## Supplementary material for "Comprehensive profiling of the human intestinal DNA virome and prediction of disease-associated bacterial hosts in severe Myalgic Encephalomyelitis/Chronic Fatigue Syndrome (ME/CFS)": S Table 2 VLP UViGs

| contig_id | checkv_quality |
| --- | --- |
| S17_extracted_k141_110830 | Not-determined |
| S17_extracted_k141_36947 | Not-determined |
| S17_extracted_k141_29558 | Not-determined |
| S17_extracted_k141_36949 | Not-determined |
| S17_extracted_k141_162550 | Not-determined |
| S17_extracted_k141_7390 | Low-quality |
| S17_extracted_k141_88672 | Not-determined |
| S17_extracted_k141_36953 | Low-quality |
| S17_extracted_k141_147775 | Not-determined |
| S17_extracted_k141_88673 | Low-quality |
| S17_extracted_k141_88674 | Not-determined |
| S17_extracted_k141_7395 | Not-determined |
| S17_extracted_k141_81289 | Not-determined |
| S17_extracted_k141_29567 | Not-determined |
| S17_extracted_k141_118228 | Not-determined |
| S17_extracted_k141_3 | Not-determined |
| S17_extracted_k141_103455 | Not-determined |
| S17_extracted_k141_36962 | Not-determined |
| S17_extracted_k141_59117 | Not-determined |
| S17_extracted_k141_9 | Not-determined |
| S17_extracted_k141_81299 | Not-determined |
| S17_extracted_k141_103460 | Not-determined |
| S17_extracted_k141_96073 | Not-determined |
| S17_extracted_k141_118236 | Low-quality |
| S17_extracted_k141_118237 | Not-determined |
| S17_extracted_k141_18 | Not-determined |
| S17_extracted_k141_125623 | Not-determined |
| S17_extracted_k141_169946 | Not-determined |
| S17_extracted_k141_66519 | Not-determined |
| S17_extracted_k141_7410 | Not-determined |
| S17_extracted_k141_73905 | Not-determined |
| S17_extracted_k141_118242 | Not-determined |
| S17_extracted_k141_44341 | Not-determined |
| S17_extracted_k141_140401 | Not-determined |
| S17_extracted_k141_73910 | Not-determined |
| S17_extracted_k141_125625 | Not-determined |
| S17_extracted_k141_73913 | Not-determined |
| S17_extracted_k141_110856 | Low-quality |
| S17_extracted_k141_133022 | Low-quality |
| S17_extracted_k141_66530 | Not-determined |
| S17_extracted_k141_140412 | Not-determined |
| S17_extracted_k141_88694 | Not-determined |
| S17_extracted_k141_73921 | Not-determined |
| S17_extracted_k141_110860 | Low-quality |
| S17_extracted_k141_42 | Low-quality |
| S17_extracted_k141_125632 | Not-determined |
| S17_extracted_k141_66535 | Not-determined |
| S17_extracted_k141_118256 | Low-quality |
| S17_extracted_k141_66537 | Not-determined |
| S17_extracted_k141_140414 | Low-quality |
| S17_extracted_k141_118258 | Not-determined |
| S17_extracted_k141_46 | Low-quality |
| S17_extracted_k141_66541 | Not-determined |
| S17_extracted_k141_103478 | Not-determined |
| S17_extracted_k141_36973 | Low-quality |
| S17_extracted_k141_103486 | Not-determined |
| S17_extracted_k141_73940 | Low-quality |
| S17_extracted_k141_125643 | Not-determined |
| S17_extracted_k141_125644 | Not-determined |
| S17_extracted_k141_14804 | Not-determined |
| S17_extracted_k141_169959 | Low-quality |
| S17_extracted_k141_14805 | Not-determined |
| S17_extracted_k141_66547 | Not-determined |
| S17_extracted_k141_59125 | Low-quality |
| S17_extracted_k141_66548 | Not-determined |
| S17_extracted_k141_169961 | Not-determined |
| S17_extracted_k141_14811 | Low-quality |
| S17_extracted_k141_96091 | Low-quality |
| S17_extracted_k141_66551 | Not-determined |
| S17_extracted_k141_96092 | Not-determined |
| S17_extracted_k141_169965 | Not-determined |
| S17_extracted_k141_73949 | Not-determined |
| S17_extracted_k141_29599 | Not-determined |
| S17_extracted_k141_36977 | Low-quality |
| S17_extracted_k141_140423 | Not-determined |
| S17_extracted_k141_59131 | Low-quality |
| S17_extracted_k141_29604 | Not-determined |
| S17_extracted_k141_155188 | Not-determined |
| S17_extracted_k141_51761 | Low-quality |
| S17_extracted_k141_155192 | Not-determined |
| S17_extracted_k141_36984 | Not-determined |
| S17_extracted_k141_14823 | Not-determined |
| S17_extracted_k141_73958 | Not-determined |
| S17_extracted_k141_51763 | Low-quality |
| S17_extracted_k141_36988 | Low-quality |
| S17_extracted_k141_7435 | Not-determined |
| S17_extracted_k141_96096 | Not-determined |
| S17_extracted_k141_125660 | Not-determined |
| S17_extracted_k141_169975 | Not-determined |
| S17_extracted_k141_7438 | Not-determined |
| S17_extracted_k141_147798 | Not-determined |
| S17_extracted_k141_51767 | Not-determined |
| S17_extracted_k141_59144 | Not-determined |
| S17_extracted_k141_36993 | Not-determined |
| S17_extracted_k141_29615 | Not-determined |
| S17_extracted_k141_147801 | Not-determined |
| S17_extracted_k141_44352 | Not-determined |
| S17_extracted_k141_44353 | Not-determined |
| S17_extracted_k141_59147 | Not-determined |
| S17_extracted_k141_44356 | Low-quality |
| S17_extracted_k141_155205 | Not-determined |
| S17_extracted_k141_96104 | Not-determined |
| S17_extracted_k141_96107 | Not-determined |
| S17_extracted_k141_169981 | Low-quality |
| S17_extracted_k141_110881 | Low-quality |
| S17_extracted_k141_96110 | Not-determined |
| S17_extracted_k141_29622 | Not-determined |
| S17_extracted_k141_110886 | Not-determined |
| S17_extracted_k141_140433 | Low-quality |
| S17_extracted_k141_22195 | Not-determined |
| S17_extracted_k141_59162 | Not-determined |
| S17_extracted_k141_51781 | Low-quality |
| S17_extracted_k141_22198 | Not-determined |
| S17_extracted_k141_59163 | Low-quality |
| S17_extracted_k141_118273 | Not-determined |
| S17_extracted_k141_169985 | Not-determined |
| S17_extracted_k141_155215 | Low-quality |
| S17_extracted_k141_155216 | Not-determined |
| S17_extracted_k141_59168 | Low-quality |
| S17_extracted_k141_22204 | Not-determined |
| S17_extracted_k141_44367 | Not-determined |
| S17_extracted_k141_44371 | Not-determined |
| S17_extracted_k141_118280 | Not-determined |
| S17_extracted_k141_103512 | Low-quality |
| S17_extracted_k141_147823 | Not-determined |
| S17_extracted_k141_147824 | Low-quality |
| S17_extracted_k141_59180 | Not-determined |
| S17_extracted_k141_51785 | Not-determined |
| S17_extracted_k141_51792 | Not-determined |
| S17_extracted_k141_44378 | Not-determined |
| S17_extracted_k141_7483 | Not-determined |
| S17_extracted_k141_44383 | Low-quality |
| S17_extracted_k141_118293 | Not-determined |
| S17_extracted_k141_118294 | Not-determined |
| S17_extracted_k141_73988 | Not-determined |
| S17_extracted_k141_88718 | Not-determined |
| S17_extracted_k141_73994 | Low-quality |
| S17_extracted_k141_96125 | Not-determined |
| S17_extracted_k141_73995 | Not-determined |
| S17_extracted_k141_73996 | Not-determined |
| S17_extracted_k141_73997 | Not-determined |
| S17_extracted_k141_162601 | Low-quality |
| S17_extracted_k141_147828 | Low-quality |
| S17_extracted_k141_133058 | Not-determined |
| S17_extracted_k141_66571 | Not-determined |
| S17_extracted_k141_133060 | Not-determined |
| S17_extracted_k141_88727 | Not-determined |
| S17_extracted_k141_96132 | Not-determined |
| S17_extracted_k141_96133 | Low-quality |
| S17_extracted_k141_29656 | Low-quality |
| S17_extracted_k141_118307 | Not-determined |
| S17_extracted_k141_96135 | Low-quality |
| S17_extracted_k141_162613 | Not-determined |
| S17_extracted_k141_162616 | Low-quality |
| S17_extracted_k141_59191 | Not-determined |
| S17_extracted_k141_155230 | Low-quality |
| S17_extracted_k141_155232 | Not-determined |
| S17_extracted_k141_133069 | Not-determined |
| S17_extracted_k141_59195 | Not-determined |
| S17_extracted_k141_133070 | Not-determined |
| S17_extracted_k141_29668 | Not-determined |
| S17_extracted_k141_96 | Not-determined |
| S17_extracted_k141_118309 | Not-determined |
| S17_extracted_k141_162626 | Not-determined |
| S17_extracted_k141_147846 | Not-determined |
| S17_extracted_k141_155238 | Not-determined |
| S17_extracted_k141_96141 | Not-determined |
| S17_extracted_k141_66582 | Not-determined |
| S17_extracted_k141_59201 | Not-determined |
| S17_extracted_k141_74011 | Low-quality |
| S17_extracted_k141_147849 | Not-determined |
| S17_extracted_k141_101 | Not-determined |
| S17_extracted_k141_7485 | Not-determined |
| S17_extracted_k141_29669 | Not-determined |
| S17_extracted_k141_88737 | Not-determined |
| S17_extracted_k141_118314 | Not-determined |
| S17_extracted_k141_118316 | Not-determined |
| S17_extracted_k141_147852 | Not-determined |
| S17_extracted_k141_81309 | Not-determined |
| S17_extracted_k141_155243 | Not-determined |
| S17_extracted_k141_29672 | Not-determined |
| S17_extracted_k141_81313 | Not-determined |
| S17_extracted_k141_118324 | Not-determined |
| S17_extracted_k141_103544 | Not-determined |
| S17_extracted_k141_37023 | Not-determined |
| S17_extracted_k141_104 | Not-determined |
| S17_extracted_k141_103545 | Not-determined |
| S17_extracted_k141_66592 | Low-quality |
| S17_extracted_k141_29676 | Not-determined |
| S17_extracted_k141_103548 | Not-determined |
| S17_extracted_k141_147867 | Low-quality |
| S17_extracted_k141_81319 | Not-determined |
| S17_extracted_k141_133078 | Not-determined |
| S17_extracted_k141_74025 | Not-determined |
| S17_extracted_k141_133079 | Not-determined |
| S17_extracted_k141_133083 | Not-determined |
| S17_extracted_k141_51814 | Low-quality |
| S17_extracted_k141_51815 | Not-determined |
| S17_extracted_k141_133087 | Not-determined |
| S17_extracted_k141_147878 | Not-determined |
| S17_extracted_k141_147882 | Not-determined |
| S17_extracted_k141_7519 | Not-determined |
| S17_extracted_k141_107 | Not-determined |
| S17_extracted_k141_66610 | Not-determined |
| S17_extracted_k141_140466 | Not-determined |
| S17_extracted_k141_162641 | Not-determined |
| S17_extracted_k141_7524 | Not-determined |
| S17_extracted_k141_59222 | Not-determined |
| S17_extracted_k141_29694 | Not-determined |
| S17_extracted_k141_118335 | Low-quality |
| S17_extracted_k141_59223 | Not-determined |
| S17_extracted_k141_96154 | Not-determined |
| S17_extracted_k141_118338 | Not-determined |
| S17_extracted_k141_59226 | Not-determined |
| S17_extracted_k141_51822 | Low-quality |
| S17_extracted_k141_96156 | Not-determined |
| S17_extracted_k141_133093 | Not-determined |
| S17_extracted_k141_66616 | Low-quality |
| S17_extracted_k141_96160 | Not-determined |
| S17_extracted_k141_81326 | Not-determined |
| S17_extracted_k141_7534 | Not-determined |
| S17_extracted_k141_14837 | Not-determined |
| S17_extracted_k141_37052 | Not-determined |
| S17_extracted_k141_37054 | Low-quality |
| S17_extracted_k141_29703 | Not-determined |
| S17_extracted_k141_81327 | Not-determined |
| S17_extracted_k141_7540 | Not-determined |
| S17_extracted_k141_133101 | Not-determined |
| S17_extracted_k141_14842 | Not-determined |
| S17_extracted_k141_29708 | Not-determined |
| S17_extracted_k141_14843 | Not-determined |
| S17_extracted_k141_44392 | Not-determined |
| S17_extracted_k141_125676 | Not-determined |
| S17_extracted_k141_44398 | Not-determined |
| S17_extracted_k141_140474 | Not-determined |
| S17_extracted_k141_96170 | Low-quality |
| S17_extracted_k141_29713 | Not-determined |
| S17_extracted_k141_14853 | Low-quality |
| S17_extracted_k141_66627 | Not-determined |
| S17_extracted_k141_59244 | Not-determined |
| S17_extracted_k141_29718 | Not-determined |
| S17_extracted_k141_44406 | Not-determined |
| S17_extracted_k141_74031 | Not-determined |
| S17_extracted_k141_66633 | Not-determined |
| S17_extracted_k141_14859 | Not-determined |
| S17_extracted_k141_66635 | Not-determined |
| S17_extracted_k141_118352 | Not-determined |
| S17_extracted_k141_14865 | Not-determined |
| S17_extracted_k141_59251 | Not-determined |
| S17_extracted_k141_51828 | Not-determined |
| S17_extracted_k141_37062 | Not-determined |
| S17_extracted_k141_133124 | Not-determined |
| S17_extracted_k141_96191 | Not-determined |
| S17_extracted_k141_96192 | Not-determined |
| S17_extracted_k141_140488 | Not-determined |
| S17_extracted_k141_29724 | Not-determined |
| S17_extracted_k141_162663 | Not-determined |
| S17_extracted_k141_140493 | Not-determined |
| S17_extracted_k141_22223 | Not-determined |
| S17_extracted_k141_22225 | Not-determined |
| S17_extracted_k141_96197 | Not-determined |
| S17_extracted_k141_162664 | Not-determined |
| S17_extracted_k141_125684 | Not-determined |
| S17_extracted_k141_14870 | Not-determined |
| S17_extracted_k141_22228 | Not-determined |
| S17_extracted_k141_96199 | Not-determined |
| S17_extracted_k141_59276 | Not-determined |
| S17_extracted_k141_162668 | Not-determined |
| S17_extracted_k141_14871 | Not-determined |
| S17_extracted_k141_44417 | Not-determined |
| S17_extracted_k141_96209 | Not-determined |
| S17_extracted_k141_125691 | Not-determined |
| S17_extracted_k141_29737 | Not-determined |
| S17_extracted_k141_22232 | Not-determined |
| S17_extracted_k141_125694 | Not-determined |
| S17_extracted_k141_96211 | Not-determined |
| S17_extracted_k141_162675 | Not-determined |
| S17_extracted_k141_140507 | Not-determined |
| S17_extracted_k141_88768 | Not-determined |
| S17_extracted_k141_125697 | Not-determined |
| S17_extracted_k141_147902 | Not-determined |
| S17_extracted_k141_74065 | Not-determined |
| S17_extracted_k141_147908 | Not-determined |
| S17_extracted_k141_96215 | Not-determined |
| S17_extracted_k141_44430 | Not-determined |
| S17_extracted_k141_140514 | Not-determined |
| S17_extracted_k141_103586 | Not-determined |
| S17_extracted_k141_44432 | Not-determined |
| S17_extracted_k141_133136 | Not-determined |
| S17_extracted_k141_88778 | Not-determined |
| S17_extracted_k141_74068 | Not-determined |
| S17_extracted_k141_66654 | Not-determined |
| S17_extracted_k141_103592 | Not-determined |
| S17_extracted_k141_133142 | Low-quality |
| S17_extracted_k141_96219 | Not-determined |
| S17_extracted_k141_44437 | Not-determined |
| S17_extracted_k141_103593 | Not-determined |
| S17_extracted_k141_96223 | Not-determined |
| S17_extracted_k141_66660 | Not-determined |
| S17_extracted_k141_88784 | Not-determined |
| S17_extracted_k141_59286 | Low-quality |
| S17_extracted_k141_103596 | Not-determined |
| S17_extracted_k141_51854 | Not-determined |
| S17_extracted_k141_51855 | Not-determined |
| S17_extracted_k141_51856 | Not-determined |
| S17_extracted_k141_29751 | Not-determined |
| S17_extracted_k141_145 | Not-determined |
| S17_extracted_k141_7555 | Not-determined |
| S17_extracted_k141_140526 | Low-quality |
| S17_extracted_k141_96229 | Low-quality |
| S17_extracted_k141_74084 | Not-determined |
| S17_extracted_k141_149 | Not-determined |
| S17_extracted_k141_147919 | Not-determined |
| S17_extracted_k141_147922 | Not-determined |
| S17_extracted_k141_59293 | Not-determined |
| S17_extracted_k141_37084 | Not-determined |
| S17_extracted_k141_162684 | Not-determined |
| S17_extracted_k141_133148 | Not-determined |
| S17_extracted_k141_7566 | Not-determined |
| S17_extracted_k141_59298 | Not-determined |
| S17_extracted_k141_88803 | Low-quality |
| S17_extracted_k141_22244 | Not-determined |
| S17_extracted_k141_162686 | Not-determined |
| S17_extracted_k141_14901 | Not-determined |
| S17_extracted_k141_147931 | Low-quality |
| S17_extracted_k141_29765 | Not-determined |
| S17_extracted_k141_140544 | Not-determined |
| S17_extracted_k141_147933 | Not-determined |
| S17_extracted_k141_37087 | Low-quality |
| S17_extracted_k141_59304 | Not-determined |
| S17_extracted_k141_22250 | Not-determined |
| S17_extracted_k141_14906 | Not-determined |
| S17_extracted_k141_14907 | Not-determined |
| S17_extracted_k141_7567 | Low-quality |
| S17_extracted_k141_14909 | Not-determined |
| S17_extracted_k141_133152 | Not-determined |
| S17_extracted_k141_133153 | Not-determined |
| S17_extracted_k141_59310 | Not-determined |
| S17_extracted_k141_51871 | Low-quality |
| S17_extracted_k141_88810 | Not-determined |
| S17_extracted_k141_22264 | Not-determined |
| S17_extracted_k141_7583 | Not-determined |
| S17_extracted_k141_88815 | Not-determined |
| S17_extracted_k141_37104 | Not-determined |
| S17_extracted_k141_96263 | Not-determined |
| S17_extracted_k141_140563 | Not-determined |
| S17_extracted_k141_103622 | Not-determined |
| S17_extracted_k141_29783 | Low-quality |
| S17_extracted_k141_96272 | Not-determined |
| S17_extracted_k141_37116 | Not-determined |
| S17_extracted_k141_29785 | Not-determined |
| S17_extracted_k141_167 | Not-determined |
| S17_extracted_k141_168 | Low-quality |
| S17_extracted_k141_51879 | Low-quality |
| S17_extracted_k141_44466 | Not-determined |
| S17_extracted_k141_51880 | Not-determined |
| S17_extracted_k141_140573 | Not-determined |
| S17_extracted_k141_29791 | Not-determined |
| S17_extracted_k141_88821 | Not-determined |
| S17_extracted_k141_59314 | Low-quality |
| S17_extracted_k141_169999 | Low-quality |
| S17_extracted_k141_29794 | Not-determined |
| S17_extracted_k141_29801 | Not-determined |
| S17_extracted_k141_125748 | Low-quality |
| S17_extracted_k141_37127 | Low-quality |
| S17_extracted_k141_125749 | Not-determined |
| S17_extracted_k141_37130 | Not-determined |
| S17_extracted_k141_37131 | Not-determined |
| S17_extracted_k141_29806 | Not-determined |
| S17_extracted_k141_51888 | Not-determined |
| S17_extracted_k141_125753 | Not-determined |
| S17_extracted_k141_22268 | Low-quality |
| S17_extracted_k141_51890 | Not-determined |
| S17_extracted_k141_37135 | Not-determined |
| S17_extracted_k141_51891 | Not-determined |
| S17_extracted_k141_103639 | Not-determined |
| S17_extracted_k141_96293 | Not-determined |
| S17_extracted_k141_162700 | Not-determined |
| S17_extracted_k141_118382 | Not-determined |
| S17_extracted_k141_170019 | Not-determined |
| S17_extracted_k141_29811 | Not-determined |
| S17_extracted_k141_140582 | Not-determined |
| S17_extracted_k141_51896 | Not-determined |
| S17_extracted_k141_103645 | Not-determined |
| S17_extracted_k141_29814 | Not-determined |
| S17_extracted_k141_51901 | Not-determined |
| S17_extracted_k141_7604 | Not-determined |
| S17_extracted_k141_103647 | Not-determined |
| S17_extracted_k141_140586 | Not-determined |
| S17_extracted_k141_103649 | Not-determined |
| S17_extracted_k141_162709 | Not-determined |
| S17_extracted_k141_51909 | Not-determined |
| S17_extracted_k141_37153 | Not-determined |
| S17_extracted_k141_7606 | Low-quality |
| S17_extracted_k141_155306 | Not-determined |
| S17_extracted_k141_103655 | Not-determined |
| S17_extracted_k141_37156 | Not-determined |
| S17_extracted_k141_103657 | Not-determined |
| S17_extracted_k141_37157 | Not-determined |
| S17_extracted_k141_59329 | Not-determined |
| S17_extracted_k141_7609 | Not-determined |
| S17_extracted_k141_96300 | Low-quality |
| S17_extracted_k141_155311 | Not-determined |
| S17_extracted_k141_37161 | Not-determined |
| S17_extracted_k141_37163 | Not-determined |
| S17_extracted_k141_88837 | Low-quality |
| S17_extracted_k141_155315 | Not-determined |
| S17_extracted_k141_147961 | Not-determined |
| S17_extracted_k141_147962 | Low-quality |
| S17_extracted_k141_162718 | Not-determined |
| S17_extracted_k141_147963 | Not-determined |
| S17_extracted_k141_96306 | Low-quality |
| S17_extracted_k141_22291 | Not-determined |
| S17_extracted_k141_118402 | Not-determined |
| S17_extracted_k141_37168 | Not-determined |
| S17_extracted_k141_162720 | Not-determined |
| S17_extracted_k141_22294 | Not-determined |
| S17_extracted_k141_103661 | Low-quality |
| S17_extracted_k141_22297 | Not-determined |
| S17_extracted_k141_7623 | Not-determined |
| S17_extracted_k141_51926 | Not-determined |
| S17_extracted_k141_37173 | Low-quality |
| S17_extracted_k141_170040 | Not-determined |
| S17_extracted_k141_96312 | Not-determined |
| S17_extracted_k141_37174 | Low-quality |
| S17_extracted_k141_96313 | Not-determined |
| S17_extracted_k141_37177 | Not-determined |
| S17_extracted_k141_133187 | Not-determined |
| S17_extracted_k141_162731 | Not-determined |
| S17_extracted_k141_96321 | Low-quality |
| S17_extracted_k141_22300 | Not-determined |
| S17_extracted_k141_14933 | Not-determined |
| S17_extracted_k141_96323 | Not-determined |
| S17_extracted_k141_37184 | Not-determined |
| S17_extracted_k141_140590 | Low-quality |
| S17_extracted_k141_14944 | Not-determined |
| S17_extracted_k141_162740 | Low-quality |
| S17_extracted_k141_7647 | Not-determined |
| S17_extracted_k141_133195 | Not-determined |
| S17_extracted_k141_162742 | Not-determined |
| S17_extracted_k141_14947 | Not-determined |
| S17_extracted_k141_44485 | Not-determined |
| S17_extracted_k141_44486 | Not-determined |
| S17_extracted_k141_88852 | Not-determined |
| S17_extracted_k141_37193 | Not-determined |
| S17_extracted_k141_147972 | Not-determined |
| S17_extracted_k141_147973 | Not-determined |
| S17_extracted_k141_7652 | Not-determined |
| S17_extracted_k141_125772 | Not-determined |
| S17_extracted_k141_37198 | Low-quality |
| S17_extracted_k141_44494 | Not-determined |
| S17_extracted_k141_162753 | Not-determined |
| S17_extracted_k141_125777 | Not-determined |
| S17_extracted_k141_7659 | Not-determined |
| S17_extracted_k141_88858 | Not-determined |
| S17_extracted_k141_147985 | Not-determined |
| S17_extracted_k141_22312 | Not-determined |
| S17_extracted_k141_59364 | Not-determined |
| S17_extracted_k141_103677 | Not-determined |
| S17_extracted_k141_103682 | Not-determined |
| S17_extracted_k141_7663 | Not-determined |
| S17_extracted_k141_37218 | Low-quality |
| S17_extracted_k141_81330 | Not-determined |
| S17_extracted_k141_22328 | Not-determined |
| S17_extracted_k141_140605 | Not-determined |
| S17_extracted_k141_7670 | Low-quality |
| S17_extracted_k141_103684 | Not-determined |
| S17_extracted_k141_170062 | Not-determined |
| S17_extracted_k141_7673 | Not-determined |
| S17_extracted_k141_162765 | Not-determined |
| S17_extracted_k141_7676 | Not-determined |
| S17_extracted_k141_7682 | Not-determined |
| S17_extracted_k141_200 | Not-determined |
| S17_extracted_k141_29841 | Not-determined |
| S17_extracted_k141_96357 | Low-quality |
| S17_extracted_k141_118435 | Not-determined |
| S17_extracted_k141_22346 | Low-quality |
| S17_extracted_k141_44510 | Low-quality |
| S17_extracted_k141_14954 | Not-determined |
| S17_extracted_k141_37232 | Low-quality |
| S17_extracted_k141_37233 | Not-determined |
| S17_extracted_k141_22350 | Not-determined |
| S17_extracted_k141_96362 | Not-determined |
| S17_extracted_k141_22352 | Not-determined |
| S17_extracted_k141_118438 | Not-determined |
| S17_extracted_k141_170075 | Not-determined |
| S17_extracted_k141_155320 | Not-determined |
| S17_extracted_k141_81344 | Not-determined |
| S17_extracted_k141_44515 | Not-determined |
| S17_extracted_k141_155322 | Low-quality |
| S17_extracted_k141_29851 | Not-determined |
| S17_extracted_k141_162767 | Low-quality |
| S17_extracted_k141_96364 | Low-quality |
| S17_extracted_k141_103696 | Not-determined |
| S17_extracted_k141_96367 | Not-determined |
| S17_extracted_k141_29856 | Not-determined |
| S17_extracted_k141_103700 | Not-determined |
| S17_extracted_k141_155333 | Not-determined |
| S17_extracted_k141_103701 | Not-determined |
| S17_extracted_k141_155335 | Not-determined |
| S17_extracted_k141_140630 | Not-determined |
| S17_extracted_k141_29864 | Low-quality |
| S17_extracted_k141_81347 | Not-determined |
| S17_extracted_k141_170087 | Not-determined |
| S17_extracted_k141_133232 | Not-determined |
| S17_extracted_k141_7697 | Not-determined |
| S17_extracted_k141_133236 | Not-determined |
| S17_extracted_k141_81355 | Not-determined |
| S17_extracted_k141_96379 | Not-determined |
| S17_extracted_k141_29872 | Not-determined |
| S17_extracted_k141_51940 | Not-determined |
| S17_extracted_k141_162779 | Not-determined |
| S17_extracted_k141_37244 | Not-determined |
| S17_extracted_k141_103703 | Low-quality |
| S17_extracted_k141_110916 | Not-determined |
| S17_extracted_k141_103704 | Low-quality |
| S17_extracted_k141_51942 | Low-quality |
| S17_extracted_k141_118454 | Not-determined |
| S17_extracted_k141_162784 | Not-determined |
| S17_extracted_k141_170098 | Not-determined |
| S17_extracted_k141_81364 | Not-determined |
| S17_extracted_k141_162788 | Not-determined |
| S17_extracted_k141_29882 | Not-determined |
| S17_extracted_k141_103710 | Not-determined |
| S17_extracted_k141_29884 | Not-determined |
| S17_extracted_k141_110928 | Not-determined |
| S17_extracted_k141_148024 | Not-determined |
| S17_extracted_k141_96385 | Not-determined |
| S17_extracted_k141_162791 | Low-quality |
| S17_extracted_k141_162792 | Not-determined |
| S17_extracted_k141_162794 | Not-determined |
| S17_extracted_k141_140636 | Low-quality |
| S17_extracted_k141_162795 | Not-determined |
| S17_extracted_k141_148027 | Not-determined |
| S17_extracted_k141_162797 | Low-quality |
| S17_extracted_k141_14971 | Low-quality |
| S17_extracted_k141_14972 | Not-determined |
| S17_extracted_k141_148031 | Low-quality |
| S17_extracted_k141_7705 | Low-quality |
| S17_extracted_k141_148032 | Not-determined |
| S17_extracted_k141_103721 | Not-determined |
| S17_extracted_k141_140640 | Low-quality |
| S17_extracted_k141_140641 | Not-determined |
| S17_extracted_k141_81375 | Not-determined |
| S17_extracted_k141_148042 | Not-determined |
| S17_extracted_k141_103727 | Not-determined |
| S17_extracted_k141_162808 | Not-determined |
| S17_extracted_k141_162811 | Not-determined |
| S17_extracted_k141_59403 | Low-quality |
| S17_extracted_k141_88861 | Not-determined |
| S17_extracted_k141_88863 | Not-determined |
| S17_extracted_k141_59404 | Not-determined |
| S17_extracted_k141_29905 | Not-determined |
| S17_extracted_k141_74093 | Not-determined |
| S17_extracted_k141_125784 | Not-determined |
| S17_extracted_k141_7709 | Not-determined |
| S17_extracted_k141_59405 | Low-quality |
| S17_extracted_k141_214 | Not-determined |
| S17_extracted_k141_74099 | Not-determined |
| S17_extracted_k141_51946 | Not-determined |
| S17_extracted_k141_218 | Not-determined |
| S17_extracted_k141_222 | Not-determined |
| S17_extracted_k141_140646 | Not-determined |
| S17_extracted_k141_170132 | Not-determined |
| S17_extracted_k141_133274 | Not-determined |
| S17_extracted_k141_118457 | Not-determined |
| S17_extracted_k141_88868 | Low-quality |
| S17_extracted_k141_155376 | Not-determined |
| S17_extracted_k141_59424 | Not-determined |
| S17_extracted_k141_74117 | Not-determined |
| S17_extracted_k141_51953 | Low-quality |
| S17_extracted_k141_44546 | Low-quality |
| S17_extracted_k141_133279 | Not-determined |
| S17_extracted_k141_74120 | Not-determined |
| S17_extracted_k141_170139 | Not-determined |
| S17_extracted_k141_148064 | Not-determined |
| S17_extracted_k141_44548 | Low-quality |
| S17_extracted_k141_74123 | Not-determined |
| S17_extracted_k141_44552 | Not-determined |
| S17_extracted_k141_29915 | Not-determined |
| S17_extracted_k141_125796 | Low-quality |
| S17_extracted_k141_96421 | Not-determined |
| S17_extracted_k141_88873 | Not-determined |
| S17_extracted_k141_125799 | Not-determined |
| S17_extracted_k141_44559 | Not-determined |
| S17_extracted_k141_7717 | Not-determined |
| S17_extracted_k141_74128 | Not-determined |
| S17_extracted_k141_51959 | Not-determined |

| | |
|---|---|
| S17_extracted_k141_133285 | Not-determined |
| S17_extracted_k141_14995 | Low-quality |
| S17_extracted_k141_44562 | Low-quality |
| S17_extracted_k141_148078 | Not-determined |
| S17_extracted_k141_51965 | Not-determined |
| S17_extracted_k141_133287 | Not-determined |
| S17_extracted_k141_37268 | Not-determined |
| S17_extracted_k141_37269 | Not-determined |
| S17_extracted_k141_74138 | Low-quality |
| S17_extracted_k141_81384 | Not-determined |
| S17_extracted_k141_118475 | Not-determined |
| S17_extracted_k141_44570 | Not-determined |
| S17_extracted_k141_103755 | Not-determined |
| S17_extracted_k141_118476 | Low-quality |
| S17_extracted_k141_118477 | Not-determined |
| S17_extracted_k141_81386 | Low-quality |
| S17_extracted_k141_155401 | Not-determined |
| S17_extracted_k141_125816 | Not-determined |
| S17_extracted_k141_155402 | Not-determined |
| S17_extracted_k141_133293 | Not-determined |
| S17_extracted_k141_44576 | Not-determined |
| S17_extracted_k141_7719 | Not-determined |
| S17_extracted_k141_155405 | Not-determined |
| S17_extracted_k141_96427 | Not-determined |
| S17_extracted_k141_51978 | Not-determined |
| S17_extracted_k141_74143 | Not-determined |
| S17_extracted_k141_88890 | Not-determined |
| S17_extracted_k141_37284 | Not-determined |
| S17_extracted_k141_88892 | Not-determined |
| S17_extracted_k141_103768 | Not-determined |
| S17_extracted_k141_44584 | Not-determined |
| S17_extracted_k141_148086 | Low-quality |
| S17_extracted_k141_37287 | Not-determined |
| S17_extracted_k141_37288 | Not-determined |
| S17_extracted_k141_15008 | Low-quality |
| S17_extracted_k141_74147 | Not-determined |
| S17_extracted_k141_51986 | Not-determined |
| S13_vs_57267\|\|full | Medium-quality |
| S17_extracted_k141_81389 | Low-quality |
| S17_extracted_k141_66686 | Not-determined |
| S17_extracted_k141_140656 | Not-determined |
| S17_extracted_k141_66687 | Not-determined |
| S17_extracted_k141_140657 | Low-quality |
| S17_extracted_k141_103778 | Not-determined |
| S17_extracted_k141_66690 | Not-determined |
| S17_extracted_k141_74152 | Low-quality |
| S17_extracted_k141_96454 | Not-determined |
| S17_extracted_k141_66696 | Low-quality |
| S17_extracted_k141_44598 | Not-determined |
| S17_extracted_k141_81397 | Not-determined |
| S17_extracted_k141_29936 | Not-determined |
| S17_extracted_k141_74158 | Not-determined |
| S17_extracted_k141_103787 | Low-quality |
| S17_extracted_k141_81403 | Not-determined |
| S17_extracted_k141_140668 | Low-quality |
| S17_extracted_k141_37292 | Not-determined |
| S17_extracted_k141_140670 | Not-determined |
| S17_extracted_k141_29940 | Low-quality |
| S17_extracted_k141_140674 | Not-determined |
| S17_extracted_k141_110961 | Not-determined |
| S17_extracted_k141_66702 | Low-quality |
| S17_extracted_k141_140676 | Not-determined |
| S17_extracted_k141_103792 | Not-determined |
| S17_extracted_k141_110963 | Not-determined |
| S17_extracted_k141_66705 | Not-determined |
| S17_extracted_k141_29949 | Not-determined |
| S17_extracted_k141_15023 | Not-determined |
| S17_extracted_k141_125839 | Not-determined |
| S17_extracted_k141_66710 | Low-quality |
| S17_extracted_k141_37295 | Not-determined |
| S17_extracted_k141_81408 | Not-determined |
| S17_extracted_k141_74171 | Low-quality |
| S17_extracted_k141_170159 | Not-determined |
| S17_extracted_k141_66712 | Low-quality |
| S17_extracted_k141_110977 | Not-determined |
| S17_extracted_k141_44612 | Not-determined |
| S17_extracted_k141_15030 | Not-determined |
| S17_extracted_k141_37304 | Not-determined |
| S17_extracted_k141_37305 | Not-determined |
| S17_extracted_k141_148097 | Low-quality |
| S17_extracted_k141_44619 | Not-determined |
| S17_extracted_k141_59453 | Low-quality |
| S17_extracted_k141_44620 | Not-determined |
| S17_extracted_k141_88922 | Not-determined |
| S17_extracted_k141_170169 | Not-determined |
| S17_extracted_k141_59454 | Not-determined |
| S17_extracted_k141_125851 | Not-determined |
| S17_extracted_k141_59455 | Low-quality |
| S17_extracted_k141_125855 | Not-determined |
| S17_extracted_k141_29958 | Not-determined |
| S17_extracted_k141_7721 | Not-determined |
| S17_extracted_k141_170176 | Not-determined |
| S17_extracted_k141_37310 | Not-determined |
| S17_extracted_k141_37312 | Not-determined |
| S17_extracted_k141_88927 | Not-determined |
| S17_extracted_k141_29962 | Not-determined |
| S17_extracted_k141_74177 | Not-determined |
| S17_extracted_k141_103796 | Not-determined |
| S17_extracted_k141_110997 | Not-determined |
| S17_extracted_k141_133306 | Not-determined |
| S17_extracted_k141_125863 | Not-determined |
| S17_extracted_k141_44632 | Not-determined |
| S17_extracted_k141_59467 | Not-determined |
| S17_extracted_k141_133311 | Low-quality |
| S17_extracted_k141_59470 | Not-determined |
| S17_extracted_k141_29971 | Not-determined |
| S17_extracted_k141_133316 | Low-quality |
| S17_extracted_k141_44639 | Not-determined |
| S17_extracted_k141_7731 | Not-determined |
| S17_extracted_k141_140702 | Not-determined |
| S17_extracted_k141_59478 | Not-determined |
| S17_extracted_k141_133328 | Low-quality |
| S17_extracted_k141_44648 | Not-determined |
| S17_extracted_k141_96462 | Not-determined |
| S17_extracted_k141_125885 | Not-determined |
| S17_extracted_k141_74186 | Not-determined |
| S17_extracted_k141_162817 | Not-determined |
| S17_extracted_k141_111008 | Low-quality |
| S17_extracted_k141_111009 | Not-determined |
| S17_extracted_k141_118507 | Not-determined |
| S17_extracted_k141_74190 | Not-determined |
| S17_extracted_k141_118508 | Low-quality |
| S17_extracted_k141_133337 | Not-determined |
| S17_extracted_k141_148103 | Not-determined |
| S17_extracted_k141_52043 | Not-determined |
| S17_extracted_k141_170187 | Not-determined |
| S17_extracted_k141_74194 | Not-determined |
| S17_extracted_k141_133340 | Not-determined |
| S17_extracted_k141_74197 | Not-determined |
| S17_extracted_k141_66746 | Not-determined |
| S17_extracted_k141_7736 | Not-determined |
| S17_extracted_k141_148113 | Not-determined |
| S17_extracted_k141_148116 | Low-quality |
| S17_extracted_k141_44670 | Not-determined |
| S17_extracted_k141_59497 | Not-determined |
| S17_extracted_k141_125894 | Not-determined |
| S17_extracted_k141_111015 | Low-quality |
| S17_extracted_k141_37316 | Not-determined |
| S17_extracted_k141_29990 | Not-determined |
| S17_extracted_k141_52053 | Not-determined |
| S17_extracted_k141_170195 | Not-determined |
| S17_extracted_k141_254 | Not-determined |
| S17_extracted_k141_44673 | Low-quality |
| S17_extracted_k141_52055 | Not-determined |
| S17_extracted_k141_59501 | Not-determined |
| S17_extracted_k141_125898 | Not-determined |
| S17_extracted_k141_29993 | Not-determined |
| S17_extracted_k141_148126 | Not-determined |
| S17_extracted_k141_118523 | Not-determined |
| S17_extracted_k141_44676 | Low-quality |
| S17_extracted_k141_29997 | Not-determined |
| S17_extracted_k141_44679 | Not-determined |
| S17_extracted_k141_44680 | Low-quality |
| S17_extracted_k141_37327 | Not-determined |
| S17_extracted_k141_81413 | Not-determined |
| S17_extracted_k141_44682 | Not-determined |
| S17_extracted_k141_66755 | Not-determined |
| S17_extracted_k141_7744 | Not-determined |
| S17_extracted_k141_118531 | Not-determined |
| S17_extracted_k141_162834 | Low-quality |
| S17_extracted_k141_81419 | Low-quality |
| S17_extracted_k141_81420 | Not-determined |
| S17_extracted_k141_66757 | Not-determined |
| S17_extracted_k141_118539 | Not-determined |
| S17_extracted_k141_44690 | Low-quality |
| S17_extracted_k141_170200 | Not-determined |
| S17_extracted_k141_81425 | Low-quality |
| S17_extracted_k141_162839 | Not-determined |
| S17_extracted_k141_125909 | Not-determined |
| S17_extracted_k141_74217 | Not-determined |
| S17_extracted_k141_170205 | Not-determined |
| S17_extracted_k141_74219 | Not-determined |
| S17_extracted_k141_74223 | Low-quality |
| S17_extracted_k141_52071 | Not-determined |
| S17_extracted_k141_66760 | Not-determined |
| S17_extracted_k141_170211 | Not-determined |
| S17_extracted_k141_59527 | Not-determined |
| S17_extracted_k141_59528 | Not-determined |
| S17_extracted_k141_37331 | Not-determined |
| S17_extracted_k141_44699 | Not-determined |
| S17_extracted_k141_52075 | Low-quality |
| S17_extracted_k141_148156 | Not-determined |
| S17_extracted_k141_66762 | Not-determined |
| S17_extracted_k141_111032 | Not-determined |
| S17_extracted_k141_96481 | Low-quality |
| S17_extracted_k141_140720 | Not-determined |
| S17_extracted_k141_66764 | Not-determined |
| S17_extracted_k141_140721 | Not-determined |
| S17_extracted_k141_59536 | Low-quality |
| S17_extracted_k141_140724 | Not-determined |
| S17_extracted_k141_118553 | Not-determined |
| S17_extracted_k141_111039 | Not-determined |
| S17_extracted_k141_140727 | Not-determined |
| S17_extracted_k141_37341 | Not-determined |
| S17_extracted_k141_278 | Not-determined |
| S17_extracted_k141_148160 | Not-determined |
| S17_extracted_k141_52082 | Not-determined |
| S17_extracted_k141_281 | Low-quality |
| S17_extracted_k141_170225 | Not-determined |
| S17_extracted_k141_282 | Low-quality |
| S17_extracted_k141_37346 | Not-determined |
| S17_extracted_k141_103839 | Not-determined |
| S17_extracted_k141_88929 | Not-determined |
| S17_extracted_k141_111048 | Not-determined |
| S17_extracted_k141_37355 | Not-determined |
| S17_extracted_k141_66778 | Not-determined |
| S17_extracted_k141_111056 | Not-determined |
| S17_extracted_k141_44720 | Not-determined |
| S17_extracted_k141_289 | Not-determined |
| S17_extracted_k141_111061 | Not-determined |
| S17_extracted_k141_162861 | Not-determined |
| S17_extracted_k141_22370 | Not-determined |
| S17_extracted_k141_111062 | Not-determined |
| S17_extracted_k141_125919 | Not-determined |
| S17_extracted_k141_111063 | Not-determined |
| S17_extracted_k141_52092 | Not-determined |
| S17_extracted_k141_37361 | Not-determined |
| S17_extracted_k141_7755 | Not-determined |
| S17_extracted_k141_88937 | Not-determined |
| S17_extracted_k141_292 | Not-determined |
| S17_extracted_k141_140750 | Not-determined |
| S17_extracted_k141_103850 | Not-determined |
| S17_extracted_k141_140751 | Not-determined |
| S17_extracted_k141_44726 | Not-determined |
| S17_extracted_k141_103851 | Not-determined |
| S17_extracted_k141_170243 | Not-determined |
| S17_extracted_k141_300 | Not-determined |
| S17_extracted_k141_170247 | Not-determined |
| S17_extracted_k141_103857 | Not-determined |
| S17_extracted_k141_74262 | Low-quality |
| S17_extracted_k141_125937 | Not-determined |
| S17_extracted_k141_96511 | Not-determined |
| S17_extracted_k141_111078 | Not-determined |
| S17_extracted_k141_81464 | Not-determined |
| S17_extracted_k141_74273 | Not-determined |
| S17_extracted_k141_111081 | Low-quality |
| S17_extracted_k141_15055 | Not-determined |
| S17_extracted_k141_125941 | Not-determined |
| S17_extracted_k141_81466 | Not-determined |
| S17_extracted_k141_306 | Not-determined |
| S17_extracted_k141_111083 | Not-determined |
| S17_extracted_k141_140766 | Not-determined |
| S17_extracted_k141_15060 | Not-determined |
| S17_extracted_k141_81467 | Low-quality |
| S17_extracted_k141_44736 | Not-determined |
| S17_extracted_k141_140770 | Not-determined |
| S17_extracted_k141_118567 | Not-determined |
| S17_extracted_k141_59542 | Low-quality |
| S17_extracted_k141_7764 | Not-determined |
| S17_extracted_k141_59545 | Not-determined |
| S17_extracted_k141_37370 | Not-determined |
| S17_extracted_k141_308 | Not-determined |
| S17_extracted_k141_310 | Low-quality |
| S17_extracted_k141_37373 | Not-determined |
| S17_extracted_k141_96522 | Low-quality |
| S17_extracted_k141_103867 | Not-determined |
| S17_extracted_k141_59550 | Not-determined |
| S17_extracted_k141_59551 | Low-quality |
| S17_extracted_k141_125953 | Not-determined |
| S17_extracted_k141_44744 | Not-determined |
| S17_extracted_k141_96525 | Not-determined |
| S17_extracted_k141_140777 | Not-determined |
| S17_extracted_k141_103873 | Not-determined |
| S17_extracted_k141_125960 | Low-quality |
| S17_extracted_k141_111096 | Not-determined |
| S17_extracted_k141_37382 | Low-quality |
| S17_extracted_k141_7778 | Not-determined |
| S17_extracted_k141_140779 | Low-quality |
| S17_extracted_k141_7780 | Not-determined |
| S17_extracted_k141_74288 | Not-determined |
| S17_extracted_k141_74289 | Not-determined |
| S17_extracted_k141_125966 | Not-determined |
| S17_extracted_k141_7782 | Not-determined |
| S17_extracted_k141_170275 | Not-determined |
| S17_extracted_k141_22397 | Not-determined |
| S17_extracted_k141_74290 | Low-quality |
| S17_extracted_k141_111105 | Not-determined |
| S17_extracted_k141_74292 | Not-determined |
| S17_extracted_k141_140783 | Not-determined |
| S17_extracted_k141_66803 | Low-quality |
| S17_extracted_k141_7787 | Not-determined |
| S17_extracted_k141_59559 | Not-determined |
| S17_extracted_k141_162867 | Not-determined |
| S17_extracted_k141_66808 | Low-quality |
| S17_extracted_k141_125972 | Not-determined |
| S17_extracted_k141_59560 | Not-determined |
| S17_extracted_k141_325 | Not-determined |
| S17_extracted_k141_66811 | Not-determined |
| S17_extracted_k141_59565 | Not-determined |
| S17_extracted_k141_59569 | Not-determined |
| S17_extracted_k141_37392 | Low-quality |
| S17_extracted_k141_170281 | Not-determined |
| S17_extracted_k141_96534 | Low-quality |
| S17_extracted_k141_103878 | Not-determined |
| S17_extracted_k141_140793 | Not-determined |
| S17_extracted_k141_103879 | Not-determined |
| S17_extracted_k141_162872 | Not-determined |
| S17_extracted_k141_96540 | Not-determined |
| S17_extracted_k141_7802 | Not-determined |
| S17_extracted_k141_81490 | Not-determined |
| S17_extracted_k141_338 | Not-determined |
| S17_extracted_k141_59582 | Not-determined |
| S17_extracted_k141_111121 | Not-determined |
| S17_extracted_k141_140802 | Not-determined |
| S17_extracted_k141_341 | Low-quality |
| S17_extracted_k141_140804 | Low-quality |
| S17_extracted_k141_133354 | Not-determined |
| S17_extracted_k141_140807 | Not-determined |
| S17_extracted_k141_103888 | Not-determined |
| S17_extracted_k141_133356 | Not-determined |
| S17_extracted_k141_66824 | Not-determined |
| S17_extracted_k141_88947 | Not-determined |
| S17_extracted_k141_140811 | Not-determined |
| S17_extracted_k141_111127 | Not-determined |
| S17_extracted_k141_7812 | Low-quality |
| S17_extracted_k141_44767 | Not-determined |
| S17_extracted_k141_103892 | Not-determined |
| S17_extracted_k141_52116 | Not-determined |
| S17_extracted_k141_44771 | Low-quality |
| S17_extracted_k141_133365 | Not-determined |
| S17_extracted_k141_148205 | Not-determined |
| S17_extracted_k141_118590 | Not-determined |
| S17_extracted_k141_52119 | Not-determined |
| S17_extracted_k141_52120 | Not-determined |
| S17_extracted_k141_170292 | Low-quality |
| S17_extracted_k141_74315 | Not-determined |
| S17_extracted_k141_103897 | Not-determined |
| S17_extracted_k141_81509 | Not-determined |
| S17_extracted_k141_103900 | Not-determined |
| S17_extracted_k141_44780 | Not-determined |
| S17_extracted_k141_30018 | Not-determined |
| S17_extracted_k141_22409 | Not-determined |
| S17_extracted_k141_59597 | Not-determined |
| S17_extracted_k141_30020 | Not-determined |
| S17_extracted_k141_88956 | Not-determined |
| S17_extracted_k141_155445 | Not-determined |
| S17_extracted_k141_7828 | Not-determined |
| S17_extracted_k141_148222 | Low-quality |
| S17_extracted_k141_148223 | Not-determined |
| S17_extracted_k141_59604 | Not-determined |
| S17_extracted_k141_59606 | Not-determined |
| S17_extracted_k141_52139 | Not-determined |
| S17_extracted_k141_140830 | Low-quality |
| S17_extracted_k141_88961 | Not-determined |
| S17_extracted_k141_111153 | Not-determined |
| S17_extracted_k141_59610 | Not-determined |
| S17_extracted_k141_118607 | Not-determined |
| S17_extracted_k141_118608 | Not-determined |
| S17_extracted_k141_148228 | Low-quality |
| S17_extracted_k141_111156 | Not-determined |
| S17_extracted_k141_133382 | Not-determined |
| S17_extracted_k141_66835 | Not-determined |
| S17_extracted_k141_30028 | Not-determined |
| S17_extracted_k141_37424 | Not-determined |
| S17_extracted_k141_155455 | Not-determined |
| S17_extracted_k141_88968 | Not-determined |
| S17_extracted_k141_155456 | Not-determined |
| S17_extracted_k141_37427 | Not-determined |
| S17_extracted_k141_15078 | Not-determined |
| S17_extracted_k141_162900 | Not-determined |
| S17_extracted_k141_88970 | Not-determined |
| S17_extracted_k141_88971 | Low-quality |
| S17_extracted_k141_111161 | Not-determined |
| S17_extracted_k141_155460 | Not-determined |
| S17_extracted_k141_22418 | Not-determined |
| S17_extracted_k141_66837 | Not-determined |
| S17_extracted_k141_118612 | Low-quality |
| S17_extracted_k141_30032 | Not-determined |
| S17_extracted_k141_66839 | Not-determined |
| S17_extracted_k141_66840 | Not-determined |
| S17_extracted_k141_355 | Low-quality |
| S17_extracted_k141_170298 | Not-determined |
| S17_extracted_k141_140839 | Not-determined |
| S17_extracted_k141_66841 | Not-determined |
| S17_extracted_k141_356 | Not-determined |
| S17_extracted_k141_22421 | Not-determined |
| S17_extracted_k141_103925 | Not-determined |
| S17_extracted_k141_111163 | Low-quality |
| S17_extracted_k141_74318 | Not-determined |
| S17_extracted_k141_140843 | Low-quality |
| S17_extracted_k141_74323 | Not-determined |
| S17_extracted_k141_360 | Not-determined |
| S17_extracted_k141_74324 | Not-determined |
| S17_extracted_k141_103928 | Not-determined |
| S17_extracted_k141_22425 | Not-determined |
| S17_extracted_k141_66851 | Not-determined |
| S17_extracted_k141_15098 | Not-determined |
| S17_extracted_k141_37432 | Not-determined |
| S17_extracted_k141_59629 | Not-determined |
| S17_extracted_k141_140852 | Not-determined |
| S17_extracted_k141_363 | Not-determined |
| S17_extracted_k141_170316 | Not-determined |
| S17_extracted_k141_103932 | Not-determined |
| S17_extracted_k141_365 | Not-determined |
| S17_extracted_k141_103934 | Not-determined |
| S17_extracted_k141_15105 | Not-determined |
| S17_extracted_k141_118620 | Not-determined |
| S17_extracted_k141_170317 | Low-quality |
| S17_extracted_k141_81512 | Not-determined |
| S17_extracted_k141_37435 | Not-determined |
| S17_extracted_k141_103948 | Not-determined |
| S17_extracted_k141_52157 | Not-determined |
| S17_extracted_k141_30059 | Low-quality |
| S17_extracted_k141_155480 | Not-determined |
| S17_extracted_k141_103951 | Not-determined |
| S17_extracted_k141_162906 | Not-determined |
| S17_extracted_k141_162907 | Not-determined |
| S17_extracted_k141_37439 | Not-determined |
| S17_extracted_k141_140865 | Not-determined |
| S17_extracted_k141_140866 | Low-quality |
| S17_extracted_k141_118634 | Low-quality |
| S17_extracted_k141_37442 | Not-determined |
| S17_extracted_k141_59635 | Not-determined |
| S17_extracted_k141_162911 | Not-determined |
| S17_extracted_k141_111182 | Low-quality |
| S17_extracted_k141_88984 | Low-quality |
| S17_extracted_k141_148243 | Not-determined |
| S17_extracted_k141_30070 | Not-determined |
| S17_extracted_k141_37451 | Low-quality |
| S17_extracted_k141_15117 | Not-determined |
| S17_extracted_k141_162922 | Not-determined |
| S17_extracted_k141_15119 | Not-determined |
| S17_extracted_k141_103955 | Not-determined |
| S17_extracted_k141_118649 | Not-determined |
| S17_extracted_k141_15122 | Not-determined |
| S17_extracted_k141_103956 | Not-determined |
| S17_extracted_k141_44819 | Not-determined |
| S17_extracted_k141_52175 | Not-determined |
| S17_extracted_k141_118651 | Not-determined |
| S17_extracted_k141_37459 | Low-quality |
| S17_extracted_k141_148250 | Not-determined |
| S17_extracted_k141_148251 | Not-determined |
| S17_extracted_k141_148252 | Low-quality |
| S17_extracted_k141_111195 | Not-determined |
| S17_extracted_k141_74336 | Not-determined |
| S17_extracted_k141_30077 | Not-determined |
| S17_extracted_k141_52179 | Not-determined |
| S17_extracted_k141_22444 | Not-determined |
| S17_extracted_k141_133411 | Not-determined |
| S17_extracted_k141_133413 | Not-determined |
| S17_extracted_k141_118658 | Low-quality |
| S17_extracted_k141_103964 | Not-determined |
| S17_extracted_k141_81544 | Not-determined |
| S17_extracted_k141_118660 | Not-determined |
| S17_extracted_k141_133421 | Not-determined |
| S17_extracted_k141_133424 | Not-determined |
| S17_extracted_k141_37481 | Not-determined |
| S17_extracted_k141_133425 | Not-determined |
| S17_extracted_k141_30083 | Low-quality |
| S17_extracted_k141_375 | Not-determined |
| S17_extracted_k141_118666 | Not-determined |
| S17_extracted_k141_377 | Not-determined |
| S17_extracted_k141_30087 | Not-determined |
| S17_extracted_k141_103966 | Not-determined |
| S17_extracted_k141_15130 | Not-determined |
| S17_extracted_k141_74351 | Not-determined |
| S17_extracted_k141_380 | Not-determined |
| S17_extracted_k141_37489 | Not-determined |
| S17_extracted_k141_37490 | Not-determined |
| S17_extracted_k141_22466 | Low-quality |
| S17_extracted_k141_103970 | Not-determined |
| S17_extracted_k141_118675 | Not-determined |
| S17_extracted_k141_118678 | Not-determined |
| S17_extracted_k141_133440 | Not-determined |
| S17_extracted_k141_22475 | Not-determined |
| S17_extracted_k141_162935 | Low-quality |
| S17_extracted_k141_22478 | Not-determined |
| S17_extracted_k141_389 | Not-determined |
| S17_extracted_k141_133448 | Not-determined |
| S17_extracted_k141_52191 | Not-determined |
| S17_extracted_k141_52192 | Not-determined |
| S17_extracted_k141_133449 | Not-determined |
| S17_extracted_k141_15136 | Not-determined |
| S17_extracted_k141_37510 | Not-determined |
| S17_extracted_k141_133451 | Not-determined |
| S17_extracted_k141_7843 | Not-determined |
| S17_extracted_k141_140877 | Not-determined |
| S17_extracted_k141_52197 | Not-determined |
| S17_extracted_k141_30090 | Not-determined |
| S17_extracted_k141_170321 | Not-determined |
| S17_extracted_k141_30091 | Not-determined |
| S17_extracted_k141_74379 | Not-determined |
| S17_extracted_k141_103995 | Not-determined |
| S17_extracted_k141_140883 | Not-determined |
| S17_extracted_k141_403 | Not-determined |
| S17_extracted_k141_103998 | Not-determined |
| S17_extracted_k141_59642 | Low-quality |
| S17_extracted_k141_22486 | Not-determined |
| S17_extracted_k141_140886 | Not-determined |
| S17_extracted_k141_30095 | Not-determined |
| S17_extracted_k141_89011 | Not-determined |
| S17_extracted_k141_52200 | Not-determined |
| S17_extracted_k141_405 | Low-quality |
| S17_extracted_k141_133460 | Not-determined |
| S17_extracted_k141_133461 | Not-determined |
| S17_extracted_k141_407 | Not-determined |
| S17_extracted_k141_52204 | Low-quality |
| S17_extracted_k141_140894 | Not-determined |
| S17_extracted_k141_409 | Low-quality |
| S17_extracted_k141_104007 | Not-determined |
| S17_extracted_k141_52205 | Not-determined |
| S17_extracted_k141_52207 | Not-determined |
| S17_extracted_k141_413 | Not-determined |
| S17_extracted_k141_140898 | Not-determined |
| S17_extracted_k141_155499 | Not-determined |
| S17_extracted_k141_7857 | Not-determined |
| S17_extracted_k141_89018 | Not-determined |
| S17_extracted_k141_52213 | Not-determined |
| S17_extracted_k141_419 | Not-determined |
| S17_extracted_k141_118694 | Not-determined |
| S17_extracted_k141_59650 | Low-quality |
| S17_extracted_k141_155502 | Not-determined |
| S17_extracted_k141_7863 | Not-determined |
| S17_extracted_k141_15156 | Not-determined |
| S17_extracted_k141_118701 | Not-determined |
| S17_extracted_k141_89020 | Not-determined |
| S17_extracted_k141_59657 | Low-quality |
| S17_extracted_k141_89022 | Not-determined |
| S17_extracted_k141_52217 | Low-quality |
| S17_extracted_k141_52222 | Not-determined |
| S17_extracted_k141_425 | Not-determined |
| S17_extracted_k141_148293 | Not-determined |
| S17_extracted_k141_140917 | Not-determined |
| S17_extracted_k141_59670 | Low-quality |
| S17_extracted_k141_155518 | Low-quality |
| S17_extracted_k141_111205 | Not-determined |
| S17_extracted_k141_74394 | Not-determined |
| S17_extracted_k141_162959 | Low-quality |
| S17_extracted_k141_111208 | Not-determined |
| S17_extracted_k141_162962 | Not-determined |
| S17_extracted_k141_118714 | Not-determined |
| S17_extracted_k141_140928 | Not-determined |
| S17_extracted_k141_37534 | Not-determined |
| S17_extracted_k141_52237 | Not-determined |
| S17_extracted_k141_37536 | Not-determined |
| S17_extracted_k141_52239 | Not-determined |
| S17_extracted_k141_140930 | Not-determined |
| S17_extracted_k141_22496 | Not-determined |
| S17_extracted_k141_162965 | Low-quality |
| S17_extracted_k141_74409 | Not-determined |
| S17_extracted_k141_22498 | Not-determined |
| S17_extracted_k141_15173 | Not-determined |
| S17_extracted_k141_155535 | Low-quality |
| S17_extracted_k141_170347 | Not-determined |
| S17_extracted_k141_162969 | Low-quality |
| S17_extracted_k141_89045 | Not-determined |
| S17_extracted_k141_89047 | Not-determined |
| S17_extracted_k141_22505 | Not-determined |
| S17_extracted_k141_162973 | Not-determined |
| S17_extracted_k141_30139 | Low-quality |
| S17_extracted_k141_7897 | Low-quality |
| S17_extracted_k141_7899 | Not-determined |
| S17_extracted_k141_118725 | Not-determined |
| S17_extracted_k141_452 | Not-determined |
| S17_extracted_k141_155546 | Not-determined |
| S17_extracted_k141_118728 | Low-quality |
| S17_extracted_k141_104036 | Not-determined |
| S17_extracted_k141_104037 | Not-determined |
| S17_extracted_k141_140937 | Not-determined |
| S17_extracted_k141_59692 | Not-determined |
| S17_extracted_k141_125990 | Not-determined |
| S17_extracted_k141_7908 | Low-quality |
| S17_extracted_k141_22507 | Not-determined |
| S17_extracted_k141_463 | Low-quality |
| S17_extracted_k141_81560 | Not-determined |
| S17_extracted_k141_52242 | Not-determined |
| S17_extracted_k141_140948 | Not-determined |
| S17_extracted_k141_81567 | Low-quality |
| S17_extracted_k141_89053 | Not-determined |
| S17_extracted_k141_104055 | Not-determined |
| S17_extracted_k141_22514 | Not-determined |
| S17_extracted_k141_74420 | Not-determined |
| S17_extracted_k141_7919 | Not-determined |
| S17_extracted_k141_52251 | Not-determined |
| S17_extracted_k141_126003 | Not-determined |
| S17_extracted_k141_140956 | Low-quality |
| S17_extracted_k141_104064 | Not-determined |
| S17_extracted_k141_7925 | Not-determined |
| S14_vs_k141_74067\|\|full | Medium-quality |
| S17_extracted_k141_22518 | Not-determined |
| S17_extracted_k141_140957 | Not-determined |
| S17_extracted_k141_170364 | Not-determined |
| S17_extracted_k141_126009 | Not-determined |
| S17_extracted_k141_140960 | Not-determined |
| S17_extracted_k141_22526 | Not-determined |
| S5_vs_k141_40848\|\|full | High-quality |
| S17_extracted_k141_111211 | Not-determined |
| S17_extracted_k141_170373 | Not-determined |
| S17_extracted_k141_37563 | Not-determined |

| | |
|---|---|
| S17_extracted_k141_81585 | Not-determined |
| S17_extracted_k141_15188 | Not-determined |
| S17_extracted_k141_74430 | Not-determined |
| S17_extracted_k141_74432 | Low-quality |
| S17_extracted_k141_37568 | Not-determined |
| S17_extracted_k141_7943 | Not-determined |
| S17_extracted_k141_170381 | Not-determined |
| S17_extracted_k141_155563 | Low-quality |
| S17_extracted_k141_37572 | Not-determined |
| S17_extracted_k141_111227 | Not-determined |
| S17_extracted_k141_7946 | Not-determined |
| S17_extracted_k141_81593 | Not-determined |
| S17_extracted_k141_15195 | Not-determined |
| S17_extracted_k141_89058 | Not-determined |
| S17_extracted_k141_37577 | Not-determined |
| S17_extracted_k141_155571 | Not-determined |
| S17_extracted_k141_81594 | Low-quality |
| S17_extracted_k141_15200 | Low-quality |
| S17_extracted_k141_81596 | Low-quality |
| S17_extracted_k141_118751 | Not-determined |
| S17_extracted_k141_491 | Not-determined |
| S17_extracted_k141_22531 | Not-determined |
| S17_extracted_k141_118754 | Not-determined |
| S17_extracted_k141_163019 | Not-determined |
| S17_extracted_k141_15207 | Not-determined |
| S17_extracted_k141_163020 | Not-determined |
| S17_extracted_k141_111236 | Not-determined |
| S17_extracted_k141_163022 | Not-determined |
| S17_extracted_k141_22536 | Not-determined |
| S17_extracted_k141_118759 | Not-determined |
| S17_extracted_k141_37590 | Not-determined |
| S17_extracted_k141_52281 | Not-determined |
| S17_extracted_k141_37592 | Not-determined |
| S17_extracted_k141_66866 | Low-quality |
| S17_extracted_k141_15216 | Low-quality |
| S17_extracted_k141_111237 | Low-quality |
| S17_extracted_k141_111238 | Not-determined |
| S17_extracted_k141_111240 | Not-determined |
| S17_extracted_k141_22539 | Not-determined |
| S17_extracted_k141_15221 | Not-determined |
| S17_extracted_k141_163029 | Low-quality |
| S17_extracted_k141_148319 | Low-quality |
| S17_extracted_k141_118766 | Not-determined |
| S17_extracted_k141_104080 | Not-determined |
| S17_extracted_k141_44849 | Not-determined |
| S17_extracted_k141_74451 | Not-determined |
| S17_extracted_k141_15224 | Not-determined |
| S17_extracted_k141_111243 | Low-quality |
| S17_extracted_k141_15227 | Not-determined |
| S17_extracted_k141_502 | Not-determined |
| S17_extracted_k141_15228 | Not-determined |
| S17_extracted_k141_59712 | Not-determined |
| S17_extracted_k141_52289 | Low-quality |
| S17_extracted_k141_504 | Not-determined |
| S17_extracted_k141_163031 | Not-determined |
| S17_extracted_k141_505 | Not-determined |
| S17_extracted_k141_118771 | Not-determined |
| S17_extracted_k141_59714 | Not-determined |
| S17_extracted_k141_44853 | Not-determined |
| S17_extracted_k141_22542 | Not-determined |
| S17_extracted_k141_15231 | Not-determined |
| S17_extracted_k141_148329 | Not-determined |
| S17_extracted_k141_111256 | Not-determined |
| S17_extracted_k141_507 | Not-determined |
| S17_extracted_k141_37609 | Low-quality |
| S17_extracted_k141_15240 | Not-determined |
| S17_extracted_k141_66871 | Not-determined |
| S17_extracted_k141_513 | Not-determined |
| S17_extracted_k141_118776 | Low-quality |
| S17_extracted_k141_89073 | Not-determined |
| S17_extracted_k141_44858 | Not-determined |
| S17_extracted_k141_148346 | Not-determined |
| S17_extracted_k141_515 | Not-determined |
| S17_extracted_k141_66876 | Not-determined |
| S17_extracted_k141_155596 | Not-determined |
| S17_extracted_k141_520 | Not-determined |
| S17_extracted_k141_44862 | Not-determined |
| S17_extracted_k141_524 | Not-determined |
| S17_extracted_k141_66881 | Low-quality |
| S17_extracted_k141_52301 | Low-quality |
| S17_extracted_k141_163051 | Not-determined |
| S17_extracted_k141_74461 | Not-determined |
| S17_extracted_k141_155603 | Not-determined |
| S17_extracted_k141_118788 | Not-determined |
| S17_extracted_k141_22560 | Not-determined |
| S17_extracted_k141_155604 | Low-quality |
| S17_extracted_k141_15248 | Not-determined |
| S17_extracted_k141_155608 | Not-determined |
| S17_extracted_k141_133487 | Low-quality |
| S17_extracted_k141_7974 | Not-determined |
| S17_extracted_k141_52317 | Not-determined |
| S17_extracted_k141_52318 | Not-determined |
| S17_extracted_k141_118794 | Not-determined |
| S17_extracted_k141_133489 | Not-determined |
| S17_extracted_k141_140981 | Not-determined |
| S17_extracted_k141_133493 | Not-determined |
| S17_extracted_k141_7982 | Not-determined |
| S17_extracted_k141_163063 | Not-determined |
| S17_extracted_k141_7984 | Not-determined |
| S17_extracted_k141_111266 | Not-determined |
| S17_extracted_k141_30221 | Not-determined |
| S17_extracted_k141_148368 | Not-determined |
| S17_extracted_k141_537 | Not-determined |
| S17_extracted_k141_111270 | Not-determined |
| S17_extracted_k141_15259 | Not-determined |
| S17_extracted_k141_155620 | Not-determined |
| S17_extracted_k141_133497 | Not-determined |
| S17_extracted_k141_133498 | Not-determined |
| S17_extracted_k141_126039 | Not-determined |
| S17_extracted_k141_155622 | Not-determined |
| S17_extracted_k141_89103 | Low-quality |
| S17_extracted_k141_148374 | Not-determined |
| S17_extracted_k141_118801 | Not-determined |
| S17_extracted_k141_30229 | Not-determined |
| S17_extracted_k141_545 | Not-determined |
| S17_extracted_k141_22574 | Not-determined |
| S17_extracted_k141_140990 | Not-determined |
| S17_extracted_k141_22577 | Low-quality |
| S17_extracted_k141_111275 | Low-quality |
| S17_extracted_k141_148381 | Not-determined |
| S17_extracted_k141_126044 | Not-determined |
| S17_extracted_k141_15271 | Not-determined |
| S17_extracted_k141_118805 | Not-determined |
| S17_extracted_k141_44884 | Not-determined |
| S17_extracted_k141_126055 | Not-determined |
| S17_extracted_k141_8006 | Not-determined |
| S17_extracted_k141_37633 | Not-determined |
| S17_extracted_k141_111282 | Not-determined |
| S17_extracted_k141_118810 | Not-determined |
| S17_extracted_k141_44886 | Not-determined |
| S17_extracted_k141_44889 | Low-quality |
| S17_extracted_k141_15275 | Low-quality |
| S17_extracted_k141_22590 | Not-determined |
| S17_extracted_k141_8010 | Not-determined |
| S17_extracted_k141_22593 | Not-determined |
| S17_extracted_k141_170421 | Not-determined |
| S17_extracted_k141_15278 | Not-determined |
| S17_extracted_k141_15279 | Not-determined |
| S17_extracted_k141_111287 | Not-determined |
| S17_extracted_k141_170424 | Not-determined |
| S17_extracted_k141_37640 | Not-determined |
| S17_extracted_k141_22604 | Not-determined |
| S17_extracted_k141_52346 | Low-quality |
| S17_extracted_k141_552 | Not-determined |
| S17_extracted_k141_118826 | Not-determined |
| S17_extracted_k141_30249 | Not-determined |
| S17_extracted_k141_30250 | Not-determined |
| S17_extracted_k141_8018 | Not-determined |
| S17_extracted_k141_44897 | Not-determined |
| S17_extracted_k141_8019 | Not-determined |
| S17_extracted_k141_118828 | Not-determined |
| S17_extracted_k141_163073 | Not-determined |
| S17_extracted_k141_133520 | Not-determined |
| S17_extracted_k141_74493 | Not-determined |
| S17_extracted_k141_133525 | Not-determined |
| S17_extracted_k141_111302 | Not-determined |
| S17_extracted_k141_118838 | Not-determined |
| S17_extracted_k141_558 | Low-quality |
| S17_extracted_k141_133527 | Low-quality |
| S17_extracted_k141_74498 | Not-determined |
| S17_extracted_k141_30265 | Not-determined |
| S17_extracted_k141_104089 | Not-determined |
| S17_extracted_k141_133530 | Not-determined |
| S17_extracted_k141_118841 | Not-determined |
| S17_extracted_k141_30268 | Not-determined |
| S17_extracted_k141_133531 | Not-determined |
| S17_extracted_k141_155649 | Not-determined |
| S17_extracted_k141_44904 | Not-determined |
| S17_extracted_k141_74505 | Not-determined |
| S17_extracted_k141_37660 | Not-determined |
| S17_extracted_k141_148398 | Not-determined |
| S17_extracted_k141_148399 | Not-determined |
| S17_extracted_k141_66914 | Not-determined |
| S17_extracted_k141_170444 | Not-determined |
| S17_extracted_k141_66916 | Not-determined |
| S17_extracted_k141_30282 | Not-determined |
| S17_extracted_k141_579 | Not-determined |
| S17_extracted_k141_37667 | Low-quality |
| S17_extracted_k141_581 | Not-determined |
| S17_extracted_k141_74516 | Not-determined |
| S17_extracted_k141_37671 | Low-quality |
| S17_extracted_k141_37675 | Not-determined |
| S17_extracted_k141_37676 | Not-determined |
| S17_extracted_k141_133538 | Not-determined |
| S17_extracted_k141_170446 | Not-determined |
| S17_extracted_k141_66924 | Not-determined |
| S17_extracted_k141_22624 | Not-determined |
| S17_extracted_k141_8050 | Not-determined |
| S17_extracted_k141_584 | Not-determined |
| S17_extracted_k141_133541 | Not-determined |
| S17_extracted_k141_37680 | Not-determined |
| S17_extracted_k141_22626 | Not-determined |
| S17_extracted_k141_586 | Not-determined |
| S17_extracted_k141_8053 | Not-determined |
| S17_extracted_k141_22629 | Not-determined |
| S17_extracted_k141_133543 | Not-determined |
| S17_extracted_k141_170448 | Not-determined |
| S17_extracted_k141_111319 | Not-determined |
| S17_extracted_k141_52363 | Not-determined |
| S17_extracted_k141_111322 | Not-determined |
| S17_extracted_k141_104116 | Not-determined |
| S17_extracted_k141_126080 | Low-quality |
| S17_extracted_k141_133548 | Not-determined |
| S17_extracted_k141_126083 | Not-determined |
| S17_extracted_k141_96543 | Not-determined |
| S17_extracted_k141_133550 | Not-determined |
| S17_extracted_k141_126084 | Not-determined |
| S17_extracted_k141_22633 | Not-determined |
| S17_extracted_k141_52371 | Not-determined |
| S17_extracted_k141_170455 | Low-quality |
| S17_extracted_k141_170457 | Not-determined |
| S17_extracted_k141_170458 | Low-quality |
| S17_extracted_k141_111332 | Not-determined |
| S17_extracted_k141_22641 | Low-quality |
| S17_extracted_k141_66930 | Not-determined |
| S17_extracted_k141_52377 | Not-determined |
| S17_extracted_k141_126097 | Not-determined |
| S17_extracted_k141_96549 | Low-quality |
| S17_extracted_k141_22646 | Not-determined |
| S17_extracted_k141_44924 | Not-determined |
| S17_extracted_k141_22647 | Not-determined |
| S17_extracted_k141_66932 | Not-determined |
| S17_extracted_k141_22648 | Not-determined |
| S17_extracted_k141_15284 | Not-determined |
| S17_extracted_k141_52383 | Not-determined |
| S17_extracted_k141_126103 | Not-determined |
| S17_extracted_k141_96553 | Low-quality |
| S17_extracted_k141_66934 | Not-determined |
| S17_extracted_k141_44926 | Not-determined |
| S17_extracted_k141_104125 | Low-quality |
| S17_extracted_k141_52385 | Not-determined |
| S17_extracted_k141_66936 | Not-determined |
| S17_extracted_k141_104127 | Not-determined |
| S17_extracted_k141_126106 | Low-quality |
| S17_extracted_k141_104133 | Not-determined |
| S17_extracted_k141_15290 | Low-quality |
| S17_extracted_k141_96560 | Not-determined |
| S17_extracted_k141_96562 | Not-determined |
| S17_extracted_k141_44938 | Low-quality |
| S17_extracted_k141_118857 | Not-determined |
| S17_extracted_k141_133576 | Not-determined |
| S17_extracted_k141_170472 | Not-determined |
| S17_extracted_k141_170473 | Not-determined |
| S17_extracted_k141_155683 | Low-quality |
| S17_extracted_k141_37701 | Low-quality |
| S17_extracted_k141_30320 | Not-determined |
| S17_extracted_k141_59740 | Low-quality |
| S17_extracted_k141_163095 | Not-determined |
| S17_extracted_k141_74525 | Not-determined |
| S17_extracted_k141_163096 | Low-quality |
| S17_extracted_k141_89114 | Not-determined |
| S17_extracted_k141_30324 | Not-determined |
| S17_extracted_k141_74529 | Not-determined |
| S17_extracted_k141_59747 | Not-determined |
| S17_extracted_k141_89122 | Not-determined |
| S17_extracted_k141_133594 | Not-determined |
| S17_extracted_k141_59748 | Not-determined |
| S17_extracted_k141_126114 | Not-determined |
| S17_extracted_k141_37713 | Not-determined |
| S17_extracted_k141_631 | Not-determined |
| S17_extracted_k141_74536 | Low-quality |
| S17_extracted_k141_141017 | Not-determined |
| S17_extracted_k141_104146 | Not-determined |
| S17_extracted_k141_37716 | Not-determined |
| S17_extracted_k141_52412 | Not-determined |
| S17_extracted_k141_81608 | Not-determined |
| S17_extracted_k141_155710 | Not-determined |
| S17_extracted_k141_155714 | Not-determined |
| S17_extracted_k141_96569 | Not-determined |
| S17_extracted_k141_96570 | Not-determined |
| S17_extracted_k141_74548 | Not-determined |
| S17_extracted_k141_74549 | Not-determined |
| S17_extracted_k141_74550 | Not-determined |
| S17_extracted_k141_96572 | Not-determined |
| S17_extracted_k141_104160 | Low-quality |
| S17_extracted_k141_96573 | Not-determined |
| S17_extracted_k141_104162 | Not-determined |
| S17_extracted_k141_634 | Low-quality |
| S17_extracted_k141_155720 | Low-quality |
| S17_extracted_k141_639 | Not-determined |
| S17_extracted_k141_155722 | Not-determined |
| S17_extracted_k141_640 | Not-determined |
| S17_extracted_k141_641 | Not-determined |
| S17_extracted_k141_74562 | Not-determined |
| S17_extracted_k141_44952 | Not-determined |
| S17_extracted_k141_81624 | Low-quality |
| S17_extracted_k141_155729 | Not-determined |
| S17_extracted_k141_648 | Not-determined |
| S17_extracted_k141_74568 | Not-determined |
| S17_extracted_k141_52422 | Not-determined |
| S17_extracted_k141_96585 | Not-determined |
| S17_extracted_k141_148423 | Not-determined |
| S17_extracted_k141_652 | Low-quality |
| S17_extracted_k141_30343 | Not-determined |
| S17_extracted_k141_52427 | Not-determined |
| S17_extracted_k141_96588 | Not-determined |
| S17_extracted_k141_59781 | Not-determined |
| S17_extracted_k141_163112 | Not-determined |
| S17_extracted_k141_66959 | Low-quality |
| S17_extracted_k141_163113 | Low-quality |
| S17_extracted_k141_155747 | Not-determined |
| S17_extracted_k141_155748 | Not-determined |
| S17_extracted_k141_81638 | Low-quality |
| S17_extracted_k141_126117 | Not-determined |
| S17_extracted_k141_89156 | Not-determined |
| S17_extracted_k141_118877 | Not-determined |
| S17_extracted_k141_59791 | Not-determined |
| S17_extracted_k141_148441 | Not-determined |
| S17_extracted_k141_59795 | Low-quality |
| S17_extracted_k141_118882 | Not-determined |
| S17_extracted_k141_15302 | Low-quality |
| S17_extracted_k141_155752 | Not-determined |
| S17_extracted_k141_104189 | Low-quality |
| S17_extracted_k141_118885 | Not-determined |
| S17_extracted_k141_118886 | Not-determined |
| S17_extracted_k141_141053 | Not-determined |
| S17_extracted_k141_104192 | Not-determined |
| S17_extracted_k141_148448 | Not-determined |
| S17_extracted_k141_15308 | Not-determined |
| S17_extracted_k141_111379 | Not-determined |
| S17_extracted_k141_59799 | Not-determined |
| S17_extracted_k141_163119 | Not-determined |
| S17_extracted_k141_163120 | Low-quality |
| S17_extracted_k141_163122 | Not-determined |
| S17_extracted_k141_59803 | Not-determined |
| S17_extracted_k141_141060 | Not-determined |
| S17_extracted_k141_111381 | Low-quality |
| S17_extracted_k141_81650 | Low-quality |
| S17_extracted_k141_81651 | Not-determined |
| S17_extracted_k141_59808 | Not-determined |
| S17_extracted_k141_81653 | Not-determined |
| S17_extracted_k141_15324 | Not-determined |
| S17_extracted_k141_15325 | Not-determined |
| S17_extracted_k141_126133 | Not-determined |
| S17_extracted_k141_66968 | Not-determined |
| S17_extracted_k141_59811 | Low-quality |
| S17_extracted_k141_30356 | Low-quality |
| S17_extracted_k141_126140 | Not-determined |
| S17_extracted_k141_81661 | Not-determined |
| S17_extracted_k141_96591 | Not-determined |
| S17_extracted_k141_118902 | Not-determined |
| S17_extracted_k141_141076 | Not-determined |
| S17_extracted_k141_118904 | Not-determined |
| S17_extracted_k141_37728 | Low-quality |
| S17_extracted_k141_163138 | Not-determined |
| S17_extracted_k141_59822 | Not-determined |
| S17_extracted_k141_104215 | Not-determined |
| S17_extracted_k141_74574 | Not-determined |
| S17_extracted_k141_155759 | Not-determined |
| S17_extracted_k141_52455 | Not-determined |
| S17_extracted_k141_37733 | Not-determined |
| S17_extracted_k141_30360 | Not-determined |
| S17_extracted_k141_170529 | Low-quality |
| S17_extracted_k141_96596 | Low-quality |
| S17_extracted_k141_15341 | Not-determined |
| S17_extracted_k141_74584 | Not-determined |
| S17_extracted_k141_659 | Low-quality |
| S17_extracted_k141_111402 | Low-quality |
| S17_extracted_k141_74585 | Not-determined |
| S17_extracted_k141_163147 | Not-determined |
| S17_extracted_k141_74586 | Low-quality |
| S17_extracted_k141_155775 | Not-determined |
| S17_extracted_k141_74591 | Not-determined |
| S17_extracted_k141_81669 | Not-determined |
| S17_extracted_k141_15346 | Not-determined |
| S17_extracted_k141_81670 | Not-determined |
| S17_extracted_k141_148477 | Not-determined |
| S17_extracted_k141_44977 | Low-quality |
| S17_extracted_k141_141091 | Not-determined |
| S17_extracted_k141_89166 | Not-determined |
| S17_extracted_k141_170541 | Not-determined |
| S17_extracted_k141_15349 | Low-quality |
| S17_extracted_k141_74601 | Not-determined |
| S17_extracted_k141_170548 | Not-determined |
| S17_extracted_k141_96608 | Not-determined |
| S17_extracted_k141_155785 | Not-determined |
| S17_extracted_k141_37739 | Not-determined |
| S17_extracted_k141_37740 | Not-determined |
| S17_extracted_k141_89176 | Not-determined |
| S17_extracted_k141_15358 | Low-quality |
| S17_extracted_k141_170552 | Not-determined |
| S17_extracted_k141_81681 | Not-determined |
| S17_extracted_k141_44987 | Not-determined |
| S17_extracted_k141_30368 | Not-determined |
| S17_extracted_k141_155794 | Not-determined |
| S17_extracted_k141_44989 | Not-determined |
| S17_extracted_k141_30369 | Not-determined |
| S17_extracted_k141_89182 | Not-determined |
| S17_extracted_k141_89184 | Not-determined |
| S17_extracted_k141_111415 | Not-determined |
| S17_extracted_k141_37746 | Low-quality |
| S17_extracted_k141_37749 | Not-determined |
| S17_extracted_k141_141105 | Not-determined |
| S17_extracted_k141_111421 | Not-determined |
| S17_extracted_k141_141111 | Not-determined |
| S17_extracted_k141_74620 | Low-quality |
| S17_extracted_k141_126169 | Not-determined |
| S17_extracted_k141_170562 | Not-determined |
| S17_extracted_k141_15379 | Not-determined |
| S17_extracted_k141_148503 | Not-determined |
| S17_extracted_k141_155801 | Not-determined |
| S17_extracted_k141_74627 | Not-determined |
| S17_extracted_k141_126172 | Low-quality |
| S17_extracted_k141_30375 | Not-determined |
| S17_extracted_k141_37752 | Not-determined |
| S17_extracted_k141_89199 | Not-determined |
| S17_extracted_k141_30380 | Low-quality |
| S17_extracted_k141_30381 | Not-determined |
| S17_extracted_k141_170567 | Not-determined |
| S17_extracted_k141_155815 | Not-determined |
| S17_extracted_k141_89202 | Low-quality |
| S17_extracted_k141_104245 | Not-determined |
| S17_extracted_k141_126185 | Not-determined |
| S17_extracted_k141_126186 | Not-determined |
| S17_extracted_k141_683 | Not-determined |
| S17_extracted_k141_155818 | Not-determined |
| S17_extracted_k141_155819 | Not-determined |
| S17_extracted_k141_155820 | Not-determined |
| S17_extracted_k141_89212 | Not-determined |
| S17_extracted_k141_148522 | Not-determined |
| S17_extracted_k141_66986 | Not-determined |
| S17_extracted_k141_104252 | Not-determined |
| S17_extracted_k141_170582 | Not-determined |
| S17_extracted_k141_148526 | Not-determined |
| S17_extracted_k141_163164 | Not-determined |
| S17_extracted_k141_141122 | Not-determined |
| S17_extracted_k141_74656 | Not-determined |
| S17_extracted_k141_104258 | Low-quality |
| S17_extracted_k141_148531 | Low-quality |
| S17_extracted_k141_148532 | Not-determined |
| S17_extracted_k141_148533 | Low-quality |
| S17_extracted_k141_126203 | Not-determined |
| S17_extracted_k141_15393 | Not-determined |
| S17_extracted_k141_81691 | Not-determined |
| S17_extracted_k141_163173 | Not-determined |
| S17_extracted_k141_126205 | Low-quality |
| S17_extracted_k141_170598 | Low-quality |
| S17_extracted_k141_141125 | Low-quality |
| S17_extracted_k141_15398 | Not-determined |
| S17_extracted_k141_141130 | Not-determined |
| S17_extracted_k141_96618 | Not-determined |
| S17_extracted_k141_30400 | Not-determined |
| S17_extracted_k141_163177 | Not-determined |
| S17_extracted_k141_111436 | Not-determined |
| S17_extracted_k141_8066 | Low-quality |
| S17_extracted_k141_30405 | Low-quality |
| S17_extracted_k141_96627 | Low-quality |
| S17_extracted_k141_141139 | Not-determined |
| S17_extracted_k141_74663 | Not-determined |
| S17_extracted_k141_163185 | Not-determined |
| S17_extracted_k141_30416 | Low-quality |
| S17_extracted_k141_141142 | Low-quality |
| S17_extracted_k141_74666 | Not-determined |
| S17_extracted_k141_141143 | Not-determined |
| S17_extracted_k141_118915 | Not-determined |
| S17_extracted_k141_67004 | Not-determined |
| S17_extracted_k141_118916 | Not-determined |
| S17_extracted_k141_37782 | Not-determined |
| S17_extracted_k141_96638 | Not-determined |
| S17_extracted_k141_37783 | Not-determined |
| S17_extracted_k141_67011 | Not-determined |
| S17_extracted_k141_8082 | Not-determined |
| S17_extracted_k141_37788 | Not-determined |
| S17_extracted_k141_133619 | Not-determined |
| S17_extracted_k141_148544 | Not-determined |
| S17_extracted_k141_148545 | Not-determined |
| S17_extracted_k141_96644 | Not-determined |
| S17_extracted_k141_133623 | Not-determined |
| S17_extracted_k141_141158 | Not-determined |
| S17_extracted_k141_148550 | Not-determined |
| S17_extracted_k141_30430 | Not-determined |
| S17_extracted_k141_170602 | Low-quality |
| S17_extracted_k141_118924 | Not-determined |
| S17_extracted_k141_67020 | Low-quality |
| S17_extracted_k141_148557 | Not-determined |
| S17_extracted_k141_89237 | Low-quality |
| S17_extracted_k141_30444 | Not-determined |
| S17_extracted_k141_104293 | Not-determined |
| S17_extracted_k141_89239 | Not-determined |
| S17_extracted_k141_8094 | Not-determined |
| S17_extracted_k141_74685 | Not-determined |
| S17_extracted_k141_118934 | Not-determined |
| S17_extracted_k141_52464 | Low-quality |
| S17_extracted_k141_74686 | Not-determined |
| S17_extracted_k141_148561 | Not-determined |
| S17_extracted_k141_37799 | Not-determined |
| S17_extracted_k141_155842 | Not-determined |
| S17_extracted_k141_148566 | Not-determined |
| S17_extracted_k141_37802 | Not-determined |
| S17_extracted_k141_67035 | Not-determined |
| S17_extracted_k141_15412 | Low-quality |
| S17_extracted_k141_104300 | Not-determined |
| S17_extracted_k141_15418 | Low-quality |
| S17_extracted_k141_163198 | Low-quality |
| S17_extracted_k141_15419 | Not-determined |
| S17_extracted_k141_163203 | Not-determined |
| S17_extracted_k141_37815 | Not-determined |
| S17_extracted_k141_163207 | Low-quality |
| S17_extracted_k141_118943 | Not-determined |
| S17_extracted_k141_702 | Not-determined |
| S17_extracted_k141_118945 | Low-quality |
| S17_extracted_k141_163211 | Not-determined |
| S17_extracted_k141_141169 | Not-determined |
| S17_extracted_k141_705 | Not-determined |
| S17_extracted_k141_155863 | Not-determined |
| S17_extracted_k141_37823 | Not-determined |
| S17_extracted_k141_148576 | Not-determined |
| S17_extracted_k141_104315 | Not-determined |
| S17_extracted_k141_163221 | Not-determined |
| S17_extracted_k141_30460 | Not-determined |
| S17_extracted_k141_30461 | Not-determined |
| S17_extracted_k141_148581 | Not-determined |
| S17_extracted_k141_104324 | Not-determined |
| S17_extracted_k141_163231 | Not-determined |
| S17_extracted_k141_148584 | Not-determined |
| S17_extracted_k141_45011 | Not-determined |
| S17_extracted_k141_89253 | Not-determined |
| S17_extracted_k141_148586 | Not-determined |
| S17_extracted_k141_30469 | Not-determined |
| S17_extracted_k141_111467 | Low-quality |
| S17_extracted_k141_111468 | Not-determined |
| S17_extracted_k141_155880 | Not-determined |
| S17_extracted_k141_163241 | Not-determined |
| S17_extracted_k141_155882 | Not-determined |
| S17_extracted_k141_163243 | Not-determined |
| S17_extracted_k141_111470 | Not-determined |
| S17_extracted_k141_163247 | Not-determined |
| S17_extracted_k141_8107 | Low-quality |
| S9_extracted_k141_13507 | Complete |
| S17_extracted_k141_163248 | Not-determined |
| S17_extracted_k141_163250 | Not-determined |
| S17_extracted_k141_170608 | Not-determined |
| S17_extracted_k141_104335 | Not-determined |
| S17_extracted_k141_170610 | Not-determined |
| S17_extracted_k141_67045 | Not-determined |
| S17_extracted_k141_22665 | Not-determined |
| S17_extracted_k141_30483 | Not-determined |
| S17_extracted_k141_163256 | Not-determined |
| S17_extracted_k141_148601 | Low-quality |
| S17_extracted_k141_45022 | Low-quality |
| S17_extracted_k141_67049 | Not-determined |
| S17_extracted_k141_170618 | Not-determined |
| S17_extracted_k141_170620 | Not-determined |
| S17_extracted_k141_37847 | Low-quality |
| S17_extracted_k141_45028 | Not-determined |
| S17_extracted_k141_45029 | Low-quality |
| S17_extracted_k141_163267 | Not-determined |
| S17_extracted_k141_163268 | Not-determined |
| S17_extracted_k141_45032 | Not-determined |
| S17_extracted_k141_30500 | Not-determined |
| S17_extracted_k141_141174 | Not-determined |
| S17_extracted_k141_170625 | Low-quality |
| S17_extracted_k141_67059 | Not-determined |
| S17_extracted_k141_45033 | Not-determined |
| S17_extracted_k141_8116 | Not-determined |
| S17_extracted_k141_141179 | Not-determined |
| S17_extracted_k141_104350 | Not-determined |
| S17_extracted_k141_37859 | Not-determined |
| S17_extracted_k141_163282 | Not-determined |
| S17_extracted_k141_37864 | Not-determined |
| S17_extracted_k141_163284 | Not-determined |
| S17_extracted_k141_163285 | Not-determined |
| S17_extracted_k141_52488 | Low-quality |
| S17_extracted_k141_45037 | Not-determined |
| S17_extracted_k141_52489 | Not-determined |
| S17_extracted_k141_148615 | Low-quality |
| S17_extracted_k141_37870 | Not-determined |
| S17_extracted_k141_163290 | Not-determined |
| S17_extracted_k141_89278 | Low-quality |
| S17_extracted_k141_163291 | Not-determined |
| S17_extracted_k141_104359 | Not-determined |
| S17_extracted_k141_104361 | Not-determined |
| S17_extracted_k141_8128 | Low-quality |
| S17_extracted_k141_89281 | Not-determined |
| S17_extracted_k141_8132 | Not-determined |
| S17_extracted_k141_45046 | Not-determined |
| S17_extracted_k141_155905 | Not-determined |
| S17_extracted_k141_155908 | Not-determined |
| S17_extracted_k141_37884 | Not-determined |
| S17_extracted_k141_118948 | Not-determined |
| S17_extracted_k141_104369 | Not-determined |
| S17_extracted_k141_141200 | Not-determined |
| S17_extracted_k141_118954 | Not-determined |
| S17_extracted_k141_141206 | Not-determined |
| S17_extracted_k141_81748 | Low-quality |
| S17_extracted_k141_141207 | Not-determined |
| S17_extracted_k141_170641 | Not-determined |
| S17_extracted_k141_141208 | Not-determined |
| S17_extracted_k141_126229 | Not-determined |

| | |
|---|---|
| S17_extracted_k141_118958 | Not-determined |
| S17_extracted_k141_141209 | Not-determined |
| S17_extracted_k141_111478 | Not-determined |
| S17_extracted_k141_118960 | Not-determined |
| S17_extracted_k141_118961 | Not-determined |
| S17_extracted_k141_141214 | Not-determined |
| S17_extracted_k141_155914 | Low-quality |
| S17_extracted_k141_155916 | Low-quality |
| S17_extracted_k141_141216 | Not-determined |
| S17_extracted_k141_52531 | Low-quality |
| S17_extracted_k141_104376 | Not-determined |
| S17_extracted_k141_45067 | Not-determined |
| S17_extracted_k141_59853 | Not-determined |
| S17_extracted_k141_141222 | Not-determined |
| S17_extracted_k141_163306 | Low-quality |
| S17_extracted_k141_59857 | Not-determined |
| S17_extracted_k141_52542 | Not-determined |
| S17_extracted_k141_37895 | Not-determined |
| S17_extracted_k141_170662 | Not-determined |
| S17_extracted_k141_155932 | Not-determined |
| S17_extracted_k141_104389 | Not-determined |
| S17_extracted_k141_81769 | Not-determined |
| S17_extracted_k141_104390 | Not-determined |
| S17_extracted_k141_126234 | Low-quality |
| S17_extracted_k141_118967 | Not-determined |
| S17_extracted_k141_52559 | Not-determined |
| S17_extracted_k141_155947 | Not-determined |
| S17_extracted_k141_81776 | Not-determined |
| S17_extracted_k141_104399 | Not-determined |
| S17_extracted_k141_74698 | Not-determined |
| S17_extracted_k141_37910 | Low-quality |
| S17_extracted_k141_89303 | Low-quality |
| S17_extracted_k141_170680 | Not-determined |
| S17_extracted_k141_126238 | Low-quality |
| S17_extracted_k141_67107 | Not-determined |
| S17_extracted_k141_74703 | Not-determined |
| S17_extracted_k141_67110 | Not-determined |
| S17_extracted_k141_74705 | Not-determined |
| S17_extracted_k141_104408 | Not-determined |
| S17_extracted_k141_89312 | Not-determined |
| S17_extracted_k141_74706 | Not-determined |
| S17_extracted_k141_67111 | Not-determined |
| S17_extracted_k141_8178 | Low-quality |
| S17_extracted_k141_96660 | Not-determined |
| S17_extracted_k141_81780 | Not-determined |
| [illegible] | [illegible] |

| | |
|---|---|
| S17_extracted_k141_67396 | Not-determined |
| S17_extracted_k141_156265 | Not-determined |
| S17_extracted_k141_22937 | Not-determined |
| S17_extracted_k141_89492 | Not-determined |
| S17_extracted_k141_148926 | Not-determined |
| S17_extracted_k141_148927 | Low-quality |
| S17_extracted_k141_148928 | Not-determined |
| S17_extracted_k141_60072 | Not-determined |
| S17_extracted_k141_38194 | Low-quality |
| S17_extracted_k141_170940 | Not-determined |
| S17_extracted_k141_8415 | Not-determined |
| S17_extracted_k141_74921 | Low-quality |
| S17_extracted_k141_38200 | Not-determined |
| S17_extracted_k141_148932 | Not-determined |
| S17_extracted_k141_30641 | Not-determined |
| S17_extracted_k141_126401 | Not-determined |
| S17_extracted_k141_30643 | Low-quality |
| S17_extracted_k141_60079 | Low-quality |
| S17_extracted_k141_45310 | Not-determined |
| S17_extracted_k141_126403 | Not-determined |
| S17_extracted_k141_8419 | Not-determined |
| S17_extracted_k141_45314 | Low-quality |
| S17_extracted_k141_60087 | Not-determined |
| S17_extracted_k141_38209 | Not-determined |
| S17_extracted_k141_976 | Low-quality |
| S17_extracted_k141_141346 | Low-quality |
| S17_extracted_k141_170950 | Not-determined |
| S17_extracted_k141_979 | Low-quality |
| S17_extracted_k141_89504 | Not-determined |
| S17_extracted_k141_156290 | Low-quality |
| S17_extracted_k141_170956 | Low-quality |
| S17_extracted_k141_156292 | Not-determined |
| S17_extracted_k141_15537 | Low-quality |
| S17_extracted_k141_148943 | Not-determined |
| S17_extracted_k141_148944 | Not-determined |
| S17_extracted_k141_89511 | Not-determined |
| S17_extracted_k141_133840 | Not-determined |
| S17_extracted_k141_104712 | Not-determined |
| S17_extracted_k141_133842 | Not-determined |
| S17_extracted_k141_119221 | Low-quality |
| S17_extracted_k141_133844 | Not-determined |
| S17_extracted_k141_15555 | Not-determined |
| S17_extracted_k141_133846 | Not-determined |
| S17_extracted_k141_156302 | Not-determined |
| S17_extracted_k141_96948 | Not-determined |
| S17_extracted_k141_38224 | Not-determined |
| S17_extracted_k141_89514 | Not-determined |
| S17_extracted_k141_60111 | Not-determined |
| S17_extracted_k141_96950 | Not-determined |
| S17_extracted_k141_170968 | Not-determined |
| S17_extracted_k141_170969 | Low-quality |
| S17_extracted_k141_89516 | Not-determined |
| S17_extracted_k141_96954 | Not-determined |
| S17_extracted_k141_60116 | Not-determined |
| S17_extracted_k141_126408 | Not-determined |
| S17_extracted_k141_126410 | Not-determined |
| S17_extracted_k141_30661 | Not-determined |
| S17_extracted_k141_104722 | Not-determined |
| S17_extracted_k141_89521 | Not-determined |
| S17_extracted_k141_156312 | Not-determined |
| S17_extracted_k141_60117 | Low-quality |
| S17_extracted_k141_148964 | Not-determined |
| S17_extracted_k141_81912 | Not-determined |
| S17_extracted_k141_96961 | Not-determined |
| S17_extracted_k141_156317 | Not-determined |
| S17_extracted_k141_45342 | Low-quality |
| S17_extracted_k141_81917 | Not-determined |
| S17_extracted_k141_60125 | Not-determined |
| S17_extracted_k141_126423 | Not-determined |
| S17_extracted_k141_170981 | Not-determined |
| S17_extracted_k141_15570 | Low-quality |
| S17_extracted_k141_156322 | Not-determined |
| S17_extracted_k141_30673 | Not-determined |
| S17_extracted_k141_170986 | Low-quality |
| S17_extracted_k141_141361 | Not-determined |
| S17_extracted_k141_15574 | Not-determined |
| S17_extracted_k141_15575 | Not-determined |
| S17_extracted_k141_8440 | Not-determined |
| S17_extracted_k141_141366 | Not-determined |
| S17_extracted_k141_119239 | Not-determined |
| S17_extracted_k141_38232 | Not-determined |
| S17_extracted_k141_119240 | Not-determined |
| S17_extracted_k141_126428 | Not-determined |
| S17_extracted_k141_126431 | Not-determined |
| S17_extracted_k141_133856 | Low-quality |
| S17_extracted_k141_30684 | Not-determined |
| S17_extracted_k141_38238 | Not-determined |
| S17_extracted_k141_133858 | Not-determined |
| S17_extracted_k141_38239 | Low-quality |
| S17_extracted_k141_171002 | Not-determined |
| S17_extracted_k141_30690 | Not-determined |
| S17_extracted_k141_141374 | Not-determined |
| S17_extracted_k141_1000 | Not-determined |
| S17_extracted_k141_30695 | Not-determined |
| S17_extracted_k141_119252 | Not-determined |
| S17_extracted_k141_30699 | Low-quality |
| S17_extracted_k141_15592 | Not-determined |
| S17_extracted_k141_119254 | Not-determined |
| S17_extracted_k141_141384 | Not-determined |
| S17_extracted_k141_119256 | Not-determined |
| S17_extracted_k141_89548 | Not-determined |
| S17_extracted_k141_148981 | Not-determined |
| S17_extracted_k141_45353 | Not-determined |
| S17_extracted_k141_15599 | Not-determined |
| S17_extracted_k141_156339 | Not-determined |
| S17_extracted_k141_45355 | Not-determined |
| S17_extracted_k141_171015 | Not-determined |
| S17_extracted_k141_1007 | Not-determined |
| S17_extracted_k141_8462 | Not-determined |
| S17_extracted_k141_141401 | Low-quality |
| S17_extracted_k141_141402 | Not-determined |
| S17_extracted_k141_67418 | Not-determined |
| S17_extracted_k141_141404 | Not-determined |
| S17_extracted_k141_111701 | Not-determined |
| S17_extracted_k141_111702 | Not-determined |
| S17_extracted_k141_8469 | Not-determined |
| S17_extracted_k141_133878 | Not-determined |
| S17_extracted_k141_89561 | Not-determined |
| S17_extracted_k141_171023 | Low-quality |
| S17_extracted_k141_171024 | Not-determined |
| S17_extracted_k141_148996 | Not-determined |
| S17_extracted_k141_163533 | Low-quality |
| S17_extracted_k141_67427 | Not-determined |
| S17_extracted_k141_104756 | Not-determined |
| S17_extracted_k141_133882 | Not-determined |
| S17_extracted_k141_156353 | Not-determined |
| S17_extracted_k141_171027 | Not-determined |
| S17_extracted_k141_67430 | Not-determined |
| S17_extracted_k141_1027 | Not-determined |
| S17_extracted_k141_171035 | Not-determined |
| S17_extracted_k141_119266 | Not-determined |
| S17_extracted_k141_67432 | Not-determined |
| S17_extracted_k141_67434 | Not-determined |
| S17_extracted_k141_171041 | Not-determined |
| S17_extracted_k141_149006 | Not-determined |
| S17_extracted_k141_8479 | Not-determined |
| S17_extracted_k141_81934 | Not-determined |
| S17_extracted_k141_67436 | Not-determined |
| S17_extracted_k141_104758 | Not-determined |
| S17_extracted_k141_89569 | Not-determined |
| S17_extracted_k141_52893 | Not-determined |
| S17_extracted_k141_52894 | Low-quality |
| S17_extracted_k141_52895 | Not-determined |
| S17_extracted_k141_60157 | Not-determined |
| S17_extracted_k141_163541 | Low-quality |
| S17_extracted_k141_1038 | Not-determined |
| S17_extracted_k141_1040 | Not-determined |
| S17_extracted_k141_52897 | Not-determined |
| S17_extracted_k141_67442 | Not-determined |
| S17_extracted_k141_96972 | Not-determined |
| S17_extracted_k141_163546 | Low-quality |
| S17_extracted_k141_1042 | Not-determined |
| S17_extracted_k141_133886 | Low-quality |
| S17_extracted_k141_133888 | Not-determined |
| S17_extracted_k141_96976 | Not-determined |
| S17_extracted_k141_45376 | Not-determined |
| S17_extracted_k141_8496 | Not-determined |
| S17_extracted_k141_111734 | Not-determined |
| S17_extracted_k141_163553 | Not-determined |
| S17_extracted_k141_89576 | Not-determined |
| S17_extracted_k141_149020 | Not-determined |
| S17_extracted_k141_133893 | Not-determined |
| S17_extracted_k141_141409 | Low-quality |
| S17_extracted_k141_60166 | Not-determined |
| S17_extracted_k141_111743 | Not-determined |
| S17_extracted_k141_96979 | Not-determined |
| S17_extracted_k141_96980 | Not-determined |
| S17_extracted_k141_22976 | Not-determined |
| S17_extracted_k141_89580 | Not-determined |
| S17_extracted_k141_96985 | Not-determined |
| S17_extracted_k141_89585 | Not-determined |
| S17_extracted_k141_22979 | Not-determined |
| S17_extracted_k141_22981 | Not-determined |
| S17_extracted_k141_89590 | Low-quality |
| S17_extracted_k141_111760 | Low-quality |
| S17_extracted_k141_149025 | Not-determined |
| S17_extracted_k141_30716 | Not-determined |
| S17_extracted_k141_52907 | Not-determined |
| S17_extracted_k141_163560 | Not-determined |
| S17_extracted_k141_104769 | Not-determined |
| S17_extracted_k141_30718 | Not-determined |
| S17_extracted_k141_163562 | Not-determined |
| S17_extracted_k141_111764 | Low-quality |
| S17_extracted_k141_156375 | Not-determined |
| S17_extracted_k141_133906 | Not-determined |
| S17_extracted_k141_149032 | Not-determined |
| S17_extracted_k141_149034 | Not-determined |
| S17_extracted_k141_133911 | Not-determined |
| S17_extracted_k141_74935 | Low-quality |
| S17_extracted_k141_60175 | Not-determined |
| S17_extracted_k141_30730 | Not-determined |
| S17_extracted_k141_104771 | Low-quality |
| S17_extracted_k141_52915 | Not-determined |
| S17_extracted_k141_60180 | Not-determined |
| S17_extracted_k141_97004 | Not-determined |
| S17_extracted_k141_111782 | Not-determined |
| S17_extracted_k141_133916 | Not-determined |
| S17_extracted_k141_163581 | Not-determined |
| S17_extracted_k141_30735 | Not-determined |
| S17_extracted_k141_163583 | Low-quality |
| S17_extracted_k141_89611 | Not-determined |
| S17_extracted_k141_133917 | Not-determined |
| S17_extracted_k141_89613 | Low-quality |
| S17_extracted_k141_52925 | Not-determined |
| S17_extracted_k141_111789 | Low-quality |
| S17_extracted_k141_45396 | Not-determined |
| S17_extracted_k141_52927 | Not-determined |
| S17_extracted_k141_45397 | Not-determined |
| S17_extracted_k141_74943 | Low-quality |
| S17_extracted_k141_67453 | Low-quality |
| S17_extracted_k141_52929 | Low-quality |
| S17_extracted_k141_38262 | Not-determined |
| S17_extracted_k141_8533 | Low-quality |
| S17_extracted_k141_38263 | Not-determined |
| S17_extracted_k141_133924 | Not-determined |
| S17_extracted_k141_38265 | Not-determined |
| S17_extracted_k141_104781 | Low-quality |
| S17_extracted_k141_74945 | Not-determined |
| S17_extracted_k141_45402 | Low-quality |
| S17_extracted_k141_8537 | Not-determined |
| S17_extracted_k141_171083 | Not-determined |
| S17_extracted_k141_104784 | Not-determined |
| S17_extracted_k141_38270 | Low-quality |
| S17_extracted_k141_30738 | Low-quality |
| S17_extracted_k141_52935 | Not-determined |
| S17_extracted_k141_141417 | Not-determined |
| S17_extracted_k141_149056 | Low-quality |
| S17_extracted_k141_52938 | Not-determined |
| S17_extracted_k141_89633 | Not-determined |
| S17_extracted_k141_156391 | Not-determined |
| S17_extracted_k141_156392 | Not-determined |
| S17_extracted_k141_60192 | Not-determined |
| S17_extracted_k141_45423 | Low-quality |
| S17_extracted_k141_141422 | Not-determined |
| S17_extracted_k141_133930 | Not-determined |
| S17_extracted_k141_163607 | Low-quality |
| S17_extracted_k141_104789 | Not-determined |
| S17_extracted_k141_52944 | Not-determined |
| S17_extracted_k141_163610 | Low-quality |
| S17_extracted_k141_22989 | Not-determined |
| S17_extracted_k141_52945 | Not-determined |
| S17_extracted_k141_67470 | Low-quality |
| S17_extracted_k141_67473 | Not-determined |
| S17_extracted_k141_67476 | Not-determined |
| S17_extracted_k141_67477 | Not-determined |
| S17_extracted_k141_97017 | Not-determined |
| S17_extracted_k141_163616 | Not-determined |
| S17_extracted_k141_52948 | Low-quality |
| S17_extracted_k141_89649 | Not-determined |
| S17_extracted_k141_52949 | Not-determined |
| S17_extracted_k141_133940 | Not-determined |
| S17_extracted_k141_156408 | Low-quality |
| S17_extracted_k141_45441 | Not-determined |
| S17_extracted_k141_156411 | Not-determined |
| S17_extracted_k141_89655 | Low-quality |
| S17_extracted_k141_89656 | Not-determined |
| S17_extracted_k141_104803 | Low-quality |
| S17_extracted_k141_30748 | Not-determined |
| S17_extracted_k141_1047 | Not-determined |
| S17_extracted_k141_45444 | Not-determined |
| S17_extracted_k141_89659 | Not-determined |
| S17_extracted_k141_149063 | Not-determined |
| S17_extracted_k141_163621 | Not-determined |
| S17_extracted_k141_119294 | Not-determined |
| S17_extracted_k141_104808 | Not-determined |
| S17_extracted_k141_45449 | Not-determined |
| S17_extracted_k141_119296 | Not-determined |
| S17_extracted_k141_45452 | Not-determined |
| S17_extracted_k141_149066 | Not-determined |
| S17_extracted_k141_97037 | Not-determined |
| S17_extracted_k141_111802 | Not-determined |
| S17_extracted_k141_30754 | Not-determined |
| S17_extracted_k141_22999 | Not-determined |
| S17_extracted_k141_119298 | Not-determined |
| S17_extracted_k141_81959 | Not-determined |
| S17_extracted_k141_23000 | Not-determined |
| S17_extracted_k141_97040 | Not-determined |
| S17_extracted_k141_141454 | Not-determined |
| S17_extracted_k141_38287 | Low-quality |
| S17_extracted_k141_89667 | Not-determined |
| S17_extracted_k141_156416 | Not-determined |
| S17_extracted_k141_38292 | Not-determined |
| S17_extracted_k141_38293 | Low-quality |
| S17_extracted_k141_30757 | Not-determined |
| S17_extracted_k141_60220 | Not-determined |
| S17_extracted_k141_111809 | Not-determined |
| S17_extracted_k141_60222 | Not-determined |
| S17_extracted_k141_149084 | Not-determined |
| S17_extracted_k141_111811 | Not-determined |
| S17_extracted_k141_141465 | Not-determined |
| S17_extracted_k141_163639 | Not-determined |
| S17_extracted_k141_163642 | Not-determined |
| S17_extracted_k141_104827 | Not-determined |
| S17_extracted_k141_60225 | Not-determined |
| S17_extracted_k141_38298 | Not-determined |
| S17_extracted_k141_149089 | Not-determined |
| S17_extracted_k141_104828 | Not-determined |
| S17_extracted_k141_45462 | Low-quality |
| S17_extracted_k141_171097 | Not-determined |
| S17_extracted_k141_111817 | Not-determined |
| S17_extracted_k141_111818 | Not-determined |
| S17_extracted_k141_45464 | Not-determined |
| S17_extracted_k141_67483 | Not-determined |
| S17_extracted_k141_38301 | Not-determined |
| S17_extracted_k141_156430 | Not-determined |
| S17_extracted_k141_30767 | Not-determined |
| S17_extracted_k141_38305 | Not-determined |
| S17_extracted_k141_30769 | Not-determined |
| S17_extracted_k141_60231 | Not-determined |
| S17_extracted_k141_89677 | Not-determined |
| S17_extracted_k141_119317 | Low-quality |
| S17_extracted_k141_163648 | Not-determined |
| S17_extracted_k141_38306 | Not-determined |
| S17_extracted_k141_8558 | Not-determined |
| S17_extracted_k141_119320 | Not-determined |
| S17_extracted_k141_111825 | Not-determined |
| S17_extracted_k141_81977 | Not-determined |
| S17_extracted_k141_60234 | Not-determined |
| S17_extracted_k141_38309 | Not-determined |
| S17_extracted_k141_171103 | Not-determined |
| S17_extracted_k141_97047 | Not-determined |
| S17_extracted_k141_60237 | Not-determined |
| S17_extracted_k141_38311 | Not-determined |
| S17_extracted_k141_89682 | Not-determined |
| S17_extracted_k141_111836 | Low-quality |
| S17_extracted_k141_163651 | Not-determined |
| S17_extracted_k141_81980 | Not-determined |
| S17_extracted_k141_38317 | Not-determined |
| S17_extracted_k141_15645 | Not-determined |
| S17_extracted_k141_97052 | Not-determined |
| S17_extracted_k141_141480 | Not-determined |
| S17_extracted_k141_81983 | Not-determined |
| S17_extracted_k141_171112 | Not-determined |
| S17_extracted_k141_81985 | Not-determined |
| S17_extracted_k141_45490 | Not-determined |
| S17_extracted_k141_171113 | Not-determined |
| S17_extracted_k141_23010 | Not-determined |
| S17_extracted_k141_119329 | Not-determined |
| S17_extracted_k141_1053 | Not-determined |
| S17_extracted_k141_141485 | Not-determined |
| S17_extracted_k141_1055 | Not-determined |
| S17_extracted_k141_23012 | Not-determined |
| S17_extracted_k141_60240 | Not-determined |
| S17_extracted_k141_23013 | Not-determined |
| S17_extracted_k141_104850 | Not-determined |
| S17_extracted_k141_23015 | Not-determined |
| S17_extracted_k141_156444 | Not-determined |
| S17_extracted_k141_156448 | Not-determined |
| S17_extracted_k141_97059 | Not-determined |
| S17_extracted_k141_30774 | Not-determined |
| S17_extracted_k141_60246 | Not-determined |
| S17_extracted_k141_30775 | Not-determined |
| S17_extracted_k141_141493 | Not-determined |
| S17_extracted_k141_149104 | Not-determined |
| S17_extracted_k141_30776 | Not-determined |
| S17_extracted_k141_156452 | Not-determined |
| S17_extracted_k141_81991 | Not-determined |
| S17_extracted_k141_141496 | Not-determined |
| S17_extracted_k141_133959 | Not-determined |
| S17_extracted_k141_97061 | Not-determined |
| S17_extracted_k141_111854 | Not-determined |
| S17_extracted_k141_81993 | Not-determined |
| S17_extracted_k141_149110 | Not-determined |
| S17_extracted_k141_1063 | Not-determined |
| S17_extracted_k141_141502 | Not-determined |
| S17_extracted_k141_8584 | Not-determined |
| S17_extracted_k141_67505 | Not-determined |
| S17_extracted_k141_45514 | Not-determined |
| S17_extracted_k141_8587 | Low-quality |
| S17_extracted_k141_104867 | Not-determined |
| S17_extracted_k141_141506 | Not-determined |
| S17_extracted_k141_104868 | Not-determined |
| S17_extracted_k141_67509 | Not-determined |
| S17_extracted_k141_23031 | Not-determined |
| S17_extracted_k141_81998 | Not-determined |
| S17_extracted_k141_67514 | Low-quality |
| S17_extracted_k141_119350 | Not-determined |
| S17_extracted_k141_82001 | Not-determined |
| S17_extracted_k141_23034 | Not-determined |
| S17_extracted_k141_97072 | Not-determined |
| S17_extracted_k141_67520 | Not-determined |
| S17_extracted_k141_38334 | Not-determined |
| S17_extracted_k141_149116 | Not-determined |
| S17_extracted_k141_149119 | Not-determined |
| S17_extracted_k141_8602 | Not-determined |
| S17_extracted_k141_38337 | Not-determined |
| S17_extracted_k141_52981 | Not-determined |
| S17_extracted_k141_52983 | Not-determined |
| S17_extracted_k141_38342 | Not-determined |
| S17_extracted_k141_45529 | Not-determined |
| S17_extracted_k141_97074 | Not-determined |
| S17_extracted_k141_67530 | Not-determined |
| S17_extracted_k141_60259 | Not-determined |
| S17_extracted_k141_119365 | Not-determined |
| S17_extracted_k141_67531 | Not-determined |
| S17_extracted_k141_156475 | Not-determined |
| S17_extracted_k141_67533 | Not-determined |
| S17_extracted_k141_38345 | Not-determined |
| S17_extracted_k141_67534 | Low-quality |
| S17_extracted_k141_97078 | Not-determined |
| S17_extracted_k141_23053 | Not-determined |
| S17_extracted_k141_67540 | Not-determined |
| S17_extracted_k141_156478 | Not-determined |
| S17_extracted_k141_119370 | Not-determined |
| S17_extracted_k141_111863 | Low-quality |
| S17_extracted_k141_67544 | Low-quality |
| S17_extracted_k141_67545 | Not-determined |
| S17_extracted_k141_1082 | Not-determined |
| S17_extracted_k141_8614 | Not-determined |
| S17_extracted_k141_8615 | Low-quality |
| S17_extracted_k141_149126 | Not-determined |
| S17_extracted_k141_67549 | Not-determined |
| S17_extracted_k141_30794 | Not-determined |
| S17_extracted_k141_30796 | Not-determined |
| S17_extracted_k141_53004 | Low-quality |
| S17_extracted_k141_163676 | Not-determined |
| S17_extracted_k141_30800 | Not-determined |
| S17_extracted_k141_8625 | Not-determined |
| S17_extracted_k141_82039 | Not-determined |
| S17_extracted_k141_171139 | Not-determined |
| S17_extracted_k141_149137 | Not-determined |
| S17_extracted_k141_38368 | Low-quality |
| S17_extracted_k141_149140 | Not-determined |
| S17_extracted_k141_163684 | Not-determined |
| S17_extracted_k141_53014 | Not-determined |
| S17_extracted_k141_97093 | Not-determined |
| S17_extracted_k141_1094 | Not-determined |
| S17_extracted_k141_97095 | Not-determined |
| S17_extracted_k141_30806 | Not-determined |
| S17_extracted_k141_1095 | Not-determined |
| S17_extracted_k141_38373 | Not-determined |
| S17_extracted_k141_163691 | Not-determined |
| S17_extracted_k141_89706 | Low-quality |
| S17_extracted_k141_111881 | Not-determined |
| S17_extracted_k141_8633 | Not-determined |
| S17_extracted_k141_45561 | Not-determined |
| S17_extracted_k141_8635 | Not-determined |
| S17_extracted_k141_163695 | Not-determined |
| S17_extracted_k141_141519 | Not-determined |
| S17_extracted_k141_149145 | Not-determined |
| S17_extracted_k141_97104 | Low-quality |
| S17_extracted_k141_171142 | Not-determined |
| S17_extracted_k141_141521 | Not-determined |
| S17_extracted_k141_104907 | Not-determined |
| S17_extracted_k141_97106 | Not-determined |
| S17_extracted_k141_104909 | Not-determined |
| S17_extracted_k141_97110 | Low-quality |
| S17_extracted_k141_97111 | Not-determined |
| S17_extracted_k141_15699 | Not-determined |
| S17_extracted_k141_60285 | Not-determined |
| S17_extracted_k141_104913 | Not-determined |
| S17_extracted_k141_149148 | Not-determined |
| S17_extracted_k141_30821 | Not-determined |
| S17_extracted_k141_30822 | Low-quality |
| S17_extracted_k141_104919 | Not-determined |
| S17_extracted_k141_133985 | Not-determined |
| S17_extracted_k141_133989 | Not-determined |
| S17_extracted_k141_23080 | Not-determined |
| S17_extracted_k141_141531 | Not-determined |
| S17_extracted_k141_141532 | Not-determined |
| S17_extracted_k141_171149 | Not-determined |
| S17_extracted_k141_38376 | Not-determined |
| S17_extracted_k141_23081 | Not-determined |
| S17_extracted_k141_53032 | Not-determined |
| S17_extracted_k141_133992 | Not-determined |
| S17_extracted_k141_23082 | Not-determined |
| S17_extracted_k141_67583 | Not-determined |
| S17_extracted_k141_1108 | Not-determined |
| S17_extracted_k141_171155 | Not-determined |
| S17_extracted_k141_23087 | Not-determined |
| S17_extracted_k141_111905 | Not-determined |
| S17_extracted_k141_141541 | Not-determined |
| S17_extracted_k141_97129 | Not-determined |
| S17_extracted_k141_149160 | Not-determined |
| S17_extracted_k141_171162 | Not-determined |
| S17_extracted_k141_111908 | Not-determined |
| S17_extracted_k141_30833 | Not-determined |
| S17_extracted_k141_45585 | Not-determined |
| S17_extracted_k141_141544 | Low-quality |
| S17_extracted_k141_53046 | Not-determined |
| S17_extracted_k141_141545 | Not-determined |
| S17_extracted_k141_89721 | Not-determined |
| S17_extracted_k141_82062 | Not-determined |
| S17_extracted_k141_141549 | Low-quality |
| S17_extracted_k141_30836 | Not-determined |
| S17_extracted_k141_156493 | Not-determined |
| S17_extracted_k141_156494 | Not-determined |
| S17_extracted_k141_38388 | Low-quality |
| S17_extracted_k141_97148 | Low-quality |
| S17_extracted_k141_67601 | Not-determined |
| S17_extracted_k141_141554 | Not-determined |
| S17_extracted_k141_97152 | Not-determined |
| S17_extracted_k141_104942 | Not-determined |
| S17_extracted_k141_133994 | Not-determined |
| S17_extracted_k141_38390 | Not-determined |
| S17_extracted_k141_156498 | Not-determined |
| S17_extracted_k141_104945 | Not-determined |
| S17_extracted_k141_133998 | Not-determined |
| S17_extracted_k141_8650 | Not-determined |
| S17_extracted_k141_67605 | Not-determined |
| S17_extracted_k141_30847 | Low-quality |
| S17_extracted_k141_149179 | Not-determined |
| S17_extracted_k141_149181 | Not-determined |
| S17_extracted_k141_171184 | Not-determined |
| S17_extracted_k141_89739 | Low-quality |
| S17_extracted_k141_156508 | Not-determined |
| S17_extracted_k141_89740 | Not-determined |
| S17_extracted_k141_38399 | Not-determined |
| S17_extracted_k141_8658 | Not-determined |
| S17_extracted_k141_171188 | Not-determined |
| S17_extracted_k141_74955 | Not-determined |
| S17_extracted_k141_141566 | Not-determined |
| S17_extracted_k141_60291 | Not-determined |
| S17_extracted_k141_89750 | Not-determined |
| S17_extracted_k141_171192 | Not-determined |
| S17_extracted_k141_53072 | Not-determined |
| S17_extracted_k141_104961 | Low-quality |
| S17_extracted_k141_89752 | Not-determined |
| S17_extracted_k141_156514 | Not-determined |
| S17_extracted_k141_156515 | Low-quality |
| S17_extracted_k141_38409 | Not-determined |
| S17_extracted_k141_45600 | Low-quality |
| S17_extracted_k141_74960 | Low-quality |
| S17_extracted_k141_141574 | Not-determined |
| S17_extracted_k141_149184 | Not-determined |
| S17_extracted_k141_149185 | Low-quality |
| S17_extracted_k141_53078 | Not-determined |
| S17_extracted_k141_104970 | Not-determined |
| S17_extracted_k141_104971 | Not-determined |
| S17_extracted_k141_156521 | Not-determined |
| S17_extracted_k141_149189 | Not-determined |
| S17_extracted_k141_141584 | Low-quality |
| S17_extracted_k141_134010 | Not-determined |
| S17_extracted_k141_171202 | Not-determined |
| S17_extracted_k141_45611 | Not-determined |
| S17_extracted_k141_156526 | Not-determined |
| S17_extracted_k141_53083 | Not-determined |
| S17_extracted_k141_134013 | Not-determined |
| S17_extracted_k141_141590 | Not-determined |
| S17_extracted_k141_1116 | Not-determined |
| S17_extracted_k141_8672 | Low-quality |
| S17_extracted_k141_1117 | Not-determined |
| S17_extracted_k141_8675 | Not-determined |
| S17_extracted_k141_8676 | Not-determined |
| S17_extracted_k141_23093 | Not-determined |
| S17_extracted_k141_23094 | Not-determined |
| S17_extracted_k141_134020 | Not-determined |
| S17_extracted_k141_119398 | Not-determined |
| S17_extracted_k141_149196 | Not-determined |
| S17_extracted_k141_45620 | Low-quality |
| S17_extracted_k141_8683 | Not-determined |
| S17_extracted_k141_89776 | Not-determined |
| S17_extracted_k141_1128 | Low-quality |
| S17_extracted_k141_149197 | Low-quality |
| S17_extracted_k141_141607 | Not-determined |
| S17_extracted_k141_104990 | Not-determined |
| S17_extracted_k141_23109 | Not-determined |
| S17_extracted_k141_163715 | Not-determined |
| S17_extracted_k141_119404 | Low-quality |
| S17_extracted_k141_53094 | Not-determined |
| S17_extracted_k141_60313 | Not-determined |
| S17_extracted_k141_53095 | Low-quality |
| S17_extracted_k141_156536 | Not-determined |
| S17_extracted_k141_134033 | Not-determined |
| S17_extracted_k141_149204 | Low-quality |
| S17_extracted_k141_119408 | Low-quality |
| S17_extracted_k141_134034 | Not-determined |
| S17_extracted_k141_141611 | Not-determined |
| S17_extracted_k141_111917 | Not-determined |
| S17_extracted_k141_53099 | Not-determined |
| S17_extracted_k141_134035 | Not-determined |
| S17_extracted_k141_156540 | Not-determined |
| S17_extracted_k141_30866 | Not-determined |
| S17_extracted_k141_119413 | Not-determined |
| S17_extracted_k141_97171 | Not-determined |
| S17_extracted_k141_45631 | Not-determined |
| S17_extracted_k141_60317 | Low-quality |
| S17_extracted_k141_38434 | Not-determined |
| S17_extracted_k141_163728 | Not-determined |
| S17_extracted_k141_104992 | Not-determined |
| S17_extracted_k141_163729 | Not-determined |
| S17_extracted_k141_134037 | Not-determined |
| S17_extracted_k141_53105 | Not-determined |
| S17_extracted_k141_111927 | Not-determined |
| S17_extracted_k141_156550 | Not-determined |
| S17_extracted_k141_171223 | Low-quality |
| S17_extracted_k141_119419 | Not-determined |
| S17_extracted_k141_111930 | Low-quality |
| S17_extracted_k141_45640 | Low-quality |

| | |
|---|---|
| S17_extracted_k141_171225 | Not-determined |
| S17_extracted_k141_45644 | Not-determined |
| S17_extracted_k141_67610 | Not-determined |
| S17_extracted_k141_53114 | Not-determined |
| S17_extracted_k141_105004 | Not-determined |
| S17_extracted_k141_67613 | Not-determined |
| S17_extracted_k141_163740 | Not-determined |
| S17_extracted_k141_163743 | Not-determined |
| S17_extracted_k141_134041 | Low-quality |
| S17_extracted_k141_134045 | Not-determined |
| S17_extracted_k141_67619 | Low-quality |
| S17_extracted_k141_141638 | Not-determined |
| S17_extracted_k141_141642 | Not-determined |
| S17_extracted_k141_30880 | Not-determined |
| S17_extracted_k141_53130 | Not-determined |
| S17_extracted_k141_74989 | Low-quality |
| S17_extracted_k141_30886 | Not-determined |
| S17_extracted_k141_156576 | Not-determined |
| S17_extracted_k141_67623 | Not-determined |
| S17_extracted_k141_38456 | Not-determined |
| S17_extracted_k141_97187 | Not-determined |
| S17_extracted_k141_74996 | Not-determined |
| S17_extracted_k141_23118 | Not-determined |
| S17_extracted_k141_141649 | Not-determined |
| S17_extracted_k141_67627 | Not-determined |
| S17_extracted_k141_97188 | Not-determined |
| S17_extracted_k141_30894 | Not-determined |
| S17_extracted_k141_141652 | Not-determined |
| S9_vs_k141_13507\|\|full | Complete |
| S17_extracted_k141_97192 | Low-quality |
| S17_extracted_k141_111932 | Not-determined |
| S17_extracted_k141_97194 | Not-determined |
| S17_extracted_k141_30899 | Low-quality |
| S17_extracted_k141_119441 | Low-quality |
| S17_extracted_k141_105023 | Not-determined |
| S17_extracted_k141_105024 | Not-determined |
| S17_extracted_k141_30902 | Not-determined |
| S17_extracted_k141_97197 | Not-determined |
| S17_extracted_k141_119445 | Not-determined |
| S17_extracted_k141_1151 | Not-determined |
| S17_extracted_k141_163770 | Not-determined |
| S17_extracted_k141_53145 | Not-determined |
| S17_extracted_k141_67633 | Not-determined |
| S17_extracted_k141_149230 | Low-quality |
| S17_extracted_k141_163771 | Low-quality |
| S17_extracted_k141_67634 | Not-determined |
| S17_extracted_k141_163772 | Not-determined |
| S17_extracted_k141_134059 | Low-quality |
| S17_extracted_k141_149234 | Not-determined |
| S17_extracted_k141_23138 | Not-determined |
| S17_extracted_k141_156595 | Not-determined |
| S17_extracted_k141_23139 | Low-quality |
| S17_extracted_k141_97203 | Not-determined |
| S17_extracted_k141_156597 | Not-determined |
| S17_extracted_k141_156598 | Not-determined |
| S17_extracted_k141_105033 | Low-quality |
| S17_extracted_k141_156599 | Low-quality |
| S17_extracted_k141_105037 | Low-quality |
| S17_extracted_k141_105038 | Not-determined |
| S17_extracted_k141_119454 | Not-determined |
| S17_extracted_k141_149239 | Not-determined |
| S17_extracted_k141_30908 | Not-determined |
| S17_extracted_k141_119459 | Not-determined |
| S17_extracted_k141_75013 | Low-quality |
| S17_extracted_k141_156609 | Not-determined |
| S17_extracted_k141_45666 | Not-determined |
| S17_extracted_k141_149242 | Not-determined |
| S17_extracted_k141_23152 | Not-determined |
| S17_extracted_k141_105049 | Not-determined |
| S17_extracted_k141_97217 | Not-determined |
| S17_extracted_k141_1165 | Not-determined |
| S17_extracted_k141_171250 | Not-determined |
| S17_extracted_k141_53163 | Not-determined |
| S17_extracted_k141_134073 | Not-determined |
| S17_extracted_k141_82064 | Not-determined |
| S17_extracted_k141_45674 | Not-determined |
| S17_extracted_k141_67664 | Not-determined |
| S17_extracted_k141_45677 | Not-determined |
| S17_extracted_k141_75022 | Not-determined |
| S17_extracted_k141_141661 | Not-determined |
| S17_extracted_k141_141663 | Not-determined |
| S17_extracted_k141_67674 | Not-determined |
| S17_extracted_k141_141668 | Low-quality |
| S17_extracted_k141_149258 | Not-determined |
| S17_extracted_k141_171257 | Not-determined |
| S17_extracted_k141_1171 | Not-determined |
| S17_extracted_k141_1172 | Not-determined |
| S17_extracted_k141_60331 | Not-determined |
| S17_extracted_k141_23166 | Not-determined |
| S17_extracted_k141_23167 | Not-determined |
| S17_extracted_k141_38477 | Low-quality |
| S17_extracted_k141_89821 | Low-quality |
| S17_extracted_k141_23171 | Not-determined |
| S17_extracted_k141_141676 | Not-determined |
| S17_extracted_k141_171264 | Not-determined |
| S17_extracted_k141_23173 | Not-determined |
| S17_extracted_k141_171265 | Not-determined |
| S17_extracted_k141_1173 | Not-determined |
| S17_extracted_k141_149272 | Not-determined |
| S17_extracted_k141_149273 | Not-determined |
| S17_extracted_k141_8696 | Low-quality |
| S17_extracted_k141_30920 | Not-determined |
| S17_extracted_k141_75034 | Low-quality |
| S17_extracted_k141_119476 | Not-determined |
| S17_extracted_k141_60334 | Not-determined |
| S17_extracted_k141_1178 | Not-determined |
| S17_extracted_k141_89827 | Not-determined |
| S17_extracted_k141_60340 | Not-determined |
| S17_extracted_k141_15733 | Not-determined |
| S17_extracted_k141_30931 | Low-quality |
| S17_extracted_k141_149284 | Not-determined |
| S17_extracted_k141_141688 | Not-determined |
| S17_extracted_k141_8701 | Not-determined |
| S17_extracted_k141_23176 | Low-quality |
| S17_extracted_k141_105074 | Not-determined |
| S17_extracted_k141_60348 | Not-determined |
| S17_extracted_k141_119485 | Not-determined |
| S17_extracted_k141_171276 | Not-determined |
| S17_extracted_k141_8705 | Not-determined |
| S17_extracted_k141_111950 | Not-determined |
| S17_extracted_k141_119490 | Not-determined |
| S17_extracted_k141_75039 | Not-determined |
| S17_extracted_k141_149288 | Low-quality |
| S17_extracted_k141_1183 | Not-determined |
| S17_extracted_k141_149290 | Not-determined |
| S17_extracted_k141_23181 | Not-determined |
| S17_extracted_k141_111959 | Not-determined |
| S17_extracted_k141_53200 | Not-determined |
| S17_extracted_k141_60362 | Not-determined |
| S17_extracted_k141_82072 | Low-quality |
| S17_extracted_k141_163784 | Low-quality |
| S17_extracted_k141_163785 | Not-determined |
| S17_extracted_k141_67701 | Low-quality |
| S17_extracted_k141_171285 | Not-determined |
| S17_extracted_k141_67702 | Not-determined |
| S17_extracted_k141_89847 | Not-determined |
| S17_extracted_k141_75049 | Low-quality |
| S17_extracted_k141_119500 | Not-determined |
| S17_extracted_k141_134107 | Not-determined |
| S17_extracted_k141_82082 | Not-determined |
| S17_extracted_k141_67707 | Not-determined |
| S17_extracted_k141_119503 | Not-determined |
| S17_extracted_k141_8726 | Not-determined |
| S17_extracted_k141_30951 | Not-determined |
| S17_extracted_k141_163788 | Not-determined |
| S17_extracted_k141_89855 | Not-determined |
| S17_extracted_k141_119504 | Low-quality |
| S17_extracted_k141_97243 | Not-determined |
| S17_extracted_k141_119507 | Not-determined |
| S17_extracted_k141_67713 | Not-determined |
| S17_extracted_k141_97252 | Not-determined |
| S17_extracted_k141_75062 | Not-determined |
| S17_extracted_k141_97253 | Not-determined |
| S17_extracted_k141_89865 | Not-determined |
| S17_extracted_k141_75065 | Not-determined |
| S17_extracted_k141_134113 | Not-determined |
| S17_extracted_k141_75066 | Not-determined |
| S17_extracted_k141_1192 | Not-determined |
| S17_extracted_k141_156645 | Not-determined |
| S17_extracted_k141_30959 | Low-quality |
| S17_extracted_k141_89867 | Not-determined |
| S17_extracted_k141_134117 | Not-determined |
| S17_extracted_k141_53209 | Not-determined |
| S17_extracted_k141_149315 | Not-determined |
| S17_extracted_k141_163797 | Not-determined |
| S17_extracted_k141_82105 | Not-determined |
| S17_extracted_k141_8742 | Not-determined |
| S17_extracted_k141_126450 | Not-determined |
| S17_extracted_k141_97270 | Not-determined |
| S17_extracted_k141_75076 | Not-determined |
| S17_extracted_k141_97274 | Not-determined |
| S17_extracted_k141_30960 | Not-determined |
| S17_extracted_k141_156657 | Not-determined |
| S17_extracted_k141_134131 | Not-determined |
| S17_extracted_k141_119518 | Not-determined |
| S17_extracted_k141_126459 | Not-determined |
| S17_extracted_k141_15748 | Low-quality |
| S17_extracted_k141_126460 | Not-determined |
| S17_extracted_k141_105110 | Not-determined |
| S17_extracted_k141_67737 | Not-determined |
| S17_extracted_k141_30967 | Not-determined |
| S17_extracted_k141_89889 | Not-determined |
| S17_extracted_k141_1208 | Not-determined |
| S17_extracted_k141_15750 | Not-determined |
| S17_extracted_k141_149321 | Not-determined |
| S17_extracted_k141_15752 | Not-determined |
| S17_extracted_k141_171299 | Not-determined |
| S17_extracted_k141_1209 | Not-determined |
| S17_extracted_k141_111992 | Not-determined |
| S17_extracted_k141_105115 | Not-determined |
| S17_extracted_k141_1210 | Not-determined |
| S17_extracted_k141_8764 | Not-determined |
| S17_extracted_k141_82131 | Not-determined |
| S17_extracted_k141_75083 | Not-determined |
| S17_extracted_k141_134139 | Not-determined |
| S17_extracted_k141_89901 | Not-determined |
| S17_extracted_k141_105123 | Not-determined |
| S17_extracted_k141_105124 | Not-determined |
| S17_extracted_k141_134141 | Not-determined |
| S17_extracted_k141_60371 | Not-determined |
| S17_extracted_k141_75088 | Not-determined |
| S17_extracted_k141_67755 | Not-determined |
| S17_extracted_k141_105127 | Low-quality |
| S17_extracted_k141_134144 | Not-determined |
| S17_extracted_k141_134145 | Not-determined |
| S17_extracted_k141_119532 | Not-determined |
| S17_extracted_k141_163807 | Not-determined |
| S17_extracted_k141_45691 | Low-quality |
| S17_extracted_k141_97287 | Not-determined |
| S17_extracted_k141_119533 | Not-determined |
| S17_extracted_k141_45694 | Low-quality |
| S17_extracted_k141_30977 | Not-determined |
| S17_extracted_k141_1219 | Not-determined |
| S17_extracted_k141_82138 | Not-determined |
| S17_extracted_k141_30978 | Not-determined |
| S17_extracted_k141_23196 | Not-determined |
| S17_extracted_k141_171309 | Not-determined |
| S17_extracted_k141_82141 | Not-determined |
| S17_extracted_k141_53236 | Not-determined |
| S17_extracted_k141_171311 | Not-determined |
| S17_extracted_k141_82145 | Not-determined |
| S17_extracted_k141_134152 | Not-determined |
| S17_extracted_k141_1226 | Not-determined |
| S17_extracted_k141_126476 | Not-determined |
| S17_extracted_k141_82146 | Not-determined |
| S17_extracted_k141_149346 | Not-determined |
| S17_extracted_k141_134155 | Low-quality |
| S17_extracted_k141_134156 | Not-determined |
| S17_extracted_k141_97296 | Low-quality |
| S17_extracted_k141_105140 | Low-quality |
| S17_extracted_k141_15776 | Not-determined |
| S17_extracted_k141_89921 | Not-determined |
| S17_extracted_k141_134159 | Not-determined |
| S17_extracted_k141_15778 | Not-determined |
| S17_extracted_k141_23207 | Not-determined |
| S17_extracted_k141_126483 | Not-determined |
| S17_extracted_k141_82157 | Not-determined |
| S17_extracted_k141_82158 | Not-determined |
| S17_extracted_k141_149354 | Not-determined |
| S17_extracted_k141_149357 | Not-determined |
| S17_extracted_k141_53244 | Not-determined |
| S17_extracted_k141_8798 | Low-quality |
| S17_extracted_k141_60378 | Not-determined |
| S17_extracted_k141_67768 | Not-determined |
| S17_extracted_k141_75114 | Not-determined |
| S17_extracted_k141_67769 | Not-determined |
| S17_extracted_k141_75115 | Not-determined |
| S17_extracted_k141_149358 | Not-determined |
| S17_extracted_k141_67771 | Low-quality |
| S17_extracted_k141_38528 | Not-determined |
| S17_extracted_k141_1242 | Low-quality |
| S17_extracted_k141_67774 | Not-determined |
| S17_extracted_k141_15793 | Not-determined |
| S17_extracted_k141_38532 | Not-determined |
| S17_extracted_k141_163824 | Not-determined |
| S17_extracted_k141_82160 | Not-determined |
| S17_extracted_k141_1247 | Not-determined |
| S17_extracted_k141_163826 | Not-determined |
| S17_extracted_k141_53255 | Not-determined |
| S17_extracted_k141_1249 | Not-determined |
| S17_extracted_k141_97300 | Low-quality |
| S17_extracted_k141_30995 | Not-determined |
| S17_extracted_k141_82165 | Not-determined |
| S17_extracted_k141_89947 | Not-determined |
| S17_extracted_k141_97302 | Not-determined |
| S17_extracted_k141_23233 | Not-determined |
| S17_extracted_k141_8811 | Not-determined |
| S17_extracted_k141_53258 | Not-determined |
| S17_extracted_k141_67786 | Not-determined |
| S17_extracted_k141_97304 | Not-determined |
| S17_extracted_k141_45721 | Not-determined |
| S17_extracted_k141_171343 | Not-determined |
| S17_extracted_k141_82177 | Not-determined |
| S17_extracted_k141_45725 | Not-determined |
| S17_extracted_k141_31003 | Not-determined |
| S17_extracted_k141_163835 | Not-determined |
| S17_extracted_k141_53260 | Not-determined |
| S17_extracted_k141_97313 | Not-determined |
| S17_extracted_k141_31008 | Not-determined |
| S17_extracted_k141_31011 | Not-determined |
| S17_extracted_k141_8828 | Not-determined |
| S17_extracted_k141_75134 | Not-determined |
| S17_extracted_k141_31015 | Not-determined |
| S17_extracted_k141_45732 | Not-determined |
| S17_extracted_k141_31017 | Not-determined |
| S17_extracted_k141_134168 | Low-quality |
| S17_extracted_k141_163841 | Not-determined |
| S17_extracted_k141_134169 | Not-determined |
| S17_extracted_k141_45737 | Low-quality |
| S17_extracted_k141_97324 | Not-determined |
| S17_extracted_k141_45739 | Not-determined |
| S17_extracted_k141_134174 | Not-determined |
| S17_extracted_k141_82183 | Low-quality |
| S17_extracted_k141_38541 | Not-determined |
| S17_extracted_k141_45744 | Not-determined |
| S17_extracted_k141_97328 | Not-determined |
| S17_extracted_k141_38543 | Not-determined |
| S17_extracted_k141_60414 | Not-determined |
| S17_extracted_k141_171354 | Not-determined |
| S17_extracted_k141_23253 | Low-quality |
| S17_extracted_k141_171355 | Not-determined |
| S17_extracted_k141_60420 | Not-determined |
| S17_extracted_k141_163855 | Not-determined |
| S17_extracted_k141_38548 | Low-quality |
| S17_extracted_k141_163856 | Not-determined |
| S17_extracted_k141_89975 | Not-determined |
| S17_extracted_k141_8840 | Low-quality |
| S17_extracted_k141_8843 | Not-determined |
| S17_extracted_k141_134182 | Not-determined |
| S17_extracted_k141_31035 | Not-determined |
| S17_extracted_k141_1262 | Low-quality |
| S17_extracted_k141_53279 | Not-determined |
| S17_extracted_k141_1265 | Low-quality |
| S17_extracted_k141_38560 | Low-quality |
| S17_extracted_k141_171360 | Not-determined |
| S17_extracted_k141_38563 | Not-determined |
| S17_extracted_k141_67808 | Not-determined |
| S17_extracted_k141_149410 | Not-determined |
| S17_extracted_k141_38564 | Not-determined |
| S17_extracted_k141_31046 | Not-determined |
| S17_extracted_k141_1273 | Not-determined |
| S17_extracted_k141_38569 | Not-determined |
| S17_extracted_k141_171366 | Not-determined |
| S17_extracted_k141_134193 | Not-determined |
| S17_extracted_k141_171367 | Not-determined |
| S17_extracted_k141_67809 | Not-determined |
| S17_extracted_k141_97350 | Low-quality |
| S17_extracted_k141_45775 | Not-determined |
| S17_extracted_k141_82191 | Low-quality |
| S17_extracted_k141_45776 | Not-determined |
| S17_extracted_k141_82195 | Not-determined |
| S17_extracted_k141_89980 | Not-determined |
| S17_extracted_k141_134208 | Low-quality |
| S17_extracted_k141_23261 | Not-determined |
| S17_extracted_k141_60441 | Not-determined |
| S17_extracted_k141_134212 | Not-determined |
| S17_extracted_k141_105174 | Not-determined |
| S17_extracted_k141_97361 | Not-determined |
| S17_extracted_k141_134213 | Not-determined |
| S17_extracted_k141_8863 | Low-quality |
| S17_extracted_k141_149417 | Not-determined |
| S17_extracted_k141_23262 | Not-determined |
| S17_extracted_k141_38585 | Low-quality |
| S17_extracted_k141_23264 | Low-quality |
| S17_extracted_k141_105186 | Not-determined |
| S17_extracted_k141_1291 | Not-determined |
| S17_extracted_k141_149427 | Not-determined |
| S17_extracted_k141_15813 | Not-determined |
| S17_extracted_k141_53311 | Not-determined |
| S17_extracted_k141_8873 | Not-determined |
| S17_extracted_k141_60450 | Low-quality |
| S17_extracted_k141_105195 | Not-determined |
| S17_extracted_k141_8876 | Not-determined |
| S17_extracted_k141_126498 | Not-determined |
| S17_extracted_k141_38598 | Not-determined |
| S12_extracted_k141_188819 | Complete |
| S17_extracted_k141_67813 | Not-determined |
| S17_extracted_k141_45797 | Not-determined |
| S17_extracted_k141_1305 | Not-determined |
| S17_extracted_k141_67816 | Not-determined |
| S17_extracted_k141_149433 | Not-determined |
| S17_extracted_k141_38608 | Low-quality |
| S17_extracted_k141_105213 | Not-determined |
| S17_extracted_k141_38610 | Not-determined |
| S17_extracted_k141_171389 | Not-determined |
| S17_extracted_k141_90006 | Not-determined |
| S17_extracted_k141_8881 | Not-determined |
| S17_extracted_k141_23274 | Not-determined |
| S17_extracted_k141_15819 | Not-determined |
| S17_extracted_k141_119559 | Low-quality |
| S17_extracted_k141_82209 | Not-determined |
| S17_extracted_k141_82210 | Not-determined |
| S17_extracted_k141_112003 | Not-determined |
| S17_extracted_k141_119562 | Not-determined |
| S17_extracted_k141_112005 | Not-determined |
| S17_extracted_k141_53321 | Low-quality |
| S17_extracted_k141_45807 | Not-determined |
| S17_extracted_k141_149447 | Low-quality |
| S17_extracted_k141_1326 | Not-determined |
| S17_extracted_k141_119569 | Not-determined |
| S17_extracted_k141_126508 | Not-determined |
| S17_extracted_k141_31057 | Low-quality |
| S17_extracted_k141_67837 | Not-determined |
| S17_extracted_k141_82222 | Not-determined |
| S17_extracted_k141_67838 | Not-determined |
| S17_extracted_k141_60468 | Not-determined |
| S17_extracted_k141_82223 | Not-determined |
| S17_extracted_k141_60470 | Not-determined |
| S17_extracted_k141_82228 | Low-quality |
| S17_extracted_k141_82229 | Not-determined |
| S17_extracted_k141_119579 | Not-determined |
| S17_extracted_k141_171404 | Not-determined |
| S17_extracted_k141_156665 | Low-quality |
| S17_extracted_k141_171405 | Not-determined |
| S17_extracted_k141_60475 | Not-determined |
| S17_extracted_k141_126520 | Not-determined |
| S17_extracted_k141_149449 | Not-determined |
| S17_extracted_k141_126521 | Not-determined |
| S17_extracted_k141_97384 | Not-determined |
| S17_extracted_k141_119587 | Not-determined |
| S17_extracted_k141_8890 | Low-quality |
| S17_extracted_k141_8891 | Not-determined |
| S17_extracted_k141_53334 | Not-determined |
| S17_extracted_k141_97395 | Not-determined |
| S17_extracted_k141_90020 | Not-determined |
| S17_extracted_k141_38630 | Not-determined |
| S17_extracted_k141_53338 | Not-determined |
| S17_extracted_k141_171415 | Not-determined |
| S17_extracted_k141_134225 | Not-determined |
| S17_extracted_k141_90022 | Low-quality |
| S17_extracted_k141_119599 | Low-quality |
| S17_extracted_k141_156676 | Not-determined |
| S17_extracted_k141_8899 | Not-determined |
| S17_extracted_k141_171423 | Not-determined |
| S17_extracted_k141_171424 | Not-determined |
| S17_extracted_k141_8902 | Not-determined |
| S17_extracted_k141_31069 | Not-determined |
| S17_extracted_k141_23292 | Not-determined |
| S17_extracted_k141_90025 | Not-determined |
| S17_extracted_k141_119606 | Not-determined |
| S17_extracted_k141_1341 | Not-determined |
| S17_extracted_k141_60480 | Not-determined |
| S17_extracted_k141_75148 | Low-quality |
| S17_extracted_k141_31074 | Not-determined |
| S17_extracted_k141_53349 | Low-quality |
| S17_extracted_k141_45837 | Not-determined |
| S17_extracted_k141_82249 | Not-determined |
| S17_extracted_k141_112019 | Not-determined |
| S17_extracted_k141_126547 | Not-determined |
| S17_extracted_k141_126551 | Not-determined |
| S17_extracted_k141_75155 | Not-determined |
| S17_extracted_k141_171431 | Not-determined |
| S17_extracted_k141_82254 | Not-determined |
| S17_extracted_k141_82255 | Not-determined |
| S17_extracted_k141_38653 | Not-determined |
| S17_extracted_k141_134242 | Not-determined |
| S17_extracted_k141_82256 | Not-determined |
| S17_extracted_k141_1359 | Not-determined |
| S17_extracted_k141_31087 | Not-determined |
| S17_extracted_k141_38658 | Low-quality |
| S17_extracted_k141_97411 | Not-determined |
| S17_extracted_k141_38659 | Not-determined |
| S17_extracted_k141_134248 | Not-determined |
| S17_extracted_k141_45846 | Not-determined |
| S17_extracted_k141_134249 | Not-determined |
| S17_extracted_k141_60496 | Not-determined |
| S17_extracted_k141_97417 | Not-determined |
| S17_extracted_k141_97420 | Not-determined |
| S17_extracted_k141_53354 | Not-determined |
| S17_extracted_k141_90034 | Low-quality |
| S17_extracted_k141_126564 | Low-quality |
| S17_extracted_k141_8929 | Not-determined |
| S17_extracted_k141_90036 | Not-determined |
| S17_extracted_k141_38668 | Not-determined |
| S17_extracted_k141_97427 | Not-determined |
| S17_extracted_k141_156690 | Not-determined |
| S17_extracted_k141_8934 | Not-determined |
| S17_extracted_k141_75169 | Not-determined |
| S17_extracted_k141_119614 | Not-determined |
| S17_extracted_k141_82261 | Low-quality |
| S17_extracted_k141_82263 | Not-determined |
| S17_extracted_k141_126573 | Low-quality |
| S17_extracted_k141_119617 | Low-quality |
| S17_extracted_k141_149483 | Not-determined |
| S17_extracted_k141_119618 | Not-determined |
| S17_extracted_k141_8940 | Not-determined |
| S17_extracted_k141_60513 | Not-determined |
| S17_extracted_k141_112030 | Not-determined |
| S17_extracted_k141_141769 | Not-determined |
| S17_extracted_k141_97432 | Low-quality |
| S17_extracted_k141_149485 | Not-determined |
| S17_extracted_k141_75174 | Not-determined |
| S17_extracted_k141_119621 | Not-determined |
| S17_extracted_k141_8942 | Not-determined |
| S17_extracted_k141_23297 | Not-determined |
| S17_extracted_k141_8947 | Low-quality |
| S17_extracted_k141_90048 | Not-determined |
| S17_extracted_k141_31108 | Not-determined |
| S17_extracted_k141_23308 | Not-determined |
| S17_extracted_k141_8954 | Not-determined |
| S17_extracted_k141_23311 | Not-determined |
| S17_extracted_k141_141789 | Not-determined |
| S17_extracted_k141_23313 | Not-determined |
| S17_extracted_k141_67842 | Not-determined |
| S17_extracted_k141_31113 | Not-determined |
| S17_extracted_k141_60533 | Not-determined |
| S17_extracted_k141_126584 | Not-determined |
| S17_extracted_k141_90066 | Not-determined |
| S17_extracted_k141_171455 | Not-determined |
| S17_extracted_k141_134268 | Not-determined |
| S17_extracted_k141_60536 | Not-determined |
| S17_extracted_k141_38691 | Not-determined |
| S17_extracted_k141_119626 | Low-quality |
| S17_extracted_k141_53388 | Not-determined |
| S17_extracted_k141_141797 | Low-quality |
| S17_extracted_k141_97438 | Not-determined |
| S17_extracted_k141_82277 | Not-determined |
| S17_extracted_k141_8963 | Low-quality |
| S17_extracted_k141_119628 | Not-determined |
| S17_extracted_k141_112051 | Not-determined |
| S17_extracted_k141_8968 | Not-determined |
| S17_extracted_k141_105226 | Not-determined |
| S17_extracted_k141_126587 | Not-determined |
| S17_extracted_k141_82282 | Not-determined |
| S17_extracted_k141_97446 | Not-determined |
| S17_extracted_k141_171467 | Not-determined |
| S17_extracted_k141_53399 | Not-determined |
| S17_extracted_k141_53400 | Not-determined |
| S17_extracted_k141_67860 | Not-determined |
| S17_extracted_k141_31130 | Not-determined |
| S17_extracted_k141_97454 | Not-determined |
| S17_extracted_k141_67864 | Not-determined |
| S17_extracted_k141_97456 | Not-determined |
| S17_extracted_k141_105229 | Not-determined |
| S17_extracted_k141_45870 | Not-determined |
| S17_extracted_k141_171469 | Not-determined |
| S17_extracted_k141_156702 | Not-determined |
| S17_extracted_k141_23322 | Not-determined |
| S17_extracted_k141_97464 | Not-determined |
| S17_extracted_k141_156705 | Not-determined |
| S17_extracted_k141_8977 | Not-determined |
| S17_extracted_k141_149526 | Not-determined |
| S17_extracted_k141_23328 | Not-determined |
| S17_extracted_k141_163894 | Not-determined |
| S17_extracted_k141_156712 | Not-determined |
| S17_extracted_k141_90072 | Not-determined |
| S17_extracted_k141_60543 | Not-determined |
| S17_extracted_k141_97468 | Not-determined |
| S17_extracted_k141_156716 | Not-determined |
| S17_extracted_k141_112062 | Not-determined |
| S17_extracted_k141_8979 | Low-quality |
| S17_extracted_k141_119643 | Not-determined |
| S17_extracted_k141_141820 | Not-determined |
| S17_extracted_k141_23336 | Low-quality |
| S17_extracted_k141_23338 | Not-determined |
| S17_extracted_k141_97476 | Low-quality |
| S17_extracted_k141_53426 | Not-determined |
| S17_extracted_k141_105254 | Not-determined |
| S17_extracted_k141_156725 | Low-quality |
| S17_extracted_k141_97478 | Low-quality |
| S17_extracted_k141_112073 | Not-determined |
| S17_extracted_k141_163901 | Low-quality |
| S17_extracted_k141_105259 | Not-determined |
| S17_extracted_k141_75211 | Low-quality |
| S17_extracted_k141_163905 | Not-determined |
| S17_extracted_k141_60559 | Not-determined |
| S17_extracted_k141_38722 | Not-determined |
| S17_extracted_k141_60561 | Not-determined |
| S17_extracted_k141_97482 | Not-determined |
| S17_extracted_k141_75216 | Not-determined |
| S17_extracted_k141_97484 | Not-determined |
| S17_extracted_k141_53437 | Not-determined |
| S17_extracted_k141_97488 | Not-determined |
| S17_extracted_k141_75222 | Not-determined |
| S17_extracted_k141_53440 | Not-determined |
| S17_extracted_k141_31155 | Not-determined |
| S17_extracted_k141_53441 | Not-determined |
| S17_extracted_k141_119660 | Low-quality |
| S17_extracted_k141_31160 | Not-determined |
| S17_extracted_k141_156749 | Not-determined |
| S17_extracted_k141_60570 | Not-determined |
| S17_extracted_k141_45894 | Low-quality |
| S17_extracted_k141_97492 | Not-determined |
| S17_extracted_k141_9000 | Not-determined |
| S17_extracted_k141_156751 | Not-determined |
| S17_extracted_k141_38730 | Not-determined |
| S17_extracted_k141_53445 | Not-determined |
| S17_extracted_k141_23354 | Not-determined |
| S17_extracted_k141_119665 | Low-quality |
| S17_extracted_k141_23356 | Not-determined |
| S17_extracted_k141_126610 | Not-determined |
| S17_extracted_k141_156754 | Not-determined |
| S17_extracted_k141_156755 | Not-determined |
| S17_extracted_k141_53448 | Low-quality |
| S17_extracted_k141_126611 | Not-determined |
| S17_extracted_k141_23359 | Not-determined |
| S17_extracted_k141_9003 | Low-quality |
| S17_extracted_k141_23361 | Not-determined |
| S17_extracted_k141_45897 | Not-determined |
| S17_extracted_k141_156758 | Not-determined |
| S17_extracted_k141_126615 | Not-determined |
| S17_extracted_k141_53453 | Low-quality |
| S17_extracted_k141_126617 | Not-determined |
| S17_extracted_k141_23369 | Not-determined |
| S17_extracted_k141_97497 | Not-determined |
| S17_extracted_k141_75238 | Not-determined |
| S17_extracted_k141_126618 | Low-quality |
| S17_extracted_k141_31169 | Low-quality |
| S17_extracted_k141_60580 | Not-determined |

| | |
|---|---|
| S17_extracted_k141_163916 | Not-determined |
| S17_extracted_k141_45904 | Not-determined |
| S17_extracted_k141_60581 | Low-quality |
| S17_extracted_k141_75242 | Not-determined |
| S17_extracted_k141_97504 | Not-determined |
| S17_extracted_k141_171489 | Not-determined |
| S17_extracted_k141_31175 | Not-determined |
| S17_extracted_k141_45914 | Not-determined |
| S17_extracted_k141_1399 | Not-determined |
| S17_extracted_k141_38738 | Not-determined |
| S17_extracted_k141_163921 | Not-determined |
| S17_extracted_k141_60589 | Not-determined |
| S17_extracted_k141_53460 | Low-quality |
| S17_extracted_k141_97517 | Not-determined |
| S17_extracted_k141_53463 | Low-quality |
| S17_extracted_k141_1404 | Low-quality |
| S17_extracted_k141_1405 | Not-determined |
| S17_extracted_k141_171500 | Not-determined |
| S17_extracted_k141_60600 | Not-determined |
| S17_extracted_k141_149546 | Not-determined |
| S17_extracted_k141_23376 | Not-determined |
| S17_extracted_k141_31186 | Not-determined |
| S17_extracted_k141_112103 | Not-determined |
| S17_extracted_k141_31187 | Not-determined |
| S17_extracted_k141_38753 | Not-determined |
| S17_extracted_k141_163925 | Low-quality |
| S17_extracted_k141_31192 | Not-determined |
| S17_extracted_k141_53466 | Low-quality |
| S17_extracted_k141_97531 | Not-determined |
| S17_extracted_k141_149552 | Not-determined |
| S17_extracted_k141_163929 | Not-determined |
| S17_extracted_k141_53470 | Not-determined |
| S17_extracted_k141_82307 | Not-determined |
| S17_extracted_k141_126644 | Not-determined |
| S17_extracted_k141_9022 | Not-determined |
| S17_extracted_k141_15823 | Not-determined |
| S17_extracted_k141_15824 | Low-quality |
| S17_extracted_k141_67876 | Low-quality |
| S17_extracted_k141_82312 | Not-determined |
| S17_extracted_k141_163940 | Not-determined |
| S17_extracted_k141_112107 | Low-quality |
| S17_extracted_k141_171518 | Low-quality |
| S17_extracted_k141_9031 | Not-determined |
| S17_extracted_k141_67882 | Not-determined |
| S17_extracted_k141_60610 | Not-determined |
| S17_extracted_k141_9033 | Not-determined |
| S17_extracted_k141_38767 | Not-determined |
| S17_extracted_k141_75268 | Not-determined |
| S17_extracted_k141_163945 | Not-determined |
| S17_extracted_k141_112111 | Low-quality |
| S17_extracted_k141_141861 | Not-determined |
| S17_extracted_k141_171525 | Not-determined |
| S17_extracted_k141_163947 | Not-determined |
| S17_extracted_k141_15841 | Low-quality |
| S17_extracted_k141_156806 | Not-determined |
| S17_extracted_k141_75269 | Not-determined |
| S17_extracted_k141_105271 | Not-determined |
| S17_extracted_k141_171528 | Not-determined |
| S17_extracted_k141_90080 | Not-determined |
| S17_extracted_k141_105277 | Not-determined |
| S17_extracted_k141_134290 | Low-quality |
| S17_extracted_k141_163959 | Not-determined |
| S17_extracted_k141_112114 | Not-determined |
| S17_extracted_k141_134292 | Not-determined |
| S17_extracted_k141_171533 | Not-determined |
| S17_extracted_k141_45941 | Not-determined |
| S17_extracted_k141_112116 | Not-determined |
| S17_extracted_k141_9035 | Not-determined |
| S17_extracted_k141_141875 | Not-determined |
| S17_extracted_k141_171534 | Not-determined |
| S17_extracted_k141_141876 | Not-determined |
| S17_extracted_k141_60621 | Not-determined |
| S17_extracted_k141_119678 | Not-determined |
| S17_extracted_k141_15848 | Low-quality |
| S17_extracted_k141_163963 | Low-quality |
| S17_extracted_k141_105281 | Not-determined |
| S17_extracted_k141_15849 | Low-quality |
| S17_extracted_k141_60625 | Not-determined |
| S17_extracted_k141_149566 | Low-quality |
| S17_extracted_k141_105286 | Not-determined |
| S17_extracted_k141_112118 | Not-determined |
| S17_extracted_k141_149568 | Not-determined |
| S17_extracted_k141_149569 | Not-determined |
| S17_extracted_k141_60630 | Not-determined |
| S17_extracted_k141_149570 | Not-determined |
| S17_extracted_k141_134303 | Low-quality |
| S17_extracted_k141_149571 | Not-determined |
| S17_extracted_k141_149572 | Not-determined |
| S17_extracted_k141_45956 | Not-determined |
| S17_extracted_k141_119690 | Low-quality |
| S17_extracted_k141_60636 | Not-determined |
| S17_extracted_k141_119691 | Not-determined |
| S17_extracted_k141_112120 | Low-quality |
| S17_extracted_k141_119692 | Not-determined |
| S17_extracted_k141_53501 | Not-determined |
| S17_extracted_k141_23384 | Not-determined |
| S17_extracted_k141_149581 | Not-determined |
| S17_extracted_k141_97564 | Not-determined |
| S17_extracted_k141_23388 | Not-determined |
| S17_extracted_k141_119697 | Not-determined |
| S17_extracted_k141_105300 | Low-quality |
| S17_extracted_k141_105301 | Not-determined |
| S17_extracted_k141_97568 | Not-determined |
| S17_extracted_k141_53508 | Low-quality |
| S17_extracted_k141_9059 | Not-determined |
| S17_extracted_k141_67896 | Not-determined |
| S17_extracted_k141_105304 | Not-determined |
| S17_extracted_k141_90105 | Not-determined |
| S17_extracted_k141_97570 | Not-determined |
| S17_extracted_k141_97572 | Not-determined |
| S17_extracted_k141_156814 | Not-determined |
| S17_extracted_k141_156815 | Not-determined |
| S17_extracted_k141_156816 | Not-determined |
| S17_extracted_k141_141894 | Not-determined |
| S17_extracted_k141_60651 | Not-determined |
| S17_extracted_k141_163982 | Not-determined |
| S17_extracted_k141_156823 | Not-determined |
| S17_extracted_k141_156824 | Not-determined |
| S17_extracted_k141_53520 | Not-determined |
| S17_extracted_k141_53523 | Not-determined |
| S17_extracted_k141_112125 | Not-determined |
| S17_extracted_k141_112126 | Not-determined |
| S17_extracted_k141_23396 | Not-determined |
| S17_extracted_k141_23397 | Not-determined |
| S17_extracted_k141_53528 | Low-quality |
| S17_extracted_k141_112130 | Not-determined |
| S17_extracted_k141_31215 | Not-determined |
| S17_extracted_k141_105327 | Not-determined |
| S17_extracted_k141_60668 | Low-quality |
| S17_extracted_k141_60670 | Not-determined |
| S17_extracted_k141_112139 | Not-determined |
| S17_extracted_k141_90116 | Not-determined |
| S17_extracted_k141_82324 | Low-quality |
| S17_extracted_k141_9075 | Low-quality |
| S17_extracted_k141_45967 | Low-quality |
| S17_extracted_k141_60673 | Not-determined |
| S17_extracted_k141_9077 | Not-determined |
| S17_extracted_k141_82326 | Not-determined |
| S17_extracted_k141_67915 | Not-determined |
| S17_extracted_k141_119715 | Not-determined |
| S17_extracted_k141_112140 | Low-quality |
| S17_extracted_k141_156840 | Not-determined |
| S17_extracted_k141_31218 | Not-determined |
| S17_extracted_k141_163999 | Low-quality |
| S17_extracted_k141_156842 | Not-determined |
| S17_extracted_k141_82332 | Not-determined |
| S17_extracted_k141_1436 | Not-determined |
| S17_extracted_k141_82333 | Low-quality |
| S17_extracted_k141_112143 | Not-determined |
| S17_extracted_k141_38776 | Low-quality |
| S17_extracted_k141_23407 | Not-determined |
| S17_extracted_k141_82336 | Not-determined |
| S17_extracted_k141_119722 | Not-determined |
| S17_extracted_k141_141925 | Not-determined |
| S17_extracted_k141_38780 | Not-determined |
| S17_extracted_k141_15892 | Low-quality |
| S17_extracted_k141_90132 | Not-determined |
| S17_extracted_k141_82340 | Not-determined |
| S17_extracted_k141_126674 | Not-determined |
| S17_extracted_k141_105335 | Low-quality |
| S17_extracted_k141_90134 | Not-determined |
| S17_extracted_k141_82345 | Low-quality |
| S17_extracted_k141_141934 | Not-determined |
| S17_extracted_k141_31227 | Low-quality |
| S17_extracted_k141_90137 | Not-determined |
| S17_extracted_k141_126680 | Not-determined |
| S17_extracted_k141_67938 | Not-determined |
| S17_extracted_k141_149591 | Not-determined |
| S17_extracted_k141_31229 | Not-determined |
| S17_extracted_k141_149592 | Not-determined |
| S17_extracted_k141_31230 | Not-determined |
| S17_extracted_k141_67942 | Not-determined |
| S17_extracted_k141_141936 | Low-quality |
| S17_extracted_k141_9087 | Not-determined |
| S17_extracted_k141_1442 | Not-determined |
| S17_extracted_k141_156859 | Not-determined |
| S17_extracted_k141_53544 | Not-determined |
| S17_extracted_k141_45984 | Not-determined |
| S17_extracted_k141_97592 | Not-determined |
| S17_extracted_k141_105339 | Not-determined |
| S17_extracted_k141_1445 | Not-determined |
| S17_extracted_k141_119733 | Not-determined |
| S17_extracted_k141_105340 | Not-determined |
| S17_extracted_k141_9092 | Not-determined |
| S17_extracted_k141_31241 | Not-determined |
| S17_extracted_k141_164011 | Not-determined |
| S17_extracted_k141_112165 | Not-determined |
| S17_extracted_k141_9102 | Not-determined |
| S17_extracted_k141_90147 | Not-determined |
| S17_extracted_k141_97598 | Not-determined |
| S17_extracted_k141_141947 | Not-determined |
| S17_extracted_k141_90150 | Not-determined |
| S17_extracted_k141_141950 | Not-determined |
| S17_extracted_k141_1453 | Low-quality |
| S17_extracted_k141_141952 | Not-determined |
| S17_extracted_k141_119743 | Low-quality |
| S17_extracted_k141_67952 | Not-determined |
| S17_extracted_k141_105348 | Not-determined |
| S17_extracted_k141_23424 | Not-determined |
| S17_extracted_k141_105352 | Not-determined |
| S17_extracted_k141_97605 | Not-determined |
| S17_extracted_k141_45990 | Low-quality |
| S17_extracted_k141_156869 | Not-determined |
| S17_extracted_k141_1457 | Low-quality |
| S17_extracted_k141_164024 | Not-determined |
| S17_extracted_k141_9116 | Not-determined |
| S17_extracted_k141_90152 | Not-determined |
| S17_extracted_k141_23427 | Not-determined |
| S17_extracted_k141_90153 | Not-determined |
| S17_extracted_k141_164030 | Not-determined |
| S17_extracted_k141_156872 | Not-determined |
| S17_extracted_k141_97620 | Not-determined |
| S17_extracted_k141_90157 | Low-quality |
| S17_extracted_k141_67963 | Not-determined |
| S17_extracted_k141_67966 | Not-determined |
| S17_extracted_k141_90161 | Not-determined |
| S17_extracted_k141_156880 | Not-determined |
| S17_extracted_k141_53558 | Not-determined |
| S17_extracted_k141_164042 | Low-quality |
| S17_extracted_k141_15932 | Not-determined |
| S17_extracted_k141_9127 | Low-quality |
| S17_extracted_k141_119761 | Low-quality |
| S17_extracted_k141_141975 | Not-determined |
| S17_extracted_k141_119762 | Not-determined |
| S17_extracted_k141_67977 | Not-determined |
| S17_extracted_k141_31246 | Not-determined |
| S17_extracted_k141_126700 | Not-determined |
| S17_extracted_k141_46008 | Not-determined |
| S17_extracted_k141_105367 | Not-determined |
| S17_extracted_k141_97636 | Not-determined |
| S17_extracted_k141_60688 | Not-determined |
| S17_extracted_k141_90177 | Not-determined |
| S17_extracted_k141_90183 | Not-determined |
| S17_extracted_k141_31256 | Not-determined |
| S17_extracted_k141_141983 | Not-determined |
| S17_extracted_k141_9141 | Not-determined |
| S17_extracted_k141_105369 | Low-quality |
| S17_extracted_k141_126708 | Not-determined |
| S17_extracted_k141_141988 | Low-quality |
| S17_extracted_k141_15933 | Low-quality |
| S17_extracted_k141_141990 | Not-determined |
| S17_extracted_k141_9144 | Not-determined |
| S17_extracted_k141_23436 | Not-determined |
| S17_extracted_k141_90188 | Not-determined |
| S17_extracted_k141_141993 | Not-determined |
| S17_extracted_k141_67988 | Not-determined |
| S17_extracted_k141_105375 | Low-quality |
| S17_extracted_k141_97651 | Not-determined |
| S17_extracted_k141_82369 | Not-determined |
| S17_extracted_k141_126712 | Not-determined |
| S17_extracted_k141_53579 | Not-determined |
| S17_extracted_k141_9155 | Not-determined |
| S17_extracted_k141_1489 | Not-determined |
| S17_extracted_k141_134359 | Not-determined |
| S17_extracted_k141_60700 | Not-determined |
| S17_extracted_k141_97661 | Low-quality |
| S17_extracted_k141_164069 | Not-determined |
| S17_extracted_k141_23438 | Not-determined |
| S17_extracted_k141_97662 | Low-quality |
| S17_extracted_k141_53580 | Low-quality |
| S17_extracted_k141_119783 | Not-determined |
| S17_extracted_k141_1495 | Not-determined |
| S17_extracted_k141_60705 | Not-determined |
| S17_extracted_k141_134365 | Not-determined |
| S17_extracted_k141_126715 | Not-determined |
| S17_extracted_k141_126717 | Not-determined |
| S17_extracted_k141_60710 | Not-determined |
| S17_extracted_k141_53584 | Not-determined |
| S17_extracted_k141_60712 | Not-determined |
| S17_extracted_k141_142008 | Low-quality |
| S17_extracted_k141_15958 | Low-quality |
| S17_extracted_k141_142009 | Not-determined |
| S17_extracted_k141_23449 | Not-determined |
| S17_extracted_k141_31262 | Not-determined |
| S17_extracted_k141_31263 | Not-determined |
| S17_extracted_k141_105394 | Not-determined |
| S17_extracted_k141_1500 | Not-determined |
| S17_extracted_k141_68002 | Not-determined |
| S17_extracted_k141_126726 | Not-determined |
| S17_extracted_k141_142016 | Not-determined |
| S17_extracted_k141_164086 | Low-quality |
| S17_extracted_k141_60720 | Not-determined |
| S17_extracted_k141_82390 | Not-determined |
| S17_extracted_k141_105405 | Low-quality |
| S17_extracted_k141_9166 | Not-determined |
| S17_extracted_k141_156899 | Not-determined |
| S17_extracted_k141_164092 | Not-determined |
| S17_extracted_k141_1511 | Not-determined |
| S17_extracted_k141_38813 | Not-determined |
| S17_extracted_k141_60727 | Not-determined |
| S17_extracted_k141_134380 | Low-quality |
| S17_extracted_k141_1520 | Not-determined |
| S17_extracted_k141_68016 | Not-determined |
| S17_extracted_k141_38826 | Not-determined |
| S17_extracted_k141_105415 | Not-determined |
| S17_extracted_k141_105417 | Low-quality |
| S17_extracted_k141_53593 | Low-quality |
| S17_extracted_k141_53594 | Low-quality |
| S17_extracted_k141_164110 | Not-determined |
| S17_extracted_k141_31278 | Not-determined |
| S17_extracted_k141_38832 | Not-determined |
| S17_extracted_k141_46032 | Not-determined |
| S17_extracted_k141_126751 | Not-determined |
| S12_vs_k141_188819\|\|full | High-quality |
| S17_extracted_k141_134390 | Not-determined |
| S17_extracted_k141_97673 | Not-determined |
| S17_extracted_k141_9185 | Not-determined |
| S17_extracted_k141_97675 | Not-determined |
| S17_extracted_k141_9186 | Low-quality |
| S17_extracted_k141_156924 | Not-determined |
| S17_extracted_k141_119787 | Not-determined |
| S17_extracted_k141_9189 | Not-determined |
| S17_extracted_k141_31284 | Not-determined |
| S17_extracted_k141_31285 | Not-determined |
| S17_extracted_k141_46041 | Not-determined |
| S17_extracted_k141_31286 | Low-quality |
| S17_extracted_k141_9192 | Low-quality |
| S17_extracted_k141_46042 | Not-determined |
| S17_extracted_k141_1535 | Not-determined |
| S17_extracted_k141_119790 | Low-quality |
| S17_extracted_k141_1536 | Not-determined |
| S17_extracted_k141_46046 | Not-determined |
| S17_extracted_k141_46050 | Not-determined |
| S17_extracted_k141_156926 | Not-determined |
| S17_extracted_k141_164117 | Not-determined |
| S17_extracted_k141_134398 | Not-determined |
| S17_extracted_k141_105427 | Not-determined |
| S17_extracted_k141_134400 | Not-determined |
| S17_extracted_k141_46053 | Not-determined |
| S17_extracted_k141_46054 | Not-determined |
| S17_extracted_k141_68040 | Not-determined |
| S17_extracted_k141_119795 | Not-determined |
| S17_extracted_k141_46055 | Not-determined |
| S17_extracted_k141_119796 | Low-quality |
| S17_extracted_k141_1544 | Low-quality |
| S17_extracted_k141_105430 | Not-determined |
| S17_extracted_k141_112185 | Not-determined |
| S17_extracted_k141_31289 | Not-determined |
| S17_extracted_k141_142031 | Not-determined |
| S17_extracted_k141_112187 | Not-determined |
| S17_extracted_k141_31292 | Not-determined |
| S17_extracted_k141_142033 | Not-determined |
| S17_extracted_k141_53614 | Low-quality |
| S17_extracted_k141_134409 | Not-determined |
| S17_extracted_k141_112192 | Not-determined |
| S17_extracted_k141_23478 | Not-determined |
| S17_extracted_k141_156931 | Not-determined |
| S17_extracted_k141_156932 | Not-determined |
| S17_extracted_k141_134412 | Not-determined |
| S17_extracted_k141_126765 | Low-quality |
| S17_extracted_k141_1551 | Not-determined |
| S17_extracted_k141_105439 | Low-quality |
| S17_extracted_k141_46059 | Not-determined |
| S17_extracted_k141_31300 | Not-determined |
| S17_extracted_k141_126767 | Low-quality |
| S17_extracted_k141_105441 | Not-determined |
| S17_extracted_k141_149631 | Not-determined |
| S17_extracted_k141_105444 | Not-determined |
| S17_extracted_k141_46061 | Not-determined |
| S17_extracted_k141_105445 | Not-determined |
| S17_extracted_k141_46062 | Low-quality |
| S17_extracted_k141_149639 | Not-determined |
| S17_extracted_k141_97677 | Not-determined |
| S17_extracted_k141_149643 | Not-determined |
| S17_extracted_k141_97678 | Not-determined |
| S17_extracted_k141_112203 | Not-determined |
| S17_extracted_k141_112204 | Not-determined |
| S17_extracted_k141_97679 | Low-quality |
| S17_extracted_k141_82446 | Not-determined |
| S17_extracted_k141_15962 | Low-quality |
| S17_extracted_k141_82450 | Not-determined |
| S17_extracted_k141_46076 | Not-determined |
| S17_extracted_k141_9206 | Not-determined |
| S17_extracted_k141_60756 | Not-determined |
| S17_extracted_k141_15970 | Not-determined |
| S17_extracted_k141_142049 | Not-determined |
| S17_extracted_k141_53635 | Not-determined |
| S17_extracted_k141_97696 | Low-quality |
| S17_extracted_k141_97697 | Not-determined |
| S17_extracted_k141_82453 | Low-quality |
| S17_extracted_k141_46082 | Low-quality |
| S17_extracted_k141_97699 | Low-quality |
| S17_extracted_k141_134422 | Not-determined |
| S17_extracted_k141_105465 | Not-determined |
| S17_extracted_k141_119815 | Not-determined |
| S17_extracted_k141_31318 | Not-determined |
| S17_extracted_k141_112209 | Not-determined |
| S17_extracted_k141_149659 | Not-determined |
| S17_extracted_k141_82456 | Not-determined |
| S17_extracted_k141_126776 | Not-determined |
| S17_extracted_k141_53645 | Not-determined |
| S17_extracted_k141_46091 | Not-determined |
| S17_extracted_k141_119825 | Low-quality |
| S17_extracted_k141_82458 | Low-quality |
| S17_extracted_k141_97702 | Not-determined |
| S17_extracted_k141_15988 | Not-determined |
| S17_extracted_k141_156959 | Not-determined |
| S17_extracted_k141_46095 | Not-determined |
| S17_extracted_k141_1574 | Not-determined |
| S17_extracted_k141_53651 | Not-determined |
| S17_extracted_k141_1576 | Not-determined |
| S17_extracted_k141_126781 | Not-determined |
| S17_extracted_k141_134432 | Low-quality |
| S17_extracted_k141_15993 | Not-determined |
| S17_extracted_k141_126783 | Not-determined |
| S17_extracted_k141_53654 | Not-determined |
| S17_extracted_k141_46103 | Not-determined |
| S17_extracted_k141_38849 | Low-quality |
| S17_extracted_k141_134438 | Low-quality |
| S17_extracted_k141_1581 | Not-determined |
| S17_extracted_k141_126784 | Not-determined |
| S17_extracted_k141_105480 | Not-determined |
| S17_extracted_k141_134441 | Low-quality |
| S17_extracted_k141_46108 | Low-quality |
| S17_extracted_k141_38855 | Not-determined |
| S17_extracted_k141_105482 | Not-determined |
| S17_extracted_k141_46110 | Not-determined |
| S17_extracted_k141_38860 | Not-determined |
| S17_extracted_k141_82475 | Not-determined |
| S17_extracted_k141_149665 | Low-quality |
| S17_extracted_k141_112222 | Not-determined |
| S17_extracted_k141_119838 | Low-quality |
| S17_extracted_k141_1585 | Not-determined |
| S17_extracted_k141_134445 | Not-determined |
| S17_extracted_k141_119840 | Not-determined |
| S17_extracted_k141_38864 | Not-determined |
| S17_extracted_k141_53665 | Not-determined |
| S17_extracted_k141_1590 | Not-determined |
| S17_extracted_k141_112229 | Not-determined |
| S17_extracted_k141_75280 | Low-quality |
| S17_extracted_k141_156988 | Not-determined |
| S17_extracted_k141_38870 | Not-determined |
| S17_extracted_k141_97720 | Not-determined |
| S17_extracted_k141_1594 | Not-determined |
| S17_extracted_k141_126798 | Not-determined |
| S17_extracted_k141_82480 | Not-determined |
| S17_extracted_k141_53669 | Not-determined |
| S17_extracted_k141_53670 | Not-determined |
| S17_extracted_k141_82482 | Not-determined |
| S17_extracted_k141_105493 | Not-determined |
| S17_extracted_k141_23506 | Not-determined |
| S17_extracted_k141_82485 | Not-determined |
| S17_extracted_k141_23507 | Not-determined |
| S17_extracted_k141_38876 | Not-determined |
| S17_extracted_k141_119851 | Not-determined |
| S17_extracted_k141_119852 | Low-quality |
| S17_extracted_k141_112246 | Not-determined |
| S17_extracted_k141_134465 | Not-determined |
| S17_extracted_k141_119858 | Not-determined |
| S17_extracted_k141_149692 | Not-determined |
| S17_extracted_k141_97743 | Not-determined |
| S17_extracted_k141_149694 | Not-determined |
| S17_extracted_k141_149696 | Not-determined |
| S17_extracted_k141_31333 | Not-determined |
| S17_extracted_k141_149698 | Low-quality |
| S17_extracted_k141_31335 | Low-quality |
| S17_extracted_k141_105503 | Low-quality |
| S17_extracted_k141_23514 | Not-determined |
| S17_extracted_k141_75303 | Low-quality |
| S17_extracted_k141_142068 | Not-determined |
| S17_extracted_k141_97752 | Low-quality |
| S17_extracted_k141_75304 | Not-determined |
| S17_extracted_k141_126810 | Not-determined |
| S17_extracted_k141_75310 | Not-determined |
| S17_extracted_k141_1612 | Not-determined |
| S17_extracted_k141_53672 | Not-determined |
| S17_extracted_k141_1613 | Not-determined |
| S17_extracted_k141_31346 | Not-determined |
| S17_extracted_k141_134485 | Not-determined |
| S17_extracted_k141_97758 | Not-determined |
| S17_extracted_k141_112260 | Not-determined |
| S17_extracted_k141_53679 | Not-determined |
| S17_extracted_k141_75321 | Not-determined |
| S17_extracted_k141_82488 | Not-determined |
| S17_extracted_k141_75323 | Not-determined |
| S17_extracted_k141_112262 | Not-determined |
| S17_extracted_k141_112263 | Not-determined |
| S17_extracted_k141_126825 | Not-determined |
| S17_extracted_k141_9234 | Not-determined |
| S17_vs_k141_54605\|\|full | Medium-quality |
| S17_extracted_k141_75329 | Not-determined |
| S17_extracted_k141_97768 | Low-quality |
| S17_extracted_k141_75332 | Not-determined |
| S17_extracted_k141_112267 | Not-determined |
| S17_extracted_k141_97771 | Not-determined |
| S17_extracted_k141_142101 | Not-determined |
| S17_extracted_k141_16024 | Not-determined |
| S17_extracted_k141_142104 | Not-determined |
| S17_extracted_k141_97780 | Not-determined |
| S17_extracted_k141_46156 | Not-determined |
| S17_extracted_k141_97783 | Not-determined |
| S17_extracted_k141_23529 | Not-determined |
| S17_extracted_k141_53689 | Not-determined |
| S17_extracted_k141_97792 | Not-determined |
| S17_extracted_k141_46168 | Not-determined |
| S17_extracted_k141_126842 | Low-quality |
| S17_extracted_k141_60773 | Not-determined |
| S17_extracted_k141_23540 | Not-determined |
| S17_extracted_k141_53692 | Not-determined |
| S17_extracted_k141_53693 | Low-quality |
| S17_extracted_k141_23541 | Not-determined |
| S17_extracted_k141_105523 | Not-determined |
| S17_extracted_k141_75336 | Not-determined |
| S17_extracted_k141_46171 | Low-quality |
| S17_extracted_k141_119869 | Not-determined |
| S17_extracted_k141_16031 | Low-quality |
| S17_extracted_k141_46173 | Not-determined |
| S17_extracted_k141_75339 | Low-quality |
| S17_extracted_k141_105528 | Not-determined |
| S17_extracted_k141_9246 | Not-determined |
| S17_extracted_k141_119874 | Not-determined |
| S17_extracted_k141_126851 | Not-determined |
| S17_extracted_k141_105531 | Not-determined |
| S17_extracted_k141_142120 | Not-determined |
| S17_extracted_k141_68061 | Not-determined |
| S17_extracted_k141_126853 | Low-quality |
| S17_extracted_k141_105533 | Not-determined |
| S17_extracted_k141_105534 | Not-determined |
| S17_extracted_k141_105538 | Not-determined |
| S17_extracted_k141_75345 | Not-determined |
| S17_extracted_k141_23548 | Not-determined |
| S17_extracted_k141_60784 | Not-determined |
| S17_extracted_k141_119879 | Not-determined |
| S17_extracted_k141_119883 | Not-determined |
| S17_extracted_k141_31368 | Low-quality |
| S17_extracted_k141_46181 | Low-quality |
| S17_extracted_k141_134511 | Not-determined |
| S17_extracted_k141_46182 | Not-determined |
| S17_extracted_k141_68077 | Not-determined |
| S17_extracted_k141_126866 | Not-determined |
| S17_extracted_k141_46185 | Not-determined |
| S17_extracted_k141_60791 | Not-determined |
| S17_extracted_k141_60792 | Not-determined |
| S17_extracted_k141_60793 | Not-determined |
| S17_extracted_k141_134515 | Not-determined |
| S17_extracted_k141_53710 | Not-determined |
| S17_extracted_k141_60794 | Low-quality |
| S14_vs_k141_66139\|\|full | Medium-quality |
| S17_extracted_k141_53712 | Not-determined |
| S17_extracted_k141_97813 | Not-determined |
| S17_extracted_k141_31376 | Not-determined |
| S17_extracted_k141_105550 | Not-determined |
| S17_extracted_k141_97816 | Not-determined |
| S17_extracted_k141_16048 | Not-determined |
| S17_extracted_k141_1618 | Not-determined |
| S17_extracted_k141_46196 | Not-determined |
| S17_extracted_k141_134520 | Not-determined |
| S17_extracted_k141_97818 | Not-determined |
| S17_extracted_k141_126872 | Not-determined |
| S17_extracted_k141_134523 | Not-determined |
| S17_extracted_k141_38894 | Not-determined |
| S17_extracted_k141_1620 | Low-quality |
| S17_extracted_k141_46200 | Not-determined |
| S17_extracted_k141_53721 | Not-determined |
| S17_extracted_k141_46202 | Not-determined |
| S17_extracted_k141_38898 | Not-determined |
| S17_extracted_k141_31388 | Not-determined |
| S17_extracted_k141_105551 | Low-quality |
| S17_extracted_k141_38900 | Not-determined |
| S17_extracted_k141_112285 | Low-quality |
| S17_extracted_k141_119905 | Low-quality |
| S17_extracted_k141_68091 | Low-quality |
| S17_extracted_k141_1623 | Not-determined |
| S17_extracted_k141_60800 | Not-determined |
| S17_extracted_k141_1624 | Not-determined |
| S17_extracted_k141_105554 | Not-determined |
| S17_extracted_k141_68094 | Not-determined |
| S17_extracted_k141_97824 | Low-quality |
| S17_extracted_k141_1627 | Low-quality |
| S17_extracted_k141_1631 | Not-determined |
| S17_extracted_k141_23561 | Not-determined |
| S17_extracted_k141_105562 | Not-determined |
| S17_extracted_k141_134536 | Not-determined |
| S17_extracted_k141_134537 | Not-determined |
| S17_extracted_k141_119911 | Low-quality |
| S17_extracted_k141_97827 | Not-determined |
| S17_extracted_k141_126889 | Low-quality |
| S17_extracted_k141_97832 | Not-determined |
| S17_extracted_k141_68104 | Not-determined |
| S17_extracted_k141_105568 | Not-determined |
| S17_extracted_k141_157013 | Low-quality |
| S17_extracted_k141_90199 | Not-determined |
| S17_extracted_k141_142126 | Low-quality |
| S17_extracted_k141_126893 | Not-determined |
| S17_extracted_k141_149746 | Not-determined |
| S17_extracted_k141_46218 | Not-determined |
| S17_extracted_k141_23573 | Not-determined |
| S17_extracted_k141_16069 | Low-quality |
| S17_extracted_k141_16071 | Not-determined |

| | |
|---|---|
| S17_extracted_k141_1650 | Low-quality |
| S17_extracted_k141_16075 | Not-determined |
| S17_extracted_k141_46222 | Not-determined |
| S17_extracted_k141_112303 | Not-determined |
| S17_extracted_k141_157022 | Not-determined |
| S17_extracted_k141_119915 | Low-quality |
| S17_extracted_k141_157024 | Not-determined |
| S17_extracted_k141_105575 | Not-determined |
| S17_extracted_k141_75383 | Not-determined |
| S17_extracted_k141_157025 | Not-determined |
| S17_extracted_k141_119916 | Not-determined |
| S17_extracted_k141_126905 | Not-determined |
| S17_extracted_k141_157026 | Not-determined |
| S17_extracted_k141_157028 | Low-quality |
| S17_extracted_k141_119918 | Low-quality |
| S17_extracted_k141_119920 | Not-determined |
| S17_extracted_k141_149758 | Not-determined |
| S17_extracted_k141_126910 | Not-determined |
| S17_extracted_k141_149761 | Low-quality |
| S17_extracted_k141_75386 | Not-determined |
| S17_extracted_k141_31400 | Not-determined |
| S17_extracted_k141_31403 | Not-determined |
| S17_extracted_k141_53746 | Not-determined |
| S17_extracted_k141_105580 | Not-determined |
| S17_extracted_k141_142138 | Not-determined |
| S17_extracted_k141_68117 | Not-determined |
| S17_extracted_k141_119921 | Low-quality |
| S17_extracted_k141_134546 | Not-determined |
| S17_extracted_k141_149764 | Low-quality |
| S17_extracted_k141_126911 | Not-determined |
| S17_extracted_k141_126912 | Not-determined |
| S17_extracted_k141_82532 | Not-determined |
| S17_extracted_k141_1666 | Low-quality |
| S17_extracted_k141_90220 | Not-determined |
| S17_extracted_k141_164122 | Low-quality |
| S17_extracted_k141_31410 | Not-determined |
| S17_extracted_k141_82534 | Not-determined |
| S17_extracted_k141_157035 | Not-determined |
| S17_extracted_k141_31411 | Not-determined |
| S17_extracted_k141_46235 | Not-determined |
| S17_extracted_k141_90226 | Not-determined |
| S17_extracted_k141_90227 | Not-determined |
| S17_extracted_k141_149775 | Not-determined |
| S17_extracted_k141_1672 | Not-determined |
| S17_extracted_k141_23597 | Not-determined |
| S17_extracted_k141_126917 | Not-determined |
| S17_extracted_k141_90233 | Not-determined |
| S17_extracted_k141_82543 | Not-determined |
| S17_extracted_k141_75393 | Not-determined |
| S17_extracted_k141_82544 | Not-determined |
| S17_extracted_k141_9261 | Not-determined |
| S17_extracted_k141_82546 | Not-determined |
| S17_extracted_k141_46245 | Not-determined |
| S17_extracted_k141_105581 | Low-quality |
| S17_extracted_k141_53763 | Not-determined |
| S17_extracted_k141_46246 | Not-determined |
| S17_extracted_k141_1682 | Not-determined |
| S17_extracted_k141_90235 | Not-determined |
| S17_extracted_k141_105583 | Low-quality |
| S17_extracted_k141_157041 | Low-quality |
| S17_extracted_k141_53766 | Not-determined |
| S17_extracted_k141_142153 | Low-quality |
| S17_extracted_k141_105586 | Not-determined |
| S17_extracted_k141_142156 | Not-determined |
| S17_extracted_k141_157043 | Low-quality |
| S17_extracted_k141_46251 | Not-determined |
| S17_extracted_k141_90239 | Not-determined |
| S17_extracted_k141_157047 | Not-determined |
| S17_extracted_k141_46255 | Not-determined |
| S17_extracted_k141_157048 | Low-quality |
| S17_extracted_k141_126927 | Not-determined |
| S17_extracted_k141_53775 | Not-determined |
| S17_extracted_k141_68147 | Not-determined |
| S17_extracted_k141_119938 | Not-determined |
| S17_extracted_k141_105597 | Not-determined |
| S17_extracted_k141_23626 | Not-determined |
| S17_extracted_k141_53784 | Not-determined |
| S17_extracted_k141_38935 | Not-determined |
| S17_extracted_k141_38938 | Not-determined |
| S17_extracted_k141_142166 | Not-determined |
| S17_extracted_k141_31419 | Low-quality |
| S17_extracted_k141_53791 | Not-determined |
| S17_extracted_k141_157058 | Low-quality |
| S17_extracted_k141_164125 | Not-determined |
| S17_extracted_k141_38941 | Not-determined |
| S17_extracted_k141_1688 | Not-determined |
| S17_extracted_k141_46265 | Low-quality |
| S17_extracted_k141_82578 | Not-determined |
| S17_extracted_k141_164131 | Not-determined |
| S17_extracted_k141_142175 | Not-determined |
| S17_extracted_k141_38946 | Not-determined |
| S17_extracted_k141_53793 | Not-determined |
| S17_extracted_k141_82583 | Not-determined |
| S17_extracted_k141_149799 | Low-quality |
| S17_extracted_k141_112344 | Not-determined |
| S17_extracted_k141_105610 | Not-determined |
| S17_extracted_k141_126938 | Not-determined |
| S17_extracted_k141_105611 | Not-determined |
| S17_extracted_k141_142184 | Not-determined |
| S17_extracted_k141_105612 | Not-determined |
| S17_extracted_k141_16084 | Not-determined |
| S17_extracted_k141_1703 | Not-determined |
| S17_extracted_k141_142186 | Not-determined |
| S17_extracted_k141_142187 | Not-determined |
| S17_extracted_k141_46272 | Not-determined |
| S17_extracted_k141_164141 | Not-determined |
| S17_extracted_k141_164142 | Not-determined |
| S17_extracted_k141_38958 | Not-determined |
| S17_extracted_k141_38959 | Not-determined |
| S17_extracted_k141_157080 | Not-determined |
| S17_extracted_k141_38960 | Not-determined |
| S17_extracted_k141_126939 | Not-determined |
| S17_extracted_k141_46286 | Not-determined |
| S17_extracted_k141_68159 | Not-determined |
| S17_extracted_k141_31434 | Not-determined |
| S17_extracted_k141_31435 | Not-determined |
| S17_extracted_k141_164149 | Low-quality |
| S17_extracted_k141_119968 | Low-quality |
| S17_extracted_k141_23641 | Not-determined |
| S17_extracted_k141_142195 | Not-determined |
| S17_extracted_k141_75428 | Not-determined |
| S17_extracted_k141_164152 | Not-determined |
| S17_extracted_k141_164153 | Not-determined |
| S17_extracted_k141_134558 | Low-quality |
| S17_extracted_k141_68163 | Not-determined |
| S17_extracted_k141_9287 | Not-determined |
| S17_extracted_k141_126943 | Not-determined |
| S17_extracted_k141_75433 | Low-quality |
| S17_extracted_k141_164159 | Not-determined |
| S17_extracted_k141_31440 | Not-determined |
| S17_extracted_k141_16092 | Not-determined |
| S17_extracted_k141_119976 | Not-determined |
| S17_extracted_k141_105626 | Not-determined |
| S17_extracted_k141_23648 | Not-determined |
| S17_extracted_k141_105627 | Not-determined |
| S17_extracted_k141_23652 | Not-determined |
| S17_extracted_k141_16094 | Not-determined |
| S17_extracted_k141_38971 | Not-determined |
| S17_extracted_k141_46301 | Not-determined |
| S17_extracted_k141_46305 | Not-determined |
| S17_extracted_k141_38978 | Not-determined |
| S17_extracted_k141_9292 | Not-determined |
| S17_extracted_k141_134567 | Not-determined |
| S17_extracted_k141_112367 | Low-quality |
| S17_extracted_k141_142212 | Not-determined |
| S17_extracted_k141_149828 | Not-determined |
| S17_extracted_k141_112371 | Not-determined |
| S17_extracted_k141_16108 | Not-determined |
| S17_extracted_k141_31457 | Not-determined |
| S17_extracted_k141_164170 | Not-determined |
| S17_extracted_k141_119987 | Not-determined |
| S17_extracted_k141_46309 | Not-determined |
| S17_extracted_k141_31463 | Not-determined |
| S17_extracted_k141_164171 | Not-determined |
| S17_extracted_k141_38986 | Not-determined |
| S17_extracted_k141_105640 | Not-determined |
| S17_extracted_k141_97853 | Low-quality |
| S17_extracted_k141_105644 | Not-determined |
| S17_extracted_k141_16117 | Low-quality |
| S17_extracted_k141_97856 | Not-determined |
| S17_extracted_k141_105646 | Not-determined |
| S17_extracted_k141_97860 | Not-determined |
| S17_extracted_k141_134575 | Low-quality |
| S17_extracted_k141_149843 | Not-determined |
| S17_extracted_k141_126955 | Low-quality |
| S17_extracted_k141_142223 | Not-determined |
| S17_extracted_k141_82586 | Low-quality |
| S17_extracted_k141_126959 | Not-determined |
| S17_extracted_k141_134580 | Not-determined |
| S17_extracted_k141_23684 | Not-determined |
| S17_extracted_k141_23685 | Not-determined |
| S17_extracted_k141_149846 | Not-determined |
| S17_extracted_k141_68184 | Not-determined |
| S17_extracted_k141_68185 | Not-determined |
| S17_extracted_k141_97864 | Not-determined |
| S17_extracted_k141_97865 | Not-determined |
| S17_extracted_k141_16128 | Low-quality |
| S17_extracted_k141_126961 | Not-determined |
| S17_extracted_k141_149853 | Not-determined |
| S17_extracted_k141_68191 | Not-determined |
| S17_extracted_k141_157119 | Not-determined |
| S17_extracted_k141_1714 | Low-quality |
| S17_extracted_k141_39001 | Low-quality |
| S17_extracted_k141_23694 | Low-quality |
| S17_extracted_k141_23695 | Not-determined |
| S17_extracted_k141_23697 | Not-determined |
| S17_extracted_k141_23698 | Not-determined |
| S17_extracted_k141_1715 | Not-determined |
| S17_extracted_k141_9306 | Not-determined |
| S17_extracted_k141_82598 | Not-determined |
| S17_extracted_k141_1717 | Not-determined |
| S17_extracted_k141_53808 | Not-determined |
| S17_extracted_k141_157123 | Not-determined |
| S17_extracted_k141_46328 | Not-determined |
| S17_extracted_k141_97876 | Not-determined |
| S17_extracted_k141_46331 | Not-determined |
| S17_extracted_k141_1718 | Not-determined |
| S17_extracted_k141_23705 | Not-determined |
| S17_extracted_k141_23706 | Not-determined |
| S17_extracted_k141_23708 | Not-determined |
| S17_extracted_k141_1722 | Not-determined |
| S17_extracted_k141_53819 | Not-determined |
| S17_extracted_k141_68209 | Not-determined |
| S17_extracted_k141_142241 | Not-determined |
| S17_extracted_k141_16139 | Not-determined |
| S17_extracted_k141_1724 | Not-determined |
| S17_extracted_k141_105660 | Not-determined |
| S17_extracted_k141_16141 | Not-determined |
| S17_extracted_k141_120035 | Low-quality |
| S17_extracted_k141_1726 | Not-determined |
| S17_extracted_k141_120036 | Not-determined |
| S17_extracted_k141_134601 | Low-quality |
| S17_extracted_k141_68213 | Not-determined |
| S17_extracted_k141_126970 | Not-determined |
| S17_extracted_k141_9324 | Not-determined |
| S17_extracted_k141_53828 | Not-determined |
| S17_extracted_k141_157139 | Not-determined |
| S17_extracted_k141_90254 | Not-determined |
| S17_extracted_k141_157142 | Not-determined |
| S17_extracted_k141_23721 | Not-determined |
| S17_extracted_k141_23722 | Not-determined |
| S17_extracted_k141_16143 | Not-determined |
| S17_extracted_k141_39006 | Not-determined |
| S17_extracted_k141_75438 | Not-determined |
| S17_extracted_k141_157146 | Not-determined |
| S17_extracted_k141_105667 | Not-determined |
| S17_extracted_k141_90258 | Not-determined |
| S17_extracted_k141_97896 | Not-determined |
| S17_extracted_k141_134611 | Not-determined |
| S17_extracted_k141_39008 | Not-determined |
| S17_extracted_k141_46347 | Not-determined |
| S17_extracted_k141_16148 | Not-determined |
| S17_extracted_k141_164180 | Not-determined |
| S17_extracted_k141_134616 | Not-determined |
| S17_extracted_k141_9335 | Not-determined |
| S17_extracted_k141_23728 | Low-quality |
| S17_extracted_k141_46351 | Low-quality |
| S17_extracted_k141_23731 | Not-determined |
| S17_extracted_k141_23732 | Not-determined |
| S17_extracted_k141_142257 | Low-quality |
| S17_extracted_k141_16150 | Not-determined |
| S17_extracted_k141_23733 | Low-quality |
| S17_extracted_k141_16151 | Not-determined |
| S17_extracted_k141_134621 | Not-determined |
| S17_extracted_k141_134624 | Not-determined |
| S17_extracted_k141_105675 | Not-determined |
| S17_extracted_k141_134625 | Not-determined |
| S17_extracted_k141_16156 | Not-determined |
| S17_extracted_k141_134628 | Low-quality |
| S17_extracted_k141_164187 | Low-quality |
| S17_extracted_k141_16162 | Not-determined |
| S17_extracted_k141_90262 | Not-determined |
| S17_extracted_k141_23738 | Not-determined |
| S17_extracted_k141_23742 | Not-determined |
| S17_extracted_k141_16171 | Low-quality |
| S17_extracted_k141_134642 | Not-determined |
| S17_extracted_k141_16172 | Not-determined |
| S17_extracted_k141_164195 | Not-determined |
| S17_extracted_k141_120042 | Not-determined |
| S17_extracted_k141_97911 | Not-determined |
| S17_extracted_k141_82622 | Not-determined |
| S17_extracted_k141_9354 | Not-determined |
| S17_extracted_k141_134648 | Not-determined |
| S17_extracted_k141_120053 | Not-determined |
| S17_extracted_k141_134651 | Low-quality |
| S17_extracted_k141_142267 | Not-determined |
| S17_extracted_k141_23755 | Not-determined |
| S17_extracted_k141_142268 | Not-determined |
| S17_extracted_k141_126993 | Not-determined |
| S17_extracted_k141_39035 | Low-quality |
| S17_extracted_k141_82635 | Not-determined |
| S17_extracted_k141_97920 | Not-determined |
| S17_extracted_k141_120056 | Not-determined |
| S17_extracted_k141_46382 | Not-determined |
| S17_extracted_k141_53861 | Not-determined |
| S17_extracted_k141_82640 | Not-determined |
| S17_extracted_k141_126995 | Low-quality |
| S17_extracted_k141_82642 | Not-determined |
| S17_extracted_k141_53864 | Low-quality |
| S17_extracted_k141_53866 | Not-determined |
| S17_extracted_k141_120062 | Low-quality |
| S17_extracted_k141_16197 | Not-determined |
| S17_extracted_k141_53870 | Not-determined |
| S17_extracted_k141_127004 | Not-determined |
| S17_extracted_k141_171571 | Not-determined |
| S17_extracted_k141_68242 | Low-quality |
| S17_extracted_k141_171573 | Not-determined |
| S17_extracted_k141_105704 | Not-determined |
| S17_extracted_k141_157156 | Not-determined |
| S17_extracted_k141_68244 | Not-determined |
| S17_extracted_k141_171575 | Not-determined |
| S17_extracted_k141_120067 | Not-determined |
| S17_extracted_k141_157157 | Not-determined |
| S17_extracted_k141_16202 | Not-determined |
| S17_extracted_k141_105707 | Not-determined |
| S17_extracted_k141_157158 | Not-determined |
| S17_extracted_k141_105708 | Not-determined |
| S17_extracted_k141_164214 | Low-quality |
| S17_extracted_k141_23775 | Not-determined |
| S17_extracted_k141_127008 | Not-determined |
| S17_extracted_k141_171577 | Low-quality |
| S17_extracted_k141_142274 | Low-quality |
| S17_extracted_k141_53877 | Not-determined |
| S17_extracted_k141_105712 | Not-determined |
| S17_extracted_k141_164219 | Not-determined |
| S17_extracted_k141_53879 | Not-determined |
| S17_extracted_k141_9387 | Not-determined |
| S17_extracted_k141_149912 | Not-determined |
| S17_extracted_k141_53880 | Low-quality |
| S17_extracted_k141_31506 | Low-quality |
| S17_extracted_k141_31507 | Not-determined |
| S17_extracted_k141_60810 | Not-determined |
| S17_extracted_k141_142280 | Low-quality |
| S17_extracted_k141_164223 | Not-determined |
| S17_extracted_k141_97938 | Low-quality |
| S17_extracted_k141_120075 | Not-determined |
| S17_extracted_k141_171579 | Not-determined |
| S17_extracted_k141_23784 | Not-determined |
| S17_extracted_k141_105719 | Not-determined |
| S17_extracted_k141_127023 | Not-determined |
| S17_extracted_k141_127024 | Low-quality |
| S17_extracted_k141_23785 | Low-quality |
| S17_extracted_k141_97943 | Not-determined |
| S17_extracted_k141_134677 | Not-determined |
| S17_extracted_k141_53886 | Not-determined |
| S17_extracted_k141_164225 | Not-determined |
| S17_extracted_k141_68266 | Not-determined |
| S17_extracted_k141_31514 | Not-determined |
| S17_extracted_k141_127030 | Low-quality |
| S17_extracted_k141_31515 | Not-determined |
| S17_extracted_k141_23790 | Not-determined |
| S17_extracted_k141_82674 | Not-determined |
| S17_extracted_k141_105731 | Not-determined |
| S17_extracted_k141_68271 | Not-determined |
| S17_extracted_k141_142293 | Not-determined |
| S17_extracted_k141_142295 | Not-determined |
| S17_extracted_k141_105735 | Not-determined |
| S17_extracted_k141_90270 | Low-quality |
| S17_extracted_k141_31519 | Not-determined |
| S17_extracted_k141_90273 | Not-determined |
| S17_extracted_k141_60815 | Low-quality |
| S17_extracted_k141_97949 | Not-determined |
| S17_extracted_k141_60817 | Not-determined |
| S17_extracted_k141_9414 | Not-determined |
| S17_extracted_k141_16218 | Not-determined |
| S17_extracted_k141_149925 | Not-determined |
| S17_extracted_k141_97953 | Not-determined |
| S17_extracted_k141_82683 | Low-quality |
| S17_extracted_k141_120097 | Not-determined |
| S17_extracted_k141_127046 | Not-determined |
| S17_extracted_k141_142297 | Not-determined |
| S17_extracted_k141_60823 | Not-determined |
| S17_extracted_k141_31526 | Not-determined |
| S17_extracted_k141_53896 | Not-determined |
| S17_extracted_k141_39061 | Not-determined |
| S8_vs_k141_59736\|\|full | High-quality |
| S17_extracted_k141_39063 | Not-determined |
| S17_extracted_k141_142302 | Not-determined |
| S17_extracted_k141_112383 | Low-quality |
| S17_extracted_k141_23810 | Not-determined |
| S17_extracted_k141_149936 | Not-determined |
| S17_extracted_k141_23812 | Not-determined |
| S17_extracted_k141_142303 | Low-quality |
| S17_extracted_k141_60829 | Not-determined |
| S17_extracted_k141_112391 | Not-determined |
| S17_extracted_k141_31536 | Not-determined |
| S17_extracted_k141_105750 | Not-determined |
| S17_extracted_k141_112395 | Not-determined |
| S17_extracted_k141_23816 | Not-determined |
| S17_extracted_k141_16232 | Not-determined |
| S17_extracted_k141_157161 | Not-determined |
| S17_extracted_k141_82695 | Low-quality |
| S17_extracted_k141_23819 | Low-quality |
| S17_extracted_k141_46400 | Not-determined |
| S17_extracted_k141_134706 | Not-determined |
| S17_extracted_k141_142309 | Low-quality |
| S17_extracted_k141_149941 | Not-determined |
| S17_extracted_k141_97976 | Low-quality |
| S17_extracted_k141_82701 | Not-determined |
| S17_extracted_k141_127068 | Not-determined |
| S17_extracted_k141_39078 | Not-determined |
| S17_extracted_k141_16236 | Not-determined |
| S17_extracted_k141_120125 | Not-determined |
| S17_extracted_k141_164255 | Low-quality |
| S17_extracted_k141_82706 | Low-quality |
| S17_extracted_k141_134712 | Not-determined |
| S17_extracted_k141_39083 | Not-determined |
| S17_extracted_k141_16240 | Low-quality |
| S17_extracted_k141_142324 | Not-determined |
| S17_extracted_k141_97983 | Low-quality |
| S17_extracted_k141_127076 | Not-determined |
| S17_extracted_k141_1765 | Not-determined |
| S17_extracted_k141_149951 | Not-determined |
| S17_extracted_k141_142330 | Not-determined |
| S17_extracted_k141_127080 | Not-determined |
| S17_extracted_k141_16248 | Not-determined |
| S17_extracted_k141_97991 | Low-quality |
| S17_extracted_k141_82716 | Low-quality |
| S17_extracted_k141_120135 | Not-determined |
| S17_extracted_k141_134726 | Not-determined |
| S17_extracted_k141_171617 | Not-determined |
| S17_extracted_k141_46405 | Not-determined |
| S17_extracted_k141_120138 | Not-determined |
| S17_extracted_k141_53928 | Not-determined |
| S17_extracted_k141_120140 | Not-determined |
| S17_extracted_k141_112403 | Not-determined |
| S17_extracted_k141_16252 | Not-determined |
| S17_extracted_k141_39091 | Low-quality |
| S17_extracted_k141_53930 | Not-determined |
| S17_extracted_k141_1772 | Not-determined |
| S17_extracted_k141_1773 | Not-determined |
| S17_extracted_k141_105766 | Low-quality |
| S17_extracted_k141_171624 | Not-determined |
| S17_extracted_k141_157179 | Not-determined |
| S17_extracted_k141_23838 | Not-determined |
| S17_extracted_k141_31555 | Not-determined |
| S17_extracted_k141_120143 | Not-determined |
| S17_extracted_k141_157180 | Not-determined |
| S17_extracted_k141_149960 | Not-determined |
| S17_extracted_k141_82725 | Low-quality |
| S17_extracted_k141_60849 | Not-determined |
| S17_extracted_k141_31560 | Not-determined |
| S17_extracted_k141_46416 | Low-quality |
| S17_extracted_k141_31561 | Not-determined |
| S17_extracted_k141_171632 | Not-determined |
| S17_extracted_k141_120155 | Low-quality |
| S17_extracted_k141_127095 | Not-determined |
| S17_extracted_k141_98006 | Not-determined |
| S17_extracted_k141_164268 | Not-determined |
| S17_extracted_k141_60854 | Not-determined |
| S17_extracted_k141_112413 | Not-determined |
| S17_extracted_k141_60856 | Not-determined |
| S17_extracted_k141_112414 | Not-determined |
| S17_extracted_k141_75477 | Not-determined |
| S17_extracted_k141_68275 | Not-determined |
| S17_extracted_k141_1780 | Not-determined |
| S17_extracted_k141_98011 | Low-quality |
| S17_extracted_k141_164274 | Not-determined |
| S17_extracted_k141_157193 | Not-determined |
| S17_extracted_k141_31573 | Not-determined |
| S17_extracted_k141_164280 | Not-determined |
| S17_extracted_k141_75485 | Not-determined |
| S17_extracted_k141_157200 | Not-determined |
| S17_extracted_k141_60867 | Low-quality |
| S17_extracted_k141_60868 | Not-determined |
| S17_extracted_k141_1789 | Not-determined |
| S17_extracted_k141_127108 | Not-determined |
| S17_extracted_k141_82734 | Not-determined |
| S17_extracted_k141_53967 | Low-quality |
| S17_extracted_k141_39121 | Not-determined |
| S17_extracted_k141_134742 | Not-determined |
| S17_extracted_k141_149971 | Not-determined |
| S17_extracted_k141_112430 | Not-determined |
| S17_extracted_k141_23847 | Not-determined |
| S17_extracted_k141_127114 | Not-determined |
| S17_extracted_k141_75493 | Not-determined |
| S17_extracted_k141_127115 | Not-determined |
| S17_extracted_k141_112433 | Not-determined |
| S17_extracted_k141_157205 | Not-determined |
| S17_extracted_k141_82743 | Not-determined |
| S17_extracted_k141_39127 | Not-determined |
| S17_extracted_k141_53976 | Not-determined |
| S17_extracted_k141_46426 | Not-determined |
| S17_extracted_k141_112435 | Not-determined |
| S17_extracted_k141_149980 | Not-determined |
| S17_extracted_k141_75499 | Not-determined |
| S17_extracted_k141_134749 | Not-determined |
| S17_extracted_k141_112438 | Low-quality |
| S17_extracted_k141_134750 | Not-determined |
| S17_extracted_k141_112440 | Not-determined |
| S17_extracted_k141_23855 | Not-determined |
| S17_extracted_k141_134752 | Not-determined |
| S17_extracted_k141_112441 | Not-determined |
| S17_extracted_k141_23857 | Not-determined |
| S17_extracted_k141_164289 | Not-determined |
| S17_extracted_k141_82749 | Low-quality |
| S17_extracted_k141_68293 | Not-determined |
| S17_extracted_k141_23861 | Not-determined |
| S17_extracted_k141_23862 | Not-determined |
| S17_extracted_k141_120182 | Low-quality |
| S17_extracted_k141_142340 | Not-determined |
| S17_extracted_k141_60891 | Not-determined |
| S17_extracted_k141_120183 | Not-determined |
| S17_extracted_k141_98028 | Not-determined |
| S17_extracted_k141_127121 | Not-determined |
| S17_extracted_k141_68301 | Low-quality |
| S17_extracted_k141_134764 | Not-determined |
| S17_extracted_k141_68304 | Not-determined |
| S17_extracted_k141_134766 | Not-determined |
| S17_extracted_k141_134767 | Not-determined |
| S17_extracted_k141_60896 | Not-determined |
| S17_extracted_k141_105786 | Not-determined |
| S17_extracted_k141_16284 | Not-determined |
| S17_extracted_k141_23870 | Not-determined |
| S17_extracted_k141_164302 | Not-determined |
| S17_extracted_k141_98030 | Not-determined |
| S17_extracted_k141_120193 | Not-determined |
| S17_extracted_k141_105789 | Not-determined |
| S17_extracted_k141_105790 | Not-determined |
| S17_extracted_k141_150000 | Not-determined |
| S17_extracted_k141_23873 | Low-quality |
| S17_extracted_k141_60901 | Not-determined |
| S17_extracted_k141_105796 | Not-determined |
| S17_extracted_k141_98043 | Not-determined |
| S17_extracted_k141_127136 | Not-determined |
| S17_extracted_k141_23878 | Not-determined |
| S17_extracted_k141_68310 | Not-determined |
| S17_extracted_k141_150003 | Not-determined |
| S17_extracted_k141_31582 | Not-determined |
| S17_extracted_k141_120203 | Not-determined |
| S17_extracted_k141_90302 | Not-determined |
| S17_extracted_k141_112454 | Not-determined |
| S17_extracted_k141_90307 | Not-determined |
| S17_extracted_k141_39148 | Not-determined |
| S17_extracted_k141_171647 | Not-determined |
| S17_extracted_k141_150009 | Not-determined |
| S17_extracted_k141_98054 | Not-determined |
| S17_extracted_k141_112461 | Not-determined |
| S17_extracted_k141_134781 | Not-determined |
| S17_extracted_k141_134782 | Low-quality |
| S17_extracted_k141_120212 | Not-determined |
| S17_extracted_k141_120213 | Not-determined |
| S17_extracted_k141_171653 | Low-quality |
| S17_extracted_k141_134786 | Not-determined |
| S17_extracted_k141_150016 | Not-determined |
| S17_extracted_k141_46432 | Low-quality |
| S17_extracted_k141_150018 | Not-determined |
| S17_extracted_k141_150019 | Not-determined |
| S17_extracted_k141_112473 | Not-determined |
| S17_extracted_k141_68319 | Low-quality |
| S17_extracted_k141_112477 | Not-determined |
| S17_extracted_k141_134790 | Not-determined |
| S17_extracted_k141_90327 | Not-determined |
| S17_extracted_k141_31596 | Not-determined |
| S17_extracted_k141_46438 | Low-quality |
| S17_extracted_k141_16303 | Not-determined |
| S17_extracted_k141_68321 | Not-determined |
| S17_extracted_k141_105806 | Low-quality |
| S17_extracted_k141_90329 | Not-determined |
| S17_extracted_k141_46440 | Not-determined |
| S17_extracted_k141_46446 | Not-determined |
| S17_extracted_k141_134793 | Low-quality |
| S17_extracted_k141_46449 | Not-determined |
| S17_extracted_k141_46450 | Not-determined |
| S17_extracted_k141_75535 | Not-determined |
| S17_extracted_k141_46453 | Not-determined |
| S17_extracted_k141_1833 | Not-determined |
| S17_extracted_k141_105818 | Not-determined |
| S17_extracted_k141_105820 | Not-determined |
| S17_extracted_k141_75539 | Low-quality |
| S17_extracted_k141_31607 | Not-determined |
| S17_extracted_k141_46460 | Not-determined |
| S17_extracted_k141_98059 | Not-determined |
| S17_extracted_k141_31613 | Low-quality |
| S17_extracted_k141_31616 | Not-determined |
| S17_extracted_k141_105826 | Not-determined |
| S17_extracted_k141_75542 | Not-determined |
| S17_extracted_k141_150060 | Not-determined |
| S17_extracted_k141_31619 | Not-determined |
| S17_extracted_k141_150061 | Not-determined |
| S17_extracted_k141_90352 | Not-determined |
| S17_extracted_k141_31620 | Not-determined |
| S17_extracted_k141_46467 | Not-determined |
| S17_extracted_k141_39172 | Not-determined |
| S17_extracted_k141_75546 | Not-determined |
| S17_extracted_k141_98067 | Low-quality |
| S17_extracted_k141_150063 | Low-quality |
| S17_extracted_k141_164324 | Not-determined |
| S17_extracted_k141_105833 | Not-determined |
| S17_extracted_k141_39177 | Not-determined |
| S17_extracted_k141_98071 | Not-determined |
| S17_extracted_k141_31629 | Not-determined |
| S17_extracted_k141_16327 | Not-determined |
| S17_extracted_k141_90356 | Low-quality |
| S17_extracted_k141_68345 | Low-quality |
| S17_extracted_k141_127164 | Not-determined |
| S17_extracted_k141_82763 | Not-determined |
| S17_extracted_k141_75553 | Not-determined |
| S17_extracted_k141_31632 | Not-determined |
| S17_extracted_k141_46470 | Not-determined |
| S17_extracted_k141_1857 | Not-determined |
| S17_extracted_k141_39183 | Not-determined |
| S17_extracted_k141_171662 | Not-determined |
| S17_extracted_k141_171663 | Low-quality |
| S17_extracted_k141_68351 | Not-determined |
| S17_extracted_k141_16333 | Not-determined |
| S17_extracted_k141_68354 | Low-quality |
| S17_extracted_k141_46476 | Not-determined |
| S17_extracted_k141_46480 | Not-determined |

| | |
|---|---|
| S17_extracted_k141_171669 | Low-quality |
| S17_extracted_k141_90368 | Not-determined |
| S17_extracted_k141_164335 | Not-determined |
| S17_extracted_k141_171672 | Not-determined |
| S17_extracted_k141_127178 | Not-determined |
| S17_extracted_k141_98092 | Not-determined |
| S17_extracted_k141_68367 | Not-determined |
| S17_extracted_k141_112488 | Not-determined |
| S17_extracted_k141_31647 | Not-determined |
| S17_extracted_k141_142403 | Not-determined |
| S17_extracted_k141_134799 | Not-determined |
| S17_extracted_k141_1872 | Not-determined |
| S17_extracted_k141_171678 | Not-determined |
| S17_extracted_k141_171679 | Not-determined |
| S17_extracted_k141_46492 | Not-determined |
| S17_extracted_k141_31653 | Not-determined |
| S17_extracted_k141_164346 | Not-determined |
| S17_extracted_k141_134801 | Not-determined |
| S17_extracted_k141_112498 | Not-determined |
| S17_extracted_k141_98097 | Not-determined |
| S17_extracted_k141_1874 | Low-quality |
| S17_extracted_k141_142409 | Not-determined |
| S17_extracted_k141_105860 | Low-quality |
| S17_extracted_k141_39200 | Not-determined |
| S17_extracted_k141_75573 | Not-determined |
| S17_extracted_k141_142416 | Not-determined |
| S17_extracted_k141_75575 | Not-determined |
| S17_extracted_k141_112506 | Not-determined |
| S17_extracted_k141_60934 | Not-determined |
| S17_extracted_k141_150067 | Not-determined |
| S17_extracted_k141_164359 | Not-determined |
| S17_extracted_k141_60936 | Not-determined |
| S17_extracted_k141_90376 | Not-determined |
| S17_extracted_k141_164363 | Low-quality |
| S17_extracted_k141_150069 | Not-determined |
| S17_extracted_k141_60942 | Not-determined |
| S17_extracted_k141_90384 | Not-determined |
| S17_extracted_k141_157227 | Not-determined |
| S17_extracted_k141_164368 | Low-quality |
| S17_extracted_k141_23892 | Not-determined |
| S17_extracted_k141_150074 | Not-determined |
| S17_extracted_k141_142431 | Not-determined |
| S17_extracted_k141_142432 | Not-determined |
| S17_extracted_k141_150076 | Not-determined |
| S17_extracted_k141_75586 | Low-quality |
| S17_extracted_k141_54007 | Low-quality |
| S17_extracted_k141_150077 | Not-determined |
| S17_extracted_k141_23895 | Not-determined |
| S17_extracted_k141_75588 | Not-determined |
| S17_extracted_k141_75593 | Not-determined |
| S17_extracted_k141_39229 | Not-determined |
| S17_extracted_k141_112518 | Low-quality |
| S17_extracted_k141_142436 | Not-determined |
| S17_extracted_k141_150078 | Not-determined |
| S17_extracted_k141_54011 | Not-determined |
| S17_extracted_k141_105878 | Low-quality |
| S17_extracted_k141_105882 | Not-determined |
| S17_extracted_k141_105884 | Not-determined |
| S17_extracted_k141_75598 | Not-determined |
| S17_extracted_k141_39237 | Not-determined |
| S17_extracted_k141_171699 | Not-determined |
| S17_extracted_k141_75600 | Not-determined |
| S17_extracted_k141_1893 | Not-determined |
| S17_extracted_k141_75602 | Not-determined |
| S17_extracted_k141_68409 | Not-determined |
| S17_extracted_k141_157235 | Not-determined |
| S17_extracted_k141_39241 | Not-determined |
| S17_extracted_k141_112532 | Not-determined |
| S17_extracted_k141_31659 | Not-determined |
| S17_extracted_k141_98108 | Not-determined |
| S17_extracted_k141_90392 | Not-determined |
| S17_extracted_k141_112535 | Not-determined |
| S17_extracted_k141_9435 | Not-determined |
| S17_extracted_k141_68417 | Not-determined |
| S17_extracted_k141_1902 | Not-determined |
| S17_extracted_k141_112537 | Low-quality |
| S17_extracted_k141_54033 | Not-determined |
| S17_extracted_k141_90397 | Not-determined |
| S17_extracted_k141_68422 | Not-determined |
| S17_extracted_k141_54036 | Not-determined |
| S17_extracted_k141_134849 | Low-quality |
| S17_extracted_k141_54039 | Not-determined |
| S17_extracted_k141_90400 | Not-determined |
| S17_extracted_k141_1907 | Not-determined |
| S17_extracted_k141_98119 | Low-quality |
| S17_extracted_k141_134855 | Not-determined |
| S17_extracted_k141_171716 | Not-determined |
| S17_extracted_k141_171717 | Not-determined |
| S17_extracted_k141_82773 | Low-quality |
| S17_extracted_k141_54045 | Not-determined |
| S17_extracted_k141_171720 | Not-determined |
| S17_extracted_k141_164399 | Not-determined |
| S17_extracted_k141_54046 | Not-determined |
| S17_extracted_k141_90406 | Low-quality |
| S17_extracted_k141_60969 | Low-quality |
| S17_extracted_k141_39262 | Not-determined |
| S17_extracted_k141_134862 | Low-quality |
| S17_extracted_k141_150100 | Not-determined |
| S17_extracted_k141_60970 | Not-determined |
| S17_extracted_k141_150101 | Not-determined |
| S17_extracted_k141_127194 | Not-determined |
| S17_extracted_k141_1917 | Not-determined |
| S17_extracted_k141_39267 | Not-determined |
| S17_extracted_k141_90411 | Low-quality |
| S17_extracted_k141_134866 | Not-determined |
| S17_extracted_k141_112549 | Not-determined |
| S17_extracted_k141_60976 | Not-determined |
| S17_extracted_k141_46501 | Not-determined |
| S17_extracted_k141_82783 | Not-determined |
| S17_extracted_k141_157254 | Not-determined |
| S17_extracted_k141_31661 | Not-determined |
| S17_extracted_k141_171729 | Not-determined |
| S17_extracted_k141_39276 | Not-determined |
| S17_extracted_k141_1931 | Not-determined |
| S17_extracted_k141_60986 | Not-determined |
| S17_extracted_k141_171734 | Not-determined |
| S17_extracted_k141_127203 | Not-determined |
| S17_extracted_k141_171735 | Not-determined |
| S17_extracted_k141_164423 | Not-determined |
| S17_extracted_k141_98141 | Not-determined |
| S17_extracted_k141_157261 | Not-determined |
| S17_extracted_k141_157262 | Not-determined |
| S17_extracted_k141_134881 | Not-determined |
| S17_extracted_k141_82794 | Not-determined |
| S17_extracted_k141_82795 | Not-determined |
| S17_extracted_k141_54057 | Not-determined |
| S17_extracted_k141_1940 | Not-determined |
| S17_extracted_k141_171739 | Not-determined |
| S17_extracted_k141_82798 | Not-determined |
| S17_extracted_k141_171743 | Not-determined |
| S17_extracted_k141_1943 | Not-determined |
| S17_extracted_k141_39296 | Not-determined |
| S17_extracted_k141_54065 | Not-determined |
| S17_extracted_k141_127216 | Not-determined |
| S17_extracted_k141_39298 | Not-determined |
| S17_extracted_k141_98150 | Not-determined |
| S17_extracted_k141_54068 | Not-determined |
| S17_extracted_k141_112561 | Not-determined |
| S17_extracted_k141_98151 | Not-determined |
| S17_extracted_k141_82812 | Low-quality |
| S17_extracted_k141_60991 | Not-determined |
| S17_extracted_k141_1954 | Not-determined |
| S17_extracted_k141_164443 | Low-quality |
| S17_extracted_k141_60995 | Not-determined |
| S17_extracted_k141_164446 | Not-determined |
| S17_extracted_k141_134882 | Low-quality |
| S17_extracted_k141_164448 | Not-determined |
| S17_extracted_k141_82820 | Not-determined |
| S17_extracted_k141_112568 | Not-determined |
| S17_extracted_k141_31671 | Not-determined |
| S17_extracted_k141_23923 | Not-determined |
| S17_extracted_k141_82822 | Low-quality |
| S17_extracted_k141_61000 | Not-determined |
| S17_extracted_k141_23924 | Low-quality |
| S17_extracted_k141_68464 | Not-determined |
| S17_extracted_k141_134888 | Not-determined |
| S17_extracted_k141_150133 | Not-determined |
| S17_extracted_k141_171749 | Not-determined |
| S17_extracted_k141_134892 | Not-determined |
| S17_extracted_k141_68467 | Low-quality |
| S17_extracted_k141_82834 | Not-determined |
| S17_extracted_k141_164458 | Not-determined |
| S17_extracted_k141_68471 | Not-determined |
| S17_extracted_k141_164459 | Low-quality |
| S17_extracted_k141_23935 | Not-determined |
| S17_extracted_k141_164461 | Not-determined |
| S17_extracted_k141_150141 | Not-determined |
| S17_extracted_k141_112578 | Not-determined |
| S17_extracted_k141_16354 | Not-determined |
| S17_extracted_k141_23937 | Low-quality |
| S17_extracted_k141_112581 | Not-determined |
| S17_extracted_k141_61003 | Not-determined |
| S17_extracted_k141_120228 | Not-determined |
| S17_extracted_k141_16356 | Low-quality |
| S17_extracted_k141_134901 | Not-determined |
| S17_extracted_k141_23943 | Not-determined |
| S17_extracted_k141_23944 | Low-quality |
| S17_extracted_k141_134903 | Not-determined |
| S17_extracted_k141_16357 | Not-determined |
| S17_extracted_k141_120231 | Low-quality |
| S17_extracted_k141_112590 | Not-determined |
| S17_extracted_k141_112591 | Not-determined |
| S17_extracted_k141_82838 | Low-quality |
| S17_extracted_k141_46507 | Not-determined |
| S17_extracted_k141_46508 | Not-determined |
| S17_extracted_k141_134904 | Not-determined |
| S17_extracted_k141_134905 | Low-quality |
| S17_extracted_k141_68484 | Not-determined |
| S17_extracted_k141_23951 | Not-determined |
| S17_extracted_k141_68485 | Not-determined |
| S17_extracted_k141_134907 | Not-determined |
| S17_extracted_k141_68486 | Not-determined |
| S17_extracted_k141_16370 | Not-determined |
| S17_extracted_k141_46513 | Low-quality |
| S17_extracted_k141_46516 | Not-determined |
| S17_extracted_k141_150158 | Not-determined |
| S17_extracted_k141_120239 | Not-determined |
| S17_extracted_k141_112600 | Not-determined |
| S17_extracted_k141_46520 | Not-determined |
| S17_extracted_k141_150161 | Not-determined |
| S17_extracted_k141_112603 | Not-determined |
| S17_extracted_k141_46524 | Not-determined |
| S17_extracted_k141_46525 | Not-determined |
| S17_extracted_k141_31702 | Not-determined |
| S17_extracted_k141_16379 | Not-determined |
| S17_extracted_k141_150169 | Not-determined |
| S17_extracted_k141_142495 | Not-determined |
| S17_extracted_k141_157318 | Not-determined |
| S17_extracted_k141_112610 | Not-determined |
| S17_extracted_k141_75637 | Not-determined |
| S17_extracted_k141_112612 | Not-determined |
| S17_extracted_k141_157322 | Not-determined |
| S17_extracted_k141_112613 | Not-determined |
| S17_extracted_k141_171763 | Not-determined |
| S17_extracted_k141_157323 | Low-quality |
| S17_extracted_k141_171764 | Not-determined |
| S17_extracted_k141_120249 | Low-quality |
| S17_extracted_k141_171765 | Not-determined |
| S17_extracted_k141_112616 | Not-determined |
| S17_extracted_k141_75641 | Not-determined |
| S17_extracted_k141_142504 | Not-determined |
| S17_extracted_k141_171768 | Low-quality |
| S17_extracted_k141_142506 | Low-quality |
| S17_extracted_k141_120252 | Not-determined |
| S17_extracted_k141_171770 | Low-quality |
| S17_extracted_k141_134923 | Not-determined |
| S17_extracted_k141_31712 | Not-determined |
| S17_extracted_k141_9463 | Not-determined |
| S17_extracted_k141_142517 | Not-determined |
| S17_extracted_k141_39336 | Not-determined |
| S17_extracted_k141_120257 | Not-determined |
| S17_extracted_k141_16384 | Not-determined |
| S17_extracted_k141_134930 | Not-determined |
| S17_extracted_k141_142522 | Not-determined |
| S17_extracted_k141_157327 | Low-quality |
| S17_extracted_k141_127223 | Not-determined |
| S17_extracted_k141_23973 | Not-determined |
| S17_extracted_k141_150177 | Not-determined |
| S17_extracted_k141_46545 | Not-determined |
| S17_extracted_k141_39338 | Not-determined |
| S17_extracted_k141_68498 | Not-determined |
| S17_extracted_k141_134934 | Low-quality |
| S17_extracted_k141_82859 | Not-determined |
| S17_extracted_k141_127229 | Not-determined |
| S17_extracted_k141_157333 | Not-determined |
| S17_extracted_k141_98168 | Not-determined |
| S17_extracted_k141_16392 | Not-determined |
| S17_extracted_k141_105938 | Not-determined |
| S17_extracted_k141_82864 | Not-determined |
| S17_extracted_k141_75650 | Not-determined |
| S17_extracted_k141_171782 | Not-determined |
| S17_extracted_k141_46551 | Not-determined |
| S17_extracted_k141_82869 | Not-determined |
| S17_extracted_k141_171783 | Not-determined |
| S17_extracted_k141_98176 | Not-determined |
| S17_extracted_k141_9478 | Low-quality |
| S17_extracted_k141_171785 | Not-determined |
| S17_extracted_k141_98178 | Not-determined |
| S17_extracted_k141_39353 | Not-determined |
| S17_extracted_k141_23980 | Low-quality |
| S17_extracted_k141_1963 | Not-determined |
| S17_extracted_k141_134952 | Low-quality |
| S17_extracted_k141_39357 | Not-determined |
| S17_extracted_k141_75658 | Not-determined |
| S17_extracted_k141_171787 | Not-determined |
| S17_extracted_k141_1974 | Low-quality |
| S17_extracted_k141_9486 | Low-quality |
| S17_extracted_k141_9487 | Not-determined |
| S17_extracted_k141_105950 | Not-determined |
| S17_extracted_k141_1976 | Not-determined |
| S17_extracted_k141_9489 | Low-quality |
| S17_extracted_k141_68510 | Not-determined |
| S17_extracted_k141_134956 | Not-determined |
| S17_extracted_k141_39364 | Not-determined |
| S17_extracted_k141_164469 | Not-determined |
| S17_extracted_k141_157343 | Not-determined |
| S17_extracted_k141_134958 | Not-determined |
| S17_extracted_k141_9494 | Not-determined |
| S17_extracted_k141_46557 | Low-quality |
| S17_extracted_k141_16400 | Not-determined |
| S17_extracted_k141_39366 | Low-quality |
| S17_extracted_k141_150188 | Not-determined |
| S17_extracted_k141_150193 | Not-determined |
| S17_extracted_k141_46564 | Not-determined |
| S17_extracted_k141_75666 | Not-determined |
| S17_extracted_k141_75667 | Low-quality |
| S17_extracted_k141_39371 | Low-quality |
| S17_extracted_k141_98213 | Low-quality |
| S17_extracted_k141_112620 | Not-determined |
| S17_extracted_k141_157346 | Not-determined |
| S17_extracted_k141_112622 | Low-quality |
| S17_extracted_k141_82878 | Not-determined |
| S17_extracted_k141_98218 | Not-determined |
| S17_extracted_k141_9507 | Not-determined |
| S17_extracted_k141_134970 | Not-determined |
| S17_extracted_k141_75679 | Not-determined |
| S17_extracted_k141_1997 | Not-determined |
| S17_extracted_k141_75680 | Not-determined |
| S17_extracted_k141_134973 | Not-determined |
| S17_extracted_k141_68539 | Not-determined |
| S17_extracted_k141_75682 | Not-determined |
| S17_extracted_k141_98228 | Not-determined |
| S17_extracted_k141_112629 | Not-determined |
| S17_extracted_k141_31741 | Not-determined |
| S17_extracted_k141_164490 | Not-determined |
| S17_extracted_k141_46582 | Not-determined |
| S17_extracted_k141_31743 | Not-determined |
| S17_extracted_k141_82883 | Not-determined |
| S17_extracted_k141_75688 | Not-determined |
| S17_extracted_k141_98233 | Not-determined |
| S17_extracted_k141_127265 | Not-determined |
| S17_extracted_k141_68541 | Low-quality |
| S17_extracted_k141_16408 | Not-determined |
| S17_extracted_k141_142544 | Not-determined |
| S17_extracted_k141_39383 | Not-determined |
| S17_extracted_k141_39386 | Not-determined |
| S17_extracted_k141_39387 | Not-determined |
| S17_extracted_k141_98239 | Not-determined |
| S17_extracted_k141_112641 | Low-quality |
| S17_extracted_k141_75697 | Not-determined |
| S17_extracted_k141_16412 | Not-determined |
| S17_extracted_k141_112644 | Not-determined |
| S17_extracted_k141_98241 | Not-determined |
| S17_extracted_k141_54103 | Not-determined |
| S17_extracted_k141_112650 | Not-determined |
| S17_extracted_k141_98246 | Not-determined |
| S17_extracted_k141_54105 | Not-determined |
| S17_extracted_k141_164499 | Not-determined |
| S17_extracted_k141_54108 | Not-determined |
| S17_extracted_k141_2004 | Not-determined |
| S17_extracted_k141_39400 | Not-determined |
| S17_extracted_k141_31753 | Not-determined |
| S17_extracted_k141_2005 | Not-determined |
| S17_extracted_k141_68553 | Not-determined |
| S17_extracted_k141_9516 | Not-determined |
| S17_extracted_k141_39401 | Not-determined |
| S17_extracted_k141_112658 | Not-determined |
| S17_extracted_k141_31760 | Not-determined |
| S17_extracted_k141_134986 | Not-determined |
| S17_extracted_k141_16425 | Not-determined |
| S17_extracted_k141_127289 | Low-quality |
| S17_extracted_k141_105974 | Not-determined |
| S17_extracted_k141_98267 | Not-determined |
| S17_extracted_k141_120281 | Low-quality |
| S17_extracted_k141_105977 | Not-determined |
| S17_extracted_k141_142567 | Not-determined |
| S17_extracted_k141_31768 | Low-quality |
| S17_extracted_k141_54132 | Not-determined |
| S17_extracted_k141_120289 | Not-determined |
| S17_extracted_k141_112668 | Not-determined |
| S17_extracted_k141_112669 | Low-quality |
| S17_extracted_k141_142573 | Not-determined |
| S17_extracted_k141_90434 | Not-determined |
| S17_extracted_k141_142574 | Not-determined |
| S17_extracted_k141_127299 | Not-determined |
| S17_extracted_k141_142577 | Not-determined |
| S17_extracted_k141_150227 | Low-quality |
| S17_extracted_k141_142580 | Not-determined |
| S17_extracted_k141_2019 | Not-determined |
| S17_extracted_k141_112677 | Not-determined |
| S17_extracted_k141_31779 | Low-quality |
| S17_extracted_k141_164520 | Not-determined |
| S17_extracted_k141_68571 | Not-determined |
| S17_extracted_k141_112678 | Not-determined |
| S17_extracted_k141_9537 | Not-determined |
| S17_extracted_k141_150232 | Low-quality |
| S17_extracted_k141_164522 | Not-determined |
| S17_extracted_k141_112680 | Not-determined |
| S17_extracted_k141_127306 | Not-determined |
| S17_extracted_k141_150235 | Not-determined |
| S17_extracted_k141_105981 | Not-determined |
| S17_extracted_k141_150237 | Not-determined |
| S17_extracted_k141_24016 | Not-determined |
| S17_extracted_k141_2027 | Not-determined |
| S17_extracted_k141_31785 | Not-determined |
| S17_extracted_k141_24018 | Not-determined |
| S17_extracted_k141_120300 | Not-determined |
| S17_extracted_k141_75728 | Not-determined |
| S17_extracted_k141_112682 | Not-determined |
| S17_extracted_k141_164527 | Not-determined |
| S17_extracted_k141_127313 | Not-determined |
| S17_extracted_k141_31790 | Not-determined |
| S17_extracted_k141_120306 | Not-determined |
| S17_extracted_k141_24021 | Not-determined |
| S17_extracted_k141_54140 | Not-determined |
| S17_extracted_k141_120311 | Not-determined |
| S17_extracted_k141_2034 | Not-determined |
| S17_extracted_k141_31795 | Not-determined |
| S17_extracted_k141_24025 | Not-determined |
| S17_extracted_k141_142586 | Not-determined |
| S17_extracted_k141_16445 | Not-determined |
| S17_extracted_k141_9551 | Low-quality |
| S17_extracted_k141_105995 | Not-determined |
| S17_extracted_k141_75732 | Not-determined |
| S17_extracted_k141_105996 | Low-quality |
| S17_extracted_k141_2038 | Not-determined |
| S17_extracted_k141_16450 | Not-determined |
| S17_extracted_k141_164530 | Not-determined |
| S17_extracted_k141_127316 | Not-determined |
| S17_extracted_k141_142593 | Not-determined |
| S17_extracted_k141_90452 | Low-quality |
| S17_extracted_k141_164532 | Not-determined |
| S17_extracted_k141_24032 | Not-determined |
| S17_extracted_k141_31801 | Not-determined |
| S17_extracted_k141_127320 | Not-determined |
| S17_extracted_k141_150252 | Not-determined |
| S17_extracted_k141_24037 | Not-determined |
| S17_extracted_k141_75742 | Not-determined |
| S17_extracted_k141_106005 | Not-determined |
| S17_extracted_k141_164537 | Not-determined |
| S17_extracted_k141_24039 | Not-determined |
| S17_extracted_k141_16454 | Not-determined |
| S17_extracted_k141_120325 | Not-determined |
| S17_extracted_k141_54162 | Not-determined |
| S17_extracted_k141_54163 | Not-determined |
| S17_extracted_k141_9564 | Not-determined |
| S17_extracted_k141_127327 | Not-determined |
| S17_extracted_k141_16455 | Not-determined |
| S17_extracted_k141_135015 | Not-determined |
| S17_extracted_k141_75747 | Not-determined |
| S17_extracted_k141_135017 | Not-determined |
| S17_extracted_k141_16459 | Not-determined |
| S17_extracted_k141_90461 | Not-determined |
| S17_extracted_k141_90462 | Not-determined |
| S17_extracted_k141_16461 | Not-determined |
| S17_extracted_k141_39424 | Not-determined |
| S17_extracted_k141_135024 | Not-determined |
| S17_extracted_k141_54180 | Not-determined |
| S17_extracted_k141_150278 | Not-determined |
| S17_extracted_k141_54183 | Not-determined |
| S17_extracted_k141_127330 | Not-determined |
| S17_extracted_k141_150280 | Not-determined |
| S17_extracted_k141_54186 | Not-determined |
| S17_extracted_k141_106021 | Not-determined |
| S17_extracted_k141_150282 | Not-determined |
| S17_extracted_k141_90472 | Not-determined |
| S17_extracted_k141_150284 | Not-determined |
| S17_extracted_k141_2049 | Not-determined |
| S17_extracted_k141_164547 | Not-determined |
| S17_extracted_k141_39436 | Not-determined |
| S17_extracted_k141_106024 | Low-quality |
| S17_extracted_k141_68596 | Not-determined |
| S17_extracted_k141_106026 | Low-quality |
| S17_extracted_k141_90479 | Not-determined |
| S17_extracted_k141_75758 | Not-determined |
| S17_extracted_k141_54194 | Not-determined |
| S17_extracted_k141_90485 | Not-determined |
| S17_extracted_k141_98270 | Not-determined |
| S17_extracted_k141_75762 | Not-determined |
| S17_extracted_k141_90486 | Low-quality |
| S17_extracted_k141_39445 | Not-determined |
| S17_extracted_k141_24064 | Low-quality |
| S17_extracted_k141_16467 | Low-quality |
| S17_extracted_k141_39449 | Not-determined |
| S17_extracted_k141_127344 | Low-quality |
| S17_extracted_k141_24067 | Not-determined |
| S17_extracted_k141_112692 | Not-determined |
| S17_extracted_k141_2066 | Not-determined |
| S17_extracted_k141_142621 | Not-determined |
| S17_extracted_k141_24071 | Not-determined |
| S17_extracted_k141_39457 | Not-determined |
| S17_extracted_k141_127349 | Low-quality |
| S17_extracted_k141_98276 | Not-determined |
| S17_extracted_k141_39463 | Not-determined |
| S17_extracted_k141_98283 | Not-determined |
| S17_extracted_k141_31813 | Not-determined |
| S17_extracted_k141_16482 | Low-quality |
| S17_extracted_k141_157415 | Not-determined |
| S17_extracted_k141_164571 | Not-determined |
| S17_extracted_k141_127356 | Not-determined |
| S17_extracted_k141_142629 | Not-determined |
| S17_extracted_k141_98289 | Not-determined |
| S17_extracted_k141_142630 | Low-quality |
| S17_extracted_k141_157422 | Not-determined |
| S17_extracted_k141_75772 | Low-quality |
| S17_extracted_k141_142633 | Low-quality |
| S17_extracted_k141_9598 | Not-determined |
| S17_extracted_k141_164574 | Not-determined |
| S17_extracted_k141_46595 | Low-quality |
| S17_extracted_k141_164575 | Not-determined |
| S17_extracted_k141_68616 | Not-determined |
| S17_extracted_k141_157426 | Not-determined |
| S17_extracted_k141_39472 | Not-determined |
| S17_extracted_k141_157436 | Not-determined |
| S17_extracted_k141_164578 | Not-determined |
| S17_extracted_k141_164579 | Not-determined |
| S17_extracted_k141_164581 | Not-determined |
| S17_extracted_k141_135038 | Not-determined |
| S17_extracted_k141_9614 | Not-determined |
| S17_extracted_k141_112702 | Low-quality |
| S17_extracted_k141_98299 | Not-determined |
| S17_extracted_k141_164584 | Low-quality |
| S17_extracted_k141_46602 | Not-determined |
| S17_extracted_k141_112714 | Not-determined |
| S17_extracted_k141_16505 | Low-quality |
| S17_extracted_k141_9626 | Not-determined |
| S17_extracted_k141_24079 | Not-determined |
| S17_extracted_k141_46608 | Not-determined |
| S17_extracted_k141_31832 | Not-determined |
| S17_extracted_k141_9628 | Not-determined |
| S17_extracted_k141_9630 | Not-determined |
| S17_extracted_k141_157441 | Low-quality |
| S17_extracted_k141_68635 | Not-determined |
| S17_extracted_k141_157442 | Not-determined |
| S17_extracted_k141_150330 | Not-determined |
| S17_extracted_k141_120336 | Not-determined |
| S17_extracted_k141_142657 | Not-determined |
| S17_extracted_k141_31839 | Not-determined |
| S17_extracted_k141_106069 | Not-determined |
| S17_extracted_k141_39481 | Not-determined |
| S17_extracted_k141_16509 | Low-quality |
| S17_extracted_k141_135047 | Not-determined |
| S17_extracted_k141_135049 | Not-determined |
| S17_extracted_k141_120342 | Low-quality |
| S17_extracted_k141_142659 | Low-quality |
| S17_extracted_k141_98312 | Not-determined |
| S17_extracted_k141_164594 | Not-determined |
| S17_extracted_k141_142660 | Not-determined |
| S17_extracted_k141_31845 | Not-determined |
| S17_extracted_k141_112727 | Low-quality |
| S17_extracted_k141_31846 | Not-determined |
| S17_extracted_k141_90517 | Not-determined |
| S17_extracted_k141_68646 | Not-determined |
| S17_extracted_k141_16514 | Low-quality |
| S17_extracted_k141_120343 | Not-determined |
| S17_extracted_k141_112731 | Not-determined |
| S17_extracted_k141_164604 | Not-determined |
| S17_extracted_k141_16517 | Low-quality |
| S17_extracted_k141_98314 | Not-determined |
| S17_extracted_k141_164606 | Not-determined |
| S17_extracted_k141_164608 | Not-determined |
| S17_extracted_k141_135058 | Not-determined |
| S17_extracted_k141_90524 | Not-determined |
| S17_extracted_k141_82893 | Low-quality |
| S17_extracted_k141_106080 | Low-quality |
| S17_extracted_k141_82896 | Not-determined |
| S17_extracted_k141_75792 | Not-determined |
| S17_extracted_k141_120349 | Not-determined |
| S17_extracted_k141_24093 | Not-determined |
| S17_extracted_k141_112739 | Not-determined |
| S17_extracted_k141_106085 | Not-determined |
| S17_extracted_k141_120354 | Not-determined |
| S17_extracted_k141_150350 | Not-determined |
| S17_extracted_k141_39487 | Not-determined |
| S17_extracted_k141_164614 | Not-determined |
| S17_extracted_k141_90526 | Low-quality |
| S17_extracted_k141_39488 | Not-determined |
| S17_extracted_k141_75799 | Not-determined |
| S17_extracted_k141_120358 | Low-quality |
| S17_extracted_k141_112742 | Not-determined |
| S17_extracted_k141_9648 | Not-determined |
| S17_extracted_k141_135066 | Low-quality |
| S17_extracted_k141_157450 | Low-quality |
| S17_extracted_k141_90528 | Low-quality |
| S17_extracted_k141_150357 | Not-determined |
| S17_extracted_k141_142669 | Not-determined |
| S17_extracted_k141_98330 | Not-determined |
| S17_extracted_k141_2084 | Low-quality |
| S17_extracted_k141_39490 | Low-quality |
| S17_extracted_k141_135072 | Low-quality |
| S17_extracted_k141_120366 | Not-determined |
| S17_extracted_k141_142674 | Not-determined |
| S17_extracted_k141_16535 | Not-determined |
| S17_extracted_k141_142677 | Not-determined |
| S17_extracted_k141_112756 | Not-determined |
| S17_extracted_k141_150363 | Not-determined |
| S17_extracted_k141_106099 | Not-determined |
| S17_extracted_k141_31868 | Not-determined |
| S17_extracted_k141_106102 | Not-determined |
| S17_extracted_k141_150370 | Not-determined |
| S17_extracted_k141_24112 | Not-determined |
| S17_extracted_k141_39495 | Not-determined |
| S17_extracted_k141_2091 | Not-determined |
| S17_extracted_k141_106108 | Not-determined |
| S17_extracted_k141_16547 | Not-determined |
| S17_extracted_k141_164635 | Not-determined |
| S17_extracted_k141_9662 | Not-determined |
| S17_extracted_k141_164640 | Not-determined |
| S17_extracted_k141_9666 | Not-determined |
| S17_extracted_k141_90539 | Not-determined |
| S17_extracted_k141_9667 | Not-determined |
| S17_extracted_k141_90540 | Low-quality |
| S17_extracted_k141_106117 | Not-determined |
| S17_extracted_k141_142688 | Not-determined |
| S17_extracted_k141_75825 | Not-determined |
| S17_extracted_k141_68673 | Not-determined |
| S17_extracted_k141_75828 | Not-determined |
| S17_extracted_k141_68675 | Not-determined |
| S17_extracted_k141_106120 | Not-determined |

| | |
|---|---|
| S17_extracted_k141_75830 | Low-quality |
| S17_extracted_k141_106121 | Not-determined |
| S17_extracted_k141_171817 | Not-determined |
| S17_extracted_k141_142693 | Not-determined |
| S17_extracted_k141_75831 | Not-determined |
| S17_extracted_k141_164649 | Not-determined |
| S17_extracted_k141_16555 | Not-determined |
| S17_extracted_k141_171818 | Not-determined |
| S17_extracted_k141_106126 | Not-determined |
| S17_extracted_k141_112769 | Not-determined |
| S17_extracted_k141_112770 | Low-quality |
| S17_extracted_k141_157481 | Not-determined |
| S17_extracted_k141_98340 | Not-determined |
| S17_extracted_k141_171820 | Low-quality |
| S17_extracted_k141_39497 | Not-determined |
| S17_extracted_k141_68684 | Not-determined |
| S17_extracted_k141_171821 | Not-determined |
| S17_extracted_k141_39498 | Not-determined |
| S17_extracted_k141_46632 | Not-determined |
| S17_extracted_k141_171822 | Not-determined |
| S17_extracted_k141_46633 | Not-determined |
| S17_extracted_k141_112775 | Low-quality |
| S17_extracted_k141_142703 | Not-determined |
| S17_extracted_k141_46637 | Not-determined |
| S17_extracted_k141_24119 | Not-determined |
| S17_extracted_k141_16566 | Not-determined |
| S17_extracted_k141_68689 | Low-quality |
| S17_extracted_k141_164656 | Not-determined |
| S17_extracted_k141_46639 | Not-determined |
| S17_extracted_k141_39501 | Not-determined |
| S17_extracted_k141_46641 | Not-determined |
| S17_extracted_k141_142708 | Not-determined |
| S17_extracted_k141_98349 | Low-quality |
| S17_extracted_k141_120391 | Not-determined |
| S17_extracted_k141_164659 | Not-determined |
| S17_extracted_k141_75836 | Not-determined |
| S17_extracted_k141_39508 | Not-determined |
| S17_extracted_k141_157486 | Not-determined |
| S17_extracted_k141_68695 | Not-determined |
| S17_extracted_k141_16575 | Not-determined |
| S17_extracted_k141_157487 | Not-determined |
| S17_extracted_k141_75839 | Not-determined |
| S17_extracted_k141_157488 | Low-quality |
| S17_extracted_k141_68697 | Not-determined |
| S17_extracted_k141_142713 | Not-determined |
| S17_extracted_k141_68700 | Not-determined |
| S17_extracted_k141_68701 | Not-determined |
| S17_extracted_k141_16578 | Not-determined |
| S17_extracted_k141_142718 | Not-determined |
| S17_extracted_k141_150392 | Not-determined |
| S17_extracted_k141_16582 | Not-determined |
| S17_extracted_k141_135096 | Low-quality |
| S17_extracted_k141_75843 | Not-determined |
| S17_extracted_k141_75845 | Not-determined |
| S17_extracted_k141_142723 | Not-determined |
| S17_extracted_k141_68709 | Not-determined |
| S17_extracted_k141_68710 | Not-determined |
| S17_extracted_k141_75846 | Not-determined |
| S17_extracted_k141_16589 | Not-determined |
| S17_extracted_k141_98364 | Not-determined |
| S17_extracted_k141_135099 | Not-determined |
| S17_extracted_k141_90571 | Not-determined |
| S17_extracted_k141_16590 | Not-determined |
| S17_extracted_k141_164667 | Low-quality |
| S17_extracted_k141_68720 | Not-determined |
| S17_extracted_k141_98367 | Not-determined |
| S17_extracted_k141_90575 | Not-determined |
| S17_extracted_k141_112796 | Not-determined |
| S17_extracted_k141_16597 | Low-quality |
| S17_extracted_k141_75856 | Not-determined |
| S17_extracted_k141_75859 | Not-determined |
| S17_extracted_k141_164672 | Not-determined |
| S17_extracted_k141_171860 | Not-determined |
| S17_extracted_k141_171861 | Low-quality |
| S17_extracted_k141_75862 | Not-determined |
| S17_extracted_k141_75863 | Not-determined |
| S17_extracted_k141_171867 | Not-determined |
| S17_extracted_k141_98381 | Low-quality |
| S17_extracted_k141_157514 | Low-quality |
| S17_extracted_k141_9717 | Not-determined |
| S17_extracted_k141_127393 | Not-determined |
| S17_extracted_k141_106163 | Not-determined |
| S17_extracted_k141_9719 | Not-determined |
| S17_extracted_k141_150408 | Not-determined |
| S17_extracted_k141_9723 | Not-determined |
| S17_extracted_k141_98385 | Not-determined |
| S17_extracted_k141_98388 | Not-determined |
| S17_extracted_k141_98389 | Not-determined |
| S17_extracted_k141_98393 | Not-determined |
| S17_extracted_k141_46666 | Not-determined |
| S17_extracted_k141_157523 | Not-determined |
| S17_extracted_k141_142739 | Not-determined |
| S17_extracted_k141_90586 | Not-determined |
| S17_extracted_k141_171887 | Not-determined |
| S17_extracted_k141_127405 | Not-determined |
| S17_extracted_k141_90588 | Low-quality |
| S17_extracted_k141_90590 | Not-determined |
| S17_extracted_k141_171889 | Not-determined |
| S17_extracted_k141_127408 | Not-determined |
| S17_extracted_k141_157530 | Not-determined |
| S17_extracted_k141_24139 | Not-determined |
| S17_extracted_k141_68737 | Low-quality |
| S17_extracted_k141_127412 | Not-determined |
| S17_extracted_k141_75891 | Not-determined |
| S17_extracted_k141_46681 | Not-determined |
| S17_extracted_k141_120433 | Low-quality |
| S17_extracted_k141_112822 | Not-determined |
| S17_extracted_k141_68739 | Not-determined |
| S17_extracted_k141_98399 | Not-determined |
| S17_extracted_k141_75893 | Not-determined |
| S17_extracted_k141_112823 | Not-determined |
| S17_extracted_k141_127417 | Not-determined |
| S17_extracted_k141_98403 | Not-determined |
| S17_extracted_k141_2129 | Not-determined |
| S17_extracted_k141_68746 | Not-determined |
| S17_extracted_k141_98407 | Not-determined |
| S17_extracted_k141_46684 | Not-determined |
| S17_extracted_k141_9748 | Not-determined |
| S17_extracted_k141_164692 | Not-determined |
| S17_extracted_k141_142745 | Not-determined |
| S17_extracted_k141_9750 | Not-determined |
| S17_extracted_k141_164696 | Not-determined |
| S17_extracted_k141_46688 | Not-determined |
| S17_extracted_k141_68752 | Not-determined |
| S17_extracted_k141_157535 | Not-determined |
| S17_extracted_k141_9761 | Not-determined |
| S17_extracted_k141_120441 | Not-determined |
| S17_extracted_k141_75905 | Not-determined |
| S17_extracted_k141_9763 | Not-determined |
| S17_extracted_k141_127424 | Not-determined |
| S17_extracted_k141_127425 | Not-determined |
| S17_extracted_k141_142758 | Low-quality |
| S17_extracted_k141_157540 | Low-quality |
| S17_extracted_k141_112825 | Not-determined |
| S17_extracted_k141_127430 | Low-quality |
| S17_extracted_k141_142762 | Not-determined |
| S17_extracted_k141_98424 | Not-determined |
| S17_extracted_k141_16610 | Low-quality |
| S17_extracted_k141_16613 | Not-determined |
| S17_extracted_k141_75915 | Not-determined |
| S17_extracted_k141_112829 | Low-quality |
| S17_extracted_k141_2145 | Not-determined |
| S17_extracted_k141_106178 | Not-determined |
| S17_extracted_k141_2147 | Not-determined |
| S17_extracted_k141_75920 | Not-determined |
| S17_extracted_k141_90603 | Not-determined |
| S17_extracted_k141_106184 | Not-determined |
| S17_extracted_k141_39535 | Not-determined |
| S17_extracted_k141_75923 | Not-determined |
| S17_extracted_k141_150433 | Not-determined |
| S17_extracted_k141_54241 | Not-determined |
| S17_extracted_k141_112841 | Not-determined |
| S17_extracted_k141_31880 | Not-determined |
| S17_extracted_k141_24145 | Not-determined |
| S17_extracted_k141_24147 | Not-determined |
| S17_extracted_k141_150441 | Not-determined |
| S17_extracted_k141_24153 | Not-determined |
| S17_extracted_k141_106199 | Not-determined |
| S17_extracted_k141_46709 | Not-determined |
| S17_extracted_k141_54244 | Low-quality |
| S17_extracted_k141_75934 | Not-determined |
| S17_extracted_k141_75936 | Not-determined |
| S17_extracted_k141_98437 | Not-determined |
| S17_extracted_k141_39551 | Not-determined |
| S17_extracted_k141_39552 | Not-determined |
| S17_extracted_k141_9788 | Not-determined |
| S17_extracted_k141_61028 | Not-determined |
| S17_extracted_k141_98442 | Not-determined |
| S17_extracted_k141_16636 | Not-determined |
| S17_extracted_k141_61032 | Not-determined |
| S17_extracted_k141_157564 | Not-determined |
| S17_extracted_k141_16639 | Low-quality |
| S17_extracted_k141_112849 | Low-quality |
| S17_extracted_k141_9792 | Not-determined |
| S17_extracted_k141_127449 | Not-determined |
| S17_extracted_k141_68786 | Not-determined |
| S17_extracted_k141_16642 | Not-determined |
| S17_extracted_k141_24161 | Not-determined |
| S17_extracted_k141_61037 | Low-quality |
| S17_extracted_k141_54253 | Not-determined |
| S17_extracted_k141_9800 | Low-quality |
| S17_extracted_k141_31888 | Not-determined |
| S17_extracted_k141_150457 | Not-determined |
| S17_extracted_k141_82914 | Not-determined |
| S17_extracted_k141_24162 | Not-determined |
| S17_extracted_k141_54257 | Not-determined |
| S17_extracted_k141_39565 | Not-determined |
| S17_extracted_k141_164715 | Low-quality |
| S17_extracted_k141_46729 | Low-quality |
| S17_extracted_k141_31891 | Not-determined |
| S17_extracted_k141_106206 | Not-determined |
| S17_extracted_k141_112853 | Not-determined |
| S17_extracted_k141_157576 | Low-quality |
| S17_extracted_k141_164718 | Low-quality |
| S17_extracted_k141_31897 | Not-determined |
| S17_extracted_k141_120477 | Not-determined |
| S17_extracted_k141_46737 | Not-determined |
| S17_extracted_k141_54269 | Not-determined |
| S17_extracted_k141_31902 | Low-quality |
| S17_extracted_k141_31904 | Low-quality |
| S17_extracted_k141_82924 | Low-quality |
| S17_extracted_k141_98459 | Not-determined |
| S17_extracted_k141_24176 | Not-determined |
| S17_extracted_k141_90634 | Not-determined |
| S17_extracted_k141_39576 | Low-quality |
| S17_extracted_k141_171935 | Not-determined |
| S17_extracted_k141_120489 | Not-determined |
| S17_extracted_k141_75969 | Not-determined |
| S17_extracted_k141_16657 | Not-determined |
| S17_extracted_k141_112869 | Low-quality |
| S17_extracted_k141_46745 | Not-determined |
| S17_extracted_k141_106215 | Not-determined |
| S17_extracted_k141_9812 | Not-determined |
| S17_extracted_k141_75972 | Not-determined |
| S17_extracted_k141_16662 | Not-determined |
| S17_extracted_k141_127470 | Not-determined |
| S17_extracted_k141_75978 | Not-determined |
| S17_extracted_k141_120502 | Not-determined |
| S17_extracted_k141_171942 | Not-determined |
| S17_extracted_k141_164731 | Low-quality |
| S17_extracted_k141_46752 | Low-quality |
| S17_extracted_k141_112874 | Not-determined |
| S17_extracted_k141_127482 | Not-determined |
| S17_extracted_k141_142779 | Not-determined |
| S17_extracted_k141_127483 | Not-determined |
| S17_extracted_k141_54284 | Not-determined |
| S17_extracted_k141_98480 | Low-quality |
| S17_extracted_k141_164733 | Not-determined |
| S17_extracted_k141_98481 | Not-determined |
| S17_extracted_k141_142783 | Not-determined |
| S17_extracted_k141_31918 | Not-determined |
| S17_extracted_k141_98482 | Not-determined |
| S17_extracted_k141_142784 | Not-determined |
| S17_extracted_k141_46758 | Not-determined |
| S17_extracted_k141_61057 | Not-determined |
| S17_extracted_k141_9831 | Low-quality |
| S17_extracted_k141_39580 | Not-determined |
| S17_extracted_k141_142786 | Not-determined |
| S17_extracted_k141_31922 | Not-determined |
| S17_extracted_k141_112885 | Not-determined |
| S17_extracted_k141_164740 | Not-determined |
| S17_extracted_k141_61066 | Not-determined |
| S17_extracted_k141_112887 | Not-determined |
| S17_extracted_k141_2179 | Low-quality |
| S17_extracted_k141_61071 | Not-determined |
| S17_extracted_k141_31929 | Not-determined |
| S17_extracted_k141_90645 | Low-quality |
| S17_extracted_k141_112892 | Not-determined |
| S17_extracted_k141_90646 | Not-determined |
| S17_extracted_k141_98494 | Not-determined |
| S17_extracted_k141_24201 | Not-determined |
| S17_extracted_k141_98497 | Not-determined |
| S17_extracted_k141_157612 | Not-determined |
| S17_extracted_k141_98499 | Not-determined |
| S17_extracted_k141_82932 | Not-determined |
| S17_extracted_k141_142802 | Not-determined |
| S17_extracted_k141_54306 | Low-quality |
| S17_extracted_k141_90649 | Not-determined |
| S17_extracted_k141_171964 | Not-determined |
| S17_extracted_k141_61083 | Low-quality |
| S17_extracted_k141_112899 | Not-determined |
| S17_extracted_k141_54308 | Not-determined |
| S17_extracted_k141_171967 | Not-determined |
| S17_extracted_k141_164750 | Low-quality |
| S17_extracted_k141_127498 | Not-determined |
| S17_extracted_k141_54311 | Not-determined |
| S17_extracted_k141_157623 | Not-determined |
| S17_extracted_k141_82950 | Not-determined |
| S17_extracted_k141_112905 | Not-determined |
| S17_extracted_k141_142811 | Not-determined |
| S17_extracted_k141_31943 | Not-determined |
| S17_extracted_k141_24215 | Not-determined |
| S17_extracted_k141_150475 | Not-determined |
| S17_extracted_k141_82957 | Not-determined |
| S17_extracted_k141_75995 | Not-determined |
| S17_extracted_k141_90665 | Not-determined |
| S17_extracted_k141_39616 | Low-quality |
| S17_extracted_k141_135118 | Not-determined |
| S17_extracted_k141_82960 | Not-determined |
| S17_extracted_k141_39620 | Not-determined |
| S17_extracted_k141_164756 | Not-determined |
| S17_extracted_k141_9854 | Not-determined |
| S17_extracted_k141_164759 | Not-determined |
| S17_extracted_k141_90671 | Low-quality |
| S17_extracted_k141_16677 | Not-determined |
| S17_extracted_k141_31951 | Not-determined |
| S17_extracted_k141_171977 | Not-determined |
| S17_extracted_k141_150486 | Low-quality |
| S17_extracted_k141_157646 | Not-determined |
| S17_extracted_k141_76005 | Not-determined |
| S17_extracted_k141_31953 | Not-determined |
| S17_extracted_k141_157648 | Not-determined |
| S17_extracted_k141_135121 | Low-quality |
| S17_extracted_k141_120529 | Not-determined |
| S17_extracted_k141_68803 | Low-quality |
| S17_extracted_k141_135123 | Not-determined |
| S17_extracted_k141_157656 | Not-determined |
| S17_extracted_k141_164772 | Low-quality |
| S17_extracted_k141_68810 | Low-quality |
| S17_extracted_k141_39641 | Not-determined |
| S17_extracted_k141_98502 | Not-determined |
| S17_extracted_k141_112919 | Not-determined |
| S17_extracted_k141_90685 | Not-determined |
| S17_extracted_k141_16684 | Not-determined |
| S17_extracted_k141_16687 | Low-quality |
| S17_extracted_k141_54324 | Low-quality |
| S17_extracted_k141_98506 | Not-determined |
| S17_extracted_k141_82971 | Not-determined |
| S17_extracted_k141_39647 | Not-determined |
| S17_extracted_k141_76021 | Low-quality |
| S17_extracted_k141_150500 | Not-determined |
| S17_extracted_k141_135129 | Low-quality |
| S17_extracted_k141_142818 | Not-determined |
| S17_extracted_k141_90696 | Low-quality |
| S17_extracted_k141_90697 | Not-determined |
| S17_extracted_k141_61107 | Not-determined |
| S17_extracted_k141_24231 | Not-determined |
| S17_extracted_k141_135135 | Not-determined |
| S17_extracted_k141_135136 | Not-determined |
| S17_extracted_k141_112929 | Not-determined |
| S17_extracted_k141_150508 | Not-determined |
| S17_extracted_k141_24234 | Not-determined |
| S17_extracted_k141_157675 | Low-quality |
| S17_extracted_k141_31974 | Not-determined |
| S17_extracted_k141_135143 | Not-determined |
| S17_extracted_k141_157680 | Not-determined |
| S17_extracted_k141_171991 | Not-determined |
| S17_extracted_k141_24242 | Not-determined |
| S17_extracted_k141_61114 | Not-determined |
| S17_extracted_k141_76032 | Not-determined |
| S17_extracted_k141_68828 | Not-determined |
| S17_extracted_k141_24243 | Not-determined |
| S17_extracted_k141_9863 | Not-determined |
| S17_extracted_k141_76035 | Not-determined |
| S17_extracted_k141_164793 | Not-determined |
| S17_extracted_k141_24246 | Low-quality |
| S17_extracted_k141_164794 | Not-determined |
| S17_extracted_k141_171998 | Not-determined |
| S17_extracted_k141_171999 | Not-determined |
| S17_extracted_k141_172000 | Not-determined |
| S17_extracted_k141_2207 | Not-determined |
| S17_extracted_k141_39659 | Not-determined |
| S17_extracted_k141_112931 | Low-quality |
| S17_extracted_k141_135153 | Not-determined |
| S17_extracted_k141_164799 | Not-determined |
| S17_extracted_k141_150524 | Not-determined |
| S17_extracted_k141_82994 | Not-determined |
| S17_extracted_k141_39662 | Not-determined |
| S17_extracted_k141_135158 | Not-determined |
| S17_extracted_k141_83005 | Not-determined |
| S17_extracted_k141_120567 | Not-determined |
| S17_extracted_k141_24258 | Not-determined |
| S17_extracted_k141_68850 | Not-determined |
| S17_extracted_k141_76046 | Not-determined |
| S17_extracted_k141_16716 | Low-quality |
| S17_extracted_k141_9875 | Low-quality |
| S17_extracted_k141_142845 | Not-determined |
| S17_extracted_k141_112940 | Not-determined |
| S17_extracted_k141_142848 | Not-determined |
| S17_extracted_k141_90716 | Not-determined |
| S17_extracted_k141_9880 | Not-determined |
| S17_extracted_k141_135166 | Not-determined |
| S17_extracted_k141_16719 | Not-determined |
| S17_extracted_k141_157704 | Not-determined |
| S17_extracted_k141_112946 | Not-determined |
| S17_extracted_k141_61137 | Not-determined |
| S17_extracted_k141_39677 | Not-determined |
| S17_extracted_k141_83020 | Not-determined |
| S17_extracted_k141_31996 | Not-determined |
| S17_extracted_k141_135173 | Not-determined |
| S17_extracted_k141_39679 | Not-determined |
| S17_extracted_k141_31999 | Not-determined |
| S17_extracted_k141_90730 | Not-determined |
| S17_extracted_k141_90731 | Not-determined |
| S17_extracted_k141_39685 | Low-quality |
| S17_extracted_k141_135179 | Not-determined |
| S17_extracted_k141_90734 | Not-determined |
| S17_extracted_k141_54339 | Low-quality |
| S17_extracted_k141_150538 | Not-determined |
| S17_extracted_k141_61145 | Not-determined |
| S17_extracted_k141_90736 | Not-determined |
| S17_extracted_k141_90737 | Not-determined |
| S17_extracted_k141_142866 | Not-determined |
| S17_extracted_k141_32004 | Not-determined |
| S17_extracted_k141_16725 | Not-determined |
| S17_extracted_k141_83029 | Not-determined |
| S17_extracted_k141_142868 | Not-determined |
| S17_extracted_k141_24279 | Not-determined |
| S17_extracted_k141_9883 | Not-determined |
| S17_extracted_k141_16727 | Not-determined |
| S17_extracted_k141_39694 | Not-determined |
| S17_extracted_k141_61156 | Not-determined |
| S17_extracted_k141_83038 | Low-quality |
| S17_extracted_k141_157718 | Not-determined |
| S17_extracted_k141_157720 | Not-determined |
| S17_extracted_k141_32012 | Low-quality |
| S17_extracted_k141_61157 | Not-determined |
| S17_extracted_k141_157721 | Not-determined |
| S17_extracted_k141_83042 | Not-determined |
| S17_extracted_k141_127520 | Not-determined |
| S17_extracted_k141_46786 | Low-quality |
| S17_extracted_k141_76051 | Not-determined |
| S17_extracted_k141_112958 | Not-determined |
| S17_extracted_k141_164809 | Not-determined |
| S17_extracted_k141_112961 | Not-determined |
| S17_extracted_k141_120597 | Not-determined |
| S17_extracted_k141_120598 | Low-quality |
| S17_extracted_k141_90748 | Low-quality |
| S17_extracted_k141_76055 | Not-determined |
| S17_extracted_k141_127523 | Not-determined |
| S17_extracted_k141_9886 | Not-determined |
| S17_extracted_k141_61169 | Low-quality |
| S17_extracted_k141_76057 | Not-determined |
| S17_extracted_k141_2215 | Not-determined |
| S17_extracted_k141_112967 | Not-determined |
| S17_extracted_k141_106225 | Not-determined |
| S17_extracted_k141_106227 | Not-determined |
| S17_extracted_k141_32024 | Not-determined |
| S17_extracted_k141_46796 | Not-determined |
| S17_extracted_k141_61175 | Not-determined |
| S17_extracted_k141_150553 | Not-determined |
| S17_extracted_k141_68883 | Low-quality |
| S17_extracted_k141_76060 | Not-determined |
| S17_extracted_k141_54351 | Not-determined |
| S17_extracted_k141_54352 | Low-quality |
| S17_extracted_k141_150558 | Not-determined |
| S17_extracted_k141_142878 | Not-determined |
| S17_extracted_k141_120613 | Not-determined |
| S17_extracted_k141_61182 | Not-determined |
| S17_extracted_k141_120614 | Not-determined |
| S17_extracted_k141_142879 | Not-determined |
| S17_extracted_k141_39705 | Not-determined |
| S17_extracted_k141_2219 | Not-determined |
| S17_extracted_k141_142884 | Low-quality |
| S17_extracted_k141_120618 | Not-determined |
| S17_extracted_k141_112983 | Not-determined |
| S17_extracted_k141_135186 | Not-determined |
| S17_extracted_k141_76064 | Low-quality |
| S17_extracted_k141_2224 | Not-determined |
| S17_extracted_k141_157735 | Not-determined |
| S17_extracted_k141_32032 | Not-determined |
| S17_extracted_k141_39714 | Not-determined |
| S17_extracted_k141_39719 | Not-determined |
| S17_extracted_k141_135191 | Not-determined |
| S17_extracted_k141_172013 | Not-determined |
| S17_extracted_k141_76075 | Not-determined |
| S17_extracted_k141_54370 | Not-determined |
| S17_extracted_k141_106240 | Not-determined |
| S17_extracted_k141_24298 | Not-determined |
| S17_extracted_k141_120626 | Not-determined |
| S17_extracted_k141_142888 | Not-determined |
| S17_extracted_k141_120628 | Not-determined |
| S17_extracted_k141_150570 | Not-determined |
| S17_extracted_k141_39727 | Not-determined |
| S17_extracted_k141_172015 | Not-determined |
| S17_extracted_k141_157745 | Not-determined |
| S17_extracted_k141_142892 | Not-determined |
| S17_extracted_k141_9890 | Not-determined |
| S17_extracted_k141_16744 | Not-determined |
| S17_extracted_k141_68904 | Not-determined |
| S17_extracted_k141_61204 | Not-determined |
| S17_extracted_k141_76082 | Low-quality |
| S17_extracted_k141_68907 | Not-determined |
| S17_extracted_k141_142899 | Not-determined |
| S17_extracted_k141_172018 | Not-determined |
| S17_extracted_k141_172021 | Not-determined |
| S17_extracted_k141_46808 | Not-determined |
| S17_extracted_k141_142905 | Not-determined |
| S17_extracted_k141_83055 | Not-determined |
| S17_extracted_k141_46811 | Not-determined |
| S17_extracted_k141_142906 | Not-determined |
| S17_extracted_k141_164828 | Not-determined |
| S17_extracted_k141_142908 | Not-determined |
| S17_extracted_k141_98552 | Not-determined |
| S17_extracted_k141_112997 | Not-determined |
| S17_extracted_k141_76092 | Not-determined |
| S17_extracted_k141_120642 | Not-determined |
| S17_extracted_k141_106248 | Not-determined |
| S17_extracted_k141_76093 | Not-determined |
| S17_extracted_k141_164832 | Not-determined |
| S17_extracted_k141_76094 | Not-determined |
| S17_extracted_k141_46813 | Not-determined |
| S17_extracted_k141_106250 | Not-determined |
| S17_extracted_k141_113000 | Not-determined |
| S17_extracted_k141_106254 | Not-determined |
| S17_extracted_k141_83068 | Not-determined |
| S17_extracted_k141_83070 | Not-determined |
| S17_extracted_k141_61212 | Not-determined |
| S17_extracted_k141_83072 | Not-determined |
| S17_extracted_k141_68927 | Not-determined |
| S17_extracted_k141_142924 | Not-determined |
| S17_extracted_k141_127573 | Not-determined |
| S17_extracted_k141_106256 | Low-quality |
| S17_extracted_k141_142927 | Not-determined |
| S17_extracted_k141_54383 | Not-determined |
| S17_extracted_k141_127576 | Not-determined |
| S17_extracted_k141_113014 | Not-determined |
| S17_extracted_k141_106260 | Not-determined |
| S17_extracted_k141_39750 | Not-determined |
| S17_extracted_k141_76112 | Not-determined |
| S17_extracted_k141_142931 | Not-determined |
| S17_extracted_k141_39754 | Low-quality |
| S17_extracted_k141_68932 | Not-determined |
| S17_extracted_k141_90769 | Not-determined |
| S17_extracted_k141_120660 | Not-determined |
| S17_extracted_k141_90770 | Not-determined |
| S17_extracted_k141_120661 | Not-determined |
| S17_extracted_k141_54395 | Not-determined |
| S17_extracted_k141_83080 | Not-determined |
| S17_extracted_k141_120663 | Not-determined |
| S17_extracted_k141_61219 | Not-determined |
| S17_extracted_k141_39760 | Low-quality |
| S17_extracted_k141_142940 | Not-determined |
| S17_extracted_k141_68939 | Not-determined |
| S17_extracted_k141_150595 | Not-determined |
| S17_extracted_k141_46832 | Not-determined |
| S17_extracted_k141_90775 | Not-determined |
| S17_extracted_k141_39765 | Not-determined |
| S17_extracted_k141_172025 | Not-determined |
| S17_extracted_k141_106274 | Not-determined |
| S17_extracted_k141_90780 | Not-determined |
| S17_extracted_k141_120668 | Not-determined |
| S17_extracted_k141_106275 | Low-quality |
| S17_extracted_k141_61228 | Not-determined |
| S17_extracted_k141_90782 | Not-determined |
| S17_extracted_k141_98575 | Low-quality |
| S17_extracted_k141_106278 | Not-determined |
| S17_extracted_k141_157775 | Not-determined |
| S17_extracted_k141_32056 | Not-determined |
| S17_extracted_k141_142945 | Not-determined |
| S17_extracted_k141_39771 | Not-determined |
| S17_extracted_k141_157776 | Not-determined |
| S17_extracted_k141_142947 | Not-determined |
| S17_extracted_k141_120671 | Not-determined |
| S17_extracted_k141_61232 | Not-determined |
| S17_extracted_k141_76120 | Not-determined |
| S17_extracted_k141_142949 | Not-determined |
| S17_extracted_k141_61233 | Not-determined |
| S17_extracted_k141_46843 | Not-determined |
| S17_extracted_k141_76122 | Not-determined |
| S17_extracted_k141_142951 | Not-determined |
| S17_extracted_k141_39776 | Not-determined |
| S17_extracted_k141_90787 | Not-determined |
| S17_extracted_k141_90789 | Not-determined |
| S17_extracted_k141_24329 | Low-quality |
| S17_extracted_k141_76127 | Not-determined |
| S17_extracted_k141_164857 | Low-quality |
| S17_extracted_k141_172030 | Not-determined |
| S17_extracted_k141_106287 | Not-determined |
| S17_extracted_k141_90794 | Low-quality |
| S17_extracted_k141_90795 | Not-determined |
| S17_extracted_k141_90798 | Not-determined |
| S17_extracted_k141_120681 | Not-determined |
| S17_extracted_k141_120682 | Not-determined |
| S17_extracted_k141_150610 | Not-determined |
| S17_extracted_k141_83091 | Not-determined |
| S17_extracted_k141_46852 | Not-determined |
| S17_extracted_k141_120683 | Low-quality |
| S17_extracted_k141_16761 | Low-quality |
| S17_extracted_k141_46853 | Not-determined |
| S17_extracted_k141_90803 | Not-determined |
| S17_extracted_k141_127583 | Not-determined |
| S17_extracted_k141_32066 | Not-determined |
| S17_extracted_k141_98591 | Low-quality |
| S17_extracted_k141_150619 | Low-quality |
| S17_extracted_k141_150620 | Not-determined |
| S17_extracted_k141_90804 | Not-determined |
| S17_extracted_k141_90805 | Not-determined |
| S17_extracted_k141_150622 | Not-determined |
| S17_extracted_k141_54399 | Low-quality |
| S17_extracted_k141_120690 | Not-determined |
| S17_extracted_k141_32069 | Not-determined |
| S17_extracted_k141_24341 | Not-determined |
| S17_extracted_k141_106303 | Not-determined |
| S17_extracted_k141_120695 | Low-quality |
| S17_extracted_k141_46859 | Not-determined |
| S17_extracted_k141_16770 | Not-determined |
| S17_extracted_k141_142963 | Not-determined |
| S17_extracted_k141_157797 | Low-quality |
| S17_extracted_k141_98601 | Not-determined |
| S17_extracted_k141_16772 | Not-determined |
| S17_extracted_k141_157799 | Not-determined |
| S17_extracted_k141_68964 | Not-determined |
| S17_extracted_k141_142966 | Not-determined |
| S17_extracted_k141_32082 | Not-determined |
| S17_extracted_k141_24352 | Not-determined |
| S17_extracted_k141_150629 | Low-quality |
| S17_extracted_k141_16774 | Low-quality |
| S17_extracted_k141_54411 | Not-determined |

| | |
|---|---|
| S17_extracted_k141_127590 | Not-determined |
| S17_extracted_k141_113045 | Not-determined |
| S17_extracted_k141_9926 | Low-quality |
| S17_extracted_k141_46862 | Low-quality |
| S17_extracted_k141_54415 | Not-determined |
| S17_extracted_k141_46863 | Not-determined |
| S17_extracted_k141_61258 | Not-determined |
| S17_extracted_k141_90816 | Not-determined |
| S17_extracted_k141_83119 | Not-determined |
| S17_extracted_k141_120712 | Not-determined |
| S17_extracted_k141_24362 | Not-determined |
| S17_extracted_k141_76153 | Not-determined |
| S17_extracted_k141_24363 | Low-quality |
| S17_extracted_k141_9933 | Not-determined |
| S17_extracted_k141_127600 | Not-determined |
| S17_extracted_k141_150644 | Not-determined |
| S17_extracted_k141_113052 | Not-determined |
| S17_extracted_k141_127603 | Not-determined |
| S17_extracted_k141_142989 | Not-determined |
| S17_extracted_k141_98619 | Not-determined |
| S17_extracted_k141_142990 | Low-quality |
| S17_extracted_k141_16780 | Not-determined |
| S17_extracted_k141_68972 | Not-determined |
| S17_extracted_k141_90831 | Not-determined |
| S17_extracted_k141_76167 | Not-determined |
| S17_extracted_k141_98624 | Low-quality |
| S17_extracted_k141_68976 | Not-determined |
| S17_extracted_k141_142996 | Not-determined |
| S17_extracted_k141_61268 | Not-determined |
| S17_extracted_k141_61269 | Low-quality |
| S17_extracted_k141_9944 | Not-determined |
| S17_extracted_k141_16787 | Low-quality |
| S17_extracted_k141_46886 | Not-determined |
| S17_extracted_k141_39787 | Not-determined |
| S17_extracted_k141_143000 | Not-determined |
| S17_extracted_k141_143002 | Not-determined |
| S17_extracted_k141_157833 | Not-determined |
| S17_extracted_k141_9948 | Not-determined |
| S17_extracted_k141_113064 | Not-determined |
| S17_extracted_k141_68985 | Not-determined |
| S17_extracted_k141_16789 | Not-determined |
| S17_extracted_k141_61271 | Not-determined |
| S17_extracted_k141_127619 | Not-determined |
| S17_extracted_k141_127621 | Not-determined |
| S17_extracted_k141_39793 | Low-quality |
| S17_extracted_k141_143008 | Low-quality |
| S17_extracted_k141_32114 | Not-determined |
| S17_extracted_k141_150674 | Not-determined |
| S17_extracted_k141_16791 | Not-determined |
| S17_extracted_k141_150676 | Not-determined |
| S17_extracted_k141_157843 | Not-determined |
| S17_extracted_k141_150677 | Not-determined |
| S17_extracted_k141_16795 | Not-determined |
| S17_extracted_k141_2259 | Not-determined |
| S17_extracted_k141_113069 | Not-determined |
| S17_extracted_k141_120721 | Not-determined |
| S17_extracted_k141_150683 | Not-determined |
| S17_extracted_k141_16800 | Not-determined |
| S17_extracted_k141_32120 | Low-quality |
| S17_extracted_k141_16801 | Not-determined |
| S17_extracted_k141_39801 | Not-determined |
| S17_extracted_k141_46909 | Not-determined |
| S17_extracted_k141_54433 | Not-determined |
| S17_extracted_k141_150690 | Not-determined |
| S17_extracted_k141_83138 | Not-determined |
| S17_extracted_k141_143018 | Low-quality |
| S17_extracted_k141_150693 | Not-determined |
| S17_extracted_k141_83142 | Not-determined |
| S17_extracted_k141_54439 | Not-determined |
| S17_extracted_k141_90842 | Not-determined |
| S17_extracted_k141_54443 | Not-determined |
| S17_extracted_k141_143024 | Not-determined |
| S17_extracted_k141_39807 | Not-determined |
| S17_extracted_k141_54445 | Low-quality |
| S17_extracted_k141_32134 | Not-determined |
| S17_extracted_k141_150703 | Not-determined |
| S17_extracted_k141_46923 | Not-determined |
| S17_extracted_k141_32137 | Not-determined |
| S17_extracted_k141_127641 | Not-determined |
| S17_extracted_k141_39815 | Not-determined |
| S17_extracted_k141_2268 | Not-determined |
| S17_extracted_k141_106326 | Not-determined |
| S17_extracted_k141_143031 | Not-determined |
| S17_extracted_k141_24371 | Not-determined |
| S17_extracted_k141_143032 | Not-determined |
| S17_extracted_k141_32142 | Low-quality |
| S17_extracted_k141_106332 | Not-determined |
| S17_extracted_k141_24373 | Not-determined |
| S17_extracted_k141_39817 | Not-determined |
| S17_extracted_k141_127644 | Not-determined |
| S17_extracted_k141_113091 | Not-determined |
| S17_extracted_k141_32146 | Not-determined |
| S17_extracted_k141_61296 | Not-determined |
| S17_extracted_k141_54462 | Not-determined |
| S17_extracted_k141_2278 | Not-determined |
| S17_extracted_k141_54465 | Not-determined |
| S17_extracted_k141_113098 | Not-determined |
| S17_extracted_k141_83173 | Not-determined |
| S17_extracted_k141_143047 | Not-determined |
| S17_extracted_k141_106341 | Not-determined |
| S17_extracted_k141_39830 | Low-quality |
| S17_extracted_k141_143051 | Not-determined |
| S17_extracted_k141_113104 | Low-quality |
| S17_extracted_k141_113106 | Not-determined |
| S17_extracted_k141_106346 | Not-determined |
| S17_extracted_k141_106347 | Not-determined |
| S17_extracted_k141_113110 | Not-determined |
| S17_extracted_k141_2292 | Low-quality |
| S17_extracted_k141_39839 | Not-determined |
| S17_extracted_k141_32152 | Not-determined |
| S17_extracted_k141_46936 | Not-determined |
| S17_extracted_k141_46939 | Not-determined |
| S17_extracted_k141_39846 | Not-determined |
| S17_extracted_k141_135243 | Not-determined |
| S17_extracted_k141_98644 | Low-quality |
| S17_extracted_k141_9978 | Not-determined |
| S17_extracted_k141_135250 | Low-quality |
| S17_extracted_k141_143068 | Not-determined |
| S17_extracted_k141_98647 | Not-determined |
| S17_extracted_k141_32163 | Low-quality |
| S17_extracted_k141_32165 | Not-determined |
| S17_extracted_k141_113118 | Not-determined |
| S17_extracted_k141_54470 | Not-determined |
| S17_extracted_k141_98649 | Low-quality |
| S17_extracted_k141_9981 | Not-determined |
| S17_extracted_k141_150729 | Not-determined |
| S17_extracted_k141_32170 | Not-determined |
| S17_extracted_k141_172044 | Not-determined |
| S17_extracted_k141_143072 | Not-determined |
| S17_extracted_k141_39854 | Low-quality |
| S17_extracted_k141_106359 | Not-determined |
| S17_extracted_k141_135255 | Not-determined |
| S17_extracted_k141_150733 | Not-determined |
| S17_extracted_k141_83205 | Not-determined |
| S17_extracted_k141_69027 | Not-determined |
| S17_extracted_k141_69029 | Not-determined |
| S17_extracted_k141_46952 | Low-quality |
| S17_extracted_k141_135262 | Not-determined |
| S17_extracted_k141_127679 | Not-determined |
| S17_extracted_k141_172055 | Low-quality |
| S17_extracted_k141_150740 | Low-quality |
| S17_extracted_k141_83209 | Not-determined |
| S17_extracted_k141_54479 | Low-quality |
| S17_extracted_k141_135268 | Not-determined |
| S17_extracted_k141_61311 | Low-quality |
| S17_extracted_k141_16845 | Not-determined |
| S17_extracted_k141_83213 | Low-quality |
| S17_extracted_k141_113141 | Low-quality |
| S17_extracted_k141_46961 | Not-determined |
| S17_extracted_k141_135275 | Not-determined |
| S17_extracted_k141_90867 | Low-quality |
| S17_extracted_k141_32185 | Not-determined |
| S17_extracted_k141_172066 | Not-determined |
| S17_extracted_k141_90868 | Not-determined |
| S17_extracted_k141_172067 | Not-determined |
| S17_extracted_k141_46967 | Not-determined |
| S17_extracted_k141_83214 | Not-determined |
| S17_extracted_k141_172069 | Not-determined |
| S17_extracted_k141_32191 | Not-determined |
| S17_extracted_k141_135285 | Not-determined |
| S17_extracted_k141_32194 | Not-determined |
| S17_extracted_k141_32195 | Not-determined |
| S17_extracted_k141_10000 | Not-determined |
| S17_extracted_k141_10002 | Not-determined |
| S17_extracted_k141_39876 | Not-determined |
| S17_extracted_k141_46977 | Not-determined |
| S17_extracted_k141_83222 | Not-determined |
| S17_extracted_k141_69034 | Not-determined |
| S17_extracted_k141_46979 | Low-quality |
| S17_extracted_k141_150762 | Not-determined |
| S17_extracted_k141_39880 | Not-determined |
| S17_extracted_k141_172085 | Not-determined |
| S17_extracted_k141_90878 | Not-determined |
| S17_extracted_k141_150763 | Not-determined |
| S17_extracted_k141_172086 | Not-determined |
| S17_extracted_k141_39881 | Not-determined |
| S17_extracted_k141_32200 | Not-determined |
| S17_extracted_k141_32201 | Not-determined |
| S17_extracted_k141_46981 | Not-determined |
| S17_extracted_k141_127702 | Not-determined |
| S17_extracted_k141_90880 | Not-determined |
| S17_extracted_k141_54493 | Not-determined |
| S17_extracted_k141_150765 | Low-quality |
| S17_extracted_k141_172087 | Low-quality |
| S17_extracted_k141_106400 | Not-determined |
| S17_extracted_k141_83227 | Not-determined |
| S17_extracted_k141_69039 | Not-determined |
| S17_extracted_k141_76207 | Not-determined |
| S17_extracted_k141_39885 | Not-determined |
| S17_extracted_k141_10010 | Not-determined |
| S17_extracted_k141_83232 | Low-quality |
| S17_extracted_k141_127708 | Not-determined |
| S17_extracted_k141_83235 | Low-quality |
| S17_extracted_k141_61333 | Low-quality |
| S17_extracted_k141_69050 | Not-determined |
| S17_extracted_k141_61334 | Not-determined |
| S17_extracted_k141_98675 | Not-determined |
| S17_extracted_k141_39895 | Not-determined |
| S17_extracted_k141_10013 | Not-determined |
| S17_extracted_k141_54502 | Not-determined |
| S17_extracted_k141_164870 | Low-quality |
| S17_extracted_k141_98682 | Low-quality |
| S17_extracted_k141_83237 | Low-quality |
| S17_extracted_k141_164875 | Not-determined |
| S17_extracted_k141_127717 | Not-determined |
| S17_extracted_k141_69057 | Not-determined |
| S17_extracted_k141_143085 | Not-determined |
| S17_extracted_k141_69058 | Not-determined |
| S17_extracted_k141_106421 | Low-quality |
| S17_extracted_k141_127722 | Not-determined |
| S17_extracted_k141_47008 | Not-determined |
| S17_extracted_k141_69062 | Not-determined |
| S17_extracted_k141_172103 | Not-determined |
| S17_extracted_k141_127724 | Not-determined |
| S17_extracted_k141_98694 | Not-determined |
| S17_extracted_k141_172106 | Not-determined |
| S17_extracted_k141_127726 | Not-determined |
| S17_extracted_k141_54515 | Low-quality |
| S17_extracted_k141_16849 | Not-determined |
| S17_extracted_k141_143099 | Not-determined |
| S17_extracted_k141_69068 | Not-determined |
| S17_extracted_k141_90886 | Not-determined |
| S17_extracted_k141_39912 | Not-determined |
| S17_extracted_k141_135314 | Not-determined |
| S17_extracted_k141_54521 | Not-determined |
| S17_extracted_k141_164888 | Not-determined |
| S17_extracted_k141_135316 | Not-determined |
| S17_extracted_k141_83248 | Not-determined |
| S17_extracted_k141_135318 | Not-determined |
| S17_extracted_k141_164893 | Not-determined |
| S17_extracted_k141_69074 | Not-determined |
| S17_extracted_k141_135321 | Not-determined |
| S17_extracted_k141_76233 | Not-determined |
| S17_extracted_k141_32215 | Not-determined |
| S17_extracted_k141_54526 | Not-determined |
| S17_extracted_k141_32217 | Not-determined |
| S17_extracted_k141_172119 | Low-quality |
| S17_extracted_k141_54530 | Not-determined |
| S17_extracted_k141_127740 | Not-determined |
| S17_extracted_k141_90891 | Not-determined |
| S17_extracted_k141_69077 | Not-determined |
| S17_extracted_k141_76238 | Not-determined |
| S17_extracted_k141_47020 | Not-determined |
| S17_extracted_k141_143114 | Not-determined |
| S17_extracted_k141_164898 | Not-determined |
| S17_extracted_k141_32224 | Not-determined |
| S17_extracted_k141_143115 | Low-quality |
| S17_extracted_k141_61364 | Low-quality |
| S17_extracted_k141_32226 | Not-determined |
| S17_extracted_k141_164902 | Low-quality |
| S17_extracted_k141_113147 | Not-determined |
| S17_extracted_k141_164903 | Not-determined |
| S17_extracted_k141_83253 | Not-determined |
| S17_extracted_k141_32228 | Low-quality |
| S17_extracted_k141_113148 | Not-determined |
| S17_extracted_k141_76243 | Low-quality |
| S17_extracted_k141_39921 | Not-determined |
| S17_extracted_k141_135334 | Not-determined |
| S17_extracted_k141_47024 | Not-determined |
| S17_extracted_k141_83257 | Not-determined |
| S17_extracted_k141_90903 | Not-determined |
| S17_extracted_k141_90904 | Not-determined |
| S17_extracted_k141_90908 | Not-determined |
| S17_extracted_k141_54541 | Not-determined |
| S17_extracted_k141_2304 | Not-determined |
| S17_extracted_k141_69090 | Not-determined |
| S17_extracted_k141_143126 | Not-determined |
| S17_extracted_k141_2307 | Not-determined |
| S17_extracted_k141_157878 | Not-determined |
| S17_extracted_k141_90915 | Not-determined |
| S17_extracted_k141_10049 | Not-determined |
| S17_extracted_k141_10050 | Not-determined |
| S17_extracted_k141_157883 | Not-determined |
| S17_extracted_k141_90919 | Not-determined |
| S17_extracted_k141_143131 | Low-quality |
| S17_extracted_k141_54552 | Not-determined |
| S17_extracted_k141_143132 | Not-determined |
| S17_extracted_k141_157889 | Low-quality |
| S17_extracted_k141_10059 | Not-determined |
| S17_extracted_k141_2317 | Not-determined |
| S17_extracted_k141_10061 | Not-determined |
| S17_extracted_k141_135352 | Not-determined |
| S17_extracted_k141_106472 | Not-determined |
| S17_extracted_k141_164927 | Not-determined |
| S17_extracted_k141_164928 | Not-determined |
| S17_extracted_k141_10065 | Not-determined |
| S17_extracted_k141_164931 | Low-quality |
| S17_extracted_k141_32267 | Low-quality |
| S17_extracted_k141_47026 | Not-determined |
| S17_extracted_k141_54559 | Low-quality |
| S17_extracted_k141_39934 | Not-determined |
| S17_extracted_k141_2326 | Not-determined |
| S17_extracted_k141_76262 | Low-quality |
| S17_extracted_k141_143144 | Not-determined |
| S15_vs_42043\|\|full | High-quality |
| S17_extracted_k141_16860 | Not-determined |
| S17_extracted_k141_106486 | Not-determined |
| S17_extracted_k141_54562 | Not-determined |
| S17_extracted_k141_172135 | Not-determined |
| S17_extracted_k141_90930 | Low-quality |
| S17_extracted_k141_90931 | Not-determined |
| S17_extracted_k141_47033 | Not-determined |
| S17_extracted_k141_47036 | Not-determined |
| S17_extracted_k141_2333 | Not-determined |
| S17_extracted_k141_143149 | Low-quality |
| S17_extracted_k141_16863 | Low-quality |
| S17_extracted_k141_172138 | Not-determined |
| S17_extracted_k141_83282 | Not-determined |
| S17_extracted_k141_150795 | Not-determined |
| S17_extracted_k141_127748 | Low-quality |
| S17_extracted_k141_2338 | Not-determined |
| S17_extracted_k141_90933 | Not-determined |
| S17_extracted_k141_32287 | Not-determined |
| S17_extracted_k141_143153 | Not-determined |
| S17_extracted_k141_54569 | Not-determined |
| S17_extracted_k141_83287 | Not-determined |
| S17_extracted_k141_24395 | Not-determined |
| S17_extracted_k141_10073 | Not-determined |
| S17_extracted_k141_127750 | Not-determined |
| S17_extracted_k141_16869 | Not-determined |
| S17_extracted_k141_39941 | Low-quality |
| S17_extracted_k141_24398 | Not-determined |
| S17_extracted_k141_61385 | Not-determined |
| S17_extracted_k141_39945 | Not-determined |
| S17_extracted_k141_150807 | Not-determined |
| S17_extracted_k141_90943 | Not-determined |
| S17_extracted_k141_127753 | Not-determined |
| S17_extracted_k141_172148 | Not-determined |
| S17_extracted_k141_54576 | Not-determined |
| S17_extracted_k141_69108 | Not-determined |
| S17_extracted_k141_61388 | Low-quality |
| S17_extracted_k141_150808 | Not-determined |
| S17_extracted_k141_98741 | Not-determined |
| S17_extracted_k141_32293 | Not-determined |
| S17_extracted_k141_164961 | Not-determined |
| S17_extracted_k141_172154 | Not-determined |
| S17_extracted_k141_98745 | Not-determined |
| S17_extracted_k141_16876 | Not-determined |
| S17_extracted_k141_150812 | Low-quality |
| S17_extracted_k141_61391 | Not-determined |
| S17_extracted_k141_2353 | Not-determined |
| S17_extracted_k141_69115 | Not-determined |
| S17_extracted_k141_76274 | Not-determined |
| S17_extracted_k141_90955 | Not-determined |
| S17_extracted_k141_143168 | Not-determined |
| S17_extracted_k141_32304 | Not-determined |
| S17_extracted_k141_76280 | Not-determined |
| S17_extracted_k141_32305 | Not-determined |
| S17_extracted_k141_106494 | Not-determined |
| S17_extracted_k141_164967 | Not-determined |
| S17_extracted_k141_143175 | Low-quality |
| S17_extracted_k141_172158 | Low-quality |
| S17_extracted_k141_76285 | Not-determined |
| S17_extracted_k141_172159 | Not-determined |
| S17_extracted_k141_54583 | Not-determined |
| S17_extracted_k141_76286 | Low-quality |
| S17_extracted_k141_83295 | Not-determined |
| S17_extracted_k141_76287 | Low-quality |
| S17_extracted_k141_106509 | Not-determined |
| S17_extracted_k141_98760 | Not-determined |
| S17_extracted_k141_69124 | Not-determined |
| S17_extracted_k141_61402 | Not-determined |
| S17_extracted_k141_83302 | Not-determined |
| S17_extracted_k141_61403 | Low-quality |
| S17_extracted_k141_98763 | Low-quality |
| S17_extracted_k141_164981 | Not-determined |
| S17_extracted_k141_98765 | Not-determined |
| S17_extracted_k141_150826 | Not-determined |
| S17_extracted_k141_90972 | Not-determined |
| S17_extracted_k141_150827 | Not-determined |
| S17_extracted_k141_32328 | Not-determined |
| S17_extracted_k141_16901 | Not-determined |
| S17_extracted_k141_143191 | Not-determined |
| S17_extracted_k141_54590 | Not-determined |
| S17_extracted_k141_16903 | Not-determined |
| S17_extracted_k141_61414 | Not-determined |
| S17_extracted_k141_98776 | Not-determined |
| S17_extracted_k141_150832 | Not-determined |
| S17_extracted_k141_135372 | Not-determined |
| S17_extracted_k141_98781 | Not-determined |
| S17_extracted_k141_2368 | Not-determined |
| S17_extracted_k141_135375 | Not-determined |
| S17_extracted_k141_172181 | Not-determined |
| S17_extracted_k141_106522 | Not-determined |
| S17_extracted_k141_83315 | Not-determined |
| S17_extracted_k141_69129 | Low-quality |
| S17_extracted_k141_47047 | Low-quality |
| S17_extracted_k141_47049 | Not-determined |
| S17_extracted_k141_157908 | Not-determined |
| S17_extracted_k141_76304 | Not-determined |
| S17_extracted_k141_39970 | Not-determined |
| S17_extracted_k141_76306 | Not-determined |
| S17_extracted_k141_164983 | Not-determined |
| S17_extracted_k141_32347 | Not-determined |
| S17_extracted_k141_2376 | Not-determined |
| S17_extracted_k141_164986 | Not-determined |
| S17_extracted_k141_98788 | Not-determined |
| S17_extracted_k141_164987 | Not-determined |
| S17_extracted_k141_172192 | Not-determined |
| S17_extracted_k141_47056 | Not-determined |
| S17_extracted_k141_47058 | Low-quality |
| S17_extracted_k141_164989 | Not-determined |
| S17_extracted_k141_106535 | Not-determined |
| S17_extracted_k141_143199 | Not-determined |
| S17_extracted_k141_47059 | Low-quality |
| S17_extracted_k141_164991 | Not-determined |
| S17_extracted_k141_98798 | Not-determined |
| S17_extracted_k141_32352 | Not-determined |
| S17_extracted_k141_24429 | Not-determined |
| S17_extracted_k141_47063 | Not-determined |
| S17_extracted_k141_106546 | Not-determined |
| S17_extracted_k141_135392 | Not-determined |
| S17_extracted_k141_106549 | Not-determined |
| S17_extracted_k141_90996 | Not-determined |
| S17_extracted_k141_135395 | Not-determined |
| S17_extracted_k141_165000 | Not-determined |
| S17_extracted_k141_47073 | Not-determined |
| S17_extracted_k141_47074 | Not-determined |
| S17_extracted_k141_165002 | Not-determined |
| S17_extracted_k141_39985 | Not-determined |
| S17_extracted_k141_98811 | Not-determined |
| S17_extracted_k141_165003 | Not-determined |
| S17_extracted_k141_39986 | Low-quality |
| S17_extracted_k141_143211 | Not-determined |
| S17_extracted_k141_135402 | Not-determined |
| S17_extracted_k141_91007 | Not-determined |
| S17_extracted_k141_106559 | Not-determined |
| S17_extracted_k141_2382 | Low-quality |
| S17_extracted_k141_91008 | Not-determined |
| S17_extracted_k141_135409 | Not-determined |
| S17_extracted_k141_24446 | Not-determined |
| S17_extracted_k141_47078 | Not-determined |
| S17_extracted_k141_24447 | Low-quality |
| S17_extracted_k141_2387 | Not-determined |
| S17_extracted_k141_120761 | Low-quality |
| S17_extracted_k141_157927 | Not-determined |
| S17_extracted_k141_120764 | Not-determined |
| S17_extracted_k141_98813 | Not-determined |
| S17_extracted_k141_2391 | Not-determined |
| S17_extracted_k141_157931 | Not-determined |
| S17_extracted_k141_98815 | Not-determined |
| S17_extracted_k141_39993 | Low-quality |
| S17_extracted_k141_2395 | Not-determined |
| S17_extracted_k141_157937 | Not-determined |
| S17_extracted_k141_39997 | Not-determined |
| S17_extracted_k141_165021 | Not-determined |
| S17_extracted_k141_10112 | Not-determined |
| S17_extracted_k141_98818 | Not-determined |
| S17_extracted_k141_91019 | Not-determined |
| S17_extracted_k141_10115 | Low-quality |
| S17_extracted_k141_157945 | Low-quality |
| S17_extracted_k141_91022 | Not-determined |
| S17_extracted_k141_76311 | Not-determined |
| S17_extracted_k141_24467 | Not-determined |
| S17_extracted_k141_40007 | Not-determined |
| S17_extracted_k141_32367 | Not-determined |
| S17_extracted_k141_10117 | Not-determined |
| S17_extracted_k141_2407 | Not-determined |
| S17_extracted_k141_150851 | Not-determined |
| S17_extracted_k141_40011 | Not-determined |
| S17_extracted_k141_150854 | Not-determined |
| S17_extracted_k141_40013 | Not-determined |
| S17_extracted_k141_91030 | Not-determined |
| S17_extracted_k141_10120 | Low-quality |
| S17_extracted_k141_135432 | Low-quality |
| S17_extracted_k141_91034 | Not-determined |
| S17_extracted_k141_165022 | Not-determined |
| S17_extracted_k141_40019 | Not-determined |
| S17_extracted_k141_150859 | Not-determined |
| S17_extracted_k141_40021 | Not-determined |
| S17_extracted_k141_16927 | Not-determined |
| S17_extracted_k141_150861 | Not-determined |
| S17_extracted_k141_120792 | Not-determined |
| S17_extracted_k141_10125 | Low-quality |
| S17_extracted_k141_165026 | Not-determined |
| S17_extracted_k141_69176 | Not-determined |
| S17_extracted_k141_143228 | Low-quality |
| S17_extracted_k141_91048 | Not-determined |
| S17_extracted_k141_150867 | Not-determined |
| S17_extracted_k141_10128 | Not-determined |
| S17_extracted_k141_135440 | Not-determined |
| S17_extracted_k141_120800 | Not-determined |
| S17_extracted_k141_157966 | Low-quality |
| S17_extracted_k141_10133 | Not-determined |
| S17_extracted_k141_113155 | Not-determined |
| S17_extracted_k141_69183 | Not-determined |
| S17_extracted_k141_165029 | Not-determined |
| S17_extracted_k141_16942 | Not-determined |
| S17_extracted_k141_69185 | Not-determined |
| S17_extracted_k141_157972 | Not-determined |
| S17_extracted_k141_76340 | Not-determined |
| S17_extracted_k141_47110 | Not-determined |
| S17_extracted_k141_98843 | Not-determined |
| S17_extracted_k141_91062 | Not-determined |
| S17_extracted_k141_135444 | Not-determined |
| S17_extracted_k141_91063 | Not-determined |
| S17_extracted_k141_91064 | Not-determined |
| S17_extracted_k141_91067 | Not-determined |
| S17_extracted_k141_127791 | Low-quality |
| S17_extracted_k141_91071 | Not-determined |
| S17_extracted_k141_40035 | Not-determined |
| S17_extracted_k141_98853 | Not-determined |
| S17_extracted_k141_135456 | Not-determined |
| S17_extracted_k141_24488 | Not-determined |
| S17_extracted_k141_24490 | Low-quality |
| S17_extracted_k141_120809 | Not-determined |
| S17_extracted_k141_150873 | Low-quality |
| S17_extracted_k141_40043 | Not-determined |
| S17_extracted_k141_91078 | Low-quality |
| S17_extracted_k141_40046 | Low-quality |
| S17_extracted_k141_106590 | Not-determined |
| S17_extracted_k141_83326 | Not-determined |
| S17_extracted_k141_91080 | Low-quality |
| S17_extracted_k141_76360 | Low-quality |
| S17_extracted_k141_150878 | Not-determined |
| S17_extracted_k141_158002 | Not-determined |
| S17_extracted_k141_113170 | Not-determined |
| S17_extracted_k141_172231 | Not-determined |
| S17_extracted_k141_40050 | Low-quality |
| S17_extracted_k141_76364 | Not-determined |
| S17_extracted_k141_16961 | Low-quality |
| S17_extracted_k141_69196 | Not-determined |
| S17_extracted_k141_127810 | Not-determined |
| S17_extracted_k141_83332 | Not-determined |
| S17_extracted_k141_91087 | Not-determined |
| S17_extracted_k141_113174 | Not-determined |
| S17_extracted_k141_91088 | Low-quality |
| S17_extracted_k141_24497 | Not-determined |
| S17_extracted_k141_150892 | Not-determined |
| S17_extracted_k141_150894 | Not-determined |
| S17_extracted_k141_40065 | Not-determined |
| S17_extracted_k141_158009 | Not-determined |
| S17_extracted_k141_91092 | Not-determined |
| S17_extracted_k141_16972 | Not-determined |
| S17_extracted_k141_143258 | Not-determined |
| S17_extracted_k141_47132 | Not-determined |
| S17_extracted_k141_69203 | Not-determined |
| S17_extracted_k141_24506 | Not-determined |
| S17_extracted_k141_106606 | Not-determined |
| S17_extracted_k141_76376 | Not-determined |
| S17_extracted_k141_32411 | Not-determined |
| S17_extracted_k141_172249 | Low-quality |
| S17_extracted_k141_113183 | Not-determined |
| S17_extracted_k141_16983 | Not-determined |
| S17_extracted_k141_47144 | Not-determined |
| S17_extracted_k141_10169 | Low-quality |
| S17_extracted_k141_127838 | Low-quality |
| S17_extracted_k141_10174 | Not-determined |
| S17_extracted_k141_47149 | Not-determined |
| S17_extracted_k141_16999 | Not-determined |
| S17_extracted_k141_47151 | Not-determined |
| S17_extracted_k141_17000 | Not-determined |
| S17_extracted_k141_158029 | Not-determined |
| S17_extracted_k141_61431 | Low-quality |
| S17_extracted_k141_127845 | Not-determined |
| S17_extracted_k141_69222 | Not-determined |
| S17_extracted_k141_91109 | Not-determined |
| S17_extracted_k141_2410 | Not-determined |
| S17_extracted_k141_32419 | Not-determined |
| S17_extracted_k141_127849 | Low-quality |
| S17_extracted_k141_17004 | Not-determined |
| S17_extracted_k141_24516 | Low-quality |
| S17_extracted_k141_120832 | Not-determined |
| S17_extracted_k141_17009 | Not-determined |
| S17_extracted_k141_150915 | Low-quality |
| S17_extracted_k141_17013 | Not-determined |
| S17_extracted_k141_76401 | Not-determined |
| S17_extracted_k141_127857 | Not-determined |
| S17_extracted_k141_10183 | Low-quality |
| S17_extracted_k141_47160 | Not-determined |
| S17_extracted_k141_76406 | Not-determined |
| S17_extracted_k141_143280 | Not-determined |
| S17_extracted_k141_10185 | Not-determined |
| S17_extracted_k141_2416 | Not-determined |
| S17_extracted_k141_17021 | Not-determined |
| S17_extracted_k141_150921 | Not-determined |
| S17_extracted_k141_143283 | Not-determined |
| S17_extracted_k141_135495 | Not-determined |
| S17_extracted_k141_127868 | Low-quality |
| S17_extracted_k141_32435 | Not-determined |
| S17_extracted_k141_135497 | Not-determined |
| S17_extracted_k141_143289 | Not-determined |
| S17_extracted_k141_10201 | Not-determined |
| S17_extracted_k141_2430 | Not-determined |
| S17_extracted_k141_150926 | Not-determined |
| S17_extracted_k141_2433 | Not-determined |
| S17_extracted_k141_40076 | Not-determined |
| S17_extracted_k141_47176 | Not-determined |
| S17_extracted_k141_2435 | Not-determined |

| | |
|---|---|
| S17_extracted_k141_127876 | Not-determined |
| S17_extracted_k141_61448 | Not-determined |
| S17_extracted_k141_32446 | Not-determined |
| S17_extracted_k141_2437 | Not-determined |
| S17_extracted_k141_91129 | Low-quality |
| S17_extracted_k141_106638 | Not-determined |
| S17_extracted_k141_106641 | Not-determined |
| S17_extracted_k141_40079 | Not-determined |
| S17_extracted_k141_91133 | Not-determined |
| S17_extracted_k141_172291 | Not-determined |
| S17_extracted_k141_106645 | Not-determined |
| S17_extracted_k141_172294 | Not-determined |
| S17_extracted_k141_120859 | Not-determined |
| S17_extracted_k141_76414 | Not-determined |
| S17_extracted_k141_113195 | Low-quality |
| S17_extracted_k141_40085 | Low-quality |
| S17_extracted_k141_32453 | Not-determined |
| S17_extracted_k141_76420 | Not-determined |
| S17_extracted_k141_172298 | Low-quality |
| S17_extracted_k141_10207 | Not-determined |
| S17_extracted_k141_76422 | Not-determined |
| S17_extracted_k141_150938 | Not-determined |
| S17_extracted_k141_10209 | Not-determined |
| S17_extracted_k141_2456 | Not-determined |
| S17_extracted_k141_2458 | Not-determined |
| S17_extracted_k141_106649 | Not-determined |
| S17_extracted_k141_106650 | Not-determined |
| S17_extracted_k141_91151 | Not-determined |
| S17_extracted_k141_2466 | Not-determined |
| S17_extracted_k141_127897 | Not-determined |
| S17_extracted_k141_17053 | Not-determined |
| S17_extracted_k141_120870 | Not-determined |
| S17_extracted_k141_47197 | Not-determined |
| S17_extracted_k141_47201 | Not-determined |
| S17_extracted_k141_69235 | Not-determined |
| S17_extracted_k141_47205 | Not-determined |
| S17_extracted_k141_40099 | Low-quality |
| S17_extracted_k141_113215 | Not-determined |
| S17_extracted_k141_17063 | Not-determined |
| S17_extracted_k141_40100 | Not-determined |
| S17_extracted_k141_165050 | Not-determined |
| S17_extracted_k141_47206 | Not-determined |
| S17_extracted_k141_69239 | Low-quality |
| S17_extracted_k141_2471 | Low-quality |
| S17_extracted_k141_69240 | Not-determined |
| S17_extracted_k141_76441 | Not-determined |
| S17_extracted_k141_135506 | Not-determined |
| S17_extracted_k141_69242 | Not-determined |
| S17_extracted_k141_17066 | Not-determined |
| S17_extracted_k141_2474 | Not-determined |
| S17_extracted_k141_98922 | Not-determined |
| S17_extracted_k141_158053 | Not-determined |
| S17_extracted_k141_158054 | Not-determined |
| S17_extracted_k141_17073 | Not-determined |
| S17_extracted_k141_32463 | Not-determined |
| S17_extracted_k141_47216 | Not-determined |
| S17_extracted_k141_32465 | Not-determined |
| S17_extracted_k141_158059 | Not-determined |
| S17_extracted_k141_24548 | Low-quality |
| S17_extracted_k141_2484 | Not-determined |
| S17_extracted_k141_69254 | Low-quality |
| S17_extracted_k141_135516 | Not-determined |
| S17_extracted_k141_47224 | Not-determined |
| S17_extracted_k141_98929 | Not-determined |
| S17_extracted_k141_98931 | Not-determined |
| S17_extracted_k141_98932 | Low-quality |
| S17_extracted_k141_61469 | Low-quality |
| S17_extracted_k141_61470 | Not-determined |
| S17_extracted_k141_165060 | Not-determined |
| S17_extracted_k141_135530 | Not-determined |
| S17_extracted_k141_165061 | Not-determined |
| S17_extracted_k141_143295 | Not-determined |
| S17_extracted_k141_113217 | Low-quality |
| S17_extracted_k141_2498 | Not-determined |
| S17_extracted_k141_135535 | Not-determined |
| S17_extracted_k141_61476 | Not-determined |
| S17_extracted_k141_150949 | Not-determined |
| S17_extracted_k141_61477 | Not-determined |
| S17_extracted_k141_135536 | Not-determined |
| S17_extracted_k141_120896 | Not-determined |
| S17_extracted_k141_120900 | Not-determined |
| S17_extracted_k141_24557 | Not-determined |
| S17_extracted_k141_69260 | Not-determined |
| S17_extracted_k141_135538 | Not-determined |
| S17_extracted_k141_40126 | Not-determined |
| S17_extracted_k141_98936 | Not-determined |
| S17_extracted_k141_98937 | Low-quality |
| S17_extracted_k141_172320 | Not-determined |
| S17_extracted_k141_40129 | Not-determined |
| S17_extracted_k141_120908 | Not-determined |
| S17_extracted_k141_47250 | Not-determined |
| S17_extracted_k141_165074 | Not-determined |
| S17_extracted_k141_165075 | Not-determined |
| S17_extracted_k141_172322 | Not-determined |
| S17_extracted_k141_32497 | Not-determined |
| S17_extracted_k141_83355 | Not-determined |
| S17_extracted_k141_113223 | Not-determined |
| S17_extracted_k141_76462 | Not-determined |
| S17_extracted_k141_91185 | Not-determined |
| S17_extracted_k141_76463 | Not-determined |
| S17_extracted_k141_135549 | Not-determined |
| S17_extracted_k141_172326 | Not-determined |
| S17_extracted_k141_69272 | Not-determined |
| S17_extracted_k141_2524 | Low-quality |
| S17_extracted_k141_98948 | Not-determined |
| S17_extracted_k141_98949 | Not-determined |
| S17_extracted_k141_61491 | Not-determined |
| S17_extracted_k141_32511 | Low-quality |
| S17_extracted_k141_61494 | Not-determined |
| S17_extracted_k141_91193 | Not-determined |
| S17_extracted_k141_2530 | Low-quality |
| S17_extracted_k141_2531 | Not-determined |
| S17_extracted_k141_76473 | Not-determined |
| S17_extracted_k141_120926 | Not-determined |
| S17_extracted_k141_69283 | Not-determined |
| S17_extracted_k141_76479 | Not-determined |
| S17_extracted_k141_69286 | Not-determined |
| S17_extracted_k141_120931 | Not-determined |
| S17_extracted_k141_83367 | Not-determined |
| S17_extracted_k141_172339 | Not-determined |
| S17_extracted_k141_98962 | Not-determined |
| S17_extracted_k141_32516 | Not-determined |
| S17_extracted_k141_76486 | Not-determined |
| S17_extracted_k141_24589 | Low-quality |
| S17_extracted_k141_172344 | Not-determined |
| S17_extracted_k141_91199 | Not-determined |
| S17_extracted_k141_24591 | Not-determined |
| S17_extracted_k141_172347 | Not-determined |
| S17_extracted_k141_158074 | Not-determined |
| S17_extracted_k141_113245 | Not-determined |
| S17_extracted_k141_113246 | Not-determined |
| S17_extracted_k141_61513 | Not-determined |
| S17_extracted_k141_61514 | Not-determined |
| S17_extracted_k141_113249 | Not-determined |
| S17_extracted_k141_76497 | Not-determined |
| S17_extracted_k141_69302 | Not-determined |
| S17_extracted_k141_158080 | Not-determined |
| S17_extracted_k141_17092 | Not-determined |
| S17_extracted_k141_120944 | Not-determined |
| S17_extracted_k141_69305 | Not-determined |
| S17_extracted_k141_69306 | Not-determined |
| S17_extracted_k141_32526 | Low-quality |
| S17_extracted_k141_2549 | Not-determined |
| S17_extracted_k141_120949 | Not-determined |
| S17_extracted_k141_76507 | Low-quality |
| S17_extracted_k141_165077 | Not-determined |
| S17_extracted_k141_127941 | Not-determined |
| S17_extracted_k141_61524 | Not-determined |
| S17_extracted_k141_10219 | Low-quality |
| S17_extracted_k141_98970 | Not-determined |
| S17_extracted_k141_98972 | Not-determined |
| S17_extracted_k141_120951 | Not-determined |
| S17_extracted_k141_61526 | Not-determined |
| S17_extracted_k141_47291 | Not-determined |
| S17_extracted_k141_91209 | Not-determined |
| S17_extracted_k141_165083 | Not-determined |
| S17_extracted_k141_106673 | Not-determined |
| S17_extracted_k141_61530 | Not-determined |
| S17_extracted_k141_76518 | Low-quality |
| S17_extracted_k141_172356 | Not-determined |
| S17_extracted_k141_113265 | Not-determined |
| S17_extracted_k141_113266 | Low-quality |
| S17_extracted_k141_10229 | Not-determined |
| S17_extracted_k141_83376 | Not-determined |
| S17_extracted_k141_113268 | Not-determined |
| S17_extracted_k141_83378 | Not-determined |
| S17_extracted_k141_83380 | Not-determined |
| S17_extracted_k141_40174 | Not-determined |
| S17_extracted_k141_106680 | Not-determined |
| S17_extracted_k141_17106 | Low-quality |
| S17_extracted_k141_32540 | Not-determined |
| S17_extracted_k141_91226 | Not-determined |
| S17_extracted_k141_83383 | Not-determined |
| S17_extracted_k141_2566 | Low-quality |
| S17_extracted_k141_143343 | Not-determined |
| S17_extracted_k141_83384 | Low-quality |
| S17_extracted_k141_17109 | Low-quality |
| S17_extracted_k141_76526 | Low-quality |
| S17_extracted_k141_158117 | Not-determined |
| S17_extracted_k141_69316 | Low-quality |
| S17_extracted_k141_127958 | Not-determined |
| S17_extracted_k141_165096 | Low-quality |
| S17_extracted_k141_17114 | Not-determined |
| S17_extracted_k141_76530 | Not-determined |
| S17_extracted_k141_24613 | Low-quality |
| S17_extracted_k141_24616 | Low-quality |
| S17_extracted_k141_2578 | Not-determined |
| S17_extracted_k141_127961 | Not-determined |
| S17_extracted_k141_2581 | Not-determined |
| S17_extracted_k141_76534 | Not-determined |
| S17_extracted_k141_172376 | Not-determined |
| S17_extracted_k141_165098 | Not-determined |
| S17_extracted_k141_143347 | Not-determined |
| S17_extracted_k141_127966 | Not-determined |
| S17_extracted_k141_165099 | Low-quality |
| S17_extracted_k141_76536 | Low-quality |
| S17_extracted_k141_106687 | Not-determined |
| S17_extracted_k141_158131 | Not-determined |
| S17_extracted_k141_76539 | Low-quality |
| S17_extracted_k141_143349 | Not-determined |
| S17_extracted_k141_61542 | Not-determined |
| S17_extracted_k141_127970 | Not-determined |
| S17_extracted_k141_143351 | Not-determined |
| S17_extracted_k141_127971 | Not-determined |
| S17_extracted_k141_172382 | Low-quality |
| S17_extracted_k141_127974 | Not-determined |
| S17_extracted_k141_165107 | Not-determined |
| S17_extracted_k141_40193 | Not-determined |
| S17_extracted_k141_83395 | Not-determined |
| S17_extracted_k141_69329 | Not-determined |
| S17_extracted_k141_172390 | Not-determined |
| S17_extracted_k141_165110 | Low-quality |
| S17_extracted_k141_172395 | Not-determined |
| S17_extracted_k141_172396 | Not-determined |
| S17_extracted_k141_91234 | Low-quality |
| S17_extracted_k141_61555 | Not-determined |
| S17_extracted_k141_76540 | Not-determined |
| S17_extracted_k141_143357 | Not-determined |
| S17_extracted_k141_40195 | Not-determined |
| S17_extracted_k141_24626 | Not-determined |
| S17_extracted_k141_127989 | Not-determined |
| S17_extracted_k141_47302 | Not-determined |
| S17_extracted_k141_17133 | Not-determined |
| S17_extracted_k141_83412 | Not-determined |
| S17_extracted_k141_143360 | Not-determined |
| S17_extracted_k141_106703 | Not-determined |
| S17_extracted_k141_158148 | Not-determined |
| S17_extracted_k141_165123 | Not-determined |
| S17_extracted_k141_47305 | Not-determined |
| S17_extracted_k141_83414 | Not-determined |
| S17_extracted_k141_76544 | Not-determined |
| S17_extracted_k141_113298 | Not-determined |
| S17_extracted_k141_76545 | Low-quality |
| S17_extracted_k141_106707 | Not-determined |
| S17_extracted_k141_24632 | Not-determined |
| S17_extracted_k141_165126 | Low-quality |
| S17_extracted_k141_113302 | Not-determined |
| S17_extracted_k141_40200 | Not-determined |
| S17_extracted_k141_76547 | Not-determined |
| S17_extracted_k141_47309 | Not-determined |
| S17_extracted_k141_113304 | Not-determined |
| S17_extracted_k141_24637 | Not-determined |
| S17_extracted_k141_172401 | Not-determined |
| S17_extracted_k141_120958 | Not-determined |
| S17_extracted_k141_47311 | Not-determined |
| S17_extracted_k141_76550 | Not-determined |
| S17_extracted_k141_158159 | Not-determined |
| S17_extracted_k141_17138 | Low-quality |
| S17_extracted_k141_61563 | Not-determined |
| S17_extracted_k141_83425 | Not-determined |
| S17_extracted_k141_61564 | Not-determined |
| S17_extracted_k141_17141 | Not-determined |
| S17_extracted_k141_76553 | Not-determined |
| S17_extracted_k141_113311 | Not-determined |
| S17_extracted_k141_120961 | Low-quality |
| S17_extracted_k141_61565 | Not-determined |
| S17_extracted_k141_2588 | Not-determined |
| S17_extracted_k141_61567 | Not-determined |
| S17_extracted_k141_158165 | Not-determined |
| S17_extracted_k141_76555 | Low-quality |
| S17_extracted_k141_76556 | Not-determined |
| S17_extracted_k141_2590 | Not-determined |
| S17_extracted_k141_143371 | Not-determined |
| S17_extracted_k141_91256 | Not-determined |
| S17_extracted_k141_32582 | Not-determined |
| S17_extracted_k141_47320 | Not-determined |
| S17_extracted_k141_24645 | Not-determined |
| S17_extracted_k141_83438 | Low-quality |
| S17_extracted_k141_83439 | Not-determined |
| S17_extracted_k141_120966 | Not-determined |
| S17_extracted_k141_24649 | Not-determined |
| S17_extracted_k141_10246 | Not-determined |
| S17_extracted_k141_47330 | Not-determined |
| S17_extracted_k141_17151 | Not-determined |
| S17_extracted_k141_10250 | Not-determined |
| S17_extracted_k141_17153 | Not-determined |
| S17_extracted_k141_98986 | Not-determined |
| S17_extracted_k141_40222 | Not-determined |
| S17_extracted_k141_98987 | Not-determined |
| S17_extracted_k141_40225 | Not-determined |
| S17_extracted_k141_106736 | Not-determined |
| S17_extracted_k141_98989 | Not-determined |
| S17_extracted_k141_98991 | Not-determined |
| S17_extracted_k141_135584 | Not-determined |
| S17_extracted_k141_76567 | Not-determined |
| S17_extracted_k141_2608 | Not-determined |
| S17_extracted_k141_32604 | Low-quality |
| S17_extracted_k141_172404 | Not-determined |
| S17_extracted_k141_128015 | Not-determined |
| S17_extracted_k141_120987 | Not-determined |
| S17_extracted_k141_2612 | Not-determined |
| S17_extracted_k141_172406 | Low-quality |
| S17_extracted_k141_106739 | Not-determined |
| S17_extracted_k141_32608 | Not-determined |
| S17_extracted_k141_2618 | Low-quality |
| S17_extracted_k141_61588 | Not-determined |
| S17_extracted_k141_91273 | Not-determined |
| S17_extracted_k141_165138 | Not-determined |
| S17_extracted_k141_32615 | Not-determined |
| S17_extracted_k141_54613 | Not-determined |
| S17_extracted_k141_83451 | Low-quality |
| S17_extracted_k141_32617 | Not-determined |
| S17_extracted_k141_91274 | Not-determined |
| S17_extracted_k141_47344 | Not-determined |
| S17_extracted_k141_172412 | Low-quality |
| S17_extracted_k141_17171 | Not-determined |
| S17_extracted_k141_47345 | Not-determined |
| S17_extracted_k141_172413 | Not-determined |
| S17_extracted_k141_99011 | Not-determined |
| S17_extracted_k141_47347 | Not-determined |
| S17_extracted_k141_143414 | Not-determined |
| S17_extracted_k141_17174 | Not-determined |
| S17_extracted_k141_172417 | Not-determined |
| S17_extracted_k141_143418 | Not-determined |
| S17_extracted_k141_121000 | Not-determined |
| S17_extracted_k141_32623 | Not-determined |
| S17_extracted_k141_143420 | Not-determined |
| S17_extracted_k141_40240 | Not-determined |
| S17_extracted_k141_24654 | Low-quality |
| S17_extracted_k141_99019 | Not-determined |
| S17_extracted_k141_40243 | Not-determined |
| S17_extracted_k141_99020 | Not-determined |
| S17_extracted_k141_2629 | Not-determined |
| S17_extracted_k141_32628 | Not-determined |
| S17_extracted_k141_32629 | Not-determined |
| S17_extracted_k141_143426 | Not-determined |
| S17_extracted_k141_172426 | Low-quality |
| S17_extracted_k141_135608 | Not-determined |
| S17_extracted_k141_165144 | Not-determined |
| S17_extracted_k141_24655 | Low-quality |
| S17_extracted_k141_106764 | Not-determined |
| S17_extracted_k141_172429 | Not-determined |
| S17_extracted_k141_17179 | Low-quality |
| S17_extracted_k141_17181 | Not-determined |
| S17_extracted_k141_143430 | Not-determined |
| S17_extracted_k141_17183 | Not-determined |
| S17_extracted_k141_172432 | Not-determined |
| S17_extracted_k141_32633 | Low-quality |
| S17_extracted_k141_91289 | Not-determined |
| S17_extracted_k141_172436 | Not-determined |
| S17_extracted_k141_165154 | Not-determined |
| S17_extracted_k141_40252 | Not-determined |
| S17_extracted_k141_32642 | Not-determined |
| S17_extracted_k141_172439 | Not-determined |
| S17_extracted_k141_76589 | Not-determined |
| S17_extracted_k141_151034 | Low-quality |
| S17_extracted_k141_10260 | Not-determined |
| S17_extracted_k141_99038 | Not-determined |
| S17_extracted_k141_24669 | Not-determined |
| S17_extracted_k141_143440 | Not-determined |
| S17_extracted_k141_91298 | Not-determined |
| S17_extracted_k141_143441 | Not-determined |
| S17_extracted_k141_17196 | Not-determined |
| S17_extracted_k141_2636 | Low-quality |
| S17_extracted_k141_40258 | Not-determined |
| S17_extracted_k141_165161 | Not-determined |
| S17_extracted_k141_40259 | Not-determined |
| S17_extracted_k141_172447 | Low-quality |
| S17_extracted_k141_69387 | Not-determined |
| S17_extracted_k141_47365 | Not-determined |
| S17_extracted_k141_54637 | Not-determined |
| S17_extracted_k141_121012 | Not-determined |
| S17_extracted_k141_121013 | Not-determined |
| S17_extracted_k141_121014 | Not-determined |
| S17_extracted_k141_83482 | Low-quality |
| S17_extracted_k141_69389 | Not-determined |
| S17_extracted_k141_54639 | Not-determined |
| S17_extracted_k141_165169 | Not-determined |
| S17_extracted_k141_172452 | Not-determined |
| S17_extracted_k141_76601 | Low-quality |
| S17_extracted_k141_165173 | Not-determined |
| S17_extracted_k141_165174 | Not-determined |
| S17_extracted_k141_99046 | Not-determined |
| S17_extracted_k141_99047 | Not-determined |
| S17_extracted_k141_54646 | Not-determined |
| S17_extracted_k141_106794 | Not-determined |
| S17_extracted_k141_172459 | Not-determined |
| S17_extracted_k141_54649 | Not-determined |
| S17_extracted_k141_76608 | Not-determined |
| S17_extracted_k141_69402 | Not-determined |
| S17_extracted_k141_61605 | Not-determined |
| S17_extracted_k141_91307 | Not-determined |
| S17_extracted_k141_61606 | Not-determined |
| S17_extracted_k141_40266 | Low-quality |
| S17_extracted_k141_128049 | Not-determined |
| S17_extracted_k141_76613 | Not-determined |
| S17_extracted_k141_54652 | Not-determined |
| S17_extracted_k141_32660 | Not-determined |
| S17_extracted_k141_54653 | Not-determined |
| S17_extracted_k141_24676 | Not-determined |
| S17_extracted_k141_121024 | Not-determined |
| S17_extracted_k141_32663 | Not-determined |
| S17_extracted_k141_99055 | Not-determined |
| S17_extracted_k141_76617 | Not-determined |
| S17_extracted_k141_151069 | Not-determined |
| S17_extracted_k141_24681 | Not-determined |
| S17_extracted_k141_17202 | Low-quality |
| S17_extracted_k141_121031 | Not-determined |
| S17_extracted_k141_151072 | Not-determined |
| S17_extracted_k141_165189 | Low-quality |
| S17_extracted_k141_2658 | Not-determined |
| S17_extracted_k141_47390 | Not-determined |
| S17_extracted_k141_24687 | Not-determined |
| S17_extracted_k141_121035 | Not-determined |
| S17_extracted_k141_172479 | Not-determined |
| S17_extracted_k141_17204 | Low-quality |
| S17_extracted_k141_47394 | Not-determined |
| S17_extracted_k141_54663 | Not-determined |
| S17_extracted_k141_165198 | Not-determined |
| S17_extracted_k141_165200 | Not-determined |
| S17_extracted_k141_40279 | Not-determined |
| S17_extracted_k141_76622 | Low-quality |
| S17_extracted_k141_76623 | Not-determined |
| S17_extracted_k141_165202 | Not-determined |
| S17_extracted_k141_2664 | Low-quality |
| S17_extracted_k141_76624 | Not-determined |
| S17_extracted_k141_2665 | Not-determined |
| S17_extracted_k141_165205 | Not-determined |
| S17_extracted_k141_24702 | Not-determined |
| S17_extracted_k141_165206 | Not-determined |
| S17_extracted_k141_99067 | Not-determined |
| S17_extracted_k141_47398 | Not-determined |
| S17_extracted_k141_2670 | Not-determined |
| S17_extracted_k141_172488 | Not-determined |
| S17_extracted_k141_172489 | Not-determined |
| S17_extracted_k141_61626 | Low-quality |
| S17_extracted_k141_128073 | Not-determined |
| S17_extracted_k141_69415 | Not-determined |
| S17_extracted_k141_47402 | Not-determined |
| S17_extracted_k141_135658 | Not-determined |
| S17_extracted_k141_99071 | Not-determined |
| S17_extracted_k141_47404 | Not-determined |
| S17_extracted_k141_54664 | Low-quality |
| S17_extracted_k141_106818 | Not-determined |
| S17_extracted_k141_172497 | Not-determined |
| S17_extracted_k141_24706 | Not-determined |
| S17_extracted_k141_61633 | Not-determined |
| S17_extracted_k141_165216 | Not-determined |
| S17_extracted_k141_91320 | Not-determined |
| S17_extracted_k141_47413 | Low-quality |
| S17_extracted_k141_165218 | Not-determined |
| S17_extracted_k141_76630 | Not-determined |
| S17_extracted_k141_61639 | Not-determined |
| S17_extracted_k141_47416 | Not-determined |
| S17_extracted_k141_69422 | Low-quality |
| S17_extracted_k141_69425 | Not-determined |
| S17_extracted_k141_10276 | Not-determined |
| S17_extracted_k141_106824 | Low-quality |
| S17_extracted_k141_135672 | Not-determined |
| S17_extracted_k141_2680 | Low-quality |
| S17_extracted_k141_76640 | Not-determined |
| S17_extracted_k141_135675 | Not-determined |
| S17_extracted_k141_172500 | Not-determined |
| S17_extracted_k141_10277 | Not-determined |
| S17_extracted_k141_76643 | Not-determined |
| S17_extracted_k141_128091 | Not-determined |
| S17_extracted_k141_24713 | Low-quality |
| S17_extracted_k141_24715 | Not-determined |
| S17_extracted_k141_69434 | Not-determined |
| S17_extracted_k141_24718 | Not-determined |
| S17_extracted_k141_17216 | Not-determined |
| S17_extracted_k141_99085 | Not-determined |
| S17_extracted_k141_2685 | Low-quality |
| S17_extracted_k141_24719 | Low-quality |
| S17_extracted_k141_47418 | Low-quality |
| S17_extracted_k141_128099 | Not-determined |
| S17_extracted_k141_135681 | Not-determined |
| S17_extracted_k141_91328 | Not-determined |
| S17_extracted_k141_172510 | Not-determined |
| S17_extracted_k141_99094 | Not-determined |
| S17_extracted_k141_2689 | Not-determined |
| S17_extracted_k141_24727 | Low-quality |
| S17_extracted_k141_172515 | Low-quality |
| S17_extracted_k141_61657 | Not-determined |
| S17_extracted_k141_47428 | Not-determined |
| S17_extracted_k141_135686 | Not-determined |
| S17_extracted_k141_61659 | Not-determined |
| S17_extracted_k141_76650 | Not-determined |
| S17_extracted_k141_91335 | Low-quality |
| S17_extracted_k141_61661 | Not-determined |
| S17_extracted_k141_76653 | Not-determined |
| S17_extracted_k141_135692 | Not-determined |
| S17_extracted_k141_47439 | Not-determined |
| S17_extracted_k141_165239 | Not-determined |
| S17_extracted_k141_76658 | Not-determined |
| S17_extracted_k141_47444 | Not-determined |
| S17_extracted_k141_2698 | Not-determined |
| S17_extracted_k141_47445 | Not-determined |
| S17_extracted_k141_10293 | Not-determined |
| S17_extracted_k141_172529 | Not-determined |
| S17_extracted_k141_172530 | Not-determined |
| S17_extracted_k141_143469 | Not-determined |
| S17_extracted_k141_47450 | Not-determined |
| S17_extracted_k141_47451 | Not-determined |
| S17_extracted_k141_165247 | Not-determined |
| S17_extracted_k141_143471 | Low-quality |
| S17_extracted_k141_172533 | Not-determined |
| S17_extracted_k141_54685 | Not-determined |
| S17_extracted_k141_17238 | Not-determined |
| S17_extracted_k141_165254 | Not-determined |
| S17_extracted_k141_17239 | Not-determined |
| S17_extracted_k141_99110 | Not-determined |
| S17_extracted_k141_61665 | Low-quality |
| S17_extracted_k141_99113 | Not-determined |
| S17_extracted_k141_165257 | Not-determined |
| S17_extracted_k141_76679 | Not-determined |
| S17_extracted_k141_99115 | Low-quality |
| S17_extracted_k141_54693 | Not-determined |
| S17_extracted_k141_165259 | Not-determined |
| S17_extracted_k141_99117 | Not-determined |
| S17_extracted_k141_113391 | Not-determined |
| S17_extracted_k141_99118 | Low-quality |
| S17_extracted_k141_91347 | Not-determined |
| S17_extracted_k141_17246 | Not-determined |
| S17_extracted_k141_76683 | Not-determined |
| S17_extracted_k141_76686 | Not-determined |
| S17_extracted_k141_69458 | Not-determined |
| S17_extracted_k141_91350 | Not-determined |
| S17_extracted_k141_165268 | Not-determined |
| S17_extracted_k141_47459 | Low-quality |
| S17_extracted_k141_91352 | Low-quality |
| S17_extracted_k141_69460 | Not-determined |
| S17_extracted_k141_99121 | Not-determined |
| S17_extracted_k141_47461 | Not-determined |
| S17_extracted_k141_172539 | Not-determined |
| S17_extracted_k141_83488 | Not-determined |
| S17_extracted_k141_24742 | Not-determined |
| S17_extracted_k141_158204 | Not-determined |
| S17_extracted_k141_17257 | Not-determined |
| S17_extracted_k141_128119 | Low-quality |
| S17_extracted_k141_76696 | Not-determined |
| S17_extracted_k141_2717 | Not-determined |
| S17_extracted_k141_106847 | Not-determined |
| S17_extracted_k141_99128 | Not-determined |
| S17_extracted_k141_99129 | Not-determined |
| S17_extracted_k141_40297 | Not-determined |
| S17_extracted_k141_143477 | Not-determined |
| S17_extracted_k141_10316 | Not-determined |
| S17_extracted_k141_172552 | Not-determined |
| S17_extracted_k141_165289 | Not-determined |
| S17_extracted_k141_61673 | Low-quality |
| S17_extracted_k141_69478 | Not-determined |
| S17_extracted_k141_158214 | Not-determined |
| S17_extracted_k141_24758 | Not-determined |
| S17_extracted_k141_121049 | Not-determined |
| S17_extracted_k141_83513 | Low-quality |
| S17_extracted_k141_40315 | Low-quality |
| S17_extracted_k141_135706 | Not-determined |
| S17_extracted_k141_2729 | Low-quality |
| S17_extracted_k141_40318 | Not-determined |
| S17_extracted_k141_17275 | Not-determined |
| S17_extracted_k141_99138 | Low-quality |
| S17_extracted_k141_76718 | Low-quality |
| S17_extracted_k141_83515 | Not-determined |
| S17_extracted_k141_106865 | Not-determined |
| S17_extracted_k141_143483 | Low-quality |
| S17_extracted_k141_24772 | Not-determined |
| S17_extracted_k141_76723 | Not-determined |
| S17_extracted_k141_172562 | Low-quality |
| S17_extracted_k141_83519 | Not-determined |
| S17_extracted_k141_10322 | Not-determined |
| S17_extracted_k141_106870 | Not-determined |
| S17_extracted_k141_76725 | Not-determined |
| S17_extracted_k141_158223 | Not-determined |
| S17_extracted_k141_69494 | Not-determined |
| S17_extracted_k141_69495 | Low-quality |
| S17_extracted_k141_76732 | Low-quality |
| S17_extracted_k141_143495 | Low-quality |
| S17_extracted_k141_106878 | Not-determined |
| S17_extracted_k141_2738 | Low-quality |
| S17_extracted_k141_17280 | Not-determined |
| S17_extracted_k141_40331 | Not-determined |
| S17_extracted_k141_113411 | Low-quality |
| S17_extracted_k141_165309 | Not-determined |
| S17_extracted_k141_17282 | Not-determined |
| S17_extracted_k141_40333 | Not-determined |
| S17_extracted_k141_135713 | Not-determined |
| S17_extracted_k141_135714 | Not-determined |
| S17_extracted_k141_2741 | Not-determined |
| S17_extracted_k141_2743 | Not-determined |
| S17_extracted_k141_69504 | Not-determined |
| S17_extracted_k141_61693 | Not-determined |
| S17_extracted_k141_69505 | Not-determined |
| S17_extracted_k141_24776 | Not-determined |
| S17_extracted_k141_69506 | Low-quality |
| S17_extracted_k141_172578 | Low-quality |
| S17_extracted_k141_83536 | Not-determined |
| S17_extracted_k141_40337 | Low-quality |
| S17_extracted_k141_135720 | Low-quality |

| | |
|---|---|
| S17_extracted_k141_54699 | Low-quality |
| S17_extracted_k141_151130 | Not-determined |
| S17_extracted_k141_165325 | Not-determined |
| S17_extracted_k141_83542 | Not-determined |
| S17_extracted_k141_91360 | Not-determined |
| S17_extracted_k141_47492 | Not-determined |
| S17_extracted_k141_99151 | Not-determined |
| S17_extracted_k141_83545 | Not-determined |
| S17_extracted_k141_24790 | Not-determined |
| S17_extracted_k141_83546 | Not-determined |
| S17_extracted_k141_17288 | Not-determined |
| S17_extracted_k141_165329 | Not-determined |
| S17_extracted_k141_61706 | Not-determined |
| S17_extracted_k141_99155 | Not-determined |
| S17_extracted_k141_40343 | Not-determined |
| S17_extracted_k141_69512 | Not-determined |
| S17_extracted_k141_106889 | Not-determined |
| S17_extracted_k141_128129 | Not-determined |
| S17_extracted_k141_24802 | Not-determined |
| S17_extracted_k141_91370 | Low-quality |
| S17_extracted_k141_10334 | Not-determined |
| S17_extracted_k141_135729 | Not-determined |
| S17_extracted_k141_69524 | Not-determined |
| S17_extracted_k141_106899 | Not-determined |
| S17_extracted_k141_2762 | Low-quality |
| S17_extracted_k141_99170 | Not-determined |
| S17_extracted_k141_10337 | Not-determined |
| S17_extracted_k141_17299 | Not-determined |
| S17_extracted_k141_61721 | Not-determined |
| S17_extracted_k141_128138 | Low-quality |
| S17_extracted_k141_113433 | Not-determined |
| S17_extracted_k141_165335 | Not-determined |
| S17_extracted_k141_10341 | Not-determined |
| S17_extracted_k141_151156 | Not-determined |
| S17_extracted_k141_83564 | Low-quality |
| S17_extracted_k141_172598 | Not-determined |
| S17_extracted_k141_143530 | Not-determined |
| S17_extracted_k141_83565 | Not-determined |
| S17_extracted_k141_99183 | Not-determined |
| S17_extracted_k141_143532 | Not-determined |
| S17_extracted_k141_47515 | Low-quality |
| S17_extracted_k141_99187 | Not-determined |
| S17_extracted_k141_106914 | Low-quality |
| S17_extracted_k141_17315 | Low-quality |
| S17_extracted_k141_47518 | Not-determined |
| S17_extracted_k141_143534 | Not-determined |
| S17_extracted_k141_121069 | Not-determined |
| S17_extracted_k141_40347 | Low-quality |
| S17_extracted_k141_99194 | Not-determined |
| S17_extracted_k141_151166 | Not-determined |
| S17_extracted_k141_121073 | Not-determined |
| S17_extracted_k141_47523 | Low-quality |
| S17_extracted_k141_172612 | Not-determined |
| S5_extracted_k141_145579 | Complete |
| S17_extracted_k141_128149 | Not-determined |
| S17_extracted_k141_128150 | Not-determined |
| S17_extracted_k141_40360 | Not-determined |
| S17_extracted_k141_135749 | Not-determined |
| S17_extracted_k141_40361 | Not-determined |
| S17_extracted_k141_172622 | Not-determined |
| S17_extracted_k141_54729 | Not-determined |
| S17_extracted_k141_61743 | Not-determined |
| S17_extracted_k141_128159 | Not-determined |
| S17_extracted_k141_99206 | Not-determined |
| S17_extracted_k141_91383 | Not-determined |
| S17_extracted_k141_91384 | Not-determined |
| S17_extracted_k141_54734 | Not-determined |
| S17_extracted_k141_10351 | Not-determined |
| S17_extracted_k141_61748 | Not-determined |
| S17_extracted_k141_172628 | Not-determined |
| S17_extracted_k141_54738 | Not-determined |
| S17_extracted_k141_106936 | Not-determined |
| S17_extracted_k141_61749 | Not-determined |
| S17_extracted_k141_76753 | Not-determined |
| S17_extracted_k141_99212 | Not-determined |
| S17_extracted_k141_47545 | Not-determined |
| S17_extracted_k141_47549 | Low-quality |
| S17_extracted_k141_106942 | Not-determined |
| S17_extracted_k141_113448 | Not-determined |
| S17_extracted_k141_143557 | Not-determined |
| S17_extracted_k141_113449 | Not-determined |
| S17_extracted_k141_76756 | Not-determined |
| S17_extracted_k141_106944 | Not-determined |
| S17_extracted_k141_47552 | Not-determined |
| S17_extracted_k141_99217 | Not-determined |
| S17_extracted_k141_54749 | Not-determined |
| S17_extracted_k141_61753 | Low-quality |
| S17_extracted_k141_61754 | Not-determined |
| S17_extracted_k141_99221 | Low-quality |
| S17_extracted_k141_135759 | Not-determined |
| S17_extracted_k141_10362 | Low-quality |
| S17_extracted_k141_24818 | Not-determined |
| S17_extracted_k141_76763 | Not-determined |
| S17_extracted_k141_32692 | Not-determined |
| S17_extracted_k141_128167 | Not-determined |
| S17_extracted_k141_61761 | Not-determined |
| S17_extracted_k141_151186 | Not-determined |
| S17_extracted_k141_61764 | Not-determined |
| S17_extracted_k141_69562 | Not-determined |
| S17_extracted_k141_54755 | Low-quality |
| S17_extracted_k141_113457 | Not-determined |
| S17_extracted_k141_2816 | Not-determined |
| S17_extracted_k141_61766 | Not-determined |
| S17_extracted_k141_32701 | Not-determined |
| S17_extracted_k141_113460 | Not-determined |
| S17_extracted_k141_151192 | Not-determined |
| S17_extracted_k141_106953 | Low-quality |
| S17_extracted_k141_135764 | Not-determined |
| S17_extracted_k141_113462 | Not-determined |
| S17_extracted_k141_113463 | Not-determined |
| S17_extracted_k141_40386 | Not-determined |
| S17_extracted_k141_24830 | Not-determined |
| S17_extracted_k141_151196 | Not-determined |
| S17_extracted_k141_99232 | Low-quality |
| S17_extracted_k141_24832 | Not-determined |
| S17_extracted_k141_76775 | Not-determined |
| S17_extracted_k141_151199 | Not-determined |
| S17_extracted_k141_172645 | Low-quality |
| S17_extracted_k141_2819 | Low-quality |
| S17_extracted_k141_47568 | Not-determined |
| S17_extracted_k141_151206 | Low-quality |
| S17_extracted_k141_135777 | Not-determined |
| S17_extracted_k141_172650 | Not-determined |
| S17_extracted_k141_165371 | Low-quality |
| S17_extracted_k141_128178 | Not-determined |
| S17_extracted_k141_32717 | Not-determined |
| S17_extracted_k141_32718 | Low-quality |
| S17_extracted_k141_113480 | Not-determined |
| S17_extracted_k141_172653 | Not-determined |
| S17_extracted_k141_128181 | Low-quality |
| S17_extracted_k141_10381 | Not-determined |
| S17_extracted_k141_32723 | Not-determined |
| S17_extracted_k141_2831 | Not-determined |
| S17_extracted_k141_143582 | Low-quality |
| S17_extracted_k141_113485 | Not-determined |
| S17_extracted_k141_17339 | Low-quality |
| S17_extracted_k141_165373 | Not-determined |
| S17_extracted_k141_99239 | Not-determined |
| S17_extracted_k141_61780 | Not-determined |
| S17_extracted_k141_76782 | Low-quality |
| S17_extracted_k141_106955 | Not-determined |
| S17_extracted_k141_143589 | Low-quality |
| S17_extracted_k141_69576 | Low-quality |
| S17_extracted_k141_143590 | Not-determined |
| S17_extracted_k141_106958 | Not-determined |
| S17_extracted_k141_91420 | Not-determined |
| S17_extracted_k141_32729 | Not-determined |
| S17_extracted_k141_10394 | Not-determined |
| S17_extracted_k141_61786 | Not-determined |
| S17_extracted_k141_158230 | Low-quality |
| S17_extracted_k141_113502 | Low-quality |
| S17_extracted_k141_165393 | Not-determined |
| S17_extracted_k141_54776 | Not-determined |
| S17_extracted_k141_151214 | Low-quality |
| S17_extracted_k141_172665 | Not-determined |
| S17_extracted_k141_10398 | Low-quality |
| S17_extracted_k141_121084 | Not-determined |
| S17_extracted_k141_10399 | Not-determined |
| S17_extracted_k141_47587 | Not-determined |
| S17_extracted_k141_165397 | Not-determined |
| S17_extracted_k141_10400 | Not-determined |
| S17_extracted_k141_113507 | Not-determined |
| S17_extracted_k141_69582 | Not-determined |
| S17_extracted_k141_151219 | Low-quality |
| S17_extracted_k141_47594 | Not-determined |
| S17_extracted_k141_172666 | Not-determined |
| S17_extracted_k141_61793 | Not-determined |
| S17_extracted_k141_113518 | Not-determined |
| S17_extracted_k141_151227 | Not-determined |
| S17_extracted_k141_165399 | Low-quality |
| S17_extracted_k141_40406 | Not-determined |
| S17_extracted_k141_99254 | Not-determined |
| S17_extracted_k141_158241 | Not-determined |
| S17_extracted_k141_99256 | Not-determined |
| S17_extracted_k141_76801 | Not-determined |
| S17_extracted_k141_76802 | Not-determined |
| S17_extracted_k141_91433 | Not-determined |
| S17_extracted_k141_165406 | Not-determined |
| S17_extracted_k141_10412 | Not-determined |
| S17_extracted_k141_2846 | Not-determined |
| S17_extracted_k141_143611 | Not-determined |
| S17_extracted_k141_99266 | Not-determined |
| S17_extracted_k141_2848 | Not-determined |
| S17_extracted_k141_151238 | Not-determined |
| S17_extracted_k141_54794 | Not-determined |
| S17_extracted_k141_143614 | Not-determined |
| S17_extracted_k141_128199 | Not-determined |
| S17_extracted_k141_40414 | Not-determined |
| S17_extracted_k141_10416 | Low-quality |
| S17_extracted_k141_113531 | Not-determined |
| S17_extracted_k141_158251 | Not-determined |
| S17_extracted_k141_151243 | Not-determined |
| S17_extracted_k141_24850 | Not-determined |
| S17_extracted_k141_17348 | Low-quality |
| S17_extracted_k141_17349 | Not-determined |
| S17_extracted_k141_17350 | Not-determined |
| S17_extracted_k141_158256 | Not-determined |
| S17_extracted_k141_91441 | Not-determined |
| S17_extracted_k141_143620 | Not-determined |
| S17_extracted_k141_24855 | Not-determined |
| S17_extracted_k141_143621 | Not-determined |
| S17_extracted_k141_61806 | Not-determined |
| S17_extracted_k141_2850 | Low-quality |
| S17_extracted_k141_91446 | Not-determined |
| S17_extracted_k141_143623 | Not-determined |
| S17_extracted_k141_69608 | Not-determined |
| S17_extracted_k141_172674 | Low-quality |
| S17_extracted_k141_91449 | Low-quality |
| S17_extracted_k141_91451 | Low-quality |
| S17_extracted_k141_143630 | Not-determined |
| S17_extracted_k141_165417 | Not-determined |
| S17_extracted_k141_24859 | Not-determined |
| S17_extracted_k141_76815 | Low-quality |
| S17_extracted_k141_113535 | Not-determined |
| S17_extracted_k141_61818 | Not-determined |
| S17_extracted_k141_113538 | Not-determined |
| S17_extracted_k141_17362 | Low-quality |
| S17_extracted_k141_151254 | Not-determined |
| S17_extracted_k141_54819 | Low-quality |
| S17_extracted_k141_17363 | Not-determined |
| S17_extracted_k141_113540 | Not-determined |
| S17_extracted_k141_2858 | Low-quality |
| S17_extracted_k141_106983 | Not-determined |
| S17_extracted_k141_61822 | Not-determined |
| S17_extracted_k141_106985 | Not-determined |
| S17_extracted_k141_158273 | Not-determined |
| S17_extracted_k141_158274 | Not-determined |
| S17_extracted_k141_10432 | Not-determined |
| S17_extracted_k141_91462 | Low-quality |
| S17_extracted_k141_61825 | Not-determined |
| S17_extracted_k141_165426 | Not-determined |
| S17_extracted_k141_17370 | Not-determined |
| S17_extracted_k141_69622 | Not-determined |
| S17_extracted_k141_158279 | Not-determined |
| S17_extracted_k141_91465 | Low-quality |
| S17_extracted_k141_69623 | Low-quality |
| S17_extracted_k141_99296 | Not-determined |
| S17_extracted_k141_135812 | Not-determined |
| S17_extracted_k141_54828 | Not-determined |
| S17_extracted_k141_135814 | Low-quality |
| S17_extracted_k141_158283 | Not-determined |
| S17_extracted_k141_17376 | Not-determined |
| S17_extracted_k141_128233 | Not-determined |
| S17_extracted_k141_69627 | Not-determined |
| S17_extracted_k141_10435 | Not-determined |
| S17_extracted_k141_165435 | Not-determined |
| S17_extracted_k141_10436 | Low-quality |
| S17_extracted_k141_158288 | Not-determined |
| S17_extracted_k141_17382 | Not-determined |
| S17_extracted_k141_128237 | Not-determined |
| S17_extracted_k141_69630 | Not-determined |
| S17_extracted_k141_113552 | Not-determined |
| S17_extracted_k141_99306 | Not-determined |
| S17_extracted_k141_69631 | Low-quality |
| S17_extracted_k141_10442 | Not-determined |
| S17_extracted_k141_128243 | Not-determined |
| S17_extracted_k141_121113 | Low-quality |
| S17_extracted_k141_135816 | Not-determined |
| S17_extracted_k141_106994 | Not-determined |
| S17_extracted_k141_69636 | Not-determined |
| S17_extracted_k141_128247 | Not-determined |
| S17_extracted_k141_113559 | Not-determined |
| S17_extracted_k141_106996 | Not-determined |
| S17_extracted_k141_165441 | Not-determined |
| S17_extracted_k141_106998 | Not-determined |
| S17_extracted_k141_54836 | Not-determined |
| S17_extracted_k141_69638 | Low-quality |
| S17_extracted_k141_10445 | Not-determined |
| S17_extracted_k141_32743 | Not-determined |
| S17_extracted_k141_121114 | Not-determined |
| S17_extracted_k141_135821 | Low-quality |
| S17_extracted_k141_10449 | Not-determined |
| S17_extracted_k141_40446 | Low-quality |
| S17_extracted_k141_10451 | Not-determined |
| S17_extracted_k141_54843 | Not-determined |
| S17_extracted_k141_107004 | Not-determined |
| S17_extracted_k141_158306 | Not-determined |
| S17_extracted_k141_40452 | Not-determined |
| S17_extracted_k141_165449 | Not-determined |
| S17_extracted_k141_158307 | Not-determined |
| S17_extracted_k141_165451 | Not-determined |
| S17_extracted_k141_47622 | Not-determined |
| S17_extracted_k141_47623 | Not-determined |
| S17_extracted_k141_2874 | Low-quality |
| S17_extracted_k141_40460 | Low-quality |
| S17_extracted_k141_91488 | Not-determined |
| S17_extracted_k141_61831 | Not-determined |
| S17_extracted_k141_40462 | Not-determined |
| S17_extracted_k141_99309 | Not-determined |
| S17_extracted_k141_172701 | Not-determined |
| S17_extracted_k141_17398 | Low-quality |
| S17_extracted_k141_91492 | Low-quality |
| S17_extracted_k141_99310 | Low-quality |
| S17_extracted_k141_76836 | Low-quality |
| S17_extracted_k141_165458 | Low-quality |
| S17_extracted_k141_113568 | Not-determined |
| S17_extracted_k141_165459 | Not-determined |
| S17_extracted_k141_17401 | Not-determined |
| S17_extracted_k141_17405 | Not-determined |
| S17_extracted_k141_135833 | Not-determined |
| S17_extracted_k141_143648 | Not-determined |
| S17_extracted_k141_17406 | Not-determined |
| S17_extracted_k141_113570 | Low-quality |
| S17_extracted_k141_40470 | Low-quality |
| S17_extracted_k141_151265 | Not-determined |
| S17_extracted_k141_99318 | Low-quality |
| S17_extracted_k141_40473 | Not-determined |
| S17_extracted_k141_151267 | Not-determined |
| S17_extracted_k141_17410 | Not-determined |
| S17_extracted_k141_99320 | Not-determined |
| S17_extracted_k141_61848 | Not-determined |
| S17_extracted_k141_143653 | Low-quality |
| S17_extracted_k141_10478 | Not-determined |
| S17_extracted_k141_91498 | Not-determined |
| S17_extracted_k141_91502 | Not-determined |
| S17_extracted_k141_99326 | Not-determined |
| S17_extracted_k141_135834 | Not-determined |
| S17_extracted_k141_91503 | Not-determined |
| S17_extracted_k141_135835 | Not-determined |
| S17_extracted_k141_113580 | Not-determined |
| S17_extracted_k141_121127 | Low-quality |
| S17_extracted_k141_143656 | Low-quality |
| S17_extracted_k141_76842 | Low-quality |
| S17_extracted_k141_99334 | Not-determined |
| S17_extracted_k141_47650 | Not-determined |
| S17_extracted_k141_121135 | Low-quality |
| S17_extracted_k141_172719 | Not-determined |
| S17_extracted_k141_121137 | Not-determined |
| S17_extracted_k141_17424 | Not-determined |
| S17_extracted_k141_121138 | Not-determined |
| S17_extracted_k141_143662 | Not-determined |
| S17_extracted_k141_99342 | Not-determined |
| S17_extracted_k141_10495 | Not-determined |
| S17_extracted_k141_121141 | Not-determined |
| S17_extracted_k141_2905 | Not-determined |
| S17_extracted_k141_61851 | Not-determined |
| S17_extracted_k141_99344 | Not-determined |
| S17_extracted_k141_2909 | Not-determined |
| S17_extracted_k141_54862 | Low-quality |
| S17_extracted_k141_151287 | Not-determined |
| S17_extracted_k141_32755 | Low-quality |
| S17_extracted_k141_135859 | Not-determined |
| S17_extracted_k141_128273 | Not-determined |
| S17_extracted_k141_2912 | Not-determined |
| S17_extracted_k141_121144 | Not-determined |
| S17_extracted_k141_121145 | Not-determined |
| S17_extracted_k141_91525 | Not-determined |
| S17_extracted_k141_2913 | Not-determined |
| S17_extracted_k141_40489 | Not-determined |
| S17_extracted_k141_143670 | Not-determined |
| S17_extracted_k141_54871 | Not-determined |
| S17_extracted_k141_113600 | Not-determined |
| S17_extracted_k141_91529 | Low-quality |
| S17_extracted_k141_143671 | Not-determined |
| S17_extracted_k141_32758 | Not-determined |
| S17_extracted_k141_47663 | Not-determined |
| S17_extracted_k141_61861 | Low-quality |
| S17_extracted_k141_113603 | Not-determined |
| S17_extracted_k141_165493 | Low-quality |
| S17_extracted_k141_107026 | Not-determined |
| S17_extracted_k141_172729 | Not-determined |
| S17_extracted_k141_165495 | Not-determined |
| S17_extracted_k141_76855 | Not-determined |
| S17_extracted_k141_17441 | Not-determined |
| S17_extracted_k141_17443 | Not-determined |
| S17_extracted_k141_99357 | Not-determined |
| S17_extracted_k141_2921 | Not-determined |
| S17_extracted_k141_107029 | Not-determined |
| S17_extracted_k141_40497 | Not-determined |
| S17_extracted_k141_113610 | Not-determined |
| S17_extracted_k141_47671 | Not-determined |
| S17_extracted_k141_32762 | Not-determined |
| S17_extracted_k141_47672 | Not-determined |
| S17_extracted_k141_107030 | Low-quality |
| S17_extracted_k141_32763 | Low-quality |
| S17_extracted_k141_135861 | Not-determined |
| S17_extracted_k141_113613 | Not-determined |
| S17_extracted_k141_121159 | Not-determined |
| S17_extracted_k141_128289 | Not-determined |
| S17_extracted_k141_165503 | Not-determined |
| S17_extracted_k141_128290 | Not-determined |
| S17_extracted_k141_10512 | Low-quality |
| S17_extracted_k141_10513 | Low-quality |
| S17_extracted_k141_165505 | Low-quality |
| S17_extracted_k141_24926 | Not-determined |
| S17_extracted_k141_61874 | Not-determined |
| S17_extracted_k141_2934 | Not-determined |
| S17_extracted_k141_40499 | Not-determined |
| S17_extracted_k141_121165 | Not-determined |
| S17_extracted_k141_107038 | Not-determined |
| S17_extracted_k141_40500 | Not-determined |
| S17_extracted_k141_121169 | Not-determined |
| S17_extracted_k141_32767 | Not-determined |
| S17_extracted_k141_24929 | Not-determined |
| S17_extracted_k141_32768 | Not-determined |
| S17_extracted_k141_91544 | Not-determined |
| S17_extracted_k141_76864 | Low-quality |
| S17_extracted_k141_61880 | Not-determined |
| S17_extracted_k141_2939 | Not-determined |
| S17_extracted_k141_143688 | Low-quality |
| S17_extracted_k141_83574 | Not-determined |
| S17_extracted_k141_17457 | Low-quality |
| S17_extracted_k141_40505 | Not-determined |
| S17_extracted_k141_24936 | Not-determined |
| S17_extracted_k141_40507 | Not-determined |
| S17_extracted_k141_91549 | Not-determined |
| S17_extracted_k141_91551 | Not-determined |
| S17_extracted_k141_17461 | Not-determined |
| S17_extracted_k141_83577 | Not-determined |
| S17_extracted_k141_54884 | Not-determined |
| S17_extracted_k141_10524 | Not-determined |
| S17_extracted_k141_165520 | Not-determined |
| S17_extracted_k141_99372 | Not-determined |
| S17_extracted_k141_40512 | Not-determined |
| S17_extracted_k141_24940 | Not-determined |
| S17_extracted_k141_61889 | Not-determined |
| S17_extracted_k141_99377 | Not-determined |
| S17_extracted_k141_40517 | Not-determined |
| S17_extracted_k141_10529 | Not-determined |
| S17_extracted_k141_172745 | Low-quality |
| S17_extracted_k141_107045 | Low-quality |
| S17_extracted_k141_17471 | Not-determined |
| S17_extracted_k141_99382 | Not-determined |
| S17_extracted_k141_17472 | Not-determined |
| S17_extracted_k141_143697 | Not-determined |
| S17_extracted_k141_69656 | Low-quality |
| S17_extracted_k141_40522 | Low-quality |
| S17_extracted_k141_91560 | Low-quality |
| S17_extracted_k141_61894 | Low-quality |
| S17_extracted_k141_143700 | Not-determined |
| S17_extracted_k141_40523 | Not-determined |
| S17_extracted_k141_143701 | Not-determined |
| S17_extracted_k141_24944 | Not-determined |
| S17_extracted_k141_54897 | Not-determined |
| S17_extracted_k141_40525 | Low-quality |
| S17_extracted_k141_158349 | Not-determined |
| S17_extracted_k141_128310 | Not-determined |
| S17_extracted_k141_2952 | Not-determined |
| S17_extracted_k141_107051 | Not-determined |
| S17_extracted_k141_99391 | Low-quality |
| S17_extracted_k141_83591 | Not-determined |
| S17_extracted_k141_47684 | Not-determined |
| S17_extracted_k141_99395 | Low-quality |
| S17_extracted_k141_69667 | Not-determined |
| S17_extracted_k141_172760 | Not-determined |
| S17_extracted_k141_121177 | Not-determined |
| S17_extracted_k141_172761 | Not-determined |
| S17_extracted_k141_143708 | Not-determined |
| S17_extracted_k141_69668 | Not-determined |
| S17_extracted_k141_113634 | Not-determined |
| S17_extracted_k141_69669 | Not-determined |
| S17_extracted_k141_40536 | Not-determined |
| S17_extracted_k141_121181 | Not-determined |
| S17_extracted_k141_47691 | Not-determined |
| S17_extracted_k141_121184 | Not-determined |
| S17_extracted_k141_40539 | Not-determined |
| S17_extracted_k141_107061 | Not-determined |
| S17_extracted_k141_143713 | Not-determined |
| S17_extracted_k141_24951 | Not-determined |
| S17_extracted_k141_143715 | Not-determined |
| S17_extracted_k141_151327 | Not-determined |
| S17_extracted_k141_76879 | Not-determined |
| S17_extracted_k141_113641 | Not-determined |
| S17_extracted_k141_91571 | Not-determined |
| S17_extracted_k141_158365 | Not-determined |
| S17_extracted_k141_107064 | Not-determined |
| S17_extracted_k141_143718 | Not-determined |
| S17_extracted_k141_158366 | Low-quality |
| S17_extracted_k141_47705 | Not-determined |
| S17_extracted_k141_143721 | Not-determined |
| S17_extracted_k141_40547 | Low-quality |
| S17_extracted_k141_17492 | Not-determined |
| S17_extracted_k141_91577 | Not-determined |
| S17_extracted_k141_158371 | Not-determined |
| S17_extracted_k141_172775 | Not-determined |
| S17_extracted_k141_10556 | Not-determined |
| S17_extracted_k141_47712 | Not-determined |
| S17_extracted_k141_143725 | Not-determined |
| S17_extracted_k141_91580 | Not-determined |
| S17_extracted_k141_83608 | Not-determined |
| S17_extracted_k141_47714 | Not-determined |
| S17_extracted_k141_91582 | Not-determined |
| S17_extracted_k141_151339 | Not-determined |
| S17_extracted_k141_2956 | Not-determined |
| S17_extracted_k141_76895 | Not-determined |
| S17_extracted_k141_17495 | Not-determined |
| S17_extracted_k141_17497 | Not-determined |
| S17_extracted_k141_128326 | Not-determined |
| S17_extracted_k141_91585 | Low-quality |
| S17_extracted_k141_61916 | Not-determined |
| S17_extracted_k141_107073 | Not-determined |
| S17_extracted_k141_17499 | Not-determined |
| S17_extracted_k141_76900 | Not-determined |
| S17_extracted_k141_128329 | Not-determined |
| S17_extracted_k141_17501 | Not-determined |
| S17_extracted_k141_91591 | Not-determined |
| S17_extracted_k141_83613 | Not-determined |
| S17_extracted_k141_113664 | Not-determined |
| S17_extracted_k141_83616 | Not-determined |
| S17_extracted_k141_151344 | Low-quality |
| S17_extracted_k141_83618 | Not-determined |
| S17_extracted_k141_113668 | Not-determined |
| S17_extracted_k141_158388 | Not-determined |
| S17_extracted_k141_40555 | Not-determined |
| S17_extracted_k141_40556 | Not-determined |
| S17_extracted_k141_17503 | Not-determined |
| S17_extracted_k141_47729 | Not-determined |
| S17_extracted_k141_2962 | Not-determined |
| S17_extracted_k141_32792 | Not-determined |
| S17_extracted_k141_47731 | Not-determined |
| S17_extracted_k141_76913 | Not-determined |
| S17_extracted_k141_2964 | Not-determined |
| S17_extracted_k141_151351 | Not-determined |
| S17_extracted_k141_24971 | Not-determined |
| S17_extracted_k141_151352 | Not-determined |
| S17_extracted_k141_128340 | Not-determined |
| S17_extracted_k141_2965 | Not-determined |
| S17_extracted_k141_76916 | Not-determined |
| S17_extracted_k141_61930 | Not-determined |
| S17_extracted_k141_151354 | Low-quality |
| S17_extracted_k141_151355 | Not-determined |
| S17_extracted_k141_121219 | Not-determined |
| S17_extracted_k141_40563 | Low-quality |
| S17_extracted_k141_143737 | Low-quality |
| S17_extracted_k141_61932 | Not-determined |
| S17_extracted_k141_83623 | Low-quality |
| S17_extracted_k141_99406 | Not-determined |
| S17_extracted_k141_61938 | Not-determined |
| S17_extracted_k141_83627 | Not-determined |
| S17_extracted_k141_158396 | Not-determined |
| S17_extracted_k141_10574 | Low-quality |
| S17_extracted_k141_128348 | Not-determined |
| S17_extracted_k141_47749 | Not-determined |
| S17_extracted_k141_121226 | Low-quality |
| S17_extracted_k141_99409 | Not-determined |
| S17_extracted_k141_107094 | Not-determined |
| S17_extracted_k141_76929 | Low-quality |
| S17_extracted_k141_76931 | Not-determined |
| S17_extracted_k141_32802 | Low-quality |
| S17_extracted_k141_76933 | Not-determined |
| S17_extracted_k141_99416 | Not-determined |
| S17_extracted_k141_151366 | Not-determined |
| S17_extracted_k141_99419 | Not-determined |
| S17_extracted_k141_107104 | Low-quality |
| S17_extracted_k141_172791 | Not-determined |
| S17_extracted_k141_135884 | Not-determined |
| S17_extracted_k141_40577 | Low-quality |
| S17_extracted_k141_24990 | Not-determined |
| S17_extracted_k141_69691 | Not-determined |
| S17_extracted_k141_69693 | Not-determined |
| S17_extracted_k141_99427 | Not-determined |
| S17_extracted_k141_121235 | Not-determined |
| S17_extracted_k141_47760 | Not-determined |
| S17_extracted_k141_128365 | Not-determined |
| S17_extracted_k141_32811 | Not-determined |
| S17_extracted_k141_91597 | Not-determined |
| S17_extracted_k141_54933 | Not-determined |
| S17_extracted_k141_121240 | Not-determined |
| S17_extracted_k141_91600 | Not-determined |
| S17_extracted_k141_107112 | Not-determined |
| S17_extracted_k141_158405 | Not-determined |
| S17_extracted_k141_69704 | Not-determined |
| S17_extracted_k141_2986 | Not-determined |
| S17_extracted_k141_107117 | Not-determined |
| S17_extracted_k141_128373 | Not-determined |
| S17_extracted_k141_172810 | Not-determined |
| S17_extracted_k141_172811 | Not-determined |
| S17_extracted_k141_10597 | Not-determined |
| S17_extracted_k141_113687 | Not-determined |
| S17_extracted_k141_172813 | Not-determined |
| S17_extracted_k141_69712 | Not-determined |
| S17_extracted_k141_69713 | Not-determined |
| S17_extracted_k141_113694 | Not-determined |
| S17_extracted_k141_69717 | Not-determined |
| S17_extracted_k141_47768 | Not-determined |
| S17_extracted_k141_69718 | Not-determined |
| S17_extracted_k141_135896 | Not-determined |
| S17_extracted_k141_54948 | Not-determined |
| S17_extracted_k141_91613 | Low-quality |
| S17_extracted_k141_83642 | Not-determined |
| S17_extracted_k141_47771 | Low-quality |
| S17_extracted_k141_69723 | Not-determined |
| S17_extracted_k141_128382 | Not-determined |
| S17_extracted_k141_135901 | Not-determined |
| S17_extracted_k141_10604 | Not-determined |
| S17_extracted_k141_135904 | Not-determined |
| S17_extracted_k141_128386 | Not-determined |
| S17_extracted_k141_172823 | Low-quality |
| S17_extracted_k141_76967 | Not-determined |

| | |
|---|---|
| S17_extracted_k141_172824 | Not-determined |
| S17_extracted_k141_47783 | Not-determined |
| S17_extracted_k141_40586 | Not-determined |
| S17_extracted_k141_172826 | Low-quality |
| S17_extracted_k141_47786 | Not-determined |
| S17_extracted_k141_40590 | Not-determined |
| S17_extracted_k141_121260 | Not-determined |
| S17_extracted_k141_135908 | Not-determined |
| S17_extracted_k141_40595 | Not-determined |
| S17_extracted_k141_158423 | Not-determined |
| S17_extracted_k141_91626 | Not-determined |
| S17_extracted_k141_10616 | Not-determined |
| S17_extracted_k141_83656 | Not-determined |
| S17_extracted_k141_91627 | Not-determined |
| S17_extracted_k141_32846 | Not-determined |
| S17_extracted_k141_10619 | Not-determined |
| S17_extracted_k141_76979 | Not-determined |
| S17_extracted_k141_143750 | Not-determined |
| S17_extracted_k141_17579 | Not-determined |
| S17_extracted_k141_91631 | Not-determined |
| S17_extracted_k141_165544 | Not-determined |
| S17_extracted_k141_3007 | Not-determined |
| S17_extracted_k141_40607 | Not-determined |
| S17_extracted_k141_91636 | Not-determined |
| S17_extracted_k141_91637 | Not-determined |
| S17_extracted_k141_128408 | Low-quality |
| S17_extracted_k141_25004 | Low-quality |
| S17_extracted_k141_17587 | Not-determined |
| S17_extracted_k141_83664 | Not-determined |
| S17_extracted_k141_91641 | Not-determined |
| S17_extracted_k141_83669 | Not-determined |
| S17_extracted_k141_107159 | Not-determined |
| S17_extracted_k141_107160 | Not-determined |
| S17_extracted_k141_107161 | Not-determined |
| S17_extracted_k141_83672 | Not-determined |
| S17_extracted_k141_128414 | Low-quality |
| S17_extracted_k141_40616 | Not-determined |
| S17_extracted_k141_113722 | Not-determined |
| S17_extracted_k141_165546 | Low-quality |
| S17_extracted_k141_143770 | Not-determined |
| S17_extracted_k141_143771 | Not-determined |
| S17_extracted_k141_128417 | Not-determined |
| S17_extracted_k141_121278 | Not-determined |
| S17_extracted_k141_113725 | Not-determined |
| S17_extracted_k141_121279 | Not-determined |
| S17_extracted_k141_76995 | Not-determined |
| S17_extracted_k141_165550 | Not-determined |
| S17_extracted_k141_40619 | Not-determined |
| S17_extracted_k141_128421 | Not-determined |
| S17_extracted_k141_61947 | Not-determined |
| S17_extracted_k141_3013 | Not-determined |
| S17_extracted_k141_113730 | Not-determined |
| S17_extracted_k141_83680 | Low-quality |
| S17_extracted_k141_113732 | Not-determined |
| S17_extracted_k141_25018 | Not-determined |
| S17_extracted_k141_113733 | Not-determined |
| S17_extracted_k141_3019 | Not-determined |
| S17_extracted_k141_165557 | Not-determined |
| S17_extracted_k141_107169 | Not-determined |
| S17_extracted_k141_107176 | Not-determined |
| S17_extracted_k141_113738 | Not-determined |
| S17_extracted_k141_113739 | Not-determined |
| S17_extracted_k141_143783 | Not-determined |
| S17_extracted_k141_143784 | Not-determined |
| S17_extracted_k141_99451 | Not-determined |
| S17_extracted_k141_77014 | Not-determined |
| S17_extracted_k141_54966 | Not-determined |
| S17_extracted_k141_25027 | Not-determined |
| S17_extracted_k141_25028 | Low-quality |
| S17_extracted_k141_54969 | Not-determined |
| S17_extracted_k141_99455 | Not-determined |
| S17_extracted_k141_54970 | Low-quality |
| S17_extracted_k141_69754 | Not-determined |
| S17_extracted_k141_121288 | Not-determined |
| S17_extracted_k141_83693 | Not-determined |
| S17_extracted_k141_107188 | Not-determined |
| S17_extracted_k141_69758 | Not-determined |
| S17_extracted_k141_77022 | Not-determined |
| S17_extracted_k141_158439 | Not-determined |
| S17_extracted_k141_165568 | Low-quality |
| S17_extracted_k141_17618 | Not-determined |
| S17_extracted_k141_113750 | Not-determined |
| S17_extracted_k141_151370 | Not-determined |
| S17_extracted_k141_17620 | Not-determined |
| S17_extracted_k141_17621 | Not-determined |
| S17_extracted_k141_143796 | Not-determined |
| S17_extracted_k141_121297 | Not-determined |
| S17_extracted_k141_113758 | Not-determined |
| S17_extracted_k141_32875 | Not-determined |
| S17_extracted_k141_143798 | Low-quality |
| S17_extracted_k141_113760 | Not-determined |
| S17_extracted_k141_135915 | Not-determined |
| S17_extracted_k141_61965 | Not-determined |
| S17_extracted_k141_40620 | Low-quality |
| S17_extracted_k141_113766 | Not-determined |
| S17_extracted_k141_99469 | Not-determined |
| S17_extracted_k141_17629 | Not-determined |
| S17_extracted_k141_128444 | Not-determined |
| S17_extracted_k141_113769 | Not-determined |
| S17_extracted_k141_143809 | Not-determined |
| S17_extracted_k141_91655 | Not-determined |
| S17_extracted_k141_107190 | Not-determined |
| S17_extracted_k141_151382 | Not-determined |
| S17_extracted_k141_69762 | Low-quality |
| S17_extracted_k141_128449 | Not-determined |
| S17_extracted_k141_77028 | Not-determined |
| S17_extracted_k141_3029 | Not-determined |
| S17_extracted_k141_165588 | Not-determined |
| S17_extracted_k141_151386 | Not-determined |
| S17_extracted_k141_151387 | Not-determined |
| S17_extracted_k141_158455 | Not-determined |
| S17_extracted_k141_32892 | Not-determined |
| S17_extracted_k141_128455 | Not-determined |
| S17_extracted_k141_143819 | Low-quality |
| S17_extracted_k141_121302 | Not-determined |
| S17_extracted_k141_143820 | Not-determined |
| S17_extracted_k141_77039 | Not-determined |
| S17_extracted_k141_32898 | Not-determined |
| S17_extracted_k141_17635 | Not-determined |
| S17_extracted_k141_3044 | Not-determined |
| S17_extracted_k141_143823 | Low-quality |
| S17_extracted_k141_77043 | Not-determined |
| S17_extracted_k141_107199 | Not-determined |
| S17_extracted_k141_107200 | Not-determined |
| S17_extracted_k141_107201 | Not-determined |
| S17_extracted_k141_32900 | Low-quality |
| S17_extracted_k141_107203 | Not-determined |
| S17_extracted_k141_77049 | Not-determined |
| S17_extracted_k141_25040 | Not-determined |
| S17_extracted_k141_77051 | Not-determined |
| S17_extracted_k141_69785 | Not-determined |
| S17_extracted_k141_135947 | Not-determined |
| S17_extracted_k141_47798 | Not-determined |
| S17_extracted_k141_55003 | Not-determined |
| S17_extracted_k141_151393 | Not-determined |
| S17_extracted_k141_77054 | Not-determined |
| S17_extracted_k141_113781 | Not-determined |
| S17_extracted_k141_151396 | Not-determined |
| S17_extracted_k141_135952 | Not-determined |
| S17_extracted_k141_40638 | Not-determined |
| S17_extracted_k141_113786 | Not-determined |
| S17_extracted_k141_172840 | Not-determined |
| S17_extracted_k141_172841 | Low-quality |
| S17_extracted_k141_143832 | Not-determined |
| S17_extracted_k141_3058 | Not-determined |
| S17_extracted_k141_10667 | Not-determined |
| S17_extracted_k141_107217 | Not-determined |
| S17_extracted_k141_165596 | Not-determined |
| S17_extracted_k141_121321 | Not-determined |
| S17_extracted_k141_121322 | Not-determined |
| S17_extracted_k141_55010 | Not-determined |
| S17_extracted_k141_143842 | Not-determined |
| S17_extracted_k141_40644 | Not-determined |
| S17_extracted_k141_69787 | Low-quality |
| S17_extracted_k141_151403 | Not-determined |
| S17_extracted_k141_158483 | Not-determined |
| S17_extracted_k141_165601 | Not-determined |
| S17_extracted_k141_55017 | Not-determined |
| S17_extracted_k141_151406 | Not-determined |
| S17_extracted_k141_3066 | Not-determined |
| S17_extracted_k141_10679 | Low-quality |
| S17_extracted_k141_165604 | Not-determined |
| S17_extracted_k141_61998 | Not-determined |
| S17_extracted_k141_3068 | Not-determined |
| S17_extracted_k141_61999 | Not-determined |
| S17_extracted_k141_32916 | Low-quality |
| S17_extracted_k141_69794 | Not-determined |
| S17_extracted_k141_3072 | Not-determined |
| S17_extracted_k141_32919 | Not-determined |
| S17_extracted_k141_113809 | Not-determined |
| S17_extracted_k141_91673 | Not-determined |
| S17_extracted_k141_62006 | Not-determined |
| S17_extracted_k141_40663 | Not-determined |
| S17_extracted_k141_113812 | Low-quality |
| S17_extracted_k141_113814 | Not-determined |
| S17_extracted_k141_113815 | Not-determined |
| S17_extracted_k141_158489 | Not-determined |
| S17_extracted_k141_91676 | Low-quality |
| S17_extracted_k141_165614 | Not-determined |
| S17_extracted_k141_107234 | Not-determined |
| S17_extracted_k141_40665 | Not-determined |
| S17_extracted_k141_69801 | Not-determined |
| S17_extracted_k141_32924 | Low-quality |
| S17_extracted_k141_3080 | Not-determined |
| S17_extracted_k141_121332 | Not-determined |
| S17_extracted_k141_69808 | Not-determined |
| S17_extracted_k141_62010 | Not-determined |
| S17_extracted_k141_55024 | Low-quality |
| S17_extracted_k141_10689 | Not-determined |
| S17_extracted_k141_55028 | Not-determined |
| S17_extracted_k141_62015 | Not-determined |
| S17_extracted_k141_55031 | Low-quality |
| S17_extracted_k141_10697 | Not-determined |
| S17_extracted_k141_3091 | Not-determined |
| S17_extracted_k141_47831 | Not-determined |
| S17_extracted_k141_172869 | Not-determined |
| S17_extracted_k141_47832 | Not-determined |
| S17_extracted_k141_47833 | Not-determined |
| S17_extracted_k141_62020 | Not-determined |
| S17_extracted_k141_25071 | Not-determined |
| S5_vs_k141_145579\|\|full | High-quality |
| S17_extracted_k141_62022 | Not-determined |
| S17_extracted_k141_40672 | Low-quality |
| S17_extracted_k141_128479 | Not-determined |
| S17_extracted_k141_158506 | Not-determined |
| S17_extracted_k141_32939 | Not-determined |
| S17_extracted_k141_69817 | Not-determined |
| S17_extracted_k141_55037 | Not-determined |
| S17_extracted_k141_10702 | Not-determined |
| S17_extracted_k141_77064 | Not-determined |
| S17_extracted_k141_3104 | Not-determined |
| S17_extracted_k141_135978 | Not-determined |
| S17_extracted_k141_165628 | Not-determined |
| S17_extracted_k141_158510 | Low-quality |
| S17_extracted_k141_83726 | Low-quality |
| S17_extracted_k141_69821 | Not-determined |
| S17_extracted_k141_158513 | Not-determined |
| S17_extracted_k141_83729 | Not-determined |
| S17_extracted_k141_151430 | Not-determined |
| S17_extracted_k141_25080 | Not-determined |
| S17_extracted_k141_165634 | Not-determined |
| S17_extracted_k141_128487 | Not-determined |
| S17_extracted_k141_77067 | Not-determined |
| S17_extracted_k141_172874 | Low-quality |
| S17_extracted_k141_165636 | Not-determined |
| S17_extracted_k141_121339 | Not-determined |
| S17_extracted_k141_107251 | Not-determined |
| S17_extracted_k141_77071 | Not-determined |
| S17_extracted_k141_113837 | Not-determined |
| S17_extracted_k141_128489 | Not-determined |
| S17_extracted_k141_107254 | Not-determined |
| S17_extracted_k141_165643 | Not-determined |
| S17_extracted_k141_165645 | Not-determined |
| S17_extracted_k141_17687 | Not-determined |
| S17_extracted_k141_32956 | Not-determined |
| S17_extracted_k141_158519 | Not-determined |
| S17_extracted_k141_47853 | Low-quality |
| S17_extracted_k141_17690 | Not-determined |
| S17_extracted_k141_47856 | Not-determined |
| S17_extracted_k141_3116 | Not-determined |
| S17_extracted_k141_55050 | Not-determined |
| S17_extracted_k141_158530 | Not-determined |
| S17_extracted_k141_69837 | Not-determined |
| S17_extracted_k141_172892 | Not-determined |
| S17_extracted_k141_69839 | Low-quality |
| S17_extracted_k141_121347 | Low-quality |
| S17_extracted_k141_47861 | Not-determined |
| S17_extracted_k141_32966 | Not-determined |
| S17_extracted_k141_62048 | Low-quality |
| S17_extracted_k141_17701 | Low-quality |
| S17_extracted_k141_107260 | Not-determined |
| S17_extracted_k141_69841 | Not-determined |
| S17_extracted_k141_77078 | Low-quality |
| S17_extracted_k141_25088 | Not-determined |
| S17_extracted_k141_99527 | Not-determined |
| S17_extracted_k141_107261 | Not-determined |
| S17_extracted_k141_121352 | Not-determined |
| S17_extracted_k141_17703 | Not-determined |
| S17_extracted_k141_107262 | Not-determined |
| S17_extracted_k141_128502 | Not-determined |
| S17_extracted_k141_107264 | Not-determined |
| S17_extracted_k141_77083 | Not-determined |
| S17_extracted_k141_165659 | Not-determined |
| S17_extracted_k141_47869 | Not-determined |
| S17_extracted_k141_91687 | Not-determined |
| S17_extracted_k141_128510 | Low-quality |
| S17_extracted_k141_91688 | Not-determined |
| S17_extracted_k141_17706 | Low-quality |
| S17_extracted_k141_47870 | Not-determined |
| S17_extracted_k141_158539 | Not-determined |
| S17_extracted_k141_10734 | Not-determined |
| S17_extracted_k141_121361 | Low-quality |
| S17_extracted_k141_17709 | Not-determined |
| S17_extracted_k141_10738 | Not-determined |
| S17_extracted_k141_136003 | Not-determined |
| S17_extracted_k141_32980 | Not-determined |
| S17_extracted_k141_10740 | Low-quality |
| S17_extracted_k141_55065 | Not-determined |
| S17_extracted_k141_17718 | Not-determined |
| S17_extracted_k141_128519 | Not-determined |
| S17_extracted_k141_165678 | Not-determined |
| S17_extracted_k141_55069 | Not-determined |
| S17_extracted_k141_32987 | Not-determined |
| S17_extracted_k141_69853 | Not-determined |
| S17_extracted_k141_10747 | Not-determined |
| S17_extracted_k141_3137 | Not-determined |
| S17_extracted_k141_77102 | Low-quality |
| S17_extracted_k141_128521 | Not-determined |
| S17_extracted_k141_25106 | Not-determined |
| S17_extracted_k141_62061 | Not-determined |
| S17_extracted_k141_128523 | Low-quality |
| S17_extracted_k141_62062 | Not-determined |
| S17_extracted_k141_55075 | Low-quality |
| S17_extracted_k141_91696 | Not-determined |
| S17_extracted_k141_83766 | Not-determined |
| S17_extracted_k141_32997 | Not-determined |
| S17_extracted_k141_10751 | Not-determined |
| S17_extracted_k141_158555 | Low-quality |
| S17_extracted_k141_3147 | Not-determined |
| S17_extracted_k141_99544 | Not-determined |
| S17_extracted_k141_17727 | Not-determined |
| S17_extracted_k141_128527 | Low-quality |
| S17_extracted_k141_77113 | Not-determined |
| S17_extracted_k141_62072 | Not-determined |
| S17_extracted_k141_33001 | Not-determined |
| S17_extracted_k141_136019 | Not-determined |
| S17_extracted_k141_47897 | Low-quality |
| S17_extracted_k141_62073 | Not-determined |
| S17_extracted_k141_10756 | Not-determined |
| S17_extracted_k141_33004 | Low-quality |
| S17_extracted_k141_99549 | Not-determined |
| S17_extracted_k141_165688 | Not-determined |
| S17_extracted_k141_99552 | Not-determined |
| S17_extracted_k141_10757 | Not-determined |
| S17_extracted_k141_77119 | Not-determined |
| S17_extracted_k141_107299 | Not-determined |
| S17_extracted_k141_128541 | Not-determined |
| S17_extracted_k141_33009 | Not-determined |
| S17_extracted_k141_128543 | Low-quality |
| S17_extracted_k141_143879 | Not-determined |
| S17_extracted_k141_107301 | Low-quality |
| S17_extracted_k141_107302 | Not-determined |
| S17_extracted_k141_25115 | Not-determined |
| S17_extracted_k141_62085 | Not-determined |
| S17_extracted_k141_33014 | Not-determined |
| S17_extracted_k141_17742 | Not-determined |
| S17_extracted_k141_3161 | Low-quality |
| S17_extracted_k141_77127 | Not-determined |
| S17_extracted_k141_47903 | Not-determined |
| S17_extracted_k141_3163 | Not-determined |
| S17_extracted_k141_121380 | Not-determined |
| S17_extracted_k141_99553 | Low-quality |
| S17_extracted_k141_77129 | Not-determined |
| S17_extracted_k141_128550 | Not-determined |
| S17_extracted_k141_77130 | Not-determined |
| S17_extracted_k141_10766 | Not-determined |
| S17_extracted_k141_3164 | Not-determined |
| S17_extracted_k141_10768 | Not-determined |
| S17_extracted_k141_25120 | Not-determined |
| S17_extracted_k141_40706 | Not-determined |
| S17_extracted_k141_77135 | Not-determined |
| S17_extracted_k141_151464 | Not-determined |
| S17_extracted_k141_62090 | Not-determined |
| S17_extracted_k141_128556 | Not-determined |
| S17_extracted_k141_40710 | Not-determined |
| S17_extracted_k141_121386 | Low-quality |
| S17_extracted_k141_172899 | Not-determined |
| S17_extracted_k141_10774 | Low-quality |
| S17_extracted_k141_128558 | Not-determined |
| S17_extracted_k141_25125 | Not-determined |
| S17_extracted_k141_165690 | Not-determined |
| S17_extracted_k141_40717 | Low-quality |
| S17_extracted_k141_10777 | Not-determined |
| S17_extracted_k141_121391 | Low-quality |
| S17_extracted_k141_62095 | Not-determined |
| S17_extracted_k141_172905 | Not-determined |
| S17_extracted_k141_33031 | Not-determined |
| S17_extracted_k141_25128 | Not-determined |
| S17_extracted_k141_113847 | Low-quality |
| S17_extracted_k141_107314 | Low-quality |
| S17_extracted_k141_113848 | Not-determined |
| S17_extracted_k141_62098 | Not-determined |
| S17_extracted_k141_151472 | Not-determined |
| S17_extracted_k141_69868 | Not-determined |
| S17_extracted_k141_10781 | Not-determined |
| S17_extracted_k141_151475 | Low-quality |
| S17_extracted_k141_91730 | Not-determined |
| S17_extracted_k141_107316 | Not-determined |
| S17_extracted_k141_121396 | Not-determined |
| S17_extracted_k141_62101 | Low-quality |
| S17_extracted_k141_121397 | Not-determined |
| S17_extracted_k141_121398 | Not-determined |
| S17_extracted_k141_113853 | Not-determined |
| S17_extracted_k141_25136 | Low-quality |
| S17_extracted_k141_10785 | Not-determined |
| S17_extracted_k141_83785 | Not-determined |
| S17_extracted_k141_172914 | Not-determined |
| S17_extracted_k141_151480 | Not-determined |
| S17_extracted_k141_143895 | Not-determined |
| S17_extracted_k141_83786 | Not-determined |
| S17_extracted_k141_62108 | Not-determined |
| S17_extracted_k141_10790 | Not-determined |
| S17_extracted_k141_33043 | Not-determined |
| S17_extracted_k141_91737 | Not-determined |
| S17_extracted_k141_91738 | Not-determined |
| S17_extracted_k141_55096 | Not-determined |
| S17_extracted_k141_172921 | Not-determined |
| S17_extracted_k141_91745 | Not-determined |
| S17_extracted_k141_165699 | Not-determined |
| S17_extracted_k141_33051 | Not-determined |
| S17_extracted_k141_17755 | Not-determined |
| S17_extracted_k141_69883 | Low-quality |
| S17_extracted_k141_107324 | Not-determined |
| S17_extracted_k141_91749 | Not-determined |
| S17_extracted_k141_69885 | Low-quality |
| S17_extracted_k141_83794 | Low-quality |
| S17_extracted_k141_107325 | Low-quality |
| S17_extracted_k141_83795 | Not-determined |
| S17_extracted_k141_69887 | Not-determined |
| S17_extracted_k141_165705 | Low-quality |
| S17_extracted_k141_158571 | Not-determined |
| S17_extracted_k141_77148 | Not-determined |
| S17_extracted_k141_158572 | Not-determined |
| S17_extracted_k141_151489 | Not-determined |
| S17_extracted_k141_121406 | Low-quality |
| S17_extracted_k141_77152 | Not-determined |
| S17_extracted_k141_33059 | Not-determined |
| S17_extracted_k141_165707 | Not-determined |
| S17_extracted_k141_25152 | Not-determined |
| S17_extracted_k141_40733 | Not-determined |
| S17_extracted_k141_143902 | Not-determined |
| S17_extracted_k141_91765 | Not-determined |
| S17_extracted_k141_172926 | Low-quality |
| S17_extracted_k141_69897 | Not-determined |
| S17_extracted_k141_25155 | Not-determined |
| S17_extracted_k141_91769 | Low-quality |
| S17_extracted_k141_10794 | Not-determined |
| S17_extracted_k141_33062 | Not-determined |
| S17_extracted_k141_17770 | Not-determined |
| S17_extracted_k141_172931 | Not-determined |
| S17_extracted_k141_69902 | Not-determined |
| S17_extracted_k141_40737 | Low-quality |
| S17_extracted_k141_165713 | Low-quality |
| S17_extracted_k141_77159 | Not-determined |
| S17_extracted_k141_121414 | Not-determined |
| S17_extracted_k141_158581 | Low-quality |
| S17_extracted_k141_136061 | Not-determined |
| S17_extracted_k141_3178 | Not-determined |
| S17_extracted_k141_143908 | Not-determined |
| S17_extracted_k141_158584 | Not-determined |
| S17_extracted_k141_62127 | Not-determined |
| S17_extracted_k141_47920 | Not-determined |
| S17_extracted_k141_33071 | Not-determined |
| S17_extracted_k141_151503 | Not-determined |
| S17_extracted_k141_143915 | Low-quality |
| S17_extracted_k141_165720 | Not-determined |
| S17_extracted_k141_128587 | Not-determined |
| S17_extracted_k141_143919 | Not-determined |
| S17_extracted_k141_128588 | Not-determined |
| S17_extracted_k141_40743 | Not-determined |
| S17_extracted_k141_107332 | Not-determined |
| S17_extracted_k141_77162 | Low-quality |
| S17_extracted_k141_77164 | Not-determined |
| S17_extracted_k141_40745 | Not-determined |
| S17_extracted_k141_40746 | Not-determined |
| S17_extracted_k141_113898 | Not-determined |
| S17_extracted_k141_143923 | Not-determined |
| S17_extracted_k141_113899 | Not-determined |
| S17_extracted_k141_47933 | Low-quality |
| S17_extracted_k141_69906 | Not-determined |
| S17_extracted_k141_33082 | Not-determined |
| S17_extracted_k141_17772 | Low-quality |
| S17_extracted_k141_33084 | Not-determined |
| S17_extracted_k141_113902 | Low-quality |
| S17_extracted_k141_3194 | Not-determined |
| S17_extracted_k141_121428 | Low-quality |
| S17_extracted_k141_77171 | Not-determined |
| S17_extracted_k141_91779 | Not-determined |
| S17_extracted_k141_143931 | Not-determined |
| S17_extracted_k141_128599 | Not-determined |
| S17_extracted_k141_172941 | Not-determined |
| S17_extracted_k141_121434 | Low-quality |
| S17_extracted_k141_172943 | Not-determined |
| S17_extracted_k141_107356 | Not-determined |
| S17_extracted_k141_128602 | Not-determined |
| S17_extracted_k141_33095 | Not-determined |
| S17_extracted_k141_77176 | Not-determined |
| S17_extracted_k141_113912 | Not-determined |
| S17_extracted_k141_165744 | Not-determined |
| S17_extracted_k141_121439 | Not-determined |
| S17_extracted_k141_69922 | Not-determined |
| S17_extracted_k141_143943 | Not-determined |
| S17_extracted_k141_33101 | Low-quality |
| S17_extracted_k141_47957 | Not-determined |
| S17_extracted_k141_158609 | Low-quality |
| S17_extracted_k141_165747 | Not-determined |
| S17_extracted_k141_91791 | Not-determined |
| S17_extracted_k141_17776 | Not-determined |
| S17_extracted_k141_69929 | Not-determined |
| S17_extracted_k141_165749 | Low-quality |
| S17_extracted_k141_121442 | Not-determined |
| S17_extracted_k141_158611 | Not-determined |
| S17_extracted_k141_165750 | Not-determined |
| S17_extracted_k141_158612 | Not-determined |
| S17_extracted_k141_62139 | Not-determined |
| S17_extracted_k141_62141 | Not-determined |
| S17_extracted_k141_62142 | Not-determined |
| S17_extracted_k141_83799 | Not-determined |
| S17_extracted_k141_172947 | Not-determined |
| S17_extracted_k141_47961 | Not-determined |
| S17_extracted_k141_3206 | Low-quality |
| S17_extracted_k141_69937 | Not-determined |
| S17_extracted_k141_3208 | Not-determined |
| S17_extracted_k141_33107 | Not-determined |
| S17_extracted_k141_113927 | Not-determined |
| S17_extracted_k141_62152 | Low-quality |
| S17_extracted_k141_91803 | Not-determined |
| S17_extracted_k141_172956 | Not-determined |
| S17_extracted_k141_136093 | Not-determined |
| S17_extracted_k141_40779 | Not-determined |
| S17_extracted_k141_172957 | Not-determined |
| S17_extracted_k141_158620 | Not-determined |
| S17_extracted_k141_91804 | Low-quality |
| S17_extracted_k141_83810 | Not-determined |
| S17_extracted_k141_55155 | Not-determined |
| S17_extracted_k141_99561 | Low-quality |
| S17_extracted_k141_113935 | Not-determined |
| S17_extracted_k141_136104 | Not-determined |
| S17_extracted_k141_40787 | Not-determined |
| S17_extracted_k141_172965 | Low-quality |
| S17_extracted_k141_40792 | Not-determined |
| S17_extracted_k141_47974 | Not-determined |
| S17_extracted_k141_113941 | Not-determined |
| S17_extracted_k141_40795 | Not-determined |
| S17_extracted_k141_136106 | Not-determined |
| S17_extracted_k141_136107 | Not-determined |
| S17_extracted_k141_62157 | Not-determined |
| S17_extracted_k141_25165 | Low-quality |
| S17_extracted_k141_91815 | Not-determined |
| S17_extracted_k141_128632 | Not-determined |
| S17_extracted_k141_62163 | Low-quality |
| S17_extracted_k141_17803 | Low-quality |
| S17_extracted_k141_83822 | Not-determined |
| S17_extracted_k141_107375 | Not-determined |
| S17_extracted_k141_91818 | Not-determined |
| S17_extracted_k141_136119 | Not-determined |
| S17_extracted_k141_91819 | Not-determined |
| S17_extracted_k141_25170 | Not-determined |
| S17_extracted_k141_83824 | Not-determined |
| S17_extracted_k141_62166 | Not-determined |
| S17_extracted_k141_91823 | Not-determined |
| S17_extracted_k141_55172 | Not-determined |
| S17_extracted_k141_136124 | Not-determined |
| S17_extracted_k141_121451 | Low-quality |
| S17_extracted_k141_121452 | Not-determined |
| S17_extracted_k141_121453 | Not-determined |
| S17_extracted_k141_165779 | Not-determined |
| S17_extracted_k141_62170 | Low-quality |
| S17_extracted_k141_121457 | Not-determined |
| S17_extracted_k141_69956 | Not-determined |
| S17_extracted_k141_158635 | Not-determined |
| S17_extracted_k141_107389 | Not-determined |
| S17_extracted_k141_17815 | Not-determined |
| S17_extracted_k141_121462 | Not-determined |
| S17_extracted_k141_121464 | Not-determined |
| S17_extracted_k141_17817 | Not-determined |
| S17_extracted_k141_47995 | Not-determined |
| S17_extracted_k141_62182 | Not-determined |
| S17_extracted_k141_47999 | Not-determined |
| S17_extracted_k141_158642 | Not-determined |
| S17_extracted_k141_121471 | Low-quality |
| S17_extracted_k141_99589 | Not-determined |
| S17_extracted_k141_136130 | Not-determined |
| S17_extracted_k141_69961 | Not-determined |
| S17_extracted_k141_17822 | Not-determined |
| S17_extracted_k141_91830 | Not-determined |
| S17_extracted_k141_158644 | Low-quality |
| S17_extracted_k141_91833 | Not-determined |
| S17_extracted_k141_99599 | Not-determined |
| S17_extracted_k141_3262 | Not-determined |
| S17_extracted_k141_136134 | Not-determined |
| S17_extracted_k141_3263 | Not-determined |
| S17_extracted_k141_158649 | Low-quality |
| S17_extracted_k141_91837 | Not-determined |
| S17_extracted_k141_3265 | Not-determined |
| S17_extracted_k141_55200 | Not-determined |
| S17_extracted_k141_77205 | Not-determined |
| S17_extracted_k141_121476 | Not-determined |
| S17_extracted_k141_158652 | Not-determined |
| S17_extracted_k141_83830 | Low-quality |
| S17_extracted_k141_62195 | Not-determined |
| S17_extracted_k141_40811 | Not-determined |
| S17_extracted_k141_83835 | Not-determined |
| S17_extracted_k141_77208 | Not-determined |
| S17_extracted_k141_33135 | Not-determined |
| S17_extracted_k141_165798 | Not-determined |
| S17_extracted_k141_33136 | Not-determined |
| S17_extracted_k141_40814 | Low-quality |
| S17_extracted_k141_91847 | Low-quality |
| S17_extracted_k141_17836 | Not-determined |
| S17_extracted_k141_91848 | Not-determined |
| S17_extracted_k141_173001 | Not-determined |
| S17_extracted_k141_55217 | Not-determined |
| S17_extracted_k141_121490 | Not-determined |
| S17_extracted_k141_136154 | Not-determined |
| S17_extracted_k141_113961 | Not-determined |
| S17_extracted_k141_173005 | Not-determined |
| S17_extracted_k141_136155 | Not-determined |
| S17_extracted_k141_91849 | Not-determined |

| | |
|---|---|
| S17_extracted_k141_173008 | Not-determined |
| S17_extracted_k141_69988 | Not-determined |
| S17_extracted_k141_99623 | Not-determined |
| S17_extracted_k141_107401 | Not-determined |
| S17_extracted_k141_25182 | Not-determined |
| S17_extracted_k141_25185 | Not-determined |
| S17_extracted_k141_136158 | Not-determined |
| S17_extracted_k141_69992 | Not-determined |
| S17_extracted_k141_91861 | Not-determined |
| S17_extracted_k141_113966 | Not-determined |
| S17_extracted_k141_136164 | Not-determined |
| S17_extracted_k141_33155 | Low-quality |
| S17_extracted_k141_99633 | Low-quality |
| S17_extracted_k141_173013 | Not-determined |
| S17_extracted_k141_33156 | Not-determined |
| S17_extracted_k141_3279 | Low-quality |
| S17_extracted_k141_143979 | Low-quality |
| S17_extracted_k141_158671 | Low-quality |
| S17_extracted_k141_143981 | Not-determined |
| S17_extracted_k141_158674 | Not-determined |
| S17_extracted_k141_48002 | Not-determined |
| S17_extracted_k141_25196 | Not-determined |
| S17_extracted_k141_173019 | Not-determined |
| S17_extracted_k141_113973 | Not-determined |
| S17_extracted_k141_69998 | Low-quality |
| S17_extracted_k141_173020 | Low-quality |
| S17_extracted_k141_69999 | Not-determined |
| S17_extracted_k141_77220 | Not-determined |
| S17_extracted_k141_48005 | Not-determined |
| S17_extracted_k141_77221 | Not-determined |
| S17_extracted_k141_136174 | Not-determined |
| S17_extracted_k141_3284 | Not-determined |
| S17_extracted_k141_40834 | Not-determined |
| S17_extracted_k141_99643 | Not-determined |
| S17_extracted_k141_33161 | Not-determined |
| S17_extracted_k141_136180 | Not-determined |
| S17_extracted_k141_77224 | Not-determined |
| S17_extracted_k141_48012 | Not-determined |
| S17_extracted_k141_121506 | Not-determined |
| S17_extracted_k141_91878 | Not-determined |
| S17_extracted_k141_33168 | Not-determined |
| S17_extracted_k141_165815 | Not-determined |
| S17_extracted_k141_173030 | Not-determined |
| S17_extracted_k141_48014 | Not-determined |
| S17_extracted_k141_91880 | Low-quality |
| S17_extracted_k141_144003 | Not-determined |
| S17_extracted_k141_165819 | Not-determined |
| S17_extracted_k141_40836 | Low-quality |
| S17_extracted_k141_40837 | Not-determined |
| S17_extracted_k141_17864 | Not-determined |
| S17_extracted_k141_55252 | Low-quality |
| S17_extracted_k141_3300 | Not-determined |
| S17_extracted_k141_70006 | Not-determined |
| S17_extracted_k141_151547 | Low-quality |
| S17_extracted_k141_165826 | Not-determined |
| S17_extracted_k141_48028 | Not-determined |
| S17_extracted_k141_113987 | Not-determined |
| S17_extracted_k141_17869 | Low-quality |
| S17_extracted_k141_40843 | Not-determined |
| S17_extracted_k141_48030 | Not-determined |
| S17_extracted_k141_70012 | Not-determined |
| S17_extracted_k141_10821 | Not-determined |
| S17_extracted_k141_55258 | Not-determined |
| S17_extracted_k141_70013 | Not-determined |
| S17_extracted_k141_3310 | Not-determined |
| S17_extracted_k141_113993 | Not-determined |
| S17_extracted_k141_83867 | Not-determined |
| S17_extracted_k141_55263 | Not-determined |
| S17_extracted_k141_165831 | Not-determined |
| S17_extracted_k141_173041 | Not-determined |
| S17_extracted_k141_77242 | Not-determined |
| S17_extracted_k141_10827 | Not-determined |
| S17_extracted_k141_113996 | Not-determined |
| S17_extracted_k141_83870 | Not-determined |
| S17_extracted_k141_151557 | Not-determined |
| S17_extracted_k141_33184 | Not-determined |
| S17_extracted_k141_128640 | Not-determined |
| S17_extracted_k141_121521 | Not-determined |
| S17_extracted_k141_99655 | Low-quality |
| S17_extracted_k141_33187 | Not-determined |
| S17_extracted_k141_77247 | Not-determined |
| S17_extracted_k141_55267 | Not-determined |
| S17_extracted_k141_121525 | Not-determined |
| S17_extracted_k141_99658 | Not-determined |
| S17_extracted_k141_114000 | Not-determined |
| S17_extracted_k141_77251 | Not-determined |
| S17_extracted_k141_40856 | Not-determined |
| S17_extracted_k141_128643 | Not-determined |
| S17_extracted_k141_128647 | Not-determined |
| S17_extracted_k141_77254 | Not-determined |
| S17_extracted_k141_128648 | Not-determined |
| S17_extracted_k141_48037 | Not-determined |
| S17_extracted_k141_3324 | Not-determined |
| S17_extracted_k141_165841 | Not-determined |
| S17_extracted_k141_121530 | Not-determined |
| S17_extracted_k141_33193 | Not-determined |
| S17_extracted_k141_128655 | Not-determined |
| S17_extracted_k141_151567 | Low-quality |
| S17_extracted_k141_99677 | Not-determined |
| S17_extracted_k141_173064 | Not-determined |
| S17_extracted_k141_91892 | Not-determined |
| S17_extracted_k141_165845 | Not-determined |
| S17_extracted_k141_128660 | Not-determined |
| S17_extracted_k141_48043 | Not-determined |
| S17_extracted_k141_48044 | Not-determined |
| S17_extracted_k141_165847 | Not-determined |
| S17_extracted_k141_91898 | Not-determined |
| S17_extracted_k141_83886 | Not-determined |
| S17_extracted_k141_158699 | Not-determined |
| S17_extracted_k141_99687 | Not-determined |
| S17_extracted_k141_3339 | Not-determined |
| S17_extracted_k141_144014 | Low-quality |
| S17_extracted_k141_55291 | Not-determined |
| S17_extracted_k141_158703 | Not-determined |
| S17_extracted_k141_114013 | Not-determined |
| S17_extracted_k141_128666 | Not-determined |
| S17_extracted_k141_33205 | Not-determined |
| S17_extracted_k141_33207 | Not-determined |
| S17_extracted_k141_173078 | Not-determined |
| S17_extracted_k141_10850 | Not-determined |
| S17_extracted_k141_128675 | Not-determined |
| S17_extracted_k141_144021 | Not-determined |
| S17_extracted_k141_77273 | Not-determined |
| S17_extracted_k141_114019 | Low-quality |
| S17_extracted_k141_77275 | Not-determined |
| S17_extracted_k141_33210 | Low-quality |
| S17_extracted_k141_99696 | Not-determined |
| S17_extracted_k141_10861 | Not-determined |
| S17_extracted_k141_136201 | Not-determined |
| S17_extracted_k141_121533 | Not-determined |
| S17_extracted_k141_107415 | Not-determined |
| S17_extracted_k141_17895 | Not-determined |
| S17_extracted_k141_121536 | Not-determined |
| S17_extracted_k141_128689 | Not-determined |
| S17_extracted_k141_165852 | Low-quality |
| S17_extracted_k141_3353 | Not-determined |
| S17_extracted_k141_48049 | Not-determined |
| S17_extracted_k141_10864 | Not-determined |
| S17_extracted_k141_136204 | Not-determined |
| S17_extracted_k141_77284 | Not-determined |
| S17_extracted_k141_3355 | Not-determined |
| S17_extracted_k141_99702 | Not-determined |
| S17_extracted_k141_70028 | Not-determined |
| S17_extracted_k141_144027 | Not-determined |
| S17_extracted_k141_83903 | Not-determined |
| S17_extracted_k141_17903 | Not-determined |
| S17_extracted_k141_33217 | Not-determined |
| S17_extracted_k141_99706 | Not-determined |
| S17_extracted_k141_158715 | Not-determined |
| S17_extracted_k141_136217 | Not-determined |
| S17_extracted_k141_136218 | Not-determined |
| S17_extracted_k141_70039 | Low-quality |
| S17_extracted_k141_70040 | Not-determined |
| S17_extracted_k141_99713 | Not-determined |
| S17_extracted_k141_121550 | Low-quality |
| S17_extracted_k141_10871 | Not-determined |
| S17_extracted_k141_165863 | Not-determined |
| S17_extracted_k141_121554 | Not-determined |
| S17_extracted_k141_136226 | Not-determined |
| S17_extracted_k141_70044 | Not-determined |
| S17_extracted_k141_3365 | Not-determined |
| S17_extracted_k141_158725 | Not-determined |
| S17_extracted_k141_40874 | Low-quality |
| S17_extracted_k141_158728 | Low-quality |
| S17_extracted_k141_99720 | Not-determined |
| S17_extracted_k141_158729 | Not-determined |
| S17_extracted_k141_3367 | Not-determined |
| S17_extracted_k141_114027 | Not-determined |
| S17_extracted_k141_158730 | Not-determined |
| S17_extracted_k141_17912 | Not-determined |
| S17_extracted_k141_136232 | Not-determined |
| S17_extracted_k141_25205 | Not-determined |
| S17_extracted_k141_10879 | Not-determined |
| S17_extracted_k141_107432 | Not-determined |
| S17_extracted_k141_3370 | Low-quality |
| S17_extracted_k141_128701 | Not-determined |
| S17_extracted_k141_158735 | Not-determined |
| S17_extracted_k141_3372 | Not-determined |
| S17_extracted_k141_121560 | Low-quality |
| S17_extracted_k141_158736 | Not-determined |
| S17_extracted_k141_17915 | Not-determined |
| S17_extracted_k141_70049 | Low-quality |
| S17_extracted_k141_136241 | Low-quality |
| S17_extracted_k141_3375 | Not-determined |
| S17_extracted_k141_136243 | Not-determined |
| S17_extracted_k141_77308 | Not-determined |
| S17_extracted_k141_33237 | Low-quality |
| S17_extracted_k141_40880 | Not-determined |
| S17_extracted_k141_77310 | Not-determined |
| S17_extracted_k141_3382 | Not-determined |
| S17_extracted_k141_33240 | Low-quality |
| S17_extracted_k141_48082 | Not-determined |
| S17_extracted_k141_70059 | Not-determined |
| S17_extracted_k141_25216 | Not-determined |
| S17_extracted_k141_158744 | Not-determined |
| S17_extracted_k141_158746 | Not-determined |
| S17_extracted_k141_121563 | Not-determined |
| S17_extracted_k141_40886 | Not-determined |
| S17_extracted_k141_33247 | Not-determined |
| S17_extracted_k141_121564 | Not-determined |
| S17_extracted_k141_107450 | Not-determined |
| S17_extracted_k141_121565 | Not-determined |
| S17_extracted_k141_91928 | Low-quality |
| S17_extracted_k141_121566 | Not-determined |
| S17_extracted_k141_62209 | Not-determined |
| S17_extracted_k141_107452 | Not-determined |
| S17_extracted_k141_3390 | Not-determined |
| S17_extracted_k141_33252 | Not-determined |
| S17_extracted_k141_40891 | Not-determined |
| S17_extracted_k141_107456 | Low-quality |
| S17_extracted_k141_10893 | Not-determined |
| S17_extracted_k141_158753 | Not-determined |
| S17_extracted_k141_17929 | Not-determined |
| S17_extracted_k141_62216 | Not-determined |
| S17_extracted_k141_62218 | Not-determined |
| S17_extracted_k141_70068 | Not-determined |
| S17_extracted_k141_48097 | Not-determined |
| S17_extracted_k141_3401 | Not-determined |
| S17_extracted_k141_40900 | Not-determined |
| S17_extracted_k141_48099 | Not-determined |
| S17_extracted_k141_40901 | Not-determined |
| S17_extracted_k141_165873 | Low-quality |
| S17_extracted_k141_136255 | Not-determined |
| S17_extracted_k141_136257 | Not-determined |
| S17_extracted_k141_70074 | Not-determined |
| S17_extracted_k141_136258 | Not-determined |
| S17_extracted_k141_144046 | Low-quality |
| S17_extracted_k141_144047 | Not-determined |
| S17_extracted_k141_70076 | Not-determined |
| S17_extracted_k141_99750 | Not-determined |
| S17_extracted_k141_158765 | Not-determined |
| S17_extracted_k141_33268 | Not-determined |
| S17_extracted_k141_70083 | Low-quality |
| S17_extracted_k141_165881 | Not-determined |
| S17_extracted_k141_158767 | Not-determined |
| S17_extracted_k141_128721 | Not-determined |
| S17_extracted_k141_3420 | Not-determined |
| S17_extracted_k141_25239 | Not-determined |
| S17_extracted_k141_70090 | Not-determined |
| S17_extracted_k141_165889 | Not-determined |
| S17_extracted_k141_3426 | Not-determined |
| S17_extracted_k141_17948 | Low-quality |
| S17_extracted_k141_99757 | Low-quality |
| S17_extracted_k141_99759 | Not-determined |
| S17_extracted_k141_151575 | Low-quality |
| S17_extracted_k141_128739 | Not-determined |
| S17_extracted_k141_91960 | Not-determined |
| S17_extracted_k141_83940 | Not-determined |
| S17_extracted_k141_121596 | Not-determined |
| S17_extracted_k141_114063 | Not-determined |
| S17_extracted_k141_114064 | Not-determined |
| S17_extracted_k141_40918 | Not-determined |
| S17_extracted_k141_33288 | Not-determined |
| S17_extracted_k141_99761 | Not-determined |
| S17_extracted_k141_70100 | Low-quality |
| S17_extracted_k141_77323 | Not-determined |
| S17_extracted_k141_121598 | Not-determined |
| S17_extracted_k141_99764 | Not-determined |
| S17_extracted_k141_107470 | Not-determined |
| S17_extracted_k141_55324 | Not-determined |
| S17_extracted_k141_55325 | Not-determined |
| S17_extracted_k141_33290 | Not-determined |
| S17_extracted_k141_158798 | Not-determined |
| S17_extracted_k141_25258 | Not-determined |
| S17_extracted_k141_77325 | Low-quality |
| S17_extracted_k141_48123 | Not-determined |
| S17_extracted_k141_25260 | Not-determined |
| S17_extracted_k141_128754 | Not-determined |
| S17_extracted_k141_107478 | Not-determined |
| S17_extracted_k141_77328 | Not-determined |
| S17_extracted_k141_128755 | Not-determined |
| S17_extracted_k141_128756 | Not-determined |
| S17_extracted_k141_128758 | Not-determined |
| S17_extracted_k141_107479 | Low-quality |
| S17_extracted_k141_91963 | Low-quality |
| S17_extracted_k141_158812 | Not-determined |
| S17_extracted_k141_99778 | Not-determined |
| S17_extracted_k141_173107 | Not-determined |
| S17_extracted_k141_151594 | Not-determined |
| S17_extracted_k141_128767 | Not-determined |
| S17_extracted_k141_121609 | Low-quality |
| S17_extracted_k141_151596 | Not-determined |
| S17_extracted_k141_151597 | Not-determined |
| S17_extracted_k141_83961 | Not-determined |
| S17_extracted_k141_3455 | Not-determined |
| S17_extracted_k141_114073 | Not-determined |
| S17_extracted_k141_33300 | Not-determined |
| S17_extracted_k141_83967 | Not-determined |
| S17_extracted_k141_77342 | Not-determined |
| S17_extracted_k141_91979 | Not-determined |
| S17_extracted_k141_173115 | Low-quality |
| S17_extracted_k141_3464 | Low-quality |
| S17_extracted_k141_158824 | Low-quality |
| S17_extracted_k141_151608 | Not-determined |
| S17_extracted_k141_151609 | Not-determined |
| S17_extracted_k141_70109 | Not-determined |
| S17_extracted_k141_70110 | Not-determined |
| S17_extracted_k141_173117 | Not-determined |
| S17_extracted_k141_70111 | Not-determined |
| S17_extracted_k141_114077 | Low-quality |
| S17_extracted_k141_3470 | Not-determined |
| S17_extracted_k141_3472 | Not-determined |
| S17_extracted_k141_151613 | Low-quality |
| S17_extracted_k141_55350 | Not-determined |
| S17_extracted_k141_99790 | Low-quality |
| S17_extracted_k141_62243 | Not-determined |
| S17_extracted_k141_173124 | Not-determined |
| S17_extracted_k141_55351 | Low-quality |
| S17_extracted_k141_107500 | Not-determined |
| S17_extracted_k141_99794 | Not-determined |
| S17_extracted_k141_99796 | Not-determined |
| S17_extracted_k141_173126 | Not-determined |
| S17_extracted_k141_62247 | Not-determined |
| S17_extracted_k141_40931 | Not-determined |
| S17_extracted_k141_128783 | Not-determined |
| S17_extracted_k141_107507 | Not-determined |
| S17_extracted_k141_17961 | Not-determined |
| S17_extracted_k141_70128 | Not-determined |
| S17_extracted_k141_17962 | Not-determined |
| S17_extracted_k141_91989 | Not-determined |
| S17_extracted_k141_17963 | Not-determined |
| S17_extracted_k141_62252 | Not-determined |
| S17_extracted_k141_77353 | Not-determined |
| S17_extracted_k141_83986 | Not-determined |
| S17_extracted_k141_17965 | Low-quality |
| S17_extracted_k141_70132 | Not-determined |
| S17_extracted_k141_107508 | Not-determined |
| S17_extracted_k141_3486 | Not-determined |
| S17_extracted_k141_114093 | Not-determined |
| S17_extracted_k141_70135 | Not-determined |
| S17_extracted_k141_151625 | Not-determined |
| S17_extracted_k141_70136 | Not-determined |
| S17_extracted_k141_70138 | Not-determined |
| S17_extracted_k141_62259 | Not-determined |
| S17_extracted_k141_33308 | Not-determined |
| S17_extracted_k141_114097 | Not-determined |
| S17_extracted_k141_151630 | Not-determined |
| S17_extracted_k141_70139 | Not-determined |
| S17_extracted_k141_17972 | Not-determined |
| S17_extracted_k141_62266 | Not-determined |
| S17_extracted_k141_91998 | Low-quality |
| S17_extracted_k141_107517 | Not-determined |
| S17_extracted_k141_158850 | Not-determined |
| S17_extracted_k141_70147 | Not-determined |
| S17_extracted_k141_40935 | Not-determined |
| S17_extracted_k141_173144 | Not-determined |
| S17_extracted_k141_62275 | Not-determined |
| S17_extracted_k141_33322 | Not-determined |
| S17_extracted_k141_84003 | Not-determined |
| S17_extracted_k141_17983 | Not-determined |
| S17_extracted_k141_55375 | Not-determined |
| S17_extracted_k141_40948 | Not-determined |
| S17_extracted_k141_99804 | Not-determined |
| S17_extracted_k141_25302 | Low-quality |
| S17_extracted_k141_99809 | Not-determined |
| S17_extracted_k141_99810 | Not-determined |
| S17_extracted_k141_40956 | Low-quality |
| S17_extracted_k141_99812 | Not-determined |
| S17_extracted_k141_17995 | Not-determined |
| S17_extracted_k141_55386 | Not-determined |
| S17_extracted_k141_92013 | Not-determined |
| S17_extracted_k141_55388 | Not-determined |
| S17_extracted_k141_17997 | Not-determined |
| S17_extracted_k141_55389 | Not-determined |
| S17_extracted_k141_158860 | Not-determined |
| S17_extracted_k141_25310 | Not-determined |
| S17_extracted_k141_25311 | Not-determined |
| S17_extracted_k141_77391 | Not-determined |
| S17_extracted_k141_99816 | Not-determined |
| S17_extracted_k141_77393 | Not-determined |
| S17_extracted_k141_55390 | Not-determined |
| S17_extracted_k141_151660 | Low-quality |
| S17_extracted_k141_99818 | Not-determined |
| S17_extracted_k141_151661 | Not-determined |
| S17_extracted_k141_48131 | Low-quality |
| S17_extracted_k141_33334 | Not-determined |
| S17_extracted_k141_84017 | Not-determined |
| S17_extracted_k141_173161 | Not-determined |
| S17_extracted_k141_144061 | Low-quality |
| S17_extracted_k141_70173 | Not-determined |
| S17_extracted_k141_40959 | Not-determined |
| S17_extracted_k141_173166 | Not-determined |
| S17_extracted_k141_40960 | Not-determined |
| S17_extracted_k141_48140 | Low-quality |
| S17_extracted_k141_114104 | Not-determined |
| S17_extracted_k141_25324 | Low-quality |
| S17_extracted_k141_136278 | Not-determined |
| S17_extracted_k141_33347 | Not-determined |
| S17_extracted_k141_33348 | Not-determined |
| S17_extracted_k141_136282 | Low-quality |
| S17_extracted_k141_18013 | Not-determined |
| S17_extracted_k141_18015 | Not-determined |
| S17_extracted_k141_25328 | Not-determined |
| S17_extracted_k141_62291 | Not-determined |
| S17_extracted_k141_84030 | Not-determined |
| S17_extracted_k141_136285 | Not-determined |
| S17_extracted_k141_136286 | Not-determined |
| S17_extracted_k141_77414 | Not-determined |
| S17_extracted_k141_70184 | Not-determined |
| S17_extracted_k141_136293 | Not-determined |
| S17_extracted_k141_173175 | Not-determined |
| S17_extracted_k141_173176 | Not-determined |
| S17_extracted_k141_128819 | Not-determined |
| S17_extracted_k141_165917 | Not-determined |
| S17_extracted_k141_165919 | Not-determined |
| S17_extracted_k141_92035 | Not-determined |
| S17_extracted_k141_165922 | Not-determined |
| S17_extracted_k141_70190 | Not-determined |
| S17_extracted_k141_55396 | Not-determined |
| S17_extracted_k141_70192 | Not-determined |
| S17_extracted_k141_144068 | Not-determined |
| S17_extracted_k141_121614 | Not-determined |
| S17_extracted_k141_33355 | Not-determined |
| S17_extracted_k141_40981 | Not-determined |
| S17_extracted_k141_25343 | Not-determined |
| S17_extracted_k141_144077 | Not-determined |
| S17_extracted_k141_173186 | Not-determined |
| S17_extracted_k141_128827 | Not-determined |
| S17_extracted_k141_18032 | Not-determined |
| S17_extracted_k141_48154 | Not-determined |
| S17_extracted_k141_128829 | Not-determined |
| S17_extracted_k141_18036 | Not-determined |
| S17_extracted_k141_114120 | Not-determined |
| S17_extracted_k141_18038 | Not-determined |
| S17_extracted_k141_158886 | Not-determined |
| S17_extracted_k141_33364 | Not-determined |
| S17_extracted_k141_40991 | Not-determined |
| S17_extracted_k141_165929 | Not-determined |
| S17_extracted_k141_18045 | Not-determined |
| S17_extracted_k141_114126 | Not-determined |
| S17_extracted_k141_114127 | Not-determined |
| S17_extracted_k141_62315 | Not-determined |
| S17_extracted_k141_48165 | Not-determined |
| S17_extracted_k141_158895 | Not-determined |
| S17_extracted_k141_84050 | Not-determined |
| S17_extracted_k141_25353 | Not-determined |
| S17_extracted_k141_158899 | Not-determined |
| S17_extracted_k141_55405 | Not-determined |
| S17_extracted_k141_18058 | Low-quality |
| S17_extracted_k141_165933 | Not-determined |
| S17_extracted_k141_121630 | Not-determined |
| S17_extracted_k141_3516 | Not-determined |
| S17_extracted_k141_55409 | Not-determined |
| S17_extracted_k141_144088 | Not-determined |
| S17_extracted_k141_144089 | Not-determined |
| S17_extracted_k141_158903 | Low-quality |
| S17_extracted_k141_173202 | Not-determined |
| S17_extracted_k141_3520 | Not-determined |
| S17_extracted_k141_77424 | Not-determined |
| S17_extracted_k141_84055 | Not-determined |
| S17_extracted_k141_173204 | Not-determined |
| S17_extracted_k141_165935 | Low-quality |
| S17_extracted_k141_62324 | Not-determined |
| S17_extracted_k141_173206 | Not-determined |
| S17_extracted_k141_173207 | Not-determined |
| S17_extracted_k141_77426 | Not-determined |
| S17_extracted_k141_128843 | Not-determined |
| S17_extracted_k141_55419 | Not-determined |
| S17_extracted_k141_3523 | Not-determined |
| S17_extracted_k141_114148 | Not-determined |
| S17_extracted_k141_77427 | Low-quality |
| S17_extracted_k141_3527 | Not-determined |
| S17_extracted_k141_25366 | Low-quality |
| S17_extracted_k141_33368 | Not-determined |
| S17_extracted_k141_173212 | Not-determined |
| S17_extracted_k141_25369 | Not-determined |
| S17_extracted_k141_99844 | Not-determined |
| S17_extracted_k141_128848 | Not-determined |
| S17_extracted_k141_158912 | Not-determined |
| S17_extracted_k141_48181 | Not-determined |
| S17_extracted_k141_99845 | Not-determined |
| S17_extracted_k141_55425 | Low-quality |
| S17_extracted_k141_114154 | Not-determined |
| S17_extracted_k141_33376 | Not-determined |
| S17_extracted_k141_114155 | Not-determined |
| S17_extracted_k141_48186 | Not-determined |
| S17_extracted_k141_114156 | Not-determined |
| S17_extracted_k141_114160 | Low-quality |
| S17_extracted_k141_144107 | Low-quality |
| S17_extracted_k141_62339 | Not-determined |
| S17_extracted_k141_33379 | Not-determined |
| S17_extracted_k141_136329 | Not-determined |
| S17_extracted_k141_33381 | Not-determined |
| S17_extracted_k141_25378 | Low-quality |
| S17_extracted_k141_173230 | Not-determined |
| S17_extracted_k141_25379 | Not-determined |
| S17_extracted_k141_84069 | Not-determined |
| S17_extracted_k141_144117 | Not-determined |
| S17_extracted_k141_70226 | Not-determined |
| S17_extracted_k141_136341 | Low-quality |
| S17_extracted_k141_70227 | Not-determined |
| S17_extracted_k141_114172 | Not-determined |
| S17_extracted_k141_114175 | Not-determined |
| S17_extracted_k141_18094 | Not-determined |
| S17_extracted_k141_114177 | Not-determined |
| S17_extracted_k141_84070 | Not-determined |
| S17_extracted_k141_55442 | Not-determined |
| S17_extracted_k141_173238 | Low-quality |
| S17_extracted_k141_62349 | Not-determined |
| S17_extracted_k141_114179 | Low-quality |
| S17_extracted_k141_70234 | Not-determined |
| S17_extracted_k141_165953 | Not-determined |
| S17_extracted_k141_144125 | Not-determined |
| S17_extracted_k141_77439 | Not-determined |
| S17_extracted_k141_165954 | Not-determined |
| S17_extracted_k141_121637 | Low-quality |
| S17_extracted_k141_41005 | Low-quality |
| S17_extracted_k141_121639 | Not-determined |
| S17_extracted_k141_62357 | Not-determined |
| S17_extracted_k141_121640 | Not-determined |
| S17_extracted_k141_128868 | Not-determined |
| S17_extracted_k141_92043 | Not-determined |
| S17_extracted_k141_33391 | Not-determined |
| S17_extracted_k141_92044 | Not-determined |
| S17_extracted_k141_99865 | Low-quality |
| S17_extracted_k141_173251 | Low-quality |
| S17_extracted_k141_165959 | Not-determined |
| S17_extracted_k141_158942 | Not-determined |
| S17_extracted_k141_165960 | Not-determined |
| S17_extracted_k141_158944 | Not-determined |
| S17_extracted_k141_114188 | Not-determined |
| S17_extracted_k141_158949 | Not-determined |
| S17_extracted_k141_41010 | Not-determined |
| S17_extracted_k141_41011 | Not-determined |
| S17_extracted_k141_107575 | Not-determined |
| S17_extracted_k141_18103 | Not-determined |
| S17_extracted_k141_55456 | Not-determined |
| S17_extracted_k141_10923 | Not-determined |
| S17_extracted_k141_136370 | Not-determined |
| S17_extracted_k141_92052 | Not-determined |
| S17_extracted_k141_77454 | Not-determined |
| S17_extracted_k141_136373 | Not-determined |
| S17_extracted_k141_121643 | Not-determined |
| S17_extracted_k141_107584 | Low-quality |
| S17_extracted_k141_114195 | Not-determined |
| S17_extracted_k141_144131 | Not-determined |
| S17_extracted_k141_173263 | Low-quality |
| S17_extracted_k141_144132 | Not-determined |
| S17_extracted_k141_25400 | Not-determined |
| S17_extracted_k141_62369 | Not-determined |
| S17_extracted_k141_92057 | Not-determined |
| S17_extracted_k141_92058 | Not-determined |
| S17_extracted_k141_92061 | Not-determined |
| S17_extracted_k141_25404 | Low-quality |
| S17_extracted_k141_173271 | Not-determined |
| S17_extracted_k141_55467 | Low-quality |
| S17_extracted_k141_107589 | Not-determined |
| S17_extracted_k141_62376 | Not-determined |
| S17_extracted_k141_121649 | Not-determined |
| S17_extracted_k141_121651 | Low-quality |
| S17_extracted_k141_55469 | Not-determined |
| S17_extracted_k141_121654 | Not-determined |
| S17_extracted_k141_18110 | Not-determined |
| S17_extracted_k141_62380 | Not-determined |
| S17_extracted_k141_107595 | Not-determined |
| S17_extracted_k141_10934 | Not-determined |
| S17_extracted_k141_18115 | Not-determined |
| S17_extracted_k141_18119 | Not-determined |
| S17_extracted_k141_158968 | Not-determined |
| S17_extracted_k141_48215 | Low-quality |
| S17_extracted_k141_62387 | Not-determined |
| S17_extracted_k141_128878 | Not-determined |
| S17_extracted_k141_62388 | Low-quality |
| S17_extracted_k141_84112 | Not-determined |
| S17_extracted_k141_18123 | Not-determined |
| S17_extracted_k141_48217 | Not-determined |
| S17_extracted_k141_10947 | Low-quality |
| S17_extracted_k141_77467 | Low-quality |
| S17_extracted_k141_77469 | Not-determined |
| S17_extracted_k141_121666 | Not-determined |
| S17_extracted_k141_136387 | Not-determined |
| S17_extracted_k141_41022 | Low-quality |
| S17_extracted_k141_33416 | Low-quality |
| S17_extracted_k141_84121 | Not-determined |
| S17_extracted_k141_173294 | Not-determined |
| S17_extracted_k141_48226 | Not-determined |
| S17_extracted_k141_158982 | Not-determined |
| S17_extracted_k141_48228 | Not-determined |
| S17_extracted_k141_158984 | Not-determined |
| S17_extracted_k141_151728 | Not-determined |
| S17_extracted_k141_107605 | Not-determined |
| S17_extracted_k141_121672 | Not-determined |
| S17_extracted_k141_41030 | Not-determined |
| S17_extracted_k141_144148 | Not-determined |
| S17_extracted_k141_107609 | Not-determined |
| S17_extracted_k141_107610 | Not-determined |
| S17_extracted_k141_121678 | Not-determined |
| S17_extracted_k141_84133 | Not-determined |
| S17_extracted_k141_151734 | Not-determined |

| | |
|---|---|
| S17_extracted_k141_70284 | Not-determined |
| S17_extracted_k141_70287 | Not-determined |
| S17_extracted_k141_84134 | Not-determined |
| S17_extracted_k141_107622 | Not-determined |
| S17_extracted_k141_70293 | Not-determined |
| S17_extracted_k141_10957 | Low-quality |
| S17_extracted_k141_128897 | Not-determined |
| S17_extracted_k141_25423 | Not-determined |
| S17_extracted_k141_107628 | Low-quality |
| S17_extracted_k141_3534 | Not-determined |
| S17_extracted_k141_151748 | Low-quality |
| S17_extracted_k141_121683 | Not-determined |
| S17_extracted_k141_25426 | Not-determined |
| S17_extracted_k141_25427 | Not-determined |
| S17_extracted_k141_107636 | Low-quality |
| S17_extracted_k141_107637 | Not-determined |
| S17_extracted_k141_84151 | Low-quality |
| S17_extracted_k141_18135 | Not-determined |
| S17_extracted_k141_25430 | Not-determined |
| S17_extracted_k141_33437 | Not-determined |
| S17_extracted_k141_173312 | Not-determined |
| S17_extracted_k141_3540 | Low-quality |
| S17_extracted_k141_18145 | Not-determined |
| S17_extracted_k141_173314 | Not-determined |
| S17_extracted_k141_128915 | Not-determined |
| S17_extracted_k141_107643 | Not-determined |
| S17_extracted_k141_77500 | Not-determined |
| S17_extracted_k141_121691 | Not-determined |
| S17_extracted_k141_3543 | Not-determined |
| S17_extracted_k141_121693 | Not-determined |
| S17_extracted_k141_136417 | Not-determined |
| S17_extracted_k141_3546 | Not-determined |
| S17_extracted_k141_136419 | Not-determined |
| S17_extracted_k141_3548 | Low-quality |
| S17_extracted_k141_48244 | Not-determined |
| S17_extracted_k141_107648 | Not-determined |
| S17_extracted_k141_18153 | Not-determined |
| S17_extracted_k141_18154 | Not-determined |
| S17_extracted_k141_128918 | Not-determined |
| S17_extracted_k141_136422 | Not-determined |
| S17_extracted_k141_25446 | Not-determined |
| S17_extracted_k141_128921 | Not-determined |
| S17_extracted_k141_25451 | Not-determined |
| S17_extracted_k141_173325 | Not-determined |
| S17_extracted_k141_55475 | Not-determined |
| S17_extracted_k141_151763 | Not-determined |
| S17_extracted_k141_62419 | Not-determined |
| S17_extracted_k141_84167 | Not-determined |
| S17_extracted_k141_173327 | Not-determined |
| S17_extracted_k141_173329 | Not-determined |
| S17_extracted_k141_25455 | Not-determined |
| S17_extracted_k141_151768 | Not-determined |
| S17_extracted_k141_84172 | Not-determined |
| S17_extracted_k141_25458 | Low-quality |
| S17_extracted_k141_48254 | Not-determined |
| S17_extracted_k141_128926 | Not-determined |
| S17_extracted_k141_33457 | Not-determined |
| S17_extracted_k141_92095 | Not-determined |
| S17_extracted_k141_158997 | Not-determined |
| S17_extracted_k141_158998 | Not-determined |
| S17_extracted_k141_173334 | Not-determined |
| S17_extracted_k141_151774 | Low-quality |
| S17_extracted_k141_92097 | Not-determined |
| S17_extracted_k141_114219 | Not-determined |
| S17_extracted_k141_92098 | Not-determined |
| S17_extracted_k141_70306 | Low-quality |
| S17_extracted_k141_84181 | Not-determined |
| S17_extracted_k141_62426 | Low-quality |
| S17_extracted_k141_165980 | Not-determined |
| S17_extracted_k141_10965 | Not-determined |
| S17_extracted_k141_10967 | Not-determined |
| S17_extracted_k141_151776 | Not-determined |
| S17_extracted_k141_3554 | Not-determined |
| S17_extracted_k141_70310 | Not-determined |
| S17_extracted_k141_3556 | Not-determined |
| S17_extracted_k141_144169 | Not-determined |
| S17_extracted_k141_41043 | Not-determined |
| S17_extracted_k141_41044 | Not-determined |
| S17_extracted_k141_144175 | Not-determined |
| S17_extracted_k141_159001 | Not-determined |
| S17_extracted_k141_165989 | Not-determined |
| S17_extracted_k141_48261 | Not-determined |
| S17_extracted_k141_173337 | Low-quality |
| S17_extracted_k141_165990 | Low-quality |
| S17_extracted_k141_70317 | Not-determined |
| S17_extracted_k141_10973 | Not-determined |
| S17_extracted_k141_144180 | Not-determined |
| S17_extracted_k141_77518 | Not-determined |
| S17_extracted_k141_151783 | Not-determined |
| S17_extracted_k141_136439 | Not-determined |
| S17_extracted_k141_18180 | Not-determined |
| S17_extracted_k141_33482 | Not-determined |
| S17_extracted_k141_92109 | Low-quality |
| S17_extracted_k141_159012 | Not-determined |
| S17_extracted_k141_92112 | Not-determined |
| S17_extracted_k141_33488 | Not-determined |
| S17_extracted_k141_144187 | Not-determined |
| S17_extracted_k141_128947 | Low-quality |
| S17_extracted_k141_166002 | Not-determined |
| S17_extracted_k141_55501 | Not-determined |
| S17_extracted_k141_33491 | Not-determined |
| S17_extracted_k141_18187 | Low-quality |
| S17_extracted_k141_33494 | Not-determined |
| S17_extracted_k141_159020 | Not-determined |
| S17_extracted_k141_77525 | Not-determined |
| S17_extracted_k141_48274 | Not-determined |
| S17_extracted_k141_10996 | Low-quality |
| S17_extracted_k141_41056 | Not-determined |
| S17_extracted_k141_10997 | Not-determined |
| S17_extracted_k141_10999 | Not-determined |
| S17_extracted_k141_151786 | Low-quality |
| S17_extracted_k141_41062 | Not-determined |
| S17_extracted_k141_92125 | Not-determined |
| S17_extracted_k141_41067 | Not-determined |
| S17_extracted_k141_173347 | Not-determined |
| S17_extracted_k141_41072 | Not-determined |
| S17_extracted_k141_151792 | Not-determined |
| S17_extracted_k141_92128 | Not-determined |
| S17_extracted_k141_11010 | Low-quality |
| S17_extracted_k141_3563 | Not-determined |
| S17_extracted_k141_84208 | Not-determined |
| S17_extracted_k141_3564 | Not-determined |
| S17_extracted_k141_114225 | Not-determined |
| S17_extracted_k141_18212 | Not-determined |
| S17_extracted_k141_70331 | Not-determined |
| S17_extracted_k141_99880 | Not-determined |
| S17_extracted_k141_55525 | Not-determined |
| S17_extracted_k141_77533 | Not-determined |
| S17_extracted_k141_77534 | Not-determined |
| S17_extracted_k141_62468 | Not-determined |
| S17_extracted_k141_70335 | Not-determined |
| S17_extracted_k141_48302 | Not-determined |
| S17_extracted_k141_70341 | Not-determined |
| S17_extracted_k141_33513 | Low-quality |
| S17_extracted_k141_55528 | Low-quality |
| S17_extracted_k141_3574 | Not-determined |
| S17_extracted_k141_159045 | Not-determined |
| S17_extracted_k141_33517 | Not-determined |
| S17_extracted_k141_128977 | Not-determined |
| S17_extracted_k141_3577 | Not-determined |
| S17_extracted_k141_41076 | Not-determined |
| S17_extracted_k141_114242 | Not-determined |
| S17_extracted_k141_84216 | Low-quality |
| S17_extracted_k141_33520 | Not-determined |
| S17_extracted_k141_128979 | Low-quality |
| S17_extracted_k141_18230 | Not-determined |
| S17_extracted_k141_114247 | Not-determined |
| S17_extracted_k141_114249 | Not-determined |
| S17_extracted_k141_18234 | Not-determined |
| S17_extracted_k141_92152 | Not-determined |
| S17_extracted_k141_173369 | Not-determined |
| S17_extracted_k141_33525 | Not-determined |
| S17_extracted_k141_114251 | Not-determined |
| S17_extracted_k141_114252 | Not-determined |
| S17_extracted_k141_33527 | Not-determined |
| S17_extracted_k141_33528 | Not-determined |
| S17_extracted_k141_41082 | Not-determined |
| S17_extracted_k141_114256 | Not-determined |
| S17_extracted_k141_114258 | Not-determined |
| S17_extracted_k141_173374 | Low-quality |
| S17_extracted_k141_33533 | Not-determined |
| S17_extracted_k141_48321 | Not-determined |
| S17_extracted_k141_151823 | Not-determined |
| S17_extracted_k141_144234 | Not-determined |
| S17_extracted_k141_25486 | Low-quality |
| S17_extracted_k141_55534 | Not-determined |
| S17_extracted_k141_33538 | Not-determined |
| S17_extracted_k141_33539 | Not-determined |
| S17_extracted_k141_144241 | Not-determined |
| S17_extracted_k141_48328 | Not-determined |
| S17_extracted_k141_48329 | Not-determined |
| S17_extracted_k141_136470 | Not-determined |
| S17_extracted_k141_136471 | Not-determined |
| S17_extracted_k141_136472 | Low-quality |
| S17_extracted_k141_11026 | Not-determined |
| S17_extracted_k141_92163 | Not-determined |
| S17_extracted_k141_70360 | Not-determined |
| S17_extracted_k141_166021 | Not-determined |
| S17_extracted_k141_173379 | Not-determined |
| S17_extracted_k141_173380 | Not-determined |
| S17_extracted_k141_92166 | Low-quality |
| S17_extracted_k141_166024 | Not-determined |
| S17_extracted_k141_114279 | Not-determined |
| S17_extracted_k141_25496 | Not-determined |
| S17_extracted_k141_41104 | Not-determined |
| S17_extracted_k141_128991 | Not-determined |
| S17_extracted_k141_173390 | Not-determined |
| S17_extracted_k141_144246 | Low-quality |
| S17_extracted_k141_48339 | Not-determined |
| S17_extracted_k141_166029 | Not-determined |
| S17_extracted_k141_18240 | Low-quality |
| S17_extracted_k141_70361 | Low-quality |
| S17_extracted_k141_18241 | Not-determined |
| S17_extracted_k141_99905 | Not-determined |
| S17_extracted_k141_173392 | Not-determined |
| S17_extracted_k141_33543 | Not-determined |
| S17_extracted_k141_114288 | Not-determined |
| S17_extracted_k141_173395 | Not-determined |
| S17_extracted_k141_99910 | Not-determined |
| S17_extracted_k141_77566 | Not-determined |
| S17_extracted_k141_128997 | Not-determined |
| S17_extracted_k141_41109 | Not-determined |
| S17_extracted_k141_41110 | Not-determined |
| S17_extracted_k141_3606 | Not-determined |
| S17_extracted_k141_166036 | Not-determined |
| S17_extracted_k141_70370 | Not-determined |
| S17_extracted_k141_70372 | Not-determined |
| S17_extracted_k141_11050 | Not-determined |
| S17_extracted_k141_114302 | Not-determined |
| S17_extracted_k141_18252 | Not-determined |
| S17_extracted_k141_129007 | Not-determined |
| S17_extracted_k141_129010 | Not-determined |
| S17_extracted_k141_129011 | Not-determined |
| S17_extracted_k141_99914 | Not-determined |
| S17_extracted_k141_33556 | Not-determined |
| S17_extracted_k141_41119 | Not-determined |
| S17_extracted_k141_173411 | Not-determined |
| S17_extracted_k141_33559 | Not-determined |
| S17_extracted_k141_18261 | Low-quality |
| S17_extracted_k141_129014 | Not-determined |
| S17_extracted_k141_18264 | Not-determined |
| S17_extracted_k141_33561 | Not-determined |
| S17_extracted_k141_77573 | Not-determined |
| S17_extracted_k141_99920 | Not-determined |
| S17_extracted_k141_151843 | Not-determined |
| S17_extracted_k141_3622 | Not-determined |
| S17_extracted_k141_144257 | Not-determined |
| S17_extracted_k141_129018 | Not-determined |
| S17_extracted_k141_62480 | Not-determined |
| S17_extracted_k141_77580 | Not-determined |
| S17_extracted_k141_33566 | Not-determined |
| S17_extracted_k141_3630 | Not-determined |
| S17_extracted_k141_25515 | Not-determined |
| S17_extracted_k141_121738 | Not-determined |
| S17_extracted_k141_114316 | Not-determined |
| S17_extracted_k141_144267 | Not-determined |
| S17_extracted_k141_166058 | Not-determined |
| S17_extracted_k141_92191 | Not-determined |
| S17_extracted_k141_166059 | Not-determined |
| S17_extracted_k141_151853 | Not-determined |
| S17_extracted_k141_159088 | Not-determined |
| S17_extracted_k141_92194 | Not-determined |
| S17_extracted_k141_159090 | Not-determined |
| S17_extracted_k141_99928 | Not-determined |
| S17_extracted_k141_136478 | Not-determined |
| S17_extracted_k141_77589 | Not-determined |
| S17_extracted_k141_114320 | Not-determined |
| S17_extracted_k141_144268 | Low-quality |
| S17_extracted_k141_3644 | Not-determined |
| S17_extracted_k141_166065 | Not-determined |
| S17_extracted_k141_136483 | Not-determined |
| S17_extracted_k141_129028 | Low-quality |
| S17_extracted_k141_144269 | Not-determined |
| S17_extracted_k141_11066 | Not-determined |
| S17_extracted_k141_33578 | Not-determined |
| S17_extracted_k141_144273 | Not-determined |
| S17_extracted_k141_33581 | Not-determined |
| S17_extracted_k141_121747 | Not-determined |
| S17_extracted_k141_84242 | Not-determined |
| S17_extracted_k141_92201 | Not-determined |
| S17_extracted_k141_151864 | Low-quality |
| S17_extracted_k141_33584 | Not-determined |
| S17_extracted_k141_159105 | Not-determined |
| S17_extracted_k141_107680 | Low-quality |
| S17_extracted_k141_70388 | Not-determined |
| S17_extracted_k141_166068 | Not-determined |
| S17_extracted_k141_55556 | Low-quality |
| S17_extracted_k141_84247 | Not-determined |
| S17_extracted_k141_99945 | Not-determined |
| S17_extracted_k141_129032 | Not-determined |
| S17_extracted_k141_144278 | Not-determined |
| S17_extracted_k141_3652 | Not-determined |
| S17_extracted_k141_136489 | Not-determined |
| S17_extracted_k141_84248 | Not-determined |
| S17_extracted_k141_173431 | Low-quality |
| S17_extracted_k141_107684 | Low-quality |
| S17_extracted_k141_18286 | Not-determined |
| S17_extracted_k141_166073 | Not-determined |
| S17_extracted_k141_129037 | Not-determined |
| S17_extracted_k141_129038 | Not-determined |
| S17_extracted_k141_18289 | Not-determined |
| S17_extracted_k141_70392 | Not-determined |
| S17_extracted_k141_77597 | Not-determined |
| S17_extracted_k141_70395 | Low-quality |
| S17_extracted_k141_114337 | Not-determined |
| S17_extracted_k141_70397 | Not-determined |
| S17_extracted_k141_129045 | Not-determined |
| S17_extracted_k141_33590 | Not-determined |
| S17_extracted_k141_77602 | Not-determined |
| S17_extracted_k141_70400 | Not-determined |
| S17_extracted_k141_92214 | Not-determined |
| S17_extracted_k141_121761 | Not-determined |
| S17_extracted_k141_70404 | Not-determined |
| S17_extracted_k141_144284 | Not-determined |
| S17_extracted_k141_151873 | Low-quality |
| S17_extracted_k141_114346 | Not-determined |
| S17_extracted_k141_114349 | Low-quality |
| S17_extracted_k141_33592 | Not-determined |
| S17_extracted_k141_92217 | Not-determined |
| S17_extracted_k141_173444 | Not-determined |
| S17_extracted_k141_55576 | Low-quality |
| S17_extracted_k141_114352 | Low-quality |
| S17_extracted_k141_33599 | Low-quality |
| S17_extracted_k141_70409 | Not-determined |
| S17_extracted_k141_33601 | Not-determined |
| S17_extracted_k141_92221 | Not-determined |
| S17_extracted_k141_11085 | Not-determined |
| S17_extracted_k141_107690 | Not-determined |
| S17_extracted_k141_3670 | Not-determined |
| S17_extracted_k141_3671 | Not-determined |
| S17_extracted_k141_129068 | Not-determined |
| S17_extracted_k141_151885 | Low-quality |
| S17_extracted_k141_55586 | Not-determined |
| S17_extracted_k141_55587 | Not-determined |
| S17_extracted_k141_55590 | Not-determined |
| S17_extracted_k141_173456 | Low-quality |
| S17_extracted_k141_41145 | Not-determined |
| S17_extracted_k141_151890 | Not-determined |
| S17_extracted_k141_107699 | Low-quality |
| S17_extracted_k141_77619 | Low-quality |
| S17_extracted_k141_77623 | Not-determined |
| S17_extracted_k141_62496 | Not-determined |
| S17_extracted_k141_107706 | Not-determined |
| S17_extracted_k141_11103 | Low-quality |
| S17_extracted_k141_114368 | Not-determined |
| S17_extracted_k141_159128 | Not-determined |
| S17_extracted_k141_159129 | Not-determined |
| S17_extracted_k141_62500 | Not-determined |
| S17_extracted_k141_159132 | Low-quality |
| S17_extracted_k141_33607 | Low-quality |
| S17_extracted_k141_92225 | Not-determined |
| S17_extracted_k141_92226 | Low-quality |
| S17_extracted_k141_84273 | Not-determined |
| S17_extracted_k141_144308 | Not-determined |
| S17_extracted_k141_55608 | Not-determined |
| S17_extracted_k141_92232 | Not-determined |
| S17_extracted_k141_70416 | Low-quality |
| S17_extracted_k141_33617 | Low-quality |
| S17_extracted_k141_107712 | Not-determined |
| S17_extracted_k141_159139 | Not-determined |
| S17_extracted_k141_144314 | Not-determined |
| S17_extracted_k141_151900 | Not-determined |
| S17_extracted_k141_92236 | Not-determined |
| S17_extracted_k141_92237 | Not-determined |
| S17_extracted_k141_144318 | Not-determined |
| S17_extracted_k141_144320 | Not-determined |
| S17_extracted_k141_159145 | Not-determined |
| S17_extracted_k141_3704 | Not-determined |
| S17_extracted_k141_166103 | Not-determined |
| S17_extracted_k141_3705 | Low-quality |
| S17_extracted_k141_92238 | Not-determined |
| S17_extracted_k141_92242 | Not-determined |
| S17_extracted_k141_70425 | Not-determined |
| S17_extracted_k141_144326 | Not-determined |
| S17_extracted_k141_3708 | Not-determined |
| S17_extracted_k141_55617 | Not-determined |
| S17_extracted_k141_129111 | Not-determined |
| S17_extracted_k141_166110 | Not-determined |
| S17_extracted_k141_33629 | Not-determined |
| S17_extracted_k141_41175 | Low-quality |
| S17_extracted_k141_159152 | Not-determined |
| S17_extracted_k141_3713 | Low-quality |
| S17_extracted_k141_3715 | Not-determined |
| S17_extracted_k141_121777 | Not-determined |
| S17_extracted_k141_18303 | Not-determined |
| S17_extracted_k141_151907 | Low-quality |
| S17_extracted_k141_151908 | Not-determined |
| S17_extracted_k141_41178 | Not-determined |
| S17_extracted_k141_166119 | Not-determined |
| S17_extracted_k141_70439 | Not-determined |
| S17_extracted_k141_33636 | Not-determined |
| S17_extracted_k141_3720 | Not-determined |
| S17_extracted_k141_121786 | Not-determined |
| S17_extracted_k141_3721 | Not-determined |
| S17_extracted_k141_151912 | Low-quality |
| S17_extracted_k141_173493 | Not-determined |
| S17_extracted_k141_3725 | Not-determined |
| S17_extracted_k141_55628 | Not-determined |
| S17_extracted_k141_55630 | Not-determined |
| S17_extracted_k141_114404 | Not-determined |
| S17_extracted_k141_41187 | Not-determined |
| S17_extracted_k141_70447 | Not-determined |
| S17_extracted_k141_41188 | Not-determined |
| S17_extracted_k141_70451 | Low-quality |
| S17_extracted_k141_151916 | Low-quality |
| S17_extracted_k141_33644 | Low-quality |
| S17_extracted_k141_77661 | Not-determined |
| S17_extracted_k141_41194 | Not-determined |
| S17_extracted_k141_114412 | Not-determined |
| S17_extracted_k141_166122 | Not-determined |
| S17_extracted_k141_92252 | Not-determined |
| S17_extracted_k141_166123 | Not-determined |
| S17_extracted_k141_92253 | Not-determined |
| S17_extracted_k141_77666 | Not-determined |
| S17_extracted_k141_55645 | Not-determined |
| S17_extracted_k141_70456 | Not-determined |
| S17_extracted_k141_70457 | Not-determined |
| S17_extracted_k141_11142 | Not-determined |
| S17_extracted_k141_99973 | Not-determined |
| S17_extracted_k141_55651 | Not-determined |
| S17_extracted_k141_41201 | Not-determined |
| S17_extracted_k141_11147 | Not-determined |
| S17_extracted_k141_62525 | Low-quality |
| S17_extracted_k141_11149 | Not-determined |
| S17_extracted_k141_151919 | Not-determined |
| S17_extracted_k141_70464 | Not-determined |
| S17_extracted_k141_151920 | Not-determined |
| S17_extracted_k141_107739 | Not-determined |
| S17_extracted_k141_114427 | Not-determined |
| S17_extracted_k141_62527 | Not-determined |
| S17_extracted_k141_107740 | Low-quality |
| S17_extracted_k141_129139 | Not-determined |
| S17_extracted_k141_114428 | Not-determined |
| S17_extracted_k141_11155 | Not-determined |
| S17_extracted_k141_129140 | Not-determined |
| S17_extracted_k141_129143 | Not-determined |
| S17_extracted_k141_41214 | Not-determined |
| S17_extracted_k141_166141 | Not-determined |
| S17_extracted_k141_33666 | Not-determined |
| S17_extracted_k141_99985 | Not-determined |
| S17_extracted_k141_41216 | Not-determined |
| S17_extracted_k141_129147 | Low-quality |
| S17_extracted_k141_166144 | Not-determined |
| S17_extracted_k141_159153 | Low-quality |
| S17_extracted_k141_151931 | Not-determined |
| S17_extracted_k141_92274 | Not-determined |
| S17_extracted_k141_151935 | Not-determined |
| S17_extracted_k141_159155 | Not-determined |
| S17_extracted_k141_70481 | Not-determined |
| S17_extracted_k141_18337 | Not-determined |
| S17_extracted_k141_129155 | Not-determined |
| S17_extracted_k141_41224 | Not-determined |
| S17_extracted_k141_166152 | Not-determined |
| S17_extracted_k141_41225 | Not-determined |
| S17_extracted_k141_41227 | Not-determined |
| S17_extracted_k141_129159 | Not-determined |
| S17_extracted_k141_18342 | Not-determined |
| S17_extracted_k141_11168 | Not-determined |
| S17_extracted_k141_99996 | Not-determined |
| S17_extracted_k141_129163 | Not-determined |
| S17_extracted_k141_99997 | Not-determined |
| S17_extracted_k141_48391 | Low-quality |
| S17_extracted_k141_41231 | Low-quality |
| S17_extracted_k141_3748 | Low-quality |
| S17_extracted_k141_3749 | Low-quality |
| S17_extracted_k141_70494 | Not-determined |
| S17_extracted_k141_18353 | Not-determined |
| S17_extracted_k141_62548 | Not-determined |
| S17_extracted_k141_55671 | Low-quality |
| S17_extracted_k141_77675 | Not-determined |
| S17_extracted_k141_18357 | Not-determined |
| S17_extracted_k141_114460 | Not-determined |
| S17_extracted_k141_48401 | Not-determined |
| S17_extracted_k141_129173 | Not-determined |
| S17_extracted_k141_151951 | Not-determined |
| S17_extracted_k141_70499 | Not-determined |
| S17_extracted_k141_55675 | Not-determined |
| S17_extracted_k141_129176 | Not-determined |
| S17_extracted_k141_100008 | Not-determined |
| S17_extracted_k141_151954 | Not-determined |
| S17_extracted_k141_33682 | Not-determined |
| S17_extracted_k141_41240 | Not-determined |
| S17_extracted_k141_166164 | Not-determined |
| S17_extracted_k141_77679 | Not-determined |
| S17_extracted_k141_100015 | Not-determined |
| S17_extracted_k141_18368 | Not-determined |
| S17_extracted_k141_11181 | Not-determined |
| S17_extracted_k141_129185 | Not-determined |
| S17_extracted_k141_3764 | Low-quality |
| S17_extracted_k141_18371 | Not-determined |
| S17_extracted_k141_129188 | Not-determined |
| S17_extracted_k141_114476 | Not-determined |
| S17_extracted_k141_151962 | Not-determined |
| S17_extracted_k141_41246 | Not-determined |
| S17_extracted_k141_41247 | Not-determined |
| S17_extracted_k141_121805 | Not-determined |
| S17_extracted_k141_41248 | Not-determined |
| S17_extracted_k141_114481 | Not-determined |
| S17_extracted_k141_33698 | Not-determined |
| S17_extracted_k141_151969 | Not-determined |
| S17_extracted_k141_41253 | Low-quality |
| S17_extracted_k141_129200 | Not-determined |
| S17_extracted_k141_77689 | Not-determined |
| S17_extracted_k141_166174 | Low-quality |
| S17_extracted_k141_41254 | Not-determined |
| S15_extracted_75924 | Complete |
| S17_extracted_k141_33703 | Not-determined |
| S17_extracted_k141_151972 | Not-determined |
| S17_extracted_k141_41255 | Not-determined |
| S17_extracted_k141_144352 | Not-determined |
| S17_extracted_k141_11195 | Not-determined |
| S17_extracted_k141_114488 | Not-determined |
| S17_extracted_k141_107744 | Not-determined |
| S17_extracted_k141_77692 | Low-quality |
| S17_extracted_k141_107747 | Not-determined |
| S17_extracted_k141_114492 | Not-determined |
| S17_extracted_k141_159157 | Low-quality |
| S17_extracted_k141_129208 | Not-determined |
| S17_extracted_k141_107749 | Not-determined |
| S17_extracted_k141_33707 | Not-determined |
| S17_extracted_k141_151974 | Not-determined |
| S17_extracted_k141_166185 | Not-determined |
| S17_extracted_k141_151978 | Not-determined |
| S17_extracted_k141_166187 | Not-determined |
| S17_extracted_k141_3772 | Low-quality |
| S17_extracted_k141_159162 | Not-determined |
| S17_extracted_k141_107755 | Not-determined |
| S17_extracted_k141_55693 | Low-quality |
| S17_extracted_k141_114499 | Not-determined |
| S17_extracted_k141_114500 | Not-determined |
| S17_extracted_k141_18388 | Not-determined |
| S17_extracted_k141_33714 | Not-determined |
| S17_extracted_k141_55696 | Not-determined |
| S17_extracted_k141_166195 | Not-determined |
| S17_extracted_k141_107762 | Not-determined |
| S17_extracted_k141_107763 | Not-determined |
| S17_extracted_k141_129222 | Not-determined |
| S17_extracted_k141_77708 | Low-quality |
| S17_extracted_k141_70504 | Not-determined |
| S17_extracted_k141_41272 | Not-determined |
| S17_extracted_k141_70505 | Not-determined |
| S17_extracted_k141_166203 | Not-determined |
| S17_extracted_k141_70506 | Not-determined |
| S17_extracted_k141_129226 | Not-determined |
| S17_extracted_k141_77714 | Not-determined |
| S17_extracted_k141_151981 | Low-quality |
| S17_extracted_k141_121835 | Not-determined |
| S17_extracted_k141_100047 | Low-quality |
| S17_extracted_k141_11218 | Not-determined |
| S17_extracted_k141_55712 | Not-determined |
| S17_extracted_k141_151986 | Not-determined |
| S17_extracted_k141_129231 | Not-determined |
| S17_extracted_k141_11221 | Not-determined |
| S17_extracted_k141_41284 | Not-determined |
| S17_extracted_k141_136496 | Not-determined |
| S17_extracted_k141_33725 | Not-determined |
| S17_extracted_k141_41288 | Not-determined |
| S17_extracted_k141_159180 | Not-determined |
| S17_extracted_k141_129240 | Not-determined |
| S17_extracted_k141_84282 | Low-quality |
| S17_extracted_k141_41293 | Low-quality |
| S17_extracted_k141_121844 | Not-determined |
| S17_extracted_k141_129242 | Not-determined |
| S17_extracted_k141_159183 | Not-determined |
| S17_extracted_k141_33737 | Not-determined |
| S17_extracted_k141_100049 | Not-determined |
| S17_extracted_k141_33739 | Not-determined |
| S17_extracted_k141_114505 | Not-determined |
| S17_extracted_k141_129250 | Low-quality |
| S17_extracted_k141_3802 | Not-determined |
| S17_extracted_k141_100052 | Not-determined |
| S17_extracted_k141_11226 | Not-determined |
| S17_extracted_k141_144370 | Not-determined |
| S17_extracted_k141_144372 | Not-determined |
| S17_extracted_k141_18413 | Not-determined |
| S17_extracted_k141_114507 | Low-quality |
| S17_extracted_k141_129253 | Not-determined |
| S17_extracted_k141_77741 | Not-determined |
| S17_extracted_k141_114510 | Not-determined |
| S17_extracted_k141_55722 | Not-determined |
| S17_extracted_k141_70517 | Not-determined |
| S17_extracted_k141_121853 | Not-determined |
| S17_extracted_k141_151992 | Not-determined |
| S17_extracted_k141_114515 | Not-determined |
| S17_extracted_k141_129256 | Not-determined |
| S17_extracted_k141_18423 | Not-determined |
| S17_extracted_k141_55727 | Not-determined |
| S17_extracted_k141_136520 | Not-determined |
| S17_extracted_k141_25525 | Not-determined |
| S17_extracted_k141_166231 | Not-determined |
| S17_extracted_k141_77752 | Not-determined |
| S17_extracted_k141_84286 | Not-determined |
| S17_extracted_k141_18425 | Not-determined |
| S17_extracted_k141_77754 | Not-determined |
| S17_extracted_k141_3830 | Not-determined |
| S17_extracted_k141_3831 | Not-determined |
| S17_extracted_k141_77761 | Not-determined |
| S17_extracted_k141_77763 | Not-determined |
| S17_extracted_k141_18434 | Low-quality |
| S17_extracted_k141_3833 | Not-determined |
| S17_extracted_k141_33758 | Low-quality |
| S17_extracted_k141_136536 | Low-quality |
| S17_extracted_k141_25541 | Not-determined |
| S17_extracted_k141_84294 | Not-determined |
| S17_extracted_k141_41314 | Not-determined |
| S17_extracted_k141_151996 | Not-determined |
| S17_extracted_k141_25543 | Not-determined |
| S17_extracted_k141_18441 | Not-determined |
| S17_extracted_k141_77771 | Low-quality |

| | |
|---|---|
| S17_extracted_k141_25546 | Not-determined |
| S17_extracted_k141_166240 | Not-determined |
| S17_extracted_k141_25548 | Not-determined |
| S17_extracted_k141_129268 | Low-quality |
| S17_extracted_k141_77775 | Not-determined |
| S17_extracted_k141_129269 | Not-determined |
| S17_extracted_k141_25549 | Low-quality |
| S17_extracted_k141_129270 | Not-determined |
| S17_extracted_k141_100079 | Not-determined |
| S17_extracted_k141_18445 | Low-quality |
| S17_extracted_k141_114531 | Not-determined |
| S17_extracted_k141_152009 | Not-determined |
| S17_extracted_k141_114532 | Not-determined |
| S17_extracted_k141_173531 | Not-determined |
| S17_extracted_k141_129273 | Not-determined |
| S17_extracted_k141_114533 | Not-determined |
| S17_extracted_k141_84299 | Not-determined |
| S17_extracted_k141_25555 | Not-determined |
| S17_extracted_k141_70526 | Low-quality |
| S17_extracted_k141_55767 | Low-quality |
| S17_extracted_k141_48443 | Not-determined |
| S17_extracted_k141_107795 | Not-determined |
| S17_extracted_k141_152013 | Not-determined |
| S17_extracted_k141_18455 | Not-determined |
| S17_extracted_k141_129279 | Low-quality |
| S17_extracted_k141_152014 | Not-determined |
| S17_extracted_k141_41322 | Not-determined |
| S17_extracted_k141_107799 | Not-determined |
| S17_extracted_k141_152016 | Not-determined |
| S17_extracted_k141_114541 | Not-determined |
| S17_extracted_k141_84308 | Not-determined |
| S17_extracted_k141_152017 | Not-determined |
| S17_extracted_k141_3854 | Not-determined |
| S17_extracted_k141_70536 | Not-determined |
| S17_extracted_k141_33778 | Low-quality |
| S17_extracted_k141_84311 | Not-determined |
| S17_extracted_k141_107810 | Not-determined |
| S17_extracted_k141_55776 | Not-determined |
| S17_extracted_k141_136551 | Not-determined |
| S17_extracted_k141_92284 | Not-determined |
| S17_extracted_k141_136552 | Not-determined |
| S17_extracted_k141_114547 | Not-determined |
| S17_extracted_k141_25563 | Low-quality |
| S17_extracted_k141_18471 | Not-determined |
| S17_extracted_k141_92287 | Not-determined |
| S17_extracted_k141_136555 | Not-determined |
| S17_extracted_k141_18472 | Not-determined |
| S17_extracted_k141_121889 | Not-determined |
| S17_extracted_k141_18473 | Not-determined |
| S17_extracted_k141_77797 | Not-determined |
| S17_extracted_k141_33780 | Not-determined |
| S17_extracted_k141_166250 | Not-determined |
| S17_extracted_k141_92291 | Not-determined |
| S17_extracted_k141_166257 | Not-determined |
| S17_extracted_k141_33788 | Low-quality |
| S17_extracted_k141_55780 | Low-quality |
| S17_extracted_k141_33789 | Not-determined |
| S17_extracted_k141_18479 | Not-determined |
| S17_extracted_k141_55781 | Not-determined |
| S17_extracted_k141_48458 | Low-quality |
| S17_extracted_k141_114561 | Not-determined |
| S17_extracted_k141_3863 | Not-determined |
| S17_extracted_k141_114564 | Low-quality |
| S17_extracted_k141_33794 | Not-determined |
| S17_extracted_k141_11257 | Not-determined |
| S17_extracted_k141_114569 | Low-quality |
| S17_extracted_k141_114570 | Not-determined |
| S17_extracted_k141_100097 | Not-determined |
| S17_extracted_k141_48466 | Not-determined |
| S17_extracted_k141_11263 | Not-determined |
| S17_extracted_k141_152042 | Not-determined |
| S17_extracted_k141_48468 | Low-quality |
| S17_extracted_k141_70552 | Low-quality |
| S17_extracted_k141_166273 | Not-determined |
| S17_extracted_k141_55784 | Not-determined |
| S17_extracted_k141_55786 | Not-determined |
| S17_extracted_k141_70555 | Not-determined |
| S17_extracted_k141_3875 | Not-determined |
| S17_extracted_k141_136583 | Low-quality |
| S17_extracted_k141_18488 | Not-determined |
| S17_extracted_k141_33807 | Low-quality |
| S17_extracted_k141_121910 | Not-determined |
| S17_extracted_k141_152045 | Not-determined |
| S17_extracted_k141_48475 | Not-determined |
| S17_extracted_k141_48476 | Low-quality |
| S17_extracted_k141_144407 | Not-determined |
| S17_extracted_k141_84332 | Not-determined |
| S17_extracted_k141_121918 | Not-determined |
| S17_extracted_k141_84337 | Not-determined |
| S17_extracted_k141_55798 | Not-determined |
| S17_extracted_k141_100118 | Not-determined |
| S17_extracted_k141_84342 | Not-determined |
| S17_extracted_k141_18495 | Not-determined |
| S17_extracted_k141_3888 | Not-determined |
| S17_extracted_k141_173561 | Not-determined |
| S17_extracted_k141_136597 | Not-determined |
| S17_extracted_k141_144421 | Not-determined |
| S17_extracted_k141_100120 | Not-determined |
| S17_extracted_k141_136599 | Not-determined |
| S17_extracted_k141_152062 | Not-determined |
| S17_extracted_k141_70564 | Low-quality |
| S17_extracted_k141_100122 | Not-determined |
| S17_extracted_k141_3895 | Low-quality |
| S17_extracted_k141_129300 | Low-quality |
| S17_extracted_k141_33825 | Not-determined |
| S17_extracted_k141_173569 | Not-determined |
| S17_extracted_k141_55803 | Not-determined |
| S17_extracted_k141_129303 | Not-determined |
| S17_extracted_k141_144425 | Low-quality |
| S17_extracted_k141_129304 | Not-determined |
| S17_extracted_k141_11269 | Not-determined |
| S17_extracted_k141_173576 | Not-determined |
| S17_extracted_k141_33830 | Not-determined |
| S17_extracted_k141_55809 | Not-determined |
| S17_extracted_k141_55810 | Not-determined |
| S17_extracted_k141_152066 | Not-determined |
| S17_extracted_k141_11272 | Not-determined |
| S17_extracted_k141_11273 | Not-determined |
| S17_extracted_k141_144432 | Not-determined |
| S17_extracted_k141_3905 | Not-determined |
| S17_extracted_k141_77832 | Not-determined |
| S17_extracted_k141_166294 | Low-quality |
| S17_extracted_k141_55813 | Not-determined |
| S17_extracted_k141_84357 | Not-determined |
| S17_extracted_k141_33834 | Not-determined |
| S17_extracted_k141_107870 | Not-determined |
| S17_extracted_k141_173585 | Not-determined |
| S17_extracted_k141_55820 | Not-determined |
| S17_extracted_k141_114594 | Low-quality |
| S17_extracted_k141_55823 | Not-determined |
| S17_extracted_k141_144437 | Low-quality |
| S17_extracted_k141_152086 | Not-determined |
| S17_extracted_k141_152087 | Not-determined |
| S17_extracted_k141_11279 | Low-quality |
| S17_extracted_k141_144439 | Not-determined |
| S17_extracted_k141_33841 | Not-determined |
| S17_extracted_k141_11282 | Not-determined |
| S17_extracted_k141_136609 | Not-determined |
| S17_extracted_k141_11283 | Low-quality |
| S17_extracted_k141_100143 | Low-quality |
| S17_extracted_k141_3919 | Not-determined |
| S17_extracted_k141_55827 | Not-determined |
| S17_extracted_k141_100146 | Not-determined |
| S17_extracted_k141_25570 | Not-determined |
| S17_extracted_k141_100147 | Not-determined |
| S17_extracted_k141_92326 | Low-quality |
| S17_extracted_k141_152098 | Not-determined |
| S17_extracted_k141_166312 | Not-determined |
| S17_extracted_k141_33845 | Not-determined |
| S17_extracted_k141_33846 | Not-determined |
| S17_extracted_k141_114606 | Not-determined |
| S17_extracted_k141_136617 | Not-determined |
| S17_extracted_k141_144443 | Low-quality |
| S17_extracted_k141_114610 | Not-determined |
| S17_extracted_k141_144444 | Not-determined |
| S17_extracted_k141_92331 | Not-determined |
| S17_extracted_k141_129319 | Not-determined |
| S17_extracted_k141_144447 | Not-determined |
| S17_extracted_k141_11292 | Not-determined |
| S17_extracted_k141_25577 | Not-determined |
| S17_extracted_k141_33855 | Not-determined |
| S17_extracted_k141_25578 | Low-quality |
| S17_extracted_k141_136622 | Not-determined |
| S17_extracted_k141_144454 | Not-determined |
| S17_extracted_k141_70596 | Not-determined |
| S17_extracted_k141_136624 | Low-quality |
| S17_extracted_k141_3940 | Not-determined |
| S17_extracted_k141_48519 | Not-determined |
| S17_extracted_k141_11299 | Not-determined |
| S17_extracted_k141_129329 | Not-determined |
| S17_extracted_k141_77852 | Not-determined |
| S17_extracted_k141_107892 | Low-quality |
| S17_extracted_k141_77855 | Not-determined |
| S17_extracted_k141_48521 | Not-determined |
| S17_extracted_k141_25585 | Not-determined |
| S17_extracted_k141_3948 | Not-determined |
| S17_extracted_k141_152114 | Not-determined |
| S17_extracted_k141_84372 | Not-determined |
| S17_extracted_k141_129334 | Not-determined |
| S17_extracted_k141_70604 | Not-determined |
| S17_extracted_k141_166326 | Not-determined |
| S17_extracted_k141_107903 | Not-determined |
| S17_extracted_k141_92342 | Not-determined |
| S17_extracted_k141_70605 | Not-determined |
| S17_extracted_k141_166328 | Not-determined |
| S17_extracted_k141_41339 | Low-quality |
| S17_extracted_k141_114633 | Not-determined |
| S17_extracted_k141_70608 | Not-determined |
| S17_extracted_k141_77861 | Low-quality |
| S17_extracted_k141_3956 | Not-determined |
| S17_extracted_k141_121936 | Not-determined |
| S17_extracted_k141_129337 | Not-determined |
| S17_extracted_k141_33860 | Not-determined |
| S17_extracted_k141_121937 | Not-determined |
| S17_extracted_k141_3957 | Not-determined |
| S17_extracted_k141_84384 | Not-determined |
| S17_extracted_k141_114636 | Not-determined |
| S17_extracted_k141_152124 | Not-determined |
| S17_extracted_k141_77864 | Not-determined |
| S17_extracted_k141_3960 | Not-determined |
| S17_extracted_k141_18513 | Not-determined |
| S17_extracted_k141_136632 | Not-determined |
| S17_extracted_k141_48531 | Not-determined |
| S17_extracted_k141_33865 | Not-determined |
| S17_extracted_k141_48533 | Not-determined |
| S17_extracted_k141_114641 | Not-determined |
| S17_extracted_k141_41348 | Low-quality |
| S17_extracted_k141_33870 | Not-determined |
| S17_extracted_k141_77869 | Not-determined |
| S17_extracted_k141_48538 | Not-determined |
| S17_extracted_k141_173604 | Not-determined |
| S17_extracted_k141_3968 | Not-determined |
| S17_extracted_k141_11307 | Low-quality |
| S17_extracted_k141_18524 | Not-determined |
| S17_extracted_k141_18526 | Not-determined |
| S17_extracted_k141_48545 | Low-quality |
| S17_extracted_k141_107910 | Low-quality |
| S17_extracted_k141_92362 | Low-quality |
| S17_extracted_k141_3976 | Not-determined |
| S17_extracted_k141_77877 | Not-determined |
| S17_extracted_k141_121951 | Not-determined |
| S17_extracted_k141_121952 | Low-quality |
| S17_extracted_k141_92367 | Not-determined |
| S17_extracted_k141_41360 | Not-determined |
| S17_extracted_k141_92369 | Not-determined |
| S17_extracted_k141_100174 | Not-determined |
| S17_extracted_k141_129354 | Not-determined |
| S17_extracted_k141_136652 | Not-determined |
| S17_extracted_k141_136653 | Not-determined |
| S17_extracted_k141_25603 | Not-determined |
| S17_extracted_k141_136654 | Not-determined |
| S17_extracted_k141_77883 | Not-determined |
| S17_extracted_k141_100179 | Not-determined |
| S17_extracted_k141_173612 | Not-determined |
| S17_extracted_k141_136660 | Not-determined |
| S17_extracted_k141_107925 | Not-determined |
| S17_extracted_k141_41371 | Low-quality |
| S17_extracted_k141_136665 | Not-determined |
| S17_extracted_k141_129364 | Not-determined |
| S17_extracted_k141_84395 | Not-determined |
| S17_extracted_k141_129365 | Not-determined |
| S17_extracted_k141_3993 | Not-determined |
| S17_extracted_k141_136670 | Not-determined |
| S17_extracted_k141_159211 | Not-determined |
| S17_extracted_k141_129368 | Not-determined |
| S17_extracted_k141_41379 | Not-determined |
| S17_extracted_k141_41382 | Not-determined |
| S17_extracted_k141_48563 | Not-determined |
| S17_extracted_k141_100193 | Low-quality |
| S17_extracted_k141_166338 | Low-quality |
| S17_extracted_k141_48566 | Not-determined |
| S17_extracted_k141_100195 | Not-determined |
| S17_extracted_k141_25620 | Not-determined |
| S17_extracted_k141_48567 | Not-determined |
| S17_extracted_k141_100197 | Not-determined |
| S17_extracted_k141_25621 | Not-determined |
| S17_extracted_k141_48570 | Not-determined |
| S17_extracted_k141_25624 | Not-determined |
| S17_extracted_k141_11329 | Not-determined |
| S17_extracted_k141_48572 | Not-determined |
| S17_extracted_k141_114673 | Not-determined |
| S17_extracted_k141_48574 | Not-determined |
| S17_extracted_k141_55872 | Not-determined |
| S17_extracted_k141_173616 | Low-quality |
| S17_extracted_k141_166341 | Not-determined |
| S17_extracted_k141_25627 | Not-determined |
| S17_extracted_k141_11333 | Not-determined |
| S17_extracted_k141_100206 | Not-determined |
| S17_extracted_k141_173621 | Low-quality |
| S17_extracted_k141_77899 | Not-determined |
| S17_extracted_k141_114682 | Low-quality |
| S17_extracted_k141_159221 | Not-determined |
| S17_extracted_k141_159223 | Not-determined |
| S17_extracted_k141_25631 | Low-quality |
| S17_extracted_k141_107943 | Not-determined |
| S17_extracted_k141_121975 | Not-determined |
| S17_extracted_k141_77907 | Not-determined |
| S17_extracted_k141_129375 | Not-determined |
| S17_extracted_k141_159224 | Not-determined |
| S17_extracted_k141_114690 | Not-determined |
| S17_extracted_k141_84413 | Not-determined |
| S17_extracted_k141_121976 | Not-determined |
| S17_extracted_k141_166351 | Not-determined |
| S17_extracted_k141_114692 | Not-determined |
| S17_extracted_k141_166352 | Not-determined |
| S17_extracted_k141_121981 | Not-determined |
| S17_extracted_k141_18567 | Not-determined |
| S17_extracted_k141_70616 | Not-determined |
| S17_extracted_k141_136685 | Not-determined |
| S17_extracted_k141_25641 | Not-determined |
| S17_extracted_k141_152132 | Not-determined |
| S17_extracted_k141_129379 | Low-quality |
| S17_extracted_k141_84420 | Not-determined |
| S17_extracted_k141_129380 | Low-quality |
| S17_extracted_k141_70623 | Not-determined |
| S17_extracted_k141_152136 | Not-determined |
| S17_extracted_k141_25648 | Low-quality |
| S17_extracted_k141_70626 | Not-determined |
| S17_extracted_k141_159238 | Not-determined |
| S17_extracted_k141_173633 | Not-determined |
| S17_extracted_k141_25651 | Not-determined |
| S17_extracted_k141_41414 | Not-determined |
| S17_extracted_k141_70628 | Not-determined |
| S17_extracted_k141_48602 | Low-quality |
| S17_extracted_k141_166363 | Not-determined |
| S17_extracted_k141_107963 | Not-determined |
| S17_extracted_k141_159242 | Not-determined |
| S17_extracted_k141_136694 | Not-determined |
| S17_extracted_k141_159245 | Not-determined |
| S17_extracted_k141_107966 | Not-determined |
| S17_extracted_k141_166368 | Not-determined |
| S17_extracted_k141_121987 | Low-quality |
| S17_extracted_k141_48609 | Not-determined |
| S17_extracted_k141_48611 | Not-determined |
| S17_extracted_k141_166373 | Not-determined |
| S17_extracted_k141_77927 | Not-determined |
| S17_extracted_k141_55883 | Not-determined |
| S17_extracted_k141_70635 | Not-determined |
| S17_extracted_k141_25667 | Not-determined |
| S17_extracted_k141_159252 | Low-quality |
| S17_extracted_k141_11341 | Not-determined |
| S17_extracted_k141_121990 | Not-determined |
| S17_extracted_k141_159254 | Not-determined |
| S17_extracted_k141_129390 | Not-determined |
| S17_extracted_k141_107970 | Not-determined |
| S17_extracted_k141_159255 | Not-determined |
| S17_extracted_k141_84432 | Low-quality |
| S17_extracted_k141_136708 | Not-determined |
| S17_extracted_k141_159258 | Not-determined |
| S17_extracted_k141_136711 | Low-quality |
| S17_extracted_k141_173649 | Not-determined |
| S17_extracted_k141_121993 | Not-determined |
| S17_extracted_k141_77934 | Low-quality |
| S17_extracted_k141_152155 | Not-determined |
| S17_extracted_k141_152156 | Not-determined |
| S17_extracted_k141_107974 | Not-determined |
| S17_extracted_k141_41438 | Not-determined |
| S17_extracted_k141_166381 | Low-quality |
| S17_extracted_k141_41439 | Low-quality |
| S17_extracted_k141_92405 | Not-determined |
| S17_extracted_k141_129398 | Not-determined |
| S17_extracted_k141_25676 | Not-determined |
| S17_extracted_k141_136721 | Not-determined |
| S17_extracted_k141_173658 | Not-determined |
| S17_extracted_k141_114704 | Low-quality |
| S17_extracted_k141_173664 | Not-determined |
| S17_extracted_k141_33904 | Low-quality |
| S17_extracted_k141_70660 | Not-determined |
| S17_extracted_k141_166395 | Not-determined |
| S17_extracted_k141_152167 | Not-determined |
| S17_extracted_k141_159273 | Not-determined |
| S17_extracted_k141_11355 | Not-determined |
| S17_extracted_k141_48621 | Not-determined |
| S17_extracted_k141_11357 | Low-quality |
| S17_extracted_k141_114713 | Not-determined |
| S17_extracted_k141_122013 | Not-determined |
| S17_extracted_k141_70665 | Not-determined |
| S17_extracted_k141_129407 | Not-determined |
| S17_extracted_k141_129408 | Not-determined |
| S17_extracted_k141_92420 | Not-determined |
| S17_extracted_k141_107983 | Not-determined |
| S17_extracted_k141_107985 | Low-quality |
| S17_extracted_k141_77951 | Not-determined |
| S17_extracted_k141_129413 | Not-determined |
| S17_extracted_k141_152171 | Not-determined |
| S17_extracted_k141_41458 | Not-determined |
| S17_extracted_k141_122025 | Not-determined |
| S17_extracted_k141_100237 | Not-determined |
| S17_extracted_k141_70670 | Not-determined |
| S17_extracted_k141_107991 | Not-determined |
| S17_extracted_k141_92429 | Low-quality |
| S17_extracted_k141_48633 | Not-determined |
| S17_extracted_k141_77957 | Low-quality |
| S17_extracted_k141_152177 | Not-determined |
| S17_extracted_k141_107992 | Not-determined |
| S17_extracted_k141_41465 | Not-determined |
| S17_extracted_k141_129416 | Not-determined |
| S17_extracted_k141_159286 | Not-determined |
| S17_extracted_k141_108002 | Not-determined |
| S17_extracted_k141_108003 | Not-determined |
| S17_extracted_k141_122035 | Not-determined |
| S17_extracted_k141_70676 | Not-determined |
| S17_extracted_k141_48636 | Low-quality |
| S17_extracted_k141_122036 | Not-determined |
| S17_extracted_k141_114735 | Not-determined |
| S17_extracted_k141_122037 | Not-determined |
| S17_extracted_k141_159290 | Not-determined |
| S17_extracted_k141_129421 | Not-determined |
| S17_extracted_k141_18609 | Not-determined |
| S17_extracted_k141_159292 | Low-quality |
| S17_extracted_k141_33919 | Low-quality |
| S17_extracted_k141_114738 | Not-determined |
| S17_extracted_k141_84453 | Not-determined |
| S17_extracted_k141_166409 | Not-determined |
| S17_extracted_k141_173674 | Not-determined |
| S17_extracted_k141_114744 | Not-determined |
| S17_extracted_k141_33927 | Not-determined |
| S17_extracted_k141_48644 | Not-determined |
| S17_extracted_k141_41478 | Not-determined |
| S17_extracted_k141_173675 | Not-determined |
| S17_extracted_k141_100247 | Not-determined |
| S17_extracted_k141_48647 | Not-determined |
| S17_extracted_k141_41481 | Not-determined |
| S17_extracted_k141_152197 | Not-determined |
| S17_extracted_k141_166420 | Not-determined |
| S17_extracted_k141_70690 | Low-quality |
| S17_extracted_k141_166423 | Low-quality |
| S17_extracted_k141_129434 | Low-quality |
| S17_extracted_k141_159299 | Not-determined |
| S17_extracted_k141_41484 | Not-determined |
| S17_extracted_k141_114748 | Not-determined |
| S17_extracted_k141_92448 | Not-determined |
| S17_extracted_k141_152201 | Not-determined |
| S17_extracted_k141_166427 | Not-determined |
| S17_extracted_k141_18626 | Not-determined |
| S17_extracted_k141_173693 | Not-determined |
| S17_extracted_k141_159305 | Not-determined |
| S17_extracted_k141_114751 | Not-determined |
| S17_extracted_k141_159310 | Not-determined |
| S17_extracted_k141_55914 | Low-quality |
| S17_extracted_k141_48654 | Not-determined |
| S17_extracted_k141_55918 | Not-determined |
| S17_extracted_k141_114756 | Not-determined |
| S17_extracted_k141_129446 | Not-determined |
| S17_extracted_k141_136742 | Not-determined |
| S17_extracted_k141_25697 | Not-determined |
| S17_extracted_k141_108019 | Not-determined |
| S17_extracted_k141_55922 | Not-determined |
| S17_extracted_k141_48656 | Not-determined |
| S17_extracted_k141_122046 | Not-determined |
| S17_extracted_k141_108024 | Not-determined |
| S17_extracted_k141_159320 | Not-determined |
| S17_extracted_k141_129448 | Not-determined |
| S17_extracted_k141_77978 | Not-determined |
| S17_extracted_k141_114760 | Not-determined |
| S17_extracted_k141_48660 | Not-determined |
| S17_extracted_k141_152224 | Low-quality |
| S17_extracted_k141_100264 | Low-quality |
| S17_extracted_k141_18638 | Not-determined |
| S17_extracted_k141_100265 | Not-determined |
| S17_extracted_k141_25705 | Not-determined |
| S17_extracted_k141_108032 | Not-determined |
| S17_extracted_k141_159325 | Not-determined |
| S17_extracted_k141_77982 | Not-determined |
| S17_extracted_k141_114767 | Low-quality |
| S17_extracted_k141_77983 | Not-determined |
| S17_extracted_k141_41506 | Not-determined |
| S17_extracted_k141_122061 | Not-determined |
| S17_extracted_k141_122063 | Not-determined |
| S17_extracted_k141_92474 | Not-determined |
| S17_extracted_k141_84464 | Not-determined |
| S17_extracted_k141_11398 | Low-quality |
| S17_extracted_k141_11403 | Low-quality |
| S17_extracted_k141_152228 | Not-determined |
| S17_extracted_k141_166452 | Not-determined |
| S17_extracted_k141_136750 | Low-quality |
| S17_extracted_k141_122074 | Not-determined |
| S17_extracted_k141_136751 | Low-quality |
| S17_extracted_k141_25707 | Not-determined |
| S17_extracted_k141_122075 | Not-determined |
| S17_extracted_k141_100284 | Not-determined |
| S17_extracted_k141_55930 | Not-determined |
| S17_extracted_k141_84472 | Not-determined |
| S17_extracted_k141_173728 | Not-determined |
| S17_extracted_k141_152237 | Not-determined |
| S17_extracted_k141_33974 | Not-determined |
| S17_extracted_k141_108045 | Not-determined |
| S17_extracted_k141_152238 | Not-determined |
| S17_extracted_k141_100287 | Not-determined |
| S17_extracted_k141_166462 | Not-determined |
| S17_extracted_k141_122077 | Not-determined |
| S17_extracted_k141_152239 | Not-determined |
| S17_extracted_k141_166464 | Not-determined |
| S17_extracted_k141_33976 | Not-determined |
| S17_extracted_k141_25713 | Not-determined |
| S17_extracted_k141_77987 | Not-determined |
| S17_extracted_k141_152240 | Not-determined |
| S17_extracted_k141_77988 | Not-determined |
| S17_extracted_k141_48671 | Not-determined |
| S17_extracted_k141_33978 | Low-quality |
| S17_extracted_k141_77991 | Low-quality |
| S17_extracted_k141_84482 | Not-determined |
| S17_extracted_k141_100288 | Low-quality |
| S17_extracted_k141_41530 | Not-determined |
| S17_extracted_k141_84485 | Not-determined |
| S17_extracted_k141_33982 | Not-determined |
| S17_extracted_k141_152245 | Not-determined |
| S17_extracted_k141_159342 | Not-determined |
| S17_extracted_k141_48677 | Not-determined |
| S17_extracted_k141_92487 | Not-determined |
| S17_extracted_k141_159346 | Not-determined |
| S17_extracted_k141_77998 | Not-determined |
| S17_extracted_k141_129478 | Not-determined |
| S17_extracted_k141_55936 | Low-quality |
| S17_extracted_k141_92491 | Low-quality |
| S17_extracted_k141_55937 | Not-determined |
| S17_extracted_k141_33988 | Low-quality |
| S17_extracted_k141_48680 | Not-determined |
| S17_extracted_k141_41539 | Low-quality |
| S17_extracted_k141_129481 | Not-determined |
| S17_extracted_k141_159350 | Not-determined |
| S17_extracted_k141_114790 | Low-quality |
| S17_extracted_k141_41542 | Low-quality |
| S17_extracted_k141_114791 | Not-determined |
| S17_extracted_k141_25726 | Not-determined |
| S17_extracted_k141_55944 | Not-determined |
| S17_extracted_k141_108060 | Not-determined |
| S17_extracted_k141_129484 | Low-quality |
| S17_extracted_k141_55946 | Not-determined |
| S17_extracted_k141_152250 | Not-determined |
| S17_extracted_k141_84494 | Not-determined |
| S17_extracted_k141_84497 | Not-determined |
| S17_extracted_k141_84498 | Not-determined |
| S17_extracted_k141_166489 | Not-determined |
| S17_extracted_k141_78009 | Not-determined |
| S17_extracted_k141_78010 | Not-determined |
| S17_extracted_k141_70712 | Not-determined |
| S17_extracted_k141_84504 | Not-determined |
| S17_extracted_k141_34000 | Not-determined |
| S17_extracted_k141_34003 | Not-determined |
| S17_extracted_k141_25735 | Not-determined |
| S17_extracted_k141_159366 | Not-determined |
| S17_extracted_k141_92505 | Not-determined |
| S17_extracted_k141_129493 | Not-determined |
| S17_extracted_k141_152255 | Not-determined |
| S17_extracted_k141_152257 | Not-determined |
| S17_extracted_k141_84512 | Not-determined |
| S17_extracted_k141_159371 | Not-determined |
| S17_extracted_k141_108074 | Not-determined |
| S17_extracted_k141_122085 | Not-determined |
| S17_extracted_k141_136786 | Not-determined |
| S17_extracted_k141_70714 | Low-quality |
| S17_extracted_k141_114819 | Not-determined |
| S17_extracted_k141_159373 | Low-quality |
| S17_extracted_k141_136788 | Not-determined |
| S17_extracted_k141_41559 | Not-determined |
| S17_extracted_k141_152261 | Not-determined |
| S17_extracted_k141_100309 | Not-determined |
| S17_extracted_k141_48696 | Not-determined |
| S17_extracted_k141_48697 | Not-determined |
| S17_extracted_k141_41561 | Not-determined |
| S17_extracted_k141_108078 | Not-determined |
| S17_extracted_k141_92516 | Not-determined |
| S17_extracted_k141_129501 | Not-determined |
| S17_extracted_k141_70719 | Low-quality |
| S17_extracted_k141_84520 | Not-determined |
| S17_extracted_k141_108081 | Not-determined |
| S17_extracted_k141_48703 | Low-quality |
| S17_extracted_k141_48706 | Not-determined |
| S17_extracted_k141_114834 | Not-determined |
| S17_extracted_k141_166508 | Not-determined |
| S17_extracted_k141_129507 | Low-quality |
| S17_extracted_k141_25756 | Not-determined |
| S17_extracted_k141_55973 | Not-determined |
| S17_extracted_k141_122098 | Low-quality |
| S17_extracted_k141_159389 | Not-determined |
| S17_extracted_k141_114839 | Not-determined |
| S17_extracted_k141_114844 | Low-quality |
| S17_extracted_k141_34021 | Not-determined |
| S17_extracted_k141_78031 | Not-determined |
| S17_extracted_k141_70727 | Not-determined |
| S17_extracted_k141_48715 | Not-determined |
| S17_extracted_k141_41565 | Not-determined |
| S17_extracted_k141_70729 | Not-determined |
| S17_extracted_k141_55981 | Not-determined |
| S17_extracted_k141_108096 | Not-determined |
| S17_extracted_k141_78033 | Not-determined |
| S17_extracted_k141_129512 | Not-determined |
| S17_extracted_k141_108100 | Not-determined |
| S17_extracted_k141_70734 | Not-determined |
| S17_extracted_k141_84534 | Not-determined |
| S17_extracted_k141_108104 | Not-determined |
| S17_extracted_k141_114849 | Not-determined |
| S17_extracted_k141_34033 | Not-determined |
| S17_extracted_k141_41574 | Not-determined |
| S17_extracted_k141_152276 | Not-determined |
| S17_extracted_k141_108109 | Not-determined |
| S17_extracted_k141_78044 | Not-determined |
| S17_extracted_k141_152280 | Not-determined |
| S17_extracted_k141_152281 | Not-determined |
| S17_extracted_k141_78047 | Not-determined |
| S17_extracted_k141_55994 | Not-determined |
| S17_extracted_k141_114856 | Low-quality |
| S17_extracted_k141_108119 | Not-determined |
| S17_extracted_k141_129527 | Not-determined |
| S17_extracted_k141_129531 | Low-quality |
| S17_extracted_k141_92559 | Not-determined |
| S17_extracted_k141_56001 | Not-determined |
| S17_extracted_k141_34048 | Not-determined |

| | |
|---|---|
| S17_extracted_k141_48744 | Not-determined |
| S17_extracted_k141_144492 | Not-determined |
| S17_extracted_k141_129538 | Not-determined |
| S17_extracted_k141_41595 | Not-determined |
| S17_extracted_k141_25779 | Low-quality |
| S17_extracted_k141_100338 | Not-determined |
| S17_extracted_k141_18668 | Not-determined |
| S17_extracted_k141_114864 | Not-determined |
| S17_extracted_k141_78064 | Not-determined |
| S17_extracted_k141_166532 | Low-quality |
| S17_extracted_k141_48750 | Not-determined |
| S17_extracted_k141_108128 | Not-determined |
| S17_extracted_k141_152309 | Not-determined |
| S17_extracted_k141_48753 | Not-determined |
| S17_extracted_k141_129547 | Not-determined |
| S17_extracted_k141_78074 | Not-determined |
| S17_extracted_k141_152311 | Not-determined |
| S17_extracted_k141_92573 | Not-determined |
| S17_extracted_k141_70744 | Not-determined |
| S17_extracted_k141_92576 | Not-determined |
| S17_extracted_k141_34057 | Not-determined |
| S17_extracted_k141_122111 | Not-determined |
| S17_extracted_k141_92577 | Not-determined |
| S17_extracted_k141_92578 | Not-determined |
| S17_extracted_k141_122114 | Not-determined |
| S17_extracted_k141_92579 | Not-determined |
| S17_extracted_k141_34059 | Low-quality |
| S17_extracted_k141_152320 | Not-determined |
| S17_extracted_k141_159406 | Not-determined |
| S17_extracted_k141_159407 | Not-determined |
| S17_extracted_k141_41615 | Not-determined |
| S17_extracted_k141_4040 | Not-determined |
| S17_extracted_k141_70750 | Not-determined |
| S17_extracted_k141_84547 | Not-determined |
| S17_extracted_k141_84549 | Not-determined |
| S17_extracted_k141_41625 | Not-determined |
| S17_extracted_k141_78094 | Not-determined |
| S17_extracted_k141_18688 | Not-determined |
| S17_extracted_k141_11414 | Not-determined |
| S17_extracted_k141_122129 | Not-determined |
| S17_extracted_k141_78096 | Not-determined |
| S17_extracted_k141_166545 | Not-determined |
| S17_extracted_k141_84556 | Not-determined |
| S17_extracted_k141_159422 | Not-determined |
| S17_extracted_k141_129569 | Not-determined |
| S17_extracted_k141_144514 | Not-determined |
| S17_extracted_k141_108154 | Not-determined |
| S17_extracted_k141_114899 | Not-determined |
| S17_extracted_k141_108155 | Not-determined |
| S17_extracted_k141_11423 | Not-determined |
| S17_extracted_k141_129577 | Not-determined |
| S17_extracted_k141_92588 | Low-quality |
| S17_extracted_k141_144520 | Not-determined |
| S17_extracted_k141_100351 | Low-quality |
| S17_extracted_k141_152340 | Not-determined |
| S17_extracted_k141_166556 | Low-quality |
| S17_extracted_k141_70752 | Low-quality |
| S17_extracted_k141_92594 | Not-determined |
| S17_extracted_k141_129583 | Not-determined |
| S17_extracted_k141_92595 | Not-determined |
| S17_extracted_k141_70754 | Not-determined |
| S17_extracted_k141_144525 | Low-quality |
| S17_extracted_k141_78103 | Low-quality |
| S17_extracted_k141_11431 | Not-determined |
| S17_extracted_k141_166559 | Not-determined |
| S17_extracted_k141_108166 | Not-determined |
| S17_extracted_k141_144529 | Not-determined |
| S17_extracted_k141_78109 | Not-determined |
| S17_extracted_k141_144532 | Not-determined |
| S17_extracted_k141_108171 | Not-determined |
| S17_extracted_k141_166562 | Not-determined |
| S17_extracted_k141_70759 | Not-determined |
| S17_extracted_k141_78113 | Not-determined |
| S17_extracted_k141_34078 | Not-determined |
| S17_extracted_k141_56036 | Low-quality |
| S17_extracted_k141_108174 | Not-determined |
| S17_extracted_k141_166572 | Not-determined |
| S17_extracted_k141_166573 | Not-determined |
| S17_extracted_k141_48763 | Not-determined |
| S17_extracted_k141_108176 | Not-determined |
| S17_extracted_k141_136866 | Not-determined |
| S17_extracted_k141_108180 | Not-determined |
| S17_extracted_k141_108181 | Not-determined |
| S17_extracted_k141_129595 | Not-determined |
| S17_extracted_k141_92605 | Low-quality |
| S17_extracted_k141_166577 | Not-determined |
| S17_extracted_k141_48768 | Not-determined |
| S17_extracted_k141_136868 | Not-determined |
| S17_extracted_k141_129596 | Not-determined |
| S17_extracted_k141_48769 | Not-determined |
| S17_extracted_k141_122150 | Not-determined |
| S17_extracted_k141_34085 | Low-quality |
| S17_extracted_k141_136871 | Not-determined |
| S17_extracted_k141_136872 | Low-quality |
| S17_extracted_k141_144547 | Not-determined |
| S17_extracted_k141_152369 | Not-determined |
| S17_extracted_k141_18702 | Not-determined |
| S17_extracted_k141_152373 | Not-determined |
| S17_extracted_k141_100374 | Not-determined |
| S17_extracted_k141_122160 | Low-quality |
| S17_extracted_k141_136877 | Not-determined |
| S17_extracted_k141_56061 | Low-quality |
| S17_extracted_k141_25811 | Not-determined |
| S17_extracted_k141_56062 | Not-determined |
| S17_extracted_k141_152381 | Not-determined |
| S17_extracted_k141_18713 | Not-determined |
| S17_extracted_k141_114935 | Not-determined |
| S17_extracted_k141_25814 | Not-determined |
| S17_extracted_k141_56065 | Not-determined |
| S17_extracted_k141_78133 | Not-determined |
| S17_extracted_k141_152383 | Low-quality |
| S17_extracted_k141_152384 | Low-quality |
| S17_extracted_k141_48781 | Not-determined |
| S17_extracted_k141_122167 | Low-quality |
| S17_extracted_k141_114940 | Not-determined |
| S17_extracted_k141_56071 | Not-determined |
| S17_extracted_k141_70779 | Low-quality |
| S17_extracted_k141_159445 | Not-determined |
| S17_extracted_k141_108191 | Not-determined |
| S17_extracted_k141_25818 | Not-determined |
| S17_extracted_k141_100383 | Not-determined |
| S17_extracted_k141_108193 | Not-determined |
| S17_extracted_k141_136885 | Not-determined |
| S17_extracted_k141_129598 | Not-determined |
| S17_extracted_k141_100388 | Low-quality |
| S17_extracted_k141_152385 | Not-determined |
| S17_extracted_k141_152386 | Not-determined |
| S17_extracted_k141_129602 | Not-determined |
| S17_extracted_k141_144559 | Not-determined |
| S17_extracted_k141_18728 | Not-determined |
| S17_extracted_k141_78135 | Not-determined |
| S17_extracted_k141_41637 | Not-determined |
| S17_extracted_k141_100391 | Low-quality |
| S17_extracted_k141_114948 | Not-determined |
| S17_extracted_k141_70792 | Not-determined |
| S17_extracted_k141_144563 | Not-determined |
| S17_extracted_k141_108201 | Low-quality |
| S17_extracted_k141_48796 | Low-quality |
| S17_extracted_k141_18732 | Not-determined |
| S17_extracted_k141_41646 | Not-determined |
| S17_extracted_k141_114956 | Not-determined |
| S17_extracted_k141_48801 | Not-determined |
| S17_extracted_k141_144566 | Not-determined |
| S17_extracted_k141_136893 | Low-quality |
| S17_extracted_k141_108205 | Not-determined |
| S17_extracted_k141_108207 | Not-determined |
| S17_extracted_k141_11451 | Not-determined |
| S17_extracted_k141_166586 | Low-quality |
| S17_extracted_k141_152394 | Not-determined |
| S17_extracted_k141_136898 | Low-quality |
| S17_extracted_k141_48807 | Not-determined |
| S17_extracted_k141_11455 | Not-determined |
| S17_extracted_k141_108213 | Not-determined |
| S17_extracted_k141_92621 | Not-determined |
| S17_extracted_k141_144571 | Low-quality |
| S17_extracted_k141_166589 | Low-quality |
| S17_extracted_k141_34100 | Not-determined |
| S17_extracted_k141_166592 | Not-determined |
| S17_extracted_k141_11460 | Not-determined |
| S17_extracted_k141_159455 | Not-determined |
| S17_extracted_k141_11461 | Not-determined |
| S17_extracted_k141_92630 | Not-determined |
| S17_extracted_k141_108222 | Not-determined |
| S17_extracted_k141_152410 | Not-determined |
| S17_extracted_k141_41658 | Not-determined |
| S17_extracted_k141_136910 | Not-determined |
| S17_extracted_k141_48823 | Not-determined |
| S17_extracted_k141_41661 | Not-determined |
| S17_extracted_k141_41663 | Not-determined |
| S17_extracted_k141_129614 | Not-determined |
| S17_extracted_k141_70806 | Not-determined |
| S17_extracted_k141_78150 | Not-determined |
| S17_extracted_k141_48825 | Not-determined |
| S17_extracted_k141_34115 | Not-determined |
| S17_extracted_k141_78152 | Low-quality |
| S17_extracted_k141_11476 | Not-determined |
| S17_extracted_k141_18750 | Not-determined |
| S17_extracted_k141_92638 | Low-quality |
| S17_extracted_k141_152423 | Not-determined |
| S17_extracted_k141_92640 | Not-determined |
| S17_extracted_k141_84586 | Not-determined |
| S17_extracted_k141_129623 | Not-determined |
| S17_extracted_k141_136915 | Not-determined |
| S17_extracted_k141_136918 | Not-determined |
| S17_extracted_k141_136919 | Not-determined |
| S17_extracted_k141_136921 | Not-determined |
| S17_extracted_k141_70826 | Not-determined |
| S17_extracted_k141_18757 | Not-determined |
| S17_extracted_k141_92644 | Low-quality |
| S17_extracted_k141_41684 | Not-determined |
| S17_extracted_k141_108238 | Not-determined |
| S17_extracted_k141_18759 | Low-quality |
| S17_extracted_k141_78167 | Not-determined |
| S17_extracted_k141_108242 | Not-determined |
| S17_extracted_k141_11483 | Not-determined |
| S17_extracted_k141_11484 | Not-determined |
| S17_extracted_k141_70832 | Not-determined |
| S17_extracted_k141_152429 | Not-determined |
| S17_extracted_k141_78172 | Not-determined |
| S17_extracted_k141_92653 | Not-determined |
| S17_extracted_k141_11490 | Low-quality |
| S17_extracted_k141_78175 | Not-determined |
| S17_extracted_k141_84590 | Not-determined |
| S17_extracted_k141_108249 | Not-determined |
| S17_extracted_k141_144576 | Not-determined |
| S17_extracted_k141_84592 | Not-determined |
| S17_extracted_k141_92658 | Not-determined |
| S17_extracted_k141_144582 | Not-determined |
| S17_extracted_k141_100418 | Not-determined |
| S17_extracted_k141_129634 | Not-determined |
| S17_extracted_k141_129635 | Not-determined |
| S17_extracted_k141_129636 | Low-quality |
| S17_extracted_k141_18780 | Not-determined |
| S17_extracted_k141_78180 | Not-determined |
| S17_extracted_k141_18783 | Not-determined |
| S17_extracted_k141_84594 | Not-determined |
| S17_extracted_k141_48859 | Not-determined |
| S17_extracted_k141_84597 | Low-quality |
| S17_extracted_k141_129642 | Low-quality |
| S17_extracted_k141_18785 | Not-determined |
| S17_extracted_k141_108253 | Low-quality |
| S17_extracted_k141_78187 | Not-determined |
| S17_extracted_k141_70841 | Low-quality |
| S17_extracted_k141_18788 | Not-determined |
| S17_extracted_k141_70844 | Low-quality |
| S17_extracted_k141_108257 | Not-determined |
| S17_extracted_k141_25847 | Low-quality |
| S17_extracted_k141_84607 | Not-determined |
| S17_extracted_k141_159468 | Not-determined |
| S17_extracted_k141_129644 | Not-determined |
| S17_extracted_k141_159469 | Not-determined |
| S17_extracted_k141_159470 | Not-determined |
| S17_extracted_k141_48864 | Not-determined |
| S17_extracted_k141_78191 | Not-determined |
| S17_extracted_k141_70847 | Not-determined |
| S17_extracted_k141_129646 | Not-determined |
| S17_extracted_k141_152438 | Not-determined |
| S17_extracted_k141_48868 | Not-determined |
| S17_extracted_k141_78195 | Not-determined |
| S17_extracted_k141_56108 | Not-determined |
| S17_extracted_k141_166604 | Not-determined |
| S17_extracted_k141_92677 | Low-quality |
| S17_extracted_k141_18798 | Not-determined |
| S17_extracted_k141_166607 | Not-determined |
| S17_extracted_k141_56109 | Not-determined |
| S17_extracted_k141_100435 | Not-determined |
| S17_extracted_k141_100436 | Not-determined |
| S17_extracted_k141_18802 | Not-determined |
| S17_extracted_k141_144587 | Not-determined |
| S17_extracted_k141_144591 | Not-determined |
| S17_extracted_k141_129655 | Not-determined |
| S17_extracted_k141_48874 | Not-determined |
| S17_extracted_k141_48875 | Not-determined |
| S17_extracted_k141_92685 | Low-quality |
| S17_extracted_k141_41706 | Not-determined |
| S17_extracted_k141_152442 | Not-determined |
| S17_extracted_k141_100442 | Not-determined |
| S17_extracted_k141_48879 | Not-determined |
| S17_extracted_k141_56116 | Not-determined |
| S17_extracted_k141_48883 | Not-determined |
| S17_extracted_k141_144594 | Not-determined |
| S17_extracted_k141_144596 | Not-determined |
| S17_extracted_k141_166613 | Not-determined |
| S17_extracted_k141_41712 | Not-determined |
| S17_extracted_k141_152445 | Not-determined |
| S17_extracted_k141_56119 | Not-determined |
| S17_extracted_k141_114976 | Not-determined |
| S17_extracted_k141_100447 | Not-determined |
| S17_extracted_k141_11506 | Not-determined |
| S17_extracted_k141_173754 | Not-determined |
| S17_extracted_k141_84628 | Not-determined |
| S17_extracted_k141_25871 | Not-determined |
| S17_extracted_k141_108289 | Not-determined |
| S17_extracted_k141_166621 | Not-determined |
| S17_extracted_k141_48893 | Not-determined |
| S17_extracted_k141_108293 | Not-determined |
| S17_extracted_k141_78208 | Not-determined |
| S17_extracted_k141_100458 | Not-determined |
| S17_extracted_k141_56132 | Not-determined |
| S17_extracted_k141_56133 | Not-determined |
| S17_extracted_k141_78214 | Not-determined |
| S17_extracted_k141_25881 | Not-determined |
| S17_extracted_k141_122173 | Low-quality |
| S17_extracted_k141_122174 | Not-determined |
| S17_extracted_k141_114993 | Not-determined |
| S17_extracted_k141_84644 | Not-determined |
| S17_extracted_k141_78218 | Not-determined |
| S17_extracted_k141_70881 | Not-determined |
| S17_extracted_k141_18823 | Not-determined |
| S17_extracted_k141_92719 | Not-determined |
| S17_extracted_k141_11521 | Not-determined |
| S17_extracted_k141_100466 | Not-determined |
| S17_extracted_k141_144612 | Not-determined |
| S17_extracted_k141_92722 | Not-determined |
| S17_extracted_k141_18825 | Not-determined |
| S17_extracted_k141_25892 | Not-determined |
| S17_extracted_k141_115002 | Not-determined |
| S17_extracted_k141_92728 | Not-determined |
| S17_extracted_k141_70890 | Not-determined |
| S17_extracted_k141_108320 | Not-determined |
| S17_extracted_k141_115005 | Not-determined |
| S17_extracted_k141_4054 | Low-quality |
| S17_extracted_k141_4055 | Low-quality |
| S17_extracted_k141_4058 | Not-determined |
| S17_extracted_k141_18840 | Not-determined |
| S17_extracted_k141_152466 | Not-determined |
| S17_extracted_k141_41731 | Not-determined |
| S17_extracted_k141_152467 | Not-determined |
| S17_extracted_k141_144627 | Not-determined |
| S17_extracted_k141_166643 | Not-determined |
| S17_extracted_k141_56147 | Not-determined |
| S17_extracted_k141_92737 | Not-determined |
| S17_extracted_k141_78229 | Low-quality |
| S17_extracted_k141_41738 | Low-quality |
| S17_extracted_k141_144628 | Not-determined |
| S17_extracted_k141_92738 | Not-determined |
| S17_extracted_k141_152475 | Not-determined |
| S17_extracted_k141_78233 | Low-quality |
| S17_extracted_k141_56153 | Not-determined |
| S17_extracted_k141_56154 | Not-determined |
| S17_extracted_k141_144632 | Not-determined |
| S17_extracted_k141_48931 | Not-determined |
| S17_extracted_k141_108329 | Not-determined |
| S17_extracted_k141_152481 | Not-determined |
| S17_extracted_k141_48935 | Not-determined |
| S17_extracted_k141_78240 | Not-determined |
| S17_extracted_k141_173777 | Not-determined |
| S17_extracted_k141_152484 | Not-determined |
| S17_extracted_k141_115022 | Not-determined |
| S17_extracted_k141_25904 | Not-determined |
| S17_extracted_k141_34135 | Not-determined |
| S17_extracted_k141_173782 | Not-determined |
| S17_extracted_k141_18863 | Not-determined |
| S17_extracted_k141_166653 | Not-determined |
| S17_extracted_k141_78244 | Not-determined |
| S17_extracted_k141_70911 | Not-determined |
| S17_extracted_k141_173788 | Not-determined |
| S17_extracted_k141_4073 | Not-determined |
| S17_extracted_k141_129665 | Not-determined |
| S17_extracted_k141_56158 | Not-determined |
| S17_extracted_k141_56160 | Low-quality |
| S17_extracted_k141_173790 | Not-determined |
| S17_extracted_k141_4083 | Not-determined |
| S17_extracted_k141_159497 | Not-determined |
| S17_extracted_k141_70920 | Not-determined |
| S17_extracted_k141_78245 | Not-determined |
| S17_extracted_k141_100489 | Not-determined |
| S17_extracted_k141_48947 | Not-determined |
| S17_extracted_k141_115033 | Low-quality |
| S17_extracted_k141_4086 | Not-determined |
| S17_extracted_k141_56166 | Not-determined |
| S17_extracted_k141_41760 | Not-determined |
| S17_extracted_k141_48948 | Not-determined |
| S17_extracted_k141_108339 | Not-determined |
| S17_extracted_k141_100496 | Not-determined |
| S17_extracted_k141_136950 | Not-determined |
| S17_extracted_k141_92752 | Not-determined |
| S17_extracted_k141_25918 | Not-determined |
| S17_extracted_k141_18881 | Low-quality |
| S17_extracted_k141_25922 | Low-quality |
| S17_extracted_k141_11556 | Low-quality |
| S17_extracted_k141_48954 | Not-determined |
| S17_extracted_k141_84666 | Not-determined |
| S17_extracted_k141_56176 | Not-determined |
| S17_extracted_k141_92757 | Not-determined |
| S17_extracted_k141_78250 | Not-determined |
| S17_extracted_k141_48959 | Not-determined |
| S17_extracted_k141_78251 | Not-determined |
| S17_extracted_k141_115036 | Not-determined |
| S17_extracted_k141_41766 | Not-determined |
| S17_extracted_k141_108354 | Not-determined |
| S17_extracted_k141_129684 | Not-determined |
| S17_extracted_k141_152512 | Not-determined |
| S17_extracted_k141_115039 | Not-determined |
| S17_extracted_k141_108356 | Not-determined |
| S17_extracted_k141_136954 | Not-determined |
| S17_extracted_k141_108357 | Not-determined |
| S17_extracted_k141_136957 | Not-determined |
| S17_extracted_k141_159511 | Not-determined |
| S17_extracted_k141_108360 | Not-determined |
| S17_extracted_k141_25933 | Not-determined |
| S17_extracted_k141_136958 | Not-determined |
| S17_extracted_k141_48967 | Not-determined |
| S17_extracted_k141_173796 | Not-determined |
| S17_extracted_k141_18895 | Not-determined |
| S17_extracted_k141_11567 | Not-determined |
| S17_extracted_k141_159515 | Not-determined |
| S17_extracted_k141_144657 | Not-determined |
| S17_extracted_k141_136964 | Not-determined |
| S17_extracted_k141_84677 | Low-quality |
| S17_extracted_k141_34173 | Not-determined |
| S17_extracted_k141_34175 | Not-determined |
| S17_extracted_k141_115048 | Not-determined |
| S17_extracted_k141_115050 | Not-determined |
| S17_extracted_k141_70935 | Low-quality |
| S17_extracted_k141_41785 | Not-determined |
| S17_extracted_k141_4107 | Not-determined |
| S17_extracted_k141_78264 | Not-determined |
| S17_extracted_k141_41789 | Not-determined |
| S17_extracted_k141_173809 | Not-determined |
| S17_extracted_k141_70940 | Not-determined |
| S17_extracted_k141_4110 | Not-determined |
| S17_extracted_k141_56198 | Not-determined |
| S17_extracted_k141_166673 | Not-determined |
| S17_extracted_k141_173810 | Low-quality |
| S17_extracted_k141_4115 | Not-determined |
| S17_extracted_k141_166675 | Not-determined |
| S17_extracted_k141_136971 | Not-determined |
| S17_extracted_k141_84686 | Not-determined |
| S17_extracted_k141_144663 | Not-determined |
| S17_extracted_k141_56201 | Low-quality |
| S17_extracted_k141_144664 | Not-determined |
| S17_extracted_k141_173814 | Low-quality |
| S17_extracted_k141_84688 | Not-determined |
| S17_extracted_k141_41795 | Not-determined |
| S17_extracted_k141_18900 | Not-determined |
| S17_extracted_k141_84690 | Not-determined |
| S17_extracted_k141_84691 | Not-determined |
| S17_extracted_k141_56204 | Not-determined |
| S17_extracted_k141_129701 | Not-determined |
| S17_extracted_k141_56206 | Not-determined |
| S17_extracted_k141_136979 | Not-determined |
| S17_extracted_k141_41798 | Not-determined |
| S17_extracted_k141_136980 | Not-determined |
| S17_extracted_k141_100513 | Not-determined |
| S17_extracted_k141_136984 | Not-determined |
| S17_extracted_k141_115062 | Not-determined |
| S17_extracted_k141_78280 | Not-determined |
| S17_extracted_k141_166682 | Not-determined |
| S17_extracted_k141_173828 | Not-determined |
| S17_extracted_k141_173829 | Low-quality |
| S17_extracted_k141_173832 | Low-quality |
| S17_extracted_k141_34195 | Not-determined |
| S17_extracted_k141_34197 | Not-determined |
| S17_extracted_k141_115068 | Not-determined |
| S17_extracted_k141_122184 | Not-determined |
| S17_extracted_k141_34198 | Low-quality |
| S17_extracted_k141_144675 | Low-quality |
| S17_extracted_k141_136993 | Not-determined |
| S17_extracted_k141_173835 | Not-determined |
| S17_extracted_k141_70947 | Low-quality |
| S17_extracted_k141_84701 | Low-quality |
| S17_extracted_k141_34202 | Not-determined |
| S17_extracted_k141_84702 | Not-determined |
| S17_extracted_k141_34205 | Not-determined |
| S17_extracted_k141_78288 | Not-determined |
| S17_extracted_k141_56220 | Low-quality |
| S17_extracted_k141_78292 | Not-determined |
| S17_extracted_k141_166693 | Not-determined |
| S15_vs_75924\|\|full | High-quality |
| S17_extracted_k141_152531 | Low-quality |
| S17_extracted_k141_108381 | Low-quality |
| S17_extracted_k141_100520 | Not-determined |
| S17_extracted_k141_108383 | Not-determined |
| S17_extracted_k141_108385 | Not-determined |
| S17_extracted_k141_173841 | Not-determined |
| S17_extracted_k141_78293 | Not-determined |
| S17_extracted_k141_92786 | Not-determined |
| S17_extracted_k141_108390 | Not-determined |
| S17_extracted_k141_129722 | Not-determined |
| S17_extracted_k141_11576 | Not-determined |
| S17_extracted_k141_49000 | Not-determined |
| S17_extracted_k141_34220 | Not-determined |
| S17_extracted_k141_18933 | Not-determined |
| S17_extracted_k141_173847 | Not-determined |
| S17_extracted_k141_144690 | Not-determined |
| S17_extracted_k141_159545 | Not-determined |
| S17_extracted_k141_41820 | Not-determined |
| S17_extracted_k141_92789 | Low-quality |
| S17_extracted_k141_144693 | Low-quality |
| S17_extracted_k141_70967 | Not-determined |
| S17_extracted_k141_25961 | Low-quality |
| S17_extracted_k141_25962 | Not-determined |
| S17_extracted_k141_70972 | Not-determined |
| S17_extracted_k141_122195 | Not-determined |
| S17_extracted_k141_108406 | Not-determined |
| S17_extracted_k141_144698 | Not-determined |
| S17_extracted_k141_92793 | Not-determined |
| S17_extracted_k141_159553 | Not-determined |
| S17_extracted_k141_152554 | Not-determined |
| S17_extracted_k141_129733 | Low-quality |
| S17_extracted_k141_129736 | Not-determined |
| S17_extracted_k141_78307 | Low-quality |
| S17_extracted_k141_173861 | Not-determined |
| S17_extracted_k141_92799 | Low-quality |
| S17_extracted_k141_56237 | Not-determined |
| S17_extracted_k141_137026 | Not-determined |
| S17_extracted_k141_92801 | Not-determined |
| S17_extracted_k141_144707 | Not-determined |
| S17_extracted_k141_34230 | Low-quality |
| S17_extracted_k141_34231 | Not-determined |
| S17_extracted_k141_18951 | Not-determined |
| S17_extracted_k141_159562 | Not-determined |
| S17_extracted_k141_152566 | Not-determined |
| S17_extracted_k141_100549 | Low-quality |
| S17_extracted_k141_144715 | Not-determined |
| S17_extracted_k141_159566 | Not-determined |
| S17_extracted_k141_78313 | Not-determined |
| S17_extracted_k141_84709 | Not-determined |
| S17_extracted_k141_159567 | Low-quality |
| S17_extracted_k141_41836 | Not-determined |
| S17_extracted_k141_115102 | Not-determined |
| S17_extracted_k141_34241 | Not-determined |
| S17_extracted_k141_166713 | Not-determined |
| S17_extracted_k141_56252 | Not-determined |
| S17_extracted_k141_108420 | Not-determined |
| S17_extracted_k141_84714 | Not-determined |
| S17_extracted_k141_56254 | Not-determined |
| S17_extracted_k141_34243 | Low-quality |
| S17_extracted_k141_108423 | Not-determined |
| S17_extracted_k141_159570 | Not-determined |
| S17_extracted_k141_84724 | Low-quality |
| S17_extracted_k141_70978 | Not-determined |
| S17_extracted_k141_84725 | Not-determined |
| S17_extracted_k141_159573 | Low-quality |
| S17_extracted_k141_4146 | Not-determined |
| S17_extracted_k141_159574 | Low-quality |
| S17_extracted_k141_152578 | Not-determined |
| S17_extracted_k141_108432 | Not-determined |
| S17_extracted_k141_11605 | Low-quality |
| S17_extracted_k141_159577 | Low-quality |
| S17_extracted_k141_18972 | Low-quality |
| S17_extracted_k141_122209 | Not-determined |
| S17_extracted_k141_159580 | Not-determined |
| S17_extracted_k141_108435 | Not-determined |
| S17_extracted_k141_84732 | Not-determined |
| S17_extracted_k141_159582 | Not-determined |
| S17_extracted_k141_129776 | Not-determined |
| S17_extracted_k141_159583 | Not-determined |
| S17_extracted_k141_11609 | Not-determined |
| S17_extracted_k141_49026 | Not-determined |
| S17_extracted_k141_100570 | Not-determined |
| S17_extracted_k141_18984 | Low-quality |
| S17_extracted_k141_129780 | Not-determined |
| S17_extracted_k141_62559 | Not-determined |
| S17_extracted_k141_4153 | Not-determined |
| S17_extracted_k141_34263 | Not-determined |
| S17_extracted_k141_4155 | Not-determined |
| S17_extracted_k141_108439 | Not-determined |
| S17_extracted_k141_4156 | Not-determined |
| S17_extracted_k141_56282 | Not-determined |
| S17_extracted_k141_70994 | Not-determined |
| S17_extracted_k141_122223 | Not-determined |
| S17_extracted_k141_173895 | Not-determined |
| S17_extracted_k141_159588 | Not-determined |
| S17_extracted_k141_34264 | Low-quality |
| S17_extracted_k141_173897 | Not-determined |
| S17_extracted_k141_173898 | Not-determined |
| S17_extracted_k141_70997 | Not-determined |
| S17_extracted_k141_4162 | Not-determined |
| S17_extracted_k141_62564 | Not-determined |
| S17_extracted_k141_25989 | Not-determined |
| S17_extracted_k141_152598 | Not-determined |
| S17_extracted_k141_18988 | Low-quality |
| S17_extracted_k141_166732 | Not-determined |
| S17_extracted_k141_108453 | Not-determined |
| S17_extracted_k141_11622 | Not-determined |
| S17_extracted_k141_11623 | Low-quality |
| S17_extracted_k141_62567 | Low-quality |
| S17_extracted_k141_137047 | Not-determined |
| S17_extracted_k141_4170 | Not-determined |
| S17_extracted_k141_137049 | Low-quality |
| S17_extracted_k141_11626 | Not-determined |
| S17_extracted_k141_71010 | Not-determined |
| S17_extracted_k141_18998 | Not-determined |
| S17_extracted_k141_152605 | Not-determined |
| S17_extracted_k141_62575 | Not-determined |
| S17_extracted_k141_19000 | Not-determined |
| S17_extracted_k141_100588 | Low-quality |
| S17_extracted_k141_159598 | Not-determined |
| S17_extracted_k141_166748 | Not-determined |
| S17_extracted_k141_159599 | Not-determined |
| S17_extracted_k141_173911 | Low-quality |
| S17_extracted_k141_166750 | Low-quality |
| S17_extracted_k141_108461 | Not-determined |
| S17_extracted_k141_144746 | Not-determined |
| S17_extracted_k141_84754 | Low-quality |
| S17_extracted_k141_71021 | Not-determined |
| S17_extracted_k141_137055 | Not-determined |
| S17_extracted_k141_34279 | Not-determined |
| S17_extracted_k141_137057 | Not-determined |
| S17_extracted_k141_144750 | Not-determined |
| S17_extracted_k141_159606 | Not-determined |
| S17_extracted_k141_92843 | Not-determined |
| S17_extracted_k141_84761 | Not-determined |
| S17_extracted_k141_137063 | Not-determined |
| S17_extracted_k141_129796 | Low-quality |
| S17_extracted_k141_122249 | Not-determined |
| S17_extracted_k141_144754 | Not-determined |

| | |
|---|---|
| S17_extracted_k141_129798 | Not-determined |
| S17_extracted_k141_137066 | Not-determined |
| S17_extracted_k141_115125 | Not-determined |
| S17_extracted_k141_34284 | Not-determined |
| S17_extracted_k141_11651 | Not-determined |
| S17_extracted_k141_173919 | Low-quality |
| S17_extracted_k141_115127 | Not-determined |
| S17_extracted_k141_173921 | Low-quality |
| S17_extracted_k141_56314 | Not-determined |
| S17_extracted_k141_152617 | Not-determined |
| S17_extracted_k141_49063 | Not-determined |
| S17_extracted_k141_62589 | Not-determined |
| S17_extracted_k141_108471 | Not-determined |
| S17_extracted_k141_26007 | Not-determined |
| S17_extracted_k141_62592 | Not-determined |
| S17_extracted_k141_49069 | Not-determined |
| S17_extracted_k141_137077 | Not-determined |
| S17_extracted_k141_122257 | Not-determined |
| S17_extracted_k141_92855 | Low-quality |
| S17_extracted_k141_92858 | Not-determined |
| S17_extracted_k141_84774 | Not-determined |
| S17_extracted_k141_26013 | Not-determined |
| S17_extracted_k141_122260 | Not-determined |
| S17_extracted_k141_49077 | Not-determined |
| S17_extracted_k141_34297 | Not-determined |
| S17_extracted_k141_115135 | Not-determined |
| S17_extracted_k141_152636 | Not-determined |
| S17_extracted_k141_49081 | Not-determined |
| S17_extracted_k141_115139 | Not-determined |
| S17_extracted_k141_144763 | Not-determined |
| S17_extracted_k141_4207 | Not-determined |
| S17_extracted_k141_115142 | Not-determined |
| S17_extracted_k141_115145 | Not-determined |
| S17_extracted_k141_166770 | Low-quality |
| S17_extracted_k141_115148 | Not-determined |
| S17_extracted_k141_108479 | Not-determined |
| S17_extracted_k141_49089 | Low-quality |
| S17_extracted_k141_71049 | Not-determined |
| S17_extracted_k141_137093 | Not-determined |
| S17_extracted_k141_137094 | Not-determined |
| S17_extracted_k141_71051 | Not-determined |
| S17_extracted_k141_100605 | Not-determined |
| S17_extracted_k141_115153 | Low-quality |
| S17_extracted_k141_144770 | Not-determined |
| S17_extracted_k141_62608 | Low-quality |
| S17_extracted_k141_137096 | Low-quality |
| S17_extracted_k141_159627 | Not-determined |
| S17_extracted_k141_34311 | Not-determined |
| S17_extracted_k141_56338 | Not-determined |
| S17_extracted_k141_108480 | Not-determined |
| S17_extracted_k141_62612 | Not-determined |
| S17_extracted_k141_137102 | Not-determined |
| S17_extracted_k141_62615 | Not-determined |
| S17_extracted_k141_62616 | Not-determined |
| S17_extracted_k141_56341 | Not-determined |
| S17_extracted_k141_4223 | Low-quality |
| S17_extracted_k141_19046 | Not-determined |
| S17_extracted_k141_152658 | Not-determined |
| S17_extracted_k141_129819 | Not-determined |
| S17_extracted_k141_71068 | Not-determined |
| S17_extracted_k141_56342 | Low-quality |
| S17_extracted_k141_11679 | Low-quality |
| S17_extracted_k141_84800 | Not-determined |
| S17_extracted_k141_71070 | Not-determined |
| S17_extracted_k141_108493 | Not-determined |
| S17_extracted_k141_115172 | Not-determined |
| S17_extracted_k141_19050 | Not-determined |
| S17_extracted_k141_19052 | Low-quality |
| S17_extracted_k141_108497 | Not-determined |
| S17_extracted_k141_129824 | Not-determined |
| S17_extracted_k141_71076 | Not-determined |
| S17_extracted_k141_173941 | Not-determined |
| S17_extracted_k141_152662 | Not-determined |
| S17_extracted_k141_159635 | Not-determined |
| S17_extracted_k141_159636 | Not-determined |
| S17_extracted_k141_49110 | Not-determined |
| S17_extracted_k141_173945 | Low-quality |
| S17_extracted_k141_56346 | Low-quality |
| S17_extracted_k141_11685 | Not-determined |
| S17_extracted_k141_152666 | Not-determined |
| S17_extracted_k141_11686 | Not-determined |
| S17_extracted_k141_49117 | Not-determined |
| S17_extracted_k141_49118 | Not-determined |
| S17_extracted_k141_129831 | Not-determined |
| S17_extracted_k141_71083 | Not-determined |
| S17_extracted_k141_49120 | Not-determined |
| S17_extracted_k141_19065 | Not-determined |
| S17_extracted_k141_26044 | Not-determined |
| S17_extracted_k141_84804 | Not-determined |
| S17_extracted_k141_144787 | Not-determined |
| S17_extracted_k141_26047 | Not-determined |
| S17_extracted_k141_19069 | Not-determined |
| S17_extracted_k141_159649 | Not-determined |
| S17_extracted_k141_159650 | Not-determined |
| S17_extracted_k141_129836 | Not-determined |
| S17_extracted_k141_166799 | Not-determined |
| S17_extracted_k141_137128 | Not-determined |
| S17_extracted_k141_41872 | Not-determined |
| S17_extracted_k141_144792 | Not-determined |
| S17_extracted_k141_129840 | Not-determined |
| S17_extracted_k141_41874 | Not-determined |
| S17_extracted_k141_56352 | Not-determined |
| S17_extracted_k141_115189 | Not-determined |
| S17_extracted_k141_129843 | Not-determined |
| S17_extracted_k141_173963 | Low-quality |
| S17_extracted_k141_166809 | Not-determined |
| S17_extracted_k141_144796 | Not-determined |
| S17_extracted_k141_115195 | Not-determined |
| S17_extracted_k141_11702 | Not-determined |
| S17_extracted_k141_62646 | Not-determined |
| S17_extracted_k141_166812 | Not-determined |
| S17_extracted_k141_115199 | Not-determined |
| S17_extracted_k141_41882 | Not-determined |
| S17_extracted_k141_78351 | Not-determined |
| S17_extracted_k141_108504 | Not-determined |
| S17_extracted_k141_49129 | Not-determined |
| S17_extracted_k141_78352 | Not-determined |
| S17_extracted_k141_71096 | Not-determined |
| S17_extracted_k141_62654 | Not-determined |
| S17_extracted_k141_78358 | Not-determined |
| S17_extracted_k141_62655 | Not-determined |
| S17_extracted_k141_137140 | Low-quality |
| S17_extracted_k141_84809 | Not-determined |
| S17_extracted_k141_100627 | Not-determined |
| S17_extracted_k141_62658 | Not-determined |
| S17_extracted_k141_108508 | Not-determined |
| S17_extracted_k141_78364 | Not-determined |
| S17_extracted_k141_108509 | Not-determined |
| S17_extracted_k141_71101 | Not-determined |
| S17_extracted_k141_100631 | Not-determined |
| S17_extracted_k141_62659 | Not-determined |
| S17_extracted_k141_26054 | Not-determined |
| S17_extracted_k141_144802 | Not-determined |
| S17_extracted_k141_41892 | Not-determined |
| S17_extracted_k141_108513 | Not-determined |
| S17_extracted_k141_26056 | Not-determined |
| S17_extracted_k141_26057 | Not-determined |
| S17_extracted_k141_159676 | Not-determined |
| S17_extracted_k141_41894 | Not-determined |
| S17_extracted_k141_100638 | Not-determined |
| S17_extracted_k141_152671 | Not-determined |
| S17_extracted_k141_115217 | Not-determined |
| S17_extracted_k141_166823 | Not-determined |
| S17_extracted_k141_41901 | Not-determined |
| S17_extracted_k141_144812 | Not-determined |
| S17_extracted_k141_173981 | Not-determined |
| S17_extracted_k141_173982 | Not-determined |
| S17_extracted_k141_62671 | Not-determined |
| S17_extracted_k141_26062 | Low-quality |
| S17_extracted_k141_26064 | Not-determined |
| S17_extracted_k141_41910 | Low-quality |
| S17_extracted_k141_166828 | Not-determined |
| S17_extracted_k141_100641 | Not-determined |
| S17_extracted_k141_137157 | Not-determined |
| S17_extracted_k141_78372 | Low-quality |
| S17_extracted_k141_4256 | Low-quality |
| S17_extracted_k141_173993 | Low-quality |
| S17_extracted_k141_62674 | Not-determined |
| S17_extracted_k141_159691 | Not-determined |
| S17_extracted_k141_115224 | Low-quality |
| S17_extracted_k141_152689 | Not-determined |
| S17_extracted_k141_71115 | Not-determined |
| S17_extracted_k141_71116 | Not-determined |
| S17_extracted_k141_26082 | Not-determined |
| S17_extracted_k141_78381 | Not-determined |
| S17_extracted_k141_137165 | Not-determined |
| S17_extracted_k141_129865 | Low-quality |
| S17_extracted_k141_78384 | Not-determined |
| S17_extracted_k141_92872 | Not-determined |
| S17_extracted_k141_26086 | Low-quality |
| S17_extracted_k141_174008 | Not-determined |
| S17_extracted_k141_62688 | Low-quality |
| S17_extracted_k141_166838 | Not-determined |
| S17_extracted_k141_78387 | Not-determined |
| S17_extracted_k141_100664 | Not-determined |
| S17_extracted_k141_56374 | Not-determined |
| S17_extracted_k141_26088 | Not-determined |
| S17_extracted_k141_159706 | Not-determined |
| S17_extracted_k141_49139 | Low-quality |
| S17_extracted_k141_78392 | Not-determined |
| S17_extracted_k141_100670 | Not-determined |
| S17_extracted_k141_26096 | Not-determined |
| S17_extracted_k141_159713 | Low-quality |
| S17_extracted_k141_129875 | Not-determined |
| S17_extracted_k141_137181 | Not-determined |
| S17_extracted_k141_26098 | Not-determined |
| S17_extracted_k141_49146 | Low-quality |
| S17_extracted_k141_144818 | Not-determined |
| S17_extracted_k141_49147 | Not-determined |
| S17_extracted_k141_144819 | Not-determined |
| S17_extracted_k141_159720 | Not-determined |
| S17_extracted_k141_4288 | Not-determined |
| S17_extracted_k141_166863 | Low-quality |
| S17_extracted_k141_49150 | Not-determined |
| S17_extracted_k141_174012 | Not-determined |
| S17_extracted_k141_92883 | Not-determined |
| S17_extracted_k141_100683 | Not-determined |
| S17_extracted_k141_159724 | Not-determined |
| S17_extracted_k141_49151 | Not-determined |
| S17_extracted_k141_84833 | Not-determined |
| S17_extracted_k141_100685 | Low-quality |
| S17_extracted_k141_56385 | Not-determined |
| S17_extracted_k141_174015 | Not-determined |
| S17_extracted_k141_78400 | Not-determined |
| S17_extracted_k141_4297 | Not-determined |
| S17_extracted_k141_159726 | Not-determined |
| S17_extracted_k141_174019 | Not-determined |
| S17_extracted_k141_4298 | Not-determined |
| S17_extracted_k141_26105 | Not-determined |
| S17_extracted_k141_78406 | Low-quality |
| S17_extracted_k141_78408 | Not-determined |
| S17_extracted_k141_41935 | Low-quality |
| S17_extracted_k141_49161 | Not-determined |
| S17_extracted_k141_100695 | Not-determined |
| S17_extracted_k141_84844 | Low-quality |
| S17_extracted_k141_108518 | Not-determined |
| S17_extracted_k141_166871 | Not-determined |
| S17_extracted_k141_11731 | Not-determined |
| S17_extracted_k141_71136 | Low-quality |
| S17_extracted_k141_129898 | Not-determined |
| S17_extracted_k141_26113 | Low-quality |
| S17_extracted_k141_100700 | Low-quality |
| S17_extracted_k141_78414 | Not-determined |
| S17_extracted_k141_78415 | Not-determined |
| S17_extracted_k141_71139 | Low-quality |
| S17_extracted_k141_34316 | Low-quality |
| S17_extracted_k141_174026 | Not-determined |
| S17_extracted_k141_174027 | Not-determined |
| S17_extracted_k141_166875 | Not-determined |
| S17_extracted_k141_100704 | Not-determined |
| S17_extracted_k141_78419 | Low-quality |
| S17_extracted_k141_115240 | Not-determined |
| S17_extracted_k141_56399 | Not-determined |
| S17_extracted_k141_152730 | Low-quality |
| S17_extracted_k141_62710 | Low-quality |
| S17_extracted_k141_56401 | Not-determined |
| S17_extracted_k141_41947 | Not-determined |
| S17_extracted_k141_34321 | Not-determined |
| S17_extracted_k141_166878 | Not-determined |
| S17_extracted_k141_115246 | Not-determined |
| S17_extracted_k141_129909 | Not-determined |
| S17_extracted_k141_129910 | Not-determined |
| S17_extracted_k141_49168 | Not-determined |
| S17_extracted_k141_49169 | Not-determined |
| S17_extracted_k141_129913 | Not-determined |
| S17_extracted_k141_129914 | Not-determined |
| S17_extracted_k141_34326 | Not-determined |
| S17_extracted_k141_34327 | Not-determined |
| S17_extracted_k141_41950 | Not-determined |
| S17_extracted_k141_26116 | Not-determined |
| S17_extracted_k141_78427 | Not-determined |
| S17_extracted_k141_92908 | Not-determined |
| S17_extracted_k141_159752 | Not-determined |
| S17_extracted_k141_34332 | Not-determined |
| S17_extracted_k141_115257 | Not-determined |
| S17_extracted_k141_49173 | Not-determined |
| S17_extracted_k141_62719 | Not-determined |
| S17_extracted_k141_62720 | Not-determined |
| S17_extracted_k141_62721 | Not-determined |
| S17_extracted_k141_92913 | Not-determined |
| S17_extracted_k141_92914 | Not-determined |
| S17_extracted_k141_78432 | Not-determined |
| S17_extracted_k141_144842 | Low-quality |
| S17_extracted_k141_100719 | Low-quality |
| S17_extracted_k141_92916 | Not-determined |
| S17_extracted_k141_84863 | Not-determined |
| S17_extracted_k141_4318 | Not-determined |
| S17_extracted_k141_34337 | Not-determined |
| S17_extracted_k141_4321 | Not-determined |
| S17_extracted_k141_62727 | Not-determined |
| S17_extracted_k141_137189 | Low-quality |
| S17_extracted_k141_78447 | Not-determined |
| S17_extracted_k141_34341 | Low-quality |
| S17_extracted_k141_92921 | Not-determined |
| S17_extracted_k141_56424 | Not-determined |
| S17_extracted_k141_144851 | Not-determined |
| S17_extracted_k141_159762 | Low-quality |
| S17_extracted_k141_56427 | Not-determined |
| S17_extracted_k141_92927 | Not-determined |
| S17_extracted_k141_137194 | Low-quality |
| S17_extracted_k141_100725 | Low-quality |
| S17_extracted_k141_41969 | Not-determined |
| S17_extracted_k141_152742 | Not-determined |
| S17_extracted_k141_115272 | Not-determined |
| S17_extracted_k141_26128 | Not-determined |
| S17_extracted_k141_159767 | Not-determined |
| S17_extracted_k141_41973 | Not-determined |
| S17_extracted_k141_92929 | Not-determined |
| S17_extracted_k141_92930 | Not-determined |
| S17_extracted_k141_152746 | Not-determined |
| S17_extracted_k141_78454 | Not-determined |
| S17_extracted_k141_108529 | Not-determined |
| S17_extracted_k141_84868 | Not-determined |
| S17_extracted_k141_92932 | Not-determined |
| S17_extracted_k141_84870 | Not-determined |
| S17_extracted_k141_159770 | Not-determined |
| S17_extracted_k141_49193 | Not-determined |
| S17_extracted_k141_92933 | Not-determined |
| S17_extracted_k141_100733 | Not-determined |
| S17_extracted_k141_49194 | Low-quality |
| S17_extracted_k141_115276 | Not-determined |
| S17_extracted_k141_49196 | Not-determined |
| S17_extracted_k141_49197 | Not-determined |
| S17_extracted_k141_62733 | Low-quality |
| S17_extracted_k141_166903 | Not-determined |
| S17_extracted_k141_56432 | Not-determined |
| S17_extracted_k141_166904 | Not-determined |
| S17_extracted_k141_84876 | Not-determined |
| S17_extracted_k141_152749 | Not-determined |
| S17_extracted_k141_137198 | Not-determined |
| S17_extracted_k141_152752 | Not-determined |
| S17_extracted_k141_62735 | Not-determined |
| S17_extracted_k141_166907 | Not-determined |
| S17_extracted_k141_4335 | Not-determined |
| S17_extracted_k141_115286 | Not-determined |
| S17_extracted_k141_166909 | Not-determined |
| S17_extracted_k141_159776 | Not-determined |
| S17_extracted_k141_78463 | Not-determined |
| S17_extracted_k141_152756 | Low-quality |
| S17_extracted_k141_78464 | Not-determined |
| S17_extracted_k141_159777 | Not-determined |
| S17_extracted_k141_62743 | Not-determined |
| S17_extracted_k141_159780 | Low-quality |
| S17_extracted_k141_115296 | Low-quality |
| S17_extracted_k141_26145 | Not-determined |
| S17_extracted_k141_26146 | Not-determined |
| S17_extracted_k141_152768 | Not-determined |
| S17_extracted_k141_34366 | Low-quality |
| S17_extracted_k141_62753 | Not-determined |
| S17_extracted_k141_49214 | Not-determined |
| S17_extracted_k141_34368 | Not-determined |
| S17_extracted_k141_62756 | Not-determined |
| S17_extracted_k141_159784 | Low-quality |
| S17_extracted_k141_152775 | Not-determined |
| S17_extracted_k141_78468 | Not-determined |
| S17_extracted_k141_34371 | Not-determined |
| S17_extracted_k141_34372 | Not-determined |
| S17_extracted_k141_41995 | Low-quality |
| S17_extracted_k141_56455 | Not-determined |
| S17_extracted_k141_71146 | Not-determined |
| S17_extracted_k141_159789 | Low-quality |
| S17_extracted_k141_115306 | Not-determined |
| S17_extracted_k141_159793 | Not-determined |
| S17_extracted_k141_62763 | Not-determined |
| S17_extracted_k141_152784 | Not-determined |
| S17_extracted_k141_166915 | Not-determined |
| S17_extracted_k141_71156 | Not-determined |
| S17_extracted_k141_49219 | Not-determined |
| S17_extracted_k141_144880 | Not-determined |
| S17_extracted_k141_49222 | Low-quality |
| S17_extracted_k141_4347 | Not-determined |
| S17_extracted_k141_92956 | Not-determined |
| S17_extracted_k141_56462 | Not-determined |
| S17_extracted_k141_159803 | Not-determined |
| S17_extracted_k141_144887 | Low-quality |
| S17_extracted_k141_26167 | Not-determined |
| S17_extracted_k141_26168 | Not-determined |
| S17_extracted_k141_144891 | Not-determined |
| S17_extracted_k141_144892 | Not-determined |
| S17_extracted_k141_137224 | Not-determined |
| S17_extracted_k141_26170 | Not-determined |
| S17_extracted_k141_152793 | Not-determined |
| S17_extracted_k141_26172 | Not-determined |
| S17_extracted_k141_56469 | Not-determined |
| S17_extracted_k141_71172 | Low-quality |
| S17_extracted_k141_137229 | Not-determined |
| S17_extracted_k141_56475 | Not-determined |
| S17_extracted_k141_49237 | Not-determined |
| S17_extracted_k141_137232 | Not-determined |
| S17_extracted_k141_71177 | Not-determined |
| S17_extracted_k141_159811 | Not-determined |
| S17_extracted_k141_152805 | Low-quality |
| S17_extracted_k141_137234 | Not-determined |
| S17_extracted_k141_152806 | Low-quality |
| S17_extracted_k141_166941 | Not-determined |
| S17_extracted_k141_115320 | Low-quality |
| S17_extracted_k141_42004 | Not-determined |
| S17_extracted_k141_152811 | Low-quality |
| S17_extracted_k141_4357 | Low-quality |
| S17_extracted_k141_144900 | Not-determined |
| S17_extracted_k141_152815 | Not-determined |
| S17_extracted_k141_152816 | Not-determined |
| S17_extracted_k141_78493 | Not-determined |
| S17_extracted_k141_26195 | Low-quality |
| S17_extracted_k141_56484 | Not-determined |
| S17_extracted_k141_84922 | Not-determined |
| S17_extracted_k141_166953 | Not-determined |
| S17_extracted_k141_34391 | Low-quality |
| S17_extracted_k141_100760 | Not-determined |
| S17_extracted_k141_159821 | Not-determined |
| S17_extracted_k141_115328 | Not-determined |
| S17_extracted_k141_100764 | Not-determined |
| S17_extracted_k141_62782 | Not-determined |
| S17_extracted_k141_4366 | Not-determined |
| S17_extracted_k141_62784 | Not-determined |
| S17_extracted_k141_152823 | Low-quality |
| S17_extracted_k141_108538 | Not-determined |
| S17_extracted_k141_115333 | Not-determined |
| S17_extracted_k141_115336 | Not-determined |
| S17_extracted_k141_4371 | Not-determined |
| S17_extracted_k141_62786 | Low-quality |
| S17_extracted_k141_56497 | Not-determined |
| S17_extracted_k141_137255 | Not-determined |
| S17_extracted_k141_62787 | Not-determined |
| S17_extracted_k141_144914 | Not-determined |
| S17_extracted_k141_62789 | Low-quality |
| S17_extracted_k141_11757 | Not-determined |
| S17_extracted_k141_115344 | Not-determined |
| S17_extracted_k141_71198 | Not-determined |
| S17_extracted_k141_100775 | Not-determined |
| S17_extracted_k141_115348 | Low-quality |
| S17_extracted_k141_100780 | Not-determined |
| S17_extracted_k141_152834 | Not-determined |
| S17_extracted_k141_100781 | Low-quality |
| S17_extracted_k141_42026 | Not-determined |
| S17_extracted_k141_71205 | Not-determined |
| S17_extracted_k141_152838 | Not-determined |
| S17_extracted_k141_49262 | Not-determined |
| S17_extracted_k141_62798 | Not-determined |
| S17_extracted_k141_42030 | Not-determined |
| S17_extracted_k141_115356 | Low-quality |
| S17_extracted_k141_115357 | Not-determined |
| S17_extracted_k141_84939 | Not-determined |
| S17_extracted_k141_144918 | Not-determined |
| S17_extracted_k141_56515 | Not-determined |
| S6_extracted_k141_11228 | Complete |
| S17_extracted_k141_56519 | Not-determined |
| S17_extracted_k141_144923 | Not-determined |
| S17_extracted_k141_174034 | Not-determined |
| S17_extracted_k141_49275 | Not-determined |
| S17_extracted_k141_174037 | Not-determined |
| S17_extracted_k141_115369 | Not-determined |
| S17_extracted_k141_115370 | Low-quality |
| S17_extracted_k141_42036 | Not-determined |
| S17_extracted_k141_84947 | Not-determined |
| S17_extracted_k141_49278 | Low-quality |
| S17_extracted_k141_115375 | Not-determined |
| S17_extracted_k141_84948 | Not-determined |
| S17_extracted_k141_11771 | Not-determined |
| S17_extracted_k141_115377 | Not-determined |
| S17_extracted_k141_137264 | Not-determined |
| S17_extracted_k141_115378 | Not-determined |
| S17_extracted_k141_49285 | Not-determined |
| S17_extracted_k141_34414 | Low-quality |
| S17_extracted_k141_42040 | Not-determined |
| S17_extracted_k141_56533 | Not-determined |
| S17_extracted_k141_56535 | Not-determined |
| S17_extracted_k141_11776 | Not-determined |
| S17_extracted_k141_108550 | Low-quality |
| S17_extracted_k141_11778 | Not-determined |
| S17_extracted_k141_11779 | Not-determined |
| S17_extracted_k141_34420 | Low-quality |
| S17_extracted_k141_100801 | Not-determined |
| S17_extracted_k141_42044 | Not-determined |
| S17_extracted_k141_115389 | Low-quality |
| S17_extracted_k141_115390 | Not-determined |
| S17_extracted_k141_115391 | Not-determined |
| S17_extracted_k141_100808 | Low-quality |
| S17_extracted_k141_152843 | Not-determined |
| S17_extracted_k141_78528 | Not-determined |
| S17_extracted_k141_11795 | Low-quality |
| S17_extracted_k141_42052 | Not-determined |
| S17_extracted_k141_26227 | Not-determined |
| S17_extracted_k141_78534 | Not-determined |
| S17_extracted_k141_42054 | Not-determined |
| S17_extracted_k141_49305 | Not-determined |
| S17_extracted_k141_49307 | Not-determined |
| S17_extracted_k141_115398 | Not-determined |
| S17_extracted_k141_49309 | Not-determined |
| S17_extracted_k141_137273 | Not-determined |
| S17_extracted_k141_159828 | Not-determined |
| S17_extracted_k141_144942 | Not-determined |
| S17_extracted_k141_84955 | Not-determined |
| S17_extracted_k141_137279 | Low-quality |
| S17_extracted_k141_78538 | Not-determined |
| S17_extracted_k141_42065 | Low-quality |
| S17_extracted_k141_84960 | Not-determined |
| S17_extracted_k141_100826 | Not-determined |
| S17_extracted_k141_84962 | Not-determined |
| S17_extracted_k141_56560 | Not-determined |
| S17_extracted_k141_115408 | Not-determined |
| S17_extracted_k141_100831 | Not-determined |
| S17_extracted_k141_56563 | Not-determined |
| S17_extracted_k141_100832 | Not-determined |
| S17_extracted_k141_56566 | Not-determined |
| S17_extracted_k141_159838 | Not-determined |
| S17_extracted_k141_11817 | Not-determined |
| S17_extracted_k141_108569 | Not-determined |
| S17_extracted_k141_11818 | Not-determined |
| S17_extracted_k141_42075 | Not-determined |
| S17_extracted_k141_144952 | Not-determined |
| S17_extracted_k141_34437 | Not-determined |
| S17_extracted_k141_115416 | Not-determined |
| S17_extracted_k141_71213 | Not-determined |
| S17_extracted_k141_174067 | Not-determined |
| S17_extracted_k141_167019 | Not-determined |
| S17_extracted_k141_84975 | Not-determined |
| S17_extracted_k141_71216 | Not-determined |
| S17_extracted_k141_42082 | Not-determined |
| S17_extracted_k141_159847 | Not-determined |
| S17_extracted_k141_108580 | Not-determined |
| S17_extracted_k141_129955 | Not-determined |
| S17_extracted_k141_92981 | Not-determined |
| S17_extracted_k141_167025 | Not-determined |
| S17_extracted_k141_71220 | Not-determined |
| S17_extracted_k141_108583 | Not-determined |
| S17_extracted_k141_137301 | Not-determined |
| S17_extracted_k141_71222 | Not-determined |
| S17_extracted_k141_71223 | Not-determined |
| S17_extracted_k141_137302 | Not-determined |
| S6_vs_k141_11228\|\|full | High-quality |
| S17_extracted_k141_108584 | Not-determined |
| S17_extracted_k141_49327 | Not-determined |
| S17_extracted_k141_174073 | Not-determined |
| S17_extracted_k141_144963 | Not-determined |
| S17_extracted_k141_174075 | Not-determined |
| S17_extracted_k141_49331 | Not-determined |
| S17_extracted_k141_122266 | Low-quality |
| S17_extracted_k141_34453 | Not-determined |
| S17_extracted_k141_34454 | Not-determined |
| S17_extracted_k141_26237 | Low-quality |
| S17_extracted_k141_100848 | Not-determined |
| S17_extracted_k141_174080 | Not-determined |
| S17_extracted_k141_167032 | Not-determined |
| S17_extracted_k141_19091 | Not-determined |
| S17_extracted_k141_19093 | Not-determined |
| S17_extracted_k141_56586 | Not-determined |
| S17_extracted_k141_152866 | Not-determined |
| S17_extracted_k141_144966 | Not-determined |
| S17_extracted_k141_49335 | Not-determined |
| S17_extracted_k141_71230 | Not-determined |
| S17_extracted_k141_4390 | Not-determined |
| S17_extracted_k141_56587 | Not-determined |
| S17_extracted_k141_137310 | Low-quality |
| S17_extracted_k141_174084 | Not-determined |
| S17_extracted_k141_115433 | Not-determined |
| S17_extracted_k141_174088 | Not-determined |
| S17_extracted_k141_71238 | Not-determined |
| S17_extracted_k141_71239 | Not-determined |
| S17_extracted_k141_167040 | Not-determined |
| S17_extracted_k141_4396 | Low-quality |
| S17_extracted_k141_137315 | Not-determined |
| S17_extracted_k141_167042 | Not-determined |
| S17_extracted_k141_4397 | Not-determined |
| S17_extracted_k141_115441 | Not-determined |
| S17_extracted_k141_26242 | Not-determined |
| S17_extracted_k141_19103 | Not-determined |
| S17_extracted_k141_137320 | Not-determined |
| S17_extracted_k141_78552 | Low-quality |
| S17_extracted_k141_78553 | Not-determined |
| S17_extracted_k141_108601 | Not-determined |
| S17_extracted_k141_137322 | Not-determined |
| S17_extracted_k141_129969 | Not-determined |
| S17_extracted_k141_56601 | Low-quality |
| S17_extracted_k141_115445 | Not-determined |
| S17_extracted_k141_167051 | Not-determined |
| S17_extracted_k141_49344 | Not-determined |
| S17_extracted_k141_62839 | Not-determined |
| S17_extracted_k141_174097 | Not-determined |
| S17_extracted_k141_4407 | Not-determined |
| S17_extracted_k141_115448 | Not-determined |
| S17_extracted_k141_4409 | Not-determined |
| S17_extracted_k141_71247 | Not-determined |
| S17_extracted_k141_78560 | Not-determined |
| S17_extracted_k141_26251 | Low-quality |
| S17_extracted_k141_42095 | Not-determined |
| S17_extracted_k141_174102 | Not-determined |
| S17_extracted_k141_56608 | Not-determined |
| S17_extracted_k141_122287 | Not-determined |
| S17_extracted_k141_56612 | Not-determined |
| S17_extracted_k141_174104 | Not-determined |
| S17_extracted_k141_62850 | Not-determined |
| S17_extracted_k141_115454 | Not-determined |
| S17_extracted_k141_137339 | Not-determined |
| S17_extracted_k141_71251 | Low-quality |
| S17_extracted_k141_56619 | Not-determined |
| S17_extracted_k141_122290 | Not-determined |
| S17_extracted_k141_42101 | Not-determined |
| S17_extracted_k141_4420 | Low-quality |
| S17_extracted_k141_42102 | Not-determined |
| S17_extracted_k141_144999 | Not-determined |
| S17_extracted_k141_174114 | Not-determined |
| S17_extracted_k141_122293 | Not-determined |
| S17_extracted_k141_34482 | Not-determined |
| S17_extracted_k141_137347 | Not-determined |
| S17_extracted_k141_129986 | Not-determined |
| S17_extracted_k141_137349 | Not-determined |
| S17_extracted_k141_159875 | Low-quality |
| S17_extracted_k141_42108 | Not-determined |

| | |
|---|---|
| S17_extracted_k141_42109 | Not-determined |
| S17_extracted_k141_137353 | Not-determined |
| S17_extracted_k141_42111 | Not-determined |
| S17_extracted_k141_145003 | Not-determined |
| S17_extracted_k141_108623 | Low-quality |
| S17_extracted_k141_92996 | Not-determined |
| S17_extracted_k141_115464 | Low-quality |
| S17_extracted_k141_92997 | Not-determined |
| S17_extracted_k141_159880 | Not-determined |
| S17_extracted_k141_34485 | Low-quality |
| S17_extracted_k141_4432 | Not-determined |
| S17_extracted_k141_159885 | Not-determined |
| S17_extracted_k141_93000 | Not-determined |
| S17_extracted_k141_4439 | Not-determined |
| S17_extracted_k141_78571 | Not-determined |
| S17_extracted_k141_129990 | Not-determined |
| S17_extracted_k141_137358 | Not-determined |
| S17_extracted_k141_122304 | Not-determined |
| S17_extracted_k141_62862 | Not-determined |
| S17_extracted_k141_129993 | Not-determined |
| S17_extracted_k141_34488 | Not-determined |
| S17_extracted_k141_11867 | Not-determined |
| S17_extracted_k141_122306 | Not-determined |
| S17_extracted_k141_174125 | Not-determined |
| S17_extracted_k141_159888 | Not-determined |
| S17_extracted_k141_129996 | Not-determined |
| S17_extracted_k141_93005 | Not-determined |
| S17_extracted_k141_122309 | Not-determined |
| S17_extracted_k141_129998 | Not-determined |
| S17_extracted_k141_137362 | Not-determined |
| S17_extracted_k141_115467 | Not-determined |
| S17_extracted_k141_71267 | Not-determined |
| S17_extracted_k141_78581 | Not-determined |
| S17_extracted_k141_78582 | Not-determined |
| S17_extracted_k141_122311 | Not-determined |
| S17_extracted_k141_130002 | Low-quality |
| S17_extracted_k141_122313 | Not-determined |
| S17_extracted_k141_85009 | Low-quality |
| S17_extracted_k141_42130 | Not-determined |
| S17_extracted_k141_130005 | Not-determined |
| S17_extracted_k141_62871 | Not-determined |
| S17_extracted_k141_85012 | Not-determined |
| S17_extracted_k141_56637 | Not-determined |
| S17_extracted_k141_71272 | Low-quality |
| S17_extracted_k141_122314 | Not-determined |
| S17_extracted_k141_115475 | Not-determined |
| S17_extracted_k141_159897 | Not-determined |
| S17_extracted_k141_115477 | Not-determined |
| S17_extracted_k141_11876 | Not-determined |
| S17_extracted_k141_85013 | Not-determined |
| S17_extracted_k141_78590 | Not-determined |
| S17_extracted_k141_93008 | Low-quality |
| S17_extracted_k141_137376 | Not-determined |
| S17_extracted_k141_115484 | Not-determined |
| S17_extracted_k141_100855 | Not-determined |
| S17_extracted_k141_62877 | Not-determined |
| S17_extracted_k141_42144 | Not-determined |
| S17_extracted_k141_167067 | Not-determined |
| S17_extracted_k141_19145 | Low-quality |
| S17_extracted_k141_167068 | Not-determined |
| S17_extracted_k141_62880 | Not-determined |
| S17_extracted_k141_115491 | Not-determined |
| S17_extracted_k141_49362 | Not-determined |
| S17_extracted_k141_93014 | Not-determined |
| S17_extracted_k141_100860 | Not-determined |
| S17_extracted_k141_56651 | Not-determined |
| S17_extracted_k141_71277 | Not-determined |
| S17_extracted_k141_34500 | Not-determined |
| S17_extracted_k141_62888 | Not-determined |
| S17_extracted_k141_71279 | Not-determined |
| S17_extracted_k141_62891 | Not-determined |
| S17_extracted_k141_34504 | Not-determined |
| S17_extracted_k141_56655 | Not-determined |
| S17_extracted_k141_42154 | Low-quality |
| S17_extracted_k141_122334 | Not-determined |
| S17_extracted_k141_62895 | Not-determined |
| S17_extracted_k141_62896 | Not-determined |
| S17_extracted_k141_100871 | Not-determined |
| S17_extracted_k141_49370 | Not-determined |
| S17_extracted_k141_34509 | Not-determined |
| S17_extracted_k141_93025 | Not-determined |
| S17_extracted_k141_122338 | Not-determined |
| S17_extracted_k141_42160 | Not-determined |
| S17_extracted_k141_145006 | Low-quality |
| S17_extracted_k141_152904 | Not-determined |
| S17_extracted_k141_11883 | Not-determined |
| S17_extracted_k141_100877 | Low-quality |
| S17_extracted_k141_42162 | Not-determined |
| S17_extracted_k141_167084 | Not-determined |
| S17_extracted_k141_78613 | Not-determined |
| S17_extracted_k141_49375 | Not-determined |
| S17_extracted_k141_152911 | Not-determined |
| S17_extracted_k141_145010 | Not-determined |
| S17_extracted_k141_49377 | Not-determined |
| S17_extracted_k141_159908 | Low-quality |
| S17_extracted_k141_26268 | Not-determined |
| S17_extracted_k141_56659 | Not-determined |
| S17_extracted_k141_42166 | Not-determined |
| S17_extracted_k141_174136 | Low-quality |
| S17_extracted_k141_42169 | Not-determined |
| S17_extracted_k141_137381 | Low-quality |
| S17_extracted_k141_71299 | Not-determined |
| S17_extracted_k141_78619 | Not-determined |
| S17_extracted_k141_174141 | Low-quality |
| S17_extracted_k141_159918 | Not-determined |
| S17_extracted_k141_93038 | Not-determined |
| S17_extracted_k141_56664 | Not-determined |
| S17_extracted_k141_152921 | Not-determined |
| S17_extracted_k141_137386 | Not-determined |
| S17_extracted_k141_174145 | Not-determined |
| S17_extracted_k141_122343 | Not-determined |
| S17_extracted_k141_159921 | Low-quality |
| S17_extracted_k141_122346 | Not-determined |
| S17_extracted_k141_152924 | Not-determined |
| S17_extracted_k141_159925 | Low-quality |
| S17_extracted_k141_174152 | Low-quality |
| S17_extracted_k141_93045 | Low-quality |
| S17_extracted_k141_174155 | Not-determined |
| S17_extracted_k141_93046 | Not-determined |
| S17_extracted_k141_11893 | Not-determined |
| S17_extracted_k141_122349 | Not-determined |
| S17_extracted_k141_49389 | Not-determined |
| S17_extracted_k141_122351 | Low-quality |
| S17_extracted_k141_93050 | Low-quality |
| S17_extracted_k141_26286 | Not-determined |
| S17_extracted_k141_26287 | Not-determined |
| S17_extracted_k141_4454 | Not-determined |
| S17_extracted_k141_100894 | Not-determined |
| S17_extracted_k141_115507 | Not-determined |
| S17_extracted_k141_11906 | Not-determined |
| S17_extracted_k141_115508 | Not-determined |
| S17_extracted_k141_42175 | Low-quality |
| S17_extracted_k141_42177 | Not-determined |
| S17_extracted_k141_85027 | Not-determined |
| S17_extracted_k141_85028 | Not-determined |
| S17_extracted_k141_56673 | Not-determined |
| S17_extracted_k141_56674 | Low-quality |
| S17_extracted_k141_145040 | Low-quality |
| S17_extracted_k141_137395 | Not-determined |
| S17_extracted_k141_4460 | Not-determined |
| S17_extracted_k141_56675 | Not-determined |
| S17_extracted_k141_4461 | Not-determined |
| S17_extracted_k141_56676 | Low-quality |
| S17_extracted_k141_34545 | Not-determined |
| S17_extracted_k141_19166 | Low-quality |
| S17_extracted_k141_56678 | Not-determined |
| S17_extracted_k141_115510 | Not-determined |
| S17_extracted_k141_49399 | Not-determined |
| S17_extracted_k141_115514 | Not-determined |
| S17_extracted_k141_4469 | Not-determined |
| S17_extracted_k141_159938 | Not-determined |
| S17_extracted_k141_11922 | Low-quality |
| S17_extracted_k141_62923 | Not-determined |
| S17_extracted_k141_145046 | Not-determined |
| S17_extracted_k141_145048 | Not-determined |
| S17_extracted_k141_11926 | Not-determined |
| S17_extracted_k141_49410 | Not-determined |
| S17_extracted_k141_71323 | Not-determined |
| S17_extracted_k141_78635 | Not-determined |
| S17_extracted_k141_34556 | Not-determined |
| S17_extracted_k141_34557 | Not-determined |
| S17_extracted_k141_115519 | Not-determined |
| S17_extracted_k141_100911 | Not-determined |
| S17_extracted_k141_174179 | Not-determined |
| S17_extracted_k141_49414 | Not-determined |
| S17_extracted_k141_167123 | Not-determined |
| S17_extracted_k141_152939 | Not-determined |
| S17_extracted_k141_145053 | Not-determined |
| S17_extracted_k141_137415 | Not-determined |
| S17_extracted_k141_145055 | Not-determined |
| S17_extracted_k141_115524 | Not-determined |
| S17_extracted_k141_11933 | Not-determined |
| S17_extracted_k141_4481 | Not-determined |
| S17_extracted_k141_71328 | Not-determined |
| S17_extracted_k141_34564 | Not-determined |
| S17_extracted_k141_174186 | Not-determined |
| S17_extracted_k141_11934 | Low-quality |
| S17_extracted_k141_49419 | Low-quality |
| S17_extracted_k141_145063 | Not-determined |
| S17_extracted_k141_78638 | Not-determined |
| S17_extracted_k141_34567 | Low-quality |
| S17_extracted_k141_34568 | Low-quality |
| S17_extracted_k141_19187 | Low-quality |
| S17_extracted_k141_100921 | Not-determined |
| S17_extracted_k141_78641 | Not-determined |
| S17_extracted_k141_4488 | Not-determined |
| S17_extracted_k141_93089 | Not-determined |
| S17_extracted_k141_145068 | Not-determined |
| S17_extracted_k141_108663 | Not-determined |
| S17_extracted_k141_71341 | Not-determined |
| S17_extracted_k141_115537 | Not-determined |
| S17_extracted_k141_145073 | Not-determined |
| S17_extracted_k141_115539 | Not-determined |
| S17_extracted_k141_159953 | Not-determined |
| S17_extracted_k141_42205 | Not-determined |
| S17_extracted_k141_100929 | Not-determined |
| S17_extracted_k141_4493 | Not-determined |
| S17_extracted_k141_122358 | Not-determined |
| S17_extracted_k141_115551 | Not-determined |
| S17_extracted_k141_78648 | Not-determined |
| S17_extracted_k141_108678 | Not-determined |
| S17_extracted_k141_42212 | Not-determined |
| S17_extracted_k141_100940 | Not-determined |
| S17_extracted_k141_19198 | Not-determined |
| S17_extracted_k141_34587 | Not-determined |
| S17_extracted_k141_115560 | Not-determined |
| S17_extracted_k141_145085 | Not-determined |
| S17_extracted_k141_11953 | Not-determined |
| S17_extracted_k141_115566 | Not-determined |
| S17_extracted_k141_137429 | Not-determined |
| S17_extracted_k141_42221 | Not-determined |
| S17_extracted_k141_78653 | Not-determined |
| S17_extracted_k141_115571 | Not-determined |
| S17_extracted_k141_34592 | Low-quality |
| S17_extracted_k141_4506 | Not-determined |
| S17_extracted_k141_19202 | Low-quality |
| S17_extracted_k141_167125 | Not-determined |
| S17_extracted_k141_78659 | Not-determined |
| S17_extracted_k141_137438 | Not-determined |
| S17_extracted_k141_152959 | Low-quality |
| S17_extracted_k141_26334 | Not-determined |
| S17_extracted_k141_56710 | Not-determined |
| S17_extracted_k141_56713 | Not-determined |
| S17_extracted_k141_19208 | Low-quality |
| S17_extracted_k141_167136 | Not-determined |
| S17_extracted_k141_78672 | Not-determined |
| S17_extracted_k141_26347 | Low-quality |
| S17_extracted_k141_137446 | Not-determined |
| S17_extracted_k141_85064 | Not-determined |
| S17_extracted_k141_115586 | Low-quality |
| S17_extracted_k141_122375 | Not-determined |
| S17_extracted_k141_167138 | Not-determined |
| S17_extracted_k141_42236 | Not-determined |
| S17_extracted_k141_85071 | Not-determined |
| S17_extracted_k141_26352 | Low-quality |
| S17_extracted_k141_100949 | Low-quality |
| S17_extracted_k141_85074 | Not-determined |
| S17_extracted_k141_78682 | Not-determined |
| S17_extracted_k141_4526 | Not-determined |
| S17_extracted_k141_85076 | Not-determined |
| S17_extracted_k141_159959 | Not-determined |
| S17_extracted_k141_42239 | Not-determined |
| S17_extracted_k141_34618 | Low-quality |
| S17_extracted_k141_159964 | Not-determined |
| S17_extracted_k141_152979 | Low-quality |
| S17_extracted_k141_93124 | Not-determined |
| S17_extracted_k141_4528 | Not-determined |
| S17_extracted_k141_85081 | Not-determined |
| S17_extracted_k141_42241 | Not-determined |
| S17_extracted_k141_93125 | Low-quality |
| S17_extracted_k141_108713 | Not-determined |
| S17_extracted_k141_4530 | Not-determined |
| S17_extracted_k141_115588 | Low-quality |
| S17_extracted_k141_85084 | Not-determined |
| S17_extracted_k141_56733 | Not-determined |
| S17_extracted_k141_34625 | Not-determined |
| S17_extracted_k141_137460 | Not-determined |
| S17_extracted_k141_34627 | Not-determined |
| S17_extracted_k141_56736 | Low-quality |
| S17_extracted_k141_100961 | Not-determined |
| S17_extracted_k141_42249 | Not-determined |
| S17_extracted_k141_4539 | Not-determined |
| S17_extracted_k141_115597 | Not-determined |
| S17_extracted_k141_100963 | Low-quality |
| S17_extracted_k141_159971 | Not-determined |
| S17_extracted_k141_26371 | Not-determined |
| S17_extracted_k141_42254 | Not-determined |
| S17_extracted_k141_42256 | Not-determined |
| S17_extracted_k141_85090 | Not-determined |
| S17_extracted_k141_11996 | Not-determined |
| S17_extracted_k141_78694 | Not-determined |
| S17_extracted_k141_115604 | Not-determined |
| S17_extracted_k141_62963 | Not-determined |
| S17_extracted_k141_78696 | Not-determined |
| S17_extracted_k141_42263 | Not-determined |
| S17_extracted_k141_71384 | Not-determined |
| S17_extracted_k141_62967 | Low-quality |
| S17_extracted_k141_108735 | Not-determined |
| S17_extracted_k141_108736 | Not-determined |
| S17_extracted_k141_122402 | Not-determined |
| S17_extracted_k141_108746 | Low-quality |
| S17_extracted_k141_115612 | Low-quality |
| S17_extracted_k141_93141 | Not-determined |
| S17_extracted_k141_62975 | Low-quality |
| S17_extracted_k141_93145 | Not-determined |
| S17_extracted_k141_34642 | Not-determined |
| S17_extracted_k141_122406 | Not-determined |
| S17_extracted_k141_42270 | Not-determined |
| S17_extracted_k141_4554 | Low-quality |
| S17_extracted_k141_115615 | Not-determined |
| S17_extracted_k141_78708 | Not-determined |
| S17_extracted_k141_122410 | Not-determined |
| S17_extracted_k141_167187 | Not-determined |
| S17_extracted_k141_122411 | Not-determined |
| S17_extracted_k141_93153 | Not-determined |
| S17_extracted_k141_137494 | Not-determined |
| S17_extracted_k141_78712 | Not-determined |
| S17_extracted_k141_137495 | Not-determined |
| S17_extracted_k141_108754 | Not-determined |
| S17_extracted_k141_71406 | Not-determined |
| S17_extracted_k141_42281 | Not-determined |
| S17_extracted_k141_34651 | Not-determined |
| S17_extracted_k141_62981 | Not-determined |
| S17_extracted_k141_108756 | Not-determined |
| S17_extracted_k141_174199 | Not-determined |
| S17_extracted_k141_159985 | Not-determined |
| S17_extracted_k141_93158 | Not-determined |
| S17_extracted_k141_100986 | Not-determined |
| S17_extracted_k141_167191 | Not-determined |
| S17_extracted_k141_78716 | Low-quality |
| S17_extracted_k141_159990 | Not-determined |
| S17_extracted_k141_4571 | Not-determined |
| S17_extracted_k141_78717 | Not-determined |
| S17_extracted_k141_71416 | Not-determined |
| S17_extracted_k141_62983 | Not-determined |
| S17_extracted_k141_56739 | Not-determined |
| S17_extracted_k141_42290 | Not-determined |
| S17_extracted_k141_85106 | Not-determined |
| S17_extracted_k141_152999 | Not-determined |
| S17_extracted_k141_34659 | Not-determined |
| S17_extracted_k141_71420 | Not-determined |
| S17_extracted_k141_108766 | Not-determined |
| S17_extracted_k141_78722 | Low-quality |
| S17_extracted_k141_4575 | Not-determined |
| S17_extracted_k141_4576 | Not-determined |
| S17_extracted_k141_115632 | Not-determined |
| S17_extracted_k141_115633 | Not-determined |
| S17_extracted_k141_56746 | Low-quality |
| S17_extracted_k141_93164 | Not-determined |
| S17_extracted_k141_12011 | Not-determined |
| S17_extracted_k141_115634 | Not-determined |
| S17_extracted_k141_108768 | Not-determined |
| S17_extracted_k141_137514 | Not-determined |
| S17_extracted_k141_49456 | Not-determined |
| S17_extracted_k141_93170 | Not-determined |
| S17_extracted_k141_62997 | Not-determined |
| S17_extracted_k141_137521 | Not-determined |
| S17_extracted_k141_115640 | Not-determined |
| S17_extracted_k141_71424 | Not-determined |
| S17_extracted_k141_115641 | Not-determined |
| S17_extracted_k141_63000 | Not-determined |
| S17_extracted_k141_63001 | Not-determined |
| S17_extracted_k141_108778 | Not-determined |
| S17_extracted_k141_108780 | Not-determined |
| S17_extracted_k141_26380 | Not-determined |
| S17_extracted_k141_145103 | Not-determined |
| S17_extracted_k141_26382 | Not-determined |
| S17_extracted_k141_122417 | Not-determined |
| S17_extracted_k141_42303 | Not-determined |
| S17_extracted_k141_71433 | Not-determined |
| S17_extracted_k141_145104 | Not-determined |
| S17_extracted_k141_49467 | Not-determined |
| S17_extracted_k141_49469 | Not-determined |
| S17_extracted_k141_78733 | Not-determined |
| S17_extracted_k141_4589 | Not-determined |
| S16_extracted_42706 | High-quality |
| S17_extracted_k141_108789 | Not-determined |
| S17_extracted_k141_115660 | Not-determined |
| S17_extracted_k141_42316 | Not-determined |
| S17_extracted_k141_153012 | Not-determined |
| S17_extracted_k141_19243 | Not-determined |
| S17_extracted_k141_145112 | Not-determined |
| S17_extracted_k141_26394 | Not-determined |
| S17_extracted_k141_93190 | Not-determined |
| S17_extracted_k141_19246 | Not-determined |
| S17_extracted_k141_137544 | Not-determined |
| S17_extracted_k141_145114 | Not-determined |
| S17_extracted_k141_93192 | Not-determined |
| S17_extracted_k141_42325 | Not-determined |
| S17_extracted_k141_12029 | Not-determined |
| S17_extracted_k141_153017 | Not-determined |
| S17_extracted_k141_42330 | Not-determined |
| S17_extracted_k141_34680 | Not-determined |
| S17_extracted_k141_167206 | Not-determined |
| S17_extracted_k141_137550 | Not-determined |
| S17_extracted_k141_42331 | Not-determined |
| S17_extracted_k141_101011 | Not-determined |
| S17_extracted_k141_137552 | Not-determined |
| S17_extracted_k141_153025 | Not-determined |
| S17_extracted_k141_78748 | Not-determined |
| S17_extracted_k141_63026 | Not-determined |
| S17_extracted_k141_4602 | Not-determined |
| S17_extracted_k141_108802 | Not-determined |
| S17_extracted_k141_4603 | Not-determined |
| S17_extracted_k141_93206 | Not-determined |
| S17_extracted_k141_4604 | Not-determined |
| S17_extracted_k141_93207 | Not-determined |
| S17_extracted_k141_63032 | Not-determined |
| S17_extracted_k141_4606 | Not-determined |
| S17_extracted_k141_145129 | Not-determined |
| S17_extracted_k141_137560 | Not-determined |
| S17_extracted_k141_108806 | Not-determined |
| S17_extracted_k141_167220 | Not-determined |
| S17_extracted_k141_167221 | Not-determined |
| S17_extracted_k141_101021 | Low-quality |
| S17_extracted_k141_159997 | Not-determined |
| S17_extracted_k141_63035 | Not-determined |
| S17_extracted_k141_137563 | Not-determined |
| S17_extracted_k141_153033 | Not-determined |
| S17_extracted_k141_159998 | Not-determined |
| S17_extracted_k141_153035 | Not-determined |
| S17_extracted_k141_34684 | Not-determined |
| S17_extracted_k141_122440 | Not-determined |
| S17_extracted_k141_174228 | Not-determined |
| S17_extracted_k141_115692 | Not-determined |
| S17_extracted_k141_115694 | Not-determined |
| S17_extracted_k141_174233 | Not-determined |
| S17_extracted_k141_93214 | Not-determined |
| S17_extracted_k141_34691 | Not-determined |
| S17_extracted_k141_115696 | Not-determined |
| S17_extracted_k141_115697 | Low-quality |
| S17_extracted_k141_160014 | Not-determined |
| S17_extracted_k141_4619 | Not-determined |
| S17_extracted_k141_153047 | Not-determined |
| S17_extracted_k141_167232 | Not-determined |
| S17_extracted_k141_101038 | Not-determined |
| S17_extracted_k141_115706 | Not-determined |
| S17_extracted_k141_85143 | Low-quality |
| S17_extracted_k141_101040 | Not-determined |
| S17_extracted_k141_34705 | Not-determined |
| S17_extracted_k141_26424 | Not-determined |
| S17_extracted_k141_34707 | Not-determined |
| S17_extracted_k141_34710 | Low-quality |
| S17_extracted_k141_12050 | Not-determined |
| S17_extracted_k141_34711 | Not-determined |
| S17_extracted_k141_153055 | Not-determined |
| S17_extracted_k141_85152 | Not-determined |
| S17_extracted_k141_101046 | Not-determined |
| S17_extracted_k141_71464 | Low-quality |
| S17_extracted_k141_71466 | Not-determined |
| S17_extracted_k141_4632 | Not-determined |
| S17_extracted_k141_49490 | Not-determined |
| S17_extracted_k141_101049 | Not-determined |
| S17_extracted_k141_160029 | Not-determined |
| S17_extracted_k141_4635 | Not-determined |
| S17_extracted_k141_85156 | Not-determined |
| S17_extracted_k141_122464 | Not-determined |
| S17_extracted_k141_122465 | Not-determined |
| S17_extracted_k141_26436 | Not-determined |
| S17_extracted_k141_122467 | Not-determined |
| S17_extracted_k141_63054 | Not-determined |
| S17_extracted_k141_122469 | Low-quality |
| S17_extracted_k141_85161 | Not-determined |
| S17_extracted_k141_115725 | Not-determined |
| S17_extracted_k141_63057 | Low-quality |
| S17_extracted_k141_12058 | Not-determined |
| S17_extracted_k141_63060 | Not-determined |
| S17_extracted_k141_71472 | Not-determined |
| S17_extracted_k141_153065 | Not-determined |
| S17_extracted_k141_174249 | Not-determined |
| S17_extracted_k141_34726 | Not-determined |
| S17_extracted_k141_71473 | Not-determined |
| S17_extracted_k141_71475 | Not-determined |
| S17_extracted_k141_85169 | Low-quality |
| S17_extracted_k141_71481 | Not-determined |
| S17_extracted_k141_63065 | Low-quality |
| S17_extracted_k141_71486 | Low-quality |
| S17_extracted_k141_78768 | Not-determined |
| S17_extracted_k141_153070 | Not-determined |
| S17_extracted_k141_71487 | Not-determined |
| S17_extracted_k141_12066 | Not-determined |
| S17_extracted_k141_137609 | Not-determined |
| S17_extracted_k141_12067 | Not-determined |
| S17_extracted_k141_160047 | Not-determined |
| S17_extracted_k141_174263 | Not-determined |
| S17_extracted_k141_174265 | Not-determined |
| S17_extracted_k141_4658 | Not-determined |
| S17_extracted_k141_49513 | Not-determined |
| S17_extracted_k141_167248 | Not-determined |
| S17_extracted_k141_108829 | Low-quality |
| S17_extracted_k141_153075 | Not-determined |
| S17_extracted_k141_56792 | Not-determined |
| S17_extracted_k141_145168 | Low-quality |
| S17_extracted_k141_174272 | Not-determined |
| S17_extracted_k141_174274 | Low-quality |
| S17_extracted_k141_160051 | Not-determined |
| S17_extracted_k141_93236 | Not-determined |
| S17_extracted_k141_153079 | Not-determined |
| S17_extracted_k141_174277 | Not-determined |
| S17_extracted_k141_71492 | Not-determined |
| S17_extracted_k141_56796 | Low-quality |
| S17_extracted_k141_93237 | Not-determined |
| S17_extracted_k141_71494 | Not-determined |
| S17_extracted_k141_78782 | Not-determined |
| S17_extracted_k141_34746 | Low-quality |
| S17_extracted_k141_56799 | Low-quality |
| S17_extracted_k141_49525 | Not-determined |
| S17_extracted_k141_145175 | Not-determined |
| S17_extracted_k141_153089 | Not-determined |
| S17_extracted_k141_145178 | Not-determined |
| S17_extracted_k141_174281 | Not-determined |
| S17_extracted_k141_34748 | Not-determined |
| S17_extracted_k141_49530 | Low-quality |
| S17_extracted_k141_160057 | Not-determined |
| S17_extracted_k141_145180 | Not-determined |
| S17_extracted_k141_137624 | Not-determined |
| S17_extracted_k141_56803 | Low-quality |
| S17_extracted_k141_12075 | Not-determined |
| S17_extracted_k141_4675 | Not-determined |
| S17_extracted_k141_56804 | Low-quality |
| S17_extracted_k141_137627 | Low-quality |
| S17_extracted_k141_12080 | Not-determined |
| S17_extracted_k141_153097 | Not-determined |
| S17_extracted_k141_12081 | Low-quality |
| S17_extracted_k141_12083 | Low-quality |
| S17_extracted_k141_34757 | Not-determined |
| S17_extracted_k141_34758 | Not-determined |
| S17_extracted_k141_174286 | Not-determined |
| S17_extracted_k141_71513 | Not-determined |
| S17_extracted_k141_12086 | Not-determined |
| S17_extracted_k141_63094 | Not-determined |
| S17_extracted_k141_71515 | Low-quality |
| S17_extracted_k141_101086 | Not-determined |
| S17_extracted_k141_101087 | Not-determined |
| S17_extracted_k141_78790 | Not-determined |
| S17_extracted_k141_137640 | Low-quality |
| S17_extracted_k141_174290 | Low-quality |
| S17_extracted_k141_122498 | Not-determined |
| S17_extracted_k141_78792 | Not-determined |
| S17_extracted_k141_174291 | Not-determined |
| S17_extracted_k141_101091 | Not-determined |
| S17_extracted_k141_42349 | Not-determined |
| S17_extracted_k141_93251 | Not-determined |
| S17_extracted_k141_153102 | Not-determined |
| S17_extracted_k141_56815 | Not-determined |
| S17_extracted_k141_4688 | Not-determined |
| S17_extracted_k141_167274 | Low-quality |
| S17_extracted_k141_160075 | Low-quality |
| S17_extracted_k141_122502 | Low-quality |
| S17_extracted_k141_42353 | Low-quality |
| S17_extracted_k141_174297 | Not-determined |
| S17_extracted_k141_153107 | Not-determined |
| S17_extracted_k141_56820 | Low-quality |
| S17_extracted_k141_4694 | Low-quality |
| S17_extracted_k141_56824 | Not-determined |
| S17_extracted_k141_160077 | Not-determined |
| S17_extracted_k141_115745 | Not-determined |
| S17_extracted_k141_145198 | Low-quality |
| S17_extracted_k141_63118 | Not-determined |
| S17_extracted_k141_34783 | Not-determined |
| S17_extracted_k141_137659 | Not-determined |
| S17_extracted_k141_34784 | Not-determined |
| S17_extracted_k141_160083 | Low-quality |
| S17_extracted_k141_115749 | Not-determined |
| S17_extracted_k141_174309 | Not-determined |
| S17_extracted_k141_42360 | Low-quality |
| S17_extracted_k141_34789 | Not-determined |
| S17_extracted_k141_63121 | Not-determined |
| S17_extracted_k141_174311 | Not-determined |
| S17_extracted_k141_122512 | Low-quality |
| S17_extracted_k141_122513 | Not-determined |
| S17_extracted_k141_12098 | Not-determined |
| S17_extracted_k141_137664 | Not-determined |
| S17_extracted_k141_4712 | Not-determined |
| S17_extracted_k141_115755 | Not-determined |
| S17_extracted_k141_93267 | Not-determined |
| S17_extracted_k141_160089 | Low-quality |
| S17_extracted_k141_42364 | Not-determined |
| S17_extracted_k141_85212 | Low-quality |
| S17_extracted_k141_34794 | Not-determined |
| S17_extracted_k141_115757 | Not-determined |
| S17_extracted_k141_160092 | Not-determined |
| S17_extracted_k141_130021 | Low-quality |
| S17_extracted_k141_160094 | Not-determined |
| S17_extracted_k141_85218 | Not-determined |
| S17_extracted_k141_34797 | Not-determined |
| S17_extracted_k141_56836 | Not-determined |
| S17_extracted_k141_167295 | Not-determined |
| S17_extracted_k141_145210 | Not-determined |
| S17_extracted_k141_34800 | Not-determined |
| S17_extracted_k141_160097 | Not-determined |
| S17_extracted_k141_12110 | Not-determined |
| S17_extracted_k141_122523 | Low-quality |
| S17_extracted_k141_34802 | Not-determined |
| S17_extracted_k141_108847 | Not-determined |
| S17_extracted_k141_137673 | Low-quality |
| S17_extracted_k141_160100 | Not-determined |
| S17_extracted_k141_122528 | Not-determined |
| S17_extracted_k141_93271 | Low-quality |
| S17_extracted_k141_115761 | Not-determined |
| S17_extracted_k141_49557 | Low-quality |
| S17_extracted_k141_145219 | Not-determined |
| S17_extracted_k141_42369 | Low-quality |
| S17_extracted_k141_115765 | Not-determined |
| S17_extracted_k141_130034 | Not-determined |
| S17_extracted_k141_137676 | Not-determined |
| S17_extracted_k141_49559 | Not-determined |
| S17_extracted_k141_49560 | Not-determined |
| S17_extracted_k141_145224 | Not-determined |

| | |
|---|---|
| S17_extracted_k141_85230 | Not-determined |
| S17_extracted_k141_34809 | Not-determined |
| S17_extracted_k141_12122 | Not-determined |
| S17_extracted_k141_93274 | Not-determined |
| S17_extracted_k141_101105 | Not-determined |
| S17_extracted_k141_78813 | Low-quality |
| S17_extracted_k141_93280 | Not-determined |
| S17_extracted_k141_108849 | Not-determined |
| S17_extracted_k141_56851 | Low-quality |
| S17_extracted_k141_108852 | Not-determined |
| S17_extracted_k141_85236 | Not-determined |
| S17_extracted_k141_71548 | Not-determined |
| S17_extracted_k141_34818 | Not-determined |
| S17_extracted_k141_137684 | Not-determined |
| S17_extracted_k141_4720 | Low-quality |
| S17_extracted_k141_122545 | Not-determined |
| S17_extracted_k141_108861 | Not-determined |
| S17_extracted_k141_78827 | Not-determined |
| S17_extracted_k141_122546 | Not-determined |
| S17_extracted_k141_101118 | Not-determined |
| S17_extracted_k141_93289 | Not-determined |
| S17_extracted_k141_34828 | Low-quality |
| S17_extracted_k141_71561 | Not-determined |
| S17_extracted_k141_85249 | Not-determined |
| S17_extracted_k141_137692 | Not-determined |
| S17_extracted_k141_101122 | Not-determined |
| S17_extracted_k141_93293 | Not-determined |
| S17_extracted_k141_34834 | Not-determined |
| S17_extracted_k141_93294 | Not-determined |
| S17_extracted_k141_34836 | Not-determined |
| S17_extracted_k141_160112 | Low-quality |
| S17_extracted_k141_78832 | Not-determined |
| S17_extracted_k141_34840 | Not-determined |
| S17_extracted_k141_122556 | Not-determined |
| S17_extracted_k141_4730 | Not-determined |
| S17_extracted_k141_4732 | Not-determined |
| S17_extracted_k141_4733 | Not-determined |
| S17_extracted_k141_108869 | Not-determined |
| S17_extracted_k141_63132 | Low-quality |
| S17_extracted_k141_71567 | Not-determined |
| S17_extracted_k141_12134 | Not-determined |
| S17_extracted_k141_93300 | Not-determined |
| S17_extracted_k141_153151 | Low-quality |
| S17_extracted_k141_49579 | Not-determined |
| S17_extracted_k141_115781 | Not-determined |
| S17_extracted_k141_19280 | Not-determined |
| S17_extracted_k141_115783 | Low-quality |
| S17_extracted_k141_108879 | Not-determined |
| S17_extracted_k141_167335 | Not-determined |
| S17_extracted_k141_153154 | Not-determined |
| S17_extracted_k141_160116 | Not-determined |
| S17_extracted_k141_85261 | Low-quality |
| S17_extracted_k141_145248 | Low-quality |
| S17_extracted_k141_115787 | Low-quality |
| S17_extracted_k141_93304 | Not-determined |
| S17_extracted_k141_93305 | Not-determined |
| S17_extracted_k141_167341 | Low-quality |
| S17_extracted_k141_167342 | Not-determined |
| S17_extracted_k141_115793 | Not-determined |
| S17_extracted_k141_4752 | Not-determined |
| S17_extracted_k141_85269 | Not-determined |
| S17_extracted_k141_115794 | Not-determined |
| S17_extracted_k141_49586 | Not-determined |
| S17_extracted_k141_108889 | Not-determined |
| S17_extracted_k141_56884 | Not-determined |
| S17_extracted_k141_108890 | Not-determined |
| S17_extracted_k141_137705 | Not-determined |
| S17_extracted_k141_137707 | Not-determined |
| S17_extracted_k141_115798 | Not-determined |
| S17_extracted_k141_122577 | Not-determined |
| S17_extracted_k141_145257 | Not-determined |
| S17_extracted_k141_137710 | Low-quality |
| S17_extracted_k141_108896 | Not-determined |
| S17_extracted_k141_71586 | Not-determined |
| S17_extracted_k141_34861 | Not-determined |
| S17_extracted_k141_101126 | Low-quality |
| S17_extracted_k141_145261 | Not-determined |
| S17_extracted_k141_145262 | Not-determined |
| S17_extracted_k141_78850 | Not-determined |
| S17_extracted_k141_34865 | Not-determined |
| S17_extracted_k141_71588 | Not-determined |
| S17_extracted_k141_108900 | Not-determined |
| S17_extracted_k141_56898 | Not-determined |
| S17_extracted_k141_78853 | Not-determined |
| S17_extracted_k141_145269 | Not-determined |
| S17_extracted_k141_19296 | Not-determined |
| S17_extracted_k141_85293 | Not-determined |
| S17_extracted_k141_85294 | Low-quality |
| S17_extracted_k141_19300 | Not-determined |
| S17_extracted_k141_34874 | Not-determined |
| S17_extracted_k141_108908 | Not-determined |
| S17_extracted_k141_108909 | Not-determined |
| S17_extracted_k141_63143 | Not-determined |
| S17_extracted_k141_108910 | Not-determined |
| S17_extracted_k141_49603 | Not-determined |
| S17_extracted_k141_85305 | Not-determined |
| S17_extracted_k141_167356 | Not-determined |
| S17_extracted_k141_174318 | Not-determined |
| S17_extracted_k141_115815 | Not-determined |
| S17_extracted_k141_115817 | Not-determined |
| S17_extracted_k141_174321 | Not-determined |
| S17_extracted_k141_145278 | Not-determined |
| S17_extracted_k141_145279 | Low-quality |
| S17_extracted_k141_34877 | Not-determined |
| S17_extracted_k141_19307 | Not-determined |
| S17_extracted_k141_34878 | Not-determined |
| S17_extracted_k141_49607 | Not-determined |
| S17_extracted_k141_137729 | Not-determined |
| S17_extracted_k141_63152 | Not-determined |
| S17_extracted_k141_153179 | Not-determined |
| S17_extracted_k141_56912 | Not-determined |
| S17_extracted_k141_167363 | Not-determined |
| S17_extracted_k141_115823 | Not-determined |
| S17_extracted_k141_71601 | Not-determined |
| S17_extracted_k141_71603 | Not-determined |
| S17_extracted_k141_85313 | Not-determined |
| S17_extracted_k141_42409 | Not-determined |
| S17_extracted_k141_174332 | Low-quality |
| S17_extracted_k141_122599 | Not-determined |
| S17_extracted_k141_160125 | Not-determined |
| S17_extracted_k141_108916 | Not-determined |
| S17_extracted_k141_19314 | Not-determined |
| S17_extracted_k141_167374 | Not-determined |
| S17_extracted_k141_101143 | Low-quality |
| S17_extracted_k141_42411 | Not-determined |
| S17_extracted_k141_56920 | Not-determined |
| S17_extracted_k141_56921 | Not-determined |
| S17_extracted_k141_108919 | Not-determined |
| S17_extracted_k141_56922 | Not-determined |
| S17_extracted_k141_101145 | Not-determined |
| S17_extracted_k141_85325 | Not-determined |
| S17_extracted_k141_93323 | Not-determined |
| S17_extracted_k141_63168 | Not-determined |
| S17_extracted_k141_49618 | Not-determined |
| S17_extracted_k141_160126 | Not-determined |
| S17_extracted_k141_63171 | Not-determined |
| S17_extracted_k141_19320 | Not-determined |
| S17_extracted_k141_101152 | Not-determined |
| S17_extracted_k141_49625 | Not-determined |
| S17_extracted_k141_122608 | Not-determined |
| S17_extracted_k141_160129 | Not-determined |
| S17_extracted_k141_153198 | Not-determined |
| S17_extracted_k141_122610 | Not-determined |
| S17_extracted_k141_93332 | Not-determined |
| S17_extracted_k141_93333 | Not-determined |
| S17_extracted_k141_122612 | Not-determined |
| S17_extracted_k141_167387 | Not-determined |
| S17_extracted_k141_153205 | Not-determined |
| S17_extracted_k141_153207 | Not-determined |
| S17_extracted_k141_160139 | Not-determined |
| S17_extracted_k141_130090 | Not-determined |
| S17_extracted_k141_137743 | Not-determined |
| S17_extracted_k141_4803 | Not-determined |
| S17_extracted_k141_63175 | Low-quality |
| S17_extracted_k141_130093 | Not-determined |
| S17_extracted_k141_71607 | Not-determined |
| S17_extracted_k141_71608 | Not-determined |
| S17_extracted_k141_101162 | Not-determined |
| S17_extracted_k141_137750 | Not-determined |
| S17_extracted_k141_63181 | Low-quality |
| S17_extracted_k141_145309 | Not-determined |
| S17_extracted_k141_26483 | Not-determined |
| S17_extracted_k141_4809 | Not-determined |
| S17_extracted_k141_26484 | Not-determined |
| S17_extracted_k141_153217 | Not-determined |
| S17_extracted_k141_85344 | Low-quality |
| S17_extracted_k141_153219 | Not-determined |
| S17_extracted_k141_115837 | Not-determined |
| S17_extracted_k141_19338 | Not-determined |
| S17_extracted_k141_85349 | Not-determined |
| S17_extracted_k141_85352 | Not-determined |
| S17_extracted_k141_63187 | Not-determined |
| S17_extracted_k141_78856 | Not-determined |
| S17_extracted_k141_56940 | Not-determined |
| S17_extracted_k141_56943 | Not-determined |
| S17_extracted_k141_160143 | Low-quality |
| S17_extracted_k141_101167 | Not-determined |
| S17_extracted_k141_153224 | Not-determined |
| S17_extracted_k141_93360 | Not-determined |
| S17_extracted_k141_4817 | Not-determined |
| S17_extracted_k141_85360 | Low-quality |
| S17_extracted_k141_34933 | Not-determined |
| S17_extracted_k141_71624 | Not-determined |
| S17_extracted_k141_130116 | Not-determined |
| S17_extracted_k141_130119 | Not-determined |
| S17_extracted_k141_130121 | Not-determined |
| S17_extracted_k141_93367 | Not-determined |
| S17_extracted_k141_130123 | Not-determined |
| S17_extracted_k141_42450 | Not-determined |
| S17_extracted_k141_34945 | Not-determined |
| S17_extracted_k141_4828 | Not-determined |
| S17_extracted_k141_4829 | Not-determined |
| S17_extracted_k141_42454 | Not-determined |
| S17_extracted_k141_71630 | Not-determined |
| S17_extracted_k141_137770 | Not-determined |
| S17_extracted_k141_108948 | Not-determined |
| S17_extracted_k141_63209 | Not-determined |
| S17_extracted_k141_4836 | Low-quality |
| S17_extracted_k141_167405 | Not-determined |
| S17_extracted_k141_49653 | Low-quality |
| S17_extracted_k141_34949 | Not-determined |
| S17_extracted_k141_34952 | Not-determined |
| S17_extracted_k141_130138 | Not-determined |
| S17_extracted_k141_160164 | Not-determined |
| S17_extracted_k141_108951 | Not-determined |
| S17_extracted_k141_160166 | Not-determined |
| S17_extracted_k141_101188 | Not-determined |
| S17_extracted_k141_153250 | Not-determined |
| S17_extracted_k141_63218 | Not-determined |
| S17_extracted_k141_130142 | Not-determined |
| S17_extracted_k141_130143 | Not-determined |
| S17_extracted_k141_160167 | Not-determined |
| S17_extracted_k141_19346 | Not-determined |
| S17_extracted_k141_85380 | Not-determined |
| S17_extracted_k141_4843 | Low-quality |
| S17_extracted_k141_108962 | Not-determined |
| S17_extracted_k141_137782 | Not-determined |
| S17_extracted_k141_101193 | Not-determined |
| S17_extracted_k141_42473 | Not-determined |
| S17_extracted_k141_167412 | Not-determined |
| S17_extracted_k141_153256 | Not-determined |
| S17_extracted_k141_153257 | Not-determined |
| S17_extracted_k141_4845 | Not-determined |
| S17_extracted_k141_108966 | Not-determined |
| S17_extracted_k141_49660 | Not-determined |
| S17_extracted_k141_93391 | Not-determined |
| S17_extracted_k141_78880 | Not-determined |
| S17_extracted_k141_115871 | Not-determined |
| S17_extracted_k141_108968 | Not-determined |
| S17_extracted_k141_4848 | Not-determined |
| S17_extracted_k141_63231 | Not-determined |
| S17_extracted_k141_4849 | Not-determined |
| S17_extracted_k141_153261 | Not-determined |
| S17_extracted_k141_101198 | Not-determined |
| S17_extracted_k141_85389 | Low-quality |
| S17_extracted_k141_56965 | Not-determined |
| S17_extracted_k141_101199 | Low-quality |
| S17_extracted_k141_153265 | Not-determined |
| S17_extracted_k141_49666 | Not-determined |
| S17_extracted_k141_19357 | Not-determined |
| S17_extracted_k141_56968 | Not-determined |
| S17_extracted_k141_137793 | Not-determined |
| S17_extracted_k141_71643 | Not-determined |
| S17_extracted_k141_78886 | Not-determined |
| S17_extracted_k141_153269 | Not-determined |
| S17_extracted_k141_71646 | Not-determined |
| S17_extracted_k141_115878 | Not-determined |
| S17_extracted_k141_108976 | Not-determined |
| S17_extracted_k141_160185 | Low-quality |
| S17_extracted_k141_130155 | Not-determined |
| S17_extracted_k141_26492 | Not-determined |
| S17_extracted_k141_56974 | Not-determined |
| S17_extracted_k141_160188 | Not-determined |
| S17_extracted_k141_145314 | Not-determined |
| S17_extracted_k141_56976 | Not-determined |
| S17_extracted_k141_130160 | Low-quality |
| S17_extracted_k141_19360 | Low-quality |
| S17_extracted_k141_26496 | Not-determined |
| S17_extracted_k141_19362 | Not-determined |
| S17_extracted_k141_26498 | Not-determined |
| S17_extracted_k141_71655 | Not-determined |
| S17_extracted_k141_93405 | Not-determined |
| S17_extracted_k141_26499 | Not-determined |
| S17_extracted_k141_63251 | Not-determined |
| S17_extracted_k141_71657 | Not-determined |
| S17_extracted_k141_115886 | Low-quality |
| S17_extracted_k141_56990 | Not-determined |
| S17_extracted_k141_93412 | Not-determined |
| S17_extracted_k141_122640 | Low-quality |
| S17_extracted_k141_93413 | Not-determined |
| S17_extracted_k141_71663 | Low-quality |
| S17_extracted_k141_137814 | Not-determined |
| S17_extracted_k141_137815 | Low-quality |
| S17_extracted_k141_160204 | Not-determined |
| S17_extracted_k141_137822 | Not-determined |
| S17_extracted_k141_85407 | Not-determined |
| S17_extracted_k141_42486 | Low-quality |
| S17_extracted_k141_108995 | Not-determined |
| S17_extracted_k141_115894 | Not-determined |
| S17_extracted_k141_137827 | Not-determined |
| S17_extracted_k141_115898 | Not-determined |
| S17_extracted_k141_85414 | Not-determined |
| S17_extracted_k141_130178 | Not-determined |
| S17_extracted_k141_78914 | Not-determined |
| S17_extracted_k141_85415 | Not-determined |
| S17_extracted_k141_57007 | Not-determined |
| S17_extracted_k141_130180 | Low-quality |
| S17_extracted_k141_93420 | Not-determined |
| S17_extracted_k141_34992 | Not-determined |
| S17_extracted_k141_122654 | Not-determined |
| S17_extracted_k141_160226 | Not-determined |
| S17_extracted_k141_57014 | Not-determined |
| S17_extracted_k141_35001 | Not-determined |
| S17_extracted_k141_115917 | Not-determined |
| S17_extracted_k141_42497 | Not-determined |
| S17_extracted_k141_145336 | Not-determined |
| S17_extracted_k141_19380 | Not-determined |
| S17_extracted_k141_115922 | Not-determined |
| S17_extracted_k141_137846 | Not-determined |
| S17_extracted_k141_71671 | Not-determined |
| S17_extracted_k141_160234 | Not-determined |
| S17_extracted_k141_137848 | Not-determined |
| S17_extracted_k141_145340 | Not-determined |
| S17_extracted_k141_19384 | Not-determined |
| S17_extracted_k141_122661 | Not-determined |
| S17_extracted_k141_57028 | Not-determined |
| S17_extracted_k141_78922 | Not-determined |
| S17_extracted_k141_160242 | Not-determined |
| S17_extracted_k141_101218 | Not-determined |
| S17_extracted_k141_42506 | Not-determined |
| S17_extracted_k141_122673 | Not-determined |
| S17_extracted_k141_93435 | Not-determined |
| S17_extracted_k141_153282 | Not-determined |
| S17_extracted_k141_167426 | Low-quality |
| S17_extracted_k141_167428 | Not-determined |
| S17_extracted_k141_19394 | Not-determined |
| S17_extracted_k141_122676 | Not-determined |
| S17_extracted_k141_130193 | Not-determined |
| S17_extracted_k141_174340 | Low-quality |
| S17_extracted_k141_4897 | Not-determined |
| S17_extracted_k141_167433 | Not-determined |
| S17_extracted_k141_85437 | Low-quality |
| S17_extracted_k141_4901 | Not-determined |
| S17_extracted_k141_57040 | Not-determined |
| S17_extracted_k141_4902 | Not-determined |
| S17_extracted_k141_4903 | Not-determined |
| S17_extracted_k141_101225 | Not-determined |
| S17_extracted_k141_160257 | Not-determined |
| S17_extracted_k141_4907 | Not-determined |
| S17_extracted_k141_160258 | Not-determined |
| S17_extracted_k141_122690 | Not-determined |
| S17_extracted_k141_167440 | Not-determined |
| S17_extracted_k141_137862 | Not-determined |
| S17_extracted_k141_167442 | Not-determined |
| S17_extracted_k141_174345 | Not-determined |
| S17_extracted_k141_42533 | Not-determined |
| S17_extracted_k141_101232 | Not-determined |
| S17_extracted_k141_115940 | Not-determined |
| S17_extracted_k141_130202 | Not-determined |
| S17_extracted_k141_174348 | Not-determined |
| S17_extracted_k141_115943 | Not-determined |
| S17_extracted_k141_153301 | Low-quality |
| S17_extracted_k141_12138 | Not-determined |
| S17_extracted_k141_130206 | Not-determined |
| S17_extracted_k141_115946 | Not-determined |
| S17_extracted_k141_85442 | Not-determined |
| S17_extracted_k141_153302 | Not-determined |
| S17_extracted_k141_85443 | Not-determined |
| S17_extracted_k141_42539 | Not-determined |
| S17_extracted_k141_153303 | Not-determined |
| S17_extracted_k141_4913 | Low-quality |
| S17_extracted_k141_42541 | Not-determined |
| S17_extracted_k141_35034 | Not-determined |
| S17_extracted_k141_12146 | Not-determined |
| S17_extracted_k141_63309 | Not-determined |
| S17_extracted_k141_122706 | Not-determined |
| S17_extracted_k141_160275 | Not-determined |
| S17_extracted_k141_122707 | Not-determined |
| S17_extracted_k141_35044 | Not-determined |
| S17_extracted_k141_115953 | Not-determined |
| S17_extracted_k141_167451 | Not-determined |
| S17_extracted_k141_101240 | Not-determined |
| S17_extracted_k141_122711 | Not-determined |
| S17_extracted_k141_85460 | Not-determined |
| S17_extracted_k141_115957 | Not-determined |
| S17_extracted_k141_174365 | Not-determined |
| S17_extracted_k141_12151 | Not-determined |
| S17_extracted_k141_153311 | Low-quality |
| S17_extracted_k141_63314 | Not-determined |
| S17_extracted_k141_93449 | Not-determined |
| S17_extracted_k141_109029 | Not-determined |
| S17_extracted_k141_109030 | Not-determined |
| S17_extracted_k141_145348 | Not-determined |
| S17_extracted_k141_101246 | Not-determined |
| S17_extracted_k141_12158 | Not-determined |
| S17_extracted_k141_71688 | Not-determined |
| S17_extracted_k141_109038 | Not-determined |
| S17_extracted_k141_115970 | Not-determined |
| S17_extracted_k141_122731 | Not-determined |
| S17_extracted_k141_153321 | Not-determined |
| S17_extracted_k141_78964 | Not-determined |
| S17_extracted_k141_12164 | Not-determined |
| S17_extracted_k141_71693 | Not-determined |
| S17_extracted_k141_42561 | Not-determined |
| S17_extracted_k141_78967 | Not-determined |
| S17_extracted_k141_167461 | Low-quality |
| S17_extracted_k141_12168 | Not-determined |
| S17_extracted_k141_57048 | Not-determined |
| S17_extracted_k141_12169 | Not-determined |
| S17_extracted_k141_109044 | Not-determined |
| S17_extracted_k141_174379 | Not-determined |
| S17_extracted_k141_115971 | Low-quality |
| S17_extracted_k141_174383 | Not-determined |
| S17_extracted_k141_93466 | Not-determined |
| S17_extracted_k141_85484 | Not-determined |
| S17_extracted_k141_93468 | Low-quality |
| S17_extracted_k141_42572 | Not-determined |
| S17_extracted_k141_101264 | Not-determined |
| S17_extracted_k141_42573 | Not-determined |
| S17_extracted_k141_85488 | Not-determined |
| S17_extracted_k141_42574 | Not-determined |
| S17_extracted_k141_4926 | Low-quality |
| S17_extracted_k141_26522 | Not-determined |
| S17_extracted_k141_93471 | Not-determined |
| S17_extracted_k141_174387 | Not-determined |
| S17_extracted_k141_145376 | Not-determined |
| S17_extracted_k141_167471 | Not-determined |
| S17_extracted_k141_115977 | Not-determined |
| S17_extracted_k141_160297 | Not-determined |
| S17_extracted_k141_4931 | Not-determined |
| S17_extracted_k141_167474 | Low-quality |
| S17_extracted_k141_57057 | Not-determined |
| S17_extracted_k141_93475 | Not-determined |
| S17_extracted_k141_12188 | Not-determined |
| S17_extracted_k141_26527 | Low-quality |
| S17_extracted_k141_71701 | Not-determined |
| S17_extracted_k141_85494 | Low-quality |
| S17_extracted_k141_174392 | Not-determined |
| S17_extracted_k141_93480 | Not-determined |
| S17_extracted_k141_57061 | Not-determined |
| S17_extracted_k141_137868 | Low-quality |
| S17_extracted_k141_57062 | Not-determined |
| S17_extracted_k141_71704 | Not-determined |
| S17_extracted_k141_78985 | Not-determined |
| S17_extracted_k141_57065 | Not-determined |
| S17_extracted_k141_26532 | Not-determined |
| S17_extracted_k141_35077 | Not-determined |
| S17_extracted_k141_137870 | Not-determined |
| S17_extracted_k141_174394 | Not-determined |
| S17_extracted_k141_130222 | Not-determined |
| S17_extracted_k141_137872 | Not-determined |
| S17_extracted_k141_130223 | Low-quality |
| S17_extracted_k141_85497 | Not-determined |
| S17_extracted_k141_78988 | Not-determined |
| S17_extracted_k141_12197 | Not-determined |
| S17_extracted_k141_78989 | Not-determined |
| S17_extracted_k141_174397 | Not-determined |
| S17_extracted_k141_4942 | Not-determined |
| S17_extracted_k141_12200 | Not-determined |
| S17_extracted_k141_71711 | Not-determined |
| S17_extracted_k141_35087 | Not-determined |
| S17_extracted_k141_63329 | Not-determined |
| S17_extracted_k141_63331 | Not-determined |
| S17_extracted_k141_101288 | Not-determined |
| S17_extracted_k141_109073 | Not-determined |
| S17_extracted_k141_71713 | Not-determined |
| S17_extracted_k141_85504 | Not-determined |
| S17_extracted_k141_35091 | Not-determined |
| S17_extracted_k141_63334 | Not-determined |
| S17_extracted_k141_57080 | Not-determined |
| S17_extracted_k141_101291 | Not-determined |
| S17_extracted_k141_4951 | Not-determined |
| S17_extracted_k141_57084 | Not-determined |
| S17_extracted_k141_35095 | Not-determined |
| S17_extracted_k141_4952 | Not-determined |
| S17_extracted_k141_85514 | Not-determined |
| S17_extracted_k141_122763 | Not-determined |
| S17_extracted_k141_4953 | Not-determined |
| S17_extracted_k141_71720 | Not-determined |
| S17_extracted_k141_12207 | Not-determined |
| S17_extracted_k141_63342 | Not-determined |
| S17_extracted_k141_12208 | Not-determined |
| S17_extracted_k141_167495 | Not-determined |
| S17_extracted_k141_12209 | Not-determined |
| S17_extracted_k141_71728 | Not-determined |
| S17_extracted_k141_174408 | Not-determined |
| S17_extracted_k141_130234 | Not-determined |
| S17_extracted_k141_71731 | Not-determined |
| S17_extracted_k141_145397 | Not-determined |
| S17_extracted_k141_153327 | Not-determined |
| S17_extracted_k141_130237 | Low-quality |
| S17_extracted_k141_153329 | Not-determined |
| S17_extracted_k141_130238 | Low-quality |
| S17_extracted_k141_19424 | Low-quality |
| S17_extracted_k141_63346 | Not-determined |
| S17_extracted_k141_122774 | Not-determined |
| S17_extracted_k141_115997 | Not-determined |
| S17_extracted_k141_145400 | Not-determined |
| S17_extracted_k141_35099 | Not-determined |
| S17_extracted_k141_19433 | Not-determined |
| S17_extracted_k141_93490 | Not-determined |
| S17_extracted_k141_35102 | Not-determined |
| S17_extracted_k141_26560 | Not-determined |
| S17_extracted_k141_167506 | Not-determined |
| S17_extracted_k141_35107 | Not-determined |
| S17_extracted_k141_167509 | Low-quality |
| S17_extracted_k141_63350 | Not-determined |
| S17_extracted_k141_57102 | Not-determined |
| S17_extracted_k141_174419 | Not-determined |
| S17_extracted_k141_19440 | Not-determined |
| S17_extracted_k141_57104 | Not-determined |
| S17_extracted_k141_71751 | Not-determined |
| S17_extracted_k141_12225 | Not-determined |
| S17_extracted_k141_71752 | Not-determined |
| S17_extracted_k141_130245 | Not-determined |
| S17_extracted_k141_101316 | Not-determined |
| S17_extracted_k141_167513 | Not-determined |
| S17_extracted_k141_26570 | Not-determined |
| S17_extracted_k141_153345 | Low-quality |
| S17_extracted_k141_71758 | Not-determined |
| S17_extracted_k141_145411 | Not-determined |
| S17_extracted_k141_145412 | Not-determined |
| S17_extracted_k141_26571 | Not-determined |
| S17_extracted_k141_174427 | Not-determined |
| S17_extracted_k141_153353 | Not-determined |
| S17_extracted_k141_101319 | Not-determined |
| S17_extracted_k141_122783 | Not-determined |
| S17_extracted_k141_12232 | Not-determined |
| S17_extracted_k141_35121 | Not-determined |
| S17_extracted_k141_35122 | Not-determined |
| S17_extracted_k141_109093 | Not-determined |
| S17_extracted_k141_109095 | Not-determined |
| S17_extracted_k141_12235 | Not-determined |
| S17_extracted_k141_78997 | Not-determined |
| S17_extracted_k141_12237 | Not-determined |
| S17_extracted_k141_35128 | Not-determined |
| S17_extracted_k141_26579 | Not-determined |
| S17_extracted_k141_109098 | Not-determined |
| S17_extracted_k141_153361 | Not-determined |
| S17_extracted_k141_116011 | Not-determined |
| S17_extracted_k141_145426 | Not-determined |
| S17_extracted_k141_153362 | Not-determined |
| S17_extracted_k141_85555 | Not-determined |
| S17_extracted_k141_167523 | Not-determined |
| S17_extracted_k141_4977 | Not-determined |
| S17_extracted_k141_4978 | Not-determined |
| S17_extracted_k141_174443 | Not-determined |
| S17_extracted_k141_122793 | Not-determined |
| S17_extracted_k141_79005 | Not-determined |
| S17_extracted_k141_26583 | Not-determined |
| S17_extracted_k141_137915 | Low-quality |
| S17_extracted_k141_26584 | Low-quality |
| S17_extracted_k141_137916 | Not-determined |
| S17_extracted_k141_63366 | Not-determined |
| S17_extracted_k141_137917 | Not-determined |
| S17_extracted_k141_93510 | Not-determined |
| S17_extracted_k141_19459 | Not-determined |
| S17_extracted_k141_145430 | Not-determined |
| S17_extracted_k141_101325 | Not-determined |
| S17_extracted_k141_57117 | Not-determined |
| S17_extracted_k141_130259 | Not-determined |
| S17_extracted_k141_137921 | Not-determined |
| S17_extracted_k141_130261 | Not-determined |
| S17_extracted_k141_167534 | Low-quality |
| S17_extracted_k141_153373 | Not-determined |
| S17_extracted_k141_26594 | Not-determined |
| S17_extracted_k141_137922 | Low-quality |
| S17_extracted_k141_19465 | Not-determined |
| S17_extracted_k141_12248 | Not-determined |
| S17_extracted_k141_79019 | Not-determined |
| S17_extracted_k141_63378 | Not-determined |
| S17_extracted_k141_12252 | Not-determined |
| S17_extracted_k141_57118 | Low-quality |
| S17_extracted_k141_57119 | Not-determined |
| S17_extracted_k141_19468 | Not-determined |
| S17_extracted_k141_26597 | Not-determined |
| S17_extracted_k141_26599 | Not-determined |
| S17_extracted_k141_71776 | Low-quality |
| S17_extracted_k141_4986 | Not-determined |
| S17_extracted_k141_71778 | Not-determined |
| S17_extracted_k141_4987 | Not-determined |
| S17_extracted_k141_57126 | Not-determined |
| S17_extracted_k141_19470 | Low-quality |
| S17_extracted_k141_26605 | Not-determined |
| S17_extracted_k141_160321 | Not-determined |
| S17_extracted_k141_160322 | Low-quality |
| S17_extracted_k141_101337 | Not-determined |
| S17_extracted_k141_19475 | Not-determined |
| S17_extracted_k141_26611 | Not-determined |
| S17_extracted_k141_130275 | Not-determined |
| S17_extracted_k141_26615 | Not-determined |
| S17_extracted_k141_101341 | Not-determined |
| S17_extracted_k141_101342 | Not-determined |
| S17_extracted_k141_160327 | Not-determined |
| S17_extracted_k141_122817 | Not-determined |
| S17_extracted_k141_79037 | Low-quality |
| S17_extracted_k141_153393 | Not-determined |
| S17_extracted_k141_145476 | Not-determined |
| S17_extracted_k141_116032 | Not-determined |
| S17_extracted_k141_4994 | Not-determined |
| S17_extracted_k141_57146 | Low-quality |
| S17_extracted_k141_160335 | Not-determined |
| S17_extracted_k141_153402 | Not-determined |
| S17_extracted_k141_19486 | Low-quality |
| S17_extracted_k141_4998 | Not-determined |
| S17_extracted_k141_19488 | Not-determined |
| S17_extracted_k141_167557 | Low-quality |
| S17_extracted_k141_160340 | Not-determined |
| S17_extracted_k141_42615 | Not-determined |

| | |
|---|---|
| S17_extracted_k141_160342 | Not-determined |
| S17_extracted_k141_167559 | Not-determined |
| S17_extracted_k141_19495 | Not-determined |
| S17_extracted_k141_42618 | Low-quality |
| S17_extracted_k141_122822 | Low-quality |
| S17_extracted_k141_145487 | Not-determined |
| S17_extracted_k141_5006 | Not-determined |
| S17_extracted_k141_5008 | Not-determined |
| S17_extracted_k141_42622 | Not-determined |
| S17_extracted_k141_71796 | Not-determined |
| S17_extracted_k141_42625 | Not-determined |
| S17_extracted_k141_49688 | Low-quality |
| S17_extracted_k141_145493 | Not-determined |
| S17_extracted_k141_137951 | Not-determined |
| S17_extracted_k141_57168 | Low-quality |
| S17_extracted_k141_57169 | Not-determined |
| S17_extracted_k141_79047 | Not-determined |
| S17_extracted_k141_49692 | Not-determined |
| S17_extracted_k141_122832 | Low-quality |
| S17_extracted_k141_79050 | Not-determined |
| S17_extracted_k141_79051 | Not-determined |
| S17_extracted_k141_42633 | Not-determined |
| S17_extracted_k141_93554 | Low-quality |
| S17_extracted_k141_116040 | Low-quality |
| S17_extracted_k141_5019 | Low-quality |
| S17_extracted_k141_167569 | Not-determined |
| S17_extracted_k141_130282 | Not-determined |
| S17_extracted_k141_49697 | Low-quality |
| S17_extracted_k141_122834 | Not-determined |
| S17_extracted_k141_5022 | Not-determined |
| S17_extracted_k141_57177 | Not-determined |
| S17_extracted_k141_145504 | Not-determined |
| S17_extracted_k141_71809 | Not-determined |
| S17_extracted_k141_167574 | Not-determined |
| S17_extracted_k141_57179 | Low-quality |
| S17_extracted_k141_137960 | Not-determined |
| S17_extracted_k141_101357 | Low-quality |
| S17_extracted_k141_5028 | Not-determined |
| S17_extracted_k141_101359 | Low-quality |
| S17_extracted_k141_19518 | Not-determined |
| S17_extracted_k141_93558 | Not-determined |
| S17_extracted_k141_145513 | Not-determined |
| S17_extracted_k141_101362 | Not-determined |
| S17_extracted_k141_71816 | Low-quality |
| S17_extracted_k141_79067 | Not-determined |
| S17_extracted_k141_19522 | Not-determined |
| S17_extracted_k141_160363 | Not-determined |
| S17_extracted_k141_145518 | Not-determined |
| S17_extracted_k141_160364 | Not-determined |
| S17_extracted_k141_101366 | Not-determined |
| S17_extracted_k141_19525 | Not-determined |
| S17_extracted_k141_153414 | Not-determined |
| S17_extracted_k141_122842 | Low-quality |
| S17_extracted_k141_122843 | Not-determined |
| S17_extracted_k141_116050 | Low-quality |
| S17_extracted_k141_79077 | Not-determined |
| S17_extracted_k141_130288 | Not-determined |
| S17_extracted_k141_137966 | Low-quality |
| S17_extracted_k141_93567 | Not-determined |
| S17_extracted_k141_137968 | Not-determined |
| S17_extracted_k141_130291 | Not-determined |
| S17_extracted_k141_130292 | Not-determined |
| S17_extracted_k141_71821 | Not-determined |
| S17_extracted_k141_19531 | Not-determined |
| S17_extracted_k141_153417 | Not-determined |
| S17_extracted_k141_116055 | Low-quality |
| S17_extracted_k141_5045 | Not-determined |
| S17_extracted_k141_5046 | Not-determined |
| S17_extracted_k141_19534 | Not-determined |
| S17_extracted_k141_49721 | Low-quality |
| S17_extracted_k141_19535 | Not-determined |
| S17_extracted_k141_5047 | Not-determined |
| S17_extracted_k141_145528 | Not-determined |
| S17_extracted_k141_130294 | Not-determined |
| S17_extracted_k141_160371 | Not-determined |
| S17_extracted_k141_19537 | Not-determined |
| S17_extracted_k141_5049 | Not-determined |
| S17_extracted_k141_145530 | Not-determined |
| S17_extracted_k141_93572 | Not-determined |
| S17_extracted_k141_5050 | Not-determined |
| S17_extracted_k141_85563 | Not-determined |
| S17_extracted_k141_116063 | Low-quality |
| S17_extracted_k141_93577 | Not-determined |
| S17_extracted_k141_145537 | Not-determined |
| S17_extracted_k141_153432 | Not-determined |
| S17_extracted_k141_153435 | Not-determined |
| S17_extracted_k141_116069 | Not-determined |
| S17_extracted_k141_174462 | Not-determined |
| S17_extracted_k141_49729 | Low-quality |
| S17_extracted_k141_93584 | Not-determined |
| S17_extracted_k141_93585 | Not-determined |
| S17_extracted_k141_153438 | Low-quality |
| S17_extracted_k141_130302 | Not-determined |
| S17_extracted_k141_174466 | Low-quality |
| S16_vs_42706\|\|full | High-quality |
| S17_extracted_k141_130304 | Not-determined |
| S17_extracted_k141_160385 | Not-determined |
| S17_extracted_k141_174468 | Not-determined |
| S17_extracted_k141_35141 | Not-determined |
| S17_extracted_k141_109118 | Not-determined |
| S17_extracted_k141_5076 | Not-determined |
| S17_extracted_k141_116078 | Not-determined |
| S17_extracted_k141_63386 | Not-determined |
| S17_extracted_k141_63388 | Not-determined |
| S17_extracted_k141_93596 | Not-determined |
| S17_extracted_k141_153455 | Not-determined |
| S17_extracted_k141_109121 | Not-determined |
| S17_extracted_k141_63391 | Not-determined |
| S17_extracted_k141_35158 | Not-determined |
| S17_extracted_k141_109126 | Not-determined |
| S17_extracted_k141_85587 | Not-determined |
| S17_extracted_k141_153463 | Not-determined |
| S17_extracted_k141_49756 | Not-determined |
| S17_extracted_k141_42638 | Not-determined |
| S17_extracted_k141_160402 | Low-quality |
| S17_extracted_k141_109133 | Not-determined |
| S17_extracted_k141_49760 | Not-determined |
| S17_extracted_k141_122879 | Not-determined |
| S17_extracted_k141_79107 | Not-determined |
| S17_extracted_k141_42641 | Low-quality |
| S17_extracted_k141_57206 | Not-determined |
| S17_extracted_k141_42643 | Low-quality |
| S17_extracted_k141_85598 | Low-quality |
| S17_extracted_k141_153469 | Low-quality |
| S17_extracted_k141_116099 | Not-determined |
| S17_extracted_k141_116101 | Low-quality |
| S17_extracted_k141_130315 | Not-determined |
| S17_extracted_k141_130316 | Not-determined |
| S17_extracted_k141_137987 | Not-determined |
| S17_extracted_k141_122889 | Not-determined |
| S17_extracted_k141_5100 | Not-determined |
| S17_extracted_k141_116102 | Not-determined |
| S17_extracted_k141_93622 | Not-determined |
| S17_extracted_k141_63406 | Not-determined |
| S17_extracted_k141_71823 | Not-determined |
| S17_extracted_k141_93624 | Not-determined |
| S17_extracted_k141_153477 | Not-determined |
| S17_extracted_k141_93625 | Not-determined |
| S17_extracted_k141_122896 | Not-determined |
| S17_extracted_k141_116109 | Not-determined |
| S17_extracted_k141_5104 | Not-determined |
| S17_extracted_k141_5105 | Not-determined |
| S17_extracted_k141_153481 | Not-determined |
| S17_extracted_k141_93633 | Not-determined |
| S17_extracted_k141_109153 | Not-determined |
| S17_extracted_k141_153482 | Not-determined |
| S17_extracted_k141_93634 | Low-quality |
| S17_extracted_k141_49776 | Not-determined |
| S17_extracted_k141_5111 | Low-quality |
| S17_extracted_k141_109160 | Not-determined |
| S17_extracted_k141_85614 | Not-determined |
| S17_extracted_k141_49779 | Not-determined |
| S17_extracted_k141_85616 | Not-determined |
| S17_extracted_k141_122899 | Not-determined |
| S17_extracted_k141_138004 | Not-determined |
| S17_extracted_k141_49785 | Not-determined |
| S17_extracted_k141_35184 | Not-determined |
| S17_extracted_k141_109170 | Low-quality |
| S17_extracted_k141_26630 | Not-determined |
| S17_extracted_k141_49787 | Not-determined |
| S17_extracted_k141_26632 | Not-determined |
| S17_extracted_k141_12258 | Not-determined |
| S17_extracted_k141_85621 | Not-determined |
| S17_extracted_k141_85622 | Not-determined |
| S17_extracted_k141_5124 | Not-determined |
| S17_extracted_k141_49788 | Not-determined |
| S17_extracted_k141_160419 | Not-determined |
| S17_extracted_k141_5127 | Not-determined |
| S17_extracted_k141_122909 | Not-determined |
| S17_extracted_k141_12268 | Not-determined |
| S17_extracted_k141_12269 | Low-quality |
| S17_extracted_k141_79128 | Not-determined |
| S17_extracted_k141_93658 | Not-determined |
| S17_extracted_k141_116127 | Not-determined |
| S17_extracted_k141_12274 | Not-determined |
| S17_extracted_k141_93659 | Low-quality |
| S17_extracted_k141_79132 | Not-determined |
| S17_extracted_k141_35196 | Not-determined |
| S17_extracted_k141_35197 | Not-determined |
| S17_extracted_k141_79133 | Not-determined |
| S17_extracted_k141_85623 | Low-quality |
| S17_extracted_k141_167594 | Not-determined |
| S17_extracted_k141_5133 | Low-quality |
| S17_extracted_k141_35200 | Not-determined |
| S17_extracted_k141_85627 | Not-determined |
| S17_extracted_k141_109194 | Not-determined |
| S17_extracted_k141_130326 | Low-quality |
| S17_extracted_k141_130327 | Not-determined |
| S17_extracted_k141_85630 | Not-determined |
| S17_extracted_k141_116130 | Not-determined |
| S17_extracted_k141_49810 | Not-determined |
| S17_extracted_k141_130329 | Not-determined |
| S17_extracted_k141_93668 | Low-quality |
| S17_extracted_k141_26633 | Low-quality |
| S17_extracted_k141_12283 | Not-determined |
| S17_extracted_k141_71848 | Not-determined |
| S17_extracted_k141_71849 | Not-determined |
| S17_extracted_k141_42693 | Not-determined |
| S17_extracted_k141_42694 | Not-determined |
| S17_extracted_k141_160434 | Not-determined |
| S17_extracted_k141_167607 | Not-determined |
| S17_extracted_k141_160437 | Low-quality |
| S17_extracted_k141_145540 | Low-quality |
| S17_extracted_k141_35214 | Not-determined |
| S17_extracted_k141_160438 | Not-determined |
| S17_extracted_k141_49816 | Low-quality |
| S17_extracted_k141_12300 | Not-determined |
| S17_extracted_k141_160440 | Not-determined |
| S17_extracted_k141_63430 | Not-determined |
| S17_extracted_k141_116142 | Not-determined |
| S17_extracted_k141_122914 | Not-determined |
| S17_extracted_k141_63434 | Not-determined |
| S17_extracted_k141_109208 | Not-determined |
| S17_extracted_k141_63435 | Not-determined |
| S17_extracted_k141_42708 | Not-determined |
| S17_extracted_k141_130339 | Not-determined |
| S17_extracted_k141_63436 | Not-determined |
| S17_extracted_k141_35225 | Low-quality |
| S17_extracted_k141_174493 | Not-determined |
| S17_extracted_k141_138035 | Not-determined |
| S17_extracted_k141_5156 | Not-determined |
| S17_extracted_k141_19560 | Not-determined |
| S17_extracted_k141_145547 | Not-determined |
| S17_extracted_k141_42712 | Not-determined |
| S17_extracted_k141_174501 | Not-determined |
| S17_extracted_k141_35231 | Low-quality |
| S17_extracted_k141_93682 | Not-determined |
| S17_extracted_k141_174502 | Not-determined |
| S17_extracted_k141_79140 | Not-determined |
| S17_extracted_k141_12308 | Not-determined |
| S17_extracted_k141_122927 | Not-determined |
| S17_extracted_k141_35233 | Not-determined |
| S17_extracted_k141_79144 | Not-determined |
| S17_extracted_k141_130354 | Not-determined |
| S17_extracted_k141_85660 | Not-determined |
| S17_extracted_k141_79145 | Not-determined |
| S17_extracted_k141_85661 | Not-determined |
| S17_extracted_k141_71876 | Not-determined |
| S17_extracted_k141_26654 | Low-quality |
| S17_extracted_k141_122931 | Not-determined |
| S17_extracted_k141_26657 | Not-determined |
| S17_extracted_k141_160450 | Low-quality |
| S17_extracted_k141_42716 | Not-determined |
| S17_extracted_k141_19574 | Not-determined |
| S17_extracted_k141_160451 | Not-determined |
| S17_extracted_k141_71879 | Not-determined |
| S17_extracted_k141_19575 | Not-determined |
| S17_extracted_k141_145554 | Not-determined |
| S17_extracted_k141_130361 | Not-determined |
| S17_extracted_k141_153524 | Not-determined |
| S17_extracted_k141_42720 | Not-determined |
| S17_extracted_k141_42721 | Low-quality |
| S17_extracted_k141_160461 | Not-determined |
| S17_extracted_k141_57240 | Not-determined |
| S17_extracted_k141_5184 | Not-determined |
| S17_extracted_k141_19587 | Not-determined |
| S17_extracted_k141_63460 | Not-determined |
| S17_extracted_k141_42723 | Low-quality |
| S17_extracted_k141_160470 | Not-determined |
| S17_extracted_k141_71892 | Not-determined |
| S17_extracted_k141_42724 | Not-determined |
| S17_extracted_k141_71893 | Low-quality |
| S17_extracted_k141_19591 | Not-determined |
| S17_extracted_k141_26661 | Low-quality |
| S17_extracted_k141_12334 | Not-determined |
| S17_extracted_k141_71896 | Not-determined |
| S17_extracted_k141_116178 | Not-determined |
| S17_extracted_k141_122950 | Not-determined |
| S17_extracted_k141_5198 | Not-determined |
| S17_extracted_k141_85678 | Not-determined |
| S17_extracted_k141_5199 | Low-quality |
| S17_extracted_k141_71899 | Not-determined |
| S17_extracted_k141_109243 | Not-determined |
| S17_extracted_k141_57248 | Not-determined |
| S17_extracted_k141_174528 | Not-determined |
| S17_extracted_k141_5201 | Low-quality |
| S17_extracted_k141_71902 | Not-determined |
| S17_extracted_k141_12340 | Not-determined |
| S17_extracted_k141_35264 | Low-quality |
| S17_extracted_k141_42733 | Low-quality |
| S17_extracted_k141_109248 | Not-determined |
| S17_extracted_k141_19600 | Not-determined |
| S17_extracted_k141_35267 | Not-determined |
| S17_extracted_k141_63465 | Low-quality |
| S17_extracted_k141_122962 | Not-determined |
| S11_extracted_k141_104068 | Complete |
| S17_extracted_k141_153553 | Not-determined |
| S17_extracted_k141_12350 | Low-quality |
| S17_extracted_k141_153554 | Not-determined |
| S17_extracted_k141_85687 | Not-determined |
| S17_extracted_k141_49853 | Not-determined |
| S17_extracted_k141_5217 | Not-determined |
| S17_extracted_k141_42744 | Not-determined |
| S17_extracted_k141_167617 | Low-quality |
| S17_extracted_k141_138048 | Low-quality |
| S17_extracted_k141_26673 | Not-determined |
| S17_extracted_k141_167618 | Not-determined |
| S17_extracted_k141_57266 | Not-determined |
| S17_extracted_k141_35279 | Not-determined |
| S17_extracted_k141_167620 | Not-determined |
| S17_extracted_k141_130392 | Not-determined |
| S17_extracted_k141_130393 | Not-determined |
| S17_extracted_k141_26678 | Not-determined |
| S17_extracted_k141_101383 | Not-determined |
| S17_extracted_k141_26679 | Not-determined |
| S17_extracted_k141_109263 | Not-determined |
| S17_extracted_k141_79156 | Not-determined |
| S17_extracted_k141_57274 | Low-quality |
| S17_extracted_k141_85695 | Not-determined |
| S17_extracted_k141_116194 | Not-determined |
| S17_extracted_k141_57275 | Not-determined |
| S17_extracted_k141_116196 | Not-determined |
| S17_extracted_k141_160498 | Low-quality |
| S17_extracted_k141_71927 | Not-determined |
| S17_extracted_k141_85699 | Not-determined |
| S17_extracted_k141_71928 | Not-determined |
| S17_extracted_k141_160500 | Not-determined |
| S17_extracted_k141_26691 | Not-determined |
| S17_extracted_k141_116202 | Not-determined |
| S17_extracted_k141_138054 | Not-determined |
| S17_extracted_k141_116204 | Not-determined |
| S17_extracted_k141_116206 | Not-determined |
| S17_extracted_k141_116207 | Not-determined |
| S17_extracted_k141_174540 | Low-quality |
| S17_extracted_k141_19616 | Not-determined |
| S17_extracted_k141_63490 | Not-determined |
| S17_extracted_k141_116209 | Not-determined |
| S17_extracted_k141_167640 | Not-determined |
| S17_extracted_k141_19619 | Not-determined |
| S17_extracted_k141_26697 | Not-determined |
| S17_extracted_k141_145559 | Not-determined |
| S17_extracted_k141_167644 | Low-quality |
| S17_extracted_k141_57287 | Not-determined |
| S17_extracted_k141_71934 | Not-determined |
| S17_extracted_k141_167645 | Not-determined |
| S17_extracted_k141_116217 | Not-determined |
| S17_extracted_k141_42761 | Low-quality |
| S17_extracted_k141_116218 | Not-determined |
| S17_extracted_k141_79161 | Not-determined |
| S17_extracted_k141_79162 | Not-determined |
| S17_extracted_k141_116219 | Not-determined |
| S17_extracted_k141_101402 | Low-quality |
| S17_extracted_k141_63497 | Not-determined |
| S17_extracted_k141_63498 | Not-determined |
| S17_extracted_k141_49875 | Not-determined |
| S17_extracted_k141_130409 | Not-determined |
| S17_extracted_k141_145569 | Not-determined |
| S17_extracted_k141_101409 | Not-determined |
| S17_extracted_k141_130411 | Low-quality |
| S17_extracted_k141_167653 | Not-determined |
| S17_extracted_k141_145573 | Not-determined |
| S17_extracted_k141_109295 | Not-determined |
| S17_extracted_k141_109296 | Not-determined |
| S17_extracted_k141_101412 | Not-determined |
| S17_extracted_k141_138070 | Not-determined |
| S17_extracted_k141_85714 | Low-quality |
| S17_extracted_k141_12359 | Not-determined |
| S17_extracted_k141_71941 | Not-determined |
| S17_extracted_k141_79174 | Not-determined |
| S17_extracted_k141_101416 | Not-determined |
| S17_extracted_k141_145581 | Not-determined |
| S17_extracted_k141_12362 | Not-determined |
| S17_extracted_k141_167664 | Not-determined |
| S17_extracted_k141_109307 | Not-determined |
| S17_extracted_k141_12364 | Not-determined |
| S17_extracted_k141_26707 | Not-determined |
| S17_extracted_k141_12365 | Not-determined |
| S17_extracted_k141_85721 | Not-determined |
| S17_extracted_k141_71948 | Not-determined |
| S17_extracted_k141_138081 | Not-determined |
| S17_extracted_k141_109309 | Low-quality |
| S17_extracted_k141_138089 | Not-determined |
| S17_extracted_k141_35323 | Low-quality |
| S17_extracted_k141_79187 | Not-determined |
| S17_extracted_k141_42781 | Not-determined |
| S17_extracted_k141_109314 | Not-determined |
| S17_extracted_k141_109316 | Not-determined |
| S17_extracted_k141_116240 | Low-quality |
| S17_extracted_k141_49905 | Not-determined |
| S17_extracted_k141_35333 | Not-determined |
| S17_extracted_k141_12374 | Not-determined |
| S17_extracted_k141_130426 | Low-quality |
| S17_extracted_k141_35336 | Not-determined |
| S17_extracted_k141_101431 | Not-determined |
| S17_extracted_k141_85729 | Low-quality |
| S17_extracted_k141_123003 | Not-determined |
| S17_extracted_k141_57297 | Not-determined |
| S17_extracted_k141_145594 | Not-determined |
| S17_extracted_k141_12378 | Not-determined |
| S17_extracted_k141_19640 | Not-determined |
| S17_extracted_k141_116249 | Not-determined |
| S17_extracted_k141_12381 | Not-determined |
| S17_extracted_k141_57303 | Low-quality |
| S17_extracted_k141_71961 | Not-determined |
| S17_extracted_k141_79203 | Not-determined |
| S17_extracted_k141_57305 | Not-determined |
| S17_extracted_k141_109331 | Low-quality |
| S17_extracted_k141_145602 | Not-determined |
| S17_extracted_k141_101444 | Not-determined |
| S17_extracted_k141_174549 | Not-determined |
| S17_extracted_k141_71969 | Not-determined |
| S17_extracted_k141_57311 | Not-determined |
| S17_extracted_k141_101448 | Low-quality |
| S17_extracted_k141_145608 | Not-determined |
| S17_extracted_k141_145609 | Not-determined |
| S17_extracted_k141_109340 | Not-determined |
| S17_extracted_k141_130435 | Not-determined |
| S17_extracted_k141_145613 | Low-quality |
| S17_extracted_k141_167680 | Not-determined |
| S17_extracted_k141_130437 | Not-determined |
| S17_extracted_k141_35359 | Not-determined |
| S17_extracted_k141_57317 | Not-determined |
| S17_extracted_k141_130439 | Not-determined |
| S17_extracted_k141_71974 | Not-determined |
| S17_extracted_k141_35365 | Not-determined |
| S17_extracted_k141_167688 | Not-determined |
| S17_extracted_k141_174555 | Not-determined |
| S17_extracted_k141_35368 | Not-determined |
| S17_extracted_k141_79213 | Not-determined |
| S17_extracted_k141_49919 | Not-determined |
| S17_extracted_k141_101459 | Not-determined |
| S17_extracted_k141_71980 | Not-determined |
| S17_extracted_k141_71981 | Not-determined |
| S17_extracted_k141_123024 | Not-determined |
| S17_extracted_k141_167696 | Not-determined |
| S17_extracted_k141_35372 | Not-determined |
| S17_extracted_k141_57329 | Not-determined |
| S17_extracted_k141_116256 | Low-quality |
| S17_extracted_k141_145626 | Not-determined |
| S17_extracted_k141_138125 | Not-determined |
| S17_extracted_k141_79223 | Not-determined |
| S17_extracted_k141_49927 | Not-determined |
| S17_extracted_k141_49929 | Not-determined |
| S17_extracted_k141_101469 | Not-determined |
| S17_extracted_k141_130453 | Not-determined |
| S17_extracted_k141_109360 | Low-quality |
| S17_extracted_k141_63534 | Low-quality |
| S17_extracted_k141_130456 | Not-determined |
| S17_extracted_k141_49934 | Not-determined |
| S17_extracted_k141_49935 | Not-determined |
| S17_extracted_k141_35384 | Not-determined |
| S17_extracted_k141_167706 | Not-determined |
| S17_extracted_k141_145631 | Not-determined |
| S17_extracted_k141_49940 | Not-determined |
| S17_extracted_k141_79235 | Not-determined |
| S17_extracted_k141_116268 | Not-determined |
| S17_extracted_k141_79237 | Not-determined |
| S17_extracted_k141_116273 | Not-determined |
| S17_extracted_k141_138133 | Not-determined |
| S17_extracted_k141_79240 | Not-determined |
| S17_extracted_k141_138139 | Not-determined |
| S17_extracted_k141_26733 | Low-quality |
| S17_extracted_k141_160516 | Not-determined |
| S17_extracted_k141_101485 | Not-determined |
| S17_extracted_k141_35398 | Not-determined |
| S17_extracted_k141_63547 | Not-determined |
| S17_extracted_k141_63548 | Not-determined |
| S17_extracted_k141_85759 | Not-determined |
| S17_extracted_k141_174574 | Not-determined |
| S17_extracted_k141_72004 | Not-determined |
| S17_extracted_k141_174578 | Not-determined |
| S17_extracted_k141_145639 | Not-determined |
| S17_extracted_k141_167722 | Not-determined |
| S17_extracted_k141_109380 | Not-determined |
| S17_extracted_k141_123048 | Not-determined |
| S17_extracted_k141_116282 | Not-determined |
| S17_extracted_k141_123052 | Not-determined |
| S17_extracted_k141_35412 | Not-determined |
| S17_extracted_k141_35413 | Low-quality |
| S17_extracted_k141_101487 | Low-quality |
| S17_extracted_k141_63559 | Not-determined |
| S17_extracted_k141_174587 | Not-determined |
| S17_extracted_k141_49963 | Not-determined |
| S17_extracted_k141_109391 | Not-determined |
| S17_extracted_k141_167737 | Not-determined |
| S17_extracted_k141_19678 | Not-determined |
| S17_extracted_k141_49973 | Not-determined |
| S17_extracted_k141_49974 | Not-determined |
| S17_extracted_k141_19681 | Not-determined |
| S17_extracted_k141_145647 | Not-determined |
| S17_extracted_k141_12443 | Not-determined |
| S17_extracted_k141_12444 | Not-determined |
| S17_extracted_k141_174594 | Low-quality |
| S17_extracted_k141_72024 | Not-determined |
| S17_extracted_k141_12447 | Not-determined |
| S17_extracted_k141_138158 | Not-determined |
| S17_extracted_k141_85774 | Low-quality |
| S17_extracted_k141_63572 | Not-determined |
| S17_extracted_k141_42808 | Not-determined |
| S17_extracted_k141_42809 | Not-determined |
| S17_extracted_k141_138161 | Not-determined |
| S17_extracted_k141_72026 | Low-quality |
| S17_extracted_k141_85778 | Low-quality |
| S17_extracted_k141_130457 | Low-quality |
| S17_extracted_k141_130458 | Not-determined |
| S17_extracted_k141_79260 | Not-determined |
| S17_extracted_k141_93707 | Not-determined |
| S17_extracted_k141_26742 | Not-determined |
| S17_extracted_k141_5237 | Not-determined |
| S17_extracted_k141_109411 | Not-determined |
| S17_extracted_k141_160542 | Low-quality |
| S17_extracted_k141_5239 | Not-determined |
| S17_extracted_k141_130462 | Not-determined |
| S17_extracted_k141_167755 | Not-determined |
| S17_extracted_k141_12461 | Not-determined |
| S17_extracted_k141_123074 | Not-determined |
| S17_extracted_k141_79263 | Not-determined |
| S17_extracted_k141_160548 | Low-quality |
| S17_extracted_k141_63584 | Not-determined |
| S17_extracted_k141_174609 | Not-determined |
| S17_extracted_k141_49994 | Not-determined |
| S17_extracted_k141_116305 | Not-determined |
| S17_extracted_k141_12467 | Not-determined |
| S17_extracted_k141_5242 | Not-determined |
| S17_extracted_k141_12470 | Not-determined |
| S17_extracted_k141_174610 | Not-determined |
| S17_extracted_k141_116310 | Not-determined |
| S17_extracted_k141_85794 | Not-determined |
| S17_extracted_k141_63589 | Not-determined |
| S17_extracted_k141_130471 | Not-determined |
| S17_extracted_k141_5244 | Not-determined |
| S17_extracted_k141_123082 | Not-determined |
| S17_extracted_k141_57364 | Not-determined |
| S17_extracted_k141_57365 | Not-determined |
| S17_extracted_k141_109430 | Not-determined |
| S17_extracted_k141_72042 | Not-determined |
| S17_extracted_k141_5248 | Not-determined |
| S17_extracted_k141_123085 | Not-determined |
| S17_extracted_k141_167758 | Not-determined |
| S17_extracted_k141_116318 | Low-quality |
| S17_extracted_k141_50000 | Not-determined |
| S17_extracted_k141_57371 | Not-determined |
| S17_extracted_k141_72047 | Not-determined |
| S17_extracted_k141_160558 | Not-determined |
| S17_extracted_k141_130477 | Not-determined |
| S17_extracted_k141_79272 | Not-determined |
| S17_extracted_k141_116322 | Not-determined |
| S17_extracted_k141_167768 | Not-determined |
| S17_extracted_k141_85806 | Not-determined |
| S17_extracted_k141_123093 | Not-determined |
| S17_extracted_k141_42833 | Not-determined |
| S17_extracted_k141_160566 | Not-determined |
| S17_extracted_k141_42834 | Not-determined |
| S17_extracted_k141_50006 | Low-quality |
| S17_extracted_k141_160567 | Not-determined |
| S17_extracted_k141_5265 | Not-determined |
| S17_extracted_k141_19706 | Not-determined |
| S17_extracted_k141_85816 | Not-determined |
| S17_extracted_k141_101523 | Not-determined |
| S17_extracted_k141_174622 | Not-determined |
| S17_extracted_k141_130488 | Not-determined |
| S17_extracted_k141_19709 | Not-determined |
| S17_extracted_k141_174624 | Low-quality |
| S17_extracted_k141_130490 | Low-quality |
| S17_extracted_k141_174625 | Not-determined |
| S17_extracted_k141_85821 | Not-determined |
| S17_extracted_k141_63601 | Low-quality |
| S17_extracted_k141_5269 | Not-determined |
| S17_extracted_k141_145664 | Not-determined |
| S17_extracted_k141_101527 | Not-determined |
| S17_extracted_k141_19710 | Not-determined |
| S17_extracted_k141_63604 | Not-determined |
| S17_extracted_k141_167779 | Not-determined |
| S17_extracted_k141_101529 | Not-determined |
| S17_extracted_k141_123096 | Not-determined |
| S17_extracted_k141_85826 | Not-determined |
| S17_extracted_k141_72064 | Low-quality |
| S17_extracted_k141_116328 | Not-determined |
| S17_extracted_k141_123097 | Not-determined |
| S17_extracted_k141_138176 | Not-determined |
| S17_extracted_k141_174629 | Low-quality |
| S17_extracted_k141_85829 | Low-quality |
| S17_extracted_k141_5279 | Not-determined |
| S17_extracted_k141_19715 | Low-quality |
| S17_extracted_k141_123103 | Not-determined |
| S17_extracted_k141_85832 | Low-quality |
| S17_extracted_k141_109441 | Not-determined |
| S17_extracted_k141_116334 | Not-determined |
| S17_extracted_k141_50020 | Not-determined |
| S17_extracted_k141_79286 | Not-determined |
| S17_extracted_k141_123106 | Not-determined |
| S17_extracted_k141_138181 | Not-determined |
| S17_extracted_k141_85838 | Not-determined |
| S17_extracted_k141_50024 | Not-determined |
| S17_extracted_k141_123111 | Not-determined |
| S17_extracted_k141_145680 | Not-determined |
| S17_extracted_k141_130503 | Low-quality |
| S17_extracted_k141_101547 | Not-determined |
| S17_extracted_k141_12486 | Not-determined |

| | |
|---|---|
| S17_extracted_k141_63625 | Low-quality |
| S17_extracted_k141_160587 | Not-determined |
| S17_extracted_k141_167794 | Not-determined |
| S17_extracted_k141_109447 | Not-determined |
| S17_extracted_k141_160589 | Not-determined |
| S17_extracted_k141_85844 | Not-determined |
| S17_extracted_k141_145685 | Not-determined |
| S17_extracted_k141_167797 | Low-quality |
| S17_extracted_k141_138188 | Not-determined |
| S17_extracted_k141_101554 | Not-determined |
| S17_extracted_k141_130512 | Not-determined |
| S17_extracted_k141_5296 | Not-determined |
| S17_extracted_k141_130514 | Not-determined |
| S17_extracted_k141_145687 | Not-determined |
| S17_extracted_k141_85852 | Not-determined |
| S17_extracted_k141_79290 | Low-quality |
| S17_extracted_k141_85853 | Low-quality |
| S17_extracted_k141_19727 | Not-determined |
| S17_extracted_k141_138190 | Low-quality |
| S17_extracted_k141_123124 | Low-quality |
| S17_extracted_k141_101558 | Not-determined |
| S17_extracted_k141_138192 | Not-determined |
| S17_extracted_k141_116346 | Not-determined |
| S17_extracted_k141_79296 | Not-determined |
| S17_extracted_k141_109451 | Not-determined |
| S17_extracted_k141_130522 | Not-determined |
| S17_extracted_k141_50047 | Not-determined |
| S17_extracted_k141_57381 | Not-determined |
| S17_extracted_k141_79298 | Low-quality |
| S17_extracted_k141_93720 | Low-quality |
| S17_extracted_k141_160596 | Not-determined |
| S17_extracted_k141_93723 | Not-determined |
| S17_extracted_k141_79300 | Not-determined |
| S17_extracted_k141_101565 | Not-determined |
| S17_extracted_k141_138198 | Not-determined |
| S17_extracted_k141_5310 | Not-determined |
| S17_extracted_k141_109459 | Not-determined |
| S17_extracted_k141_57388 | Low-quality |
| S17_extracted_k141_93730 | Not-determined |
| S17_extracted_k141_12509 | Not-determined |
| S17_extracted_k141_138204 | Not-determined |
| S17_extracted_k141_101570 | Not-determined |
| S17_extracted_k141_93736 | Not-determined |
| S17_extracted_k141_5323 | Not-determined |
| S17_extracted_k141_167800 | Not-determined |
| S17_extracted_k141_5324 | Not-determined |
| S17_extracted_k141_167803 | Low-quality |
| S17_extracted_k141_57400 | Not-determined |
| S17_extracted_k141_101573 | Low-quality |
| S17_extracted_k141_174656 | Not-determined |
| S17_extracted_k141_5327 | Not-determined |
| S17_extracted_k141_174657 | Low-quality |
| S17_extracted_k141_42871 | Not-determined |
| S17_extracted_k141_85873 | Not-determined |
| S17_extracted_k141_12517 | Not-determined |
| S17_extracted_k141_79311 | Not-determined |
| S17_extracted_k141_5332 | Not-determined |
| S17_extracted_k141_35419 | Not-determined |
| S17_extracted_k141_85874 | Not-determined |
| S17_extracted_k141_116380 | Not-determined |
| S17_extracted_k141_57408 | Low-quality |
| S17_extracted_k141_79316 | Not-determined |
| S17_extracted_k141_12520 | Not-determined |
| S17_extracted_k141_167811 | Not-determined |
| S17_extracted_k141_85877 | Not-determined |
| S17_extracted_k141_42877 | Not-determined |
| S17_extracted_k141_79320 | Not-determined |
| S17_extracted_k141_5345 | Not-determined |
| S17_extracted_k141_5347 | Not-determined |
| S17_extracted_k141_63653 | Not-determined |
| S17_extracted_k141_35425 | Not-determined |
| S17_extracted_k141_138218 | Not-determined |
| S17_extracted_k141_116382 | Not-determined |
| S17_extracted_k141_174661 | Not-determined |
| S17_extracted_k141_85887 | Not-determined |
| S17_extracted_k141_138222 | Not-determined |
| S17_extracted_k141_167822 | Not-determined |
| S17_extracted_k141_79325 | Not-determined |
| S17_extracted_k141_130553 | Not-determined |
| S17_extracted_k141_63661 | Not-determined |
| S17_extracted_k141_160619 | Not-determined |
| S17_extracted_k141_174665 | Not-determined |
| S17_extracted_k141_26775 | Not-determined |
| S17_extracted_k141_63664 | Not-determined |
| S17_extracted_k141_116387 | Not-determined |
| S17_extracted_k141_153606 | Not-determined |
| S17_extracted_k141_35433 | Not-determined |
| S17_extracted_k141_79329 | Not-determined |
| S17_extracted_k141_19736 | Low-quality |
| S17_extracted_k141_174667 | Low-quality |
| S17_extracted_k141_167830 | Low-quality |
| S17_extracted_k141_116393 | Not-determined |
| S17_extracted_k141_130566 | Not-determined |
| S17_extracted_k141_101594 | Not-determined |
| S17_extracted_k141_85895 | Not-determined |
| S17_extracted_k141_101595 | Not-determined |
| S17_extracted_k141_26778 | Not-determined |
| S17_extracted_k141_101596 | Not-determined |
| S17_extracted_k141_79332 | Not-determined |
| S17_extracted_k141_145709 | Not-determined |
| S17_extracted_k141_174673 | Not-determined |
| S17_extracted_k141_79336 | Not-determined |
| S17_extracted_k141_145711 | Not-determined |
| S17_extracted_k141_130571 | Not-determined |
| S17_extracted_k141_12532 | Low-quality |
| S17_extracted_k141_174678 | Low-quality |
| S17_extracted_k141_174679 | Low-quality |
| S17_extracted_k141_79341 | Not-determined |
| S17_extracted_k141_26782 | Not-determined |
| S17_extracted_k141_79342 | Not-determined |
| S17_extracted_k141_174682 | Not-determined |
| S17_extracted_k141_160627 | Not-determined |
| S17_extracted_k141_79344 | Not-determined |
| S17_extracted_k141_109468 | Not-determined |
| S17_extracted_k141_153614 | Not-determined |
| S17_extracted_k141_174684 | Not-determined |
| S17_extracted_k141_116408 | Not-determined |
| S17_extracted_k141_12540 | Not-determined |
| S17_extracted_k141_63670 | Not-determined |
| S17_extracted_k141_12542 | Not-determined |
| S17_extracted_k141_116410 | Low-quality |
| S17_extracted_k141_35445 | Not-determined |
| S17_extracted_k141_50088 | Not-determined |
| S17_extracted_k141_116413 | Not-determined |
| S17_extracted_k141_85913 | Not-determined |
| S17_extracted_k141_160634 | Low-quality |
| S17_extracted_k141_50093 | Low-quality |
| S17_extracted_k141_123159 | Not-determined |
| S17_extracted_k141_145724 | Not-determined |
| S17_extracted_k141_174688 | Low-quality |
| S17_extracted_k141_5379 | Not-determined |
| S17_extracted_k141_145729 | Low-quality |
| S17_extracted_k141_93743 | Not-determined |
| S17_extracted_k141_174692 | Not-determined |
| S17_extracted_k141_153624 | Not-determined |
| S17_extracted_k141_12550 | Not-determined |
| S17_extracted_k141_109488 | Not-determined |
| S17_extracted_k141_57437 | Low-quality |
| S17_extracted_k141_26787 | Not-determined |
| S17_extracted_k141_101620 | Not-determined |
| S17_extracted_k141_35460 | Not-determined |
| S17_extracted_k141_167859 | Not-determined |
| S17_extracted_k141_57440 | Not-determined |
| S17_extracted_k141_35463 | Not-determined |
| S17_extracted_k141_109494 | Low-quality |
| S17_extracted_k141_145741 | Not-determined |
| S17_extracted_k141_109495 | Not-determined |
| S17_extracted_k141_116426 | Low-quality |
| S17_extracted_k141_174696 | Not-determined |
| S17_extracted_k141_79353 | Not-determined |
| S17_extracted_k141_153634 | Not-determined |
| S17_extracted_k141_138235 | Not-determined |
| S17_extracted_k141_109503 | Not-determined |
| S17_extracted_k141_93758 | Low-quality |
| S17_extracted_k141_93760 | Not-determined |
| S17_extracted_k141_35468 | Not-determined |
| S17_extracted_k141_153637 | Not-determined |
| S17_extracted_k141_26800 | Not-determined |
| S17_extracted_k141_167867 | Not-determined |
| S17_extracted_k141_101626 | Not-determined |
| S17_extracted_k141_63695 | Not-determined |
| S17_extracted_k141_63697 | Not-determined |
| S17_extracted_k141_35473 | Not-determined |
| S17_extracted_k141_167869 | Not-determined |
| S17_extracted_k141_50110 | Not-determined |
| S17_extracted_k141_160648 | Not-determined |
| S17_extracted_k141_50112 | Not-determined |
| S17_extracted_k141_79363 | Not-determined |
| S17_extracted_k141_42906 | Not-determined |
| S17_extracted_k141_116441 | Not-determined |
| S17_extracted_k141_101636 | Not-determined |
| S17_extracted_k141_12557 | Not-determined |
| S17_extracted_k141_5398 | Not-determined |
| S17_extracted_k141_63704 | Low-quality |
| S17_extracted_k141_50122 | Low-quality |
| S17_extracted_k141_12559 | Not-determined |
| S17_extracted_k141_116445 | Not-determined |
| S17_extracted_k141_109512 | Low-quality |
| S17_extracted_k141_50125 | Not-determined |
| S17_extracted_k141_153653 | Not-determined |
| S17_extracted_k141_42916 | Not-determined |
| S17_extracted_k141_79368 | Not-determined |
| S17_extracted_k141_174703 | Not-determined |
| S17_extracted_k141_57459 | Not-determined |
| S17_extracted_k141_153657 | Not-determined |
| S17_extracted_k141_160657 | Low-quality |
| S17_extracted_k141_93772 | Not-determined |
| S17_extracted_k141_79372 | Not-determined |
| S17_extracted_k141_93775 | Not-determined |
| S17_extracted_k141_101648 | Not-determined |
| S17_extracted_k141_5407 | Not-determined |
| S17_extracted_k141_79374 | Not-determined |
| S17_extracted_k141_5408 | Not-determined |
| S17_extracted_k141_109519 | Not-determined |
| S17_extracted_k141_63706 | Not-determined |
| S17_extracted_k141_123173 | Not-determined |
| S17_extracted_k141_116449 | Not-determined |
| S17_extracted_k141_50136 | Low-quality |
| S17_extracted_k141_101649 | Not-determined |
| S17_extracted_k141_116452 | Not-determined |
| S17_extracted_k141_63712 | Low-quality |
| S17_extracted_k141_145759 | Not-determined |
| S17_extracted_k141_174712 | Not-determined |
| S17_extracted_k141_167883 | Not-determined |
| S17_extracted_k141_109530 | Not-determined |
| S17_extracted_k141_35476 | Low-quality |
| S17_extracted_k141_101654 | Low-quality |
| S17_extracted_k141_35477 | Low-quality |
| S17_extracted_k141_101655 | Not-determined |
| S17_extracted_k141_63717 | Not-determined |
| S17_extracted_k141_85934 | Not-determined |
| S17_extracted_k141_116461 | Not-determined |
| S17_extracted_k141_50141 | Not-determined |
| S17_extracted_k141_85939 | Not-determined |
| S17_extracted_k141_50142 | Not-determined |
| S17_extracted_k141_50143 | Not-determined |
| S17_extracted_k141_12591 | Not-determined |
| S17_extracted_k141_85940 | Not-determined |
| S17_extracted_k141_42934 | Not-determined |
| S17_extracted_k141_63729 | Not-determined |
| S17_extracted_k141_145773 | Not-determined |
| S17_extracted_k141_123190 | Not-determined |
| S17_extracted_k141_5418 | Not-determined |
| S17_extracted_k141_167897 | Not-determined |
| S17_extracted_k141_63732 | Low-quality |
| S17_extracted_k141_50147 | Not-determined |
| S17_extracted_k141_153681 | Low-quality |
| S17_extracted_k141_167901 | Not-determined |
| S17_extracted_k141_174730 | Not-determined |
| S17_extracted_k141_93781 | Not-determined |
| S17_extracted_k141_109542 | Not-determined |
| S17_extracted_k141_174734 | Not-determined |
| S17_extracted_k141_174738 | Not-determined |
| S17_extracted_k141_116475 | Not-determined |
| S17_extracted_k141_153693 | Not-determined |
| S17_extracted_k141_57487 | Low-quality |
| S17_extracted_k141_79386 | Not-determined |
| S17_extracted_k141_153697 | Not-determined |
| S17_extracted_k141_79389 | Low-quality |
| S17_extracted_k141_101669 | Not-determined |
| S17_extracted_k141_12612 | Low-quality |
| S17_extracted_k141_160697 | Not-determined |
| S17_extracted_k141_153701 | Not-determined |
| S17_extracted_k141_123210 | Not-determined |
| S17_extracted_k141_138246 | Not-determined |
| S17_extracted_k141_79392 | Low-quality |
| S17_extracted_k141_109562 | Not-determined |
| S17_extracted_k141_79394 | Not-determined |
| S17_extracted_k141_123211 | Not-determined |
| S17_extracted_k141_123212 | Not-determined |
| S17_extracted_k141_57501 | Not-determined |
| S17_extracted_k141_63755 | Not-determined |
| S17_extracted_k141_160707 | Not-determined |
| S17_extracted_k141_123217 | Not-determined |
| S17_extracted_k141_79400 | Not-determined |
| S17_extracted_k141_50167 | Not-determined |
| S17_extracted_k141_35497 | Not-determined |
| S17_extracted_k141_50168 | Not-determined |
| S17_extracted_k141_5427 | Low-quality |
| S17_extracted_k141_35498 | Not-determined |
| S17_extracted_k141_93801 | Not-determined |
| S17_extracted_k141_145807 | Not-determined |
| S17_extracted_k141_57508 | Not-determined |
| S17_extracted_k141_85960 | Not-determined |
| S17_extracted_k141_153715 | Not-determined |
| S17_extracted_k141_138252 | Not-determined |
| S17_extracted_k141_57510 | Not-determined |
| S17_extracted_k141_123225 | Low-quality |
| S17_extracted_k141_42954 | Not-determined |
| S17_extracted_k141_160715 | Not-determined |
| S17_extracted_k141_101685 | Not-determined |
| S17_extracted_k141_93807 | Not-determined |
| S17_extracted_k141_160720 | Not-determined |
| S17_extracted_k141_79403 | Low-quality |
| S17_extracted_k141_42957 | Low-quality |
| S17_extracted_k141_167919 | Not-determined |
| S17_extracted_k141_145812 | Not-determined |
| S17_extracted_k141_101692 | Low-quality |
| S17_extracted_k141_72097 | Low-quality |
| S17_extracted_k141_93817 | Not-determined |
| S17_extracted_k141_101695 | Not-determined |
| S17_extracted_k141_160725 | Low-quality |
| S17_extracted_k141_145818 | Not-determined |
| S17_extracted_k141_50183 | Not-determined |
| S17_extracted_k141_138270 | Not-determined |
| S17_extracted_k141_109581 | Not-determined |
| S17_extracted_k141_26817 | Not-determined |
| S17_extracted_k141_42974 | Not-determined |
| S17_extracted_k141_145827 | Not-determined |
| S17_extracted_k141_123236 | Not-determined |
| S17_extracted_k141_153730 | Not-determined |
| S17_extracted_k141_160733 | Not-determined |
| S17_extracted_k141_138274 | Not-determined |
| S17_extracted_k141_79408 | Not-determined |
| S17_extracted_k141_42978 | Not-determined |
| S17_extracted_k141_145829 | Low-quality |
| S17_extracted_k141_5452 | Not-determined |
| S17_extracted_k141_109588 | Not-determined |
| S17_extracted_k141_85963 | Low-quality |
| S17_extracted_k141_123237 | Not-determined |
| S17_extracted_k141_42983 | Not-determined |
| S17_extracted_k141_63774 | Not-determined |
| S17_extracted_k141_35511 | Not-determined |
| S17_extracted_k141_145831 | Not-determined |
| S17_extracted_k141_12642 | Not-determined |
| S17_extracted_k141_63776 | Not-determined |
| S17_extracted_k141_123242 | Not-determined |
| S17_extracted_k141_109596 | Not-determined |
| S17_extracted_k141_85970 | Not-determined |
| S17_extracted_k141_57530 | Low-quality |
| S17_extracted_k141_79413 | Not-determined |
| S17_extracted_k141_109597 | Not-determined |
| S17_extracted_k141_85972 | Not-determined |
| S17_extracted_k141_145834 | Not-determined |
| S17_extracted_k141_116491 | Not-determined |
| S17_extracted_k141_153744 | Not-determined |
| S17_extracted_k141_5458 | Not-determined |
| S17_extracted_k141_101713 | Not-determined |
| S17_extracted_k141_174755 | Not-determined |
| S17_extracted_k141_85977 | Not-determined |
| S17_extracted_k141_85980 | Low-quality |
| S17_extracted_k141_160747 | Low-quality |
| S17_extracted_k141_63784 | Not-determined |
| S17_extracted_k141_93837 | Not-determined |
| S17_extracted_k141_101716 | Not-determined |
| S17_extracted_k141_123247 | Not-determined |
| S17_extracted_k141_116498 | Not-determined |
| S17_extracted_k141_5463 | Not-determined |
| S17_extracted_k141_63788 | Not-determined |
| S17_extracted_k141_93838 | Not-determined |
| S17_extracted_k141_116500 | Low-quality |
| S17_extracted_k141_79420 | Not-determined |
| S17_extracted_k141_116501 | Not-determined |
| S17_extracted_k141_35523 | Not-determined |
| S17_extracted_k141_145844 | Not-determined |
| S17_extracted_k141_79425 | Not-determined |
| S17_extracted_k141_101722 | Not-determined |
| S17_extracted_k141_123255 | Not-determined |
| S17_extracted_k141_123257 | Not-determined |
| S17_extracted_k141_145849 | Not-determined |
| S17_extracted_k141_123258 | Not-determined |
| S17_extracted_k141_63800 | Not-determined |
| S17_extracted_k141_167946 | Not-determined |
| S17_extracted_k141_93848 | Not-determined |
| S17_extracted_k141_145855 | Not-determined |
| S17_extracted_k141_50200 | Not-determined |
| S17_extracted_k141_123267 | Not-determined |
| S17_extracted_k141_5480 | Not-determined |
| S17_extracted_k141_42999 | Not-determined |
| S17_extracted_k141_101737 | Not-determined |
| S17_extracted_k141_109619 | Not-determined |
| S17_extracted_k141_123268 | Low-quality |
| S17_extracted_k141_26831 | Not-determined |
| S17_extracted_k141_12668 | Not-determined |
| S17_extracted_k141_174776 | Not-determined |
| S17_extracted_k141_5486 | Not-determined |
| S17_extracted_k141_12671 | Low-quality |
| S17_extracted_k141_5488 | Not-determined |
| S17_extracted_k141_12672 | Low-quality |
| S17_extracted_k141_86000 | Not-determined |
| S17_extracted_k141_12675 | Low-quality |
| S17_extracted_k141_26838 | Low-quality |
| S17_extracted_k141_116508 | Not-determined |
| S17_extracted_k141_86003 | Not-determined |
| S17_extracted_k141_101743 | Low-quality |
| S17_extracted_k141_5494 | Not-determined |
| S17_extracted_k141_101744 | Not-determined |
| S17_extracted_k141_116509 | Not-determined |
| S17_extracted_k141_123275 | Not-determined |
| S17_extracted_k141_145867 | Not-determined |
| S17_extracted_k141_130584 | Low-quality |
| S17_extracted_k141_101747 | Low-quality |
| S17_extracted_k141_174787 | Not-determined |
| S17_extracted_k141_145872 | Low-quality |
| S17_extracted_k141_167953 | Not-determined |
| S17_extracted_k141_116516 | Not-determined |
| S17_extracted_k141_174792 | Not-determined |
| S17_extracted_k141_123279 | Low-quality |
| S17_extracted_k141_5505 | Not-determined |
| S17_extracted_k141_174794 | Not-determined |
| S17_extracted_k141_26849 | Not-determined |
| S17_extracted_k141_26852 | Low-quality |
| S17_extracted_k141_109635 | Not-determined |
| S17_extracted_k141_35565 | Not-determined |
| S17_extracted_k141_167961 | Not-determined |
| S17_extracted_k141_93869 | Not-determined |
| S17_extracted_k141_35567 | Not-determined |
| S17_extracted_k141_160752 | Not-determined |
| S17_extracted_k141_160753 | Not-determined |
| S17_extracted_k141_93871 | Not-determined |
| S17_extracted_k141_116528 | Not-determined |
| S17_extracted_k141_35569 | Not-determined |
| S17_extracted_k141_167967 | Not-determined |
| S17_extracted_k141_93876 | Not-determined |
| S17_extracted_k141_35571 | Low-quality |
| S17_extracted_k141_79461 | Not-determined |
| S17_extracted_k141_86029 | Not-determined |
| S17_extracted_k141_160756 | Not-determined |
| S17_extracted_k141_50233 | Low-quality |
| S17_extracted_k141_86030 | Not-determined |
| S17_extracted_k141_109651 | Not-determined |
| S17_extracted_k141_167976 | Not-determined |
| S17_extracted_k141_138301 | Not-determined |
| S17_extracted_k141_167977 | Not-determined |
| S17_extracted_k141_86032 | Not-determined |
| S17_extracted_k141_167980 | Not-determined |
| S17_extracted_k141_79470 | Not-determined |
| S17_extracted_k141_101772 | Not-determined |
| S17_extracted_k141_50241 | Not-determined |
| S17_extracted_k141_50242 | Not-determined |
| S17_extracted_k141_145892 | Not-determined |
| S17_extracted_k141_116547 | Not-determined |
| S17_extracted_k141_93884 | Low-quality |
| S17_extracted_k141_93885 | Low-quality |
| S17_extracted_k141_116549 | Not-determined |
| S17_extracted_k141_72160 | Not-determined |
| S17_extracted_k141_123295 | Not-determined |
| S17_extracted_k141_50247 | Not-determined |
| S17_extracted_k141_109663 | Low-quality |
| S17_extracted_k141_123297 | Not-determined |
| S17_extracted_k141_167989 | Not-determined |
| S17_extracted_k141_174813 | Not-determined |
| S17_extracted_k141_5536 | Not-determined |
| S17_extracted_k141_101782 | Not-determined |
| S17_extracted_k141_109666 | Not-determined |
| S17_extracted_k141_72164 | Not-determined |
| S17_extracted_k141_50253 | Not-determined |
| S17_extracted_k141_50254 | Not-determined |
| S17_extracted_k141_86037 | Not-determined |
| S17_extracted_k141_72167 | Not-determined |
| S17_extracted_k141_79484 | Not-determined |
| S17_extracted_k141_86040 | Not-determined |
| S17_extracted_k141_153803 | Low-quality |
| S17_extracted_k141_109673 | Not-determined |
| S17_extracted_k141_86041 | Low-quality |
| S17_extracted_k141_160783 | Not-determined |
| S17_extracted_k141_79490 | Not-determined |
| S17_extracted_k141_174828 | Not-determined |
| S17_extracted_k141_5544 | Not-determined |
| S17_extracted_k141_101787 | Not-determined |
| S17_extracted_k141_123306 | Not-determined |
| S17_extracted_k141_86047 | Not-determined |
| S17_extracted_k141_168003 | Low-quality |
| S17_extracted_k141_86049 | Low-quality |
| S17_extracted_k141_72176 | Not-determined |
| S17_extracted_k141_153813 | Not-determined |
| S17_extracted_k141_116566 | Not-determined |
| S17_extracted_k141_160793 | Not-determined |
| S17_extracted_k141_116571 | Not-determined |
| S17_extracted_k141_79501 | Not-determined |
| S17_extracted_k141_116577 | Not-determined |
| S17_extracted_k141_123311 | Not-determined |
| S17_extracted_k141_72181 | Not-determined |
| S17_extracted_k141_72183 | Not-determined |
| S17_extracted_k141_63824 | Not-determined |
| S17_extracted_k141_123313 | Not-determined |
| S17_extracted_k141_93903 | Not-determined |
| S17_extracted_k141_101805 | Not-determined |
| S17_extracted_k141_101806 | Low-quality |
| S17_extracted_k141_43010 | Not-determined |
| S17_extracted_k141_101808 | Not-determined |
| S17_extracted_k141_43011 | Not-determined |
| S17_extracted_k141_79511 | Not-determined |
| S17_extracted_k141_35597 | Not-determined |
| S17_extracted_k141_160805 | Not-determined |
| S17_extracted_k141_93908 | Not-determined |
| S17_extracted_k141_138322 | Not-determined |
| S17_extracted_k141_109685 | Low-quality |
| S17_extracted_k141_153819 | Not-determined |
| S17_extracted_k141_160807 | Not-determined |
| S17_extracted_k141_72188 | Not-determined |
| S17_extracted_k141_93910 | Not-determined |
| S17_extracted_k141_101815 | Not-determined |
| S17_extracted_k141_168014 | Low-quality |
| S17_extracted_k141_123323 | Not-determined |
| S17_extracted_k141_116583 | Not-determined |
| S17_extracted_k141_26870 | Not-determined |
| S17_extracted_k141_50272 | Not-determined |
| S17_extracted_k141_101822 | Not-determined |
| S17_extracted_k141_5550 | Not-determined |
| S17_extracted_k141_35610 | Low-quality |
| S17_extracted_k141_50275 | Not-determined |
| S17_extracted_k141_43027 | Not-determined |
| S17_extracted_k141_50276 | Not-determined |
| S17_extracted_k141_160818 | Not-determined |
| S17_extracted_k141_123328 | Not-determined |
| S17_extracted_k141_26876 | Not-determined |
| S17_extracted_k141_123330 | Not-determined |
| S17_extracted_k141_160824 | Not-determined |
| S17_extracted_k141_101824 | Not-determined |
| S17_extracted_k141_35621 | Not-determined |
| S17_extracted_k141_26883 | Not-determined |
| S17_extracted_k141_93922 | Not-determined |
| S17_extracted_k141_101826 | Not-determined |
| S17_extracted_k141_35626 | Not-determined |
| S17_extracted_k141_35627 | Not-determined |
| S17_extracted_k141_86060 | Not-determined |
| S17_extracted_k141_57544 | Not-determined |
| S17_extracted_k141_101829 | Not-determined |
| S17_extracted_k141_168020 | Not-determined |
| S17_extracted_k141_35629 | Not-determined |
| S17_extracted_k141_116601 | Not-determined |
| S17_extracted_k141_35639 | Not-determined |
| S17_extracted_k141_145924 | Not-determined |
| S17_extracted_k141_35640 | Not-determined |
| S17_extracted_k141_19769 | Not-determined |
| S17_extracted_k141_63845 | Not-determined |
| S17_extracted_k141_153835 | Not-determined |
| S17_extracted_k141_72209 | Low-quality |
| S17_extracted_k141_138347 | Not-determined |
| S17_extracted_k141_43039 | Not-determined |
| S17_extracted_k141_26888 | Low-quality |
| S17_extracted_k141_93932 | Not-determined |
| S17_extracted_k141_116613 | Not-determined |
| S17_extracted_k141_93935 | Low-quality |
| S17_extracted_k141_174836 | Not-determined |
| S17_extracted_k141_101848 | Not-determined |
| S17_extracted_k141_5576 | Not-determined |
| S17_extracted_k141_26892 | Not-determined |
| S17_extracted_k141_101850 | Not-determined |
| S17_extracted_k141_5577 | Not-determined |
| S17_extracted_k141_63848 | Not-determined |
| S17_extracted_k141_35655 | Not-determined |
| S17_extracted_k141_123340 | Not-determined |
| S17_extracted_k141_5580 | Not-determined |
| S17_extracted_k141_168029 | Not-determined |
| S17_extracted_k141_35656 | Not-determined |
| S17_extracted_k141_116619 | Not-determined |
| S17_extracted_k141_43055 | Not-determined |
| S17_extracted_k141_63850 | Not-determined |
| S17_extracted_k141_109722 | Not-determined |
| S17_extracted_k141_63853 | Not-determined |
| S17_extracted_k141_130604 | Not-determined |
| S17_extracted_k141_35663 | Not-determined |
| S17_extracted_k141_138362 | Low-quality |
| S17_extracted_k141_174844 | Low-quality |
| S17_extracted_k141_79518 | Not-determined |
| S17_extracted_k141_153855 | Not-determined |
| S17_extracted_k141_109727 | Not-determined |
| S17_extracted_k141_63857 | Not-determined |
| S17_extracted_k141_160862 | Low-quality |
| S17_extracted_k141_174847 | Not-determined |
| S17_extracted_k141_72230 | Low-quality |
| S17_extracted_k141_174850 | Not-determined |
| S17_extracted_k141_101861 | Not-determined |
| S17_extracted_k141_160865 | Not-determined |
| S17_extracted_k141_35677 | Not-determined |
| S17_extracted_k141_130620 | Not-determined |
| S17_extracted_k141_174852 | Not-determined |
| S17_extracted_k141_101866 | Low-quality |
| S17_extracted_k141_35681 | Low-quality |
| S17_extracted_k141_138375 | Not-determined |
| S17_extracted_k141_26914 | Not-determined |
| S17_extracted_k141_43066 | Not-determined |
| S17_extracted_k141_43069 | Not-determined |
| S17_extracted_k141_116637 | Not-determined |
| S17_extracted_k141_79524 | Not-determined |
| S17_extracted_k141_50305 | Not-determined |
| S17_extracted_k141_43071 | Not-determined |
| S17_extracted_k141_174857 | Not-determined |
| S17_extracted_k141_79527 | Not-determined |
| S17_extracted_k141_174859 | Not-determined |
| S17_extracted_k141_101876 | Not-determined |
| S17_extracted_k141_138377 | Not-determined |
| S17_extracted_k141_153861 | Not-determined |
| S17_extracted_k141_101877 | Not-determined |
| S17_extracted_k141_72242 | Low-quality |
| S17_extracted_k141_79535 | Not-determined |
| S17_extracted_k141_138383 | Not-determined |
| S17_extracted_k141_63870 | Not-determined |
| S17_extracted_k141_160882 | Not-determined |
| S17_extracted_k141_57548 | Not-determined |
| S17_extracted_k141_109745 | Not-determined |
| S17_extracted_k141_123360 | Low-quality |
| S17_extracted_k141_153867 | Low-quality |
| S17_extracted_k141_19771 | Not-determined |
| S17_extracted_k141_19772 | Not-determined |
| S17_extracted_k141_63871 | Not-determined |
| S17_extracted_k141_109748 | Not-determined |
| S17_extracted_k141_57551 | Not-determined |
| S17_extracted_k141_168058 | Not-determined |
| S17_extracted_k141_50315 | Not-determined |
| S17_extracted_k141_109750 | Not-determined |
| S17_extracted_k141_19774 | Not-determined |
| S17_extracted_k141_168065 | Not-determined |
| S17_extracted_k141_79542 | Low-quality |
| S17_extracted_k141_19783 | Not-determined |
| S17_extracted_k141_79545 | Not-determined |
| S17_extracted_k141_86068 | Not-determined |

| | |
|---|---|
| S17_extracted_k141_72249 | Not-determined |
| S17_extracted_k141_130641 | Not-determined |
| S17_extracted_k141_138396 | Not-determined |
| S17_extracted_k141_86071 | Not-determined |
| S17_extracted_k141_160895 | Not-determined |
| S17_extracted_k141_86072 | Low-quality |
| S17_extracted_k141_26932 | Not-determined |
| S17_extracted_k141_130642 | Not-determined |
| S17_extracted_k141_86073 | Not-determined |
| S17_extracted_k141_43086 | Not-determined |
| S17_extracted_k141_109758 | Not-determined |
| S17_extracted_k141_63886 | Not-determined |
| S17_extracted_k141_43087 | Not-determined |
| S17_extracted_k141_19789 | Not-determined |
| S17_extracted_k141_109759 | Not-determined |
| S17_extracted_k141_19790 | Not-determined |
| S17_extracted_k141_35706 | Not-determined |
| S17_extracted_k141_101891 | Not-determined |
| S17_extracted_k141_50329 | Not-determined |
| S17_extracted_k141_35713 | Not-determined |
| S17_extracted_k141_35714 | Low-quality |
| S17_extracted_k141_57565 | Not-determined |
| S17_extracted_k141_160914 | Not-determined |
| S17_extracted_k141_174870 | Not-determined |
| S17_extracted_k141_160918 | Low-quality |
| S17_extracted_k141_26948 | Not-determined |
| S17_extracted_k141_123385 | Not-determined |
| S17_extracted_k141_86094 | Not-determined |
| S17_extracted_k141_19791 | Low-quality |
| S17_extracted_k141_138412 | Not-determined |
| S17_extracted_k141_174871 | Not-determined |
| S17_extracted_k141_50334 | Not-determined |
| S17_extracted_k141_72276 | Not-determined |
| S17_extracted_k141_26950 | Not-determined |
| S17_extracted_k141_160922 | Not-determined |
| S17_extracted_k141_153887 | Not-determined |
| S17_extracted_k141_43105 | Not-determined |
| S17_extracted_k141_101895 | Not-determined |
| S17_extracted_k141_79551 | Not-determined |
| S17_extracted_k141_72282 | Not-determined |
| S17_extracted_k141_101897 | Not-determined |
| S17_extracted_k141_26952 | Not-determined |
| S17_extracted_k141_86100 | Not-determined |
| S17_extracted_k141_123393 | Not-determined |
| S17_extracted_k141_5615 | Not-determined |
| S17_extracted_k141_130671 | Not-determined |
| S17_extracted_k141_168097 | Not-determined |
| S17_extracted_k141_50338 | Not-determined |
| S17_extracted_k141_109775 | Not-determined |
| S17_extracted_k141_50339 | Low-quality |
| S17_extracted_k141_116647 | Not-determined |
| S17_extracted_k141_19800 | Not-determined |
| S17_extracted_k141_86106 | Not-determined |
| S17_extracted_k141_116650 | Not-determined |
| S17_extracted_k141_138417 | Not-determined |
| S17_extracted_k141_72291 | Not-determined |
| S17_extracted_k141_174876 | Low-quality |
| S17_extracted_k141_116654 | Low-quality |
| S17_extracted_k141_43116 | Not-determined |
| S17_extracted_k141_101899 | Not-determined |
| S17_extracted_k141_50346 | Not-determined |
| S17_extracted_k141_19807 | Not-determined |
| S17_extracted_k141_109783 | Not-determined |
| S17_extracted_k141_168108 | Not-determined |
| S17_extracted_k141_153899 | Not-determined |
| S17_extracted_k141_50348 | Not-determined |
| S17_extracted_k141_43123 | Not-determined |
| S17_extracted_k141_116659 | Not-determined |
| S17_extracted_k141_153901 | Not-determined |
| S17_extracted_k141_116660 | Not-determined |
| S17_extracted_k141_174881 | Not-determined |
| S17_extracted_k141_109789 | Not-determined |
| S17_extracted_k141_174885 | Not-determined |
| S17_extracted_k141_109791 | Not-determined |
| S17_extracted_k141_174886 | Not-determined |
| S17_extracted_k141_86124 | Not-determined |
| S17_extracted_k141_160937 | Not-determined |
| S17_extracted_k141_19821 | Low-quality |
| S17_extracted_k141_109795 | Not-determined |
| S17_extracted_k141_86128 | Low-quality |
| S17_extracted_k141_72310 | Not-determined |
| S17_extracted_k141_160943 | Not-determined |
| S17_extracted_k141_116668 | Not-determined |
| S17_extracted_k141_145949 | Not-determined |
| S17_extracted_k141_116670 | Not-determined |
| S17_extracted_k141_50355 | Low-quality |
| S17_extracted_k141_19828 | Low-quality |
| S17_extracted_k141_145951 | Not-determined |
| S17_extracted_k141_12685 | Not-determined |
| S17_extracted_k141_101920 | Not-determined |
| S17_extracted_k141_43136 | Not-determined |
| S17_extracted_k141_86131 | Not-determined |
| S17_extracted_k141_153908 | Low-quality |
| S17_extracted_k141_86132 | Not-determined |
| S17_extracted_k141_145956 | Not-determined |
| S17_extracted_k141_12689 | Not-determined |
| S17_extracted_k141_123412 | Not-determined |
| S17_extracted_k141_153911 | Not-determined |
| S17_extracted_k141_79578 | Low-quality |
| S17_extracted_k141_168112 | Not-determined |
| S17_extracted_k141_168114 | Not-determined |
| S17_extracted_k141_79580 | Not-determined |
| S17_extracted_k141_168115 | Not-determined |
| S17_extracted_k141_43145 | Not-determined |
| S17_extracted_k141_72327 | Not-determined |
| S17_extracted_k141_145965 | Not-determined |
| S17_extracted_k141_145966 | Not-determined |
| S17_extracted_k141_145968 | Not-determined |
| S17_extracted_k141_79586 | Not-determined |
| S17_extracted_k141_145971 | Not-determined |
| S17_extracted_k141_86147 | Not-determined |
| S17_extracted_k141_145972 | Not-determined |
| S17_extracted_k141_35724 | Not-determined |
| S17_extracted_k141_63912 | Not-determined |
| S17_extracted_k141_35725 | Low-quality |
| S17_extracted_k141_116678 | Not-determined |
| S17_extracted_k141_109809 | Not-determined |
| S17_extracted_k141_86152 | Not-determined |
| S17_extracted_k141_86153 | Not-determined |
| S17_extracted_k141_153922 | Not-determined |
| S17_extracted_k141_116682 | Not-determined |
| S17_extracted_k141_35731 | Not-determined |
| S17_extracted_k141_116684 | Not-determined |
| S17_extracted_k141_86157 | Not-determined |
| S17_extracted_k141_145981 | Low-quality |
| S17_extracted_k141_43151 | Not-determined |
| S17_extracted_k141_174906 | Not-determined |
| S17_extracted_k141_123427 | Not-determined |
| S17_extracted_k141_5647 | Not-determined |
| S17_extracted_k141_101935 | Not-determined |
| S17_extracted_k141_79591 | Not-determined |
| S17_extracted_k141_123430 | Not-determined |
| S17_extracted_k141_79592 | Not-determined |
| S17_extracted_k141_5651 | Low-quality |
| S17_extracted_k141_27005 | Not-determined |
| S17_extracted_k141_109820 | Not-determined |
| S17_extracted_k141_5653 | Not-determined |
| S17_extracted_k141_86168 | Not-determined |
| S17_extracted_k141_63914 | Not-determined |
| S17_extracted_k141_50383 | Not-determined |
| S17_extracted_k141_43158 | Not-determined |
| S17_extracted_k141_27008 | Not-determined |
| S17_extracted_k141_174913 | Not-determined |
| S17_extracted_k141_86170 | Not-determined |
| S17_extracted_k141_153939 | Not-determined |
| S17_extracted_k141_130694 | Not-determined |
| S17_extracted_k141_57585 | Not-determined |
| S17_extracted_k141_12721 | Not-determined |
| S17_extracted_k141_130696 | Not-determined |
| S17_extracted_k141_153942 | Low-quality |
| S17_extracted_k141_160968 | Low-quality |
| S17_extracted_k141_86174 | Low-quality |
| S17_extracted_k141_174917 | Low-quality |
| S17_extracted_k141_168142 | Not-determined |
| S17_extracted_k141_72334 | Low-quality |
| S17_extracted_k141_86177 | Not-determined |
| S17_extracted_k141_43169 | Not-determined |
| S17_extracted_k141_123439 | Low-quality |
| S17_extracted_k141_109831 | Not-determined |
| S17_extracted_k141_72339 | Not-determined |
| S17_extracted_k141_72343 | Not-determined |
| S17_extracted_k141_168147 | Not-determined |
| S17_extracted_k141_19858 | Low-quality |
| S17_extracted_k141_35761 | Not-determined |
| S17_extracted_k141_86183 | Not-determined |
| S17_extracted_k141_116707 | Not-determined |
| S17_extracted_k141_168151 | Not-determined |
| S17_extracted_k141_146010 | Not-determined |
| S17_extracted_k141_109844 | Not-determined |
| S17_extracted_k141_116714 | Not-determined |
| S17_extracted_k141_27029 | Not-determined |
| S17_extracted_k141_116718 | Not-determined |
| S17_extracted_k141_27032 | Not-determined |
| S17_extracted_k141_27033 | Not-determined |
| S17_extracted_k141_146015 | Not-determined |
| S17_extracted_k141_116721 | Not-determined |
| S17_extracted_k141_63945 | Not-determined |
| S17_extracted_k141_101949 | Low-quality |
| S17_extracted_k141_146020 | Not-determined |
| S17_extracted_k141_57613 | Not-determined |
| S17_extracted_k141_86196 | Not-determined |
| S17_extracted_k141_57616 | Not-determined |
| S17_extracted_k141_160993 | Not-determined |
| S17_extracted_k141_12742 | Not-determined |
| S17_extracted_k141_146029 | Not-determined |
| S17_extracted_k141_86199 | Not-determined |
| S17_extracted_k141_116737 | Not-determined |
| S17_extracted_k141_79620 | Not-determined |
| S17_extracted_k141_101954 | Low-quality |
| S17_extracted_k141_63959 | Not-determined |
| S17_extracted_k141_79624 | Not-determined |
| S17_extracted_k141_12752 | Not-determined |
| S17_extracted_k141_123452 | Low-quality |
| S17_extracted_k141_109867 | Not-determined |
| S17_extracted_k141_109868 | Not-determined |
| S17_extracted_k141_123455 | Low-quality |
| S17_extracted_k141_130713 | Not-determined |
| S17_extracted_k141_63961 | Not-determined |
| S17_extracted_k141_101956 | Not-determined |
| S17_extracted_k141_35784 | Not-determined |
| S17_extracted_k141_161003 | Not-determined |
| S17_extracted_k141_101959 | Low-quality |
| S17_extracted_k141_130716 | Not-determined |
| S17_extracted_k141_174932 | Not-determined |
| S17_extracted_k141_161004 | Not-determined |
| S17_extracted_k141_109870 | Not-determined |
| S17_extracted_k141_146031 | Not-determined |
| S17_extracted_k141_63964 | Not-determined |
| S17_extracted_k141_63965 | Not-determined |
| S17_extracted_k141_57627 | Not-determined |
| S17_extracted_k141_109875 | Not-determined |
| S17_extracted_k141_101965 | Not-determined |
| S17_extracted_k141_63968 | Not-determined |
| S17_extracted_k141_27054 | Not-determined |
| S17_extracted_k141_130723 | Not-determined |
| S17_extracted_k141_168175 | Not-determined |
| S17_extracted_k141_123465 | Not-determined |
| S17_extracted_k141_19890 | Low-quality |
| S17_extracted_k141_50413 | Not-determined |
| S17_extracted_k141_130724 | Not-determined |
| S17_extracted_k141_168177 | Not-determined |
| S17_extracted_k141_19892 | Not-determined |
| S17_extracted_k141_116745 | Low-quality |
| S17_extracted_k141_57632 | Not-determined |
| S17_extracted_k141_161019 | Not-determined |
| S17_extracted_k141_19895 | Not-determined |
| S17_extracted_k141_109882 | Low-quality |
| S17_extracted_k141_153972 | Low-quality |
| S17_extracted_k141_63982 | Not-determined |
| S17_extracted_k141_116756 | Not-determined |
| S17_extracted_k141_5668 | Not-determined |
| S17_extracted_k141_43215 | Not-determined |
| S17_extracted_k141_12771 | Not-determined |
| S17_extracted_k141_161028 | Not-determined |
| S17_extracted_k141_130729 | Not-determined |
| S17_extracted_k141_109887 | Not-determined |
| S17_extracted_k141_27066 | Low-quality |
| S17_extracted_k141_123481 | Not-determined |
| S17_extracted_k141_57644 | Not-determined |
| S17_extracted_k141_123483 | Not-determined |
| S17_extracted_k141_174947 | Not-determined |
| S17_extracted_k141_101983 | Low-quality |
| S17_extracted_k141_79648 | Not-determined |
| S17_extracted_k141_86230 | Not-determined |
| S17_extracted_k141_50419 | Not-determined |
| S17_extracted_k141_5674 | Not-determined |
| S17_extracted_k141_86232 | Not-determined |
| S17_extracted_k141_72375 | Not-determined |
| S17_extracted_k141_93960 | Not-determined |
| S17_extracted_k141_161033 | Not-determined |
| S17_extracted_k141_35797 | Low-quality |
| S17_extracted_k141_79657 | Low-quality |
| S17_extracted_k141_161035 | Not-determined |
| S17_extracted_k141_123493 | Not-determined |
| S17_extracted_k141_146046 | Not-determined |
| S17_extracted_k141_27082 | Not-determined |
| S17_extracted_k141_130741 | Not-determined |
| S17_extracted_k141_19906 | Not-determined |
| S17_extracted_k141_161041 | Not-determined |
| S17_extracted_k141_43234 | Not-determined |
| S17_extracted_k141_35808 | Not-determined |
| S17_extracted_k141_153987 | Not-determined |
| S17_extracted_k141_50432 | Not-determined |
| S17_extracted_k141_161045 | Not-determined |
| S17_extracted_k141_109893 | Not-determined |
| S17_extracted_k141_123499 | Not-determined |
| S17_extracted_k141_19913 | Not-determined |
| S17_extracted_k141_35809 | Low-quality |
| S17_extracted_k141_50433 | Not-determined |
| S17_extracted_k141_146057 | Not-determined |
| S17_extracted_k141_19915 | Not-determined |
| S17_extracted_k141_35816 | Not-determined |
| S17_extracted_k141_153993 | Not-determined |
| S17_extracted_k141_57660 | Not-determined |
| S17_extracted_k141_50440 | Not-determined |
| S17_extracted_k141_79664 | Not-determined |
| S17_extracted_k141_146058 | Not-determined |
| S17_extracted_k141_161054 | Not-determined |
| S17_extracted_k141_146059 | Not-determined |
| S17_extracted_k141_161055 | Not-determined |
| S17_extracted_k141_57662 | Not-determined |
| S17_extracted_k141_109904 | Not-determined |
| S17_extracted_k141_138476 | Not-determined |
| S17_extracted_k141_50442 | Not-determined |
| S17_extracted_k141_5696 | Not-determined |
| S17_extracted_k141_146063 | Not-determined |
| S17_extracted_k141_5698 | Not-determined |
| S17_extracted_k141_161059 | Not-determined |
| S17_extracted_k141_57667 | Not-determined |
| S17_extracted_k141_12810 | Not-determined |
| S17_extracted_k141_43257 | Not-determined |
| S17_extracted_k141_168209 | Low-quality |
| S17_extracted_k141_5700 | Not-determined |
| S17_extracted_k141_35826 | Not-determined |
| S17_extracted_k141_35827 | Not-determined |
| S17_extracted_k141_146074 | Not-determined |
| S17_extracted_k141_79672 | Not-determined |
| S17_extracted_k141_174973 | Not-determined |
| S17_extracted_k141_174976 | Not-determined |
| S17_extracted_k141_109920 | Not-determined |
| S17_extracted_k141_168214 | Not-determined |
| S17_extracted_k141_5703 | Not-determined |
| S17_extracted_k141_174977 | Not-determined |
| S17_extracted_k141_168215 | Not-determined |
| S17_extracted_k141_93979 | Not-determined |
| S17_extracted_k141_123516 | Not-determined |
| S17_extracted_k141_168217 | Not-determined |
| S17_extracted_k141_168218 | Low-quality |
| S17_extracted_k141_130763 | Not-determined |
| S17_extracted_k141_93983 | Not-determined |
| S17_extracted_k141_43265 | Not-determined |
| S17_extracted_k141_130764 | Low-quality |
| S17_extracted_k141_154009 | Not-determined |
| S17_extracted_k141_27110 | Not-determined |
| S17_extracted_k141_138498 | Low-quality |
| S17_extracted_k141_109923 | Not-determined |
| S17_extracted_k141_5714 | Not-determined |
| S17_extracted_k141_168223 | Not-determined |
| S17_extracted_k141_27116 | Not-determined |
| S17_extracted_k141_93995 | Not-determined |
| S17_extracted_k141_12828 | Low-quality |
| S17_extracted_k141_93997 | Low-quality |
| S17_extracted_k141_43274 | Not-determined |
| S17_extracted_k141_146087 | Not-determined |
| S17_extracted_k141_35848 | Not-determined |
| S17_extracted_k141_116791 | Not-determined |
| S17_extracted_k141_5717 | Not-determined |
| S17_extracted_k141_79689 | Not-determined |
| S17_extracted_k141_116792 | Not-determined |
| S17_extracted_k141_79690 | Not-determined |
| S17_extracted_k141_27124 | Not-determined |
| S17_extracted_k141_50466 | Not-determined |
| S17_extracted_k141_50468 | Low-quality |
| S17_extracted_k141_130786 | Low-quality |
| S17_extracted_k141_138509 | Not-determined |
| S17_extracted_k141_168233 | Not-determined |
| S17_extracted_k141_146101 | Not-determined |
| S17_extracted_k141_109933 | Not-determined |
| S17_extracted_k141_130791 | Not-determined |
| S17_extracted_k141_72418 | Not-determined |
| S17_extracted_k141_109939 | Not-determined |
| S17_extracted_k141_109940 | Low-quality |
| S17_extracted_k141_5732 | Not-determined |
| S17_extracted_k141_130797 | Not-determined |
| S17_extracted_k141_161084 | Not-determined |
| S17_extracted_k141_12846 | Low-quality |
| S17_extracted_k141_123538 | Not-determined |
| S17_extracted_k141_12848 | Not-determined |
| S17_extracted_k141_27137 | Not-determined |
| S17_extracted_k141_86267 | Not-determined |
| S17_extracted_k141_146110 | Not-determined |
| S17_extracted_k141_50483 | Low-quality |
| S17_extracted_k141_130807 | Not-determined |
| S17_extracted_k141_86271 | Not-determined |
| S17_extracted_k141_102005 | Not-determined |
| S17_extracted_k141_27141 | Not-determined |
| S17_extracted_k141_35860 | Not-determined |
| S17_extracted_k141_116812 | Not-determined |
| S17_extracted_k141_64006 | Low-quality |
| S17_extracted_k141_5744 | Not-determined |
| S17_extracted_k141_64007 | Not-determined |
| S17_extracted_k141_146117 | Not-determined |
| S17_extracted_k141_102009 | Not-determined |
| S17_extracted_k141_19930 | Not-determined |
| S17_extracted_k141_64013 | Not-determined |
| S17_extracted_k141_109957 | Not-determined |
| S17_extracted_k141_154032 | Not-determined |
| S17_extracted_k141_94002 | Low-quality |
| S17_extracted_k141_102015 | Low-quality |
| S17_extracted_k141_123545 | Low-quality |
| S17_extracted_k141_50494 | Not-determined |
| S17_extracted_k141_72436 | Not-determined |
| S17_extracted_k141_161091 | Not-determined |
| S17_extracted_k141_138541 | Not-determined |
| S17_extracted_k141_123546 | Low-quality |
| S17_extracted_k141_19933 | Not-determined |
| S17_extracted_k141_161095 | Not-determined |
| S17_extracted_k141_146131 | Not-determined |
| S17_extracted_k141_123552 | Not-determined |
| S17_extracted_k141_35873 | Not-determined |
| S17_extracted_k141_19938 | Not-determined |
| S17_extracted_k141_123554 | Not-determined |
| S17_extracted_k141_72444 | Not-determined |
| S17_extracted_k141_72445 | Not-determined |
| S17_extracted_k141_50502 | Low-quality |
| S17_extracted_k141_116826 | Low-quality |
| S17_extracted_k141_102028 | Not-determined |
| S17_extracted_k141_94014 | Not-determined |
| S17_extracted_k141_154045 | Not-determined |
| S17_extracted_k141_146135 | Not-determined |
| S17_extracted_k141_123561 | Not-determined |
| S17_extracted_k141_138550 | Not-determined |
| S17_extracted_k141_79700 | Not-determined |
| S17_extracted_k141_116830 | Low-quality |
| S17_extracted_k141_146137 | Not-determined |
| S17_extracted_k141_35876 | Not-determined |
| S17_extracted_k141_146138 | Not-determined |
| S17_extracted_k141_138552 | Not-determined |
| S17_extracted_k141_146139 | Not-determined |
| S17_extracted_k141_123567 | Not-determined |
| S17_extracted_k141_5763 | Not-determined |
| S17_extracted_k141_86289 | Low-quality |
| S17_extracted_k141_109977 | Not-determined |
| S17_extracted_k141_19945 | Not-determined |
| S17_extracted_k141_116838 | Not-determined |
| S17_extracted_k141_72450 | Not-determined |
| S17_extracted_k141_12872 | Not-determined |
| S17_extracted_k141_109979 | Not-determined |
| S17_extracted_k141_130824 | Not-determined |
| S17_extracted_k141_109984 | Not-determined |
| S17_extracted_k141_161116 | Not-determined |
| S17_extracted_k141_19952 | Low-quality |
| S17_extracted_k141_64032 | Not-determined |
| S17_extracted_k141_102034 | Not-determined |
| S17_extracted_k141_146147 | Not-determined |
| S17_extracted_k141_146149 | Not-determined |
| S17_extracted_k141_154062 | Not-determined |
| S17_extracted_k141_146150 | Low-quality |
| S17_extracted_k141_64033 | Low-quality |
| S17_extracted_k141_64034 | Not-determined |
| S17_extracted_k141_35899 | Not-determined |
| S17_extracted_k141_72455 | Not-determined |
| S17_extracted_k141_116852 | Not-determined |
| S17_extracted_k141_86307 | Not-determined |
| S17_extracted_k141_43284 | Not-determined |
| S17_extracted_k141_5778 | Not-determined |
| S17_extracted_k141_86309 | Low-quality |
| S17_extracted_k141_138562 | Not-determined |
| S17_extracted_k141_138563 | Not-determined |
| S17_extracted_k141_5782 | Not-determined |
| S17_extracted_k141_123579 | Not-determined |
| S17_extracted_k141_12878 | Not-determined |
| S17_extracted_k141_50535 | Not-determined |
| S17_extracted_k141_161135 | Not-determined |
| S17_extracted_k141_146154 | Not-determined |
| S17_extracted_k141_116855 | Low-quality |
| S17_extracted_k141_146155 | Not-determined |
| S17_extracted_k141_138570 | Not-determined |
| S17_extracted_k141_138571 | Not-determined |
| S17_extracted_k141_19964 | Not-determined |
| S17_extracted_k141_94035 | Not-determined |
| S17_extracted_k141_5787 | Not-determined |
| S17_extracted_k141_161139 | Not-determined |
| S17_extracted_k141_94039 | Not-determined |
| S17_extracted_k141_64047 | Not-determined |
| S17_extracted_k141_123585 | Not-determined |
| S17_extracted_k141_161142 | Not-determined |
| S17_extracted_k141_43298 | Low-quality |
| S17_extracted_k141_5796 | Not-determined |
| S17_extracted_k141_79731 | Not-determined |
| S17_extracted_k141_116870 | Not-determined |
| S17_extracted_k141_123589 | Not-determined |
| S17_extracted_k141_27161 | Not-determined |
| S17_extracted_k141_64055 | Not-determined |
| S17_extracted_k141_79735 | Not-determined |
| S17_extracted_k141_27163 | Not-determined |
| S17_extracted_k141_50547 | Not-determined |
| S17_extracted_k141_79740 | Not-determined |
| S17_extracted_k141_138584 | Not-determined |
| S17_extracted_k141_72472 | Not-determined |
| S17_extracted_k141_146179 | Not-determined |
| S17_extracted_k141_94053 | Not-determined |
| S17_extracted_k141_146181 | Not-determined |
| S17_extracted_k141_72475 | Not-determined |
| S17_extracted_k141_79747 | Not-determined |
| S17_extracted_k141_79749 | Low-quality |
| S17_extracted_k141_72480 | Not-determined |
| S17_extracted_k141_79751 | Low-quality |
| S17_extracted_k141_79752 | Not-determined |
| S17_extracted_k141_154101 | Low-quality |
| S17_extracted_k141_94062 | Not-determined |
| S17_extracted_k141_64062 | Low-quality |
| S17_extracted_k141_146190 | Not-determined |
| S17_extracted_k141_154103 | Not-determined |
| S17_extracted_k141_5811 | Not-determined |
| S17_extracted_k141_154104 | Not-determined |
| S17_extracted_k141_27171 | Not-determined |
| S17_extracted_k141_72483 | Not-determined |
| S17_extracted_k141_79755 | Low-quality |
| S17_extracted_k141_79760 | Not-determined |
| S17_extracted_k141_154110 | Not-determined |
| S17_extracted_k141_146198 | Not-determined |
| S17_extracted_k141_27177 | Not-determined |
| S17_extracted_k141_138597 | Not-determined |
| S17_extracted_k141_146200 | Not-determined |
| S17_extracted_k141_110019 | Not-determined |
| S17_extracted_k141_5816 | Low-quality |
| S17_extracted_k141_123605 | Low-quality |
| S17_extracted_k141_138602 | Not-determined |
| S17_extracted_k141_94069 | Not-determined |
| S17_extracted_k141_102047 | Low-quality |
| S17_extracted_k141_19997 | Not-determined |
| S17_extracted_k141_5821 | Not-determined |
| S17_extracted_k141_50562 | Not-determined |
| S17_extracted_k141_102049 | Not-determined |
| S17_extracted_k141_79769 | Not-determined |
| S17_extracted_k141_64076 | Low-quality |
| S17_extracted_k141_27185 | Not-determined |
| S17_extracted_k141_12916 | Not-determined |
| S17_extracted_k141_161183 | Not-determined |
| S17_extracted_k141_50567 | Not-determined |
| S17_extracted_k141_5825 | Not-determined |
| S17_extracted_k141_5826 | Not-determined |
| S17_extracted_k141_94075 | Not-determined |
| S17_extracted_k141_102058 | Not-determined |
| S17_extracted_k141_138605 | Not-determined |
| S17_extracted_k141_154126 | Low-quality |
| S17_extracted_k141_110031 | Not-determined |
| S17_extracted_k141_50574 | Not-determined |
| S17_extracted_k141_50575 | Not-determined |
| S17_extracted_k141_20001 | Low-quality |
| S17_extracted_k141_102062 | Not-determined |
| S17_extracted_k141_64085 | Not-determined |
| S17_extracted_k141_35920 | Not-determined |
| S17_extracted_k141_64086 | Not-determined |
| S17_extracted_k141_138608 | Not-determined |
| S17_extracted_k141_27200 | Not-determined |
| S17_extracted_k141_123625 | Not-determined |
| S17_extracted_k141_79777 | Low-quality |
| S17_extracted_k141_123628 | Not-determined |
| S17_extracted_k141_79778 | Not-determined |
| S17_extracted_k141_57708 | Low-quality |
| S17_extracted_k141_20007 | Not-determined |
| S17_extracted_k141_12927 | Not-determined |
| S17_extracted_k141_5838 | Not-determined |
| S17_extracted_k141_79783 | Not-determined |
| S17_extracted_k141_123637 | Not-determined |
| S17_extracted_k141_57711 | Not-determined |
| S17_extracted_k141_110042 | Not-determined |
| S17_extracted_k141_138623 | Not-determined |
| S17_extracted_k141_50584 | Not-determined |
| S17_extracted_k141_57718 | Not-determined |
| S17_extracted_k141_79789 | Not-determined |
| S17_extracted_k141_27209 | Not-determined |
| S17_extracted_k141_94101 | Not-determined |
| S17_extracted_k141_161199 | Low-quality |
| S17_extracted_k141_79795 | Not-determined |
| S17_extracted_k141_50587 | Low-quality |
| S17_extracted_k141_64095 | Not-determined |
| S17_extracted_k141_130914 | Not-determined |
| S17_extracted_k141_27212 | Not-determined |
| S17_extracted_k141_154150 | Not-determined |
| S17_extracted_k141_110055 | Not-determined |
| S17_extracted_k141_20018 | Not-determined |
| S17_extracted_k141_161205 | Not-determined |
| S17_extracted_k141_110056 | Not-determined |
| S17_extracted_k141_57728 | Not-determined |
| S17_extracted_k141_110060 | Not-determined |
| S17_extracted_k141_138629 | Not-determined |
| S17_extracted_k141_110061 | Not-determined |
| S17_extracted_k141_79804 | Not-determined |
| S17_extracted_k141_161207 | Not-determined |
| S17_extracted_k141_138631 | Not-determined |
| S17_extracted_k141_12958 | Low-quality |
| S17_extracted_k141_5843 | Not-determined |
| S17_extracted_k141_79807 | Not-determined |
| S17_extracted_k141_110064 | Not-determined |
| S17_extracted_k141_12961 | Not-determined |
| S17_extracted_k141_110065 | Not-determined |
| S17_extracted_k141_174998 | Low-quality |
| S17_extracted_k141_123652 | Not-determined |
| S17_extracted_k141_72511 | Not-determined |
| S17_extracted_k141_130922 | Low-quality |
| S17_extracted_k141_130923 | Not-determined |
| S17_extracted_k141_20026 | Low-quality |
| S17_extracted_k141_130925 | Low-quality |
| S17_extracted_k141_110066 | Low-quality |
| S17_extracted_k141_123655 | Not-determined |
| S17_extracted_k141_146209 | Low-quality |
| S17_extracted_k141_130927 | Low-quality |
| S17_extracted_k141_175005 | Low-quality |
| S17_extracted_k141_79817 | Not-determined |
| S17_extracted_k141_27229 | Not-determined |
| S17_extracted_k141_5859 | Low-quality |
| S17_extracted_k141_72517 | Not-determined |
| S17_extracted_k141_94117 | Not-determined |
| S17_extracted_k141_5860 | Not-determined |
| S17_extracted_k141_64103 | Not-determined |
| S17_extracted_k141_123659 | Not-determined |
| S17_extracted_k141_12974 | Not-determined |
| S17_extracted_k141_116922 | Not-determined |
| S17_extracted_k141_27230 | Not-determined |
| S17_extracted_k141_161224 | Not-determined |
| S17_extracted_k141_72520 | Low-quality |
| S17_extracted_k141_57734 | Not-determined |
| S17_extracted_k141_123661 | Not-determined |
| S17_extracted_k141_57736 | Not-determined |
| S17_extracted_k141_27234 | Low-quality |
| S17_extracted_k141_35936 | Low-quality |

| | |
|---|---|
| S17_extracted_k141_86368 | Not-determined |
| S17_extracted_k141_5866 | Not-determined |
| S17_extracted_k141_116926 | Not-determined |
| S17_extracted_k141_146217 | Not-determined |
| S17_extracted_k141_154173 | Not-determined |
| S17_extracted_k141_20038 | Not-determined |
| S17_extracted_k141_57744 | Low-quality |
| S17_extracted_k141_154174 | Not-determined |
| S17_extracted_k141_154177 | Low-quality |
| S17_extracted_k141_5873 | Not-determined |
| S17_extracted_k141_72532 | Not-determined |
| S17_extracted_k141_175009 | Not-determined |
| S17_extracted_k141_154180 | Not-determined |
| S17_extracted_k141_79826 | Not-determined |
| S17_extracted_k141_64116 | Not-determined |
| S17_extracted_k141_72538 | Not-determined |
| S17_extracted_k141_94124 | Not-determined |
| S17_extracted_k141_110076 | Not-determined |
| S17_extracted_k141_138654 | Not-determined |
| S17_extracted_k141_146225 | Not-determined |
| S17_extracted_k141_27243 | Not-determined |
| S17_extracted_k141_110079 | Not-determined |
| S17_extracted_k141_175014 | Not-determined |
| S17_extracted_k141_35955 | Not-determined |
| S17_extracted_k141_102075 | Not-determined |
| S17_extracted_k141_86378 | Not-determined |
| S17_extracted_k141_27245 | Not-determined |
| S17_extracted_k141_5881 | Not-determined |
| S17_extracted_k141_43309 | Not-determined |
| S17_extracted_k141_86382 | Not-determined |
| S17_extracted_k141_20049 | Not-determined |
| S17_extracted_k141_110082 | Low-quality |
| S17_extracted_k141_64125 | Not-determined |
| S17_extracted_k141_161235 | Not-determined |
| S17_extracted_k141_175025 | Not-determined |
| S17_extracted_k141_110085 | Not-determined |
| S17_extracted_k141_86386 | Not-determined |
| S17_extracted_k141_175026 | Not-determined |
| S17_extracted_k141_161237 | Not-determined |
| S17_extracted_k141_5886 | Low-quality |
| S17_extracted_k141_20055 | Not-determined |
| S17_extracted_k141_161238 | Not-determined |
| S17_extracted_k141_102087 | Not-determined |
| S17_extracted_k141_102088 | Not-determined |
| S17_extracted_k141_175029 | Not-determined |
| S17_extracted_k141_27256 | Not-determined |
| S17_extracted_k141_161239 | Not-determined |
| S17_extracted_k141_79829 | Not-determined |
| S17_extracted_k141_94137 | Not-determined |
| S17_extracted_k141_154191 | Not-determined |
| S17_extracted_k141_161243 | Not-determined |
| S17_extracted_k141_27258 | Not-determined |
| S17_extracted_k141_86394 | Not-determined |
| S17_extracted_k141_86395 | Not-determined |
| S17_extracted_k141_175034 | Not-determined |
| S17_extracted_k141_102091 | Not-determined |
| S17_extracted_k141_20061 | Not-determined |
| S17_extracted_k141_123667 | Not-determined |
| S17_extracted_k141_86396 | Low-quality |
| S17_extracted_k141_175036 | Not-determined |
| S17_extracted_k141_27263 | Not-determined |
| S17_extracted_k141_64132 | Low-quality |
| S17_extracted_k141_146240 | Not-determined |
| S17_extracted_k141_102095 | Not-determined |
| S17_extracted_k141_123672 | Not-determined |
| S17_extracted_k141_154198 | Low-quality |
| S17_extracted_k141_35961 | Low-quality |
| S17_extracted_k141_154202 | Not-determined |
| S17_extracted_k141_13000 | Not-determined |
| S17_extracted_k141_20070 | Not-determined |
| S17_extracted_k141_102101 | Not-determined |
| S17_extracted_k141_110095 | Not-determined |
| S17_extracted_k141_35966 | Not-determined |
| S17_extracted_k141_64141 | Not-determined |
| S17_extracted_k141_175039 | Not-determined |
| S17_extracted_k141_35967 | Not-determined |
| S17_extracted_k141_154211 | Not-determined |
| S17_extracted_k141_27277 | Not-determined |
| S17_extracted_k141_175040 | Not-determined |
| S17_extracted_k141_161255 | Not-determined |
| S17_extracted_k141_175044 | Not-determined |
| S17_extracted_k141_168244 | Not-determined |
| S17_extracted_k141_20075 | Not-determined |
| S17_extracted_k141_116941 | Not-determined |
| S17_extracted_k141_64147 | Not-determined |
| S17_extracted_k141_116942 | Not-determined |
| S17_extracted_k141_35974 | Low-quality |
| S17_extracted_k141_123675 | Low-quality |
| S17_extracted_k141_64150 | Low-quality |
| S17_extracted_k141_13013 | Not-determined |
| S17_extracted_k141_20079 | Not-determined |
| S17_extracted_k141_94160 | Not-determined |
| S17_extracted_k141_5896 | Not-determined |
| S17_extracted_k141_102110 | Low-quality |
| S17_extracted_k141_102111 | Not-determined |
| S17_extracted_k141_35977 | Not-determined |
| S17_extracted_k141_161263 | Not-determined |
| S17_extracted_k141_130942 | Not-determined |
| S17_extracted_k141_64155 | Low-quality |
| S17_extracted_k141_94162 | Low-quality |
| S17_extracted_k141_43334 | Not-determined |
| S17_extracted_k141_27284 | Low-quality |
| S17_extracted_k141_168255 | Not-determined |
| S17_extracted_k141_123682 | Not-determined |
| S17_extracted_k141_43338 | Not-determined |
| S17_extracted_k141_123685 | Not-determined |
| S17_extracted_k141_123687 | Not-determined |
| S17_extracted_k141_64162 | Low-quality |
| S17_extracted_k141_57793 | Not-determined |
| S17_extracted_k141_123688 | Low-quality |
| S17_extracted_k141_161273 | Not-determined |
| S17_extracted_k141_20089 | Not-determined |
| S17_extracted_k141_175053 | Not-determined |
| S17_extracted_k141_5908 | Not-determined |
| S17_extracted_k141_57795 | Not-determined |
| S17_extracted_k141_64165 | Not-determined |
| S17_extracted_k141_50658 | Low-quality |
| S17_extracted_k141_175056 | Low-quality |
| S17_extracted_k141_123697 | Not-determined |
| S17_extracted_k141_79855 | Not-determined |
| S17_extracted_k141_5912 | Not-determined |
| S17_extracted_k141_123698 | Not-determined |
| S17_extracted_k141_79857 | Not-determined |
| S17_extracted_k141_123699 | Not-determined |
| S17_extracted_k141_116962 | Not-determined |
| S17_extracted_k141_161283 | Not-determined |
| S17_extracted_k141_86414 | Not-determined |
| S17_extracted_k141_102133 | Not-determined |
| S17_extracted_k141_35992 | Not-determined |
| S17_extracted_k141_110113 | Not-determined |
| S17_extracted_k141_161287 | Not-determined |
| S17_extracted_k141_57799 | Not-determined |
| S17_extracted_k141_5918 | Not-determined |
| S17_extracted_k141_20096 | Low-quality |
| S17_extracted_k141_110117 | Not-determined |
| S17_extracted_k141_154245 | Low-quality |
| S17_extracted_k141_130952 | Low-quality |
| S17_extracted_k141_102139 | Not-determined |
| S17_extracted_k141_102140 | Not-determined |
| S17_extracted_k141_86420 | Not-determined |
| S17_extracted_k141_27301 | Not-determined |
| S17_extracted_k141_13030 | Low-quality |
| S17_extracted_k141_27302 | Low-quality |
| S17_extracted_k141_123707 | Low-quality |
| S17_extracted_k141_154250 | Not-determined |
| S17_extracted_k141_168263 | Not-determined |
| S17_extracted_k141_116970 | Not-determined |
| S17_extracted_k141_13034 | Not-determined |
| S17_extracted_k141_161298 | Not-determined |
| S17_extracted_k141_116972 | Not-determined |
| S17_extracted_k141_161299 | Not-determined |
| S17_extracted_k141_154254 | Not-determined |
| S17_extracted_k141_116973 | Not-determined |
| S17_extracted_k141_123710 | Low-quality |
| S17_extracted_k141_154257 | Not-determined |
| S17_extracted_k141_20104 | Not-determined |
| S17_extracted_k141_20105 | Not-determined |
| S17_extracted_k141_123714 | Not-determined |
| S17_extracted_k141_27312 | Not-determined |
| S17_extracted_k141_146283 | Not-determined |
| S17_extracted_k141_102152 | Not-determined |
| S17_extracted_k141_5926 | Not-determined |
| S17_extracted_k141_168271 | Not-determined |
| S17_extracted_k141_138688 | Not-determined |
| S17_extracted_k141_161305 | Not-determined |
| S17_extracted_k141_130966 | Low-quality |
| S17_extracted_k141_146286 | Low-quality |
| S17_extracted_k141_154263 | Not-determined |
| S17_extracted_k141_175063 | Not-determined |
| S17_extracted_k141_79870 | Low-quality |
| S17_extracted_k141_64190 | Not-determined |
| S17_extracted_k141_5931 | Not-determined |
| S17_extracted_k141_130968 | Not-determined |
| S17_extracted_k141_146289 | Not-determined |
| S17_extracted_k141_110134 | Not-determined |
| S17_extracted_k141_130970 | Not-determined |
| S17_extracted_k141_130971 | Not-determined |
| S17_extracted_k141_168275 | Not-determined |
| S17_extracted_k141_102160 | Not-determined |
| S17_extracted_k141_5934 | Not-determined |
| S17_extracted_k141_154269 | Low-quality |
| S17_extracted_k141_168278 | Not-determined |
| S17_extracted_k141_154271 | Not-determined |
| S17_extracted_k141_20111 | Not-determined |
| S17_extracted_k141_161311 | Not-determined |
| S17_extracted_k141_57812 | Not-determined |
| S17_extracted_k141_27327 | Low-quality |
| S17_extracted_k141_20117 | Not-determined |
| S17_extracted_k141_27332 | Not-determined |
| S17_extracted_k141_116982 | Not-determined |
| S17_extracted_k141_175083 | Not-determined |
| S17_extracted_k141_116984 | Low-quality |
| S17_extracted_k141_161317 | Low-quality |
| S17_extracted_k141_175084 | Not-determined |
| S17_extracted_k141_161319 | Not-determined |
| S17_extracted_k141_168286 | Not-determined |
| S17_extracted_k141_175085 | Not-determined |
| S17_extracted_k141_102172 | Not-determined |
| S17_extracted_k141_116987 | Low-quality |
| S17_extracted_k141_146297 | Not-determined |
| S17_extracted_k141_161324 | Not-determined |
| S17_extracted_k141_146300 | Not-determined |
| S17_extracted_k141_168287 | Not-determined |
| S17_extracted_k141_20125 | Not-determined |
| S17_extracted_k141_146303 | Not-determined |
| S17_extracted_k141_27341 | Not-determined |
| S17_extracted_k141_146304 | Not-determined |
| S17_extracted_k141_130992 | Low-quality |
| S17_extracted_k141_138704 | Not-determined |
| S17_extracted_k141_102180 | Not-determined |
| S17_extracted_k141_116992 | Low-quality |
| S17_extracted_k141_138705 | Not-determined |
| S17_extracted_k141_57817 | Not-determined |
| S17_extracted_k141_130996 | Low-quality |
| S17_extracted_k141_102183 | Not-determined |
| S17_extracted_k141_138708 | Low-quality |
| S17_extracted_k141_64222 | Not-determined |
| S17_extracted_k141_116998 | Low-quality |
| S17_extracted_k141_123746 | Not-determined |
| S17_extracted_k141_146309 | Not-determined |
| S17_extracted_k141_117000 | Not-determined |
| S17_extracted_k141_20135 | Not-determined |
| S17_extracted_k141_110156 | Not-determined |
| S17_extracted_k141_123750 | Not-determined |
| S17_extracted_k141_117003 | Not-determined |
| S17_extracted_k141_79891 | Not-determined |
| S17_extracted_k141_36006 | Not-determined |
| S17_extracted_k141_161329 | Not-determined |
| S17_extracted_k141_117006 | Not-determined |
| S17_extracted_k141_110158 | Not-determined |
| S17_extracted_k141_57829 | Not-determined |
| S17_extracted_k141_110161 | Not-determined |
| S17_extracted_k141_146316 | Not-determined |
| S17_extracted_k141_36009 | Not-determined |
| S17_extracted_k141_79897 | Not-determined |
| S17_extracted_k141_138723 | Not-determined |
| S17_extracted_k141_110167 | Not-determined |
| S17_extracted_k141_27349 | Not-determined |
| S17_extracted_k141_57837 | Low-quality |
| S17_extracted_k141_138726 | Not-determined |
| S17_extracted_k141_131011 | Not-determined |
| S17_extracted_k141_50677 | Not-determined |
| S17_extracted_k141_146322 | Not-determined |
| S17_extracted_k141_43371 | Low-quality |
| S17_extracted_k141_154307 | Not-determined |
| S17_extracted_k141_102185 | Not-determined |
| S17_extracted_k141_154309 | Low-quality |
| S17_extracted_k141_154310 | Not-determined |
| S17_extracted_k141_131013 | Not-determined |
| S17_extracted_k141_110173 | Low-quality |
| S17_extracted_k141_57844 | Not-determined |
| S17_extracted_k141_5950 | Not-determined |
| S17_extracted_k141_36022 | Not-determined |
| S17_extracted_k141_5953 | Not-determined |
| S17_extracted_k141_154316 | Not-determined |
| S17_extracted_k141_64242 | Not-determined |
| S17_extracted_k141_94173 | Low-quality |
| S17_extracted_k141_57852 | Low-quality |
| S17_extracted_k141_20155 | Low-quality |
| S17_extracted_k141_117015 | Not-determined |
| S17_extracted_k141_5957 | Not-determined |
| S17_extracted_k141_117016 | Not-determined |
| S17_extracted_k141_36025 | Not-determined |
| S17_extracted_k141_102194 | Low-quality |
| S17_extracted_k141_57855 | Low-quality |
| S17_extracted_k141_138736 | Low-quality |
| S17_extracted_k141_36027 | Not-determined |
| S17_extracted_k141_27372 | Not-determined |
| S17_extracted_k141_123781 | Low-quality |
| S17_extracted_k141_168300 | Not-determined |
| S17_extracted_k141_131021 | Low-quality |
| S17_extracted_k141_57861 | Not-determined |
| S17_extracted_k141_154327 | Not-determined |
| S17_extracted_k141_154328 | Not-determined |
| S17_extracted_k141_94184 | Not-determined |
| S17_extracted_k141_72564 | Not-determined |
| S17_extracted_k141_117026 | Not-determined |
| S17_extracted_k141_79912 | Not-determined |
| S17_extracted_k141_131026 | Not-determined |
| S17_extracted_k141_86467 | Not-determined |
| S17_extracted_k141_94192 | Not-determined |
| S17_extracted_k141_27385 | Low-quality |
| S17_extracted_k141_154338 | Not-determined |
| S17_extracted_k141_43383 | Not-determined |
| S17_extracted_k141_27387 | Not-determined |
| S17_extracted_k141_154340 | Not-determined |
| S17_extracted_k141_146344 | Not-determined |
| S17_extracted_k141_64255 | Not-determined |
| S17_extracted_k141_117035 | Not-determined |
| S17_extracted_k141_79922 | Low-quality |
| S17_extracted_k141_79924 | Low-quality |
| S17_extracted_k141_110192 | Not-determined |
| S17_extracted_k141_138753 | Not-determined |
| S17_extracted_k141_50700 | Not-determined |
| S17_extracted_k141_154348 | Not-determined |
| S17_extracted_k141_161354 | Low-quality |
| S17_extracted_k141_64263 | Not-determined |
| S17_extracted_k141_117040 | Not-determined |
| S17_extracted_k141_86479 | Not-determined |
| S17_extracted_k141_117044 | Not-determined |
| S17_extracted_k141_161358 | Not-determined |
| S17_extracted_k141_5972 | Not-determined |
| S17_extracted_k141_5973 | Not-determined |
| S17_extracted_k141_146355 | Not-determined |
| S17_extracted_k141_50706 | Not-determined |
| S17_extracted_k141_50707 | Not-determined |
| S17_extracted_k141_102206 | Not-determined |
| S17_extracted_k141_102208 | Not-determined |
| S17_extracted_k141_5977 | Not-determined |
| S17_extracted_k141_161361 | Low-quality |
| S17_extracted_k141_146357 | Not-determined |
| S17_extracted_k141_50714 | Not-determined |
| S17_extracted_k141_138762 | Not-determined |
| S17_extracted_k141_43393 | Not-determined |
| S17_extracted_k141_117058 | Low-quality |
| S17_extracted_k141_5982 | Not-determined |
| S17_extracted_k141_168314 | Not-determined |
| S17_extracted_k141_102219 | Not-determined |
| S17_extracted_k141_102223 | Not-determined |
| S17_extracted_k141_110206 | Not-determined |
| S17_extracted_k141_36051 | Not-determined |
| S17_extracted_k141_79934 | Low-quality |
| S17_extracted_k141_102228 | Not-determined |
| S17_extracted_k141_154368 | Not-determined |
| S17_extracted_k141_5986 | Low-quality |
| S17_extracted_k141_72586 | Not-determined |
| S17_extracted_k141_79936 | Not-determined |
| S17_extracted_k141_79937 | Not-determined |
| S17_extracted_k141_43403 | Not-determined |
| S17_extracted_k141_36053 | Not-determined |
| S17_extracted_k141_64287 | Not-determined |
| S17_extracted_k141_146379 | Not-determined |
| S17_extracted_k141_72589 | Low-quality |
| S17_extracted_k141_154370 | Low-quality |
| S17_extracted_k141_138776 | Not-determined |
| S17_extracted_k141_154372 | Not-determined |
| S17_extracted_k141_131053 | Not-determined |
| S17_extracted_k141_102231 | Not-determined |
| S17_extracted_k141_110217 | Not-determined |
| S17_extracted_k141_110218 | Not-determined |
| S17_extracted_k141_102233 | Not-determined |
| S17_extracted_k141_161364 | Not-determined |
| S17_extracted_k141_102235 | Not-determined |
| S17_extracted_k141_50738 | Not-determined |
| S17_extracted_k141_146385 | Not-determined |
| S17_extracted_k141_13070 | Not-determined |
| S17_extracted_k141_86488 | Not-determined |
| S17_extracted_k141_6004 | Not-determined |
| S17_extracted_k141_102240 | Not-determined |
| S17_extracted_k141_6005 | Not-determined |
| S17_extracted_k141_154381 | Low-quality |
| S17_extracted_k141_131059 | Not-determined |
| S17_extracted_k141_27389 | Not-determined |
| S17_extracted_k141_117067 | Not-determined |
| S17_extracted_k141_117070 | Not-determined |
| S17_extracted_k141_50752 | Not-determined |
| S17_extracted_k141_138782 | Not-determined |
| S17_extracted_k141_154387 | Not-determined |
| S17_extracted_k141_168332 | Not-determined |
| S17_extracted_k141_123816 | Not-determined |
| S17_extracted_k141_110224 | Not-determined |
| S17_extracted_k141_117076 | Not-determined |
| S17_extracted_k141_168334 | Not-determined |
| S17_extracted_k141_117078 | Not-determined |
| S17_extracted_k141_168335 | Not-determined |
| S17_extracted_k141_154392 | Not-determined |
| S17_extracted_k141_43427 | Not-determined |
| S17_extracted_k141_27398 | Not-determined |
| S17_extracted_k141_131073 | Not-determined |
| S17_extracted_k141_94241 | Not-determined |
| S17_extracted_k141_161390 | Not-determined |
| S17_extracted_k141_161391 | Not-determined |
| S17_extracted_k141_168339 | Not-determined |
| S17_extracted_k141_86505 | Not-determined |
| S17_extracted_k141_27404 | Not-determined |
| S17_extracted_k141_94246 | Not-determined |
| S17_extracted_k141_117086 | Low-quality |
| S17_extracted_k141_123825 | Not-determined |
| S17_extracted_k141_154398 | Not-determined |
| S17_extracted_k141_57914 | Not-determined |
| S17_extracted_k141_86508 | Low-quality |
| S17_extracted_k141_102256 | Low-quality |
| S17_extracted_k141_57916 | Low-quality |
| S17_extracted_k141_131085 | Not-determined |
| S17_extracted_k141_27411 | Not-determined |
| S17_extracted_k141_102259 | Low-quality |
| S17_extracted_k141_117095 | Not-determined |
| S17_extracted_k141_168348 | Not-determined |
| S17_extracted_k141_110237 | Not-determined |
| S17_extracted_k141_123828 | Not-determined |
| S17_extracted_k141_20175 | Not-determined |
| S17_extracted_k141_20176 | Not-determined |
| S17_extracted_k141_168350 | Not-determined |
| S17_extracted_k141_123833 | Not-determined |
| S17_extracted_k141_64298 | Not-determined |
| S7_extracted_6664 | Complete |
| S17_extracted_k141_123835 | Not-determined |
| S17_extracted_k141_94260 | Not-determined |
| S17_extracted_k141_13093 | Not-determined |
| S17_extracted_k141_110238 | Not-determined |
| S17_extracted_k141_146407 | Not-determined |
| S17_extracted_k141_43447 | Not-determined |
| S17_extracted_k141_161404 | Not-determined |
| S17_extracted_k141_6032 | Not-determined |
| S17_extracted_k141_64304 | Not-determined |
| S17_extracted_k141_102266 | Not-determined |
| S17_extracted_k141_102269 | Not-determined |
| S17_extracted_k141_94264 | Not-determined |
| S17_extracted_k141_20190 | Not-determined |
| S17_extracted_k141_110244 | Not-determined |
| S17_extracted_k141_175111 | Not-determined |
| S17_extracted_k141_146416 | Not-determined |
| S17_extracted_k141_168357 | Not-determined |
| S17_extracted_k141_168358 | Not-determined |
| S17_extracted_k141_102276 | Not-determined |
| S17_extracted_k141_20193 | Not-determined |
| S17_extracted_k141_6040 | Not-determined |
| S17_extracted_k141_27430 | Low-quality |
| S17_extracted_k141_20195 | Not-determined |
| S17_extracted_k141_161417 | Not-determined |
| S17_extracted_k141_146422 | Not-determined |
| S17_extracted_k141_43454 | Not-determined |
| S17_extracted_k141_64311 | Not-determined |
| S17_extracted_k141_102281 | Not-determined |
| S17_extracted_k141_94273 | Not-determined |
| S17_extracted_k141_86517 | Not-determined |
| S17_extracted_k141_146429 | Not-determined |
| S17_extracted_k141_13114 | Not-determined |
| S17_extracted_k141_175119 | Not-determined |
| S17_extracted_k141_154423 | Not-determined |
| S17_extracted_k141_13116 | Low-quality |
| S17_extracted_k141_168365 | Not-determined |
| S17_extracted_k141_13117 | Not-determined |
| S17_extracted_k141_175121 | Not-determined |
| S17_extracted_k141_94278 | Not-determined |
| S17_extracted_k141_154427 | Not-determined |
| S17_extracted_k141_117122 | Not-determined |
| S17_extracted_k141_57925 | Not-determined |
| S17_extracted_k141_175124 | Low-quality |
| S17_extracted_k141_102284 | Not-determined |
| S17_extracted_k141_117124 | Not-determined |
| S17_extracted_k141_13122 | Not-determined |
| S17_extracted_k141_13124 | Not-determined |
| S17_extracted_k141_6058 | Not-determined |
| S17_extracted_k141_175125 | Not-determined |
| S17_extracted_k141_57930 | Not-determined |
| S17_extracted_k141_86522 | Not-determined |
| S17_extracted_k141_6060 | Not-determined |
| S17_extracted_k141_146432 | Not-determined |
| S17_extracted_k141_154432 | Not-determined |
| S17_extracted_k141_131114 | Not-determined |
| S17_extracted_k141_138830 | Not-determined |
| S17_extracted_k141_110261 | Not-determined |
| S17_extracted_k141_20208 | Not-determined |
| S17_extracted_k141_57937 | Not-determined |
| S17_extracted_k141_123852 | Low-quality |
| S17_extracted_k141_57938 | Not-determined |
| S17_extracted_k141_146434 | Low-quality |
| S17_extracted_k141_161431 | Not-determined |
| S17_extracted_k141_102293 | Not-determined |
| S17_extracted_k141_168367 | Not-determined |
| S17_extracted_k141_102294 | Not-determined |
| S17_extracted_k141_117134 | Not-determined |
| S17_extracted_k141_161434 | Not-determined |
| S17_extracted_k141_43465 | Not-determined |
| S17_extracted_k141_27433 | Not-determined |
| S17_extracted_k141_102297 | Not-determined |
| S17_extracted_k141_27435 | Not-determined |
| S17_extracted_k141_13135 | Not-determined |
| S17_extracted_k141_57939 | Not-determined |
| S17_extracted_k141_154443 | Not-determined |
| S17_extracted_k141_117141 | Low-quality |
| S17_extracted_k141_13136 | Not-determined |
| S17_extracted_k141_13138 | Not-determined |
| S17_extracted_k141_131125 | Not-determined |
| S17_extracted_k141_57942 | Not-determined |
| S17_extracted_k141_161442 | Not-determined |
| S17_extracted_k141_86535 | Not-determined |
| S17_extracted_k141_117148 | Not-determined |
| S17_extracted_k141_117149 | Low-quality |
| S17_extracted_k141_175144 | Not-determined |
| S17_extracted_k141_86538 | Low-quality |
| S17_extracted_k141_110273 | Low-quality |
| S17_extracted_k141_154445 | Low-quality |
| S17_extracted_k141_168377 | Not-determined |
| S17_extracted_k141_57952 | Low-quality |
| S17_extracted_k141_64334 | Not-determined |
| S17_extracted_k141_168378 | Not-determined |
| S17_extracted_k141_20219 | Low-quality |
| S17_extracted_k141_161452 | Not-determined |
| S17_extracted_k141_20220 | Not-determined |
| S17_extracted_k141_20224 | Low-quality |
| S17_extracted_k141_110284 | Not-determined |
| S17_extracted_k141_117164 | Not-determined |
| S17_extracted_k141_102305 | Not-determined |
| S17_extracted_k141_57958 | Not-determined |
| S17_extracted_k141_102308 | Not-determined |
| S17_extracted_k141_64335 | Low-quality |
| S17_extracted_k141_102310 | Not-determined |
| S17_extracted_k141_64337 | Not-determined |
| S17_extracted_k141_94296 | Low-quality |
| S17_extracted_k141_146450 | Not-determined |
| S17_extracted_k141_20231 | Not-determined |
| S17_extracted_k141_64343 | Not-determined |
| S17_extracted_k141_154457 | Not-determined |
| S17_extracted_k141_154460 | Not-determined |
| S17_extracted_k141_154461 | Not-determined |
| S17_extracted_k141_50807 | Not-determined |
| S17_extracted_k141_168389 | Not-determined |
| S17_extracted_k141_175159 | Low-quality |
| S17_extracted_k141_168390 | Not-determined |
| S17_extracted_k141_110294 | Not-determined |
| S17_extracted_k141_13148 | Not-determined |
| S17_extracted_k141_13149 | Not-determined |
| S17_extracted_k141_94303 | Low-quality |
| S17_extracted_k141_175165 | Not-determined |
| S17_extracted_k141_110297 | Not-determined |
| S17_extracted_k141_13152 | Low-quality |
| S17_extracted_k141_86560 | Not-determined |
| S17_extracted_k141_138867 | Low-quality |
| S17_extracted_k141_146470 | Not-determined |
| S17_extracted_k141_86565 | Not-determined |
| S17_extracted_k141_86566 | Not-determined |
| S17_extracted_k141_94312 | Not-determined |
| S17_extracted_k141_20249 | Not-determined |
| S17_extracted_k141_6083 | Not-determined |
| S17_extracted_k141_102330 | Not-determined |
| S17_extracted_k141_13159 | Low-quality |
| S17_extracted_k141_110315 | Not-determined |
| S17_extracted_k141_168409 | Not-determined |
| S17_extracted_k141_27463 | Not-determined |
| S17_extracted_k141_6091 | Not-determined |
| S17_extracted_k141_6092 | Not-determined |
| S17_extracted_k141_27470 | Not-determined |
| S17_extracted_k141_154467 | Not-determined |
| S17_extracted_k141_146482 | Not-determined |
| S17_extracted_k141_94329 | Low-quality |
| S17_extracted_k141_64375 | Not-determined |
| S17_extracted_k141_6098 | Not-determined |
| S17_extracted_k141_20257 | Not-determined |
| S17_extracted_k141_175181 | Not-determined |
| S17_extracted_k141_79964 | Not-determined |
| S17_extracted_k141_110322 | Not-determined |
| S17_extracted_k141_110323 | Not-determined |
| S17_extracted_k141_64379 | Not-determined |
| S17_extracted_k141_6103 | Not-determined |
| S17_extracted_k141_154472 | Not-determined |
| S17_extracted_k141_154473 | Not-determined |
| S17_extracted_k141_64382 | Not-determined |
| S17_extracted_k141_110327 | Not-determined |
| S17_extracted_k141_27482 | Low-quality |
| S17_extracted_k141_64384 | Not-determined |
| S17_extracted_k141_86585 | Low-quality |
| S17_extracted_k141_117192 | Not-determined |
| S17_extracted_k141_20264 | Not-determined |
| S17_extracted_k141_154477 | Not-determined |
| S17_extracted_k141_27485 | Not-determined |
| S17_extracted_k141_27486 | Not-determined |
| S17_extracted_k141_117195 | Not-determined |
| S17_extracted_k141_110331 | Not-determined |
| S17_extracted_k141_123916 | Not-determined |
| S17_extracted_k141_13180 | Not-determined |
| S17_extracted_k141_146489 | Not-determined |
| S17_extracted_k141_168429 | Not-determined |
| S17_extracted_k141_154482 | Not-determined |
| S17_extracted_k141_64392 | Not-determined |
| S17_extracted_k141_43480 | Not-determined |
| S17_extracted_k141_72622 | Not-determined |
| S17_extracted_k141_86590 | Not-determined |
| S17_extracted_k141_154485 | Not-determined |
| S17_extracted_k141_13185 | Not-determined |
| S17_extracted_k141_146491 | Not-determined |
| S17_extracted_k141_79979 | Not-determined |
| S17_extracted_k141_123921 | Not-determined |
| S17_extracted_k141_94343 | Not-determined |
| S17_extracted_k141_138901 | Low-quality |
| S17_extracted_k141_123922 | Low-quality |
| S17_extracted_k141_138902 | Not-determined |
| S17_extracted_k141_154489 | Not-determined |
| S17_extracted_k141_123923 | Not-determined |
| S17_extracted_k141_79984 | Not-determined |
| S17_extracted_k141_146497 | Not-determined |
| S17_extracted_k141_146498 | Not-determined |
| S17_extracted_k141_110342 | Not-determined |
| S17_extracted_k141_117200 | Not-determined |
| S17_extracted_k141_20272 | Not-determined |
| S17_extracted_k141_13192 | Not-determined |
| S17_extracted_k141_72629 | Not-determined |
| S17_extracted_k141_146500 | Not-determined |

| | |
|---|---|
| S17_extracted_k141_110347 | Not-determined |
| S17_extracted_k141_123932 | Not-determined |
| S17_extracted_k141_161482 | Not-determined |
| S17_extracted_k141_86596 | Not-determined |
| S17_extracted_k141_72634 | Not-determined |
| S17_extracted_k141_72635 | Not-determined |
| S17_extracted_k141_6122 | Low-quality |
| S17_extracted_k141_154494 | Low-quality |
| S17_extracted_k141_86599 | Not-determined |
| S17_extracted_k141_72640 | Not-determined |
| S17_extracted_k141_146512 | Not-determined |
| S17_extracted_k141_86600 | Not-determined |
| S17_extracted_k141_13205 | Not-determined |
| S17_extracted_k141_154497 | Not-determined |
| S17_extracted_k141_64403 | Not-determined |
| S17_extracted_k141_154498 | Not-determined |
| S17_extracted_k141_64405 | Not-determined |
| S17_extracted_k141_154499 | Not-determined |
| S17_extracted_k141_57998 | Not-determined |
| S17_extracted_k141_110356 | Not-determined |
| S17_extracted_k141_117215 | Not-determined |
| S17_extracted_k141_58002 | Not-determined |
| S17_extracted_k141_64413 | Low-quality |
| S17_extracted_k141_175206 | Not-determined |
| S17_extracted_k141_154507 | Not-determined |
| S17_extracted_k141_64417 | Not-determined |
| S17_extracted_k141_58004 | Not-determined |
| S17_extracted_k141_43501 | Not-determined |
| S17_extracted_k141_86614 | Not-determined |
| S17_extracted_k141_168445 | Low-quality |
| S17_extracted_k141_168446 | Not-determined |
| S17_extracted_k141_161500 | Not-determined |
| S17_extracted_k141_43506 | Not-determined |
| S17_extracted_k141_117222 | Not-determined |
| S17_extracted_k141_6142 | Not-determined |
| S17_extracted_k141_86619 | Not-determined |
| S17_extracted_k141_168448 | Not-determined |
| S17_extracted_k141_64423 | Not-determined |
| S17_extracted_k141_123949 | Low-quality |
| S17_extracted_k141_117225 | Not-determined |
| S17_extracted_k141_123950 | Not-determined |
| S17_extracted_k141_58012 | Not-determined |
| S17_extracted_k141_6150 | Not-determined |
| S17_extracted_k141_36095 | Low-quality |
| S17_extracted_k141_64427 | Low-quality |
| S17_extracted_k141_20296 | Not-determined |
| S17_extracted_k141_138924 | Not-determined |
| S17_extracted_k141_43515 | Not-determined |
| S17_extracted_k141_64429 | Not-determined |
| S17_extracted_k141_86626 | Not-determined |
| S17_extracted_k141_43522 | Not-determined |
| S17_extracted_k141_138932 | Not-determined |
| S17_extracted_k141_64437 | Not-determined |
| S17_extracted_k141_36109 | Low-quality |
| S17_extracted_k141_86630 | Not-determined |
| S17_extracted_k141_110386 | Not-determined |
| S17_extracted_k141_146556 | Not-determined |
| S17_extracted_k141_43525 | Not-determined |
| S17_extracted_k141_72674 | Not-determined |
| S17_extracted_k141_102357 | Not-determined |
| S17_extracted_k141_86636 | Not-determined |
| S17_extracted_k141_64439 | Not-determined |
| S17_extracted_k141_36112 | Not-determined |
| S17_extracted_k141_27513 | Not-determined |
| S17_extracted_k141_131192 | Not-determined |
| S17_extracted_k141_36115 | Low-quality |
| S17_extracted_k141_110395 | Not-determined |
| S17_extracted_k141_117234 | Not-determined |
| S17_extracted_k141_138946 | Not-determined |
| S17_extracted_k141_72677 | Not-determined |
| S17_extracted_k141_123966 | Not-determined |
| S17_extracted_k141_102361 | Low-quality |
| S17_extracted_k141_72681 | Not-determined |
| S17_extracted_k141_102364 | Low-quality |
| S17_extracted_k141_43542 | Not-determined |
| S17_extracted_k141_36127 | Not-determined |
| S17_extracted_k141_6185 | Not-determined |
| S17_extracted_k141_102365 | Low-quality |
| S17_extracted_k141_36130 | Not-determined |
| S17_extracted_k141_175226 | Not-determined |
| S17_extracted_k141_146578 | Not-determined |
| S17_extracted_k141_154549 | Not-determined |
| S17_extracted_k141_123975 | Not-determined |
| S17_extracted_k141_138962 | Not-determined |
| S17_extracted_k141_102373 | Low-quality |
| S17_extracted_k141_154556 | Low-quality |
| S17_extracted_k141_138964 | Not-determined |
| S17_extracted_k141_86652 | Not-determined |
| S17_extracted_k141_6195 | Not-determined |
| S17_extracted_k141_27521 | Not-determined |
| S17_extracted_k141_86653 | Not-determined |
| S17_extracted_k141_36139 | Not-determined |
| S17_extracted_k141_117245 | Low-quality |
| S17_extracted_k141_27524 | Not-determined |
| S17_extracted_k141_110404 | Not-determined |
| S17_extracted_k141_36143 | Not-determined |
| S17_extracted_k141_138970 | Not-determined |
| S17_extracted_k141_80015 | Not-determined |
| S17_extracted_k141_138973 | Not-determined |
| S17_extracted_k141_13258 | Not-determined |
| S17_extracted_k141_138974 | Not-determined |
| S17_extracted_k141_102376 | Not-determined |
| S17_extracted_k141_138976 | Not-determined |
| S17_extracted_k141_117251 | Low-quality |
| S17_extracted_k141_110413 | Not-determined |
| S17_extracted_k141_43570 | Not-determined |
| S17_extracted_k141_64467 | Low-quality |
| S17_extracted_k141_13263 | Not-determined |
| S17_extracted_k141_86663 | Not-determined |
| S17_extracted_k141_154572 | Not-determined |
| S17_extracted_k141_27532 | Low-quality |
| S17_extracted_k141_6207 | Low-quality |
| S17_extracted_k141_13270 | Not-determined |
| S17_extracted_k141_27536 | Not-determined |
| S17_extracted_k141_43577 | Not-determined |
| S17_extracted_k141_36161 | Not-determined |
| S17_extracted_k141_27538 | Not-determined |
| S17_extracted_k141_6209 | Not-determined |
| S17_extracted_k141_138984 | Not-determined |
| S17_extracted_k141_43581 | Not-determined |
| S17_extracted_k141_123984 | Not-determined |
| S17_extracted_k141_102390 | Not-determined |
| S17_extracted_k141_80029 | Not-determined |
| S17_extracted_k141_138991 | Not-determined |
| S17_extracted_k141_43584 | Not-determined |
| S17_extracted_k141_58021 | Not-determined |
| S17_extracted_k141_80032 | Not-determined |
| S17_extracted_k141_131196 | Not-determined |
| S17_extracted_k141_154587 | Not-determined |
| S17_extracted_k141_20327 | Not-determined |
| S17_extracted_k141_36170 | Not-determined |
| S17_extracted_k141_146584 | Not-determined |
| S17_extracted_k141_161560 | Not-determined |
| S17_extracted_k141_146587 | Not-determined |
| S17_extracted_k141_110418 | Not-determined |
| S17_extracted_k141_6210 | Low-quality |
| S17_extracted_k141_6211 | Low-quality |
| S17_extracted_k141_146589 | Not-determined |
| S17_extracted_k141_146590 | Not-determined |
| S17_extracted_k141_110421 | Not-determined |
| S17_extracted_k141_146592 | Low-quality |
| S17_extracted_k141_146593 | Not-determined |
| S17_extracted_k141_117286 | Not-determined |
| S17_extracted_k141_154598 | Not-determined |
| S17_extracted_k141_154599 | Not-determined |
| S17_extracted_k141_27556 | Not-determined |
| S17_extracted_k141_6217 | Not-determined |
| S17_extracted_k141_146598 | Not-determined |
| S17_extracted_k141_117290 | Not-determined |
| S17_extracted_k141_175249 | Not-determined |
| S17_extracted_k141_117293 | Not-determined |
| S17_extracted_k141_27559 | Not-determined |
| S17_extracted_k141_117295 | Not-determined |
| S17_extracted_k141_102400 | Low-quality |
| S17_extracted_k141_175255 | Not-determined |
| S17_extracted_k141_102402 | Not-determined |
| S17_extracted_k141_110429 | Not-determined |
| S17_extracted_k141_117296 | Low-quality |
| S17_extracted_k141_6223 | Low-quality |
| S17_extracted_k141_175256 | Not-determined |
| S17_extracted_k141_80044 | Not-determined |
| S17_extracted_k141_102405 | Low-quality |
| S17_extracted_k141_72698 | Low-quality |
| S17_extracted_k141_131206 | Not-determined |
| S17_extracted_k141_27571 | Not-determined |
| S17_extracted_k141_72699 | Low-quality |
| S17_extracted_k141_123992 | Not-determined |
| S17_extracted_k141_64497 | Not-determined |
| S17_extracted_k141_36183 | Not-determined |
| S17_extracted_k141_131207 | Not-determined |
| S17_extracted_k141_154613 | Not-determined |
| S17_extracted_k141_20359 | Not-determined |
| S17_extracted_k141_64500 | Low-quality |
| S17_extracted_k141_27576 | Not-determined |
| S17_extracted_k141_13293 | Not-determined |
| S17_extracted_k141_64502 | Not-determined |
| S17_extracted_k141_139028 | Not-determined |
| S17_extracted_k141_175265 | Not-determined |
| S17_extracted_k141_154616 | Not-determined |
| S17_extracted_k141_27582 | Not-determined |
| S17_extracted_k141_86685 | Not-determined |
| S17_extracted_k141_86686 | Not-determined |
| S17_extracted_k141_27584 | Low-quality |
| S17_extracted_k141_175267 | Not-determined |
| S17_extracted_k141_124006 | Not-determined |
| S17_extracted_k141_117311 | Not-determined |
| S17_extracted_k141_154621 | Not-determined |
| S17_extracted_k141_175268 | Low-quality |
| S17_extracted_k141_117313 | Not-determined |
| S17_extracted_k141_154623 | Not-determined |
| S17_extracted_k141_117316 | Not-determined |
| S17_extracted_k141_36195 | Not-determined |
| S17_extracted_k141_80062 | Not-determined |
| S17_extracted_k141_36198 | Not-determined |
| S17_extracted_k141_146631 | Not-determined |
| S17_extracted_k141_102427 | Not-determined |
| S17_extracted_k141_27592 | Not-determined |
| S17_extracted_k141_161583 | Not-determined |
| S17_extracted_k141_36205 | Not-determined |
| S17_extracted_k141_175276 | Not-determined |
| S17_extracted_k141_102431 | Not-determined |
| S17_extracted_k141_36206 | Not-determined |
| S17_extracted_k141_117328 | Not-determined |
| S17_extracted_k141_124014 | Not-determined |
| S17_extracted_k141_110438 | Not-determined |
| S17_extracted_k141_117331 | Not-determined |
| S17_extracted_k141_110439 | Not-determined |
| S17_extracted_k141_86701 | Not-determined |
| S17_extracted_k141_72731 | Not-determined |
| S17_extracted_k141_131225 | Low-quality |
| S17_extracted_k141_86704 | Not-determined |
| S17_extracted_k141_131226 | Not-determined |
| S17_extracted_k141_72738 | Not-determined |
| S17_extracted_k141_161596 | Not-determined |
| S17_extracted_k141_43609 | Low-quality |
| S17_extracted_k141_131230 | Not-determined |
| S17_extracted_k141_27609 | Low-quality |
| S17_extracted_k141_43610 | Not-determined |
| S17_extracted_k141_146638 | Low-quality |
| S17_extracted_k141_36213 | Not-determined |
| S17_extracted_k141_102445 | Not-determined |
| S17_extracted_k141_6237 | Not-determined |
| S17_extracted_k141_13318 | Not-determined |
| S17_extracted_k141_146642 | Not-determined |
| S17_extracted_k141_27612 | Low-quality |
| S17_extracted_k141_80076 | Low-quality |
| S17_extracted_k141_58059 | Not-determined |
| S17_extracted_k141_161601 | Not-determined |
| S17_extracted_k141_64531 | Not-determined |
| S17_extracted_k141_131235 | Not-determined |
| S17_extracted_k141_64533 | Low-quality |
| S17_extracted_k141_13321 | Low-quality |
| S17_extracted_k141_175305 | Not-determined |
| S17_extracted_k141_94363 | Low-quality |
| S17_extracted_k141_36224 | Not-determined |
| S17_extracted_k141_80084 | Not-determined |
| S17_extracted_k141_43614 | Low-quality |
| S17_extracted_k141_102454 | Not-determined |
| S17_extracted_k141_124018 | Not-determined |
| S17_extracted_k141_36228 | Not-determined |
| S17_extracted_k141_175310 | Not-determined |
| S17_extracted_k141_64541 | Not-determined |
| S17_extracted_k141_124020 | Low-quality |
| S17_extracted_k141_36233 | Not-determined |
| S17_extracted_k141_20385 | Not-determined |
| S17_extracted_k141_131244 | Not-determined |
| S17_extracted_k141_36234 | Not-determined |
| S17_extracted_k141_86718 | Not-determined |
| S17_extracted_k141_20387 | Not-determined |
| S17_extracted_k141_131246 | Low-quality |
| S17_extracted_k141_6252 | Not-determined |
| S17_extracted_k141_139052 | Not-determined |
| S17_extracted_k141_124023 | Low-quality |
| S17_extracted_k141_175316 | Not-determined |
| S17_extracted_k141_102463 | Low-quality |
| S17_extracted_k141_124029 | Not-determined |
| S17_extracted_k141_43623 | Not-determined |
| S17_extracted_k141_36238 | Not-determined |
| S17_extracted_k141_43625 | Not-determined |
| S17_extracted_k141_94373 | Low-quality |
| S17_extracted_k141_36240 | Not-determined |
| S17_extracted_k141_117357 | Not-determined |
| S17_extracted_k141_6257 | Low-quality |
| S17_extracted_k141_13339 | Not-determined |
| S17_extracted_k141_6258 | Not-determined |
| S17_extracted_k141_117358 | Not-determined |
| S17_extracted_k141_110462 | Low-quality |
| S17_extracted_k141_110463 | Not-determined |
| S17_extracted_k141_131260 | Low-quality |
| S17_extracted_k141_43631 | Not-determined |
| S17_extracted_k141_110465 | Not-determined |
| S17_extracted_k141_175325 | Low-quality |
| S17_extracted_k141_86737 | Not-determined |
| S17_extracted_k141_43635 | Not-determined |
| S17_extracted_k141_175327 | Low-quality |
| S17_extracted_k141_154660 | Not-determined |
| S17_extracted_k141_43639 | Not-determined |
| S17_extracted_k141_58074 | Not-determined |
| S17_extracted_k141_161619 | Not-determined |
| S17_extracted_k141_110473 | Not-determined |
| S17_extracted_k141_117369 | Low-quality |
| S17_extracted_k141_131274 | Not-determined |
| S17_extracted_k141_64546 | Low-quality |
| S17_extracted_k141_110482 | Not-determined |
| S17_extracted_k141_110483 | Not-determined |
| S17_extracted_k141_86748 | Not-determined |
| S17_extracted_k141_58076 | Not-determined |
| S17_extracted_k141_110485 | Not-determined |
| S17_extracted_k141_6265 | Not-determined |
| S17_extracted_k141_94387 | Low-quality |
| S17_extracted_k141_161625 | Low-quality |
| S17_extracted_k141_64551 | Not-determined |
| S17_extracted_k141_72761 | Not-determined |
| S17_extracted_k141_72763 | Not-determined |
| S17_extracted_k141_94388 | Not-determined |
| S17_extracted_k141_27642 | Not-determined |
| S17_extracted_k141_131286 | Not-determined |
| S17_extracted_k141_161632 | Not-determined |
| S17_extracted_k141_168481 | Not-determined |
| S17_extracted_k141_50836 | Not-determined |
| S17_extracted_k141_175350 | Not-determined |
| S17_extracted_k141_146684 | Not-determined |
| S17_extracted_k141_168487 | Not-determined |
| S17_extracted_k141_43655 | Not-determined |
| S17_extracted_k141_72769 | Low-quality |
| S17_extracted_k141_146686 | Not-determined |
| S17_extracted_k141_146689 | Not-determined |
| S17_extracted_k141_117384 | Low-quality |
| S17_extracted_k141_80120 | Not-determined |
| S17_extracted_k141_58091 | Not-determined |
| S17_extracted_k141_20413 | Not-determined |
| S17_extracted_k141_72773 | Not-determined |
| S17_extracted_k141_27647 | Not-determined |
| S17_extracted_k141_154670 | Not-determined |
| S17_extracted_k141_27649 | Not-determined |
| S17_extracted_k141_64571 | Not-determined |
| S17_extracted_k141_43664 | Not-determined |
| S17_extracted_k141_124051 | Not-determined |
| S17_extracted_k141_20417 | Not-determined |
| S17_extracted_k141_20418 | Not-determined |
| S17_extracted_k141_161642 | Not-determined |
| S17_extracted_k141_72785 | Not-determined |
| S17_extracted_k141_117398 | Not-determined |
| S17_extracted_k141_110500 | Not-determined |
| S17_extracted_k141_50852 | Not-determined |
| S17_extracted_k141_50853 | Not-determined |
| S17_extracted_k141_161647 | Not-determined |
| S17_extracted_k141_117405 | Not-determined |
| S17_extracted_k141_27658 | Not-determined |
| S17_extracted_k141_72793 | Not-determined |
| S17_extracted_k141_27663 | Not-determined |
| S17_extracted_k141_154678 | Low-quality |
| S17_extracted_k141_43673 | Not-determined |
| S17_extracted_k141_86767 | Not-determined |
| S17_extracted_k141_36263 | Not-determined |
| S17_extracted_k141_94409 | Not-determined |
| S17_extracted_k141_102497 | Not-determined |
| S17_extracted_k141_36264 | Not-determined |
| S17_extracted_k141_43677 | Not-determined |
| S17_extracted_k141_72799 | Not-determined |
| S17_extracted_k141_72802 | Not-determined |
| S17_extracted_k141_146710 | Low-quality |
| S17_extracted_k141_117414 | Low-quality |
| S17_extracted_k141_36276 | Not-determined |
| S17_extracted_k141_124061 | Not-determined |
| S17_extracted_k141_161655 | Not-determined |
| S17_extracted_k141_161656 | Not-determined |
| S17_extracted_k141_80139 | Not-determined |
| S17_extracted_k141_124062 | Not-determined |
| S17_extracted_k141_146716 | Not-determined |
| S17_extracted_k141_124063 | Not-determined |
| S17_extracted_k141_94418 | Not-determined |
| S17_extracted_k141_175367 | Not-determined |
| S17_extracted_k141_80142 | Not-determined |
| S17_extracted_k141_43682 | Not-determined |
| S17_extracted_k141_146717 | Not-determined |
| S17_extracted_k141_117419 | Not-determined |
| S17_extracted_k141_102511 | Not-determined |
| S17_extracted_k141_161661 | Low-quality |
| S17_extracted_k141_117422 | Not-determined |
| S17_extracted_k141_86783 | Not-determined |
| S17_extracted_k141_72817 | Low-quality |
| S17_extracted_k141_131321 | Not-determined |
| S17_extracted_k141_139064 | Not-determined |
| S17_extracted_k141_64599 | Not-determined |
| S17_extracted_k141_6297 | Not-determined |
| S17_extracted_k141_58102 | Not-determined |
| S17_extracted_k141_117425 | Not-determined |
| S17_extracted_k141_64601 | Not-determined |
| S17_extracted_k141_161666 | Not-determined |
| S17_extracted_k141_131324 | Not-determined |
| S17_extracted_k141_6300 | Not-determined |
| S17_extracted_k141_102519 | Low-quality |
| S17_extracted_k141_175380 | Not-determined |
| S17_extracted_k141_175381 | Not-determined |
| S17_extracted_k141_124072 | Not-determined |
| S17_extracted_k141_80156 | Not-determined |
| S17_extracted_k141_124075 | Not-determined |
| S17_extracted_k141_86796 | Not-determined |
| S17_extracted_k141_146731 | Not-determined |
| S17_extracted_k141_154681 | Not-determined |
| S17_extracted_k141_146732 | Not-determined |
| S17_extracted_k141_154682 | Not-determined |
| S17_extracted_k141_36283 | Low-quality |
| S17_extracted_k141_72831 | Low-quality |
| S17_extracted_k141_131327 | Not-determined |
| S17_extracted_k141_124078 | Not-determined |
| S17_extracted_k141_27688 | Not-determined |
| S17_extracted_k141_102531 | Not-determined |
| S17_extracted_k141_161677 | Low-quality |
| S17_extracted_k141_27690 | Not-determined |
| S17_extracted_k141_154688 | Not-determined |
| S17_extracted_k141_58112 | Not-determined |
| S17_extracted_k141_58113 | Low-quality |
| S17_extracted_k141_27691 | Not-determined |
| S17_extracted_k141_146742 | Not-determined |
| S17_extracted_k141_102532 | Not-determined |
| S17_extracted_k141_43689 | Not-determined |
| S17_extracted_k141_86809 | Not-determined |
| S17_extracted_k141_146748 | Not-determined |
| S17_extracted_k141_161689 | Not-determined |
| S17_extracted_k141_102538 | Not-determined |
| S17_extracted_k141_110518 | Low-quality |
| S17_extracted_k141_175389 | Not-determined |
| S17_extracted_k141_80178 | Low-quality |
| S17_extracted_k141_36303 | Not-determined |
| S17_extracted_k141_110520 | Not-determined |
| S17_extracted_k141_131340 | Not-determined |
| S17_extracted_k141_154697 | Not-determined |
| S17_extracted_k141_131341 | Not-determined |
| S17_extracted_k141_131342 | Low-quality |
| S17_extracted_k141_154699 | Not-determined |
| S17_extracted_k141_131343 | Not-determined |
| S17_extracted_k141_102543 | Not-determined |
| S17_extracted_k141_72838 | Low-quality |
| S17_extracted_k141_131345 | Not-determined |
| S17_extracted_k141_86820 | Not-determined |
| S17_extracted_k141_86821 | Not-determined |
| S17_extracted_k141_102551 | Not-determined |
| S17_extracted_k141_175397 | Not-determined |
| S17_extracted_k141_72845 | Not-determined |
| S17_extracted_k141_50886 | Not-determined |
| S17_extracted_k141_80187 | Low-quality |
| S17_extracted_k141_175400 | Not-determined |
| S17_extracted_k141_50887 | Not-determined |
| S17_extracted_k141_20474 | Not-determined |
| S17_extracted_k141_110529 | Not-determined |
| S17_extracted_k141_13355 | Not-determined |
| S17_extracted_k141_139089 | Not-determined |
| S17_extracted_k141_110531 | Not-determined |
| S17_extracted_k141_139090 | Not-determined |
| S17_extracted_k141_110532 | Not-determined |
| S17_extracted_k141_139093 | Not-determined |
| S17_extracted_k141_117441 | Not-determined |
| S17_extracted_k141_175404 | Low-quality |
| S17_extracted_k141_110537 | Not-determined |
| S17_extracted_k141_175406 | Not-determined |
| S17_extracted_k141_27716 | Not-determined |
| S17_extracted_k141_139099 | Low-quality |
| S17_extracted_k141_6335 | Not-determined |
| S17_extracted_k141_175411 | Not-determined |
| S17_extracted_k141_154711 | Not-determined |
| S17_extracted_k141_43705 | Not-determined |
| S17_extracted_k141_175412 | Not-determined |
| S17_extracted_k141_43708 | Not-determined |
| S17_extracted_k141_117453 | Not-determined |
| S17_extracted_k141_146782 | Low-quality |
| S17_extracted_k141_80199 | Not-determined |
| S17_extracted_k141_102565 | Low-quality |
| S17_extracted_k141_80202 | Not-determined |
| S17_extracted_k141_102566 | Not-determined |
| S17_extracted_k141_43710 | Not-determined |
| S17_extracted_k141_58149 | Not-determined |
| S17_extracted_k141_58150 | Not-determined |
| S17_extracted_k141_146789 | Not-determined |
| S17_extracted_k141_139111 | Not-determined |
| S17_extracted_k141_139112 | Low-quality |
| S17_extracted_k141_80208 | Not-determined |
| S17_extracted_k141_146793 | Not-determined |
| S17_extracted_k141_124125 | Not-determined |
| S17_extracted_k141_6350 | Not-determined |
| S17_extracted_k141_139119 | Low-quality |
| S17_extracted_k141_146795 | Not-determined |
| S17_extracted_k141_139122 | Not-determined |
| S17_extracted_k141_154724 | Not-determined |
| S17_extracted_k141_139123 | Not-determined |
| S17_extracted_k141_72875 | Not-determined |
| S17_extracted_k141_80211 | Low-quality |
| S17_extracted_k141_6354 | Not-determined |
| S17_extracted_k141_43716 | Low-quality |
| S17_extracted_k141_124130 | Not-determined |
| S17_extracted_k141_110551 | Not-determined |
| S17_extracted_k141_6355 | Not-determined |
| S17_extracted_k141_20488 | Low-quality |
| S17_extracted_k141_20490 | Not-determined |
| S17_extracted_k141_6356 | Not-determined |
| S17_extracted_k141_72881 | Not-determined |
| S17_extracted_k141_50907 | Not-determined |
| S17_extracted_k141_6359 | Not-determined |
| S17_extracted_k141_13382 | Not-determined |
| S17_extracted_k141_43722 | Not-determined |
| S17_extracted_k141_80217 | Not-determined |
| S17_extracted_k141_124137 | Low-quality |
| S17_extracted_k141_124140 | Not-determined |
| S17_extracted_k141_175431 | Low-quality |
| S17_extracted_k141_27724 | Not-determined |
| S17_extracted_k141_80219 | Not-determined |
| S17_extracted_k141_110556 | Not-determined |
| S17_extracted_k141_43725 | Not-determined |
| S17_extracted_k141_131378 | Not-determined |
| S17_extracted_k141_110558 | Low-quality |
| S17_extracted_k141_72885 | Not-determined |
| S17_extracted_k141_43728 | Not-determined |
| S17_extracted_k141_86873 | Not-determined |
| S17_extracted_k141_146808 | Not-determined |
| S17_extracted_k141_72888 | Not-determined |
| S17_extracted_k141_102583 | Not-determined |
| S17_extracted_k141_72890 | Not-determined |
| S17_extracted_k141_58177 | Not-determined |
| S17_extracted_k141_124149 | Not-determined |
| S17_extracted_k141_72892 | Not-determined |
| S17_extracted_k141_72893 | Not-determined |
| S17_extracted_k141_13393 | Low-quality |
| S17_extracted_k141_72894 | Not-determined |
| S17_extracted_k141_80228 | Not-determined |
| S17_extracted_k141_175434 | Not-determined |
| S17_extracted_k141_72895 | Not-determined |
| S17_extracted_k141_13396 | Low-quality |
| S17_extracted_k141_6382 | Not-determined |
| S17_extracted_k141_94434 | Low-quality |
| S17_extracted_k141_27734 | Not-determined |
| S17_extracted_k141_50918 | Not-determined |
| S17_extracted_k141_110567 | Not-determined |
| S17_extracted_k141_13400 | Not-determined |
| S17_extracted_k141_168499 | Not-determined |
| S17_extracted_k141_72901 | Not-determined |
| S17_extracted_k141_168500 | Not-determined |
| S17_extracted_k141_36325 | Not-determined |
| S17_extracted_k141_13405 | Low-quality |
| S17_extracted_k141_124159 | Not-determined |
| S17_extracted_k141_20501 | Not-determined |
| S17_extracted_k141_146814 | Not-determined |
| S17_extracted_k141_168503 | Not-determined |
| S17_extracted_k141_131380 | Not-determined |
| S17_extracted_k141_168505 | Not-determined |
| S17_extracted_k141_86888 | Not-determined |
| S17_extracted_k141_27740 | Not-determined |
| S17_extracted_k141_36330 | Not-determined |
| S17_extracted_k141_50921 | Not-determined |
| S17_extracted_k141_36332 | Not-determined |
| S17_extracted_k141_43734 | Not-determined |
| S17_extracted_k141_50925 | Not-determined |
| S17_extracted_k141_6391 | Low-quality |
| S17_extracted_k141_72902 | Low-quality |
| S17_extracted_k141_43742 | Not-determined |
| S17_extracted_k141_102593 | Low-quality |
| S17_extracted_k141_50937 | Not-determined |
| S17_extracted_k141_20508 | Low-quality |
| S17_extracted_k141_80254 | Not-determined |
| S17_extracted_k141_154754 | Not-determined |
| S17_extracted_k141_168517 | Not-determined |
| S17_extracted_k141_175449 | Not-determined |
| S17_extracted_k141_13428 | Not-determined |
| S17_extracted_k141_168523 | Not-determined |
| S17_extracted_k141_86898 | Not-determined |
| S17_extracted_k141_6393 | Low-quality |
| S17_extracted_k141_117487 | Not-determined |
| S17_extracted_k141_175451 | Not-determined |
| S17_extracted_k141_131406 | Not-determined |
| S17_extracted_k141_36347 | Not-determined |
| S17_extracted_k141_72918 | Not-determined |
| S17_extracted_k141_6401 | Low-quality |
| S17_extracted_k141_154757 | Not-determined |
| S17_extracted_k141_146837 | Low-quality |
| S17_extracted_k141_154759 | Not-determined |
| S17_extracted_k141_146838 | Not-determined |
| S17_extracted_k141_175463 | Not-determined |
| S17_extracted_k141_139134 | Low-quality |
| S17_extracted_k141_102607 | Low-quality |
| S17_extracted_k141_86912 | Not-determined |
| S17_extracted_k141_168537 | Not-determined |
| S17_extracted_k141_146844 | Not-determined |
| S17_extracted_k141_6410 | Not-determined |
| S17_extracted_k141_175465 | Not-determined |
| S17_extracted_k141_58200 | Not-determined |
| S17_extracted_k141_13445 | Not-determined |
| S17_extracted_k141_110591 | Not-determined |
| S17_extracted_k141_50964 | Not-determined |
| S17_extracted_k141_72928 | Not-determined |
| S17_extracted_k141_50967 | Not-determined |
| S17_extracted_k141_175474 | Not-determined |
| S17_extracted_k141_139138 | Not-determined |
| S17_extracted_k141_168544 | Not-determined |
| S17_extracted_k141_86919 | Not-determined |
| S17_extracted_k141_154761 | Not-determined |
| S17_extracted_k141_6414 | Not-determined |
| S17_extracted_k141_80279 | Not-determined |
| S17_extracted_k141_20525 | Not-determined |
| S17_extracted_k141_20526 | Not-determined |
| S17_extracted_k141_50972 | Not-determined |
| S17_extracted_k141_20527 | Not-determined |
| S17_extracted_k141_27767 | Low-quality |
| S17_extracted_k141_27768 | Not-determined |
| S17_extracted_k141_13454 | Not-determined |
| S17_extracted_k141_168554 | Not-determined |
| S17_extracted_k141_94451 | Low-quality |
| S17_extracted_k141_110601 | Not-determined |
| S17_extracted_k141_6418 | Not-determined |
| S17_extracted_k141_175489 | Not-determined |

| | |
|---|---|
| S17_extracted_k141_117503 | Not-determined |
| S17_extracted_k141_154771 | Not-determined |
| S17_extracted_k141_117505 | Not-determined |
| S17_extracted_k141_131424 | Not-determined |
| S17_extracted_k141_117506 | Not-determined |
| S17_extracted_k141_94458 | Not-determined |
| S17_extracted_k141_139147 | Not-determined |
| S17_extracted_k141_86930 | Not-determined |
| S17_extracted_k141_139148 | Not-determined |
| S17_extracted_k141_94461 | Not-determined |
| S17_extracted_k141_27771 | Not-determined |
| S17_extracted_k141_43761 | Not-determined |
| S17_extracted_k141_43762 | Not-determined |
| S17_extracted_k141_58216 | Not-determined |
| S17_extracted_k141_102636 | Not-determined |
| S17_extracted_k141_20531 | Not-determined |
| S17_extracted_k141_161750 | Not-determined |
| S17_extracted_k141_43764 | Not-determined |
| S17_extracted_k141_139153 | Not-determined |
| S17_extracted_k141_80291 | Not-determined |
| S17_extracted_k141_36367 | Not-determined |
| S17_extracted_k141_102641 | Not-determined |
| S17_extracted_k141_94468 | Not-determined |
| S17_extracted_k141_154782 | Not-determined |
| S17_extracted_k141_13466 | Not-determined |
| S17_extracted_k141_94470 | Not-determined |
| S17_extracted_k141_124205 | Not-determined |
| S17_extracted_k141_43769 | Not-determined |
| S17_extracted_k141_80296 | Not-determined |
| S17_extracted_k141_13470 | Not-determined |
| S17_extracted_k141_124211 | Low-quality |
| S17_extracted_k141_58227 | Not-determined |
| S17_extracted_k141_6436 | Not-determined |
| S17_extracted_k141_80305 | Not-determined |
| S17_extracted_k141_102651 | Not-determined |
| S17_extracted_k141_50993 | Not-determined |
| S17_extracted_k141_146882 | Low-quality |
| S17_extracted_k141_80308 | Not-determined |
| S17_extracted_k141_58229 | Low-quality |
| S17_extracted_k141_86938 | Not-determined |
| S17_extracted_k141_13476 | Not-determined |
| S17_extracted_k141_13477 | Not-determined |
| S17_extracted_k141_80310 | Not-determined |
| S17_extracted_k141_161764 | Not-determined |
| S17_extracted_k141_20541 | Not-determined |
| S17_extracted_k141_86940 | Not-determined |
| S17_extracted_k141_13480 | Not-determined |
| S17_extracted_k141_86942 | Not-determined |
| S17_extracted_k141_43779 | Low-quality |
| S17_extracted_k141_36377 | Not-determined |
| S17_extracted_k141_43784 | Not-determined |
| S17_extracted_k141_13486 | Not-determined |
| S17_extracted_k141_27790 | Not-determined |
| S17_extracted_k141_146890 | Not-determined |
| S17_extracted_k141_58235 | Not-determined |
| S17_extracted_k141_161775 | Not-determined |
| S17_extracted_k141_27795 | Low-quality |
| S17_extracted_k141_131451 | Not-determined |
| S17_extracted_k141_27796 | Not-determined |
| S17_extracted_k141_20552 | Low-quality |
| S17_extracted_k141_50996 | Not-determined |
| S17_extracted_k141_139165 | Not-determined |
| S17_extracted_k141_50998 | Not-determined |
| S17_extracted_k141_146901 | Not-determined |
| S17_extracted_k141_58241 | Not-determined |
| S17_extracted_k141_110613 | Low-quality |
| S17_extracted_k141_13493 | Not-determined |
| S17_extracted_k141_58242 | Not-determined |
| S17_extracted_k141_43795 | Not-determined |
| S17_extracted_k141_131457 | Not-determined |
| S17_extracted_k141_27805 | Not-determined |
| S17_extracted_k141_161783 | Not-determined |
| S17_extracted_k141_94481 | Not-determined |
| S17_extracted_k141_131461 | Not-determined |
| S17_extracted_k141_175519 | Not-determined |
| S17_extracted_k141_161785 | Not-determined |
| S17_extracted_k141_110616 | Not-determined |
| S17_extracted_k141_110617 | Not-determined |
| S17_extracted_k141_6463 | Not-determined |
| S17_extracted_k141_94486 | Not-determined |
| S17_extracted_k141_13500 | Not-determined |
| S17_extracted_k141_131469 | Not-determined |
| S17_extracted_k141_110621 | Not-determined |
| S17_extracted_k141_124233 | Not-determined |
| S17_extracted_k141_51007 | Not-determined |
| S17_extracted_k141_72968 | Not-determined |
| S17_extracted_k141_124234 | Not-determined |
| S17_extracted_k141_72969 | Not-determined |
| S17_extracted_k141_102671 | Not-determined |
| S17_extracted_k141_51008 | Not-determined |
| S17_extracted_k141_20565 | Not-determined |
| S17_extracted_k141_27809 | Low-quality |
| S17_extracted_k141_80339 | Not-determined |
| S17_extracted_k141_154807 | Not-determined |
| S17_extracted_k141_110623 | Not-determined |
| S17_extracted_k141_43811 | Not-determined |
| S17_extracted_k141_94494 | Low-quality |
| S17_extracted_k141_175529 | Not-determined |
| S17_extracted_k141_161794 | Not-determined |
| S17_extracted_k141_161795 | Not-determined |
| S17_extracted_k141_124243 | Not-determined |
| S17_extracted_k141_110630 | Not-determined |
| S17_extracted_k141_72979 | Not-determined |
| S17_extracted_k141_86972 | Not-determined |
| S17_extracted_k141_86973 | Not-determined |
| S17_extracted_k141_94502 | Not-determined |
| S17_extracted_k141_146922 | Not-determined |
| S17_extracted_k141_72980 | Low-quality |
| S17_extracted_k141_139176 | Low-quality |
| S17_extracted_k141_175543 | Not-determined |
| S17_extracted_k141_72983 | Not-determined |
| S17_extracted_k141_43823 | Not-determined |
| S17_extracted_k141_168581 | Not-determined |
| S17_extracted_k141_13522 | Not-determined |
| S17_extracted_k141_80355 | Low-quality |
| S17_extracted_k141_110638 | Not-determined |
| S17_extracted_k141_146926 | Not-determined |
| S17_extracted_k141_58259 | Low-quality |
| S17_extracted_k141_161809 | Not-determined |
| S17_extracted_k141_131484 | Not-determined |
| S17_extracted_k141_131485 | Not-determined |
| S17_extracted_k141_80356 | Not-determined |
| S17_extracted_k141_13524 | Low-quality |
| S17_extracted_k141_154812 | Low-quality |
| S17_extracted_k141_58261 | Low-quality |
| S17_extracted_k141_36385 | Not-determined |
| S17_extracted_k141_51027 | Not-determined |
| S17_extracted_k141_110641 | Low-quality |
| S17_extracted_k141_117519 | Not-determined |
| S17_extracted_k141_161815 | Not-determined |
| S17_extracted_k141_36388 | Not-determined |
| S17_extracted_k141_43834 | Low-quality |
| S17_extracted_k141_168592 | Not-determined |
| S17_extracted_k141_20577 | Not-determined |
| S17_extracted_k141_110642 | Not-determined |
| S17_extracted_k141_51032 | Not-determined |
| S17_extracted_k141_117523 | Not-determined |
| S17_extracted_k141_154821 | Not-determined |
| S17_extracted_k141_154824 | Not-determined |
| S17_extracted_k141_86989 | Not-determined |
| S17_extracted_k141_102696 | Not-determined |
| S17_extracted_k141_154826 | Not-determined |
| S17_extracted_k141_161826 | Not-determined |
| S17_extracted_k141_161827 | Low-quality |
| S17_extracted_k141_146940 | Not-determined |
| S17_extracted_k141_154830 | Not-determined |
| S17_extracted_k141_102699 | Low-quality |
| S17_extracted_k141_80360 | Not-determined |
| S17_extracted_k141_43847 | Not-determined |
| S17_extracted_k141_80362 | Not-determined |
| S17_extracted_k141_20584 | Not-determined |
| S17_extracted_k141_86994 | Not-determined |
| S17_extracted_k141_117531 | Not-determined |
| S17_extracted_k141_86995 | Low-quality |
| S17_extracted_k141_20585 | Not-determined |
| S17_extracted_k141_51044 | Not-determined |
| S17_extracted_k141_124254 | Not-determined |
| S17_extracted_k141_6477 | Not-determined |
| S17_extracted_k141_36399 | Not-determined |
| S17_extracted_k141_51045 | Not-determined |
| S17_extracted_k141_124259 | Not-determined |
| S17_extracted_k141_72999 | Not-determined |
| S17_extracted_k141_102708 | Not-determined |
| S17_extracted_k141_6480 | Not-determined |
| S17_extracted_k141_51046 | Not-determined |
| S17_extracted_k141_51048 | Not-determined |
| S17_extracted_k141_13545 | Not-determined |
| S17_extracted_k141_117540 | Not-determined |
| S17_extracted_k141_36409 | Not-determined |
| S17_extracted_k141_13551 | Not-determined |
| S17_extracted_k141_86999 | Not-determined |
| S17_extracted_k141_94523 | Not-determined |
| S17_extracted_k141_36412 | Not-determined |
| S17_extracted_k141_110645 | Not-determined |
| S17_extracted_k141_110646 | Not-determined |
| S17_extracted_k141_6489 | Not-determined |
| S17_extracted_k141_102719 | Low-quality |
| S17_extracted_k141_124273 | Not-determined |
| S17_extracted_k141_51059 | Low-quality |
| S17_extracted_k141_146945 | Not-determined |
| S17_extracted_k141_87000 | Not-determined |
| S17_extracted_k141_94528 | Not-determined |
| S17_extracted_k141_43865 | Not-determined |
| S17_extracted_k141_87004 | Not-determined |
| S17_extracted_k141_20591 | Low-quality |
| S17_extracted_k141_6496 | Not-determined |
| S17_extracted_k141_6497 | Not-determined |
| S17_extracted_k141_20592 | Low-quality |
| S17_extracted_k141_20595 | Not-determined |
| S17_extracted_k141_124281 | Not-determined |
| S17_extracted_k141_110654 | Not-determined |
| S17_extracted_k141_80389 | Not-determined |
| S17_extracted_k141_6505 | Not-determined |
| S17_extracted_k141_102730 | Low-quality |
| S17_extracted_k141_117553 | Low-quality |
| S17_extracted_k141_27845 | Not-determined |
| S17_extracted_k141_117555 | Low-quality |
| S17_extracted_k141_80394 | Not-determined |
| S17_extracted_k141_117558 | Not-determined |
| S17_extracted_k141_27848 | Not-determined |
| S17_extracted_k141_27849 | Not-determined |
| S17_extracted_k141_168623 | Not-determined |
| S17_extracted_k141_27850 | Not-determined |
| S17_extracted_k141_161857 | Not-determined |
| S17_extracted_k141_13557 | Not-determined |
| S17_extracted_k141_146964 | Not-determined |
| S17_extracted_k141_161860 | Not-determined |
| S17_extracted_k141_27855 | Not-determined |
| S17_extracted_k141_6523 | Not-determined |
| S17_extracted_k141_13561 | Not-determined |
| S17_extracted_k141_110664 | Not-determined |
| S17_extracted_k141_117563 | Low-quality |
| S17_extracted_k141_27857 | Not-determined |
| S17_extracted_k141_20616 | Not-determined |
| S17_extracted_k141_117569 | Not-determined |
| S17_extracted_k141_102747 | Not-determined |
| S17_extracted_k141_13564 | Not-determined |
| S17_extracted_k141_161865 | Not-determined |
| S17_extracted_k141_13565 | Not-determined |
| S17_extracted_k141_168640 | Not-determined |
| S17_extracted_k141_58279 | Not-determined |
| S17_extracted_k141_102751 | Low-quality |
| S17_extracted_k141_43877 | Not-determined |
| S17_extracted_k141_146982 | Not-determined |
| S17_extracted_k141_80398 | Low-quality |
| S17_extracted_k141_58285 | Not-determined |
| S17_extracted_k141_131534 | Not-determined |
| S17_extracted_k141_139198 | Not-determined |
| S17_extracted_k141_131535 | Not-determined |
| S17_extracted_k141_20624 | Not-determined |
| S17_extracted_k141_58289 | Not-determined |
| S17_extracted_k141_117574 | Not-determined |
| S17_extracted_k141_117576 | Not-determined |
| S17_extracted_k141_73031 | Not-determined |
| S17_extracted_k141_27862 | Not-determined |
| S17_extracted_k141_6538 | Not-determined |
| S17_extracted_k141_161871 | Not-determined |
| S17_extracted_k141_80408 | Not-determined |
| S17_extracted_k141_73036 | Not-determined |
| S17_extracted_k141_117581 | Not-determined |
| S17_extracted_k141_51086 | Low-quality |
| S17_extracted_k141_58295 | Not-determined |
| S17_extracted_k141_43896 | Low-quality |
| S17_extracted_k141_58297 | Not-determined |
| S17_extracted_k141_73041 | Low-quality |
| S17_extracted_k141_161882 | Not-determined |
| S17_extracted_k141_27867 | Low-quality |
| S17_extracted_k141_13587 | Low-quality |
| S17_extracted_k141_43901 | Not-determined |
| S17_extracted_k141_139211 | Not-determined |
| S17_extracted_k141_27876 | Not-determined |
| S17_extracted_k141_139213 | Low-quality |
| S17_extracted_k141_80423 | Not-determined |
| S17_extracted_k141_51095 | Not-determined |
| S17_extracted_k141_27879 | Not-determined |
| S17_extracted_k141_102768 | Low-quality |
| S17_extracted_k141_161893 | Not-determined |
| S17_extracted_k141_147019 | Not-determined |
| S17_extracted_k141_161894 | Not-determined |
| S17_extracted_k141_147020 | Not-determined |
| S17_extracted_k141_102771 | Not-determined |
| S17_extracted_k141_168655 | Not-determined |
| S17_extracted_k141_124319 | Not-determined |
| S17_extracted_k141_161897 | Not-determined |
| S17_extracted_k141_27886 | Low-quality |
| S17_extracted_k141_147026 | Not-determined |
| S17_extracted_k141_117606 | Not-determined |
| S17_extracted_k141_168660 | Not-determined |
| S17_extracted_k141_168661 | Not-determined |
| S17_extracted_k141_117607 | Not-determined |
| S17_extracted_k141_27890 | Not-determined |
| S17_extracted_k141_94535 | Not-determined |
| S17_extracted_k141_161904 | Not-determined |
| S17_extracted_k141_168671 | Not-determined |
| S17_extracted_k141_124325 | Not-determined |
| S17_extracted_k141_124326 | Not-determined |
| S17_extracted_k141_94543 | Not-determined |
| S17_extracted_k141_36443 | Not-determined |
| S17_extracted_k141_117615 | Not-determined |
| S17_extracted_k141_80441 | Not-determined |
| S17_extracted_k141_43919 | Not-determined |
| S17_extracted_k141_94547 | Low-quality |
| S17_extracted_k141_168672 | Not-determined |
| S17_extracted_k141_58324 | Not-determined |
| S17_extracted_k141_51110 | Not-determined |
| S17_extracted_k141_58325 | Not-determined |
| S17_extracted_k141_36450 | Not-determined |
| S17_extracted_k141_102788 | Not-determined |
| S17_extracted_k141_154887 | Not-determined |
| S17_extracted_k141_161916 | Not-determined |
| S17_extracted_k141_94549 | Not-determined |
| S17_extracted_k141_94550 | Not-determined |
| S17_extracted_k141_51119 | Not-determined |
| S17_extracted_k141_124336 | Not-determined |
| S17_extracted_k141_80450 | Not-determined |
| S17_extracted_k141_161922 | Not-determined |
| S17_extracted_k141_80451 | Not-determined |
| S17_extracted_k141_131552 | Not-determined |
| S17_extracted_k141_147049 | Not-determined |
| S17_extracted_k141_124342 | Not-determined |
| S17_extracted_k141_43923 | Not-determined |
| S17_extracted_k141_94559 | Not-determined |
| S17_extracted_k141_147053 | Not-determined |
| S17_extracted_k141_131557 | Not-determined |
| S17_extracted_k141_168688 | Not-determined |
| S17_extracted_k141_102805 | Not-determined |
| S17_extracted_k141_124346 | Not-determined |
| S17_extracted_k141_124348 | Not-determined |
| S17_extracted_k141_43931 | Low-quality |
| S17_extracted_k141_43932 | Not-determined |
| S17_extracted_k141_131559 | Not-determined |
| S17_extracted_k141_154901 | Not-determined |
| S17_extracted_k141_51134 | Not-determined |
| S17_extracted_k141_51135 | Not-determined |
| S17_extracted_k141_168693 | Low-quality |
| S17_extracted_k141_94567 | Not-determined |
| S17_extracted_k141_94569 | Not-determined |
| S17_extracted_k141_43938 | Not-determined |
| S17_extracted_k141_147063 | Not-determined |
| S17_extracted_k141_147064 | Not-determined |
| S17_extracted_k141_27912 | Low-quality |
| S17_extracted_k141_124349 | Not-determined |
| S11_vs_k141_104068\|\|full | High-quality |
| S17_extracted_k141_43941 | Not-determined |
| S17_extracted_k141_13601 | Not-determined |
| S17_extracted_k141_6569 | Not-determined |
| S17_extracted_k141_51143 | Not-determined |
| S17_extracted_k141_13603 | Not-determined |
| S17_extracted_k141_51144 | Not-determined |
| S17_extracted_k141_161946 | Not-determined |
| S17_extracted_k141_13605 | Not-determined |
| S17_extracted_k141_73060 | Not-determined |
| S17_extracted_k141_36460 | Not-determined |
| S17_extracted_k141_6572 | Not-determined |
| S17_extracted_k141_27925 | Not-determined |
| S17_extracted_k141_102814 | Low-quality |
| S17_extracted_k141_27927 | Low-quality |
| S17_extracted_k141_27928 | Not-determined |
| S17_extracted_k141_147078 | Not-determined |
| S17_extracted_k141_51157 | Not-determined |
| S17_extracted_k141_27929 | Not-determined |
| S17_extracted_k141_94584 | Not-determined |
| S17_extracted_k141_147083 | Not-determined |
| S17_extracted_k141_168710 | Not-determined |
| S17_extracted_k141_102816 | Low-quality |
| S17_extracted_k141_94588 | Not-determined |
| S17_extracted_k141_13616 | Not-determined |
| S17_extracted_k141_94590 | Not-determined |
| S17_extracted_k141_147092 | Not-determined |
| S17_extracted_k141_147093 | Not-determined |
| S17_extracted_k141_102823 | Not-determined |
| S17_extracted_k141_154921 | Not-determined |
| S17_extracted_k141_6584 | Low-quality |
| S17_extracted_k141_94594 | Not-determined |
| S17_extracted_k141_27941 | Not-determined |
| S17_extracted_k141_175557 | Not-determined |
| S17_extracted_k141_139242 | Not-determined |
| S17_extracted_k141_131578 | Low-quality |
| S17_extracted_k141_139245 | Not-determined |
| S17_extracted_k141_80481 | Not-determined |
| S17_extracted_k141_102830 | Not-determined |
| S17_extracted_k141_131581 | Not-determined |
| S17_extracted_k141_131584 | Not-determined |
| S17_extracted_k141_154934 | Not-determined |
| S17_extracted_k141_80486 | Not-determined |
| S17_extracted_k141_51171 | Low-quality |
| S17_extracted_k141_139254 | Not-determined |
| S17_extracted_k141_175562 | Not-determined |
| S17_extracted_k141_102840 | Not-determined |
| S17_extracted_k141_94597 | Not-determined |
| S17_extracted_k141_13637 | Not-determined |
| S17_extracted_k141_175564 | Not-determined |
| S17_extracted_k141_36478 | Low-quality |
| S17_extracted_k141_80488 | Not-determined |
| S17_extracted_k141_94599 | Not-determined |
| S17_extracted_k141_102842 | Low-quality |
| S17_extracted_k141_51175 | Not-determined |
| S17_extracted_k141_51176 | Not-determined |
| S17_extracted_k141_161961 | Not-determined |
| S17_extracted_k141_147118 | Not-determined |
| S17_extracted_k141_27958 | Low-quality |
| S17_extracted_k141_87086 | Not-determined |
| S17_extracted_k141_147119 | Not-determined |
| S17_extracted_k141_161963 | Not-determined |
| S17_extracted_k141_139261 | Not-determined |
| S17_extracted_k141_110674 | Not-determined |
| S17_extracted_k141_147125 | Not-determined |
| S17_extracted_k141_73091 | Not-determined |
| S17_extracted_k141_131592 | Not-determined |
| S17_extracted_k141_168736 | Not-determined |
| S17_extracted_k141_161971 | Not-determined |
| S17_extracted_k141_13651 | Not-determined |
| S17_extracted_k141_102855 | Not-determined |
| S17_extracted_k141_175575 | Not-determined |
| S17_extracted_k141_168738 | Not-determined |
| S17_extracted_k141_175578 | Not-determined |
| S17_extracted_k141_154964 | Not-determined |
| S17_extracted_k141_13654 | Not-determined |
| S17_extracted_k141_102861 | Not-determined |
| S17_extracted_k141_80497 | Not-determined |
| S17_extracted_k141_6597 | Low-quality |
| S17_extracted_k141_94614 | Not-determined |
| S17_extracted_k141_110681 | Not-determined |
| S17_extracted_k141_102864 | Not-determined |
| S17_extracted_k141_102865 | Not-determined |
| S17_extracted_k141_94618 | Not-determined |
| S17_extracted_k141_102868 | Not-determined |
| S17_extracted_k141_94619 | Not-determined |
| S17_extracted_k141_87097 | Not-determined |
| S17_extracted_k141_175589 | Not-determined |
| S17_extracted_k141_147138 | Not-determined |
| S17_extracted_k141_102870 | Not-determined |
| S17_extracted_k141_102871 | Not-determined |
| S17_extracted_k141_87098 | Low-quality |
| S17_extracted_k141_43986 | Not-determined |
| S17_extracted_k141_43988 | Not-determined |
| S17_extracted_k141_175592 | Not-determined |
| S17_extracted_k141_110684 | Not-determined |
| S17_extracted_k141_110685 | Not-determined |
| S17_extracted_k141_175594 | Not-determined |
| S17_extracted_k141_110686 | Not-determined |
| S17_extracted_k141_51193 | Not-determined |
| S17_extracted_k141_102881 | Low-quality |
| S17_extracted_k141_87102 | Not-determined |
| S17_extracted_k141_147144 | Not-determined |
| S17_extracted_k141_73103 | Not-determined |
| S17_extracted_k141_131611 | Not-determined |
| S17_extracted_k141_168745 | Not-determined |
| S17_extracted_k141_43990 | Not-determined |
| S17_extracted_k141_124373 | Not-determined |
| S17_extracted_k141_27966 | Not-determined |
| S17_extracted_k141_43994 | Not-determined |
| S17_extracted_k141_27967 | Low-quality |
| S17_extracted_k141_154972 | Not-determined |
| S17_extracted_k141_139277 | Not-determined |
| S17_extracted_k141_175601 | Not-determined |
| S17_extracted_k141_13674 | Low-quality |
| S17_extracted_k141_154975 | Not-determined |
| S17_extracted_k141_43999 | Not-determined |
| S17_extracted_k141_73113 | Not-determined |
| S17_extracted_k141_73114 | Low-quality |
| S17_extracted_k141_139286 | Not-determined |
| S17_extracted_k141_6603 | Not-determined |
| S17_extracted_k141_87110 | Not-determined |
| S17_extracted_k141_6605 | Not-determined |
| S17_extracted_k141_147152 | Not-determined |
| S17_extracted_k141_110697 | Not-determined |
| S17_extracted_k141_110698 | Not-determined |
| S17_extracted_k141_51206 | Not-determined |
| S17_extracted_k141_27982 | Not-determined |
| S17_extracted_k141_168763 | Not-determined |
| S17_extracted_k141_147159 | Not-determined |
| S17_extracted_k141_73127 | Not-determined |
| S17_extracted_k141_124383 | Not-determined |
| S17_extracted_k141_51210 | Not-determined |
| S17_extracted_k141_124385 | Not-determined |
| S17_extracted_k141_36491 | Low-quality |
| S17_extracted_k141_73130 | Not-determined |
| S17_extracted_k141_36493 | Not-determined |
| S17_extracted_k141_6610 | Not-determined |
| S17_extracted_k141_124388 | Not-determined |
| S17_extracted_k141_6611 | Not-determined |
| S17_extracted_k141_175610 | Not-determined |
| S17_extracted_k141_175611 | Not-determined |
| S17_extracted_k141_131629 | Low-quality |
| S17_extracted_k141_6613 | Low-quality |
| S17_extracted_k141_139298 | Not-determined |
| S17_extracted_k141_44012 | Not-determined |
| S17_extracted_k141_124392 | Not-determined |
| S17_extracted_k141_58354 | Not-determined |
| S17_extracted_k141_58359 | Low-quality |
| S17_extracted_k141_102909 | Not-determined |
| S17_extracted_k141_36506 | Not-determined |
| S17_extracted_k141_139303 | Not-determined |
| S17_extracted_k141_80537 | Not-determined |
| S17_extracted_k141_154996 | Not-determined |
| S17_extracted_k141_28005 | Not-determined |
| S17_extracted_k141_168775 | Not-determined |
| S17_extracted_k141_168777 | Not-determined |
| S17_extracted_k141_28006 | Not-determined |
| S17_extracted_k141_168782 | Low-quality |
| S17_extracted_k141_175628 | Not-determined |
| S17_extracted_k141_147174 | Not-determined |
| S17_extracted_k141_168783 | Not-determined |
| S17_extracted_k141_175629 | Not-determined |
| S17_extracted_k141_147178 | Not-determined |
| S17_extracted_k141_58371 | Not-determined |
| S17_extracted_k141_131642 | Not-determined |
| S17_extracted_k141_168786 | Not-determined |
| S17_extracted_k141_36516 | Not-determined |
| S17_extracted_k141_87111 | Not-determined |
| S17_extracted_k141_44022 | Not-determined |
| S17_extracted_k141_155006 | Not-determined |
| S17_extracted_k141_162002 | Not-determined |
| S17_extracted_k141_175637 | Not-determined |
| S17_extracted_k141_36522 | Not-determined |
| S17_extracted_k141_168793 | Not-determined |
| S17_extracted_k141_73150 | Not-determined |
| S17_extracted_k141_155008 | Not-determined |
| S17_extracted_k141_28017 | Not-determined |
| S17_extracted_k141_175638 | Not-determined |
| S17_extracted_k141_6625 | Not-determined |
| S17_extracted_k141_155009 | Not-determined |
| S17_extracted_k141_155010 | Not-determined |
| S17_extracted_k141_44028 | Not-determined |
| S17_extracted_k141_87116 | Not-determined |
| S17_extracted_k141_13706 | Not-determined |
| S17_extracted_k141_168799 | Not-determined |
| S17_extracted_k141_80545 | Not-determined |
| S17_extracted_k141_102915 | Not-determined |
| S17_extracted_k141_131657 | Low-quality |
| S17_extracted_k141_168800 | Not-determined |
| S17_extracted_k141_124400 | Low-quality |
| S17_extracted_k141_175646 | Not-determined |
| S17_extracted_k141_6631 | Not-determined |
| S17_extracted_k141_175648 | Not-determined |
| S17_extracted_k141_139323 | Not-determined |
| S17_extracted_k141_175649 | Not-determined |
| S17_extracted_k141_102917 | Not-determined |
| S17_extracted_k141_175650 | Not-determined |
| S17_extracted_k141_147190 | Not-determined |
| S17_extracted_k141_147195 | Not-determined |
| S17_extracted_k141_51225 | Not-determined |
| S17_extracted_k141_44035 | Not-determined |
| S17_extracted_k141_147197 | Not-determined |
| S17_extracted_k141_51227 | Not-determined |
| S17_extracted_k141_175658 | Low-quality |
| S17_extracted_k141_6636 | Not-determined |
| S17_extracted_k141_102924 | Not-determined |
| S17_extracted_k141_51230 | Not-determined |
| S17_extracted_k141_73162 | Not-determined |
| S17_extracted_k141_73167 | Not-determined |
| S17_extracted_k141_80561 | Not-determined |
| S17_extracted_k141_162022 | Not-determined |
| S17_extracted_k141_73173 | Not-determined |
| S17_extracted_k141_13724 | Not-determined |
| S17_extracted_k141_13725 | Not-determined |
| S17_extracted_k141_110700 | Low-quality |
| S17_extracted_k141_80568 | Not-determined |
| S17_extracted_k141_162024 | Not-determined |
| S17_extracted_k141_175670 | Not-determined |
| S17_extracted_k141_58410 | Not-determined |
| S17_extracted_k141_175671 | Not-determined |
| S17_extracted_k141_51246 | Not-determined |
| S17_extracted_k141_168812 | Low-quality |
| S17_extracted_k141_147212 | Not-determined |
| S17_extracted_k141_51249 | Low-quality |
| S17_extracted_k141_117629 | Low-quality |
| S17_extracted_k141_13733 | Not-determined |
| S17_extracted_k141_80574 | Not-determined |
| S17_extracted_k141_117632 | Low-quality |
| S17_extracted_k141_73175 | Not-determined |
| S17_extracted_k141_117634 | Low-quality |
| S17_extracted_k141_58417 | Not-determined |
| S17_extracted_k141_110712 | Not-determined |
| S17_extracted_k141_87147 | Not-determined |
| S17_extracted_k141_155030 | Not-determined |
| S17_extracted_k141_175679 | Not-determined |
| S17_extracted_k141_87149 | Not-determined |
| S17_extracted_k141_51261 | Not-determined |
| S17_extracted_k141_80579 | Low-quality |
| S17_extracted_k141_131691 | Not-determined |
| S17_extracted_k141_73187 | Not-determined |
| S17_extracted_k141_155034 | Not-determined |
| S17_extracted_k141_73188 | Not-determined |
| S17_extracted_k141_94631 | Low-quality |
| S17_extracted_k141_102951 | Not-determined |
| S17_extracted_k141_131694 | Not-determined |
| S17_extracted_k141_102952 | Not-determined |
| S17_extracted_k141_6659 | Not-determined |
| S17_extracted_k141_175685 | Not-determined |
| S17_extracted_k141_162029 | Not-determined |
| S17_extracted_k141_80581 | Not-determined |
| S17_extracted_k141_139352 | Not-determined |
| S17_extracted_k141_73192 | Not-determined |
| S17_extracted_k141_80583 | Not-determined |
| S17_extracted_k141_73193 | Not-determined |
| S17_extracted_k141_102960 | Not-determined |
| S17_extracted_k141_147222 | Not-determined |
| S17_extracted_k141_73197 | Not-determined |
| S17_extracted_k141_175693 | Low-quality |
| S17_extracted_k141_168830 | Not-determined |
| S17_extracted_k141_87164 | Low-quality |
| S17_extracted_k141_51270 | Not-determined |
| S17_extracted_k141_80590 | Not-determined |
| S17_extracted_k141_58434 | Not-determined |
| S17_extracted_k141_162043 | Not-determined |
| S17_extracted_k141_155044 | Not-determined |
| S17_extracted_k141_58438 | Not-determined |

| | |
|---|---|
| S17_extracted_k141_13741 | Not-determined |
| S17_extracted_k141_58441 | Not-determined |
| S17_extracted_k141_73209 | Not-determined |
| S17_extracted_k141_73210 | Not-determined |
| S17_extracted_k141_155047 | Not-determined |
| S17_extracted_k141_73211 | Not-determined |
| S17_extracted_k141_80597 | Not-determined |
| S17_extracted_k141_73214 | Low-quality |
| S17_extracted_k141_13743 | Not-determined |
| S17_extracted_k141_13745 | Not-determined |
| S17_extracted_k141_87173 | Low-quality |
| S17_extracted_k141_117671 | Not-determined |
| S17_extracted_k141_20652 | Not-determined |
| S17_extracted_k141_87177 | Not-determined |
| S17_extracted_k141_51284 | Not-determined |
| S17_extracted_k141_102976 | Not-determined |
| S17_extracted_k141_87180 | Not-determined |
| S17_extracted_k141_131724 | Not-determined |
| S17_extracted_k141_94646 | Not-determined |
| S17_extracted_k141_102983 | Low-quality |
| S17_extracted_k141_73224 | Low-quality |
| S17_extracted_k141_162053 | Low-quality |
| S17_extracted_k141_147233 | Low-quality |
| S17_extracted_k141_168848 | Not-determined |
| S17_extracted_k141_13755 | Not-determined |
| S17_extracted_k141_117683 | Not-determined |
| S17_extracted_k141_139373 | Not-determined |
| S17_extracted_k141_44056 | Not-determined |
| S17_extracted_k141_168852 | Not-determined |
| S17_extracted_k141_110731 | Not-determined |
| S17_extracted_k141_80620 | Not-determined |
| S17_extracted_k141_117687 | Not-determined |
| S17_extracted_k141_175719 | Not-determined |
| S17_extracted_k141_110738 | Not-determined |
| S17_extracted_k141_175723 | Not-determined |
| S17_extracted_k141_6684 | Not-determined |
| S17_extracted_k141_87196 | Low-quality |
| S17_extracted_k141_124438 | Not-determined |
| S17_extracted_k141_168859 | Not-determined |
| S17_extracted_k141_73226 | Not-determined |
| S17_extracted_k141_58462 | Not-determined |
| S17_extracted_k141_80629 | Not-determined |
| S17_extracted_k141_73228 | Low-quality |
| S17_extracted_k141_58463 | Not-determined |
| S17_extracted_k141_44060 | Not-determined |
| S17_extracted_k141_124442 | Not-determined |
| S17_extracted_k141_58465 | Not-determined |
| S17_extracted_k141_175731 | Not-determined |
| S17_extracted_k141_73232 | Not-determined |
| S17_extracted_k141_51294 | Not-determined |
| S17_extracted_k141_139388 | Not-determined |
| S17_extracted_k141_117696 | Low-quality |
| S17_extracted_k141_175735 | Not-determined |
| S17_extracted_k141_13765 | Not-determined |
| S17_extracted_k141_94666 | Not-determined |
| S17_extracted_k141_58472 | Not-determined |
| S17_extracted_k141_162064 | Not-determined |
| S17_extracted_k141_94669 | Low-quality |
| S17_extracted_k141_87208 | Not-determined |
| S17_extracted_k141_124455 | Not-determined |
| S17_extracted_k141_162072 | Not-determined |
| S17_extracted_k141_175750 | Not-determined |
| S17_extracted_k141_162074 | Not-determined |
| S17_extracted_k141_110742 | Not-determined |
| S17_extracted_k141_124456 | Not-determined |
| S17_extracted_k141_110744 | Low-quality |
| S17_extracted_k141_13772 | Not-determined |
| S17_extracted_k141_73249 | Not-determined |
| S17_extracted_k141_94678 | Not-determined |
| S17_extracted_k141_13775 | Not-determined |
| S17_extracted_k141_13776 | Not-determined |
| S17_extracted_k141_73250 | Not-determined |
| S17_extracted_k141_94680 | Low-quality |
| S17_extracted_k141_94681 | Not-determined |
| S17_extracted_k141_162082 | Not-determined |
| S17_extracted_k141_44073 | Not-determined |
| S17_extracted_k141_36532 | Not-determined |
| S17_extracted_k141_36533 | Not-determined |
| S17_extracted_k141_73254 | Not-determined |
| S17_extracted_k141_131751 | Not-determined |
| S17_extracted_k141_162088 | Not-determined |
| S17_extracted_k141_36536 | Not-determined |
| S17_extracted_k141_58485 | Low-quality |
| S17_extracted_k141_51319 | Not-determined |
| S17_extracted_k141_20687 | Not-determined |
| S17_extracted_k141_94688 | Not-determined |
| S17_extracted_k141_20689 | Not-determined |
| S17_extracted_k141_124472 | Not-determined |
| S17_extracted_k141_58495 | Not-determined |
| S17_extracted_k141_44080 | Low-quality |
| S17_extracted_k141_131755 | Not-determined |
| S17_extracted_k141_131756 | Not-determined |
| S17_extracted_k141_20696 | Not-determined |
| S17_extracted_k141_6715 | Not-determined |
| S17_extracted_k141_6716 | Not-determined |
| S17_extracted_k141_139410 | Low-quality |
| S17_extracted_k141_168887 | Not-determined |
| S17_extracted_k141_117709 | Not-determined |
| S17_extracted_k141_44089 | Not-determined |
| S17_extracted_k141_44090 | Not-determined |
| S17_extracted_k141_155079 | Low-quality |
| S17_extracted_k141_110754 | Not-determined |
| S17_extracted_k141_28031 | Not-determined |
| S17_extracted_k141_139414 | Not-determined |
| S17_extracted_k141_117714 | Not-determined |
| S17_extracted_k141_51332 | Not-determined |
| S17_extracted_k141_6720 | Not-determined |
| S17_extracted_k141_36542 | Not-determined |
| S17_extracted_k141_13792 | Not-determined |
| S17_extracted_k141_175754 | Not-determined |
| S17_extracted_k141_87233 | Not-determined |
| S17_extracted_k141_155083 | Not-determined |
| S17_extracted_k141_131769 | Not-determined |
| S17_extracted_k141_73275 | Not-determined |
| S17_extracted_k141_147262 | Not-determined |
| S17_extracted_k141_139420 | Not-determined |
| S17_extracted_k141_87240 | Not-determined |
| S17_extracted_k141_51337 | Not-determined |
| S17_extracted_k141_13796 | Not-determined |
| S17_extracted_k141_124487 | Not-determined |
| S17_extracted_k141_87244 | Not-determined |
| S17_extracted_k141_44102 | Not-determined |
| S17_extracted_k141_110767 | Not-determined |
| S17_extracted_k141_13800 | Not-determined |
| S17_extracted_k141_73279 | Not-determined |
| S17_extracted_k141_175767 | Not-determined |
| S17_extracted_k141_13802 | Low-quality |
| S17_extracted_k141_28039 | Not-determined |
| S17_extracted_k141_28040 | Not-determined |
| S17_extracted_k141_20705 | Not-determined |
| S17_extracted_k141_94717 | Not-determined |
| S17_extracted_k141_73283 | Not-determined |
| S17_extracted_k141_73284 | Low-quality |
| S17_extracted_k141_6736 | Not-determined |
| S17_extracted_k141_51345 | Not-determined |
| S17_extracted_k141_147278 | Not-determined |
| S17_extracted_k141_58523 | Not-determined |
| S17_extracted_k141_175773 | Low-quality |
| S17_extracted_k141_131793 | Not-determined |
| S17_extracted_k141_139430 | Not-determined |
| S17_extracted_k141_147284 | Not-determined |
| S17_extracted_k141_147285 | Not-determined |
| S17_extracted_k141_139433 | Not-determined |
| S17_extracted_k141_155089 | Low-quality |
| S17_extracted_k141_155091 | Not-determined |
| S17_extracted_k141_28051 | Not-determined |
| S17_extracted_k141_110776 | Not-determined |
| S17_extracted_k141_44110 | Not-determined |
| S17_extracted_k141_131798 | Not-determined |
| S17_extracted_k141_87262 | Not-determined |
| S17_extracted_k141_36559 | Not-determined |
| S17_extracted_k141_28055 | Not-determined |
| S17_extracted_k141_44114 | Not-determined |
| S17_extracted_k141_58531 | Low-quality |
| S17_extracted_k141_139438 | Not-determined |
| S17_extracted_k141_117724 | Not-determined |
| S17_extracted_k141_131806 | Low-quality |
| S17_extracted_k141_175780 | Not-determined |
| S17_extracted_k141_131808 | Not-determined |
| S17_extracted_k141_28062 | Not-determined |
| S17_extracted_k141_94746 | Low-quality |
| S17_extracted_k141_6759 | Not-determined |
| S17_extracted_k141_64632 | Low-quality |
| S17_extracted_k141_124501 | Low-quality |
| S17_extracted_k141_13821 | Not-determined |
| S17_extracted_k141_13822 | Not-determined |
| S17_extracted_k141_6764 | Low-quality |
| S17_extracted_k141_36565 | Not-determined |
| S17_extracted_k141_44122 | Low-quality |
| S17_extracted_k141_131818 | Not-determined |
| S17_extracted_k141_36566 | Low-quality |
| S17_extracted_k141_13826 | Not-determined |
| S17_extracted_k141_58555 | Not-determined |
| S17_extracted_k141_168903 | Not-determined |
| S17_extracted_k141_28079 | Not-determined |
| S17_extracted_k141_64644 | Not-determined |
| S17_extracted_k141_168906 | Not-determined |
| S17_extracted_k141_73308 | Low-quality |
| S17_extracted_k141_58558 | Not-determined |
| S17_extracted_k141_6769 | Not-determined |
| S17_extracted_k141_168908 | Not-determined |
| S17_extracted_k141_124507 | Not-determined |
| S17_extracted_k141_64650 | Not-determined |
| S17_extracted_k141_6773 | Not-determined |
| S17_extracted_k141_87296 | Not-determined |
| S17_extracted_k141_58566 | Not-determined |
| S17_extracted_k141_58568 | Not-determined |
| S17_extracted_k141_124510 | Not-determined |
| S17_extracted_k141_44132 | Low-quality |
| S17_extracted_k141_117735 | Not-determined |
| S17_extracted_k141_36586 | Not-determined |
| S17_extracted_k141_6779 | Not-determined |
| S17_extracted_k141_44138 | Not-determined |
| S17_extracted_k141_51358 | Not-determined |
| S17_extracted_k141_124521 | Not-determined |
| S17_extracted_k141_94769 | Not-determined |
| S17_extracted_k141_139473 | Not-determined |
| S17_extracted_k141_73322 | Not-determined |
| S17_extracted_k141_175805 | Not-determined |
| S17_extracted_k141_124526 | Low-quality |
| S17_extracted_k141_168916 | Not-determined |
| S17_extracted_k141_28093 | Not-determined |
| S17_extracted_k141_87309 | Not-determined |
| S17_extracted_k141_139480 | Not-determined |
| S17_extracted_k141_51363 | Not-determined |
| S17_extracted_k141_44149 | Not-determined |
| S17_extracted_k141_73328 | Not-determined |
| S17_extracted_k141_64655 | Not-determined |
| S17_extracted_k141_162103 | Not-determined |
| S17_extracted_k141_147331 | Not-determined |
| S17_extracted_k141_147332 | Not-determined |
| S17_extracted_k141_147333 | Not-determined |
| S17_extracted_k141_103027 | Not-determined |
| S17_extracted_k141_6794 | Not-determined |
| S17_extracted_k141_175812 | Low-quality |
| S17_extracted_k141_6795 | Low-quality |
| S17_extracted_k141_131853 | Not-determined |
| S17_extracted_k141_124537 | Not-determined |
| S17_extracted_k141_131854 | Not-determined |
| S17_extracted_k141_64665 | Not-determined |
| S17_extracted_k141_168930 | Not-determined |
| S17_extracted_k141_87320 | Low-quality |
| S17_extracted_k141_94785 | Low-quality |
| S17_extracted_k141_36610 | Not-determined |
| S17_extracted_k141_103035 | Not-determined |
| S17_extracted_k141_103036 | Not-determined |
| S17_extracted_k141_131861 | Not-determined |
| S17_extracted_k141_117766 | Not-determined |
| S17_extracted_k141_20772 | Not-determined |
| S17_extracted_k141_36613 | Low-quality |
| S17_extracted_k141_168938 | Not-determined |
| S17_extracted_k141_94787 | Not-determined |
| S17_extracted_k141_36616 | Low-quality |
| S17_extracted_k141_51383 | Low-quality |
| S17_extracted_k141_6799 | Not-determined |
| S17_extracted_k141_51384 | Not-determined |
| S17_extracted_k141_87328 | Not-determined |
| S17_extracted_k141_20776 | Not-determined |
| S17_extracted_k141_117769 | Low-quality |
| S17_extracted_k141_94790 | Not-determined |
| S17_extracted_k141_36619 | Not-determined |
| S17_extracted_k141_147344 | Not-determined |
| S17_extracted_k141_147346 | Not-determined |
| S17_extracted_k141_168947 | Not-determined |
| S17_extracted_k141_28102 | Low-quality |
| S17_extracted_k141_87332 | Not-determined |
| S17_extracted_k141_131878 | Not-determined |
| S17_extracted_k141_6803 | Not-determined |
| S17_extracted_k141_117773 | Not-determined |
| S17_extracted_k141_58600 | Not-determined |
| S17_extracted_k141_110808 | Low-quality |
| S17_extracted_k141_175818 | Not-determined |
| S17_extracted_k141_94796 | Not-determined |
| S17_extracted_k141_117775 | Not-determined |
| S17_extracted_k141_44156 | Not-determined |
| S17_extracted_k141_87337 | Not-determined |
| S17_extracted_k141_162120 | Not-determined |
| S17_extracted_k141_73349 | Not-determined |
| S17_extracted_k141_87339 | Not-determined |
| S17_extracted_k141_20793 | Not-determined |
| S17_extracted_k141_87341 | Not-determined |
| S17_extracted_k141_51393 | Not-determined |
| S17_extracted_k141_20794 | Not-determined |
| S17_extracted_k141_147357 | Not-determined |
| S17_extracted_k141_51395 | Not-determined |
| S17_extracted_k141_103061 | Not-determined |
| S17_extracted_k141_6807 | Not-determined |
| S17_extracted_k141_80659 | Not-determined |
| S17_extracted_k141_168960 | Not-determined |
| S17_extracted_k141_139508 | Not-determined |
| S17_extracted_k141_36627 | Not-determined |
| S17_extracted_k141_162128 | Not-determined |
| S17_extracted_k141_110825 | Not-determined |
| S17_extracted_k141_94810 | Not-determined |
| S17_extracted_k141_94811 | Not-determined |
| S17_extracted_k141_162129 | Not-determined |
| S17_extracted_k141_103067 | Not-determined |
| S17_extracted_k141_36629 | Low-quality |
| S17_extracted_k141_20803 | Not-determined |
| S17_extracted_k141_147365 | Not-determined |
| S17_extracted_k141_162131 | Not-determined |
| S17_extracted_k141_94813 | Not-determined |
| S17_extracted_k141_162132 | Not-determined |
| S17_extracted_k141_94816 | Not-determined |
| S17_extracted_k141_139516 | Not-determined |
| S17_extracted_k141_6808 | Low-quality |
| S17_extracted_k141_147370 | Low-quality |
| S17_extracted_k141_117786 | Low-quality |
| S17_extracted_k141_110829 | Not-determined |
| S17_extracted_k141_139519 | Not-determined |
| S17_extracted_k141_117790 | Not-determined |
| S17_extracted_k141_36635 | Not-determined |
| S17_extracted_k141_87356 | Not-determined |
| S17_extracted_k141_64687 | Not-determined |
| S17_extracted_k141_51405 | Not-determined |
| S17_extracted_k141_44163 | Not-determined |
| S17_extracted_k141_28133 | Not-determined |
| S17_extracted_k141_168978 | Not-determined |
| S17_extracted_k141_94823 | Not-determined |
| S17_extracted_k141_124562 | Not-determined |
| S17_extracted_k141_117793 | Not-determined |
| S17_extracted_k141_103084 | Low-quality |
| S17_extracted_k141_117795 | Not-determined |
| S17_extracted_k141_51417 | Not-determined |
| S17_extracted_k141_139527 | Not-determined |
| S17_extracted_k141_73374 | Not-determined |
| S17_extracted_k141_6822 | Not-determined |
| S17_extracted_k141_36641 | Not-determined |
| S17_extracted_k141_64697 | Not-determined |
| S17_extracted_k141_87367 | Not-determined |
| S17_extracted_k141_28136 | Not-determined |
| S17_extracted_k141_175834 | Not-determined |
| S17_extracted_k141_80679 | Not-determined |
| S17_extracted_k141_147392 | Not-determined |
| S17_extracted_k141_13846 | Low-quality |
| S17_extracted_k141_36647 | Not-determined |
| S17_extracted_k141_73380 | Low-quality |
| S17_extracted_k141_162146 | Not-determined |
| S17_extracted_k141_64711 | Low-quality |
| S17_extracted_k141_103096 | Not-determined |
| S17_extracted_k141_64712 | Not-determined |
| S17_extracted_k141_87379 | Low-quality |
| S17_extracted_k141_64714 | Not-determined |
| S17_extracted_k141_117808 | Not-determined |
| S17_extracted_k141_147397 | Not-determined |
| S17_extracted_k141_44173 | Not-determined |
| S17_extracted_k141_73386 | Not-determined |
| S17_extracted_k141_147401 | Not-determined |
| S17_extracted_k141_175850 | Not-determined |
| S17_extracted_k141_175851 | Not-determined |
| S17_extracted_k141_28154 | Not-determined |
| S17_extracted_k141_175852 | Not-determined |
| S17_extracted_k141_28155 | Not-determined |
| S17_extracted_k141_80689 | Not-determined |
| S17_extracted_k141_20821 | Not-determined |
| S17_extracted_k141_155119 | Low-quality |
| S17_extracted_k141_155120 | Not-determined |
| S17_extracted_k141_155125 | Not-determined |
| S17_extracted_k141_51435 | Not-determined |
| S17_extracted_k141_169009 | Not-determined |
| S17_extracted_k141_36650 | Not-determined |
| S17_extracted_k141_73390 | Not-determined |
| S17_extracted_k141_155129 | Not-determined |
| S17_extracted_k141_80703 | Not-determined |
| S17_extracted_k141_51438 | Low-quality |
| S17_extracted_k141_20823 | Low-quality |
| S17_extracted_k141_155130 | Not-determined |
| S17_extracted_k141_175865 | Low-quality |
| S17_extracted_k141_80704 | Low-quality |
| S17_extracted_k141_155133 | Not-determined |
| S17_extracted_k141_51442 | Not-determined |
| S17_extracted_k141_64726 | Not-determined |
| S17_extracted_k141_73396 | Not-determined |
| S17_extracted_k141_20828 | Not-determined |
| S17_extracted_k141_139544 | Not-determined |
| S17_extracted_k141_13859 | Not-determined |
| S17_extracted_k141_64731 | Low-quality |
| S17_extracted_k141_64732 | Not-determined |
| S17_extracted_k141_36660 | Low-quality |
| S17_extracted_k141_28173 | Not-determined |
| S17_extracted_k141_36662 | Not-determined |
| S17_extracted_k141_6835 | Not-determined |
| S17_extracted_k141_87397 | Not-determined |
| S17_extracted_k141_117825 | Not-determined |
| S17_extracted_k141_87398 | Low-quality |
| S17_extracted_k141_6837 | Low-quality |
| S17_extracted_k141_139549 | Not-determined |
| S17_extracted_k141_20834 | Not-determined |
| S17_extracted_k141_155137 | Not-determined |
| S17_extracted_k141_103117 | Low-quality |
| S17_extracted_k141_28181 | Not-determined |
| S17_extracted_k141_169024 | Not-determined |
| S17_extracted_k141_169027 | Not-determined |
| S17_extracted_k141_103122 | Low-quality |
| S17_extracted_k141_87405 | Low-quality |
| S17_extracted_k141_6846 | Not-determined |
| S17_extracted_k141_51451 | Not-determined |
| S17_extracted_k141_13872 | Low-quality |
| S17_extracted_k141_162180 | Not-determined |
| S17_extracted_k141_139559 | Not-determined |
| S17_extracted_k141_87408 | Low-quality |
| S17_extracted_k141_44205 | Not-determined |
| S17_extracted_k141_80706 | Not-determined |
| S17_extracted_k141_87409 | Not-determined |
| S17_extracted_k141_28186 | Not-determined |
| S17_extracted_k141_103126 | Not-determined |
| S17_extracted_k141_20846 | Not-determined |
| S17_extracted_k141_13880 | Not-determined |
| S17_extracted_k141_103137 | Not-determined |
| S17_extracted_k141_155146 | Not-determined |
| S17_extracted_k141_13884 | Not-determined |
| S17_extracted_k141_80716 | Not-determined |
| S17_extracted_k141_169034 | Not-determined |
| S17_extracted_k141_28192 | Low-quality |
| S17_extracted_k141_80717 | Not-determined |
| S17_extracted_k141_131932 | Not-determined |
| S17_extracted_k141_64749 | Not-determined |
| S17_extracted_k141_80720 | Not-determined |
| S17_extracted_k141_103139 | Not-determined |
| S17_extracted_k141_6853 | Not-determined |
| S17_extracted_k141_36672 | Not-determined |
| S17_extracted_k141_147431 | Not-determined |
| S17_extracted_k141_131935 | Not-determined |
| S17_extracted_k141_20859 | Low-quality |
| S17_extracted_k141_73420 | Not-determined |
| S17_extracted_k141_131937 | Not-determined |
| S17_extracted_k141_6857 | Not-determined |
| S17_extracted_k141_80726 | Not-determined |
| S17_extracted_k141_147437 | Not-determined |
| S17_extracted_k141_13885 | Not-determined |
| S17_extracted_k141_131943 | Low-quality |
| S17_extracted_k141_13886 | Not-determined |
| S17_extracted_k141_51466 | Low-quality |
| S17_extracted_k141_20866 | Not-determined |
| S17_extracted_k141_131947 | Not-determined |
| S17_extracted_k141_155153 | Low-quality |
| S17_extracted_k141_131957 | Not-determined |
| S17_extracted_k141_131959 | Not-determined |
| S17_extracted_k141_131960 | Not-determined |
| S17_extracted_k141_131961 | Not-determined |
| S17_extracted_k141_139579 | Low-quality |
| S17_extracted_k141_131962 | Not-determined |
| S17_extracted_k141_64764 | Not-determined |
| S17_extracted_k141_36687 | Low-quality |
| S17_extracted_k141_87421 | Low-quality |
| S17_extracted_k141_6872 | Not-determined |
| S17_extracted_k141_80735 | Not-determined |
| S17_extracted_k141_44215 | Low-quality |
| S17_extracted_k141_162200 | Not-determined |
| S17_extracted_k141_20881 | Not-determined |
| S17_extracted_k141_64767 | Not-determined |
| S17_extracted_k141_175890 | Not-determined |
| S17_extracted_k141_94842 | Not-determined |
| S17_extracted_k141_175894 | Not-determined |
| S17_extracted_k141_80746 | Not-determined |
| S17_extracted_k141_64775 | Not-determined |
| S17_extracted_k141_36694 | Not-determined |
| S17_extracted_k141_36695 | Not-determined |
| S17_extracted_k141_51481 | Low-quality |
| S17_extracted_k141_73443 | Not-determined |
| S17_extracted_k141_175895 | Not-determined |
| S17_extracted_k141_124612 | Low-quality |
| S17_extracted_k141_175897 | Low-quality |
| S17_extracted_k141_64780 | Not-determined |
| S17_extracted_k141_13900 | Not-determined |
| S17_extracted_k141_20889 | Not-determined |
| S17_extracted_k141_51486 | Not-determined |
| S17_extracted_k141_147446 | Not-determined |
| S17_extracted_k141_80750 | Not-determined |
| S17_extracted_k141_147447 | Low-quality |
| S17_extracted_k141_175903 | Not-determined |
| S17_extracted_k141_73446 | Not-determined |
| S17_extracted_k141_94848 | Not-determined |
| S17_extracted_k141_175906 | Not-determined |
| S17_extracted_k141_131977 | Not-determined |
| S17_extracted_k141_175908 | Low-quality |
| S17_extracted_k141_131979 | Not-determined |
| S17_extracted_k141_36708 | Low-quality |
| S17_extracted_k141_36709 | Not-determined |
| S17_extracted_k141_147453 | Not-determined |
| S7_vs_6664\|\|full | High-quality |
| S17_extracted_k141_36712 | Low-quality |
| S17_extracted_k141_80757 | Not-determined |
| S17_extracted_k141_124628 | Low-quality |
| S17_extracted_k141_6886 | Low-quality |
| S17_extracted_k141_6887 | Not-determined |
| S17_extracted_k141_87443 | Not-determined |
| S17_extracted_k141_28214 | Not-determined |
| S17_extracted_k141_36714 | Not-determined |
| S17_extracted_k141_169056 | Not-determined |
| S17_extracted_k141_28215 | Not-determined |
| S17_extracted_k141_87444 | Not-determined |
| S17_extracted_k141_147458 | Not-determined |
| S17_extracted_k141_147459 | Not-determined |
| S17_extracted_k141_6889 | Not-determined |
| S17_extracted_k141_147460 | Not-determined |
| S17_extracted_k141_175915 | Not-determined |
| S17_extracted_k141_20895 | Not-determined |
| S17_extracted_k141_175917 | Low-quality |
| S17_extracted_k141_28223 | Not-determined |
| S17_extracted_k141_20899 | Not-determined |
| S17_extracted_k141_87451 | Not-determined |
| S17_extracted_k141_51501 | Low-quality |
| S17_extracted_k141_36727 | Not-determined |
| S17_extracted_k141_80767 | Not-determined |
| S17_extracted_k141_103188 | Not-determined |
| S17_extracted_k141_94867 | Low-quality |
| S17_extracted_k141_28229 | Not-determined |
| S17_extracted_k141_103191 | Not-determined |
| S17_extracted_k141_87461 | Not-determined |
| S17_extracted_k141_103195 | Not-determined |
| S17_extracted_k141_87464 | Not-determined |
| S17_extracted_k141_87465 | Not-determined |
| S17_extracted_k141_28231 | Not-determined |
| S17_extracted_k141_28232 | Not-determined |
| S17_extracted_k141_131991 | Not-determined |
| S17_extracted_k141_87466 | Low-quality |
| S17_extracted_k141_6904 | Not-determined |
| S17_extracted_k141_169067 | Not-determined |
| S17_extracted_k141_103203 | Not-determined |
| S17_extracted_k141_73468 | Not-determined |
| S17_extracted_k141_103204 | Not-determined |
| S17_extracted_k141_147477 | Not-determined |
| S17_extracted_k141_169069 | Not-determined |
| S17_extracted_k141_94875 | Not-determined |
| S17_extracted_k141_162236 | Not-determined |
| S17_extracted_k141_28235 | Not-determined |
| S17_extracted_k141_87470 | Not-determined |
| S17_extracted_k141_44267 | Not-determined |
| S17_extracted_k141_80773 | Not-determined |
| S17_extracted_k141_124635 | Not-determined |
| S17_extracted_k141_131997 | Not-determined |
| S17_extracted_k141_162237 | Not-determined |
| S17_extracted_k141_175942 | Not-determined |
| S17_extracted_k141_94879 | Not-determined |
| S17_extracted_k141_36740 | Low-quality |
| S17_extracted_k141_162240 | Not-determined |
| S17_extracted_k141_162241 | Not-determined |
| S17_extracted_k141_73476 | Not-determined |
| S17_extracted_k141_162246 | Not-determined |
| S17_extracted_k141_20906 | Not-determined |
| S17_extracted_k141_124642 | Low-quality |
| S17_extracted_k141_28244 | Not-determined |
| S17_extracted_k141_169077 | Low-quality |
| S17_extracted_k141_162249 | Not-determined |
| S17_extracted_k141_124643 | Not-determined |
| S17_extracted_k141_94888 | Not-determined |
| S17_extracted_k141_175947 | Not-determined |
| S17_extracted_k141_20912 | Low-quality |
| S17_extracted_k141_6918 | Not-determined |
| S17_extracted_k141_73484 | Low-quality |
| S17_extracted_k141_147494 | Not-determined |
| S17_extracted_k141_94891 | Not-determined |
| S17_extracted_k141_139601 | Low-quality |
| S17_extracted_k141_124648 | Not-determined |
| S17_extracted_k141_103221 | Not-determined |
| S17_extracted_k141_124650 | Not-determined |
| S17_extracted_k141_117887 | Not-determined |
| S17_extracted_k141_80788 | Not-determined |
| S17_extracted_k141_36752 | Not-determined |
| S17_extracted_k141_139603 | Low-quality |
| S17_extracted_k141_132008 | Not-determined |
| S17_extracted_k141_87492 | Not-determined |
| S17_extracted_k141_175952 | Not-determined |
| S17_extracted_k141_147505 | Not-determined |
| S17_extracted_k141_73499 | Not-determined |
| S17_extracted_k141_44283 | Low-quality |
| S17_extracted_k141_20923 | Not-determined |
| S17_extracted_k141_13927 | Not-determined |
| S17_extracted_k141_87499 | Not-determined |
| S17_extracted_k141_80796 | Not-determined |
| S17_extracted_k141_175959 | Not-determined |
| S17_extracted_k141_175962 | Not-determined |
| S17_extracted_k141_51531 | Not-determined |
| S17_extracted_k141_132021 | Not-determined |
| S17_extracted_k141_175965 | Not-determined |
| S17_extracted_k141_117898 | Not-determined |
| S17_extracted_k141_169092 | Not-determined |
| S17_extracted_k141_103239 | Not-determined |
| S17_extracted_k141_117900 | Not-determined |
| S17_extracted_k141_117901 | Not-determined |
| S17_extracted_k141_36758 | Not-determined |
| S17_extracted_k141_162266 | Not-determined |
| S17_extracted_k141_132027 | Not-determined |
| S17_extracted_k141_51536 | Not-determined |
| S17_extracted_k141_175968 | Not-determined |
| S17_extracted_k141_28279 | Not-determined |
| S17_extracted_k141_175969 | Not-determined |
| S17_extracted_k141_175970 | Not-determined |
| S17_extracted_k141_147509 | Not-determined |
| S17_extracted_k141_175971 | Not-determined |
| S17_extracted_k141_147511 | Not-determined |
| S17_extracted_k141_36766 | Not-determined |
| S17_extracted_k141_36767 | Not-determined |
| S17_extracted_k141_117909 | Low-quality |
| S17_extracted_k141_162274 | Not-determined |
| S17_extracted_k141_73515 | Not-determined |
| S17_extracted_k141_147513 | Low-quality |
| S17_extracted_k141_132039 | Not-determined |
| S17_extracted_k141_87515 | Not-determined |
| S17_extracted_k141_13938 | Low-quality |
| S17_extracted_k141_169109 | Not-determined |
| S17_extracted_k141_103257 | Not-determined |
| S17_extracted_k141_162282 | Not-determined |
| S17_extracted_k141_20950 | Not-determined |
| S17_extracted_k141_20952 | Not-determined |
| S17_extracted_k141_169113 | Not-determined |
| S17_extracted_k141_117916 | Not-determined |
| S17_extracted_k141_147521 | Not-determined |

| | |
|---|---|
| S17_extracted_k141_28289 | Not-determined |
| S17_extracted_k141_147523 | Not-determined |
| S17_extracted_k141_175979 | Not-determined |
| S17_extracted_k141_13941 | Not-determined |
| S17_extracted_k141_103265 | Not-determined |
| S17_extracted_k141_169122 | Not-determined |
| S17_extracted_k141_51559 | Not-determined |
| S17_extracted_k141_147525 | Not-determined |
| S17_extracted_k141_117923 | Low-quality |
| S17_extracted_k141_20959 | Not-determined |
| S17_extracted_k141_175983 | Not-determined |
| S17_extracted_k141_94913 | Low-quality |
| S17_extracted_k141_162287 | Not-determined |
| S17_extracted_k141_20964 | Low-quality |
| S17_extracted_k141_73525 | Low-quality |
| S17_extracted_k141_13948 | Not-determined |
| S17_extracted_k141_94917 | Not-determined |
| S17_extracted_k141_147531 | Not-determined |
| S17_extracted_k141_13950 | Not-determined |
| S17_extracted_k141_87526 | Low-quality |
| S17_extracted_k141_132056 | Not-determined |
| S17_extracted_k141_175992 | Not-determined |
| S17_extracted_k141_51566 | Not-determined |
| S17_extracted_k141_117933 | Low-quality |
| S17_extracted_k141_175995 | Low-quality |
| S17_extracted_k141_175997 | Not-determined |
| S17_extracted_k141_87532 | Not-determined |
| S17_extracted_k141_103274 | Not-determined |
| S17_extracted_k141_147541 | Not-determined |
| S17_extracted_k141_147542 | Not-determined |
| S17_extracted_k141_162300 | Not-determined |
| S17_extracted_k141_28306 | Low-quality |
| S17_extracted_k141_73536 | Not-determined |
| S17_extracted_k141_176001 | Not-determined |
| S17_extracted_k141_87538 | Not-determined |
| S17_extracted_k141_132066 | Not-determined |
| S17_extracted_k141_87539 | Not-determined |
| S17_extracted_k141_28309 | Not-determined |
| S17_extracted_k141_132068 | Not-determined |
| S17_extracted_k141_87542 | Not-determined |
| S17_extracted_k141_44308 | Low-quality |
| S17_extracted_k141_139626 | Not-determined |
| S17_extracted_k141_103285 | Not-determined |
| S17_extracted_k141_139631 | Not-determined |
| S17_extracted_k141_169135 | Low-quality |
| S17_extracted_k141_124653 | Not-determined |
| S17_extracted_k141_13960 | Not-determined |
| S17_extracted_k141_139635 | Not-determined |
| S17_extracted_k141_20974 | Not-determined |
| S17_extracted_k141_103291 | Low-quality |
| S17_extracted_k141_6957 | Not-determined |
| S17_extracted_k141_132078 | Low-quality |
| S17_extracted_k141_169142 | Not-determined |
| S17_extracted_k141_6959 | Not-determined |
| S17_extracted_k141_80827 | Not-determined |
| S17_extracted_k141_147560 | Not-determined |
| S17_extracted_k141_44319 | Not-determined |
| S17_extracted_k141_80828 | Not-determined |
| S17_extracted_k141_132081 | Not-determined |
| S17_extracted_k141_51585 | Not-determined |
| S17_extracted_k141_80829 | Not-determined |
| S17_extracted_k141_28324 | Low-quality |
| S17_extracted_k141_147566 | Not-determined |
| S17_extracted_k141_103299 | Not-determined |
| S17_extracted_k141_147568 | Not-determined |
| S17_extracted_k141_51587 | Not-determined |
| S17_extracted_k141_44322 | Not-determined |
| S17_extracted_k141_6965 | Low-quality |
| S17_extracted_k141_132087 | Not-determined |
| S17_extracted_k141_73559 | Not-determined |
| S17_extracted_k141_73560 | Not-determined |
| S17_extracted_k141_6968 | Not-determined |
| S17_extracted_k141_13975 | Not-determined |
| S17_extracted_k141_6969 | Not-determined |
| S17_extracted_k141_13979 | Low-quality |
| S17_extracted_k141_80841 | Not-determined |
| S17_extracted_k141_147573 | Not-determined |
| S17_extracted_k141_51598 | Not-determined |
| S17_extracted_k141_64798 | Low-quality |
| S17_extracted_k141_51599 | Not-determined |
| S17_extracted_k141_147575 | Not-determined |
| S17_extracted_k141_176006 | Not-determined |
| S17_extracted_k141_132096 | Not-determined |
| S17_extracted_k141_169153 | Not-determined |
| S17_extracted_k141_64800 | Not-determined |
| S17_extracted_k141_139646 | Not-determined |
| S17_extracted_k141_162326 | Not-determined |
| S17_extracted_k141_21001 | Not-determined |
| S17_extracted_k141_28329 | Low-quality |
| S17_extracted_k141_6983 | Not-determined |
| S17_extracted_k141_73572 | Not-determined |
| S17_extracted_k141_64802 | Not-determined |
| S17_extracted_k141_73573 | Low-quality |
| S17_extracted_k141_117949 | Not-determined |
| S17_extracted_k141_73575 | Not-determined |
| S17_extracted_k141_162329 | Not-determined |
| S17_extracted_k141_162332 | Not-determined |
| S17_extracted_k141_13986 | Not-determined |
| S17_extracted_k141_162334 | Not-determined |
| S17_extracted_k141_51601 | Not-determined |
| S17_extracted_k141_162337 | Low-quality |
| S17_extracted_k141_28336 | Not-determined |
| S17_extracted_k141_169164 | Low-quality |
| S17_extracted_k141_169165 | Low-quality |
| S17_extracted_k141_117957 | Not-determined |
| S17_extracted_k141_64813 | Not-determined |
| S17_extracted_k141_87566 | Not-determined |
| S17_extracted_k141_28344 | Not-determined |
| S17_extracted_k141_21027 | Low-quality |
| S17_extracted_k141_80858 | Not-determined |
| S17_extracted_k141_21028 | Not-determined |
| S17_extracted_k141_94946 | Not-determined |
| S17_extracted_k141_87581 | Not-determined |
| S17_extracted_k141_117977 | Not-determined |
| S17_extracted_k141_147610 | Not-determined |
| S17_extracted_k141_117978 | Not-determined |
| S17_extracted_k141_147612 | Not-determined |
| S17_extracted_k141_94952 | Not-determined |
| S17_extracted_k141_14000 | Not-determined |
| S17_extracted_k141_162357 | Not-determined |
| S17_extracted_k141_21034 | Low-quality |
| S17_extracted_k141_132117 | Not-determined |
| S17_extracted_k141_132118 | Not-determined |
| S17_extracted_k141_14004 | Low-quality |
| S17_extracted_k141_21037 | Low-quality |
| S17_extracted_k141_162364 | Low-quality |
| S17_extracted_k141_14007 | Not-determined |
| S17_extracted_k141_87590 | Not-determined |
| S17_extracted_k141_132123 | Not-determined |
| S17_extracted_k141_73593 | Low-quality |
| S17_extracted_k141_117985 | Low-quality |
| S17_extracted_k141_80874 | Not-determined |
| S17_extracted_k141_28358 | Not-determined |
| S17_extracted_k141_132124 | Not-determined |
| S17_extracted_k141_73595 | Not-determined |
| S17_extracted_k141_117987 | Not-determined |
| S17_extracted_k141_124691 | Not-determined |
| S17_extracted_k141_51613 | Not-determined |
| S17_extracted_k141_94962 | Not-determined |
| S17_extracted_k141_87595 | Not-determined |
| S17_extracted_k141_14016 | Not-determined |
| S17_extracted_k141_169171 | Not-determined |
| S17_extracted_k141_7005 | Not-determined |
| S17_extracted_k141_80884 | Not-determined |
| S17_extracted_k141_64844 | Not-determined |
| S17_extracted_k141_80885 | Not-determined |
| S17_extracted_k141_73602 | Low-quality |
| S17_extracted_k141_21055 | Not-determined |
| S17_extracted_k141_169181 | Not-determined |
| S17_extracted_k141_64847 | Not-determined |
| S17_extracted_k141_64848 | Not-determined |
| S17_extracted_k141_21058 | Not-determined |
| S17_extracted_k141_169182 | Not-determined |
| S17_extracted_k141_73609 | Not-determined |
| S17_extracted_k141_87601 | Not-determined |
| S17_extracted_k141_73613 | Not-determined |
| S17_extracted_k141_21064 | Not-determined |
| S17_extracted_k141_176045 | Not-determined |
| S17_extracted_k141_162391 | Not-determined |
| S17_extracted_k141_124705 | Not-determined |
| S17_extracted_k141_169193 | Not-determined |
| S17_extracted_k141_36776 | Not-determined |
| S17_extracted_k141_51621 | Not-determined |
| S17_extracted_k141_169195 | Not-determined |
| S17_extracted_k141_176054 | Not-determined |
| S17_extracted_k141_124708 | Not-determined |
| S17_extracted_k141_7026 | Not-determined |
| S17_extracted_k141_73623 | Not-determined |
| S17_extracted_k141_58655 | Low-quality |
| S17_extracted_k141_28377 | Not-determined |
| S17_extracted_k141_80906 | Not-determined |
| S17_extracted_k141_7031 | Not-determined |
| S17_extracted_k141_147630 | Not-determined |
| S17_extracted_k141_58657 | Low-quality |
| S17_extracted_k141_162403 | Not-determined |
| S17_extracted_k141_94971 | Not-determined |
| S17_extracted_k141_147634 | Low-quality |
| S17_extracted_k141_162405 | Not-determined |
| S17_extracted_k141_169207 | Not-determined |
| S17_extracted_k141_51634 | Not-determined |
| S17_extracted_k141_147636 | Not-determined |
| S17_extracted_k141_51635 | Not-determined |
| S17_extracted_k141_94976 | Low-quality |
| S17_extracted_k141_169208 | Not-determined |
| S17_extracted_k141_118004 | Not-determined |
| S17_extracted_k141_7046 | Not-determined |
| S17_extracted_k141_124715 | Low-quality |
| S17_extracted_k141_21084 | Not-determined |
| S17_extracted_k141_28392 | Not-determined |
| S17_extracted_k141_58665 | Not-determined |
| S17_extracted_k141_7049 | Not-determined |
| S17_extracted_k141_58667 | Not-determined |
| S17_extracted_k141_147639 | Low-quality |
| S17_extracted_k141_7054 | Not-determined |
| S17_extracted_k141_64861 | Low-quality |
| S17_extracted_k141_147641 | Low-quality |
| S17_extracted_k141_64863 | Not-determined |
| S17_extracted_k141_118016 | Not-determined |
| S17_extracted_k141_28397 | Not-determined |
| S17_extracted_k141_169221 | Not-determined |
| S17_extracted_k141_139655 | Not-determined |
| S17_extracted_k141_132170 | Not-determined |
| S17_extracted_k141_14030 | Not-determined |
| S17_extracted_k141_147647 | Not-determined |
| S17_extracted_k141_7065 | Not-determined |
| S17_extracted_k141_28399 | Not-determined |
| S17_extracted_k141_21100 | Not-determined |
| S17_extracted_k141_124729 | Not-determined |
| S17_extracted_k141_162425 | Not-determined |
| S17_extracted_k141_7068 | Not-determined |
| S17_extracted_k141_51652 | Not-determined |
| S17_extracted_k141_21102 | Not-determined |
| S17_extracted_k141_21104 | Not-determined |
| S17_extracted_k141_169227 | Not-determined |
| S17_extracted_k141_7070 | Not-determined |
| S17_extracted_k141_169228 | Not-determined |
| S17_extracted_k141_124734 | Not-determined |
| S17_extracted_k141_95000 | Not-determined |
| S17_extracted_k141_14042 | Not-determined |
| S17_extracted_k141_51658 | Not-determined |
| S17_extracted_k141_58670 | Not-determined |
| S17_extracted_k141_64880 | Not-determined |
| S17_extracted_k141_124745 | Not-determined |
| S17_extracted_k141_169235 | Not-determined |
| S17_extracted_k141_7082 | Not-determined |
| S17_extracted_k141_176087 | Not-determined |
| S17_extracted_k141_14050 | Not-determined |
| S17_extracted_k141_7084 | Not-determined |
| S17_extracted_k141_103359 | Not-determined |
| S17_extracted_k141_147668 | Not-determined |
| S17_extracted_k141_7085 | Not-determined |
| S17_extracted_k141_64893 | Not-determined |
| S17_extracted_k141_176089 | Low-quality |
| S17_extracted_k141_103361 | Low-quality |
| S17_extracted_k141_14056 | Not-determined |
| S17_extracted_k141_64896 | Not-determined |
| S17_extracted_k141_124751 | Not-determined |
| S17_extracted_k141_7092 | Not-determined |
| S17_extracted_k141_73654 | Not-determined |
| S17_extracted_k141_103367 | Not-determined |
| S17_extracted_k141_51671 | Not-determined |
| S17_extracted_k141_176096 | Not-determined |
| S17_extracted_k141_14065 | Not-determined |
| S17_extracted_k141_139682 | Not-determined |
| S17_extracted_k141_51676 | Not-determined |
| S17_extracted_k141_176102 | Not-determined |
| S17_extracted_k141_21127 | Not-determined |
| S17_extracted_k141_7109 | Not-determined |
| S17_extracted_k141_147682 | Low-quality |
| S17_extracted_k141_64909 | Not-determined |
| S17_extracted_k141_103373 | Not-determined |
| S17_extracted_k141_73667 | Not-determined |
| S17_extracted_k141_28417 | Not-determined |
| S17_extracted_k141_64911 | Not-determined |
| S17_extracted_k141_95010 | Not-determined |
| S17_extracted_k141_80965 | Not-determined |
| S17_extracted_k141_176109 | Not-determined |
| S17_extracted_k141_51685 | Not-determined |
| S17_extracted_k141_64912 | Low-quality |
| S17_extracted_k141_95011 | Not-determined |
| S17_extracted_k141_103378 | Not-determined |
| S17_extracted_k141_28423 | Not-determined |
| S17_extracted_k141_80967 | Not-determined |
| S17_extracted_k141_28424 | Not-determined |
| S17_extracted_k141_21143 | Not-determined |
| S17_extracted_k141_162436 | Not-determined |
| S17_extracted_k141_124776 | Not-determined |
| S17_extracted_k141_58709 | Not-determined |
| S17_extracted_k141_162442 | Not-determined |
| S17_extracted_k141_73682 | Not-determined |
| S17_extracted_k141_64915 | Not-determined |
| S17_extracted_k141_87633 | Not-determined |
| S17_extracted_k141_64916 | Not-determined |
| S17_extracted_k141_95026 | Not-determined |
| S17_extracted_k141_80974 | Not-determined |
| S17_extracted_k141_80975 | Not-determined |
| S17_extracted_k141_147703 | Not-determined |
| S17_extracted_k141_87634 | Not-determined |
| S17_extracted_k141_64920 | Not-determined |
| S17_extracted_k141_21152 | Low-quality |
| S17_extracted_k141_169240 | Not-determined |
| S17_extracted_k141_87638 | Not-determined |
| S17_extracted_k141_73684 | Not-determined |
| S17_extracted_k141_132213 | Not-determined |
| S17_extracted_k141_7131 | Low-quality |
| S17_extracted_k141_124789 | Not-determined |
| S17_extracted_k141_21155 | Not-determined |
| S17_extracted_k141_58719 | Not-determined |
| S17_extracted_k141_103395 | Not-determined |
| S17_extracted_k141_87642 | Not-determined |
| S17_extracted_k141_132218 | Not-determined |
| S17_extracted_k141_124791 | Not-determined |
| S17_extracted_k141_7133 | Not-determined |
| S17_extracted_k141_73688 | Not-determined |
| S17_extracted_k141_139697 | Not-determined |
| S17_extracted_k141_36791 | Low-quality |
| S17_extracted_k141_64923 | Not-determined |
| S17_extracted_k141_139701 | Low-quality |
| S17_extracted_k141_162449 | Low-quality |
| S17_extracted_k141_162450 | Not-determined |
| S17_extracted_k141_21163 | Low-quality |
| S17_extracted_k141_64926 | Not-determined |
| S17_extracted_k141_58724 | Low-quality |
| S17_extracted_k141_21166 | Not-determined |
| S17_extracted_k141_147714 | Not-determined |
| S17_extracted_k141_162453 | Not-determined |
| S17_extracted_k141_64928 | Not-determined |
| S17_extracted_k141_176119 | Not-determined |
| S17_extracted_k141_169248 | Not-determined |
| S17_extracted_k141_36794 | Not-determined |
| S17_extracted_k141_58727 | Not-determined |
| S17_extracted_k141_51713 | Not-determined |
| S17_extracted_k141_21169 | Low-quality |
| S17_extracted_k141_169250 | Low-quality |
| S17_extracted_k141_162459 | Low-quality |
| S17_extracted_k141_87647 | Low-quality |
| S17_extracted_k141_36802 | Not-determined |
| S17_extracted_k141_169256 | Not-determined |
| S17_extracted_k141_21170 | Not-determined |
| S17_extracted_k141_64934 | Not-determined |
| S17_extracted_k141_36804 | Not-determined |
| S17_extracted_k141_28462 | Not-determined |
| S17_extracted_k141_21171 | Not-determined |
| S17_extracted_k141_169261 | Not-determined |
| S17_extracted_k141_87656 | Not-determined |
| S17_extracted_k141_7152 | Not-determined |
| S17_extracted_k141_36811 | Not-determined |
| S17_extracted_k141_7153 | Not-determined |
| S17_extracted_k141_162470 | Not-determined |
| S17_extracted_k141_176135 | Not-determined |
| S17_extracted_k141_73695 | Not-determined |
| S17_extracted_k141_64942 | Not-determined |
| S17_extracted_k141_73698 | Not-determined |
| S17_extracted_k141_176138 | Not-determined |
| S17_extracted_k141_36814 | Not-determined |
| S17_extracted_k141_162475 | Not-determined |
| S17_extracted_k141_176142 | Not-determined |
| S17_extracted_k141_21190 | Low-quality |
| S17_extracted_k141_176143 | Not-determined |
| S17_extracted_k141_14092 | Not-determined |
| S17_extracted_k141_64951 | Not-determined |
| S17_extracted_k141_21192 | Not-determined |
| S17_extracted_k141_64953 | Not-determined |
| S17_extracted_k141_169276 | Not-determined |
| S17_extracted_k141_14094 | Low-quality |
| S17_extracted_k141_169279 | Not-determined |
| S17_extracted_k141_21196 | Not-determined |
| S17_extracted_k141_73703 | Not-determined |
| S17_extracted_k141_162485 | Low-quality |
| S17_extracted_k141_103412 | Not-determined |
| S17_extracted_k141_176147 | Not-determined |
| S17_extracted_k141_58748 | Low-quality |
| S17_extracted_k141_73708 | Not-determined |
| S17_extracted_k141_64959 | Not-determined |
| S17_extracted_k141_87686 | Not-determined |
| S17_extracted_k141_14102 | Not-determined |
| S17_extracted_k141_118033 | Not-determined |
| S17_extracted_k141_176151 | Not-determined |
| S17_extracted_k141_7168 | Not-determined |
| S17_extracted_k141_7169 | Low-quality |
| S17_extracted_k141_21209 | Not-determined |
| S17_extracted_k141_14109 | Low-quality |
| S17_extracted_k141_73716 | Not-determined |
| S17_extracted_k141_147762 | Not-determined |
| S17_extracted_k141_176158 | Not-determined |
| S17_extracted_k141_7179 | Not-determined |
| S17_extracted_k141_176159 | Not-determined |
| S17_extracted_k141_147767 | Not-determined |
| S17_extracted_k141_73724 | Low-quality |
| S17_extracted_k141_124805 | Not-determined |
| S17_extracted_k141_124808 | Not-determined |
| S17_extracted_k141_147769 | Not-determined |
| S17_extracted_k141_73726 | Low-quality |
| S17_extracted_k141_73727 | Not-determined |
| S17_extracted_k141_21229 | Not-determined |
| S17_extracted_k141_118052 | Low-quality |
| S17_extracted_k141_7190 | Not-determined |
| S17_extracted_k141_169307 | Not-determined |
| S17_extracted_k141_176170 | Not-determined |
| S17_extracted_k141_176171 | Not-determined |
| S17_extracted_k141_169309 | Not-determined |
| S17_extracted_k141_139715 | Not-determined |
| S17_extracted_k141_7192 | Low-quality |
| S17_extracted_k141_7195 | Not-determined |
| S17_extracted_k141_21233 | Not-determined |
| S17_extracted_k141_124815 | Low-quality |
| S17_extracted_k141_132244 | Not-determined |
| S17_extracted_k141_81031 | Not-determined |
| S17_extracted_k141_124820 | Low-quality |
| S17_extracted_k141_28487 | Not-determined |
| S17_extracted_k141_81035 | Not-determined |
| S17_extracted_k141_176177 | Not-determined |
| S17_extracted_k141_162510 | Not-determined |
| S17_extracted_k141_58769 | Not-determined |
| S17_extracted_k141_103426 | Not-determined |
| S17_extracted_k141_73744 | Low-quality |
| S17_extracted_k141_28488 | Not-determined |
| S17_extracted_k141_7201 | Low-quality |
| S17_extracted_k141_103428 | Not-determined |
| S17_extracted_k141_64989 | Not-determined |
| S17_extracted_k141_124826 | Not-determined |
| S17_extracted_k141_169319 | Low-quality |
| S17_extracted_k141_162513 | Not-determined |
| S17_extracted_k141_103431 | Not-determined |
| S17_extracted_k141_95071 | Not-determined |
| S17_extracted_k141_103432 | Not-determined |
| S17_extracted_k141_118070 | Not-determined |
| S17_extracted_k141_169321 | Not-determined |
| S17_extracted_k141_28494 | Not-determined |
| S17_extracted_k141_132260 | Not-determined |
| S17_extracted_k141_73754 | Not-determined |
| S17_extracted_k141_87716 | Not-determined |
| S17_extracted_k141_65006 | Not-determined |
| S17_extracted_k141_58783 | Low-quality |
| S17_extracted_k141_28500 | Not-determined |
| S17_extracted_k141_28501 | Not-determined |
| S17_extracted_k141_124837 | Not-determined |
| S17_extracted_k141_21249 | Low-quality |
| S17_extracted_k141_21251 | Not-determined |
| S17_extracted_k141_118078 | Not-determined |
| S17_extracted_k141_103436 | Not-determined |
| S17_extracted_k141_132266 | Not-determined |
| S17_extracted_k141_65013 | Not-determined |
| S17_extracted_k141_21254 | Not-determined |
| S17_extracted_k141_103439 | Not-determined |
| S17_extracted_k141_21255 | Not-determined |
| S17_extracted_k141_162522 | Not-determined |
| S17_extracted_k141_132268 | Not-determined |
| S17_extracted_k141_28513 | Not-determined |
| S17_extracted_k141_87727 | Not-determined |
| S17_extracted_k141_176180 | Low-quality |
| S17_extracted_k141_162527 | Not-determined |
| S17_extracted_k141_81045 | Not-determined |
| S17_extracted_k141_169342 | Not-determined |
| S17_extracted_k141_87731 | Low-quality |
| S17_extracted_k141_176186 | Not-determined |
| S17_extracted_k141_169344 | Not-determined |
| S17_extracted_k141_132274 | Not-determined |
| S17_extracted_k141_95079 | Low-quality |
| S17_extracted_k141_139740 | Not-determined |
| S17_extracted_k141_95084 | Not-determined |
| S17_extracted_k141_7226 | Not-determined |
| S17_extracted_k141_21270 | Not-determined |
| S17_extracted_k141_21271 | Not-determined |
| S17_extracted_k141_176192 | Low-quality |
| S17_extracted_k141_28518 | Not-determined |
| S17_extracted_k141_118085 | Not-determined |
| S17_extracted_k141_95090 | Not-determined |
| S17_extracted_k141_176196 | Not-determined |
| S17_extracted_k141_176197 | Not-determined |
| S17_extracted_k141_162537 | Not-determined |
| S17_extracted_k141_118090 | Not-determined |
| S17_extracted_k141_124866 | Not-determined |
| S17_extracted_k141_81067 | Not-determined |
| S17_extracted_k141_95097 | Low-quality |
| S17_extracted_k141_58787 | Not-determined |
| S17_extracted_k141_95100 | Not-determined |
| S17_extracted_k141_139757 | Not-determined |
| S17_extracted_k141_21286 | Not-determined |
| S17_extracted_k141_65033 | Not-determined |
| S17_extracted_k141_169366 | Not-determined |
| S17_extracted_k141_73799 | Not-determined |
| S17_extracted_k141_81071 | Not-determined |
| S17_extracted_k141_95107 | Not-determined |
| S17_extracted_k141_176200 | Not-determined |
| S17_extracted_k141_21291 | Not-determined |
| S17_extracted_k141_132283 | Low-quality |
| S17_extracted_k141_58790 | Low-quality |
| S17_extracted_k141_21295 | Not-determined |
| S17_extracted_k141_7231 | Not-determined |
| S17_extracted_k141_132289 | Not-determined |
| S17_extracted_k141_124874 | Not-determined |
| S17_extracted_k141_73806 | Not-determined |
| S17_extracted_k141_65042 | Not-determined |
| S17_extracted_k141_169372 | Not-determined |
| S17_extracted_k141_118100 | Not-determined |
| S17_extracted_k141_169374 | Not-determined |
| S17_extracted_k141_14165 | Not-determined |
| S17_extracted_k141_65048 | Not-determined |
| S17_extracted_k141_139769 | Not-determined |
| S17_extracted_k141_87766 | Not-determined |
| S17_extracted_k141_95115 | Not-determined |
| S17_extracted_k141_73813 | Not-determined |
| S17_extracted_k141_169379 | Not-determined |
| S17_extracted_k141_132298 | Low-quality |
| S17_extracted_k141_139770 | Not-determined |
| S17_extracted_k141_73815 | Not-determined |
| S17_extracted_k141_73816 | Low-quality |
| S17_extracted_k141_73820 | Not-determined |
| S17_extracted_k141_169386 | Not-determined |
| S17_extracted_k141_73824 | Not-determined |
| S17_extracted_k141_81085 | Low-quality |
| S17_extracted_k141_169391 | Not-determined |
| S17_extracted_k141_95122 | Not-determined |
| S17_extracted_k141_58805 | Low-quality |
| S17_extracted_k141_124887 | Low-quality |
| S17_extracted_k141_95126 | Not-determined |
| S17_extracted_k141_118112 | Low-quality |
| S17_extracted_k141_73829 | Low-quality |
| S17_extracted_k141_169395 | Not-determined |
| S17_extracted_k141_28533 | Not-determined |
| S17_extracted_k141_65067 | Not-determined |
| S17_extracted_k141_132306 | Not-determined |
| S17_extracted_k141_14185 | Not-determined |
| S17_extracted_k141_21316 | Not-determined |
| S17_extracted_k141_132310 | Not-determined |
| S17_extracted_k141_95130 | Not-determined |
| S17_extracted_k141_21317 | Not-determined |
| S17_extracted_k141_124889 | Low-quality |
| S17_extracted_k141_95131 | Not-determined |
| S17_extracted_k141_176221 | Not-determined |
| S17_extracted_k141_95132 | Not-determined |
| S17_extracted_k141_169403 | Low-quality |
| S17_extracted_k141_65077 | Not-determined |
| S17_extracted_k141_81097 | Low-quality |
| S17_extracted_k141_28542 | Not-determined |
| S17_extracted_k141_95134 | Not-determined |
| S17_extracted_k141_14195 | Not-determined |
| S17_extracted_k141_169406 | Not-determined |
| S17_extracted_k141_169408 | Not-determined |
| S17_extracted_k141_7252 | Not-determined |
| S17_extracted_k141_21327 | Low-quality |
| S17_extracted_k141_176232 | Not-determined |
| S17_extracted_k141_28551 | Not-determined |
| S17_extracted_k141_28553 | Not-determined |
| S17_extracted_k141_118127 | Low-quality |
| S17_extracted_k141_176237 | Not-determined |
| S17_extracted_k141_65085 | Not-determined |
| S17_extracted_k141_65086 | Not-determined |
| S17_extracted_k141_65089 | Not-determined |
| S17_extracted_k141_87794 | Not-determined |
| S17_extracted_k141_58816 | Not-determined |
| S17_extracted_k141_118137 | Not-determined |
| S17_extracted_k141_14208 | Not-determined |
| S17_extracted_k141_36832 | Low-quality |
| S17_extracted_k141_81110 | Not-determined |
| S17_extracted_k141_124914 | Not-determined |
| S17_extracted_k141_81114 | Not-determined |
| S17_extracted_k141_28567 | Not-determined |
| S17_extracted_k141_139789 | Not-determined |
| S17_extracted_k141_132339 | Not-determined |
| S17_extracted_k141_28568 | Not-determined |
| S17_extracted_k141_139790 | Not-determined |
| S17_extracted_k141_87809 | Low-quality |
| S17_extracted_k141_36842 | Low-quality |
| S17_extracted_k141_124921 | Not-determined |
| S17_extracted_k141_87814 | Not-determined |
| S17_extracted_k141_139797 | Not-determined |
| S17_extracted_k141_81124 | Not-determined |
| S17_extracted_k141_139799 | Not-determined |
| S17_extracted_k141_36847 | Not-determined |
| S17_extracted_k141_58831 | Not-determined |
| S17_extracted_k141_58832 | Not-determined |
| S17_extracted_k141_28577 | Not-determined |
| S17_extracted_k141_14220 | Low-quality |
| S17_extracted_k141_81129 | Not-determined |
| S17_extracted_k141_87818 | Not-determined |
| S17_extracted_k141_132361 | Not-determined |
| S17_extracted_k141_132362 | Not-determined |
| S17_extracted_k141_118153 | Not-determined |
| S17_extracted_k141_139809 | Low-quality |
| S17_extracted_k141_95146 | Not-determined |
| S17_extracted_k141_118155 | Low-quality |
| S17_extracted_k141_36856 | Not-determined |
| S17_extracted_k141_176262 | Not-determined |
| S17_extracted_k141_21349 | Low-quality |
| S17_extracted_k141_36858 | Not-determined |
| S17_extracted_k141_139812 | Not-determined |
| S17_extracted_k141_118157 | Not-determined |
| S17_extracted_k141_65105 | Not-determined |
| S17_extracted_k141_28590 | Not-determined |
| S17_extracted_k141_28592 | Not-determined |
| S17_extracted_k141_176265 | Not-determined |
| S17_extracted_k141_87829 | Not-determined |
| S17_extracted_k141_21354 | Low-quality |
| S17_extracted_k141_21355 | Low-quality |
| S17_extracted_k141_176268 | Not-determined |
| S17_extracted_k141_87835 | Not-determined |
| S17_extracted_k141_118167 | Not-determined |
| S17_extracted_k141_73844 | Not-determined |
| S17_extracted_k141_176270 | Not-determined |
| S17_extracted_k141_36875 | Not-determined |
| S17_extracted_k141_21366 | Not-determined |
| S17_extracted_k141_176278 | Not-determined |
| S17_extracted_k141_124950 | Not-determined |
| S17_extracted_k141_21368 | Not-determined |
| S17_extracted_k141_87845 | Not-determined |
| S17_extracted_k141_73849 | Not-determined |
| S17_extracted_k141_36882 | Not-determined |

| | |
|---|---|
| S17_extracted_k141_21376 | Not-determined |
| S17_extracted_k141_139841 | Low-quality |
| S17_extracted_k141_28606 | Low-quality |
| S17_extracted_k141_87851 | Not-determined |
| S17_extracted_k141_169445 | Not-determined |
| S17_extracted_k141_124955 | Not-determined |
| S17_extracted_k141_87855 | Not-determined |
| S17_extracted_k141_95171 | Not-determined |
| S17_extracted_k141_139847 | Not-determined |
| S17_extracted_k141_124959 | Not-determined |
| S17_extracted_k141_118191 | Not-determined |
| S17_extracted_k141_87860 | Not-determined |
| S17_extracted_k141_36894 | Not-determined |
| S17_extracted_k141_21396 | Not-determined |
| S17_extracted_k141_58877 | Low-quality |
| S17_extracted_k141_124971 | Not-determined |
| S17_extracted_k141_65146 | Not-determined |
| S17_extracted_k141_139854 | Not-determined |
| S17_extracted_k141_65149 | Low-quality |
| S17_extracted_k141_95174 | Not-determined |
| S17_extracted_k141_65150 | Low-quality |
| S17_extracted_k141_36907 | Not-determined |
| S17_extracted_k141_81160 | Not-determined |
| S17_extracted_k141_139859 | Not-determined |
| S17_extracted_k141_28632 | Not-determined |
| S17_extracted_k141_95183 | Not-determined |
| S17_extracted_k141_28633 | Not-determined |
| S17_extracted_k141_124975 | Not-determined |
| S17_extracted_k141_169477 | Not-determined |
| S17_extracted_k141_169481 | Not-determined |
| S17_extracted_k141_118208 | Not-determined |
| S17_extracted_k141_36913 | Not-determined |
| S17_extracted_k141_81168 | Low-quality |
| S17_extracted_k141_7301 | Low-quality |
| S17_extracted_k141_36917 | Not-determined |
| S17_extracted_k141_73874 | Not-determined |
| S17_extracted_k141_65154 | Not-determined |
| S17_extracted_k141_81177 | Not-determined |
| S17_extracted_k141_81178 | Not-determined |
| S17_extracted_k141_81179 | Not-determined |
| S17_extracted_k141_176282 | Low-quality |
| S17_extracted_k141_87897 | Not-determined |
| S17_extracted_k141_73878 | Not-determined |
| S17_extracted_k141_58893 | Not-determined |
| S17_extracted_k141_169500 | Not-determined |
| S17_extracted_k141_73883 | Low-quality |
| S17_extracted_k141_87905 | Not-determined |
| S17_extracted_k141_65168 | Not-determined |
| S17_extracted_k141_21435 | Not-determined |
| S17_extracted_k141_176285 | Low-quality |
| S17_extracted_k141_132392 | Not-determined |
| S17_extracted_k141_132397 | Not-determined |
| S17_extracted_k141_28659 | Not-determined |
| S17_extracted_k141_81196 | Not-determined |
| S17_extracted_k141_21449 | Not-determined |
| S17_extracted_k141_95203 | Not-determined |
| S17_extracted_k141_21454 | Not-determined |
| S17_extracted_k141_176296 | Low-quality |
| S17_extracted_k141_169505 | Not-determined |
| S17_extracted_k141_28668 | Not-determined |
| S17_extracted_k141_65178 | Low-quality |
| S17_extracted_k141_139862 | Not-determined |
| S17_extracted_k141_21460 | Not-determined |
| S17_extracted_k141_95209 | Not-determined |
| S17_extracted_k141_28674 | Not-determined |
| S17_extracted_k141_95211 | Not-determined |
| S17_extracted_k141_132398 | Low-quality |
| S17_extracted_k141_132399 | Not-determined |
| S17_extracted_k141_7325 | Not-determined |
| S17_extracted_k141_87917 | Not-determined |
| S17_extracted_k141_21468 | Not-determined |
| S17_extracted_k141_132404 | Not-determined |
| S17_extracted_k141_7330 | Not-determined |
| S17_extracted_k141_28685 | Not-determined |
| S17_extracted_k141_95222 | Not-determined |
| S17_extracted_k141_36922 | Not-determined |
| S17_extracted_k141_7334 | Not-determined |
| S17_extracted_k141_169517 | Not-determined |
| S17_extracted_k141_87926 | Not-determined |
| S17_extracted_k141_169519 | Not-determined |
| S17_extracted_k141_21481 | Not-determined |
| S17_extracted_k141_132412 | Not-determined |
| S17_extracted_k141_139876 | Not-determined |
| S17_extracted_k141_7335 | Not-determined |
| S17_extracted_k141_36927 | Low-quality |
| S17_extracted_k141_169522 | Not-determined |
| S17_extracted_k141_169523 | Not-determined |
| S17_extracted_k141_21492 | Not-determined |
| S17_extracted_k141_28700 | Not-determined |
| S17_extracted_k141_169526 | Not-determined |
| S17_extracted_k141_169529 | Not-determined |
| S17_extracted_k141_139886 | Not-determined |
| S17_extracted_k141_28707 | Low-quality |
| S17_extracted_k141_139890 | Not-determined |
| S17_extracted_k141_169533 | Low-quality |
| S17_extracted_k141_176330 | Not-determined |
| S17_extracted_k141_36933 | Low-quality |
| S17_extracted_k141_65210 | Not-determined |
| S17_extracted_k141_176332 | Not-determined |
| S17_extracted_k141_65214 | Not-determined |
| S17_extracted_k141_21505 | Not-determined |
| S17_extracted_k141_132421 | Low-quality |
| S17_extracted_k141_36943 | Not-determined |
| S17_extracted_k141_169550 | Not-determined |
| S17_extracted_k141_169551 | Not-determined |
| S17_extracted_k141_21513 | Not-determined |
| S17_extracted_k141_81214 | Not-determined |
| S17_extracted_k141_132431 | Not-determined |
| S17_extracted_k141_21517 | Not-determined |
| S17_extracted_k141_21518 | Not-determined |
| S17_extracted_k141_139904 | Not-determined |
| S17_extracted_k141_169558 | Not-determined |
| S17_extracted_k141_176351 | Not-determined |
| S17_extracted_k141_28732 | Not-determined |
| S17_extracted_k141_14239 | Not-determined |
| S17_extracted_k141_14240 | Not-determined |
| S17_extracted_k141_139918 | Not-determined |
| S17_extracted_k141_28736 | Not-determined |
| S17_extracted_k141_28740 | Low-quality |
| S17_extracted_k141_169570 | Not-determined |
| S17_extracted_k141_65232 | Not-determined |
| S17_extracted_k141_139922 | Not-determined |
| S17_extracted_k141_132450 | Not-determined |
| S17_extracted_k141_139924 | Not-determined |
| S17_extracted_k141_65236 | Not-determined |
| S17_extracted_k141_176358 | Not-determined |
| S17_extracted_k141_28746 | Not-determined |
| S17_extracted_k141_176362 | Not-determined |
| S17_extracted_k141_176367 | Low-quality |
| S17_extracted_k141_14262 | Not-determined |
| S17_extracted_k141_28754 | Not-determined |
| S17_extracted_k141_139936 | Not-determined |
| S17_extracted_k141_139937 | Not-determined |
| S17_extracted_k141_132468 | Not-determined |
| S17_extracted_k141_169590 | Not-determined |
| S17_extracted_k141_21541 | Not-determined |
| S17_extracted_k141_28767 | Not-determined |
| S17_extracted_k141_124988 | Not-determined |
| S17_extracted_k141_132481 | Not-determined |
| S17_extracted_k141_14285 | Not-determined |
| S17_extracted_k141_65272 | Not-determined |
| S17_extracted_k141_14286 | Not-determined |
| S17_extracted_k141_124995 | Low-quality |
| S17_extracted_k141_132492 | Low-quality |
| S17_extracted_k141_65274 | Not-determined |
| S17_extracted_k141_139962 | Not-determined |
| S17_extracted_k141_132497 | Not-determined |
| S17_extracted_k141_95271 | Not-determined |
| S17_extracted_k141_21566 | Not-determined |
| S17_extracted_k141_176389 | Not-determined |
| S17_extracted_k141_21569 | Not-determined |
| S17_extracted_k141_14295 | Not-determined |
| S17_extracted_k141_28783 | Not-determined |
| S17_extracted_k141_87990 | Not-determined |
| S17_extracted_k141_14300 | Not-determined |
| S17_extracted_k141_169641 | Not-determined |
| S17_extracted_k141_14304 | Not-determined |
| S17_extracted_k141_95278 | Not-determined |
| S17_extracted_k141_21579 | Not-determined |
| S17_extracted_k141_21580 | Low-quality |
| S17_extracted_k141_28793 | Not-determined |
| S17_extracted_k141_28794 | Not-determined |
| S17_extracted_k141_125015 | Not-determined |
| S17_extracted_k141_125017 | Not-determined |
| S17_extracted_k141_169645 | Not-determined |
| S17_extracted_k141_139982 | Not-determined |
| S17_extracted_k141_176396 | Not-determined |
| S17_extracted_k141_139983 | Not-determined |
| S17_extracted_k141_95286 | Not-determined |
| S17_extracted_k141_28801 | Not-determined |
| S17_extracted_k141_58929 | Not-determined |
| S17_extracted_k141_176400 | Not-determined |
| S17_extracted_k141_65288 | Not-determined |
| S17_extracted_k141_132503 | Not-determined |
| S17_extracted_k141_58937 | Low-quality |
| S17_extracted_k141_139997 | Not-determined |
| S17_extracted_k141_176406 | Low-quality |
| S17_extracted_k141_169660 | Low-quality |
| S17_extracted_k141_87997 | Not-determined |
| S17_extracted_k141_58942 | Low-quality |
| S17_extracted_k141_88000 | Not-determined |
| S17_extracted_k141_140002 | Low-quality |
| S17_extracted_k141_140006 | Not-determined |
| S17_extracted_k141_14331 | Not-determined |
| S17_extracted_k141_140011 | Low-quality |
| S17_extracted_k141_132526 | Not-determined |
| S17_extracted_k141_14338 | Not-determined |
| S17_extracted_k141_125045 | Not-determined |
| S17_extracted_k141_95312 | Not-determined |
| S17_extracted_k141_14342 | Not-determined |
| S17_extracted_k141_81220 | Not-determined |
| S17_extracted_k141_21611 | Not-determined |
| S17_extracted_k141_28825 | Not-determined |
| S17_extracted_k141_169679 | Not-determined |
| S17_extracted_k141_81224 | Not-determined |
| S17_extracted_k141_125052 | Not-determined |
| S17_extracted_k141_81226 | Not-determined |
| S17_extracted_k141_95319 | Not-determined |
| S17_extracted_k141_28829 | Low-quality |
| S17_extracted_k141_95323 | Not-determined |
| S17_extracted_k141_88028 | Not-determined |
| S17_extracted_k141_169686 | Not-determined |
| S17_extracted_k141_21627 | Not-determined |
| S17_extracted_k141_21628 | Not-determined |
| S17_extracted_k141_14359 | Not-determined |
| S17_extracted_k141_21630 | Not-determined |
| S17_extracted_k141_176440 | Low-quality |
| S17_extracted_k141_81231 | Not-determined |
| S17_extracted_k141_88037 | Not-determined |
| S17_extracted_k141_125074 | Not-determined |
| S17_extracted_k141_65321 | Not-determined |
| S17_extracted_k141_65323 | Not-determined |
| S17_extracted_k141_176441 | Not-determined |
| S17_extracted_k141_176445 | Not-determined |
| S17_extracted_k141_14368 | Low-quality |
| S17_extracted_k141_65330 | Not-determined |
| S17_extracted_k141_14370 | Not-determined |
| S17_extracted_k141_21643 | Not-determined |
| S17_extracted_k141_95342 | Not-determined |
| S17_extracted_k141_95343 | Low-quality |
| S17_extracted_k141_58970 | Not-determined |
| S17_extracted_k141_65333 | Not-determined |
| S17_extracted_k141_28841 | Not-determined |
| S17_extracted_k141_125086 | Not-determined |
| S17_extracted_k141_58974 | Not-determined |
| S17_extracted_k141_95351 | Not-determined |
| S17_extracted_k141_132550 | Not-determined |
| S17_extracted_k141_28849 | Not-determined |
| S17_extracted_k141_58981 | Not-determined |
| S17_extracted_k141_21656 | Not-determined |
| S17_extracted_k141_169717 | Not-determined |
| S17_extracted_k141_65344 | Not-determined |
| S17_extracted_k141_125097 | Not-determined |
| S17_extracted_k141_132555 | Not-determined |
| S17_extracted_k141_95354 | Not-determined |
| S17_extracted_k141_28853 | Not-determined |
| S17_extracted_k141_21659 | Not-determined |
| S17_extracted_k141_125103 | Not-determined |
| S17_extracted_k141_169722 | Not-determined |
| S17_extracted_k141_65348 | Not-determined |
| S17_extracted_k141_95359 | Not-determined |
| S17_extracted_k141_58985 | Low-quality |
| S17_extracted_k141_125108 | Low-quality |
| S17_extracted_k141_21666 | Not-determined |
| S17_extracted_k141_58989 | Not-determined |
| S17_extracted_k141_28860 | Not-determined |
| S17_extracted_k141_132565 | Not-determined |
| S17_extracted_k141_21670 | Not-determined |
| S17_extracted_k141_140034 | Not-determined |
| S17_extracted_k141_14383 | Not-determined |
| S17_extracted_k141_140035 | Low-quality |
| S17_extracted_k141_132575 | Not-determined |
| S17_extracted_k141_28869 | Not-determined |
| S17_extracted_k141_125121 | Low-quality |
| S17_extracted_k141_125122 | Low-quality |
| S17_extracted_k141_169731 | Not-determined |
| S17_extracted_k141_140045 | Not-determined |
| S17_extracted_k141_65364 | Not-determined |
| S17_extracted_k141_140046 | Not-determined |
| S17_extracted_k141_125128 | Low-quality |
| S17_extracted_k141_169737 | Not-determined |
| S17_extracted_k141_176475 | Not-determined |
| S17_extracted_k141_169739 | Not-determined |
| S17_extracted_k141_14396 | Not-determined |
| S17_extracted_k141_28881 | Low-quality |
| S17_extracted_k141_169746 | Not-determined |
| S17_extracted_k141_169748 | Not-determined |
| S17_extracted_k141_125134 | Low-quality |
| S17_extracted_k141_169756 | Not-determined |
| S17_extracted_k141_14403 | Low-quality |
| S17_extracted_k141_125142 | Not-determined |
| S17_extracted_k141_132604 | Not-determined |
| S17_extracted_k141_125147 | Not-determined |
| S17_extracted_k141_65385 | Not-determined |
| S17_extracted_k141_176489 | Not-determined |
| S17_extracted_k141_125150 | Not-determined |
| S17_extracted_k141_21704 | Not-determined |
| S17_extracted_k141_28883 | Not-determined |
| S17_extracted_k141_125151 | Not-determined |
| S17_extracted_k141_21707 | Low-quality |
| S17_extracted_k141_125153 | Not-determined |
| S17_extracted_k141_14408 | Not-determined |
| S17_extracted_k141_176496 | Not-determined |
| S17_extracted_k141_14414 | Not-determined |
| S17_extracted_k141_132612 | Low-quality |
| S17_extracted_k141_88042 | Not-determined |
| S17_extracted_k141_28895 | Not-determined |
| S17_extracted_k141_132624 | Not-determined |
| S17_extracted_k141_7361 | Low-quality |
| S17_extracted_k141_28898 | Low-quality |
| S17_extracted_k141_59003 | Not-determined |
| S17_extracted_k141_132631 | Not-determined |
| S17_extracted_k141_176508 | Not-determined |
| S17_extracted_k141_88052 | Not-determined |
| S17_extracted_k141_88058 | Not-determined |
| S17_extracted_k141_169784 | Not-determined |
| S17_extracted_k141_59008 | Low-quality |
| S17_extracted_k141_59014 | Not-determined |
| S17_extracted_k141_140066 | Not-determined |
| S17_extracted_k141_176518 | Not-determined |
| S17_extracted_k141_59016 | Not-determined |
| S17_extracted_k141_59017 | Not-determined |
| S17_extracted_k141_59021 | Not-determined |
| S17_extracted_k141_140071 | Not-determined |
| S17_extracted_k141_132647 | Not-determined |
| S17_extracted_k141_88066 | Not-determined |
| S17_extracted_k141_169787 | Not-determined |
| S17_extracted_k141_7367 | Not-determined |
| S17_extracted_k141_14423 | Not-determined |
| S17_extracted_k141_65392 | Not-determined |
| S17_extracted_k141_132648 | Not-determined |
| S17_extracted_k141_59026 | Not-determined |
| S17_extracted_k141_176535 | Not-determined |
| S17_extracted_k141_59027 | Low-quality |
| S17_extracted_k141_125161 | Not-determined |
| S17_extracted_k141_59031 | Not-determined |
| S17_extracted_k141_132657 | Not-determined |
| S17_extracted_k141_59035 | Low-quality |
| S17_extracted_k141_95369 | Not-determined |
| S17_extracted_k141_95371 | Not-determined |
| S17_extracted_k141_28924 | Low-quality |
| S17_extracted_k141_125173 | Not-determined |
| S17_extracted_k141_59041 | Not-determined |
| S17_extracted_k141_140072 | Low-quality |
| S17_extracted_k141_14436 | Not-determined |
| S17_extracted_k141_140074 | Not-determined |
| S17_extracted_k141_125175 | Not-determined |
| S17_extracted_k141_125177 | Low-quality |
| S17_extracted_k141_65411 | Not-determined |
| S17_extracted_k141_65412 | Not-determined |
| S17_extracted_k141_125179 | Not-determined |
| S17_extracted_k141_14439 | Not-determined |
| S17_extracted_k141_59045 | Not-determined |
| S17_extracted_k141_65415 | Not-determined |
| S17_extracted_k141_28934 | Not-determined |
| S17_extracted_k141_125185 | Not-determined |
| S17_extracted_k141_28936 | Not-determined |
| S17_extracted_k141_88092 | Not-determined |
| S17_extracted_k141_140087 | Not-determined |
| S17_extracted_k141_125189 | Low-quality |
| S17_extracted_k141_28938 | Not-determined |
| S17_extracted_k141_14449 | Not-determined |
| S17_extracted_k141_65423 | Not-determined |
| S17_extracted_k141_14452 | Low-quality |
| S17_extracted_k141_65424 | Not-determined |
| S17_extracted_k141_95388 | Not-determined |
| S17_extracted_k141_65430 | Not-determined |
| S17_extracted_k141_28947 | Not-determined |
| S17_extracted_k141_132688 | Not-determined |
| S17_extracted_k141_169800 | Low-quality |
| S17_extracted_k141_169802 | Not-determined |
| S17_extracted_k141_169805 | Not-determined |
| S17_extracted_k141_169811 | Low-quality |
| S17_extracted_k141_59065 | Low-quality |
| S17_extracted_k141_59067 | Not-determined |
| S17_extracted_k141_125207 | Not-determined |
| S17_extracted_k141_28957 | Not-determined |
| S17_extracted_k141_59071 | Not-determined |
| S17_extracted_k141_65449 | Low-quality |
| S17_extracted_k141_132696 | Not-determined |
| S17_extracted_k141_176562 | Low-quality |
| S17_extracted_k141_132697 | Not-determined |
| S17_extracted_k141_28958 | Not-determined |
| S17_extracted_k141_7376 | Not-determined |
| S17_extracted_k141_95398 | Not-determined |
| S17_extracted_k141_14477 | Not-determined |
| S17_extracted_k141_95399 | Not-determined |
| S17_extracted_k141_28961 | Not-determined |
| S17_extracted_k141_122263 | High-quality |
| S17_extracted_k141_169824 | Not-determined |
| S17_extracted_k141_140094 | Not-determined |
| S17_extracted_k141_59081 | Not-determined |
| S17_extracted_k141_169825 | Not-determined |
| S17_extracted_k141_7379 | Low-quality |
| S17_extracted_k141_14478 | Low-quality |
| S17_extracted_k141_7381 | Not-determined |
| S17_extracted_k141_140099 | Not-determined |
| S17_extracted_k141_7382 | Not-determined |
| S17_extracted_k141_95402 | Low-quality |
| S17_extracted_k141_59082 | Not-determined |
| S17_extracted_k141_59083 | Not-determined |
| S17_extracted_k141_169833 | Not-determined |
| S17_extracted_k141_140103 | Not-determined |
| S17_extracted_k141_21721 | Low-quality |
| S17_extracted_k141_88117 | Not-determined |
| S17_vs_k141_122263\|\|full | High-quality |
| S17_extracted_k141_28969 | Not-determined |
| S17_extracted_k141_140108 | Not-determined |
| S17_extracted_k141_81234 | Low-quality |
| S17_extracted_k141_88123 | Low-quality |
| S17_extracted_k141_7388 | Low-quality |
| S17_extracted_k141_169843 | Low-quality |
| S17_extracted_k141_28977 | Not-determined |
| S17_extracted_k141_176577 | Not-determined |
| S17_extracted_k141_132711 | Not-determined |
| S17_extracted_k141_65455 | Not-determined |
| S17_extracted_k141_169848 | Not-determined |
| S17_extracted_k141_81245 | Not-determined |
| S17_extracted_k141_95413 | Not-determined |
| S17_extracted_k141_140119 | Not-determined |
| S17_extracted_k141_21731 | Low-quality |
| S17_extracted_k141_140120 | Not-determined |
| S17_extracted_k141_140121 | Not-determined |
| S17_extracted_k141_140124 | Not-determined |
| S17_extracted_k141_132724 | Not-determined |
| S17_extracted_k141_169859 | Low-quality |
| S17_extracted_k141_95424 | Not-determined |
| S17_extracted_k141_140126 | Not-determined |
| S17_extracted_k141_65469 | Not-determined |
| S17_extracted_k141_95427 | Not-determined |
| S17_extracted_k141_132728 | Low-quality |
| S17_extracted_k141_28994 | Not-determined |
| S17_extracted_k141_81250 | Not-determined |
| S17_extracted_k141_169867 | Low-quality |
| S17_extracted_k141_65475 | Not-determined |
| S17_extracted_k141_81252 | Not-determined |
| S17_extracted_k141_132740 | Not-determined |
| S17_extracted_k141_132744 | Not-determined |
| S17_extracted_k141_14513 | Low-quality |
| S17_extracted_k141_88157 | Low-quality |
| S17_extracted_k141_81258 | Not-determined |
| S17_extracted_k141_176593 | Not-determined |
| S17_extracted_k141_81266 | Low-quality |
| S17_extracted_k141_176599 | Not-determined |
| S17_extracted_k141_21737 | Not-determined |
| S17_extracted_k141_176602 | Not-determined |
| S17_extracted_k141_176603 | Not-determined |
| S17_extracted_k141_88168 | Not-determined |
| S17_extracted_k141_14523 | Not-determined |
| S17_extracted_k141_14525 | Not-determined |
| S17_extracted_k141_88176 | Not-determined |
| S17_extracted_k141_21744 | Not-determined |
| S17_extracted_k141_125209 | Low-quality |
| S17_extracted_k141_81272 | Not-determined |
| S17_extracted_k141_81273 | Not-determined |
| S17_extracted_k141_169885 | Not-determined |
| S17_extracted_k141_140147 | Low-quality |
| S17_extracted_k141_81275 | Not-determined |
| S17_extracted_k141_29003 | Not-determined |
| S17_extracted_k141_169891 | Not-determined |
| S17_extracted_k141_14535 | Not-determined |
| S17_extracted_k141_29011 | Not-determined |
| S17_extracted_k141_95462 | Not-determined |
| S17_extracted_k141_169895 | Not-determined |
| S17_extracted_k141_95464 | Low-quality |
| S17_extracted_k141_88186 | Not-determined |
| S17_extracted_k141_88190 | Low-quality |
| S17_extracted_k141_14543 | Not-determined |
| S17_extracted_k141_21751 | Not-determined |
| S17_extracted_k141_21752 | Not-determined |
| S17_extracted_k141_169899 | Not-determined |
| S17_extracted_k141_169900 | Not-determined |
| S17_extracted_k141_65503 | Not-determined |
| S17_extracted_k141_140164 | Not-determined |
| S17_extracted_k141_65510 | Not-determined |
| S17_extracted_k141_125234 | Low-quality |
| S17_extracted_k141_95487 | Low-quality |
| S17_extracted_k141_14555 | Not-determined |
| S17_extracted_k141_65525 | Not-determined |
| S17_extracted_k141_95493 | Not-determined |
| S17_extracted_k141_169924 | Low-quality |
| S17_extracted_k141_125242 | Not-determined |
| S17_extracted_k141_140183 | Not-determined |
| S17_extracted_k141_140184 | Not-determined |
| S17_extracted_k141_95507 | Not-determined |
| S17_extracted_k141_140189 | Low-quality |
| S17_extracted_k141_21786 | Not-determined |
| S17_extracted_k141_14565 | Not-determined |
| S17_extracted_k141_132769 | Not-determined |
| S17_extracted_k141_125256 | Not-determined |
| S17_extracted_k141_95518 | Not-determined |
| S17_extracted_k141_21787 | Not-determined |
| S17_extracted_k141_125264 | Not-determined |
| S17_extracted_k141_95521 | Not-determined |
| S17_extracted_k141_125266 | Not-determined |
| S17_extracted_k141_125267 | Not-determined |
| S17_extracted_k141_95523 | Not-determined |
| S17_extracted_k141_14572 | Not-determined |
| S17_extracted_k141_14576 | Not-determined |
| S17_extracted_k141_29040 | Not-determined |
| S17_extracted_k141_21795 | Not-determined |
| S17_extracted_k141_95535 | Not-determined |
| S17_extracted_k141_65566 | Not-determined |
| S17_extracted_k141_125271 | Not-determined |
| S17_extracted_k141_132786 | Not-determined |
| S17_extracted_k141_95541 | Not-determined |
| S17_extracted_k141_140212 | Not-determined |
| S17_extracted_k141_132790 | Low-quality |
| S17_extracted_k141_65573 | Low-quality |
| S17_extracted_k141_132793 | Not-determined |
| S17_extracted_k141_132795 | Not-determined |
| S17_extracted_k141_95546 | Not-determined |
| S17_extracted_k141_95547 | Low-quality |
| S17_extracted_k141_176609 | Not-determined |
| S17_extracted_k141_88252 | Not-determined |
| S17_extracted_k141_14589 | Not-determined |
| S17_extracted_k141_65578 | Not-determined |
| S17_extracted_k141_95551 | Not-determined |
| S17_extracted_k141_176614 | Not-determined |
| S17_extracted_k141_132802 | Not-determined |
| S17_extracted_k141_125285 | Not-determined |
| S17_extracted_k141_95559 | Not-determined |
| S17_extracted_k141_176620 | Not-determined |
| S17_extracted_k141_176621 | Not-determined |
| S17_extracted_k141_14598 | Not-determined |
| S17_extracted_k141_14601 | Low-quality |
| S17_extracted_k141_176624 | Not-determined |
| S17_extracted_k141_65592 | Not-determined |
| S17_extracted_k141_14602 | Not-determined |
| S17_extracted_k141_176625 | Not-determined |
| S17_extracted_k141_132808 | Not-determined |
| S17_extracted_k141_29053 | Not-determined |
| S17_extracted_k141_176627 | Not-determined |
| S17_extracted_k141_125289 | Low-quality |
| S17_extracted_k141_65598 | Not-determined |
| S17_extracted_k141_88266 | Not-determined |
| S17_extracted_k141_140233 | Not-determined |
| S17_extracted_k141_88268 | Not-determined |
| S17_extracted_k141_21809 | Low-quality |
| S17_extracted_k141_65603 | Not-determined |
| S17_extracted_k141_176634 | Not-determined |
| S17_extracted_k141_29059 | Not-determined |
| S17_extracted_k141_140243 | Not-determined |
| S17_extracted_k141_95582 | Low-quality |
| S17_extracted_k141_21815 | Low-quality |
| S17_extracted_k141_21816 | Low-quality |
| S17_extracted_k141_21821 | Not-determined |
| S17_extracted_k141_65611 | Low-quality |
| S17_extracted_k141_65612 | Not-determined |
| S17_extracted_k141_14613 | Not-determined |
| S17_extracted_k141_65617 | Not-determined |
| S17_extracted_k141_132829 | Not-determined |
| S17_extracted_k141_140245 | Not-determined |
| S17_extracted_k141_95597 | Not-determined |
| S17_extracted_k141_132832 | Not-determined |
| S17_extracted_k141_21831 | Not-determined |
| S17_extracted_k141_65628 | Not-determined |
| S17_extracted_k141_125306 | Not-determined |
| S17_extracted_k141_176646 | Low-quality |
| S17_extracted_k141_132836 | Not-determined |
| S17_extracted_k141_176647 | Not-determined |
| S17_extracted_k141_125311 | Not-determined |
| S17_extracted_k141_132838 | Not-determined |
| S17_extracted_k141_14624 | Not-determined |
| S17_extracted_k141_140267 | Not-determined |
| S17_extracted_k141_140269 | Not-determined |
| S17_extracted_k141_132842 | Not-determined |
| S17_extracted_k141_132843 | Not-determined |
| S17_extracted_k141_140272 | Not-determined |
| S17_extracted_k141_176660 | Not-determined |
| S17_extracted_k141_176661 | Not-determined |
| S17_extracted_k141_132848 | Low-quality |
| S17_extracted_k141_125327 | Not-determined |
| S17_extracted_k141_65647 | Not-determined |
| S17_extracted_k141_29090 | Not-determined |
| S17_extracted_k141_132856 | Not-determined |
| S17_extracted_k141_125337 | Not-determined |
| S17_extracted_k141_29095 | Not-determined |
| S17_extracted_k141_14643 | Not-determined |
| S17_extracted_k141_29100 | Not-determined |
| S17_extracted_k141_29103 | Not-determined |
| S17_extracted_k141_125351 | Not-determined |
| S17_extracted_k141_132862 | Not-determined |
| S17_extracted_k141_125354 | Not-determined |
| S17_extracted_k141_14648 | Not-determined |
| S17_extracted_k141_125355 | Not-determined |
| S17_extracted_k141_95616 | Not-determined |
| S17_extracted_k141_29109 | Not-determined |
| S17_extracted_k141_176672 | Not-determined |
| S17_extracted_k141_176676 | Not-determined |
| S17_extracted_k141_140285 | Not-determined |
| S17_extracted_k141_125359 | Low-quality |
| S17_extracted_k141_140286 | Low-quality |
| S17_extracted_k141_29111 | Not-determined |
| S17_extracted_k141_95633 | Not-determined |
| S17_extracted_k141_14660 | Not-determined |
| S17_extracted_k141_95641 | Not-determined |
| S17_extracted_k141_14666 | Not-determined |
| S17_extracted_k141_140295 | Not-determined |
| S17_extracted_k141_14667 | Not-determined |
| S17_extracted_k141_140302 | Low-quality |
| S17_extracted_k141_132877 | Not-determined |
| S17_extracted_k141_140306 | Not-determined |
| S17_extracted_k141_29131 | Not-determined |
| S17_extracted_k141_125374 | Not-determined |
| S17_extracted_k141_132880 | Not-determined |
| S17_extracted_k141_176702 | Not-determined |
| S17_extracted_k141_29135 | Not-determined |
| S17_extracted_k141_132882 | Not-determined |

| | |
|---|---|
| S17_extracted_k141_176703 | Not-determined |
| S17_extracted_k141_125392 | Not-determined |
| S17_extracted_k141_176707 | Not-determined |
| S17_extracted_k141_125394 | Low-quality |
| S17_extracted_k141_14675 | Not-determined |
| S17_extracted_k141_88290 | Not-determined |
| S17_extracted_k141_29149 | Not-determined |
| S17_extracted_k141_14684 | Not-determined |
| S17_extracted_k141_88293 | Not-determined |
| S17_extracted_k141_88295 | Not-determined |
| S17_extracted_k141_140318 | Not-determined |
| S17_extracted_k141_95680 | Not-determined |
| S17_extracted_k141_176723 | Not-determined |
| S17_extracted_k141_176725 | Not-determined |
| S17_extracted_k141_140327 | Low-quality |
| S17_extracted_k141_176728 | Not-determined |
| S17_extracted_k141_176729 | Not-determined |
| S17_extracted_k141_88308 | Not-determined |
| S17_extracted_k141_21842 | Not-determined |
| S17_extracted_k141_176742 | Not-determined |
| S17_extracted_k141_132915 | Not-determined |
| S17_extracted_k141_95701 | Not-determined |
| S17_extracted_k141_132919 | Not-determined |
| S17_extracted_k141_140342 | Not-determined |
| S17_extracted_k141_29165 | Not-determined |
| S17_extracted_k141_140348 | Not-determined |
| S17_extracted_k141_125406 | Not-determined |
| S17_extracted_k141_88330 | Not-determined |
| S17_extracted_k141_21852 | Low-quality |
| S17_extracted_k141_21853 | Not-determined |
| S17_extracted_k141_125410 | Not-determined |
| S17_extracted_k141_125411 | Not-determined |
| S17_extracted_k141_29178 | Not-determined |
| S17_extracted_k141_176751 | Not-determined |
| S17_extracted_k141_29179 | Not-determined |
| S17_extracted_k141_21864 | Not-determined |
| S17_extracted_k141_125414 | Not-determined |
| S17_extracted_k141_21866 | Not-determined |
| S17_extracted_k141_29183 | Not-determined |
| S17_extracted_k141_125418 | Low-quality |
| S17_extracted_k141_125424 | Not-determined |
| S17_extracted_k141_132939 | Not-determined |
| S17_extracted_k141_88347 | Not-determined |
| S17_extracted_k141_21872 | Not-determined |
| S17_extracted_k141_21877 | Not-determined |
| S17_extracted_k141_140349 | Low-quality |
| S17_extracted_k141_140352 | Not-determined |
| S17_extracted_k141_65668 | Not-determined |
| S17_extracted_k141_140360 | Not-determined |
| S17_extracted_k141_125442 | Low-quality |
| S17_extracted_k141_29192 | Not-determined |
| S17_extracted_k141_21882 | Low-quality |
| S17_extracted_k141_95717 | Not-determined |
| S17_extracted_k141_95718 | Low-quality |
| S17_extracted_k141_132941 | Not-determined |
| S17_extracted_k141_125447 | Low-quality |
| S17_extracted_k141_95727 | Not-determined |
| S17_extracted_k141_88358 | Not-determined |
| S17_extracted_k141_88359 | Not-determined |
| S17_extracted_k141_29195 | Low-quality |
| S17_extracted_k141_88364 | Not-determined |
| S17_extracted_k141_88365 | Not-determined |
| S17_extracted_k141_65688 | Not-determined |
| S17_extracted_k141_88366 | Not-determined |
| S17_extracted_k141_65691 | Not-determined |
| S17_extracted_k141_132955 | Not-determined |
| S17_extracted_k141_176760 | Not-determined |
| S17_extracted_k141_176762 | Not-determined |
| S17_extracted_k141_140363 | Low-quality |
| S17_extracted_k141_140364 | Low-quality |
| S17_extracted_k141_65704 | Not-determined |
| S17_extracted_k141_29215 | Not-determined |
| S17_extracted_k141_125471 | Not-determined |
| S17_extracted_k141_132964 | Not-determined |
| S17_extracted_k141_176771 | Not-determined |
| S17_extracted_k141_140375 | Low-quality |
| S17_extracted_k141_88378 | Not-determined |
| S17_extracted_k141_65715 | Not-determined |
| S17_extracted_k141_125482 | Not-determined |
| S17_extracted_k141_132970 | Not-determined |
| S17_extracted_k141_14716 | Not-determined |
| S17_extracted_k141_125484 | Not-determined |
| S17_extracted_k141_88380 | Low-quality |
| S17_extracted_k141_14720 | Not-determined |
| S17_extracted_k141_14722 | Not-determined |
| S17_extracted_k141_132973 | Not-determined |
| S17_extracted_k141_65726 | Low-quality |
| S17_extracted_k141_140380 | Not-determined |
| S17_extracted_k141_176777 | Not-determined |
| S17_extracted_k141_176780 | Not-determined |
| S17_extracted_k141_132987 | Not-determined |
| S17_extracted_k141_125498 | Not-determined |
| S17_extracted_k141_88395 | Not-determined |
| S17_extracted_k141_132991 | Not-determined |
| S17_extracted_k141_176787 | Not-determined |
| S17_extracted_k141_21921 | Not-determined |
| S17_extracted_k141_95772 | Not-determined |
| S17_extracted_k141_125501 | Not-determined |
| S17_extracted_k141_14731 | Not-determined |
| S17_extracted_k141_95777 | Not-determined |
| S17_extracted_k141_95780 | Not-determined |
| S17_extracted_k141_29229 | Not-determined |
| S17_extracted_k141_176798 | Low-quality |
| S17_extracted_k141_65738 | Not-determined |
| S17_extracted_k141_65741 | Not-determined |
| S17_extracted_k141_29234 | Not-determined |
| S17_extracted_k141_21932 | Not-determined |
| S17_extracted_k141_14750 | Low-quality |
| S17_extracted_k141_125509 | Not-determined |
| S17_extracted_k141_65748 | Low-quality |
| S17_extracted_k141_29244 | Not-determined |
| S17_extracted_k141_176819 | Not-determined |
| S17_extracted_k141_88431 | Not-determined |
| S17_extracted_k141_65761 | Not-determined |
| S17_extracted_k141_176821 | Not-determined |
| S17_extracted_k141_95808 | Not-determined |
| S17_extracted_k141_29260 | Not-determined |
| S17_extracted_k141_88440 | Not-determined |
| S17_extracted_k141_14757 | Low-quality |
| S17_extracted_k141_14760 | Not-determined |
| S17_extracted_k141_176830 | Not-determined |
| S17_extracted_k141_176833 | Not-determined |
| S17_extracted_k141_65774 | Not-determined |
| S17_extracted_k141_176845 | Not-determined |
| S17_extracted_k141_125541 | Not-determined |
| S17_extracted_k141_95826 | Not-determined |
| S17_extracted_k141_21948 | Not-determined |
| S17_extracted_k141_14772 | Not-determined |
| S17_extracted_k141_125547 | Not-determined |
| S17_extracted_k141_14776 | Not-determined |
| S17_extracted_k141_176853 | Not-determined |
| S17_extracted_k141_65791 | Not-determined |
| S17_extracted_k141_125553 | Not-determined |
| S17_extracted_k141_65793 | Not-determined |
| S17_extracted_k141_65794 | Not-determined |
| S17_extracted_k141_65795 | Not-determined |
| S17_extracted_k141_21951 | Not-determined |
| S17_extracted_k141_29283 | Not-determined |
| S17_extracted_k141_176864 | Not-determined |
| S17_extracted_k141_125561 | Not-determined |
| S17_extracted_k141_29290 | Not-determined |
| S17_extracted_k141_65800 | Not-determined |
| S17_extracted_k141_29295 | Not-determined |
| S17_extracted_k141_65806 | Not-determined |
| S17_extracted_k141_29300 | Not-determined |
| S17_extracted_k141_88464 | Not-determined |
| S17_extracted_k141_176870 | Low-quality |
| S17_extracted_k141_21972 | Not-determined |
| S17_extracted_k141_21975 | Low-quality |
| S17_extracted_k141_176883 | Not-determined |
| S17_extracted_k141_88473 | Low-quality |
| S17_extracted_k141_88474 | Not-determined |
| S17_extracted_k141_176891 | Not-determined |
| S17_extracted_k141_65817 | Not-determined |
| S17_extracted_k141_125580 | Not-determined |
| S17_extracted_k141_88479 | Low-quality |
| S17_extracted_k141_176903 | Not-determined |
| S17_extracted_k141_176904 | Not-determined |
| S17_extracted_k141_176910 | Not-determined |
| S17_extracted_k141_176912 | Not-determined |
| S17_extracted_k141_176927 | Not-determined |
| S17_extracted_k141_176929 | Not-determined |
| S17_extracted_k141_65846 | Not-determined |
| S17_extracted_k141_65852 | Not-determined |
| S17_extracted_k141_65855 | Not-determined |
| S17_extracted_k141_65857 | Low-quality |
| S17_extracted_k141_29317 | Not-determined |
| S17_extracted_k141_65866 | Not-determined |
| S17_extracted_k141_29319 | Not-determined |
| S17_extracted_k141_22010 | Not-determined |
| S17_extracted_k141_29322 | Not-determined |
| S17_extracted_k141_22012 | Not-determined |
| S17_extracted_k141_29350 | Not-determined |
| S17_extracted_k141_29356 | Not-determined |
| S17_extracted_k141_65909 | Not-determined |
| S17_extracted_k141_88508 | Not-determined |
| S17_extracted_k141_29365 | Not-determined |
| S17_extracted_k141_29370 | Not-determined |
| S17_extracted_k141_29371 | Not-determined |
| S17_extracted_k141_65919 | Not-determined |
| S17_extracted_k141_29377 | Not-determined |
| S17_extracted_k141_29381 | Not-determined |
| S17_extracted_k141_29382 | Not-determined |
| S17_extracted_k141_29387 | Not-determined |
| S17_extracted_k141_65935 | Not-determined |
| S17_extracted_k141_29393 | Low-quality |
| S17_extracted_k141_29396 | Not-determined |
| S17_extracted_k141_29399 | Not-determined |
| S17_extracted_k141_65943 | Low-quality |
| S17_extracted_k141_88536 | Not-determined |
| S17_extracted_k141_88544 | Low-quality |
| S17_extracted_k141_29426 | Not-determined |
| S17_extracted_k141_176946 | Not-determined |
| S17_extracted_k141_95849 | Low-quality |
| S17_extracted_k141_176963 | Not-determined |
| S17_extracted_k141_88565 | Not-determined |
| S17_extracted_k141_29441 | Not-determined |
| S17_extracted_k141_176969 | Not-determined |
| S17_extracted_k141_95856 | Not-determined |
| S17_extracted_k141_176974 | Not-determined |
| S17_extracted_k141_176975 | Not-determined |
| S17_extracted_k141_22019 | Not-determined |
| S17_extracted_k141_176976 | Not-determined |
| S17_extracted_k141_88577 | Not-determined |
| S17_extracted_k141_29447 | Not-determined |
| S17_extracted_k141_88578 | Not-determined |
| S17_extracted_k141_176978 | Not-determined |
| S17_extracted_k141_88580 | Not-determined |
| S17_extracted_k141_22033 | Not-determined |
| S17_extracted_k141_29459 | Not-determined |
| S17_extracted_k141_176992 | Not-determined |
| S17_extracted_k141_22040 | Not-determined |
| S17_extracted_k141_88596 | Not-determined |
| S17_extracted_k141_88602 | Low-quality |
| S17_extracted_k141_88604 | Not-determined |
| S17_extracted_k141_95902 | Not-determined |
| S17_extracted_k141_95903 | Not-determined |
| S17_extracted_k141_95904 | Not-determined |
| S17_extracted_k141_95906 | Not-determined |
| S17_extracted_k141_88620 | Not-determined |
| S17_extracted_k141_95915 | Not-determined |
| S17_extracted_k141_29476 | Not-determined |
| S17_extracted_k141_177024 | Not-determined |
| S17_extracted_k141_29478 | Low-quality |
| S17_extracted_k141_65969 | Low-quality |
| S17_extracted_k141_22067 | Not-determined |
| S17_extracted_k141_29484 | Low-quality |
| S17_extracted_k141_95918 | Not-determined |
| S17_extracted_k141_65980 | Not-determined |
| S17_extracted_k141_88630 | Not-determined |
| S17_extracted_k141_177041 | Not-determined |
| S17_extracted_k141_95926 | Low-quality |
| S17_extracted_k141_95931 | Not-determined |
| S17_extracted_k141_29493 | Not-determined |
| S17_extracted_k141_177050 | Not-determined |
| S17_extracted_k141_88642 | Not-determined |
| S17_extracted_k141_22080 | Not-determined |
| S17_extracted_k141_29499 | Not-determined |
| S17_extracted_k141_29500 | Not-determined |
| S17_extracted_k141_29502 | Not-determined |
| S17_extracted_k141_177061 | Not-determined |
| S17_extracted_k141_177062 | Not-determined |
| S17_extracted_k141_29512 | Not-determined |
| S17_extracted_k141_29513 | Not-determined |
| S17_extracted_k141_95948 | Not-determined |
| S17_extracted_k141_95949 | Not-determined |
| S17_extracted_k141_177070 | Not-determined |
| S17_extracted_k141_66008 | Not-determined |
| S17_extracted_k141_177072 | Not-determined |
| S17_extracted_k141_95955 | Not-determined |
| S17_extracted_k141_177077 | Not-determined |
| S17_extracted_k141_177079 | Not-determined |
| S17_extracted_k141_177081 | Not-determined |
| S17_extracted_k141_88665 | Not-determined |
| S17_extracted_k141_177088 | Not-determined |
| S17_extracted_k141_177090 | Low-quality |
| S17_extracted_k141_66018 | Low-quality |
| S17_extracted_k141_22094 | Not-determined |
| S17_extracted_k141_22095 | Not-determined |
| S17_extracted_k141_29525 | Not-determined |
| S17_extracted_k141_66038 | Not-determined |
| S17_extracted_k141_177107 | Not-determined |
| S17_extracted_k141_177109 | Not-determined |
| S17_extracted_k141_177114 | Not-determined |
| S17_extracted_k141_29528 | Not-determined |
| S17_extracted_k141_95964 | Not-determined |
| S17_extracted_k141_66042 | Not-determined |
| S17_extracted_k141_95966 | Not-determined |
| S17_extracted_k141_177125 | Not-determined |
| S17_extracted_k141_66054 | Not-determined |
| S17_extracted_k141_29533 | Not-determined |
| S17_extracted_k141_29534 | Not-determined |
| S17_extracted_k141_95976 | Not-determined |
| S17_extracted_k141_66066 | Not-determined |
| S17_extracted_k141_177145 | Not-determined |
| S17_extracted_k141_66071 | Not-determined |
| S17_extracted_k141_177147 | Not-determined |
| S17_extracted_k141_29547 | Low-quality |
| S17_extracted_k141_177157 | Low-quality |
| S17_extracted_k141_177163 | Not-determined |
| S17_extracted_k141_95991 | Not-determined |
| S17_extracted_k141_95993 | Not-determined |
| S17_extracted_k141_95998 | Not-determined |
| S17_extracted_k141_177167 | Not-determined |
| S17_extracted_k141_66099 | Not-determined |
| S17_extracted_k141_29555 | Not-determined |
| S17_extracted_k141_66107 | Not-determined |
| S17_extracted_k141_66110 | Not-determined |
| S17_extracted_k141_96010 | Not-determined |
| S17_extracted_k141_177178 | Not-determined |
| S17_extracted_k141_96015 | Not-determined |
| S17_extracted_k141_66119 | Not-determined |
| S17_extracted_k141_177183 | Not-determined |
| S17_extracted_k141_177189 | Low-quality |
| S17_extracted_k141_96024 | Not-determined |
| S17_extracted_k141_66131 | Not-determined |
| S17_extracted_k141_96034 | Low-quality |
| S17_extracted_k141_66139 | Not-determined |
| S17_extracted_k141_66146 | Not-determined |
| S17_extracted_k141_66147 | Not-determined |
| S17_extracted_k141_66148 | Not-determined |
| S17_extracted_k141_66159 | Low-quality |
| S17_extracted_k141_22125 | Not-determined |
| S17_extracted_k141_96042 | Not-determined |
| S17_extracted_k141_66182 | Not-determined |
| S17_extracted_k141_22153 | Not-determined |
| S17_extracted_k141_96050 | Not-determined |
| S17_extracted_k141_66194 | Not-determined |
| S17_extracted_k141_22165 | Not-determined |
| S17_extracted_k141_66208 | Not-determined |
| S17_extracted_k141_66209 | Low-quality |
| S17_extracted_k141_66239 | Not-determined |
| S17_extracted_k141_66250 | Not-determined |
| S12_extracted_k141_173572 | Complete |
| S17_extracted_k141_66281 | Not-determined |
| S17_extracted_k141_66285 | Low-quality |
| S17_extracted_k141_66286 | Not-determined |
| S17_extracted_k141_66290 | Not-determined |
| S17_extracted_k141_66304 | Low-quality |
| S17_extracted_k141_66307 | Not-determined |
| S12_vs_k141_173572\|\|full | High-quality |
| S17_extracted_k141_66320 | Not-determined |
| S17_extracted_k141_66321 | Not-determined |
| S17_extracted_k141_66326 | Not-determined |
| S17_extracted_k141_66329 | Not-determined |
| S17_extracted_k141_66340 | Not-determined |
| S5_vs_k141_97574\|\|full | High-quality |
| S17_extracted_k141_66343 | Not-determined |
| S17_extracted_k141_66345 | Not-determined |
| S17_extracted_k141_66346 | Not-determined |
| S17_extracted_k141_66347 | Not-determined |
| S17_extracted_k141_66356 | Not-determined |
| S17_extracted_k141_66357 | Not-determined |
| S17_extracted_k141_66370 | Not-determined |
| S17_extracted_k141_66375 | Not-determined |
| S17_extracted_k141_66376 | Not-determined |
| S17_extracted_k141_66378 | Not-determined |
| S17_extracted_k141_66382 | Not-determined |
| S17_extracted_k141_66403 | Not-determined |
| S17_extracted_k141_66409 | Not-determined |
| S17_extracted_k141_177221 | Not-determined |
| S17_extracted_k141_177226 | Low-quality |
| S17_extracted_k141_66447 | Not-determined |
| S17_extracted_k141_177231 | Not-determined |
| S17_extracted_k141_177241 | Not-determined |
| S17_extracted_k141_177242 | Not-determined |
| S17_extracted_k141_177243 | Low-quality |
| S17_extracted_k141_177246 | Not-determined |
| S17_extracted_k141_177254 | Not-determined |
| S17_extracted_k141_66459 | Low-quality |
| S17_extracted_k141_66461 | Not-determined |
| S17_extracted_k141_177257 | Not-determined |
| S17_extracted_k141_177259 | Not-determined |
| S17_extracted_k141_177265 | Not-determined |
| S17_extracted_k141_177274 | Not-determined |
| S17_extracted_k141_177275 | Low-quality |
| S17_extracted_k141_177276 | Not-determined |
| S17_extracted_k141_177277 | Not-determined |
| S17_extracted_k141_177282 | Not-determined |
| S17_extracted_k141_177285 | Not-determined |
| S17_extracted_k141_177287 | Not-determined |
| S17_extracted_k141_177288 | Not-determined |
| S17_extracted_k141_177289 | Low-quality |
| S17_extracted_k141_177290 | Not-determined |
| S17_extracted_k141_177296 | Not-determined |
| S17_extracted_k141_177298 | Not-determined |
| S17_extracted_k141_177299 | Not-determined |
| S17_extracted_k141_177301 | Not-determined |
| S17_extracted_k141_177309 | Not-determined |
| S17_extracted_k141_177312 | Not-determined |
| S17_extracted_k141_177315 | Not-determined |
| S17_extracted_k141_177317 | Not-determined |
| S17_extracted_k141_177320 | Not-determined |
| S17_extracted_k141_66488 | Not-determined |
| S17_extracted_k141_66493 | Not-determined |
| S17_extracted_k141_66494 | Not-determined |
| S17_extracted_k141_66499 | Not-determined |
| S17_vs_k141_15243\|\|full | Not-determined |
| S10_extracted_29265 | High-quality |
| S10_vs_29265\|\|full | High-quality |
| S14_vs_k141_85765\|\|full | High-quality |
| S17_vs_k141_61673\|\|full | Low-quality |
| S17_vs_k141_48258\|\|full | Not-determined |
| S17_vs_k141_160104\|\|full | Not-determined |
| S17_vs_k141_153490\|\|full | Not-determined |
| S12_extracted_k141_32615 | Complete |
| S17_vs_k141_57885\|\|full | Not-determined |
| S12_vs_k141_32615\|\|full | High-quality |
| S17_vs_k141_169681\|\|full | Not-determined |
| S17_vs_k141_7367\|\|full | Not-determined |
| S15_extracted_33505 | Complete |
| S15_vs_33505\|\|full | High-quality |
| S9_vs_k141_90727\|\|full | High-quality |
| S3_extracted_k141_99244 | High-quality |
| S3_vs_k141_99244\|\|full | High-quality |
| S11_vs_k141_45658\|\|full | Medium-quality |
| S8_vs_k141_49154\|\|full | High-quality |
| S14_vs_k141_26320\|\|full | High-quality |
| S17_vs_k141_177222\|\|full | Not-determined |
| S17_vs_k141_132997\|\|full | Not-determined |
| S17_vs_k141_96058\|\|full | Low-quality |
| S17_vs_k141_66506\|\|full | Not-determined |
| S17_vs_k141_81294\|\|full | Low-quality |
| S17_vs_k141_29580\|\|full | Low-quality |
| S17_vs_k141_103464\|\|full | Not-determined |
| S17_vs_k141_133010\|\|full | Not-determined |
| S17_vs_k141_125625\|\|full | Not-determined |
| S17_vs_k141_118249\|\|full | Not-determined |
| S17_vs_k141_29590\|\|full | Not-determined |
| S17_vs_k141_96090\|\|full | Low-quality |
| S17_vs_k141_133028\|\|full | Low-quality |
| S17_vs_k141_125634\|\|full | Not-determined |
| S17_vs_k141_140415\|\|full | Low-quality |
| S17_vs_k141_73944\|\|full | Not-determined |
| S17_vs_k141_59125\|\|full | Low-quality |
| S17_vs_k141_14811\|\|full | Low-quality |
| S17_vs_k141_96091\|\|full | Low-quality |
| S17_vs_k141_125657\|\|full | Not-determined |
| S17_vs_k141_51763\|\|full | Low-quality |
| S17_vs_k141_155200\|\|full | Low-quality |
| S17_vs_k141_110876\|\|full | Not-determined |
| S17_vs_k141_155207\|\|full | Not-determined |
| S17_vs_k141_59152\|\|full | Not-determined |
| S17_vs_k141_66560\|\|full | Low-quality |
| S17_vs_k141_169981\|\|full | Low-quality |
| S17_vs_k141_147812\|\|full | Low-quality |
| S17_vs_k141_51781\|\|full | Low-quality |
| S17_vs_k141_162569\|\|full | Low-quality |
| S17_vs_k141_155215\|\|full | Low-quality |
| S17_vs_k141_162570\|\|full | Low-quality |
| S17_vs_k141_110896\|\|full | Low-quality |
| S17_vs_k141_59168\|\|full | Low-quality |
| S17_vs_k141_147818\|\|full | Low-quality |
| S17_vs_k141_110897\|\|full | Low-quality |
| S17_vs_k141_44366\|\|full | Not-determined |
| S17_vs_k141_140445\|\|full | Not-determined |
| S17_vs_k141_110903\|\|full | Not-determined |
| S17_vs_k141_59181\|\|full | Not-determined |
| S17_vs_k141_110905\|\|full | Not-determined |
| S17_vs_k141_118290\|\|full | Not-determined |
| S17_vs_k141_7479\|\|full | Low-quality |
| S17_vs_k141_29652\|\|full | Low-quality |
| S17_vs_k141_73994\|\|full | Low-quality |
| S17_vs_k141_44386\|\|full | Not-determined |
| S17_vs_k141_162600\|\|full | Not-determined |
| S17_vs_k141_147828\|\|full | Low-quality |
| S17_vs_k141_96133\|\|full | Low-quality |
| S17_vs_k141_29656\|\|full | Low-quality |
| S17_vs_k141_162612\|\|full | Not-determined |
| S17_vs_k141_37008\|\|full | Not-determined |
| S17_vs_k141_96137\|\|full | Low-quality |
| S17_vs_k141_155230\|\|full | Low-quality |
| S17_vs_k141_118308\|\|full | Not-determined |
| S17_vs_k141_37011\|\|full | Low-quality |
| S17_vs_k141_51806\|\|full | Low-quality |
| S17_vs_k141_88736\|\|full | Low-quality |
| S17_vs_k141_37017\|\|full | Not-determined |
| S17_vs_k141_96146\|\|full | Low-quality |
| S17_vs_k141_59208\|\|full | Not-determined |
| S17_vs_k141_66592\|\|full | Low-quality |
| S17_vs_k141_133074\|\|full | Not-determined |
| S17_vs_k141_140464\|\|full | Not-determined |
| S17_vs_k141_88753\|\|full | Not-determined |
| S17_vs_k141_59211\|\|full | Low-quality |
| S17_vs_k141_29686\|\|full | Low-quality |
| S17_vs_k141_66605\|\|full | Low-quality |
| S17_vs_k141_7525\|\|full | Not-determined |
| S17_vs_k141_162643\|\|full | Not-determined |
| S17_vs_k141_51822\|\|full | Low-quality |
| S17_vs_k141_29696\|\|full | Low-quality |
| S17_vs_k141_66616\|\|full | Low-quality |
| S17_vs_k141_114\|\|full | Not-determined |
| S17_vs_k141_29706\|\|full | Not-determined |
| S17_vs_k141_59236\|\|full | Not-determined |
| S17_vs_k141_96170\|\|full | Low-quality |
| S17_vs_k141_96176\|\|full | Low-quality |
| S17_vs_k141_51825\|\|full | Not-determined |
| S17_vs_k141_140481\|\|full | Not-determined |
| S17_vs_k141_44414\|\|full | Low-quality |
| S17_vs_k141_125686\|\|full | Low-quality |
| S17_vs_k141_88760\|\|full | Low-quality |
| S17_vs_k141_88761\|\|full | Low-quality |
| S17_vs_k141_14875\|\|full | Low-quality |
| S17_vs_k141_29739\|\|full | Low-quality |
| S17_vs_k141_103581\|\|full | Not-determined |
| S17_vs_k141_7549\|\|full | Not-determined |
| S17_vs_k141_74066\|\|full | Not-determined |
| S17_vs_k141_51847\|\|full | Not-determined |
| S17_vs_k141_44436\|\|full | Low-quality |
| S17_vs_k141_14886\|\|full | Not-determined |
| S17_vs_k141_51851\|\|full | Low-quality |
| S17_vs_k141_140526\|\|full | Low-quality |
| S17_vs_k141_22238\|\|full | Not-determined |
| S17_vs_k141_103599\|\|full | Not-determined |
| S17_vs_k141_37082\|\|full | Low-quality |
| S17_vs_k141_22240\|\|full | Not-determined |
| S17_vs_k141_51866\|\|full | Low-quality |
| S17_vs_k141_96230\|\|full | Not-determined |
| S17_vs_k141_147926\|\|full | Low-quality |
| S17_vs_k141_14899\|\|full | Low-quality |
| S17_vs_k141_51869\|\|full | Low-quality |
| S17_vs_k141_147931\|\|full | Low-quality |
| S17_vs_k141_66672\|\|full | Not-determined |
| S17_vs_k141_37087\|\|full | Low-quality |
| S17_vs_k141_96243\|\|full | Low-quality |
| S17_vs_k141_14905\|\|full | Not-determined |
| S17_vs_k141_7567\|\|full | Low-quality |
| S17_vs_k141_51871\|\|full | Low-quality |
| S17_vs_k141_88810\|\|full | Not-determined |
| S17_vs_k141_160\|\|full | Low-quality |
| S17_vs_k141_125736\|\|full | Low-quality |
| S17_vs_k141_96271\|\|full | Not-determined |
| S17_vs_k141_59312\|\|full | Not-determined |
| S17_vs_k141_44467\|\|full | Low-quality |
| S17_vs_k141_59315\|\|full | Low-quality |
| S17_vs_k141_118363\|\|full | Not-determined |
| S17_vs_k141_133175\|\|full | Not-determined |
| S17_vs_k141_59322\|\|full | Not-determined |
| S17_vs_k141_22275\|\|full | Low-quality |
| S17_vs_k141_88837\|\|full | Low-quality |
| S17_vs_k141_44479\|\|full | Low-quality |
| S17_vs_k141_96326\|\|full | Low-quality |
| S17_vs_k141_133190\|\|full | Not-determined |
| S17_vs_k141_29833\|\|full | Low-quality |
| S17_vs_k141_7647\|\|full | Not-determined |
| S17_vs_k141_37189\|\|full | Low-quality |
| S17_vs_k141_140597\|\|full | Not-determined |
| S17_vs_k141_44495\|\|full | Not-determined |
| S17_vs_k141_44496\|\|full | Not-determined |
| S17_vs_k141_7661\|\|full | Not-determined |
| S17_vs_k141_22328\|\|full | Not-determined |
| S17_vs_k141_118426\|\|full | Not-determined |
| S17_vs_k141_190\|\|full | Low-quality |
| S17_vs_k141_118428\|\|full | Low-quality |
| S17_vs_k141_81335\|\|full | Not-determined |
| S17_vs_k141_7683\|\|full | Low-quality |
| S17_vs_k141_147999\|\|full | Low-quality |
| S17_vs_k141_22346\|\|full | Low-quality |
| S17_vs_k141_170070\|\|full | Low-quality |
| S17_vs_k141_140615\|\|full | Not-determined |
| S17_vs_k141_7690\|\|full | Not-determined |
| S17_vs_k141_59377\|\|full | Low-quality |
| S17_vs_k141_162767\|\|full | Low-quality |
| S17_vs_k141_96364\|\|full | Low-quality |
| S17_vs_k141_148004\|\|full | Not-determined |
| S7_vs_16344\|\|full | Medium-quality |
| S17_vs_k141_96376\|\|full | Not-determined |
| S17_vs_k141_162777\|\|full | Low-quality |
| S17_vs_k141_103703\|\|full | Low-quality |
| S17_vs_k141_162783\|\|full | Not-determined |
| S17_vs_k141_44534\|\|full | Not-determined |
| S17_vs_k141_148022\|\|full | Not-determined |
| S17_vs_k141_155347\|\|full | Low-quality |
| S17_vs_k141_14971\|\|full | Low-quality |
| S17_vs_k141_140639\|\|full | Low-quality |
| S17_vs_k141_162801\|\|full | Low-quality |
| S17_vs_k141_210\|\|full | Low-quality |
| S17_vs_k141_59405\|\|full | Low-quality |
| S17_vs_k141_88865\|\|full | Not-determined |
| S17_vs_k141_133260\|\|full | Not-determined |
| S17_vs_k141_7712\|\|full | Not-determined |
| S17_vs_k141_215\|\|full | Low-quality |
| S17_vs_k141_14983\|\|full | Low-quality |
| S17_vs_k141_51947\|\|full | Not-determined |
| S17_vs_k141_155372\|\|full | Not-determined |
| S17_vs_k141_148060\|\|full | Not-determined |
| S17_vs_k141_44546\|\|full | Low-quality |
| S17_vs_k141_44548\|\|full | Low-quality |
| S17_vs_k141_125796\|\|full | Low-quality |
| S17_vs_k141_96422\|\|full | Not-determined |
| S17_vs_k141_14995\|\|full | Low-quality |
| S17_vs_k141_103755\|\|full | Not-determined |
| S17_vs_k141_118476\|\|full | Low-quality |
| S17_vs_k141_14999\|\|full | Low-quality |
| S17_vs_k141_155407\|\|full | Not-determined |
| S17_vs_k141_96433\|\|full | Low-quality |
| S17_vs_k141_148086\|\|full | Low-quality |
| S17_vs_k141_96439\|\|full | Not-determined |
| S17_vs_k141_88905\|\|full | Low-quality |
| S17_vs_k141_81387\|\|full | Low-quality |
| S17_vs_k141_88906\|\|full | Not-determined |
| S12_vs_k141_172379\|\|full | Medium-quality |
| S17_vs_k141_81389\|\|full | Low-quality |
| S17_vs_k141_110953\|\|full | Not-determined |
| S17_vs_k141_51995\|\|full | Low-quality |
| S17_vs_k141_125825\|\|full | Low-quality |
| S17_vs_k141_133303\|\|full | Low-quality |
| S17_vs_k141_88918\|\|full | Not-determined |
| S17_vs_k141_37289\|\|full | Not-determined |

| | |
|---|---|
| S17_vs_k141_81404\|\|full | Not-determined |
| S17_vs_k141_59445\|\|full | Low-quality |
| S17_vs_k141_74165\|\|full | Low-quality |
| S17_vs_k141_37297\|\|full | Low-quality |
| S17_vs_k141_66712\|\|full | Low-quality |
| S17_vs_k141_59453\|\|full | Low-quality |
| S17_vs_k141_170173\|\|full | Not-determined |
| S17_vs_k141_170180\|\|full | Low-quality |
| S17_vs_k141_125870\|\|full | Not-determined |
| S17_vs_k141_133322\|\|full | Not-determined |
| S17_vs_k141_111001\|\|full | Low-quality |
| S17_vs_k141_7730\|\|full | Low-quality |
| S17_vs_k141_125875\|\|full | Low-quality |
| S17_vs_k141_59474\|\|full | Low-quality |
| S17_vs_k141_52037\|\|full | Low-quality |
| S17_vs_k141_44657\|\|full | Low-quality |
| S17_vs_k141_162817\|\|full | Not-determined |
| S17_vs_k141_118515\|\|full | Not-determined |
| S17_vs_k141_252\|\|full | Low-quality |
| S17_vs_k141_59494\|\|full | Low-quality |
| S17_vs_k141_96476\|\|full | Not-determined |
| S17_vs_k141_111014\|\|full | Not-determined |
| S17_vs_k141_44676\|\|full | Low-quality |
| S17_vs_k141_111016\|\|full | Not-determined |
| S17_vs_k141_162831\|\|full | Not-determined |
| S17_vs_k141_140709\|\|full | Low-quality |
| S17_vs_k141_103828\|\|full | Not-determined |
| S17_vs_k141_74215\|\|full | Not-determined |
| S17_vs_k141_140711\|\|full | Low-quality |
| S17_vs_k141_170208\|\|full | Low-quality |
| S17_vs_k141_264\|\|full | Not-determined |
| S17_vs_k141_44697\|\|full | Not-determined |
| S17_vs_k141_22356\|\|full | Not-determined |
| S17_vs_k141_37340\|\|full | Low-quality |
| S17_vs_k141_96491\|\|full | Low-quality |
| S17_vs_k141_148164\|\|full | Not-determined |
| S17_vs_k141_81443\|\|full | Not-determined |
| S17_vs_k141_30007\|\|full | Not-determined |
| S17_vs_k141_111052\|\|full | Not-determined |
| S17_vs_k141_170236\|\|full | Low-quality |
| S17_vs_k141_74255\|\|full | Not-determined |
| S17_vs_k141_66781\|\|full | Low-quality |
| S17_vs_k141_44728\|\|full | Low-quality |
| S17_vs_k141_22382\|\|full | Low-quality |
| S17_vs_k141_74266\|\|full | Low-quality |
| S17_vs_k141_44730\|\|full | Low-quality |
| S17_vs_k141_111081\|\|full | Low-quality |
| S17_vs_k141_59539\|\|full | Low-quality |
| S17_vs_k141_103864\|\|full | Low-quality |
| S17_vs_k141_66798\|\|full | Low-quality |
| S17_vs_k141_96522\|\|full | Low-quality |
| S17_vs_k141_66802\|\|full | Not-determined |
| S17_vs_k141_37382\|\|full | Low-quality |
| S17_vs_k141_66803\|\|full | Low-quality |
| S17_vs_k141_96529\|\|full | Not-determined |
| S17_vs_k141_170279\|\|full | Not-determined |
| S17_vs_k141_140792\|\|full | Low-quality |
| S17_vs_k141_59573\|\|full | Low-quality |
| S17_vs_k141_74304\|\|full | Low-quality |
| S17_vs_k141_111116\|\|full | Low-quality |
| S17_vs_k141_81488\|\|full | Low-quality |
| S17_vs_k141_170284\|\|full | Not-determined |
| S17_vs_k141_148197\|\|full | Low-quality |
| S17_vs_k141_7807\|\|full | Not-determined |
| S17_vs_k141_88942\|\|full | Not-determined |
| S17_vs_k141_170290\|\|full | Not-determined |
| S17_vs_k141_140809\|\|full | Low-quality |
| S17_vs_k141_111126\|\|full | Low-quality |
| S17_vs_k141_118579\|\|full | Not-determined |
| S17_vs_k141_52117\|\|full | Low-quality |
| S17_vs_k141_74311\|\|full | Low-quality |
| S17_vs_k141_170292\|\|full | Low-quality |
| S17_vs_k141_44782\|\|full | Low-quality |
| S17_vs_k141_162880\|\|full | Not-determined |
| S17_vs_k141_52135\|\|full | Low-quality |
| S17_vs_k141_66827\|\|full | Not-determined |
| S17_vs_k141_7831\|\|full | Low-quality |
| S17_vs_k141_133375\|\|full | Not-determined |
| S17_vs_k141_111149\|\|full | Not-determined |
| S17_vs_k141_140830\|\|full | Low-quality |
| S17_vs_k141_140837\|\|full | Low-quality |
| S17_vs_k141_155461\|\|full | Not-determined |
| S17_vs_k141_111162\|\|full | Not-determined |
| S17_vs_k141_59618\|\|full | Low-quality |
| S17_vs_k141_355\|\|full | Low-quality |
| S17_vs_k141_358\|\|full | Low-quality |
| S17_vs_k141_140843\|\|full | Low-quality |
| S17_vs_k141_155465\|\|full | Low-quality |
| S17_vs_k141_111165\|\|full | Low-quality |
| S17_vs_k141_59625\|\|full | Not-determined |
| S17_vs_k141_88979\|\|full | Not-determined |
| S17_vs_k141_59631\|\|full | Not-determined |
| S17_vs_k141_37433\|\|full | Not-determined |
| S17_vs_k141_162902\|\|full | Low-quality |
| S17_vs_k141_369\|\|full | Not-determined |
| S17_vs_k141_103950\|\|full | Not-determined |
| S17_vs_k141_111180\|\|full | Low-quality |
| S17_vs_k141_81521\|\|full | Not-determined |
| S17_vs_k141_111182\|\|full | Low-quality |
| S17_vs_k141_103952\|\|full | Low-quality |
| S17_vs_k141_37451\|\|full | Low-quality |
| S17_vs_k141_44818\|\|full | Not-determined |
| S17_vs_k141_148250\|\|full | Not-determined |
| S17_vs_k141_22442\|\|full | Not-determined |
| S17_vs_k141_22444\|\|full | Not-determined |
| S17_vs_k141_118658\|\|full | Low-quality |
| S17_vs_k141_81545\|\|full | Not-determined |
| S17_vs_k141_37491\|\|full | Not-determined |
| S17_vs_k141_89008\|\|full | Not-determined |
| S17_vs_k141_118676\|\|full | Low-quality |
| S17_vs_k141_162935\|\|full | Low-quality |
| S17_vs_k141_15131\|\|full | Not-determined |
| S17_vs_k141_388\|\|full | Not-determined |
| S17_vs_k141_162938\|\|full | Low-quality |
| S17_vs_k141_103986\|\|full | Low-quality |
| S17_vs_k141_148266\|\|full | Low-quality |
| S17_vs_k141_399\|\|full | Low-quality |
| S17_vs_k141_59642\|\|full | Low-quality |
| S17_vs_k141_405\|\|full | Low-quality |
| S17_vs_k141_59645\|\|full | Low-quality |
| S17_vs_k141_170328\|\|full | Not-determined |
| S17_vs_k141_7862\|\|full | Not-determined |
| S17_vs_k141_7868\|\|full | Low-quality |
| S17_vs_k141_89020\|\|full | Not-determined |
| S17_vs_k141_52217\|\|full | Low-quality |
| S17_vs_k141_155512\|\|full | Not-determined |
| S17_vs_k141_89026\|\|full | Not-determined |
| S17_vs_k141_426\|\|full | Not-determined |
| S17_vs_k141_59670\|\|full | Low-quality |
| S17_vs_k141_15165\|\|full | Not-determined |
| S17_vs_k141_59674\|\|full | Not-determined |
| S17_vs_k141_140922\|\|full | Not-determined |
| S17_vs_k141_170340\|\|full | Low-quality |
| S17_vs_k141_118710\|\|full | Low-quality |
| S17_vs_k141_59678\|\|full | Not-determined |
| S17_vs_k141_170343\|\|full | Not-determined |
| S17_vs_k141_74407\|\|full | Low-quality |
| S17_vs_k141_104025\|\|full | Low-quality |
| S17_vs_k141_170347\|\|full | Not-determined |
| S17_vs_k141_30134\|\|full | Not-determined |
| S17_vs_k141_162969\|\|full | Low-quality |
| S17_vs_k141_118719\|\|full | Not-determined |
| S17_vs_k141_59683\|\|full | Low-quality |
| S17_vs_k141_7897\|\|full | Low-quality |
| S17_vs_k141_118728\|\|full | Low-quality |
| S17_vs_k141_104037\|\|full | Not-determined |
| S17_vs_k141_118731\|\|full | Not-determined |
| S17_vs_k141_7908\|\|full | Low-quality |
| S5_vs_k141_131510\|\|full | Complete |
| S17_vs_k141_126005\|\|full | Low-quality |
| S12_extracted_k141_145044 | High-quality |
| S17_vs_k141_81581\|\|full | Not-determined |
| S17_vs_k141_37565\|\|full | Not-determined |
| S17_vs_k141_111224\|\|full | Not-determined |
| S17_vs_k141_155563\|\|full | Low-quality |
| S17_vs_k141_30171\|\|full | Low-quality |
| S17_vs_k141_81594\|\|full | Low-quality |
| S17_vs_k141_37581\|\|full | Low-quality |
| S17_vs_k141_126012\|\|full | Low-quality |
| S17_vs_k141_111237\|\|full | Low-quality |
| S17_vs_k141_148319\|\|full | Low-quality |
| S17_vs_k141_118768\|\|full | Not-determined |
| S17_vs_k141_52289\|\|full | Low-quality |
| S17_vs_k141_22540\|\|full | Not-determined |
| S17_vs_k141_155576\|\|full | Low-quality |
| S17_vs_k141_52299\|\|full | Low-quality |
| S17_vs_k141_148335\|\|full | Low-quality |
| S17_vs_k141_37608\|\|full | Not-determined |
| S17_vs_k141_155589\|\|full | Low-quality |
| S17_vs_k141_37609\|\|full | Low-quality |
| S17_vs_k141_37615\|\|full | Not-determined |
| S17_vs_k141_22555\|\|full | Low-quality |
| S17_vs_k141_30203\|\|full | Not-determined |
| S17_vs_k141_155606\|\|full | Low-quality |
| S17_vs_k141_118790\|\|full | Low-quality |
| S17_vs_k141_74468\|\|full | Low-quality |
| S17_vs_k141_59724\|\|full | Not-determined |
| S17_vs_k141_22563\|\|full | Not-determined |
| S17_vs_k141_89094\|\|full | Not-determined |
| S17_vs_k141_111266\|\|full | Not-determined |
| S17_vs_k141_22568\|\|full | Low-quality |
| S17_vs_k141_170402\|\|full | Not-determined |
| S17_vs_k141_111268\|\|full | Not-determined |
| S17_vs_k141_133494\|\|full | Not-determined |
| S17_vs_k141_163064\|\|full | Low-quality |
| S17_vs_k141_89103\|\|full | Low-quality |
| S17_vs_k141_7990\|\|full | Low-quality |
| S17_vs_k141_74485\|\|full | Not-determined |
| S17_vs_k141_126040\|\|full | Low-quality |
| S17_vs_k141_8002\|\|full | Low-quality |
| S17_vs_k141_170412\|\|full | Low-quality |
| S17_vs_k141_52328\|\|full | Not-determined |
| S17_vs_k141_155625\|\|full | Low-quality |
| S17_vs_k141_66907\|\|full | Low-quality |
| S17_vs_k141_15274\|\|full | Not-determined |
| S17_vs_k141_118812\|\|full | Not-determined |
| S17_vs_k141_37635\|\|full | Not-determined |
| S17_vs_k141_170428\|\|full | Not-determined |
| S17_vs_k141_74490\|\|full | Not-determined |
| S17_vs_k141_163072\|\|full | Low-quality |
| S17_vs_k141_30258\|\|full | Low-quality |
| S17_vs_k141_44901\|\|full | Low-quality |
| S17_vs_k141_37655\|\|full | Low-quality |
| S17_vs_k141_44910\|\|full | Low-quality |
| S17_vs_k141_170447\|\|full | Low-quality |
| S17_vs_k141_59727\|\|full | Not-determined |
| S17_vs_k141_126079\|\|full | Not-determined |
| S17_vs_k141_44916\|\|full | Low-quality |
| S17_vs_k141_155672\|\|full | Low-quality |
| S17_vs_k141_15281\|\|full | Low-quality |
| S17_vs_k141_170455\|\|full | Low-quality |
| S17_vs_k141_96548\|\|full | Low-quality |
| S17_vs_k141_118848\|\|full | Low-quality |
| S17_vs_k141_59730\|\|full | Low-quality |
| S17_vs_k141_44929\|\|full | Low-quality |
| S17_vs_k141_59733\|\|full | Not-determined |
| S17_vs_k141_15290\|\|full | Low-quality |
| S17_vs_k141_44934\|\|full | Not-determined |
| S17_vs_k141_37695\|\|full | Not-determined |
| S17_vs_k141_52393\|\|full | Not-determined |
| S17_vs_k141_37701\|\|full | Low-quality |
| S17_vs_k141_170476\|\|full | Low-quality |
| S17_vs_k141_155691\|\|full | Low-quality |
| S17_vs_k141_111351\|\|full | Low-quality |
| S17_vs_k141_126112\|\|full | Not-determined |
| S17_vs_k141_59749\|\|full | Not-determined |
| S17_vs_k141_118864\|\|full | Not-determined |
| S17_vs_k141_170506\|\|full | Not-determined |
| S17_vs_k141_111354\|\|full | Not-determined |
| S17_vs_k141_37717\|\|full | Low-quality |
| S17_vs_k141_96574\|\|full | Not-determined |
| S17_vs_k141_104163\|\|full | Not-determined |
| S17_vs_k141_15298\|\|full | Low-quality |
| S17_vs_k141_111359\|\|full | Low-quality |
| S17_vs_k141_89147\|\|full | Low-quality |
| S17_vs_k141_37725\|\|full | Not-determined |
| S17_vs_k141_52426\|\|full | Not-determined |
| S17_vs_k141_141040\|\|full | Low-quality |
| S17_vs_k141_66951\|\|full | Not-determined |
| S17_vs_k141_141043\|\|full | Low-quality |
| S17_vs_k141_104189\|\|full | Low-quality |
| S17_vs_k141_52436\|\|full | Not-determined |
| S17_vs_k141_163119\|\|full | Not-determined |
| S17_vs_k141_118887\|\|full | Low-quality |
| S17_vs_k141_111380\|\|full | Not-determined |
| S17_vs_k141_37726\|\|full | Low-quality |
| S17_vs_k141_37729\|\|full | Low-quality |
| S17_vs_k141_74577\|\|full | Low-quality |
| S17_vs_k141_163142\|\|full | Not-determined |
| S17_vs_k141_118906\|\|full | Not-determined |
| S17_vs_k141_74580\|\|full | Not-determined |
| S17_vs_k141_44971\|\|full | Not-determined |
| S17_vs_k141_111402\|\|full | Low-quality |
| S17_vs_k141_66970\|\|full | Not-determined |
| S17_vs_k141_96600\|\|full | Not-determined |
| S17_vs_k141_15349\|\|full | Low-quality |
| S17_vs_k141_155779\|\|full | Low-quality |
| S17_vs_k141_148482\|\|full | Low-quality |
| S17_vs_k141_104226\|\|full | Low-quality |
| S17_vs_k141_74611\|\|full | Low-quality |
| S17_vs_k141_163150\|\|full | Low-quality |
| S17_vs_k141_670\|\|full | Not-determined |
| S17_vs_k141_74637\|\|full | Low-quality |
| S17_vs_k141_679\|\|full | Low-quality |
| S17_vs_k141_148516\|\|full | Low-quality |
| S17_vs_k141_66986\|\|full | Not-determined |
| S17_vs_k141_89213\|\|full | Not-determined |
| S17_vs_k141_104257\|\|full | Not-determined |
| S17_vs_k141_133604\|\|full | Not-determined |
| S17_vs_k141_104258\|\|full | Low-quality |
| S17_vs_k141_89214\|\|full | Low-quality |
| S17_vs_k141_141124\|\|full | Not-determined |
| S17_vs_k141_148534\|\|full | Low-quality |
| S17_vs_k141_141125\|\|full | Low-quality |
| S17_vs_k141_81694\|\|full | Not-determined |
| S17_vs_k141_170599\|\|full | Not-determined |
| S17_vs_k141_163176\|\|full | Not-determined |
| S17_vs_k141_15400\|\|full | Not-determined |
| S17_vs_k141_74660\|\|full | Low-quality |
| S17_vs_k141_96627\|\|full | Low-quality |
| S17_vs_k141_104279\|\|full | Not-determined |
| S17_vs_k141_141142\|\|full | Low-quality |
| S17_vs_k141_67003\|\|full | Not-determined |
| S17_vs_k141_170602\|\|full | Low-quality |
| S17_vs_k141_8088\|\|full | Not-determined |
| S17_vs_k141_52464\|\|full | Low-quality |
| S17_vs_k141_52467\|\|full | Low-quality |
| S17_vs_k141_74691\|\|full | Low-quality |
| S17_vs_k141_67038\|\|full | Low-quality |
| S17_vs_k141_81707\|\|full | Not-determined |
| S17_vs_k141_111445\|\|full | Not-determined |
| S17_vs_k141_163207\|\|full | Low-quality |
| S17_vs_k141_89241\|\|full | Not-determined |
| S17_vs_k141_89251\|\|full | Low-quality |
| S17_vs_k141_111467\|\|full | Low-quality |
| S17_vs_k141_148589\|\|full | Not-determined |
| S17_vs_k141_163244\|\|full | Low-quality |
| S17_vs_k141_8107\|\|full | Low-quality |
| S12_vs_k141_145044\|\|full | High-quality |
| S17_vs_k141_111473\|\|full | Low-quality |
| S17_vs_k141_170614\|\|full | Not-determined |
| S17_vs_k141_8115\|\|full | Not-determined |
| S17_vs_k141_141176\|\|full | Not-determined |
| S17_vs_k141_22680\|\|full | Low-quality |
| S17_vs_k141_89278\|\|full | Low-quality |
| S17_vs_k141_81728\|\|full | Not-determined |
| S17_vs_k141_30522\|\|full | Low-quality |
| S17_vs_k141_45041\|\|full | Not-determined |
| S17_vs_k141_155903\|\|full | Low-quality |
| S17_vs_k141_141192\|\|full | Not-determined |
| S17_vs_k141_37880\|\|full | Low-quality |
| S17_vs_k141_141204\|\|full | Not-determined |
| S17_vs_k141_89290\|\|full | Low-quality |
| S17_vs_k141_155914\|\|full | Low-quality |
| S17_vs_k141_67093\|\|full | Low-quality |
| S17_vs_k141_52538\|\|full | Not-determined |
| S17_vs_k141_81764\|\|full | Low-quality |
| S17_vs_k141_163309\|\|full | Low-quality |
| S17_vs_k141_170661\|\|full | Low-quality |
| S17_vs_k141_141223\|\|full | Low-quality |
| S17_vs_k141_126231\|\|full | Not-determined |
| S17_vs_k141_37900\|\|full | Not-determined |
| S17_vs_k141_126234\|\|full | Low-quality |
| S17_vs_k141_89302\|\|full | Low-quality |
| S17_vs_k141_37910\|\|full | Low-quality |
| S17_vs_k141_126238\|\|full | Low-quality |
| S17_vs_k141_148621\|\|full | Low-quality |
| S17_vs_k141_118977\|\|full | Not-determined |
| S17_vs_k141_37924\|\|full | Not-determined |
| S17_vs_k141_155955\|\|full | Low-quality |
| S17_vs_k141_45072\|\|full | Low-quality |
| S17_vs_k141_133632\|\|full | Low-quality |
| S17_vs_k141_37932\|\|full | Low-quality |
| S17_vs_k141_74723\|\|full | Not-determined |
| S17_vs_k141_170699\|\|full | Not-determined |
| S17_vs_k141_133634\|\|full | Not-determined |
| S17_vs_k141_155961\|\|full | Not-determined |
| S17_vs_k141_96669\|\|full | Not-determined |
| S17_vs_k141_155977\|\|full | Not-determined |
| S17_vs_k141_163319\|\|full | Low-quality |
| S17_vs_k141_133659\|\|full | Low-quality |
| S17_vs_k141_104459\|\|full | Low-quality |
| S17_vs_k141_148653\|\|full | Low-quality |
| S17_vs_k141_22742\|\|full | Not-determined |
| S17_vs_k141_119013\|\|full | Low-quality |
| S17_vs_k141_89339\|\|full | Not-determined |
| S17_vs_k141_96710\|\|full | Low-quality |
| S17_vs_k141_89345\|\|full | Low-quality |
| S17_vs_k141_148675\|\|full | Low-quality |
| S17_vs_k141_45107\|\|full | Low-quality |
| S17_vs_k141_148684\|\|full | Not-determined |
| S17_vs_k141_52640\|\|full | Not-determined |
| S17_vs_k141_89360\|\|full | Low-quality |
| S17_vs_k141_170745\|\|full | Low-quality |
| S17_vs_k141_67155\|\|full | Not-determined |
| S17_vs_k141_133695\|\|full | Low-quality |
| S17_vs_k141_111539\|\|full | Not-determined |
| S17_vs_k141_170747\|\|full | Not-determined |
| S17_vs_k141_104488\|\|full | Not-determined |
| S17_vs_k141_170748\|\|full | Low-quality |
| S17_vs_k141_156037\|\|full | Low-quality |
| S17_vs_k141_89372\|\|full | Low-quality |
| S17_vs_k141_170755\|\|full | Low-quality |
| S17_vs_k141_104501\|\|full | Low-quality |
| S17_vs_k141_119048\|\|full | Low-quality |
| S5_extracted_k141_67622 | High-quality |
| S17_vs_k141_74802\|\|full | Low-quality |
| S17_vs_k141_111556\|\|full | Low-quality |
| S17_vs_k141_74807\|\|full | Low-quality |
| S17_vs_k141_170767\|\|full | Not-determined |
| S17_vs_k141_119057\|\|full | Not-determined |
| S17_vs_k141_59882\|\|full | Low-quality |
| S17_vs_k141_163359\|\|full | Not-determined |
| S17_vs_k141_45115\|\|full | Not-determined |
| S17_vs_k141_163368\|\|full | Low-quality |
| S17_vs_k141_30553\|\|full | Low-quality |
| S17_vs_k141_170782\|\|full | Not-determined |
| S17_vs_k141_96788\|\|full | Not-determined |
| S17_vs_k141_148712\|\|full | Low-quality |
| S17_vs_k141_30560\|\|full | Low-quality |
| S17_vs_k141_170793\|\|full | Low-quality |
| S17_vs_k141_148714\|\|full | Not-determined |
| S17_vs_k141_67196\|\|full | Not-determined |
| S17_vs_k141_22820\|\|full | Not-determined |
| S17_vs_k141_52677\|\|full | Low-quality |
| S17_vs_k141_148723\|\|full | Low-quality |
| S17_vs_k141_156085\|\|full | Low-quality |
| S17_vs_k141_126331\|\|full | Low-quality |
| S17_vs_k141_163394\|\|full | Not-determined |
| S17_vs_k141_22832\|\|full | Not-determined |
| S17_vs_k141_104552\|\|full | Not-determined |
| S17_vs_k141_89379\|\|full | Not-determined |
| S17_vs_k141_45152\|\|full | Low-quality |
| S17_vs_k141_148741\|\|full | Low-quality |
| S17_vs_k141_89384\|\|full | Not-determined |
| S17_vs_k141_8288\|\|full | Not-determined |
| S17_vs_k141_119072\|\|full | Low-quality |
| S17_vs_k141_104562\|\|full | Low-quality |
| S17_vs_k141_59931\|\|full | Low-quality |
| S17_vs_k141_74821\|\|full | Low-quality |
| S17_vs_k141_163418\|\|full | Not-determined |
| S17_vs_k141_170830\|\|full | Not-determined |
| S17_vs_k141_22852\|\|full | Low-quality |
| S17_vs_k141_22857\|\|full | Low-quality |
| S17_vs_k141_170840\|\|full | Not-determined |
| S17_vs_k141_67243\|\|full | Not-determined |
| S17_vs_k141_8302\|\|full | Low-quality |
| S17_vs_k141_8304\|\|full | Low-quality |
| S17_vs_k141_38033\|\|full | Not-determined |
| S17_vs_k141_96831\|\|full | Not-determined |
| S17_vs_k141_104572\|\|full | Not-determined |
| S17_vs_k141_22873\|\|full | Not-determined |
| S17_vs_k141_156128\|\|full | Low-quality |
| S17_vs_k141_126346\|\|full | Not-determined |
| S17_vs_k141_38040\|\|full | Low-quality |
| S17_vs_k141_8319\|\|full | Low-quality |
| S17_vs_k141_163439\|\|full | Low-quality |
| S17_vs_k141_45179\|\|full | Not-determined |
| S17_vs_k141_156138\|\|full | Not-determined |
| S17_vs_k141_163440\|\|full | Low-quality |
| S17_vs_k141_67253\|\|full | Low-quality |
| S17_vs_k141_45183\|\|full | Not-determined |
| S17_vs_k141_163445\|\|full | Low-quality |
| S17_vs_k141_844\|\|full | Low-quality |
| S17_vs_k141_156145\|\|full | Low-quality |
| S17_vs_k141_38063\|\|full | Not-determined |
| S17_vs_k141_59945\|\|full | Low-quality |
| S17_vs_k141_59946\|\|full | Low-quality |
| S17_vs_k141_74850\|\|full | Low-quality |
| S17_vs_k141_89425\|\|full | Low-quality |
| S17_vs_k141_67269\|\|full | Low-quality |
| S17_vs_k141_148787\|\|full | Low-quality |
| S17_vs_k141_59956\|\|full | Low-quality |
| S17_vs_k141_119090\|\|full | Low-quality |
| S17_vs_k141_8356\|\|full | Low-quality |
| S17_vs_k141_163469\|\|full | Not-determined |
| S17_vs_k141_148795\|\|full | Not-determined |
| S17_vs_k141_96868\|\|full | Low-quality |
| S17_vs_k141_148803\|\|full | Low-quality |
| S17_vs_k141_81839\|\|full | Low-quality |
| S17_vs_k141_52764\|\|full | Low-quality |
| S17_vs_k141_45223\|\|full | Low-quality |
| S17_vs_k141_119097\|\|full | Not-determined |
| S17_vs_k141_89454\|\|full | Low-quality |
| S17_vs_k141_52775\|\|full | Low-quality |
| S17_vs_k141_15464\|\|full | Low-quality |
| S17_vs_k141_74889\|\|full | Not-determined |
| S17_vs_k141_52789\|\|full | Not-determined |
| S17_vs_k141_163474\|\|full | Low-quality |
| S17_vs_k141_163480\|\|full | Low-quality |
| S17_vs_k141_892\|\|full | Low-quality |
| S17_vs_k141_148834\|\|full | Not-determined |
| S17_vs_k141_111597\|\|full | Not-determined |
| S17_vs_k141_897\|\|full | Not-determined |
| S17_vs_k141_38130\|\|full | Low-quality |
| S17_vs_k141_96906\|\|full | Low-quality |
| S17_vs_k141_52806\|\|full | Not-determined |
| S17_vs_k141_119139\|\|full | Low-quality |
| S17_vs_k141_111618\|\|full | Not-determined |
| S17_vs_k141_920\|\|full | Low-quality |
| S17_vs_k141_133785\|\|full | Not-determined |
| S17_vs_k141_67359\|\|full | Not-determined |
| S17_vs_k141_133787\|\|full | Low-quality |
| S17_vs_k141_96922\|\|full | Not-determined |
| S17_vs_k141_74918\|\|full | Not-determined |
| S17_vs_k141_119149\|\|full | Not-determined |
| S17_vs_k141_30597\|\|full | Not-determined |
| S17_vs_k141_15486\|\|full | Low-quality |
| S17_vs_k141_67361\|\|full | Not-determined |
| S17_vs_k141_133797\|\|full | Low-quality |
| S17_vs_k141_163494\|\|full | Not-determined |
| S17_vs_k141_22900\|\|full | Not-determined |
| S17_vs_k141_96940\|\|full | Low-quality |
| S17_vs_k141_111640\|\|full | Low-quality |
| S17_vs_k141_52838\|\|full | Not-determined |
| S17_vs_k141_45303\|\|full | Low-quality |
| S17_vs_k141_45306\|\|full | Low-quality |
| S17_vs_k141_104678\|\|full | Low-quality |
| S17_vs_k141_81899\|\|full | Not-determined |
| S17_vs_k141_22912\|\|full | Not-determined |
| S17_vs_k141_156237\|\|full | Not-determined |
| S17_vs_k141_111663\|\|full | Not-determined |
| S17_vs_k141_89468\|\|full | Not-determined |
| S17_vs_k141_148910\|\|full | Low-quality |
| S17_vs_k141_170932\|\|full | Low-quality |
| S17_vs_k141_15531\|\|full | Not-determined |
| S17_vs_k141_67401\|\|full | Not-determined |
| S17_vs_k141_126402\|\|full | Low-quality |
| S17_vs_k141_30648\|\|full | Not-determined |
| S17_vs_k141_8424\|\|full | Not-determined |
| S17_vs_k141_15537\|\|full | Low-quality |
| S17_vs_k141_15541\|\|full | Low-quality |
| S17_vs_k141_170960\|\|full | Not-determined |
| S17_vs_k141_89510\|\|full | Low-quality |
| S17_vs_k141_8430\|\|full | Low-quality |
| S17_vs_k141_8433\|\|full | Low-quality |
| S17_vs_k141_119226\|\|full | Low-quality |
| S17_vs_k141_38225\|\|full | Not-determined |
| S17_vs_k141_45336\|\|full | Not-determined |
| S17_vs_k141_81910\|\|full | Not-determined |
| S17_vs_k141_60117\|\|full | Low-quality |
| S17_vs_k141_133847\|\|full | Low-quality |
| S17_vs_k141_170974\|\|full | Not-determined |
| S17_vs_k141_984\|\|full | Not-determined |
| S17_vs_k141_15570\|\|full | Low-quality |
| S17_vs_k141_15577\|\|full | Not-determined |
| S17_vs_k141_133856\|\|full | Low-quality |
| S17_vs_k141_119243\|\|full | Low-quality |
| S17_vs_k141_141371\|\|full | Not-determined |
| S17_vs_k141_38239\|\|full | Low-quality |
| S17_vs_k141_111698\|\|full | Low-quality |
| S17_vs_k141_30699\|\|full | Low-quality |
| S17_vs_k141_148982\|\|full | Low-quality |
| S17_vs_k141_171018\|\|full | Low-quality |
| S17_vs_k141_148996\|\|full | Not-determined |
| S6_extracted_k141_43812 | High-quality |
| S17_vs_k141_163533\|\|full | Low-quality |
| S17_vs_k141_52891\|\|full | Low-quality |
| S17_vs_k141_81929\|\|full | Not-determined |
| S17_vs_k141_52892\|\|full | Low-quality |
| S17_vs_k141_52894\|\|full | Low-quality |
| S17_vs_k141_163541\|\|full | Low-quality |
| S17_vs_k141_119269\|\|full | Not-determined |
| S17_vs_k141_60159\|\|full | Low-quality |
| S17_vs_k141_133886\|\|full | Low-quality |
| S17_vs_k141_156356\|\|full | Not-determined |
| S17_vs_k141_8497\|\|full | Not-determined |
| S17_vs_k141_111739\|\|full | Not-determined |
| S17_vs_k141_81943\|\|full | Not-determined |
| S17_vs_k141_60169\|\|full | Low-quality |
| S17_vs_k141_96988\|\|full | Low-quality |
| S17_vs_k141_96991\|\|full | Not-determined |
| S17_vs_k141_60173\|\|full | Low-quality |
| S17_vs_k141_38246\|\|full | Low-quality |
| S17_vs_k141_52916\|\|full | Low-quality |
| S17_vs_k141_111781\|\|full | Low-quality |
| S17_vs_k141_163580\|\|full | Low-quality |
| S17_vs_k141_74937\|\|full | Not-determined |
| S17_vs_k141_89609\|\|full | Low-quality |
| S17_vs_k141_111789\|\|full | Low-quality |
| S17_vs_k141_74943\|\|full | Low-quality |
| S17_vs_k141_156381\|\|full | Low-quality |
| S17_vs_k141_149053\|\|full | Not-determined |
| S17_vs_k141_30738\|\|full | Low-quality |
| S17_vs_k141_30739\|\|full | Not-determined |
| S17_vs_k141_156388\|\|full | Low-quality |
| S17_vs_k141_133930\|\|full | Not-determined |
| S17_vs_k141_8553\|\|full | Low-quality |
| S17_vs_k141_104792\|\|full | Low-quality |
| S17_vs_k141_52948\|\|full | Low-quality |
| S17_vs_k141_141435\|\|full | Low-quality |
| S17_vs_k141_156408\|\|full | Low-quality |
| S17_vs_k141_97028\|\|full | Not-determined |
| S17_vs_k141_60196\|\|full | Low-quality |
| S17_vs_k141_38278\|\|full | Not-determined |
| S17_vs_k141_104804\|\|full | Low-quality |
| S17_vs_k141_149067\|\|full | Low-quality |
| S17_vs_k141_133955\|\|full | Not-determined |
| S17_vs_k141_104812\|\|full | Not-determined |
| S17_vs_k141_81960\|\|full | Not-determined |
| S17_vs_k141_141462\|\|full | Low-quality |
| S17_vs_k141_156425\|\|full | Not-determined |
| S17_vs_k141_45462\|\|full | Low-quality |
| S17_vs_k141_104840\|\|full | Not-determined |
| S17_vs_k141_97048\|\|full | Not-determined |
| S17_vs_k141_1050\|\|full | Not-determined |
| S17_vs_k141_104847\|\|full | Not-determined |
| S17_vs_k141_38321\|\|full | Low-quality |
| S17_vs_k141_97056\|\|full | Low-quality |
| S17_vs_k141_60240\|\|full | Not-determined |
| S17_vs_k141_156442\|\|full | Low-quality |
| S17_vs_k141_119332\|\|full | Not-determined |
| S17_vs_k141_111843\|\|full | Not-determined |
| S17_vs_k141_163659\|\|full | Not-determined |
| S17_vs_k141_15652\|\|full | Low-quality |
| S17_vs_k141_104856\|\|full | Low-quality |
| S17_vs_k141_60250\|\|full | Low-quality |
| S17_vs_k141_30780\|\|full | Not-determined |
| S17_vs_k141_8586\|\|full | Low-quality |
| S17_vs_k141_111855\|\|full | Not-determined |
| S17_vs_k141_104866\|\|full | Low-quality |
| S17_vs_k141_97067\|\|full | Not-determined |
| S17_vs_k141_133967\|\|full | Not-determined |
| S17_vs_k141_119353\|\|full | Not-determined |
| S17_vs_k141_67523\|\|full | Not-determined |
| S17_vs_k141_149121\|\|full | Low-quality |
| S17_vs_k141_60257\|\|full | Low-quality |
| S17_vs_k141_97077\|\|full | Low-quality |
| S17_vs_k141_8606\|\|full | Low-quality |

| | |
|---|---|
| S17_vs_k141_38346\|\|full | Low-quality |
| S17_vs_k141_119367\|\|full | Not-determined |
| S17_vs_k141_15682\|\|full | Low-quality |
| S17_vs_k141_67544\|\|full | Low-quality |
| S17_vs_k141_67549\|\|full | Not-determined |
| S17_vs_k141_163670\|\|full | Not-determined |
| S17_vs_k141_52998\|\|full | Low-quality |
| S17_vs_k141_23064\|\|full | Low-quality |
| S17_vs_k141_104899\|\|full | Low-quality |
| S17_vs_k141_163680\|\|full | Not-determined |
| S17_vs_k141_38368\|\|full | Low-quality |
| S17_vs_k141_1087\|\|full | Not-determined |
| S17_vs_k141_89705\|\|full | Low-quality |
| S17_vs_k141_111875\|\|full | Not-determined |
| S17_vs_k141_8631\|\|full | Not-determined |
| S17_vs_k141_53018\|\|full | Low-quality |
| S17_vs_k141_89706\|\|full | Low-quality |
| S17_vs_k141_141519\|\|full | Not-determined |
| S17_vs_k141_149144\|\|full | Not-determined |
| S17_vs_k141_111887\|\|full | Not-determined |
| S17_vs_k141_111889\|\|full | Not-determined |
| S17_vs_k141_67579\|\|full | Not-determined |
| S17_vs_k141_67581\|\|full | Low-quality |
| S17_vs_k141_53033\|\|full | Low-quality |
| S17_vs_k141_141549\|\|full | Low-quality |
| S17_vs_k141_104940\|\|full | Not-determined |
| S17_vs_k141_133993\|\|full | Not-determined |
| S17_vs_k141_30845\|\|full | Low-quality |
| S17_vs_k141_89741\|\|full | Low-quality |
| S17_vs_k141_119394\|\|full | Not-determined |
| S17_vs_k141_30859\|\|full | Low-quality |
| S17_vs_k141_156519\|\|full | Not-determined |
| S17_vs_k141_171198\|\|full | Not-determined |
| S17_vs_k141_60305\|\|full | Not-determined |
| S17_vs_k141_60306\|\|full | Not-determined |
| S17_vs_k141_1115\|\|full | Low-quality |
| S17_vs_k141_8672\|\|full | Low-quality |
| S17_vs_k141_171212\|\|full | Not-determined |
| S17_vs_k141_149197\|\|full | Low-quality |
| S17_vs_k141_53092\|\|full | Not-determined |
| S17_vs_k141_149201\|\|full | Low-quality |
| S17_vs_k141_119408\|\|full | Low-quality |
| S17_vs_k141_141612\|\|full | Not-determined |
| S17_vs_k141_38429\|\|full | Not-determined |
| S17_vs_k141_163726\|\|full | Low-quality |
| S17_vs_k141_97177\|\|full | Low-quality |
| S17_vs_k141_134039\|\|full | Low-quality |
| S17_vs_k141_171223\|\|full | Low-quality |
| S17_vs_k141_141622\|\|full | Low-quality |
| S17_vs_k141_163733\|\|full | Not-determined |
| S17_vs_k141_45646\|\|full | Not-determined |
| S17_vs_k141_163736\|\|full | Not-determined |
| S17_vs_k141_156557\|\|full | Low-quality |
| S17_vs_k141_74975\|\|full | Not-determined |
| S17_vs_k141_67617\|\|full | Low-quality |
| S17_vs_k141_134044\|\|full | Not-determined |
| S17_vs_k141_67619\|\|full | Low-quality |
| S17_vs_k141_45650\|\|full | Low-quality |
| S17_vs_k141_149224\|\|full | Low-quality |
| S17_vs_k141_119426\|\|full | Not-determined |
| S17_vs_k141_171234\|\|full | Not-determined |
| S17_vs_k141_156571\|\|full | Not-determined |
| S17_vs_k141_134052\|\|full | Low-quality |
| S17_vs_k141_163760\|\|full | Not-determined |
| S5_vs_k141_67622\|\|full | High-quality |
| S17_vs_k141_38464\|\|full | Not-determined |
| S17_vs_k141_163771\|\|full | Low-quality |
| S17_vs_k141_134059\|\|full | Low-quality |
| S17_vs_k141_163773\|\|full | Low-quality |
| S17_vs_k141_119449\|\|full | Not-determined |
| S17_vs_k141_23139\|\|full | Low-quality |
| S17_vs_k141_1156\|\|full | Low-quality |
| S17_vs_k141_1160\|\|full | Not-determined |
| S17_vs_k141_89792\|\|full | Low-quality |
| S17_vs_k141_23145\|\|full | Not-determined |
| S17_vs_k141_67649\|\|full | Low-quality |
| S17_vs_k141_119457\|\|full | Not-determined |
| S17_vs_k141_149240\|\|full | Low-quality |
| S17_vs_k141_171244\|\|full | Low-quality |
| S17_vs_k141_23157\|\|full | Low-quality |
| S17_vs_k141_105050\|\|full | Low-quality |
| S17_vs_k141_53167\|\|full | Not-determined |
| S17_vs_k141_45678\|\|full | Not-determined |
| S17_vs_k141_149255\|\|full | Not-determined |
| S17_vs_k141_141668\|\|full | Low-quality |
| S17_vs_k141_23163\|\|full | Not-determined |
| S17_vs_k141_105060\|\|full | Low-quality |
| S17_vs_k141_67682\|\|full | Low-quality |
| S17_vs_k141_1173\|\|full | Not-determined |
| S17_vs_k141_119476\|\|full | Not-determined |
| S17_vs_k141_141681\|\|full | Low-quality |
| S17_vs_k141_67686\|\|full | Not-determined |
| S17_vs_k141_141684\|\|full | Not-determined |
| S17_vs_k141_89829\|\|full | Not-determined |
| S17_vs_k141_23176\|\|full | Low-quality |
| S17_vs_k141_89832\|\|full | Not-determined |
| S17_vs_k141_149288\|\|full | Low-quality |
| S17_vs_k141_23181\|\|full | Not-determined |
| S17_vs_k141_134098\|\|full | Low-quality |
| S17_vs_k141_75040\|\|full | Not-determined |
| S17_vs_k141_82072\|\|full | Low-quality |
| S17_vs_k141_141701\|\|full | Not-determined |
| S17_vs_k141_171286\|\|full | Low-quality |
| S17_vs_k141_75049\|\|full | Low-quality |
| S17_vs_k141_89851\|\|full | Low-quality |
| S17_vs_k141_119504\|\|full | Low-quality |
| S17_vs_k141_75057\|\|full | Low-quality |
| S17_vs_k141_134110\|\|full | Not-determined |
| S17_vs_k141_82095\|\|full | Low-quality |
| S17_vs_k141_171294\|\|full | Not-determined |
| S17_vs_k141_30959\|\|full | Low-quality |
| S17_vs_k141_163794\|\|full | Not-determined |
| S17_vs_k141_82106\|\|full | Low-quality |
| S17_vs_k141_134127\|\|full | Low-quality |
| S17_vs_k141_111982\|\|full | Not-determined |
| S17_vs_k141_119517\|\|full | Not-determined |
| S17_vs_k141_1206\|\|full | Not-determined |
| S17_vs_k141_15748\|\|full | Low-quality |
| S17_vs_k141_119522\|\|full | Not-determined |
| S17_vs_k141_8760\|\|full | Not-determined |
| S17_vs_k141_53227\|\|full | Not-determined |
| S17_vs_k141_126467\|\|full | Low-quality |
| S17_vs_k141_97282\|\|full | Not-determined |
| S17_vs_k141_89907\|\|full | Low-quality |
| S17_vs_k141_89908\|\|full | Low-quality |
| S17_vs_k141_8774\|\|full | Not-determined |
| S17_vs_k141_171307\|\|full | Not-determined |
| S17_vs_k141_15772\|\|full | Low-quality |
| S17_vs_k141_23201\|\|full | Not-determined |
| S17_vs_k141_30984\|\|full | Low-quality |
| S17_vs_k141_105140\|\|full | Low-quality |
| S17_vs_k141_126480\|\|full | Low-quality |
| S17_vs_k141_134166\|\|full | Low-quality |
| S17_vs_k141_8800\|\|full | Low-quality |
| S17_vs_k141_1240\|\|full | Not-determined |
| S17_vs_k141_89937\|\|full | Low-quality |
| S17_vs_k141_97300\|\|full | Low-quality |
| S17_vs_k141_82166\|\|full | Low-quality |
| S17_vs_k141_53256\|\|full | Low-quality |
| S17_vs_k141_75118\|\|full | Not-determined |
| S17_vs_k141_23234\|\|full | Low-quality |
| S17_vs_k141_89952\|\|full | Not-determined |
| S17_vs_k141_8823\|\|full | Not-determined |
| S17_vs_k141_134168\|\|full | Low-quality |
| S17_vs_k141_134176\|\|full | Not-determined |
| S17_vs_k141_134184\|\|full | Low-quality |
| S17_vs_k141_1262\|\|full | Low-quality |
| S17_vs_k141_53279\|\|full | Not-determined |
| S17_vs_k141_31041\|\|full | Not-determined |
| S17_vs_k141_89976\|\|full | Low-quality |
| S17_vs_k141_97347\|\|full | Low-quality |
| S17_vs_k141_53283\|\|full | Low-quality |
| S17_vs_k141_97350\|\|full | Low-quality |
| S17_vs_k141_134208\|\|full | Low-quality |
| S17_vs_k141_1276\|\|full | Not-determined |
| S17_vs_k141_8860\|\|full | Low-quality |
| S17_vs_k141_53302\|\|full | Low-quality |
| S17_vs_k141_89992\|\|full | Not-determined |
| S17_vs_k141_53310\|\|full | Not-determined |
| S17_vs_k141_60450\|\|full | Low-quality |
| S6_vs_k141_43812\|\|full | High-quality |
| S17_vs_k141_171380\|\|full | Low-quality |
| S17_vs_k141_171381\|\|full | Low-quality |
| S17_vs_k141_45799\|\|full | Low-quality |
| S17_vs_k141_82207\|\|full | Low-quality |
| S17_vs_k141_111999\|\|full | Low-quality |
| S17_vs_k141_90009\|\|full | Not-determined |
| S17_vs_k141_119559\|\|full | Low-quality |
| S17_vs_k141_171396\|\|full | Not-determined |
| S17_vs_k141_149447\|\|full | Low-quality |
| S17_vs_k141_1331\|\|full | Low-quality |
| S17_vs_k141_45818\|\|full | Low-quality |
| S17_vs_k141_31064\|\|full | Low-quality |
| S17_vs_k141_126521\|\|full | Not-determined |
| S17_vs_k141_97389\|\|full | Not-determined |
| S17_vs_k141_171409\|\|full | Low-quality |
| S17_vs_k141_156670\|\|full | Not-determined |
| S17_vs_k141_134219\|\|full | Not-determined |
| S17_vs_k141_97397\|\|full | Low-quality |
| S17_vs_k141_134221\|\|full | Not-determined |
| S17_vs_k141_90022\|\|full | Low-quality |
| S17_vs_k141_141732\|\|full | Not-determined |
| S17_vs_k141_75148\|\|full | Low-quality |
| S17_vs_k141_90026\|\|full | Low-quality |
| S17_vs_k141_31081\|\|full | Not-determined |
| S17_vs_k141_97406\|\|full | Not-determined |
| S17_vs_k141_90029\|\|full | Low-quality |
| S17_vs_k141_8913\|\|full | Not-determined |
| S17_vs_k141_90033\|\|full | Not-determined |
| S17_vs_k141_90034\|\|full | Low-quality |
| S17_vs_k141_38666\|\|full | Low-quality |
| S17_vs_k141_31099\|\|full | Not-determined |
| S17_vs_k141_82261\|\|full | Low-quality |
| S17_vs_k141_31100\|\|full | Low-quality |
| S17_vs_k141_60509\|\|full | Low-quality |
| S17_vs_k141_97431\|\|full | Not-determined |
| S17_vs_k141_8941\|\|full | Low-quality |
| S17_vs_k141_112032\|\|full | Low-quality |
| S17_vs_k141_171446\|\|full | Low-quality |
| S17_vs_k141_149491\|\|full | Low-quality |
| S17_vs_k141_60521\|\|full | Not-determined |
| S17_vs_k141_171449\|\|full | Not-determined |
| S17_vs_k141_38685\|\|full | Low-quality |
| S17_vs_k141_126583\|\|full | Low-quality |
| S17_vs_k141_119624\|\|full | Low-quality |
| S17_vs_k141_134269\|\|full | Low-quality |
| S17_vs_k141_141797\|\|full | Low-quality |
| S17_vs_k141_119627\|\|full | Not-determined |
| S17_vs_k141_1377\|\|full | Not-determined |
| S17_vs_k141_75187\|\|full | Not-determined |
| S17_vs_k141_82284\|\|full | Low-quality |
| S17_vs_k141_75192\|\|full | Low-quality |
| S17_vs_k141_126592\|\|full | Not-determined |
| S17_vs_k141_23322\|\|full | Not-determined |
| S17_vs_k141_45877\|\|full | Not-determined |
| S17_vs_k141_1383\|\|full | Low-quality |
| S17_vs_k141_156708\|\|full | Not-determined |
| S17_vs_k141_38706\|\|full | Low-quality |
| S17_vs_k141_53421\|\|full | Low-quality |
| S17_vs_k141_97469\|\|full | Low-quality |
| S17_vs_k141_38710\|\|full | Not-determined |
| S17_vs_k141_105249\|\|full | Low-quality |
| S17_vs_k141_23336\|\|full | Low-quality |
| S17_vs_k141_141823\|\|full | Low-quality |
| S17_vs_k141_75205\|\|full | Not-determined |
| S17_vs_k141_23341\|\|full | Not-determined |
| S17_vs_k141_31144\|\|full | Low-quality |
| S17_vs_k141_75215\|\|full | Low-quality |
| S17_vs_k141_31152\|\|full | Low-quality |
| S17_vs_k141_149532\|\|full | Low-quality |
| S17_vs_k141_156747\|\|full | Low-quality |
| S17_vs_k141_60574\|\|full | Low-quality |
| S17_vs_k141_149541\|\|full | Low-quality |
| S17_vs_k141_38734\|\|full | Not-determined |
| S17_vs_k141_31169\|\|full | Low-quality |
| S17_vs_k141_163919\|\|full | Low-quality |
| S17_vs_k141_163922\|\|full | Low-quality |
| S17_vs_k141_156772\|\|full | Low-quality |
| S17_vs_k141_53460\|\|full | Low-quality |
| S17_vs_k141_53461\|\|full | Not-determined |
| S17_vs_k141_75256\|\|full | Not-determined |
| S17_vs_k141_141847\|\|full | Not-determined |
| S17_vs_k141_163925\|\|full | Low-quality |
| S17_vs_k141_53466\|\|full | Low-quality |
| S17_vs_k141_60602\|\|full | Low-quality |
| S17_vs_k141_15824\|\|full | Low-quality |
| S17_vs_k141_67876\|\|full | Low-quality |
| S17_vs_k141_38763\|\|full | Low-quality |
| S17_vs_k141_15841\|\|full | Low-quality |
| S17_vs_k141_141866\|\|full | Low-quality |
| S17_vs_k141_134290\|\|full | Low-quality |
| S17_vs_k141_9036\|\|full | Not-determined |
| S17_vs_k141_15848\|\|full | Low-quality |
| S17_vs_k141_105280\|\|full | Not-determined |
| S17_vs_k141_15849\|\|full | Low-quality |
| S17_vs_k141_134303\|\|full | Low-quality |
| S17_vs_k141_67887\|\|full | Not-determined |
| S17_vs_k141_67889\|\|full | Not-determined |
| S17_vs_k141_156808\|\|full | Not-determined |
| S17_vs_k141_53508\|\|full | Low-quality |
| S17_vs_k141_163977\|\|full | Not-determined |
| S17_vs_k141_97569\|\|full | Low-quality |
| S17_vs_k141_105308\|\|full | Low-quality |
| S17_vs_k141_163980\|\|full | Low-quality |
| S17_vs_k141_163987\|\|full | Not-determined |
| S17_vs_k141_53528\|\|full | Low-quality |
| S17_vs_k141_82319\|\|full | Low-quality |
| S17_vs_k141_67911\|\|full | Low-quality |
| S17_vs_k141_82324\|\|full | Low-quality |
| S17_vs_k141_45968\|\|full | Low-quality |
| S17_vs_k141_112140\|\|full | Low-quality |
| S17_vs_k141_31218\|\|full | Not-determined |
| S17_vs_k141_38776\|\|full | Low-quality |
| S17_vs_k141_15892\|\|full | Low-quality |
| S17_vs_k141_105335\|\|full | Low-quality |
| S17_vs_k141_141932\|\|full | Low-quality |
| S17_vs_k141_82345\|\|full | Low-quality |
| S17_vs_k141_97588\|\|full | Low-quality |
| S17_vs_k141_119727\|\|full | Low-quality |
| S17_vs_k141_164013\|\|full | Low-quality |
| S17_vs_k141_97596\|\|full | Not-determined |
| S17_vs_k141_105343\|\|full | Not-determined |
| S17_vs_k141_119743\|\|full | Low-quality |
| S17_vs_k141_45987\|\|full | Low-quality |
| S17_vs_k141_1457\|\|full | Low-quality |
| S17_vs_k141_119745\|\|full | Not-determined |
| S17_vs_k141_67959\|\|full | Low-quality |
| S17_vs_k141_15928\|\|full | Not-determined |
| S17_vs_k141_53561\|\|full | Not-determined |
| S17_vs_k141_90168\|\|full | Not-determined |
| S17_vs_k141_126693\|\|full | Not-determined |
| S17_vs_k141_67973\|\|full | Not-determined |
| S17_vs_k141_82360\|\|full | Low-quality |
| S17_vs_k141_1478\|\|full | Low-quality |
| S17_vs_k141_141981\|\|full | Low-quality |
| S17_vs_k141_9143\|\|full | Not-determined |
| S17_vs_k141_46016\|\|full | Low-quality |
| S17_vs_k141_23436\|\|full | Not-determined |
| S17_vs_k141_82367\|\|full | Not-determined |
| S17_vs_k141_141999\|\|full | Low-quality |
| S17_vs_k141_97658\|\|full | Low-quality |
| S17_vs_k141_90192\|\|full | Not-determined |
| S17_vs_k141_164073\|\|full | Low-quality |
| S17_vs_k141_142008\|\|full | Low-quality |
| S17_vs_k141_105389\|\|full | Low-quality |
| S17_vs_k141_82386\|\|full | Not-determined |
| S17_vs_k141_134372\|\|full | Low-quality |
| S17_vs_k141_68010\|\|full | Not-determined |
| S17_vs_k141_134380\|\|full | Low-quality |
| S17_vs_k141_142025\|\|full | Not-determined |
| S17_vs_k141_105412\|\|full | Not-determined |
| S17_vs_k141_68018\|\|full | Low-quality |
| S17_vs_k141_38825\|\|full | Low-quality |
| S17_vs_k141_82403\|\|full | Not-determined |
| S17_vs_k141_156913\|\|full | Low-quality |
| S17_vs_k141_105417\|\|full | Low-quality |
| S13_extracted_138153 | Complete |
| S17_vs_k141_60737\|\|full | Not-determined |
| S17_vs_k141_53601\|\|full | Not-determined |
| S17_vs_k141_171548\|\|full | Low-quality |
| S17_vs_k141_68037\|\|full | Not-determined |
| S17_vs_k141_23465\|\|full | Not-determined |
| S17_vs_k141_142027\|\|full | Low-quality |
| S17_vs_k141_53605\|\|full | Not-determined |
| S17_vs_k141_134405\|\|full | Not-determined |
| S17_vs_k141_126765\|\|full | Low-quality |
| S17_vs_k141_53617\|\|full | Not-determined |
| S17_vs_k141_82435\|\|full | Not-determined |
| S17_vs_k141_149637\|\|full | Not-determined |
| S17_vs_k141_156938\|\|full | Not-determined |
| S17_vs_k141_1564\|\|full | Low-quality |
| S17_vs_k141_46072\|\|full | Not-determined |
| S17_vs_k141_9201\|\|full | Low-quality |
| S17_vs_k141_149647\|\|full | Low-quality |
| S17_vs_k141_53634\|\|full | Low-quality |
| S17_vs_k141_31315\|\|full | Low-quality |
| S17_vs_k141_82453\|\|full | Low-quality |
| S17_vs_k141_105470\|\|full | Not-determined |
| S17_vs_k141_156961\|\|full | Not-determined |
| S17_vs_k141_46108\|\|full | Low-quality |
| S17_vs_k141_149665\|\|full | Low-quality |
| S17_vs_k141_23496\|\|full | Not-determined |
| S17_vs_k141_23498\|\|full | Low-quality |
| S17_vs_k141_38871\|\|full | Low-quality |
| S17_vs_k141_105493\|\|full | Not-determined |
| S17_vs_k141_38878\|\|full | Not-determined |
| S17_vs_k141_142064\|\|full | Low-quality |
| S17_vs_k141_149698\|\|full | Low-quality |
| S17_vs_k141_105503\|\|full | Low-quality |
| S17_vs_k141_126807\|\|full | Low-quality |
| S17_vs_k141_9210\|\|full | Not-determined |
| S17_vs_k141_97755\|\|full | Not-determined |
| S17_vs_k141_1611\|\|full | Low-quality |
| S17_vs_k141_23524\|\|full | Low-quality |
| S17_vs_k141_126823\|\|full | Low-quality |
| S2_extracted_k141_21631 | Complete |
| S17_vs_k141_97768\|\|full | Low-quality |
| S17_vs_k141_149719\|\|full | Low-quality |
| S17_vs_k141_31363\|\|full | Not-determined |
| S17_vs_k141_126839\|\|full | Not-determined |
| S17_vs_k141_126848\|\|full | Low-quality |
| S17_vs_k141_82499\|\|full | Not-determined |
| S17_vs_k141_60781\|\|full | Low-quality |
| S17_vs_k141_31368\|\|full | Low-quality |
| S17_vs_k141_60786\|\|full | Not-determined |
| S17_vs_k141_134513\|\|full | Not-determined |
| S17_vs_k141_68079\|\|full | Low-quality |
| S17_vs_k141_46188\|\|full | Not-determined |
| S13_vs_138153\|\|full | High-quality |
| S17_vs_k141_31376\|\|full | Not-determined |
| S17_vs_k141_68082\|\|full | Low-quality |
| S17_vs_k141_126869\|\|full | Not-determined |
| S17_vs_k141_126871\|\|full | Not-determined |
| S17_vs_k141_126873\|\|full | Low-quality |
| S17_vs_k141_1620\|\|full | Low-quality |
| S17_vs_k141_126875\|\|full | Not-determined |
| S17_vs_k141_1621\|\|full | Not-determined |
| S17_vs_k141_60795\|\|full | Low-quality |
| S17_vs_k141_16054\|\|full | Not-determined |
| S17_vs_k141_119907\|\|full | Low-quality |
| S17_vs_k141_68091\|\|full | Low-quality |
| S17_vs_k141_97824\|\|full | Low-quality |
| S17_vs_k141_53727\|\|full | Low-quality |
| S17_vs_k141_46212\|\|full | Not-determined |
| S17_vs_k141_112297\|\|full | Low-quality |
| S17_vs_k141_157013\|\|full | Low-quality |
| S17_vs_k141_142126\|\|full | Low-quality |
| S17_vs_k141_1653\|\|full | Low-quality |
| S17_vs_k141_23577\|\|full | Low-quality |
| S17_vs_k141_119915\|\|full | Low-quality |
| S17_vs_k141_149756\|\|full | Low-quality |
| S17_vs_k141_75384\|\|full | Not-determined |
| S17_vs_k141_31399\|\|full | Low-quality |
| S17_vs_k141_68114\|\|full | Not-determined |
| S17_vs_k141_75386\|\|full | Not-determined |
| S17_vs_k141_149762\|\|full | Not-determined |
| S17_vs_k141_68115\|\|full | Low-quality |
| S17_vs_k141_119921\|\|full | Low-quality |
| S17_vs_k141_149764\|\|full | Low-quality |
| S17_vs_k141_68119\|\|full | Not-determined |
| S17_vs_k141_1666\|\|full | Low-quality |
| S17_vs_k141_134548\|\|full | Low-quality |
| S17_vs_k141_31409\|\|full | Not-determined |
| S17_vs_k141_46233\|\|full | Low-quality |
| S17_vs_k141_97840\|\|full | Low-quality |
| S17_vs_k141_53760\|\|full | Low-quality |
| S17_vs_k141_105581\|\|full | Low-quality |
| S17_vs_k141_157043\|\|full | Low-quality |
| S17_vs_k141_46248\|\|full | Not-determined |
| S17_vs_k141_157044\|\|full | Low-quality |
| S17_vs_k141_75406\|\|full | Not-determined |
| S17_vs_k141_157053\|\|full | Low-quality |
| S17_vs_k141_149793\|\|full | Low-quality |
| S17_vs_k141_31423\|\|full | Low-quality |
| S17_vs_k141_38949\|\|full | Not-determined |
| S17_vs_k141_31426\|\|full | Not-determined |
| S17_vs_k141_105617\|\|full | Not-determined |
| S17_vs_k141_1705\|\|full | Low-quality |
| S17_vs_k141_38955\|\|full | Not-determined |
| S17_vs_k141_112359\|\|full | Not-determined |
| S17_vs_k141_134558\|\|full | Low-quality |
| S17_vs_k141_164156\|\|full | Low-quality |
| S17_vs_k141_126945\|\|full | Not-determined |
| S17_vs_k141_142202\|\|full | Not-determined |
| S17_vs_k141_112367\|\|full | Low-quality |
| S17_vs_k141_68175\|\|full | Not-determined |
| S17_vs_k141_16108\|\|full | Not-determined |
| S17_vs_k141_164173\|\|full | Low-quality |
| S17_vs_k141_31467\|\|full | Low-quality |
| S17_vs_k141_97854\|\|full | Low-quality |
| S17_vs_k141_16117\|\|full | Low-quality |
| S17_vs_k141_82586\|\|full | Low-quality |
| S17_vs_k141_149844\|\|full | Not-determined |
| S17_vs_k141_16125\|\|full | Low-quality |
| S17_vs_k141_157108\|\|full | Low-quality |
| S17_vs_k141_157111\|\|full | Low-quality |
| S17_vs_k141_164175\|\|full | Not-determined |
| S17_vs_k141_97866\|\|full | Low-quality |
| S17_vs_k141_1714\|\|full | Low-quality |
| S17_vs_k141_39001\|\|full | Low-quality |
| S17_vs_k141_46327\|\|full | Low-quality |
| S17_vs_k141_68201\|\|full | Not-determined |
| S17_vs_k141_134599\|\|full | Low-quality |
| S17_vs_k141_120029\|\|full | Low-quality |
| S17_vs_k141_97887\|\|full | Not-determined |
| S17_vs_k141_134601\|\|full | Low-quality |
| S17_vs_k141_82601\|\|full | Not-determined |
| S17_vs_k141_126974\|\|full | Not-determined |
| S17_vs_k141_75441\|\|full | Not-determined |
| S17_vs_k141_105671\|\|full | Low-quality |
| S17_vs_k141_142256\|\|full | Low-quality |
| S17_vs_k141_142257\|\|full | Low-quality |
| S17_vs_k141_164187\|\|full | Low-quality |
| S17_vs_k141_9344\|\|full | Low-quality |
| S17_vs_k141_82616\|\|full | Not-determined |
| S17_vs_k141_126985\|\|full | Low-quality |
| S17_vs_k141_39022\|\|full | Low-quality |
| S17_vs_k141_16177\|\|full | Low-quality |
| S17_vs_k141_120044\|\|full | Not-determined |
| S17_vs_k141_105688\|\|full | Low-quality |
| S17_vs_k141_164200\|\|full | Low-quality |
| S17_vs_k141_134651\|\|full | Low-quality |
| S17_vs_k141_53858\|\|full | Low-quality |
| S17_vs_k141_126995\|\|full | Low-quality |
| S17_vs_k141_97924\|\|full | Low-quality |
| S17_vs_k141_9374\|\|full | Low-quality |
| S17_vs_k141_97928\|\|full | Low-quality |
| S17_vs_k141_105703\|\|full | Not-determined |
| S17_vs_k141_60802\|\|full | Low-quality |
| S17_vs_k141_60805\|\|full | Not-determined |
| S17_vs_k141_23778\|\|full | Low-quality |
| S17_vs_k141_97938\|\|full | Low-quality |
| S17_vs_k141_9398\|\|full | Low-quality |
| S17_vs_k141_90265\|\|full | Low-quality |
| S17_vs_k141_68264\|\|full | Not-determined |
| S17_vs_k141_82679\|\|full | Low-quality |
| S17_vs_k141_90270\|\|full | Low-quality |
| S17_vs_k141_16213\|\|full | Low-quality |
| S17_vs_k141_134681\|\|full | Not-determined |
| S17_vs_k141_164233\|\|full | Not-determined |
| S17_vs_k141_120096\|\|full | Low-quality |
| S17_vs_k141_82685\|\|full | Low-quality |
| S2_vs_k141_21631\|\|full | High-quality |
| S17_vs_k141_164243\|\|full | Low-quality |
| S17_vs_k141_39064\|\|full | Not-determined |
| S17_vs_k141_53900\|\|full | Low-quality |
| S17_vs_k141_82692\|\|full | Not-determined |
| S17_vs_k141_16228\|\|full | Not-determined |
| S17_vs_k141_16231\|\|full | Low-quality |
| S17_vs_k141_75464\|\|full | Not-determined |
| S17_vs_k141_82695\|\|full | Low-quality |
| S17_vs_k141_164253\|\|full | Not-determined |
| S17_vs_k141_60838\|\|full | Not-determined |
| S17_vs_k141_16240\|\|full | Low-quality |
| S17_vs_k141_149950\|\|full | Low-quality |
| S17_vs_k141_134716\|\|full | Not-determined |
| S17_vs_k141_127082\|\|full | Not-determined |
| S17_vs_k141_82714\|\|full | Not-determined |
| S17_vs_k141_82716\|\|full | Low-quality |
| S17_vs_k141_1769\|\|full | Low-quality |
| S17_vs_k141_157170\|\|full | Low-quality |
| S17_vs_k141_23832\|\|full | Not-determined |
| S17_vs_k141_39091\|\|full | Low-quality |
| S17_vs_k141_105766\|\|full | Low-quality |
| S17_vs_k141_16253\|\|full | Low-quality |
| S17_vs_k141_157178\|\|full | Low-quality |
| S17_vs_k141_60847\|\|full | Not-determined |
| S17_vs_k141_82725\|\|full | Low-quality |
| S17_vs_k141_164262\|\|full | Not-determined |
| S17_vs_k141_60850\|\|full | Not-determined |
| S17_vs_k141_46423\|\|full | Low-quality |
| S17_vs_k141_164273\|\|full | Low-quality |
| S17_vs_k141_157195\|\|full | Low-quality |
| S17_vs_k141_60861\|\|full | Not-determined |
| S17_vs_k141_164283\|\|full | Low-quality |
| S17_vs_k141_23842\|\|full | Not-determined |
| S17_vs_k141_149970\|\|full | Low-quality |
| S17_vs_k141_23845\|\|full | Not-determined |
| S17_vs_k141_157204\|\|full | Not-determined |
| S17_vs_k141_112438\|\|full | Low-quality |
| S17_vs_k141_149985\|\|full | Not-determined |
| S17_vs_k141_23859\|\|full | Not-determined |
| S17_vs_k141_75506\|\|full | Not-determined |
| S17_vs_k141_60896\|\|full | Not-determined |
| S17_vs_k141_134769\|\|full | Not-determined |
| S17_vs_k141_120192\|\|full | Low-quality |
| S17_vs_k141_142349\|\|full | Low-quality |
| S17_vs_k141_23873\|\|full | Low-quality |
| S17_vs_k141_134773\|\|full | Low-quality |
| S17_vs_k141_120199\|\|full | Not-determined |
| S17_vs_k141_120201\|\|full | Low-quality |
| S17_vs_k141_127139\|\|full | Not-determined |
| S17_vs_k141_16296\|\|full | Low-quality |
| S17_vs_k141_120216\|\|full | Not-determined |
| S17_vs_k141_68319\|\|full | Low-quality |
| S17_vs_k141_1820\|\|full | Not-determined |
| S17_vs_k141_90330\|\|full | Low-quality |
| S17_vs_k141_164322\|\|full | Low-quality |
| S17_vs_k141_68325\|\|full | Low-quality |
| S17_vs_k141_134793\|\|full | Low-quality |
| S17_vs_k141_105815\|\|full | Low-quality |
| S17_vs_k141_98067\|\|full | Low-quality |
| S17_vs_k141_75548\|\|full | Low-quality |
| S11_extracted_k141_104046 | Complete |
| S17_vs_k141_1851\|\|full | Not-determined |
| S17_vs_k141_31634\|\|full | Low-quality |
| S17_vs_k141_16330\|\|full | Not-determined |
| S17_vs_k141_1866\|\|full | Low-quality |
| S17_vs_k141_134798\|\|full | Not-determined |
| S17_vs_k141_164349\|\|full | Low-quality |
| S17_vs_k141_1874\|\|full | Low-quality |
| S17_vs_k141_90369\|\|full | Low-quality |
| S17_vs_k141_112507\|\|full | Low-quality |
| S17_vs_k141_23889\|\|full | Not-determined |
| S17_vs_k141_60940\|\|full | Not-determined |
| S17_vs_k141_75582\|\|full | Not-determined |
| S17_vs_k141_75586\|\|full | Low-quality |
| S17_vs_k141_54007\|\|full | Low-quality |
| S17_vs_k141_112517\|\|full | Low-quality |
| S17_vs_k141_134830\|\|full | Low-quality |
| S17_vs_k141_98101\|\|full | Low-quality |
| S17_vs_k141_112524\|\|full | Not-determined |
| S11_vs_k141_104046\|\|full | High-quality |
| S17_vs_k141_39240\|\|full | Not-determined |
| S17_vs_k141_157240\|\|full | Not-determined |
| S17_vs_k141_75610\|\|full | Low-quality |
| S17_vs_k141_82768\|\|full | Low-quality |
| S17_vs_k141_54041\|\|full | Not-determined |
| S17_vs_k141_98119\|\|full | Low-quality |
| S17_vs_k141_82770\|\|full | Not-determined |
| S17_vs_k141_171718\|\|full | Low-quality |
| S17_vs_k141_23901\|\|full | Not-determined |
| S17_vs_k141_150099\|\|full | Low-quality |
| S17_vs_k141_82775\|\|full | Not-determined |
| S17_vs_k141_112544\|\|full | Not-determined |
| S17_vs_k141_171722\|\|full | Not-determined |
| S17_vs_k141_68431\|\|full | Not-determined |
| S17_vs_k141_90411\|\|full | Low-quality |
| S17_vs_k141_90413\|\|full | Not-determined |
| S17_vs_k141_127200\|\|full | Not-determined |
| S17_vs_k141_164416\|\|full | Low-quality |
| S17_vs_k141_90414\|\|full | Low-quality |
| S17_vs_k141_82794\|\|full | Not-determined |
| S17_vs_k141_142454\|\|full | Not-determined |
| S17_vs_k141_31662\|\|full | Low-quality |
| S17_vs_k141_112565\|\|full | Low-quality |
| S17_vs_k141_134882\|\|full | Low-quality |
| S17_vs_k141_82822\|\|full | Low-quality |
| S17_vs_k141_16344\|\|full | Low-quality |

S17_vs_k141_68465||full Low-quality
S17_vs_k141_134895||full Low-quality
S17_vs_k141_150136||full Not-determined
S17_vs_k141_23937||full Low-quality
S17_vs_k141_98155||full Low-quality
S17_vs_k141_61003||full Not-determined
S17_vs_k141_16356||full Low-quality
S17_vs_k141_171756||full Not-determined
S17_vs_k141_82838||full Low-quality
S17_vs_k141_134905||full Low-quality
S17_vs_k141_68483||full Not-determined
S17_vs_k141_16364||full Not-determined
S17_vs_k141_105909||full Low-quality
S17_vs_k141_120238||full Not-determined
S17_vs_k141_23958||full Low-quality
S17_vs_k141_134911||full Not-determined
S17_vs_k141_157316||full Low-quality
S17_vs_k141_120249||full Low-quality
S17_vs_k141_54095||full Low-quality
S17_vs_k141_112617||full Not-determined
S17_vs_k141_9459||full Not-determined
S17_vs_k141_31716||full Not-determined
S17_vs_k141_157327||full Low-quality
S17_vs_k141_105933||full Low-quality
S17_vs_k141_134934||full Low-quality
S17_vs_k141_150181||full Not-determined
S17_vs_k141_157335||full Not-determined
S17_vs_k141_75654||full Low-quality
S17_vs_k141_23980||full Low-quality
S17_vs_k141_46553||full Low-quality
S17_vs_k141_75657||full Low-quality
S17_vs_k141_105944||full Low-quality
S17_vs_k141_75659||full Not-determined
S17_vs_k141_98183||full Not-determined
S17_vs_k141_134954||full Not-determined
S17_vs_k141_9483||full Low-quality
S17_vs_k141_31718||full Low-quality
S17_vs_k141_31721||full Not-determined
S17_vs_k141_9492||full Low-quality
S17_vs_k141_46557||full Low-quality
S17_vs_k141_39366||full Low-quality
S17_vs_k141_98200||full Not-determined
S17_vs_k141_134961||full Low-quality
S17_vs_k141_75667||full Low-quality
S17_vs_k141_39371||full Low-quality
S17_vs_k141_164475||full Not-determined
S17_vs_k141_23998||full Low-quality
S17_vs_k141_98219||full Low-quality
S17_vs_k141_171795||full Low-quality
S17_vs_k141_82881||full Low-quality
S17_vs_k141_112633||full Low-quality
S17_vs_k141_68541||full Low-quality
S17_vs_k141_171804||full Low-quality
S17_vs_k141_112634||full Not-determined
S17_vs_k141_150216||full Not-determined
S17_vs_k141_75709||full Low-quality
S17_vs_k141_112654||full Low-quality
S17_vs_k141_9514||full Low-quality
S17_vs_k141_75711||full Not-determined
S17_vs_k141_24008||full Low-quality
S17_vs_k141_112669||full Low-quality
S17_vs_k141_105979||full Low-quality
S17_vs_k141_127297||full Low-quality
S17_vs_k141_24009||full Not-determined
S17_vs_k141_68565||full Low-quality
S17_vs_k141_150227||full Low-quality
S17_vs_k141_24010||full Low-quality
S17_vs_k141_24011||full Low-quality
S17_vs_k141_134992||full Not-determined
S17_vs_k141_90439||full Low-quality
S17_vs_k141_105983||full Not-determined
S17_vs_k141_112681||full Not-determined
S17_vs_k141_142586||full Not-determined
S17_vs_k141_24027||full Not-determined
S17_vs_k141_54149||full Low-quality
S17_vs_k141_90450||full Low-quality
S17_vs_k141_106000||full Not-determined
S17_vs_k141_90452||full Low-quality
S17_vs_k141_68587||full Low-quality
S17_vs_k141_90453||full Not-determined
S17_vs_k141_24033||full Not-determined
S17_vs_k141_39414||full Low-quality
S17_vs_k141_150251||full Not-determined
S17_vs_k141_16453||full Low-quality
S17_vs_k141_135015||full Not-determined
S17_vs_k141_39421||full Not-determined
S17_vs_k141_68589||full Not-determined
S17_vs_k141_90467||full Low-quality
S17_vs_k141_2044||full Not-determined
S17_vs_k141_2046||full Low-quality
S17_vs_k141_164546||full Low-quality
S17_vs_k141_9575||full Low-quality
S17_vs_k141_150286||full Not-determined
S17_vs_k141_127333||full Not-determined
S17_vs_k141_164549||full Low-quality
S17_vs_k141_75757||full Not-determined
S17_vs_k141_142610||full Not-determined
S17_vs_k141_75760||full Low-quality
S17_vs_k141_135031||full Low-quality
S17_vs_k141_16467||full Low-quality
S17_vs_k141_112693||full Low-quality
S17_vs_k141_127349||full Low-quality
S17_vs_k141_90499||full Not-determined
S17_vs_k141_16482||full Low-quality
S17_vs_k141_127356||full Not-determined
S17_vs_k141_157418||full Low-quality
S17_vs_k141_31819||full Low-quality
S17_vs_k141_9602||full Not-determined
S17_vs_k141_39473||full Not-determined
S17_vs_k141_112702||full Low-quality
S17_vs_k141_164584||full Low-quality
S17_vs_k141_68628||full Not-determined
S17_vs_k141_164589||full Not-determined
S17_vs_k141_68634||full Low-quality
S17_vs_k141_24083||full Not-determined
S17_vs_k141_54212||full Not-determined
S17_vs_k141_142659||full Low-quality
S17_vs_k141_68641||full Not-determined
S17_vs_k141_106072||full Low-quality
S17_vs_k141_82893||full Low-quality
S17_vs_k141_150348||full Low-quality
S13_extracted_150365 Complete
S17_vs_k141_135064||full Not-determined
S17_vs_k141_157450||full Low-quality
S17_vs_k141_135069||full Low-quality
S17_vs_k141_2089||full Low-quality
S17_vs_k141_157464||full Low-quality
S17_vs_k141_150372||full Not-determined
S17_vs_k141_106106||full Low-quality
S17_vs_k141_164625||full Not-determined
S17_vs_k141_135083||full Low-quality
S17_vs_k141_150375||full Low-quality
S17_vs_k141_16552||full Not-determined
S17_vs_k141_98335||full Not-determined
S17_vs_k141_9675||full Not-determined
S17_vs_k141_39496||full Not-determined
S17_vs_k141_171820||full Low-quality
S17_vs_k141_2095||full Low-quality
S17_vs_k141_68689||full Low-quality
S17_vs_k141_157485||full Low-quality
S17_vs_k141_171834||full Not-determined
S17_vs_k141_164661||full Low-quality
S17_vs_k141_106140||full Low-quality
S17_vs_k141_157491||full Low-quality
S17_vs_k141_68706||full Not-determined
S17_vs_k141_135096||full Low-quality
S17_vs_k141_164667||full Low-quality
S17_vs_k141_157510||full Not-determined
S17_vs_k141_112799||full Low-quality
S17_vs_k141_157513||full Low-quality
S17_vs_k141_98384||full Low-quality
S17_vs_k141_150412||full Not-determined
S17_vs_k141_68732||full Low-quality
S17_vs_k141_157531||full Not-determined
S17_vs_k141_171894||full Not-determined
S17_vs_k141_90597||full Not-determined
S13_vs_150365||full High-quality
S17_vs_k141_46692||full Not-determined
S17_vs_k141_112829||full Low-quality
S17_vs_k141_54223||full Low-quality
S17_vs_k141_9771||full Not-determined
S17_vs_k141_171906||full Not-determined
S17_vs_k141_157551||full Not-determined
S17_vs_k141_61018||full Low-quality
S17_vs_k141_98434||full Not-determined
S17_vs_k141_98435||full Not-determined
S17_vs_k141_150440||full Not-determined
S17_vs_k141_171912||full Not-determined
S17_vs_k141_9783||full Low-quality
S17_vs_k141_68775||full Not-determined
S17_vs_k141_127444||full Not-determined
S17_vs_k141_157563||full Not-determined
S17_vs_k141_112849||full Low-quality
S17_vs_k141_82906||full Low-quality
S17_vs_k141_16641||full Not-determined
S17_vs_k141_75950||full Low-quality
S17_vs_k141_120473||full Low-quality
S17_vs_k141_90625||full Low-quality
S17_vs_k141_157575||full Low-quality
S17_vs_k141_24165||full Not-determined
S17_vs_k141_164718||full Low-quality
S17_vs_k141_61044||full Not-determined
S17_vs_k141_24174||full Low-quality
S17_vs_k141_112863||full Not-determined
S17_vs_k141_82924||full Low-quality
S17_vs_k141_106209||full Low-quality
S17_vs_k141_61049||full Not-determined
S17_vs_k141_16654||full Not-determined
S17_vs_k141_9811||full Not-determined
S17_vs_k141_127471||full Not-determined
S17_vs_k141_157585||full Low-quality
S17_vs_k141_142776||full Not-determined
S17_vs_k141_164731||full Low-quality
S17_vs_k141_171944||full Not-determined
S17_vs_k141_112886||full Low-quality
S17_vs_k141_157606||full Low-quality
S17_vs_k141_90642||full Not-determined
S17_vs_k141_54294||full Low-quality
S17_vs_k141_2180||full Not-determined
S17_vs_k141_90645||full Low-quality
S17_vs_k141_112893||full Low-quality
S17_vs_k141_61079||full Not-determined
S17_vs_k141_90647||full Low-quality
S17_vs_k141_24204||full Not-determined
S17_vs_k141_61083||full Low-quality
S17_vs_k141_82943||full Not-determined
S17_vs_k141_54314||full Low-quality
S17_vs_k141_82953||full Not-determined
S17_vs_k141_16667||full Not-determined
S17_vs_k141_24218||full Low-quality
S17_vs_k141_75996||full Low-quality
S17_vs_k141_120527||full Not-determined
S17_vs_k141_135121||full Low-quality
S17_vs_k141_61104||full Not-determined
S17_vs_k141_16684||full Not-determined
S17_vs_k141_54326||full Low-quality
S17_vs_k141_16695||full Low-quality
S17_vs_k141_39651||full Not-determined
S17_vs_k141_142823||full Low-quality
S17_vs_k141_127517||full Not-determined
S17_vs_k141_90698||full Low-quality
S17_vs_k141_54328||full Low-quality
S17_vs_k141_68824||full Not-determined
S17_vs_k141_98515||full Low-quality
S17_vs_k141_112931||full Low-quality
S17_vs_k141_76038||full Low-quality
S17_vs_k141_142838||full Not-determined
S17_vs_k141_135154||full Low-quality
S17_vs_k141_112932||full Low-quality
S17_vs_k141_31982||full Low-quality
S17_vs_k141_90714||full Not-determined
S17_vs_k141_24259||full Low-quality
S17_vs_k141_61131||full Not-determined
S17_vs_k141_32000||full Not-determined
S17_vs_k141_24273||full Low-quality
S17_vs_k141_157711||full Low-quality
S17_vs_k141_54340||full Not-determined
S17_vs_k141_24280||full Low-quality
S17_vs_k141_46783||full Low-quality
S17_vs_k141_68872||full Low-quality
S17_vs_k141_32008||full Low-quality
S17_vs_k141_150547||full Low-quality
S17_vs_k141_76048||full Not-determined
S14_vs_k141_70985||full Medium-quality
S17_vs_k141_90744||full Not-determined
S17_vs_k141_106218||full Low-quality
S17_vs_k141_127533||full Not-determined
S17_vs_k141_135182||full Low-quality
S17_vs_k141_127535||full Low-quality
S17_vs_k141_54355||full Not-determined
S17_vs_k141_157729||full Not-determined
S17_vs_k141_76064||full Low-quality
S17_vs_k141_16738||full Low-quality
S17_vs_k141_106234||full Low-quality
S17_vs_k141_98529||full Low-quality
S17_vs_k141_76071||full Low-quality
S17_vs_k141_106238||full Not-determined
S17_vs_k141_142888||full Not-determined
S17_vs_k141_164814||full Low-quality
S17_vs_k141_90752||full Not-determined
S17_vs_k141_172016||full Not-determined
S17_vs_k141_24305||full Not-determined
S17_vs_k141_16748||full Low-quality
S17_vs_k141_9894||full Low-quality
S17_vs_k141_68913||full Low-quality
S17_vs_k141_157755||full Not-determined
S17_vs_k141_24311||full Not-determined
S17_vs_k141_83055||full Not-determined
S17_vs_k141_120642||full Not-determined
S17_vs_k141_54375||full Low-quality
S17_vs_k141_106259||full Low-quality
S17_vs_k141_106261||full Low-quality
S17_vs_k141_142933||full Low-quality
S17_vs_k141_90767||full Low-quality
S17_vs_k141_54395||full Not-determined
S17_vs_k141_106268||full Low-quality
S17_vs_k141_120665||full Not-determined
S17_vs_k141_164848||full Low-quality
S17_vs_k141_106271||full Low-quality
S17_vs_k141_142949||full Not-determined
S17_vs_k141_16758||full Low-quality
S17_vs_k141_172029||full Low-quality
S17_vs_k141_150606||full Not-determined
S17_vs_k141_135206||full Low-quality
S17_vs_k141_164861||full Low-quality
S17_vs_k141_39780||full Low-quality
S17_vs_k141_142956||full Not-determined
S17_vs_k141_157786||full Low-quality
S17_vs_k141_142957||full Low-quality
S17_vs_k141_54399||full Low-quality
S17_vs_k141_46855||full Low-quality
S17_vs_k141_16767||full Not-determined
S17_vs_k141_46857||full Not-determined
S17_vs_k141_16774||full Low-quality
S17_vs_k141_61252||full Not-determined
S17_vs_k141_46862||full Low-quality
S17_vs_k141_32085||full Low-quality
S17_vs_k141_113050||full Not-determined
S17_vs_k141_83122||full Not-determined
S17_vs_k141_142985||full Not-determined
S17_vs_k141_127606||full Low-quality
S17_vs_k141_98620||full Low-quality
S17_vs_k141_61263||full Low-quality
S17_vs_k141_9943||full Low-quality
S17_vs_k141_39786||full Not-determined
S17_vs_k141_46887||full Not-determined
S17_vs_k141_113061||full Not-determined
S17_vs_k141_83134||full Low-quality
S17_vs_k141_143008||full Low-quality
S17_vs_k141_39794||full Not-determined
S17_vs_k141_150694||full Low-quality
S17_vs_k141_9952||full Low-quality
S17_vs_k141_46917||full Not-determined
S17_vs_k141_54451||full Not-determined
S17_vs_k141_54454||full Not-determined
S17_vs_k141_2270||full Not-determined
S17_vs_k141_32145||full Low-quality
S17_vs_k141_54465||full Not-determined
S17_vs_k141_113104||full Low-quality
S17_vs_k141_2292||full Low-quality
S17_vs_k141_98643||full Low-quality
S17_vs_k141_32155||full Low-quality
S17_vs_k141_127663||full Not-determined
S17_vs_k141_98644||full Low-quality
S17_vs_k141_135244||full Low-quality
S17_vs_k141_32161||full Low-quality
S17_vs_k141_83189||full Not-determined
S17_vs_k141_135251||full Not-determined
S17_vs_k141_54469||full Low-quality
S17_vs_k141_16837||full Low-quality
S17_vs_k141_135253||full Low-quality
S17_vs_k141_83204||full Low-quality
S17_vs_k141_150732||full Low-quality
S17_vs_k141_9983||full Low-quality
S17_vs_k141_54483||full Not-determined
S17_vs_k141_32186||full Low-quality
S17_vs_k141_172071||full Not-determined
S17_vs_k141_98661||full Not-determined
S17_vs_k141_106406||full Not-determined
S17_vs_k141_39891||full Low-quality
S17_vs_k141_76211||full Low-quality
S17_vs_k141_83232||full Low-quality
S17_vs_k141_127706||full Low-quality
S17_vs_k141_61333||full Low-quality
S17_vs_k141_164870||full Low-quality
S17_vs_k141_127721||full Low-quality
S17_vs_k141_61343||full Low-quality
S17_vs_k141_76223||full Not-determined
S17_vs_k141_143094||full Low-quality
S17_vs_k141_47012||full Low-quality
S17_vs_k141_90885||full Not-determined
S17_vs_k141_76224||full Low-quality
S17_vs_k141_83245||full Low-quality
S17_vs_k141_76229||full Not-determined
S17_vs_k141_54525||full Low-quality
S17_vs_k141_98699||full Not-determined
S17_vs_k141_127736||full Low-quality
S17_vs_k141_143115||full Low-quality
S17_vs_k141_61364||full Low-quality
S17_vs_k141_2294||full Low-quality
S17_vs_k141_61371||full Not-determined
S17_vs_k141_164906||full Low-quality
S17_vs_k141_47025||full Not-determined
S17_vs_k141_2314||full Low-quality
S17_vs_k141_164923||full Low-quality
S17_vs_k141_32255||full Low-quality
S17_vs_k141_143133||full Low-quality
S17_vs_k141_10063||full Low-quality
S17_vs_k141_90924||full Not-determined
S17_vs_k141_10067||full Not-determined
S17_vs_k141_47026||full Not-determined
S17_vs_k141_150788||full Low-quality
S5_vs_k141_21778||full High-quality
S17_vs_k141_2327||full Low-quality
S17_vs_k141_90930||full Low-quality
S17_vs_k141_76264||full Low-quality
S17_vs_k141_83280||full Low-quality
S17_vs_k141_135364||full Low-quality
S17_vs_k141_2332||full Low-quality
S17_vs_k141_16863||full Low-quality
S17_vs_k141_127748||full Low-quality
S17_vs_k141_172140||full Low-quality
S17_vs_k141_24394||full Not-determined
S17_vs_k141_90941||full Low-quality
S17_vs_k141_24401||full Low-quality
S17_vs_k141_69105||full Low-quality
S17_vs_k141_90942||full Not-determined
S17_vs_k141_54577||full Low-quality
S17_vs_k141_54578||full Not-determined
S17_vs_k141_69118||full Low-quality
S17_vs_k141_172158||full Low-quality
S17_vs_k141_76286||full Low-quality
S17_vs_k141_61397||full Not-determined
S17_vs_k141_76287||full Low-quality
S17_vs_k141_157905||full Not-determined
S17_vs_k141_90970||full Not-determined
S17_vs_k141_47043||full Not-determined
S17_vs_k141_150829||full Low-quality
S17_vs_k141_90984||full Not-determined
S17_vs_k141_106522||full Not-determined
S17_vs_k141_98783||full Low-quality
S17_vs_k141_2369||full Not-determined
S17_vs_k141_90988||full Low-quality
S17_vs_k141_39975||full Not-determined
S17_vs_k141_90993||full Low-quality
S17_vs_k141_47059||full Low-quality
S17_vs_k141_172195||full Low-quality
S17_vs_k141_157914||full Not-determined
S17_vs_k141_120752||full Not-determined
S17_vs_k141_47076||full Low-quality
S17_vs_k141_2382||full Low-quality
S17_vs_k141_98816||full Not-determined
S17_vs_k141_16918||full Low-quality
S17_vs_k141_10115||full Low-quality
S17_vs_k141_157945||full Low-quality
S17_vs_k141_150849||full Low-quality
S17_vs_k141_135432||full Low-quality
S17_vs_k141_165022||full Not-determined
S17_vs_k141_40026||full Low-quality
S17_vs_k141_157957||full Not-determined
S17_vs_k141_10125||full Low-quality
S17_vs_k141_16931||full Not-determined
S17_vs_k141_98832||full Low-quality
S17_vs_k141_143228||full Low-quality
S17_vs_k141_135436||full Not-determined
S17_vs_k141_120801||full Low-quality
S17_vs_k141_91054||full Low-quality
S17_vs_k141_120805||full Not-determined
S17_vs_k141_32390||full Low-quality
S17_vs_k141_10139||full Not-determined
S17_vs_k141_32391||full Low-quality
S17_vs_k141_165035||full Not-determined
S17_vs_k141_16943||full Low-quality
S17_vs_k141_157983||full Not-determined
S17_vs_k141_113158||full Low-quality
S17_vs_k141_150872||full Low-quality
S17_vs_k141_135458||full Not-determined
S17_vs_k141_40046||full Low-quality
S17_vs_k141_113165||full Not-determined
S17_vs_k141_120820||full Low-quality
S17_vs_k141_76362||full Not-determined
S17_vs_k141_10160||full Low-quality
S17_vs_k141_172234||full Low-quality
S17_vs_k141_158005||full Low-quality
S17_vs_k141_47126||full Not-determined
S17_vs_k141_32407||full Not-determined
S17_vs_k141_135473||full Low-quality
S17_vs_k141_172240||full Not-determined
S17_vs_k141_10162||full Low-quality
S17_vs_k141_47136||full Not-determined
S17_vs_k141_76379||full Low-quality
S17_vs_k141_127837||full Not-determined
S17_vs_k141_32414||full Not-determined
S17_vs_k141_91107||full Low-quality
S17_vs_k141_127849||full Low-quality
S17_vs_k141_24516||full Low-quality
S17_vs_k141_32423||full Not-determined
S17_vs_k141_61435||full Low-quality
S17_vs_k141_10183||full Low-quality
S17_vs_k141_17016||full Not-determined
S17_vs_k141_172265||full Low-quality
S17_vs_k141_91118||full Low-quality
S17_vs_k141_32429||full Low-quality
S17_vs_k141_24527||full Low-quality
S17_vs_k141_120849||full Not-determined
S17_vs_k141_47167||full Low-quality
S17_vs_k141_32440||full Low-quality
S17_vs_k141_17043||full Low-quality
S17_vs_k141_91135||full Not-determined
S17_vs_k141_47193||full Not-determined
S17_vs_k141_120869||full Low-quality
S17_vs_k141_127900||full Low-quality
S17_vs_k141_76435||full Low-quality
S17_vs_k141_135505||full Low-quality
S17_vs_k141_24545||full Low-quality
S17_vs_k141_40099||full Low-quality
S17_vs_k141_17064||full Low-quality
S17_vs_k141_40103||full Not-determined
S17_vs_k141_32462||full Low-quality
S17_vs_k141_2483||full Low-quality
S17_vs_k141_127918||full Low-quality
S17_vs_k141_120886||full Low-quality
S17_vs_k141_120892||full Low-quality
S17_vs_k141_32472||full Not-determined
S17_vs_k141_113217||full Low-quality
S17_vs_k141_150947||full Not-determined
S17_vs_k141_150951||full Low-quality
S17_vs_k141_76458||full Low-quality
S17_vs_k141_143303||full Low-quality
S17_vs_k141_98933||full Not-determined
S17_vs_k141_120904||full Low-quality
S17_vs_k141_69262||full Not-determined
S17_vs_k141_143306||full Not-determined
S17_vs_k141_24563||full Not-determined
S17_vs_k141_2514||full Not-determined
S17_vs_k141_40130||full Not-determined
S17_vs_k141_24566||full Low-quality
S17_vs_k141_83354||full Low-quality
S17_vs_k141_135548||full Low-quality
S17_vs_k141_61485||full Low-quality
S17_vs_k141_69269||full Not-determined
S17_vs_k141_135552||full Low-quality
S17_vs_k141_98946||full Low-quality
S17_vs_k141_24576||full Not-determined
S17_vs_k141_91195||full Not-determined
S17_vs_k141_2530||full Low-quality
S17_vs_k141_113232||full Not-determined
S17_vs_k141_69287||full Low-quality
S17_vs_k141_158069||full Low-quality
S17_vs_k141_91199||full Not-determined
S17_vs_k141_98964||full Low-quality
S17_vs_k141_120935||full Low-quality
S17_vs_k141_61513||full Not-determined
S17_vs_k141_83373||full Not-determined
S17_vs_k141_32525||full Not-determined
S17_vs_k141_69307||full Low-quality
S17_vs_k141_98969||full Low-quality
S17_vs_k141_143334||full Low-quality
S17_vs_k141_91211||full Not-determined
S17_vs_k141_17105||full Low-quality
S17_vs_k141_17106||full Low-quality
S17_vs_k141_2566||full Low-quality
S17_vs_k141_106681||full Not-determined
S17_vs_k141_69314||full Low-quality
S17_vs_k141_17109||full Low-quality
S17_vs_k141_172372||full Low-quality
S17_vs_k141_69317||full Low-quality
S17_vs_k141_76536||full Low-quality
S17_vs_k141_61543||full Not-determined
S17_vs_k141_158141||full Low-quality
S17_vs_k141_91234||full Low-quality
S17_vs_k141_32561||full Not-determined
S17_vs_k141_158148||full Not-determined
S17_vs_k141_158157||full Not-determined
S17_vs_k141_32568||full Not-determined
S17_vs_k141_47310||full Low-quality
S17_vs_k141_17137||full Not-determined
S17_vs_k141_47316||full Not-determined
S17_vs_k141_17144||full Low-quality
S17_vs_k141_98977||full Low-quality
S17_vs_k141_24648||full Low-quality
S17_vs_k141_106728||full Low-quality
S17_vs_k141_76563||full Not-determined
S17_vs_k141_32599||full Not-determined
S17_vs_k141_135583||full Low-quality
S7_extracted_28429 Complete
S17_vs_k141_106743||full Not-determined
S17_vs_k141_61585||full Not-determined
S17_vs_k141_61589||full Low-quality
S17_vs_k141_158174||full Not-determined
S17_vs_k141_2625||full Not-determined
S17_vs_k141_128025||full Low-quality
S17_vs_k141_143433||full Low-quality
S17_vs_k141_83471||full Not-determined
S17_vs_k141_40253||full Low-quality
S17_vs_k141_54632||full Low-quality
S17_vs_k141_151034||full Low-quality
S17_vs_k141_2636||full Low-quality
S17_vs_k141_32647||full Not-determined
S17_vs_k141_99040||full Not-determined
S17_vs_k141_172447||full Low-quality
S17_vs_k141_106787||full Not-determined
S17_vs_k141_76601||full Low-quality
S17_vs_k141_99045||full Not-determined
S17_vs_k141_76606||full Low-quality
S17_vs_k141_69400||full Low-quality
S17_vs_k141_143453||full Low-quality
S17_vs_k141_76609||full Not-determined
S17_vs_k141_113337||full Not-determined
S17_vs_k141_61606||full Not-determined
S17_vs_k141_76615||full Low-quality
S17_vs_k141_165181||full Low-quality
S17_vs_k141_24679||full Low-quality
S17_vs_k141_61609||full Not-determined
S17_vs_k141_40268||full Not-determined
S17_vs_k141_47387||full Low-quality
S17_vs_k141_17204||full Low-quality
S17_vs_k141_40277||full Not-determined
S17_vs_k141_76622||full Low-quality
S17_vs_k141_2664||full Low-quality
S17_vs_k141_54664||full Low-quality
S17_vs_k141_69420||full Not-determined
S17_vs_k141_106822||full Not-determined
S17_vs_k141_2680||full Low-quality
S17_vs_k141_113378||full Not-determined
S17_vs_k141_99083||full Low-quality
S17_vs_k141_10277||full Not-determined
S17_vs_k141_128089||full Low-quality
S17_vs_k141_24713||full Low-quality

S17_vs_k141_2686||full Low-quality
S17_vs_k141_47418||full Low-quality
S17_vs_k141_10278||full Low-quality
S17_vs_k141_106833||full Low-quality
S17_vs_k141_165235||full Low-quality
S17_vs_k141_69453||full Low-quality
S17_vs_k141_54682||full Not-determined
S17_vs_k141_113389||full Not-determined
S17_vs_k141_61665||full Low-quality
S17_vs_k141_128103||full Not-determined
S17_vs_k141_10304||full Not-determined
S17_vs_k141_47459||full Low-quality
S17_vs_k141_172539||full Not-determined
S7_vs_28429||full High-quality
S17_vs_k141_121047||full Not-determined
S17_vs_k141_165295||full Not-determined
S17_vs_k141_83513||full Low-quality
S17_vs_k141_24764||full Not-determined
S17_vs_k141_106862||full Low-quality
S17_vs_k141_76720||full Not-determined
S17_vs_k141_172562||full Low-quality
S17_vs_k141_165302||full Low-quality
S17_vs_k141_135708||full Not-determined
S17_vs_k141_172567||full Low-quality
S17_vs_k141_10324||full Not-determined
S17_vs_k141_99139||full Not-determined
S17_vs_k141_2738||full Low-quality
S17_vs_k141_17280||full Not-determined
S17_vs_k141_76739||full Not-determined
S17_vs_k141_135717||full Low-quality
S17_vs_k141_61694||full Low-quality
S17_vs_k141_40337||full Low-quality
S17_vs_k141_47490||full Low-quality
S17_vs_k141_24788||full Not-determined
S17_vs_k141_83550||full Not-determined
S17_vs_k141_106890||full Low-quality
S17_vs_k141_24807||full Low-quality
S17_vs_k141_2762||full Low-quality
S17_vs_k141_10336||full Low-quality
S17_vs_k141_61720||full Not-determined
S17_vs_k141_128138||full Low-quality
S17_vs_k141_17307||full Not-determined
S17_vs_k141_47511||full Low-quality
S17_vs_k141_61726||full Not-determined
S17_vs_k141_2777||full Low-quality
S17_vs_k141_40347||full Low-quality
S17_vs_k141_47523||full Low-quality
S17_vs_k141_121074||full Low-quality
S12_vs_k141_8204||full Medium-quality
S17_vs_k141_143540||full Low-quality
S17_vs_k141_113436||full Not-determined
S17_vs_k141_172626||full Low-quality
S17_vs_k141_99213||full Low-quality
S17_vs_k141_61752||full Not-determined
S17_vs_k141_165348||full Not-determined
S17_vs_k141_143566||full Low-quality
S17_vs_k141_69562||full Not-determined
S17_vs_k141_113454||full Low-quality
S17_vs_k141_54755||full Low-quality
S17_vs_k141_2814||full Not-determined
S17_vs_k141_61765||full Not-determined
S17_vs_k141_47560||full Not-determined
S17_vs_k141_24828||full Not-determined
S17_vs_k141_99232||full Low-quality
S17_vs_k141_24833||full Not-determined
S17_vs_k141_2819||full Low-quality
S17_vs_k141_151206||full Low-quality
S17_vs_k141_135783||full Not-determined
S17_vs_k141_143581||full Not-determined
S17_vs_k141_76782||full Low-quality
S17_vs_k141_69576||full Low-quality
S17_vs_k141_158228||full Low-quality
S17_vs_k141_47579||full Low-quality
S17_vs_k141_32731||full Not-determined
S17_vs_k141_158232||full Low-quality
S17_vs_k141_99249||full Not-determined
S17_vs_k141_2837||full Low-quality
S17_vs_k141_54784||full Not-determined
S17_vs_k141_113520||full Low-quality
S17_vs_k141_165399||full Low-quality
S17_vs_k141_172670||full Not-determined
S17_vs_k141_2845||full Low-quality
S17_vs_k141_10416||full Low-quality
S17_vs_k141_99270||full Low-quality
S17_vs_k141_106962||full Low-quality
S17_vs_k141_91450||full Low-quality
S17_vs_k141_76815||full Low-quality
S17_vs_k141_2858||full Low-quality
S17_vs_k141_135810||full Low-quality
S17_vs_k141_10434||full Low-quality
S17_vs_k141_121107||full Not-determined
S17_vs_k141_10438||full Low-quality
S17_vs_k141_121113||full Low-quality
S17_vs_k141_135821||full Low-quality
S17_vs_k141_128249||full Low-quality
S17_vs_k141_158305||full Low-quality
S17_vs_k141_135822||full Not-determined
S17_vs_k141_47625||full Low-quality
S17_vs_k141_143643||full Not-determined
S17_vs_k141_99310||full Low-quality
S17_vs_k141_135832||full Low-quality
S17_vs_k141_40470||full Low-quality
S17_vs_k141_121119||full Not-determined
S17_vs_k141_40477||full Not-determined
S17_vs_k141_113577||full Low-quality
S17_vs_k141_113578||full Not-determined
S17_vs_k141_143656||full Low-quality
S17_vs_k141_24905||full Low-quality
S17_vs_k141_10485||full Not-determined
S17_vs_k141_24909||full Low-quality
S17_vs_k141_143657||full Not-determined
S17_vs_k141_99341||full Not-determined
S17_vs_k141_107014||full Low-quality
S17_vs_k141_128268||full Low-quality
S17_vs_k141_143665||full Not-determined
S17_vs_k141_99346||full Not-determined
S17_vs_k141_107017||full Not-determined
S17_vs_k141_143671||full Not-determined
S17_vs_k141_165493||full Low-quality
S17_vs_k141_91533||full Not-determined
S17_vs_k141_128285||full Not-determined
S17_vs_k141_107030||full Low-quality
S17_vs_k141_172731||full Not-determined
S17_vs_k141_128287||full Not-determined
S17_vs_k141_17449||full Not-determined
S17_vs_k141_151304||full Low-quality
S17_vs_k141_17455||full Low-quality
S17_vs_k141_107041||full Low-quality
S17_vs_k141_61882||full Low-quality
S17_vs_k141_151310||full Low-quality
S17_vs_k141_107045||full Low-quality
S17_vs_k141_143696||full Low-quality
S17_vs_k141_83583||full Low-quality
S17_vs_k141_61894||full Low-quality
S17_vs_k141_151316||full Low-quality
S17_vs_k141_24943||full Not-determined
S17_vs_k141_158356||full Not-determined
S17_vs_k141_151326||full Low-quality
S17_vs_k141_69671||full Not-determined
S17_vs_k141_17493||full Not-determined
S17_vs_k141_24957||full Not-determined
S17_vs_k141_61915||full Not-determined
S17_vs_k141_172783||full Not-determined
S17_vs_k141_54915||full Not-determined
S17_vs_k141_17503||full Not-determined
S17_vs_k141_121215||full Low-quality
S17_vs_k141_40563||full Low-quality
S17_vs_k141_143737||full Low-quality
S17_vs_k141_128345||full Low-quality
S17_vs_k141_10574||full Low-quality
S17_vs_k141_113670||full Not-determined
S17_vs_k141_107104||full Low-quality
S17_vs_k141_128358||full Low-quality
S17_vs_k141_135879||full Not-determined
S17_vs_k141_76946||full Low-quality
S17_vs_k141_24989||full Not-determined
S17_vs_k141_113677||full Low-quality
S17_vs_k141_99429||full Low-quality
S17_vs_k141_47764||full Low-quality
S17_vs_k141_128369||full Not-determined
S17_vs_k141_121244||full Low-quality
S17_vs_k141_54940||full Low-quality
S17_vs_k141_17544||full Low-quality
S17_vs_k141_47771||full Low-quality
S17_vs_k141_135898||full Low-quality
S17_vs_k141_128382||full Not-determined
S17_vs_k141_172823||full Low-quality
S17_vs_k141_76967||full Low-quality
S17_vs_k141_76972||full Low-quality
S8_vs_k141_182288||full High-quality
S17_vs_k141_107147||full Not-determined
S17_vs_k141_17594||full Low-quality
S17_vs_k141_61947||full Not-determined
S17_vs_k141_113735||full Low-quality
S17_vs_k141_54964||full Low-quality
S17_vs_k141_113746||full Not-determined
S17_vs_k141_151374||full Not-determined
S17_vs_k141_113762||full Low-quality
S17_vs_k141_40620||full Low-quality
S17_vs_k141_128446||full Low-quality
S17_vs_k141_69762||full Low-quality
S17_vs_k141_158451||full Low-quality
S17_vs_k141_3031||full Not-determined
S17_vs_k141_151390||full Low-quality
S17_vs_k141_121302||full Not-determined
S17_vs_k141_69776||full Low-quality
S17_vs_k141_83701||full Low-quality
S17_vs_k141_32900||full Low-quality
S17_vs_k141_3053||full Not-determined
S17_vs_k141_47807||full Low-quality
S17_vs_k141_151398||full Low-quality
S17_vs_k141_61990||full Not-determined
S17_vs_k141_61991||full Not-determined
S17_vs_k141_10669||full Not-determined
S17_vs_k141_158480||full Not-determined
S17_vs_k141_69787||full Low-quality
S17_vs_k141_113800||full Low-quality
S17_vs_k141_172852||full Low-quality
S17_vs_k141_10679||full Low-quality
S17_vs_k141_32916||full Low-quality
S17_vs_k141_107231||full Not-determined
S17_vs_k141_128460||full Low-quality
S17_vs_k141_99512||full Low-quality
S17_vs_k141_91677||full Low-quality
S17_vs_k141_158494||full Not-determined
S17_vs_k141_165621||full Not-determined
S17_vs_k141_128474||full Low-quality
S17_vs_k141_113820||full Not-determined
S17_vs_k141_32938||full Not-determined
S13_vs_15645||full High-quality
S17_vs_k141_55035||full Low-quality
S17_vs_k141_113827||full Not-determined
S17_vs_k141_121337||full Not-determined
S17_vs_k141_158510||full Low-quality
S17_vs_k141_40675||full Low-quality
S17_vs_k141_47841||full Not-determined
S17_vs_k141_62032||full Not-determined
S17_vs_k141_55043||full Low-quality
S17_vs_k141_3109||full Not-determined
S17_vs_k141_165641||full Low-quality
S17_vs_k141_3112||full Low-quality
S17_vs_k141_121345||full Not-determined
S17_vs_k141_47859||full Low-quality
S17_vs_k141_62048||full Low-quality
S17_vs_k141_62049||full Low-quality
S17_vs_k141_10723||full Not-determined
S17_vs_k141_165656||full Not-determined
S17_vs_k141_32974||full Low-quality
S17_vs_k141_99533||full Low-quality
S17_vs_k141_25099||full Low-quality
S17_vs_k141_55059||full Low-quality
S17_vs_k141_158544||full Low-quality
S17_vs_k141_69850||full Not-determined
S17_vs_k141_77096||full Not-determined
S17_vs_k141_121365||full Low-quality
S17_vs_k141_10740||full Low-quality
S17_vs_k141_165673||full Not-determined
S17_vs_k141_25100||full Not-determined
S17_vs_k141_47886||full Not-determined
S17_vs_k141_128520||full Not-determined
S17_vs_k141_55075||full Low-quality
S17_vs_k141_10749||full Low-quality
S17_vs_k141_151453||full Not-determined
S17_vs_k141_158555||full Low-quality
S17_vs_k141_91703||full Low-quality
S17_vs_k141_165686||full Low-quality
S17_vs_k141_107298||full Not-determined
S17_vs_k141_121379||full Low-quality
S17_vs_k141_25117||full Not-determined
S17_vs_k141_107307||full Not-determined
S17_vs_k141_172896||full Low-quality
S17_vs_k141_25121||full Not-determined
S17_vs_k141_136040||full Not-determined
S17_vs_k141_77138||full Not-determined
S17_vs_k141_128564||full Low-quality
S17_vs_k141_83788||full Not-determined
S17_vs_k141_17752||full Low-quality
S17_vs_k141_151484||full Low-quality
S17_vs_k141_128574||full Not-determined
S17_vs_k141_113869||full Low-quality
S17_vs_k141_40728||full Low-quality
S17_vs_k141_172925||full Not-determined
S17_vs_k141_136057||full Not-determined
S17_vs_k141_128581||full Low-quality
S17_vs_k141_165712||full Low-quality
S17_vs_k141_10796||full Not-determined
S17_vs_k141_143915||full Low-quality
S17_vs_k141_3183||full Not-determined
S17_vs_k141_143922||full Low-quality
S17_vs_k141_17772||full Low-quality
S17_vs_k141_121428||full Low-quality
S17_vs_k141_107352||full Low-quality
S17_vs_k141_69926||full Low-quality
S17_vs_k141_62152||full Low-quality
S17_vs_k141_33113||full Not-determined
S17_vs_k141_99561||full Not-determined
S17_vs_k141_165753||full Low-quality
S17_vs_k141_91805||full Not-determined
S17_vs_k141_17787||full Not-determined
S17_vs_k141_17800||full Low-quality
S17_vs_k141_62163||full Low-quality
S17_vs_k141_121450||full Low-quality
S17_vs_k141_91826||full Not-determined
S17_vs_k141_158637||full Not-determined
S17_vs_k141_47991||full Low-quality
S17_vs_k141_165782||full Low-quality
S17_vs_k141_47996||full Low-quality
S17_vs_k141_113946||full Low-quality
S17_vs_k141_143965||full Low-quality
S17_vs_k141_158644||full Low-quality
S17_vs_k141_136132||full Not-determined
S17_vs_k141_113953||full Low-quality
S17_vs_k141_17828||full Not-determined
S17_vs_k141_77207||full Low-quality
S17_vs_k141_33135||full Not-determined
S17_vs_k141_121486||full Not-determined
S17_vs_k141_77210||full Low-quality
S17_vs_k141_99620||full Low-quality
S17_vs_k141_55221||full Low-quality
S17_vs_k141_158657||full Not-determined
S17_vs_k141_143972||full Low-quality
S17_vs_k141_173012||full Low-quality
S17_vs_k141_91863||full Low-quality
S17_vs_k141_83851||full Not-determined
S17_vs_k141_48007||full Not-determined
S17_vs_k141_25199||full Low-quality
S17_vs_k141_113983||full Not-determined
S17_vs_k141_40836||full Low-quality
S17_vs_k141_55252||full Low-quality
S17_vs_k141_113987||full Not-determined
S17_vs_k141_128635||full Not-determined
S17_vs_k141_55265||full Low-quality
S17_vs_k141_83871||full Low-quality
S17_vs_k141_33192||full Low-quality
S17_vs_k141_77261||full Low-quality
S17_vs_k141_3326||full Not-determined
S17_vs_k141_83885||full Low-quality
S17_vs_k141_128661||full Not-determined
S17_vs_k141_144014||full Low-quality
S17_vs_k141_99689||full Low-quality
S17_vs_k141_33204||full Low-quality
S17_vs_k141_55292||full Not-determined
S17_vs_k141_33210||full Low-quality
S17_vs_k141_10860||full Not-determined
S17_vs_k141_91912||full Not-determined
S17_vs_k141_144023||full Not-determined
S17_vs_k141_33216||full Low-quality
S17_vs_k141_91917||full Low-quality
S17_vs_k141_121542||full Not-determined
S17_vs_k141_121543||full Not-determined
S17_vs_k141_165857||full Low-quality
S17_vs_k141_121550||full Low-quality
S17_vs_k141_114024||full Not-determined
S17_vs_k141_3366||full Low-quality
S17_vs_k141_77305||full Not-determined
S17_vs_k141_33237||full Low-quality
S17_vs_k141_136245||full Not-determined
S17_vs_k141_25211||full Low-quality
S17_vs_k141_77311||full Low-quality
S17_vs_k141_33240||full Low-quality
S17_vs_k141_91930||full Low-quality
S17_vs_k141_48093||full Not-determined
S17_vs_k141_173094||full Low-quality
S17_vs_k141_25234||full Not-determined
S17_vs_k141_99736||full Not-determined
S17_vs_k141_144042||full Not-determined
S17_vs_k141_10900||full Low-quality
S17_vs_k141_70075||full Low-quality
S17_vs_k141_10903||full Low-quality
S17_vs_k141_83918||full Low-quality
S17_vs_k141_3415||full Not-determined
S17_vs_k141_107460||full Low-quality
S17_vs_k141_165888||full Not-determined
S17_vs_k141_17948||full Low-quality
S17_vs_k141_99757||full Low-quality
S17_vs_k141_33285||full Not-determined
S17_vs_k141_40918||full Not-determined
S17_vs_k141_151576||full Not-determined
S17_vs_k141_77325||full Low-quality
S17_vs_k141_83949||full Not-determined
S17_vs_k141_99768||full Low-quality
S17_vs_k141_77328||full Not-determined
S17_vs_k141_70102||full Low-quality
S17_vs_k141_107479||full Low-quality
S17_vs_k141_91963||full Low-quality
S17_vs_k141_114072||full Not-determined
S17_vs_k141_99780||full Low-quality
S17_vs_k141_107490||full Not-determined
S17_vs_k141_114077||full Low-quality
S17_vs_k141_107493||full Low-quality
S17_vs_k141_70113||full Not-determined
S17_vs_k141_70118||full Low-quality
S17_vs_k141_17958||full Low-quality
S17_vs_k141_128784||full Not-determined
S17_vs_k141_99797||full Low-quality
S17_vs_k141_151625||full Low-quality
S17_vs_k141_83993||full Low-quality
S17_vs_k141_3490||full Low-quality
S17_vs_k141_77358||full Low-quality
S17_vs_k141_70145||full Not-determined
S17_vs_k141_91999||full Low-quality
S17_vs_k141_84002||full Low-quality
S17_vs_k141_158851||full Low-quality
S17_vs_k141_151642||full Low-quality
S17_vs_k141_99801||full Low-quality
S17_vs_k141_55385||full Low-quality
S17_vs_k141_33329||full Not-determined
S17_vs_k141_173157||full Low-quality
S17_vs_k141_99815||full Not-determined
S17_vs_k141_55392||full Not-determined
S17_vs_k141_84016||full Not-determined
S17_vs_k141_144061||full Low-quality
S17_vs_k141_77403||full Not-determined
S17_vs_k141_25322||full Not-determined
S17_vs_k141_33342||full Not-determined
S17_vs_k141_92025||full Low-quality
S17_vs_k141_92027||full Low-quality
S17_vs_k141_40965||full Not-determined
S17_vs_k141_40966||full Low-quality
S17_vs_k141_18017||full Not-determined
S17_vs_k141_158876||full Not-determined
S17_vs_k141_128820||full Not-determined
S17_vs_k141_121617||full Not-determined
S17_vs_k141_84047||full Low-quality
S17_vs_k141_48174||full Low-quality
S17_vs_k141_165936||full Low-quality
S17_vs_k141_48178||full Low-quality
S17_vs_k141_158911||full Not-determined
S17_vs_k141_158913||full Low-quality
S17_vs_k141_70217||full Low-quality
S17_vs_k141_128857||full Low-quality
S17_vs_k141_144107||full Low-quality
S17_vs_k141_107544||full Low-quality
S17_vs_k141_25377||full Not-determined
S17_vs_k141_18087||full Not-determined
S17_vs_k141_18091||full Low-quality
S17_vs_k141_25382||full Low-quality
S17_vs_k141_136343||full Low-quality
S17_vs_k141_107559||full Low-quality
S17_vs_k141_25387||full Not-determined
S17_vs_k141_114181||full Not-determined
S17_vs_k141_121637||full Low-quality
S17_vs_k141_107567||full Low-quality
S17_vs_k141_18096||full Low-quality
S17_vs_k141_48200||full Not-determined
S17_vs_k141_107579||full Low-quality
S17_vs_k141_62365||full Low-quality
S17_vs_k141_107584||full Low-quality
S17_vs_k141_173263||full Low-quality
S17_vs_k141_10926||full Low-quality
S17_vs_k141_151693||full Not-determined
S17_vs_k141_25404||full Low-quality
S17_vs_k141_18107||full Low-quality
S17_vs_k141_158961||full Low-quality
S17_vs_k141_62378||full Not-determined
S17_vs_k141_121656||full Low-quality
S17_vs_k141_84104||full Low-quality
S17_vs_k141_151702||full Not-determined
S17_vs_k141_62385||full Not-determined
S17_vs_k141_92067||full Not-determined
S17_vs_k141_70268||full Low-quality
S17_vs_k141_10946||full Low-quality
S17_vs_k141_173285||full Not-determined
S17_vs_k141_77467||full Low-quality
S17_vs_k141_41022||full Low-quality
S17_vs_k141_33416||full Low-quality
S17_vs_k141_41027||full Low-quality
S17_vs_k141_77472||full Not-determined
S17_vs_k141_173298||full Low-quality
S17_vs_k141_62399||full Not-determined
S17_vs_k141_151733||full Not-determined
S17_vs_k141_55472||full Not-determined
S17_vs_k141_173300||full Not-determined
S17_vs_k141_92087||full Low-quality
S17_vs_k141_62406||full Not-determined
S17_vs_k141_173304||full Low-quality
S17_vs_k141_10957||full Low-quality
S17_vs_k141_107636||full Low-quality
S17_vs_k141_173313||full Low-quality
S17_vs_k141_33450||full Not-determined
S17_vs_k141_107650||full Low-quality
S17_vs_k141_121700||full Not-determined
S17_vs_k141_121701||full Not-determined
S17_vs_k141_114214||full Not-determined
S17_vs_k141_121703||full Low-quality
S17_vs_k141_18160||full Low-quality
S17_vs_k141_48255||full Not-determined
S17_vs_k141_25462||full Not-determined
S17_vs_k141_77512||full Not-determined
S17_vs_k141_158999||full Not-determined
S17_vs_k141_62429||full Not-determined
S17_vs_k141_92104||full Low-quality
S17_vs_k141_136436||full Not-determined
S17_vs_k141_77515||full Low-quality
S17_vs_k141_165992||full Not-determined
S17_vs_k141_33479||full Low-quality
S17_vs_k141_165993||full Not-determined
S17_vs_k141_128947||full Low-quality
S17_vs_k141_10985||full Low-quality
S17_vs_k141_18187||full Low-quality
S17_vs_k141_77525||full Not-determined
S17_vs_k141_84197||full Not-determined
S17_vs_k141_159026||full Low-quality
S17_vs_k141_144198||full Low-quality
S17_vs_k141_33501||full Not-determined
S17_vs_k141_92121||full Low-quality
S17_vs_k141_92127||full Not-determined
S17_vs_k141_114220||full Not-determined
S17_vs_k141_18206||full Low-quality
S17_vs_k141_11010||full Low-quality
S17_vs_k141_173348||full Low-quality
S17_vs_k141_166017||full Not-determined
S17_vs_k141_62461||full Not-determined
S17_vs_k141_92130||full Not-determined
S17_vs_k141_48290||full Not-determined
S17_vs_k141_3566||full Not-determined
S17_vs_k141_159039||full Low-quality
S17_vs_k141_92134||full Low-quality
S17_vs_k141_159042||full Low-quality
S17_vs_k141_92137||full Low-quality
S17_vs_k141_33513||full Low-quality
S17_vs_k141_144224||full Low-quality
S17_vs_k141_84214||full Not-determined
S17_vs_k141_70345||full Low-quality
S17_vs_k141_128979||full Low-quality
S17_vs_k141_136462||full Low-quality
S17_vs_k141_173369||full Not-determined
S17_vs_k141_11011||full Not-determined
S17_vs_k141_55534||full Not-determined
S17_vs_k141_128983||full Not-determined
S17_vs_k141_3592||full Low-quality
S17_vs_k141_55538||full Not-determined
S17_vs_k141_41099||full Not-determined
S17_vs_k141_128985||full Not-determined
S17_vs_k141_128992||full Not-determined
S17_vs_k141_173398||full Low-quality
S17_vs_k141_33544||full Low-quality
S17_vs_k141_3609||full Not-determined
S17_vs_k141_48352||full Not-determined
S17_vs_k141_25502||full Not-determined
S17_vs_k141_151843||full Not-determined
S17_vs_k141_166050||full Not-determined
S17_vs_k141_114311||full Not-determined
S17_vs_k141_151849||full Not-determined
S17_vs_k141_18271||full Low-quality
S17_vs_k141_33571||full Low-quality
S17_vs_k141_25516||full Low-quality
S17_vs_k141_18279||full Low-quality
S17_vs_k141_41136||full Low-quality
S17_vs_k141_151865||full Low-quality
S17_vs_k141_84245||full Low-quality
S17_vs_k141_144275||full Low-quality
S17_vs_k141_107681||full Not-determined
S17_vs_k141_173431||full Low-quality
S17_vs_k141_77593||full Low-quality
S17_vs_k141_144280||full Low-quality
S17_vs_k141_121763||full Not-determined
S17_vs_k141_173456||full Low-quality
S17_vs_k141_107699||full Low-quality
S17_vs_k141_114365||full Not-determined
S17_vs_k141_107704||full Low-quality
S17_vs_k141_114367||full Not-determined
S17_vs_k141_107707||full Low-quality
S17_vs_k141_33607||full Low-quality
S17_vs_k141_159136||full Low-quality
S17_vs_k141_159137||full Low-quality
S17_vs_k141_151897||full Low-quality
S17_vs_k141_129089||full Not-determined
S17_vs_k141_62501||full Not-determined
S17_vs_k141_33617||full Low-quality
S17_vs_k141_77640||full Not-determined
S17_vs_k141_166099||full Low-quality
S17_vs_k141_18300||full Not-determined
S17_vs_k141_41175||full Low-quality
S17_vs_k141_166118||full Low-quality
S17_vs_k141_70437||full Low-quality
S17_vs_k141_33641||full Not-determined
S17_vs_k141_3728||full Not-determined
S17_vs_k141_33644||full Low-quality
S17_vs_k141_99972||full Low-quality
S17_vs_k141_129135||full Low-quality
S17_vs_k141_62525||full Low-quality
S17_vs_k141_151919||full Not-determined
S17_vs_k141_55655||full Not-determined
S17_vs_k141_99979||full Not-determined
S17_vs_k141_159153||full Low-quality
S17_vs_k141_11165||full Not-determined
S17_vs_k141_55664||full Not-determined
S17_vs_k141_48391||full Low-quality
S17_vs_k141_18346||full Low-quality
S17_vs_k141_62546||full Low-quality
S17_vs_k141_55671||full Low-quality
S17_vs_k141_166157||full Not-determined
S17_vs_k141_33677||full Not-determined
S17_vs_k141_33682||full Not-determined
S17_vs_k141_166163||full Low-quality
S17_vs_k141_77679||full Not-determined
S17_vs_k141_11180||full Low-quality
S17_vs_k141_114473||full Low-quality
S17_vs_k141_151959||full Not-determined
S17_vs_k141_11187||full Not-determined
S17_vs_k141_121806||full Low-quality
S17_vs_k141_41250||full Low-quality
S5_vs_k141_80496||full Medium-quality
S17_vs_k141_144351||full Low-quality
S17_vs_k141_3771||full Not-determined
S17_vs_k141_166179||full Low-quality
S17_vs_k141_100033||full Low-quality
S17_vs_k141_159157||full Low-quality

| | |
|---|---|
| S17_vs_k141_11203\|\|full | Low-quality |
| S17_vs_k141_166185\|\|full | Not-determined |
| S17_vs_k141_55692\|\|full | Low-quality |
| S17_vs_k141_3772\|\|full | Low-quality |
| S17_vs_k141_107758\|\|full | Low-quality |
| S17_vs_k141_11210\|\|full | Low-quality |
| S17_vs_k141_41270\|\|full | Low-quality |
| S17_vs_k141_77708\|\|full | Low-quality |
| S17_vs_k141_3780\|\|full | Not-determined |
| S17_vs_k141_55706\|\|full | Low-quality |
| S17_vs_k141_151981\|\|full | Low-quality |
| S17_vs_k141_77718\|\|full | Not-determined |
| S17_vs_k141_136494\|\|full | Low-quality |
| S17_vs_k141_18398\|\|full | Low-quality |
| S17_vs_k141_84282\|\|full | Low-quality |
| S17_vs_k141_41300\|\|full | Not-determined |
| S17_vs_k141_107770\|\|full | Low-quality |
| S17_vs_k141_114507\|\|full | Low-quality |
| S17_vs_k141_3810\|\|full | Low-quality |
| S17_vs_k141_33745\|\|full | Low-quality |
| S17_vs_k141_25528\|\|full | Low-quality |
| S17_vs_k141_136524\|\|full | Not-determined |
| S17_vs_k141_70522\|\|full | Low-quality |
| S17_vs_k141_25534\|\|full | Not-determined |
| S17_vs_k141_3843\|\|full | Low-quality |
| S17_vs_k141_114531\|\|full | Not-determined |
| S17_vs_k141_25550\|\|full | Not-determined |
| S17_vs_k141_48439\|\|full | Not-determined |
| S17_vs_k141_121868\|\|full | Not-determined |
| S17_vs_k141_166245\|\|full | Low-quality |
| S17_vs_k141_77779\|\|full | Not-determined |
| S17_vs_k141_70526\|\|full | Low-quality |
| S17_vs_k141_55767\|\|full | Low-quality |
| S17_vs_k141_33775\|\|full | Low-quality |
| S17_vs_k141_159199\|\|full | Low-quality |
| S17_vs_k141_129283\|\|full | Low-quality |
| S17_vs_k141_33778\|\|full | Low-quality |
| S17_vs_k141_114543\|\|full | Not-determined |
| S17_vs_k141_55780\|\|full | Low-quality |
| S17_vs_k141_33790\|\|full | Low-quality |
| S17_vs_k141_33791\|\|full | Not-determined |
| S17_vs_k141_107819\|\|full | Low-quality |
| S17_vs_k141_107820\|\|full | Low-quality |
| S17_vs_k141_121906\|\|full | Not-determined |
| S17_vs_k141_48468\|\|full | Low-quality |
| S17_vs_k141_70552\|\|full | Low-quality |
| S17_vs_k141_166279\|\|full | Not-determined |
| S17_vs_k141_152043\|\|full | Low-quality |
| S17_vs_k141_107832\|\|full | Not-determined |
| S17_vs_k141_3879\|\|full | Not-determined |
| S17_vs_k141_70561\|\|full | Low-quality |
| S17_vs_k141_77818\|\|full | Not-determined |
| S17_vs_k141_136591\|\|full | Low-quality |
| S17_vs_k141_48480\|\|full | Low-quality |
| S17_vs_k141_70564\|\|full | Low-quality |
| S17_vs_k141_152063\|\|full | Not-determined |
| S17_vs_k141_33819\|\|full | Low-quality |
| S17_vs_k141_84347\|\|full | Low-quality |
| S17_vs_k141_18502\|\|full | Not-determined |
| S17_vs_k141_107860\|\|full | Low-quality |
| S17_vs_k141_33828\|\|full | Low-quality |
| S17_vs_k141_166291\|\|full | Low-quality |
| S17_vs_k141_152065\|\|full | Not-determined |
| S17_vs_k141_166294\|\|full | Low-quality |
| S17_vs_k141_70575\|\|full | Low-quality |
| S17_vs_k141_166296\|\|full | Not-determined |
| S17_vs_k141_144437\|\|full | Low-quality |
| S17_vs_k141_77836\|\|full | Not-determined |
| S17_vs_k141_100143\|\|full | Low-quality |
| S17_vs_k141_55825\|\|full | Low-quality |
| S17_vs_k141_136612\|\|full | Not-determined |
| S17_vs_k141_100148\|\|full | Low-quality |
| S17_vs_k141_55828\|\|full | Not-determined |
| S17_vs_k141_70597\|\|full | Low-quality |
| S17_vs_k141_107891\|\|full | Low-quality |
| S17_vs_k141_3945\|\|full | Low-quality |
| S17_vs_k141_107892\|\|full | Low-quality |
| S17_vs_k141_55841\|\|full | Low-quality |
| S17_vs_k141_41339\|\|full | Low-quality |
| S17_vs_k141_173595\|\|full | Low-quality |
| S17_vs_k141_18509\|\|full | Not-determined |
| S17_vs_k141_48536\|\|full | Low-quality |
| S17_vs_k141_100161\|\|full | Low-quality |
| S17_vs_k141_107910\|\|full | Low-quality |
| S17_vs_k141_121950\|\|full | Not-determined |
| S17_vs_k141_100169\|\|full | Not-determined |
| S17_vs_k141_173606\|\|full | Low-quality |
| S17_vs_k141_25601\|\|full | Low-quality |
| S17_vs_k141_33879\|\|full | Not-determined |
| S17_vs_k141_159210\|\|full | Low-quality |
| S17_vs_k141_25618\|\|full | Low-quality |
| S17_vs_k141_114668\|\|full | Not-determined |
| S17_vs_k141_55871\|\|full | Low-quality |
| S17_vs_k141_173616\|\|full | Low-quality |
| S17_vs_k141_92383\|\|full | Low-quality |
| S17_vs_k141_173621\|\|full | Low-quality |
| S17_vs_k141_41391\|\|full | Low-quality |
| S17_vs_k141_159219\|\|full | Not-determined |
| S17_vs_k141_100210\|\|full | Not-determined |
| S17_vs_k141_25631\|\|full | Low-quality |
| S17_vs_k141_84413\|\|full | Not-determined |
| S17_vs_k141_129380\|\|full | Low-quality |
| S17_vs_k141_84421\|\|full | Not-determined |
| S17_vs_k141_152137\|\|full | Low-quality |
| S17_vs_k141_84428\|\|full | Low-quality |
| S17_vs_k141_159252\|\|full | Low-quality |
| S17_vs_k141_11341\|\|full | Not-determined |
| S17_vs_k141_48618\|\|full | Not-determined |
| S17_vs_k141_77934\|\|full | Low-quality |
| S17_vs_k141_166381\|\|full | Low-quality |
| S17_vs_k141_136714\|\|full | Low-quality |
| S17_vs_k141_48619\|\|full | Low-quality |
| S17_vs_k141_11352\|\|full | Not-determined |
| S17_vs_k141_100224\|\|full | Not-determined |
| S17_vs_k141_100230\|\|full | Not-determined |
| S17_vs_k141_114717\|\|full | Low-quality |
| S17_vs_k141_166400\|\|full | Low-quality |
| S17_vs_k141_114725\|\|full | Not-determined |
| S17_vs_k141_18603\|\|full | Low-quality |
| S17_vs_k141_100238\|\|full | Not-determined |
| S17_vs_k141_70672\|\|full | Not-determined |
| S17_vs_k141_25690\|\|full | Low-quality |
| S17_vs_k141_114748\|\|full | Not-determined |
| S17_vs_k141_152199\|\|full | Low-quality |
| S17_vs_k141_41492\|\|full | Not-determined |
| S17_vs_k141_108017\|\|full | Not-determined |
| S17_vs_k141_48656\|\|full | Not-determined |
| S14_vs_k141_15228\|\|full | High-quality |
| S17_vs_k141_108031\|\|full | Low-quality |
| S17_vs_k141_84463\|\|full | Not-determined |
| S17_vs_k141_129460\|\|full | Low-quality |
| S17_vs_k141_114777\|\|full | Low-quality |
| S17_vs_k141_136750\|\|full | Low-quality |
| S17_vs_k141_100288\|\|full | Low-quality |
| S17_vs_k141_84483\|\|full | Not-determined |
| S17_vs_k141_77993\|\|full | Not-determined |
| S17_vs_k141_136760\|\|full | Not-determined |
| S17_vs_k141_41534\|\|full | Not-determined |
| S17_vs_k141_55936\|\|full | Low-quality |
| S17_vs_k141_92496\|\|full | Low-quality |
| S17_vs_k141_159356\|\|full | Not-determined |
| S17_vs_k141_129484\|\|full | Low-quality |
| S17_vs_k141_108064\|\|full | Low-quality |
| S17_vs_k141_25729\|\|full | Low-quality |
| S17_vs_k141_84502\|\|full | Low-quality |
| S17_vs_k141_129488\|\|full | Not-determined |
| S17_vs_k141_84503\|\|full | Low-quality |
| S17_vs_k141_114814\|\|full | Low-quality |
| S17_vs_k141_159373\|\|full | Low-quality |
| S17_vs_k141_34008\|\|full | Low-quality |
| S17_vs_k141_122091\|\|full | Not-determined |
| S17_vs_k141_100319\|\|full | Not-determined |
| S17_vs_k141_122101\|\|full | Low-quality |
| S17_vs_k141_92542\|\|full | Low-quality |
| S17_vs_k141_34022\|\|full | Not-determined |
| S17_vs_k141_41573\|\|full | Not-determined |
| S17_vs_k141_78052\|\|full | Low-quality |
| S17_vs_k141_41591\|\|full | Low-quality |
| S17_vs_k141_122103\|\|full | Low-quality |
| S17_vs_k141_4027\|\|full | Low-quality |
| S17_vs_k141_41597\|\|full | Not-determined |
| S17_vs_k141_152303\|\|full | Low-quality |
| S17_vs_k141_166532\|\|full | Low-quality |
| S17_vs_k141_166534\|\|full | Not-determined |
| S17_vs_k141_108135\|\|full | Low-quality |
| S17_vs_k141_34059\|\|full | Low-quality |
| S17_vs_k141_25798\|\|full | Not-determined |
| S17_vs_k141_56009\|\|full | Not-determined |
| S17_vs_k141_114882\|\|full | Not-determined |
| S17_vs_k141_4041\|\|full | Low-quality |
| S17_vs_k141_108149\|\|full | Low-quality |
| S17_vs_k141_56012\|\|full | Not-determined |
| S17_vs_k141_18689\|\|full | Low-quality |
| S17_vs_k141_129567\|\|full | Low-quality |
| S17_vs_k141_11420\|\|full | Not-determined |
| S17_vs_k141_114902\|\|full | Not-determined |
| S17_vs_k141_11424\|\|full | Not-determined |
| S17_vs_k141_11428\|\|full | Low-quality |
| S17_vs_k141_78103\|\|full | Low-quality |
| S17_vs_k141_122141\|\|full | Low-quality |
| S17_vs_k141_84558\|\|full | Not-determined |
| S17_vs_k141_56036\|\|full | Low-quality |
| S17_vs_k141_78115\|\|full | Low-quality |
| S17_vs_k141_56040\|\|full | Low-quality |
| S17_vs_k141_92601\|\|full | Low-quality |
| S17_vs_k141_166574\|\|full | Not-determined |
| S17_vs_k141_56044\|\|full | Low-quality |
| S17_vs_k141_34085\|\|full | Low-quality |
| S17_vs_k141_18702\|\|full | Not-determined |
| S17_vs_k141_34091\|\|full | Low-quality |
| S17_vs_k141_48771\|\|full | Low-quality |
| S17_vs_k141_152377\|\|full | Not-determined |
| S17_vs_k141_122165\|\|full | Not-determined |
| S17_vs_k141_122167\|\|full | Low-quality |
| S17_vs_k141_70775\|\|full | Low-quality |
| S17_vs_k141_159443\|\|full | Low-quality |
| S17_vs_k141_11440\|\|full | Low-quality |
| S17_vs_k141_144555\|\|full | Low-quality |
| S17_vs_k141_48789\|\|full | Not-determined |
| S17_vs_k141_41633\|\|full | Not-determined |
| S17_vs_k141_11445\|\|full | Low-quality |
| S17_vs_k141_56074\|\|full | Not-determined |
| S17_vs_k141_78137\|\|full | Not-determined |
| S17_vs_k141_144567\|\|full | Low-quality |
| S17_vs_k141_166586\|\|full | Low-quality |
| S17_vs_k141_92618\|\|full | Low-quality |
| S17_vs_k141_166588\|\|full | Not-determined |
| S17_vs_k141_166589\|\|full | Low-quality |
| S17_vs_k141_34100\|\|full | Not-determined |
| S17_vs_k141_48817\|\|full | Not-determined |
| S17_vs_k141_78152\|\|full | Low-quality |
| S17_vs_k141_11475\|\|full | Low-quality |
| S17_vs_k141_78154\|\|full | Not-determined |
| S17_vs_k141_152422\|\|full | Low-quality |
| S17_vs_k141_92643\|\|full | Not-determined |
| S17_vs_k141_41689\|\|full | Not-determined |
| S17_vs_k141_70836\|\|full | Not-determined |
| S17_vs_k141_48858\|\|full | Low-quality |
| S17_vs_k141_144583\|\|full | Not-determined |
| S17_vs_k141_108252\|\|full | Not-determined |
| S17_vs_k141_108253\|\|full | Low-quality |
| S17_vs_k141_100427\|\|full | Not-determined |
| S17_vs_k141_152433\|\|full | Low-quality |
| S17_vs_k141_152434\|\|full | Low-quality |
| S17_vs_k141_78194\|\|full | Not-determined |
| S17_vs_k141_18793\|\|full | Not-determined |
| S17_vs_k141_166610\|\|full | Not-determined |
| S17_vs_k141_48870\|\|full | Low-quality |
| S17_vs_k141_48882\|\|full | Low-quality |
| S17_vs_k141_144593\|\|full | Not-determined |
| S17_vs_k141_41708\|\|full | Low-quality |
| S17_vs_k141_56119\|\|full | Not-determined |
| S17_vs_k141_92698\|\|full | Not-determined |
| S17_vs_k141_48893\|\|full | Not-determined |
| S17_vs_k141_18812\|\|full | Low-quality |
| S17_vs_k141_152455\|\|full | Not-determined |
| S17_vs_k141_70872\|\|full | Not-determined |
| S17_vs_k141_166626\|\|full | Not-determined |
| S17_vs_k141_144605\|\|full | Low-quality |
| S17_vs_k141_144607\|\|full | Low-quality |
| S17_vs_k141_56136\|\|full | Low-quality |
| S17_vs_k141_173767\|\|full | Not-determined |
| S17_vs_k141_78221\|\|full | Low-quality |
| S17_vs_k141_152459\|\|full | Low-quality |
| S17_vs_k141_11527\|\|full | Low-quality |
| S17_vs_k141_70893\|\|full | Not-determined |
| S17_vs_k141_78229\|\|full | Low-quality |
| S17_vs_k141_115013\|\|full | Low-quality |
| S17_vs_k141_78233\|\|full | Low-quality |
| S17_vs_k141_152479\|\|full | Not-determined |
| S17_vs_k141_115020\|\|full | Not-determined |
| S17_vs_k141_11539\|\|full | Low-quality |
| S17_vs_k141_11547\|\|full | Not-determined |
| S17_vs_k141_100478\|\|full | Not-determined |
| S17_vs_k141_129667\|\|full | Not-determined |
| S17_vs_k141_4085\|\|full | Low-quality |
| S17_vs_k141_11556\|\|full | Low-quality |
| S17_vs_k141_92758\|\|full | Low-quality |
| S17_vs_k141_100503\|\|full | Not-determined |
| S17_vs_k141_56185\|\|full | Not-determined |
| S17_vs_k141_108362\|\|full | Low-quality |
| S17_vs_k141_159516\|\|full | Low-quality |
| S17_vs_k141_34173\|\|full | Not-determined |
| S17_vs_k141_166667\|\|full | Low-quality |
| S17_vs_k141_129687\|\|full | Not-determined |
| S17_vs_k141_136968\|\|full | Not-determined |
| S17_vs_k141_129691\|\|full | Low-quality |
| S17_vs_k141_92770\|\|full | Not-determined |
| S17_vs_k141_173813\|\|full | Low-quality |
| S17_vs_k141_25940\|\|full | Not-determined |
| S17_vs_k141_25947\|\|full | Not-determined |
| S17_vs_k141_78281\|\|full | Low-quality |
| S17_vs_k141_108372\|\|full | Not-determined |
| S17_vs_k141_84701\|\|full | Low-quality |
| S17_vs_k141_18921\|\|full | Not-determined |
| S17_vs_k141_129715\|\|full | Not-determined |
| S17_vs_k141_166693\|\|full | Not-determined |
| S6_vs_k141_53748\|\|full | High-quality |
| S17_vs_k141_136996\|\|full | Not-determined |
| S17_vs_k141_92785\|\|full | Low-quality |
| S17_vs_k141_56227\|\|full | Not-determined |
| S17_vs_k141_48996\|\|full | Not-determined |
| S17_vs_k141_11577\|\|full | Not-determined |
| S17_vs_k141_144691\|\|full | Low-quality |
| S17_vs_k141_25961\|\|full | Low-quality |
| S17_vs_k141_122194\|\|full | Not-determined |
| S17_vs_k141_11585\|\|full | Low-quality |
| S17_vs_k141_159554\|\|full | Low-quality |
| S17_vs_k141_41825\|\|full | Not-determined |
| S17_vs_k141_159558\|\|full | Low-quality |
| S17_vs_k141_100549\|\|full | Low-quality |
| S17_vs_k141_78314\|\|full | Low-quality |
| S17_vs_k141_34243\|\|full | Low-quality |
| S17_vs_k141_115107\|\|full | Not-determined |
| S17_vs_k141_4142\|\|full | Low-quality |
| S17_vs_k141_144720\|\|full | Not-determined |
| S17_vs_k141_159577\|\|full | Low-quality |
| S17_vs_k141_137039\|\|full | Not-determined |
| S17_vs_k141_108444\|\|full | Not-determined |
| S17_vs_k141_84733\|\|full | Not-determined |
| S17_vs_k141_100573\|\|full | Low-quality |
| S17_vs_k141_122233\|\|full | Low-quality |
| S17_vs_k141_62568\|\|full | Not-determined |
| S17_vs_k141_34276\|\|full | Low-quality |
| S17_vs_k141_100592\|\|full | Low-quality |
| S17_vs_k141_56308\|\|full | Low-quality |
| S17_vs_k141_129788\|\|full | Low-quality |
| S17_vs_k141_122245\|\|full | Low-quality |
| S17_vs_k141_166752\|\|full | Not-determined |
| S17_vs_k141_173917\|\|full | Not-determined |
| S17_vs_k141_71032\|\|full | Not-determined |
| S17_vs_k141_152618\|\|full | Not-determined |
| S17_vs_k141_144760\|\|full | Low-quality |
| S17_vs_k141_100596\|\|full | Low-quality |
| S17_vs_k141_4190\|\|full | Low-quality |
| S17_vs_k141_92855\|\|full | Low-quality |
| S17_vs_k141_11655\|\|full | Low-quality |
| S17_vs_k141_34293\|\|full | Not-determined |
| S17_vs_k141_92861\|\|full | Low-quality |
| S3_vs_k141_80151\|\|full | High-quality |
| S17_vs_k141_152632\|\|full | Not-determined |
| S17_vs_k141_115136\|\|full | Not-determined |
| S17_vs_k141_166770\|\|full | Low-quality |
| S17_vs_k141_137088\|\|full | Not-determined |
| S17_vs_k141_49089\|\|full | Low-quality |
| S17_vs_k141_4217\|\|full | Low-quality |
| S17_vs_k141_11661\|\|full | Low-quality |
| S17_vs_k141_137096\|\|full | Low-quality |
| S17_vs_k141_144773\|\|full | Not-determined |
| S17_vs_k141_137101\|\|full | Low-quality |
| S17_vs_k141_4223\|\|full | Low-quality |
| S17_vs_k141_159630\|\|full | Low-quality |
| S17_vs_k141_173938\|\|full | Not-determined |
| S17_vs_k141_62626\|\|full | Low-quality |
| S17_vs_k141_78325\|\|full | Low-quality |
| S17_vs_k141_152663\|\|full | Not-determined |
| S17_vs_k141_56346\|\|full | Low-quality |
| S17_vs_k141_49114\|\|full | Low-quality |
| S17_vs_k141_144787\|\|full | Not-determined |
| S17_vs_k141_62635\|\|full | Not-determined |
| S17_vs_k141_11693\|\|full | Low-quality |
| S17_vs_k141_71089\|\|full | Not-determined |
| S17_vs_k141_166801\|\|full | Low-quality |
| S17_vs_k141_56351\|\|full | Low-quality |
| S17_vs_k141_71094\|\|full | Low-quality |
| S17_vs_k141_115200\|\|full | Not-determined |
| S17_vs_k141_108507\|\|full | Low-quality |
| S17_vs_k141_62663\|\|full | Not-determined |
| S17_vs_k141_84815\|\|full | Low-quality |
| S17_vs_k141_62665\|\|full | Low-quality |
| S17_vs_k141_129862\|\|full | Not-determined |
| S17_vs_k141_159680\|\|full | Low-quality |
| S17_vs_k141_26065\|\|full | Low-quality |
| S9_extracted_k141_90847 | Complete |
| S17_vs_k141_26070\|\|full | Not-determined |
| S17_vs_k141_56361\|\|full | Not-determined |
| S17_vs_k141_4260\|\|full | Low-quality |
| S17_vs_k141_115224\|\|full | Low-quality |
| S17_vs_k141_166833\|\|full | Low-quality |
| S17_vs_k141_129865\|\|full | Low-quality |
| S17_vs_k141_78385\|\|full | Low-quality |
| S17_vs_k141_92876\|\|full | Low-quality |
| S17_vs_k141_152700\|\|full | Low-quality |
| S17_vs_k141_49140\|\|full | Low-quality |
| S17_vs_k141_56377\|\|full | Not-determined |
| S17_vs_k141_159713\|\|full | Low-quality |
| S17_vs_k141_78399\|\|full | Low-quality |
| S17_vs_k141_26098\|\|full | Not-determined |
| S17_vs_k141_49146\|\|full | Low-quality |
| S17_vs_k141_62706\|\|full | Not-determined |
| S17_vs_k141_56390\|\|full | Low-quality |
| S17_vs_k141_41935\|\|full | Low-quality |
| S17_vs_k141_71132\|\|full | Low-quality |
| S17_vs_k141_92886\|\|full | Low-quality |
| S17_vs_k141_144822\|\|full | Not-determined |
| S17_vs_k141_26113\|\|full | Low-quality |
| S17_vs_k141_129900\|\|full | Not-determined |
| S17_vs_k141_78421\|\|full | Low-quality |
| S17_vs_k141_34324\|\|full | Low-quality |
| S17_vs_k141_84860\|\|full | Low-quality |
| S17_vs_k141_115262\|\|full | Not-determined |
| S17_vs_k141_62725\|\|full | Low-quality |
| S17_vs_k141_108523\|\|full | Not-determined |
| S17_vs_k141_152734\|\|full | Low-quality |
| S17_vs_k141_4326\|\|full | Low-quality |
| S17_vs_k141_137194\|\|full | Low-quality |
| S17_vs_k141_78457\|\|full | Low-quality |
| S17_vs_k141_56434\|\|full | Not-determined |
| S17_vs_k141_92942\|\|full | Not-determined |
| S17_vs_k141_84883\|\|full | Not-determined |
| S17_vs_k141_92949\|\|full | Not-determined |
| S17_vs_k141_41995\|\|full | Low-quality |
| S17_vs_k141_56458\|\|full | Not-determined |
| S17_vs_k141_92955\|\|full | Not-determined |
| S17_vs_k141_159801\|\|full | Not-determined |
| S17_vs_k141_49228\|\|full | Not-determined |
| S17_vs_k141_26169\|\|full | Low-quality |
| S17_vs_k141_159806\|\|full | Not-determined |
| S17_vs_k141_84910\|\|full | Not-determined |
| S17_vs_k141_78481\|\|full | Low-quality |
| S17_vs_k141_115320\|\|full | Low-quality |
| S17_vs_k141_26183\|\|full | Not-determined |
| S17_vs_k141_4357\|\|full | Low-quality |
| S17_vs_k141_71183\|\|full | Low-quality |
| S17_vs_k141_159812\|\|full | Not-determined |
| S17_vs_k141_49247\|\|full | Not-determined |
| S17_vs_k141_4361\|\|full | Low-quality |
| S17_vs_k141_115327\|\|full | Not-determined |
| S17_vs_k141_71191\|\|full | Not-determined |
| S17_vs_k141_56500\|\|full | Low-quality |
| S17_vs_k141_78507\|\|full | Low-quality |
| S17_vs_k141_49253\|\|full | Low-quality |
| S17_vs_k141_11759\|\|full | Not-determined |
| S17_vs_k141_26212\|\|full | Low-quality |
| S17_vs_k141_56515\|\|full | Not-determined |
| S17_vs_k141_56520\|\|full | Low-quality |
| S17_vs_k141_49278\|\|full | Low-quality |
| S17_vs_k141_62814\|\|full | Not-determined |
| S17_vs_k141_166981\|\|full | Not-determined |
| S17_vs_k141_108547\|\|full | Low-quality |
| S17_vs_k141_115381\|\|full | Not-determined |
| S17_vs_k141_166992\|\|full | Not-determined |
| S17_vs_k141_137270\|\|full | Not-determined |
| S17_vs_k141_167005\|\|full | Low-quality |
| S17_vs_k141_115404\|\|full | Low-quality |
| S17_vs_k141_34431\|\|full | Not-determined |
| S17_vs_k141_11809\|\|full | Not-determined |
| S17_vs_k141_174065\|\|full | Low-quality |
| S17_vs_k141_100834\|\|full | Not-determined |
| S17_vs_k141_167018\|\|full | Low-quality |
| S17_vs_k141_92978\|\|full | Low-quality |
| S17_vs_k141_100836\|\|full | Low-quality |
| S17_vs_k141_49319\|\|full | Low-quality |
| S9_vs_k141_90847\|\|full | Medium-quality |
| S17_vs_k141_49330\|\|full | Low-quality |
| S17_vs_k141_122266\|\|full | Low-quality |
| S17_vs_k141_137310\|\|full | Low-quality |
| S17_vs_k141_159857\|\|full | Not-determined |
| S17_vs_k141_115430\|\|full | Low-quality |
| S17_vs_k141_4391\|\|full | Low-quality |
| S17_vs_k141_11842\|\|full | Low-quality |
| S17_vs_k141_26239\|\|full | Not-determined |
| S17_vs_k141_108601\|\|full | Not-determined |
| S17_vs_k141_62837\|\|full | Low-quality |
| S17_vs_k141_26244\|\|full | Not-determined |
| S17_vs_k141_122281\|\|full | Not-determined |
| S17_vs_k141_84999\|\|full | Low-quality |
| S17_vs_k141_174098\|\|full | Not-determined |
| S17_vs_k141_62849\|\|full | Not-determined |
| S17_vs_k141_108622\|\|full | Low-quality |
| S17_vs_k141_78567\|\|full | Low-quality |
| S17_vs_k141_34485\|\|full | Low-quality |
| S17_vs_k141_56626\|\|full | Not-determined |
| S17_vs_k141_78574\|\|full | Low-quality |
| S17_vs_k141_137360\|\|full | Not-determined |
| S17_vs_k141_42127\|\|full | Low-quality |
| S17_vs_k141_130002\|\|full | Low-quality |
| S17_vs_k141_85009\|\|full | Low-quality |
| S17_vs_k141_122314\|\|full | Not-determined |
| S17_vs_k141_34493\|\|full | Low-quality |
| S17_vs_k141_93008\|\|full | Low-quality |
| S17_vs_k141_19135\|\|full | Not-determined |
| S17_vs_k141_56643\|\|full | Low-quality |
| S17_vs_k141_19145\|\|full | Low-quality |
| S17_vs_k141_174131\|\|full | Low-quality |
| S17_vs_k141_122333\|\|full | Low-quality |
| S17_vs_k141_108639\|\|full | Not-determined |
| S17_vs_k141_145006\|\|full | Low-quality |
| S17_vs_k141_174134\|\|full | Not-determined |
| S17_vs_k141_42164\|\|full | Not-determined |
| S17_vs_k141_159908\|\|full | Low-quality |
| S17_vs_k141_174136\|\|full | Low-quality |
| S17_vs_k141_174141\|\|full | Low-quality |
| S17_vs_k141_93037\|\|full | Low-quality |
| S17_vs_k141_174148\|\|full | Low-quality |
| S17_vs_k141_71306\|\|full | Low-quality |
| S17_vs_k141_167094\|\|full | Not-determined |
| S17_vs_k141_56669\|\|full | Low-quality |
| S17_vs_k141_26285\|\|full | Low-quality |
| S17_vs_k141_152935\|\|full | Not-determined |
| S17_vs_k141_56674\|\|full | Low-quality |
| S17_vs_k141_159929\|\|full | Low-quality |
| S17_vs_k141_56676\|\|full | Low-quality |
| S17_vs_k141_19166\|\|full | Low-quality |
| S17_vs_k141_93064\|\|full | Low-quality |
| S17_vs_k141_49399\|\|full | Not-determined |
| S17_vs_k141_159934\|\|full | Low-quality |
| S17_vs_k141_85039\|\|full | Low-quality |
| S17_vs_k141_137407\|\|full | Low-quality |
| S17_vs_k141_174179\|\|full | Not-determined |
| S17_vs_k141_71326\|\|full | Low-quality |
| S17_vs_k141_19181\|\|full | Not-determined |
| S17_vs_k141_11934\|\|full | Low-quality |
| S17_vs_k141_49419\|\|full | Low-quality |
| S17_vs_k141_152943\|\|full | Not-determined |
| S17_vs_k141_34565\|\|full | Not-determined |
| S17_vs_k141_115530\|\|full | Low-quality |
| S17_vs_k141_71331\|\|full | Not-determined |
| S17_vs_k141_34567\|\|full | Low-quality |
| S17_vs_k141_159949\|\|full | Not-determined |
| S17_vs_k141_100928\|\|full | Low-quality |
| S17_vs_k141_26315\|\|full | Not-determined |
| S17_vs_k141_71342\|\|full | Not-determined |
| S17_vs_k141_26316\|\|full | Not-determined |
| S17_vs_k141_42214\|\|full | Not-determined |
| S17_vs_k141_56703\|\|full | Low-quality |
| S17_vs_k141_11951\|\|full | Low-quality |
| S17_vs_k141_137428\|\|full | Low-quality |
| S17_vs_k141_56704\|\|full | Low-quality |
| S17_vs_k141_34590\|\|full | Low-quality |
| S17_vs_k141_26333\|\|full | Low-quality |
| S17_vs_k141_56717\|\|full | Low-quality |
| S17_vs_k141_11966\|\|full | Low-quality |
| S17_vs_k141_152962\|\|full | Low-quality |
| S17_vs_k141_62944\|\|full | Low-quality |
| S17_vs_k141_26347\|\|full | Low-quality |
| S17_vs_k141_167139\|\|full | Not-determined |
| S17_vs_k141_100949\|\|full | Low-quality |
| S17_vs_k141_34618\|\|full | Low-quality |
| S17_vs_k141_62954\|\|full | Low-quality |
| S17_vs_k141_93125\|\|full | Low-quality |
| S17_vs_k141_71371\|\|full | Low-quality |
| S17_vs_k141_56736\|\|full | Low-quality |
| S17_vs_k141_62957\|\|full | Not-determined |
| S17_vs_k141_159968\|\|full | Low-quality |
| S17_vs_k141_11988\|\|full | Low-quality |
| S17_vs_k141_71375\|\|full | Not-determined |
| S17_vs_k141_4540\|\|full | Low-quality |
| S17_vs_k141_108722\|\|full | Low-quality |
| S17_vs_k141_34634\|\|full | Low-quality |
| S17_vs_k141_49424\|\|full | Not-determined |
| S17_vs_k141_93135\|\|full | Low-quality |
| S17_vs_k141_137479\|\|full | Not-determined |
| S17_vs_k141_93151\|\|full | Not-determined |
| S17_vs_k141_62978\|\|full | Low-quality |
| S17_vs_k141_174201\|\|full | Not-determined |
| S17_vs_k141_122413\|\|full | Not-determined |
| S17_vs_k141_49446\|\|full | Not-determined |
| S17_vs_k141_78722\|\|full | Low-quality |
| S17_vs_k141_42296\|\|full | Low-quality |
| S17_vs_k141_71422\|\|full | Low-quality |
| S17_vs_k141_115639\|\|full | Not-determined |
| S17_vs_k141_145096\|\|full | Low-quality |
| S17_vs_k141_63005\|\|full | Not-determined |
| S17_vs_k141_153010\|\|full | Low-quality |
| S3_vs_k141_82895\|\|full | High-quality |
| S17_vs_k141_4590\|\|full | Not-determined |
| S17_vs_k141_101004\|\|full | Low-quality |
| S17_vs_k141_137547\|\|full | Not-determined |
| S17_vs_k141_174210\|\|full | Not-determined |
| S17_vs_k141_167207\|\|full | Not-determined |
| S17_vs_k141_26407\|\|full | Low-quality |
| S17_vs_k141_167213\|\|full | Low-quality |
| S17_vs_k141_145129\|\|full | Not-determined |
| S17_vs_k141_4607\|\|full | Low-quality |
| S17_vs_k141_85133\|\|full | Not-determined |
| S17_vs_k141_49472\|\|full | Low-quality |
| S17_vs_k141_93211\|\|full | Low-quality |
| S17_vs_k141_63043\|\|full | Low-quality |
| S17_vs_k141_153054\|\|full | Low-quality |
| S17_vs_k141_137580\|\|full | Low-quality |
| S17_vs_k141_26433\|\|full | Low-quality |
| S17_vs_k141_71467\|\|full | Not-determined |
| S17_vs_k141_137587\|\|full | Not-determined |
| S17_vs_k141_160037\|\|full | Not-determined |
| S17_vs_k141_71471\|\|full | Low-quality |
| S17_vs_k141_34725\|\|full | Not-determined |
| S17_vs_k141_26451\|\|full | Low-quality |
| S17_vs_k141_101066\|\|full | Not-determined |
| S17_vs_k141_63065\|\|full | Low-quality |
| S17_vs_k141_160045\|\|full | Not-determined |
| S17_vs_k141_167244\|\|full | Not-determined |
| S17_vs_k141_34736\|\|full | Low-quality |
| S17_vs_k141_4657\|\|full | Low-quality |
| S17_vs_k141_85173\|\|full | Low-quality |
| S17_vs_k141_167258\|\|full | Not-determined |
| S17_vs_k141_137623\|\|full | Low-quality |
| S17_vs_k141_4674\|\|full | Not-determined |
| S17_vs_k141_56802\|\|full | Low-quality |
| S17_vs_k141_26460\|\|full | Low-quality |
| S17_vs_k141_153099\|\|full | Low-quality |
| S17_vs_k141_85178\|\|full | Not-determined |
| S17_vs_k141_4687\|\|full | Not-determined |
| S17_vs_k141_26473\|\|full | Low-quality |
| S17_vs_k141_167274\|\|full | Low-quality |
| S17_vs_k141_167283\|\|full | Low-quality |
| S17_vs_k141_12091\|\|full | Not-determined |
| S17_vs_k141_42360\|\|full | Low-quality |
| S17_vs_k141_122512\|\|full | Low-quality |

| | |
|---|---|
| S17_vs_k141_160086\|\|full | Not-determined |
| S17_vs_k141_167294\|\|full | Not-determined |
| S17_vs_k141_108845\|\|full | Not-determined |
| S17_vs_k141_122523\|\|full | Low-quality |
| S17_vs_k141_85226\|\|full | Low-quality |
| S17_vs_k141_93271\|\|full | Low-quality |
| S17_vs_k141_49557\|\|full | Low-quality |
| S17_vs_k141_42369\|\|full | Low-quality |
| S17_vs_k141_34808\|\|full | Low-quality |
| S17_vs_k141_145224\|\|full | Not-determined |
| S17_vs_k141_42371\|\|full | Not-determined |
| S17_vs_k141_49566\|\|full | Low-quality |
| S17_vs_k141_4720\|\|full | Low-quality |
| S17_vs_k141_42384\|\|full | Not-determined |
| S17_vs_k141_137690\|\|full | Not-determined |
| S17_vs_k141_153138\|\|full | Not-determined |
| S17_vs_k141_160112\|\|full | Low-quality |
| S17_vs_k141_63132\|\|full | Low-quality |
| S17_vs_k141_78836\|\|full | Not-determined |
| S17_vs_k141_160115\|\|full | Not-determined |
| S17_vs_k141_145248\|\|full | Low-quality |
| S17_vs_k141_160117\|\|full | Not-determined |
| S17_vs_k141_122565\|\|full | Low-quality |
| S17_vs_k141_167341\|\|full | Low-quality |
| S17_vs_k141_122569\|\|full | Low-quality |
| S17_vs_k141_115796\|\|full | Not-determined |
| S17_vs_k141_167347\|\|full | Low-quality |
| S17_vs_k141_56892\|\|full | Not-determined |
| S17_vs_k141_71587\|\|full | Not-determined |
| S17_vs_k141_78851\|\|full | Low-quality |
| S17_vs_k141_78852\|\|full | Low-quality |
| S17_vs_k141_19293\|\|full | Low-quality |
| S17_vs_k141_63142\|\|full | Not-determined |
| S17_vs_k141_167354\|\|full | Low-quality |
| S17_vs_k141_174324\|\|full | Low-quality |
| S17_vs_k141_56916\|\|full | Not-determined |
| S17_vs_k141_101143\|\|full | Low-quality |
| S17_vs_k141_93321\|\|full | Low-quality |
| S17_vs_k141_101146\|\|full | Not-determined |
| S17_vs_k141_42418\|\|full | Low-quality |
| S17_vs_k141_19327\|\|full | Low-quality |
| S17_vs_k141_34910\|\|full | Not-determined |
| S17_vs_k141_93338\|\|full | Not-determined |
| S17_vs_k141_49634\|\|full | Not-determined |
| S17_vs_k141_19335\|\|full | Low-quality |
| S17_vs_k141_85344\|\|full | Low-quality |
| S17_vs_k141_167392\|\|full | Low-quality |
| S17_vs_k141_93360\|\|full | Not-determined |
| S17_vs_k141_130110\|\|full | Not-determined |
| S17_vs_k141_115847\|\|full | Not-determined |
| S17_vs_k141_85360\|\|full | Low-quality |
| S17_vs_k141_160147\|\|full | Low-quality |
| S17_vs_k141_115848\|\|full | Low-quality |
| S17_vs_k141_49636\|\|full | Low-quality |
| S17_vs_k141_34940\|\|full | Not-determined |
| S17_vs_k141_78864\|\|full | Not-determined |
| S17_vs_k141_93369\|\|full | Low-quality |
| S17_vs_k141_130127\|\|full | Low-quality |
| S17_vs_k141_4839\|\|full | Low-quality |
| S17_vs_k141_78870\|\|full | Not-determined |
| S17_vs_k141_153249\|\|full | Low-quality |
| S17_vs_k141_56953\|\|full | Not-determined |
| S17_vs_k141_4843\|\|full | Low-quality |
| S17_vs_k141_56964\|\|full | Not-determined |
| S17_vs_k141_115873\|\|full | Not-determined |
| S17_vs_k141_49665\|\|full | Not-determined |
| S17_vs_k141_4851\|\|full | Not-determined |
| S17_vs_k141_85402\|\|full | Not-determined |
| S17_vs_k141_122637\|\|full | Not-determined |
| S17_vs_k141_26500\|\|full | Not-determined |
| S17_vs_k141_93411\|\|full | Not-determined |
| S17_vs_k141_49675\|\|full | Not-determined |
| S17_vs_k141_42486\|\|full | Low-quality |
| S17_vs_k141_130178\|\|full | Not-determined |
| S17_vs_k141_130180\|\|full | Low-quality |
| S17_vs_k141_34992\|\|full | Not-determined |
| S17_vs_k141_93423\|\|full | Low-quality |
| S17_vs_k141_130189\|\|full | Not-determined |
| S17_vs_k141_63277\|\|full | Low-quality |
| S17_vs_k141_57026\|\|full | Low-quality |
| S17_vs_k141_42503\|\|full | Not-determined |
| S17_vs_k141_19388\|\|full | Low-quality |
| S17_vs_k141_130191\|\|full | Not-determined |
| S17_vs_k141_35008\|\|full | Not-determined |
| S17_vs_k141_78926\|\|full | Low-quality |
| S17_vs_k141_174340\|\|full | Low-quality |
| S17_vs_k141_42515\|\|full | Not-determined |
| S17_vs_k141_109012\|\|full | Low-quality |
| S17_vs_k141_137856\|\|full | Low-quality |
| S17_vs_k141_109014\|\|full | Low-quality |
| S17_vs_k141_109015\|\|full | Low-quality |
| S17_vs_k141_160264\|\|full | Low-quality |
| S17_vs_k141_153301\|\|full | Low-quality |
| S17_vs_k141_4913\|\|full | Low-quality |
| S17_vs_k141_101233\|\|full | Not-determined |
| S17_vs_k141_101234\|\|full | Not-determined |
| S17_vs_k141_122704\|\|full | Low-quality |
| S17_vs_k141_93443\|\|full | Low-quality |
| S17_vs_k141_153305\|\|full | Not-determined |
| S17_vs_k141_78957\|\|full | Not-determined |
| S17_vs_k141_63314\|\|full | Not-determined |
| S17_vs_k141_145357\|\|full | Low-quality |
| S17_vs_k141_85469\|\|full | Low-quality |
| S17_vs_k141_35065\|\|full | Not-determined |
| S17_vs_k141_115971\|\|full | Low-quality |
| S17_vs_k141_167464\|\|full | Low-quality |
| S17_vs_k141_57053\|\|full | Not-determined |
| S17_vs_k141_78973\|\|full | Not-determined |
| S17_vs_k141_115976\|\|full | Not-determined |
| S17_vs_k141_57056\|\|full | Low-quality |
| S17_vs_k141_85494\|\|full | Low-quality |
| S17_vs_k141_122758\|\|full | Low-quality |
| S17_vs_k141_12206\|\|full | Not-determined |
| S17_vs_k141_63341\|\|full | Not-determined |
| S17_vs_k141_145392\|\|full | Not-determined |
| S17_vs_k141_85523\|\|full | Low-quality |
| S17_vs_k141_19424\|\|full | Low-quality |
| S17_vs_k141_35097\|\|full | Low-quality |
| S17_vs_k141_116000\|\|full | Not-determined |
| S17_vs_k141_116001\|\|full | Not-determined |
| S17_vs_k141_145407\|\|full | Not-determined |
| S17_vs_k141_167510\|\|full | Not-determined |
| S17_vs_k141_19445\|\|full | Low-quality |
| S17_vs_k141_63358\|\|full | Low-quality |
| S17_vs_k141_174433\|\|full | Low-quality |
| S17_vs_k141_109097\|\|full | Not-determined |
| S17_vs_k141_137910\|\|full | Not-determined |
| S17_vs_k141_12240\|\|full | Low-quality |
| S17_vs_k141_42587\|\|full | Low-quality |
| S17_vs_k141_153372\|\|full | Not-determined |
| S17_vs_k141_137922\|\|full | Low-quality |
| S17_vs_k141_63375\|\|full | Not-determined |
| S17_vs_k141_42598\|\|full | Low-quality |
| S17_vs_k141_71772\|\|full | Not-determined |
| S17_vs_k141_4982\|\|full | Not-determined |
| S17_vs_k141_57118\|\|full | Low-quality |
| S17_vs_k141_79024\|\|full | Not-determined |
| S17_vs_k141_167536\|\|full | Low-quality |
| S17_vs_k141_19470\|\|full | Low-quality |
| S17_vs_k141_167550\|\|full | Low-quality |
| S17_vs_k141_57142\|\|full | Not-determined |
| S17_vs_k141_145479\|\|full | Not-determined |
| S17_vs_k141_160343\|\|full | Not-determined |
| S17_vs_k141_19499\|\|full | Not-determined |
| S17_vs_k141_57168\|\|full | Low-quality |
| S17_vs_k141_145497\|\|full | Not-determined |
| S17_vs_k141_116044\|\|full | Low-quality |
| S17_vs_k141_101354\|\|full | Not-determined |
| S17_vs_k141_71816\|\|full | Low-quality |
| S17_vs_k141_5030\|\|full | Low-quality |
| S17_vs_k141_122842\|\|full | Low-quality |
| S17_vs_k141_116050\|\|full | Low-quality |
| S17_vs_k141_19527\|\|full | Not-determined |
| S17_vs_k141_5044\|\|full | Low-quality |
| S17_vs_k141_49721\|\|full | Low-quality |
| S17_vs_k141_145529\|\|full | Not-determined |
| S17_vs_k141_153422\|\|full | Not-determined |
| S17_vs_k141_49723\|\|full | Not-determined |
| S17_vs_k141_85569\|\|full | Low-quality |
| S5_vs_k141_135486\|\|full | High-quality |
| S17_vs_k141_122864\|\|full | Not-determined |
| S17_vs_k141_79095\|\|full | Low-quality |
| S17_vs_k141_93589\|\|full | Not-determined |
| S17_vs_k141_63384\|\|full | Low-quality |
| S17_vs_k141_85575\|\|full | Low-quality |
| S17_vs_k141_63387\|\|full | Low-quality |
| S17_vs_k141_93605\|\|full | Low-quality |
| S17_vs_k141_79101\|\|full | Low-quality |
| S17_vs_k141_85588\|\|full | Low-quality |
| S17_vs_k141_42638\|\|full | Not-determined |
| S17_vs_k141_57204\|\|full | Low-quality |
| S17_vs_k141_42643\|\|full | Low-quality |
| S17_vs_k141_153469\|\|full | Low-quality |
| S17_vs_k141_93621\|\|full | Low-quality |
| S17_vs_k141_49773\|\|full | Low-quality |
| S17_vs_k141_153484\|\|full | Low-quality |
| S17_vs_k141_5112\|\|full | Low-quality |
| S17_vs_k141_42661\|\|full | Not-determined |
| S17_vs_k141_138008\|\|full | Not-determined |
| S17_vs_k141_116127\|\|full | Not-determined |
| S9_vs_k141_12761\|\|full | High-quality |
| S17_vs_k141_93659\|\|full | Low-quality |
| S17_vs_k141_5130\|\|full | Not-determined |
| S17_vs_k141_85623\|\|full | Low-quality |
| S17_vs_k141_174479\|\|full | Not-determined |
| S17_vs_k141_138013\|\|full | Not-determined |
| S17_vs_k141_42678\|\|full | Not-determined |
| S17_vs_k141_160424\|\|full | Not-determined |
| S17_vs_k141_167601\|\|full | Not-determined |
| S17_vs_k141_19539\|\|full | Low-quality |
| S17_vs_k141_116129\|\|full | Not-determined |
| S17_vs_k141_167606\|\|full | Not-determined |
| S17_vs_k141_116134\|\|full | Low-quality |
| S17_vs_k141_138021\|\|full | Not-determined |
| S17_vs_k141_145540\|\|full | Low-quality |
| S17_vs_k141_49816\|\|full | Low-quality |
| S17_vs_k141_63424\|\|full | Low-quality |
| S17_vs_k141_71864\|\|full | Not-determined |
| S17_vs_k141_174492\|\|full | Not-determined |
| S17_vs_k141_71866\|\|full | Not-determined |
| S17_vs_k141_49824\|\|full | Low-quality |
| S17_vs_k141_57230\|\|full | Low-quality |
| S17_vs_k141_19568\|\|full | Low-quality |
| S17_vs_k141_26649\|\|full | Low-quality |
| S17_vs_k141_116158\|\|full | Not-determined |
| S17_vs_k141_35233\|\|full | Not-determined |
| S17_vs_k141_57234\|\|full | Not-determined |
| S17_vs_k141_145552\|\|full | Low-quality |
| S17_vs_k141_42715\|\|full | Not-determined |
| S17_vs_k141_153520\|\|full | Not-determined |
| S17_vs_k141_49836\|\|full | Not-determined |
| S17_vs_k141_71883\|\|full | Low-quality |
| S17_vs_k141_122945\|\|full | Low-quality |
| S17_vs_k141_116175\|\|full | Not-determined |
| S17_vs_k141_85673\|\|full | Not-determined |
| S17_vs_k141_19595\|\|full | Low-quality |
| S17_vs_k141_160473\|\|full | Low-quality |
| S17_vs_k141_57247\|\|full | Low-quality |
| S17_vs_k141_42726\|\|full | Low-quality |
| S17_vs_k141_122952\|\|full | Low-quality |
| S17_vs_k141_49843\|\|full | Low-quality |
| S17_vs_k141_35265\|\|full | Low-quality |
| S17_vs_k141_63465\|\|full | Low-quality |
| S17_vs_k141_138044\|\|full | Low-quality |
| S7_vs_137611\|\|full | Medium-quality |
| S17_vs_k141_5216\|\|full | Low-quality |
| S17_vs_k141_85688\|\|full | Low-quality |
| S17_vs_k141_153564\|\|full | Not-determined |
| S17_vs_k141_42746\|\|full | Not-determined |
| S17_vs_k141_160491\|\|full | Low-quality |
| S17_vs_k141_63481\|\|full | Low-quality |
| S17_vs_k141_19610\|\|full | Low-quality |
| S17_vs_k141_101390\|\|full | Low-quality |
| S17_vs_k141_145557\|\|full | Low-quality |
| S17_vs_k141_71934\|\|full | Not-determined |
| S17_vs_k141_19620\|\|full | Low-quality |
| S17_vs_k141_79162\|\|full | Not-determined |
| S17_vs_k141_153585\|\|full | Not-determined |
| S17_vs_k141_35299\|\|full | Not-determined |
| S17_vs_k141_167653\|\|full | Not-determined |
| S17_vs_k141_116231\|\|full | Low-quality |
| S17_vs_k141_109309\|\|full | Low-quality |
| S17_vs_k141_109312\|\|full | Low-quality |
| S17_vs_k141_138095\|\|full | Low-quality |
| S17_vs_k141_130426\|\|full | Low-quality |
| S17_vs_k141_63511\|\|full | Not-determined |
| S17_vs_k141_85729\|\|full | Low-quality |
| S17_vs_k141_116247\|\|full | Not-determined |
| S17_vs_k141_167672\|\|full | Low-quality |
| S17_vs_k141_12377\|\|full | Low-quality |
| S17_vs_k141_109327\|\|full | Not-determined |
| S17_vs_k141_63513\|\|full | Low-quality |
| S17_vs_k141_26714\|\|full | Low-quality |
| S17_vs_k141_49909\|\|full | Low-quality |
| S17_vs_k141_63516\|\|full | Not-determined |
| S17_vs_k141_101445\|\|full | Low-quality |
| S17_vs_k141_79211\|\|full | Not-determined |
| S17_vs_k141_85735\|\|full | Not-determined |
| S17_vs_k141_167679\|\|full | Not-determined |
| S17_vs_k141_109342\|\|full | Not-determined |
| S17_vs_k141_63520\|\|full | Low-quality |
| S17_vs_k141_138118\|\|full | Low-quality |
| S17_vs_k141_71972\|\|full | Not-determined |
| S5_vs_k141_109527\|\|full | Medium-quality |
| S17_vs_k141_101455\|\|full | Low-quality |
| S17_vs_k141_123018\|\|full | Low-quality |
| S17_vs_k141_35367\|\|full | Low-quality |
| S17_vs_k141_145620\|\|full | Low-quality |
| S17_vs_k141_101462\|\|full | Low-quality |
| S17_vs_k141_12413\|\|full | Low-quality |
| S17_vs_k141_85751\|\|full | Low-quality |
| S17_vs_k141_167712\|\|full | Low-quality |
| S17_vs_k141_85756\|\|full | Low-quality |
| S17_vs_k141_19661\|\|full | Low-quality |
| S17_vs_k141_71998\|\|full | Low-quality |
| S17_vs_k141_123050\|\|full | Not-determined |
| S17_vs_k141_101487\|\|full | Low-quality |
| S17_vs_k141_63567\|\|full | Not-determined |
| S17_vs_k141_5228\|\|full | Low-quality |
| S17_vs_k141_174594\|\|full | Low-quality |
| S17_vs_k141_85774\|\|full | Low-quality |
| S17_vs_k141_72026\|\|full | Low-quality |
| S17_vs_k141_19686\|\|full | Low-quality |
| S17_vs_k141_130457\|\|full | Low-quality |
| S17_vs_k141_63579\|\|full | Low-quality |
| S17_vs_k141_123073\|\|full | Low-quality |
| S17_vs_k141_49993\|\|full | Low-quality |
| S17_vs_k141_123077\|\|full | Not-determined |
| S17_vs_k141_130470\|\|full | Not-determined |
| S17_vs_k141_42822\|\|full | Low-quality |
| S17_vs_k141_167758\|\|full | Not-determined |
| S17_vs_k141_116318\|\|full | Low-quality |
| S17_vs_k141_72046\|\|full | Low-quality |
| S17_vs_k141_174617\|\|full | Not-determined |
| S17_vs_k141_19700\|\|full | Low-quality |
| S17_vs_k141_19701\|\|full | Low-quality |
| S17_vs_k141_167773\|\|full | Low-quality |
| S17_vs_k141_26758\|\|full | Not-determined |
| S17_vs_k141_79284\|\|full | Not-determined |
| S17_vs_k141_12484\|\|full | Low-quality |
| S17_vs_k141_101546\|\|full | Low-quality |
| S17_vs_k141_42848\|\|full | Low-quality |
| S17_vs_k141_63625\|\|full | Low-quality |
| S17_vs_k141_85840\|\|full | Low-quality |
| S17_vs_k141_123118\|\|full | Not-determined |
| S17_vs_k141_160590\|\|full | Not-determined |
| S17_vs_k141_145686\|\|full | Low-quality |
| S17_vs_k141_101555\|\|full | Low-quality |
| S17_vs_k141_130516\|\|full | Low-quality |
| S17_vs_k141_50042\|\|full | Not-determined |
| S17_vs_k141_79290\|\|full | Low-quality |
| S17_vs_k141_85853\|\|full | Low-quality |
| S17_vs_k141_138190\|\|full | Low-quality |
| S17_vs_k141_145688\|\|full | Not-determined |
| S17_vs_k141_42860\|\|full | Not-determined |
| S17_vs_k141_101569\|\|full | Low-quality |
| S17_vs_k141_12513\|\|full | Low-quality |
| S17_vs_k141_101576\|\|full | Low-quality |
| S17_vs_k141_167808\|\|full | Not-determined |
| S17_vs_k141_12520\|\|full | Not-determined |
| S17_vs_k141_79322\|\|full | Low-quality |
| S17_vs_k141_116381\|\|full | Not-determined |
| S17_vs_k141_26774\|\|full | Not-determined |
| S17_vs_k141_26776\|\|full | Low-quality |
| S17_vs_k141_12521\|\|full | Not-determined |
| S17_vs_k141_174667\|\|full | Low-quality |
| S17_vs_k141_145708\|\|full | Low-quality |
| S17_vs_k141_12528\|\|full | Not-determined |
| S17_vs_k141_101599\|\|full | Low-quality |
| S17_vs_k141_153613\|\|full | Not-determined |
| S17_vs_k141_138232\|\|full | Low-quality |
| S17_vs_k141_79345\|\|full | Low-quality |
| S17_vs_k141_12536\|\|full | Not-determined |
| S17_vs_k141_167845\|\|full | Low-quality |
| S17_vs_k141_116410\|\|full | Low-quality |
| S17_vs_k141_160634\|\|full | Low-quality |
| S17_vs_k141_85914\|\|full | Not-determined |
| S17_vs_k141_145727\|\|full | Low-quality |
| S17_vs_k141_153627\|\|full | Low-quality |
| S17_vs_k141_116422\|\|full | Low-quality |
| S17_vs_k141_50103\|\|full | Not-determined |
| S17_vs_k141_57443\|\|full | Not-determined |
| S17_vs_k141_116428\|\|full | Low-quality |
| S17_vs_k141_93758\|\|full | Low-quality |
| S17_vs_k141_42905\|\|full | Low-quality |
| S17_vs_k141_153647\|\|full | Low-quality |
| S17_vs_k141_93770\|\|full | Not-determined |
| S17_vs_k141_101638\|\|full | Low-quality |
| S17_vs_k141_5401\|\|full | Not-determined |
| S17_vs_k141_35476\|\|full | Low-quality |
| S17_vs_k141_167884\|\|full | Low-quality |
| S17_vs_k141_63716\|\|full | Not-determined |
| S17_vs_k141_50138\|\|full | Low-quality |
| S17_vs_k141_160675\|\|full | Low-quality |
| S17_vs_k141_160676\|\|full | Not-determined |
| S17_vs_k141_109540\|\|full | Low-quality |
| S17_vs_k141_93781\|\|full | Not-determined |
| S17_vs_k141_42937\|\|full | Low-quality |
| S17_vs_k141_123195\|\|full | Not-determined |
| S17_vs_k141_145784\|\|full | Not-determined |
| S17_vs_k141_79382\|\|full | Low-quality |
| S17_vs_k141_63744\|\|full | Low-quality |
| S17_vs_k141_35486\|\|full | Low-quality |
| S17_vs_k141_109558\|\|full | Not-determined |
| S17_vs_k141_160710\|\|full | Low-quality |
| S17_vs_k141_93799\|\|full | Low-quality |
| S17_vs_k141_85955\|\|full | Low-quality |
| S17_vs_k141_72087\|\|full | Not-determined |
| S17_vs_k141_167916\|\|full | Low-quality |
| S17_vs_k141_145811\|\|full | Not-determined |
| S17_vs_k141_174740\|\|full | Low-quality |
| S17_vs_k141_160725\|\|full | Low-quality |
| S17_vs_k141_50182\|\|full | Low-quality |
| S17_vs_k141_93825\|\|full | Low-quality |
| S17_vs_k141_72104\|\|full | Not-determined |
| S17_vs_k141_50185\|\|full | Low-quality |
| S17_vs_k141_145829\|\|full | Low-quality |
| S17_vs_k141_85962\|\|full | Not-determined |
| S17_vs_k141_35500\|\|full | Low-quality |
| S17_vs_k141_138281\|\|full | Not-determined |
| S17_vs_k141_160741\|\|full | Not-determined |
| S17_vs_k141_109594\|\|full | Low-quality |
| S17_vs_k141_101710\|\|full | Low-quality |
| S17_vs_k141_57530\|\|full | Low-quality |
| S17_vs_k141_174752\|\|full | Not-determined |
| S17_vs_k141_109602\|\|full | Low-quality |
| S17_vs_k141_174758\|\|full | Not-determined |
| S17_vs_k141_145848\|\|full | Low-quality |
| S17_vs_k141_50195\|\|full | Low-quality |
| S17_vs_k141_109611\|\|full | Not-determined |
| S17_vs_k141_12665\|\|full | Not-determined |
| S17_vs_k141_145859\|\|full | Low-quality |
| S17_vs_k141_79436\|\|full | Not-determined |
| S17_vs_k141_93855\|\|full | Not-determined |
| S17_vs_k141_174782\|\|full | Low-quality |
| S17_vs_k141_130584\|\|full | Low-quality |
| S17_vs_k141_86007\|\|full | Low-quality |
| S17_vs_k141_35559\|\|full | Low-quality |
| S17_vs_k141_5507\|\|full | Not-determined |
| S17_vs_k141_93872\|\|full | Low-quality |
| S17_vs_k141_174798\|\|full | Not-determined |
| S17_vs_k141_93873\|\|full | Not-determined |
| S17_vs_k141_50233\|\|full | Low-quality |
| S17_vs_k141_101764\|\|full | Low-quality |
| S17_vs_k141_93877\|\|full | Low-quality |
| S17_vs_k141_79472\|\|full | Not-determined |
| S17_vs_k141_167981\|\|full | Low-quality |
| S17_vs_k141_5525\|\|full | Not-determined |
| S17_vs_k141_174807\|\|full | Low-quality |
| S17_vs_k141_93884\|\|full | Low-quality |
| S17_vs_k141_101776\|\|full | Low-quality |
| S17_vs_k141_138306\|\|full | Not-determined |
| S17_vs_k141_160776\|\|full | Low-quality |
| S17_vs_k141_116559\|\|full | Not-determined |
| S17_vs_k141_86041\|\|full | Low-quality |
| S17_vs_k141_160787\|\|full | Low-quality |
| S17_vs_k141_116562\|\|full | Not-determined |
| S17_vs_k141_86049\|\|full | Low-quality |
| S17_vs_k141_72176\|\|full | Not-determined |
| S17_vs_k141_72180\|\|full | Low-quality |
| S17_vs_k141_138320\|\|full | Not-determined |
| S17_vs_k141_35594\|\|full | Not-determined |
| S17_vs_k141_86053\|\|full | Low-quality |
| S17_vs_k141_35596\|\|full | Low-quality |
| S17_vs_k141_138322\|\|full | Not-determined |
| S17_vs_k141_109685\|\|full | Low-quality |
| S17_vs_k141_50267\|\|full | Low-quality |
| S17_vs_k141_116583\|\|full | Not-determined |
| S17_vs_k141_116589\|\|full | Low-quality |
| S17_vs_k141_93920\|\|full | Low-quality |
| S17_vs_k141_130588\|\|full | Not-determined |
| S17_vs_k141_5565\|\|full | Low-quality |
| S17_vs_k141_43038\|\|full | Not-determined |
| S17_vs_k141_26888\|\|full | Low-quality |
| S17_vs_k141_138348\|\|full | Low-quality |
| S17_vs_k141_168026\|\|full | Low-quality |
| S17_vs_k141_109712\|\|full | Low-quality |
| S17_vs_k141_160844\|\|full | Not-determined |
| S17_vs_k141_63846\|\|full | Not-determined |
| S17_vs_k141_93949\|\|full | Low-quality |
| S17_vs_k141_160861\|\|full | Low-quality |
| S17_vs_k141_160862\|\|full | Low-quality |
| S17_vs_k141_101858\|\|full | Low-quality |
| S17_vs_k141_160863\|\|full | Low-quality |
| S17_vs_k141_138367\|\|full | Low-quality |
| S17_vs_k141_26913\|\|full | Not-determined |
| S17_vs_k141_116634\|\|full | Not-determined |
| S17_vs_k141_168049\|\|full | Not-determined |
| S17_vs_k141_43072\|\|full | Low-quality |
| S17_vs_k141_130633\|\|full | Not-determined |
| S17_vs_k141_57559\|\|full | Not-determined |
| S17_vs_k141_50321\|\|full | Low-quality |
| S17_vs_k141_123372\|\|full | Not-determined |
| S17_vs_k141_130642\|\|full | Not-determined |
| S17_vs_k141_26941\|\|full | Low-quality |
| S17_vs_k141_130653\|\|full | Low-quality |
| S17_vs_k141_79547\|\|full | Low-quality |
| S17_vs_k141_26945\|\|full | Not-determined |
| S17_vs_k141_138410\|\|full | Low-quality |
| S10_extracted_120738 | Complete |
| S17_vs_k141_19791\|\|full | Low-quality |
| S17_vs_k141_50334\|\|full | Not-determined |
| S17_vs_k141_116644\|\|full | Not-determined |
| S17_vs_k141_153891\|\|full | Not-determined |
| S17_vs_k141_19800\|\|full | Not-determined |
| S17_vs_k141_123394\|\|full | Low-quality |
| S17_vs_k141_174879\|\|full | Low-quality |
| S17_vs_k141_72311\|\|full | Low-quality |
| S17_vs_k141_50352\|\|full | Low-quality |
| S17_vs_k141_26970\|\|full | Low-quality |
| S17_vs_k141_43140\|\|full | Low-quality |
| S17_vs_k141_153913\|\|full | Low-quality |
| S17_vs_k141_19837\|\|full | Low-quality |
| S17_vs_k141_12692\|\|full | Not-determined |
| S17_vs_k141_123414\|\|full | Low-quality |
| S17_vs_k141_168129\|\|full | Low-quality |
| S17_vs_k141_19843\|\|full | Not-determined |
| S17_vs_k141_79589\|\|full | Not-determined |
| S17_vs_k141_116691\|\|full | Low-quality |
| S17_vs_k141_130684\|\|full | Not-determined |
| S17_vs_k141_160966\|\|full | Not-determined |
| S17_vs_k141_43157\|\|full | Low-quality |
| S17_vs_k141_27007\|\|full | Not-determined |
| S17_vs_k141_35744\|\|full | Not-determined |
| S17_vs_k141_43162\|\|full | Low-quality |
| S17_vs_k141_160968\|\|full | Low-quality |
| S17_vs_k141_43165\|\|full | Not-determined |
| S17_vs_k141_123439\|\|full | Low-quality |
| S17_vs_k141_79599\|\|full | Not-determined |
| S17_vs_k141_35758\|\|full | Low-quality |
| S17_vs_k141_19858\|\|full | Low-quality |
| S17_vs_k141_153951\|\|full | Low-quality |
| S17_vs_k141_123449\|\|full | Low-quality |
| S17_vs_k141_160987\|\|full | Not-determined |
| S17_vs_k141_72353\|\|full | Not-determined |
| S17_vs_k141_27032\|\|full | Not-determined |
| S17_vs_k141_35766\|\|full | Not-determined |
| S17_vs_k141_168159\|\|full | Not-determined |
| S17_vs_k141_101950\|\|full | Low-quality |
| S17_vs_k141_160991\|\|full | Low-quality |
| S17_vs_k141_50400\|\|full | Not-determined |
| S17_vs_k141_19877\|\|full | Not-determined |
| S17_vs_k141_101954\|\|full | Low-quality |
| S17_vs_k141_109864\|\|full | Low-quality |
| S17_vs_k141_27041\|\|full | Not-determined |
| S17_vs_k141_130712\|\|full | Not-determined |
| S17_vs_k141_35783\|\|full | Not-determined |
| S17_vs_k141_161002\|\|full | Not-determined |
| S17_vs_k141_50403\|\|full | Not-determined |
| S17_vs_k141_123457\|\|full | Low-quality |
| S17_vs_k141_79628\|\|full | Not-determined |
| S17_vs_k141_138439\|\|full | Low-quality |
| S17_vs_k141_130721\|\|full | Not-determined |
| S17_vs_k141_79632\|\|full | Not-determined |
| S17_vs_k141_174936\|\|full | Not-determined |
| S17_vs_k141_19890\|\|full | Low-quality |
| S10_vs_120738\|\|full | High-quality |
| S17_vs_k141_116745\|\|full | Low-quality |
| S17_vs_k141_93952\|\|full | Low-quality |
| S17_vs_k141_35788\|\|full | Low-quality |
| S17_vs_k141_93957\|\|full | Not-determined |
| S17_vs_k141_168190\|\|full | Low-quality |
| S17_vs_k141_101979\|\|full | Not-determined |
| S17_vs_k141_27066\|\|full | Low-quality |
| S17_vs_k141_101986\|\|full | Low-quality |
| S17_vs_k141_116759\|\|full | Low-quality |
| S17_vs_k141_35797\|\|full | Low-quality |
| S17_vs_k141_153979\|\|full | Low-quality |
| S17_vs_k141_161038\|\|full | Not-determined |
| S17_vs_k141_57650\|\|full | Not-determined |
| S17_vs_k141_43236\|\|full | Low-quality |
| S17_vs_k141_93970\|\|full | Low-quality |
| S17_vs_k141_116769\|\|full | Not-determined |
| S17_vs_k141_174964\|\|full | Low-quality |
| S17_vs_k141_161064\|\|full | Low-quality |
| S17_vs_k141_93978\|\|full | Low-quality |
| S17_vs_k141_72398\|\|full | Low-quality |
| S17_vs_k141_5702\|\|full | Low-quality |
| S17_vs_k141_27108\|\|full | Not-determined |
| S17_vs_k141_146081\|\|full | Low-quality |
| S17_vs_k141_138498\|\|full | Low-quality |
| S17_vs_k141_161070\|\|full | Not-determined |
| S17_vs_k141_12828\|\|full | Low-quality |
| S17_vs_k141_63990\|\|full | Low-quality |
| S17_vs_k141_109926\|\|full | Low-quality |
| S17_vs_k141_50464\|\|full | Low-quality |
| S17_vs_k141_116791\|\|full | Not-determined |
| S17_vs_k141_146092\|\|full | Not-determined |
| S17_vs_k141_5725\|\|full | Not-determined |
| S17_vs_k141_63999\|\|full | Not-determined |
| S17_vs_k141_94000\|\|full | Low-quality |
| S17_vs_k141_12846\|\|full | Low-quality |
| S17_vs_k141_138528\|\|full | Low-quality |
| S17_vs_k141_161090\|\|full | Not-determined |
| S17_vs_k141_35864\|\|full | Not-determined |
| S17_vs_k141_138532\|\|full | Low-quality |
| S17_vs_k141_50491\|\|full | Low-quality |
| S17_vs_k141_57689\|\|full | Low-quality |
| S17_vs_k141_19933\|\|full | Not-determined |
| S17_vs_k141_50495\|\|full | Low-quality |
| S17_vs_k141_5749\|\|full | Low-quality |
| S17_vs_k141_27147\|\|full | Low-quality |
| S17_vs_k141_138542\|\|full | Low-quality |
| S17_vs_k141_50502\|\|full | Low-quality |
| S17_vs_k141_161105\|\|full | Low-quality |
| S17_vs_k141_5753\|\|full | Not-determined |
| S17_vs_k141_116830\|\|full | Low-quality |
| S17_vs_k141_5761\|\|full | Not-determined |
| S17_vs_k141_86289\|\|full | Low-quality |
| S17_vs_k141_50520\|\|full | Low-quality |
| S17_vs_k141_64032\|\|full | Not-determined |
| S17_vs_k141_94025\|\|full | Low-quality |
| S17_vs_k141_5772\|\|full | Low-quality |
| S17_vs_k141_123573\|\|full | Not-determined |
| S17_vs_k141_12878\|\|full | Not-determined |
| S17_vs_k141_116855\|\|full | Low-quality |
| S17_vs_k141_19964\|\|full | Not-determined |
| S17_vs_k141_79720\|\|full | Low-quality |
| S17_vs_k141_123582\|\|full | Not-determined |
| S17_vs_k141_12881\|\|full | Not-determined |
| S17_vs_k141_138575\|\|full | Not-determined |
| S17_vs_k141_146163\|\|full | Low-quality |
| S17_vs_k141_43298\|\|full | Low-quality |
| S17_vs_k141_64049\|\|full | Low-quality |
| S17_vs_k141_138580\|\|full | Low-quality |
| S17_vs_k141_43300\|\|full | Not-determined |
| S17_vs_k141_19974\|\|full | Not-determined |
| S17_vs_k141_5803\|\|full | Low-quality |
| S17_vs_k141_50552\|\|full | Low-quality |

| | |
|---|---|
| S17_vs_k141_146187\|\|full | Not-determined |
| S17_vs_k141_50559\|\|full | Low-quality |
| S17_vs_k141_5812\|\|full | Low-quality |
| S17_vs_k141_138596\|\|full | Low-quality |
| S17_vs_k141_116885\|\|full | Low-quality |
| S17_vs_k141_102049\|\|full | Not-determined |
| S17_vs_k141_79768\|\|full | Low-quality |
| S17_vs_k141_168235\|\|full | Not-determined |
| S17_vs_k141_27183\|\|full | Not-determined |
| S17_vs_k141_154126\|\|full | Low-quality |
| S17_vs_k141_57702\|\|full | Low-quality |
| S17_vs_k141_79771\|\|full | Low-quality |
| S17_vs_k141_50578\|\|full | Not-determined |
| S17_vs_k141_27206\|\|full | Low-quality |
| S17_vs_k141_110041\|\|full | Low-quality |
| S17_vs_k141_161199\|\|full | Low-quality |
| S17_vs_k141_174992\|\|full | Not-determined |
| S17_vs_k141_123642\|\|full | Low-quality |
| S17_vs_k141_110056\|\|full | Not-determined |
| S17_vs_k141_130920\|\|full | Not-determined |
| S17_vs_k141_94108\|\|full | Low-quality |
| S17_vs_k141_138634\|\|full | Not-determined |
| S17_vs_k141_94112\|\|full | Low-quality |
| S17_vs_k141_12966\|\|full | Low-quality |
| S17_vs_k141_102068\|\|full | Not-determined |
| S17_vs_k141_110068\|\|full | Not-determined |
| S17_vs_k141_20038\|\|full | Not-determined |
| S17_vs_k141_86370\|\|full | Not-determined |
| S17_vs_k141_94121\|\|full | Low-quality |
| S17_vs_k141_146222\|\|full | Not-determined |
| S17_vs_k141_86377\|\|full | Not-determined |
| S9_extracted_k141_47365 | Complete |
| S17_vs_k141_5881\|\|full | Not-determined |
| S17_vs_k141_138661\|\|full | Low-quality |
| S17_vs_k141_27246\|\|full | Low-quality |
| S17_vs_k141_35957\|\|full | Not-determined |
| S17_vs_k141_154186\|\|full | Not-determined |
| S17_vs_k141_43314\|\|full | Low-quality |
| S17_vs_k141_138662\|\|full | Low-quality |
| S17_vs_k141_57762\|\|full | Not-determined |
| S17_vs_k141_57765\|\|full | Not-determined |
| S17_vs_k141_94150\|\|full | Low-quality |
| S17_vs_k141_13005\|\|full | Low-quality |
| S17_vs_k141_110100\|\|full | Not-determined |
| S17_vs_k141_5891\|\|full | Low-quality |
| S17_vs_k141_168246\|\|full | Low-quality |
| S17_vs_k141_35974\|\|full | Low-quality |
| S17_vs_k141_154218\|\|full | Low-quality |
| S17_vs_k141_64155\|\|full | Low-quality |
| S17_vs_k141_130943\|\|full | Low-quality |
| S17_vs_k141_110104\|\|full | Not-determined |
| S17_vs_k141_27284\|\|full | Low-quality |
| S17_vs_k141_154222\|\|full | Low-quality |
| S17_vs_k141_27285\|\|full | Not-determined |
| S17_vs_k141_102122\|\|full | Not-determined |
| S17_vs_k141_35986\|\|full | Not-determined |
| S17_vs_k141_50658\|\|full | Low-quality |
| S17_vs_k141_175056\|\|full | Low-quality |
| S17_vs_k141_20095\|\|full | Not-determined |
| S17_vs_k141_123703\|\|full | Not-determined |
| S17_vs_k141_20096\|\|full | Low-quality |
| S17_vs_k141_79861\|\|full | Not-determined |
| S17_vs_k141_123707\|\|full | Low-quality |
| S17_vs_k141_116969\|\|full | Low-quality |
| S17_vs_k141_20101\|\|full | Not-determined |
| S17_vs_k141_64184\|\|full | Not-determined |
| S17_vs_k141_146279\|\|full | Not-determined |
| S17_vs_k141_123710\|\|full | Low-quality |
| S17_vs_k141_130966\|\|full | Low-quality |
| S17_vs_k141_161306\|\|full | Not-determined |
| S17_vs_k141_154269\|\|full | Low-quality |
| S17_vs_k141_146291\|\|full | Not-determined |
| S17_vs_k141_161317\|\|full | Low-quality |
| S17_vs_k141_130979\|\|full | Low-quality |
| S17_vs_k141_168286\|\|full | Not-determined |
| S17_vs_k141_20120\|\|full | Low-quality |
| S17_vs_k141_154285\|\|full | Low-quality |
| S17_vs_k141_79875\|\|full | Not-determined |
| S17_vs_k141_138703\|\|full | Low-quality |
| S17_vs_k141_20129\|\|full | Not-determined |
| S17_vs_k141_79878\|\|full | Low-quality |
| S17_vs_k141_146308\|\|full | Not-determined |
| S17_vs_k141_161329\|\|full | Not-determined |
| S17_vs_k141_131012\|\|full | Not-determined |
| S17_vs_k141_102187\|\|full | Not-determined |
| S17_vs_k141_110173\|\|full | Not-determined |
| S17_vs_k141_57850\|\|full | Low-quality |
| S17_vs_k141_50680\|\|full | Low-quality |
| S17_vs_k141_94174\|\|full | Low-quality |
| S17_vs_k141_102193\|\|full | Not-determined |
| S17_vs_k141_131017\|\|full | Not-determined |
| S17_vs_k141_161338\|\|full | Not-determined |
| S17_vs_k141_146333\|\|full | Not-determined |
| S17_vs_k141_131021\|\|full | Low-quality |
| S17_vs_k141_50691\|\|full | Not-determined |
| S17_vs_k141_5960\|\|full | Low-quality |
| S17_vs_k141_72566\|\|full | Low-quality |
| S17_vs_k141_36028\|\|full | Not-determined |
| S17_vs_k141_50693\|\|full | Low-quality |
| S17_vs_k141_117030\|\|full | Low-quality |
| S17_vs_k141_110198\|\|full | Low-quality |
| S17_vs_k141_86481\|\|full | Low-quality |
| S17_vs_k141_146356\|\|full | Not-determined |
| S17_vs_k141_117058\|\|full | Low-quality |
| S17_vs_k141_79934\|\|full | Low-quality |
| S17_vs_k141_168323\|\|full | Low-quality |
| S17_vs_k141_5988\|\|full | Low-quality |
| S17_vs_k141_161362\|\|full | Low-quality |
| S17_vs_k141_123807\|\|full | Low-quality |
| S17_vs_k141_146384\|\|full | Not-determined |
| S17_vs_k141_72594\|\|full | Low-quality |
| S17_vs_k141_57897\|\|full | Low-quality |
| S17_vs_k141_72599\|\|full | Not-determined |
| S17_vs_k141_117074\|\|full | Low-quality |
| S17_vs_k141_123814\|\|full | Low-quality |
| S17_vs_k141_86500\|\|full | Not-determined |
| S17_vs_k141_161388\|\|full | Not-determined |
| S17_vs_k141_27400\|\|full | Low-quality |
| S17_vs_k141_86509\|\|full | Not-determined |
| S17_vs_k141_50768\|\|full | Low-quality |
| S17_vs_k141_102259\|\|full | Low-quality |
| S17_vs_k141_20175\|\|full | Not-determined |
| S9_vs_k141_47365\|\|full | High-quality |
| S17_vs_k141_13096\|\|full | Not-determined |
| S17_vs_k141_43451\|\|full | Not-determined |
| S17_vs_k141_50785\|\|full | Low-quality |
| S17_vs_k141_123845\|\|full | Not-determined |
| S17_vs_k141_168359\|\|full | Low-quality |
| S17_vs_k141_154424\|\|full | Not-determined |
| S17_vs_k141_131104\|\|full | Low-quality |
| S17_vs_k141_20204\|\|full | Low-quality |
| S17_vs_k141_154434\|\|full | Low-quality |
| S17_vs_k141_43465\|\|full | Not-determined |
| S17_vs_k141_123858\|\|full | Not-determined |
| S17_vs_k141_123862\|\|full | Low-quality |
| S17_vs_k141_154445\|\|full | Low-quality |
| S17_vs_k141_102301\|\|full | Not-determined |
| S17_vs_k141_86543\|\|full | Low-quality |
| S17_vs_k141_64335\|\|full | Low-quality |
| S17_vs_k141_161456\|\|full | Low-quality |
| S17_vs_k141_94296\|\|full | Low-quality |
| S17_vs_k141_102315\|\|full | Not-determined |
| S17_vs_k141_20238\|\|full | Low-quality |
| S17_vs_k141_94303\|\|full | Low-quality |
| S17_vs_k141_64350\|\|full | Not-determined |
| S17_vs_k141_146471\|\|full | Low-quality |
| S17_vs_k141_102326\|\|full | Low-quality |
| S17_vs_k141_131151\|\|full | Low-quality |
| S17_vs_k141_110303\|\|full | Not-determined |
| S17_vs_k141_13155\|\|full | Not-determined |
| S17_vs_k141_123891\|\|full | Low-quality |
| S17_vs_k141_138876\|\|full | Not-determined |
| S17_vs_k141_110316\|\|full | Low-quality |
| S17_vs_k141_20254\|\|full | Low-quality |
| S17_vs_k141_13168\|\|full | Not-determined |
| S17_vs_k141_86581\|\|full | Not-determined |
| S17_vs_k141_27482\|\|full | Low-quality |
| S17_vs_k141_72620\|\|full | Not-determined |
| S17_vs_k141_175185\|\|full | Not-determined |
| S17_vs_k141_138897\|\|full | Not-determined |
| S17_vs_k141_161481\|\|full | Not-determined |
| S17_vs_k141_57995\|\|full | Not-determined |
| S17_vs_k141_110349\|\|full | Low-quality |
| S17_vs_k141_131168\|\|full | Not-determined |
| S17_vs_k141_79996\|\|full | Not-determined |
| S17_vs_k141_168442\|\|full | Not-determined |
| S17_vs_k141_36090\|\|full | Not-determined |
| S17_vs_k141_86622\|\|full | Not-determined |
| S17_vs_k141_72666\|\|full | Not-determined |
| S17_vs_k141_20302\|\|full | Not-determined |
| S17_vs_k141_154524\|\|full | Not-determined |
| S17_vs_k141_64441\|\|full | Not-determined |
| S17_vs_k141_146564\|\|full | Not-determined |
| S17_vs_k141_131194\|\|full | Not-determined |
| S17_vs_k141_110398\|\|full | Low-quality |
| S17_vs_k141_64447\|\|full | Not-determined |
| S17_vs_k141_80008\|\|full | Low-quality |
| S17_vs_k141_146571\|\|full | Low-quality |
| S17_vs_k141_102370\|\|full | Not-determined |
| S17_vs_k141_161530\|\|full | Low-quality |
| S17_vs_k141_64456\|\|full | Low-quality |
| S17_vs_k141_6194\|\|full | Low-quality |
| S17_vs_k141_64458\|\|full | Low-quality |
| S17_vs_k141_117245\|\|full | Low-quality |
| S17_vs_k141_175234\|\|full | Not-determined |
| S17_vs_k141_86662\|\|full | Low-quality |
| S17_vs_k141_27532\|\|full | Low-quality |
| S17_vs_k141_20317\|\|full | Not-determined |
| S17_vs_k141_64484\|\|full | Low-quality |
| S17_vs_k141_64489\|\|full | Low-quality |
| S17_vs_k141_6210\|\|full | Low-quality |
| S17_vs_k141_175242\|\|full | Not-determined |
| S17_vs_k141_110426\|\|full | Not-determined |
| S17_vs_k141_102400\|\|full | Low-quality |
| S17_vs_k141_123986\|\|full | Low-quality |
| S17_vs_k141_117297\|\|full | Low-quality |
| S17_vs_k141_139013\|\|full | Low-quality |
| S17_vs_k141_154607\|\|full | Low-quality |
| S8_extracted_k141_208110 | Complete |
| S17_vs_k141_102412\|\|full | Not-determined |
| S17_vs_k141_139021\|\|full | Low-quality |
| S17_vs_k141_64500\|\|full | Low-quality |
| S17_vs_k141_124001\|\|full | Low-quality |
| S17_vs_k141_146620\|\|full | Low-quality |
| S17_vs_k141_36188\|\|full | Not-determined |
| S17_vs_k141_58036\|\|full | Not-determined |
| S17_vs_k141_13301\|\|full | Low-quality |
| S17_vs_k141_117314\|\|full | Not-determined |
| S17_vs_k141_27588\|\|full | Not-determined |
| S17_vs_k141_110435\|\|full | Not-determined |
| S17_vs_k141_13310\|\|full | Low-quality |
| S17_vs_k141_64516\|\|full | Not-determined |
| S17_vs_k141_102440\|\|full | Low-quality |
| S17_vs_k141_146638\|\|full | Low-quality |
| S17_vs_k141_36213\|\|full | Not-determined |
| S17_vs_k141_139038\|\|full | Not-determined |
| S17_vs_k141_110448\|\|full | Low-quality |
| S17_vs_k141_94359\|\|full | Low-quality |
| S17_vs_k141_161600\|\|full | Not-determined |
| S17_vs_k141_175302\|\|full | Not-determined |
| S17_vs_k141_13321\|\|full | Low-quality |
| S17_vs_k141_58065\|\|full | Not-determined |
| S17_vs_k141_102458\|\|full | Not-determined |
| S17_vs_k141_124023\|\|full | Low-quality |
| S17_vs_k141_117355\|\|full | Not-determined |
| S17_vs_k141_64543\|\|full | Not-determined |
| S17_vs_k141_117369\|\|full | Low-quality |
| S17_vs_k141_102465\|\|full | Low-quality |
| S17_vs_k141_64546\|\|full | Low-quality |
| S17_vs_k141_6263\|\|full | Not-determined |
| S17_vs_k141_161625\|\|full | Low-quality |
| S17_vs_k141_64553\|\|full | Low-quality |
| S17_vs_k141_6280\|\|full | Low-quality |
| S17_vs_k141_117388\|\|full | Not-determined |
| S17_vs_k141_175357\|\|full | Low-quality |
| S17_vs_k141_94401\|\|full | Not-determined |
| S17_vs_k141_64581\|\|full | Not-determined |
| S17_vs_k141_43668\|\|full | Not-determined |
| S17_vs_k141_6284\|\|full | Low-quality |
| S17_vs_k141_146706\|\|full | Low-quality |
| S17_vs_k141_64584\|\|full | Low-quality |
| S17_vs_k141_124057\|\|full | Low-quality |
| S17_vs_k141_27666\|\|full | Low-quality |
| S17_vs_k141_72808\|\|full | Low-quality |
| S17_vs_k141_72810\|\|full | Not-determined |
| S17_vs_k141_6297\|\|full | Not-determined |
| S17_vs_k141_64602\|\|full | Not-determined |
| S17_vs_k141_102519\|\|full | Low-quality |
| S17_vs_k141_161671\|\|full | Low-quality |
| S17_vs_k141_64607\|\|full | Not-determined |
| S17_vs_k141_72831\|\|full | Low-quality |
| S17_vs_k141_131327\|\|full | Not-determined |
| S17_vs_k141_64611\|\|full | Not-determined |
| S17_vs_k141_6316\|\|full | Low-quality |
| S17_vs_k141_50876\|\|full | Not-determined |
| S17_vs_k141_124083\|\|full | Low-quality |
| S17_vs_k141_27691\|\|full | Not-determined |
| S17_vs_k141_102532\|\|full | Not-determined |
| S17_vs_k141_154693\|\|full | Low-quality |
| S17_vs_k141_139078\|\|full | Not-determined |
| S17_vs_k141_43697\|\|full | Not-determined |
| S17_vs_k141_58122\|\|full | Low-quality |
| S17_vs_k141_175407\|\|full | Not-determined |
| S17_vs_k141_58144\|\|full | Not-determined |
| S17_vs_k141_139103\|\|full | Not-determined |
| S17_vs_k141_50896\|\|full | Not-determined |
| S17_vs_k141_175417\|\|full | Not-determined |
| S17_vs_k141_139108\|\|full | Low-quality |
| S17_vs_k141_154719\|\|full | Not-determined |
| S17_vs_k141_43711\|\|full | Low-quality |
| S17_vs_k141_154721\|\|full | Low-quality |
| S17_vs_k141_50901\|\|full | Low-quality |
| S17_vs_k141_72874\|\|full | Not-determined |
| S17_vs_k141_20488\|\|full | Low-quality |
| S17_vs_k141_80217\|\|full | Not-determined |
| S17_vs_k141_27722\|\|full | Low-quality |
| S17_vs_k141_20493\|\|full | Not-determined |
| S17_vs_k141_6364\|\|full | Low-quality |
| S17_vs_k141_175431\|\|full | Low-quality |
| S17_vs_k141_50914\|\|full | Low-quality |
| S17_vs_k141_86873\|\|full | Not-determined |
| S17_vs_k141_154736\|\|full | Not-determined |
| S17_vs_k141_175432\|\|full | Low-quality |
| S17_vs_k141_168494\|\|full | Not-determined |
| S17_vs_k141_86876\|\|full | Low-quality |
| S17_vs_k141_168496\|\|full | Not-determined |
| S17_vs_k141_13393\|\|full | Low-quality |
| S17_vs_k141_117455\|\|full | Low-quality |
| S17_vs_k141_110565\|\|full | Not-determined |
| S17_vs_k141_94434\|\|full | Low-quality |
| S17_vs_k141_154742\|\|full | Not-determined |
| S17_vs_k141_161732\|\|full | Not-determined |
| S17_vs_k141_6388\|\|full | Not-determined |
| S17_vs_k141_43733\|\|full | Low-quality |
| S17_vs_k141_168507\|\|full | Not-determined |
| S17_vs_k141_72902\|\|full | Low-quality |
| S17_vs_k141_146824\|\|full | Low-quality |
| S17_vs_k141_102593\|\|full | Low-quality |
| S17_vs_k141_20508\|\|full | Low-quality |
| S17_vs_k141_20511\|\|full | Not-determined |
| S17_vs_k141_6393\|\|full | Low-quality |
| S17_vs_k141_50944\|\|full | Low-quality |
| S17_vs_k141_58188\|\|full | Low-quality |
| S17_vs_k141_175451\|\|full | Not-determined |
| S17_vs_k141_94449\|\|full | Low-quality |
| S17_vs_k141_86903\|\|full | Not-determined |
| S17_vs_k141_36350\|\|full | Low-quality |
| S17_vs_k141_50957\|\|full | Low-quality |
| S17_vs_k141_139134\|\|full | Low-quality |
| S17_vs_k141_102607\|\|full | Low-quality |
| S17_vs_k141_146853\|\|full | Not-determined |
| S17_vs_k141_94451\|\|full | Low-quality |
| S17_vs_k141_168555\|\|full | Low-quality |
| S17_vs_k141_146865\|\|full | Low-quality |
| S17_vs_k141_161749\|\|full | Not-determined |
| S17_vs_k141_131428\|\|full | Not-determined |
| S17_vs_k141_154783\|\|full | Low-quality |
| S17_vs_k141_124211\|\|full | Low-quality |
| S17_vs_k141_161757\|\|full | Low-quality |
| S17_vs_k141_27778\|\|full | Low-quality |
| S17_vs_k141_175501\|\|full | Not-determined |
| S17_vs_k141_175502\|\|full | Not-determined |
| S17_vs_k141_175504\|\|full | Not-determined |
| S17_vs_k141_139161\|\|full | Not-determined |
| S17_vs_k141_161768\|\|full | Low-quality |
| S17_vs_k141_175507\|\|full | Not-determined |
| S17_vs_k141_20542\|\|full | Not-determined |
| S17_vs_k141_168565\|\|full | Not-determined |
| S17_vs_k141_154785\|\|full | Not-determined |
| S17_vs_k141_168567\|\|full | Low-quality |
| S17_vs_k141_110613\|\|full | Low-quality |
| S17_vs_k141_110614\|\|full | Not-determined |
| S17_vs_k141_58249\|\|full | Low-quality |
| S8_vs_k141_208110\|\|full | High-quality |
| S17_vs_k141_51008\|\|full | Not-determined |
| S17_vs_k141_27809\|\|full | Low-quality |
| S17_vs_k141_86963\|\|full | Low-quality |
| S17_vs_k141_124236\|\|full | Not-determined |
| S17_vs_k141_131474\|\|full | Low-quality |
| S17_vs_k141_80348\|\|full | Not-determined |
| S17_vs_k141_139176\|\|full | Low-quality |
| S17_vs_k141_13519\|\|full | Low-quality |
| S17_vs_k141_58259\|\|full | Low-quality |
| S17_vs_k141_58260\|\|full | Not-determined |
| S17_vs_k141_13523\|\|full | Low-quality |
| S17_vs_k141_58261\|\|full | Low-quality |
| S17_vs_k141_20575\|\|full | Low-quality |
| S17_vs_k141_146935\|\|full | Low-quality |
| S17_vs_k141_51030\|\|full | Low-quality |
| S17_vs_k141_161825\|\|full | Low-quality |
| S17_vs_k141_20580\|\|full | Not-determined |
| S17_vs_k141_102703\|\|full | Low-quality |
| S17_vs_k141_36398\|\|full | Not-determined |
| S17_vs_k141_102707\|\|full | Low-quality |
| S17_vs_k141_80369\|\|full | Low-quality |
| S17_vs_k141_73008\|\|full | Not-determined |
| S17_vs_k141_20592\|\|full | Low-quality |
| S17_vs_k141_146946\|\|full | Low-quality |
| S17_vs_k141_124278\|\|full | Not-determined |
| S17_vs_k141_51069\|\|full | Low-quality |
| S17_vs_k141_51071\|\|full | Not-determined |
| S17_vs_k141_36418\|\|full | Not-determined |
| S17_vs_k141_146958\|\|full | Low-quality |
| S17_vs_k141_87026\|\|full | Low-quality |
| S17_vs_k141_13570\|\|full | Not-determined |
| S17_vs_k141_80398\|\|full | Low-quality |
| S17_vs_k141_43882\|\|full | Low-quality |
| S17_vs_k141_73028\|\|full | Low-quality |
| S17_vs_k141_36437\|\|full | Not-determined |
| S17_vs_k141_124312\|\|full | Low-quality |
| S17_vs_k141_58303\|\|full | Not-determined |
| S17_vs_k141_139213\|\|full | Low-quality |
| S17_vs_k141_80423\|\|full | Not-determined |
| S17_vs_k141_161889\|\|full | Not-determined |
| S17_vs_k141_102768\|\|full | Low-quality |
| S17_vs_k141_58306\|\|full | Not-determined |
| S17_vs_k141_27887\|\|full | Low-quality |
| S17_vs_k141_51104\|\|full | Not-determined |
| S17_vs_k141_94533\|\|full | Low-quality |
| S17_vs_k141_124322\|\|full | Not-determined |
| S17_vs_k141_117611\|\|full | Low-quality |
| S17_vs_k141_58318\|\|full | Not-determined |
| S17_vs_k141_51108\|\|full | Not-determined |
| S17_vs_k141_94545\|\|full | Not-determined |
| S17_vs_k141_94547\|\|full | Low-quality |
| S17_vs_k141_168677\|\|full | Low-quality |
| S17_vs_k141_51117\|\|full | Low-quality |
| S17_vs_k141_102796\|\|full | Low-quality |
| S17_vs_k141_80458\|\|full | Low-quality |
| S17_vs_k141_102805\|\|full | Not-determined |
| S17_vs_k141_51136\|\|full | Low-quality |
| S8_vs_k141_115958\|\|full | High-quality |
| S17_vs_k141_27912\|\|full | Low-quality |
| S17_vs_k141_124349\|\|full | Not-determined |
| S5_vs_k141_77593\|\|full | Medium-quality |
| S17_vs_k141_131563\|\|full | Low-quality |
| S17_vs_k141_27913\|\|full | Low-quality |
| S17_vs_k141_154910\|\|full | Low-quality |
| S17_vs_k141_168702\|\|full | Not-determined |
| S17_vs_k141_154915\|\|full | Low-quality |
| S17_vs_k141_102816\|\|full | Low-quality |
| S17_vs_k141_147088\|\|full | Low-quality |
| S17_vs_k141_147095\|\|full | Not-determined |
| S17_vs_k141_87067\|\|full | Low-quality |
| S17_vs_k141_139241\|\|full | Not-determined |
| S17_vs_k141_175559\|\|full | Not-determined |
| S8_extracted_k141_208105 | Complete |
| S17_vs_k141_94597\|\|full | Not-determined |
| S17_vs_k141_51172\|\|full | Not-determined |
| S17_vs_k141_6588\|\|full | Low-quality |
| S17_vs_k141_168727\|\|full | Low-quality |
| S17_vs_k141_110669\|\|full | Low-quality |
| S17_vs_k141_27958\|\|full | Low-quality |
| S17_vs_k141_139260\|\|full | Low-quality |
| S17_vs_k141_175571\|\|full | Low-quality |
| S17_vs_k141_43968\|\|full | Not-determined |
| S17_vs_k141_102852\|\|full | Not-determined |
| S17_vs_k141_13650\|\|full | Low-quality |
| S17_vs_k141_175574\|\|full | Not-determined |
| S17_vs_k141_94612\|\|full | Not-determined |
| S17_vs_k141_80501\|\|full | Not-determined |
| S17_vs_k141_154968\|\|full | Low-quality |
| S17_vs_k141_87098\|\|full | Low-quality |
| S17_vs_k141_175591\|\|full | Not-determined |
| S17_vs_k141_73104\|\|full | Not-determined |
| S17_vs_k141_73105\|\|full | Low-quality |
| S17_vs_k141_168748\|\|full | Low-quality |
| S17_vs_k141_124387\|\|full | Not-determined |
| S17_vs_k141_131629\|\|full | Low-quality |
| S17_vs_k141_139299\|\|full | Not-determined |
| S17_vs_k141_102902\|\|full | Low-quality |
| S17_vs_k141_131637\|\|full | Low-quality |
| S17_vs_k141_168769\|\|full | Not-determined |
| S17_vs_k141_139301\|\|full | Low-quality |
| S17_vs_k141_139302\|\|full | Not-determined |
| S17_vs_k141_51213\|\|full | Low-quality |
| S17_vs_k141_147165\|\|full | Not-determined |
| S17_vs_k141_28009\|\|full | Low-quality |
| S17_vs_k141_73148\|\|full | Low-quality |
| S17_vs_k141_44024\|\|full | Not-determined |
| S17_vs_k141_147187\|\|full | Not-determined |
| S17_vs_k141_117627\|\|full | Not-determined |
| S17_vs_k141_155014\|\|full | Low-quality |
| S17_vs_k141_6635\|\|full | Not-determined |
| S17_vs_k141_147198\|\|full | Not-determined |
| S17_vs_k141_6639\|\|full | Low-quality |
| S17_vs_k141_102934\|\|full | Low-quality |
| S17_vs_k141_110700\|\|full | Low-quality |
| S17_vs_k141_147203\|\|full | Not-determined |
| S17_vs_k141_110702\|\|full | Not-determined |
| S17_vs_k141_73174\|\|full | Not-determined |
| S17_vs_k141_117629\|\|full | Low-quality |
| S17_vs_k141_131679\|\|full | Not-determined |
| S17_vs_k141_139339\|\|full | Not-determined |
| S17_vs_k141_102941\|\|full | Low-quality |
| S16_extracted_124668 | High-quality |
| S17_vs_k141_131689\|\|full | Low-quality |
| S17_vs_k141_80579\|\|full | Low-quality |
| S17_vs_k141_51265\|\|full | Not-determined |
| S17_vs_k141_168824\|\|full | Not-determined |
| S17_vs_k141_87173\|\|full | Low-quality |
| S17_vs_k141_94642\|\|full | Low-quality |
| S17_vs_k141_162053\|\|full | Not-determined |
| S17_vs_k141_80606\|\|full | Not-determined |
| S17_vs_k141_162058\|\|full | Low-quality |
| S17_vs_k141_139373\|\|full | Not-determined |
| S17_vs_k141_51291\|\|full | Not-determined |
| S17_vs_k141_131733\|\|full | Not-determined |
| S17_vs_k141_175727\|\|full | Not-determined |
| S17_vs_k141_87206\|\|full | Not-determined |
| S17_vs_k141_6697\|\|full | Not-determined |
| S17_vs_k141_124452\|\|full | Low-quality |
| S17_vs_k141_73244\|\|full | Not-determined |
| S17_vs_k141_44069\|\|full | Not-determined |
| S17_vs_k141_110744\|\|full | Low-quality |
| S17_vs_k141_155066\|\|full | Not-determined |
| S17_vs_k141_73257\|\|full | Not-determined |
| S17_vs_k141_80638\|\|full | Not-determined |
| S17_vs_k141_117707\|\|full | Not-determined |
| S17_vs_k141_58490\|\|full | Low-quality |
| S17_vs_k141_44076\|\|full | Not-determined |
| S17_vs_k141_51330\|\|full | Not-determined |
| S17_vs_k141_73267\|\|full | Low-quality |
| S17_vs_k141_139415\|\|full | Low-quality |
| S17_vs_k141_110756\|\|full | Low-quality |
| S17_vs_k141_87236\|\|full | Low-quality |
| S17_vs_k141_58513\|\|full | Low-quality |
| S17_vs_k141_124485\|\|full | Low-quality |
| S17_vs_k141_139423\|\|full | Not-determined |
| S17_vs_k141_20703\|\|full | Not-determined |
| S17_vs_k141_73288\|\|full | Not-determined |
| S17_vs_k141_36550\|\|full | Low-quality |
| S17_vs_k141_139429\|\|full | Low-quality |
| S17_vs_k141_175773\|\|full | Low-quality |
| S17_vs_k141_44103\|\|full | Not-determined |
| S17_vs_k141_155089\|\|full | Low-quality |
| S17_vs_k141_175775\|\|full | Not-determined |
| S17_vs_k141_175777\|\|full | Low-quality |
| S17_vs_k141_13813\|\|full | Low-quality |
| S17_vs_k141_117722\|\|full | Not-determined |
| S17_vs_k141_131806\|\|full | Low-quality |
| S17_vs_k141_64628\|\|full | Low-quality |
| S17_vs_k141_117725\|\|full | Low-quality |
| S17_vs_k141_64629\|\|full | Low-quality |
| S17_vs_k141_124501\|\|full | Low-quality |
| S17_vs_k141_58546\|\|full | Low-quality |
| S17_vs_k141_168898\|\|full | Not-determined |
| S17_vs_k141_13823\|\|full | Not-determined |
| S17_vs_k141_44122\|\|full | Low-quality |
| S17_vs_k141_36566\|\|full | Low-quality |
| S17_vs_k141_147312\|\|full | Low-quality |
| S17_vs_k141_73308\|\|full | Low-quality |
| S17_vs_k141_73309\|\|full | Low-quality |
| S17_vs_k141_139463\|\|full | Not-determined |
| S17_vs_k141_44138\|\|full | Not-determined |
| S17_vs_k141_44139\|\|full | Low-quality |
| S17_vs_k141_175802\|\|full | Low-quality |
| S17_vs_k141_124526\|\|full | Low-quality |
| S17_vs_k141_110805\|\|full | Not-determined |
| S17_vs_k141_139487\|\|full | Low-quality |
| S17_vs_k141_175812\|\|full | Low-quality |
| S17_vs_k141_73330\|\|full | Not-determined |
| S17_vs_k141_94785\|\|full | Low-quality |
| S17_vs_k141_20768\|\|full | Not-determined |
| S17_vs_k141_51377\|\|full | Low-quality |
| S17_vs_k141_147350\|\|full | Low-quality |
| S17_vs_k141_94801\|\|full | Low-quality |
| S17_vs_k141_64684\|\|full | Not-determined |
| S17_vs_k141_51396\|\|full | Low-quality |
| S17_vs_k141_117786\|\|full | Low-quality |
| S17_vs_k141_175826\|\|full | Low-quality |
| S17_vs_k141_64688\|\|full | Low-quality |
| S17_vs_k141_80680\|\|full | Low-quality |
| S17_vs_k141_6827\|\|full | Low-quality |
| S17_vs_k141_117802\|\|full | Not-determined |
| S17_vs_k141_73382\|\|full | Low-quality |
| S17_vs_k141_13847\|\|full | Low-quality |
| S17_vs_k141_124583\|\|full | Low-quality |
| S17_vs_k141_87391\|\|full | Not-determined |
| S17_vs_k141_169010\|\|full | Not-determined |
| S17_vs_k141_64731\|\|full | Not-determined |
| S17_vs_k141_117824\|\|full | Not-determined |
| S17_vs_k141_87398\|\|full | Low-quality |
| S17_vs_k141_28182\|\|full | Low-quality |
| S17_vs_k141_13868\|\|full | Low-quality |
| S17_vs_k141_117830\|\|full | Not-determined |
| S17_vs_k141_131917\|\|full | Not-determined |
| S17_vs_k141_6849\|\|full | Low-quality |
| S17_vs_k141_147428\|\|full | Low-quality |
| S17_vs_k141_51459\|\|full | Low-quality |
| S17_vs_k141_80727\|\|full | Not-determined |
| S17_vs_k141_131943\|\|full | Low-quality |
| S17_vs_k141_51471\|\|full | Not-determined |
| S17_vs_k141_44213\|\|full | Low-quality |
| S17_vs_k141_87421\|\|full | Low-quality |
| S17_vs_k141_103150\|\|full | Not-determined |
| S17_vs_k141_175884\|\|full | Low-quality |
| S17_vs_k141_51477\|\|full | Low-quality |
| S17_vs_k141_175888\|\|full | Low-quality |
| S17_vs_k141_80743\|\|full | Low-quality |
| S17_vs_k141_131970\|\|full | Low-quality |
| S17_vs_k141_117862\|\|full | Not-determined |
| S17_vs_k141_147447\|\|full | Low-quality |
| S17_vs_k141_131974\|\|full | Not-determined |
| S17_vs_k141_175908\|\|full | Low-quality |
| S17_vs_k141_36708\|\|full | Low-quality |
| S8_vs_k141_208105\|\|full | High-quality |
| S17_vs_k141_6886\|\|full | Low-quality |
| S17_vs_k141_103177\|\|full | Low-quality |
| S17_vs_k141_51493\|\|full | Low-quality |
| S17_vs_k141_175917\|\|full | Low-quality |
| S17_vs_k141_94860\|\|full | Low-quality |
| S17_vs_k141_44245\|\|full | Low-quality |
| S17_vs_k141_51503\|\|full | Low-quality |
| S17_vs_k141_28227\|\|full | Not-determined |
| S17_vs_k141_20902\|\|full | Low-quality |
| S17_vs_k141_169063\|\|full | Low-quality |
| S17_vs_k141_103193\|\|full | Not-determined |
| S17_vs_k141_169065\|\|full | Low-quality |
| S17_vs_k141_28233\|\|full | Low-quality |
| S17_vs_k141_80771\|\|full | Not-determined |
| S17_vs_k141_94881\|\|full | Low-quality |
| S17_vs_k141_169079\|\|full | Not-determined |
| S17_vs_k141_73484\|\|full | Low-quality |
| S17_vs_k141_80783\|\|full | Low-quality |
| S17_vs_k141_117885\|\|full | Not-determined |
| S17_vs_k141_139602\|\|full | Low-quality |
| S17_vs_k141_139603\|\|full | Low-quality |
| S17_vs_k141_6932\|\|full | Low-quality |

| | |
|---|---|
| S17_vs_k141_13929\|\|full | Not-determined |
| S17_vs_k141_175966\|\|full | Not-determined |
| S17_vs_k141_13938\|\|full | Low-quality |
| S17_vs_k141_175974\|\|full | Not-determined |
| S17_vs_k141_132049\|\|full | Not-determined |
| S17_vs_k141_87526\|\|full | Low-quality |
| S17_vs_k141_13953\|\|full | Not-determined |
| S17_vs_k141_87536\|\|full | Low-quality |
| S17_vs_k141_28306\|\|full | Low-quality |
| S17_vs_k141_94924\|\|full | Not-determined |
| S17_vs_k141_162305\|\|full | Not-determined |
| S17_vs_k141_139628\|\|full | Not-determined |
| S17_vs_k141_169135\|\|full | Low-quality |
| S17_vs_k141_44313\|\|full | Low-quality |
| S17_vs_k141_162310\|\|full | Not-determined |
| S17_vs_k141_169142\|\|full | Not-determined |
| S17_vs_k141_28324\|\|full | Low-quality |
| S17_vs_k141_139642\|\|full | Not-determined |
| S17_vs_k141_28327\|\|full | Not-determined |
| S17_vs_k141_147571\|\|full | Low-quality |
| S17_vs_k141_162319\|\|full | Not-determined |
| S17_vs_k141_139644\|\|full | Low-quality |
| S17_vs_k141_87548\|\|full | Not-determined |
| S17_vs_k141_28329\|\|full | Low-quality |
| S17_vs_k141_169156\|\|full | Low-quality |
| S17_vs_k141_80851\|\|full | Not-determined |
| S17_vs_k141_28335\|\|full | Not-determined |
| S17_vs_k141_103311\|\|full | Not-determined |
| S17_vs_k141_73581\|\|full | Low-quality |
| S17_vs_k141_80858\|\|full | Not-determined |
| S17_vs_k141_147602\|\|full | Low-quality |
| S17_vs_k141_73585\|\|full | Not-determined |
| S17_vs_k141_21034\|\|full | Low-quality |
| S17_vs_k141_80867\|\|full | Low-quality |
| S17_vs_k141_147618\|\|full | Low-quality |
| S17_vs_k141_64834\|\|full | Not-determined |
| S17_vs_k141_176033\|\|full | Not-determined |
| S17_vs_k141_6999\|\|full | Not-determined |
| S17_vs_k141_94961\|\|full | Low-quality |
| S17_vs_k141_28362\|\|full | Low-quality |
| S17_vs_k141_73602\|\|full | Low-quality |
| S17_vs_k141_7008\|\|full | Not-determined |
| S17_vs_k141_87604\|\|full | Low-quality |
| S17_vs_k141_14017\|\|full | Low-quality |
| S17_vs_k141_117991\|\|full | Not-determined |
| S17_vs_k141_147634\|\|full | Low-quality |
| S17_vs_k141_132155\|\|full | Low-quality |
| S17_vs_k141_64850\|\|full | Low-quality |
| S17_vs_k141_58663\|\|full | Low-quality |
| S17_vs_k141_132160\|\|full | Low-quality |
| S17_vs_k141_147639\|\|full | Low-quality |
| S17_vs_k141_162415\|\|full | Not-determined |
| S17_vs_k141_21099\|\|full | Not-determined |
| S17_vs_k141_139660\|\|full | Not-determined |
| S17_vs_k141_139661\|\|full | Not-determined |
| S17_vs_k141_169230\|\|full | Not-determined |
| S17_vs_k141_124738\|\|full | Not-determined |
| S17_vs_k141_103356\|\|full | Low-quality |
| S17_vs_k141_21113\|\|full | Not-determined |
| S17_vs_k141_80943\|\|full | Not-determined |
| S17_vs_k141_176089\|\|full | Low-quality |
| S17_vs_k141_124757\|\|full | Not-determined |
| S17_vs_k141_124771\|\|full | Low-quality |
| S17_vs_k141_51688\|\|full | Low-quality |
| S17_vs_k141_124775\|\|full | Low-quality |
| S17_vs_k141_87629\|\|full | Low-quality |
| S17_vs_k141_21152\|\|full | Low-quality |
| S17_vs_k141_132211\|\|full | Low-quality |
| S17_vs_k141_73685\|\|full | Low-quality |
| S17_vs_k141_58724\|\|full | Low-quality |
| S17_vs_k141_36792\|\|full | Not-determined |
| S17_vs_k141_103398\|\|full | Not-determined |
| S17_vs_k141_21168\|\|full | Low-quality |
| S17_vs_k141_21169\|\|full | Low-quality |
| S17_vs_k141_169250\|\|full | Low-quality |
| S17_vs_k141_7140\|\|full | Not-determined |
| S17_vs_k141_36801\|\|full | Low-quality |
| S17_vs_k141_87651\|\|full | Low-quality |
| S17_vs_k141_36809\|\|full | Low-quality |
| S17_vs_k141_162470\|\|full | Not-determined |
| S17_vs_k141_118022\|\|full | Not-determined |
| S17_vs_k141_14094\|\|full | Low-quality |
| S17_vs_k141_95044\|\|full | Low-quality |
| S17_vs_k141_64955\|\|full | Not-determined |
| S17_vs_k141_162490\|\|full | Not-determined |
| S17_vs_k141_176156\|\|full | Low-quality |
| S17_vs_k141_169304\|\|full | Not-determined |
| S17_vs_k141_14110\|\|full | Low-quality |
| S17_vs_k141_64974\|\|full | Not-determined |
| S17_vs_k141_118050\|\|full | Not-determined |
| S17_vs_k141_81020\|\|full | Low-quality |
| S17_vs_k141_7189\|\|full | Not-determined |
| S17_vs_k141_58752\|\|full | Not-determined |
| S17_vs_k141_21232\|\|full | Not-determined |
| S17_vs_k141_95055\|\|full | Low-quality |
| S17_vs_k141_132238\|\|full | Not-determined |
| S17_vs_k141_124818\|\|full | Low-quality |
| S17_vs_k141_64984\|\|full | Not-determined |
| S17_vs_k141_7201\|\|full | Low-quality |
| S17_vs_k141_64998\|\|full | Low-quality |
| S17_vs_k141_28491\|\|full | Low-quality |
| S17_vs_k141_118070\|\|full | Not-determined |
| S17_vs_k141_95075\|\|full | Low-quality |
| S17_vs_k141_124828\|\|full | Not-determined |
| S17_vs_k141_81041\|\|full | Low-quality |
| S17_vs_k141_118076\|\|full | Not-determined |
| S17_vs_k141_28505\|\|full | Not-determined |
| S17_vs_k141_65013\|\|full | Not-determined |
| S17_vs_k141_132268\|\|full | Not-determined |
| S17_vs_k141_169351\|\|full | Not-determined |
| S17_vs_k141_81059\|\|full | Low-quality |
| S17_vs_k141_176193\|\|full | Low-quality |
| S17_vs_k141_65027\|\|full | Low-quality |
| S17_vs_k141_139750\|\|full | Low-quality |
| S17_vs_k141_118094\|\|full | Not-determined |
| S17_vs_k141_81067\|\|full | Not-determined |
| S17_vs_k141_65031\|\|full | Low-quality |
| S17_vs_k141_58789\|\|full | Low-quality |
| S17_vs_k141_139759\|\|full | Not-determined |
| S17_vs_k141_176200\|\|full | Not-determined |
| S17_vs_k141_7232\|\|full | Low-quality |
| S17_vs_k141_14165\|\|full | Not-determined |
| S17_vs_k141_132295\|\|full | Not-determined |
| S17_vs_k141_169384\|\|full | Not-determined |
| S17_vs_k141_28531\|\|full | Low-quality |
| S17_vs_k141_118111\|\|full | Not-determined |
| S17_vs_k141_132303\|\|full | Low-quality |
| S17_vs_k141_14181\|\|full | Low-quality |
| S17_vs_k141_28534\|\|full | Not-determined |
| S17_vs_k141_124889\|\|full | Low-quality |
| S17_vs_k141_95133\|\|full | Low-quality |
| S17_vs_k141_87780\|\|full | Low-quality |
| S17_vs_k141_14197\|\|full | Not-determined |
| S17_vs_k141_118127\|\|full | Low-quality |
| S17_vs_k141_14201\|\|full | Not-determined |
| S17_vs_k141_14202\|\|full | Not-determined |
| S17_vs_k141_118133\|\|full | Low-quality |
| S17_vs_k141_124906\|\|full | Not-determined |
| S17_vs_k141_176255\|\|full | Not-determined |
| S17_vs_k141_14222\|\|full | Low-quality |
| S17_vs_k141_132360\|\|full | Low-quality |
| S11_extracted_k141_104043 | Complete |
| S17_vs_k141_95143\|\|full | Low-quality |
| S17_vs_k141_65098\|\|full | Low-quality |
| S17_vs_k141_118152\|\|full | Low-quality |
| S17_vs_k141_73832\|\|full | Low-quality |
| S17_vs_k141_139809\|\|full | Low-quality |
| S17_vs_k141_21347\|\|full | Low-quality |
| S17_vs_k141_169440\|\|full | Not-determined |
| S17_vs_k141_124939\|\|full | Not-determined |
| S17_vs_k141_81137\|\|full | Not-determined |
| S17_vs_k141_36879\|\|full | Low-quality |
| S17_vs_k141_65125\|\|full | Not-determined |
| S17_vs_k141_132376\|\|full | Not-determined |
| S17_vs_k141_118182\|\|full | Not-determined |
| S17_vs_k141_118186\|\|full | Not-determined |
| S17_vs_k141_28614\|\|full | Not-determined |
| S17_vs_k141_65138\|\|full | Low-quality |
| S17_vs_k141_21395\|\|full | Low-quality |
| S17_vs_k141_95172\|\|full | Low-quality |
| S17_vs_k141_65150\|\|full | Low-quality |
| S17_vs_k141_7302\|\|full | Low-quality |
| S17_vs_k141_124979\|\|full | Low-quality |
| S17_vs_k141_169490\|\|full | Low-quality |
| S17_vs_k141_95188\|\|full | Low-quality |
| S17_vs_k141_176282\|\|full | Low-quality |
| S17_vs_k141_73883\|\|full | Low-quality |
| S17_vs_k141_176285\|\|full | Low-quality |
| S17_vs_k141_95200\|\|full | Not-determined |
| S17_vs_k141_7313\|\|full | Not-determined |
| S17_vs_k141_176301\|\|full | Low-quality |
| S17_vs_k141_95208\|\|full | Low-quality |
| S17_vs_k141_7321\|\|full | Low-quality |
| S17_vs_k141_95211\|\|full | Not-determined |
| S17_vs_k141_132398\|\|full | Low-quality |
| S17_vs_k141_21462\|\|full | Low-quality |
| S17_vs_k141_81202\|\|full | Not-determined |
| S17_vs_k141_36924\|\|full | Not-determined |
| S17_vs_k141_21484\|\|full | Not-determined |
| S17_vs_k141_28698\|\|full | Low-quality |
| S17_vs_k141_36929\|\|full | Not-determined |
| S17_vs_k141_58920\|\|full | Not-determined |
| S17_vs_k141_21505\|\|full | Not-determined |
| S17_vs_k141_65219\|\|full | Low-quality |
| S17_vs_k141_21523\|\|full | Not-determined |
| S17_vs_k141_28740\|\|full | Low-quality |
| S17_vs_k141_65231\|\|full | Not-determined |
| S17_vs_k141_139926\|\|full | Low-quality |
| S17_vs_k141_176367\|\|full | Low-quality |
| S17_vs_k141_28753\|\|full | Low-quality |
| S17_vs_k141_87973\|\|full | Low-quality |
| S17_vs_k141_14270\|\|full | Not-determined |
| S17_vs_k141_124990\|\|full | Not-determined |
| S17_vs_k141_176388\|\|full | Not-determined |
| S17_vs_k141_125004\|\|full | Not-determined |
| S17_vs_k141_14290\|\|full | Not-determined |
| S17_vs_k141_21577\|\|full | Not-determined |
| S17_vs_k141_14304\|\|full | Not-determined |
| S17_vs_k141_95278\|\|full | Not-determined |
| S17_vs_k141_95284\|\|full | Low-quality |
| S17_vs_k141_65286\|\|full | Not-determined |
| S17_vs_k141_176410\|\|full | Not-determined |
| S17_vs_k141_176419\|\|full | Low-quality |
| S17_vs_k141_14341\|\|full | Not-determined |
| S17_vs_k141_58946\|\|full | Not-determined |
| S17_vs_k141_65320\|\|full | Low-quality |
| S17_vs_k141_14365\|\|full | Not-determined |
| S17_vs_k141_176443\|\|full | Low-quality |
| S17_vs_k141_14368\|\|full | Low-quality |
| S17_vs_k141_7360\|\|full | Not-determined |
| S17_vs_k141_14373\|\|full | Not-determined |
| S17_vs_k141_95349\|\|full | Low-quality |
| S17_vs_k141_132543\|\|full | Low-quality |
| S17_vs_k141_21652\|\|full | Not-determined |
| S17_vs_k141_95363\|\|full | Not-determined |
| S17_vs_k141_132591\|\|full | Not-determined |
| S17_vs_k141_14399\|\|full | Low-quality |
| S17_vs_k141_169752\|\|full | Low-quality |
| S17_vs_k141_14405\|\|full | Not-determined |
| S17_vs_k141_28888\|\|full | Low-quality |
| S17_vs_k141_14413\|\|full | Not-determined |
| S17_vs_k141_132612\|\|full | Low-quality |
| S17_vs_k141_132613\|\|full | Not-determined |
| S17_vs_k141_58999\|\|full | Not-determined |
| S17_vs_k141_7361\|\|full | Low-quality |
| S17_vs_k141_28907\|\|full | Low-quality |
| S17_vs_k141_169786\|\|full | Not-determined |
| S17_vs_k141_59022\|\|full | Not-determined |
| S17_vs_k141_125155\|\|full | Low-quality |
| S17_vs_k141_28919\|\|full | Not-determined |
| S17_vs_k141_132658\|\|full | Low-quality |
| S17_vs_k141_59037\|\|full | Low-quality |
| S17_vs_k141_28924\|\|full | Low-quality |
| S17_vs_k141_125172\|\|full | Low-quality |
| S17_vs_k141_95372\|\|full | Not-determined |
| S17_vs_k141_95373\|\|full | Low-quality |
| S17_vs_k141_140072\|\|full | Low-quality |
| S17_vs_k141_28929\|\|full | Low-quality |
| S17_vs_k141_95381\|\|full | Low-quality |
| S17_vs_k141_88099\|\|full | Low-quality |
| S17_vs_k141_169811\|\|full | Low-quality |
| S17_vs_k141_88106\|\|full | Low-quality |
| S17_vs_k141_125206\|\|full | Not-determined |
| S17_vs_k141_176562\|\|full | Low-quality |
| S17_vs_k141_176564\|\|full | Low-quality |
| S17_vs_k141_14478\|\|full | Low-quality |
| S17_vs_k141_169855\|\|full | Low-quality |
| S17_vs_k141_65466\|\|full | Low-quality |
| S17_vs_k141_132728\|\|full | Low-quality |
| S17_vs_k141_14510\|\|full | Not-determined |
| S17_vs_k141_169875\|\|full | Not-determined |
| S17_vs_k141_81255\|\|full | Not-determined |
| S17_vs_k141_125209\|\|full | Low-quality |
| S17_vs_k141_65485\|\|full | Not-determined |
| S17_vs_k141_14532\|\|full | Low-quality |
| S17_vs_k141_125223\|\|full | Not-determined |
| S17_vs_k141_65502\|\|full | Not-determined |
| S17_vs_k141_140170\|\|full | Low-quality |
| S17_vs_k141_21763\|\|full | Not-determined |
| S17_vs_k141_169921\|\|full | Not-determined |
| S17_vs_k141_65530\|\|full | Low-quality |
| S17_vs_k141_95504\|\|full | Not-determined |
| S17_vs_k141_88218\|\|full | Not-determined |
| S17_vs_k141_65551\|\|full | Low-quality |
| S17_vs_k141_14569\|\|full | Not-determined |
| S17_vs_k141_132777\|\|full | Low-quality |
| S17_vs_k141_21790\|\|full | Not-determined |
| S17_vs_k141_95544\|\|full | Low-quality |
| S17_vs_k141_176606\|\|full | Low-quality |
| S17_vs_k141_140215\|\|full | Low-quality |
| S17_vs_k141_125289\|\|full | Low-quality |
| S17_vs_k141_88265\|\|full | Low-quality |
| S17_vs_k141_95567\|\|full | Low-quality |
| S17_vs_k141_88269\|\|full | Not-determined |
| S17_vs_k141_14607\|\|full | Not-determined |
| S17_vs_k141_21816\|\|full | Low-quality |
| S17_vs_k141_140244\|\|full | Low-quality |
| S17_vs_k141_125312\|\|full | Not-determined |
| S17_vs_k141_29084\|\|full | Low-quality |
| S17_vs_k141_95618\|\|full | Not-determined |
| S17_vs_k141_140286\|\|full | Low-quality |
| S17_vs_k141_125361\|\|full | Not-determined |
| S17_vs_k141_176686\|\|full | Not-determined |
| S17_vs_k141_176709\|\|full | Low-quality |
| S17_vs_k141_140316\|\|full | Not-determined |
| S17_vs_k141_132895\|\|full | Not-determined |
| S17_vs_k141_88305\|\|full | Not-determined |
| S17_vs_k141_125407\|\|full | Low-quality |
| S17_vs_k141_21860\|\|full | Low-quality |
| S17_vs_k141_125413\|\|full | Low-quality |
| S17_vs_k141_88354\|\|full | Low-quality |
| S17_vs_k141_125458\|\|full | Not-determined |
| S17_vs_k141_140363\|\|full | Low-quality |
| S17_vs_k141_95757\|\|full | Low-quality |
| S17_vs_k141_65721\|\|full | Not-determined |
| S17_vs_k141_29234\|\|full | Not-determined |
| S17_vs_k141_14750\|\|full | Low-quality |
| S17_vs_k141_14753\|\|full | Low-quality |
| S17_vs_k141_65746\|\|full | Not-determined |
| S17_vs_k141_95797\|\|full | Low-quality |
| S17_vs_k141_29249\|\|full | Low-quality |
| S17_vs_k141_125556\|\|full | Not-determined |
| S17_vs_k141_21975\|\|full | Low-quality |
| S17_vs_k141_88466\|\|full | Not-determined |
| S17_vs_k141_176884\|\|full | Not-determined |
| S17_vs_k141_21993\|\|full | Not-determined |
| S17_vs_k141_176922\|\|full | Not-determined |
| S17_vs_k141_176929\|\|full | Not-determined |
| S17_vs_k141_29323\|\|full | Low-quality |
| S17_vs_k141_65882\|\|full | Not-determined |
| S17_vs_k141_29357\|\|full | Low-quality |
| S17_vs_k141_29361\|\|full | Not-determined |
| S17_vs_k141_88522\|\|full | Not-determined |
| S17_vs_k141_65943\|\|full | Low-quality |
| S17_vs_k141_29413\|\|full | Not-determined |
| S17_vs_k141_176943\|\|full | Low-quality |
| S17_vs_k141_29434\|\|full | Low-quality |
| S17_vs_k141_95855\|\|full | Not-determined |
| S17_vs_k141_176976\|\|full | Not-determined |
| S17_vs_k141_22026\|\|full | Low-quality |
| S17_vs_k141_95866\|\|full | Not-determined |
| S17_vs_k141_88602\|\|full | Low-quality |
| S17_vs_k141_177009\|\|full | Low-quality |
| S17_vs_k141_29467\|\|full | Low-quality |
| S17_vs_k141_88622\|\|full | Low-quality |
| S17_vs_k141_65969\|\|full | Low-quality |
| S17_vs_k141_29489\|\|full | Low-quality |
| S17_vs_k141_88632\|\|full | Not-determined |
| S17_vs_k141_22071\|\|full | Not-determined |
| S17_vs_k141_177075\|\|full | Low-quality |
| S17_vs_k141_177090\|\|full | Low-quality |
| S17_vs_k141_177092\|\|full | Low-quality |
| S17_vs_k141_22103\|\|full | Low-quality |
| S17_vs_k141_177097\|\|full | Low-quality |
| S17_vs_k141_177101\|\|full | Low-quality |
| S17_vs_k141_95960\|\|full | Not-determined |
| S17_vs_k141_29528\|\|full | Not-determined |
| S17_vs_k141_95962\|\|full | Low-quality |
| S17_vs_k141_177118\|\|full | Not-determined |
| S17_vs_k141_66040\|\|full | Low-quality |
| S17_vs_k141_177134\|\|full | Not-determined |
| S17_vs_k141_29541\|\|full | Low-quality |
| S17_vs_k141_177146\|\|full | Low-quality |
| S17_vs_k141_66070\|\|full | Low-quality |
| S17_vs_k141_29544\|\|full | Low-quality |
| S17_vs_k141_66075\|\|full | Low-quality |
| S17_vs_k141_95978\|\|full | Not-determined |
| S17_vs_k141_29547\|\|full | Low-quality |
| S17_vs_k141_177157\|\|full | Low-quality |
| S17_vs_k141_29551\|\|full | Low-quality |
| S17_vs_k141_96007\|\|full | Not-determined |
| S17_vs_k141_66121\|\|full | Not-determined |
| S17_vs_k141_177189\|\|full | Low-quality |
| S17_vs_k141_66201\|\|full | Low-quality |
| S17_vs_k141_66209\|\|full | Low-quality |
| S17_vs_k141_66261\|\|full | Low-quality |
| S17_vs_k141_66263\|\|full | Low-quality |
| S17_vs_k141_66285\|\|full | Low-quality |
| S17_vs_k141_66298\|\|full | Not-determined |
| S17_vs_k141_66317\|\|full | Low-quality |
| S17_vs_k141_66330\|\|full | Not-determined |
| S17_vs_k141_66388\|\|full | Not-determined |
| S17_vs_k141_66423\|\|full | Not-determined |
| S17_vs_k141_81304\|\|0_partial | Low-quality |
| S17_vs_k141_147886\|\|0_partial | Not-determined |
| S17_vs_k141_141233\|\|0_partial | Not-determined |
| S17_vs_k141_111575\|\|0_partial | Not-determined |
| S17_vs_k141_45389\|\|0_partial | Not-determined |
| S11_vs_k141_104043\|\|full | High-quality |
| S17_vs_k141_82185\|\|0_partial | Not-determined |
| S17_vs_k141_134275\|\|0_partial | Low-quality |
| S17_vs_k141_83317\|\|0_partial | Low-quality |
| S17_vs_k141_165534\|\|0_partial | Low-quality |
| S17_vs_k141_91924\|\|0_partial | Not-determined |
| S17_vs_k141_11405\|\|0_partial | Not-determined |
| S17_vs_k141_62552\|\|0_partial | Not-determined |
| S17_vs_k141_64619\|\|0_partial | Low-quality |
| S17_vs_k141_139583\|\|0_partial | Not-determined |
| S17_vs_k141_147770\|\|lt2gene | Low-quality |
| S17_vs_k141_118220\|\|lt2gene | Not-determined |
| S17_vs_k141_7390\|\|lt2gene | Low-quality |
| S17_vs_k141_73894\|\|lt2gene | Low-quality |
| S17_vs_k141_96071\|\|lt2gene | Low-quality |
| S17_vs_k141_133011\|\|lt2gene | Low-quality |
| S17_vs_k141_29583\|\|lt2gene | Not-determined |
| S17_vs_k141_133013\|\|lt2gene | Low-quality |
| S17_vs_k141_42\|\|lt2gene | Low-quality |
| S17_vs_k141_46\|\|lt2gene | Low-quality |
| S17_vs_k141_36988\|\|lt2gene | Low-quality |
| S17_vs_k141_36991\|\|lt2gene | Not-determined |
| S17_vs_k141_22185\|\|lt2gene | Low-quality |
| S17_vs_k141_125666\|\|lt2gene | Low-quality |
| S17_vs_k141_110881\|\|lt2gene | Low-quality |
| S17_vs_k141_88\|\|lt2gene | Low-quality |
| S17_vs_k141_51789\|\|lt2gene | Low-quality |
| S17_vs_k141_29651\|\|lt2gene | Low-quality |
| S17_vs_k141_44383\|\|lt2gene | Low-quality |
| S17_vs_k141_147832\|\|lt2gene | Low-quality |
| S17_vs_k141_155234\|\|lt2gene | Low-quality |
| S17_vs_k141_22214\|\|lt2gene | Low-quality |
| S17_vs_k141_37053\|\|lt2gene | Low-quality |
| S17_vs_k141_14853\|\|lt2gene | Low-quality |
| S17_vs_k141_59286\|\|lt2gene | Low-quality |
| S17_vs_k141_66665\|\|lt2gene | Low-quality |
| S17_vs_k141_14894\|\|lt2gene | Low-quality |
| S17_vs_k141_140539\|\|lt2gene | Low-quality |
| S17_vs_k141_88803\|\|lt2gene | Low-quality |
| S17_vs_k141_147930\|\|lt2gene | Low-quality |
| S17_vs_k141_96253\|\|lt2gene | Not-determined |
| S17_vs_k141_140550\|\|lt2gene | Low-quality |
| S17_vs_k141_168\|\|lt2gene | Low-quality |
| S17_vs_k141_125740\|\|lt2gene | Low-quality |
| S17_vs_k141_29798\|\|lt2gene | Low-quality |
| S17_vs_k141_125746\|\|lt2gene | Not-determined |
| S17_vs_k141_118380\|\|lt2gene | Not-determined |
| S17_vs_k141_88824\|\|lt2gene | Not-determined |
| S17_vs_k141_51908\|\|lt2gene | Not-determined |
| S17_vs_k141_29820\|\|lt2gene | Low-quality |
| S17_vs_k141_37153\|\|lt2gene | Not-determined |
| S17_vs_k141_14917\|\|lt2gene | Low-quality |
| S17_vs_k141_162726\|\|lt2gene | Low-quality |
| S17_vs_k141_96321\|\|lt2gene | Low-quality |
| S17_vs_k141_125779\|\|lt2gene | Low-quality |
| S17_vs_k141_37222\|\|lt2gene | Not-determined |
| S17_vs_k141_7670\|\|lt2gene | Low-quality |
| S17_vs_k141_118432\|\|lt2gene | Not-determined |
| S17_vs_k141_118435\|\|lt2gene | Not-determined |
| S17_vs_k141_37233\|\|lt2gene | Not-determined |
| S17_vs_k141_170087\|\|lt2gene | Not-determined |
| S17_vs_k141_81352\|\|lt2gene | Low-quality |
| S17_vs_k141_51937\|\|lt2gene | Low-quality |
| S17_vs_k141_29895\|\|lt2gene | Not-determined |
| S17_vs_k141_59399\|\|lt2gene | Not-determined |
| S17_vs_k141_96407\|\|lt2gene | Low-quality |
| S17_vs_k141_51953\|\|lt2gene | Low-quality |
| S17_vs_k141_118461\|\|lt2gene | Not-determined |
| S17_vs_k141_118468\|\|lt2gene | Low-quality |
| S17_vs_k141_96424\|\|lt2gene | Not-determined |
| S17_vs_k141_44562\|\|lt2gene | Low-quality |
| S17_vs_k141_81385\|\|lt2gene | Low-quality |
| S17_vs_k141_103757\|\|lt2gene | Not-determined |
| S17_vs_k141_51972\|\|lt2gene | Low-quality |
| S17_vs_k141_140657\|\|lt2gene | Low-quality |
| S17_vs_k141_66688\|\|lt2gene | Low-quality |
| S17_vs_k141_66689\|\|lt2gene | Low-quality |
| S17_vs_k141_66695\|\|lt2gene | Low-quality |
| S17_vs_k141_66697\|\|lt2gene | Low-quality |
| S17_vs_k141_140668\|\|lt2gene | Low-quality |
| S17_vs_k141_110963\|\|lt2gene | Not-determined |
| S17_vs_k141_125837\|\|lt2gene | Not-determined |
| S17_vs_k141_66710\|\|lt2gene | Low-quality |
| S17_vs_k141_44606\|\|lt2gene | Low-quality |
| S17_vs_k141_59455\|\|lt2gene | Low-quality |
| S17_vs_k141_103805\|\|lt2gene | Low-quality |
| S17_vs_k141_111002\|\|lt2gene | Low-quality |
| S17_vs_k141_125877\|\|lt2gene | Low-quality |
| S17_vs_k141_44647\|\|lt2gene | Low-quality |
| S17_vs_k141_59501\|\|lt2gene | Not-determined |
| S17_vs_k141_66756\|\|lt2gene | Not-determined |
| S17_vs_k141_148134\|\|lt2gene | Low-quality |
| S17_vs_k141_261\|\|lt2gene | Not-determined |
| S17_vs_k141_74223\|\|lt2gene | Low-quality |
| S17_vs_k141_59536\|\|lt2gene | Low-quality |
| S17_vs_k141_282\|\|lt2gene | Low-quality |
| S17_vs_k141_74246\|\|lt2gene | Low-quality |
| S17_vs_k141_52091\|\|lt2gene | Low-quality |
| S17_vs_k141_22379\|\|lt2gene | Low-quality |
| S17_vs_k141_140759\|\|lt2gene | Not-determined |
| S17_vs_k141_118555\|\|lt2gene | Not-determined |
| S17_vs_k141_96515\|\|lt2gene | Low-quality |
| S17_vs_k141_59542\|\|lt2gene | Low-quality |
| S17_vs_k141_125948\|\|lt2gene | Low-quality |
| S17_vs_k141_44750\|\|lt2gene | Low-quality |
| S17_vs_k141_111106\|\|lt2gene | Not-determined |
| S17_vs_k141_96534\|\|lt2gene | Low-quality |
| S17_vs_k141_81505\|\|lt2gene | Not-determined |
| S17_vs_k141_37417\|\|lt2gene | Low-quality |
| S17_vs_k141_44803\|\|lt2gene | Low-quality |
| S17_vs_k141_111163\|\|lt2gene | Low-quality |
| S17_vs_k141_59622\|\|lt2gene | Low-quality |
| S17_vs_k141_155467\|\|lt2gene | Not-determined |
| S17_vs_k141_148240\|\|lt2gene | Low-quality |
| S17_vs_k141_111178\|\|lt2gene | Low-quality |
| S17_vs_k141_118634\|\|lt2gene | Low-quality |
| S17_vs_k141_155489\|\|lt2gene | Not-determined |
| S17_vs_k141_15117\|\|lt2gene | Not-determined |
| S17_vs_k141_15124\|\|lt2gene | Low-quality |
| S17_vs_k141_89007\|\|lt2gene | Low-quality |
| S17_vs_k141_52194\|\|lt2gene | Low-quality |
| S17_vs_k141_140896\|\|lt2gene | Low-quality |
| S17_vs_k141_155507\|\|lt2gene | Low-quality |
| S17_vs_k141_59669\|\|lt2gene | Not-determined |
| S17_vs_k141_52227\|\|lt2gene | Low-quality |
| S17_vs_k141_89036\|\|lt2gene | Not-determined |
| S17_vs_k141_170344\|\|lt2gene | Low-quality |
| S17_vs_k141_155538\|\|lt2gene | Low-quality |
| S17_vs_k141_30139\|\|lt2gene | Low-quality |
| S17_vs_k141_118729\|\|lt2gene | Low-quality |
| S17_vs_k141_7910\|\|lt2gene | Low-quality |
| S17_vs_k141_140943\|\|lt2gene | Low-quality |
| S17_vs_k141_118736\|\|lt2gene | Low-quality |
| S17_vs_k141_155556\|\|lt2gene | Not-determined |
| S17_vs_k141_37568\|\|lt2gene | Not-determined |
| S17_vs_k141_7943\|\|lt2gene | Not-determined |
| S17_vs_k141_37571\|\|lt2gene | Low-quality |
| S17_vs_k141_7946\|\|lt2gene | Not-determined |
| S17_vs_k141_15196\|\|lt2gene | Low-quality |
| S17_vs_k141_15200\|\|lt2gene | Low-quality |
| S17_vs_k141_81596\|\|lt2gene | Low-quality |
| S17_vs_k141_81598\|\|lt2gene | Low-quality |
| S17_vs_k141_22536\|\|lt2gene | Not-determined |
| S17_vs_k141_52282\|\|lt2gene | Low-quality |
| S17_vs_k141_163029\|\|lt2gene | Low-quality |
| S17_vs_k141_44852\|\|lt2gene | Low-quality |
| S17_vs_k141_111252\|\|lt2gene | Low-quality |
| S17_vs_k141_7969\|\|lt2gene | Low-quality |
| S17_vs_k141_148336\|\|lt2gene | Not-determined |
| S17_vs_k141_155591\|\|lt2gene | Low-quality |
| S17_vs_k141_44857\|\|lt2gene | Not-determined |
| S17_vs_k141_66889\|\|lt2gene | Low-quality |
| S17_vs_k141_15264\|\|lt2gene | Not-determined |
| S17_vs_k141_22577\|\|lt2gene | Low-quality |
| S17_vs_k141_22604\|\|lt2gene | Not-determined |
| S17_vs_k141_52346\|\|lt2gene | Low-quality |
| S17_vs_k141_104085\|\|lt2gene | Low-quality |
| S17_vs_k141_577\|\|lt2gene | Not-determined |
| S17_vs_k141_66916\|\|lt2gene | Not-determined |
| S17_vs_k141_37671\|\|lt2gene | Low-quality |
| S17_vs_k141_141002\|\|lt2gene | Low-quality |
| S17_vs_k141_155671\|\|lt2gene | Low-quality |
| S17_vs_k141_126086\|\|lt2gene | Low-quality |
| S17_vs_k141_44919\|\|lt2gene | Low-quality |
| S17_vs_k141_37684\|\|lt2gene | Not-determined |
| S17_vs_k141_96549\|\|lt2gene | Low-quality |
| S17_vs_k141_133585\|\|lt2gene | Not-determined |
| S17_vs_k141_170499\|\|lt2gene | Low-quality |
| S17_vs_k141_66959\|\|lt2gene | Low-quality |
| S17_vs_k141_52431\|\|lt2gene | Low-quality |
| S17_vs_k141_148442\|\|lt2gene | Not-determined |
| S17_vs_k141_59795\|\|lt2gene | Low-quality |
| S17_vs_k141_163120\|\|lt2gene | Low-quality |
| S17_vs_k141_141074\|\|lt2gene | Low-quality |
| S17_vs_k141_59819\|\|lt2gene | Low-quality |
| S17_vs_k141_74574\|\|lt2gene | Not-determined |
| S17_vs_k141_74613\|\|lt2gene | Low-quality |
| S17_vs_k141_15368\|\|lt2gene | Low-quality |
| S17_vs_k141_74619\|\|lt2gene | Not-determined |
| S17_vs_k141_126174\|\|lt2gene | Low-quality |
| S17_vs_k141_155810\|\|lt2gene | Low-quality |
| S17_vs_k141_163159\|\|lt2gene | Low-quality |
| S17_vs_k141_104265\|\|lt2gene | Low-quality |
| S17_vs_k141_126205\|\|lt2gene | Low-quality |
| S17_vs_k141_170598\|\|lt2gene | Low-quality |
| S17_vs_k141_67004\|\|lt2gene | Not-determined |
| S17_vs_k141_8095\|\|lt2gene | Not-determined |
| S17_vs_k141_163198\|\|lt2gene | Low-quality |
| S17_vs_k141_89261\|\|lt2gene | Not-determined |
| S17_vs_k141_52480\|\|lt2gene | Low-quality |
| S17_vs_k141_30480\|\|lt2gene | Low-quality |
| S17_vs_k141_8114\|\|lt2gene | Low-quality |
| S17_vs_k141_37855\|\|lt2gene | Low-quality |
| S17_vs_k141_141178\|\|lt2gene | Low-quality |
| S17_vs_k141_163278\|\|lt2gene | Low-quality |
| S17_vs_k141_163292\|\|lt2gene | Low-quality |
| S17_vs_k141_89284\|\|lt2gene | Low-quality |
| S17_vs_k141_81744\|\|lt2gene | Low-quality |
| S17_vs_k141_81748\|\|lt2gene | Low-quality |
| S17_vs_k141_141211\|\|lt2gene | Low-quality |
| S17_vs_k141_155916\|\|lt2gene | Low-quality |
| S17_vs_k141_141217\|\|lt2gene | Low-quality |
| S17_vs_k141_59854\|\|lt2gene | Low-quality |
| S17_vs_k141_155938\|\|lt2gene | Low-quality |
| S17_vs_k141_8160\|\|lt2gene | Low-quality |
| S17_vs_k141_37906\|\|lt2gene | Low-quality |
| S17_vs_k141_118964\|\|lt2gene | Low-quality |
| S17_vs_k141_89304\|\|lt2gene | Not-determined |
| S17_vs_k141_118972\|\|lt2gene | Low-quality |
| S17_vs_k141_170700\|\|lt2gene | Low-quality |
| S17_vs_k141_22725\|\|lt2gene | Not-determined |
| S17_vs_k141_45087\|\|lt2gene | Not-determined |
| S17_vs_k141_89337\|\|lt2gene | Low-quality |
| S17_vs_k141_119014\|\|lt2gene | Low-quality |
| S17_vs_k141_89349\|\|lt2gene | Low-quality |
| S17_vs_k141_170726\|\|lt2gene | Not-determined |
| S17_vs_k141_170728\|\|lt2gene | Not-determined |
| S17_vs_k141_156020\|\|lt2gene | Low-quality |
| S17_vs_k141_8228\|\|lt2gene | Low-quality |
| S17_vs_k141_133697\|\|lt2gene | Low-quality |
| S17_vs_k141_133705\|\|lt2gene | Low-quality |

S17_vs_k141_111541||lt2gene Not-determined
S17_vs_k141_133714||lt2gene Low-quality
S17_vs_k141_37973||lt2gene Low-quality
S17_vs_k141_8242||lt2gene Low-quality
S17_vs_k141_774||lt2gene Low-quality
S17_vs_k141_52658||lt2gene Low-quality
S17_vs_k141_45113||lt2gene Not-determined
S17_vs_k141_67186||lt2gene Low-quality
S17_vs_k141_8258||lt2gene Low-quality
S17_vs_k141_156067||lt2gene Not-determined
S17_vs_k141_170803||lt2gene Not-determined
S17_vs_k141_45127||lt2gene Low-quality
S17_vs_k141_96807||lt2gene Low-quality
S17_vs_k141_59913||lt2gene Not-determined
S17_vs_k141_119076||lt2gene Low-quality
S17_vs_k141_52697||lt2gene Low-quality
S17_vs_k141_59941||lt2gene Low-quality
S17_vs_k141_141291||lt2gene Not-determined
S17_vs_k141_860||lt2gene Low-quality
S17_vs_k141_74856||lt2gene Not-determined
S17_vs_k141_864||lt2gene Low-quality
S17_vs_k141_15444||lt2gene Low-quality
S17_vs_k141_96898||lt2gene Not-determined
S17_vs_k141_163481||lt2gene Low-quality
S17_vs_k141_104627||lt2gene Low-quality
S17_vs_k141_45254||lt2gene Low-quality
S17_vs_k141_156183||lt2gene Low-quality
S17_vs_k141_148868||lt2gene Not-determined
S17_vs_k141_141322||lt2gene Not-determined
S17_vs_k141_22923||lt2gene Low-quality
S17_vs_k141_126370||lt2gene Not-determined
S17_vs_k141_119177||lt2gene Low-quality
S17_vs_k141_38182||lt2gene Low-quality
S17_vs_k141_163521||lt2gene Low-quality
S17_vs_k141_119187||lt2gene Not-determined
S17_vs_k141_30643||lt2gene Low-quality
S17_vs_k141_8422||lt2gene Low-quality
S17_vs_k141_976||lt2gene Low-quality
S17_vs_k141_979||lt2gene Low-quality
S17_vs_k141_156290||lt2gene Low-quality
S17_vs_k141_119221||lt2gene Low-quality
S17_vs_k141_119231||lt2gene Low-quality
S17_vs_k141_30658||lt2gene Low-quality
S17_vs_k141_30663||lt2gene Low-quality
S17_vs_k141_156313||lt2gene Low-quality
S17_vs_k141_45342||lt2gene Low-quality
S17_vs_k141_126423||lt2gene Not-determined
S17_vs_k141_133863||lt2gene Not-determined
S17_vs_k141_133868||lt2gene Not-determined
S17_vs_k141_89542||lt2gene Not-determined
S17_vs_k141_8456||lt2gene Not-determined
S17_vs_k141_67420||lt2gene Not-determined
S17_vs_k141_156350||lt2gene Low-quality
S17_vs_k141_1031||lt2gene Low-quality
S17_vs_k141_15619||lt2gene Low-quality
S17_vs_k141_163546||lt2gene Low-quality
S17_vs_k141_45382||lt2gene Low-quality
S17_vs_k141_111740||lt2gene Not-determined
S17_vs_k141_171048||lt2gene Not-determined
S17_vs_k141_96980||lt2gene Not-determined
S17_vs_k141_171051||lt2gene Low-quality
S17_vs_k141_171056||lt2gene Low-quality
S17_vs_k141_111764||lt2gene Low-quality
S17_vs_k141_89600||lt2gene Low-quality
S17_vs_k141_111772||lt2gene Low-quality
S17_vs_k141_74938||lt2gene Not-determined
S17_vs_k141_67450||lt2gene Low-quality
S17_vs_k141_45395||lt2gene Not-determined
S17_vs_k141_52929||lt2gene Low-quality
S17_vs_k141_45402||lt2gene Low-quality
S17_vs_k141_52939||lt2gene Low-quality
S17_vs_k141_163607||lt2gene Low-quality
S17_vs_k141_156397||lt2gene Low-quality
S17_vs_k141_97017||lt2gene Not-determined
S17_vs_k141_141432||lt2gene Low-quality
S17_vs_k141_89647||lt2gene Not-determined
S17_vs_k141_89649||lt2gene Not-determined
S17_vs_k141_156409||lt2gene Not-determined
S17_vs_k141_149069||lt2gene Low-quality
S17_vs_k141_38291||lt2gene Low-quality
S17_vs_k141_163627||lt2gene Low-quality
S17_vs_k141_119310||lt2gene Not-determined
S17_vs_k141_149090||lt2gene Not-determined
S17_vs_k141_156431||lt2gene Low-quality
S17_vs_k141_8560||lt2gene Not-determined
S17_vs_k141_149104||lt2gene Not-determined
S17_vs_k141_23032||lt2gene Not-determined
S17_vs_k141_156469||lt2gene Low-quality
S17_vs_k141_119364||lt2gene Not-determined
S17_vs_k141_52988||lt2gene Low-quality
S17_vs_k141_53002||lt2gene Low-quality
S17_vs_k141_53008||lt2gene Low-quality
S17_vs_k141_156491||lt2gene Low-quality
S17_vs_k141_134003||lt2gene Low-quality
S17_vs_k141_156530||lt2gene Low-quality
S17_vs_k141_60309||lt2gene Not-determined
S17_vs_k141_141593||lt2gene Low-quality
S17_vs_k141_8685||lt2gene Not-determined
S17_vs_k141_171213||lt2gene Low-quality
S17_vs_k141_163711||lt2gene Not-determined
S17_vs_k141_97164||lt2gene Low-quality
S17_vs_k141_53095||lt2gene Low-quality
S17_vs_k141_149204||lt2gene Low-quality
S17_vs_k141_171221||lt2gene Low-quality
S17_vs_k141_163763||lt2gene Low-quality
S17_vs_k141_30901||lt2gene Low-quality
S17_vs_k141_75003||lt2gene Low-quality
S17_vs_k141_119459||lt2gene Not-determined
S17_vs_k141_45670||lt2gene Low-quality
S17_vs_k141_45675||lt2gene Low-quality
S17_vs_k141_163784||lt2gene Low-quality
S17_vs_k141_8718||lt2gene Not-determined
S17_vs_k141_171285||lt2gene Not-determined
S17_vs_k141_171289||lt2gene Low-quality
S17_vs_k141_67716||lt2gene Not-determined
S17_vs_k141_38488||lt2gene Low-quality
S17_vs_k141_105110||lt2gene Not-determined
S17_vs_k141_53225||lt2gene Low-quality
S17_vs_k141_15751||lt2gene Low-quality
S17_vs_k141_75086||lt2gene Low-quality
S17_vs_k141_45691||lt2gene Low-quality
S17_vs_k141_45694||lt2gene Low-quality
S17_vs_k141_30976||lt2gene Not-determined
S17_vs_k141_119537||lt2gene Low-quality
S17_vs_k141_75095||lt2gene Low-quality
S17_vs_k141_149342||lt2gene Not-determined
S17_vs_k141_1229||lt2gene Low-quality
S17_vs_k141_8798||lt2gene Low-quality
S17_vs_k141_23219||lt2gene Low-quality
S17_vs_k141_97308||lt2gene Not-determined
S17_vs_k141_67793||lt2gene Low-quality
S17_vs_k141_105151||lt2gene Low-quality
S17_vs_k141_97325||lt2gene Not-determined
S17_vs_k141_149395||lt2gene Low-quality
S17_vs_k141_8840||lt2gene Low-quality
S17_vs_k141_1265||lt2gene Low-quality
S17_vs_k141_141717||lt2gene Low-quality
S17_vs_k141_82191||lt2gene Low-quality
S17_vs_k141_163872||lt2gene Low-quality
S17_vs_k141_38585||lt2gene Low-quality
S17_vs_k141_105194||lt2gene Low-quality
S17_vs_k141_90002||lt2gene Low-quality
S17_vs_k141_23269||lt2gene Low-quality
S17_vs_k141_15817||lt2gene Low-quality
S17_vs_k141_82228||lt2gene Low-quality
S17_vs_k141_156665||lt2gene Low-quality
S17_vs_k141_171407||lt2gene Low-quality
S17_vs_k141_75155||lt2gene Not-determined
S17_vs_k141_8911||lt2gene Not-determined
S17_vs_k141_53352||lt2gene Not-determined
S17_vs_k141_126564||lt2gene Low-quality
S17_vs_k141_141782||lt2gene Not-determined
S17_vs_k141_171455||lt2gene Not-determined
S17_vs_k141_8961||lt2gene Low-quality
S17_vs_k141_8963||lt2gene Low-quality
S17_vs_k141_75201||lt2gene Low-quality
S17_vs_k141_163897||lt2gene Low-quality
S17_vs_k141_97478||lt2gene Low-quality
S17_vs_k141_97481||lt2gene Low-quality
S17_vs_k141_75211||lt2gene Low-quality
S17_vs_k141_112078||lt2gene Not-determined
S17_vs_k141_9003||lt2gene Low-quality
S17_vs_k141_156761||lt2gene Low-quality
S17_vs_k141_97499||lt2gene Not-determined
S17_vs_k141_38748||lt2gene Low-quality
S17_vs_k141_53463||lt2gene Low-quality
S17_vs_k141_9014||lt2gene Low-quality
S17_vs_k141_171510||lt2gene Low-quality
S17_vs_k141_15827||lt2gene Not-determined
S17_vs_k141_126653||lt2gene Low-quality
S17_vs_k141_149569||lt2gene Not-determined
S17_vs_k141_45955||lt2gene Low-quality
S17_vs_k141_105300||lt2gene Low-quality
S17_vs_k141_134314||lt2gene Low-quality
S17_vs_k141_23394||lt2gene Not-determined
S17_vs_k141_23395||lt2gene Not-determined
S17_vs_k141_45967||lt2gene Low-quality
S17_vs_k141_90124||lt2gene Low-quality
S17_vs_k141_9084||lt2gene Not-determined
S17_vs_k141_141931||lt2gene Low-quality
S17_vs_k141_15905||lt2gene Low-quality
S17_vs_k141_164019||lt2gene Not-determined
S17_vs_k141_45990||lt2gene Low-quality
S17_vs_k141_164045||lt2gene Not-determined
S17_vs_k141_9131||lt2gene Low-quality
S17_vs_k141_164065||lt2gene Not-determined
S17_vs_k141_46020||lt2gene Low-quality
S17_vs_k141_134351||lt2gene Low-quality
S17_vs_k141_38803||lt2gene Low-quality
S17_vs_k141_134357||lt2gene Low-quality
S17_vs_k141_126731||lt2gene Low-quality
S17_vs_k141_105405||lt2gene Low-quality
S17_vs_k141_164097||lt2gene Low-quality
S17_vs_k141_31268||lt2gene Low-quality
S17_vs_k141_105414||lt2gene Low-quality
S17_vs_k141_1523||lt2gene Low-quality
S17_vs_k141_9177||lt2gene Not-determined
S17_vs_k141_9192||lt2gene Low-quality
S17_vs_k141_119790||lt2gene Low-quality
S17_vs_k141_119796||lt2gene Low-quality
S17_vs_k141_156930||lt2gene Not-determined
S17_vs_k141_15962||lt2gene Low-quality
S17_vs_k141_149651||lt2gene Not-determined
S17_vs_k141_156963||lt2gene Low-quality
S17_vs_k141_38849||lt2gene Low-quality
S17_vs_k141_82471||lt2gene Low-quality
S17_vs_k141_38855||lt2gene Not-determined
S17_vs_k141_82475||lt2gene Not-determined
S17_vs_k141_149694||lt2gene Not-determined
S17_vs_k141_16012||lt2gene Low-quality
S17_vs_k141_97761||lt2gene Low-quality
S17_vs_k141_126842||lt2gene Low-quality
S17_vs_k141_53693||lt2gene Low-quality
S17_vs_k141_75338||lt2gene Low-quality
S17_vs_k141_75339||lt2gene Low-quality
S17_vs_k141_53705||lt2gene Low-quality
S17_vs_k141_46181||lt2gene Low-quality
S17_vs_k141_119905||lt2gene Low-quality
S17_vs_k141_16056||lt2gene Low-quality
S17_vs_k141_119911||lt2gene Low-quality
S17_vs_k141_142124||lt2gene Low-quality
S17_vs_k141_16069||lt2gene Low-quality
S17_vs_k141_112303||lt2gene Not-determined
S17_vs_k141_1655||lt2gene Not-determined
S17_vs_k141_1659||lt2gene Low-quality
S17_vs_k141_119918||lt2gene Low-quality
S17_vs_k141_38908||lt2gene Low-quality
S17_vs_k141_157035||lt2gene Not-determined
S17_vs_k141_97845||lt2gene Low-quality
S17_vs_k141_9270||lt2gene Low-quality
S17_vs_k141_82575||lt2gene Low-quality
S17_vs_k141_142174||lt2gene Low-quality
S17_vs_k141_142181||lt2gene Low-quality
S17_vs_k141_157074||lt2gene Not-determined
S17_vs_k141_119962||lt2gene Low-quality
S17_vs_k141_75432||lt2gene Not-determined
S17_vs_k141_75433||lt2gene Low-quality
S17_vs_k141_149819||lt2gene Not-determined
S17_vs_k141_23651||lt2gene Low-quality
S17_vs_k141_16095||lt2gene Low-quality
S17_vs_k141_142206||lt2gene Low-quality
S17_vs_k141_105642||lt2gene Not-determined
S17_vs_k141_126955||lt2gene Low-quality
S17_vs_k141_120010||lt2gene Low-quality
S17_vs_k141_120017||lt2gene Low-quality
S17_vs_k141_120018||lt2gene Low-quality
S17_vs_k141_134600||lt2gene Low-quality
S17_vs_k141_120035||lt2gene Low-quality
S17_vs_k141_157138||lt2gene Low-quality
S17_vs_k141_149874||lt2gene Low-quality
S17_vs_k141_16171||lt2gene Low-quality
S17_vs_k141_9352||lt2gene Low-quality
S17_vs_k141_82635||lt2gene Not-determined
S17_vs_k141_157152||lt2gene Not-determined
S17_vs_k141_120062||lt2gene Low-quality
S17_vs_k141_23766||lt2gene Low-quality
S17_vs_k141_31495||lt2gene Not-determined
S17_vs_k141_171577||lt2gene Low-quality
S17_vs_k141_31506||lt2gene Low-quality
S17_vs_k141_60815||lt2gene Low-quality
S17_vs_k141_9420||lt2gene Low-quality
S17_vs_k141_31529||lt2gene Low-quality
S17_vs_k141_31531||lt2gene Not-determined
S17_vs_k141_53904||lt2gene Not-determined
S17_vs_k141_171594||lt2gene Not-determined
S17_vs_k141_23819||lt2gene Low-quality
S17_vs_k141_23820||lt2gene Not-determined
S17_vs_k141_164255||lt2gene Low-quality
S17_vs_k141_127077||lt2gene Not-determined
S17_vs_k141_97994||lt2gene Not-determined
S17_vs_k141_82722||lt2gene Low-quality
S17_vs_k141_39103||lt2gene Not-determined
S17_vs_k141_98011||lt2gene Low-quality
S17_vs_k141_23844||lt2gene Low-quality
S17_vs_k141_53967||lt2gene Low-quality
S17_vs_k141_39121||lt2gene Not-determined
S17_vs_k141_127118||lt2gene Low-quality
S17_vs_k141_68298||lt2gene Low-quality
S17_vs_k141_68300||lt2gene Low-quality
S17_vs_k141_68301||lt2gene Low-quality
S17_vs_k141_68306||lt2gene Not-determined
S17_vs_k141_39135||lt2gene Low-quality
S17_vs_k141_112449||lt2gene Low-quality
S17_vs_k141_149999||lt2gene Not-determined
S17_vs_k141_98050||lt2gene Not-determined
S17_vs_k141_142357||lt2gene Low-quality
S17_vs_k141_134780||lt2gene Low-quality
S17_vs_k141_31590||lt2gene Not-determined
S17_vs_k141_46432||lt2gene Low-quality
S17_vs_k141_112470||lt2gene Not-determined
S17_vs_k141_90331||lt2gene Low-quality
S17_vs_k141_98058||lt2gene Low-quality
S17_vs_k141_150042||lt2gene Low-quality
S17_vs_k141_105824||lt2gene Low-quality
S17_vs_k141_75543||lt2gene Low-quality
S17_vs_k141_1849||lt2gene Not-determined
S17_vs_k141_68345||lt2gene Low-quality
S17_vs_k141_82762||lt2gene Low-quality
S17_vs_k141_68354||lt2gene Low-quality
S17_vs_k141_1865||lt2gene Low-quality
S17_vs_k141_164337||lt2gene Low-quality
S17_vs_k141_112487||lt2gene Low-quality
S17_vs_k141_16340||lt2gene Not-determined
S17_vs_k141_127188||lt2gene Low-quality
S17_vs_k141_164363||lt2gene Low-quality
S17_vs_k141_60941||lt2gene Not-determined
S17_vs_k141_75588||lt2gene Not-determined
S17_vs_k141_39227||lt2gene Low-quality
S17_vs_k141_134849||lt2gene Low-quality
S17_vs_k141_112540||lt2gene Low-quality
S17_vs_k141_134862||lt2gene Low-quality
S17_vs_k141_112559||lt2gene Low-quality
S17_vs_k141_82812||lt2gene Low-quality
S17_vs_k141_1957||lt2gene Low-quality
S17_vs_k141_1960||lt2gene Low-quality
S17_vs_k141_82835||lt2gene Not-determined
S17_vs_k141_75626||lt2gene Low-quality
S17_vs_k141_54083||lt2gene Low-quality
S17_vs_k141_150153||lt2gene Low-quality
S17_vs_k141_68485||lt2gene Not-determined
S17_vs_k141_142506||lt2gene Low-quality
S17_vs_k141_46543||lt2gene Low-quality
S17_vs_k141_120263||lt2gene Low-quality
S17_vs_k141_150180||lt2gene Low-quality
S17_vs_k141_9478||lt2gene Low-quality
S17_vs_k141_1974||lt2gene Low-quality
S17_vs_k141_9486||lt2gene Low-quality
S17_vs_k141_98205||lt2gene Low-quality
S17_vs_k141_82875||lt2gene Low-quality
S17_vs_k141_98213||lt2gene Low-quality
S17_vs_k141_98243||lt2gene Low-quality
S17_vs_k141_127284||lt2gene Low-quality
S17_vs_k141_31779||lt2gene Low-quality
S17_vs_k141_134997||lt2gene Not-determined
S17_vs_k141_9551||lt2gene Low-quality
S17_vs_k141_142596||lt2gene Not-determined
S17_vs_k141_54159||lt2gene Low-quality
S17_vs_k141_16455||lt2gene Not-determined
S17_vs_k141_54165||lt2gene Not-determined
S17_vs_k141_75746||lt2gene Not-determined
S17_vs_k141_106014||lt2gene Low-quality
S17_vs_k141_39437||lt2gene Low-quality
S17_vs_k141_2052||lt2gene Not-determined
S17_vs_k141_54195||lt2gene Not-determined
S17_vs_k141_127359||lt2gene Low-quality
S17_vs_k141_46595||lt2gene Low-quality
S17_vs_k141_68615||lt2gene Low-quality
S17_vs_k141_68626||lt2gene Low-quality
S17_vs_k141_112717||lt2gene Low-quality
S17_vs_k141_46610||lt2gene Low-quality
S17_vs_k141_112737||lt2gene Low-quality
S17_vs_k141_112740||lt2gene Low-quality
S17_vs_k141_150355||lt2gene Low-quality
S17_vs_k141_112749||lt2gene Low-quality
S17_vs_k141_112750||lt2gene Low-quality
S17_vs_k141_39490||lt2gene Low-quality
S17_vs_k141_112754||lt2gene Low-quality
S17_vs_k141_142676||lt2gene Not-determined
S17_vs_k141_90536||lt2gene Low-quality
S17_vs_k141_90540||lt2gene Low-quality
S17_vs_k141_142698||lt2gene Low-quality
S17_vs_k141_16586||lt2gene Low-quality
S17_vs_k141_142724||lt2gene Low-quality
S17_vs_k141_157514||lt2gene Low-quality
S17_vs_k141_150404||lt2gene Low-quality
S17_vs_k141_157520||lt2gene Low-quality
S17_vs_k141_2115||lt2gene Not-determined
S17_vs_k141_120433||lt2gene Low-quality
S17_vs_k141_164695||lt2gene Not-determined
S17_vs_k141_75900||lt2gene Low-quality
S17_vs_k141_142758||lt2gene Low-quality
S17_vs_k141_90601||lt2gene Low-quality
S17_vs_k141_75914||lt2gene Low-quality
S17_vs_k141_112838||lt2gene Low-quality
S17_vs_k141_150446||lt2gene Not-determined
S17_vs_k141_46729||lt2gene Low-quality
S17_vs_k141_157576||lt2gene Low-quality
S17_vs_k141_112858||lt2gene Not-determined
S17_vs_k141_31902||lt2gene Low-quality
S17_vs_k141_142774||lt2gene Not-determined
S17_vs_k141_127486||lt2gene Low-quality
S17_vs_k141_9832||lt2gene Low-quality
S17_vs_k141_142790||lt2gene Low-quality
S17_vs_k141_157605||lt2gene Low-quality
S17_vs_k141_39590||lt2gene Not-determined
S17_vs_k141_164750||lt2gene Low-quality
S17_vs_k141_127499||lt2gene Low-quality
S17_vs_k141_9849||lt2gene Low-quality
S17_vs_k141_90671||lt2gene Low-quality
S17_vs_k141_150486||lt2gene Low-quality
S17_vs_k141_76017||lt2gene Low-quality
S17_vs_k141_135125||lt2gene Low-quality
S17_vs_k141_135129||lt2gene Low-quality
S17_vs_k141_171985||lt2gene Not-determined
S17_vs_k141_164801||lt2gene Low-quality
S17_vs_k141_164802||lt2gene Low-quality
S17_vs_k141_90709||lt2gene Low-quality
S17_vs_k141_31979||lt2gene Low-quality
S17_vs_k141_54336||lt2gene Low-quality
S17_vs_k141_32012||lt2gene Low-quality
S17_vs_k141_120598||lt2gene Low-quality
S17_vs_k141_157724||lt2gene Not-determined
S17_vs_k141_39702||lt2gene Low-quality
S17_vs_k141_142884||lt2gene Low-quality
S17_vs_k141_39712||lt2gene Low-quality
S17_vs_k141_172012||lt2gene Low-quality
S17_vs_k141_157747||lt2gene Low-quality
S17_vs_k141_61208||lt2gene Low-quality
S17_vs_k141_46810||lt2gene Low-quality
S17_vs_k141_98555||lt2gene Not-determined
S17_vs_k141_46816||lt2gene Not-determined
S17_vs_k141_68925||lt2gene Low-quality
S17_vs_k141_39760||lt2gene Low-quality
S17_vs_k141_90773||lt2gene Low-quality
S17_vs_k141_150599||lt2gene Low-quality
S17_vs_k141_24336||lt2gene Low-quality
S17_vs_k141_83116||lt2gene Low-quality
S17_vs_k141_113047||lt2gene Low-quality
S17_vs_k141_46868||lt2gene Low-quality
S17_vs_k141_135220||lt2gene Not-determined
S17_vs_k141_61269||lt2gene Low-quality
S17_vs_k141_46894||lt2gene Low-quality
S17_vs_k141_16792||lt2gene Low-quality
S17_vs_k141_143010||lt2gene Low-quality
S17_vs_k141_157846||lt2gene Low-quality
S17_vs_k141_32120||lt2gene Low-quality
S17_vs_k141_127636||lt2gene Not-determined
S17_vs_k141_90848||lt2gene Not-determined
S17_vs_k141_32142||lt2gene Low-quality
S17_vs_k141_68990||lt2gene Not-determined
S17_vs_k141_2281||lt2gene Low-quality
S17_vs_k141_39830||lt2gene Low-quality
S17_vs_k141_83175||lt2gene Low-quality
S17_vs_k141_113106||lt2gene Not-determined
S17_vs_k141_39847||lt2gene Low-quality
S17_vs_k141_98649||lt2gene Low-quality
S17_vs_k141_135255||lt2gene Not-determined
S17_vs_k141_150735||lt2gene Not-determined
S17_vs_k141_39856||lt2gene Low-quality
S17_vs_k141_9993||lt2gene Not-determined
S17_vs_k141_76209||lt2gene Low-quality
S17_vs_k141_76214||lt2gene Low-quality
S17_vs_k141_16850||lt2gene Low-quality
S17_vs_k141_39912||lt2gene Not-determined
S17_vs_k141_76227||lt2gene Low-quality
S17_vs_k141_135321||lt2gene Not-determined
S17_vs_k141_32221||lt2gene Not-determined
S17_vs_k141_143112||lt2gene Not-determined
S17_vs_k141_172126||lt2gene Low-quality
S17_vs_k141_135328||lt2gene Low-quality
S17_vs_k141_157889||lt2gene Low-quality
S17_vs_k141_164931||lt2gene Low-quality
S17_vs_k141_32284||lt2gene Low-quality
S17_vs_k141_127749||lt2gene Low-quality
S17_vs_k141_90934||lt2gene Low-quality
S17_vs_k141_98745||lt2gene Not-determined
S17_vs_k141_2354||lt2gene Low-quality
S17_vs_k141_143175||lt2gene Low-quality
S17_vs_k141_61403||lt2gene Low-quality
S17_vs_k141_83307||lt2gene Not-determined
S17_vs_k141_90979||lt2gene Low-quality
S17_vs_k141_150835||lt2gene Not-determined
S17_vs_k141_143193||lt2gene Low-quality
S17_vs_k141_172190||lt2gene Low-quality
S17_vs_k141_32348||lt2gene Low-quality
S17_vs_k141_47058||lt2gene Low-quality
S17_vs_k141_106538||lt2gene Low-quality
S17_vs_k141_143212||lt2gene Low-quality
S17_vs_k141_39993||lt2gene Low-quality
S17_vs_k141_150842||lt2gene Low-quality
S17_vs_k141_150847||lt2gene Not-determined
S17_vs_k141_91052||lt2gene Not-determined
S17_vs_k141_91071||lt2gene Not-determined
S17_vs_k141_106591||lt2gene Not-determined
S17_vs_k141_76360||lt2gene Low-quality
S17_vs_k141_120819||lt2gene Not-determined
S17_vs_k141_91084||lt2gene Low-quality
S17_vs_k141_172233||lt2gene Low-quality
S17_vs_k141_32400||lt2gene Low-quality
S17_vs_k141_16962||lt2gene Low-quality
S17_vs_k141_150884||lt2gene Low-quality
S17_vs_k141_76369||lt2gene Low-quality
S17_vs_k141_69222||lt2gene Not-determined
S17_vs_k141_91114||lt2gene Not-determined
S17_vs_k141_98892||lt2gene Low-quality
S17_vs_k141_127862||lt2gene Not-determined
S17_vs_k141_91129||lt2gene Low-quality
S17_vs_k141_24533||lt2gene Low-quality
S17_vs_k141_47181||lt2gene Not-determined
S17_vs_k141_113195||lt2gene Low-quality
S17_vs_k141_172297||lt2gene Low-quality
S17_vs_k141_127881||lt2gene Not-determined
S17_vs_k141_172298||lt2gene Low-quality
S17_vs_k141_91144||lt2gene Low-quality
S17_vs_k141_106654||lt2gene Low-quality
S17_vs_k141_69239||lt2gene Low-quality
S17_vs_k141_135511||lt2gene Low-quality
S17_vs_k141_120908||lt2gene Not-determined
S17_vs_k141_32489||lt2gene Low-quality
S17_vs_k141_91187||lt2gene Not-determined
S17_vs_k141_32511||lt2gene Low-quality
S17_vs_k141_150985||lt2gene Not-determined
S17_vs_k141_172352||lt2gene Not-determined
S17_vs_k141_165096||lt2gene Low-quality
S17_vs_k141_61536||lt2gene Low-quality
S17_vs_k141_113276||lt2gene Low-quality
S17_vs_k141_76539||lt2gene Low-quality
S17_vs_k141_47300||lt2gene Low-quality
S17_vs_k141_76545||lt2gene Low-quality
S17_vs_k141_106711||lt2gene Low-quality
S17_vs_k141_128004||lt2gene Low-quality
S17_vs_k141_83438||lt2gene Low-quality
S17_vs_k141_47323||lt2gene Low-quality
S17_vs_k141_61572||lt2gene Low-quality
S17_vs_k141_10248||lt2gene Low-quality
S17_vs_k141_98986||lt2gene Not-determined
S17_vs_k141_135589||lt2gene Low-quality
S17_vs_k141_32604||lt2gene Low-quality
S17_vs_k141_2615||lt2gene Low-quality
S17_vs_k141_106745||lt2gene Not-determined
S17_vs_k141_2618||lt2gene Low-quality
S17_vs_k141_135593||lt2gene Not-determined
S17_vs_k141_47352||lt2gene Low-quality
S17_vs_k141_24656||lt2gene Low-quality
S17_vs_k141_17183||lt2gene Not-determined
S17_vs_k141_99026||lt2gene Low-quality
S17_vs_k141_32643||lt2gene Low-quality
S17_vs_k141_151045||lt2gene Low-quality
S17_vs_k141_69390||lt2gene Low-quality
S17_vs_k141_151050||lt2gene Not-determined
S17_vs_k141_54654||lt2gene Not-determined
S17_vs_k141_135633||lt2gene Low-quality
S17_vs_k141_61612||lt2gene Low-quality
S17_vs_k141_32670||lt2gene Not-determined
S17_vs_k141_172488||lt2gene Not-determined
S17_vs_k141_54670||lt2gene Low-quality
S17_vs_k141_47413||lt2gene Low-quality
S17_vs_k141_17211||lt2gene Not-determined
S17_vs_k141_128092||lt2gene Low-quality
S17_vs_k141_17220||lt2gene Low-quality
S17_vs_k141_24727||lt2gene Low-quality
S17_vs_k141_61659||lt2gene Not-determined
S17_vs_k141_91335||lt2gene Low-quality
S17_vs_k141_24734||lt2gene Low-quality
S17_vs_k141_143471||lt2gene Low-quality
S17_vs_k141_172534||lt2gene Not-determined
S17_vs_k141_158208||lt2gene Low-quality
S17_vs_k141_121043||lt2gene Low-quality
S17_vs_k141_99138||lt2gene Low-quality
S17_vs_k141_76718||lt2gene Low-quality
S17_vs_k141_172559||lt2gene Not-determined
S17_vs_k141_83522||lt2gene Low-quality
S17_vs_k141_76732||lt2gene Low-quality
S17_vs_k141_106878||lt2gene Not-determined
S17_vs_k141_83530||lt2gene Low-quality
S17_vs_k141_10328||lt2gene Low-quality
S17_vs_k141_99156||lt2gene Low-quality
S17_vs_k141_128131||lt2gene Not-determined
S17_vs_k141_83553||lt2gene Low-quality
S17_vs_k141_106896||lt2gene Low-quality
S17_vs_k141_17306||lt2gene Not-determined
S17_vs_k141_83564||lt2gene Low-quality
S17_vs_k141_165341||lt2gene Low-quality
S17_vs_k141_143538||lt2gene Not-determined
S17_vs_k141_135747||lt2gene Not-determined
S17_vs_k141_61740||lt2gene Low-quality
S17_vs_k141_54740||lt2gene Low-quality
S17_vs_k141_128167||lt2gene Not-determined
S17_vs_k141_106953||lt2gene Low-quality
S17_vs_k141_32705||lt2gene Not-determined
S17_vs_k141_61773||lt2gene Low-quality
S17_vs_k141_135785||lt2gene Not-determined
S17_vs_k141_10382||lt2gene Not-determined
S17_vs_k141_17339||lt2gene Low-quality
S17_vs_k141_158230||lt2gene Low-quality
S17_vs_k141_151214||lt2gene Low-quality
S17_vs_k141_10398||lt2gene Low-quality
S17_vs_k141_69593||lt2gene Not-determined
S17_vs_k141_47605||lt2gene Not-determined
S17_vs_k141_10424||lt2gene Not-determined
S17_vs_k141_47609||lt2gene Not-determined
S17_vs_k141_172682||lt2gene Low-quality
S17_vs_k141_17362||lt2gene Low-quality
S17_vs_k141_91462||lt2gene Low-quality
S17_vs_k141_54826||lt2gene Low-quality
S17_vs_k141_113557||lt2gene Not-determined
S17_vs_k141_69638||lt2gene Low-quality
S17_vs_k141_91492||lt2gene Low-quality
S17_vs_k141_165458||lt2gene Low-quality

| | |
|---|---|
| S17_vs_k141_158313||lt2gene | Not-determined |
| S17_vs_k141_165472||lt2gene | Low-quality |
| S17_vs_k141_121135||lt2gene | Low-quality |
| S17_vs_k141_17423||lt2gene | Low-quality |
| S17_vs_k141_172720||lt2gene | Low-quality |
| S17_vs_k141_151283||lt2gene | Not-determined |
| S17_vs_k141_99344||lt2gene | Not-determined |
| S17_vs_k141_61857||lt2gene | Low-quality |
| S17_vs_k141_61859||lt2gene | Not-determined |
| S17_vs_k141_54869||lt2gene | Low-quality |
| S17_vs_k141_32763||lt2gene | Low-quality |
| S17_vs_k141_165500||lt2gene | Low-quality |
| S17_vs_k141_121159||lt2gene | Not-determined |
| S17_vs_k141_10512||lt2gene | Low-quality |
| S17_vs_k141_10513||lt2gene | Low-quality |
| S17_vs_k141_143688||lt2gene | Low-quality |
| S17_vs_k141_151306||lt2gene | Low-quality |
| S17_vs_k141_151308||lt2gene | Low-quality |
| S17_vs_k141_17464||lt2gene | Low-quality |
| S17_vs_k141_151311||lt2gene | Low-quality |
| S17_vs_k141_10530||lt2gene | Not-determined |
| S17_vs_k141_121173||lt2gene | Not-determined |
| S17_vs_k141_158349||lt2gene | Not-determined |
| S17_vs_k141_99391||lt2gene | Low-quality |
| S17_vs_k141_99395||lt2gene | Low-quality |
| S17_vs_k141_143713||lt2gene | Not-determined |
| S17_vs_k141_91571||lt2gene | Not-determined |
| S17_vs_k141_47705||lt2gene | Not-determined |
| S17_vs_k141_40546||lt2gene | Not-determined |
| S17_vs_k141_128332||lt2gene | Not-determined |
| S17_vs_k141_61920||lt2gene | Not-determined |
| S17_vs_k141_76912||lt2gene | Low-quality |
| S17_vs_k141_54925||lt2gene | Not-determined |
| S17_vs_k141_107085||lt2gene | Low-quality |
| S17_vs_k141_151357||lt2gene | Low-quality |
| S17_vs_k141_143740||lt2gene | Low-quality |
| S17_vs_k141_10579||lt2gene | Not-determined |
| S17_vs_k141_32801||lt2gene | Low-quality |
| S17_vs_k141_99414||lt2gene | Low-quality |
| S17_vs_k141_10587||lt2gene | Low-quality |
| S17_vs_k141_40577||lt2gene | Low-quality |
| S17_vs_k141_17530||lt2gene | Not-determined |
| S17_vs_k141_128367||lt2gene | Low-quality |
| S17_vs_k141_128372||lt2gene | Low-quality |
| S17_vs_k141_32822||lt2gene | Low-quality |
| S17_vs_k141_128376||lt2gene | Not-determined |
| S17_vs_k141_69710||lt2gene | Low-quality |
| S17_vs_k141_172813||lt2gene | Not-determined |
| S17_vs_k141_17558||lt2gene | Low-quality |
| S17_vs_k141_17566||lt2gene | Low-quality |
| S17_vs_k141_10617||lt2gene | Low-quality |
| S17_vs_k141_17576||lt2gene | Low-quality |
| S17_vs_k141_17578||lt2gene | Low-quality |
| S17_vs_k141_32854||lt2gene | Low-quality |
| S17_vs_k141_17588||lt2gene | Low-quality |
| S17_vs_k141_107152||lt2gene | Low-quality |
| S17_vs_k141_128412||lt2gene | Low-quality |
| S17_vs_k141_83674||lt2gene | Low-quality |
| S17_vs_k141_40615||lt2gene | Low-quality |
| S17_vs_k141_128416||lt2gene | Low-quality |
| S17_vs_k141_128426||lt2gene | Not-determined |
| S17_vs_k141_107174||lt2gene | Low-quality |
| S17_vs_k141_17614||lt2gene | Low-quality |
| S17_vs_k141_151377||lt2gene | Not-determined |
| S17_vs_k141_158468||lt2gene | Not-determined |
| S17_vs_k141_121306||lt2gene | Low-quality |
| S17_vs_k141_32907||lt2gene | Low-quality |
| S17_vs_k141_40670||lt2gene | Low-quality |
| S17_vs_k141_77059||lt2gene | Low-quality |
| S17_vs_k141_62026||lt2gene | Low-quality |
| S17_vs_k141_77065||lt2gene | Low-quality |
| S17_vs_k141_77068||lt2gene | Low-quality |
| S17_vs_k141_62035||lt2gene | Low-quality |
| S17_vs_k141_69839||lt2gene | Low-quality |
| S17_vs_k141_62046||lt2gene | Low-quality |
| S17_vs_k141_17706||lt2gene | Low-quality |
| S17_vs_k141_121361||lt2gene | Low-quality |
| S17_vs_k141_47878||lt2gene | Low-quality |
| S17_vs_k141_32986||lt2gene | Low-quality |
| S17_vs_k141_3136||lt2gene | Low-quality |
| S17_vs_k141_77102||lt2gene | Low-quality |
| S17_vs_k141_83768||lt2gene | Low-quality |
| S17_vs_k141_55086||lt2gene | Low-quality |
| S17_vs_k141_47897||lt2gene | Low-quality |
| S17_vs_k141_143879||lt2gene | Not-determined |
| S17_vs_k141_33015||lt2gene | Low-quality |
| S17_vs_k141_143883||lt2gene | Low-quality |
| S17_vs_k141_121386||lt2gene | Low-quality |
| S17_vs_k141_91720||lt2gene | Low-quality |
| S17_vs_k141_33031||lt2gene | Not-determined |
| S17_vs_k141_107314||lt2gene | Low-quality |
| S17_vs_k141_151475||lt2gene | Low-quality |
| S17_vs_k141_91736||lt2gene | Low-quality |
| S17_vs_k141_128569||lt2gene | Not-determined |
| S17_vs_k141_69878||lt2gene | Low-quality |
| S17_vs_k141_113861||lt2gene | Low-quality |
| S17_vs_k141_172922||lt2gene | Low-quality |
| S17_vs_k141_33057||lt2gene | Not-determined |
| S17_vs_k141_121406||lt2gene | Low-quality |
| S17_vs_k141_172926||lt2gene | Low-quality |
| S17_vs_k141_113878||lt2gene | Not-determined |
| S17_vs_k141_165713||lt2gene | Low-quality |
| S17_vs_k141_158581||lt2gene | Low-quality |
| S17_vs_k141_143907||lt2gene | Low-quality |
| S17_vs_k141_128586||lt2gene | Low-quality |
| S17_vs_k141_151508||lt2gene | Low-quality |
| S17_vs_k141_55136||lt2gene | Low-quality |
| S17_vs_k141_121434||lt2gene | Low-quality |
| S17_vs_k141_33101||lt2gene | Low-quality |
| S17_vs_k141_62138||lt2gene | Not-determined |
| S17_vs_k141_47963||lt2gene | Low-quality |
| S17_vs_k141_136090||lt2gene | Low-quality |
| S17_vs_k141_69941||lt2gene | Low-quality |
| S17_vs_k141_143951||lt2gene | Low-quality |
| S17_vs_k141_136107||lt2gene | Not-determined |
| S17_vs_k141_91820||lt2gene | Low-quality |
| S17_vs_k141_136121||lt2gene | Not-determined |
| S17_vs_k141_55170||lt2gene | Low-quality |
| S17_vs_k141_62170||lt2gene | Low-quality |
| S17_vs_k141_77202||lt2gene | Not-determined |
| S17_vs_k141_83829||lt2gene | Low-quality |
| S17_vs_k141_83830||lt2gene | Low-quality |
| S17_vs_k141_165802||lt2gene | Low-quality |
| S17_vs_k141_25178||lt2gene | Low-quality |
| S17_vs_k141_107401||lt2gene | Not-determined |
| S17_vs_k141_69990||lt2gene | Low-quality |
| S17_vs_k141_99633||lt2gene | Low-quality |
| S17_vs_k141_3279||lt2gene | Low-quality |
| S17_vs_k141_40829||lt2gene | Low-quality |
| S17_vs_k141_55235||lt2gene | Low-quality |
| S17_vs_k141_48028||lt2gene | Not-determined |
| S17_vs_k141_17869||lt2gene | Low-quality |
| S17_vs_k141_48030||lt2gene | Not-determined |
| S17_vs_k141_40846||lt2gene | Not-determined |
| S17_vs_k141_62199||lt2gene | Low-quality |
| S17_vs_k141_128643||lt2gene | Not-determined |
| S17_vs_k141_151564||lt2gene | Not-determined |
| S17_vs_k141_151567||lt2gene | Low-quality |
| S17_vs_k141_3336||lt2gene | Not-determined |
| S17_vs_k141_114019||lt2gene | Low-quality |
| S17_vs_k141_114023||lt2gene | Low-quality |
| S17_vs_k141_70039||lt2gene | Low-quality |
| S17_vs_k141_83908||lt2gene | Low-quality |
| S17_vs_k141_158721||lt2gene | Low-quality |
| S17_vs_k141_10873||lt2gene | Low-quality |
| S17_vs_k141_70044||lt2gene | Not-determined |
| S17_vs_k141_99719||lt2gene | Not-determined |
| S17_vs_k141_99722||lt2gene | Low-quality |
| S17_vs_k141_158731||lt2gene | Not-determined |
| S17_vs_k141_10881||lt2gene | Low-quality |
| S17_vs_k141_136243||lt2gene | Not-determined |
| S17_vs_k141_173089||lt2gene | Not-determined |
| S17_vs_k141_91928||lt2gene | Low-quality |
| S17_vs_k141_77316||lt2gene | Low-quality |
| S17_vs_k141_40891||lt2gene | Not-determined |
| S17_vs_k141_144044||lt2gene | Low-quality |
| S17_vs_k141_10905||lt2gene | Low-quality |
| S17_vs_k141_33267||lt2gene | Not-determined |
| S17_vs_k141_165889||lt2gene | Not-determined |
| S17_vs_k141_128741||lt2gene | Not-determined |
| S17_vs_k141_3436||lt2gene | Low-quality |
| S17_vs_k141_17953||lt2gene | Not-determined |
| S17_vs_k141_70100||lt2gene | Low-quality |
| S17_vs_k141_128746||lt2gene | Not-determined |
| S17_vs_k141_128762||lt2gene | Low-quality |
| S17_vs_k141_128767||lt2gene | Not-determined |
| S17_vs_k141_3459||lt2gene | Not-determined |
| S17_vs_k141_83967||lt2gene | Not-determined |
| S17_vs_k141_128771||lt2gene | Low-quality |
| S17_vs_k141_173115||lt2gene | Low-quality |
| S17_vs_k141_158824||lt2gene | Low-quality |
| S17_vs_k141_25276||lt2gene | Not-determined |
| S17_vs_k141_151613||lt2gene | Low-quality |
| S17_vs_k141_55351||lt2gene | Low-quality |
| S17_vs_k141_91985||lt2gene | Low-quality |
| S17_vs_k141_173127||lt2gene | Low-quality |
| S17_vs_k141_17965||lt2gene | Low-quality |
| S17_vs_k141_107518||lt2gene | Low-quality |
| S17_vs_k141_107520||lt2gene | Not-determined |
| S17_vs_k141_107522||lt2gene | Not-determined |
| S17_vs_k141_40942||lt2gene | Not-determined |
| S17_vs_k141_151650||lt2gene | Low-quality |
| S17_vs_k141_136273||lt2gene | Low-quality |
| S17_vs_k141_165906||lt2gene | Low-quality |
| S17_vs_k141_136282||lt2gene | Low-quality |
| S17_vs_k141_173173||lt2gene | Low-quality |
| S17_vs_k141_40975||lt2gene | Low-quality |
| S17_vs_k141_40978||lt2gene | Low-quality |
| S17_vs_k141_121630||lt2gene | Not-determined |
| S17_vs_k141_158903||lt2gene | Low-quality |
| S17_vs_k141_114145||lt2gene | Low-quality |
| S17_vs_k141_77425||lt2gene | Not-determined |
| S17_vs_k141_77427||lt2gene | Low-quality |
| S17_vs_k141_114149||lt2gene | Low-quality |
| S17_vs_k141_128845||lt2gene | Low-quality |
| S17_vs_k141_84063||lt2gene | Low-quality |
| S17_vs_k141_99857||lt2gene | Low-quality |
| S17_vs_k141_70227||lt2gene | Not-determined |
| S17_vs_k141_114171||lt2gene | Low-quality |
| S17_vs_k141_173238||lt2gene | Low-quality |
| S17_vs_k141_114179||lt2gene | Low-quality |
| S17_vs_k141_158931||lt2gene | Low-quality |
| S17_vs_k141_99865||lt2gene | Low-quality |
| S17_vs_k141_136370||lt2gene | Not-determined |
| S17_vs_k141_136382||lt2gene | Not-determined |
| S17_vs_k141_144156||lt2gene | Low-quality |
| S17_vs_k141_33434||lt2gene | Not-determined |
| S17_vs_k141_70298||lt2gene | Low-quality |
| S17_vs_k141_151748||lt2gene | Low-quality |
| S17_vs_k141_107639||lt2gene | Low-quality |
| S17_vs_k141_128908||lt2gene | Low-quality |
| S17_vs_k141_33439||lt2gene | Low-quality |
| S17_vs_k141_3540||lt2gene | Low-quality |
| S17_vs_k141_121693||lt2gene | Not-determined |
| S17_vs_k141_3548||lt2gene | Low-quality |
| S17_vs_k141_84176||lt2gene | Low-quality |
| S17_vs_k141_128957||lt2gene | Low-quality |
| S17_vs_k141_151786||lt2gene | Low-quality |
| S17_vs_k141_11007||lt2gene | Low-quality |
| S17_vs_k141_48291||lt2gene | Low-quality |
| S17_vs_k141_77544||lt2gene | Not-determined |
| S17_vs_k141_33537||lt2gene | Low-quality |
| S17_vs_k141_48324||lt2gene | Not-determined |
| S17_vs_k141_151839||lt2gene | Low-quality |
| S17_vs_k141_114300||lt2gene | Low-quality |
| S17_vs_k141_114308||lt2gene | Low-quality |
| S17_vs_k141_3627||lt2gene | Low-quality |
| S17_vs_k141_55547||lt2gene | Low-quality |
| S17_vs_k141_3642||lt2gene | Low-quality |
| S17_vs_k141_151864||lt2gene | Low-quality |
| S17_vs_k141_129041||lt2gene | Low-quality |
| S17_vs_k141_151873||lt2gene | Low-quality |
| S17_vs_k141_77606||lt2gene | Low-quality |
| S17_vs_k141_151885||lt2gene | Low-quality |
| S17_vs_k141_107692||lt2gene | Low-quality |
| S17_vs_k141_129070||lt2gene | Low-quality |
| S17_vs_k141_77619||lt2gene | Low-quality |
| S17_vs_k141_92226||lt2gene | Low-quality |
| S17_vs_k141_11111||lt2gene | Low-quality |
| S17_vs_k141_114389||lt2gene | Low-quality |
| S17_vs_k141_33622||lt2gene | Low-quality |
| S17_vs_k141_11130||lt2gene | Low-quality |
| S17_vs_k141_129106||lt2gene | Low-quality |
| S17_vs_k141_62516||lt2gene | Not-determined |
| S17_vs_k141_55627||lt2gene | Low-quality |
| S17_vs_k141_151912||lt2gene | Low-quality |
| S17_vs_k141_151916||lt2gene | Low-quality |
| S17_vs_k141_3732||lt2gene | Low-quality |
| S17_vs_k141_77664||lt2gene | Low-quality |
| S17_vs_k141_129147||lt2gene | Low-quality |
| S17_vs_k141_114442||lt2gene | Low-quality |
| S17_vs_k141_41231||lt2gene | Low-quality |
| S17_vs_k141_3748||lt2gene | Low-quality |
| S17_vs_k141_3749||lt2gene | Low-quality |
| S17_vs_k141_3750||lt2gene | Low-quality |
| S17_vs_k141_3763||lt2gene | Low-quality |
| S17_vs_k141_166174||lt2gene | Low-quality |
| S17_vs_k141_107752||lt2gene | Low-quality |
| S17_vs_k141_129213||lt2gene | Low-quality |
| S17_vs_k141_107757||lt2gene | Not-determined |
| S17_vs_k141_41303||lt2gene | Low-quality |
| S17_vs_k141_33746||lt2gene | Low-quality |
| S17_vs_k141_107782||lt2gene | Low-quality |
| S17_vs_k141_55739||lt2gene | Low-quality |
| S17_vs_k141_18434||lt2gene | Low-quality |
| S17_vs_k141_159197||lt2gene | Low-quality |
| S17_vs_k141_107810||lt2gene | Not-determined |
| S17_vs_k141_84320||lt2gene | Low-quality |
| S17_vs_k141_18484||lt2gene | Low-quality |
| S17_vs_k141_114573||lt2gene | Not-determined |
| S17_vs_k141_70554||lt2gene | Not-determined |
| S17_vs_k141_3876||lt2gene | Not-determined |
| S17_vs_k141_100109||lt2gene | Low-quality |
| S17_vs_k141_144425||lt2gene | Low-quality |
| S17_vs_k141_107863||lt2gene | Low-quality |
| S17_vs_k141_77826||lt2gene | Not-determined |
| S17_vs_k141_55813||lt2gene | Not-determined |
| S17_vs_k141_48504||lt2gene | Low-quality |
| S17_vs_k141_3912||lt2gene | Not-determined |
| S17_vs_k141_136624||lt2gene | Low-quality |
| S17_vs_k141_41348||lt2gene | Low-quality |
| S17_vs_k141_11307||lt2gene | Low-quality |
| S17_vs_k141_48545||lt2gene | Low-quality |
| S17_vs_k141_121952||lt2gene | Low-quality |
| S17_vs_k141_48551||lt2gene | Low-quality |
| S17_vs_k141_18534||lt2gene | Low-quality |
| S17_vs_k141_3987||lt2gene | Not-determined |
| S17_vs_k141_121959||lt2gene | Low-quality |
| S17_vs_k141_166338||lt2gene | Low-quality |
| S17_vs_k141_25648||lt2gene | Low-quality |
| S17_vs_k141_92397||lt2gene | Low-quality |
| S17_vs_k141_48609||lt2gene | Not-determined |
| S17_vs_k141_136704||lt2gene | Low-quality |
| S17_vs_k141_84432||lt2gene | Low-quality |
| S17_vs_k141_11346||lt2gene | Not-determined |
| S17_vs_k141_41446||lt2gene | Low-quality |
| S17_vs_k141_55894||lt2gene | Not-determined |
| S17_vs_k141_11357||lt2gene | Low-quality |
| S17_vs_k141_129404||lt2gene | Low-quality |
| S17_vs_k141_159280||lt2gene | Low-quality |
| S17_vs_k141_77956||lt2gene | Not-determined |
| S17_vs_k141_11373||lt2gene | Low-quality |
| S17_vs_k141_122033||lt2gene | Low-quality |
| S17_vs_k141_159292||lt2gene | Low-quality |
| S17_vs_k141_25692||lt2gene | Low-quality |
| S17_vs_k141_166422||lt2gene | Low-quality |
| S17_vs_k141_77966||lt2gene | Low-quality |
| S17_vs_k141_92444||lt2gene | Not-determined |
| S17_vs_k141_159309||lt2gene | Low-quality |
| S17_vs_k141_55914||lt2gene | Low-quality |
| S17_vs_k141_55923||lt2gene | Low-quality |
| S17_vs_k141_159318||lt2gene | Not-determined |
| S17_vs_k141_152224||lt2gene | Low-quality |
| S17_vs_k141_114767||lt2gene | Low-quality |
| S17_vs_k141_122060||lt2gene | Not-determined |
| S17_vs_k141_108038||lt2gene | Low-quality |
| S17_vs_k141_129457||lt2gene | Low-quality |
| S17_vs_k141_18656||lt2gene | Low-quality |
| S17_vs_k141_152240||lt2gene | Not-determined |
| S17_vs_k141_33978||lt2gene | Low-quality |
| S17_vs_k141_77991||lt2gene | Low-quality |
| S17_vs_k141_166466||lt2gene | Low-quality |
| S17_vs_k141_33989||lt2gene | Low-quality |
| S17_vs_k141_41542||lt2gene | Low-quality |
| S17_vs_k141_159358||lt2gene | Low-quality |
| S17_vs_k141_136775||lt2gene | Not-determined |
| S17_vs_k141_25737||lt2gene | Low-quality |
| S17_vs_k141_55965||lt2gene | Low-quality |
| S17_vs_k141_100311||lt2gene | Low-quality |
| S17_vs_k141_129504||lt2gene | Not-determined |
| S17_vs_k141_108081||lt2gene | Not-determined |
| S17_vs_k141_56003||lt2gene | Low-quality |
| S17_vs_k141_129538||lt2gene | Not-determined |
| S17_vs_k141_122117||lt2gene | Low-quality |
| S17_vs_k141_56006||lt2gene | Low-quality |
| S17_vs_k141_11425||lt2gene | Low-quality |
| S17_vs_k141_129580||lt2gene | Low-quality |
| S17_vs_k141_56027||lt2gene | Not-determined |
| S17_vs_k141_11429||lt2gene | Low-quality |
| S17_vs_k141_100363||lt2gene | Low-quality |
| S17_vs_k141_100367||lt2gene | Low-quality |
| S17_vs_k141_70768||lt2gene | Low-quality |
| S17_vs_k141_122160||lt2gene | Low-quality |
| S17_vs_k141_114931||lt2gene | Low-quality |
| S17_vs_k141_108201||lt2gene | Low-quality |
| S17_vs_k141_41645||lt2gene | Not-determined |
| S17_vs_k141_152410||lt2gene | Not-determined |
| S17_vs_k141_34109||lt2gene | Low-quality |
| S17_vs_k141_18759||lt2gene | Low-quality |
| S17_vs_k141_25839||lt2gene | Not-determined |
| S17_vs_k141_78198||lt2gene | Low-quality |
| S17_vs_k141_25865||lt2gene | Not-determined |
| S17_vs_k141_100441||lt2gene | Low-quality |
| S17_vs_k141_100445||lt2gene | Low-quality |
| S17_vs_k141_84645||lt2gene | Not-determined |
| S17_vs_k141_144612||lt2gene | Not-determined |
| S17_vs_k141_11524||lt2gene | Low-quality |
| S17_vs_k141_18833||lt2gene | Low-quality |
| S17_vs_k141_4054||lt2gene | Low-quality |
| S17_vs_k141_41738||lt2gene | Low-quality |
| S17_vs_k141_18857||lt2gene | Low-quality |
| S17_vs_k141_48936||lt2gene | Low-quality |
| S17_vs_k141_41746||lt2gene | Not-determined |
| S17_vs_k141_115028||lt2gene | Low-quality |
| S17_vs_k141_115033||lt2gene | Low-quality |
| S17_vs_k141_159501||lt2gene | Low-quality |
| S17_vs_k141_56174||lt2gene | Low-quality |
| S17_vs_k141_25923||lt2gene | Low-quality |
| S17_vs_k141_84677||lt2gene | Low-quality |
| S17_vs_k141_166673||lt2gene | Not-determined |
| S17_vs_k141_129693||lt2gene | Low-quality |
| S17_vs_k141_173832||lt2gene | Low-quality |
| S17_vs_k141_144675||lt2gene | Low-quality |
| S17_vs_k141_129713||lt2gene | Not-determined |
| S17_vs_k141_70962||lt2gene | Low-quality |
| S17_vs_k141_152550||lt2gene | Low-quality |
| S17_vs_k141_100542||lt2gene | Low-quality |
| S17_vs_k141_115103||lt2gene | Low-quality |
| S17_vs_k141_84722||lt2gene | Low-quality |
| S17_vs_k141_129763||lt2gene | Low-quality |
| S17_vs_k141_56260||lt2gene | Low-quality |
| S17_vs_k141_129777||lt2gene | Low-quality |
| S17_vs_k141_100567||lt2gene | Low-quality |
| S17_vs_k141_122217||lt2gene | Low-quality |
| S17_vs_k141_34264||lt2gene | Low-quality |
| S17_vs_k141_166733||lt2gene | Low-quality |
| S17_vs_k141_173911||lt2gene | Low-quality |
| S17_vs_k141_159606||lt2gene | Not-determined |
| S17_vs_k141_92843||lt2gene | Not-determined |
| S17_vs_k141_56315||lt2gene | Low-quality |
| S17_vs_k141_4198||lt2gene | Low-quality |
| S17_vs_k141_144777||lt2gene | Not-determined |
| S17_vs_k141_19052||lt2gene | Low-quality |
| S17_vs_k141_137114||lt2gene | Low-quality |
| S17_vs_k141_11685||lt2gene | Not-determined |
| S17_vs_k141_41869||lt2gene | Not-determined |
| S17_vs_k141_173963||lt2gene | Low-quality |
| S17_vs_k141_78348||lt2gene | Low-quality |
| S17_vs_k141_78349||lt2gene | Low-quality |
| S17_vs_k141_49126||lt2gene | Not-determined |
| S17_vs_k141_115205||lt2gene | Not-determined |
| S17_vs_k141_41910||lt2gene | Low-quality |
| S17_vs_k141_152683||lt2gene | Not-determined |
| S17_vs_k141_137158||lt2gene | Not-determined |
| S17_vs_k141_78372||lt2gene | Low-quality |
| S17_vs_k141_4256||lt2gene | Low-quality |
| S17_vs_k141_166834||lt2gene | Low-quality |
| S17_vs_k141_56365||lt2gene | Low-quality |
| S17_vs_k141_26090||lt2gene | Not-determined |
| S17_vs_k141_78395||lt2gene | Low-quality |
| S17_vs_k141_78406||lt2gene | Low-quality |
| S17_vs_k141_26108||lt2gene | Low-quality |
| S17_vs_k141_129891||lt2gene | Low-quality |
| S17_vs_k141_84843||lt2gene | Low-quality |
| S17_vs_k141_71139||lt2gene | Low-quality |
| S17_vs_k141_174024||lt2gene | Low-quality |
| S17_vs_k141_144826||lt2gene | Not-determined |
| S17_vs_k141_100705||lt2gene | Not-determined |
| S17_vs_k141_100707||lt2gene | Low-quality |
| S17_vs_k141_56401||lt2gene | Not-determined |
| S17_vs_k141_49168||lt2gene | Not-determined |
| S17_vs_k141_100712||lt2gene | Low-quality |
| S17_vs_k141_34341||lt2gene | Low-quality |
| S17_vs_k141_159762||lt2gene | Low-quality |
| S17_vs_k141_108529||lt2gene | Not-determined |
| S17_vs_k141_49192||lt2gene | Low-quality |
| S17_vs_k141_62737||lt2gene | Not-determined |
| S17_vs_k141_34361||lt2gene | Low-quality |
| S17_vs_k141_34366||lt2gene | Low-quality |
| S17_vs_k141_56451||lt2gene | Low-quality |
| S17_vs_k141_84888||lt2gene | Not-determined |
| S17_vs_k141_152779||lt2gene | Low-quality |
| S17_vs_k141_137215||lt2gene | Low-quality |
| S17_vs_k141_137218||lt2gene | Low-quality |
| S17_vs_k141_4352||lt2gene | Not-determined |
| S17_vs_k141_84906||lt2gene | Not-determined |
| S17_vs_k141_71178||lt2gene | Low-quality |
| S17_vs_k141_166941||lt2gene | Not-determined |
| S17_vs_k141_42005||lt2gene | Low-quality |
| S17_vs_k141_11740||lt2gene | Low-quality |
| S17_vs_k141_49245||lt2gene | Low-quality |
| S17_vs_k141_84931||lt2gene | Not-determined |
| S17_vs_k141_78515||lt2gene | Not-determined |
| S17_vs_k141_166980||lt2gene | Low-quality |
| S17_vs_k141_100808||lt2gene | Low-quality |
| S17_vs_k141_152843||lt2gene | Not-determined |
| S17_vs_k141_11795||lt2gene | Low-quality |
| S17_vs_k141_152846||lt2gene | Low-quality |
| S17_vs_k141_11801||lt2gene | Low-quality |
| S17_vs_k141_167001||lt2gene | Low-quality |
| S17_vs_k141_100824||lt2gene | Low-quality |
| S17_vs_k141_92974||lt2gene | Low-quality |
| S17_vs_k141_108573||lt2gene | Low-quality |
| S17_vs_k141_4384||lt2gene | Low-quality |
| S17_vs_k141_34451||lt2gene | Low-quality |
| S17_vs_k141_100846||lt2gene | Not-determined |
| S17_vs_k141_56601||lt2gene | Low-quality |
| S17_vs_k141_62844||lt2gene | Not-determined |
| S17_vs_k141_137335||lt2gene | Not-determined |
| S17_vs_k141_108619||lt2gene | Low-quality |
| S17_vs_k141_4420||lt2gene | Low-quality |
| S17_vs_k141_11846||lt2gene | Low-quality |
| S17_vs_k141_71264||lt2gene | Not-determined |
| S17_vs_k141_4430||lt2gene | Low-quality |
| S17_vs_k141_115464||lt2gene | Low-quality |
| S17_vs_k141_85018||lt2gene | Low-quality |
| S17_vs_k141_115500||lt2gene | Low-quality |
| S17_vs_k141_93045||lt2gene | Low-quality |
| S17_vs_k141_122351||lt2gene | Low-quality |
| S17_vs_k141_49391||lt2gene | Low-quality |
| S17_vs_k141_115552||lt2gene | Low-quality |
| S17_vs_k141_34592||lt2gene | Low-quality |
| S17_vs_k141_19202||lt2gene | Low-quality |
| S17_vs_k141_4511||lt2gene | Not-determined |
| S17_vs_k141_115577||lt2gene | Not-determined |
| S17_vs_k141_11970||lt2gene | Low-quality |
| S17_vs_k141_26352||lt2gene | Low-quality |
| S17_vs_k141_93122||lt2gene | Low-quality |
| S17_vs_k141_34627||lt2gene | Not-determined |
| S17_vs_k141_122392||lt2gene | Not-determined |
| S17_vs_k141_42261||lt2gene | Low-quality |
| S17_vs_k141_4543||lt2gene | Low-quality |
| S17_vs_k141_62967||lt2gene | Low-quality |
| S17_vs_k141_108746||lt2gene | Low-quality |
| S17_vs_k141_49433||lt2gene | Low-quality |
| S17_vs_k141_78705||lt2gene | Not-determined |
| S17_vs_k141_34653||lt2gene | Low-quality |
| S17_vs_k141_78716||lt2gene | Low-quality |
| S17_vs_k141_93161||lt2gene | Not-determined |
| S17_vs_k141_42290||lt2gene | Not-determined |
| S17_vs_k141_108767||lt2gene | Low-quality |
| S17_vs_k141_108768||lt2gene | Not-determined |
| S17_vs_k141_108788||lt2gene | Low-quality |
| S17_vs_k141_34678||lt2gene | Low-quality |
| S17_vs_k141_174212||lt2gene | Low-quality |
| S17_vs_k141_63027||lt2gene | Low-quality |
| S17_vs_k141_85135||lt2gene | Low-quality |
| S17_vs_k141_101034||lt2gene | Low-quality |
| S17_vs_k141_63046||lt2gene | Low-quality |
| S17_vs_k141_137575||lt2gene | Low-quality |
| S17_vs_k141_26425||lt2gene | Low-quality |
| S17_vs_k141_34710||lt2gene | Low-quality |
| S17_vs_k141_19256||lt2gene | Low-quality |
| S17_vs_k141_115729||lt2gene | Low-quality |
| S17_vs_k141_78761||lt2gene | Low-quality |
| S17_vs_k141_174256||lt2gene | Not-determined |
| S17_vs_k141_12064||lt2gene | Low-quality |
| S17_vs_k141_34735||lt2gene | Low-quality |
| S17_vs_k141_145168||lt2gene | Low-quality |
| S17_vs_k141_4663||lt2gene | Low-quality |
| S17_vs_k141_174274||lt2gene | Low-quality |
| S17_vs_k141_34746||lt2gene | Low-quality |
| S17_vs_k141_56799||lt2gene | Low-quality |
| S17_vs_k141_56803||lt2gene | Low-quality |
| S17_vs_k141_56811||lt2gene | Low-quality |
| S17_vs_k141_71525||lt2gene | Low-quality |
| S17_vs_k141_145198||lt2gene | Low-quality |
| S17_vs_k141_160083||lt2gene | Low-quality |
| S17_vs_k141_93262||lt2gene | Low-quality |
| S17_vs_k141_137671||lt2gene | Low-quality |
| S17_vs_k141_160100||lt2gene | Not-determined |
| S17_vs_k141_78813||lt2gene | Low-quality |
| S17_vs_k141_56851||lt2gene | Low-quality |
| S17_vs_k141_34824||lt2gene | Low-quality |
| S17_vs_k141_153143||lt2gene | Not-determined |
| S17_vs_k141_78831||lt2gene | Low-quality |
| S17_vs_k141_122562||lt2gene | Not-determined |
| S17_vs_k141_115783||lt2gene | Low-quality |
| S17_vs_k141_115787||lt2gene | Low-quality |
| S17_vs_k141_49587||lt2gene | Low-quality |
| S17_vs_k141_108903||lt2gene | Low-quality |
| S17_vs_k141_56913||lt2gene | Low-quality |
| S17_vs_k141_115825||lt2gene | Low-quality |
| S17_vs_k141_174332||lt2gene | Low-quality |
| S17_vs_k141_85323||lt2gene | Not-determined |
| S17_vs_k141_85336||lt2gene | Low-quality |
| S17_vs_k141_130099||lt2gene | Low-quality |
| S17_vs_k141_78856||lt2gene | Not-determined |
| S17_vs_k141_56947||lt2gene | Not-determined |
| S17_vs_k141_42455||lt2gene | Low-quality |
| S17_vs_k141_63216||lt2gene | Low-quality |
| S17_vs_k141_153253||lt2gene | Low-quality |
| S17_vs_k141_63223||lt2gene | Low-quality |
| S17_vs_k141_85385||lt2gene | Low-quality |
| S17_vs_k141_93394||lt2gene | Low-quality |
| S17_vs_k141_153266||lt2gene | Low-quality |
| S17_vs_k141_130160||lt2gene | Low-quality |
| S17_vs_k141_71661||lt2gene | Low-quality |
| S17_vs_k141_160203||lt2gene | Not-determined |
| S17_vs_k141_57003||lt2gene | Low-quality |
| S17_vs_k141_167434||lt2gene | Low-quality |
| S17_vs_k141_122687||lt2gene | Low-quality |
| S17_vs_k141_4912||lt2gene | Low-quality |
| S17_vs_k141_115939||lt2gene | Low-quality |
| S17_vs_k141_93459||lt2gene | Not-determined |
| S17_vs_k141_12181||lt2gene | Low-quality |
| S17_vs_k141_26546||lt2gene | Not-determined |
| S17_vs_k141_160312||lt2gene | Low-quality |
| S17_vs_k141_167493||lt2gene | Low-quality |
| S17_vs_k141_12208||lt2gene | Not-determined |
| S17_vs_k141_101303||lt2gene | Low-quality |
| S17_vs_k141_57100||lt2gene | Low-quality |
| S17_vs_k141_71749||lt2gene | Not-determined |
| S17_vs_k141_35112||lt2gene | Low-quality |
| S17_vs_k141_174428||lt2gene | Not-determined |
| S17_vs_k141_174429||lt2gene | Not-determined |
| S17_vs_k141_35128||lt2gene | Not-determined |
| S17_vs_k141_153362||lt2gene | Not-determined |
| S17_vs_k141_167523||lt2gene | Not-determined |
| S17_vs_k141_26599||lt2gene | Not-determined |
| S17_vs_k141_57146||lt2gene | Low-quality |
| S17_vs_k141_26625||lt2gene | Not-determined |
| S17_vs_k141_137953||lt2gene | Low-quality |
| S17_vs_k141_116040||lt2gene | Low-quality |
| S17_vs_k141_5019||lt2gene | Low-quality |
| S17_vs_k141_49697||lt2gene | Low-quality |
| S17_vs_k141_101357||lt2gene | Low-quality |
| S17_vs_k141_101359||lt2gene | Low-quality |
| S17_vs_k141_5057||lt2gene | Not-determined |
| S17_vs_k141_93584||lt2gene | Not-determined |
| S17_vs_k141_49730||lt2gene | Not-determined |
| S17_vs_k141_174466||lt2gene | Low-quality |
| S17_vs_k141_85573||lt2gene | Low-quality |
| S17_vs_k141_122870||lt2gene | Low-quality |
| S17_vs_k141_153450||lt2gene | Low-quality |
| S17_vs_k141_153464||lt2gene | Low-quality |
| S17_vs_k141_79109||lt2gene | Low-quality |
| S17_vs_k141_137999||lt2gene | Not-determined |
| S17_vs_k141_85613||lt2gene | Not-determined |

| | |
|---|---|
| S17_vs_k141_5115\|\|lt2gene | Low-quality |
| S17_vs_k141_5124\|\|lt2gene | Not-determined |
| S17_vs_k141_12267\|\|lt2gene | Low-quality |
| S17_vs_k141_12269\|\|lt2gene | Low-quality |
| S17_vs_k141_160421\|\|lt2gene | Not-determined |
| S17_vs_k141_138017\|\|lt2gene | Low-quality |
| S17_vs_k141_42684\|\|lt2gene | Low-quality |
| S17_vs_k141_116131\|\|lt2gene | Low-quality |
| S17_vs_k141_93668\|\|lt2gene | Low-quality |
| S17_vs_k141_71859\|\|lt2gene | Low-quality |
| S17_vs_k141_85647\|\|lt2gene | Not-determined |
| S17_vs_k141_130341\|\|lt2gene | Not-determined |
| S17_vs_k141_79137\|\|lt2gene | Not-determined |
| S17_vs_k141_57235\|\|lt2gene | Not-determined |
| S17_vs_k141_79148\|\|lt2gene | Not-determined |
| S17_vs_k141_160461\|\|lt2gene | Not-determined |
| S17_vs_k141_71893\|\|lt2gene | Low-quality |
| S17_vs_k141_116184\|\|lt2gene | Not-determined |
| S17_vs_k141_12354\|\|lt2gene | Low-quality |
| S17_vs_k141_35277\|\|lt2gene | Low-quality |
| S17_vs_k141_71913\|\|lt2gene | Not-determined |
| S17_vs_k141_63477\|\|lt2gene | Low-quality |
| S17_vs_k141_160494\|\|lt2gene | Low-quality |
| S17_vs_k141_42762\|\|lt2gene | Low-quality |
| S17_vs_k141_19623\|\|lt2gene | Not-determined |
| S17_vs_k141_85714\|\|lt2gene | Low-quality |
| S17_vs_k141_63505\|\|lt2gene | Low-quality |
| S17_vs_k141_35323\|\|lt2gene | Low-quality |
| S17_vs_k141_79185\|\|lt2gene | Low-quality |
| S17_vs_k141_109328\|\|lt2gene | Low-quality |
| S17_vs_k141_174548\|\|lt2gene | Not-determined |
| S17_vs_k141_71967\|\|lt2gene | Low-quality |
| S17_vs_k141_109334\|\|lt2gene | Not-determined |
| S17_vs_k141_85733\|\|lt2gene | Not-determined |
| S17_vs_k141_63517\|\|lt2gene | Low-quality |
| S17_vs_k141_35346\|\|lt2gene | Not-determined |
| S17_vs_k141_109341\|\|lt2gene | Low-quality |
| S17_vs_k141_57328\|\|lt2gene | Low-quality |
| S17_vs_k141_174564\|\|lt2gene | Low-quality |
| S17_vs_k141_109362\|\|lt2gene | Low-quality |
| S17_vs_k141_63538\|\|lt2gene | Low-quality |
| S17_vs_k141_63544\|\|lt2gene | Not-determined |
| S17_vs_k141_109381\|\|lt2gene | Low-quality |
| S17_vs_k141_85772\|\|lt2gene | Low-quality |
| S17_vs_k141_109399\|\|lt2gene | Not-determined |
| S17_vs_k141_85778\|\|lt2gene | Low-quality |
| S17_vs_k141_160542\|\|lt2gene | Low-quality |
| S17_vs_k141_5244\|\|lt2gene | Not-determined |
| S17_vs_k141_57365\|\|lt2gene | Not-determined |
| S17_vs_k141_5248\|\|lt2gene | Not-determined |
| S17_vs_k141_72051\|\|lt2gene | Low-quality |
| S17_vs_k141_50006\|\|lt2gene | Low-quality |
| S17_vs_k141_63609\|\|lt2gene | Low-quality |
| S17_vs_k141_19715\|\|lt2gene | Low-quality |
| S17_vs_k141_50023\|\|lt2gene | Low-quality |
| S17_vs_k141_42849\|\|lt2gene | Low-quality |
| S17_vs_k141_167797\|\|lt2gene | Low-quality |
| S17_vs_k141_160591\|\|lt2gene | Not-determined |
| S17_vs_k141_57388\|\|lt2gene | Low-quality |
| S17_vs_k141_138209\|\|lt2gene | Not-determined |
| S17_vs_k141_174654\|\|lt2gene | Low-quality |
| S17_vs_k141_63647\|\|lt2gene | Low-quality |
| S17_vs_k141_167817\|\|lt2gene | Low-quality |
| S17_vs_k141_167827\|\|lt2gene | Low-quality |
| S17_vs_k141_116389\|\|lt2gene | Low-quality |
| S17_vs_k141_167830\|\|lt2gene | Low-quality |
| S17_vs_k141_12523\|\|lt2gene | Low-quality |
| S17_vs_k141_130573\|\|lt2gene | Low-quality |
| S17_vs_k141_174680\|\|lt2gene | Low-quality |
| S17_vs_k141_109468\|\|lt2gene | Not-determined |
| S17_vs_k141_5368\|\|lt2gene | Not-determined |
| S17_vs_k141_42901\|\|lt2gene | Low-quality |
| S17_vs_k141_145722\|\|lt2gene | Low-quality |
| S17_vs_k141_116419\|\|lt2gene | Not-determined |
| S17_vs_k141_116426\|\|lt2gene | Low-quality |
| S17_vs_k141_109505\|\|lt2gene | Low-quality |
| S17_vs_k141_42909\|\|lt2gene | Low-quality |
| S17_vs_k141_79368\|\|lt2gene | Not-determined |
| S17_vs_k141_93776\|\|lt2gene | Low-quality |
| S17_vs_k141_63712\|\|lt2gene | Low-quality |
| S17_vs_k141_35481\|\|lt2gene | Low-quality |
| S17_vs_k141_35483\|\|lt2gene | Low-quality |
| S17_vs_k141_145774\|\|lt2gene | Not-determined |
| S17_vs_k141_153685\|\|lt2gene | Low-quality |
| S17_vs_k141_79389\|\|lt2gene | Low-quality |
| S17_vs_k141_12612\|\|lt2gene | Low-quality |
| S17_vs_k141_160700\|\|lt2gene | Not-determined |
| S17_vs_k141_79392\|\|lt2gene | Low-quality |
| S17_vs_k141_35490\|\|lt2gene | Low-quality |
| S17_vs_k141_42949\|\|lt2gene | Low-quality |
| S17_vs_k141_109571\|\|lt2gene | Low-quality |
| S17_vs_k141_123225\|\|lt2gene | Low-quality |
| S17_vs_k141_167923\|\|lt2gene | Low-quality |
| S17_vs_k141_26816\|\|lt2gene | Low-quality |
| S17_vs_k141_12636\|\|lt2gene | Low-quality |
| S17_vs_k141_35514\|\|lt2gene | Not-determined |
| S17_vs_k141_5455\|\|lt2gene | Not-determined |
| S17_vs_k141_85980\|\|lt2gene | Low-quality |
| S17_vs_k141_160748\|\|lt2gene | Low-quality |
| S17_vs_k141_167944\|\|lt2gene | Low-quality |
| S17_vs_k141_42994\|\|lt2gene | Low-quality |
| S17_vs_k141_42995\|\|lt2gene | Low-quality |
| S17_vs_k141_12672\|\|lt2gene | Low-quality |
| S17_vs_k141_63804\|\|lt2gene | Low-quality |
| S17_vs_k141_12675\|\|lt2gene | Low-quality |
| S17_vs_k141_174780\|\|lt2gene | Not-determined |
| S17_vs_k141_79440\|\|lt2gene | Low-quality |
| S17_vs_k141_86005\|\|lt2gene | Not-determined |
| S17_vs_k141_145872\|\|lt2gene | Low-quality |
| S17_vs_k141_72139\|\|lt2gene | Low-quality |
| S17_vs_k141_116537\|\|lt2gene | Low-quality |
| S17_vs_k141_109663\|\|lt2gene | Low-quality |
| S17_vs_k141_79483\|\|lt2gene | Low-quality |
| S17_vs_k141_101786\|\|lt2gene | Low-quality |
| S17_vs_k141_116571\|\|lt2gene | Not-determined |
| S17_vs_k141_116574\|\|lt2gene | Low-quality |
| S17_vs_k141_109688\|\|lt2gene | Low-quality |
| S17_vs_k141_168014\|\|lt2gene | Low-quality |
| S17_vs_k141_35606\|\|lt2gene | Not-determined |
| S17_vs_k141_160812\|\|lt2gene | Low-quality |
| S17_vs_k141_43022\|\|lt2gene | Not-determined |
| S17_vs_k141_50276\|\|lt2gene | Not-determined |
| S17_vs_k141_35618\|\|lt2gene | Low-quality |
| S17_vs_k141_35619\|\|lt2gene | Low-quality |
| S17_vs_k141_168022\|\|lt2gene | Not-determined |
| S17_vs_k141_138353\|\|lt2gene | Not-determined |
| S17_vs_k141_138357\|\|lt2gene | Low-quality |
| S17_vs_k141_153854\|\|lt2gene | Low-quality |
| S17_vs_k141_101861\|\|lt2gene | Not-determined |
| S17_vs_k141_72232\|\|lt2gene | Not-determined |
| S17_vs_k141_35681\|\|lt2gene | Low-quality |
| S17_vs_k141_63865\|\|lt2gene | Low-quality |
| S17_vs_k141_57560\|\|lt2gene | Low-quality |
| S17_vs_k141_86072\|\|lt2gene | Low-quality |
| S17_vs_k141_72261\|\|lt2gene | Low-quality |
| S17_vs_k141_138407\|\|lt2gene | Not-determined |
| S17_vs_k141_123378\|\|lt2gene | Low-quality |
| S17_vs_k141_43110\|\|lt2gene | Low-quality |
| S17_vs_k141_5613\|\|lt2gene | Not-determined |
| S17_vs_k141_153892\|\|lt2gene | Low-quality |
| S17_vs_k141_19806\|\|lt2gene | Low-quality |
| S17_vs_k141_19807\|\|lt2gene | Not-determined |
| S17_vs_k141_86122\|\|lt2gene | Low-quality |
| S17_vs_k141_101913\|\|lt2gene | Low-quality |
| S17_vs_k141_50355\|\|lt2gene | Low-quality |
| S17_vs_k141_153908\|\|lt2gene | Low-quality |
| S17_vs_k141_153917\|\|lt2gene | Low-quality |
| S17_vs_k141_35727\|\|lt2gene | Low-quality |
| S17_vs_k141_26991\|\|lt2gene | Low-quality |
| S17_vs_k141_12706\|\|lt2gene | Low-quality |
| S17_vs_k141_5640\|\|lt2gene | Low-quality |
| S17_vs_k141_145981\|\|lt2gene | Low-quality |
| S17_vs_k141_26999\|\|lt2gene | Not-determined |
| S17_vs_k141_5651\|\|lt2gene | Low-quality |
| S17_vs_k141_72334\|\|lt2gene | Low-quality |
| S17_vs_k141_35764\|\|lt2gene | Low-quality |
| S17_vs_k141_168156\|\|lt2gene | Not-determined |
| S17_vs_k141_109854\|\|lt2gene | Low-quality |
| S17_vs_k141_101949\|\|lt2gene | Low-quality |
| S17_vs_k141_63947\|\|lt2gene | Low-quality |
| S17_vs_k141_174927\|\|lt2gene | Low-quality |
| S17_vs_k141_123452\|\|lt2gene | Low-quality |
| S17_vs_k141_153972\|\|lt2gene | Low-quality |
| S17_vs_k141_101983\|\|lt2gene | Low-quality |
| S17_vs_k141_35792\|\|lt2gene | Low-quality |
| S17_vs_k141_116765\|\|lt2gene | Low-quality |
| S17_vs_k141_130748\|\|lt2gene | Low-quality |
| S17_vs_k141_161059\|\|lt2gene | Not-determined |
| S17_vs_k141_72394\|\|lt2gene | Low-quality |
| S17_vs_k141_27104\|\|lt2gene | Not-determined |
| S17_vs_k141_63986\|\|lt2gene | Low-quality |
| S17_vs_k141_50468\|\|lt2gene | Low-quality |
| S17_vs_k141_168232\|\|lt2gene | Low-quality |
| S17_vs_k141_138512\|\|lt2gene | Low-quality |
| S17_vs_k141_27129\|\|lt2gene | Low-quality |
| S17_vs_k141_35855\|\|lt2gene | Low-quality |
| S17_vs_k141_12842\|\|lt2gene | Low-quality |
| S17_vs_k141_5732\|\|lt2gene | Not-determined |
| S17_vs_k141_12847\|\|lt2gene | Low-quality |
| S17_vs_k141_50483\|\|lt2gene | Low-quality |
| S17_vs_k141_5740\|\|lt2gene | Low-quality |
| S17_vs_k141_123542\|\|lt2gene | Low-quality |
| S17_vs_k141_35862\|\|lt2gene | Not-determined |
| S17_vs_k141_64033\|\|lt2gene | Low-quality |
| S17_vs_k141_116847\|\|lt2gene | Not-determined |
| S17_vs_k141_79714\|\|lt2gene | Low-quality |
| S17_vs_k141_86309\|\|lt2gene | Low-quality |
| S17_vs_k141_5781\|\|lt2gene | Not-determined |
| S17_vs_k141_109993\|\|lt2gene | Low-quality |
| S17_vs_k141_130847\|\|lt2gene | Not-determined |
| S17_vs_k141_116863\|\|lt2gene | Low-quality |
| S17_vs_k141_123590\|\|lt2gene | Low-quality |
| S17_vs_k141_27163\|\|lt2gene | Not-determined |
| S17_vs_k141_79749\|\|lt2gene | Low-quality |
| S17_vs_k141_50556\|\|lt2gene | Low-quality |
| S17_vs_k141_123596\|\|lt2gene | Low-quality |
| S17_vs_k141_79755\|\|lt2gene | Low-quality |
| S17_vs_k141_72487\|\|lt2gene | Low-quality |
| S17_vs_k141_64076\|\|lt2gene | Low-quality |
| S17_vs_k141_27186\|\|lt2gene | Not-determined |
| S17_vs_k141_86331\|\|lt2gene | Not-determined |
| S17_vs_k141_27192\|\|lt2gene | Low-quality |
| S17_vs_k141_20001\|\|lt2gene | Low-quality |
| S17_vs_k141_64085\|\|lt2gene | Not-determined |
| S17_vs_k141_79773\|\|lt2gene | Low-quality |
| S17_vs_k141_161187\|\|lt2gene | Low-quality |
| S17_vs_k141_12958\|\|lt2gene | Low-quality |
| S17_vs_k141_130922\|\|lt2gene | Low-quality |
| S17_vs_k141_20026\|\|lt2gene | Low-quality |
| S17_vs_k141_20031\|\|lt2gene | Low-quality |
| S17_vs_k141_146213\|\|lt2gene | Low-quality |
| S17_vs_k141_72521\|\|lt2gene | Low-quality |
| S17_vs_k141_27234\|\|lt2gene | Low-quality |
| S17_vs_k141_35936\|\|lt2gene | Low-quality |
| S17_vs_k141_12980\|\|lt2gene | Low-quality |
| S17_vs_k141_50621\|\|lt2gene | Not-determined |
| S17_vs_k141_154177\|\|lt2gene | Low-quality |
| S17_vs_k141_5873\|\|lt2gene | Not-determined |
| S17_vs_k141_86372\|\|lt2gene | Low-quality |
| S17_vs_k141_5877\|\|lt2gene | Not-determined |
| S17_vs_k141_146224\|\|lt2gene | Low-quality |
| S17_vs_k141_5880\|\|lt2gene | Low-quality |
| S17_vs_k141_20045\|\|lt2gene | Not-determined |
| S17_vs_k141_154183\|\|lt2gene | Not-determined |
| S17_vs_k141_110085\|\|lt2gene | Not-determined |
| S17_vs_k141_175026\|\|lt2gene | Not-determined |
| S17_vs_k141_5886\|\|lt2gene | Low-quality |
| S17_vs_k141_161241\|\|lt2gene | Low-quality |
| S17_vs_k141_94148\|\|lt2gene | Not-determined |
| S17_vs_k141_79839\|\|lt2gene | Low-quality |
| S17_vs_k141_154209\|\|lt2gene | Low-quality |
| S17_vs_k141_123675\|\|lt2gene | Low-quality |
| S17_vs_k141_64150\|\|lt2gene | Low-quality |
| S17_vs_k141_102110\|\|lt2gene | Low-quality |
| S17_vs_k141_154219\|\|lt2gene | Low-quality |
| S17_vs_k141_35979\|\|lt2gene | Low-quality |
| S17_vs_k141_64162\|\|lt2gene | Low-quality |
| S17_vs_k141_79852\|\|lt2gene | Low-quality |
| S17_vs_k141_154239\|\|lt2gene | Low-quality |
| S17_vs_k141_130955\|\|lt2gene | Low-quality |
| S17_vs_k141_13038\|\|lt2gene | Low-quality |
| S17_vs_k141_146284\|\|lt2gene | Low-quality |
| S17_vs_k141_146286\|\|lt2gene | Low-quality |
| S17_vs_k141_175063\|\|lt2gene | Not-determined |
| S17_vs_k141_123719\|\|lt2gene | Low-quality |
| S17_vs_k141_175066\|\|lt2gene | Low-quality |
| S17_vs_k141_116987\|\|lt2gene | Low-quality |
| S17_vs_k141_130992\|\|lt2gene | Low-quality |
| S17_vs_k141_130996\|\|lt2gene | Low-quality |
| S17_vs_k141_117002\|\|lt2gene | Low-quality |
| S17_vs_k141_117003\|\|lt2gene | Not-determined |
| S17_vs_k141_57827\|\|lt2gene | Low-quality |
| S17_vs_k141_20141\|\|lt2gene | Not-determined |
| S17_vs_k141_64236\|\|lt2gene | Low-quality |
| S17_vs_k141_5951\|\|lt2gene | Low-quality |
| S17_vs_k141_20155\|\|lt2gene | Low-quality |
| S17_vs_k141_102194\|\|lt2gene | Low-quality |
| S17_vs_k141_123781\|\|lt2gene | Low-quality |
| S17_vs_k141_161343\|\|lt2gene | Low-quality |
| S17_vs_k141_154340\|\|lt2gene | Not-determined |
| S17_vs_k141_5968\|\|lt2gene | Low-quality |
| S17_vs_k141_110197\|\|lt2gene | Low-quality |
| S17_vs_k141_154357\|\|lt2gene | Low-quality |
| S17_vs_k141_72577\|\|lt2gene | Not-determined |
| S17_vs_k141_72582\|\|lt2gene | Low-quality |
| S17_vs_k141_36049\|\|lt2gene | Low-quality |
| S17_vs_k141_57887\|\|lt2gene | Low-quality |
| S17_vs_k141_72589\|\|lt2gene | Low-quality |
| S17_vs_k141_102237\|\|lt2gene | Not-determined |
| S17_vs_k141_94226\|\|lt2gene | Low-quality |
| S17_vs_k141_43414\|\|lt2gene | Not-determined |
| S17_vs_k141_94230\|\|lt2gene | Low-quality |
| S17_vs_k141_43417\|\|lt2gene | Not-determined |
| S17_vs_k141_27395\|\|lt2gene | Low-quality |
| S17_vs_k141_6017\|\|lt2gene | Low-quality |
| S17_vs_k141_94251\|\|lt2gene | Low-quality |
| S17_vs_k141_117103\|\|lt2gene | Low-quality |
| S17_vs_k141_117105\|\|lt2gene | Low-quality |
| S17_vs_k141_138810\|\|lt2gene | Not-determined |
| S17_vs_k141_20206\|\|lt2gene | Not-determined |
| S17_vs_k141_117127\|\|lt2gene | Low-quality |
| S17_vs_k141_146434\|\|lt2gene | Low-quality |
| S17_vs_k141_94287\|\|lt2gene | Not-determined |
| S17_vs_k141_146440\|\|lt2gene | Low-quality |
| S17_vs_k141_175149\|\|lt2gene | Low-quality |
| S17_vs_k141_20224\|\|lt2gene | Low-quality |
| S17_vs_k141_168380\|\|lt2gene | Low-quality |
| S17_vs_k141_123871\|\|lt2gene | Not-determined |
| S17_vs_k141_57967\|\|lt2gene | Not-determined |
| S17_vs_k141_175159\|\|lt2gene | Low-quality |
| S17_vs_k141_94308\|\|lt2gene | Low-quality |
| S17_vs_k141_64359\|\|lt2gene | Low-quality |
| S17_vs_k141_110319\|\|lt2gene | Low-quality |
| S17_vs_k141_154468\|\|lt2gene | Not-determined |
| S17_vs_k141_13166\|\|lt2gene | Low-quality |
| S17_vs_k141_175181\|\|lt2gene | Not-determined |
| S17_vs_k141_168421\|\|lt2gene | Low-quality |
| S17_vs_k141_117188\|\|lt2gene | Low-quality |
| S17_vs_k141_86585\|\|lt2gene | Low-quality |
| S17_vs_k141_57986\|\|lt2gene | Low-quality |
| S17_vs_k141_154486\|\|lt2gene | Low-quality |
| S17_vs_k141_123921\|\|lt2gene | Not-determined |
| S17_vs_k141_123922\|\|lt2gene | Low-quality |
| S17_vs_k141_123924\|\|lt2gene | Not-determined |
| S17_vs_k141_64408\|\|lt2gene | Not-determined |
| S17_vs_k141_138910\|\|lt2gene | Not-determined |
| S17_vs_k141_64427\|\|lt2gene | Low-quality |
| S17_vs_k141_6152\|\|lt2gene | Low-quality |
| S17_vs_k141_6153\|\|lt2gene | Low-quality |
| S17_vs_k141_72667\|\|lt2gene | Low-quality |
| S17_vs_k141_36109\|\|lt2gene | Low-quality |
| S17_vs_k141_36115\|\|lt2gene | Low-quality |
| S17_vs_k141_6190\|\|lt2gene | Not-determined |
| S17_vs_k141_154556\|\|lt2gene | Low-quality |
| S17_vs_k141_110410\|\|lt2gene | Low-quality |
| S17_vs_k141_36164\|\|lt2gene | Low-quality |
| S17_vs_k141_6211\|\|lt2gene | Low-quality |
| S17_vs_k141_117283\|\|lt2gene | Low-quality |
| S17_vs_k141_27555\|\|lt2gene | Not-determined |
| S17_vs_k141_146616\|\|lt2gene | Low-quality |
| S17_vs_k141_124004\|\|lt2gene | Low-quality |
| S17_vs_k141_117311\|\|lt2gene | Not-determined |
| S17_vs_k141_110434\|\|lt2gene | Not-determined |
| S17_vs_k141_124011\|\|lt2gene | Low-quality |
| S17_vs_k141_110437\|\|lt2gene | Low-quality |
| S17_vs_k141_20372\|\|lt2gene | Low-quality |
| S17_vs_k141_117331\|\|lt2gene | Not-determined |
| S17_vs_k141_102438\|\|lt2gene | Low-quality |
| S17_vs_k141_64530\|\|lt2gene | Low-quality |
| S17_vs_k141_139042\|\|lt2gene | Low-quality |
| S17_vs_k141_13329\|\|lt2gene | Low-quality |
| S17_vs_k141_124020\|\|lt2gene | Low-quality |
| S17_vs_k141_117352\|\|lt2gene | Not-determined |
| S17_vs_k141_43617\|\|lt2gene | Low-quality |
| S17_vs_k141_20387\|\|lt2gene | Not-determined |
| S17_vs_k141_86727\|\|lt2gene | Low-quality |
| S17_vs_k141_43632\|\|lt2gene | Not-determined |
| S17_vs_k141_124032\|\|lt2gene | Low-quality |
| S17_vs_k141_58075\|\|lt2gene | Low-quality |
| S17_vs_k141_110482\|\|lt2gene | Not-determined |
| S17_vs_k141_43654\|\|lt2gene | Not-determined |
| S17_vs_k141_117384\|\|lt2gene | Low-quality |
| S17_vs_k141_43661\|\|lt2gene | Low-quality |
| S17_vs_k141_154678\|\|lt2gene | Low-quality |
| S17_vs_k141_94418\|\|lt2gene | Not-determined |
| S17_vs_k141_72817\|\|lt2gene | Low-quality |
| S17_vs_k141_86785\|\|lt2gene | Low-quality |
| S17_vs_k141_50870\|\|lt2gene | Low-quality |
| S17_vs_k141_146733\|\|lt2gene | Low-quality |
| S17_vs_k141_110507\|\|lt2gene | Not-determined |
| S17_vs_k141_80178\|\|lt2gene | Low-quality |
| S17_vs_k141_58134\|\|lt2gene | Low-quality |
| S17_vs_k141_6335\|\|lt2gene | Not-determined |
| S17_vs_k141_102565\|\|lt2gene | Low-quality |
| S17_vs_k141_13367\|\|lt2gene | Not-determined |
| S17_vs_k141_86845\|\|lt2gene | Low-quality |
| S17_vs_k141_80211\|\|lt2gene | Low-quality |
| S17_vs_k141_43716\|\|lt2gene | Low-quality |
| S17_vs_k141_50907\|\|lt2gene | Not-determined |
| S17_vs_k141_124143\|\|lt2gene | Low-quality |
| S17_vs_k141_6373\|\|lt2gene | Not-determined |
| S17_vs_k141_13405\|\|lt2gene | Low-quality |
| S17_vs_k141_13406\|\|lt2gene | Low-quality |
| S17_vs_k141_6391\|\|lt2gene | Low-quality |
| S17_vs_k141_50941\|\|lt2gene | Low-quality |
| S17_vs_k141_50947\|\|lt2gene | Low-quality |
| S17_vs_k141_154759\|\|lt2gene | Not-determined |
| S17_vs_k141_72924\|\|lt2gene | Not-determined |
| S17_vs_k141_175470\|\|lt2gene | Low-quality |
| S17_vs_k141_20527\|\|lt2gene | Not-determined |
| S17_vs_k141_131420\|\|lt2gene | Low-quality |
| S17_vs_k141_94459\|\|lt2gene | Low-quality |
| S17_vs_k141_50982\|\|lt2gene | Low-quality |
| S17_vs_k141_124201\|\|lt2gene | Low-quality |
| S17_vs_k141_146882\|\|lt2gene | Low-quality |
| S17_vs_k141_27791\|\|lt2gene | Low-quality |
| S17_vs_k141_154796\|\|lt2gene | Low-quality |
| S17_vs_k141_94494\|\|lt2gene | Low-quality |
| S17_vs_k141_117513\|\|lt2gene | Low-quality |
| S17_vs_k141_80355\|\|lt2gene | Low-quality |
| S17_vs_k141_131485\|\|lt2gene | Not-determined |
| S17_vs_k141_13524\|\|lt2gene | Low-quality |
| S17_vs_k141_154812\|\|lt2gene | Low-quality |
| S17_vs_k141_43834\|\|lt2gene | Low-quality |
| S17_vs_k141_13527\|\|lt2gene | Not-determined |
| S17_vs_k141_110645\|\|lt2gene | Not-determined |
| S17_vs_k141_36414\|\|lt2gene | Low-quality |
| S17_vs_k141_102719\|\|lt2gene | Low-quality |
| S17_vs_k141_51059\|\|lt2gene | Low-quality |
| S17_vs_k141_20591\|\|lt2gene | Low-quality |
| S17_vs_k141_51068\|\|lt2gene | Low-quality |
| S17_vs_k141_117555\|\|lt2gene | Low-quality |
| S17_vs_k141_51073\|\|lt2gene | Not-determined |
| S17_vs_k141_124292\|\|lt2gene | Low-quality |
| S17_vs_k141_124294\|\|lt2gene | Low-quality |
| S17_vs_k141_36429\|\|lt2gene | Low-quality |
| S17_vs_k141_168648\|\|lt2gene | Low-quality |
| S17_vs_k141_13576\|\|lt2gene | Not-determined |
| S17_vs_k141_43895\|\|lt2gene | Low-quality |
| S17_vs_k141_43896\|\|lt2gene | Low-quality |
| S17_vs_k141_6542\|\|lt2gene | Not-determined |
| S17_vs_k141_117589\|\|lt2gene | Not-determined |
| S17_vs_k141_102766\|\|lt2gene | Low-quality |
| S17_vs_k141_51110\|\|lt2gene | Not-determined |
| S17_vs_k141_51115\|\|lt2gene | Low-quality |
| S17_vs_k141_161915\|\|lt2gene | Low-quality |
| S17_vs_k141_124341\|\|lt2gene | Not-determined |
| S17_vs_k141_94558\|\|lt2gene | Low-quality |
| S17_vs_k141_51130\|\|lt2gene | Low-quality |
| S17_vs_k141_161939\|\|lt2gene | Not-determined |
| S17_vs_k141_102814\|\|lt2gene | Low-quality |
| S17_vs_k141_27939\|\|lt2gene | Not-determined |
| S17_vs_k141_43948\|\|lt2gene | Not-determined |
| S17_vs_k141_87074\|\|lt2gene | Not-determined |
| S17_vs_k141_102842\|\|lt2gene | Low-quality |
| S17_vs_k141_168729\|\|lt2gene | Low-quality |
| S17_vs_k141_6593\|\|lt2gene | Low-quality |
| S17_vs_k141_124362\|\|lt2gene | Low-quality |
| S17_vs_k141_110681\|\|lt2gene | Not-determined |
| S17_vs_k141_36491\|\|lt2gene | Low-quality |
| S17_vs_k141_13687\|\|lt2gene | Low-quality |
| S17_vs_k141_147184\|\|lt2gene | Not-determined |
| S17_vs_k141_13703\|\|lt2gene | Not-determined |
| S17_vs_k141_44031\|\|lt2gene | Not-determined |
| S17_vs_k141_87130\|\|lt2gene | Low-quality |
| S17_vs_k141_147199\|\|lt2gene | Not-determined |
| S17_vs_k141_51249\|\|lt2gene | Low-quality |
| S17_vs_k141_110710\|\|lt2gene | Low-quality |
| S17_vs_k141_175683\|\|lt2gene | Not-determined |
| S17_vs_k141_131698\|\|lt2gene | Low-quality |
| S17_vs_k141_131702\|\|lt2gene | Low-quality |
| S17_vs_k141_131714\|\|lt2gene | Low-quality |
| S17_vs_k141_58442\|\|lt2gene | Low-quality |
| S17_vs_k141_73216\|\|lt2gene | Low-quality |
| S17_vs_k141_94650\|\|lt2gene | Low-quality |
| S17_vs_k141_175716\|\|lt2gene | Low-quality |
| S17_vs_k141_175719\|\|lt2gene | Not-determined |
| S17_vs_k141_58467\|\|lt2gene | Not-determined |
| S17_vs_k141_6692\|\|lt2gene | Low-quality |
| S17_vs_k141_139410\|\|lt2gene | Low-quality |
| S17_vs_k141_155079\|\|lt2gene | Low-quality |
| S17_vs_k141_94700\|\|lt2gene | Low-quality |
| S17_vs_k141_36543\|\|lt2gene | Not-determined |
| S17_vs_k141_87238\|\|lt2gene | Low-quality |
| S17_vs_k141_139428\|\|lt2gene | Low-quality |
| S17_vs_k141_28049\|\|lt2gene | Not-determined |
| S17_vs_k141_44132\|\|lt2gene | Low-quality |
| S17_vs_k141_87299\|\|lt2gene | Not-determined |
| S17_vs_k141_139467\|\|lt2gene | Low-quality |
| S17_vs_k141_36596\|\|lt2gene | Low-quality |
| S17_vs_k141_168911\|\|lt2gene | Low-quality |
| S17_vs_k141_6795\|\|lt2gene | Low-quality |
| S17_vs_k141_36613\|\|lt2gene | Low-quality |
| S17_vs_k141_64669\|\|lt2gene | Not-determined |
| S17_vs_k141_162113\|\|lt2gene | Low-quality |
| S17_vs_k141_110807\|\|lt2gene | Low-quality |
| S17_vs_k141_110808\|\|lt2gene | Low-quality |
| S17_vs_k141_147352\|\|lt2gene | Low-quality |
| S17_vs_k141_87342\|\|lt2gene | Low-quality |
| S17_vs_k141_139510\|\|lt2gene | Low-quality |
| S17_vs_k141_139511\|\|lt2gene | Low-quality |
| S17_vs_k141_94819\|\|lt2gene | Not-determined |
| S17_vs_k141_175827\|\|lt2gene | Low-quality |
| S17_vs_k141_103081\|\|lt2gene | Low-quality |
| S17_vs_k141_124567\|\|lt2gene | Not-determined |
| S17_vs_k141_13846\|\|lt2gene | Low-quality |
| S17_vs_k141_28163\|\|lt2gene | Not-determined |
| S17_vs_k141_64726\|\|lt2gene | Not-determined |
| S17_vs_k141_36658\|\|lt2gene | Low-quality |
| S17_vs_k141_6835\|\|lt2gene | Not-determined |
| S17_vs_k141_103117\|\|lt2gene | Low-quality |
| S17_vs_k141_87405\|\|lt2gene | Low-quality |
| S17_vs_k141_13872\|\|lt2gene | Low-quality |
| S17_vs_k141_13873\|\|lt2gene | Low-quality |
| S17_vs_k141_28192\|\|lt2gene | Low-quality |
| S17_vs_k141_80722\|\|lt2gene | Low-quality |
| S17_vs_k141_162188\|\|lt2gene | Low-quality |
| S17_vs_k141_131939\|\|lt2gene | Low-quality |
| S17_vs_k141_13893\|\|lt2gene | Low-quality |
| S17_vs_k141_36685\|\|lt2gene | Low-quality |
| S17_vs_k141_36687\|\|lt2gene | Low-quality |
| S17_vs_k141_44214\|\|lt2gene | Low-quality |
| S17_vs_k141_175891\|\|lt2gene | Low-quality |
| S17_vs_k141_162211\|\|lt2gene | Low-quality |
| S17_vs_k141_124612\|\|lt2gene | Low-quality |
| S17_vs_k141_44264\|\|lt2gene | Low-quality |
| S17_vs_k141_103209\|\|lt2gene | Low-quality |
| S17_vs_k141_28240\|\|lt2gene | Low-quality |
| S17_vs_k141_103215\|\|lt2gene | Not-determined |
| S17_vs_k141_169077\|\|lt2gene | Low-quality |
| S17_vs_k141_36747\|\|lt2gene | Low-quality |
| S17_vs_k141_139601\|\|lt2gene | Low-quality |
| S17_vs_k141_132007\|\|lt2gene | Low-quality |
| S17_vs_k141_147513\|\|lt2gene | Low-quality |
| S17_vs_k141_51565\|\|lt2gene | Low-quality |
| S17_vs_k141_175995\|\|lt2gene | Low-quality |
| S17_vs_k141_162299\|\|lt2gene | Low-quality |
| S17_vs_k141_147549\|\|lt2gene | Not-determined |
| S17_vs_k141_20971\|\|lt2gene | Low-quality |
| S17_vs_k141_147554\|\|lt2gene | Not-determined |
| S17_vs_k141_73543\|\|lt2gene | Not-determined |
| S17_vs_k141_13979\|\|lt2gene | Low-quality |
| S17_vs_k141_139643\|\|lt2gene | Low-quality |
| S17_vs_k141_20994\|\|lt2gene | Low-quality |
| S17_vs_k141_20996\|\|lt2gene | Low-quality |
| S17_vs_k141_117954\|\|lt2gene | Low-quality |
| S17_vs_k141_162337\|\|lt2gene | Low-quality |
| S17_vs_k141_87576\|\|lt2gene | Low-quality |
| S17_vs_k141_80862\|\|lt2gene | Not-determined |
| S17_vs_k141_103330\|\|lt2gene | Low-quality |
| S17_vs_k141_176021\|\|lt2gene | Low-quality |
| S17_vs_k141_147616\|\|lt2gene | Not-determined |
| S17_vs_k141_14004\|\|lt2gene | Low-quality |
| S17_vs_k141_73589\|\|lt2gene | Low-quality |
| S17_vs_k141_162364\|\|lt2gene | Low-quality |
| S17_vs_k141_117985\|\|lt2gene | Low-quality |
| S17_vs_k141_7001\|\|lt2gene | Not-determined |
| S17_vs_k141_87594\|\|lt2gene | Low-quality |
| S17_vs_k141_80879\|\|lt2gene | Not-determined |
| S17_vs_k141_162386\|\|lt2gene | Not-determined |
| S17_vs_k141_176057\|\|lt2gene | Not-determined |
| S17_vs_k141_132151\|\|lt2gene | Not-determined |
| S17_vs_k141_21084\|\|lt2gene | Not-determined |
| S17_vs_k141_147641\|\|lt2gene | Low-quality |
| S17_vs_k141_21122\|\|lt2gene | Low-quality |
| S17_vs_k141_7093\|\|lt2gene | Low-quality |
| S17_vs_k141_124766\|\|lt2gene | Not-determined |
| S17_vs_k141_132209\|\|lt2gene | Low-quality |
| S17_vs_k141_162429\|\|lt2gene | Not-determined |
| S17_vs_k141_103382\|\|lt2gene | Low-quality |
| S17_vs_k141_103385\|\|lt2gene | Low-quality |
| S17_vs_k141_73686\|\|lt2gene | Not-determined |
| S17_vs_k141_139701\|\|lt2gene | Low-quality |
| S17_vs_k141_58726\|\|lt2gene | Low-quality |
| S17_vs_k141_147730\|\|lt2gene | Low-quality |
| S17_vs_k141_64944\|\|lt2gene | Low-quality |
| S17_vs_k141_21187\|\|lt2gene | Low-quality |
| S17_vs_k141_21190\|\|lt2gene | Low-quality |
| S17_vs_k141_147765\|\|lt2gene | Low-quality |
| S17_vs_k141_28485\|\|lt2gene | Low-quality |
| S17_vs_k141_124815\|\|lt2gene | Low-quality |
| S17_vs_k141_162512\|\|lt2gene | Low-quality |
| S17_vs_k141_14137\|\|lt2gene | Not-determined |
| S17_vs_k141_162518\|\|lt2gene | Low-quality |
| S17_vs_k141_87731\|\|lt2gene | Low-quality |
| S17_vs_k141_21278\|\|lt2gene | Not-determined |
| S17_vs_k141_118091\|\|lt2gene | Low-quality |
| S17_vs_k141_124864\|\|lt2gene | Low-quality |
| S17_vs_k141_95095\|\|lt2gene | Low-quality |
| S17_vs_k141_73790\|\|lt2gene | Not-determined |
| S17_vs_k141_169379\|\|lt2gene | Not-determined |
| S17_vs_k141_58798\|\|lt2gene | Low-quality |
| S17_vs_k141_73829\|\|lt2gene | Low-quality |
| S17_vs_k141_132320\|\|lt2gene | Low-quality |
| S17_vs_k141_95139\|\|lt2gene | Low-quality |
| S17_vs_k141_169425\|\|lt2gene | Low-quality |
| S17_vs_k141_28578\|\|lt2gene | Low-quality |
| S17_vs_k141_132356\|\|lt2gene | Low-quality |
| S17_vs_k141_28586\|\|lt2gene | Low-quality |
| S17_vs_k141_118155\|\|lt2gene | Low-quality |
| S17_vs_k141_118159\|\|lt2gene | Low-quality |
| S17_vs_k141_58843\|\|lt2gene | Low-quality |
| S17_vs_k141_87837\|\|lt2gene | Low-quality |
| S17_vs_k141_139841\|\|lt2gene | Low-quality |
| S17_vs_k141_65149\|\|lt2gene | Low-quality |
| S17_vs_k141_7300\|\|lt2gene | Low-quality |
| S17_vs_k141_28645\|\|lt2gene | Low-quality |
| S17_vs_k141_21459\|\|lt2gene | Low-quality |
| S17_vs_k141_21471\|\|lt2gene | Low-quality |
| S17_vs_k141_28695\|\|lt2gene | Low-quality |
| S17_vs_k141_28707\|\|lt2gene | Low-quality |
| S17_vs_k141_36933\|\|lt2gene | Low-quality |
| S17_vs_k141_21509\|\|lt2gene | Not-determined |
| S17_vs_k141_58923\|\|lt2gene | Low-quality |
| S17_vs_k141_95253\|\|lt2gene | Low-quality |
| S17_vs_k141_21525\|\|lt2gene | Not-determined |
| S17_vs_k141_124995\|\|lt2gene | Low-quality |
| S17_vs_k141_176386\|\|lt2gene | Low-quality |
| S17_vs_k141_95277\|\|lt2gene | Not-determined |
| S17_vs_k141_28796\|\|lt2gene | Not-determined |
| S17_vs_k141_139981\|\|lt2gene | Low-quality |
| S17_vs_k141_169657\|\|lt2gene | Low-quality |
| S17_vs_k141_140010\|\|lt2gene | Not-determined |
| S17_vs_k141_14354\|\|lt2gene | Low-quality |
| S17_vs_k141_176440\|\|lt2gene | Low-quality |
| S17_vs_k141_169711\|\|lt2gene | Low-quality |
| S17_vs_k141_65352\|\|lt2gene | Low-quality |
| S17_vs_k141_65360\|\|lt2gene | Low-quality |
| S17_vs_k141_28870\|\|lt2gene | Low-quality |

| | |
|---|---|
| S17_vs_k141_125128\|\|lt2gene | Low-quality |
| S17_vs_k141_21707\|\|lt2gene | Low-quality |
| S17_vs_k141_28899\|\|lt2gene | Low-quality |
| S17_vs_k141_59044\|\|lt2gene | Low-quality |
| S17_vs_k141_140079\|\|lt2gene | Low-quality |
| S17_vs_k141_88086\|\|lt2gene | Low-quality |
| S17_vs_k141_59055\|\|lt2gene | Low-quality |
| S17_vs_k141_132691\|\|lt2gene | Not-determined |
| S17_vs_k141_59078\|\|lt2gene | Not-determined |
| S17_vs_k141_7379\|\|lt2gene | Low-quality |
| S17_vs_k141_88123\|\|lt2gene | Low-quality |
| S17_vs_k141_7388\|\|lt2gene | Low-quality |
| S17_vs_k141_132742\|\|lt2gene | Not-determined |
| S17_vs_k141_14513\|\|lt2gene | Low-quality |
| S17_vs_k141_88183\|\|lt2gene | Low-quality |
| S17_vs_k141_29012\|\|lt2gene | Low-quality |
| S17_vs_k141_140189\|\|lt2gene | Low-quality |
| S17_vs_k141_95521\|\|lt2gene | Not-determined |
| S17_vs_k141_125277\|\|lt2gene | Low-quality |
| S17_vs_k141_29053\|\|lt2gene | Not-determined |
| S17_vs_k141_65612\|\|lt2gene | Not-determined |
| S17_vs_k141_140246\|\|lt2gene | Not-determined |
| S17_vs_k141_140256\|\|lt2gene | Not-determined |
| S17_vs_k141_132848\|\|lt2gene | Low-quality |
| S17_vs_k141_95610\|\|lt2gene | Low-quality |
| S17_vs_k141_125359\|\|lt2gene | Low-quality |
| S17_vs_k141_95633\|\|lt2gene | Not-determined |
| S17_vs_k141_125365\|\|lt2gene | Low-quality |
| S17_vs_k141_29132\|\|lt2gene | Low-quality |
| S17_vs_k141_176715\|\|lt2gene | Not-determined |
| S17_vs_k141_140337\|\|lt2gene | Low-quality |
| S17_vs_k141_140339\|\|lt2gene | Low-quality |
| S17_vs_k141_14690\|\|lt2gene | Not-determined |
| S17_vs_k141_21852\|\|lt2gene | Low-quality |
| S17_vs_k141_140359\|\|lt2gene | Low-quality |
| S17_vs_k141_125442\|\|lt2gene | Low-quality |
| S17_vs_k141_125447\|\|lt2gene | Low-quality |
| S17_vs_k141_125448\|\|lt2gene | Low-quality |
| S17_vs_k141_88364\|\|lt2gene | Not-determined |
| S17_vs_k141_132959\|\|lt2gene | Low-quality |
| S17_vs_k141_65706\|\|lt2gene | Low-quality |
| S17_vs_k141_140379\|\|lt2gene | Low-quality |
| S17_vs_k141_176803\|\|lt2gene | Low-quality |
| S17_vs_k141_65748\|\|lt2gene | Low-quality |
| S17_vs_k141_14758\|\|lt2gene | Low-quality |
| S17_vs_k141_65767\|\|lt2gene | Not-determined |
| S17_vs_k141_95825\|\|lt2gene | Low-quality |
| S17_vs_k141_88456\|\|lt2gene | Not-determined |
| S17_vs_k141_176891\|\|lt2gene | Not-determined |
| S17_vs_k141_29439\|\|lt2gene | Not-determined |
| S17_vs_k141_88567\|\|lt2gene | Low-quality |
| S17_vs_k141_22037\|\|lt2gene | Low-quality |
| S17_vs_k141_95909\|\|lt2gene | Low-quality |
| S17_vs_k141_65965\|\|lt2gene | Low-quality |
| S17_vs_k141_65997\|\|lt2gene | Low-quality |
| S17_vs_k141_66018\|\|lt2gene | Low-quality |
| S17_vs_k141_95991\|\|lt2gene | Not-determined |
| S17_vs_k141_66088\|\|lt2gene | Not-determined |
| S17_vs_k141_177170\|\|lt2gene | Not-determined |
| S17_vs_k141_177173\|\|lt2gene | Low-quality |
| S17_vs_k141_22125\|\|lt2gene | Not-determined |
| S17_vs_k141_66294\|\|lt2gene | Low-quality |
| S17_vs_k141_66304\|\|lt2gene | Low-quality |
| S17_vs_k141_66426\|\|lt2gene | Low-quality |
| S9_extracted_k141_64416 | Not-determined |
| S9_extracted_k141_56838 | Not-determined |
| S9_extracted_k141_37893 | Not-determined |
| S9_extracted_k141_18948 | Not-determined |
| S9_extracted_k141_68205 | Not-determined |
| S9_extracted_k141_4 | Not-determined |
| S9_extracted_k141_22737 | Not-determined |
| S9_extracted_k141_3793 | Not-determined |
| S9_extracted_k141_64421 | Not-determined |
| S9_extracted_k141_7584 | Not-determined |
| S9_extracted_k141_49262 | Not-determined |
| S9_extracted_k141_15162 | Not-determined |
| S9_extracted_k141_64425 | Not-determined |
| S9_extracted_k141_11 | Not-determined |
| S9_extracted_k141_26531 | Not-determined |
| S9_extracted_k141_49265 | Not-determined |
| S9_extracted_k141_75787 | Not-determined |
| S9_extracted_k141_7588 | Not-determined |
| S9_extracted_k141_3796 | Not-determined |
| S9_extracted_k141_26532 | Not-determined |
| S9_extracted_k141_56844 | Not-determined |
| S9_extracted_k141_60633 | Not-determined |
| S9_extracted_k141_34109 | Not-determined |
| S9_extracted_k141_16 | Not-determined |
| S9_extracted_k141_49271 | Low-quality |
| S9_extracted_k141_79578 | Not-determined |
| S9_extracted_k141_30320 | Not-determined |
| S9_extracted_k141_3800 | Not-determined |
| S9_extracted_k141_41687 | Not-determined |
| S9_extracted_k141_72003 | Not-determined |
| S9_extracted_k141_15171 | Not-determined |
| S9_extracted_k141_30323 | Not-determined |
| S9_extracted_k141_83368 | Not-determined |
| S9_extracted_k141_79581 | Not-determined |
| S9_extracted_k141_41688 | Low-quality |
| S9_extracted_k141_7592 | Not-determined |
| S9_extracted_k141_49275 | Low-quality |
| S9_extracted_k141_7593 | Not-determined |
| S9_extracted_k141_21 | Not-determined |
| S9_extracted_k141_26540 | Not-determined |
| S9_extracted_k141_41693 | Not-determined |
| S9_extracted_k141_83372 | Not-determined |
| S9_extracted_k141_34117 | Low-quality |
| S9_extracted_k141_60642 | Not-determined |
| S9_extracted_k141_83373 | Not-determined |
| S9_extracted_k141_34118 | Low-quality |
| S9_extracted_k141_41695 | Not-determined |
| S9_extracted_k141_60643 | Not-determined |
| S9_extracted_k141_15177 | Not-determined |
| S9_extracted_k141_22747 | Not-determined |
| S9_extracted_k141_64438 | Not-determined |
| S9_extracted_k141_53067 | Not-determined |
| S9_extracted_k141_45474 | Not-determined |
| S9_extracted_k141_87163 | Not-determined |
| S9_extracted_k141_79586 | Not-determined |
| S9_extracted_k141_41700 | Not-determined |
| S9_extracted_k141_18970 | Low-quality |
| S9_extracted_k141_22751 | Not-determined |
| S9_extracted_k141_56857 | Not-determined |
| S9_extracted_k141_34124 | Not-determined |
| S9_extracted_k141_64442 | Not-determined |
| S9_extracted_k141_49283 | Low-quality |
| S9_extracted_k141_87166 | Not-determined |
| S9_extracted_k141_11395 | Not-determined |
| S9_extracted_k141_22755 | Not-determined |
| S9_extracted_k141_79590 | Low-quality |
| S9_extracted_k141_11399 | Not-determined |
| S9_extracted_k141_22757 | Not-determined |
| S9_extracted_k141_53076 | Low-quality |
| S9_extracted_k141_87169 | Low-quality |
| S9_extracted_k141_37915 | Not-determined |
| S9_extracted_k141_22762 | Not-determined |
| S9_extracted_k141_37918 | Not-determined |
| S9_extracted_k141_87173 | Not-determined |
| S9_extracted_k141_26551 | Not-determined |
| S9_extracted_k141_72024 | Not-determined |
| S9_extracted_k141_11404 | Not-determined |
| S9_extracted_k141_75798 | Not-determined |
| S9_extracted_k141_3813 | Low-quality |
| S9_extracted_k141_41709 | Not-determined |
| S9_extracted_k141_18980 | Low-quality |
| S9_extracted_k141_75799 | Low-quality |
| S9_extracted_k141_83380 | Not-determined |
| S9_extracted_k141_11406 | Not-determined |
| S9_extracted_k141_75802 | Low-quality |
| S9_extracted_k141_83383 | Not-determined |
| S9_extracted_k141_3815 | Not-determined |
| S9_extracted_k141_72030 | Not-determined |
| S9_extracted_k141_34142 | Not-determined |
| S9_extracted_k141_51 | Not-determined |
| S9_extracted_k141_75804 | Low-quality |
| S9_extracted_k141_3818 | Not-determined |
| S9_extracted_k141_15195 | Not-determined |
| S9_extracted_k141_41718 | Not-determined |
| S9_extracted_k141_72033 | Not-determined |
| S9_extracted_k141_41719 | Low-quality |
| S9_extracted_k141_54 | Not-determined |
| S9_extracted_k141_83393 | Not-determined |
| S9_extracted_k141_87189 | Low-quality |
| S9_extracted_k141_68221 | Not-determined |
| S9_extracted_k141_62 | Not-determined |
| S9_extracted_k141_34148 | Not-determined |
| S9_extracted_k141_3819 | Low-quality |
| S9_extracted_k141_26581 | Not-determined |
| S9_extracted_k141_83402 | Not-determined |
| S9_extracted_k141_75808 | Not-determined |
| S9_extracted_k141_18988 | Low-quality |
| S9_extracted_k141_26583 | Low-quality |
| S9_extracted_k141_87193 | Not-determined |
| S9_extracted_k141_83408 | Low-quality |
| S9_extracted_k141_72047 | Not-determined |
| S9_extracted_k141_68229 | Not-determined |
| S9_extracted_k141_68231 | Not-determined |
| S9_extracted_k141_68236 | Not-determined |
| S9_extracted_k141_45491 | Not-determined |
| S9_extracted_k141_41733 | Not-determined |
| S9_extracted_k141_34159 | Not-determined |
| S9_extracted_k141_75818 | Not-determined |
| S9_extracted_k141_75819 | Not-determined |
| S9_extracted_k141_45493 | Not-determined |
| S9_extracted_k141_34161 | Not-determined |
| S9_extracted_k141_72055 | Not-determined |
| S9_extracted_k141_45495 | Not-determined |
| S9_extracted_k141_56893 | Not-determined |
| S9_extracted_k141_83420 | Not-determined |
| S9_extracted_k141_41742 | Not-determined |
| S9_extracted_k141_34167 | Not-determined |
| S9_extracted_k141_22788 | Not-determined |
| S9_extracted_k141_26585 | Low-quality |
| S9_extracted_k141_7631 | Not-determined |
| S9_extracted_k141_72058 | Not-determined |
| S9_extracted_k141_7634 | Not-determined |
| S9_extracted_k141_7635 | Not-determined |
| S9_extracted_k141_7636 | Not-determined |
| S9_extracted_k141_30348 | Not-determined |
| S9_extracted_k141_79629 | Not-determined |
| S9_extracted_k141_7637 | Not-determined |
| S9_extracted_k141_45507 | Low-quality |
| S9_extracted_k141_45510 | Low-quality |
| S9_extracted_k141_30350 | Low-quality |
| S9_extracted_k141_34177 | Not-determined |
| S9_extracted_k141_53098 | Low-quality |
| S9_extracted_k141_68250 | Not-determined |
| S9_extracted_k141_30352 | Not-determined |
| S9_extracted_k141_53102 | Low-quality |
| S9_extracted_k141_53103 | Not-determined |
| S9_extracted_k141_22800 | Not-determined |
| S9_extracted_k141_3845 | Not-determined |
| S9_extracted_k141_34181 | Not-determined |
| S9_extracted_k141_3847 | Not-determined |
| S9_extracted_k141_15230 | Not-determined |
| S9_extracted_k141_53105 | Not-determined |
| S9_extracted_k141_7641 | Not-determined |
| S9_extracted_k141_15232 | Not-determined |
| S9_extracted_k141_68257 | Not-determined |
| S9_extracted_k141_26594 | Not-determined |
| S9_extracted_k141_15233 | Low-quality |
| S9_extracted_k141_64489 | Not-determined |
| S9_extracted_k141_72067 | Not-determined |
| S9_extracted_k141_68260 | Low-quality |
| S9_extracted_k141_79642 | Not-determined |
| S9_extracted_k141_49320 | Low-quality |
| S9_extracted_k141_79643 | Not-determined |
| S9_extracted_k141_7646 | Not-determined |
| S9_extracted_k141_53112 | Not-determined |
| S9_extracted_k141_87218 | Not-determined |
| S9_extracted_k141_87219 | Not-determined |
| S9_extracted_k141_3856 | Not-determined |
| S9_extracted_k141_45528 | Not-determined |
| S9_extracted_k141_60679 | Not-determined |
| S9_extracted_k141_60681 | Not-determined |
| S9_extracted_k141_30368 | Not-determined |
| S9_extracted_k141_83446 | Not-determined |
| S9_extracted_k141_87232 | Not-determined |
| S9_extracted_k141_49333 | Not-determined |
| S9_extracted_k141_56921 | Not-determined |
| S9_extracted_k141_37935 | Not-determined |
| S9_extracted_k141_60687 | Not-determined |
| S9_extracted_k141_18994 | Not-determined |
| S9_extracted_k141_26616 | Not-determined |
| S9_extracted_k141_45537 | Not-determined |
| S9_extracted_k141_83453 | Low-quality |
| S9_extracted_k141_15250 | Not-determined |
| S9_extracted_k141_72084 | Low-quality |
| S9_extracted_k141_37940 | Not-determined |
| S9_extracted_k141_72085 | Not-determined |
| S9_extracted_k141_79664 | Low-quality |
| S9_extracted_k141_75855 | Not-determined |
| S9_extracted_k141_75856 | Not-determined |
| S9_extracted_k141_34207 | Not-determined |
| S9_extracted_k141_56931 | Not-determined |
| S9_extracted_k141_83456 | Low-quality |
| S9_extracted_k141_64509 | Not-determined |
| S9_extracted_k141_49345 | Low-quality |
| S9_extracted_k141_30376 | Not-determined |
| S9_extracted_k141_79666 | Not-determined |
| S9_extracted_k141_87242 | Low-quality |
| S9_extracted_k141_64511 | Not-determined |
| S9_extracted_k141_83459 | Not-determined |
| S9_extracted_k141_49350 | Low-quality |
| S9_extracted_k141_11416 | Not-determined |
| S9_extracted_k141_30380 | Not-determined |
| S9_extracted_k141_22826 | Low-quality |
| S9_extracted_k141_64513 | Not-determined |
| S9_extracted_k141_15259 | Not-determined |
| S9_extracted_k141_7666 | Not-determined |
| S9_extracted_k141_34214 | Not-determined |
| S9_extracted_k141_60695 | Not-determined |
| S9_extracted_k141_22831 | Not-determined |
| S9_extracted_k141_83461 | Not-determined |
| S9_extracted_k141_11425 | Not-determined |
| S9_extracted_k141_11427 | Not-determined |
| S9_extracted_k141_72099 | Not-determined |
| S9_extracted_k141_19019 | Not-determined |
| S9_extracted_k141_79678 | Not-determined |
| S9_extracted_k141_45548 | Not-determined |
| S9_extracted_k141_37942 | Not-determined |
| S9_extracted_k141_87255 | Not-determined |
| S9_extracted_k141_37943 | Not-determined |
| S9_extracted_k141_79681 | Not-determined |
| S9_extracted_k141_75875 | Not-determined |
| S9_extracted_k141_7671 | Low-quality |
| S9_extracted_k141_60700 | Not-determined |
| S9_extracted_k141_3871 | Not-determined |
| S9_extracted_k141_37945 | Not-determined |
| S9_extracted_k141_87259 | Not-determined |
| S9_extracted_k141_37947 | Not-determined |
| S9_extracted_k141_30396 | Low-quality |
| S9_extracted_k141_19027 | Low-quality |
| S9_extracted_k141_22844 | Low-quality |
| S9_extracted_k141_7674 | Low-quality |
| S9_extracted_k141_79686 | Low-quality |
| S9_extracted_k141_15273 | Not-determined |
| S9_extracted_k141_7675 | Not-determined |
| S9_extracted_k141_11439 | Not-determined |
| S9_extracted_k141_37951 | Not-determined |
| S9_extracted_k141_26628 | Low-quality |
| S9_extracted_k141_11442 | Not-determined |
| S9_extracted_k141_3882 | Not-determined |
| S9_extracted_k141_19029 | Not-determined |
| S9_extracted_k141_19030 | Low-quality |
| S9_extracted_k141_26631 | Not-determined |
| S9_extracted_k141_75883 | Not-determined |
| S9_extracted_k141_83471 | Not-determined |
| S9_extracted_k141_30403 | Low-quality |
| S9_extracted_k141_64530 | Low-quality |
| S9_extracted_k141_11448 | Not-determined |
| S9_extracted_k141_75886 | Not-determined |
| S9_extracted_k141_19034 | Low-quality |
| S9_extracted_k141_41761 | Not-determined |
| S9_extracted_k141_22852 | Not-determined |
| S9_extracted_k141_7683 | Not-determined |
| S9_extracted_k141_53150 | Not-determined |
| S9_extracted_k141_19038 | Not-determined |
| S9_extracted_k141_22855 | Low-quality |
| S9_extracted_k141_30411 | Not-determined |
| S9_extracted_k141_41769 | Not-determined |
| S9_extracted_k141_19042 | Low-quality |
| S9_extracted_k141_78 | Not-determined |
| S9_extracted_k141_53153 | Not-determined |
| S9_extracted_k141_80 | Not-determined |
| S9_extracted_k141_64536 | Not-determined |
| S9_extracted_k141_41775 | Not-determined |
| S9_extracted_k141_22860 | Not-determined |
| S9_extracted_k141_53159 | Not-determined |
| S9_extracted_k141_34227 | Not-determined |
| S9_extracted_k141_34228 | Not-determined |
| S9_extracted_k141_85 | Not-determined |
| S9_extracted_k141_87 | Low-quality |
| S9_extracted_k141_3900 | Not-determined |
| S9_extracted_k141_41780 | Not-determined |
| S9_extracted_k141_3901 | Not-determined |
| S9_extracted_k141_22863 | Not-determined |
| S9_extracted_k141_30422 | Not-determined |
| S9_extracted_k141_30424 | Not-determined |
| S9_extracted_k141_60728 | Not-determined |
| S9_extracted_k141_37975 | Not-determined |
| S9_extracted_k141_22867 | Not-determined |
| S9_extracted_k141_83488 | Low-quality |
| S9_extracted_k141_60731 | Not-determined |
| S9_extracted_k141_60732 | Not-determined |
| S9_extracted_k141_15289 | Low-quality |
| S9_extracted_k141_60733 | Not-determined |
| S9_extracted_k141_41790 | Not-determined |
| S9_extracted_k141_11475 | Not-determined |
| S9_extracted_k141_53172 | Not-determined |
| S9_extracted_k141_22877 | Not-determined |
| S9_extracted_k141_83494 | Not-determined |
| S9_extracted_k141_34237 | Not-determined |
| S9_extracted_k141_19049 | Not-determined |
| S9_extracted_k141_15295 | Not-determined |
| S9_extracted_k141_41798 | Not-determined |
| S9_extracted_k141_60744 | Not-determined |
| S9_extracted_k141_11487 | Not-determined |
| S9_extracted_k141_83498 | Low-quality |
| S9_extracted_k141_72106 | Not-determined |
| S9_extracted_k141_83501 | Not-determined |
| S9_extracted_k141_30441 | Not-determined |
| S9_extracted_k141_72107 | Low-quality |
| S9_extracted_k141_30442 | Not-determined |
| S9_extracted_k141_41812 | Not-determined |
| S9_extracted_k141_87308 | Not-determined |
| S9_extracted_k141_22890 | Not-determined |
| S9_extracted_k141_53187 | Not-determined |
| S9_extracted_k141_34251 | Not-determined |
| S9_extracted_k141_75893 | Not-determined |
| S9_extracted_k141_7697 | Not-determined |
| S9_extracted_k141_41814 | Not-determined |
| S9_extracted_k141_53189 | Not-determined |
| S9_extracted_k141_75895 | Not-determined |
| S9_extracted_k141_37999 | Low-quality |
| S9_extracted_k141_38002 | Not-determined |
| S9_extracted_k141_83516 | Not-determined |
| S9_extracted_k141_30451 | Not-determined |
| S9_extracted_k141_83517 | Not-determined |
| S9_extracted_k141_83518 | Not-determined |
| S9_extracted_k141_127 | Not-determined |
| S9_extracted_k141_75905 | Not-determined |
| S9_extracted_k141_83523 | Not-determined |
| S9_extracted_k141_87311 | Not-determined |
| S9_extracted_k141_38013 | Not-determined |
| S9_extracted_k141_30460 | Not-determined |
| S9_extracted_k141_75906 | Not-determined |
| S9_extracted_k141_56939 | Low-quality |
| S9_extracted_k141_22898 | Not-determined |
| S9_extracted_k141_30461 | Low-quality |
| S9_extracted_k141_22900 | Not-determined |
| S9_extracted_k141_30465 | Not-determined |
| S9_extracted_k141_53213 | Not-determined |
| S9_extracted_k141_38023 | Not-determined |
| S9_extracted_k141_30467 | Not-determined |
| S9_extracted_k141_83529 | Not-determined |
| S9_extracted_k141_11514 | Not-determined |
| S9_extracted_k141_56947 | Not-determined |
| S9_extracted_k141_83532 | Not-determined |
| S9_extracted_k141_22903 | Not-determined |
| S9_extracted_k141_53217 | Low-quality |
| S9_extracted_k141_147 | Not-determined |
| S9_extracted_k141_38031 | Not-determined |
| S9_extracted_k141_30475 | Not-determined |
| S9_extracted_k141_83536 | Not-determined |
| S9_extracted_k141_30477 | Not-determined |
| S9_extracted_k141_60774 | Not-determined |
| S9_extracted_k141_22907 | Not-determined |
| S9_extracted_k141_19075 | Not-determined |
| S9_extracted_k141_60775 | Low-quality |
| S9_extracted_k141_22909 | Not-determined |
| S16_vs_124668\|\|full | High-quality |
| S9_extracted_k141_53222 | Low-quality |
| S9_extracted_k141_79699 | Not-determined |
| S9_extracted_k141_75912 | Not-determined |
| S9_extracted_k141_30480 | Not-determined |
| S9_extracted_k141_3934 | Not-determined |
| S9_extracted_k141_19080 | Not-determined |
| S9_extracted_k141_11523 | Not-determined |
| S9_extracted_k141_41834 | Not-determined |
| S9_extracted_k141_152 | Not-determined |
| S9_extracted_k141_30484 | Not-determined |
| S9_extracted_k141_30485 | Low-quality |
| S9_extracted_k141_83544 | Not-determined |
| S9_extracted_k141_79704 | Not-determined |
| S9_extracted_k141_68277 | Low-quality |
| S9_extracted_k141_15321 | Low-quality |
| S9_extracted_k141_30487 | Not-determined |
| S9_extracted_k141_79705 | Low-quality |
| S9_extracted_k141_79707 | Not-determined |
| S9_extracted_k141_38045 | Low-quality |
| S9_extracted_k141_158 | Low-quality |
| S9_extracted_k141_79709 | Low-quality |
| S9_extracted_k141_19092 | Low-quality |
| S9_extracted_k141_22920 | Not-determined |
| S9_extracted_k141_68282 | Not-determined |
| S9_extracted_k141_22922 | Low-quality |
| S9_extracted_k141_68284 | Not-determined |
| S9_extracted_k141_26645 | Not-determined |
| S9_extracted_k141_41840 | Low-quality |
| S9_extracted_k141_68286 | Low-quality |
| S9_extracted_k141_53241 | Not-determined |
| S9_extracted_k141_34288 | Not-determined |
| S9_extracted_k141_38053 | Not-determined |
| S9_extracted_k141_79715 | Low-quality |
| S9_extracted_k141_11540 | Not-determined |
| S9_extracted_k141_164 | Low-quality |
| S9_extracted_k141_26649 | Not-determined |
| S9_extracted_k141_60789 | Low-quality |
| S8_extracted_k141_11794 | Complete |
| S9_extracted_k141_19102 | Low-quality |
| S9_extracted_k141_68294 | Low-quality |
| S9_extracted_k141_83558 | Low-quality |
| S9_extracted_k141_26652 | Low-quality |
| S9_extracted_k141_79719 | Low-quality |
| S9_extracted_k141_30493 | Not-determined |
| S9_extracted_k141_19106 | Not-determined |
| S9_extracted_k141_19109 | Low-quality |
| S9_extracted_k141_60799 | Not-determined |
| S9_extracted_k141_45570 | Not-determined |
| S9_extracted_k141_45571 | Not-determined |
| S9_extracted_k141_30502 | Not-determined |
| S9_extracted_k141_53244 | Low-quality |
| S9_extracted_k141_53246 | Low-quality |
| S9_extracted_k141_19114 | Not-determined |
| S9_extracted_k141_38067 | Not-determined |
| S9_extracted_k141_19115 | Not-determined |
| S9_extracted_k141_34300 | Not-determined |
| S9_extracted_k141_68306 | Not-determined |
| S9_extracted_k141_87342 | Not-determined |
| S9_extracted_k141_83566 | Not-determined |
| S9_extracted_k141_41863 | Not-determined |
| S9_extracted_k141_11566 | Not-determined |
| S9_extracted_k141_38076 | Not-determined |
| S9_extracted_k141_22928 | Low-quality |
| S9_extracted_k141_22929 | Not-determined |
| S9_extracted_k141_19124 | Not-determined |
| S9_extracted_k141_41867 | Not-determined |
| S9_extracted_k141_79729 | Not-determined |
| S9_extracted_k141_79730 | Not-determined |
| S9_extracted_k141_30520 | Not-determined |
| S9_extracted_k141_79734 | Not-determined |
| S9_extracted_k141_45586 | Not-determined |
| S9_extracted_k141_45587 | Not-determined |
| S9_extracted_k141_30523 | Not-determined |
| S9_extracted_k141_3981 | Not-determined |
| S9_extracted_k141_45591 | Not-determined |
| S9_extracted_k141_26657 | Low-quality |
| S9_extracted_k141_11573 | Low-quality |
| S9_extracted_k141_60823 | Not-determined |
| S9_extracted_k141_56954 | Not-determined |
| S9_extracted_k141_22940 | Not-determined |
| S9_extracted_k141_60825 | Not-determined |
| S9_extracted_k141_53257 | Not-determined |
| S9_extracted_k141_26662 | Low-quality |
| S9_extracted_k141_45599 | Not-determined |
| S9_extracted_k141_79747 | Not-determined |
| S9_extracted_k141_197 | Not-determined |
| S9_extracted_k141_26667 | Not-determined |
| S9_extracted_k141_198 | Low-quality |
| S9_extracted_k141_53263 | Low-quality |
| S9_extracted_k141_19135 | Not-determined |
| S9_extracted_k141_34331 | Not-determined |
| S9_extracted_k141_38096 | Not-determined |
| S9_extracted_k141_64559 | Not-determined |
| S9_extracted_k141_68327 | Not-determined |
| S9_extracted_k141_202 | Not-determined |
| S9_extracted_k141_34337 | Not-determined |
| S9_extracted_k141_53268 | Low-quality |
| S9_extracted_k141_22949 | Not-determined |
| S9_extracted_k141_87361 | Not-determined |
| S9_extracted_k141_87362 | Not-determined |
| S9_extracted_k141_53271 | Not-determined |
| S9_extracted_k141_79752 | Not-determined |
| S9_extracted_k141_11588 | Not-determined |
| S9_extracted_k141_60840 | Not-determined |
| S9_extracted_k141_53279 | Not-determined |
| S9_extracted_k141_53282 | Not-determined |
| S9_extracted_k141_60843 | Not-determined |
| S9_extracted_k141_72115 | Not-determined |
| S9_extracted_k141_22958 | Not-determined |
| S9_extracted_k141_45612 | Not-determined |
| S9_extracted_k141_60848 | Not-determined |
| S9_extracted_k141_68338 | Not-determined |
| S9_extracted_k141_220 | Not-determined |
| S9_extracted_k141_68340 | Not-determined |
| S9_extracted_k141_60851 | Not-determined |
| S9_extracted_k141_72123 | Not-determined |
| S9_extracted_k141_64570 | Not-determined |
| S9_extracted_k141_45616 | Low-quality |
| S9_extracted_k141_19149 | Not-determined |
| S9_extracted_k141_64577 | Not-determined |
| S9_extracted_k141_34362 | Not-determined |
| S9_extracted_k141_223 | Low-quality |
| S9_extracted_k141_56974 | Not-determined |
| S9_extracted_k141_30557 | Not-determined |
| S9_extracted_k141_45620 | Low-quality |
| S9_extracted_k141_68344 | Not-determined |
| S9_extracted_k141_229 | Not-determined |
| S9_extracted_k141_38120 | Not-determined |
| S9_extracted_k141_38122 | Not-determined |
| S9_extracted_k141_49361 | Not-determined |
| S9_extracted_k141_19155 | Not-determined |
| S9_extracted_k141_11614 | Not-determined |
| S9_extracted_k141_87392 | Low-quality |
| S9_extracted_k141_15353 | Not-determined |
| S9_extracted_k141_19167 | Not-determined |
| S9_extracted_k141_11618 | Low-quality |
| S9_extracted_k141_38128 | Not-determined |
| S9_extracted_k141_56986 | Low-quality |
| S9_extracted_k141_30575 | Not-determined |
| S9_extracted_k141_53300 | Low-quality |
| S9_extracted_k141_34390 | Not-determined |
| S9_extracted_k141_79781 | Not-determined |
| S9_extracted_k141_53301 | Not-determined |
| S9_extracted_k141_45637 | Not-determined |
| S9_extracted_k141_15366 | Low-quality |
| S9_extracted_k141_30578 | Not-determined |
| S9_extracted_k141_68355 | Not-determined |
| S9_extracted_k141_11628 | Not-determined |
| S9_extracted_k141_30580 | Not-determined |
| S9_extracted_k141_38138 | Not-determined |
| S9_extracted_k141_68359 | Not-determined |
| S9_extracted_k141_19176 | Low-quality |
| S9_extracted_k141_53310 | Not-determined |
| S9_extracted_k141_53312 | Not-determined |
| S9_extracted_k141_53314 | Not-determined |
| S9_extracted_k141_15375 | Low-quality |
| S9_extracted_k141_45649 | Not-determined |
| S9_extracted_k141_30584 | Not-determined |
| S9_extracted_k141_7714 | Not-determined |
| S9_extracted_k141_15379 | Not-determined |
| S9_extracted_k141_258 | Not-determined |
| S9_extracted_k141_79790 | Not-determined |
| S9_extracted_k141_45652 | Low-quality |
| S9_extracted_k141_34409 | Low-quality |
| S9_extracted_k141_38146 | Not-determined |
| S9_extracted_k141_34410 | Low-quality |
| S9_extracted_k141_11632 | Not-determined |
| S9_extracted_k141_53317 | Not-determined |
| S9_extracted_k141_11633 | Not-determined |
| S9_extracted_k141_79793 | Not-determined |
| S9_extracted_k141_19186 | Not-determined |
| S9_extracted_k141_264 | Low-quality |
| S9_extracted_k141_72161 | Not-determined |
| S9_extracted_k141_64609 | Low-quality |
| S9_extracted_k141_79796 | Not-determined |
| S9_extracted_k141_30597 | Not-determined |
| S9_extracted_k141_11641 | Not-determined |

| | |
|---|---|
| S9_extracted_k141_34419 | Not-determined |
| S9_extracted_k141_72165 | Not-determined |
| S9_extracted_k141_7718 | Low-quality |
| S9_extracted_k141_34421 | Not-determined |
| S9_extracted_k141_34425 | Not-determined |
| S9_extracted_k141_34426 | Low-quality |
| S9_extracted_k141_38166 | Not-determined |
| S9_extracted_k141_64618 | Not-determined |
| S9_extracted_k141_49388 | Low-quality |
| S9_extracted_k141_64619 | Not-determined |
| S9_extracted_k141_87411 | Not-determined |
| S9_extracted_k141_75937 | Not-determined |
| S9_extracted_k141_34432 | Low-quality |
| S9_extracted_k141_19204 | Not-determined |
| S9_extracted_k141_30611 | Not-determined |
| S9_extracted_k141_11648 | Not-determined |
| S9_extracted_k141_19207 | Not-determined |
| S9_extracted_k141_83583 | Not-determined |
| S9_extracted_k141_34435 | Not-determined |
| S9_extracted_k141_15406 | Low-quality |
| S9_extracted_k141_15407 | Not-determined |
| S9_extracted_k141_22970 | Not-determined |
| S9_extracted_k141_292 | Not-determined |
| S9_extracted_k141_68384 | Not-determined |
| S9_extracted_k141_26695 | Not-determined |
| S9_extracted_k141_57023 | Not-determined |
| S9_extracted_k141_34440 | Not-determined |
| S9_extracted_k141_19212 | Low-quality |
| S9_extracted_k141_34442 | Not-determined |
| S9_extracted_k141_83587 | Not-determined |
| S9_extracted_k141_49397 | Low-quality |
| S9_extracted_k141_11654 | Not-determined |
| S9_extracted_k141_49399 | Not-determined |
| S9_extracted_k141_79818 | Not-determined |
| S9_extracted_k141_75943 | Not-determined |
| S9_extracted_k141_72175 | Low-quality |
| S9_extracted_k141_64631 | Not-determined |
| S9_extracted_k141_79823 | Not-determined |
| S9_extracted_k141_87426 | Low-quality |
| S9_extracted_k141_26707 | Not-determined |
| S9_extracted_k141_72178 | Not-determined |
| S9_extracted_k141_30628 | Not-determined |
| S9_extracted_k141_45669 | Low-quality |
| S9_extracted_k141_68397 | Low-quality |
| S9_extracted_k141_41908 | Not-determined |
| S9_extracted_k141_45671 | Not-determined |
| S9_extracted_k141_75949 | Not-determined |
| S9_extracted_k141_26711 | Not-determined |
| S9_extracted_k141_72180 | Not-determined |
| S9_extracted_k141_45676 | Not-determined |
| S9_extracted_k141_72182 | Not-determined |
| S9_extracted_k141_15421 | Not-determined |
| S9_extracted_k141_22985 | Low-quality |
| S9_extracted_k141_79829 | Low-quality |
| S9_extracted_k141_34454 | Not-determined |
| S9_extracted_k141_64639 | Not-determined |
| S9_extracted_k141_15424 | Not-determined |
| S9_extracted_k141_68401 | Not-determined |
| S9_extracted_k141_19233 | Not-determined |
| S9_extracted_k141_79832 | Not-determined |
| S9_extracted_k141_49415 | Not-determined |
| S9_extracted_k141_79834 | Not-determined |
| S9_extracted_k141_79836 | Not-determined |
| S9_extracted_k141_45680 | Not-determined |
| S9_extracted_k141_22999 | Not-determined |
| S9_extracted_k141_15430 | Low-quality |
| S9_extracted_k141_15431 | Not-determined |
| S9_extracted_k141_53360 | Not-determined |
| S9_extracted_k141_72188 | Not-determined |
| S9_extracted_k141_64645 | Not-determined |
| S9_extracted_k141_72192 | Not-determined |
| S9_extracted_k141_72193 | Low-quality |
| S9_extracted_k141_49425 | Not-determined |
| S9_extracted_k141_53367 | Not-determined |
| S9_extracted_k141_23005 | Not-determined |
| S9_extracted_k141_49426 | Not-determined |
| S9_extracted_k141_325 | Not-determined |
| S9_extracted_k141_23007 | Not-determined |
| S9_extracted_k141_11685 | Not-determined |
| S9_extracted_k141_23008 | Not-determined |
| S9_extracted_k141_11686 | Not-determined |
| S9_extracted_k141_23009 | Not-determined |
| S9_extracted_k141_7758 | Low-quality |
| S9_extracted_k141_7760 | Not-determined |
| S9_extracted_k141_57028 | Not-determined |
| S9_extracted_k141_72195 | Not-determined |
| S9_extracted_k141_72197 | Not-determined |
| S9_extracted_k141_87446 | Low-quality |
| S9_extracted_k141_4094 | Not-determined |
| S9_extracted_k141_72200 | Not-determined |
| S9_extracted_k141_23016 | Not-determined |
| S9_extracted_k141_87449 | Not-determined |
| S9_extracted_k141_34472 | Not-determined |
| S9_extracted_k141_19259 | Not-determined |
| S9_extracted_k141_336 | Not-determined |
| S9_extracted_k141_83608 | Not-determined |
| S9_extracted_k141_72204 | Not-determined |
| S9_extracted_k141_72206 | Not-determined |
| S9_extracted_k141_15444 | Not-determined |
| S9_extracted_k141_19267 | Low-quality |
| S9_extracted_k141_26734 | Low-quality |
| S9_extracted_k141_4098 | Not-determined |
| S9_extracted_k141_7773 | Not-determined |
| S9_extracted_k141_11699 | Low-quality |
| S9_extracted_k141_30656 | Not-determined |
| S9_extracted_k141_64662 | Not-determined |
| S9_extracted_k141_38207 | Not-determined |
| S9_extracted_k141_41942 | Not-determined |
| S9_extracted_k141_30658 | Not-determined |
| S9_extracted_k141_57042 | Not-determined |
| S9_extracted_k141_30659 | Not-determined |
| S9_extracted_k141_64668 | Not-determined |
| S9_extracted_k141_4102 | Low-quality |
| S9_extracted_k141_34488 | Not-determined |
| S9_extracted_k141_64670 | Not-determined |
| S9_extracted_k141_23028 | Not-determined |
| S9_extracted_k141_19272 | Not-determined |
| S9_extracted_k141_7776 | Low-quality |
| S9_extracted_k141_68424 | Not-determined |
| S9_extracted_k141_34491 | Not-determined |
| S9_extracted_k141_38215 | Not-determined |
| S9_extracted_k141_23033 | Low-quality |
| S9_extracted_k141_4109 | Not-determined |
| S9_extracted_k141_38220 | Not-determined |
| S9_extracted_k141_79856 | Low-quality |
| S9_extracted_k141_23039 | Not-determined |
| S9_extracted_k141_34500 | Not-determined |
| S9_extracted_k141_34501 | Not-determined |
| S9_extracted_k141_87462 | Not-determined |
| S9_extracted_k141_79860 | Not-determined |
| S9_extracted_k141_23042 | Not-determined |
| S9_extracted_k141_79862 | Low-quality |
| S9_extracted_k141_79868 | Not-determined |
| S9_extracted_k141_79869 | Not-determined |
| S9_extracted_k141_26758 | Not-determined |
| S9_extracted_k141_34510 | Not-determined |
| S9_extracted_k141_7787 | Not-determined |
| S9_extracted_k141_23051 | Not-determined |
| S9_extracted_k141_49438 | Low-quality |
| S9_extracted_k141_360 | Not-determined |
| S9_extracted_k141_83630 | Not-determined |
| S9_extracted_k141_72230 | Not-determined |
| S9_extracted_k141_34515 | Not-determined |
| S9_extracted_k141_83633 | Not-determined |
| S9_extracted_k141_34518 | Not-determined |
| S9_extracted_k141_45682 | Low-quality |
| S9_extracted_k141_79882 | Not-determined |
| S9_extracted_k141_83636 | Not-determined |
| S9_extracted_k141_68439 | Not-determined |
| S9_extracted_k141_49445 | Low-quality |
| S9_extracted_k141_87482 | Not-determined |
| S9_extracted_k141_41968 | Not-determined |
| S9_extracted_k141_68443 | Not-determined |
| S9_extracted_k141_72239 | Not-determined |
| S9_extracted_k141_41970 | Low-quality |
| S9_extracted_k141_15459 | Low-quality |
| S9_extracted_k141_49448 | Not-determined |
| S9_extracted_k141_68444 | Not-determined |
| S9_extracted_k141_87487 | Not-determined |
| S9_extracted_k141_49451 | Not-determined |
| S9_extracted_k141_79888 | Low-quality |
| S9_extracted_k141_38247 | Not-determined |
| S9_extracted_k141_45691 | Not-determined |
| S9_extracted_k141_49453 | Not-determined |
| S9_extracted_k141_72244 | Not-determined |
| S9_extracted_k141_41975 | Not-determined |
| S9_extracted_k141_26772 | Not-determined |
| S9_extracted_k141_7800 | Not-determined |
| S9_extracted_k141_49455 | Low-quality |
| S9_extracted_k141_41981 | Not-determined |
| S9_extracted_k141_83643 | Not-determined |
| S9_extracted_k141_49461 | Not-determined |
| S9_extracted_k141_49463 | Not-determined |
| S9_extracted_k141_41983 | Low-quality |
| S9_extracted_k141_83651 | Not-determined |
| S9_extracted_k141_87499 | Not-determined |
| S9_extracted_k141_87501 | Not-determined |
| S9_extracted_k141_64708 | Not-determined |
| S9_extracted_k141_83657 | Low-quality |
| S9_extracted_k141_49469 | Not-determined |
| S9_extracted_k141_23082 | Not-determined |
| S9_extracted_k141_57068 | Not-determined |
| S9_extracted_k141_15481 | Not-determined |
| S9_extracted_k141_60871 | Low-quality |
| S9_extracted_k141_34525 | Low-quality |
| S9_extracted_k141_41995 | Not-determined |
| S9_extracted_k141_34527 | Not-determined |
| S9_extracted_k141_30674 | Not-determined |
| S9_extracted_k141_7819 | Not-determined |
| S9_extracted_k141_45700 | Not-determined |
| S9_extracted_k141_41999 | Not-determined |
| S9_extracted_k141_68467 | Not-determined |
| S9_extracted_k141_72267 | Low-quality |
| S9_extracted_k141_19279 | Low-quality |
| S9_extracted_k141_30683 | Not-determined |
| S9_extracted_k141_87518 | Not-determined |
| S9_extracted_k141_72271 | Not-determined |
| S8_vs_k141_11794\|\|full | High-quality |
| S9_extracted_k141_4118 | Low-quality |
| S9_extracted_k141_30689 | Low-quality |
| S9_extracted_k141_38261 | Not-determined |
| S9_extracted_k141_57090 | Not-determined |
| S9_extracted_k141_30694 | Not-determined |
| S9_extracted_k141_87533 | Not-determined |
| S9_extracted_k141_49482 | Not-determined |
| S9_extracted_k141_34536 | Not-determined |
| S9_extracted_k141_30699 | Low-quality |
| S9_extracted_k141_42011 | Not-determined |
| S9_extracted_k141_34537 | Low-quality |
| S9_extracted_k141_68478 | Not-determined |
| S9_extracted_k141_34539 | Low-quality |
| S9_extracted_k141_30707 | Not-determined |
| S12_vs_k141_133070\|\|full | Medium-quality |
| S9_extracted_k141_49492 | Low-quality |
| S9_extracted_k141_72293 | Not-determined |
| S9_extracted_k141_23106 | Low-quality |
| S9_extracted_k141_408 | Not-determined |
| S9_extracted_k141_30714 | Not-determined |
| S9_extracted_k141_49496 | Not-determined |
| S9_extracted_k141_68490 | Not-determined |
| S9_extracted_k141_38286 | Not-determined |
| S9_extracted_k141_49498 | Low-quality |
| S9_extracted_k141_42025 | Not-determined |
| S9_extracted_k141_72296 | Not-determined |
| S9_extracted_k141_15517 | Not-determined |
| S9_extracted_k141_72297 | Not-determined |
| S9_extracted_k141_23109 | Not-determined |
| S9_extracted_k141_38293 | Not-determined |
| S9_extracted_k141_23110 | Not-determined |
| S9_extracted_k141_79915 | Not-determined |
| S9_extracted_k141_4134 | Low-quality |
| S9_extracted_k141_83674 | Not-determined |
| S9_extracted_k141_15519 | Not-determined |
| S9_extracted_k141_15520 | Not-determined |
| S9_extracted_k141_4137 | Not-determined |
| S9_extracted_k141_38296 | Low-quality |
| S9_extracted_k141_38297 | Not-determined |
| S9_extracted_k141_83679 | Low-quality |
| S9_extracted_k141_75954 | Low-quality |
| S9_extracted_k141_416 | Not-determined |
| S9_extracted_k141_64761 | Not-determined |
| S9_extracted_k141_75959 | Not-determined |
| S9_extracted_k141_49515 | Not-determined |
| S9_extracted_k141_4146 | Not-determined |
| S9_extracted_k141_11706 | Low-quality |
| S9_extracted_k141_4147 | Not-determined |
| S9_extracted_k141_53396 | Not-determined |
| S9_extracted_k141_424 | Not-determined |
| S9_extracted_k141_7842 | Not-determined |
| S9_extracted_k141_45718 | Not-determined |
| S9_extracted_k141_7846 | Not-determined |
| S9_extracted_k141_30733 | Low-quality |
| S9_extracted_k141_23120 | Low-quality |
| S9_extracted_k141_60923 | Not-determined |
| S9_extracted_k141_64778 | Not-determined |
| S9_extracted_k141_23122 | Not-determined |
| S9_extracted_k141_75969 | Not-determined |
| S9_extracted_k141_7849 | Not-determined |
| S9_extracted_k141_11717 | Not-determined |
| S9_extracted_k141_30737 | Low-quality |
| S9_extracted_k141_87556 | Not-determined |
| S9_extracted_k141_42048 | Not-determined |
| S9_extracted_k141_4154 | Low-quality |
| S9_extracted_k141_434 | Not-determined |
| S9_extracted_k141_57133 | Not-determined |
| S9_extracted_k141_72300 | Low-quality |
| S9_extracted_k141_57135 | Low-quality |
| S9_extracted_k141_87561 | Not-determined |
| S9_extracted_k141_45730 | Not-determined |
| S9_extracted_k141_42053 | Low-quality |
| S9_extracted_k141_11720 | Not-determined |
| S9_extracted_k141_45733 | Not-determined |
| S9_extracted_k141_87566 | Not-determined |
| S9_extracted_k141_72313 | Low-quality |
| S9_extracted_k141_60939 | Not-determined |
| S9_extracted_k141_42065 | Not-determined |
| S9_extracted_k141_15545 | Not-determined |
| S9_extracted_k141_49543 | Not-determined |
| S9_extracted_k141_75983 | Not-determined |
| S9_extracted_k141_30747 | Not-determined |
| S9_extracted_k141_75984 | Low-quality |
| S9_extracted_k141_30748 | Not-determined |
| S9_extracted_k141_30750 | Low-quality |
| S9_extracted_k141_4172 | Not-determined |
| S9_extracted_k141_34548 | Low-quality |
| S9_extracted_k141_34549 | Not-determined |
| S9_extracted_k141_83710 | Low-quality |
| S9_extracted_k141_34552 | Not-determined |
| S9_extracted_k141_23142 | Not-determined |
| S9_extracted_k141_68542 | Low-quality |
| S9_extracted_k141_34553 | Not-determined |
| S9_extracted_k141_87578 | Not-determined |
| S9_extracted_k141_68543 | Not-determined |
| S9_extracted_k141_23143 | Not-determined |
| S9_extracted_k141_60945 | Not-determined |
| S9_extracted_k141_60946 | Not-determined |
| S9_extracted_k141_30761 | Not-determined |
| S9_extracted_k141_64796 | Not-determined |
| S9_extracted_k141_75995 | Not-determined |
| S9_extracted_k141_42075 | Not-determined |
| S12_vs_k141_121913\|\|full | Medium-quality |
| S9_extracted_k141_11743 | Not-determined |
| S9_extracted_k141_83722 | Low-quality |
| S9_extracted_k141_26824 | Not-determined |
| S9_extracted_k141_23150 | Low-quality |
| S9_extracted_k141_19288 | Not-determined |
| S9_extracted_k141_72333 | Low-quality |
| S9_extracted_k141_26827 | Not-determined |
| S9_extracted_k141_4188 | Not-determined |
| S9_extracted_k141_72335 | Not-determined |
| S9_extracted_k141_42083 | Not-determined |
| S9_extracted_k141_68555 | Not-determined |
| S9_extracted_k141_68556 | Not-determined |
| S12_extracted_k141_169796 | High-quality |
| S9_extracted_k141_75999 | Low-quality |
| S9_extracted_k141_49556 | Not-determined |
| S9_extracted_k141_57157 | Not-determined |
| S9_extracted_k141_79921 | Not-determined |
| S9_extracted_k141_68558 | Not-determined |
| S9_extracted_k141_4195 | Low-quality |
| S9_extracted_k141_64802 | Not-determined |
| S9_extracted_k141_34557 | Not-determined |
| S9_extracted_k141_26838 | Not-determined |
| S9_extracted_k141_42090 | Not-determined |
| S9_extracted_k141_30772 | Low-quality |
| S9_extracted_k141_49562 | Not-determined |
| S9_extracted_k141_49564 | Not-determined |
| S9_extracted_k141_72340 | Not-determined |
| S9_extracted_k141_42096 | Not-determined |
| S9_extracted_k141_79933 | Not-determined |
| S9_extracted_k141_19305 | Not-determined |
| S9_extracted_k141_42097 | Not-determined |
| S9_extracted_k141_83735 | Low-quality |
| S9_extracted_k141_60967 | Low-quality |
| S9_extracted_k141_68561 | Not-determined |
| S9_extracted_k141_30776 | Not-determined |
| S9_extracted_k141_72344 | Not-determined |
| S9_extracted_k141_45756 | Not-determined |
| S9_extracted_k141_76008 | Not-determined |
| S9_extracted_k141_42101 | Not-determined |
| S9_extracted_k141_83738 | Low-quality |
| S9_extracted_k141_19310 | Low-quality |
| S9_extracted_k141_465 | Not-determined |
| S9_extracted_k141_45757 | Low-quality |
| S9_extracted_k141_87589 | Not-determined |
| S9_extracted_k141_34566 | Not-determined |
| S9_extracted_k141_467 | Not-determined |
| S9_extracted_k141_87592 | Not-determined |
| S9_extracted_k141_57168 | Low-quality |
| S9_extracted_k141_76014 | Not-determined |
| S9_extracted_k141_60978 | Not-determined |
| S9_extracted_k141_49576 | Not-determined |
| S9_extracted_k141_15581 | Not-determined |
| S9_extracted_k141_475 | Not-determined |
| S9_extracted_k141_23159 | Low-quality |
| S9_extracted_k141_83746 | Not-determined |
| S9_extracted_k141_4216 | Not-determined |
| S9_extracted_k141_68570 | Not-determined |
| S9_extracted_k141_87599 | Not-determined |
| S9_extracted_k141_49581 | Not-determined |
| S9_extracted_k141_15585 | Not-determined |
| S9_extracted_k141_68572 | Not-determined |
| S9_extracted_k141_30789 | Low-quality |
| S9_extracted_k141_79942 | Not-determined |
| S9_extracted_k141_68573 | Not-determined |
| S9_extracted_k141_87601 | Not-determined |
| S9_extracted_k141_38314 | Not-determined |
| S9_extracted_k141_15590 | Not-determined |
| S9_extracted_k141_19326 | Not-determined |
| S9_extracted_k141_23164 | Not-determined |
| S9_extracted_k141_68577 | Not-determined |
| S9_extracted_k141_83754 | Not-determined |
| S9_extracted_k141_23165 | Not-determined |
| S9_extracted_k141_15595 | Not-determined |
| S9_extracted_k141_30792 | Not-determined |
| S9_extracted_k141_79945 | Not-determined |
| S9_extracted_k141_83757 | Not-determined |
| S9_extracted_k141_49587 | Low-quality |
| S9_extracted_k141_49588 | Not-determined |
| S9_extracted_k141_26862 | Not-determined |
| S9_extracted_k141_23171 | Not-determined |
| S9_extracted_k141_15600 | Not-determined |
| S9_extracted_k141_30802 | Not-determined |
| S9_extracted_k141_19332 | Not-determined |
| S9_extracted_k141_38325 | Low-quality |
| S9_extracted_k141_30805 | Not-determined |
| S9_extracted_k141_76020 | Not-determined |
| S9_extracted_k141_38327 | Not-determined |
| S9_extracted_k141_53413 | Not-determined |
| S9_extracted_k141_68579 | Low-quality |
| S9_extracted_k141_57174 | Low-quality |
| S9_extracted_k141_68580 | Not-determined |
| S9_extracted_k141_87616 | Not-determined |
| S9_extracted_k141_57176 | Not-determined |
| S9_extracted_k141_4233 | Not-determined |
| S9_extracted_k141_23177 | Not-determined |
| S9_extracted_k141_57178 | Not-determined |
| S9_extracted_k141_61006 | Not-determined |
| S9_extracted_k141_53422 | Not-determined |
| S9_extracted_k141_53423 | Not-determined |
| S9_extracted_k141_30815 | Low-quality |
| S9_extracted_k141_15612 | Not-determined |
| S9_extracted_k141_49611 | Not-determined |
| S9_extracted_k141_72363 | Low-quality |
| S9_extracted_k141_45766 | Not-determined |
| S9_extracted_k141_4244 | Not-determined |
| S9_extracted_k141_15617 | Not-determined |
| S9_extracted_k141_53432 | Not-determined |
| S9_extracted_k141_42122 | Low-quality |
| S9_extracted_k141_38341 | Not-determined |
| S9_extracted_k141_72367 | Not-determined |
| S9_extracted_k141_49614 | Not-determined |
| S9_extracted_k141_42127 | Low-quality |
| S9_extracted_k141_83769 | Not-determined |
| S9_extracted_k141_495 | Not-determined |
| S9_extracted_k141_4250 | Not-determined |
| S9_extracted_k141_49616 | Not-determined |
| S9_extracted_k141_72374 | Not-determined |
| S9_extracted_k141_15625 | Not-determined |
| S9_extracted_k141_72376 | Not-determined |
| S9_extracted_k141_76044 | Not-determined |
| S9_extracted_k141_502 | Not-determined |
| S9_extracted_k141_49623 | Not-determined |
| S9_extracted_k141_83777 | Not-determined |
| S9_extracted_k141_57196 | Not-determined |
| S9_extracted_k141_57197 | Low-quality |
| S9_extracted_k141_83780 | Not-determined |
| S9_extracted_k141_7886 | Not-determined |
| S9_extracted_k141_72380 | Not-determined |
| S9_extracted_k141_45780 | Not-determined |
| S9_extracted_k141_15630 | Not-determined |
| S9_extracted_k141_42137 | Not-determined |
| S9_extracted_k141_42138 | Not-determined |
| S9_extracted_k141_26906 | Not-determined |
| S9_extracted_k141_83789 | Not-determined |
| S9_extracted_k141_516 | Not-determined |
| S9_extracted_k141_45786 | Not-determined |
| S9_extracted_k141_61027 | Not-determined |
| S9_extracted_k141_42143 | Not-determined |
| S9_extracted_k141_15638 | Low-quality |
| S9_extracted_k141_15640 | Not-determined |
| S9_extracted_k141_38364 | Not-determined |
| S9_extracted_k141_76052 | Not-determined |
| S9_extracted_k141_7897 | Not-determined |
| S9_extracted_k141_76054 | Not-determined |
| S9_extracted_k141_53450 | Low-quality |
| S9_extracted_k141_23186 | Not-determined |
| S9_extracted_k141_34575 | Low-quality |
| S9_extracted_k141_4264 | Not-determined |
| S9_extracted_k141_19362 | Not-determined |
| S9_extracted_k141_7907 | Not-determined |
| S9_extracted_k141_61036 | Not-determined |
| S9_extracted_k141_7908 | Not-determined |
| S9_extracted_k141_524 | Not-determined |
| S9_extracted_k141_34578 | Low-quality |
| S9_extracted_k141_23188 | Low-quality |
| S9_extracted_k141_34580 | Not-determined |
| S9_extracted_k141_83801 | Not-determined |
| S9_extracted_k141_42156 | Not-determined |
| S9_extracted_k141_76066 | Low-quality |
| S9_extracted_k141_49637 | Not-determined |
| S9_extracted_k141_83803 | Low-quality |
| S9_extracted_k141_528 | Not-determined |
| S9_extracted_k141_79979 | Not-determined |
| S9_extracted_k141_83805 | Not-determined |
| S9_extracted_k141_15655 | Not-determined |
| S9_extracted_k141_57226 | Low-quality |
| S9_extracted_k141_26926 | Low-quality |
| S9_extracted_k141_4275 | Low-quality |
| S9_extracted_k141_49647 | Low-quality |
| S9_extracted_k141_7913 | Low-quality |
| S9_extracted_k141_15656 | Low-quality |
| S9_extracted_k141_4276 | Low-quality |
| S9_extracted_k141_30819 | Not-determined |
| S9_extracted_k141_38379 | Not-determined |
| S9_extracted_k141_38380 | Low-quality |
| S9_extracted_k141_76076 | Not-determined |
| S9_extracted_k141_4280 | Not-determined |
| S9_extracted_k141_72414 | Not-determined |
| S9_extracted_k141_87633 | Not-determined |
| S9_extracted_k141_23208 | Not-determined |
| S9_extracted_k141_49660 | Not-determined |
| S9_extracted_k141_79989 | Low-quality |
| S9_extracted_k141_42166 | Not-determined |
| S9_extracted_k141_7919 | Not-determined |
| S9_extracted_k141_53476 | Not-determined |
| S9_extracted_k141_79992 | Low-quality |
| S9_extracted_k141_79993 | Not-determined |
| S9_extracted_k141_87639 | Not-determined |
| S9_extracted_k141_26944 | Not-determined |
| S9_extracted_k141_15669 | Not-determined |
| S9_extracted_k141_30827 | Low-quality |
| S9_extracted_k141_53482 | Not-determined |
| S9_extracted_k141_19380 | Not-determined |
| S9_extracted_k141_38398 | Not-determined |
| S9_extracted_k141_76091 | Not-determined |
| S9_extracted_k141_79995 | Not-determined |
| S9_extracted_k141_7924 | Not-determined |
| S9_extracted_k141_15676 | Not-determined |
| S9_extracted_k141_76096 | Low-quality |
| S9_extracted_k141_79999 | Not-determined |
| S9_extracted_k141_42175 | Not-determined |
| S9_extracted_k141_53487 | Not-determined |
| S9_extracted_k141_57235 | Low-quality |
| S9_extracted_k141_61060 | Low-quality |
| S9_extracted_k141_26953 | Not-determined |
| S9_extracted_k141_23210 | Not-determined |
| S9_extracted_k141_15680 | Not-determined |
| S9_extracted_k141_538 | Low-quality |
| S9_extracted_k141_30832 | Not-determined |
| S9_extracted_k141_30833 | Not-determined |
| S9_extracted_k141_11876 | Not-determined |
| S9_extracted_k141_49668 | Not-determined |
| S9_extracted_k141_49670 | Not-determined |
| S9_extracted_k141_15690 | Not-determined |
| S9_extracted_k141_38409 | Not-determined |
| S9_extracted_k141_546 | Not-determined |
| S9_extracted_k141_548 | Not-determined |
| S9_extracted_k141_26966 | Not-determined |
| S9_extracted_k141_49676 | Low-quality |
| S9_extracted_k141_11883 | Not-determined |
| S9_extracted_k141_57248 | Low-quality |
| S9_extracted_k141_26968 | Not-determined |
| S9_extracted_k141_23222 | Not-determined |
| S9_extracted_k141_42188 | Not-determined |
| S9_extracted_k141_57250 | Not-determined |
| S9_extracted_k141_551 | Not-determined |
| S9_extracted_k141_26970 | Not-determined |
| S9_extracted_k141_76111 | Not-determined |
| S9_extracted_k141_19385 | Not-determined |
| S9_extracted_k141_83833 | Not-determined |
| S9_extracted_k141_15693 | Not-determined |
| S9_extracted_k141_11890 | Not-determined |
| S9_extracted_k141_83835 | Not-determined |
| S9_extracted_k141_26973 | Not-determined |
| S9_extracted_k141_76115 | Not-determined |
| S9_extracted_k141_15698 | Not-determined |
| S9_extracted_k141_61075 | Low-quality |
| S9_extracted_k141_42194 | Low-quality |
| S9_extracted_k141_38414 | Not-determined |
| S9_extracted_k141_23233 | Not-determined |
| S9_extracted_k141_87665 | Not-determined |
| S9_extracted_k141_38417 | Low-quality |
| S9_extracted_k141_42200 | Not-determined |
| S9_extracted_k141_76119 | Low-quality |
| S9_extracted_k141_38423 | Not-determined |
| S9_extracted_k141_23235 | Low-quality |
| S9_extracted_k141_42203 | Not-determined |
| S9_extracted_k141_61079 | Low-quality |
| S9_extracted_k141_11899 | Not-determined |
| S9_extracted_k141_38428 | Low-quality |
| S9_extracted_k141_64823 | Not-determined |
| S9_extracted_k141_76125 | Not-determined |
| S9_extracted_k141_26988 | Not-determined |
| S9_extracted_k141_38429 | Low-quality |
| S9_extracted_k141_19394 | Low-quality |
| S9_extracted_k141_15714 | Not-determined |
| S9_extracted_k141_19395 | Not-determined |
| S9_extracted_k141_19397 | Not-determined |
| S9_extracted_k141_566 | Not-determined |
| S9_extracted_k141_38437 | Not-determined |
| S9_extracted_k141_42211 | Not-determined |
| S9_extracted_k141_15721 | Not-determined |
| S9_extracted_k141_19403 | Not-determined |
| S9_extracted_k141_83846 | Not-determined |
| S9_extracted_k141_76133 | Not-determined |
| S9_extracted_k141_87680 | Not-determined |
| S9_extracted_k141_38441 | Not-determined |
| S9_extracted_k141_83848 | Not-determined |
| S9_extracted_k141_7958 | Not-determined |
| S9_extracted_k141_42217 | Not-determined |
| S9_extracted_k141_76134 | Not-determined |
| S9_extracted_k141_23240 | Low-quality |
| S9_extracted_k141_27000 | Not-determined |
| S9_extracted_k141_27001 | Not-determined |
| S9_extracted_k141_61094 | Not-determined |
| S9_extracted_k141_11908 | Low-quality |
| S9_extracted_k141_4322 | Low-quality |
| S9_extracted_k141_27004 | Not-determined |
| S9_extracted_k141_38451 | Not-determined |
| S9_extracted_k141_27005 | Not-determined |
| S9_extracted_k141_64834 | Low-quality |
| S9_extracted_k141_87692 | Not-determined |
| S9_extracted_k141_27007 | Low-quality |
| S9_extracted_k141_30857 | Not-determined |
| S9_extracted_k141_76139 | Not-determined |
| S9_extracted_k141_19417 | Not-determined |
| S9_extracted_k141_61102 | Not-determined |
| S9_extracted_k141_87697 | Not-determined |

| | |
|---|---|
| S9_extracted_k141_80036 | Low-quality |
| S9_extracted_k141_578 | Not-determined |
| S9_extracted_k141_64838 | Low-quality |
| S9_extracted_k141_64839 | Not-determined |
| S9_extracted_k141_87702 | Not-determined |
| S9_extracted_k141_19424 | Not-determined |
| S9_extracted_k141_83853 | Not-determined |
| S9_extracted_k141_80043 | Not-determined |
| S9_extracted_k141_61114 | Low-quality |
| S9_extracted_k141_72451 | Not-determined |
| S9_extracted_k141_80047 | Not-determined |
| S9_extracted_k141_83857 | Not-determined |
| S9_extracted_k141_30864 | Not-determined |
| S9_extracted_k141_76151 | Not-determined |
| S9_extracted_k141_87711 | Low-quality |
| S9_extracted_k141_11931 | Not-determined |
| S9_extracted_k141_27023 | Not-determined |
| S9_extracted_k141_83863 | Not-determined |
| S9_extracted_k141_80054 | Not-determined |
| S9_extracted_k141_30870 | Not-determined |
| S9_extracted_k141_83866 | Not-determined |
| S9_extracted_k141_30872 | Not-determined |
| S9_extracted_k141_61124 | Not-determined |
| S9_extracted_k141_53533 | Not-determined |
| S9_extracted_k141_87720 | Not-determined |
| S9_extracted_k141_27028 | Not-determined |
| S9_extracted_k141_72461 | Low-quality |
| S9_extracted_k141_53536 | Not-determined |
| S9_extracted_k141_11938 | Not-determined |
| S9_extracted_k141_7989 | Not-determined |
| S9_extracted_k141_87725 | Not-determined |
| S9_extracted_k141_53541 | Not-determined |
| S9_extracted_k141_42232 | Not-determined |
| S9_extracted_k141_87728 | Not-determined |
| S9_extracted_k141_19440 | Not-determined |
| S9_extracted_k141_597 | Not-determined |
| S9_extracted_k141_87730 | Not-determined |
| S9_extracted_k141_61131 | Not-determined |
| S9_extracted_k141_53546 | Not-determined |
| S9_extracted_k141_72473 | Not-determined |
| S9_extracted_k141_72474 | Not-determined |
| S9_extracted_k141_11951 | Not-determined |
| S9_extracted_k141_53553 | Not-determined |
| S9_extracted_k141_80071 | Not-determined |
| S9_extracted_k141_19446 | Not-determined |
| S9_extracted_k141_4336 | Not-determined |
| S9_extracted_k141_23262 | Low-quality |
| S9_extracted_k141_80074 | Not-determined |
| S9_extracted_k141_8001 | Not-determined |
| S9_extracted_k141_76172 | Not-determined |
| S9_extracted_k141_42246 | Not-determined |
| S9_extracted_k141_72481 | Not-determined |
| S9_extracted_k141_72483 | Not-determined |
| S9_extracted_k141_30893 | Not-determined |
| S9_extracted_k141_42249 | Not-determined |
| [illegible] | [illegible] |

| | |
|---|---|
| S9_extracted_k141_57421 | Not-determined |
| S9_extracted_k141_31172 | Not-determined |
| S9_extracted_k141_80373 | Not-determined |
| S9_extracted_k141_34838 | Not-determined |
| S9_extracted_k141_34839 | Low-quality |
| S9_extracted_k141_49803 | Not-determined |
| S9_extracted_k141_80375 | Low-quality |
| S9_extracted_k141_76407 | Not-determined |
| S9_extracted_k141_31175 | Not-determined |
| S9_extracted_k141_12268 | Low-quality |
| S9_extracted_k141_31176 | Not-determined |
| S9_extracted_k141_61416 | Not-determined |
| S9_extracted_k141_12269 | Not-determined |
| S9_extracted_k141_46144 | Not-determined |
| S9_extracted_k141_88024 | Not-determined |
| S9_extracted_k141_23477 | Low-quality |
| S9_extracted_k141_80377 | Not-determined |
| S9_extracted_k141_76409 | Low-quality |
| S9_extracted_k141_34840 | Low-quality |
| S9_extracted_k141_88027 | Not-determined |
| S9_extracted_k141_764 | Low-quality |
| S9_extracted_k141_38822 | Not-determined |
| S9_extracted_k141_4529 | Not-determined |
| S9_extracted_k141_76411 | Low-quality |
| S9_extracted_k141_23483 | Not-determined |
| S9_extracted_k141_768 | Not-determined |
| S9_extracted_k141_34848 | Not-determined |
| S9_extracted_k141_46154 | Not-determined |
| S9_extracted_k141_12278 | Low-quality |
| S9_extracted_k141_42565 | Not-determined |
| S9_extracted_k141_34853 | Not-determined |
| S9_extracted_k141_88037 | Not-determined |
| S9_extracted_k141_31185 | Low-quality |
| S9_extracted_k141_16062 | Not-determined |
| S9_extracted_k141_31187 | Not-determined |
| S9_extracted_k141_34858 | Not-determined |
| S9_extracted_k141_38840 | Not-determined |
| S9_extracted_k141_88042 | Not-determined |
| S9_extracted_k141_31189 | Low-quality |
| S9_extracted_k141_23500 | Not-determined |
| S9_extracted_k141_777 | Not-determined |
| S9_extracted_k141_23503 | Not-determined |
| S9_extracted_k141_4544 | Not-determined |
| S9_extracted_k141_34862 | Not-determined |
| S9_extracted_k141_49826 | Low-quality |
| S9_extracted_k141_31199 | Not-determined |
| S9_extracted_k141_57432 | Low-quality |
| S9_extracted_k141_80389 | Low-quality |
| S9_extracted_k141_38851 | Not-determined |
| S9_extracted_k141_31201 | Not-determined |
| S9_extracted_k141_34867 | Low-quality |
| S9_extracted_k141_783 | Not-determined |
| S9_extracted_k141_46176 | Not-determined |
| S9_extracted_k141_4554 | Not-determined |
| S9_extracted_k141_61444 | Not-determined |
| S9_extracted_k141_23511 | Not-determined |
| S9_extracted_k141_53917 | Not-determined |
| S9_extracted_k141_34872 | Not-determined |
| S9_extracted_k141_80404 | Not-determined |
| S9_extracted_k141_72761 | Not-determined |
| S9_extracted_k141_76423 | Low-quality |
| S9_extracted_k141_76424 | Not-determined |
| S9_extracted_k141_80408 | Not-determined |
| S9_extracted_k141_80412 | Not-determined |
| S9_extracted_k141_53926 | Not-determined |
| S9_extracted_k141_38856 | Low-quality |
| S9_extracted_k141_88068 | Not-determined |
| S9_extracted_k141_72771 | Not-determined |
| S9_extracted_k141_16090 | Low-quality |
| S9_extracted_k141_88072 | Low-quality |
| S9_extracted_k141_791 | Not-determined |
| S9_extracted_k141_61456 | Not-determined |
| S9_extracted_k141_53928 | Low-quality |
| S9_extracted_k141_46194 | Not-determined |
| S9_extracted_k141_46195 | Not-determined |
| S9_extracted_k141_798 | Not-determined |
| S9_extracted_k141_61461 | Not-determined |
| S9_extracted_k141_76431 | Not-determined |
| S9_extracted_k141_61462 | Not-determined |
| S9_extracted_k141_76432 | Not-determined |
| S9_extracted_k141_72777 | Not-determined |
| S9_extracted_k141_34880 | Not-determined |
| S9_extracted_k141_34882 | Not-determined |
| S9_extracted_k141_65090 | Not-determined |
| S9_extracted_k141_61471 | Low-quality |
| S9_extracted_k141_53940 | Not-determined |
| S9_extracted_k141_49851 | Not-determined |
| S9_extracted_k141_31206 | Not-determined |
| S9_extracted_k141_61473 | Not-determined |
| S9_extracted_k141_72786 | Not-determined |
| S9_extracted_k141_61475 | Not-determined |
| S9_extracted_k141_65093 | Not-determined |
| S9_extracted_k141_72788 | Not-determined |
| S9_extracted_k141_31214 | Not-determined |
| S9_extracted_k141_34891 | Not-determined |
| S9_extracted_k141_72792 | Not-determined |
| S9_extracted_k141_810 | Low-quality |
| S9_extracted_k141_23538 | Not-determined |
| S9_extracted_k141_31221 | Not-determined |
| S9_extracted_k141_76436 | Not-determined |
| S9_extracted_k141_76437 | Not-determined |
| S9_extracted_k141_65097 | Not-determined |
| S9_extracted_k141_31224 | Not-determined |
| S9_extracted_k141_813 | Not-determined |
| S9_extracted_k141_72797 | Low-quality |
| S9_extracted_k141_72798 | Not-determined |
| S9_extracted_k141_38883 | Not-determined |
| S9_extracted_k141_49864 | Not-determined |
| S9_extracted_k141_49865 | Not-determined |
| S9_extracted_k141_65106 | Not-determined |
| S9_extracted_k141_46222 | Not-determined |
| S9_extracted_k141_820 | Not-determined |
| S9_extracted_k141_46223 | Not-determined |
| S9_extracted_k141_65109 | Not-determined |
| S9_extracted_k141_46224 | Not-determined |
| S9_extracted_k141_823 | Not-determined |
| S9_extracted_k141_65110 | Not-determined |
| S9_extracted_k141_61491 | Not-determined |
| S9_extracted_k141_23559 | Not-determined |
| S9_extracted_k141_61494 | Low-quality |
| S9_extracted_k141_828 | Not-determined |
| S9_extracted_k141_53963 | Not-determined |
| S9_extracted_k141_88099 | Not-determined |
| S9_extracted_k141_42604 | Not-determined |
| S9_extracted_k141_38891 | Not-determined |
| S9_extracted_k141_31238 | Low-quality |
| S9_extracted_k141_19698 | Not-determined |
| S9_extracted_k141_49873 | Not-determined |
| S9_extracted_k141_42606 | Low-quality |
| S9_extracted_k141_16112 | Not-determined |
| S9_extracted_k141_38894 | Not-determined |
| S9_extracted_k141_19700 | Low-quality |
| S9_extracted_k141_12293 | Low-quality |
| S9_extracted_k141_88100 | Low-quality |
| S9_extracted_k141_72812 | Low-quality |
| S9_extracted_k141_65121 | Not-determined |
| S9_extracted_k141_53975 | Not-determined |
| S9_extracted_k141_49880 | Not-determined |
| S9_extracted_k141_80437 | Not-determined |
| S9_extracted_k141_38904 | Not-determined |
| S9_extracted_k141_19709 | Low-quality |
| S9_extracted_k141_42612 | Not-determined |
| S9_extracted_k141_72822 | Not-determined |
| S9_extracted_k141_842 | Low-quality |
| S9_extracted_k141_49893 | Not-determined |
| S9_extracted_k141_61510 | Not-determined |
| S9_extracted_k141_49894 | Not-determined |
| S9_extracted_k141_42616 | Not-determined |
| S9_extracted_k141_53993 | Not-determined |
| S9_extracted_k141_61511 | Low-quality |
| S9_extracted_k141_12302 | Low-quality |
| S9_extracted_k141_88110 | Low-quality |
| S9_extracted_k141_49899 | Not-determined |
| S9_extracted_k141_80448 | Not-determined |
| S9_extracted_k141_72834 | Not-determined |
| S9_extracted_k141_54001 | Not-determined |
| S9_extracted_k141_23565 | Low-quality |
| S9_extracted_k141_23566 | Not-determined |
| S9_extracted_k141_65138 | Not-determined |
| S9_extracted_k141_16125 | Not-determined |
| S9_extracted_k141_61516 | Not-determined |
| S9_extracted_k141_42626 | Not-determined |
| S9_extracted_k141_72841 | Not-determined |
| S9_extracted_k141_57479 | Not-determined |
| S9_extracted_k141_858 | Low-quality |
| S9_extracted_k141_72842 | Not-determined |
| S9_extracted_k141_38917 | Not-determined |
| S9_extracted_k141_65143 | Low-quality |
| S9_extracted_k141_38919 | Low-quality |
| S9_extracted_k141_861 | Not-determined |
| S9_extracted_k141_38922 | Not-determined |
| S9_extracted_k141_27364 | Not-determined |
| S9_extracted_k141_65149 | Not-determined |
| S9_extracted_k141_65150 | Not-determined |
| S9_extracted_k141_31279 | Not-determined |
| S9_extracted_k141_65151 | Low-quality |
| S9_extracted_k141_57486 | Low-quality |
| S9_extracted_k141_42634 | Low-quality |
| S9_extracted_k141_46261 | Not-determined |
| S9_extracted_k141_65153 | Low-quality |
| S9_extracted_k141_42638 | Low-quality |
| S9_extracted_k141_34899 | Low-quality |
| S9_extracted_k141_42640 | Not-determined |
| S9_extracted_k141_12317 | Not-determined |
| S9_extracted_k141_54013 | Low-quality |
| S9_extracted_k141_34907 | Not-determined |
| S9_extracted_k141_61530 | Not-determined |
| S9_extracted_k141_49923 | Not-determined |
| S9_extracted_k141_61531 | Not-determined |
| S9_extracted_k141_46272 | Not-determined |
| S9_extracted_k141_46274 | Not-determined |
| S9_extracted_k141_34910 | Not-determined |
| S9_extracted_k141_61535 | Not-determined |
| S9_extracted_k141_57492 | Not-determined |
| S9_extracted_k141_34911 | Not-determined |
| S9_extracted_k141_19747 | Not-determined |
| S9_extracted_k141_16152 | Not-determined |
| S9_extracted_k141_16153 | Not-determined |
| S9_extracted_k141_49929 | Low-quality |
| S9_extracted_k141_12324 | Not-determined |
| S9_extracted_k141_876 | Not-determined |
| S9_extracted_k141_877 | Not-determined |
| S9_extracted_k141_88126 | Low-quality |
| S9_extracted_k141_88128 | Not-determined |
| S9_extracted_k141_38947 | Low-quality |
| S9_extracted_k141_34919 | Low-quality |
| S9_extracted_k141_31291 | Low-quality |
| S9_extracted_k141_16157 | Not-determined |
| S9_extracted_k141_46286 | Not-determined |
| S9_extracted_k141_19754 | Not-determined |
| S9_extracted_k141_23571 | Not-determined |
| S9_extracted_k141_68825 | Not-determined |
| S9_extracted_k141_23575 | Low-quality |
| S9_extracted_k141_38950 | Not-determined |
| S9_extracted_k141_16164 | Not-determined |
| S9_extracted_k141_72864 | Not-determined |
| S9_extracted_k141_34931 | Low-quality |
| S9_extracted_k141_65172 | Not-determined |
| S9_extracted_k141_65173 | Not-determined |
| S9_extracted_k141_16167 | Not-determined |
| S9_extracted_k141_72868 | Not-determined |
| S9_extracted_k141_4574 | Not-determined |
| S9_extracted_k141_88153 | Not-determined |
| S9_extracted_k141_46301 | Not-determined |
| S9_extracted_k141_31305 | Not-determined |
| S9_extracted_k141_54038 | Not-determined |
| S9_extracted_k141_72878 | Not-determined |
| S9_extracted_k141_68843 | Not-determined |
| S9_extracted_k141_34943 | Not-determined |
| S9_extracted_k141_61561 | Low-quality |
| S9_extracted_k141_34944 | Not-determined |
| S9_extracted_k141_31314 | Not-determined |
| S9_extracted_k141_46313 | Not-determined |
| S9_extracted_k141_16178 | Not-determined |
| S9_extracted_k141_38963 | Low-quality |
| S9_extracted_k141_27398 | Not-determined |
| S9_extracted_k141_891 | Not-determined |
| S9_extracted_k141_31318 | Low-quality |
| S9_extracted_k141_57500 | Not-determined |
| S9_extracted_k141_54049 | Not-determined |
| S9_extracted_k141_27405 | Not-determined |
| S9_extracted_k141_34956 | Not-determined |
| S9_extracted_k141_38973 | Not-determined |
| S9_extracted_k141_31323 | Not-determined |
| S9_extracted_k141_57503 | Not-determined |
| S9_extracted_k141_88157 | Low-quality |
| S9_extracted_k141_68851 | Low-quality |
| S9_extracted_k141_31325 | Low-quality |
| S9_extracted_k141_49963 | Not-determined |
| S9_extracted_k141_61567 | Not-determined |
| S9_extracted_k141_42666 | Not-determined |
| S9_extracted_k141_49965 | Not-determined |
| S9_extracted_k141_61570 | Low-quality |
| S9_extracted_k141_16183 | Low-quality |
| S9_extracted_k141_61571 | Not-determined |
| S9_extracted_k141_57507 | Not-determined |
| S9_extracted_k141_72894 | Not-determined |
| S9_extracted_k141_4600 | Not-determined |
| S9_extracted_k141_31334 | Not-determined |
| S9_extracted_k141_903 | Not-determined |
| S9_extracted_k141_4603 | Not-determined |
| S9_extracted_k141_12350 | Not-determined |
| S9_extracted_k141_31336 | Not-determined |
| S9_extracted_k141_88172 | Not-determined |
| S9_extracted_k141_72900 | Not-determined |
| S9_extracted_k141_49982 | Not-determined |
| S9_extracted_k141_4609 | Not-determined |
| S9_extracted_k141_57515 | Not-determined |
| S9_extracted_k141_12358 | Not-determined |
| S9_extracted_k141_57517 | Not-determined |
| S9_extracted_k141_8152 | Not-determined |
| S9_extracted_k141_911 | Not-determined |
| S9_extracted_k141_34977 | Not-determined |
| S9_extracted_k141_42682 | Not-determined |
| S9_extracted_k141_65197 | Not-determined |
| S9_extracted_k141_8155 | Low-quality |
| S9_extracted_k141_61583 | Low-quality |
| S9_extracted_k141_61585 | Not-determined |
| S9_extracted_k141_54060 | Low-quality |
| S9_extracted_k141_42689 | Low-quality |
| S9_extracted_k141_31340 | Low-quality |
| S9_extracted_k141_68867 | Not-determined |
| S9_extracted_k141_42692 | Low-quality |
| S9_extracted_k141_916 | Not-determined |
| S9_extracted_k141_31341 | Not-determined |
| S9_extracted_k141_4619 | Not-determined |
| S9_extracted_k141_57526 | Low-quality |
| S9_extracted_k141_54066 | Not-determined |
| S9_extracted_k141_68874 | Not-determined |
| S9_extracted_k141_38992 | Not-determined |
| S9_extracted_k141_65206 | Not-determined |
| S9_extracted_k141_42701 | Not-determined |
| S9_extracted_k141_46345 | Not-determined |
| S9_extracted_k141_61594 | Low-quality |
| S9_extracted_k141_34996 | Not-determined |
| S9_extracted_k141_16211 | Not-determined |
| S9_extracted_k141_57529 | Not-determined |
| S9_extracted_k141_16212 | Not-determined |
| S9_extracted_k141_42705 | Not-determined |
| S9_extracted_k141_35001 | Not-determined |
| S9_extracted_k141_923 | Not-determined |
| S9_extracted_k141_31356 | Not-determined |
| S9_extracted_k141_72908 | Not-determined |
| S9_extracted_k141_16218 | Not-determined |
| S9_extracted_k141_8179 | Not-determined |
| S9_extracted_k141_35002 | Low-quality |
| S9_extracted_k141_88179 | Not-determined |
| S9_extracted_k141_12371 | Not-determined |
| S9_extracted_k141_54083 | Not-determined |
| S9_extracted_k141_16221 | Not-determined |
| S9_extracted_k141_61606 | Not-determined |
| S9_extracted_k141_42713 | Low-quality |
| S9_extracted_k141_35009 | Not-determined |
| S9_extracted_k141_12372 | Low-quality |
| S9_extracted_k141_8183 | Not-determined |
| S9_extracted_k141_42714 | Not-determined |
| S9_extracted_k141_61608 | Low-quality |
| S9_extracted_k141_61609 | Not-determined |
| S9_extracted_k141_68882 | Not-determined |
| S9_extracted_k141_12373 | Low-quality |
| S9_extracted_k141_61612 | Not-determined |
| S9_extracted_k141_39012 | Not-determined |
| S9_extracted_k141_12375 | Not-determined |
| S9_extracted_k141_16232 | Not-determined |
| S9_extracted_k141_57544 | Not-determined |
| S9_extracted_k141_19788 | Not-determined |
| S9_extracted_k141_31369 | Not-determined |
| S9_extracted_k141_88193 | Not-determined |
| S9_extracted_k141_4640 | Not-determined |
| S9_extracted_k141_42720 | Not-determined |
| S9_extracted_k141_27412 | Not-determined |
| S9_extracted_k141_4642 | Not-determined |
| S9_extracted_k141_68887 | Not-determined |
| S9_extracted_k141_57553 | Low-quality |
| S9_extracted_k141_54095 | Low-quality |
| S9_extracted_k141_88198 | Low-quality |
| S9_extracted_k141_54096 | Not-determined |
| S9_extracted_k141_35022 | Not-determined |
| S9_extracted_k141_4646 | Not-determined |
| S9_extracted_k141_88201 | Not-determined |
| S9_extracted_k141_72929 | Not-determined |
| S9_extracted_k141_4648 | Not-determined |
| S9_extracted_k141_39027 | Not-determined |
| S9_extracted_k141_88203 | Not-determined |
| S9_extracted_k141_72930 | Not-determined |
| S9_extracted_k141_50025 | Low-quality |
| S9_extracted_k141_12393 | Not-determined |
| S9_extracted_k141_54102 | Low-quality |
| S9_extracted_k141_50026 | Low-quality |
| S9_extracted_k141_88211 | Low-quality |
| S9_extracted_k141_51656 | Complete |
| S9_extracted_k141_935 | Not-determined |
| S9_extracted_k141_57570 | Not-determined |
| S9_extracted_k141_54107 | Low-quality |
| S9_extracted_k141_68891 | Not-determined |
| S9_extracted_k141_31383 | Not-determined |
| S9_extracted_k141_39035 | Not-determined |
| S9_extracted_k141_27424 | Not-determined |
| S9_extracted_k141_68896 | Not-determined |
| S9_extracted_k141_68898 | Not-determined |
| S9_extracted_k141_46351 | Not-determined |
| S9_extracted_k141_35041 | Low-quality |
| S9_extracted_k141_42726 | Low-quality |
| S9_extracted_k141_31389 | Not-determined |
| S9_extracted_k141_88218 | Not-determined |
| S9_extracted_k141_88219 | Not-determined |
| S9_extracted_k141_16262 | Not-determined |
| S9_extracted_k141_31391 | Not-determined |
| S9_extracted_k141_4651 | Not-determined |
| S9_extracted_k141_72946 | Not-determined |
| S9_extracted_k141_35048 | Not-determined |
| S9_extracted_k141_4653 | Not-determined |
| S9_extracted_k141_46360 | Not-determined |
| S9_extracted_k141_35050 | Not-determined |
| S9_extracted_k141_35052 | Not-determined |
| S9_extracted_k141_8215 | Not-determined |
| S9_extracted_k141_46362 | Not-determined |
| S9_extracted_k141_54112 | Not-determined |
| S9_extracted_k141_50035 | Not-determined |
| S9_extracted_k141_54115 | Not-determined |
| S9_extracted_k141_954 | Not-determined |
| S9_extracted_k141_4655 | Low-quality |
| S9_extracted_k141_46369 | Not-determined |
| S9_extracted_k141_4656 | Not-determined |
| S9_extracted_k141_19810 | Not-determined |
| S9_extracted_k141_50044 | Not-determined |
| S9_extracted_k141_958 | Low-quality |
| S9_extracted_k141_65239 | Not-determined |
| S9_extracted_k141_54125 | Not-determined |
| S9_extracted_k141_80464 | Low-quality |
| S9_extracted_k141_39045 | Not-determined |
| S9_extracted_k141_31403 | Not-determined |
| S9_extracted_k141_72955 | Low-quality |
| S9_extracted_k141_54128 | Not-determined |
| S9_extracted_k141_27443 | Not-determined |
| S9_extracted_k141_88240 | Not-determined |
| S9_extracted_k141_80468 | Not-determined |
| S9_extracted_k141_12415 | Not-determined |
| S9_extracted_k141_35064 | Not-determined |
| S9_extracted_k141_50055 | Not-determined |
| S9_extracted_k141_35066 | Not-determined |
| S9_extracted_k141_4667 | Not-determined |
| S9_extracted_k141_68921 | Not-determined |
| S9_extracted_k141_80470 | Low-quality |
| S9_extracted_k141_39053 | Not-determined |
| S9_extracted_k141_23590 | Low-quality |
| S9_extracted_k141_68923 | Low-quality |
| S9_extracted_k141_57599 | Not-determined |
| S9_extracted_k141_50061 | Low-quality |
| S9_extracted_k141_16284 | Not-determined |
| S9_extracted_k141_16285 | Not-determined |
| S9_extracted_k141_12422 | Not-determined |
| S9_extracted_k141_88246 | Not-determined |
| S9_extracted_k141_16289 | Not-determined |
| S9_extracted_k141_39060 | Low-quality |
| S9_extracted_k141_57605 | Not-determined |
| S9_extracted_k141_72973 | Not-determined |
| S9_extracted_k141_16291 | Not-determined |
| S9_extracted_k141_23596 | Not-determined |
| S9_extracted_k141_65246 | Not-determined |
| S9_extracted_k141_50067 | Low-quality |
| S9_extracted_k141_39066 | Not-determined |
| S9_extracted_k141_50068 | Low-quality |
| S9_extracted_k141_50070 | Not-determined |
| S9_extracted_k141_65251 | Not-determined |
| S9_extracted_k141_8229 | Not-determined |
| S9_extracted_k141_4674 | Not-determined |
| S9_extracted_k141_16299 | Not-determined |
| S9_extracted_k141_31417 | Not-determined |
| S9_extracted_k141_974 | Low-quality |
| S9_extracted_k141_50075 | Not-determined |
| S9_extracted_k141_8232 | Not-determined |
| S9_extracted_k141_4677 | Not-determined |
| S9_extracted_k141_27460 | Not-determined |
| S9_extracted_k141_8234 | Not-determined |
| S9_extracted_k141_31423 | Not-determined |
| S9_extracted_k141_57618 | Not-determined |
| S9_extracted_k141_983 | Not-determined |
| S9_extracted_k141_35091 | Not-determined |
| S9_extracted_k141_31425 | Low-quality |
| S9_extracted_k141_39070 | Not-determined |
| S9_extracted_k141_12437 | Not-determined |
| S9_extracted_k141_31428 | Not-determined |
| S9_extracted_k141_88267 | Not-determined |
| S9_extracted_k141_988 | Low-quality |
| S9_extracted_k141_31430 | Not-determined |
| S9_extracted_k141_16308 | Not-determined |
| S9_extracted_k141_12441 | Not-determined |
| S9_extracted_k141_50084 | Low-quality |
| S9_extracted_k141_12442 | Not-determined |
| S9_extracted_k141_46397 | Not-determined |
| S9_extracted_k141_42765 | Not-determined |
| S9_extracted_k141_23609 | Not-determined |
| S9_extracted_k141_23610 | Not-determined |
| S9_extracted_k141_76464 | Not-determined |
| S9_extracted_k141_72985 | Not-determined |
| S9_extracted_k141_65265 | Not-determined |
| S9_extracted_k141_76465 | Low-quality |
| S9_extracted_k141_65268 | Low-quality |
| S9_extracted_k141_72989 | Not-determined |
| S9_extracted_k141_12448 | Low-quality |
| S9_extracted_k141_8251 | Not-determined |
| S9_extracted_k141_4698 | Not-determined |
| S9_extracted_k141_57625 | Not-determined |
| S9_extracted_k141_19838 | Not-determined |
| S9_extracted_k141_65271 | Not-determined |
| S9_extracted_k141_39086 | Low-quality |
| S9_extracted_k141_50100 | Not-determined |
| S9_extracted_k141_76468 | Low-quality |
| S9_extracted_k141_76469 | Not-determined |
| S9_extracted_k141_50102 | Not-determined |
| S9_extracted_k141_57628 | Not-determined |
| S9_extracted_k141_46407 | Low-quality |
| S9_extracted_k141_12453 | Not-determined |
| S9_extracted_k141_35104 | Not-determined |
| S9_extracted_k141_50105 | Not-determined |
| S9_extracted_k141_76472 | Not-determined |
| S9_extracted_k141_27466 | Low-quality |
| S9_extracted_k141_72999 | Not-determined |
| S9_extracted_k141_46413 | Low-quality |
| S9_extracted_k141_76475 | Low-quality |
| S9_extracted_k141_23621 | Not-determined |
| S9_extracted_k141_12458 | Not-determined |
| S9_extracted_k141_50110 | Not-determined |
| S9_extracted_k141_35112 | Not-determined |
| S9_extracted_k141_16335 | Not-determined |
| S9_extracted_k141_73002 | Low-quality |
| S9_extracted_k141_31442 | Not-determined |
| S9_extracted_k141_46418 | Not-determined |
| S9_extracted_k141_39099 | Not-determined |
| S9_extracted_k141_23627 | Not-determined |
| S9_extracted_k141_35116 | Not-determined |
| S9_extracted_k141_57644 | Low-quality |
| S9_extracted_k141_19850 | Not-determined |
| S9_extracted_k141_39103 | Not-determined |
| S9_extracted_k141_84099 | Not-determined |
| S9_extracted_k141_8262 | Not-determined |
| S9_extracted_k141_1010 | Not-determined |
| S9_extracted_k141_23631 | Low-quality |
| S9_extracted_k141_84100 | Not-determined |
| S9_extracted_k141_46427 | Low-quality |
| S9_extracted_k141_8267 | Not-determined |
| S9_extracted_k141_19857 | Not-determined |
| S9_extracted_k141_65289 | Not-determined |
| S9_extracted_k141_76496 | Not-determined |
| S9_extracted_k141_61626 | Not-determined |
| S9_extracted_k141_27480 | Not-determined |
| S9_extracted_k141_84106 | Not-determined |
| S9_extracted_k141_61628 | Not-determined |
| S9_extracted_k141_42792 | Low-quality |
| S9_extracted_k141_12478 | Not-determined |
| S9_extracted_k141_27485 | Not-determined |
| S9_extracted_k141_61634 | Not-determined |
| S9_extracted_k141_61635 | Low-quality |
| S9_extracted_k141_65296 | Not-determined |
| S9_extracted_k141_16354 | Not-determined |
| S9_extracted_k141_27493 | Low-quality |
| S9_extracted_k141_8275 | Not-determined |
| S9_extracted_k141_39123 | Low-quality |
| S9_extracted_k141_39124 | Not-determined |
| S9_extracted_k141_1027 | Not-determined |
| S9_extracted_k141_1030 | Not-determined |
| S9_extracted_k141_42806 | Not-determined |
| S9_extracted_k141_12483 | Low-quality |
| S9_extracted_k141_84114 | Not-determined |
| S9_extracted_k141_19867 | Low-quality |
| S9_extracted_k141_31468 | Not-determined |
| S9_extracted_k141_31469 | Not-determined |
| S9_extracted_k141_27502 | Not-determined |
| S9_extracted_k141_27504 | Not-determined |
| S9_extracted_k141_4721 | Not-determined |
| S9_extracted_k141_84121 | Not-determined |
| S9_extracted_k141_19871 | Not-determined |
| S9_extracted_k141_54187 | Not-determined |
| S9_extracted_k141_46443 | Low-quality |
| S9_extracted_k141_35127 | Low-quality |
| S9_extracted_k141_76513 | Not-determined |
| S9_extracted_k141_76514 | Not-determined |
| S9_extracted_k141_76515 | Not-determined |
| S9_extracted_k141_57666 | Not-determined |
| S9_extracted_k141_76516 | Not-determined |
| S9_extracted_k141_16369 | Not-determined |
| S9_extracted_k141_31473 | Not-determined |
| S9_extracted_k141_31475 | Not-determined |
| S9_extracted_k141_27511 | Not-determined |
| S9_extracted_k141_50131 | Not-determined |
| S9_extracted_k141_4733 | Not-determined |
| S9_extracted_k141_4734 | Not-determined |
| S9_extracted_k141_65310 | Low-quality |
| S9_extracted_k141_35133 | Not-determined |
| S9_extracted_k141_65314 | Not-determined |
| S9_extracted_k141_84133 | Not-determined |
| S9_extracted_k141_4741 | Not-determined |
| S9_extracted_k141_12508 | Not-determined |
| S9_extracted_k141_46452 | Low-quality |
| S9_extracted_k141_27521 | Low-quality |
| S9_extracted_k141_84137 | Not-determined |
| S9_extracted_k141_76527 | Low-quality |
| S9_extracted_k141_19882 | Not-determined |
| S9_extracted_k141_50143 | Low-quality |
| S9_extracted_k141_27529 | Not-determined |
| S9_extracted_k141_12515 | Not-determined |
| S9_extracted_k141_1047 | Low-quality |
| S9_extracted_k141_19887 | Not-determined |
| S9_extracted_k141_12518 | Not-determined |
| S9_extracted_k141_31480 | Not-determined |
| S9_extracted_k141_27537 | Not-determined |
| S9_extracted_k141_31485 | Not-determined |
| S9_extracted_k141_27539 | Not-determined |
| S9_extracted_k141_12524 | Not-determined |
| S9_extracted_k141_84153 | Low-quality |
| S9_extracted_k141_16399 | Not-determined |
| S9_extracted_k141_8314 | Low-quality |
| S9_extracted_k141_73012 | Low-quality |
| S9_extracted_k141_1056 | Not-determined |
| S9_extracted_k141_16402 | Not-determined |
| S9_extracted_k141_27551 | Not-determined |
| S9_extracted_k141_46474 | Not-determined |
| S9_extracted_k141_65343 | Not-determined |
| S9_extracted_k141_31495 | Not-determined |
| S9_extracted_k141_12527 | Not-determined |
| S9_extracted_k141_84161 | Not-determined |
| S9_extracted_k141_19902 | Not-determined |
| S9_extracted_k141_54224 | Not-determined |
| S9_extracted_k141_54225 | Not-determined |
| S9_extracted_k141_68933 | Not-determined |
| S9_extracted_k141_4757 | Not-determined |
| S9_extracted_k141_39160 | Not-determined |
| S9_extracted_k141_54230 | Not-determined |

| | |
|---|---|
| S9_extracted_k141_27557 | Not-determined |
| S9_extracted_k141_50154 | Low-quality |
| S9_extracted_k141_46481 | Not-determined |
| S9_extracted_k141_50156 | Not-determined |
| S9_extracted_k141_73025 | Low-quality |
| S9_extracted_k141_31506 | Not-determined |
| S9_extracted_k141_31509 | Low-quality |
| S9_extracted_k141_39165 | Not-determined |
| S9_extracted_k141_1062 | Not-determined |
| S9_extracted_k141_12545 | Not-determined |
| S9_extracted_k141_4770 | Not-determined |
| S9_extracted_k141_42813 | Not-determined |
| S9_extracted_k141_19909 | Not-determined |
| S9_extracted_k141_1066 | Not-determined |
| S9_extracted_k141_50165 | Not-determined |
| S9_extracted_k141_19910 | Not-determined |
| S9_extracted_k141_31513 | Not-determined |
| S9_extracted_k141_27576 | Not-determined |
| S9_extracted_k141_39169 | Not-determined |
| S9_extracted_k141_42815 | Not-determined |
| S9_extracted_k141_65358 | Low-quality |
| S9_extracted_k141_31514 | Not-determined |
| S9_extracted_k141_42816 | Not-determined |
| S9_extracted_k141_8328 | Not-determined |
| S9_extracted_k141_39171 | Not-determined |
| S9_extracted_k141_19915 | Not-determined |
| S9_extracted_k141_65361 | Low-quality |
| S9_extracted_k141_8330 | Not-determined |
| S9_extracted_k141_19916 | Not-determined |
| S9_extracted_k141_50171 | Not-determined |
| S9_extracted_k141_31522 | Not-determined |
| S9_extracted_k141_8333 | Low-quality |
| S9_extracted_k141_27580 | Low-quality |
| S9_extracted_k141_19921 | Not-determined |
| S9_extracted_k141_4781 | Low-quality |
| S9_extracted_k141_16432 | Not-determined |
| S9_extracted_k141_65364 | Not-determined |
| S9_extracted_k141_46493 | Not-determined |
| S9_extracted_k141_65365 | Not-determined |
| S9_extracted_k141_42819 | Not-determined |
| S9_extracted_k141_4786 | Not-determined |
| S9_extracted_k141_73050 | Not-determined |
| S9_extracted_k141_19925 | Not-determined |
| S9_extracted_k141_54256 | Not-determined |
| S9_extracted_k141_57700 | Not-determined |
| S9_extracted_k141_54257 | Not-determined |
| S9_extracted_k141_84188 | Not-determined |
| S9_extracted_k141_12559 | Not-determined |
| S9_extracted_k141_27586 | Not-determined |
| S9_extracted_k141_1071 | Not-determined |
| S9_extracted_k141_57704 | Low-quality |
| S9_extracted_k141_68956 | Not-determined |
| S9_extracted_k141_57705 | Low-quality |
| S9_extracted_k141_12562 | Not-determined |
| S9_extracted_k141_1074 | Not-determined |
| S9_extracted_k141_16439 | Not-determined |
| S9_extracted_k141_16440 | Low-quality |
| S9_extracted_k141_27595 | Not-determined |
| S9_extracted_k141_88301 | Not-determined |
| S9_extracted_k141_31531 | Not-determined |
| S9_extracted_k141_19935 | Not-determined |
| S9_extracted_k141_31535 | Not-determined |
| S9_extracted_k141_19936 | Not-determined |
| S9_extracted_k141_12564 | Low-quality |
| S9_extracted_k141_12566 | Not-determined |
| S9_extracted_k141_84199 | Not-determined |
| S9_extracted_k141_73061 | Not-determined |
| S9_extracted_k141_4803 | Not-determined |
| S9_extracted_k141_68972 | Not-determined |
| S9_extracted_k141_1080 | Not-determined |
| S9_extracted_k141_1081 | Not-determined |
| S9_extracted_k141_27600 | Not-determined |
| S9_extracted_k141_73062 | Low-quality |
| S9_extracted_k141_65389 | Not-determined |
| S9_extracted_k141_4809 | Not-determined |
| S9_extracted_k141_42835 | Low-quality |
| S9_extracted_k141_50182 | Low-quality |
| S9_extracted_k141_84211 | Not-determined |
| S9_extracted_k141_39182 | Low-quality |
| S9_extracted_k141_57708 | Not-determined |
| S9_extracted_k141_4812 | Not-determined |
| S9_extracted_k141_12576 | Not-determined |
| S9_extracted_k141_8337 | Not-determined |
| S9_extracted_k141_57710 | Not-determined |
| S9_extracted_k141_42841 | Not-determined |
| S9_extracted_k141_61637 | Not-determined |
| S9_extracted_k141_8341 | Not-determined |
| S9_extracted_k141_1096 | Not-determined |
| S9_extracted_k141_23650 | Not-determined |
| S9_extracted_k141_68987 | Not-determined |
| S9_extracted_k141_23653 | Not-determined |
| S9_extracted_k141_27614 | Not-determined |
| S9_extracted_k141_61645 | Low-quality |
| S9_extracted_k141_35196 | Low-quality |
| S9_extracted_k141_65392 | Not-determined |
| S9_extracted_k141_31551 | Low-quality |
| S9_extracted_k141_35197 | Not-determined |
| S9_extracted_k141_50195 | Not-determined |
| S9_extracted_k141_61650 | Not-determined |
| S9_extracted_k141_57721 | Not-determined |
| S9_extracted_k141_23660 | Not-determined |
| S9_extracted_k141_50198 | Not-determined |
| S9_extracted_k141_19957 | Not-determined |
| S9_extracted_k141_16460 | Not-determined |
| S9_extracted_k141_65397 | Not-determined |
| S9_extracted_k141_23663 | Not-determined |
| S9_extracted_k141_16462 | Low-quality |
| S9_extracted_k141_31559 | Not-determined |
| S9_extracted_k141_31562 | Not-determined |
| S9_extracted_k141_19962 | Not-determined |
| S9_extracted_k141_35203 | Not-determined |
| S9_extracted_k141_12599 | Not-determined |
| S9_extracted_k141_84227 | Not-determined |
| S9_extracted_k141_1107 | Low-quality |
| S9_extracted_k141_16466 | Not-determined |
| S9_extracted_k141_42854 | Not-determined |
| S9_extracted_k141_31565 | Not-determined |
| S9_extracted_k141_54274 | Not-determined |
| S9_extracted_k141_31566 | Not-determined |
| S9_extracted_k141_54275 | Not-determined |
| S9_extracted_k141_84230 | Not-determined |
| S9_extracted_k141_42856 | Low-quality |
| S9_extracted_k141_35209 | Not-determined |
| S9_extracted_k141_8365 | Not-determined |
| S9_extracted_k141_57730 | Not-determined |
| S9_extracted_k141_42859 | Not-determined |
| S9_extracted_k141_65401 | Not-determined |
| S9_extracted_k141_16472 | Not-determined |
| S9_extracted_k141_12604 | Not-determined |
| S9_extracted_k141_39206 | Not-determined |
| S9_extracted_k141_35214 | Not-determined |
| S9_extracted_k141_8366 | Not-determined |
| S9_extracted_k141_73095 | Low-quality |
| S9_extracted_k141_73096 | Not-determined |
| S9_extracted_k141_12605 | Low-quality |
| S9_extracted_k141_84236 | Not-determined |
| S9_extracted_k141_27626 | Not-determined |
| S9_extracted_k141_42865 | Not-determined |
| S9_extracted_k141_16480 | Not-determined |
| S9_extracted_k141_69000 | Low-quality |
| S9_extracted_k141_73099 | Not-determined |
| S9_extracted_k141_65409 | Not-determined |
| S9_extracted_k141_16483 | Not-determined |
| S9_extracted_k141_35218 | Low-quality |
| S9_extracted_k141_69003 | Not-determined |
| S9_extracted_k141_88336 | Not-determined |
| S9_extracted_k141_54278 | Low-quality |
| S9_extracted_k141_31582 | Not-determined |
| S9_extracted_k141_35221 | Low-quality |
| S9_extracted_k141_31583 | Not-determined |
| S9_extracted_k141_54282 | Low-quality |
| S9_extracted_k141_46518 | Low-quality |
| S9_extracted_k141_8377 | Not-determined |
| S9_extracted_k141_1125 | Not-determined |
| S9_extracted_k141_27633 | Not-determined |
| S9_extracted_k141_73105 | Not-determined |
| S9_extracted_k141_69012 | Not-determined |
| S9_extracted_k141_42882 | Not-determined |
| S9_extracted_k141_35230 | Low-quality |
| S9_extracted_k141_61659 | Not-determined |
| S9_extracted_k141_1131 | Not-determined |
| S9_extracted_k141_16497 | Not-determined |
| S9_extracted_k141_88350 | Not-determined |
| S9_extracted_k141_1133 | Not-determined |
| S9_extracted_k141_50223 | Low-quality |
| S9_extracted_k141_1134 | Not-determined |
| S9_extracted_k141_84250 | Not-determined |
| S9_extracted_k141_35231 | Not-determined |
| S9_extracted_k141_73112 | Not-determined |
| S9_extracted_k141_73115 | Not-determined |
| S9_extracted_k141_39216 | Not-determined |
| S9_extracted_k141_50226 | Low-quality |
| S9_extracted_k141_61667 | Not-determined |
| S9_extracted_k141_42893 | Not-determined |
| S9_extracted_k141_12636 | Not-determined |
| S9_extracted_k141_35239 | Not-determined |
| S9_extracted_k141_88357 | Not-determined |
| S9_extracted_k141_54298 | Not-determined |
| S9_extracted_k141_42899 | Not-determined |
| S9_extracted_k141_39221 | Not-determined |
| S9_extracted_k141_4841 | Not-determined |
| S9_extracted_k141_54302 | Low-quality |
| S9_extracted_k141_54303 | Not-determined |
| S9_extracted_k141_19995 | Not-determined |
| S9_extracted_k141_61670 | Not-determined |
| S9_extracted_k141_57739 | Low-quality |
| S9_extracted_k141_23691 | Not-determined |
| S9_extracted_k141_50234 | Not-determined |
| S9_extracted_k141_16518 | Low-quality |
| S9_extracted_k141_27653 | Not-determined |
| S9_extracted_k141_73133 | Not-determined |
| S9_extracted_k141_57740 | Not-determined |
| S9_extracted_k141_73135 | Low-quality |
| S9_extracted_k141_88364 | Not-determined |
| S9_extracted_k141_12647 | Not-determined |
| S9_extracted_k141_39240 | Not-determined |
| S9_extracted_k141_8406 | Not-determined |
| S9_extracted_k141_4853 | Not-determined |
| S9_extracted_k141_42917 | Not-determined |
| S9_extracted_k141_69029 | Low-quality |
| S9_extracted_k141_27660 | Not-determined |
| S9_extracted_k141_16526 | Not-determined |
| S9_extracted_k141_57744 | Low-quality |
| S9_extracted_k141_12654 | Low-quality |
| S9_extracted_k141_73138 | Low-quality |
| S9_extracted_k141_42920 | Not-determined |
| S9_extracted_k141_12655 | Not-determined |
| S9_extracted_k141_4857 | Not-determined |
| S9_extracted_k141_27661 | Low-quality |
| S9_extracted_k141_69036 | Low-quality |
| S9_extracted_k141_61677 | Not-determined |
| S9_extracted_k141_57749 | Not-determined |
| S9_extracted_k141_23703 | Not-determined |
| S9_extracted_k141_42928 | Not-determined |
| S9_extracted_k141_27666 | Not-determined |
| S9_extracted_k141_57752 | Low-quality |
| S9_extracted_k141_57755 | Not-determined |
| S9_extracted_k141_27668 | Not-determined |
| S9_extracted_k141_46529 | Low-quality |
| S9_extracted_k141_73146 | Low-quality |
| S9_extracted_k141_46530 | Not-determined |
| S9_extracted_k141_4863 | Not-determined |
| S9_extracted_k141_57757 | Low-quality |
| S9_extracted_k141_54335 | Not-determined |
| S9_extracted_k141_54337 | Not-determined |
| S9_extracted_k141_80511 | Low-quality |
| S9_extracted_k141_35275 | Not-determined |
| S9_extracted_k141_61694 | Not-determined |
| S9_extracted_k141_84293 | Not-determined |
| S9_extracted_k141_20027 | Not-determined |
| S9_extracted_k141_27678 | Not-determined |
| S9_extracted_k141_61696 | Low-quality |
| S9_extracted_k141_73158 | Not-determined |
| S9_extracted_k141_27681 | Not-determined |
| S9_extracted_k141_84296 | Low-quality |
| S9_extracted_k141_61698 | Not-determined |
| S9_extracted_k141_35277 | Not-determined |
| S9_extracted_k141_80517 | Low-quality |
| S9_extracted_k141_73163 | Not-determined |
| S9_extracted_k141_80519 | Not-determined |
| S9_extracted_k141_12657 | Low-quality |
| S9_extracted_k141_54347 | Low-quality |
| S9_extracted_k141_27687 | Not-determined |
| S9_extracted_k141_27688 | Not-determined |
| S9_extracted_k141_1177 | Low-quality |
| S9_extracted_k141_16545 | Not-determined |
| S9_extracted_k141_16547 | Not-determined |
| S9_extracted_k141_39265 | Low-quality |
| S9_extracted_k141_35283 | Not-determined |
| S9_extracted_k141_54350 | Low-quality |
| S9_extracted_k141_88389 | Not-determined |
| S9_extracted_k141_8415 | Not-determined |
| S9_extracted_k141_39267 | Not-determined |
| S9_extracted_k141_76543 | Not-determined |
| S9_extracted_k141_8418 | Not-determined |
| S9_extracted_k141_16553 | Not-determined |
| S9_extracted_k141_73171 | Low-quality |
| S9_extracted_k141_88394 | Not-determined |
| S9_extracted_k141_76547 | Not-determined |
| S9_extracted_k141_35292 | Not-determined |
| S9_extracted_k141_73175 | Not-determined |
| S9_extracted_k141_88400 | Not-determined |
| S9_extracted_k141_16559 | Not-determined |
| S9_extracted_k141_69054 | Low-quality |
| S9_extracted_k141_39277 | Low-quality |
| S9_extracted_k141_4883 | Not-determined |
| S9_extracted_k141_73180 | Not-determined |
| S9_extracted_k141_88405 | Not-determined |
| S9_extracted_k141_4888 | Not-determined |
| S9_extracted_k141_31606 | Not-determined |
| S9_extracted_k141_23730 | Not-determined |
| S9_extracted_k141_4894 | Not-determined |
| S9_extracted_k141_57785 | Not-determined |
| S9_extracted_k141_54367 | Not-determined |
| S9_extracted_k141_39284 | Low-quality |
| S9_extracted_k141_84318 | Not-determined |
| S9_extracted_k141_35300 | Not-determined |
| S9_extracted_k141_57789 | Not-determined |
| S9_extracted_k141_1186 | Not-determined |
| S9_extracted_k141_57790 | Not-determined |
| S9_extracted_k141_61717 | Not-determined |
| S9_extracted_k141_31613 | Not-determined |
| S9_extracted_k141_57791 | Not-determined |
| S9_extracted_k141_35304 | Not-determined |
| S9_extracted_k141_31615 | Not-determined |
| S9_extracted_k141_39289 | Not-determined |
| S9_extracted_k141_42951 | Not-determined |
| S9_extracted_k141_61725 | Not-determined |
| S9_extracted_k141_1194 | Not-determined |
| S9_extracted_k141_69070 | Not-determined |
| S9_extracted_k141_73189 | Not-determined |
| S9_extracted_k141_35309 | Not-determined |
| S9_extracted_k141_35311 | Not-determined |
| S9_extracted_k141_20050 | Not-determined |
| S9_extracted_k141_42957 | Not-determined |
| S9_extracted_k141_61727 | Not-determined |
| S9_extracted_k141_88408 | Not-determined |
| S9_extracted_k141_73194 | Low-quality |
| S9_extracted_k141_88409 | Not-determined |
| S9_extracted_k141_12676 | Not-determined |
| S9_extracted_k141_35313 | Not-determined |
| S9_extracted_k141_69080 | Not-determined |
| S9_extracted_k141_31622 | Low-quality |
| S9_extracted_k141_8449 | Not-determined |
| S9_extracted_k141_61736 | Not-determined |
| S9_extracted_k141_73202 | Low-quality |
| S9_extracted_k141_88414 | Not-determined |
| S9_extracted_k141_39303 | Not-determined |
| S9_extracted_k141_12677 | Not-determined |
| S9_extracted_k141_16583 | Not-determined |
| S9_extracted_k141_27729 | Not-determined |
| S9_extracted_k141_69089 | Not-determined |
| S9_extracted_k141_61740 | Not-determined |
| S9_extracted_k141_1204 | Not-determined |
| S9_extracted_k141_39310 | Not-determined |
| S9_extracted_k141_16588 | Not-determined |
| S9_extracted_k141_54391 | Low-quality |
| S9_extracted_k141_20066 | Not-determined |
| S9_extracted_k141_76582 | Not-determined |
| S9_extracted_k141_12685 | Not-determined |
| S9_extracted_k141_4904 | Not-determined |
| S9_extracted_k141_80565 | Not-determined |
| S9_extracted_k141_88420 | Low-quality |
| S9_extracted_k141_42977 | Not-determined |
| S9_extracted_k141_42978 | Not-determined |
| S9_extracted_k141_73215 | Low-quality |
| S9_extracted_k141_54398 | Not-determined |
| S9_extracted_k141_80566 | Low-quality |
| S9_extracted_k141_12690 | Not-determined |
| S9_extracted_k141_88423 | Not-determined |
| S9_extracted_k141_65426 | Not-determined |
| S9_extracted_k141_35330 | Not-determined |
| S9_extracted_k141_27741 | Not-determined |
| S9_extracted_k141_88424 | Not-determined |
| S9_extracted_k141_73217 | Not-determined |
| S9_extracted_k141_27742 | Not-determined |
| S9_extracted_k141_4909 | Low-quality |
| S9_extracted_k141_27743 | Not-determined |
| S9_extracted_k141_54399 | Low-quality |
| S9_extracted_k141_35334 | Not-determined |
| S9_extracted_k141_12694 | Low-quality |
| S9_extracted_k141_39315 | Not-determined |
| S9_extracted_k141_1220 | Not-determined |
| S9_extracted_k141_61748 | Not-determined |
| S9_extracted_k141_65428 | Not-determined |
| S9_extracted_k141_16597 | Not-determined |
| S9_extracted_k141_1224 | Not-determined |
| S9_extracted_k141_4919 | Not-determined |
| S9_extracted_k141_80568 | Low-quality |
| S9_extracted_k141_65433 | Low-quality |
| S9_extracted_k141_35343 | Low-quality |
| S9_extracted_k141_1230 | Not-determined |
| S9_extracted_k141_20085 | Not-determined |
| S9_extracted_k141_1231 | Not-determined |
| S9_extracted_k141_20086 | Low-quality |
| S9_extracted_k141_39317 | Low-quality |
| S9_extracted_k141_35345 | Not-determined |
| S9_extracted_k141_1233 | Not-determined |
| S9_extracted_k141_73229 | Not-determined |
| S9_extracted_k141_80575 | Not-determined |
| S9_extracted_k141_69106 | Not-determined |
| S9_extracted_k141_12710 | Low-quality |
| S9_extracted_k141_46558 | Not-determined |
| S9_extracted_k141_31629 | Not-determined |
| S9_extracted_k141_54412 | Not-determined |
| S9_extracted_k141_69109 | Low-quality |
| S9_extracted_k141_76595 | Not-determined |
| S9_extracted_k141_31633 | Not-determined |
| S9_extracted_k141_69113 | Low-quality |
| S9_extracted_k141_43002 | Not-determined |
| S9_extracted_k141_27761 | Not-determined |
| S9_extracted_k141_12716 | Not-determined |
| S9_extracted_k141_76601 | Not-determined |
| S9_extracted_k141_27763 | Low-quality |
| S9_extracted_k141_80580 | Low-quality |
| S9_extracted_k141_31640 | Low-quality |
| S9_extracted_k141_80582 | Not-determined |
| S9_extracted_k141_1244 | Not-determined |
| S9_extracted_k141_20105 | Not-determined |
| S9_extracted_k141_76604 | Not-determined |
| S9_extracted_k141_31643 | Not-determined |
| S9_extracted_k141_31644 | Low-quality |
| S9_extracted_k141_43007 | Not-determined |
| S9_extracted_k141_27770 | Not-determined |
| S9_extracted_k141_31648 | Not-determined |
| S9_extracted_k141_80589 | Not-determined |
| S9_extracted_k141_4929 | Low-quality |
| S9_extracted_k141_80590 | Not-determined |
| S9_extracted_k141_39330 | Not-determined |
| S9_extracted_k141_61764 | Not-determined |
| S9_extracted_k141_27773 | Not-determined |
| S9_extracted_k141_23749 | Not-determined |
| S9_extracted_k141_1247 | Not-determined |
| S9_extracted_k141_39333 | Not-determined |
| S9_extracted_k141_69124 | Not-determined |
| S9_extracted_k141_1248 | Not-determined |
| S9_extracted_k141_50254 | Low-quality |
| S9_extracted_k141_20115 | Not-determined |
| S9_extracted_k141_12733 | Not-determined |
| S9_extracted_k141_73244 | Not-determined |
| S9_extracted_k141_12737 | Not-determined |
| S9_extracted_k141_61772 | Not-determined |
| S9_extracted_k141_12739 | Not-determined |
| S9_extracted_k141_46583 | Not-determined |
| S9_extracted_k141_8482 | Not-determined |
| S9_extracted_k141_35371 | Not-determined |
| S9_extracted_k141_8483 | Low-quality |
| S9_extracted_k141_4944 | Not-determined |
| S9_extracted_k141_88463 | Low-quality |
| S9_extracted_k141_73253 | Not-determined |
| S9_extracted_k141_31657 | Low-quality |
| S9_extracted_k141_61778 | Not-determined |
| S9_extracted_k141_80602 | Not-determined |
| S9_extracted_k141_43019 | Not-determined |
| S9_extracted_k141_31659 | Not-determined |
| S9_extracted_k141_43020 | Not-determined |
| S9_extracted_k141_69142 | Low-quality |
| S9_extracted_k141_61781 | Not-determined |
| S9_extracted_k141_43023 | Not-determined |
| S9_extracted_k141_88471 | Not-determined |
| S9_extracted_k141_39342 | Not-determined |
| S9_extracted_k141_50269 | Not-determined |
| S9_extracted_k141_88472 | Not-determined |
| S9_extracted_k141_69145 | Not-determined |
| S9_extracted_k141_69146 | Not-determined |
| S9_extracted_k141_80604 | Not-determined |
| S9_extracted_k141_73262 | Not-determined |
| S9_extracted_k141_35383 | Not-determined |
| S9_extracted_k141_73263 | Not-determined |
| S9_extracted_k141_23769 | Low-quality |
| S9_extracted_k141_39350 | Not-determined |
| S9_extracted_k141_8498 | Not-determined |
| S9_extracted_k141_27790 | Not-determined |
| S9_extracted_k141_39354 | Not-determined |
| S9_extracted_k141_35388 | Not-determined |
| S9_extracted_k141_73269 | Not-determined |
| S9_extracted_k141_43039 | Not-determined |
| S9_extracted_k141_23774 | Not-determined |
| S9_extracted_k141_35391 | Not-determined |
| S9_extracted_k141_88479 | Not-determined |
| S9_extracted_k141_4963 | Not-determined |
| S9_extracted_k141_4964 | Not-determined |
| S9_extracted_k141_73274 | Not-determined |
| S9_extracted_k141_39364 | Not-determined |
| S9_extracted_k141_23778 | Not-determined |
| S9_extracted_k141_61795 | Not-determined |
| S9_extracted_k141_23780 | Not-determined |
| S9_extracted_k141_43050 | Low-quality |
| S9_extracted_k141_46600 | Not-determined |
| S9_extracted_k141_76638 | Not-determined |
| S9_extracted_k141_65475 | Low-quality |
| S9_extracted_k141_23786 | Not-determined |
| S9_extracted_k141_39370 | Low-quality |
| S9_extracted_k141_80617 | Not-determined |
| S9_extracted_k141_31690 | Low-quality |
| S9_extracted_k141_39373 | Not-determined |
| S9_extracted_k141_50282 | Not-determined |
| S9_extracted_k141_8505 | Not-determined |
| S9_extracted_k141_61804 | Not-determined |
| S9_extracted_k141_76643 | Not-determined |
| S9_extracted_k141_65481 | Low-quality |
| S9_extracted_k141_73288 | Not-determined |
| S9_extracted_k141_27807 | Not-determined |
| S9_extracted_k141_80622 | Not-determined |
| S9_extracted_k141_76644 | Low-quality |
| S9_extracted_k141_31698 | Not-determined |
| S9_extracted_k141_8510 | Not-determined |
| S9_extracted_k141_43056 | Not-determined |
| S9_extracted_k141_80625 | Not-determined |
| S9_extracted_k141_43057 | Not-determined |
| S9_extracted_k141_35408 | Not-determined |
| S9_extracted_k141_65486 | Not-determined |
| S9_extracted_k141_4979 | Low-quality |
| S9_extracted_k141_57820 | Not-determined |
| S9_extracted_k141_1295 | Not-determined |
| S9_extracted_k141_35411 | Low-quality |
| S9_extracted_k141_57823 | Not-determined |
| S9_extracted_k141_65492 | Low-quality |
| S9_extracted_k141_23795 | Not-determined |
| S9_extracted_k141_65493 | Not-determined |
| S9_extracted_k141_46617 | Not-determined |
| S9_extracted_k141_8515 | Low-quality |
| S9_extracted_k141_88500 | Not-determined |
| S9_extracted_k141_76651 | Not-determined |
| S9_extracted_k141_43066 | Low-quality |
| S9_extracted_k141_65496 | Not-determined |
| S9_extracted_k141_46622 | Not-determined |
| S9_extracted_k141_69173 | Not-determined |
| S9_extracted_k141_8520 | Not-determined |
| S9_extracted_k141_80631 | Not-determined |
| S9_extracted_k141_35420 | Not-determined |
| S9_extracted_k141_46626 | Not-determined |
| S9_extracted_k141_4991 | Not-determined |
| S9_extracted_k141_35426 | Not-determined |
| S9_extracted_k141_4992 | Not-determined |
| S9_extracted_k141_80635 | Low-quality |
| S9_extracted_k141_8528 | Not-determined |
| S9_extracted_k141_76658 | Not-determined |
| S9_extracted_k141_57833 | Not-determined |
| S9_extracted_k141_73302 | Not-determined |
| S9_extracted_k141_8531 | Not-determined |
| S9_extracted_k141_80638 | Not-determined |
| S9_extracted_k141_50296 | Not-determined |
| S9_extracted_k141_69188 | Not-determined |
| S9_extracted_k141_43077 | Not-determined |
| S9_extracted_k141_73307 | Not-determined |
| S9_extracted_k141_39402 | Not-determined |
| S9_extracted_k141_50299 | Not-determined |
| S9_extracted_k141_39405 | Not-determined |
| S9_extracted_k141_88511 | Not-determined |
| S9_extracted_k141_46641 | Not-determined |
| S9_extracted_k141_61815 | Low-quality |
| S9_extracted_k141_8543 | Not-determined |
| S9_extracted_k141_23816 | Not-determined |
| S9_extracted_k141_88516 | Not-determined |
| S9_extracted_k141_80643 | Not-determined |
| S9_extracted_k141_80645 | Not-determined |
| S9_extracted_k141_8548 | Not-determined |
| S9_extracted_k141_65507 | Not-determined |
| S9_extracted_k141_31736 | Not-determined |
| S9_extracted_k141_4997 | Not-determined |
| S9_extracted_k141_39417 | Not-determined |
| S9_extracted_k141_88520 | Not-determined |
| S9_extracted_k141_5000 | Not-determined |
| S9_extracted_k141_31740 | Not-determined |
| S9_extracted_k141_61825 | Not-determined |
| S9_extracted_k141_50310 | Low-quality |
| S9_extracted_k141_61828 | Not-determined |
| S9_extracted_k141_76674 | Not-determined |
| S9_extracted_k141_31744 | Not-determined |
| S9_extracted_k141_50314 | Not-determined |
| S9_extracted_k141_76678 | Not-determined |
| S9_extracted_k141_31745 | Not-determined |
| S9_extracted_k141_20161 | Not-determined |
| S9_extracted_k141_1334 | Not-determined |
| S9_extracted_k141_35458 | Not-determined |
| S9_extracted_k141_43097 | Not-determined |
| S9_extracted_k141_23831 | Low-quality |
| S9_extracted_k141_20165 | Not-determined |
| S9_extracted_k141_39425 | Low-quality |
| S9_extracted_k141_5011 | Not-determined |
| S9_extracted_k141_88539 | Low-quality |
| S9_extracted_k141_23833 | Low-quality |
| S9_extracted_k141_65509 | Low-quality |
| S9_extracted_k141_1344 | Not-determined |
| S9_extracted_k141_35460 | Not-determined |
| S9_extracted_k141_80664 | Not-determined |
| S9_extracted_k141_31753 | Not-determined |
| S9_extracted_k141_88542 | Not-determined |
| S9_extracted_k141_20170 | Not-determined |
| S9_extracted_k141_16617 | Not-determined |
| S9_extracted_k141_46658 | Not-determined |
| S9_extracted_k141_46659 | Low-quality |
| S9_extracted_k141_39433 | Not-determined |
| S9_extracted_k141_35466 | Not-determined |
| S9_extracted_k141_23839 | Low-quality |
| S9_extracted_k141_69217 | Not-determined |
| S9_extracted_k141_76692 | Low-quality |
| S9_extracted_k141_80670 | Not-determined |
| S9_extracted_k141_69221 | Not-determined |
| S9_extracted_k141_69222 | Not-determined |
| S9_extracted_k141_80673 | Not-determined |
| S9_extracted_k141_76698 | Not-determined |
| S9_extracted_k141_20177 | Not-determined |
| S9_extracted_k141_61846 | Not-determined |
| S9_extracted_k141_61847 | Not-determined |
| S9_extracted_k141_43110 | Not-determined |
| S9_extracted_k141_57868 | Not-determined |
| S9_extracted_k141_16629 | Not-determined |
| S9_extracted_k141_1351 | Not-determined |
| S9_extracted_k141_5032 | Not-determined |
| S9_extracted_k141_57873 | Low-quality |
| S9_extracted_k141_61855 | Not-determined |
| S9_extracted_k141_5036 | Not-determined |
| S9_extracted_k141_50340 | Not-determined |
| S9_extracted_k141_23852 | Not-determined |
| S9_extracted_k141_20184 | Not-determined |
| S9_extracted_k141_5040 | Not-determined |
| S9_extracted_k141_35490 | Not-determined |
| S9_extracted_k141_88557 | Not-determined |
| S9_extracted_k141_23855 | Not-determined |
| S9_extracted_k141_50342 | Not-determined |
| S9_extracted_k141_23856 | Not-determined |
| S9_extracted_k141_31776 | Not-determined |
| S9_extracted_k141_35494 | Not-determined |
| S9_extracted_k141_65530 | Not-determined |
| S9_extracted_k141_61867 | Not-determined |
| S9_extracted_k141_27822 | Not-determined |
| S9_extracted_k141_76705 | Low-quality |

| | |
|---|---|
| S9_extracted_k141_88564 | Not-determined |
| S9_extracted_k141_27824 | Not-determined |
| S9_extracted_k141_20198 | Not-determined |
| S9_extracted_k141_31781 | Not-determined |
| S9_extracted_k141_27829 | Not-determined |
| S9_extracted_k141_69231 | Low-quality |
| S9_extracted_k141_61877 | Not-determined |
| S9_extracted_k141_27833 | Not-determined |
| S9_extracted_k141_76714 | Not-determined |
| S9_extracted_k141_43143 | Not-determined |
| S9_extracted_k141_27839 | Not-determined |
| S9_vs_k141_51656\|\|full | High-quality |
| S9_extracted_k141_31788 | Not-determined |
| S9_extracted_k141_5062 | Not-determined |
| S9_extracted_k141_27842 | Not-determined |
| S9_extracted_k141_5064 | Not-determined |
| S9_extracted_k141_35502 | Not-determined |
| S9_extracted_k141_69239 | Low-quality |
| S9_extracted_k141_50360 | Not-determined |
| S9_extracted_k141_27847 | Not-determined |
| S9_extracted_k141_73317 | Not-determined |
| S9_extracted_k141_23882 | Not-determined |
| S9_extracted_k141_23884 | Not-determined |
| S9_extracted_k141_50365 | Not-determined |
| S9_extracted_k141_57906 | Not-determined |
| S9_extracted_k141_8588 | Low-quality |
| S9_extracted_k141_84346 | Not-determined |
| S9_extracted_k141_8593 | Low-quality |
| S9_extracted_k141_57913 | Not-determined |
| S9_extracted_k141_27857 | Not-determined |
| S9_extracted_k141_31809 | Not-determined |
| S9_extracted_k141_35514 | Not-determined |
| S9_extracted_k141_31810 | Not-determined |
| S9_extracted_k141_23893 | Not-determined |
| S9_extracted_k141_31812 | Low-quality |
| S9_extracted_k141_5079 | Not-determined |
| S9_extracted_k141_65571 | Not-determined |
| S9_extracted_k141_8598 | Not-determined |
| S9_extracted_k141_20224 | Not-determined |
| S9_extracted_k141_76720 | Not-determined |
| S9_extracted_k141_8605 | Not-determined |
| S9_extracted_k141_1383 | Low-quality |
| S9_extracted_k141_50377 | Not-determined |
| S9_extracted_k141_76726 | Not-determined |
| S9_extracted_k141_76727 | Low-quality |
| S9_extracted_k141_20231 | Not-determined |
| S9_extracted_k141_23897 | Low-quality |
| S9_extracted_k141_31819 | Not-determined |
| S9_extracted_k141_35530 | Low-quality |
| S9_extracted_k141_27867 | Not-determined |
| S9_extracted_k141_20237 | Low-quality |
| S9_extracted_k141_20239 | Not-determined |
| S9_extracted_k141_43167 | Low-quality |
| S9_extracted_k141_65582 | Not-determined |
| S9_extracted_k141_76736 | Not-determined |
| S9_extracted_k141_65584 | Not-determined |
| S9_extracted_k141_80688 | Not-determined |
| S9_extracted_k141_8611 | Not-determined |
| S9_extracted_k141_80689 | Not-determined |
| S9_extracted_k141_76740 | Not-determined |
| S9_extracted_k141_35538 | Not-determined |
| S9_extracted_k141_80692 | Not-determined |
| S9_extracted_k141_39448 | Low-quality |
| S9_extracted_k141_8616 | Not-determined |
| S8_extracted_k141_187742 | High-quality |
| S9_extracted_k141_61913 | Not-determined |
| S9_extracted_k141_20253 | Not-determined |
| S9_extracted_k141_76751 | Not-determined |
| S9_extracted_k141_27882 | Not-determined |
| S9_extracted_k141_27883 | Not-determined |
| S9_extracted_k141_84380 | Not-determined |
| S9_extracted_k141_27884 | Not-determined |
| S9_extracted_k141_69269 | Not-determined |
| S9_extracted_k141_20256 | Not-determined |
| S9_extracted_k141_8625 | Not-determined |
| S9_extracted_k141_5119 | Not-determined |
| S9_extracted_k141_35549 | Not-determined |
| S9_extracted_k141_65602 | Not-determined |
| S9_extracted_k141_1402 | Not-determined |
| S9_extracted_k141_8630 | Low-quality |
| S9_extracted_k141_35553 | Not-determined |
| S9_extracted_k141_27890 | Not-determined |
| S9_extracted_k141_43174 | Not-determined |
| S9_extracted_k141_5124 | Not-determined |
| S9_extracted_k141_73360 | Not-determined |
| S9_extracted_k141_35555 | Low-quality |
| S9_extracted_k141_35556 | Not-determined |
| S9_extracted_k141_50386 | Low-quality |
| S9_extracted_k141_50387 | Not-determined |
| S9_extracted_k141_5128 | Not-determined |
| S9_extracted_k141_50390 | Not-determined |
| S9_extracted_k141_80713 | Not-determined |
| S9_extracted_k141_57953 | Not-determined |
| S9_extracted_k141_57955 | Not-determined |
| S9_extracted_k141_5131 | Not-determined |
| S9_extracted_k141_80717 | Not-determined |
| S9_extracted_k141_23921 | Not-determined |
| S9_extracted_k141_69277 | Not-determined |
| S9_extracted_k141_80720 | Low-quality |
| S9_extracted_k141_57957 | Low-quality |
| S9_extracted_k141_69279 | Not-determined |
| S9_extracted_k141_8640 | Not-determined |
| S9_extracted_k141_27905 | Not-determined |
| S9_extracted_k141_5135 | Not-determined |
| S9_extracted_k141_80722 | Not-determined |
| S9_extracted_k141_20272 | Not-determined |
| S9_extracted_k141_31858 | Not-determined |
| S9_extracted_k141_8644 | Not-determined |
| S9_extracted_k141_31862 | Low-quality |
| S9_extracted_k141_39471 | Not-determined |
| S9_extracted_k141_80729 | Not-determined |
| S9_extracted_k141_50400 | Not-determined |
| S9_extracted_k141_23932 | Not-determined |
| S9_extracted_k141_57966 | Not-determined |
| S9_extracted_k141_80733 | Not-determined |
| S9_extracted_k141_27913 | Not-determined |
| S9_extracted_k141_5146 | Not-determined |
| S9_extracted_k141_39481 | Low-quality |
| S9_extracted_k141_80738 | Not-determined |
| S9_extracted_k141_1413 | Not-determined |
| S9_extracted_k141_57976 | Not-determined |
| S9_extracted_k141_39485 | Low-quality |
| S9_extracted_k141_31877 | Not-determined |
| S9_extracted_k141_31879 | Not-determined |
| S9_extracted_k141_57981 | Low-quality |
| S9_extracted_k141_5156 | Not-determined |
| S9_extracted_k141_50413 | Not-determined |
| S9_extracted_k141_57983 | Not-determined |
| S9_extracted_k141_35584 | Not-determined |
| S9_extracted_k141_39490 | Not-determined |
| S9_extracted_k141_65618 | Not-determined |
| S9_extracted_k141_65619 | Not-determined |
| S9_extracted_k141_69290 | Not-determined |
| S9_extracted_k141_23947 | Not-determined |
| S9_extracted_k141_23949 | Not-determined |
| S9_extracted_k141_65621 | Low-quality |
| S9_extracted_k141_1422 | Low-quality |
| S9_extracted_k141_69294 | Not-determined |
| S9_extracted_k141_20288 | Not-determined |
| S9_extracted_k141_84415 | Not-determined |
| S9_extracted_k141_20289 | Low-quality |
| S9_extracted_k141_39500 | Not-determined |
| S9_extracted_k141_43178 | Not-determined |
| S9_extracted_k141_8663 | Not-determined |
| S9_extracted_k141_69297 | Low-quality |
| S9_extracted_k141_39501 | Not-determined |
| S9_extracted_k141_43179 | Not-determined |
| S9_extracted_k141_43181 | Not-determined |
| S9_extracted_k141_27938 | Not-determined |
| S9_extracted_k141_69300 | Not-determined |
| S9_extracted_k141_35594 | Not-determined |
| S9_extracted_k141_46678 | Not-determined |
| S9_extracted_k141_61957 | Not-determined |
| S9_extracted_k141_5167 | Not-determined |
| S9_extracted_k141_35597 | Not-determined |
| S9_extracted_k141_5170 | Not-determined |
| S9_extracted_k141_43187 | Not-determined |
| S9_extracted_k141_84426 | Not-determined |
| S9_extracted_k141_20293 | Low-quality |
| S9_extracted_k141_31898 | Not-determined |
| S9_extracted_k141_58006 | Not-determined |
| S9_extracted_k141_35602 | Not-determined |
| S9_extracted_k141_84431 | Not-determined |
| S9_extracted_k141_31905 | Not-determined |
| S9_extracted_k141_16655 | Low-quality |
| S9_extracted_k141_65639 | Not-determined |
| S9_extracted_k141_61967 | Not-determined |
| S9_extracted_k141_65640 | Not-determined |
| S9_extracted_k141_61970 | Not-determined |
| S9_extracted_k141_80770 | Low-quality |
| S9_extracted_k141_1441 | Not-determined |
| S9_extracted_k141_58015 | Not-determined |
| S9_extracted_k141_31911 | Not-determined |
| S9_extracted_k141_50423 | Low-quality |
| S9_extracted_k141_5185 | Not-determined |
| S9_extracted_k141_23979 | Not-determined |
| S9_extracted_k141_43203 | Not-determined |
| S9_extracted_k141_16665 | Not-determined |
| S9_extracted_k141_31918 | Not-determined |
| S9_extracted_k141_46700 | Not-determined |
| S9_extracted_k141_65650 | Low-quality |
| S9_extracted_k141_50430 | Low-quality |
| S9_extracted_k141_16667 | Not-determined |
| S9_extracted_k141_43205 | Low-quality |
| S9_extracted_k141_50431 | Not-determined |
| S9_extracted_k141_31922 | Not-determined |
| S9_extracted_k141_50433 | Not-determined |
| S9_extracted_k141_16668 | Not-determined |
| S9_extracted_k141_80779 | Not-determined |
| S9_extracted_k141_31924 | Not-determined |
| S9_extracted_k141_16669 | Not-determined |
| S9_extracted_k141_23987 | Not-determined |
| S9_extracted_k141_35619 | Low-quality |
| S9_extracted_k141_23992 | Not-determined |
| S9_extracted_k141_80791 | Not-determined |
| S9_extracted_k141_16679 | Not-determined |
| S9_extracted_k141_84456 | Not-determined |
| S9_extracted_k141_35623 | Not-determined |
| S9_extracted_k141_16682 | Not-determined |
| S9_extracted_k141_54423 | Not-determined |
| S9_extracted_k141_23999 | Low-quality |
| S9_extracted_k141_5196 | Not-determined |
| S9_extracted_k141_20317 | Low-quality |
| S9_extracted_k141_54425 | Not-determined |
| S9_extracted_k141_20320 | Low-quality |
| S9_extracted_k141_54426 | Not-determined |
| S9_extracted_k141_43218 | Low-quality |
| S9_extracted_k141_35632 | Not-determined |
| S9_extracted_k141_46722 | Not-determined |
| S9_extracted_k141_8680 | Not-determined |
| S9_extracted_k141_80800 | Low-quality |
| S9_extracted_k141_12764 | Not-determined |
| S9_extracted_k141_16688 | Low-quality |
| S9_extracted_k141_65677 | Not-determined |
| S9_extracted_k141_73365 | Not-determined |
| S9_extracted_k141_16693 | Not-determined |
| S9_extracted_k141_1470 | Not-determined |
| S9_extracted_k141_12771 | Not-determined |
| S9_extracted_k141_16695 | Not-determined |
| S9_extracted_k141_31945 | Not-determined |
| S9_extracted_k141_31947 | Not-determined |
| S9_extracted_k141_80814 | Not-determined |
| S9_extracted_k141_31948 | Not-determined |
| S9_extracted_k141_73370 | Not-determined |
| S9_extracted_k141_1472 | Not-determined |
| S9_extracted_k141_12774 | Not-determined |
| S9_extracted_k141_84482 | Not-determined |
| S9_extracted_k141_16701 | Not-determined |
| S9_extracted_k141_12775 | Not-determined |
| S9_extracted_k141_16703 | Not-determined |
| S9_extracted_k141_54432 | Low-quality |
| S9_extracted_k141_46738 | Not-determined |
| S9_extracted_k141_65683 | Not-determined |
| S9_extracted_k141_43227 | Not-determined |
| S9_extracted_k141_43228 | Not-determined |
| S9_extracted_k141_46740 | Not-determined |
| S9_extracted_k141_80822 | Not-determined |
| S9_extracted_k141_20333 | Not-determined |
| S9_extracted_k141_80824 | Low-quality |
| S9_extracted_k141_16708 | Not-determined |
| S9_extracted_k141_31956 | Not-determined |
| S9_extracted_k141_35653 | Not-determined |
| S9_extracted_k141_84490 | Not-determined |
| S9_extracted_k141_43233 | Not-determined |
| S9_extracted_k141_24015 | Not-determined |
| S9_extracted_k141_8698 | Not-determined |
| S9_extracted_k141_46746 | Not-determined |
| S9_extracted_k141_35656 | Not-determined |
| S9_extracted_k141_20339 | Not-determined |
| S9_extracted_k141_16711 | Low-quality |
| S9_extracted_k141_84496 | Not-determined |
| S9_extracted_k141_43239 | Not-determined |
| S9_extracted_k141_5227 | Not-determined |
| S9_extracted_k141_8708 | Not-determined |
| S9_extracted_k141_31964 | Not-determined |
| S9_extracted_k141_69324 | Not-determined |
| S9_extracted_k141_61999 | Low-quality |
| S9_extracted_k141_8713 | Not-determined |
| S9_extracted_k141_43248 | Not-determined |
| S9_extracted_k141_80839 | Not-determined |
| S9_extracted_k141_46758 | Not-determined |
| S9_extracted_k141_54452 | Not-determined |
| S9_extracted_k141_27957 | Low-quality |
| S9_extracted_k141_24033 | Not-determined |
| S9_extracted_k141_24036 | Not-determined |
| S9_extracted_k141_69331 | Not-determined |
| S9_extracted_k141_31975 | Not-determined |
| S9_extracted_k141_62013 | Not-determined |
| S9_extracted_k141_69332 | Not-determined |
| S9_extracted_k141_31976 | Not-determined |
| S9_extracted_k141_46767 | Not-determined |
| S9_extracted_k141_65703 | Not-determined |
| S9_extracted_k141_65704 | Not-determined |
| S9_extracted_k141_54463 | Not-determined |
| S9_extracted_k141_65709 | Not-determined |
| S9_extracted_k141_73384 | Not-determined |
| S9_extracted_k141_1508 | Low-quality |
| S9_extracted_k141_46774 | Not-determined |
| S9_extracted_k141_16731 | Not-determined |
| S9_extracted_k141_46777 | Low-quality |
| S9_extracted_k141_27969 | Not-determined |
| S9_extracted_k141_1522 | Not-determined |
| S9_extracted_k141_58027 | Not-determined |
| S9_extracted_k141_58029 | Not-determined |
| S9_extracted_k141_43269 | Not-determined |
| S9_extracted_k141_84528 | Low-quality |
| S9_extracted_k141_1524 | Not-determined |
| S9_extracted_k141_54481 | Not-determined |
| S9_extracted_k141_31996 | Not-determined |
| S9_extracted_k141_69347 | Not-determined |
| S9_extracted_k141_80840 | Low-quality |
| S9_extracted_k141_69348 | Low-quality |
| S9_extracted_k141_58032 | Not-determined |
| S9_extracted_k141_46788 | Not-determined |
| S9_extracted_k141_80844 | Not-determined |
| S9_extracted_k141_69353 | Low-quality |
| S9_extracted_k141_58034 | Not-determined |
| S9_extracted_k141_58035 | Not-determined |
| S9_extracted_k141_43281 | Not-determined |
| S9_extracted_k141_32004 | Not-determined |
| S9_extracted_k141_73392 | Not-determined |
| S9_extracted_k141_20354 | Low-quality |
| S9_extracted_k141_54489 | Not-determined |
| S9_extracted_k141_54490 | Not-determined |
| S9_extracted_k141_5263 | Low-quality |
| S9_extracted_k141_80851 | Not-determined |
| S9_extracted_k141_8752 | Not-determined |
| S9_extracted_k141_62049 | Not-determined |
| S9_extracted_k141_80854 | Not-determined |
| S9_extracted_k141_62052 | Not-determined |
| S9_extracted_k141_80857 | Not-determined |
| S9_extracted_k141_73402 | Not-determined |
| S9_extracted_k141_24072 | Not-determined |
| S9_extracted_k141_80859 | Low-quality |
| S9_extracted_k141_65727 | Low-quality |
| S9_extracted_k141_5276 | Not-determined |
| S9_extracted_k141_65730 | Not-determined |
| S9_extracted_k141_8760 | Not-determined |
| S9_extracted_k141_80869 | Not-determined |
| S9_extracted_k141_24079 | Not-determined |
| S9_extracted_k141_80870 | Not-determined |
| S9_extracted_k141_84561 | Not-determined |
| S9_extracted_k141_65736 | Not-determined |
| S9_extracted_k141_43302 | Not-determined |
| S9_extracted_k141_27993 | Low-quality |
| S9_extracted_k141_16774 | Not-determined |
| S9_extracted_k141_8766 | Not-determined |
| S8_vs_k141_187742\|\|full | High-quality |
| S9_extracted_k141_69373 | Not-determined |
| S9_extracted_k141_65744 | Not-determined |
| S9_extracted_k141_43307 | Not-determined |
| S9_extracted_k141_27999 | Not-determined |
| S9_extracted_k141_16782 | Low-quality |
| S9_extracted_k141_73413 | Not-determined |
| S9_extracted_k141_73414 | Not-determined |
| S9_extracted_k141_32031 | Not-determined |
| S9_extracted_k141_84567 | Low-quality |
| S9_extracted_k141_54517 | Not-determined |
| S9_extracted_k141_24091 | Low-quality |
| S9_extracted_k141_84569 | Not-determined |
| S9_extracted_k141_84570 | Low-quality |
| S9_extracted_k141_50442 | Not-determined |
| S9_extracted_k141_69382 | Not-determined |
| S9_extracted_k141_43317 | Not-determined |
| S9_extracted_k141_84573 | Not-determined |
| S9_extracted_k141_65751 | Not-determined |
| S9_extracted_k141_20389 | Low-quality |
| S9_extracted_k141_69385 | Not-determined |
| S9_extracted_k141_76779 | Not-determined |
| S9_extracted_k141_32039 | Low-quality |
| S9_extracted_k141_16795 | Not-determined |
| S9_extracted_k141_80878 | Not-determined |
| S9_extracted_k141_76781 | Not-determined |
| S9_extracted_k141_65753 | Not-determined |
| S9_extracted_k141_62075 | Not-determined |
| S9_extracted_k141_20394 | Not-determined |
| S9_extracted_k141_76783 | Not-determined |
| S9_extracted_k141_65757 | Not-determined |
| S9_extracted_k141_62078 | Not-determined |
| S9_extracted_k141_24097 | Low-quality |
| S9_extracted_k141_69392 | Not-determined |
| S9_extracted_k141_20397 | Low-quality |
| S9_extracted_k141_69395 | Not-determined |
| S9_extracted_k141_32049 | Not-determined |
| S9_extracted_k141_76785 | Not-determined |
| S9_extracted_k141_54527 | Not-determined |
| S9_extracted_k141_12797 | Not-determined |
| S9_extracted_k141_69397 | Low-quality |
| S9_extracted_k141_73433 | Not-determined |
| S9_extracted_k141_80888 | Not-determined |
| S9_extracted_k141_39509 | Not-determined |
| S9_extracted_k141_73434 | Not-determined |
| S9_extracted_k141_69400 | Not-determined |
| S9_extracted_k141_20400 | Not-determined |
| S9_extracted_k141_65766 | Not-determined |
| S9_extracted_k141_84588 | Not-determined |
| S9_extracted_k141_54530 | Low-quality |
| S9_extracted_k141_16811 | Not-determined |
| S9_extracted_k141_76792 | Not-determined |
| S9_extracted_k141_8792 | Not-determined |
| S9_extracted_k141_62083 | Low-quality |
| S9_extracted_k141_12810 | Not-determined |
| S9_extracted_k141_8794 | Not-determined |
| S9_extracted_k141_50468 | Low-quality |
| S9_extracted_k141_50469 | Not-determined |
| S9_extracted_k141_39521 | Not-determined |
| S9_extracted_k141_54535 | Not-determined |
| S9_extracted_k141_69411 | Not-determined |
| S9_extracted_k141_39523 | Not-determined |
| S9_extracted_k141_35678 | Low-quality |
| S9_extracted_k141_84598 | Low-quality |
| S9_extracted_k141_20410 | Not-determined |
| S9_extracted_k141_65772 | Low-quality |
| S9_extracted_k141_12811 | Low-quality |
| S9_extracted_k141_32054 | Not-determined |
| S9_extracted_k141_62086 | Low-quality |
| S9_extracted_k141_20413 | Not-determined |
| S9_extracted_k141_5313 | Not-determined |
| S9_extracted_k141_35685 | Low-quality |
| S9_extracted_k141_20415 | Low-quality |
| S9_extracted_k141_39531 | Not-determined |
| S9_extracted_k141_39532 | Not-determined |
| S9_extracted_k141_69420 | Not-determined |
| S9_extracted_k141_80905 | Low-quality |
| S9_extracted_k141_5316 | Not-determined |
| S9_extracted_k141_76804 | Not-determined |
| S9_extracted_k141_5317 | Not-determined |
| S9_extracted_k141_8809 | Not-determined |
| S9_extracted_k141_65780 | Not-determined |
| S9_extracted_k141_84608 | Not-determined |
| S9_extracted_k141_24104 | Not-determined |
| S9_extracted_k141_5321 | Low-quality |
| S9_extracted_k141_69425 | Not-determined |
| S9_extracted_k141_54551 | Not-determined |
| S9_extracted_k141_39537 | Not-determined |
| S9_extracted_k141_39538 | Not-determined |
| S9_extracted_k141_5325 | Not-determined |
| S9_extracted_k141_32067 | Not-determined |
| S9_extracted_k141_20429 | Not-determined |
| S9_extracted_k141_39543 | Not-determined |
| S9_extracted_k141_1561 | Not-determined |
| S9_extracted_k141_5329 | Not-determined |
| S9_extracted_k141_20433 | Not-determined |
| S9_extracted_k141_12826 | Not-determined |
| S9_extracted_k141_65782 | Low-quality |
| S9_extracted_k141_54564 | Not-determined |
| S9_extracted_k141_32069 | Low-quality |
| S9_extracted_k141_24112 | Not-determined |
| S9_extracted_k141_80918 | Not-determined |
| S9_extracted_k141_24113 | Low-quality |
| S9_extracted_k141_8816 | Not-determined |
| S9_extracted_k141_69437 | Not-determined |
| S9_extracted_k141_80919 | Low-quality |
| S9_extracted_k141_5333 | Not-determined |
| S9_extracted_k141_20438 | Not-determined |
| S9_extracted_k141_80920 | Not-determined |
| S9_extracted_k141_28023 | Not-determined |
| S9_extracted_k141_8823 | Not-determined |
| S9_extracted_k141_20440 | Low-quality |
| S9_extracted_k141_69442 | Not-determined |
| S9_extracted_k141_5335 | Not-determined |
| S9_extracted_k141_5336 | Not-determined |
| S9_extracted_k141_12835 | Not-determined |
| S9_extracted_k141_54568 | Not-determined |
| S9_extracted_k141_73463 | Not-determined |
| S9_extracted_k141_1574 | Low-quality |
| S9_extracted_k141_76815 | Not-determined |
| S9_extracted_k141_35697 | Low-quality |
| S9_extracted_k141_28030 | Not-determined |
| S9_extracted_k141_35698 | Not-determined |
| S9_extracted_k141_69446 | Low-quality |
| S9_extracted_k141_20447 | Not-determined |
| S9_extracted_k141_76821 | Not-determined |
| S9_extracted_k141_5343 | Not-determined |
| S9_extracted_k141_80927 | Not-determined |
| S9_extracted_k141_12841 | Not-determined |
| S9_extracted_k141_50487 | Not-determined |
| S9_extracted_k141_35706 | Not-determined |
| S9_extracted_k141_12842 | Low-quality |
| S9_extracted_k141_69454 | Low-quality |
| S9_extracted_k141_5349 | Not-determined |
| S9_extracted_k141_80934 | Not-determined |
| S9_extracted_k141_69458 | Low-quality |
| S9_extracted_k141_76827 | Not-determined |
| S9_extracted_k141_35713 | Not-determined |
| S9_extracted_k141_80937 | Not-determined |
| S9_extracted_k141_76830 | Not-determined |
| S9_extracted_k141_54578 | Not-determined |
| S9_extracted_k141_76832 | Not-determined |
| S9_extracted_k141_28045 | Not-determined |
| S9_extracted_k141_5353 | Not-determined |
| S9_extracted_k141_16827 | Not-determined |
| S9_extracted_k141_8834 | Not-determined |
| S9_extracted_k141_20451 | Low-quality |
| S9_extracted_k141_54580 | Not-determined |
| S9_extracted_k141_16829 | Low-quality |
| S9_extracted_k141_54582 | Not-determined |
| S9_extracted_k141_12851 | Low-quality |
| S9_extracted_k141_5359 | Not-determined |
| S9_extracted_k141_1591 | Low-quality |
| S9_extracted_k141_50496 | Not-determined |
| S9_extracted_k141_32098 | Not-determined |
| S9_extracted_k141_32102 | Not-determined |
| S9_extracted_k141_50500 | Low-quality |
| S9_extracted_k141_80948 | Low-quality |
| S9_extracted_k141_69473 | Not-determined |
| S9_extracted_k141_8839 | Low-quality |
| S9_extracted_k141_5369 | Not-determined |
| S9_extracted_k141_12859 | Not-determined |
| S9_extracted_k141_50506 | Not-determined |
| S9_extracted_k141_54593 | Not-determined |
| S9_extracted_k141_5372 | Not-determined |
| S9_extracted_k141_69476 | Not-determined |
| S9_extracted_k141_76856 | Not-determined |
| S9_extracted_k141_69478 | Not-determined |
| S9_extracted_k141_76858 | Not-determined |
| S9_extracted_k141_54597 | Low-quality |
| S9_extracted_k141_73489 | Low-quality |
| S9_extracted_k141_28072 | Low-quality |
| S9_extracted_k141_73490 | Not-determined |
| S9_extracted_k141_35723 | Not-determined |
| S9_extracted_k141_32120 | Not-determined |
| S9_extracted_k141_73493 | Low-quality |
| S9_extracted_k141_5380 | Low-quality |
| S9_extracted_k141_69488 | Low-quality |
| S9_extracted_k141_73495 | Not-determined |
| S9_extracted_k141_39578 | Not-determined |
| S9_extracted_k141_76867 | Not-determined |
| S9_extracted_k141_80964 | Low-quality |
| S9_extracted_k141_28079 | Low-quality |
| S9_extracted_k141_16856 | Not-determined |
| S9_extracted_k141_50518 | Not-determined |
| S9_extracted_k141_80967 | Not-determined |
| S9_extracted_k141_39583 | Not-determined |
| S9_extracted_k141_69495 | Not-determined |
| S9_extracted_k141_76873 | Low-quality |
| S9_extracted_k141_35726 | Not-determined |
| S9_extracted_k141_80973 | Not-determined |
| S9_extracted_k141_50525 | Not-determined |
| S9_extracted_k141_24128 | Not-determined |
| S9_extracted_k141_32126 | Not-determined |
| S9_extracted_k141_20484 | Not-determined |
| S9_extracted_k141_39585 | Not-determined |
| S9_extracted_k141_28089 | Not-determined |
| S9_extracted_k141_58055 | Not-determined |
| S9_extracted_k141_80979 | Not-determined |
| S9_extracted_k141_50531 | Not-determined |
| S9_extracted_k141_8860 | Not-determined |
| S9_extracted_k141_20486 | Not-determined |
| S9_extracted_k141_58057 | Low-quality |
| S9_extracted_k141_12881 | Not-determined |
| S9_extracted_k141_80983 | Not-determined |
| S9_extracted_k141_20490 | Not-determined |
| S9_extracted_k141_76884 | Low-quality |
| S9_extracted_k141_24140 | Not-determined |
| S9_extracted_k141_62141 | Not-determined |
| S9_extracted_k141_20491 | Not-determined |
| S9_extracted_k141_28093 | Not-determined |
| S9_extracted_k141_12888 | Not-determined |
| S9_extracted_k141_73520 | Not-determined |
| S9_extracted_k141_80990 | Not-determined |
| S9_extracted_k141_65794 | Not-determined |
| S9_extracted_k141_50536 | Not-determined |
| S9_extracted_k141_8869 | Not-determined |
| S9_extracted_k141_35742 | Not-determined |
| S9_extracted_k141_50540 | Not-determined |
| S9_extracted_k141_24150 | Not-determined |
| S9_extracted_k141_39600 | Not-determined |
| S9_extracted_k141_69513 | Low-quality |
| S9_extracted_k141_20498 | Not-determined |
| S9_extracted_k141_65802 | Not-determined |
| S9_extracted_k141_35746 | Not-determined |
| S9_extracted_k141_16884 | Not-determined |
| S9_extracted_k141_54622 | Low-quality |
| S9_extracted_k141_20499 | Not-determined |
| S9_extracted_k141_58065 | Not-determined |
| S9_extracted_k141_62153 | Not-determined |
| S9_extracted_k141_54625 | Not-determined |
| S9_extracted_k141_20500 | Low-quality |
| S9_extracted_k141_54626 | Not-determined |
| S9_extracted_k141_62155 | Not-determined |
| S9_extracted_k141_24154 | Low-quality |
| S9_extracted_k141_32143 | Not-determined |
| S9_extracted_k141_81001 | Not-determined |
| S9_extracted_k141_35749 | Not-determined |
| S9_extracted_k141_20503 | Not-determined |
| S9_extracted_k141_28105 | Not-determined |
| S9_extracted_k141_69519 | Not-determined |
| S9_extracted_k141_16894 | Not-determined |
| S9_extracted_k141_39615 | Low-quality |
| S9_extracted_k141_76889 | Not-determined |
| S9_extracted_k141_54630 | Not-determined |
| S9_extracted_k141_62167 | Not-determined |
| S9_extracted_k141_32152 | Low-quality |
| S9_extracted_k141_39618 | Not-determined |
| S9_extracted_k141_69524 | Low-quality |
| S9_extracted_k141_54633 | Low-quality |
| S9_extracted_k141_35756 | Not-determined |
| S9_extracted_k141_28113 | Not-determined |
| S9_extracted_k141_39620 | Not-determined |
| S9_extracted_k141_35757 | Not-determined |
| S9_extracted_k141_69528 | Not-determined |
| S9_extracted_k141_8890 | Low-quality |
| S9_extracted_k141_69530 | Not-determined |
| S9_extracted_k141_58076 | Not-determined |
| S9_extracted_k141_69531 | Not-determined |
| S9_extracted_k141_58079 | Not-determined |
| S9_extracted_k141_32159 | Not-determined |
| S9_extracted_k141_39628 | Not-determined |

| | |
|---|---|
| S9_extracted_k141_24177 | Not-determined |
| S9_extracted_k141_50562 | Not-determined |
| S9_extracted_k141_16910 | Not-determined |
| S9_extracted_k141_32164 | Low-quality |
| S9_extracted_k141_24180 | Not-determined |
| S9_extracted_k141_62174 | Low-quality |
| S9_extracted_k141_73546 | Not-determined |
| S9_extracted_k141_39631 | Not-determined |
| S9_extracted_k141_28124 | Not-determined |
| S9_extracted_k141_5396 | Not-determined |
| S9_extracted_k141_24184 | Not-determined |
| S9_extracted_k141_39634 | Low-quality |
| S9_extracted_k141_46801 | Not-determined |
| S9_extracted_k141_24187 | Not-determined |
| S9_extracted_k141_73549 | Not-determined |
| S9_extracted_k141_20519 | Low-quality |
| S9_extracted_k141_69546 | Not-determined |
| S9_extracted_k141_50571 | Low-quality |
| S9_extracted_k141_32179 | Low-quality |
| S9_extracted_k141_39642 | Not-determined |
| S9_extracted_k141_50574 | Not-determined |
| S9_extracted_k141_32180 | Not-determined |
| S9_extracted_k141_69553 | Low-quality |
| S9_extracted_k141_32184 | Not-determined |
| S9_extracted_k141_54647 | Not-determined |
| S9_extracted_k141_32187 | Not-determined |
| S9_extracted_k141_1634 | Not-determined |
| S9_extracted_k141_16918 | Not-determined |
| S9_extracted_k141_62194 | Not-determined |
| S9_extracted_k141_16919 | Not-determined |
| S9_extracted_k141_46805 | Not-determined |
| S9_extracted_k141_16920 | Not-determined |
| S9_extracted_k141_39647 | Low-quality |
| S9_extracted_k141_54655 | Not-determined |
| S9_extracted_k141_5412 | Not-determined |
| S9_extracted_k141_24202 | Not-determined |
| S9_extracted_k141_24203 | Not-determined |
| S9_extracted_k141_62197 | Not-determined |
| S9_extracted_k141_32195 | Not-determined |
| S9_extracted_k141_46808 | Low-quality |
| S9_extracted_k141_32200 | Not-determined |
| S9_extracted_k141_32201 | Not-determined |
| S9_extracted_k141_20529 | Not-determined |
| S9_extracted_k141_46813 | Not-determined |
| S9_extracted_k141_54661 | Low-quality |
| S9_extracted_k141_24212 | Not-determined |
| S9_extracted_k141_24213 | Not-determined |
| S9_extracted_k141_54662 | Not-determined |
| S9_extracted_k141_62204 | Not-determined |
| S9_extracted_k141_24214 | Not-determined |
| S9_extracted_k141_8918 | Not-determined |
| S9_extracted_k141_8919 | Not-determined |
| S9_extracted_k141_69573 | Low-quality |
| S9_extracted_k141_8920 | Not-determined |
| S9_extracted_k141_32210 | Not-determined |
| S9_extracted_k141_8922 | Not-determined |
| S9_extracted_k141_28136 | Not-determined |
| S9_extracted_k141_43338 | Not-determined |
| S9_extracted_k141_28137 | Not-determined |
| S9_extracted_k141_62210 | Not-determined |
| S9_extracted_k141_43340 | Not-determined |
| S9_extracted_k141_16931 | Not-determined |
| S9_extracted_k141_62211 | Not-determined |
| S9_extracted_k141_5430 | Not-determined |
| S9_extracted_k141_8927 | Not-determined |
| S9_extracted_k141_20544 | Not-determined |
| S9_extracted_k141_69582 | Not-determined |
| S9_extracted_k141_8929 | Not-determined |
| S9_extracted_k141_46824 | Not-determined |
| S9_extracted_k141_69584 | Low-quality |
| S9_extracted_k141_35780 | Low-quality |
| S9_extracted_k141_32221 | Not-determined |
| S9_extracted_k141_46828 | Not-determined |
| S9_extracted_k141_62222 | Not-determined |
| S9_extracted_k141_1654 | Not-determined |
| S9_extracted_k141_5431 | Not-determined |
| S9_extracted_k141_43348 | Low-quality |
| S9_extracted_k141_28151 | Not-determined |
| S9_extracted_k141_8934 | Not-determined |
| S9_extracted_k141_12921 | Not-determined |
| S9_extracted_k141_16938 | Not-determined |
| S9_extracted_k141_50610 | Not-determined |
| S9_extracted_k141_32230 | Low-quality |
| S9_extracted_k141_24225 | Low-quality |
| S9_extracted_k141_50613 | Not-determined |
| S9_extracted_k141_62226 | Not-determined |
| S9_extracted_k141_43359 | Not-determined |
| S9_extracted_k141_43361 | Low-quality |
| S9_extracted_k141_84649 | Not-determined |
| S9_extracted_k141_50621 | Not-determined |
| S9_extracted_k141_12934 | Not-determined |
| S9_extracted_k141_20568 | Not-determined |
| S9_extracted_k141_8948 | Not-determined |
| S9_extracted_k141_54680 | Not-determined |
| S9_extracted_k141_81005 | Low-quality |
| S9_extracted_k141_50626 | Not-determined |
| S9_extracted_k141_1670 | Not-determined |
| S9_extracted_k141_5448 | Not-determined |
| S9_extracted_k141_62236 | Not-determined |
| S9_extracted_k141_8953 | Not-determined |
| S9_extracted_k141_76903 | Not-determined |
| S9_extracted_k141_76904 | Not-determined |
| S9_extracted_k141_76905 | Not-determined |
| S9_extracted_k141_43373 | Not-determined |
| S9_extracted_k141_32241 | Low-quality |
| S9_extracted_k141_76907 | Low-quality |
| S9_extracted_k141_8959 | Not-determined |
| S9_extracted_k141_20579 | Not-determined |
| S9_extracted_k141_54691 | Low-quality |
| S9_extracted_k141_62244 | Low-quality |
| S9_extracted_k141_8964 | Not-determined |
| S9_extracted_k141_20582 | Not-determined |
| S9_extracted_k141_28179 | Not-determined |
| S9_extracted_k141_50642 | Low-quality |
| S9_extracted_k141_39660 | Low-quality |
| S9_extracted_k141_81019 | Not-determined |
| S9_extracted_k141_76915 | Not-determined |
| S9_extracted_k141_43386 | Not-determined |
| S9_extracted_k141_20587 | Low-quality |
| S9_extracted_k141_1690 | Not-determined |
| S9_extracted_k141_16980 | Not-determined |
| S9_extracted_k141_54700 | Low-quality |
| S9_extracted_k141_43388 | Not-determined |
| S9_extracted_k141_12952 | Not-determined |
| S9_extracted_k141_24232 | Not-determined |
| S9_extracted_k141_43389 | Not-determined |
| S9_extracted_k141_32250 | Not-determined |
| S9_extracted_k141_28189 | Not-determined |
| S9_extracted_k141_28192 | Not-determined |
| S9_extracted_k141_16989 | Low-quality |
| S9_extracted_k141_24236 | Low-quality |
| S9_extracted_k141_32254 | Not-determined |
| S9_extracted_k141_54703 | Not-determined |
| S9_extracted_k141_12959 | Not-determined |
| S9_extracted_k141_24239 | Not-determined |
| S9_extracted_k141_58090 | Not-determined |
| S9_extracted_k141_16993 | Low-quality |
| S9_extracted_k141_12961 | Not-determined |
| S9_extracted_k141_84669 | Not-determined |
| S9_extracted_k141_32260 | Not-determined |
| S9_extracted_k141_24241 | Not-determined |
| S9_extracted_k141_20591 | Low-quality |
| S9_extracted_k141_43397 | Low-quality |
| S9_extracted_k141_76920 | Low-quality |
| S9_extracted_k141_35810 | Not-determined |
| S9_extracted_k141_84671 | Not-determined |
| S9_extracted_k141_50655 | Not-determined |
| S9_extracted_k141_12965 | Not-determined |
| S9_extracted_k141_84674 | Not-determined |
| S9_extracted_k141_84675 | Not-determined |
| S9_extracted_k141_39684 | Not-determined |
| S9_extracted_k141_69601 | Not-determined |
| S9_extracted_k141_35815 | Not-determined |
| S9_extracted_k141_81040 | Not-determined |
| S9_extracted_k141_58105 | Low-quality |
| S9_extracted_k141_1717 | Low-quality |
| S9_extracted_k141_76935 | Not-determined |
| S9_extracted_k141_1718 | Not-determined |
| S9_extracted_k141_54730 | Not-determined |
| S9_extracted_k141_58115 | Not-determined |
| S9_extracted_k141_58118 | Low-quality |
| S9_extracted_k141_24262 | Low-quality |
| S9_extracted_k141_17019 | Not-determined |
| S9_extracted_k141_1727 | Not-determined |
| S9_extracted_k141_35821 | Not-determined |
| S9_extracted_k141_81065 | Not-determined |
| S9_extracted_k141_9006 | Not-determined |
| S9_extracted_k141_17024 | Not-determined |
| S9_extracted_k141_54741 | Low-quality |
| S9_extracted_k141_43429 | Not-determined |
| S9_extracted_k141_35823 | Low-quality |
| S9_extracted_k141_81066 | Not-determined |
| S9_extracted_k141_24272 | Low-quality |
| S9_extracted_k141_24273 | Low-quality |
| S9_extracted_k141_35829 | Not-determined |
| S9_extracted_k141_84695 | Low-quality |
| S9_extracted_k141_17030 | Not-determined |
| S9_extracted_k141_9010 | Low-quality |
| S9_extracted_k141_81071 | Not-determined |
| S9_extracted_k141_69626 | Not-determined |
| S9_extracted_k141_17036 | Not-determined |
| S9_extracted_k141_84701 | Not-determined |
| S9_extracted_k141_43445 | Not-determined |
| S9_extracted_k141_81076 | Low-quality |
| S9_extracted_k141_5485 | Low-quality |
| S9_extracted_k141_76958 | Not-determined |
| S9_extracted_k141_32276 | Not-determined |
| S9_extracted_k141_88589 | Not-determined |
| S9_extracted_k141_58140 | Not-determined |
| S9_extracted_k141_58142 | Not-determined |
| S9_extracted_k141_12978 | Low-quality |
| S12_vs_k141_34387\|\|full | High-quality |
| S9_extracted_k141_24284 | Not-determined |
| S9_extracted_k141_1744 | Low-quality |
| S9_extracted_k141_35843 | Not-determined |
| S9_extracted_k141_43454 | Not-determined |
| S9_extracted_k141_12981 | Low-quality |
| S9_extracted_k141_58150 | Not-determined |
| S9_extracted_k141_28201 | Not-determined |
| S9_extracted_k141_1746 | Not-determined |
| S9_extracted_k141_50668 | Not-determined |
| S9_extracted_k141_65819 | Not-determined |
| S9_extracted_k141_35846 | Not-determined |
| S9_extracted_k141_17051 | Low-quality |
| S9_extracted_k141_17052 | Not-determined |
| S9_extracted_k141_24293 | Not-determined |
| S9_extracted_k141_28205 | Not-determined |
| S9_extracted_k141_81089 | Not-determined |
| S9_extracted_k141_24294 | Not-determined |
| S9_extracted_k141_5495 | Not-determined |
| S9_extracted_k141_76971 | Low-quality |
| S9_extracted_k141_12994 | Not-determined |
| S9_extracted_k141_28212 | Not-determined |
| S9_extracted_k141_35856 | Not-determined |
| S9_extracted_k141_20620 | Not-determined |
| S9_extracted_k141_50680 | Not-determined |
| S9_extracted_k141_76974 | Not-determined |
| S9_extracted_k141_24301 | Not-determined |
| S9_extracted_k141_5498 | Not-determined |
| S9_extracted_k141_35860 | Not-determined |
| S9_extracted_k141_1763 | Not-determined |
| S9_extracted_k141_54750 | Not-determined |
| S9_extracted_k141_1764 | Not-determined |
| S9_extracted_k141_39720 | Low-quality |
| S9_extracted_k141_32297 | Low-quality |
| S9_extracted_k141_76979 | Not-determined |
| S9_extracted_k141_35865 | Not-determined |
| S9_extracted_k141_28219 | Not-determined |
| S9_extracted_k141_46835 | Not-determined |
| S9_extracted_k141_84731 | Not-determined |
| S9_extracted_k141_76991 | Not-determined |
| S9_extracted_k141_35875 | Not-determined |
| S9_extracted_k141_84735 | Low-quality |
| S9_extracted_k141_65839 | Not-determined |
| S9_extracted_k141_76998 | Not-determined |
| S9_extracted_k141_46836 | Low-quality |
| S9_extracted_k141_35882 | Not-determined |
| S9_extracted_k141_54770 | Not-determined |
| S9_extracted_k141_46838 | Low-quality |
| S9_extracted_k141_77003 | Not-determined |
| S9_extracted_k141_46840 | Not-determined |
| S9_extracted_k141_69648 | Not-determined |
| S9_extracted_k141_58180 | Not-determined |
| S9_extracted_k141_54774 | Not-determined |
| S9_extracted_k141_77006 | Low-quality |
| S9_extracted_k141_13010 | Low-quality |
| S9_extracted_k141_50701 | Not-determined |
| S9_extracted_k141_28239 | Not-determined |
| S9_extracted_k141_5516 | Not-determined |
| S9_extracted_k141_88619 | Not-determined |
| S9_extracted_k141_46845 | Not-determined |
| S9_extracted_k141_77008 | Not-determined |
| S9_extracted_k141_9035 | Not-determined |
| S9_extracted_k141_65843 | Low-quality |
| S9_extracted_k141_17088 | Not-determined |
| S9_extracted_k141_28246 | Not-determined |
| S9_extracted_k141_28248 | Not-determined |
| S9_extracted_k141_50708 | Low-quality |
| S9_extracted_k141_84747 | Not-determined |
| S9_extracted_k141_28249 | Not-determined |
| S9_extracted_k141_13015 | Not-determined |
| S9_extracted_k141_46851 | Not-determined |
| S9_extracted_k141_50711 | Not-determined |
| S9_extracted_k141_62250 | Low-quality |
| S9_extracted_k141_5524 | Not-determined |
| S9_extracted_k141_84751 | Low-quality |
| S9_extracted_k141_54786 | Not-determined |
| S9_extracted_k141_46854 | Not-determined |
| S9_extracted_k141_9044 | Not-determined |
| S9_extracted_k141_54788 | Not-determined |
| S9_extracted_k141_9045 | Not-determined |
| S9_extracted_k141_54790 | Not-determined |
| S9_extracted_k141_9046 | Not-determined |
| S9_extracted_k141_35893 | Not-determined |
| S9_extracted_k141_69662 | Not-determined |
| S9_extracted_k141_84760 | Not-determined |
| S9_extracted_k141_13024 | Not-determined |
| S9_extracted_k141_69663 | Not-determined |
| S9_extracted_k141_84761 | Not-determined |
| S9_extracted_k141_69664 | Low-quality |
| S9_extracted_k141_9051 | Not-determined |
| S9_extracted_k141_1798 | Low-quality |
| S9_extracted_k141_39751 | Not-determined |
| S9_extracted_k141_43504 | Not-determined |
| S9_extracted_k141_58189 | Not-determined |
| S9_extracted_k141_77016 | Not-determined |
| S9_extracted_k141_5536 | Not-determined |
| S9_extracted_k141_54801 | Not-determined |
| S9_extracted_k141_9058 | Low-quality |
| S9_extracted_k141_39758 | Not-determined |
| S9_extracted_k141_24331 | Not-determined |
| S9_extracted_k141_62273 | Not-determined |
| S9_extracted_k141_35902 | Low-quality |
| S9_extracted_k141_20631 | Not-determined |
| S9_extracted_k141_88633 | Low-quality |
| S9_extracted_k141_17103 | Not-determined |
| S9_extracted_k141_35907 | Not-determined |
| S9_extracted_k141_58199 | Not-determined |
| S9_extracted_k141_50737 | Not-determined |
| S9_extracted_k141_69686 | Low-quality |
| S9_extracted_k141_77030 | Not-determined |
| S9_extracted_k141_62282 | Not-determined |
| S9_extracted_k141_84782 | Not-determined |
| S9_extracted_k141_88635 | Not-determined |
| S9_extracted_k141_69692 | Not-determined |
| S9_extracted_k141_17111 | Not-determined |
| S9_extracted_k141_50748 | Low-quality |
| S9_extracted_k141_46880 | Not-determined |
| S9_extracted_k141_84786 | Not-determined |
| S9_extracted_k141_69697 | Not-determined |
| S9_extracted_k141_9082 | Not-determined |
| S9_extracted_k141_39773 | Not-determined |
| S9_extracted_k141_13042 | Not-determined |
| S9_extracted_k141_39774 | Not-determined |
| S9_extracted_k141_69700 | Not-determined |
| S9_extracted_k141_46891 | Not-determined |
| S9_extracted_k141_43532 | Not-determined |
| S9_extracted_k141_35914 | Low-quality |
| S9_extracted_k141_35915 | Not-determined |
| S9_extracted_k141_9087 | Not-determined |
| S9_extracted_k141_77046 | Not-determined |
| S9_extracted_k141_20655 | Not-determined |
| S9_extracted_k141_88647 | Not-determined |
| S9_extracted_k141_1832 | Low-quality |
| S9_extracted_k141_5564 | Not-determined |
| S9_extracted_k141_1834 | Not-determined |
| S9_extracted_k141_5565 | Not-determined |
| S9_extracted_k141_58215 | Not-determined |
| S9_extracted_k141_81134 | Not-determined |
| S9_extracted_k141_50771 | Not-determined |
| S9_extracted_k141_5568 | Not-determined |
| S9_extracted_k141_81136 | Not-determined |
| S9_extracted_k141_62302 | Not-determined |
| S9_extracted_k141_20658 | Not-determined |
| S9_extracted_k141_24373 | Not-determined |
| S9_extracted_k141_54832 | Not-determined |
| S9_extracted_k141_28289 | Not-determined |
| S9_extracted_k141_50777 | Not-determined |
| S9_extracted_k141_9097 | Not-determined |
| S9_extracted_k141_58222 | Not-determined |
| S9_extracted_k141_54838 | Not-determined |
| S9_extracted_k141_62308 | Not-determined |
| S9_extracted_k141_81141 | Not-determined |
| S9_extracted_k141_24376 | Not-determined |
| S9_extracted_k141_43544 | Low-quality |
| S9_extracted_k141_5573 | Not-determined |
| S9_extracted_k141_20665 | Not-determined |
| S9_extracted_k141_5576 | Not-determined |
| S9_extracted_k141_46911 | Not-determined |
| S9_extracted_k141_32319 | Not-determined |
| S9_extracted_k141_5582 | Not-determined |
| S9_extracted_k141_1850 | Not-determined |
| S9_extracted_k141_81151 | Not-determined |
| S9_extracted_k141_17134 | Not-determined |
| S9_extracted_k141_54847 | Not-determined |
| S9_extracted_k141_54848 | Not-determined |
| S9_extracted_k141_39779 | Low-quality |
| S9_extracted_k141_28299 | Not-determined |
| S9_extracted_k141_32322 | Not-determined |
| S9_extracted_k141_28300 | Not-determined |
| S9_extracted_k141_81156 | Not-determined |
| S9_extracted_k141_58241 | Not-determined |
| S9_extracted_k141_43558 | Low-quality |
| S9_extracted_k141_54855 | Not-determined |
| S9_extracted_k141_28304 | Not-determined |
| S9_extracted_k141_81160 | Not-determined |
| S9_extracted_k141_28305 | Low-quality |
| S9_extracted_k141_88655 | Not-determined |
| S9_extracted_k141_24390 | Low-quality |
| S9_extracted_k141_1864 | Not-determined |
| S9_extracted_k141_24395 | Not-determined |
| S9_extracted_k141_54861 | Low-quality |
| S9_extracted_k141_84820 | Not-determined |
| S9_extracted_k141_81165 | Not-determined |
| S9_extracted_k141_81166 | Not-determined |
| S9_extracted_k141_24399 | Not-determined |
| S9_extracted_k141_88662 | Not-determined |
| S9_extracted_k141_9119 | Not-determined |
| S9_extracted_k141_39795 | Not-determined |
| S9_extracted_k141_24405 | Not-determined |
| S9_extracted_k141_88667 | Not-determined |
| S9_extracted_k141_9123 | Not-determined |
| S9_extracted_k141_58255 | Low-quality |
| S9_extracted_k141_28317 | Not-determined |
| S9_extracted_k141_69736 | Not-determined |
| S9_extracted_k141_77104 | Low-quality |
| S9_extracted_k141_13089 | Not-determined |
| S9_extracted_k141_39801 | Low-quality |
| S9_extracted_k141_24412 | Not-determined |
| S9_extracted_k141_77107 | Not-determined |
| S9_extracted_k141_58259 | Not-determined |
| S9_extracted_k141_20688 | Not-determined |
| S9_extracted_k141_9129 | Not-determined |
| S9_extracted_k141_43579 | Not-determined |
| S9_extracted_k141_28328 | Low-quality |
| S9_extracted_k141_84834 | Not-determined |
| S9_extracted_k141_58264 | Not-determined |
| S9_extracted_k141_28333 | Not-determined |
| S9_extracted_k141_54880 | Not-determined |
| S9_extracted_k141_58265 | Low-quality |
| S9_extracted_k141_88673 | Not-determined |
| S9_extracted_k141_24420 | Not-determined |
| S9_extracted_k141_81183 | Not-determined |
| S9_extracted_k141_9136 | Not-determined |
| S9_extracted_k141_58268 | Not-determined |
| S9_extracted_k141_28336 | Not-determined |
| S9_extracted_k141_58271 | Low-quality |
| S9_extracted_k141_43588 | Low-quality |
| S9_extracted_k141_28340 | Not-determined |
| S9_extracted_k141_81186 | Not-determined |
| S9_extracted_k141_28341 | Not-determined |
| S9_extracted_k141_43590 | Not-determined |
| S9_extracted_k141_54886 | Not-determined |
| S9_extracted_k141_39808 | Not-determined |
| S9_extracted_k141_9143 | Not-determined |
| S9_extracted_k141_9145 | Low-quality |
| S9_extracted_k141_28347 | Not-determined |
| S9_extracted_k141_77112 | Low-quality |
| S9_extracted_k141_50832 | Not-determined |
| S9_extracted_k141_35956 | Not-determined |
| S9_extracted_k141_5617 | Not-determined |
| S9_extracted_k141_35957 | Not-determined |
| S9_extracted_k141_35958 | Not-determined |
| S9_extracted_k141_24427 | Low-quality |
| S9_extracted_k141_81196 | Not-determined |
| S9_extracted_k141_39815 | Not-determined |
| S9_extracted_k141_54894 | Not-determined |
| S9_extracted_k141_5621 | Not-determined |
| S9_extracted_k141_9154 | Not-determined |
| S9_extracted_k141_28357 | Not-determined |
| S9_extracted_k141_35967 | Not-determined |
| S9_extracted_k141_5623 | Not-determined |
| S9_extracted_k141_9157 | Not-determined |
| S9_extracted_k141_73564 | Not-determined |
| S9_extracted_k141_58281 | Not-determined |
| S9_extracted_k141_43604 | Not-determined |
| S9_extracted_k141_39821 | Not-determined |
| S9_extracted_k141_43606 | Not-determined |
| S9_extracted_k141_5632 | Not-determined |
| S9_extracted_k141_9164 | Not-determined |
| S9_extracted_k141_24437 | Not-determined |
| S9_extracted_k141_17178 | Not-determined |
| S9_extracted_k141_77123 | Not-determined |
| S9_extracted_k141_17180 | Not-determined |
| S9_extracted_k141_5634 | Low-quality |
| S9_extracted_k141_28363 | Low-quality |
| S9_extracted_k141_88697 | Not-determined |
| S9_extracted_k141_28365 | Not-determined |
| S9_extracted_k141_28368 | Not-determined |
| S9_extracted_k141_5639 | Not-determined |
| S9_extracted_k141_73581 | Not-determined |
| S9_extracted_k141_24448 | Not-determined |
| S9_extracted_k141_28374 | Not-determined |
| S9_extracted_k141_69747 | Not-determined |
| S9_extracted_k141_58295 | Not-determined |
| S9_extracted_k141_17185 | Not-determined |
| S9_extracted_k141_24451 | Not-determined |
| S9_extracted_k141_50844 | Not-determined |
| S9_extracted_k141_43620 | Not-determined |
| S9_extracted_k141_69753 | Not-determined |
| S9_extracted_k141_50848 | Not-determined |
| S9_extracted_k141_69754 | Not-determined |
| S9_extracted_k141_43623 | Not-determined |
| S9_extracted_k141_58301 | Not-determined |
| S9_extracted_k141_69758 | Not-determined |
| S9_extracted_k141_77126 | Low-quality |
| S9_extracted_k141_17196 | Low-quality |
| S9_extracted_k141_62385 | Low-quality |
| S9_extracted_k141_81215 | Not-determined |
| S9_extracted_k141_65852 | Not-determined |
| S9_extracted_k141_1883 | Not-determined |
| S9_extracted_k141_43630 | Not-determined |
| S9_extracted_k141_77130 | Not-determined |
| S9_extracted_k141_43631 | Not-determined |
| S9_extracted_k141_17202 | Not-determined |
| S9_extracted_k141_43634 | Low-quality |
| S9_extracted_k141_81221 | Not-determined |
| S9_extracted_k141_28393 | Not-determined |
| S9_extracted_k141_69769 | Not-determined |
| S9_extracted_k141_69772 | Not-determined |
| S9_extracted_k141_50860 | Not-determined |
| S9_extracted_k141_65859 | Low-quality |
| S9_extracted_k141_65860 | Not-determined |
| S9_extracted_k141_65865 | Not-determined |
| S9_extracted_k141_69775 | Not-determined |
| S9_extracted_k141_1891 | Not-determined |
| S9_extracted_k141_20701 | Low-quality |
| S9_extracted_k141_65867 | Low-quality |
| S9_extracted_k141_13099 | Not-determined |
| S9_extracted_k141_62393 | Not-determined |
| S9_extracted_k141_17215 | Not-determined |
| S9_extracted_k141_5661 | Not-determined |
| S9_extracted_k141_62398 | Not-determined |
| S9_extracted_k141_46940 | Not-determined |
| S9_extracted_k141_43640 | Not-determined |
| S9_extracted_k141_43643 | Low-quality |
| S9_extracted_k141_20707 | Not-determined |
| S9_extracted_k141_24480 | Not-determined |
| S9_extracted_k141_77145 | Not-determined |
| S9_extracted_k141_69784 | Low-quality |
| S9_extracted_k141_5666 | Not-determined |
| S9_extracted_k141_58329 | Not-determined |
| S9_extracted_k141_1901 | Not-determined |
| S9_extracted_k141_54950 | Not-determined |
| S9_extracted_k141_54952 | Not-determined |
| S9_extracted_k141_1902 | Not-determined |
| S9_extracted_k141_73618 | Low-quality |
| S9_extracted_k141_24483 | Low-quality |
| S9_extracted_k141_39869 | Not-determined |
| S9_extracted_k141_58333 | Low-quality |
| S9_extracted_k141_62406 | Not-determined |
| S9_extracted_k141_28424 | Not-determined |
| S9_extracted_k141_54956 | Not-determined |
| S9_extracted_k141_28425 | Not-determined |
| S9_extracted_k141_58337 | Not-determined |
| S9_extracted_k141_43650 | Not-determined |
| S9_extracted_k141_58338 | Not-determined |
| S9_extracted_k141_24484 | Low-quality |
| S9_extracted_k141_28432 | Low-quality |
| S9_extracted_k141_20724 | Not-determined |
| S9_extracted_k141_39879 | Not-determined |
| S9_extracted_k141_73628 | Not-determined |
| S9_extracted_k141_88749 | Not-determined |
| S9_extracted_k141_73630 | Not-determined |
| S9_extracted_k141_39882 | Not-determined |
| S9_extracted_k141_65896 | Not-determined |
| S9_extracted_k141_54963 | Low-quality |
| S9_extracted_k141_13122 | Not-determined |
| S9_extracted_k141_46971 | Not-determined |
| S9_extracted_k141_69799 | Low-quality |
| S9_extracted_k141_20731 | Not-determined |
| S9_extracted_k141_54967 | Low-quality |
| S9_extracted_k141_54969 | Not-determined |
| S9_extracted_k141_73633 | Not-determined |
| S9_extracted_k141_17243 | Low-quality |
| S9_extracted_k141_65908 | Not-determined |
| S9_extracted_k141_58344 | Not-determined |
| S9_extracted_k141_54975 | Not-determined |
| S9_extracted_k141_62421 | Not-determined |
| S9_extracted_k141_62422 | Not-determined |
| S9_extracted_k141_5673 | Not-determined |
| S9_extracted_k141_28445 | Low-quality |
| S9_extracted_k141_73641 | Not-determined |
| S9_extracted_k141_39888 | Low-quality |
| S9_extracted_k141_73642 | Not-determined |
| S9_extracted_k141_54981 | Not-determined |
| S9_extracted_k141_43672 | Not-determined |
| S9_extracted_k141_17257 | Not-determined |
| S9_extracted_k141_77175 | Not-determined |
| S9_extracted_k141_69819 | Not-determined |
| S9_extracted_k141_43677 | Not-determined |
| S9_extracted_k141_39894 | Not-determined |
| S9_extracted_k141_62436 | Not-determined |
| S9_extracted_k141_46988 | Not-determined |
| S9_extracted_k141_54987 | Not-determined |
| S9_extracted_k141_58362 | Not-determined |
| S9_extracted_k141_20753 | Low-quality |
| S9_extracted_k141_58363 | Low-quality |
| S9_extracted_k141_46991 | Not-determined |
| S9_extracted_k141_1924 | Low-quality |
| S9_extracted_k141_39902 | Not-determined |
| S9_extracted_k141_62440 | Not-determined |
| S9_extracted_k141_17265 | Not-determined |
| S9_extracted_k141_24511 | Not-determined |
| S9_extracted_k141_54994 | Not-determined |
| S9_extracted_k141_39907 | Not-determined |
| S9_extracted_k141_81239 | Not-determined |
| S9_extracted_k141_65917 | Not-determined |
| S9_extracted_k141_43686 | Not-determined |
| S9_extracted_k141_36013 | Not-determined |
| S9_extracted_k141_17272 | Not-determined |
| S9_extracted_k141_46996 | Not-determined |
| S9_extracted_k141_69840 | Not-determined |
| S9_extracted_k141_73651 | Not-determined |
| S9_extracted_k141_58374 | Not-determined |
| S9_extracted_k141_69841 | Not-determined |
| S9_extracted_k141_69845 | Not-determined |
| S9_extracted_k141_73656 | Not-determined |
| S9_extracted_k141_81247 | Low-quality |
| S9_extracted_k141_81248 | Not-determined |
| S9_extracted_k141_47005 | Not-determined |
| S9_extracted_k141_28464 | Not-determined |
| S9_extracted_k141_69851 | Not-determined |
| S9_extracted_k141_69853 | Low-quality |
| S9_extracted_k141_84855 | Low-quality |
| S9_extracted_k141_65929 | Not-determined |
| S9_extracted_k141_17281 | Not-determined |
| S9_extracted_k141_28469 | Not-determined |
| S9_extracted_k141_36025 | Not-determined |

| | |
|---|---|
| S9_extracted_k141_24520 | Not-determined |
| S9_extracted_k141_24521 | Not-determined |
| S9_extracted_k141_24522 | Not-determined |
| S9_extracted_k141_17286 | Not-determined |
| S9_extracted_k141_17289 | Low-quality |
| S9_extracted_k141_17290 | Low-quality |
| S9_extracted_k141_24523 | Not-determined |
| S9_extracted_k141_39939 | Not-determined |
| S9_extracted_k141_81263 | Not-determined |
| S9_extracted_k141_81264 | Not-determined |
| S9_extracted_k141_69861 | Not-determined |
| S9_extracted_k141_20769 | Not-determined |
| S9_extracted_k141_36036 | Low-quality |
| S9_extracted_k141_65936 | Not-determined |
| S9_extracted_k141_36037 | Not-determined |
| S9_extracted_k141_24529 | Not-determined |
| S9_extracted_k141_17296 | Not-determined |
| S9_extracted_k141_13167 | Not-determined |
| S9_extracted_k141_36038 | Not-determined |
| S9_extracted_k141_47015 | Not-determined |
| S9_extracted_k141_36039 | Not-determined |
| S9_extracted_k141_47016 | Not-determined |
| S9_extracted_k141_13168 | Not-determined |
| S9_extracted_k141_47017 | Low-quality |
| S9_extracted_k141_73675 | Not-determined |
| S9_extracted_k141_13170 | Not-determined |
| S9_extracted_k141_73676 | Not-determined |
| S9_extracted_k141_20775 | Not-determined |
| S9_extracted_k141_58391 | Not-determined |
| S9_extracted_k141_58392 | Not-determined |
| S9_extracted_k141_43710 | Not-determined |
| S9_extracted_k141_1962 | Not-determined |
| S9_extracted_k141_58393 | Not-determined |
| S9_extracted_k141_13173 | Not-determined |
| S9_extracted_k141_77198 | Not-determined |
| S9_extracted_k141_81271 | Low-quality |
| S9_extracted_k141_28482 | Not-determined |
| S9_extracted_k141_47025 | Not-determined |
| S9_extracted_k141_20778 | Low-quality |
| S9_extracted_k141_24544 | Low-quality |
| S9_extracted_k141_47027 | Low-quality |
| S9_extracted_k141_24545 | Not-determined |
| S9_extracted_k141_1965 | Not-determined |
| S9_extracted_k141_20781 | Not-determined |
| S9_extracted_k141_39951 | Not-determined |
| S9_extracted_k141_43720 | Not-determined |
| S9_extracted_k141_39952 | Low-quality |
| S9_extracted_k141_77203 | Not-determined |
| S9_extracted_k141_9170 | Low-quality |
| S9_extracted_k141_47034 | Not-determined |
| S9_extracted_k141_5677 | Not-determined |
| S9_extracted_k141_73683 | Not-determined |
| S9_extracted_k141_84882 | Low-quality |
| S9_extracted_k141_69888 | Not-determined |
| S9_extracted_k141_24551 | Not-determined |
| S9_extracted_k141_36054 | Not-determined |
| S9_extracted_k141_47038 | Low-quality |
| S9_extracted_k141_73687 | Not-determined |
| S9_extracted_k141_58401 | Not-determined |
| S9_extracted_k141_13186 | Not-determined |
| S9_extracted_k141_24556 | Low-quality |
| S9_extracted_k141_28494 | Low-quality |
| S9_extracted_k141_5683 | Low-quality |
| S9_extracted_k141_13188 | Not-determined |
| S9_extracted_k141_47041 | Not-determined |
| S9_extracted_k141_81290 | Not-determined |
| S9_extracted_k141_62483 | Not-determined |
| S9_extracted_k141_24560 | Not-determined |
| S9_extracted_k141_20796 | Not-determined |
| S9_extracted_k141_39973 | Not-determined |
| S9_extracted_k141_36059 | Low-quality |
| S9_extracted_k141_9189 | Not-determined |
| S9_extracted_k141_77217 | Not-determined |
| S9_extracted_k141_84900 | Not-determined |
| S9_extracted_k141_9190 | Not-determined |
| S9_extracted_k141_17319 | Low-quality |
| S9_extracted_k141_84904 | Not-determined |
| S9_extracted_k141_17320 | Low-quality |
| S9_extracted_k141_43750 | Low-quality |
| S9_extracted_k141_9194 | Not-determined |
| S9_extracted_k141_43751 | Not-determined |
| S9_extracted_k141_47057 | Not-determined |
| S9_extracted_k141_13210 | Not-determined |
| S9_extracted_k141_65968 | Low-quality |
| S9_extracted_k141_55044 | Not-determined |
| S9_extracted_k141_47062 | Low-quality |
| S9_extracted_k141_28508 | Not-determined |
| S9_extracted_k141_13215 | Not-determined |
| S9_extracted_k141_73701 | Not-determined |
| S9_extracted_k141_32388 | Not-determined |
| S9_extracted_k141_47064 | Not-determined |
| S9_extracted_k141_32389 | Not-determined |
| S9_extracted_k141_5700 | Not-determined |
| S9_extracted_k141_88795 | Not-determined |
| S9_extracted_k141_47067 | Not-determined |
| S9_extracted_k141_17325 | Not-determined |
| S9_extracted_k141_77222 | Not-determined |
| S9_extracted_k141_43753 | Low-quality |
| S9_extracted_k141_81312 | Not-determined |
| S9_extracted_k141_39992 | Not-determined |
| S9_extracted_k141_69915 | Not-determined |
| S9_extracted_k141_73708 | Not-determined |
| S9_extracted_k141_9201 | Not-determined |
| S9_extracted_k141_17331 | Not-determined |
| S9_extracted_k141_13222 | Not-determined |
| S9_extracted_k141_36083 | Not-determined |
| S9_extracted_k141_88801 | Not-determined |
| S9_extracted_k141_81315 | Not-determined |
| S9_extracted_k141_9206 | Not-determined |
| S9_extracted_k141_55052 | Not-determined |
| S9_extracted_k141_20809 | Low-quality |
| S9_extracted_k141_9208 | Not-determined |
| S9_extracted_k141_28517 | Not-determined |
| S9_extracted_k141_77233 | Not-determined |
| S9_extracted_k141_69918 | Not-determined |
| S9_extracted_k141_88807 | Not-determined |
| S9_extracted_k141_9211 | Not-determined |
| S9_extracted_k141_88810 | Not-determined |
| S9_extracted_k141_5709 | Not-determined |
| S9_extracted_k141_5711 | Not-determined |
| S9_extracted_k141_65983 | Not-determined |
| S9_extracted_k141_55060 | Not-determined |
| S9_extracted_k141_36088 | Not-determined |
| S9_extracted_k141_65986 | Not-determined |
| S9_extracted_k141_58404 | Not-determined |
| S9_extracted_k141_9219 | Not-determined |
| S9_extracted_k141_73724 | Not-determined |
| S9_extracted_k141_69926 | Not-determined |
| S9_extracted_k141_77244 | Not-determined |
| S9_extracted_k141_1972 | Low-quality |
| S9_extracted_k141_1973 | Not-determined |
| S9_extracted_k141_5715 | Not-determined |
| S9_extracted_k141_58412 | Not-determined |
| S9_extracted_k141_5717 | Low-quality |
| S9_extracted_k141_32410 | Not-determined |
| S9_extracted_k141_1979 | Not-determined |
| S9_extracted_k141_58417 | Not-determined |
| S9_extracted_k141_24600 | Not-determined |
| S9_extracted_k141_43773 | Low-quality |
| S9_extracted_k141_73733 | Not-determined |
| S9_extracted_k141_81333 | Not-determined |
| S9_extracted_k141_13229 | Not-determined |
| S9_extracted_k141_20825 | Low-quality |
| S9_extracted_k141_73736 | Not-determined |
| S9_extracted_k141_20826 | Not-determined |
| S9_extracted_k141_9231 | Low-quality |
| S9_extracted_k141_81335 | Not-determined |
| S9_extracted_k141_88833 | Not-determined |
| S9_extracted_k141_77254 | Not-determined |
| S9_extracted_k141_55078 | Not-determined |
| S9_extracted_k141_36103 | Not-determined |
| S9_extracted_k141_1988 | Not-determined |
| S9_extracted_k141_9239 | Not-determined |
| S9_extracted_k141_55083 | Low-quality |
| S9_extracted_k141_43778 | Not-determined |
| S9_extracted_k141_81337 | Not-determined |
| S9_extracted_k141_20833 | Not-determined |
| S9_extracted_k141_43779 | Not-determined |
| S9_extracted_k141_24609 | Low-quality |
| S9_extracted_k141_17345 | Not-determined |
| S9_extracted_k141_1995 | Not-determined |
| S9_extracted_k141_1996 | Not-determined |
| S9_extracted_k141_13237 | Low-quality |
| S9_extracted_k141_81339 | Not-determined |
| S9_extracted_k141_13238 | Not-determined |
| S9_extracted_k141_88842 | Not-determined |
| S9_extracted_k141_77262 | Low-quality |
| S9_extracted_k141_69945 | Not-determined |
| S9_extracted_k141_17353 | Not-determined |
| S9_extracted_k141_88845 | Low-quality |
| S9_extracted_k141_40012 | Not-determined |
| S9_extracted_k141_36114 | Low-quality |
| S9_extracted_k141_47102 | Not-determined |
| S9_extracted_k141_47104 | Not-determined |
| S9_extracted_k141_69950 | Not-determined |
| S9_extracted_k141_20844 | Not-determined |
| S9_extracted_k141_2005 | Not-determined |
| S9_extracted_k141_36120 | Not-determined |
| S9_extracted_k141_13249 | Not-determined |
| S9_extracted_k141_17360 | Not-determined |
| S9_extracted_k141_17362 | Low-quality |
| S9_extracted_k141_73762 | Not-determined |
| S9_extracted_k141_9259 | Not-determined |
| S9_extracted_k141_73763 | Not-determined |
| S9_extracted_k141_77276 | Not-determined |
| S9_extracted_k141_69954 | Not-determined |
| S9_extracted_k141_84927 | Low-quality |
| S9_extracted_k141_81345 | Not-determined |
| S9_extracted_k141_77279 | Not-determined |
| S9_extracted_k141_20854 | Low-quality |
| S9_extracted_k141_24626 | Not-determined |
| S9_extracted_k141_77281 | Not-determined |
| S9_extracted_k141_9262 | Not-determined |
| S9_extracted_k141_43795 | Not-determined |
| S9_extracted_k141_32420 | Not-determined |
| S9_extracted_k141_2013 | Not-determined |
| S9_extracted_k141_81348 | Not-determined |
| S9_extracted_k141_84933 | Not-determined |
| S9_extracted_k141_24627 | Not-determined |
| S9_extracted_k141_69959 | Not-determined |
| S9_extracted_k141_69960 | Not-determined |
| S9_extracted_k141_77285 | Low-quality |
| S9_extracted_k141_84934 | Not-determined |
| S9_extracted_k141_17367 | Low-quality |
| S9_extracted_k141_36125 | Not-determined |
| S9_extracted_k141_9269 | Not-determined |
| S9_extracted_k141_58446 | Not-determined |
| S9_extracted_k141_17368 | Low-quality |
| S9_extracted_k141_43801 | Low-quality |
| S9_extracted_k141_40023 | Low-quality |
| S9_extracted_k141_58447 | Not-determined |
| S9_extracted_k141_81355 | Not-determined |
| S9_extracted_k141_84938 | Not-determined |
| S9_extracted_k141_5743 | Not-determined |
| S9_extracted_k141_55094 | Not-determined |
| S9_extracted_k141_84939 | Low-quality |
| S9_extracted_k141_43804 | Not-determined |
| S9_extracted_k141_13259 | Not-determined |
| S9_extracted_k141_69972 | Not-determined |
| S9_extracted_k141_69973 | Not-determined |
| S9_extracted_k141_84942 | Not-determined |
| S9_extracted_k141_81361 | Not-determined |
| S9_extracted_k141_2026 | Not-determined |
| S9_extracted_k141_77294 | Not-determined |
| S9_extracted_k141_24639 | Low-quality |
| S9_extracted_k141_69977 | Low-quality |
| S9_extracted_k141_9273 | Not-determined |
| S9_extracted_k141_84946 | Not-determined |
| S9_extracted_k141_24641 | Not-determined |
| S9_extracted_k141_9275 | Not-determined |
| S9_extracted_k141_17375 | Not-determined |
| S9_extracted_k141_55100 | Not-determined |
| S9_extracted_k141_58458 | Not-determined |
| S9_extracted_k141_77299 | Not-determined |
| S9_extracted_k141_5755 | Not-determined |
| S9_extracted_k141_62489 | Not-determined |
| S9_extracted_k141_47127 | Not-determined |
| S9_extracted_k141_24644 | Not-determined |
| S9_extracted_k141_40034 | Not-determined |
| S9_extracted_k141_62492 | Not-determined |
| S9_extracted_k141_5760 | Not-determined |
| S9_extracted_k141_2031 | Not-determined |
| S9_extracted_k141_65991 | Not-determined |
| S9_extracted_k141_55106 | Not-determined |
| S9_extracted_k141_55109 | Not-determined |
| S9_extracted_k141_47136 | Low-quality |
| S9_extracted_k141_77305 | Not-determined |
| S9_extracted_k141_84959 | Not-determined |
| S9_extracted_k141_47137 | Not-determined |
| S9_extracted_k141_84960 | Not-determined |
| S9_extracted_k141_65996 | Not-determined |
| S9_extracted_k141_69997 | Not-determined |
| S9_extracted_k141_58465 | Not-determined |
| S9_extracted_k141_81371 | Not-determined |
| S9_extracted_k141_5768 | Not-determined |
| S9_extracted_k141_9286 | Not-determined |
| S9_extracted_k141_24658 | Not-determined |
| S9_extracted_k141_84969 | Not-determined |
| S9_extracted_k141_70002 | Not-determined |
| S9_extracted_k141_70003 | Not-determined |
| S9_extracted_k141_28553 | Not-determined |
| S9_extracted_k141_13283 | Low-quality |
| S9_extracted_k141_28554 | Not-determined |
| S9_extracted_k141_47149 | Not-determined |
| S9_extracted_k141_81378 | Not-determined |
| S9_extracted_k141_36139 | Low-quality |
| S9_extracted_k141_43814 | Not-determined |
| S9_extracted_k141_84974 | Not-determined |
| S9_extracted_k141_20878 | Not-determined |
| S9_extracted_k141_81381 | Not-determined |
| S9_extracted_k141_24666 | Not-determined |
| S9_extracted_k141_9297 | Not-determined |
| S9_extracted_k141_81382 | Not-determined |
| S9_extracted_k141_77322 | Not-determined |
| S9_extracted_k141_43817 | Not-determined |
| S9_extracted_k141_47161 | Not-determined |
| S9_extracted_k141_20880 | Not-determined |
| S9_extracted_k141_43819 | Not-determined |
| S9_extracted_k141_36145 | Not-determined |
| S9_extracted_k141_2047 | Low-quality |
| S9_extracted_k141_20882 | Low-quality |
| S9_extracted_k141_58472 | Not-determined |
| S9_extracted_k141_81386 | Low-quality |
| S9_extracted_k141_13298 | Not-determined |
| S9_extracted_k141_70017 | Low-quality |
| S9_extracted_k141_47171 | Not-determined |
| S9_extracted_k141_58478 | Not-determined |
| S9_extracted_k141_36151 | Not-determined |
| S9_extracted_k141_62520 | Not-determined |
| S9_extracted_k141_36156 | Not-determined |
| S9_extracted_k141_47178 | Not-determined |
| S9_extracted_k141_32477 | Not-determined |
| S9_extracted_k141_47182 | Not-determined |
| S9_extracted_k141_62525 | Not-determined |
| S9_extracted_k141_47183 | Not-determined |
| S9_extracted_k141_17400 | Not-determined |
| S9_extracted_k141_24688 | Not-determined |
| S9_extracted_k141_70029 | Low-quality |
| S9_extracted_k141_17402 | Not-determined |
| S9_extracted_k141_84997 | Not-determined |
| S9_extracted_k141_43840 | Not-determined |
| S9_extracted_k141_13312 | Low-quality |
| S9_extracted_k141_20906 | Not-determined |
| S9_extracted_k141_77343 | Not-determined |
| S9_extracted_k141_40084 | Not-determined |
| S9_extracted_k141_13316 | Not-determined |
| S9_extracted_k141_40086 | Not-determined |
| S9_extracted_k141_24691 | Not-determined |
| S9_extracted_k141_13318 | Low-quality |
| S9_extracted_k141_62535 | Not-determined |
| S9_extracted_k141_88859 | Not-determined |
| S9_extracted_k141_73776 | Low-quality |
| S9_extracted_k141_43846 | Not-determined |
| S9_extracted_k141_47192 | Not-determined |
| S9_extracted_k141_73778 | Not-determined |
| S9_extracted_k141_13319 | Not-determined |
| S9_extracted_k141_47194 | Not-determined |
| S9_extracted_k141_77345 | Not-determined |
| S9_extracted_k141_32494 | Low-quality |
| S9_extracted_k141_13321 | Not-determined |
| S9_extracted_k141_81417 | Not-determined |
| S9_extracted_k141_32496 | Not-determined |
| S9_extracted_k141_62538 | Not-determined |
| S9_extracted_k141_81418 | Not-determined |
| S9_extracted_k141_66027 | Not-determined |
| S9_extracted_k141_32498 | Not-determined |
| S9_extracted_k141_17409 | Not-determined |
| S9_extracted_k141_47200 | Not-determined |
| S9_extracted_k141_62541 | Not-determined |
| S9_extracted_k141_20917 | Not-determined |
| S9_extracted_k141_70049 | Not-determined |
| S9_extracted_k141_24696 | Not-determined |
| S9_extracted_k141_9323 | Not-determined |
| S9_extracted_k141_32500 | Not-determined |
| S9_extracted_k141_13326 | Low-quality |
| S9_extracted_k141_66035 | Not-determined |
| S9_extracted_k141_55146 | Not-determined |
| S9_extracted_k141_17415 | Not-determined |
| S9_extracted_k141_2053 | Low-quality |
| S9_extracted_k141_62545 | Not-determined |
| S9_extracted_k141_70052 | Not-determined |
| S9_extracted_k141_62549 | Low-quality |
| S9_extracted_k141_5816 | Not-determined |
| S9_extracted_k141_28597 | Low-quality |
| S9_extracted_k141_40111 | Not-determined |
| S9_extracted_k141_81429 | Low-quality |
| S9_extracted_k141_55152 | Not-determined |
| S9_extracted_k141_17422 | Not-determined |
| S9_extracted_k141_88874 | Not-determined |
| S9_extracted_k141_28601 | Not-determined |
| S9_extracted_k141_5821 | Not-determined |
| S9_extracted_k141_24707 | Not-determined |
| S9_extracted_k141_66045 | Not-determined |
| S9_extracted_k141_47218 | Not-determined |
| S9_extracted_k141_88881 | Not-determined |
| S9_extracted_k141_47219 | Not-determined |
| S9_extracted_k141_24712 | Not-determined |
| S9_extracted_k141_81433 | Not-determined |
| S9_extracted_k141_70066 | Not-determined |
| S9_extracted_k141_85016 | Not-determined |
| S9_extracted_k141_28606 | Not-determined |
| S9_extracted_k141_24720 | Low-quality |
| S9_extracted_k141_85019 | Not-determined |
| S9_extracted_k141_66052 | Not-determined |
| S9_extracted_k141_13341 | Low-quality |
| S9_extracted_k141_28609 | Not-determined |
| S9_extracted_k141_43873 | Low-quality |
| S9_extracted_k141_58497 | Not-determined |
| S9_extracted_k141_43875 | Not-determined |
| S9_extracted_k141_43876 | Not-determined |
| S9_extracted_k141_9355 | Not-determined |
| S9_extracted_k141_73803 | Low-quality |
| S9_extracted_k141_5826 | Not-determined |
| S9_extracted_k141_43880 | Not-determined |
| S9_extracted_k141_66054 | Low-quality |
| S9_extracted_k141_66056 | Not-determined |
| S9_extracted_k141_13348 | Low-quality |
| S9_extracted_k141_73810 | Not-determined |
| S9_extracted_k141_47228 | Not-determined |
| S9_extracted_k141_43885 | Not-determined |
| S9_extracted_k141_66060 | Low-quality |
| S9_extracted_k141_36193 | Not-determined |
| S9_extracted_k141_17441 | Not-determined |
| S9_extracted_k141_85031 | Not-determined |
| S9_extracted_k141_73815 | Not-determined |
| S9_extracted_k141_40118 | Not-determined |
| S9_extracted_k141_85036 | Not-determined |
| S9_extracted_k141_9367 | Not-determined |
| S9_extracted_k141_73816 | Not-determined |
| S9_extracted_k141_43892 | Not-determined |
| S9_extracted_k141_58514 | Not-determined |
| S9_extracted_k141_70078 | Not-determined |
| S9_extracted_k141_36199 | Not-determined |
| S9_extracted_k141_70079 | Not-determined |
| S9_extracted_k141_28615 | Not-determined |
| S9_extracted_k141_70080 | Not-determined |
| S9_extracted_k141_62582 | Not-determined |
| S9_extracted_k141_40125 | Low-quality |
| S9_extracted_k141_28619 | Not-determined |
| S9_extracted_k141_13363 | Not-determined |
| S9_extracted_k141_73820 | Not-determined |
| S9_extracted_k141_70086 | Low-quality |
| S9_extracted_k141_5842 | Low-quality |
| S9_extracted_k141_40129 | Not-determined |
| S9_extracted_k141_24746 | Low-quality |
| S9_extracted_k141_58519 | Not-determined |
| S9_extracted_k141_32523 | Low-quality |
| S9_extracted_k141_28622 | Low-quality |
| S9_extracted_k141_5845 | Low-quality |
| S9_extracted_k141_36202 | Not-determined |
| S9_extracted_k141_13370 | Not-determined |
| S9_extracted_k141_66072 | Not-determined |
| S9_extracted_k141_43897 | Not-determined |
| S9_extracted_k141_62589 | Low-quality |
| S9_extracted_k141_13372 | Low-quality |
| S9_extracted_k141_36205 | Low-quality |
| S9_extracted_k141_58524 | Low-quality |
| S9_extracted_k141_40139 | Not-determined |
| S9_extracted_k141_47246 | Not-determined |
| S9_extracted_k141_24749 | Not-determined |
| S9_extracted_k141_58529 | Low-quality |
| S9_extracted_k141_73832 | Not-determined |
| S9_extracted_k141_24753 | Not-determined |
| S9_extracted_k141_5854 | Not-determined |
| S9_extracted_k141_58531 | Not-determined |
| S9_extracted_k141_85050 | Not-determined |
| S9_extracted_k141_58535 | Not-determined |
| S9_extracted_k141_43907 | Not-determined |
| S9_extracted_k141_17464 | Low-quality |
| S9_extracted_k141_58540 | Not-determined |
| S9_extracted_k141_28637 | Low-quality |
| S9_extracted_k141_17469 | Not-determined |
| S9_extracted_k141_62599 | Low-quality |
| S9_extracted_k141_28639 | Not-determined |
| S9_extracted_k141_36220 | Not-determined |
| S9_extracted_k141_66092 | Not-determined |
| S9_extracted_k141_5861 | Not-determined |
| S9_extracted_k141_24765 | Low-quality |
| S9_extracted_k141_36221 | Not-determined |
| S9_extracted_k141_73841 | Low-quality |
| S9_extracted_k141_66093 | Not-determined |
| S9_extracted_k141_88915 | Not-determined |
| S9_extracted_k141_47261 | Low-quality |
| S9_extracted_k141_73842 | Low-quality |
| S9_extracted_k141_58547 | Low-quality |
| S9_extracted_k141_58548 | Not-determined |
| S9_extracted_k141_58549 | Not-determined |
| S9_extracted_k141_47264 | Low-quality |
| S9_extracted_k141_58552 | Not-determined |
| S9_extracted_k141_58553 | Not-determined |
| S9_extracted_k141_43913 | Low-quality |
| S9_extracted_k141_9411 | Not-determined |
| S9_extracted_k141_47268 | Not-determined |
| S9_extracted_k141_5868 | Not-determined |
| S9_extracted_k141_62603 | Low-quality |
| S9_extracted_k141_13402 | Not-determined |
| S9_extracted_k141_70107 | Not-determined |
| S9_extracted_k141_85063 | Low-quality |
| S9_extracted_k141_17475 | Not-determined |
| S9_extracted_k141_73853 | Not-determined |
| S9_extracted_k141_88924 | Low-quality |
| S9_extracted_k141_58557 | Not-determined |
| S9_extracted_k141_32544 | Not-determined |
| S9_extracted_k141_5878 | Not-determined |
| S9_extracted_k141_17480 | Not-determined |
| S9_extracted_k141_81459 | Not-determined |
| S9_extracted_k141_28654 | Not-determined |
| S9_extracted_k141_20938 | Not-determined |
| S9_extracted_k141_47276 | Not-determined |
| S9_extracted_k141_47281 | Not-determined |
| S9_extracted_k141_88928 | Not-determined |
| S9_extracted_k141_5882 | Not-determined |
| S9_extracted_k141_24783 | Not-determined |
| S9_extracted_k141_55184 | Not-determined |
| S9_extracted_k141_5887 | Not-determined |
| S9_extracted_k141_17491 | Not-determined |
| S9_extracted_k141_40176 | Low-quality |
| S9_extracted_k141_43924 | Low-quality |
| S9_extracted_k141_5889 | Not-determined |
| S9_extracted_k141_62609 | Low-quality |
| S9_extracted_k141_73866 | Not-determined |
| S9_extracted_k141_77367 | Not-determined |
| S9_extracted_k141_20951 | Not-determined |
| S9_extracted_k141_77369 | Not-determined |
| S9_extracted_k141_58568 | Not-determined |
| S9_extracted_k141_88933 | Not-determined |
| S9_extracted_k141_66105 | Not-determined |
| S9_extracted_k141_24794 | Not-determined |
| S9_extracted_k141_55187 | Not-determined |
| S9_extracted_k141_40186 | Low-quality |
| S9_extracted_k141_62611 | Not-determined |
| S9_extracted_k141_47289 | Not-determined |
| S9_extracted_k141_62612 | Not-determined |
| S9_extracted_k141_9431 | Not-determined |
| S9_extracted_k141_5900 | Not-determined |
| S9_extracted_k141_17502 | Not-determined |
| S9_extracted_k141_36242 | Not-determined |
| S9_extracted_k141_13423 | Not-determined |
| S9_extracted_k141_5902 | Not-determined |
| S9_extracted_k141_58581 | Not-determined |
| S9_extracted_k141_81471 | Not-determined |
| S9_extracted_k141_5905 | Not-determined |
| S9_extracted_k141_43936 | Not-determined |
| S9_extracted_k141_88945 | Not-determined |
| S9_extracted_k141_20965 | Not-determined |
| S9_extracted_k141_70111 | Low-quality |
| S9_extracted_k141_9441 | Not-determined |
| S9_extracted_k141_36248 | Not-determined |
| S9_extracted_k141_58587 | Low-quality |
| S9_extracted_k141_17510 | Low-quality |
| S9_extracted_k141_47294 | Not-determined |
| S9_extracted_k141_36251 | Low-quality |
| S9_extracted_k141_40200 | Not-determined |
| S9_extracted_k141_88951 | Not-determined |
| S9_extracted_k141_47303 | Not-determined |
| S9_extracted_k141_58594 | Not-determined |
| S9_extracted_k141_36257 | Not-determined |
| S9_extracted_k141_9446 | Low-quality |
| S9_extracted_k141_36259 | Low-quality |
| S9_extracted_k141_55193 | Low-quality |
| S9_extracted_k141_66116 | Not-determined |
| S9_extracted_k141_47306 | Not-determined |
| S9_extracted_k141_85075 | Not-determined |
| S9_extracted_k141_36261 | Not-determined |
| S9_extracted_k141_20984 | Not-determined |
| S9_extracted_k141_43945 | Low-quality |
| S9_extracted_k141_20986 | Not-determined |
| S9_extracted_k141_40210 | Not-determined |
| S9_extracted_k141_17526 | Not-determined |
| S9_extracted_k141_24816 | Not-determined |
| S9_extracted_k141_70127 | Low-quality |
| S9_extracted_k141_43946 | Low-quality |
| S9_extracted_k141_28688 | Not-determined |
| S9_extracted_k141_40218 | Not-determined |
| S9_extracted_k141_43948 | Not-determined |
| S9_extracted_k141_32570 | Low-quality |
| S9_extracted_k141_2086 | Not-determined |
| S9_extracted_k141_17531 | Not-determined |
| S9_extracted_k141_58601 | Not-determined |
| S9_extracted_k141_9453 | Low-quality |
| S9_extracted_k141_9454 | Not-determined |
| S9_extracted_k141_55200 | Low-quality |
| S9_extracted_k141_55201 | Low-quality |
| S9_extracted_k141_66120 | Low-quality |
| S9_extracted_k141_24826 | Not-determined |
| S9_extracted_k141_2093 | Not-determined |
| S9_extracted_k141_85091 | Not-determined |
| S9_extracted_k141_40226 | Not-determined |
| S9_extracted_k141_66123 | Not-determined |
| S9_extracted_k141_5906 | Low-quality |
| S9_extracted_k141_21002 | Not-determined |
| S9_extracted_k141_5907 | Low-quality |
| S9_extracted_k141_24830 | Not-determined |
| S9_extracted_k141_24833 | Not-determined |
| S9_extracted_k141_5912 | Low-quality |
| S9_extracted_k141_21006 | Not-determined |
| S9_extracted_k141_85101 | Not-determined |
| S9_extracted_k141_24838 | Not-determined |
| S9_extracted_k141_88987 | Low-quality |
| S9_extracted_k141_58618 | Not-determined |
| S9_extracted_k141_43965 | Not-determined |
| S9_extracted_k141_85105 | Not-determined |
| S9_extracted_k141_32575 | Low-quality |
| S9_extracted_k141_66132 | Not-determined |
| S9_extracted_k141_62647 | Not-determined |
| S9_extracted_k141_2108 | Not-determined |
| S9_extracted_k141_66133 | Not-determined |
| S9_extracted_k141_62648 | Low-quality |
| S9_extracted_k141_66134 | Low-quality |
| S9_extracted_k141_62649 | Not-determined |
| S9_extracted_k141_21011 | Not-determined |
| S9_extracted_k141_28702 | Low-quality |
| S9_extracted_k141_47337 | Low-quality |
| S9_extracted_k141_32579 | Low-quality |
| S9_extracted_k141_85110 | Not-determined |
| S9_extracted_k141_28703 | Not-determined |
| S9_extracted_k141_32581 | Not-determined |
| S9_extracted_k141_32582 | Not-determined |
| S9_extracted_k141_85113 | Not-determined |
| S9_extracted_k141_36262 | Low-quality |
| S9_extracted_k141_17558 | Not-determined |
| S9_extracted_k141_36263 | Not-determined |
| S9_extracted_k141_81478 | Low-quality |
| S9_extracted_k141_13459 | Not-determined |
| S9_extracted_k141_36265 | Not-determined |
| S9_extracted_k141_66139 | Not-determined |
| S9_extracted_k141_81482 | Not-determined |
| S9_extracted_k141_43972 | Not-determined |
| S9_extracted_k141_62657 | Not-determined |

| | |
|---|---|
| S9_extracted_k141_58637 | Not-determined |
| S9_extracted_k141_66144 | Not-determined |
| S9_extracted_k141_85118 | Not-determined |
| S9_extracted_k141_43980 | Not-determined |
| S9_extracted_k141_89000 | Not-determined |
| S9_extracted_k141_81493 | Low-quality |
| S9_extracted_k141_89001 | Not-determined |
| S9_extracted_k141_13466 | Low-quality |
| S9_extracted_k141_89004 | Not-determined |
| S9_extracted_k141_17572 | Not-determined |
| S9_extracted_k141_9474 | Not-determined |
| S9_extracted_k141_17573 | Not-determined |
| S9_extracted_k141_43989 | Not-determined |
| S9_extracted_k141_28719 | Low-quality |
| S9_extracted_k141_36277 | Not-determined |
| S9_extracted_k141_5927 | Not-determined |
| S9_extracted_k141_70141 | Not-determined |
| S9_extracted_k141_58648 | Not-determined |
| S9_extracted_k141_66153 | Low-quality |
| S9_extracted_k141_66154 | Not-determined |
| S9_extracted_k141_81505 | Not-determined |
| S9_extracted_k141_40260 | Not-determined |
| S9_extracted_k141_43994 | Not-determined |
| S9_extracted_k141_13479 | Not-determined |
| S9_extracted_k141_89016 | Not-determined |
| S9_extracted_k141_81508 | Not-determined |
| S9_extracted_k141_28723 | Not-determined |
| S9_extracted_k141_21034 | Not-determined |
| S9_extracted_k141_28724 | Not-determined |
| S9_extracted_k141_66159 | Not-determined |
| S9_extracted_k141_13483 | Low-quality |
| S9_extracted_k141_89018 | Not-determined |
| S9_extracted_k141_70149 | Not-determined |
| S9_extracted_k141_66162 | Not-determined |
| S9_extracted_k141_58658 | Not-determined |
| S9_extracted_k141_9492 | Not-determined |
| S9_extracted_k141_40268 | Not-determined |
| S9_extracted_k141_89021 | Not-determined |
| S9_extracted_k141_70155 | Not-determined |
| S9_extracted_k141_85132 | Not-determined |
| S9_extracted_k141_44008 | Not-determined |
| S9_extracted_k141_44009 | Not-determined |
| S9_extracted_k141_28735 | Not-determined |
| S9_extracted_k141_81518 | Not-determined |
| S9_extracted_k141_36292 | Not-determined |
| S9_extracted_k141_40275 | Not-determined |
| S9_extracted_k141_89028 | Not-determined |
| S9_extracted_k141_36294 | Not-determined |
| S9_extracted_k141_44013 | Not-determined |
| S9_extracted_k141_89030 | Not-determined |
| S9_extracted_k141_17598 | Not-determined |
| S9_extracted_k141_70165 | Not-determined |
| S9_extracted_k141_9498 | Not-determined |
| S9_extracted_k141_73926 | Not-determined |
| S9_extracted_k141_13498 | Not-determined |
| S9_extracted_k141_70167 | Not-determined |
| S9_extracted_k141_70168 | Low-quality |
| S9_extracted_k141_55241 | Low-quality |
| S9_extracted_k141_28745 | Not-determined |
| S9_extracted_k141_13503 | Not-determined |
| S9_extracted_k141_47349 | Low-quality |
| S9_extracted_k141_66174 | Not-determined |
| S9_extracted_k141_13504 | Not-determined |
| S9_extracted_k141_40283 | Not-determined |
| S9_extracted_k141_66176 | Not-determined |
| S9_extracted_k141_70180 | Low-quality |
| S9_extracted_k141_44018 | Low-quality |
| S9_extracted_k141_66181 | Not-determined |
| S9_extracted_k141_21044 | Low-quality |
| S9_extracted_k141_40291 | Low-quality |
| S9_extracted_k141_44023 | Not-determined |
| S9_extracted_k141_9511 | Low-quality |
| S9_extracted_k141_5957 | Not-determined |
| S9_extracted_k141_5958 | Not-determined |
| S9_extracted_k141_17609 | Not-determined |
| S9_extracted_k141_73944 | Not-determined |
| S9_extracted_k141_70186 | Not-determined |
| S9_extracted_k141_85152 | Not-determined |
| S9_extracted_k141_73946 | Not-determined |
| S9_extracted_k141_40296 | Not-determined |
| S9_extracted_k141_85155 | Low-quality |
| S9_extracted_k141_81534 | Not-determined |
| S9_extracted_k141_81535 | Not-determined |
| S9_extracted_k141_85157 | Not-determined |
| S9_extracted_k141_81536 | Not-determined |
| S9_extracted_k141_36314 | Low-quality |
| S9_extracted_k141_81538 | Low-quality |
| S9_extracted_k141_50865 | Not-determined |
| S9_extracted_k141_36317 | Not-determined |
| S9_extracted_k141_44034 | Not-determined |
| S9_extracted_k141_85159 | Not-determined |
| S9_extracted_k141_50868 | Not-determined |
| S9_extracted_k141_50869 | Low-quality |
| S9_extracted_k141_9521 | Not-determined |
| S9_extracted_k141_5963 | Not-determined |
| S9_extracted_k141_24881 | Not-determined |
| S9_extracted_k141_73955 | Not-determined |
| S9_extracted_k141_40302 | Low-quality |
| S9_extracted_k141_24883 | Not-determined |
| S9_extracted_k141_36323 | Not-determined |
| S9_extracted_k141_9524 | Not-determined |
| S9_extracted_k141_44039 | Not-determined |
| S9_extracted_k141_9527 | Not-determined |
| S9_extracted_k141_50871 | Not-determined |
| S9_extracted_k141_28771 | Not-determined |
| S13_vs_45488\|\|full | Medium-quality |
| S9_extracted_k141_5972 | Not-determined |
| S9_extracted_k141_81547 | Not-determined |
| S9_extracted_k141_24890 | Not-determined |
| S9_extracted_k141_21072 | Not-determined |
| S9_extracted_k141_50876 | Not-determined |
| S9_extracted_k141_85170 | Not-determined |
| S9_extracted_k141_21073 | Not-determined |
| S9_extracted_k141_62666 | Low-quality |
| S9_extracted_k141_44049 | Low-quality |
| S9_extracted_k141_36334 | Not-determined |
| S9_extracted_k141_44050 | Low-quality |
| S9_extracted_k141_21076 | Not-determined |
| S9_extracted_k141_36337 | Not-determined |
| S9_extracted_k141_85174 | Not-determined |
| S9_extracted_k141_85175 | Not-determined |
| S9_extracted_k141_85176 | Not-determined |
| S9_extracted_k141_9544 | Not-determined |
| S9_extracted_k141_36341 | Not-determined |
| S9_extracted_k141_44053 | Low-quality |
| S9_extracted_k141_21082 | Low-quality |
| S9_extracted_k141_85183 | Not-determined |
| S9_extracted_k141_2167 | Low-quality |
| S9_extracted_k141_81548 | Not-determined |
| S9_extracted_k141_85185 | Low-quality |
| S9_extracted_k141_21088 | Not-determined |
| S9_extracted_k141_85187 | Not-determined |
| S9_extracted_k141_40325 | Not-determined |
| S9_extracted_k141_89067 | Low-quality |
| S9_extracted_k141_36352 | Not-determined |
| S9_extracted_k141_77393 | Not-determined |
| S9_extracted_k141_89069 | Not-determined |
| S9_extracted_k141_77394 | Not-determined |
| S9_extracted_k141_21093 | Not-determined |
| S9_extracted_k141_55276 | Low-quality |
| S9_extracted_k141_6006 | Not-determined |
| S9_extracted_k141_89074 | Not-determined |
| S9_extracted_k141_36355 | Not-determined |
| S9_extracted_k141_36358 | Not-determined |
| S9_extracted_k141_28794 | Not-determined |
| S9_extracted_k141_62676 | Low-quality |
| S9_extracted_k141_62680 | Not-determined |
| S9_extracted_k141_6015 | Low-quality |
| S9_extracted_k141_62685 | Not-determined |
| S9_extracted_k141_77400 | Low-quality |
| S9_extracted_k141_28808 | Not-determined |
| S9_extracted_k141_81566 | Not-determined |
| S9_extracted_k141_2188 | Low-quality |
| S9_extracted_k141_6023 | Not-determined |
| S9_extracted_k141_32603 | Not-determined |
| S9_extracted_k141_6027 | Not-determined |
| S9_extracted_k141_44084 | Low-quality |
| S9_extracted_k141_55296 | Not-determined |
| S9_extracted_k141_2200 | Not-determined |
| S9_extracted_k141_44086 | Not-determined |
| S9_extracted_k141_55299 | Not-determined |
| S9_extracted_k141_32614 | Not-determined |
| S9_extracted_k141_28820 | Low-quality |
| S9_extracted_k141_55301 | Low-quality |
| S9_extracted_k141_81573 | Not-determined |
| S9_extracted_k141_32616 | Low-quality |
| S9_extracted_k141_36375 | Not-determined |
| S9_extracted_k141_81580 | Not-determined |
| S9_extracted_k141_36378 | Not-determined |
| S9_extracted_k141_36379 | Not-determined |
| S9_extracted_k141_55313 | Not-determined |
| S9_extracted_k141_89081 | Low-quality |
| S9_extracted_k141_2214 | Not-determined |
| S9_extracted_k141_81590 | Low-quality |
| S9_extracted_k141_36385 | Not-determined |
| S9_extracted_k141_44101 | Not-determined |
| S9_extracted_k141_36388 | Not-determined |
| S6_vs_k141_53393\|\|full | Medium-quality |
| S9_extracted_k141_81595 | Low-quality |
| S9_extracted_k141_6035 | Not-determined |
| S9_extracted_k141_6036 | Not-determined |
| S9_extracted_k141_55322 | Not-determined |
| S9_extracted_k141_2221 | Not-determined |
| S9_extracted_k141_6039 | Low-quality |
| S9_extracted_k141_32634 | Not-determined |
| S9_extracted_k141_77414 | Not-determined |
| S9_extracted_k141_21146 | Not-determined |
| S9_extracted_k141_55328 | Not-determined |
| S9_extracted_k141_2225 | Low-quality |
| S9_extracted_k141_73967 | Not-determined |
| S9_extracted_k141_85234 | Not-determined |
| S9_extracted_k141_62732 | Not-determined |
| S9_extracted_k141_55332 | Not-determined |
| S9_extracted_k141_62733 | Not-determined |
| S9_extracted_k141_21153 | Not-determined |
| S9_extracted_k141_62734 | Not-determined |
| S9_extracted_k141_55333 | Not-determined |
| S9_extracted_k141_2230 | Not-determined |
| S9_extracted_k141_6051 | Not-determined |
| S9_extracted_k141_89092 | Not-determined |
| S9_extracted_k141_62736 | Not-determined |
| S9_extracted_k141_9557 | Low-quality |
| S9_extracted_k141_55336 | Low-quality |
| S9_extracted_k141_77423 | Not-determined |
| S9_extracted_k141_36398 | Not-determined |
| S9_extracted_k141_2232 | Low-quality |
| S9_extracted_k141_62738 | Low-quality |
| S9_extracted_k141_44120 | Low-quality |
| S9_extracted_k141_55340 | Not-determined |
| S9_extracted_k141_17617 | Not-determined |
| S9_extracted_k141_36405 | Low-quality |
| S9_extracted_k141_55350 | Not-determined |
| S9_extracted_k141_17623 | Not-determined |
| S9_extracted_k141_44130 | Not-determined |
| S9_extracted_k141_55355 | Not-determined |
| S9_extracted_k141_81631 | Not-determined |
| S9_extracted_k141_40341 | Not-determined |
| S9_extracted_k141_9571 | Not-determined |
| S9_extracted_k141_70208 | Not-determined |
| S9_extracted_k141_9572 | Not-determined |
| S9_extracted_k141_77441 | Not-determined |
| S9_extracted_k141_28862 | Low-quality |
| S9_extracted_k141_89107 | Not-determined |
| S9_extracted_k141_17626 | Low-quality |
| S9_extracted_k141_6066 | Low-quality |
| S9_extracted_k141_70211 | Not-determined |
| S9_extracted_k141_17629 | Not-determined |
| S9_extracted_k141_62749 | Not-determined |
| S9_extracted_k141_36413 | Not-determined |
| S9_extracted_k141_21170 | Not-determined |
| S9_extracted_k141_85262 | Not-determined |
| S9_extracted_k141_9580 | Not-determined |
| S9_extracted_k141_40344 | Not-determined |
| S9_extracted_k141_70218 | Not-determined |
| S9_extracted_k141_89117 | Not-determined |
| S9_extracted_k141_6075 | Not-determined |
| S9_extracted_k141_2257 | Not-determined |
| S9_extracted_k141_44147 | Not-determined |
| S9_extracted_k141_28874 | Low-quality |
| S9_extracted_k141_89120 | Not-determined |
| S9_extracted_k141_73993 | Not-determined |
| S9_extracted_k141_89123 | Not-determined |
| S9_extracted_k141_73994 | Not-determined |
| S9_extracted_k141_77461 | Not-determined |
| S9_extracted_k141_66213 | Not-determined |
| S9_extracted_k141_73996 | Low-quality |
| S9_extracted_k141_21176 | Low-quality |
| S9_extracted_k141_36430 | Not-determined |
| S9_extracted_k141_70227 | Not-determined |
| S9_extracted_k141_89127 | Not-determined |
| S9_extracted_k141_89129 | Not-determined |
| S9_extracted_k141_36433 | Not-determined |
| S9_extracted_k141_40361 | Not-determined |
| S9_extracted_k141_6080 | Low-quality |
| S9_extracted_k141_36436 | Not-determined |
| S9_extracted_k141_85282 | Not-determined |
| S9_extracted_k141_9600 | Not-determined |
| S9_extracted_k141_55386 | Low-quality |
| S9_extracted_k141_89140 | Low-quality |
| S9_extracted_k141_9604 | Not-determined |
| S9_extracted_k141_77469 | Low-quality |
| S9_extracted_k141_62767 | Low-quality |
| S9_extracted_k141_89146 | Not-determined |
| S9_extracted_k141_66226 | Low-quality |
| S9_extracted_k141_44161 | Not-determined |
| S9_extracted_k141_85296 | Not-determined |
| S9_extracted_k141_66231 | Not-determined |
| S9_extracted_k141_85298 | Not-determined |
| S9_extracted_k141_40374 | Not-determined |
| S9_extracted_k141_40375 | Not-determined |
| S9_extracted_k141_66234 | Not-determined |
| S9_extracted_k141_85300 | Not-determined |
| S9_extracted_k141_40376 | Low-quality |
| S9_extracted_k141_2259 | Low-quality |
| S9_extracted_k141_36451 | Not-determined |
| S9_extracted_k141_66237 | Not-determined |
| S9_extracted_k141_66238 | Not-determined |
| S9_extracted_k141_2264 | Not-determined |
| S9_extracted_k141_9623 | Not-determined |
| S9_extracted_k141_9624 | Not-determined |
| S9_extracted_k141_77487 | Not-determined |
| S9_extracted_k141_2268 | Not-determined |
| S9_extracted_k141_9628 | Low-quality |
| S9_extracted_k141_9630 | Not-determined |
| S9_extracted_k141_24979 | Low-quality |
| S9_extracted_k141_17657 | Low-quality |
| S9_extracted_k141_66242 | Not-determined |
| S9_extracted_k141_40387 | Not-determined |
| S9_extracted_k141_77489 | Low-quality |
| S9_extracted_k141_77492 | Not-determined |
| S9_extracted_k141_36461 | Not-determined |
| S9_extracted_k141_36462 | Not-determined |
| S9_extracted_k141_77493 | Not-determined |
| S9_extracted_k141_74029 | Not-determined |
| S9_extracted_k141_36463 | Not-determined |
| S9_extracted_k141_85311 | Not-determined |
| S9_extracted_k141_28906 | Low-quality |
| S9_extracted_k141_24985 | Low-quality |
| S9_extracted_k141_74031 | Not-determined |
| S12_extracted_k141_31630 | Medium-quality |
| S9_extracted_k141_24988 | Not-determined |
| S9_extracted_k141_2275 | Not-determined |
| S9_extracted_k141_70239 | Not-determined |
| S9_extracted_k141_2276 | Low-quality |
| S9_extracted_k141_40395 | Not-determined |
| S9_extracted_k141_66250 | Not-determined |
| S9_extracted_k141_58700 | Low-quality |
| S9_extracted_k141_28914 | Not-determined |
| S9_extracted_k141_70243 | Not-determined |
| S9_extracted_k141_85320 | Not-determined |
| S9_extracted_k141_6104 | Not-determined |
| S9_extracted_k141_89161 | Not-determined |
| S9_extracted_k141_58703 | Not-determined |
| S9_extracted_k141_77502 | Not-determined |
| S9_extracted_k141_17667 | Not-determined |
| S9_extracted_k141_58706 | Not-determined |
| S9_extracted_k141_2285 | Not-determined |
| S9_extracted_k141_28919 | Not-determined |
| S9_extracted_k141_74038 | Not-determined |
| S9_extracted_k141_77506 | Not-determined |
| S9_extracted_k141_58710 | Not-determined |
| S9_extracted_k141_89169 | Not-determined |
| S9_extracted_k141_21207 | Not-determined |
| S9_extracted_k141_28923 | Not-determined |
| S9_extracted_k141_58714 | Not-determined |
| S9_extracted_k141_6107 | Low-quality |
| S9_extracted_k141_24999 | Not-determined |
| S9_extracted_k141_40408 | Not-determined |
| S9_extracted_k141_28928 | Not-determined |
| S9_extracted_k141_58718 | Not-determined |
| S9_extracted_k141_21215 | Not-determined |
| S9_extracted_k141_70260 | Not-determined |
| S9_extracted_k141_70262 | Not-determined |
| S9_extracted_k141_70265 | Not-determined |
| S9_extracted_k141_21219 | Not-determined |
| S9_extracted_k141_58726 | Low-quality |
| S9_extracted_k141_74058 | Not-determined |
| S9_extracted_k141_17676 | Not-determined |
| S9_extracted_k141_44203 | Not-determined |
| S9_extracted_k141_36498 | Not-determined |
| S9_extracted_k141_25014 | Not-determined |
| S9_extracted_k141_21228 | Low-quality |
| S9_extracted_k141_58734 | Not-determined |
| S9_extracted_k141_85351 | Low-quality |
| S9_extracted_k141_40419 | Not-determined |
| S9_extracted_k141_44211 | Not-determined |
| S9_extracted_k141_81654 | Not-determined |
| S9_extracted_k141_40420 | Not-determined |
| S9_extracted_k141_28941 | Low-quality |
| S9_extracted_k141_44214 | Not-determined |
| S9_extracted_k141_17686 | Low-quality |
| S9_extracted_k141_25020 | Not-determined |
| S9_extracted_k141_70282 | Not-determined |
| S9_extracted_k141_66288 | Not-determined |
| S9_extracted_k141_66289 | Not-determined |
| S9_extracted_k141_28948 | Low-quality |
| S9_extracted_k141_25022 | Not-determined |
| S9_extracted_k141_28951 | Low-quality |
| S9_extracted_k141_25025 | Not-determined |
| S9_extracted_k141_25027 | Not-determined |
| S9_extracted_k141_2316 | Not-determined |
| S9_extracted_k141_17690 | Low-quality |
| S9_extracted_k141_66296 | Low-quality |
| S9_extracted_k141_81668 | Not-determined |
| S9_extracted_k141_6135 | Not-determined |
| S9_extracted_k141_17693 | Low-quality |
| S9_extracted_k141_17695 | Not-determined |
| S9_extracted_k141_74063 | Not-determined |
| S9_extracted_k141_25034 | Not-determined |
| S9_extracted_k141_9644 | Not-determined |
| S9_extracted_k141_58746 | Low-quality |
| S9_extracted_k141_17701 | Not-determined |
| S9_extracted_k141_2322 | Low-quality |
| S9_extracted_k141_36525 | Low-quality |
| S9_extracted_k141_6141 | Not-determined |
| S9_extracted_k141_9650 | Not-determined |
| S9_extracted_k141_17704 | Not-determined |
| S9_extracted_k141_21257 | Not-determined |
| S9_extracted_k141_36528 | Not-determined |
| S9_extracted_k141_81682 | Not-determined |
| S9_extracted_k141_44224 | Low-quality |
| S9_extracted_k141_81684 | Not-determined |
| S9_extracted_k141_44227 | Low-quality |
| S9_extracted_k141_21266 | Not-determined |
| S9_extracted_k141_85369 | Not-determined |
| S9_extracted_k141_28971 | Not-determined |
| S9_extracted_k141_70304 | Not-determined |
| S9_extracted_k141_21272 | Not-determined |
| S9_extracted_k141_77526 | Not-determined |
| S9_extracted_k141_25062 | Not-determined |
| S9_extracted_k141_55400 | Not-determined |
| S9_extracted_k141_36544 | Not-determined |
| S9_extracted_k141_25067 | Not-determined |
| S9_extracted_k141_9674 | Not-determined |
| S9_extracted_k141_74079 | Not-determined |
| S9_extracted_k141_21279 | Not-determined |
| S12_vs_k141_31630\|\|full | Medium-quality |
| S9_extracted_k141_9683 | Not-determined |
| S9_extracted_k141_66318 | Not-determined |
| S9_extracted_k141_58789 | Not-determined |
| S9_extracted_k141_74087 | Not-determined |
| S9_extracted_k141_66322 | Not-determined |
| S9_extracted_k141_74091 | Not-determined |
| S9_extracted_k141_28990 | Not-determined |
| S9_extracted_k141_21288 | Not-determined |
| S9_extracted_k141_44256 | Low-quality |
| S9_extracted_k141_74100 | Not-determined |
| S9_extracted_k141_66331 | Not-determined |
| S9_extracted_k141_58803 | Not-determined |
| S9_extracted_k141_25081 | Not-determined |
| S9_extracted_k141_25082 | Not-determined |
| S9_extracted_k141_66336 | Low-quality |
| S9_extracted_k141_89182 | Not-determined |
| S9_extracted_k141_81703 | Low-quality |
| S9_extracted_k141_50888 | Not-determined |
| S9_extracted_k141_66341 | Not-determined |
| S9_extracted_k141_21304 | Low-quality |
| S9_extracted_k141_40485 | Not-determined |
| S9_extracted_k141_44264 | Low-quality |
| S9_extracted_k141_50891 | Not-determined |
| S9_extracted_k141_44266 | Not-determined |
| S9_extracted_k141_6168 | Not-determined |
| S9_extracted_k141_17721 | Not-determined |
| S9_extracted_k141_40487 | Not-determined |
| S9_extracted_k141_6172 | Not-determined |
| S9_extracted_k141_25094 | Not-determined |
| S9_extracted_k141_17723 | Not-determined |
| S9_extracted_k141_21317 | Not-determined |
| S9_extracted_k141_17724 | Not-determined |
| S9_extracted_k141_32650 | Not-determined |
| S9_extracted_k141_9708 | Not-determined |
| S9_extracted_k141_74117 | Not-determined |
| S9_extracted_k141_21320 | Not-determined |
| S9_extracted_k141_47398 | Not-determined |
| S9_extracted_k141_44272 | Not-determined |
| S9_extracted_k141_81723 | Not-determined |
| S9_extracted_k141_17728 | Not-determined |
| S9_extracted_k141_66352 | Low-quality |
| S9_extracted_k141_36576 | Not-determined |
| S9_extracted_k141_62787 | Not-determined |
| S9_extracted_k141_21326 | Not-determined |
| S9_extracted_k141_62788 | Not-determined |
| S9_extracted_k141_9718 | Not-determined |
| S9_extracted_k141_17730 | Low-quality |
| S9_extracted_k141_62795 | Not-determined |
| S9_extracted_k141_29000 | Low-quality |
| S9_extracted_k141_66365 | Not-determined |
| S9_extracted_k141_21330 | Low-quality |
| S9_extracted_k141_47412 | Not-determined |
| S9_extracted_k141_17737 | Not-determined |
| S9_extracted_k141_55433 | Not-determined |
| S9_extracted_k141_17741 | Not-determined |
| S9_extracted_k141_58822 | Not-determined |
| S9_extracted_k141_40498 | Not-determined |
| S9_extracted_k141_40500 | Not-determined |
| S9_extracted_k141_25111 | Not-determined |
| S9_extracted_k141_17742 | Low-quality |
| S9_extracted_k141_66380 | Not-determined |
| S9_extracted_k141_58828 | Not-determined |
| S9_extracted_k141_58830 | Not-determined |
| S9_extracted_k141_58831 | Not-determined |
| S9_extracted_k141_47421 | Low-quality |
| S9_extracted_k141_32658 | Not-determined |
| S9_extracted_k141_40513 | Not-determined |
| S9_extracted_k141_47425 | Not-determined |
| S9_extracted_k141_58838 | Not-determined |
| S9_extracted_k141_47429 | Not-determined |
| S9_extracted_k141_66390 | Not-determined |
| S9_extracted_k141_32664 | Not-determined |
| S9_extracted_k141_9738 | Not-determined |
| S9_extracted_k141_29008 | Not-determined |
| S9_extracted_k141_9739 | Not-determined |
| S9_extracted_k141_29010 | Not-determined |
| S9_extracted_k141_29011 | Not-determined |
| S9_extracted_k141_40519 | Not-determined |
| S9_extracted_k141_9741 | Not-determined |
| S9_extracted_k141_25132 | Not-determined |
| S9_extracted_k141_47439 | Low-quality |
| S9_extracted_k141_74155 | Not-determined |
| S9_extracted_k141_58845 | Not-determined |
| S9_extracted_k141_21335 | Low-quality |
| S9_extracted_k141_74157 | Not-determined |
| S9_extracted_k141_25141 | Not-determined |
| S9_extracted_k141_25142 | Not-determined |
| S9_extracted_k141_2385 | Not-determined |
| S9_extracted_k141_21337 | Not-determined |
| S9_extracted_k141_44299 | Not-determined |
| S9_extracted_k141_25143 | Not-determined |
| S9_extracted_k141_6180 | Not-determined |
| S9_extracted_k141_17770 | Not-determined |
| S9_extracted_k141_40533 | Low-quality |
| S9_extracted_k141_85384 | Not-determined |
| S9_extracted_k141_17773 | Not-determined |
| S9_extracted_k141_47453 | Not-determined |
| S9_extracted_k141_40537 | Not-determined |
| S9_extracted_k141_66414 | Low-quality |
| S9_extracted_k141_81751 | Not-determined |
| S9_extracted_k141_36619 | Low-quality |
| S9_extracted_k141_29028 | Not-determined |
| S9_extracted_k141_81753 | Not-determined |
| S9_extracted_k141_81754 | Not-determined |
| S9_extracted_k141_29036 | Not-determined |
| S9_extracted_k141_40543 | Low-quality |
| S9_extracted_k141_32675 | Not-determined |
| S9_extracted_k141_6189 | Not-determined |
| S9_extracted_k141_36623 | Not-determined |
| S9_extracted_k141_25161 | Not-determined |
| S9_extracted_k141_36625 | Low-quality |
| S9_extracted_k141_58866 | Not-determined |
| S9_extracted_k141_47466 | Not-determined |
| S9_extracted_k141_66428 | Low-quality |
| S9_extracted_k141_25166 | Low-quality |
| S9_extracted_k141_29042 | Not-determined |
| S9_extracted_k141_55474 | Not-determined |
| S9_extracted_k141_85404 | Not-determined |
| S9_extracted_k141_29046 | Not-determined |
| S9_extracted_k141_2403 | Not-determined |
| S9_extracted_k141_21360 | Not-determined |
| S9_extracted_k141_6205 | Not-determined |
| S9_extracted_k141_6206 | Not-determined |
| S9_extracted_k141_74176 | Low-quality |
| S9_extracted_k141_29049 | Low-quality |
| S9_extracted_k141_6216 | Not-determined |
| S9_extracted_k141_21369 | Not-determined |
| S9_extracted_k141_85415 | Low-quality |
| S9_extracted_k141_2412 | Low-quality |
| S9_extracted_k141_58878 | Not-determined |
| S9_extracted_k141_81770 | Not-determined |
| S9_extracted_k141_58881 | Not-determined |
| S9_extracted_k141_62807 | Not-determined |
| S9_extracted_k141_62808 | Not-determined |
| S9_extracted_k141_85420 | Not-determined |
| S9_extracted_k141_29056 | Not-determined |
| S9_extracted_k141_55494 | Not-determined |
| S9_extracted_k141_6224 | Not-determined |
| S9_extracted_k141_36644 | Low-quality |
| S9_extracted_k141_21378 | Not-determined |
| S9_extracted_k141_29063 | Low-quality |
| S9_extracted_k141_85428 | Not-determined |
| S9_extracted_k141_55498 | Low-quality |
| S9_extracted_k141_17784 | Low-quality |
| S9_extracted_k141_81783 | Not-determined |
| S9_extracted_k141_36648 | Not-determined |
| S9_extracted_k141_29069 | Not-determined |
| S9_extracted_k141_47495 | Not-determined |
| S9_extracted_k141_81789 | Not-determined |
| S9_extracted_k141_55504 | Low-quality |
| S9_extracted_k141_6229 | Not-determined |
| S9_extracted_k141_47497 | Not-determined |
| S9_extracted_k141_29075 | Not-determined |
| S4_extracted_31973 | Complete |
| S9_extracted_k141_47500 | Not-determined |
| S9_extracted_k141_62811 | Not-determined |
| S9_extracted_k141_21392 | Not-determined |
| S9_extracted_k141_13511 | Not-determined |
| S9_extracted_k141_32712 | Not-determined |
| S9_extracted_k141_44323 | Low-quality |
| S9_extracted_k141_25181 | Low-quality |
| S9_extracted_k141_32714 | Low-quality |
| S9_extracted_k141_25182 | Not-determined |
| S9_extracted_k141_6237 | Low-quality |
| S9_extracted_k141_9773 | Low-quality |
| S9_extracted_k141_62817 | Not-determined |
| S9_extracted_k141_13516 | Not-determined |
| S9_extracted_k141_66439 | Not-determined |
| S9_extracted_k141_66440 | Not-determined |
| S9_extracted_k141_81798 | Low-quality |
| S9_extracted_k141_2439 | Not-determined |
| S9_extracted_k141_40566 | Not-determined |
| S9_extracted_k141_58905 | Not-determined |
| S9_extracted_k141_85451 | Low-quality |
| S9_extracted_k141_85452 | Not-determined |
| S9_extracted_k141_25192 | Not-determined |
| S9_extracted_k141_21398 | Not-determined |
| S9_extracted_k141_62820 | Not-determined |
| S9_extracted_k141_6248 | Not-determined |
| S9_extracted_k141_9780 | Not-determined |
| S9_extracted_k141_81799 | Not-determined |
| S9_extracted_k141_44329 | Not-determined |
| S9_extracted_k141_40572 | Not-determined |
| S9_extracted_k141_85456 | Low-quality |
| S9_extracted_k141_32719 | Not-determined |
| S9_extracted_k141_81802 | Not-determined |
| S9_extracted_k141_62829 | Not-determined |
| S9_extracted_k141_36662 | Not-determined |
| S9_extracted_k141_17791 | Not-determined |
| S9_extracted_k141_9783 | Not-determined |
| S9_extracted_k141_13526 | Not-determined |
| S9_extracted_k141_58911 | Not-determined |

| | |
|---|---|
| S9_extracted_k141_32723 | Not-determined |
| S9_extracted_k141_29088 | Low-quality |
| S9_extracted_k141_47507 | Low-quality |
| S9_extracted_k141_21409 | Not-determined |
| S9_extracted_k141_9787 | Not-determined |
| S9_extracted_k141_13531 | Not-determined |
| S9_extracted_k141_77562 | Not-determined |
| S9_extracted_k141_58916 | Not-determined |
| S9_extracted_k141_55517 | Low-quality |
| S9_extracted_k141_58917 | Low-quality |
| S9_extracted_k141_66454 | Not-determined |
| S9_extracted_k141_25199 | Not-determined |
| S9_extracted_k141_36665 | Not-determined |
| S9_extracted_k141_47511 | Not-determined |
| S9_extracted_k141_2451 | Not-determined |
| S9_extracted_k141_74199 | Not-determined |
| S9_extracted_k141_58919 | Low-quality |
| S9_extracted_k141_9790 | Not-determined |
| S9_extracted_k141_17801 | Low-quality |
| S9_extracted_k141_29095 | Not-determined |
| S9_extracted_k141_21423 | Not-determined |
| S9_extracted_k141_25203 | Not-determined |
| S9_extracted_k141_81809 | Not-determined |
| S9_extracted_k141_58927 | Not-determined |
| S9_extracted_k141_29101 | Not-determined |
| S9_extracted_k141_58930 | Not-determined |
| S9_extracted_k141_50922 | Not-determined |
| S9_extracted_k141_17812 | Not-determined |
| S9_extracted_k141_13543 | Not-determined |
| S9_extracted_k141_6282 | Not-determined |
| S9_extracted_k141_50924 | Not-determined |
| S9_extracted_k141_74222 | Low-quality |
| S9_extracted_k141_29107 | Not-determined |
| S9_extracted_k141_85463 | Not-determined |
| S9_extracted_k141_55532 | Not-determined |
| S9_extracted_k141_13546 | Not-determined |
| S9_extracted_k141_77580 | Not-determined |
| S9_extracted_k141_17821 | Not-determined |
| S9_extracted_k141_85471 | Not-determined |
| S9_extracted_k141_36680 | Not-determined |
| S9_extracted_k141_25208 | Not-determined |
| S9_extracted_k141_36682 | Low-quality |
| S9_extracted_k141_13556 | Not-determined |
| S9_extracted_k141_9821 | Not-determined |
| S9_extracted_k141_21444 | Not-determined |
| S9_extracted_k141_77589 | Low-quality |
| S9_extracted_k141_50939 | Not-determined |
| S9_extracted_k141_6287 | Not-determined |
| S9_extracted_k141_32757 | Not-determined |
| S9_extracted_k141_25217 | Not-determined |
| S9_extracted_k141_55546 | Not-determined |
| S9_extracted_k141_62840 | Not-determined |
| S9_extracted_k141_58947 | Low-quality |
| S9_extracted_k141_50942 | Not-determined |
| S9_extracted_k141_81819 | Not-determined |
| S9_extracted_k141_50943 | Not-determined |
| S9_extracted_k141_29118 | Low-quality |
| S9_extracted_k141_32762 | Not-determined |
| S9_extracted_k141_2479 | Not-determined |
| S9_extracted_k141_21450 | Low-quality |
| S9_extracted_k141_21452 | Not-determined |
| S9_extracted_k141_81825 | Low-quality |
| S9_extracted_k141_47539 | Not-determined |
| S9_extracted_k141_9839 | Not-determined |
| S9_extracted_k141_55553 | Not-determined |
| S9_extracted_k141_2490 | Not-determined |
| S9_extracted_k141_77597 | Not-determined |
| S9_extracted_k141_62850 | Not-determined |
| S9_extracted_k141_58958 | Not-determined |
| S9_extracted_k141_85491 | Not-determined |
| S9_extracted_k141_2498 | Not-determined |
| S9_extracted_k141_21459 | Not-determined |
| S9_extracted_k141_58963 | Not-determined |
| S9_extracted_k141_81835 | Not-determined |
| S9_extracted_k141_29127 | Low-quality |
| S9_extracted_k141_44334 | Low-quality |
| S9_extracted_k141_29129 | Not-determined |
| S9_extracted_k141_66492 | Not-determined |
| S9_extracted_k141_21465 | Not-determined |
| S9_extracted_k141_58969 | Not-determined |
| S9_extracted_k141_77609 | Not-determined |
| S9_extracted_k141_58971 | Not-determined |
| S9_extracted_k141_36708 | Low-quality |
| S9_extracted_k141_85492 | Low-quality |
| S9_extracted_k141_47556 | Not-determined |
| S9_extracted_k141_32779 | Not-determined |
| S9_extracted_k141_36712 | Low-quality |
| S9_extracted_k141_29135 | Not-determined |
| S9_extracted_k141_17857 | Not-determined |
| S9_extracted_k141_2513 | Not-determined |
| S9_extracted_k141_21472 | Not-determined |
| S9_extracted_k141_36717 | Not-determined |
| S9_extracted_k141_32785 | Not-determined |
| S9_extracted_k141_13584 | Not-determined |
| S9_extracted_k141_21473 | Not-determined |
| S9_extracted_k141_2519 | Not-determined |
| S9_extracted_k141_66502 | Not-determined |
| S9_extracted_k141_13591 | Not-determined |
| S9_extracted_k141_47572 | Not-determined |
| S9_extracted_k141_44344 | Not-determined |
| S9_extracted_k141_77621 | Not-determined |
| S9_extracted_k141_13595 | Low-quality |
| S9_extracted_k141_40587 | Not-determined |
| S9_extracted_k141_36736 | Not-determined |
| S9_extracted_k141_77626 | Low-quality |
| S9_extracted_k141_66510 | Not-determined |
| S9_extracted_k141_40590 | Not-determined |
| S9_extracted_k141_77629 | Not-determined |
| S9_extracted_k141_47580 | Low-quality |
| S9_extracted_k141_40593 | Not-determined |
| S9_extracted_k141_6319 | Not-determined |
| S9_extracted_k141_44356 | Low-quality |
| S9_extracted_k141_29159 | Not-determined |
| S9_extracted_k141_6324 | Low-quality |
| S9_extracted_k141_44361 | Low-quality |
| S3_extracted_k141_127402 | Complete |
| S9_extracted_k141_32799 | Not-determined |
| S9_extracted_k141_36752 | Not-determined |
| S9_extracted_k141_81853 | Not-determined |
| S9_extracted_k141_32801 | Not-determined |
| S9_extracted_k141_50986 | Not-determined |
| S9_extracted_k141_66523 | Low-quality |
| S9_extracted_k141_40610 | Not-determined |
| S9_extracted_k141_9875 | Not-determined |
| S9_extracted_k141_13616 | Not-determined |
| S9_extracted_k141_62883 | Not-determined |
| S9_extracted_k141_13617 | Not-determined |
| S9_extracted_k141_21497 | Low-quality |
| S9_extracted_k141_40612 | Not-determined |
| S9_extracted_k141_81856 | Not-determined |
| S9_extracted_k141_44370 | Not-determined |
| S9_extracted_k141_62886 | Not-determined |
| S9_extracted_k141_21503 | Low-quality |
| S9_extracted_k141_21504 | Not-determined |
| S9_extracted_k141_50999 | Not-determined |
| S9_extracted_k141_77645 | Not-determined |
| S9_extracted_k141_9887 | Not-determined |
| S9_extracted_k141_44374 | Not-determined |
| S9_extracted_k141_9890 | Not-determined |
| S9_extracted_k141_66540 | Not-determined |
| S9_extracted_k141_25234 | Not-determined |
| S9_extracted_k141_47602 | Not-determined |
| S9_extracted_k141_36771 | Not-determined |
| S9_extracted_k141_9892 | Not-determined |
| S9_extracted_k141_25235 | Not-determined |
| S9_extracted_k141_36774 | Not-determined |
| S9_extracted_k141_9896 | Not-determined |
| S9_extracted_k141_62901 | Not-determined |
| S9_extracted_k141_21511 | Not-determined |
| S9_extracted_k141_62905 | Not-determined |
| S9_extracted_k141_9900 | Not-determined |
| S9_extracted_k141_32813 | Not-determined |
| S9_extracted_k141_66547 | Not-determined |
| S9_extracted_k141_81874 | Not-determined |
| S9_extracted_k141_2574 | Not-determined |
| S9_extracted_k141_21520 | Not-determined |
| S9_extracted_k141_62910 | Not-determined |
| S9_extracted_k141_81875 | Not-determined |
| S9_extracted_k141_66552 | Not-determined |
| S9_extracted_k141_81880 | Not-determined |
| S9_extracted_k141_6361 | Not-determined |
| S9_extracted_k141_9906 | Not-determined |
| S9_extracted_k141_2580 | Not-determined |
| S9_extracted_k141_89204 | Low-quality |
| S9_extracted_k141_21525 | Not-determined |
| S9_extracted_k141_47613 | Not-determined |
| S9_extracted_k141_55617 | Not-determined |
| S9_extracted_k141_81884 | Not-determined |
| S9_extracted_k141_55618 | Low-quality |
| S9_extracted_k141_17886 | Low-quality |
| S9_extracted_k141_85506 | Not-determined |
| S9_extracted_k141_17891 | Not-determined |
| S9_extracted_k141_59019 | Low-quality |
| S9_extracted_k141_62925 | Not-determined |
| S9_extracted_k141_66556 | Not-determined |
| S9_extracted_k141_9921 | Not-determined |
| S9_extracted_k141_6365 | Low-quality |
| S9_extracted_k141_66559 | Not-determined |
| S9_extracted_k141_44379 | Not-determined |
| S9_extracted_k141_62928 | Not-determined |
| S9_extracted_k141_62929 | Low-quality |
| S9_extracted_k141_47627 | Not-determined |
| S9_extracted_k141_32827 | Not-determined |
| S9_extracted_k141_59021 | Not-determined |
| S9_extracted_k141_59022 | Low-quality |
| S9_extracted_k141_89215 | Low-quality |
| S9_extracted_k141_89216 | Not-determined |
| S3_vs_k141_127402\|\|full | High-quality |
| S9_extracted_k141_47632 | Not-determined |
| S9_extracted_k141_21544 | Not-determined |
| S9_extracted_k141_74238 | Not-determined |
| S9_extracted_k141_85516 | Not-determined |
| S9_extracted_k141_89218 | Not-determined |
| S9_extracted_k141_77661 | Not-determined |
| S9_extracted_k141_55625 | Not-determined |
| S9_extracted_k141_6376 | Not-determined |
| S9_extracted_k141_59030 | Not-determined |
| S9_extracted_k141_36802 | Not-determined |
| S9_extracted_k141_66567 | Not-determined |
| S9_extracted_k141_29205 | Not-determined |
| S9_extracted_k141_77665 | Not-determined |
| S9_extracted_k141_62944 | Not-determined |
| S9_extracted_k141_66570 | Not-determined |
| S9_extracted_k141_62946 | Not-determined |
| S9_extracted_k141_66573 | Not-determined |
| S9_extracted_k141_13643 | Low-quality |
| S9_extracted_k141_81899 | Not-determined |
| S9_extracted_k141_66576 | Not-determined |
| S9_extracted_k141_66577 | Not-determined |
| S9_extracted_k141_6391 | Low-quality |
| S9_extracted_k141_29215 | Not-determined |
| S9_extracted_k141_21561 | Low-quality |
| S9_extracted_k141_85537 | Not-determined |
| S9_extracted_k141_59039 | Not-determined |
| S9_extracted_k141_47639 | Not-determined |
| S9_extracted_k141_9947 | Not-determined |
| S9_extracted_k141_21567 | Not-determined |
| S9_extracted_k141_9950 | Not-determined |
| S9_extracted_k141_29218 | Low-quality |
| S9_extracted_k141_13656 | Not-determined |
| S9_extracted_k141_29220 | Not-determined |
| S9_extracted_k141_62960 | Low-quality |
| S9_extracted_k141_55637 | Not-determined |
| S9_extracted_k141_66585 | Not-determined |
| S9_extracted_k141_29222 | Low-quality |
| S9_extracted_k141_6397 | Low-quality |
| S9_extracted_k141_62962 | Not-determined |
| S9_extracted_k141_6399 | Not-determined |
| S9_extracted_k141_29225 | Not-determined |
| S9_extracted_k141_77688 | Not-determined |
| S9_extracted_k141_6402 | Not-determined |
| S9_extracted_k141_17922 | Low-quality |
| S9_extracted_k141_21576 | Not-determined |
| S9_extracted_k141_6403 | Not-determined |
| S9_extracted_k141_36823 | Low-quality |
| S9_extracted_k141_44414 | Not-determined |
| S9_extracted_k141_55648 | Not-determined |
| S9_extracted_k141_44415 | Not-determined |
| S9_extracted_k141_62967 | Not-determined |
| S9_extracted_k141_36830 | Not-determined |
| S9_extracted_k141_17928 | Not-determined |
| S9_extracted_k141_6410 | Not-determined |
| S9_extracted_k141_89242 | Not-determined |
| S9_extracted_k141_36835 | Not-determined |
| S9_extracted_k141_6413 | Not-determined |
| S9_extracted_k141_21584 | Not-determined |
| S9_extracted_k141_29242 | Not-determined |
| S9_extracted_k141_44424 | Not-determined |
| S9_extracted_k141_74265 | Not-determined |
| S9_extracted_k141_55656 | Low-quality |
| S9_extracted_k141_13680 | Not-determined |
| S9_extracted_k141_36839 | Not-determined |
| S9_extracted_k141_74267 | Not-determined |
| S9_extracted_k141_6417 | Not-determined |
| S9_extracted_k141_89246 | Not-determined |
| S9_extracted_k141_9968 | Not-determined |
| S9_extracted_k141_62970 | Not-determined |
| S9_extracted_k141_89249 | Not-determined |
| S9_extracted_k141_59056 | Not-determined |
| S9_extracted_k141_6421 | Not-determined |
| S9_extracted_k141_77705 | Not-determined |
| S9_extracted_k141_77706 | Not-determined |
| S9_extracted_k141_29251 | Not-determined |
| S9_extracted_k141_59058 | Not-determined |
| S9_extracted_k141_85546 | Low-quality |
| S9_extracted_k141_74277 | Not-determined |
| S9_extracted_k141_44428 | Not-determined |
| S9_extracted_k141_21595 | Not-determined |
| S9_extracted_k141_2620 | Not-determined |
| S9_extracted_k141_89257 | Not-determined |
| S9_extracted_k141_6427 | Not-determined |
| S9_extracted_k141_21596 | Not-determined |
| S9_extracted_k141_2621 | Not-determined |
| S9_extracted_k141_36850 | Low-quality |
| S9_extracted_k141_21598 | Not-determined |
| S9_extracted_k141_47660 | Not-determined |
| S9_extracted_k141_59068 | Not-determined |
| S9_extracted_k141_29256 | Not-determined |
| S9_extracted_k141_81918 | Not-determined |
| S9_extracted_k141_36852 | Not-determined |
| S9_extracted_k141_89263 | Not-determined |
| S9_extracted_k141_47666 | Low-quality |
| S9_extracted_k141_89264 | Not-determined |
| S9_extracted_k141_62980 | Not-determined |
| S9_extracted_k141_36857 | Not-determined |
| S9_extracted_k141_89266 | Not-determined |
| S9_extracted_k141_21603 | Low-quality |
| S9_extracted_k141_89269 | Not-determined |
| S9_extracted_k141_36863 | Not-determined |
| S9_extracted_k141_85559 | Not-determined |
| S9_extracted_k141_59080 | Not-determined |
| S9_extracted_k141_36865 | Not-determined |
| S9_extracted_k141_62987 | Low-quality |
| S9_extracted_k141_85563 | Low-quality |
| S9_extracted_k141_85565 | Not-determined |
| S9_extracted_k141_21612 | Not-determined |
| S9_extracted_k141_9992 | Not-determined |
| S9_extracted_k141_9993 | Not-determined |
| S9_extracted_k141_9998 | Not-determined |
| S9_extracted_k141_74299 | Not-determined |
| S9_extracted_k141_55687 | Not-determined |
| S9_extracted_k141_25240 | Not-determined |
| S9_extracted_k141_63002 | Not-determined |
| S9_extracted_k141_85579 | Not-determined |
| S9_extracted_k141_81942 | Not-determined |
| S9_extracted_k141_6451 | Not-determined |
| S9_extracted_k141_36878 | Not-determined |
| S9_extracted_k141_59098 | Not-determined |
| S9_extracted_k141_10010 | Not-determined |
| S9_extracted_k141_13705 | Not-determined |
| S9_extracted_k141_29261 | Not-determined |
| S9_extracted_k141_63008 | Not-determined |
| S9_extracted_k141_29265 | Not-determined |
| S9_extracted_k141_47694 | Not-determined |
| S9_extracted_k141_59102 | Not-determined |
| S9_extracted_k141_74315 | Not-determined |
| S9_extracted_k141_29268 | Not-determined |
| S9_extracted_k141_47700 | Not-determined |
| S9_extracted_k141_81952 | Low-quality |
| S9_extracted_k141_6463 | Not-determined |
| S9_extracted_k141_25248 | Not-determined |
| S9_extracted_k141_6464 | Not-determined |
| S9_extracted_k141_44463 | Not-determined |
| S9_extracted_k141_13711 | Not-determined |
| S9_extracted_k141_89291 | Not-determined |
| S9_extracted_k141_85597 | Not-determined |
| S9_extracted_k141_6468 | Not-determined |
| S9_extracted_k141_89292 | Not-determined |
| S9_extracted_k141_63020 | Not-determined |
| S9_extracted_k141_21640 | Low-quality |
| S9_extracted_k141_81960 | Low-quality |
| S9_extracted_k141_89295 | Not-determined |
| S9_extracted_k141_21642 | Not-determined |
| S9_extracted_k141_6470 | Not-determined |
| S9_extracted_k141_2641 | Not-determined |
| S9_extracted_k141_17955 | Not-determined |
| S9_extracted_k141_81962 | Low-quality |
| S9_extracted_k141_32859 | Not-determined |
| S9_extracted_k141_47706 | Not-determined |
| S9_extracted_k141_32860 | Not-determined |
| S9_extracted_k141_89301 | Not-determined |
| S9_extracted_k141_63031 | Not-determined |
| S9_extracted_k141_85602 | Not-determined |
| S9_extracted_k141_47708 | Low-quality |
| S9_extracted_k141_32862 | Not-determined |
| S9_extracted_k141_10026 | Not-determined |
| S9_extracted_k141_59114 | Low-quality |
| S9_extracted_k141_25262 | Not-determined |
| S9_extracted_k141_74328 | Low-quality |
| S9_extracted_k141_85607 | Not-determined |
| S9_extracted_k141_89304 | Not-determined |
| S9_extracted_k141_47710 | Low-quality |
| S9_extracted_k141_74331 | Not-determined |
| S9_extracted_k141_74333 | Not-determined |
| S9_extracted_k141_32868 | Not-determined |
| S9_extracted_k141_2647 | Low-quality |
| S9_extracted_k141_47714 | Not-determined |
| S9_extracted_k141_44476 | Not-determined |
| S9_extracted_k141_17959 | Not-determined |
| S9_extracted_k141_40616 | Not-determined |
| S9_extracted_k141_44477 | Not-determined |
| S9_extracted_k141_21656 | Low-quality |
| S9_extracted_k141_2650 | Not-determined |
| S9_extracted_k141_2652 | Low-quality |
| S9_extracted_k141_10040 | Not-determined |
| S9_extracted_k141_40623 | Not-determined |
| S9_extracted_k141_74338 | Not-determined |
| S9_extracted_k141_55700 | Low-quality |
| S9_extracted_k141_74342 | Not-determined |
| S9_extracted_k141_59126 | Not-determined |
| S9_extracted_k141_6479 | Low-quality |
| S9_extracted_k141_2657 | Not-determined |
| S9_extracted_k141_40627 | Low-quality |
| S9_extracted_k141_89308 | Not-determined |
| S9_extracted_k141_13735 | Not-determined |
| S9_extracted_k141_77760 | Not-determined |
| S9_extracted_k141_17963 | Not-determined |
| S9_extracted_k141_77761 | Not-determined |
| S9_extracted_k141_25278 | Not-determined |
| S9_extracted_k141_40631 | Low-quality |
| S9_extracted_k141_13739 | Not-determined |
| S9_extracted_k141_25281 | Low-quality |
| S9_extracted_k141_44484 | Not-determined |
| S9_extracted_k141_25282 | Not-determined |
| S9_extracted_k141_81974 | Not-determined |
| S9_extracted_k141_66605 | Not-determined |
| S9_extracted_k141_44488 | Not-determined |
| S9_extracted_k141_10059 | Low-quality |
| S9_extracted_k141_13744 | Not-determined |
| S9_extracted_k141_77769 | Not-determined |
| S9_extracted_k141_44490 | Not-determined |
| S9_extracted_k141_59135 | Not-determined |
| S9_extracted_k141_66607 | Not-determined |
| S9_extracted_k141_85622 | Not-determined |
| S9_extracted_k141_66608 | Not-determined |
| S9_extracted_k141_85624 | Not-determined |
| S9_extracted_k141_55711 | Not-determined |
| S9_extracted_k141_85626 | Low-quality |
| S9_extracted_k141_44495 | Not-determined |
| S9_extracted_k141_44496 | Not-determined |
| S9_extracted_k141_17978 | Not-determined |
| S9_extracted_k141_70326 | Not-determined |
| S9_extracted_k141_77775 | Not-determined |
| S9_extracted_k141_13753 | Not-determined |
| S9_extracted_k141_59142 | Low-quality |
| S9_extracted_k141_6487 | Not-determined |
| S9_extracted_k141_74353 | Not-determined |
| S9_extracted_k141_13756 | Not-determined |
| S9_extracted_k141_21667 | Low-quality |
| S9_extracted_k141_85630 | Low-quality |
| S9_extracted_k141_21668 | Low-quality |
| S9_extracted_k141_70330 | Not-determined |
| S9_extracted_k141_81986 | Not-determined |
| S9_extracted_k141_55718 | Not-determined |
| S9_extracted_k141_32890 | Not-determined |
| S9_extracted_k141_59144 | Low-quality |
| S9_extracted_k141_81987 | Not-determined |
| S9_extracted_k141_74357 | Not-determined |
| S9_extracted_k141_6490 | Low-quality |
| S9_extracted_k141_25295 | Not-determined |
| S9_extracted_k141_59146 | Low-quality |
| S9_extracted_k141_13758 | Not-determined |
| S9_extracted_k141_13759 | Not-determined |
| S9_extracted_k141_32893 | Not-determined |
| S9_extracted_k141_40645 | Low-quality |
| S9_extracted_k141_40646 | Not-determined |
| S9_extracted_k141_63063 | Low-quality |
| S9_extracted_k141_13762 | Not-determined |
| S9_extracted_k141_40649 | Not-determined |
| S9_extracted_k141_81991 | Low-quality |
| S9_extracted_k141_55731 | Low-quality |
| S9_extracted_k141_47743 | Not-determined |
| S9_extracted_k141_81994 | Not-determined |
| S9_extracted_k141_17990 | Not-determined |
| S9_extracted_k141_2682 | Not-determined |
| S9_extracted_k141_6503 | Not-determined |
| S9_extracted_k141_63070 | Low-quality |
| S9_extracted_k141_59159 | Not-determined |
| S9_extracted_k141_74365 | Not-determined |
| S9_extracted_k141_51004 | Low-quality |
| S9_extracted_k141_74367 | Low-quality |
| S9_extracted_k141_85640 | Not-determined |
| S9_extracted_k141_74368 | Not-determined |
| S9_extracted_k141_85642 | Not-determined |
| S9_extracted_k141_17999 | Not-determined |
| S9_extracted_k141_13774 | Not-determined |
| S9_extracted_k141_2690 | Not-determined |
| S9_extracted_k141_32913 | Not-determined |
| S9_extracted_k141_10076 | Not-determined |
| S9_extracted_k141_2692 | Low-quality |
| S9_extracted_k141_85647 | Not-determined |
| S9_extracted_k141_32915 | Low-quality |
| S9_extracted_k141_74371 | Not-determined |
| S9_extracted_k141_74373 | Not-determined |
| S9_extracted_k141_32918 | Not-determined |
| S9_extracted_k141_2697 | Not-determined |
| S9_extracted_k141_25316 | Not-determined |
| S9_extracted_k141_6516 | Not-determined |
| S9_extracted_k141_59169 | Low-quality |
| S9_extracted_k141_18008 | Not-determined |
| S9_extracted_k141_51014 | Not-determined |
| S9_extracted_k141_59171 | Low-quality |
| S9_extracted_k141_21675 | Not-determined |
| S9_extracted_k141_13782 | Not-determined |
| S9_extracted_k141_25318 | Not-determined |
| S9_extracted_k141_25320 | Not-determined |
| S9_extracted_k141_13784 | Not-determined |
| S9_extracted_k141_66619 | Low-quality |
| S9_extracted_k141_10083 | Not-determined |
| S9_extracted_k141_25323 | Not-determined |
| S9_extracted_k141_32924 | Not-determined |
| S9_extracted_k141_59178 | Not-determined |
| S9_extracted_k141_77799 | Not-determined |
| S9_extracted_k141_40662 | Not-determined |
| S9_extracted_k141_40663 | Low-quality |
| S9_extracted_k141_44519 | Not-determined |
| S9_extracted_k141_81998 | Low-quality |
| S9_extracted_k141_59180 | Low-quality |
| S9_extracted_k141_70349 | Not-determined |
| S9_extracted_k141_66629 | Not-determined |
| S9_extracted_k141_70351 | Not-determined |
| S9_extracted_k141_32928 | Not-determined |
| S9_extracted_k141_10085 | Not-determined |
| S9_extracted_k141_25329 | Not-determined |
| S9_extracted_k141_59185 | Not-determined |
| S9_extracted_k141_44526 | Not-determined |
| S9_extracted_k141_21687 | Not-determined |
| S9_extracted_k141_32931 | Not-determined |
| S9_extracted_k141_32932 | Low-quality |
| S9_extracted_k141_74383 | Not-determined |
| S9_extracted_k141_29313 | Not-determined |
| S9_extracted_k141_59192 | Not-determined |
| S9_extracted_k141_63100 | Not-determined |
| S9_extracted_k141_2707 | Not-determined |
| S9_extracted_k141_77804 | Not-determined |
| S9_extracted_k141_66637 | Not-determined |
| S9_extracted_k141_63105 | Not-determined |
| S9_extracted_k141_40676 | Not-determined |
| S9_extracted_k141_6535 | Not-determined |
| S9_extracted_k141_40679 | Not-determined |
| S9_extracted_k141_51020 | Not-determined |
| S9_extracted_k141_70367 | Not-determined |
| S9_extracted_k141_70368 | Not-determined |
| S9_extracted_k141_40684 | Not-determined |
| S9_extracted_k141_32948 | Not-determined |
| S9_extracted_k141_66644 | Not-determined |
| S9_extracted_k141_40687 | Not-determined |
| S9_extracted_k141_74400 | Not-determined |
| S9_extracted_k141_89326 | Low-quality |
| S9_extracted_k141_21699 | Not-determined |
| S9_extracted_k141_77818 | Low-quality |
| S9_extracted_k141_51025 | Not-determined |
| S9_extracted_k141_74402 | Low-quality |
| S9_extracted_k141_63123 | Not-determined |
| S9_extracted_k141_66651 | Not-determined |
| S9_extracted_k141_51027 | Not-determined |
| S9_extracted_k141_29326 | Not-determined |
| S9_extracted_k141_51029 | Not-determined |
| S9_extracted_k141_6546 | Not-determined |
| S9_extracted_k141_32962 | Not-determined |
| S9_extracted_k141_10108 | Not-determined |
| S9_extracted_k141_89330 | Not-determined |
| S9_extracted_k141_10112 | Not-determined |
| S9_extracted_k141_18043 | Not-determined |
| S9_extracted_k141_18045 | Not-determined |
| S9_extracted_k141_51035 | Not-determined |
| S9_extracted_k141_85681 | Not-determined |
| S9_extracted_k141_25341 | Not-determined |
| S9_extracted_k141_10116 | Not-determined |
| S9_extracted_k141_36906 | Not-determined |
| S9_extracted_k141_21710 | Not-determined |
| S9_extracted_k141_25344 | Low-quality |
| S9_extracted_k141_25345 | Not-determined |
| S9_extracted_k141_70383 | Not-determined |
| S9_extracted_k141_70385 | Not-determined |
| S9_extracted_k141_74407 | Not-determined |
| S9_extracted_k141_32973 | Not-determined |
| S9_extracted_k141_44555 | Not-determined |
| S9_extracted_k141_74409 | Not-determined |
| S9_extracted_k141_21717 | Not-determined |
| S9_extracted_k141_59215 | Not-determined |
| S9_extracted_k141_40713 | Not-determined |
| S9_extracted_k141_29344 | Not-determined |
| S9_extracted_k141_44560 | Low-quality |
| S9_extracted_k141_29345 | Not-determined |
| S9_extracted_k141_59218 | Low-quality |
| S9_extracted_k141_29348 | Not-determined |
| S9_extracted_k141_70392 | Not-determined |
| S9_extracted_k141_21725 | Not-determined |
| S9_extracted_k141_59222 | Not-determined |
| S9_extracted_k141_25361 | Low-quality |
| S9_extracted_k141_85701 | Low-quality |
| S9_extracted_k141_82005 | Not-determined |
| S9_extracted_k141_32980 | Not-determined |
| S9_extracted_k141_29352 | Not-determined |
| S9_extracted_k141_85705 | Not-determined |
| S9_extracted_k141_36922 | Not-determined |
| S9_extracted_k141_77832 | Not-determined |
| S9_extracted_k141_77834 | Not-determined |
| S9_extracted_k141_85710 | Not-determined |
| S9_extracted_k141_36929 | Not-determined |
| S9_extracted_k141_74423 | Not-determined |
| S9_extracted_k141_63129 | Low-quality |
| S9_extracted_k141_74426 | Not-determined |
| S9_extracted_k141_59234 | Low-quality |
| S9_extracted_k141_63133 | Not-determined |
| S9_extracted_k141_36934 | Not-determined |
| S9_extracted_k141_66674 | Not-determined |
| S9_extracted_k141_44572 | Not-determined |
| S9_extracted_k141_10139 | Low-quality |
| S9_extracted_k141_2741 | Not-determined |
| S9_extracted_k141_10142 | Low-quality |
| S9_extracted_k141_6554 | Not-determined |
| S9_extracted_k141_63139 | Not-determined |
| S9_extracted_k141_40732 | Not-determined |
| S9_extracted_k141_32991 | Low-quality |
| S9_extracted_k141_66679 | Not-determined |
| S9_extracted_k141_18084 | Low-quality |
| S9_extracted_k141_66680 | Not-determined |
| S9_extracted_k141_2746 | Not-determined |
| S9_extracted_k141_21739 | Not-determined |
| S9_extracted_k141_18086 | Low-quality |
| S9_extracted_k141_74440 | Not-determined |
| S9_extracted_k141_74441 | Not-determined |
| S9_extracted_k141_85726 | Low-quality |
| S9_extracted_k141_18089 | Not-determined |
| S9_extracted_k141_10150 | Low-quality |
| S9_extracted_k141_85727 | Not-determined |
| S9_extracted_k141_32999 | Not-determined |

| | |
|---|---|
| S9_extracted_k141_33000 | Low-quality |
| S9_extracted_k141_66683 | Not-determined |
| S9_extracted_k141_25389 | Not-determined |
| S9_extracted_k141_2748 | Low-quality |
| S9_extracted_k141_6566 | Not-determined |
| S9_extracted_k141_63149 | Not-determined |
| S9_extracted_k141_21746 | Not-determined |
| S9_extracted_k141_63152 | Not-determined |
| S9_extracted_k141_82026 | Not-determined |
| S9_extracted_k141_55756 | Not-determined |
| S9_extracted_k141_18095 | Not-determined |
| S9_extracted_k141_77856 | Not-determined |
| S9_extracted_k141_85731 | Not-determined |
| S9_extracted_k141_33007 | Not-determined |
| S9_extracted_k141_63154 | Not-determined |
| S9_extracted_k141_6573 | Not-determined |
| S9_extracted_k141_77859 | Low-quality |
| S9_extracted_k141_10156 | Not-determined |
| S9_extracted_k141_10158 | Not-determined |
| S9_extracted_k141_13843 | Low-quality |
| S9_extracted_k141_44594 | Not-determined |
| S9_extracted_k141_36958 | Not-determined |
| S9_extracted_k141_10160 | Not-determined |
| S9_extracted_k141_29376 | Not-determined |
| S9_extracted_k141_29377 | Not-determined |
| S9_extracted_k141_85736 | Not-determined |
| S9_extracted_k141_44600 | Not-determined |
| S9_extracted_k141_33014 | Not-determined |
| S9_extracted_k141_29381 | Not-determined |
| S9_extracted_k141_33016 | Not-determined |
| S9_extracted_k141_29383 | Not-determined |
| S9_extracted_k141_47755 | Not-determined |
| S9_extracted_k141_82035 | Not-determined |
| S9_extracted_k141_47757 | Not-determined |
| S9_extracted_k141_66702 | Not-determined |
| S9_extracted_k141_25404 | Not-determined |
| S9_extracted_k141_33023 | Not-determined |
| S9_extracted_k141_82041 | Not-determined |
| S9_extracted_k141_63163 | Not-determined |
| S9_extracted_k141_63166 | Not-determined |
| S9_extracted_k141_82043 | Not-determined |
| S9_extracted_k141_70432 | Low-quality |
| S9_extracted_k141_70433 | Not-determined |
| S9_extracted_k141_18103 | Low-quality |
| S9_extracted_k141_70435 | Not-determined |
| S9_extracted_k141_47766 | Not-determined |
| S9_extracted_k141_70436 | Low-quality |
| S9_extracted_k141_82047 | Not-determined |
| S9_extracted_k141_21773 | Not-determined |
| S9_extracted_k141_70439 | Not-determined |
| S9_extracted_k141_21776 | Low-quality |
| S9_extracted_k141_36970 | Not-determined |
| S9_extracted_k141_47772 | Not-determined |
| S9_extracted_k141_47777 | Not-determined |
| S9_extracted_k141_47779 | Not-determined |
| S9_extracted_k141_33034 | Low-quality |
| S9_extracted_k141_55790 | Not-determined |
| S9_extracted_k141_13871 | Not-determined |
| S9_extracted_k141_10180 | Low-quality |
| S9_extracted_k141_47782 | Not-determined |
| S9_extracted_k141_10181 | Not-determined |
| S9_extracted_k141_13873 | Not-determined |
| S9_extracted_k141_6603 | Not-determined |
| S9_extracted_k141_82067 | Not-determined |
| S9_extracted_k141_29415 | Not-determined |
| S9_extracted_k141_47787 | Not-determined |
| S9_extracted_k141_18123 | Not-determined |
| S9_extracted_k141_36980 | Not-determined |
| S9_extracted_k141_2779 | Not-determined |
| S9_extracted_k141_47788 | Low-quality |
| S9_extracted_k141_2780 | Not-determined |
| S9_extracted_k141_33043 | Low-quality |
| S9_extracted_k141_2782 | Not-determined |
| S9_extracted_k141_36986 | Low-quality |
| S9_extracted_k141_70463 | Low-quality |
| S9_extracted_k141_63192 | Not-determined |
| S9_extracted_k141_10195 | Not-determined |
| S9_extracted_k141_25422 | Low-quality |
| S9_extracted_k141_47794 | Low-quality |
| S9_extracted_k141_63193 | Low-quality |
| S9_extracted_k141_85768 | Not-determined |
| S9_extracted_k141_10196 | Low-quality |
| S9_extracted_k141_51055 | Not-determined |
| S9_extracted_k141_82076 | Not-determined |
| S9_extracted_k141_89355 | Not-determined |
| S9_extracted_k141_70468 | Not-determined |
| S9_extracted_k141_18132 | Not-determined |
| S9_extracted_k141_82079 | Not-determined |
| S9_extracted_k141_36996 | Not-determined |
| S9_extracted_k141_10200 | Not-determined |
| S9_extracted_k141_89358 | Not-determined |
| S9_extracted_k141_2790 | Not-determined |
| S9_extracted_k141_25430 | Not-determined |
| S9_extracted_k141_18135 | Not-determined |
| S9_extracted_k141_37000 | Low-quality |
| S9_extracted_k141_21796 | Low-quality |
| S9_extracted_k141_18140 | Not-determined |
| S9_extracted_k141_33051 | Not-determined |
| S9_extracted_k141_13892 | Not-determined |
| S9_extracted_k141_10210 | Not-determined |
| S9_extracted_k141_66722 | Not-determined |
| S9_extracted_k141_2795 | Not-determined |
| S9_extracted_k141_37003 | Low-quality |
| S9_extracted_k141_51061 | Not-determined |
| S9_extracted_k141_82092 | Not-determined |
| S9_extracted_k141_70478 | Not-determined |
| S9_extracted_k141_6627 | Not-determined |
| S9_extracted_k141_47812 | Not-determined |
| S9_extracted_k141_70482 | Not-determined |
| S9_extracted_k141_77917 | Not-determined |
| S9_extracted_k141_37008 | Not-determined |
| S9_extracted_k141_89367 | Not-determined |
| S9_extracted_k141_25443 | Not-determined |
| S9_extracted_k141_70486 | Low-quality |
| S9_extracted_k141_25446 | Not-determined |
| S9_extracted_k141_13901 | Not-determined |
| S9_extracted_k141_66735 | Not-determined |
| S9_extracted_k141_55821 | Not-determined |
| S9_extracted_k141_77921 | Not-determined |
| S9_extracted_k141_55823 | Not-determined |
| S9_extracted_k141_89369 | Low-quality |
| S9_extracted_k141_37012 | Not-determined |
| S9_extracted_k141_77926 | Not-determined |
| S9_extracted_k141_10224 | Not-determined |
| S9_extracted_k141_37015 | Not-determined |
| S9_extracted_k141_47817 | Not-determined |
| S9_extracted_k141_82105 | Not-determined |
| S9_extracted_k141_37017 | Not-determined |
| S9_extracted_k141_89381 | Not-determined |
| S9_extracted_k141_18152 | Not-determined |
| S9_extracted_k141_66749 | Not-determined |
| S9_extracted_k141_70494 | Low-quality |
| S9_extracted_k141_85796 | Not-determined |
| S9_extracted_k141_25458 | Not-determined |
| S9_extracted_k141_21807 | Low-quality |
| S9_extracted_k141_89384 | Not-determined |
| S9_extracted_k141_21810 | Not-determined |
| S9_extracted_k141_6640 | Not-determined |
| S9_extracted_k141_13913 | Low-quality |
| S9_extracted_k141_18164 | Not-determined |
| S9_extracted_k141_89390 | Not-determined |
| S9_extracted_k141_37029 | Not-determined |
| S9_extracted_k141_37032 | Not-determined |
| S9_extracted_k141_85804 | Not-determined |
| S9_extracted_k141_70503 | Not-determined |
| S9_extracted_k141_51092 | Not-determined |
| S9_extracted_k141_63226 | Low-quality |
| S9_extracted_k141_63227 | Not-determined |
| S9_extracted_k141_82111 | Not-determined |
| S9_extracted_k141_82112 | Not-determined |
| S9_extracted_k141_18175 | Not-determined |
| S9_extracted_k141_18178 | Low-quality |
| S9_extracted_k141_77948 | Low-quality |
| S9_extracted_k141_37036 | Not-determined |
| S9_extracted_k141_37037 | Not-determined |
| S9_extracted_k141_77951 | Not-determined |
| S9_extracted_k141_63231 | Not-determined |
| S9_extracted_k141_82118 | Not-determined |
| S9_extracted_k141_6658 | Not-determined |
| S9_extracted_k141_63233 | Not-determined |
| S9_extracted_k141_21829 | Not-determined |
| S9_extracted_k141_6659 | Not-determined |
| S9_extracted_k141_77954 | Not-determined |
| S9_extracted_k141_51100 | Low-quality |
| S9_extracted_k141_66769 | Low-quality |
| S9_extracted_k141_63234 | Not-determined |
| S9_extracted_k141_85807 | Not-determined |
| S9_extracted_k141_29455 | Not-determined |
| S9_extracted_k141_33087 | Not-determined |
| S9_extracted_k141_47843 | Not-determined |
| S9_extracted_k141_44615 | Not-determined |
| S9_extracted_k141_66781 | Not-determined |
| S9_extracted_k141_21839 | Not-determined |
| S9_extracted_k141_25482 | Not-determined |
| S9_extracted_k141_70523 | Not-determined |
| S9_extracted_k141_85820 | Not-determined |
| S9_extracted_k141_37054 | Not-determined |
| S9_extracted_k141_66782 | Not-determined |
| S9_extracted_k141_47848 | Not-determined |
| S9_extracted_k141_47849 | Not-determined |
| S9_extracted_k141_82126 | Not-determined |
| S9_extracted_k141_63244 | Low-quality |
| S9_extracted_k141_85828 | Not-determined |
| S9_extracted_k141_85829 | Not-determined |
| S9_extracted_k141_47853 | Not-determined |
| S9_extracted_k141_51113 | Not-determined |
| S9_extracted_k141_29458 | Not-determined |
| S9_extracted_k141_82128 | Not-determined |
| S9_extracted_k141_59249 | Not-determined |
| S9_extracted_k141_82131 | Low-quality |
| S9_extracted_k141_37065 | Not-determined |
| S9_extracted_k141_66791 | Low-quality |
| S9_extracted_k141_13963 | Not-determined |
| S9_extracted_k141_59252 | Not-determined |
| S9_extracted_k141_29465 | Not-determined |
| S9_extracted_k141_59253 | Low-quality |
| S9_extracted_k141_44635 | Not-determined |
| S9_extracted_k141_29466 | Not-determined |
| S9_extracted_k141_6678 | Not-determined |
| S9_extracted_k141_59256 | Not-determined |
| S9_extracted_k141_6680 | Not-determined |
| S9_extracted_k141_47868 | Not-determined |
| S9_extracted_k141_82134 | Low-quality |
| S9_extracted_k141_55835 | Not-determined |
| S9_extracted_k141_59258 | Low-quality |
| S9_extracted_k141_85836 | Low-quality |
| S9_extracted_k141_40774 | Not-determined |
| S9_extracted_k141_77974 | Not-determined |
| S9_extracted_k141_33108 | Not-determined |
| S9_extracted_k141_51124 | Low-quality |
| S9_extracted_k141_47872 | Not-determined |
| S9_extracted_k141_77976 | Not-determined |
| S9_extracted_k141_47874 | Not-determined |
| S9_extracted_k141_82136 | Not-determined |
| S9_extracted_k141_44640 | Not-determined |
| S9_extracted_k141_40777 | Not-determined |
| S9_extracted_k141_51135 | Not-determined |
| S9_extracted_k141_10254 | Not-determined |
| S9_extracted_k141_47883 | Not-determined |
| S9_extracted_k141_55848 | Not-determined |
| S9_extracted_k141_10257 | Not-determined |
| S9_extracted_k141_66805 | Not-determined |
| S9_extracted_k141_44649 | Not-determined |
| S9_extracted_k141_37079 | Low-quality |
| S9_extracted_k141_40783 | Not-determined |
| S9_extracted_k141_85847 | Not-determined |
| S9_extracted_k141_82147 | Low-quality |
| S9_extracted_k141_66808 | Not-determined |
| S9_extracted_k141_10267 | Not-determined |
| S9_extracted_k141_66810 | Not-determined |
| S9_extracted_k141_37085 | Not-determined |
| S9_extracted_k141_89416 | Not-determined |
| S9_extracted_k141_77993 | Low-quality |
| S9_extracted_k141_33124 | Not-determined |
| S9_extracted_k141_40789 | Not-determined |
| S9_extracted_k141_10271 | Low-quality |
| S9_extracted_k141_89423 | Not-determined |
| S9_extracted_k141_70530 | Not-determined |
| S9_extracted_k141_47888 | Not-determined |
| S9_extracted_k141_44651 | Low-quality |
| S9_extracted_k141_89426 | Not-determined |
| S9_extracted_k141_10276 | Not-determined |
| S9_extracted_k141_10277 | Not-determined |
| S9_extracted_k141_47893 | Not-determined |
| S9_extracted_k141_66824 | Not-determined |
| S9_extracted_k141_78000 | Low-quality |
| S9_extracted_k141_40800 | Low-quality |
| S9_extracted_k141_33134 | Not-determined |
| S9_extracted_k141_55856 | Low-quality |
| S9_extracted_k141_55858 | Not-determined |
| S9_extracted_k141_10282 | Not-determined |
| S9_extracted_k141_89440 | Not-determined |
| S9_extracted_k141_59290 | Low-quality |
| S9_extracted_k141_89444 | Low-quality |
| S9_extracted_k141_66837 | Low-quality |
| S9_extracted_k141_63282 | Not-determined |
| S9_extracted_k141_55865 | Low-quality |
| S9_extracted_k141_40812 | Not-determined |
| S9_extracted_k141_2802 | Not-determined |
| S9_extracted_k141_14000 | Not-determined |
| S9_extracted_k141_85864 | Not-determined |
| S9_extracted_k141_40815 | Not-determined |
| S9_extracted_k141_40816 | Not-determined |
| S9_extracted_k141_29504 | Not-determined |
| S9_extracted_k141_66847 | Not-determined |
| S9_extracted_k141_59297 | Low-quality |
| S9_extracted_k141_89453 | Not-determined |
| S9_extracted_k141_33146 | Not-determined |
| S9_extracted_k141_2807 | Not-determined |
| S9_extracted_k141_44669 | Low-quality |
| S9_extracted_k141_14006 | Not-determined |
| S9_extracted_k141_63289 | Low-quality |
| S9_extracted_k141_89457 | Low-quality |
| S9_extracted_k141_40822 | Not-determined |
| S9_extracted_k141_40823 | Not-determined |
| S9_extracted_k141_2812 | Not-determined |
| S9_extracted_k141_89460 | Not-determined |
| S9_extracted_k141_2813 | Not-determined |
| S9_extracted_k141_29516 | Not-determined |
| S9_extracted_k141_47910 | Not-determined |
| S9_extracted_k141_14012 | Not-determined |
| S9_extracted_k141_10306 | Not-determined |
| S9_extracted_k141_89466 | Not-determined |
| S9_extracted_k141_2818 | Not-determined |
| S9_extracted_k141_63296 | Not-determined |
| S9_extracted_k141_33157 | Not-determined |
| S9_extracted_k141_10312 | Not-determined |
| S9_extracted_k141_33158 | Not-determined |
| S9_extracted_k141_25535 | Low-quality |
| S9_extracted_k141_40834 | Not-determined |
| S9_extracted_k141_40836 | Not-determined |
| S9_extracted_k141_89470 | Not-determined |
| S9_extracted_k141_25537 | Not-determined |
| S9_extracted_k141_2825 | Not-determined |
| S9_extracted_k141_25538 | Not-determined |
| S9_extracted_k141_89472 | Not-determined |
| S9_extracted_k141_59312 | Low-quality |
| S9_extracted_k141_33167 | Not-determined |
| S9_extracted_k141_21854 | Not-determined |
| S9_extracted_k141_14013 | Low-quality |
| S9_extracted_k141_89477 | Not-determined |
| S9_extracted_k141_14015 | Not-determined |
| S9_extracted_k141_55884 | Not-determined |
| S9_extracted_k141_59316 | Not-determined |
| S9_extracted_k141_14016 | Not-determined |
| S9_extracted_k141_70567 | Not-determined |
| S9_extracted_k141_85887 | Not-determined |
| S9_extracted_k141_29533 | Not-determined |
| S9_extracted_k141_63307 | Low-quality |
| S9_extracted_k141_66867 | Not-determined |
| S9_extracted_k141_33174 | Not-determined |
| S9_extracted_k141_55889 | Not-determined |
| S9_extracted_k141_18183 | Not-determined |
| S9_extracted_k141_29538 | Not-determined |
| S9_extracted_k141_33177 | Not-determined |
| S9_extracted_k141_33178 | Low-quality |
| S9_extracted_k141_21872 | Low-quality |
| S9_extracted_k141_59321 | Not-determined |
| S9_extracted_k141_59322 | Not-determined |
| S9_extracted_k141_2837 | Not-determined |
| S9_extracted_k141_89493 | Not-determined |
| S9_extracted_k141_47930 | Not-determined |
| S9_extracted_k141_59325 | Not-determined |
| S9_extracted_k141_40858 | Low-quality |
| S9_extracted_k141_2840 | Not-determined |
| S9_extracted_k141_10323 | Not-determined |
| S9_extracted_k141_37099 | Not-determined |
| S9_extracted_k141_59328 | Not-determined |
| S9_extracted_k141_18190 | Not-determined |
| S9_extracted_k141_29541 | Low-quality |
| S9_extracted_k141_40864 | Not-determined |
| S9_extracted_k141_18193 | Not-determined |
| S9_extracted_k141_89500 | Low-quality |
| S9_extracted_k141_70576 | Not-determined |
| S9_extracted_k141_85893 | Not-determined |
| S9_extracted_k141_59331 | Not-determined |
| S9_extracted_k141_82155 | Low-quality |
| S9_extracted_k141_82156 | Not-determined |
| S9_extracted_k141_14025 | Low-quality |
| S9_extracted_k141_47939 | Not-determined |
| S9_extracted_k141_2850 | Low-quality |
| S9_extracted_k141_70582 | Not-determined |
| S9_extracted_k141_29547 | Not-determined |
| S9_extracted_k141_85899 | Not-determined |
| S9_extracted_k141_10337 | Not-determined |
| S9_extracted_k141_70586 | Not-determined |
| S9_extracted_k141_70587 | Not-determined |
| S9_extracted_k141_89510 | Not-determined |
| S9_extracted_k141_70588 | Not-determined |
| S9_extracted_k141_14031 | Low-quality |
| S9_extracted_k141_47948 | Not-determined |
| S9_extracted_k141_10341 | Not-determined |
| S9_extracted_k141_63333 | Not-determined |
| S9_extracted_k141_29556 | Not-determined |
| S9_extracted_k141_37116 | Not-determined |
| S9_extracted_k141_47953 | Not-determined |
| S9_extracted_k141_33193 | Low-quality |
| S9_extracted_k141_70593 | Low-quality |
| S9_extracted_k141_29559 | Not-determined |
| S9_extracted_k141_33197 | Not-determined |
| S9_extracted_k141_10343 | Not-determined |
| S9_extracted_k141_29563 | Not-determined |
| S9_extracted_k141_25565 | Not-determined |
| S9_extracted_k141_37121 | Not-determined |
| S9_extracted_k141_85911 | Not-determined |
| S9_extracted_k141_6694 | Not-determined |
| S9_extracted_k141_89519 | Not-determined |
| S9_extracted_k141_6695 | Low-quality |
| S9_extracted_k141_10351 | Not-determined |
| S9_extracted_k141_82173 | Not-determined |
| S9_extracted_k141_47968 | Low-quality |
| S9_extracted_k141_10353 | Not-determined |
| S9_extracted_k141_37134 | Not-determined |
| S9_extracted_k141_51187 | Low-quality |
| S9_extracted_k141_10354 | Not-determined |
| S9_extracted_k141_29574 | Low-quality |
| S9_extracted_k141_63346 | Not-determined |
| S9_extracted_k141_51193 | Not-determined |
| S9_extracted_k141_89532 | Not-determined |
| S9_extracted_k141_82180 | Not-determined |
| S9_extracted_k141_21905 | Not-determined |
| S9_extracted_k141_33216 | Not-determined |
| S9_extracted_k141_63353 | Not-determined |
| S9_extracted_k141_37145 | Not-determined |
| S9_extracted_k141_55912 | Not-determined |
| S9_extracted_k141_44688 | Not-determined |
| S9_extracted_k141_51205 | Low-quality |
| S9_extracted_k141_82184 | Not-determined |
| S9_extracted_k141_10367 | Not-determined |
| S9_extracted_k141_44692 | Not-determined |
| S9_extracted_k141_85933 | Low-quality |
| S9_extracted_k141_40884 | Not-determined |
| S9_extracted_k141_6714 | Not-determined |
| S9_extracted_k141_14049 | Not-determined |
| S9_extracted_k141_33223 | Not-determined |
| S9_extracted_k141_47985 | Low-quality |
| S9_extracted_k141_47986 | Low-quality |
| S9_extracted_k141_63363 | Not-determined |
| S9_extracted_k141_18220 | Not-determined |
| S9_extracted_k141_21920 | Not-determined |
| S9_extracted_k141_6717 | Not-determined |
| S9_extracted_k141_14052 | Not-determined |
| S9_extracted_k141_6718 | Not-determined |
| S9_extracted_k141_59353 | Not-determined |
| S9_extracted_k141_14054 | Not-determined |
| S9_extracted_k141_21923 | Not-determined |
| S9_extracted_k141_47992 | Low-quality |
| S9_extracted_k141_74463 | Not-determined |
| S9_extracted_k141_74464 | Low-quality |
| S9_extracted_k141_82194 | Not-determined |
| S9_extracted_k141_70622 | Not-determined |
| S9_extracted_k141_6725 | Not-determined |
| S9_extracted_k141_70626 | Not-determined |
| S9_extracted_k141_55923 | Low-quality |
| S9_extracted_k141_89557 | Low-quality |
| S9_extracted_k141_85946 | Not-determined |
| S9_extracted_k141_2878 | Not-determined |
| S9_extracted_k141_2879 | Not-determined |
| S9_extracted_k141_44706 | Not-determined |
| S9_extracted_k141_89560 | Not-determined |
| S9_extracted_k141_6729 | Not-determined |
| S9_extracted_k141_21934 | Low-quality |
| S9_extracted_k141_40898 | Not-determined |
| S9_extracted_k141_82204 | Not-determined |
| S9_extracted_k141_59366 | Not-determined |
| S9_extracted_k141_70633 | Not-determined |
| S9_extracted_k141_82207 | Not-determined |
| S9_extracted_k141_14066 | Not-determined |
| S9_extracted_k141_89563 | Low-quality |
| S9_extracted_k141_6733 | Not-determined |
| S9_extracted_k141_33245 | Not-determined |
| S9_extracted_k141_63372 | Not-determined |
| S9_extracted_k141_10390 | Not-determined |
| S9_extracted_k141_89566 | Not-determined |
| S9_extracted_k141_40903 | Not-determined |
| S9_extracted_k141_74478 | Low-quality |
| S9_extracted_k141_74479 | Not-determined |
| S9_extracted_k141_37154 | Low-quality |
| S9_extracted_k141_78023 | Not-determined |
| S9_extracted_k141_85953 | Not-determined |
| S9_extracted_k141_74481 | Not-determined |
| S9_extracted_k141_63376 | Low-quality |
| S9_extracted_k141_70636 | Not-determined |
| S9_extracted_k141_82217 | Not-determined |
| S9_extracted_k141_18227 | Not-determined |
| S9_extracted_k141_14074 | Not-determined |
| S9_extracted_k141_85955 | Not-determined |
| S9_extracted_k141_55936 | Low-quality |
| S9_extracted_k141_89572 | Low-quality |
| S9_extracted_k141_2896 | Not-determined |
| S9_extracted_k141_10402 | Not-determined |
| S9_extracted_k141_55939 | Not-determined |
| S9_extracted_k141_10404 | Not-determined |
| S9_extracted_k141_2903 | Not-determined |
| S9_extracted_k141_70645 | Not-determined |
| S9_extracted_k141_18237 | Not-determined |
| S9_extracted_k141_70647 | Not-determined |
| S9_extracted_k141_89575 | Not-determined |
| S9_extracted_k141_89577 | Not-determined |
| S9_extracted_k141_70653 | Not-determined |
| S9_extracted_k141_78031 | Not-determined |
| S9_extracted_k141_2906 | Not-determined |
| S9_extracted_k141_78033 | Not-determined |
| S9_extracted_k141_55945 | Not-determined |
| S9_extracted_k141_55946 | Not-determined |
| S9_extracted_k141_82237 | Not-determined |
| S9_extracted_k141_63388 | Low-quality |
| S9_extracted_k141_70657 | Not-determined |
| S9_extracted_k141_78037 | Low-quality |
| S9_extracted_k141_2911 | Not-determined |
| S9_extracted_k141_44726 | Low-quality |
| S9_extracted_k141_66896 | Not-determined |
| S9_extracted_k141_18247 | Not-determined |
| S9_extracted_k141_44732 | Not-determined |
| S9_extracted_k141_85975 | Not-determined |
| S9_extracted_k141_18253 | Not-determined |
| S9_extracted_k141_55955 | Not-determined |
| S9_extracted_k141_37173 | Low-quality |
| S9_extracted_k141_51248 | Not-determined |
| S9_extracted_k141_85978 | Not-determined |
| S9_extracted_k141_2916 | Not-determined |
| S9_extracted_k141_18255 | Not-determined |
| S9_extracted_k141_44734 | Low-quality |
| S9_extracted_k141_70664 | Not-determined |
| S9_extracted_k141_44735 | Not-determined |
| S9_extracted_k141_55959 | Not-determined |
| S9_extracted_k141_51253 | Low-quality |
| S9_extracted_k141_89588 | Not-determined |
| S9_extracted_k141_44736 | Not-determined |
| S9_extracted_k141_85986 | Not-determined |
| S9_extracted_k141_29601 | Not-determined |
| S9_extracted_k141_63404 | Not-determined |
| S9_extracted_k141_82248 | Not-determined |
| S9_extracted_k141_18260 | Not-determined |
| S9_extracted_k141_55967 | Not-determined |
| S9_extracted_k141_2920 | Low-quality |
| S9_extracted_k141_89592 | Not-determined |
| S9_extracted_k141_55969 | Not-determined |
| S9_extracted_k141_82251 | Not-determined |
| S9_extracted_k141_82254 | Not-determined |
| S9_extracted_k141_2925 | Not-determined |
| S9_extracted_k141_51257 | Low-quality |
| S9_extracted_k141_18267 | Not-determined |
| S9_extracted_k141_18268 | Not-determined |
| S9_extracted_k141_63412 | Not-determined |
| S9_extracted_k141_85994 | Not-determined |
| S9_extracted_k141_40929 | Not-determined |
| S9_extracted_k141_33283 | Not-determined |
| S9_extracted_k141_85996 | Not-determined |
| S9_extracted_k141_48045 | Not-determined |
| S9_extracted_k141_29612 | Not-determined |
| S9_extracted_k141_25615 | Not-determined |
| S9_extracted_k141_6761 | Not-determined |
| S9_extracted_k141_89596 | Not-determined |
| S9_extracted_k141_40935 | Not-determined |
| S9_extracted_k141_63415 | Not-determined |
| S9_extracted_k141_74507 | Not-determined |
| S9_extracted_k141_66910 | Not-determined |
| S9_extracted_k141_25619 | Not-determined |
| S9_extracted_k141_82260 | Not-determined |
| S9_extracted_k141_86005 | Not-determined |
| S9_extracted_k141_25621 | Not-determined |
| S9_extracted_k141_25622 | Not-determined |
| S9_extracted_k141_48053 | Not-determined |
| S9_extracted_k141_2932 | Not-determined |
| S9_extracted_k141_89603 | Low-quality |
| S9_extracted_k141_6767 | Low-quality |
| S9_extracted_k141_89606 | Not-determined |
| S9_extracted_k141_89607 | Not-determined |
| S9_extracted_k141_29628 | Not-determined |
| S9_extracted_k141_89609 | Not-determined |
| S9_extracted_k141_18289 | Not-determined |
| S9_extracted_k141_18290 | Not-determined |
| S9_extracted_k141_89611 | Not-determined |
| S9_extracted_k141_59397 | Low-quality |
| S9_extracted_k141_74514 | Not-determined |
| S9_extracted_k141_29632 | Not-determined |
| S9_extracted_k141_21975 | Low-quality |
| S9_extracted_k141_2945 | Low-quality |
| S9_extracted_k141_66916 | Low-quality |
| S9_extracted_k141_25633 | Not-determined |
| S9_extracted_k141_33302 | Low-quality |
| S9_extracted_k141_29640 | Low-quality |
| S9_extracted_k141_78071 | Not-determined |
| S9_extracted_k141_6778 | Not-determined |
| S9_extracted_k141_25637 | Not-determined |
| S9_extracted_k141_66922 | Low-quality |
| S9_extracted_k141_78076 | Not-determined |
| S9_extracted_k141_29647 | Not-determined |
| S9_extracted_k141_29650 | Not-determined |
| S9_extracted_k141_78079 | Not-determined |
| S9_extracted_k141_78080 | Not-determined |
| S9_extracted_k141_86028 | Not-determined |
| S9_extracted_k141_86031 | Not-determined |
| S9_extracted_k141_63424 | Not-determined |
| S9_extracted_k141_74530 | Not-determined |
| S9_extracted_k141_44780 | Low-quality |
| S9_extracted_k141_74533 | Not-determined |
| S9_extracted_k141_63429 | Not-determined |
| S9_extracted_k141_44783 | Not-determined |
| S9_extracted_k141_2959 | Not-determined |
| S9_extracted_k141_33322 | Not-determined |
| S9_extracted_k141_10425 | Not-determined |
| S9_extracted_k141_74541 | Not-determined |
| S9_extracted_k141_29661 | Not-determined |
| S9_extracted_k141_25650 | Low-quality |
| S9_extracted_k141_78090 | Not-determined |
| S9_extracted_k141_25651 | Not-determined |
| S9_extracted_k141_74547 | Not-determined |
| S9_extracted_k141_40952 | Not-determined |
| S9_extracted_k141_63434 | Not-determined |
| S9_extracted_k141_56011 | Not-determined |
| S9_extracted_k141_56012 | Not-determined |
| S9_extracted_k141_59419 | Not-determined |
| S9_extracted_k141_89614 | Low-quality |
| S9_extracted_k141_2966 | Not-determined |
| S9_extracted_k141_10437 | Not-determined |
| S9_extracted_k141_33323 | Not-determined |
| S9_extracted_k141_10438 | Not-determined |
| S9_extracted_k141_63443 | Not-determined |
| S9_extracted_k141_59423 | Not-determined |
| S9_extracted_k141_37208 | Not-determined |
| S9_extracted_k141_56019 | Not-determined |
| S9_extracted_k141_74552 | Not-determined |
| S9_extracted_k141_29677 | Not-determined |
| S9_extracted_k141_48089 | Not-determined |
| S9_extracted_k141_78098 | Not-determined |
| S9_extracted_k141_48090 | Not-determined |
| S9_extracted_k141_29679 | Low-quality |

| | |
|---|---|
| S9_extracted_k141_51282 | Not-determined |
| S9_extracted_k141_29682 | Not-determined |
| S9_extracted_k141_18315 | Not-determined |
| S9_extracted_k141_10446 | Not-determined |
| S9_extracted_k141_44804 | Not-determined |
| S9_extracted_k141_74558 | Low-quality |
| S9_extracted_k141_86041 | Low-quality |
| S9_extracted_k141_6789 | Not-determined |
| S9_extracted_k141_25667 | Not-determined |
| S9_extracted_k141_63449 | Not-determined |
| S9_extracted_k141_63452 | Not-determined |
| S9_extracted_k141_48096 | Not-determined |
| S9_extracted_k141_86048 | Low-quality |
| S9_extracted_k141_51293 | Low-quality |
| S9_extracted_k141_74566 | Not-determined |
| S9_extracted_k141_63456 | Not-determined |
| S9_extracted_k141_63457 | Not-determined |
| S9_extracted_k141_29695 | Not-determined |
| S9_extracted_k141_48099 | Not-determined |
| S9_extracted_k141_89637 | Not-determined |
| S9_extracted_k141_51296 | Not-determined |
| S9_extracted_k141_51297 | Not-determined |
| S9_extracted_k141_37228 | Low-quality |
| S9_extracted_k141_86052 | Low-quality |
| S9_extracted_k141_51298 | Low-quality |
| S9_extracted_k141_25679 | Not-determined |
| S9_extracted_k141_70681 | Not-determined |
| S9_extracted_k141_10457 | Low-quality |
| S9_extracted_k141_74570 | Not-determined |
| S9_extracted_k141_86055 | Low-quality |
| S9_extracted_k141_10459 | Not-determined |
| S9_extracted_k141_74573 | Low-quality |
| S9_extracted_k141_74574 | Not-determined |
| S9_extracted_k141_29701 | Low-quality |
| S9_extracted_k141_25685 | Low-quality |
| S9_extracted_k141_48100 | Low-quality |
| S9_extracted_k141_59456 | Not-determined |
| S9_extracted_k141_37234 | Not-determined |
| S9_extracted_k141_86059 | Low-quality |
| S9_extracted_k141_37235 | Not-determined |
| S9_extracted_k141_2978 | Not-determined |
| S9_extracted_k141_59459 | Not-determined |
| S9_extracted_k141_18341 | Not-determined |
| S9_extracted_k141_78126 | Not-determined |
| S9_extracted_k141_18342 | Not-determined |
| S9_extracted_k141_66976 | Not-determined |
| S9_extracted_k141_78128 | Not-determined |
| S9_extracted_k141_48106 | Low-quality |
| S9_extracted_k141_63462 | Not-determined |
| S9_extracted_k141_51317 | Not-determined |
| S9_extracted_k141_25695 | Not-determined |
| S9_extracted_k141_29713 | Not-determined |
| S9_extracted_k141_25696 | Not-determined |
| S9_extracted_k141_29715 | Not-determined |
| S9_extracted_k141_59468 | Not-determined |
| S9_extracted_k141_18346 | Not-determined |
| S9_extracted_k141_51324 | Not-determined |
| S9_extracted_k141_6818 | Not-determined |
| S9_extracted_k141_86070 | Not-determined |
| S9_extracted_k141_40979 | Not-determined |
| S9_extracted_k141_40980 | Not-determined |
| S9_extracted_k141_18350 | Not-determined |
| S9_extracted_k141_18351 | Not-determined |
| S9_extracted_k141_74595 | Not-determined |
| S9_extracted_k141_18353 | Not-determined |
| S9_extracted_k141_51331 | Not-determined |
| S9_extracted_k141_89670 | Not-determined |
| S9_extracted_k141_21993 | Not-determined |
| S9_extracted_k141_21994 | Not-determined |
| S9_extracted_k141_37250 | Low-quality |
| S9_extracted_k141_89673 | Low-quality |
| S9_extracted_k141_51338 | Not-determined |
| S9_extracted_k141_40984 | Not-determined |
| S9_extracted_k141_51339 | Not-determined |
| S9_extracted_k141_59485 | Low-quality |
| S9_extracted_k141_18360 | Not-determined |
| S9_extracted_k141_3002 | Not-determined |
| S9_extracted_k141_33373 | Not-determined |
| S9_extracted_k141_59490 | Not-determined |
| S9_extracted_k141_78137 | Low-quality |
| S9_extracted_k141_78142 | Not-determined |
| S9_extracted_k141_51344 | Low-quality |
| S9_extracted_k141_89684 | Not-determined |
| S9_extracted_k141_86089 | Low-quality |
| S9_extracted_k141_33379 | Not-determined |
| S9_extracted_k141_3012 | Not-determined |
| S9_extracted_k141_6846 | Not-determined |
| S9_extracted_k141_59499 | Not-determined |
| S9_extracted_k141_25721 | Not-determined |
| S9_extracted_k141_89688 | Not-determined |
| S9_extracted_k141_59501 | Not-determined |
| S9_extracted_k141_25723 | Not-determined |
| S9_extracted_k141_25726 | Not-determined |
| S9_extracted_k141_51350 | Not-determined |
| S9_extracted_k141_59505 | Not-determined |
| S9_extracted_k141_82302 | Not-determined |
| S9_extracted_k141_56057 | Not-determined |
| S9_extracted_k141_48149 | Not-determined |
| S9_extracted_k141_37270 | Not-determined |
| S9_extracted_k141_22019 | Not-determined |
| S9_extracted_k141_66983 | Not-determined |
| S9_extracted_k141_3017 | Low-quality |
| S9_extracted_k141_37272 | Not-determined |
| S9_extracted_k141_25734 | Not-determined |
| S9_extracted_k141_86109 | Low-quality |
| S9_extracted_k141_82305 | Not-determined |
| S9_extracted_k141_66986 | Not-determined |
| S9_extracted_k141_51356 | Low-quality |
| S9_extracted_k141_78152 | Not-determined |
| S9_extracted_k141_78153 | Not-determined |
| S9_extracted_k141_10488 | Low-quality |
| S9_extracted_k141_78156 | Not-determined |
| S9_extracted_k141_40999 | Not-determined |
| S9_extracted_k141_48162 | Not-determined |
| S9_extracted_k141_3021 | Low-quality |
| S9_extracted_k141_70719 | Not-determined |
| S9_extracted_k141_82314 | Not-determined |
| S9_extracted_k141_56066 | Not-determined |
| S9_extracted_k141_82315 | Low-quality |
| S9_extracted_k141_29741 | Not-determined |
| S9_extracted_k141_56068 | Not-determined |
| S9_extracted_k141_18391 | Not-determined |
| S9_extracted_k141_56070 | Not-determined |
| S9_extracted_k141_18393 | Not-determined |
| S9_extracted_k141_33404 | Not-determined |
| S9_extracted_k141_82320 | Low-quality |
| S9_extracted_k141_22034 | Not-determined |
| S9_extracted_k141_6864 | Not-determined |
| S9_extracted_k141_37280 | Not-determined |
| S9_extracted_k141_6867 | Not-determined |
| S9_extracted_k141_41004 | Not-determined |
| S9_extracted_k141_78168 | Not-determined |
| S9_extracted_k141_59528 | Low-quality |
| S9_extracted_k141_89703 | Not-determined |
| S9_extracted_k141_22037 | Not-determined |
| S9_extracted_k141_3032 | Not-determined |
| S9_extracted_k141_82326 | Not-determined |
| S9_extracted_k141_86120 | Not-determined |
| S9_extracted_k141_22039 | Not-determined |
| S9_extracted_k141_37287 | Not-determined |
| S9_extracted_k141_78171 | Not-determined |
| S9_extracted_k141_82332 | Not-determined |
| S9_extracted_k141_70734 | Not-determined |
| S9_extracted_k141_48171 | Not-determined |
| S9_extracted_k141_18401 | Not-determined |
| S9_extracted_k141_10506 | Low-quality |
| S9_extracted_k141_44847 | Not-determined |
| S9_extracted_k141_89713 | Not-determined |
| S9_extracted_k141_82335 | Not-determined |
| S9_extracted_k141_44850 | Not-determined |
| S9_extracted_k141_10515 | Not-determined |
| S9_extracted_k141_10517 | Not-determined |
| S9_extracted_k141_66996 | Not-determined |
| S9_extracted_k141_86131 | Not-determined |
| S9_extracted_k141_66999 | Low-quality |
| S9_extracted_k141_22046 | Not-determined |
| S9_extracted_k141_74637 | Not-determined |
| S9_extracted_k141_82343 | Not-determined |
| S9_extracted_k141_37298 | Not-determined |
| S9_extracted_k141_78181 | Not-determined |
| S9_extracted_k141_18416 | Not-determined |
| S9_extracted_k141_89719 | Not-determined |
| S9_extracted_k141_25767 | Not-determined |
| S9_extracted_k141_10524 | Not-determined |
| S9_extracted_k141_89721 | Low-quality |
| S9_extracted_k141_78183 | Not-determined |
| S9_extracted_k141_10527 | Not-determined |
| S9_extracted_k141_44857 | Not-determined |
| S9_extracted_k141_6883 | Low-quality |
| S9_extracted_k141_22052 | Low-quality |
| S9_extracted_k141_25772 | Not-determined |
| S9_extracted_k141_67009 | Low-quality |
| S9_extracted_k141_67010 | Low-quality |
| S9_extracted_k141_86134 | Low-quality |
| S9_extracted_k141_37306 | Not-determined |
| S9_extracted_k141_56098 | Not-determined |
| S9_extracted_k141_29758 | Not-determined |
| S9_extracted_k141_29760 | Not-determined |
| S9_extracted_k141_44866 | Low-quality |
| S9_extracted_k141_86138 | Not-determined |
| S9_extracted_k141_89725 | Low-quality |
| S9_extracted_k141_44869 | Not-determined |
| S9_extracted_k141_10536 | Not-determined |
| S9_extracted_k141_44873 | Not-determined |
| S9_extracted_k141_56104 | Not-determined |
| S9_extracted_k141_18425 | Not-determined |
| S9_extracted_k141_25790 | Not-determined |
| S9_extracted_k141_70756 | Not-determined |
| S9_extracted_k141_33430 | Not-determined |
| S9_extracted_k141_41029 | Not-determined |
| S9_extracted_k141_56109 | Not-determined |
| S9_extracted_k141_63486 | Not-determined |
| S9_extracted_k141_63487 | Not-determined |
| S9_extracted_k141_63489 | Not-determined |
| S9_extracted_k141_67031 | Not-determined |
| S9_extracted_k141_10539 | Not-determined |
| S9_extracted_k141_70761 | Not-determined |
| S9_extracted_k141_67034 | Not-determined |
| S9_extracted_k141_56115 | Not-determined |
| S9_extracted_k141_18431 | Low-quality |
| S9_extracted_k141_56117 | Not-determined |
| S9_extracted_k141_59553 | Not-determined |
| S9_extracted_k141_89742 | Low-quality |
| S9_extracted_k141_6907 | Not-determined |
| S9_extracted_k141_29771 | Not-determined |
| S9_extracted_k141_29772 | Not-determined |
| S9_extracted_k141_89746 | Low-quality |
| S9_extracted_k141_89747 | Not-determined |
| S9_extracted_k141_70768 | Not-determined |
| S9_extracted_k141_25812 | Not-determined |
| S9_extracted_k141_3086 | Not-determined |
| S9_extracted_k141_56123 | Not-determined |
| S9_extracted_k141_48190 | Not-determined |
| S9_extracted_k141_6917 | Not-determined |
| S9_extracted_k141_44891 | Low-quality |
| S9_extracted_k141_82379 | Not-determined |
| S9_extracted_k141_44893 | Not-determined |
| S9_extracted_k141_10543 | Not-determined |
| S9_extracted_k141_22075 | Low-quality |
| S9_extracted_k141_3093 | Not-determined |
| S9_extracted_k141_63504 | Not-determined |
| S9_extracted_k141_70774 | Low-quality |
| S9_extracted_k141_63505 | Not-determined |
| S9_extracted_k141_89754 | Not-determined |
| S9_extracted_k141_10547 | Not-determined |
| S9_extracted_k141_63506 | Not-determined |
| S9_extracted_k141_56130 | Low-quality |
| S9_extracted_k141_6921 | Not-determined |
| S9_extracted_k141_63507 | Low-quality |
| S9_extracted_k141_6923 | Not-determined |
| S9_extracted_k141_22082 | Not-determined |
| S9_extracted_k141_59573 | Not-determined |
| S9_extracted_k141_44904 | Not-determined |
| S9_extracted_k141_29788 | Not-determined |
| S9_extracted_k141_18442 | Not-determined |
| S9_extracted_k141_51378 | Not-determined |
| S9_extracted_k141_59575 | Not-determined |
| S9_extracted_k141_33448 | Not-determined |
| S9_extracted_k141_44909 | Not-determined |
| S9_extracted_k141_74678 | Not-determined |
| S9_extracted_k141_10557 | Low-quality |
| S9_extracted_k141_70789 | Not-determined |
| S9_extracted_k141_6927 | Low-quality |
| S9_extracted_k141_37339 | Low-quality |
| S9_extracted_k141_48204 | Not-determined |
| S9_extracted_k141_6929 | Not-determined |
| S9_extracted_k141_18445 | Not-determined |
| S9_extracted_k141_18449 | Not-determined |
| S9_extracted_k141_18451 | Not-determined |
| S9_extracted_k141_18452 | Not-determined |
| S9_extracted_k141_74685 | Low-quality |
| S9_extracted_k141_48208 | Not-determined |
| S9_extracted_k141_18455 | Not-determined |
| S9_extracted_k141_74686 | Not-determined |
| S9_extracted_k141_37347 | Not-determined |
| S9_extracted_k141_48211 | Not-determined |
| S9_extracted_k141_22095 | Not-determined |
| S9_extracted_k141_37348 | Not-determined |
| S9_extracted_k141_33460 | Not-determined |
| S9_extracted_k141_10564 | Not-determined |
| S9_extracted_k141_56146 | Low-quality |
| S9_extracted_k141_63516 | Not-determined |
| S9_extracted_k141_33462 | Not-determined |
| S9_extracted_k141_59584 | Not-determined |
| S9_extracted_k141_63518 | Not-determined |
| S9_extracted_k141_74689 | Not-determined |
| S9_extracted_k141_56149 | Not-determined |
| S9_extracted_k141_70805 | Not-determined |
| S9_extracted_k141_82399 | Not-determined |
| S9_extracted_k141_56150 | Not-determined |
| S9_extracted_k141_70810 | Not-determined |
| S9_extracted_k141_25827 | Not-determined |
| S9_extracted_k141_82405 | Not-determined |
| S9_extracted_k141_86187 | Not-determined |
| S9_extracted_k141_82406 | Not-determined |
| S9_extracted_k141_29808 | Not-determined |
| S9_extracted_k141_78215 | Not-determined |
| S9_extracted_k141_41072 | Not-determined |
| S9_extracted_k141_37353 | Not-determined |
| S9_extracted_k141_25830 | Not-determined |
| S9_extracted_k141_59592 | Not-determined |
| S9_extracted_k141_89775 | Not-determined |
| S9_extracted_k141_59593 | Not-determined |
| S9_extracted_k141_33472 | Low-quality |
| S9_extracted_k141_44920 | Low-quality |
| S9_extracted_k141_44922 | Not-determined |
| S9_extracted_k141_74701 | Not-determined |
| S9_extracted_k141_29812 | Not-determined |
| S9_extracted_k141_59596 | Not-determined |
| S9_extracted_k141_48219 | Not-determined |
| S9_extracted_k141_37357 | Not-determined |
| S9_extracted_k141_37358 | Not-determined |
| S9_extracted_k141_10574 | Not-determined |
| S9_extracted_k141_82413 | Not-determined |
| S9_extracted_k141_18470 | Not-determined |
| S9_extracted_k141_10575 | Not-determined |
| S9_extracted_k141_6952 | Not-determined |
| S9_extracted_k141_37361 | Not-determined |
| S9_extracted_k141_63531 | Not-determined |
| S9_extracted_k141_56159 | Low-quality |
| S9_extracted_k141_51403 | Not-determined |
| S9_extracted_k141_37363 | Not-determined |
| S9_extracted_k141_59599 | Low-quality |
| S9_extracted_k141_44928 | Not-determined |
| S9_extracted_k141_89779 | Low-quality |
| S9_extracted_k141_51407 | Not-determined |
| S9_extracted_k141_70818 | Not-determined |
| S9_extracted_k141_48224 | Not-determined |
| S9_extracted_k141_6957 | Not-determined |
| S9_extracted_k141_78227 | Not-determined |
| S9_extracted_k141_44934 | Low-quality |
| S9_extracted_k141_41089 | Low-quality |
| S9_extracted_k141_37369 | Not-determined |
| S9_extracted_k141_10588 | Not-determined |
| S9_extracted_k141_25838 | Not-determined |
| S9_extracted_k141_14105 | Not-determined |
| S9_extracted_k141_89786 | Not-determined |
| S9_extracted_k141_29820 | Not-determined |
| S9_extracted_k141_89787 | Not-determined |
| S9_extracted_k141_37375 | Not-determined |
| S9_extracted_k141_41096 | Not-determined |
| S9_extracted_k141_89789 | Not-determined |
| S9_extracted_k141_37377 | Not-determined |
| S9_extracted_k141_67097 | Not-determined |
| S9_extracted_k141_44942 | Not-determined |
| S9_extracted_k141_14114 | Low-quality |
| S9_extracted_k141_29827 | Not-determined |
| S9_extracted_k141_3113 | Not-determined |
| S9_extracted_k141_59618 | Not-determined |
| S9_extracted_k141_25848 | Not-determined |
| S9_extracted_k141_59620 | Not-determined |
| S9_extracted_k141_29831 | Not-determined |
| S9_extracted_k141_86190 | Low-quality |
| S9_extracted_k141_48239 | Low-quality |
| S9_extracted_k141_33489 | Not-determined |
| S9_extracted_k141_74726 | Not-determined |
| S9_extracted_k141_63550 | Low-quality |
| S9_extracted_k141_70837 | Not-determined |
| S9_extracted_k141_6969 | Not-determined |
| S9_extracted_k141_37385 | Low-quality |
| S9_extracted_k141_86194 | Not-determined |
| S9_extracted_k141_25851 | Low-quality |
| S9_extracted_k141_6972 | Not-determined |
| S9_extracted_k141_10606 | Not-determined |
| S9_extracted_k141_67104 | Not-determined |
| S9_extracted_k141_56165 | Low-quality |
| S9_extracted_k141_51431 | Not-determined |
| S9_extracted_k141_63556 | Low-quality |
| S9_extracted_k141_56167 | Not-determined |
| S9_extracted_k141_56169 | Not-determined |
| S9_extracted_k141_63559 | Not-determined |
| S9_extracted_k141_74734 | Not-determined |
| S9_extracted_k141_74738 | Not-determined |
| S9_extracted_k141_78245 | Not-determined |
| S9_extracted_k141_29839 | Low-quality |
| S9_extracted_k141_74739 | Not-determined |
| S9_extracted_k141_70842 | Not-determined |
| S9_extracted_k141_18494 | Not-determined |
| S9_extracted_k141_6979 | Not-determined |
| S9_extracted_k141_74743 | Not-determined |
| S9_extracted_k141_44950 | Not-determined |
| S9_extracted_k141_63562 | Not-determined |
| S9_extracted_k141_22113 | Not-determined |
| S9_extracted_k141_67115 | Not-determined |
| S9_extracted_k141_78248 | Not-determined |
| S9_extracted_k141_33498 | Low-quality |
| S9_extracted_k141_29847 | Not-determined |
| S9_extracted_k141_86213 | Low-quality |
| S9_extracted_k141_78251 | Not-determined |
| S9_extracted_k141_67119 | Not-determined |
| S9_extracted_k141_25866 | Low-quality |
| S9_extracted_k141_37397 | Not-determined |
| S9_extracted_k141_33502 | Low-quality |
| S9_extracted_k141_25869 | Low-quality |
| S9_extracted_k141_70852 | Not-determined |
| S9_extracted_k141_59639 | Not-determined |
| S9_extracted_k141_86217 | Not-determined |
| S9_extracted_k141_74754 | Not-determined |
| S9_extracted_k141_74755 | Low-quality |
| S9_extracted_k141_44961 | Not-determined |
| S9_extracted_k141_37404 | Low-quality |
| S9_extracted_k141_10635 | Not-determined |
| S9_extracted_k141_22115 | Not-determined |
| S9_extracted_k141_67129 | Not-determined |
| S9_extracted_k141_18509 | Low-quality |
| S9_extracted_k141_29863 | Not-determined |
| S9_extracted_k141_18510 | Not-determined |
| S9_extracted_k141_89821 | Not-determined |
| S9_extracted_k141_22119 | Not-determined |
| S9_extracted_k141_89823 | Not-determined |
| S9_extracted_k141_7000 | Not-determined |
| S9_extracted_k141_63570 | Not-determined |
| S9_extracted_k141_29867 | Not-determined |
| S9_extracted_k141_74768 | Not-determined |
| S9_extracted_k141_70869 | Not-determined |
| S9_extracted_k141_78266 | Not-determined |
| S9_extracted_k141_86228 | Not-determined |
| S9_extracted_k141_29872 | Not-determined |
| S9_extracted_k141_89829 | Not-determined |
| S9_extracted_k141_7005 | Low-quality |
| S9_extracted_k141_41128 | Not-determined |
| S9_extracted_k141_89832 | Not-determined |
| S9_extracted_k141_59660 | Not-determined |
| S9_extracted_k141_44979 | Not-determined |
| S9_extracted_k141_18514 | Low-quality |
| S9_extracted_k141_74775 | Not-determined |
| S9_extracted_k141_37418 | Not-determined |
| S9_extracted_k141_18517 | Not-determined |
| S9_extracted_k141_37419 | Not-determined |
| S9_extracted_k141_89839 | Not-determined |
| S9_extracted_k141_33514 | Not-determined |
| S9_extracted_k141_89840 | Not-determined |
| S9_extracted_k141_70881 | Low-quality |
| S9_extracted_k141_67145 | Low-quality |
| S9_extracted_k141_14123 | Not-determined |
| S9_extracted_k141_25888 | Not-determined |
| S9_extracted_k141_67151 | Not-determined |
| S9_extracted_k141_25894 | Not-determined |
| S9_extracted_k141_25895 | Low-quality |
| S9_extracted_k141_44985 | Not-determined |
| S9_extracted_k141_44988 | Not-determined |
| S9_extracted_k141_89853 | Not-determined |
| S9_extracted_k141_44989 | Not-determined |
| S9_extracted_k141_67161 | Not-determined |
| S9_extracted_k141_44992 | Not-determined |
| S9_extracted_k141_18540 | Not-determined |
| S9_extracted_k141_82415 | Not-determined |
| S9_extracted_k141_82416 | Not-determined |
| S9_extracted_k141_25901 | Low-quality |
| S9_extracted_k141_10662 | Not-determined |
| S9_extracted_k141_48241 | Not-determined |
| S9_extracted_k141_67167 | Not-determined |
| S9_extracted_k141_82418 | Low-quality |
| S9_extracted_k141_25902 | Not-determined |
| S9_extracted_k141_18544 | Low-quality |
| S9_extracted_k141_14135 | Not-determined |
| S9_extracted_k141_33535 | Not-determined |
| S9_extracted_k141_70900 | Not-determined |
| S9_extracted_k141_59680 | Not-determined |
| S9_extracted_k141_41143 | Not-determined |
| S9_extracted_k141_7035 | Not-determined |
| S9_extracted_k141_10666 | Not-determined |
| S9_extracted_k141_41144 | Low-quality |
| S9_extracted_k141_70903 | Not-determined |
| S9_extracted_k141_89866 | Not-determined |
| S9_extracted_k141_56210 | Not-determined |
| S9_extracted_k141_29910 | Not-determined |
| S9_extracted_k141_33543 | Not-determined |
| S9_extracted_k141_7038 | Not-determined |
| S9_extracted_k141_33546 | Not-determined |
| S9_extracted_k141_25910 | Not-determined |
| S9_extracted_k141_48248 | Not-determined |
| S9_extracted_k141_48249 | Not-determined |
| S9_extracted_k141_74800 | Low-quality |
| S9_extracted_k141_45010 | Low-quality |
| S9_extracted_k141_18556 | Not-determined |
| S9_extracted_k141_25915 | Not-determined |
| S9_extracted_k141_82428 | Low-quality |
| S9_extracted_k141_7046 | Not-determined |
| S9_extracted_k141_7047 | Not-determined |
| S9_extracted_k141_7048 | Low-quality |
| S9_extracted_k141_33551 | Not-determined |
| S9_extracted_k141_45015 | Not-determined |
| S9_extracted_k141_7049 | Low-quality |
| S9_extracted_k141_70911 | Not-determined |
| S9_extracted_k141_56223 | Not-determined |
| S9_extracted_k141_70912 | Not-determined |
| S9_extracted_k141_78288 | Not-determined |
| S9_extracted_k141_67189 | Not-determined |
| S9_extracted_k141_67190 | Not-determined |
| S9_extracted_k141_29923 | Not-determined |
| S9_extracted_k141_89882 | Low-quality |
| S9_extracted_k141_59691 | Not-determined |
| S9_extracted_k141_33555 | Not-determined |
| S9_extracted_k141_59692 | Not-determined |
| S9_extracted_k141_82436 | Low-quality |
| S9_extracted_k141_45018 | Not-determined |
| S9_extracted_k141_89883 | Not-determined |
| S9_extracted_k141_14142 | Not-determined |
| S9_extracted_k141_70918 | Low-quality |
| S9_extracted_k141_59693 | Low-quality |
| S9_extracted_k141_45022 | Not-determined |
| S9_extracted_k141_70919 | Not-determined |
| S9_extracted_k141_59695 | Not-determined |
| S9_extracted_k141_29932 | Not-determined |
| S9_extracted_k141_89892 | Not-determined |
| S9_extracted_k141_70924 | Not-determined |
| S9_extracted_k141_56230 | Not-determined |
| S9_extracted_k141_89894 | Low-quality |
| S9_extracted_k141_67196 | Not-determined |
| S9_extracted_k141_74824 | Not-determined |
| S9_extracted_k141_70925 | Low-quality |
| S9_extracted_k141_82445 | Not-determined |
| S9_extracted_k141_33567 | Low-quality |
| S9_extracted_k141_29939 | Low-quality |
| S9_extracted_k141_45028 | Not-determined |
| S9_extracted_k141_70928 | Not-determined |
| S9_extracted_k141_41169 | Not-determined |
| S9_extracted_k141_78301 | Not-determined |
| S9_extracted_k141_29942 | Not-determined |
| S9_extracted_k141_14151 | Not-determined |
| S9_extracted_k141_74828 | Not-determined |
| S9_extracted_k141_33576 | Not-determined |
| S9_extracted_k141_7076 | Not-determined |
| S9_extracted_k141_82449 | Not-determined |
| S9_extracted_k141_33578 | Low-quality |
| S9_extracted_k141_48274 | Not-determined |
| S9_extracted_k141_89908 | Low-quality |
| S9_extracted_k141_89909 | Not-determined |
| S9_extracted_k141_89910 | Not-determined |
| S9_extracted_k141_86295 | Not-determined |
| S9_extracted_k141_78311 | Not-determined |
| S9_extracted_k141_82454 | Not-determined |
| S9_extracted_k141_59705 | Not-determined |
| S9_extracted_k141_70937 | Not-determined |
| S9_extracted_k141_74833 | Not-determined |
| S9_extracted_k141_45037 | Not-determined |
| S9_extracted_k141_7084 | Not-determined |
| S9_extracted_k141_29952 | Not-determined |
| S9_extracted_k141_25939 | Not-determined |
| S9_extracted_k141_29958 | Not-determined |
| S9_extracted_k141_45042 | Not-determined |
| S9_extracted_k141_18581 | Not-determined |
| S9_extracted_k141_86300 | Not-determined |
| S9_extracted_k141_59710 | Not-determined |
| S9_extracted_k141_29961 | Low-quality |
| S9_extracted_k141_22166 | Not-determined |
| S9_extracted_k141_25945 | Not-determined |
| S9_extracted_k141_14172 | Low-quality |
| S9_extracted_k141_29964 | Not-determined |
| S9_extracted_k141_86303 | Not-determined |
| S9_extracted_k141_74841 | Not-determined |
| S9_extracted_k141_48281 | Not-determined |
| S9_extracted_k141_82468 | Not-determined |
| S9_extracted_k141_18592 | Not-determined |
| S9_extracted_k141_14179 | Not-determined |
| S10_vs_39822\|\|full | High-quality |
| S9_extracted_k141_41203 | Low-quality |
| S9_extracted_k141_56239 | Not-determined |
| S9_extracted_k141_14181 | Low-quality |
| S9_extracted_k141_10696 | Not-determined |
| S9_extracted_k141_45051 | Not-determined |
| S9_extracted_k141_14182 | Low-quality |
| S9_extracted_k141_3127 | Not-determined |
| S9_extracted_k141_70953 | Not-determined |
| S9_extracted_k141_18597 | Not-determined |
| S9_extracted_k141_10699 | Not-determined |
| S9_extracted_k141_78337 | Not-determined |
| S9_extracted_k141_45059 | Low-quality |
| S9_extracted_k141_70957 | Not-determined |
| S9_extracted_k141_14185 | Not-determined |
| S9_extracted_k141_45060 | Not-determined |
| S9_extracted_k141_29983 | Not-determined |
| S9_extracted_k141_78344 | Not-determined |
| S9_extracted_k141_78346 | Low-quality |
| S9_extracted_k141_3138 | Low-quality |
| S9_extracted_k141_25959 | Low-quality |
| S9_extracted_k141_56254 | Not-determined |
| S9_extracted_k141_10708 | Not-determined |
| S9_extracted_k141_56256 | Not-determined |
| S9_extracted_k141_10709 | Low-quality |
| S9_extracted_k141_82488 | Not-determined |
| S9_extracted_k141_33593 | Not-determined |
| S9_extracted_k141_18616 | Not-determined |
| S9_extracted_k141_3143 | Not-determined |
| S9_extracted_k141_89934 | Not-determined |
| S9_extracted_k141_78351 | Not-determined |
| S9_extracted_k141_45070 | Not-determined |
| S9_extracted_k141_10717 | Not-determined |
| S9_extracted_k141_45071 | Not-determined |
| S9_extracted_k141_33598 | Not-determined |
| S9_extracted_k141_74868 | Not-determined |
| S9_extracted_k141_48297 | Not-determined |
| S9_extracted_k141_74870 | Not-determined |
| S9_extracted_k141_25965 | Not-determined |
| S9_extracted_k141_59724 | Not-determined |
| S9_extracted_k141_3148 | Not-determined |
| S9_extracted_k141_18623 | Not-determined |
| S9_extracted_k141_30001 | Not-determined |
| S9_extracted_k141_10725 | Not-determined |
| S9_extracted_k141_37428 | Not-determined |
| S9_extracted_k141_82495 | Not-determined |
| S9_extracted_k141_78361 | Not-determined |
| S9_extracted_k141_14204 | Low-quality |
| S9_extracted_k141_41228 | Not-determined |
| S9_extracted_k141_14205 | Not-determined |
| S9_extracted_k141_74875 | Not-determined |
| S9_extracted_k141_48304 | Low-quality |
| S9_extracted_k141_78366 | Not-determined |

| | |
|---|---|
| S9_extracted_k141_10734 | Not-determined |
| S9_extracted_k141_22188 | Not-determined |
| S9_extracted_k141_70968 | Not-determined |
| S9_extracted_k141_25973 | Not-determined |
| S9_extracted_k141_74883 | Not-determined |
| S9_extracted_k141_25974 | Not-determined |
| S9_extracted_k141_22190 | Not-determined |
| S9_extracted_k141_7127 | Not-determined |
| S9_extracted_k141_33618 | Not-determined |
| S9_extracted_k141_30006 | Not-determined |
| S9_extracted_k141_30009 | Not-determined |
| S9_extracted_k141_22194 | Not-determined |
| S9_extracted_k141_18642 | Not-determined |
| S9_extracted_k141_3166 | Not-determined |
| S9_extracted_k141_70973 | Not-determined |
| S9_extracted_k141_22201 | Not-determined |
| S9_extracted_k141_56272 | Low-quality |
| S9_extracted_k141_56273 | Not-determined |
| S9_extracted_k141_70978 | Not-determined |
| S9_extracted_k141_25982 | Not-determined |
| S9_extracted_k141_70984 | Low-quality |
| S9_extracted_k141_74897 | Not-determined |
| S9_extracted_k141_74898 | Not-determined |
| S9_extracted_k141_41244 | Not-determined |
| S9_extracted_k141_10747 | Not-determined |
| S9_extracted_k141_78380 | Not-determined |
| S9_extracted_k141_33629 | Low-quality |
| S9_extracted_k141_25985 | Not-determined |
| S9_extracted_k141_56278 | Not-determined |
| S9_extracted_k141_41248 | Not-determined |
| S9_extracted_k141_33633 | Not-determined |
| S9_extracted_k141_78383 | Not-determined |
| S9_extracted_k141_74899 | Low-quality |
| S9_extracted_k141_82516 | Not-determined |
| S9_extracted_k141_78384 | Not-determined |
| S9_extracted_k141_22208 | Not-determined |
| S9_extracted_k141_45090 | Not-determined |
| S9_extracted_k141_30025 | Not-determined |
| S9_extracted_k141_41252 | Not-determined |
| S9_extracted_k141_30026 | Not-determined |
| S9_extracted_k141_3188 | Low-quality |
| S9_extracted_k141_18659 | Not-determined |
| S9_extracted_k141_82519 | Not-determined |
| S9_extracted_k141_59751 | Not-determined |
| S9_extracted_k141_59753 | Low-quality |
| S9_extracted_k141_18663 | Not-determined |
| S9_extracted_k141_78392 | Not-determined |
| S9_extracted_k141_78394 | Not-determined |
| S9_extracted_k141_41262 | Not-determined |
| S9_extracted_k141_59758 | Not-determined |
| S9_extracted_k141_7144 | Not-determined |
| S9_extracted_k141_30034 | Not-determined |
| S9_extracted_k141_22223 | Not-determined |
| S9_extracted_k141_30040 | Not-determined |
| S9_extracted_k141_41271 | Not-determined |
| S9_extracted_k141_26007 | Not-determined |
| S9_extracted_k141_86326 | Not-determined |
| S9_extracted_k141_45106 | Low-quality |
| S9_extracted_k141_78409 | Not-determined |
| S9_extracted_k141_30047 | Not-determined |
| S9_extracted_k141_74913 | Not-determined |
| S9_extracted_k141_56314 | Low-quality |
| S9_extracted_k141_30049 | Not-determined |
| S9_extracted_k141_74914 | Not-determined |
| S9_extracted_k141_30050 | Not-determined |
| S9_extracted_k141_45115 | Not-determined |
| S9_extracted_k141_45117 | Low-quality |
| S9_extracted_k141_26017 | Not-determined |
| S9_extracted_k141_86335 | Not-determined |
| S9_extracted_k141_3209 | Low-quality |
| S9_extracted_k141_26021 | Not-determined |
| S9_extracted_k141_59766 | Low-quality |
| S9_extracted_k141_30056 | Not-determined |
| S9_extracted_k141_48343 | Not-determined |
| S9_extracted_k141_86338 | Not-determined |
| S9_extracted_k141_71008 | Low-quality |
| S9_extracted_k141_51468 | Not-determined |
| S9_extracted_k141_78419 | Not-determined |
| S9_extracted_k141_26026 | Not-determined |
| S9_extracted_k141_78421 | Not-determined |
| S9_extracted_k141_30062 | Not-determined |
| S9_extracted_k141_26027 | Not-determined |
| S9_extracted_k141_74917 | Low-quality |
| S9_extracted_k141_22237 | Not-determined |
| S9_extracted_k141_56320 | Not-determined |
| S9_extracted_k141_18697 | Not-determined |
| S9_extracted_k141_56321 | Not-determined |
| S9_extracted_k141_56322 | Not-determined |
| S9_extracted_k141_86343 | Not-determined |
| S9_extracted_k141_26031 | Not-determined |
| S9_extracted_k141_78426 | Not-determined |
| S9_extracted_k141_45129 | Not-determined |
| S9_extracted_k141_56325 | Not-determined |
| S9_extracted_k141_78427 | Not-determined |
| S9_extracted_k141_45131 | Low-quality |
| S9_extracted_k141_26033 | Low-quality |
| S9_extracted_k141_51477 | Not-determined |
| S9_extracted_k141_26034 | Low-quality |
| S9_extracted_k141_59780 | Not-determined |
| S9_extracted_k141_30069 | Not-determined |
| S9_extracted_k141_3227 | Not-determined |
| S9_extracted_k141_18701 | Low-quality |
| S9_extracted_k141_74923 | Not-determined |
| S9_extracted_k141_3230 | Not-determined |
| S9_extracted_k141_59785 | Not-determined |
| S9_extracted_k141_74925 | Not-determined |
| S9_extracted_k141_22248 | Not-determined |
| S9_extracted_k141_59787 | Not-determined |
| S9_extracted_k141_22250 | Low-quality |
| S9_extracted_k141_78432 | Low-quality |
| S9_extracted_k141_59788 | Not-determined |
| S9_extracted_k141_22251 | Not-determined |
| S9_extracted_k141_26044 | Not-determined |
| S9_extracted_k141_18705 | Not-determined |
| S9_extracted_k141_7182 | Not-determined |
| S9_extracted_k141_59790 | Not-determined |
| S9_extracted_k141_78433 | Low-quality |
| S9_extracted_k141_37438 | Not-determined |
| S9_extracted_k141_37440 | Not-determined |
| S9_extracted_k141_14226 | Not-determined |
| S9_extracted_k141_22256 | Not-determined |
| S9_extracted_k141_78436 | Low-quality |
| S9_extracted_k141_22257 | Not-determined |
| S9_extracted_k141_10792 | Not-determined |
| S9_extracted_k141_30076 | Not-determined |
| S9_extracted_k141_45146 | Not-determined |
| S9_extracted_k141_22261 | Low-quality |
| S9_extracted_k141_67198 | Not-determined |
| S9_extracted_k141_14231 | Not-determined |
| S9_extracted_k141_14232 | Not-determined |
| S9_extracted_k141_37448 | Not-determined |
| S9_extracted_k141_10803 | Low-quality |
| S9_extracted_k141_51504 | Not-determined |
| S9_extracted_k141_51505 | Not-determined |
| S9_extracted_k141_59802 | Not-determined |
| S9_extracted_k141_67204 | Low-quality |
| S9_extracted_k141_22275 | Not-determined |
| S9_extracted_k141_41297 | Not-determined |
| S9_extracted_k141_74943 | Not-determined |
| S9_extracted_k141_22279 | Not-determined |
| S9_extracted_k141_33667 | Low-quality |
| S9_extracted_k141_51512 | Not-determined |
| S9_extracted_k141_18713 | Not-determined |
| S9_extracted_k141_22283 | Not-determined |
| S9_extracted_k141_89954 | Low-quality |
| S9_extracted_k141_18717 | Not-determined |
| S9_extracted_k141_51516 | Not-determined |
| S9_extracted_k141_7206 | Not-determined |
| S9_extracted_k141_22291 | Not-determined |
| S9_extracted_k141_67216 | Not-determined |
| S9_extracted_k141_30085 | Not-determined |
| S9_extracted_k141_71049 | Not-determined |
| S9_extracted_k141_51517 | Not-determined |
| S9_extracted_k141_18720 | Not-determined |
| S9_extracted_k141_18721 | Not-determined |
| S9_extracted_k141_59819 | Low-quality |
| S9_extracted_k141_18726 | Low-quality |
| S9_extracted_k141_22292 | Not-determined |
| S9_extracted_k141_33672 | Not-determined |
| S9_extracted_k141_78445 | Low-quality |
| S9_extracted_k141_18727 | Not-determined |
| S9_extracted_k141_7212 | Not-determined |
| S9_extracted_k141_30089 | Not-determined |
| S9_extracted_k141_10826 | Low-quality |
| S9_extracted_k141_3264 | Not-determined |
| S9_extracted_k141_45172 | Not-determined |
| S9_extracted_k141_78450 | Not-determined |
| S9_extracted_k141_67224 | Not-determined |
| S9_extracted_k141_45173 | Not-determined |
| S9_extracted_k141_30090 | Low-quality |
| S9_extracted_k141_30092 | Not-determined |
| S9_extracted_k141_71054 | Low-quality |
| S9_extracted_k141_67228 | Not-determined |
| S9_extracted_k141_74967 | Not-determined |
| S9_extracted_k141_45180 | Not-determined |
| S9_extracted_k141_30100 | Not-determined |
| S9_extracted_k141_45184 | Not-determined |
| S9_extracted_k141_71062 | Low-quality |
| S9_extracted_k141_71063 | Low-quality |
| S9_extracted_k141_67233 | Low-quality |
| S9_extracted_k141_30105 | Not-determined |
| S9_extracted_k141_67234 | Not-determined |
| S9_extracted_k141_48351 | Not-determined |
| S9_extracted_k141_41328 | Not-determined |
| S9_extracted_k141_74975 | Not-determined |
| S9_extracted_k141_59832 | Not-determined |
| S9_extracted_k141_22311 | Not-determined |
| S9_extracted_k141_86356 | Not-determined |
| S9_extracted_k141_71071 | Not-determined |
| S9_extracted_k141_78467 | Not-determined |
| S9_extracted_k141_45195 | Low-quality |
| S9_extracted_k141_51534 | Not-determined |
| S9_extracted_k141_41333 | Not-determined |
| S9_extracted_k141_59842 | Not-determined |
| S9_extracted_k141_3286 | Not-determined |
| S9_extracted_k141_86363 | Not-determined |
| S9_extracted_k141_37465 | Not-determined |
| S9_extracted_k141_86364 | Not-determined |
| S9_extracted_k141_86365 | Not-determined |
| S9_extracted_k141_89986 | Low-quality |
| S9_extracted_k141_41337 | Not-determined |
| S9_extracted_k141_71078 | Not-determined |
| S9_extracted_k141_7235 | Not-determined |
| S9_extracted_k141_86368 | Not-determined |
| S9_extracted_k141_7236 | Not-determined |
| S9_extracted_k141_7237 | Not-determined |
| S9_extracted_k141_67245 | Not-determined |
| S9_extracted_k141_51540 | Not-determined |
| S9_extracted_k141_78478 | Not-determined |
| S9_extracted_k141_10848 | Not-determined |
| S9_extracted_k141_45208 | Not-determined |
| S9_extracted_k141_89990 | Not-determined |
| S9_extracted_k141_10851 | Not-determined |
| S9_extracted_k141_78482 | Not-determined |
| S9_extracted_k141_67251 | Not-determined |
| S9_extracted_k141_89993 | Not-determined |
| S9_extracted_k141_18758 | Not-determined |
| S9_extracted_k141_45213 | Not-determined |
| S9_extracted_k141_37472 | Low-quality |
| S9_extracted_k141_7241 | Low-quality |
| S9_extracted_k141_14243 | Low-quality |
| S9_extracted_k141_10857 | Not-determined |
| S9_extracted_k141_22338 | Not-determined |
| S9_extracted_k141_26069 | Low-quality |
| S9_extracted_k141_82522 | Low-quality |
| S9_extracted_k141_78494 | Not-determined |
| S9_extracted_k141_89994 | Not-determined |
| S9_extracted_k141_10862 | Not-determined |
| S9_extracted_k141_33703 | Not-determined |
| S9_extracted_k141_10865 | Not-determined |
| S9_extracted_k141_41351 | Not-determined |
| S9_extracted_k141_48376 | Not-determined |
| S9_extracted_k141_22345 | Low-quality |
| S9_extracted_k141_45223 | Not-determined |
| S9_extracted_k141_7253 | Not-determined |
| S9_extracted_k141_59863 | Not-determined |
| S9_extracted_k141_18770 | Not-determined |
| S9_extracted_k141_86386 | Low-quality |
| S9_extracted_k141_59864 | Not-determined |
| S9_extracted_k141_82538 | Low-quality |
| S9_extracted_k141_90005 | Not-determined |
| S9_extracted_k141_33713 | Not-determined |
| S9_extracted_k141_33714 | Not-determined |
| S9_extracted_k141_48385 | Not-determined |
| S9_extracted_k141_82540 | Not-determined |
| S9_extracted_k141_37492 | Low-quality |
| S9_extracted_k141_3294 | Not-determined |
| S9_extracted_k141_86391 | Not-determined |
| S9_extracted_k141_48387 | Low-quality |
| S9_extracted_k141_82545 | Not-determined |
| S9_extracted_k141_86394 | Not-determined |
| S9_extracted_k141_67275 | Not-determined |
| S9_extracted_k141_51563 | Not-determined |
| S9_extracted_k141_75020 | Not-determined |
| S9_extracted_k141_10887 | Not-determined |
| S9_extracted_k141_26086 | Not-determined |
| S9_extracted_k141_82547 | Not-determined |
| S9_extracted_k141_26091 | Not-determined |
| S9_extracted_k141_3296 | Not-determined |
| S9_extracted_k141_22362 | Not-determined |
| S9_extracted_k141_56357 | Not-determined |
| S9_extracted_k141_75027 | Not-determined |
| S9_extracted_k141_10890 | Not-determined |
| S9_extracted_k141_75028 | Not-determined |
| S9_extracted_k141_86408 | Not-determined |
| S9_extracted_k141_90015 | Low-quality |
| S9_extracted_k141_18791 | Low-quality |
| S9_extracted_k141_90016 | Low-quality |
| S9_extracted_k141_10897 | Not-determined |
| S9_extracted_k141_59880 | Not-determined |
| S9_extracted_k141_14270 | Not-determined |
| S9_extracted_k141_26098 | Not-determined |
| S9_extracted_k141_26099 | Not-determined |
| S9_extracted_k141_75032 | Not-determined |
| S9_extracted_k141_90018 | Low-quality |
| S9_extracted_k141_3307 | Not-determined |
| S9_extracted_k141_18795 | Not-determined |
| S9_extracted_k141_18797 | Not-determined |
| S9_extracted_k141_59883 | Low-quality |
| S9_extracted_k141_71122 | Not-determined |
| S9_extracted_k141_56372 | Not-determined |
| S9_extracted_k141_90019 | Not-determined |
| S9_extracted_k141_86413 | Not-determined |
| S9_extracted_k141_7269 | Not-determined |
| S9_extracted_k141_30145 | Not-determined |
| S9_extracted_k141_90023 | Not-determined |
| S9_extracted_k141_18804 | Not-determined |
| S9_extracted_k141_56379 | Low-quality |
| S9_extracted_k141_10906 | Not-determined |
| S9_extracted_k141_3319 | Low-quality |
| S9_extracted_k141_14285 | Not-determined |
| S9_extracted_k141_3324 | Not-determined |
| S9_extracted_k141_56384 | Not-determined |
| S9_extracted_k141_7277 | Not-determined |
| S9_extracted_k141_56385 | Not-determined |
| S9_extracted_k141_37525 | Not-determined |
| S9_extracted_k141_30154 | Low-quality |
| S9_extracted_k141_3327 | Not-determined |
| S9_extracted_k141_86419 | Low-quality |
| S9_extracted_k141_26109 | Low-quality |
| S9_extracted_k141_51594 | Not-determined |
| S9_extracted_k141_14294 | Not-determined |
| S9_extracted_k141_78545 | Not-determined |
| S9_extracted_k141_82557 | Not-determined |
| S9_extracted_k141_18815 | Low-quality |
| S9_extracted_k141_90047 | Low-quality |
| S9_extracted_k141_90048 | Not-determined |
| S9_extracted_k141_90049 | Not-determined |
| S9_extracted_k141_86429 | Not-determined |
| S9_extracted_k141_78550 | Not-determined |
| S9_extracted_k141_45261 | Not-determined |
| S9_extracted_k141_86431 | Not-determined |
| S9_extracted_k141_45262 | Low-quality |
| S9_extracted_k141_45263 | Not-determined |
| S9_extracted_k141_51603 | Not-determined |
| S9_extracted_k141_71151 | Not-determined |
| S9_extracted_k141_90054 | Not-determined |
| S9_extracted_k141_86437 | Not-determined |
| S9_extracted_k141_90057 | Not-determined |
| S9_extracted_k141_90058 | Not-determined |
| S9_extracted_k141_30173 | Not-determined |
| S9_extracted_k141_51607 | Not-determined |
| S9_extracted_k141_71157 | Not-determined |
| S9_extracted_k141_37548 | Low-quality |
| S9_extracted_k141_90064 | Not-determined |
| S9_extracted_k141_56387 | Low-quality |
| S9_extracted_k141_3350 | Not-determined |
| S9_extracted_k141_86446 | Not-determined |
| S9_extracted_k141_26116 | Not-determined |
| S9_extracted_k141_86450 | Not-determined |
| S9_extracted_k141_7308 | Not-determined |
| S9_extracted_k141_41389 | Not-determined |
| S9_extracted_k141_26118 | Not-determined |
| S9_extracted_k141_78560 | Not-determined |
| S9_extracted_k141_22377 | Not-determined |
| S9_extracted_k141_51619 | Low-quality |
| S9_extracted_k141_90071 | Low-quality |
| S9_extracted_k141_51621 | Not-determined |
| S9_extracted_k141_90072 | Not-determined |
| S9_extracted_k141_18833 | Not-determined |
| S9_extracted_k141_67313 | Not-determined |
| S9_extracted_k141_14322 | Not-determined |
| S9_extracted_k141_14326 | Low-quality |
| S9_extracted_k141_18840 | Not-determined |
| S9_extracted_k141_26125 | Not-determined |
| S9_extracted_k141_30193 | Low-quality |
| S9_extracted_k141_45283 | Low-quality |
| S9_extracted_k141_30194 | Not-determined |
| S9_extracted_k141_67316 | Not-determined |
| S9_extracted_k141_14330 | Not-determined |
| S9_extracted_k141_14332 | Not-determined |
| S9_extracted_k141_51624 | Low-quality |
| S9_extracted_k141_67319 | Not-determined |
| S9_extracted_k141_67320 | Not-determined |
| S9_extracted_k141_18847 | Not-determined |
| S9_extracted_k141_67321 | Not-determined |
| S9_extracted_k141_33742 | Not-determined |
| S9_extracted_k141_3367 | Not-determined |
| S9_extracted_k141_67324 | Not-determined |
| S9_extracted_k141_67325 | Not-determined |
| S9_extracted_k141_3368 | Not-determined |
| S9_extracted_k141_14333 | Low-quality |
| S9_extracted_k141_51634 | Not-determined |
| S9_extracted_k141_33745 | Low-quality |
| S9_extracted_k141_67326 | Not-determined |
| S9_extracted_k141_67327 | Not-determined |
| S9_extracted_k141_33748 | Not-determined |
| S9_extracted_k141_71175 | Not-determined |
| S9_extracted_k141_3371 | Low-quality |
| S9_extracted_k141_71176 | Not-determined |
| S9_extracted_k141_3372 | Not-determined |
| S9_extracted_k141_33754 | Not-determined |
| S9_extracted_k141_59938 | Not-determined |
| S9_extracted_k141_78579 | Low-quality |
| S9_extracted_k141_71181 | Not-determined |
| S9_extracted_k141_51641 | Not-determined |
| S9_extracted_k141_41409 | Not-determined |
| S9_extracted_k141_30207 | Not-determined |
| S9_extracted_k141_90094 | Low-quality |
| S9_extracted_k141_45288 | Not-determined |
| S9_extracted_k141_67340 | Not-determined |
| S9_extracted_k141_90098 | Not-determined |
| S9_extracted_k141_51647 | Not-determined |
| S9_extracted_k141_14344 | Not-determined |
| S9_extracted_k141_30212 | Low-quality |
| S9_extracted_k141_56413 | Not-determined |
| S9_extracted_k141_22392 | Not-determined |
| S9_extracted_k141_78592 | Low-quality |
| S9_extracted_k141_75056 | Not-determined |
| S9_extracted_k141_33766 | Not-determined |
| S9_extracted_k141_26158 | Not-determined |
| S9_extracted_k141_56420 | Not-determined |
| S9_extracted_k141_33769 | Not-determined |
| S9_extracted_k141_45294 | Low-quality |
| S9_extracted_k141_51655 | Low-quality |
| S9_extracted_k141_14345 | Not-determined |
| S9_extracted_k141_56423 | Not-determined |
| S9_extracted_k141_67350 | Not-determined |
| S9_extracted_k141_3383 | Not-determined |
| S9_extracted_k141_26166 | Not-determined |
| S9_extracted_k141_45297 | Low-quality |
| S9_extracted_k141_18874 | Not-determined |
| S9_extracted_k141_71190 | Not-determined |
| S9_extracted_k141_26169 | Not-determined |
| S9_extracted_k141_75065 | Not-determined |
| S9_extracted_k141_33776 | Not-determined |
| S9_extracted_k141_59953 | Low-quality |
| S9_extracted_k141_71198 | Not-determined |
| S9_extracted_k141_14349 | Not-determined |
| S9_extracted_k141_41421 | Not-determined |
| S9_extracted_k141_75070 | Not-determined |
| S9_extracted_k141_14351 | Not-determined |
| S9_extracted_k141_71203 | Not-determined |
| S9_extracted_k141_26178 | Not-determined |
| S9_extracted_k141_75073 | Not-determined |
| S9_extracted_k141_78602 | Low-quality |
| S9_extracted_k141_45309 | Not-determined |
| S9_extracted_k141_14355 | Low-quality |
| S9_extracted_k141_75079 | Not-determined |
| S9_extracted_k141_30226 | Not-determined |
| S9_extracted_k141_41430 | Not-determined |
| S9_extracted_k141_75081 | Not-determined |
| S9_extracted_k141_78611 | Not-determined |
| S9_extracted_k141_18881 | Low-quality |
| S9_extracted_k141_30228 | Not-determined |
| S9_extracted_k141_78618 | Not-determined |
| S9_extracted_k141_67368 | Not-determined |
| S9_extracted_k141_3393 | Not-determined |
| S9_extracted_k141_30234 | Not-determined |
| S9_extracted_k141_33799 | Not-determined |
| S9_extracted_k141_67371 | Not-determined |
| S9_extracted_k141_59981 | Not-determined |
| S9_extracted_k141_67374 | Not-determined |
| S9_extracted_k141_71212 | Low-quality |
| S9_extracted_k141_75094 | Not-determined |
| S9_extracted_k141_3396 | Not-determined |
| S9_extracted_k141_37578 | Not-determined |
| S9_extracted_k141_56450 | Not-determined |
| S9_extracted_k141_37582 | Not-determined |
| S9_extracted_k141_71217 | Not-determined |
| S9_extracted_k141_7336 | Not-determined |
| S9_extracted_k141_14381 | Not-determined |
| S9_extracted_k141_7337 | Not-determined |
| S9_extracted_k141_37590 | Low-quality |
| S9_extracted_k141_33818 | Not-determined |
| S9_extracted_k141_3402 | Not-determined |
| S9_extracted_k141_37593 | Not-determined |
| S9_extracted_k141_90146 | Not-determined |
| S9_extracted_k141_71224 | Low-quality |
| S9_extracted_k141_71226 | Not-determined |
| S9_extracted_k141_3410 | Not-determined |
| S9_extracted_k141_78640 | Not-determined |
| S9_extracted_k141_18895 | Low-quality |
| S9_extracted_k141_22419 | Not-determined |
| S9_extracted_k141_90151 | Not-determined |
| S9_extracted_k141_10972 | Not-determined |
| S9_extracted_k141_75113 | Low-quality |
| S9_extracted_k141_3412 | Not-determined |
| S9_extracted_k141_3413 | Not-determined |
| S9_extracted_k141_14390 | Low-quality |
| S9_extracted_k141_30251 | Not-determined |
| S9_extracted_k141_78642 | Not-determined |
| S9_extracted_k141_37601 | Not-determined |
| S9_extracted_k141_75117 | Not-determined |
| S9_extracted_k141_33829 | Not-determined |
| S9_extracted_k141_45348 | Low-quality |
| S9_extracted_k141_30256 | Not-determined |
| S9_extracted_k141_67402 | Not-determined |
| S9_extracted_k141_33835 | Not-determined |
| S9_extracted_k141_30261 | Not-determined |
| S9_extracted_k141_7359 | Not-determined |
| S9_extracted_k141_71237 | Not-determined |
| S9_extracted_k141_22430 | Not-determined |
| S9_extracted_k141_7363 | Not-determined |
| S9_extracted_k141_71240 | Not-determined |
| S9_extracted_k141_78648 | Not-determined |
| S9_extracted_k141_18906 | Not-determined |
| S9_extracted_k141_18909 | Not-determined |
| S9_extracted_k141_67409 | Not-determined |
| S9_extracted_k141_56480 | Not-determined |
| S9_extracted_k141_41451 | Not-determined |
| S9_extracted_k141_22437 | Not-determined |
| S9_extracted_k141_67411 | Not-determined |
| S9_extracted_k141_33851 | Not-determined |
| S9_extracted_k141_14413 | Not-determined |
| S9_extracted_k141_56484 | Not-determined |
| S9_extracted_k141_45364 | Not-determined |
| S9_extracted_k141_22446 | Not-determined |
| S9_extracted_k141_56487 | Not-determined |
| S9_extracted_k141_56488 | Not-determined |
| S9_extracted_k141_22449 | Not-determined |
| S9_extracted_k141_11002 | Not-determined |
| S9_extracted_k141_22453 | Low-quality |
| S9_extracted_k141_26194 | Low-quality |
| S9_extracted_k141_30282 | Not-determined |
| S9_extracted_k141_56492 | Not-determined |
| S9_extracted_k141_33862 | Not-determined |
| S9_extracted_k141_45370 | Not-determined |
| S9_extracted_k141_71256 | Not-determined |
| S9_extracted_k141_67421 | Not-determined |
| S9_extracted_k141_26199 | Not-determined |
| S9_extracted_k141_67422 | Not-determined |
| S9_extracted_k141_18926 | Not-determined |
| S9_extracted_k141_11011 | Not-determined |
| S9_extracted_k141_67428 | Not-determined |
| S9_extracted_k141_7389 | Not-determined |
| S9_extracted_k141_90161 | Low-quality |
| S9_extracted_k141_45386 | Low-quality |
| S9_extracted_k141_33876 | Not-determined |
| S9_extracted_k141_45387 | Not-determined |
| S9_extracted_k141_26215 | Not-determined |
| S9_extracted_k141_14431 | Not-determined |
| S9_extracted_k141_90166 | Not-determined |
| S9_extracted_k141_75134 | Not-determined |
| S9_extracted_k141_60005 | Not-determined |
| S9_extracted_k141_14434 | Not-determined |
| S9_extracted_k141_41476 | Not-determined |
| S9_extracted_k141_60006 | Not-determined |
| S9_extracted_k141_78674 | Not-determined |
| S9_extracted_k141_60007 | Not-determined |
| S9_extracted_k141_45389 | Not-determined |
| S9_extracted_k141_14437 | Not-determined |
| S9_extracted_k141_60009 | Not-determined |
| S9_extracted_k141_7400 | Low-quality |
| S9_extracted_k141_60010 | Not-determined |
| S9_extracted_k141_18943 | Not-determined |
| S9_extracted_k141_67438 | Not-determined |
| S9_extracted_k141_41483 | Not-determined |
| S9_extracted_k141_26227 | Low-quality |
| S9_extracted_k141_33888 | Not-determined |
| S9_extracted_k141_41488 | Not-determined |
| S9_extracted_k141_67443 | Not-determined |
| S9_extracted_k141_37652 | Not-determined |
| S9_extracted_k141_86459 | Not-determined |
| S9_extracted_k141_3435 | Not-determined |
| S9_extracted_k141_26236 | Not-determined |
| S9_extracted_k141_82575 | Not-determined |
| S9_extracted_k141_37658 | Low-quality |
| S9_extracted_k141_78680 | Low-quality |
| S9_extracted_k141_22474 | Not-determined |
| S9_extracted_k141_86468 | Not-determined |
| S9_extracted_k141_67454 | Not-determined |
| S9_extracted_k141_22480 | Not-determined |
| S9_extracted_k141_86472 | Not-determined |
| S9_extracted_k141_48429 | Not-determined |
| S9_extracted_k141_30308 | Not-determined |
| S9_extracted_k141_37667 | Not-determined |
| S9_extracted_k141_26247 | Not-determined |
| S9_extracted_k141_22486 | Low-quality |
| S9_extracted_k141_82585 | Not-determined |
| S9_extracted_k141_71296 | Not-determined |
| S9_extracted_k141_60040 | Low-quality |
| S9_extracted_k141_45415 | Low-quality |
| S9_extracted_k141_3445 | Not-determined |
| S9_extracted_k141_41496 | Not-determined |
| S9_extracted_k141_48439 | Not-determined |
| S9_extracted_k141_86483 | Not-determined |
| S9_extracted_k141_22493 | Not-determined |
| S9_extracted_k141_14465 | Not-determined |
| S9_extracted_k141_14467 | Not-determined |
| S9_extracted_k141_71305 | Not-determined |
| S9_extracted_k141_3448 | Low-quality |
| S9_extracted_k141_7417 | Not-determined |
| S9_extracted_k141_14469 | Not-determined |
| S9_extracted_k141_11045 | Not-determined |
| S9_extracted_k141_41498 | Not-determined |
| S9_extracted_k141_22497 | Not-determined |
| S9_extracted_k141_7419 | Not-determined |
| S9_extracted_k141_11049 | Not-determined |
| S9_extracted_k141_26262 | Not-determined |
| S9_extracted_k141_60046 | Not-determined |
| S9_extracted_k141_33917 | Not-determined |
| S9_extracted_k141_11052 | Not-determined |
| S9_extracted_k141_82596 | Not-determined |
| S9_extracted_k141_37681 | Not-determined |
| S9_extracted_k141_26267 | Not-determined |
| S9_extracted_k141_22510 | Not-determined |
| S9_extracted_k141_86490 | Not-determined |
| S9_extracted_k141_82599 | Not-determined |
| S9_extracted_k141_90195 | Not-determined |
| S9_extracted_k141_67476 | Not-determined |
| S9_extracted_k141_3460 | Not-determined |
| S9_extracted_k141_45428 | Not-determined |
| S9_extracted_k141_33925 | Not-determined |
| S9_extracted_k141_82601 | Not-determined |
| S9_extracted_k141_45429 | Not-determined |
| S9_extracted_k141_14478 | Not-determined |
| S9_extracted_k141_48449 | Not-determined |
| S9_extracted_k141_60064 | Not-determined |

| | |
|---|---|
| S9_extracted_k141_67479 | Low-quality |
| S9_extracted_k141_82604 | Not-determined |
| S9_extracted_k141_78685 | Low-quality |
| S9_extracted_k141_71319 | Not-determined |
| S9_extracted_k141_86501 | Not-determined |
| S9_extracted_k141_75177 | Not-determined |
| S9_extracted_k141_60067 | Not-determined |
| S9_extracted_k141_33934 | Not-determined |
| S9_extracted_k141_56523 | Not-determined |
| S9_extracted_k141_78688 | Low-quality |
| S9_extracted_k141_45437 | Not-determined |
| S9_extracted_k141_26277 | Not-determined |
| S9_extracted_k141_75181 | Not-determined |
| S9_extracted_k141_67485 | Not-determined |
| S9_extracted_k141_11069 | Not-determined |
| S9_extracted_k141_37692 | Not-determined |
| S9_extracted_k141_22524 | Not-determined |
| S9_extracted_k141_86507 | Not-determined |
| S9_extracted_k141_37697 | Not-determined |
| S9_extracted_k141_22528 | Not-determined |
| S9_extracted_k141_67500 | Not-determined |
| S9_extracted_k141_78698 | Not-determined |
| S9_extracted_k141_60085 | Not-determined |
| S9_extracted_k141_11076 | Not-determined |
| S9_extracted_k141_71334 | Not-determined |
| S9_extracted_k141_78700 | Not-determined |
| S9_extracted_k141_48471 | Not-determined |
| S9_extracted_k141_3483 | Not-determined |
| S9_extracted_k141_11081 | Not-determined |
| S9_extracted_k141_71339 | Not-determined |
| S9_extracted_k141_11082 | Low-quality |
| S9_extracted_k141_14498 | Not-determined |
| S9_extracted_k141_7452 | Not-determined |
| S9_extracted_k141_90225 | Low-quality |
| S9_extracted_k141_75196 | Not-determined |
| S9_extracted_k141_26288 | Low-quality |
| S9_extracted_k141_71343 | Not-determined |
| S9_extracted_k141_63583 | Not-determined |
| S9_extracted_k141_86524 | Not-determined |
| S9_extracted_k141_71344 | Not-determined |
| S9_extracted_k141_86525 | Not-determined |
| S9_extracted_k141_71346 | Not-determined |
| S9_extracted_k141_56542 | Low-quality |
| S9_extracted_k141_56543 | Not-determined |
| S9_extracted_k141_86527 | Not-determined |
| S9_extracted_k141_22538 | Not-determined |
| S9_extracted_k141_37704 | Not-determined |
| S9_extracted_k141_67516 | Not-determined |
| S9_extracted_k141_75201 | Low-quality |
| S9_extracted_k141_78722 | Not-determined |
| S9_extracted_k141_45461 | Not-determined |
| S9_extracted_k141_67518 | Not-determined |
| S9_extracted_k141_78724 | Low-quality |
| S9_extracted_k141_56551 | Not-determined |
| S9_extracted_k141_11089 | Low-quality |
| S9_extracted_k141_14508 | Low-quality |
| S9_extracted_k141_56552 | Not-determined |
| S9_extracted_k141_37709 | Not-determined |
| S9_extracted_k141_86534 | Not-determined |
| S9_extracted_k141_67521 | Not-determined |
| S9_extracted_k141_48486 | Not-determined |
| S9_extracted_k141_45467 | Not-determined |
| S9_extracted_k141_48488 | Low-quality |
| S9_extracted_k141_26314 | Not-determined |
| S9_extracted_k141_33975 | Not-determined |
| S9_extracted_k141_37720 | Not-determined |
| S9_extracted_k141_82608 | Low-quality |
| S9_extracted_k141_67529 | Not-determined |
| S9_extracted_k141_33977 | Not-determined |
| S9_extracted_k141_82610 | Not-determined |
| S9_extracted_k141_14521 | Not-determined |
| S9_extracted_k141_33981 | Not-determined |
| S9_extracted_k141_37726 | Not-determined |
| S9_extracted_k141_75211 | Not-determined |
| S9_extracted_k141_71353 | Not-determined |
| S9_extracted_k141_78733 | Not-determined |
| S9_extracted_k141_48493 | Low-quality |
| S9_extracted_k141_37731 | Not-determined |
| S9_extracted_k141_22563 | Not-determined |
| S9_extracted_k141_71356 | Not-determined |
| S9_extracted_k141_22565 | Not-determined |
| S9_extracted_k141_41507 | Low-quality |
| S9_extracted_k141_26325 | Not-determined |
| S9_extracted_k141_26326 | Not-determined |
| S9_extracted_k141_14528 | Not-determined |
| S9_extracted_k141_26327 | Not-determined |
| S9_extracted_k141_37741 | Not-determined |
| S9_extracted_k141_7460 | Not-determined |
| S9_extracted_k141_71366 | Not-determined |
| S9_extracted_k141_71367 | Not-determined |
| S9_extracted_k141_14532 | Not-determined |
| S9_extracted_k141_41509 | Low-quality |
| S9_extracted_k141_14533 | Low-quality |
| S9_extracted_k141_71370 | Not-determined |
| S9_extracted_k141_22580 | Not-determined |
| S9_extracted_k141_67530 | Not-determined |
| S9_extracted_k141_75221 | Low-quality |
| S9_extracted_k141_71372 | Not-determined |
| S9_extracted_k141_75225 | Not-determined |
| S9_extracted_k141_41514 | Not-determined |
| S9_extracted_k141_86547 | Low-quality |
| S9_extracted_k141_78754 | Not-determined |
| S9_extracted_k141_22588 | Not-determined |
| S9_extracted_k141_78755 | Not-determined |
| S12_extracted_k141_18902 | Complete |
| S9_extracted_k141_41520 | Not-determined |
| S9_extracted_k141_51660 | Low-quality |
| S9_extracted_k141_67540 | Not-determined |
| S9_extracted_k141_82631 | Not-determined |
| S9_extracted_k141_75233 | Not-determined |
| S9_extracted_k141_51661 | Not-determined |
| S9_extracted_k141_75234 | Low-quality |
| S9_extracted_k141_37757 | Not-determined |
| S9_extracted_k141_34010 | Not-determined |
| S9_extracted_k141_51665 | Not-determined |
| S9_extracted_k141_75235 | Not-determined |
| S9_extracted_k141_51667 | Not-determined |
| S9_extracted_k141_51668 | Not-determined |
| S9_extracted_k141_71388 | Not-determined |
| S9_extracted_k141_51671 | Not-determined |
| S9_extracted_k141_75240 | Not-determined |
| S9_extracted_k141_78765 | Not-determined |
| S9_extracted_k141_67548 | Not-determined |
| S9_extracted_k141_48524 | Not-determined |
| S9_extracted_k141_22596 | Not-determined |
| S9_extracted_k141_14556 | Not-determined |
| S9_extracted_k141_78767 | Low-quality |
| S9_extracted_k141_75243 | Not-determined |
| S9_extracted_k141_11102 | Not-determined |
| S9_extracted_k141_34019 | Not-determined |
| S9_extracted_k141_82636 | Not-determined |
| S9_extracted_k141_22602 | Not-determined |
| S9_extracted_k141_34020 | Low-quality |
| S9_extracted_k141_75247 | Not-determined |
| S9_extracted_k141_22604 | Not-determined |
| S9_extracted_k141_48528 | Not-determined |
| S9_extracted_k141_75250 | Not-determined |
| S9_extracted_k141_56594 | Low-quality |
| S9_extracted_k141_51681 | Not-determined |
| S9_extracted_k141_14561 | Not-determined |
| S9_extracted_k141_51683 | Not-determined |
| S9_extracted_k141_34026 | Low-quality |
| S9_extracted_k141_34027 | Not-determined |
| S9_extracted_k141_51685 | Not-determined |
| S9_extracted_k141_78776 | Not-determined |
| S9_extracted_k141_41533 | Not-determined |
| S9_extracted_k141_56596 | Not-determined |
| S9_extracted_k141_71406 | Not-determined |
| S9_extracted_k141_71408 | Low-quality |
| S9_extracted_k141_22614 | Not-determined |
| S9_extracted_k141_3504 | Not-determined |
| S9_extracted_k141_75258 | Low-quality |
| S9_extracted_k141_71411 | Not-determined |
| S9_extracted_k141_51693 | Not-determined |
| S9_extracted_k141_41541 | Not-determined |
| S9_extracted_k141_14568 | Not-determined |
| S9_extracted_k141_75262 | Not-determined |
| S9_extracted_k141_75263 | Not-determined |
| S9_extracted_k141_37772 | Not-determined |
| S9_extracted_k141_48539 | Not-determined |
| S9_extracted_k141_56605 | Not-determined |
| S9_extracted_k141_75270 | Not-determined |
| S9_extracted_k141_48545 | Not-determined |
| S9_extracted_k141_37781 | Low-quality |
| S9_extracted_k141_56607 | Low-quality |
| S9_extracted_k141_26345 | Not-determined |
| S9_extracted_k141_86572 | Not-determined |
| S9_extracted_k141_56614 | Not-determined |
| S9_extracted_k141_86574 | Not-determined |
| S9_extracted_k141_41554 | Low-quality |
| S9_extracted_k141_37787 | Not-determined |
| S9_extracted_k141_67550 | Low-quality |
| S9_extracted_k141_37789 | Not-determined |
| S9_extracted_k141_86576 | Not-determined |
| S9_extracted_k141_37791 | Not-determined |
| S9_extracted_k141_22621 | Not-determined |
| S9_extracted_k141_22622 | Not-determined |
| S9_extracted_k141_71433 | Not-determined |
| S9_extracted_k141_51706 | Low-quality |
| S9_extracted_k141_78799 | Not-determined |
| S9_extracted_k141_14596 | Not-determined |
| S9_extracted_k141_78800 | Not-determined |
| S9_extracted_k141_71441 | Not-determined |
| S9_extracted_k141_14598 | Not-determined |
| S9_extracted_k141_41561 | Not-determined |
| S9_extracted_k141_86578 | Not-determined |
| S9_extracted_k141_48562 | Not-determined |
| S9_extracted_k141_48563 | Not-determined |
| S9_extracted_k141_41564 | Not-determined |
| S9_extracted_k141_75284 | Not-determined |
| S9_extracted_k141_90270 | Not-determined |
| S9_extracted_k141_3529 | Not-determined |
| S9_extracted_k141_37800 | Not-determined |
| S9_extracted_k141_60090 | Low-quality |
| S9_extracted_k141_41571 | Not-determined |
| S9_extracted_k141_37803 | Low-quality |
| S9_extracted_k141_14613 | Not-determined |
| S9_extracted_k141_37805 | Not-determined |
| S9_extracted_k141_22636 | Not-determined |
| S9_extracted_k141_37806 | Not-determined |
| S9_extracted_k141_3532 | Not-determined |
| S9_extracted_k141_22638 | Not-determined |
| S9_extracted_k141_48571 | Not-determined |
| S9_extracted_k141_67561 | Not-determined |
| S9_extracted_k141_41576 | Not-determined |
| S9_extracted_k141_60095 | Low-quality |
| S9_extracted_k141_26359 | Not-determined |
| S9_extracted_k141_78813 | Not-determined |
| S9_extracted_k141_48574 | Not-determined |
| S9_extracted_k141_26360 | Not-determined |
| S9_extracted_k141_71448 | Not-determined |
| S9_extracted_k141_11123 | Not-determined |
| S9_extracted_k141_67570 | Not-determined |
| S9_extracted_k141_26364 | Not-determined |
| S9_extracted_k141_56633 | Not-determined |
| S9_extracted_k141_60099 | Not-determined |
| S9_extracted_k141_67572 | Not-determined |
| S9_extracted_k141_67575 | Not-determined |
| S9_extracted_k141_56636 | Not-determined |
| S9_extracted_k141_86591 | Low-quality |
| S9_extracted_k141_26368 | Not-determined |
| S9_extracted_k141_41588 | Low-quality |
| S9_extracted_k141_86592 | Not-determined |
| S9_extracted_k141_60101 | Not-determined |
| S9_extracted_k141_51727 | Not-determined |
| S9_extracted_k141_51728 | Low-quality |
| S9_extracted_k141_3545 | Not-determined |
| S9_extracted_k141_14631 | Not-determined |
| S9_extracted_k141_22654 | Low-quality |
| S9_extracted_k141_3549 | Not-determined |
| S9_extracted_k141_51735 | Not-determined |
| S9_extracted_k141_14634 | Low-quality |
| S9_extracted_k141_78829 | Low-quality |
| S9_extracted_k141_78830 | Not-determined |
| S9_extracted_k141_48578 | Not-determined |
| S9_extracted_k141_60107 | Low-quality |
| S9_extracted_k141_51738 | Low-quality |
| S9_extracted_k141_82647 | Not-determined |
| S9_extracted_k141_78836 | Not-determined |
| S9_extracted_k141_67597 | Not-determined |
| S9_extracted_k141_86602 | Not-determined |
| S9_extracted_k141_56645 | Not-determined |
| S9_extracted_k141_82651 | Not-determined |
| S9_extracted_k141_11138 | Not-determined |
| S9_extracted_k141_26384 | Not-determined |
| S9_extracted_k141_37832 | Low-quality |
| S9_extracted_k141_86606 | Low-quality |
| S9_extracted_k141_3558 | Not-determined |
| S9_extracted_k141_60115 | Not-determined |
| S9_extracted_k141_26389 | Not-determined |
| S9_extracted_k141_51746 | Not-determined |
| S9_extracted_k141_51749 | Not-determined |
| S9_extracted_k141_86608 | Low-quality |
| S9_extracted_k141_51750 | Not-determined |
| S9_extracted_k141_60119 | Not-determined |
| S9_extracted_k141_90293 | Not-determined |
| S9_extracted_k141_71473 | Not-determined |
| S9_extracted_k141_37841 | Not-determined |
| S9_extracted_k141_11158 | Not-determined |
| S9_extracted_k141_41614 | Not-determined |
| S9_extracted_k141_86614 | Not-determined |
| S9_extracted_k141_78847 | Not-determined |
| S9_extracted_k141_71479 | Low-quality |
| S9_extracted_k141_11161 | Not-determined |
| S9_extracted_k141_14661 | Not-determined |
| S9_extracted_k141_56667 | Not-determined |
| S9_extracted_k141_78852 | Not-determined |
| S9_extracted_k141_22672 | Not-determined |
| S9_extracted_k141_56671 | Not-determined |
| S9_extracted_k141_71488 | Not-determined |
| S9_extracted_k141_60133 | Not-determined |
| S9_extracted_k141_26412 | Low-quality |
| S9_extracted_k141_82672 | Low-quality |
| S9_extracted_k141_86624 | Not-determined |
| S9_extracted_k141_60136 | Not-determined |
| S9_extracted_k141_51771 | Low-quality |
| S9_extracted_k141_82677 | Not-determined |
| S9_extracted_k141_67626 | Not-determined |
| S9_extracted_k141_78859 | Not-determined |
| S9_extracted_k141_67627 | Low-quality |
| S9_extracted_k141_3575 | Not-determined |
| S9_extracted_k141_51773 | Not-determined |
| S9_extracted_k141_14673 | Not-determined |
| S9_extracted_k141_14674 | Low-quality |
| S9_extracted_k141_3578 | Not-determined |
| S9_extracted_k141_78862 | Not-determined |
| S9_extracted_k141_22682 | Not-determined |
| S9_extracted_k141_37860 | Not-determined |
| S9_extracted_k141_60146 | Not-determined |
| S9_extracted_k141_60149 | Not-determined |
| S9_extracted_k141_86639 | Low-quality |
| S9_extracted_k141_90307 | Not-determined |
| S9_extracted_k141_78867 | Not-determined |
| S9_extracted_k141_78868 | Not-determined |
| S9_extracted_k141_41644 | Not-determined |
| S9_extracted_k141_90308 | Not-determined |
| S9_extracted_k141_41645 | Low-quality |
| S9_extracted_k141_78872 | Not-determined |
| S9_extracted_k141_51791 | Not-determined |
| S9_extracted_k141_82689 | Not-determined |
| S9_extracted_k141_86644 | Not-determined |
| S9_extracted_k141_86645 | Not-determined |
| S9_extracted_k141_26428 | Not-determined |
| S9_extracted_k141_78873 | Low-quality |
| S9_extracted_k141_14682 | Not-determined |
| S9_extracted_k141_22694 | Not-determined |
| S9_extracted_k141_22696 | Not-determined |
| S9_extracted_k141_26435 | Not-determined |
| S9_extracted_k141_90323 | Not-determined |
| S9_extracted_k141_51798 | Not-determined |
| S9_extracted_k141_90328 | Not-determined |
| S9_extracted_k141_26440 | Not-determined |
| S9_extracted_k141_60163 | Low-quality |
| S9_extracted_k141_86664 | Not-determined |
| S9_extracted_k141_86666 | Not-determined |
| S9_extracted_k141_67654 | Not-determined |
| S9_extracted_k141_56686 | Not-determined |
| S9_extracted_k141_60167 | Not-determined |
| S9_extracted_k141_37880 | Not-determined |
| S9_extracted_k141_51809 | Not-determined |
| S9_extracted_k141_22712 | Low-quality |
| S9_extracted_k141_86672 | Not-determined |
| S9_extracted_k141_3598 | Not-determined |
| S9_extracted_k141_7510 | Not-determined |
| S9_extracted_k141_41658 | Low-quality |
| S9_extracted_k141_90335 | Not-determined |
| S9_extracted_k141_41659 | Not-determined |
| S9_extracted_k141_67658 | Not-determined |
| S9_extracted_k141_67659 | Not-determined |
| S9_extracted_k141_86677 | Not-determined |
| S9_extracted_k141_75312 | Not-determined |
| S9_extracted_k141_3603 | Not-determined |
| S9_extracted_k141_3607 | Not-determined |
| S9_extracted_k141_26444 | Not-determined |
| S9_extracted_k141_22727 | Not-determined |
| S9_extracted_k141_51826 | Not-determined |
| S9_extracted_k141_63590 | Low-quality |
| S9_extracted_k141_22733 | Not-determined |
| S9_extracted_k141_22734 | Not-determined |
| S9_extracted_k141_51828 | Not-determined |
| S9_extracted_k141_63592 | Not-determined |
| S9_extracted_k141_51830 | Low-quality |
| S9_extracted_k141_14699 | Not-determined |
| S9_extracted_k141_3613 | Not-determined |
| S9_extracted_k141_7531 | Not-determined |
| S9_extracted_k141_71527 | Not-determined |
| S9_extracted_k141_26454 | Not-determined |
| S9_extracted_k141_11202 | Not-determined |
| S9_extracted_k141_51840 | Not-determined |
| S9_extracted_k141_14708 | Not-determined |
| S9_extracted_k141_26458 | Not-determined |
| S9_extracted_k141_82694 | Not-determined |
| S9_extracted_k141_7541 | Not-determined |
| S9_extracted_k141_14712 | Not-determined |
| S9_extracted_k141_14713 | Not-determined |
| S9_extracted_k141_26463 | Not-determined |
| S9_extracted_k141_3625 | Not-determined |
| S9_extracted_k141_3626 | Not-determined |
| S9_extracted_k141_63608 | Not-determined |
| S9_extracted_k141_82702 | Not-determined |
| S9_extracted_k141_67679 | Not-determined |
| S9_extracted_k141_7550 | Not-determined |
| S9_extracted_k141_14716 | Not-determined |
| S9_extracted_k141_78909 | Not-determined |
| S9_extracted_k141_3632 | Low-quality |
| S9_extracted_k141_63610 | Low-quality |
| S9_extracted_k141_7553 | Not-determined |
| S9_extracted_k141_11217 | Not-determined |
| S9_extracted_k141_78913 | Not-determined |
| S9_extracted_k141_11220 | Low-quality |
| S9_extracted_k141_48596 | Not-determined |
| S9_extracted_k141_63614 | Not-determined |
| S9_extracted_k141_51856 | Low-quality |
| S9_extracted_k141_86699 | Not-determined |
| S9_extracted_k141_48599 | Not-determined |
| S9_extracted_k141_11224 | Not-determined |
| S9_extracted_k141_26476 | Not-determined |
| S9_extracted_k141_48602 | Not-determined |
| S9_extracted_k141_75340 | Not-determined |
| S9_extracted_k141_14724 | Not-determined |
| S9_extracted_k141_78923 | Not-determined |
| S9_extracted_k141_56733 | Low-quality |
| S9_extracted_k141_60192 | Not-determined |
| S9_extracted_k141_26484 | Not-determined |
| S9_extracted_k141_82716 | Not-determined |
| S9_extracted_k141_86706 | Not-determined |
| S9_extracted_k141_26485 | Not-determined |
| S9_extracted_k141_51863 | Not-determined |
| S9_extracted_k141_26486 | Not-determined |
| S9_extracted_k141_78930 | Not-determined |
| S9_extracted_k141_82721 | Not-determined |
| S9_extracted_k141_78932 | Not-determined |
| S9_extracted_k141_67700 | Not-determined |
| S9_extracted_k141_63630 | Not-determined |
| S9_extracted_k141_67701 | Not-determined |
| S9_extracted_k141_75350 | Not-determined |
| S9_extracted_k141_48607 | Not-determined |
| S9_extracted_k141_75351 | Not-determined |
| S9_extracted_k141_82724 | Not-determined |
| S9_extracted_k141_48609 | Not-determined |
| S9_extracted_k141_86713 | Not-determined |
| S9_extracted_k141_90393 | Low-quality |
| S9_extracted_k141_48613 | Not-determined |
| S9_extracted_k141_78936 | Not-determined |
| S9_extracted_k141_7575 | Not-determined |
| S9_extracted_k141_67719 | Not-determined |
| S9_extracted_k141_26499 | Not-determined |
| S9_extracted_k141_48615 | Low-quality |
| S9_extracted_k141_90400 | Not-determined |
| S9_extracted_k141_14737 | Not-determined |
| S9_extracted_k141_90401 | Not-determined |
| S9_extracted_k141_67721 | Not-determined |
| S9_extracted_k141_63642 | Low-quality |
| S9_extracted_k141_71548 | Not-determined |
| S9_extracted_k141_71549 | Not-determined |
| S9_extracted_k141_86730 | Not-determined |
| S9_extracted_k141_14746 | Not-determined |
| S9_extracted_k141_90409 | Not-determined |
| S9_extracted_k141_90412 | Not-determined |
| S9_extracted_k141_71559 | Not-determined |
| S9_extracted_k141_78947 | Low-quality |
| S9_extracted_k141_71561 | Not-determined |
| S9_extracted_k141_63651 | Low-quality |
| S9_extracted_k141_51878 | Not-determined |
| S9_extracted_k141_86735 | Not-determined |
| S9_extracted_k141_71562 | Not-determined |
| S9_extracted_k141_78949 | Not-determined |
| S9_extracted_k141_56750 | Not-determined |
| S9_extracted_k141_48628 | Not-determined |
| S9_extracted_k141_56751 | Not-determined |
| S9_extracted_k141_86738 | Low-quality |
| S9_extracted_k141_82753 | Not-determined |
| S9_extracted_k141_82754 | Low-quality |
| S9_extracted_k141_51887 | Not-determined |
| S9_extracted_k141_90425 | Low-quality |
| S9_extracted_k141_63658 | Not-determined |
| S9_extracted_k141_67732 | Not-determined |
| S9_extracted_k141_56754 | Not-determined |
| S9_extracted_k141_71572 | Not-determined |
| S9_extracted_k141_60237 | Not-determined |
| S9_extracted_k141_86749 | Not-determined |
| S9_extracted_k141_14756 | Not-determined |
| S9_extracted_k141_78956 | Low-quality |
| S9_extracted_k141_86753 | Low-quality |
| S9_extracted_k141_60243 | Not-determined |
| S9_extracted_k141_75357 | Not-determined |
| S9_extracted_k141_78962 | Not-determined |
| S9_extracted_k141_51905 | Low-quality |
| S9_extracted_k141_86760 | Not-determined |
| S9_extracted_k141_78966 | Not-determined |
| S9_extracted_k141_14771 | Not-determined |
| S9_extracted_k141_51908 | Not-determined |
| S9_extracted_k141_51909 | Not-determined |
| S9_extracted_k141_82767 | Not-determined |
| S9_extracted_k141_14772 | Not-determined |
| S9_extracted_k141_60249 | Not-determined |
| S9_extracted_k141_51911 | Low-quality |
| S9_extracted_k141_71597 | Not-determined |
| S9_extracted_k141_78970 | Not-determined |
| S9_extracted_k141_3655 | Not-determined |
| S9_extracted_k141_63683 | Low-quality |
| S9_extracted_k141_56775 | Low-quality |
| S9_extracted_k141_82774 | Not-determined |
| S9_extracted_k141_51914 | Not-determined |
| S9_extracted_k141_71601 | Not-determined |
| S9_extracted_k141_48656 | Low-quality |
| S9_extracted_k141_71603 | Not-determined |
| S9_extracted_k141_67755 | Not-determined |
| S9_extracted_k141_78974 | Not-determined |
| S9_extracted_k141_71606 | Not-determined |
| S9_extracted_k141_63687 | Not-determined |
| S9_extracted_k141_56782 | Not-determined |
| S9_extracted_k141_82779 | Not-determined |
| S9_extracted_k141_51923 | Not-determined |
| S9_extracted_k141_63690 | Not-determined |
| S9_extracted_k141_63691 | Not-determined |
| S9_extracted_k141_41678 | Low-quality |
| S9_extracted_k141_71609 | Not-determined |
| S9_extracted_k141_11252 | Not-determined |
| S9_extracted_k141_3668 | Not-determined |
| S9_extracted_k141_48670 | Low-quality |
| S9_extracted_k141_67770 | Not-determined |
| S9_extracted_k141_67772 | Not-determined |
| S9_extracted_k141_51935 | Not-determined |
| S12_vs_k141_18902\|\|full | High-quality |
| S9_extracted_k141_51937 | Not-determined |
| S9_extracted_k141_51938 | Not-determined |
| S9_extracted_k141_67775 | Low-quality |
| S9_extracted_k141_82789 | Not-determined |
| S9_extracted_k141_3671 | Low-quality |
| S9_extracted_k141_14794 | Not-determined |
| S9_extracted_k141_67779 | Low-quality |
| S9_extracted_k141_78986 | Not-determined |
| S9_extracted_k141_67781 | Not-determined |
| S9_extracted_k141_63697 | Not-determined |
| S9_extracted_k141_71622 | Not-determined |
| S9_extracted_k141_51945 | Not-determined |
| S9_extracted_k141_56789 | Low-quality |
| S9_extracted_k141_78989 | Low-quality |
| S9_extracted_k141_82793 | Not-determined |
| S9_extracted_k141_71628 | Not-determined |
| S9_extracted_k141_63707 | Low-quality |
| S9_extracted_k141_86781 | Not-determined |
| S9_extracted_k141_86782 | Not-determined |
| S9_extracted_k141_75376 | Not-determined |
| S9_extracted_k141_60281 | Not-determined |
| S9_extracted_k141_86783 | Not-determined |
| S9_extracted_k141_48690 | Not-determined |
| S9_extracted_k141_56791 | Not-determined |
| S9_extracted_k141_48691 | Not-determined |
| S9_extracted_k141_75381 | Not-determined |
| S9_extracted_k141_75382 | Low-quality |
| S9_extracted_k141_78994 | Not-determined |
| S9_extracted_k141_11286 | Not-determined |
| S9_extracted_k141_86796 | Not-determined |
| S9_extracted_k141_48704 | Not-determined |
| S9_extracted_k141_71638 | Not-determined |
| S9_extracted_k141_3688 | Low-quality |
| S9_extracted_k141_48705 | Not-determined |
| S9_extracted_k141_51973 | Not-determined |
| S9_extracted_k141_56803 | Low-quality |
| S9_extracted_k141_51975 | Not-determined |
| S9_extracted_k141_71640 | Not-determined |
| S9_extracted_k141_51976 | Not-determined |
| S9_extracted_k141_82808 | Not-determined |
| S9_extracted_k141_86800 | Not-determined |
| S9_extracted_k141_56808 | Not-determined |
| S9_extracted_k141_79009 | Not-determined |
| S9_extracted_k141_71646 | Not-determined |
| S9_extracted_k141_71647 | Not-determined |
| S9_extracted_k141_79011 | Not-determined |
| S9_extracted_k141_51989 | Not-determined |
| S9_extracted_k141_56819 | Not-determined |
| S9_extracted_k141_56821 | Not-determined |
| S9_extracted_k141_3711 | Not-determined |
| S9_extracted_k141_71656 | Low-quality |
| S9_extracted_k141_51995 | Low-quality |
| S9_extracted_k141_51996 | Not-determined |
| S9_extracted_k141_56828 | Not-determined |
| S9_extracted_k141_11309 | Not-determined |
| S9_extracted_k141_79025 | Low-quality |
| S9_extracted_k141_11316 | Not-determined |
| S9_extracted_k141_3714 | Not-determined |
| S9_extracted_k141_86818 | Not-determined |
| S9_extracted_k141_3716 | Not-determined |
| S9_extracted_k141_82837 | Not-determined |
| S9_extracted_k141_3722 | Not-determined |
| S9_extracted_k141_86825 | Low-quality |
| S9_extracted_k141_86826 | Not-determined |
| S9_extracted_k141_11327 | Not-determined |
| S9_extracted_k141_60322 | Low-quality |
| S9_extracted_k141_82845 | Not-determined |
| S9_extracted_k141_63750 | Low-quality |
| S9_extracted_k141_86829 | Not-determined |
| S9_extracted_k141_60324 | Not-determined |
| S9_extracted_k141_71667 | Not-determined |
| S9_extracted_k141_63757 | Not-determined |
| S9_extracted_k141_48728 | Not-determined |
| S9_extracted_k141_52031 | Not-determined |
| S9_extracted_k141_52032 | Not-determined |
| S9_extracted_k141_63761 | Not-determined |
| S9_extracted_k141_48730 | Not-determined |
| S9_extracted_k141_52034 | Not-determined |
| S9_extracted_k141_79045 | Not-determined |
| S9_extracted_k141_11337 | Not-determined |
| S9_extracted_k141_71674 | Not-determined |
| S9_extracted_k141_63770 | Not-determined |
| S9_extracted_k141_71676 | Not-determined |
| S9_extracted_k141_48737 | Not-determined |
| S9_extracted_k141_60336 | Low-quality |
| S9_extracted_k141_11340 | Not-determined |
| S9_extracted_k141_75434 | Not-determined |
| S9_extracted_k141_52045 | Not-determined |
| S9_extracted_k141_14822 | Low-quality |
| S9_extracted_k141_63775 | Low-quality |
| S9_extracted_k141_48743 | Low-quality |
| S9_extracted_k141_82867 | Not-determined |
| S9_extracted_k141_48744 | Not-determined |
| S9_extracted_k141_86846 | Not-determined |
| S9_extracted_k141_71683 | Not-determined |
| S9_extracted_k141_79062 | Low-quality |
| S9_extracted_k141_82872 | Not-determined |
| S9_extracted_k141_14827 | Not-determined |
| S9_extracted_k141_52055 | Not-determined |
| S9_extracted_k141_82879 | Not-determined |
| S9_extracted_k141_63795 | Not-determined |
| S9_extracted_k141_79073 | Not-determined |
| S9_extracted_k141_52061 | Not-determined |
| S9_extracted_k141_71695 | Low-quality |

| | |
|---|---|
| S9_extracted_k141_86860 | Not-determined |
| S9_extracted_k141_11359 | Not-determined |
| S9_extracted_k141_48765 | Not-determined |
| S9_extracted_k141_82885 | Not-determined |
| S9_extracted_k141_86863 | Low-quality |
| S9_extracted_k141_86864 | Not-determined |
| S9_extracted_k141_82890 | Not-determined |
| S9_extracted_k141_71701 | Not-determined |
| S9_extracted_k141_60357 | Not-determined |
| S9_extracted_k141_14847 | Not-determined |
| S9_extracted_k141_52072 | Not-determined |
| S9_extracted_k141_86875 | Not-determined |
| S9_extracted_k141_75457 | Not-determined |
| S9_extracted_k141_63814 | Not-determined |
| S9_extracted_k141_86882 | Not-determined |
| S9_extracted_k141_52081 | Not-determined |
| S9_extracted_k141_86885 | Not-determined |
| S9_extracted_k141_14856 | Not-determined |
| S9_extracted_k141_90428 | Not-determined |
| S9_extracted_k141_52083 | Not-determined |
| S9_extracted_k141_48779 | Not-determined |
| S9_extracted_k141_86890 | Not-determined |
| S9_extracted_k141_90432 | Not-determined |
| S9_extracted_k141_48784 | Low-quality |
| S9_extracted_k141_48785 | Low-quality |
| S9_extracted_k141_48786 | Not-determined |
| S9_extracted_k141_48788 | Not-determined |
| S9_extracted_k141_75468 | Not-determined |
| S9_extracted_k141_79101 | Not-determined |
| S9_extracted_k141_63834 | Not-determined |
| S9_extracted_k141_86901 | Low-quality |
| S9_extracted_k141_86904 | Not-determined |
| S9_extracted_k141_82920 | Not-determined |
| S9_extracted_k141_75481 | Not-determined |
| S9_extracted_k141_3740 | Low-quality |
| S9_extracted_k141_86910 | Low-quality |
| S9_extracted_k141_90450 | Not-determined |
| S9_extracted_k141_71738 | Not-determined |
| S9_extracted_k141_86917 | Not-determined |
| S9_extracted_k141_86918 | Low-quality |
| S9_extracted_k141_90455 | Not-determined |
| S9_extracted_k141_86920 | Not-determined |
| S9_extracted_k141_71743 | Not-determined |
| S9_extracted_k141_82928 | Not-determined |
| S9_extracted_k141_71745 | Not-determined |
| S9_extracted_k141_67795 | Low-quality |
| S9_extracted_k141_48820 | Not-determined |
| S9_extracted_k141_82929 | Not-determined |
| S9_extracted_k141_71746 | Low-quality |
| S9_extracted_k141_79129 | Not-determined |
| S9_extracted_k141_79130 | Not-determined |
| S9_extracted_k141_82932 | Low-quality |
| S9_extracted_k141_90468 | Not-determined |
| S9_extracted_k141_75494 | Not-determined |
| S9_extracted_k141_90470 | Low-quality |
| S9_extracted_k141_86931 | Low-quality |
| S9_extracted_k141_90472 | Not-determined |
| S9_extracted_k141_67803 | Not-determined |
| S9_extracted_k141_86933 | Not-determined |
| S9_extracted_k141_48824 | Not-determined |
| S9_extracted_k141_86934 | Not-determined |
| S9_extracted_k141_86935 | Not-determined |
| S9_extracted_k141_14896 | Not-determined |
| S9_extracted_k141_67808 | Not-determined |
| S9_extracted_k141_71753 | Not-determined |
| S9_extracted_k141_82941 | Not-determined |
| S9_extracted_k141_3763 | Not-determined |
| S9_extracted_k141_82945 | Not-determined |
| S9_extracted_k141_3764 | Low-quality |
| S9_extracted_k141_90489 | Low-quality |
| S9_extracted_k141_75505 | Not-determined |
| S9_extracted_k141_71757 | Low-quality |
| S9_extracted_k141_67819 | Not-determined |
| S9_extracted_k141_82952 | Not-determined |
| S9_extracted_k141_86948 | Low-quality |
| S9_extracted_k141_71764 | Not-determined |
| S9_extracted_k141_82955 | Not-determined |
| S9_extracted_k141_52133 | Not-determined |
| S9_extracted_k141_86952 | Not-determined |
| S9_extracted_k141_75518 | Low-quality |
| S9_extracted_k141_67827 | Not-determined |
| S9_extracted_k141_67828 | Not-determined |
| S9_extracted_k141_86956 | Not-determined |
| S9_extracted_k141_75522 | Not-determined |
| S9_extracted_k141_52138 | Low-quality |
| S9_extracted_k141_71773 | Not-determined |
| S9_extracted_k141_67833 | Not-determined |
| S9_extracted_k141_90501 | Not-determined |
| S9_extracted_k141_86960 | Not-determined |
| S9_extracted_k141_75527 | Not-determined |
| S9_extracted_k141_75528 | Not-determined |
| S9_extracted_k141_3786 | Not-determined |
| S9_extracted_k141_86965 | Not-determined |
| S9_extracted_k141_67838 | Not-determined |
| S9_extracted_k141_52146 | Not-determined |
| S9_extracted_k141_52147 | Not-determined |
| S9_extracted_k141_82966 | Not-determined |
| S9_extracted_k141_71782 | Not-determined |
| S9_extracted_k141_48851 | Not-determined |
| S9_extracted_k141_90507 | Not-determined |
| S9_extracted_k141_75541 | Not-determined |
| S9_extracted_k141_75543 | Not-determined |
| S9_extracted_k141_71789 | Not-determined |
| S9_extracted_k141_75545 | Low-quality |
| S9_extracted_k141_71792 | Not-determined |
| S9_extracted_k141_86970 | Not-determined |
| S9_extracted_k141_90514 | Not-determined |
| S9_extracted_k141_71796 | Not-determined |
| S9_extracted_k141_48865 | Not-determined |
| S9_extracted_k141_71797 | Low-quality |
| S9_extracted_k141_71799 | Not-determined |
| S9_extracted_k141_79168 | Not-determined |
| S9_extracted_k141_82986 | Not-determined |
| S9_extracted_k141_90519 | Not-determined |
| S9_extracted_k141_75556 | Not-determined |
| S9_extracted_k141_71800 | Low-quality |
| S9_extracted_k141_71801 | Not-determined |
| S9_extracted_k141_48868 | Low-quality |
| S9_extracted_k141_71803 | Not-determined |
| S9_extracted_k141_90523 | Not-determined |
| S9_extracted_k141_75568 | Not-determined |
| S9_extracted_k141_14939 | Not-determined |
| S9_extracted_k141_67864 | Not-determined |
| S9_extracted_k141_67868 | Not-determined |
| S9_extracted_k141_67871 | Not-determined |
| S9_extracted_k141_75584 | Not-determined |
| S9_extracted_k141_83003 | Not-determined |
| S9_extracted_k141_75587 | Not-determined |
| S9_extracted_k141_90532 | Not-determined |
| S9_extracted_k141_90533 | Not-determined |
| S9_extracted_k141_14953 | Not-determined |
| S9_extracted_k141_63881 | Not-determined |
| S9_extracted_k141_79189 | Not-determined |
| S9_extracted_k141_71831 | Not-determined |
| S9_extracted_k141_90538 | Not-determined |
| S9_extracted_k141_75592 | Low-quality |
| S9_extracted_k141_14958 | Not-determined |
| S9_extracted_k141_79192 | Not-determined |
| S9_extracted_k141_90541 | Not-determined |
| S9_extracted_k141_79197 | Not-determined |
| S9_extracted_k141_79200 | Low-quality |
| S9_extracted_k141_67883 | Low-quality |
| S9_extracted_k141_86997 | Not-determined |
| S9_extracted_k141_14963 | Low-quality |
| S9_extracted_k141_83016 | Not-determined |
| S9_extracted_k141_75606 | Low-quality |
| S9_extracted_k141_87000 | Low-quality |
| S9_extracted_k141_75607 | Not-determined |
| S9_extracted_k141_52162 | Low-quality |
| S9_extracted_k141_67888 | Not-determined |
| S9_extracted_k141_63889 | Not-determined |
| S9_extracted_k141_75608 | Not-determined |
| S9_extracted_k141_83022 | Low-quality |
| S9_extracted_k141_90548 | Not-determined |
| S9_extracted_k141_14976 | Low-quality |
| S9_extracted_k141_52170 | Not-determined |
| S9_extracted_k141_63898 | Not-determined |
| S9_extracted_k141_90553 | Not-determined |
| S9_extracted_k141_14981 | Not-determined |
| S9_extracted_k141_83037 | Not-determined |
| S9_extracted_k141_87020 | Not-determined |
| S9_extracted_k141_83038 | Not-determined |
| S9_extracted_k141_87021 | Not-determined |
| S9_extracted_k141_87023 | Not-determined |
| S9_extracted_k141_83044 | Not-determined |
| S9_extracted_k141_67910 | Not-determined |
| S9_extracted_k141_87026 | Not-determined |
| S9_extracted_k141_83048 | Not-determined |
| S9_extracted_k141_67914 | Not-determined |
| S9_extracted_k141_34031 | Not-determined |
| S9_extracted_k141_34032 | Not-determined |
| S9_extracted_k141_87029 | Not-determined |
| S9_extracted_k141_71859 | Not-determined |
| S9_extracted_k141_79232 | Not-determined |
| S9_extracted_k141_67920 | Not-determined |
| S9_extracted_k141_79235 | Not-determined |
| S9_extracted_k141_83057 | Low-quality |
| S9_extracted_k141_79236 | Not-determined |
| S9_extracted_k141_71863 | Not-determined |
| S9_extracted_k141_75639 | Low-quality |
| S9_extracted_k141_34041 | Not-determined |
| S9_extracted_k141_75641 | Not-determined |
| S9_extracted_k141_87036 | Low-quality |
| S9_extracted_k141_75642 | Not-determined |
| S9_extracted_k141_15008 | Not-determined |
| S9_extracted_k141_79245 | Not-determined |
| S9_extracted_k141_79246 | Not-determined |
| S9_extracted_k141_79249 | Not-determined |
| S9_extracted_k141_63922 | Not-determined |
| S9_extracted_k141_83062 | Not-determined |
| S9_extracted_k141_67929 | Not-determined |
| S9_extracted_k141_79254 | Not-determined |
| S9_extracted_k141_67935 | Low-quality |
| S9_extracted_k141_79255 | Not-determined |
| S9_extracted_k141_71886 | Not-determined |
| S9_extracted_k141_75657 | Low-quality |
| S9_extracted_k141_71889 | Not-determined |
| S9_extracted_k141_67945 | Not-determined |
| S9_extracted_k141_79264 | Low-quality |
| S9_extracted_k141_34058 | Low-quality |
| S9_extracted_k141_67947 | Low-quality |
| S9_extracted_k141_75664 | Not-determined |
| S9_extracted_k141_75665 | Low-quality |
| S9_extracted_k141_83081 | Low-quality |
| S9_extracted_k141_63932 | Not-determined |
| S9_extracted_k141_79269 | Not-determined |
| S9_extracted_k141_79270 | Not-determined |
| S9_extracted_k141_48938 | Low-quality |
| S9_extracted_k141_67955 | Not-determined |
| S9_extracted_k141_83091 | Low-quality |
| S9_extracted_k141_67961 | Not-determined |
| S9_extracted_k141_63948 | Not-determined |
| S9_extracted_k141_67964 | Not-determined |
| S9_extracted_k141_63951 | Not-determined |
| S9_extracted_k141_83098 | Not-determined |
| S9_extracted_k141_87057 | Not-determined |
| S9_extracted_k141_90599 | Not-determined |
| S9_extracted_k141_63957 | Not-determined |
| S9_extracted_k141_90603 | Low-quality |
| S9_extracted_k141_34084 | Not-determined |
| S9_extracted_k141_90605 | Not-determined |
| S9_extracted_k141_71912 | Not-determined |
| S9_extracted_k141_71917 | Not-determined |
| S9_extracted_k141_60387 | Not-determined |
| S9_extracted_k141_83104 | Not-determined |
| S9_extracted_k141_71920 | Not-determined |
| S9_extracted_k141_83107 | Not-determined |
| S5_vs_k141_113397\|\|full | Medium-quality |
| S9_extracted_k141_90621 | Not-determined |
| S9_extracted_k141_60398 | Not-determined |
| S9_extracted_k141_52223 | Low-quality |
| S9_extracted_k141_48958 | Not-determined |
| S9_extracted_k141_15017 | Low-quality |
| S9_extracted_k141_52228 | Not-determined |
| S9_extracted_k141_87088 | Not-determined |
| S9_extracted_k141_63973 | Low-quality |
| S9_extracted_k141_52233 | Not-determined |
| S9_extracted_k141_63975 | Not-determined |
| S9_extracted_k141_15028 | Not-determined |
| S9_extracted_k141_90635 | Not-determined |
| S9_extracted_k141_60412 | Low-quality |
| S9_extracted_k141_71943 | Not-determined |
| S9_extracted_k141_87099 | Not-determined |
| S9_extracted_k141_79299 | Not-determined |
| S9_extracted_k141_68003 | Low-quality |
| S9_extracted_k141_71948 | Not-determined |
| S9_extracted_k141_63985 | Not-determined |
| S9_extracted_k141_90645 | Not-determined |
| S9_extracted_k141_75688 | Not-determined |
| S9_extracted_k141_71955 | Low-quality |
| S9_extracted_k141_52249 | Not-determined |
| S9_extracted_k141_90653 | Not-determined |
| S9_extracted_k141_64002 | Not-determined |
| S9_extracted_k141_60429 | Low-quality |
| S9_extracted_k141_90655 | Not-determined |
| S9_extracted_k141_90657 | Low-quality |
| S9_extracted_k141_64006 | Not-determined |
| S9_extracted_k141_79300 | Low-quality |
| S9_extracted_k141_60431 | Not-determined |
| S9_extracted_k141_79311 | Not-determined |
| S9_extracted_k141_90666 | Low-quality |
| S9_extracted_k141_90667 | Not-determined |
| S9_extracted_k141_79313 | Not-determined |
| S9_extracted_k141_87115 | Not-determined |
| S9_extracted_k141_64009 | Low-quality |
| S9_extracted_k141_64011 | Not-determined |
| S9_extracted_k141_64018 | Not-determined |
| S9_extracted_k141_87125 | Low-quality |
| S9_extracted_k141_75711 | Not-determined |
| S9_extracted_k141_87126 | Not-determined |
| S9_extracted_k141_79324 | Not-determined |
| S9_extracted_k141_48999 | Not-determined |
| S9_extracted_k141_87129 | Not-determined |
| S9_extracted_k141_87130 | Not-determined |
| S9_extracted_k141_90683 | Not-determined |
| S9_extracted_k141_49003 | Not-determined |
| S9_extracted_k141_90685 | Low-quality |
| S9_extracted_k141_49004 | Not-determined |
| S9_extracted_k141_68011 | Not-determined |
| S9_extracted_k141_49008 | Not-determined |
| S9_extracted_k141_90689 | Not-determined |
| S9_extracted_k141_64034 | Not-determined |
| S9_extracted_k141_64037 | Low-quality |
| S9_extracted_k141_68017 | Not-determined |
| S9_extracted_k141_64039 | Not-determined |
| S9_extracted_k141_75720 | Low-quality |
| S9_extracted_k141_90696 | Not-determined |
| S9_extracted_k141_75725 | Not-determined |
| S9_extracted_k141_64045 | Not-determined |
| S9_extracted_k141_87148 | Not-determined |
| S9_extracted_k141_79348 | Not-determined |
| S9_extracted_k141_64049 | Not-determined |
| S9_extracted_k141_68032 | Not-determined |
| S9_extracted_k141_15085 | Not-determined |
| S9_extracted_k141_71990 | Not-determined |
| S9_extracted_k141_15087 | Not-determined |
| S9_extracted_k141_15094 | Low-quality |
| S9_extracted_k141_15095 | Low-quality |
| S9_extracted_k141_64061 | Not-determined |
| S9_extracted_k141_75747 | Low-quality |
| S9_extracted_k141_60457 | Not-determined |
| S9_extracted_k141_49034 | Low-quality |
| S9_extracted_k141_68047 | Not-determined |
| S9_extracted_k141_64067 | Not-determined |
| S9_extracted_k141_75753 | Not-determined |
| S9_extracted_k141_64071 | Not-determined |
| S9_extracted_k141_60460 | Low-quality |
| S9_extracted_k141_15109 | Low-quality |
| S9_extracted_k141_75768 | Not-determined |
| S9_extracted_k141_90708 | Not-determined |
| S9_extracted_k141_90709 | Not-determined |
| S9_extracted_k141_75771 | Low-quality |
| S9_extracted_k141_90710 | Not-determined |
| S9_extracted_k141_64081 | Not-determined |
| S9_extracted_k141_79389 | Not-determined |
| S9_extracted_k141_15125 | Not-determined |
| S9_extracted_k141_75773 | Not-determined |
| S9_extracted_k141_75781 | Not-determined |
| S9_extracted_k141_64086 | Not-determined |
| S9_extracted_k141_68074 | Low-quality |
| S9_extracted_k141_90721 | Low-quality |
| S9_extracted_k141_68082 | Low-quality |
| S9_extracted_k141_60480 | Not-determined |
| S9_extracted_k141_60483 | Not-determined |
| S9_extracted_k141_83116 | Not-determined |
| S9_extracted_k141_60487 | Low-quality |
| S9_extracted_k141_79418 | Not-determined |
| S9_extracted_k141_68095 | Not-determined |
| S9_extracted_k141_83121 | Not-determined |
| S9_extracted_k141_79424 | Not-determined |
| S9_extracted_k141_79427 | Not-determined |
| S9_extracted_k141_83130 | Not-determined |
| S9_extracted_k141_83132 | Not-determined |
| S9_extracted_k141_79434 | Not-determined |
| S9_extracted_k141_83134 | Not-determined |
| S9_extracted_k141_79437 | Not-determined |
| S9_extracted_k141_49086 | Not-determined |
| S9_extracted_k141_52277 | Low-quality |
| S9_extracted_k141_68100 | Not-determined |
| S9_extracted_k141_68103 | Not-determined |
| S9_extracted_k141_83140 | Not-determined |
| S9_extracted_k141_60501 | Not-determined |
| S9_extracted_k141_60503 | Not-determined |
| S9_extracted_k141_79453 | Not-determined |
| S9_extracted_k141_64112 | Low-quality |
| S9_extracted_k141_68114 | Not-determined |
| S9_extracted_k141_52283 | Low-quality |
| S9_extracted_k141_68117 | Not-determined |
| S9_extracted_k141_79456 | Low-quality |
| S9_extracted_k141_64119 | Not-determined |
| S9_extracted_k141_68121 | Not-determined |
| S9_extracted_k141_60519 | Not-determined |
| S9_extracted_k141_83156 | Not-determined |
| S9_extracted_k141_83159 | Not-determined |
| S9_extracted_k141_68136 | Not-determined |
| S9_extracted_k141_68137 | Not-determined |
| S9_extracted_k141_64130 | Not-determined |
| S9_extracted_k141_79492 | Low-quality |
| S9_extracted_k141_64137 | Low-quality |
| S9_extracted_k141_64140 | Low-quality |
| S9_extracted_k141_68149 | Low-quality |
| S9_extracted_k141_79496 | Low-quality |
| S9_extracted_k141_79500 | Not-determined |
| S9_extracted_k141_64149 | Not-determined |
| S9_extracted_k141_52314 | Not-determined |
| S9_extracted_k141_68156 | Not-determined |
| S9_extracted_k141_79512 | Low-quality |
| S9_extracted_k141_64158 | Not-determined |
| S9_extracted_k141_68163 | Not-determined |
| S9_extracted_k141_64165 | Low-quality |
| S9_extracted_k141_68167 | Not-determined |
| S9_extracted_k141_49113 | Low-quality |
| S9_extracted_k141_68175 | Not-determined |
| S9_extracted_k141_68180 | Not-determined |
| S9_extracted_k141_64175 | Low-quality |
| S9_extracted_k141_68184 | Low-quality |
| S9_extracted_k141_49126 | Not-determined |
| S9_extracted_k141_64182 | Not-determined |
| S9_extracted_k141_64183 | Not-determined |
| S9_extracted_k141_68191 | Not-determined |
| S9_extracted_k141_79539 | Not-determined |
| S9_extracted_k141_83219 | Not-determined |
| S9_extracted_k141_68193 | Not-determined |
| S9_extracted_k141_83220 | Not-determined |
| S9_extracted_k141_64188 | Low-quality |
| S9_extracted_k141_52336 | Not-determined |
| S9_extracted_k141_64196 | Not-determined |
| S9_extracted_k141_83226 | Not-determined |
| S9_extracted_k141_83231 | Not-determined |
| S9_extracted_k141_64199 | Not-determined |
| S9_extracted_k141_64200 | Low-quality |
| S9_extracted_k141_52348 | Low-quality |
| S9_extracted_k141_52350 | Not-determined |
| S9_extracted_k141_52351 | Not-determined |
| S9_extracted_k141_49138 | Not-determined |
| S9_extracted_k141_83236 | Not-determined |
| S9_extracted_k141_83242 | Low-quality |
| S9_extracted_k141_79553 | Not-determined |
| S9_extracted_k141_79564 | Not-determined |
| S9_extracted_k141_52380 | Not-determined |
| S9_extracted_k141_52381 | Not-determined |
| S9_extracted_k141_52388 | Not-determined |
| S9_extracted_k141_83269 | Not-determined |
| S9_extracted_k141_60577 | Not-determined |
| S9_extracted_k141_64216 | Low-quality |
| S9_extracted_k141_64221 | Not-determined |
| S9_extracted_k141_60584 | Low-quality |
| S9_extracted_k141_60587 | Not-determined |
| S9_extracted_k141_49173 | Not-determined |
| S9_extracted_k141_52394 | Not-determined |
| S9_extracted_k141_49174 | Not-determined |
| S9_extracted_k141_83283 | Not-determined |
| S9_extracted_k141_60592 | Not-determined |
| S9_extracted_k141_83284 | Not-determined |
| S9_extracted_k141_60593 | Not-determined |
| S9_extracted_k141_52397 | Not-determined |
| S9_extracted_k141_64235 | Low-quality |
| S9_extracted_k141_83293 | Not-determined |
| S9_extracted_k141_52404 | Not-determined |
| S9_extracted_k141_52405 | Not-determined |
| S9_extracted_k141_49183 | Not-determined |
| S9_extracted_k141_64245 | Not-determined |
| S9_extracted_k141_64246 | Not-determined |
| S9_extracted_k141_49195 | Not-determined |
| S9_extracted_k141_64256 | Not-determined |
| S9_extracted_k141_83314 | Not-determined |
| S9_extracted_k141_64257 | Not-determined |
| S9_extracted_k141_52432 | Not-determined |
| S9_extracted_k141_52435 | Not-determined |
| S9_extracted_k141_49212 | Not-determined |
| S9_extracted_k141_52443 | Not-determined |
| S9_extracted_k141_49219 | Not-determined |
| S9_extracted_k141_64276 | Not-determined |
| S9_extracted_k141_64279 | Not-determined |
| S9_extracted_k141_49221 | Not-determined |
| S9_extracted_k141_49224 | Not-determined |
| S9_extracted_k141_49225 | Not-determined |
| S9_extracted_k141_52451 | Not-determined |
| S9_extracted_k141_49238 | Not-determined |
| S9_extracted_k141_83353 | Not-determined |
| S9_extracted_k141_49239 | Not-determined |
| S9_extracted_k141_52464 | Not-determined |
| S9_extracted_k141_64310 | Low-quality |
| S9_extracted_k141_52478 | Not-determined |
| S9_extracted_k141_52497 | Not-determined |
| S9_extracted_k141_52504 | Not-determined |
| S9_extracted_k141_90738 | Not-determined |
| S9_extracted_k141_90739 | Low-quality |
| S9_extracted_k141_90753 | Not-determined |
| S9_extracted_k141_90761 | Low-quality |
| S9_extracted_k141_90763 | Not-determined |
| S9_extracted_k141_64331 | Not-determined |
| S9_extracted_k141_52584 | Not-determined |
| S9_extracted_k141_64345 | Not-determined |
| S9_extracted_k141_52589 | Not-determined |
| S9_extracted_k141_90789 | Not-determined |
| S9_extracted_k141_52591 | Not-determined |
| S9_extracted_k141_90790 | Low-quality |
| S9_extracted_k141_64360 | Not-determined |
| S9_extracted_k141_64361 | Not-determined |
| S9_extracted_k141_90797 | Not-determined |
| S9_extracted_k141_90802 | Not-determined |
| S9_extracted_k141_90804 | Not-determined |
| S9_extracted_k141_90807 | Not-determined |
| S9_extracted_k141_90808 | Not-determined |
| S9_extracted_k141_90810 | Not-determined |
| S9_extracted_k141_52611 | Not-determined |
| S9_extracted_k141_90818 | Not-determined |
| S9_extracted_k141_52619 | Not-determined |
| S9_extracted_k141_90830 | Low-quality |
| S9_extracted_k141_90839 | Not-determined |
| S9_extracted_k141_52629 | Not-determined |
| S9_extracted_k141_52630 | Low-quality |
| S9_extracted_k141_90844 | Low-quality |
| S9_extracted_k141_60625 | Not-determined |
| S9_extracted_k141_64398 | Not-determined |
| S9_extracted_k141_64399 | Low-quality |
| S9_extracted_k141_52651 | Not-determined |
| S9_extracted_k141_52653 | Not-determined |
| S9_extracted_k141_64406 | Low-quality |
| S9_extracted_k141_52661 | Not-determined |
| S9_extracted_k141_52707 | Not-determined |
| S9_extracted_k141_52710 | Not-determined |
| S9_extracted_k141_52713 | Not-determined |
| S9_extracted_k141_52715 | Not-determined |
| S9_extracted_k141_52722 | Not-determined |
| S9_extracted_k141_52738 | Not-determined |
| S9_extracted_k141_52756 | Not-determined |
| S9_extracted_k141_52787 | Not-determined |
| S9_extracted_k141_52792 | Not-determined |
| S9_extracted_k141_52798 | Not-determined |
| S9_extracted_k141_52803 | Low-quality |
| S9_extracted_k141_52813 | Not-determined |
| S9_extracted_k141_52826 | Low-quality |
| S9_extracted_k141_52828 | Not-determined |
| S9_extracted_k141_52834 | Not-determined |
| S9_extracted_k141_52836 | Not-determined |
| S9_extracted_k141_52850 | Low-quality |
| S9_extracted_k141_52853 | Not-determined |
| S9_extracted_k141_52862 | Not-determined |
| S9_extracted_k141_52877 | Low-quality |
| S9_extracted_k141_52880 | Not-determined |
| S9_extracted_k141_52887 | Not-determined |
| S9_extracted_k141_52893 | Not-determined |
| S9_extracted_k141_52895 | Not-determined |
| S9_extracted_k141_52897 | Not-determined |
| S9_extracted_k141_52929 | Not-determined |
| S9_extracted_k141_52937 | Not-determined |
| S9_extracted_k141_52938 | Not-determined |
| S9_extracted_k141_52942 | Not-determined |
| S9_extracted_k141_52943 | Not-determined |
| S10_extracted_80712 | Medium-quality |
| S9_extracted_k141_52949 | Low-quality |
| S9_extracted_k141_52965 | Low-quality |
| S9_extracted_k141_52967 | Not-determined |
| S9_extracted_k141_52974 | Not-determined |
| S9_extracted_k141_52982 | Not-determined |
| S9_extracted_k141_52996 | Low-quality |
| S9_extracted_k141_53022 | Not-determined |
| S9_extracted_k141_53030 | Not-determined |
| S10_vs_80712\|\|full | Medium-quality |
| S9_extracted_k141_90853 | Low-quality |
| S12_extracted_k141_67909 | Medium-quality |
| S12_vs_k141_123620\|\|full | Complete |
| S12_vs_k141_67909\|\|full | Medium-quality |
| S9_extracted_k141_90880 | Not-determined |
| S9_extracted_k141_90883 | Not-determined |
| S9_extracted_k141_90885 | Not-determined |
| S9_extracted_k141_90888 | Low-quality |
| S9_extracted_k141_90895 | Not-determined |
| S9_extracted_k141_90897 | Not-determined |
| S9_extracted_k141_90901 | Not-determined |
| S9_extracted_k141_90902 | Not-determined |
| S9_extracted_k141_90907 | Not-determined |
| S9_extracted_k141_90915 | Low-quality |
| S9_extracted_k141_90922 | Not-determined |
| S9_extracted_k141_90929 | Not-determined |
| S9_extracted_k141_90931 | Not-determined |
| S9_extracted_k141_90932 | Not-determined |
| S9_extracted_k141_90935 | Not-determined |
| S16_vs_82116\|\|full | High-quality |
| S13_vs_30101\|\|full | Medium-quality |
| S9_vs_k141_49529\|\|full | Not-determined |
| S6_vs_k141_46039\|\|full | Medium-quality |
| S16_extracted_161663 | Medium-quality |
| S10_vs_37379\|\|full | Medium-quality |
| S16_vs_161663\|\|full | Medium-quality |
| S9_vs_k141_49943\|\|full | Not-determined |
| S8_vs_k141_179208\|\|full | Medium-quality |
| S5_vs_k141_117319\|\|full | Complete |
| S14_vs_k141_25852\|\|full | Medium-quality |
| S6_extracted_k141_56486 | Complete |
| S9_vs_k141_16826\|\|full | Low-quality |
| S5_extracted_k141_145580 | Complete |
| S6_vs_k141_56486\|\|full | High-quality |
| S9_vs_k141_55092\|\|full | Not-determined |
| S5_vs_k141_145580\|\|full | High-quality |
| S11_extracted_k141_104033 | Complete |
| S11_vs_k141_104033\|\|full | High-quality |
| S5_vs_k141_124677\|\|full | Medium-quality |
| S3_extracted_k141_25163 | Complete |
| S3_vs_k141_17168\|\|full | Complete |
| S3_vs_k141_25163\|\|full | High-quality |
| S10_extracted_3644 | High-quality |
| S10_vs_3644\|\|full | High-quality |
| S8_extracted_k141_161703 | High-quality |
| S8_vs_k141_161703\|\|full | High-quality |
| S9_vs_k141_90853\|\|full | Low-quality |
| S9_vs_k141_90854\|\|full | Not-determined |
| S14_vs_k141_9162\|\|full | Medium-quality |
| S17_extracted_k141_81232 | Complete |
| S7_extracted_136803 | Complete |
| S17_vs_k141_81232\|\|full | High-quality |
| S9_vs_k141_90862\|\|full | Not-determined |
| S7_vs_136803\|\|full | High-quality |
| S9_vs_k141_90864\|\|full | Low-quality |
| S9_vs_k141_90865\|\|full | Low-quality |
| S13_extracted_40252 | Complete |
| S9_vs_k141_90868\|\|full | Low-quality |
| S1_extracted_k141_159951 | Complete |
| S9_vs_k141_90875\|\|full | Low-quality |
| S9_vs_k141_90894\|\|full | Not-determined |
| S9_vs_k141_90915\|\|full | Low-quality |
| S9_vs_k141_41681\|\|full | Not-determined |
| S9_vs_k141_64417\|\|full | Low-quality |
| S9_vs_k141_7581\|\|full | Not-determined |
| S9_vs_k141_7583\|\|full | Low-quality |
| S9_vs_k141_30316\|\|full | Low-quality |
| S9_vs_k141_56843\|\|full | Low-quality |

| | |
|---|---|
| S9_vs_k141_41688\|\|full | Low-quality |
| S9_vs_k141_83371\|\|full | Low-quality |
| S9_vs_k141_34117\|\|full | Low-quality |
| S9_vs_k141_53066\|\|full | Low-quality |
| S9_vs_k141_3803\|\|full | Low-quality |
| S9_vs_k141_3810\|\|full | Not-determined |
| S9_vs_k141_3814\|\|full | Low-quality |
| S9_vs_k141_75802\|\|full | Low-quality |
| S9_vs_k141_18983\|\|full | Low-quality |
| S9_vs_k141_26568\|\|full | Low-quality |
| S9_vs_k141_53086\|\|full | Not-determined |
| S9_vs_k141_75804\|\|full | Low-quality |
| S9_vs_k141_87184\|\|full | Not-determined |
| S9_vs_k141_3819\|\|full | Low-quality |
| S9_vs_k141_22773\|\|full | Not-determined |
| S9_vs_k141_34149\|\|full | Not-determined |
| S9_vs_k141_7617\|\|full | Low-quality |
| S9_vs_k141_83412\|\|full | Low-quality |
| S9_vs_k141_30339\|\|full | Low-quality |
| S9_vs_k141_83413\|\|full | Not-determined |
| S9_vs_k141_68233\|\|full | Low-quality |
| S9_vs_k141_72051\|\|full | Low-quality |
| S9_vs_k141_75816\|\|full | Not-determined |
| S9_vs_k141_68239\|\|full | Low-quality |
| S9_vs_k141_49309\|\|full | Low-quality |
| S9_vs_k141_60649\|\|full | Low-quality |
| S9_vs_k141_26585\|\|full | Low-quality |
| S9_vs_k141_79625\|\|full | Not-determined |
| S9_vs_k141_53092\|\|full | Not-determined |
| S9_vs_k141_30347\|\|full | Not-determined |
| S9_vs_k141_83430\|\|full | Not-determined |
| S9_vs_k141_75836\|\|full | Not-determined |
| S9_vs_k141_15232\|\|full | Not-determined |
| S9_vs_k141_22804\|\|full | Low-quality |
| S9_vs_k141_37919\|\|full | Low-quality |
| S9_vs_k141_83431\|\|full | Not-determined |
| S9_vs_k141_26597\|\|full | Low-quality |
| S9_vs_k141_45526\|\|full | Low-quality |
| S9_vs_k141_34191\|\|full | Not-determined |
| S9_vs_k141_83436\|\|full | Low-quality |
| S9_vs_k141_22811\|\|full | Not-determined |
| S9_vs_k141_79657\|\|full | Low-quality |
| S9_vs_k141_49330\|\|full | Low-quality |
| S9_vs_k141_15247\|\|full | Not-determined |
| S9_vs_k141_37933\|\|full | Low-quality |
| S9_vs_k141_18991\|\|full | Not-determined |
| S9_vs_k141_34201\|\|full | Low-quality |
| S9_vs_k141_7655\|\|full | Not-determined |
| S9_vs_k141_22823\|\|full | Not-determined |
| S9_vs_k141_64509\|\|full | Not-determined |
| S9_vs_k141_49347\|\|full | Low-quality |
| S9_vs_k141_75859\|\|full | Low-quality |
| S9_vs_k141_60688\|\|full | Low-quality |
| S9_vs_k141_83457\|\|full | Not-determined |
| S9_vs_k141_87242\|\|full | Low-quality |
| S9_vs_k141_64510\|\|full | Not-determined |
| S9_vs_k141_68267\|\|full | Low-quality |
| S9_vs_k141_22825\|\|full | Not-determined |
| S9_vs_k141_83459\|\|full | Not-determined |
| S9_vs_k141_68270\|\|full | Not-determined |
| S9_vs_k141_83460\|\|full | Not-determined |
| S9_vs_k141_60696\|\|full | Not-determined |
| S9_vs_k141_3870\|\|full | Not-determined |
| S9_vs_k141_60700\|\|full | Not-determined |
| S9_vs_k141_3871\|\|full | Not-determined |
| S9_vs_k141_37944\|\|full | Not-determined |
| S9_vs_k141_15268\|\|full | Not-determined |
| S9_vs_k141_83469\|\|full | Not-determined |
| S9_vs_k141_7676\|\|full | Low-quality |
| S9_vs_k141_30401\|\|full | Not-determined |
| S9_vs_k141_83472\|\|full | Low-quality |
| S9_vs_k141_19034\|\|full | Low-quality |
| S9_vs_k141_19039\|\|full | Not-determined |
| S9_vs_k141_19046\|\|full | Low-quality |
| S9_vs_k141_22868\|\|full | Low-quality |
| S9_vs_k141_15289\|\|full | Low-quality |
| S9_vs_k141_53171\|\|full | Not-determined |
| S9_vs_k141_3911\|\|full | Not-determined |
| S9_vs_k141_87293\|\|full | Low-quality |
| S9_vs_k141_41792\|\|full | Low-quality |
| S9_vs_k141_97\|\|full | Not-determined |
| S9_vs_k141_34246\|\|full | Not-determined |
| S9_vs_k141_83498\|\|full | Low-quality |
| S9_vs_k141_60749\|\|full | Low-quality |
| S9_vs_k141_15310\|\|full | Low-quality |
| S9_vs_k141_60754\|\|full | Low-quality |
| S9_vs_k141_38001\|\|full | Not-determined |
| S9_vs_k141_30449\|\|full | Low-quality |
| S9_vs_k141_83517\|\|full | Not-determined |
| S9_vs_k141_19064\|\|full | Not-determined |
| S9_vs_k141_53204\|\|full | Not-determined |
| S9_vs_k141_34264\|\|full | Not-determined |
| S9_vs_k141_75906\|\|full | Not-determined |
| S9_vs_k141_11521\|\|full | Low-quality |
| S9_vs_k141_79697\|\|full | Not-determined |
| S1_vs_k141_159951\|\|full | High-quality |
| S9_vs_k141_11522\|\|full | Not-determined |
| S9_vs_k141_87322\|\|full | Low-quality |
| S9_vs_k141_53222\|\|full | Low-quality |
| S9_vs_k141_11523\|\|full | Not-determined |
| S9_vs_k141_79703\|\|full | Low-quality |
| S9_vs_k141_38045\|\|full | Low-quality |
| S9_vs_k141_22918\|\|full | Not-determined |
| S9_vs_k141_79709\|\|full | Low-quality |
| S9_vs_k141_34287\|\|full | Low-quality |
| S9_vs_k141_26647\|\|full | Low-quality |
| S9_vs_k141_30489\|\|full | Not-determined |
| S9_vs_k141_164\|\|full | Low-quality |
| S9_vs_k141_87330\|\|full | Not-determined |
| S9_vs_k141_68296\|\|full | Low-quality |
| S9_vs_k141_87334\|\|full | Low-quality |
| S9_vs_k141_53244\|\|full | Low-quality |
| S9_vs_k141_68298\|\|full | Not-determined |
| S9_vs_k141_45573\|\|full | Not-determined |
| S9_vs_k141_60804\|\|full | Low-quality |
| S9_vs_k141_87340\|\|full | Low-quality |
| S9_vs_k141_175\|\|full | Not-determined |
| S9_vs_k141_83562\|\|full | Low-quality |
| S9_vs_k141_22925\|\|full | Not-determined |
| S9_vs_k141_45576\|\|full | Not-determined |
| S9_vs_k141_60813\|\|full | Low-quality |
| S9_vs_k141_22928\|\|full | Low-quality |
| S9_vs_k141_19122\|\|full | Low-quality |
| S9_vs_k141_60816\|\|full | Low-quality |
| S9_vs_k141_22932\|\|full | Low-quality |
| S9_vs_k141_11573\|\|full | Low-quality |
| S9_vs_k141_30531\|\|full | Low-quality |
| S9_vs_k141_38093\|\|full | Not-determined |
| S9_vs_k141_15329\|\|full | Not-determined |
| S9_vs_k141_45601\|\|full | Not-determined |
| S9_vs_k141_199\|\|full | Low-quality |
| S9_vs_k141_34333\|\|full | Not-determined |
| S9_vs_k141_45602\|\|full | Low-quality |
| S9_vs_k141_56959\|\|full | Low-quality |
| S9_vs_k141_11586\|\|full | Not-determined |
| S9_vs_k141_79752\|\|full | Not-determined |
| S9_vs_k141_87368\|\|full | Low-quality |
| S9_vs_k141_38100\|\|full | Low-quality |
| S9_vs_k141_72127\|\|full | Low-quality |
| S9_vs_k141_30556\|\|full | Not-determined |
| S9_vs_k141_19154\|\|full | Low-quality |
| S9_vs_k141_11606\|\|full | Not-determined |
| S9_vs_k141_45624\|\|full | Low-quality |
| S9_vs_k141_34379\|\|full | Low-quality |
| S9_vs_k141_68351\|\|full | Low-quality |
| S9_vs_k141_19157\|\|full | Low-quality |
| S9_vs_k141_56986\|\|full | Low-quality |
| S9_vs_k141_34388\|\|full | Not-determined |
| S9_vs_k141_64591\|\|full | Low-quality |
| S9_vs_k141_19176\|\|full | Low-quality |
| S9_vs_k141_7716\|\|full | Low-quality |
| S9_vs_k141_45651\|\|full | Not-determined |
| S9_vs_k141_38148\|\|full | Low-quality |
| S9_vs_k141_57003\|\|full | Not-determined |
| S9_vs_k141_64608\|\|full | Not-determined |
| S9_vs_k141_4043\|\|full | Not-determined |
| S9_vs_k141_49380\|\|full | Not-determined |
| S9_vs_k141_68371\|\|full | Not-determined |
| S9_vs_k141_11646\|\|full | Low-quality |
| S9_vs_k141_7718\|\|full | Low-quality |
| S9_vs_k141_83574\|\|full | Not-determined |
| S9_vs_k141_276\|\|full | Low-quality |
| S9_vs_k141_53342\|\|full | Not-determined |
| S9_vs_k141_19208\|\|full | Low-quality |
| S9_vs_k141_83584\|\|full | Low-quality |
| S9_vs_k141_19212\|\|full | Low-quality |
| S9_vs_k141_49397\|\|full | Low-quality |
| S9_vs_k141_30624\|\|full | Low-quality |
| S9_vs_k141_72175\|\|full | Low-quality |
| S9_vs_k141_15416\|\|full | Low-quality |
| S9_vs_k141_68394\|\|full | Low-quality |
| S9_vs_k141_45669\|\|full | Low-quality |
| S9_vs_k141_45670\|\|full | Low-quality |
| S9_vs_k141_83593\|\|full | Low-quality |
| S9_vs_k141_4068\|\|full | Low-quality |
| S9_vs_k141_53355\|\|full | Not-determined |
| S9_vs_k141_45678\|\|full | Low-quality |
| S9_vs_k141_49422\|\|full | Not-determined |
| S9_vs_k141_34461\|\|full | Not-determined |
| S9_vs_k141_322\|\|full | Not-determined |
| S9_vs_k141_26724\|\|full | Low-quality |
| S9_vs_k141_34462\|\|full | Not-determined |
| S9_vs_k141_41929\|\|full | Low-quality |
| S9_vs_k141_79846\|\|full | Low-quality |
| S9_vs_k141_64649\|\|full | Low-quality |
| S9_vs_k141_38192\|\|full | Not-determined |
| S9_vs_k141_41933\|\|full | Not-determined |
| S9_vs_k141_83605\|\|full | Low-quality |
| S9_vs_k141_68415\|\|full | Not-determined |
| S9_vs_k141_30647\|\|full | Low-quality |
| S9_vs_k141_38198\|\|full | Not-determined |
| S9_vs_k141_4101\|\|full | Not-determined |
| S9_vs_k141_53379\|\|full | Low-quality |
| S9_vs_k141_68417\|\|full | Low-quality |
| S9_vs_k141_53383\|\|full | Not-determined |
| S9_vs_k141_4102\|\|full | Low-quality |
| S9_vs_k141_53388\|\|full | Low-quality |
| S9_vs_k141_87452\|\|full | Not-determined |
| S9_vs_k141_7776\|\|full | Low-quality |
| S9_vs_k141_64671\|\|full | Not-determined |
| S9_vs_k141_7778\|\|full | Low-quality |
| S9_vs_k141_72212\|\|full | Low-quality |
| S9_vs_k141_64675\|\|full | Not-determined |
| S9_vs_k141_34494\|\|full | Low-quality |
| S9_vs_k141_57044\|\|full | Not-determined |
| S9_vs_k141_7779\|\|full | Low-quality |
| S9_vs_k141_23043\|\|full | Low-quality |
| S9_vs_k141_7784\|\|full | Low-quality |
| S9_vs_k141_72223\|\|full | Low-quality |
| S9_vs_k141_34504\|\|full | Low-quality |
| S9_vs_k141_57053\|\|full | Not-determined |
| S9_vs_k141_83625\|\|full | Low-quality |
| S9_vs_k141_64690\|\|full | Low-quality |
| S9_vs_k141_72232\|\|full | Low-quality |
| S9_vs_k141_64696\|\|full | Low-quality |
| S9_vs_k141_49443\|\|full | Not-determined |
| S9_vs_k141_45682\|\|full | Low-quality |
| S9_vs_k141_15459\|\|full | Low-quality |
| S9_vs_k141_60854\|\|full | Low-quality |
| S9_vs_k141_23060\|\|full | Not-determined |
| S9_vs_k141_45690\|\|full | Low-quality |
| S9_vs_k141_49455\|\|full | Low-quality |
| S9_vs_k141_79890\|\|full | Low-quality |
| S9_vs_k141_38250\|\|full | Not-determined |
| S9_vs_k141_49466\|\|full | Not-determined |
| S9_vs_k141_79894\|\|full | Low-quality |
| S9_vs_k141_60867\|\|full | Not-determined |
| S9_vs_k141_83657\|\|full | Low-quality |
| S9_vs_k141_38259\|\|full | Not-determined |
| S9_vs_k141_49470\|\|full | Low-quality |
| S9_vs_k141_60871\|\|full | Low-quality |
| S9_vs_k141_34525\|\|full | Low-quality |
| S9_vs_k141_23088\|\|full | Not-determined |
| S9_vs_k141_7821\|\|full | Low-quality |
| S13_vs_40252\|\|full | High-quality |
| S9_vs_k141_57083\|\|full | Low-quality |
| S9_vs_k141_49473\|\|full | Not-determined |
| S9_vs_k141_15495\|\|full | Low-quality |
| S9_vs_k141_60878\|\|full | Not-determined |
| S9_vs_k141_49478\|\|full | Low-quality |
| S9_vs_k141_395\|\|full | Low-quality |
| S9_vs_k141_34535\|\|full | Low-quality |
| S9_vs_k141_34537\|\|full | Low-quality |
| S9_vs_k141_7829\|\|full | Low-quality |
| S9_vs_k141_15509\|\|full | Not-determined |
| S9_vs_k141_42020\|\|full | Not-determined |
| S9_vs_k141_15510\|\|full | Low-quality |
| S9_vs_k141_7834\|\|full | Not-determined |
| S9_vs_k141_83670\|\|full | Low-quality |
| S9_vs_k141_72295\|\|full | Low-quality |
| S9_vs_k141_4131\|\|full | Low-quality |
| S9_vs_k141_64753\|\|full | Not-determined |
| S9_vs_k141_57111\|\|full | Low-quality |
| S9_vs_k141_4137\|\|full | Not-determined |
| S9_vs_k141_49504\|\|full | Low-quality |
| S9_vs_k141_15521\|\|full | Low-quality |
| S9_vs_k141_415\|\|full | Low-quality |
| S9_vs_k141_30719\|\|full | Low-quality |
| S9_vs_k141_83685\|\|full | Not-determined |
| S7_vs_137915\|\|full | High-quality |
| S9_vs_k141_60911\|\|full | Low-quality |
| S9_vs_k141_23120\|\|full | Low-quality |
| S9_vs_k141_87556\|\|full | Not-determined |
| S9_vs_k141_436\|\|full | Low-quality |
| S9_vs_k141_72300\|\|full | Low-quality |
| S9_vs_k141_57135\|\|full | Low-quality |
| S9_vs_k141_23126\|\|full | Low-quality |
| S9_vs_k141_15536\|\|full | Low-quality |
| S9_vs_k141_30738\|\|full | Low-quality |
| S9_vs_k141_60928\|\|full | Low-quality |
| S9_vs_k141_57141\|\|full | Low-quality |
| S9_vs_k141_68530\|\|full | Not-determined |
| S9_vs_k141_11724\|\|full | Not-determined |
| S9_vs_k141_64786\|\|full | Not-determined |
| S9_vs_k141_34548\|\|full | Low-quality |
| S9_vs_k141_4173\|\|full | Low-quality |
| S9_vs_k141_83710\|\|full | Low-quality |
| S9_vs_k141_42069\|\|full | Low-quality |
| S9_vs_k141_72322\|\|full | Low-quality |
| S9_vs_k141_26819\|\|full | Low-quality |
| S9_vs_k141_83718\|\|full | Low-quality |
| S9_vs_k141_49551\|\|full | Low-quality |
| S9_vs_k141_7878\|\|full | Low-quality |
| S9_vs_k141_64797\|\|full | Not-determined |
| S9_vs_k141_64798\|\|full | Low-quality |
| S9_vs_k141_30768\|\|full | Not-determined |
| S9_vs_k141_68558\|\|full | Not-determined |
| S9_vs_k141_4192\|\|full | Low-quality |
| S9_vs_k141_76000\|\|full | Low-quality |
| S9_vs_k141_87586\|\|full | Low-quality |
| S9_vs_k141_72340\|\|full | Not-determined |
| S9_vs_k141_463\|\|full | Not-determined |
| S9_vs_k141_26844\|\|full | Low-quality |
| S9_vs_k141_64809\|\|full | Not-determined |
| S9_vs_k141_23157\|\|full | Not-determined |
| S9_vs_k141_19310\|\|full | Low-quality |
| S9_vs_k141_34564\|\|full | Low-quality |
| S9_vs_k141_57168\|\|full | Low-quality |
| S9_vs_k141_72350\|\|full | Not-determined |
| S9_vs_k141_76015\|\|full | Not-determined |
| S9_vs_k141_23159\|\|full | Low-quality |
| S9_vs_k141_19323\|\|full | Low-quality |
| S9_vs_k141_30792\|\|full | Not-determined |
| S9_vs_k141_23168\|\|full | Low-quality |
| S9_vs_k141_30795\|\|full | Not-determined |
| S9_vs_k141_19331\|\|full | Low-quality |
| S9_vs_k141_38325\|\|full | Low-quality |
| S9_vs_k141_23174\|\|full | Low-quality |
| S9_vs_k141_68579\|\|full | Low-quality |
| S9_vs_k141_57174\|\|full | Low-quality |
| S9_vs_k141_26873\|\|full | Low-quality |
| S9_vs_k141_45763\|\|full | Low-quality |
| S9_vs_k141_19338\|\|full | Low-quality |
| S9_vs_k141_38333\|\|full | Low-quality |
| S9_vs_k141_26879\|\|full | Low-quality |
| S9_vs_k141_61011\|\|full | Low-quality |
| S9_vs_k141_76029\|\|full | Not-determined |
| S9_vs_k141_492\|\|full | Not-determined |
| S9_vs_k141_72364\|\|full | Not-determined |
| S9_vs_k141_49612\|\|full | Low-quality |
| S9_vs_k141_72370\|\|full | Low-quality |
| S9_vs_k141_26888\|\|full | Low-quality |
| S3_extracted_k141_127447 | Complete |
| S9_vs_k141_61025\|\|full | Not-determined |
| S9_vs_k141_76046\|\|full | Low-quality |
| S9_vs_k141_45783\|\|full | Low-quality |
| S9_vs_k141_87627\|\|full | Low-quality |
| S9_vs_k141_23181\|\|full | Low-quality |
| S9_vs_k141_53442\|\|full | Not-determined |
| S9_vs_k141_15638\|\|full | Low-quality |
| S9_vs_k141_72385\|\|full | Not-determined |
| S9_vs_k141_23184\|\|full | Not-determined |
| S9_vs_k141_57212\|\|full | Low-quality |
| S9_vs_k141_15647\|\|full | Not-determined |
| S9_vs_k141_34578\|\|full | Low-quality |
| S9_vs_k141_23189\|\|full | Low-quality |
| S9_vs_k141_76066\|\|full | Low-quality |
| S9_vs_k141_23194\|\|full | Low-quality |
| S9_vs_k141_53457\|\|full | Low-quality |
| S9_vs_k141_19367\|\|full | Not-determined |
| S9_vs_k141_15656\|\|full | Low-quality |
| S9_vs_k141_4276\|\|full | Low-quality |
| S9_vs_k141_26934\|\|full | Not-determined |
| S9_vs_k141_76079\|\|full | Low-quality |
| S9_vs_k141_34601\|\|full | Not-determined |
| S9_vs_k141_30827\|\|full | Low-quality |
| S9_vs_k141_11866\|\|full | Low-quality |
| S9_vs_k141_76096\|\|full | Low-quality |
| S9_vs_k141_83828\|\|full | Low-quality |
| S9_vs_k141_76102\|\|full | Low-quality |
| S9_vs_k141_11881\|\|full | Not-determined |
| S9_vs_k141_49677\|\|full | Low-quality |
| S9_vs_k141_30841\|\|full | Low-quality |
| S9_vs_k141_76114\|\|full | Not-determined |
| S9_vs_k141_83838\|\|full | Low-quality |
| S9_vs_k141_87663\|\|full | Low-quality |
| S9_vs_k141_61075\|\|full | Low-quality |
| S9_vs_k141_42194\|\|full | Low-quality |
| S9_vs_k141_72431\|\|full | Not-determined |
| S9_vs_k141_61077\|\|full | Not-determined |
| S9_vs_k141_72435\|\|full | Low-quality |
| S9_vs_k141_564\|\|full | Low-quality |
| S9_vs_k141_64825\|\|full | Low-quality |
| S9_vs_k141_568\|\|full | Not-determined |
| S9_vs_k141_80026\|\|full | Not-determined |
| S9_vs_k141_87678\|\|full | Low-quality |
| S9_vs_k141_30850\|\|full | Not-determined |
| S9_vs_k141_23240\|\|full | Low-quality |
| S9_vs_k141_11911\|\|full | Low-quality |
| S9_vs_k141_64834\|\|full | Low-quality |
| S9_vs_k141_64837\|\|full | Low-quality |
| S9_vs_k141_38455\|\|full | Low-quality |
| S9_vs_k141_30862\|\|full | Low-quality |
| S9_vs_k141_11921\|\|full | Low-quality |
| S9_vs_k141_61112\|\|full | Low-quality |
| S9_vs_k141_19428\|\|full | Low-quality |
| S9_vs_k141_83857\|\|full | Not-determined |
| S9_vs_k141_80048\|\|full | Not-determined |
| S9_vs_k141_87711\|\|full | Low-quality |
| S9_vs_k141_72461\|\|full | Low-quality |
| S9_vs_k141_7988\|\|full | Low-quality |
| S9_vs_k141_38468\|\|full | Not-determined |
| S9_vs_k141_72464\|\|full | Low-quality |
| S9_vs_k141_19439\|\|full | Low-quality |
| S9_vs_k141_11942\|\|full | Low-quality |
| S9_vs_k141_7993\|\|full | Not-determined |
| S9_vs_k141_30888\|\|full | Not-determined |
| S9_vs_k141_72477\|\|full | Low-quality |
| S9_vs_k141_8000\|\|full | Low-quality |
| S9_vs_k141_61145\|\|full | Low-quality |
| S9_vs_k141_23267\|\|full | Not-determined |
| S9_vs_k141_27057\|\|full | Not-determined |
| S9_vs_k141_38472\|\|full | Not-determined |
| S9_vs_k141_76178\|\|full | Low-quality |
| S9_vs_k141_15732\|\|full | Low-quality |
| S9_vs_k141_11969\|\|full | Low-quality |
| S9_vs_k141_38483\|\|full | Low-quality |
| S9_vs_k141_8020\|\|full | Low-quality |
| S9_vs_k141_15743\|\|full | Low-quality |
| S9_vs_k141_27077\|\|full | Not-determined |
| S9_vs_k141_23279\|\|full | Low-quality |
| S9_vs_k141_4342\|\|full | Low-quality |
| S9_vs_k141_87752\|\|full | Low-quality |
| S9_vs_k141_87753\|\|full | Low-quality |
| S9_vs_k141_53572\|\|full | Low-quality |
| S9_vs_k141_30926\|\|full | Low-quality |
| S9_vs_k141_42276\|\|full | Low-quality |
| S9_vs_k141_80079\|\|full | Low-quality |
| S9_vs_k141_30929\|\|full | Low-quality |
| S9_vs_k141_15768\|\|full | Low-quality |
| S9_vs_k141_38516\|\|full | Low-quality |
| S9_vs_k141_12008\|\|full | Low-quality |
| S9_vs_k141_61202\|\|full | Low-quality |
| S9_vs_k141_80085\|\|full | Low-quality |
| S9_vs_k141_640\|\|full | Low-quality |
| S9_vs_k141_53586\|\|full | Low-quality |
| S9_vs_k141_12010\|\|full | Low-quality |
| S9_vs_k141_30956\|\|full | Not-determined |
| S9_vs_k141_8062\|\|full | Not-determined |
| S9_vs_k141_15794\|\|full | Low-quality |
| S9_vs_k141_72526\|\|full | Low-quality |
| S9_vs_k141_34648\|\|full | Low-quality |
| S9_vs_k141_30963\|\|full | Not-determined |
| S9_vs_k141_61223\|\|full | Not-determined |
| S9_vs_k141_72531\|\|full | Low-quality |
| S9_vs_k141_49695\|\|full | Low-quality |
| S9_vs_k141_68606\|\|full | Low-quality |
| S9_vs_k141_38540\|\|full | Low-quality |
| S9_vs_k141_30968\|\|full | Low-quality |
| S9_vs_k141_76216\|\|full | Low-quality |
| S9_vs_k141_45858\|\|full | Low-quality |
| S9_vs_k141_87793\|\|full | Low-quality |
| S9_vs_k141_76220\|\|full | Not-determined |
| S9_vs_k141_45865\|\|full | Not-determined |
| S9_vs_k141_15823\|\|full | Low-quality |
| S3_vs_k141_127447\|\|full | High-quality |
| S9_vs_k141_666\|\|full | Low-quality |
| S9_vs_k141_34670\|\|full | Low-quality |
| S9_vs_k141_61245\|\|full | Low-quality |
| S9_vs_k141_45889\|\|full | Low-quality |
| S9_vs_k141_61251\|\|full | Not-determined |
| S9_vs_k141_80135\|\|full | Low-quality |
| S9_vs_k141_31007\|\|full | Low-quality |
| S9_vs_k141_34678\|\|full | Not-determined |
| S9_vs_k141_72581\|\|full | Not-determined |
| S9_vs_k141_34682\|\|full | Not-determined |
| S3_extracted_k141_115304 | Complete |
| S9_vs_k141_45910\|\|full | Low-quality |
| S9_vs_k141_68651\|\|full | Low-quality |
| S9_vs_k141_15835\|\|full | Not-determined |
| S9_vs_k141_87845\|\|full | Not-determined |
| S9_vs_k141_68658\|\|full | Not-determined |
| S9_vs_k141_8096\|\|full | Low-quality |
| S9_vs_k141_68666\|\|full | Low-quality |
| S9_vs_k141_72605\|\|full | Low-quality |
| S9_vs_k141_87855\|\|full | Low-quality |
| S9_vs_k141_61287\|\|full | Not-determined |
| S9_vs_k141_27161\|\|full | Low-quality |
| S9_vs_k141_4360\|\|full | Not-determined |
| S9_vs_k141_23302\|\|full | Low-quality |
| S9_vs_k141_80189\|\|full | Low-quality |
| S9_vs_k141_45938\|\|full | Low-quality |
| S9_vs_k141_19593\|\|full | Low-quality |
| S9_vs_k141_87864\|\|full | Low-quality |
| S9_vs_k141_68688\|\|full | Low-quality |
| S9_vs_k141_42419\|\|full | Low-quality |
| S9_vs_k141_87865\|\|full | Low-quality |
| S9_vs_k141_27192\|\|full | Not-determined |
| S9_vs_k141_19613\|\|full | Low-quality |
| S9_vs_k141_53736\|\|full | Low-quality |
| S9_vs_k141_64929\|\|full | Low-quality |
| S9_vs_k141_38613\|\|full | Low-quality |
| S9_vs_k141_23349\|\|full | Not-determined |
| S9_vs_k141_80211\|\|full | Not-determined |
| S9_vs_k141_53738\|\|full | Low-quality |
| S9_vs_k141_27194\|\|full | Low-quality |
| S9_vs_k141_61326\|\|full | Not-determined |
| S9_vs_k141_68694\|\|full | Low-quality |
| S9_vs_k141_80226\|\|full | Low-quality |
| S9_vs_k141_64939\|\|full | Not-determined |
| S9_vs_k141_15895\|\|full | Not-determined |
| S9_vs_k141_80231\|\|full | Low-quality |
| S9_vs_k141_68710\|\|full | Not-determined |
| S9_vs_k141_83981\|\|full | Low-quality |
| S9_vs_k141_87901\|\|full | Not-determined |
| S9_vs_k141_4386\|\|full | Low-quality |
| S9_vs_k141_49724\|\|full | Low-quality |
| S9_vs_k141_15920\|\|full | Low-quality |
| S9_vs_k141_83995\|\|full | Not-determined |
| S9_vs_k141_61343\|\|full | Low-quality |
| S4_extracted_2441 | Complete |
| S9_vs_k141_12124\|\|full | Low-quality |
| S9_vs_k141_68738\|\|full | Low-quality |
| S9_vs_k141_87917\|\|full | Low-quality |
| S9_vs_k141_27240\|\|full | Not-determined |
| S9_vs_k141_12128\|\|full | Low-quality |
| S9_vs_k141_42454\|\|full | Low-quality |
| S9_vs_k141_38675\|\|full | Not-determined |
| S9_vs_k141_87923\|\|full | Not-determined |
| S9_vs_k141_27246\|\|full | Low-quality |
| S9_vs_k141_46031\|\|full | Low-quality |
| S9_vs_k141_72683\|\|full | Low-quality |
| S9_vs_k141_84011\|\|full | Not-determined |
| S9_vs_k141_64976\|\|full | Not-determined |
| S9_vs_k141_57298\|\|full | Low-quality |
| S9_vs_k141_49747\|\|full | Not-determined |
| S9_vs_k141_15949\|\|full | Not-determined |
| S9_vs_k141_34769\|\|full | Low-quality |
| S9_vs_k141_68758\|\|full | Low-quality |
| S9_vs_k141_31059\|\|full | Low-quality |
| S9_vs_k141_87940\|\|full | Low-quality |
| S9_vs_k141_87941\|\|full | Low-quality |
| S9_vs_k141_76314\|\|full | Low-quality |
| S9_vs_k141_27274\|\|full | Not-determined |
| S9_vs_k141_76319\|\|full | Not-determined |
| S9_vs_k141_12174\|\|full | Low-quality |
| S9_vs_k141_4422\|\|full | Not-determined |
| S9_vs_k141_15967\|\|full | Not-determined |
| S9_vs_k141_76326\|\|full | Low-quality |
| S9_vs_k141_42510\|\|full | Not-determined |
| S9_vs_k141_31077\|\|full | Low-quality |
| S9_vs_k141_53808\|\|full | Not-determined |
| S9_vs_k141_72693\|\|full | Low-quality |
| S9_vs_k141_15972\|\|full | Low-quality |
| S9_vs_k141_87953\|\|full | Low-quality |
| S9_vs_k141_31082\|\|full | Low-quality |
| S9_vs_k141_80322\|\|full | Not-determined |
| S9_vs_k141_23412\|\|full | Not-determined |
| S9_vs_k141_4441\|\|full | Low-quality |
| S9_vs_k141_87958\|\|full | Not-determined |
| S9_vs_k141_46081\|\|full | Low-quality |
| S9_vs_k141_27295\|\|full | Low-quality |
| S9_vs_k141_87962\|\|full | Low-quality |
| S9_vs_k141_57345\|\|full | Not-determined |
| S9_vs_k141_76351\|\|full | Not-determined |
| S9_vs_k141_23417\|\|full | Not-determined |
| S9_vs_k141_42534\|\|full | Low-quality |
| S9_vs_k141_61358\|\|full | Not-determined |
| S9_vs_k141_72712\|\|full | Not-determined |
| S9_vs_k141_68803\|\|full | Not-determined |
| S9_vs_k141_12219\|\|full | Low-quality |
| S9_vs_k141_12223\|\|full | Low-quality |
| S9_vs_k141_76359\|\|full | Low-quality |
| S9_vs_k141_15985\|\|full | Low-quality |
| S9_vs_k141_61361\|\|full | Low-quality |
| S9_vs_k141_49761\|\|full | Low-quality |
| S9_vs_k141_38768\|\|full | Low-quality |
| S9_vs_k141_705\|\|full | Low-quality |
| S9_vs_k141_4472\|\|full | Low-quality |
| S9_vs_k141_27315\|\|full | Not-determined |
| S3_vs_k141_115304\|\|full | High-quality |
| S9_vs_k141_42547\|\|full | Low-quality |
| S9_vs_k141_23437\|\|full | Low-quality |
| S9_vs_k141_49763\|\|full | Not-determined |
| S9_vs_k141_4483\|\|full | Low-quality |
| S9_vs_k141_34801\|\|full | Low-quality |
| S9_vs_k141_76374\|\|full | Not-determined |
| S9_vs_k141_16018\|\|full | Low-quality |
| S9_vs_k141_57388\|\|full | Not-determined |
| S9_vs_k141_12228\|\|full | Low-quality |
| S9_vs_k141_65082\|\|full | Not-determined |
| S9_vs_k141_53854\|\|full | Low-quality |
| S9_vs_k141_16023\|\|full | Low-quality |
| S9_vs_k141_4506\|\|full | Low-quality |
| S9_vs_k141_23450\|\|full | Not-determined |
| S9_vs_k141_12233\|\|full | Low-quality |
| S9_vs_k141_31151\|\|full | Low-quality |
| S9_vs_k141_34821\|\|full | Low-quality |
| S9_vs_k141_38798\|\|full | Low-quality |
| S9_vs_k141_87990\|\|full | Not-determined |
| S9_vs_k141_23454\|\|full | Not-determined |
| S9_vs_k141_76383\|\|full | Low-quality |
| S9_vs_k141_84065\|\|full | Low-quality |
| S9_vs_k141_38799\|\|full | Not-determined |
| S9_vs_k141_19649\|\|full | Not-determined |
| S9_vs_k141_57411\|\|full | Low-quality |
| S9_vs_k141_34830\|\|full | Not-determined |
| S9_vs_k141_751\|\|full | Not-determined |
| S9_vs_k141_42551\|\|full | Low-quality |
| S9_vs_k141_46141\|\|full | Not-determined |
| S9_vs_k141_4520\|\|full | Low-quality |
| S9_vs_k141_12268\|\|full | Low-quality |
| S9_vs_k141_49807\|\|full | Not-determined |
| S9_vs_k141_23477\|\|full | Low-quality |
| S9_vs_k141_57424\|\|full | Not-determined |
| S9_vs_k141_84085\|\|full | Not-determined |
| S9_vs_k141_76409\|\|full | Low-quality |
| S9_vs_k141_34840\|\|full | Low-quality |
| S9_vs_k141_4529\|\|full | Not-determined |

S9_vs_k141_53888||full Low-quality
S9_vs_k141_34851||full Low-quality
S9_vs_k141_80382||full Low-quality
S9_vs_k141_42568||full Low-quality
S9_vs_k141_61431||full Low-quality
S9_vs_k141_19675||full Low-quality
S9_vs_k141_31185||full Low-quality
S9_vs_k141_774||full Low-quality
S9_vs_k141_4541||full Low-quality
S9_vs_k141_16064||full Not-determined
S9_vs_k141_49825||full Low-quality
S9_vs_k141_23505||full Low-quality
S9_vs_k141_80389||full Low-quality
S9_vs_k141_46171||full Not-determined
S9_vs_k141_34867||full Low-quality
S9_vs_k141_38853||full Not-determined
S9_vs_k141_23517||full Low-quality
S9_vs_k141_23518||full Not-determined
S9_vs_k141_72767||full Low-quality
S9_vs_k141_88072||full Low-quality
S9_vs_k141_34875||full Low-quality
S9_vs_k141_46200||full Not-determined
S11_extracted_k141_104044 Complete
S9_vs_k141_31202||full Low-quality
S9_vs_k141_80425||full Low-quality
S9_vs_k141_76434||full Low-quality
S9_vs_k141_23532||full Low-quality
S9_vs_k141_88083||full Low-quality
S9_vs_k141_53943||full Low-quality
S9_vs_k141_34895||full Not-determined
S9_vs_k141_65094||full Low-quality
S9_vs_k141_38879||full Low-quality
S9_vs_k141_72795||full Low-quality
S9_vs_k141_53956||full Not-determined
S11_vs_k141_104044||full High-quality
S9_vs_k141_23555||full Not-determined
S9_vs_k141_832||full Low-quality
S9_vs_k141_80427||full Low-quality
S9_vs_k141_88100||full Low-quality
S9_vs_k141_49877||full Low-quality
S9_vs_k141_835||full Low-quality
S9_vs_k141_46231||full Low-quality
S9_vs_k141_61504||full Not-determined
S9_vs_k141_80435||full Not-determined
S9_vs_k141_65132||full Not-determined
S9_vs_k141_27348||full Not-determined
S9_vs_k141_12304||full Low-quality
S9_vs_k141_42619||full Not-determined
S9_vs_k141_27356||full Low-quality
S9_vs_k141_23565||full Low-quality
S9_vs_k141_72837||full Not-determined
S9_vs_k141_12307||full Not-determined
S9_vs_k141_49905||full Low-quality
S9_vs_k141_858||full Low-quality
S12_extracted_k141_128327 Complete
S9_vs_k141_65151||full Low-quality
S9_vs_k141_72853||full Not-determined
S9_vs_k141_34899||full Low-quality
S9_vs_k141_42641||full Low-quality
S9_vs_k141_27371||full Not-determined
S9_vs_k141_65155||full Not-determined
S9_vs_k141_61525||full Not-determined
S9_vs_k141_46270||full Low-quality
S9_vs_k141_34909||full Low-quality
S9_vs_k141_57492||full Not-determined
S9_vs_k141_38942||full Not-determined
S9_vs_k141_12323||full Low-quality
S9_vs_k141_88126||full Low-quality
S12_vs_k141_128327||full High-quality
S9_vs_k141_19751||full Low-quality
S9_vs_k141_46285||full Not-determined
S9_vs_k141_16156||full Low-quality
S9_vs_k141_38948||full Not-determined
S9_vs_k141_61546||full Low-quality
S9_vs_k141_23579||full Low-quality
S9_vs_k141_4564||full Low-quality
S9_vs_k141_72863||full Low-quality
S9_vs_k141_31299||full Not-determined
S9_vs_k141_38958||full Low-quality
S9_vs_k141_27396||full Low-quality
S9_vs_k141_38963||full Low-quality
S9_vs_k141_46315||full Low-quality
S9_vs_k141_68847||full Not-determined
S9_vs_k141_61563||full Low-quality
S9_vs_k141_88157||full Low-quality
S9_vs_k141_57504||full Low-quality
S9_vs_k141_61570||full Low-quality
S9_vs_k141_34964||full Low-quality
S9_vs_k141_42668||full Not-determined
S9_vs_k141_46328||full Low-quality
S9_vs_k141_16192||full Not-determined
S9_vs_k141_34976||full Low-quality
S9_vs_k141_4617||full Low-quality
S9_vs_k141_65203||full Not-determined
S9_vs_k141_31341||full Not-determined
S9_vs_k141_46337||full Not-determined
S9_vs_k141_57526||full Low-quality
S9_vs_k141_31342||full Not-determined
S9_vs_k141_8163||full Low-quality
S9_vs_k141_917||full Low-quality
S9_vs_k141_4626||full Low-quality
S9_vs_k141_19777||full Not-determined
S9_vs_k141_39005||full Not-determined
S9_vs_k141_12372||full Low-quality
S9_vs_k141_88183||full Low-quality
S9_vs_k141_12373||full Low-quality
S9_vs_k141_8187||full Low-quality
S9_vs_k141_42715||full Not-determined
S9_vs_k141_4637||full Low-quality
S9_vs_k141_4638||full Low-quality
S9_vs_k141_54095||full Low-quality
S9_vs_k141_12385||full Low-quality
S9_vs_k141_88198||full Low-quality
S9_vs_k141_8199||full Low-quality
S9_vs_k141_50026||full Low-quality
S9_vs_k141_4649||full Low-quality
S9_vs_k141_8201||full Low-quality
S2_extracted_k141_4130 High-quality
S9_vs_k141_54104||full Low-quality
S9_vs_k141_65231||full Low-quality
S9_vs_k141_19803||full Low-quality
S9_vs_k141_42726||full Low-quality
S9_vs_k141_16260||full Low-quality
S9_vs_k141_88217||full Low-quality
S9_vs_k141_19804||full Low-quality
S9_vs_k141_35043||full Not-determined
S9_vs_k141_46358||full Low-quality
S9_vs_k141_35049||full Low-quality
S9_vs_k141_88221||full Not-determined
S9_vs_k141_8212||full Low-quality
S9_vs_k141_12402||full Not-determined
S9_vs_k141_19806||full Low-quality
S9_vs_k141_4655||full Low-quality
S9_vs_k141_955||full Low-quality
S9_vs_k141_956||full Low-quality
S9_vs_k141_16270||full Not-determined
S9_vs_k141_80464||full Low-quality
S9_vs_k141_12408||full Low-quality
S9_vs_k141_961||full Low-quality
S9_vs_k141_57593||full Not-determined
S9_vs_k141_54131||full Not-determined
S9_vs_k141_57595||full Low-quality
S9_vs_k141_19817||full Not-determined
S9_vs_k141_80470||full Low-quality
S9_vs_k141_23590||full Low-quality
S9_vs_k141_27444||full Not-determined
S9_vs_k141_88245||full Low-quality
S9_vs_k141_50067||full Low-quality
S9_vs_k141_80483||full Not-determined
S9_vs_k141_27455||full Low-quality
S9_vs_k141_39069||full Low-quality
S9_vs_k141_31425||full Low-quality
S9_vs_k141_16310||full Not-determined
S9_vs_k141_46397||full Not-determined
S9_vs_k141_989||full Low-quality
S9_vs_k141_76465||full Low-quality
S9_vs_k141_35095||full Low-quality
S9_vs_k141_39086||full Low-quality
S9_vs_k141_27463||full Low-quality
S9_vs_k141_35106||full Low-quality
S9_vs_k141_16329||full Low-quality
S9_vs_k141_19846||full Not-determined
S9_vs_k141_31440||full Low-quality
S9_vs_k141_8260||full Not-determined
S9_vs_k141_42784||full Low-quality
S9_vs_k141_73001||full Low-quality
S9_vs_k141_39096||full Low-quality
S9_vs_k141_8262||full Not-determined
S9_vs_k141_54163||full Low-quality
S9_vs_k141_27479||full Low-quality
S9_vs_k141_50124||full Low-quality
S9_vs_k141_65292||full Low-quality
S9_vs_k141_35117||full Not-determined
S9_vs_k141_23643||full Not-determined
S9_vs_k141_19862||full Low-quality
S9_vs_k141_8272||full Low-quality
S9_vs_k141_16353||full Low-quality
S9_vs_k141_8279||full Not-determined
S9_vs_k141_19864||full Low-quality
S9_vs_k141_84111||full Low-quality
S9_vs_k141_65301||full Low-quality
S9_vs_k141_19867||full Low-quality
S9_vs_k141_23648||full Not-determined
S9_vs_k141_57660||full Not-determined
S9_vs_k141_16363||full Not-determined
S9_vs_k141_65307||full Low-quality
S9_vs_k141_46443||full Low-quality
S9_vs_k141_19872||full Not-determined
S9_vs_k141_27506||full Not-determined
S9_vs_k141_31471||full Low-quality
S9_vs_k141_65310||full Low-quality
S9_vs_k141_76517||full Low-quality
S9_vs_k141_76522||full Low-quality
S9_vs_k141_46452||full Low-quality
S9_vs_k141_35138||full Low-quality
S9_vs_k141_76527||full Low-quality
S9_vs_k141_19882||full Not-determined
S9_vs_k141_50143||full Low-quality
S9_vs_k141_57678||full Low-quality
S9_vs_k141_84149||full Not-determined
S9_vs_k141_31486||full Low-quality
S9_vs_k141_84153||full Low-quality
S9_vs_k141_8314||full Low-quality
S9_vs_k141_50152||full Not-determined
S9_vs_k141_39152||full Not-determined
S9_vs_k141_54219||full Not-determined
S9_vs_k141_73025||full Low-quality
S9_vs_k141_27562||full Not-determined
S9_vs_k141_31509||full Low-quality
S9_vs_k141_50157||full Not-determined
S9_vs_k141_54241||full Low-quality
S9_vs_k141_27572||full Low-quality
S9_vs_k141_12548||full Low-quality
S9_vs_k141_68947||full Not-determined
S9_vs_k141_65361||full Low-quality
S9_vs_k141_4781||full Low-quality
S9_vs_k141_46491||full Low-quality
S9_vs_k141_73048||full Low-quality
S9_vs_k141_68949||full Not-determined
S9_vs_k141_31526||full Not-determined
S9_vs_k141_27584||full Low-quality
S9_vs_k141_57702||full Not-determined
S9_vs_k141_1071||full Not-determined
S9_vs_k141_65368||full Low-quality
S9_vs_k141_4796||full Low-quality
S9_vs_k141_12564||full Low-quality
S9_vs_k141_73062||full Low-quality
S9_vs_k141_50182||full Low-quality
S9_vs_k141_39181||full Low-quality
S9_vs_k141_16452||full Low-quality
S9_vs_k141_84214||full Not-determined
S9_vs_k141_1092||full Not-determined
S9_vs_k141_12581||full Low-quality
S9_vs_k141_42843||full Low-quality
S9_vs_k141_73082||full Low-quality
S9_vs_k141_88318||full Not-determined
S9_vs_k141_65390||full Low-quality
S9_vs_k141_8351||full Low-quality
S9_vs_k141_12590||full Low-quality
S9_vs_k141_88324||full Low-quality
S9_vs_k141_73088||full Low-quality
S8_vs_k141_208103||full High-quality
S9_vs_k141_68988||full Low-quality
S9_vs_k141_23664||full Low-quality
S9_vs_k141_88332||full Low-quality
S9_vs_k141_73095||full Low-quality
S9_vs_k141_31572||full Low-quality
S9_vs_k141_12605||full Low-quality
S9_vs_k141_4824||full Not-determined
S9_vs_k141_46511||full Not-determined
S9_vs_k141_69000||full Low-quality
S9_vs_k141_54278||full Low-quality
S9_vs_k141_65410||full Low-quality
S9_vs_k141_23670||full Low-quality
S9_vs_k141_1123||full Not-determined
S9_vs_k141_42873||full Not-determined
S9_vs_k141_54282||full Low-quality
S9_vs_k141_31584||full Not-determined
S9_vs_k141_88345||full Low-quality
S9_vs_k141_54286||full Low-quality
S9_vs_k141_69014||full Low-quality
S9_vs_k141_35231||full Not-determined
S9_vs_k141_23683||full Low-quality
S9_vs_k141_61664||full Low-quality
S9_vs_k141_88353||full Not-determined
S9_vs_k141_8388||full Low-quality
S9_vs_k141_50232||full Not-determined
S9_vs_k141_54302||full Low-quality
S9_vs_k141_23687||full Low-quality
S9_vs_k141_42905||full Low-quality
S9_vs_k141_16516||full Not-determined
S9_vs_k141_57739||full Low-quality
S9_vs_k141_16518||full Low-quality
S9_vs_k141_8401||full Low-quality
S9_vs_k141_35255||full Low-quality
S9_vs_k141_20008||full Low-quality
S9_vs_k141_1163||full Not-determined
S9_vs_k141_88372||full Low-quality
S9_vs_k141_54325||full Low-quality
S9_vs_k141_69039||full Not-determined
S9_vs_k141_46529||full Low-quality
S9_vs_k141_35270||full Not-determined
S9_vs_k141_4865||full Low-quality
S9_vs_k141_57763||full Not-determined
S9_vs_k141_61696||full Low-quality
S9_vs_k141_88382||full Not-determined
S9_vs_k141_31590||full Low-quality
S9_vs_k141_69040||full Low-quality
S9_vs_k141_35276||full Low-quality
S9_vs_k141_23711||full Not-determined
S9_vs_k141_88385||full Not-determined
S9_vs_k141_27686||full Not-determined
S9_vs_k141_12657||full Low-quality
S9_vs_k141_73164||full Low-quality
S9_vs_k141_39265||full Low-quality
S9_vs_k141_54350||full Low-quality
S9_vs_k141_20036||full Low-quality
S9_vs_k141_12660||full Not-determined
S9_vs_k141_35289||full Low-quality
S9_vs_k141_8420||full Low-quality
S9_vs_k141_61710||full Not-determined
S9_vs_k141_20047||full Not-determined
S9_vs_k141_57781||full Not-determined
S9_vs_k141_57782||full Low-quality
S9_vs_k141_27704||full Not-determined
S9_vs_k141_76554||full Low-quality
S9_vs_k141_16562||full Not-determined
S9_vs_k141_61722||full Low-quality
S9_vs_k141_54372||full Low-quality
S9_vs_k141_27714||full Not-determined
S9_vs_k141_31616||full Low-quality
S9_vs_k141_65414||full Low-quality
S9_vs_k141_54375||full Not-determined
S9_vs_k141_46532||full Low-quality
S9_vs_k141_61732||full Not-determined
S9_vs_k141_20061||full Low-quality
S9_vs_k141_42968||full Low-quality
S9_vs_k141_61739||full Not-determined
S9_vs_k141_65418||full Low-quality
S9_vs_k141_84342||full Low-quality
S9_vs_k141_1212||full Not-determined
S9_vs_k141_80566||full Low-quality
S9_vs_k141_4909||full Low-quality
S9_vs_k141_76587||full Not-determined
S9_vs_k141_80568||full Low-quality
S9_vs_k141_54408||full Low-quality
S9_vs_k141_12704||full Not-determined
S9_vs_k141_39317||full Low-quality
S9_vs_k141_73228||full Low-quality
S9_vs_k141_54410||full Not-determined
S9_vs_k141_88443||full Low-quality
S9_vs_k141_27764||full Not-determined
S9_vs_k141_50249||full Not-determined
S9_vs_k141_88450||full Not-determined
S9_vs_k141_4929||full Low-quality
S9_vs_k141_88453||full Low-quality
S9_vs_k141_50254||full Low-quality
S9_vs_k141_35366||full Not-determined
S9_vs_k141_80599||full Not-determined
S9_vs_k141_65448||full Not-determined
S9_vs_k141_76622||full Low-quality
S9_vs_k141_35376||full Low-quality
S9_vs_k141_8486||full Low-quality
S9_vs_k141_61779||full Not-determined
S9_vs_k141_46588||full Low-quality
S9_vs_k141_1256||full Not-determined
S9_vs_k141_4950||full Low-quality
S9_vs_k141_27783||full Low-quality
S9_vs_k141_61786||full Not-determined
S9_vs_k141_69148||full Low-quality
S9_vs_k141_80608||full Low-quality
S9_vs_k141_61788||full Low-quality
S9_vs_k141_23769||full Low-quality
S9_vs_k141_76630||full Low-quality
S9_vs_k141_12757||full Low-quality
S9_vs_k141_80611||full Low-quality
S9_vs_k141_46596||full Not-determined
S9_vs_k141_8499||full Low-quality
S9_vs_k141_46597||full Low-quality
S9_vs_k141_1280||full Not-determined
S9_vs_k141_65475||full Low-quality
S9_vs_k141_69154||full Not-determined
S9_vs_k141_27805||full Low-quality
S9_vs_k141_1288||full Low-quality
S9_vs_k141_76644||full Low-quality
S9_vs_k141_61809||full Low-quality
S9_vs_k141_4979||full Low-quality
S9_vs_k141_35410||full Not-determined
S9_vs_k141_57822||full Low-quality
S9_vs_k141_23800||full Not-determined
S9_vs_k141_39388||full Not-determined
S9_vs_k141_23801||full Not-determined
S9_vs_k141_65498||full Low-quality
S9_vs_k141_39399||full Not-determined
S9_vs_k141_88507||full Low-quality
S9_vs_k141_46633||full Not-determined
S9_vs_k141_65499||full Low-quality
S9_vs_k141_76664||full Low-quality
S9_vs_k141_46643||full Low-quality
S9_vs_k141_88516||full Not-determined
S9_vs_k141_23821||full Low-quality
S9_vs_k141_88522||full Low-quality
S9_vs_k141_43089||full Not-determined
S9_vs_k141_50312||full Low-quality
S9_vs_k141_88533||full Low-quality
S9_vs_k141_23831||full Low-quality
S9_vs_k141_69207||full Not-determined
S9_vs_k141_23833||full Low-quality
S9_vs_k141_65509||full Low-quality
S9_vs_k141_31753||full Not-determined
S9_vs_k141_23839||full Low-quality
S9_vs_k141_20175||full Low-quality
S9_vs_k141_16628||full Low-quality
S9_vs_k141_8552||full Low-quality
S9_vs_k141_8553||full Low-quality
S9_vs_k141_5034||full Low-quality
S4_extracted_4838 Complete
S9_vs_k141_5041||full Low-quality
S9_vs_k141_88561||full Not-determined
S9_vs_k141_57881||full Not-determined
S9_vs_k141_76712||full Not-determined
S9_vs_k141_5055||full Not-determined
S9_vs_k141_69232||full Low-quality
S9_vs_k141_88571||full Not-determined
S9_vs_k141_69235||full Low-quality
S9_vs_k141_43148||full Low-quality
S9_vs_k141_27850||full Low-quality
S9_vs_k141_8593||full Low-quality
S9_vs_k141_5070||full Not-determined
S9_vs_k141_27856||full Low-quality
S9_vs_k141_8597||full Low-quality
S9_vs_k141_69251||full Low-quality
S9_vs_k141_31818||full Low-quality
S9_vs_k141_23897||full Low-quality
S9_vs_k141_27863||full Low-quality
S9_vs_k141_43165||full Low-quality
S9_vs_k141_43167||full Low-quality
S9_vs_k141_8609||full Low-quality
S9_vs_k141_20242||full Low-quality
S9_vs_k141_84375||full Low-quality
S9_vs_k141_35544||full Low-quality
S9_vs_k141_1400||full Not-determined
S9_vs_k141_80704||full Low-quality
S9_vs_k141_43175||full Low-quality
S9_vs_k141_35555||full Low-quality
S9_vs_k141_50386||full Low-quality
S9_vs_k141_23914||full Not-determined
S9_vs_k141_1407||full Not-determined
S9_vs_k141_39459||full Not-determined
S9_vs_k141_57958||full Low-quality
S9_vs_k141_1408||full Low-quality
S9_vs_k141_20273||full Low-quality
S9_vs_k141_16635||full Low-quality
S9_vs_k141_35573||full Low-quality
S9_vs_k141_8652||full Low-quality
S9_vs_k141_23945||full Not-determined
S9_vs_k141_20284||full Not-determined
S9_vs_k141_20288||full Not-determined
S9_vs_k141_43177||full Low-quality
S9_vs_k141_58002||full Low-quality
S9_vs_k141_58003||full Not-determined
S9_vs_k141_20293||full Low-quality
S9_vs_k141_1435||full Not-determined
S9_vs_k141_69304||full Low-quality
S9_vs_k141_8674||full Low-quality
S9_vs_k141_16655||full Low-quality
S9_vs_k141_43191||full Low-quality
S9_vs_k141_43200||full Not-determined
S9_vs_k141_69315||full Not-determined
S9_vs_k141_35610||full Low-quality
S9_vs_k141_31917||full Low-quality
S9_vs_k141_43205||full Low-quality
S9_vs_k141_84446||full Low-quality
S9_vs_k141_23985||full Not-determined
S9_vs_k141_61984||full Low-quality
S9_vs_k141_23999||full Low-quality
S9_vs_k141_20317||full Low-quality
S9_vs_k141_80792||full Low-quality
S9_vs_k141_54426||full Not-determined
S9_vs_k141_31931||full Low-quality
S9_vs_k141_31932||full Low-quality
S11_extracted_k141_104035 Complete
S9_vs_k141_8681||full Not-determined
S9_vs_k141_1466||full Low-quality
S9_vs_k141_31939||full Low-quality
S9_vs_k141_20326||full Low-quality
S9_vs_k141_1474||full Low-quality
S9_vs_k141_46735||full Not-determined
S9_vs_k141_65682||full Low-quality
S9_vs_k141_65684||full Not-determined
S9_vs_k141_31961||full Not-determined
S6_extracted_k141_23629 Complete
S9_vs_k141_61999||full Low-quality
S9_vs_k141_80834||full Low-quality
S9_vs_k141_8717||full Not-determined
S9_vs_k141_54452||full Not-determined
S9_vs_k141_84512||full Not-determined
S9_vs_k141_43255||full Not-determined
S9_vs_k141_16723||full Low-quality
S9_vs_k141_62021||full Low-quality
S9_vs_k141_46777||full Low-quality
S9_vs_k141_27970||full Low-quality
S9_vs_k141_80840||full Low-quality
S9_vs_k141_24067||full Not-determined
S9_vs_k141_54485||full Low-quality
S9_vs_k141_1531||full Not-determined
S9_vs_k141_8758||full Low-quality
S9_vs_k141_24073||full Low-quality
S9_vs_k141_65734||full Low-quality
S9_vs_k141_69366||full Low-quality
S9_vs_k141_73408||full Low-quality
S4_vs_VIRSorter_4838-circular-cat_ High-quality
S9_vs_k141_16778||full Low-quality
S9_vs_k141_54512||full Not-determined
S9_vs_k141_69378||full Not-determined
S9_vs_k141_16791||full Low-quality
S9_vs_k141_50447||full Not-determined
S9_vs_k141_20391||full Not-determined
S9_vs_k141_76779||full Not-determined
S9_vs_k141_84577||full Low-quality
S9_vs_k141_80877||full Low-quality
S9_vs_k141_65752||full Not-determined
S9_vs_k141_16796||full Low-quality
S9_vs_k141_24095||full Low-quality
S9_vs_k141_24098||full Low-quality
S9_vs_k141_8782||full Not-determined
S9_vs_k141_73435||full Low-quality
S9_vs_k141_65771||full Not-determined
S9_vs_k141_62086||full Low-quality
S9_vs_k141_65776||full Low-quality
S9_vs_k141_76801||full Not-determined
S9_vs_k141_69421||full Low-quality
S9_vs_k141_54548||full Low-quality
S9_vs_k141_8811||full Not-determined
S9_vs_k141_80913||full Not-determined
S9_vs_k141_65782||full Low-quality
S9_vs_k141_28017||full Not-determined
S9_vs_k141_24113||full Low-quality
S9_vs_k141_65784||full Low-quality
S9_vs_k141_80919||full Low-quality
S9_vs_k141_73457||full Not-determined
S9_vs_k141_32076||full Low-quality
S9_vs_k141_24118||full Low-quality
S9_vs_k141_12840||full Not-determined
S9_vs_k141_12842||full Low-quality
S9_vs_k141_5350||full Low-quality
S9_vs_k141_69464||full Not-determined
S9_vs_k141_20451||full Low-quality
S9_vs_k141_80939||full Not-determined
S9_vs_k141_28050||full Not-determined
S9_vs_k141_5357||full Low-quality
S9_vs_k141_69466||full Not-determined
S9_vs_k141_20456||full Not-determined
S9_vs_k141_32104||full Low-quality
S9_vs_k141_8839||full Low-quality
S9_vs_k141_28067||full Not-determined
S9_vs_k141_24121||full Low-quality
S9_vs_k141_54594||full Not-determined
S9_vs_k141_62110||full Not-determined
S9_vs_k141_50508||full Low-quality
S9_vs_k141_73489||full Low-quality
S9_vs_k141_28072||full Low-quality
S9_vs_k141_50513||full Low-quality
S9_vs_k141_12864||full Not-determined
S9_vs_k141_54603||full Low-quality
S9_vs_k141_50515||full Low-quality
S9_vs_k141_35724||full Low-quality
S9_vs_k141_80965||full Not-determined
S9_vs_k141_12870||full Low-quality
S9_vs_k141_54614||full Low-quality
S9_vs_k141_62138||full Not-determined
S9_vs_k141_58057||full Low-quality
S9_vs_k141_20494||full Low-quality
S9_vs_k141_54622||full Low-quality
S9_vs_k141_73523||full Not-determined
S9_vs_k141_20502||full Low-quality
S9_vs_k141_32145||full Low-quality
S9_vs_k141_62162||full Low-quality
S9_vs_k141_32151||full Not-determined
S9_vs_k141_62172||full Low-quality
S9_vs_k141_58080||full Low-quality
S9_vs_k141_50562||full Low-quality
S9_vs_k141_73543||full Low-quality
S9_vs_k141_62174||full Low-quality
S9_vs_k141_20518||full Not-determined
S9_vs_k141_50565||full Not-determined
S9_vs_k141_69538||full Low-quality
S9_vs_k141_39634||full Low-quality
S9_vs_k141_39635||full Low-quality
S9_vs_k141_12900||full Not-determined
S9_vs_k141_20519||full Low-quality
S9_vs_k141_50571||full Low-quality
S9_vs_k141_32179||full Low-quality
S9_vs_k141_54643||full Low-quality
S9_vs_k141_24193||full Low-quality
S9_vs_k141_69553||full Low-quality
S9_vs_k141_1632||full Low-quality
S9_vs_k141_8906||full Low-quality
S9_vs_k141_46804||full Low-quality
S9_vs_k141_39647||full Low-quality
S9_vs_k141_24201||full Not-determined
S9_vs_k141_54656||full Not-determined
S9_vs_k141_62200||full Low-quality
S9_vs_k141_54660||full Not-determined
S9_vs_k141_46812||full Low-quality
S9_vs_k141_16927||full Low-quality
S9_vs_k141_8921||full Low-quality
S9_vs_k141_32213||full Low-quality
S9_vs_k141_50601||full Low-quality

| | |
|---|---|
| S9_vs_k141_12913||full | Low-quality |
| S9_vs_k141_16933||full | Not-determined |
| S9_vs_k141_20550||full | Low-quality |
| S9_vs_k141_1659||full | Low-quality |
| S6_vs_k141_23629||full | High-quality |
| S9_vs_k141_28152||full | Low-quality |
| S9_vs_k141_69592||full | Low-quality |
| S9_vs_k141_62228||full | Low-quality |
| S9_vs_k141_32237||full | Not-determined |
| S9_vs_k141_43361||full | Low-quality |
| S9_vs_k141_81005||full | Low-quality |
| S9_vs_k141_12937||full | Low-quality |
| S9_vs_k141_32241||full | Low-quality |
| S9_vs_k141_12944||full | Low-quality |
| S9_vs_k141_1678||full | Not-determined |
| S9_vs_k141_50632||full | Low-quality |
| S9_vs_k141_76912||full | Low-quality |
| S9_vs_k141_28176||full | Low-quality |
| S9_vs_k141_81015||full | Low-quality |
| S9_vs_k141_76913||full | Low-quality |
| S9_vs_k141_39660||full | Low-quality |
| S9_vs_k141_81020||full | Not-determined |
| S9_vs_k141_54700||full | Low-quality |
| S9_vs_k141_54701||full | Low-quality |
| S9_vs_k141_24236||full | Low-quality |
| S9_vs_k141_32256||full | Not-determined |
| S9_vs_k141_84666||full | Low-quality |
| S9_vs_k141_16993||full | Low-quality |
| S9_vs_k141_39674||full | Not-determined |
| S9_vs_k141_76918||full | Not-determined |
| S9_vs_k141_35808||full | Not-determined |
| S9_vs_k141_84672||full | Not-determined |
| S9_vs_k141_39685||full | Low-quality |
| S9_vs_k141_84678||full | Not-determined |
| S9_vs_k141_84680||full | Low-quality |
| S9_vs_k141_35818||full | Low-quality |
| S9_vs_k141_84681||full | Low-quality |
| S9_vs_k141_84683||full | Low-quality |
| S9_vs_k141_24262||full | Low-quality |
| S9_vs_k141_35820||full | Not-determined |
| S9_vs_k141_5475||full | Not-determined |
| S9_vs_k141_88579||full | Low-quality |
| S9_vs_k141_69620||full | Low-quality |
| S9_vs_k141_81068||full | Low-quality |
| S9_vs_k141_35830||full | Not-determined |
| S9_vs_k141_5482||full | Low-quality |
| S9_vs_k141_43446||full | Low-quality |
| S9_vs_k141_12974||full | Low-quality |
| S11_vs_k141_104035||full | High-quality |
| S9_vs_k141_20608||full | Low-quality |
| S9_vs_k141_1748||full | Not-determined |
| S9_vs_k141_1750||full | Low-quality |
| S9_vs_k141_5494||full | Not-determined |
| S9_vs_k141_20616||full | Not-determined |
| S9_vs_k141_17055||full | Low-quality |
| S9_vs_k141_43469||full | Low-quality |
| S9_vs_k141_35857||full | Not-determined |
| S9_vs_k141_54746||full | Low-quality |
| S9_vs_k141_43475||full | Low-quality |
| S9_vs_k141_24303||full | Low-quality |
| S9_vs_k141_54753||full | Not-determined |
| S9_vs_k141_39723||full | Not-determined |
| S9_vs_k141_65837||full | Not-determined |
| S9_vs_k141_46836||full | Low-quality |
| S9_vs_k141_43488||full | Not-determined |
| S9_vs_k141_84739||full | Low-quality |
| S9_vs_k141_46847||full | Not-determined |
| S9_vs_k141_69658||full | Low-quality |
| S9_vs_k141_39742||full | Not-determined |
| S9_vs_k141_24319||full | Low-quality |
| S9_vs_k141_84757||full | Low-quality |
| S9_vs_k141_1797||full | Low-quality |
| S9_vs_k141_81100||full | Not-determined |
| S9_vs_k141_28252||full | Low-quality |
| S9_vs_k141_46866||full | Not-determined |
| S9_vs_k141_39761||full | Low-quality |
| S9_vs_k141_88633||full | Low-quality |
| S9_vs_k141_58197||full | Not-determined |
| S9_vs_k141_50735||full | Not-determined |
| S9_vs_k141_17106||full | Not-determined |
| S9_vs_k141_13038||full | Low-quality |
| S9_vs_k141_46875||full | Not-determined |
| S9_vs_k141_5553||full | Low-quality |
| S9_vs_k141_58202||full | Low-quality |
| S9_vs_k141_77037||full | Not-determined |
| S9_vs_k141_28269||full | Low-quality |
| S9_vs_k141_62288||full | Low-quality |
| S9_vs_k141_17112||full | Low-quality |
| S9_vs_k141_35910||full | Low-quality |
| S9_vs_k141_28276||full | Low-quality |
| S9_vs_k141_9087||full | Not-determined |
| S9_vs_k141_46896||full | Low-quality |
| S9_vs_k141_1832||full | Low-quality |
| S9_vs_k141_69707||full | Low-quality |
| S9_vs_k141_43544||full | Low-quality |
| S9_vs_k141_46908||full | Not-determined |
| S9_vs_k141_62311||full | Not-determined |
| S9_vs_k141_1853||full | Low-quality |
| S9_vs_k141_9103||full | Low-quality |
| S9_vs_k141_62315||full | Low-quality |
| S9_vs_k141_39779||full | Low-quality |
| S9_vs_k141_20673||full | Low-quality |
| S9_vs_k141_20674||full | Low-quality |
| S9_vs_k141_28299||full | Not-determined |
| S9_vs_k141_43558||full | Low-quality |
| S9_vs_k141_17140||full | Not-determined |
| S9_vs_k141_9113||full | Not-determined |
| S9_vs_k141_62323||full | Low-quality |
| S9_vs_k141_32331||full | Low-quality |
| S9_vs_k141_13080||full | Low-quality |
| S9_vs_k141_43567||full | Low-quality |
| S9_vs_k141_81169||full | Not-determined |
| S9_vs_k141_5600||full | Not-determined |
| S9_vs_k141_54876||full | Low-quality |
| S9_vs_k141_62334||full | Not-determined |
| S9_vs_k141_58259||full | Not-determined |
| S9_vs_k141_84831||full | Low-quality |
| S9_vs_k141_54878||full | Not-determined |
| S9_vs_k141_28331||full | Not-determined |
| S9_vs_k141_54879||full | Low-quality |
| S9_vs_k141_1879||full | Low-quality |
| S9_vs_k141_20698||full | Low-quality |
| S9_vs_k141_24425||full | Not-determined |
| S9_vs_k141_54891||full | Not-determined |
| S9_vs_k141_32343||full | Low-quality |
| S9_vs_k141_9149||full | Low-quality |
| S9_vs_k141_62353||full | Not-determined |
| S9_vs_k141_88690||full | Low-quality |
| S9_vs_k141_17175||full | Not-determined |
| S9_vs_k141_54903||full | Not-determined |
| S9_vs_k141_32350||full | Low-quality |
| S9_vs_k141_28360||full | Low-quality |
| S9_vs_k141_77121||full | Low-quality |
| S9_vs_k141_35978||full | Low-quality |
| S9_vs_k141_5634||full | Low-quality |
| S9_vs_k141_28363||full | Low-quality |
| S9_vs_k141_88696||full | Low-quality |
| S9_vs_k141_28370||full | Low-quality |
| S9_vs_k141_54917||full | Low-quality |
| S9_vs_k141_54920||full | Low-quality |
| S9_vs_k141_81209||full | Not-determined |
| S9_vs_k141_5645||full | Not-determined |
| S9_vs_k141_77126||full | Low-quality |
| S9_vs_k141_73597||full | Low-quality |
| S9_vs_k141_43634||full | Low-quality |
| S9_vs_k141_5653||full | Low-quality |
| S9_vs_k141_46927||full | Not-determined |
| S9_vs_k141_73603||full | Low-quality |
| S9_vs_k141_69772||full | Not-determined |
| S9_vs_k141_20701||full | Low-quality |
| S9_vs_k141_5657||full | Low-quality |
| S9_vs_k141_43638||full | Low-quality |
| S9_vs_k141_24472||full | Not-determined |
| S9_vs_k141_20704||full | Not-determined |
| S9_vs_k141_81230||full | Low-quality |
| S9_vs_k141_1894||full | Not-determined |
| S9_vs_k141_77141||full | Low-quality |
| S9_vs_k141_28411||full | Low-quality |
| S9_vs_k141_28417||full | Low-quality |
| S9_vs_k141_73618||full | Low-quality |
| S9_vs_k141_1904||full | Low-quality |
| S9_vs_k141_58333||full | Low-quality |
| S9_vs_k141_17230||full | Low-quality |
| S9_vs_k141_65890||full | Not-determined |
| S9_vs_k141_24484||full | Low-quality |
| S9_vs_k141_28432||full | Low-quality |
| S9_vs_k141_62410||full | Not-determined |
| S9_vs_k141_88751||full | Low-quality |
| S9_vs_k141_69799||full | Low-quality |
| S9_vs_k141_69800||full | Low-quality |
| S9_vs_k141_65910||full | Not-determined |
| S9_vs_k141_20745||full | Low-quality |
| S9_vs_k141_28445||full | Low-quality |
| S9_vs_k141_28450||full | Not-determined |
| S9_vs_k141_1924||full | Low-quality |
| S9_vs_k141_39904||full | Low-quality |
| S9_vs_k141_1930||full | Not-determined |
| S9_vs_k141_39914||full | Low-quality |
| S9_vs_k141_54996||full | Low-quality |
| S9_vs_k141_81246||full | Not-determined |
| S9_vs_k141_69853||full | Low-quality |
| S9_vs_k141_43696||full | Low-quality |
| S9_vs_k141_28470||full | Low-quality |
| S9_vs_k141_77195||full | Not-determined |
| S9_vs_k141_65934||full | Not-determined |
| S9_vs_k141_17293||full | Low-quality |
| S9_vs_k141_55008||full | Low-quality |
| S9_vs_k141_58390||full | Low-quality |
| S9_vs_k141_1964||full | Low-quality |
| S9_vs_k141_20778||full | Low-quality |
| S9_vs_k141_58395||full | Low-quality |
| S9_vs_k141_73679||full | Low-quality |
| S9_vs_k141_24546||full | Not-determined |
| S9_vs_k141_55023||full | Low-quality |
| S9_vs_k141_88781||full | Low-quality |
| S9_vs_k141_20790||full | Low-quality |
| S9_vs_k141_9176||full | Not-determined |
| S9_vs_k141_17303||full | Not-determined |
| S9_vs_k141_24556||full | Low-quality |
| S9_vs_k141_39963||full | Low-quality |
| S9_vs_k141_36059||full | Low-quality |
| S9_vs_k141_13202||full | Low-quality |
| S9_vs_k141_65960||full | Low-quality |
| S9_vs_k141_13205||full | Not-determined |
| S9_vs_k141_17319||full | Low-quality |
| S9_vs_k141_36066||full | Low-quality |
| S9_vs_k141_17320||full | Low-quality |
| S9_vs_k141_9193||full | Low-quality |
| S9_vs_k141_43750||full | Low-quality |
| S9_vs_k141_88789||full | Not-determined |
| S9_vs_k141_28505||full | Low-quality |
| S9_vs_k141_5696||full | Low-quality |
| S9_vs_k141_81303||full | Low-quality |
| S9_vs_k141_73700||full | Not-determined |
| S9_vs_k141_88793||full | Low-quality |
| S9_vs_k141_84909||full | Low-quality |
| S9_vs_k141_43753||full | Low-quality |
| S9_vs_k141_47071||full | Not-determined |
| S9_vs_k141_13219||full | Low-quality |
| S9_vs_k141_24586||full | Low-quality |
| S9_vs_k141_73712||full | Low-quality |
| S2_vs_k141_4130||full | High-quality |
| S9_vs_k141_9215||full | Low-quality |
| S8_extracted_k141_49282 | High-quality |
| S9_vs_k141_9220||full | Low-quality |
| S9_vs_k141_55068||full | Low-quality |
| S9_vs_k141_58408||full | Low-quality |
| S9_vs_k141_1973||full | Not-determined |
| S9_vs_k141_47091||full | Low-quality |
| S9_vs_k141_55072||full | Not-determined |
| S9_vs_k141_81333||full | Low-quality |
| S9_vs_k141_43774||full | Not-determined |
| S9_vs_k141_24609||full | Low-quality |
| S9_vs_k141_47097||full | Not-determined |
| S9_vs_k141_9250||full | Not-determined |
| S9_vs_k141_77266||full | Low-quality |
| S9_vs_k141_5730||full | Low-quality |
| S9_vs_k141_84927||full | Low-quality |
| S9_vs_k141_32419||full | Low-quality |
| S9_vs_k141_81347||full | Not-determined |
| S9_vs_k141_36121||full | Not-determined |
| S9_vs_k141_77285||full | Low-quality |
| S9_vs_k141_58446||full | Not-determined |
| S9_vs_k141_17368||full | Low-quality |
| S9_vs_k141_40023||full | Low-quality |
| S9_vs_k141_55096||full | Not-determined |
| S9_vs_k141_69974||full | Not-determined |
| S9_vs_k141_47124||full | Low-quality |
| S9_vs_k141_13279||full | Low-quality |
| S9_vs_k141_5764||full | Not-determined |
| S9_vs_k141_20869||full | Not-determined |
| S9_vs_k141_40043||full | Not-determined |
| S9_vs_k141_36131||full | Low-quality |
| S9_vs_k141_81370||full | Not-determined |
| S9_vs_k141_5767||full | Low-quality |
| S9_vs_k141_9285||full | Low-quality |
| S9_vs_k141_36133||full | Low-quality |
| S9_vs_k141_36142||full | Not-determined |
| S9_vs_k141_77323||full | Low-quality |
| S9_vs_k141_2047||full | Low-quality |
| S9_vs_k141_5779||full | Not-determined |
| S9_vs_k141_81386||full | Low-quality |
| S8_extracted_k141_131802 | Medium-quality |
| S9_vs_k141_20884||full | Low-quality |
| S9_vs_k141_70017||full | Low-quality |
| S8_vs_k141_131802||full | Medium-quality |
| S9_vs_k141_55122||full | Low-quality |
| S9_vs_k141_5787||full | Low-quality |
| S9_vs_k141_20892||full | Low-quality |
| S9_vs_k141_88850||full | Low-quality |
| S9_vs_k141_32477||full | Not-determined |
| S9_vs_k141_77340||full | Not-determined |
| S9_vs_k141_70029||full | Low-quality |
| S9_vs_k141_73775||full | Low-quality |
| S9_vs_k141_66022||full | Low-quality |
| S9_vs_k141_62534||full | Not-determined |
| S9_vs_k141_43844||full | Not-determined |
| S9_vs_k141_36162||full | Not-determined |
| S9_vs_k141_13318||full | Low-quality |
| S9_vs_k141_73776||full | Low-quality |
| S9_vs_k141_55136||full | Low-quality |
| S9_vs_k141_13319||full | Not-determined |
| S9_vs_k141_43849||full | Low-quality |
| S9_vs_k141_24693||full | Not-determined |
| S9_vs_k141_55144||full | Not-determined |
| S9_vs_k141_9328||full | Not-determined |
| S9_vs_k141_24702||full | Low-quality |
| S9_vs_k141_32504||full | Low-quality |
| S9_vs_k141_66044||full | Low-quality |
| S9_vs_k141_36181||full | Low-quality |
| S9_vs_k141_62557||full | Not-determined |
| S9_vs_k141_13335||full | Low-quality |
| S9_vs_k141_32506||full | Not-determined |
| S9_vs_k141_77360||full | Not-determined |
| S9_vs_k141_40115||full | Not-determined |
| S9_vs_k141_17434||full | Not-determined |
| S9_vs_k141_13341||full | Low-quality |
| S9_vs_k141_43872||full | Low-quality |
| S9_vs_k141_85024||full | Low-quality |
| S9_vs_k141_13344||full | Low-quality |
| S9_vs_k141_13348||full | Low-quality |
| S9_vs_k141_62572||full | Not-determined |
| S9_vs_k141_32520||full | Not-determined |
| S9_vs_k141_5838||full | Low-quality |
| S9_vs_k141_88895||full | Not-determined |
| S9_vs_k141_47233||full | Not-determined |
| S9_vs_k141_2057||full | Low-quality |
| S9_vs_k141_32523||full | Low-quality |
| S9_vs_k141_17448||full | Low-quality |
| S9_vs_k141_62589||full | Low-quality |
| S9_vs_k141_28625||full | Low-quality |
| S9_vs_k141_13372||full | Low-quality |
| S9_vs_k141_70092||full | Low-quality |
| S9_vs_k141_24749||full | Not-determined |
| S9_vs_k141_58529||full | Low-quality |
| S9_vs_k141_13377||full | Not-determined |
| S9_vs_k141_66081||full | Not-determined |
| S9_vs_k141_13380||full | Not-determined |
| S9_vs_k141_88910||full | Not-determined |
| S9_vs_k141_28637||full | Low-quality |
| S9_vs_k141_47261||full | Low-quality |
| S9_vs_k141_58547||full | Low-quality |
| S9_vs_k141_47264||full | Low-quality |
| S9_vs_k141_40161||full | Not-determined |
| S9_vs_k141_43913||full | Low-quality |
| S9_vs_k141_17471||full | Low-quality |
| S9_vs_k141_20932||full | Low-quality |
| S9_vs_k141_81453||full | Low-quality |
| S9_vs_k141_85064||full | Low-quality |
| S9_vs_k141_58556||full | Low-quality |
| S9_vs_k141_88924||full | Low-quality |
| S9_vs_k141_85067||full | Low-quality |
| S9_vs_k141_70109||full | Low-quality |
| S9_vs_k141_36233||full | Low-quality |
| S9_vs_k141_43924||full | Not-determined |
| S9_vs_k141_58564||full | Low-quality |
| S9_vs_k141_24791||full | Not-determined |
| S9_vs_k141_43931||full | Low-quality |
| S9_vs_k141_73875||full | Not-determined |
| S9_vs_k141_88942||full | Low-quality |
| S9_vs_k141_70111||full | Low-quality |
| S9_vs_k141_58587||full | Low-quality |
| S9_vs_k141_17510||full | Low-quality |
| S9_vs_k141_9442||full | Not-determined |
| S9_vs_k141_77381||full | Low-quality |
| S9_vs_k141_58596||full | Not-determined |
| S9_vs_k141_43945||full | Low-quality |
| S9_vs_k141_62629||full | Low-quality |
| S9_vs_k141_17526||full | Not-determined |
| S9_vs_k141_62632||full | Low-quality |
| S9_vs_k141_28687||full | Low-quality |
| S9_vs_k141_43946||full | Low-quality |
| S9_vs_k141_32570||full | Low-quality |
| S9_vs_k141_28691||full | Low-quality |
| S9_vs_k141_9453||full | Low-quality |
| S9_vs_k141_55200||full | Low-quality |
| S9_vs_k141_66120||full | Low-quality |
| S9_vs_k141_5906||full | Low-quality |
| S9_vs_k141_24828||full | Not-determined |
| S9_vs_k141_58610||full | Not-determined |
| S9_vs_k141_85100||full | Not-determined |
| S9_vs_k141_73892||full | Low-quality |
| S9_vs_k141_70132||full | Low-quality |
| S9_vs_k141_32575||full | Low-quality |
| S9_vs_k141_17555||full | Low-quality |
| S9_vs_k141_73896||full | Low-quality |
| S9_vs_k141_81478||full | Low-quality |
| S9_vs_k141_88997||full | Not-determined |
| S9_vs_k141_13466||full | Low-quality |
| S9_vs_k141_70135||full | Low-quality |
| S9_vs_k141_36273||full | Not-determined |
| S9_vs_k141_40253||full | Low-quality |
| S9_vs_k141_9481||full | Not-determined |
| S9_vs_k141_66152||full | Not-determined |
| S9_vs_k141_89011||full | Low-quality |
| S9_vs_k141_17578||full | Not-determined |
| S9_vs_k141_36286||full | Low-quality |
| S9_vs_k141_13487||full | Not-determined |
| S9_vs_k141_36290||full | Not-determined |
| S9_vs_k141_2136||full | Low-quality |
| S9_vs_k141_47341||full | Low-quality |
| S9_vs_k141_47350||full | Low-quality |
| S9_vs_k141_28749||full | Not-determined |
| S9_vs_k141_9508||full | Not-determined |
| S9_vs_k141_44018||full | Low-quality |
| S9_vs_k141_21044||full | Low-quality |
| S9_vs_k141_9511||full | Low-quality |
| S9_vs_k141_24869||full | Low-quality |
| S9_vs_k141_5960||full | Not-determined |
| S9_vs_k141_36314||full | Low-quality |
| S9_vs_k141_21058||full | Not-determined |
| S9_vs_k141_5965||full | Low-quality |
| S9_vs_k141_28771||full | Not-determined |
| S11_extracted_k141_28515 | Medium-quality |
| S9_vs_k141_81544||full | Low-quality |
| S9_vs_k141_89052||full | Low-quality |
| S9_vs_k141_24885||full | Not-determined |
| S9_vs_k141_62666||full | Low-quality |
| S9_vs_k141_44052||full | Low-quality |
| S9_vs_k141_40322||full | Low-quality |
| S9_vs_k141_44053||full | Low-quality |
| S9_vs_k141_21092||full | Low-quality |
| S9_vs_k141_55276||full | Low-quality |
| S9_vs_k141_21094||full | Low-quality |
| S5_extracted_k141_55415 | High-quality |
| S9_vs_k141_28793||full | Low-quality |
| S9_vs_k141_62686||full | Low-quality |
| S9_vs_k141_55292||full | Low-quality |
| S9_vs_k141_62699||full | Not-determined |
| S9_vs_k141_81572||full | Not-determined |
| S9_vs_k141_28820||full | Low-quality |
| S9_vs_k141_62707||full | Low-quality |
| S9_vs_k141_32623||full | Not-determined |
| S9_vs_k141_89081||full | Low-quality |
| S6_extracted_k141_39623 | High-quality |
| S9_vs_k141_77407||full | Not-determined |
| S9_vs_k141_62719||full | Low-quality |
| S9_vs_k141_44103||full | Not-determined |
| S9_vs_k141_89083||full | Not-determined |
| S9_vs_k141_36392||full | Not-determined |
| S9_vs_k141_85236||full | Low-quality |
| S9_vs_k141_73972||full | Low-quality |
| S9_vs_k141_2231||full | Not-determined |
| S9_vs_k141_6056||full | Low-quality |
| S9_vs_k141_2235||full | Not-determined |
| S9_vs_k141_85259||full | Low-quality |
| S9_vs_k141_62746||full | Not-determined |
| S9_vs_k141_36411||full | Low-quality |
| S9_vs_k141_81633||full | Not-determined |
| S9_vs_k141_55364||full | Low-quality |
| S9_vs_k141_70216||full | Low-quality |
| S9_vs_k141_66204||full | Low-quality |
| S9_vs_k141_62755||full | Low-quality |
| S9_vs_k141_73990||full | Not-determined |
| S9_vs_k141_21171||full | Low-quality |
| S9_vs_k141_9585||full | Low-quality |
| S9_vs_k141_70221||full | Not-determined |
| S9_vs_k141_73992||full | Low-quality |
| S9_vs_k141_85272||full | Not-determined |
| S9_vs_k141_89126||full | Low-quality |
| S9_vs_k141_66219||full | Low-quality |
| S9_vs_k141_9594||full | Low-quality |
| S9_vs_k141_44154||full | Low-quality |
| S9_vs_k141_77467||full | Low-quality |
| S9_vs_k141_89140||full | Low-quality |
| S9_vs_k141_77469||full | Low-quality |
| S9_vs_k141_66226||full | Low-quality |
| S9_vs_k141_9612||full | Not-determined |
| S9_vs_k141_28894||full | Low-quality |
| S9_vs_k141_2259||full | Low-quality |
| S9_vs_k141_2263||full | Low-quality |
| S9_vs_k141_24979||full | Low-quality |
| S9_vs_k141_28906||full | Low-quality |
| S4_extracted_6168 | High-quality |
| S9_vs_k141_58700||full | Low-quality |
| S9_vs_k141_77505||full | Not-determined |
| S9_vs_k141_36480||full | Not-determined |
| S9_vs_k141_40397||full | Not-determined |
| S9_vs_k141_6107||full | Low-quality |
| S9_vs_k141_70255||full | Low-quality |
| S9_vs_k141_70256||full | Low-quality |
| S9_vs_k141_77509||full | Not-determined |
| S9_vs_k141_2296||full | Low-quality |
| S9_vs_k141_25010||full | Low-quality |
| S9_vs_k141_58730||full | Low-quality |
| S9_vs_k141_28939||full | Low-quality |
| S9_vs_k141_28948||full | Low-quality |
| S9_vs_k141_21238||full | Low-quality |
| S9_vs_k141_66290||full | Low-quality |
| S9_vs_k141_40430||full | Low-quality |
| S9_vs_k141_17690||full | Low-quality |
| S9_vs_k141_66299||full | Low-quality |
| S9_vs_k141_74067||full | Low-quality |
| S9_vs_k141_85361||full | Low-quality |
| S9_vs_k141_2322||full | Low-quality |
| S9_vs_k141_28964||full | Low-quality |
| S9_vs_k141_70303||full | Low-quality |
| S9_vs_k141_74068||full | Low-quality |
| S9_vs_k141_44236||full | Low-quality |
| S9_vs_k141_70308||full | Not-determined |
| S9_vs_k141_40468||full | Low-quality |
| S9_vs_k141_28982||full | Low-quality |
| S9_vs_k141_21276||full | Not-determined |
| S9_vs_k141_25068||full | Low-quality |
| S9_vs_k141_40472||full | Not-determined |
| S9_vs_k141_2346||full | Not-determined |
| S9_vs_k141_77539||full | Low-quality |
| S9_vs_k141_28986||full | Not-determined |
| S9_vs_k141_44251||full | Low-quality |
| S9_vs_k141_40475||full | Not-determined |
| S9_vs_k141_9689||full | Low-quality |
| S9_vs_k141_44257||full | Low-quality |
| S9_vs_k141_55416||full | Not-determined |
| S9_vs_k141_44258||full | Low-quality |
| S9_vs_k141_74111||full | Low-quality |
| S9_vs_k141_66336||full | Low-quality |
| S9_vs_k141_81703||full | Low-quality |
| S9_vs_k141_44260||full | Low-quality |
| S9_vs_k141_21304||full | Low-quality |
| S9_vs_k141_55422||full | Not-determined |
| S9_vs_k141_32646||full | Not-determined |
| S9_vs_k141_21315||full | Low-quality |
| S9_vs_k141_58805||full | Not-determined |
| S9_vs_k141_9709||full | Low-quality |
| S9_vs_k141_9712||full | Low-quality |
| S9_vs_k141_58816||full | Not-determined |
| S9_vs_k141_47405||full | Not-determined |
| S9_vs_k141_17730||full | Low-quality |
| S9_vs_k141_47411||full | Low-quality |
| S9_vs_k141_9725||full | Low-quality |
| S9_vs_k141_17742||full | Low-quality |
| S9_vs_k141_32651||full | Low-quality |
| S9_vs_k141_81734||full | Not-determined |
| S9_vs_k141_55437||full | Not-determined |
| S9_vs_k141_58834||full | Low-quality |
| S9_vs_k141_81744||full | Low-quality |
| S9_vs_k141_74149||full | Low-quality |
| S9_vs_k141_21334||full | Not-determined |
| S9_vs_k141_21335||full | Low-quality |
| S9_vs_k141_17768||full | Low-quality |
| S9_vs_k141_81747||full | Low-quality |
| S9_vs_k141_25149||full | Low-quality |
| S9_vs_k141_66414||full | Low-quality |
| S9_vs_k141_85389||full | Low-quality |
| S9_vs_k141_2387||full | Low-quality |
| S9_vs_k141_85395||full | Low-quality |
| S9_vs_k141_6187||full | Not-determined |
| S9_vs_k141_40551||full | Not-determined |
| S9_vs_k141_66428||full | Low-quality |
| S9_vs_k141_25166||full | Low-quality |
| S9_vs_k141_29049||full | Low-quality |
| S9_vs_k141_2408||full | Low-quality |
| S4_vs_VIRSorter_6168-cat_3 | High-quality |
| S9_vs_k141_85415||full | Low-quality |
| S9_vs_k141_55481||full | Not-determined |
| S9_vs_k141_58883||full | Not-determined |
| S9_vs_k141_32695||full | Low-quality |
| S9_vs_k141_25171||full | Low-quality |
| S9_vs_k141_36645||full | Low-quality |
| S9_vs_k141_29062||full | Not-determined |
| S9_vs_k141_17784||full | Low-quality |
| S9_vs_k141_36646||full | Not-determined |
| S9_vs_k141_2425||full | Low-quality |
| S9_vs_k141_32702||full | Low-quality |
| S9_vs_k141_44321||full | Low-quality |
| S9_vs_k141_2430||full | Low-quality |
| S4_vs_VIRSorter_13190-cat_2 | Medium-quality |
| S9_vs_k141_62810||full | Not-determined |
| S9_vs_k141_25178||full | Low-quality |
| S9_vs_k141_74182||full | Low-quality |
| S9_vs_k141_40559||full | Low-quality |
| S9_vs_k141_9773||full | Low-quality |
| S9_vs_k141_66437||full | Not-determined |
| S9_vs_k141_47505||full | Not-determined |
| S9_vs_k141_81798||full | Low-quality |
| S9_vs_k141_62818||full | Not-determined |
| S9_vs_k141_74184||full | Not-determined |
| S9_vs_k141_17785||full | Low-quality |
| S9_vs_k141_2442||full | Low-quality |
| S9_vs_k141_47507||full | Low-quality |
| S9_vs_k141_9790||full | Not-determined |
| S9_vs_k141_32727||full | Low-quality |
| S9_vs_k141_29094||full | Low-quality |
| S9_vs_k141_25200||full | Low-quality |
| S9_vs_k141_77567||full | Low-quality |
| S9_vs_k141_55518||full | Not-determined |
| S9_vs_k141_66463||full | Not-determined |
| S9_vs_k141_6279||full | Not-determined |
| S9_vs_k141_74222||full | Low-quality |
| S9_vs_k141_77579||full | Not-determined |
| S9_vs_k141_77581||full | Not-determined |
| S9_vs_k141_36682||full | Low-quality |
| S9_vs_k141_85472||full | Not-determined |
| S9_vs_k141_17823||full | Low-quality |
| S9_vs_k141_66473||full | Low-quality |
| S9_vs_k141_13557||full | Low-quality |
| S9_vs_k141_55540||full | Not-determined |
| S9_vs_k141_74227||full | Not-determined |
| S9_vs_k141_66477||full | Not-determined |
| S9_vs_k141_47537||full | Low-quality |
| S9_vs_k141_17830||full | Low-quality |
| S9_vs_k141_2482||full | Low-quality |
| S9_vs_k141_21450||full | Low-quality |
| S9_vs_k141_81825||full | Low-quality |
| S9_vs_k141_66481||full | Low-quality |
| S9_vs_k141_32768||full | Not-determined |
| S11_vs_k141_28515||full | Medium-quality |
| S9_vs_k141_21456||full | Low-quality |
| S9_vs_k141_32769||full | Low-quality |

S9_vs_k141_32772||full Low-quality
S9_vs_k141_77604||full Not-determined
S9_vs_k141_44334||full Low-quality
S9_vs_k141_13577||full Low-quality
S9_vs_k141_36708||full Low-quality
S9_vs_k141_2504||full Low-quality
S9_vs_k141_32782||full Low-quality
S9_vs_k141_58973||full Low-quality
S9_vs_k141_58975||full Low-quality
S9_vs_k141_13590||full Low-quality
S9_vs_k141_2520||full Low-quality
S9_vs_k141_77619||full Low-quality
S9_vs_k141_66508||full Not-determined
S9_vs_k141_6318||full Low-quality
S9_vs_k141_77626||full Low-quality
S9_vs_k141_13599||full Low-quality
S9_vs_k141_2527||full Low-quality
S9_vs_k141_47582||full Low-quality
S9_vs_k141_44356||full Low-quality
S9_vs_k141_50979||full Not-determined
S9_vs_k141_77633||full Low-quality
S9_extracted_k141_76769 High-quality
S9_vs_k141_40606||full Not-determined
S9_vs_k141_17865||full Not-determined
S9_vs_k141_36757||full Low-quality
S9_vs_k141_36758||full Not-determined
S9_vs_k141_77639||full Not-determined
S9_vs_k141_55596||full Low-quality
S9_vs_k141_59008||full Not-determined
S9_vs_k141_29176||full Not-determined
S9_vs_k141_17873||full Low-quality
S9_vs_k141_36780||full Not-determined
S9_vs_k141_17886||full Low-quality
S9_vs_k141_89205||full Not-determined
S9_vs_k141_29195||full Low-quality
S9_vs_k141_47628||full Low-quality
S9_vs_k141_66561||full Low-quality
S9_vs_k141_59026||full Low-quality
S9_vs_k141_76769||full High-quality
S9_vs_k141_77658||full Not-determined
S9_vs_k141_47631||full Not-determined
S9_vs_k141_66562||full Low-quality
S9_vs_k141_85519||full Low-quality
S9_vs_k141_9939||full Not-determined
S9_vs_k141_17911||full Low-quality
S9_vs_k141_74250||full Not-determined
S9_vs_k141_55634||full Low-quality
S9_vs_k141_44394||full Low-quality
S9_vs_k141_47634||full Not-determined
S9_vs_k141_77679||full Low-quality
S9_vs_k141_29216||full Not-determined
S9_vs_k141_2600||full Not-determined
S9_vs_k141_17917||full Low-quality
S9_vs_k141_55636||full Low-quality
S9_vs_k141_62960||full Low-quality
S9_vs_k141_29222||full Low-quality
S9_vs_k141_66587||full Low-quality
S9_vs_k141_17919||full Low-quality
S9_vs_k141_74262||full Not-determined
S9_vs_k141_6406||full Low-quality
S9_vs_k141_9961||full Low-quality
S9_vs_k141_85541||full Not-determined
S9_vs_k141_36842||full Low-quality
S9_vs_k141_55660||full Not-determined
S9_vs_k141_21590||full Not-determined
S9_vs_k141_9972||full Low-quality
S9_vs_k141_47657||full Low-quality
S9_vs_k141_9974||full Low-quality
S9_vs_k141_36850||full Low-quality
S9_vs_k141_29255||full Low-quality
S9_vs_k141_59069||full Not-determined
S9_vs_k141_47666||full Low-quality
S9_vs_k141_44434||full Not-determined
S9_vs_k141_6434||full Low-quality
S9_vs_k141_85560||full Not-determined
S9_vs_k141_13692||full Low-quality
S9_vs_k141_81932||full Low-quality
S9_vs_k141_62993||full Not-determined
S9_vs_k141_6440||full Low-quality
S9_vs_k141_6443||full Low-quality
S9_vs_k141_81934||full Not-determined
S9_vs_k141_6445||full Low-quality
S9_vs_k141_25242||full Low-quality
S8_extracted_k141_173225 High-quality
S9_vs_k141_21629||full Low-quality
S9_vs_k141_81945||full Low-quality
S9_vs_k141_32844||full Low-quality
S9_vs_k141_25243||full Low-quality
S9_vs_k141_17947||full Low-quality
S9_vs_k141_17948||full Not-determined
S9_vs_k141_10015||full Not-determined
S9_vs_k141_55698||full Not-determined
S9_vs_k141_59105||full Not-determined
S9_vs_k141_81960||full Low-quality
S9_vs_k141_85600||full Low-quality
S9_vs_k141_59106||full Not-determined
S9_vs_k141_81962||full Low-quality
S9_vs_k141_32862||full Not-determined
S9_vs_k141_44469||full Low-quality
S9_vs_k141_32867||full Not-determined
S9_vs_k141_2647||full Low-quality
S9_vs_k141_6474||full Low-quality
S9_vs_k141_13727||full Not-determined
S9_vs_k141_74336||full Not-determined
S9_vs_k141_55700||full Low-quality
S9_vs_k141_40627||full Low-quality
S9_vs_k141_44478||full Low-quality
S9_vs_k141_21657||full Low-quality
S9_vs_k141_25279||full Low-quality
S9_vs_k141_77773||full Low-quality
S9_vs_k141_66609||full Low-quality
S9_vs_k141_21662||full Low-quality
S9_vs_k141_2672||full Not-determined
S9_vs_k141_32888||full Not-determined
S9_vs_k141_59144||full Low-quality
S9_vs_k141_6490||full Low-quality
S9_vs_k141_10064||full Not-determined
S9_vs_k141_17983||full Not-determined
S9_vs_k141_81991||full Low-quality
S9_vs_k141_2680||full Low-quality
S9_vs_k141_2681||full Low-quality
S9_vs_k141_17991||full Low-quality
S9_vs_k141_63070||full Low-quality
S9_vs_k141_70343||full Low-quality
S9_vs_k141_77784||full Low-quality
S9_vs_k141_81996||full Low-quality
S9_vs_k141_10071||full Low-quality
S9_vs_k141_29282||full Low-quality
S9_vs_k141_44505||full Low-quality
S9_vs_k141_51004||full Low-quality
S9_vs_k141_10074||full Low-quality
S9_vs_k141_59167||full Low-quality
S9_vs_k141_25315||full Not-determined
S9_vs_k141_21671||full Low-quality
S9_vs_k141_32919||full Not-determined
S9_vs_k141_66619||full Low-quality
S9_vs_k141_21678||full Not-determined
S9_vs_k141_81998||full Low-quality
S9_vs_k141_59180||full Low-quality
S9_vs_k141_70348||full Not-determined
S9_vs_k141_63089||full Not-determined
S9_vs_k141_32932||full Low-quality
S9_vs_k141_18017||full Low-quality
S9_vs_k141_32934||full Not-determined
S9_vs_k141_66635||full Not-determined
S9_vs_k141_10086||full Low-quality
S9_vs_k141_18020||full Not-determined
S9_vs_k141_85665||full Low-quality
S9_vs_k141_10092||full Low-quality
S9_vs_k141_44539||full Not-determined
S9_vs_k141_29319||full Not-determined
S9_vs_k141_6541||full Low-quality
S9_vs_k141_6544||full Low-quality
S9_vs_k141_89328||full Not-determined
S9_vs_k141_74403||full Low-quality
S9_vs_k141_89331||full Low-quality
S9_vs_k141_25340||full Low-quality
S9_vs_k141_36903||full Low-quality
S9_vs_k141_59209||full Not-determined
S9_vs_k141_10117||full Not-determined
S9_vs_k141_2713||full Not-determined
S9_vs_k141_10119||full Not-determined
S9_vs_k141_21713||full Not-determined
S9_vs_k141_13824||full Not-determined
S9_vs_k141_21720||full Not-determined
S9_vs_k141_63127||full Not-determined
S9_vs_k141_82002||full Low-quality
S9_vs_k141_74415||full Low-quality
S9_vs_k141_44562||full Not-determined
S9_vs_k141_36917||full Low-quality
S9_vs_k141_40722||full Not-determined
S9_vs_k141_21726||full Low-quality
S9_vs_k141_77830||full Not-determined
S9_vs_k141_70400||full Low-quality
S9_vs_k141_44568||full Low-quality
S9_vs_k141_85711||full Low-quality
S9_vs_k141_63129||full Low-quality
S9_vs_k141_32984||full Low-quality
S4_extracted_31825 Complete
S9_vs_k141_10139||full Low-quality
S9_vs_k141_44575||full Low-quality
S9_vs_k141_44576||full Not-determined
S9_vs_k141_6555||full Not-determined
S9_vs_k141_18084||full Low-quality
S9_vs_k141_77844||full Not-determined
S9_vs_k141_2747||full Low-quality
S9_vs_k141_63142||full Low-quality
S9_vs_k141_44581||full Low-quality
S9_vs_k141_85728||full Low-quality
S9_vs_k141_2748||full Low-quality
S9_vs_k141_74444||full Low-quality
S9_vs_k141_70418||full Low-quality
S9_vs_k141_77867||full Low-quality
S9_vs_k141_47758||full Low-quality
S9_vs_k141_13859||full Low-quality
S9_vs_k141_21769||full Low-quality
S9_vs_k141_18103||full Low-quality
S9_vs_k141_47769||full Low-quality
S9_vs_k141_55777||full Low-quality
S9_vs_k141_6597||full Not-determined
S9_vs_k141_70448||full Not-determined
S9_vs_k141_18113||full Not-determined
S9_vs_k141_6602||full Low-quality
S9_vs_k141_85757||full Low-quality
S9_vs_k141_6604||full Low-quality
S9_vs_k141_29415||full Not-determined
S9_vs_k141_33043||full Low-quality
S9_vs_k141_25420||full Not-determined
S9_vs_k141_36985||full Low-quality
S9_vs_k141_70463||full Low-quality
S9_vs_k141_21791||full Not-determined
S9_vs_k141_47794||full Low-quality
S9_vs_k141_10196||full Low-quality
S9_vs_k141_13880||full Not-determined
S9_vs_k141_36991||full Not-determined
S9_vs_k141_25426||full Low-quality
S9_vs_k141_37003||full Low-quality
S9_vs_k141_70482||full Not-determined
S9_vs_k141_25441||full Low-quality
S9_vs_k141_37010||full Not-determined
S9_vs_k141_47813||full Not-determined
S9_vs_k141_63215||full Low-quality
S9_vs_k141_21800||full Low-quality
S9_vs_k141_89369||full Low-quality
S9_vs_k141_85790||full Low-quality
S9_vs_k141_47816||full Low-quality
S9_vs_k141_6633||full Not-determined
S8_vs_k141_173225||full High-quality
S9_vs_k141_70490||full Low-quality
S9_vs_k141_25455||full Low-quality
S9_vs_k141_66748||full Low-quality
S9_vs_k141_21806||full Not-determined
S9_vs_k141_37023||full Low-quality
S9_vs_k141_6639||full Low-quality
S9_vs_k141_66751||full Not-determined
S9_vs_k141_33063||full Low-quality
S9_vs_k141_18162||full Low-quality
S9_vs_k141_77937||full Low-quality
S9_vs_k141_13920||full Low-quality
S9_vs_k141_10242||full Low-quality
S9_vs_k141_51100||full Low-quality
S9_vs_k141_66769||full Low-quality
S9_vs_k141_10243||full Not-determined
S9_vs_k141_51101||full Not-determined
S9_vs_k141_10246||full Not-determined
S9_vs_k141_47843||full Not-determined
S9_vs_k141_59237||full Low-quality
S9_vs_k141_47846||full Low-quality
S9_vs_k141_37052||full Low-quality
S9_vs_k141_40765||full Low-quality
S9_vs_k141_29457||full Low-quality
S9_vs_k141_59247||full Low-quality
S9_vs_k141_40767||full Not-determined
S9_vs_k141_21848||full Not-determined
S9_vs_k141_85831||full Low-quality
S9_vs_k141_33105||full Not-determined
S9_vs_k141_59258||full Low-quality
S9_vs_k141_47875||full Low-quality
S9_vs_k141_82136||full Not-determined
S9_vs_k141_66802||full Low-quality
S9_vs_k141_40778||full Not-determined
S9_vs_k141_44647||full Low-quality
S9_vs_k141_13975||full Low-quality
S9_vs_k141_37079||full Low-quality
S9_vs_k141_33115||full Not-determined
S9_vs_k141_66808||full Not-determined
S9_vs_k141_63264||full Not-determined
S9_vs_k141_47885||full Low-quality
S9_vs_k141_10271||full Low-quality
S9_vs_k141_29483||full Not-determined
S9_vs_k141_44651||full Low-quality
S9_vs_k141_44652||full Low-quality
S9_vs_k141_55856||full Low-quality
S9_vs_k141_63271||full Low-quality
S9_vs_k141_66830||full Low-quality
S9_vs_k141_40810||full Not-determined
S5_vs_k141_55415||full High-quality
S9_vs_k141_44663||full Not-determined
S9_vs_k141_47897||full Low-quality
S9_vs_k141_44669||full Low-quality
S9_vs_k141_14011||full Low-quality
S9_vs_k141_40828||full Low-quality
S9_vs_k141_55880||full Not-determined
S9_vs_k141_63299||full Low-quality
S8_vs_k141_49282||full High-quality
S9_vs_k141_66858||full Low-quality
S9_vs_k141_55882||full Low-quality
S9_vs_k141_14013||full Low-quality
S9_vs_k141_63304||full Not-determined
S9_vs_k141_33173||full Low-quality
S9_vs_k141_21872||full Low-quality
S9_vs_k141_47928||full Not-determined
S9_vs_k141_89494||full Low-quality
S9_vs_k141_29540||full Low-quality
S9_vs_k141_70570||full Low-quality
S9_vs_k141_59326||full Low-quality
S9_vs_k141_63319||full Low-quality
S9_vs_k141_89500||full Low-quality
S9_vs_k141_14023||full Low-quality
S9_vs_k141_59331||full Not-determined
S9_vs_k141_2850||full Low-quality
S9_vs_k141_63331||full Low-quality
S9_vs_k141_82161||full Not-determined
S9_vs_k141_33193||full Low-quality
S9_vs_k141_82175||full Low-quality
S9_vs_k141_47977||full Low-quality
S9_vs_k141_51204||full Low-quality
S9_vs_k141_85933||full Low-quality
S9_vs_k141_82185||full Low-quality
S9_vs_k141_55915||full Low-quality
S9_vs_k141_14052||full Low-quality
S9_vs_k141_89547||full Low-quality
S9_vs_k141_44698||full Low-quality
S9_vs_k141_40891||full Low-quality
S9_vs_k141_33231||full Not-determined
S9_vs_k141_6726||full Not-determined
S9_vs_k141_51223||full Low-quality
S9_vs_k141_51225||full Low-quality
S9_vs_k141_48006||full Not-determined
S9_vs_k141_18227||full Not-determined
S9_vs_k141_37160||full Low-quality
S9_vs_k141_89572||full Low-quality
S9_vs_k141_18228||full Not-determined
S9_vs_k141_70641||full Not-determined
S9_vs_k141_59377||full Low-quality
S9_vs_k141_55942||full Low-quality
S9_vs_k141_85961||full Not-determined
S9_vs_k141_10412||full Low-quality
S9_vs_k141_63388||full Low-quality
S9_vs_k141_44726||full Low-quality
S9_vs_k141_25597||full Low-quality
S9_vs_k141_85975||full Not-determined
S9_vs_k141_37173||full Low-quality
S9_vs_k141_70662||full Not-determined
S9_vs_k141_44734||full Low-quality
S9_vs_k141_44737||full Low-quality
S9_vs_k141_25607||full Not-determined
S9_vs_k141_6746||full Low-quality
S9_vs_k141_2920||full Low-quality
S9_vs_k141_85988||full Not-determined
S9_vs_k141_78054||full Low-quality
S9_vs_k141_25610||full Low-quality
S9_vs_k141_48044||full Low-quality
S9_vs_k141_6756||full Not-determined
S9_vs_k141_74505||full Low-quality
S9_vs_k141_25616||full Low-quality
S9_vs_k141_25625||full Low-quality
S9_vs_k141_6769||full Low-quality
S9_vs_k141_59397||full Low-quality
S9_vs_k141_74514||full Not-determined
S9_vs_k141_25629||full Low-quality
S9_vs_k141_70678||full Low-quality
S9_vs_k141_33297||full Low-quality
S9_vs_k141_25631||full Low-quality
S9_vs_k141_78069||full Low-quality
S9_vs_k141_44770||full Not-determined
S9_vs_k141_66920||full Low-quality
S9_vs_k141_48067||full Not-determined
S9_vs_k141_48071||full Low-quality
S9_vs_k141_37191||full Not-determined
S9_vs_k141_29653||full Not-determined
S9_vs_k141_44780||full Low-quality
S9_vs_k141_40944||full Low-quality
S9_vs_k141_29654||full Not-determined
S9_vs_k141_59411||full Not-determined
S9_vs_k141_48075||full Low-quality
S6_vs_k141_39623||full High-quality
S9_vs_k141_78088||full Low-quality
S9_vs_k141_25650||full Low-quality
S9_vs_k141_78091||full Low-quality
S9_vs_k141_48082||full Not-determined
S9_vs_k141_37200||full Not-determined
S9_vs_k141_89614||full Low-quality
S9_vs_k141_44798||full Low-quality
S9_vs_k141_51275||full Low-quality
S9_vs_k141_40959||full Not-determined
S9_vs_k141_2970||full Not-determined
S9_vs_k141_56021||full Not-determined
S9_vs_k141_6787||full Low-quality
S9_vs_k141_74558||full Low-quality
S9_vs_k141_48095||full Low-quality
S9_vs_k141_78111||full Low-quality
S9_vs_k141_25676||full Low-quality
S9_vs_k141_33342||full Low-quality
S9_vs_k141_6801||full Not-determined
S9_vs_k141_86055||full Low-quality
S9_vs_k141_25680||full Low-quality
S9_vs_k141_18337||full Low-quality
S9_vs_k141_51309||full Low-quality
S9_vs_k141_48100||full Low-quality
S9_vs_k141_40976||full Low-quality
S9_vs_k141_86066||full Low-quality
S9_vs_k141_21986||full Low-quality
S9_vs_k141_2986||full Low-quality
S9_vs_k141_21987||full Low-quality
S9_vs_k141_18346||full Not-determined
S9_vs_k141_2989||full Low-quality
S9_vs_k141_86070||full Not-determined
S9_vs_k141_56029||full Low-quality
S9_vs_k141_25699||full Not-determined
S9_vs_k141_18355||full Low-quality
S9_vs_k141_89673||full Low-quality
S9_vs_k141_33372||full Not-determined
S9_vs_k141_63478||full Not-determined
S9_vs_k141_78137||full Low-quality
S9_vs_k141_51342||full Low-quality
S9_vs_k141_40992||full Not-determined
S9_vs_k141_33375||full Not-determined
S9_vs_k141_56047||full Not-determined
S9_vs_k141_18370||full Not-determined
S9_vs_k141_51348||full Not-determined
S9_vs_k141_74607||full Low-quality
S9_vs_k141_66977||full Low-quality
S9_vs_k141_6852||full Low-quality
S9_vs_k141_82303||full Low-quality
S9_vs_k141_3017||full Low-quality
S9_vs_k141_22020||full Not-determined
S9_vs_k141_89695||full Low-quality
S9_vs_k141_3021||full Low-quality
S9_vs_k141_22028||full Low-quality
S9_vs_k141_25742||full Low-quality
S9_vs_k141_82320||full Low-quality
S9_vs_k141_89698||full Not-determined
S9_vs_k141_29745||full Low-quality
S9_vs_k141_6870||full Not-determined
S9_vs_k141_41007||full Not-determined
S9_vs_k141_78170||full Low-quality
S9_vs_k141_29746||full Low-quality
S9_vs_k141_3036||full Not-determined
S9_vs_k141_10506||full Low-quality
S9_vs_k141_51367||full Low-quality
S9_vs_k141_89717||full Not-determined
S9_vs_k141_41009||full Not-determined
S9_vs_k141_89719||full Not-determined
S9_vs_k141_41011||full Low-quality
S9_vs_k141_89721||full Low-quality
S9_vs_k141_56089||full Not-determined
S9_vs_k141_29751||full Low-quality
S9_vs_k141_59549||full Not-determined
S9_vs_k141_18424||full Not-determined
S9_vs_k141_89728||full Low-quality
S9_vs_k141_74653||full Not-determined
S9_vs_k141_37314||full Low-quality
S9_vs_k141_56109||full Not-determined
S11_vs_k141_82331||full Complete
S9_vs_k141_82359||full Low-quality
S9_vs_k141_67031||full Not-determined
S9_vs_k141_25798||full Low-quality
S9_vs_k141_48179||full Low-quality
S9_vs_k141_37315||full Not-determined
S9_vs_k141_22070||full Not-determined
S9_vs_k141_18431||full Low-quality
S9_vs_k141_67040||full Low-quality
S9_vs_k141_44891||full Low-quality
S9_vs_k141_70773||full Low-quality
S9_vs_k141_67052||full Not-determined
S9_vs_k141_25816||full Not-determined
S9_vs_k141_6927||full Low-quality
S9_vs_k141_70790||full Not-determined
S9_vs_k141_41057||full Low-quality
S9_vs_k141_14089||full Not-determined
S9_vs_k141_33457||full Not-determined
S9_vs_k141_67059||full Not-determined
S9_vs_k141_86178||full Not-determined
S9_vs_k141_37350||full Low-quality
S9_vs_k141_6937||full Not-determined
S9_vs_k141_48212||full Low-quality
S9_vs_k141_22101||full Low-quality
S9_vs_k141_56154||full Not-determined
S9_vs_k141_44920||full Low-quality
S9_vs_k141_14098||full Not-determined
S9_vs_k141_82411||full Not-determined
S9_vs_k141_74705||full Not-determined
S9_vs_k141_89776||full Not-determined
S9_vs_k141_70815||full Not-determined
S9_vs_k141_89779||full Low-quality
S9_vs_k141_14100||full Low-quality
S9_vs_k141_6961||full Not-determined
S9_vs_k141_3111||full Low-quality
S9_vs_k141_33485||full Low-quality
S9_vs_k141_89793||full Not-determined
S9_vs_k141_86190||full Low-quality
S9_vs_k141_44943||full Low-quality
S9_vs_k141_63550||full Low-quality
S9_vs_k141_56161||full Low-quality
S9_vs_k141_25851||full Low-quality
S9_vs_k141_25852||full Low-quality
S9_vs_k141_18488||full Not-determined
S9_vs_k141_22106||full Low-quality
S9_vs_k141_33492||full Not-determined
S9_vs_k141_25853||full Not-determined
S9_vs_k141_51432||full Low-quality
S9_vs_k141_70838||full Not-determined
S9_vs_k141_10612||full Not-determined
S9_vs_k141_63560||full Not-determined
S9_vs_k141_56170||full Not-determined
S9_vs_k141_78246||full Not-determined
S9_vs_k141_33498||full Low-quality
S9_vs_k141_74746||full Low-quality
S9_vs_k141_25866||full Low-quality
S9_vs_k141_18502||full Low-quality
S9_vs_k141_59640||full Not-determined
S9_vs_k141_18505||full Low-quality
S9_vs_k141_86218||full Not-determined
S9_vs_k141_70866||full Not-determined
S9_vs_k141_70868||full Low-quality
S9_vs_k141_41123||full Low-quality
S9_vs_k141_22123||full Not-determined
S9_vs_k141_86242||full Low-quality
S9_vs_k141_7024||full Low-quality
S9_vs_k141_41136||full Low-quality
S9_vs_k141_56196||full Low-quality
S12_extracted_k141_4027 High-quality
S9_vs_k141_82418||full Low-quality
S9_vs_k141_22136||full Not-determined
S9_vs_k141_45010||full Low-quality
S9_vs_k141_10676||full Not-determined
S9_vs_k141_48256||full Low-quality
S9_vs_k141_10682||full Low-quality
S9_vs_k141_82436||full Low-quality
S9_vs_k141_41157||full Not-determined
S9_vs_k141_59693||full Low-quality
S9_vs_k141_7060||full Not-determined
S9_vs_k141_67194||full Not-determined
S9_vs_k141_86282||full Low-quality
S9_vs_k141_7071||full Low-quality
S9_vs_k141_25935||full Not-determined
S9_vs_k141_18577||full Not-determined
S9_vs_k141_56234||full Not-determined
S9_vs_k141_14166||full Not-determined
S9_vs_k141_59709||full Not-determined
S9_vs_k141_29961||full Low-quality
S9_vs_k141_74839||full Low-quality
S13_vs_151510||full High-quality
S9_vs_k141_41203||full Low-quality
S9_vs_k141_70952||full Not-determined
S9_vs_k141_56241||full Not-determined
S9_vs_k141_7102||full Low-quality
S9_vs_k141_89924||full Low-quality
S9_vs_k141_10703||full Not-determined
S9_vs_k141_74858||full Low-quality
S9_vs_k141_89926||full Not-determined
S9_vs_k141_70958||full Low-quality
S9_vs_k141_56262||full Low-quality
S9_vs_k141_78358||full Not-determined
S9_vs_k141_45073||full Not-determined
S9_vs_k141_45074||full Low-quality
S9_vs_k141_22184||full Not-determined
S9_vs_k141_82496||full Low-quality
S9_vs_k141_48304||full Low-quality
S9_vs_k141_48309||full Not-determined
S9_vs_k141_59739||full Low-quality
S9_vs_k141_41235||full Low-quality
S9_vs_k141_33625||full Not-determined
S9_vs_k141_3173||full Not-determined
S9_vs_k141_3174||full Not-determined
S9_vs_k141_82514||full Low-quality
S9_vs_k141_30019||full Not-determined
S9_vs_k141_74899||full Low-quality
S9_vs_k141_7134||full Low-quality
S9_vs_k141_70998||full Low-quality
S9_vs_k141_30030||full Not-determined
S9_vs_k141_18668||full Low-quality
S9_vs_k141_48325||full Not-determined
S9_vs_k141_59760||full Not-determined
S9_vs_k141_78410||full Low-quality
S9_vs_k141_74917||full Low-quality
S9_vs_k141_45133||full Low-quality
S9_vs_k141_71018||full Low-quality
S9_vs_k141_18701||full Low-quality
S9_vs_k141_3229||full Not-determined
S9_vs_k141_78432||full Low-quality
S9_vs_k141_78433||full Low-quality
S10_vs_24907||full High-quality
S9_vs_k141_59792||full Low-quality
S9_vs_k141_45143||full Low-quality
S9_vs_k141_30075||full Not-determined
S9_vs_k141_3241||full Not-determined
S9_vs_k141_7192||full Low-quality
S9_vs_k141_71037||full Not-determined
S9_vs_k141_37449||full Not-determined
S9_vs_k141_67204||full Low-quality
S9_vs_k141_18711||full Low-quality
S9_vs_k141_74947||full Low-quality
S9_vs_k141_45161||full Low-quality
S9_vs_k141_89957||full Not-determined
S9_vs_k141_78445||full Low-quality
S9_vs_k141_3263||full Low-quality
S9_vs_k141_10826||full Low-quality

| | |
|---|---|
| S9_vs_k141_41316\|\|full | Low-quality |
| S9_vs_k141_30090\|\|full | Low-quality |
| S9_vs_k141_71054\|\|full | Low-quality |
| S9_vs_k141_30094\|\|full | Low-quality |
| S9_vs_k141_18739\|\|full | Low-quality |
| S9_vs_k141_3273\|\|full | Not-determined |
| S9_vs_k141_89971\|\|full | Not-determined |
| S9_vs_k141_51524\|\|full | Not-determined |
| S12_vs_k141_4027\|\|full | High-quality |
| S9_vs_k141_41327\|\|full | Low-quality |
| S9_vs_k141_51527\|\|full | Low-quality |
| S9_vs_k141_37462\|\|full | Low-quality |
| S9_vs_k141_18744\|\|full | Not-determined |
| S3_extracted_k141_127399 | Complete |
| S9_vs_k141_51532\|\|full | Not-determined |
| S9_vs_k141_74984\|\|full | Low-quality |
| S9_vs_k141_41332\|\|full | Low-quality |
| S9_vs_k141_37463\|\|full | Low-quality |
| S9_vs_k141_22322\|\|full | Not-determined |
| S9_vs_k141_67241\|\|full | Low-quality |
| S9_vs_k141_41337\|\|full | Not-determined |
| S9_vs_k141_48360\|\|full | Low-quality |
| S9_vs_k141_18750\|\|full | Low-quality |
| S9_vs_k141_59844\|\|full | Low-quality |
| S9_vs_k141_51543\|\|full | Not-determined |
| S9_vs_k141_33700\|\|full | Not-determined |
| S9_vs_k141_86383\|\|full | Not-determined |
| S9_vs_k141_59855\|\|full | Low-quality |
| S9_vs_k141_10867\|\|full | Not-determined |
| S9_vs_k141_3292\|\|full | Low-quality |
| S9_vs_k141_41361\|\|full | Not-determined |
| S9_vs_k141_37492\|\|full | Low-quality |
| S9_vs_k141_78502\|\|full | Not-determined |
| S9_vs_k141_48387\|\|full | Low-quality |
| S13_extracted_11246 | Complete |
| S9_vs_k141_30134\|\|full | Low-quality |
| S9_vs_k141_45230\|\|full | Low-quality |
| S9_vs_k141_86405\|\|full | Not-determined |
| S9_vs_k141_82552\|\|full | Low-quality |
| S9_vs_k141_75033\|\|full | Not-determined |
| S9_vs_k141_90018\|\|full | Low-quality |
| S9_vs_k141_78509\|\|full | Low-quality |
| S9_vs_k141_7263\|\|full | Not-determined |
| S9_vs_k141_59883\|\|full | Low-quality |
| S9_vs_k141_37515\|\|full | Low-quality |
| S9_vs_k141_30144\|\|full | Not-determined |
| S9_vs_k141_33736\|\|full | Low-quality |
| S9_vs_k141_48414\|\|full | Not-determined |
| S9_vs_k141_10906\|\|full | Not-determined |
| S9_vs_k141_14283\|\|full | Low-quality |
| S9_vs_k141_59891\|\|full | Low-quality |
| S9_vs_k141_37523\|\|full | Low-quality |
| S9_vs_k141_67291\|\|full | Not-determined |
| S9_vs_k141_59895\|\|full | Low-quality |
| S9_vs_k141_90036\|\|full | Low-quality |
| S9_vs_k141_86419\|\|full | Low-quality |
| S9_vs_k141_51593\|\|full | Low-quality |
| S9_vs_k141_26109\|\|full | Low-quality |
| S9_vs_k141_90037\|\|full | Low-quality |
| S9_vs_k141_37532\|\|full | Low-quality |
| S9_vs_k141_18815\|\|full | Low-quality |
| S9_vs_k141_26114\|\|full | Not-determined |
| S9_vs_k141_82559\|\|full | Low-quality |
| S9_vs_k141_7291\|\|full | Not-determined |
| S9_vs_k141_78552\|\|full | Low-quality |
| S14_extracted_k141_113008 | Complete |
| S9_vs_k141_37548\|\|full | Low-quality |
| S9_vs_k141_59910\|\|full | Low-quality |
| S9_vs_k141_10915\|\|full | Low-quality |
| S9_vs_k141_30178\|\|full | Not-determined |
| S9_vs_k141_3349\|\|full | Low-quality |
| S9_vs_k141_56387\|\|full | Low-quality |
| S9_vs_k141_26115\|\|full | Low-quality |
| S9_vs_k141_7307\|\|full | Low-quality |
| S9_vs_k141_59919\|\|full | Not-determined |
| S9_vs_k141_26122\|\|full | Low-quality |
| S9_vs_k141_18835\|\|full | Low-quality |
| S9_vs_k141_3357\|\|full | Low-quality |
| S9_vs_k141_41397\|\|full | Not-determined |
| S9_vs_k141_3358\|\|full | Low-quality |
| S9_vs_k141_37560\|\|full | Not-determined |
| S9_vs_k141_56398\|\|full | Low-quality |
| S9_vs_k141_71174\|\|full | Not-determined |
| S9_vs_k141_78576\|\|full | Not-determined |
| S9_vs_k141_3373\|\|full | Low-quality |
| S9_vs_k141_41408\|\|full | Low-quality |
| S9_vs_k141_30206\|\|full | Not-determined |
| S9_vs_k141_18866\|\|full | Low-quality |
| S9_vs_k141_45294\|\|full | Low-quality |
| S9_vs_k141_26162\|\|full | Low-quality |
| S9_vs_k141_56424\|\|full | Low-quality |
| S9_vs_k141_18873\|\|full | Low-quality |
| S9_vs_k141_3385\|\|full | Low-quality |
| S9_vs_k141_56427\|\|full | Not-determined |
| S9_vs_k141_71203\|\|full | Not-determined |
| S9_vs_k141_33780\|\|full | Low-quality |
| S9_vs_k141_14352\|\|full | Not-determined |
| S9_vs_k141_71206\|\|full | Not-determined |
| S9_vs_k141_33790\|\|full | Low-quality |
| S9_vs_k141_41431\|\|full | Low-quality |
| S9_vs_k141_71207\|\|full | Low-quality |
| S9_vs_k141_78619\|\|full | Not-determined |
| S9_vs_k141_18890\|\|full | Not-determined |
| S9_vs_k141_71212\|\|full | Low-quality |
| S9_vs_k141_41444\|\|full | Low-quality |
| S9_vs_k141_90131\|\|full | Low-quality |
| S9_vs_k141_71216\|\|full | Not-determined |
| S9_vs_k141_3398\|\|full | Low-quality |
| S9_vs_k141_30243\|\|full | Not-determined |
| S9_vs_k141_75105\|\|full | Low-quality |
| S9_vs_k141_22413\|\|full | Low-quality |
| S9_vs_k141_75109\|\|full | Not-determined |
| S9_vs_k141_45345\|\|full | Low-quality |
| S9_vs_k141_14390\|\|full | Low-quality |
| S9_vs_k141_37600\|\|full | Low-quality |
| S2_extracted_k141_1399 | Complete |
| S9_vs_k141_22421\|\|full | Low-quality |
| S9_vs_k141_45348\|\|full | Low-quality |
| S9_vs_k141_75120\|\|full | Low-quality |
| S9_vs_k141_59994\|\|full | Low-quality |
| S9_vs_k141_45352\|\|full | Low-quality |
| S9_vs_k141_3414\|\|full | Low-quality |
| S9_vs_k141_33839\|\|full | Not-determined |
| S9_vs_k141_45353\|\|full | Low-quality |
| S9_vs_k141_3417\|\|full | Not-determined |
| S9_vs_k141_60000\|\|full | Low-quality |
| S9_vs_k141_67414\|\|full | Not-determined |
| S9_vs_k141_22441\|\|full | Low-quality |
| S9_vs_k141_33855\|\|full | Low-quality |
| S9_vs_k141_56486\|\|full | Low-quality |
| S9_vs_k141_45364\|\|full | Not-determined |
| S9_vs_k141_14415\|\|full | Low-quality |
| S9_vs_k141_14419\|\|full | Low-quality |
| S9_vs_k141_41462\|\|full | Not-determined |
| S9_vs_k141_67419\|\|full | Not-determined |
| S9_vs_k141_22454\|\|full | Low-quality |
| S9_vs_k141_7379\|\|full | Low-quality |
| S9_vs_k141_41467\|\|full | Not-determined |
| S9_vs_k141_14427\|\|full | Low-quality |
| S9_vs_k141_75129\|\|full | Low-quality |
| S9_vs_k141_90161\|\|full | Low-quality |
| S9_vs_k141_14429\|\|full | Not-determined |
| S9_vs_k141_37637\|\|full | Low-quality |
| S9_vs_k141_26217\|\|full | Low-quality |
| S9_vs_k141_45392\|\|full | Low-quality |
| S9_vs_k141_33886\|\|full | Not-determined |
| S9_vs_k141_3429\|\|full | Low-quality |
| S9_vs_k141_60016\|\|full | Low-quality |
| S9_vs_k141_22470\|\|full | Low-quality |
| S9_vs_k141_86457\|\|full | Not-determined |
| S9_vs_k141_90168\|\|full | Low-quality |
| S9_vs_k141_37654\|\|full | Low-quality |
| S9_vs_k141_14448\|\|full | Not-determined |
| S9_vs_k141_78680\|\|full | Low-quality |
| S9_vs_k141_14450\|\|full | Not-determined |
| S9_vs_k141_71291\|\|full | Low-quality |
| S9_vs_k141_75151\|\|full | Low-quality |
| S9_vs_k141_45412\|\|full | Low-quality |
| S9_vs_k141_90185\|\|full | Not-determined |
| S9_vs_k141_3443\|\|full | Low-quality |
| S9_vs_k141_45415\|\|full | Low-quality |
| S9_vs_k141_60041\|\|full | Low-quality |
| S9_vs_k141_11044\|\|full | Not-determined |
| S9_vs_k141_90190\|\|full | Not-determined |
| S9_vs_k141_60050\|\|full | Not-determined |
| S9_vs_k141_26264\|\|full | Low-quality |
| S9_vs_k141_75168\|\|full | Not-determined |
| S9_vs_k141_60056\|\|full | Low-quality |
| S9_vs_k141_3459\|\|full | Low-quality |
| S9_vs_k141_7433\|\|full | Low-quality |
| S9_vs_k141_75175\|\|full | Not-determined |
| S9_vs_k141_3467\|\|full | Not-determined |
| S9_vs_k141_67484\|\|full | Low-quality |
| S9_vs_k141_22518\|\|full | Not-determined |
| S9_vs_k141_78689\|\|full | Not-determined |
| S9_vs_k141_7440\|\|full | Not-determined |
| S9_vs_k141_26279\|\|full | Low-quality |
| S9_vs_k141_7444\|\|full | Low-quality |
| S9_vs_k141_48466\|\|full | Not-determined |
| S9_vs_k141_11078\|\|full | Not-determined |
| S9_vs_k141_75195\|\|full | Not-determined |
| S9_vs_k141_11082\|\|full | Low-quality |
| S9_vs_k141_26288\|\|full | Low-quality |
| S9_vs_k141_3484\|\|full | Low-quality |
| S9_vs_k141_90228\|\|full | Not-determined |
| S9_vs_k141_14508\|\|full | Low-quality |
| S9_vs_k141_86533\|\|full | Not-determined |
| S9_vs_k141_22543\|\|full | Low-quality |
| S9_vs_k141_45463\|\|full | Low-quality |
| S9_vs_k141_56556\|\|full | Not-determined |
| S9_vs_k141_45469\|\|full | Low-quality |
| S9_vs_k141_48489\|\|full | Low-quality |
| S9_vs_k141_48491\|\|full | Low-quality |
| S9_vs_k141_90254\|\|full | Low-quality |
| S9_vs_k141_37738\|\|full | Not-determined |
| S9_vs_k141_26324\|\|full | Not-determined |
| S9_vs_k141_41507\|\|full | Low-quality |
| S9_vs_k141_75216\|\|full | Low-quality |
| S9_vs_k141_41509\|\|full | Low-quality |
| S9_vs_k141_86547\|\|full | Low-quality |
| S9_vs_k141_67538\|\|full | Not-determined |
| S9_vs_k141_26336\|\|full | Low-quality |
| S9_vs_k141_82632\|\|full | Low-quality |
| S9_vs_k141_82634\|\|full | Low-quality |
| S9_vs_k141_51676\|\|full | Not-determined |
| S9_vs_k141_56594\|\|full | Low-quality |
| S9_vs_k141_14562\|\|full | Low-quality |
| S9_vs_k141_78779\|\|full | Not-determined |
| S9_vs_k141_37768\|\|full | Not-determined |
| S9_vs_k141_51694\|\|full | Not-determined |
| S9_vs_k141_48538\|\|full | Not-determined |
| S9_vs_k141_3511\|\|full | Not-determined |
| S9_vs_k141_51704\|\|full | Low-quality |
| S9_vs_k141_26343\|\|full | Low-quality |
| S9_vs_k141_3515\|\|full | Low-quality |
| S9_vs_k141_14588\|\|full | Not-determined |
| S9_vs_k141_78796\|\|full | Not-determined |
| S9_vs_k141_51706\|\|full | Low-quality |
| S9_vs_k141_22626\|\|full | Low-quality |
| S9_vs_k141_67561\|\|full | Not-determined |
| S9_vs_k141_75287\|\|full | Low-quality |
| S9_vs_k141_11129\|\|full | Low-quality |
| S9_vs_k141_86591\|\|full | Low-quality |
| S9_vs_k141_22649\|\|full | Not-determined |
| S9_vs_k141_90277\|\|full | Low-quality |
| S9_vs_k141_22654\|\|full | Low-quality |
| S9_vs_k141_78829\|\|full | Low-quality |
| S9_vs_k141_11138\|\|full | Not-determined |
| S9_vs_k141_37832\|\|full | Low-quality |
| S9_vs_k141_3557\|\|full | Not-determined |
| S9_vs_k141_86606\|\|full | Low-quality |
| S3_vs_k141_127399\|\|full | High-quality |
| S9_vs_k141_86608\|\|full | Low-quality |
| S9_vs_k141_51751\|\|full | Low-quality |
| S9_vs_k141_56667\|\|full | Not-determined |
| S9_vs_k141_51759\|\|full | Low-quality |
| S9_vs_k141_14662\|\|full | Not-determined |
| S9_vs_k141_26410\|\|full | Low-quality |
| S9_vs_k141_67628\|\|full | Low-quality |
| S9_vs_k141_90304\|\|full | Not-determined |
| S9_vs_k141_14677\|\|full | Low-quality |
| S9_vs_k141_7485\|\|full | Low-quality |
| S9_vs_k141_26428\|\|full | Not-determined |
| S9_vs_k141_3590\|\|full | Low-quality |
| S9_vs_k141_67642\|\|full | Low-quality |
| S9_vs_k141_75298\|\|full | Low-quality |
| S9_vs_k141_51796\|\|full | Not-determined |
| S9_vs_k141_11174\|\|full | Low-quality |
| S9_vs_k141_56685\|\|full | Low-quality |
| S9_vs_k141_14688\|\|full | Low-quality |
| S9_vs_k141_41658\|\|full | Low-quality |
| S9_vs_k141_78881\|\|full | Low-quality |
| S9_vs_k141_67662\|\|full | Not-determined |
| S9_vs_k141_86690\|\|full | Low-quality |
| S9_vs_k141_14698\|\|full | Not-determined |
| S9_vs_k141_63602\|\|full | Low-quality |
| S9_vs_k141_82696\|\|full | Low-quality |
| S9_vs_k141_90348\|\|full | Not-determined |
| S9_vs_k141_51847\|\|full | Not-determined |
| S9_vs_k141_63610\|\|full | Low-quality |
| S9_vs_k141_67686\|\|full | Not-determined |
| S9_vs_k141_82711\|\|full | Not-determined |
| S9_vs_k141_63617\|\|full | Low-quality |
| S9_vs_k141_63618\|\|full | Not-determined |
| S9_vs_k141_90376\|\|full | Low-quality |
| S9_vs_k141_75341\|\|full | Not-determined |
| S9_vs_k141_51862\|\|full | Low-quality |
| S9_vs_k141_48604\|\|full | Low-quality |
| S9_vs_k141_56737\|\|full | Not-determined |
| S9_vs_k141_26486\|\|full | Not-determined |
| S9_vs_k141_67696\|\|full | Not-determined |
| S9_vs_k141_60197\|\|full | Low-quality |
| S9_vs_k141_14734\|\|full | Not-determined |
| S9_vs_k141_51869\|\|full | Not-determined |
| S9_vs_k141_26506\|\|full | Low-quality |
| S9_vs_k141_56747\|\|full | Not-determined |
| S9_vs_k141_78947\|\|full | Low-quality |
| S9_vs_k141_63651\|\|full | Low-quality |
| S9_vs_k141_51880\|\|full | Low-quality |
| S9_vs_k141_14748\|\|full | Low-quality |
| S9_vs_k141_71578\|\|full | Low-quality |
| S9_vs_k141_78956\|\|full | Low-quality |
| S9_vs_k141_75356\|\|full | Low-quality |
| S9_vs_k141_56769\|\|full | Low-quality |
| S9_vs_k141_3657\|\|full | Not-determined |
| S9_vs_k141_60254\|\|full | Low-quality |
| S9_vs_k141_3664\|\|full | Low-quality |
| S9_vs_k141_60256\|\|full | Not-determined |
| S9_vs_k141_41678\|\|full | Low-quality |
| S9_vs_k141_48669\|\|full | Low-quality |
| S9_vs_k141_71612\|\|full | Not-determined |
| S9_vs_k141_48670\|\|full | Low-quality |
| S9_vs_k141_71615\|\|full | Low-quality |
| S13_vs_11246\|\|full | High-quality |
| S9_vs_k141_3671\|\|full | Low-quality |
| S9_vs_k141_56789\|\|full | Low-quality |
| S9_vs_k141_3678\|\|full | Not-determined |
| S9_vs_k141_60276\|\|full | Low-quality |
| S9_vs_k141_11271\|\|full | Low-quality |
| S9_vs_k141_56791\|\|full | Not-determined |
| S9_vs_k141_48692\|\|full | Not-determined |
| S9_vs_k141_78993\|\|full | Not-determined |
| S9_vs_k141_75386\|\|full | Not-determined |
| S9_vs_k141_79004\|\|full | Low-quality |
| S9_vs_k141_63732\|\|full | Not-determined |
| S9_vs_k141_51989\|\|full | Not-determined |
| S9_vs_k141_51997\|\|full | Low-quality |
| S9_vs_k141_75406\|\|full | Not-determined |
| S9_vs_k141_82829\|\|full | Not-determined |
| S9_vs_k141_75418\|\|full | Not-determined |
| S9_vs_k141_14806\|\|full | Low-quality |
| S9_vs_k141_60322\|\|full | Low-quality |
| S9_vs_k141_82847\|\|full | Not-determined |
| S9_vs_k141_14816\|\|full | Not-determined |
| S9_vs_k141_60325\|\|full | Low-quality |
| S9_vs_k141_71669\|\|full | Not-determined |
| S9_vs_k141_82860\|\|full | Low-quality |
| S9_vs_k141_60335\|\|full | Low-quality |
| S9_vs_k141_75432\|\|full | Low-quality |
| S9_vs_k141_11339\|\|full | Not-determined |
| S9_vs_k141_60336\|\|full | Low-quality |
| S9_vs_k141_63774\|\|full | Low-quality |
| S9_vs_k141_11342\|\|full | Low-quality |
| S9_vs_k141_63775\|\|full | Low-quality |
| S9_vs_k141_86844\|\|full | Not-determined |
| S9_vs_k141_71681\|\|full | Not-determined |
| S9_vs_k141_3731\|\|full | Low-quality |
| S9_vs_k141_71695\|\|full | Low-quality |
| S9_vs_k141_60353\|\|full | Low-quality |
| S9_vs_k141_82888\|\|full | Low-quality |
| S9_vs_k141_52069\|\|full | Not-determined |
| S9_vs_k141_71705\|\|full | Low-quality |
| S9_vs_k141_82896\|\|full | Low-quality |
| S9_vs_k141_63810\|\|full | Not-determined |
| S9_vs_k141_52075\|\|full | Not-determined |
| S9_vs_k141_60368\|\|full | Not-determined |
| S9_vs_k141_52089\|\|full | Not-determined |
| S9_vs_k141_86892\|\|full | Not-determined |
| S9_vs_k141_3740\|\|full | Low-quality |
| S9_vs_k141_71737\|\|full | Low-quality |
| S14_vs_k141_113008\|\|full | High-quality |
| S9_vs_k141_48818\|\|full | Not-determined |
| S9_vs_k141_90476\|\|full | Not-determined |
| S9_vs_k141_75498\|\|full | Low-quality |
| S9_vs_k141_71754\|\|full | Not-determined |
| S9_vs_k141_75502\|\|full | Low-quality |
| S9_vs_k141_71756\|\|full | Not-determined |
| S9_vs_k141_82948\|\|full | Not-determined |
| S9_vs_k141_86943\|\|full | Not-determined |
| S9_vs_k141_71761\|\|full | Not-determined |
| S9_vs_k141_67830\|\|full | Not-determined |
| S9_vs_k141_3778\|\|full | Low-quality |
| S9_vs_k141_82962\|\|full | Not-determined |
| S9_vs_k141_90504\|\|full | Low-quality |
| S9_vs_k141_67844\|\|full | Not-determined |
| S9_vs_k141_75545\|\|full | Low-quality |
| S9_vs_k141_71794\|\|full | Low-quality |
| S9_vs_k141_71797\|\|full | Low-quality |
| S9_vs_k141_63856\|\|full | Low-quality |
| S9_vs_k141_52161\|\|full | Low-quality |
| S9_vs_k141_48868\|\|full | Low-quality |
| S9_vs_k141_67853\|\|full | Low-quality |
| S9_vs_k141_63859\|\|full | Not-determined |
| S9_vs_k141_14940\|\|full | Low-quality |
| S9_vs_k141_48874\|\|full | Not-determined |
| S9_vs_k141_63863\|\|full | Low-quality |
| S9_vs_k141_48876\|\|full | Not-determined |
| S9_vs_k141_14951\|\|full | Low-quality |
| S9_vs_k141_90535\|\|full | Low-quality |
| S9_vs_k141_75591\|\|full | Not-determined |
| S9_vs_k141_71837\|\|full | Not-determined |
| S9_vs_k141_14963\|\|full | Low-quality |
| S9_vs_k141_63884\|\|full | Low-quality |
| S9_vs_k141_87000\|\|full | Low-quality |
| S9_vs_k141_87001\|\|full | Low-quality |
| S9_vs_k141_14967\|\|full | Not-determined |
| S9_vs_k141_67899\|\|full | Low-quality |
| S9_vs_k141_87013\|\|full | Not-determined |
| S9_vs_k141_67901\|\|full | Low-quality |
| S9_vs_k141_79217\|\|full | Not-determined |
| S9_vs_k141_63903\|\|full | Low-quality |
| S9_vs_k141_90565\|\|full | Low-quality |
| S9_vs_k141_71857\|\|full | Low-quality |
| S9_vs_k141_83057\|\|full | Low-quality |
| S9_vs_k141_52192\|\|full | Low-quality |
| S9_vs_k141_34056\|\|full | Low-quality |
| S9_vs_k141_83072\|\|full | Low-quality |
| S9_vs_k141_79263\|\|full | Low-quality |
| S9_vs_k141_79264\|\|full | Low-quality |
| S9_vs_k141_34058\|\|full | Low-quality |
| S9_vs_k141_71907\|\|full | Low-quality |
| S9_vs_k141_83099\|\|full | Low-quality |
| S9_vs_k141_87071\|\|full | Low-quality |
| S9_vs_k141_34092\|\|full | Not-determined |
| S3_extracted_k141_91904 | High-quality |
| S9_vs_k141_52222\|\|full | Not-determined |
| S9_vs_k141_15014\|\|full | Not-determined |
| S9_vs_k141_79283\|\|full | Not-determined |
| S9_vs_k141_79284\|\|full | Low-quality |
| S9_vs_k141_63973\|\|full | Low-quality |
| S9_vs_k141_71939\|\|full | Not-determined |
| S9_vs_k141_87096\|\|full | Not-determined |
| S9_vs_k141_60412\|\|full | Low-quality |
| S9_vs_k141_90638\|\|full | Low-quality |
| S9_vs_k141_60417\|\|full | Low-quality |
| S9_vs_k141_71955\|\|full | Low-quality |
| S9_vs_k141_68005\|\|full | Not-determined |
| S9_vs_k141_60422\|\|full | Low-quality |
| S9_vs_k141_87104\|\|full | Low-quality |
| S9_vs_k141_64004\|\|full | Not-determined |
| S9_vs_k141_60430\|\|full | Not-determined |
| S9_vs_k141_15047\|\|full | Low-quality |
| S9_vs_k141_90670\|\|full | Low-quality |
| S9_vs_k141_64009\|\|full | Low-quality |
| S9_vs_k141_90674\|\|full | Low-quality |
| S9_vs_k141_90675\|\|full | Low-quality |
| S9_vs_k141_87125\|\|full | Low-quality |
| S9_vs_k141_79328\|\|full | Low-quality |
| S9_vs_k141_90687\|\|full | Not-determined |
| S9_vs_k141_49006\|\|full | Not-determined |
| S9_vs_k141_52264\|\|full | Low-quality |
| S9_vs_k141_87143\|\|full | Not-determined |
| S9_vs_k141_79341\|\|full | Not-determined |
| S9_vs_k141_87147\|\|full | Not-determined |
| S9_vs_k141_79357\|\|full | Low-quality |
| S9_vs_k141_15086\|\|full | Not-determined |
| S9_vs_k141_75741\|\|full | Low-quality |
| S9_vs_k141_79367\|\|full | Low-quality |
| S9_vs_k141_15096\|\|full | Not-determined |
| S9_vs_k141_49034\|\|full | Low-quality |
| S9_vs_k141_79377\|\|full | Not-determined |
| S9_vs_k141_68053\|\|full | Low-quality |
| S9_vs_k141_60460\|\|full | Low-quality |
| S9_vs_k141_15112\|\|full | Low-quality |
| S9_vs_k141_79380\|\|full | Low-quality |
| S9_vs_k141_49060\|\|full | Low-quality |
| S9_vs_k141_49063\|\|full | Not-determined |
| S9_vs_k141_90716\|\|full | Low-quality |
| S9_vs_k141_64090\|\|full | Low-quality |
| S9_vs_k141_90721\|\|full | Low-quality |
| S9_vs_k141_64095\|\|full | Low-quality |
| S3_vs_k141_91904\|\|full | High-quality |
| S9_vs_k141_68095\|\|full | Not-determined |
| S9_vs_k141_79419\|\|full | Low-quality |
| S9_vs_k141_83137\|\|full | Not-determined |
| S9_vs_k141_52277\|\|full | Low-quality |
| S9_vs_k141_52280\|\|full | Not-determined |
| S9_vs_k141_49106\|\|full | Low-quality |
| S9_vs_k141_79456\|\|full | Low-quality |
| S9_vs_k141_64122\|\|full | Low-quality |
| S9_vs_k141_83148\|\|full | Low-quality |
| S9_vs_k141_83158\|\|full | Low-quality |
| S9_vs_k141_64132\|\|full | Not-determined |
| S9_vs_k141_79492\|\|full | Low-quality |
| S9_vs_k141_79493\|\|full | Low-quality |
| S9_vs_k141_68149\|\|full | Low-quality |
| S9_vs_k141_79496\|\|full | Low-quality |
| S9_vs_k141_64167\|\|full | Low-quality |
| S9_vs_k141_79520\|\|full | Low-quality |
| S9_vs_k141_52322\|\|full | Not-determined |
| S9_vs_k141_49113\|\|full | Low-quality |
| S9_vs_k141_83203\|\|full | Low-quality |
| S9_vs_k141_68182\|\|full | Low-quality |
| S9_vs_k141_52327\|\|full | Low-quality |
| S9_vs_k141_52332\|\|full | Low-quality |
| S9_vs_k141_52358\|\|full | Low-quality |
| S9_vs_k141_83258\|\|full | Low-quality |
| S9_vs_k141_49152\|\|full | Not-determined |
| S9_vs_k141_64213\|\|full | Low-quality |
| S9_vs_k141_60584\|\|full | Low-quality |
| S9_vs_k141_64223\|\|full | Low-quality |
| S9_vs_k141_49174\|\|full | Low-quality |
| S9_vs_k141_83290\|\|full | Not-determined |
| S9_vs_k141_64237\|\|full | Not-determined |
| S9_vs_k141_64247\|\|full | Not-determined |
| S9_vs_k141_52434\|\|full | Not-determined |
| S9_vs_k141_49214\|\|full | Low-quality |
| S9_vs_k141_52445\|\|full | Low-quality |
| S9_vs_k141_49234\|\|full | Low-quality |
| S9_vs_k141_49246\|\|full | Not-determined |
| S9_vs_k141_64310\|\|full | Low-quality |
| S3_extracted_k141_17748 | Complete |
| S9_vs_k141_90739\|\|full | Low-quality |
| S9_vs_k141_52543\|\|full | Not-determined |
| S9_vs_k141_52550\|\|full | Not-determined |
| S9_vs_k141_90761\|\|full | Low-quality |
| S9_vs_k141_52582\|\|full | Low-quality |
| S9_vs_k141_64345\|\|full | Not-determined |
| S9_vs_k141_90785\|\|full | Low-quality |
| S9_vs_k141_90790\|\|full | Low-quality |
| S9_vs_k141_90794\|\|full | Low-quality |
| S9_vs_k141_90813\|\|full | Low-quality |
| S9_vs_k141_64390\|\|full | Low-quality |
| S9_vs_k141_52629\|\|full | Not-determined |
| S9_vs_k141_52630\|\|full | Low-quality |
| S9_vs_k141_90844\|\|full | Low-quality |
| S9_vs_k141_52631\|\|full | Low-quality |
| S9_vs_k141_64399\|\|full | Low-quality |
| S9_vs_k141_52650\|\|full | Low-quality |
| S9_vs_k141_52704\|\|full | Low-quality |
| S9_vs_k141_52757\|\|full | Not-determined |
| S9_vs_k141_52881\|\|full | Low-quality |
| S9_vs_k141_52896\|\|full | Low-quality |
| S9_vs_k141_52922\|\|full | Low-quality |
| S9_vs_k141_52924\|\|full | Low-quality |
| S9_vs_k141_52949\|\|full | Low-quality |
| S9_vs_k141_52978\|\|full | Low-quality |
| S9_vs_k141_52995\|\|full | Low-quality |
| S9_vs_k141_53017\|\|full | Low-quality |
| S9_vs_k141_23649\|\|0_partial | Not-determined |
| S9_vs_k141_32352\|\|0_partial | Not-determined |
| S9_vs_k141_51168\|\|0_partial | Low-quality |
| S9_vs_k141_82414\|\|0_partial | Low-quality |
| S9_vs_k141_3795\|\|lt2gene | Low-quality |
| S9_vs_k141_49266\|\|lt2gene | Low-quality |
| S9_vs_k141_49267\|\|lt2gene | Not-determined |
| S9_vs_k141_37900\|\|lt2gene | Not-determined |
| S9_vs_k141_18\|\|lt2gene | Low-quality |
| S9_vs_k141_75791\|\|lt2gene | Low-quality |
| S9_vs_k141_30326\|\|lt2gene | Low-quality |
| S9_vs_k141_37907\|\|lt2gene | Low-quality |
| S9_vs_k141_18970\|\|lt2gene | Low-quality |
| S9_vs_k141_72010\|\|lt2gene | Low-quality |
| S9_vs_k141_64451\|\|lt2gene | Low-quality |
| S9_vs_k141_18980\|\|lt2gene | Low-quality |
| S9_vs_k141_75799\|\|lt2gene | Low-quality |
| S9_vs_k141_26583\|\|lt2gene | Low-quality |
| S9_vs_k141_56881\|\|lt2gene | Low-quality |
| S9_vs_k141_83408\|\|lt2gene | Low-quality |
| S9_vs_k141_34155\|\|lt2gene | Not-determined |
| S9_vs_k141_3825\|\|lt2gene | Low-quality |
| S9_vs_k141_72058\|\|lt2gene | Not-determined |
| S9_vs_k141_26586\|\|lt2gene | Low-quality |
| S9_vs_k141_30348\|\|lt2gene | Not-determined |
| S9_vs_k141_53102\|\|lt2gene | Low-quality |
| S9_vs_k141_30353\|\|lt2gene | Low-quality |
| S9_vs_k141_15229\|\|lt2gene | Low-quality |
| S9_vs_k141_15233\|\|lt2gene | Low-quality |
| S9_vs_k141_30359\|\|lt2gene | Low-quality |
| S9_vs_k141_34192\|\|lt2gene | Not-determined |
| S9_vs_k141_87231\|\|lt2gene | Low-quality |
| S9_vs_k141_87232\|\|lt2gene | Not-determined |
| S9_vs_k141_49334\|\|lt2gene | Low-quality |
| S9_vs_k141_87235\|\|lt2gene | Low-quality |
| S9_vs_k141_72084\|\|lt2gene | Low-quality |
| S9_vs_k141_79664\|\|lt2gene | Low-quality |
| S9_vs_k141_75861\|\|lt2gene | Not-determined |
| S9_vs_k141_22826\|\|lt2gene | Low-quality |
| S9_vs_k141_72096\|\|lt2gene | Low-quality |
| S9_vs_k141_87250\|\|lt2gene | Low-quality |
| S9_vs_k141_30388\|\|lt2gene | Not-determined |
| S9_vs_k141_79683\|\|lt2gene | Low-quality |
| S9_vs_k141_30399\|\|lt2gene | Not-determined |
| S9_vs_k141_79686\|\|lt2gene | Low-quality |
| S9_vs_k141_53143\|\|lt2gene | Low-quality |
| S9_vs_k141_22877\|\|lt2gene | Not-determined |
| S9_vs_k141_37988\|\|lt2gene | Not-determined |
| S9_vs_k141_34237\|\|lt2gene | Not-determined |
| S9_vs_k141_53186\|\|lt2gene | Low-quality |
| S9_vs_k141_53197\|\|lt2gene | Low-quality |
| S9_vs_k141_87311\|\|lt2gene | Not-determined |
| S9_vs_k141_30475\|\|lt2gene | Not-determined |
| S9_vs_k141_60775\|\|lt2gene | Low-quality |
| S9_vs_k141_68274\|\|lt2gene | Low-quality |
| S9_vs_k141_34278\|\|lt2gene | Not-determined |
| S9_vs_k141_158\|\|lt2gene | Low-quality |
| S9_vs_k141_53236\|\|lt2gene | Not-determined |
| S9_vs_k141_19092\|\|lt2gene | Low-quality |
| S9_vs_k141_34283\|\|lt2gene | Not-determined |
| S9_vs_k141_60784\|\|lt2gene | Low-quality |
| S9_vs_k141_11537\|\|lt2gene | Not-determined |
| S9_vs_k141_68286\|\|lt2gene | Low-quality |
| S9_vs_k141_34290\|\|lt2gene | Not-determined |
| S9_vs_k141_83555\|\|lt2gene | Low-quality |
| S9_vs_k141_83558\|\|lt2gene | Low-quality |
| S9_vs_k141_68300\|\|lt2gene | Low-quality |
| S9_vs_k141_30504\|\|lt2gene | Not-determined |
| S9_vs_k141_60806\|\|lt2gene | Not-determined |
| S9_vs_k141_3965\|\|lt2gene | Low-quality |
| S9_vs_k141_41864\|\|lt2gene | Low-quality |
| S9_vs_k141_79729\|\|lt2gene | Not-determined |
| S9_vs_k141_79735\|\|lt2gene | Low-quality |
| S9_vs_k141_38083\|\|lt2gene | Not-determined |
| S9_vs_k141_38095\|\|lt2gene | Not-determined |
| S9_vs_k141_87359\|\|lt2gene | Not-determined |
| S9_vs_k141_30553\|\|lt2gene | Low-quality |
| S9_vs_k141_60849\|\|lt2gene | Not-determined |
| S9_vs_k141_4004\|\|lt2gene | Low-quality |
| S9_vs_k141_30561\|\|lt2gene | Low-quality |

| | |
|---|---|
| S9_vs_k141_30565\|\|lt2gene | Low-quality |
| S9_vs_k141_45628\|\|lt2gene | Not-determined |
| S9_vs_k141_30578\|\|lt2gene | Not-determined |
| S9_vs_k141_56992\|\|lt2gene | Low-quality |
| S9_vs_k141_15375\|\|lt2gene | Low-quality |
| S9_vs_k141_68362\|\|lt2gene | Low-quality |
| S9_vs_k141_45652\|\|lt2gene | Low-quality |
| S9_vs_k141_34410\|\|lt2gene | Low-quality |
| S9_vs_k141_34411\|\|lt2gene | Low-quality |
| S9_vs_k141_19187\|\|lt2gene | Low-quality |
| S9_vs_k141_269\|\|lt2gene | Low-quality |
| S9_vs_k141_15394\|\|lt2gene | Low-quality |
| S9_vs_k141_34426\|\|lt2gene | Low-quality |
| S9_vs_k141_4051\|\|lt2gene | Not-determined |
| S9_vs_k141_34432\|\|lt2gene | Low-quality |
| S9_vs_k141_34442\|\|lt2gene | Not-determined |
| S9_vs_k141_68397\|\|lt2gene | Low-quality |
| S9_vs_k141_30631\|\|lt2gene | Low-quality |
| S9_vs_k141_79829\|\|lt2gene | Low-quality |
| S9_vs_k141_79832\|\|lt2gene | Not-determined |
| S9_vs_k141_22994\|\|lt2gene | Low-quality |
| S9_vs_k141_45681\|\|lt2gene | Low-quality |
| S9_vs_k141_19246\|\|lt2gene | Low-quality |
| S9_vs_k141_72193\|\|lt2gene | Low-quality |
| S9_vs_k141_7755\|\|lt2gene | Low-quality |
| S9_vs_k141_7757\|\|lt2gene | Low-quality |
| S9_vs_k141_19253\|\|lt2gene | Low-quality |
| S9_vs_k141_7758\|\|lt2gene | Low-quality |
| S9_vs_k141_87446\|\|lt2gene | Low-quality |
| S9_vs_k141_38201\|\|lt2gene | Low-quality |
| S9_vs_k141_11699\|\|lt2gene | Low-quality |
| S9_vs_k141_64669\|\|lt2gene | Low-quality |
| S9_vs_k141_23033\|\|lt2gene | Low-quality |
| S9_vs_k141_30669\|\|lt2gene | Low-quality |
| S9_vs_k141_34502\|\|lt2gene | Low-quality |
| S9_vs_k141_79862\|\|lt2gene | Low-quality |
| S9_vs_k141_79886\|\|lt2gene | Low-quality |
| S9_vs_k141_79888\|\|lt2gene | Low-quality |
| S9_vs_k141_26773\|\|lt2gene | Low-quality |
| S9_vs_k141_41983\|\|lt2gene | Low-quality |
| S9_vs_k141_7808\|\|lt2gene | Low-quality |
| S9_vs_k141_57067\|\|lt2gene | Low-quality |
| S9_vs_k141_41997\|\|lt2gene | Low-quality |
| S9_vs_k141_87514\|\|lt2gene | Low-quality |
| S9_vs_k141_4118\|\|lt2gene | Low-quality |
| S9_vs_k141_38265\|\|lt2gene | Low-quality |
| S9_vs_k141_64735\|\|lt2gene | Low-quality |
| S9_vs_k141_4124\|\|lt2gene | Low-quality |
| S9_vs_k141_4125\|\|lt2gene | Not-determined |
| S9_vs_k141_49486\|\|lt2gene | Low-quality |
| S9_vs_k141_49488\|\|lt2gene | Low-quality |
| S9_vs_k141_64747\|\|lt2gene | Low-quality |
| S9_vs_k141_23106\|\|lt2gene | Low-quality |
| S9_vs_k141_15512\|\|lt2gene | Low-quality |
| S9_vs_k141_30715\|\|lt2gene | Low-quality |
| S9_vs_k141_83671\|\|lt2gene | Low-quality |
| S9_vs_k141_38296\|\|lt2gene | Low-quality |
| S9_vs_k141_7844\|\|lt2gene | Not-determined |
| S9_vs_k141_30733\|\|lt2gene | Low-quality |
| S9_vs_k141_11713\|\|lt2gene | Low-quality |
| S9_vs_k141_30737\|\|lt2gene | Low-quality |
| S9_vs_k141_23134\|\|lt2gene | Low-quality |
| S9_vs_k141_30750\|\|lt2gene | Low-quality |
| S9_vs_k141_30767\|\|lt2gene | Not-determined |
| S9_vs_k141_23150\|\|lt2gene | Low-quality |
| S9_vs_k141_30771\|\|lt2gene | Low-quality |
| S9_vs_k141_45747\|\|lt2gene | Low-quality |
| S9_vs_k141_30773\|\|lt2gene | Low-quality |
| S9_vs_k141_60967\|\|lt2gene | Low-quality |
| S9_vs_k141_49568\|\|lt2gene | Low-quality |
| S9_vs_k141_19308\|\|lt2gene | Low-quality |
| S9_vs_k141_42103\|\|lt2gene | Low-quality |
| S9_vs_k141_83742\|\|lt2gene | Low-quality |
| S9_vs_k141_49576\|\|lt2gene | Not-determined |
| S9_vs_k141_19325\|\|lt2gene | Low-quality |
| S9_vs_k141_68576\|\|lt2gene | Low-quality |
| S9_vs_k141_87605\|\|lt2gene | Not-determined |
| S9_vs_k141_49587\|\|lt2gene | Low-quality |
| S9_vs_k141_60995\|\|lt2gene | Low-quality |
| S9_vs_k141_4227\|\|lt2gene | Low-quality |
| S9_vs_k141_79949\|\|lt2gene | Low-quality |
| S9_vs_k141_49607\|\|lt2gene | Low-quality |
| S9_vs_k141_26881\|\|lt2gene | Low-quality |
| S9_vs_k141_11803\|\|lt2gene | Low-quality |
| S9_vs_k141_76033\|\|lt2gene | Not-determined |
| S9_vs_k141_49623\|\|lt2gene | Not-determined |
| S9_vs_k141_42140\|\|lt2gene | Low-quality |
| S9_vs_k141_53444\|\|lt2gene | Not-determined |
| S9_vs_k141_26913\|\|lt2gene | Low-quality |
| S9_vs_k141_57209\|\|lt2gene | Low-quality |
| S9_vs_k141_11835\|\|lt2gene | Low-quality |
| S9_vs_k141_4268\|\|lt2gene | Low-quality |
| S9_vs_k141_57226\|\|lt2gene | Low-quality |
| S9_vs_k141_26926\|\|lt2gene | Low-quality |
| S9_vs_k141_49650\|\|lt2gene | Low-quality |
| S9_vs_k141_23202\|\|lt2gene | Low-quality |
| S9_vs_k141_72411\|\|lt2gene | Not-determined |
| S9_vs_k141_4282\|\|lt2gene | Low-quality |
| S9_vs_k141_87640\|\|lt2gene | Not-determined |
| S9_vs_k141_57235\|\|lt2gene | Low-quality |
| S9_vs_k141_26953\|\|lt2gene | Not-determined |
| S9_vs_k141_76100\|\|lt2gene | Low-quality |
| S9_vs_k141_87652\|\|lt2gene | Low-quality |
| S9_vs_k141_49676\|\|lt2gene | Low-quality |
| S9_vs_k141_42195\|\|lt2gene | Low-quality |
| S9_vs_k141_19393\|\|lt2gene | Low-quality |
| S9_vs_k141_61079\|\|lt2gene | Low-quality |
| S9_vs_k141_87672\|\|lt2gene | Low-quality |
| S9_vs_k141_38429\|\|lt2gene | Low-quality |
| S9_vs_k141_19394\|\|lt2gene | Low-quality |
| S9_vs_k141_53505\|\|lt2gene | Low-quality |
| S9_vs_k141_23242\|\|lt2gene | Low-quality |
| S9_vs_k141_61105\|\|lt2gene | Low-quality |
| S9_vs_k141_7972\|\|lt2gene | Low-quality |
| S9_vs_k141_580\|\|lt2gene | Low-quality |
| S9_vs_k141_80047\|\|lt2gene | Not-determined |
| S9_vs_k141_27022\|\|lt2gene | Not-determined |
| S9_vs_k141_61123\|\|lt2gene | Low-quality |
| S9_vs_k141_61124\|\|lt2gene | Not-determined |
| S9_vs_k141_27028\|\|lt2gene | Not-determined |
| S9_vs_k141_27034\|\|lt2gene | Low-quality |
| S9_vs_k141_11951\|\|lt2gene | Not-determined |
| S9_vs_k141_42243\|\|lt2gene | Low-quality |
| S9_vs_k141_76169\|\|lt2gene | Low-quality |
| S9_vs_k141_19452\|\|lt2gene | Low-quality |
| S9_vs_k141_76188\|\|lt2gene | Not-determined |
| S9_vs_k141_83912\|\|lt2gene | Not-determined |
| S9_vs_k141_8037\|\|lt2gene | Not-determined |
| S9_vs_k141_15759\|\|lt2gene | Low-quality |
| S9_vs_k141_87761\|\|lt2gene | Not-determined |
| S9_vs_k141_15763\|\|lt2gene | Low-quality |
| S9_vs_k141_8040\|\|lt2gene | Low-quality |
| S9_vs_k141_27101\|\|lt2gene | Low-quality |
| S9_vs_k141_45820\|\|lt2gene | Low-quality |
| S9_vs_k141_15767\|\|lt2gene | Not-determined |
| S9_vs_k141_53578\|\|lt2gene | Low-quality |
| S9_vs_k141_42300\|\|lt2gene | Low-quality |
| S9_vs_k141_34637\|\|lt2gene | Low-quality |
| S9_vs_k141_38532\|\|lt2gene | Not-determined |
| S9_vs_k141_53593\|\|lt2gene | Low-quality |
| S9_vs_k141_83938\|\|lt2gene | Low-quality |
| S9_vs_k141_53599\|\|lt2gene | Low-quality |
| S9_vs_k141_30962\|\|lt2gene | Low-quality |
| S9_vs_k141_80099\|\|lt2gene | Low-quality |
| S9_vs_k141_8066\|\|lt2gene | Low-quality |
| S9_vs_k141_76215\|\|lt2gene | Low-quality |
| S9_vs_k141_19503\|\|lt2gene | Low-quality |
| S9_vs_k141_80112\|\|lt2gene | Not-determined |
| S9_vs_k141_72548\|\|lt2gene | Low-quality |
| S9_vs_k141_38549\|\|lt2gene | Low-quality |
| S9_vs_k141_68625\|\|lt2gene | Low-quality |
| S9_vs_k141_80128\|\|lt2gene | Low-quality |
| S9_vs_k141_80155\|\|lt2gene | Low-quality |
| S9_vs_k141_64858\|\|lt2gene | Not-determined |
| S9_vs_k141_15845\|\|lt2gene | Low-quality |
| S9_vs_k141_72600\|\|lt2gene | Not-determined |
| S9_vs_k141_61278\|\|lt2gene | Low-quality |
| S9_vs_k141_64887\|\|lt2gene | Low-quality |
| S9_vs_k141_45932\|\|lt2gene | Not-determined |
| S9_vs_k141_34713\|\|lt2gene | Low-quality |
| S9_vs_k141_23299\|\|lt2gene | Low-quality |
| S9_vs_k141_53686\|\|lt2gene | Low-quality |
| S9_vs_k141_8122\|\|lt2gene | Low-quality |
| S9_vs_k141_8124\|\|lt2gene | Low-quality |
| S9_vs_k141_72632\|\|lt2gene | Low-quality |
| S9_vs_k141_72633\|\|lt2gene | Low-quality |
| S9_vs_k141_23311\|\|lt2gene | Not-determined |
| S9_vs_k141_64912\|\|lt2gene | Not-determined |
| S9_vs_k141_42400\|\|lt2gene | Low-quality |
| S9_vs_k141_19606\|\|lt2gene | Low-quality |
| S9_vs_k141_72668\|\|lt2gene | Low-quality |
| S9_vs_k141_53732\|\|lt2gene | Low-quality |
| S9_vs_k141_64925\|\|lt2gene | Low-quality |
| S9_vs_k141_19621\|\|lt2gene | Low-quality |
| S9_vs_k141_15869\|\|lt2gene | Not-determined |
| S9_vs_k141_72672\|\|lt2gene | Low-quality |
| S9_vs_k141_34721\|\|lt2gene | Low-quality |
| S9_vs_k141_12095\|\|lt2gene | Low-quality |
| S9_vs_k141_23353\|\|lt2gene | Low-quality |
| S9_vs_k141_80232\|\|lt2gene | Not-determined |
| S9_vs_k141_15900\|\|lt2gene | Not-determined |
| S9_vs_k141_42450\|\|lt2gene | Not-determined |
| S9_vs_k141_76296\|\|lt2gene | Low-quality |
| S9_vs_k141_64943\|\|lt2gene | Not-determined |
| S9_vs_k141_23369\|\|lt2gene | Low-quality |
| S9_vs_k141_46011\|\|lt2gene | Low-quality |
| S9_vs_k141_64961\|\|lt2gene | Low-quality |
| S9_vs_k141_12130\|\|lt2gene | Low-quality |
| S9_vs_k141_15934\|\|lt2gene | Low-quality |
| S9_vs_k141_64966\|\|lt2gene | Low-quality |
| S9_vs_k141_53784\|\|lt2gene | Low-quality |
| S9_vs_k141_12147\|\|lt2gene | Low-quality |
| S9_vs_k141_57301\|\|lt2gene | Not-determined |
| S9_vs_k141_27265\|\|lt2gene | Low-quality |
| S9_vs_k141_68754\|\|lt2gene | Low-quality |
| S9_vs_k141_23381\|\|lt2gene | Low-quality |
| S9_vs_k141_42497\|\|lt2gene | Low-quality |
| S9_vs_k141_57333\|\|lt2gene | Low-quality |
| S9_vs_k141_65005\|\|lt2gene | Low-quality |
| S9_vs_k141_76323\|\|lt2gene | Low-quality |
| S9_vs_k141_53805\|\|lt2gene | Not-determined |
| S9_vs_k141_23410\|\|lt2gene | Low-quality |
| S9_vs_k141_23415\|\|lt2gene | Low-quality |
| S9_vs_k141_61351\|\|lt2gene | Low-quality |
| S9_vs_k141_15979\|\|lt2gene | Low-quality |
| S9_vs_k141_65032\|\|lt2gene | Low-quality |
| S9_vs_k141_12208\|\|lt2gene | Low-quality |
| S9_vs_k141_72710\|\|lt2gene | Not-determined |
| S9_vs_k141_23426\|\|lt2gene | Low-quality |
| S9_vs_k141_72713\|\|lt2gene | Low-quality |
| S9_vs_k141_34792\|\|lt2gene | Low-quality |
| S9_vs_k141_23434\|\|lt2gene | Low-quality |
| S9_vs_k141_31126\|\|lt2gene | Low-quality |
| S9_vs_k141_80341\|\|lt2gene | Low-quality |
| S9_vs_k141_80343\|\|lt2gene | Low-quality |
| S9_vs_k141_80344\|\|lt2gene | Low-quality |
| S9_vs_k141_16014\|\|lt2gene | Low-quality |
| S9_vs_k141_84055\|\|lt2gene | Low-quality |
| S9_vs_k141_84058\|\|lt2gene | Low-quality |
| S9_vs_k141_31158\|\|lt2gene | Low-quality |
| S9_vs_k141_80360\|\|lt2gene | Low-quality |
| S9_vs_k141_61407\|\|lt2gene | Low-quality |
| S9_vs_k141_12258\|\|lt2gene | Low-quality |
| S9_vs_k141_88017\|\|lt2gene | Low-quality |
| S9_vs_k141_76411\|\|lt2gene | Low-quality |
| S9_vs_k141_19672\|\|lt2gene | Low-quality |
| S9_vs_k141_19678\|\|lt2gene | Low-quality |
| S9_vs_k141_16066\|\|lt2gene | Low-quality |
| S9_vs_k141_72754\|\|lt2gene | Low-quality |
| S9_vs_k141_57443\|\|lt2gene | Not-determined |
| S9_vs_k141_49835\|\|lt2gene | Low-quality |
| S9_vs_k141_16090\|\|lt2gene | Low-quality |
| S9_vs_k141_23524\|\|lt2gene | Low-quality |
| S9_vs_k141_53928\|\|lt2gene | Low-quality |
| S9_vs_k141_72774\|\|lt2gene | Low-quality |
| S9_vs_k141_61473\|\|lt2gene | Not-determined |
| S9_vs_k141_810\|\|lt2gene | Low-quality |
| S9_vs_k141_76435\|\|lt2gene | Low-quality |
| S9_vs_k141_61494\|\|lt2gene | Low-quality |
| S9_vs_k141_31238\|\|lt2gene | Low-quality |
| S9_vs_k141_42606\|\|lt2gene | Low-quality |
| S9_vs_k141_19700\|\|lt2gene | Low-quality |
| S3_vs_k141_17748\|\|full | High-quality |
| S9_vs_k141_16114\|\|lt2gene | Low-quality |
| S9_vs_k141_53981\|\|lt2gene | Low-quality |
| S9_vs_k141_72817\|\|lt2gene | Low-quality |
| S9_vs_k141_72823\|\|lt2gene | Low-quality |
| S9_vs_k141_12302\|\|lt2gene | Low-quality |
| S9_vs_k141_54000\|\|lt2gene | Low-quality |
| S9_vs_k141_31261\|\|lt2gene | Not-determined |
| S9_vs_k141_72840\|\|lt2gene | Low-quality |
| S9_vs_k141_65143\|\|lt2gene | Low-quality |
| S9_vs_k141_38932\|\|lt2gene | Not-determined |
| S9_vs_k141_72856\|\|lt2gene | Low-quality |
| S9_vs_k141_34908\|\|lt2gene | Low-quality |
| S9_vs_k141_46275\|\|lt2gene | Low-quality |
| S9_vs_k141_34919\|\|lt2gene | Low-quality |
| S9_vs_k141_80461\|\|lt2gene | Low-quality |
| S9_vs_k141_23575\|\|lt2gene | Low-quality |
| S9_vs_k141_31298\|\|lt2gene | Low-quality |
| S9_vs_k141_61561\|\|lt2gene | Low-quality |
| S9_vs_k141_890\|\|lt2gene | Low-quality |
| S9_vs_k141_893\|\|lt2gene | Low-quality |
| S9_vs_k141_72891\|\|lt2gene | Low-quality |
| S9_vs_k141_31325\|\|lt2gene | Low-quality |
| S9_vs_k141_4594\|\|lt2gene | Not-determined |
| S9_vs_k141_16183\|\|lt2gene | Low-quality |
| S9_vs_k141_16185\|\|lt2gene | Low-quality |
| S9_vs_k141_8150\|\|lt2gene | Low-quality |
| S9_vs_k141_57525\|\|lt2gene | Low-quality |
| S9_vs_k141_54060\|\|lt2gene | Low-quality |
| S9_vs_k141_34993\|\|lt2gene | Low-quality |
| S9_vs_k141_46342\|\|lt2gene | Low-quality |
| S9_vs_k141_61594\|\|lt2gene | Low-quality |
| S9_vs_k141_46346\|\|lt2gene | Low-quality |
| S9_vs_k141_4629\|\|lt2gene | Not-determined |
| S9_vs_k141_924\|\|lt2gene | Low-quality |
| S9_vs_k141_35002\|\|lt2gene | Low-quality |
| S9_vs_k141_929\|\|lt2gene | Not-determined |
| S9_vs_k141_88190\|\|lt2gene | Low-quality |
| S9_vs_k141_57559\|\|lt2gene | Low-quality |
| S9_vs_k141_88211\|\|lt2gene | Low-quality |
| S9_vs_k141_936\|\|lt2gene | Not-determined |
| S9_vs_k141_54107\|\|lt2gene | Low-quality |
| S9_vs_k141_35055\|\|lt2gene | Low-quality |
| S9_vs_k141_50045\|\|lt2gene | Low-quality |
| S9_vs_k141_960\|\|lt2gene | Low-quality |
| S9_vs_k141_16278\|\|lt2gene | Low-quality |
| S9_vs_k141_50057\|\|lt2gene | Low-quality |
| S9_vs_k141_68922\|\|lt2gene | Low-quality |
| S9_vs_k141_50061\|\|lt2gene | Low-quality |
| S9_vs_k141_27450\|\|lt2gene | Not-determined |
| S9_vs_k141_31418\|\|lt2gene | Low-quality |
| S9_vs_k141_988\|\|lt2gene | Low-quality |
| S9_vs_k141_12448\|\|lt2gene | Low-quality |
| S9_vs_k141_999\|\|lt2gene | Low-quality |
| S9_vs_k141_46407\|\|lt2gene | Low-quality |
| S9_vs_k141_73002\|\|lt2gene | Low-quality |
| S9_vs_k141_39099\|\|lt2gene | Not-determined |
| S9_vs_k141_57644\|\|lt2gene | Low-quality |
| S9_vs_k141_39103\|\|lt2gene | Not-determined |
| S9_vs_k141_1009\|\|lt2gene | Low-quality |
| S9_vs_k141_8266\|\|lt2gene | Low-quality |
| S9_vs_k141_42792\|\|lt2gene | Low-quality |
| S9_vs_k141_27488\|\|lt2gene | Low-quality |
| S9_vs_k141_46436\|\|lt2gene | Low-quality |
| S9_vs_k141_39126\|\|lt2gene | Low-quality |
| S9_vs_k141_35127\|\|lt2gene | Low-quality |
| S9_vs_k141_27509\|\|lt2gene | Low-quality |
| S9_vs_k141_35128\|\|lt2gene | Not-determined |
| S9_vs_k141_1040\|\|lt2gene | Low-quality |
| S9_vs_k141_27521\|\|lt2gene | Low-quality |
| S9_vs_k141_4742\|\|lt2gene | Low-quality |
| S9_vs_k141_1047\|\|lt2gene | Low-quality |
| S9_vs_k141_65335\|\|lt2gene | Low-quality |
| S9_vs_k141_1054\|\|lt2gene | Not-determined |
| S9_vs_k141_73012\|\|lt2gene | Low-quality |
| S9_vs_k141_19899\|\|lt2gene | Low-quality |
| S9_vs_k141_46484\|\|lt2gene | Low-quality |
| S9_vs_k141_27567\|\|lt2gene | Low-quality |
| S9_vs_k141_19909\|\|lt2gene | Not-determined |
| S9_vs_k141_46489\|\|lt2gene | Not-determined |
| S9_vs_k141_19915\|\|lt2gene | Not-determined |
| S9_vs_k141_57704\|\|lt2gene | Low-quality |
| S9_vs_k141_4798\|\|lt2gene | Low-quality |
| S9_vs_k141_19940\|\|lt2gene | Low-quality |
| S9_vs_k141_65380\|\|lt2gene | Not-determined |
| S9_vs_k141_12571\|\|lt2gene | Low-quality |
| S9_vs_k141_57710\|\|lt2gene | Not-determined |
| S9_vs_k141_27610\|\|lt2gene | Not-determined |
| S9_vs_k141_1097\|\|lt2gene | Low-quality |
| S9_vs_k141_46518\|\|lt2gene | Low-quality |
| S9_vs_k141_27642\|\|lt2gene | Not-determined |
| S9_vs_k141_16505\|\|lt2gene | Low-quality |
| S9_vs_k141_50226\|\|lt2gene | Low-quality |
| S9_vs_k141_39218\|\|lt2gene | Low-quality |
| S9_vs_k141_1143\|\|lt2gene | Low-quality |
| S9_vs_k141_8389\|\|lt2gene | Low-quality |
| S9_vs_k141_88356\|\|lt2gene | Not-determined |
| S9_vs_k141_42901\|\|lt2gene | Low-quality |
| S9_vs_k141_16515\|\|lt2gene | Not-determined |
| S9_vs_k141_19999\|\|lt2gene | Low-quality |
| S9_vs_k141_54316\|\|lt2gene | Low-quality |
| S9_vs_k141_12654\|\|lt2gene | Low-quality |
| S9_vs_k141_73138\|\|lt2gene | Low-quality |
| S9_vs_k141_35273\|\|lt2gene | Low-quality |
| S9_vs_k141_12658\|\|lt2gene | Low-quality |
| S9_vs_k141_54351\|\|lt2gene | Not-determined |
| S9_vs_k141_73181\|\|lt2gene | Low-quality |
| S9_vs_k141_4889\|\|lt2gene | Low-quality |
| S9_vs_k141_80538\|\|lt2gene | Low-quality |
| S9_vs_k141_16564\|\|lt2gene | Low-quality |
| S9_vs_k141_39284\|\|lt2gene | Low-quality |
| S9_vs_k141_80542\|\|lt2gene | Low-quality |
| S9_vs_k141_73194\|\|lt2gene | Low-quality |
| S9_vs_k141_27717\|\|lt2gene | Low-quality |
| S9_vs_k141_20074\|\|lt2gene | Low-quality |
| S9_vs_k141_42989\|\|lt2gene | Low-quality |
| S9_vs_k141_12701\|\|lt2gene | Low-quality |
| S9_vs_k141_8467\|\|lt2gene | Low-quality |
| S9_vs_k141_20086\|\|lt2gene | Low-quality |
| S9_vs_k141_69112\|\|lt2gene | Not-determined |
| S9_vs_k141_27760\|\|lt2gene | Low-quality |
| S9_vs_k141_76609\|\|lt2gene | Low-quality |
| S9_vs_k141_27773\|\|lt2gene | Not-determined |
| S9_vs_k141_1248\|\|lt2gene | Not-determined |
| S9_vs_k141_8483\|\|lt2gene | Low-quality |
| S9_vs_k141_69142\|\|lt2gene | Low-quality |
| S9_vs_k141_23759\|\|lt2gene | Low-quality |
| S9_vs_k141_39342\|\|lt2gene | Not-determined |
| S9_vs_k141_27796\|\|lt2gene | Not-determined |
| S9_vs_k141_73278\|\|lt2gene | Low-quality |
| S9_vs_k141_88486\|\|lt2gene | Low-quality |
| S9_vs_k141_39370\|\|lt2gene | Low-quality |
| S9_vs_k141_35398\|\|lt2gene | Low-quality |
| S9_vs_k141_35399\|\|lt2gene | Low-quality |
| S9_vs_k141_35411\|\|lt2gene | Low-quality |
| S9_vs_k141_8515\|\|lt2gene | Low-quality |
| S9_vs_k141_43068\|\|lt2gene | Low-quality |
| S9_vs_k141_69175\|\|lt2gene | Low-quality |
| S9_vs_k141_23806\|\|lt2gene | Low-quality |
| S9_vs_k141_61827\|\|lt2gene | Low-quality |
| S9_vs_k141_39419\|\|lt2gene | Low-quality |
| S9_vs_k141_88536\|\|lt2gene | Low-quality |
| S9_vs_k141_39425\|\|lt2gene | Low-quality |
| S9_vs_k141_43109\|\|lt2gene | Low-quality |
| S9_vs_k141_61857\|\|lt2gene | Not-determined |
| S9_vs_k141_35499\|\|lt2gene | Low-quality |
| S9_vs_k141_69227\|\|lt2gene | Low-quality |
| S9_vs_k141_69230\|\|lt2gene | Low-quality |
| S9_vs_k141_8588\|\|lt2gene | Low-quality |
| S9_vs_k141_8596\|\|lt2gene | Low-quality |
| S9_vs_k141_35514\|\|lt2gene | Not-determined |
| S9_vs_k141_57917\|\|lt2gene | Low-quality |
| S9_vs_k141_8600\|\|lt2gene | Low-quality |
| S9_vs_k141_73328\|\|lt2gene | Low-quality |
| S9_vs_k141_65576\|\|lt2gene | Low-quality |
| S9_vs_k141_84381\|\|lt2gene | Low-quality |
| S9_vs_k141_69269\|\|lt2gene | Not-determined |
| S9_vs_k141_31842\|\|lt2gene | Low-quality |
| S9_vs_k141_23913\|\|lt2gene | Not-determined |
| S9_vs_k141_65611\|\|lt2gene | Low-quality |
| S9_vs_k141_39468\|\|lt2gene | Low-quality |
| S9_vs_k141_31862\|\|lt2gene | Low-quality |
| S9_vs_k141_50402\|\|lt2gene | Low-quality |
| S9_vs_k141_80737\|\|lt2gene | Low-quality |
| S9_vs_k141_39481\|\|lt2gene | Low-quality |
| S9_vs_k141_39487\|\|lt2gene | Low-quality |
| S9_vs_k141_27925\|\|lt2gene | Not-determined |
| S9_vs_k141_57981\|\|lt2gene | Low-quality |
| S9_vs_k141_1419\|\|lt2gene | Not-determined |
| S9_vs_k141_20289\|\|lt2gene | Low-quality |
| S9_vs_k141_1425\|\|lt2gene | Not-determined |
| S9_vs_k141_69297\|\|lt2gene | Low-quality |
| S9_vs_k141_43185\|\|lt2gene | Low-quality |
| S9_vs_k141_1431\|\|lt2gene | Low-quality |
| S9_vs_k141_61961\|\|lt2gene | Low-quality |
| S9_vs_k141_65635\|\|lt2gene | Not-determined |
| S9_vs_k141_16657\|\|lt2gene | Low-quality |
| S9_vs_k141_50419\|\|lt2gene | Low-quality |
| S9_vs_k141_84441\|\|lt2gene | Low-quality |
| S9_vs_k141_35615\|\|lt2gene | Low-quality |
| S9_vs_k141_61981\|\|lt2gene | Not-determined |
| S9_vs_k141_46713\|\|lt2gene | Low-quality |
| S9_vs_k141_46715\|\|lt2gene | Low-quality |
| S9_vs_k141_16680\|\|lt2gene | Not-determined |
| S9_vs_k141_16688\|\|lt2gene | Low-quality |
| S9_vs_k141_12769\|\|lt2gene | Low-quality |
| S9_vs_k141_35650\|\|lt2gene | Not-determined |
| S9_vs_k141_80824\|\|lt2gene | Low-quality |
| S9_vs_k141_16708\|\|lt2gene | Not-determined |
| S9_vs_k141_16711\|\|lt2gene | Low-quality |
| S9_vs_k141_1508\|\|lt2gene | Low-quality |
| S9_vs_k141_46774\|\|lt2gene | Not-determined |
| S9_vs_k141_65710\|\|lt2gene | Low-quality |
| S9_vs_k141_84527\|\|lt2gene | Low-quality |
| S9_vs_k141_35671\|\|lt2gene | Not-determined |
| S9_vs_k141_32022\|\|lt2gene | Low-quality |
| S9_vs_k141_65735\|\|lt2gene | Low-quality |
| S9_vs_k141_27996\|\|lt2gene | Low-quality |
| S9_vs_k141_5287\|\|lt2gene | Not-determined |
| S9_vs_k141_5289\|\|lt2gene | Not-determined |
| S9_vs_k141_84567\|\|lt2gene | Low-quality |
| S9_vs_k141_50441\|\|lt2gene | Low-quality |
| S9_vs_k141_20387\|\|lt2gene | Low-quality |
| S9_vs_k141_32039\|\|lt2gene | Low-quality |
| S9_vs_k141_80880\|\|lt2gene | Low-quality |
| S9_vs_k141_24097\|\|lt2gene | Low-quality |
| S9_vs_k141_20397\|\|lt2gene | Low-quality |
| S9_vs_k141_84585\|\|lt2gene | Not-determined |
| S9_vs_k141_28005\|\|lt2gene | Low-quality |
| S9_vs_k141_50464\|\|lt2gene | Not-determined |
| S9_vs_k141_50468\|\|lt2gene | Low-quality |
| S9_vs_k141_80897\|\|lt2gene | Low-quality |
| S9_vs_k141_35678\|\|lt2gene | Low-quality |
| S9_vs_k141_84608\|\|lt2gene | Not-determined |
| S9_vs_k141_5321\|\|lt2gene | Low-quality |
| S9_vs_k141_84618\|\|lt2gene | Not-determined |
| S9_vs_k141_24120\|\|lt2gene | Low-quality |
| S9_vs_k141_32078\|\|lt2gene | Low-quality |
| S9_vs_k141_1587\|\|lt2gene | Not-determined |
| S9_vs_k141_76828\|\|lt2gene | Not-determined |
| S9_vs_k141_28060\|\|lt2gene | Low-quality |
| S9_vs_k141_1597\|\|lt2gene | Low-quality |
| S9_vs_k141_62111\|\|lt2gene | Low-quality |
| S9_vs_k141_80964\|\|lt2gene | Low-quality |
| S9_vs_k141_73501\|\|lt2gene | Low-quality |
| S9_vs_k141_12874\|\|lt2gene | Low-quality |
| S9_vs_k141_32127\|\|lt2gene | Low-quality |
| S9_vs_k141_54617\|\|lt2gene | Low-quality |
| S9_vs_k141_76884\|\|lt2gene | Low-quality |
| S9_vs_k141_16881\|\|lt2gene | Low-quality |
| S9_vs_k141_24149\|\|lt2gene | Low-quality |
| S9_vs_k141_54626\|\|lt2gene | Not-determined |
| S9_vs_k141_39606\|\|lt2gene | Not-determined |
| S9_vs_k141_39615\|\|lt2gene | Low-quality |
| S9_vs_k141_32152\|\|lt2gene | Low-quality |
| S9_vs_k141_73550\|\|lt2gene | Low-quality |
| S9_vs_k141_54644\|\|lt2gene | Low-quality |
| S9_vs_k141_24222\|\|lt2gene | Not-determined |
| S9_vs_k141_35780\|\|lt2gene | Low-quality |
| S9_vs_k141_62220\|\|lt2gene | Low-quality |
| S9_vs_k141_16937\|\|lt2gene | Low-quality |
| S9_vs_k141_32230\|\|lt2gene | Low-quality |
| S9_vs_k141_54692\|\|lt2gene | Low-quality |
| S9_vs_k141_8972\|\|lt2gene | Not-determined |
| S9_vs_k141_20587\|\|lt2gene | Low-quality |
| S9_vs_k141_5459\|\|lt2gene | Low-quality |
| S9_vs_k141_16992\|\|lt2gene | Low-quality |
| S9_vs_k141_81029\|\|lt2gene | Low-quality |
| S9_vs_k141_81030\|\|lt2gene | Low-quality |
| S9_vs_k141_43397\|\|lt2gene | Low-quality |
| S9_vs_k141_39680\|\|lt2gene | Low-quality |
| S9_vs_k141_58115\|\|lt2gene | Not-determined |
| S9_vs_k141_54737\|\|lt2gene | Low-quality |
| S9_vs_k141_76947\|\|lt2gene | Not-determined |
| S9_vs_k141_24272\|\|lt2gene | Low-quality |
| S9_vs_k141_24273\|\|lt2gene | Low-quality |
| S9_vs_k141_9010\|\|lt2gene | Low-quality |
| S9_vs_k141_69622\|\|lt2gene | Low-quality |
| S9_vs_k141_12969\|\|lt2gene | Low-quality |
| S9_vs_k141_32270\|\|lt2gene | Low-quality |
| S9_vs_k141_5485\|\|lt2gene | Low-quality |
| S9_vs_k141_88588\|\|lt2gene | Low-quality |
| S9_vs_k141_84706\|\|lt2gene | Low-quality |
| S9_vs_k141_12981\|\|lt2gene | Low-quality |
| S9_vs_k141_32297\|\|lt2gene | Low-quality |
| S9_vs_k141_46835\|\|lt2gene | Not-determined |
| S9_vs_k141_84735\|\|lt2gene | Low-quality |
| S9_vs_k141_65838\|\|lt2gene | Low-quality |
| S9_vs_k141_46838\|\|lt2gene | Low-quality |
| S9_vs_k141_77007\|\|lt2gene | Low-quality |
| S9_vs_k141_1785\|\|lt2gene | Low-quality |
| S9_vs_k141_17086\|\|lt2gene | Low-quality |
| S9_vs_k141_28246\|\|lt2gene | Not-determined |
| S9_vs_k141_5519\|\|lt2gene | Low-quality |
| S9_vs_k141_46851\|\|lt2gene | Not-determined |
| S9_vs_k141_46854\|\|lt2gene | Not-determined |
| S9_vs_k141_5527\|\|lt2gene | Low-quality |
| S9_vs_k141_54788\|\|lt2gene | Not-determined |
| S9_vs_k141_54790\|\|lt2gene | Not-determined |
| S9_vs_k141_1798\|\|lt2gene | Low-quality |
| S9_vs_k141_77019\|\|lt2gene | Low-quality |
| S9_vs_k141_50727\|\|lt2gene | Low-quality |
| S9_vs_k141_46871\|\|lt2gene | Low-quality |
| S9_vs_k141_28254\|\|lt2gene | Not-determined |
| S9_vs_k141_13037\|\|lt2gene | Low-quality |
| S9_vs_k141_50748\|\|lt2gene | Low-quality |
| S9_vs_k141_50757\|\|lt2gene | Low-quality |
| S9_vs_k141_35915\|\|lt2gene | Not-determined |
| S9_vs_k141_5568\|\|lt2gene | Not-determined |
| S9_vs_k141_58218\|\|lt2gene | Low-quality |
| S9_vs_k141_50776\|\|lt2gene | Low-quality |
| S9_vs_k141_58219\|\|lt2gene | Not-determined |
| S9_vs_k141_24381\|\|lt2gene | Low-quality |
| S9_vs_k141_39784\|\|lt2gene | Low-quality |
| S9_vs_k141_35931\|\|lt2gene | Not-determined |
| S9_vs_k141_77097\|\|lt2gene | Not-determined |
| S9_vs_k141_58255\|\|lt2gene | Low-quality |
| S9_vs_k141_77104\|\|lt2gene | Low-quality |
| S9_vs_k141_13090\|\|lt2gene | Low-quality |
| S9_vs_k141_43588\|\|lt2gene | Low-quality |
| S9_vs_k141_9142\|\|lt2gene | Not-determined |
| S9_vs_k141_43591\|\|lt2gene | Low-quality |
| S9_vs_k141_9145\|\|lt2gene | Low-quality |
| S9_vs_k141_77117\|\|lt2gene | Low-quality |
| S9_vs_k141_88691\|\|lt2gene | Not-determined |
| S9_vs_k141_28371\|\|lt2gene | Low-quality |
| S9_vs_k141_62374\|\|lt2gene | Low-quality |
| S9_vs_k141_54922\|\|lt2gene | Low-quality |
| S9_vs_k141_58305\|\|lt2gene | Low-quality |
| S9_vs_k141_62385\|\|lt2gene | Low-quality |
| S9_vs_k141_88721\|\|lt2gene | Not-determined |
| S9_vs_k141_28399\|\|lt2gene | Low-quality |
| S9_vs_k141_28401\|\|lt2gene | Low-quality |
| S9_vs_k141_17209\|\|lt2gene | Not-determined |
| S9_vs_k141_13101\|\|lt2gene | Low-quality |
| S9_vs_k141_43643\|\|lt2gene | Low-quality |
| S9_vs_k141_46960\|\|lt2gene | Low-quality |
| S9_vs_k141_54960\|\|lt2gene | Low-quality |
| S9_vs_k141_62418\|\|lt2gene | Low-quality |
| S9_vs_k141_35993\|\|lt2gene | Low-quality |
| S9_vs_k141_62430\|\|lt2gene | Not-determined |
| S9_vs_k141_20753\|\|lt2gene | Low-quality |
| S9_vs_k141_58363\|\|lt2gene | Low-quality |
| S9_vs_k141_1928\|\|lt2gene | Low-quality |
| S9_vs_k141_69851\|\|lt2gene | Not-determined |
| S9_vs_k141_77191\|\|lt2gene | Low-quality |
| S9_vs_k141_13165\|\|lt2gene | Not-determined |
| S9_vs_k141_43717\|\|lt2gene | Low-quality |
| S9_vs_k141_39952\|\|lt2gene | Low-quality |
| S9_vs_k141_9189\|\|lt2gene | Not-determined |
| S9_vs_k141_9195\|\|lt2gene | Not-determined |
| S9_vs_k141_39983\|\|lt2gene | Low-quality |
| S9_vs_k141_20812\|\|lt2gene | Low-quality |
| S9_vs_k141_20813\|\|lt2gene | Low-quality |
| S9_vs_k141_81321\|\|lt2gene | Not-determined |
| S9_vs_k141_17339\|\|lt2gene | Low-quality |
| S9_vs_k141_55059\|\|lt2gene | Not-determined |
| S9_vs_k141_55061\|\|lt2gene | Not-determined |
| S9_vs_k141_5717\|\|lt2gene | Low-quality |
| S9_vs_k141_43771\|\|lt2gene | Low-quality |
| S9_vs_k141_58422\|\|lt2gene | Low-quality |
| S9_vs_k141_9231\|\|lt2gene | Low-quality |
| S9_vs_k141_47095\|\|lt2gene | Not-determined |
| S9_vs_k141_36108\|\|lt2gene | Low-quality |
| S9_vs_k141_13241\|\|lt2gene | Low-quality |
| S9_vs_k141_2003\|\|lt2gene | Low-quality |
| S9_vs_k141_47105\|\|lt2gene | Low-quality |

S9_vs_k141_17362||lt2gene Low-quality
S9_vs_k141_24629||lt2gene Not-determined
S9_vs_k141_24639||lt2gene Low-quality
S9_vs_k141_32437||lt2gene Not-determined
S9_vs_k141_13273||lt2gene Low-quality
S9_vs_k141_40033||lt2gene Low-quality
S9_vs_k141_9284||lt2gene Low-quality
S9_vs_k141_24656||lt2gene Low-quality
S9_vs_k141_13283||lt2gene Low-quality
S9_vs_k141_28555||lt2gene Low-quality
S9_vs_k141_32452||lt2gene Low-quality
S9_vs_k141_47150||lt2gene Low-quality
S9_vs_k141_20882||lt2gene Low-quality
S9_vs_k141_77334||lt2gene Low-quality
S9_vs_k141_40071||lt2gene Low-quality
S9_vs_k141_20896||lt2gene Not-determined
S9_vs_k141_20903||lt2gene Low-quality
S9_vs_k141_20909||lt2gene Low-quality
S9_vs_k141_85002||lt2gene Low-quality
S9_vs_k141_9318||lt2gene Low-quality
S9_vs_k141_17428||lt2gene Not-determined
S9_vs_k141_81433||lt2gene Not-determined
S9_vs_k141_43865||lt2gene Low-quality
S9_vs_k141_58494||lt2gene Low-quality
S9_vs_k141_43873||lt2gene Low-quality
S9_vs_k141_9358||lt2gene Low-quality
S9_vs_k141_40125||lt2gene Low-quality
S9_vs_k141_28622||lt2gene Low-quality
S9_vs_k141_36205||lt2gene Low-quality
S9_vs_k141_58524||lt2gene Low-quality
S9_vs_k141_73832||lt2gene Not-determined
S9_vs_k141_66080||lt2gene Low-quality
S9_vs_k141_28631||lt2gene Not-determined
S9_vs_k141_9394||lt2gene Low-quality
S9_vs_k141_17462||lt2gene Low-quality
S9_vs_k141_28644||lt2gene Low-quality
S9_vs_k141_9409||lt2gene Low-quality
S9_vs_k141_36226||lt2gene Low-quality
S9_vs_k141_47275||lt2gene Low-quality
S9_vs_k141_20943||lt2gene Low-quality
S9_vs_k141_28669||lt2gene Not-determined
S9_vs_k141_32561||lt2gene Low-quality
S9_vs_k141_36251||lt2gene Low-quality
S9_vs_k141_62621||lt2gene Low-quality
S9_vs_k141_20982||lt2gene Low-quality
S9_vs_k141_85078||lt2gene Low-quality
S9_vs_k141_47317||lt2gene Not-determined
S9_vs_k141_47325||lt2gene Low-quality
S9_vs_k141_66123||lt2gene Not-determined
S9_vs_k141_66134||lt2gene Low-quality
S9_vs_k141_32579||lt2gene Low-quality
S9_vs_k141_85110||lt2gene Not-determined
S9_vs_k141_28704||lt2gene Low-quality
S9_vs_k141_17559||lt2gene Low-quality
S9_vs_k141_81485||lt2gene Not-determined
S9_vs_k141_13464||lt2gene Low-quality
S9_vs_k141_88998||lt2gene Low-quality
S9_vs_k141_17576||lt2gene Low-quality
S9_vs_k141_89009||lt2gene Low-quality
S9_vs_k141_73901||lt2gene Low-quality
S9_vs_k141_66153||lt2gene Low-quality
S9_vs_k141_58650||lt2gene Low-quality
S9_vs_k141_17597||lt2gene Not-determined
S9_vs_k141_36296||lt2gene Low-quality
S9_vs_k141_85141||lt2gene Low-quality
S9_vs_k141_9499||lt2gene Low-quality
S9_vs_k141_21040||lt2gene Low-quality
S9_vs_k141_85156||lt2gene Not-determined
S9_vs_k141_70190||lt2gene Low-quality
S9_vs_k141_70191||lt2gene Low-quality
S9_vs_k141_44034||lt2gene Not-determined
S9_vs_k141_40302||lt2gene Low-quality
S9_vs_k141_9523||lt2gene Low-quality
S9_vs_k141_66191||lt2gene Low-quality
S9_vs_k141_44049||lt2gene Low-quality
S9_vs_k141_24896||lt2gene Low-quality
S9_vs_k141_36337||lt2gene Not-determined
S9_vs_k141_36339||lt2gene Low-quality
S9_vs_k141_36341||lt2gene Not-determined
S9_vs_k141_21080||lt2gene Low-quality
S9_vs_k141_28781||lt2gene Low-quality
S9_vs_k141_2174||lt2gene Low-quality
S9_vs_k141_36357||lt2gene Low-quality
S9_vs_k141_55295||lt2gene Low-quality
S9_vs_k141_55301||lt2gene Low-quality
S9_vs_k141_62703||lt2gene Low-quality
S9_vs_k141_36382||lt2gene Low-quality
S9_vs_k141_89085||lt2gene Not-determined
S9_vs_k141_9557||lt2gene Low-quality
S9_vs_k141_2232||lt2gene Low-quality
S9_vs_k141_44120||lt2gene Low-quality
S9_vs_k141_73976||lt2gene Low-quality
S9_vs_k141_55344||lt2gene Low-quality
S9_vs_k141_85250||lt2gene Not-determined
S9_vs_k141_40329||lt2gene Low-quality
S9_vs_k141_44135||lt2gene Not-determined
S9_vs_k141_70212||lt2gene Low-quality
S9_vs_k141_36413||lt2gene Not-determined
S9_vs_k141_66203||lt2gene Low-quality
S9_vs_k141_17633||lt2gene Not-determined
S9_vs_k141_89125||lt2gene Low-quality
S9_vs_k141_24954||lt2gene Not-determined
S9_vs_k141_55381||lt2gene Low-quality
S9_vs_k141_55386||lt2gene Low-quality
S9_vs_k141_9608||lt2gene Low-quality
S9_vs_k141_89151||lt2gene Low-quality
S9_vs_k141_66236||lt2gene Low-quality
S9_vs_k141_66237||lt2gene Not-determined
S9_vs_k141_9628||lt2gene Low-quality
S9_vs_k141_9631||lt2gene Low-quality
S9_vs_k141_77489||lt2gene Low-quality
S9_vs_k141_85335||lt2gene Not-determined
S9_vs_k141_85337||lt2gene Low-quality
S9_vs_k141_36492||lt2gene Low-quality
S9_vs_k141_6112||lt2gene Low-quality
S9_vs_k141_74055||lt2gene Low-quality
S9_vs_k141_85342||lt2gene Not-determined
S9_vs_k141_2300||lt2gene Low-quality
S9_vs_k141_85351||lt2gene Low-quality
S9_vs_k141_17693||lt2gene Low-quality
S9_vs_k141_2321||lt2gene Low-quality
S9_vs_k141_36521||lt2gene Not-determined
S9_vs_k141_28959||lt2gene Low-quality
S9_vs_k141_58746||lt2gene Low-quality
S9_vs_k141_9652||lt2gene Low-quality
S9_vs_k141_44227||lt2gene Low-quality
S9_vs_k141_85372||lt2gene Low-quality
S9_vs_k141_9668||lt2gene Low-quality
S9_vs_k141_28981||lt2gene Low-quality
S9_vs_k141_2344||lt2gene Low-quality
S9_vs_k141_21283||lt2gene Not-determined
S9_vs_k141_28987||lt2gene Low-quality
S9_vs_k141_55411||lt2gene Low-quality
S9_vs_k141_2355||lt2gene Low-quality
S9_vs_k141_21302||lt2gene Low-quality
S9_vs_k141_32640||lt2gene Not-determined
S9_vs_k141_44264||lt2gene Low-quality
S9_vs_k141_58807||lt2gene Low-quality
S9_vs_k141_36577||lt2gene Low-quality
S9_vs_k141_62793||lt2gene Not-determined
S9_vs_k141_21330||lt2gene Low-quality
S9_vs_k141_17743||lt2gene Low-quality
S9_vs_k141_66396||lt2gene Low-quality
S9_vs_k141_66404||lt2gene Not-determined
S9_vs_k141_74158||lt2gene Low-quality
S9_vs_k141_40533||lt2gene Low-quality
S9_vs_k141_85386||lt2gene Low-quality
S9_vs_k141_40543||lt2gene Low-quality
S9_vs_k141_6191||lt2gene Not-determined
S9_vs_k141_66427||lt2gene Not-determined
S9_vs_k141_2390||lt2gene Low-quality
S9_vs_k141_25164||lt2gene Not-determined
S9_vs_k141_74176||lt2gene Low-quality
S9_vs_k141_2412||lt2gene Low-quality
S9_vs_k141_36651||lt2gene Low-quality
S9_vs_k141_81787||lt2gene Not-determined
S9_vs_k141_25181||lt2gene Low-quality
S9_vs_k141_74183||lt2gene Low-quality
S9_vs_k141_17801||lt2gene Low-quality
S9_vs_k141_2456||lt2gene Low-quality
S9_vs_k141_29099||lt2gene Low-quality
S9_vs_k141_29105||lt2gene Low-quality
S9_vs_k141_6282||lt2gene Not-determined
S9_vs_k141_9808||lt2gene Low-quality
S9_vs_k141_13546||lt2gene Not-determined
S9_vs_k141_58963||lt2gene Not-determined
S9_vs_k141_13574||lt2gene Low-quality
S9_vs_k141_58969||lt2gene Not-determined
S9_vs_k141_55567||lt2gene Low-quality
S9_vs_k141_29136||lt2gene Low-quality
S9_vs_k141_21474||lt2gene Low-quality
S9_vs_k141_44339||lt2gene Low-quality
S9_vs_k141_47585||lt2gene Low-quality
S9_vs_k141_66523||lt2gene Low-quality
S9_vs_k141_40612||lt2gene Not-determined
S9_vs_k141_59005||lt2gene Low-quality
S9_vs_k141_13633||lt2gene Low-quality
S9_vs_k141_6347||lt2gene Low-quality
S9_vs_k141_17874||lt2gene Low-quality
S9_vs_k141_32815||lt2gene Low-quality
S9_vs_k141_77654||lt2gene Low-quality
S9_vs_k141_6363||lt2gene Low-quality
S9_vs_k141_6365||lt2gene Low-quality
S9_vs_k141_47622||lt2gene Low-quality
S9_vs_k141_62929||lt2gene Low-quality
S9_vs_k141_85509||lt2gene Low-quality
S9_vs_k141_77674||lt2gene Not-determined
S9_vs_k141_81899||lt2gene Not-determined
S9_vs_k141_21558||lt2gene Not-determined
S9_vs_k141_85531||lt2gene Low-quality
S9_vs_k141_13656||lt2gene Not-determined
S9_vs_k141_6397||lt2gene Low-quality
S9_vs_k141_62965||lt2gene Low-quality
S9_vs_k141_55642||lt2gene Low-quality
S9_vs_k141_55644||lt2gene Not-determined
S9_vs_k141_44411||lt2gene Low-quality
S9_vs_k141_47655||lt2gene Low-quality
S9_vs_k141_59060||lt2gene Low-quality
S9_vs_k141_59078||lt2gene Not-determined
S9_vs_k141_62987||lt2gene Low-quality
S9_vs_k141_77717||lt2gene Low-quality
S9_vs_k141_21618||lt2gene Low-quality
S9_vs_k141_85582||lt2gene Low-quality
S9_vs_k141_10008||lt2gene Low-quality
S9_vs_k141_59098||lt2gene Not-determined
S9_vs_k141_10010||lt2gene Not-determined
S9_vs_k141_36880||lt2gene Low-quality
S9_vs_k141_13709||lt2gene Low-quality
S9_vs_k141_47709||lt2gene Low-quality
S9_vs_k141_63041||lt2gene Not-determined
S9_vs_k141_40629||lt2gene Not-determined
S9_vs_k141_10050||lt2gene Low-quality
S9_vs_k141_25281||lt2gene Low-quality
S9_vs_k141_2664||lt2gene Low-quality
S9_vs_k141_10059||lt2gene Low-quality
S9_vs_k141_17974||lt2gene Not-determined
S9_vs_k141_55710||lt2gene Low-quality
S9_vs_k141_66611||lt2gene Not-determined
S9_vs_k141_21667||lt2gene Low-quality
S9_vs_k141_40645||lt2gene Low-quality
S9_vs_k141_63063||lt2gene Low-quality
S9_vs_k141_63066||lt2gene Low-quality
S9_vs_k141_55731||lt2gene Low-quality
S9_vs_k141_47744||lt2gene Low-quality
S9_vs_k141_17990||lt2gene Not-determined
S9_vs_k141_74367||lt2gene Low-quality
S9_vs_k141_32915||lt2gene Low-quality
S9_vs_k141_29292||lt2gene Low-quality
S9_vs_k141_10080||lt2gene Low-quality
S9_vs_k141_66627||lt2gene Not-determined
S9_vs_k141_70352||lt2gene Not-determined
S9_vs_k141_66642||lt2gene Low-quality
S9_vs_k141_32944||lt2gene Low-quality
S9_vs_k141_89326||lt2gene Low-quality
S9_vs_k141_40688||lt2gene Low-quality
S9_vs_k141_63122||lt2gene Low-quality
S9_vs_k141_85676||lt2gene Low-quality
S9_vs_k141_70385||lt2gene Not-determined
S9_vs_k141_36914||lt2gene Low-quality
S9_vs_k141_63128||lt2gene Not-determined
S9_vs_k141_74420||lt2gene Low-quality
S9_vs_k141_66676||lt2gene Low-quality
S9_vs_k141_85726||lt2gene Low-quality
S9_vs_k141_10150||lt2gene Low-quality
S9_vs_k141_63143||lt2gene Low-quality
S9_vs_k141_85731||lt2gene Not-determined
S9_vs_k141_63154||lt2gene Not-determined
S9_vs_k141_29377||lt2gene Not-determined
S9_vs_k141_40758||lt2gene Not-determined
S9_vs_k141_55763||lt2gene Low-quality
S9_vs_k141_66707||lt2gene Low-quality
S9_vs_k141_70432||lt2gene Low-quality
S9_vs_k141_70436||lt2gene Low-quality
S9_vs_k141_25414||lt2gene Not-determined
S9_vs_k141_21776||lt2gene Low-quality
S9_vs_k141_47778||lt2gene Not-determined
S9_vs_k141_13868||lt2gene Low-quality
S9_vs_k141_10180||lt2gene Low-quality
S9_vs_k141_2781||lt2gene Low-quality
S9_vs_k141_63193||lt2gene Low-quality
S9_vs_k141_47797||lt2gene Low-quality
S9_vs_k141_25431||lt2gene Not-determined
S9_vs_k141_47808||lt2gene Low-quality
S9_vs_k141_77913||lt2gene Not-determined
S9_vs_k141_70486||lt2gene Low-quality
S9_vs_k141_70487||lt2gene Not-determined
S9_vs_k141_66747||lt2gene Low-quality
S9_vs_k141_25457||lt2gene Low-quality
S9_vs_k141_70494||lt2gene Low-quality
S9_vs_k141_21807||lt2gene Low-quality
S9_vs_k141_13913||lt2gene Low-quality
S9_vs_k141_21815||lt2gene Low-quality
S9_vs_k141_70504||lt2gene Low-quality
S9_vs_k141_25468||lt2gene Not-determined
S9_vs_k141_66768||lt2gene Low-quality
S9_vs_k141_18178||lt2gene Low-quality
S9_vs_k141_10249||lt2gene Not-determined
S9_vs_k141_82131||lt2gene Low-quality
S9_vs_k141_66791||lt2gene Low-quality
S9_vs_k141_59253||lt2gene Low-quality
S9_vs_k141_37068||lt2gene Low-quality
S9_vs_k141_77972||lt2gene Low-quality
S9_vs_k141_66795||lt2gene Low-quality
S9_vs_k141_85836||lt2gene Low-quality
S9_vs_k141_51124||lt2gene Low-quality
S9_vs_k141_44644||lt2gene Low-quality
S9_vs_k141_51141||lt2gene Low-quality
S9_vs_k141_82145||lt2gene Low-quality
S9_vs_k141_59270||lt2gene Low-quality
S9_vs_k141_51144||lt2gene Low-quality
S9_vs_k141_77993||lt2gene Low-quality
S9_vs_k141_66813||lt2gene Low-quality
S9_vs_k141_89434||lt2gene Not-determined
S9_vs_k141_10281||lt2gene Low-quality
S9_vs_k141_44666||lt2gene Low-quality
S9_vs_k141_85873||lt2gene Low-quality
S9_vs_k141_2817||lt2gene Low-quality
S9_vs_k141_63295||lt2gene Low-quality
S9_vs_k141_29530||lt2gene Low-quality
S9_vs_k141_21862||lt2gene Low-quality
S9_vs_k141_29541||lt2gene Low-quality
S9_vs_k141_47933||lt2gene Low-quality
S9_vs_k141_21884||lt2gene Low-quality
S9_vs_k141_47956||lt2gene Low-quality
S9_vs_k141_89517||lt2gene Low-quality
S9_vs_k141_82170||lt2gene Not-determined
S9_vs_k141_51187||lt2gene Low-quality
S9_vs_k141_44692||lt2gene Not-determined
S9_vs_k141_74454||lt2gene Not-determined
S9_vs_k141_14051||lt2gene Not-determined
S9_vs_k141_47986||lt2gene Low-quality
S9_vs_k141_82188||lt2gene Not-determined
S9_vs_k141_85939||lt2gene Low-quality
S9_vs_k141_14067||lt2gene Low-quality
S9_vs_k141_74476||lt2gene Not-determined
S9_vs_k141_55931||lt2gene Low-quality
S9_vs_k141_14070||lt2gene Low-quality
S9_vs_k141_21938||lt2gene Not-determined
S9_vs_k141_55936||lt2gene Low-quality
S9_vs_k141_55938||lt2gene Low-quality
S9_vs_k141_70644||lt2gene Low-quality
S9_vs_k141_48023||lt2gene Low-quality
S9_vs_k141_78039||lt2gene Low-quality
S9_vs_k141_21955||lt2gene Low-quality
S9_vs_k141_18249||lt2gene Low-quality
S9_vs_k141_21959||lt2gene Low-quality
S9_vs_k141_29601||lt2gene Not-determined
S9_vs_k141_29603||lt2gene Low-quality
S9_vs_k141_21967||lt2gene Low-quality
S9_vs_k141_6757||lt2gene Low-quality
S9_vs_k141_51261||lt2gene Low-quality
S9_vs_k141_82267||lt2gene Low-quality
S9_vs_k141_44762||lt2gene Low-quality
S9_vs_k141_33302||lt2gene Low-quality
S9_vs_k141_66922||lt2gene Low-quality
S9_vs_k141_78076||lt2gene Not-determined
S9_vs_k141_18306||lt2gene Low-quality
S9_vs_k141_56015||lt2gene Low-quality
S9_vs_k141_10435||lt2gene Low-quality
S9_vs_k141_59423||lt2gene Not-determined
S9_vs_k141_74557||lt2gene Low-quality
S9_vs_k141_63451||lt2gene Low-quality
S9_vs_k141_89631||lt2gene Low-quality
S9_vs_k141_51293||lt2gene Low-quality
S9_vs_k141_25678||lt2gene Low-quality
S9_vs_k141_78122||lt2gene Not-determined
S9_vs_k141_29701||lt2gene Low-quality
S9_vs_k141_29702||lt2gene Low-quality
S9_vs_k141_86059||lt2gene Low-quality
S9_vs_k141_33349||lt2gene Low-quality
S9_vs_k141_29720||lt2gene Low-quality
S9_vs_k141_56036||lt2gene Low-quality
S9_vs_k141_37250||lt2gene Low-quality
S9_vs_k141_22005||lt2gene Low-quality
S9_vs_k141_86112||lt2gene Low-quality
S9_vs_k141_48159||lt2gene Low-quality
S9_vs_k141_41002||lt2gene Low-quality
S9_vs_k141_44840||lt2gene Low-quality
S9_vs_k141_44843||lt2gene Not-determined
S9_vs_k141_18408||lt2gene Low-quality
S9_vs_k141_44860||lt2gene Low-quality
S9_vs_k141_3054||lt2gene Low-quality
S9_vs_k141_10532||lt2gene Low-quality
S9_vs_k141_44866||lt2gene Low-quality
S9_vs_k141_78191||lt2gene Low-quality
S9_vs_k141_25792||lt2gene Low-quality
S9_vs_k141_3068||lt2gene Not-determined
S9_vs_k141_33441||lt2gene Not-determined
S9_vs_k141_59556||lt2gene Not-determined
S9_vs_k141_89747||lt2gene Not-determined
S9_vs_k141_70767||lt2gene Not-determined
S9_vs_k141_22075||lt2gene Low-quality
S9_vs_k141_44896||lt2gene Not-determined
S9_vs_k141_10545||lt2gene Not-determined
S9_vs_k141_56130||lt2gene Low-quality
S9_vs_k141_33445||lt2gene Low-quality
S9_vs_k141_10557||lt2gene Low-quality
S9_vs_k141_89762||lt2gene Low-quality
S9_vs_k141_37345||lt2gene Low-quality
S9_vs_k141_18456||lt2gene Low-quality
S9_vs_k141_70801||lt2gene Low-quality
S9_vs_k141_33468||lt2gene Not-determined
S9_vs_k141_33472||lt2gene Low-quality
S9_vs_k141_48221||lt2gene Low-quality
S9_vs_k141_59599||lt2gene Low-quality
S9_vs_k141_18474||lt2gene Low-quality
S9_vs_k141_41089||lt2gene Low-quality
S9_vs_k141_18480||lt2gene Low-quality
S9_vs_k141_14108||lt2gene Low-quality
S9_vs_k141_70827||lt2gene Not-determined
S9_vs_k141_14111||lt2gene Low-quality
S9_vs_k141_14114||lt2gene Low-quality
S9_vs_k141_37385||lt2gene Low-quality
S9_vs_k141_56165||lt2gene Low-quality
S9_vs_k141_29846||lt2gene Low-quality
S9_vs_k141_86213||lt2gene Low-quality
S9_vs_k141_56175||lt2gene Not-determined
S9_vs_k141_74754||lt2gene Not-determined
S9_vs_k141_74755||lt2gene Low-quality
S9_vs_k141_89819||lt2gene Low-quality
S9_vs_k141_59649||lt2gene Low-quality
S9_vs_k141_10640||lt2gene Low-quality
S9_vs_k141_78268||lt2gene Low-quality
S9_vs_k141_29877||lt2gene Low-quality
S9_vs_k141_70880||lt2gene Low-quality
S9_vs_k141_67145||lt2gene Low-quality
S9_vs_k141_14123||lt2gene Not-determined
S9_vs_k141_78278||lt2gene Low-quality
S9_vs_k141_25895||lt2gene Low-quality
S9_vs_k141_29894||lt2gene Low-quality
S9_vs_k141_44992||lt2gene Not-determined
S9_vs_k141_29896||lt2gene Low-quality
S9_vs_k141_18544||lt2gene Low-quality
S9_vs_k141_7049||lt2gene Low-quality
S9_vs_k141_70912||lt2gene Not-determined
S9_vs_k141_82435||lt2gene Low-quality
S9_vs_k141_7062||lt2gene Low-quality
S9_vs_k141_78304||lt2gene Low-quality
S9_vs_k141_33578||lt2gene Low-quality
S9_vs_k141_70934||lt2gene Low-quality
S9_vs_k141_89908||lt2gene Low-quality
S9_vs_k141_45040||lt2gene Not-determined
S9_vs_k141_22161||lt2gene Low-quality
S9_vs_k141_18580||lt2gene Low-quality
S9_vs_k141_82472||lt2gene Low-quality
S9_vs_k141_25952||lt2gene Low-quality
S9_vs_k141_3131||lt2gene Not-determined
S9_vs_k141_78337||lt2gene Not-determined
S9_vs_k141_78346||lt2gene Low-quality
S9_vs_k141_22183||lt2gene Low-quality
S9_vs_k141_78362||lt2gene Low-quality
S9_vs_k141_33614||lt2gene Low-quality
S9_vs_k141_3154||lt2gene Not-determined
S9_vs_k141_82501||lt2gene Low-quality
S9_vs_k141_3162||lt2gene Low-quality
S9_vs_k141_22201||lt2gene Not-determined
S9_vs_k141_22207||lt2gene Low-quality
S9_vs_k141_59753||lt2gene Low-quality
S9_vs_k141_33642||lt2gene Low-quality
S9_vs_k141_26006||lt2gene Low-quality
S9_vs_k141_7153||lt2gene Low-quality
S9_vs_k141_30055||lt2gene Low-quality
S9_vs_k141_56317||lt2gene Low-quality
S9_vs_k141_51475||lt2gene Low-quality
S9_vs_k141_26033||lt2gene Low-quality
S9_vs_k141_45136||lt2gene Low-quality
S9_vs_k141_56335||lt2gene Low-quality
S9_vs_k141_7191||lt2gene Low-quality
S9_vs_k141_41290||lt2gene Not-determined
S9_vs_k141_41299||lt2gene Low-quality
S9_vs_k141_33667||lt2gene Low-quality
S9_vs_k141_22298||lt2gene Low-quality
S9_vs_k141_71062||lt2gene Low-quality
S9_vs_k141_45190||lt2gene Not-determined
S9_vs_k141_7225||lt2gene Low-quality
S9_vs_k141_71072||lt2gene Low-quality
S9_vs_k141_45195||lt2gene Low-quality
S9_vs_k141_7241||lt2gene Low-quality
S9_vs_k141_59859||lt2gene Not-determined
S9_vs_k141_59862||lt2gene Not-determined
S9_vs_k141_82538||lt2gene Low-quality
S9_vs_k141_37494||lt2gene Low-quality
S9_vs_k141_82546||lt2gene Low-quality
S9_vs_k141_59873||lt2gene Not-determined
S9_vs_k141_82548||lt2gene Low-quality
S9_vs_k141_30136||lt2gene Low-quality
S9_vs_k141_90015||lt2gene Low-quality
S9_vs_k141_22369||lt2gene Low-quality
S9_vs_k141_59886||lt2gene Low-quality
S9_vs_k141_51578||lt2gene Low-quality
S9_vs_k141_30146||lt2gene Not-determined
S9_vs_k141_90027||lt2gene Not-determined
S9_vs_k141_51592||lt2gene Low-quality
S9_vs_k141_82557||lt2gene Not-determined
S9_vs_k141_86426||lt2gene Low-quality
S9_vs_k141_90047||lt2gene Low-quality
S9_vs_k141_7293||lt2gene Low-quality
S9_vs_k141_7295||lt2gene Not-determined
S9_vs_k141_67315||lt2gene Low-quality
S9_vs_k141_90078||lt2gene Not-determined
S9_vs_k141_67320||lt2gene Not-determined
S9_vs_k141_71172||lt2gene Low-quality
S9_vs_k141_78572||lt2gene Not-determined
S9_vs_k141_3371||lt2gene Low-quality
S9_vs_k141_56402||lt2gene Low-quality
S9_vs_k141_18865||lt2gene Low-quality
S9_vs_k141_26177||lt2gene Low-quality
S9_vs_k141_90114||lt2gene Low-quality
S9_vs_k141_56433||lt2gene Low-quality
S9_vs_k141_59978||lt2gene Not-determined
S9_vs_k141_90135||lt2gene Low-quality
S9_vs_k141_78634||lt2gene Low-quality
S9_vs_k141_37604||lt2gene Low-quality
S9_vs_k141_14394||lt2gene Low-quality
S9_vs_k141_56473||lt2gene Not-determined
S9_vs_k141_22434||lt2gene Low-quality
S9_vs_k141_71243||lt2gene Low-quality
S9_vs_k141_71254||lt2gene Low-quality
S9_vs_k141_14425||lt2gene Low-quality
S9_vs_k141_75137||lt2gene Low-quality
S9_vs_k141_71282||lt2gene Low-quality
S9_vs_k141_75149||lt2gene Not-determined
S9_vs_k141_14455||lt2gene Not-determined
S9_vs_k141_30309||lt2gene Low-quality
S9_vs_k141_26257||lt2gene Low-quality
S9_vs_k141_37684||lt2gene Not-determined
S9_vs_k141_67479||lt2gene Low-quality
S9_vs_k141_33933||lt2gene Not-determined
S9_vs_k141_45436||lt2gene Low-quality
S9_vs_k141_56526||lt2gene Not-determined
S9_vs_k141_78702||lt2gene Not-determined
S9_vs_k141_7452||lt2gene Not-determined
S9_vs_k141_14499||lt2gene Low-quality
S9_vs_k141_67515||lt2gene Low-quality
S9_vs_k141_78724||lt2gene Low-quality
S9_vs_k141_11089||lt2gene Low-quality
S9_vs_k141_86536||lt2gene Low-quality
S9_vs_k141_37727||lt2gene Low-quality
S9_vs_k141_71358||lt2gene Low-quality
S9_vs_k141_33991||lt2gene Low-quality
S9_vs_k141_7462||lt2gene Low-quality
S9_vs_k141_14533||lt2gene Low-quality
S9_vs_k141_75221||lt2gene Low-quality
S9_vs_k141_67531||lt2gene Low-quality
S9_vs_k141_48507||lt2gene Not-determined
S9_vs_k141_51660||lt2gene Low-quality
S9_vs_k141_75234||lt2gene Low-quality
S9_vs_k141_78767||lt2gene Low-quality
S9_vs_k141_34020||lt2gene Low-quality
S9_vs_k141_34026||lt2gene Low-quality
S9_vs_k141_75255||lt2gene Not-determined
S9_vs_k141_75261||lt2gene Low-quality
S9_vs_k141_51702||lt2gene Low-quality
S9_vs_k141_78792||lt2gene Low-quality
S9_vs_k141_14594||lt2gene Low-quality
S9_vs_k141_37798||lt2gene Low-quality
S9_vs_k141_14612||lt2gene Not-determined
S9_vs_k141_48573||lt2gene Low-quality
S9_vs_k141_26362||lt2gene Low-quality
S9_vs_k141_67572||lt2gene Not-determined
S9_vs_k141_11131||lt2gene Low-quality
S9_vs_k141_26371||lt2gene Not-determined
S9_vs_k141_67601||lt2gene Not-determined
S9_vs_k141_11140||lt2gene Low-quality
S9_vs_k141_14650||lt2gene Low-quality
S9_vs_k141_86612||lt2gene Low-quality
S9_vs_k141_71479||lt2gene Low-quality
S9_vs_k141_26412||lt2gene Low-quality
S9_vs_k141_82672||lt2gene Low-quality
S9_vs_k141_56673||lt2gene Low-quality
S9_vs_k141_82676||lt2gene Low-quality
S9_vs_k141_51771||lt2gene Low-quality
S9_vs_k141_14674||lt2gene Low-quality
S9_vs_k141_86639||lt2gene Low-quality
S9_vs_k141_41645||lt2gene Low-quality
S9_vs_k141_7491||lt2gene Low-quality
S9_vs_k141_71507||lt2gene Low-quality
S9_vs_k141_67650||lt2gene Low-quality
S9_vs_k141_26444||lt2gene Not-determined
S9_vs_k141_63590||lt2gene Low-quality
S9_vs_k141_7529||lt2gene Low-quality
S9_vs_k141_14699||lt2gene Not-determined
S9_vs_k141_14700||lt2gene Low-quality
S9_vs_k141_26455||lt2gene Not-determined
S9_vs_k141_14709||lt2gene Low-quality
S9_vs_k141_75327||lt2gene Low-quality
S9_vs_k141_3632||lt2gene Low-quality
S9_vs_k141_82709||lt2gene Low-quality
S9_vs_k141_51856||lt2gene Low-quality
S9_vs_k141_26474||lt2gene Low-quality
S9_vs_k141_56733||lt2gene Low-quality
S9_vs_k141_14726||lt2gene Low-quality
S9_vs_k141_90401||lt2gene Not-determined
S9_vs_k141_67723||lt2gene Low-quality
S9_vs_k141_71550||lt2gene Low-quality
S9_vs_k141_51886||lt2gene Not-determined
S9_vs_k141_82754||lt2gene Low-quality
S9_vs_k141_48636||lt2gene Low-quality
S9_vs_k141_67745||lt2gene Not-determined
S9_vs_k141_63683||lt2gene Low-quality
S9_vs_k141_56775||lt2gene Low-quality
S9_vs_k141_67751||lt2gene Low-quality
S9_vs_k141_63685||lt2gene Low-quality
S9_vs_k141_67771||lt2gene Low-quality
S9_vs_k141_82789||lt2gene Not-determined
S9_vs_k141_78987||lt2gene Low-quality
S9_vs_k141_75382||lt2gene Low-quality
S9_vs_k141_11280||lt2gene Not-determined
S9_vs_k141_48695||lt2gene Not-determined

| | |
|---|---|
| S9_vs_k141_71656\|\|lt2gene | Low-quality |
| S9_vs_k141_63750\|\|lt2gene | Low-quality |
| S9_vs_k141_82855\|\|lt2gene | Low-quality |
| S9_vs_k141_63767\|\|lt2gene | Low-quality |
| S9_vs_k141_11340\|\|lt2gene | Not-determined |
| S9_vs_k141_52047\|\|lt2gene | Low-quality |
| S9_vs_k141_48743\|\|lt2gene | Low-quality |
| S9_vs_k141_71683\|\|lt2gene | Not-determined |
| S9_vs_k141_3735\|\|lt2gene | Low-quality |
| S9_vs_k141_86863\|\|lt2gene | Low-quality |
| S9_vs_k141_86869\|\|lt2gene | Low-quality |
| S9_vs_k141_79088\|\|lt2gene | Low-quality |
| S9_vs_k141_48785\|\|lt2gene | Low-quality |
| S9_vs_k141_14867\|\|lt2gene | Not-determined |
| S9_vs_k141_86898\|\|lt2gene | Not-determined |
| S9_vs_k141_86910\|\|lt2gene | Low-quality |
| S9_vs_k141_67795\|\|lt2gene | Low-quality |
| S9_vs_k141_71746\|\|lt2gene | Low-quality |
| S9_vs_k141_52119\|\|lt2gene | Low-quality |
| S9_vs_k141_86948\|\|lt2gene | Low-quality |
| S9_vs_k141_75518\|\|lt2gene | Low-quality |
| S9_vs_k141_52136\|\|lt2gene | Low-quality |
| S9_vs_k141_90501\|\|lt2gene | Not-determined |
| S9_vs_k141_48841\|\|lt2gene | Not-determined |
| S9_vs_k141_71784\|\|lt2gene | Low-quality |
| S9_vs_k141_86995\|\|lt2gene | Low-quality |
| S9_vs_k141_48888\|\|lt2gene | Not-determined |
| S9_vs_k141_52163\|\|lt2gene | Not-determined |
| S9_vs_k141_48896\|\|lt2gene | Low-quality |
| S9_vs_k141_63896\|\|lt2gene | Low-quality |
| S9_vs_k141_67897\|\|lt2gene | Low-quality |
| S9_vs_k141_14994\|\|lt2gene | Low-quality |
| S9_vs_k141_75628\|\|lt2gene | Low-quality |
| S9_vs_k141_75639\|\|lt2gene | Low-quality |
| S9_vs_k141_34041\|\|lt2gene | Not-determined |
| S9_vs_k141_79242\|\|lt2gene | Not-determined |
| S9_vs_k141_75650\|\|lt2gene | Not-determined |
| S9_vs_k141_75657\|\|lt2gene | Low-quality |
| S9_vs_k141_67947\|\|lt2gene | Low-quality |
| S9_vs_k141_48938\|\|lt2gene | Low-quality |
| S9_vs_k141_90603\|\|lt2gene | Low-quality |
| S9_vs_k141_67978\|\|lt2gene | Low-quality |
| S9_vs_k141_60392\|\|lt2gene | Not-determined |
| S9_vs_k141_60398\|\|lt2gene | Not-determined |
| S9_vs_k141_87082\|\|lt2gene | Low-quality |
| S9_vs_k141_34101\|\|lt2gene | Low-quality |
| S9_vs_k141_63987\|\|lt2gene | Low-quality |
| S9_vs_k141_90644\|\|lt2gene | Low-quality |
| S9_vs_k141_48989\|\|lt2gene | Not-determined |
| S9_vs_k141_87122\|\|lt2gene | Low-quality |
| S9_vs_k141_90695\|\|lt2gene | Not-determined |
| S9_vs_k141_60437\|\|lt2gene | Not-determined |
| S9_vs_k141_15103\|\|lt2gene | Not-determined |
| S9_vs_k141_15109\|\|lt2gene | Low-quality |
| S9_vs_k141_64079\|\|lt2gene | Low-quality |
| S9_vs_k141_68074\|\|lt2gene | Low-quality |
| S9_vs_k141_15145\|\|lt2gene | Low-quality |
| S9_vs_k141_52285\|\|lt2gene | Low-quality |
| S9_vs_k141_83160\|\|lt2gene | Low-quality |
| S9_vs_k141_64175\|\|lt2gene | Low-quality |
| S9_vs_k141_52328\|\|lt2gene | Low-quality |
| S9_vs_k141_49161\|\|lt2gene | Not-determined |
| S9_vs_k141_64235\|\|lt2gene | Low-quality |
| S9_vs_k141_52411\|\|lt2gene | Low-quality |
| S9_vs_k141_49208\|\|lt2gene | Low-quality |
| S9_vs_k141_52526\|\|lt2gene | Low-quality |
| S9_vs_k141_52539\|\|lt2gene | Low-quality |
| S9_vs_k141_52620\|\|lt2gene | Not-determined |
| S9_vs_k141_52765\|\|lt2gene | Not-determined |
| S9_vs_k141_52824\|\|lt2gene | Not-determined |
| S9_vs_k141_52826\|\|lt2gene | Low-quality |
| S9_vs_k141_52994\|\|lt2gene | Not-determined |
| S9_vs_k141_53040\|\|lt2gene | Low-quality |
| S11_extracted_k141_32540 | Low-quality |
| S11_extracted_k141_97619 | Not-determined |
| S11_extracted_k141_5 | Not-determined |
| S11_extracted_k141_91115 | Not-determined |
| S11_extracted_k141_19528 | Not-determined |
| S11_extracted_k141_45562 | Not-determined |
| S11_extracted_k141_84609 | Not-determined |
| S11_extracted_k141_97625 | Not-determined |
| S11_extracted_k141_84611 | Low-quality |
| S11_extracted_k141_97626 | Not-determined |
| S11_extracted_k141_91116 | Not-determined |
| S11_extracted_k141_13023 | Low-quality |
| S11_extracted_k141_97628 | Not-determined |
| S11_extracted_k141_78106 | Not-determined |
| S11_extracted_k141_39057 | Not-determined |
| S11_extracted_k141_26045 | Not-determined |
| S11_extracted_k141_52076 | Low-quality |
| S11_extracted_k141_39060 | Not-determined |
| S11_extracted_k141_65099 | Not-determined |
| S11_extracted_k141_65100 | Not-determined |
| S11_extracted_k141_39062 | Not-determined |
| S11_extracted_k141_52081 | Not-determined |
| S11_extracted_k141_21 | Not-determined |
| S11_extracted_k141_58581 | Not-determined |
| S11_extracted_k141_52082 | Not-determined |
| S11_extracted_k141_65103 | Low-quality |
| S11_extracted_k141_65104 | Low-quality |
| S11_extracted_k141_65105 | Low-quality |
| S11_extracted_k141_52086 | Low-quality |
| S11_extracted_k141_32561 | Not-determined |
| S11_extracted_k141_29 | Not-determined |
| S11_extracted_k141_52089 | Not-determined |
| S11_extracted_k141_45576 | Not-determined |
| S11_extracted_k141_71598 | Not-determined |
| S11_extracted_k141_65108 | Not-determined |
| S11_extracted_k141_65109 | Not-determined |
| S11_extracted_k141_6526 | Not-determined |
| S11_extracted_k141_65110 | Not-determined |
| S11_extracted_k141_26053 | Not-determined |
| S11_extracted_k141_32566 | Low-quality |
| S11_extracted_k141_32567 | Not-determined |
| S11_extracted_k141_52092 | Not-determined |
| S11_extracted_k141_91131 | Not-determined |
| S11_extracted_k141_78129 | Not-determined |
| S11_extracted_k141_91133 | Not-determined |
| S11_extracted_k141_52095 | Not-determined |
| S11_extracted_k141_52098 | Low-quality |
| S11_extracted_k141_26059 | Not-determined |
| S11_extracted_k141_26060 | Not-determined |
| S11_extracted_k141_91142 | Not-determined |
| S11_extracted_k141_6537 | Not-determined |
| S11_extracted_k141_97642 | Low-quality |
| S11_extracted_k141_97643 | Not-determined |
| S11_extracted_k141_65124 | Not-determined |
| S11_extracted_k141_19546 | Not-determined |
| S11_extracted_k141_97646 | Not-determined |
| S11_extracted_k141_6539 | Not-determined |
| S11_extracted_k141_65131 | Not-determined |
| S11_extracted_k141_45587 | Not-determined |
| S11_extracted_k141_71618 | Not-determined |
| S11_extracted_k141_65134 | Not-determined |
| S11_extracted_k141_91155 | Not-determined |
| S11_extracted_k141_32572 | Not-determined |
| S2_vs_k141_1399\|\|full | High-quality |
| S11_extracted_k141_91159 | Not-determined |
| S11_extracted_k141_65136 | Low-quality |
| S11_extracted_k141_77 | Not-determined |
| S11_extracted_k141_52115 | Not-determined |
| S11_extracted_k141_78143 | Low-quality |
| S11_extracted_k141_45603 | Low-quality |
| S11_extracted_k141_58596 | Not-determined |
| S11_extracted_k141_78146 | Not-determined |
| S11_extracted_k141_52120 | Not-determined |
| S11_extracted_k141_71633 | Not-determined |
| S11_extracted_k141_65140 | Not-determined |
| S11_extracted_k141_91168 | Low-quality |
| S11_extracted_k141_58601 | Not-determined |
| S11_extracted_k141_32586 | Low-quality |
| S11_extracted_k141_58604 | Not-determined |
| S11_extracted_k141_26090 | Not-determined |
| S11_extracted_k141_65145 | Low-quality |
| S11_extracted_k141_32590 | Not-determined |
| S11_extracted_k141_45605 | Not-determined |
| S11_extracted_k141_97663 | Not-determined |
| S11_extracted_k141_65151 | Not-determined |
| S11_extracted_k141_45610 | Not-determined |
| S11_extracted_k141_45611 | Not-determined |
| S11_extracted_k141_26098 | Not-determined |
| S11_extracted_k141_52125 | Not-determined |
| S11_extracted_k141_65155 | Not-determined |
| S11_extracted_k141_52127 | Not-determined |
| S11_extracted_k141_52128 | Not-determined |
| S11_extracted_k141_19568 | Not-determined |
| S11_extracted_k141_52131 | Not-determined |
| S11_extracted_k141_65160 | Not-determined |
| S11_extracted_k141_58624 | Not-determined |
| S11_extracted_k141_91181 | Low-quality |
| S11_extracted_k141_39084 | Not-determined |
| S11_extracted_k141_65167 | Not-determined |
| S11_extracted_k141_91190 | Not-determined |
| S11_extracted_k141_58630 | Not-determined |
| S11_extracted_k141_84628 | Not-determined |
| S11_extracted_k141_91193 | Not-determined |
| S11_extracted_k141_71642 | Not-determined |
| S11_extracted_k141_45624 | Not-determined |
| S11_extracted_k141_71657 | Not-determined |
| S11_extracted_k141_65183 | Not-determined |
| S11_extracted_k141_58642 | Not-determined |
| S11_extracted_k141_19591 | Not-determined |
| S11_extracted_k141_71663 | Not-determined |
| S11_extracted_k141_84647 | Not-determined |
| S11_extracted_k141_19596 | Not-determined |
| S11_extracted_k141_32596 | Not-determined |
| S11_extracted_k141_45633 | Not-determined |
| S11_extracted_k141_65187 | Low-quality |
| S11_extracted_k141_39102 | Not-determined |
| S11_extracted_k141_19600 | Not-determined |
| S11_extracted_k141_93 | Not-determined |
| S11_extracted_k141_32606 | Not-determined |
| S11_extracted_k141_97 | Not-determined |
| S11_extracted_k141_32610 | Not-determined |
| S11_extracted_k141_19616 | Not-determined |
| S11_extracted_k141_26146 | Low-quality |
| S16_extracted_173095 | Complete |
| S11_extracted_k141_52174 | Low-quality |
| S11_extracted_k141_103 | Not-determined |
| S11_extracted_k141_39117 | Not-determined |
| S11_extracted_k141_19631 | Not-determined |
| S11_extracted_k141_58675 | Not-determined |
| S11_extracted_k141_97672 | Not-determined |
| S11_extracted_k141_113 | Not-determined |
| S11_extracted_k141_58676 | Not-determined |
| S11_extracted_k141_91206 | Not-determined |
| S11_extracted_k141_97677 | Not-determined |
| S11_extracted_k141_84670 | Not-determined |
| S11_extracted_k141_91207 | Not-determined |
| S11_extracted_k141_65204 | Not-determined |
| S11_extracted_k141_84672 | Not-determined |
| S11_extracted_k141_19638 | Low-quality |
| S11_extracted_k141_13056 | Not-determined |
| S11_extracted_k141_97679 | Not-determined |
| S11_extracted_k141_126 | Not-determined |
| S11_extracted_k141_19640 | Not-determined |
| S11_extracted_k141_45640 | Low-quality |
| S11_extracted_k141_133 | Not-determined |
| S11_extracted_k141_84681 | Not-determined |
| S11_extracted_k141_97683 | Low-quality |
| S11_extracted_k141_84683 | Not-determined |
| S11_extracted_k141_13065 | Not-determined |
| S11_extracted_k141_32623 | Not-determined |
| S11_extracted_k141_13066 | Low-quality |
| S11_extracted_k141_45655 | Not-determined |
| S11_extracted_k141_45657 | Not-determined |
| S11_extracted_k141_84691 | Low-quality |
| S11_extracted_k141_39145 | Not-determined |
| S11_extracted_k141_13074 | Low-quality |
| S11_extracted_k141_65229 | Not-determined |
| S11_extracted_k141_135 | Low-quality |
| S11_extracted_k141_52218 | Not-determined |
| S11_extracted_k141_39150 | Not-determined |
| S11_extracted_k141_13079 | Not-determined |
| S11_extracted_k141_52221 | Not-determined |
| S11_extracted_k141_65236 | Low-quality |
| S11_extracted_k141_32625 | Low-quality |
| S11_extracted_k141_91243 | Low-quality |
| S11_extracted_k141_32628 | Not-determined |
| S11_extracted_k141_91250 | Not-determined |
| S11_extracted_k141_142 | Low-quality |
| S11_extracted_k141_32636 | Low-quality |
| S11_extracted_k141_52229 | Not-determined |
| S11_extracted_k141_52235 | Low-quality |
| S11_extracted_k141_13100 | Not-determined |
| S11_extracted_k141_150 | Not-determined |
| S11_extracted_k141_65255 | Not-determined |
| S11_extracted_k141_13104 | Not-determined |
| S11_extracted_k141_84717 | Not-determined |
| S11_extracted_k141_52250 | Not-determined |
| S11_extracted_k141_65274 | Not-determined |
| S11_extracted_k141_71699 | Not-determined |
| S11_extracted_k141_32645 | Not-determined |
| S11_extracted_k141_169 | Not-determined |
| S11_extracted_k141_182 | Not-determined |
| S11_extracted_k141_97697 | Not-determined |
| S11_extracted_k141_91277 | Not-determined |
| S11_extracted_k141_26166 | Not-determined |
| S11_extracted_k141_84752 | Not-determined |
| S11_extracted_k141_189 | Not-determined |
| S11_extracted_k141_84754 | Not-determined |
| S11_extracted_k141_97704 | Not-determined |
| S11_extracted_k141_19665 | Not-determined |
| S11_extracted_k141_19668 | Not-determined |
| S11_extracted_k141_52276 | Low-quality |
| S11_extracted_k141_32655 | Not-determined |
| S11_extracted_k141_52280 | Not-determined |
| S11_extracted_k141_204 | Not-determined |
| S11_extracted_k141_39167 | Low-quality |
| S11_extracted_k141_97719 | Not-determined |
| S11_extracted_k141_97722 | Low-quality |
| S11_extracted_k141_39172 | Not-determined |
| S11_extracted_k141_97724 | Not-determined |
| S11_extracted_k141_97728 | Not-determined |
| S11_extracted_k141_221 | Not-determined |
| S11_extracted_k141_223 | Not-determined |
| S11_extracted_k141_52287 | Not-determined |
| S11_extracted_k141_26197 | Not-determined |
| S11_extracted_k141_13118 | Not-determined |
| S11_extracted_k141_26199 | Not-determined |
| S11_extracted_k141_26200 | Not-determined |
| S11_extracted_k141_97734 | Not-determined |
| S11_extracted_k141_19682 | Not-determined |
| S11_extracted_k141_39180 | Not-determined |
| S11_extracted_k141_26203 | Not-determined |
| S11_extracted_k141_13124 | Not-determined |
| S11_extracted_k141_26205 | Not-determined |
| S11_extracted_k141_13130 | Not-determined |
| S11_extracted_k141_26209 | Not-determined |
| S11_extracted_k141_238 | Not-determined |
| S11_extracted_k141_6562 | Low-quality |
| S11_extracted_k141_97744 | Low-quality |
| S11_extracted_k141_97747 | Not-determined |
| S11_extracted_k141_65285 | Not-determined |
| S11_extracted_k141_97753 | Not-determined |
| S11_extracted_k141_19694 | Not-determined |
| S11_extracted_k141_19696 | Low-quality |
| S11_extracted_k141_13140 | Not-determined |
| S11_extracted_k141_91303 | Not-determined |
| S11_extracted_k141_52313 | Not-determined |
| S11_extracted_k141_13142 | Not-determined |
| S11_extracted_k141_13147 | Not-determined |
| S11_extracted_k141_52315 | Not-determined |
| S11_extracted_k141_6577 | Not-determined |
| S11_extracted_k141_19708 | Low-quality |
| S11_extracted_k141_259 | Low-quality |
| S11_extracted_k141_13155 | Not-determined |
| S11_extracted_k141_19712 | Not-determined |
| S11_extracted_k141_262 | Low-quality |
| S11_extracted_k141_91310 | Low-quality |
| S11_extracted_k141_84777 | Low-quality |
| S11_extracted_k141_71708 | Low-quality |
| S11_extracted_k141_71709 | Not-determined |
| S11_extracted_k141_266 | Not-determined |
| S11_extracted_k141_19721 | Not-determined |
| S11_extracted_k141_52321 | Low-quality |
| S11_extracted_k141_91317 | Not-determined |
| S11_extracted_k141_78191 | Not-determined |
| S11_extracted_k141_78192 | Not-determined |
| S11_extracted_k141_278 | Not-determined |
| S11_extracted_k141_71715 | Not-determined |
| S11_extracted_k141_284 | Not-determined |
| S11_extracted_k141_71720 | Not-determined |
| S11_extracted_k141_58708 | Not-determined |
| S11_extracted_k141_84799 | Not-determined |
| S11_extracted_k141_39194 | Not-determined |
| S11_extracted_k141_84802 | Low-quality |
| S11_extracted_k141_6601 | Not-determined |
| S11_extracted_k141_32693 | Low-quality |
| S11_extracted_k141_6603 | Not-determined |
| S11_extracted_k141_6604 | Not-determined |
| S11_extracted_k141_6605 | Low-quality |
| S11_extracted_k141_13170 | Low-quality |
| S11_extracted_k141_91332 | Not-determined |
| S11_extracted_k141_84807 | Not-determined |
| S11_extracted_k141_84810 | Not-determined |
| S11_extracted_k141_39211 | Not-determined |
| S11_extracted_k141_71734 | Not-determined |
| S11_extracted_k141_58714 | Low-quality |
| S11_extracted_k141_13183 | Not-determined |
| S11_extracted_k141_71736 | Not-determined |
| S11_extracted_k141_58720 | Not-determined |
| S11_extracted_k141_78208 | Not-determined |
| S11_extracted_k141_32718 | Not-determined |
| S11_extracted_k141_58724 | Not-determined |
| S11_extracted_k141_91335 | Low-quality |
| S11_extracted_k141_71744 | Not-determined |
| S11_extracted_k141_58727 | Not-determined |
| S11_extracted_k141_32723 | Low-quality |
| S11_extracted_k141_71753 | Not-determined |
| S11_extracted_k141_97794 | Low-quality |
| S11_extracted_k141_32728 | Not-determined |
| S11_extracted_k141_91343 | Not-determined |
| S11_extracted_k141_6614 | Not-determined |
| S11_extracted_k141_71757 | Not-determined |
| S11_extracted_k141_6618 | Low-quality |
| S11_extracted_k141_78224 | Not-determined |
| S11_extracted_k141_320 | Not-determined |
| S11_extracted_k141_39221 | Not-determined |
| S11_extracted_k141_78227 | Low-quality |
| S11_extracted_k141_6621 | Not-determined |
| S11_extracted_k141_6622 | Not-determined |
| S11_extracted_k141_13194 | Low-quality |
| S11_extracted_k141_78234 | Not-determined |
| S11_extracted_k141_84821 | Low-quality |
| S11_extracted_k141_71768 | Not-determined |
| S11_extracted_k141_91359 | Not-determined |
| S11_extracted_k141_13203 | Not-determined |
| S11_extracted_k141_71769 | Low-quality |
| S11_extracted_k141_78243 | Low-quality |
| S11_extracted_k141_71770 | Low-quality |
| S11_extracted_k141_78244 | Not-determined |
| S11_extracted_k141_32744 | Not-determined |
| S11_extracted_k141_58738 | Not-determined |
| S11_extracted_k141_91366 | Not-determined |
| S11_extracted_k141_71776 | Not-determined |
| S11_extracted_k141_78249 | Not-determined |
| S11_extracted_k141_78250 | Not-determined |
| S11_extracted_k141_338 | Not-determined |
| S11_extracted_k141_58744 | Low-quality |
| S11_extracted_k141_39225 | Not-determined |
| S11_extracted_k141_52327 | Not-determined |
| S11_extracted_k141_71779 | Low-quality |
| S11_extracted_k141_52330 | Not-determined |
| S11_extracted_k141_52331 | Not-determined |
| S11_extracted_k141_97822 | Low-quality |
| S11_extracted_k141_84847 | Not-determined |
| S11_extracted_k141_19759 | Not-determined |
| S11_extracted_k141_32759 | Not-determined |
| S11_extracted_k141_97826 | Not-determined |
| S11_extracted_k141_65302 | Not-determined |
| S11_extracted_k141_32763 | Not-determined |
| S11_extracted_k141_65303 | Low-quality |
| S11_extracted_k141_52347 | Low-quality |
| S11_extracted_k141_91397 | Not-determined |
| S11_extracted_k141_78296 | Not-determined |
| S11_extracted_k141_52354 | Not-determined |
| S11_extracted_k141_19778 | Not-determined |
| S11_extracted_k141_91400 | Not-determined |
| S11_extracted_k141_19779 | Low-quality |
| S11_extracted_k141_78302 | Low-quality |
| S11_extracted_k141_32776 | Not-determined |
| S11_extracted_k141_19783 | Not-determined |
| S11_extracted_k141_52362 | Low-quality |
| S11_extracted_k141_71803 | Not-determined |
| S11_extracted_k141_375 | Low-quality |
| S11_extracted_k141_71805 | Not-determined |
| S11_extracted_k141_377 | Not-determined |
| S11_extracted_k141_71806 | Not-determined |
| S11_extracted_k141_58779 | Not-determined |
| S11_extracted_k141_97842 | Low-quality |
| S11_extracted_k141_52367 | Not-determined |
| S11_extracted_k141_58780 | Not-determined |
| S11_extracted_k141_52369 | Low-quality |
| S11_extracted_k141_71810 | Not-determined |
| S11_extracted_k141_71812 | Low-quality |
| S11_extracted_k141_19791 | Not-determined |
| S11_extracted_k141_52371 | Not-determined |
| S11_extracted_k141_32786 | Not-determined |
| S11_extracted_k141_52372 | Not-determined |
| S11_extracted_k141_6679 | Not-determined |
| S11_extracted_k141_32789 | Not-determined |
| S11_extracted_k141_6682 | Not-determined |
| S11_extracted_k141_78331 | Low-quality |
| S11_extracted_k141_52383 | Not-determined |
| S11_extracted_k141_65313 | Not-determined |
| S11_extracted_k141_399 | Not-determined |
| S11_extracted_k141_45661 | Not-determined |
| S11_extracted_k141_6691 | Not-determined |
| S11_extracted_k141_6693 | Low-quality |
| S11_extracted_k141_402 | Not-determined |
| S11_extracted_k141_52389 | Not-determined |
| S11_extracted_k141_78332 | Low-quality |
| S11_extracted_k141_404 | Not-determined |
| S11_extracted_k141_32808 | Low-quality |
| S11_extracted_k141_407 | Not-determined |
| S11_extracted_k141_408 | Low-quality |
| S11_extracted_k141_78336 | Low-quality |
| S11_extracted_k141_413 | Low-quality |
| S11_extracted_k141_97872 | Not-determined |
| S11_extracted_k141_19807 | Low-quality |
| S11_extracted_k141_45666 | Not-determined |
| S11_extracted_k141_417 | Not-determined |
| S11_extracted_k141_71838 | Not-determined |
| S11_extracted_k141_84856 | Not-determined |
| S11_extracted_k141_6707 | Not-determined |
| S11_extracted_k141_45673 | Not-determined |
| S11_extracted_k141_32823 | Low-quality |
| S11_extracted_k141_78344 | Not-determined |
| S11_extracted_k141_426 | Not-determined |
| S11_extracted_k141_45679 | Not-determined |
| S11_extracted_k141_26263 | Not-determined |
| S11_extracted_k141_65339 | Not-determined |
| S11_extracted_k141_78350 | Not-determined |
| S11_extracted_k141_39258 | Not-determined |
| S11_extracted_k141_45684 | Not-determined |
| S11_extracted_k141_65347 | Not-determined |
| S11_extracted_k141_45690 | Not-determined |
| S11_extracted_k141_45692 | Not-determined |
| S11_extracted_k141_444 | Not-determined |
| S11_extracted_k141_91419 | Not-determined |
| S11_extracted_k141_447 | Not-determined |
| S11_extracted_k141_65350 | Not-determined |
| S11_extracted_k141_84871 | Not-determined |
| S11_extracted_k141_26277 | Not-determined |
| S11_extracted_k141_39277 | Not-determined |
| S11_extracted_k141_65366 | Not-determined |
| S11_extracted_k141_454 | Low-quality |
| S11_extracted_k141_45707 | Not-determined |
| S11_extracted_k141_91451 | Not-determined |
| S11_extracted_k141_32857 | Not-determined |
| S11_extracted_k141_84893 | Not-determined |
| S11_extracted_k141_84896 | Not-determined |
| S11_extracted_k141_26290 | Low-quality |
| S11_extracted_k141_26292 | Not-determined |
| S11_extracted_k141_58799 | Not-determined |
| S11_extracted_k141_91455 | Not-determined |
| S11_extracted_k141_26297 | Not-determined |
| S11_extracted_k141_32863 | Not-determined |
| S11_extracted_k141_26298 | Not-determined |
| S11_extracted_k141_97905 | Not-determined |
| S11_extracted_k141_78374 | Not-determined |
| S11_extracted_k141_97908 | Low-quality |
| S11_extracted_k141_26308 | Not-determined |
| S11_extracted_k141_6724 | Not-determined |
| S11_extracted_k141_58810 | Not-determined |
| S11_extracted_k141_97917 | Not-determined |
| S11_extracted_k141_91473 | Not-determined |
| S11_extracted_k141_19820 | Not-determined |
| S11_extracted_k141_478 | Not-determined |
| S11_extracted_k141_97925 | Not-determined |
| S11_extracted_k141_58822 | Low-quality |
| S11_extracted_k141_97928 | Not-determined |
| S11_extracted_k141_84920 | Low-quality |
| S11_extracted_k141_39299 | Not-determined |
| S11_extracted_k141_91483 | Low-quality |
| S11_extracted_k141_84928 | Not-determined |
| S11_extracted_k141_71858 | Not-determined |
| S11_extracted_k141_91485 | Not-determined |
| S11_extracted_k141_84931 | Not-determined |
| S11_extracted_k141_91488 | Low-quality |
| S11_extracted_k141_497 | Not-determined |
| S11_extracted_k141_78402 | Low-quality |
| S11_extracted_k141_26329 | Not-determined |
| S11_extracted_k141_78408 | Not-determined |
| S11_extracted_k141_78409 | Not-determined |
| S11_extracted_k141_84940 | Not-determined |
| S11_extracted_k141_84942 | Not-determined |
| S11_extracted_k141_39321 | Low-quality |
| S11_extracted_k141_84943 | Not-determined |
| S11_extracted_k141_97943 | Not-determined |
| S11_extracted_k141_78413 | Low-quality |
| S11_extracted_k141_39325 | Not-determined |
| S11_extracted_k141_97948 | Not-determined |
| S11_extracted_k141_39326 | Not-determined |
| S11_extracted_k141_39328 | Low-quality |
| S11_extracted_k141_78418 | Not-determined |
| S11_extracted_k141_6757 | Not-determined |
| S11_extracted_k141_39332 | Not-determined |
| S11_extracted_k141_26352 | Not-determined |
| S11_extracted_k141_91499 | Low-quality |
| S11_extracted_k141_39337 | Not-determined |
| S11_extracted_k141_91501 | Not-determined |
| S11_extracted_k141_97959 | Not-determined |
| S11_extracted_k141_32880 | Low-quality |
| S11_extracted_k141_71875 | Not-determined |
| S11_extracted_k141_84967 | Not-determined |
| S11_extracted_k141_97962 | Not-determined |
| S11_extracted_k141_32885 | Not-determined |
| S11_extracted_k141_84970 | Not-determined |
| S11_extracted_k141_510 | Not-determined |
| S11_extracted_k141_32887 | Not-determined |
| S11_extracted_k141_97968 | Not-determined |
| S11_extracted_k141_52397 | Not-determined |
| S11_extracted_k141_97970 | Not-determined |
| S11_extracted_k141_39340 | Low-quality |
| S11_extracted_k141_32896 | Not-determined |
| S11_extracted_k141_78437 | Not-determined |
| S11_extracted_k141_19840 | Not-determined |
| S11_extracted_k141_84982 | Not-determined |
| S11_extracted_k141_26367 | Not-determined |
| S11_extracted_k141_26368 | Not-determined |
| S11_extracted_k141_91513 | Not-determined |
| S11_extracted_k141_78443 | Low-quality |
| S11_extracted_k141_91514 | Not-determined |
| S11_extracted_k141_39357 | Low-quality |
| S11_extracted_k141_78445 | Not-determined |
| S11_extracted_k141_58826 | Not-determined |
| S11_extracted_k141_52409 | Not-determined |
| S11_extracted_k141_71887 | Low-quality |
| S11_extracted_k141_52410 | Not-determined |
| S11_extracted_k141_65385 | Not-determined |
| S11_extracted_k141_26374 | Not-determined |
| S11_extracted_k141_39364 | Low-quality |
| S11_extracted_k141_45751 | Not-determined |
| S11_extracted_k141_78449 | Low-quality |
| S11_extracted_k141_71889 | Not-determined |
| S11_extracted_k141_530 | Not-determined |
| S11_extracted_k141_84995 | Not-determined |
| S11_extracted_k141_78458 | Not-determined |
| S11_extracted_k141_91528 | Not-determined |
| S11_extracted_k141_39372 | Not-determined |
| S11_extracted_k141_91530 | Not-determined |
| S11_extracted_k141_13229 | Not-determined |
| S11_extracted_k141_78463 | Not-determined |
| S11_extracted_k141_19868 | Low-quality |
| S11_extracted_k141_71903 | Not-determined |
| S11_extracted_k141_85003 | Not-determined |
| S11_extracted_k141_85005 | Not-determined |
| S11_extracted_k141_58836 | Low-quality |
| S11_extracted_k141_91537 | Not-determined |
| S11_extracted_k141_19874 | Not-determined |
| S11_extracted_k141_19876 | Not-determined |
| S11_extracted_k141_39376 | Not-determined |
| S11_extracted_k141_58841 | Not-determined |
| S11_extracted_k141_91541 | Low-quality |
| S11_extracted_k141_19878 | Low-quality |
| S11_extracted_k141_26384 | Not-determined |
| S11_extracted_k141_71906 | Low-quality |
| S11_extracted_k141_39378 | Not-determined |
| S11_extracted_k141_91547 | Not-determined |
| S11_extracted_k141_26387 | Not-determined |
| S11_extracted_k141_26388 | Not-determined |
| S11_extracted_k141_26389 | Low-quality |
| S11_extracted_k141_71910 | Not-determined |
| S11_extracted_k141_52443 | Not-determined |
| S11_extracted_k141_71911 | Not-determined |
| S11_extracted_k141_45776 | Not-determined |
| S11_extracted_k141_19884 | Not-determined |

| | |
|---|---|
| S11_extracted_k141_26398 | Low-quality |
| S11_extracted_k141_85036 | Not-determined |
| S11_extracted_k141_85037 | Not-determined |
| S11_extracted_k141_71925 | Not-determined |
| S11_extracted_k141_71926 | Not-determined |
| S11_extracted_k141_26403 | Not-determined |
| S11_extracted_k141_78475 | Not-determined |
| S11_extracted_k141_566 | Low-quality |
| S11_extracted_k141_568 | Not-determined |
| S11_extracted_k141_85043 | Low-quality |
| S11_extracted_k141_39399 | Not-determined |
| S11_extracted_k141_71933 | Low-quality |
| S11_extracted_k141_71935 | Not-determined |
| S11_extracted_k141_39400 | Not-determined |
| S11_extracted_k141_85051 | Not-determined |
| S11_extracted_k141_78490 | Not-determined |
| S11_extracted_k141_91559 | Not-determined |
| S11_extracted_k141_78492 | Not-determined |
| S11_extracted_k141_39405 | Not-determined |
| S11_extracted_k141_26406 | Low-quality |
| S11_extracted_k141_26408 | Not-determined |
| S11_extracted_k141_97988 | Low-quality |
| S11_extracted_k141_6819 | Not-determined |
| S11_extracted_k141_85056 | Low-quality |
| S11_extracted_k141_52467 | Not-determined |
| S11_extracted_k141_97991 | Not-determined |
| S11_extracted_k141_45802 | Not-determined |
| S11_extracted_k141_39408 | Not-determined |
| S11_extracted_k141_26418 | Not-determined |
| S11_extracted_k141_71941 | Not-determined |
| S11_extracted_k141_78504 | Not-determined |
| S11_extracted_k141_6824 | Not-determined |
| S11_extracted_k141_85067 | Not-determined |
| S11_extracted_k141_71948 | Not-determined |
| S11_extracted_k141_26427 | Not-determined |
| S11_extracted_k141_26434 | Not-determined |
| S11_extracted_k141_39412 | Not-determined |
| S11_extracted_k141_71959 | Not-determined |
| S11_extracted_k141_85079 | Not-determined |
| S11_extracted_k141_78519 | Not-determined |
| S11_extracted_k141_6835 | Not-determined |
| S11_extracted_k141_45810 | Not-determined |
| S11_extracted_k141_6837 | Low-quality |
| S11_extracted_k141_91592 | Not-determined |
| S11_extracted_k141_58871 | Low-quality |
| S11_extracted_k141_98009 | Not-determined |
| S11_extracted_k141_71972 | Not-determined |
| S11_extracted_k141_19897 | Not-determined |
| S11_extracted_k141_71977 | Not-determined |
| S11_extracted_k141_71979 | Not-determined |
| S11_extracted_k141_19901 | Not-determined |
| S11_extracted_k141_91599 | Not-determined |
| S11_extracted_k141_78535 | Low-quality |
| S11_extracted_k141_19909 | Not-determined |
| S11_extracted_k141_6846 | Not-determined |
| S11_extracted_k141_19915 | Not-determined |
| S11_extracted_k141_32930 | Low-quality |
| S11_extracted_k141_6857 | Not-determined |
| S11_extracted_k141_32936 | Not-determined |
| S11_extracted_k141_32938 | Not-determined |
| S11_extracted_k141_58883 | Not-determined |
| S11_extracted_k141_58886 | Not-determined |
| S11_extracted_k141_19923 | Low-quality |
| S11_extracted_k141_91611 | Low-quality |
| S11_extracted_k141_6861 | Low-quality |
| S11_extracted_k141_6864 | Low-quality |
| S11_extracted_k141_98029 | Not-determined |
| S11_extracted_k141_98030 | Low-quality |
| S11_extracted_k141_58892 | Low-quality |
| S11_extracted_k141_58896 | Not-determined |
| S11_extracted_k141_6872 | Not-determined |
| S11_extracted_k141_71986 | Not-determined |
| S11_extracted_k141_91622 | Not-determined |
| S11_extracted_k141_6875 | Low-quality |
| S11_extracted_k141_19937 | Not-determined |
| S11_extracted_k141_32951 | Not-determined |
| S11_extracted_k141_91625 | Not-determined |
| S11_extracted_k141_58901 | Low-quality |
| S11_extracted_k141_45828 | Not-determined |
| S11_extracted_k141_45829 | Low-quality |
| S11_extracted_k141_65410 | Low-quality |
| S11_extracted_k141_65412 | Not-determined |
| S11_extracted_k141_65413 | Not-determined |
| S11_extracted_k141_624 | Not-determined |
| S11_extracted_k141_26438 | Not-determined |
| S11_extracted_k141_627 | Not-determined |
| S11_extracted_k141_91636 | Low-quality |
| S11_extracted_k141_45843 | Low-quality |
| S11_extracted_k141_45844 | Not-determined |
| S11_extracted_k141_26444 | Not-determined |
| S11_extracted_k141_65418 | Not-determined |
| S11_extracted_k141_65420 | Not-determined |
| S11_extracted_k141_91644 | Not-determined |
| S11_extracted_k141_98060 | Not-determined |
| S11_extracted_k141_26452 | Not-determined |
| S11_extracted_k141_633 | Not-determined |
| S11_extracted_k141_78593 | Not-determined |
| S11_extracted_k141_65428 | Not-determined |
| S11_extracted_k141_91649 | Not-determined |
| S11_extracted_k141_45862 | Not-determined |
| S11_extracted_k141_78602 | Not-determined |
| S11_extracted_k141_91653 | Not-determined |
| S11_extracted_k141_71993 | Not-determined |
| S11_extracted_k141_26468 | Low-quality |
| S11_extracted_k141_45867 | Not-determined |
| S11_extracted_k141_52488 | Not-determined |
| S11_extracted_k141_78606 | Not-determined |
| S11_extracted_k141_39453 | Not-determined |
| S11_extracted_k141_91659 | Not-determined |
| S11_extracted_k141_78610 | Not-determined |
| S11_extracted_k141_45871 | Not-determined |
| S11_extracted_k141_71998 | Not-determined |
| S11_extracted_k141_78612 | Not-determined |
| S11_extracted_k141_91663 | Not-determined |
| S11_extracted_k141_72001 | Not-determined |
| S11_extracted_k141_643 | Not-determined |
| S11_extracted_k141_91668 | Not-determined |
| S11_extracted_k141_19968 | Not-determined |
| S11_extracted_k141_32969 | Low-quality |
| S11_extracted_k141_19971 | Not-determined |
| S11_extracted_k141_19972 | Low-quality |
| S11_extracted_k141_19974 | Not-determined |
| S11_extracted_k141_32974 | Low-quality |
| S11_extracted_k141_65441 | Not-determined |
| S11_extracted_k141_19979 | Not-determined |
| S11_extracted_k141_19981 | Low-quality |
| S11_extracted_k141_26480 | Low-quality |
| S11_extracted_k141_39478 | Not-determined |
| S11_extracted_k141_19985 | Low-quality |
| S11_extracted_k141_19987 | Not-determined |
| S11_extracted_k141_65452 | Not-determined |
| S11_extracted_k141_39484 | Not-determined |
| S11_extracted_k141_52504 | Not-determined |
| S11_extracted_k141_32983 | Not-determined |
| S11_extracted_k141_39488 | Low-quality |
| S11_extracted_k141_32985 | Not-determined |
| S11_extracted_k141_72024 | Not-determined |
| S11_extracted_k141_19996 | Not-determined |
| S11_extracted_k141_39489 | Low-quality |
| S11_extracted_k141_72029 | Not-determined |
| S11_extracted_k141_65458 | Low-quality |
| S11_extracted_k141_72033 | Not-determined |
| S11_extracted_k141_26498 | Not-determined |
| S11_extracted_k141_6881 | Not-determined |
| S11_extracted_k141_26500 | Not-determined |
| S11_extracted_k141_26501 | Not-determined |
| S11_extracted_k141_78632 | Low-quality |
| S11_extracted_k141_39497 | Not-determined |
| S11_extracted_k141_6885 | Not-determined |
| S11_extracted_k141_6887 | Not-determined |
| S11_extracted_k141_45905 | Not-determined |
| S11_extracted_k141_6888 | Not-determined |
| S11_extracted_k141_45907 | Not-determined |
| S11_extracted_k141_39501 | Not-determined |
| S11_extracted_k141_39502 | Low-quality |
| S11_extracted_k141_6895 | Not-determined |
| S11_extracted_k141_78637 | Not-determined |
| S11_extracted_k141_72047 | Not-determined |
| S11_extracted_k141_58941 | Not-determined |
| S11_extracted_k141_98099 | Not-determined |
| S11_extracted_k141_13236 | Not-determined |
| S11_extracted_k141_52524 | Not-determined |
| S11_extracted_k141_58945 | Not-determined |
| S11_extracted_k141_98107 | Low-quality |
| S11_extracted_k141_52530 | Not-determined |
| S11_extracted_k141_45926 | Not-determined |
| S11_extracted_k141_78651 | Not-determined |
| S11_extracted_k141_33012 | Low-quality |
| S11_extracted_k141_78652 | Not-determined |
| S11_extracted_k141_72056 | Not-determined |
| S11_extracted_k141_39518 | Not-determined |
| S11_extracted_k141_72058 | Not-determined |
| S11_extracted_k141_65486 | Not-determined |
| S11_extracted_k141_13246 | Not-determined |
| S11_extracted_k141_52533 | Low-quality |
| S11_extracted_k141_6920 | Not-determined |
| S11_extracted_k141_65492 | Not-determined |
| S11_extracted_k141_72072 | Not-determined |
| S11_extracted_k141_65495 | Not-determined |
| S11_extracted_k141_667 | Not-determined |
| S11_extracted_k141_98114 | Low-quality |
| S11_extracted_k141_669 | Not-determined |
| S11_extracted_k141_72078 | Not-determined |
| S11_extracted_k141_72079 | Not-determined |
| S11_extracted_k141_65502 | Not-determined |
| S11_extracted_k141_78660 | Not-determined |
| S11_extracted_k141_72087 | Not-determined |
| S11_extracted_k141_72088 | Not-determined |
| S11_extracted_k141_65509 | Low-quality |
| S11_extracted_k141_98129 | Not-determined |
| S11_extracted_k141_52550 | Not-determined |
| S11_extracted_k141_13267 | Not-determined |
| S11_extracted_k141_78670 | Low-quality |
| S11_extracted_k141_20022 | Not-determined |
| S11_extracted_k141_85100 | Not-determined |
| S11_extracted_k141_52563 | Not-determined |
| S11_extracted_k141_45939 | Not-determined |
| S11_extracted_k141_39554 | Not-determined |
| S11_extracted_k141_6931 | Not-determined |
| S11_extracted_k141_98140 | Not-determined |
| S11_extracted_k141_52571 | Not-determined |
| S11_extracted_k141_39557 | Not-determined |
| S11_extracted_k141_39560 | Not-determined |
| S11_extracted_k141_78694 | Low-quality |
| S11_extracted_k141_33034 | Not-determined |
| S11_extracted_k141_45947 | Low-quality |
| S11_extracted_k141_39563 | Not-determined |
| S11_extracted_k141_6944 | Not-determined |
| S11_extracted_k141_98157 | Low-quality |
| S11_extracted_k141_677 | Low-quality |
| S11_extracted_k141_13306 | Not-determined |
| S11_extracted_k141_13308 | Not-determined |
| S11_extracted_k141_678 | Not-determined |
| S11_extracted_k141_98161 | Not-determined |
| S11_extracted_k141_33043 | Not-determined |
| S11_extracted_k141_679 | Not-determined |
| S11_extracted_k141_13310 | Low-quality |
| S11_extracted_k141_687 | Not-determined |
| S11_extracted_k141_85136 | Not-determined |
| S11_extracted_k141_45956 | Low-quality |
| S11_extracted_k141_691 | Not-determined |
| S11_extracted_k141_98176 | Not-determined |
| S11_extracted_k141_65532 | Not-determined |
| S11_extracted_k141_98177 | Not-determined |
| S11_extracted_k141_45962 | Low-quality |
| S11_extracted_k141_65542 | Not-determined |
| S11_extracted_k141_72118 | Not-determined |
| S11_extracted_k141_85140 | Low-quality |
| S11_extracted_k141_72126 | Not-determined |
| S11_extracted_k141_45967 | Not-determined |
| S11_extracted_k141_72132 | Low-quality |
| S11_extracted_k141_703 | Not-determined |
| S11_extracted_k141_13332 | Not-determined |
| S11_extracted_k141_72137 | Not-determined |
| S11_extracted_k141_39580 | Not-determined |
| S11_extracted_k141_6979 | Low-quality |
| S16_vs_173095\|\|full | High-quality |
| S11_extracted_k141_711 | Low-quality |
| S11_extracted_k141_45981 | Not-determined |
| S11_extracted_k141_45982 | Not-determined |
| S11_extracted_k141_39585 | Not-determined |
| S11_extracted_k141_45985 | Not-determined |
| S11_extracted_k141_52578 | Not-determined |
| S11_extracted_k141_39587 | Not-determined |
| S11_extracted_k141_52580 | Not-determined |
| S11_extracted_k141_85156 | Not-determined |
| S11_extracted_k141_52584 | Not-determined |
| S11_extracted_k141_91681 | Not-determined |
| S11_extracted_k141_91683 | Low-quality |
| S11_extracted_k141_85162 | Not-determined |
| S11_extracted_k141_723 | Not-determined |
| S11_extracted_k141_39596 | Not-determined |
| S11_extracted_k141_33068 | Not-determined |
| S11_extracted_k141_85163 | Not-determined |
| S11_extracted_k141_727 | Low-quality |
| S11_extracted_k141_728 | Not-determined |
| S11_extracted_k141_91691 | Not-determined |
| S11_extracted_k141_45995 | Not-determined |
| S11_extracted_k141_91692 | Not-determined |
| S11_extracted_k141_65565 | Not-determined |
| S11_extracted_k141_45999 | Low-quality |
| S11_extracted_k141_20083 | Low-quality |
| S11_extracted_k141_65568 | Not-determined |
| S11_extracted_k141_85171 | Not-determined |
| S11_extracted_k141_46002 | Not-determined |
| S11_extracted_k141_26521 | Not-determined |
| S11_extracted_k141_58954 | Low-quality |
| S11_extracted_k141_6991 | Low-quality |
| S11_extracted_k141_26534 | Not-determined |
| S11_extracted_k141_13338 | Not-determined |
| S11_extracted_k141_20095 | Not-determined |
| S11_extracted_k141_6994 | Not-determined |
| S11_extracted_k141_58967 | Low-quality |
| S11_extracted_k141_13342 | Not-determined |
| S11_extracted_k141_20107 | Not-determined |
| S11_extracted_k141_733 | Not-determined |
| S11_extracted_k141_13351 | Not-determined |
| S11_extracted_k141_78700 | Low-quality |
| S11_extracted_k141_78702 | Not-determined |
| S11_extracted_k141_78704 | Not-determined |
| S11_extracted_k141_78707 | Not-determined |
| S11_extracted_k141_52609 | Not-determined |
| S11_extracted_k141_20119 | Not-determined |
| S11_extracted_k141_52610 | Not-determined |
| S11_extracted_k141_46036 | Not-determined |
| S11_extracted_k141_743 | Not-determined |
| S11_extracted_k141_78714 | Not-determined |
| S11_extracted_k141_26558 | Low-quality |
| S11_extracted_k141_26559 | Not-determined |
| S11_extracted_k141_13362 | Not-determined |
| S11_extracted_k141_91735 | Low-quality |
| S11_extracted_k141_20125 | Not-determined |
| S11_extracted_k141_98199 | Low-quality |
| S11_extracted_k141_78717 | Not-determined |
| S11_extracted_k141_98200 | Not-determined |
| S11_extracted_k141_26565 | Not-determined |
| S11_extracted_k141_13367 | Not-determined |
| S11_extracted_k141_91740 | Not-determined |
| S11_extracted_k141_98201 | Low-quality |
| S11_extracted_k141_20127 | Not-determined |
| S11_extracted_k141_98202 | Not-determined |
| S11_extracted_k141_78723 | Not-determined |
| S11_extracted_k141_98203 | Not-determined |
| S11_extracted_k141_20133 | Not-determined |
| S11_extracted_k141_13381 | Not-determined |
| S11_extracted_k141_52616 | Low-quality |
| S11_extracted_k141_52617 | Not-determined |
| S11_extracted_k141_78729 | Not-determined |
| S11_extracted_k141_78730 | Not-determined |
| S11_extracted_k141_65577 | Not-determined |
| S11_extracted_k141_52622 | Low-quality |
| S11_extracted_k141_52624 | Not-determined |
| S11_extracted_k141_85199 | Not-determined |
| S11_extracted_k141_767 | Not-determined |
| S11_extracted_k141_65580 | Not-determined |
| S11_extracted_k141_39636 | Not-determined |
| S11_extracted_k141_78749 | Not-determined |
| S11_extracted_k141_85205 | Not-determined |
| S11_extracted_k141_39639 | Not-determined |
| S11_extracted_k141_13393 | Not-determined |
| S11_extracted_k141_39640 | Not-determined |
| S11_extracted_k141_39641 | Not-determined |
| S11_extracted_k141_20140 | Low-quality |
| S11_extracted_k141_20141 | Not-determined |
| S11_extracted_k141_98220 | Low-quality |
| S11_extracted_k141_20142 | Low-quality |
| S11_extracted_k141_13397 | Not-determined |
| S11_extracted_k141_78751 | Low-quality |
| S11_extracted_k141_13405 | Not-determined |
| S11_extracted_k141_78755 | Not-determined |
| S11_extracted_k141_20164 | Not-determined |
| S11_extracted_k141_39660 | Low-quality |
| S11_extracted_k141_20165 | Not-determined |
| S11_extracted_k141_78761 | Not-determined |
| S11_extracted_k141_39662 | Not-determined |
| S11_extracted_k141_91744 | Low-quality |
| S11_extracted_k141_13413 | Not-determined |
| S11_extracted_k141_785 | Not-determined |
| S11_extracted_k141_65612 | Not-determined |
| S11_extracted_k141_20177 | Not-determined |
| S11_extracted_k141_98240 | Low-quality |
| S11_extracted_k141_13418 | Not-determined |
| S11_extracted_k141_20178 | Low-quality |
| S11_extracted_k141_39672 | Not-determined |
| S11_extracted_k141_20179 | Not-determined |
| S11_extracted_k141_46049 | Not-determined |
| S11_extracted_k141_39673 | Not-determined |
| S11_extracted_k141_85253 | Not-determined |
| S13_extracted_4901 | Medium-quality |
| S11_extracted_k141_65623 | Not-determined |
| S11_extracted_k141_91760 | Not-determined |
| S11_extracted_k141_26596 | Low-quality |
| S11_extracted_k141_799 | Not-determined |
| S11_extracted_k141_98255 | Not-determined |
| S11_extracted_k141_52646 | Low-quality |
| S11_extracted_k141_58979 | Not-determined |
| S11_extracted_k141_65628 | Not-determined |
| S11_extracted_k141_98257 | Not-determined |
| S11_extracted_k141_46062 | Not-determined |
| S11_extracted_k141_39676 | Not-determined |
| S11_extracted_k141_52651 | Not-determined |
| S11_extracted_k141_91767 | Not-determined |
| S11_extracted_k141_78789 | Not-determined |
| S11_extracted_k141_20207 | Not-determined |
| S11_extracted_k141_13438 | Not-determined |
| S11_extracted_k141_20208 | Not-determined |
| S11_extracted_k141_52658 | Not-determined |
| S11_extracted_k141_72208 | Low-quality |
| S11_extracted_k141_98264 | Not-determined |
| S11_extracted_k141_58999 | Not-determined |
| S11_extracted_k141_59000 | Not-determined |
| S11_extracted_k141_72217 | Not-determined |
| S11_extracted_k141_26603 | Not-determined |
| S13_extracted_24158 | Complete |
| S11_extracted_k141_26607 | Not-determined |
| S11_extracted_k141_72224 | Not-determined |
| S11_extracted_k141_46088 | Not-determined |
| S11_extracted_k141_91777 | Not-determined |
| S11_extracted_k141_59006 | Not-determined |
| S11_extracted_k141_91779 | Not-determined |
| S11_extracted_k141_91781 | Not-determined |
| S11_extracted_k141_85263 | Not-determined |
| S11_extracted_k141_46093 | Not-determined |
| S11_extracted_k141_20224 | Not-determined |
| S11_extracted_k141_33129 | Not-determined |
| S11_extracted_k141_46096 | Not-determined |
| S11_extracted_k141_59016 | Not-determined |
| S11_extracted_k141_91789 | Not-determined |
| S11_extracted_k141_72240 | Not-determined |
| S11_extracted_k141_91793 | Low-quality |
| S11_extracted_k141_26622 | Not-determined |
| S11_extracted_k141_72243 | Not-determined |
| S11_extracted_k141_59020 | Low-quality |
| S11_extracted_k141_13453 | Low-quality |
| S11_extracted_k141_52666 | Not-determined |
| S11_extracted_k141_91796 | Not-determined |
| S11_extracted_k141_33145 | Not-determined |
| S11_extracted_k141_91797 | Not-determined |
| S11_extracted_k141_46109 | Not-determined |
| S11_extracted_k141_98287 | Not-determined |
| S11_extracted_k141_26633 | Not-determined |
| S11_extracted_k141_13461 | Not-determined |
| S11_extracted_k141_72254 | Not-determined |
| S11_extracted_k141_98290 | Low-quality |
| S11_extracted_k141_52676 | Not-determined |
| S11_extracted_k141_52683 | Not-determined |
| S11_extracted_k141_78815 | Not-determined |
| S11_extracted_k141_52685 | Not-determined |
| S11_extracted_k141_13469 | Low-quality |
| S11_extracted_k141_85288 | Not-determined |
| S11_extracted_k141_65642 | Not-determined |
| S11_extracted_k141_85291 | Not-determined |
| S11_extracted_k141_39690 | Not-determined |
| S11_extracted_k141_85294 | Not-determined |
| S11_extracted_k141_78822 | Not-determined |
| S11_extracted_k141_13474 | Not-determined |
| S11_extracted_k141_78824 | Not-determined |
| S11_extracted_k141_78827 | Low-quality |
| S11_extracted_k141_26661 | Not-determined |
| S11_extracted_k141_85302 | Not-determined |
| S11_extracted_k141_85303 | Not-determined |
| S11_extracted_k141_819 | Not-determined |
| S11_extracted_k141_65649 | Low-quality |
| S11_extracted_k141_65650 | Not-determined |
| S11_extracted_k141_46127 | Not-determined |
| S11_extracted_k141_33175 | Not-determined |
| S11_extracted_k141_46128 | Not-determined |
| S11_extracted_k141_46130 | Not-determined |
| S11_extracted_k141_26680 | Not-determined |
| S11_extracted_k141_85312 | Not-determined |
| S11_extracted_k141_72277 | Not-determined |
| S11_extracted_k141_91819 | Not-determined |
| S11_extracted_k141_13486 | Not-determined |
| S11_extracted_k141_46135 | Not-determined |
| S11_extracted_k141_85313 | Not-determined |
| S11_extracted_k141_98315 | Not-determined |
| S11_extracted_k141_85319 | Not-determined |
| S11_extracted_k141_85320 | Not-determined |
| S11_extracted_k141_26689 | Low-quality |
| S11_extracted_k141_85321 | Not-determined |
| S11_extracted_k141_85322 | Not-determined |
| S11_extracted_k141_78844 | Not-determined |
| S11_extracted_k141_842 | Not-determined |
| S11_extracted_k141_91829 | Not-determined |
| S11_extracted_k141_91830 | Not-determined |
| S11_extracted_k141_33192 | Not-determined |
| S11_extracted_k141_848 | Low-quality |
| S11_extracted_k141_59028 | Low-quality |
| S11_extracted_k141_72293 | Not-determined |
| S11_extracted_k141_46151 | Not-determined |
| S11_extracted_k141_46152 | Low-quality |
| S11_extracted_k141_98339 | Not-determined |
| S11_extracted_k141_39720 | Low-quality |
| S11_extracted_k141_46154 | Not-determined |
| S11_extracted_k141_85329 | Not-determined |
| S11_extracted_k141_39723 | Not-determined |
| S11_extracted_k141_59035 | Not-determined |
| S11_extracted_k141_85332 | Not-determined |
| S11_extracted_k141_91838 | Not-determined |
| S11_extracted_k141_46157 | Low-quality |
| S11_extracted_k141_46158 | Not-determined |
| S11_extracted_k141_59040 | Not-determined |
| S11_extracted_k141_78850 | Low-quality |
| S11_extracted_k141_91844 | Not-determined |
| S11_extracted_k141_859 | Low-quality |
| S11_extracted_k141_65686 | Not-determined |
| S11_extracted_k141_98354 | Not-determined |
| S11_extracted_k141_13508 | Low-quality |
| S11_extracted_k141_98355 | Not-determined |
| S11_extracted_k141_39729 | Low-quality |
| S11_extracted_k141_91857 | Not-determined |
| S11_extracted_k141_91862 | Low-quality |
| S11_extracted_k141_65695 | Not-determined |
| S11_extracted_k141_65696 | Low-quality |
| S11_extracted_k141_85349 | Not-determined |
| S11_extracted_k141_878 | Not-determined |
| S11_extracted_k141_20244 | Not-determined |
| S11_extracted_k141_65700 | Low-quality |
| S11_extracted_k141_98375 | Not-determined |
| S11_extracted_k141_98378 | Not-determined |
| S11_extracted_k141_98380 | Not-determined |
| S11_extracted_k141_85353 | Low-quality |
| S11_extracted_k141_26735 | Not-determined |
| S11_extracted_k141_98385 | Not-determined |
| S11_extracted_k141_46185 | Low-quality |
| S11_extracted_k141_7029 | Not-determined |
| S11_extracted_k141_98388 | Low-quality |
| S11_extracted_k141_98389 | Low-quality |
| S11_extracted_k141_13519 | Not-determined |
| S11_extracted_k141_78882 | Not-determined |
| S11_extracted_k141_20256 | Not-determined |
| S11_extracted_k141_59061 | Not-determined |
| S11_extracted_k141_85355 | Not-determined |
| S11_extracted_k141_26751 | Not-determined |
| S11_extracted_k141_46191 | Not-determined |
| S11_extracted_k141_33221 | Not-determined |
| S11_extracted_k141_85362 | Not-determined |
| S11_extracted_k141_20269 | Not-determined |
| S11_extracted_k141_26759 | Low-quality |
| S11_extracted_k141_65735 | Not-determined |
| S11_extracted_k141_33227 | Low-quality |
| S11_extracted_k141_65737 | Not-determined |
| S11_extracted_k141_909 | Not-determined |
| S11_extracted_k141_65741 | Not-determined |
| S11_extracted_k141_85370 | Not-determined |
| S11_extracted_k141_65742 | Not-determined |
| S11_extracted_k141_912 | Not-determined |
| S11_extracted_k141_65743 | Low-quality |
| S11_extracted_k141_59071 | Not-determined |
| S11_extracted_k141_33236 | Not-determined |
| S11_extracted_k141_98405 | Not-determined |
| S11_extracted_k141_33237 | Not-determined |
| S11_extracted_k141_20287 | Not-determined |
| S11_extracted_k141_13539 | Not-determined |
| S11_extracted_k141_98415 | Low-quality |
| S11_extracted_k141_59080 | Not-determined |
| S11_extracted_k141_916 | Not-determined |
| S11_extracted_k141_13543 | Low-quality |
| S11_extracted_k141_98418 | Not-determined |
| S11_extracted_k141_72342 | Not-determined |
| S11_extracted_k141_921 | Not-determined |
| S11_extracted_k141_85374 | Not-determined |
| S11_extracted_k141_78910 | Not-determined |
| S11_extracted_k141_98419 | Low-quality |
| S11_extracted_k141_59088 | Not-determined |
| S11_extracted_k141_52707 | Not-determined |
| S11_extracted_k141_98426 | Low-quality |
| S11_extracted_k141_46220 | Not-determined |
| S11_extracted_k141_46226 | Not-determined |
| S11_extracted_k141_85381 | Low-quality |
| S11_extracted_k141_91886 | Low-quality |
| S11_extracted_k141_91890 | Not-determined |
| S11_extracted_k141_98440 | Not-determined |
| S11_extracted_k141_52717 | Low-quality |
| S11_extracted_k141_927 | Low-quality |
| S11_extracted_k141_98441 | Not-determined |
| S11_extracted_k141_72368 | Low-quality |
| S11_extracted_k141_98445 | Not-determined |
| S11_extracted_k141_59105 | Not-determined |
| S11_extracted_k141_26769 | Not-determined |
| S11_extracted_k141_59106 | Not-determined |
| S11_extracted_k141_91897 | Not-determined |
| S11_extracted_k141_91900 | Low-quality |
| S11_extracted_k141_13559 | Not-determined |
| S11_extracted_k141_98455 | Not-determined |
| S11_extracted_k141_85409 | Not-determined |
| S11_extracted_k141_78949 | Not-determined |
| S11_extracted_k141_98456 | Low-quality |
| S11_extracted_k141_33253 | Not-determined |
| S11_extracted_k141_944 | Not-determined |
| S11_extracted_k141_33256 | Not-determined |
| S11_extracted_k141_91915 | Not-determined |
| S11_extracted_k141_98460 | Not-determined |
| S11_extracted_k141_85415 | Low-quality |
| S11_extracted_k141_85416 | Not-determined |
| S11_extracted_k141_52738 | Not-determined |
| S11_extracted_k141_59122 | Not-determined |
| S11_extracted_k141_26799 | Not-determined |
| S11_extracted_k141_46264 | Not-determined |
| S11_extracted_k141_78961 | Not-determined |
| S11_extracted_k141_72382 | Not-determined |
| S11_extracted_k141_91927 | Not-determined |
| S11_extracted_k141_78964 | Not-determined |
| S11_extracted_k141_91929 | Not-determined |
| S11_extracted_k141_947 | Not-determined |
| S11_extracted_k141_20346 | Not-determined |
| S11_extracted_k141_85437 | Not-determined |
| S11_extracted_k141_52750 | Not-determined |
| S11_extracted_k141_52752 | Not-determined |
| S11_extracted_k141_98472 | Not-determined |
| S11_extracted_k141_33282 | Not-determined |
| S11_extracted_k141_46278 | Not-determined |
| S11_extracted_k141_72396 | Not-determined |
| S11_extracted_k141_98474 | Not-determined |
| S11_extracted_k141_39753 | Low-quality |
| S11_extracted_k141_72403 | Low-quality |
| S11_extracted_k141_72405 | Not-determined |
| S11_extracted_k141_91945 | Not-determined |
| S11_extracted_k141_72407 | Not-determined |
| S11_extracted_k141_20361 | Not-determined |
| S11_extracted_k141_52764 | Not-determined |
| S11_extracted_k141_46289 | Not-determined |
| S11_extracted_k141_65753 | Not-determined |
| S11_extracted_k141_20365 | Not-determined |
| S11_extracted_k141_26822 | Not-determined |
| S11_extracted_k141_39763 | Not-determined |
| S11_extracted_k141_72418 | Not-determined |
| S11_extracted_k141_7050 | Not-determined |
| S11_extracted_k141_65764 | Low-quality |

| | |
|---|---|
| S11_extracted_k141_39773 | Not-determined |
| S11_extracted_k141_91950 | Not-determined |
| S11_extracted_k141_72423 | Not-determined |
| S11_extracted_k141_33302 | Not-determined |
| S11_extracted_k141_33303 | Not-determined |
| S11_extracted_k141_33304 | Not-determined |
| S11_extracted_k141_26831 | Low-quality |
| S11_extracted_k141_13582 | Not-determined |
| S11_extracted_k141_33308 | Not-determined |
| S11_extracted_k141_72430 | Not-determined |
| S11_extracted_k141_46307 | Not-determined |
| S11_extracted_k141_46308 | Not-determined |
| S11_extracted_k141_98494 | Not-determined |
| S11_extracted_k141_26837 | Not-determined |
| S11_extracted_k141_65770 | Not-determined |
| S11_extracted_k141_26839 | Not-determined |
| S11_extracted_k141_13586 | Not-determined |
| S11_extracted_k141_72437 | Not-determined |
| S11_extracted_k141_39792 | Not-determined |
| S11_extracted_k141_72441 | Not-determined |
| S11_extracted_k141_7071 | Not-determined |
| S11_extracted_k141_13595 | Not-determined |
| S11_extracted_k141_46317 | Not-determined |
| S11_extracted_k141_85456 | Not-determined |
| S11_extracted_k141_46318 | Not-determined |
| S11_extracted_k141_46319 | Not-determined |
| S11_extracted_k141_91969 | Not-determined |
| S11_extracted_k141_85462 | Not-determined |
| S11_extracted_k141_33316 | Not-determined |
| S11_extracted_k141_91976 | Not-determined |
| S11_extracted_k141_13601 | Not-determined |
| S11_extracted_k141_85469 | Not-determined |
| S11_extracted_k141_7090 | Not-determined |
| S11_extracted_k141_91978 | Not-determined |
| S11_extracted_k141_72452 | Low-quality |
| S11_extracted_k141_91980 | Not-determined |
| S11_extracted_k141_72454 | Not-determined |
| S11_extracted_k141_85473 | Not-determined |
| S11_extracted_k141_13604 | Not-determined |
| S11_extracted_k141_85476 | Not-determined |
| S11_extracted_k141_7096 | Low-quality |
| S11_extracted_k141_52798 | Not-determined |
| S11_extracted_k141_39810 | Not-determined |
| S11_extracted_k141_52803 | Low-quality |
| S11_extracted_k141_85484 | Low-quality |
| S11_extracted_k141_85487 | Not-determined |
| S11_extracted_k141_72457 | Not-determined |
| S11_extracted_k141_39815 | Not-determined |
| S11_extracted_k141_91984 | Not-determined |
| S11_extracted_k141_65788 | Low-quality |
| S11_extracted_k141_59147 | Not-determined |
| S11_extracted_k141_59148 | Not-determined |
| S11_extracted_k141_79028 | Not-determined |
| S11_extracted_k141_85492 | Not-determined |
| S11_extracted_k141_20380 | Not-determined |
| S11_extracted_k141_91989 | Not-determined |
| S11_extracted_k141_20385 | Not-determined |
| S11_extracted_k141_65793 | Not-determined |
| S11_extracted_k141_98523 | Not-determined |
| S11_extracted_k141_91999 | Not-determined |
| S11_extracted_k141_26885 | Not-determined |
| S11_extracted_k141_65795 | Low-quality |
| S11_extracted_k141_59169 | Not-determined |
| S11_extracted_k141_72465 | Low-quality |
| S11_extracted_k141_65798 | Low-quality |
| S11_extracted_k141_26894 | Not-determined |
| S11_extracted_k141_85504 | Not-determined |
| S11_extracted_k141_7109 | Low-quality |
| S11_extracted_k141_39837 | Not-determined |
| S11_extracted_k141_85508 | Low-quality |
| S11_extracted_k141_46334 | Not-determined |
| S11_extracted_k141_92011 | Not-determined |
| S11_extracted_k141_20390 | Not-determined |
| S11_extracted_k141_39839 | Not-determined |
| S11_extracted_k141_72475 | Not-determined |
| S11_extracted_k141_20392 | Low-quality |
| S11_extracted_k141_98532 | Not-determined |
| S11_extracted_k141_46338 | Not-determined |
| S11_extracted_k141_7115 | Not-determined |
| S11_extracted_k141_1014 | Low-quality |
| S11_extracted_k141_65804 | Not-determined |
| S11_extracted_k141_46341 | Not-determined |
| S11_extracted_k141_52827 | Not-determined |
| S11_extracted_k141_7121 | Not-determined |
| S11_extracted_k141_92026 | Low-quality |
| S11_extracted_k141_20400 | Not-determined |
| S11_extracted_k141_26910 | Not-determined |
| S11_extracted_k141_26911 | Not-determined |
| S11_extracted_k141_65814 | Not-determined |
| S11_extracted_k141_39843 | Low-quality |
| S11_extracted_k141_98542 | Low-quality |
| S11_extracted_k141_20407 | Not-determined |
| S11_extracted_k141_26913 | Not-determined |
| S11_extracted_k141_20410 | Not-determined |
| S11_extracted_k141_92028 | Low-quality |
| S11_extracted_k141_1027 | Not-determined |
| S11_extracted_k141_20411 | Not-determined |
| S11_extracted_k141_26916 | Low-quality |
| S11_extracted_k141_85518 | Not-determined |
| S11_extracted_k141_46352 | Not-determined |
| S11_extracted_k141_52837 | Not-determined |
| S11_extracted_k141_46353 | Not-determined |
| S11_extracted_k141_85520 | Not-determined |
| S11_extracted_k141_46355 | Not-determined |
| S11_extracted_k141_98553 | Not-determined |
| S11_extracted_k141_20420 | Not-determined |
| S11_extracted_k141_1035 | Not-determined |
| S11_extracted_k141_98557 | Not-determined |
| S11_extracted_k141_39858 | Not-determined |
| S11_extracted_k141_20427 | Not-determined |
| S11_extracted_k141_85529 | Not-determined |
| S11_extracted_k141_20428 | Not-determined |
| S11_extracted_k141_33350 | Low-quality |
| S11_extracted_k141_46370 | Not-determined |
| S11_extracted_k141_20432 | Not-determined |
| S11_extracted_k141_20433 | Not-determined |
| S11_extracted_k141_65833 | Low-quality |
| S11_extracted_k141_46375 | Not-determined |
| S11_extracted_k141_79067 | Not-determined |
| S11_extracted_k141_46379 | Not-determined |
| S11_extracted_k141_79071 | Not-determined |
| S11_extracted_k141_65839 | Not-determined |
| S11_extracted_k141_33368 | Not-determined |
| S11_extracted_k141_46384 | Not-determined |
| S11_extracted_k141_46385 | Not-determined |
| S11_extracted_k141_7143 | Not-determined |
| S11_extracted_k141_7145 | Not-determined |
| S11_extracted_k141_1059 | Not-determined |
| S11_extracted_k141_46390 | Not-determined |
| S11_extracted_k141_52849 | Not-determined |
| S11_extracted_k141_7147 | Not-determined |
| S11_extracted_k141_46392 | Not-determined |
| S11_extracted_k141_7148 | Low-quality |
| S11_extracted_k141_65844 | Low-quality |
| S11_extracted_k141_92052 | Not-determined |
| S11_extracted_k141_72531 | Not-determined |
| S11_extracted_k141_85539 | Not-determined |
| S11_extracted_k141_72532 | Low-quality |
| S11_extracted_k141_33379 | Low-quality |
| S13_vs_24158\|\|full | High-quality |
| S11_extracted_k141_92055 | Not-determined |
| S11_extracted_k141_59230 | Not-determined |
| S11_extracted_k141_46399 | Not-determined |
| S11_extracted_k141_39869 | Not-determined |
| S11_extracted_k141_33386 | Not-determined |
| S11_extracted_k141_52862 | Not-determined |
| S11_extracted_k141_65854 | Not-determined |
| S11_extracted_k141_1069 | Not-determined |
| S11_extracted_k141_92072 | Not-determined |
| S11_extracted_k141_98578 | Not-determined |
| S11_extracted_k141_39882 | Not-determined |
| S11_extracted_k141_85554 | Low-quality |
| S11_extracted_k141_52871 | Not-determined |
| S11_extracted_k141_85555 | Not-determined |
| S11_extracted_k141_85558 | Not-determined |
| S11_extracted_k141_59241 | Low-quality |
| S11_extracted_k141_98586 | Not-determined |
| S11_extracted_k141_52875 | Low-quality |
| S11_extracted_k141_39891 | Not-determined |
| S11_extracted_k141_85559 | Low-quality |
| S11_extracted_k141_7162 | Not-determined |
| S11_extracted_k141_52878 | Not-determined |
| S11_extracted_k141_39894 | Low-quality |
| S11_extracted_k141_59246 | Not-determined |
| S11_extracted_k141_65871 | Not-determined |
| S11_extracted_k141_59250 | Not-determined |
| S11_extracted_k141_52883 | Not-determined |
| S11_extracted_k141_7167 | Not-determined |
| S11_extracted_k141_33408 | Not-determined |
| S11_extracted_k141_65877 | Not-determined |
| S11_extracted_k141_52888 | Not-determined |
| S11_extracted_k141_59260 | Not-determined |
| S11_extracted_k141_33410 | Not-determined |
| S11_extracted_k141_52893 | Not-determined |
| S11_extracted_k141_52894 | Not-determined |
| S11_extracted_k141_98594 | Not-determined |
| S11_extracted_k141_52895 | Not-determined |
| S11_extracted_k141_46421 | Not-determined |
| S11_extracted_k141_98596 | Not-determined |
| S11_extracted_k141_98597 | Not-determined |
| S11_extracted_k141_20485 | Not-determined |
| S11_extracted_k141_92076 | Not-determined |
| S11_extracted_k141_7178 | Low-quality |
| S11_extracted_k141_59272 | Not-determined |
| S11_extracted_k141_52903 | Not-determined |
| S11_extracted_k141_92079 | Not-determined |
| S11_extracted_k141_85579 | Not-determined |
| S11_extracted_k141_20493 | Not-determined |
| S11_extracted_k141_20494 | Not-determined |
| S11_extracted_k141_85584 | Low-quality |
| S11_extracted_k141_98610 | Not-determined |
| S11_extracted_k141_85585 | Not-determined |
| S11_extracted_k141_92087 | Not-determined |
| S11_extracted_k141_98614 | Not-determined |
| S11_extracted_k141_85589 | Low-quality |
| S11_extracted_k141_7187 | Not-determined |
| S11_extracted_k141_85590 | Low-quality |
| S11_extracted_k141_1095 | Low-quality |
| S11_extracted_k141_72556 | Not-determined |
| S11_extracted_k141_52912 | Not-determined |
| S11_extracted_k141_98618 | Not-determined |
| S11_extracted_k141_85593 | Not-determined |
| S11_extracted_k141_72560 | Not-determined |
| S11_extracted_k141_7193 | Not-determined |
| S11_extracted_k141_85596 | Not-determined |
| S11_extracted_k141_39899 | Not-determined |
| S11_extracted_k141_1101 | Not-determined |
| S11_extracted_k141_92095 | Not-determined |
| S11_extracted_k141_85598 | Not-determined |
| S11_extracted_k141_85599 | Low-quality |
| S11_extracted_k141_98626 | Not-determined |
| S11_extracted_k141_92098 | Low-quality |
| S11_extracted_k141_92101 | Not-determined |
| S11_extracted_k141_85608 | Low-quality |
| S11_extracted_k141_20508 | Not-determined |
| S11_extracted_k141_85612 | Not-determined |
| S11_extracted_k141_20510 | Not-determined |
| S11_extracted_k141_92107 | Low-quality |
| S11_extracted_k141_7209 | Not-determined |
| S11_extracted_k141_7210 | Not-determined |
| S11_extracted_k141_98636 | Not-determined |
| S11_extracted_k141_20513 | Low-quality |
| S11_extracted_k141_85620 | Not-determined |
| S11_extracted_k141_52939 | Not-determined |
| S11_extracted_k141_72564 | Not-determined |
| S11_extracted_k141_79114 | Not-determined |
| S11_extracted_k141_46426 | Low-quality |
| S11_extracted_k141_52941 | Low-quality |
| S11_extracted_k141_52943 | Not-determined |
| S11_extracted_k141_46429 | Not-determined |
| S11_extracted_k141_1121 | Not-determined |
| S11_extracted_k141_98645 | Low-quality |
| S11_extracted_k141_59291 | Low-quality |
| S11_extracted_k141_92116 | Not-determined |
| S11_extracted_k141_59298 | Low-quality |
| S11_extracted_k141_52954 | Not-determined |
| S11_extracted_k141_52955 | Not-determined |
| S11_extracted_k141_85629 | Not-determined |
| S11_extracted_k141_85630 | Not-determined |
| S11_extracted_k141_7230 | Not-determined |
| S11_extracted_k141_39929 | Low-quality |
| S11_extracted_k141_85635 | Not-determined |
| S11_extracted_k141_59312 | Low-quality |
| S11_extracted_k141_59316 | Not-determined |
| S11_extracted_k141_46447 | Low-quality |
| S11_extracted_k141_52958 | Not-determined |
| S11_extracted_k141_46449 | Not-determined |
| S11_extracted_k141_92126 | Not-determined |
| S11_extracted_k141_7239 | Not-determined |
| S11_extracted_k141_7240 | Not-determined |
| S11_extracted_k141_52961 | Not-determined |
| S11_extracted_k141_98670 | Not-determined |
| S11_extracted_k141_46454 | Not-determined |
| S11_extracted_k141_7243 | Low-quality |
| S11_extracted_k141_79141 | Low-quality |
| S11_extracted_k141_1137 | Low-quality |
| S11_extracted_k141_59325 | Not-determined |
| S11_extracted_k141_7248 | Not-determined |
| S11_extracted_k141_1141 | Not-determined |
| S11_extracted_k141_79149 | Not-determined |
| S11_extracted_k141_59337 | Not-determined |
| S11_extracted_k141_65889 | Not-determined |
| S11_extracted_k141_72600 | Not-determined |
| S11_extracted_k141_39945 | Not-determined |
| S11_extracted_k141_65899 | Not-determined |
| S11_extracted_k141_59343 | Not-determined |
| S11_extracted_k141_59345 | Not-determined |
| S11_extracted_k141_7263 | Not-determined |
| S11_extracted_k141_7264 | Not-determined |
| S11_extracted_k141_52977 | Not-determined |
| S11_extracted_k141_1165 | Not-determined |
| S11_extracted_k141_7273 | Not-determined |
| S11_extracted_k141_85663 | Not-determined |
| S11_extracted_k141_65919 | Not-determined |
| S11_extracted_k141_7276 | Not-determined |
| S11_extracted_k141_72606 | Not-determined |
| S11_extracted_k141_52988 | Not-determined |
| S11_extracted_k141_79183 | Low-quality |
| S11_extracted_k141_39953 | Low-quality |
| S11_extracted_k141_72608 | Not-determined |
| S11_extracted_k141_39956 | Not-determined |
| S11_extracted_k141_7282 | Not-determined |
| S11_extracted_k141_79188 | Not-determined |
| S11_extracted_k141_79197 | Not-determined |
| S11_extracted_k141_39964 | Not-determined |
| S11_extracted_k141_98695 | Low-quality |
| S11_extracted_k141_85675 | Low-quality |
| S11_extracted_k141_7289 | Not-determined |
| S11_extracted_k141_85678 | Not-determined |
| S11_extracted_k141_53010 | Not-determined |
| S11_extracted_k141_7300 | Not-determined |
| S11_extracted_k141_53011 | Not-determined |
| S11_extracted_k141_79209 | Not-determined |
| S11_extracted_k141_98706 | Not-determined |
| S13_vs_4901\|\|full | Medium-quality |
| S11_extracted_k141_20515 | Not-determined |
| S11_extracted_k141_85688 | Not-determined |
| S11_extracted_k141_79216 | Not-determined |
| S11_extracted_k141_85690 | Not-determined |
| S11_extracted_k141_7307 | Not-determined |
| S11_extracted_k141_85694 | Low-quality |
| S11_extracted_k141_1197 | Not-determined |
| S11_extracted_k141_1198 | Not-determined |
| S11_extracted_k141_26939 | Not-determined |
| S11_extracted_k141_20523 | Not-determined |
| S11_extracted_k141_98717 | Not-determined |
| S11_extracted_k141_79223 | Low-quality |
| S11_extracted_k141_65962 | Not-determined |
| S11_extracted_k141_59363 | Not-determined |
| S11_extracted_k141_26946 | Low-quality |
| S11_extracted_k141_53025 | Low-quality |
| S11_extracted_k141_1207 | Low-quality |
| S11_extracted_k141_72641 | Not-determined |
| S11_extracted_k141_46482 | Not-determined |
| S11_extracted_k141_20535 | Not-determined |
| S11_extracted_k141_46484 | Not-determined |
| S11_extracted_k141_33459 | Not-determined |
| S11_extracted_k141_92141 | Not-determined |
| S11_extracted_k141_59366 | Not-determined |
| S11_extracted_k141_26953 | Not-determined |
| S11_extracted_k141_26955 | Not-determined |
| S11_extracted_k141_39973 | Not-determined |
| S11_extracted_k141_39974 | Low-quality |
| S11_extracted_k141_72652 | Not-determined |
| S11_extracted_k141_72653 | Not-determined |
| S11_extracted_k141_85705 | Not-determined |
| S11_extracted_k141_65971 | Low-quality |
| S11_extracted_k141_46497 | Not-determined |
| S11_extracted_k141_20550 | Not-determined |
| S11_extracted_k141_46502 | Not-determined |
| S11_extracted_k141_98734 | Not-determined |
| S11_extracted_k141_33475 | Not-determined |
| S11_extracted_k141_39990 | Low-quality |
| S11_extracted_k141_26972 | Not-determined |
| S11_extracted_k141_39996 | Not-determined |
| S11_extracted_k141_26979 | Not-determined |
| S11_extracted_k141_40000 | Not-determined |
| S11_extracted_k141_20562 | Not-determined |
| S11_extracted_k141_46510 | Low-quality |
| S11_extracted_k141_20564 | Not-determined |
| S11_extracted_k141_65990 | Low-quality |
| S11_extracted_k141_85718 | Not-determined |
| S11_extracted_k141_7335 | Not-determined |
| S11_extracted_k141_85721 | Low-quality |
| S11_extracted_k141_72665 | Not-determined |
| S11_extracted_k141_20572 | Low-quality |
| S11_extracted_k141_40007 | Not-determined |
| S11_extracted_k141_20576 | Not-determined |
| S11_extracted_k141_92164 | Not-determined |
| S11_extracted_k141_1219 | Not-determined |
| S11_extracted_k141_46521 | Not-determined |
| S11_extracted_k141_72670 | Not-determined |
| S11_extracted_k141_40015 | Not-determined |
| S11_extracted_k141_20580 | Low-quality |
| S11_extracted_k141_20582 | Not-determined |
| S11_extracted_k141_33490 | Not-determined |
| S11_extracted_k141_92175 | Not-determined |
| S11_extracted_k141_1230 | Not-determined |
| S11_extracted_k141_53044 | Not-determined |
| S11_extracted_k141_1232 | Low-quality |
| S11_extracted_k141_20589 | Not-determined |
| S11_extracted_k141_33506 | Low-quality |
| S11_extracted_k141_79240 | Low-quality |
| S11_extracted_k141_85737 | Low-quality |
| S11_extracted_k141_79243 | Not-determined |
| S11_extracted_k141_66001 | Not-determined |
| S11_extracted_k141_85743 | Not-determined |
| S11_extracted_k141_72691 | Not-determined |
| S11_extracted_k141_53069 | Not-determined |
| S11_extracted_k141_46540 | Low-quality |
| S11_extracted_k141_13610 | Low-quality |
| S11_extracted_k141_53073 | Not-determined |
| S11_extracted_k141_7381 | Not-determined |
| S11_extracted_k141_46544 | Not-determined |
| S11_extracted_k141_53076 | Not-determined |
| S11_extracted_k141_33521 | Not-determined |
| S11_extracted_k141_79259 | Not-determined |
| S11_extracted_k141_79262 | Not-determined |
| S11_extracted_k141_40040 | Not-determined |
| S11_extracted_k141_13616 | Low-quality |
| S11_extracted_k141_40042 | Not-determined |
| S11_extracted_k141_66025 | Not-determined |
| S11_extracted_k141_13617 | Not-determined |
| S11_extracted_k141_40045 | Low-quality |
| S11_extracted_k141_92196 | Not-determined |
| S11_extracted_k141_92197 | Low-quality |
| S11_extracted_k141_79267 | Not-determined |
| S11_extracted_k141_7398 | Not-determined |
| S11_extracted_k141_53090 | Not-determined |
| S11_extracted_k141_92200 | Not-determined |
| S11_extracted_k141_79271 | Not-determined |
| S11_extracted_k141_72717 | Low-quality |
| S11_extracted_k141_7400 | Not-determined |
| S11_extracted_k141_85766 | Not-determined |
| S11_extracted_k141_13623 | Not-determined |
| S11_extracted_k141_59401 | Not-determined |
| S11_extracted_k141_13625 | Not-determined |
| S11_extracted_k141_1236 | Not-determined |
| S11_extracted_k141_13627 | Low-quality |
| S11_extracted_k141_85769 | Not-determined |
| S11_extracted_k141_1238 | Not-determined |
| S11_extracted_k141_20618 | Not-determined |
| S11_extracted_k141_92213 | Not-determined |
| S11_extracted_k141_20620 | Not-determined |
| S11_extracted_k141_85772 | Not-determined |
| S11_extracted_k141_53099 | Not-determined |
| S11_extracted_k141_85773 | Not-determined |
| S11_extracted_k141_53101 | Not-determined |
| S11_extracted_k141_92215 | Not-determined |
| S11_extracted_k141_33538 | Not-determined |
| S11_extracted_k141_72725 | Not-determined |
| S11_extracted_k141_72727 | Not-determined |
| S11_extracted_k141_13642 | Not-determined |
| S11_extracted_k141_59416 | Not-determined |
| S11_extracted_k141_33542 | Low-quality |
| S11_extracted_k141_33543 | Low-quality |
| S11_extracted_k141_72732 | Not-determined |
| S11_extracted_k141_53106 | Not-determined |
| S11_extracted_k141_1249 | Not-determined |
| S11_extracted_k141_7428 | Low-quality |
| S11_extracted_k141_59422 | Not-determined |
| S11_extracted_k141_40069 | Not-determined |
| S11_extracted_k141_59424 | Low-quality |
| S11_extracted_k141_59426 | Not-determined |
| S11_extracted_k141_85785 | Not-determined |
| S11_extracted_k141_40073 | Not-determined |
| S11_extracted_k141_27004 | Not-determined |
| S11_extracted_k141_33547 | Not-determined |
| S11_extracted_k141_33548 | Not-determined |
| S11_extracted_k141_53110 | Not-determined |
| S11_extracted_k141_66049 | Not-determined |
| S11_extracted_k141_66051 | Not-determined |
| S11_extracted_k141_1264 | Low-quality |
| S11_extracted_k141_27020 | Not-determined |
| S11_extracted_k141_72745 | Not-determined |
| S11_extracted_k141_27023 | Low-quality |
| S11_extracted_k141_27024 | Not-determined |
| S11_extracted_k141_72747 | Not-determined |
| S11_extracted_k141_27026 | Not-determined |
| S11_extracted_k141_33552 | Low-quality |
| S11_extracted_k141_72752 | Not-determined |
| S11_extracted_k141_53126 | Not-determined |
| S11_extracted_k141_66060 | Not-determined |
| S11_extracted_k141_20630 | Not-determined |
| S11_extracted_k141_72754 | Not-determined |
| S11_extracted_k141_33554 | Not-determined |
| S11_extracted_k141_20632 | Not-determined |
| S11_extracted_k141_53129 | Not-determined |
| S11_extracted_k141_59441 | Not-determined |
| S11_extracted_k141_72757 | Not-determined |
| S11_extracted_k141_72758 | Not-determined |
| S11_extracted_k141_33559 | Not-determined |
| S11_extracted_k141_33561 | Not-determined |
| S11_extracted_k141_33562 | Not-determined |
| S11_extracted_k141_20637 | Not-determined |
| S11_extracted_k141_20638 | Not-determined |
| S11_extracted_k141_33565 | Not-determined |
| S11_extracted_k141_7442 | Not-determined |
| S11_extracted_k141_7443 | Not-determined |
| S11_extracted_k141_40083 | Not-determined |
| S11_extracted_k141_53142 | Not-determined |
| S11_extracted_k141_27035 | Not-determined |
| S11_extracted_k141_66078 | Not-determined |
| S11_extracted_k141_40093 | Not-determined |
| S11_extracted_k141_53151 | Not-determined |
| S11_extracted_k141_72763 | Not-determined |
| S11_extracted_k141_20654 | Not-determined |
| S11_extracted_k141_59454 | Low-quality |
| S11_extracted_k141_53152 | Not-determined |
| S11_extracted_k141_13654 | Not-determined |
| S11_extracted_k141_13658 | Not-determined |
| S11_extracted_k141_92224 | Not-determined |
| S11_extracted_k141_72773 | Not-determined |
| S11_extracted_k141_72777 | Not-determined |
| S11_extracted_k141_13664 | Not-determined |
| S11_extracted_k141_33587 | Not-determined |
| S11_extracted_k141_53165 | Not-determined |
| S11_extracted_k141_33589 | Not-determined |
| S11_extracted_k141_40101 | Not-determined |
| S11_extracted_k141_20671 | Not-determined |
| S11_extracted_k141_92230 | Low-quality |
| S11_extracted_k141_53169 | Not-determined |
| S11_extracted_k141_27059 | Not-determined |
| S11_extracted_k141_53177 | Not-determined |
| S11_extracted_k141_79306 | Not-determined |
| S11_extracted_k141_53181 | Not-determined |
| S11_extracted_k141_33605 | Not-determined |
| S11_extracted_k141_66096 | Not-determined |
| S11_extracted_k141_59467 | Not-determined |
| S11_extracted_k141_79318 | Not-determined |
| S11_extracted_k141_98766 | Not-determined |
| S11_extracted_k141_53186 | Not-determined |
| S11_extracted_k141_59478 | Not-determined |
| S11_extracted_k141_33614 | Not-determined |
| S11_extracted_k141_27080 | Not-determined |
| S11_extracted_k141_53189 | Not-determined |
| S11_extracted_k141_72804 | Not-determined |
| S11_extracted_k141_33620 | Not-determined |
| S11_extracted_k141_1280 | Low-quality |
| S11_extracted_k141_59487 | Not-determined |
| S11_extracted_k141_40108 | Not-determined |
| S11_extracted_k141_53197 | Low-quality |
| S11_extracted_k141_46574 | Not-determined |
| S11_extracted_k141_53200 | Not-determined |
| S11_extracted_k141_40115 | Low-quality |
| S11_extracted_k141_98774 | Not-determined |
| S11_extracted_k141_59494 | Not-determined |
| S11_extracted_k141_98775 | Not-determined |
| S11_extracted_k141_72812 | Not-determined |
| S11_extracted_k141_98776 | Not-determined |
| S11_extracted_k141_53202 | Not-determined |
| S11_extracted_k141_59497 | Not-determined |
| S11_extracted_k141_98778 | Not-determined |
| S11_extracted_k141_13672 | Not-determined |
| S11_extracted_k141_59500 | Low-quality |
| S11_extracted_k141_46582 | Not-determined |
| S11_extracted_k141_79333 | Not-determined |
| S11_extracted_k141_7463 | Not-determined |
| S11_extracted_k141_53205 | Not-determined |
| S11_extracted_k141_13678 | Low-quality |
| S11_extracted_k141_33632 | Not-determined |
| S11_extracted_k141_27084 | Low-quality |
| S11_extracted_k141_72828 | Not-determined |
| S11_extracted_k141_7470 | Low-quality |
| S11_extracted_k141_40125 | Not-determined |
| S11_extracted_k141_1294 | Not-determined |
| S11_extracted_k141_13688 | Not-determined |
| S11_extracted_k141_79342 | Not-determined |
| S11_extracted_k141_85837 | Not-determined |
| S11_extracted_k141_79344 | Not-determined |
| S11_extracted_k141_33644 | Not-determined |
| S11_extracted_k141_59508 | Not-determined |
| S11_extracted_k141_79346 | Not-determined |
| S11_extracted_k141_46596 | Not-determined |
| S11_extracted_k141_7483 | Not-determined |
| S11_extracted_k141_72838 | Not-determined |
| S11_extracted_k141_59514 | Not-determined |
| S11_extracted_k141_40129 | Not-determined |
| S11_extracted_k141_33648 | Not-determined |
| S11_extracted_k141_98786 | Not-determined |
| S11_extracted_k141_72844 | Not-determined |
| S11_extracted_k141_46609 | Low-quality |
| S11_extracted_k141_85856 | Low-quality |
| S11_extracted_k141_85858 | Not-determined |
| S11_extracted_k141_53222 | Not-determined |
| S11_extracted_k141_66110 | Not-determined |
| S11_extracted_k141_66113 | Low-quality |
| S11_extracted_k141_59515 | Low-quality |
| S11_extracted_k141_40140 | Not-determined |
| S11_extracted_k141_13701 | Not-determined |
| S11_extracted_k141_85867 | Not-determined |
| S11_extracted_k141_59518 | Not-determined |
| S11_extracted_k141_13706 | Not-determined |
| S11_extracted_k141_85868 | Low-quality |
| S11_extracted_k141_27097 | Not-determined |
| S11_extracted_k141_92250 | Not-determined |
| S11_extracted_k141_85873 | Not-determined |
| S11_extracted_k141_53233 | Not-determined |
| S11_extracted_k141_85874 | Not-determined |
| S11_extracted_k141_85876 | Not-determined |
| S11_extracted_k141_79358 | Not-determined |
| S11_extracted_k141_27105 | Not-determined |
| S11_extracted_k141_66134 | Not-determined |
| S11_extracted_k141_59538 | Not-determined |
| S11_extracted_k141_59541 | Not-determined |
| S11_extracted_k141_13721 | Not-determined |
| S11_extracted_k141_13722 | Not-determined |
| S11_extracted_k141_66141 | Not-determined |
| S11_extracted_k141_33658 | Not-determined |
| S11_extracted_k141_59545 | Not-determined |
| S11_extracted_k141_33660 | Not-determined |
| S11_extracted_k141_79371 | Low-quality |
| S11_extracted_k141_33661 | Not-determined |
| S11_extracted_k141_79373 | Not-determined |
| S11_extracted_k141_85899 | Low-quality |
| S11_extracted_k141_27127 | Not-determined |
| S11_extracted_k141_7512 | Not-determined |
| S11_extracted_k141_72879 | Not-determined |
| S11_extracted_k141_72883 | Not-determined |
| S11_extracted_k141_98812 | Not-determined |
| S11_extracted_k141_53272 | Not-determined |
| S11_extracted_k141_53273 | Not-determined |
| S11_extracted_k141_66148 | Not-determined |
| S11_extracted_k141_79387 | Not-determined |
| S11_extracted_k141_79389 | Not-determined |
| S11_extracted_k141_27136 | Not-determined |
| S11_extracted_k141_66152 | Not-determined |
| S11_extracted_k141_46648 | Not-determined |
| S11_extracted_k141_72897 | Not-determined |

| | |
|---|---|
| S11_extracted_k141_92252 | Not-determined |
| S11_extracted_k141_98825 | Not-determined |
| S11_extracted_k141_27146 | Low-quality |
| S11_extracted_k141_98827 | Not-determined |
| S11_extracted_k141_7523 | Not-determined |
| S11_extracted_k141_85922 | Not-determined |
| S11_extracted_k141_85923 | Not-determined |
| S11_extracted_k141_27155 | Not-determined |
| S11_extracted_k141_46660 | Not-determined |
| S11_extracted_k141_85927 | Not-determined |
| S11_extracted_k141_85929 | Low-quality |
| S11_extracted_k141_98832 | Low-quality |
| S11_extracted_k141_98836 | Low-quality |
| S11_extracted_k141_1303 | Not-determined |
| S11_extracted_k141_1305 | Not-determined |
| S11_extracted_k141_79416 | Not-determined |
| S11_extracted_k141_92268 | Not-determined |
| S11_extracted_k141_85933 | Low-quality |
| S11_extracted_k141_1306 | Low-quality |
| S11_extracted_k141_98846 | Not-determined |
| S11_extracted_k141_33671 | Not-determined |
| S11_extracted_k141_7534 | Not-determined |
| S11_extracted_k141_72906 | Not-determined |
| S11_extracted_k141_20748 | Not-determined |
| S11_extracted_k141_7536 | Not-determined |
| S11_extracted_k141_92270 | Low-quality |
| S11_extracted_k141_1313 | Not-determined |
| S11_extracted_k141_98853 | Not-determined |
| S11_extracted_k141_98855 | Not-determined |
| S11_extracted_k141_1317 | Not-determined |
| S11_extracted_k141_20756 | Not-determined |
| S11_extracted_k141_7546 | Low-quality |
| S11_extracted_k141_53284 | Low-quality |
| S11_extracted_k141_85959 | Not-determined |
| S11_extracted_k141_33686 | Not-determined |
| S11_extracted_k141_20760 | Low-quality |
| S11_extracted_k141_7552 | Not-determined |
| S11_extracted_k141_46678 | Not-determined |
| S11_extracted_k141_53287 | Not-determined |
| S11_extracted_k141_59607 | Not-determined |
| S11_extracted_k141_59609 | Not-determined |
| S11_extracted_k141_13784 | Low-quality |
| S11_extracted_k141_59611 | Not-determined |
| S11_extracted_k141_85967 | Not-determined |
| S11_extracted_k141_59612 | Not-determined |
| S11_extracted_k141_1336 | Not-determined |
| S11_extracted_k141_7561 | Not-determined |
| S11_extracted_k141_46686 | Not-determined |
| S11_extracted_k141_7565 | Not-determined |
| S11_extracted_k141_46692 | Not-determined |
| S11_extracted_k141_72919 | Low-quality |
| S11_extracted_k141_1346 | Low-quality |
| S11_extracted_k141_59625 | Not-determined |
| S11_extracted_k141_53302 | Low-quality |
| S11_extracted_k141_20778 | Not-determined |
| S11_extracted_k141_59639 | Not-determined |
| S11_extracted_k141_53308 | Not-determined |
| S11_extracted_k141_13813 | Not-determined |
| S11_extracted_k141_92279 | Not-determined |
| S11_extracted_k141_33713 | Not-determined |
| S11_extracted_k141_1362 | Not-determined |
| S11_extracted_k141_1372 | Low-quality |
| S11_extracted_k141_1375 | Not-determined |
| S11_extracted_k141_33729 | Not-determined |
| S11_extracted_k141_20789 | Low-quality |
| S11_extracted_k141_1377 | Not-determined |
| S11_extracted_k141_1378 | Not-determined |
| S11_extracted_k141_13831 | Not-determined |
| S11_extracted_k141_59651 | Not-determined |
| S11_extracted_k141_33735 | Low-quality |
| S11_extracted_k141_1383 | Not-determined |
| S11_extracted_k141_1385 | Low-quality |
| S11_extracted_k141_53346 | Not-determined |
| S11_extracted_k141_7602 | Not-determined |
| S11_extracted_k141_7605 | Not-determined |
| S11_extracted_k141_7606 | Low-quality |
| S11_extracted_k141_13841 | Not-determined |
| S11_extracted_k141_1399 | Not-determined |
| S11_extracted_k141_72929 | Low-quality |
| S11_extracted_k141_7612 | Not-determined |
| S11_extracted_k141_1401 | Not-determined |
| S11_extracted_k141_98867 | Not-determined |
| S11_extracted_k141_13852 | Not-determined |
| S11_extracted_k141_98872 | Not-determined |
| S11_extracted_k141_59677 | Not-determined |
| S16_extracted_151450 | Complete |
| S11_extracted_k141_98877 | Low-quality |
| S11_extracted_k141_92307 | Low-quality |
| S11_extracted_k141_40152 | Not-determined |
| S11_extracted_k141_13862 | Not-determined |
| S11_extracted_k141_20801 | Not-determined |
| S11_extracted_k141_85999 | Not-determined |
| S11_extracted_k141_7630 | Not-determined |
| S11_extracted_k141_92315 | Not-determined |
| S11_extracted_k141_13865 | Not-determined |
| S11_extracted_k141_33736 | Low-quality |
| S11_extracted_k141_13866 | Not-determined |
| S11_extracted_k141_72951 | Not-determined |
| S11_extracted_k141_40165 | Not-determined |
| S11_extracted_k141_86008 | Not-determined |
| S11_extracted_k141_1420 | Not-determined |
| S11_extracted_k141_72970 | Not-determined |
| S11_extracted_k141_72976 | Not-determined |
| S11_extracted_k141_20813 | Not-determined |
| S11_extracted_k141_92319 | Not-determined |
| S11_extracted_k141_20817 | Not-determined |
| S11_extracted_k141_40180 | Not-determined |
| S11_extracted_k141_72985 | Not-determined |
| S11_extracted_k141_72989 | Not-determined |
| S11_extracted_k141_92335 | Not-determined |
| S11_extracted_k141_59708 | Not-determined |
| S11_extracted_k141_20830 | Not-determined |
| S11_extracted_k141_92336 | Not-determined |
| S11_extracted_k141_27199 | Not-determined |
| S11_extracted_k141_79418 | Low-quality |
| S11_extracted_k141_20833 | Not-determined |
| S11_extracted_k141_59713 | Not-determined |
| S11_extracted_k141_79420 | Low-quality |
| S11_extracted_k141_92343 | Low-quality |
| S11_extracted_k141_59719 | Low-quality |
| S11_extracted_k141_72998 | Not-determined |
| S11_extracted_k141_7657 | Not-determined |
| S11_extracted_k141_7658 | Not-determined |
| S11_extracted_k141_13909 | Not-determined |
| S11_extracted_k141_98891 | Not-determined |
| S11_extracted_k141_40197 | Not-determined |
| S11_extracted_k141_40199 | Not-determined |
| S11_extracted_k141_98896 | Not-determined |
| S11_extracted_k141_86033 | Not-determined |
| S11_extracted_k141_13914 | Not-determined |
| S11_extracted_k141_92356 | Not-determined |
| S11_extracted_k141_86037 | Not-determined |
| S11_extracted_k141_79427 | Not-determined |
| S11_extracted_k141_7662 | Low-quality |
| S11_extracted_k141_98899 | Low-quality |
| S11_extracted_k141_86046 | Not-determined |
| S11_extracted_k141_98902 | Not-determined |
| S11_extracted_k141_20857 | Not-determined |
| S11_extracted_k141_92367 | Not-determined |
| S11_extracted_k141_13918 | Not-determined |
| S11_extracted_k141_40202 | Low-quality |
| S11_extracted_k141_1473 | Low-quality |
| S11_extracted_k141_53394 | Low-quality |
| S11_extracted_k141_73019 | Low-quality |
| S11_extracted_k141_53399 | Not-determined |
| S11_extracted_k141_20867 | Not-determined |
| S11_extracted_k141_73023 | Not-determined |
| S11_extracted_k141_73024 | Not-determined |
| S11_extracted_k141_40214 | Not-determined |
| S11_extracted_k141_73028 | Not-determined |
| S11_extracted_k141_73029 | Low-quality |
| S11_extracted_k141_86066 | Low-quality |
| S11_extracted_k141_20878 | Not-determined |
| S11_extracted_k141_27236 | Not-determined |
| S11_extracted_k141_1490 | Not-determined |
| S11_extracted_k141_40220 | Not-determined |
| S11_extracted_k141_98910 | Not-determined |
| S11_extracted_k141_53413 | Low-quality |
| S11_extracted_k141_66216 | Not-determined |
| S11_extracted_k141_53414 | Not-determined |
| S11_extracted_k141_40221 | Not-determined |
| S11_extracted_k141_86072 | Not-determined |
| S11_extracted_k141_98913 | Not-determined |
| S11_extracted_k141_27241 | Not-determined |
| S11_extracted_k141_73040 | Not-determined |
| S11_extracted_k141_86078 | Low-quality |
| S11_extracted_k141_92387 | Not-determined |
| S11_extracted_k141_98917 | Not-determined |
| S11_extracted_k141_7693 | Low-quality |
| S11_extracted_k141_98921 | Not-determined |
| S11_extracted_k141_1494 | Not-determined |
| S11_extracted_k141_66222 | Not-determined |
| S11_extracted_k141_7695 | Not-determined |
| S11_extracted_k141_73047 | Not-determined |
| S11_extracted_k141_1497 | Not-determined |
| S11_extracted_k141_7699 | Not-determined |
| S11_extracted_k141_20892 | Low-quality |
| S11_extracted_k141_1501 | Not-determined |
| S11_extracted_k141_73054 | Not-determined |
| S11_extracted_k141_7702 | Not-determined |
| S11_extracted_k141_20897 | Not-determined |
| S11_extracted_k141_27262 | Not-determined |
| S11_extracted_k141_98929 | Not-determined |
| S11_extracted_k141_33769 | Not-determined |
| S11_extracted_k141_98936 | Not-determined |
| S11_extracted_k141_98937 | Not-determined |
| S11_extracted_k141_86087 | Not-determined |
| S11_extracted_k141_7713 | Not-determined |
| S11_extracted_k141_86093 | Not-determined |
| S11_extracted_k141_33774 | Not-determined |
| S11_extracted_k141_33775 | Not-determined |
| S11_extracted_k141_20906 | Not-determined |
| S7_extracted_58751 | Medium-quality |
| S11_extracted_k141_27276 | Not-determined |
| S11_extracted_k141_20907 | Not-determined |
| S11_extracted_k141_66238 | Not-determined |
| S11_extracted_k141_59749 | Not-determined |
| S11_extracted_k141_66240 | Not-determined |
| S11_extracted_k141_86100 | Not-determined |
| S11_extracted_k141_73075 | Not-determined |
| S11_extracted_k141_92396 | Not-determined |
| S11_extracted_k141_7721 | Not-determined |
| S11_extracted_k141_33787 | Not-determined |
| S11_extracted_k141_13938 | Not-determined |
| S11_extracted_k141_59756 | Low-quality |
| S11_extracted_k141_13939 | Not-determined |
| S11_extracted_k141_20913 | Not-determined |
| S11_extracted_k141_1522 | Low-quality |
| S11_extracted_k141_59762 | Not-determined |
| S11_extracted_k141_92412 | Not-determined |
| S11_extracted_k141_92415 | Not-determined |
| S11_extracted_k141_98955 | Not-determined |
| S11_extracted_k141_20922 | Not-determined |
| S11_extracted_k141_79449 | Not-determined |
| S11_extracted_k141_92419 | Not-determined |
| S11_extracted_k141_20925 | Not-determined |
| S11_extracted_k141_13948 | Low-quality |
| S11_extracted_k141_46742 | Not-determined |
| S11_extracted_k141_20932 | Not-determined |
| S11_extracted_k141_66256 | Not-determined |
| S11_extracted_k141_73103 | Not-determined |
| S11_extracted_k141_59769 | Low-quality |
| S11_extracted_k141_59774 | Not-determined |
| S11_extracted_k141_66263 | Not-determined |
| S11_extracted_k141_59777 | Not-determined |
| S11_extracted_k141_59780 | Not-determined |
| S11_extracted_k141_92432 | Not-determined |
| S11_extracted_k141_59782 | Not-determined |
| S11_extracted_k141_59784 | Not-determined |
| S11_extracted_k141_86115 | Low-quality |
| S11_extracted_k141_7750 | Not-determined |
| S11_extracted_k141_1541 | Not-determined |
| S11_extracted_k141_7753 | Low-quality |
| S11_extracted_k141_1545 | Low-quality |
| S11_extracted_k141_27292 | Not-determined |
| S11_extracted_k141_7758 | Not-determined |
| S11_extracted_k141_13962 | Low-quality |
| S11_extracted_k141_7761 | Not-determined |
| S11_extracted_k141_27296 | Not-determined |
| S11_extracted_k141_27297 | Not-determined |
| S11_extracted_k141_79468 | Low-quality |
| S11_extracted_k141_66272 | Not-determined |
| S11_extracted_k141_1555 | Low-quality |
| S11_extracted_k141_13972 | Not-determined |
| S11_extracted_k141_86127 | Not-determined |
| S11_extracted_k141_27302 | Not-determined |
| S11_extracted_k141_7774 | Low-quality |
| S11_extracted_k141_13975 | Not-determined |
| S11_extracted_k141_7776 | Not-determined |
| S11_extracted_k141_98987 | Not-determined |
| S11_extracted_k141_13979 | Not-determined |
| S11_extracted_k141_7779 | Not-determined |
| S11_extracted_k141_7780 | Not-determined |
| S11_extracted_k141_27311 | Not-determined |
| S11_extracted_k141_86137 | Not-determined |
| S11_extracted_k141_73138 | Not-determined |
| S11_extracted_k141_66287 | Not-determined |
| S11_extracted_k141_66288 | Not-determined |
| S11_extracted_k141_73140 | Not-determined |
| S11_extracted_k141_46758 | Not-determined |
| S11_extracted_k141_79477 | Low-quality |
| S11_extracted_k141_79478 | Not-determined |
| S11_extracted_k141_27320 | Not-determined |
| S11_extracted_k141_46762 | Not-determined |
| S11_extracted_k141_79481 | Not-determined |
| S11_extracted_k141_20964 | Not-determined |
| S11_extracted_k141_13985 | Not-determined |
| S11_extracted_k141_79482 | Not-determined |
| S11_extracted_k141_59791 | Not-determined |
| S11_extracted_k141_66300 | Not-determined |
| S11_extracted_k141_79485 | Not-determined |
| S11_extracted_k141_79487 | Not-determined |
| S11_extracted_k141_99014 | Not-determined |
| S11_extracted_k141_7795 | Not-determined |
| S11_extracted_k141_73154 | Not-determined |
| S11_extracted_k141_79488 | Low-quality |
| S11_extracted_k141_79490 | Not-determined |
| S11_extracted_k141_86153 | Not-determined |
| S11_extracted_k141_86155 | Not-determined |
| S11_extracted_k141_1587 | Low-quality |
| S11_extracted_k141_79494 | Not-determined |
| S11_extracted_k141_59809 | Low-quality |
| S11_extracted_k141_20970 | Not-determined |
| S11_extracted_k141_79497 | Not-determined |
| S11_extracted_k141_66303 | Not-determined |
| S11_extracted_k141_73165 | Not-determined |
| S11_extracted_k141_1593 | Not-determined |
| S11_extracted_k141_99022 | Not-determined |
| S11_extracted_k141_86165 | Low-quality |
| S11_extracted_k141_79500 | Low-quality |
| S11_extracted_k141_73171 | Not-determined |
| S11_extracted_k141_20981 | Not-determined |
| S11_extracted_k141_99027 | Not-determined |
| S11_extracted_k141_86170 | Not-determined |
| S11_extracted_k141_27331 | Not-determined |
| S11_extracted_k141_86171 | Low-quality |
| S11_extracted_k141_66314 | Not-determined |
| S11_extracted_k141_1605 | Not-determined |
| S11_extracted_k141_40240 | Not-determined |
| S11_extracted_k141_99035 | Not-determined |
| S11_extracted_k141_14013 | Not-determined |
| S11_extracted_k141_27335 | Not-determined |
| S11_extracted_k141_66322 | Low-quality |
| S11_extracted_k141_46787 | Not-determined |
| S11_extracted_k141_27343 | Not-determined |
| S11_extracted_k141_79515 | Not-determined |
| S11_extracted_k141_14023 | Low-quality |
| S11_extracted_k141_27345 | Not-determined |
| S11_extracted_k141_14025 | Not-determined |
| S11_extracted_k141_79518 | Not-determined |
| S11_extracted_k141_73176 | Not-determined |
| S11_extracted_k141_86192 | Not-determined |
| S11_extracted_k141_79523 | Not-determined |
| S11_extracted_k141_46796 | Low-quality |
| S11_extracted_k141_99042 | Low-quality |
| S11_extracted_k141_27356 | Not-determined |
| S11_extracted_k141_14037 | Not-determined |
| S11_extracted_k141_20994 | Low-quality |
| S11_extracted_k141_99045 | Not-determined |
| S11_extracted_k141_14040 | Not-determined |
| S11_extracted_k141_40263 | Low-quality |
| S11_extracted_k141_7843 | Not-determined |
| S11_extracted_k141_21003 | Not-determined |
| S11_extracted_k141_14053 | Low-quality |
| S11_extracted_k141_14054 | Not-determined |
| S11_extracted_k141_40281 | Not-determined |
| S11_extracted_k141_1628 | Not-determined |
| S11_extracted_k141_86211 | Not-determined |
| S11_extracted_k141_86213 | Not-determined |
| S11_extracted_k141_79549 | Not-determined |
| S11_extracted_k141_79552 | Not-determined |
| S11_extracted_k141_21016 | Low-quality |
| S11_extracted_k141_79555 | Not-determined |
| S11_extracted_k141_86219 | Not-determined |
| S11_extracted_k141_1650 | Not-determined |
| S11_extracted_k141_86221 | Not-determined |
| S11_extracted_k141_53477 | Not-determined |
| S11_extracted_k141_53478 | Not-determined |
| S11_extracted_k141_86225 | Low-quality |
| S11_extracted_k141_66341 | Not-determined |
| S11_extracted_k141_53479 | Not-determined |
| S11_extracted_k141_53481 | Not-determined |
| S11_extracted_k141_33801 | Not-determined |
| S11_extracted_k141_14072 | Not-determined |
| S11_extracted_k141_79564 | Not-determined |
| S11_extracted_k141_53485 | Not-determined |
| S11_extracted_k141_66347 | Not-determined |
| S11_extracted_k141_14075 | Not-determined |
| S11_extracted_k141_27391 | Not-determined |
| S11_extracted_k141_14081 | Not-determined |
| S11_extracted_k141_1674 | Low-quality |
| S11_extracted_k141_27395 | Not-determined |
| S11_extracted_k141_7860 | Low-quality |
| S11_extracted_k141_40309 | Not-determined |
| S11_extracted_k141_40311 | Not-determined |
| S11_extracted_k141_86230 | Low-quality |
| S11_extracted_k141_53499 | Not-determined |
| S11_extracted_k141_33822 | Not-determined |
| S11_extracted_k141_33823 | Not-determined |
| S11_extracted_k141_33825 | Low-quality |
| S11_extracted_k141_73186 | Low-quality |
| S11_extracted_k141_86240 | Not-determined |
| S11_extracted_k141_53509 | Low-quality |
| S11_extracted_k141_40326 | Not-determined |
| S11_extracted_k141_73189 | Low-quality |
| S11_extracted_k141_53513 | Not-determined |
| S11_extracted_k141_73195 | Low-quality |
| S11_extracted_k141_21034 | Not-determined |
| S11_extracted_k141_21037 | Low-quality |
| S11_extracted_k141_27407 | Not-determined |
| S11_extracted_k141_86252 | Not-determined |
| S11_extracted_k141_86253 | Not-determined |
| S11_extracted_k141_27409 | Not-determined |
| S11_extracted_k141_53524 | Not-determined |
| S11_extracted_k141_79582 | Not-determined |
| S11_extracted_k141_86259 | Not-determined |
| S11_extracted_k141_21043 | Low-quality |
| S11_extracted_k141_86267 | Not-determined |
| S11_extracted_k141_14093 | Not-determined |
| S11_extracted_k141_40339 | Not-determined |
| S11_extracted_k141_33843 | Low-quality |
| S11_extracted_k141_33848 | Not-determined |
| S11_extracted_k141_33850 | Not-determined |
| S11_extracted_k141_53544 | Not-determined |
| S11_extracted_k141_21051 | Not-determined |
| S11_extracted_k141_86277 | Not-determined |
| S11_extracted_k141_7894 | Not-determined |
| S11_extracted_k141_21056 | Not-determined |
| S11_extracted_k141_40350 | Not-determined |
| S11_extracted_k141_21059 | Not-determined |
| S11_extracted_k141_21060 | Not-determined |
| S11_extracted_k141_53550 | Not-determined |
| S11_extracted_k141_53551 | Not-determined |
| S11_extracted_k141_27433 | Not-determined |
| S11_extracted_k141_86292 | Low-quality |
| S11_extracted_k141_73227 | Low-quality |
| S11_extracted_k141_7900 | Not-determined |
| S11_extracted_k141_14110 | Not-determined |
| S11_extracted_k141_14111 | Not-determined |
| S11_extracted_k141_40352 | Low-quality |
| S11_extracted_k141_40354 | Not-determined |
| S11_extracted_k141_86300 | Not-determined |
| S11_extracted_k141_86306 | Not-determined |
| S11_extracted_k141_40362 | Not-determined |
| S11_extracted_k141_86310 | Not-determined |
| S11_extracted_k141_14129 | Not-determined |
| S11_extracted_k141_14132 | Not-determined |
| S11_extracted_k141_86314 | Not-determined |
| S11_extracted_k141_53568 | Not-determined |
| S11_extracted_k141_14134 | Not-determined |
| S11_extracted_k141_53571 | Not-determined |
| S11_extracted_k141_14140 | Not-determined |
| S11_extracted_k141_99068 | Not-determined |
| S11_extracted_k141_14142 | Not-determined |
| S11_extracted_k141_14143 | Not-determined |
| S11_extracted_k141_40367 | Low-quality |
| S11_extracted_k141_40368 | Not-determined |
| S11_extracted_k141_40369 | Not-determined |
| S11_extracted_k141_14146 | Low-quality |
| S11_extracted_k141_27457 | Not-determined |
| S11_extracted_k141_21063 | Low-quality |
| S11_extracted_k141_33868 | Not-determined |
| S11_extracted_k141_73237 | Not-determined |
| S11_extracted_k141_7902 | Not-determined |
| S11_extracted_k141_59821 | Low-quality |
| S11_extracted_k141_7903 | Not-determined |
| S11_extracted_k141_73241 | Not-determined |
| S11_extracted_k141_99076 | Not-determined |
| S11_extracted_k141_53577 | Not-determined |
| S11_extracted_k141_7908 | Not-determined |
| S11_extracted_k141_53578 | Not-determined |
| S11_extracted_k141_46848 | Low-quality |
| S11_extracted_k141_1704 | Not-determined |
| S11_extracted_k141_14151 | Low-quality |
| S11_extracted_k141_79611 | Not-determined |
| S11_extracted_k141_46854 | Not-determined |
| S11_extracted_k141_46855 | Not-determined |
| S11_extracted_k141_21073 | Low-quality |
| S11_extracted_k141_1711 | Not-determined |
| S11_extracted_k141_40378 | Low-quality |
| S11_extracted_k141_53586 | Not-determined |
| S11_extracted_k141_27484 | Not-determined |
| S11_extracted_k141_40382 | Not-determined |
| S11_extracted_k141_1716 | Not-determined |
| S11_extracted_k141_27488 | Not-determined |
| S11_extracted_k141_73252 | Not-determined |
| S11_extracted_k141_27491 | Not-determined |
| S11_extracted_k141_59838 | Not-determined |
| S11_extracted_k141_27495 | Not-determined |
| S11_extracted_k141_14161 | Not-determined |
| S11_extracted_k141_59843 | Not-determined |
| S11_extracted_k141_73258 | Not-determined |
| S11_extracted_k141_40395 | Not-determined |
| S11_extracted_k141_59844 | Not-determined |
| S11_extracted_k141_59845 | Not-determined |
| S11_extracted_k141_7927 | Not-determined |
| S11_extracted_k141_53591 | Not-determined |
| S11_extracted_k141_7930 | Not-determined |
| S11_extracted_k141_73260 | Not-determined |
| S11_extracted_k141_7933 | Not-determined |
| S11_extracted_k141_40401 | Low-quality |
| S11_extracted_k141_1735 | Not-determined |
| S11_extracted_k141_53599 | Not-determined |
| S11_extracted_k141_40403 | Not-determined |
| S11_extracted_k141_86349 | Not-determined |
| S11_extracted_k141_73266 | Not-determined |
| S11_extracted_k141_1739 | Not-determined |
| S11_extracted_k141_73268 | Not-determined |
| S11_extracted_k141_40407 | Not-determined |
| S11_extracted_k141_99085 | Not-determined |
| S11_extracted_k141_40408 | Not-determined |
| S11_extracted_k141_73271 | Not-determined |
| S11_extracted_k141_59858 | Not-determined |
| S11_extracted_k141_92483 | Not-determined |
| S11_extracted_k141_27515 | Not-determined |
| S11_extracted_k141_7944 | Not-determined |
| S11_extracted_k141_27518 | Not-determined |
| S11_extracted_k141_7947 | Not-determined |
| S11_extracted_k141_27519 | Not-determined |
| S11_extracted_k141_99089 | Not-determined |
| S11_extracted_k141_27520 | Not-determined |
| S11_extracted_k141_21106 | Not-determined |
| S11_extracted_k141_46871 | Not-determined |
| S11_extracted_k141_27526 | Not-determined |
| S11_extracted_k141_21109 | Not-determined |
| S11_extracted_k141_21110 | Not-determined |
| S11_extracted_k141_46876 | Not-determined |
| S11_extracted_k141_14176 | Not-determined |
| S11_extracted_k141_79638 | Not-determined |
| S11_extracted_k141_7960 | Low-quality |
| S11_extracted_k141_14183 | Not-determined |
| S11_extracted_k141_79641 | Low-quality |
| S11_extracted_k141_46885 | Not-determined |
| S11_extracted_k141_46887 | Not-determined |
| S11_extracted_k141_59868 | Not-determined |
| S11_extracted_k141_66409 | Not-determined |
| S11_extracted_k141_59870 | Not-determined |
| S11_extracted_k141_27540 | Not-determined |
| S11_extracted_k141_59872 | Not-determined |
| S11_extracted_k141_79651 | Not-determined |
| S11_extracted_k141_40431 | Low-quality |
| S11_extracted_k141_14200 | Not-determined |
| S11_extracted_k141_79653 | Not-determined |
| S11_extracted_k141_59877 | Not-determined |
| S11_extracted_k141_40437 | Not-determined |
| S11_extracted_k141_79660 | Not-determined |
| S11_extracted_k141_46902 | Not-determined |
| S11_extracted_k141_79663 | Not-determined |
| S11_extracted_k141_7978 | Not-determined |
| S11_extracted_k141_86357 | Low-quality |
| S11_extracted_k141_46906 | Not-determined |
| S11_extracted_k141_53616 | Low-quality |
| S11_extracted_k141_79667 | Low-quality |
| S11_extracted_k141_46908 | Low-quality |
| S11_extracted_k141_66426 | Not-determined |
| S11_extracted_k141_7984 | Low-quality |
| S11_extracted_k141_1779 | Not-determined |
| S11_extracted_k141_40450 | Not-determined |
| S11_extracted_k141_14218 | Not-determined |
| S11_extracted_k141_14220 | Not-determined |
| S11_extracted_k141_40454 | Not-determined |
| S11_extracted_k141_7990 | Not-determined |
| S11_extracted_k141_86368 | Not-determined |
| S11_extracted_k141_40456 | Not-determined |
| S11_extracted_k141_86369 | Not-determined |
| S11_extracted_k141_66440 | Not-determined |
| S11_extracted_k141_27571 | Not-determined |
| S11_extracted_k141_1786 | Not-determined |
| S11_extracted_k141_59893 | Not-determined |
| S11_extracted_k141_53627 | Not-determined |
| S11_extracted_k141_1787 | Not-determined |
| S11_extracted_k141_1788 | Not-determined |
| S11_extracted_k141_46924 | Not-determined |
| S11_extracted_k141_14227 | Low-quality |
| S11_extracted_k141_1800 | Low-quality |
| S11_extracted_k141_59907 | Not-determined |
| S11_extracted_k141_53639 | Not-determined |
| S11_extracted_k141_21136 | Not-determined |
| S11_extracted_k141_14230 | Not-determined |
| S11_extracted_k141_27578 | Not-determined |
| S11_extracted_k141_27579 | Not-determined |
| S11_extracted_k141_1807 | Not-determined |
| S11_extracted_k141_59913 | Not-determined |
| S11_extracted_k141_21139 | Not-determined |
| S11_extracted_k141_8007 | Not-determined |
| S11_extracted_k141_53646 | Not-determined |
| S11_extracted_k141_46935 | Low-quality |
| S11_extracted_k141_79683 | Low-quality |
| S11_extracted_k141_99104 | Not-determined |
| S11_extracted_k141_79685 | Not-determined |
| S11_extracted_k141_8014 | Not-determined |
| S11_extracted_k141_79689 | Not-determined |
| S11_extracted_k141_79691 | Not-determined |
| S11_extracted_k141_79693 | Not-determined |
| S11_extracted_k141_99107 | Not-determined |
| S11_extracted_k141_79696 | Not-determined |
| S11_extracted_k141_99110 | Low-quality |
| S11_extracted_k141_99111 | Not-determined |
| S11_extracted_k141_86402 | Not-determined |
| S11_extracted_k141_14248 | Not-determined |
| S11_extracted_k141_79703 | Low-quality |
| S11_extracted_k141_86404 | Not-determined |
| S11_extracted_k141_14251 | Low-quality |
| S11_extracted_k141_99114 | Low-quality |
| S11_extracted_k141_99115 | Not-determined |
| S11_extracted_k141_66456 | Not-determined |
| S11_extracted_k141_79709 | Not-determined |
| S11_extracted_k141_21148 | Not-determined |
| S11_extracted_k141_99119 | Not-determined |
| S11_extracted_k141_27602 | Low-quality |
| S11_extracted_k141_14262 | Not-determined |
| S11_extracted_k141_99122 | Not-determined |
| S11_extracted_k141_99124 | Not-determined |
| S11_extracted_k141_46962 | Not-determined |
| S11_extracted_k141_73330 | Not-determined |
| S11_extracted_k141_73331 | Low-quality |
| S11_extracted_k141_79724 | Not-determined |
| S11_extracted_k141_46969 | Not-determined |
| S11_extracted_k141_27614 | Not-determined |
| S11_extracted_k141_14270 | Low-quality |
| S11_extracted_k141_99134 | Not-determined |
| S11_extracted_k141_79729 | Low-quality |
| S11_extracted_k141_1840 | Not-determined |
| S11_extracted_k141_21166 | Not-determined |
| S11_extracted_k141_8030 | Not-determined |
| S11_extracted_k141_8032 | Low-quality |
| S11_extracted_k141_99136 | Not-determined |
| S11_extracted_k141_46975 | Not-determined |
| S11_extracted_k141_27626 | Not-determined |

| | |
|---|---|
| S11_extracted_k141_8040 | Not-determined |
| S11_extracted_k141_99141 | Low-quality |
| S11_extracted_k141_27628 | Not-determined |
| S11_extracted_k141_73348 | Not-determined |
| S11_extracted_k141_46978 | Not-determined |
| S11_extracted_k141_66459 | Not-determined |
| S11_extracted_k141_27631 | Not-determined |
| S11_extracted_k141_73351 | Low-quality |
| S11_extracted_k141_66462 | Not-determined |
| S11_extracted_k141_59928 | Low-quality |
| S11_extracted_k141_59929 | Not-determined |
| S11_extracted_k141_59934 | Not-determined |
| S11_extracted_k141_21204 | Not-determined |
| S11_extracted_k141_66467 | Low-quality |
| S11_extracted_k141_21211 | Not-determined |
| S11_extracted_k141_59945 | Not-determined |
| S11_extracted_k141_14299 | Not-determined |
| S11_extracted_k141_59946 | Not-determined |
| S11_extracted_k141_59948 | Not-determined |
| S11_extracted_k141_8055 | Not-determined |
| S11_extracted_k141_99162 | Not-determined |
| S11_extracted_k141_46984 | Not-determined |
| S11_extracted_k141_99166 | Low-quality |
| S11_extracted_k141_99167 | Not-determined |
| S11_extracted_k141_8060 | Not-determined |
| S11_extracted_k141_99172 | Not-determined |
| S11_extracted_k141_14315 | Not-determined |
| S11_extracted_k141_14323 | Not-determined |
| S11_extracted_k141_66485 | Low-quality |
| S11_extracted_k141_53652 | Not-determined |
| S11_extracted_k141_79774 | Not-determined |
| S11_extracted_k141_8076 | Not-determined |
| S11_extracted_k141_27685 | Not-determined |
| S11_extracted_k141_1851 | Low-quality |
| S11_extracted_k141_46997 | Not-determined |
| S11_extracted_k141_79777 | Not-determined |
| S11_extracted_k141_66491 | Not-determined |
| S11_extracted_k141_8080 | Not-determined |
| S11_extracted_k141_27695 | Not-determined |
| S11_extracted_k141_47007 | Not-determined |
| S11_extracted_k141_8088 | Not-determined |
| S11_extracted_k141_1865 | Not-determined |
| S11_extracted_k141_47009 | Not-determined |
| S11_extracted_k141_47011 | Not-determined |
| S11_extracted_k141_73386 | Low-quality |
| S11_extracted_k141_53666 | Not-determined |
| S11_extracted_k141_59955 | Low-quality |
| S11_extracted_k141_59956 | Not-determined |
| S11_extracted_k141_66513 | Not-determined |
| S11_extracted_k141_1883 | Not-determined |
| S11_extracted_k141_79790 | Low-quality |
| S11_extracted_k141_79792 | Not-determined |
| S11_extracted_k141_99189 | Not-determined |
| S11_extracted_k141_1886 | Not-determined |
| S11_extracted_k141_66518 | Not-determined |
| S11_extracted_k141_21249 | Not-determined |
| S11_extracted_k141_99194 | Not-determined |
| S11_extracted_k141_27700 | Not-determined |
| S11_extracted_k141_8110 | Not-determined |
| S11_extracted_k141_14349 | Low-quality |
| S11_extracted_k141_14354 | Low-quality |
| S11_extracted_k141_47024 | Low-quality |
| S11_extracted_k141_92531 | Not-determined |
| S11_extracted_k141_92532 | Not-determined |
| S11_extracted_k141_59975 | Not-determined |
| S11_extracted_k141_47030 | Not-determined |
| S11_extracted_k141_21258 | Low-quality |
| S11_extracted_k141_14360 | Not-determined |
| S11_extracted_k141_27709 | Low-quality |
| S11_extracted_k141_8125 | Not-determined |
| S11_extracted_k141_8126 | Low-quality |
| S11_extracted_k141_1903 | Not-determined |
| S11_extracted_k141_8131 | Low-quality |
| S11_extracted_k141_47036 | Not-determined |
| S11_extracted_k141_8133 | Not-determined |
| S11_extracted_k141_99201 | Not-determined |
| S11_extracted_k141_8136 | Not-determined |
| S11_extracted_k141_59982 | Not-determined |
| S11_extracted_k141_1906 | Not-determined |
| S11_extracted_k141_59984 | Not-determined |
| S11_extracted_k141_47046 | Not-determined |
| S11_extracted_k141_53677 | Not-determined |
| S11_extracted_k141_8145 | Low-quality |
| S11_extracted_k141_27716 | Not-determined |
| S11_extracted_k141_14378 | Not-determined |
| S11_extracted_k141_8150 | Low-quality |
| S11_extracted_k141_47054 | Not-determined |
| S11_extracted_k141_27719 | Low-quality |
| S11_extracted_k141_92548 | Not-determined |
| S11_extracted_k141_53688 | Not-determined |
| S11_extracted_k141_92551 | Not-determined |
| S11_extracted_k141_27725 | Not-determined |
| S11_extracted_k141_53689 | Not-determined |
| S11_extracted_k141_21283 | Not-determined |
| S11_extracted_k141_47060 | Not-determined |
| S11_extracted_k141_8158 | Not-determined |
| S11_extracted_k141_47062 | Not-determined |
| S16_vs_151450\|\|full | High-quality |
| S11_extracted_k141_53695 | Not-determined |
| S11_extracted_k141_27732 | Not-determined |
| S11_extracted_k141_79822 | Low-quality |
| S11_extracted_k141_79824 | Not-determined |
| S11_extracted_k141_59992 | Low-quality |
| S11_extracted_k141_92560 | Low-quality |
| S11_extracted_k141_21295 | Not-determined |
| S11_extracted_k141_8167 | Not-determined |
| S11_extracted_k141_92562 | Not-determined |
| S11_extracted_k141_59996 | Not-determined |
| S11_extracted_k141_99213 | Not-determined |
| S11_extracted_k141_66535 | Not-determined |
| S11_extracted_k141_27738 | Not-determined |
| S11_extracted_k141_47069 | Low-quality |
| S11_extracted_k141_99218 | Not-determined |
| S11_extracted_k141_92567 | Not-determined |
| S11_extracted_k141_73392 | Not-determined |
| S11_extracted_k141_92569 | Not-determined |
| S11_extracted_k141_47075 | Not-determined |
| S11_extracted_k141_47080 | Not-determined |
| S11_extracted_k141_99225 | Low-quality |
| S11_extracted_k141_8177 | Not-determined |
| S11_extracted_k141_8179 | Not-determined |
| S11_extracted_k141_92575 | Not-determined |
| S11_extracted_k141_60019 | Not-determined |
| S11_extracted_k141_27750 | Not-determined |
| S11_extracted_k141_40460 | Not-determined |
| S11_extracted_k141_99236 | Not-determined |
| S11_extracted_k141_47098 | Not-determined |
| S11_extracted_k141_14415 | Not-determined |
| S11_extracted_k141_53719 | Not-determined |
| S11_extracted_k141_14419 | Not-determined |
| S11_extracted_k141_27751 | Not-determined |
| S11_extracted_k141_60029 | Not-determined |
| S11_extracted_k141_21309 | Not-determined |
| S11_extracted_k141_40468 | Not-determined |
| S11_extracted_k141_14422 | Not-determined |
| S11_extracted_k141_60033 | Not-determined |
| S11_extracted_k141_40471 | Not-determined |
| S11_extracted_k141_14426 | Not-determined |
| S11_extracted_k141_53727 | Not-determined |
| S11_extracted_k141_27762 | Not-determined |
| S11_extracted_k141_14427 | Not-determined |
| S11_extracted_k141_14428 | Not-determined |
| S11_extracted_k141_21320 | Not-determined |
| S11_extracted_k141_92583 | Not-determined |
| S11_extracted_k141_66551 | Low-quality |
| S11_extracted_k141_66554 | Low-quality |
| S11_extracted_k141_53734 | Not-determined |
| S11_extracted_k141_14435 | Low-quality |
| S11_extracted_k141_60048 | Low-quality |
| S11_extracted_k141_40482 | Not-determined |
| S11_extracted_k141_92587 | Not-determined |
| S11_extracted_k141_53737 | Not-determined |
| S11_extracted_k141_99242 | Low-quality |
| S11_extracted_k141_99243 | Not-determined |
| S11_extracted_k141_21338 | Not-determined |
| S11_extracted_k141_14444 | Not-determined |
| S11_extracted_k141_92590 | Low-quality |
| S11_extracted_k141_60053 | Not-determined |
| S11_extracted_k141_73425 | Not-determined |
| S11_extracted_k141_14451 | Not-determined |
| S11_extracted_k141_21341 | Not-determined |
| S11_extracted_k141_47102 | Low-quality |
| S11_extracted_k141_21345 | Not-determined |
| S11_extracted_k141_47104 | Low-quality |
| S11_extracted_k141_53747 | Low-quality |
| S11_extracted_k141_60062 | Not-determined |
| S11_extracted_k141_40494 | Not-determined |
| S11_extracted_k141_66572 | Low-quality |
| S11_extracted_k141_21357 | Not-determined |
| S11_extracted_k141_60071 | Not-determined |
| S11_extracted_k141_66574 | Not-determined |
| S11_extracted_k141_47116 | Low-quality |
| S11_extracted_k141_99249 | Not-determined |
| S11_extracted_k141_47121 | Not-determined |
| S11_extracted_k141_53758 | Not-determined |
| S11_extracted_k141_92616 | Not-determined |
| S11_extracted_k141_47124 | Not-determined |
| S11_extracted_k141_40509 | Not-determined |
| S11_extracted_k141_99254 | Not-determined |
| S11_extracted_k141_92623 | Not-determined |
| S11_extracted_k141_99255 | Not-determined |
| S11_extracted_k141_99256 | Not-determined |
| S11_extracted_k141_53766 | Not-determined |
| S11_extracted_k141_21363 | Low-quality |
| S11_extracted_k141_21364 | Not-determined |
| S11_extracted_k141_99262 | Not-determined |
| S11_extracted_k141_40512 | Not-determined |
| S11_extracted_k141_99263 | Not-determined |
| S11_extracted_k141_21367 | Not-determined |
| S11_extracted_k141_66584 | Not-determined |
| S11_extracted_k141_60072 | Not-determined |
| S11_extracted_k141_73439 | Not-determined |
| S11_extracted_k141_99272 | Not-determined |
| S11_extracted_k141_27813 | Not-determined |
| S11_extracted_k141_92645 | Low-quality |
| S11_extracted_k141_60083 | Not-determined |
| S11_extracted_k141_73449 | Not-determined |
| S11_extracted_k141_40533 | Not-determined |
| S11_extracted_k141_27816 | Not-determined |
| S11_extracted_k141_40536 | Low-quality |
| S11_extracted_k141_60090 | Low-quality |
| S11_extracted_k141_92653 | Low-quality |
| S11_extracted_k141_14487 | Low-quality |
| S11_extracted_k141_14488 | Not-determined |
| S11_extracted_k141_21386 | Not-determined |
| S11_extracted_k141_14489 | Not-determined |
| S11_extracted_k141_47141 | Not-determined |
| S11_extracted_k141_27829 | Low-quality |
| S11_extracted_k141_8239 | Not-determined |
| S11_extracted_k141_99282 | Not-determined |
| S11_extracted_k141_14496 | Not-determined |
| S11_extracted_k141_1941 | Not-determined |
| S11_extracted_k141_66629 | Low-quality |
| S11_extracted_k141_14499 | Not-determined |
| S11_extracted_k141_40546 | Not-determined |
| S11_extracted_k141_99287 | Low-quality |
| S11_extracted_k141_1942 | Low-quality |
| S11_extracted_k141_73456 | Not-determined |
| S11_extracted_k141_27838 | Not-determined |
| S11_extracted_k141_73463 | Not-determined |
| S11_extracted_k141_53806 | Not-determined |
| S11_extracted_k141_1947 | Not-determined |
| S11_extracted_k141_27842 | Low-quality |
| S11_extracted_k141_27844 | Not-determined |
| S11_extracted_k141_86439 | Low-quality |
| S11_extracted_k141_66647 | Low-quality |
| S11_extracted_k141_21396 | Not-determined |
| S11_extracted_k141_92665 | Not-determined |
| S11_extracted_k141_73476 | Low-quality |
| S11_extracted_k141_92667 | Not-determined |
| S11_extracted_k141_92669 | Not-determined |
| S11_extracted_k141_86455 | Low-quality |
| S11_extracted_k141_92672 | Not-determined |
| S11_extracted_k141_14516 | Low-quality |
| S11_extracted_k141_14517 | Not-determined |
| S11_extracted_k141_79842 | Low-quality |
| S11_extracted_k141_92682 | Not-determined |
| S11_extracted_k141_14524 | Not-determined |
| S11_extracted_k141_60103 | Not-determined |
| S11_extracted_k141_60105 | Not-determined |
| S11_extracted_k141_66655 | Not-determined |
| S11_extracted_k141_92689 | Not-determined |
| S11_extracted_k141_79851 | Not-determined |
| S11_extracted_k141_53833 | Not-determined |
| S11_extracted_k141_47144 | Not-determined |
| S11_extracted_k141_21412 | Low-quality |
| S11_extracted_k141_60110 | Not-determined |
| S11_extracted_k141_40574 | Not-determined |
| S11_extracted_k141_86461 | Not-determined |
| S11_extracted_k141_8271 | Not-determined |
| S11_extracted_k141_53843 | Not-determined |
| S11_extracted_k141_73486 | Not-determined |
| S11_extracted_k141_73488 | Not-determined |
| S11_extracted_k141_92698 | Not-determined |
| S11_extracted_k141_21419 | Not-determined |
| S11_extracted_k141_21421 | Not-determined |
| S11_extracted_k141_21422 | Low-quality |
| S11_extracted_k141_86474 | Not-determined |
| S11_extracted_k141_8278 | Not-determined |
| S11_extracted_k141_21424 | Not-determined |
| S11_extracted_k141_1978 | Not-determined |
| S11_extracted_k141_40586 | Not-determined |
| S11_extracted_k141_79858 | Not-determined |
| S11_extracted_k141_1981 | Not-determined |
| S11_extracted_k141_40590 | Low-quality |
| S11_extracted_k141_40592 | Not-determined |
| S11_extracted_k141_14535 | Not-determined |
| S11_extracted_k141_1988 | Not-determined |
| S11_extracted_k141_60123 | Low-quality |
| S11_extracted_k141_8288 | Not-determined |
| S11_extracted_k141_60127 | Not-determined |
| S11_extracted_k141_60128 | Not-determined |
| S11_extracted_k141_53857 | Low-quality |
| S11_extracted_k141_79860 | Not-determined |
| S11_extracted_k141_40601 | Not-determined |
| S11_extracted_k141_47161 | Not-determined |
| S11_extracted_k141_79865 | Low-quality |
| S11_extracted_k141_60133 | Low-quality |
| S11_extracted_k141_47166 | Not-determined |
| S11_extracted_k141_99327 | Not-determined |
| S11_extracted_k141_92712 | Low-quality |
| S11_extracted_k141_53861 | Low-quality |
| S11_extracted_k141_14544 | Not-determined |
| S11_extracted_k141_53865 | Not-determined |
| S11_extracted_k141_92717 | Not-determined |
| S11_extracted_k141_79867 | Low-quality |
| S11_extracted_k141_47174 | Not-determined |
| S11_extracted_k141_53868 | Not-determined |
| S11_extracted_k141_73515 | Not-determined |
| S11_extracted_k141_1994 | Not-determined |
| S11_extracted_k141_73516 | Not-determined |
| S11_extracted_k141_1995 | Not-determined |
| S11_extracted_k141_14547 | Not-determined |
| S11_extracted_k141_33904 | Not-determined |
| S11_extracted_k141_2001 | Not-determined |
| S11_extracted_k141_47177 | Not-determined |
| S11_extracted_k141_79874 | Not-determined |
| S11_extracted_k141_79880 | Not-determined |
| S11_extracted_k141_99342 | Not-determined |
| S11_extracted_k141_21441 | Not-determined |
| S11_extracted_k141_79884 | Not-determined |
| S11_extracted_k141_79885 | Not-determined |
| S11_extracted_k141_33908 | Not-determined |
| S11_extracted_k141_21442 | Low-quality |
| S11_extracted_k141_47189 | Not-determined |
| S11_extracted_k141_53885 | Not-determined |
| S11_extracted_k141_33911 | Not-determined |
| S11_extracted_k141_40617 | Low-quality |
| S11_extracted_k141_47192 | Not-determined |
| S11_extracted_k141_40619 | Not-determined |
| S11_extracted_k141_99357 | Not-determined |
| S11_extracted_k141_99358 | Low-quality |
| S11_extracted_k141_2017 | Low-quality |
| S11_extracted_k141_79896 | Not-determined |
| S11_extracted_k141_92736 | Not-determined |
| S11_extracted_k141_86504 | Not-determined |
| S11_extracted_k141_79898 | Not-determined |
| S11_extracted_k141_21455 | Not-determined |
| S11_extracted_k141_33918 | Not-determined |
| S11_extracted_k141_33919 | Not-determined |
| S11_extracted_k141_99360 | Not-determined |
| S11_extracted_k141_53889 | Not-determined |
| S11_extracted_k141_99363 | Not-determined |
| S11_extracted_k141_40632 | Not-determined |
| S11_extracted_k141_66685 | Not-determined |
| S8_extracted_k141_49008 | Complete |
| S11_extracted_k141_33926 | Not-determined |
| S11_extracted_k141_33927 | Not-determined |
| S11_extracted_k141_2024 | Not-determined |
| S11_extracted_k141_53893 | Not-determined |
| S11_extracted_k141_86517 | Low-quality |
| S11_extracted_k141_92740 | Not-determined |
| S11_extracted_k141_86519 | Not-determined |
| S11_extracted_k141_8320 | Not-determined |
| S11_extracted_k141_27848 | Low-quality |
| S11_extracted_k141_21466 | Not-determined |
| S11_extracted_k141_53899 | Not-determined |
| S11_extracted_k141_92743 | Not-determined |
| S11_extracted_k141_21468 | Low-quality |
| S11_extracted_k141_2030 | Not-determined |
| S11_extracted_k141_86524 | Not-determined |
| S11_extracted_k141_47202 | Not-determined |
| S11_extracted_k141_53905 | Not-determined |
| S11_extracted_k141_92749 | Not-determined |
| S11_extracted_k141_73558 | Not-determined |
| S11_extracted_k141_47212 | Not-determined |
| S11_extracted_k141_27853 | Not-determined |
| S11_extracted_k141_66698 | Not-determined |
| S11_extracted_k141_27856 | Not-determined |
| S11_extracted_k141_86534 | Not-determined |
| S11_extracted_k141_2046 | Not-determined |
| S11_extracted_k141_47214 | Not-determined |
| S11_extracted_k141_53914 | Low-quality |
| S11_extracted_k141_21485 | Not-determined |
| S11_extracted_k141_66705 | Not-determined |
| S11_extracted_k141_33952 | Not-determined |
| S11_extracted_k141_92760 | Not-determined |
| S11_extracted_k141_47215 | Not-determined |
| S11_extracted_k141_21490 | Not-determined |
| S11_extracted_k141_27864 | Not-determined |
| S11_extracted_k141_33958 | Not-determined |
| S11_extracted_k141_86549 | Low-quality |
| S11_extracted_k141_27870 | Not-determined |
| S11_extracted_k141_79935 | Not-determined |
| S11_extracted_k141_92775 | Not-determined |
| S11_extracted_k141_79941 | Not-determined |
| S11_extracted_k141_53923 | Not-determined |
| S11_extracted_k141_92778 | Not-determined |
| S11_extracted_k141_21511 | Low-quality |
| S11_extracted_k141_92779 | Not-determined |
| S11_extracted_k141_79947 | Not-determined |
| S11_extracted_k141_53926 | Not-determined |
| S11_extracted_k141_40636 | Not-determined |
| S11_extracted_k141_73579 | Not-determined |
| S11_extracted_k141_79954 | Not-determined |
| S11_extracted_k141_21517 | Not-determined |
| S11_extracted_k141_53933 | Not-determined |
| S11_extracted_k141_92784 | Low-quality |
| S11_extracted_k141_99393 | Not-determined |
| S11_extracted_k141_99394 | Not-determined |
| S11_extracted_k141_21521 | Low-quality |
| S11_extracted_k141_86572 | Not-determined |
| S11_extracted_k141_8335 | Not-determined |
| S11_extracted_k141_79958 | Not-determined |
| S11_extracted_k141_21523 | Low-quality |
| S11_extracted_k141_79959 | Not-determined |
| S11_extracted_k141_99399 | Not-determined |
| S11_extracted_k141_8343 | Not-determined |
| S11_extracted_k141_8344 | Not-determined |
| S11_extracted_k141_53942 | Low-quality |
| S11_extracted_k141_86587 | Not-determined |
| S11_extracted_k141_8356 | Low-quality |
| S11_extracted_k141_53949 | Not-determined |
| S11_extracted_k141_99414 | Not-determined |
| S11_extracted_k141_8361 | Not-determined |
| S11_extracted_k141_47253 | Not-determined |
| S11_extracted_k141_79975 | Not-determined |
| S11_extracted_k141_86595 | Low-quality |
| S11_extracted_k141_8369 | Not-determined |
| S11_extracted_k141_40663 | Low-quality |
| S11_extracted_k141_8373 | Not-determined |
| S11_extracted_k141_60153 | Not-determined |
| S11_extracted_k141_2065 | Not-determined |
| S11_extracted_k141_73606 | Not-determined |
| S11_extracted_k141_99431 | Not-determined |
| S11_extracted_k141_86607 | Low-quality |
| S11_extracted_k141_79979 | Low-quality |
| S11_extracted_k141_2069 | Not-determined |
| S11_extracted_k141_73612 | Low-quality |
| S11_extracted_k141_21555 | Not-determined |
| S11_extracted_k141_2077 | Not-determined |
| S11_extracted_k141_8383 | Not-determined |
| S11_extracted_k141_53961 | Low-quality |
| S11_extracted_k141_79990 | Low-quality |
| S11_extracted_k141_53965 | Not-determined |
| S11_extracted_k141_99445 | Not-determined |
| S11_extracted_k141_21564 | Not-determined |
| S11_extracted_k141_21566 | Low-quality |
| S11_extracted_k141_40679 | Not-determined |
| S11_extracted_k141_8394 | Low-quality |
| S11_extracted_k141_21570 | Not-determined |
| S11_extracted_k141_21572 | Not-determined |
| S11_extracted_k141_53971 | Not-determined |
| S11_extracted_k141_86624 | Not-determined |
| S11_extracted_k141_2087 | Not-determined |
| S11_extracted_k141_21575 | Low-quality |
| S11_extracted_k141_80005 | Low-quality |
| S11_extracted_k141_2096 | Not-determined |
| S11_extracted_k141_40691 | Not-determined |
| S11_extracted_k141_40692 | Not-determined |
| S11_extracted_k141_33978 | Not-determined |
| S11_extracted_k141_99458 | Not-determined |
| S11_extracted_k141_53988 | Not-determined |
| S11_extracted_k141_53993 | Not-determined |
| S11_extracted_k141_53996 | Low-quality |
| S11_extracted_k141_53997 | Not-determined |
| S11_extracted_k141_8405 | Low-quality |
| S11_extracted_k141_92796 | Not-determined |
| S11_extracted_k141_8408 | Not-determined |
| S11_extracted_k141_8410 | Not-determined |
| S11_extracted_k141_33989 | Not-determined |
| S11_extracted_k141_8415 | Not-determined |
| S11_extracted_k141_92804 | Low-quality |
| S11_extracted_k141_54012 | Not-determined |
| S11_extracted_k141_92808 | Low-quality |
| S11_extracted_k141_92810 | Not-determined |
| S11_extracted_k141_54014 | Not-determined |
| S11_extracted_k141_54015 | Not-determined |
| S11_extracted_k141_80021 | Not-determined |
| S11_extracted_k141_54018 | Not-determined |
| S11_extracted_k141_80022 | Not-determined |
| S11_extracted_k141_92818 | Not-determined |
| S11_extracted_k141_14550 | Not-determined |
| S11_extracted_k141_14552 | Not-determined |
| S11_extracted_k141_2138 | Not-determined |
| S11_extracted_k141_2140 | Not-determined |
| S11_extracted_k141_86650 | Low-quality |
| S11_extracted_k141_60175 | Not-determined |
| S11_extracted_k141_14562 | Low-quality |
| S11_extracted_k141_8430 | Not-determined |
| S11_extracted_k141_8432 | Not-determined |
| S11_extracted_k141_54040 | Not-determined |
| S11_extracted_k141_54042 | Low-quality |
| S11_extracted_k141_54044 | Low-quality |
| S11_extracted_k141_14571 | Not-determined |
| S11_extracted_k141_80031 | Not-determined |
| S11_extracted_k141_8442 | Not-determined |
| S11_extracted_k141_8443 | Low-quality |
| S11_extracted_k141_80032 | Not-determined |
| S11_extracted_k141_80033 | Not-determined |
| S11_extracted_k141_99471 | Low-quality |
| S11_extracted_k141_2155 | Low-quality |
| S11_extracted_k141_86664 | Not-determined |
| S11_extracted_k141_40714 | Low-quality |
| S11_extracted_k141_40715 | Not-determined |
| S11_extracted_k141_73618 | Not-determined |
| S11_extracted_k141_99478 | Not-determined |
| S11_extracted_k141_86671 | Not-determined |
| S11_extracted_k141_99480 | Not-determined |
| S11_extracted_k141_40722 | Not-determined |
| S11_extracted_k141_73620 | Not-determined |
| S11_extracted_k141_27900 | Not-determined |
| S11_extracted_k141_99486 | Not-determined |
| S11_extracted_k141_80051 | Low-quality |
| S11_extracted_k141_73624 | Not-determined |
| S11_extracted_k141_99490 | Not-determined |
| S11_extracted_k141_2167 | Not-determined |
| S11_extracted_k141_40729 | Not-determined |
| S11_extracted_k141_2170 | Not-determined |
| S11_extracted_k141_80055 | Not-determined |
| S11_extracted_k141_80056 | Not-determined |
| S11_extracted_k141_86685 | Low-quality |
| S11_extracted_k141_99500 | Not-determined |
| S11_extracted_k141_2172 | Not-determined |
| S11_extracted_k141_54067 | Not-determined |
| S11_extracted_k141_40737 | Not-determined |
| S11_extracted_k141_40742 | Not-determined |
| S11_extracted_k141_2180 | Low-quality |
| S12_extracted_k141_134657 | High-quality |
| S11_extracted_k141_2181 | Low-quality |
| S11_extracted_k141_86690 | Not-determined |
| S11_extracted_k141_40747 | Not-determined |
| S11_extracted_k141_27923 | Not-determined |
| S11_extracted_k141_27925 | Not-determined |
| S11_extracted_k141_99509 | Not-determined |
| S11_extracted_k141_92823 | Not-determined |
| S11_extracted_k141_40755 | Low-quality |
| S11_extracted_k141_27928 | Not-determined |
| S11_extracted_k141_80065 | Not-determined |
| S11_extracted_k141_99519 | Low-quality |
| S11_extracted_k141_86695 | Not-determined |
| S11_extracted_k141_14574 | Not-determined |
| S11_extracted_k141_92829 | Low-quality |
| S11_extracted_k141_86698 | Not-determined |
| S11_extracted_k141_92831 | Low-quality |
| S11_extracted_k141_14580 | Not-determined |
| S11_extracted_k141_80080 | Not-determined |
| S11_extracted_k141_86705 | Not-determined |
| S11_extracted_k141_40765 | Not-determined |
| S11_extracted_k141_54080 | Not-determined |
| S11_extracted_k141_73655 | Not-determined |
| S11_extracted_k141_86707 | Not-determined |
| S11_extracted_k141_92837 | Low-quality |
| S11_extracted_k141_86713 | Not-determined |
| S11_extracted_k141_2205 | Not-determined |
| S11_extracted_k141_40776 | Not-determined |
| S11_extracted_k141_2206 | Not-determined |
| S11_extracted_k141_14595 | Not-determined |
| S11_extracted_k141_40780 | Not-determined |
| S11_extracted_k141_14599 | Not-determined |
| S11_extracted_k141_73665 | Low-quality |
| S11_extracted_k141_40783 | Not-determined |
| S11_extracted_k141_34016 | Not-determined |
| S11_extracted_k141_73667 | Not-determined |
| S11_extracted_k141_92849 | Not-determined |
| S11_extracted_k141_47274 | Not-determined |
| S11_extracted_k141_40788 | Not-determined |
| S11_extracted_k141_92852 | Not-determined |
| S11_extracted_k141_86719 | Low-quality |
| S11_extracted_k141_34021 | Not-determined |
| S11_extracted_k141_34024 | Not-determined |
| S11_extracted_k141_92857 | Not-determined |
| S11_extracted_k141_27953 | Not-determined |
| S11_extracted_k141_14609 | Not-determined |
| S11_extracted_k141_54098 | Not-determined |
| S11_extracted_k141_54100 | Not-determined |
| S11_extracted_k141_86731 | Low-quality |
| S11_extracted_k141_54104 | Not-determined |
| S11_extracted_k141_86733 | Not-determined |
| S11_extracted_k141_80116 | Not-determined |
| S11_extracted_k141_40811 | Not-determined |
| S11_extracted_k141_80119 | Not-determined |
| S11_extracted_k141_34039 | Not-determined |
| S11_extracted_k141_73692 | Not-determined |
| S11_extracted_k141_14618 | Not-determined |
| S11_extracted_k141_8464 | Low-quality |
| S11_extracted_k141_73693 | Not-determined |
| S11_extracted_k141_8466 | Not-determined |
| S11_extracted_k141_86736 | Not-determined |
| S11_extracted_k141_40826 | Not-determined |
| S11_extracted_k141_73696 | Not-determined |
| S11_extracted_k141_73697 | Not-determined |
| S11_extracted_k141_34048 | Low-quality |
| S11_extracted_k141_80138 | Not-determined |
| S11_extracted_k141_47288 | Low-quality |
| S11_extracted_k141_73700 | Not-determined |
| S11_extracted_k141_2224 | Low-quality |
| S11_extracted_k141_40843 | Not-determined |
| S11_extracted_k141_14632 | Not-determined |
| S11_extracted_k141_73702 | Not-determined |
| S11_extracted_k141_40845 | Low-quality |
| S11_extracted_k141_99562 | Not-determined |
| S11_extracted_k141_92887 | Not-determined |
| S11_extracted_k141_47300 | Low-quality |
| S11_extracted_k141_86756 | Low-quality |
| S11_extracted_k141_99565 | Not-determined |
| S11_extracted_k141_47304 | Not-determined |
| S11_extracted_k141_92895 | Not-determined |
| S11_extracted_k141_60203 | Not-determined |
| S11_extracted_k141_47310 | Not-determined |
| S11_extracted_k141_14644 | Not-determined |
| S11_extracted_k141_80141 | Not-determined |
| S11_extracted_k141_2237 | Not-determined |
| S11_extracted_k141_92902 | Not-determined |
| S11_extracted_k141_60208 | Not-determined |
| S11_extracted_k141_2239 | Not-determined |
| S11_extracted_k141_40855 | Not-determined |
| S11_extracted_k141_92908 | Not-determined |
| S11_extracted_k141_99578 | Not-determined |

| | |
|---|---|
| S11_extracted_k141_2244 | Not-determined |
| S11_extracted_k141_80148 | Low-quality |
| S11_extracted_k141_14656 | Not-determined |
| S11_extracted_k141_99581 | Not-determined |
| S11_extracted_k141_92915 | Not-determined |
| S11_extracted_k141_80150 | Low-quality |
| S11_extracted_k141_40862 | Not-determined |
| S11_extracted_k141_34082 | Not-determined |
| S11_extracted_k141_8499 | Not-determined |
| S11_extracted_k141_73718 | Low-quality |
| S11_extracted_k141_8501 | Not-determined |
| S11_extracted_k141_80155 | Not-determined |
| S11_extracted_k141_21631 | Not-determined |
| S11_extracted_k141_8507 | Not-determined |
| S11_extracted_k141_86766 | Not-determined |
| S11_extracted_k141_60227 | Low-quality |
| S11_extracted_k141_92927 | Low-quality |
| S11_extracted_k141_8512 | Not-determined |
| S11_extracted_k141_60233 | Not-determined |
| S11_extracted_k141_73735 | Not-determined |
| S11_extracted_k141_21643 | Not-determined |
| S11_extracted_k141_73737 | Not-determined |
| S11_extracted_k141_8514 | Not-determined |
| S11_extracted_k141_21649 | Low-quality |
| S11_extracted_k141_92938 | Not-determined |
| S11_extracted_k141_92939 | Not-determined |
| S11_extracted_k141_8521 | Not-determined |
| S11_extracted_k141_92944 | Not-determined |
| S11_extracted_k141_86777 | Low-quality |
| S11_extracted_k141_21656 | Not-determined |
| S11_extracted_k141_14680 | Not-determined |
| S11_extracted_k141_21659 | Not-determined |
| S11_extracted_k141_8533 | Not-determined |
| S11_extracted_k141_40871 | Not-determined |
| S11_extracted_k141_86790 | Low-quality |
| S11_extracted_k141_73752 | Low-quality |
| S11_extracted_k141_73755 | Not-determined |
| S11_extracted_k141_34110 | Not-determined |
| S11_extracted_k141_21665 | Low-quality |
| S11_extracted_k141_40884 | Not-determined |
| S11_extracted_k141_86798 | Low-quality |
| S11_extracted_k141_40886 | Not-determined |
| S11_extracted_k141_92971 | Not-determined |
| S11_extracted_k141_73764 | Not-determined |
| S11_extracted_k141_73765 | Not-determined |
| S11_extracted_k141_34112 | Not-determined |
| S11_extracted_k141_54147 | Not-determined |
| S11_extracted_k141_86807 | Not-determined |
| S11_extracted_k141_92974 | Not-determined |
| S11_extracted_k141_34116 | Not-determined |
| S11_extracted_k141_54152 | Not-determined |
| S11_extracted_k141_54153 | Not-determined |
| S11_extracted_k141_34119 | Not-determined |
| S11_extracted_k141_34120 | Not-determined |
| S11_extracted_k141_8554 | Not-determined |
| S11_extracted_k141_86812 | Low-quality |
| S11_extracted_k141_54156 | Not-determined |
| S11_extracted_k141_8556 | Not-determined |
| S11_extracted_k141_40899 | Not-determined |
| S11_extracted_k141_34126 | Not-determined |
| S11_extracted_k141_73771 | Not-determined |
| S11_extracted_k141_8560 | Not-determined |
| S11_extracted_k141_60253 | Not-determined |
| S11_extracted_k141_73774 | Low-quality |
| S11_extracted_k141_73775 | Not-determined |
| S11_extracted_k141_14718 | Not-determined |
| S11_extracted_k141_99584 | Not-determined |
| S11_extracted_k141_80157 | Low-quality |
| S11_extracted_k141_99585 | Not-determined |
| S11_extracted_k141_99586 | Not-determined |
| S11_extracted_k141_60264 | Not-determined |
| S11_extracted_k141_40922 | Not-determined |
| S11_extracted_k141_40927 | Not-determined |
| S11_extracted_k141_21690 | Not-determined |
| S11_extracted_k141_99591 | Low-quality |
| S11_extracted_k141_80163 | Not-determined |
| S11_extracted_k141_80165 | Not-determined |
| S11_extracted_k141_54168 | Not-determined |
| S11_extracted_k141_21692 | Not-determined |
| S11_extracted_k141_92993 | Not-determined |
| S11_extracted_k141_66727 | Not-determined |
| S11_extracted_k141_28011 | Not-determined |
| S11_extracted_k141_28013 | Not-determined |
| S11_extracted_k141_73792 | Not-determined |
| S11_extracted_k141_99595 | Not-determined |
| S11_extracted_k141_54176 | Not-determined |
| S11_extracted_k141_99597 | Not-determined |
| S11_extracted_k141_73794 | Not-determined |
| S11_extracted_k141_92998 | Not-determined |
| S11_extracted_k141_47327 | Not-determined |
| S11_extracted_k141_99607 | Not-determined |
| S11_extracted_k141_28022 | Low-quality |
| S11_extracted_k141_73798 | Low-quality |
| S11_extracted_k141_93008 | Not-determined |
| S11_extracted_k141_14743 | Not-determined |
| S11_extracted_k141_28027 | Not-determined |
| S11_extracted_k141_73805 | Not-determined |
| S11_extracted_k141_21718 | Not-determined |
| S11_extracted_k141_93015 | Not-determined |
| S11_extracted_k141_93017 | Not-determined |
| S11_extracted_k141_73811 | Not-determined |
| S11_extracted_k141_60282 | Not-determined |
| S11_extracted_k141_73814 | Not-determined |
| S11_extracted_k141_99619 | Not-determined |
| S11_extracted_k141_21724 | Not-determined |
| S11_extracted_k141_21726 | Not-determined |
| S11_extracted_k141_93026 | Low-quality |
| S11_extracted_k141_99620 | Low-quality |
| S11_extracted_k141_21727 | Not-determined |
| S11_extracted_k141_21732 | Low-quality |
| S11_extracted_k141_21733 | Not-determined |
| S11_extracted_k141_14762 | Low-quality |
| S11_extracted_k141_54204 | Not-determined |
| S11_extracted_k141_86871 | Not-determined |
| S11_extracted_k141_21737 | Low-quality |
| S11_extracted_k141_93033 | Not-determined |
| S11_extracted_k141_93034 | Not-determined |
| S11_extracted_k141_93035 | Not-determined |
| S11_extracted_k141_93037 | Not-determined |
| S11_extracted_k141_60301 | Not-determined |
| S11_extracted_k141_73827 | Not-determined |
| S11_extracted_k141_34147 | Low-quality |
| S11_extracted_k141_14772 | Not-determined |
| S11_extracted_k141_21740 | Not-determined |
| S11_extracted_k141_93047 | Not-determined |
| S11_extracted_k141_93049 | Not-determined |
| S11_extracted_k141_60309 | Not-determined |
| S11_extracted_k141_60315 | Not-determined |
| S11_extracted_k141_80175 | Not-determined |
| S11_extracted_k141_99638 | Not-determined |
| S11_extracted_k141_54207 | Low-quality |
| S11_extracted_k141_99640 | Low-quality |
| S11_extracted_k141_80183 | Not-determined |
| S11_extracted_k141_34152 | Not-determined |
| S11_extracted_k141_99646 | Not-determined |
| S11_extracted_k141_60328 | Low-quality |
| S11_extracted_k141_80189 | Not-determined |
| S11_extracted_k141_54210 | Not-determined |
| S11_extracted_k141_60333 | Not-determined |
| S11_extracted_k141_80191 | Not-determined |
| S11_extracted_k141_54214 | Not-determined |
| S11_extracted_k141_99652 | Not-determined |
| S11_extracted_k141_54216 | Low-quality |
| S11_extracted_k141_60342 | Not-determined |
| S11_extracted_k141_40947 | Not-determined |
| S11_extracted_k141_40949 | Not-determined |
| S11_extracted_k141_80207 | Not-determined |
| S11_extracted_k141_34178 | Not-determined |
| S11_extracted_k141_34180 | Not-determined |
| S11_extracted_k141_86910 | Not-determined |
| S11_extracted_k141_86913 | Low-quality |
| S11_extracted_k141_14806 | Not-determined |
| S11_extracted_k141_99675 | Not-determined |
| S11_extracted_k141_21747 | Low-quality |
| S11_extracted_k141_60348 | Not-determined |
| S11_extracted_k141_80229 | Not-determined |
| S11_extracted_k141_21754 | Not-determined |
| S11_extracted_k141_34196 | Low-quality |
| S11_extracted_k141_21758 | Low-quality |
| S11_extracted_k141_8618 | Not-determined |
| S11_extracted_k141_99685 | Not-determined |
| S11_extracted_k141_99686 | Low-quality |
| S11_extracted_k141_99690 | Not-determined |
| S11_extracted_k141_99695 | Not-determined |
| S11_extracted_k141_99696 | Not-determined |
| S11_extracted_k141_60367 | Not-determined |
| S11_extracted_k141_66752 | Not-determined |
| S11_extracted_k141_47357 | Not-determined |
| S11_extracted_k141_60373 | Not-determined |
| S11_extracted_k141_34208 | Not-determined |
| S11_extracted_k141_99707 | Not-determined |
| S11_extracted_k141_60377 | Not-determined |
| S11_extracted_k141_93059 | Not-determined |
| S11_extracted_k141_99711 | Not-determined |
| S11_extracted_k141_93061 | Not-determined |
| S11_extracted_k141_8633 | Not-determined |
| S11_extracted_k141_40959 | Low-quality |
| S11_extracted_k141_2330 | Low-quality |
| S11_extracted_k141_34215 | Not-determined |
| S11_extracted_k141_86949 | Not-determined |
| S11_extracted_k141_66784 | Not-determined |
| S11_extracted_k141_40986 | Not-determined |
| S11_extracted_k141_34218 | Low-quality |
| S11_extracted_k141_2348 | Not-determined |
| S11_extracted_k141_60405 | Not-determined |
| S11_extracted_k141_60408 | Low-quality |
| S11_extracted_k141_99723 | Low-quality |
| S11_extracted_k141_21796 | Not-determined |
| S11_extracted_k141_21797 | Not-determined |
| S11_extracted_k141_99726 | Not-determined |
| S11_extracted_k141_40995 | Not-determined |
| S11_extracted_k141_2363 | Not-determined |
| S11_extracted_k141_99730 | Not-determined |
| S11_extracted_k141_80232 | Not-determined |
| S11_extracted_k141_80233 | Not-determined |
| S11_extracted_k141_86972 | Not-determined |
| S11_extracted_k141_2371 | Low-quality |
| S11_extracted_k141_41003 | Not-determined |
| S11_extracted_k141_66801 | Not-determined |
| S11_extracted_k141_8645 | Not-determined |
| S11_extracted_k141_99734 | Not-determined |
| S11_extracted_k141_99736 | Not-determined |
| S11_extracted_k141_8650 | Low-quality |
| S11_extracted_k141_73873 | Not-determined |
| S11_extracted_k141_80239 | Not-determined |
| S11_extracted_k141_2376 | Not-determined |
| S11_extracted_k141_8654 | Not-determined |
| S11_extracted_k141_99743 | Low-quality |
| S11_extracted_k141_47397 | Not-determined |
| S11_extracted_k141_8657 | Not-determined |
| S11_extracted_k141_47399 | Not-determined |
| S11_extracted_k141_41021 | Not-determined |
| S11_extracted_k141_54226 | Not-determined |
| S11_extracted_k141_73892 | Not-determined |
| S11_extracted_k141_66831 | Not-determined |
| S11_extracted_k141_14840 | Not-determined |
| S11_extracted_k141_86998 | Not-determined |
| S11_extracted_k141_80249 | Not-determined |
| S11_extracted_k141_66837 | Low-quality |
| S11_extracted_k141_73901 | Not-determined |
| S11_extracted_k141_28085 | Not-determined |
| S11_extracted_k141_8675 | Not-determined |
| S11_extracted_k141_41025 | Not-determined |
| S11_extracted_k141_54235 | Not-determined |
| S11_extracted_k141_28092 | Not-determined |
| S11_extracted_k141_41031 | Not-determined |
| S11_extracted_k141_73906 | Not-determined |
| S11_extracted_k141_14853 | Not-determined |
| S11_extracted_k141_14854 | Not-determined |
| S11_extracted_k141_73911 | Not-determined |
| S11_extracted_k141_47421 | Low-quality |
| S11_extracted_k141_54249 | Not-determined |
| S11_extracted_k141_47424 | Not-determined |
| S11_extracted_k141_28104 | Not-determined |
| S11_extracted_k141_87020 | Not-determined |
| S11_extracted_k141_80261 | Not-determined |
| S11_extracted_k141_41043 | Not-determined |
| S12_vs_k141_134657\|\|full | High-quality |
| S11_extracted_k141_80264 | Low-quality |
| S11_extracted_k141_87026 | Not-determined |
| S11_extracted_k141_2409 | Low-quality |
| S11_extracted_k141_2411 | Low-quality |
| S11_extracted_k141_73923 | Not-determined |
| S11_extracted_k141_28106 | Low-quality |
| S11_extracted_k141_73926 | Not-determined |
| S11_extracted_k141_41050 | Not-determined |
| S11_extracted_k141_87036 | Not-determined |
| S11_extracted_k141_73932 | Not-determined |
| S11_extracted_k141_34235 | Not-determined |
| S11_extracted_k141_73936 | Not-determined |
| S11_extracted_k141_54273 | Not-determined |
| S11_extracted_k141_54274 | Not-determined |
| S11_extracted_k141_80275 | Not-determined |
| S11_extracted_k141_21822 | Not-determined |
| S11_extracted_k141_41060 | Not-determined |
| S11_extracted_k141_87045 | Not-determined |
| S11_extracted_k141_80279 | Not-determined |
| S11_extracted_k141_41066 | Not-determined |
| S11_extracted_k141_41068 | Not-determined |
| S11_extracted_k141_41069 | Not-determined |
| S11_extracted_k141_54281 | Not-determined |
| S11_extracted_k141_41073 | Low-quality |
| S11_extracted_k141_54288 | Not-determined |
| S11_extracted_k141_21837 | Not-determined |
| S11_extracted_k141_80298 | Not-determined |
| S11_extracted_k141_41084 | Not-determined |
| S11_extracted_k141_80303 | Low-quality |
| S11_extracted_k141_54299 | Not-determined |
| S11_extracted_k141_73937 | Not-determined |
| S11_extracted_k141_21846 | Not-determined |
| S11_extracted_k141_87051 | Not-determined |
| S11_extracted_k141_28148 | Not-determined |
| S11_extracted_k141_80308 | Not-determined |
| S11_extracted_k141_41093 | Not-determined |
| S11_extracted_k141_73944 | Not-determined |
| S11_extracted_k141_8699 | Not-determined |
| S11_extracted_k141_54308 | Not-determined |
| S11_extracted_k141_60437 | Not-determined |
| S11_extracted_k141_54309 | Not-determined |
| S11_extracted_k141_60438 | Not-determined |
| S11_extracted_k141_28158 | Not-determined |
| S11_extracted_k141_93111 | Low-quality |
| S11_extracted_k141_34265 | Not-determined |
| S11_extracted_k141_80318 | Not-determined |
| S11_extracted_k141_21853 | Not-determined |
| S11_extracted_k141_8711 | Low-quality |
| S11_extracted_k141_2441 | Not-determined |
| S11_extracted_k141_21860 | Low-quality |
| S11_extracted_k141_93119 | Not-determined |
| S11_extracted_k141_54327 | Not-determined |
| S11_extracted_k141_8718 | Low-quality |
| S11_extracted_k141_2445 | Not-determined |
| S11_extracted_k141_54329 | Not-determined |
| S11_extracted_k141_2446 | Not-determined |
| S11_extracted_k141_21868 | Not-determined |
| S11_extracted_k141_54330 | Not-determined |
| S11_extracted_k141_2448 | Not-determined |
| S11_extracted_k141_54334 | Low-quality |
| S11_extracted_k141_8725 | Not-determined |
| S11_extracted_k141_73969 | Not-determined |
| S11_extracted_k141_2454 | Not-determined |
| S11_extracted_k141_2455 | Not-determined |
| S11_extracted_k141_2456 | Not-determined |
| S11_extracted_k141_80333 | Low-quality |
| S11_extracted_k141_80335 | Not-determined |
| S11_extracted_k141_28183 | Not-determined |
| S11_extracted_k141_28184 | Not-determined |
| S11_extracted_k141_87082 | Not-determined |
| S11_extracted_k141_28186 | Not-determined |
| S11_extracted_k141_8732 | Not-determined |
| S11_extracted_k141_21882 | Not-determined |
| S11_extracted_k141_80344 | Low-quality |
| S11_extracted_k141_87091 | Low-quality |
| S11_extracted_k141_80347 | Not-determined |
| S11_extracted_k141_8739 | Low-quality |
| S10_extracted_61680 | Complete |
| S11_extracted_k141_28194 | Not-determined |
| S11_extracted_k141_34287 | Not-determined |
| S11_extracted_k141_73990 | Not-determined |
| S11_extracted_k141_73991 | Not-determined |
| S11_extracted_k141_66851 | Not-determined |
| S11_extracted_k141_73993 | Not-determined |
| S11_extracted_k141_73994 | Not-determined |
| S11_extracted_k141_28195 | Not-determined |
| S11_extracted_k141_8742 | Not-determined |
| S11_extracted_k141_93153 | Not-determined |
| S11_extracted_k141_21894 | Not-determined |
| S11_extracted_k141_2474 | Not-determined |
| S11_extracted_k141_93155 | Not-determined |
| S11_extracted_k141_93161 | Not-determined |
| S11_extracted_k141_99765 | Not-determined |
| S11_extracted_k141_47444 | Low-quality |
| S11_extracted_k141_87118 | Not-determined |
| S11_extracted_k141_34303 | Not-determined |
| S11_extracted_k141_93167 | Not-determined |
| S11_extracted_k141_28208 | Low-quality |
| S11_extracted_k141_54366 | Not-determined |
| S11_extracted_k141_93172 | Not-determined |
| S11_extracted_k141_34307 | Not-determined |
| S11_extracted_k141_66869 | Low-quality |
| S11_extracted_k141_2479 | Not-determined |
| S11_extracted_k141_87121 | Not-determined |
| S11_extracted_k141_87128 | Not-determined |
| S11_extracted_k141_2491 | Not-determined |
| S11_extracted_k141_54377 | Not-determined |
| S11_extracted_k141_41119 | Not-determined |
| S11_extracted_k141_54378 | Not-determined |
| S11_extracted_k141_2493 | Not-determined |
| S11_extracted_k141_54380 | Not-determined |
| S11_extracted_k141_8753 | Not-determined |
| S11_extracted_k141_8754 | Not-determined |
| S11_extracted_k141_8756 | Not-determined |
| S11_extracted_k141_54381 | Not-determined |
| S11_extracted_k141_54383 | Low-quality |
| S11_extracted_k141_74022 | Not-determined |
| S11_extracted_k141_14904 | Not-determined |
| S11_extracted_k141_14905 | Not-determined |
| S11_extracted_k141_54387 | Not-determined |
| S11_extracted_k141_8758 | Not-determined |
| S11_extracted_k141_8761 | Not-determined |
| S11_extracted_k141_8762 | Not-determined |
| S11_extracted_k141_8763 | Not-determined |
| S11_extracted_k141_8770 | Low-quality |
| S11_extracted_k141_2507 | Not-determined |
| S11_extracted_k141_2508 | Not-determined |
| S11_extracted_k141_80372 | Not-determined |
| S11_extracted_k141_34316 | Low-quality |
| S11_extracted_k141_80375 | Not-determined |
| S11_extracted_k141_87138 | Not-determined |
| S11_extracted_k141_34322 | Not-determined |
| S11_extracted_k141_8779 | Not-determined |
| S11_extracted_k141_66887 | Not-determined |
| S11_extracted_k141_66888 | Not-determined |
| S11_extracted_k141_41138 | Not-determined |
| S11_extracted_k141_87147 | Not-determined |
| S11_extracted_k141_34329 | Not-determined |
| S11_extracted_k141_14916 | Not-determined |
| S11_extracted_k141_8787 | Not-determined |
| S11_extracted_k141_14920 | Not-determined |
| S11_extracted_k141_80385 | Low-quality |
| S11_extracted_k141_21939 | Not-determined |
| S11_extracted_k141_99806 | Not-determined |
| S11_extracted_k141_87149 | Not-determined |
| S11_extracted_k141_87150 | Not-determined |
| S11_extracted_k141_34335 | Not-determined |
| S11_extracted_k141_41150 | Not-determined |
| S11_extracted_k141_34337 | Not-determined |
| S11_extracted_k141_34338 | Not-determined |
| S11_extracted_k141_47457 | Low-quality |
| S11_extracted_k141_47459 | Low-quality |
| S11_extracted_k141_54393 | Not-determined |
| S11_extracted_k141_87166 | Low-quality |
| S11_extracted_k141_14923 | Low-quality |
| S11_extracted_k141_93179 | Not-determined |
| S11_extracted_k141_93182 | Low-quality |
| S11_extracted_k141_21956 | Not-determined |
| S11_extracted_k141_87172 | Low-quality |
| S11_extracted_k141_21959 | Not-determined |
| S11_extracted_k141_34358 | Low-quality |
| S11_extracted_k141_41163 | Not-determined |
| S11_extracted_k141_99830 | Not-determined |
| S11_extracted_k141_41164 | Not-determined |
| S11_extracted_k141_8810 | Not-determined |
| S11_extracted_k141_54413 | Not-determined |
| S11_extracted_k141_14942 | Not-determined |
| S11_extracted_k141_74057 | Low-quality |
| S11_extracted_k141_14944 | Not-determined |
| S11_extracted_k141_99839 | Not-determined |
| S11_extracted_k141_41175 | Not-determined |
| S11_extracted_k141_74062 | Low-quality |
| S11_extracted_k141_99840 | Not-determined |
| S11_extracted_k141_8817 | Not-determined |
| S11_extracted_k141_8818 | Not-determined |
| S11_extracted_k141_41176 | Not-determined |
| S11_extracted_k141_74068 | Not-determined |
| S11_extracted_k141_93206 | Not-determined |
| S11_extracted_k141_99844 | Not-determined |
| S11_extracted_k141_74069 | Not-determined |
| S11_extracted_k141_87193 | Not-determined |
| S11_extracted_k141_74070 | Not-determined |
| S11_extracted_k141_47466 | Not-determined |
| S11_extracted_k141_8824 | Not-determined |
| S11_extracted_k141_93214 | Not-determined |
| S11_extracted_k141_8828 | Not-determined |
| S11_extracted_k141_47470 | Not-determined |
| S11_extracted_k141_8832 | Low-quality |
| S11_extracted_k141_47473 | Not-determined |
| S11_extracted_k141_74080 | Low-quality |
| S11_extracted_k141_2536 | Not-determined |
| S11_extracted_k141_8837 | Low-quality |
| S11_extracted_k141_14975 | Not-determined |
| S11_extracted_k141_54430 | Not-determined |
| S11_extracted_k141_41199 | Not-determined |
| S11_extracted_k141_28235 | Not-determined |
| S11_extracted_k141_66918 | Not-determined |
| S11_extracted_k141_99865 | Not-determined |
| S11_extracted_k141_41200 | Not-determined |
| S11_extracted_k141_41203 | Not-determined |
| S11_extracted_k141_66924 | Not-determined |
| S11_extracted_k141_14984 | Not-determined |
| S11_extracted_k141_47476 | Low-quality |
| S11_extracted_k141_14987 | Not-determined |
| S11_extracted_k141_28245 | Not-determined |
| S11_extracted_k141_28248 | Not-determined |
| S11_extracted_k141_2555 | Not-determined |
| S11_extracted_k141_99878 | Not-determined |
| S11_extracted_k141_14996 | Not-determined |
| S11_extracted_k141_47481 | Not-determined |
| S11_extracted_k141_14998 | Not-determined |
| S11_extracted_k141_34391 | Not-determined |
| S11_extracted_k141_34393 | Not-determined |
| S11_extracted_k141_47488 | Not-determined |
| S11_extracted_k141_41222 | Not-determined |
| S11_extracted_k141_2562 | Not-determined |
| S11_extracted_k141_54442 | Not-determined |
| S11_extracted_k141_66935 | Low-quality |
| S11_extracted_k141_41223 | Not-determined |
| S11_extracted_k141_66936 | Not-determined |
| S11_extracted_k141_21967 | Not-determined |
| S11_extracted_k141_54448 | Not-determined |
| S11_extracted_k141_93245 | Not-determined |
| S11_extracted_k141_66943 | Not-determined |
| S11_extracted_k141_15008 | Not-determined |
| S11_extracted_k141_93248 | Not-determined |
| S11_extracted_k141_41235 | Not-determined |
| S11_extracted_k141_15013 | Not-determined |
| S11_extracted_k141_34403 | Not-determined |
| S11_extracted_k141_60471 | Low-quality |
| S11_extracted_k141_66947 | Low-quality |
| S11_extracted_k141_87206 | Not-determined |
| S11_extracted_k141_28266 | Not-determined |
| S11_extracted_k141_34405 | Not-determined |
| S11_extracted_k141_93255 | Not-determined |
| S11_extracted_k141_80421 | Not-determined |
| S11_extracted_k141_99898 | Not-determined |
| S11_extracted_k141_93260 | Not-determined |
| S11_extracted_k141_34408 | Not-determined |
| S11_extracted_k141_47503 | Not-determined |
| S11_extracted_k141_28274 | Not-determined |
| S11_extracted_k141_93266 | Not-determined |
| S11_extracted_k141_93267 | Not-determined |
| S11_extracted_k141_54469 | Low-quality |
| S11_extracted_k141_47509 | Low-quality |
| S11_extracted_k141_66959 | Not-determined |
| S11_extracted_k141_2572 | Low-quality |
| S11_extracted_k141_47516 | Not-determined |
| S11_extracted_k141_87214 | Not-determined |
| S11_extracted_k141_99902 | Not-determined |
| S11_extracted_k141_60476 | Low-quality |
| S11_extracted_k141_66963 | Low-quality |
| S11_extracted_k141_66964 | Not-determined |
| S11_extracted_k141_41251 | Not-determined |
| S11_extracted_k141_54483 | Not-determined |
| S11_extracted_k141_2582 | Not-determined |
| S11_extracted_k141_2587 | Not-determined |
| S11_extracted_k141_2589 | Not-determined |
| S11_extracted_k141_54495 | Not-determined |
| S11_extracted_k141_47536 | Low-quality |
| S11_extracted_k141_2592 | Not-determined |
| S11_extracted_k141_74109 | Not-determined |
| S11_extracted_k141_41259 | Low-quality |
| S11_extracted_k141_15038 | Not-determined |
| S11_extracted_k141_41266 | Not-determined |
| S11_extracted_k141_87228 | Not-determined |
| S11_extracted_k141_28297 | Not-determined |
| S11_extracted_k141_66983 | Not-determined |
| S11_extracted_k141_28300 | Not-determined |
| S11_extracted_k141_87231 | Not-determined |
| S11_extracted_k141_99924 | Not-determined |
| S11_extracted_k141_60506 | Low-quality |
| S11_extracted_k141_66985 | Low-quality |
| S11_extracted_k141_28305 | Not-determined |
| S11_extracted_k141_93281 | Low-quality |
| S11_extracted_k141_54504 | Not-determined |
| S11_extracted_k141_28311 | Not-determined |
| S11_extracted_k141_15065 | Low-quality |
| S11_extracted_k141_2608 | Low-quality |
| S11_extracted_k141_2610 | Not-determined |
| S11_extracted_k141_93292 | Low-quality |
| S11_extracted_k141_74128 | Not-determined |
| S11_extracted_k141_54515 | Low-quality |
| S11_extracted_k141_28319 | Not-determined |
| S11_extracted_k141_15068 | Not-determined |
| S11_extracted_k141_54518 | Not-determined |
| S11_extracted_k141_67001 | Not-determined |
| S11_extracted_k141_2617 | Low-quality |
| S11_extracted_k141_41287 | Low-quality |
| S11_extracted_k141_93296 | Not-determined |
| S11_extracted_k141_60525 | Not-determined |
| S11_extracted_k141_41291 | Not-determined |
| S11_extracted_k141_54524 | Not-determined |
| S11_extracted_k141_15076 | Low-quality |
| S11_extracted_k141_47564 | Not-determined |
| S11_extracted_k141_15079 | Not-determined |
| S11_extracted_k141_28334 | Not-determined |
| S11_extracted_k141_28335 | Not-determined |
| S11_extracted_k141_54528 | Not-determined |
| S11_extracted_k141_54531 | Low-quality |
| S11_extracted_k141_74142 | Not-determined |
| S11_extracted_k141_93307 | Low-quality |
| S11_extracted_k141_74145 | Not-determined |
| S11_extracted_k141_67017 | Not-determined |
| S11_extracted_k141_21980 | Not-determined |
| S11_extracted_k141_67023 | Not-determined |
| S11_extracted_k141_67024 | Not-determined |
| S11_extracted_k141_74154 | Not-determined |
| S11_extracted_k141_74155 | Not-determined |
| S11_extracted_k141_80450 | Not-determined |
| S11_extracted_k141_80452 | Not-determined |
| S11_extracted_k141_99940 | Not-determined |
| S11_extracted_k141_93325 | Not-determined |
| S11_extracted_k141_54541 | Low-quality |
| S11_extracted_k141_47576 | Not-determined |
| S11_extracted_k141_47577 | Not-determined |
| S11_extracted_k141_8908 | Not-determined |
| S11_extracted_k141_87285 | Low-quality |
| S11_extracted_k141_99943 | Not-determined |
| S11_extracted_k141_74159 | Not-determined |
| S11_extracted_k141_99944 | Not-determined |
| S11_extracted_k141_80459 | Not-determined |
| S11_extracted_k141_67030 | Not-determined |
| S11_extracted_k141_15092 | Not-determined |
| S11_extracted_k141_54549 | Not-determined |
| S11_extracted_k141_74160 | Low-quality |
| S11_extracted_k141_93336 | Not-determined |
| S11_extracted_k141_41317 | Low-quality |
| S11_extracted_k141_54553 | Low-quality |
| S11_extracted_k141_80465 | Not-determined |
| S11_extracted_k141_54555 | Not-determined |
| S11_extracted_k141_99949 | Not-determined |
| S11_extracted_k141_41319 | Not-determined |
| S11_extracted_k141_41324 | Not-determined |
| S11_extracted_k141_34465 | Not-determined |
| S11_extracted_k141_87293 | Not-determined |
| S11_extracted_k141_41325 | Low-quality |
| S11_extracted_k141_34466 | Low-quality |
| S11_extracted_k141_28355 | Not-determined |
| S11_extracted_k141_87296 | Not-determined |
| S11_extracted_k141_80470 | Not-determined |
| S11_extracted_k141_41328 | Not-determined |
| S11_extracted_k141_99953 | Low-quality |
| S11_extracted_k141_80472 | Not-determined |
| S11_extracted_k141_99954 | Low-quality |
| S11_extracted_k141_41332 | Not-determined |
| S11_extracted_k141_34472 | Not-determined |
| S11_extracted_k141_15098 | Not-determined |
| S11_extracted_k141_41341 | Low-quality |
| S11_extracted_k141_15105 | Low-quality |
| S11_extracted_k141_67055 | Not-determined |

| | |
|---|---|
| S11_extracted_k141_93352 | Low-quality |
| S11_extracted_k141_41348 | Not-determined |
| S11_extracted_k141_74179 | Not-determined |
| S11_extracted_k141_60567 | Not-determined |
| S11_extracted_k141_54575 | Not-determined |
| S11_extracted_k141_15108 | Low-quality |
| S11_extracted_k141_93354 | Not-determined |
| S11_extracted_k141_41354 | Not-determined |
| S11_extracted_k141_93355 | Not-determined |
| S11_extracted_k141_41360 | Not-determined |
| S11_extracted_k141_8928 | Not-determined |
| S11_extracted_k141_60578 | Not-determined |
| S11_extracted_k141_41361 | Not-determined |
| S11_extracted_k141_74189 | Low-quality |
| S11_extracted_k141_54587 | Low-quality |
| S11_extracted_k141_54593 | Not-determined |
| S11_extracted_k141_28381 | Not-determined |
| S11_extracted_k141_34501 | Not-determined |
| S11_extracted_k141_8934 | Low-quality |
| S11_extracted_k141_22008 | Low-quality |
| S11_extracted_k141_74200 | Not-determined |
| S11_extracted_k141_74204 | Not-determined |
| S11_extracted_k141_8943 | Not-determined |
| S11_extracted_k141_74207 | Not-determined |
| S11_extracted_k141_93367 | Not-determined |
| S11_extracted_k141_87327 | Not-determined |
| S11_extracted_k141_93369 | Not-determined |
| S11_extracted_k141_22012 | Low-quality |
| S11_extracted_k141_41380 | Not-determined |
| S11_extracted_k141_80489 | Low-quality |
| S11_extracted_k141_34516 | Not-determined |
| S11_extracted_k141_28399 | Not-determined |
| S11_extracted_k141_8946 | Not-determined |
| S11_extracted_k141_34520 | Not-determined |
| S11_extracted_k141_99962 | Low-quality |
| S11_extracted_k141_15133 | Not-determined |
| S11_extracted_k141_99966 | Not-determined |
| S11_extracted_k141_99969 | Not-determined |
| S11_extracted_k141_99973 | Not-determined |
| S11_extracted_k141_74218 | Low-quality |
| S11_extracted_k141_28415 | Not-determined |
| S11_extracted_k141_87344 | Not-determined |
| S11_extracted_k141_22037 | Not-determined |
| S11_extracted_k141_80496 | Low-quality |
| S11_extracted_k141_99980 | Not-determined |
| S11_extracted_k141_22040 | Not-determined |
| S11_extracted_k141_74222 | Low-quality |
| S11_extracted_k141_74227 | Not-determined |
| S11_extracted_k141_8961 | Not-determined |
| S11_extracted_k141_74228 | Not-determined |
| S11_extracted_k141_15156 | Not-determined |
| S11_extracted_k141_99991 | Not-determined |
| S11_extracted_k141_22048 | Not-determined |
| S11_extracted_k141_54611 | Not-determined |
| S11_extracted_k141_93398 | Not-determined |
| S11_extracted_k141_15161 | Low-quality |
| S11_extracted_k141_74238 | Not-determined |
| S11_extracted_k141_2645 | Low-quality |
| S11_extracted_k141_60585 | Not-determined |
| S11_extracted_k141_22057 | Not-determined |
| S11_extracted_k141_8971 | Not-determined |
| S11_extracted_k141_2646 | Not-determined |
| S11_extracted_k141_87363 | Not-determined |
| S11_extracted_k141_54617 | Low-quality |
| S11_extracted_k141_2650 | Not-determined |
| S11_extracted_k141_60591 | Low-quality |
| S11_extracted_k141_87366 | Not-determined |
| S11_extracted_k141_100002 | Not-determined |
| S11_extracted_k141_15171 | Low-quality |
| S11_extracted_k141_2656 | Not-determined |
| S11_extracted_k141_2660 | Not-determined |
| S11_extracted_k141_15178 | Not-determined |
| S11_extracted_k141_54627 | Low-quality |
| S11_extracted_k141_87372 | Not-determined |
| S11_extracted_k141_93414 | Not-determined |
| S11_extracted_k141_8974 | Low-quality |
| S11_extracted_k141_54631 | Not-determined |
| S11_extracted_k141_67075 | Not-determined |
| S11_extracted_k141_2669 | Not-determined |
| S11_extracted_k141_93418 | Low-quality |
| S11_extracted_k141_100012 | Not-determined |
| S11_extracted_k141_54636 | Not-determined |
| S11_extracted_k141_60613 | Not-determined |
| S11_extracted_k141_100017 | Not-determined |
| S11_extracted_k141_67085 | Not-determined |
| S11_extracted_k141_93425 | Not-determined |
| S11_extracted_k141_8986 | Low-quality |
| S8_vs_k141_49008\|\|full | High-quality |
| S11_extracted_k141_47634 | Not-determined |
| S11_extracted_k141_60630 | Not-determined |
| S11_extracted_k141_2688 | Not-determined |
| S11_extracted_k141_60632 | Not-determined |
| S11_extracted_k141_100022 | Not-determined |
| S11_extracted_k141_87390 | Low-quality |
| S11_extracted_k141_28432 | Low-quality |
| S11_extracted_k141_15199 | Not-determined |
| S11_extracted_k141_47640 | Not-determined |
| S11_extracted_k141_60634 | Not-determined |
| S11_extracted_k141_2692 | Not-determined |
| S11_extracted_k141_60635 | Not-determined |
| S11_extracted_k141_93437 | Not-determined |
| S11_extracted_k141_67093 | Not-determined |
| S11_extracted_k141_93440 | Low-quality |
| S11_extracted_k141_60637 | Low-quality |
| S11_extracted_k141_87396 | Low-quality |
| S11_extracted_k141_28442 | Not-determined |
| S11_extracted_k141_28443 | Not-determined |
| S11_extracted_k141_41418 | Not-determined |
| S11_extracted_k141_87403 | Not-determined |
| S11_extracted_k141_87409 | Not-determined |
| S11_extracted_k141_60647 | Low-quality |
| S11_extracted_k141_87411 | Not-determined |
| S11_extracted_k141_80513 | Low-quality |
| S11_extracted_k141_15216 | Low-quality |
| S11_extracted_k141_60648 | Low-quality |
| S11_extracted_k141_60650 | Not-determined |
| S11_extracted_k141_60652 | Low-quality |
| S11_extracted_k141_28459 | Not-determined |
| S11_extracted_k141_47655 | Not-determined |
| S11_extracted_k141_74268 | Not-determined |
| S11_extracted_k141_80520 | Low-quality |
| S11_extracted_k141_74271 | Not-determined |
| S11_extracted_k141_15223 | Not-determined |
| S11_extracted_k141_41426 | Low-quality |
| S11_extracted_k141_41428 | Not-determined |
| S11_extracted_k141_28465 | Not-determined |
| S11_extracted_k141_15228 | Low-quality |
| S11_extracted_k141_15230 | Not-determined |
| S11_extracted_k141_9010 | Not-determined |
| S11_extracted_k141_22077 | Low-quality |
| S11_extracted_k141_80530 | Not-determined |
| S11_extracted_k141_22082 | Not-determined |
| S11_extracted_k141_9021 | Not-determined |
| S11_extracted_k141_41437 | Not-determined |
| S11_extracted_k141_80531 | Not-determined |
| S11_extracted_k141_22088 | Not-determined |
| S11_extracted_k141_80534 | Low-quality |
| S11_extracted_k141_100036 | Low-quality |
| S11_extracted_k141_2717 | Low-quality |
| S11_extracted_k141_28472 | Not-determined |
| S11_extracted_k141_100038 | Not-determined |
| S11_extracted_k141_87423 | Not-determined |
| S11_extracted_k141_87424 | Not-determined |
| S11_extracted_k141_100045 | Not-determined |
| S11_extracted_k141_41443 | Not-determined |
| S11_extracted_k141_67110 | Low-quality |
| S11_extracted_k141_22099 | Not-determined |
| S11_extracted_k141_80543 | Not-determined |
| S11_extracted_k141_87433 | Not-determined |
| S11_extracted_k141_87436 | Not-determined |
| S11_extracted_k141_87437 | Low-quality |
| S11_extracted_k141_100050 | Not-determined |
| S11_extracted_k141_80551 | Low-quality |
| S11_extracted_k141_100052 | Not-determined |
| S11_extracted_k141_15257 | Low-quality |
| S11_extracted_k141_87443 | Not-determined |
| S11_extracted_k141_100053 | Not-determined |
| S11_extracted_k141_9047 | Not-determined |
| S11_extracted_k141_28492 | Not-determined |
| S11_extracted_k141_100059 | Not-determined |
| S11_extracted_k141_34531 | Not-determined |
| S11_extracted_k141_67127 | Not-determined |
| S11_extracted_k141_67128 | Not-determined |
| S11_extracted_k141_100065 | Not-determined |
| S11_extracted_k141_28501 | Not-determined |
| S11_extracted_k141_87458 | Low-quality |
| S11_extracted_k141_34538 | Not-determined |
| S11_extracted_k141_74289 | Not-determined |
| S11_extracted_k141_41469 | Low-quality |
| S11_extracted_k141_67138 | Not-determined |
| S11_extracted_k141_34541 | Not-determined |
| S11_extracted_k141_100070 | Low-quality |
| S11_extracted_k141_15271 | Not-determined |
| S11_extracted_k141_2744 | Not-determined |
| S11_extracted_k141_100075 | Not-determined |
| S11_extracted_k141_15276 | Low-quality |
| S11_extracted_k141_22134 | Not-determined |
| S11_extracted_k141_47709 | Low-quality |
| S11_extracted_k141_47712 | Not-determined |
| S11_extracted_k141_87473 | Not-determined |
| S11_extracted_k141_9068 | Not-determined |
| S11_extracted_k141_15278 | Not-determined |
| S11_extracted_k141_47715 | Not-determined |
| S11_extracted_k141_100086 | Not-determined |
| S11_extracted_k141_15282 | Not-determined |
| S11_extracted_k141_87479 | Not-determined |
| S11_extracted_k141_34563 | Not-determined |
| S11_extracted_k141_74321 | Not-determined |
| S11_extracted_k141_87486 | Not-determined |
| S11_extracted_k141_74325 | Not-determined |
| S11_extracted_k141_93490 | Not-determined |
| S11_extracted_k141_93491 | Low-quality |
| S11_extracted_k141_87488 | Not-determined |
| S11_extracted_k141_15291 | Not-determined |
| S11_extracted_k141_93500 | Not-determined |
| S11_extracted_k141_34574 | Not-determined |
| S11_extracted_k141_2768 | Not-determined |
| S11_extracted_k141_9091 | Not-determined |
| S11_extracted_k141_100104 | Not-determined |
| S11_extracted_k141_34576 | Low-quality |
| S11_extracted_k141_100105 | Not-determined |
| S11_extracted_k141_87499 | Not-determined |
| S11_extracted_k141_2778 | Not-determined |
| S11_extracted_k141_93508 | Low-quality |
| S11_extracted_k141_22172 | Not-determined |
| S11_extracted_k141_93515 | Not-determined |
| S11_extracted_k141_9106 | Not-determined |
| S11_extracted_k141_74348 | Low-quality |
| S11_extracted_k141_74349 | Not-determined |
| S11_extracted_k141_87507 | Not-determined |
| S11_extracted_k141_60685 | Low-quality |
| S11_extracted_k141_100113 | Not-determined |
| S11_extracted_k141_80569 | Not-determined |
| S11_extracted_k141_9110 | Not-determined |
| S11_extracted_k141_100115 | Not-determined |
| S11_extracted_k141_93521 | Not-determined |
| S11_extracted_k141_80571 | Not-determined |
| S11_extracted_k141_87514 | Low-quality |
| S11_extracted_k141_100120 | Not-determined |
| S11_extracted_k141_93526 | Not-determined |
| S11_extracted_k141_100124 | Not-determined |
| S11_extracted_k141_34600 | Not-determined |
| S11_extracted_k141_74358 | Not-determined |
| S11_extracted_k141_93534 | Not-determined |
| S11_extracted_k141_80583 | Not-determined |
| S11_extracted_k141_80587 | Not-determined |
| S11_extracted_k141_100135 | Not-determined |
| S11_extracted_k141_100136 | Low-quality |
| S11_extracted_k141_87526 | Not-determined |
| S11_extracted_k141_9123 | Low-quality |
| S11_extracted_k141_80610 | Not-determined |
| S11_extracted_k141_47725 | Not-determined |
| S11_extracted_k141_22183 | Not-determined |
| S11_extracted_k141_60702 | Not-determined |
| S11_extracted_k141_34615 | Not-determined |
| S11_extracted_k141_80614 | Not-determined |
| S11_extracted_k141_22185 | Not-determined |
| S11_extracted_k141_87535 | Low-quality |
| S11_extracted_k141_22186 | Not-determined |
| S11_extracted_k141_60706 | Low-quality |
| S11_extracted_k141_9128 | Low-quality |
| S11_extracted_k141_41490 | Low-quality |
| S11_extracted_k141_41491 | Not-determined |
| S11_extracted_k141_22188 | Low-quality |
| S11_extracted_k141_87540 | Low-quality |
| S11_extracted_k141_41493 | Not-determined |
| S11_extracted_k141_15329 | Not-determined |
| S11_extracted_k141_15334 | Not-determined |
| S11_extracted_k141_41500 | Not-determined |
| S11_extracted_k141_87548 | Not-determined |
| S11_extracted_k141_87550 | Not-determined |
| S11_extracted_k141_9139 | Not-determined |
| S11_extracted_k141_22197 | Low-quality |
| S11_extracted_k141_9142 | Low-quality |
| S11_extracted_k141_74397 | Not-determined |
| S11_extracted_k141_74398 | Not-determined |
| S11_extracted_k141_9144 | Not-determined |
| S11_extracted_k141_22201 | Not-determined |
| S11_extracted_k141_60732 | Not-determined |
| S11_extracted_k141_60737 | Not-determined |
| S11_extracted_k141_22207 | Not-determined |
| S11_extracted_k141_2848 | Not-determined |
| S11_extracted_k141_74408 | Not-determined |
| S11_extracted_k141_34630 | Low-quality |
| S11_extracted_k141_60741 | Not-determined |
| S11_extracted_k141_47746 | Not-determined |
| S11_extracted_k141_47747 | Low-quality |
| S11_extracted_k141_47750 | Not-determined |
| S11_extracted_k141_9151 | Not-determined |
| S11_extracted_k141_2859 | Not-determined |
| S11_extracted_k141_2861 | Not-determined |
| S11_extracted_k141_34642 | Not-determined |
| S11_extracted_k141_60746 | Not-determined |
| S11_extracted_k141_41521 | Not-determined |
| S11_extracted_k141_34645 | Not-determined |
| S11_extracted_k141_34647 | Not-determined |
| S11_extracted_k141_87569 | Not-determined |
| S11_extracted_k141_15359 | Not-determined |
| S11_extracted_k141_22215 | Not-determined |
| S11_extracted_k141_60752 | Not-determined |
| S11_extracted_k141_87572 | Not-determined |
| S11_extracted_k141_22221 | Low-quality |
| S11_extracted_k141_2879 | Not-determined |
| S11_extracted_k141_93557 | Not-determined |
| S11_extracted_k141_87581 | Not-determined |
| S11_extracted_k141_87582 | Not-determined |
| S11_extracted_k141_2884 | Low-quality |
| S11_extracted_k141_93559 | Not-determined |
| S11_extracted_k141_47768 | Not-determined |
| S11_extracted_k141_93564 | Not-determined |
| S11_extracted_k141_41550 | Not-determined |
| S11_extracted_k141_93574 | Not-determined |
| S11_extracted_k141_80669 | Not-determined |
| S11_extracted_k141_60766 | Low-quality |
| S11_extracted_k141_80670 | Not-determined |
| S11_extracted_k141_41554 | Low-quality |
| S11_extracted_k141_60767 | Not-determined |
| S11_extracted_k141_93576 | Not-determined |
| S11_extracted_k141_67169 | Not-determined |
| S11_extracted_k141_60769 | Not-determined |
| S11_extracted_k141_87585 | Not-determined |
| S11_extracted_k141_34666 | Not-determined |
| S11_extracted_k141_34668 | Low-quality |
| S11_extracted_k141_87593 | Not-determined |
| S11_extracted_k141_34669 | Not-determined |
| S11_extracted_k141_60775 | Not-determined |
| S11_extracted_k141_2908 | Not-determined |
| S11_extracted_k141_9178 | Not-determined |
| S11_extracted_k141_41566 | Not-determined |
| S11_extracted_k141_9180 | Not-determined |
| S7_vs_58751\|\|full | Medium-quality |
| S11_extracted_k141_15367 | Not-determined |
| S11_extracted_k141_47787 | Not-determined |
| S11_extracted_k141_80685 | Low-quality |
| S11_extracted_k141_15370 | Not-determined |
| S11_extracted_k141_41572 | Low-quality |
| S11_extracted_k141_93589 | Low-quality |
| S11_extracted_k141_47793 | Not-determined |
| S11_extracted_k141_9191 | Not-determined |
| S11_extracted_k141_47794 | Low-quality |
| S11_extracted_k141_74444 | Low-quality |
| S11_extracted_k141_87601 | Low-quality |
| S11_extracted_k141_87602 | Not-determined |
| S11_extracted_k141_87605 | Not-determined |
| S11_extracted_k141_87611 | Low-quality |
| S11_extracted_k141_15383 | Low-quality |
| S11_extracted_k141_93599 | Low-quality |
| S11_extracted_k141_41584 | Not-determined |
| S11_extracted_k141_2929 | Not-determined |
| S11_extracted_k141_93600 | Not-determined |
| S11_extracted_k141_87614 | Low-quality |
| S11_extracted_k141_67202 | Not-determined |
| S11_extracted_k141_47806 | Low-quality |
| S11_extracted_k141_74454 | Not-determined |
| S11_extracted_k141_2936 | Low-quality |
| S11_extracted_k141_2938 | Low-quality |
| S11_extracted_k141_74456 | Not-determined |
| S11_extracted_k141_74457 | Not-determined |
| S11_extracted_k141_87623 | Low-quality |
| S11_extracted_k141_74459 | Not-determined |
| S11_extracted_k141_74461 | Not-determined |
| S11_extracted_k141_93606 | Not-determined |
| S11_extracted_k141_2943 | Not-determined |
| S11_extracted_k141_9196 | Low-quality |
| S11_extracted_k141_60791 | Not-determined |
| S11_extracted_k141_80701 | Low-quality |
| S11_extracted_k141_47816 | Not-determined |
| S11_extracted_k141_9202 | Not-determined |
| S11_extracted_k141_34671 | Low-quality |
| S11_extracted_k141_60797 | Low-quality |
| S11_extracted_k141_93610 | Low-quality |
| S11_extracted_k141_93611 | Not-determined |
| S11_extracted_k141_22250 | Not-determined |
| S11_extracted_k141_28517 | Not-determined |
| S11_extracted_k141_9210 | Not-determined |
| S11_extracted_k141_93624 | Not-determined |
| S11_extracted_k141_9219 | Not-determined |
| S11_extracted_k141_41604 | Not-determined |
| S11_extracted_k141_67208 | Not-determined |
| S11_extracted_k141_100141 | Not-determined |
| S11_extracted_k141_100142 | Not-determined |
| S11_extracted_k141_2951 | Not-determined |
| S11_extracted_k141_74478 | Not-determined |
| S11_extracted_k141_15394 | Not-determined |
| S11_extracted_k141_100148 | Not-determined |
| S11_extracted_k141_100149 | Not-determined |
| S11_extracted_k141_41612 | Not-determined |
| S11_extracted_k141_60814 | Not-determined |
| S11_extracted_k141_74485 | Not-determined |
| S11_extracted_k141_93634 | Not-determined |
| S11_extracted_k141_67213 | Not-determined |
| S11_extracted_k141_41617 | Not-determined |
| S11_extracted_k141_100156 | Low-quality |
| S11_extracted_k141_93639 | Not-determined |
| S11_extracted_k141_60820 | Not-determined |
| S11_extracted_k141_93644 | Not-determined |
| S11_extracted_k141_80720 | Not-determined |
| S11_extracted_k141_9231 | Not-determined |
| S11_extracted_k141_100169 | Not-determined |
| S11_extracted_k141_9235 | Low-quality |
| S11_extracted_k141_60832 | Not-determined |
| S11_extracted_k141_67233 | Not-determined |
| S11_extracted_k141_60833 | Not-determined |
| S11_extracted_k141_22272 | Not-determined |
| S11_extracted_k141_22278 | Not-determined |
| S11_extracted_k141_74513 | Low-quality |
| S11_extracted_k141_74516 | Not-determined |
| S11_extracted_k141_41632 | Not-determined |
| S11_extracted_k141_15414 | Not-determined |
| S11_extracted_k141_41633 | Not-determined |
| S11_extracted_k141_74518 | Not-determined |
| S11_extracted_k141_41640 | Not-determined |
| S11_extracted_k141_74522 | Not-determined |
| S11_extracted_k141_47847 | Not-determined |
| S11_extracted_k141_93673 | Not-determined |
| S11_extracted_k141_93675 | Not-determined |
| S11_extracted_k141_47853 | Not-determined |
| S11_extracted_k141_67244 | Not-determined |
| S11_extracted_k141_47856 | Low-quality |
| S11_extracted_k141_74536 | Not-determined |
| S11_extracted_k141_41654 | Not-determined |
| S11_extracted_k141_41655 | Not-determined |
| S11_extracted_k141_41662 | Not-determined |
| S11_extracted_k141_67252 | Low-quality |
| S11_extracted_k141_60858 | Low-quality |
| S11_extracted_k141_41664 | Low-quality |
| S11_extracted_k141_47872 | Not-determined |
| S11_extracted_k141_100178 | Not-determined |
| S11_extracted_k141_41666 | Not-determined |
| S11_extracted_k141_41667 | Low-quality |
| S11_extracted_k141_93694 | Not-determined |
| S11_extracted_k141_67261 | Not-determined |
| S11_extracted_k141_67262 | Low-quality |
| S11_extracted_k141_41674 | Not-determined |
| S11_extracted_k141_60864 | Not-determined |
| S11_extracted_k141_9275 | Not-determined |
| S11_extracted_k141_60866 | Not-determined |
| S11_extracted_k141_9276 | Low-quality |
| S11_extracted_k141_100183 | Not-determined |
| S11_extracted_k141_74556 | Not-determined |
| S11_extracted_k141_22310 | Low-quality |
| S11_extracted_k141_100185 | Low-quality |
| S11_extracted_k141_9278 | Not-determined |
| S11_extracted_k141_100186 | Not-determined |
| S11_extracted_k141_100188 | Low-quality |
| S11_extracted_k141_60868 | Low-quality |
| S11_extracted_k141_22318 | Not-determined |
| S11_extracted_k141_41690 | Not-determined |
| S11_extracted_k141_93708 | Not-determined |
| S11_extracted_k141_41691 | Not-determined |
| S11_extracted_k141_41692 | Not-determined |
| S11_extracted_k141_93710 | Not-determined |
| S11_extracted_k141_9286 | Not-determined |
| S11_extracted_k141_22323 | Not-determined |
| S11_extracted_k141_9288 | Not-determined |
| S11_extracted_k141_67278 | Not-determined |
| S11_extracted_k141_47902 | Not-determined |
| S11_extracted_k141_22330 | Not-determined |
| S11_extracted_k141_2954 | Not-determined |
| S11_extracted_k141_15442 | Not-determined |
| S11_extracted_k141_60874 | Low-quality |
| S11_extracted_k141_93721 | Not-determined |
| S11_extracted_k141_2957 | Not-determined |
| S11_extracted_k141_93725 | Low-quality |
| S11_extracted_k141_34695 | Low-quality |
| S11_extracted_k141_100205 | Not-determined |
| S11_extracted_k141_60882 | Not-determined |
| S10_vs_61680\|\|full | Medium-quality |
| S11_extracted_k141_2966 | Not-determined |
| S11_extracted_k141_93731 | Not-determined |
| S11_extracted_k141_87664 | Low-quality |
| S11_extracted_k141_2967 | Not-determined |
| S11_extracted_k141_22345 | Low-quality |
| S11_extracted_k141_22346 | Not-determined |
| S11_extracted_k141_93734 | Not-determined |
| S11_extracted_k141_93735 | Not-determined |
| S11_extracted_k141_34707 | Not-determined |
| S11_extracted_k141_22348 | Not-determined |
| S11_extracted_k141_22349 | Not-determined |
| S11_extracted_k141_60890 | Not-determined |
| S11_extracted_k141_28523 | Not-determined |
| S11_extracted_k141_28526 | Not-determined |
| S11_extracted_k141_74594 | Not-determined |
| S11_extracted_k141_22355 | Not-determined |
| S11_extracted_k141_22357 | Not-determined |
| S11_extracted_k141_60894 | Not-determined |
| S11_extracted_k141_74598 | Not-determined |
| S11_extracted_k141_100215 | Low-quality |
| S11_extracted_k141_22361 | Not-determined |
| S11_extracted_k141_100216 | Low-quality |
| S11_extracted_k141_54685 | Not-determined |
| S11_extracted_k141_34717 | Not-determined |
| S11_extracted_k141_9305 | Low-quality |
| S11_extracted_k141_54688 | Not-determined |
| S11_extracted_k141_93761 | Not-determined |
| S11_extracted_k141_9308 | Not-determined |
| S11_extracted_k141_54690 | Low-quality |
| S11_extracted_k141_9310 | Not-determined |
| S11_extracted_k141_2982 | Not-determined |
| S11_extracted_k141_2987 | Not-determined |
| S11_extracted_k141_74607 | Not-determined |
| S11_extracted_k141_54698 | Not-determined |
| S11_extracted_k141_100231 | Not-determined |
| S11_extracted_k141_74609 | Low-quality |
| S11_extracted_k141_60896 | Not-determined |
| S11_extracted_k141_74610 | Low-quality |
| S11_extracted_k141_60897 | Low-quality |
| S11_extracted_k141_2990 | Low-quality |
| S11_extracted_k141_74614 | Not-determined |
| S11_extracted_k141_22378 | Not-determined |
| S11_extracted_k141_74617 | Low-quality |
| S11_extracted_k141_60909 | Not-determined |
| S11_extracted_k141_87681 | Not-determined |
| S11_extracted_k141_15478 | Not-determined |
| S11_extracted_k141_87682 | Not-determined |
| S11_extracted_k141_22386 | Not-determined |
| S11_extracted_k141_74620 | Not-determined |
| S11_extracted_k141_60911 | Low-quality |
| S11_extracted_k141_74621 | Not-determined |
| S11_extracted_k141_34734 | Not-determined |
| S11_extracted_k141_22391 | Not-determined |
| S11_extracted_k141_47935 | Not-determined |
| S11_extracted_k141_80743 | Not-determined |
| S11_extracted_k141_87689 | Low-quality |
| S11_extracted_k141_15482 | Low-quality |
| S11_extracted_k141_34740 | Not-determined |
| S11_extracted_k141_9320 | Low-quality |
| S11_extracted_k141_41709 | Low-quality |
| S11_extracted_k141_100240 | Not-determined |
| S11_extracted_k141_9321 | Not-determined |
| S11_extracted_k141_34745 | Not-determined |
| S11_extracted_k141_100244 | Low-quality |
| S11_extracted_k141_100245 | Not-determined |
| S11_extracted_k141_80750 | Not-determined |
| S11_extracted_k141_100248 | Not-determined |
| S11_extracted_k141_74638 | Not-determined |
| S11_extracted_k141_80753 | Not-determined |
| S11_extracted_k141_87701 | Not-determined |
| S11_extracted_k141_100249 | Not-determined |
| S11_extracted_k141_80754 | Not-determined |
| S11_extracted_k141_22408 | Not-determined |
| S11_extracted_k141_67322 | Low-quality |
| S11_extracted_k141_93782 | Not-determined |
| S11_extracted_k141_41720 | Low-quality |
| S11_extracted_k141_93784 | Not-determined |
| S11_extracted_k141_67325 | Not-determined |
| S11_extracted_k141_41722 | Not-determined |
| S11_extracted_k141_47964 | Not-determined |
| S11_extracted_k141_41723 | Not-determined |
| S11_extracted_k141_47966 | Not-determined |
| S11_extracted_k141_34754 | Not-determined |
| S11_extracted_k141_54736 | Not-determined |
| S11_extracted_k141_67326 | Not-determined |
| S11_extracted_k141_15494 | Not-determined |
| S11_extracted_k141_34757 | Not-determined |
| S11_extracted_k141_87710 | Not-determined |
| S11_extracted_k141_74650 | Not-determined |
| S11_extracted_k141_34759 | Not-determined |
| S11_extracted_k141_47975 | Not-determined |
| S11_extracted_k141_22419 | Not-determined |
| S11_extracted_k141_60935 | Not-determined |
| S11_extracted_k141_87714 | Not-determined |
| S11_extracted_k141_87716 | Not-determined |
| S11_extracted_k141_60937 | Not-determined |
| S11_extracted_k141_93794 | Not-determined |
| S11_extracted_k141_87717 | Not-determined |
| S11_extracted_k141_74657 | Not-determined |
| S11_extracted_k141_74658 | Not-determined |
| S11_extracted_k141_100268 | Low-quality |
| S11_extracted_k141_34764 | Not-determined |
| S11_extracted_k141_3002 | Not-determined |
| S11_extracted_k141_80765 | Not-determined |
| S11_extracted_k141_34766 | Low-quality |
| S11_extracted_k141_67335 | Not-determined |
| S11_extracted_k141_93801 | Not-determined |
| S11_extracted_k141_54755 | Not-determined |
| S11_extracted_k141_100278 | Low-quality |
| S11_extracted_k141_93806 | Not-determined |
| S11_extracted_k141_87726 | Not-determined |
| S11_extracted_k141_93809 | Not-determined |
| S11_extracted_k141_74663 | Not-determined |
| S11_extracted_k141_100280 | Low-quality |
| S11_extracted_k141_100284 | Low-quality |
| S11_extracted_k141_80773 | Not-determined |
| S11_extracted_k141_67350 | Not-determined |
| S11_extracted_k141_87733 | Not-determined |
| S11_extracted_k141_80784 | Not-determined |
| S11_extracted_k141_60962 | Not-determined |
| S11_extracted_k141_67354 | Low-quality |
| S11_extracted_k141_15515 | Low-quality |
| S11_extracted_k141_60968 | Not-determined |
| S11_extracted_k141_9357 | Not-determined |
| S11_extracted_k141_22435 | Not-determined |
| S11_extracted_k141_93815 | Low-quality |
| S11_extracted_k141_74667 | Not-determined |
| S11_extracted_k141_74668 | Not-determined |
| S11_extracted_k141_60974 | Not-determined |
| S11_extracted_k141_80791 | Not-determined |
| S11_extracted_k141_80794 | Not-determined |
| S11_extracted_k141_87747 | Not-determined |
| S11_extracted_k141_22448 | Not-determined |
| S11_extracted_k141_80799 | Not-determined |
| S11_extracted_k141_67374 | Not-determined |
| S11_extracted_k141_15520 | Not-determined |
| S11_extracted_k141_87756 | Not-determined |
| S11_extracted_k141_93829 | Not-determined |
| S11_extracted_k141_15522 | Not-determined |
| S11_extracted_k141_22454 | Low-quality |
| S11_extracted_k141_80809 | Not-determined |

| | |
|---|---|
| S11_extracted_k141_60983 | Not-determined |
| S11_extracted_k141_87764 | Not-determined |
| S11_extracted_k141_93841 | Not-determined |
| S11_extracted_k141_87770 | Not-determined |
| S11_extracted_k141_80819 | Not-determined |
| S11_extracted_k141_28531 | Low-quality |
| S11_extracted_k141_60993 | Low-quality |
| S11_extracted_k141_60994 | Not-determined |
| S11_extracted_k141_100315 | Low-quality |
| S11_extracted_k141_22468 | Not-determined |
| S11_extracted_k141_100316 | Not-determined |
| S11_extracted_k141_87784 | Not-determined |
| S11_extracted_k141_3013 | Not-determined |
| S11_extracted_k141_100321 | Not-determined |
| S11_extracted_k141_41772 | Not-determined |
| S11_extracted_k141_61007 | Not-determined |
| S11_extracted_k141_67404 | Not-determined |
| S11_extracted_k141_87790 | Not-determined |
| S11_extracted_k141_80840 | Not-determined |
| S11_extracted_k141_80841 | Not-determined |
| S11_extracted_k141_41774 | Not-determined |
| S11_extracted_k141_87796 | Not-determined |
| S11_extracted_k141_80846 | Not-determined |
| S11_extracted_k141_100327 | Not-determined |
| S11_extracted_k141_93848 | Not-determined |
| S11_extracted_k141_100330 | Not-determined |
| S11_extracted_k141_61014 | Not-determined |
| S11_extracted_k141_22482 | Not-determined |
| S11_extracted_k141_80850 | Not-determined |
| S11_extracted_k141_41788 | Not-determined |
| S11_extracted_k141_93851 | Not-determined |
| S11_extracted_k141_48008 | Not-determined |
| S11_extracted_k141_28559 | Not-determined |
| S11_extracted_k141_22489 | Low-quality |
| S11_extracted_k141_9391 | Not-determined |
| S11_extracted_k141_3018 | Low-quality |
| S11_extracted_k141_22491 | Not-determined |
| S11_extracted_k141_100337 | Not-determined |
| S11_extracted_k141_74694 | Not-determined |
| S11_extracted_k141_80863 | Not-determined |
| S11_extracted_k141_100339 | Not-determined |
| S11_extracted_k141_87815 | Not-determined |
| S11_extracted_k141_15553 | Not-determined |
| S11_extracted_k141_100340 | Low-quality |
| S11_extracted_k141_9393 | Not-determined |
| S11_extracted_k141_28570 | Not-determined |
| S11_extracted_k141_74700 | Not-determined |
| S11_extracted_k141_3032 | Not-determined |
| S11_extracted_k141_3035 | Not-determined |
| S11_extracted_k141_3036 | Not-determined |
| S11_extracted_k141_61033 | Not-determined |
| S11_extracted_k141_3047 | Not-determined |
| S11_extracted_k141_22514 | Not-determined |
| S11_extracted_k141_15572 | Not-determined |
| S11_extracted_k141_61036 | Not-determined |
| S11_extracted_k141_74710 | Not-determined |
| S11_extracted_k141_54768 | Not-determined |
| S11_extracted_k141_80889 | Not-determined |
| S11_extracted_k141_9415 | Not-determined |
| S11_extracted_k141_3053 | Low-quality |
| S11_extracted_k141_93878 | Not-determined |
| S11_extracted_k141_61041 | Low-quality |
| S11_extracted_k141_80896 | Not-determined |
| S11_extracted_k141_54777 | Not-determined |
| S11_extracted_k141_61049 | Not-determined |
| S11_extracted_k141_54781 | Not-determined |
| S11_extracted_k141_34776 | Not-determined |
| S11_extracted_k141_80905 | Low-quality |
| S11_extracted_k141_41790 | Not-determined |
| S11_extracted_k141_93899 | Low-quality |
| S11_extracted_k141_48017 | Not-determined |
| S11_extracted_k141_67414 | Low-quality |
| S11_extracted_k141_67415 | Low-quality |
| S11_extracted_k141_80912 | Not-determined |
| S11_extracted_k141_67418 | Not-determined |
| S11_extracted_k141_80913 | Not-determined |
| S11_extracted_k141_34783 | Not-determined |
| S11_extracted_k141_3076 | Low-quality |
| S11_extracted_k141_67424 | Low-quality |
| S11_extracted_k141_100384 | Not-determined |
| S11_extracted_k141_100385 | Low-quality |
| S11_extracted_k141_15588 | Not-determined |
| S11_extracted_k141_61065 | Not-determined |
| S11_extracted_k141_100387 | Not-determined |
| S11_extracted_k141_34800 | Not-determined |
| S11_extracted_k141_41810 | Not-determined |
| S11_extracted_k141_28608 | Low-quality |
| S11_extracted_k141_93917 | Not-determined |
| S11_extracted_k141_22552 | Not-determined |
| S11_extracted_k141_54792 | Not-determined |
| S11_extracted_k141_3084 | Low-quality |
| S11_extracted_k141_22553 | Low-quality |
| S11_extracted_k141_41814 | Not-determined |
| S11_extracted_k141_22557 | Not-determined |
| S11_extracted_k141_22560 | Not-determined |
| S11_extracted_k141_41822 | Not-determined |
| S11_extracted_k141_54799 | Not-determined |
| S11_extracted_k141_34807 | Not-determined |
| S11_extracted_k141_54801 | Not-determined |
| S11_extracted_k141_100400 | Not-determined |
| S11_extracted_k141_61070 | Not-determined |
| S11_extracted_k141_100401 | Not-determined |
| S11_extracted_k141_100402 | Not-determined |
| S11_extracted_k141_28625 | Not-determined |
| S11_extracted_k141_3102 | Not-determined |
| S11_extracted_k141_3104 | Not-determined |
| S11_extracted_k141_100410 | Not-determined |
| S11_extracted_k141_22580 | Low-quality |
| S11_extracted_k141_15607 | Not-determined |
| S11_extracted_k141_34824 | Not-determined |
| S11_extracted_k141_87845 | Not-determined |
| S11_extracted_k141_34825 | Not-determined |
| S11_extracted_k141_41836 | Not-determined |
| S11_extracted_k141_61082 | Not-determined |
| S11_extracted_k141_100417 | Not-determined |
| S11_extracted_k141_41852 | Low-quality |
| S11_extracted_k141_15619 | Low-quality |
| S11_extracted_k141_9441 | Not-determined |
| S11_extracted_k141_34835 | Low-quality |
| S11_extracted_k141_61087 | Not-determined |
| S11_extracted_k141_67482 | Not-determined |
| S11_extracted_k141_61089 | Low-quality |
| S11_extracted_k141_61090 | Not-determined |
| S11_extracted_k141_34840 | Low-quality |
| S11_extracted_k141_34843 | Low-quality |
| S11_extracted_k141_3128 | Not-determined |
| S11_extracted_k141_9446 | Not-determined |
| S11_extracted_k141_61098 | Not-determined |
| S11_extracted_k141_93965 | Not-determined |
| S11_extracted_k141_61099 | Not-determined |
| S10_extracted_43964 | Complete |
| S11_extracted_k141_48049 | Not-determined |
| S11_extracted_k141_9449 | Not-determined |
| S11_extracted_k141_34851 | Not-determined |
| S11_extracted_k141_28647 | Low-quality |
| S11_extracted_k141_61105 | Not-determined |
| S11_extracted_k141_28651 | Not-determined |
| S11_extracted_k141_48054 | Low-quality |
| S11_extracted_k141_61108 | Not-determined |
| S11_extracted_k141_48056 | Not-determined |
| S11_extracted_k141_41859 | Low-quality |
| S11_extracted_k141_100442 | Low-quality |
| S11_extracted_k141_9451 | Low-quality |
| S11_extracted_k141_34856 | Not-determined |
| S11_extracted_k141_3146 | Not-determined |
| S11_extracted_k141_67498 | Not-determined |
| S11_extracted_k141_100450 | Not-determined |
| S11_extracted_k141_61113 | Not-determined |
| S11_extracted_k141_3148 | Not-determined |
| S11_extracted_k141_67502 | Not-determined |
| S11_extracted_k141_3149 | Not-determined |
| S11_extracted_k141_34862 | Not-determined |
| S11_extracted_k141_22601 | Not-determined |
| S11_extracted_k141_67506 | Not-determined |
| S11_extracted_k141_9460 | Not-determined |
| S11_extracted_k141_22604 | Not-determined |
| S11_extracted_k141_48070 | Not-determined |
| S11_extracted_k141_34863 | Low-quality |
| S11_extracted_k141_67514 | Not-determined |
| S11_extracted_k141_54806 | Not-determined |
| S11_extracted_k141_93972 | Not-determined |
| S11_extracted_k141_22611 | Not-determined |
| S11_extracted_k141_67516 | Not-determined |
| S11_extracted_k141_93974 | Not-determined |
| S11_extracted_k141_67518 | Not-determined |
| S11_extracted_k141_3164 | Low-quality |
| S11_extracted_k141_34871 | Low-quality |
| S11_extracted_k141_61124 | Not-determined |
| S11_extracted_k141_67519 | Low-quality |
| S11_extracted_k141_100477 | Not-determined |
| S11_extracted_k141_28657 | Not-determined |
| S11_extracted_k141_3165 | Not-determined |
| S11_extracted_k141_48072 | Low-quality |
| S11_extracted_k141_9473 | Not-determined |
| S11_extracted_k141_41875 | Not-determined |
| S11_extracted_k141_22619 | Not-determined |
| S11_extracted_k141_54813 | Not-determined |
| S11_extracted_k141_15628 | Not-determined |
| S11_extracted_k141_15629 | Not-determined |
| S11_extracted_k141_22620 | Not-determined |
| S11_extracted_k141_54816 | Low-quality |
| S11_extracted_k141_48074 | Not-determined |
| S11_extracted_k141_28669 | Not-determined |
| S11_extracted_k141_28670 | Not-determined |
| S11_extracted_k141_34880 | Not-determined |
| S11_extracted_k141_61128 | Not-determined |
| S11_extracted_k141_15636 | Not-determined |
| S11_extracted_k141_15637 | Not-determined |
| S11_extracted_k141_61130 | Low-quality |
| S11_extracted_k141_48077 | Not-determined |
| S11_extracted_k141_54823 | Not-determined |
| S11_extracted_k141_100488 | Not-determined |
| S11_extracted_k141_9494 | Not-determined |
| S11_extracted_k141_48078 | Not-determined |
| S11_extracted_k141_34886 | Low-quality |
| S11_extracted_k141_22631 | Not-determined |
| S11_extracted_k141_41885 | Not-determined |
| S11_extracted_k141_54827 | Low-quality |
| S11_extracted_k141_67540 | Low-quality |
| S11_extracted_k141_100493 | Not-determined |
| S11_extracted_k141_100494 | Low-quality |
| S11_extracted_k141_61142 | Not-determined |
| S11_extracted_k141_22638 | Not-determined |
| S11_extracted_k141_15655 | Not-determined |
| S11_extracted_k141_100498 | Not-determined |
| S11_extracted_k141_54842 | Not-determined |
| S11_extracted_k141_15658 | Not-determined |
| S11_extracted_k141_3194 | Not-determined |
| S11_extracted_k141_48094 | Not-determined |
| S11_extracted_k141_100504 | Not-determined |
| S11_extracted_k141_48101 | Not-determined |
| S11_extracted_k141_61147 | Low-quality |
| S11_extracted_k141_3199 | Not-determined |
| S11_extracted_k141_61150 | Not-determined |
| S11_extracted_k141_100511 | Low-quality |
| S11_extracted_k141_9502 | Not-determined |
| S11_extracted_k141_74752 | Not-determined |
| S11_extracted_k141_67546 | Not-determined |
| S11_extracted_k141_9503 | Low-quality |
| S11_extracted_k141_54851 | Low-quality |
| S11_extracted_k141_34909 | Low-quality |
| S11_extracted_k141_74756 | Not-determined |
| S11_extracted_k141_100518 | Not-determined |
| S11_extracted_k141_54855 | Low-quality |
| S11_extracted_k141_74762 | Not-determined |
| S11_extracted_k141_74766 | Not-determined |
| S11_extracted_k141_28687 | Low-quality |
| S11_extracted_k141_28688 | Not-determined |
| S11_extracted_k141_54863 | Not-determined |
| S11_extracted_k141_100528 | Not-determined |
| S11_extracted_k141_100529 | Low-quality |
| S11_extracted_k141_28692 | Not-determined |
| S11_extracted_k141_15671 | Not-determined |
| S11_extracted_k141_15672 | Not-determined |
| S11_extracted_k141_22673 | Not-determined |
| S11_extracted_k141_100530 | Low-quality |
| S11_extracted_k141_93996 | Low-quality |
| S11_extracted_k141_54870 | Not-determined |
| S11_extracted_k141_54871 | Not-determined |
| S11_extracted_k141_48105 | Not-determined |
| S11_extracted_k141_61175 | Not-determined |
| S11_extracted_k141_22678 | Low-quality |
| S11_extracted_k141_22679 | Not-determined |
| S11_extracted_k141_94000 | Low-quality |
| S11_extracted_k141_9523 | Not-determined |
| S11_extracted_k141_22680 | Low-quality |
| S11_extracted_k141_34923 | Not-determined |
| S11_extracted_k141_74784 | Not-determined |
| S11_extracted_k141_15675 | Not-determined |
| S11_extracted_k141_9525 | Not-determined |
| S11_extracted_k141_28701 | Not-determined |
| S11_extracted_k141_22683 | Not-determined |
| S11_extracted_k141_61176 | Not-determined |
| S11_extracted_k141_3220 | Low-quality |
| S11_extracted_k141_100545 | Not-determined |
| S11_extracted_k141_94009 | Not-determined |
| S11_extracted_k141_9530 | Not-determined |
| S11_extracted_k141_15677 | Not-determined |
| S11_extracted_k141_100553 | Not-determined |
| S11_extracted_k141_15682 | Not-determined |
| S11_extracted_k141_34929 | Low-quality |
| S11_extracted_k141_41916 | Not-determined |
| S11_extracted_k141_41920 | Not-determined |
| S11_extracted_k141_100558 | Not-determined |
| S11_extracted_k141_87856 | Not-determined |
| S11_extracted_k141_34932 | Low-quality |
| S11_extracted_k141_74795 | Not-determined |
| S11_extracted_k141_15691 | Not-determined |
| S11_extracted_k141_9540 | Not-determined |
| S11_extracted_k141_61192 | Low-quality |
| S11_extracted_k141_94022 | Not-determined |
| S11_extracted_k141_80965 | Low-quality |
| S11_extracted_k141_74804 | Not-determined |
| S11_extracted_k141_80969 | Not-determined |
| S11_extracted_k141_100574 | Not-determined |
| S11_extracted_k141_9549 | Not-determined |
| S11_extracted_k141_80976 | Not-determined |
| S11_extracted_k141_41935 | Not-determined |
| S11_extracted_k141_80978 | Not-determined |
| S11_extracted_k141_28707 | Not-determined |
| S11_extracted_k141_15698 | Low-quality |
| S11_extracted_k141_74806 | Not-determined |
| S11_extracted_k141_74808 | Not-determined |
| S11_extracted_k141_3251 | Not-determined |
| S11_extracted_k141_80983 | Not-determined |
| S11_extracted_k141_94033 | Not-determined |
| S11_extracted_k141_100581 | Low-quality |
| S11_extracted_k141_28717 | Not-determined |
| S11_extracted_k141_22714 | Low-quality |
| S11_extracted_k141_94041 | Not-determined |
| S11_extracted_k141_74821 | Not-determined |
| S11_extracted_k141_3258 | Not-determined |
| S11_extracted_k141_28722 | Not-determined |
| S11_extracted_k141_74826 | Not-determined |
| S11_extracted_k141_9570 | Not-determined |
| S11_extracted_k141_74832 | Not-determined |
| S11_extracted_k141_3267 | Not-determined |
| S11_extracted_k141_22718 | Low-quality |
| S11_extracted_k141_3269 | Not-determined |
| S11_extracted_k141_3271 | Not-determined |
| S11_extracted_k141_74838 | Not-determined |
| S11_extracted_k141_100607 | Not-determined |
| S11_extracted_k141_81011 | Not-determined |
| S11_extracted_k141_61202 | Not-determined |
| S11_extracted_k141_41955 | Not-determined |
| S11_extracted_k141_41956 | Not-determined |
| S11_extracted_k141_54923 | Not-determined |
| S11_extracted_k141_61204 | Low-quality |
| S11_extracted_k141_74852 | Not-determined |
| S11_extracted_k141_34949 | Not-determined |
| S11_extracted_k141_15729 | Not-determined |
| S11_extracted_k141_15732 | Not-determined |
| S11_extracted_k141_61211 | Not-determined |
| S11_extracted_k141_41966 | Low-quality |
| S11_extracted_k141_61212 | Not-determined |
| S11_extracted_k141_9583 | Not-determined |
| S11_extracted_k141_100621 | Not-determined |
| S11_extracted_k141_94068 | Low-quality |
| S11_extracted_k141_61223 | Not-determined |
| S11_extracted_k141_61224 | Not-determined |
| S11_extracted_k141_61226 | Not-determined |
| S11_extracted_k141_9598 | Low-quality |
| S11_extracted_k141_74871 | Not-determined |
| S11_extracted_k141_61229 | Not-determined |
| S11_extracted_k141_54934 | Not-determined |
| S11_extracted_k141_9602 | Low-quality |
| S11_extracted_k141_54942 | Not-determined |
| S11_extracted_k141_81036 | Not-determined |
| S11_extracted_k141_94075 | Not-determined |
| S11_extracted_k141_15753 | Not-determined |
| S11_extracted_k141_22731 | Low-quality |
| S11_extracted_k141_28750 | Not-determined |
| S11_extracted_k141_100631 | Not-determined |
| S11_extracted_k141_94086 | Not-determined |
| S11_extracted_k141_94093 | Low-quality |
| S11_extracted_k141_28757 | Low-quality |
| S11_extracted_k141_34970 | Not-determined |
| S11_extracted_k141_28761 | Not-determined |
| S11_extracted_k141_54958 | Not-determined |
| S11_extracted_k141_22733 | Not-determined |
| S11_extracted_k141_28768 | Not-determined |
| S11_extracted_k141_9625 | Low-quality |
| S11_extracted_k141_28772 | Not-determined |
| S11_extracted_k141_34978 | Not-determined |
| S11_extracted_k141_74900 | Not-determined |
| S11_extracted_k141_74902 | Not-determined |
| S11_extracted_k141_22737 | Not-determined |
| S11_extracted_k141_61263 | Not-determined |
| S11_extracted_k141_54971 | Not-determined |
| S11_extracted_k141_28778 | Low-quality |
| S11_extracted_k141_67598 | Not-determined |
| S11_extracted_k141_74909 | Low-quality |
| S11_extracted_k141_81064 | Not-determined |
| S11_extracted_k141_15778 | Not-determined |
| S11_extracted_k141_3278 | Not-determined |
| S11_extracted_k141_15783 | Not-determined |
| S11_extracted_k141_61274 | Not-determined |
| S11_extracted_k141_67607 | Not-determined |
| S11_extracted_k141_22740 | Not-determined |
| S11_extracted_k141_74919 | Not-determined |
| S11_extracted_k141_15789 | Not-determined |
| S11_extracted_k141_81072 | Not-determined |
| S11_extracted_k141_9642 | Not-determined |
| S11_extracted_k141_54986 | Low-quality |
| S11_extracted_k141_81075 | Not-determined |
| S11_extracted_k141_94111 | Not-determined |
| S11_extracted_k141_67616 | Not-determined |
| S11_extracted_k141_3287 | Low-quality |
| S11_extracted_k141_48124 | Not-determined |
| S11_extracted_k141_54992 | Low-quality |
| S11_extracted_k141_3291 | Not-determined |
| S11_extracted_k141_61288 | Low-quality |
| S11_extracted_k141_61292 | Not-determined |
| S11_extracted_k141_54994 | Not-determined |
| S11_extracted_k141_61296 | Not-determined |
| S11_extracted_k141_61299 | Low-quality |
| S11_extracted_k141_74940 | Not-determined |
| S11_extracted_k141_74941 | Not-determined |
| S11_extracted_k141_22762 | Not-determined |
| S11_extracted_k141_9668 | Not-determined |
| S11_extracted_k141_15801 | Not-determined |
| S11_extracted_k141_74943 | Not-determined |
| S11_extracted_k141_61304 | Not-determined |
| S11_extracted_k141_3304 | Not-determined |
| S11_extracted_k141_48140 | Not-determined |
| S11_extracted_k141_41988 | Not-determined |
| S11_extracted_k141_67641 | Not-determined |
| S11_extracted_k141_41994 | Not-determined |
| S11_extracted_k141_35024 | Not-determined |
| S11_extracted_k141_9677 | Low-quality |
| S11_extracted_k141_74946 | Not-determined |
| S11_extracted_k141_55018 | Low-quality |
| S11_extracted_k141_48154 | Low-quality |
| S11_extracted_k141_22785 | Not-determined |
| S11_extracted_k141_9690 | Not-determined |
| S11_extracted_k141_35028 | Not-determined |
| S11_extracted_k141_22793 | Not-determined |
| S11_extracted_k141_67666 | Not-determined |
| S11_extracted_k141_61322 | Low-quality |
| S11_extracted_k141_100654 | Not-determined |
| S11_extracted_k141_61324 | Not-determined |
| S11_extracted_k141_3332 | Not-determined |
| S11_extracted_k141_15828 | Not-determined |
| S11_extracted_k141_42022 | Low-quality |
| S11_extracted_k141_55029 | Low-quality |
| S11_extracted_k141_42030 | Not-determined |
| S11_extracted_k141_94160 | Not-determined |
| S11_extracted_k141_100677 | Not-determined |
| S11_extracted_k141_67687 | Low-quality |
| S11_extracted_k141_94162 | Not-determined |
| S11_extracted_k141_15841 | Not-determined |
| S11_extracted_k141_94168 | Not-determined |
| S11_extracted_k141_22821 | Not-determined |
| S11_extracted_k141_28813 | Not-determined |
| S11_extracted_k141_35064 | Not-determined |
| S11_extracted_k141_42033 | Not-determined |
| S11_extracted_k141_94170 | Not-determined |
| S11_extracted_k141_15848 | Not-determined |
| S11_extracted_k141_94172 | Not-determined |
| S11_extracted_k141_42037 | Low-quality |
| S11_extracted_k141_48176 | Not-determined |
| S11_extracted_k141_48177 | Not-determined |
| S11_extracted_k141_22838 | Not-determined |
| S11_extracted_k141_28822 | Not-determined |
| S11_extracted_k141_15858 | Not-determined |
| S11_extracted_k141_15859 | Not-determined |
| S11_extracted_k141_94177 | Not-determined |
| S11_extracted_k141_15861 | Low-quality |
| S11_extracted_k141_22844 | Not-determined |
| S11_extracted_k141_87910 | Not-determined |
| S11_extracted_k141_22852 | Not-determined |
| S11_extracted_k141_94193 | Not-determined |
| S11_extracted_k141_87917 | Not-determined |
| S11_extracted_k141_42050 | Low-quality |
| S11_extracted_k141_87920 | Not-determined |
| S11_extracted_k141_42054 | Not-determined |
| S11_extracted_k141_87926 | Not-determined |
| S11_extracted_k141_22861 | Not-determined |
| S11_extracted_k141_28848 | Not-determined |
| S11_extracted_k141_42060 | Not-determined |
| S11_extracted_k141_94201 | Not-determined |
| S11_extracted_k141_55080 | Not-determined |
| S11_extracted_k141_74986 | Not-determined |
| S11_extracted_k141_55082 | Not-determined |
| S11_extracted_k141_74989 | Not-determined |
| S11_extracted_k141_22869 | Not-determined |
| S11_extracted_k141_55083 | Not-determined |
| S11_extracted_k141_9712 | Not-determined |
| S11_extracted_k141_28862 | Not-determined |
| S11_extracted_k141_74993 | Low-quality |
| S11_extracted_k141_94207 | Not-determined |
| S11_extracted_k141_87938 | Low-quality |
| S11_extracted_k141_55086 | Not-determined |
| S11_extracted_k141_94209 | Low-quality |
| S11_extracted_k141_22880 | Not-determined |
| S11_extracted_k141_35083 | Not-determined |
| S11_extracted_k141_22881 | Not-determined |
| S11_extracted_k141_75001 | Not-determined |
| S11_extracted_k141_94213 | Not-determined |
| S11_extracted_k141_42082 | Low-quality |
| S11_extracted_k141_75002 | Not-determined |
| S11_extracted_k141_48199 | Not-determined |
| S11_extracted_k141_9720 | Low-quality |
| S11_extracted_k141_75014 | Not-determined |
| S11_extracted_k141_100699 | Not-determined |
| S11_extracted_k141_61354 | Not-determined |
| S11_extracted_k141_15895 | Not-determined |
| S11_extracted_k141_28886 | Not-determined |
| S11_extracted_k141_35093 | Low-quality |
| S11_extracted_k141_22892 | Not-determined |
| S11_extracted_k141_100704 | Not-determined |
| S11_extracted_k141_94228 | Not-determined |
| S11_extracted_k141_55093 | Not-determined |
| S11_extracted_k141_35096 | Not-determined |
| S11_extracted_k141_28894 | Not-determined |
| S11_extracted_k141_94230 | Not-determined |
| S11_extracted_k141_35097 | Not-determined |
| S11_extracted_k141_100707 | Not-determined |
| S11_extracted_k141_48208 | Not-determined |
| S11_extracted_k141_55096 | Not-determined |
| S11_extracted_k141_42095 | Low-quality |
| S11_extracted_k141_15905 | Low-quality |
| S11_extracted_k141_87960 | Not-determined |
| S11_extracted_k141_35107 | Not-determined |
| S11_extracted_k141_35109 | Not-determined |
| S11_extracted_k141_55101 | Not-determined |
| S11_extracted_k141_48220 | Not-determined |
| S11_extracted_k141_94238 | Not-determined |
| S11_extracted_k141_94239 | Not-determined |
| S11_extracted_k141_94240 | Not-determined |
| S11_extracted_k141_15909 | Not-determined |
| S11_extracted_k141_48226 | Not-determined |
| S11_extracted_k141_22905 | Low-quality |
| S11_extracted_k141_75031 | Not-determined |
| S11_extracted_k141_55111 | Not-determined |
| S11_extracted_k141_48233 | Not-determined |
| S11_extracted_k141_87966 | Not-determined |
| S11_extracted_k141_87968 | Not-determined |
| S11_extracted_k141_75037 | Low-quality |
| S11_extracted_k141_42104 | Not-determined |
| S11_extracted_k141_75038 | Not-determined |
| S11_extracted_k141_28896 | Not-determined |
| S11_extracted_k141_22915 | Not-determined |
| S11_extracted_k141_22917 | Not-determined |
| S11_extracted_k141_67708 | Not-determined |
| S11_extracted_k141_55123 | Not-determined |
| S11_extracted_k141_55124 | Not-determined |
| S11_extracted_k141_100718 | Not-determined |
| S11_extracted_k141_67712 | Not-determined |
| S11_extracted_k141_48240 | Not-determined |
| S11_extracted_k141_42124 | Not-determined |
| S11_extracted_k141_15943 | Low-quality |
| S11_extracted_k141_55131 | Not-determined |
| S11_extracted_k141_100721 | Not-determined |
| S11_extracted_k141_15945 | Not-determined |
| S11_extracted_k141_15947 | Not-determined |
| S11_extracted_k141_42129 | Not-determined |
| S11_extracted_k141_94250 | Not-determined |
| S11_extracted_k141_42132 | Low-quality |
| S11_extracted_k141_75068 | Low-quality |
| S11_extracted_k141_75069 | Not-determined |
| S11_extracted_k141_48253 | Not-determined |
| S11_extracted_k141_48254 | Not-determined |
| S11_extracted_k141_100728 | Not-determined |
| S11_extracted_k141_100729 | Not-determined |
| S11_extracted_k141_94261 | Not-determined |
| S11_extracted_k141_75074 | Low-quality |
| S11_extracted_k141_35140 | Not-determined |
| S11_extracted_k141_75080 | Not-determined |
| S11_extracted_k141_67724 | Not-determined |
| S11_extracted_k141_48260 | Not-determined |
| S11_extracted_k141_94270 | Low-quality |
| S11_extracted_k141_75084 | Not-determined |
| S11_extracted_k141_67728 | Low-quality |
| S11_extracted_k141_100743 | Not-determined |
| S11_extracted_k141_42143 | Low-quality |
| S11_extracted_k141_22947 | Not-determined |
| S11_extracted_k141_15979 | Not-determined |
| S11_extracted_k141_22950 | Not-determined |
| S11_extracted_k141_94280 | Not-determined |
| S11_extracted_k141_94281 | Low-quality |
| S11_extracted_k141_42146 | Low-quality |
| S11_extracted_k141_35157 | Not-determined |
| S11_extracted_k141_100752 | Low-quality |
| S11_extracted_k141_28919 | Not-determined |
| S11_extracted_k141_28921 | Not-determined |
| S11_extracted_k141_42150 | Not-determined |
| S11_extracted_k141_81113 | Low-quality |
| S11_extracted_k141_100758 | Low-quality |
| S11_extracted_k141_42152 | Low-quality |
| S11_extracted_k141_9756 | Not-determined |
| S11_extracted_k141_9757 | Low-quality |
| S11_extracted_k141_9758 | Not-determined |
| S11_extracted_k141_28927 | Not-determined |
| S11_extracted_k141_28930 | Not-determined |
| S11_extracted_k141_42159 | Not-determined |
| S11_extracted_k141_100772 | Not-determined |
| S11_extracted_k141_67737 | Not-determined |
| S11_extracted_k141_100777 | Not-determined |
| S11_extracted_k141_15990 | Not-determined |
| S11_extracted_k141_67743 | Not-determined |
| S11_extracted_k141_22980 | Not-determined |
| S11_extracted_k141_42162 | Low-quality |
| S11_extracted_k141_48274 | Not-determined |
| S11_extracted_k141_22983 | Not-determined |
| S11_extracted_k141_35182 | Low-quality |
| S11_extracted_k141_81135 | Not-determined |
| S11_extracted_k141_75102 | Low-quality |
| S11_extracted_k141_28939 | Not-determined |
| S11_extracted_k141_42165 | Not-determined |
| S11_extracted_k141_81138 | Not-determined |
| S11_extracted_k141_48278 | Not-determined |
| S11_extracted_k141_42166 | Not-determined |
| S11_extracted_k141_42167 | Not-determined |
| S11_extracted_k141_100788 | Low-quality |
| S11_extracted_k141_15997 | Not-determined |
| S11_extracted_k141_100791 | Not-determined |
| S11_extracted_k141_16000 | Not-determined |
| S11_extracted_k141_9777 | Not-determined |
| S11_extracted_k141_67750 | Not-determined |
| S11_extracted_k141_42170 | Low-quality |
| S11_extracted_k141_28951 | Not-determined |
| S11_extracted_k141_28953 | Not-determined |
| S11_extracted_k141_22995 | Not-determined |
| S11_extracted_k141_35192 | Not-determined |
| S11_extracted_k141_67753 | Low-quality |
| S11_extracted_k141_100797 | Not-determined |
| S11_extracted_k141_100799 | Low-quality |
| S11_extracted_k141_35197 | Not-determined |

| | |
|---|---|
| S11_extracted_k141_22997 | Not-determined |
| S11_extracted_k141_23000 | Not-determined |
| S11_extracted_k141_100806 | Not-determined |
| S11_extracted_k141_35208 | Not-determined |
| S11_extracted_k141_67757 | Not-determined |
| S11_extracted_k141_48292 | Not-determined |
| S11_extracted_k141_48293 | Not-determined |
| S11_extracted_k141_35211 | Not-determined |
| S11_extracted_k141_48296 | Not-determined |
| S11_extracted_k141_42184 | Not-determined |
| S11_extracted_k141_67763 | Not-determined |
| S11_extracted_k141_28957 | Not-determined |
| S11_extracted_k141_16022 | Not-determined |
| S11_extracted_k141_55151 | Not-determined |
| S11_extracted_k141_55152 | Not-determined |
| S11_extracted_k141_67772 | Not-determined |
| S11_extracted_k141_67773 | Not-determined |
| S11_extracted_k141_61378 | Not-determined |
| S11_extracted_k141_9792 | Not-determined |
| S11_extracted_k141_28967 | Not-determined |
| S11_extracted_k141_28969 | Not-determined |
| S11_extracted_k141_100831 | Not-determined |
| S11_extracted_k141_48305 | Not-determined |
| S11_extracted_k141_3375 | Not-determined |
| S11_extracted_k141_100836 | Low-quality |
| S11_extracted_k141_81149 | Not-determined |
| S11_extracted_k141_9798 | Not-determined |
| S11_extracted_k141_67780 | Not-determined |
| S11_extracted_k141_3376 | Not-determined |
| S11_extracted_k141_100841 | Not-determined |
| S11_extracted_k141_61389 | Not-determined |
| S11_extracted_k141_23037 | Low-quality |
| S11_extracted_k141_28980 | Low-quality |
| S11_extracted_k141_94317 | Not-determined |
| S11_extracted_k141_3379 | Not-determined |
| S11_extracted_k141_28985 | Low-quality |
| S11_extracted_k141_35237 | Not-determined |
| S11_extracted_k141_94319 | Not-determined |
| S11_extracted_k141_94320 | Low-quality |
| S11_extracted_k141_55177 | Not-determined |
| S11_extracted_k141_81152 | Not-determined |
| S11_extracted_k141_23042 | Not-determined |
| S11_extracted_k141_42213 | Not-determined |
| S11_extracted_k141_75122 | Low-quality |
| S11_extracted_k141_67797 | Not-determined |
| S11_extracted_k141_9811 | Low-quality |
| S11_extracted_k141_48320 | Low-quality |
| S11_extracted_k141_100863 | Not-determined |
| S11_extracted_k141_94334 | Not-determined |
| S11_extracted_k141_9816 | Not-determined |
| S11_extracted_k141_100864 | Not-determined |
| S11_extracted_k141_16049 | Not-determined |
| S11_extracted_k141_29000 | Not-determined |
| S11_extracted_k141_100866 | Not-determined |
| S11_extracted_k141_35256 | Low-quality |
| S11_extracted_k141_48327 | Not-determined |
| S11_extracted_k141_61393 | Not-determined |
| S11_extracted_k141_42221 | Not-determined |
| S11_extracted_k141_35262 | Not-determined |
| S11_extracted_k141_16060 | Not-determined |
| S11_extracted_k141_42223 | Not-determined |
| S11_extracted_k141_35265 | Not-determined |
| S11_extracted_k141_23053 | Not-determined |
| S11_extracted_k141_3395 | Low-quality |
| S11_extracted_k141_42229 | Low-quality |
| S11_extracted_k141_61403 | Not-determined |
| S11_extracted_k141_35273 | Not-determined |
| S11_extracted_k141_61405 | Not-determined |
| S11_extracted_k141_29018 | Not-determined |
| S11_extracted_k141_29020 | Not-determined |
| S11_extracted_k141_55191 | Not-determined |
| S11_extracted_k141_81166 | Not-determined |
| S11_extracted_k141_3402 | Not-determined |
| S11_extracted_k141_81168 | Not-determined |
| S11_extracted_k141_3403 | Not-determined |
| S11_extracted_k141_94341 | Not-determined |
| S11_extracted_k141_9837 | Not-determined |
| S11_extracted_k141_48340 | Not-determined |
| S11_extracted_k141_35287 | Not-determined |
| S11_extracted_k141_94343 | Not-determined |
| S11_extracted_k141_29030 | Not-determined |
| S11_extracted_k141_35289 | Not-determined |
| S11_extracted_k141_94346 | Low-quality |
| S11_extracted_k141_42247 | Not-determined |
| S11_extracted_k141_61418 | Not-determined |
| S11_extracted_k141_23073 | Not-determined |
| S11_extracted_k141_48347 | Not-determined |
| S11_extracted_k141_94351 | Not-determined |
| S11_extracted_k141_42257 | Low-quality |
| S11_extracted_k141_55203 | Not-determined |
| S11_extracted_k141_48355 | Not-determined |
| S11_extracted_k141_48358 | Not-determined |
| S11_extracted_k141_94357 | Not-determined |
| S11_extracted_k141_55208 | Not-determined |
| S11_extracted_k141_29036 | Low-quality |
| S11_extracted_k141_42271 | Low-quality |
| S11_extracted_k141_48369 | Not-determined |
| S11_extracted_k141_29041 | Not-determined |
| S11_extracted_k141_48370 | Not-determined |
| S11_extracted_k141_67816 | Not-determined |
| S11_extracted_k141_42275 | Not-determined |
| S11_extracted_k141_16070 | Low-quality |
| S11_extracted_k141_29046 | Low-quality |
| S11_extracted_k141_81190 | Not-determined |
| S11_extracted_k141_42282 | Not-determined |
| S11_extracted_k141_29055 | Not-determined |
| S11_extracted_k141_29056 | Not-determined |
| S11_extracted_k141_16074 | Not-determined |
| S11_extracted_k141_61439 | Not-determined |
| S11_extracted_k141_16082 | Not-determined |
| S11_extracted_k141_23089 | Low-quality |
| S11_extracted_k141_75139 | Low-quality |
| S11_extracted_k141_9858 | Not-determined |
| S11_extracted_k141_42290 | Not-determined |
| S11_extracted_k141_94373 | Not-determined |
| S11_extracted_k141_75141 | Low-quality |
| S11_extracted_k141_35325 | Not-determined |
| S11_extracted_k141_75146 | Not-determined |
| S11_extracted_k141_23099 | Not-determined |
| S11_extracted_k141_23101 | Not-determined |
| S11_extracted_k141_23103 | Not-determined |
| S11_extracted_k141_9868 | Not-determined |
| S11_extracted_k141_9869 | Not-determined |
| S11_extracted_k141_61455 | Not-determined |
| S11_extracted_k141_75159 | Not-determined |
| S11_extracted_k141_9875 | Not-determined |
| S11_extracted_k141_29077 | Low-quality |
| S11_extracted_k141_16102 | Low-quality |
| S11_extracted_k141_48380 | Not-determined |
| S11_extracted_k141_16103 | Not-determined |
| S11_extracted_k141_3469 | Not-determined |
| S11_extracted_k141_9881 | Not-determined |
| S11_extracted_k141_61461 | Not-determined |
| S11_extracted_k141_35342 | Not-determined |
| S11_extracted_k141_3473 | Not-determined |
| S11_extracted_k141_16106 | Not-determined |
| S11_extracted_k141_16107 | Not-determined |
| S11_extracted_k141_55237 | Not-determined |
| S11_extracted_k141_42309 | Not-determined |
| S11_extracted_k141_23117 | Not-determined |
| S11_extracted_k141_3477 | Low-quality |
| S11_extracted_k141_100879 | Not-determined |
| S11_extracted_k141_16111 | Not-determined |
| S11_extracted_k141_87971 | Not-determined |
| S11_extracted_k141_67826 | Not-determined |
| S11_extracted_k141_48387 | Not-determined |
| S11_extracted_k141_35349 | Not-determined |
| S11_extracted_k141_42317 | Not-determined |
| S11_extracted_k141_75172 | Not-determined |
| S11_extracted_k141_67832 | Not-determined |
| S11_extracted_k141_67833 | Not-determined |
| S11_extracted_k141_87976 | Not-determined |
| S11_extracted_k141_61466 | Not-determined |
| S11_extracted_k141_9900 | Low-quality |
| S11_extracted_k141_61468 | Not-determined |
| S11_extracted_k141_42320 | Low-quality |
| S11_extracted_k141_16127 | Not-determined |
| S11_extracted_k141_75178 | Not-determined |
| S11_extracted_k141_16129 | Not-determined |
| S11_extracted_k141_81220 | Not-determined |
| S11_extracted_k141_61474 | Not-determined |
| S11_extracted_k141_23134 | Low-quality |
| S11_extracted_k141_81222 | Not-determined |
| S11_extracted_k141_16131 | Low-quality |
| S11_extracted_k141_29104 | Not-determined |
| S11_extracted_k141_100891 | Not-determined |
| S11_extracted_k141_87985 | Not-determined |
| S11_extracted_k141_48403 | Not-determined |
| S11_extracted_k141_9913 | Not-determined |
| S11_extracted_k141_16138 | Not-determined |
| S11_extracted_k141_55254 | Not-determined |
| S11_extracted_k141_16140 | Not-determined |
| S11_extracted_k141_35369 | Not-determined |
| S11_extracted_k141_61481 | Not-determined |
| S11_extracted_k141_87994 | Not-determined |
| S11_extracted_k141_81229 | Not-determined |
| S11_extracted_k141_61486 | Not-determined |
| S11_extracted_k141_23141 | Not-determined |
| S11_extracted_k141_42333 | Not-determined |
| S11_extracted_k141_55257 | Low-quality |
| S11_extracted_k141_55258 | Not-determined |
| S11_extracted_k141_35378 | Low-quality |
| S11_extracted_k141_29120 | Not-determined |
| S11_extracted_k141_42337 | Not-determined |
| S11_extracted_k141_61497 | Not-determined |
| S11_extracted_k141_48415 | Not-determined |
| S11_extracted_k141_81231 | Not-determined |
| S11_extracted_k141_48420 | Not-determined |
| S11_extracted_k141_9924 | Low-quality |
| S11_extracted_k141_35383 | Not-determined |
| S11_extracted_k141_55267 | Not-determined |
| S11_extracted_k141_100905 | Not-determined |
| S11_extracted_k141_100906 | Not-determined |
| S11_extracted_k141_48425 | Not-determined |
| S11_extracted_k141_55269 | Not-determined |
| S11_extracted_k141_29128 | Not-determined |
| S11_extracted_k141_16148 | Not-determined |
| S11_extracted_k141_61507 | Not-determined |
| S11_extracted_k141_48429 | Low-quality |
| S11_extracted_k141_3524 | Not-determined |
| S11_extracted_k141_16151 | Low-quality |
| S11_extracted_k141_29134 | Low-quality |
| S11_extracted_k141_75208 | Not-determined |
| S11_extracted_k141_81244 | Not-determined |
| S11_extracted_k141_16154 | Not-determined |
| S11_extracted_k141_23155 | Not-determined |
| S11_extracted_k141_9926 | Low-quality |
| S11_extracted_k141_35396 | Not-determined |
| S11_extracted_k141_29139 | Not-determined |
| S11_extracted_k141_94381 | Low-quality |
| S11_extracted_k141_94383 | Not-determined |
| S11_extracted_k141_3531 | Not-determined |
| S11_extracted_k141_75212 | Low-quality |
| S11_extracted_k141_48441 | Not-determined |
| S11_extracted_k141_35401 | Not-determined |
| S11_extracted_k141_75214 | Not-determined |
| S11_extracted_k141_9936 | Low-quality |
| S11_extracted_k141_9937 | Not-determined |
| S11_extracted_k141_23164 | Not-determined |
| S11_extracted_k141_81256 | Not-determined |
| S11_extracted_k141_35405 | Not-determined |
| S11_extracted_k141_81258 | Not-determined |
| S11_extracted_k141_88025 | Not-determined |
| S11_extracted_k141_42358 | Not-determined |
| S11_extracted_k141_88028 | Not-determined |
| S11_extracted_k141_100923 | Low-quality |
| S11_extracted_k141_94394 | Not-determined |
| S11_extracted_k141_94395 | Not-determined |
| S11_extracted_k141_55289 | Low-quality |
| S11_extracted_k141_29156 | Low-quality |
| S11_extracted_k141_94401 | Not-determined |
| S11_extracted_k141_23170 | Low-quality |
| S11_extracted_k141_55295 | Not-determined |
| S11_extracted_k141_29159 | Not-determined |
| S11_extracted_k141_55296 | Not-determined |
| S11_extracted_k141_88038 | Not-determined |
| S11_extracted_k141_75229 | Not-determined |
| S11_extracted_k141_35418 | Not-determined |
| S11_extracted_k141_100929 | Not-determined |
| S11_extracted_k141_88043 | Not-determined |
| S11_extracted_k141_61525 | Not-determined |
| S11_extracted_k141_88045 | Not-determined |
| S11_extracted_k141_100933 | Low-quality |
| S11_extracted_k141_88046 | Not-determined |
| S11_extracted_k141_9942 | Not-determined |
| S11_extracted_k141_55302 | Not-determined |
| S11_extracted_k141_88051 | Not-determined |
| S11_extracted_k141_23185 | Not-determined |
| S11_extracted_k141_81283 | Not-determined |
| S11_extracted_k141_81286 | Not-determined |
| S11_extracted_k141_55314 | Low-quality |
| S11_extracted_k141_81292 | Not-determined |
| S11_extracted_k141_94420 | Low-quality |
| S11_extracted_k141_88058 | Not-determined |
| S11_extracted_k141_35436 | Not-determined |
| S11_extracted_k141_35441 | Not-determined |
| S11_extracted_k141_75249 | Not-determined |
| S11_extracted_k141_55320 | Not-determined |
| S11_extracted_k141_81300 | Low-quality |
| S11_extracted_k141_29177 | Low-quality |
| S11_extracted_k141_55329 | Low-quality |
| S11_extracted_k141_16196 | Not-determined |
| S11_extracted_k141_75254 | Not-determined |
| S11_extracted_k141_35451 | Not-determined |
| S11_extracted_k141_88064 | Not-determined |
| S11_extracted_k141_61544 | Not-determined |
| S11_extracted_k141_35455 | Not-determined |
| S11_extracted_k141_16202 | Low-quality |
| S11_extracted_k141_16203 | Not-determined |
| S11_extracted_k141_16207 | Not-determined |
| S11_extracted_k141_9980 | Not-determined |
| S11_extracted_k141_100965 | Not-determined |
| S11_extracted_k141_67863 | Not-determined |
| S11_extracted_k141_29186 | Not-determined |
| S11_extracted_k141_81314 | Not-determined |
| S11_extracted_k141_81316 | Not-determined |
| S11_extracted_k141_61550 | Low-quality |
| S11_extracted_k141_61551 | Not-determined |
| S11_extracted_k141_35466 | Not-determined |
| S11_extracted_k141_67874 | Not-determined |
| S11_extracted_k141_55344 | Not-determined |
| S11_extracted_k141_42394 | Not-determined |
| S11_extracted_k141_42395 | Not-determined |
| S11_extracted_k141_35469 | Not-determined |
| S11_extracted_k141_67878 | Not-determined |
| S11_extracted_k141_29192 | Not-determined |
| S11_extracted_k141_67887 | Not-determined |
| S11_extracted_k141_61561 | Low-quality |
| S11_extracted_k141_16220 | Low-quality |
| S11_extracted_k141_61564 | Not-determined |
| S11_extracted_k141_75272 | Not-determined |
| S11_extracted_k141_61567 | Low-quality |
| S11_extracted_k141_61569 | Low-quality |
| S11_extracted_k141_35485 | Low-quality |
| S11_extracted_k141_55358 | Low-quality |
| S11_extracted_k141_94427 | Low-quality |
| S11_extracted_k141_94428 | Not-determined |
| S11_extracted_k141_67898 | Not-determined |
| S11_extracted_k141_35492 | Low-quality |
| S11_extracted_k141_81329 | Not-determined |
| S11_extracted_k141_23189 | Not-determined |
| S11_extracted_k141_48462 | Not-determined |
| S11_extracted_k141_10007 | Low-quality |
| S11_extracted_k141_35493 | Low-quality |
| S11_extracted_k141_88076 | Not-determined |
| S11_extracted_k141_81339 | Not-determined |
| S11_extracted_k141_94438 | Not-determined |
| S11_extracted_k141_67910 | Not-determined |
| S11_extracted_k141_16237 | Not-determined |
| S11_extracted_k141_16238 | Not-determined |
| S11_extracted_k141_10016 | Low-quality |
| S11_extracted_k141_42415 | Not-determined |
| S11_extracted_k141_23197 | Not-determined |
| S11_extracted_k141_42419 | Low-quality |
| S11_extracted_k141_42420 | Not-determined |
| S11_extracted_k141_100982 | Not-determined |
| S11_extracted_k141_67911 | Not-determined |
| S11_extracted_k141_75295 | Not-determined |
| S11_extracted_k141_29236 | Not-determined |
| S11_extracted_k141_23201 | Low-quality |
| S11_extracted_k141_29237 | Not-determined |
| S11_extracted_k141_67916 | Low-quality |
| S11_extracted_k141_42429 | Not-determined |
| S11_extracted_k141_67920 | Not-determined |
| S11_extracted_k141_29240 | Not-determined |
| S11_extracted_k141_100987 | Not-determined |
| S11_extracted_k141_35515 | Not-determined |
| S11_extracted_k141_16245 | Not-determined |
| S11_extracted_k141_42430 | Not-determined |
| S11_extracted_k141_23207 | Not-determined |
| S11_extracted_k141_23208 | Not-determined |
| S11_extracted_k141_81346 | Low-quality |
| S11_extracted_k141_16249 | Not-determined |
| S11_extracted_k141_23214 | Not-determined |
| S11_extracted_k141_75304 | Low-quality |
| S11_extracted_k141_67932 | Not-determined |
| S11_extracted_k141_48477 | Not-determined |
| S11_extracted_k141_48479 | Low-quality |
| S11_extracted_k141_88078 | Not-determined |
| S11_extracted_k141_101005 | Not-determined |
| S11_extracted_k141_29250 | Not-determined |
| S11_extracted_k141_88081 | Not-determined |
| S11_extracted_k141_67949 | Not-determined |
| S11_extracted_k141_23217 | Not-determined |
| S11_extracted_k141_35539 | Low-quality |
| S11_extracted_k141_48484 | Low-quality |
| S11_extracted_k141_101013 | Not-determined |
| S11_extracted_k141_55387 | Not-determined |
| S11_extracted_k141_55388 | Not-determined |
| S11_extracted_k141_75326 | Not-determined |
| S11_extracted_k141_75330 | Not-determined |
| S11_extracted_k141_75331 | Low-quality |
| S11_extracted_k141_23223 | Not-determined |
| S11_extracted_k141_10035 | Not-determined |
| S11_extracted_k141_94467 | Not-determined |
| S11_extracted_k141_94468 | Not-determined |
| S11_extracted_k141_23226 | Low-quality |
| S11_extracted_k141_67953 | Low-quality |
| S11_extracted_k141_35542 | Not-determined |
| S11_extracted_k141_61585 | Not-determined |
| S11_extracted_k141_10037 | Not-determined |
| S11_extracted_k141_16269 | Not-determined |
| S11_extracted_k141_94472 | Not-determined |
| S11_extracted_k141_67956 | Not-determined |
| S11_extracted_k141_35545 | Not-determined |
| S11_extracted_k141_67957 | Not-determined |
| S11_extracted_k141_75338 | Low-quality |
| S11_extracted_k141_35550 | Not-determined |
| S11_extracted_k141_101025 | Not-determined |
| S11_extracted_k141_101026 | Not-determined |
| S11_extracted_k141_48504 | Not-determined |
| S11_extracted_k141_42437 | Not-determined |
| S11_extracted_k141_55401 | Not-determined |
| S11_extracted_k141_42438 | Not-determined |
| S11_extracted_k141_81373 | Not-determined |
| S11_extracted_k141_42442 | Not-determined |
| S11_extracted_k141_61602 | Not-determined |
| S11_extracted_k141_101032 | Not-determined |
| S11_extracted_k141_101034 | Not-determined |
| S11_extracted_k141_67972 | Low-quality |
| S11_extracted_k141_61605 | Low-quality |
| S11_extracted_k141_94487 | Not-determined |
| S11_extracted_k141_23232 | Not-determined |
| S11_extracted_k141_94488 | Not-determined |
| S11_extracted_k141_35560 | Not-determined |
| S11_extracted_k141_61607 | Not-determined |
| S11_extracted_k141_81381 | Not-determined |
| S11_extracted_k141_75353 | Not-determined |
| S11_extracted_k141_88110 | Low-quality |
| S11_extracted_k141_61611 | Not-determined |
| S11_extracted_k141_55406 | Not-determined |
| S11_extracted_k141_42456 | Not-determined |
| S11_extracted_k141_48519 | Low-quality |
| S11_extracted_k141_10052 | Low-quality |
| S11_extracted_k141_67979 | Not-determined |
| S11_extracted_k141_101041 | Not-determined |
| S11_extracted_k141_61621 | Low-quality |
| S11_extracted_k141_48520 | Low-quality |
| S11_extracted_k141_67981 | Not-determined |
| S11_extracted_k141_101042 | Not-determined |
| S11_extracted_k141_42463 | Not-determined |
| S11_extracted_k141_101045 | Low-quality |
| S11_extracted_k141_94493 | Not-determined |
| S11_extracted_k141_10058 | Not-determined |
| S11_extracted_k141_48531 | Not-determined |
| S11_extracted_k141_88123 | Not-determined |
| S11_extracted_k141_94499 | Low-quality |
| S11_extracted_k141_48535 | Not-determined |
| S11_extracted_k141_94500 | Not-determined |
| S11_extracted_k141_101046 | Low-quality |
| S11_extracted_k141_10061 | Not-determined |
| S11_extracted_k141_101048 | Low-quality |
| S11_extracted_k141_75356 | Not-determined |
| S11_extracted_k141_61631 | Not-determined |
| S11_extracted_k141_16303 | Low-quality |
| S11_extracted_k141_88134 | Low-quality |
| S11_extracted_k141_48541 | Not-determined |
| S11_extracted_k141_10071 | Not-determined |
| S11_extracted_k141_48542 | Not-determined |
| S11_extracted_k141_42468 | Not-determined |
| S11_extracted_k141_10073 | Not-determined |
| S11_extracted_k141_16304 | Not-determined |
| S11_extracted_k141_61639 | Low-quality |
| S11_extracted_k141_61640 | Not-determined |
| S11_extracted_k141_88137 | Not-determined |
| S11_extracted_k141_10076 | Not-determined |
| S11_extracted_k141_75378 | Not-determined |
| S11_extracted_k141_10077 | Not-determined |
| S11_extracted_k141_23250 | Not-determined |
| S11_extracted_k141_75383 | Not-determined |
| S11_extracted_k141_88142 | Low-quality |
| S11_extracted_k141_81415 | Not-determined |
| S11_extracted_k141_35598 | Not-determined |
| S11_extracted_k141_35599 | Not-determined |
| S11_extracted_k141_81418 | Not-determined |
| S11_extracted_k141_35606 | Not-determined |
| S11_extracted_k141_101074 | Low-quality |
| S11_extracted_k141_10083 | Not-determined |
| S11_extracted_k141_55418 | Low-quality |
| S11_extracted_k141_88146 | Low-quality |
| S11_extracted_k141_101077 | Not-determined |
| S11_extracted_k141_42483 | Low-quality |
| S11_extracted_k141_55421 | Not-determined |
| S11_extracted_k141_42487 | Not-determined |
| S11_extracted_k141_23257 | Not-determined |
| S11_extracted_k141_67995 | Not-determined |
| S11_extracted_k141_16321 | Not-determined |
| S11_extracted_k141_10093 | Not-determined |
| S11_extracted_k141_23261 | Not-determined |
| S11_extracted_k141_88155 | Not-determined |
| S11_extracted_k141_55426 | Not-determined |
| S11_extracted_k141_55428 | Low-quality |
| S11_extracted_k141_10097 | Low-quality |
| S11_extracted_k141_68004 | Not-determined |
| S11_extracted_k141_10099 | Not-determined |
| S11_extracted_k141_75392 | Not-determined |
| S11_extracted_k141_68009 | Not-determined |
| S11_extracted_k141_68012 | Not-determined |
| S11_extracted_k141_10104 | Low-quality |
| S11_extracted_k141_42499 | Not-determined |
| S11_extracted_k141_101087 | Not-determined |
| S11_extracted_k141_68020 | Not-determined |
| S11_extracted_k141_23275 | Not-determined |
| S11_extracted_k141_101089 | Not-determined |
| S11_extracted_k141_23278 | Not-determined |
| S11_extracted_k141_42503 | Not-determined |
| S11_extracted_k141_48573 | Not-determined |
| S11_extracted_k141_88171 | Not-determined |
| S11_extracted_k141_68033 | Not-determined |
| S11_extracted_k141_75410 | Not-determined |
| S11_extracted_k141_42508 | Not-determined |
| S11_extracted_k141_68038 | Not-determined |
| S11_extracted_k141_16352 | Low-quality |
| S11_extracted_k141_88177 | Not-determined |
| S11_extracted_k141_10111 | Not-determined |
| S11_extracted_k141_55455 | Not-determined |
| S11_extracted_k141_23292 | Not-determined |
| S11_extracted_k141_16360 | Not-determined |
| S11_extracted_k141_23293 | Not-determined |
| S11_extracted_k141_42521 | Not-determined |
| S11_extracted_k141_94543 | Not-determined |
| S11_extracted_k141_48590 | Not-determined |
| S11_extracted_k141_75413 | Low-quality |
| S11_extracted_k141_55460 | Not-determined |
| S11_extracted_k141_35643 | Not-determined |
| S11_extracted_k141_94551 | Not-determined |
| S11_extracted_k141_101107 | Low-quality |
| S11_extracted_k141_81420 | Not-determined |
| S11_extracted_k141_29282 | Not-determined |
| S11_extracted_k141_61658 | Not-determined |
| S11_extracted_k141_42538 | Not-determined |
| S11_extracted_k141_29285 | Low-quality |
| S11_extracted_k141_48601 | Not-determined |
| S11_extracted_k141_81423 | Not-determined |
| S11_extracted_k141_88200 | Not-determined |
| S11_extracted_k141_23309 | Low-quality |
| S11_extracted_k141_75426 | Low-quality |
| S11_extracted_k141_61663 | Not-determined |
| S11_extracted_k141_23310 | Not-determined |
| S11_extracted_k141_68044 | Low-quality |
| S11_extracted_k141_16366 | Not-determined |
| S11_extracted_k141_88205 | Not-determined |
| S11_extracted_k141_101113 | Low-quality |
| S11_extracted_k141_68049 | Not-determined |
| S11_extracted_k141_81429 | Not-determined |
| S11_extracted_k141_42544 | Not-determined |
| S11_extracted_k141_29292 | Low-quality |
| S11_extracted_k141_81435 | Not-determined |
| S11_extracted_k141_75437 | Not-determined |
| S11_extracted_k141_68052 | Not-determined |
| S11_extracted_k141_101125 | Not-determined |
| S11_extracted_k141_101126 | Not-determined |
| S11_extracted_k141_16378 | Low-quality |
| S11_extracted_k141_42548 | Low-quality |
| S11_extracted_k141_16379 | Not-determined |
| S11_extracted_k141_55474 | Not-determined |
| S11_extracted_k141_35659 | Not-determined |
| S11_extracted_k141_88214 | Not-determined |
| S11_extracted_k141_88215 | Not-determined |
| S11_extracted_k141_42551 | Not-determined |
| S11_extracted_k141_16383 | Not-determined |
| S11_extracted_k141_94571 | Not-determined |
| S11_extracted_k141_55479 | Not-determined |
| S11_extracted_k141_42556 | Not-determined |
| S11_extracted_k141_23318 | Not-determined |
| S11_extracted_k141_75448 | Not-determined |
| S11_extracted_k141_35663 | Not-determined |
| S11_extracted_k141_101135 | Not-determined |
| S11_extracted_k141_75452 | Not-determined |
| S11_extracted_k141_75455 | Not-determined |
| S11_extracted_k141_10137 | Not-determined |
| S11_extracted_k141_101139 | Not-determined |
| S11_extracted_k141_23325 | Not-determined |
| S11_extracted_k141_94584 | Not-determined |
| S11_extracted_k141_94588 | Not-determined |
| S11_extracted_k141_23330 | Not-determined |
| S11_extracted_k141_29294 | Not-determined |
| S11_extracted_k141_23334 | Not-determined |
| S11_extracted_k141_23336 | Not-determined |
| S11_extracted_k141_61680 | Low-quality |
| S11_extracted_k141_29301 | Not-determined |
| S11_extracted_k141_48630 | Not-determined |
| S11_extracted_k141_75466 | Low-quality |
| S11_extracted_k141_35670 | Not-determined |
| S11_extracted_k141_16402 | Low-quality |
| S11_extracted_k141_68072 | Not-determined |
| S11_extracted_k141_35676 | Not-determined |
| S11_extracted_k141_75475 | Low-quality |
| S11_extracted_k141_10152 | Not-determined |
| S11_extracted_k141_81460 | Not-determined |
| S11_extracted_k141_94601 | Not-determined |
| S11_extracted_k141_16404 | Not-determined |
| S11_extracted_k141_94604 | Not-determined |
| S11_extracted_k141_55497 | Low-quality |
| S11_extracted_k141_68083 | Not-determined |
| S11_extracted_k141_23355 | Not-determined |
| S11_extracted_k141_75482 | Not-determined |
| S11_extracted_k141_10168 | Low-quality |
| S11_extracted_k141_16410 | Not-determined |
| S11_extracted_k141_81474 | Not-determined |
| S11_extracted_k141_75488 | Low-quality |
| S11_extracted_k141_16414 | Not-determined |
| S11_extracted_k141_55512 | Not-determined |
| S11_extracted_k141_68095 | Low-quality |
| S11_extracted_k141_75498 | Not-determined |
| S11_extracted_k141_55519 | Not-determined |
| S11_extracted_k141_35705 | Not-determined |
| S11_extracted_k141_48659 | Not-determined |
| S11_extracted_k141_48663 | Not-determined |
| S11_extracted_k141_16419 | Not-determined |
| S11_extracted_k141_42582 | Low-quality |
| S11_extracted_k141_101180 | Not-determined |
| S11_extracted_k141_35714 | Not-determined |
| S11_extracted_k141_55533 | Low-quality |
| S11_extracted_k141_16434 | Not-determined |
| S11_extracted_k141_61709 | Not-determined |
| S11_extracted_k141_94634 | Not-determined |
| S11_extracted_k141_35720 | Low-quality |
| S11_extracted_k141_55538 | Not-determined |
| S11_extracted_k141_35721 | Not-determined |
| S11_extracted_k141_55540 | Not-determined |
| S11_extracted_k141_42585 | Not-determined |
| S11_extracted_k141_35726 | Not-determined |
| S11_extracted_k141_55542 | Not-determined |
| S11_extracted_k141_42588 | Not-determined |
| S11_extracted_k141_42590 | Low-quality |
| S11_extracted_k141_75522 | Not-determined |
| S11_extracted_k141_55546 | Not-determined |
| S11_extracted_k141_68115 | Not-determined |

| | |
|---|---|
| S11_extracted_k141_75524 | Not-determined |
| S11_extracted_k141_94644 | Low-quality |
| S11_extracted_k141_55548 | Low-quality |
| S11_extracted_k141_42598 | Not-determined |
| S11_extracted_k141_94655 | Not-determined |
| S11_extracted_k141_101191 | Low-quality |
| S11_extracted_k141_101192 | Not-determined |
| S11_extracted_k141_101193 | Not-determined |
| S11_extracted_k141_42603 | Not-determined |
| S11_extracted_k141_68127 | Not-determined |
| S11_extracted_k141_35745 | Low-quality |
| S11_extracted_k141_55566 | Not-determined |
| S11_extracted_k141_42613 | Not-determined |
| S11_extracted_k141_61721 | Not-determined |
| S11_extracted_k141_55569 | Not-determined |
| S11_extracted_k141_3537 | Low-quality |
| S11_extracted_k141_75543 | Not-determined |
| S11_extracted_k141_3540 | Not-determined |
| S11_extracted_k141_94671 | Not-determined |
| S11_extracted_k141_94674 | Low-quality |
| S11_extracted_k141_42618 | Low-quality |
| S11_extracted_k141_75547 | Not-determined |
| S11_extracted_k141_55574 | Not-determined |
| S11_extracted_k141_42624 | Not-determined |
| S11_extracted_k141_42625 | Not-determined |
| S11_extracted_k141_81480 | Not-determined |
| S11_extracted_k141_3552 | Not-determined |
| S11_extracted_k141_23358 | Not-determined |
| S11_extracted_k141_94682 | Not-determined |
| S11_extracted_k141_94689 | Not-determined |
| S11_extracted_k141_10175 | Not-determined |
| S11_extracted_k141_55583 | Low-quality |
| S11_extracted_k141_94692 | Not-determined |
| S11_extracted_k141_94697 | Not-determined |
| S11_extracted_k141_81493 | Not-determined |
| S11_extracted_k141_61737 | Not-determined |
| S11_extracted_k141_23367 | Not-determined |
| S11_extracted_k141_10182 | Not-determined |
| S11_extracted_k141_94702 | Not-determined |
| S11_extracted_k141_94703 | Low-quality |
| S11_extracted_k141_101211 | Low-quality |
| S11_extracted_k141_55601 | Not-determined |
| S11_extracted_k141_94709 | Low-quality |
| S11_extracted_k141_101213 | Not-determined |
| S11_extracted_k141_23377 | Not-determined |
| S11_extracted_k141_61748 | Not-determined |
| S11_extracted_k141_101218 | Not-determined |
| S11_extracted_k141_101221 | Not-determined |
| S11_extracted_k141_23383 | Not-determined |
| S11_extracted_k141_10200 | Not-determined |
| S11_extracted_k141_23385 | Not-determined |
| S11_extracted_k141_16468 | Not-determined |
| S11_extracted_k141_61755 | Not-determined |
| S11_extracted_k141_29329 | Not-determined |
| S11_extracted_k141_29330 | Not-determined |
| S11_extracted_k141_48698 | Not-determined |
| S11_extracted_k141_75558 | Not-determined |
| S11_extracted_k141_61757 | Not-determined |
| S11_extracted_k141_29334 | Low-quality |
| S11_extracted_k141_75559 | Not-determined |
| S11_extracted_k141_55613 | Low-quality |
| S11_extracted_k141_88245 | Not-determined |
| S11_extracted_k141_61766 | Not-determined |
| S11_extracted_k141_48705 | Not-determined |
| S11_extracted_k141_75567 | Not-determined |
| S11_extracted_k141_29339 | Not-determined |
| S11_extracted_k141_16476 | Not-determined |
| S11_extracted_k141_94741 | Not-determined |
| S11_extracted_k141_29341 | Not-determined |
| S11_extracted_k141_55622 | Not-determined |
| S11_extracted_k141_48717 | Low-quality |
| S11_extracted_k141_29345 | Not-determined |
| S11_extracted_k141_48720 | Not-determined |
| S11_extracted_k141_94745 | Not-determined |
| S11_extracted_k141_81504 | Not-determined |
| S11_extracted_k141_61772 | Not-determined |
| S11_extracted_k141_61774 | Low-quality |
| S11_extracted_k141_61775 | Not-determined |
| S11_extracted_k141_29353 | Not-determined |
| S11_extracted_k141_55633 | Not-determined |
| S11_extracted_k141_61778 | Not-determined |
| S11_extracted_k141_61781 | Not-determined |
| S11_extracted_k141_61783 | Not-determined |
| S11_extracted_k141_35789 | Not-determined |
| S11_extracted_k141_61784 | Not-determined |
| S11_extracted_k141_88255 | Low-quality |
| S11_extracted_k141_94751 | Not-determined |
| S11_extracted_k141_35791 | Not-determined |
| S11_extracted_k141_16495 | Not-determined |
| S11_extracted_k141_55642 | Not-determined |
| S11_extracted_k141_75578 | Not-determined |
| S11_extracted_k141_23411 | Not-determined |
| S11_extracted_k141_16497 | Not-determined |
| S11_extracted_k141_55645 | Not-determined |
| S11_extracted_k141_3572 | Not-determined |
| S11_extracted_k141_16500 | Not-determined |
| S11_extracted_k141_75583 | Not-determined |
| S11_extracted_k141_10230 | Not-determined |
| S11_extracted_k141_88266 | Not-determined |
| S11_extracted_k141_3581 | Not-determined |
| S11_extracted_k141_16514 | Not-determined |
| S11_extracted_k141_23415 | Not-determined |
| S11_extracted_k141_88273 | Not-determined |
| S11_extracted_k141_23420 | Not-determined |
| S11_extracted_k141_23421 | Not-determined |
| S11_extracted_k141_23424 | Not-determined |
| S11_extracted_k141_55662 | Not-determined |
| S11_extracted_k141_81517 | Low-quality |
| S11_extracted_k141_94778 | Low-quality |
| S11_extracted_k141_10251 | Not-determined |
| S11_extracted_k141_10252 | Not-determined |
| S11_extracted_k141_81519 | Not-determined |
| S11_extracted_k141_3592 | Low-quality |
| S11_extracted_k141_10254 | Not-determined |
| S11_extracted_k141_101260 | Not-determined |
| S11_extracted_k141_94784 | Low-quality |
| S11_extracted_k141_10258 | Not-determined |
| S11_extracted_k141_61820 | Low-quality |
| S11_extracted_k141_81522 | Low-quality |
| S11_extracted_k141_55677 | Not-determined |
| S11_extracted_k141_88294 | Not-determined |
| S11_extracted_k141_88295 | Low-quality |
| S11_extracted_k141_3603 | Not-determined |
| S11_extracted_k141_88296 | Not-determined |
| S11_extracted_k141_35841 | Not-determined |
| S11_extracted_k141_3606 | Not-determined |
| S11_extracted_k141_10267 | Not-determined |
| S11_extracted_k141_35843 | Not-determined |
| S11_extracted_k141_101270 | Not-determined |
| S11_extracted_k141_29379 | Not-determined |
| S10_vs_43964\|\|full | Complete |
| S11_extracted_k141_35848 | Not-determined |
| S11_extracted_k141_75603 | Not-determined |
| S11_extracted_k141_29385 | Low-quality |
| S11_extracted_k141_101274 | Not-determined |
| S11_extracted_k141_42638 | Not-determined |
| S11_extracted_k141_10277 | Not-determined |
| S11_extracted_k141_29393 | Not-determined |
| S11_extracted_k141_75618 | Not-determined |
| S11_extracted_k141_61838 | Low-quality |
| S11_extracted_k141_88299 | Not-determined |
| S11_extracted_k141_55697 | Not-determined |
| S11_extracted_k141_88301 | Not-determined |
| S11_extracted_k141_81536 | Not-determined |
| S11_extracted_k141_88303 | Not-determined |
| S11_extracted_k141_55699 | Low-quality |
| S11_extracted_k141_101282 | Not-determined |
| S11_extracted_k141_81542 | Not-determined |
| S11_extracted_k141_3628 | Low-quality |
| S11_extracted_k141_81545 | Not-determined |
| S11_extracted_k141_101285 | Not-determined |
| S11_extracted_k141_35856 | Not-determined |
| S11_extracted_k141_68149 | Not-determined |
| S11_extracted_k141_88314 | Not-determined |
| S11_extracted_k141_42650 | Not-determined |
| S11_extracted_k141_81551 | Not-determined |
| S11_extracted_k141_10292 | Low-quality |
| S11_extracted_k141_88317 | Low-quality |
| S11_extracted_k141_3640 | Not-determined |
| S11_extracted_k141_10297 | Not-determined |
| S11_extracted_k141_48738 | Low-quality |
| S11_extracted_k141_81557 | Not-determined |
| S11_extracted_k141_94817 | Not-determined |
| S11_extracted_k141_75642 | Not-determined |
| S11_extracted_k141_94822 | Not-determined |
| S11_extracted_k141_88329 | Not-determined |
| S11_extracted_k141_75645 | Low-quality |
| S11_extracted_k141_48741 | Not-determined |
| S11_extracted_k141_3642 | Low-quality |
| S11_extracted_k141_10311 | Not-determined |
| S11_extracted_k141_35870 | Low-quality |
| S11_extracted_k141_55719 | Not-determined |
| S11_extracted_k141_88338 | Not-determined |
| S11_extracted_k141_3652 | Not-determined |
| S11_extracted_k141_35876 | Not-determined |
| S11_extracted_k141_42669 | Low-quality |
| S11_extracted_k141_101316 | Not-determined |
| S11_extracted_k141_3656 | Not-determined |
| S11_extracted_k141_88345 | Not-determined |
| S11_extracted_k141_81575 | Not-determined |
| S11_extracted_k141_61870 | Not-determined |
| S11_extracted_k141_101319 | Not-determined |
| S11_extracted_k141_94831 | Not-determined |
| S11_extracted_k141_10320 | Not-determined |
| S11_extracted_k141_10321 | Not-determined |
| S11_extracted_k141_88349 | Not-determined |
| S11_extracted_k141_94835 | Not-determined |
| S11_extracted_k141_75658 | Low-quality |
| S11_extracted_k141_55736 | Not-determined |
| S11_extracted_k141_3673 | Low-quality |
| S11_extracted_k141_42678 | Low-quality |
| S11_extracted_k141_75661 | Not-determined |
| S11_extracted_k141_10329 | Not-determined |
| S11_extracted_k141_42683 | Not-determined |
| S11_extracted_k141_42686 | Low-quality |
| S11_extracted_k141_42688 | Not-determined |
| S11_extracted_k141_3679 | Not-determined |
| S11_extracted_k141_3680 | Not-determined |
| S11_extracted_k141_55745 | Low-quality |
| S11_extracted_k141_94845 | Not-determined |
| S11_extracted_k141_94847 | Not-determined |
| S11_extracted_k141_3682 | Low-quality |
| S11_extracted_k141_55752 | Not-determined |
| S11_extracted_k141_94848 | Not-determined |
| S11_extracted_k141_35893 | Not-determined |
| S11_extracted_k141_61882 | Not-determined |
| S11_extracted_k141_94853 | Not-determined |
| S11_extracted_k141_101331 | Not-determined |
| S11_extracted_k141_35903 | Low-quality |
| S11_extracted_k141_101332 | Not-determined |
| S11_extracted_k141_55758 | Not-determined |
| S11_extracted_k141_94860 | Not-determined |
| S11_extracted_k141_3689 | Not-determined |
| S11_extracted_k141_94866 | Not-determined |
| S11_extracted_k141_88359 | Not-determined |
| S11_extracted_k141_61891 | Not-determined |
| S11_extracted_k141_61897 | Not-determined |
| S11_extracted_k141_61898 | Low-quality |
| S11_extracted_k141_48755 | Not-determined |
| S11_extracted_k141_75699 | Not-determined |
| S11_extracted_k141_61902 | Low-quality |
| S11_extracted_k141_55776 | Low-quality |
| S11_extracted_k141_10349 | Not-determined |
| S11_extracted_k141_42707 | Low-quality |
| S11_extracted_k141_48765 | Not-determined |
| S11_extracted_k141_35922 | Not-determined |
| S11_extracted_k141_94879 | Not-determined |
| S11_extracted_k141_55780 | Not-determined |
| S11_extracted_k141_61910 | Not-determined |
| S11_extracted_k141_42713 | Not-determined |
| S11_extracted_k141_3710 | Not-determined |
| S11_extracted_k141_48779 | Not-determined |
| S11_extracted_k141_10353 | Low-quality |
| S11_extracted_k141_94895 | Not-determined |
| S11_extracted_k141_94897 | Not-determined |
| S11_extracted_k141_3720 | Not-determined |
| S11_extracted_k141_10355 | Not-determined |
| S11_extracted_k141_94902 | Not-determined |
| S11_extracted_k141_35930 | Not-determined |
| S11_extracted_k141_61920 | Not-determined |
| S11_extracted_k141_3724 | Not-determined |
| S11_extracted_k141_61924 | Not-determined |
| S11_extracted_k141_3727 | Not-determined |
| S11_extracted_k141_3728 | Not-determined |
| S11_extracted_k141_3729 | Not-determined |
| S11_extracted_k141_55783 | Not-determined |
| S11_extracted_k141_75707 | Not-determined |
| S11_extracted_k141_55786 | Not-determined |
| S11_extracted_k141_61933 | Not-determined |
| S11_extracted_k141_23463 | Not-determined |
| S11_extracted_k141_61938 | Low-quality |
| S11_extracted_k141_101350 | Low-quality |
| S11_extracted_k141_42734 | Not-determined |
| S11_extracted_k141_42736 | Low-quality |
| S11_extracted_k141_101360 | Not-determined |
| S11_extracted_k141_35956 | Not-determined |
| S11_extracted_k141_55804 | Not-determined |
| S11_extracted_k141_42744 | Not-determined |
| S11_extracted_k141_88367 | Not-determined |
| S11_extracted_k141_55810 | Not-determined |
| S11_extracted_k141_68170 | Low-quality |
| S11_extracted_k141_10364 | Not-determined |
| S11_extracted_k141_68171 | Not-determined |
| S11_extracted_k141_23487 | Not-determined |
| S11_extracted_k141_55818 | Not-determined |
| S11_extracted_k141_101374 | Not-determined |
| S11_extracted_k141_35964 | Low-quality |
| S11_extracted_k141_55824 | Not-determined |
| S11_extracted_k141_68180 | Not-determined |
| S11_extracted_k141_55827 | Not-determined |
| S11_extracted_k141_23498 | Not-determined |
| S11_extracted_k141_10370 | Not-determined |
| S11_extracted_k141_88382 | Not-determined |
| S11_extracted_k141_23501 | Not-determined |
| S11_extracted_k141_10372 | Not-determined |
| S10_vs_14557\|\|full | Complete |
| S11_extracted_k141_68189 | Not-determined |
| S11_extracted_k141_23503 | Not-determined |
| S11_extracted_k141_68191 | Not-determined |
| S11_extracted_k141_23504 | Not-determined |
| S11_extracted_k141_61974 | Not-determined |
| S11_extracted_k141_75716 | Not-determined |
| S11_extracted_k141_29428 | Not-determined |
| S11_extracted_k141_61976 | Low-quality |
| S11_extracted_k141_42762 | Not-determined |
| S11_extracted_k141_75720 | Not-determined |
| S11_extracted_k141_23508 | Low-quality |
| S11_extracted_k141_75721 | Low-quality |
| S11_extracted_k141_61981 | Not-determined |
| S11_extracted_k141_10380 | Not-determined |
| S11_extracted_k141_88395 | Not-determined |
| S11_extracted_k141_88398 | Low-quality |
| S11_extracted_k141_55833 | Not-determined |
| S11_extracted_k141_23515 | Not-determined |
| S11_extracted_k141_55835 | Not-determined |
| S11_extracted_k141_101390 | Not-determined |
| S11_extracted_k141_81588 | Not-determined |
| S11_extracted_k141_88400 | Not-determined |
| S11_extracted_k141_10387 | Low-quality |
| S11_extracted_k141_68203 | Not-determined |
| S11_extracted_k141_68204 | Not-determined |
| S11_extracted_k141_55840 | Not-determined |
| S11_extracted_k141_61993 | Not-determined |
| S11_extracted_k141_81595 | Not-determined |
| S11_extracted_k141_35979 | Not-determined |
| S11_extracted_k141_35980 | Not-determined |
| S11_extracted_k141_10394 | Not-determined |
| S11_extracted_k141_10396 | Not-determined |
| S11_extracted_k141_61997 | Not-determined |
| S11_extracted_k141_61998 | Not-determined |
| S11_extracted_k141_81599 | Not-determined |
| S11_extracted_k141_68209 | Low-quality |
| S11_extracted_k141_3767 | Not-determined |
| S11_extracted_k141_62002 | Not-determined |
| S11_extracted_k141_81604 | Not-determined |
| S11_extracted_k141_42781 | Not-determined |
| S11_extracted_k141_35990 | Not-determined |
| S11_extracted_k141_23531 | Low-quality |
| S11_extracted_k141_42784 | Not-determined |
| S11_extracted_k141_42785 | Low-quality |
| S11_extracted_k141_10402 | Not-determined |
| S11_extracted_k141_75743 | Not-determined |
| S11_extracted_k141_10407 | Not-determined |
| S11_extracted_k141_101418 | Not-determined |
| S11_extracted_k141_36000 | Not-determined |
| S10_extracted_36880 | Complete |
| S11_extracted_k141_75748 | Not-determined |
| S11_extracted_k141_29461 | Not-determined |
| S11_extracted_k141_3781 | Not-determined |
| S11_extracted_k141_81616 | Not-determined |
| S11_extracted_k141_10411 | Not-determined |
| S11_extracted_k141_81622 | Not-determined |
| S11_extracted_k141_10416 | Not-determined |
| S11_extracted_k141_3787 | Not-determined |
| S11_extracted_k141_3789 | Low-quality |
| S11_extracted_k141_81629 | Not-determined |
| S11_extracted_k141_81630 | Not-determined |
| S11_extracted_k141_94936 | Not-determined |
| S11_extracted_k141_94938 | Not-determined |
| S11_extracted_k141_94939 | Not-determined |
| S11_extracted_k141_94941 | Low-quality |
| S11_extracted_k141_94943 | Not-determined |
| S11_extracted_k141_16531 | Not-determined |
| S11_extracted_k141_68227 | Not-determined |
| S11_extracted_k141_62014 | Not-determined |
| S11_extracted_k141_42814 | Not-determined |
| S11_extracted_k141_16533 | Not-determined |
| S11_extracted_k141_29485 | Not-determined |
| S11_extracted_k141_94947 | Not-determined |
| S11_extracted_k141_94948 | Not-determined |
| S11_extracted_k141_88437 | Not-determined |
| S11_extracted_k141_68234 | Low-quality |
| S11_extracted_k141_101440 | Not-determined |
| S11_extracted_k141_94952 | Not-determined |
| S11_extracted_k141_101442 | Not-determined |
| S11_extracted_k141_42824 | Not-determined |
| S11_extracted_k141_16542 | Not-determined |
| S11_extracted_k141_94957 | Not-determined |
| S11_extracted_k141_16545 | Not-determined |
| S11_extracted_k141_81642 | Low-quality |
| S11_extracted_k141_29494 | Low-quality |
| S11_extracted_k141_48794 | Not-determined |
| S11_extracted_k141_88447 | Not-determined |
| S11_extracted_k141_42834 | Not-determined |
| S11_extracted_k141_48798 | Not-determined |
| S11_extracted_k141_81651 | Not-determined |
| S11_extracted_k141_42841 | Low-quality |
| S11_extracted_k141_29503 | Low-quality |
| S11_extracted_k141_101449 | Low-quality |
| S11_extracted_k141_94969 | Not-determined |
| S11_extracted_k141_42844 | Low-quality |
| S11_extracted_k141_81653 | Not-determined |
| S11_extracted_k141_81656 | Low-quality |
| S11_extracted_k141_16548 | Not-determined |
| S11_extracted_k141_42847 | Not-determined |
| S11_extracted_k141_29511 | Not-determined |
| S11_extracted_k141_81660 | Not-determined |
| S11_extracted_k141_16552 | Not-determined |
| S11_extracted_k141_16554 | Not-determined |
| S11_extracted_k141_101455 | Low-quality |
| S11_extracted_k141_48811 | Not-determined |
| S11_extracted_k141_48813 | Not-determined |
| S11_extracted_k141_36012 | Not-determined |
| S11_extracted_k141_88466 | Low-quality |
| S11_extracted_k141_94985 | Not-determined |
| S11_extracted_k141_75763 | Not-determined |
| S11_extracted_k141_68248 | Not-determined |
| S11_extracted_k141_75765 | Not-determined |
| S11_extracted_k141_36021 | Not-determined |
| S11_extracted_k141_36024 | Not-determined |
| S11_extracted_k141_88480 | Not-determined |
| S11_extracted_k141_55886 | Not-determined |
| S11_extracted_k141_42862 | Low-quality |
| S11_extracted_k141_75779 | Not-determined |
| S11_extracted_k141_55893 | Not-determined |
| S11_extracted_k141_88485 | Not-determined |
| S11_extracted_k141_42869 | Not-determined |
| S11_extracted_k141_23541 | Not-determined |
| S11_extracted_k141_68261 | Low-quality |
| S11_extracted_k141_55897 | Not-determined |
| S11_extracted_k141_88490 | Not-determined |
| S11_extracted_k141_29543 | Not-determined |
| S11_extracted_k141_48852 | Not-determined |
| S11_extracted_k141_48854 | Not-determined |
| S11_extracted_k141_62062 | Not-determined |
| S11_extracted_k141_23547 | Low-quality |
| S11_extracted_k141_55902 | Low-quality |
| S11_extracted_k141_36030 | Not-determined |
| S11_extracted_k141_29549 | Not-determined |
| S11_extracted_k141_75799 | Not-determined |
| S11_extracted_k141_42880 | Not-determined |
| S11_extracted_k141_81695 | Not-determined |
| S11_extracted_k141_36039 | Not-determined |
| S11_extracted_k141_29554 | Not-determined |
| S11_extracted_k141_75806 | Not-determined |
| S11_extracted_k141_101477 | Not-determined |
| S11_extracted_k141_29557 | Not-determined |
| S11_extracted_k141_36041 | Not-determined |
| S11_extracted_k141_75809 | Not-determined |
| S11_extracted_k141_75810 | Not-determined |
| S11_extracted_k141_81702 | Not-determined |
| S11_extracted_k141_81703 | Not-determined |
| S11_extracted_k141_62088 | Not-determined |
| S11_extracted_k141_101488 | Low-quality |
| S11_extracted_k141_81708 | Not-determined |
| S11_extracted_k141_62089 | Not-determined |
| S11_extracted_k141_88513 | Not-determined |
| S11_extracted_k141_23559 | Not-determined |
| S11_extracted_k141_81714 | Not-determined |
| S11_extracted_k141_55907 | Not-determined |
| S11_extracted_k141_81717 | Not-determined |
| S11_extracted_k141_29566 | Not-determined |
| S11_extracted_k141_55912 | Not-determined |
| S9_extracted_k141_64844 | High-quality |
| S11_extracted_k141_81721 | Not-determined |
| S11_extracted_k141_88527 | Not-determined |
| S11_extracted_k141_68266 | Low-quality |
| S11_extracted_k141_23574 | Not-determined |
| S11_extracted_k141_81725 | Low-quality |
| S11_extracted_k141_81726 | Low-quality |
| S11_extracted_k141_88536 | Not-determined |
| S11_extracted_k141_55933 | Not-determined |
| S11_extracted_k141_101512 | Not-determined |
| S11_extracted_k141_68275 | Low-quality |
| S11_extracted_k141_16600 | Not-determined |
| S11_extracted_k141_23586 | Not-determined |
| S11_extracted_k141_29582 | Low-quality |
| S11_extracted_k141_55934 | Low-quality |
| S11_extracted_k141_101521 | Not-determined |
| S11_extracted_k141_101524 | Not-determined |
| S11_extracted_k141_68278 | Low-quality |
| S11_extracted_k141_101526 | Not-determined |
| S11_extracted_k141_88557 | Not-determined |
| S11_extracted_k141_10423 | Not-determined |
| S11_extracted_k141_36063 | Low-quality |
| S11_extracted_k141_29593 | Not-determined |
| S11_extracted_k141_55939 | Not-determined |
| S11_extracted_k141_10424 | Not-determined |
| S11_extracted_k141_3815 | Not-determined |
| S11_extracted_k141_3818 | Not-determined |
| S11_extracted_k141_94992 | Not-determined |
| S11_extracted_k141_36069 | Not-determined |
| S11_extracted_k141_75824 | Not-determined |
| S11_extracted_k141_75825 | Not-determined |
| S11_extracted_k141_3822 | Not-determined |
| S11_extracted_k141_68293 | Not-determined |
| S11_extracted_k141_16619 | Not-determined |
| S11_extracted_k141_36074 | Not-determined |
| S11_extracted_k141_68295 | Not-determined |
| S11_extracted_k141_81734 | Low-quality |
| S11_extracted_k141_29603 | Not-determined |
| S11_extracted_k141_23606 | Not-determined |
| S11_extracted_k141_75829 | Not-determined |
| S11_extracted_k141_88565 | Not-determined |
| S11_extracted_k141_42891 | Not-determined |
| S11_extracted_k141_95001 | Low-quality |
| S11_extracted_k141_95002 | Not-determined |
| S11_extracted_k141_23615 | Not-determined |
| S11_extracted_k141_16626 | Not-determined |
| S11_extracted_k141_23618 | Low-quality |
| S11_extracted_k141_16628 | Not-determined |
| S11_extracted_k141_16629 | Not-determined |
| S11_extracted_k141_95011 | Not-determined |
| S11_extracted_k141_36088 | Not-determined |
| S11_extracted_k141_55967 | Not-determined |
| S11_extracted_k141_62115 | Not-determined |
| S11_extracted_k141_75849 | Not-determined |
| S11_extracted_k141_81736 | Not-determined |
| S11_extracted_k141_101540 | Not-determined |
| S11_extracted_k141_88582 | Low-quality |
| S11_extracted_k141_88583 | Not-determined |
| S11_extracted_k141_101542 | Not-determined |
| S11_extracted_k141_23642 | Not-determined |
| S11_extracted_k141_36098 | Low-quality |
| S11_extracted_k141_88588 | Not-determined |
| S11_extracted_k141_68313 | Not-determined |
| S11_extracted_k141_42913 | Not-determined |
| S11_extracted_k141_62125 | Not-determined |
| S11_extracted_k141_3837 | Not-determined |
| S11_extracted_k141_23650 | Not-determined |
| S11_extracted_k141_36104 | Not-determined |
| S11_extracted_k141_36106 | Not-determined |
| S11_extracted_k141_55986 | Not-determined |
| S11_extracted_k141_3845 | Low-quality |
| S11_extracted_k141_62133 | Not-determined |
| S11_extracted_k141_88602 | Low-quality |
| S11_extracted_k141_23658 | Low-quality |
| S11_extracted_k141_95049 | Not-determined |
| S11_extracted_k141_48868 | Low-quality |
| S11_extracted_k141_88607 | Not-determined |
| S11_extracted_k141_88609 | Not-determined |
| S11_extracted_k141_3854 | Not-determined |
| S11_extracted_k141_16655 | Not-determined |
| S11_extracted_k141_3856 | Low-quality |
| S11_extracted_k141_36112 | Not-determined |
| S11_extracted_k141_88614 | Not-determined |
| S11_extracted_k141_68324 | Low-quality |
| S11_extracted_k141_75861 | Not-determined |
| S11_extracted_k141_3859 | Low-quality |
| S11_extracted_k141_36115 | Not-determined |
| S11_extracted_k141_16657 | Not-determined |
| S11_extracted_k141_88618 | Not-determined |
| S11_extracted_k141_68328 | Not-determined |
| S11_extracted_k141_101577 | Not-determined |
| S11_extracted_k141_68329 | Low-quality |
| S11_extracted_k141_16661 | Not-determined |
| S11_extracted_k141_3865 | Not-determined |
| S11_extracted_k141_16664 | Not-determined |
| S11_extracted_k141_36122 | Not-determined |
| S11_extracted_k141_68333 | Not-determined |
| S11_extracted_k141_101587 | Low-quality |
| S11_extracted_k141_101589 | Low-quality |
| S11_extracted_k141_68338 | Not-determined |
| S11_extracted_k141_75875 | Not-determined |
| S11_extracted_k141_101601 | Not-determined |
| S11_extracted_k141_101602 | Not-determined |
| S11_extracted_k141_88644 | Not-determined |
| S11_extracted_k141_62149 | Low-quality |
| S11_extracted_k141_16681 | Not-determined |
| S11_extracted_k141_56027 | Not-determined |
| S11_extracted_k141_88645 | Low-quality |
| S11_extracted_k141_16685 | Not-determined |
| S11_extracted_k141_48900 | Low-quality |
| S11_extracted_k141_23671 | Not-determined |
| S11_extracted_k141_48909 | Low-quality |
| S11_extracted_k141_75888 | Not-determined |
| S11_extracted_k141_23674 | Low-quality |
| S11_extracted_k141_16695 | Not-determined |
| S11_extracted_k141_36135 | Not-determined |
| S11_extracted_k141_16697 | Not-determined |
| S11_extracted_k141_48913 | Not-determined |
| S11_extracted_k141_16700 | Low-quality |
| S11_extracted_k141_48915 | Not-determined |
| S11_extracted_k141_16706 | Not-determined |
| S11_extracted_k141_62166 | Not-determined |
| S11_extracted_k141_16710 | Low-quality |
| S11_extracted_k141_36147 | Not-determined |
| S11_extracted_k141_88668 | Not-determined |
| S11_extracted_k141_10453 | Not-determined |
| S11_extracted_k141_3884 | Low-quality |
| S11_extracted_k141_48922 | Not-determined |
| S11_extracted_k141_48923 | Low-quality |
| S11_extracted_k141_16719 | Not-determined |
| S11_extracted_k141_23685 | Not-determined |
| S11_extracted_k141_56048 | Not-determined |
| S11_extracted_k141_88676 | Not-determined |
| S11_extracted_k141_75909 | Not-determined |
| S11_extracted_k141_88677 | Not-determined |
| S11_extracted_k141_3892 | Not-determined |
| S11_extracted_k141_56049 | Not-determined |
| S11_extracted_k141_3895 | Not-determined |
| S11_extracted_k141_36148 | Not-determined |
| S11_extracted_k141_29623 | Not-determined |
| S11_extracted_k141_3897 | Not-determined |
| S11_extracted_k141_3899 | Not-determined |
| S11_extracted_k141_56054 | Not-determined |
| S11_extracted_k141_3904 | Not-determined |
| S11_extracted_k141_88680 | Not-determined |
| S11_extracted_k141_48929 | Low-quality |
| S11_extracted_k141_10464 | Not-determined |
| S11_extracted_k141_3912 | Not-determined |
| S11_extracted_k141_56061 | Not-determined |
| S11_extracted_k141_88691 | Not-determined |
| S11_extracted_k141_10468 | Not-determined |
| S11_extracted_k141_29631 | Not-determined |
| S11_extracted_k141_16725 | Not-determined |
| S11_extracted_k141_16728 | Low-quality |
| S11_extracted_k141_29634 | Not-determined |
| S11_extracted_k141_3925 | Low-quality |

| | |
|---|---|
| S11_extracted_k141_88693 | Low-quality |
| S11_extracted_k141_3927 | Low-quality |
| S11_extracted_k141_56074 | Not-determined |
| S11_extracted_k141_3929 | Low-quality |
| S11_extracted_k141_81777 | Not-determined |
| S11_extracted_k141_36168 | Not-determined |
| S11_extracted_k141_3933 | Not-determined |
| S11_extracted_k141_10483 | Not-determined |
| S11_extracted_k141_29642 | Not-determined |
| S11_extracted_k141_62195 | Not-determined |
| S11_extracted_k141_23716 | Not-determined |
| S11_extracted_k141_88705 | Not-determined |
| S11_extracted_k141_88706 | Not-determined |
| S11_extracted_k141_88709 | Not-determined |
| S11_extracted_k141_3935 | Not-determined |
| S11_extracted_k141_56081 | Not-determined |
| S11_extracted_k141_16752 | Not-determined |
| S11_extracted_k141_16755 | Not-determined |
| S11_extracted_k141_10491 | Not-determined |
| S11_extracted_k141_29649 | Not-determined |
| S11_extracted_k141_36182 | Not-determined |
| S11_extracted_k141_68353 | Not-determined |
| S11_extracted_k141_10494 | Not-determined |
| S11_extracted_k141_36184 | Not-determined |
| S11_extracted_k141_88714 | Not-determined |
| S11_extracted_k141_29654 | Low-quality |
| S11_extracted_k141_81795 | Low-quality |
| S11_extracted_k141_42929 | Not-determined |
| S11_extracted_k141_62205 | Not-determined |
| S11_extracted_k141_23731 | Not-determined |
| S11_extracted_k141_23733 | Not-determined |
| S11_extracted_k141_81796 | Low-quality |
| S11_extracted_k141_36193 | Not-determined |
| S11_extracted_k141_62208 | Not-determined |
| S11_extracted_k141_16768 | Not-determined |
| S11_extracted_k141_36196 | Not-determined |
| S11_extracted_k141_3943 | Not-determined |
| S11_extracted_k141_3945 | Not-determined |
| S11_extracted_k141_3946 | Not-determined |
| S11_extracted_k141_23740 | Not-determined |
| S11_extracted_k141_16775 | Low-quality |
| S11_extracted_k141_36201 | Not-determined |
| S11_extracted_k141_95066 | Not-determined |
| S11_extracted_k141_62213 | Not-determined |
| S11_extracted_k141_56103 | Low-quality |
| S11_extracted_k141_23750 | Not-determined |
| S11_extracted_k141_68365 | Not-determined |
| S11_extracted_k141_3960 | Not-determined |
| S11_extracted_k141_16793 | Low-quality |
| S11_extracted_k141_23755 | Not-determined |
| S11_extracted_k141_23760 | Low-quality |
| S11_extracted_k141_81817 | Not-determined |
| S11_extracted_k141_16806 | Not-determined |
| S11_extracted_k141_88728 | Not-determined |
| S11_extracted_k141_23766 | Not-determined |
| S11_extracted_k141_81822 | Not-determined |
| S11_extracted_k141_3974 | Not-determined |
| S11_extracted_k141_48968 | Low-quality |
| S11_extracted_k141_62220 | Not-determined |
| S11_extracted_k141_48970 | Not-determined |
| S11_extracted_k141_36208 | Not-determined |
| S11_extracted_k141_36209 | Not-determined |
| S11_extracted_k141_62222 | Not-determined |
| S11_extracted_k141_3977 | Not-determined |
| S11_extracted_k141_48972 | Not-determined |
| S11_extracted_k141_36214 | Low-quality |
| S11_extracted_k141_3980 | Not-determined |
| S11_extracted_k141_3983 | Not-determined |
| S11_extracted_k141_62226 | Not-determined |
| S11_extracted_k141_3985 | Not-determined |
| S11_extracted_k141_88736 | Not-determined |
| S11_extracted_k141_101629 | Not-determined |
| S11_extracted_k141_101631 | Not-determined |
| S11_extracted_k141_56124 | Not-determined |
| S11_extracted_k141_56125 | Not-determined |
| S11_extracted_k141_36218 | Not-determined |
| S11_extracted_k141_10507 | Not-determined |
| S11_extracted_k141_88743 | Low-quality |
| S11_extracted_k141_68387 | Not-determined |
| S11_extracted_k141_10510 | Not-determined |
| S11_extracted_k141_88748 | Not-determined |
| S11_extracted_k141_62238 | Not-determined |
| S11_extracted_k141_42970 | Not-determined |
| S11_extracted_k141_36232 | Not-determined |
| S11_extracted_k141_16834 | Not-determined |
| S11_extracted_k141_88756 | Not-determined |
| S11_extracted_k141_81841 | Low-quality |
| S11_extracted_k141_48996 | Not-determined |
| S11_extracted_k141_68402 | Not-determined |
| S11_extracted_k141_88759 | Not-determined |
| S11_extracted_k141_101645 | Not-determined |
| S11_extracted_k141_56146 | Low-quality |
| S11_extracted_k141_68404 | Not-determined |
| S11_extracted_k141_23797 | Not-determined |
| S11_extracted_k141_23798 | Not-determined |
| S11_extracted_k141_36239 | Not-determined |
| S11_extracted_k141_36240 | Not-determined |
| S11_extracted_k141_10525 | Not-determined |
| S11_extracted_k141_3993 | Low-quality |
| S11_extracted_k141_10527 | Not-determined |
| S11_extracted_k141_62251 | Not-determined |
| S11_extracted_k141_75911 | Not-determined |
| S11_extracted_k141_75912 | Not-determined |
| S11_extracted_k141_23809 | Not-determined |
| S11_extracted_k141_36252 | Not-determined |
| S11_extracted_k141_16851 | Low-quality |
| S11_extracted_k141_49005 | Not-determined |
| S11_extracted_k141_62266 | Not-determined |
| S10_vs_36880\|\|full | High-quality |
| S11_extracted_k141_23826 | Not-determined |
| S11_extracted_k141_101658 | Not-determined |
| S11_extracted_k141_23827 | Not-determined |
| S11_extracted_k141_36266 | Not-determined |
| S11_extracted_k141_16861 | Not-determined |
| S11_extracted_k141_43009 | Not-determined |
| S11_extracted_k141_49023 | Not-determined |
| S11_extracted_k141_75922 | Not-determined |
| S11_extracted_k141_16871 | Not-determined |
| S11_extracted_k141_10562 | Not-determined |
| S11_extracted_k141_56160 | Low-quality |
| S11_extracted_k141_81882 | Low-quality |
| S11_extracted_k141_81883 | Not-determined |
| S11_extracted_k141_23834 | Low-quality |
| S11_extracted_k141_36277 | Not-determined |
| S11_extracted_k141_43020 | Not-determined |
| S11_extracted_k141_16882 | Not-determined |
| S11_extracted_k141_23838 | Not-determined |
| S11_extracted_k141_101661 | Not-determined |
| S11_extracted_k141_81889 | Not-determined |
| S11_extracted_k141_56178 | Not-determined |
| S11_extracted_k141_43030 | Not-determined |
| S11_extracted_k141_81895 | Not-determined |
| S11_extracted_k141_62296 | Not-determined |
| S11_extracted_k141_49043 | Not-determined |
| S11_extracted_k141_4013 | Not-determined |
| S11_extracted_k141_56192 | Not-determined |
| S11_extracted_k141_23858 | Not-determined |
| S11_extracted_k141_68470 | Not-determined |
| S11_extracted_k141_62300 | Not-determined |
| S11_extracted_k141_68473 | Not-determined |
| S11_extracted_k141_101672 | Low-quality |
| S11_extracted_k141_36285 | Not-determined |
| S11_extracted_k141_101674 | Not-determined |
| S11_extracted_k141_49048 | Not-determined |
| S11_extracted_k141_62306 | Low-quality |
| S11_extracted_k141_36291 | Not-determined |
| S11_extracted_k141_81899 | Not-determined |
| S11_extracted_k141_23871 | Not-determined |
| S11_extracted_k141_81906 | Low-quality |
| S11_extracted_k141_101678 | Not-determined |
| S11_extracted_k141_68499 | Not-determined |
| S11_extracted_k141_4030 | Not-determined |
| S11_extracted_k141_68505 | Not-determined |
| S11_extracted_k141_101682 | Not-determined |
| S11_extracted_k141_81914 | Not-determined |
| S11_extracted_k141_49065 | Not-determined |
| S11_extracted_k141_49066 | Not-determined |
| S11_extracted_k141_36302 | Low-quality |
| S11_extracted_k141_43051 | Not-determined |
| S11_extracted_k141_81919 | Not-determined |
| S11_extracted_k141_95096 | Not-determined |
| S11_extracted_k141_16896 | Not-determined |
| S11_extracted_k141_23891 | Not-determined |
| S11_extracted_k141_23896 | Not-determined |
| S11_extracted_k141_4050 | Not-determined |
| S11_extracted_k141_23900 | Not-determined |
| S11_extracted_k141_16913 | Not-determined |
| S11_extracted_k141_23901 | Low-quality |
| S11_extracted_k141_68518 | Not-determined |
| S11_extracted_k141_68522 | Not-determined |
| S11_extracted_k141_16919 | Not-determined |
| S11_extracted_k141_95115 | Low-quality |
| S11_extracted_k141_23908 | Not-determined |
| S11_extracted_k141_95118 | Not-determined |
| S11_extracted_k141_95121 | Not-determined |
| S11_extracted_k141_68528 | Low-quality |
| S11_extracted_k141_23913 | Not-determined |
| S11_extracted_k141_62339 | Not-determined |
| S11_extracted_k141_36320 | Low-quality |
| S11_extracted_k141_36322 | Not-determined |
| S11_extracted_k141_95127 | Not-determined |
| S11_extracted_k141_4066 | Low-quality |
| S11_extracted_k141_81949 | Low-quality |
| S11_extracted_k141_10583 | Not-determined |
| S11_extracted_k141_62344 | Not-determined |
| S11_extracted_k141_68532 | Not-determined |
| S11_extracted_k141_81952 | Not-determined |
| S11_extracted_k141_43079 | Not-determined |
| S11_extracted_k141_62354 | Not-determined |
| S11_extracted_k141_95142 | Not-determined |
| S11_extracted_k141_10587 | Low-quality |
| S11_extracted_k141_10588 | Low-quality |
| S11_extracted_k141_4072 | Not-determined |
| S11_extracted_k141_36333 | Not-determined |
| S11_extracted_k141_23925 | Not-determined |
| S11_extracted_k141_56259 | Not-determined |
| S11_extracted_k141_62363 | Not-determined |
| S11_extracted_k141_4077 | Low-quality |
| S11_extracted_k141_43097 | Not-determined |
| S11_extracted_k141_4083 | Not-determined |
| S11_extracted_k141_88781 | Low-quality |
| S11_extracted_k141_56262 | Not-determined |
| S11_extracted_k141_36352 | Not-determined |
| S11_extracted_k141_62371 | Low-quality |
| S11_extracted_k141_62373 | Not-determined |
| S11_extracted_k141_88784 | Not-determined |
| S11_extracted_k141_43100 | Low-quality |
| S11_extracted_k141_62375 | Not-determined |
| S11_extracted_k141_36355 | Not-determined |
| S11_extracted_k141_95158 | Not-determined |
| S11_extracted_k141_62381 | Not-determined |
| S11_extracted_k141_95161 | Not-determined |
| S11_extracted_k141_4093 | Not-determined |
| S11_extracted_k141_68553 | Low-quality |
| S11_extracted_k141_43101 | Low-quality |
| S11_extracted_k141_62384 | Low-quality |
| S11_extracted_k141_36360 | Low-quality |
| S11_extracted_k141_95163 | Low-quality |
| S11_extracted_k141_88793 | Not-determined |
| S11_extracted_k141_36361 | Not-determined |
| S11_extracted_k141_4100 | Not-determined |
| S11_extracted_k141_75929 | Low-quality |
| S11_extracted_k141_81969 | Not-determined |
| S11_extracted_k141_95166 | Not-determined |
| S11_extracted_k141_68559 | Not-determined |
| S11_extracted_k141_36369 | Not-determined |
| S11_extracted_k141_81971 | Not-determined |
| S11_extracted_k141_88798 | Low-quality |
| S11_extracted_k141_68562 | Not-determined |
| S11_extracted_k141_81973 | Not-determined |
| S11_extracted_k141_75936 | Not-determined |
| S11_extracted_k141_56277 | Not-determined |
| S11_extracted_k141_36373 | Not-determined |
| S11_extracted_k141_68565 | Low-quality |
| S11_extracted_k141_81974 | Not-determined |
| S11_extracted_k141_75944 | Low-quality |
| S11_extracted_k141_75951 | Not-determined |
| S11_extracted_k141_29686 | Not-determined |
| S11_extracted_k141_16921 | Not-determined |
| S11_extracted_k141_43120 | Not-determined |
| S11_extracted_k141_62401 | Low-quality |
| S11_extracted_k141_43122 | Not-determined |
| S11_extracted_k141_101706 | Not-determined |
| S11_extracted_k141_43127 | Low-quality |
| S11_extracted_k141_56290 | Not-determined |
| S11_extracted_k141_43130 | Low-quality |
| S11_extracted_k141_4112 | Not-determined |
| S11_extracted_k141_56294 | Not-determined |
| S11_extracted_k141_62407 | Not-determined |
| S11_extracted_k141_81996 | Not-determined |
| S11_extracted_k141_56303 | Not-determined |
| S11_extracted_k141_29708 | Not-determined |
| S11_extracted_k141_56309 | Not-determined |
| S11_extracted_k141_23968 | Not-determined |
| S11_extracted_k141_23969 | Not-determined |
| S11_extracted_k141_56315 | Not-determined |
| S11_extracted_k141_101727 | Not-determined |
| S11_extracted_k141_62426 | Not-determined |
| S11_extracted_k141_62429 | Not-determined |
| S11_extracted_k141_23976 | Low-quality |
| S11_extracted_k141_23980 | Not-determined |
| S11_extracted_k141_16956 | Not-determined |
| S11_extracted_k141_4121 | Not-determined |
| S11_extracted_k141_29736 | Low-quality |
| S11_extracted_k141_4130 | Not-determined |
| S11_extracted_k141_56334 | Not-determined |
| S11_extracted_k141_56335 | Not-determined |
| S11_extracted_k141_4134 | Not-determined |
| S11_extracted_k141_62448 | Not-determined |
| S11_extracted_k141_4136 | Not-determined |
| S11_extracted_k141_101745 | Not-determined |
| S11_extracted_k141_10601 | Not-determined |
| S11_extracted_k141_101747 | Not-determined |
| S11_extracted_k141_101748 | Not-determined |
| S11_extracted_k141_4138 | Not-determined |
| S11_extracted_k141_56346 | Not-determined |
| S11_extracted_k141_29739 | Low-quality |
| S11_extracted_k141_101750 | Not-determined |
| S11_extracted_k141_29740 | Not-determined |
| S11_extracted_k141_101753 | Not-determined |
| S11_extracted_k141_43171 | Not-determined |
| S11_extracted_k141_43172 | Not-determined |
| S11_extracted_k141_56358 | Not-determined |
| S11_extracted_k141_10611 | Not-determined |
| S11_extracted_k141_10612 | Not-determined |
| S11_extracted_k141_36404 | Low-quality |
| S11_extracted_k141_43179 | Low-quality |
| S11_extracted_k141_43185 | Low-quality |
| S11_extracted_k141_88848 | Not-determined |
| S11_extracted_k141_4153 | Not-determined |
| S11_extracted_k141_36418 | Not-determined |
| S11_extracted_k141_29745 | Not-determined |
| S11_extracted_k141_24005 | Not-determined |
| S11_extracted_k141_82038 | Not-determined |
| S11_extracted_k141_62460 | Not-determined |
| S11_extracted_k141_95204 | Not-determined |
| S11_extracted_k141_10618 | Not-determined |
| S11_extracted_k141_82044 | Low-quality |
| S11_extracted_k141_43189 | Not-determined |
| S11_extracted_k141_10620 | Not-determined |
| S11_extracted_k141_29750 | Not-determined |
| S11_extracted_k141_62466 | Not-determined |
| S11_extracted_k141_24018 | Not-determined |
| S11_extracted_k141_43191 | Not-determined |
| S11_extracted_k141_82048 | Not-determined |
| S11_extracted_k141_95211 | Not-determined |
| S11_extracted_k141_49085 | Not-determined |
| S11_extracted_k141_68600 | Not-determined |
| S11_extracted_k141_10627 | Not-determined |
| S11_extracted_k141_56381 | Not-determined |
| S11_extracted_k141_62479 | Not-determined |
| S11_extracted_k141_88865 | Low-quality |
| S11_extracted_k141_10634 | Low-quality |
| S11_extracted_k141_95224 | Not-determined |
| S11_extracted_k141_95225 | Not-determined |
| S11_extracted_k141_49093 | Not-determined |
| S11_extracted_k141_36434 | Not-determined |
| S11_extracted_k141_24027 | Not-determined |
| S11_extracted_k141_10641 | Not-determined |
| S11_extracted_k141_88871 | Not-determined |
| S11_extracted_k141_88873 | Not-determined |
| S11_extracted_k141_29763 | Not-determined |
| S11_extracted_k141_68610 | Not-determined |
| S11_extracted_k141_49097 | Low-quality |
| S11_extracted_k141_36441 | Not-determined |
| S11_extracted_k141_82062 | Low-quality |
| S11_extracted_k141_95238 | Not-determined |
| S11_extracted_k141_36446 | Not-determined |
| S11_extracted_k141_49105 | Low-quality |
| S11_extracted_k141_62491 | Not-determined |
| S11_extracted_k141_43211 | Low-quality |
| S11_extracted_k141_82069 | Not-determined |
| S11_extracted_k141_68621 | Not-determined |
| S11_extracted_k141_56401 | Not-determined |
| S11_extracted_k141_95243 | Low-quality |
| S11_extracted_k141_49107 | Not-determined |
| S11_extracted_k141_49109 | Not-determined |
| S11_extracted_k141_36453 | Not-determined |
| S11_extracted_k141_95247 | Not-determined |
| S11_extracted_k141_43216 | Not-determined |
| S11_extracted_k141_101775 | Not-determined |
| S11_extracted_k141_49115 | Not-determined |
| S11_extracted_k141_95256 | Not-determined |
| S11_extracted_k141_62498 | Not-determined |
| S11_extracted_k141_62499 | Not-determined |
| S11_extracted_k141_49119 | Not-determined |
| S11_extracted_k141_24049 | Low-quality |
| S11_extracted_k141_56417 | Not-determined |
| S11_extracted_k141_24053 | Not-determined |
| S11_extracted_k141_24055 | Low-quality |
| S11_extracted_k141_95263 | Not-determined |
| S11_extracted_k141_62513 | Not-determined |
| S11_extracted_k141_88882 | Low-quality |
| S11_extracted_k141_24060 | Low-quality |
| S11_extracted_k141_24061 | Not-determined |
| S11_extracted_k141_56425 | Low-quality |
| S11_extracted_k141_88885 | Not-determined |
| S11_extracted_k141_36464 | Not-determined |
| S11_extracted_k141_56426 | Not-determined |
| S11_extracted_k141_4160 | Not-determined |
| S11_extracted_k141_95276 | Not-determined |
| S11_extracted_k141_10672 | Not-determined |
| S11_extracted_k141_16964 | Not-determined |
| S11_extracted_k141_75959 | Not-determined |
| S11_extracted_k141_43225 | Low-quality |
| S11_extracted_k141_95281 | Not-determined |
| S11_extracted_k141_43227 | Not-determined |
| S11_extracted_k141_101799 | Not-determined |
| S11_extracted_k141_29785 | Not-determined |
| S11_extracted_k141_43229 | Not-determined |
| S11_extracted_k141_4180 | Not-determined |
| S11_extracted_k141_95287 | Not-determined |
| S11_extracted_k141_95288 | Not-determined |
| S11_extracted_k141_43235 | Not-determined |
| S11_extracted_k141_36485 | Not-determined |
| S11_extracted_k141_75972 | Not-determined |
| S11_extracted_k141_95291 | Not-determined |
| S11_extracted_k141_29792 | Low-quality |
| S11_extracted_k141_88908 | Not-determined |
| S11_extracted_k141_88911 | Low-quality |
| S11_extracted_k141_36497 | Low-quality |
| S11_extracted_k141_56439 | Not-determined |
| S11_extracted_k141_75976 | Low-quality |
| S11_extracted_k141_88915 | Not-determined |
| S11_extracted_k141_36503 | Not-determined |
| S11_extracted_k141_36508 | Low-quality |
| S11_extracted_k141_43247 | Not-determined |
| S11_extracted_k141_75990 | Not-determined |
| S11_extracted_k141_24105 | Not-determined |
| S11_extracted_k141_24107 | Not-determined |
| S11_extracted_k141_95313 | Not-determined |
| S11_extracted_k141_101831 | Not-determined |
| S11_extracted_k141_56457 | Not-determined |
| S11_extracted_k141_56458 | Not-determined |
| S11_extracted_k141_36516 | Not-determined |
| S11_extracted_k141_43257 | Low-quality |
| S11_extracted_k141_36519 | Not-determined |
| S11_extracted_k141_43260 | Not-determined |
| S11_extracted_k141_101835 | Low-quality |
| S11_extracted_k141_16987 | Not-determined |
| S11_extracted_k141_95319 | Not-determined |
| S11_extracted_k141_4194 | Not-determined |
| S11_extracted_k141_76001 | Not-determined |
| S11_extracted_k141_88923 | Low-quality |
| S11_extracted_k141_36523 | Not-determined |
| S11_extracted_k141_16990 | Not-determined |
| S11_extracted_k141_68648 | Not-determined |
| S11_extracted_k141_36527 | Not-determined |
| S11_extracted_k141_43264 | Not-determined |
| S11_extracted_k141_68654 | Not-determined |
| S11_extracted_k141_24119 | Not-determined |
| S11_extracted_k141_16994 | Not-determined |
| S11_extracted_k141_29827 | Low-quality |
| S11_extracted_k141_43269 | Not-determined |
| S11_extracted_k141_62569 | Not-determined |
| S11_extracted_k141_24125 | Not-determined |
| S11_extracted_k141_95332 | Not-determined |
| S11_extracted_k141_76016 | Not-determined |
| S11_extracted_k141_68662 | Not-determined |
| S11_extracted_k141_36536 | Not-determined |
| S11_extracted_k141_62570 | Low-quality |
| S11_extracted_k141_88939 | Not-determined |
| S11_extracted_k141_101844 | Not-determined |
| S11_extracted_k141_76022 | Not-determined |
| S11_extracted_k141_68670 | Not-determined |
| S11_extracted_k141_16998 | Not-determined |
| S11_extracted_k141_43282 | Low-quality |
| S11_extracted_k141_17000 | Low-quality |
| S11_extracted_k141_36551 | Not-determined |
| S11_extracted_k141_17002 | Not-determined |
| S11_extracted_k141_10734 | Not-determined |
| S11_extracted_k141_24130 | Not-determined |
| S11_extracted_k141_95356 | Not-determined |
| S11_extracted_k141_68681 | Not-determined |
| S11_extracted_k141_36557 | Low-quality |
| S11_extracted_k141_4204 | Not-determined |
| S11_extracted_k141_43294 | Not-determined |
| S11_extracted_k141_56493 | Not-determined |
| S11_extracted_k141_56494 | Not-determined |
| S11_extracted_k141_43295 | Not-determined |
| S11_extracted_k141_95360 | Not-determined |
| S11_extracted_k141_76037 | Not-determined |
| S11_extracted_k141_95364 | Not-determined |
| S11_extracted_k141_43299 | Not-determined |
| S11_extracted_k141_88963 | Not-determined |
| S11_extracted_k141_4213 | Not-determined |
| S11_extracted_k141_43300 | Not-determined |
| S11_extracted_k141_17017 | Not-determined |
| S11_extracted_k141_10750 | Low-quality |
| S11_extracted_k141_36571 | Not-determined |
| S11_extracted_k141_4216 | Not-determined |
| S11_extracted_k141_24148 | Not-determined |
| S11_extracted_k141_88973 | Low-quality |
| S11_extracted_k141_43307 | Not-determined |
| S11_extracted_k141_10753 | Not-determined |
| S11_extracted_k141_4224 | Not-determined |
| S11_extracted_k141_29848 | Not-determined |
| S11_extracted_k141_4230 | Not-determined |
| S11_extracted_k141_17026 | Not-determined |
| S11_extracted_k141_43320 | Not-determined |
| S11_extracted_k141_88980 | Not-determined |
| S11_extracted_k141_88983 | Not-determined |
| S11_extracted_k141_82088 | Not-determined |
| S11_extracted_k141_95389 | Not-determined |
| S11_extracted_k141_88984 | Not-determined |
| S11_extracted_k141_56509 | Not-determined |
| S11_extracted_k141_43326 | Not-determined |
| S11_extracted_k141_56510 | Not-determined |
| S11_extracted_k141_95392 | Low-quality |
| S11_extracted_k141_56515 | Not-determined |
| S11_extracted_k141_17038 | Not-determined |
| S11_extracted_k141_88992 | Not-determined |
| S11_extracted_k141_82103 | Not-determined |
| S11_extracted_k141_36596 | Not-determined |
| S11_extracted_k141_82105 | Not-determined |
| S11_extracted_k141_36598 | Not-determined |
| S11_extracted_k141_29865 | Not-determined |
| S11_extracted_k141_29866 | Not-determined |
| S11_extracted_k141_95402 | Not-determined |
| S11_extracted_k141_36601 | Not-determined |
| S11_extracted_k141_95405 | Low-quality |
| S11_extracted_k141_10770 | Not-determined |
| S11_extracted_k141_82110 | Not-determined |
| S11_extracted_k141_29873 | Low-quality |
| S11_extracted_k141_29876 | Not-determined |
| S11_extracted_k141_10772 | Not-determined |
| S11_extracted_k141_95412 | Low-quality |
| S11_extracted_k141_95413 | Not-determined |
| S11_extracted_k141_17055 | Not-determined |
| S11_extracted_k141_82116 | Not-determined |
| S11_extracted_k141_10774 | Low-quality |
| S11_extracted_k141_17060 | Not-determined |
| S11_extracted_k141_10777 | Not-determined |
| S11_extracted_k141_82118 | Not-determined |
| S11_extracted_k141_76058 | Not-determined |
| S11_extracted_k141_76063 | Not-determined |
| S11_extracted_k141_29886 | Not-determined |
| S11_extracted_k141_10784 | Not-determined |
| S11_extracted_k141_82121 | Not-determined |
| S11_extracted_k141_4251 | Not-determined |
| S11_extracted_k141_10786 | Not-determined |
| S11_extracted_k141_4253 | Not-determined |
| S11_extracted_k141_29888 | Not-determined |
| S11_extracted_k141_82123 | Not-determined |
| S11_extracted_k141_10789 | Not-determined |
| S11_extracted_k141_101902 | Not-determined |
| S11_extracted_k141_24193 | Not-determined |
| S11_extracted_k141_17076 | Not-determined |
| S11_extracted_k141_95432 | Low-quality |
| S11_extracted_k141_82137 | Not-determined |
| S11_extracted_k141_24199 | Not-determined |
| S11_extracted_k141_4268 | Low-quality |
| S11_extracted_k141_82139 | Low-quality |
| S11_extracted_k141_76078 | Low-quality |
| S11_extracted_k141_56517 | Not-determined |
| S11_extracted_k141_24206 | Low-quality |
| S11_extracted_k141_10805 | Low-quality |
| S11_extracted_k141_101916 | Not-determined |
| S13_extracted_3728 | Complete |
| S11_extracted_k141_62576 | Low-quality |
| S11_extracted_k141_101918 | Not-determined |
| S11_extracted_k141_76083 | Not-determined |
| S11_extracted_k141_10808 | Not-determined |
| S11_extracted_k141_56527 | Not-determined |
| S11_extracted_k141_62578 | Low-quality |
| S11_extracted_k141_89032 | Low-quality |
| S11_extracted_k141_68697 | Not-determined |
| S11_extracted_k141_24219 | Not-determined |
| S11_extracted_k141_43334 | Not-determined |
| S11_extracted_k141_49144 | Not-determined |
| S11_extracted_k141_4276 | Not-determined |
| S11_extracted_k141_4278 | Not-determined |
| S11_extracted_k141_43340 | Not-determined |
| S11_extracted_k141_76093 | Not-determined |
| S11_extracted_k141_82155 | Not-determined |
| S11_extracted_k141_82156 | Not-determined |
| S11_extracted_k141_101936 | Not-determined |
| S11_extracted_k141_10826 | Not-determined |
| S11_extracted_k141_56543 | Not-determined |
| S11_extracted_k141_56546 | Not-determined |
| S11_extracted_k141_29912 | Low-quality |
| S11_extracted_k141_68718 | Not-determined |
| S11_extracted_k141_68721 | Not-determined |
| S11_extracted_k141_4283 | Low-quality |
| S11_extracted_k141_17105 | Not-determined |
| S11_extracted_k141_24245 | Low-quality |
| S11_extracted_k141_82163 | Not-determined |
| S11_extracted_k141_62591 | Low-quality |
| S11_extracted_k141_89060 | Not-determined |
| S11_extracted_k141_17108 | Not-determined |
| S11_extracted_k141_4293 | Not-determined |
| S11_extracted_k141_36640 | Not-determined |
| S11_extracted_k141_10848 | Not-determined |
| S11_extracted_k141_10850 | Not-determined |
| S11_extracted_k141_56561 | Not-determined |
| S11_extracted_k141_36645 | Not-determined |
| S11_extracted_k141_49172 | Not-determined |
| S11_extracted_k141_101960 | Not-determined |
| S11_extracted_k141_56565 | Not-determined |
| S11_extracted_k141_36651 | Not-determined |
| S11_extracted_k141_24258 | Low-quality |
| S11_extracted_k141_17120 | Not-determined |
| S11_extracted_k141_82173 | Not-determined |
| S11_extracted_k141_17122 | Low-quality |
| S11_extracted_k141_49178 | Not-determined |
| S11_extracted_k141_4308 | Not-determined |
| S11_extracted_k141_4309 | Not-determined |
| S11_extracted_k141_24265 | Not-determined |
| S11_extracted_k141_24266 | Not-determined |
| S11_extracted_k141_56571 | Low-quality |
| S11_extracted_k141_101970 | Not-determined |
| S11_extracted_k141_29937 | Not-determined |
| S11_extracted_k141_4316 | Not-determined |
| S11_extracted_k141_29940 | Not-determined |
| S11_extracted_k141_10865 | Not-determined |
| S11_extracted_k141_62614 | Not-determined |
| S11_extracted_k141_95461 | Not-determined |
| S11_extracted_k141_24275 | Not-determined |
| S11_extracted_k141_101974 | Not-determined |
| S11_extracted_k141_17125 | Low-quality |
| S11_extracted_k141_10869 | Not-determined |
| S11_extracted_k141_95469 | Not-determined |
| S11_extracted_k141_36664 | Low-quality |
| S11_extracted_k141_10871 | Not-determined |
| S11_extracted_k141_101979 | Not-determined |
| S11_extracted_k141_43382 | Not-determined |
| S11_extracted_k141_49188 | Not-determined |
| S11_extracted_k141_49192 | Not-determined |
| S11_extracted_k141_101985 | Not-determined |

| | |
|---|---|
| S11_extracted_k141_95476 | Not-determined |
| S11_extracted_k141_24292 | Not-determined |
| S11_extracted_k141_36675 | Not-determined |
| S11_extracted_k141_95480 | Not-determined |
| S11_extracted_k141_10886 | Not-determined |
| S11_extracted_k141_24295 | Not-determined |
| S11_extracted_k141_49202 | Low-quality |
| S11_extracted_k141_49204 | Not-determined |
| S11_extracted_k141_43394 | Low-quality |
| S11_extracted_k141_17133 | Not-determined |
| S11_extracted_k141_17134 | Low-quality |
| S11_extracted_k141_43400 | Not-determined |
| S11_extracted_k141_101993 | Not-determined |
| S11_extracted_k141_4343 | Low-quality |
| S11_extracted_k141_29970 | Not-determined |
| S11_extracted_k141_17135 | Low-quality |
| S11_extracted_k141_43406 | Not-determined |
| S11_extracted_k141_43409 | Not-determined |
| S11_extracted_k141_95497 | Not-determined |
| S11_extracted_k141_24311 | Not-determined |
| S11_extracted_k141_49222 | Not-determined |
| S11_extracted_k141_24315 | Not-determined |
| S11_extracted_k141_24317 | Not-determined |
| S11_extracted_k141_62656 | Not-determined |
| S11_extracted_k141_95503 | Low-quality |
| S11_extracted_k141_95504 | Not-determined |
| S11_extracted_k141_43421 | Not-determined |
| S11_extracted_k141_4350 | Not-determined |
| S11_extracted_k141_62662 | Not-determined |
| S11_extracted_k141_76137 | Not-determined |
| S11_extracted_k141_43427 | Not-determined |
| S11_extracted_k141_95509 | Low-quality |
| S11_extracted_k141_43431 | Not-determined |
| S11_extracted_k141_95510 | Not-determined |
| S11_extracted_k141_102015 | Low-quality |
| S11_extracted_k141_4359 | Not-determined |
| S11_extracted_k141_29991 | Not-determined |
| S11_extracted_k141_102021 | Not-determined |
| S11_extracted_k141_4362 | Not-determined |
| S11_extracted_k141_4364 | Not-determined |
| S11_extracted_k141_49247 | Not-determined |
| S11_extracted_k141_17162 | Not-determined |
| S11_extracted_k141_24347 | Not-determined |
| S11_extracted_k141_30007 | Not-determined |
| S11_extracted_k141_30010 | Low-quality |
| S11_extracted_k141_95524 | Not-determined |
| S11_extracted_k141_36727 | Not-determined |
| S11_extracted_k141_24354 | Low-quality |
| S11_extracted_k141_24357 | Not-determined |
| S11_extracted_k141_95534 | Not-determined |
| S11_extracted_k141_62681 | Not-determined |
| S11_extracted_k141_102058 | Not-determined |
| S11_extracted_k141_4386 | Not-determined |
| S11_extracted_k141_24362 | Low-quality |
| S11_extracted_k141_62688 | Not-determined |
| S11_extracted_k141_36741 | Not-determined |
| S11_extracted_k141_4393 | Not-determined |
| S11_extracted_k141_24369 | Not-determined |
| S11_extracted_k141_4394 | Not-determined |
| S11_extracted_k141_102070 | Not-determined |
| S11_extracted_k141_43450 | Not-determined |
| S11_extracted_k141_4396 | Not-determined |
| S11_extracted_k141_4397 | Not-determined |
| S11_extracted_k141_62696 | Not-determined |
| S2_extracted_k141_112065 | Complete |
| S11_extracted_k141_102073 | Not-determined |
| S11_extracted_k141_30023 | Low-quality |
| S11_extracted_k141_30024 | Not-determined |
| S11_extracted_k141_30025 | Not-determined |
| S11_extracted_k141_10909 | Not-determined |
| S11_extracted_k141_68739 | Not-determined |
| S11_extracted_k141_62701 | Not-determined |
| S11_extracted_k141_62702 | Not-determined |
| S11_extracted_k141_10915 | Not-determined |
| S11_extracted_k141_49267 | Low-quality |
| S11_extracted_k141_95553 | Not-determined |
| S11_extracted_k141_62703 | Not-determined |
| S11_extracted_k141_68747 | Low-quality |
| S11_extracted_k141_4414 | Not-determined |
| S11_extracted_k141_56583 | Low-quality |
| S11_extracted_k141_68751 | Not-determined |
| S11_extracted_k141_56585 | Not-determined |
| S11_extracted_k141_56588 | Not-determined |
| S11_extracted_k141_68757 | Not-determined |
| S11_extracted_k141_68759 | Low-quality |
| S11_extracted_k141_24389 | Low-quality |
| S11_extracted_k141_49276 | Not-determined |
| S11_extracted_k141_68762 | Low-quality |
| S11_extracted_k141_82191 | Not-determined |
| S11_extracted_k141_49280 | Not-determined |
| S11_extracted_k141_4429 | Not-determined |
| S11_extracted_k141_49281 | Not-determined |
| S11_extracted_k141_4431 | Not-determined |
| S11_extracted_k141_95569 | Not-determined |
| S11_extracted_k141_68766 | Not-determined |
| S11_extracted_k141_30036 | Not-determined |
| S11_extracted_k141_68768 | Not-determined |
| S11_extracted_k141_24399 | Low-quality |
| S11_extracted_k141_95574 | Not-determined |
| S11_extracted_k141_43462 | Not-determined |
| S11_extracted_k141_102106 | Not-determined |
| S11_extracted_k141_82198 | Low-quality |
| S11_extracted_k141_10931 | Not-determined |
| S11_extracted_k141_49286 | Not-determined |
| S11_extracted_k141_49288 | Not-determined |
| S11_extracted_k141_102118 | Not-determined |
| S11_extracted_k141_4448 | Not-determined |
| S11_extracted_k141_4453 | Not-determined |
| S11_extracted_k141_62735 | Not-determined |
| S11_extracted_k141_102128 | Low-quality |
| S11_extracted_k141_102130 | Not-determined |
| S11_extracted_k141_4456 | Not-determined |
| S11_extracted_k141_17222 | Not-determined |
| S11_extracted_k141_102132 | Low-quality |
| S11_extracted_k141_43476 | Not-determined |
| S11_extracted_k141_30054 | Not-determined |
| S11_extracted_k141_82214 | Not-determined |
| S11_extracted_k141_10945 | Not-determined |
| S11_extracted_k141_17226 | Not-determined |
| S11_extracted_k141_49306 | Not-determined |
| S11_extracted_k141_17229 | Not-determined |
| S11_extracted_k141_62750 | Not-determined |
| S11_extracted_k141_82224 | Not-determined |
| S11_extracted_k141_82227 | Not-determined |
| S11_extracted_k141_102145 | Not-determined |
| S11_extracted_k141_82228 | Not-determined |
| S11_extracted_k141_62754 | Not-determined |
| S11_extracted_k141_49316 | Low-quality |
| S11_extracted_k141_102151 | Low-quality |
| S11_extracted_k141_24430 | Not-determined |
| S11_extracted_k141_68774 | Not-determined |
| S11_extracted_k141_49324 | Not-determined |
| S11_extracted_k141_24436 | Not-determined |
| S11_extracted_k141_24438 | Not-determined |
| S11_extracted_k141_10968 | Not-determined |
| S11_extracted_k141_10970 | Not-determined |
| S11_extracted_k141_68783 | Not-determined |
| S11_extracted_k141_102158 | Not-determined |
| S11_extracted_k141_10975 | Not-determined |
| S11_extracted_k141_24442 | Not-determined |
| S11_extracted_k141_95630 | Not-determined |
| S11_extracted_k141_10977 | Not-determined |
| S11_extracted_k141_89103 | Not-determined |
| S11_extracted_k141_24454 | Not-determined |
| S11_extracted_k141_24458 | Not-determined |
| S11_extracted_k141_95641 | Not-determined |
| S11_extracted_k141_89116 | Not-determined |
| S11_extracted_k141_68801 | Not-determined |
| S11_extracted_k141_56619 | Not-determined |
| S11_extracted_k141_102175 | Not-determined |
| S11_extracted_k141_95650 | Not-determined |
| S11_extracted_k141_89120 | Not-determined |
| S11_extracted_k141_95651 | Low-quality |
| S11_extracted_k141_89122 | Not-determined |
| S11_extracted_k141_102178 | Not-determined |
| S11_extracted_k141_17247 | Not-determined |
| S11_extracted_k141_95653 | Low-quality |
| S11_extracted_k141_62777 | Not-determined |
| S11_extracted_k141_62778 | Not-determined |
| S11_extracted_k141_49342 | Not-determined |
| S11_extracted_k141_82266 | Not-determined |
| S11_extracted_k141_49346 | Not-determined |
| S11_extracted_k141_36771 | Not-determined |
| S11_extracted_k141_10999 | Not-determined |
| S11_extracted_k141_56633 | Not-determined |
| S11_extracted_k141_89127 | Not-determined |
| S11_extracted_k141_36773 | Not-determined |
| S11_extracted_k141_68821 | Not-determined |
| S11_extracted_k141_82273 | Not-determined |
| S11_extracted_k141_82274 | Not-determined |
| S11_extracted_k141_62788 | Not-determined |
| S11_extracted_k141_56645 | Not-determined |
| S11_extracted_k141_17255 | Not-determined |
| S11_extracted_k141_36780 | Not-determined |
| S11_extracted_k141_95664 | Low-quality |
| S11_extracted_k141_24487 | Not-determined |
| S11_extracted_k141_56650 | Low-quality |
| S11_extracted_k141_82286 | Not-determined |
| S11_extracted_k141_24491 | Not-determined |
| S11_extracted_k141_62792 | Low-quality |
| S11_extracted_k141_11013 | Not-determined |
| S11_extracted_k141_17262 | Not-determined |
| S11_extracted_k141_11014 | Not-determined |
| S11_extracted_k141_102188 | Not-determined |
| S11_extracted_k141_24496 | Not-determined |
| S11_extracted_k141_36790 | Not-determined |
| S11_extracted_k141_24498 | Not-determined |
| S11_extracted_k141_62804 | Low-quality |
| S11_extracted_k141_11023 | Not-determined |
| S11_extracted_k141_68835 | Not-determined |
| S11_extracted_k141_4463 | Not-determined |
| S11_extracted_k141_24509 | Low-quality |
| S11_extracted_k141_24510 | Not-determined |
| S11_extracted_k141_95685 | Low-quality |
| S11_extracted_k141_17273 | Not-determined |
| S11_extracted_k141_17274 | Not-determined |
| S11_extracted_k141_17276 | Not-determined |
| S11_extracted_k141_24517 | Low-quality |
| S11_extracted_k141_4470 | Not-determined |
| S11_extracted_k141_76146 | Low-quality |
| S11_extracted_k141_56671 | Not-determined |
| S11_extracted_k141_36803 | Not-determined |
| S11_extracted_k141_4473 | Not-determined |
| S11_extracted_k141_4475 | Not-determined |
| S11_extracted_k141_89158 | Not-determined |
| S11_extracted_k141_4477 | Not-determined |
| S11_extracted_k141_102208 | Low-quality |
| S11_extracted_k141_43490 | Not-determined |
| S11_extracted_k141_24522 | Not-determined |
| S11_extracted_k141_68858 | Not-determined |
| S11_extracted_k141_11040 | Not-determined |
| S11_extracted_k141_89168 | Not-determined |
| S11_extracted_k141_49370 | Not-determined |
| S11_extracted_k141_43500 | Not-determined |
| S11_extracted_k141_95700 | Not-determined |
| S11_extracted_k141_24538 | Not-determined |
| S11_extracted_k141_102215 | Not-determined |
| S11_extracted_k141_11048 | Not-determined |
| S11_extracted_k141_24542 | Not-determined |
| S11_extracted_k141_62822 | Not-determined |
| S11_extracted_k141_24544 | Not-determined |
| S11_extracted_k141_102220 | Low-quality |
| S11_extracted_k141_68873 | Not-determined |
| S11_extracted_k141_102222 | Not-determined |
| S11_extracted_k141_82301 | Not-determined |
| S11_extracted_k141_24551 | Low-quality |
| S11_extracted_k141_76166 | Low-quality |
| S11_extracted_k141_62834 | Not-determined |
| S11_extracted_k141_4492 | Not-determined |
| S11_extracted_k141_11059 | Not-determined |
| S11_extracted_k141_4493 | Not-determined |
| S11_extracted_k141_68879 | Not-determined |
| S11_extracted_k141_95720 | Not-determined |
| S11_extracted_k141_11064 | Not-determined |
| S11_extracted_k141_95726 | Not-determined |
| S11_extracted_k141_24564 | Not-determined |
| S11_extracted_k141_11069 | Low-quality |
| S11_extracted_k141_24568 | Not-determined |
| S11_extracted_k141_11073 | Not-determined |
| S11_extracted_k141_24573 | Not-determined |
| S11_extracted_k141_36845 | Not-determined |
| S11_extracted_k141_24577 | Low-quality |
| S11_extracted_k141_68897 | Not-determined |
| S11_extracted_k141_11081 | Not-determined |
| S11_extracted_k141_102235 | Not-determined |
| S11_extracted_k141_43535 | Low-quality |
| S11_extracted_k141_102236 | Low-quality |
| S11_extracted_k141_4513 | Not-determined |
| S11_extracted_k141_76183 | Not-determined |
| S11_extracted_k141_24586 | Not-determined |
| S11_extracted_k141_82326 | Not-determined |
| S11_extracted_k141_4524 | Not-determined |
| S11_extracted_k141_82329 | Not-determined |
| S11_extracted_k141_95753 | Not-determined |
| S11_extracted_k141_95754 | Low-quality |
| S11_extracted_k141_49405 | Not-determined |
| S11_extracted_k141_102248 | Low-quality |
| S11_extracted_k141_11096 | Not-determined |
| S11_extracted_k141_24593 | Not-determined |
| S11_extracted_k141_76194 | Not-determined |
| S11_extracted_k141_49416 | Low-quality |
| S11_extracted_k141_49420 | Not-determined |
| S11_extracted_k141_49422 | Not-determined |
| S11_extracted_k141_24605 | Low-quality |
| S11_extracted_k141_76200 | Not-determined |
| S11_extracted_k141_49424 | Not-determined |
| S11_extracted_k141_11108 | Not-determined |
| S11_extracted_k141_17295 | Not-determined |
| S11_extracted_k141_43550 | Not-determined |
| S11_extracted_k141_95770 | Not-determined |
| S11_extracted_k141_49429 | Not-determined |
| S11_extracted_k141_49430 | Low-quality |
| S11_extracted_k141_76208 | Not-determined |
| S11_extracted_k141_11109 | Not-determined |
| S11_extracted_k141_76209 | Not-determined |
| S11_extracted_k141_24616 | Not-determined |
| S11_extracted_k141_11110 | Not-determined |
| S11_extracted_k141_11115 | Not-determined |
| S11_extracted_k141_36881 | Not-determined |
| S11_extracted_k141_102262 | Not-determined |
| S11_extracted_k141_68932 | Not-determined |
| S11_extracted_k141_24624 | Not-determined |
| S11_extracted_k141_17317 | Not-determined |
| S11_extracted_k141_11122 | Not-determined |
| S11_extracted_k141_56733 | Low-quality |
| S11_extracted_k141_102265 | Not-determined |
| S11_extracted_k141_102269 | Low-quality |
| S11_extracted_k141_49445 | Not-determined |
| S11_extracted_k141_24631 | Not-determined |
| S11_extracted_k141_4557 | Low-quality |
| S11_extracted_k141_102279 | Low-quality |
| S11_extracted_k141_102280 | Not-determined |
| S11_extracted_k141_36893 | Not-determined |
| S11_extracted_k141_49451 | Not-determined |
| S11_extracted_k141_11136 | Low-quality |
| S11_extracted_k141_11138 | Low-quality |
| S11_extracted_k141_4564 | Not-determined |
| S11_extracted_k141_68934 | Low-quality |
| S11_extracted_k141_49457 | Low-quality |
| S11_extracted_k141_4571 | Not-determined |
| S11_extracted_k141_36898 | Not-determined |
| S11_extracted_k141_17344 | Not-determined |
| S11_extracted_k141_95794 | Low-quality |
| S11_extracted_k141_17347 | Low-quality |
| S11_extracted_k141_95795 | Not-determined |
| S11_extracted_k141_24652 | Not-determined |
| S11_extracted_k141_36903 | Not-determined |
| S11_extracted_k141_95800 | Low-quality |
| S11_extracted_k141_56744 | Not-determined |
| S11_extracted_k141_17357 | Not-determined |
| S11_extracted_k141_43573 | Not-determined |
| S11_extracted_k141_95809 | Not-determined |
| S11_extracted_k141_36906 | Low-quality |
| S11_extracted_k141_24655 | Not-determined |
| S11_extracted_k141_49473 | Low-quality |
| S11_extracted_k141_30069 | Not-determined |
| S11_extracted_k141_30070 | Low-quality |
| S11_extracted_k141_49474 | Not-determined |
| S11_extracted_k141_68952 | Not-determined |
| S11_extracted_k141_11143 | Low-quality |
| S11_extracted_k141_4600 | Not-determined |
| S11_extracted_k141_24658 | Low-quality |
| S11_extracted_k141_62873 | Not-determined |
| S11_extracted_k141_24663 | Not-determined |
| S11_extracted_k141_49481 | Not-determined |
| S11_extracted_k141_4609 | Not-determined |
| S11_extracted_k141_62875 | Not-determined |
| S11_extracted_k141_95830 | Not-determined |
| S11_extracted_k141_68960 | Not-determined |
| S11_extracted_k141_43584 | Low-quality |
| S11_extracted_k141_76212 | Low-quality |
| S11_extracted_k141_30081 | Low-quality |
| S11_extracted_k141_36921 | Not-determined |
| S11_extracted_k141_56760 | Not-determined |
| S11_extracted_k141_76214 | Not-determined |
| S11_extracted_k141_89174 | Not-determined |
| S11_extracted_k141_36929 | Not-determined |
| S11_extracted_k141_4617 | Not-determined |
| S11_extracted_k141_49488 | Not-determined |
| S11_extracted_k141_17388 | Low-quality |
| S11_extracted_k141_89177 | Not-determined |
| S11_extracted_k141_95842 | Not-determined |
| S11_extracted_k141_95844 | Not-determined |
| S11_extracted_k141_102315 | Low-quality |
| S11_extracted_k141_68966 | Not-determined |
| S11_extracted_k141_68969 | Not-determined |
| S11_extracted_k141_62888 | Not-determined |
| S11_extracted_k141_89182 | Not-determined |
| S11_extracted_k141_62889 | Not-determined |
| S11_extracted_k141_4627 | Not-determined |
| S11_extracted_k141_89186 | Not-determined |
| S11_extracted_k141_11164 | Low-quality |
| S11_extracted_k141_68972 | Not-determined |
| S11_extracted_k141_68974 | Not-determined |
| S11_extracted_k141_95854 | Not-determined |
| S11_extracted_k141_68977 | Not-determined |
| S11_extracted_k141_95859 | Not-determined |
| S11_extracted_k141_17399 | Not-determined |
| S11_extracted_k141_24678 | Not-determined |
| S11_extracted_k141_17403 | Low-quality |
| S11_extracted_k141_95865 | Low-quality |
| S11_extracted_k141_95869 | Not-determined |
| S11_extracted_k141_30109 | Low-quality |
| S11_extracted_k141_49500 | Low-quality |
| S11_extracted_k141_76234 | Low-quality |
| S11_extracted_k141_62909 | Not-determined |
| S11_extracted_k141_24687 | Low-quality |
| S11_extracted_k141_89198 | Not-determined |
| S11_extracted_k141_4644 | Not-determined |
| S11_extracted_k141_4645 | Not-determined |
| S11_extracted_k141_62914 | Not-determined |
| S11_extracted_k141_76242 | Not-determined |
| S11_extracted_k141_76244 | Low-quality |
| S11_extracted_k141_62915 | Not-determined |
| S11_extracted_k141_30115 | Not-determined |
| S11_extracted_k141_17407 | Not-determined |
| S11_extracted_k141_24697 | Not-determined |
| S11_extracted_k141_24699 | Not-determined |
| S11_extracted_k141_36950 | Not-determined |
| S11_extracted_k141_76250 | Low-quality |
| S11_extracted_k141_17412 | Low-quality |
| S11_extracted_k141_24701 | Not-determined |
| S11_extracted_k141_49509 | Not-determined |
| S11_extracted_k141_76252 | Not-determined |
| S11_extracted_k141_95886 | Low-quality |
| S11_extracted_k141_4660 | Not-determined |
| S11_extracted_k141_43612 | Not-determined |
| S11_extracted_k141_76257 | Not-determined |
| S11_extracted_k141_30128 | Not-determined |
| S11_extracted_k141_56766 | Not-determined |
| S11_extracted_k141_95893 | Not-determined |
| S11_extracted_k141_95894 | Not-determined |
| S11_extracted_k141_56768 | Not-determined |
| S11_extracted_k141_56770 | Not-determined |
| S11_extracted_k141_17421 | Not-determined |
| S11_extracted_k141_89226 | Not-determined |
| S11_extracted_k141_17423 | Not-determined |
| S11_extracted_k141_89228 | Not-determined |
| S11_extracted_k141_62944 | Not-determined |
| S11_extracted_k141_49520 | Not-determined |
| S11_extracted_k141_56775 | Not-determined |
| S11_extracted_k141_76276 | Not-determined |
| S11_extracted_k141_76277 | Not-determined |
| S11_extracted_k141_4674 | Not-determined |
| S11_extracted_k141_102325 | Not-determined |
| S11_extracted_k141_30144 | Not-determined |
| S11_extracted_k141_49521 | Low-quality |
| S11_extracted_k141_89237 | Not-determined |
| S11_extracted_k141_4677 | Low-quality |
| S11_extracted_k141_102333 | Not-determined |
| S11_extracted_k141_30148 | Not-determined |
| S11_extracted_k141_76283 | Not-determined |
| S11_extracted_k141_95909 | Not-determined |
| S11_extracted_k141_36970 | Not-determined |
| S11_extracted_k141_30153 | Not-determined |
| S11_extracted_k141_95911 | Low-quality |
| S11_extracted_k141_69006 | Low-quality |
| S11_extracted_k141_69008 | Not-determined |
| S11_extracted_k141_36973 | Not-determined |
| S11_extracted_k141_49529 | Low-quality |
| S11_extracted_k141_56789 | Low-quality |
| S11_extracted_k141_82334 | Not-determined |
| S11_extracted_k141_49533 | Low-quality |
| S11_extracted_k141_17458 | Not-determined |
| S11_extracted_k141_82335 | Not-determined |
| S11_extracted_k141_36979 | Not-determined |
| S11_extracted_k141_49538 | Not-determined |
| S11_extracted_k141_36984 | Not-determined |
| S11_extracted_k141_89252 | Not-determined |
| S11_extracted_k141_89253 | Not-determined |
| S11_extracted_k141_36986 | Not-determined |
| S11_extracted_k141_69010 | Low-quality |
| S11_extracted_k141_102344 | Not-determined |
| S11_extracted_k141_36992 | Not-determined |
| S11_extracted_k141_24725 | Not-determined |
| S11_extracted_k141_82344 | Not-determined |
| S11_extracted_k141_24729 | Not-determined |
| S11_extracted_k141_102358 | Not-determined |
| S11_extracted_k141_89262 | Low-quality |
| S11_extracted_k141_49544 | Not-determined |
| S11_extracted_k141_76315 | Not-determined |
| S11_extracted_k141_17480 | Not-determined |
| S11_extracted_k141_56809 | Not-determined |
| S11_extracted_k141_17481 | Not-determined |
| S11_extracted_k141_4699 | Low-quality |
| S11_extracted_k141_89271 | Low-quality |
| S11_extracted_k141_49553 | Not-determined |
| S11_extracted_k141_37007 | Not-determined |
| S11_extracted_k141_49554 | Low-quality |
| S11_extracted_k141_37010 | Low-quality |
| S11_extracted_k141_17489 | Not-determined |
| S11_extracted_k141_95929 | Low-quality |
| S11_extracted_k141_95933 | Not-determined |
| S11_extracted_k141_30188 | Not-determined |
| S11_extracted_k141_24754 | Low-quality |
| S11_extracted_k141_82359 | Not-determined |
| S11_extracted_k141_62960 | Not-determined |
| S11_extracted_k141_49563 | Not-determined |
| S11_extracted_k141_76325 | Not-determined |
| S11_extracted_k141_69043 | Not-determined |
| S11_extracted_k141_24757 | Not-determined |
| S11_extracted_k141_95944 | Not-determined |
| S11_extracted_k141_24759 | Not-determined |
| S11_extracted_k141_56828 | Not-determined |
| S11_extracted_k141_49566 | Not-determined |
| S11_extracted_k141_49567 | Not-determined |
| S11_extracted_k141_24763 | Not-determined |
| S11_extracted_k141_56834 | Not-determined |
| S11_extracted_k141_49571 | Not-determined |
| S11_extracted_k141_24766 | Low-quality |
| S11_extracted_k141_49572 | Not-determined |
| S11_extracted_k141_24767 | Not-determined |
| S11_extracted_k141_24769 | Not-determined |
| S11_extracted_k141_24774 | Not-determined |
| S11_extracted_k141_95957 | Low-quality |
| S11_extracted_k141_37032 | Low-quality |
| S11_extracted_k141_30213 | Not-determined |
| S11_extracted_k141_37036 | Not-determined |
| S11_extracted_k141_56843 | Not-determined |
| S11_extracted_k141_37039 | Not-determined |
| S11_extracted_k141_49585 | Not-determined |
| S11_extracted_k141_37040 | Not-determined |
| S11_extracted_k141_56847 | Not-determined |
| S11_extracted_k141_95962 | Not-determined |
| S11_extracted_k141_89294 | Not-determined |
| S11_extracted_k141_11184 | Low-quality |
| S11_extracted_k141_43655 | Low-quality |
| S11_extracted_k141_102371 | Not-determined |
| S11_extracted_k141_49590 | Not-determined |
| S11_extracted_k141_56851 | Not-determined |
| S11_extracted_k141_56856 | Not-determined |
| S11_extracted_k141_49593 | Not-determined |
| S11_extracted_k141_102375 | Not-determined |
| S11_extracted_k141_37057 | Not-determined |
| S11_extracted_k141_17522 | Not-determined |
| S11_extracted_k141_43670 | Not-determined |
| S11_extracted_k141_95979 | Not-determined |
| S11_extracted_k141_76355 | Not-determined |
| S11_extracted_k141_24791 | Not-determined |
| S11_extracted_k141_56861 | Not-determined |
| S11_extracted_k141_43677 | Not-determined |
| S11_extracted_k141_37067 | Not-determined |
| S11_extracted_k141_56870 | Low-quality |
| S11_extracted_k141_17541 | Not-determined |
| S11_extracted_k141_17542 | Not-determined |
| S11_extracted_k141_56874 | Not-determined |
| S11_extracted_k141_95986 | Not-determined |
| S11_extracted_k141_37076 | Not-determined |
| S11_extracted_k141_37079 | Not-determined |
| S11_extracted_k141_30245 | Not-determined |
| S11_extracted_k141_11212 | Not-determined |
| S11_extracted_k141_37083 | Not-determined |
| S11_extracted_k141_95990 | Not-determined |
| S11_extracted_k141_37084 | Not-determined |
| S11_extracted_k141_43689 | Not-determined |
| S11_extracted_k141_43690 | Not-determined |
| S11_extracted_k141_63017 | Not-determined |
| S11_extracted_k141_4735 | Not-determined |
| S11_extracted_k141_4737 | Low-quality |
| S11_extracted_k141_63022 | Not-determined |
| S11_extracted_k141_30255 | Not-determined |
| S11_extracted_k141_4739 | Not-determined |
| S11_extracted_k141_56882 | Not-determined |
| S11_extracted_k141_24812 | Not-determined |
| S11_extracted_k141_24814 | Low-quality |
| S11_extracted_k141_43703 | Not-determined |
| S11_extracted_k141_4750 | Not-determined |
| S11_extracted_k141_102406 | Low-quality |
| S11_extracted_k141_63029 | Low-quality |
| S11_extracted_k141_102409 | Not-determined |
| S11_extracted_k141_43705 | Low-quality |
| S11_extracted_k141_30268 | Low-quality |
| S11_extracted_k141_43709 | Not-determined |
| S11_extracted_k141_63042 | Not-determined |
| S11_extracted_k141_76380 | Not-determined |
| S11_extracted_k141_102418 | Not-determined |
| S11_extracted_k141_4768 | Low-quality |
| S11_extracted_k141_30280 | Not-determined |
| S11_extracted_k141_102420 | Not-determined |
| S11_extracted_k141_49631 | Low-quality |
| S11_extracted_k141_63049 | Not-determined |
| S11_extracted_k141_43712 | Not-determined |
| S11_extracted_k141_56907 | Not-determined |
| S11_extracted_k141_37096 | Not-determined |
| S11_extracted_k141_56912 | Not-determined |
| S11_extracted_k141_24822 | Not-determined |
| S11_extracted_k141_82383 | Not-determined |
| S11_extracted_k141_63060 | Not-determined |
| S11_extracted_k141_17571 | Not-determined |
| S11_extracted_k141_82389 | Not-determined |
| S11_extracted_k141_4783 | Not-determined |
| S11_extracted_k141_69067 | Not-determined |
| S11_extracted_k141_56922 | Not-determined |
| S11_extracted_k141_102436 | Not-determined |
| S11_extracted_k141_4788 | Low-quality |
| S11_extracted_k141_63066 | Not-determined |
| S11_extracted_k141_95998 | Not-determined |
| S11_extracted_k141_24825 | Not-determined |
| S11_extracted_k141_4790 | Not-determined |
| S11_extracted_k141_69075 | Not-determined |
| S11_extracted_k141_96001 | Low-quality |
| S11_extracted_k141_69076 | Not-determined |
| S11_extracted_k141_96002 | Not-determined |
| S11_extracted_k141_96005 | Not-determined |
| S11_extracted_k141_96006 | Not-determined |
| S11_extracted_k141_43725 | Not-determined |
| S11_extracted_k141_43728 | Not-determined |
| S13_vs_3728\|\|full | High-quality |
| S11_extracted_k141_30287 | Not-determined |
| S11_extracted_k141_56931 | Not-determined |
| S11_extracted_k141_30289 | Not-determined |
| S11_extracted_k141_69080 | Not-determined |
| S11_extracted_k141_89316 | Not-determined |
| S11_extracted_k141_102445 | Not-determined |
| S11_extracted_k141_37118 | Low-quality |
| S11_extracted_k141_24844 | Not-determined |
| S11_extracted_k141_30294 | Not-determined |
| S11_extracted_k141_96019 | Not-determined |
| S11_extracted_k141_37120 | Low-quality |
| S11_extracted_k141_56940 | Not-determined |
| S11_extracted_k141_37124 | Not-determined |
| S11_extracted_k141_37125 | Not-determined |
| S11_extracted_k141_30300 | Not-determined |
| S11_extracted_k141_37127 | Not-determined |
| S11_extracted_k141_69091 | Not-determined |
| S11_extracted_k141_89321 | Low-quality |
| S11_extracted_k141_69095 | Not-determined |
| S11_extracted_k141_69096 | Not-determined |
| S11_extracted_k141_30309 | Not-determined |

| | |
|---|---|
| S11_extracted_k141_89327 | Not-determined |
| S11_extracted_k141_24856 | Low-quality |
| S11_extracted_k141_30311 | Not-determined |
| S11_extracted_k141_43744 | Not-determined |
| S11_extracted_k141_30312 | Not-determined |
| S11_extracted_k141_89329 | Low-quality |
| S11_extracted_k141_89331 | Not-determined |
| S11_extracted_k141_4799 | Not-determined |
| S11_extracted_k141_24864 | Not-determined |
| S11_extracted_k141_76399 | Low-quality |
| S11_extracted_k141_69104 | Not-determined |
| S11_extracted_k141_89340 | Not-determined |
| S11_extracted_k141_63087 | Low-quality |
| S11_extracted_k141_82422 | Not-determined |
| S11_extracted_k141_30320 | Low-quality |
| S11_extracted_k141_4811 | Not-determined |
| S11_extracted_k141_76408 | Not-determined |
| S11_extracted_k141_89344 | Not-determined |
| S11_extracted_k141_96046 | Not-determined |
| S11_extracted_k141_37145 | Not-determined |
| S11_extracted_k141_102464 | Not-determined |
| S11_extracted_k141_69111 | Low-quality |
| S11_extracted_k141_37147 | Not-determined |
| S11_extracted_k141_76414 | Not-determined |
| S11_extracted_k141_37150 | Not-determined |
| S11_extracted_k141_4818 | Not-determined |
| S11_extracted_k141_4819 | Not-determined |
| S11_extracted_k141_102467 | Not-determined |
| S11_extracted_k141_49667 | Not-determined |
| S11_extracted_k141_37151 | Not-determined |
| S11_extracted_k141_102470 | Not-determined |
| S11_extracted_k141_4828 | Not-determined |
| S11_extracted_k141_4830 | Low-quality |
| S11_extracted_k141_76428 | Not-determined |
| S11_extracted_k141_69130 | Not-determined |
| S11_extracted_k141_56972 | Not-determined |
| S11_extracted_k141_63107 | Not-determined |
| S11_extracted_k141_82428 | Not-determined |
| S11_extracted_k141_43750 | Not-determined |
| S11_extracted_k141_49681 | Not-determined |
| S11_extracted_k141_63109 | Not-determined |
| S11_extracted_k141_56974 | Not-determined |
| S11_extracted_k141_96062 | Low-quality |
| S11_extracted_k141_30348 | Not-determined |
| S11_extracted_k141_30350 | Not-determined |
| S11_extracted_k141_63112 | Not-determined |
| S11_extracted_k141_76446 | Not-determined |
| S11_extracted_k141_76447 | Not-determined |
| S11_extracted_k141_96070 | Low-quality |
| S11_extracted_k141_49695 | Not-determined |
| S11_extracted_k141_49701 | Not-determined |
| S11_extracted_k141_82449 | Not-determined |
| S11_extracted_k141_49705 | Not-determined |
| S11_extracted_k141_102491 | Not-determined |
| S11_extracted_k141_30359 | Not-determined |
| S11_extracted_k141_69161 | Not-determined |
| S11_extracted_k141_63120 | Not-determined |
| S11_extracted_k141_102494 | Not-determined |
| S11_extracted_k141_82455 | Not-determined |
| S11_extracted_k141_89349 | Not-determined |
| S11_extracted_k141_82456 | Low-quality |
| S11_extracted_k141_4848 | Not-determined |
| S11_extracted_k141_63127 | Not-determined |
| S11_extracted_k141_69162 | Low-quality |
| S11_extracted_k141_63130 | Not-determined |
| S11_extracted_k141_30375 | Not-determined |
| S11_extracted_k141_49715 | Not-determined |
| S11_extracted_k141_89365 | Low-quality |
| S11_extracted_k141_49723 | Not-determined |
| S11_extracted_k141_63148 | Not-determined |
| S11_extracted_k141_30378 | Not-determined |
| S11_extracted_k141_76469 | Not-determined |
| S11_extracted_k141_69174 | Not-determined |
| S11_extracted_k141_43782 | Not-determined |
| S11_extracted_k141_96109 | Not-determined |
| S11_extracted_k141_49728 | Not-determined |
| S11_extracted_k141_69178 | Not-determined |
| S11_extracted_k141_43784 | Not-determined |
| S11_extracted_k141_69180 | Not-determined |
| S11_extracted_k141_24900 | Not-determined |
| S11_extracted_k141_57010 | Low-quality |
| S11_extracted_k141_4869 | Not-determined |
| S11_extracted_k141_4870 | Not-determined |
| S11_extracted_k141_57014 | Not-determined |
| S11_extracted_k141_69184 | Not-determined |
| S11_extracted_k141_49741 | Not-determined |
| S11_extracted_k141_57019 | Not-determined |
| S11_extracted_k141_43791 | Not-determined |
| S11_extracted_k141_102516 | Not-determined |
| S11_extracted_k141_76480 | Not-determined |
| S11_extracted_k141_49745 | Not-determined |
| S11_extracted_k141_96124 | Not-determined |
| S11_extracted_k141_57022 | Not-determined |
| S11_extracted_k141_11238 | Not-determined |
| S11_extracted_k141_24917 | Not-determined |
| S11_extracted_k141_24919 | Low-quality |
| S11_extracted_k141_69192 | Not-determined |
| S11_extracted_k141_57025 | Low-quality |
| S11_extracted_k141_49749 | Not-determined |
| S11_extracted_k141_76489 | Low-quality |
| S11_extracted_k141_4879 | Not-determined |
| S11_extracted_k141_69202 | Not-determined |
| S11_extracted_k141_49754 | Not-determined |
| S11_extracted_k141_30397 | Not-determined |
| S11_extracted_k141_69204 | Not-determined |
| S11_extracted_k141_96131 | Not-determined |
| S11_extracted_k141_69211 | Low-quality |
| S11_extracted_k141_89376 | Not-determined |
| S11_extracted_k141_96137 | Not-determined |
| S11_extracted_k141_11254 | Not-determined |
| S11_extracted_k141_30406 | Not-determined |
| S11_extracted_k141_30407 | Not-determined |
| S11_extracted_k141_49768 | Not-determined |
| S11_extracted_k141_69217 | Not-determined |
| S11_extracted_k141_76508 | Not-determined |
| S11_extracted_k141_37205 | Low-quality |
| S11_extracted_k141_49777 | Not-determined |
| S11_extracted_k141_37210 | Not-determined |
| S11_extracted_k141_37212 | Not-determined |
| S11_extracted_k141_76513 | Not-determined |
| S11_extracted_k141_57054 | Not-determined |
| S11_extracted_k141_89398 | Not-determined |
| S11_extracted_k141_43810 | Low-quality |
| S11_extracted_k141_37217 | Low-quality |
| S11_extracted_k141_17615 | Not-determined |
| S11_extracted_k141_89402 | Low-quality |
| S11_extracted_k141_63178 | Low-quality |
| S11_extracted_k141_17617 | Not-determined |
| S11_extracted_k141_57064 | Not-determined |
| S11_extracted_k141_49788 | Low-quality |
| S11_extracted_k141_24935 | Not-determined |
| S11_extracted_k141_43815 | Not-determined |
| S11_extracted_k141_89410 | Not-determined |
| S11_extracted_k141_17626 | Not-determined |
| S11_extracted_k141_89412 | Not-determined |
| S11_extracted_k141_76516 | Not-determined |
| S11_extracted_k141_63186 | Not-determined |
| S11_extracted_k141_24940 | Low-quality |
| S11_extracted_k141_63188 | Low-quality |
| S11_extracted_k141_96171 | Low-quality |
| S11_extracted_k141_4900 | Not-determined |
| S11_extracted_k141_63194 | Not-determined |
| S11_extracted_k141_43827 | Low-quality |
| S11_extracted_k141_82484 | Not-determined |
| S11_extracted_k141_43828 | Not-determined |
| S11_extracted_k141_57079 | Not-determined |
| S11_extracted_k141_69233 | Not-determined |
| S11_extracted_k141_63199 | Not-determined |
| S11_extracted_k141_57086 | Not-determined |
| S11_extracted_k141_49791 | Not-determined |
| S11_extracted_k141_69239 | Not-determined |
| S11_extracted_k141_49794 | Not-determined |
| S11_extracted_k141_57092 | Not-determined |
| S11_extracted_k141_76536 | Not-determined |
| S11_extracted_k141_89438 | Low-quality |
| S11_extracted_k141_24962 | Not-determined |
| S11_extracted_k141_11284 | Not-determined |
| S11_extracted_k141_49804 | Low-quality |
| S11_extracted_k141_11287 | Not-determined |
| S11_extracted_k141_11288 | Low-quality |
| S11_extracted_k141_11289 | Not-determined |
| S11_extracted_k141_57104 | Low-quality |
| S11_extracted_k141_4926 | Not-determined |
| S11_extracted_k141_17662 | Not-determined |
| S11_extracted_k141_17666 | Not-determined |
| S11_extracted_k141_57110 | Low-quality |
| S11_extracted_k141_17667 | Not-determined |
| S11_extracted_k141_89443 | Not-determined |
| S11_extracted_k141_102536 | Not-determined |
| S11_extracted_k141_4929 | Not-determined |
| S11_extracted_k141_102537 | Not-determined |
| S11_extracted_k141_89448 | Not-determined |
| S11_extracted_k141_17673 | Not-determined |
| S11_extracted_k141_69268 | Not-determined |
| S11_extracted_k141_30434 | Not-determined |
| S11_extracted_k141_4940 | Not-determined |
| S11_extracted_k141_76571 | Not-determined |
| S11_extracted_k141_17679 | Not-determined |
| S11_extracted_k141_82500 | Not-determined |
| S11_extracted_k141_76573 | Not-determined |
| S11_extracted_k141_11307 | Not-determined |
| S11_extracted_k141_11308 | Not-determined |
| S11_extracted_k141_37254 | Not-determined |
| S11_extracted_k141_24981 | Not-determined |
| S11_extracted_k141_11312 | Not-determined |
| S11_extracted_k141_37256 | Not-determined |
| S11_extracted_k141_82508 | Not-determined |
| S11_extracted_k141_96206 | Not-determined |
| S11_extracted_k141_96207 | Low-quality |
| S11_extracted_k141_96210 | Not-determined |
| S11_extracted_k141_17696 | Not-determined |
| S11_extracted_k141_4957 | Not-determined |
| S11_extracted_k141_49841 | Not-determined |
| S11_extracted_k141_69278 | Not-determined |
| S11_extracted_k141_96213 | Not-determined |
| S11_extracted_k141_49843 | Not-determined |
| S11_extracted_k141_82517 | Low-quality |
| S11_extracted_k141_57120 | Not-determined |
| S11_extracted_k141_76586 | Not-determined |
| S11_extracted_k141_96217 | Not-determined |
| S11_extracted_k141_76587 | Not-determined |
| S11_extracted_k141_4964 | Not-determined |
| S11_extracted_k141_69284 | Not-determined |
| S11_extracted_k141_96221 | Not-determined |
| S11_extracted_k141_57125 | Low-quality |
| S11_extracted_k141_30441 | Not-determined |
| S11_extracted_k141_69287 | Not-determined |
| S11_extracted_k141_76591 | Not-determined |
| S11_extracted_k141_96224 | Not-determined |
| S11_extracted_k141_49852 | Not-determined |
| S11_extracted_k141_82529 | Not-determined |
| S11_extracted_k141_82531 | Not-determined |
| S11_extracted_k141_57131 | Low-quality |
| S11_extracted_k141_69289 | Not-determined |
| S11_extracted_k141_49857 | Not-determined |
| S11_extracted_k141_69294 | Not-determined |
| S11_extracted_k141_82540 | Not-determined |
| S11_extracted_k141_96236 | Not-determined |
| S11_extracted_k141_11331 | Not-determined |
| S11_extracted_k141_76596 | Low-quality |
| S11_extracted_k141_96243 | Not-determined |
| S11_extracted_k141_4989 | Low-quality |
| S11_extracted_k141_25027 | Not-determined |
| S11_extracted_k141_11336 | Low-quality |
| S11_extracted_k141_76599 | Not-determined |
| S11_extracted_k141_49880 | Not-determined |
| S11_extracted_k141_57150 | Not-determined |
| S11_extracted_k141_11340 | Not-determined |
| S11_extracted_k141_4993 | Not-determined |
| S11_extracted_k141_76603 | Not-determined |
| S11_extracted_k141_57154 | Not-determined |
| S11_extracted_k141_11343 | Not-determined |
| S11_extracted_k141_37286 | Low-quality |
| S11_extracted_k141_25034 | Not-determined |
| S11_extracted_k141_4996 | Low-quality |
| S11_extracted_k141_25044 | Not-determined |
| S11_extracted_k141_49894 | Not-determined |
| S11_extracted_k141_25050 | Not-determined |
| S11_extracted_k141_57170 | Not-determined |
| S11_extracted_k141_17731 | Not-determined |
| S11_extracted_k141_82563 | Not-determined |
| S11_extracted_k141_11364 | Not-determined |
| S11_extracted_k141_5010 | Not-determined |
| S11_extracted_k141_43882 | Not-determined |
| S11_extracted_k141_82571 | Low-quality |
| S11_extracted_k141_11373 | Not-determined |
| S11_extracted_k141_37306 | Not-determined |
| S11_extracted_k141_96253 | Low-quality |
| S11_extracted_k141_37308 | Not-determined |
| S11_extracted_k141_57183 | Low-quality |
| S11_extracted_k141_57184 | Not-determined |
| S11_extracted_k141_82577 | Not-determined |
| S11_extracted_k141_82581 | Not-determined |
| S11_extracted_k141_57190 | Not-determined |
| S11_extracted_k141_76619 | Not-determined |
| S11_extracted_k141_76620 | Not-determined |
| S11_extracted_k141_25069 | Not-determined |
| S11_extracted_k141_76623 | Not-determined |
| S11_extracted_k141_69310 | Not-determined |
| S11_extracted_k141_37313 | Not-determined |
| S11_extracted_k141_37314 | Not-determined |
| S11_extracted_k141_69315 | Not-determined |
| S11_extracted_k141_43899 | Not-determined |
| S11_extracted_k141_30477 | Not-determined |
| S11_extracted_k141_37318 | Not-determined |
| S11_extracted_k141_30478 | Not-determined |
| S11_extracted_k141_57199 | Low-quality |
| S11_extracted_k141_49916 | Not-determined |
| S11_extracted_k141_25071 | Low-quality |
| S11_extracted_k141_96263 | Not-determined |
| S11_extracted_k141_76633 | Not-determined |
| S11_extracted_k141_43905 | Not-determined |
| S11_extracted_k141_30481 | Low-quality |
| S11_extracted_k141_25078 | Not-determined |
| S11_extracted_k141_25079 | Not-determined |
| S11_extracted_k141_57205 | Not-determined |
| S11_extracted_k141_17746 | Not-determined |
| S11_extracted_k141_25082 | Low-quality |
| S11_extracted_k141_43910 | Not-determined |
| S11_extracted_k141_25084 | Not-determined |
| S11_extracted_k141_69326 | Not-determined |
| S11_extracted_k141_11385 | Low-quality |
| S11_extracted_k141_76639 | Low-quality |
| S11_extracted_k141_57215 | Not-determined |
| S11_extracted_k141_49933 | Not-determined |
| S11_extracted_k141_96272 | Not-determined |
| S11_extracted_k141_17757 | Not-determined |
| S11_extracted_k141_37338 | Not-determined |
| S11_extracted_k141_30502 | Not-determined |
| S11_extracted_k141_25098 | Not-determined |
| S11_extracted_k141_17759 | Low-quality |
| S11_extracted_k141_49936 | Not-determined |
| S11_extracted_k141_30504 | Not-determined |
| S11_extracted_k141_37339 | Not-determined |
| S11_extracted_k141_25100 | Not-determined |
| S11_extracted_k141_30509 | Not-determined |
| S11_extracted_k141_57217 | Low-quality |
| S11_extracted_k141_30510 | Not-determined |
| S11_extracted_k141_30511 | Not-determined |
| S11_extracted_k141_11389 | Not-determined |
| S11_extracted_k141_11391 | Not-determined |
| S11_extracted_k141_49943 | Not-determined |
| S11_extracted_k141_96292 | Not-determined |
| S11_extracted_k141_43926 | Not-determined |
| S11_extracted_k141_25106 | Not-determined |
| S11_extracted_k141_43931 | Not-determined |
| S11_extracted_k141_76652 | Not-determined |
| S11_extracted_k141_43935 | Not-determined |
| S11_extracted_k141_43937 | Not-determined |
| S11_extracted_k141_11402 | Not-determined |
| S11_extracted_k141_49955 | Not-determined |
| S11_extracted_k141_82620 | Not-determined |
| S11_extracted_k141_57230 | Low-quality |
| S11_extracted_k141_17770 | Not-determined |
| S11_extracted_k141_43942 | Not-determined |
| S11_extracted_k141_43944 | Not-determined |
| S11_extracted_k141_11408 | Not-determined |
| S11_extracted_k141_11409 | Not-determined |
| S11_extracted_k141_30524 | Not-determined |
| S11_extracted_k141_11410 | Not-determined |
| S11_extracted_k141_96304 | Not-determined |
| S11_extracted_k141_11411 | Not-determined |
| S11_extracted_k141_30525 | Low-quality |
| S11_extracted_k141_82625 | Not-determined |
| S11_extracted_k141_82626 | Not-determined |
| S11_extracted_k141_25114 | Not-determined |
| S11_extracted_k141_57239 | Not-determined |
| S11_extracted_k141_69363 | Not-determined |
| S11_extracted_k141_30530 | Not-determined |
| S11_extracted_k141_69364 | Not-determined |
| S11_extracted_k141_69365 | Not-determined |
| S11_extracted_k141_37354 | Not-determined |
| S11_extracted_k141_57244 | Not-determined |
| S11_extracted_k141_25118 | Low-quality |
| S11_extracted_k141_69372 | Not-determined |
| S11_extracted_k141_76658 | Not-determined |
| S11_extracted_k141_17785 | Not-determined |
| S11_extracted_k141_37360 | Not-determined |
| S11_extracted_k141_76662 | Not-determined |
| S11_extracted_k141_11427 | Not-determined |
| S11_extracted_k141_69381 | Not-determined |
| S11_extracted_k141_25124 | Not-determined |
| S11_extracted_k141_25125 | Not-determined |
| S11_extracted_k141_37371 | Not-determined |
| S11_extracted_k141_11431 | Not-determined |
| S11_extracted_k141_96325 | Not-determined |
| S11_extracted_k141_17796 | Not-determined |
| S11_extracted_k141_49976 | Not-determined |
| S11_extracted_k141_76673 | Not-determined |
| S11_extracted_k141_57263 | Not-determined |
| S11_extracted_k141_43975 | Not-determined |
| S11_extracted_k141_43977 | Not-determined |
| S11_extracted_k141_96330 | Low-quality |
| S11_extracted_k141_11440 | Not-determined |
| S11_extracted_k141_57269 | Not-determined |
| S11_extracted_k141_82659 | Low-quality |
| S11_extracted_k141_49977 | Not-determined |
| S11_extracted_k141_96341 | Not-determined |
| S11_extracted_k141_82665 | Not-determined |
| S11_extracted_k141_30544 | Not-determined |
| S11_extracted_k141_37385 | Not-determined |
| S11_extracted_k141_43992 | Not-determined |
| S11_extracted_k141_76680 | Low-quality |
| S11_extracted_k141_43994 | Not-determined |
| S11_extracted_k141_37389 | Not-determined |
| S11_extracted_k141_17813 | Not-determined |
| S11_extracted_k141_30554 | Not-determined |
| S11_extracted_k141_37395 | Not-determined |
| S11_extracted_k141_82673 | Not-determined |
| S11_extracted_k141_82674 | Not-determined |
| S11_extracted_k141_30556 | Low-quality |
| S11_extracted_k141_17818 | Not-determined |
| S11_extracted_k141_96351 | Not-determined |
| S11_extracted_k141_17819 | Not-determined |
| S11_extracted_k141_17821 | Not-determined |
| S11_extracted_k141_25136 | Not-determined |
| S11_extracted_k141_69400 | Not-determined |
| S11_extracted_k141_57287 | Not-determined |
| S11_extracted_k141_17824 | Not-determined |
| S11_extracted_k141_49992 | Not-determined |
| S11_extracted_k141_25139 | Not-determined |
| S11_extracted_k141_17827 | Not-determined |
| S11_extracted_k141_89489 | Not-determined |
| S11_extracted_k141_49995 | Not-determined |
| S11_extracted_k141_89491 | Not-determined |
| S11_extracted_k141_11458 | Not-determined |
| S11_extracted_k141_49997 | Low-quality |
| S11_extracted_k141_69408 | Not-determined |
| S11_extracted_k141_69409 | Not-determined |
| S11_extracted_k141_69411 | Not-determined |
| S11_extracted_k141_11467 | Low-quality |
| S11_extracted_k141_25154 | Not-determined |
| S11_extracted_k141_76711 | Not-determined |
| S11_extracted_k141_11478 | Not-determined |
| S11_extracted_k141_37421 | Not-determined |
| S11_extracted_k141_69421 | Low-quality |
| S11_extracted_k141_89505 | Not-determined |
| S11_extracted_k141_30579 | Not-determined |
| S11_extracted_k141_89516 | Not-determined |
| S11_extracted_k141_30583 | Low-quality |
| S11_extracted_k141_50011 | Not-determined |
| S11_extracted_k141_82714 | Not-determined |
| S11_extracted_k141_96368 | Not-determined |
| S11_extracted_k141_76717 | Low-quality |
| S11_extracted_k141_89520 | Not-determined |
| S11_extracted_k141_37441 | Not-determined |
| S11_extracted_k141_63242 | Not-determined |
| S11_extracted_k141_37442 | Not-determined |
| S11_extracted_k141_89524 | Not-determined |
| S11_extracted_k141_30594 | Not-determined |
| S11_extracted_k141_76721 | Low-quality |
| S11_extracted_k141_102572 | Low-quality |
| S11_extracted_k141_76722 | Not-determined |
| S11_extracted_k141_96375 | Low-quality |
| S11_extracted_k141_76726 | Not-determined |
| S11_extracted_k141_82728 | Not-determined |
| S11_extracted_k141_76729 | Not-determined |
| S11_extracted_k141_30599 | Low-quality |
| S11_extracted_k141_96379 | Not-determined |
| S11_extracted_k141_17837 | Low-quality |
| S11_extracted_k141_57291 | Not-determined |
| S11_extracted_k141_96380 | Low-quality |
| S11_extracted_k141_57292 | Not-determined |
| S11_extracted_k141_57296 | Not-determined |
| S11_extracted_k141_57297 | Not-determined |
| S11_extracted_k141_17841 | Low-quality |
| S11_extracted_k141_63251 | Not-determined |
| S11_extracted_k141_96388 | Not-determined |
| S11_extracted_k141_17843 | Not-determined |
| S11_extracted_k141_11503 | Not-determined |
| S11_extracted_k141_50027 | Not-determined |
| S11_extracted_k141_63254 | Low-quality |
| S11_extracted_k141_50029 | Not-determined |
| S11_extracted_k141_30609 | Low-quality |
| S11_extracted_k141_50032 | Not-determined |
| S11_extracted_k141_11509 | Not-determined |
| S11_extracted_k141_96400 | Low-quality |
| S11_extracted_k141_11511 | Low-quality |
| S11_extracted_k141_25180 | Not-determined |
| S11_extracted_k141_25181 | Not-determined |
| S11_extracted_k141_89530 | Not-determined |
| S11_extracted_k141_37470 | Not-determined |
| S11_extracted_k141_25186 | Not-determined |
| S11_extracted_k141_96408 | Not-determined |
| S11_extracted_k141_25190 | Not-determined |
| S11_extracted_k141_50040 | Not-determined |
| S11_extracted_k141_82753 | Not-determined |
| S11_extracted_k141_89535 | Not-determined |
| S11_extracted_k141_102595 | Not-determined |
| S11_extracted_k141_96415 | Not-determined |
| S11_extracted_k141_89545 | Not-determined |
| S11_extracted_k141_96421 | Not-determined |
| S11_extracted_k141_57310 | Not-determined |
| S11_extracted_k141_89553 | Low-quality |
| S11_extracted_k141_11521 | Not-determined |
| S11_extracted_k141_57313 | Not-determined |
| S11_extracted_k141_89554 | Low-quality |
| S11_extracted_k141_102613 | Not-determined |
| S11_extracted_k141_89556 | Low-quality |
| S11_extracted_k141_76761 | Not-determined |
| S11_extracted_k141_89560 | Low-quality |
| S11_extracted_k141_30638 | Not-determined |
| S9_vs_k141_64844\|\|full | High-quality |
| S11_extracted_k141_76763 | Not-determined |
| S11_extracted_k141_50062 | Not-determined |
| S11_extracted_k141_102624 | Not-determined |
| S11_extracted_k141_89571 | Not-determined |
| S11_extracted_k141_30647 | Not-determined |
| S11_extracted_k141_50067 | Not-determined |
| S11_extracted_k141_82762 | Not-determined |
| S11_extracted_k141_76778 | Not-determined |
| S11_extracted_k141_102627 | Not-determined |
| S11_extracted_k141_50070 | Not-determined |
| S11_extracted_k141_50071 | Not-determined |
| S11_extracted_k141_50074 | Not-determined |
| S11_extracted_k141_17872 | Not-determined |
| S11_extracted_k141_30649 | Not-determined |
| S11_extracted_k141_102635 | Not-determined |
| S11_extracted_k141_17875 | Not-determined |
| S11_extracted_k141_96453 | Not-determined |
| S11_extracted_k141_96456 | Not-determined |
| S11_extracted_k141_37495 | Low-quality |
| S11_extracted_k141_17880 | Not-determined |
| S11_extracted_k141_63282 | Not-determined |
| S11_extracted_k141_63286 | Not-determined |
| S11_extracted_k141_30654 | Not-determined |
| S11_extracted_k141_37497 | Not-determined |
| S11_extracted_k141_30656 | Not-determined |
| S11_extracted_k141_69433 | Not-determined |
| S11_extracted_k141_17886 | Not-determined |
| S11_extracted_k141_30665 | Not-determined |
| S11_extracted_k141_96470 | Not-determined |
| S11_extracted_k141_69439 | Not-determined |
| S11_extracted_k141_57332 | Not-determined |
| S11_extracted_k141_96472 | Not-determined |
| S11_extracted_k141_44020 | Not-determined |
| S11_extracted_k141_96475 | Not-determined |
| S11_extracted_k141_82775 | Not-determined |
| S11_extracted_k141_17890 | Not-determined |
| S11_extracted_k141_57340 | Not-determined |
| S11_extracted_k141_63313 | Not-determined |
| S11_extracted_k141_50094 | Not-determined |
| S11_extracted_k141_50095 | Not-determined |
| S11_extracted_k141_63317 | Not-determined |
| S11_extracted_k141_96484 | Not-determined |
| S11_extracted_k141_57344 | Not-determined |
| S11_extracted_k141_82782 | Not-determined |
| S11_extracted_k141_69446 | Not-determined |
| S11_extracted_k141_96489 | Low-quality |
| S11_extracted_k141_30684 | Low-quality |
| S11_extracted_k141_57349 | Not-determined |
| S11_extracted_k141_63325 | Not-determined |
| S11_extracted_k141_37531 | Not-determined |
| S11_extracted_k141_17912 | Not-determined |
| S11_extracted_k141_63330 | Not-determined |
| S11_extracted_k141_44044 | Not-determined |
| S11_extracted_k141_57352 | Low-quality |
| S11_extracted_k141_69466 | Not-determined |
| S11_extracted_k141_96504 | Low-quality |
| S11_extracted_k141_44050 | Not-determined |
| S11_extracted_k141_57359 | Not-determined |
| S11_extracted_k141_44057 | Low-quality |
| S11_extracted_k141_89573 | Not-determined |
| S11_extracted_k141_57362 | Low-quality |
| S11_extracted_k141_96515 | Not-determined |
| S11_extracted_k141_44061 | Low-quality |
| S11_extracted_k141_89578 | Not-determined |
| S11_extracted_k141_57370 | Low-quality |
| S11_extracted_k141_96523 | Low-quality |
| S11_extracted_k141_69475 | Low-quality |
| S11_extracted_k141_44070 | Low-quality |
| S11_extracted_k141_89586 | Not-determined |
| S11_extracted_k141_102644 | Not-determined |
| S11_extracted_k141_69480 | Not-determined |
| S11_extracted_k141_5071 | Not-determined |
| S11_extracted_k141_89587 | Not-determined |
| S11_extracted_k141_5072 | Not-determined |
| S11_extracted_k141_76785 | Low-quality |
| S11_extracted_k141_69484 | Not-determined |
| S11_extracted_k141_82790 | Not-determined |
| S11_extracted_k141_57383 | Not-determined |
| S11_extracted_k141_37546 | Not-determined |
| S11_extracted_k141_96536 | Not-determined |
| S11_extracted_k141_30734 | Not-determined |
| S11_extracted_k141_102655 | Not-determined |
| S11_extracted_k141_96539 | Not-determined |
| S11_extracted_k141_37550 | Not-determined |
| S11_extracted_k141_11565 | Not-determined |
| S11_extracted_k141_17947 | Not-determined |
| S11_extracted_k141_89612 | Not-determined |
| S11_extracted_k141_44104 | Not-determined |
| S11_extracted_k141_25202 | Not-determined |
| S11_extracted_k141_89616 | Not-determined |
| S11_extracted_k141_102684 | Not-determined |
| S11_extracted_k141_69503 | Not-determined |
| S11_extracted_k141_96559 | Not-determined |
| S11_extracted_k141_69505 | Not-determined |
| S11_extracted_k141_82809 | Not-determined |
| S11_extracted_k141_102691 | Not-determined |
| S11_extracted_k141_82812 | Not-determined |
| S11_extracted_k141_57426 | Low-quality |
| S11_extracted_k141_11587 | Low-quality |
| S11_extracted_k141_102695 | Low-quality |
| S11_extracted_k141_37582 | Low-quality |
| S11_extracted_k141_25217 | Low-quality |
| S11_extracted_k141_44122 | Not-determined |
| S11_extracted_k141_96567 | Low-quality |
| S11_extracted_k141_57441 | Low-quality |
| S11_extracted_k141_5079 | Not-determined |
| S11_extracted_k141_82820 | Low-quality |
| S11_extracted_k141_17957 | Not-determined |
| S11_extracted_k141_25225 | Low-quality |
| S11_extracted_k141_82821 | Not-determined |
| S11_extracted_k141_25229 | Not-determined |
| S11_extracted_k141_37593 | Not-determined |
| S11_extracted_k141_17961 | Not-determined |
| S11_extracted_k141_89643 | Not-determined |
| S11_extracted_k141_37599 | Not-determined |
| S11_extracted_k141_89646 | Not-determined |
| S11_extracted_k141_96576 | Not-determined |
| S11_extracted_k141_76800 | Not-determined |
| S11_extracted_k141_25240 | Not-determined |
| S11_extracted_k141_96579 | Not-determined |
| S11_extracted_k141_5091 | Low-quality |
| S11_extracted_k141_44144 | Not-determined |
| S11_extracted_k141_89657 | Not-determined |
| S11_extracted_k141_25252 | Not-determined |
| S11_extracted_k141_96592 | Not-determined |
| S11_extracted_k141_89665 | Not-determined |

| | |
|---|---|
| S11_extracted_k141_25256 | Not-determined |
| S11_extracted_k141_69541 | Not-determined |
| S11_extracted_k141_82835 | Not-determined |
| S11_extracted_k141_96596 | Not-determined |
| S11_extracted_k141_44150 | Low-quality |
| S11_extracted_k141_25259 | Not-determined |
| S11_extracted_k141_82841 | Not-determined |
| S11_extracted_k141_82845 | Not-determined |
| S11_extracted_k141_96603 | Not-determined |
| S11_extracted_k141_89677 | Not-determined |
| S11_extracted_k141_69545 | Not-determined |
| S11_extracted_k141_82849 | Not-determined |
| S11_extracted_k141_76805 | Not-determined |
| S11_extracted_k141_82851 | Low-quality |
| S11_extracted_k141_25268 | Not-determined |
| S11_extracted_k141_30750 | Low-quality |
| S11_extracted_k141_96613 | Not-determined |
| S11_extracted_k141_89684 | Low-quality |
| S11_extracted_k141_37622 | Not-determined |
| S11_extracted_k141_89685 | Low-quality |
| S11_extracted_k141_37623 | Not-determined |
| S11_extracted_k141_82858 | Not-determined |
| S11_extracted_k141_57448 | Not-determined |
| S11_extracted_k141_37625 | Not-determined |
| S11_extracted_k141_30758 | Not-determined |
| S11_extracted_k141_69553 | Low-quality |
| S11_extracted_k141_69556 | Not-determined |
| S11_extracted_k141_89695 | Not-determined |
| S11_extracted_k141_96619 | Not-determined |
| S11_extracted_k141_82866 | Not-determined |
| S11_extracted_k141_44169 | Low-quality |
| S11_extracted_k141_37635 | Not-determined |
| S11_extracted_k141_17993 | Not-determined |
| S11_extracted_k141_37637 | Not-determined |
| S11_extracted_k141_5101 | Not-determined |
| S11_extracted_k141_57453 | Not-determined |
| S11_extracted_k141_96625 | Not-determined |
| S11_extracted_k141_76823 | Not-determined |
| S11_extracted_k141_37645 | Not-determined |
| S11_extracted_k141_69563 | Not-determined |
| S11_extracted_k141_5114 | Not-determined |
| S11_extracted_k141_69567 | Not-determined |
| S11_extracted_k141_25296 | Low-quality |
| S11_extracted_k141_17995 | Low-quality |
| S11_extracted_k141_17998 | Not-determined |
| S11_extracted_k141_69574 | Not-determined |
| S11_extracted_k141_69576 | Not-determined |
| S11_extracted_k141_18008 | Not-determined |
| S11_extracted_k141_18009 | Not-determined |
| S11_extracted_k141_5137 | Not-determined |
| S11_extracted_k141_37659 | Not-determined |
| S11_extracted_k141_76848 | Not-determined |
| S11_extracted_k141_76849 | Not-determined |
| S11_extracted_k141_30790 | Not-determined |
| S11_extracted_k141_18013 | Not-determined |
| S11_extracted_k141_69583 | Not-determined |
| S11_extracted_k141_82898 | Not-determined |
| S11_extracted_k141_25310 | Not-determined |
| S11_extracted_k141_76853 | Not-determined |
| S11_extracted_k141_82900 | Not-determined |
| S11_extracted_k141_82903 | Not-determined |
| S11_extracted_k141_63361 | Not-determined |
| S11_extracted_k141_30798 | Not-determined |
| S11_extracted_k141_82904 | Not-determined |
| S11_extracted_k141_82905 | Not-determined |
| S11_extracted_k141_63362 | Not-determined |
| S11_extracted_k141_5139 | Not-determined |
| S11_extracted_k141_25317 | Not-determined |
| S11_extracted_k141_76860 | Not-determined |
| S11_extracted_k141_96649 | Low-quality |
| S11_extracted_k141_69584 | Not-determined |
| S11_extracted_k141_82911 | Not-determined |
| S11_extracted_k141_5142 | Not-determined |
| S11_extracted_k141_18021 | Low-quality |
| S11_extracted_k141_96653 | Not-determined |
| S11_extracted_k141_30808 | Not-determined |
| S11_extracted_k141_30809 | Low-quality |
| S11_extracted_k141_76865 | Not-determined |
| S11_extracted_k141_44177 | Not-determined |
| S11_extracted_k141_5151 | Not-determined |
| S11_extracted_k141_69599 | Not-determined |
| S11_extracted_k141_44189 | Not-determined |
| S11_extracted_k141_82926 | Not-determined |
| S11_extracted_k141_76878 | Not-determined |
| S11_extracted_k141_69609 | Not-determined |
| S11_extracted_k141_69613 | Not-determined |
| S11_extracted_k141_25342 | Not-determined |
| S11_extracted_k141_44202 | Not-determined |
| S11_extracted_k141_82939 | Not-determined |
| S11_extracted_k141_5163 | Not-determined |
| S11_extracted_k141_18036 | Not-determined |
| S11_extracted_k141_44207 | Not-determined |
| S11_extracted_k141_76887 | Low-quality |
| S11_extracted_k141_63387 | Not-determined |
| S11_extracted_k141_44212 | Not-determined |
| S11_extracted_k141_18045 | Not-determined |
| S11_extracted_k141_96667 | Not-determined |
| S11_extracted_k141_44219 | Not-determined |
| S11_extracted_k141_76899 | Not-determined |
| S11_extracted_k141_96670 | Not-determined |
| S11_extracted_k141_96673 | Low-quality |
| S11_extracted_k141_37676 | Not-determined |
| S11_extracted_k141_96676 | Not-determined |
| S11_extracted_k141_76902 | Not-determined |
| S11_extracted_k141_30838 | Low-quality |
| S11_extracted_k141_76903 | Not-determined |
| S11_extracted_k141_18056 | Not-determined |
| S11_extracted_k141_44224 | Low-quality |
| S11_extracted_k141_63406 | Low-quality |
| S11_extracted_k141_30842 | Not-determined |
| S11_extracted_k141_18061 | Not-determined |
| S11_extracted_k141_5192 | Not-determined |
| S11_extracted_k141_30845 | Not-determined |
| S11_extracted_k141_82954 | Low-quality |
| S11_extracted_k141_69627 | Not-determined |
| S11_extracted_k141_37687 | Not-determined |
| S11_extracted_k141_44233 | Not-determined |
| S11_extracted_k141_69633 | Not-determined |
| S11_extracted_k141_18074 | Not-determined |
| S11_extracted_k141_69635 | Not-determined |
| S11_extracted_k141_37691 | Low-quality |
| S11_extracted_k141_69639 | Not-determined |
| S11_extracted_k141_5208 | Not-determined |
| S11_extracted_k141_37694 | Not-determined |
| S11_extracted_k141_82960 | Not-determined |
| S11_extracted_k141_82961 | Not-determined |
| S11_extracted_k141_30853 | Not-determined |
| S11_extracted_k141_44236 | Not-determined |
| S11_extracted_k141_30854 | Not-determined |
| S11_extracted_k141_44237 | Not-determined |
| S11_extracted_k141_63416 | Not-determined |
| S11_extracted_k141_5218 | Not-determined |
| S11_extracted_k141_30858 | Not-determined |
| S11_extracted_k141_25363 | Not-determined |
| S11_extracted_k141_69647 | Low-quality |
| S11_extracted_k141_25366 | Not-determined |
| S11_extracted_k141_69656 | Not-determined |
| S11_extracted_k141_96682 | Not-determined |
| S11_extracted_k141_63427 | Not-determined |
| S11_extracted_k141_63428 | Not-determined |
| S11_extracted_k141_69669 | Low-quality |
| S11_extracted_k141_37722 | Not-determined |
| S11_extracted_k141_69673 | Not-determined |
| S11_extracted_k141_37723 | Not-determined |
| S11_extracted_k141_96687 | Not-determined |
| S11_extracted_k141_63439 | Not-determined |
| S11_extracted_k141_63440 | Not-determined |
| S11_extracted_k141_44263 | Not-determined |
| S11_extracted_k141_63444 | Not-determined |
| S11_extracted_k141_11600 | Not-determined |
| S11_extracted_k141_30889 | Low-quality |
| S11_extracted_k141_11603 | Not-determined |
| S11_extracted_k141_44266 | Not-determined |
| S11_extracted_k141_37737 | Low-quality |
| S11_extracted_k141_63448 | Not-determined |
| S11_extracted_k141_30894 | Not-determined |
| S11_extracted_k141_44270 | Not-determined |
| S11_extracted_k141_69681 | Not-determined |
| S11_extracted_k141_37739 | Not-determined |
| S11_extracted_k141_25390 | Low-quality |
| S11_extracted_k141_37743 | Low-quality |
| S11_extracted_k141_18088 | Not-determined |
| S11_extracted_k141_83001 | Not-determined |
| S11_extracted_k141_83003 | Not-determined |
| S11_extracted_k141_5247 | Not-determined |
| S11_extracted_k141_44276 | Not-determined |
| S11_extracted_k141_5248 | Not-determined |
| S11_extracted_k141_30903 | Not-determined |
| S11_extracted_k141_11621 | Not-determined |
| S11_extracted_k141_69694 | Not-determined |
| S11_extracted_k141_5253 | Not-determined |
| S11_extracted_k141_63469 | Not-determined |
| S11_extracted_k141_76969 | Not-determined |
| S11_extracted_k141_11630 | Not-determined |
| S11_extracted_k141_63473 | Low-quality |
| S11_extracted_k141_76971 | Not-determined |
| S11_extracted_k141_44289 | Not-determined |
| S11_extracted_k141_30912 | Low-quality |
| S11_extracted_k141_30914 | Not-determined |
| S11_extracted_k141_25406 | Low-quality |
| S11_extracted_k141_30917 | Not-determined |
| S11_extracted_k141_63477 | Low-quality |
| S11_extracted_k141_37758 | Not-determined |
| S11_extracted_k141_76977 | Not-determined |
| S11_extracted_k141_76978 | Not-determined |
| S11_extracted_k141_44294 | Not-determined |
| S11_extracted_k141_18108 | Not-determined |
| S11_extracted_k141_69710 | Not-determined |
| S11_extracted_k141_30924 | Not-determined |
| S11_extracted_k141_44300 | Low-quality |
| S11_extracted_k141_30926 | Not-determined |
| S11_extracted_k141_30929 | Not-determined |
| S11_extracted_k141_30934 | Not-determined |
| S11_extracted_k141_18122 | Not-determined |
| S11_extracted_k141_89713 | Low-quality |
| S11_extracted_k141_44309 | Not-determined |
| S11_extracted_k141_89716 | Not-determined |
| S11_extracted_k141_18125 | Not-determined |
| S11_extracted_k141_89718 | Not-determined |
| S11_extracted_k141_18126 | Not-determined |
| S11_extracted_k141_96732 | Not-determined |
| S11_extracted_k141_63496 | Not-determined |
| S11_extracted_k141_11639 | Low-quality |
| S11_extracted_k141_44322 | Low-quality |
| S11_extracted_k141_25417 | Low-quality |
| S11_extracted_k141_63500 | Not-determined |
| S11_extracted_k141_37784 | Not-determined |
| S11_extracted_k141_63504 | Not-determined |
| S11_extracted_k141_96735 | Not-determined |
| S11_extracted_k141_25422 | Not-determined |
| S11_extracted_k141_63513 | Not-determined |
| S11_extracted_k141_57529 | Low-quality |
| S11_extracted_k141_83015 | Low-quality |
| S11_extracted_k141_5282 | Not-determined |
| S11_extracted_k141_63516 | Not-determined |
| S11_extracted_k141_83018 | Not-determined |
| S11_extracted_k141_18140 | Low-quality |
| S11_extracted_k141_25429 | Not-determined |
| S11_extracted_k141_89728 | Not-determined |
| S11_extracted_k141_37801 | Not-determined |
| S11_extracted_k141_11657 | Not-determined |
| S11_extracted_k141_83020 | Not-determined |
| S11_extracted_k141_5291 | Not-determined |
| S11_extracted_k141_18145 | Not-determined |
| S11_extracted_k141_5293 | Low-quality |
| S11_extracted_k141_18146 | Not-determined |
| S11_extracted_k141_37806 | Low-quality |
| S11_extracted_k141_63524 | Not-determined |
| S11_extracted_k141_11663 | Not-determined |
| S11_extracted_k141_69748 | Not-determined |
| S11_extracted_k141_57548 | Low-quality |
| S11_extracted_k141_50116 | Not-determined |
| S11_extracted_k141_37812 | Not-determined |
| S11_extracted_k141_63538 | Low-quality |
| S11_extracted_k141_69772 | Low-quality |
| S11_extracted_k141_69774 | Not-determined |
| S11_extracted_k141_50128 | Not-determined |
| S11_extracted_k141_63549 | Not-determined |
| S11_extracted_k141_25448 | Not-determined |
| S11_extracted_k141_96761 | Not-determined |
| S11_extracted_k141_102758 | Low-quality |
| S11_extracted_k141_37826 | Not-determined |
| S11_extracted_k141_83043 | Not-determined |
| S11_extracted_k141_25459 | Not-determined |
| S11_extracted_k141_89756 | Low-quality |
| S11_extracted_k141_11689 | Not-determined |
| S11_extracted_k141_102773 | Not-determined |
| S11_extracted_k141_11691 | Not-determined |
| S11_extracted_k141_96773 | Not-determined |
| S11_extracted_k141_69791 | Not-determined |
| S11_extracted_k141_5307 | Not-determined |
| S11_extracted_k141_77055 | Not-determined |
| S11_extracted_k141_57572 | Not-determined |
| S11_extracted_k141_57576 | Not-determined |
| S11_extracted_k141_37841 | Not-determined |
| S11_extracted_k141_57577 | Low-quality |
| S11_extracted_k141_37843 | Not-determined |
| S11_extracted_k141_5314 | Not-determined |
| S11_extracted_k141_77062 | Not-determined |
| S11_extracted_k141_11699 | Low-quality |
| S11_extracted_k141_77063 | Low-quality |
| S11_extracted_k141_89768 | Not-determined |
| S11_extracted_k141_50154 | Low-quality |
| S11_extracted_k141_57582 | Not-determined |
| S11_extracted_k141_11701 | Low-quality |
| S11_extracted_k141_31004 | Not-determined |
| S11_extracted_k141_25467 | Not-determined |
| S11_extracted_k141_11704 | Not-determined |
| S11_extracted_k141_18171 | Low-quality |
| S11_extracted_k141_69811 | Not-determined |
| S11_extracted_k141_57591 | Not-determined |
| S11_extracted_k141_96777 | Not-determined |
| S11_extracted_k141_44350 | Not-determined |
| S11_extracted_k141_11712 | Not-determined |
| S11_extracted_k141_57594 | Not-determined |
| S11_extracted_k141_25477 | Not-determined |
| S11_extracted_k141_44354 | Not-determined |
| S11_extracted_k141_63580 | Not-determined |
| S11_extracted_k141_57600 | Not-determined |
| S11_extracted_k141_57601 | Not-determined |
| S11_extracted_k141_25481 | Not-determined |
| S11_extracted_k141_18183 | Not-determined |
| S11_extracted_k141_102793 | Not-determined |
| S11_extracted_k141_102794 | Not-determined |
| S11_extracted_k141_63590 | Not-determined |
| S11_extracted_k141_69818 | Low-quality |
| S11_extracted_k141_96782 | Not-determined |
| S11_extracted_k141_63594 | Not-determined |
| S11_extracted_k141_31021 | Not-determined |
| S11_extracted_k141_77086 | Low-quality |
| S11_extracted_k141_57605 | Not-determined |
| S11_extracted_k141_37855 | Low-quality |
| S11_extracted_k141_44359 | Not-determined |
| S11_extracted_k141_44360 | Not-determined |
| S11_extracted_k141_25490 | Not-determined |
| S11_extracted_k141_77088 | Not-determined |
| S11_extracted_k141_63600 | Not-determined |
| S11_extracted_k141_44366 | Not-determined |
| S11_extracted_k141_44367 | Not-determined |
| S11_extracted_k141_31026 | Not-determined |
| S11_extracted_k141_44368 | Not-determined |
| S11_extracted_k141_31027 | Not-determined |
| S11_extracted_k141_44370 | Not-determined |
| S11_extracted_k141_5328 | Not-determined |
| S11_extracted_k141_77092 | Not-determined |
| S11_extracted_k141_96786 | Not-determined |
| S11_extracted_k141_96788 | Not-determined |
| S11_extracted_k141_83068 | Low-quality |
| S11_extracted_k141_83070 | Not-determined |
| S11_extracted_k141_37871 | Low-quality |
| S11_extracted_k141_31034 | Not-determined |
| S11_extracted_k141_18192 | Low-quality |
| S11_extracted_k141_83084 | Not-determined |
| S11_extracted_k141_83085 | Not-determined |
| S11_extracted_k141_50172 | Low-quality |
| S11_extracted_k141_63619 | Not-determined |
| S11_extracted_k141_96795 | Not-determined |
| S11_extracted_k141_31046 | Low-quality |
| S11_extracted_k141_83099 | Not-determined |
| S11_extracted_k141_69839 | Low-quality |
| S11_extracted_k141_89785 | Not-determined |
| S11_extracted_k141_63632 | Not-determined |
| S11_extracted_k141_37886 | Low-quality |
| S11_extracted_k141_102808 | Low-quality |
| S11_extracted_k141_5336 | Not-determined |
| S11_extracted_k141_37887 | Not-determined |
| S11_extracted_k141_37888 | Not-determined |
| S11_extracted_k141_102814 | Low-quality |
| S11_extracted_k141_18207 | Not-determined |
| S11_extracted_k141_5345 | Not-determined |
| S11_extracted_k141_37892 | Not-determined |
| S11_extracted_k141_96803 | Not-determined |
| S11_extracted_k141_63641 | Not-determined |
| S11_extracted_k141_57618 | Low-quality |
| S11_extracted_k141_77119 | Not-determined |
| S11_extracted_k141_44404 | Not-determined |
| S11_extracted_k141_31057 | Not-determined |
| S11_extracted_k141_37898 | Not-determined |
| S11_extracted_k141_11726 | Not-determined |
| S11_extracted_k141_57619 | Not-determined |
| S11_extracted_k141_11727 | Not-determined |
| S11_extracted_k141_96806 | Not-determined |
| S11_extracted_k141_77125 | Not-determined |
| S11_extracted_k141_50188 | Not-determined |
| S11_extracted_k141_50189 | Not-determined |
| S11_extracted_k141_63649 | Not-determined |
| S11_extracted_k141_25510 | Not-determined |
| S11_extracted_k141_5359 | Not-determined |
| S11_extracted_k141_50194 | Low-quality |
| S11_extracted_k141_11736 | Not-determined |
| S11_extracted_k141_57624 | Not-determined |
| S11_extracted_k141_37901 | Not-determined |
| S11_extracted_k141_63656 | Not-determined |
| S11_extracted_k141_25523 | Not-determined |
| S11_extracted_k141_37907 | Not-determined |
| S11_extracted_k141_83124 | Not-determined |
| S11_extracted_k141_25529 | Not-determined |
| S11_extracted_k141_96825 | Not-determined |
| S11_extracted_k141_69850 | Not-determined |
| S11_extracted_k141_50202 | Not-determined |
| S11_extracted_k141_77134 | Not-determined |
| S11_extracted_k141_50205 | Not-determined |
| S11_extracted_k141_31079 | Not-determined |
| S11_extracted_k141_57644 | Not-determined |
| S11_extracted_k141_44430 | Not-determined |
| S11_extracted_k141_96833 | Not-determined |
| S11_extracted_k141_83130 | Not-determined |
| S11_extracted_k141_57655 | Not-determined |
| S11_extracted_k141_5382 | Not-determined |
| S11_extracted_k141_83134 | Not-determined |
| S11_extracted_k141_96835 | Low-quality |
| S11_extracted_k141_102857 | Not-determined |
| S11_extracted_k141_69864 | Not-determined |
| S11_extracted_k141_57663 | Not-determined |
| S11_extracted_k141_63677 | Not-determined |
| S11_extracted_k141_57665 | Not-determined |
| S11_extracted_k141_69872 | Not-determined |
| S11_extracted_k141_57667 | Low-quality |
| S11_extracted_k141_96846 | Not-determined |
| S11_extracted_k141_5388 | Not-determined |
| S11_extracted_k141_89818 | Not-determined |
| S11_extracted_k141_89820 | Not-determined |
| S11_extracted_k141_50219 | Not-determined |
| S11_extracted_k141_5394 | Not-determined |
| S11_extracted_k141_96851 | Not-determined |
| S11_extracted_k141_31103 | Not-determined |
| S11_extracted_k141_96852 | Not-determined |
| S11_extracted_k141_31105 | Not-determined |
| S11_extracted_k141_63692 | Not-determined |
| S11_extracted_k141_5395 | Low-quality |
| S11_extracted_k141_77147 | Not-determined |
| S11_extracted_k141_96856 | Not-determined |
| S11_extracted_k141_77148 | Not-determined |
| S11_extracted_k141_63694 | Low-quality |
| S11_extracted_k141_63695 | Not-determined |
| S11_extracted_k141_69880 | Not-determined |
| S11_extracted_k141_25547 | Not-determined |
| S11_extracted_k141_96858 | Not-determined |
| S11_extracted_k141_31112 | Low-quality |
| S11_extracted_k141_31113 | Not-determined |
| S11_extracted_k141_96861 | Low-quality |
| S11_extracted_k141_102873 | Not-determined |
| S11_extracted_k141_50228 | Low-quality |
| S11_extracted_k141_5400 | Not-determined |
| S11_extracted_k141_25556 | Not-determined |
| S11_extracted_k141_25557 | Not-determined |
| S11_extracted_k141_5403 | Low-quality |
| S11_extracted_k141_5404 | Not-determined |
| S11_extracted_k141_96871 | Not-determined |
| S11_extracted_k141_44455 | Not-determined |
| S11_extracted_k141_25561 | Not-determined |
| S11_extracted_k141_31122 | Not-determined |
| S11_extracted_k141_25563 | Not-determined |
| S11_extracted_k141_5410 | Not-determined |
| S11_extracted_k141_102887 | Not-determined |
| S11_extracted_k141_50243 | Not-determined |
| S11_extracted_k141_11769 | Not-determined |
| S11_extracted_k141_63708 | Not-determined |
| S11_extracted_k141_18255 | Not-determined |
| S11_extracted_k141_25573 | Low-quality |
| S11_extracted_k141_31129 | Not-determined |
| S11_extracted_k141_31130 | Not-determined |
| S11_extracted_k141_96879 | Low-quality |
| S11_extracted_k141_57691 | Not-determined |
| S11_extracted_k141_50249 | Not-determined |
| S11_extracted_k141_57694 | Not-determined |
| S11_extracted_k141_25576 | Not-determined |
| S11_extracted_k141_63716 | Not-determined |
| S11_extracted_k141_5418 | Low-quality |
| S11_extracted_k141_102899 | Low-quality |
| S11_extracted_k141_69900 | Not-determined |
| S11_extracted_k141_102904 | Not-determined |
| S11_extracted_k141_69904 | Low-quality |
| S11_extracted_k141_83177 | Not-determined |
| S11_extracted_k141_63722 | Not-determined |
| S11_extracted_k141_77159 | Not-determined |
| S11_extracted_k141_18263 | Not-determined |
| S11_extracted_k141_5427 | Not-determined |
| S11_extracted_k141_57704 | Not-determined |
| S11_extracted_k141_83181 | Low-quality |
| S11_extracted_k141_37917 | Not-determined |
| S11_extracted_k141_5434 | Not-determined |
| S11_extracted_k141_69917 | Not-determined |
| S11_extracted_k141_25592 | Not-determined |
| S11_extracted_k141_69921 | Not-determined |
| S11_extracted_k141_89830 | Not-determined |
| S11_extracted_k141_18274 | Not-determined |
| S11_extracted_k141_18275 | Not-determined |
| S11_extracted_k141_83187 | Not-determined |
| S11_extracted_k141_96893 | Low-quality |
| S11_extracted_k141_77167 | Not-determined |
| S11_extracted_k141_18277 | Low-quality |
| S11_extracted_k141_77168 | Low-quality |
| S11_extracted_k141_37923 | Not-determined |
| S11_extracted_k141_31150 | Low-quality |
| S11_extracted_k141_63734 | Not-determined |
| S11_extracted_k141_96898 | Not-determined |
| S11_extracted_k141_89840 | Not-determined |
| S11_extracted_k141_83197 | Not-determined |
| S11_extracted_k141_44464 | Not-determined |
| S11_extracted_k141_44465 | Low-quality |
| S11_extracted_k141_57719 | Low-quality |
| S11_extracted_k141_96910 | Not-determined |
| S11_extracted_k141_18298 | Not-determined |
| S11_extracted_k141_57723 | Not-determined |
| S11_extracted_k141_44472 | Not-determined |
| S11_extracted_k141_5451 | Not-determined |
| S11_extracted_k141_50275 | Not-determined |
| S11_extracted_k141_89845 | Not-determined |
| S11_extracted_k141_83206 | Low-quality |
| S11_extracted_k141_63749 | Not-determined |
| S11_extracted_k141_50276 | Not-determined |
| S11_extracted_k141_5454 | Not-determined |
| S11_extracted_k141_37938 | Not-determined |
| S11_extracted_k141_63750 | Low-quality |
| S11_extracted_k141_69923 | Low-quality |
| S11_extracted_k141_50279 | Not-determined |
| S11_extracted_k141_63754 | Not-determined |
| S11_extracted_k141_89848 | Not-determined |
| S11_extracted_k141_89850 | Not-determined |
| S11_extracted_k141_50283 | Low-quality |
| S11_extracted_k141_69927 | Low-quality |
| S11_extracted_k141_18306 | Not-determined |
| S11_extracted_k141_18307 | Not-determined |
| S11_extracted_k141_25609 | Not-determined |
| S11_extracted_k141_25610 | Not-determined |
| S11_extracted_k141_18311 | Low-quality |
| S11_extracted_k141_31176 | Not-determined |
| S11_extracted_k141_77180 | Not-determined |
| S11_extracted_k141_18312 | Low-quality |
| S11_extracted_k141_25619 | Not-determined |
| S11_extracted_k141_44484 | Not-determined |
| S11_extracted_k141_37947 | Not-determined |
| S11_extracted_k141_44486 | Not-determined |
| S11_extracted_k141_44487 | Not-determined |
| S11_extracted_k141_44488 | Not-determined |
| S11_extracted_k141_25625 | Not-determined |
| S11_extracted_k141_57741 | Not-determined |
| S11_extracted_k141_5474 | Not-determined |
| S11_extracted_k141_31191 | Not-determined |
| S11_extracted_k141_5475 | Not-determined |
| S11_extracted_k141_57743 | Not-determined |
| S11_extracted_k141_5476 | Not-determined |
| S11_extracted_k141_25629 | Not-determined |
| S11_extracted_k141_44492 | Not-determined |
| S11_extracted_k141_96919 | Low-quality |
| S11_extracted_k141_77197 | Not-determined |
| S11_extracted_k141_37963 | Not-determined |
| S11_extracted_k141_18322 | Not-determined |
| S11_extracted_k141_63769 | Not-determined |
| S11_extracted_k141_50305 | Not-determined |
| S11_extracted_k141_63770 | Not-determined |
| S11_extracted_k141_31197 | Not-determined |
| S11_extracted_k141_96926 | Low-quality |
| S11_extracted_k141_83211 | Not-determined |
| S11_extracted_k141_96928 | Not-determined |
| S11_extracted_k141_25649 | Not-determined |
| S11_extracted_k141_44504 | Not-determined |
| S11_extracted_k141_25652 | Not-determined |
| S11_extracted_k141_63779 | Not-determined |
| S11_extracted_k141_11826 | Not-determined |
| S11_extracted_k141_83220 | Not-determined |
| S11_extracted_k141_11829 | Not-determined |
| S11_extracted_k141_5493 | Low-quality |
| S11_extracted_k141_37971 | Not-determined |
| S11_extracted_k141_44507 | Not-determined |
| S11_extracted_k141_11836 | Not-determined |
| S11_extracted_k141_89853 | Low-quality |
| S11_extracted_k141_44508 | Not-determined |
| S11_extracted_k141_37979 | Not-determined |
| S11_extracted_k141_89856 | Low-quality |
| S11_extracted_k141_63796 | Not-determined |
| S11_extracted_k141_96941 | Not-determined |
| S11_extracted_k141_37983 | Not-determined |
| S11_extracted_k141_77212 | Not-determined |
| S11_extracted_k141_25656 | Low-quality |
| S11_extracted_k141_83227 | Not-determined |
| S11_extracted_k141_44510 | Not-determined |
| S11_extracted_k141_25658 | Not-determined |
| S11_extracted_k141_96946 | Not-determined |
| S11_extracted_k141_18333 | Not-determined |
| S11_extracted_k141_5498 | Low-quality |
| S11_extracted_k141_37989 | Not-determined |
| S11_extracted_k141_96953 | Not-determined |
| S11_extracted_k141_5501 | Low-quality |
| S11_extracted_k141_102912 | Not-determined |
| S11_extracted_k141_102914 | Not-determined |
| S11_extracted_k141_102915 | Not-determined |
| S11_extracted_k141_44522 | Not-determined |
| S11_extracted_k141_50330 | Not-determined |
| S11_extracted_k141_25672 | Not-determined |
| S11_extracted_k141_31224 | Not-determined |
| S11_extracted_k141_96961 | Low-quality |
| S11_extracted_k141_102921 | Not-determined |
| S11_extracted_k141_89870 | Not-determined |
| S11_extracted_k141_31229 | Not-determined |
| S11_extracted_k141_83244 | Not-determined |
| S11_extracted_k141_31231 | Not-determined |
| S11_extracted_k141_18344 | Not-determined |
| S11_extracted_k141_38009 | Not-determined |
| S11_extracted_k141_63835 | Not-determined |
| S11_extracted_k141_89878 | Not-determined |
| S11_extracted_k141_83247 | Not-determined |
| S11_extracted_k141_77228 | Not-determined |
| S11_extracted_k141_83248 | Low-quality |
| S11_extracted_k141_5517 | Not-determined |
| S11_extracted_k141_50340 | Not-determined |
| S11_extracted_k141_89882 | Not-determined |
| S11_extracted_k141_96967 | Not-determined |
| S11_extracted_k141_96968 | Not-determined |
| S11_extracted_k141_50345 | Not-determined |
| S11_extracted_k141_77239 | Not-determined |
| S11_extracted_k141_44538 | Low-quality |
| S11_extracted_k141_83253 | Not-determined |
| S11_extracted_k141_63853 | Not-determined |
| S11_extracted_k141_63854 | Not-determined |
| S11_extracted_k141_18363 | Not-determined |
| S11_extracted_k141_44541 | Not-determined |
| S11_extracted_k141_44542 | Not-determined |
| S11_extracted_k141_102930 | Not-determined |
| S11_extracted_k141_11852 | Not-determined |
| S11_extracted_k141_89898 | Not-determined |
| S11_extracted_k141_5536 | Not-determined |
| S11_extracted_k141_5538 | Not-determined |
| S11_extracted_k141_89902 | Low-quality |
| S11_extracted_k141_89903 | Not-determined |
| S11_extracted_k141_5541 | Low-quality |
| S11_extracted_k141_11858 | Not-determined |
| S11_extracted_k141_5542 | Not-determined |

| | |
|---|---|
| S11_extracted_k141_69944 | Not-determined |
| S11_extracted_k141_18373 | Low-quality |
| S11_extracted_k141_102940 | Not-determined |
| S11_extracted_k141_11862 | Not-determined |
| S11_extracted_k141_77254 | Low-quality |
| S11_extracted_k141_18378 | Not-determined |
| S11_extracted_k141_44553 | Not-determined |
| S11_extracted_k141_102946 | Not-determined |
| S11_extracted_k141_83266 | Low-quality |
| S11_extracted_k141_50380 | Not-determined |
| S11_extracted_k141_18386 | Low-quality |
| S11_extracted_k141_38048 | Not-determined |
| S11_extracted_k141_31256 | Not-determined |
| S11_extracted_k141_18387 | Low-quality |
| S11_extracted_k141_96973 | Low-quality |
| S11_extracted_k141_89909 | Not-determined |
| S11_extracted_k141_18389 | Low-quality |
| S11_extracted_k141_18390 | Not-determined |
| S11_extracted_k141_31263 | Not-determined |
| S11_extracted_k141_44568 | Not-determined |
| S11_extracted_k141_63867 | Not-determined |
| S11_extracted_k141_11872 | Not-determined |
| S11_extracted_k141_69947 | Low-quality |
| S11_extracted_k141_44573 | Not-determined |
| S11_extracted_k141_102963 | Not-determined |
| S11_extracted_k141_69950 | Not-determined |
| S11_extracted_k141_96980 | Not-determined |
| S11_extracted_k141_5560 | Not-determined |
| S11_extracted_k141_102968 | Not-determined |
| S11_extracted_k141_102969 | Not-determined |
| S11_extracted_k141_11875 | Not-determined |
| S11_extracted_k141_38059 | Not-determined |
| S11_extracted_k141_63876 | Not-determined |
| S11_extracted_k141_77284 | Low-quality |
| S11_extracted_k141_18408 | Not-determined |
| S11_extracted_k141_77288 | Not-determined |
| S11_extracted_k141_77289 | Not-determined |
| S11_extracted_k141_18414 | Low-quality |
| S11_extracted_k141_18419 | Not-determined |
| S11_extracted_k141_102983 | Not-determined |
| S11_extracted_k141_102984 | Not-determined |
| S11_extracted_k141_83295 | Low-quality |
| S11_extracted_k141_5577 | Not-determined |
| S11_extracted_k141_50388 | Low-quality |
| S11_extracted_k141_69964 | Not-determined |
| S11_extracted_k141_44592 | Not-determined |
| S11_extracted_k141_5582 | Not-determined |
| S11_extracted_k141_44595 | Not-determined |
| S11_extracted_k141_5587 | Not-determined |
| S11_extracted_k141_11890 | Not-determined |
| S11_extracted_k141_5591 | Not-determined |
| S11_extracted_k141_50395 | Not-determined |
| S11_extracted_k141_102996 | Not-determined |
| S11_extracted_k141_103000 | Not-determined |
| S11_extracted_k141_11897 | Not-determined |
| S11_extracted_k141_11899 | Not-determined |
| S11_extracted_k141_97010 | Not-determined |
| S11_extracted_k141_97011 | Not-determined |
| S11_extracted_k141_50403 | Not-determined |
| S11_extracted_k141_63904 | Low-quality |
| S11_extracted_k141_11903 | Low-quality |
| S11_extracted_k141_89913 | Not-determined |
| S11_extracted_k141_89914 | Not-determined |
| S11_extracted_k141_11907 | Not-determined |
| S11_extracted_k141_89917 | Not-determined |
| S11_extracted_k141_103010 | Low-quality |
| S11_extracted_k141_11910 | Not-determined |
| S11_extracted_k141_63912 | Low-quality |
| S11_extracted_k141_5597 | Not-determined |
| S11_extracted_k141_89926 | Not-determined |
| S11_extracted_k141_97016 | Not-determined |
| S11_extracted_k141_38071 | Low-quality |
| S11_extracted_k141_44617 | Not-determined |
| S11_extracted_k141_38077 | Not-determined |
| S11_extracted_k141_83315 | Low-quality |
| S11_extracted_k141_89930 | Not-determined |
| S11_extracted_k141_5611 | Not-determined |
| S11_extracted_k141_38082 | Low-quality |
| S11_extracted_k141_89931 | Not-determined |
| S11_extracted_k141_44626 | Not-determined |
| S11_extracted_k141_103030 | Low-quality |
| S11_extracted_k141_44629 | Not-determined |
| S11_extracted_k141_38086 | Not-determined |
| S11_extracted_k141_83325 | Not-determined |
| S11_extracted_k141_83330 | Not-determined |
| S11_extracted_k141_5617 | Not-determined |
| S11_extracted_k141_83332 | Not-determined |
| S11_extracted_k141_11928 | Not-determined |
| S11_extracted_k141_5618 | Not-determined |
| S11_extracted_k141_11931 | Not-determined |
| S11_extracted_k141_69999 | Not-determined |
| S11_extracted_k141_18425 | Not-determined |
| S11_extracted_k141_83342 | Not-determined |
| S11_extracted_k141_77325 | Not-determined |
| S11_extracted_k141_83343 | Not-determined |
| S11_extracted_k141_5628 | Not-determined |
| S11_extracted_k141_70008 | Not-determined |
| S11_extracted_k141_11947 | Not-determined |
| S11_extracted_k141_83348 | Not-determined |
| S11_extracted_k141_97049 | Not-determined |
| S11_extracted_k141_77332 | Not-determined |
| S11_extracted_k141_11955 | Not-determined |
| S11_extracted_k141_50424 | Not-determined |
| S11_extracted_k141_18431 | Not-determined |
| S11_extracted_k141_83354 | Not-determined |
| S11_extracted_k141_38113 | Low-quality |
| S11_extracted_k141_18433 | Not-determined |
| S11_extracted_k141_38116 | Low-quality |
| S11_extracted_k141_50433 | Not-determined |
| S11_extracted_k141_38117 | Not-determined |
| S11_extracted_k141_83357 | Not-determined |
| S11_extracted_k141_103050 | Low-quality |
| S11_extracted_k141_44652 | Not-determined |
| S11_extracted_k141_38119 | Not-determined |
| S11_extracted_k141_38121 | Not-determined |
| S11_extracted_k141_18445 | Not-determined |
| S11_extracted_k141_18446 | Not-determined |
| S11_extracted_k141_11971 | Low-quality |
| S11_extracted_k141_5643 | Not-determined |
| S11_extracted_k141_50447 | Not-determined |
| S11_extracted_k141_103061 | Not-determined |
| S11_extracted_k141_31312 | Not-determined |
| S11_extracted_k141_5649 | Not-determined |
| S11_extracted_k141_77356 | Not-determined |
| S11_extracted_k141_97061 | Low-quality |
| S11_extracted_k141_38131 | Not-determined |
| S11_extracted_k141_31315 | Not-determined |
| S11_extracted_k141_89969 | Not-determined |
| S11_extracted_k141_44664 | Not-determined |
| S11_extracted_k141_25694 | Not-determined |
| S11_extracted_k141_25695 | Not-determined |
| S11_extracted_k141_89974 | Not-determined |
| S11_extracted_k141_18455 | Not-determined |
| S11_extracted_k141_25698 | Not-determined |
| S11_extracted_k141_18457 | Not-determined |
| S11_extracted_k141_103070 | Low-quality |
| S11_extracted_k141_31322 | Not-determined |
| S11_extracted_k141_97064 | Not-determined |
| S11_extracted_k141_11979 | Low-quality |
| S11_extracted_k141_83379 | Not-determined |
| S11_extracted_k141_83380 | Not-determined |
| S11_extracted_k141_18461 | Not-determined |
| S11_extracted_k141_31323 | Not-determined |
| S11_extracted_k141_5656 | Not-determined |
| S11_extracted_k141_31324 | Not-determined |
| S11_extracted_k141_31327 | Not-determined |
| S11_extracted_k141_11988 | Not-determined |
| S11_extracted_k141_83387 | Not-determined |
| S11_extracted_k141_77372 | Not-determined |
| S11_extracted_k141_18475 | Not-determined |
| S11_extracted_k141_50453 | Not-determined |
| S11_extracted_k141_25704 | Low-quality |
| S11_extracted_k141_50455 | Not-determined |
| S11_extracted_k141_89988 | Not-determined |
| S11_extracted_k141_50459 | Not-determined |
| S11_extracted_k141_18480 | Not-determined |
| S11_extracted_k141_83397 | Low-quality |
| S11_extracted_k141_83399 | Not-determined |
| S11_extracted_k141_89993 | Not-determined |
| S11_extracted_k141_38149 | Not-determined |
| S11_extracted_k141_103079 | Not-determined |
| S11_extracted_k141_103081 | Low-quality |
| S11_extracted_k141_38154 | Not-determined |
| S11_extracted_k141_5671 | Not-determined |
| S11_extracted_k141_50465 | Not-determined |
| S11_extracted_k141_90002 | Not-determined |
| S11_extracted_k141_50467 | Not-determined |
| S11_extracted_k141_5673 | Not-determined |
| S11_extracted_k141_5674 | Not-determined |
| S11_extracted_k141_31357 | Not-determined |
| S11_extracted_k141_103092 | Low-quality |
| S11_extracted_k141_25732 | Not-determined |
| S11_extracted_k141_5676 | Low-quality |
| S11_extracted_k141_38163 | Not-determined |
| S11_extracted_k141_83410 | Not-determined |
| S11_extracted_k141_90007 | Not-determined |
| S11_extracted_k141_77398 | Not-determined |
| S11_extracted_k141_83416 | Not-determined |
| S11_extracted_k141_31368 | Not-determined |
| S11_extracted_k141_38164 | Low-quality |
| S11_extracted_k141_50480 | Not-determined |
| S11_extracted_k141_38169 | Not-determined |
| S11_extracted_k141_90018 | Not-determined |
| S11_extracted_k141_103110 | Not-determined |
| S11_extracted_k141_44702 | Not-determined |
| S11_extracted_k141_77406 | Not-determined |
| S11_extracted_k141_5681 | Not-determined |
| S11_extracted_k141_18506 | Low-quality |
| S11_extracted_k141_31373 | Not-determined |
| S11_extracted_k141_77411 | Not-determined |
| S11_extracted_k141_90026 | Not-determined |
| S11_extracted_k141_83428 | Low-quality |
| S11_extracted_k141_83430 | Low-quality |
| S11_extracted_k141_77418 | Not-determined |
| S11_extracted_k141_31380 | Not-determined |
| S11_extracted_k141_103125 | Not-determined |
| S11_extracted_k141_83433 | Low-quality |
| S11_extracted_k141_103127 | Not-determined |
| S11_extracted_k141_31387 | Not-determined |
| S11_extracted_k141_83437 | Not-determined |
| S11_extracted_k141_44712 | Not-determined |
| S8_extracted_k141_166677 | High-quality |
| S11_extracted_k141_103141 | Not-determined |
| S11_extracted_k141_70019 | Not-determined |
| S11_extracted_k141_50487 | Not-determined |
| S11_extracted_k141_25770 | Not-determined |
| S11_extracted_k141_44717 | Not-determined |
| S11_extracted_k141_83438 | Low-quality |
| S11_extracted_k141_44722 | Not-determined |
| S11_extracted_k141_5729 | Not-determined |
| S11_extracted_k141_38197 | Not-determined |
| S11_extracted_k141_25779 | Not-determined |
| S11_extracted_k141_70035 | Low-quality |
| S11_extracted_k141_77439 | Not-determined |
| S11_extracted_k141_44733 | Low-quality |
| S11_extracted_k141_25788 | Not-determined |
| S11_extracted_k141_50503 | Not-determined |
| S11_extracted_k141_63971 | Low-quality |
| S11_extracted_k141_83450 | Not-determined |
| S11_extracted_k141_18539 | Not-determined |
| S11_extracted_k141_63973 | Not-determined |
| S11_extracted_k141_63975 | Not-determined |
| S11_extracted_k141_83453 | Low-quality |
| S11_extracted_k141_31402 | Not-determined |
| S11_extracted_k141_18553 | Not-determined |
| S11_extracted_k141_25800 | Not-determined |
| S11_extracted_k141_31409 | Not-determined |
| S11_extracted_k141_90053 | Not-determined |
| S11_extracted_k141_63984 | Not-determined |
| S11_extracted_k141_97075 | Not-determined |
| S11_extracted_k141_50524 | Not-determined |
| S11_extracted_k141_18560 | Not-determined |
| S11_extracted_k141_18562 | Low-quality |
| S11_extracted_k141_90062 | Not-determined |
| S11_extracted_k141_50528 | Low-quality |
| S11_extracted_k141_97084 | Not-determined |
| S11_extracted_k141_97086 | Not-determined |
| S11_extracted_k141_57761 | Not-determined |
| S11_extracted_k141_12015 | Not-determined |
| S11_extracted_k141_63999 | Low-quality |
| S11_extracted_k141_90071 | Not-determined |
| S11_extracted_k141_64001 | Not-determined |
| S11_extracted_k141_44766 | Not-determined |
| S11_extracted_k141_44767 | Not-determined |
| S11_extracted_k141_18572 | Not-determined |
| S11_extracted_k141_25810 | Not-determined |
| S11_extracted_k141_50537 | Not-determined |
| S11_extracted_k141_83471 | Not-determined |
| S11_extracted_k141_25811 | Not-determined |
| S11_extracted_k141_25812 | Not-determined |
| S11_extracted_k141_25813 | Not-determined |
| S11_extracted_k141_38224 | Not-determined |
| S11_extracted_k141_12025 | Low-quality |
| S11_extracted_k141_50542 | Low-quality |
| S11_extracted_k141_77470 | Not-determined |
| S11_extracted_k141_25816 | Not-determined |
| S11_extracted_k141_38228 | Not-determined |
| S11_extracted_k141_70078 | Not-determined |
| S11_extracted_k141_70079 | Not-determined |
| S11_extracted_k141_44771 | Not-determined |
| S11_extracted_k141_90079 | Not-determined |
| S11_extracted_k141_38233 | Not-determined |
| S11_extracted_k141_12031 | Not-determined |
| S11_extracted_k141_5752 | Not-determined |
| S11_extracted_k141_31414 | Not-determined |
| S11_extracted_k141_64013 | Not-determined |
| S11_extracted_k141_31416 | Not-determined |
| S11_extracted_k141_44778 | Not-determined |
| S11_extracted_k141_44780 | Not-determined |
| S11_extracted_k141_31419 | Low-quality |
| S11_extracted_k141_12032 | Not-determined |
| S11_extracted_k141_90092 | Not-determined |
| S11_extracted_k141_31421 | Not-determined |
| S11_extracted_k141_25833 | Not-determined |
| S11_extracted_k141_50562 | Not-determined |
| S11_extracted_k141_25837 | Not-determined |
| S11_extracted_k141_77480 | Not-determined |
| S11_extracted_k141_83504 | Low-quality |
| S11_extracted_k141_83505 | Not-determined |
| S11_extracted_k141_57792 | Not-determined |
| S11_extracted_k141_18600 | Low-quality |
| S11_extracted_k141_31424 | Not-determined |
| S11_extracted_k141_44790 | Not-determined |
| S11_extracted_k141_25840 | Low-quality |
| S11_extracted_k141_57795 | Not-determined |
| S11_extracted_k141_31428 | Not-determined |
| S11_extracted_k141_64016 | Not-determined |
| S11_extracted_k141_31429 | Not-determined |
| S11_extracted_k141_38255 | Not-determined |
| S11_extracted_k141_38259 | Not-determined |
| S11_extracted_k141_57805 | Not-determined |
| S11_extracted_k141_57806 | Not-determined |
| S11_extracted_k141_90115 | Not-determined |
| S11_extracted_k141_5767 | Not-determined |
| S11_extracted_k141_44795 | Not-determined |
| S11_extracted_k141_90118 | Not-determined |
| S11_extracted_k141_83521 | Low-quality |
| S11_extracted_k141_25855 | Not-determined |
| S11_extracted_k141_18627 | Not-determined |
| S11_extracted_k141_70105 | Not-determined |
| S11_extracted_k141_83530 | Not-determined |
| S11_extracted_k141_38272 | Not-determined |
| S11_extracted_k141_90132 | Not-determined |
| S11_extracted_k141_12058 | Not-determined |
| S11_extracted_k141_38275 | Not-determined |
| S11_extracted_k141_5778 | Not-determined |
| S11_extracted_k141_90140 | Not-determined |
| S11_extracted_k141_77486 | Not-determined |
| S11_extracted_k141_70110 | Not-determined |
| S11_extracted_k141_64029 | Not-determined |
| S11_extracted_k141_64030 | Not-determined |
| S11_extracted_k141_70111 | Low-quality |
| S11_extracted_k141_18635 | Not-determined |
| S11_extracted_k141_18640 | Not-determined |
| S11_extracted_k141_25869 | Not-determined |
| S11_extracted_k141_12062 | Not-determined |
| S11_extracted_k141_12067 | Not-determined |
| S11_extracted_k141_18648 | Not-determined |
| S11_extracted_k141_83555 | Not-determined |
| S11_extracted_k141_18654 | Low-quality |
| S11_extracted_k141_38301 | Low-quality |
| S11_extracted_k141_38302 | Not-determined |
| S11_extracted_k141_57834 | Not-determined |
| S11_extracted_k141_97114 | Low-quality |
| S11_extracted_k141_50569 | Not-determined |
| S11_extracted_k141_83560 | Not-determined |
| S11_extracted_k141_77511 | Not-determined |
| S11_extracted_k141_50571 | Not-determined |
| S11_extracted_k141_50572 | Low-quality |
| S11_extracted_k141_12090 | Not-determined |
| S11_extracted_k141_64055 | Low-quality |
| S11_extracted_k141_5799 | Low-quality |
| S11_extracted_k141_18666 | Not-determined |
| S11_extracted_k141_50576 | Not-determined |
| S11_extracted_k141_64059 | Not-determined |
| S11_extracted_k141_12093 | Not-determined |
| S11_extracted_k141_83567 | Not-determined |
| S11_extracted_k141_12094 | Low-quality |
| S11_extracted_k141_18669 | Low-quality |
| S11_extracted_k141_97124 | Not-determined |
| S11_extracted_k141_5804 | Not-determined |
| S11_extracted_k141_83570 | Not-determined |
| S11_extracted_k141_57848 | Not-determined |
| S11_extracted_k141_70133 | Not-determined |
| S11_extracted_k141_12100 | Not-determined |
| S11_extracted_k141_18673 | Not-determined |
| S11_extracted_k141_70135 | Low-quality |
| S11_extracted_k141_83576 | Not-determined |
| S11_extracted_k141_97131 | Not-determined |
| S11_extracted_k141_64072 | Not-determined |
| S11_extracted_k141_50584 | Not-determined |
| S11_extracted_k141_50585 | Low-quality |
| S11_extracted_k141_25893 | Low-quality |
| S11_extracted_k141_83579 | Not-determined |
| S11_extracted_k141_25894 | Not-determined |
| S11_extracted_k141_90172 | Not-determined |
| S11_extracted_k141_90174 | Not-determined |
| S11_extracted_k141_25896 | Not-determined |
| S11_extracted_k141_57853 | Not-determined |
| S11_extracted_k141_90179 | Not-determined |
| S11_extracted_k141_31475 | Not-determined |
| S11_extracted_k141_25907 | Not-determined |
| S11_extracted_k141_18691 | Not-determined |
| S11_extracted_k141_18692 | Not-determined |
| S11_extracted_k141_83590 | Not-determined |
| S11_extracted_k141_12112 | Not-determined |
| S11_extracted_k141_18693 | Not-determined |
| S11_extracted_k141_50602 | Not-determined |
| S11_extracted_k141_97144 | Not-determined |
| S11_extracted_k141_97145 | Not-determined |
| S11_extracted_k141_70159 | Not-determined |
| S11_extracted_k141_103160 | Not-determined |
| S11_extracted_k141_57868 | Low-quality |
| S11_extracted_k141_18699 | Low-quality |
| S11_extracted_k141_57869 | Not-determined |
| S11_extracted_k141_70164 | Not-determined |
| S11_extracted_k141_18701 | Not-determined |
| S11_extracted_k141_70166 | Not-determined |
| S11_extracted_k141_12121 | Not-determined |
| S11_extracted_k141_90196 | Not-determined |
| S11_extracted_k141_12125 | Not-determined |
| S11_extracted_k141_97157 | Not-determined |
| S11_extracted_k141_12132 | Not-determined |
| S11_extracted_k141_90204 | Not-determined |
| S11_extracted_k141_50627 | Not-determined |
| S11_extracted_k141_77550 | Not-determined |
| S11_extracted_k141_70179 | Not-determined |
| S11_extracted_k141_18712 | Low-quality |
| S11_extracted_k141_90206 | Not-determined |
| S11_extracted_k141_83592 | Low-quality |
| S11_extracted_k141_64105 | Not-determined |
| S11_extracted_k141_12140 | Not-determined |
| S11_extracted_k141_83596 | Not-determined |
| S11_extracted_k141_70180 | Low-quality |
| S11_extracted_k141_90211 | Low-quality |
| S11_extracted_k141_38319 | Not-determined |
| S11_extracted_k141_83599 | Not-determined |
| S11_extracted_k141_31505 | Low-quality |
| S11_extracted_k141_18719 | Not-determined |
| S11_extracted_k141_25945 | Low-quality |
| S11_extracted_k141_97171 | Not-determined |
| S11_extracted_k141_57892 | Not-determined |
| S11_extracted_k141_97173 | Not-determined |
| S11_extracted_k141_57895 | Not-determined |
| S11_extracted_k141_12152 | Not-determined |
| S11_extracted_k141_70194 | Low-quality |
| S11_extracted_k141_70196 | Not-determined |
| S11_extracted_k141_90228 | Not-determined |
| S11_extracted_k141_44798 | Not-determined |
| S11_extracted_k141_83604 | Not-determined |
| S11_extracted_k141_5840 | Not-determined |
| S11_extracted_k141_70198 | Not-determined |
| S11_extracted_k141_83606 | Not-determined |
| S11_extracted_k141_25958 | Not-determined |
| S11_extracted_k141_18731 | Not-determined |
| S11_extracted_k141_57900 | Not-determined |
| S11_extracted_k141_12156 | Not-determined |
| S11_extracted_k141_25959 | Low-quality |
| S11_extracted_k141_25961 | Not-determined |
| S11_extracted_k141_50656 | Not-determined |
| S11_extracted_k141_57905 | Not-determined |
| S11_extracted_k141_83611 | Not-determined |
| S11_extracted_k141_83612 | Not-determined |
| S11_extracted_k141_5848 | Not-determined |
| S11_extracted_k141_57906 | Not-determined |
| S11_extracted_k141_90235 | Low-quality |
| S11_extracted_k141_38335 | Not-determined |
| S11_extracted_k141_50660 | Not-determined |
| S11_extracted_k141_12163 | Not-determined |
| S11_extracted_k141_50661 | Low-quality |
| S11_extracted_k141_103168 | Not-determined |
| S11_extracted_k141_38338 | Not-determined |
| S11_extracted_k141_90241 | Low-quality |
| S11_extracted_k141_44808 | Not-determined |
| S11_extracted_k141_5854 | Not-determined |
| S11_extracted_k141_12173 | Not-determined |
| S11_extracted_k141_31530 | Not-determined |
| S11_extracted_k141_50674 | Not-determined |
| S11_extracted_k141_12177 | Low-quality |
| S11_extracted_k141_44817 | Low-quality |
| S11_extracted_k141_50676 | Not-determined |
| S11_extracted_k141_57914 | Not-determined |
| S11_extracted_k141_83625 | Not-determined |
| S11_extracted_k141_5865 | Not-determined |
| S11_extracted_k141_44823 | Not-determined |
| S11_extracted_k141_50682 | Not-determined |
| S11_extracted_k141_38346 | Low-quality |
| S11_extracted_k141_90252 | Low-quality |
| S11_extracted_k141_12185 | Not-determined |
| S11_extracted_k141_25988 | Low-quality |
| S11_extracted_k141_77575 | Low-quality |
| S11_extracted_k141_83629 | Low-quality |
| S11_extracted_k141_77576 | Not-determined |
| S11_extracted_k141_83630 | Not-determined |
| S11_extracted_k141_90254 | Not-determined |
| S11_extracted_k141_44833 | Not-determined |
| S11_extracted_k141_5873 | Not-determined |
| S11_extracted_k141_97199 | Not-determined |
| S11_extracted_k141_12194 | Not-determined |
| S11_extracted_k141_25997 | Not-determined |
| S11_extracted_k141_50693 | Not-determined |
| S11_extracted_k141_70212 | Not-determined |
| S11_extracted_k141_38352 | Not-determined |
| S11_extracted_k141_26004 | Not-determined |
| S11_extracted_k141_97208 | Not-determined |
| S11_extracted_k141_12202 | Not-determined |
| S11_extracted_k141_18762 | Not-determined |
| S11_extracted_k141_18764 | Not-determined |
| S11_extracted_k141_38359 | Not-determined |
| S11_extracted_k141_77603 | Not-determined |
| S11_extracted_k141_77608 | Not-determined |
| S11_extracted_k141_64139 | Not-determined |
| S11_extracted_k141_77610 | Not-determined |
| S11_extracted_k141_90276 | Not-determined |
| S11_extracted_k141_97221 | Not-determined |
| S11_extracted_k141_90278 | Not-determined |
| S11_extracted_k141_64141 | Not-determined |
| S11_extracted_k141_97222 | Not-determined |
| S11_extracted_k141_38370 | Not-determined |
| S11_extracted_k141_18780 | Not-determined |
| S11_extracted_k141_83656 | Not-determined |
| S11_extracted_k141_44868 | Not-determined |
| S11_extracted_k141_97226 | Not-determined |
| S11_extracted_k141_18781 | Low-quality |
| S11_extracted_k141_77620 | Not-determined |
| S11_extracted_k141_26023 | Low-quality |
| S11_extracted_k141_97227 | Not-determined |
| S11_extracted_k141_90288 | Not-determined |
| S11_extracted_k141_38379 | Not-determined |
| S11_extracted_k141_97232 | Low-quality |
| S11_extracted_k141_31550 | Not-determined |
| S11_extracted_k141_70228 | Not-determined |
| S11_extracted_k141_97235 | Not-determined |
| S11_extracted_k141_64147 | Low-quality |
| S11_extracted_k141_97237 | Not-determined |
| S11_extracted_k141_26028 | Low-quality |
| S11_extracted_k141_97239 | Not-determined |
| S11_extracted_k141_70231 | Not-determined |
| S11_extracted_k141_12206 | Low-quality |
| S11_extracted_k141_5891 | Not-determined |
| S11_extracted_k141_64152 | Not-determined |
| S11_extracted_k141_57943 | Not-determined |
| S11_extracted_k141_90299 | Not-determined |
| S11_extracted_k141_90300 | Not-determined |
| S11_extracted_k141_70237 | Not-determined |
| S11_extracted_k141_70238 | Not-determined |
| S11_extracted_k141_77639 | Not-determined |
| S11_extracted_k141_12212 | Low-quality |
| S11_extracted_k141_31562 | Not-determined |
| S11_extracted_k141_38386 | Not-determined |
| S11_extracted_k141_12217 | Not-determined |
| S11_extracted_k141_64163 | Not-determined |
| S11_extracted_k141_97254 | Not-determined |
| S11_extracted_k141_64167 | Not-determined |
| S11_extracted_k141_97257 | Not-determined |
| S11_extracted_k141_57954 | Not-determined |
| S11_extracted_k141_97261 | Not-determined |
| S11_extracted_k141_5904 | Not-determined |
| S11_extracted_k141_12222 | Not-determined |
| S11_extracted_k141_12223 | Not-determined |
| S11_extracted_k141_57960 | Not-determined |
| S11_extracted_k141_57964 | Not-determined |
| S11_extracted_k141_18816 | Not-determined |
| S11_extracted_k141_77660 | Not-determined |
| S11_extracted_k141_18819 | Low-quality |
| S11_extracted_k141_64186 | Not-determined |
| S11_extracted_k141_12235 | Not-determined |
| S11_extracted_k141_50740 | Not-determined |
| S11_extracted_k141_97267 | Not-determined |
| S11_extracted_k141_12236 | Not-determined |
| S11_extracted_k141_50741 | Not-determined |
| S11_extracted_k141_64188 | Not-determined |
| S11_extracted_k141_97268 | Not-determined |
| S11_extracted_k141_31577 | Not-determined |
| S11_extracted_k141_57972 | Not-determined |
| S11_extracted_k141_57973 | Not-determined |
| S11_extracted_k141_77667 | Low-quality |
| S11_extracted_k141_44914 | Not-determined |
| S11_extracted_k141_5917 | Not-determined |
| S11_extracted_k141_64195 | Not-determined |
| S11_extracted_k141_44916 | Not-determined |
| S11_extracted_k141_64196 | Low-quality |
| S11_extracted_k141_64205 | Not-determined |
| S11_extracted_k141_12250 | Not-determined |
| S11_extracted_k141_77679 | Not-determined |
| S11_extracted_k141_18830 | Low-quality |
| S11_extracted_k141_77685 | Not-determined |
| S11_extracted_k141_12262 | Not-determined |
| S11_extracted_k141_77687 | Not-determined |
| S11_extracted_k141_38394 | Not-determined |
| S11_extracted_k141_103192 | Not-determined |
| S11_extracted_k141_83676 | Not-determined |
| S11_extracted_k141_64216 | Not-determined |
| S11_extracted_k141_38396 | Not-determined |
| S11_extracted_k141_5924 | Not-determined |
| S11_extracted_k141_50757 | Low-quality |
| S11_extracted_k141_50765 | Not-determined |
| S11_extracted_k141_12268 | Low-quality |
| S11_extracted_k141_64221 | Not-determined |
| S11_extracted_k141_64223 | Not-determined |
| S11_extracted_k141_38403 | Low-quality |
| S11_extracted_k141_97288 | Not-determined |
| S11_extracted_k141_38408 | Low-quality |
| S11_extracted_k141_90311 | Low-quality |
| S11_extracted_k141_5935 | Low-quality |
| S11_extracted_k141_103200 | Not-determined |
| S11_extracted_k141_12281 | Low-quality |
| S11_extracted_k141_83700 | Low-quality |
| S11_extracted_k141_90318 | Low-quality |
| S11_extracted_k141_90319 | Not-determined |
| S11_extracted_k141_12288 | Low-quality |
| S11_extracted_k141_103203 | Low-quality |
| S11_extracted_k141_58013 | Not-determined |
| S11_extracted_k141_18852 | Not-determined |
| S11_extracted_k141_103205 | Not-determined |
| S11_extracted_k141_12292 | Not-determined |
| S11_extracted_k141_70258 | Not-determined |
| S11_extracted_k141_103209 | Not-determined |
| S11_extracted_k141_58021 | Not-determined |
| S11_extracted_k141_83711 | Not-determined |
| S11_extracted_k141_103213 | Not-determined |
| S11_extracted_k141_103214 | Not-determined |
| S11_extracted_k141_58025 | Low-quality |
| S11_extracted_k141_58027 | Not-determined |
| S11_extracted_k141_18856 | Not-determined |
| S11_extracted_k141_83718 | Not-determined |
| S11_extracted_k141_64248 | Not-determined |
| S11_extracted_k141_97305 | Not-determined |
| S11_extracted_k141_18860 | Not-determined |

| | |
|---|---|
| S11_extracted_k141_83724 | Low-quality |
| S11_extracted_k141_97308 | Low-quality |
| S11_extracted_k141_31603 | Not-determined |
| S11_extracted_k141_12310 | Not-determined |
| S11_extracted_k141_97309 | Low-quality |
| S11_extracted_k141_70273 | Not-determined |
| S11_extracted_k141_12312 | Not-determined |
| S11_extracted_k141_18866 | Not-determined |
| S11_extracted_k141_38421 | Low-quality |
| S11_extracted_k141_97312 | Not-determined |
| S11_extracted_k141_58038 | Low-quality |
| S11_extracted_k141_97316 | Not-determined |
| S11_extracted_k141_97318 | Not-determined |
| S11_extracted_k141_70285 | Low-quality |
| S11_extracted_k141_5955 | Not-determined |
| S11_extracted_k141_18877 | Not-determined |
| S11_extracted_k141_5957 | Low-quality |
| S11_extracted_k141_31608 | Not-determined |
| S11_extracted_k141_12319 | Low-quality |
| S11_extracted_k141_83742 | Low-quality |
| S11_extracted_k141_12323 | Not-determined |
| S11_extracted_k141_18886 | Low-quality |
| S11_extracted_k141_5965 | Not-determined |
| S11_extracted_k141_12328 | Not-determined |
| S11_extracted_k141_97323 | Low-quality |
| S11_extracted_k141_83752 | Not-determined |
| S11_extracted_k141_103240 | Not-determined |
| S11_extracted_k141_70294 | Low-quality |
| S11_extracted_k141_12334 | Not-determined |
| S11_extracted_k141_12335 | Not-determined |
| S11_extracted_k141_18895 | Not-determined |
| S11_extracted_k141_31615 | Not-determined |
| S11_extracted_k141_103246 | Low-quality |
| S11_extracted_k141_5967 | Low-quality |
| S11_extracted_k141_90359 | Not-determined |
| S11_extracted_k141_38448 | Not-determined |
| S11_extracted_k141_50769 | Not-determined |
| S11_extracted_k141_70302 | Not-determined |
| S11_extracted_k141_70303 | Not-determined |
| S11_extracted_k141_12339 | Not-determined |
| S11_extracted_k141_58054 | Not-determined |
| S11_extracted_k141_103250 | Not-determined |
| S11_extracted_k141_58055 | Not-determined |
| S11_extracted_k141_77723 | Not-determined |
| S11_extracted_k141_64288 | Not-determined |
| S11_extracted_k141_77727 | Not-determined |
| S11_extracted_k141_5976 | Not-determined |
| S11_extracted_k141_58060 | Low-quality |
| S11_extracted_k141_58061 | Not-determined |
| S11_extracted_k141_77730 | Low-quality |
| S11_extracted_k141_64290 | Not-determined |
| S11_extracted_k141_5979 | Low-quality |
| S11_extracted_k141_70312 | Not-determined |
| S11_extracted_k141_83777 | Not-determined |
| S11_extracted_k141_83778 | Not-determined |
| S11_extracted_k141_58069 | Not-determined |
| S11_extracted_k141_83781 | Not-determined |
| S11_extracted_k141_103262 | Not-determined |
| S11_extracted_k141_58072 | Not-determined |
| S11_extracted_k141_64296 | Not-determined |
| S11_extracted_k141_64297 | Not-determined |
| S11_extracted_k141_64299 | Not-determined |
| S11_extracted_k141_58075 | Not-determined |
| S11_extracted_k141_5987 | Not-determined |
| S11_extracted_k141_83791 | Not-determined |
| S11_extracted_k141_97351 | Low-quality |
| S11_extracted_k141_18913 | Not-determined |
| S11_extracted_k141_97355 | Not-determined |
| S11_extracted_k141_77736 | Low-quality |
| S11_extracted_k141_44955 | Not-determined |
| S11_extracted_k141_50800 | Not-determined |
| S11_extracted_k141_44957 | Not-determined |
| S11_extracted_k141_97363 | Not-determined |
| S11_extracted_k141_64309 | Not-determined |
| S11_extracted_k141_12349 | Not-determined |
| S11_extracted_k141_70344 | Not-determined |
| S11_extracted_k141_97369 | Not-determined |
| S11_extracted_k141_64313 | Not-determined |
| S11_extracted_k141_44960 | Low-quality |
| S11_extracted_k141_90396 | Not-determined |
| S11_extracted_k141_77752 | Not-determined |
| S11_extracted_k141_5997 | Not-determined |
| S11_extracted_k141_18928 | Not-determined |
| S11_extracted_k141_77753 | Not-determined |
| S11_extracted_k141_44967 | Low-quality |
| S11_extracted_k141_31655 | Not-determined |
| S11_extracted_k141_90403 | Low-quality |
| S11_extracted_k141_6001 | Not-determined |
| S11_extracted_k141_90404 | Low-quality |
| S11_extracted_k141_58094 | Low-quality |
| S11_extracted_k141_70347 | Low-quality |
| S11_extracted_k141_38453 | Low-quality |
| S11_extracted_k141_44972 | Not-determined |
| S11_extracted_k141_18938 | Not-determined |
| S11_extracted_k141_6009 | Low-quality |
| S11_extracted_k141_44976 | Not-determined |
| S11_extracted_k141_31664 | Not-determined |
| S11_extracted_k141_44978 | Not-determined |
| S11_extracted_k141_38464 | Not-determined |
| S11_extracted_k141_64321 | Low-quality |
| S11_extracted_k141_64322 | Not-determined |
| S11_extracted_k141_50829 | Not-determined |
| S11_extracted_k141_31678 | Not-determined |
| S11_extracted_k141_90409 | Low-quality |
| S11_extracted_k141_97384 | Low-quality |
| S11_extracted_k141_18957 | Not-determined |
| S11_extracted_k141_50834 | Low-quality |
| S11_extracted_k141_90415 | Not-determined |
| S11_extracted_k141_58113 | Not-determined |
| S11_extracted_k141_18961 | Low-quality |
| S11_extracted_k141_77763 | Not-determined |
| S11_extracted_k141_77764 | Not-determined |
| S11_extracted_k141_18963 | Not-determined |
| S11_extracted_k141_18965 | Not-determined |
| S11_extracted_k141_6030 | Not-determined |
| S11_extracted_k141_38481 | Low-quality |
| S11_extracted_k141_6033 | Not-determined |
| S11_extracted_k141_6034 | Not-determined |
| S11_extracted_k141_31690 | Not-determined |
| S11_extracted_k141_12393 | Not-determined |
| S11_extracted_k141_97386 | Not-determined |
| S11_extracted_k141_6039 | Not-determined |
| S11_extracted_k141_90422 | Not-determined |
| S11_extracted_k141_58127 | Low-quality |
| S11_extracted_k141_6043 | Low-quality |
| S11_extracted_k141_70374 | Not-determined |
| S11_extracted_k141_45012 | Not-determined |
| S11_extracted_k141_70375 | Not-determined |
| S11_extracted_k141_45014 | Not-determined |
| S11_extracted_k141_18975 | Not-determined |
| S11_extracted_k141_12404 | Not-determined |
| S11_extracted_k141_90425 | Not-determined |
| S11_extracted_k141_6049 | Not-determined |
| S11_extracted_k141_77781 | Not-determined |
| S11_extracted_k141_45018 | Not-determined |
| S11_extracted_k141_38500 | Not-determined |
| S11_extracted_k141_45020 | Not-determined |
| S11_extracted_k141_31704 | Not-determined |
| S11_extracted_k141_97394 | Not-determined |
| S11_extracted_k141_90432 | Not-determined |
| S11_extracted_k141_38506 | Not-determined |
| S11_extracted_k141_90433 | Low-quality |
| S11_extracted_k141_97396 | Not-determined |
| S11_extracted_k141_103284 | Not-determined |
| S11_extracted_k141_6057 | Not-determined |
| S11_extracted_k141_83812 | Not-determined |
| S11_extracted_k141_90447 | Not-determined |
| S11_extracted_k141_18987 | Not-determined |
| S11_extracted_k141_38514 | Not-determined |
| S11_extracted_k141_64330 | Low-quality |
| S11_extracted_k141_12422 | Not-determined |
| S11_extracted_k141_83823 | Not-determined |
| S11_extracted_k141_70403 | Not-determined |
| S11_extracted_k141_90457 | Not-determined |
| S11_extracted_k141_77795 | Low-quality |
| S11_extracted_k141_64337 | Not-determined |
| S11_extracted_k141_38518 | Not-determined |
| S11_extracted_k141_12430 | Not-determined |
| S11_extracted_k141_31722 | Not-determined |
| S11_extracted_k141_64340 | Not-determined |
| S11_extracted_k141_31723 | Not-determined |
| S11_extracted_k141_83832 | Low-quality |
| S11_extracted_k141_83834 | Not-determined |
| S11_extracted_k141_83838 | Not-determined |
| S11_extracted_k141_70411 | Not-determined |
| S11_extracted_k141_31730 | Not-determined |
| S11_extracted_k141_12439 | Not-determined |
| S11_extracted_k141_77809 | Not-determined |
| S11_extracted_k141_12440 | Not-determined |
| S11_extracted_k141_31734 | Not-determined |
| S11_extracted_k141_12442 | Not-determined |
| S11_extracted_k141_12443 | Low-quality |
| S11_extracted_k141_83849 | Not-determined |
| S11_extracted_k141_90472 | Low-quality |
| S11_extracted_k141_90475 | Low-quality |
| S11_extracted_k141_77816 | Not-determined |
| S11_extracted_k141_103295 | Not-determined |
| S11_extracted_k141_31750 | Not-determined |
| S11_extracted_k141_97436 | Not-determined |
| S11_extracted_k141_83856 | Not-determined |
| S11_extracted_k141_64342 | Not-determined |
| S11_extracted_k141_77822 | Not-determined |
| S11_extracted_k141_77823 | Not-determined |
| S11_extracted_k141_97439 | Not-determined |
| S11_extracted_k141_90487 | Low-quality |
| S11_extracted_k141_90488 | Not-determined |
| S11_extracted_k141_45046 | Not-determined |
| S11_extracted_k141_83859 | Not-determined |
| S11_extracted_k141_50840 | Not-determined |
| S11_extracted_k141_90490 | Not-determined |
| S11_extracted_k141_64347 | Not-determined |
| S11_extracted_k141_31764 | Not-determined |
| S11_extracted_k141_103299 | Not-determined |
| S11_extracted_k141_6088 | Not-determined |
| S11_extracted_k141_77836 | Not-determined |
| S11_extracted_k141_103303 | Low-quality |
| S11_extracted_k141_50851 | Not-determined |
| S11_extracted_k141_97445 | Not-determined |
| S11_extracted_k141_83868 | Low-quality |
| S11_extracted_k141_97446 | Not-determined |
| S11_extracted_k141_31772 | Not-determined |
| S11_extracted_k141_97449 | Not-determined |
| S11_extracted_k141_64361 | Not-determined |
| S11_extracted_k141_45057 | Low-quality |
| S11_extracted_k141_103320 | Not-determined |
| S11_extracted_k141_50864 | Not-determined |
| S11_extracted_k141_103321 | Not-determined |
| S11_extracted_k141_31776 | Not-determined |
| S11_extracted_k141_97464 | Not-determined |
| S11_extracted_k141_19024 | Not-determined |
| S11_extracted_k141_103323 | Not-determined |
| S11_extracted_k141_50877 | Not-determined |
| S11_extracted_k141_45072 | Not-determined |
| S11_extracted_k141_50881 | Low-quality |
| S11_extracted_k141_50883 | Not-determined |
| S11_extracted_k141_19033 | Not-determined |
| S11_extracted_k141_6105 | Not-determined |
| S11_extracted_k141_6106 | Not-determined |
| S11_extracted_k141_70435 | Low-quality |
| S11_extracted_k141_50889 | Not-determined |
| S11_extracted_k141_45077 | Not-determined |
| S11_extracted_k141_83891 | Not-determined |
| S11_extracted_k141_45078 | Not-determined |
| S11_extracted_k141_31779 | Not-determined |
| S11_extracted_k141_19038 | Low-quality |
| S11_extracted_k141_50895 | Not-determined |
| S11_extracted_k141_83896 | Not-determined |
| S11_extracted_k141_50897 | Not-determined |
| S11_extracted_k141_50898 | Not-determined |
| S11_extracted_k141_19042 | Low-quality |
| S11_extracted_k141_12492 | Low-quality |
| S11_extracted_k141_12497 | Not-determined |
| S11_extracted_k141_19046 | Not-determined |
| S11_extracted_k141_58192 | Not-determined |
| S11_extracted_k141_97498 | Not-determined |
| S11_extracted_k141_97500 | Not-determined |
| S11_extracted_k141_50910 | Not-determined |
| S11_extracted_k141_31805 | Low-quality |
| S11_extracted_k141_103346 | Not-determined |
| S11_extracted_k141_90524 | Not-determined |
| S11_extracted_k141_90525 | Not-determined |
| S11_extracted_k141_45097 | Not-determined |
| S11_extracted_k141_77865 | Not-determined |
| S11_extracted_k141_31806 | Not-determined |
| S11_extracted_k141_31807 | Not-determined |
| S11_extracted_k141_70454 | Not-determined |
| S11_extracted_k141_77870 | Not-determined |
| S11_extracted_k141_31810 | Not-determined |
| S11_extracted_k141_70458 | Not-determined |
| S11_extracted_k141_83920 | Low-quality |
| S11_extracted_k141_83922 | Not-determined |
| S11_extracted_k141_45111 | Not-determined |
| S11_extracted_k141_70463 | Not-determined |
| S11_extracted_k141_97510 | Not-determined |
| S11_extracted_k141_12516 | Not-determined |
| S11_extracted_k141_45112 | Low-quality |
| S11_extracted_k141_83934 | Not-determined |
| S11_extracted_k141_31814 | Low-quality |
| S11_extracted_k141_58215 | Not-determined |
| S11_extracted_k141_77889 | Not-determined |
| S11_extracted_k141_83946 | Low-quality |
| S11_extracted_k141_50921 | Not-determined |
| S11_extracted_k141_12528 | Not-determined |
| S11_extracted_k141_83955 | Not-determined |
| S11_extracted_k141_64401 | Not-determined |
| S11_extracted_k141_45124 | Not-determined |
| S11_extracted_k141_45127 | Not-determined |
| S11_extracted_k141_83957 | Not-determined |
| S11_extracted_k141_12534 | Not-determined |
| S11_extracted_k141_83964 | Not-determined |
| S11_extracted_k141_64414 | Not-determined |
| S11_extracted_k141_12545 | Not-determined |
| S11_extracted_k141_83972 | Not-determined |
| S11_extracted_k141_77905 | Not-determined |
| S11_extracted_k141_50940 | Low-quality |
| S11_extracted_k141_83976 | Not-determined |
| S11_extracted_k141_77911 | Not-determined |
| S11_extracted_k141_58239 | Low-quality |
| S11_extracted_k141_45149 | Not-determined |
| S11_extracted_k141_83983 | Not-determined |
| S11_extracted_k141_45152 | Not-determined |
| S11_extracted_k141_45153 | Not-determined |
| S11_extracted_k141_31838 | Not-determined |
| S11_extracted_k141_50959 | Not-determined |
| S11_extracted_k141_50962 | Not-determined |
| S11_extracted_k141_77941 | Not-determined |
| S11_extracted_k141_83999 | Not-determined |
| S11_extracted_k141_97549 | Not-determined |
| S11_extracted_k141_97553 | Not-determined |
| S11_extracted_k141_45174 | Not-determined |
| S11_extracted_k141_64437 | Not-determined |
| S11_extracted_k141_31859 | Low-quality |
| S11_extracted_k141_50980 | Not-determined |
| S11_extracted_k141_50987 | Low-quality |
| S11_extracted_k141_50991 | Not-determined |
| S11_extracted_k141_84014 | Not-determined |
| S11_extracted_k141_50993 | Not-determined |
| S11_extracted_k141_58256 | Not-determined |
| S11_extracted_k141_77958 | Not-determined |
| S11_extracted_k141_77959 | Low-quality |
| S11_extracted_k141_31863 | Low-quality |
| S11_extracted_k141_12586 | Not-determined |
| S11_extracted_k141_97570 | Not-determined |
| S11_extracted_k141_77962 | Low-quality |
| S11_extracted_k141_51004 | Low-quality |
| S11_extracted_k141_51005 | Low-quality |
| S11_extracted_k141_77967 | Low-quality |
| S11_extracted_k141_64458 | Not-determined |
| S11_extracted_k141_51010 | Low-quality |
| S11_extracted_k141_84015 | Low-quality |
| S11_extracted_k141_64461 | Not-determined |
| S11_extracted_k141_51014 | Not-determined |
| S11_extracted_k141_58265 | Not-determined |
| S11_extracted_k141_12602 | Not-determined |
| S11_extracted_k141_12603 | Not-determined |
| S11_extracted_k141_45194 | Not-determined |
| S11_extracted_k141_84021 | Not-determined |
| S11_extracted_k141_45197 | Not-determined |
| S11_extracted_k141_12608 | Low-quality |
| S11_extracted_k141_97593 | Not-determined |
| S11_extracted_k141_12611 | Not-determined |
| S11_extracted_k141_77988 | Not-determined |
| S11_extracted_k141_12613 | Not-determined |
| S11_extracted_k141_38545 | Not-determined |
| S11_extracted_k141_64478 | Not-determined |
| S11_extracted_k141_84042 | Not-determined |
| S11_extracted_k141_51036 | Not-determined |
| S11_extracted_k141_78001 | Not-determined |
| S11_extracted_k141_45221 | Not-determined |
| S11_extracted_k141_103354 | Not-determined |
| S11_extracted_k141_97615 | Low-quality |
| S11_extracted_k141_51041 | Not-determined |
| S11_extracted_k141_64499 | Low-quality |
| S11_extracted_k141_45226 | Low-quality |
| S11_extracted_k141_45232 | Not-determined |
| S11_extracted_k141_84054 | Not-determined |
| S11_extracted_k141_51055 | Not-determined |
| S11_extracted_k141_78018 | Not-determined |
| S11_extracted_k141_12627 | Not-determined |
| S11_extracted_k141_64515 | Not-determined |
| S11_extracted_k141_12629 | Not-determined |
| S11_extracted_k141_6117 | Not-determined |
| S11_extracted_k141_38567 | Low-quality |
| S11_extracted_k141_78021 | Not-determined |
| S11_extracted_k141_6120 | Low-quality |
| S11_extracted_k141_6121 | Not-determined |
| S11_extracted_k141_90580 | Not-determined |
| S11_extracted_k141_90581 | Not-determined |
| S11_extracted_k141_64534 | Not-determined |
| S11_extracted_k141_12649 | Not-determined |
| S11_extracted_k141_51062 | Not-determined |
| S11_extracted_k141_6127 | Not-determined |
| S11_extracted_k141_64537 | Not-determined |
| S11_extracted_k141_84075 | Not-determined |
| S11_extracted_k141_58281 | Not-determined |
| S11_extracted_k141_103387 | Not-determined |
| S11_extracted_k141_45261 | Low-quality |
| S11_extracted_k141_45264 | Not-determined |
| S11_extracted_k141_31893 | Not-determined |
| S11_extracted_k141_90601 | Low-quality |
| S11_extracted_k141_12658 | Low-quality |
| S11_extracted_k141_45266 | Not-determined |
| S11_extracted_k141_58287 | Not-determined |
| S11_extracted_k141_45269 | Not-determined |
| S11_extracted_k141_38592 | Not-determined |
| S11_extracted_k141_12664 | Not-determined |
| S11_extracted_k141_12665 | Not-determined |
| S11_extracted_k141_38595 | Not-determined |
| S11_extracted_k141_84088 | Not-determined |
| S11_extracted_k141_38601 | Not-determined |
| S11_extracted_k141_103405 | Not-determined |
| S11_extracted_k141_84092 | Not-determined |
| S11_extracted_k141_38606 | Not-determined |
| S11_extracted_k141_90620 | Not-determined |
| S11_extracted_k141_45282 | Not-determined |
| S11_extracted_k141_78037 | Low-quality |
| S11_extracted_k141_19105 | Not-determined |
| S11_extracted_k141_45287 | Not-determined |
| S11_extracted_k141_38613 | Not-determined |
| S11_extracted_k141_84101 | Low-quality |
| S11_extracted_k141_45299 | Low-quality |
| S11_extracted_k141_58295 | Not-determined |
| S11_extracted_k141_19115 | Not-determined |
| S11_extracted_k141_19119 | Low-quality |
| S11_extracted_k141_51073 | Not-determined |
| S11_extracted_k141_31903 | Not-determined |
| S11_extracted_k141_64548 | Not-determined |
| S11_extracted_k141_38619 | Not-determined |
| S11_extracted_k141_6140 | Not-determined |
| S11_extracted_k141_84106 | Not-determined |
| S11_extracted_k141_38621 | Not-determined |
| S11_extracted_k141_38622 | Low-quality |
| S11_extracted_k141_78050 | Not-determined |
| S11_extracted_k141_31905 | Not-determined |
| S11_extracted_k141_19123 | Not-determined |
| S11_extracted_k141_84107 | Not-determined |
| S11_extracted_k141_12689 | Not-determined |
| S11_extracted_k141_31907 | Not-determined |
| S11_extracted_k141_64557 | Not-determined |
| S11_extracted_k141_45303 | Not-determined |
| S11_extracted_k141_84110 | Low-quality |
| S11_extracted_k141_19128 | Not-determined |
| S11_extracted_k141_90642 | Not-determined |
| S11_extracted_k141_19130 | Low-quality |
| S11_extracted_k141_51081 | Not-determined |
| S11_extracted_k141_58307 | Not-determined |
| S11_extracted_k141_51085 | Not-determined |
| S11_extracted_k141_64563 | Low-quality |
| S11_extracted_k141_58310 | Not-determined |
| S11_extracted_k141_64566 | Not-determined |
| S11_extracted_k141_31916 | Not-determined |
| S11_extracted_k141_38632 | Not-determined |
| S11_extracted_k141_6154 | Not-determined |
| S11_extracted_k141_58314 | Not-determined |
| S11_extracted_k141_19139 | Not-determined |
| S11_extracted_k141_64571 | Low-quality |
| S11_extracted_k141_64574 | Not-determined |
| S11_extracted_k141_64575 | Not-determined |
| S11_extracted_k141_64576 | Not-determined |
| S11_extracted_k141_38639 | Not-determined |
| S11_extracted_k141_64577 | Low-quality |
| S11_extracted_k141_38642 | Not-determined |
| S11_extracted_k141_51090 | Not-determined |
| S11_extracted_k141_38646 | Not-determined |
| S11_extracted_k141_58322 | Low-quality |
| S11_extracted_k141_6167 | Not-determined |
| S11_extracted_k141_84129 | Not-determined |
| S11_extracted_k141_45316 | Not-determined |
| S11_extracted_k141_6170 | Not-determined |
| S11_extracted_k141_90652 | Low-quality |
| S11_extracted_k141_12702 | Low-quality |
| S11_extracted_k141_45321 | Not-determined |
| S11_extracted_k141_45326 | Not-determined |
| S11_extracted_k141_12710 | Not-determined |
| S11_extracted_k141_90657 | Not-determined |
| S11_extracted_k141_103415 | Not-determined |
| S11_extracted_k141_45334 | Not-determined |
| S11_extracted_k141_51116 | Not-determined |
| S11_extracted_k141_103419 | Not-determined |
| S11_extracted_k141_31932 | Not-determined |
| S11_extracted_k141_78070 | Not-determined |
| S11_extracted_k141_90659 | Low-quality |
| S11_extracted_k141_70472 | Not-determined |
| S11_extracted_k141_70478 | Not-determined |
| S11_extracted_k141_51130 | Low-quality |
| S11_extracted_k141_103424 | Not-determined |
| S11_extracted_k141_12724 | Not-determined |
| S11_extracted_k141_31936 | Low-quality |
| S11_extracted_k141_45346 | Not-determined |
| S11_extracted_k141_103428 | Not-determined |
| S11_extracted_k141_51140 | Not-determined |
| S11_extracted_k141_6199 | Not-determined |
| S11_extracted_k141_70495 | Not-determined |
| S11_extracted_k141_84132 | Low-quality |
| S11_extracted_k141_31947 | Not-determined |
| S11_extracted_k141_90666 | Not-determined |
| S11_extracted_k141_70500 | Not-determined |
| S11_extracted_k141_6204 | Low-quality |
| S11_extracted_k141_84135 | Low-quality |
| S11_extracted_k141_31952 | Not-determined |
| S11_extracted_k141_103433 | Not-determined |
| S11_extracted_k141_6209 | Not-determined |
| S11_extracted_k141_70508 | Not-determined |
| S11_extracted_k141_103434 | Not-determined |
| S11_extracted_k141_38684 | Low-quality |
| S11_extracted_k141_84140 | Not-determined |
| S11_extracted_k141_58343 | Not-determined |
| S11_extracted_k141_45359 | Not-determined |
| S11_extracted_k141_84144 | Not-determined |
| S11_extracted_k141_90678 | Not-determined |
| S11_extracted_k141_19161 | Not-determined |
| S11_extracted_k141_45363 | Not-determined |
| S11_extracted_k141_12743 | Not-determined |
| S11_extracted_k141_90682 | Not-determined |
| S11_extracted_k141_12745 | Not-determined |
| S11_extracted_k141_19166 | Not-determined |
| S11_extracted_k141_19167 | Not-determined |
| S11_extracted_k141_58346 | Not-determined |
| S11_extracted_k141_38692 | Not-determined |
| S11_extracted_k141_12749 | Not-determined |
| S11_extracted_k141_6217 | Not-determined |
| S11_extracted_k141_45370 | Not-determined |
| S11_extracted_k141_70523 | Not-determined |
| S11_extracted_k141_12754 | Not-determined |
| S11_extracted_k141_103446 | Not-determined |
| S11_extracted_k141_19175 | Low-quality |
| S11_extracted_k141_51153 | Not-determined |
| S11_extracted_k141_64597 | Not-determined |
| S11_extracted_k141_103447 | Not-determined |
| S11_extracted_k141_64598 | Not-determined |
| S11_extracted_k141_64599 | Not-determined |
| S11_extracted_k141_31956 | Not-determined |
| S11_extracted_k141_103450 | Low-quality |
| S11_extracted_k141_90689 | Not-determined |
| S11_extracted_k141_90690 | Low-quality |
| S11_extracted_k141_64600 | Not-determined |
| S11_extracted_k141_84150 | Not-determined |
| S11_extracted_k141_6228 | Not-determined |
| S11_extracted_k141_31962 | Not-determined |
| S11_extracted_k141_51159 | Not-determined |
| S11_extracted_k141_45384 | Not-determined |
| S11_extracted_k141_6235 | Low-quality |
| S11_extracted_k141_103460 | Not-determined |
| S11_extracted_k141_38708 | Not-determined |
| S11_extracted_k141_31965 | Not-determined |
| S11_extracted_k141_84158 | Not-determined |
| S11_extracted_k141_45391 | Low-quality |
| S11_extracted_k141_38710 | Not-determined |
| S11_extracted_k141_19186 | Not-determined |
| S11_extracted_k141_103465 | Not-determined |
| S11_extracted_k141_38714 | Not-determined |
| S11_extracted_k141_90693 | Not-determined |
| S11_extracted_k141_12775 | Not-determined |
| S11_extracted_k141_78080 | Low-quality |
| S11_extracted_k141_19201 | Not-determined |
| S11_extracted_k141_38724 | Not-determined |
| S11_extracted_k141_90696 | Not-determined |
| S11_extracted_k141_78085 | Not-determined |
| S11_extracted_k141_64605 | Not-determined |
| S11_extracted_k141_90697 | Not-determined |
| S11_extracted_k141_78088 | Not-determined |
| S11_extracted_k141_90699 | Not-determined |
| S11_extracted_k141_64611 | Low-quality |
| S11_extracted_k141_45411 | Not-determined |
| S11_extracted_k141_6256 | Not-determined |
| S11_extracted_k141_38733 | Not-determined |
| S11_extracted_k141_12789 | Not-determined |
| S11_extracted_k141_19214 | Not-determined |
| S11_extracted_k141_58372 | Not-determined |
| S11_extracted_k141_64614 | Low-quality |
| S11_extracted_k141_19217 | Not-determined |
| S11_extracted_k141_6259 | Not-determined |
| S11_extracted_k141_84170 | Not-determined |
| S11_extracted_k141_19221 | Not-determined |
| S11_extracted_k141_103473 | Low-quality |
| S11_extracted_k141_90713 | Low-quality |
| S11_extracted_k141_38751 | Not-determined |
| S11_extracted_k141_90716 | Not-determined |
| S11_extracted_k141_103478 | Not-determined |
| S11_extracted_k141_19225 | Not-determined |
| S11_extracted_k141_51183 | Low-quality |
| S11_extracted_k141_84172 | Not-determined |
| S11_extracted_k141_64621 | Not-determined |
| S11_extracted_k141_70546 | Not-determined |
| S11_extracted_k141_90719 | Low-quality |
| S11_extracted_k141_64623 | Not-determined |
| S11_extracted_k141_70551 | Not-determined |
| S11_extracted_k141_103486 | Not-determined |
| S11_extracted_k141_12804 | Not-determined |
| S11_extracted_k141_19228 | Not-determined |
| S11_extracted_k141_19230 | Not-determined |
| S11_extracted_k141_70556 | Not-determined |
| S11_extracted_k141_103491 | Not-determined |
| S11_extracted_k141_70557 | Not-determined |
| S11_extracted_k141_12813 | Not-determined |
| S11_extracted_k141_12815 | Not-determined |
| S11_extracted_k141_84191 | Not-determined |
| S11_extracted_k141_58387 | Low-quality |
| S11_extracted_k141_103495 | Low-quality |
| S11_extracted_k141_70563 | Not-determined |
| S11_extracted_k141_19256 | Not-determined |
| S11_extracted_k141_6289 | Not-determined |
| S11_extracted_k141_70571 | Not-determined |
| S11_extracted_k141_6292 | Not-determined |
| S11_extracted_k141_64640 | Low-quality |
| S11_extracted_k141_12837 | Not-determined |
| S11_extracted_k141_12838 | Low-quality |
| S11_extracted_k141_70578 | Not-determined |
| S11_extracted_k141_64645 | Low-quality |
| S11_extracted_k141_84213 | Not-determined |
| S11_extracted_k141_12846 | Not-determined |
| S11_extracted_k141_19276 | Not-determined |
| S11_extracted_k141_19280 | Not-determined |
| S11_extracted_k141_84222 | Not-determined |
| S11_extracted_k141_70594 | Low-quality |
| S11_extracted_k141_84225 | Not-determined |
| S11_extracted_k141_12855 | Not-determined |
| S11_extracted_k141_51205 | Not-determined |
| S11_extracted_k141_64656 | Not-determined |
| S11_extracted_k141_51207 | Not-determined |
| S11_extracted_k141_90761 | Not-determined |
| S11_extracted_k141_38787 | Low-quality |
| S11_extracted_k141_90762 | Not-determined |
| S11_extracted_k141_51216 | Low-quality |
| S11_extracted_k141_103519 | Not-determined |
| S11_extracted_k141_6303 | Low-quality |
| S11_extracted_k141_64662 | Not-determined |
| S11_extracted_k141_38795 | Not-determined |
| S11_extracted_k141_103524 | Not-determined |

| | |
|---|---|
| S11_extracted_k141_6312 | Not-determined |
| S11_extracted_k141_64676 | Not-determined |
| S11_extracted_k141_84253 | Not-determined |
| S11_extracted_k141_70614 | Low-quality |
| S11_extracted_k141_90780 | Low-quality |
| S11_extracted_k141_64678 | Not-determined |
| S11_extracted_k141_64679 | Not-determined |
| S11_extracted_k141_70620 | Not-determined |
| S11_extracted_k141_6326 | Low-quality |
| S11_extracted_k141_64684 | Not-determined |
| S11_extracted_k141_64685 | Not-determined |
| S11_extracted_k141_31999 | Low-quality |
| S11_extracted_k141_64688 | Not-determined |
| S11_extracted_k141_58426 | Not-determined |
| S11_extracted_k141_103543 | Low-quality |
| S11_extracted_k141_12894 | Not-determined |
| S11_extracted_k141_38816 | Low-quality |
| S11_extracted_k141_58430 | Low-quality |
| S11_extracted_k141_58431 | Not-determined |
| S11_extracted_k141_38819 | Not-determined |
| S11_extracted_k141_64706 | Not-determined |
| S11_extracted_k141_19312 | Low-quality |
| S11_extracted_k141_70640 | Not-determined |
| S11_extracted_k141_6330 | Not-determined |
| S11_extracted_k141_6331 | Not-determined |
| S11_extracted_k141_64711 | Not-determined |
| S11_extracted_k141_51256 | Not-determined |
| S11_extracted_k141_70645 | Not-determined |
| S11_extracted_k141_38833 | Not-determined |
| S11_extracted_k141_19320 | Low-quality |
| S11_extracted_k141_103553 | Not-determined |
| S11_extracted_k141_84270 | Not-determined |
| S11_extracted_k141_58449 | Not-determined |
| S11_extracted_k141_19323 | Not-determined |
| S11_extracted_k141_12913 | Not-determined |
| S11_extracted_k141_19324 | Low-quality |
| S11_extracted_k141_19326 | Low-quality |
| S11_extracted_k141_103559 | Not-determined |
| S11_extracted_k141_58452 | Not-determined |
| S11_extracted_k141_103561 | Not-determined |
| S11_extracted_k141_51263 | Not-determined |
| S11_extracted_k141_70662 | Not-determined |
| S11_extracted_k141_38844 | Not-determined |
| S11_extracted_k141_70663 | Not-determined |
| S11_extracted_k141_51266 | Not-determined |
| S11_extracted_k141_103565 | Not-determined |
| S11_extracted_k141_19332 | Low-quality |
| S11_extracted_k141_90802 | Not-determined |
| S11_extracted_k141_90803 | Not-determined |
| S11_extracted_k141_84280 | Not-determined |
| S11_extracted_k141_58456 | Low-quality |
| S11_extracted_k141_103567 | Low-quality |
| S11_extracted_k141_12918 | Not-determined |
| S11_extracted_k141_64724 | Low-quality |
| S11_extracted_k141_12919 | Not-determined |
| S11_extracted_k141_84288 | Not-determined |
| S11_extracted_k141_103569 | Not-determined |
| S11_extracted_k141_84294 | Not-determined |
| S11_extracted_k141_64731 | Not-determined |
| S11_extracted_k141_84296 | Not-determined |
| S11_extracted_k141_12927 | Not-determined |
| S11_extracted_k141_84301 | Not-determined |
| S11_extracted_k141_19346 | Not-determined |
| S11_extracted_k141_58473 | Not-determined |
| S11_extracted_k141_32021 | Low-quality |
| S11_extracted_k141_32022 | Not-determined |
| S11_extracted_k141_58479 | Not-determined |
| S11_extracted_k141_12938 | Not-determined |
| S11_extracted_k141_64746 | Low-quality |
| S11_extracted_k141_90829 | Not-determined |
| S11_extracted_k141_12944 | Low-quality |
| S11_extracted_k141_64750 | Not-determined |
| S11_extracted_k141_64751 | Not-determined |
| S11_extracted_k141_64752 | Not-determined |
| S11_extracted_k141_51293 | Not-determined |
| S11_extracted_k141_64759 | Not-determined |
| S11_extracted_k141_45433 | Not-determined |
| S11_extracted_k141_12950 | Not-determined |
| S11_extracted_k141_103590 | Low-quality |
| S11_extracted_k141_70685 | Not-determined |
| S11_extracted_k141_6369 | Not-determined |
| S11_extracted_k141_58485 | Low-quality |
| S11_extracted_k141_32043 | Low-quality |
| S11_extracted_k141_32044 | Not-determined |
| S11_extracted_k141_32045 | Not-determined |
| S11_extracted_k141_51306 | Low-quality |
| S11_extracted_k141_6372 | Not-determined |
| S11_extracted_k141_32047 | Not-determined |
| S11_extracted_k141_6378 | Not-determined |
| S11_extracted_k141_70694 | Not-determined |
| S11_extracted_k141_51311 | Not-determined |
| S11_extracted_k141_90850 | Not-determined |
| S11_extracted_k141_6380 | Not-determined |
| S11_extracted_k141_58502 | Not-determined |
| S11_extracted_k141_90854 | Not-determined |
| S11_extracted_k141_103607 | Not-determined |
| S11_extracted_k141_58508 | Not-determined |
| S11_extracted_k141_58509 | Not-determined |
| S11_extracted_k141_12982 | Not-determined |
| S11_extracted_k141_103611 | Low-quality |
| S11_extracted_k141_12984 | Not-determined |
| S11_extracted_k141_19381 | Not-determined |
| S11_extracted_k141_70718 | Not-determined |
| S11_extracted_k141_45468 | Low-quality |
| S11_extracted_k141_51326 | Not-determined |
| S11_extracted_k141_103619 | Not-determined |
| S11_extracted_k141_64769 | Not-determined |
| S11_extracted_k141_90866 | Not-determined |
| S11_extracted_k141_90867 | Not-determined |
| S11_extracted_k141_19391 | Not-determined |
| S11_extracted_k141_51335 | Low-quality |
| S11_extracted_k141_12995 | Not-determined |
| S11_extracted_k141_12996 | Not-determined |
| S11_extracted_k141_38867 | Not-determined |
| S11_extracted_k141_64783 | Not-determined |
| S11_extracted_k141_70732 | Not-determined |
| S11_extracted_k141_45481 | Not-determined |
| S11_extracted_k141_70733 | Not-determined |
| S11_extracted_k141_90875 | Low-quality |
| S11_extracted_k141_6409 | Not-determined |
| S11_extracted_k141_103628 | Low-quality |
| S8_vs_k141_166677\|\|full | High-quality |
| S11_extracted_k141_70736 | Low-quality |
| S11_extracted_k141_6415 | Not-determined |
| S11_extracted_k141_6416 | Not-determined |
| S11_extracted_k141_90885 | Not-determined |
| S11_extracted_k141_103637 | Not-determined |
| S11_extracted_k141_51349 | Low-quality |
| S11_extracted_k141_64795 | Not-determined |
| S11_extracted_k141_6422 | Not-determined |
| S11_extracted_k141_90887 | Not-determined |
| S11_extracted_k141_51350 | Not-determined |
| S11_extracted_k141_51351 | Not-determined |
| S11_extracted_k141_19409 | Not-determined |
| S11_extracted_k141_58533 | Not-determined |
| S11_extracted_k141_51354 | Not-determined |
| S11_extracted_k141_6428 | Low-quality |
| S11_extracted_k141_51356 | Not-determined |
| S11_extracted_k141_45486 | Low-quality |
| S11_extracted_k141_45491 | Not-determined |
| S11_extracted_k141_70759 | Not-determined |
| S11_extracted_k141_58544 | Low-quality |
| S11_extracted_k141_6440 | Not-determined |
| S11_extracted_k141_103655 | Not-determined |
| S11_extracted_k141_6445 | Not-determined |
| S11_extracted_k141_58548 | Not-determined |
| S11_extracted_k141_84342 | Not-determined |
| S11_extracted_k141_84345 | Not-determined |
| S11_extracted_k141_19419 | Not-determined |
| S11_extracted_k141_19420 | Not-determined |
| S11_extracted_k141_58557 | Not-determined |
| S11_extracted_k141_19425 | Not-determined |
| S11_extracted_k141_19428 | Not-determined |
| S11_extracted_k141_90909 | Not-determined |
| S11_extracted_k141_70773 | Not-determined |
| S11_extracted_k141_84361 | Not-determined |
| S11_extracted_k141_90910 | Low-quality |
| S11_extracted_k141_38911 | Not-determined |
| S11_extracted_k141_45503 | Low-quality |
| S11_extracted_k141_70779 | Not-determined |
| S11_extracted_k141_84369 | Not-determined |
| S11_extracted_k141_45506 | Not-determined |
| S11_extracted_k141_103662 | Not-determined |
| S11_extracted_k141_84372 | Low-quality |
| S11_extracted_k141_70782 | Not-determined |
| S11_extracted_k141_51381 | Not-determined |
| S11_extracted_k141_51384 | Not-determined |
| S11_extracted_k141_84373 | Not-determined |
| S11_extracted_k141_45513 | Not-determined |
| S11_extracted_k141_38924 | Not-determined |
| S11_extracted_k141_38925 | Not-determined |
| S11_extracted_k141_84379 | Not-determined |
| S11_extracted_k141_19437 | Low-quality |
| S11_extracted_k141_38929 | Not-determined |
| S11_extracted_k141_38931 | Low-quality |
| S11_extracted_k141_38932 | Not-determined |
| S11_extracted_k141_103667 | Not-determined |
| S11_extracted_k141_70784 | Low-quality |
| S11_extracted_k141_103669 | Not-determined |
| S11_extracted_k141_45530 | Not-determined |
| S11_extracted_k141_84396 | Not-determined |
| S11_extracted_k141_51396 | Not-determined |
| S11_extracted_k141_19453 | Not-determined |
| S11_extracted_k141_84403 | Not-determined |
| S11_extracted_k141_45534 | Not-determined |
| S11_extracted_k141_84404 | Not-determined |
| S11_extracted_k141_19458 | Not-determined |
| S11_extracted_k141_70791 | Low-quality |
| S11_extracted_k141_45540 | Not-determined |
| S11_extracted_k141_103677 | Not-determined |
| S11_extracted_k141_45543 | Not-determined |
| S11_extracted_k141_70793 | Not-determined |
| S11_extracted_k141_51409 | Not-determined |
| S11_extracted_k141_38948 | Not-determined |
| S11_extracted_k141_84412 | Not-determined |
| S11_extracted_k141_90926 | Not-determined |
| S11_extracted_k141_45547 | Not-determined |
| S11_extracted_k141_19467 | Not-determined |
| S11_extracted_k141_45549 | Not-determined |
| S11_extracted_k141_19468 | Not-determined |
| S11_extracted_k141_38955 | Not-determined |
| S11_extracted_k141_90935 | Not-determined |
| S11_extracted_k141_70806 | Low-quality |
| S11_extracted_k141_38960 | Low-quality |
| S11_extracted_k141_84427 | Not-determined |
| S11_extracted_k141_38965 | Low-quality |
| S11_extracted_k141_38968 | Not-determined |
| S11_extracted_k141_51419 | Not-determined |
| S11_extracted_k141_70814 | Not-determined |
| S11_extracted_k141_103696 | Not-determined |
| S11_extracted_k141_38976 | Not-determined |
| S11_extracted_k141_38978 | Not-determined |
| S11_extracted_k141_38979 | Not-determined |
| S11_extracted_k141_19479 | Not-determined |
| S11_extracted_k141_70819 | Not-determined |
| S11_extracted_k141_70820 | Not-determined |
| S11_extracted_k141_6464 | Not-determined |
| S11_extracted_k141_38988 | Not-determined |
| S11_extracted_k141_6468 | Not-determined |
| S11_extracted_k141_103699 | Not-determined |
| S11_extracted_k141_103701 | Not-determined |
| S11_extracted_k141_51429 | Not-determined |
| S11_extracted_k141_38998 | Not-determined |
| S11_extracted_k141_103703 | Not-determined |
| S11_extracted_k141_6484 | Not-determined |
| S11_extracted_k141_103707 | Not-determined |
| S11_extracted_k141_103708 | Not-determined |
| S11_extracted_k141_70836 | Low-quality |
| S11_extracted_k141_90966 | Low-quality |
| S11_extracted_k141_103711 | Not-determined |
| S11_extracted_k141_103714 | Low-quality |
| S11_extracted_k141_90976 | Not-determined |
| S11_extracted_k141_39014 | Not-determined |
| S11_extracted_k141_32110 | Not-determined |
| S11_extracted_k141_39018 | Not-determined |
| S11_extracted_k141_64848 | Not-determined |
| S11_extracted_k141_51444 | Not-determined |
| S11_extracted_k141_64849 | Not-determined |
| S11_extracted_k141_90992 | Not-determined |
| S11_extracted_k141_45551 | Low-quality |
| S11_extracted_k141_51451 | Not-determined |
| S11_extracted_k141_64858 | Low-quality |
| S11_extracted_k141_32120 | Not-determined |
| S11_extracted_k141_51453 | Not-determined |
| S11_extracted_k141_51454 | Not-determined |
| S11_extracted_k141_39022 | Not-determined |
| S11_extracted_k141_32124 | Not-determined |
| S11_extracted_k141_90999 | Not-determined |
| S11_extracted_k141_19507 | Not-determined |
| S11_extracted_k141_103730 | Low-quality |
| S11_extracted_k141_32134 | Low-quality |
| S11_extracted_k141_32135 | Not-determined |
| S11_extracted_k141_32141 | Not-determined |
| S11_extracted_k141_19514 | Not-determined |
| S11_extracted_k141_64875 | Not-determined |
| S11_extracted_k141_70884 | Not-determined |
| S11_extracted_k141_32143 | Low-quality |
| S11_extracted_k141_103737 | Low-quality |
| S11_extracted_k141_51468 | Not-determined |
| S11_extracted_k141_32148 | Not-determined |
| S11_extracted_k141_32149 | Not-determined |
| S11_extracted_k141_103740 | Not-determined |
| S11_extracted_k141_64879 | Not-determined |
| S11_extracted_k141_103742 | Not-determined |
| S11_extracted_k141_70893 | Not-determined |
| S11_extracted_k141_19520 | Not-determined |
| S11_extracted_k141_91016 | Not-determined |
| S11_extracted_k141_70896 | Not-determined |
| S11_extracted_k141_39043 | Not-determined |
| S11_extracted_k141_103761 | Not-determined |
| S11_extracted_k141_64891 | Low-quality |
| S11_extracted_k141_51476 | Not-determined |
| S11_extracted_k141_103770 | Not-determined |
| S11_extracted_k141_64898 | Not-determined |
| S11_extracted_k141_32156 | Not-determined |
| S11_extracted_k141_51480 | Not-determined |
| S11_extracted_k141_70921 | Low-quality |
| S11_extracted_k141_64906 | Not-determined |
| S11_extracted_k141_32163 | Not-determined |
| S11_extracted_k141_64913 | Not-determined |
| S11_extracted_k141_51495 | Not-determined |
| S11_extracted_k141_32169 | Not-determined |
| S11_extracted_k141_70944 | Not-determined |
| S11_extracted_k141_70947 | Not-determined |
| S11_extracted_k141_70949 | Not-determined |
| S11_extracted_k141_70958 | Not-determined |
| S11_extracted_k141_91065 | Not-determined |
| S11_extracted_k141_91069 | Low-quality |
| S11_extracted_k141_51525 | Not-determined |
| S11_extracted_k141_70968 | Not-determined |
| S11_extracted_k141_70971 | Not-determined |
| S11_extracted_k141_91073 | Not-determined |
| S11_extracted_k141_91088 | Not-determined |
| S11_extracted_k141_70981 | Not-determined |
| S11_extracted_k141_64944 | Not-determined |
| S11_extracted_k141_51547 | Not-determined |
| S11_extracted_k141_91093 | Not-determined |
| S11_extracted_k141_51548 | Not-determined |
| S11_extracted_k141_91094 | Not-determined |
| S11_extracted_k141_70986 | Low-quality |
| S11_extracted_k141_103810 | Not-determined |
| S11_extracted_k141_64947 | Not-determined |
| S11_extracted_k141_51560 | Not-determined |
| S11_extracted_k141_51561 | Not-determined |
| S11_extracted_k141_84448 | Not-determined |
| S11_extracted_k141_103823 | Low-quality |
| S11_extracted_k141_71006 | Not-determined |
| S11_extracted_k141_71009 | Not-determined |
| S11_extracted_k141_71012 | Not-determined |
| S11_extracted_k141_64963 | Low-quality |
| S11_extracted_k141_32179 | Not-determined |
| S11_extracted_k141_32180 | Not-determined |
| S11_extracted_k141_64972 | Not-determined |
| S11_extracted_k141_51570 | Low-quality |
| S11_extracted_k141_84472 | Not-determined |
| S11_extracted_k141_103837 | Not-determined |
| S11_extracted_k141_103844 | Not-determined |
| S11_extracted_k141_51581 | Not-determined |
| S11_extracted_k141_51586 | Not-determined |
| S11_extracted_k141_51590 | Low-quality |
| S11_extracted_k141_103858 | Low-quality |
| S11_extracted_k141_51604 | Not-determined |
| S11_extracted_k141_64996 | Not-determined |
| S11_extracted_k141_64997 | Not-determined |
| S11_extracted_k141_103873 | Not-determined |
| S11_extracted_k141_71047 | Not-determined |
| S11_extracted_k141_65005 | Not-determined |
| S11_extracted_k141_51616 | Not-determined |
| S11_extracted_k141_71052 | Low-quality |
| S11_extracted_k141_65007 | Not-determined |
| S11_extracted_k141_65008 | Not-determined |
| S11_extracted_k141_65010 | Not-determined |
| S11_extracted_k141_51626 | Not-determined |
| S11_extracted_k141_51627 | Not-determined |
| S11_extracted_k141_65018 | Not-determined |
| S11_extracted_k141_65020 | Not-determined |
| S11_extracted_k141_84490 | Not-determined |
| S11_extracted_k141_84492 | Not-determined |
| S11_extracted_k141_71071 | Not-determined |
| S11_extracted_k141_103886 | Not-determined |
| S11_extracted_k141_84498 | Low-quality |
| S11_extracted_k141_84502 | Not-determined |
| S11_extracted_k141_103897 | Not-determined |
| S11_extracted_k141_84516 | Not-determined |
| S11_extracted_k141_65036 | Not-determined |
| S11_extracted_k141_103906 | Not-determined |
| S11_extracted_k141_103910 | Not-determined |
| S11_extracted_k141_84523 | Not-determined |
| S11_extracted_k141_65041 | Not-determined |
| S11_extracted_k141_51654 | Not-determined |
| S11_extracted_k141_51656 | Not-determined |
| S11_extracted_k141_103916 | Not-determined |
| S11_extracted_k141_103917 | Not-determined |
| S11_extracted_k141_65054 | Not-determined |
| S11_extracted_k141_51660 | Not-determined |
| S11_extracted_k141_103925 | Not-determined |
| S11_extracted_k141_103935 | Not-determined |
| S11_extracted_k141_103938 | Not-determined |
| S11_extracted_k141_32203 | Low-quality |
| S11_extracted_k141_51677 | Not-determined |
| S11_extracted_k141_103948 | Not-determined |
| S11_extracted_k141_71084 | Not-determined |
| S11_extracted_k141_103954 | Not-determined |
| S11_extracted_k141_103964 | Not-determined |
| S11_extracted_k141_32223 | Low-quality |
| S11_extracted_k141_65075 | Not-determined |
| S11_extracted_k141_65077 | Low-quality |
| S11_extracted_k141_103967 | Not-determined |
| S11_extracted_k141_32232 | Low-quality |
| S11_extracted_k141_103970 | Not-determined |
| S11_extracted_k141_103975 | Not-determined |
| S11_extracted_k141_103979 | Not-determined |
| S11_extracted_k141_71105 | Low-quality |
| S11_extracted_k141_32238 | Not-determined |
| S11_extracted_k141_71114 | Not-determined |
| S11_extracted_k141_84592 | Not-determined |
| S11_extracted_k141_32250 | Not-determined |
| S11_extracted_k141_71119 | Low-quality |
| S11_extracted_k141_103993 | Not-determined |
| S11_extracted_k141_103996 | Not-determined |
| S11_extracted_k141_103999 | Not-determined |
| S11_extracted_k141_71129 | Not-determined |
| S11_extracted_k141_32259 | Not-determined |
| S11_extracted_k141_104001 | Not-determined |
| S11_extracted_k141_71132 | Low-quality |
| S11_extracted_k141_104005 | Low-quality |
| S11_extracted_k141_71140 | Not-determined |
| S11_extracted_k141_32272 | Not-determined |
| S11_extracted_k141_51691 | Not-determined |
| S11_extracted_k141_32274 | Low-quality |
| S11_extracted_k141_51694 | Not-determined |
| S11_extracted_k141_71142 | Low-quality |
| S11_extracted_k141_71158 | Not-determined |
| S11_extracted_k141_51721 | Low-quality |
| S11_extracted_k141_71167 | Not-determined |
| S11_extracted_k141_71168 | Not-determined |
| S11_extracted_k141_51724 | Not-determined |
| S11_extracted_k141_71170 | Not-determined |
| S11_extracted_k141_51728 | Not-determined |
| S11_extracted_k141_71175 | Not-determined |
| S11_extracted_k141_104015 | Not-determined |
| S11_extracted_k141_104018 | Not-determined |
| S11_extracted_k141_104022 | Not-determined |
| S11_extracted_k141_32288 | Not-determined |
| S11_extracted_k141_71206 | Not-determined |
| S11_extracted_k141_32290 | Low-quality |
| S11_extracted_k141_32291 | Low-quality |
| S11_extracted_k141_104029 | Not-determined |
| S11_extracted_k141_32296 | Low-quality |
| S11_extracted_k141_71213 | Not-determined |
| S11_extracted_k141_51778 | Not-determined |
| S11_extracted_k141_71232 | Not-determined |
| S11_extracted_k141_51779 | Not-determined |
| S11_extracted_k141_32307 | Not-determined |
| S11_extracted_k141_71237 | Not-determined |
| S11_extracted_k141_32309 | Not-determined |
| S11_extracted_k141_51793 | Not-determined |
| S11_extracted_k141_32331 | Low-quality |
| S11_extracted_k141_32332 | Not-determined |
| S11_extracted_k141_32336 | Not-determined |
| S11_extracted_k141_32339 | Not-determined |
| S11_extracted_k141_71260 | Not-determined |
| S11_extracted_k141_71267 | Not-determined |
| S11_extracted_k141_32345 | Not-determined |
| S11_extracted_k141_32347 | Low-quality |
| S11_extracted_k141_32363 | Not-determined |
| S11_extracted_k141_32364 | Not-determined |
| S11_extracted_k141_71282 | Not-determined |
| S11_extracted_k141_32366 | Low-quality |
| S11_extracted_k141_71297 | Low-quality |
| S11_extracted_k141_71299 | Not-determined |
| S11_extracted_k141_32390 | Not-determined |
| S11_extracted_k141_32394 | Not-determined |
| S11_extracted_k141_32397 | Not-determined |
| S11_extracted_k141_71314 | Not-determined |
| S11_extracted_k141_32407 | Not-determined |
| S11_extracted_k141_32410 | Not-determined |
| S11_extracted_k141_71318 | Low-quality |
| S11_extracted_k141_71320 | Not-determined |
| S11_extracted_k141_32414 | Not-determined |
| S11_extracted_k141_71325 | Not-determined |
| S11_extracted_k141_32423 | Not-determined |
| S11_extracted_k141_51845 | Low-quality |
| S11_extracted_k141_32429 | Not-determined |
| S11_extracted_k141_71336 | Not-determined |
| S11_extracted_k141_71341 | Low-quality |
| S11_extracted_k141_51852 | Low-quality |
| S11_extracted_k141_32434 | Not-determined |
| S11_extracted_k141_51857 | Not-determined |
| S11_extracted_k141_32441 | Low-quality |
| S11_extracted_k141_51864 | Low-quality |
| S11_extracted_k141_71355 | Not-determined |
| S11_extracted_k141_51865 | Not-determined |
| S11_extracted_k141_32450 | Not-determined |
| S11_extracted_k141_51890 | Not-determined |
| S11_extracted_k141_71364 | Low-quality |
| S11_extracted_k141_51901 | Not-determined |
| S11_extracted_k141_71370 | Not-determined |
| S11_extracted_k141_71377 | Low-quality |
| S11_extracted_k141_51918 | Low-quality |
| S11_extracted_k141_71383 | Not-determined |
| S11_extracted_k141_32476 | Not-determined |
| S11_extracted_k141_32480 | Low-quality |
| S11_extracted_k141_32484 | Low-quality |
| S11_extracted_k141_32491 | Not-determined |
| S11_extracted_k141_51931 | Not-determined |
| S3_vs_k141_112065\|\|full | High-quality |
| S11_extracted_k141_71392 | Not-determined |
| S11_extracted_k141_32502 | Not-determined |
| S11_extracted_k141_32507 | Not-determined |
| S11_extracted_k141_71400 | Not-determined |
| S11_extracted_k141_71405 | Not-determined |
| S11_extracted_k141_32536 | Not-determined |
| S11_extracted_k141_32537 | Not-determined |
| S11_extracted_k141_71425 | Not-determined |
| S11_extracted_k141_71442 | Not-determined |
| S7_extracted_13952 | Complete |
| S11_extracted_k141_71447 | Low-quality |
| S11_extracted_k141_71451 | Not-determined |
| S11_extracted_k141_52012 | Not-determined |
| S11_extracted_k141_71463 | Not-determined |
| S11_extracted_k141_71479 | Not-determined |
| S11_extracted_k141_71488 | Not-determined |
| S11_extracted_k141_52053 | Not-determined |
| S11_extracted_k141_52055 | Not-determined |
| S11_extracted_k141_71517 | Not-determined |
| S11_extracted_k141_71520 | Not-determined |
| S11_extracted_k141_71522 | Not-determined |
| S11_extracted_k141_71530 | Not-determined |
| S11_extracted_k141_71546 | Not-determined |
| S7_vs_13952\|\|full | High-quality |
| S11_extracted_k141_71554 | Not-determined |
| S11_extracted_k141_71560 | Not-determined |
| S11_extracted_k141_71565 | Not-determined |
| S11_extracted_k141_71566 | Not-determined |
| S8_extracted_k141_23955 | High-quality |
| S8_vs_k141_23955\|\|full | High-quality |
| S9_extracted_k141_65811 | Complete |
| S9_vs_k141_65811\|\|full | High-quality |
| S4_extracted_31981 | Complete |
| S15_extracted_58467 | High-quality |
| S15_vs_58467\|\|full | High-quality |
| S4_vs_VIRSorter_31981-circular-cat | High-quality |
| S11_extracted_k141_104066 | Not-determined |
| S11_extracted_k141_104059 | Complete |
| S13_vs_67429\|\|full | High-quality |
| S11_vs_k141_104059\|\|full | High-quality |
| S11_extracted_k141_104086 | Not-determined |
| S11_extracted_k141_104088 | Not-determined |
| S11_extracted_k141_104097 | Not-determined |
| S11_extracted_k141_104098 | Low-quality |
| S11_extracted_k141_104099 | Low-quality |
| S11_extracted_k141_104100 | Not-determined |
| S11_extracted_k141_104102 | Not-determined |
| S11_extracted_k141_104104 | Not-determined |
| S11_extracted_k141_104105 | Not-determined |
| S11_extracted_k141_104113 | Not-determined |
| S11_extracted_k141_104114 | Not-determined |
| S11_extracted_k141_104115 | Not-determined |
| S11_extracted_k141_104122 | Not-determined |
| S11_extracted_k141_104125 | Not-determined |
| S7_vs_101751\|\|full | High-quality |
| S15_extracted_11823 | High-quality |
| S11_vs_k141_7049\|\|full | Low-quality |
| S15_vs_11823\|\|full | High-quality |
| S8_extracted_k141_158225 | Complete |
| S8_vs_k141_158225\|\|full | High-quality |
| S11_vs_k141_20912\|\|full | Not-determined |
| S11_vs_k141_1684\|\|full | Not-determined |
| S11_vs_k141_79743\|\|full | Not-determined |
| S12_extracted_k141_32668 | Complete |
| S12_vs_k141_32668\|\|full | High-quality |
| S14_vs_k141_65503\|\|full | High-quality |
| S5_vs_k141_74071\|\|full | High-quality |
| S11_vs_k141_47716\|\|full | Low-quality |
| S11_vs_k141_15294\|\|full | Not-determined |
| S3_vs_k141_45147\|\|full | High-quality |
| S16_vs_7630\|\|full | Medium-quality |
| S6_extracted_k141_36011 | Complete |
| S5_extracted_k141_144326 | High-quality |
| S5_vs_k141_144326\|\|full | High-quality |
| S6_vs_k141_36011\|\|full | High-quality |
| S11_vs_k141_49071\|\|full | High-quality |
| S8_extracted_k141_74113 | High-quality |
| S8_vs_k141_74113\|\|full | High-quality |
| S14_vs_k141_2870\|\|full | Medium-quality |
| S9_vs_k141_83108\|\|full | High-quality |
| S12_extracted_k141_188824 | Complete |
| S12_vs_k141_188824\|\|full | High-quality |
| S7_vs_137998\|\|full | High-quality |
| S4_extracted_27861 | High-quality |
| S4_vs_VIRSorter_27861-circular-cat | High-quality |
| S4_vs_VIRSorter_30222-cat_2 | High-quality |
| S3_extracted_k141_18829 | Complete |
| S3_vs_k141_18829\|\|full | High-quality |
| S5_vs_k141_123256\|\|full | Medium-quality |
| S12_vs_k141_14927\|\|full | High-quality |
| S13_extracted_151045 | Complete |
| S13_vs_33947\|\|full | Medium-quality |
| S13_vs_151045\|\|full | High-quality |
| S16_extracted_169985 | Complete |
| S11_vs_k141_104050\|\|full | Low-quality |
| S11_vs_k141_104051\|\|full | Low-quality |
| S16_vs_169985\|\|full | High-quality |
| S12_vs_k141_92868\|\|full | High-quality |
| S15_extracted_74586 | Complete |
| S4_extracted_22219 | Complete |
| S11_vs_k141_104061\|\|full | Not-determined |
| S3_extracted_k141_25983 | Complete |
| S5_vs_k141_142195\|\|full | Medium-quality |
| S15_vs_74586\|\|full | High-quality |
| S4_vs_VIRSorter_22219-circular-cat | High-quality |
| S3_vs_k141_25983\|\|full | High-quality |
| S16_vs_89511\|\|full | High-quality |
| S11_vs_k141_104077\|\|full | Low-quality |
| S11_vs_k141_104103\|\|full | Low-quality |
| S11_vs_k141_71588\|\|full | Low-quality |
| S11_vs_k141_13031\|\|full | Low-quality |
| S11_vs_k141_45573\|\|full | Low-quality |
| S11_vs_k141_97632\|\|full | Not-determined |
| S11_vs_k141_91127\|\|full | Low-quality |
| S11_vs_k141_71599\|\|full | Low-quality |
| S11_vs_k141_39064\|\|full | Low-quality |
| S11_vs_k141_91129\|\|full | Not-determined |
| S11_vs_k141_52091\|\|full | Not-determined |
| S11_vs_k141_38\|\|full | Not-determined |
| S11_vs_k141_26054\|\|full | Not-determined |
| S11_vs_k141_52098\|\|full | Low-quality |
| S11_vs_k141_26058\|\|full | Not-determined |

| | |
|---|---|
| S11_vs_k141_97641\|\|full | Low-quality |
| S11_vs_k141_97642\|\|full | Low-quality |
| S11_vs_k141_6538\|\|full | Low-quality |
| S11_vs_k141_52102\|\|full | Low-quality |
| S11_vs_k141_45586\|\|full | Low-quality |
| S5_vs_k141_56998\|\|full | High-quality |
| S11_vs_k141_91160\|\|full | Not-determined |
| S11_vs_k141_65136\|\|full | Low-quality |
| S11_vs_k141_52116\|\|full | Not-determined |
| S11_vs_k141_97653\|\|full | Low-quality |
| S11_vs_k141_26084\|\|full | Low-quality |
| S11_vs_k141_32581\|\|full | Low-quality |
| S11_vs_k141_32582\|\|full | Low-quality |
| S11_vs_k141_58607\|\|full | Low-quality |
| S11_vs_k141_19558\|\|full | Not-determined |
| S11_vs_k141_65147\|\|full | Low-quality |
| S11_vs_k141_78148\|\|full | Not-determined |
| S11_vs_k141_26096\|\|full | Low-quality |
| S11_vs_k141_91178\|\|full | Not-determined |
| S11_vs_k141_91180\|\|full | Low-quality |
| S11_vs_k141_19572\|\|full | Low-quality |
| S11_vs_k141_52135\|\|full | Low-quality |
| S11_vs_k141_71645\|\|full | Low-quality |
| S11_vs_k141_71649\|\|full | Low-quality |
| S11_vs_k141_65181\|\|full | Low-quality |
| S11_vs_k141_39092\|\|full | Not-determined |
| S11_vs_k141_19592\|\|full | Low-quality |
| S11_vs_k141_65186\|\|full | Not-determined |
| S11_vs_k141_89\|\|full | Not-determined |
| S11_vs_k141_65187\|\|full | Low-quality |
| S11_vs_k141_58652\|\|full | Not-determined |
| S11_vs_k141_32599\|\|full | Not-determined |
| S11_vs_k141_52165\|\|full | Not-determined |
| S11_vs_k141_58656\|\|full | Not-determined |
| S6_vs_k141_22318\|\|full | High-quality |
| S11_vs_k141_52174\|\|full | Low-quality |
| S11_vs_k141_104\|\|full | Low-quality |
| S11_vs_k141_111\|\|full | Low-quality |
| S11_vs_k141_65200\|\|full | Low-quality |
| S11_vs_k141_19638\|\|full | Low-quality |
| S11_vs_k141_97678\|\|full | Not-determined |
| S11_vs_k141_58686\|\|full | Not-determined |
| S11_vs_k141_45640\|\|full | Low-quality |
| S11_vs_k141_97680\|\|full | Not-determined |
| S11_vs_k141_13062\|\|full | Low-quality |
| S11_vs_k141_13065\|\|full | Not-determined |
| S11_vs_k141_135\|\|full | Low-quality |
| S11_vs_k141_97686\|\|full | Not-determined |
| S11_vs_k141_58692\|\|full | Low-quality |
| S11_vs_k141_91243\|\|full | Low-quality |
| S11_vs_k141_32630\|\|full | Not-determined |
| S11_vs_k141_141\|\|full | Not-determined |
| S11_vs_k141_65242\|\|full | Low-quality |
| S11_vs_k141_13096\|\|full | Low-quality |
| S11_vs_k141_65249\|\|full | Low-quality |
| S11_vs_k141_13105\|\|full | Low-quality |
| S11_vs_k141_164\|\|full | Not-determined |
| S11_vs_k141_39161\|\|full | Not-determined |
| S11_vs_k141_97691\|\|full | Low-quality |
| S11_vs_k141_52268\|\|full | Not-determined |
| S11_vs_k141_52269\|\|full | Not-determined |
| S11_vs_k141_84750\|\|full | Low-quality |
| S11_vs_k141_52274\|\|full | Not-determined |
| S11_vs_k141_52276\|\|full | Low-quality |
| S11_vs_k141_198\|\|full | Not-determined |
| S11_vs_k141_19676\|\|full | Not-determined |
| S11_vs_k141_39167\|\|full | Low-quality |
| S11_vs_k141_26189\|\|full | Low-quality |
| S11_vs_k141_6553\|\|full | Not-determined |
| S11_vs_k141_32679\|\|full | Low-quality |
| S11_vs_k141_84755\|\|full | Low-quality |
| S11_vs_k141_78168\|\|full | Not-determined |
| S11_vs_k141_6560\|\|full | Low-quality |
| S11_vs_k141_241\|\|full | Not-determined |
| S11_vs_k141_13132\|\|full | Low-quality |
| S11_vs_k141_19696\|\|full | Low-quality |
| S11_vs_k141_91303\|\|full | Not-determined |
| S11_vs_k141_97757\|\|full | Low-quality |
| S11_vs_k141_13150\|\|full | Low-quality |
| S11_vs_k141_91307\|\|full | Not-determined |
| S11_vs_k141_39184\|\|full | Low-quality |
| S11_vs_k141_259\|\|full | Low-quality |
| S11_vs_k141_52317\|\|full | Low-quality |
| S11_vs_k141_26233\|\|full | Not-determined |
| S11_vs_k141_91310\|\|full | Low-quality |
| S11_vs_k141_84774\|\|full | Low-quality |
| S11_vs_k141_78182\|\|full | Not-determined |
| S11_vs_k141_52321\|\|full | Low-quality |
| S11_vs_k141_71710\|\|full | Not-determined |
| S4_extracted_26417 | High-quality |
| S11_vs_k141_84792\|\|full | Low-quality |
| S11_vs_k141_39191\|\|full | Low-quality |
| S11_vs_k141_6594\|\|full | Low-quality |
| S11_vs_k141_97777\|\|full | Not-determined |
| S11_vs_k141_91331\|\|full | Not-determined |
| S11_vs_k141_39201\|\|full | Low-quality |
| S11_vs_k141_58713\|\|full | Low-quality |
| S11_vs_k141_91335\|\|full | Low-quality |
| S11_vs_k141_32721\|\|full | Low-quality |
| S11_vs_k141_32722\|\|full | Low-quality |
| S11_vs_k141_78211\|\|full | Low-quality |
| S11_vs_k141_32723\|\|full | Low-quality |
| S11_vs_k141_97794\|\|full | Low-quality |
| S11_vs_k141_97796\|\|full | Low-quality |
| S11_vs_k141_71754\|\|full | Not-determined |
| S11_vs_k141_39217\|\|full | Not-determined |
| S11_vs_k141_97804\|\|full | Low-quality |
| S11_vs_k141_97806\|\|full | Low-quality |
| S11_vs_k141_84820\|\|full | Low-quality |
| S11_vs_k141_325\|\|full | Low-quality |
| S11_vs_k141_84824\|\|full | Not-determined |
| S11_vs_k141_71769\|\|full | Low-quality |
| S11_vs_k141_91363\|\|full | Low-quality |
| S11_vs_k141_6632\|\|full | Not-determined |
| S4_vs_VIRSorter_26417-cat_3 | High-quality |
| S11_vs_k141_71779\|\|full | Low-quality |
| S11_vs_k141_91371\|\|full | Low-quality |
| S11_vs_k141_52329\|\|full | Low-quality |
| S11_vs_k141_6644\|\|full | Not-determined |
| S11_vs_k141_19751\|\|full | Low-quality |
| S11_vs_k141_78267\|\|full | Low-quality |
| S11_vs_k141_32758\|\|full | Not-determined |
| S11_vs_k141_32762\|\|full | Low-quality |
| S11_vs_k141_78276\|\|full | Low-quality |
| S11_vs_k141_91384\|\|full | Low-quality |
| S11_vs_k141_65303\|\|full | Low-quality |
| S11_vs_k141_65304\|\|full | Not-determined |
| S11_vs_k141_52347\|\|full | Low-quality |
| S11_vs_k141_58772\|\|full | Low-quality |
| S11_vs_k141_65306\|\|full | Low-quality |
| S11_vs_k141_78305\|\|full | Not-determined |
| S11_vs_k141_78307\|\|full | Not-determined |
| S11_vs_k141_71801\|\|full | Low-quality |
| S11_vs_k141_91408\|\|full | Not-determined |
| S11_vs_k141_65307\|\|full | Not-determined |
| S11_vs_k141_32783\|\|full | Low-quality |
| S11_vs_k141_375\|\|full | Low-quality |
| S11_vs_k141_58776\|\|full | Not-determined |
| S11_vs_k141_97842\|\|full | Low-quality |
| S11_vs_k141_52369\|\|full | Low-quality |
| S11_vs_k141_6672\|\|full | Low-quality |
| S11_vs_k141_71812\|\|full | Low-quality |
| S6_extracted_k141_1829 | Complete |
| S11_vs_k141_78333\|\|full | Low-quality |
| S11_vs_k141_78336\|\|full | Low-quality |
| S11_vs_k141_413\|\|full | Low-quality |
| S6_vs_k141_1829\|\|full | High-quality |
| S11_vs_k141_78341\|\|full | Low-quality |
| S11_vs_k141_39254\|\|full | Not-determined |
| S11_vs_k141_97879\|\|full | Low-quality |
| S11_vs_k141_71849\|\|full | Low-quality |
| S11_vs_k141_65342\|\|full | Low-quality |
| S11_vs_k141_71854\|\|full | Low-quality |
| S11_vs_k141_84866\|\|full | Low-quality |
| S11_vs_k141_32832\|\|full | Not-determined |
| S11_vs_k141_454\|\|full | Low-quality |
| S11_vs_k141_45707\|\|full | Not-determined |
| S11_vs_k141_45708\|\|full | Not-determined |
| S11_vs_k141_78365\|\|full | Low-quality |
| S11_vs_k141_39279\|\|full | Not-determined |
| S11_vs_k141_91465\|\|full | Low-quality |
| S11_vs_k141_78389\|\|full | Low-quality |
| S11_vs_k141_39289\|\|full | Not-determined |
| S11_vs_k141_84917\|\|full | Not-determined |
| S11_vs_k141_91478\|\|full | Not-determined |
| S11_vs_k141_84920\|\|full | Low-quality |
| S11_vs_k141_78395\|\|full | Low-quality |
| S11_vs_k141_6747\|\|full | Low-quality |
| S11_vs_k141_84932\|\|full | Low-quality |
| S11_vs_k141_91488\|\|full | Low-quality |
| S11_vs_k141_78402\|\|full | Low-quality |
| S11_vs_k141_97937\|\|full | Low-quality |
| S11_vs_k141_39317\|\|full | Low-quality |
| S11_vs_k141_19834\|\|full | Low-quality |
| S11_vs_k141_19836\|\|full | Low-quality |
| S11_vs_k141_97955\|\|full | Low-quality |
| S11_vs_k141_32880\|\|full | Low-quality |
| S11_vs_k141_506\|\|full | Low-quality |
| S11_vs_k141_39340\|\|full | Low-quality |
| S11_vs_k141_97971\|\|full | Not-determined |
| S11_vs_k141_45741\|\|full | Low-quality |
| S11_vs_k141_52403\|\|full | Low-quality |
| S11_vs_k141_78443\|\|full | Low-quality |
| S11_vs_k141_6776\|\|full | Not-determined |
| S11_vs_k141_19851\|\|full | Not-determined |
| S11_vs_k141_26372\|\|full | Not-determined |
| S11_vs_k141_65387\|\|full | Not-determined |
| S11_vs_k141_78449\|\|full | Low-quality |
| S11_vs_k141_58832\|\|full | Low-quality |
| S11_vs_k141_522\|\|full | Not-determined |
| S11_vs_k141_65390\|\|full | Low-quality |
| S11_vs_k141_71889\|\|full | Not-determined |
| S11_vs_k141_78454\|\|full | Low-quality |
| S11_vs_k141_13220\|\|full | Not-determined |
| S11_vs_k141_26376\|\|full | Low-quality |
| S11_vs_k141_71899\|\|full | Not-determined |
| S11_vs_k141_19868\|\|full | Low-quality |
| S11_vs_k141_52430\|\|full | Not-determined |
| S11_vs_k141_78466\|\|full | Low-quality |
| S11_vs_k141_58836\|\|full | Low-quality |
| S11_vs_k141_91541\|\|full | Low-quality |
| S11_vs_k141_26383\|\|full | Not-determined |
| S11_vs_k141_71906\|\|full | Low-quality |
| S11_vs_k141_6796\|\|full | Not-determined |
| S11_vs_k141_85022\|\|full | Not-determined |
| S11_vs_k141_32907\|\|full | Low-quality |
| S11_vs_k141_71913\|\|full | Low-quality |
| S11_vs_k141_58848\|\|full | Low-quality |
| S11_vs_k141_39387\|\|full | Not-determined |
| S11_vs_k141_78467\|\|full | Low-quality |
| S11_vs_k141_26400\|\|full | Low-quality |
| S11_vs_k141_6809\|\|full | Low-quality |
| S11_vs_k141_566\|\|full | Low-quality |
| S11_vs_k141_45782\|\|full | Low-quality |
| S11_vs_k141_78484\|\|full | Not-determined |
| S11_vs_k141_78490\|\|full | Not-determined |
| S11_vs_k141_26406\|\|full | Low-quality |
| S11_vs_k141_45793\|\|full | Low-quality |
| S11_vs_k141_91564\|\|full | Low-quality |
| S11_vs_k141_85056\|\|full | Low-quality |
| S11_vs_k141_26421\|\|full | Low-quality |
| S11_vs_k141_6824\|\|full | Not-determined |
| S11_vs_k141_58870\|\|full | Low-quality |
| S11_vs_k141_39411\|\|full | Low-quality |
| S11_vs_k141_71952\|\|full | Low-quality |
| S11_vs_k141_78516\|\|full | Not-determined |
| S11_vs_k141_45807\|\|full | Low-quality |
| S6_vs_k141_10314\|\|full | Complete |
| S11_vs_k141_78527\|\|full | Not-determined |
| S11_vs_k141_58871\|\|full | Low-quality |
| S11_vs_k141_6842\|\|full | Low-quality |
| S11_vs_k141_19895\|\|full | Low-quality |
| S11_vs_k141_19905\|\|full | Low-quality |
| S11_vs_k141_78534\|\|full | Not-determined |
| S11_vs_k141_32930\|\|full | Low-quality |
| S16_extracted_48131 | Complete |
| S11_vs_k141_19919\|\|full | Low-quality |
| S11_vs_k141_91611\|\|full | Low-quality |
| S11_vs_k141_98028\|\|full | Not-determined |
| S11_vs_k141_58892\|\|full | Low-quality |
| S11_vs_k141_78555\|\|full | Low-quality |
| S11_vs_k141_98032\|\|full | Low-quality |
| S11_vs_k141_32947\|\|full | Not-determined |
| S11_vs_k141_85089\|\|full | Low-quality |
| S11_vs_k141_58901\|\|full | Low-quality |
| S11_vs_k141_19942\|\|full | Low-quality |
| S11_vs_k141_6877\|\|full | Low-quality |
| S11_vs_k141_32954\|\|full | Not-determined |
| S11_vs_k141_58905\|\|full | Low-quality |
| S11_vs_k141_633\|\|full | Not-determined |
| S11_vs_k141_58913\|\|full | Low-quality |
| S11_vs_k141_52477\|\|full | Low-quality |
| S11_vs_k141_52479\|\|full | Low-quality |
| S11_vs_k141_635\|\|full | Low-quality |
| S11_vs_k141_52481\|\|full | Not-determined |
| S11_vs_k141_636\|\|full | Low-quality |
| S11_vs_k141_78607\|\|full | Not-determined |
| S11_vs_k141_45869\|\|full | Not-determined |
| S11_vs_k141_645\|\|full | Not-determined |
| S11_vs_k141_91665\|\|full | Low-quality |
| S11_vs_k141_19972\|\|full | Low-quality |
| S11_vs_k141_91676\|\|full | Low-quality |
| S11_vs_k141_52494\|\|full | Not-determined |
| S11_vs_k141_26480\|\|full | Low-quality |
| S11_vs_k141_19985\|\|full | Low-quality |
| S11_vs_k141_39485\|\|full | Low-quality |
| S11_vs_k141_32984\|\|full | Low-quality |
| S11_vs_k141_65454\|\|full | Not-determined |
| S11_vs_k141_72026\|\|full | Low-quality |
| S16_vs_48131\|\|full | High-quality |
| S11_vs_k141_72034\|\|full | Not-determined |
| S11_vs_k141_6884\|\|full | Low-quality |
| S12_extracted_k141_150961 | High-quality |
| S11_vs_k141_52518\|\|full | Low-quality |
| S11_vs_k141_39502\|\|full | Low-quality |
| S11_vs_k141_98098\|\|full | Not-determined |
| S11_vs_k141_72045\|\|full | Low-quality |
| S11_vs_k141_13239\|\|full | Not-determined |
| S11_vs_k141_660\|\|full | Not-determined |
| S11_vs_k141_65477\|\|full | Low-quality |
| S11_vs_k141_33012\|\|full | Low-quality |
| S11_vs_k141_52533\|\|full | Low-quality |
| S11_vs_k141_52540\|\|full | Low-quality |
| S11_vs_k141_39532\|\|full | Low-quality |
| S11_vs_k141_673\|\|full | Low-quality |
| S11_vs_k141_65509\|\|full | Low-quality |
| S11_vs_k141_20005\|\|full | Low-quality |
| S11_vs_k141_20007\|\|full | Low-quality |
| S11_vs_k141_85097\|\|full | Low-quality |
| S11_vs_k141_45935\|\|full | Low-quality |
| S11_vs_k141_78682\|\|full | Not-determined |
| S11_vs_k141_85113\|\|full | Low-quality |
| S11_vs_k141_85114\|\|full | Low-quality |
| S11_vs_k141_85116\|\|full | Not-determined |
| S11_vs_k141_39557\|\|full | Not-determined |
| S11_vs_k141_78694\|\|full | Low-quality |
| S11_vs_k141_85120\|\|full | Low-quality |
| S11_vs_k141_20035\|\|full | Low-quality |
| S11_vs_k141_85123\|\|full | Low-quality |
| S11_vs_k141_20041\|\|full | Low-quality |
| S11_vs_k141_13310\|\|full | Low-quality |
| S11_vs_k141_45954\|\|full | Low-quality |
| S11_vs_k141_72109\|\|full | Not-determined |
| S11_vs_k141_6957\|\|full | Low-quality |
| S11_vs_k141_85140\|\|full | Low-quality |
| S11_vs_k141_45970\|\|full | Not-determined |
| S11_vs_k141_45974\|\|full | Low-quality |
| S12_vs_k141_150961\|\|full | High-quality |
| S16_vs_142561\|\|full | High-quality |
| S11_vs_k141_33063\|\|full | Low-quality |
| S11_vs_k141_33065\|\|full | Not-determined |
| S11_vs_k141_33069\|\|full | Not-determined |
| S11_vs_k141_20079\|\|full | Not-determined |
| S11_vs_k141_72161\|\|full | Not-determined |
| S11_vs_k141_20084\|\|full | Low-quality |
| S11_vs_k141_91702\|\|full | Not-determined |
| S11_vs_k141_13340\|\|full | Low-quality |
| S11_vs_k141_20106\|\|full | Low-quality |
| S11_vs_k141_731\|\|full | Low-quality |
| S11_vs_k141_72186\|\|full | Low-quality |
| S11_vs_k141_33102\|\|full | Low-quality |
| S11_vs_k141_78700\|\|full | Low-quality |
| S11_vs_k141_13355\|\|full | Not-determined |
| S2_extracted_k141_8253 | High-quality |
| S11_vs_k141_741\|\|full | Not-determined |
| S11_vs_k141_26558\|\|full | Low-quality |
| S11_vs_k141_98194\|\|full | Not-determined |
| S11_vs_k141_6998\|\|full | Low-quality |
| S11_vs_k141_98201\|\|full | Low-quality |
| S11_vs_k141_78726\|\|full | Not-determined |
| S11_vs_k141_26580\|\|full | Low-quality |
| S11_vs_k141_20140\|\|full | Low-quality |
| S11_vs_k141_774\|\|full | Low-quality |
| S11_vs_k141_20145\|\|full | Low-quality |
| S11_vs_k141_20147\|\|full | Not-determined |
| S11_vs_k141_65586\|\|full | Low-quality |
| S11_vs_k141_46044\|\|full | Low-quality |
| S11_vs_k141_7012\|\|full | Not-determined |
| S11_vs_k141_91744\|\|full | Low-quality |
| S11_vs_k141_13413\|\|full | Not-determined |
| S11_vs_k141_65603\|\|full | Low-quality |
| S11_vs_k141_52627\|\|full | Low-quality |
| S11_vs_k141_789\|\|full | Not-determined |
| S2_vs_k141_8253\|\|full | High-quality |
| S11_vs_k141_39674\|\|full | Low-quality |
| S11_vs_k141_52654\|\|full | Low-quality |
| S11_vs_k141_72208\|\|full | Low-quality |
| S11_vs_k141_39684\|\|full | Not-determined |
| S11_vs_k141_26600\|\|full | Low-quality |
| S11_vs_k141_13439\|\|full | Low-quality |
| S11_vs_k141_59002\|\|full | Low-quality |
| S11_vs_k141_72229\|\|full | Low-quality |
| S11_vs_k141_85267\|\|full | Low-quality |
| S11_vs_k141_98284\|\|full | Not-determined |
| S11_vs_k141_52665\|\|full | Low-quality |
| S11_vs_k141_85274\|\|full | Low-quality |
| S11_vs_k141_72248\|\|full | Low-quality |
| S11_vs_k141_91802\|\|full | Low-quality |
| S11_vs_k141_72257\|\|full | Low-quality |
| S11_vs_k141_13468\|\|full | Low-quality |
| S11_vs_k141_78817\|\|full | Low-quality |
| S11_vs_k141_33160\|\|full | Low-quality |
| S11_vs_k141_46119\|\|full | Low-quality |
| S11_vs_k141_13475\|\|full | Not-determined |
| S11_vs_k141_85300\|\|full | Not-determined |
| S11_vs_k141_816\|\|full | Not-determined |
| S11_vs_k141_825\|\|full | Low-quality |
| S11_vs_k141_13482\|\|full | Low-quality |
| S11_vs_k141_65651\|\|full | Low-quality |
| S11_vs_k141_832\|\|full | Low-quality |
| S11_vs_k141_33184\|\|full | Low-quality |
| S11_vs_k141_39705\|\|full | Low-quality |
| S11_vs_k141_13488\|\|full | Not-determined |
| S11_vs_k141_65657\|\|full | Low-quality |
| S11_vs_k141_46142\|\|full | Low-quality |
| S16_vs_127669\|\|full | High-quality |
| S11_vs_k141_46148\|\|full | Not-determined |
| S11_vs_k141_848\|\|full | Low-quality |
| S11_vs_k141_33198\|\|full | Low-quality |
| S11_vs_k141_78850\|\|full | Low-quality |
| S11_vs_k141_13505\|\|full | Low-quality |
| S11_vs_k141_13508\|\|full | Low-quality |
| S11_vs_k141_65688\|\|full | Not-determined |
| S11_vs_k141_39728\|\|full | Not-determined |
| S11_vs_k141_26714\|\|full | Low-quality |
| S11_vs_k141_39729\|\|full | Low-quality |
| S11_vs_k141_65695\|\|full | Not-determined |
| S11_vs_k141_72309\|\|full | Low-quality |
| S11_vs_k141_39741\|\|full | Low-quality |
| S11_vs_k141_65700\|\|full | Low-quality |
| S11_vs_k141_65704\|\|full | Low-quality |
| S11_vs_k141_39744\|\|full | Low-quality |
| S11_vs_k141_59044\|\|full | Low-quality |
| S11_vs_k141_13515\|\|full | Low-quality |
| S11_vs_k141_85353\|\|full | Low-quality |
| S11_vs_k141_91868\|\|full | Not-determined |
| S11_vs_k141_898\|\|full | Low-quality |
| S11_vs_k141_26737\|\|full | Low-quality |
| S11_vs_k141_98389\|\|full | Low-quality |
| S11_vs_k141_91871\|\|full | Not-determined |
| S11_vs_k141_78885\|\|full | Not-determined |
| S11_vs_k141_85368\|\|full | Low-quality |
| S11_vs_k141_78888\|\|full | Not-determined |
| S11_vs_k141_26763\|\|full | Low-quality |
| S11_vs_k141_59073\|\|full | Low-quality |
| S11_vs_k141_13548\|\|full | Low-quality |
| S11_vs_k141_13551\|\|full | Low-quality |
| S11_vs_k141_85378\|\|full | Low-quality |
| S11_vs_k141_20307\|\|full | Low-quality |
| S11_vs_k141_922\|\|full | Low-quality |
| S11_vs_k141_91886\|\|full | Low-quality |
| S11_vs_k141_52717\|\|full | Low-quality |
| S11_vs_k141_927\|\|full | Low-quality |
| S11_vs_k141_72365\|\|full | Low-quality |
| S16_vs_34021\|\|full | Medium-quality |
| S11_vs_k141_13554\|\|full | Low-quality |
| S11_vs_k141_85405\|\|full | Not-determined |
| S11_vs_k141_20321\|\|full | Not-determined |
| S11_vs_k141_85411\|\|full | Not-determined |
| S11_vs_k141_91918\|\|full | Low-quality |
| S11_vs_k141_46258\|\|full | Not-determined |
| S11_vs_k141_72381\|\|full | Low-quality |
| S11_vs_k141_20337\|\|full | Low-quality |
| S11_vs_k141_85428\|\|full | Not-determined |
| S11_vs_k141_20339\|\|full | Not-determined |
| S11_vs_k141_52747\|\|full | Low-quality |
| S11_vs_k141_91928\|\|full | Low-quality |
| S11_vs_k141_33272\|\|full | Low-quality |
| S11_vs_k141_85434\|\|full | Not-determined |
| S11_vs_k141_98470\|\|full | Low-quality |
| S11_vs_k141_39753\|\|full | Low-quality |
| S11_vs_k141_72398\|\|full | Not-determined |
| S11_vs_k141_78975\|\|full | Low-quality |
| S11_vs_k141_26821\|\|full | Not-determined |
| S11_vs_k141_39758\|\|full | Not-determined |
| S11_vs_k141_965\|\|full | Not-determined |
| S11_vs_k141_65758\|\|full | Low-quality |
| S11_vs_k141_52775\|\|full | Not-determined |
| S11_vs_k141_26824\|\|full | Not-determined |
| S11_vs_k141_91947\|\|full | Not-determined |
| S11_vs_k141_78980\|\|full | Low-quality |
| S11_vs_k141_33297\|\|full | Not-determined |
| S11_vs_k141_72422\|\|full | Low-quality |
| S11_vs_k141_52777\|\|full | Not-determined |
| S11_vs_k141_98490\|\|full | Low-quality |
| S11_vs_k141_65767\|\|full | Not-determined |
| S11_vs_k141_46307\|\|full | Not-determined |
| S11_vs_k141_13585\|\|full | Low-quality |
| S11_vs_k141_65772\|\|full | Low-quality |
| S11_vs_k141_7076\|\|full | Not-determined |
| S11_vs_k141_72447\|\|full | Low-quality |
| S11_vs_k141_26850\|\|full | Low-quality |
| S11_vs_k141_39795\|\|full | Low-quality |
| S11_vs_k141_85463\|\|full | Not-determined |
| S11_vs_k141_26854\|\|full | Not-determined |
| S11_vs_k141_52789\|\|full | Not-determined |
| S11_vs_k141_72450\|\|full | Not-determined |
| S11_vs_k141_79013\|\|full | Not-determined |
| S11_vs_k141_98501\|\|full | Low-quality |
| S11_vs_k141_98506\|\|full | Low-quality |
| S11_vs_k141_79021\|\|full | Not-determined |
| S11_vs_k141_26868\|\|full | Low-quality |
| S11_vs_k141_52804\|\|full | Not-determined |
| S11_vs_k141_65787\|\|full | Low-quality |
| S11_vs_k141_65788\|\|full | Low-quality |
| S11_vs_k141_20383\|\|full | Low-quality |
| S11_vs_k141_20384\|\|full | Low-quality |
| S11_vs_k141_1000\|\|full | Low-quality |
| S11_vs_k141_85502\|\|full | Not-determined |
| S11_vs_k141_65792\|\|full | Low-quality |
| S11_vs_k141_98521\|\|full | Not-determined |
| S11_vs_k141_52820\|\|full | Low-quality |
| S11_vs_k141_39833\|\|full | Low-quality |
| S11_vs_k141_20387\|\|full | Not-determined |
| S11_vs_k141_33345\|\|full | Low-quality |
| S11_vs_k141_1011\|\|full | Low-quality |
| S11_vs_k141_26897\|\|full | Not-determined |
| S11_vs_k141_72479\|\|full | Low-quality |
| S11_vs_k141_39843\|\|full | Low-quality |
| S11_vs_k141_39845\|\|full | Low-quality |
| S11_vs_k141_92028\|\|full | Low-quality |
| S11_vs_k141_26916\|\|full | Low-quality |
| S11_vs_k141_39848\|\|full | Not-determined |
| S11_vs_k141_46352\|\|full | Not-determined |
| S11_vs_k141_33350\|\|full | Low-quality |
| S11_vs_k141_65830\|\|full | Not-determined |
| S11_vs_k141_59206\|\|full | Low-quality |
| S11_vs_k141_46373\|\|full | Not-determined |
| S11_vs_k141_65836\|\|full | Low-quality |
| S11_vs_k141_79073\|\|full | Not-determined |
| S11_vs_k141_92046\|\|full | Low-quality |
| S11_vs_k141_52845\|\|full | Low-quality |
| S11_vs_k141_65844\|\|full | Low-quality |
| S8_extracted_k141_147128 | High-quality |
| S11_vs_k141_79084\|\|full | Low-quality |
| S11_vs_k141_52860\|\|full | Not-determined |
| S11_vs_k141_39872\|\|full | Not-determined |
| S11_vs_k141_79090\|\|full | Not-determined |
| S11_vs_k141_52862\|\|full | Not-determined |
| S11_vs_k141_59235\|\|full | Not-determined |
| S11_vs_k141_20449\|\|full | Not-determined |
| S11_vs_k141_20453\|\|full | Low-quality |
| S11_vs_k141_65858\|\|full | Low-quality |
| S11_vs_k141_33396\|\|full | Low-quality |
| S11_vs_k141_52875\|\|full | Not-determined |
| S11_vs_k141_85559\|\|full | Low-quality |
| S11_vs_k141_59245\|\|full | Not-determined |
| S11_vs_k141_7173\|\|full | Low-quality |
| S11_vs_k141_52890\|\|full | Not-determined |
| S11_vs_k141_46421\|\|full | Not-determined |
| S11_vs_k141_7178\|\|full | Low-quality |
| S11_vs_k141_52907\|\|full | Not-determined |
| S11_vs_k141_72554\|\|full | Not-determined |
| S11_vs_k141_1095\|\|full | Low-quality |
| S11_vs_k141_7188\|\|full | Not-determined |
| S11_vs_k141_59277\|\|full | Low-quality |
| S11_vs_k141_20499\|\|full | Low-quality |
| S11_vs_k141_20500\|\|full | Low-quality |
| S11_vs_k141_39898\|\|full | Low-quality |
| S11_vs_k141_52920\|\|full | Not-determined |
| S11_vs_k141_39910\|\|full | Low-quality |
| S11_vs_k141_7207\|\|full | Low-quality |
| S11_vs_k141_98634\|\|full | Not-determined |
| S11_vs_k141_46426\|\|full | Low-quality |
| S11_vs_k141_1117\|\|full | Low-quality |
| S11_vs_k141_52941\|\|full | Low-quality |
| S11_vs_k141_92115\|\|full | Not-determined |
| S11_vs_k141_72569\|\|full | Not-determined |
| S11_vs_k141_92117\|\|full | Low-quality |
| S11_vs_k141_46432\|\|full | Low-quality |
| S11_vs_k141_59294\|\|full | Low-quality |
| S11_vs_k141_7223\|\|full | Not-determined |
| S11_vs_k141_79126\|\|full | Low-quality |
| S11_vs_k141_46442\|\|full | Not-determined |
| S11_vs_k141_39929\|\|full | Low-quality |
| S11_vs_k141_7233\|\|full | Low-quality |
| S11_vs_k141_79129\|\|full | Low-quality |
| S11_vs_k141_92124\|\|full | Not-determined |
| S11_vs_k141_59318\|\|full | Low-quality |
| S11_vs_k141_46452\|\|full | Not-determined |
| S11_vs_k141_79135\|\|full | Low-quality |
| S11_vs_k141_98673\|\|full | Low-quality |
| S11_vs_k141_79141\|\|full | Low-quality |
| S11_vs_k141_7248\|\|full | Not-determined |
| S11_vs_k141_7257\|\|full | Low-quality |
| S11_vs_k141_85653\|\|full | Not-determined |
| S11_vs_k141_52974\|\|full | Low-quality |
| S11_vs_k141_79168\|\|full | Low-quality |
| S11_vs_k141_72602\|\|full | Low-quality |
| S11_vs_k141_72607\|\|full | Low-quality |
| S11_vs_k141_65928\|\|full | Not-determined |
| S11_vs_k141_52995\|\|full | Not-determined |
| S11_vs_k141_1176\|\|full | Low-quality |
| S11_vs_k141_85673\|\|full | Low-quality |
| S11_vs_k141_39961\|\|full | Not-determined |
| S11_vs_k141_53001\|\|full | Low-quality |
| S11_vs_k141_98695\|\|full | Low-quality |
| S11_vs_k141_72617\|\|full | Not-determined |
| S11_vs_k141_98712\|\|full | Not-determined |
| S8_vs_k141_147128\|\|full | High-quality |
| S11_vs_k141_72635\|\|full | Low-quality |
| S11_vs_k141_79223\|\|full | Low-quality |
| S11_vs_k141_72636\|\|full | Not-determined |
| S11_vs_k141_26945\|\|full | Low-quality |
| S11_vs_k141_53025\|\|full | Low-quality |
| S11_vs_k141_72639\|\|full | Not-determined |
| S11_vs_k141_1207\|\|full | Low-quality |
| S11_vs_k141_79235\|\|full | Low-quality |
| S11_vs_k141_98726\|\|full | Low-quality |
| S11_vs_k141_33470\|\|full | Low-quality |
| S11_vs_k141_7320\|\|full | Not-determined |
| S11_vs_k141_33476\|\|full | Not-determined |
| S11_vs_k141_39994\|\|full | Low-quality |
| S11_vs_k141_33482\|\|full | Low-quality |
| S11_vs_k141_39998\|\|full | Low-quality |
| S11_vs_k141_46510\|\|full | Low-quality |
| S11_vs_k141_40002\|\|full | Low-quality |
| S11_vs_k141_65990\|\|full | Low-quality |
| S11_vs_k141_1212\|\|full | Low-quality |
| S11_vs_k141_53039\|\|full | Low-quality |
| S11_vs_k141_20580\|\|full | Low-quality |
| S11_vs_k141_72671\|\|full | Low-quality |
| S11_vs_k141_1232\|\|full | Low-quality |
| S11_vs_k141_65995\|\|full | Low-quality |
| S11_vs_k141_7346\|\|full | Low-quality |
| S11_vs_k141_72679\|\|full | Low-quality |
| S11_vs_k141_20594\|\|full | Low-quality |
| S11_vs_k141_79240\|\|full | Low-quality |
| S11_vs_k141_85737\|\|full | Low-quality |

| | |
|---|---|
| S11_vs_k141_20600\|\|full | Low-quality |
| S11_vs_k141_92188\|\|full | Not-determined |
| S11_vs_k141_79242\|\|full | Not-determined |
| S11_vs_k141_7363\|\|full | Low-quality |
| S11_vs_k141_79250\|\|full | Low-quality |
| S11_vs_k141_72692\|\|full | Low-quality |
| S11_vs_k141_46538\|\|full | Low-quality |
| S11_vs_k141_13608\|\|full | Not-determined |
| S11_vs_k141_40038\|\|full | Not-determined |
| S11_vs_k141_46549\|\|full | Low-quality |
| S11_vs_k141_40039\|\|full | Low-quality |
| S11_vs_k141_33530\|\|full | Not-determined |
| S11_vs_k141_59395\|\|full | Low-quality |
| S11_vs_k141_72718\|\|full | Not-determined |
| S11_vs_k141_92210\|\|full | Low-quality |
| S11_vs_k141_72719\|\|full | Low-quality |
| S11_vs_k141_13636\|\|full | Low-quality |
| S11_vs_k141_66035\|\|full | Not-determined |
| S11_vs_k141_26997\|\|full | Low-quality |
| S11_vs_k141_33543\|\|full | Low-quality |
| S11_vs_k141_1249\|\|full | Not-determined |
| S11_vs_k141_40070\|\|full | Not-determined |
| S11_vs_k141_59424\|\|full | Low-quality |
| S11_vs_k141_72737\|\|full | Low-quality |
| S11_vs_k141_66046\|\|full | Not-determined |
| S11_vs_k141_85787\|\|full | Low-quality |
| S11_vs_k141_72738\|\|full | Not-determined |
| S11_vs_k141_27020\|\|full | Not-determined |
| S11_vs_k141_20627\|\|full | Low-quality |
| S11_vs_k141_85799\|\|full | Low-quality |
| S11_vs_k141_27030\|\|full | Low-quality |
| S10_extracted_46282 | High-quality |
| S6_vs_k141_40557\|\|full | High-quality |
| S11_vs_k141_59445\|\|full | Low-quality |
| S11_vs_k141_33572\|\|full | Not-determined |
| S11_vs_k141_72764\|\|full | Low-quality |
| S11_vs_k141_27045\|\|full | Low-quality |
| S11_vs_k141_1269\|\|full | Low-quality |
| S11_vs_k141_27052\|\|full | Not-determined |
| S11_vs_k141_92230\|\|full | Low-quality |
| S11_vs_k141_1270\|\|full | Not-determined |
| S11_vs_k141_72783\|\|full | Low-quality |
| S11_vs_k141_27068\|\|full | Low-quality |
| S11_vs_k141_66093\|\|full | Low-quality |
| S11_vs_k141_59465\|\|full | Low-quality |
| S11_vs_k141_79315\|\|full | Not-determined |
| S11_vs_k141_33606\|\|full | Low-quality |
| S11_vs_k141_20687\|\|full | Not-determined |
| S11_vs_k141_85812\|\|full | Low-quality |
| S11_vs_k141_1280\|\|full | Low-quality |
| S11_vs_k141_53195\|\|full | Low-quality |
| S11_vs_k141_33623\|\|full | Low-quality |
| S11_vs_k141_53197\|\|full | Low-quality |
| S11_vs_k141_1282\|\|full | Low-quality |
| S11_vs_k141_1288\|\|full | Low-quality |
| S11_vs_k141_53204\|\|full | Low-quality |
| S11_vs_k141_40119\|\|full | Not-determined |
| S11_vs_k141_27084\|\|full | Low-quality |
| S11_vs_k141_72831\|\|full | Low-quality |
| S11_vs_k141_1291\|\|full | Not-determined |
| S11_vs_k141_85835\|\|full | Low-quality |
| S5_vs_k141_62515\|\|full | High-quality |
| S11_vs_k141_53210\|\|full | Low-quality |
| S11_vs_k141_46607\|\|full | Low-quality |
| S11_vs_k141_13693\|\|full | Not-determined |
| S11_vs_k141_59515\|\|full | Low-quality |
| S11_vs_k141_98793\|\|full | Not-determined |
| S11_vs_k141_85868\|\|full | Low-quality |
| S11_vs_k141_98796\|\|full | Low-quality |
| S11_vs_k141_27099\|\|full | Low-quality |
| S11_vs_k141_13721\|\|full | Not-determined |
| S11_vs_k141_7503\|\|full | Not-determined |
| S11_vs_k141_53263\|\|full | Low-quality |
| S11_vs_k141_85899\|\|full | Low-quality |
| S11_vs_k141_27126\|\|full | Low-quality |
| S11_vs_k141_59547\|\|full | Low-quality |
| S11_vs_k141_7511\|\|full | Not-determined |
| S11_vs_k141_85908\|\|full | Not-determined |
| S11_vs_k141_85911\|\|full | Low-quality |
| S11_vs_k141_27136\|\|full | Not-determined |
| S11_vs_k141_46646\|\|full | Low-quality |
| S11_vs_k141_20731\|\|full | Low-quality |
| S11_vs_k141_92254\|\|full | Low-quality |
| S11_vs_k141_98832\|\|full | Low-quality |
| S11_vs_k141_72898\|\|full | Low-quality |
| S11_vs_k141_1306\|\|full | Low-quality |
| S11_vs_k141_59584\|\|full | Low-quality |
| S11_vs_k141_13763\|\|full | Low-quality |
| S11_vs_k141_13766\|\|full | Low-quality |
| S11_vs_k141_92270\|\|full | Low-quality |
| S11_vs_k141_33677\|\|full | Low-quality |
| S11_vs_k141_13767\|\|full | Not-determined |
| S11_vs_k141_59592\|\|full | Low-quality |
| S11_vs_k141_85951\|\|full | Low-quality |
| S11_vs_k141_33683\|\|full | Low-quality |
| S11_vs_k141_92274\|\|full | Low-quality |
| S11_vs_k141_53284\|\|full | Low-quality |
| S11_vs_k141_46681\|\|full | Low-quality |
| S11_vs_k141_1334\|\|full | Low-quality |
| S11_vs_k141_7564\|\|full | Low-quality |
| S11_vs_k141_46694\|\|full | Low-quality |
| S11_vs_k141_20779\|\|full | Low-quality |
| S11_vs_k141_53311\|\|full | Not-determined |
| S11_vs_k141_33735\|\|full | Low-quality |
| S11_vs_k141_1385\|\|full | Low-quality |
| S11_vs_k141_20790\|\|full | Low-quality |
| S11_vs_k141_85982\|\|full | Low-quality |
| S11_vs_k141_1393\|\|full | Not-determined |
| S11_vs_k141_7605\|\|full | Low-quality |
| S11_vs_k141_1394\|\|full | Low-quality |
| S11_vs_k141_92299\|\|full | Low-quality |
| S11_vs_k141_59667\|\|full | Not-determined |
| S11_vs_k141_85991\|\|full | Not-determined |
| S11_vs_k141_59675\|\|full | Not-determined |
| S3_vs_k141_68031\|\|full | Medium-quality |
| S11_vs_k141_92310\|\|full | Low-quality |
| S11_vs_k141_33736\|\|full | Low-quality |
| S11_vs_k141_13873\|\|full | Low-quality |
| S11_vs_k141_40167\|\|full | Low-quality |
| S11_vs_k141_59693\|\|full | Not-determined |
| S11_vs_k141_59695\|\|full | Not-determined |
| S11_vs_k141_40183\|\|full | Low-quality |
| S11_vs_k141_13896\|\|full | Low-quality |
| S11_vs_k141_1445\|\|full | Low-quality |
| S11_vs_k141_98885\|\|full | Low-quality |
| S11_vs_k141_27207\|\|full | Low-quality |
| S11_vs_k141_73000\|\|full | Not-determined |
| S11_vs_k141_92349\|\|full | Low-quality |
| S11_vs_k141_53380\|\|full | Low-quality |
| S11_vs_k141_59724\|\|full | Low-quality |
| S11_vs_k141_79425\|\|full | Low-quality |
| S11_vs_k141_7662\|\|full | Low-quality |
| S11_vs_k141_98899\|\|full | Low-quality |
| S11_vs_k141_53391\|\|full | Low-quality |
| S11_vs_k141_40201\|\|full | Low-quality |
| S11_vs_k141_20858\|\|full | Low-quality |
| S11_vs_k141_79435\|\|full | Low-quality |
| S11_vs_k141_66204\|\|full | Low-quality |
| S11_vs_k141_13922\|\|full | Low-quality |
| S11_vs_k141_53413\|\|full | Low-quality |
| S11_vs_k141_7693\|\|full | Low-quality |
| S11_vs_k141_1493\|\|full | Not-determined |
| S11_vs_k141_73046\|\|full | Low-quality |
| S11_vs_k141_20891\|\|full | Not-determined |
| S11_vs_k141_66225\|\|full | Not-determined |
| S11_vs_k141_53425\|\|full | Not-determined |
| S11_vs_k141_20896\|\|full | Not-determined |
| S11_vs_k141_27260\|\|full | Low-quality |
| S11_vs_k141_1506\|\|full | Low-quality |
| S11_vs_k141_66230\|\|full | Not-determined |
| S11_vs_k141_73063\|\|full | Low-quality |
| S11_vs_k141_1511\|\|full | Not-determined |
| S11_vs_k141_7711\|\|full | Low-quality |
| S11_vs_k141_59745\|\|full | Low-quality |
| S11_vs_k141_59749\|\|full | Not-determined |
| S11_vs_k141_13930\|\|full | Low-quality |
| S11_vs_k141_92402\|\|full | Not-determined |
| S11_vs_k141_1522\|\|full | Low-quality |
| S11_vs_k141_7724\|\|full | Low-quality |
| S11_vs_k141_20921\|\|full | Low-quality |
| S11_vs_k141_66248\|\|full | Not-determined |
| S11_vs_k141_46742\|\|full | Not-determined |
| S11_vs_k141_59769\|\|full | Low-quality |
| S11_vs_k141_79453\|\|full | Low-quality |
| S11_vs_k141_59771\|\|full | Not-determined |
| S11_vs_k141_7744\|\|full | Low-quality |
| S11_vs_k141_20941\|\|full | Low-quality |
| S11_vs_k141_86115\|\|full | Low-quality |
| S11_vs_k141_66266\|\|full | Low-quality |
| S11_vs_k141_13962\|\|full | Low-quality |
| S11_vs_k141_7763\|\|full | Low-quality |
| S11_vs_k141_86123\|\|full | Not-determined |
| S11_vs_k141_98996\|\|full | Not-determined |
| S11_vs_k141_27313\|\|full | Not-determined |
| S11_vs_k141_86138\|\|full | Low-quality |
| S11_vs_k141_46754\|\|full | Not-determined |
| S11_vs_k141_79482\|\|full | Not-determined |
| S11_vs_k141_46770\|\|full | Not-determined |
| S11_vs_k141_79492\|\|full | Low-quality |
| S11_vs_k141_59809\|\|full | Low-quality |
| S11_vs_k141_13996\|\|full | Not-determined |
| S11_vs_k141_7804\|\|full | Low-quality |
| S11_vs_k141_86165\|\|full | Low-quality |
| S11_vs_k141_59813\|\|full | Not-determined |
| S11_vs_k141_14004\|\|full | Low-quality |
| S11_vs_k141_79505\|\|full | Low-quality |
| S11_vs_k141_86174\|\|full | Low-quality |
| S11_vs_k141_99037\|\|full | Not-determined |
| S11_vs_k141_73172\|\|full | Not-determined |
| S11_vs_k141_66322\|\|full | Low-quality |
| S11_vs_k141_66326\|\|full | Not-determined |
| S11_vs_k141_20987\|\|full | Low-quality |
| S11_vs_k141_7825\|\|full | Low-quality |
| S11_vs_k141_79518\|\|full | Not-determined |
| S11_vs_k141_7828\|\|full | Low-quality |
| S11_vs_k141_99042\|\|full | Low-quality |
| S11_vs_k141_79533\|\|full | Low-quality |
| S11_vs_k141_14040\|\|full | Not-determined |
| S11_vs_k141_40263\|\|full | Low-quality |
| S11_vs_k141_86204\|\|full | Not-determined |
| S11_vs_k141_27373\|\|full | Not-determined |
| S11_vs_k141_79540\|\|full | Low-quality |
| S11_vs_k141_86209\|\|full | Low-quality |
| S11_vs_k141_66338\|\|full | Not-determined |
| S11_vs_k141_79545\|\|full | Not-determined |
| S11_vs_k141_79556\|\|full | Low-quality |
| S11_vs_k141_27387\|\|full | Low-quality |
| S11_vs_k141_27388\|\|full | Not-determined |
| S11_vs_k141_40300\|\|full | Low-quality |
| S11_vs_k141_27393\|\|full | Low-quality |
| S11_vs_k141_66353\|\|full | Low-quality |
| S11_vs_k141_40302\|\|full | Low-quality |
| S11_vs_k141_66354\|\|full | Not-determined |
| S11_vs_k141_14082\|\|full | Low-quality |
| S11_vs_k141_7860\|\|full | Low-quality |
| S11_vs_k141_27397\|\|full | Not-determined |
| S11_vs_k141_1680\|\|full | Low-quality |
| S11_vs_k141_86230\|\|full | Low-quality |
| S11_vs_k141_7863\|\|full | Low-quality |
| S11_vs_k141_66373\|\|full | Not-determined |
| S11_vs_k141_73187\|\|full | Not-determined |
| S11_vs_k141_73189\|\|full | Low-quality |
| S11_vs_k141_40328\|\|full | Low-quality |
| S11_vs_k141_73195\|\|full | Low-quality |
| S11_vs_k141_27404\|\|full | Not-determined |
| S11_vs_k141_7871\|\|full | Low-quality |
| S11_vs_k141_21037\|\|full | Low-quality |
| S11_vs_k141_79581\|\|full | Not-determined |
| S11_vs_k141_46820\|\|full | Low-quality |
| S11_vs_k141_21043\|\|full | Low-quality |
| S11_vs_k141_73206\|\|full | Low-quality |
| S11_vs_k141_27415\|\|full | Low-quality |
| S11_vs_k141_33843\|\|full | Low-quality |
| S11_vs_k141_27420\|\|full | Low-quality |
| S11_vs_k141_86291\|\|full | Low-quality |
| S11_vs_k141_1692\|\|full | Low-quality |
| S11_vs_k141_86309\|\|full | Low-quality |
| S11_vs_k141_53567\|\|full | Not-determined |
| S11_vs_k141_86314\|\|full | Not-determined |
| S11_vs_k141_46825\|\|full | Low-quality |
| S11_vs_k141_92448\|\|full | Not-determined |
| S11_vs_k141_66378\|\|full | Low-quality |
| S11_vs_k141_92451\|\|full | Not-determined |
| S11_vs_k141_27455\|\|full | Low-quality |
| S11_vs_k141_21063\|\|full | Low-quality |
| S11_vs_k141_40372\|\|full | Low-quality |
| S11_vs_k141_86317\|\|full | Not-determined |
| S11_vs_k141_99074\|\|full | Low-quality |
| S11_vs_k141_79602\|\|full | Low-quality |
| S11_vs_k141_59830\|\|full | Low-quality |
| S11_vs_k141_21073\|\|full | Low-quality |
| S11_vs_k141_40378\|\|full | Low-quality |
| S11_vs_k141_73247\|\|full | Not-determined |
| S11_vs_k141_46860\|\|full | Low-quality |
| S11_vs_k141_92463\|\|full | Low-quality |
| S11_vs_k141_14160\|\|full | Low-quality |
| S11_vs_k141_1722\|\|full | Low-quality |
| S11_vs_k141_53591\|\|full | Not-determined |
| S11_vs_k141_40397\|\|full | Not-determined |
| S11_vs_k141_1730\|\|full | Low-quality |
| S11_vs_k141_1731\|\|full | Not-determined |
| S11_vs_k141_92472\|\|full | Low-quality |
| S11_vs_k141_99081\|\|full | Low-quality |
| S11_vs_k141_1739\|\|full | Low-quality |
| S11_vs_k141_99088\|\|full | Not-determined |
| S11_vs_k141_79630\|\|full | Low-quality |
| S11_vs_k141_21111\|\|full | Low-quality |
| S11_vs_k141_1751\|\|full | Not-determined |
| S11_vs_k141_40421\|\|full | Not-determined |
| S11_vs_k141_79643\|\|full | Not-determined |
| S11_vs_k141_92492\|\|full | Low-quality |
| S11_vs_k141_66409\|\|full | Not-determined |
| S11_vs_k141_40431\|\|full | Low-quality |
| S11_vs_k141_66411\|\|full | Not-determined |
| S11_vs_k141_14202\|\|full | Not-determined |
| S11_vs_k141_7972\|\|full | Low-quality |
| S11_vs_k141_7973\|\|full | Low-quality |
| S11_vs_k141_53616\|\|full | Low-quality |
| S11_vs_k141_7981\|\|full | Not-determined |
| S11_vs_k141_21120\|\|full | Not-determined |
| S11_vs_k141_14216\|\|full | Low-quality |
| S11_vs_k141_79668\|\|full | Low-quality |
| S11_vs_k141_8001\|\|full | Low-quality |
| S11_vs_k141_79681\|\|full | Low-quality |
| S11_vs_k141_8004\|\|full | Low-quality |
| S11_vs_k141_73308\|\|full | Low-quality |
| S11_vs_k141_46935\|\|full | Low-quality |
| S11_vs_k141_14236\|\|full | Low-quality |
| S11_vs_k141_46940\|\|full | Not-determined |
| S11_vs_k141_73316\|\|full | Not-determined |
| S11_vs_k141_79700\|\|full | Low-quality |
| S11_vs_k141_86407\|\|full | Low-quality |
| S11_vs_k141_99114\|\|full | Low-quality |
| S11_vs_k141_99116\|\|full | Low-quality |
| S11_vs_k141_99117\|\|full | Low-quality |
| S11_vs_k141_27602\|\|full | Low-quality |
| S11_vs_k141_79712\|\|full | Not-determined |
| S11_vs_k141_8015\|\|full | Low-quality |
| S11_vs_k141_79729\|\|full | Low-quality |
| S11_vs_k141_1840\|\|full | Not-determined |
| S11_vs_k141_79732\|\|full | Low-quality |
| S11_vs_k141_14280\|\|full | Low-quality |
| S11_vs_k141_73351\|\|full | Low-quality |
| S11_vs_k141_21189\|\|full | Not-determined |
| S11_vs_k141_27638\|\|full | Low-quality |
| S11_vs_k141_21200\|\|full | Not-determined |
| S11_vs_k141_99152\|\|full | Not-determined |
| S11_vs_k141_8051\|\|full | Low-quality |
| S11_vs_k141_79756\|\|full | Not-determined |
| S11_vs_k141_8069\|\|full | Not-determined |
| S11_vs_k141_73374\|\|full | Low-quality |
| S11_vs_k141_1851\|\|full | Low-quality |
| S11_vs_k141_21237\|\|full | Low-quality |
| S11_vs_k141_59955\|\|full | Low-quality |
| S11_vs_k141_8098\|\|full | Low-quality |
| S11_vs_k141_66516\|\|full | Low-quality |
| S11_vs_k141_79790\|\|full | Low-quality |
| S11_vs_k141_92522\|\|full | Low-quality |
| S11_vs_k141_59968\|\|full | Low-quality |
| S11_vs_k141_14349\|\|full | Low-quality |
| S11_vs_k141_21255\|\|full | Low-quality |
| S11_vs_k141_79811\|\|full | Not-determined |
| S11_vs_k141_27705\|\|full | Not-determined |
| S11_vs_k141_14357\|\|full | Not-determined |
| S11_vs_k141_1899\|\|full | Not-determined |
| S11_vs_k141_21258\|\|full | Low-quality |
| S11_vs_k141_59977\|\|full | Low-quality |
| S11_vs_k141_8141\|\|full | Low-quality |
| S12_vs_k141_142625\|\|full | Complete |
| S11_vs_k141_8145\|\|full | Low-quality |
| S11_vs_k141_27716\|\|full | Not-determined |
| S11_vs_k141_27717\|\|full | Not-determined |
| S11_vs_k141_14383\|\|full | Not-determined |
| S11_vs_k141_21281\|\|full | Not-determined |
| S10_vs_46282\|\|full | High-quality |
| S11_vs_k141_21289\|\|full | Low-quality |
| S11_vs_k141_8170\|\|full | Low-quality |
| S11_vs_k141_47069\|\|full | Low-quality |
| S11_vs_k141_53702\|\|full | Not-determined |
| S11_vs_k141_66539\|\|full | Low-quality |
| S11_vs_k141_99225\|\|full | Low-quality |
| S11_vs_k141_53711\|\|full | Low-quality |
| S11_vs_k141_92574\|\|full | Low-quality |
| S11_vs_k141_14404\|\|full | Low-quality |
| S11_vs_k141_40461\|\|full | Low-quality |
| S11_vs_k141_60028\|\|full | Low-quality |
| S11_vs_k141_27751\|\|full | Not-determined |
| S11_vs_k141_40471\|\|full | Not-determined |
| S11_vs_k141_66551\|\|full | Low-quality |
| S11_vs_k141_99242\|\|full | Low-quality |
| S11_vs_k141_60051\|\|full | Not-determined |
| S11_vs_k141_14445\|\|full | Low-quality |
| S11_vs_k141_99246\|\|full | Low-quality |
| S11_vs_k141_66572\|\|full | Low-quality |
| S11_vs_k141_99247\|\|full | Low-quality |
| S11_vs_k141_53755\|\|full | Low-quality |
| S11_vs_k141_99248\|\|full | Low-quality |
| S11_vs_k141_47117\|\|full | Low-quality |
| S11_vs_k141_53762\|\|full | Low-quality |
| S11_vs_k141_40511\|\|full | Not-determined |
| S11_vs_k141_66583\|\|full | Low-quality |
| S11_vs_k141_92635\|\|full | Low-quality |
| S11_vs_k141_47130\|\|full | Low-quality |
| S11_vs_k141_47131\|\|full | Not-determined |
| S11_vs_k141_53780\|\|full | Low-quality |
| S7_extracted_84974 | Medium-quality |
| S11_vs_k141_8230\|\|full | Low-quality |
| S7_vs_84974\|\|full | Medium-quality |
| S11_vs_k141_60090\|\|full | Low-quality |
| S11_vs_k141_14487\|\|full | Low-quality |
| S11_vs_k141_92655\|\|full | Low-quality |
| S11_vs_k141_14493\|\|full | Low-quality |
| S11_vs_k141_53793\|\|full | Low-quality |
| S11_vs_k141_1942\|\|full | Low-quality |
| S11_vs_k141_27836\|\|full | Low-quality |
| S11_vs_k141_66637\|\|full | Not-determined |
| S11_vs_k141_21394\|\|full | Not-determined |
| S11_vs_k141_1953\|\|full | Low-quality |
| S11_vs_k141_8243\|\|full | Low-quality |
| S11_vs_k141_73476\|\|full | Low-quality |
| S11_vs_k141_53818\|\|full | Not-determined |
| S11_vs_k141_66652\|\|full | Not-determined |
| S11_vs_k141_14519\|\|full | Low-quality |
| S11_vs_k141_14523\|\|full | Low-quality |
| S11_vs_k141_1963\|\|full | Low-quality |
| S11_vs_k141_40568\|\|full | Not-determined |
| S11_vs_k141_53829\|\|full | Low-quality |
| S11_vs_k141_21408\|\|full | Low-quality |
| S11_vs_k141_60106\|\|full | Not-determined |
| S11_vs_k141_21412\|\|full | Low-quality |
| S11_vs_k141_1973\|\|full | Low-quality |
| S11_vs_k141_1975\|\|full | Low-quality |
| S11_vs_k141_86468\|\|full | Not-determined |
| S11_vs_k141_53846\|\|full | Low-quality |
| S11_vs_k141_40580\|\|full | Low-quality |
| S11_vs_k141_1976\|\|full | Not-determined |
| S11_vs_k141_79853\|\|full | Low-quality |
| S11_vs_k141_73489\|\|full | Not-determined |
| S11_vs_k141_40590\|\|full | Low-quality |
| S11_vs_k141_73494\|\|full | Not-determined |
| S11_vs_k141_1987\|\|full | Low-quality |
| S11_vs_k141_60123\|\|full | Low-quality |
| S11_vs_k141_79860\|\|full | Not-determined |
| S11_vs_k141_8292\|\|full | Not-determined |
| S11_vs_k141_60132\|\|full | Low-quality |
| S11_vs_k141_21431\|\|full | Low-quality |
| S11_vs_k141_66673\|\|full | Not-determined |
| S11_vs_k141_86484\|\|full | Not-determined |
| S11_vs_k141_53861\|\|full | Low-quality |
| S11_vs_k141_40610\|\|full | Not-determined |
| S11_vs_k141_79867\|\|full | Low-quality |
| S11_vs_k141_86489\|\|full | Low-quality |
| S11_vs_k141_99330\|\|full | Not-determined |
| S11_vs_k141_53878\|\|full | Not-determined |
| S11_vs_k141_92722\|\|full | Not-determined |
| S11_vs_k141_21442\|\|full | Low-quality |
| S11_vs_k141_60142\|\|full | Low-quality |
| S11_vs_k141_47188\|\|full | Not-determined |
| S11_vs_k141_66678\|\|full | Low-quality |
| S11_vs_k141_2017\|\|full | Low-quality |
| S11_vs_k141_53886\|\|full | Not-determined |
| S11_vs_k141_73531\|\|full | Low-quality |
| S11_vs_k141_92738\|\|full | Low-quality |
| S7_extracted_37279 | Complete |
| S11_vs_k141_86517\|\|full | Low-quality |
| S11_vs_k141_21461\|\|full | Not-determined |
| S11_vs_k141_92740\|\|full | Not-determined |
| S11_vs_k141_33941\|\|full | Low-quality |
| S11_vs_k141_53903\|\|full | Low-quality |
| S11_vs_k141_2034\|\|full | Not-determined |
| S11_vs_k141_86527\|\|full | Low-quality |
| S11_vs_k141_92749\|\|full | Not-determined |
| S11_vs_k141_33950\|\|full | Not-determined |
| S11_vs_k141_79921\|\|full | Not-determined |
| S11_vs_k141_47215\|\|full | Not-determined |
| S11_vs_k141_33963\|\|full | Low-quality |
| S11_vs_k141_92769\|\|full | Not-determined |
| S11_vs_k141_86552\|\|full | Low-quality |
| S11_vs_k141_27875\|\|full | Low-quality |
| S11_vs_k141_73572\|\|full | Low-quality |
| S11_vs_k141_21511\|\|full | Low-quality |
| S11_vs_k141_73576\|\|full | Not-determined |
| S11_vs_k141_53931\|\|full | Not-determined |
| S11_vs_k141_79955\|\|full | Low-quality |
| S11_vs_k141_99398\|\|full | Low-quality |
| S11_vs_k141_86583\|\|full | Not-determined |
| S11_vs_k141_40648\|\|full | Low-quality |
| S11_vs_k141_8347\|\|full | Not-determined |
| S11_vs_k141_47250\|\|full | Low-quality |
| S11_vs_k141_53949\|\|full | Not-determined |
| S11_vs_k141_21540\|\|full | Low-quality |
| S11_vs_k141_86599\|\|full | Not-determined |
| S11_vs_k141_79976\|\|full | Low-quality |
| S11_vs_k141_21552\|\|full | Not-determined |
| S11_vs_k141_73616\|\|full | Low-quality |
| S11_vs_k141_53961\|\|full | Low-quality |
| S11_vs_k141_86617\|\|full | Not-determined |
| S11_vs_k141_21573\|\|full | Low-quality |
| S11_vs_k141_79993\|\|full | Low-quality |
| S11_vs_k141_86624\|\|full | Not-determined |
| S11_vs_k141_86626\|\|full | Not-determined |
| S11_vs_k141_80009\|\|full | Not-determined |
| S11_vs_k141_53996\|\|full | Low-quality |
| S11_vs_k141_8405\|\|full | Low-quality |
| S11_vs_k141_86634\|\|full | Low-quality |
| S11_vs_k141_54007\|\|full | Not-determined |
| S11_vs_k141_60165\|\|full | Low-quality |
| S11_vs_k141_8421\|\|full | Low-quality |
| S11_vs_k141_2136\|\|full | Not-determined |
| S11_vs_k141_86652\|\|full | Low-quality |
| S11_vs_k141_27891\|\|full | Low-quality |
| S11_vs_k141_2150\|\|full | Low-quality |
| S11_vs_k141_54044\|\|full | Low-quality |
| S11_vs_k141_2153\|\|full | Low-quality |
| S11_vs_k141_80040\|\|full | Low-quality |
| S11_vs_k141_2159\|\|full | Low-quality |
| S11_vs_k141_8454\|\|full | Low-quality |
| S11_vs_k141_73621\|\|full | Not-determined |
| S11_vs_k141_80047\|\|full | Low-quality |
| S11_vs_k141_80053\|\|full | Low-quality |
| S11_vs_k141_99495\|\|full | Low-quality |
| S11_vs_k141_86682\|\|full | Not-determined |
| S7_vs_37279\|\|full | High-quality |
| S11_vs_k141_80057\|\|full | Not-determined |
| S11_vs_k141_34010\|\|full | Low-quality |
| S11_vs_k141_99505\|\|full | Not-determined |
| S11_vs_k141_80061\|\|full | Low-quality |
| S11_vs_k141_92829\|\|full | Low-quality |
| S11_vs_k141_40762\|\|full | Not-determined |
| S11_vs_k141_92831\|\|full | Low-quality |
| S11_vs_k141_92833\|\|full | Low-quality |
| S11_vs_k141_40768\|\|full | Low-quality |
| S11_vs_k141_2200\|\|full | Not-determined |
| S11_vs_k141_92838\|\|full | Low-quality |
| S11_vs_k141_99531\|\|full | Low-quality |
| S11_vs_k141_92845\|\|full | Not-determined |
| S11_vs_k141_80092\|\|full | Low-quality |
| S11_vs_k141_54093\|\|full | Low-quality |
| S14_vs_k141_32499\|\|full | Complete |
| S11_vs_k141_86721\|\|full | Low-quality |
| S11_vs_k141_54096\|\|full | Not-determined |
| S11_vs_k141_34030\|\|full | Low-quality |
| S11_vs_k141_92865\|\|full | Low-quality |
| S11_vs_k141_14610\|\|full | Not-determined |
| S11_vs_k141_47286\|\|full | Low-quality |
| S11_vs_k141_54108\|\|full | Low-quality |
| S11_vs_k141_86736\|\|full | Not-determined |
| S11_vs_k141_86737\|\|full | Low-quality |
| S11_vs_k141_54113\|\|full | Low-quality |
| S11_vs_k141_34049\|\|full | Not-determined |
| S11_vs_k141_47288\|\|full | Low-quality |
| S11_vs_k141_14629\|\|full | Not-determined |
| S11_vs_k141_2226\|\|full | Not-determined |
| S11_vs_k141_21611\|\|full | Low-quality |
| S11_vs_k141_40846\|\|full | Low-quality |
| S11_vs_k141_8485\|\|full | Low-quality |
| S11_vs_k141_14655\|\|full | Not-determined |
| S11_vs_k141_80148\|\|full | Low-quality |
| S11_vs_k141_8498\|\|full | Not-determined |
| S11_vs_k141_34082\|\|full | Not-determined |
| S11_vs_k141_92917\|\|full | Not-determined |
| S11_vs_k141_73741\|\|full | Low-quality |
| S11_vs_k141_60236\|\|full | Low-quality |
| S11_vs_k141_40869\|\|full | Not-determined |
| S11_vs_k141_34100\|\|full | Not-determined |
| S11_vs_k141_86785\|\|full | Not-determined |
| S11_vs_k141_34107\|\|full | Low-quality |
| S11_vs_k141_14690\|\|full | Not-determined |
| S11_vs_k141_86792\|\|full | Not-determined |
| S11_vs_k141_21665\|\|full | Low-quality |
| S11_vs_k141_60242\|\|full | Low-quality |
| S11_vs_k141_47316\|\|full | Low-quality |
| S11_vs_k141_21672\|\|full | Low-quality |
| S3_vs_k141_6604\|\|full | Medium-quality |
| S11_vs_k141_73774\|\|full | Low-quality |
| S11_vs_k141_86827\|\|full | Low-quality |
| S11_vs_k141_86830\|\|full | Not-determined |
| S11_vs_k141_40916\|\|full | Not-determined |
| S11_vs_k141_14723\|\|full | Not-determined |
| S11_vs_k141_80157\|\|full | Low-quality |
| S11_vs_k141_86831\|\|full | Not-determined |
| S11_vs_k141_80158\|\|full | Not-determined |
| S11_vs_k141_80160\|\|full | Low-quality |
| S11_vs_k141_99590\|\|full | Low-quality |
| S11_vs_k141_92998\|\|full | Not-determined |
| S11_vs_k141_40939\|\|full | Not-determined |
| S11_vs_k141_86847\|\|full | Low-quality |
| S11_vs_k141_73798\|\|full | Low-quality |
| S11_vs_k141_66736\|\|full | Not-determined |
| S11_vs_k141_86849\|\|full | Not-determined |
| S11_vs_k141_93019\|\|full | Low-quality |
| S11_vs_k141_99619\|\|full | Not-determined |
| S11_vs_k141_60283\|\|full | Low-quality |
| S11_vs_k141_99620\|\|full | Low-quality |
| S11_vs_k141_60285\|\|full | Low-quality |
| S11_vs_k141_28038\|\|full | Low-quality |
| S11_vs_k141_21732\|\|full | Low-quality |
| S11_vs_k141_93031\|\|full | Low-quality |
| S11_vs_k141_14762\|\|full | Low-quality |
| S11_vs_k141_86870\|\|full | Low-quality |
| S11_vs_k141_54203\|\|full | Low-quality |
| S13_extracted_30221 | Medium-quality |
| S11_vs_k141_8584\|\|full | Low-quality |
| S11_vs_k141_99624\|\|full | Low-quality |
| S11_vs_k141_8588\|\|full | Low-quality |
| S11_vs_k141_86885\|\|full | Not-determined |
| S11_vs_k141_34149\|\|full | Low-quality |
| S11_vs_k141_99640\|\|full | Low-quality |
| S11_vs_k141_28050\|\|full | Low-quality |
| S11_vs_k141_8605\|\|full | Not-determined |
| S11_vs_k141_86895\|\|full | Low-quality |
| S11_vs_k141_14780\|\|full | Low-quality |
| S11_vs_k141_14783\|\|full | Not-determined |
| S11_vs_k141_54216\|\|full | Low-quality |
| S11_vs_k141_86909\|\|full | Low-quality |
| S11_vs_k141_34188\|\|full | Not-determined |
| S11_vs_k141_21747\|\|full | Low-quality |
| S11_vs_k141_60347\|\|full | Not-determined |
| S11_vs_k141_60351\|\|full | Low-quality |
| S11_vs_k141_34196\|\|full | Low-quality |
| S11_vs_k141_99697\|\|full | Low-quality |
| S11_vs_k141_34204\|\|full | Low-quality |
| S11_vs_k141_8633\|\|full | Not-determined |
| S11_vs_k141_40959\|\|full | Low-quality |
| S11_vs_k141_21765\|\|full | Not-determined |
| S11_vs_k141_60382\|\|full | Low-quality |
| S11_vs_k141_86938\|\|full | Not-determined |
| S11_vs_k141_2330\|\|full | Low-quality |
| S11_vs_k141_73858\|\|full | Not-determined |

| | |
|---|---|
| S11_vs_k141_21782\|\|full | Not-determined |
| S11_vs_k141_34218\|\|full | Low-quality |
| S11_vs_k141_47377\|\|full | Not-determined |
| S11_vs_k141_60404\|\|full | Low-quality |
| S11_vs_k141_60408\|\|full | Low-quality |
| S11_vs_k141_93100\|\|full | Low-quality |
| S11_vs_k141_47395\|\|full | Low-quality |
| S11_vs_k141_86979\|\|full | Low-quality |
| S11_vs_k141_86991\|\|full | Not-determined |
| S11_vs_k141_28075\|\|full | Not-determined |
| S11_vs_k141_73889\|\|full | Low-quality |
| S11_vs_k141_47406\|\|full | Low-quality |
| S11_vs_k141_28084\|\|full | Low-quality |
| S11_vs_k141_66837\|\|full | Not-determined |
| S11_vs_k141_73902\|\|full | Not-determined |
| S11_vs_k141_2394\|\|full | Not-determined |
| S11_vs_k141_28094\|\|full | Low-quality |
| S11_vs_k141_73917\|\|full | Low-quality |
| S11_vs_k141_2409\|\|full | Low-quality |
| S11_vs_k141_54260\|\|full | Low-quality |
| S11_vs_k141_28106\|\|full | Low-quality |
| S11_vs_k141_34230\|\|full | Low-quality |
| S11_vs_k141_28108\|\|full | Low-quality |
| S11_vs_k141_34231\|\|full | Low-quality |
| S11_vs_k141_2427\|\|full | Not-determined |
| S11_vs_k141_34245\|\|full | Low-quality |
| S11_vs_k141_60428\|\|full | Low-quality |
| S11_vs_k141_80303\|\|full | Low-quality |
| S11_vs_k141_41088\|\|full | Not-determined |
| S11_vs_k141_41091\|\|full | Low-quality |
| S11_vs_k141_87057\|\|full | Not-determined |
| S11_vs_k141_60435\|\|full | Low-quality |
| S11_vs_k141_54310\|\|full | Low-quality |
| S11_vs_k141_60438\|\|full | Not-determined |
| S11_vs_k141_87060\|\|full | Low-quality |
| S11_vs_k141_73951\|\|full | Low-quality |
| S11_vs_k141_93113\|\|full | Not-determined |
| S11_vs_k141_87066\|\|full | Low-quality |
| S11_vs_k141_21871\|\|full | Not-determined |
| S11_vs_k141_87074\|\|full | Low-quality |
| S11_vs_k141_87086\|\|full | Low-quality |
| S11_vs_k141_21883\|\|full | Low-quality |
| S11_vs_k141_93139\|\|full | Not-determined |
| S11_vs_k141_93141\|\|full | Low-quality |
| S13_vs_30221\|\|full | Medium-quality |
| S11_vs_k141_47433\|\|full | Not-determined |
| S11_vs_k141_21886\|\|full | Low-quality |
| S11_vs_k141_87095\|\|full | Low-quality |
| S11_vs_k141_34288\|\|full | Low-quality |
| S3_extracted_k141_127406 | Complete |
| S11_vs_k141_80357\|\|full | Low-quality |
| S11_vs_k141_34301\|\|full | Not-determined |
| S11_vs_k141_54364\|\|full | Not-determined |
| S11_vs_k141_54368\|\|full | Not-determined |
| S11_vs_k141_66869\|\|full | Low-quality |
| S11_vs_k141_74005\|\|full | Low-quality |
| S3_vs_k141_127406\|\|full | High-quality |
| S11_vs_k141_87127\|\|full | Low-quality |
| S11_vs_k141_8752\|\|full | Not-determined |
| S11_vs_k141_54381\|\|full | Not-determined |
| S11_vs_k141_99781\|\|full | Low-quality |
| S11_vs_k141_41129\|\|full | Not-determined |
| S11_vs_k141_34312\|\|full | Low-quality |
| S11_vs_k141_34313\|\|full | Not-determined |
| S11_vs_k141_34316\|\|full | Low-quality |
| S11_vs_k141_41131\|\|full | Not-determined |
| S11_vs_k141_80385\|\|full | Low-quality |
| S11_vs_k141_66897\|\|full | Low-quality |
| S11_vs_k141_21944\|\|full | Not-determined |
| S11_vs_k141_47459\|\|full | Low-quality |
| S11_vs_k141_87166\|\|full | Low-quality |
| S11_vs_k141_14923\|\|full | Low-quality |
| S11_vs_k141_66901\|\|full | Low-quality |
| S11_vs_k141_14926\|\|full | Low-quality |
| S11_vs_k141_14928\|\|full | Low-quality |
| S11_vs_k141_87172\|\|full | Low-quality |
| S11_vs_k141_99827\|\|full | Low-quality |
| S11_vs_k141_14933\|\|full | Not-determined |
| S11_vs_k141_99836\|\|full | Low-quality |
| S11_vs_k141_80399\|\|full | Not-determined |
| S11_vs_k141_28222\|\|full | Low-quality |
| S11_vs_k141_93204\|\|full | Not-determined |
| S11_vs_k141_41180\|\|full | Low-quality |
| S11_vs_k141_99845\|\|full | Not-determined |
| S11_vs_k141_41187\|\|full | Low-quality |
| S11_vs_k141_8837\|\|full | Low-quality |
| S11_vs_k141_60470\|\|full | Low-quality |
| S11_vs_k141_28238\|\|full | Low-quality |
| S11_vs_k141_47476\|\|full | Low-quality |
| S11_vs_k141_66926\|\|full | Not-determined |
| S11_vs_k141_93221\|\|full | Low-quality |
| S11_vs_k141_14987\|\|full | Not-determined |
| S11_vs_k141_93225\|\|full | Low-quality |
| S11_vs_k141_66930\|\|full | Low-quality |
| S11_vs_k141_80408\|\|full | Not-determined |
| S11_vs_k141_2553\|\|full | Not-determined |
| S11_vs_k141_41217\|\|full | Low-quality |
| S11_vs_k141_28250\|\|full | Low-quality |
| S11_vs_k141_34389\|\|full | Not-determined |
| S11_vs_k141_28254\|\|full | Low-quality |
| S11_vs_k141_14999\|\|full | Low-quality |
| S11_vs_k141_47490\|\|full | Not-determined |
| S11_vs_k141_47494\|\|full | Not-determined |
| S11_vs_k141_74087\|\|full | Low-quality |
| S11_vs_k141_60471\|\|full | Low-quality |
| S11_vs_k141_80417\|\|full | Not-determined |
| S11_vs_k141_47498\|\|full | Low-quality |
| S11_vs_k141_47499\|\|full | Not-determined |
| S11_vs_k141_2565\|\|full | Low-quality |
| S11_vs_k141_41243\|\|full | Low-quality |
| S11_vs_k141_28276\|\|full | Low-quality |
| S11_vs_k141_93269\|\|full | Not-determined |
| S11_vs_k141_2572\|\|full | Low-quality |
| S11_vs_k141_60476\|\|full | Low-quality |
| S11_vs_k141_66963\|\|full | Low-quality |
| S11_vs_k141_47536\|\|full | Low-quality |
| S11_vs_k141_8867\|\|full | Low-quality |
| S11_vs_k141_54498\|\|full | Low-quality |
| S11_vs_k141_80427\|\|full | Not-determined |
| S11_vs_k141_87228\|\|full | Not-determined |
| S11_vs_k141_41268\|\|full | Low-quality |
| S11_vs_k141_74115\|\|full | Low-quality |
| S11_vs_k141_66984\|\|full | Low-quality |
| S11_vs_k141_60506\|\|full | Low-quality |
| S11_vs_k141_47545\|\|full | Low-quality |
| S11_vs_k141_74118\|\|full | Low-quality |
| S11_vs_k141_15052\|\|full | Not-determined |
| S11_vs_k141_54503\|\|full | Low-quality |
| S11_vs_k141_2608\|\|full | Low-quality |
| S11_vs_k141_47559\|\|full | Low-quality |
| S11_vs_k141_87244\|\|full | Not-determined |
| S11_vs_k141_87246\|\|full | Low-quality |
| S11_vs_k141_2617\|\|full | Low-quality |
| S11_vs_k141_47561\|\|full | Not-determined |
| S11_vs_k141_93299\|\|full | Low-quality |
| S11_vs_k141_8884\|\|full | Not-determined |
| S11_vs_k141_74136\|\|full | Low-quality |
| S11_vs_k141_67015\|\|full | Not-determined |
| S11_vs_k141_8889\|\|full | Low-quality |
| S11_vs_k141_87260\|\|full | Low-quality |
| S11_vs_k141_99934\|\|full | Low-quality |
| S11_vs_k141_41299\|\|full | Low-quality |
| S11_vs_k141_60537\|\|full | Low-quality |
| S11_vs_k141_74151\|\|full | Low-quality |
| S11_vs_k141_80446\|\|full | Low-quality |
| S11_vs_k141_21984\|\|full | Low-quality |
| S11_vs_k141_74153\|\|full | Low-quality |
| S11_vs_k141_60540\|\|full | Not-determined |
| S11_vs_k141_54541\|\|full | Low-quality |
| S11_vs_k141_47579\|\|full | Low-quality |
| S11_vs_k141_2628\|\|full | Low-quality |
| S11_vs_k141_80460\|\|full | Not-determined |
| S11_vs_k141_41325\|\|full | Low-quality |
| S11_vs_k141_60551\|\|full | Low-quality |
| S11_vs_k141_99953\|\|full | Low-quality |
| S11_vs_k141_54563\|\|full | Not-determined |
| S11_vs_k141_15094\|\|full | Low-quality |
| S11_vs_k141_67052\|\|full | Not-determined |
| S11_vs_k141_93352\|\|full | Low-quality |
| S11_vs_k141_15108\|\|full | Low-quality |
| S11_vs_k141_93354\|\|full | Not-determined |
| S11_vs_k141_47609\|\|full | Low-quality |
| S11_vs_k141_8928\|\|full | Not-determined |
| S11_vs_k141_15112\|\|full | Low-quality |
| S11_vs_k141_74189\|\|full | Low-quality |
| S11_vs_k141_80473\|\|full | Low-quality |
| S11_vs_k141_34497\|\|full | Low-quality |
| S11_vs_k141_22001\|\|full | Not-determined |
| S11_vs_k141_34501\|\|full | Not-determined |
| S11_vs_k141_60579\|\|full | Low-quality |
| S11_vs_k141_93357\|\|full | Low-quality |
| S11_vs_k141_54594\|\|full | Low-quality |
| S11_vs_k141_54595\|\|full | Low-quality |
| S11_vs_k141_54596\|\|full | Low-quality |
| S11_vs_k141_22012\|\|full | Low-quality |
| S11_vs_k141_87334\|\|full | Low-quality |
| S11_vs_k141_22021\|\|full | Low-quality |
| S11_vs_k141_47628\|\|full | Not-determined |
| S11_vs_k141_87339\|\|full | Low-quality |
| S11_vs_k141_87340\|\|full | Low-quality |
| S11_vs_k141_80501\|\|full | Low-quality |
| S11_vs_k141_15150\|\|full | Low-quality |
| S11_vs_k141_8954\|\|full | Low-quality |
| S11_vs_k141_74226\|\|full | Low-quality |
| S11_vs_k141_8963\|\|full | Low-quality |
| S11_vs_k141_80508\|\|full | Not-determined |
| S11_vs_k141_87360\|\|full | Low-quality |
| S11_vs_k141_2645\|\|full | Low-quality |
| S11_vs_k141_15163\|\|full | Low-quality |
| S11_vs_k141_2653\|\|full | Low-quality |
| S11_vs_k141_15171\|\|full | Low-quality |
| S11_vs_k141_8974\|\|full | Low-quality |
| S11_vs_k141_100010\|\|full | Low-quality |
| S11_vs_k141_87375\|\|full | Low-quality |
| S11_vs_k141_41393\|\|full | Low-quality |
| S11_vs_k141_54639\|\|full | Not-determined |
| S11_vs_k141_8986\|\|full | Low-quality |
| S11_vs_k141_54643\|\|full | Not-determined |
| S11_vs_k141_15194\|\|full | Low-quality |
| S11_vs_k141_93428\|\|full | Low-quality |
| S11_vs_k141_100019\|\|full | Low-quality |
| S11_vs_k141_15198\|\|full | Low-quality |
| S11_vs_k141_87390\|\|full | Low-quality |
| S11_vs_k141_93440\|\|full | Low-quality |
| S11_vs_k141_74256\|\|full | Low-quality |
| S11_vs_k141_15209\|\|full | Not-determined |
| S11_vs_k141_60640\|\|full | Not-determined |
| S11_vs_k141_2696\|\|full | Low-quality |
| S11_vs_k141_60649\|\|full | Low-quality |
| S11_vs_k141_41422\|\|full | Low-quality |
| S11_vs_k141_47659\|\|full | Low-quality |
| S11_vs_k141_80522\|\|full | Low-quality |
| S11_vs_k141_2704\|\|full | Low-quality |
| S11_vs_k141_28463\|\|full | Low-quality |
| S11_vs_k141_74273\|\|full | Not-determined |
| S11_vs_k141_93472\|\|full | Low-quality |
| S11_vs_k141_47673\|\|full | Not-determined |
| S11_vs_k141_100036\|\|full | Low-quality |
| S11_vs_k141_28471\|\|full | Low-quality |
| S11_vs_k141_41441\|\|full | Not-determined |
| S11_vs_k141_80539\|\|full | Not-determined |
| S11_vs_k141_28479\|\|full | Not-determined |
| S11_vs_k141_87437\|\|full | Low-quality |
| S11_vs_k141_41448\|\|full | Not-determined |
| S11_vs_k141_80550\|\|full | Low-quality |
| S11_vs_k141_9044\|\|full | Low-quality |
| S11_vs_k141_28487\|\|full | Low-quality |
| S11_vs_k141_87445\|\|full | Not-determined |
| S11_vs_k141_47699\|\|full | Low-quality |
| S11_vs_k141_34539\|\|full | Not-determined |
| S11_vs_k141_22122\|\|full | Not-determined |
| S11_vs_k141_9057\|\|full | Not-determined |
| S11_vs_k141_28504\|\|full | Not-determined |
| S11_vs_k141_87461\|\|full | Low-quality |
| S11_vs_k141_34548\|\|full | Not-determined |
| S11_vs_k141_74297\|\|full | Low-quality |
| S11_vs_k141_22135\|\|full | Not-determined |
| S11_vs_k141_34550\|\|full | Low-quality |
| S11_vs_k141_100083\|\|full | Not-determined |
| S11_vs_k141_22143\|\|full | Not-determined |
| S11_vs_k141_2758\|\|full | Low-quality |
| S11_vs_k141_100097\|\|full | Not-determined |
| S11_vs_k141_87487\|\|full | Low-quality |
| S11_vs_k141_47723\|\|full | Not-determined |
| S11_vs_k141_93491\|\|full | Low-quality |
| S11_vs_k141_34567\|\|full | Low-quality |
| S11_vs_k141_34571\|\|full | Not-determined |
| S11_vs_k141_22162\|\|full | Low-quality |
| S11_vs_k141_9090\|\|full | Low-quality |
| S11_vs_k141_60677\|\|full | Low-quality |
| S11_vs_k141_34576\|\|full | Low-quality |
| S11_vs_k141_67157\|\|full | Not-determined |
| S11_vs_k141_93506\|\|full | Low-quality |
| S11_vs_k141_22168\|\|full | Not-determined |
| S11_vs_k141_74348\|\|full | Low-quality |
| S11_vs_k141_34587\|\|full | Low-quality |
| S11_vs_k141_93520\|\|full | Low-quality |
| S11_vs_k141_15311\|\|full | Not-determined |
| S11_vs_k141_2805\|\|full | Low-quality |
| S11_vs_k141_87524\|\|full | Not-determined |
| S11_vs_k141_41478\|\|full | Not-determined |
| S11_vs_k141_34610\|\|full | Not-determined |
| S11_vs_k141_87531\|\|full | Low-quality |
| S11_vs_k141_34611\|\|full | Not-determined |
| S11_vs_k141_47728\|\|full | Low-quality |
| S11_vs_k141_60706\|\|full | Low-quality |
| S11_vs_k141_22187\|\|full | Low-quality |
| S11_vs_k141_15327\|\|full | Low-quality |
| S11_vs_k141_87540\|\|full | Low-quality |
| S11_vs_k141_87544\|\|full | Not-determined |
| S11_vs_k141_15330\|\|full | Not-determined |
| S11_vs_k141_9132\|\|full | Not-determined |
| S11_vs_k141_22197\|\|full | Low-quality |
| S11_vs_k141_2839\|\|full | Not-determined |
| S11_vs_k141_15341\|\|full | Low-quality |
| S11_vs_k141_47754\|\|full | Not-determined |
| S11_vs_k141_93547\|\|full | Low-quality |
| S11_vs_k141_87570\|\|full | Not-determined |
| S11_vs_k141_15363\|\|full | Not-determined |
| S11_vs_k141_93551\|\|full | Low-quality |
| S11_vs_k141_60756\|\|full | Not-determined |
| S11_vs_k141_34659\|\|full | Not-determined |
| S11_vs_k141_67166\|\|full | Low-quality |
| S11_vs_k141_80669\|\|full | Not-determined |
| S11_vs_k141_87590\|\|full | Low-quality |
| S11_vs_k141_74427\|\|full | Low-quality |
| S11_vs_k141_41558\|\|full | Low-quality |
| S11_vs_k141_47781\|\|full | Not-determined |
| S11_vs_k141_22228\|\|full | Not-determined |
| S11_vs_k141_74430\|\|full | Not-determined |
| S11_vs_k141_2913\|\|full | Not-determined |
| S11_extracted_k141_33433 | Complete |
| S11_vs_k141_93590\|\|full | Low-quality |
| S11_vs_k141_74444\|\|full | Low-quality |
| S11_vs_k141_87601\|\|full | Low-quality |
| S11_vs_k141_15374\|\|full | Low-quality |
| S11_vs_k141_47802\|\|full | Low-quality |
| S11_vs_k141_80699\|\|full | Low-quality |
| S11_vs_k141_15383\|\|full | Low-quality |
| S11_vs_k141_93602\|\|full | Low-quality |
| S11_vs_k141_74454\|\|full | Not-determined |
| S11_vs_k141_87623\|\|full | Low-quality |
| S11_vs_k141_87624\|\|full | Not-determined |
| S11_vs_k141_9196\|\|full | Low-quality |
| S11_vs_k141_80701\|\|full | Low-quality |
| S11_vs_k141_34671\|\|full | Low-quality |
| S11_vs_k141_87632\|\|full | Low-quality |
| S11_vs_k141_22246\|\|full | Low-quality |
| S11_vs_k141_47825\|\|full | Low-quality |
| S11_vs_k141_80708\|\|full | Low-quality |
| S11_vs_k141_60805\|\|full | Low-quality |
| S11_vs_k141_60807\|\|full | Low-quality |
| S11_vs_k141_93625\|\|full | Low-quality |
| S11_vs_k141_80713\|\|full | Not-determined |
| S11_vs_k141_22260\|\|full | Low-quality |
| S11_vs_k141_87649\|\|full | Low-quality |
| S11_vs_k141_60814\|\|full | Not-determined |
| S11_vs_k141_22264\|\|full | Not-determined |
| S11_vs_k141_93644\|\|full | Not-determined |
| S11_vs_k141_74495\|\|full | Low-quality |
| S11_vs_k141_15410\|\|full | Not-determined |
| S11_vs_k141_60824\|\|full | Low-quality |
| S11_vs_k141_74510\|\|full | Not-determined |
| S11_vs_k141_74517\|\|full | Low-quality |
| S11_vs_k141_93671\|\|full | Not-determined |
| S11_vs_k141_15418\|\|full | Low-quality |
| S11_vs_k141_9262\|\|full | Low-quality |
| S11_vs_k141_47866\|\|full | Low-quality |
| S11_vs_k141_67252\|\|full | Low-quality |
| S11_vs_k141_60858\|\|full | Low-quality |
| S11_vs_k141_9270\|\|full | Not-determined |
| S11_vs_k141_67259\|\|full | Low-quality |
| S11_vs_k141_47874\|\|full | Not-determined |
| S11_vs_k141_100180\|\|full | Not-determined |
| S11_vs_k141_74555\|\|full | Low-quality |
| S11_vs_k141_100185\|\|full | Low-quality |
| S11_vs_k141_60867\|\|full | Not-determined |
| S11_vs_k141_22322\|\|full | Low-quality |
| S11_vs_k141_54666\|\|full | Not-determined |
| S11_vs_k141_74571\|\|full | Low-quality |
| S11_vs_k141_54670\|\|full | Low-quality |
| S11_vs_k141_60870\|\|full | Low-quality |
| S11_vs_k141_34695\|\|full | Low-quality |
| S13_extracted_56074 | Complete |
| S11_vs_k141_22345\|\|full | Low-quality |
| S11_vs_k141_87670\|\|full | Not-determined |
| S11_vs_k141_2975\|\|full | Low-quality |
| S11_vs_k141_9300\|\|full | Low-quality |
| S11_vs_k141_100215\|\|full | Low-quality |
| S11_vs_k141_9305\|\|full | Low-quality |
| S11_vs_k141_34720\|\|full | Low-quality |
| S11_vs_k141_87673\|\|full | Low-quality |
| S11_vs_k141_9311\|\|full | Low-quality |
| S11_vs_k141_34727\|\|full | Not-determined |
| S11_vs_k141_74617\|\|full | Low-quality |
| S11_vs_k141_9313\|\|full | Low-quality |
| S11_vs_k141_34729\|\|full | Not-determined |
| S11_vs_k141_74619\|\|full | Not-determined |
| S11_vs_k141_22388\|\|full | Low-quality |
| S11_vs_k141_60913\|\|full | Low-quality |
| S11_vs_k141_54710\|\|full | Not-determined |
| S11_vs_k141_15479\|\|full | Low-quality |
| S11_vs_k141_34736\|\|full | Low-quality |
| S11_vs_k141_9314\|\|full | Not-determined |
| S11_vs_k141_15482\|\|full | Low-quality |
| S11_vs_k141_22398\|\|full | Low-quality |
| S11_vs_k141_9318\|\|full | Not-determined |
| S11_vs_k141_87692\|\|full | Low-quality |
| S11_vs_k141_41709\|\|full | Low-quality |
| S11_vs_k141_54717\|\|full | Low-quality |
| S11_vs_k141_2993\|\|full | Low-quality |
| S11_vs_k141_93775\|\|full | Low-quality |
| S11_vs_k141_22406\|\|full | Low-quality |
| S11_vs_k141_67322\|\|full | Low-quality |
| S11_vs_k141_15490\|\|full | Low-quality |
| S11_vs_k141_41720\|\|full | Low-quality |
| S11_vs_k141_54740\|\|full | Low-quality |
| S11_vs_k141_60933\|\|full | Not-determined |
| S11_vs_k141_67327\|\|full | Low-quality |
| S11_vs_k141_22418\|\|full | Not-determined |
| S11_vs_k141_100280\|\|full | Low-quality |
| S11_vs_k141_87727\|\|full | Low-quality |
| S11_vs_k141_22422\|\|full | Not-determined |
| S11_vs_k141_15510\|\|full | Not-determined |
| S11_vs_k141_22430\|\|full | Not-determined |
| S11_vs_k141_15514\|\|full | Low-quality |
| S11_vs_k141_67354\|\|full | Low-quality |
| S11_vs_k141_15515\|\|full | Low-quality |
| S11_vs_k141_9356\|\|full | Low-quality |
| S11_vs_k141_22434\|\|full | Low-quality |
| S11_vs_k141_93825\|\|full | Not-determined |
| S11_vs_k141_67375\|\|full | Low-quality |
| S11_vs_k141_93838\|\|full | Not-determined |
| S11_vs_k141_47993\|\|full | Low-quality |
| S11_vs_k141_22461\|\|full | Low-quality |
| S11_vs_k141_28537\|\|full | Low-quality |
| S11_vs_k141_67390\|\|full | Not-determined |
| S11_vs_k141_3009\|\|full | Low-quality |
| S11_vs_k141_61010\|\|full | Low-quality |
| S11_vs_k141_93847\|\|full | Low-quality |
| S11_vs_k141_28556\|\|full | Low-quality |
| S11_vs_k141_22489\|\|full | Low-quality |
| S11_vs_k141_15548\|\|full | Not-determined |
| S11_vs_k141_3023\|\|full | Low-quality |
| S11_vs_k141_80866\|\|full | Not-determined |
| S11_vs_k141_61027\|\|full | Not-determined |
| S11_vs_k141_61032\|\|full | Not-determined |
| S11_vs_k141_87827\|\|full | Low-quality |
| S11_vs_k141_54769\|\|full | Low-quality |
| S11_vs_k141_28574\|\|full | Low-quality |
| S11_vs_k141_54773\|\|full | Not-determined |
| S11_vs_k141_34778\|\|full | Low-quality |
| S11_vs_k141_15578\|\|full | Low-quality |
| S11_vs_k141_80914\|\|full | Not-determined |
| S11_vs_k141_3076\|\|full | Low-quality |
| S11_vs_k141_93905\|\|full | Low-quality |
| S11_vs_k141_67424\|\|full | Low-quality |
| S11_vs_k141_41802\|\|full | Not-determined |
| S11_vs_k141_48029\|\|full | Low-quality |
| S11_vs_k141_3084\|\|full | Low-quality |
| S11_vs_k141_22553\|\|full | Low-quality |
| S11_vs_k141_34803\|\|full | Not-determined |
| S11_vs_k141_22558\|\|full | Low-quality |
| S11_vs_k141_100396\|\|full | Low-quality |
| S11_vs_k141_28617\|\|full | Not-determined |
| S11_vs_k141_34808\|\|full | Low-quality |
| S11_vs_k141_61070\|\|full | Not-determined |
| S11_vs_k141_3102\|\|full | Not-determined |
| S11_vs_k141_41825\|\|full | Not-determined |
| S11_vs_k141_34825\|\|full | Not-determined |
| S11_vs_k141_15617\|\|full | Low-quality |
| S11_vs_k141_67468\|\|full | Low-quality |
| S11_vs_k141_22583\|\|full | Low-quality |
| S11_vs_k141_48039\|\|full | Low-quality |
| S11_vs_k141_41852\|\|full | Low-quality |
| S11_vs_k141_15619\|\|full | Low-quality |
| S11_vs_k141_15621\|\|full | Low-quality |
| S11_vs_k141_9442\|\|full | Not-determined |
| S11_vs_k141_93964\|\|full | Low-quality |
| S11_vs_k141_28647\|\|full | Not-determined |
| S11_vs_k141_48052\|\|full | Low-quality |
| S11_vs_k141_34852\|\|full | Not-determined |
| S11_vs_k141_100445\|\|full | Low-quality |
| S11_vs_k141_9451\|\|full | Low-quality |
| S11_vs_k141_9454\|\|full | Low-quality |
| S11_vs_k141_93968\|\|full | Low-quality |
| S11_vs_k141_34863\|\|full | Low-quality |
| S11_vs_k141_22610\|\|full | Low-quality |
| S11_vs_k141_67519\|\|full | Low-quality |
| S11_vs_k141_48072\|\|full | Low-quality |
| S11_vs_k141_9471\|\|full | Not-determined |
| S11_vs_k141_3170\|\|full | Low-quality |
| S11_vs_k141_41883\|\|full | Low-quality |
| S11_vs_k141_34886\|\|full | Low-quality |
| S11_vs_k141_54829\|\|full | Low-quality |
| S11_vs_k141_67540\|\|full | Low-quality |
| S11_vs_k141_100494\|\|full | Low-quality |
| S11_vs_k141_54840\|\|full | Not-determined |
| S11_vs_k141_48091\|\|full | Low-quality |
| S11_vs_k141_61147\|\|full | Low-quality |
| S11_vs_k141_34900\|\|full | Low-quality |
| S11_vs_k141_28678\|\|full | Not-determined |
| S11_vs_k141_93990\|\|full | Low-quality |
| S11_vs_k141_9506\|\|full | Not-determined |
| S11_vs_k141_100524\|\|full | Low-quality |
| S11_vs_k141_22668\|\|full | Low-quality |
| S11_vs_k141_61162\|\|full | Not-determined |
| S11_vs_k141_54865\|\|full | Low-quality |
| S11_vs_k141_93995\|\|full | Not-determined |
| S11_vs_k141_100530\|\|full | Low-quality |
| S11_vs_k141_74777\|\|full | Low-quality |
| S11_vs_k141_67572\|\|full | Low-quality |
| S11_vs_k141_22678\|\|full | Low-quality |
| S11_vs_k141_94000\|\|full | Low-quality |
| S11_vs_k141_3220\|\|full | Low-quality |
| S11_vs_k141_61179\|\|full | Low-quality |
| S11_vs_k141_15676\|\|full | Low-quality |
| S11_vs_k141_9534\|\|full | Low-quality |
| S11_vs_k141_34927\|\|full | Not-determined |
| S11_vs_k141_34929\|\|full | Low-quality |
| S11_vs_k141_22687\|\|full | Low-quality |
| S11_vs_k141_41919\|\|full | Low-quality |
| S11_vs_k141_34931\|\|full | Not-determined |
| S11_vs_k141_9542\|\|full | Low-quality |
| S11_vs_k141_80965\|\|full | Low-quality |
| S11_vs_k141_41934\|\|full | Not-determined |
| S11_vs_k141_15698\|\|full | Low-quality |
| S11_vs_k141_33433\|\|full | High-quality |
| S11_vs_k141_80990\|\|full | Low-quality |
| S11_vs_k141_22714\|\|full | Low-quality |
| S11_vs_k141_80992\|\|full | Not-determined |
| S11_vs_k141_9562\|\|full | Low-quality |
| S11_vs_k141_67575\|\|full | Low-quality |
| S11_vs_k141_74827\|\|full | Low-quality |
| S11_vs_k141_54904\|\|full | Low-quality |
| S11_vs_k141_41949\|\|full | Not-determined |
| S11_vs_k141_3274\|\|full | Low-quality |
| S11_vs_k141_28734\|\|full | Low-quality |
| S11_vs_k141_15720\|\|full | Low-quality |
| S11_vs_k141_54914\|\|full | Not-determined |
| S11_vs_k141_54915\|\|full | Low-quality |
| S11_vs_k141_34944\|\|full | Low-quality |
| S11_vs_k141_9572\|\|full | Low-quality |
| S11_vs_k141_61204\|\|full | Low-quality |
| S11_vs_k141_15728\|\|full | Low-quality |
| S11_vs_k141_41966\|\|full | Low-quality |
| S11_vs_k141_74867\|\|full | Low-quality |
| S11_vs_k141_34954\|\|full | Not-determined |
| S11_vs_k141_9598\|\|full | Low-quality |
| S11_vs_k141_61233\|\|full | Low-quality |
| S11_vs_k141_28747\|\|full | Low-quality |
| S11_vs_k141_22731\|\|full | Low-quality |
| S11_vs_k141_74885\|\|full | Low-quality |
| S11_vs_k141_34964\|\|full | Not-determined |
| S11_vs_k141_94093\|\|full | Low-quality |
| S11_vs_k141_67590\|\|full | Not-determined |
| S11_vs_k141_74895\|\|full | Low-quality |
| S13_vs_56074\|\|full | High-quality |
| S11_vs_k141_81053\|\|full | Low-quality |
| S11_vs_k141_54965\|\|full | Not-determined |
| S11_vs_k141_48112\|\|full | Low-quality |
| S11_vs_k141_74908\|\|full | Low-quality |
| S11_vs_k141_28778\|\|full | Low-quality |
| S16_extracted_90048 | Medium-quality |
| S11_vs_k141_15787\|\|full | Low-quality |
| S11_vs_k141_67611\|\|full | Low-quality |
| S11_vs_k141_74923\|\|full | Not-determined |
| S11_vs_k141_54988\|\|full | Low-quality |
| S11_vs_k141_3287\|\|full | Low-quality |
| S11_vs_k141_15792\|\|full | Low-quality |
| S11_vs_k141_15793\|\|full | Low-quality |
| S11_vs_k141_3292\|\|full | Not-determined |
| S11_vs_k141_22758\|\|full | Low-quality |
| S11_vs_k141_54995\|\|full | Low-quality |
| S11_vs_k141_3294\|\|full | Low-quality |
| S11_vs_k141_94123\|\|full | Low-quality |
| S11_vs_k141_15815\|\|full | Low-quality |
| S11_vs_k141_67648\|\|full | Low-quality |
| S11_vs_k141_9677\|\|full | Low-quality |
| S11_vs_k141_81101\|\|full | Not-determined |
| S11_vs_k141_22782\|\|full | Low-quality |
| S11_vs_k141_61318\|\|full | Low-quality |
| S11_vs_k141_67659\|\|full | Low-quality |
| S11_vs_k141_100645\|\|full | Low-quality |
| S11_vs_k141_55019\|\|full | Not-determined |
| S11_vs_k141_3326\|\|full | Not-determined |
| S11_vs_k141_48160\|\|full | Low-quality |
| S11_vs_k141_67662\|\|full | Not-determined |
| S11_vs_k141_35040\|\|full | Not-determined |
| S11_vs_k141_42022\|\|full | Low-quality |
| S11_vs_k141_55029\|\|full | Low-quality |
| S11_vs_k141_42027\|\|full | Low-quality |
| S11_vs_k141_94159\|\|full | Not-determined |
| S11_vs_k141_28806\|\|full | Low-quality |
| S11_vs_k141_15834\|\|full | Low-quality |
| S11_vs_k141_55044\|\|full | Low-quality |
| S11_vs_k141_42031\|\|full | Not-determined |
| S11_vs_k141_48171\|\|full | Low-quality |
| S11_vs_k141_42037\|\|full | Low-quality |
| S11_vs_k141_48175\|\|full | Low-quality |
| S15_extracted_20644 | High-quality |
| S11_vs_k141_74972\|\|full | Low-quality |
| S11_vs_k141_87914\|\|full | Low-quality |
| S11_vs_k141_42050\|\|full | Low-quality |
| S11_vs_k141_9702\|\|full | Low-quality |
| S11_vs_k141_15876\|\|full | Low-quality |
| S11_vs_k141_48187\|\|full | Not-determined |
| S11_vs_k141_94199\|\|full | Not-determined |
| S11_vs_k141_74993\|\|full | Low-quality |
| S11_vs_k141_87940\|\|full | Low-quality |
| S11_vs_k141_35085\|\|full | Low-quality |
| S11_vs_k141_48199\|\|full | Not-determined |
| S11_vs_k141_9718\|\|full | Low-quality |
| S11_vs_k141_9720\|\|full | Low-quality |
| S11_vs_k141_48202\|\|full | Low-quality |
| S11_vs_k141_28883\|\|full | Low-quality |
| S11_vs_k141_35091\|\|full | Low-quality |
| S11_vs_k141_94223\|\|full | Not-determined |
| S11_vs_k141_42090\|\|full | Low-quality |
| S11_vs_k141_15896\|\|full | Not-determined |
| S11_vs_k141_87954\|\|full | Low-quality |
| S11_vs_k141_35100\|\|full | Low-quality |
| S11_vs_k141_42095\|\|full | Low-quality |
| S11_vs_k141_94232\|\|full | Low-quality |
| S11_vs_k141_75029\|\|full | Low-quality |
| S11_vs_k141_22903\|\|full | Low-quality |
| S11_vs_k141_35113\|\|full | Low-quality |
| S11_vs_k141_9725\|\|full | Not-determined |
| S11_vs_k141_42098\|\|full | Low-quality |

| | |
|---|---|
| S11_vs_k141_100710\|\|full | Not-determined |
| S11_vs_k141_75037\|\|full | Low-quality |
| S11_vs_k141_22912\|\|full | Not-determined |
| S11_vs_k141_100715\|\|full | Low-quality |
| S11_vs_k141_55120\|\|full | Not-determined |
| S11_vs_k141_9740\|\|full | Low-quality |
| S11_vs_k141_75065\|\|full | Low-quality |
| S11_vs_k141_15948\|\|full | Not-determined |
| S11_vs_k141_28897\|\|full | Low-quality |
| S11_vs_k141_42137\|\|full | Not-determined |
| S11_vs_k141_28901\|\|full | Low-quality |
| S11_vs_k141_100737\|\|full | Not-determined |
| S11_vs_k141_67725\|\|full | Low-quality |
| S11_vs_k141_15980\|\|full | Low-quality |
| S11_vs_k141_100752\|\|full | Low-quality |
| S11_vs_k141_81112\|\|full | Low-quality |
| S11_vs_k141_42152\|\|full | Low-quality |
| S11_vs_k141_15985\|\|full | Not-determined |
| S11_vs_k141_35162\|\|full | Low-quality |
| S11_vs_k141_48262\|\|full | Low-quality |
| S11_vs_k141_67735\|\|full | Low-quality |
| S11_vs_k141_28933\|\|full | Low-quality |
| S11_vs_k141_15990\|\|full | Not-determined |
| S11_vs_k141_42162\|\|full | Low-quality |
| S11_vs_k141_75102\|\|full | Low-quality |
| S11_vs_k141_15994\|\|full | Low-quality |
| S11_vs_k141_42168\|\|full | Not-determined |
| S11_vs_k141_22991\|\|full | Low-quality |
| S11_vs_k141_35189\|\|full | Low-quality |
| S11_vs_k141_16014\|\|full | Low-quality |
| S11_vs_k141_16017\|\|full | Not-determined |
| S11_vs_k141_42181\|\|full | Low-quality |
| S16_extracted_173099 | Complete |
| S11_vs_k141_61365\|\|full | Not-determined |
| S11_vs_k141_61366\|\|full | Not-determined |
| S11_vs_k141_28962\|\|full | Not-determined |
| S11_vs_k141_67778\|\|full | Not-determined |
| S11_vs_k141_61383\|\|full | Not-determined |
| S11_vs_k141_3377\|\|full | Not-determined |
| S11_vs_k141_28980\|\|full | Low-quality |
| S11_vs_k141_42200\|\|full | Low-quality |
| S11_vs_k141_94318\|\|full | Not-determined |
| S11_vs_k141_16034\|\|full | Not-determined |
| S11_vs_k141_100848\|\|full | Low-quality |
| S11_vs_k141_3379\|\|full | Not-determined |
| S11_vs_k141_48312\|\|full | Not-determined |
| S11_vs_k141_55173\|\|full | Not-determined |
| S11_vs_k141_94320\|\|full | Low-quality |
| S11_vs_k141_23040\|\|full | Low-quality |
| S11_vs_k141_81150\|\|full | Not-determined |
| S11_vs_k141_75122\|\|full | Low-quality |
| S11_vs_k141_48320\|\|full | Low-quality |
| S11_vs_k141_23045\|\|full | Low-quality |
| S11_vs_k141_61393\|\|full | Not-determined |
| S11_vs_k141_42220\|\|full | Low-quality |
| S11_vs_k141_9819\|\|full | Low-quality |
| S11_vs_k141_81157\|\|full | Low-quality |
| S11_vs_k141_29014\|\|full | Low-quality |
| S11_vs_k141_29021\|\|full | Not-determined |
| S11_vs_k141_48339\|\|full | Not-determined |
| S11_vs_k141_9839\|\|full | Low-quality |
| S11_vs_k141_81174\|\|full | Low-quality |
| S11_vs_k141_94346\|\|full | Low-quality |
| S11_vs_k141_42246\|\|full | Low-quality |
| S11_vs_k141_55201\|\|full | Low-quality |
| S11_vs_k141_29036\|\|full | Low-quality |
| S11_vs_k141_81185\|\|full | Not-determined |
| S11_vs_k141_29045\|\|full | Not-determined |
| S11_vs_k141_16070\|\|full | Low-quality |
| S11_vs_k141_29046\|\|full | Low-quality |
| S11_vs_k141_42277\|\|full | Low-quality |
| S11_vs_k141_81191\|\|full | Low-quality |
| S11_vs_k141_23089\|\|full | Low-quality |
| S11_vs_k141_3442\|\|full | Not-determined |
| S11_vs_k141_3444\|\|full | Not-determined |
| S11_vs_k141_75141\|\|full | Low-quality |
| S11_vs_k141_81192\|\|full | Not-determined |
| S11_vs_k141_29066\|\|full | Not-determined |
| S11_vs_k141_3462\|\|full | Low-quality |
| S11_vs_k141_9874\|\|full | Low-quality |
| S11_vs_k141_29077\|\|full | Low-quality |
| S11_vs_k141_3470\|\|full | Not-determined |
| S11_vs_k141_35341\|\|full | Low-quality |
| S11_vs_k141_48382\|\|full | Low-quality |
| S11_vs_k141_61464\|\|full | Low-quality |
| S11_vs_k141_29080\|\|full | Not-determined |
| S11_vs_k141_100876\|\|full | Low-quality |
| S11_vs_k141_67826\|\|full | Not-determined |
| S11_vs_k141_35350\|\|full | Low-quality |
| S11_vs_k141_87972\|\|full | Low-quality |
| S11_vs_k141_81213\|\|full | Low-quality |
| S11_vs_k141_3484\|\|full | Not-determined |
| S11_vs_k141_35352\|\|full | Low-quality |
| S11_vs_k141_100885\|\|full | Low-quality |
| S11_vs_k141_100886\|\|full | Not-determined |
| S11_vs_k141_55246\|\|full | Not-determined |
| S11_vs_k141_9904\|\|full | Low-quality |
| S11_vs_k141_23134\|\|full | Low-quality |
| S11_vs_k141_67844\|\|full | Low-quality |
| S11_vs_k141_23139\|\|full | Low-quality |
| S11_vs_k141_100894\|\|full | Not-determined |
| S11_vs_k141_87998\|\|full | Not-determined |
| S11_vs_k141_42337\|\|full | Not-determined |
| S11_vs_k141_48414\|\|full | Not-determined |
| S11_vs_k141_23151\|\|full | Low-quality |
| S11_vs_k141_67858\|\|full | Not-determined |
| S11_vs_k141_9924\|\|full | Low-quality |
| S11_vs_k141_61502\|\|full | Low-quality |
| S11_vs_k141_3516\|\|full | Not-determined |
| S11_vs_k141_23153\|\|full | Not-determined |
| S11_vs_k141_88004\|\|full | Low-quality |
| S11_vs_k141_48429\|\|full | Low-quality |
| S11_vs_k141_55272\|\|full | Low-quality |
| S11_vs_k141_9926\|\|full | Low-quality |
| S11_vs_k141_61510\|\|full | Not-determined |
| S11_vs_k141_75210\|\|full | Low-quality |
| S11_vs_k141_23162\|\|full | Low-quality |
| S11_vs_k141_23165\|\|full | Low-quality |
| S11_vs_k141_35403\|\|full | Low-quality |
| S11_vs_k141_23167\|\|full | Low-quality |
| S11_vs_k141_55283\|\|full | Low-quality |
| S11_vs_k141_23170\|\|full | Low-quality |
| S11_vs_k141_61520\|\|full | Low-quality |
| S11_vs_k141_75231\|\|full | Not-determined |
| S11_vs_k141_100933\|\|full | Low-quality |
| S11_vs_k141_23180\|\|full | Not-determined |
| S11_vs_k141_42363\|\|full | Not-determined |
| S11_vs_k141_75241\|\|full | Not-determined |
| S11_vs_k141_35437\|\|full | Low-quality |
| S11_vs_k141_9951\|\|full | Low-quality |
| S11_vs_k141_94423\|\|full | Not-determined |
| S11_vs_k141_75246\|\|full | Low-quality |
| S11_vs_k141_100954\|\|full | Low-quality |
| S11_vs_k141_61540\|\|full | Not-determined |
| S11_vs_k141_16190\|\|full | Low-quality |
| S11_vs_k141_9957\|\|full | Not-determined |
| S11_vs_k141_35443\|\|full | Low-quality |
| S11_vs_k141_29177\|\|full | Low-quality |
| S11_vs_k141_16198\|\|full | Not-determined |
| S11_vs_k141_55331\|\|full | Not-determined |
| S11_vs_k141_61541\|\|full | Not-determined |
| S11_vs_k141_100960\|\|full | Not-determined |
| S11_vs_k141_55334\|\|full | Low-quality |
| S11_vs_k141_67862\|\|full | Low-quality |
| S11_vs_k141_100964\|\|full | Low-quality |
| S11_vs_k141_29189\|\|full | Low-quality |
| S11_vs_k141_61550\|\|full | Low-quality |
| S11_vs_k141_61552\|\|full | Not-determined |
| S11_vs_k141_75257\|\|full | Low-quality |
| S11_vs_k141_61554\|\|full | Not-determined |
| S11_vs_k141_35473\|\|full | Low-quality |
| S11_vs_k141_81322\|\|full | Low-quality |
| S11_vs_k141_61561\|\|full | Low-quality |
| S11_vs_k141_16218\|\|full | Not-determined |
| S11_vs_k141_61567\|\|full | Low-quality |
| S11_vs_k141_16223\|\|full | Low-quality |
| S11_vs_k141_81327\|\|full | Not-determined |
| S11_vs_k141_48460\|\|full | Not-determined |
| S16_vs_173099\|\|full | High-quality |
| S11_vs_k141_94430\|\|full | Not-determined |
| S11_vs_k141_10007\|\|full | Low-quality |
| S11_vs_k141_35493\|\|full | Low-quality |
| S11_vs_k141_42408\|\|full | Not-determined |
| S11_vs_k141_23192\|\|full | Low-quality |
| S11_vs_k141_55376\|\|full | Low-quality |
| S11_vs_k141_29249\|\|full | Not-determined |
| S11_vs_k141_94458\|\|full | Low-quality |
| S11_vs_k141_55382\|\|full | Low-quality |
| S11_vs_k141_48484\|\|full | Low-quality |
| S11_vs_k141_94464\|\|full | Low-quality |
| S11_vs_k141_67951\|\|full | Low-quality |
| S11_vs_k141_101017\|\|full | Low-quality |
| S11_vs_k141_75334\|\|full | Low-quality |
| S11_vs_k141_75337\|\|full | Low-quality |
| S11_vs_k141_61589\|\|full | Low-quality |
| S11_vs_k141_55396\|\|full | Low-quality |
| S11_vs_k141_88095\|\|full | Not-determined |
| S11_vs_k141_88097\|\|full | Not-determined |
| S11_vs_k141_23228\|\|full | Low-quality |
| S11_vs_k141_16282\|\|full | Low-quality |
| S11_vs_k141_81380\|\|full | Not-determined |
| S11_vs_k141_48519\|\|full | Low-quality |
| S11_vs_k141_10052\|\|full | Low-quality |
| S11_vs_k141_35575\|\|full | Not-determined |
| S11_vs_k141_101046\|\|full | Low-quality |
| S11_vs_k141_61632\|\|full | Not-determined |
| S11_vs_k141_67983\|\|full | Low-quality |
| S11_vs_k141_75379\|\|full | Low-quality |
| S11_vs_k141_67986\|\|full | Low-quality |
| S11_vs_k141_10080\|\|full | Low-quality |
| S11_vs_k141_88146\|\|full | Low-quality |
| S11_vs_k141_101076\|\|full | Low-quality |
| S11_vs_k141_88150\|\|full | Low-quality |
| S11_vs_k141_55425\|\|full | Low-quality |
| S11_vs_k141_94520\|\|full | Low-quality |
| S11_vs_k141_55428\|\|full | Low-quality |
| S11_vs_k141_16325\|\|full | Low-quality |
| S11_vs_k141_10097\|\|full | Low-quality |
| S11_vs_k141_10103\|\|full | Not-determined |
| S11_vs_k141_42496\|\|full | Not-determined |
| S11_vs_k141_16336\|\|full | Not-determined |
| S11_vs_k141_10104\|\|full | Low-quality |
| S11_vs_k141_23271\|\|full | Low-quality |
| S11_vs_k141_101094\|\|full | Not-determined |
| S11_vs_k141_94531\|\|full | Low-quality |
| S11_vs_k141_35627\|\|full | Low-quality |
| S11_vs_k141_88172\|\|full | Low-quality |
| S11_vs_k141_88175\|\|full | Low-quality |
| S11_vs_k141_10118\|\|full | Not-determined |
| S11_vs_k141_101104\|\|full | Low-quality |
| S11_vs_k141_61652\|\|full | Not-determined |
| S11_vs_k141_61662\|\|full | Not-determined |
| S11_vs_k141_88201\|\|full | Not-determined |
| S11_vs_k141_23311\|\|full | Low-quality |
| S11_vs_k141_94560\|\|full | Low-quality |
| S11_vs_k141_42543\|\|full | Not-determined |
| S11_vs_k141_29292\|\|full | Low-quality |
| S11_vs_k141_94561\|\|full | Low-quality |
| S11_vs_k141_16373\|\|full | Not-determined |
| S11_vs_k141_42548\|\|full | Low-quality |
| S11_vs_k141_68056\|\|full | Not-determined |
| S11_vs_k141_81441\|\|full | Low-quality |
| S11_vs_k141_88227\|\|full | Low-quality |
| S11_vs_k141_81449\|\|full | Not-determined |
| S11_vs_k141_23338\|\|full | Not-determined |
| S11_vs_k141_75466\|\|full | Low-quality |
| S11_vs_k141_29310\|\|full | Low-quality |
| S11_vs_k141_75480\|\|full | Low-quality |
| S11_vs_k141_35681\|\|full | Low-quality |
| S11_vs_k141_10161\|\|full | Low-quality |
| S11_vs_k141_55497\|\|full | Low-quality |
| S11_vs_k141_75483\|\|full | Low-quality |
| S11_vs_k141_55509\|\|full | Low-quality |
| S11_vs_k141_61682\|\|full | Low-quality |
| S11_vs_k141_42570\|\|full | Low-quality |
| S11_vs_k141_61688\|\|full | Low-quality |
| S11_vs_k141_16415\|\|full | Low-quality |
| S11_vs_k141_35706\|\|full | Not-determined |
| S11_vs_k141_35714\|\|full | Not-determined |
| S11_vs_k141_68099\|\|full | Not-determined |
| S11_vs_k141_55533\|\|full | Low-quality |
| S11_vs_k141_35728\|\|full | Low-quality |
| S11_vs_k141_94644\|\|full | Low-quality |
| S11_vs_k141_42598\|\|full | Not-determined |
| S11_vs_k141_101191\|\|full | Low-quality |
| S11_vs_k141_61712\|\|full | Low-quality |
| S11_vs_k141_101198\|\|full | Low-quality |
| S11_vs_k141_75542\|\|full | Not-determined |
| S11_vs_k141_61724\|\|full | Low-quality |
| S11_vs_k141_42616\|\|full | Not-determined |
| S11_vs_k141_48690\|\|full | Not-determined |
| S11_vs_k141_75547\|\|full | Not-determined |
| S8_vs_k141_53614\|\|full | Complete |
| S11_vs_k141_35755\|\|full | Low-quality |
| S11_vs_k141_55579\|\|full | Low-quality |
| S16_vs_90048\|\|full | Medium-quality |
| S11_vs_k141_94690\|\|full | Not-determined |
| S11_vs_k141_55583\|\|full | Low-quality |
| S11_vs_k141_16438\|\|full | Not-determined |
| S11_vs_k141_10181\|\|full | Not-determined |
| S11_vs_k141_94699\|\|full | Low-quality |
| S11_vs_k141_81494\|\|full | Low-quality |
| S11_vs_k141_94703\|\|full | Low-quality |
| S11_vs_k141_61748\|\|full | Not-determined |
| S11_vs_k141_35780\|\|full | Low-quality |
| S11_vs_k141_23381\|\|full | Not-determined |
| S11_vs_k141_29327\|\|full | Low-quality |
| S11_vs_k141_88233\|\|full | Low-quality |
| S11_vs_k141_88236\|\|full | Low-quality |
| S11_vs_k141_75559\|\|full | Not-determined |
| S11_vs_k141_55613\|\|full | Low-quality |
| S11_vs_k141_29335\|\|full | Not-determined |
| S11_vs_k141_35784\|\|full | Low-quality |
| S11_vs_k141_48706\|\|full | Not-determined |
| S11_vs_k141_81497\|\|full | Low-quality |
| S11_vs_k141_94746\|\|full | Low-quality |
| S11_vs_k141_88248\|\|full | Low-quality |
| S11_vs_k141_23404\|\|full | Not-determined |
| S11_vs_k141_81508\|\|full | Low-quality |
| S11_vs_k141_55638\|\|full | Low-quality |
| S11_vs_k141_75576\|\|full | Not-determined |
| S11_vs_k141_3569\|\|full | Not-determined |
| S11_vs_k141_88257\|\|full | Low-quality |
| S11_vs_k141_23410\|\|full | Low-quality |
| S11_vs_k141_94754\|\|full | Low-quality |
| S11_vs_k141_68133\|\|full | Low-quality |
| S11_vs_k141_23412\|\|full | Low-quality |
| S11_vs_k141_10240\|\|full | Not-determined |
| S11_vs_k141_94765\|\|full | Not-determined |
| S11_vs_k141_29364\|\|full | Low-quality |
| S11_vs_k141_10249\|\|full | Low-quality |
| S11_vs_k141_81513\|\|full | Not-determined |
| S11_vs_k141_75588\|\|full | Low-quality |
| S11_vs_k141_94784\|\|full | Not-determined |
| S11_vs_k141_10256\|\|full | Low-quality |
| S11_vs_k141_61817\|\|full | Low-quality |
| S11_vs_k141_55670\|\|full | Low-quality |
| S11_vs_k141_10263\|\|full | Low-quality |
| S11_vs_k141_81522\|\|full | Low-quality |
| S4_vs_VIRSorter_31973-circular_ge | Medium-quality |
| S11_vs_k141_42638\|\|full | Not-determined |
| S11_vs_k141_10276\|\|full | Not-determined |
| S11_vs_k141_55695\|\|full | Low-quality |
| S11_vs_k141_35855\|\|full | Not-determined |
| S11_vs_k141_48729\|\|full | Low-quality |
| S11_vs_k141_94808\|\|full | Not-determined |
| S11_vs_k141_101290\|\|full | Low-quality |
| S11_vs_k141_88318\|\|full | Not-determined |
| S11_vs_k141_94815\|\|full | Low-quality |
| S11_vs_k141_61853\|\|full | Low-quality |
| S11_vs_k141_42656\|\|full | Low-quality |
| S11_vs_k141_101308\|\|full | Low-quality |
| S11_vs_k141_10313\|\|full | Low-quality |
| S11_vs_k141_81576\|\|full | Not-determined |
| S11_vs_k141_55731\|\|full | Not-determined |
| S11_vs_k141_3666\|\|full | Low-quality |
| S11_vs_k141_61873\|\|full | Not-determined |
| S11_vs_k141_101322\|\|full | Low-quality |
| S11_vs_k141_35881\|\|full | Low-quality |
| S11_vs_k141_75658\|\|full | Low-quality |
| S11_vs_k141_42677\|\|full | Not-determined |
| S11_vs_k141_88354\|\|full | Not-determined |
| S11_vs_k141_42686\|\|full | Low-quality |
| S11_vs_k141_101326\|\|full | Low-quality |
| S11_vs_k141_61877\|\|full | Not-determined |
| S11_vs_k141_3682\|\|full | Low-quality |
| S11_vs_k141_88356\|\|full | Low-quality |
| S11_vs_k141_75682\|\|full | Not-determined |
| S11_vs_k141_61885\|\|full | Low-quality |
| S11_vs_k141_101341\|\|full | Low-quality |
| S11_vs_k141_55768\|\|full | Not-determined |
| S11_vs_k141_94868\|\|full | Not-determined |
| S11_vs_k141_61901\|\|full | Low-quality |
| S11_vs_k141_35911\|\|full | Not-determined |
| S11_vs_k141_55776\|\|full | Low-quality |
| S11_vs_k141_75701\|\|full | Low-quality |
| S11_vs_k141_101345\|\|full | Low-quality |
| S11_vs_k141_61908\|\|full | Low-quality |
| S11_vs_k141_42707\|\|full | Low-quality |
| S11_vs_k141_35923\|\|full | Low-quality |
| S11_vs_k141_3715\|\|full | Low-quality |
| S11_vs_k141_35927\|\|full | Not-determined |
| S8_vs_k141_47265\|\|full | Complete |
| S11_vs_k141_61937\|\|full | Not-determined |
| S11_vs_k141_35944\|\|full | Not-determined |
| S11_vs_k141_68161\|\|full | Low-quality |
| S11_vs_k141_35949\|\|full | Low-quality |
| S11_vs_k141_35952\|\|full | Not-determined |
| S11_vs_k141_61954\|\|full | Not-determined |
| S11_vs_k141_29410\|\|full | Low-quality |
| S11_vs_k141_68170\|\|full | Low-quality |
| S11_vs_k141_35960\|\|full | Low-quality |
| S11_vs_k141_88378\|\|full | Low-quality |
| S3_vs_k141_127445\|\|full | High-quality |
| S11_vs_k141_3750\|\|full | Low-quality |
| S11_vs_k141_75720\|\|full | Not-determined |
| S11_vs_k141_55830\|\|full | Low-quality |
| S11_vs_k141_35975\|\|full | Low-quality |
| S11_vs_k141_75722\|\|full | Low-quality |
| S11_vs_k141_88398\|\|full | Low-quality |
| S11_vs_k141_68202\|\|full | Low-quality |
| S11_vs_k141_35977\|\|full | Low-quality |
| S11_vs_k141_23523\|\|full | Low-quality |
| S11_vs_k141_81598\|\|full | Not-determined |
| S11_vs_k141_101401\|\|full | Not-determined |
| S11_vs_k141_62000\|\|full | Low-quality |
| S11_vs_k141_42778\|\|full | Low-quality |
| S11_vs_k141_68209\|\|full | Low-quality |
| S11_vs_k141_35988\|\|full | Low-quality |
| S11_vs_k141_10401\|\|full | Low-quality |
| S11_vs_k141_3771\|\|full | Not-determined |
| S15_vs_20644\|\|full | High-quality |
| S11_vs_k141_94925\|\|full | Not-determined |
| S11_vs_k141_42800\|\|full | Not-determined |
| S11_vs_k141_3789\|\|full | Low-quality |
| S11_vs_k141_68220\|\|full | Low-quality |
| S11_vs_k141_62008\|\|full | Low-quality |
| S11_vs_k141_81635\|\|full | Not-determined |
| S11_vs_k141_68224\|\|full | Low-quality |
| S11_vs_k141_16529\|\|full | Not-determined |
| S11_vs_k141_42813\|\|full | Low-quality |
| S11_vs_k141_48783\|\|full | Low-quality |
| S11_vs_k141_16539\|\|full | Low-quality |
| S11_vs_k141_68235\|\|full | Low-quality |
| S11_vs_k141_42825\|\|full | Not-determined |
| S11_vs_k141_88443\|\|full | Low-quality |
| S11_vs_k141_81642\|\|full | Low-quality |
| S11_vs_k141_55875\|\|full | Not-determined |
| S11_vs_k141_48803\|\|full | Low-quality |
| S11_vs_k141_88451\|\|full | Low-quality |
| S11_vs_k141_81653\|\|full | Not-determined |
| S11_vs_k141_81656\|\|full | Low-quality |
| S11_vs_k141_48807\|\|full | Low-quality |
| S11_vs_k141_48810\|\|full | Low-quality |
| S11_vs_k141_62043\|\|full | Not-determined |
| S11_vs_k141_55880\|\|full | Low-quality |
| S11_vs_k141_101466\|\|full | Not-determined |
| S11_vs_k141_101467\|\|full | Low-quality |
| S11_vs_k141_29527\|\|full | Low-quality |
| S11_vs_k141_16567\|\|full | Not-determined |
| S11_vs_k141_29529\|\|full | Not-determined |
| S11_vs_k141_88477\|\|full | Low-quality |
| S11_vs_k141_62049\|\|full | Low-quality |
| S11_vs_k141_42862\|\|full | Low-quality |
| S11_vs_k141_48838\|\|full | Low-quality |
| S11_vs_k141_16571\|\|full | Not-determined |
| S11_vs_k141_68261\|\|full | Low-quality |
| S11_vs_k141_62058\|\|full | Low-quality |
| S11_vs_k141_81677\|\|full | Not-determined |
| S11_vs_k141_23547\|\|full | Low-quality |
| S11_vs_k141_101471\|\|full | Low-quality |
| S11_vs_k141_55901\|\|full | Low-quality |
| S11_vs_k141_48861\|\|full | Low-quality |
| S11_vs_k141_36043\|\|full | Not-determined |
| S11_vs_k141_23558\|\|full | Low-quality |
| S11_vs_k141_81712\|\|full | Low-quality |
| S8_extracted_k141_113806 | Medium-quality |
| S11_vs_k141_101500\|\|full | Low-quality |
| S11_vs_k141_36046\|\|full | Low-quality |
| S11_vs_k141_101501\|\|full | Not-determined |
| S11_vs_k141_29571\|\|full | Low-quality |
| S11_vs_k141_23581\|\|full | Not-determined |
| S11_vs_k141_36060\|\|full | Low-quality |
| S11_vs_k141_55934\|\|full | Low-quality |
| S11_vs_k141_75811\|\|full | Low-quality |
| S11_vs_k141_68276\|\|full | Low-quality |
| S11_vs_k141_68278\|\|full | Low-quality |
| S11_vs_k141_101525\|\|full | Not-determined |
| S11_vs_k141_36063\|\|full | Low-quality |
| S11_vs_k141_23595\|\|full | Low-quality |
| S11_vs_k141_101529\|\|full | Low-quality |
| S11_vs_k141_81732\|\|full | Low-quality |
| S11_vs_k141_88564\|\|full | Low-quality |
| S11_vs_k141_3826\|\|full | Low-quality |
| S11_vs_k141_23618\|\|full | Low-quality |
| S11_vs_k141_23620\|\|full | Low-quality |
| S11_vs_k141_68306\|\|full | Low-quality |
| S11_vs_k141_81735\|\|full | Low-quality |
| S11_vs_k141_75850\|\|full | Not-determined |
| S11_vs_k141_88582\|\|full | Low-quality |
| S11_vs_k141_42909\|\|full | Low-quality |
| S11_vs_k141_3836\|\|full | Not-determined |
| S11_vs_k141_62127\|\|full | Not-determined |
| S11_vs_k141_3840\|\|full | Low-quality |
| S11_vs_k141_36105\|\|full | Not-determined |
| S11_vs_k141_62130\|\|full | Low-quality |
| S11_vs_k141_55982\|\|full | Not-determined |
| S11_vs_k141_42921\|\|full | Not-determined |
| S11_vs_k141_48868\|\|full | Low-quality |
| S11_vs_k141_56002\|\|full | Not-determined |
| S11_vs_k141_36120\|\|full | Low-quality |
| S11_vs_k141_81768\|\|full | Not-determined |
| S11_vs_k141_16667\|\|full | Not-determined |
| S11_vs_k141_81771\|\|full | Low-quality |
| S11_vs_k141_23661\|\|full | Low-quality |
| S11_vs_k141_88633\|\|full | Not-determined |
| S11_vs_k141_75868\|\|full | Not-determined |
| S11_vs_k141_56021\|\|full | Not-determined |
| S11_vs_k141_88643\|\|full | Low-quality |
| S11_vs_k141_88645\|\|full | Low-quality |
| S11_vs_k141_62150\|\|full | Low-quality |
| S11_vs_k141_88646\|\|full | Low-quality |
| S11_vs_k141_48903\|\|full | Low-quality |
| S11_vs_k141_48909\|\|full | Low-quality |
| S11_vs_k141_23674\|\|full | Low-quality |
| S11_vs_k141_48912\|\|full | Low-quality |
| S11_vs_k141_101603\|\|full | Low-quality |
| S11_vs_k141_56040\|\|full | Low-quality |
| S11_vs_k141_88667\|\|full | Not-determined |
| S11_vs_k141_3884\|\|full | Low-quality |
| S11_vs_k141_48927\|\|full | Low-quality |
| S11_vs_k141_36152\|\|full | Low-quality |
| S11_vs_k141_62190\|\|full | Not-determined |
| S11_vs_k141_16721\|\|full | Low-quality |
| S11_vs_k141_3914\|\|full | Not-determined |
| S11_vs_k141_36158\|\|full | Low-quality |
| S11_vs_k141_48936\|\|full | Not-determined |
| S11_vs_k141_88692\|\|full | Not-determined |
| S8_vs_k141_113806\|\|full | Medium-quality |
| S11_vs_k141_95059\|\|full | Not-determined |
| S11_vs_k141_62198\|\|full | Low-quality |
| S11_vs_k141_23719\|\|full | Low-quality |
| S11_vs_k141_3934\|\|full | Not-determined |
| S3_vs_k141_64424\|\|full | High-quality |
| S11_vs_k141_81787\|\|full | Not-determined |
| S8_extracted_k141_108709 | Medium-quality |
| S11_vs_k141_23724\|\|full | Not-determined |
| S11_vs_k141_62204\|\|full | Not-determined |
| S11_vs_k141_29656\|\|full | Low-quality |
| S11_vs_k141_68355\|\|full | Not-determined |
| S11_vs_k141_81796\|\|full | Low-quality |
| S11_vs_k141_23738\|\|full | Not-determined |
| S11_vs_k141_62210\|\|full | Low-quality |
| S11_vs_k141_62212\|\|full | Low-quality |
| S11_vs_k141_29659\|\|full | Low-quality |
| S11_vs_k141_42946\|\|full | Not-determined |
| S11_vs_k141_23751\|\|full | Low-quality |
| S11_vs_k141_88723\|\|full | Low-quality |
| S11_vs_k141_42950\|\|full | Not-determined |
| S11_vs_k141_42953\|\|full | Not-determined |
| S11_vs_k141_88741\|\|full | Low-quality |
| S11_vs_k141_48979\|\|full | Low-quality |
| S11_vs_k141_68390\|\|full | Low-quality |
| S11_vs_k141_88753\|\|full | Low-quality |
| S11_vs_k141_81841\|\|full | Low-quality |
| S11_vs_k141_42973\|\|full | Low-quality |
| S11_vs_k141_81845\|\|full | Not-determined |
| S11_vs_k141_56146\|\|full | Low-quality |
| S11_vs_k141_49002\|\|full | Not-determined |
| S11_vs_k141_88764\|\|full | Low-quality |
| S11_vs_k141_62253\|\|full | Low-quality |
| S11_vs_k141_62259\|\|full | Low-quality |
| S11_vs_k141_16852\|\|full | Not-determined |
| S11_vs_k141_62260\|\|full | Not-determined |
| S11_vs_k141_10531\|\|full | Not-determined |
| S11_vs_k141_68418\|\|full | Low-quality |
| S11_vs_k141_62262\|\|full | Not-determined |
| S11_vs_k141_10535\|\|full | Low-quality |
| S11_vs_k141_10537\|\|full | Not-determined |
| S11_vs_k141_62267\|\|full | Low-quality |
| S11_vs_k141_10540\|\|full | Low-quality |
| S11_vs_k141_36258\|\|full | Low-quality |
| S11_vs_k141_81870\|\|full | Low-quality |
| S11_vs_k141_68442\|\|full | Low-quality |
| S11_vs_k141_56160\|\|full | Low-quality |
| S11_vs_k141_23832\|\|full | Low-quality |
| S11_vs_k141_81882\|\|full | Low-quality |
| S11_vs_k141_23834\|\|full | Low-quality |
| S11_vs_k141_95080\|\|full | Low-quality |
| S11_vs_k141_68451\|\|full | Low-quality |
| S11_vs_k141_36281\|\|full | Low-quality |
| S11_vs_k141_68455\|\|full | Not-determined |
| S11_vs_k141_101660\|\|full | Low-quality |
| S11_vs_k141_4007\|\|full | Low-quality |
| S11_vs_k141_81891\|\|full | Low-quality |
| S11_vs_k141_62288\|\|full | Low-quality |
| S11_vs_k141_4009\|\|full | Low-quality |
| S11_vs_k141_16887\|\|full | Low-quality |
| S11_vs_k141_101672\|\|full | Low-quality |
| S11_vs_k141_10578\|\|full | Low-quality |
| S11_vs_k141_36288\|\|full | Not-determined |
| S11_vs_k141_43031\|\|full | Low-quality |
| S11_vs_k141_56210\|\|full | Not-determined |
| S11_vs_k141_4040\|\|full | Not-determined |
| S11_vs_k141_56219\|\|full | Low-quality |
| S11_vs_k141_68513\|\|full | Low-quality |
| S11_vs_k141_43058\|\|full | Low-quality |
| S11_vs_k141_36310\|\|full | Low-quality |
| S11_vs_k141_81949\|\|full | Low-quality |
| S11_vs_k141_68533\|\|full | Low-quality |
| S11_vs_k141_10586\|\|full | Low-quality |
| S11_vs_k141_23923\|\|full | Not-determined |
| S11_vs_k141_23924\|\|full | Not-determined |
| S11_vs_k141_62361\|\|full | Low-quality |
| S11_vs_k141_4077\|\|full | Low-quality |
| S11_vs_k141_62365\|\|full | Not-determined |
| S11_vs_k141_4084\|\|full | Low-quality |
| S11_vs_k141_95151\|\|full | Low-quality |
| S11_vs_k141_88781\|\|full | Low-quality |
| S11_vs_k141_36352\|\|full | Not-determined |
| S11_vs_k141_36360\|\|full | Low-quality |
| S11_vs_k141_81966\|\|full | Not-determined |
| S11_vs_k141_43115\|\|full | Low-quality |
| S11_vs_k141_68565\|\|full | Low-quality |
| S11_vs_k141_23935\|\|full | Low-quality |
| S11_vs_k141_23936\|\|full | Low-quality |
| S11_vs_k141_81976\|\|full | Low-quality |
| S11_vs_k141_88810\|\|full | Low-quality |
| S11_vs_k141_43117\|\|full | Low-quality |
| S11_vs_k141_29683\|\|full | Not-determined |
| S11_vs_k141_23943\|\|full | Not-determined |
| S11_vs_k141_4104\|\|full | Not-determined |
| S11_vs_k141_101707\|\|full | Low-quality |
| S11_vs_k141_36386\|\|full | Low-quality |
| S11_vs_k141_95186\|\|full | Low-quality |
| S11_vs_k141_68582\|\|full | Not-determined |
| S11_vs_k141_95190\|\|full | Low-quality |
| S11_vs_k141_43158\|\|full | Not-determined |
| S11_vs_k141_62439\|\|full | Low-quality |
| S11_vs_k141_36397\|\|full | Low-quality |
| S11_vs_k141_4135\|\|full | Low-quality |
| S11_vs_k141_29739\|\|full | Low-quality |
| S11_vs_k141_88836\|\|full | Low-quality |
| S11_vs_k141_36404\|\|full | Low-quality |
| S11_vs_k141_88844\|\|full | Low-quality |
| S11_vs_k141_43179\|\|full | Low-quality |
| S11_vs_k141_36408\|\|full | Low-quality |
| S11_vs_k141_10613\|\|full | Not-determined |
| S11_vs_k141_68589\|\|full | Low-quality |

| | |
|---|---|
| S11_vs_k141_56370\|\|full | Low-quality |
| S11_vs_k141_62458\|\|full | Low-quality |
| S11_vs_k141_49082\|\|full | Not-determined |
| S11_vs_k141_29746\|\|full | Low-quality |
| S11_vs_k141_68597\|\|full | Not-determined |
| S11_vs_k141_24019\|\|full | Low-quality |
| S11_vs_k141_10633\|\|full | Low-quality |
| S11_vs_k141_36432\|\|full | Low-quality |
| S11_vs_k141_88865\|\|full | Low-quality |
| S11_vs_k141_101768\|\|full | Not-determined |
| S11_vs_k141_68606\|\|full | Low-quality |
| S11_vs_k141_43201\|\|full | Not-determined |
| S11_vs_k141_49097\|\|full | Low-quality |
| S11_vs_k141_29765\|\|full | Not-determined |
| S11_vs_k141_62486\|\|full | Not-determined |
| S11_vs_k141_82062\|\|full | Low-quality |
| S11_vs_k141_49101\|\|full | Low-quality |
| S11_vs_k141_10650\|\|full | Not-determined |
| S11_vs_k141_43211\|\|full | Low-quality |
| S11_vs_k141_101783\|\|full | Low-quality |
| S11_vs_k141_24055\|\|full | Low-quality |
| S11_vs_k141_88882\|\|full | Low-quality |
| S11_vs_k141_56425\|\|full | Low-quality |
| S11_vs_k141_95267\|\|full | Low-quality |
| S11_vs_k141_24066\|\|full | Low-quality |
| S11_vs_k141_88890\|\|full | Not-determined |
| S11_vs_k141_10661\|\|full | Low-quality |
| S11_vs_k141_29773\|\|full | Low-quality |
| S11_vs_k141_24067\|\|full | Not-determined |
| S11_vs_k141_62526\|\|full | Not-determined |
| S11_vs_k141_4181\|\|full | Low-quality |
| S11_vs_k141_43230\|\|full | Low-quality |
| S11_vs_k141_36484\|\|full | Not-determined |
| S11_vs_k141_62536\|\|full | Low-quality |
| S11_vs_k141_24082\|\|full | Not-determined |
| S11_vs_k141_75976\|\|full | Low-quality |
| S11_vs_k141_62540\|\|full | Not-determined |
| S11_vs_k141_36508\|\|full | Low-quality |
| S11_vs_k141_43246\|\|full | Low-quality |
| S11_vs_k141_62548\|\|full | Low-quality |
| S11_vs_k141_43257\|\|full | Low-quality |
| S11_vs_k141_16986\|\|full | Low-quality |
| S11_vs_k141_101836\|\|full | Low-quality |
| S11_vs_k141_95328\|\|full | Not-determined |
| S11_vs_k141_10725\|\|full | Low-quality |
| S11_vs_k141_101846\|\|full | Low-quality |
| S11_vs_k141_36541\|\|full | Low-quality |
| S11_vs_k141_29831\|\|full | Not-determined |
| S11_vs_k141_16997\|\|full | Low-quality |
| S11_vs_k141_56481\|\|full | Not-determined |
| S11_vs_k141_4195\|\|full | Not-determined |
| S11_vs_k141_56483\|\|full | Low-quality |
| S11_vs_k141_24128\|\|full | Not-determined |
| S11_vs_k141_17014\|\|full | Low-quality |
| S11_vs_k141_36568\|\|full | Low-quality |
| S11_vs_k141_43300\|\|full | Not-determined |
| S11_vs_k141_88973\|\|full | Low-quality |
| S11_vs_k141_24154\|\|full | Not-determined |
| S11_vs_k141_43316\|\|full | Low-quality |
| S11_vs_k141_36577\|\|full | Not-determined |
| S11_vs_k141_36578\|\|full | Not-determined |
| S11_vs_k141_56504\|\|full | Not-determined |
| S11_vs_k141_29855\|\|full | Low-quality |
| S11_vs_k141_17031\|\|full | Not-determined |
| S11_vs_k141_24165\|\|full | Low-quality |
| S11_vs_k141_10765\|\|full | Low-quality |
| S11_vs_k141_95401\|\|full | Low-quality |
| S11_vs_k141_88997\|\|full | Low-quality |
| S11_vs_k141_101873\|\|full | Low-quality |
| S11_vs_k141_10770\|\|full | Not-determined |
| S11_vs_k141_10774\|\|full | Low-quality |
| S11_vs_k141_101884\|\|full | Not-determined |
| S11_vs_k141_76061\|\|full | Low-quality |
| S11_vs_k141_89010\|\|full | Low-quality |
| S11_vs_k141_76073\|\|full | Low-quality |
| S11_vs_k141_89019\|\|full | Low-quality |
| S11_vs_k141_89023\|\|full | Low-quality |
| S11_vs_k141_76078\|\|full | Low-quality |
| S11_vs_k141_76079\|\|full | Not-determined |
| S8_vs_k141_108709\|\|full | Medium-quality |
| S11_vs_k141_89026\|\|full | Low-quality |
| S7_vs_97737\|\|full | High-quality |
| S11_vs_k141_89027\|\|full | Low-quality |
| S11_vs_k141_10807\|\|full | Low-quality |
| S11_vs_k141_101924\|\|full | Not-determined |
| S11_vs_k141_68697\|\|full | Not-determined |
| S11_vs_k141_29912\|\|full | Low-quality |
| S11_vs_k141_4283\|\|full | Low-quality |
| S11_vs_k141_76094\|\|full | Low-quality |
| S11_vs_k141_24243\|\|full | Not-determined |
| S11_vs_k141_10842\|\|full | Not-determined |
| S11_vs_k141_89068\|\|full | Not-determined |
| S11_vs_k141_29929\|\|full | Low-quality |
| S11_vs_k141_62599\|\|full | Low-quality |
| S11_vs_k141_24258\|\|full | Low-quality |
| S11_vs_k141_49176\|\|full | Low-quality |
| S11_vs_k141_17120\|\|full | Not-determined |
| S11_vs_k141_101966\|\|full | Low-quality |
| S11_vs_k141_82176\|\|full | Not-determined |
| S11_vs_k141_101974\|\|full | Not-determined |
| S11_vs_k141_17125\|\|full | Low-quality |
| S11_vs_k141_76120\|\|full | Low-quality |
| S11_vs_k141_43383\|\|full | Low-quality |
| S11_vs_k141_56577\|\|full | Low-quality |
| S11_vs_k141_29955\|\|full | Low-quality |
| S11_vs_k141_43389\|\|full | Not-determined |
| S11_vs_k141_76125\|\|full | Not-determined |
| S11_vs_k141_4332\|\|full | Low-quality |
| S11_vs_k141_10895\|\|full | Low-quality |
| S11_vs_k141_95490\|\|full | Low-quality |
| S11_vs_k141_24299\|\|full | Low-quality |
| S11_vs_k141_10901\|\|full | Not-determined |
| S11_vs_k141_4343\|\|full | Low-quality |
| S11_vs_k141_17135\|\|full | Low-quality |
| S11_vs_k141_43404\|\|full | Not-determined |
| S11_vs_k141_49220\|\|full | Low-quality |
| S11_vs_k141_101999\|\|full | Low-quality |
| S11_vs_k141_43412\|\|full | Low-quality |
| S11_vs_k141_17139\|\|full | Not-determined |
| S11_vs_k141_43419\|\|full | Low-quality |
| S11_vs_k141_95503\|\|full | Low-quality |
| S11_vs_k141_17154\|\|full | Not-determined |
| S11_vs_k141_36714\|\|full | Not-determined |
| S11_vs_k141_49241\|\|full | Not-determined |
| S11_vs_k141_4371\|\|full | Not-determined |
| S11_vs_k141_17164\|\|full | Not-determined |
| S11_vs_k141_49252\|\|full | Low-quality |
| S11_vs_k141_62673\|\|full | Not-determined |
| S11_vs_k141_24354\|\|full | Low-quality |
| S11_vs_k141_4381\|\|full | Low-quality |
| S11_vs_k141_17187\|\|full | Not-determined |
| S11_vs_k141_95537\|\|full | Low-quality |
| S11_vs_k141_68729\|\|full | Not-determined |
| S11_vs_k141_43449\|\|full | Not-determined |
| S11_vs_k141_62694\|\|full | Low-quality |
| S11_vs_k141_49264\|\|full | Low-quality |
| S11_vs_k141_24377\|\|full | Low-quality |
| S11_vs_k141_102076\|\|full | Low-quality |
| S11_vs_k141_49267\|\|full | Low-quality |
| S11_vs_k141_49270\|\|full | Low-quality |
| S11_vs_k141_56583\|\|full | Low-quality |
| S11_vs_k141_95559\|\|full | Low-quality |
| S11_vs_k141_49275\|\|full | Not-determined |
| S11_vs_k141_24389\|\|full | Low-quality |
| S11_vs_k141_10924\|\|full | Not-determined |
| S11_vs_k141_68763\|\|full | Low-quality |
| S11_vs_k141_24396\|\|full | Low-quality |
| S11_vs_k141_82198\|\|full | Low-quality |
| S11_vs_k141_102134\|\|full | Low-quality |
| S11_vs_k141_24416\|\|full | Not-determined |
| S11_vs_k141_49307\|\|full | Low-quality |
| S11_vs_k141_89074\|\|full | Low-quality |
| S11_vs_k141_24423\|\|full | Low-quality |
| S11_vs_k141_89082\|\|full | Low-quality |
| S11_vs_k141_102147\|\|full | Not-determined |
| S11_vs_k141_82238\|\|full | Low-quality |
| S11_vs_k141_89088\|\|full | Low-quality |
| S11_vs_k141_24442\|\|full | Not-determined |
| S11_vs_k141_89100\|\|full | Low-quality |
| S11_vs_k141_102166\|\|full | Low-quality |
| S11_vs_k141_17242\|\|full | Not-determined |
| S11_vs_k141_102176\|\|full | Low-quality |
| S11_vs_k141_62773\|\|full | Not-determined |
| S11_vs_k141_95653\|\|full | Low-quality |
| S11_vs_k141_82267\|\|full | Low-quality |
| S11_vs_k141_36775\|\|full | Not-determined |
| S11_vs_k141_62787\|\|full | Not-determined |
| S11_vs_k141_17254\|\|full | Low-quality |
| S11_vs_k141_62790\|\|full | Not-determined |
| S11_vs_k141_24493\|\|full | Low-quality |
| S11_vs_k141_102190\|\|full | Not-determined |
| S11_vs_k141_49361\|\|full | Low-quality |
| S11_vs_k141_95685\|\|full | Low-quality |
| S11_vs_k141_68842\|\|full | Not-determined |
| S11_vs_k141_76146\|\|full | Low-quality |
| S11_vs_k141_43482\|\|full | Not-determined |
| S11_vs_k141_4474\|\|full | Low-quality |
| S11_vs_k141_24520\|\|full | Low-quality |
| S11_vs_k141_49362\|\|full | Not-determined |
| S11_vs_k141_89161\|\|full | Low-quality |
| S11_vs_k141_102208\|\|full | Low-quality |
| S11_vs_k141_95696\|\|full | Low-quality |
| S11_vs_k141_43493\|\|full | Not-determined |
| S11_vs_k141_102210\|\|full | Low-quality |
| S11_vs_k141_102220\|\|full | Low-quality |
| S11_vs_k141_68876\|\|full | Low-quality |
| S11_vs_k141_76168\|\|full | Low-quality |
| S11_vs_k141_11059\|\|full | Not-determined |
| S11_vs_k141_4498\|\|full | Low-quality |
| S11_vs_k141_102226\|\|full | Low-quality |
| S11_vs_k141_68886\|\|full | Not-determined |
| S11_vs_k141_11069\|\|full | Low-quality |
| S11_vs_k141_56713\|\|full | Low-quality |
| S11_vs_k141_102230\|\|full | Low-quality |
| S11_vs_k141_56715\|\|full | Low-quality |
| S11_vs_k141_68893\|\|full | Low-quality |
| S11_vs_k141_4509\|\|full | Low-quality |
| S11_vs_k141_43533\|\|full | Not-determined |
| S11_vs_k141_102241\|\|full | Not-determined |
| S11_vs_k141_49416\|\|full | Low-quality |
| S11_vs_k141_49418\|\|full | Not-determined |
| S11_vs_k141_68913\|\|full | Not-determined |
| S11_vs_k141_102250\|\|full | Low-quality |
| S11_vs_k141_49426\|\|full | Low-quality |
| S11_vs_k141_95771\|\|full | Low-quality |
| S11_vs_k141_17306\|\|full | Low-quality |
| S11_vs_k141_49433\|\|full | Low-quality |
| S11_vs_k141_24617\|\|full | Low-quality |
| S11_vs_k141_4539\|\|full | Low-quality |
| S11_vs_k141_43557\|\|full | Not-determined |
| S11_vs_k141_24620\|\|full | Low-quality |
| S11_vs_k141_24621\|\|full | Low-quality |
| S11_vs_k141_102261\|\|full | Low-quality |
| S11_vs_k141_49437\|\|full | Low-quality |
| S11_vs_k141_56732\|\|full | Low-quality |
| S11_vs_k141_24625\|\|full | Low-quality |
| S11_vs_k141_56733\|\|full | Low-quality |
| S11_vs_k141_95781\|\|full | Low-quality |
| S11_vs_k141_43567\|\|full | Not-determined |
| S11_vs_k141_11129\|\|full | Low-quality |
| S11_vs_k141_102275\|\|full | Not-determined |
| S11_vs_k141_102279\|\|full | Low-quality |
| S11_vs_k141_11136\|\|full | Low-quality |
| S11_vs_k141_102284\|\|full | Low-quality |
| S11_vs_k141_68934\|\|full | Low-quality |
| S11_vs_k141_49455\|\|full | Not-determined |
| S11_vs_k141_36895\|\|full | Not-determined |
| S11_vs_k141_24648\|\|full | Not-determined |
| S11_vs_k141_49464\|\|full | Not-determined |
| S11_vs_k141_43569\|\|full | Low-quality |
| S11_vs_k141_49473\|\|full | Low-quality |
| S11_vs_k141_95814\|\|full | Low-quality |
| S11_vs_k141_4600\|\|full | Not-determined |
| S11_vs_k141_49476\|\|full | Low-quality |
| S11_vs_k141_30073\|\|full | Low-quality |
| S11_vs_k141_76211\|\|full | Low-quality |
| S11_vs_k141_89173\|\|full | Low-quality |
| S11_vs_k141_24666\|\|full | Low-quality |
| S11_vs_k141_95839\|\|full | Low-quality |
| S11_vs_k141_62884\|\|full | Not-determined |
| S11_vs_k141_76220\|\|full | Low-quality |
| S11_vs_k141_24669\|\|full | Low-quality |
| S11_vs_k141_11164\|\|full | Low-quality |
| S11_vs_k141_68972\|\|full | Not-determined |
| S11_vs_k141_95849\|\|full | Not-determined |
| S11_vs_k141_17397\|\|full | Low-quality |
| S11_vs_k141_4637\|\|full | Not-determined |
| S11_vs_k141_4638\|\|full | Low-quality |
| S11_vs_k141_49500\|\|full | Low-quality |
| S11_vs_k141_49502\|\|full | Low-quality |
| S11_vs_k141_24696\|\|full | Low-quality |
| S11_vs_k141_76250\|\|full | Low-quality |
| S11_vs_k141_62923\|\|full | Low-quality |
| S11_vs_k141_17416\|\|full | Low-quality |
| S11_vs_k141_30122\|\|full | Low-quality |
| S11_vs_k141_36954\|\|full | Low-quality |
| S11_vs_k141_43610\|\|full | Low-quality |
| S3_vs_k141_81793\|\|full | High-quality |
| S11_vs_k141_69001\|\|full | Low-quality |
| S11_vs_k141_56764\|\|full | Low-quality |
| S11_vs_k141_56767\|\|full | Low-quality |
| S11_vs_k141_62946\|\|full | Low-quality |
| S11_vs_k141_30137\|\|full | Low-quality |
| S11_vs_k141_24717\|\|full | Not-determined |
| S11_vs_k141_43626\|\|full | Low-quality |
| S11_vs_k141_49521\|\|full | Low-quality |
| S11_vs_k141_4677\|\|full | Low-quality |
| S11_vs_k141_95908\|\|full | Not-determined |
| S11_vs_k141_56781\|\|full | Low-quality |
| S11_vs_k141_76284\|\|full | Low-quality |
| S11_vs_k141_36967\|\|full | Not-determined |
| S11_vs_k141_56786\|\|full | Low-quality |
| S11_vs_k141_69006\|\|full | Low-quality |
| S3_vs_k141_39774\|\|full | Medium-quality |
| S11_vs_k141_30155\|\|full | Low-quality |
| S11_vs_k141_49529\|\|full | Low-quality |
| S11_vs_k141_56789\|\|full | Low-quality |
| S11_vs_k141_69010\|\|full | Low-quality |
| S11_vs_k141_24722\|\|full | Low-quality |
| S11_vs_k141_4686\|\|full | Low-quality |
| S11_vs_k141_82343\|\|full | Not-determined |
| S11_vs_k141_69019\|\|full | Low-quality |
| S11_vs_k141_89262\|\|full | Low-quality |
| S11_vs_k141_24736\|\|full | Low-quality |
| S11_vs_k141_49554\|\|full | Low-quality |
| S11_vs_k141_37010\|\|full | Low-quality |
| S11_vs_k141_95929\|\|full | Low-quality |
| S11_vs_k141_30184\|\|full | Low-quality |
| S11_vs_k141_37022\|\|full | Low-quality |
| S11_vs_k141_56831\|\|full | Not-determined |
| S11_vs_k141_37026\|\|full | Low-quality |
| S11_vs_k141_43650\|\|full | Low-quality |
| S11_vs_k141_24777\|\|full | Low-quality |
| S11_vs_k141_37052\|\|full | Not-determined |
| S11_vs_k141_102376\|\|full | Low-quality |
| S11_vs_k141_43665\|\|full | Low-quality |
| S11_vs_k141_30223\|\|full | Low-quality |
| S11_vs_k141_89308\|\|full | Not-determined |
| S11_vs_k141_95982\|\|full | Not-determined |
| S11_vs_k141_11205\|\|full | Low-quality |
| S11_vs_k141_56867\|\|full | Low-quality |
| S11_vs_k141_30237\|\|full | Low-quality |
| S11_vs_k141_43684\|\|full | Low-quality |
| S11_vs_k141_69049\|\|full | Low-quality |
| S11_vs_k141_30253\|\|full | Not-determined |
| S11_vs_k141_11219\|\|full | Low-quality |
| S11_vs_k141_76373\|\|full | Low-quality |
| S11_vs_k141_24817\|\|full | Not-determined |
| S11_vs_k141_4768\|\|full | Low-quality |
| S11_vs_k141_102419\|\|full | Low-quality |
| S11_vs_k141_63050\|\|full | Low-quality |
| S11_vs_k141_17555\|\|full | Not-determined |
| S11_vs_k141_4776\|\|full | Not-determined |
| S11_vs_k141_63060\|\|full | Not-determined |
| S11_vs_k141_56916\|\|full | Low-quality |
| S11_vs_k141_102435\|\|full | Not-determined |
| S11_vs_k141_69073\|\|full | Not-determined |
| S11_vs_k141_43717\|\|full | Low-quality |
| S11_vs_k141_24825\|\|full | Not-determined |
| S11_vs_k141_56928\|\|full | Low-quality |
| S11_vs_k141_37109\|\|full | Low-quality |
| S11_vs_k141_37118\|\|full | Low-quality |
| S11_vs_k141_89317\|\|full | Low-quality |
| S11_vs_k141_37120\|\|full | Low-quality |
| S11_vs_k141_89320\|\|full | Not-determined |
| S11_vs_k141_102454\|\|full | Low-quality |
| S11_vs_k141_49648\|\|full | Low-quality |
| S11_vs_k141_96031\|\|full | Not-determined |
| S11_vs_k141_30313\|\|full | Low-quality |
| S11_vs_k141_76399\|\|full | Low-quality |
| S11_vs_k141_63087\|\|full | Low-quality |
| S11_vs_k141_37137\|\|full | Low-quality |
| S11_vs_k141_30320\|\|full | Low-quality |
| S11_vs_k141_76406\|\|full | Low-quality |
| S11_vs_k141_69107\|\|full | Not-determined |
| S11_vs_k141_69109\|\|full | Not-determined |
| S11_vs_k141_102463\|\|full | Low-quality |
| S11_vs_k141_96048\|\|full | Not-determined |
| S11_vs_k141_69111\|\|full | Low-quality |
| S11_vs_k141_30324\|\|full | Low-quality |
| S11_vs_k141_24880\|\|full | Low-quality |
| S11_vs_k141_49674\|\|full | Not-determined |
| S11_vs_k141_69127\|\|full | Not-determined |
| S11_vs_k141_96059\|\|full | Not-determined |
| S11_vs_k141_63109\|\|full | Not-determined |
| S11_vs_k141_30349\|\|full | Low-quality |
| S11_vs_k141_43757\|\|full | Low-quality |
| S11_vs_k141_63113\|\|full | Low-quality |
| S11_vs_k141_37171\|\|full | Not-determined |
| S11_vs_k141_96072\|\|full | Low-quality |
| S11_vs_k141_4842\|\|full | Not-determined |
| S11_vs_k141_56984\|\|full | Low-quality |
| S11_vs_k141_37179\|\|full | Low-quality |
| S11_vs_k141_82456\|\|full | Low-quality |
| S11_vs_k141_56994\|\|full | Not-determined |
| S11_vs_k141_69162\|\|full | Low-quality |
| S11_vs_k141_56995\|\|full | Not-determined |
| S11_vs_k141_4853\|\|full | Low-quality |
| S11_vs_k141_89365\|\|full | Low-quality |
| S11_vs_k141_89366\|\|full | Not-determined |
| S11_vs_k141_57009\|\|full | Not-determined |
| S11_vs_k141_4868\|\|full | Not-determined |
| S11_vs_k141_37189\|\|full | Low-quality |
| S11_vs_k141_11234\|\|full | Low-quality |
| S11_vs_k141_37193\|\|full | Not-determined |
| S11_vs_k141_49746\|\|full | Low-quality |
| S11_vs_k141_69191\|\|full | Not-determined |
| S11_vs_k141_96127\|\|full | Low-quality |
| S11_vs_k141_82477\|\|full | Not-determined |
| S11_vs_k141_57030\|\|full | Not-determined |
| S11_vs_k141_69213\|\|full | Low-quality |
| S11_vs_k141_63162\|\|full | Low-quality |
| S11_vs_k141_37205\|\|full | Low-quality |
| S11_vs_k141_43801\|\|full | Not-determined |
| S11_vs_k141_30414\|\|full | Low-quality |
| S11_vs_k141_17611\|\|full | Not-determined |
| S11_vs_k141_57048\|\|full | Low-quality |
| S11_vs_k141_43808\|\|full | Low-quality |
| S11_vs_k141_89402\|\|full | Low-quality |
| S11_vs_k141_57061\|\|full | Low-quality |
| S11_vs_k141_37219\|\|full | Low-quality |
| S11_vs_k141_89403\|\|full | Not-determined |
| S11_vs_k141_89411\|\|full | Low-quality |
| S11_vs_k141_96164\|\|full | Not-determined |
| S11_vs_k141_69219\|\|full | Low-quality |
| S11_vs_k141_17628\|\|full | Not-determined |
| S11_vs_k141_24940\|\|full | Low-quality |
| S11_vs_k141_96171\|\|full | Low-quality |
| S11_vs_k141_37237\|\|full | Low-quality |
| S11_vs_k141_24944\|\|full | Low-quality |
| S2_extracted_k141_21628 | Complete |
| S11_vs_k141_17649\|\|full | Not-determined |
| S11_vs_k141_49796\|\|full | Low-quality |
| S11_vs_k141_102531\|\|full | Not-determined |
| S11_vs_k141_102533\|\|full | Low-quality |
| S11_vs_k141_11280\|\|full | Low-quality |
| S11_vs_k141_11281\|\|full | Not-determined |
| S11_vs_k141_96195\|\|full | Not-determined |
| S11_vs_k141_89441\|\|full | Low-quality |
| S11_vs_k141_102534\|\|full | Low-quality |
| S11_vs_k141_17667\|\|full | Not-determined |
| S11_vs_k141_11294\|\|full | Low-quality |
| S11_vs_k141_49813\|\|full | Not-determined |
| S11_vs_k141_57111\|\|full | Not-determined |
| S11_vs_k141_37241\|\|full | Not-determined |
| S11_vs_k141_102543\|\|full | Low-quality |
| S11_vs_k141_82497\|\|full | Not-determined |
| S11_vs_k141_49828\|\|full | Low-quality |
| S11_vs_k141_30436\|\|full | Low-quality |
| S11_vs_k141_49831\|\|full | Low-quality |
| S11_vs_k141_96207\|\|full | Low-quality |
| S11_vs_k141_43841\|\|full | Low-quality |
| S11_vs_k141_49850\|\|full | Low-quality |
| S11_vs_k141_11319\|\|full | Low-quality |
| S11_vs_k141_82536\|\|full | Low-quality |
| S11_vs_k141_96232\|\|full | Low-quality |
| S11_vs_k141_57136\|\|full | Low-quality |
| S11_vs_k141_11326\|\|full | Low-quality |
| S11_vs_k141_76596\|\|full | Low-quality |
| S11_vs_k141_37276\|\|full | Low-quality |
| S11_vs_k141_4989\|\|full | Low-quality |
| S11_vs_k141_76600\|\|full | Low-quality |
| S11_vs_k141_17723\|\|full | Low-quality |
| S11_vs_k141_4996\|\|full | Low-quality |
| S11_vs_k141_37293\|\|full | Not-determined |
| S11_vs_k141_43875\|\|full | Low-quality |
| S11_vs_k141_57174\|\|full | Low-quality |
| S11_vs_k141_57176\|\|full | Low-quality |
| S11_vs_k141_37298\|\|full | Low-quality |
| S11_vs_k141_96253\|\|full | Low-quality |
| S11_vs_k141_49905\|\|full | Not-determined |
| S11_vs_k141_37311\|\|full | Low-quality |
| S11_vs_k141_17743\|\|full | Low-quality |
| S11_vs_k141_69309\|\|full | Low-quality |
| S11_vs_k141_43895\|\|full | Low-quality |
| S11_vs_k141_69314\|\|full | Not-determined |
| S11_vs_k141_57199\|\|full | Low-quality |
| S11_vs_k141_25071\|\|full | Low-quality |
| S11_vs_k141_11374\|\|full | Low-quality |
| S11_vs_k141_30481\|\|full | Low-quality |
| S11_vs_k141_25080\|\|full | Low-quality |
| S11_vs_k141_96267\|\|full | Low-quality |
| S11_vs_k141_96269\|\|full | Not-determined |
| S11_vs_k141_30487\|\|full | Not-determined |
| S11_vs_k141_37327\|\|full | Low-quality |
| S11_vs_k141_69329\|\|full | Low-quality |
| S11_vs_k141_25098\|\|full | Not-determined |
| S11_vs_k141_49935\|\|full | Low-quality |
| S11_vs_k141_11386\|\|full | Low-quality |
| S11_vs_k141_37345\|\|full | Not-determined |
| S11_vs_k141_11394\|\|full | Low-quality |
| S11_vs_k141_69339\|\|full | Low-quality |
| S11_vs_k141_25106\|\|full | Not-determined |
| S11_vs_k141_82619\|\|full | Not-determined |
| S11_vs_k141_57230\|\|full | Low-quality |
| S11_vs_k141_49956\|\|full | Not-determined |
| S11_vs_k141_82621\|\|full | Low-quality |
| S11_vs_k141_30525\|\|full | Low-quality |
| S11_vs_k141_17774\|\|full | Low-quality |
| S11_vs_k141_37351\|\|full | Low-quality |
| S11_vs_k141_76655\|\|full | Not-determined |
| S11_vs_k141_82638\|\|full | Low-quality |
| S11_vs_k141_43956\|\|full | Low-quality |
| S11_vs_k141_96315\|\|full | Low-quality |
| S11_vs_k141_49961\|\|full | Low-quality |
| S11_vs_k141_30533\|\|full | Low-quality |
| S11_vs_k141_96321\|\|full | Low-quality |
| S11_vs_k141_82647\|\|full | Low-quality |
| S11_vs_k141_11431\|\|full | Not-determined |
| S11_vs_k141_57263\|\|full | Not-determined |
| S11_vs_k141_11437\|\|full | Not-determined |
| S11_vs_k141_69387\|\|full | Low-quality |
| S11_vs_k141_30540\|\|full | Low-quality |
| S11_vs_k141_17808\|\|full | Not-determined |
| S11_vs_k141_11444\|\|full | Low-quality |
| S11_vs_k141_25132\|\|full | Not-determined |
| S11_vs_k141_76680\|\|full | Low-quality |
| S11_vs_k141_30549\|\|full | Not-determined |
| S11_vs_k141_30551\|\|full | Not-determined |
| S11_vs_k141_57282\|\|full | Low-quality |
| S11_vs_k141_82677\|\|full | Low-quality |
| S11_vs_k141_57283\|\|full | Not-determined |
| S11_vs_k141_49988\|\|full | Low-quality |
| S11_vs_k141_49993\|\|full | Low-quality |
| S11_vs_k141_69409\|\|full | Not-determined |
| S11_vs_k141_37408\|\|full | Low-quality |
| S11_vs_k141_63229\|\|full | Not-determined |
| S11_vs_k141_82705\|\|full | Not-determined |
| S11_vs_k141_50010\|\|full | Low-quality |
| S11_vs_k141_69425\|\|full | Not-determined |
| S11_vs_k141_89517\|\|full | Low-quality |
| S11_vs_k141_76717\|\|full | Low-quality |
| S11_vs_k141_50017\|\|full | Low-quality |
| S11_vs_k141_96375\|\|full | Low-quality |
| S11_vs_k141_17837\|\|full | Low-quality |
| S11_vs_k141_50018\|\|full | Low-quality |
| S11_vs_k141_5037\|\|full | Not-determined |
| S11_vs_k141_30609\|\|full | Low-quality |
| S11_vs_k141_37466\|\|full | Not-determined |
| S11_vs_k141_11512\|\|full | Not-determined |
| S11_vs_k141_102588\|\|full | Low-quality |
| S11_vs_k141_30614\|\|full | Low-quality |
| S11_vs_k141_76745\|\|full | Not-determined |
| S11_vs_k141_17857\|\|full | Low-quality |
| S11_vs_k141_102607\|\|full | Low-quality |
| S11_vs_k141_89554\|\|full | Low-quality |
| S11_vs_k141_63264\|\|full | Not-determined |
| S2_vs_k141_21628\|\|full | High-quality |
| S11_vs_k141_102619\|\|full | Low-quality |
| S11_vs_k141_76768\|\|full | Not-determined |
| S11_vs_k141_30648\|\|full | Low-quality |
| S11_vs_k141_37489\|\|full | Low-quality |
| S11_vs_k141_96454\|\|full | Low-quality |
| S11_vs_k141_37495\|\|full | Low-quality |
| S11_vs_k141_30652\|\|full | Not-determined |
| S11_vs_k141_96458\|\|full | Not-determined |
| S11_vs_k141_17884\|\|full | Low-quality |
| S11_vs_k141_63298\|\|full | Not-determined |
| S11_vs_k141_50087\|\|full | Not-determined |
| S11_vs_k141_17895\|\|full | Not-determined |
| S11_vs_k141_30683\|\|full | Low-quality |
| S11_vs_k141_69453\|\|full | Not-determined |
| S11_vs_k141_57351\|\|full | Low-quality |
| S11_vs_k141_11540\|\|full | Low-quality |
| S11_vs_k141_57357\|\|full | Low-quality |
| S11_vs_k141_44057\|\|full | Low-quality |
| S11_vs_k141_11554\|\|full | Not-determined |
| S11_vs_k141_89577\|\|full | Not-determined |
| S11_vs_k141_57370\|\|full | Low-quality |
| S11_extracted_k141_99751 | Medium-quality |
| S11_vs_k141_57373\|\|full | Not-determined |
| S11_vs_k141_76785\|\|full | Low-quality |
| S11_vs_k141_44072\|\|full | Low-quality |
| S11_vs_k141_11556\|\|full | Low-quality |
| S11_vs_k141_17934\|\|full | Not-determined |
| S11_vs_k141_69492\|\|full | Not-determined |
| S11_vs_k141_11563\|\|full | Not-determined |
| S11_vs_k141_102662\|\|full | Low-quality |
| S11_vs_k141_69493\|\|full | Low-quality |
| S11_vs_k141_17935\|\|full | Not-determined |
| S11_vs_k141_37561\|\|full | Not-determined |
| S11_vs_k141_82797\|\|full | Low-quality |
| S11_vs_k141_82798\|\|full | Low-quality |
| S11_vs_k141_17947\|\|full | Not-determined |
| S11_vs_k141_57410\|\|full | Not-determined |
| S11_vs_k141_25206\|\|full | Not-determined |
| S11_vs_k141_96564\|\|full | Low-quality |
| S11_vs_k141_89620\|\|full | Not-determined |
| S11_vs_k141_102695\|\|full | Low-quality |
| S11_vs_k141_44117\|\|full | Low-quality |
| S11_vs_k141_37581\|\|full | Not-determined |
| S11_vs_k141_25217\|\|full | Low-quality |
| S11_vs_k141_99751\|\|full | Medium-quality |
| S11_vs_k141_82820\|\|full | Low-quality |
| S11_vs_k141_37589\|\|full | Low-quality |
| S11_vs_k141_44126\|\|full | Not-determined |
| S11_vs_k141_37592\|\|full | Low-quality |
| S11_vs_k141_17963\|\|full | Low-quality |
| S11_vs_k141_82825\|\|full | Not-determined |
| S11_vs_k141_102705\|\|full | Low-quality |
| S11_vs_k141_5093\|\|full | Low-quality |
| S9_vs_k141_67792\|\|full | Complete |
| S11_vs_k141_82833\|\|full | Low-quality |
| S11_vs_k141_17967\|\|full | Low-quality |
| S11_vs_k141_57446\|\|full | Not-determined |
| S11_vs_k141_82851\|\|full | Low-quality |
| S11_vs_k141_30750\|\|full | Low-quality |
| S11_vs_k141_89685\|\|full | Low-quality |
| S11_vs_k141_76807\|\|full | Low-quality |
| S11_vs_k141_57449\|\|full | Low-quality |
| S11_vs_k141_69553\|\|full | Low-quality |
| S11_vs_k141_82863\|\|full | Low-quality |
| S11_vs_k141_69557\|\|full | Low-quality |
| S11_vs_k141_30764\|\|full | Not-determined |
| S11_vs_k141_17991\|\|full | Low-quality |
| S11_vs_k141_30770\|\|full | Low-quality |
| S11_vs_k141_69566\|\|full | Not-determined |
| S11_vs_k141_17995\|\|full | Low-quality |
| S11_vs_k141_25300\|\|full | Low-quality |
| S11_vs_k141_30784\|\|full | Low-quality |
| S11_vs_k141_30788\|\|full | Low-quality |
| S11_vs_k141_18011\|\|full | Low-quality |
| S11_vs_k141_30797\|\|full | Low-quality |
| S11_vs_k141_63361\|\|full | Not-determined |
| S11_vs_k141_96649\|\|full | Low-quality |
| S11_vs_k141_5140\|\|full | Not-determined |
| S11_vs_k141_18021\|\|full | Low-quality |
| S11_vs_k141_76863\|\|full | Low-quality |
| S11_vs_k141_44177\|\|full | Not-determined |

| | |
|---|---|
| S11_vs_k141_63370||full | Not-determined |
| S11_vs_k141_69597||full | Not-determined |
| S11_vs_k141_96657||full | Low-quality |
| S11_vs_k141_44187||full | Low-quality |
| S11_vs_k141_44193||full | Not-determined |
| S11_vs_k141_25347||full | Not-determined |
| S11_vs_k141_30834||full | Low-quality |
| S11_vs_k141_76887||full | Low-quality |
| S11_vs_k141_82944||full | Not-determined |
| S7_vs_35914||full | High-quality |
| S11_vs_k141_63390||full | Low-quality |
| S11_vs_k141_18049||full | Low-quality |
| S11_vs_k141_76900||full | Not-determined |
| S11_vs_k141_44221||full | Not-determined |
| S11_vs_k141_69626||full | Not-determined |
| S11_vs_k141_44224||full | Low-quality |
| S11_vs_k141_82954||full | Low-quality |
| S11_vs_k141_18072||full | Low-quality |
| S11_vs_k141_25358||full | Low-quality |
| S11_vs_k141_44241||full | Low-quality |
| S11_vs_k141_5224||full | Low-quality |
| S11_vs_k141_96678||full | Low-quality |
| S11_vs_k141_76923||full | Low-quality |
| S11_vs_k141_25368||full | Not-determined |
| S11_vs_k141_37716||full | Low-quality |
| S11_vs_k141_82964||full | Low-quality |
| S11_vs_k141_44255||full | Not-determined |
| S11_vs_k141_30879||full | Low-quality |
| S11_vs_k141_44259||full | Not-determined |
| S11_vs_k141_5242||full | Low-quality |
| S11_vs_k141_37737||full | Low-quality |
| S11_vs_k141_30895||full | Low-quality |
| S11_vs_k141_18084||full | Low-quality |
| S11_vs_k141_76958||full | Low-quality |
| S11_vs_k141_76965||full | Low-quality |
| S11_vs_k141_44283||full | Not-determined |
| S11_vs_k141_25406||full | Low-quality |
| S11_vs_k141_63477||full | Low-quality |
| S11_vs_k141_37764||full | Low-quality |
| S11_vs_k141_69717||full | Low-quality |
| S11_vs_k141_37778||full | Low-quality |
| S11_vs_k141_96733||full | Low-quality |
| S11_vs_k141_44322||full | Not-determined |
| S11_vs_k141_25417||full | Low-quality |
| S11_vs_k141_44323||full | Low-quality |
| S11_vs_k141_44324||full | Low-quality |
| S11_vs_k141_96737||full | Low-quality |
| S11_vs_k141_96738||full | Low-quality |
| S11_vs_k141_37795||full | Low-quality |
| S11_vs_k141_18140||full | Low-quality |
| S11_vs_k141_69740||full | Not-determined |
| S11_vs_k141_57535||full | Low-quality |
| S11_vs_k141_63523||full | Low-quality |
| S11_vs_k141_96739||full | Not-determined |
| S11_vs_k141_77026||full | Not-determined |
| S11_vs_k141_25436||full | Low-quality |
| S11_vs_k141_50108||full | Not-determined |
| S11_vs_k141_89736||full | Low-quality |
| S11_vs_k141_18152||full | Low-quality |
| S11_vs_k141_57548||full | Low-quality |
| S11_vs_k141_69759||full | Low-quality |
| S11_vs_k141_77034||full | Not-determined |
| S11_vs_k141_63538||full | Low-quality |
| S11_vs_k141_77036||full | Low-quality |
| S11_vs_k141_89748||full | Not-determined |
| S11_vs_k141_25447||full | Not-determined |
| S11_vs_k141_11680||full | Low-quality |
| S11_vs_k141_57563||full | Low-quality |
| S11_vs_k141_77050||full | Low-quality |
| S11_vs_k141_77053||full | Low-quality |
| S11_vs_k141_63563||full | Not-determined |
| S11_vs_k141_57577||full | Low-quality |
| S11_vs_k141_44342||full | Not-determined |
| S11_vs_k141_57578||full | Not-determined |
| S11_vs_k141_77063||full | Low-quality |
| S11_vs_k141_102780||full | Low-quality |
| S11_vs_k141_11701||full | Low-quality |
| S11_vs_k141_57584||full | Low-quality |
| S11_vs_k141_77067||full | Low-quality |
| S11_vs_k141_69815||full | Low-quality |
| S11_vs_k141_102790||full | Low-quality |
| S11_vs_k141_83060||full | Low-quality |
| S11_vs_k141_31012||full | Low-quality |
| S11_vs_k141_69816||full | Not-determined |
| S11_vs_k141_69818||full | Low-quality |
| S11_vs_k141_25485||full | Not-determined |
| S11_vs_k141_37855||full | Low-quality |
| S11_vs_k141_25492||full | Low-quality |
| S11_vs_k141_77087||full | Not-determined |
| S11_vs_k141_69823||full | Low-quality |
| S11_vs_k141_5329||full | Low-quality |
| S11_vs_k141_83066||full | Low-quality |
| S11_vs_k141_102807||full | Low-quality |
| S11_vs_k141_50161||full | Low-quality |
| S11_vs_k141_96790||full | Low-quality |
| S11_vs_k141_18191||full | Not-determined |
| S11_vs_k141_77101||full | Not-determined |
| S11_vs_k141_50171||full | Not-determined |
| S11_vs_k141_50172||full | Low-quality |
| S11_vs_k141_44383||full | Not-determined |
| S11_vs_k141_37881||full | Not-determined |
| S11_vs_k141_31046||full | Low-quality |
| S11_vs_k141_102808||full | Low-quality |
| S11_vs_k141_96797||full | Low-quality |
| S11_vs_k141_83115||full | Low-quality |
| S11_vs_k141_77117||full | Not-determined |
| S11_vs_k141_57618||full | Low-quality |
| S11_vs_k141_5350||full | Low-quality |
| S11_vs_k141_25509||full | Not-determined |
| S11_vs_k141_11735||full | Not-determined |
| S11_vs_k141_18219||full | Low-quality |
| S11_vs_k141_50200||full | Not-determined |
| S11_vs_k141_96829||full | Low-quality |
| S11_vs_k141_5369||full | Low-quality |
| S11_vs_k141_102841||full | Not-determined |
| S11_vs_k141_69853||full | Low-quality |
| S11_vs_k141_57649||full | Low-quality |
| S11_vs_k141_31087||full | Not-determined |
| S11_vs_k141_31088||full | Low-quality |
| S11_vs_k141_57664||full | Not-determined |
| S11_vs_k141_5387||full | Low-quality |
| S11_vs_k141_102860||full | Low-quality |
| S11_vs_k141_77139||full | Low-quality |
| S11_vs_k141_69875||full | Low-quality |
| S11_vs_k141_11756||full | Low-quality |
| S11_vs_k141_18241||full | Low-quality |
| S11_vs_k141_31103||full | Not-determined |
| S11_vs_k141_5395||full | Low-quality |
| S11_vs_k141_63694||full | Low-quality |
| S11_vs_k141_18242||full | Low-quality |
| S11_vs_k141_25548||full | Low-quality |
| S11_vs_k141_31112||full | Low-quality |
| S11_vs_k141_50227||full | Not-determined |
| S11_vs_k141_69882||full | Low-quality |
| S11_vs_k141_57675||full | Low-quality |
| S11_vs_k141_50228||full | Low-quality |
| S11_vs_k141_96864||full | Not-determined |
| S11_vs_k141_5403||full | Low-quality |
| S11_vs_k141_5408||full | Low-quality |
| S11_vs_k141_63707||full | Low-quality |
| S11_vs_k141_96876||full | Not-determined |
| S11_vs_k141_18256||full | Not-determined |
| S11_vs_k141_83167||full | Low-quality |
| S11_vs_k141_69897||full | Not-determined |
| S11_vs_k141_77151||full | Low-quality |
| S11_vs_k141_5419||full | Low-quality |
| S11_vs_k141_5424||full | Low-quality |
| S11_vs_k141_37914||full | Low-quality |
| S11_vs_k141_63727||full | Low-quality |
| S11_vs_k141_31140||full | Not-determined |
| S11_vs_k141_11781||full | Low-quality |
| S11_vs_k141_57710||full | Low-quality |
| S11_vs_k141_89833||full | Low-quality |
| S11_vs_k141_18277||full | Low-quality |
| S11_vs_k141_37922||full | Low-quality |
| S11_vs_k141_57715||full | Low-quality |
| S11_vs_k141_96904||full | Low-quality |
| S11_vs_k141_37929||full | Not-determined |
| S11_vs_k141_57719||full | Low-quality |
| S11_vs_k141_57720||full | Not-determined |
| S11_vs_k141_69925||full | Not-determined |
| S11_vs_k141_25604||full | Low-quality |
| S11_vs_k141_5456||full | Low-quality |
| S11_vs_k141_69927||full | Low-quality |
| S11_vs_k141_63756||full | Not-determined |
| S11_vs_k141_63757||full | Low-quality |
| S11_vs_k141_69931||full | Not-determined |
| S11_vs_k141_5462||full | Low-quality |
| S11_vs_k141_18312||full | Low-quality |
| S11_vs_k141_5463||full | Low-quality |
| S11_vs_k141_18314||full | Low-quality |
| S11_vs_k141_44489||full | Not-determined |
| S11_vs_k141_44492||full | Not-determined |
| S11_vs_k141_63764||full | Low-quality |
| S11_vs_k141_96919||full | Low-quality |
| S11_vs_k141_5488||full | Low-quality |
| S11_vs_k141_31199||full | Low-quality |
| S11_vs_k141_96926||full | Low-quality |
| S11_vs_k141_50309||full | Not-determined |
| S11_vs_k141_5493||full | Low-quality |
| S11_vs_k141_44506||full | Not-determined |
| S11_vs_k141_89853||full | Low-quality |
| S11_vs_k141_31201||full | Not-determined |
| S11_vs_k141_50313||full | Low-quality |
| S11_vs_k141_37983||full | Not-determined |
| S11_vs_k141_25656||full | Low-quality |
| S11_vs_k141_18331||full | Not-determined |
| S11_vs_k141_31209||full | Low-quality |
| S11_vs_k141_96950||full | Low-quality |
| S11_vs_k141_50328||full | Low-quality |
| S11_vs_k141_96957||full | Low-quality |
| S11_vs_k141_50330||full | Not-determined |
| S11_vs_k141_5503||full | Low-quality |
| S11_vs_k141_83241||full | Low-quality |
| S11_vs_k141_63825||full | Not-determined |
| S11_vs_k141_5511||full | Not-determined |
| S11_vs_k141_89879||full | Low-quality |
| S11_vs_k141_50337||full | Not-determined |
| S11_vs_k141_83249||full | Not-determined |
| S11_vs_k141_44538||full | Low-quality |
| S11_vs_k141_11850||full | Not-determined |
| S11_vs_k141_69942||full | Not-determined |
| S11_vs_k141_63864||full | Low-quality |
| S11_vs_k141_18373||full | Low-quality |
| S11_vs_k141_77254||full | Low-quality |
| S11_vs_k141_44550||full | Low-quality |
| S11_vs_k141_31250||full | Low-quality |
| S11_vs_k141_38049||full | Not-determined |
| S11_vs_k141_69947||full | Low-quality |
| S11_vs_k141_44574||full | Not-determined |
| S11_vs_k141_38054||full | Not-determined |
| S11_vs_k141_96979||full | Low-quality |
| S11_vs_k141_5558||full | Low-quality |
| S11_vs_k141_11873||full | Low-quality |
| S11_vs_k141_69953||full | Not-determined |
| S11_vs_k141_44582||full | Low-quality |
| S11_vs_k141_77284||full | Low-quality |
| S11_vs_k141_83284||full | Low-quality |
| S11_vs_k141_69962||full | Low-quality |
| S11_vs_k141_50388||full | Low-quality |
| S11_vs_k141_44591||full | Low-quality |
| S11_vs_k141_96997||full | Low-quality |
| S11_vs_k141_83298||full | Not-determined |
| S6_vs_k141_29152||full | Complete |
| S11_vs_k141_5589||full | Low-quality |
| S11_vs_k141_11889||full | Not-determined |
| S11_vs_k141_69967||full | Low-quality |
| S11_vs_k141_83299||full | Not-determined |
| S11_vs_k141_50402||full | Not-determined |
| S11_vs_k141_89913||full | Not-determined |
| S11_vs_k141_11905||full | Low-quality |
| S11_vs_k141_11906||full | Low-quality |
| S11_vs_k141_63919||full | Not-determined |
| S11_vs_k141_63924||full | Not-determined |
| S11_vs_k141_38076||full | Not-determined |
| S11_vs_k141_5611||full | Not-determined |
| S11_vs_k141_38082||full | Low-quality |
| S11_vs_k141_11918||full | Not-determined |
| S11_vs_k141_103035||full | Not-determined |
| S11_vs_k141_38088||full | Low-quality |
| S11_vs_k141_63940||full | Low-quality |
| S11_vs_k141_97035||full | Not-determined |
| S11_vs_k141_103040||full | Low-quality |
| S11_vs_k141_18428||full | Low-quality |
| S11_vs_k141_5627||full | Low-quality |
| S11_vs_k141_83349||full | Not-determined |
| S11_vs_k141_18431||full | Not-determined |
| S11_vs_k141_38113||full | Low-quality |
| S11_vs_k141_103047||full | Low-quality |
| S11_vs_k141_38116||full | Low-quality |
| S11_vs_k141_11971||full | Low-quality |
| S11_vs_k141_5641||full | Low-quality |
| S11_vs_k141_11972||full | Low-quality |
| S11_vs_k141_89968||full | Not-determined |
| S11_vs_k141_83374||full | Not-determined |
| S11_vs_k141_103070||full | Low-quality |
| S11_vs_k141_11979||full | Low-quality |
| S11_vs_k141_38132||full | Not-determined |
| S11_vs_k141_89985||full | Low-quality |
| S11_vs_k141_25704||full | Low-quality |
| S11_vs_k141_50460||full | Low-quality |
| S11_vs_k141_77381||full | Not-determined |
| S11_vs_k141_44697||full | Not-determined |
| S11_vs_k141_38160||full | Low-quality |
| S11_vs_k141_25731||full | Low-quality |
| S11_vs_k141_90010||full | Not-determined |
| S11_vs_k141_38164||full | Low-quality |
| S11_vs_k141_18499||full | Low-quality |
| S11_vs_k141_38167||full | Low-quality |
| S11_vs_k141_83417||full | Not-determined |
| S11_vs_k141_31372||full | Not-determined |
| S11_vs_k141_97071||full | Not-determined |
| S11_vs_k141_83424||full | Low-quality |
| S11_vs_k141_5709||full | Low-quality |
| S5_vs_k141_20500||full | Complete |
| S11_vs_k141_5726||full | Not-determined |
| S11_vs_k141_83438||full | Low-quality |
| S11_vs_k141_50502||full | Not-determined |
| S11_vs_k141_25790||full | Not-determined |
| S11_vs_k141_83455||full | Not-determined |
| S11_vs_k141_25793||full | Not-determined |
| S11_vs_k141_77451||full | Low-quality |
| S7_vs_29595||full | Medium-quality |
| S11_vs_k141_97080||full | Not-determined |
| S11_vs_k141_50530||full | Not-determined |
| S11_vs_k141_50538||full | Low-quality |
| S11_vs_k141_90076||full | Not-determined |
| S11_vs_k141_90077||full | Low-quality |
| S11_vs_k141_83479||full | Low-quality |
| S11_vs_k141_57778||full | Not-determined |
| S11_vs_k141_70083||full | Low-quality |
| S11_vs_k141_64012||full | Low-quality |
| S11_vs_k141_31415||full | Low-quality |
| S11_vs_k141_31419||full | Low-quality |
| S11_vs_k141_70085||full | Low-quality |
| S11_vs_k141_38239||full | Not-determined |
| S11_vs_k141_18596||full | Not-determined |
| S11_vs_k141_64014||full | Low-quality |
| S11_vs_k141_83511||full | Low-quality |
| S11_vs_k141_83516||full | Low-quality |
| S11_vs_k141_57809||full | Not-determined |
| S11_vs_k141_70095||full | Low-quality |
| S11_vs_k141_5776||full | Low-quality |
| S11_vs_k141_83524||full | Low-quality |
| S11_vs_k141_25858||full | Not-determined |
| S11_vs_k141_18633||full | Low-quality |
| S11_vs_k141_90148||full | Low-quality |
| S11_vs_k141_57829||full | Low-quality |
| S11_vs_k141_18645||full | Low-quality |
| S11_vs_k141_38289||full | Low-quality |
| S11_vs_k141_25874||full | Low-quality |
| S11_vs_k141_5790||full | Low-quality |
| S11_vs_k141_38304||full | Not-determined |
| S11_vs_k141_5792||full | Not-determined |
| S11_vs_k141_83559||full | Low-quality |
| S11_vs_k141_50572||full | Low-quality |
| S11_vs_k141_25887||full | Low-quality |
| S11_vs_k141_70130||full | Low-quality |
| S11_vs_k141_57847||full | Not-determined |
| S11_vs_k141_57848||full | Not-determined |
| S11_vs_k141_5809||full | Not-determined |
| S11_vs_k141_50585||full | Low-quality |
| S11_vs_k141_83579||full | Not-determined |
| S11_vs_k141_31468||full | Low-quality |
| S11_vs_k141_18683||full | Not-determined |
| S11_vs_k141_31472||full | Low-quality |
| S11_vs_k141_77528||full | Not-determined |
| S11_vs_k141_18686||full | Low-quality |
| S11_vs_k141_12111||full | Low-quality |
| S11_vs_k141_50604||full | Low-quality |
| S11_vs_k141_31477||full | Not-determined |
| S11_vs_k141_5822||full | Not-determined |
| S11_vs_k141_31479||full | Not-determined |
| S11_vs_k141_31480||full | Low-quality |
| S11_vs_k141_25919||full | Not-determined |
| S11_vs_k141_64093||full | Not-determined |
| S11_vs_k141_83592||full | Low-quality |
| S11_vs_k141_38318||full | Not-determined |
| S11_vs_k141_12142||full | Low-quality |
| S11_vs_k141_31505||full | Low-quality |
| S11_vs_k141_77555||full | Low-quality |
| S11_vs_k141_25941||full | Low-quality |
| S11_vs_k141_64109||full | Not-determined |
| S11_vs_k141_57889||full | Low-quality |
| S11_vs_k141_38322||full | Low-quality |
| S11_vs_k141_18722||full | Low-quality |
| S11_vs_k141_97178||full | Low-quality |
| S11_vs_k141_83602||full | Low-quality |
| S11_vs_k141_18728||full | Not-determined |
| S11_vs_k141_38327||full | Low-quality |
| S11_vs_k141_12156||full | Not-determined |
| S11_vs_k141_38334||full | Not-determined |
| S11_vs_k141_83614||full | Not-determined |
| S11_vs_k141_90241||full | Low-quality |
| S11_vs_k141_31522||full | Not-determined |
| S11_vs_k141_97189||full | Not-determined |
| S11_vs_k141_57915||full | Low-quality |
| S11_vs_k141_44822||full | Not-determined |
| S11_vs_k141_90248||full | Low-quality |
| S11_vs_k141_38346||full | Low-quality |
| S11_vs_k141_90252||full | Low-quality |
| S11_vs_k141_83629||full | Low-quality |
| S11_vs_k141_70211||full | Not-determined |
| S11_vs_k141_64128||full | Low-quality |
| S11_vs_k141_18759||full | Not-determined |
| S11_vs_k141_38353||full | Low-quality |
| S11_vs_k141_77599||full | Low-quality |
| S11_vs_k141_26012||full | Not-determined |
| S11_vs_k141_38363||full | Low-quality |
| S11_vs_k141_83651||full | Low-quality |
| S11_vs_k141_18781||full | Low-quality |
| S11_vs_k141_97232||full | Low-quality |
| S11_vs_k141_70227||full | Not-determined |
| S11_vs_k141_64146||full | Low-quality |
| S11_vs_k141_26028||full | Low-quality |
| S11_vs_k141_31553||full | Low-quality |
| S11_vs_k141_64151||full | Not-determined |
| S11_vs_k141_12206||full | Low-quality |
| S11_vs_k141_90296||full | Low-quality |
| S11_vs_k141_12207||full | Not-determined |
| S11_vs_k141_31557||full | Not-determined |
| S11_vs_k141_50728||full | Low-quality |
| S11_vs_k141_64165||full | Low-quality |
| S11_vs_k141_18817||full | Low-quality |
| S11_vs_k141_50738||full | Low-quality |
| S11_vs_k141_18822||full | Not-determined |
| S11_vs_k141_103170||full | Not-determined |
| S11_vs_k141_31578||full | Low-quality |
| S11_vs_k141_97274||full | Low-quality |
| S11_vs_k141_77675||full | Low-quality |
| S12_extracted_k141_181024 | Medium-quality |
| S11_vs_k141_12250||full | Not-determined |
| S11_vs_k141_70253||full | Low-quality |
| S11_vs_k141_38408||full | Low-quality |
| S11_vs_k141_97292||full | Low-quality |
| S11_vs_k141_5935||full | Low-quality |
| S11_vs_k141_103201||full | Low-quality |
| S11_vs_k141_64232||full | Not-determined |
| S11_vs_k141_103203||full | Low-quality |
| S11_vs_k141_90327||full | Low-quality |
| S11_vs_k141_12299||full | Low-quality |
| S11_vs_k141_58025||full | Low-quality |
| S11_vs_k141_70263||full | Not-determined |
| S11_vs_k141_12310||full | Not-determined |
| S11_vs_k141_70281||full | Low-quality |
| S11_vs_k141_58038||full | Low-quality |
| S11_vs_k141_90335||full | Not-determined |
| S11_vs_k141_5958||full | Low-quality |
| S11_vs_k141_12318||full | Low-quality |
| S11_vs_k141_103229||full | Not-determined |
| S11_vs_k141_83742||full | Low-quality |
| S11_vs_k141_18886||full | Low-quality |
| S11_vs_k141_77704||full | Not-determined |
| S11_vs_k141_97323||full | Low-quality |
| S11_vs_k141_12329||full | Low-quality |
| S11_vs_k141_31612||full | Low-quality |
| S11_vs_k141_70294||full | Low-quality |
| S11_vs_k141_38443||full | Low-quality |
| S11_vs_k141_12337||full | Low-quality |
| S11_vs_k141_5967||full | Low-quality |
| S11_vs_k141_31621||full | Low-quality |
| S11_vs_k141_64285||full | Low-quality |
| S11_vs_k141_103252||full | Not-determined |
| S11_vs_k141_38452||full | Low-quality |
| S11_vs_k141_58060||full | Low-quality |
| S11_vs_k141_31626||full | Low-quality |
| S11_vs_k141_5987||full | Not-determined |
| S11_vs_k141_77734||full | Not-determined |
| S11_vs_k141_90379||full | Low-quality |
| S11_vs_k141_64309||full | Not-determined |
| S11_vs_k141_31649||full | Low-quality |
| S11_vs_k141_90393||full | Low-quality |
| S11_vs_k141_64317||full | Not-determined |
| S11_vs_k141_44968||full | Low-quality |
| S11_vs_k141_58094||full | Low-quality |
| S11_vs_k141_83803||full | Low-quality |
| S11_vs_k141_77759||full | Low-quality |
| S11_vs_k141_38453||full | Low-quality |
| S11_vs_k141_6009||full | Low-quality |
| S11_vs_k141_64321||full | Low-quality |
| S11_vs_k141_90408||full | Low-quality |
| S11_vs_k141_90409||full | Low-quality |
| S11_vs_k141_58119||full | Not-determined |
| S11_vs_k141_70366||full | Low-quality |
| S11_vs_k141_103273||full | Not-determined |
| S11_vs_k141_45021||full | Low-quality |
| S11_vs_k141_12406||full | Low-quality |
| S11_vs_k141_90433||full | Low-quality |
| S11_vs_k141_31707||full | Low-quality |
| S11_vs_k141_103289||full | Low-quality |
| S11_vs_k141_38508||full | Not-determined |
| S11_vs_k141_70395||full | Not-determined |
| S11_vs_k141_64329||full | Not-determined |
| S11_vs_k141_45024||full | Low-quality |
| S11_vs_k141_58146||full | Not-determined |
| S11_vs_k141_77795||full | Low-quality |
| S11_vs_k141_38522||full | Low-quality |
| S11_vs_k141_97424||full | Low-quality |
| S11_vs_k141_77812||full | Low-quality |
| S11_vs_k141_77814||full | Low-quality |
| S11_vs_k141_77815||full | Low-quality |
| S11_vs_k141_97429||full | Not-determined |
| S11_vs_k141_45044||full | Low-quality |
| S11_vs_k141_12468||full | Low-quality |
| S11_vs_k141_83868||full | Low-quality |
| S11_vs_k141_45057||full | Low-quality |
| S11_vs_k141_12476||full | Not-determined |
| S11_vs_k141_58174||full | Low-quality |
| S11_vs_k141_58178||full | Low-quality |
| S11_vs_k141_12478||full | Low-quality |
| S11_vs_k141_45063||full | Low-quality |
| S11_vs_k141_58183||full | Low-quality |
| S11_vs_k141_70434||full | Not-determined |
| S11_vs_k141_70435||full | Low-quality |
| S11_vs_k141_90510||full | Not-determined |
| S11_vs_k141_12483||full | Not-determined |
| S11_vs_k141_58184||full | Low-quality |
| S11_vs_k141_70438||full | Low-quality |
| S11_vs_k141_45092||full | Low-quality |
| S11_vs_k141_97502||full | Not-determined |
| S11_vs_k141_45103||full | Not-determined |
| S11_vs_k141_58200||full | Not-determined |
| S11_vs_k141_97511||full | Low-quality |
| S11_vs_k141_45112||full | Low-quality |
| S11_vs_k141_64382||full | Low-quality |
| S11_vs_k141_77883||full | Low-quality |
| S11_vs_k141_58215||full | Not-determined |
| S11_vs_k141_90540||full | Low-quality |
| S11_vs_k141_50916||full | Low-quality |
| S11_vs_k141_77888||full | Not-determined |
| S11_vs_k141_45120||full | Not-determined |
| S11_vs_k141_31819||full | Low-quality |
| S11_vs_k141_97526||full | Low-quality |
| S11_vs_k141_50935||full | Low-quality |
| S11_vs_k141_64419||full | Low-quality |
| S11_vs_k141_58237||full | Low-quality |
| S11_vs_k141_50940||full | Low-quality |
| S11_vs_k141_97536||full | Not-determined |
| S11_vs_k141_64430||full | Low-quality |
| S11_vs_k141_83997||full | Not-determined |
| S11_vs_k141_77945||full | Low-quality |
| S11_vs_k141_12575||full | Not-determined |
| S11_vs_k141_31859||full | Low-quality |
| S11_vs_k141_45177||full | Low-quality |
| S11_vs_k141_97573||full | Low-quality |
| S11_vs_k141_84015||full | Low-quality |
| S11_vs_k141_77970||full | Not-determined |
| S11_vs_k141_97586||full | Low-quality |
| S11_vs_k141_97591||full | Not-determined |
| S11_vs_k141_12607||full | Low-quality |
| S11_vs_k141_51026||full | Low-quality |
| S11_vs_k141_51032||full | Low-quality |
| S11_vs_k141_51036||full | Not-determined |
| S11_vs_k141_38554||full | Low-quality |
| S11_vs_k141_45223||full | Not-determined |
| S11_vs_k141_51044||full | Not-determined |
| S11_vs_k141_51048||full | Not-determined |
| S11_vs_k141_51057||full | Not-determined |
| S11_vs_k141_78020||full | Low-quality |
| S11_vs_k141_64531||full | Low-quality |
| S11_vs_k141_103377||full | Low-quality |
| S11_vs_k141_51062||full | Not-determined |
| S11_vs_k141_103384||full | Not-determined |
| S11_vs_k141_38583||full | Not-determined |
| S11_vs_k141_90601||full | Low-quality |
| S11_vs_k141_12658||full | Low-quality |
| S11_vs_k141_45270||full | Low-quality |
| S11_vs_k141_58291||full | Not-determined |
| S11_vs_k141_84090||full | Low-quality |
| S11_vs_k141_78031||full | Low-quality |
| S11_vs_k141_84093||full | Low-quality |
| S11_vs_k141_12674||full | Low-quality |
| S11_vs_k141_90625||full | Low-quality |
| S11_vs_k141_38608||full | Not-determined |
| S11_vs_k141_84096||full | Low-quality |
| S11_vs_k141_38612||full | Low-quality |
| S11_vs_k141_12685||full | Low-quality |
| S11_vs_k141_78057||full | Low-quality |
| S11_vs_k141_64577||full | Low-quality |
| S11_vs_k141_45308||full | Low-quality |
| S11_vs_k141_58322||full | Low-quality |
| S11_vs_k141_90650||full | Not-determined |
| S11_vs_k141_78062||full | Low-quality |
| S11_vs_k141_45325||full | Not-determined |
| S11_vs_k141_70469||full | Low-quality |
| S11_vs_k141_6190||full | Low-quality |
| S11_vs_k141_51127||full | Low-quality |
| S11_vs_k141_38675||full | Low-quality |
| S11_vs_k141_38682||full | Not-determined |
| S11_vs_k141_6214||full | Low-quality |
| S11_vs_k141_58347||full | Low-quality |
| S11_vs_k141_70518||full | Low-quality |
| S11_vs_k141_6219||full | Not-determined |
| S11_vs_k141_103447||full | Not-determined |
| S11_vs_k141_31956||full | Not-determined |
| S11_vs_k141_38700||full | Not-determined |
| S11_vs_k141_103452||full | Low-quality |
| S11_vs_k141_12764||full | Low-quality |
| S11_vs_k141_31963||full | Low-quality |
| S11_vs_k141_31968||full | Low-quality |
| S11_vs_k141_6246||full | Not-determined |
| S11_vs_k141_51161||full | Not-determined |
| S11_vs_k141_64606||full | Low-quality |
| S11_vs_k141_90697||full | Not-determined |
| S11_vs_k141_78089||full | Low-quality |
| S11_vs_k141_51168||full | Low-quality |
| S11_vs_k141_70533||full | Not-determined |
| S11_vs_k141_103473||full | Low-quality |
| S11_vs_k141_38749||full | Low-quality |
| S11_vs_k141_51181||full | Not-determined |
| S11_vs_k141_6265||full | Low-quality |
| S11_vs_k141_84172||full | Not-determined |
| S11_vs_k141_12798||full | Low-quality |
| S11_vs_k141_90718||full | Low-quality |
| S11_vs_k141_90719||full | Low-quality |
| S11_vs_k141_58385||full | Low-quality |
| S11_vs_k141_58386||full | Not-determined |
| S11_vs_k141_19227||full | Low-quality |
| S11_vs_k141_38773||full | Not-determined |
| S11_vs_k141_58387||full | Low-quality |
| S11_vs_k141_103495||full | Low-quality |
| S11_vs_k141_84200||full | Not-determined |
| S11_vs_k141_6290||full | Low-quality |
| S11_vs_k141_64640||full | Low-quality |
| S11_vs_k141_19261||full | Low-quality |
| S11_vs_k141_103502||full | Low-quality |
| S11_vs_k141_12838||full | Low-quality |
| S11_vs_k141_64642||full | Not-determined |
| S11_vs_k141_58397||full | Low-quality |

S11_vs_k141_70594||full Low-quality
S11_vs_k141_12852||full Low-quality
S11_vs_k141_64657||full Not-determined
S11_vs_k141_51217||full Low-quality
S11_vs_k141_6303||full Low-quality
S11_vs_k141_70606||full Low-quality
S11_vs_k141_6320||full Not-determined
S11_vs_k141_84258||full Not-determined
S11_vs_k141_19299||full Low-quality
S11_vs_k141_38812||full Low-quality
S11_vs_k141_31999||full Low-quality
S11_vs_k141_103543||full Low-quality
S11_vs_k141_58427||full Low-quality
S11_vs_k141_103545||full Low-quality
S11_vs_k141_103546||full Not-determined
S11_vs_k141_70642||full Low-quality
S11_vs_k141_6328||full Not-determined
S11_vs_k141_32019||full Low-quality
S11_vs_k141_70649||full Not-determined
S11_vs_k141_84269||full Not-determined
S11_vs_k141_38836||full Low-quality
S11_vs_k141_64715||full Not-determined
S11_vs_k141_38842||full Low-quality
S11_vs_k141_38846||full Not-determined
S11_vs_k141_84279||full Low-quality
S11_vs_k141_6350||full Not-determined
S11_vs_k141_38847||full Low-quality
S11_vs_k141_103567||full Low-quality
S11_vs_k141_12921||full Not-determined
S11_vs_k141_12925||full Low-quality
S11_vs_k141_90813||full Low-quality
S11_vs_k141_32021||full Low-quality
S11_vs_k141_19353||full Low-quality
S11_vs_k141_90823||full Low-quality
S11_vs_k141_45427||full Not-determined
S11_vs_k141_64746||full Low-quality
S11_vs_k141_103587||full Low-quality
S11_vs_k141_51290||full Low-quality
S11_vs_k141_19361||full Not-determined
S11_vs_k141_90835||full Not-determined
S11_vs_k141_51297||full Low-quality
S11_vs_k141_103590||full Low-quality
S11_vs_k141_58485||full Low-quality
S11_vs_k141_32044||full Not-determined
S11_vs_k141_6372||full Not-determined
S11_vs_k141_12969||full Not-determined
S11_vs_k141_58503||full Low-quality
S11_vs_k141_12979||full Low-quality
S11_vs_k141_103609||full Low-quality
S11_vs_k141_58510||full Low-quality
S11_vs_k141_32052||full Not-determined
S11_vs_k141_90856||full Not-determined
S11_vs_k141_70720||full Not-determined
S11_vs_k141_12988||full Not-determined
S12_extracted_k141_104517 Complete
S11_vs_k141_19386||full Low-quality
S11_vs_k141_51335||full Low-quality
S11_vs_k141_103627||full Low-quality
S11_vs_k141_51339||full Low-quality
S11_vs_k141_32067||full Not-determined
S11_vs_k141_70736||full Low-quality
S11_vs_k141_51342||full Not-determined
S11_vs_k141_51345||full Not-determined
S11_vs_k141_103636||full Low-quality
S11_vs_k141_58535||full Low-quality
S11_vs_k141_51364||full Low-quality
S11_vs_k141_70769||full Low-quality
S11_vs_k141_58558||full Low-quality
S11_vs_k141_84368||full Low-quality
S11_vs_k141_70782||full Not-determined
S11_vs_k141_58568||full Low-quality
S11_vs_k141_90913||full Not-determined
S11_vs_k141_38930||full Low-quality
S11_vs_k141_70784||full Low-quality
S11_vs_k141_38936||full Low-quality
S11_vs_k141_51414||full Low-quality
S11_vs_k141_84433||full Low-quality
S12_vs_k141_104517||full Complete
S11_vs_k141_70816||full Low-quality
S11_vs_k141_70817||full Low-quality
S11_vs_k141_103698||full Not-determined
S11_vs_k141_51420||full Low-quality
S11_vs_k141_90954||full Low-quality
S11_vs_k141_51428||full Not-determined
S11_vs_k141_39006||full Low-quality
S11_vs_k141_103714||full Low-quality
S11_vs_k141_51438||full Not-determined
S11_vs_k141_19482||full Low-quality
S11_vs_k141_39010||full Not-determined
S11_vs_k141_70856||full Low-quality
S11_vs_k141_39019||full Low-quality
S11_vs_k141_51450||full Not-determined
S11_vs_k141_45551||full Low-quality
S11_vs_k141_70860||full Not-determined
S11_vs_k141_70872||full Not-determined
S11_vs_k141_32134||full Low-quality
S11_vs_k141_32142||full Low-quality
S11_vs_k141_103736||full Low-quality
S11_vs_k141_91011||full Not-determined
S11_vs_k141_39036||full Not-determined
S11_vs_k141_70895||full Not-determined
S11_vs_k141_32155||full Low-quality
S11_vs_k141_32158||full Low-quality
S11_vs_k141_70921||full Low-quality
S11_vs_k141_32159||full Low-quality
S11_vs_k141_91047||full Not-determined
S11_vs_k141_103786||full Low-quality
S11_vs_k141_91058||full Not-determined
S11_vs_k141_70969||full Low-quality
S11_vs_k141_51531||full Not-determined
S11_vs_k141_70986||full Low-quality
S11_vs_k141_103809||full Not-determined
S11_vs_k141_51554||full Low-quality
S11_vs_k141_64947||full Not-determined
S11_vs_k141_70991||full Not-determined
S11_vs_k141_64953||full Low-quality
S11_vs_k141_91104||full Low-quality
S11_vs_k141_91106||full Not-determined
S11_vs_k141_103828||full Low-quality
S11_vs_k141_51567||full Low-quality
S11_vs_k141_84468||full Low-quality
S11_vs_k141_32189||full Not-determined
S11_vs_k141_64993||full Low-quality
S11_vs_k141_71038||full Not-determined
S11_vs_k141_84480||full Not-determined
S11_vs_k141_51614||full Not-determined
S11_vs_k141_51615||full Low-quality
S11_vs_k141_103885||full Not-determined
S11_vs_k141_71077||full Low-quality
S11_vs_k141_71082||full Low-quality
S11_vs_k141_51642||full Not-determined
S11_vs_k141_84515||full Low-quality
S11_vs_k141_103921||full Not-determined
S11_vs_k141_103924||full Low-quality
S11_vs_k141_84557||full Not-determined
S11_vs_k141_51676||full Low-quality
S11_vs_k141_32203||full Low-quality
S11_vs_k141_71084||full Not-determined
S11_vs_k141_71085||full Not-determined
S11_vs_k141_84565||full Low-quality
S11_vs_k141_71090||full Not-determined
S11_vs_k141_71094||full Not-determined
S11_vs_k141_32223||full Low-quality
S11_vs_k141_65072||full Low-quality
S11_vs_k141_32236||full Not-determined
S11_vs_k141_32245||full Low-quality
S11_vs_k141_71132||full Low-quality
S11_vs_k141_32269||full Low-quality
S11_vs_k141_51698||full Low-quality
S11_vs_k141_51701||full Low-quality
S11_vs_k141_32282||full Low-quality
S11_vs_k141_51702||full Low-quality
S11_vs_k141_32290||full Low-quality
S11_vs_k141_71208||full Not-determined
S11_vs_k141_32296||full Low-quality
S11_vs_k141_71244||full Low-quality
S11_vs_k141_51809||full Not-determined
S11_vs_k141_71262||full Not-determined
S11_vs_k141_32365||full Low-quality
S11_vs_k141_32401||full Low-quality
S11_vs_k141_51845||full Low-quality
S11_vs_k141_71339||full Low-quality
S11_vs_k141_32438||full Not-determined
S11_vs_k141_51872||full Low-quality
S11_vs_k141_51878||full Low-quality
S11_vs_k141_71365||full Not-determined
S11_vs_k141_51910||full Low-quality
S11_vs_k141_51918||full Low-quality
S11_vs_k141_51921||full Low-quality
S11_vs_k141_51932||full Not-determined
S11_vs_k141_51944||full Not-determined
S11_vs_k141_71424||full Not-determined
S11_vs_k141_51980||full Low-quality
S11_vs_k141_52049||full Low-quality
S11_vs_k141_52059||full Not-determined
S11_vs_k141_52061||full Low-quality
S11_vs_k141_71576||full Not-determined
S11_vs_k141_92442||0_partial Not-determined
S11_vs_k141_93054||0_partial Low-quality
S11_vs_k141_2527||0_partial Not-determined
S11_vs_k141_82077||0_partial Low-quality
S11_vs_k141_43481||0_partial Low-quality
S11_vs_k141_11166||0_partial Low-quality
S11_vs_k141_89309||0_partial Low-quality
S11_vs_k141_89451||0_partial Not-determined
S11_vs_k141_63209||0_partial Low-quality
S11_vs_k141_64544||0_partial Not-determined
S11_vs_k141_84609||lt2gene Not-determined
S11_vs_k141_78103||lt2gene Not-determined
S11_vs_k141_65092||lt2gene Low-quality
S11_vs_k141_78110||lt2gene Not-determined
S11_vs_k141_32554||lt2gene Not-determined
S11_vs_k141_65105||lt2gene Low-quality
S11_vs_k141_28||lt2gene Not-determined
S11_vs_k141_65106||lt2gene Low-quality
S11_vs_k141_45||lt2gene Low-quality
S11_vs_k141_6536||lt2gene Low-quality
S11_vs_k141_19550||lt2gene Not-determined
S11_vs_k141_66||lt2gene Low-quality
S11_vs_k141_71619||lt2gene Low-quality
S11_vs_k141_52112||lt2gene Low-quality
S11_vs_k141_78139||lt2gene Low-quality
S11_vs_k141_91164||lt2gene Not-determined
S11_vs_k141_83||lt2gene Low-quality
S11_vs_k141_32586||lt2gene Low-quality
S11_vs_k141_19571||lt2gene Low-quality
S11_vs_k141_26112||lt2gene Not-determined
S11_vs_k141_45628||lt2gene Not-determined
S11_vs_k141_52155||lt2gene Low-quality
S11_vs_k141_65189||lt2gene Not-determined
S11_vs_k141_45635||lt2gene Low-quality
S11_vs_k141_58671||lt2gene Low-quality
S11_vs_k141_65227||lt2gene Low-quality
S11_vs_k141_136||lt2gene Low-quality
S11_vs_k141_142||lt2gene Low-quality
S11_vs_k141_32636||lt2gene Low-quality
S11_vs_k141_65246||lt2gene Not-determined
S11_vs_k141_84711||lt2gene Low-quality
S11_vs_k141_65262||lt2gene Low-quality
S11_vs_k141_71699||lt2gene Not-determined
S11_vs_k141_52260||lt2gene Low-quality
S11_vs_k141_26161||lt2gene Not-determined
S11_vs_k141_97703||lt2gene Not-determined
S11_vs_k141_32666||lt2gene Low-quality
S11_vs_k141_97717||lt2gene Low-quality
S11_vs_k141_97721||lt2gene Low-quality
S11_vs_k141_91292||lt2gene Low-quality
S11_vs_k141_97746||lt2gene Low-quality
S11_vs_k141_246||lt2gene Low-quality
S11_vs_k141_97765||lt2gene Low-quality
S11_vs_k141_19714||lt2gene Not-determined
S11_vs_k141_26241||lt2gene Low-quality
S11_vs_k141_78194||lt2gene Low-quality
S11_vs_k141_32693||lt2gene Low-quality
S11_vs_k141_13170||lt2gene Low-quality
S11_vs_k141_58714||lt2gene Low-quality
S11_vs_k141_32716||lt2gene Low-quality
S11_vs_k141_97787||lt2gene Low-quality
S11_vs_k141_32735||lt2gene Low-quality
S11_vs_k141_13194||lt2gene Low-quality
S11_vs_k141_97808||lt2gene Not-determined
S11_vs_k141_84821||lt2gene Low-quality
S11_vs_k141_71773||lt2gene Low-quality
S11_vs_k141_91364||lt2gene Low-quality
S11_vs_k141_97822||lt2gene Low-quality
S11_vs_k141_39234||lt2gene Low-quality
S11_vs_k141_365||lt2gene Low-quality
S11_vs_k141_19780||lt2gene Not-determined
S11_vs_k141_71793||lt2gene Low-quality
S11_vs_k141_52360||lt2gene Low-quality
S11_vs_k141_78308||lt2gene Low-quality
S11_vs_k141_39245||lt2gene Low-quality
S11_vs_k141_78319||lt2gene Not-determined
S11_vs_k141_78331||lt2gene Low-quality
S11_vs_k141_78332||lt2gene Low-quality
S11_vs_k141_84849||lt2gene Low-quality
S11_vs_k141_71841||lt2gene Low-quality
S11_vs_k141_71846||lt2gene Not-determined
S11_vs_k141_32823||lt2gene Low-quality
S11_vs_k141_26252||lt2gene Low-quality
S11_vs_k141_433||lt2gene Low-quality
S11_vs_k141_78350||lt2gene Not-determined
S11_vs_k141_91412||lt2gene Low-quality
S11_vs_k141_26280||lt2gene Low-quality
S11_vs_k141_91444||lt2gene Low-quality
S11_vs_k141_91451||lt2gene Not-determined
S11_vs_k141_26289||lt2gene Low-quality
S11_vs_k141_26290||lt2gene Low-quality
S11_vs_k141_26311||lt2gene Low-quality
S11_vs_k141_78386||lt2gene Low-quality
S11_vs_k141_6733||lt2gene Low-quality
S11_vs_k141_58822||lt2gene Low-quality
S11_vs_k141_97946||lt2gene Low-quality
S11_vs_k141_78413||lt2gene Low-quality
S11_vs_k141_32875||lt2gene Not-determined
S11_vs_k141_39357||lt2gene Low-quality
S11_vs_k141_6777||lt2gene Low-quality
S11_vs_k141_71887||lt2gene Low-quality
S11_vs_k141_6782||lt2gene Low-quality
S11_vs_k141_71894||lt2gene Low-quality
S11_vs_k141_19864||lt2gene Not-determined
S11_vs_k141_26379||lt2gene Low-quality
S11_vs_k141_539||lt2gene Low-quality
S11_vs_k141_26388||lt2gene Not-determined
S11_vs_k141_6802||lt2gene Low-quality
S11_vs_k141_26398||lt2gene Low-quality
S11_vs_k141_71922||lt2gene Low-quality
S11_vs_k141_78495||lt2gene Low-quality
S11_vs_k141_97988||lt2gene Low-quality
S11_vs_k141_71951||lt2gene Not-determined
S11_vs_k141_26433||lt2gene Low-quality
S11_vs_k141_58873||lt2gene Low-quality
S11_vs_k141_58881||lt2gene Low-quality
S11_vs_k141_45829||lt2gene Low-quality
S11_vs_k141_91631||lt2gene Low-quality
S11_vs_k141_625||lt2gene Low-quality
S11_vs_k141_32963||lt2gene Not-determined
S11_vs_k141_91648||lt2gene Low-quality
S11_vs_k141_640||lt2gene Low-quality
S11_vs_k141_32969||lt2gene Low-quality
S11_vs_k141_32974||lt2gene Low-quality
S11_vs_k141_26484||lt2gene Low-quality
S11_vs_k141_39488||lt2gene Low-quality
S11_vs_k141_20000||lt2gene Low-quality
S11_vs_k141_39492||lt2gene Low-quality
S11_vs_k141_52513||lt2gene Low-quality
S11_vs_k141_65462||lt2gene Low-quality
S11_vs_k141_32997||lt2gene Low-quality
S11_vs_k141_72048||lt2gene Low-quality
S11_vs_k141_33005||lt2gene Low-quality
S11_vs_k141_98107||lt2gene Low-quality
S11_vs_k141_72055||lt2gene Low-quality
S11_vs_k141_13267||lt2gene Not-determined
S11_vs_k141_85094||lt2gene Not-determined
S11_vs_k141_45936||lt2gene Not-determined
S11_vs_k141_33025||lt2gene Low-quality
S11_vs_k141_72101||lt2gene Low-quality
S11_vs_k141_6952||lt2gene Low-quality
S11_vs_k141_13330||lt2gene Low-quality
S11_vs_k141_33062||lt2gene Low-quality
S11_vs_k141_33094||lt2gene Low-quality
S11_vs_k141_46022||lt2gene Low-quality
S11_vs_k141_58974||lt2gene Low-quality
S11_vs_k141_91726||lt2gene Low-quality
S11_vs_k141_26567||lt2gene Low-quality
S11_vs_k141_26570||lt2gene Not-determined
S11_vs_k141_52616||lt2gene Low-quality
S11_vs_k141_26576||lt2gene Low-quality
S11_vs_k141_39644||lt2gene Not-determined
S11_vs_k141_20142||lt2gene Low-quality
S11_vs_k141_65588||lt2gene Low-quality
S11_vs_k141_20159||lt2gene Low-quality
S11_vs_k141_20178||lt2gene Low-quality
S11_vs_k141_98253||lt2gene Low-quality
S11_vs_k141_26596||lt2gene Low-quality
S11_vs_k141_91764||lt2gene Low-quality
S11_vs_k141_58982||lt2gene Low-quality
S11_vs_k141_46067||lt2gene Low-quality
S11_vs_k141_52660||lt2gene Low-quality
S11_vs_k141_46091||lt2gene Low-quality
S11_vs_k141_72230||lt2gene Low-quality
S11_vs_k141_13453||lt2gene Low-quality
S11_vs_k141_59022||lt2gene Low-quality
S11_vs_k141_52666||lt2gene Not-determined
S11_vs_k141_91804||lt2gene Low-quality
S11_vs_k141_85294||lt2gene Not-determined
S11_vs_k141_91805||lt2gene Low-quality
S11_vs_k141_78823||lt2gene Low-quality
S11_vs_k141_26666||lt2gene Low-quality
S11_vs_k141_65650||lt2gene Not-determined
S11_vs_k141_26681||lt2gene Low-quality
S11_vs_k141_26688||lt2gene Low-quality
S11_vs_k141_26689||lt2gene Low-quality
S11_vs_k141_59028||lt2gene Low-quality
S11_vs_k141_46157||lt2gene Low-quality
S11_vs_k141_91864||lt2gene Low-quality
S11_vs_k141_26721||lt2gene Low-quality
S11_vs_k141_65707||lt2gene Low-quality
S11_vs_k141_65714||lt2gene Low-quality
S11_vs_k141_20252||lt2gene Not-determined
S11_vs_k141_7026||lt2gene Not-determined
S11_vs_k141_46185||lt2gene Low-quality
S11_vs_k141_78880||lt2gene Low-quality
S11_vs_k141_91876||lt2gene Low-quality
S11_vs_k141_91879||lt2gene Low-quality
S11_vs_k141_26765||lt2gene Low-quality
S11_vs_k141_98407||lt2gene Low-quality
S11_vs_k141_98415||lt2gene Low-quality
S11_vs_k141_98426||lt2gene Low-quality
S11_vs_k141_46219||lt2gene Not-determined
S11_vs_k141_46235||lt2gene Low-quality
S11_vs_k141_26776||lt2gene Low-quality
S11_vs_k141_91926||lt2gene Low-quality
S11_vs_k141_65764||lt2gene Low-quality
S11_vs_k141_13587||lt2gene Low-quality
S11_vs_k141_65773||lt2gene Low-quality
S11_vs_k141_72452||lt2gene Low-quality
S11_vs_k141_98505||lt2gene Not-determined
S11_vs_k141_7096||lt2gene Low-quality
S11_vs_k141_98510||lt2gene Not-determined
S11_vs_k141_52803||lt2gene Low-quality
S11_vs_k141_85484||lt2gene Low-quality
S11_vs_k141_39816||lt2gene Not-determined
S11_vs_k141_39817||lt2gene Low-quality
S11_vs_k141_20382||lt2gene Not-determined
S11_vs_k141_85501||lt2gene Low-quality
S11_vs_k141_65795||lt2gene Low-quality
S11_vs_k141_65798||lt2gene Low-quality
S11_vs_k141_1007||lt2gene Low-quality
S11_vs_k141_85508||lt2gene Low-quality
S11_vs_k141_79058||lt2gene Low-quality
S11_vs_k141_1014||lt2gene Low-quality
S11_vs_k141_72491||lt2gene Low-quality
S11_vs_k141_52832||lt2gene Low-quality
S11_vs_k141_1031||lt2gene Low-quality
S11_vs_k141_46366||lt2gene Low-quality
S11_vs_k141_1041||lt2gene Low-quality
S11_vs_k141_65842||lt2gene Low-quality
S11_vs_k141_46402||lt2gene Low-quality
S11_vs_k141_92066||lt2gene Low-quality
S11_vs_k141_39876||lt2gene Low-quality
S11_vs_k141_79096||lt2gene Low-quality
S11_vs_k141_39884||lt2gene Low-quality
S11_vs_k141_85554||lt2gene Low-quality
S11_vs_k141_52872||lt2gene Not-determined
S11_vs_k141_33405||lt2gene Low-quality
S11_vs_k141_52884||lt2gene Not-determined
S11_vs_k141_33409||lt2gene Low-quality
S11_vs_k141_98602||lt2gene Low-quality
S11_vs_k141_92087||lt2gene Not-determined
S11_vs_k141_33431||lt2gene Not-determined
S11_vs_k141_85590||lt2gene Low-quality
S11_vs_k141_85599||lt2gene Low-quality
S11_vs_k141_1105||lt2gene Low-quality
S11_vs_k141_92107||lt2gene Low-quality
S11_vs_k141_92110||lt2gene Low-quality
S11_vs_k141_39917||lt2gene Low-quality
S11_vs_k141_20513||lt2gene Low-quality
S11_vs_k141_79110||lt2gene Not-determined
S11_vs_k141_1118||lt2gene Not-determined
S11_vs_k141_1128||lt2gene Not-determined
S11_vs_k141_59299||lt2gene Not-determined
S11_vs_k141_59317||lt2gene Not-determined
S11_vs_k141_39935||lt2gene Low-quality
S11_vs_k141_39936||lt2gene Not-determined
S11_vs_k141_79136||lt2gene Not-determined
S11_vs_k141_79164||lt2gene Low-quality
S11_vs_k141_79180||lt2gene Low-quality
S11_vs_k141_1184||lt2gene Low-quality
S11_vs_k141_98703||lt2gene Not-determined
S11_vs_k141_7310||lt2gene Low-quality
S11_vs_k141_65961||lt2gene Low-quality
S11_vs_k141_33461||lt2gene Low-quality
S11_vs_k141_33474||lt2gene Not-determined
S11_vs_k141_65987||lt2gene Low-quality
S11_vs_k141_85721||lt2gene Low-quality
S11_vs_k141_33489||lt2gene Low-quality
S11_vs_k141_53043||lt2gene Low-quality
S11_vs_k141_92180||lt2gene Low-quality
S11_vs_k141_20595||lt2gene Low-quality
S11_vs_k141_46534||lt2gene Low-quality
S11_vs_k141_85749||lt2gene Low-quality
S11_vs_k141_53070||lt2gene Low-quality
S11_vs_k141_46540||lt2gene Low-quality
S11_vs_k141_13610||lt2gene Low-quality
S11_vs_k141_40040||lt2gene Not-determined
S11_vs_k141_13616||lt2gene Low-quality
S11_vs_k141_72706||lt2gene Low-quality
S11_vs_k141_40043||lt2gene Low-quality
S11_vs_k141_7394||lt2gene Not-determined
S11_vs_k141_72717||lt2gene Low-quality
S11_vs_k141_85765||lt2gene Not-determined
S11_vs_k141_40062||lt2gene Low-quality
S11_vs_k141_92216||lt2gene Low-quality
S11_vs_k141_7416||lt2gene Not-determined
S11_vs_k141_13642||lt2gene Not-determined
S11_vs_k141_72740||lt2gene Low-quality
S11_vs_k141_27023||lt2gene Low-quality
S11_vs_k141_53124||lt2gene Low-quality
S11_vs_k141_13655||lt2gene Low-quality
S11_vs_k141_33576||lt2gene Not-determined
S11_vs_k141_66090||lt2gene Low-quality
S11_vs_k141_33577||lt2gene Low-quality
S11_vs_k141_40103||lt2gene Low-quality
S11_vs_k141_53174||lt2gene Not-determined
S11_vs_k141_40104||lt2gene Low-quality
S11_vs_k141_79319||lt2gene Low-quality
S11_vs_k141_20690||lt2gene Low-quality
S11_vs_k141_59495||lt2gene Low-quality
S11_vs_k141_33632||lt2gene Not-determined
S11_vs_k141_13679||lt2gene Low-quality
S11_vs_k141_46589||lt2gene Low-quality
S11_vs_k141_59504||lt2gene Low-quality
S11_vs_k141_98784||lt2gene Low-quality
S11_vs_k141_72850||lt2gene Low-quality
S11_vs_k141_53225||lt2gene Low-quality
S11_vs_k141_66116||lt2gene Low-quality
S11_vs_k141_59524||lt2gene Low-quality
S11_vs_k141_59525||lt2gene Low-quality
S11_vs_k141_13717||lt2gene Not-determined
S11_vs_k141_53245||lt2gene Not-determined
S11_vs_k141_27118||lt2gene Low-quality
S11_vs_k141_79362||lt2gene Not-determined
S11_vs_k141_79365||lt2gene Low-quality
S11_vs_k141_7505||lt2gene Not-determined
S11_vs_k141_27127||lt2gene Not-determined
S11_vs_k141_79379||lt2gene Not-determined
S11_vs_k141_46652||lt2gene Low-quality
S11_vs_k141_7527||lt2gene Not-determined
S11_vs_k141_59570||lt2gene Not-determined
S11_vs_k141_79409||lt2gene Not-determined
S11_vs_k141_85933||lt2gene Low-quality
S11_vs_k141_72904||lt2gene Not-determined
S11_vs_k141_7538||lt2gene Low-quality
S11_vs_k141_33678||lt2gene Low-quality
S11_vs_k141_33681||lt2gene Low-quality
S11_vs_k141_7547||lt2gene Low-quality
S11_vs_k141_59601||lt2gene Low-quality
S11_vs_k141_46683||lt2gene Low-quality
S11_vs_k141_13786||lt2gene Low-quality
S11_vs_k141_59619||lt2gene Low-quality
S11_vs_k141_72918||lt2gene Not-determined
S11_vs_k141_13808||lt2gene Low-quality
S11_vs_k141_7571||lt2gene Low-quality
S11_vs_k141_1357||lt2gene Low-quality
S11_vs_k141_13824||lt2gene Low-quality
S11_vs_k141_1372||lt2gene Low-quality
S11_vs_k141_1375||lt2gene Not-determined
S11_vs_k141_1397||lt2gene Low-quality
S11_vs_k141_98864||lt2gene Low-quality
S11_vs_k141_13863||lt2gene Low-quality
S11_vs_k141_86002||lt2gene Not-determined
S11_vs_k141_1416||lt2gene Low-quality
S11_vs_k141_7645||lt2gene Not-determined
S11_vs_k141_40175||lt2gene Low-quality
S11_vs_k141_72982||lt2gene Low-quality
S11_vs_k141_53369||lt2gene Not-determined
S11_vs_k141_53371||lt2gene Not-determined
S11_vs_k141_53372||lt2gene Not-determined
S11_vs_k141_53373||lt2gene Not-determined
S11_vs_k141_53378||lt2gene Not-determined
S11_vs_k141_72996||lt2gene Low-quality
S11_vs_k141_20846||lt2gene Low-quality
S11_vs_k141_73005||lt2gene Low-quality
S11_vs_k141_1473||lt2gene Low-quality
S11_vs_k141_73023||lt2gene Not-determined
S11_vs_k141_86066||lt2gene Low-quality
S11_vs_k141_66214||lt2gene Not-determined
S11_vs_k141_92388||lt2gene Low-quality
S11_vs_k141_66223||lt2gene Not-determined
S11_vs_k141_53424||lt2gene Low-quality
S11_vs_k141_98926||lt2gene Low-quality
S11_vs_k141_27273||lt2gene Low-quality
S11_vs_k141_33782||lt2gene Low-quality
S11_vs_k141_73082||lt2gene Low-quality
S11_vs_k141_46736||lt2gene Not-determined
S11_vs_k141_79444||lt2gene Low-quality
S11_vs_k141_20916||lt2gene Low-quality
S11_vs_k141_92420||lt2gene Low-quality
S11_vs_k141_59782||lt2gene Not-determined
S11_vs_k141_7751||lt2gene Not-determined
S11_vs_k141_98978||lt2gene Low-quality
S11_vs_k141_79471||lt2gene Low-quality
S11_vs_k141_66281||lt2gene Not-determined
S11_vs_k141_46750||lt2gene Not-determined
S11_vs_k141_79473||lt2gene Low-quality
S11_vs_k141_13986||lt2gene Low-quality
S11_vs_k141_1587||lt2gene Low-quality
S11_vs_k141_20977||lt2gene Not-determined
S11_vs_k141_73171||lt2gene Not-determined
S11_vs_k141_14010||lt2gene Not-determined
S11_vs_k141_20983||lt2gene Low-quality
S11_vs_k141_66327||lt2gene Low-quality
S11_vs_k141_14035||lt2gene Low-quality
S11_vs_k141_1619||lt2gene Low-quality
S11_vs_k141_27370||lt2gene Not-determined
S11_vs_k141_21002||lt2gene Low-quality
S11_vs_k141_1632||lt2gene Low-quality
S11_vs_k141_79553||lt2gene Not-determined
S11_vs_k141_33798||lt2gene Low-quality
S11_vs_k141_66349||lt2gene Low-quality
S11_vs_k141_1674||lt2gene Low-quality
S11_vs_k141_33825||lt2gene Low-quality
S11_vs_k141_66375||lt2gene Low-quality
S11_vs_k141_53515||lt2gene Not-determined
S11_vs_k141_86255||lt2gene Low-quality
S11_vs_k141_7898||lt2gene Low-quality
S11_vs_k141_86292||lt2gene Low-quality
S11_vs_k141_86304||lt2gene Not-determined
S11_vs_k141_27447||lt2gene Low-quality
S11_vs_k141_99065||lt2gene Low-quality
S11_vs_k141_27451||lt2gene Low-quality
S11_vs_k141_53571||lt2gene Not-determined
S11_vs_k141_46832||lt2gene Low-quality
S11_vs_k141_40367||lt2gene Low-quality
S11_vs_k141_14146||lt2gene Low-quality
S11_vs_k141_99075||lt2gene Not-determined
S11_vs_k141_46848||lt2gene Low-quality
S11_vs_k141_27475||lt2gene Low-quality
S11_vs_k141_40389||lt2gene Low-quality
S11_vs_k141_27506||lt2gene Low-quality
S11_vs_k141_21108||lt2gene Low-quality
S11_vs_k141_1749||lt2gene Not-determined
S11_vs_k141_79633||lt2gene Not-determined
S11_vs_k141_14194||lt2gene Low-quality
S11_vs_k141_40434||lt2gene Low-quality
S11_vs_k141_46898||lt2gene Low-quality
S11_vs_k141_59885||lt2gene Not-determined
S11_vs_k141_86358||lt2gene Low-quality

| | |
|---|---|
| S11_vs_k141_86366\|\|lt2gene | Low-quality |
| S11_vs_k141_46929\|\|lt2gene | Low-quality |
| S11_vs_k141_1814\|\|lt2gene | Low-quality |
| S11_vs_k141_14237\|\|lt2gene | Low-quality |
| S11_vs_k141_46942\|\|lt2gene | Low-quality |
| S11_vs_k141_99110\|\|lt2gene | Low-quality |
| S11_vs_k141_73331\|\|lt2gene | Low-quality |
| S11_vs_k141_8032\|\|lt2gene | Low-quality |
| S11_vs_k141_21181\|\|lt2gene | Low-quality |
| S11_vs_k141_27632\|\|lt2gene | Low-quality |
| S11_vs_k141_14287\|\|lt2gene | Low-quality |
| S11_vs_k141_99156\|\|lt2gene | Low-quality |
| S11_vs_k141_46984\|\|lt2gene | Not-determined |
| S11_vs_k141_99165\|\|lt2gene | Low-quality |
| S11_vs_k141_8072\|\|lt2gene | Not-determined |
| S11_vs_k141_14328\|\|lt2gene | Not-determined |
| S11_vs_k141_8104\|\|lt2gene | Low-quality |
| S11_vs_k141_92533\|\|lt2gene | Not-determined |
| S11_vs_k141_92551\|\|lt2gene | Not-determined |
| S11_vs_k141_92554\|\|lt2gene | Low-quality |
| S11_vs_k141_53698\|\|lt2gene | Low-quality |
| S11_vs_k141_60013\|\|lt2gene | Low-quality |
| S11_vs_k141_27750\|\|lt2gene | Not-determined |
| S11_vs_k141_14408\|\|lt2gene | Low-quality |
| S11_vs_k141_1928\|\|lt2gene | Low-quality |
| S11_vs_k141_8191\|\|lt2gene | Low-quality |
| S11_vs_k141_99240\|\|lt2gene | Low-quality |
| S11_vs_k141_60035\|\|lt2gene | Low-quality |
| S11_vs_k141_79830\|\|lt2gene | Low-quality |
| S11_vs_k141_21315\|\|lt2gene | Low-quality |
| S11_vs_k141_66545\|\|lt2gene | Not-determined |
| S11_vs_k141_40478\|\|lt2gene | Low-quality |
| S11_vs_k141_60048\|\|lt2gene | Low-quality |
| S11_vs_k141_27773\|\|lt2gene | Low-quality |
| S11_vs_k141_60061\|\|lt2gene | Low-quality |
| S11_vs_k141_14472\|\|lt2gene | Low-quality |
| S11_vs_k141_92633\|\|lt2gene | Low-quality |
| S11_vs_k141_14475\|\|lt2gene | Low-quality |
| S11_vs_k141_27815\|\|lt2gene | Not-determined |
| S11_vs_k141_40534\|\|lt2gene | Not-determined |
| S11_vs_k141_40536\|\|lt2gene | Low-quality |
| S11_vs_k141_92653\|\|lt2gene | Low-quality |
| S11_vs_k141_8239\|\|lt2gene | Not-determined |
| S11_vs_k141_99291\|\|lt2gene | Low-quality |
| S11_vs_k141_73461\|\|lt2gene | Low-quality |
| S11_vs_k141_27842\|\|lt2gene | Low-quality |
| S11_vs_k141_86434\|\|lt2gene | Low-quality |
| S11_vs_k141_86436\|\|lt2gene | Low-quality |
| S11_vs_k141_86439\|\|lt2gene | Low-quality |
| S11_vs_k141_86447\|\|lt2gene | Not-determined |
| S11_vs_k141_47148\|\|lt2gene | Low-quality |
| S11_vs_k141_66666\|\|lt2gene | Low-quality |
| S11_vs_k141_14536\|\|lt2gene | Low-quality |
| S11_vs_k141_40600\|\|lt2gene | Not-determined |
| S11_vs_k141_60133\|\|lt2gene | Low-quality |
| S11_vs_k141_99323\|\|lt2gene | Low-quality |
| S11_vs_k141_1995\|\|lt2gene | Not-determined |
| S11_vs_k141_40617\|\|lt2gene | Low-quality |
| S11_vs_k141_79894\|\|lt2gene | Low-quality |
| S11_vs_k141_66682\|\|lt2gene | Low-quality |
| S11_vs_k141_21454\|\|lt2gene | Not-determined |
| S11_vs_k141_40628\|\|lt2gene | Low-quality |
| S11_vs_k141_86521\|\|lt2gene | Low-quality |
| S11_vs_k141_47212\|\|lt2gene | Not-determined |
| S11_vs_k141_79925\|\|lt2gene | Not-determined |
| S11_vs_k141_99369\|\|lt2gene | Not-determined |
| S11_vs_k141_53926\|\|lt2gene | Not-determined |
| S11_vs_k141_79954\|\|lt2gene | Not-determined |
| S11_vs_k141_92784\|\|lt2gene | Low-quality |
| S11_vs_k141_21521\|\|lt2gene | Low-quality |
| S11_vs_k141_8349\|\|lt2gene | Low-quality |
| S11_vs_k141_8351\|\|lt2gene | Low-quality |
| S11_vs_k141_47259\|\|lt2gene | Low-quality |
| S11_vs_k141_47264\|\|lt2gene | Low-quality |
| S11_vs_k141_86615\|\|lt2gene | Low-quality |
| S11_vs_k141_21574\|\|lt2gene | Not-determined |
| S11_vs_k141_79994\|\|lt2gene | Low-quality |
| S11_vs_k141_79995\|\|lt2gene | Low-quality |
| S11_vs_k141_53986\|\|lt2gene | Low-quality |
| S11_vs_k141_53999\|\|lt2gene | Low-quality |
| S11_vs_k141_54000\|\|lt2gene | Not-determined |
| S11_vs_k141_54010\|\|lt2gene | Low-quality |
| S11_vs_k141_2122\|\|lt2gene | Low-quality |
| S11_vs_k141_8425\|\|lt2gene | Not-determined |
| S11_vs_k141_14561\|\|lt2gene | Low-quality |
| S11_vs_k141_54042\|\|lt2gene | Low-quality |
| S11_vs_k141_2151\|\|lt2gene | Low-quality |
| S11_vs_k141_99471\|\|lt2gene | Low-quality |
| S11_vs_k141_60186\|\|lt2gene | Low-quality |
| S11_vs_k141_2158\|\|lt2gene | Not-determined |
| S11_vs_k141_80051\|\|lt2gene | Low-quality |
| S11_vs_k141_99492\|\|lt2gene | Low-quality |
| S11_vs_k141_54061\|\|lt2gene | Low-quality |
| S11_vs_k141_40741\|\|lt2gene | Low-quality |
| S11_vs_k141_2180\|\|lt2gene | Low-quality |
| S11_vs_k141_2181\|\|lt2gene | Low-quality |
| S11_vs_k141_73637\|\|lt2gene | Low-quality |
| S11_vs_k141_80100\|\|lt2gene | Low-quality |
| S11_vs_k141_80102\|\|lt2gene | Low-quality |
| S11_vs_k141_92857\|\|lt2gene | Not-determined |
| S11_vs_k141_80105\|\|lt2gene | Low-quality |
| S11_vs_k141_14606\|\|lt2gene | Low-quality |
| S11_vs_k141_73681\|\|lt2gene | Low-quality |
| S11_vs_k141_80135\|\|lt2gene | Low-quality |
| S11_vs_k141_34056\|\|lt2gene | Not-determined |
| S11_vs_k141_73702\|\|lt2gene | Not-determined |
| S11_vs_k141_8482\|\|lt2gene | Not-determined |
| S11_vs_k141_47300\|\|lt2gene | Low-quality |
| S11_vs_k141_14637\|\|lt2gene | Low-quality |
| S11_vs_k141_92896\|\|lt2gene | Low-quality |
| S11_vs_k141_40855\|\|lt2gene | Not-determined |
| S11_vs_k141_80150\|\|lt2gene | Low-quality |
| S11_vs_k141_73719\|\|lt2gene | Low-quality |
| S11_vs_k141_14662\|\|lt2gene | Not-determined |
| S11_vs_k141_8506\|\|lt2gene | Not-determined |
| S11_vs_k141_73728\|\|lt2gene | Low-quality |
| S11_vs_k141_8525\|\|lt2gene | Not-determined |
| S11_vs_k141_86777\|\|lt2gene | Low-quality |
| S11_vs_k141_86804\|\|lt2gene | Low-quality |
| S11_vs_k141_2291\|\|lt2gene | Not-determined |
| S11_vs_k141_92981\|\|lt2gene | Not-determined |
| S11_vs_k141_54167\|\|lt2gene | Low-quality |
| S11_vs_k141_54173\|\|lt2gene | Low-quality |
| S11_vs_k141_54182\|\|lt2gene | Low-quality |
| S11_vs_k141_60267\|\|lt2gene | Low-quality |
| S11_vs_k141_93006\|\|lt2gene | Low-quality |
| S11_vs_k141_54195\|\|lt2gene | Not-determined |
| S11_vs_k141_28040\|\|lt2gene | Low-quality |
| S11_vs_k141_47344\|\|lt2gene | Low-quality |
| S11_vs_k141_99638\|\|lt2gene | Not-determined |
| S11_vs_k141_8607\|\|lt2gene | Low-quality |
| S11_vs_k141_60332\|\|lt2gene | Not-determined |
| S11_vs_k141_54221\|\|lt2gene | Low-quality |
| S11_vs_k141_14799\|\|lt2gene | Low-quality |
| S11_vs_k141_60356\|\|lt2gene | Low-quality |
| S11_vs_k141_34210\|\|lt2gene | Low-quality |
| S11_vs_k141_93086\|\|lt2gene | Low-quality |
| S11_vs_k141_93090\|\|lt2gene | Low-quality |
| S11_vs_k141_60406\|\|lt2gene | Low-quality |
| S11_vs_k141_60412\|\|lt2gene | Low-quality |
| S11_vs_k141_66810\|\|lt2gene | Low-quality |
| S11_vs_k141_8655\|\|lt2gene | Low-quality |
| S11_vs_k141_99743\|\|lt2gene | Low-quality |
| S11_vs_k141_86983\|\|lt2gene | Low-quality |
| S11_vs_k141_86990\|\|lt2gene | Low-quality |
| S11_vs_k141_66825\|\|lt2gene | Low-quality |
| S11_vs_k141_73897\|\|lt2gene | Low-quality |
| S11_vs_k141_54238\|\|lt2gene | Low-quality |
| S11_vs_k141_14863\|\|lt2gene | Low-quality |
| S11_vs_k141_8718\|\|lt2gene | Low-quality |
| S11_vs_k141_28186\|\|lt2gene | Not-determined |
| S11_vs_k141_87091\|\|lt2gene | Low-quality |
| S11_vs_k141_73995\|\|lt2gene | Low-quality |
| S11_vs_k141_2473\|\|lt2gene | Not-determined |
| S11_vs_k141_34300\|\|lt2gene | Not-determined |
| S11_vs_k141_21908\|\|lt2gene | Low-quality |
| S11_vs_k141_2477\|\|lt2gene | Not-determined |
| S11_vs_k141_8771\|\|lt2gene | Low-quality |
| S11_vs_k141_34321\|\|lt2gene | Low-quality |
| S11_vs_k141_99809\|\|lt2gene | Low-quality |
| S11_vs_k141_80387\|\|lt2gene | Not-determined |
| S11_vs_k141_99814\|\|lt2gene | Low-quality |
| S11_vs_k141_87157\|\|lt2gene | Low-quality |
| S11_vs_k141_41153\|\|lt2gene | Low-quality |
| S11_vs_k141_54396\|\|lt2gene | Low-quality |
| S11_vs_k141_21950\|\|lt2gene | Low-quality |
| S11_vs_k141_41165\|\|lt2gene | Not-determined |
| S11_vs_k141_74057\|\|lt2gene | Low-quality |
| S11_vs_k141_74062\|\|lt2gene | Low-quality |
| S11_vs_k141_8819\|\|lt2gene | Low-quality |
| S11_vs_k141_74070\|\|lt2gene | Not-determined |
| S11_vs_k141_14961\|\|lt2gene | Low-quality |
| S11_vs_k141_8832\|\|lt2gene | Low-quality |
| S11_vs_k141_74080\|\|lt2gene | Low-quality |
| S11_vs_k141_2540\|\|lt2gene | Low-quality |
| S11_vs_k141_66935\|\|lt2gene | Low-quality |
| S11_vs_k141_15008\|\|lt2gene | Not-determined |
| S11_vs_k141_66947\|\|lt2gene | Low-quality |
| S11_vs_k141_34404\|\|lt2gene | Low-quality |
| S11_vs_k141_87207\|\|lt2gene | Low-quality |
| S11_vs_k141_47504\|\|lt2gene | Low-quality |
| S11_vs_k141_15022\|\|lt2gene | Low-quality |
| S11_vs_k141_87217\|\|lt2gene | Low-quality |
| S11_vs_k141_60483\|\|lt2gene | Low-quality |
| S11_vs_k141_87227\|\|lt2gene | Low-quality |
| S11_vs_k141_15034\|\|lt2gene | Low-quality |
| S11_vs_k141_60493\|\|lt2gene | Low-quality |
| S11_vs_k141_80425\|\|lt2gene | Low-quality |
| S11_vs_k141_41259\|\|lt2gene | Low-quality |
| S11_vs_k141_41274\|\|lt2gene | Low-quality |
| S11_vs_k141_2603\|\|lt2gene | Low-quality |
| S11_vs_k141_66985\|\|lt2gene | Low-quality |
| S11_vs_k141_28305\|\|lt2gene | Not-determined |
| S11_vs_k141_54515\|\|lt2gene | Low-quality |
| S11_vs_k141_34439\|\|lt2gene | Not-determined |
| S11_vs_k141_60520\|\|lt2gene | Low-quality |
| S11_vs_k141_15076\|\|lt2gene | Low-quality |
| S11_vs_k141_87252\|\|lt2gene | Low-quality |
| S11_vs_k141_80437\|\|lt2gene | Low-quality |
| S11_vs_k141_60533\|\|lt2gene | Low-quality |
| S11_vs_k141_54538\|\|lt2gene | Low-quality |
| S11_vs_k141_41302\|\|lt2gene | Not-determined |
| S11_vs_k141_67024\|\|lt2gene | Not-determined |
| S11_vs_k141_2622\|\|lt2gene | Low-quality |
| S11_vs_k141_87284\|\|lt2gene | Not-determined |
| S11_vs_k141_34460\|\|lt2gene | Not-determined |
| S11_vs_k141_54553\|\|lt2gene | Low-quality |
| S11_vs_k141_87292\|\|lt2gene | Low-quality |
| S11_vs_k141_74177\|\|lt2gene | Low-quality |
| S11_vs_k141_87304\|\|lt2gene | Low-quality |
| S11_vs_k141_34495\|\|lt2gene | Not-determined |
| S11_vs_k141_54587\|\|lt2gene | Low-quality |
| S11_vs_k141_8934\|\|lt2gene | Low-quality |
| S11_vs_k141_22006\|\|lt2gene | Not-determined |
| S11_vs_k141_87333\|\|lt2gene | Low-quality |
| S11_vs_k141_99962\|\|lt2gene | Low-quality |
| S11_vs_k141_60591\|\|lt2gene | Low-quality |
| S11_vs_k141_87368\|\|lt2gene | Not-determined |
| S11_vs_k141_93411\|\|lt2gene | Not-determined |
| S11_vs_k141_8975\|\|lt2gene | Not-determined |
| S11_vs_k141_93431\|\|lt2gene | Low-quality |
| S11_vs_k141_60632\|\|lt2gene | Not-determined |
| S11_vs_k141_74251\|\|lt2gene | Low-quality |
| S11_vs_k141_60635\|\|lt2gene | Not-determined |
| S11_vs_k141_28433\|\|lt2gene | Low-quality |
| S11_vs_k141_15221\|\|lt2gene | Low-quality |
| S11_vs_k141_2702\|\|lt2gene | Low-quality |
| S11_vs_k141_80520\|\|lt2gene | Low-quality |
| S11_vs_k141_80521\|\|lt2gene | Not-determined |
| S11_vs_k141_80534\|\|lt2gene | Low-quality |
| S11_vs_k141_67105\|\|lt2gene | Not-determined |
| S11_vs_k141_28477\|\|lt2gene | Low-quality |
| S11_vs_k141_100060\|\|lt2gene | Low-quality |
| S11_vs_k141_93481\|\|lt2gene | Not-determined |
| S11_vs_k141_47702\|\|lt2gene | Low-quality |
| S11_vs_k141_74293\|\|lt2gene | Not-determined |
| S11_vs_k141_47710\|\|lt2gene | Not-determined |
| S11_vs_k141_67153\|\|lt2gene | Not-determined |
| S11_vs_k141_80562\|\|lt2gene | Low-quality |
| S11_vs_k141_9083\|\|lt2gene | Low-quality |
| S11_vs_k141_74333\|\|lt2gene | Low-quality |
| S11_vs_k141_22173\|\|lt2gene | Not-determined |
| S11_vs_k141_2790\|\|lt2gene | Low-quality |
| S11_vs_k141_9106\|\|lt2gene | Not-determined |
| S11_vs_k141_34593\|\|lt2gene | Low-quality |
| S11_vs_k141_74361\|\|lt2gene | Low-quality |
| S11_vs_k141_22178\|\|lt2gene | Low-quality |
| S11_vs_k141_2833\|\|lt2gene | Low-quality |
| S11_vs_k141_9128\|\|lt2gene | Low-quality |
| S11_vs_k141_41490\|\|lt2gene | Low-quality |
| S11_vs_k141_41501\|\|lt2gene | Low-quality |
| S11_vs_k141_87552\|\|lt2gene | Low-quality |
| S11_vs_k141_41507\|\|lt2gene | Low-quality |
| S11_vs_k141_15354\|\|lt2gene | Not-determined |
| S11_vs_k141_22214\|\|lt2gene | Low-quality |
| S11_vs_k141_60746\|\|lt2gene | Not-determined |
| S11_vs_k141_2870\|\|lt2gene | Low-quality |
| S11_vs_k141_22221\|\|lt2gene | Low-quality |
| S11_vs_k141_2884\|\|lt2gene | Low-quality |
| S11_vs_k141_9165\|\|lt2gene | Not-determined |
| S11_vs_k141_93561\|\|lt2gene | Not-determined |
| S11_vs_k141_60766\|\|lt2gene | Low-quality |
| S11_vs_k141_93584\|\|lt2gene | Low-quality |
| S11_vs_k141_41572\|\|lt2gene | Low-quality |
| S11_vs_k141_47790\|\|lt2gene | Low-quality |
| S11_vs_k141_93589\|\|lt2gene | Low-quality |
| S11_vs_k141_47794\|\|lt2gene | Low-quality |
| S11_vs_k141_60783\|\|lt2gene | Not-determined |
| S11_vs_k141_100147\|\|lt2gene | Not-determined |
| S11_vs_k141_9234\|\|lt2gene | Not-determined |
| S11_vs_k141_9235\|\|lt2gene | Low-quality |
| S11_vs_k141_41631\|\|lt2gene | Low-quality |
| S11_vs_k141_47857\|\|lt2gene | Low-quality |
| S11_vs_k141_74540\|\|lt2gene | Not-determined |
| S11_vs_k141_93697\|\|lt2gene | Low-quality |
| S11_vs_k141_22310\|\|lt2gene | Low-quality |
| S11_vs_k141_60868\|\|lt2gene | Low-quality |
| S11_vs_k141_67272\|\|lt2gene | Low-quality |
| S11_vs_k141_60874\|\|lt2gene | Low-quality |
| S11_vs_k141_60883\|\|lt2gene | Low-quality |
| S11_vs_k141_74587\|\|lt2gene | Low-quality |
| S11_vs_k141_2970\|\|lt2gene | Not-determined |
| S11_vs_k141_47910\|\|lt2gene | Not-determined |
| S11_vs_k141_2973\|\|lt2gene | Low-quality |
| S11_vs_k141_100211\|\|lt2gene | Low-quality |
| S11_vs_k141_22389\|\|lt2gene | Low-quality |
| S11_vs_k141_67324\|\|lt2gene | Low-quality |
| S11_vs_k141_74647\|\|lt2gene | Low-quality |
| S11_vs_k141_9339\|\|lt2gene | Low-quality |
| S11_vs_k141_100268\|\|lt2gene | Low-quality |
| S11_vs_k141_34766\|\|lt2gene | Low-quality |
| S11_vs_k141_60960\|\|lt2gene | Not-determined |
| S11_vs_k141_15525\|\|lt2gene | Not-determined |
| S11_vs_k141_60993\|\|lt2gene | Low-quality |
| S11_vs_k141_100332\|\|lt2gene | Not-determined |
| S11_vs_k141_74683\|\|lt2gene | Low-quality |
| S11_vs_k141_74689\|\|lt2gene | Low-quality |
| S11_vs_k141_100340\|\|lt2gene | Low-quality |
| S11_vs_k141_9403\|\|lt2gene | Low-quality |
| S11_vs_k141_15570\|\|lt2gene | Low-quality |
| S11_vs_k141_87828\|\|lt2gene | Low-quality |
| S11_vs_k141_9411\|\|lt2gene | Low-quality |
| S11_vs_k141_22525\|\|lt2gene | Low-quality |
| S11_vs_k141_22529\|\|lt2gene | Low-quality |
| S11_vs_k141_80905\|\|lt2gene | Low-quality |
| S11_vs_k141_67414\|\|lt2gene | Low-quality |
| S11_vs_k141_15581\|\|lt2gene | Low-quality |
| S11_vs_k141_48025\|\|lt2gene | Low-quality |
| S11_vs_k141_67436\|\|lt2gene | Not-determined |
| S11_vs_k141_93926\|\|lt2gene | Low-quality |
| S11_vs_k141_100412\|\|lt2gene | Low-quality |
| S11_vs_k141_15606\|\|lt2gene | Not-determined |
| S11_vs_k141_3118\|\|lt2gene | Low-quality |
| S11_vs_k141_41840\|\|lt2gene | Not-determined |
| S11_vs_k141_22585\|\|lt2gene | Low-quality |
| S11_vs_k141_67470\|\|lt2gene | Not-determined |
| S11_vs_k141_67476\|\|lt2gene | Low-quality |
| S11_vs_k141_34843\|\|lt2gene | Low-quality |
| S11_vs_k141_3128\|\|lt2gene | Not-determined |
| S11_vs_k141_100439\|\|lt2gene | Low-quality |
| S11_vs_k141_3142\|\|lt2gene | Not-determined |
| S11_vs_k141_22593\|\|lt2gene | Low-quality |
| S11_vs_k141_9458\|\|lt2gene | Not-determined |
| S11_vs_k141_61117\|\|lt2gene | Not-determined |
| S11_vs_k141_93973\|\|lt2gene | Not-determined |
| S11_vs_k141_3169\|\|lt2gene | Low-quality |
| S11_vs_k141_34879\|\|lt2gene | Low-quality |
| S11_vs_k141_100488\|\|lt2gene | Not-determined |
| S11_vs_k141_22630\|\|lt2gene | Not-determined |
| S11_vs_k141_22635\|\|lt2gene | Low-quality |
| S11_vs_k141_34894\|\|lt2gene | Not-determined |
| S11_vs_k141_48096\|\|lt2gene | Low-quality |
| S11_vs_k141_28679\|\|lt2gene | Not-determined |
| S11_vs_k141_22665\|\|lt2gene | Not-determined |
| S11_vs_k141_93993\|\|lt2gene | Low-quality |
| S11_vs_k141_41912\|\|lt2gene | Low-quality |
| S11_vs_k141_22677\|\|lt2gene | Low-quality |
| S11_vs_k141_22681\|\|lt2gene | Not-determined |
| S11_vs_k141_41933\|\|lt2gene | Not-determined |
| S11_vs_k141_22698\|\|lt2gene | Low-quality |
| S11_vs_k141_74821\|\|lt2gene | Not-determined |
| S11_vs_k141_22718\|\|lt2gene | Low-quality |
| S11_vs_k141_81003\|\|lt2gene | Not-determined |
| S11_vs_k141_81011\|\|lt2gene | Not-determined |
| S11_vs_k141_15724\|\|lt2gene | Not-determined |
| S11_vs_k141_54924\|\|lt2gene | Low-quality |
| S11_vs_k141_61211\|\|lt2gene | Not-determined |
| S11_vs_k141_81040\|\|lt2gene | Low-quality |
| S11_vs_k141_74888\|\|lt2gene | Low-quality |
| S11_vs_k141_15758\|\|lt2gene | Low-quality |
| S11_vs_k141_28758\|\|lt2gene | Low-quality |
| S11_vs_k141_9618\|\|lt2gene | Low-quality |
| S11_vs_k141_54959\|\|lt2gene | Not-determined |
| S11_vs_k141_28786\|\|lt2gene | Not-determined |
| S11_vs_k141_48119\|\|lt2gene | Not-determined |
| S11_vs_k141_15790\|\|lt2gene | Low-quality |
| S11_vs_k141_9655\|\|lt2gene | Not-determined |
| S11_vs_k141_61299\|\|lt2gene | Low-quality |
| S11_vs_k141_22767\|\|lt2gene | Low-quality |
| S11_vs_k141_67640\|\|lt2gene | Low-quality |
| S11_vs_k141_67642\|\|lt2gene | Low-quality |
| S11_vs_k141_35025\|\|lt2gene | Low-quality |
| S11_vs_k141_9684\|\|lt2gene | Not-determined |
| S11_vs_k141_35049\|\|lt2gene | Low-quality |
| S11_vs_k141_35053\|\|lt2gene | Not-determined |
| S11_vs_k141_67687\|\|lt2gene | Low-quality |
| S11_vs_k141_22858\|\|lt2gene | Not-determined |
| S11_vs_k141_22862\|\|lt2gene | Low-quality |
| S11_vs_k141_28849\|\|lt2gene | Low-quality |
| S11_vs_k141_55081\|\|lt2gene | Low-quality |
| S11_vs_k141_55082\|\|lt2gene | Not-determined |
| S11_vs_k141_87938\|\|lt2gene | Low-quality |
| S11_vs_k141_28871\|\|lt2gene | Low-quality |
| S11_vs_k141_42082\|\|lt2gene | Low-quality |
| S11_vs_k141_94241\|\|lt2gene | Low-quality |
| S11_vs_k141_75035\|\|lt2gene | Not-determined |
| S11_vs_k141_75046\|\|lt2gene | Low-quality |
| S11_vs_k141_55128\|\|lt2gene | Not-determined |
| S11_vs_k141_75074\|\|lt2gene | Low-quality |
| S11_vs_k141_9746\|\|lt2gene | Low-quality |
| S11_vs_k141_35146\|\|lt2gene | Low-quality |
| S11_vs_k141_15962\|\|lt2gene | Low-quality |
| S11_vs_k141_94279\|\|lt2gene | Not-determined |
| S11_vs_k141_42146\|\|lt2gene | Low-quality |
| S11_vs_k141_81113\|\|lt2gene | Low-quality |
| S11_vs_k141_100758\|\|lt2gene | Low-quality |
| S11_vs_k141_28924\|\|lt2gene | Not-determined |
| S11_vs_k141_81129\|\|lt2gene | Low-quality |
| S11_vs_k141_15991\|\|lt2gene | Low-quality |
| S11_vs_k141_28936\|\|lt2gene | Low-quality |
| S11_vs_k141_100785\|\|lt2gene | Low-quality |
| S11_vs_k141_9767\|\|lt2gene | Low-quality |
| S11_vs_k141_42170\|\|lt2gene | Low-quality |
| S11_vs_k141_16010\|\|lt2gene | Not-determined |
| S11_vs_k141_67759\|\|lt2gene | Not-determined |
| S11_vs_k141_9787\|\|lt2gene | Low-quality |
| S11_vs_k141_100836\|\|lt2gene | Low-quality |
| S11_vs_k141_9797\|\|lt2gene | Low-quality |
| S11_vs_k141_100849\|\|lt2gene | Not-determined |
| S11_vs_k141_28987\|\|lt2gene | Not-determined |
| S11_vs_k141_35251\|\|lt2gene | Low-quality |
| S11_vs_k141_16052\|\|lt2gene | Low-quality |
| S11_vs_k141_67805\|\|lt2gene | Low-quality |
| S11_vs_k141_94347\|\|lt2gene | Not-determined |
| S11_vs_k141_55202\|\|lt2gene | Low-quality |
| S11_vs_k141_75139\|\|lt2gene | Low-quality |
| S11_vs_k141_75166\|\|lt2gene | Not-determined |
| S11_vs_k141_29081\|\|lt2gene | Low-quality |
| S11_vs_k141_35347\|\|lt2gene | Low-quality |
| S11_vs_k141_9896\|\|lt2gene | Low-quality |
| S11_vs_k141_9900\|\|lt2gene | Low-quality |
| S11_vs_k141_16136\|\|lt2gene | Not-determined |
| S11_vs_k141_9913\|\|lt2gene | Not-determined |
| S11_vs_k141_16141\|\|lt2gene | Low-quality |
| S11_vs_k141_42333\|\|lt2gene | Not-determined |
| S11_vs_k141_35381\|\|lt2gene | Low-quality |
| S11_vs_k141_3526\|\|lt2gene | Low-quality |
| S11_vs_k141_55277\|\|lt2gene | Low-quality |
| S11_vs_k141_88027\|\|lt2gene | Not-determined |
| S11_vs_k141_88053\|\|lt2gene | Not-determined |
| S11_vs_k141_55314\|\|lt2gene | Low-quality |
| S11_vs_k141_42378\|\|lt2gene | Not-determined |
| S11_vs_k141_88062\|\|lt2gene | Low-quality |
| S11_vs_k141_55329\|\|lt2gene | Low-quality |
| S11_vs_k141_16199\|\|lt2gene | Low-quality |
| S11_vs_k141_67876\|\|lt2gene | Low-quality |
| S11_vs_k141_75273\|\|lt2gene | Not-determined |
| S11_vs_k141_61571\|\|lt2gene | Low-quality |
| S11_vs_k141_35492\|\|lt2gene | Low-quality |
| S11_vs_k141_10008\|\|lt2gene | Low-quality |
| S11_vs_k141_29230\|\|lt2gene | Not-determined |
| S11_vs_k141_42419\|\|lt2gene | Low-quality |
| S11_vs_k141_10025\|\|lt2gene | Not-determined |
| S11_vs_k141_81346\|\|lt2gene | Low-quality |
| S11_vs_k141_48479\|\|lt2gene | Low-quality |
| S11_vs_k141_81362\|\|lt2gene | Not-determined |
| S11_vs_k141_67952\|\|lt2gene | Low-quality |
| S11_vs_k141_23226\|\|lt2gene | Low-quality |
| S11_vs_k141_23227\|\|lt2gene | Low-quality |
| S11_vs_k141_61587\|\|lt2gene | Low-quality |
| S11_vs_k141_75338\|\|lt2gene | Low-quality |
| S11_vs_k141_61600\|\|lt2gene | Low-quality |
| S11_vs_k141_42440\|\|lt2gene | Low-quality |
| S11_vs_k141_88112\|\|lt2gene | Low-quality |
| S11_vs_k141_55408\|\|lt2gene | Low-quality |
| S11_vs_k141_61621\|\|lt2gene | Low-quality |
| S11_vs_k141_16289\|\|lt2gene | Not-determined |
| S11_vs_k141_23239\|\|lt2gene | Low-quality |
| S11_vs_k141_23243\|\|lt2gene | Low-quality |
| S11_vs_k141_10057\|\|lt2gene | Low-quality |
| S11_vs_k141_48540\|\|lt2gene | Low-quality |
| S11_vs_k141_88134\|\|lt2gene | Low-quality |
| S11_vs_k141_88142\|\|lt2gene | Low-quality |
| S11_vs_k141_10096\|\|lt2gene | Low-quality |
| S11_vs_k141_88165\|\|lt2gene | Low-quality |
| S11_vs_k141_55449\|\|lt2gene | Not-determined |
| S11_vs_k141_16352\|\|lt2gene | Low-quality |
| S11_vs_k141_48580\|\|lt2gene | Low-quality |
| S11_vs_k141_101107\|\|lt2gene | Low-quality |
| S11_vs_k141_88200\|\|lt2gene | Not-determined |
| S11_vs_k141_88203\|\|lt2gene | Low-quality |
| S11_vs_k141_81426\|\|lt2gene | Low-quality |
| S11_vs_k141_101115\|\|lt2gene | Low-quality |
| S11_vs_k141_48617\|\|lt2gene | Low-quality |
| S11_vs_k141_16378\|\|lt2gene | Low-quality |
| S11_vs_k141_55483\|\|lt2gene | Low-quality |
| S11_vs_k141_23331\|\|lt2gene | Not-determined |
| S11_vs_k141_23336\|\|lt2gene | Not-determined |
| S11_vs_k141_75472\|\|lt2gene | Low-quality |
| S11_vs_k141_35676\|\|lt2gene | Not-determined |
| S11_vs_k141_10168\|\|lt2gene | Low-quality |
| S11_vs_k141_35694\|\|lt2gene | Not-determined |
| S11_vs_k141_75497\|\|lt2gene | Low-quality |
| S11_vs_k141_68095\|\|lt2gene | Low-quality |
| S11_vs_k141_61701\|\|lt2gene | Not-determined |
| S11_vs_k141_94629\|\|lt2gene | Low-quality |
| S11_vs_k141_68108\|\|lt2gene | Low-quality |
| S11_vs_k141_94649\|\|lt2gene | Low-quality |
| S11_vs_k141_23369\|\|lt2gene | Low-quality |
| S11_vs_k141_35779\|\|lt2gene | Low-quality |
| S11_vs_k141_29326\|\|lt2gene | Low-quality |
| S11_vs_k141_55609\|\|lt2gene | Low-quality |
| S11_vs_k141_23390\|\|lt2gene | Low-quality |
| S11_vs_k141_3564\|\|lt2gene | Low-quality |
| S11_vs_k141_48716\|\|lt2gene | Not-determined |
| S11_vs_k141_48718\|\|lt2gene | Low-quality |
| S11_vs_k141_55626\|\|lt2gene | Low-quality |
| S11_vs_k141_61784\|\|lt2gene | Not-determined |
| S11_vs_k141_88255\|\|lt2gene | Low-quality |
| S11_vs_k141_61785\|\|lt2gene | Low-quality |
| S11_vs_k141_94759\|\|lt2gene | Low-quality |
| S11_vs_k141_101243\|\|lt2gene | Low-quality |
| S11_vs_k141_29370\|\|lt2gene | Low-quality |
| S11_vs_k141_88282\|\|lt2gene | Low-quality |
| S11_vs_k141_23430\|\|lt2gene | Low-quality |
| S11_vs_k141_35833\|\|lt2gene | Low-quality |
| S11_vs_k141_3598\|\|lt2gene | Low-quality |
| S11_vs_k141_10261\|\|lt2gene | Not-determined |
| S11_vs_k141_101264\|\|lt2gene | Not-determined |
| S11_vs_k141_61821\|\|lt2gene | Not-determined |
| S11_vs_k141_68142\|\|lt2gene | Low-quality |
| S11_vs_k141_101271\|\|lt2gene | Low-quality |
| S11_vs_k141_3628\|\|lt2gene | Low-quality |
| S11_vs_k141_3631\|\|lt2gene | Not-determined |
| S11_vs_k141_10292\|\|lt2gene | Low-quality |
| S11_vs_k141_48738\|\|lt2gene | Low-quality |
| S11_vs_k141_10300\|\|lt2gene | Not-determined |
| S11_vs_k141_75640\|\|lt2gene | Not-determined |
| S11_vs_k141_101313\|\|lt2gene | Not-determined |
| S11_vs_k141_55726\|\|lt2gene | Low-quality |
| S11_vs_k141_42687\|\|lt2gene | Not-determined |
| S11_vs_k141_101332\|\|lt2gene | Not-determined |
| S11_vs_k141_55770\|\|lt2gene | Not-determined |
| S11_vs_k141_75695\|\|lt2gene | Low-quality |
| S11_vs_k141_48756\|\|lt2gene | Low-quality |
| S11_vs_k141_94869\|\|lt2gene | Low-quality |
| S11_vs_k141_42736\|\|lt2gene | Low-quality |
| S11_vs_k141_42741\|\|lt2gene | Low-quality |
| S11_vs_k141_35964\|\|lt2gene | Low-quality |
| S11_vs_k141_68182\|\|lt2gene | Low-quality |
| S11_vs_k141_35969\|\|lt2gene | Low-quality |
| S11_vs_k141_68188\|\|lt2gene | Low-quality |
| S11_vs_k141_75716\|\|lt2gene | Not-determined |
| S11_vs_k141_23508\|\|lt2gene | Low-quality |
| S11_vs_k141_75721\|\|lt2gene | Low-quality |
| S11_vs_k141_29435\|\|lt2gene | Low-quality |
| S11_vs_k141_23530\|\|lt2gene | Low-quality |
| S11_vs_k141_81608\|\|lt2gene | Low-quality |
| S11_vs_k141_94941\|\|lt2gene | Low-quality |
| S11_vs_k141_62024\|\|lt2gene | Not-determined |
| S11_vs_k141_29507\|\|lt2gene | Low-quality |
| S11_vs_k141_48817\|\|lt2gene | Low-quality |
| S11_vs_k141_29530\|\|lt2gene | Low-quality |
| S11_vs_k141_48837\|\|lt2gene | Low-quality |
| S11_vs_k141_55902\|\|lt2gene | Low-quality |
| S11_vs_k141_29551\|\|lt2gene | Low-quality |
| S11_vs_k141_88511\|\|lt2gene | Not-determined |
| S11_vs_k141_62087\|\|lt2gene | Low-quality |
| S11_vs_k141_101488\|\|lt2gene | Low-quality |
| S11_vs_k141_81713\|\|lt2gene | Low-quality |
| S11_vs_k141_88516\|\|lt2gene | Low-quality |
| S11_vs_k141_55917\|\|lt2gene | Not-determined |
| S11_vs_k141_23569\|\|lt2gene | Low-quality |
| S11_vs_k141_81725\|\|lt2gene | Low-quality |
| S11_vs_k141_81726\|\|lt2gene | Low-quality |
| S11_vs_k141_88540\|\|lt2gene | Low-quality |
| S11_vs_k141_29582\|\|lt2gene | Low-quality |
| S11_vs_k141_29587\|\|lt2gene | Low-quality |
| S11_vs_k141_16614\|\|lt2gene | Low-quality |
| S11_vs_k141_23627\|\|lt2gene | Not-determined |
| S11_vs_k141_36098\|\|lt2gene | Low-quality |
| S11_vs_k141_101589\|\|lt2gene | Low-quality |
| S11_vs_k141_56013\|\|lt2gene | Low-quality |
| S11_vs_k141_62149\|\|lt2gene | Low-quality |
| S11_vs_k141_75885\|\|lt2gene | Low-quality |
| S11_vs_k141_16728\|\|lt2gene | Low-quality |
| S11_vs_k141_3925\|\|lt2gene | Low-quality |
| S11_vs_k141_42928\|\|lt2gene | Low-quality |
| S11_vs_k141_23730\|\|lt2gene | Low-quality |
| S11_vs_k141_68370\|\|lt2gene | Low-quality |
| S11_vs_k141_23760\|\|lt2gene | Low-quality |
| S11_vs_k141_88725\|\|lt2gene | Low-quality |
| S11_vs_k141_10501\|\|lt2gene | Not-determined |
| S11_vs_k141_48968\|\|lt2gene | Low-quality |
| S11_vs_k141_62220\|\|lt2gene | Not-determined |
| S11_vs_k141_68384\|\|lt2gene | Not-determined |
| S11_vs_k141_56134\|\|lt2gene | Low-quality |
| S11_vs_k141_101640\|\|lt2gene | Low-quality |
| S11_vs_k141_10526\|\|lt2gene | Low-quality |
| S11_vs_k141_3993\|\|lt2gene | Low-quality |
| S11_vs_k141_16846\|\|lt2gene | Low-quality |
| S11_vs_k141_68430\|\|lt2gene | Not-determined |
| S11_vs_k141_81875\|\|lt2gene | Low-quality |
| S11_vs_k141_101665\|\|lt2gene | Low-quality |
| S11_vs_k141_4022\|\|lt2gene | Low-quality |
| S11_vs_k141_81897\|\|lt2gene | Low-quality |
| S11_vs_k141_81906\|\|lt2gene | Low-quality |
| S11_vs_k141_43041\|\|lt2gene | Low-quality |
| S11_vs_k141_23885\|\|lt2gene | Low-quality |
| S11_vs_k141_36302\|\|lt2gene | Low-quality |
| S11_vs_k141_56223\|\|lt2gene | Not-determined |
| S11_vs_k141_16908\|\|lt2gene | Not-determined |
| S11_vs_k141_56226\|\|lt2gene | Not-determined |
| S11_vs_k141_101689\|\|lt2gene | Low-quality |
| S11_vs_k141_23906\|\|lt2gene | Not-determined |
| S11_vs_k141_10587\|\|lt2gene | Low-quality |
| S11_vs_k141_68538\|\|lt2gene | Low-quality |
| S11_vs_k141_95144\|\|lt2gene | Not-determined |
| S11_vs_k141_43100\|\|lt2gene | Low-quality |

S11_vs_k141_88787||lt2gene Low-quality
S11_vs_k141_68553||lt2gene Low-quality
S11_vs_k141_75935||lt2gene Low-quality
S11_vs_k141_62395||lt2gene Low-quality
S11_vs_k141_62401||lt2gene Low-quality
S11_vs_k141_56294||lt2gene Not-determined
S11_vs_k141_81993||lt2gene Low-quality
S11_vs_k141_56303||lt2gene Not-determined
S11_vs_k141_43139||lt2gene Not-determined
S11_vs_k141_56325||lt2gene Low-quality
S11_vs_k141_16953||lt2gene Not-determined
S11_vs_k141_88833||lt2gene Low-quality
S11_vs_k141_16960||lt2gene Low-quality
S11_vs_k141_36403||lt2gene Low-quality
S11_vs_k141_4143||lt2gene Low-quality
S11_vs_k141_10609||lt2gene Low-quality
S11_vs_k141_62457||lt2gene Low-quality
S11_vs_k141_56365||lt2gene Not-determined
S11_vs_k141_43185||lt2gene Low-quality
S11_vs_k141_24003||lt2gene Low-quality
S11_vs_k141_82041||lt2gene Low-quality
S11_vs_k141_29750||lt2gene Not-determined
S11_vs_k141_24028||lt2gene Low-quality
S11_vs_k141_24052||lt2gene Low-quality
S11_vs_k141_36470||lt2gene Low-quality
S11_vs_k141_10678||lt2gene Low-quality
S11_vs_k141_36482||lt2gene Low-quality
S11_vs_k141_101806||lt2gene Not-determined
S11_vs_k141_43235||lt2gene Not-determined
S11_vs_k141_101812||lt2gene Not-determined
S11_vs_k141_29792||lt2gene Low-quality
S11_vs_k141_56436||lt2gene Low-quality
S11_vs_k141_36497||lt2gene Low-quality
S11_vs_k141_68626||lt2gene Not-determined
S11_vs_k141_88915||lt2gene Not-determined
S11_vs_k141_29797||lt2gene Not-determined
S11_vs_k141_10714||lt2gene Not-determined
S11_vs_k141_75984||lt2gene Low-quality
S11_vs_k141_16977||lt2gene Low-quality
S11_vs_k141_88919||lt2gene Not-determined
S11_vs_k141_29814||lt2gene Low-quality
S11_vs_k141_56463||lt2gene Low-quality
S11_vs_k141_43282||lt2gene Low-quality
S11_vs_k141_95354||lt2gene Low-quality
S11_vs_k141_36557||lt2gene Low-quality
S11_vs_k141_101871||lt2gene Low-quality
S11_vs_k141_82103||lt2gene Not-determined
S11_vs_k141_82104||lt2gene Low-quality
S11_vs_k141_36598||lt2gene Not-determined
S11_vs_k141_29873||lt2gene Low-quality
S11_vs_k141_76058||lt2gene Not-determined
S11_vs_k141_36609||lt2gene Not-determined
S11_vs_k141_10790||lt2gene Not-determined
S11_vs_k141_17067||lt2gene Low-quality
S11_vs_k141_82134||lt2gene Low-quality
S11_vs_k141_82139||lt2gene Low-quality
S11_vs_k141_49140||lt2gene Low-quality
S11_vs_k141_43332||lt2gene Low-quality
S11_vs_k141_17099||lt2gene Low-quality
S11_vs_k141_43350||lt2gene Low-quality
S11_vs_k141_101949||lt2gene Not-determined
S11_vs_k141_17111||lt2gene Not-determined
S11_vs_k141_49173||lt2gene Low-quality
S11_vs_k141_10858||lt2gene Low-quality
S11_vs_k141_36655||lt2gene Low-quality
S11_vs_k141_95463||lt2gene Low-quality
S11_vs_k141_49195||lt2gene Not-determined
S11_vs_k141_36671||lt2gene Low-quality
S11_vs_k141_17134||lt2gene Low-quality
S11_vs_k141_101995||lt2gene Low-quality
S11_vs_k141_102001||lt2gene Low-quality
S11_vs_k141_49235||lt2gene Not-determined
S11_vs_k141_102029||lt2gene Low-quality
S11_vs_k141_17178||lt2gene Low-quality
S11_vs_k141_17185||lt2gene Low-quality
S11_vs_k141_24362||lt2gene Low-quality
S11_vs_k141_17191||lt2gene Low-quality
S11_vs_k141_62696||lt2gene Not-determined
S11_vs_k141_30023||lt2gene Low-quality
S11_vs_k141_68744||lt2gene Not-determined
S11_vs_k141_68747||lt2gene Low-quality
S11_vs_k141_82186||lt2gene Low-quality
S11_vs_k141_68765||lt2gene Low-quality
S11_vs_k141_17220||lt2gene Low-quality
S11_vs_k141_102132||lt2gene Low-quality
S11_vs_k141_89083||lt2gene Not-determined
S11_vs_k141_89090||lt2gene Low-quality
S11_vs_k141_49323||lt2gene Not-determined
S11_vs_k141_56622||lt2gene Low-quality
S11_vs_k141_10992||lt2gene Low-quality
S11_vs_k141_36766||lt2gene Low-quality
S11_vs_k141_89126||lt2gene Low-quality
S11_vs_k141_24478||lt2gene Low-quality
S11_vs_k141_17256||lt2gene Not-determined
S11_vs_k141_17257||lt2gene Not-determined
S11_vs_k141_56654||lt2gene Low-quality
S11_vs_k141_24517||lt2gene Low-quality
S11_vs_k141_4471||lt2gene Low-quality
S11_vs_k141_68849||lt2gene Low-quality
S11_vs_k141_4482||lt2gene Not-determined
S11_vs_k141_76149||lt2gene Not-determined
S11_vs_k141_95702||lt2gene Low-quality
S11_vs_k141_62823||lt2gene Low-quality
S11_vs_k141_24547||lt2gene Not-determined
S11_vs_k141_43515||lt2gene Low-quality
S11_vs_k141_62831||lt2gene Not-determined
S11_vs_k141_62841||lt2gene Low-quality
S11_vs_k141_102236||lt2gene Low-quality
S11_vs_k141_11088||lt2gene Low-quality
S11_vs_k141_43552||lt2gene Low-quality
S11_vs_k141_43555||lt2gene Low-quality
S11_vs_k141_49434||lt2gene Low-quality
S11_vs_k141_11117||lt2gene Low-quality
S11_vs_k141_95783||lt2gene Low-quality
S11_vs_k141_49447||lt2gene Not-determined
S11_vs_k141_24631||lt2gene Not-determined
S11_vs_k141_24642||lt2gene Not-determined
S11_vs_k141_95794||lt2gene Low-quality
S11_vs_k141_36906||lt2gene Low-quality
S11_vs_k141_17363||lt2gene Low-quality
S11_vs_k141_11143||lt2gene Low-quality
S11_vs_k141_11148||lt2gene Low-quality
S11_vs_k141_76212||lt2gene Low-quality
S11_vs_k141_68967||lt2gene Low-quality
S11_vs_k141_76229||lt2gene Low-quality
S11_vs_k141_17398||lt2gene Low-quality
S11_vs_k141_30109||lt2gene Low-quality
S11_vs_k141_95873||lt2gene Low-quality
S11_vs_k141_24687||lt2gene Low-quality
S11_vs_k141_76243||lt2gene Low-quality
S11_vs_k141_17412||lt2gene Low-quality
S11_vs_k141_17415||lt2gene Low-quality
S11_vs_k141_30157||lt2gene Low-quality
S11_vs_k141_102337||lt2gene Not-determined
S11_vs_k141_102339||lt2gene Not-determined
S11_vs_k141_82339||lt2gene Not-determined
S11_vs_k141_102343||lt2gene Not-determined
S11_vs_k141_102345||lt2gene Low-quality
S11_vs_k141_36990||lt2gene Low-quality
S11_vs_k141_69018||lt2gene Not-determined
S11_vs_k141_17483||lt2gene Low-quality
S11_vs_k141_17493||lt2gene Not-determined
S11_vs_k141_24754||lt2gene Low-quality
S11_vs_k141_24766||lt2gene Low-quality
S11_vs_k141_76334||lt2gene Low-quality
S11_vs_k141_76337||lt2gene Low-quality
S11_vs_k141_95957||lt2gene Low-quality
S11_vs_k141_37032||lt2gene Low-quality
S11_vs_k141_24776||lt2gene Low-quality
S11_vs_k141_43655||lt2gene Low-quality
S11_vs_k141_56859||lt2gene Low-quality
S11_vs_k141_76356||lt2gene Low-quality
S11_vs_k141_43673||lt2gene Not-determined
S11_vs_k141_63004||lt2gene Low-quality
S11_vs_k141_43681||lt2gene Low-quality
S11_vs_k141_30235||lt2gene Low-quality
S11_vs_k141_56877||lt2gene Low-quality
S11_vs_k141_56878||lt2gene Low-quality
S11_vs_k141_102406||lt2gene Low-quality
S11_vs_k141_63029||lt2gene Low-quality
S11_vs_k141_43705||lt2gene Low-quality
S11_vs_k141_49631||lt2gene Low-quality
S11_vs_k141_24819||lt2gene Low-quality
S11_vs_k141_43713||lt2gene Low-quality
S11_vs_k141_56912||lt2gene Not-determined
S11_vs_k141_63056||lt2gene Low-quality
S11_vs_k141_24834||lt2gene Low-quality
S11_vs_k141_37127||lt2gene Not-determined
S11_vs_k141_82412||lt2gene Not-determined
S11_vs_k141_89324||lt2gene Low-quality
S11_vs_k141_56957||lt2gene Low-quality
S11_vs_k141_96070||lt2gene Low-quality
S11_vs_k141_63115||lt2gene Low-quality
S11_vs_k141_96081||lt2gene Low-quality
S11_vs_k141_11232||lt2gene Low-quality
S11_vs_k141_43770||lt2gene Not-determined
S11_vs_k141_69178||lt2gene Not-determined
S11_vs_k141_57010||lt2gene Low-quality
S11_vs_k141_57025||lt2gene Low-quality
S11_vs_k141_82479||lt2gene Not-determined
S11_vs_k141_17601||lt2gene Low-quality
S11_vs_k141_57047||lt2gene Low-quality
S11_vs_k141_89390||lt2gene Low-quality
S11_vs_k141_76511||lt2gene Low-quality
S11_vs_k141_57059||lt2gene Not-determined
S11_vs_k141_49788||lt2gene Low-quality
S11_vs_k141_17625||lt2gene Low-quality
S11_vs_k141_37225||lt2gene Not-determined
S11_vs_k141_63182||lt2gene Low-quality
S11_vs_k141_76516||lt2gene Not-determined
S11_vs_k141_96170||lt2gene Not-determined
S11_vs_k141_57075||lt2gene Low-quality
S11_vs_k141_57089||lt2gene Low-quality
S11_vs_k141_69248||lt2gene Low-quality
S11_vs_k141_49804||lt2gene Low-quality
S11_vs_k141_11288||lt2gene Low-quality
S11_vs_k141_17677||lt2gene Not-determined
S11_vs_k141_69278||lt2gene Not-determined
S11_vs_k141_4971||lt2gene Not-determined
S11_vs_k141_82531||lt2gene Not-determined
S11_vs_k141_57130||lt2gene Low-quality
S11_vs_k141_57131||lt2gene Low-quality
S11_vs_k141_11325||lt2gene Low-quality
S11_vs_k141_25007||lt2gene Not-determined
S11_vs_k141_96237||lt2gene Low-quality
S11_vs_k141_43865||lt2gene Low-quality
S11_vs_k141_4988||lt2gene Low-quality
S11_vs_k141_57171||lt2gene Low-quality
S11_vs_k141_102554||lt2gene Low-quality
S11_vs_k141_69311||lt2gene Low-quality
S11_vs_k141_49922||lt2gene Low-quality
S11_vs_k141_25082||lt2gene Low-quality
S11_vs_k141_82600||lt2gene Not-determined
S11_vs_k141_37347||lt2gene Low-quality
S11_vs_k141_49946||lt2gene Not-determined
S11_vs_k141_96295||lt2gene Low-quality
S11_vs_k141_76669||lt2gene Low-quality
S11_vs_k141_96328||lt2gene Low-quality
S11_vs_k141_37397||lt2gene Low-quality
S11_vs_k141_96358||lt2gene Low-quality
S11_vs_k141_11466||lt2gene Low-quality
S11_vs_k141_11467||lt2gene Low-quality
S11_vs_k141_76716||lt2gene Not-determined
S11_vs_k141_30578||lt2gene Not-determined
S11_vs_k141_63240||lt2gene Not-determined
S11_vs_k141_96362||lt2gene Low-quality
S11_vs_k141_102562||lt2gene Low-quality
S11_vs_k141_37441||lt2gene Not-determined
S11_vs_k141_30599||lt2gene Low-quality
S11_vs_k141_17841||lt2gene Low-quality
S11_vs_k141_25166||lt2gene Low-quality
S11_vs_k141_17847||lt2gene Low-quality
S11_vs_k141_96443||lt2gene Not-determined
S11_vs_k141_96451||lt2gene Not-determined
S11_vs_k141_17878||lt2gene Low-quality
S11_vs_k141_63303||lt2gene Not-determined
S11_vs_k141_37508||lt2gene Low-quality
S11_vs_k141_63318||lt2gene Low-quality
S11_vs_k141_69448||lt2gene Low-quality
S11_vs_k141_37525||lt2gene Low-quality
S11_vs_k141_57352||lt2gene Low-quality
S11_vs_k141_57362||lt2gene Low-quality
S11_vs_k141_30709||lt2gene Low-quality
S11_vs_k141_44061||lt2gene Low-quality
S11_vs_k141_44070||lt2gene Low-quality
S11_vs_k141_57378||lt2gene Low-quality
S11_vs_k141_102647||lt2gene Low-quality
S11_vs_k141_30726||lt2gene Low-quality
S11_vs_k141_30730||lt2gene Low-quality
S11_vs_k141_57388||lt2gene Low-quality
S11_vs_k141_57426||lt2gene Low-quality
S11_vs_k141_102694||lt2gene Not-determined
S11_vs_k141_11587||lt2gene Low-quality
S11_vs_k141_69518||lt2gene Low-quality
S11_vs_k141_57441||lt2gene Low-quality
S11_vs_k141_25223||lt2gene Low-quality
S11_vs_k141_96568||lt2gene Low-quality
S11_vs_k141_25226||lt2gene Low-quality
S11_vs_k141_44134||lt2gene Low-quality
S11_vs_k141_89650||lt2gene Low-quality
S11_vs_k141_25256||lt2gene Not-determined
S11_vs_k141_96593||lt2gene Low-quality
S11_vs_k141_37627||lt2gene Not-determined
S11_vs_k141_69560||lt2gene Low-quality
S11_vs_k141_25296||lt2gene Low-quality
S11_vs_k141_96644||lt2gene Low-quality
S11_vs_k141_76850||lt2gene Not-determined
S11_vs_k141_69592||lt2gene Low-quality
S11_vs_k141_69593||lt2gene Low-quality
S11_vs_k141_76882||lt2gene Not-determined
S11_vs_k141_25344||lt2gene Low-quality
S11_vs_k141_18035||lt2gene Low-quality
S11_vs_k141_82943||lt2gene Low-quality
S11_vs_k141_82953||lt2gene Low-quality
S11_vs_k141_5185||lt2gene Low-quality
S11_vs_k141_30844||lt2gene Low-quality
S11_vs_k141_18065||lt2gene Low-quality
S11_vs_k141_44235||lt2gene Low-quality
S11_vs_k141_30885||lt2gene Low-quality
S11_vs_k141_37733||lt2gene Low-quality
S11_vs_k141_25384||lt2gene Low-quality
S11_vs_k141_44268||lt2gene Not-determined
S11_vs_k141_63456||lt2gene Low-quality
S11_vs_k141_76956||lt2gene Low-quality
S11_vs_k141_11622||lt2gene Low-quality
S11_vs_k141_18092||lt2gene Not-determined
S11_vs_k141_30905||lt2gene Low-quality
S11_vs_k141_11625||lt2gene Not-determined
S11_vs_k141_30909||lt2gene Low-quality
S11_vs_k141_37755||lt2gene Low-quality
S11_vs_k141_44314||lt2gene Low-quality
S11_vs_k141_11639||lt2gene Low-quality
S11_vs_k141_37804||lt2gene Low-quality
S11_vs_k141_18151||lt2gene Low-quality
S11_vs_k141_18158||lt2gene Low-quality
S11_vs_k141_69772||lt2gene Low-quality
S11_vs_k141_63548||lt2gene Low-quality
S11_vs_k141_25456||lt2gene Low-quality
S11_vs_k141_50142||lt2gene Low-quality
S11_vs_k141_102777||lt2gene Low-quality
S11_vs_k141_57581||lt2gene Low-quality
S11_vs_k141_18171||lt2gene Low-quality
S11_vs_k141_63574||lt2gene Low-quality
S11_vs_k141_5322||lt2gene Low-quality
S11_vs_k141_31022||lt2gene Not-determined
S11_vs_k141_44377||lt2gene Low-quality
S11_vs_k141_89786||lt2gene Low-quality
S11_vs_k141_50191||lt2gene Not-determined
S11_vs_k141_37902||lt2gene Not-determined
S11_vs_k141_57630||lt2gene Low-quality
S11_vs_k141_25534||lt2gene Low-quality
S11_vs_k141_83133||lt2gene Low-quality
S11_vs_k141_69868||lt2gene Low-quality
S11_vs_k141_63677||lt2gene Not-determined
S11_vs_k141_69883||lt2gene Low-quality
S11_vs_k141_44448||lt2gene Low-quality
S11_vs_k141_11773||lt2gene Low-quality
S11_vs_k141_102906||lt2gene Low-quality
S11_vs_k141_77157||lt2gene Low-quality
S11_vs_k141_57703||lt2gene Low-quality
S11_vs_k141_18271||lt2gene Low-quality
S11_vs_k141_83183||lt2gene Low-quality
S11_vs_k141_57714||lt2gene Low-quality
S11_vs_k141_31150||lt2gene Low-quality
S11_vs_k141_5452||lt2gene Low-quality
S11_vs_k141_83205||lt2gene Not-determined
S11_vs_k141_69930||lt2gene Low-quality
S11_vs_k141_5461||lt2gene Low-quality
S11_vs_k141_25616||lt2gene Low-quality
S11_vs_k141_37961||lt2gene Low-quality
S11_vs_k141_77196||lt2gene Low-quality
S11_vs_k141_89856||lt2gene Low-quality
S11_vs_k141_5498||lt2gene Low-quality
S11_vs_k141_37989||lt2gene Not-determined
S11_vs_k141_50333||lt2gene Low-quality
S11_vs_k141_77221||lt2gene Low-quality
S11_vs_k141_83248||lt2gene Low-quality
S11_vs_k141_50342||lt2gene Low-quality
S11_vs_k141_44537||lt2gene Low-quality
S11_vs_k141_83273||lt2gene Low-quality
S11_vs_k141_5562||lt2gene Low-quality
S11_vs_k141_44593||lt2gene Low-quality
S11_vs_k141_63882||lt2gene Low-quality
S11_vs_k141_69969||lt2gene Low-quality
S11_vs_k141_11903||lt2gene Low-quality
S11_vs_k141_69975||lt2gene Low-quality
S11_vs_k141_103017||lt2gene Not-determined
S11_vs_k141_44618||lt2gene Not-determined
S11_vs_k141_18430||lt2gene Low-quality
S11_vs_k141_38106||lt2gene Low-quality
S11_vs_k141_77330||lt2gene Not-determined
S11_vs_k141_44643||lt2gene Low-quality
S11_vs_k141_50433||lt2gene Not-determined
S11_vs_k141_103050||lt2gene Low-quality
S11_vs_k141_103051||lt2gene Low-quality
S11_vs_k141_11966||lt2gene Low-quality
S11_vs_k141_38122||lt2gene Not-determined
S11_vs_k141_44664||lt2gene Not-determined
S11_vs_k141_77360||lt2gene Low-quality
S11_vs_k141_77368||lt2gene Low-quality
S11_vs_k141_103076||lt2gene Low-quality
S11_vs_k141_44682||lt2gene Low-quality
S11_vs_k141_31331||lt2gene Not-determined
S11_vs_k141_83393||lt2gene Low-quality
S11_vs_k141_25729||lt2gene Low-quality
S11_vs_k141_77390||lt2gene Low-quality
S11_vs_k141_25749||lt2gene Not-determined
S11_vs_k141_77413||lt2gene Not-determined
S11_vs_k141_5687||lt2gene Low-quality
S11_vs_k141_5693||lt2gene Low-quality
S11_vs_k141_18525||lt2gene Low-quality
S11_vs_k141_63953||lt2gene Low-quality
S11_vs_k141_5719||lt2gene Not-determined
S11_vs_k141_83440||lt2gene Not-determined
S11_vs_k141_70032||lt2gene Not-determined
S11_vs_k141_70040||lt2gene Low-quality
S11_vs_k141_18543||lt2gene Low-quality
S11_vs_k141_31401||lt2gene Low-quality
S11_vs_k141_77454||lt2gene Low-quality
S11_vs_k141_50521||lt2gene Low-quality
S11_vs_k141_57753||lt2gene Low-quality
S11_vs_k141_77462||lt2gene Low-quality
S11_vs_k141_63994||lt2gene Low-quality
S11_vs_k141_63999||lt2gene Low-quality
S11_vs_k141_12020||lt2gene Low-quality
S11_vs_k141_83476||lt2gene Low-quality
S11_vs_k141_5749||lt2gene Low-quality
S11_vs_k141_64013||lt2gene Not-determined
S11_vs_k141_25826||lt2gene Low-quality
S11_vs_k141_5756||lt2gene Low-quality
S11_vs_k141_12038||lt2gene Low-quality
S11_vs_k141_5759||lt2gene Not-determined
S11_vs_k141_38246||lt2gene Low-quality
S11_vs_k141_12049||lt2gene Low-quality
S11_vs_k141_57802||lt2gene Low-quality
S11_vs_k141_5772||lt2gene Low-quality
S11_vs_k141_25855||lt2gene Not-determined
S11_vs_k141_25864||lt2gene Low-quality
S11_vs_k141_25867||lt2gene Low-quality
S11_vs_k141_97114||lt2gene Low-quality
S11_vs_k141_90161||lt2gene Not-determined
S11_vs_k141_64055||lt2gene Low-quality
S11_vs_k141_97119||lt2gene Low-quality
S11_vs_k141_18669||lt2gene Low-quality
S11_vs_k141_97132||lt2gene Low-quality
S11_vs_k141_97152||lt2gene Low-quality
S11_vs_k141_50615||lt2gene Not-determined
S11_vs_k141_90203||lt2gene Low-quality
S11_vs_k141_70178||lt2gene Low-quality
S11_vs_k141_50634||lt2gene Low-quality
S11_vs_k141_18717||lt2gene Not-determined
S11_vs_k141_25945||lt2gene Low-quality
S11_vs_k141_50652||lt2gene Low-quality
S11_vs_k141_50659||lt2gene Low-quality
S11_vs_k141_50661||lt2gene Low-quality
S11_vs_k141_31521||lt2gene Low-quality
S11_vs_k141_5866||lt2gene Low-quality
S11_vs_k141_25988||lt2gene Low-quality
S11_vs_k141_50696||lt2gene Low-quality
S11_vs_k141_44856||lt2gene Low-quality
S11_vs_k141_38366||lt2gene Low-quality
S11_vs_k141_18775||lt2gene Not-determined
S11_vs_k141_50715||lt2gene Low-quality
S11_vs_k141_44877||lt2gene Low-quality
S11_vs_k141_31562||lt2gene Not-determined
S11_vs_k141_64174||lt2gene Low-quality
S11_vs_k141_44900||lt2gene Low-quality
S11_vs_k141_18819||lt2gene Low-quality
S11_vs_k141_5916||lt2gene Low-quality
S11_vs_k141_57987||lt2gene Low-quality
S11_vs_k141_57992||lt2gene Not-determined
S11_vs_k141_12268||lt2gene Low-quality
S11_vs_k141_12271||lt2gene Low-quality
S11_vs_k141_31604||lt2gene Low-quality
S11_vs_k141_12327||lt2gene Not-determined
S11_vs_k141_18889||lt2gene Low-quality
S11_vs_k141_77715||lt2gene Not-determined
S11_vs_k141_90367||lt2gene Low-quality
S11_vs_k141_5979||lt2gene Low-quality
S11_vs_k141_18900||lt2gene Low-quality
S11_vs_k141_50787||lt2gene Low-quality
S11_vs_k141_97351||lt2gene Low-quality
S11_vs_k141_77736||lt2gene Low-quality
S11_vs_k141_83800||lt2gene Low-quality
S11_vs_k141_31646||lt2gene Low-quality
S11_vs_k141_97359||lt2gene Not-determined
S11_vs_k141_12348||lt2gene Low-quality
S11_vs_k141_90386||lt2gene Not-determined
S11_vs_k141_90388||lt2gene Low-quality
S11_vs_k141_97376||lt2gene Low-quality
S11_vs_k141_12370||lt2gene Not-determined
S11_vs_k141_18943||lt2gene Low-quality
S11_vs_k141_18946||lt2gene Low-quality
S11_vs_k141_6026||lt2gene Low-quality
S11_vs_k141_45004||lt2gene Low-quality
S11_vs_k141_38481||lt2gene Low-quality
S11_vs_k141_12401||lt2gene Not-determined
S11_vs_k141_58127||lt2gene Low-quality
S11_vs_k141_18974||lt2gene Low-quality
S11_vs_k141_6046||lt2gene Low-quality
S11_vs_k141_18989||lt2gene Not-determined
S11_vs_k141_64330||lt2gene Low-quality
S11_vs_k141_83826||lt2gene Low-quality
S11_vs_k141_58147||lt2gene Low-quality
S11_vs_k141_45031||lt2gene Not-determined
S11_vs_k141_83835||lt2gene Low-quality
S11_vs_k141_58154||lt2gene Low-quality
S11_vs_k141_58158||lt2gene Low-quality
S11_vs_k141_12453||lt2gene Low-quality
S11_vs_k141_103303||lt2gene Low-quality
S11_vs_k141_64360||lt2gene Low-quality
S11_vs_k141_64376||lt2gene Low-quality
S11_vs_k141_50883||lt2gene Not-determined
S11_vs_k141_31778||lt2gene Low-quality
S11_vs_k141_12492||lt2gene Low-quality
S11_vs_k141_31793||lt2gene Low-quality
S11_vs_k141_58187||lt2gene Low-quality
S11_vs_k141_97495||lt2gene Low-quality
S11_vs_k141_31805||lt2gene Low-quality
S11_vs_k141_83920||lt2gene Low-quality
S11_vs_k141_77885||lt2gene Not-determined
S11_vs_k141_31814||lt2gene Low-quality
S11_vs_k141_64395||lt2gene Not-determined
S11_vs_k141_64401||lt2gene Not-determined
S11_vs_k141_45136||lt2gene Low-quality
S11_vs_k141_90558||lt2gene Low-quality
S11_vs_k141_50952||lt2gene Low-quality
S11_vs_k141_12573||lt2gene Low-quality
S11_vs_k141_77950||lt2gene Low-quality
S11_vs_k141_64441||lt2gene Low-quality
S11_vs_k141_58255||lt2gene Not-determined
S11_vs_k141_77959||lt2gene Low-quality
S11_vs_k141_31863||lt2gene Low-quality
S11_vs_k141_77967||lt2gene Low-quality
S11_vs_k141_12608||lt2gene Low-quality
S11_vs_k141_38543||lt2gene Low-quality
S11_vs_k141_64488||lt2gene Not-determined
S11_vs_k141_64489||lt2gene Low-quality
S11_vs_k141_78010||lt2gene Not-determined
S11_vs_k141_78018||lt2gene Not-determined
S11_vs_k141_90571||lt2gene Low-quality
S11_vs_k141_6120||lt2gene Low-quality
S11_vs_k141_19073||lt2gene Low-quality
S11_vs_k141_103387||lt2gene Not-determined
S11_vs_k141_31887||lt2gene Low-quality
S11_vs_k141_38585||lt2gene Low-quality
S11_vs_k141_38593||lt2gene Not-determined
S11_vs_k141_90614||lt2gene Low-quality
S11_vs_k141_19107||lt2gene Low-quality
S11_vs_k141_19124||lt2gene Low-quality
S11_vs_k141_84109||lt2gene Low-quality
S11_vs_k141_84110||lt2gene Low-quality
S11_vs_k141_19131||lt2gene Not-determined
S11_vs_k141_78061||lt2gene Low-quality
S11_vs_k141_90647||lt2gene Low-quality
S11_vs_k141_12702||lt2gene Low-quality
S11_vs_k141_103414||lt2gene Not-determined
S11_vs_k141_51130||lt2gene Low-quality
S11_vs_k141_84135||lt2gene Low-quality
S11_vs_k141_6216||lt2gene Low-quality
S11_vs_k141_70520||lt2gene Not-determined
S11_vs_k141_64592||lt2gene Low-quality
S11_vs_k141_19175||lt2gene Low-quality
S11_vs_k141_51156||lt2gene Low-quality
S11_vs_k141_12765||lt2gene Low-quality
S11_vs_k141_103458||lt2gene Not-determined
S11_vs_k141_6235||lt2gene Low-quality
S11_vs_k141_45396||lt2gene Low-quality
S11_vs_k141_78081||lt2gene Low-quality
S11_vs_k141_45404||lt2gene Not-determined
S11_vs_k141_70530||lt2gene Low-quality
S11_vs_k141_38734||lt2gene Low-quality
S11_vs_k141_38740||lt2gene Low-quality
S11_vs_k141_90709||lt2gene Not-determined
S11_vs_k141_51183||lt2gene Low-quality
S11_vs_k141_12812||lt2gene Low-quality
S11_vs_k141_90730||lt2gene Low-quality
S11_vs_k141_19239||lt2gene Low-quality
S11_vs_k141_58389||lt2gene Low-quality
S11_vs_k141_84199||lt2gene Low-quality
S11_vs_k141_90741||lt2gene Not-determined
S11_vs_k141_84220||lt2gene Low-quality
S11_vs_k141_19287||lt2gene Low-quality
S11_vs_k141_103515||lt2gene Low-quality
S11_vs_k141_64679||lt2gene Not-determined
S11_vs_k141_6326||lt2gene Low-quality
S11_vs_k141_32007||lt2gene Low-quality
S11_vs_k141_38829||lt2gene Not-determined
S11_vs_k141_19322||lt2gene Low-quality
S11_vs_k141_19332||lt2gene Low-quality
S11_vs_k141_6354||lt2gene Not-determined
S11_vs_k141_64724||lt2gene Low-quality
S11_vs_k141_64732||lt2gene Not-determined
S11_vs_k141_90816||lt2gene Not-determined
S11_vs_k141_51279||lt2gene Low-quality
S11_vs_k141_90833||lt2gene Low-quality
S11_vs_k141_6366||lt2gene Not-determined
S11_vs_k141_12970||lt2gene Not-determined
S11_vs_k141_103611||lt2gene Low-quality
S11_vs_k141_45467||lt2gene Low-quality
S11_vs_k141_32057||lt2gene Not-determined
S11_vs_k141_64782||lt2gene Low-quality
S11_vs_k141_58522||lt2gene Low-quality
S11_vs_k141_6412||lt2gene Low-quality
S11_vs_k141_90895||lt2gene Not-determined
S11_vs_k141_45488||lt2gene Not-determined
S11_vs_k141_103656||lt2gene Low-quality
S11_vs_k141_51379||lt2gene Low-quality
S11_vs_k141_38939||lt2gene Low-quality
S11_vs_k141_51409||lt2gene Not-determined
S11_vs_k141_103680||lt2gene Low-quality
S11_vs_k141_38952||lt2gene Low-quality
S11_vs_k141_103694||lt2gene Not-determined
S11_vs_k141_90966||lt2gene Low-quality
S11_vs_k141_32115||lt2gene Low-quality
S11_vs_k141_64874||lt2gene Low-quality
S11_vs_k141_70892||lt2gene Low-quality
S11_vs_k141_64900||lt2gene Low-quality
S11_vs_k141_51487||lt2gene Low-quality
S11_vs_k141_91043||lt2gene Low-quality
S11_vs_k141_51498||lt2gene Low-quality
S11_vs_k141_70977||lt2gene Low-quality
S11_vs_k141_91088||lt2gene Not-determined
S11_vs_k141_71024||lt2gene Low-quality
S11_vs_k141_71032||lt2gene Not-determined
S11_vs_k141_84496||lt2gene Low-quality
S11_vs_k141_65038||lt2gene Low-quality
S11_vs_k141_65039||lt2gene Not-determined
S11_vs_k141_51653||lt2gene Not-determined

| | |
|---|---|
| S11_vs_k141_84547\|\|lt2gene | Low-quality |
| S11_vs_k141_32194\|\|lt2gene | Not-determined |
| S11_vs_k141_65059\|\|lt2gene | Low-quality |
| S11_vs_k141_84582\|\|lt2gene | Low-quality |
| S11_vs_k141_71119\|\|lt2gene | Low-quality |
| S11_vs_k141_71158\|\|lt2gene | Not-determined |
| S11_vs_k141_51727\|\|lt2gene | Low-quality |
| S11_vs_k141_51732\|\|lt2gene | Not-determined |
| S11_vs_k141_51735\|\|lt2gene | Low-quality |
| S11_vs_k141_104019\|\|lt2gene | Not-determined |
| S11_vs_k141_71207\|\|lt2gene | Low-quality |
| S11_vs_k141_71217\|\|lt2gene | Low-quality |
| S11_vs_k141_32329\|\|lt2gene | Not-determined |
| S11_vs_k141_32375\|\|lt2gene | Not-determined |
| S11_vs_k141_32385\|\|lt2gene | Low-quality |
| S11_vs_k141_71318\|\|lt2gene | Low-quality |
| S11_vs_k141_51831\|\|lt2gene | Low-quality |
| S11_vs_k141_32421\|\|lt2gene | Low-quality |
| S11_vs_k141_51877\|\|lt2gene | Low-quality |
| S11_vs_k141_32480\|\|lt2gene | Low-quality |
| S11_vs_k141_32484\|\|lt2gene | Low-quality |
| S11_vs_k141_51924\|\|lt2gene | Low-quality |
| S11_vs_k141_51995\|\|lt2gene | Low-quality |
| S11_vs_k141_71449\|\|lt2gene | Low-quality |
| S11_vs_k141_71468\|\|lt2gene | Low-quality |
| S11_vs_k141_71482\|\|lt2gene | Not-determined |
| S11_vs_k141_52063\|\|lt2gene | Low-quality |
| S11_vs_k141_71573\|\|lt2gene | Low-quality |
| S13_extracted_8 | Not-determined |
| S13_extracted_17 | Not-determined |
| S13_extracted_19 | Not-determined |
| S13_extracted_20 | Low-quality |
| S13_extracted_33 | Not-determined |
| S13_extracted_36 | Not-determined |
| S13_extracted_49 | Not-determined |
| S13_extracted_52 | Not-determined |
| S13_extracted_64 | Not-determined |
| S13_extracted_68 | Not-determined |
| S13_extracted_71 | Not-determined |
| S13_extracted_74 | Low-quality |
| S13_extracted_80 | Not-determined |
| S13_extracted_87 | Not-determined |
| S13_extracted_88 | Not-determined |
| S13_extracted_92 | Not-determined |
| S13_extracted_98 | Low-quality |
| S13_extracted_100 | Low-quality |
| S13_extracted_105 | Not-determined |
| S13_extracted_118 | Not-determined |
| S13_extracted_133 | Low-quality |
| S13_extracted_134 | Not-determined |
| S13_extracted_146 | Low-quality |
| S13_extracted_153 | Not-determined |
| S13_extracted_167 | Not-determined |
| S13_extracted_179 | Low-quality |
| S13_extracted_191 | Not-determined |
| S13_extracted_206 | Not-determined |
| S13_extracted_207 | Not-determined |
| S13_extracted_213 | Not-determined |
| S13_extracted_225 | Not-determined |
| S13_extracted_226 | Not-determined |
| S13_extracted_230 | Not-determined |
| S13_extracted_238 | Not-determined |
| S13_extracted_253 | Not-determined |
| S13_extracted_255 | Not-determined |
| S13_extracted_270 | Not-determined |
| S13_extracted_285 | Not-determined |
| S13_extracted_289 | Not-determined |
| S13_extracted_309 | Low-quality |
| S13_extracted_327 | Not-determined |
| S13_extracted_339 | Low-quality |
| S13_extracted_342 | Not-determined |
| S13_extracted_353 | Not-determined |
| S13_extracted_359 | Not-determined |
| S13_extracted_361 | Not-determined |
| S13_extracted_369 | Not-determined |
| S13_extracted_378 | Not-determined |
| S13_extracted_379 | Low-quality |
| S13_extracted_389 | Not-determined |
| S13_extracted_404 | Not-determined |
| S13_extracted_407 | Not-determined |
| S13_extracted_412 | Not-determined |
| S13_extracted_413 | Not-determined |
| S13_extracted_456 | Not-determined |
| S13_extracted_465 | Not-determined |
| S13_extracted_470 | Not-determined |
| S13_extracted_471 | Not-determined |
| S13_extracted_473 | Not-determined |
| S13_extracted_474 | Not-determined |
| S13_extracted_483 | Not-determined |
| S13_extracted_488 | Not-determined |
| S13_extracted_496 | Not-determined |
| S13_extracted_507 | Not-determined |
| S13_extracted_517 | Not-determined |
| S13_extracted_523 | Low-quality |
| S13_extracted_524 | Not-determined |
| S13_extracted_533 | Low-quality |
| S13_extracted_540 | Not-determined |
| S13_extracted_543 | Not-determined |
| S13_extracted_566 | Not-determined |
| S13_extracted_567 | Not-determined |
| S13_extracted_569 | Not-determined |
| S13_extracted_592 | Not-determined |
| S13_extracted_596 | Not-determined |
| S13_extracted_601 | Not-determined |
| S13_extracted_617 | Not-determined |
| S13_extracted_620 | Not-determined |
| S13_extracted_640 | Not-determined |
| S13_extracted_644 | Not-determined |
| S13_extracted_646 | Not-determined |
| S13_extracted_647 | Not-determined |
| S13_extracted_648 | Not-determined |
| S13_extracted_651 | Not-determined |
| S13_extracted_658 | Not-determined |
| S13_extracted_661 | Not-determined |
| S13_extracted_679 | Not-determined |
| S13_extracted_681 | Not-determined |
| S13_extracted_694 | Not-determined |
| S13_extracted_696 | Not-determined |
| S13_extracted_698 | Not-determined |
| S13_extracted_712 | Not-determined |
| S13_extracted_720 | Not-determined |
| S13_extracted_721 | Not-determined |
| S13_extracted_738 | Not-determined |
| S13_extracted_743 | Not-determined |
| S13_extracted_746 | Not-determined |
| S13_extracted_751 | Not-determined |
| S13_extracted_758 | Not-determined |
| S13_extracted_766 | Not-determined |
| S13_extracted_780 | Not-determined |
| S13_extracted_783 | Not-determined |
| S13_extracted_790 | Not-determined |
| S13_extracted_794 | Not-determined |
| S13_extracted_802 | Not-determined |
| S13_extracted_816 | Not-determined |
| S13_extracted_824 | Low-quality |
| S13_extracted_830 | Not-determined |
| S13_extracted_834 | Not-determined |
| S13_extracted_835 | Low-quality |
| S13_extracted_841 | Not-determined |
| S13_extracted_853 | Not-determined |
| S13_extracted_859 | Not-determined |
| S13_extracted_863 | Not-determined |
| S13_extracted_895 | Not-determined |
| S13_extracted_902 | Not-determined |
| S13_extracted_911 | Not-determined |
| S13_extracted_918 | Not-determined |
| S13_extracted_920 | Not-determined |
| S13_extracted_940 | Low-quality |
| S13_extracted_945 | Not-determined |
| S13_extracted_955 | Not-determined |
| S13_extracted_966 | Not-determined |
| S13_extracted_968 | Not-determined |
| S13_extracted_972 | Low-quality |
| S13_extracted_977 | Not-determined |
| S13_extracted_983 | Not-determined |
| S13_extracted_986 | Not-determined |
| S13_extracted_1000 | Low-quality |
| S13_extracted_1019 | Not-determined |
| S13_extracted_1048 | Not-determined |
| S13_extracted_1054 | Not-determined |
| S13_extracted_1061 | Not-determined |
| S13_extracted_1075 | Not-determined |
| S13_extracted_1076 | Not-determined |
| S13_extracted_1095 | Not-determined |
| S13_extracted_1111 | Not-determined |
| S13_extracted_1117 | Low-quality |
| S13_extracted_1136 | Low-quality |
| S13_extracted_1137 | Not-determined |
| S13_extracted_1143 | Not-determined |
| S13_extracted_1145 | Low-quality |
| S13_extracted_1159 | Not-determined |
| S13_extracted_1165 | Not-determined |
| S13_extracted_1168 | Not-determined |
| S13_extracted_1172 | Not-determined |
| S13_extracted_1177 | Not-determined |
| S13_extracted_1181 | Not-determined |
| S13_extracted_1187 | Not-determined |
| S13_extracted_1188 | Not-determined |
| S13_extracted_1189 | Not-determined |
| S13_extracted_1205 | Not-determined |
| S13_extracted_1214 | Not-determined |
| S13_extracted_1225 | Not-determined |
| S13_extracted_1226 | Low-quality |
| S13_extracted_1241 | Not-determined |
| S13_extracted_1251 | Not-determined |
| S13_extracted_1257 | Not-determined |
| S13_extracted_1264 | Not-determined |
| S13_extracted_1271 | Not-determined |
| S13_extracted_1293 | Not-determined |
| S13_extracted_1301 | Not-determined |
| S13_extracted_1311 | Not-determined |
| S13_extracted_1326 | Not-determined |
| S13_extracted_1335 | Not-determined |
| S13_extracted_1341 | Not-determined |
| S13_extracted_1347 | Not-determined |
| S13_extracted_1376 | Not-determined |
| S13_extracted_1406 | Not-determined |
| S13_extracted_1409 | Not-determined |
| S13_extracted_1412 | Not-determined |
| S13_extracted_1420 | Not-determined |
| S13_extracted_1435 | Low-quality |
| S13_extracted_1469 | Not-determined |
| S13_extracted_1487 | Not-determined |
| S13_extracted_1492 | Not-determined |
| S13_extracted_1494 | Not-determined |
| S13_extracted_1497 | Not-determined |
| S13_extracted_1507 | Not-determined |
| S13_extracted_1522 | Not-determined |
| S13_extracted_1524 | Not-determined |
| S13_extracted_1527 | Not-determined |
| S13_extracted_1538 | Not-determined |
| S13_extracted_1546 | Not-determined |
| S13_extracted_1549 | Not-determined |
| S13_extracted_1561 | Not-determined |
| S13_extracted_1579 | Not-determined |
| S13_extracted_1583 | Not-determined |
| S13_extracted_1604 | Not-determined |
| S13_extracted_1628 | Not-determined |
| S13_extracted_1631 | Not-determined |
| S13_extracted_1632 | Not-determined |
| S13_extracted_1648 | Not-determined |
| S13_extracted_1649 | Not-determined |
| S13_extracted_1655 | Low-quality |
| S13_extracted_1660 | Not-determined |
| S13_extracted_1677 | Not-determined |
| S13_extracted_1684 | Not-determined |
| S13_extracted_1694 | Not-determined |
| S13_extracted_1709 | Low-quality |
| S13_extracted_1714 | Not-determined |
| S13_extracted_1715 | Not-determined |
| S13_extracted_1720 | Not-determined |
| S13_extracted_1724 | Not-determined |
| S13_extracted_1731 | Not-determined |
| S13_extracted_1739 | Not-determined |
| S13_extracted_1750 | Not-determined |
| S13_extracted_1757 | Not-determined |
| S13_extracted_1759 | Not-determined |
| S13_extracted_1765 | Not-determined |
| S13_extracted_1768 | Not-determined |
| S12_extracted_k141_2226 | Medium-quality |
| S13_extracted_1780 | Not-determined |
| S13_extracted_1795 | Not-determined |
| S13_extracted_1807 | Not-determined |
| S13_extracted_1818 | Not-determined |
| S13_extracted_1820 | Not-determined |
| S13_extracted_1828 | Not-determined |
| S13_extracted_1831 | Not-determined |
| S13_extracted_1841 | Not-determined |
| S13_extracted_1848 | Not-determined |
| S13_extracted_1881 | Not-determined |
| S13_extracted_1888 | Not-determined |
| S13_extracted_1889 | Not-determined |
| S13_extracted_1896 | Not-determined |
| S13_extracted_1899 | Not-determined |
| S13_extracted_1912 | Not-determined |
| S13_extracted_1924 | Not-determined |
| S13_extracted_1933 | Not-determined |
| S13_extracted_1936 | Not-determined |
| S13_extracted_1947 | Not-determined |
| S13_extracted_1968 | Not-determined |
| S13_extracted_1972 | Not-determined |
| S13_extracted_1982 | Not-determined |
| S13_extracted_1986 | Not-determined |
| S13_extracted_2007 | Not-determined |
| S13_extracted_2052 | Not-determined |
| S13_extracted_2053 | Not-determined |
| S13_extracted_2054 | Not-determined |
| S13_extracted_2060 | Not-determined |
| S13_extracted_2066 | Low-quality |
| S13_extracted_2074 | Low-quality |
| S13_extracted_2080 | Not-determined |
| S13_extracted_2083 | Low-quality |
| S13_extracted_2091 | Not-determined |
| S13_extracted_2097 | Not-determined |
| S13_extracted_2105 | Not-determined |
| S13_extracted_2117 | Not-determined |
| S13_extracted_2118 | Not-determined |
| S13_extracted_2133 | Low-quality |
| S13_extracted_2134 | Not-determined |
| S13_extracted_2135 | Not-determined |
| S13_extracted_2139 | Not-determined |
| S13_extracted_2153 | Not-determined |
| S13_extracted_2155 | Not-determined |
| S13_extracted_2158 | Not-determined |
| S13_extracted_2165 | Not-determined |
| S13_extracted_2172 | Not-determined |
| S13_extracted_2184 | Not-determined |
| S13_extracted_2186 | Not-determined |
| S13_extracted_2189 | Not-determined |
| S13_extracted_2194 | Not-determined |
| S13_extracted_2209 | Not-determined |
| S13_extracted_2225 | Not-determined |
| S13_extracted_2229 | Not-determined |
| S13_extracted_2236 | Low-quality |
| S13_extracted_2239 | Not-determined |
| S13_extracted_2285 | Not-determined |
| S13_extracted_2292 | Low-quality |
| S13_extracted_2299 | Not-determined |
| S13_extracted_2307 | Not-determined |
| S13_extracted_2316 | Not-determined |
| S13_extracted_2322 | Not-determined |
| S13_extracted_2323 | Not-determined |
| S13_extracted_2332 | Not-determined |
| S13_extracted_2334 | Not-determined |
| S13_extracted_2338 | Not-determined |
| S13_extracted_2339 | Not-determined |
| S13_extracted_2348 | Not-determined |
| S13_extracted_2349 | Not-determined |
| S13_extracted_2372 | Not-determined |
| S13_extracted_2384 | Low-quality |
| S13_extracted_2403 | Not-determined |
| S13_extracted_2411 | Not-determined |
| S13_extracted_2419 | Not-determined |
| S13_extracted_2432 | Not-determined |
| S13_extracted_2436 | Not-determined |
| S13_extracted_2442 | Not-determined |
| S13_extracted_2463 | Not-determined |
| S13_extracted_2464 | Not-determined |
| S13_extracted_2473 | Not-determined |
| S13_extracted_2474 | Not-determined |
| S13_extracted_2475 | Not-determined |
| S13_extracted_2495 | Not-determined |
| S13_extracted_2514 | Not-determined |
| S13_extracted_2522 | Not-determined |
| S13_extracted_2523 | Not-determined |
| S13_extracted_2524 | Not-determined |
| S13_extracted_2526 | Not-determined |
| S13_extracted_2551 | Not-determined |
| S13_extracted_2568 | Not-determined |
| S13_extracted_2570 | Not-determined |
| S13_extracted_2591 | Not-determined |
| S13_extracted_2617 | Not-determined |
| S13_extracted_2627 | Low-quality |
| S13_extracted_2635 | Not-determined |
| S13_extracted_2644 | Low-quality |
| S13_extracted_2645 | Not-determined |
| S13_extracted_2646 | Not-determined |
| S13_extracted_2647 | Not-determined |
| S13_extracted_2652 | Not-determined |
| S13_extracted_2671 | Not-determined |
| S13_extracted_2692 | Not-determined |
| S13_extracted_2697 | Not-determined |
| S13_extracted_2698 | Not-determined |
| S13_extracted_2723 | Low-quality |
| S13_extracted_2755 | Not-determined |
| S13_extracted_2763 | Not-determined |
| S13_extracted_2772 | Not-determined |
| S13_extracted_2774 | Not-determined |
| S13_extracted_2777 | Not-determined |
| S13_extracted_2795 | Not-determined |
| S13_extracted_2810 | Not-determined |
| S13_extracted_2839 | Not-determined |
| S13_extracted_2853 | Not-determined |
| S13_extracted_2859 | Not-determined |
| S13_extracted_2865 | Not-determined |
| S13_extracted_2901 | Not-determined |
| S13_extracted_2902 | Not-determined |
| S13_extracted_2907 | Not-determined |
| S13_extracted_2925 | Low-quality |
| S13_extracted_2954 | Not-determined |
| S13_extracted_2972 | Not-determined |
| S13_extracted_2974 | Not-determined |
| S13_extracted_2987 | Not-determined |
| S13_extracted_2991 | Not-determined |
| S13_extracted_3009 | Not-determined |
| S13_extracted_3016 | Not-determined |
| S13_extracted_3028 | Low-quality |
| S13_extracted_3037 | Not-determined |
| S13_extracted_3047 | Not-determined |
| S13_extracted_3049 | Not-determined |
| S13_extracted_3059 | Not-determined |
| S13_extracted_3075 | Not-determined |
| S13_extracted_3092 | Not-determined |
| S13_extracted_3113 | Not-determined |
| S13_extracted_3124 | Not-determined |
| S13_extracted_3136 | Low-quality |
| S13_extracted_3140 | Not-determined |
| S13_extracted_3161 | Not-determined |
| S13_extracted_3180 | Not-determined |
| S13_extracted_3186 | Not-determined |
| S13_extracted_3206 | Not-determined |
| S13_extracted_3216 | Not-determined |
| S13_extracted_3219 | Not-determined |
| S13_extracted_3222 | Not-determined |
| S13_extracted_3248 | Not-determined |
| S13_extracted_3254 | Not-determined |
| S13_extracted_3255 | Not-determined |
| S13_extracted_3258 | Not-determined |
| S13_extracted_3263 | Not-determined |
| S13_extracted_3270 | Not-determined |
| S13_extracted_3296 | Not-determined |
| S13_extracted_3315 | Not-determined |
| S13_extracted_3323 | Not-determined |
| S13_extracted_3349 | Not-determined |
| S13_extracted_3351 | Low-quality |
| S13_extracted_3371 | Not-determined |
| S13_extracted_3384 | Not-determined |
| S13_extracted_3408 | Not-determined |
| S13_extracted_3416 | Not-determined |
| S13_extracted_3427 | Not-determined |
| S13_extracted_3450 | Low-quality |
| S13_extracted_3468 | Low-quality |
| S13_extracted_3469 | Not-determined |
| S13_extracted_3470 | Not-determined |
| S13_extracted_3472 | Not-determined |
| S13_extracted_3478 | Not-determined |
| S13_extracted_3482 | Low-quality |
| S13_extracted_3507 | Not-determined |
| S13_extracted_3527 | Not-determined |
| S13_extracted_3533 | Not-determined |
| S13_extracted_3552 | Low-quality |
| S13_extracted_3571 | Not-determined |
| S13_extracted_3590 | Not-determined |
| S13_extracted_3605 | Not-determined |
| S13_extracted_3606 | Not-determined |
| S13_extracted_3614 | Not-determined |
| S13_extracted_3615 | Low-quality |
| S13_extracted_3618 | Not-determined |
| S13_extracted_3629 | Not-determined |
| S13_extracted_3633 | Not-determined |
| S13_extracted_3637 | Not-determined |
| S13_extracted_3639 | Not-determined |
| S13_extracted_3645 | Not-determined |
| S13_extracted_3648 | Not-determined |
| S13_extracted_3679 | Not-determined |
| S13_extracted_3681 | Not-determined |
| S13_extracted_3701 | Not-determined |
| S13_extracted_3703 | Not-determined |
| S13_extracted_3723 | Low-quality |
| S13_extracted_3724 | Not-determined |
| S12_vs_k141_2226\|\|full | Medium-quality |
| S13_extracted_3749 | Not-determined |
| S13_extracted_3757 | Not-determined |
| S13_extracted_3759 | Not-determined |
| S13_extracted_3770 | Not-determined |
| S13_extracted_3781 | Not-determined |
| S13_extracted_3787 | Not-determined |
| S13_extracted_3792 | Not-determined |
| S13_extracted_3799 | Not-determined |
| S13_extracted_3801 | Not-determined |
| S13_extracted_3819 | Not-determined |
| S13_extracted_3820 | Not-determined |
| S13_extracted_3826 | Not-determined |
| S13_extracted_3828 | Not-determined |
| S13_extracted_3831 | Not-determined |
| S13_extracted_3837 | Not-determined |
| S13_extracted_3845 | Not-determined |
| S13_extracted_3847 | Not-determined |
| S13_extracted_3860 | Not-determined |
| S13_extracted_3861 | Not-determined |
| S13_extracted_3871 | Not-determined |
| S13_extracted_3891 | Not-determined |
| S13_extracted_3892 | Low-quality |
| S13_extracted_3893 | Not-determined |
| S13_extracted_3898 | Not-determined |
| S13_extracted_3911 | Not-determined |
| S13_extracted_3926 | Not-determined |
| S13_extracted_3948 | Low-quality |
| S13_extracted_3951 | Not-determined |
| S13_extracted_3967 | Not-determined |
| S13_extracted_3979 | Not-determined |
| S13_extracted_4031 | Low-quality |
| S13_extracted_4037 | Not-determined |
| S13_extracted_4044 | Not-determined |
| S13_extracted_4045 | Not-determined |
| S13_extracted_4064 | Not-determined |
| S13_extracted_4067 | Not-determined |
| S13_extracted_4074 | Not-determined |
| S13_extracted_4076 | Low-quality |
| S13_extracted_4095 | Not-determined |
| S13_extracted_4108 | Not-determined |
| S13_extracted_4113 | Not-determined |
| S13_extracted_4129 | Not-determined |
| S13_extracted_4146 | Not-determined |
| S13_extracted_4150 | Not-determined |
| S13_extracted_4154 | Not-determined |
| S13_extracted_4163 | Not-determined |
| S13_extracted_4177 | Not-determined |
| S13_extracted_4178 | Low-quality |
| S13_extracted_4181 | Not-determined |
| S13_extracted_4182 | Not-determined |
| S13_extracted_4192 | Not-determined |
| S13_extracted_4211 | Low-quality |
| S13_extracted_4212 | Low-quality |
| S13_extracted_4218 | Not-determined |
| S13_extracted_4219 | Not-determined |
| S13_extracted_4224 | Not-determined |
| S13_extracted_4233 | Not-determined |
| S13_extracted_4234 | Low-quality |
| S13_extracted_4238 | Not-determined |
| S13_extracted_4247 | Not-determined |
| S13_extracted_4267 | Not-determined |
| S13_extracted_4271 | Not-determined |
| S13_extracted_4272 | Not-determined |
| S13_extracted_4294 | Not-determined |
| S13_extracted_4301 | Not-determined |
| S13_extracted_4304 | Not-determined |
| S13_extracted_4314 | Not-determined |
| S13_extracted_4328 | Not-determined |
| S13_extracted_4354 | Not-determined |
| S13_extracted_4364 | Low-quality |
| S13_extracted_4375 | Not-determined |
| S13_extracted_4386 | Not-determined |
| S13_extracted_4396 | Not-determined |
| S13_extracted_4416 | Not-determined |
| S13_extracted_4420 | Not-determined |
| S13_extracted_4430 | Not-determined |
| S13_extracted_4436 | Not-determined |
| S13_extracted_4446 | Not-determined |
| S13_extracted_4448 | Not-determined |
| S13_extracted_4452 | Not-determined |
| S13_extracted_4463 | Not-determined |
| S13_extracted_4464 | Not-determined |
| S13_extracted_4468 | Not-determined |
| S13_extracted_4474 | Low-quality |
| S13_extracted_4503 | Not-determined |
| S13_extracted_4504 | Not-determined |
| S13_extracted_4520 | Not-determined |
| S13_extracted_4532 | Not-determined |
| S13_extracted_4536 | Low-quality |
| S13_extracted_4553 | Not-determined |
| S13_extracted_4554 | Low-quality |
| S13_extracted_4558 | Not-determined |
| S13_extracted_4574 | Low-quality |
| S13_extracted_4577 | Not-determined |
| S13_extracted_4580 | Not-determined |
| S13_extracted_4589 | Low-quality |
| S13_extracted_4593 | Not-determined |
| S13_extracted_4598 | Not-determined |
| S13_extracted_4599 | Not-determined |
| S13_extracted_4610 | Not-determined |
| S13_extracted_4622 | Low-quality |
| S13_extracted_4623 | Not-determined |
| S13_extracted_4637 | Not-determined |
| S13_extracted_4641 | Not-determined |
| S13_extracted_4642 | Not-determined |
| S13_extracted_4656 | Low-quality |
| S13_extracted_4657 | Not-determined |
| S13_extracted_4659 | Not-determined |
| S13_extracted_4674 | Not-determined |
| S13_extracted_4685 | Not-determined |
| S13_extracted_4703 | Not-determined |
| S13_extracted_4710 | Not-determined |
| S13_extracted_4713 | Not-determined |
| S13_extracted_4717 | Not-determined |
| S13_extracted_4744 | Not-determined |
| S13_extracted_4752 | Not-determined |
| S13_extracted_4758 | Not-determined |
| S13_extracted_4771 | Not-determined |
| S13_extracted_4777 | Not-determined |
| S13_extracted_4780 | Not-determined |
| S13_extracted_4788 | Low-quality |
| S13_extracted_4793 | Not-determined |
| S13_extracted_4798 | Not-determined |
| S13_extracted_4802 | Not-determined |
| S13_extracted_4809 | Not-determined |
| S13_extracted_4824 | Not-determined |
| S13_extracted_4831 | Not-determined |
| S13_extracted_4856 | Not-determined |
| S13_extracted_4862 | Not-determined |
| S13_extracted_4867 | Not-determined |
| S13_extracted_4868 | Not-determined |
| S13_extracted_4870 | Not-determined |
| S13_extracted_4877 | Not-determined |
| S17_vs_k141_110666\|\|full | High-quality |
| S13_extracted_4902 | Not-determined |
| S13_extracted_4908 | Not-determined |
| S13_extracted_4923 | Not-determined |
| S13_extracted_4945 | Not-determined |
| S13_extracted_4973 | Not-determined |
| S13_extracted_4974 | Not-determined |
| S13_extracted_4981 | Not-determined |
| S13_extracted_4989 | Not-determined |
| S13_extracted_4998 | Not-determined |
| S13_extracted_5007 | Not-determined |
| S13_extracted_5018 | Not-determined |
| S13_extracted_5037 | Not-determined |
| S13_extracted_5042 | Not-determined |
| S13_extracted_5043 | Not-determined |
| S13_extracted_5057 | Not-determined |
| S13_extracted_5061 | Not-determined |
| S13_extracted_5074 | Low-quality |
| S13_extracted_5117 | Not-determined |
| S13_extracted_5119 | Not-determined |
| S13_extracted_5120 | Not-determined |
| S13_extracted_5139 | Not-determined |
| S13_extracted_5154 | Not-determined |
| S13_extracted_5170 | Not-determined |
| S13_extracted_5172 | Low-quality |

| | |
|---|---|
| S13_extracted_5173 | Low-quality |
| S13_extracted_5184 | Not-determined |
| S13_extracted_5195 | Not-determined |
| S13_extracted_5203 | Not-determined |
| S13_extracted_5213 | Not-determined |
| S13_extracted_5215 | Not-determined |
| S13_extracted_5217 | Not-determined |
| S13_extracted_5225 | Not-determined |
| S13_extracted_5238 | Not-determined |
| S13_extracted_5253 | Not-determined |
| S13_extracted_5273 | Not-determined |
| S13_extracted_5275 | Not-determined |
| S13_extracted_5284 | Not-determined |
| S13_extracted_5294 | Not-determined |
| S13_extracted_5300 | Not-determined |
| S13_extracted_5305 | Not-determined |
| S13_extracted_5306 | Not-determined |
| S13_extracted_5319 | Not-determined |
| S13_extracted_5322 | Not-determined |
| S13_extracted_5334 | Not-determined |
| S13_extracted_5346 | Not-determined |
| S13_extracted_5373 | Not-determined |
| S13_extracted_5376 | Not-determined |
| S13_extracted_5380 | Not-determined |
| S13_extracted_5387 | Not-determined |
| S13_extracted_5407 | Not-determined |
| S13_extracted_5409 | Not-determined |
| S13_extracted_5410 | Not-determined |
| S13_extracted_5430 | Not-determined |
| S13_extracted_5434 | Not-determined |
| S13_extracted_5435 | Not-determined |
| S13_extracted_5437 | Not-determined |
| S13_extracted_5447 | Not-determined |
| S13_extracted_5451 | Not-determined |
| S13_extracted_5456 | Not-determined |
| S13_extracted_5483 | Not-determined |
| S13_extracted_5491 | Not-determined |
| S13_extracted_5498 | Not-determined |
| S13_extracted_5505 | Not-determined |
| S13_extracted_5511 | Not-determined |
| S13_extracted_5512 | Not-determined |
| S13_extracted_5533 | Not-determined |
| S13_extracted_5541 | Not-determined |
| S13_extracted_5545 | Low-quality |
| S13_extracted_5551 | Not-determined |
| S13_extracted_5558 | Not-determined |
| S13_extracted_5570 | Not-determined |
| S13_extracted_5578 | Not-determined |
| S13_extracted_5579 | Not-determined |
| S13_extracted_5597 | Not-determined |
| S13_extracted_5611 | Low-quality |
| S13_extracted_5612 | Not-determined |
| S13_extracted_5613 | Not-determined |
| S13_extracted_5636 | Not-determined |
| S13_extracted_5641 | Not-determined |
| S13_extracted_5649 | Not-determined |
| S13_extracted_5654 | Not-determined |
| S13_extracted_5658 | Not-determined |
| S13_extracted_5661 | Not-determined |
| S13_extracted_5676 | Not-determined |
| S13_extracted_5677 | Not-determined |
| S13_extracted_5683 | Low-quality |
| S13_extracted_5693 | Low-quality |
| S13_extracted_5707 | Not-determined |
| S13_extracted_5717 | Not-determined |
| S13_extracted_5723 | Not-determined |
| S13_extracted_5724 | Low-quality |
| S13_extracted_5726 | Not-determined |
| S13_extracted_5733 | Not-determined |
| S13_extracted_5735 | Not-determined |
| S13_extracted_5737 | Not-determined |
| S13_extracted_5738 | Not-determined |
| S13_extracted_5748 | Not-determined |
| S13_extracted_5757 | Not-determined |
| S13_extracted_5759 | Not-determined |
| S13_extracted_5762 | Low-quality |
| S13_extracted_5768 | Not-determined |
| S13_extracted_5777 | Not-determined |
| S13_extracted_5793 | Not-determined |
| S13_extracted_5795 | Not-determined |
| S13_extracted_5797 | Not-determined |
| S13_extracted_5812 | Not-determined |
| S13_extracted_5831 | Not-determined |
| S13_extracted_5838 | Not-determined |
| S13_extracted_5854 | Not-determined |
| S13_extracted_5859 | Not-determined |
| S13_extracted_5881 | Not-determined |
| S13_extracted_5888 | Not-determined |
| S13_extracted_5890 | Low-quality |
| S13_extracted_5891 | Low-quality |
| S13_extracted_5905 | Not-determined |
| S13_extracted_5906 | Not-determined |
| S13_extracted_5908 | Not-determined |
| S13_extracted_5917 | Not-determined |
| S13_extracted_5923 | Not-determined |
| S13_extracted_5930 | Not-determined |
| S13_extracted_5947 | Not-determined |
| S13_extracted_5959 | Low-quality |
| S13_extracted_5967 | Not-determined |
| S13_extracted_5968 | Not-determined |
| S13_extracted_5970 | Not-determined |
| S13_extracted_5974 | Not-determined |
| S13_extracted_5977 | Not-determined |
| S13_extracted_5989 | Not-determined |
| S13_extracted_5994 | Not-determined |
| S13_extracted_5996 | Not-determined |
| S13_extracted_6001 | Not-determined |
| S13_extracted_6002 | Not-determined |
| S13_extracted_6034 | Not-determined |
| S13_extracted_6036 | Not-determined |
| S13_extracted_6051 | Not-determined |
| S13_extracted_6069 | Not-determined |
| S13_extracted_6071 | Not-determined |
| S13_extracted_6076 | Not-determined |
| S13_extracted_6085 | Not-determined |
| S13_extracted_6105 | Not-determined |
| S13_extracted_6107 | Not-determined |
| S13_extracted_6118 | Not-determined |
| S13_extracted_6125 | Not-determined |
| S13_extracted_6138 | Low-quality |
| S13_extracted_6161 | Not-determined |
| S13_extracted_6164 | Not-determined |
| S13_extracted_6168 | Not-determined |
| S13_extracted_6169 | Not-determined |
| S13_extracted_6177 | Not-determined |
| S13_extracted_6220 | Low-quality |
| S13_extracted_6223 | Not-determined |
| S13_extracted_6224 | Not-determined |
| S13_extracted_6228 | Not-determined |
| S13_extracted_6235 | Not-determined |
| S13_extracted_6237 | Not-determined |
| S13_extracted_6272 | Not-determined |
| S13_extracted_6294 | Not-determined |
| S13_extracted_6327 | Not-determined |
| S13_extracted_6332 | Not-determined |
| S13_extracted_6348 | Not-determined |
| S13_extracted_6352 | Not-determined |
| S13_extracted_6365 | Low-quality |
| S13_extracted_6376 | Not-determined |
| S13_extracted_6379 | Low-quality |
| S13_extracted_6387 | Not-determined |
| S13_extracted_6405 | Not-determined |
| S13_extracted_6430 | Not-determined |
| S13_extracted_6435 | Not-determined |
| S13_extracted_6438 | Low-quality |
| S13_extracted_6476 | Not-determined |
| S13_extracted_6480 | Not-determined |
| S13_extracted_6488 | Not-determined |
| S13_extracted_6517 | Not-determined |
| S13_extracted_6533 | Not-determined |
| S13_extracted_6537 | Not-determined |
| S13_extracted_6544 | Not-determined |
| S13_extracted_6564 | Not-determined |
| S13_extracted_6568 | Not-determined |
| S13_extracted_6574 | Not-determined |
| S13_extracted_6602 | Low-quality |
| S13_extracted_6606 | Not-determined |
| S13_extracted_6607 | Not-determined |
| S13_extracted_6612 | Low-quality |
| S13_extracted_6614 | Low-quality |
| S13_extracted_6628 | Not-determined |
| S13_extracted_6633 | Not-determined |
| S13_extracted_6634 | Not-determined |
| S13_extracted_6637 | Not-determined |
| S13_extracted_6654 | Not-determined |
| S13_extracted_6663 | Not-determined |
| S13_extracted_6674 | Not-determined |
| S13_extracted_6681 | Not-determined |
| S13_extracted_6683 | Not-determined |
| S13_extracted_6693 | Not-determined |
| S13_extracted_6704 | Not-determined |
| S13_extracted_6719 | Not-determined |
| S13_extracted_6735 | Not-determined |
| S13_extracted_6750 | Not-determined |
| S13_extracted_6753 | Not-determined |
| S13_extracted_6764 | Not-determined |
| S13_extracted_6782 | Not-determined |
| S13_extracted_6785 | Not-determined |
| S13_extracted_6791 | Low-quality |
| S13_extracted_6796 | Not-determined |
| S13_extracted_6808 | Not-determined |
| S13_extracted_6810 | Low-quality |
| S13_extracted_6814 | Not-determined |
| S13_extracted_6817 | Low-quality |
| S13_extracted_6821 | Not-determined |
| S13_extracted_6835 | Not-determined |
| S13_extracted_6847 | Not-determined |
| S13_extracted_6872 | Not-determined |
| S13_extracted_6915 | Not-determined |
| S13_extracted_6922 | Not-determined |
| S13_extracted_6926 | Not-determined |
| S13_extracted_6933 | Not-determined |
| S13_extracted_6935 | Not-determined |
| S13_extracted_6936 | Not-determined |
| S13_extracted_6940 | Not-determined |
| S13_extracted_6941 | Not-determined |
| S13_extracted_6945 | Not-determined |
| S13_extracted_6947 | Low-quality |
| S13_extracted_6962 | Not-determined |
| S13_extracted_6967 | Not-determined |
| S13_extracted_6969 | Not-determined |
| S13_extracted_6980 | Not-determined |
| S13_extracted_6989 | Not-determined |
| S13_extracted_6998 | Not-determined |
| S13_extracted_7020 | Not-determined |
| S13_extracted_7024 | Not-determined |
| S13_extracted_7031 | Not-determined |
| S13_extracted_7043 | Not-determined |
| S13_extracted_7055 | Not-determined |
| S13_extracted_7062 | Low-quality |
| S13_extracted_7074 | Not-determined |
| S13_extracted_7081 | Not-determined |
| S13_extracted_7082 | Not-determined |
| S13_extracted_7091 | Not-determined |
| S13_extracted_7094 | Not-determined |
| S13_extracted_7100 | Not-determined |
| S13_extracted_7105 | Not-determined |
| S13_extracted_7109 | Low-quality |
| S13_extracted_7110 | Not-determined |
| S13_extracted_7111 | Not-determined |
| S13_extracted_7127 | Not-determined |
| S13_extracted_7133 | Not-determined |
| S13_extracted_7136 | Not-determined |
| S13_extracted_7140 | Low-quality |
| S13_extracted_7143 | Low-quality |
| S13_extracted_7147 | Not-determined |
| S13_extracted_7150 | Not-determined |
| S13_extracted_7161 | Low-quality |
| S13_extracted_7163 | Not-determined |
| S13_extracted_7166 | Not-determined |
| S13_extracted_7174 | Not-determined |
| S13_extracted_7177 | Not-determined |
| S13_extracted_7178 | Not-determined |
| S13_extracted_7179 | Not-determined |
| S13_extracted_7186 | Not-determined |
| S13_extracted_7221 | Not-determined |
| S13_extracted_7232 | Not-determined |
| S13_extracted_7243 | Not-determined |
| S13_extracted_7245 | Not-determined |
| S13_extracted_7251 | Not-determined |
| S13_extracted_7253 | Not-determined |
| S13_extracted_7257 | Not-determined |
| S13_extracted_7265 | Not-determined |
| S13_extracted_7273 | Not-determined |
| S13_extracted_7275 | Not-determined |
| S13_extracted_7281 | Not-determined |
| S13_extracted_7293 | Low-quality |
| S13_extracted_7296 | Not-determined |
| S13_extracted_7300 | Not-determined |
| S13_extracted_7322 | Not-determined |
| S13_extracted_7323 | Not-determined |
| S13_extracted_7335 | Low-quality |
| S13_extracted_7337 | Not-determined |
| S13_extracted_7340 | Not-determined |
| S13_extracted_7345 | Not-determined |
| S13_extracted_7362 | Low-quality |
| S13_extracted_7367 | Not-determined |
| S13_extracted_7394 | Not-determined |
| S13_extracted_7395 | Not-determined |
| S13_extracted_7401 | Not-determined |
| S13_extracted_7414 | Not-determined |
| S13_extracted_7427 | Not-determined |
| S13_extracted_7433 | Not-determined |
| S13_extracted_7435 | Not-determined |
| S13_extracted_7443 | Not-determined |
| S13_extracted_7458 | Not-determined |
| S13_extracted_7473 | Not-determined |
| S13_extracted_7479 | Not-determined |
| S13_extracted_7482 | Not-determined |
| S13_extracted_7483 | Not-determined |
| S13_extracted_7486 | Not-determined |
| S13_extracted_7516 | Not-determined |
| S13_extracted_7522 | Not-determined |
| S13_extracted_7527 | Not-determined |
| S13_extracted_7543 | Not-determined |
| S13_extracted_7561 | Not-determined |
| S13_extracted_7588 | Not-determined |
| S13_extracted_7600 | Not-determined |
| S13_extracted_7604 | Not-determined |
| S13_extracted_7625 | Not-determined |
| S13_extracted_7636 | Not-determined |
| S13_extracted_7638 | Not-determined |
| S13_extracted_7641 | Not-determined |
| S13_extracted_7642 | Not-determined |
| S13_extracted_7647 | Not-determined |
| S13_extracted_7652 | Not-determined |
| S13_extracted_7663 | Not-determined |
| S13_extracted_7669 | Not-determined |
| S8_vs_k141_149700\|\|full | Medium-quality |
| S13_extracted_7671 | Low-quality |
| S13_extracted_7676 | Not-determined |
| S13_extracted_7689 | Low-quality |
| S13_extracted_7695 | Not-determined |
| S13_extracted_7702 | Not-determined |
| S13_extracted_7703 | Low-quality |
| S13_extracted_7719 | Not-determined |
| S13_extracted_7720 | Not-determined |
| S13_extracted_7728 | Not-determined |
| S13_extracted_7749 | Not-determined |
| S13_extracted_7766 | Not-determined |
| S13_extracted_7768 | Not-determined |
| S13_extracted_7771 | Not-determined |
| S13_extracted_7780 | Not-determined |
| S13_extracted_7787 | Not-determined |
| S13_extracted_7814 | Not-determined |
| S13_extracted_7838 | Not-determined |
| S13_extracted_7853 | Low-quality |
| S13_extracted_7854 | Not-determined |
| S13_extracted_7859 | Not-determined |
| S13_extracted_7871 | Not-determined |
| S13_extracted_7877 | Not-determined |
| S13_extracted_7883 | Not-determined |
| S13_extracted_7884 | Not-determined |
| S13_extracted_7885 | Not-determined |
| S13_extracted_7887 | Not-determined |
| S13_extracted_7889 | Not-determined |
| S13_extracted_7900 | Not-determined |
| S13_extracted_7905 | Not-determined |
| S13_extracted_7908 | Not-determined |
| S13_extracted_7912 | Not-determined |
| S13_extracted_7921 | Not-determined |
| S13_extracted_7923 | Not-determined |
| S13_extracted_7924 | Not-determined |
| S13_extracted_7930 | Not-determined |
| S13_extracted_7952 | Not-determined |
| S13_extracted_7954 | Not-determined |
| S13_extracted_7973 | Low-quality |
| S13_extracted_7974 | Low-quality |
| S13_extracted_7988 | Low-quality |
| S13_extracted_7996 | Not-determined |
| S13_extracted_8005 | Not-determined |
| S13_extracted_8007 | Not-determined |
| S13_extracted_8017 | Not-determined |
| S13_extracted_8034 | Not-determined |
| S13_extracted_8039 | Not-determined |
| S13_extracted_8040 | Not-determined |
| S13_extracted_8048 | Not-determined |
| S13_extracted_8050 | Not-determined |
| S13_extracted_8051 | Not-determined |
| S13_extracted_8053 | Not-determined |
| S13_extracted_8055 | Not-determined |
| S13_extracted_8063 | Not-determined |
| S13_extracted_8082 | Not-determined |
| S13_extracted_8088 | Not-determined |
| S13_extracted_8091 | Not-determined |
| S13_extracted_8100 | Not-determined |
| S13_extracted_8102 | Not-determined |
| S13_extracted_8164 | Not-determined |
| S13_extracted_8165 | Not-determined |
| S13_extracted_8173 | Low-quality |
| S13_extracted_8179 | Not-determined |
| S13_extracted_8199 | Not-determined |
| S13_extracted_8201 | Not-determined |
| S13_extracted_8202 | Not-determined |
| S13_extracted_8205 | Not-determined |
| S13_extracted_8210 | Not-determined |
| S13_extracted_8215 | Not-determined |
| S13_extracted_8232 | Not-determined |
| S13_extracted_8235 | Not-determined |
| S13_extracted_8239 | Not-determined |
| S13_extracted_8269 | Not-determined |
| S13_extracted_8275 | Not-determined |
| S13_extracted_8285 | Not-determined |
| S13_extracted_8294 | Not-determined |
| S13_extracted_8296 | Not-determined |
| S13_extracted_8303 | Not-determined |
| S13_extracted_8309 | Not-determined |
| S13_extracted_8312 | Not-determined |
| S13_extracted_8315 | Low-quality |
| S13_extracted_8317 | Not-determined |
| S13_extracted_8321 | Not-determined |
| S13_extracted_8330 | Not-determined |
| S13_extracted_8331 | Not-determined |
| S13_extracted_8332 | Not-determined |
| S13_extracted_8344 | Not-determined |
| S13_extracted_8346 | Low-quality |
| S13_extracted_8349 | Not-determined |
| S13_extracted_8350 | Not-determined |
| S13_extracted_8362 | Not-determined |
| S13_extracted_8378 | Not-determined |
| S13_extracted_8392 | Not-determined |
| S13_extracted_8417 | Not-determined |
| S13_extracted_8419 | Not-determined |
| S13_extracted_8427 | Not-determined |
| S13_extracted_8430 | Low-quality |
| S13_extracted_8435 | Not-determined |
| S13_extracted_8450 | Not-determined |
| S13_extracted_8493 | Not-determined |
| S13_extracted_8496 | Not-determined |
| S13_extracted_8501 | Not-determined |
| S13_extracted_8502 | Not-determined |
| S13_extracted_8504 | Not-determined |
| S13_extracted_8519 | Not-determined |
| S13_extracted_8526 | Not-determined |
| S13_extracted_8527 | Not-determined |
| S13_extracted_8532 | Not-determined |
| S13_extracted_8535 | Low-quality |
| S13_extracted_8538 | Not-determined |
| S13_extracted_8551 | Not-determined |
| S13_extracted_8568 | Not-determined |
| S13_extracted_8570 | Not-determined |
| S13_extracted_8575 | Not-determined |
| S13_extracted_8578 | Not-determined |
| S13_extracted_8580 | Low-quality |
| S13_extracted_8583 | Not-determined |
| S13_extracted_8585 | Not-determined |
| S13_extracted_8587 | Not-determined |
| S13_extracted_8589 | Low-quality |
| S13_extracted_8591 | Not-determined |
| S13_extracted_8592 | Not-determined |
| S13_extracted_8604 | Not-determined |
| S13_extracted_8623 | Not-determined |
| S13_extracted_8624 | Not-determined |
| S13_extracted_8636 | Not-determined |
| S13_extracted_8645 | Not-determined |
| S13_extracted_8646 | Not-determined |
| S13_extracted_8662 | Not-determined |
| S13_extracted_8667 | Not-determined |
| S13_extracted_8684 | Not-determined |
| S13_extracted_8685 | Not-determined |
| S13_extracted_8686 | Low-quality |
| S13_extracted_8697 | Not-determined |
| S13_extracted_8708 | Not-determined |
| S13_extracted_8712 | Not-determined |
| S13_extracted_8719 | Not-determined |
| S13_extracted_8725 | Not-determined |
| S13_extracted_8728 | Not-determined |
| S13_extracted_8730 | Not-determined |
| S13_extracted_8731 | Not-determined |
| S13_extracted_8734 | Not-determined |
| S13_extracted_8754 | Not-determined |
| S13_extracted_8760 | Not-determined |
| S13_extracted_8761 | Not-determined |
| S13_extracted_8763 | Not-determined |
| S13_extracted_8764 | Not-determined |
| S13_extracted_8782 | Not-determined |
| S13_extracted_8793 | Not-determined |
| S13_extracted_8806 | Not-determined |
| S13_extracted_8808 | Low-quality |
| S13_extracted_8830 | Not-determined |
| S13_extracted_8843 | Not-determined |
| S13_extracted_8848 | Low-quality |
| S13_extracted_8850 | Not-determined |
| S13_extracted_8854 | Not-determined |
| S13_extracted_8857 | Not-determined |
| S13_extracted_8858 | Not-determined |
| S13_extracted_8859 | Not-determined |
| S13_extracted_8871 | Not-determined |
| S13_extracted_8874 | Not-determined |
| S13_extracted_8875 | Not-determined |
| S13_extracted_8885 | Not-determined |
| S13_extracted_8899 | Not-determined |
| S13_extracted_8926 | Not-determined |
| S13_extracted_8931 | Not-determined |
| S13_extracted_8939 | Not-determined |
| S13_extracted_8943 | Not-determined |
| S13_extracted_8945 | Not-determined |
| S13_extracted_8950 | Not-determined |
| S13_extracted_8952 | Not-determined |
| S13_extracted_8955 | Not-determined |
| S13_extracted_8958 | Not-determined |
| S13_extracted_8959 | Not-determined |
| S13_extracted_8965 | Low-quality |
| S13_extracted_8969 | Not-determined |
| S13_extracted_8971 | Not-determined |
| S13_extracted_8996 | Not-determined |
| S13_extracted_9002 | Not-determined |
| S13_extracted_9005 | Not-determined |
| S13_extracted_9007 | Not-determined |
| S13_extracted_9014 | Not-determined |
| S13_extracted_9023 | Not-determined |
| S13_extracted_9024 | Not-determined |
| S13_extracted_9031 | Not-determined |
| S13_extracted_9042 | Low-quality |
| S13_extracted_9046 | Not-determined |
| S13_extracted_9050 | Not-determined |
| S13_extracted_9054 | Not-determined |
| S13_extracted_9082 | Not-determined |
| S13_extracted_9086 | Low-quality |
| S13_extracted_9094 | Not-determined |
| S13_extracted_9102 | Not-determined |
| S13_extracted_9106 | Low-quality |
| S13_extracted_9119 | Not-determined |
| S13_extracted_9121 | Not-determined |
| S13_extracted_9146 | Low-quality |
| S13_extracted_9154 | Low-quality |
| S13_extracted_9159 | Low-quality |
| S13_extracted_9170 | Not-determined |
| S13_extracted_9171 | Not-determined |
| S13_extracted_9173 | Not-determined |
| S13_extracted_9198 | Not-determined |
| S13_extracted_9210 | Low-quality |
| S13_extracted_9249 | Not-determined |
| S13_extracted_9253 | Not-determined |
| S13_extracted_9259 | Not-determined |
| S13_extracted_9260 | Not-determined |
| S13_extracted_9264 | Not-determined |
| S13_extracted_9281 | Not-determined |
| S13_extracted_9282 | Not-determined |
| S13_extracted_9285 | Not-determined |
| S13_extracted_9294 | Not-determined |
| S13_extracted_9296 | Not-determined |
| S13_extracted_9301 | Low-quality |
| S13_extracted_9318 | Not-determined |
| S13_extracted_9324 | Not-determined |
| S13_extracted_9329 | Not-determined |
| S13_extracted_9339 | Not-determined |
| S13_extracted_9349 | Not-determined |
| S13_extracted_9354 | Not-determined |
| S13_extracted_9367 | Not-determined |
| S13_extracted_9372 | Not-determined |
| S13_extracted_9377 | Not-determined |
| S13_extracted_9388 | Not-determined |
| S13_extracted_9391 | Low-quality |
| S13_extracted_9398 | Not-determined |
| S13_extracted_9407 | Low-quality |
| S13_extracted_9412 | Not-determined |
| S13_extracted_9419 | Not-determined |
| S13_extracted_9425 | Not-determined |
| S13_extracted_9429 | Not-determined |
| S13_extracted_9437 | Not-determined |
| S13_extracted_9440 | Not-determined |
| S13_extracted_9463 | Not-determined |
| S13_extracted_9470 | Not-determined |
| S13_extracted_9475 | Not-determined |
| S13_extracted_9477 | Not-determined |
| S13_extracted_9483 | Not-determined |
| S13_extracted_9486 | Not-determined |
| S13_extracted_9494 | Not-determined |
| S13_extracted_9499 | Not-determined |
| S13_extracted_9511 | Low-quality |
| S13_extracted_9520 | Not-determined |
| S13_extracted_9530 | Not-determined |
| S13_extracted_9537 | Not-determined |
| S13_extracted_9544 | Low-quality |
| S13_extracted_9545 | Not-determined |
| S13_extracted_9546 | Not-determined |
| S13_extracted_9547 | Not-determined |
| S13_extracted_9549 | Not-determined |
| S13_extracted_9562 | Not-determined |
| S13_extracted_9580 | Low-quality |
| S13_extracted_9588 | Not-determined |
| S13_extracted_9602 | Not-determined |
| S13_extracted_9608 | Not-determined |
| S13_extracted_9618 | Not-determined |
| S13_extracted_9622 | Not-determined |
| S13_extracted_9634 | Not-determined |
| S13_extracted_9638 | Low-quality |
| S13_extracted_9642 | Not-determined |
| S13_extracted_9648 | Not-determined |
| S13_extracted_9665 | Not-determined |
| S13_extracted_9672 | Not-determined |
| S13_extracted_9673 | Not-determined |
| S13_extracted_9689 | Not-determined |
| S13_extracted_9691 | Not-determined |
| S13_extracted_9695 | Not-determined |
| S13_extracted_9699 | Not-determined |
| S13_extracted_9706 | Not-determined |
| S13_extracted_9709 | Low-quality |
| S13_extracted_9738 | Not-determined |
| S13_extracted_9750 | Not-determined |
| S13_extracted_9753 | Not-determined |
| S13_extracted_9762 | Not-determined |
| S13_extracted_9767 | Not-determined |
| S13_extracted_9774 | Not-determined |
| S13_extracted_9788 | Not-determined |
| S13_extracted_9802 | Low-quality |
| S13_extracted_9806 | Low-quality |
| S13_extracted_9811 | Not-determined |
| S13_extracted_9819 | Not-determined |
| S13_extracted_9825 | Not-determined |
| S13_extracted_9842 | Not-determined |
| S13_extracted_9849 | Not-determined |
| S13_extracted_9855 | Not-determined |
| S13_extracted_9857 | Low-quality |
| S13_extracted_9864 | Low-quality |
| S13_extracted_9865 | Not-determined |
| S13_extracted_9875 | Not-determined |
| S13_extracted_9876 | Not-determined |
| S13_extracted_9884 | Not-determined |
| S13_extracted_9886 | Not-determined |
| S13_extracted_9890 | Not-determined |
| S13_extracted_9899 | Not-determined |
| S13_extracted_9908 | Low-quality |
| S13_extracted_9914 | Not-determined |
| S13_extracted_9924 | Not-determined |
| S13_extracted_9929 | Not-determined |
| S13_extracted_9954 | Not-determined |
| S13_extracted_9970 | Not-determined |

| | |
|---|---|
| S13_extracted_9982 | Not-determined |
| S13_extracted_9984 | Not-determined |
| S13_extracted_9991 | Not-determined |
| S13_extracted_9993 | Not-determined |
| S13_extracted_9997 | Not-determined |
| S13_extracted_10006 | Not-determined |
| S13_extracted_10016 | Not-determined |
| S13_extracted_10019 | Not-determined |
| S13_extracted_10039 | Not-determined |
| S13_extracted_10040 | Not-determined |
| S13_extracted_10055 | Not-determined |
| S13_extracted_10058 | Not-determined |
| S13_extracted_10059 | Not-determined |
| S13_extracted_10063 | Not-determined |
| S13_extracted_10072 | Not-determined |
| S13_extracted_10079 | Not-determined |
| S13_extracted_10082 | Not-determined |
| S13_extracted_10090 | Not-determined |
| S13_extracted_10094 | Not-determined |
| S13_extracted_10103 | Low-quality |
| S13_extracted_10116 | Low-quality |
| S13_extracted_10117 | Not-determined |
| S13_extracted_10119 | Not-determined |
| S13_extracted_10127 | Not-determined |
| S13_extracted_10146 | Not-determined |
| S13_extracted_10156 | Low-quality |
| S13_extracted_10157 | Not-determined |
| S13_extracted_10183 | Low-quality |
| S13_extracted_10184 | Not-determined |
| S13_extracted_10187 | Not-determined |
| S13_extracted_10188 | Not-determined |
| S13_extracted_10195 | Not-determined |
| S13_extracted_10201 | Not-determined |
| S13_extracted_10205 | Not-determined |
| S13_extracted_10209 | Not-determined |
| S13_extracted_10217 | Not-determined |
| S13_extracted_10226 | Not-determined |
| S13_extracted_10240 | Not-determined |
| S13_extracted_10244 | Not-determined |
| S13_extracted_10251 | Low-quality |
| S13_extracted_10252 | Not-determined |
| S13_extracted_10260 | Not-determined |
| S13_extracted_10280 | Low-quality |
| S13_extracted_10286 | Not-determined |
| S13_extracted_10296 | Not-determined |
| S13_extracted_10302 | Not-determined |
| S13_extracted_10310 | Not-determined |
| S13_extracted_10313 | Not-determined |
| S13_extracted_10322 | Not-determined |
| S13_extracted_10326 | Low-quality |
| S13_extracted_10330 | Low-quality |
| S13_extracted_10344 | Not-determined |
| S13_extracted_10367 | Not-determined |
| S13_extracted_10372 | Not-determined |
| S13_extracted_10382 | Not-determined |
| S13_extracted_10389 | Not-determined |
| S13_extracted_10415 | Not-determined |
| S13_extracted_10425 | Not-determined |
| S13_extracted_10436 | Not-determined |
| S13_extracted_10441 | Not-determined |
| S13_extracted_10443 | Not-determined |
| S13_extracted_10448 | Not-determined |
| S13_extracted_10449 | Not-determined |
| S13_extracted_10450 | Not-determined |
| S13_extracted_10460 | Low-quality |
| S13_extracted_10468 | Not-determined |
| S13_extracted_10473 | Not-determined |
| S13_extracted_10480 | Not-determined |
| S13_extracted_10482 | Not-determined |
| S13_extracted_10522 | Low-quality |
| S13_extracted_10533 | Not-determined |
| S13_extracted_10536 | Low-quality |
| S13_extracted_10539 | Not-determined |
| S13_extracted_10556 | Not-determined |
| S13_extracted_10557 | Not-determined |
| S13_extracted_10560 | Not-determined |
| S13_extracted_10564 | Not-determined |
| S13_extracted_10568 | Not-determined |
| S13_extracted_10572 | Not-determined |
| S13_extracted_10576 | Low-quality |
| S13_extracted_10578 | Not-determined |
| S13_extracted_10589 | Not-determined |
| S13_extracted_10602 | Low-quality |
| S13_extracted_10615 | Low-quality |
| S13_extracted_10619 | Low-quality |
| S13_extracted_10620 | Not-determined |
| S13_extracted_10622 | Not-determined |
| S13_extracted_10635 | Low-quality |
| S13_extracted_10637 | Not-determined |
| S13_extracted_10641 | Not-determined |
| S13_extracted_10646 | Not-determined |
| S13_extracted_10661 | Not-determined |
| S13_extracted_10662 | Not-determined |
| S13_extracted_10665 | Not-determined |
| S13_extracted_10671 | Not-determined |
| S13_extracted_10674 | Low-quality |
| S13_extracted_10684 | Low-quality |
| S13_extracted_10690 | Not-determined |
| S13_extracted_10698 | Not-determined |
| S13_extracted_10700 | Not-determined |
| S13_extracted_10715 | Not-determined |
| S13_extracted_10717 | Not-determined |
| S13_extracted_10741 | Not-determined |
| S13_extracted_10760 | Not-determined |
| S13_extracted_10766 | Low-quality |
| S13_extracted_10767 | Not-determined |
| S13_extracted_10770 | Not-determined |
| S13_extracted_10791 | Not-determined |
| S13_extracted_10797 | Not-determined |
| S13_extracted_10807 | Low-quality |
| S13_extracted_10808 | Low-quality |
| S13_extracted_10833 | Not-determined |
| S13_extracted_10843 | Not-determined |
| S13_extracted_10846 | Not-determined |
| S13_extracted_10853 | Not-determined |
| S13_extracted_10859 | Not-determined |
| S13_extracted_10864 | Low-quality |
| S13_extracted_10871 | Not-determined |
| S13_extracted_10878 | Not-determined |
| S13_extracted_10880 | Not-determined |
| S13_extracted_10891 | Not-determined |
| S13_extracted_10900 | Not-determined |
| S13_extracted_10919 | Not-determined |
| S13_extracted_10920 | Low-quality |
| S13_extracted_10931 | Not-determined |
| S13_extracted_10932 | Not-determined |
| S13_extracted_10938 | Not-determined |
| S13_extracted_10942 | Low-quality |
| S13_extracted_10966 | Not-determined |
| S13_extracted_10970 | Not-determined |
| S13_extracted_10997 | Low-quality |
| S13_extracted_11015 | Not-determined |
| S13_extracted_11028 | Not-determined |
| S13_extracted_11034 | Not-determined |
| S13_extracted_11047 | Not-determined |
| S13_extracted_11054 | Low-quality |
| S13_extracted_11060 | Not-determined |
| S13_extracted_11067 | Not-determined |
| S13_extracted_11070 | Not-determined |
| S13_extracted_11080 | Not-determined |
| S13_extracted_11087 | Not-determined |
| S13_extracted_11090 | Not-determined |
| S13_extracted_11114 | Not-determined |
| S13_extracted_11145 | Low-quality |
| S13_extracted_11159 | Not-determined |
| S13_extracted_11160 | Not-determined |
| S13_extracted_11165 | Not-determined |
| S13_extracted_11167 | Not-determined |
| S13_extracted_11175 | Low-quality |
| S13_extracted_11187 | Not-determined |
| S13_extracted_11189 | Not-determined |
| S13_extracted_11193 | Not-determined |
| S13_extracted_11206 | Not-determined |
| S13_extracted_11215 | Not-determined |
| S13_extracted_11219 | Not-determined |
| S13_extracted_11242 | Not-determined |
| S4_vs_VIRSorter_27316-cat_2 | High-quality |
| S13_extracted_11274 | Low-quality |
| S13_extracted_11275 | Not-determined |
| S13_extracted_11285 | Not-determined |
| S13_extracted_11290 | Low-quality |
| S13_extracted_11294 | Not-determined |
| S13_extracted_11305 | Not-determined |
| S13_extracted_11310 | Not-determined |
| S13_extracted_11314 | Not-determined |
| S13_extracted_11316 | Not-determined |
| S13_extracted_11324 | Not-determined |
| S13_extracted_11340 | Not-determined |
| S13_extracted_11352 | Not-determined |
| S13_extracted_11353 | Not-determined |
| S13_extracted_11359 | Not-determined |
| S13_extracted_11366 | Not-determined |
| S13_extracted_11392 | Not-determined |
| S13_extracted_11432 | Not-determined |
| S13_extracted_11438 | Low-quality |
| S13_extracted_11442 | Not-determined |
| S13_extracted_11444 | Not-determined |
| S13_extracted_11446 | Not-determined |
| S13_extracted_11453 | Low-quality |
| S13_extracted_11457 | Not-determined |
| S13_extracted_11487 | Not-determined |
| S13_extracted_11490 | Not-determined |
| S13_extracted_11523 | Low-quality |
| S13_extracted_11538 | Not-determined |
| S13_extracted_11551 | Not-determined |
| S13_extracted_11565 | Not-determined |
| S13_extracted_11568 | Not-determined |
| S13_extracted_11597 | Not-determined |
| S13_extracted_11606 | Not-determined |
| S13_extracted_11611 | Not-determined |
| S13_extracted_11615 | Not-determined |
| S13_extracted_11621 | Not-determined |
| S13_extracted_11625 | Not-determined |
| S13_extracted_11629 | Not-determined |
| S13_extracted_11630 | Low-quality |
| S13_extracted_11639 | Not-determined |
| S13_extracted_11641 | Not-determined |
| S13_extracted_11678 | Not-determined |
| S13_extracted_11684 | Not-determined |
| S13_extracted_11689 | Not-determined |
| S13_extracted_11701 | Not-determined |
| S13_extracted_11702 | Not-determined |
| S13_extracted_11723 | Not-determined |
| S13_extracted_11724 | Low-quality |
| S13_extracted_11727 | Not-determined |
| S13_extracted_11735 | Not-determined |
| S13_extracted_11736 | Not-determined |
| S13_extracted_11740 | Not-determined |
| S13_extracted_11747 | Not-determined |
| S13_extracted_11748 | Low-quality |
| S13_extracted_11755 | Not-determined |
| S13_extracted_11758 | Not-determined |
| S13_extracted_11763 | Not-determined |
| S13_extracted_11801 | Not-determined |
| S13_extracted_11817 | Not-determined |
| S13_extracted_11820 | Not-determined |
| S13_extracted_11826 | Not-determined |
| S13_extracted_11827 | Not-determined |
| S13_extracted_11850 | Not-determined |
| S13_extracted_11867 | Not-determined |
| S13_extracted_11872 | Low-quality |
| S13_extracted_11877 | Not-determined |
| S13_extracted_11878 | Not-determined |
| S13_extracted_11891 | Not-determined |
| S13_extracted_11898 | Not-determined |
| S13_extracted_11904 | Not-determined |
| S13_extracted_11911 | Low-quality |
| S13_extracted_11924 | Not-determined |
| S13_extracted_11925 | Not-determined |
| S13_extracted_11929 | Not-determined |
| S13_extracted_11933 | Not-determined |
| S13_extracted_11940 | Not-determined |
| S13_extracted_11943 | Low-quality |
| S13_extracted_11950 | Not-determined |
| S13_extracted_11951 | Not-determined |
| S13_extracted_11969 | Not-determined |
| S13_extracted_11971 | Not-determined |
| S13_extracted_11978 | Not-determined |
| S13_extracted_11983 | Not-determined |
| S13_extracted_11992 | Low-quality |
| S13_extracted_11997 | Not-determined |
| S13_extracted_11999 | Not-determined |
| S13_extracted_12005 | Low-quality |
| S13_extracted_12007 | Not-determined |
| S13_extracted_12012 | Low-quality |
| S13_extracted_12026 | Not-determined |
| S13_extracted_12028 | Not-determined |
| S13_extracted_12031 | Not-determined |
| S13_extracted_12033 | Not-determined |
| S13_extracted_12042 | Low-quality |
| S13_extracted_12050 | Not-determined |
| S13_extracted_12051 | Not-determined |
| S13_extracted_12061 | Not-determined |
| S13_extracted_12070 | Not-determined |
| S13_extracted_12073 | Low-quality |
| S13_extracted_12080 | Not-determined |
| S13_extracted_12088 | Not-determined |
| S13_extracted_12089 | Not-determined |
| S13_extracted_12095 | Not-determined |
| S13_extracted_12100 | Not-determined |
| S13_extracted_12111 | Low-quality |
| S13_extracted_12122 | Not-determined |
| S13_extracted_12131 | Not-determined |
| S13_extracted_12154 | Low-quality |
| S13_extracted_12159 | Not-determined |
| S13_extracted_12165 | Not-determined |
| S13_extracted_12177 | Not-determined |
| S13_extracted_12194 | Not-determined |
| S13_extracted_12197 | Not-determined |
| S13_extracted_12200 | Not-determined |
| S13_extracted_12201 | Not-determined |
| S13_extracted_12209 | Not-determined |
| S13_extracted_12217 | Not-determined |
| S13_extracted_12219 | Low-quality |
| S13_extracted_12220 | Not-determined |
| S13_extracted_12226 | Low-quality |
| S13_extracted_12236 | Low-quality |
| S13_extracted_12241 | Not-determined |
| S13_extracted_12243 | Not-determined |
| S13_extracted_12266 | Not-determined |
| S13_extracted_12297 | Not-determined |
| S13_extracted_12301 | Low-quality |
| S13_extracted_12308 | Not-determined |
| S13_extracted_12316 | Not-determined |
| S13_extracted_12317 | Not-determined |
| S13_extracted_12323 | Not-determined |
| S13_extracted_12330 | Low-quality |
| S13_extracted_12334 | Not-determined |
| S13_extracted_12338 | Not-determined |
| S13_extracted_12343 | Not-determined |
| S13_extracted_12351 | Not-determined |
| S13_extracted_12352 | Not-determined |
| S13_extracted_12355 | Not-determined |
| S13_extracted_12358 | Not-determined |
| S13_extracted_12363 | Low-quality |
| S13_extracted_12367 | Low-quality |
| S13_extracted_12399 | Not-determined |
| S13_extracted_12407 | Not-determined |
| S13_extracted_12408 | Not-determined |
| S13_extracted_12415 | Not-determined |
| S13_extracted_12432 | Not-determined |
| S13_extracted_12436 | Not-determined |
| S13_extracted_12447 | Not-determined |
| S13_extracted_12449 | Not-determined |
| S13_extracted_12459 | Not-determined |
| S13_extracted_12463 | Not-determined |
| S13_extracted_12474 | Not-determined |
| S13_extracted_12477 | Low-quality |
| S13_extracted_12492 | Not-determined |
| S13_extracted_12504 | Low-quality |
| S13_extracted_12513 | Not-determined |
| S13_extracted_12525 | Not-determined |
| S13_extracted_12531 | Low-quality |
| S13_extracted_12534 | Not-determined |
| S13_extracted_12552 | Not-determined |
| S13_extracted_12557 | Not-determined |
| S13_extracted_12563 | Not-determined |
| S13_extracted_12571 | Not-determined |
| S13_extracted_12572 | Not-determined |
| S13_extracted_12573 | Not-determined |
| S13_extracted_12578 | Not-determined |
| S13_extracted_12588 | Not-determined |
| S13_extracted_12589 | Not-determined |
| S13_extracted_12594 | Not-determined |
| S13_extracted_12595 | Not-determined |
| S13_extracted_12599 | Not-determined |
| S13_extracted_12602 | Low-quality |
| S13_extracted_12613 | Not-determined |
| S13_extracted_12618 | Not-determined |
| S13_extracted_12620 | Not-determined |
| S13_extracted_12623 | Not-determined |
| S13_extracted_12624 | Not-determined |
| S13_extracted_12633 | Not-determined |
| S13_extracted_12641 | Low-quality |
| S13_extracted_12644 | Not-determined |
| S13_extracted_12653 | Not-determined |
| S13_extracted_12656 | Not-determined |
| S13_extracted_12661 | Not-determined |
| S13_extracted_12675 | Not-determined |
| S13_extracted_12679 | Not-determined |
| S13_extracted_12683 | Not-determined |
| S13_extracted_12711 | Not-determined |
| S13_extracted_12719 | Not-determined |
| S13_extracted_12722 | Not-determined |
| S13_extracted_12732 | Low-quality |
| S13_extracted_12739 | Not-determined |
| S13_extracted_12758 | Not-determined |
| S13_extracted_12761 | Not-determined |
| S13_extracted_12775 | Not-determined |
| S13_extracted_12787 | Not-determined |
| S13_extracted_12788 | Low-quality |
| S13_extracted_12796 | Not-determined |
| S13_extracted_12803 | Not-determined |
| S13_extracted_12811 | Not-determined |
| S13_extracted_12813 | Low-quality |
| S13_extracted_12815 | Not-determined |
| S13_extracted_12824 | Not-determined |
| S13_extracted_12826 | Not-determined |
| S13_extracted_12829 | Not-determined |
| S13_extracted_12831 | Not-determined |
| S13_extracted_12832 | Not-determined |
| S13_extracted_12833 | Not-determined |
| S13_extracted_12838 | Not-determined |
| S13_extracted_12862 | Not-determined |
| S13_extracted_12872 | Not-determined |
| S13_extracted_12885 | Not-determined |
| S13_extracted_12886 | Low-quality |
| S13_extracted_12893 | Not-determined |
| S13_extracted_12920 | Low-quality |
| S13_extracted_12921 | Not-determined |
| S13_extracted_12934 | Not-determined |
| S13_extracted_12962 | Low-quality |
| S13_extracted_12969 | Not-determined |
| S13_extracted_12973 | Not-determined |
| S13_extracted_12980 | Low-quality |
| S13_extracted_13002 | Low-quality |
| S13_extracted_13004 | Not-determined |
| S13_extracted_13008 | Not-determined |
| S13_extracted_13010 | Not-determined |
| S13_extracted_13013 | Low-quality |
| S13_extracted_13031 | Not-determined |
| S13_extracted_13034 | Not-determined |
| S13_extracted_13059 | Not-determined |
| S13_extracted_13061 | Not-determined |
| S13_extracted_13063 | Not-determined |
| S13_extracted_13068 | Not-determined |
| S13_extracted_13070 | Low-quality |
| S13_extracted_13075 | Not-determined |
| S13_extracted_13076 | Not-determined |
| S13_extracted_13107 | Low-quality |
| S13_extracted_13110 | Not-determined |
| S13_extracted_13127 | Not-determined |
| S13_extracted_13128 | Not-determined |
| S13_extracted_13140 | Low-quality |
| S13_extracted_13142 | Not-determined |
| S13_extracted_13155 | Not-determined |
| S13_extracted_13161 | Not-determined |
| S13_extracted_13168 | Not-determined |
| S13_extracted_13179 | Low-quality |
| S13_extracted_13195 | Not-determined |
| S13_extracted_13209 | Not-determined |
| S13_extracted_13219 | Not-determined |
| S13_extracted_13220 | Not-determined |
| S13_extracted_13236 | Not-determined |
| S13_extracted_13245 | Not-determined |
| S13_extracted_13251 | Not-determined |
| S13_extracted_13256 | Low-quality |
| S13_extracted_13259 | Not-determined |
| S13_extracted_13267 | Not-determined |
| S13_extracted_13280 | Not-determined |
| S13_extracted_13283 | Not-determined |
| S13_extracted_13285 | Not-determined |
| S13_extracted_13290 | Not-determined |
| S13_extracted_13303 | Low-quality |
| S13_extracted_13317 | Not-determined |
| S13_extracted_13338 | Not-determined |
| S13_extracted_13354 | Not-determined |
| S13_extracted_13375 | Not-determined |
| S13_extracted_13380 | Not-determined |
| S13_extracted_13383 | Not-determined |
| S13_extracted_13388 | Not-determined |
| S13_extracted_13394 | Not-determined |
| S13_extracted_13402 | Not-determined |
| S13_extracted_13411 | Not-determined |
| S13_extracted_13423 | Not-determined |
| S13_extracted_13443 | Not-determined |
| S13_extracted_13456 | Not-determined |
| S13_extracted_13484 | Low-quality |
| S13_extracted_13489 | Low-quality |
| S13_extracted_13492 | Not-determined |
| S13_extracted_13504 | Not-determined |
| S13_extracted_13507 | Low-quality |
| S13_extracted_13531 | Not-determined |
| S13_extracted_13537 | Not-determined |
| S13_extracted_13539 | Not-determined |
| S13_extracted_13540 | Not-determined |
| S13_extracted_13548 | Not-determined |
| S13_extracted_13599 | Not-determined |
| S13_extracted_13600 | Not-determined |
| S13_extracted_13607 | Not-determined |
| S13_extracted_13611 | Not-determined |
| S13_extracted_13613 | Not-determined |
| S13_extracted_13618 | Low-quality |
| S13_extracted_13633 | Not-determined |
| S13_extracted_13669 | Not-determined |
| S13_extracted_13670 | Not-determined |
| S13_extracted_13674 | Not-determined |
| S13_extracted_13691 | Not-determined |
| S13_extracted_13714 | Not-determined |
| S13_extracted_13724 | Low-quality |
| S13_extracted_13729 | Not-determined |
| S13_extracted_13735 | Not-determined |
| S13_extracted_13758 | Not-determined |
| S13_extracted_13763 | Not-determined |
| S13_extracted_13764 | Low-quality |
| S13_extracted_13767 | Not-determined |
| S13_extracted_13788 | Not-determined |
| S13_extracted_13807 | Not-determined |
| S13_extracted_13829 | Not-determined |
| S13_extracted_13830 | Not-determined |
| S13_extracted_13851 | Not-determined |
| S13_extracted_13854 | Not-determined |
| S13_extracted_13860 | Not-determined |
| S13_extracted_13873 | Not-determined |
| S13_extracted_13876 | Not-determined |
| S13_extracted_13883 | Not-determined |
| S13_extracted_13902 | Not-determined |
| S13_extracted_13908 | Not-determined |
| S13_extracted_13913 | Not-determined |
| S13_extracted_13919 | Not-determined |
| S13_extracted_13920 | Not-determined |
| S13_extracted_13937 | Not-determined |
| S13_extracted_13944 | Not-determined |
| S13_extracted_13946 | Not-determined |
| S13_extracted_13949 | Not-determined |
| S13_extracted_13951 | Not-determined |
| S13_extracted_13956 | Not-determined |
| S13_extracted_14002 | Not-determined |
| S13_extracted_14008 | Low-quality |
| S13_extracted_14015 | Not-determined |
| S13_extracted_14022 | Not-determined |
| S13_extracted_14043 | Not-determined |
| S13_extracted_14047 | Not-determined |
| S13_extracted_14052 | Not-determined |
| S13_extracted_14062 | Not-determined |
| S13_extracted_14067 | Not-determined |
| S13_extracted_14074 | Not-determined |
| S13_extracted_14081 | Not-determined |
| S13_extracted_14085 | Not-determined |
| S13_extracted_14097 | Not-determined |
| S13_extracted_14130 | Not-determined |
| S13_extracted_14131 | Low-quality |
| S13_extracted_14141 | Not-determined |
| S13_extracted_14149 | Low-quality |
| S13_extracted_14173 | Not-determined |
| S13_extracted_14181 | Not-determined |
| S13_extracted_14187 | Not-determined |
| S13_extracted_14195 | Not-determined |
| S13_extracted_14198 | Not-determined |
| S13_extracted_14212 | Not-determined |
| S13_extracted_14232 | Low-quality |
| S13_extracted_14241 | Not-determined |
| S13_extracted_14246 | Not-determined |
| S13_extracted_14252 | Not-determined |
| S13_extracted_14269 | Not-determined |
| S13_extracted_14274 | Not-determined |
| S13_extracted_14278 | Not-determined |
| S13_extracted_14284 | Low-quality |
| S13_extracted_14294 | Not-determined |
| S13_extracted_14299 | Not-determined |
| S13_extracted_14307 | Not-determined |
| S13_extracted_14308 | Not-determined |
| S13_extracted_14323 | Not-determined |
| S13_extracted_14325 | Low-quality |
| S13_extracted_14328 | Not-determined |
| S13_extracted_14345 | Not-determined |
| S13_extracted_14361 | Not-determined |
| S13_extracted_14365 | Not-determined |
| S13_extracted_14367 | Low-quality |
| S13_extracted_14388 | Low-quality |
| S13_extracted_14397 | Not-determined |
| S13_extracted_14404 | Not-determined |
| S13_extracted_14458 | Low-quality |
| S13_extracted_14467 | Not-determined |
| S13_extracted_14492 | Not-determined |
| S13_extracted_14496 | Not-determined |
| S13_extracted_14524 | Not-determined |
| S13_extracted_14531 | Low-quality |
| S13_extracted_14545 | Low-quality |
| S13_extracted_14546 | Not-determined |
| S13_extracted_14552 | Not-determined |
| S13_extracted_14560 | Not-determined |
| S13_extracted_14568 | Not-determined |
| S13_extracted_14573 | Not-determined |
| S13_extracted_14581 | Not-determined |
| S13_extracted_14586 | Not-determined |
| S13_extracted_14591 | Not-determined |
| S13_extracted_14599 | Low-quality |
| S13_extracted_14617 | Not-determined |
| S13_extracted_14620 | Not-determined |
| S13_extracted_14623 | Not-determined |
| S13_extracted_14637 | Not-determined |
| S13_extracted_14647 | Not-determined |
| S13_extracted_14649 | Not-determined |
| S13_extracted_14654 | Not-determined |
| S13_extracted_14670 | Not-determined |
| S13_extracted_14684 | Not-determined |
| S13_extracted_14694 | Not-determined |
| S13_extracted_14696 | Low-quality |
| S13_extracted_14698 | Low-quality |
| S13_extracted_14699 | Not-determined |
| S13_extracted_14710 | Not-determined |
| S13_extracted_14711 | Not-determined |
| S13_extracted_14717 | Not-determined |
| S13_extracted_14733 | Not-determined |
| S13_extracted_14736 | Not-determined |
| S13_extracted_14737 | Not-determined |
| S13_extracted_14743 | Not-determined |
| S13_extracted_14749 | Not-determined |
| S13_extracted_14756 | Not-determined |
| S13_extracted_14765 | Not-determined |
| S13_extracted_14766 | Not-determined |
| S13_extracted_14768 | Not-determined |
| S13_extracted_14776 | Not-determined |
| S13_extracted_14783 | Not-determined |
| S13_extracted_14795 | Not-determined |
| S13_extracted_14801 | Not-determined |
| S13_extracted_14807 | Not-determined |
| S13_extracted_14811 | Not-determined |
| S13_extracted_14829 | Not-determined |
| S13_extracted_14833 | Not-determined |
| S13_extracted_14841 | Not-determined |
| S13_extracted_14847 | Not-determined |
| S13_extracted_14852 | Low-quality |
| S13_extracted_14870 | Not-determined |
| S13_extracted_14893 | Not-determined |
| S13_extracted_14901 | Low-quality |
| S13_extracted_14908 | Not-determined |
| S13_extracted_14910 | Not-determined |
| S13_extracted_14911 | Not-determined |
| S13_extracted_14929 | Not-determined |
| S13_extracted_14935 | Not-determined |
| S13_extracted_14944 | Not-determined |
| S13_extracted_14955 | Not-determined |
| S13_extracted_14966 | Not-determined |
| S13_extracted_14967 | Low-quality |
| S13_extracted_14993 | Not-determined |
| S13_extracted_14998 | Low-quality |
| S13_extracted_15001 | Not-determined |

| | |
|---|---|
| S13_extracted_15002 | Not-determined |
| S13_extracted_15003 | Not-determined |
| S13_extracted_15007 | Not-determined |
| S13_extracted_15043 | Low-quality |
| S13_extracted_15051 | Low-quality |
| S13_extracted_15054 | Not-determined |
| S13_extracted_15075 | Low-quality |
| S13_extracted_15101 | Not-determined |
| S13_extracted_15110 | Not-determined |
| S13_extracted_15112 | Not-determined |
| S13_extracted_15115 | Low-quality |
| S13_extracted_15128 | Not-determined |
| S13_extracted_15129 | Not-determined |
| S13_extracted_15132 | Not-determined |
| S13_extracted_15173 | Not-determined |
| S13_extracted_15184 | Not-determined |
| S13_extracted_15189 | Not-determined |
| S13_extracted_15194 | Not-determined |
| S13_extracted_15195 | Not-determined |
| S13_extracted_15200 | Not-determined |
| S13_extracted_15204 | Not-determined |
| S13_extracted_15206 | Not-determined |
| S13_extracted_15209 | Not-determined |
| S13_extracted_15212 | Low-quality |
| S13_extracted_15219 | Not-determined |
| S13_extracted_15230 | Not-determined |
| S13_extracted_15240 | Low-quality |
| S13_extracted_15245 | Not-determined |
| S13_extracted_15260 | Not-determined |
| S13_extracted_15266 | Not-determined |
| S13_extracted_15291 | Not-determined |
| S13_extracted_15296 | Low-quality |
| S13_extracted_15300 | Not-determined |
| S13_extracted_15301 | Not-determined |
| S13_extracted_15304 | Not-determined |
| S13_extracted_15306 | Not-determined |
| S13_extracted_15316 | Not-determined |
| S13_extracted_15318 | Not-determined |
| S13_extracted_15321 | Not-determined |
| S13_extracted_15322 | Not-determined |
| S13_extracted_15331 | Low-quality |
| S13_extracted_15335 | Not-determined |
| S13_extracted_15349 | Not-determined |
| S13_extracted_15350 | Not-determined |
| S13_extracted_15353 | Not-determined |
| S13_extracted_15369 | Not-determined |
| S13_extracted_15373 | Not-determined |
| S13_extracted_15384 | Not-determined |
| S13_extracted_15394 | Not-determined |
| S13_extracted_15398 | Not-determined |
| S13_extracted_15401 | Not-determined |
| S13_extracted_15408 | Not-determined |
| S13_extracted_15413 | Not-determined |
| S13_extracted_15414 | Not-determined |
| S13_extracted_15419 | Not-determined |
| S13_extracted_15425 | Not-determined |
| S13_extracted_15426 | Not-determined |
| S13_extracted_15463 | Not-determined |
| S13_extracted_15469 | Not-determined |
| S13_extracted_15470 | Not-determined |
| S13_extracted_15474 | Low-quality |
| S13_extracted_15488 | Not-determined |
| S13_extracted_15494 | Low-quality |
| S13_extracted_15500 | Not-determined |
| S13_extracted_15501 | Not-determined |
| S13_extracted_15507 | Not-determined |
| S13_extracted_15523 | Not-determined |
| S13_extracted_15526 | Not-determined |
| S13_extracted_15528 | Not-determined |
| S13_extracted_15532 | Not-determined |
| S13_extracted_15534 | Not-determined |
| S13_extracted_15536 | Low-quality |
| S13_extracted_15539 | Not-determined |
| S13_extracted_15543 | Not-determined |
| S13_extracted_15545 | Low-quality |
| S13_extracted_15558 | Low-quality |
| S13_extracted_15560 | Not-determined |
| S13_extracted_15562 | Not-determined |
| S13_extracted_15570 | Not-determined |
| S13_extracted_15574 | Not-determined |
| S13_extracted_15577 | Low-quality |
| S13_extracted_15585 | Not-determined |
| S13_extracted_15586 | Not-determined |
| S13_extracted_15591 | Not-determined |
| S13_extracted_15595 | Not-determined |
| S13_extracted_15618 | Not-determined |
| S13_extracted_15619 | Not-determined |
| S13_extracted_15623 | Not-determined |
| S13_extracted_15630 | Not-determined |
| S13_extracted_15640 | Not-determined |
| S13_extracted_15643 | Not-determined |
| S13_extracted_15656 | Not-determined |
| S13_extracted_15671 | Not-determined |
| S13_extracted_15674 | Not-determined |
| S13_extracted_15684 | Not-determined |
| S13_extracted_15692 | Not-determined |
| S13_extracted_15712 | Low-quality |
| S13_extracted_15725 | Not-determined |
| S13_extracted_15729 | Not-determined |
| S13_extracted_15741 | Not-determined |
| S13_extracted_15750 | Not-determined |
| S13_extracted_15757 | Not-determined |
| S13_extracted_15760 | Not-determined |
| S13_extracted_15777 | Not-determined |
| S13_extracted_15784 | Not-determined |
| S13_extracted_15791 | Low-quality |
| S13_extracted_15793 | Not-determined |
| S13_extracted_15806 | Not-determined |
| S13_extracted_15807 | Not-determined |
| S13_extracted_15812 | Not-determined |
| S13_extracted_15813 | Not-determined |
| S13_extracted_15814 | Not-determined |
| S13_extracted_15815 | Not-determined |
| S13_extracted_15817 | Not-determined |
| S13_extracted_15826 | Not-determined |
| S13_extracted_15829 | Not-determined |
| S13_extracted_15830 | Not-determined |
| S13_extracted_15838 | Not-determined |
| S13_extracted_15860 | Not-determined |
| S13_extracted_15879 | Not-determined |
| S13_extracted_15910 | Not-determined |
| S13_extracted_15916 | Not-determined |
| S13_extracted_15919 | Not-determined |
| S13_extracted_15934 | Not-determined |
| S13_extracted_15942 | Not-determined |
| S13_extracted_15954 | Not-determined |
| S13_extracted_15977 | Low-quality |
| S13_extracted_15986 | Not-determined |
| S13_extracted_15988 | Not-determined |
| S13_extracted_15989 | Low-quality |
| S13_extracted_15993 | Not-determined |
| S13_extracted_15994 | Not-determined |
| S13_extracted_16017 | Not-determined |
| S13_extracted_16018 | Low-quality |
| S13_extracted_16025 | Not-determined |
| S13_extracted_16030 | Not-determined |
| S13_extracted_16038 | Not-determined |
| S13_extracted_16043 | Not-determined |
| S13_extracted_16045 | Not-determined |
| S13_extracted_16046 | Not-determined |
| S13_extracted_16054 | Not-determined |
| S13_extracted_16055 | Not-determined |
| S13_extracted_16073 | Not-determined |
| S13_extracted_16079 | Not-determined |
| S13_extracted_16088 | Not-determined |
| S13_extracted_16121 | Low-quality |
| S13_extracted_16132 | Not-determined |
| S13_extracted_16138 | Not-determined |
| S13_extracted_16144 | Not-determined |
| S13_extracted_16151 | Not-determined |
| S13_extracted_16167 | Not-determined |
| S13_extracted_16196 | Not-determined |
| S13_extracted_16209 | Not-determined |
| S13_extracted_16222 | Not-determined |
| S13_extracted_16223 | Not-determined |
| S13_extracted_16230 | Not-determined |
| S13_extracted_16238 | Not-determined |
| S13_extracted_16240 | Low-quality |
| S13_extracted_16265 | Not-determined |
| S13_extracted_16272 | Low-quality |
| S13_extracted_16276 | Not-determined |
| S13_extracted_16281 | Not-determined |
| S13_extracted_16289 | Not-determined |
| S13_extracted_16292 | Not-determined |
| S13_extracted_16309 | Not-determined |
| S13_extracted_16318 | Not-determined |
| S13_extracted_16320 | Not-determined |
| S13_extracted_16327 | Not-determined |
| S13_extracted_16365 | Not-determined |
| S13_extracted_16369 | Not-determined |
| S13_extracted_16370 | Low-quality |
| S13_extracted_16373 | Not-determined |
| S13_extracted_16381 | Not-determined |
| S13_extracted_16382 | Not-determined |
| S13_extracted_16416 | Not-determined |
| S13_extracted_16418 | Not-determined |
| S13_extracted_16419 | Not-determined |
| S13_extracted_16435 | Low-quality |
| S13_extracted_16442 | Not-determined |
| S13_extracted_16450 | Not-determined |
| S13_extracted_16453 | Not-determined |
| S13_extracted_16468 | Not-determined |
| S13_extracted_16469 | Not-determined |
| S13_extracted_16474 | Low-quality |
| S13_extracted_16477 | Not-determined |
| S13_extracted_16483 | Not-determined |
| S13_extracted_16486 | Not-determined |
| S13_extracted_16500 | Not-determined |
| S13_extracted_16527 | Not-determined |
| S13_extracted_16529 | Not-determined |
| S13_extracted_16543 | Low-quality |
| S13_extracted_16552 | Not-determined |
| S13_extracted_16562 | Not-determined |
| S13_extracted_16565 | Not-determined |
| S13_extracted_16573 | Not-determined |
| S13_extracted_16578 | Not-determined |
| S13_extracted_16586 | Not-determined |
| S13_extracted_16589 | Not-determined |
| S13_extracted_16591 | Not-determined |
| S13_extracted_16596 | Not-determined |
| S13_extracted_16605 | Not-determined |
| S13_extracted_16606 | Not-determined |
| S13_extracted_16633 | Not-determined |
| S13_extracted_16635 | Not-determined |
| S13_extracted_16639 | Not-determined |
| S13_extracted_16640 | Not-determined |
| S13_extracted_16654 | Not-determined |
| S13_extracted_16664 | Not-determined |
| S13_extracted_16666 | Not-determined |
| S13_extracted_16687 | Not-determined |
| S13_extracted_16696 | Not-determined |
| S13_extracted_16699 | Not-determined |
| S13_extracted_16704 | Low-quality |
| S13_extracted_16719 | Not-determined |
| S13_extracted_16720 | Not-determined |
| S13_extracted_16734 | Low-quality |
| S13_extracted_16738 | Not-determined |
| S13_extracted_16746 | Not-determined |
| S13_extracted_16753 | Low-quality |
| S13_extracted_16755 | Low-quality |
| S13_extracted_16765 | Low-quality |
| S13_extracted_16776 | Not-determined |
| S13_extracted_16780 | Not-determined |
| S13_extracted_16802 | Not-determined |
| S13_extracted_16807 | Not-determined |
| S13_extracted_16809 | Not-determined |
| S13_extracted_16834 | Not-determined |
| S13_extracted_16840 | Not-determined |
| S13_extracted_16846 | Not-determined |
| S13_extracted_16847 | Not-determined |
| S13_extracted_16856 | Not-determined |
| S13_extracted_16883 | Not-determined |
| S13_extracted_16886 | Not-determined |
| S13_extracted_16895 | Not-determined |
| S13_extracted_16913 | Not-determined |
| S13_extracted_16919 | Not-determined |
| S13_extracted_16934 | Not-determined |
| S13_extracted_16935 | Not-determined |
| S13_extracted_16948 | Not-determined |
| S13_extracted_16953 | Not-determined |
| S13_extracted_16963 | Not-determined |
| S13_extracted_16966 | Not-determined |
| S13_extracted_16979 | Low-quality |
| S13_extracted_16997 | Not-determined |
| S13_extracted_16998 | Not-determined |
| S13_extracted_17000 | Not-determined |
| S13_extracted_17027 | Not-determined |
| S13_extracted_17037 | Not-determined |
| S13_extracted_17039 | Low-quality |
| S13_extracted_17047 | Not-determined |
| S13_extracted_17058 | Not-determined |
| S13_extracted_17075 | Not-determined |
| S13_extracted_17081 | Not-determined |
| S13_extracted_17085 | Not-determined |
| S13_extracted_17093 | Not-determined |
| S13_extracted_17095 | Not-determined |
| S13_extracted_17098 | Not-determined |
| S13_extracted_17099 | Not-determined |
| S13_extracted_17106 | Not-determined |
| S13_extracted_17114 | Low-quality |
| S13_extracted_17121 | Not-determined |
| S13_extracted_17136 | Not-determined |
| S13_extracted_17140 | Not-determined |
| S13_extracted_17147 | Not-determined |
| S13_extracted_17149 | Not-determined |
| S13_extracted_17154 | Low-quality |
| S13_extracted_17169 | Not-determined |
| S13_extracted_17171 | Not-determined |
| S13_extracted_17172 | Not-determined |
| S13_extracted_17185 | Not-determined |
| S13_extracted_17188 | Not-determined |
| S13_extracted_17191 | Not-determined |
| S13_extracted_17195 | Not-determined |
| S13_extracted_17221 | Not-determined |
| S13_extracted_17230 | Not-determined |
| S13_extracted_17242 | Not-determined |
| S13_extracted_17253 | Low-quality |
| S13_extracted_17254 | Not-determined |
| S13_extracted_17260 | Not-determined |
| S13_extracted_17261 | Low-quality |
| S13_extracted_17278 | Low-quality |
| S13_extracted_17294 | Not-determined |
| S13_extracted_17297 | Not-determined |
| S13_extracted_17322 | Not-determined |
| S13_extracted_17354 | Not-determined |
| S13_extracted_17355 | Not-determined |
| S13_extracted_17356 | Not-determined |
| S13_extracted_17369 | Not-determined |
| S13_extracted_17372 | Not-determined |
| S13_extracted_17379 | Not-determined |
| S13_extracted_17381 | Not-determined |
| S13_extracted_17397 | Not-determined |
| S13_extracted_17398 | Not-determined |
| S13_extracted_17403 | Not-determined |
| S13_extracted_17414 | Not-determined |
| S13_extracted_17415 | Not-determined |
| S13_extracted_17422 | Not-determined |
| S13_extracted_17426 | Not-determined |
| S13_extracted_17428 | Not-determined |
| S13_extracted_17436 | Not-determined |
| S13_extracted_17442 | Low-quality |
| S13_extracted_17450 | Not-determined |
| S13_extracted_17451 | Not-determined |
| S13_extracted_17453 | Not-determined |
| S13_extracted_17488 | Not-determined |
| S13_extracted_17494 | Not-determined |
| S13_extracted_17499 | Not-determined |
| S13_extracted_17518 | Not-determined |
| S13_extracted_17522 | Not-determined |
| S13_extracted_17526 | Low-quality |
| S13_extracted_17534 | Not-determined |
| S13_extracted_17544 | Not-determined |
| S13_extracted_17546 | Not-determined |
| S13_extracted_17555 | Not-determined |
| S13_extracted_17563 | Not-determined |
| S13_extracted_17564 | Not-determined |
| S13_extracted_17567 | Not-determined |
| S13_extracted_17571 | Not-determined |
| S13_extracted_17590 | Not-determined |
| S13_extracted_17591 | Not-determined |
| S13_extracted_17612 | Low-quality |
| S13_extracted_17626 | Not-determined |
| S13_extracted_17634 | Low-quality |
| S13_extracted_17648 | Not-determined |
| S13_extracted_17661 | Not-determined |
| S13_extracted_17663 | Not-determined |
| S13_extracted_17691 | Not-determined |
| S13_extracted_17699 | Not-determined |
| S13_extracted_17710 | Not-determined |
| S13_extracted_17715 | Not-determined |
| S13_extracted_17724 | Not-determined |
| S13_extracted_17727 | Low-quality |
| S13_extracted_17729 | Not-determined |
| S13_extracted_17730 | Not-determined |
| S13_extracted_17733 | Not-determined |
| S13_extracted_17740 | Not-determined |
| S13_extracted_17761 | Low-quality |
| S13_extracted_17788 | Not-determined |
| S13_extracted_17793 | Not-determined |
| S13_extracted_17806 | Not-determined |
| S13_extracted_17824 | Not-determined |
| S13_extracted_17833 | Not-determined |
| S13_extracted_17845 | Not-determined |
| S13_extracted_17848 | Not-determined |
| S13_extracted_17851 | Not-determined |
| S13_extracted_17863 | Not-determined |
| S13_extracted_17864 | Not-determined |
| S13_extracted_17872 | Not-determined |
| S13_extracted_17877 | Not-determined |
| S13_extracted_17878 | Not-determined |
| S13_extracted_17883 | Not-determined |
| S13_extracted_17898 | Not-determined |
| S13_extracted_17917 | Not-determined |
| S13_extracted_17922 | Low-quality |
| S13_extracted_17928 | Low-quality |
| S13_extracted_17934 | Not-determined |
| S13_extracted_17940 | Not-determined |
| S13_extracted_17944 | Not-determined |
| S13_extracted_17946 | Not-determined |
| S13_extracted_17956 | Low-quality |
| S13_extracted_17957 | Not-determined |
| S13_extracted_17963 | Not-determined |
| S13_extracted_17981 | Not-determined |
| S13_extracted_17986 | Not-determined |
| S13_extracted_17989 | Low-quality |
| S13_extracted_17995 | Not-determined |
| S13_extracted_17997 | Not-determined |
| S13_extracted_18013 | Not-determined |
| S13_extracted_18026 | Not-determined |
| S13_extracted_18034 | Not-determined |
| S13_extracted_18038 | Not-determined |
| S13_extracted_18040 | Not-determined |
| S13_extracted_18044 | Not-determined |
| S13_extracted_18051 | Not-determined |
| S13_extracted_18056 | Not-determined |
| S13_extracted_18059 | Not-determined |
| S13_extracted_18078 | Not-determined |
| S13_extracted_18097 | Not-determined |
| S13_extracted_18100 | Not-determined |
| S13_extracted_18107 | Not-determined |
| S13_extracted_18115 | Not-determined |
| S13_extracted_18136 | Not-determined |
| S13_extracted_18139 | Not-determined |
| S13_extracted_18148 | Not-determined |
| S13_extracted_18157 | Not-determined |
| S13_extracted_18160 | Not-determined |
| S13_extracted_18162 | Not-determined |
| S13_extracted_18177 | Not-determined |
| S13_extracted_18195 | Not-determined |
| S13_extracted_18199 | Not-determined |
| S13_extracted_18201 | Not-determined |
| S13_extracted_18209 | Not-determined |
| S13_extracted_18229 | Not-determined |
| S13_extracted_18230 | Not-determined |
| S13_extracted_18247 | Not-determined |
| S13_extracted_18281 | Not-determined |
| S13_extracted_18282 | Not-determined |
| S13_extracted_18286 | Not-determined |
| S13_extracted_18301 | Not-determined |
| S13_extracted_18316 | Not-determined |
| S13_extracted_18319 | Not-determined |
| S13_extracted_18325 | Not-determined |
| S13_extracted_18330 | Not-determined |
| S13_extracted_18335 | Not-determined |
| S13_extracted_18342 | Not-determined |
| S13_extracted_18356 | Not-determined |
| S13_extracted_18363 | Not-determined |
| S13_extracted_18377 | Not-determined |
| S13_extracted_18382 | Not-determined |
| S13_extracted_18384 | Not-determined |
| S13_extracted_18394 | Not-determined |
| S13_extracted_18399 | Not-determined |
| S13_extracted_18424 | Not-determined |
| S13_extracted_18435 | Not-determined |
| S13_extracted_18455 | Not-determined |
| S13_extracted_18461 | Not-determined |
| S13_extracted_18473 | Not-determined |
| S13_extracted_18474 | Not-determined |
| S13_extracted_18480 | Not-determined |
| S13_extracted_18487 | Not-determined |
| S13_extracted_18489 | Not-determined |
| S13_extracted_18493 | Not-determined |
| S13_extracted_18500 | Low-quality |
| S13_extracted_18509 | Not-determined |
| S13_extracted_18510 | Not-determined |
| S13_extracted_18512 | Not-determined |
| S13_extracted_18518 | Not-determined |
| S13_extracted_18546 | Not-determined |
| S13_extracted_18559 | Not-determined |
| S13_extracted_18564 | Not-determined |
| S13_extracted_18570 | Not-determined |
| S13_extracted_18584 | Not-determined |
| S13_extracted_18602 | Not-determined |
| S13_extracted_18603 | Not-determined |
| S13_extracted_18606 | Not-determined |
| S13_extracted_18607 | Low-quality |
| S13_extracted_18612 | Not-determined |
| S13_extracted_18620 | Not-determined |
| S13_extracted_18624 | Low-quality |
| S13_extracted_18629 | Not-determined |
| S13_extracted_18646 | Not-determined |
| S13_extracted_18663 | Not-determined |
| S13_extracted_18677 | Not-determined |
| S13_extracted_18686 | Not-determined |
| S13_extracted_18709 | Low-quality |
| S13_extracted_18724 | Not-determined |
| S13_extracted_18742 | Not-determined |
| S13_extracted_18772 | Not-determined |
| S13_extracted_18776 | Low-quality |
| S13_extracted_18792 | Not-determined |
| S13_extracted_18797 | Not-determined |
| S13_extracted_18803 | Not-determined |
| S13_extracted_18804 | Not-determined |
| S13_extracted_18811 | Not-determined |
| S13_extracted_18832 | Not-determined |
| S13_extracted_18838 | Not-determined |
| S13_extracted_18846 | Not-determined |
| S13_extracted_18863 | Not-determined |
| S13_extracted_18870 | Not-determined |
| S13_extracted_18874 | Not-determined |
| S13_extracted_18884 | Not-determined |
| S13_extracted_18885 | Not-determined |
| S13_extracted_18900 | Not-determined |
| S13_extracted_18908 | Not-determined |
| S13_extracted_18910 | Not-determined |
| S13_extracted_18939 | Not-determined |
| S13_extracted_18963 | Not-determined |
| S13_extracted_18981 | Low-quality |
| S13_extracted_18989 | Not-determined |
| S13_extracted_18990 | Not-determined |
| S13_extracted_18993 | Not-determined |
| S13_extracted_19010 | Not-determined |
| S13_extracted_19017 | Not-determined |
| S13_extracted_19020 | Not-determined |
| S13_extracted_19021 | Not-determined |
| S13_extracted_19028 | Not-determined |
| S13_extracted_19031 | Not-determined |
| S13_extracted_19040 | Not-determined |
| S13_extracted_19041 | Low-quality |
| S13_extracted_19051 | Not-determined |
| S13_extracted_19053 | Not-determined |
| S13_extracted_19054 | Not-determined |
| S13_extracted_19064 | Not-determined |
| S13_extracted_19066 | Not-determined |
| S13_extracted_19070 | Not-determined |
| S13_extracted_19084 | Not-determined |
| S13_extracted_19091 | Not-determined |
| S13_extracted_19102 | Not-determined |
| S13_extracted_19105 | Not-determined |
| S13_extracted_19106 | Not-determined |
| S13_extracted_19114 | Not-determined |
| S13_extracted_19118 | Not-determined |
| S13_extracted_19140 | Not-determined |
| S13_extracted_19142 | Not-determined |
| S13_extracted_19143 | Not-determined |
| S13_extracted_19153 | Not-determined |
| S13_extracted_19158 | Not-determined |
| S13_extracted_19180 | Not-determined |
| S13_extracted_19187 | Not-determined |
| S13_extracted_19189 | Not-determined |
| S13_extracted_19194 | Not-determined |
| S13_extracted_19206 | Not-determined |
| S13_extracted_19207 | Not-determined |
| S13_extracted_19215 | Low-quality |
| S13_extracted_19220 | Not-determined |
| S13_extracted_19228 | Not-determined |
| S13_extracted_19236 | Not-determined |
| S13_extracted_19237 | Not-determined |
| S13_extracted_19243 | Not-determined |
| S13_extracted_19261 | Not-determined |
| S13_extracted_19263 | Low-quality |
| S13_extracted_19268 | Not-determined |
| S13_extracted_19273 | Not-determined |
| S13_extracted_19278 | Low-quality |
| S13_extracted_19279 | Not-determined |
| S13_extracted_19284 | Not-determined |
| S13_extracted_19292 | Not-determined |
| S13_extracted_19300 | Not-determined |
| S13_extracted_19303 | Not-determined |
| S13_extracted_19304 | Not-determined |
| S13_extracted_19337 | Low-quality |
| S13_extracted_19340 | Not-determined |
| S13_extracted_19343 | Not-determined |
| S13_extracted_19352 | Not-determined |
| S13_extracted_19355 | Low-quality |
| S13_extracted_19363 | Not-determined |
| S13_extracted_19367 | Not-determined |
| S13_extracted_19380 | Not-determined |
| S13_extracted_19398 | Not-determined |
| S13_extracted_19408 | Not-determined |
| S13_extracted_19409 | Not-determined |
| S13_extracted_19410 | Not-determined |
| S13_extracted_19411 | Not-determined |
| S13_extracted_19412 | Not-determined |
| S13_extracted_19413 | Not-determined |
| S13_extracted_19414 | Not-determined |
| S13_extracted_19419 | Not-determined |
| S13_extracted_19424 | Not-determined |
| S13_extracted_19433 | Not-determined |
| S13_extracted_19444 | Not-determined |
| S13_extracted_19459 | Low-quality |
| S13_extracted_19462 | Not-determined |
| S13_extracted_19481 | Low-quality |
| S13_extracted_19523 | Not-determined |
| S13_extracted_19524 | Not-determined |
| S13_extracted_19553 | Not-determined |
| S13_extracted_19557 | Not-determined |
| S13_extracted_19565 | Low-quality |
| S13_extracted_19570 | Not-determined |
| S13_extracted_19571 | Not-determined |
| S13_extracted_19575 | Not-determined |
| S13_extracted_19579 | Not-determined |
| S13_extracted_19584 | Not-determined |
| S13_extracted_19586 | Not-determined |
| S13_extracted_19588 | Not-determined |
| S13_extracted_19609 | Not-determined |
| S13_extracted_19629 | Not-determined |
| S13_extracted_19630 | Not-determined |
| S13_extracted_19635 | Not-determined |
| S13_extracted_19646 | Low-quality |
| S13_extracted_19663 | Not-determined |
| S13_extracted_19668 | Not-determined |
| S13_extracted_19702 | Not-determined |
| S13_extracted_19706 | Not-determined |
| S13_extracted_19711 | Not-determined |
| S13_extracted_19712 | Not-determined |
| S13_extracted_19717 | Not-determined |
| S13_extracted_19723 | Not-determined |
| S13_extracted_19738 | Not-determined |
| S13_extracted_19741 | Not-determined |
| S13_extracted_19749 | Not-determined |
| S13_extracted_19753 | Not-determined |
| S13_extracted_19756 | Not-determined |
| S13_extracted_19758 | Not-determined |
| S13_extracted_19761 | Not-determined |
| S13_extracted_19765 | Not-determined |
| S13_extracted_19770 | Not-determined |
| S13_extracted_19773 | Not-determined |
| S13_extracted_19774 | Not-determined |
| S13_extracted_19785 | Not-determined |
| S13_extracted_19791 | Not-determined |
| S13_extracted_19796 | Low-quality |
| S13_extracted_19810 | Not-determined |
| S13_extracted_19823 | Low-quality |
| S13_extracted_19825 | Not-determined |
| S13_extracted_19836 | Not-determined |
| S13_extracted_19838 | Not-determined |
| S13_extracted_19840 | Not-determined |
| S13_extracted_19843 | Not-determined |

| | |
|---|---|
| S13_extracted_19852 | Not-determined |
| S13_extracted_19854 | Not-determined |
| S13_extracted_19857 | Not-determined |
| S13_extracted_19860 | Not-determined |
| S13_extracted_19885 | Not-determined |
| S13_extracted_19900 | Not-determined |
| S13_extracted_19916 | Not-determined |
| S13_extracted_19926 | Not-determined |
| S13_extracted_19937 | Not-determined |
| S13_extracted_19945 | Not-determined |
| S13_extracted_19955 | Low-quality |
| S13_extracted_19956 | Not-determined |
| S13_extracted_19959 | Not-determined |
| S13_extracted_19969 | Not-determined |
| S13_extracted_19985 | Not-determined |
| S13_extracted_19989 | Not-determined |
| S13_extracted_20003 | Not-determined |
| S13_extracted_20011 | Not-determined |
| S13_extracted_20019 | Not-determined |
| S13_extracted_20027 | Not-determined |
| S13_extracted_20029 | Not-determined |
| S13_extracted_20032 | Low-quality |
| S13_extracted_20046 | Not-determined |
| S13_extracted_20047 | Not-determined |
| S13_extracted_20056 | Not-determined |
| S13_extracted_20064 | Not-determined |
| S13_extracted_20081 | Not-determined |
| S13_extracted_20083 | Not-determined |
| S13_extracted_20088 | Low-quality |
| S13_extracted_20090 | Not-determined |
| S13_extracted_20093 | Not-determined |
| S13_extracted_20112 | Not-determined |
| S13_extracted_20116 | Not-determined |
| S13_extracted_20138 | Not-determined |
| S13_extracted_20143 | Not-determined |
| S13_extracted_20150 | Not-determined |
| S13_extracted_20153 | Not-determined |
| S13_extracted_20181 | Not-determined |
| S13_extracted_20191 | Not-determined |
| S13_extracted_20215 | Not-determined |
| S13_extracted_20227 | Not-determined |
| S13_extracted_20229 | Not-determined |
| S13_extracted_20237 | Not-determined |
| S13_extracted_20238 | Not-determined |
| S13_extracted_20243 | Not-determined |
| S13_extracted_20247 | Low-quality |
| S13_extracted_20248 | Not-determined |
| S13_extracted_20290 | Not-determined |
| S13_extracted_20297 | Not-determined |
| S13_extracted_20308 | Low-quality |
| S13_extracted_20323 | Not-determined |
| S13_extracted_20324 | Not-determined |
| S13_extracted_20326 | Not-determined |
| S13_extracted_20332 | Not-determined |
| S13_extracted_20333 | Not-determined |
| S13_extracted_20335 | Not-determined |
| S13_extracted_20344 | Not-determined |
| S13_extracted_20366 | Not-determined |
| S13_extracted_20368 | Not-determined |
| S13_extracted_20377 | Not-determined |
| S13_extracted_20386 | Not-determined |
| S13_extracted_20396 | Not-determined |
| S13_extracted_20402 | Not-determined |
| S13_extracted_20403 | Not-determined |
| S13_extracted_20436 | Low-quality |
| S13_extracted_20447 | Not-determined |
| S13_extracted_20450 | Not-determined |
| S13_extracted_20476 | Low-quality |
| S13_extracted_20481 | Not-determined |
| S13_extracted_20485 | Not-determined |
| S13_extracted_20496 | Not-determined |
| S13_extracted_20510 | Not-determined |
| S13_extracted_20511 | Not-determined |
| S13_extracted_20514 | Not-determined |
| S13_extracted_20522 | Not-determined |
| S13_extracted_20527 | Not-determined |
| S13_extracted_20529 | Not-determined |
| S13_extracted_20573 | Not-determined |
| S13_extracted_20580 | Not-determined |
| S13_extracted_20590 | Not-determined |
| S13_extracted_20617 | Not-determined |
| S13_extracted_20630 | Not-determined |
| S13_extracted_20633 | Not-determined |
| S13_extracted_20647 | Not-determined |
| S13_extracted_20652 | Low-quality |
| S13_extracted_20669 | Not-determined |
| S13_extracted_20674 | Not-determined |
| S13_extracted_20679 | Not-determined |
| S13_extracted_20683 | Not-determined |
| S13_extracted_20685 | Low-quality |
| S13_extracted_20712 | Not-determined |
| S13_extracted_20714 | Not-determined |
| S13_extracted_20724 | Not-determined |
| S13_extracted_20741 | Not-determined |
| S13_extracted_20744 | Low-quality |
| S13_extracted_20752 | Not-determined |
| S13_extracted_20778 | Not-determined |
| S13_extracted_20795 | Not-determined |
| S13_extracted_20805 | Not-determined |
| S13_extracted_20806 | Not-determined |
| S13_extracted_20821 | Not-determined |
| S13_extracted_20833 | Not-determined |
| S13_extracted_20835 | Not-determined |
| S13_extracted_20840 | Not-determined |
| S13_extracted_20843 | Not-determined |
| S13_extracted_20846 | Low-quality |
| S13_extracted_20852 | Not-determined |
| S13_extracted_20866 | Not-determined |
| S13_extracted_20867 | Not-determined |
| S13_extracted_20882 | Not-determined |
| S13_extracted_20889 | Not-determined |
| S13_extracted_20897 | Low-quality |
| S13_extracted_20906 | Not-determined |
| S13_extracted_20911 | Not-determined |
| S13_extracted_20929 | Not-determined |
| S13_extracted_20944 | Not-determined |
| S13_extracted_20951 | Not-determined |
| S13_extracted_20954 | Not-determined |
| S13_extracted_20959 | Not-determined |
| S13_extracted_20964 | Low-quality |
| S13_extracted_20984 | Not-determined |
| S13_extracted_21009 | Not-determined |
| S13_extracted_21012 | Not-determined |
| S13_extracted_21014 | Low-quality |
| S13_extracted_21026 | Low-quality |
| S13_extracted_21032 | Not-determined |
| S13_extracted_21033 | Not-determined |
| S13_extracted_21044 | Not-determined |
| S13_extracted_21061 | Low-quality |
| S13_extracted_21068 | Not-determined |
| S13_extracted_21071 | Not-determined |
| S13_extracted_21074 | Not-determined |
| S13_extracted_21083 | Not-determined |
| S13_extracted_21091 | Not-determined |
| S13_extracted_21092 | Not-determined |
| S13_extracted_21098 | Not-determined |
| S13_extracted_21109 | Not-determined |
| S13_extracted_21110 | Not-determined |
| S13_extracted_21116 | Not-determined |
| S13_extracted_21123 | Not-determined |
| S13_extracted_21126 | Not-determined |
| S13_extracted_21128 | Not-determined |
| S13_extracted_21135 | Not-determined |
| S13_extracted_21144 | Not-determined |
| S13_extracted_21153 | Not-determined |
| S13_extracted_21163 | Not-determined |
| S13_extracted_21165 | Not-determined |
| S13_extracted_21174 | Not-determined |
| S13_extracted_21190 | Not-determined |
| S13_extracted_21198 | Not-determined |
| S13_extracted_21201 | Not-determined |
| S13_extracted_21203 | Low-quality |
| S13_extracted_21205 | Not-determined |
| S13_extracted_21210 | Not-determined |
| S13_extracted_21211 | Not-determined |
| S13_extracted_21212 | Low-quality |
| S13_extracted_21217 | Not-determined |
| S13_extracted_21237 | Not-determined |
| S13_extracted_21240 | Not-determined |
| S13_extracted_21251 | Not-determined |
| S13_extracted_21254 | Not-determined |
| S13_extracted_21272 | Not-determined |
| S13_extracted_21277 | Not-determined |
| S13_extracted_21279 | Not-determined |
| S13_extracted_21287 | Low-quality |
| S13_extracted_21297 | Not-determined |
| S13_extracted_21301 | Not-determined |
| S13_extracted_21310 | Not-determined |
| S13_extracted_21318 | Not-determined |
| S13_extracted_21323 | Not-determined |
| S13_extracted_21335 | Not-determined |
| S13_extracted_21343 | Not-determined |
| S13_extracted_21348 | Not-determined |
| S13_extracted_21364 | Not-determined |
| S13_extracted_21365 | Not-determined |
| S13_extracted_21366 | Not-determined |
| S13_extracted_21385 | Not-determined |
| S13_extracted_21414 | Not-determined |
| S13_extracted_21426 | Not-determined |
| S13_extracted_21442 | Not-determined |
| S13_extracted_21447 | Not-determined |
| S13_extracted_21453 | Not-determined |
| S13_extracted_21457 | Not-determined |
| S13_extracted_21459 | Not-determined |
| S13_extracted_21461 | Not-determined |
| S13_extracted_21462 | Low-quality |
| S13_extracted_21467 | Not-determined |
| S13_extracted_21471 | Not-determined |
| S13_extracted_21481 | Not-determined |
| S13_extracted_21485 | Not-determined |
| S13_extracted_21494 | Low-quality |
| S13_extracted_21517 | Not-determined |
| S13_extracted_21533 | Not-determined |
| S13_extracted_21534 | Not-determined |
| S13_extracted_21539 | Not-determined |
| S13_extracted_21541 | Not-determined |
| S13_extracted_21555 | Not-determined |
| S13_extracted_21557 | Not-determined |
| S13_extracted_21558 | Not-determined |
| S13_extracted_21559 | Not-determined |
| S13_extracted_21575 | Not-determined |
| S13_extracted_21577 | Not-determined |
| S13_extracted_21591 | Low-quality |
| S13_extracted_21592 | Low-quality |
| S13_extracted_21598 | Not-determined |
| S13_extracted_21600 | Not-determined |
| S13_extracted_21605 | Not-determined |
| S13_extracted_21616 | Low-quality |
| S13_extracted_21633 | Not-determined |
| S13_extracted_21664 | Low-quality |
| S13_extracted_21675 | Not-determined |
| S13_extracted_21692 | Not-determined |
| S13_extracted_21693 | Not-determined |
| S13_extracted_21694 | Not-determined |
| S13_extracted_21695 | Low-quality |
| S13_extracted_21710 | Low-quality |
| S13_extracted_21711 | Not-determined |
| S13_extracted_21715 | Not-determined |
| S13_extracted_21740 | Not-determined |
| S13_extracted_21750 | Not-determined |
| S13_extracted_21754 | Not-determined |
| S13_extracted_21758 | Low-quality |
| S13_extracted_21760 | Not-determined |
| S13_extracted_21770 | Not-determined |
| S13_extracted_21772 | Not-determined |
| S13_extracted_21775 | Not-determined |
| S13_extracted_21777 | Not-determined |
| S13_extracted_21779 | Not-determined |
| S13_extracted_21783 | Not-determined |
| S13_extracted_21798 | Not-determined |
| S13_extracted_21809 | Not-determined |
| S13_extracted_21816 | Not-determined |
| S13_extracted_21824 | Not-determined |
| S13_extracted_21833 | Not-determined |
| S13_extracted_21835 | Not-determined |
| S13_extracted_21836 | Not-determined |
| S13_extracted_21854 | Not-determined |
| S13_extracted_21856 | Not-determined |
| S13_extracted_21864 | Not-determined |
| S13_extracted_21874 | Not-determined |
| S13_extracted_21878 | Not-determined |
| S13_extracted_21884 | Not-determined |
| S13_extracted_21894 | Not-determined |
| S13_extracted_21898 | Not-determined |
| S13_extracted_21901 | Not-determined |
| S13_extracted_21919 | Not-determined |
| S13_extracted_21925 | Not-determined |
| S13_extracted_21938 | Not-determined |
| S13_extracted_21940 | Low-quality |
| S13_extracted_21949 | Not-determined |
| S13_extracted_21955 | Not-determined |
| S13_extracted_21958 | Low-quality |
| S13_extracted_21974 | Not-determined |
| S13_extracted_21981 | Not-determined |
| S13_extracted_21984 | Not-determined |
| S13_extracted_21991 | Not-determined |
| S13_extracted_21996 | Not-determined |
| S13_extracted_21997 | Not-determined |
| S13_extracted_22004 | Not-determined |
| S13_extracted_22013 | Not-determined |
| S13_extracted_22015 | Not-determined |
| S13_extracted_22028 | Not-determined |
| S13_extracted_22043 | Not-determined |
| S13_extracted_22045 | Not-determined |
| S13_extracted_22053 | Not-determined |
| S13_extracted_22060 | Not-determined |
| S13_extracted_22062 | Not-determined |
| S13_extracted_22070 | Not-determined |
| S13_extracted_22078 | Not-determined |
| S13_extracted_22083 | Not-determined |
| S13_extracted_22105 | Not-determined |
| S13_extracted_22115 | Low-quality |
| S13_extracted_22134 | Low-quality |
| S13_extracted_22151 | Not-determined |
| S13_extracted_22162 | Not-determined |
| S4_vs_VIRSorter_31975-circular-cat | Medium-quality |
| S13_extracted_22181 | Not-determined |
| S13_extracted_22186 | Not-determined |
| S13_extracted_22197 | Not-determined |
| S13_extracted_22200 | Not-determined |
| S13_extracted_22203 | Not-determined |
| S13_extracted_22204 | Not-determined |
| S13_extracted_22221 | Not-determined |
| S13_extracted_22227 | Not-determined |
| S13_extracted_22234 | Not-determined |
| S13_extracted_22236 | Not-determined |
| S13_extracted_22248 | Not-determined |
| S13_extracted_22255 | Not-determined |
| S13_extracted_22260 | Not-determined |
| S13_extracted_22265 | Not-determined |
| S13_extracted_22275 | Not-determined |
| S13_extracted_22289 | Not-determined |
| S13_extracted_22291 | Not-determined |
| S13_extracted_22294 | Not-determined |
| S13_extracted_22305 | Not-determined |
| S13_extracted_22312 | Not-determined |
| S13_extracted_22317 | Not-determined |
| S13_extracted_22328 | Not-determined |
| S13_extracted_22332 | Not-determined |
| S13_extracted_22338 | Low-quality |
| S13_extracted_22352 | Low-quality |
| S13_extracted_22353 | Low-quality |
| S13_extracted_22363 | Not-determined |
| S13_extracted_22368 | Not-determined |
| S13_extracted_22384 | Not-determined |
| S13_extracted_22402 | Not-determined |
| S13_extracted_22406 | Low-quality |
| S13_extracted_22409 | Not-determined |
| S13_extracted_22424 | Not-determined |
| S13_extracted_22429 | Low-quality |
| S13_extracted_22438 | Not-determined |
| S13_extracted_22468 | Not-determined |
| S13_extracted_22479 | Not-determined |
| S13_extracted_22480 | Not-determined |
| S13_extracted_22517 | Not-determined |
| S13_extracted_22542 | Not-determined |
| S13_extracted_22549 | Not-determined |
| S13_extracted_22554 | Not-determined |
| S13_extracted_22555 | Not-determined |
| S13_extracted_22561 | Not-determined |
| S13_extracted_22564 | Not-determined |
| S13_extracted_22580 | Not-determined |
| S13_extracted_22585 | Not-determined |
| S13_extracted_22588 | Not-determined |
| S13_extracted_22596 | Not-determined |
| S13_extracted_22604 | Not-determined |
| S13_extracted_22605 | Not-determined |
| S13_extracted_22626 | Not-determined |
| S13_extracted_22631 | Not-determined |
| S13_extracted_22636 | Not-determined |
| S13_extracted_22656 | Not-determined |
| S13_extracted_22658 | Not-determined |
| S13_extracted_22662 | Low-quality |
| S13_extracted_22663 | Not-determined |
| S13_extracted_22664 | Not-determined |
| S13_extracted_22665 | Not-determined |
| S13_extracted_22668 | Low-quality |
| S13_extracted_22676 | Not-determined |
| S13_extracted_22686 | Not-determined |
| S13_extracted_22691 | Low-quality |
| S13_extracted_22694 | Not-determined |
| S13_extracted_22696 | Low-quality |
| S13_extracted_22701 | Not-determined |
| S13_extracted_22704 | Not-determined |
| S13_extracted_22706 | Not-determined |
| S13_extracted_22711 | Low-quality |
| S13_extracted_22718 | Not-determined |
| S13_extracted_22723 | Not-determined |
| S13_extracted_22726 | Not-determined |
| S13_extracted_22728 | Not-determined |
| S13_extracted_22744 | Not-determined |
| S13_extracted_22745 | Not-determined |
| S13_extracted_22769 | Not-determined |
| S13_extracted_22778 | Not-determined |
| S13_extracted_22779 | Not-determined |
| S13_extracted_22785 | Not-determined |
| S13_extracted_22791 | Not-determined |
| S13_extracted_22795 | Not-determined |
| S13_extracted_22799 | Not-determined |
| S13_extracted_22800 | Not-determined |
| S13_extracted_22801 | Low-quality |
| S13_extracted_22804 | Not-determined |
| S13_extracted_22831 | Low-quality |
| S13_extracted_22841 | Not-determined |
| S13_extracted_22853 | Not-determined |
| S13_extracted_22855 | Not-determined |
| S13_extracted_22879 | Not-determined |
| S13_extracted_22880 | Not-determined |
| S13_extracted_22897 | Not-determined |
| S13_extracted_22906 | Low-quality |
| S13_extracted_22915 | Not-determined |
| S13_extracted_22921 | Not-determined |
| S13_extracted_22924 | Not-determined |
| S13_extracted_22952 | Not-determined |
| S13_extracted_22953 | Not-determined |
| S13_extracted_22964 | Not-determined |
| S13_extracted_22968 | Low-quality |
| S13_extracted_22972 | Not-determined |
| S13_extracted_22976 | Low-quality |
| S13_extracted_22990 | Not-determined |
| S13_extracted_22992 | Not-determined |
| S13_extracted_23004 | Not-determined |
| S13_extracted_23010 | Low-quality |
| S13_extracted_23011 | Not-determined |
| S13_extracted_23012 | Not-determined |
| S13_extracted_23033 | Not-determined |
| S13_extracted_23066 | Not-determined |
| S13_extracted_23068 | Not-determined |
| S13_extracted_23082 | Low-quality |
| S13_extracted_23098 | Not-determined |
| S13_extracted_23113 | Low-quality |
| S13_extracted_23132 | Not-determined |
| S13_extracted_23148 | Not-determined |
| S13_extracted_23149 | Low-quality |
| S13_extracted_23162 | Low-quality |
| S13_extracted_23166 | Not-determined |
| S13_extracted_23172 | Not-determined |
| S13_extracted_23181 | Low-quality |
| S13_extracted_23187 | Not-determined |
| S13_extracted_23192 | Not-determined |
| S13_extracted_23195 | Low-quality |
| S13_extracted_23202 | Low-quality |
| S13_extracted_23212 | Not-determined |
| S13_extracted_23217 | Not-determined |
| S13_extracted_23230 | Not-determined |
| S13_extracted_23266 | Not-determined |
| S13_extracted_23268 | Not-determined |
| S13_extracted_23272 | Not-determined |
| S13_extracted_23288 | Low-quality |
| S13_extracted_23296 | Not-determined |
| S13_extracted_23309 | Not-determined |
| S13_extracted_23324 | Not-determined |
| S13_extracted_23327 | Not-determined |
| S13_extracted_23334 | Not-determined |
| S13_extracted_23350 | Not-determined |
| S13_extracted_23355 | Not-determined |
| S13_extracted_23357 | Not-determined |
| S13_extracted_23376 | Not-determined |
| S13_extracted_23396 | Not-determined |
| S13_extracted_23402 | Not-determined |
| S13_extracted_23444 | Not-determined |
| S13_extracted_23452 | Not-determined |
| S13_extracted_23457 | Not-determined |
| S13_extracted_23465 | Not-determined |
| S13_extracted_23468 | Not-determined |
| S13_extracted_23474 | Not-determined |
| S13_extracted_23485 | Not-determined |
| S13_extracted_23488 | Not-determined |
| S13_extracted_23501 | Not-determined |
| S13_extracted_23512 | Not-determined |
| S13_extracted_23524 | Low-quality |
| S13_extracted_23533 | Not-determined |
| S13_extracted_23536 | Not-determined |
| S13_extracted_23552 | Not-determined |
| S13_extracted_23563 | Not-determined |
| S13_extracted_23568 | Not-determined |
| S13_extracted_23578 | Low-quality |
| S13_extracted_23579 | Not-determined |
| S13_extracted_23590 | Not-determined |
| S13_extracted_23591 | Not-determined |
| S13_extracted_23593 | Not-determined |
| S13_extracted_23594 | Low-quality |
| S13_extracted_23613 | Not-determined |
| S13_extracted_23621 | Not-determined |
| S13_extracted_23629 | Not-determined |
| S13_extracted_23634 | Not-determined |
| S13_extracted_23640 | Not-determined |
| S13_extracted_23641 | Not-determined |
| S13_extracted_23642 | Low-quality |
| S13_extracted_23647 | Not-determined |
| S13_extracted_23648 | Not-determined |
| S13_extracted_23665 | Not-determined |
| S13_extracted_23671 | Low-quality |
| S13_extracted_23676 | Not-determined |
| S13_extracted_23690 | Low-quality |
| S13_extracted_23697 | Not-determined |
| S13_extracted_23717 | Not-determined |
| S13_extracted_23721 | Not-determined |
| S13_extracted_23724 | Not-determined |
| S13_extracted_23727 | Not-determined |
| S13_extracted_23740 | Not-determined |
| S13_extracted_23744 | Not-determined |
| S13_extracted_23747 | Low-quality |
| S13_extracted_23757 | Not-determined |
| S13_extracted_23760 | Not-determined |
| S13_extracted_23764 | Not-determined |
| S13_extracted_23778 | Not-determined |
| S13_extracted_23799 | Not-determined |
| S13_extracted_23803 | Not-determined |
| S13_extracted_23807 | Not-determined |
| S13_extracted_23812 | Not-determined |
| S13_extracted_23815 | Low-quality |
| S13_extracted_23817 | Not-determined |
| S13_extracted_23842 | Low-quality |
| S13_extracted_23844 | Not-determined |
| S13_extracted_23852 | Not-determined |
| S13_extracted_23857 | Not-determined |
| S13_extracted_23888 | Not-determined |
| S13_extracted_23895 | Not-determined |
| S13_extracted_23900 | Not-determined |
| S13_extracted_23925 | Not-determined |
| S13_extracted_23928 | Not-determined |
| S13_extracted_23929 | Not-determined |
| S13_extracted_23932 | Not-determined |
| S13_extracted_23948 | Not-determined |
| S13_extracted_23950 | Not-determined |
| S13_extracted_23980 | Not-determined |
| S13_extracted_23987 | Low-quality |
| S13_extracted_24025 | Not-determined |
| S13_extracted_24035 | Not-determined |
| S13_extracted_24036 | Not-determined |
| S13_extracted_24039 | Low-quality |
| S13_extracted_24041 | Low-quality |
| S13_extracted_24043 | Not-determined |
| S13_extracted_24049 | Not-determined |
| S13_extracted_24052 | Not-determined |
| S13_extracted_24057 | Not-determined |
| S13_extracted_24059 | Not-determined |
| S13_extracted_24068 | Not-determined |
| S13_extracted_24070 | Not-determined |
| S13_extracted_24071 | Not-determined |
| S13_extracted_24092 | Not-determined |
| S13_extracted_24104 | Not-determined |
| S13_extracted_24118 | Not-determined |
| S13_extracted_24127 | Not-determined |
| S13_extracted_24130 | Not-determined |
| S13_extracted_24151 | Not-determined |
| S13_extracted_24157 | Not-determined |
| S12_vs_k141_181024\|\|full | Medium-quality |
| S13_extracted_24165 | Not-determined |
| S13_extracted_24167 | Not-determined |
| S13_extracted_24179 | Not-determined |
| S13_extracted_24184 | Not-determined |
| S13_extracted_24198 | Not-determined |
| S13_extracted_24201 | Not-determined |
| S13_extracted_24202 | Not-determined |
| S13_extracted_24210 | Not-determined |
| S13_extracted_24212 | Not-determined |
| S13_extracted_24214 | Not-determined |
| S13_extracted_24221 | Not-determined |
| S13_extracted_24229 | Not-determined |
| S13_extracted_24231 | Not-determined |
| S13_extracted_24234 | Low-quality |
| S13_extracted_24236 | Not-determined |
| S13_extracted_24243 | Not-determined |
| S13_extracted_24271 | Not-determined |
| S13_extracted_24273 | Not-determined |
| S13_extracted_24276 | Not-determined |
| S13_extracted_24285 | Not-determined |
| S13_extracted_24302 | Not-determined |
| S13_extracted_24308 | Not-determined |
| S13_extracted_24317 | Not-determined |
| S13_extracted_24329 | Not-determined |
| S13_extracted_24337 | Not-determined |
| S13_extracted_24363 | Not-determined |
| S13_extracted_24403 | Not-determined |
| S13_extracted_24405 | Not-determined |
| S13_extracted_24412 | Not-determined |
| S13_extracted_24418 | Not-determined |
| S13_extracted_24429 | Not-determined |
| S13_extracted_24455 | Not-determined |
| S13_extracted_24465 | Not-determined |
| S13_extracted_24473 | Not-determined |
| S13_extracted_24487 | Not-determined |
| S13_extracted_24499 | Not-determined |
| S13_extracted_24502 | Not-determined |
| S13_extracted_24514 | Not-determined |
| S13_extracted_24520 | Not-determined |
| S13_extracted_24524 | Not-determined |
| S13_extracted_24538 | Not-determined |
| S13_extracted_24540 | Not-determined |
| S13_extracted_24594 | Not-determined |
| S13_extracted_24595 | Not-determined |
| S13_extracted_24607 | Not-determined |
| S13_extracted_24608 | Not-determined |
| S13_extracted_24610 | Not-determined |
| S13_extracted_24624 | Not-determined |
| S13_extracted_24628 | Not-determined |
| S13_extracted_24630 | Not-determined |
| S13_extracted_24657 | Not-determined |
| S13_extracted_24669 | Not-determined |
| S13_extracted_24670 | Not-determined |
| S13_extracted_24673 | Not-determined |
| S13_extracted_24682 | Not-determined |
| S13_extracted_24693 | Not-determined |
| S13_extracted_24698 | Not-determined |
| S13_extracted_24699 | Not-determined |
| S13_extracted_24701 | Low-quality |
| S13_extracted_24703 | Not-determined |
| S13_extracted_24706 | Not-determined |
| S13_extracted_24712 | Low-quality |
| S13_extracted_24741 | Not-determined |
| S13_extracted_24743 | Not-determined |
| S13_extracted_24749 | Not-determined |
| S13_extracted_24757 | Not-determined |
| S13_extracted_24765 | Not-determined |
| S13_extracted_24774 | Not-determined |
| S13_extracted_24775 | Not-determined |
| S13_extracted_24776 | Not-determined |
| S13_extracted_24783 | Not-determined |
| S13_extracted_24792 | Not-determined |
| S13_extracted_24798 | Not-determined |
| S13_extracted_24804 | Low-quality |
| S13_extracted_24812 | Low-quality |
| S13_extracted_24818 | Not-determined |
| S13_extracted_24819 | Low-quality |
| S13_extracted_24827 | Not-determined |
| S13_extracted_24835 | Not-determined |
| S13_extracted_24843 | Not-determined |

| | |
|---|---|
| S13_extracted_24844 | Not-determined |
| S13_extracted_24847 | Not-determined |
| S13_extracted_24851 | Not-determined |
| S13_extracted_24867 | Not-determined |
| S13_extracted_24872 | Not-determined |
| S13_extracted_24880 | Not-determined |
| S13_extracted_24881 | Not-determined |
| S13_extracted_24884 | Not-determined |
| S13_extracted_24902 | Not-determined |
| S13_extracted_24904 | Not-determined |
| S13_extracted_24906 | Not-determined |
| S13_extracted_24920 | Not-determined |
| S13_extracted_24928 | Not-determined |
| S13_extracted_24931 | Not-determined |
| S13_extracted_24935 | Not-determined |
| S13_extracted_24937 | Not-determined |
| S13_extracted_24938 | Not-determined |
| S13_extracted_24946 | Not-determined |
| S13_extracted_24947 | Low-quality |
| S7_vs_48708\|\|full | High-quality |
| S13_extracted_24992 | Not-determined |
| S13_extracted_24995 | Not-determined |
| S13_extracted_25016 | Not-determined |
| S13_extracted_25030 | Not-determined |
| S13_extracted_25036 | Not-determined |
| S13_extracted_25040 | Not-determined |
| S13_extracted_25045 | Not-determined |
| S13_extracted_25047 | Not-determined |
| S13_extracted_25055 | Not-determined |
| S13_extracted_25058 | Not-determined |
| S13_extracted_25068 | Low-quality |
| S13_extracted_25070 | Not-determined |
| S13_extracted_25073 | Not-determined |
| S13_extracted_25077 | Not-determined |
| S13_extracted_25078 | Not-determined |
| S13_extracted_25080 | Not-determined |
| S13_extracted_25088 | Low-quality |
| S13_extracted_25089 | Not-determined |
| S13_extracted_25102 | Not-determined |
| S13_extracted_25103 | Not-determined |
| S13_extracted_25136 | Not-determined |
| S13_extracted_25137 | Not-determined |
| S13_extracted_25140 | Low-quality |
| S13_extracted_25142 | Not-determined |
| S13_extracted_25178 | Not-determined |
| S13_extracted_25221 | Not-determined |
| S13_extracted_25224 | Not-determined |
| S13_extracted_25225 | Not-determined |
| S13_extracted_25244 | Not-determined |
| S13_extracted_25265 | Not-determined |
| S13_extracted_25288 | Not-determined |
| S13_extracted_25295 | Not-determined |
| S13_extracted_25314 | Not-determined |
| S13_extracted_25330 | Not-determined |
| S13_extracted_25336 | Not-determined |
| S13_extracted_25353 | Not-determined |
| S13_extracted_25367 | Not-determined |
| S13_extracted_25373 | Not-determined |
| S13_extracted_25374 | Not-determined |
| S13_extracted_25376 | Not-determined |
| S13_extracted_25379 | Not-determined |
| S13_extracted_25383 | Not-determined |
| S13_extracted_25386 | Not-determined |
| S13_extracted_25396 | Not-determined |
| S13_extracted_25404 | Not-determined |
| S13_extracted_25412 | Not-determined |
| S13_extracted_25430 | Not-determined |
| S13_extracted_25441 | Not-determined |
| S13_extracted_25443 | Not-determined |
| S13_extracted_25446 | Not-determined |
| S13_extracted_25451 | Not-determined |
| S13_extracted_25461 | Not-determined |
| S13_extracted_25472 | Not-determined |
| S13_extracted_25474 | Not-determined |
| S13_extracted_25482 | Not-determined |
| S13_extracted_25483 | Not-determined |
| S13_extracted_25488 | Not-determined |
| S13_extracted_25491 | Not-determined |
| S13_extracted_25494 | Not-determined |
| S13_extracted_25506 | Not-determined |
| S13_extracted_25509 | Not-determined |
| S13_extracted_25518 | Low-quality |
| S13_extracted_25526 | Not-determined |
| S13_extracted_25535 | Not-determined |
| S13_extracted_25553 | Not-determined |
| S13_extracted_25565 | Not-determined |
| S13_extracted_25586 | Low-quality |
| S13_extracted_25608 | Not-determined |
| S13_extracted_25611 | Not-determined |
| S13_extracted_25612 | Not-determined |
| S13_extracted_25615 | Not-determined |
| S13_extracted_25618 | Low-quality |
| S13_extracted_25642 | Not-determined |
| S13_extracted_25643 | Low-quality |
| S13_extracted_25650 | Not-determined |
| S13_extracted_25661 | Not-determined |
| S13_extracted_25667 | Not-determined |
| S13_extracted_25672 | Not-determined |
| S13_extracted_25674 | Not-determined |
| S13_extracted_25693 | Not-determined |
| S13_extracted_25723 | Not-determined |
| S13_extracted_25740 | Not-determined |
| S13_extracted_25745 | Not-determined |
| S13_extracted_25746 | Not-determined |
| S13_extracted_25747 | Not-determined |
| S13_extracted_25764 | Not-determined |
| S13_extracted_25775 | Not-determined |
| S13_extracted_25788 | Not-determined |
| S13_extracted_25800 | Not-determined |
| S13_extracted_25810 | Not-determined |
| S13_extracted_25822 | Not-determined |
| S13_extracted_25823 | Not-determined |
| S13_extracted_25829 | Not-determined |
| S13_extracted_25830 | Not-determined |
| S13_extracted_25836 | Not-determined |
| S13_extracted_25844 | Not-determined |
| S13_extracted_25845 | Not-determined |
| S13_extracted_25863 | Not-determined |
| S13_extracted_25864 | Not-determined |
| S13_extracted_25874 | Not-determined |
| S13_extracted_25878 | Not-determined |
| S13_extracted_25895 | Not-determined |
| S13_extracted_25905 | Not-determined |
| S13_extracted_25915 | Low-quality |
| S13_extracted_25930 | Not-determined |
| S13_extracted_25932 | Not-determined |
| S13_extracted_25937 | Not-determined |
| S13_extracted_25952 | Not-determined |
| S13_extracted_25957 | Not-determined |
| S13_extracted_25969 | Not-determined |
| S13_extracted_25982 | Not-determined |
| S13_extracted_25995 | Not-determined |
| S13_extracted_26011 | Not-determined |
| S13_extracted_26013 | Not-determined |
| S13_extracted_26020 | Not-determined |
| S13_extracted_26026 | Not-determined |
| S13_extracted_26037 | Not-determined |
| S13_extracted_26040 | Not-determined |
| S13_extracted_26045 | Not-determined |
| S13_extracted_26046 | Low-quality |
| S13_extracted_26053 | Not-determined |
| S13_extracted_26074 | Not-determined |
| S13_extracted_26084 | Not-determined |
| S13_extracted_26093 | Not-determined |
| S13_extracted_26097 | Not-determined |
| S13_extracted_26106 | Not-determined |
| S13_extracted_26112 | Not-determined |
| S13_extracted_26113 | Low-quality |
| S13_extracted_26114 | Not-determined |
| S13_extracted_26115 | Not-determined |
| S13_extracted_26119 | Not-determined |
| S13_extracted_26164 | Not-determined |
| S13_extracted_26169 | Not-determined |
| S13_extracted_26207 | Not-determined |
| S13_extracted_26235 | Not-determined |
| S13_extracted_26237 | Not-determined |
| S13_extracted_26257 | Not-determined |
| S13_extracted_26261 | Not-determined |
| S13_extracted_26270 | Not-determined |
| S13_extracted_26276 | Not-determined |
| S13_extracted_26319 | Low-quality |
| S13_extracted_26324 | Low-quality |
| S13_extracted_26331 | Not-determined |
| S13_extracted_26348 | Low-quality |
| S13_extracted_26354 | Not-determined |
| S13_extracted_26358 | Not-determined |
| S13_extracted_26381 | Not-determined |
| S13_extracted_26398 | Low-quality |
| S13_extracted_26399 | Not-determined |
| S13_extracted_26402 | Not-determined |
| S13_extracted_26414 | Low-quality |
| S13_extracted_26417 | Not-determined |
| S13_extracted_26437 | Not-determined |
| S13_extracted_26446 | Not-determined |
| S13_extracted_26470 | Not-determined |
| S13_extracted_26477 | Not-determined |
| S13_extracted_26483 | Not-determined |
| S13_extracted_26486 | Not-determined |
| S13_extracted_26489 | Not-determined |
| S13_extracted_26492 | Not-determined |
| S13_extracted_26506 | Not-determined |
| S13_extracted_26508 | Not-determined |
| S13_extracted_26517 | Not-determined |
| S13_extracted_26529 | Not-determined |
| S13_extracted_26535 | Low-quality |
| S13_extracted_26536 | Not-determined |
| S13_extracted_26538 | Not-determined |
| S13_extracted_26539 | Not-determined |
| S13_extracted_26556 | Low-quality |
| S13_extracted_26562 | Low-quality |
| S13_extracted_26565 | Not-determined |
| S13_extracted_26584 | Not-determined |
| S13_extracted_26586 | Not-determined |
| S13_extracted_26607 | Not-determined |
| S13_extracted_26632 | Not-determined |
| S13_extracted_26634 | Not-determined |
| S13_extracted_26642 | Not-determined |
| S13_extracted_26646 | Not-determined |
| S13_extracted_26650 | Not-determined |
| S13_extracted_26654 | Not-determined |
| S13_extracted_26659 | Not-determined |
| S13_extracted_26663 | Not-determined |
| S13_extracted_26667 | Not-determined |
| S13_extracted_26695 | Not-determined |
| S13_extracted_26701 | Not-determined |
| S13_extracted_26708 | Not-determined |
| S13_extracted_26711 | Not-determined |
| S13_extracted_26715 | Low-quality |
| S13_extracted_26719 | Low-quality |
| S13_extracted_26721 | Low-quality |
| S13_extracted_26732 | Not-determined |
| S13_extracted_26735 | Not-determined |
| S13_extracted_26750 | Not-determined |
| S13_extracted_26770 | Not-determined |
| S13_extracted_26776 | Not-determined |
| S13_extracted_26781 | Not-determined |
| S13_extracted_26796 | Low-quality |
| S13_extracted_26805 | Not-determined |
| S13_extracted_26842 | Not-determined |
| S13_extracted_26854 | Not-determined |
| S13_extracted_26858 | Not-determined |
| S13_extracted_26859 | Not-determined |
| S13_extracted_26861 | Not-determined |
| S13_extracted_26864 | Not-determined |
| S13_extracted_26872 | Not-determined |
| S13_extracted_26873 | Not-determined |
| S13_extracted_26879 | Not-determined |
| S13_extracted_26881 | Not-determined |
| S13_extracted_26892 | Not-determined |
| S13_extracted_26899 | Not-determined |
| S13_extracted_26904 | Not-determined |
| S13_extracted_26905 | Not-determined |
| S13_extracted_26917 | Not-determined |
| S13_extracted_26925 | Not-determined |
| S13_extracted_26944 | Not-determined |
| S13_extracted_26952 | Not-determined |
| S13_extracted_26953 | Not-determined |
| S13_extracted_26957 | Low-quality |
| S13_extracted_26958 | Not-determined |
| S13_extracted_26962 | Not-determined |
| S13_extracted_26966 | Not-determined |
| S13_extracted_26991 | Not-determined |
| S13_extracted_26995 | Not-determined |
| S13_extracted_27017 | Low-quality |
| S13_extracted_27027 | Not-determined |
| S13_extracted_27042 | Not-determined |
| S13_extracted_27046 | Low-quality |
| S13_extracted_27051 | Not-determined |
| S13_extracted_27056 | Not-determined |
| S13_extracted_27079 | Not-determined |
| S13_extracted_27083 | Not-determined |
| S13_extracted_27096 | Not-determined |
| S13_extracted_27101 | Not-determined |
| S13_extracted_27129 | Low-quality |
| S13_extracted_27130 | Low-quality |
| S13_extracted_27132 | Not-determined |
| S13_extracted_27134 | Not-determined |
| S13_extracted_27137 | Not-determined |
| S13_extracted_27154 | Low-quality |
| S13_extracted_27160 | Low-quality |
| S13_extracted_27161 | Not-determined |
| S13_extracted_27178 | Not-determined |
| S13_extracted_27184 | Not-determined |
| S13_extracted_27192 | Not-determined |
| S13_extracted_27206 | Not-determined |
| S13_extracted_27216 | Not-determined |
| S13_extracted_27224 | Not-determined |
| S13_extracted_27226 | Not-determined |
| S13_extracted_27244 | Not-determined |
| S13_extracted_27262 | Low-quality |
| S13_extracted_27277 | Not-determined |
| S13_extracted_27282 | Not-determined |
| S13_extracted_27302 | Not-determined |
| S13_extracted_27304 | Low-quality |
| S13_extracted_27307 | Not-determined |
| S13_extracted_27313 | Low-quality |
| S13_extracted_27323 | Not-determined |
| S13_extracted_27326 | Not-determined |
| S13_extracted_27331 | Not-determined |
| S13_extracted_27339 | Not-determined |
| S13_extracted_27341 | Low-quality |
| S13_extracted_27347 | Not-determined |
| S13_extracted_27365 | Not-determined |
| S13_extracted_27366 | Low-quality |
| S13_extracted_27368 | Not-determined |
| S13_extracted_27380 | Low-quality |
| S13_extracted_27390 | Not-determined |
| S13_extracted_27419 | Not-determined |
| S13_extracted_27430 | Low-quality |
| S13_extracted_27450 | Not-determined |
| S13_extracted_27459 | Not-determined |
| S13_extracted_27460 | Not-determined |
| S13_extracted_27466 | Not-determined |
| S13_extracted_27468 | Low-quality |
| S13_extracted_27474 | Not-determined |
| S13_extracted_27483 | Low-quality |
| S13_extracted_27511 | Low-quality |
| S13_extracted_27512 | Not-determined |
| S13_extracted_27514 | Low-quality |
| S13_extracted_27522 | Not-determined |
| S13_extracted_27530 | Low-quality |
| S13_extracted_27531 | Not-determined |
| S13_extracted_27537 | Not-determined |
| S13_extracted_27542 | Not-determined |
| S13_extracted_27555 | Not-determined |
| S13_extracted_27563 | Low-quality |
| S13_extracted_27566 | Not-determined |
| S13_extracted_27567 | Not-determined |
| S13_extracted_27568 | Not-determined |
| S13_extracted_27586 | Not-determined |
| S13_extracted_27593 | Not-determined |
| S13_extracted_27599 | Not-determined |
| S13_extracted_27608 | Not-determined |
| S13_extracted_27621 | Not-determined |
| S13_extracted_27630 | Not-determined |
| S13_extracted_27648 | Not-determined |
| S13_extracted_27652 | Not-determined |
| S13_extracted_27687 | Not-determined |
| S13_extracted_27705 | Not-determined |
| S13_extracted_27739 | Not-determined |
| S13_extracted_27743 | Low-quality |
| S13_extracted_27756 | Not-determined |
| S13_extracted_27762 | Not-determined |
| S13_extracted_27771 | Not-determined |
| S13_extracted_27775 | Not-determined |
| S13_extracted_27789 | Not-determined |
| S13_extracted_27817 | Not-determined |
| S13_extracted_27838 | Not-determined |
| S13_extracted_27896 | Not-determined |
| S13_extracted_27900 | Not-determined |
| S13_extracted_27918 | Not-determined |
| S13_extracted_27936 | Not-determined |
| S13_extracted_27939 | Not-determined |
| S13_extracted_27958 | Low-quality |
| S13_extracted_27969 | Not-determined |
| S13_extracted_27991 | Low-quality |
| S13_extracted_27995 | Not-determined |
| S13_extracted_27997 | Not-determined |
| S13_extracted_27999 | Not-determined |
| S13_extracted_28002 | Not-determined |
| S13_extracted_28007 | Not-determined |
| S13_extracted_28027 | Low-quality |
| S13_extracted_28035 | Not-determined |
| S13_extracted_28036 | Not-determined |
| S13_extracted_28063 | Not-determined |
| S13_extracted_28072 | Not-determined |
| S13_extracted_28080 | Not-determined |
| S13_extracted_28117 | Low-quality |
| S13_extracted_28129 | Not-determined |
| S13_extracted_28132 | Not-determined |
| S13_extracted_28143 | Not-determined |
| S13_extracted_28159 | Not-determined |
| S13_extracted_28161 | Not-determined |
| S13_extracted_28168 | Not-determined |
| S13_extracted_28176 | Not-determined |
| S13_extracted_28186 | Not-determined |
| S13_extracted_28193 | Not-determined |
| S13_extracted_28209 | Not-determined |
| S13_extracted_28225 | Not-determined |
| S13_extracted_28227 | Not-determined |
| S13_extracted_28231 | Not-determined |
| S13_extracted_28233 | Not-determined |
| S13_extracted_28248 | Not-determined |
| S13_extracted_28268 | Not-determined |
| S13_extracted_28271 | Not-determined |
| S13_extracted_28275 | Low-quality |
| S13_extracted_28276 | Low-quality |
| S13_extracted_28278 | Not-determined |
| S13_extracted_28282 | Not-determined |
| S13_extracted_28288 | Not-determined |
| S13_extracted_28300 | Not-determined |
| S15_extracted_2032 | Complete |
| S13_extracted_28321 | Not-determined |
| S13_extracted_28323 | Not-determined |
| S13_extracted_28340 | Not-determined |
| S13_extracted_28352 | Not-determined |
| S13_extracted_28356 | Not-determined |
| S13_extracted_28362 | Not-determined |
| S13_extracted_28368 | Not-determined |
| S13_extracted_28382 | Not-determined |
| S13_extracted_28393 | Not-determined |
| S13_extracted_28400 | Not-determined |
| S13_extracted_28413 | Not-determined |
| S13_extracted_28415 | Low-quality |
| S13_extracted_28416 | Not-determined |
| S13_extracted_28430 | Low-quality |
| S13_extracted_28445 | Not-determined |
| S13_extracted_28470 | Not-determined |
| S13_extracted_28476 | Not-determined |
| S13_extracted_28480 | Not-determined |
| S13_extracted_28482 | Not-determined |
| S13_extracted_28484 | Not-determined |
| S13_extracted_28491 | Not-determined |
| S13_extracted_28519 | Not-determined |
| S13_extracted_28553 | Not-determined |
| S13_extracted_28554 | Not-determined |
| S13_extracted_28557 | Not-determined |
| S13_extracted_28560 | Not-determined |
| S13_extracted_28574 | Not-determined |
| S13_extracted_28577 | Not-determined |
| S13_extracted_28582 | Not-determined |
| S13_extracted_28583 | Not-determined |
| S13_extracted_28588 | Not-determined |
| S13_extracted_28601 | Not-determined |
| S13_extracted_28630 | Not-determined |
| S13_extracted_28634 | Low-quality |
| S13_extracted_28642 | Not-determined |
| S13_extracted_28647 | Not-determined |
| S13_extracted_28652 | Not-determined |
| S13_extracted_28655 | Not-determined |
| S13_extracted_28668 | Low-quality |
| S13_extracted_28678 | Not-determined |
| S13_extracted_28683 | Not-determined |
| S13_extracted_28706 | Not-determined |
| S13_extracted_28708 | Not-determined |
| S13_extracted_28733 | Not-determined |
| S13_extracted_28741 | Not-determined |
| S13_extracted_28748 | Low-quality |
| S13_extracted_28773 | Not-determined |
| S13_extracted_28778 | Not-determined |
| S13_extracted_28785 | Not-determined |
| S13_extracted_28789 | Not-determined |
| S13_extracted_28790 | Not-determined |
| S13_extracted_28791 | Not-determined |
| S13_extracted_28792 | Not-determined |
| S13_extracted_28812 | Not-determined |
| S13_extracted_28821 | Not-determined |
| S13_extracted_28827 | Not-determined |
| S13_extracted_28852 | Low-quality |
| S13_extracted_28857 | Not-determined |
| S13_extracted_28877 | Not-determined |
| S13_extracted_28880 | Not-determined |
| S13_extracted_28888 | Not-determined |
| S13_extracted_28904 | Not-determined |
| S13_extracted_28908 | Not-determined |
| S13_extracted_28918 | Low-quality |
| S13_extracted_28928 | Not-determined |
| S13_extracted_28930 | Not-determined |
| S13_extracted_28933 | Not-determined |
| S13_extracted_28934 | Not-determined |
| S13_extracted_28943 | Not-determined |
| S13_extracted_28959 | Not-determined |
| S13_extracted_28980 | Not-determined |
| S13_extracted_28984 | Not-determined |
| S13_extracted_28993 | Not-determined |
| S13_extracted_28999 | Not-determined |
| S13_extracted_29010 | Not-determined |
| S13_extracted_29016 | Not-determined |
| S13_extracted_29020 | Low-quality |
| S13_extracted_29045 | Not-determined |
| S13_extracted_29048 | Not-determined |
| S13_extracted_29052 | Not-determined |
| S13_extracted_29059 | Not-determined |
| S13_extracted_29064 | Not-determined |
| S13_extracted_29085 | Not-determined |
| S13_extracted_29091 | Not-determined |
| S13_extracted_29112 | Low-quality |
| S13_extracted_29118 | Not-determined |
| S13_extracted_29141 | Not-determined |
| S13_extracted_29149 | Not-determined |
| S13_extracted_29150 | Not-determined |
| S13_extracted_29157 | Not-determined |
| S13_extracted_29167 | Not-determined |
| S13_extracted_29182 | Not-determined |
| S13_extracted_29196 | Low-quality |
| S13_extracted_29220 | Low-quality |
| S13_extracted_29221 | Low-quality |
| S13_extracted_29252 | Not-determined |
| S13_extracted_29265 | Not-determined |
| S13_extracted_29271 | Not-determined |
| S13_extracted_29272 | Not-determined |
| S13_extracted_29275 | Not-determined |
| S13_extracted_29277 | Not-determined |
| S13_extracted_29296 | Not-determined |
| S13_extracted_29304 | Not-determined |
| S13_extracted_29309 | Low-quality |
| S13_extracted_29331 | Not-determined |
| S13_extracted_29337 | Not-determined |
| S13_extracted_29358 | Low-quality |
| S13_extracted_29360 | Not-determined |
| S13_extracted_29362 | Not-determined |
| S13_extracted_29389 | Not-determined |
| S13_extracted_29412 | Not-determined |
| S13_extracted_29415 | Not-determined |
| S13_extracted_29422 | Not-determined |
| S13_extracted_29440 | Not-determined |
| S13_extracted_29443 | Low-quality |
| S13_extracted_29444 | Not-determined |
| S13_extracted_29445 | Not-determined |
| S13_extracted_29455 | Not-determined |
| S13_extracted_29461 | Not-determined |
| S13_extracted_29466 | Not-determined |
| S13_extracted_29481 | Not-determined |
| S13_extracted_29482 | Not-determined |
| S13_extracted_29485 | Not-determined |
| S13_extracted_29487 | Not-determined |
| S13_extracted_29503 | Not-determined |
| S13_extracted_29515 | Not-determined |
| S13_extracted_29518 | Not-determined |
| S13_extracted_29559 | Not-determined |
| S13_extracted_29564 | Not-determined |
| S13_extracted_29578 | Not-determined |
| S13_extracted_29584 | Not-determined |
| S13_extracted_29593 | Not-determined |
| S13_extracted_29602 | Not-determined |
| S13_extracted_29614 | Not-determined |
| S13_extracted_29615 | Low-quality |
| S13_extracted_29617 | Not-determined |
| S13_extracted_29618 | Not-determined |
| S13_extracted_29639 | Not-determined |
| S13_extracted_29641 | Not-determined |
| S13_extracted_29649 | Not-determined |
| S13_extracted_29668 | Low-quality |
| S13_extracted_29669 | Not-determined |
| S13_extracted_29688 | Not-determined |
| S13_extracted_29700 | Not-determined |
| S13_extracted_29706 | Not-determined |
| S13_extracted_29734 | Not-determined |
| S13_extracted_29748 | Not-determined |
| S13_extracted_29753 | Not-determined |
| S13_extracted_29758 | Not-determined |
| S13_extracted_29764 | Low-quality |
| S13_extracted_29767 | Not-determined |
| S13_extracted_29783 | Not-determined |
| S13_extracted_29801 | Not-determined |
| S13_extracted_29803 | Not-determined |
| S13_extracted_29809 | Not-determined |
| S13_extracted_29824 | Not-determined |
| S13_extracted_29825 | Not-determined |
| S13_extracted_29840 | Not-determined |
| S13_extracted_29848 | Not-determined |
| S13_extracted_29849 | Not-determined |
| S13_extracted_29850 | Not-determined |
| S13_extracted_29865 | Not-determined |
| S13_extracted_29877 | Not-determined |
| S13_extracted_29889 | Not-determined |
| S13_extracted_29895 | Not-determined |
| S13_extracted_29901 | Not-determined |
| S13_extracted_29946 | Low-quality |
| S13_extracted_29947 | Low-quality |
| S13_extracted_29951 | Not-determined |
| S13_extracted_29966 | Low-quality |
| S13_extracted_29980 | Not-determined |
| S13_extracted_29994 | Not-determined |
| S13_extracted_30032 | Low-quality |
| S13_extracted_30040 | Low-quality |
| S13_extracted_30049 | Not-determined |
| S13_extracted_30055 | Low-quality |
| S13_extracted_30068 | Not-determined |
| S13_extracted_30076 | Not-determined |
| S13_extracted_30088 | Not-determined |
| S13_extracted_30130 | Not-determined |
| S13_extracted_30138 | Not-determined |
| S13_extracted_30139 | Low-quality |
| S13_extracted_30143 | Not-determined |
| S13_extracted_30144 | Low-quality |
| S13_extracted_30150 | Not-determined |
| S13_extracted_30152 | Not-determined |
| S13_extracted_30163 | Not-determined |
| S13_extracted_30177 | Not-determined |
| S13_extracted_30179 | Not-determined |
| S13_extracted_30181 | Not-determined |
| S13_extracted_30184 | Not-determined |
| S13_extracted_30192 | Not-determined |
| S13_extracted_30208 | Not-determined |
| S13_extracted_30218 | Not-determined |
| S13_extracted_30219 | Not-determined |
| S3_vs_k141_12731\|\|full | High-quality |
| S13_extracted_30225 | Not-determined |
| S13_extracted_30253 | Not-determined |
| S13_extracted_30265 | Not-determined |
| S13_extracted_30268 | Not-determined |
| S13_extracted_30273 | Not-determined |
| S13_extracted_30285 | Low-quality |
| S13_extracted_30290 | Not-determined |
| S13_extracted_30305 | Low-quality |
| S13_extracted_30321 | Not-determined |
| S13_extracted_30324 | Not-determined |
| S13_extracted_30325 | Not-determined |
| S13_extracted_30331 | Not-determined |
| S13_extracted_30333 | Not-determined |
| S13_extracted_30342 | Not-determined |
| S13_extracted_30345 | Not-determined |
| S13_extracted_30350 | Not-determined |
| S13_extracted_30353 | Not-determined |
| S13_extracted_30363 | Not-determined |
| S13_extracted_30364 | Not-determined |
| S13_extracted_30375 | Not-determined |
| S13_extracted_30379 | Low-quality |

| | |
|---|---|
| S13_extracted_30383 | Not-determined |
| S13_extracted_30391 | Not-determined |
| S13_extracted_30392 | Not-determined |
| S13_extracted_30393 | Not-determined |
| S13_extracted_30394 | Not-determined |
| S13_extracted_30395 | Not-determined |
| S13_extracted_30399 | Not-determined |
| S13_extracted_30402 | Not-determined |
| S13_extracted_30403 | Not-determined |
| S13_extracted_30419 | Low-quality |
| S13_extracted_30424 | Not-determined |
| S13_extracted_30448 | Not-determined |
| S13_extracted_30465 | Not-determined |
| S13_extracted_30474 | Not-determined |
| S13_extracted_30479 | Not-determined |
| S13_extracted_30490 | Not-determined |
| S13_extracted_30500 | Not-determined |
| S13_extracted_30524 | Not-determined |
| S13_extracted_30529 | Not-determined |
| S13_extracted_30530 | Not-determined |
| S13_extracted_30546 | Not-determined |
| S13_extracted_30549 | Not-determined |
| S13_extracted_30583 | Not-determined |
| S13_extracted_30596 | Not-determined |
| S13_extracted_30599 | Not-determined |
| S13_extracted_30602 | Not-determined |
| S13_extracted_30623 | Not-determined |
| S13_extracted_30632 | Not-determined |
| S13_extracted_30633 | Low-quality |
| S13_extracted_30635 | Not-determined |
| S13_extracted_30637 | Low-quality |
| S13_extracted_30656 | Not-determined |
| S13_extracted_30657 | Not-determined |
| S13_extracted_30670 | Low-quality |
| S13_extracted_30673 | Not-determined |
| S13_extracted_30674 | Not-determined |
| S13_extracted_30684 | Not-determined |
| S13_extracted_30697 | Not-determined |
| S13_extracted_30717 | Not-determined |
| S13_extracted_30771 | Not-determined |
| S13_extracted_30782 | Not-determined |
| S13_extracted_30787 | Not-determined |
| S13_extracted_30791 | Low-quality |
| S13_extracted_30816 | Not-determined |
| S13_extracted_30820 | Low-quality |
| S13_extracted_30825 | Not-determined |
| S13_extracted_30829 | Low-quality |
| S13_extracted_30833 | Not-determined |
| S13_extracted_30839 | Not-determined |
| S13_extracted_30844 | Not-determined |
| S13_extracted_30864 | Not-determined |
| S13_extracted_30867 | Not-determined |
| S13_extracted_30872 | Not-determined |
| S13_extracted_30882 | Not-determined |
| S13_extracted_30890 | Low-quality |
| S13_extracted_30891 | Not-determined |
| S13_extracted_30896 | Not-determined |
| S13_extracted_30911 | Not-determined |
| S13_extracted_30968 | Low-quality |
| S13_extracted_30979 | Low-quality |
| S13_extracted_30988 | Not-determined |
| S13_extracted_30990 | Not-determined |
| S13_extracted_30996 | Not-determined |
| S13_extracted_31001 | Not-determined |
| S13_extracted_31004 | Not-determined |
| S13_extracted_31007 | Not-determined |
| S13_extracted_31014 | Not-determined |
| S13_extracted_31017 | Not-determined |
| S13_extracted_31019 | Not-determined |
| S13_extracted_31022 | Not-determined |
| S13_extracted_31024 | Not-determined |
| S13_extracted_31034 | Not-determined |
| S13_extracted_31038 | Not-determined |
| S13_extracted_31041 | Not-determined |
| S13_extracted_31042 | Not-determined |
| S13_extracted_31062 | Not-determined |
| S13_extracted_31073 | Not-determined |
| S13_extracted_31079 | Not-determined |
| S13_extracted_31081 | Low-quality |
| S13_extracted_31087 | Not-determined |
| S13_extracted_31094 | Not-determined |
| S13_extracted_31105 | Not-determined |
| S13_extracted_31120 | Not-determined |
| S13_extracted_31133 | Not-determined |
| S13_extracted_31136 | Not-determined |
| S13_extracted_31139 | Low-quality |
| S13_extracted_31141 | Not-determined |
| S13_extracted_31152 | Not-determined |
| S13_extracted_31207 | Not-determined |
| S13_extracted_31217 | Not-determined |
| S13_extracted_31218 | Not-determined |
| S13_extracted_31228 | Not-determined |
| S13_extracted_31234 | Not-determined |
| S13_extracted_31235 | Low-quality |
| S13_extracted_31240 | Not-determined |
| S13_extracted_31246 | Not-determined |
| S13_extracted_31254 | Not-determined |
| S13_extracted_31258 | Not-determined |
| S13_extracted_31279 | Not-determined |
| S13_extracted_31281 | Not-determined |
| S13_extracted_31285 | Low-quality |
| S13_extracted_31288 | Not-determined |
| S13_extracted_31336 | Not-determined |
| S13_extracted_31347 | Low-quality |
| S13_extracted_31350 | Not-determined |
| S13_extracted_31352 | Not-determined |
| S13_extracted_31358 | Not-determined |
| S13_extracted_31366 | Not-determined |
| S13_extracted_31373 | Not-determined |
| S13_extracted_31379 | Not-determined |
| S13_extracted_31385 | Not-determined |
| S13_extracted_31393 | Low-quality |
| S13_extracted_31396 | Not-determined |
| S13_extracted_31405 | Not-determined |
| S13_extracted_31417 | Not-determined |
| S13_extracted_31421 | Not-determined |
| S13_extracted_31429 | Not-determined |
| S13_extracted_31430 | Not-determined |
| S13_extracted_31432 | Not-determined |
| S13_extracted_31434 | Not-determined |
| S13_extracted_31436 | Not-determined |
| S13_extracted_31437 | Not-determined |
| S13_extracted_31442 | Not-determined |
| S13_extracted_31444 | Not-determined |
| S13_extracted_31454 | Not-determined |
| S13_extracted_31466 | Not-determined |
| S13_extracted_31487 | Not-determined |
| S13_extracted_31495 | Not-determined |
| S13_extracted_31497 | Not-determined |
| S13_extracted_31499 | Not-determined |
| S13_extracted_31509 | Not-determined |
| S13_extracted_31529 | Low-quality |
| S13_extracted_31534 | Not-determined |
| S13_extracted_31535 | Not-determined |
| S13_extracted_31537 | Not-determined |
| S13_extracted_31544 | Not-determined |
| S13_extracted_31546 | Not-determined |
| S13_extracted_31554 | Not-determined |
| S13_extracted_31567 | Low-quality |
| S13_extracted_31570 | Not-determined |
| S13_extracted_31584 | Not-determined |
| S13_extracted_31593 | Not-determined |
| S13_extracted_31605 | Not-determined |
| S13_extracted_31607 | Not-determined |
| S13_extracted_31620 | Not-determined |
| S13_extracted_31631 | Low-quality |
| S13_extracted_31637 | Not-determined |
| S13_extracted_31648 | Not-determined |
| S13_extracted_31657 | Not-determined |
| S13_extracted_31672 | Low-quality |
| S13_extracted_31673 | Low-quality |
| S13_extracted_31677 | Not-determined |
| S13_extracted_31684 | Not-determined |
| S13_extracted_31685 | Not-determined |
| S13_extracted_31689 | Not-determined |
| S13_extracted_31699 | Not-determined |
| S13_extracted_31704 | Not-determined |
| S13_extracted_31711 | Not-determined |
| S13_extracted_31759 | Not-determined |
| S13_extracted_31765 | Not-determined |
| S13_extracted_31768 | Low-quality |
| S13_extracted_31770 | Low-quality |
| S13_extracted_31780 | Not-determined |
| S13_extracted_31794 | Not-determined |
| S13_extracted_31797 | Not-determined |
| S13_extracted_31802 | Not-determined |
| S13_extracted_31804 | Not-determined |
| S13_extracted_31806 | Not-determined |
| S13_extracted_31816 | Not-determined |
| S13_extracted_31819 | Not-determined |
| S13_extracted_31825 | Not-determined |
| S13_extracted_31831 | Not-determined |
| S13_extracted_31837 | Not-determined |
| S13_extracted_31841 | Not-determined |
| S13_extracted_31844 | Not-determined |
| S13_extracted_31848 | Not-determined |
| S13_extracted_31854 | Not-determined |
| S13_extracted_31860 | Not-determined |
| S13_extracted_31873 | Not-determined |
| S13_extracted_31878 | Not-determined |
| S13_extracted_31888 | Not-determined |
| S13_extracted_31908 | Not-determined |
| S13_extracted_31914 | Not-determined |
| S13_extracted_31916 | Not-determined |
| S13_extracted_31917 | Not-determined |
| S13_extracted_31934 | Not-determined |
| S13_extracted_31940 | Low-quality |
| S13_extracted_31951 | Not-determined |
| S13_extracted_31953 | Low-quality |
| S13_extracted_31954 | Not-determined |
| S13_extracted_31961 | Not-determined |
| S13_extracted_31967 | Not-determined |
| S13_extracted_31971 | Not-determined |
| S13_extracted_31982 | Not-determined |
| S13_extracted_31983 | Not-determined |
| S13_extracted_31987 | Not-determined |
| S13_extracted_31993 | Not-determined |
| S13_extracted_31999 | Not-determined |
| S13_extracted_32018 | Not-determined |
| S13_extracted_32024 | Not-determined |
| S13_extracted_32040 | Not-determined |
| S13_extracted_32044 | Not-determined |
| S13_extracted_32048 | Not-determined |
| S13_extracted_32052 | Not-determined |
| S13_extracted_32063 | Not-determined |
| S13_extracted_32068 | Not-determined |
| S13_extracted_32086 | Low-quality |
| S13_extracted_32094 | Low-quality |
| S13_extracted_32097 | Not-determined |
| S13_extracted_32104 | Low-quality |
| S13_extracted_32112 | Low-quality |
| S13_extracted_32114 | Not-determined |
| S13_extracted_32140 | Not-determined |
| S13_extracted_32148 | Not-determined |
| S13_extracted_32153 | Not-determined |
| S13_extracted_32198 | Not-determined |
| S13_extracted_32199 | Not-determined |
| S13_extracted_32206 | Not-determined |
| S13_extracted_32207 | Not-determined |
| S13_extracted_32209 | Not-determined |
| S13_extracted_32222 | Low-quality |
| S13_extracted_32225 | Not-determined |
| S13_extracted_32229 | Low-quality |
| S13_extracted_32237 | Not-determined |
| S13_extracted_32251 | Not-determined |
| S13_extracted_32268 | Not-determined |
| S13_extracted_32269 | Low-quality |
| S13_extracted_32280 | Not-determined |
| S13_extracted_32289 | Not-determined |
| S13_extracted_32296 | Not-determined |
| S13_extracted_32308 | Not-determined |
| S13_extracted_32310 | Not-determined |
| S13_extracted_32321 | Not-determined |
| S13_extracted_32323 | Not-determined |
| S13_extracted_32325 | Not-determined |
| S13_extracted_32333 | Not-determined |
| S13_extracted_32344 | Not-determined |
| S13_extracted_32371 | Not-determined |
| S13_extracted_32374 | Not-determined |
| S13_extracted_32385 | Not-determined |
| S13_extracted_32403 | Not-determined |
| S13_extracted_32408 | Low-quality |
| S13_extracted_32410 | Not-determined |
| S13_extracted_32415 | Not-determined |
| S13_extracted_32421 | Not-determined |
| S13_extracted_32439 | Not-determined |
| S13_extracted_32442 | Not-determined |
| S13_extracted_32448 | Not-determined |
| S13_extracted_32452 | Low-quality |
| S13_extracted_32455 | Low-quality |
| S13_extracted_32456 | Not-determined |
| S13_extracted_32471 | Not-determined |
| S13_extracted_32475 | Not-determined |
| S13_extracted_32478 | Not-determined |
| S13_extracted_32504 | Not-determined |
| S13_extracted_32509 | Not-determined |
| S13_extracted_32519 | Low-quality |
| S13_extracted_32523 | Not-determined |
| S13_extracted_32525 | Not-determined |
| S13_extracted_32529 | Not-determined |
| S13_extracted_32531 | Not-determined |
| S13_extracted_32534 | Not-determined |
| S13_extracted_32554 | Not-determined |
| S13_extracted_32567 | Not-determined |
| S13_extracted_32569 | Not-determined |
| S13_extracted_32573 | Low-quality |
| S13_extracted_32589 | Not-determined |
| S13_extracted_32602 | Low-quality |
| S13_extracted_32629 | Not-determined |
| S13_extracted_32632 | Not-determined |
| S13_extracted_32634 | Not-determined |
| S13_extracted_32654 | Not-determined |
| S13_extracted_32657 | Not-determined |
| S13_extracted_32658 | Not-determined |
| S13_extracted_32664 | Low-quality |
| S13_extracted_32672 | Not-determined |
| S13_extracted_32680 | Not-determined |
| S13_extracted_32685 | Low-quality |
| S13_extracted_32686 | Not-determined |
| S13_extracted_32694 | Not-determined |
| S13_extracted_32705 | Not-determined |
| S13_extracted_32707 | Low-quality |
| S13_extracted_32708 | Not-determined |
| S13_extracted_32721 | Low-quality |
| S13_extracted_32726 | Low-quality |
| S13_extracted_32760 | Not-determined |
| S13_extracted_32765 | Not-determined |
| S13_extracted_32779 | Not-determined |
| S13_extracted_32780 | Not-determined |
| S13_extracted_32789 | Not-determined |
| S13_extracted_32793 | Not-determined |
| S13_extracted_32798 | Not-determined |
| S13_extracted_32803 | Not-determined |
| S13_extracted_32810 | Not-determined |
| S13_extracted_32821 | Low-quality |
| S13_extracted_32832 | Not-determined |
| S13_extracted_32838 | Not-determined |
| S13_extracted_32846 | Not-determined |
| S13_extracted_32847 | Not-determined |
| S13_extracted_32849 | Not-determined |
| S13_extracted_32851 | Not-determined |
| S13_extracted_32852 | Not-determined |
| S13_extracted_32870 | Not-determined |
| S13_extracted_32879 | Not-determined |
| S13_extracted_32883 | Not-determined |
| S13_extracted_32891 | Not-determined |
| S13_extracted_32901 | Not-determined |
| S13_extracted_32909 | Low-quality |
| S13_extracted_32919 | Not-determined |
| S13_extracted_32950 | Not-determined |
| S13_extracted_32955 | Low-quality |
| S13_extracted_32967 | Not-determined |
| S13_extracted_32972 | Not-determined |
| S13_extracted_32975 | Not-determined |
| S13_extracted_32979 | Not-determined |
| S13_extracted_32980 | Not-determined |
| S13_extracted_32986 | Not-determined |
| S13_extracted_32987 | Not-determined |
| S13_extracted_33003 | Not-determined |
| S13_extracted_33030 | Not-determined |
| S13_extracted_33035 | Not-determined |
| S13_extracted_33044 | Not-determined |
| S13_extracted_33070 | Not-determined |
| S13_extracted_33087 | Not-determined |
| S13_extracted_33113 | Not-determined |
| S13_extracted_33122 | Not-determined |
| S13_extracted_33142 | Not-determined |
| S13_extracted_33143 | Not-determined |
| S13_extracted_33146 | Low-quality |
| S13_extracted_33147 | Low-quality |
| S13_extracted_33151 | Low-quality |
| S13_extracted_33163 | Not-determined |
| S13_extracted_33175 | Not-determined |
| S13_extracted_33176 | Not-determined |
| S13_extracted_33180 | Not-determined |
| S13_extracted_33212 | Not-determined |
| S13_extracted_33227 | Not-determined |
| S13_extracted_33231 | Not-determined |
| S13_extracted_33245 | Not-determined |
| S13_extracted_33247 | Low-quality |
| S13_extracted_33254 | Not-determined |
| S13_extracted_33262 | Not-determined |
| S13_extracted_33265 | Not-determined |
| S13_extracted_33266 | Not-determined |
| S13_extracted_33277 | Low-quality |
| S13_extracted_33301 | Not-determined |
| S13_extracted_33305 | Not-determined |
| S13_extracted_33317 | Not-determined |
| S13_extracted_33319 | Not-determined |
| S13_extracted_33337 | Not-determined |
| S13_extracted_33356 | Not-determined |
| S13_extracted_33363 | Not-determined |
| S13_extracted_33374 | Not-determined |
| S13_extracted_33375 | Low-quality |
| S13_extracted_33379 | Not-determined |
| S13_extracted_33380 | Not-determined |
| S13_extracted_33389 | Low-quality |
| S13_extracted_33398 | Not-determined |
| S13_extracted_33402 | Low-quality |
| S13_extracted_33405 | Not-determined |
| S13_extracted_33413 | Not-determined |
| S13_extracted_33420 | Not-determined |
| S13_extracted_33426 | Not-determined |
| S13_extracted_33438 | Not-determined |
| S13_extracted_33458 | Not-determined |
| S13_extracted_33465 | Not-determined |
| S13_extracted_33481 | Not-determined |
| S13_extracted_33484 | Not-determined |
| S13_extracted_33486 | Not-determined |
| S13_extracted_33498 | Not-determined |
| S13_extracted_33501 | Not-determined |
| S13_extracted_33510 | Low-quality |
| S13_extracted_33512 | Low-quality |
| S13_extracted_33519 | Low-quality |
| S13_extracted_33521 | Not-determined |
| S13_extracted_33537 | Not-determined |
| S13_extracted_33540 | Low-quality |
| S13_extracted_33543 | Not-determined |
| S13_extracted_33550 | Not-determined |
| S13_extracted_33553 | Not-determined |
| S13_extracted_33569 | Not-determined |
| S13_extracted_33580 | Not-determined |
| S13_extracted_33596 | Not-determined |
| S13_extracted_33625 | Not-determined |
| S13_extracted_33637 | Not-determined |
| S13_extracted_33653 | Not-determined |
| S13_extracted_33654 | Not-determined |
| S13_extracted_33655 | Not-determined |
| S13_extracted_33663 | Not-determined |
| S13_extracted_33671 | Not-determined |
| S13_extracted_33680 | Not-determined |
| S13_extracted_33684 | Not-determined |
| S13_extracted_33691 | Not-determined |
| S13_extracted_33731 | Not-determined |
| S13_extracted_33732 | Not-determined |
| S13_extracted_33738 | Not-determined |
| S13_extracted_33750 | Not-determined |
| S13_extracted_33760 | Low-quality |
| S13_extracted_33761 | Not-determined |
| S13_extracted_33764 | Not-determined |
| S13_extracted_33773 | Not-determined |
| S13_extracted_33774 | Not-determined |
| S13_extracted_33775 | Not-determined |
| S13_extracted_33783 | Not-determined |
| S13_extracted_33793 | Not-determined |
| S13_extracted_33794 | Not-determined |
| S13_extracted_33796 | Not-determined |
| S13_extracted_33800 | Not-determined |
| S13_extracted_33815 | Not-determined |
| S13_extracted_33842 | Not-determined |
| S13_extracted_33846 | Not-determined |
| S13_extracted_33851 | Not-determined |
| S13_extracted_33866 | Not-determined |
| S13_extracted_33899 | Not-determined |
| S13_extracted_33914 | Not-determined |
| S13_extracted_33923 | Not-determined |
| S13_extracted_33927 | Not-determined |
| S13_extracted_33942 | Not-determined |
| S13_extracted_33944 | Not-determined |
| S13_extracted_33948 | Not-determined |
| S13_extracted_33957 | Low-quality |
| S13_extracted_33960 | Low-quality |
| S13_extracted_33976 | Not-determined |
| S13_extracted_33978 | Not-determined |
| S13_extracted_33983 | Not-determined |
| S13_extracted_33992 | Not-determined |
| S13_extracted_33997 | Not-determined |
| S13_extracted_33999 | Low-quality |
| S13_extracted_34003 | Not-determined |
| S13_extracted_34030 | Not-determined |
| S13_extracted_34055 | Not-determined |
| S13_extracted_34062 | Not-determined |
| S13_extracted_34075 | Not-determined |
| S13_extracted_34099 | Not-determined |
| S13_extracted_34100 | Low-quality |
| S13_extracted_34105 | Not-determined |
| S13_extracted_34127 | Not-determined |
| S13_extracted_34142 | Low-quality |
| S13_extracted_34148 | Not-determined |
| S13_extracted_34150 | Not-determined |
| S13_extracted_34157 | Not-determined |
| S13_extracted_34167 | Not-determined |
| S13_extracted_34173 | Not-determined |
| S13_extracted_34199 | Not-determined |
| S13_extracted_34202 | Not-determined |
| S13_extracted_34203 | Not-determined |
| S13_extracted_34215 | Not-determined |
| S13_extracted_34218 | Not-determined |
| S13_extracted_34219 | Not-determined |
| S13_extracted_34223 | Not-determined |
| S13_extracted_34229 | Low-quality |
| S13_extracted_34235 | Not-determined |
| S13_extracted_34240 | Not-determined |
| S13_extracted_34244 | Not-determined |
| S13_extracted_34251 | Not-determined |
| S13_extracted_34255 | Not-determined |
| S13_extracted_34256 | Not-determined |
| S13_extracted_34261 | Not-determined |
| S13_extracted_34268 | Not-determined |
| S13_extracted_34273 | Not-determined |
| S13_extracted_34277 | Not-determined |
| S13_extracted_34294 | Not-determined |
| S13_extracted_34305 | Not-determined |
| S13_extracted_34313 | Not-determined |
| S13_extracted_34315 | Not-determined |
| S13_extracted_34349 | Low-quality |
| S13_extracted_34359 | Low-quality |
| S13_extracted_34361 | Not-determined |
| S13_extracted_34378 | Low-quality |
| S13_extracted_34385 | Not-determined |
| S13_extracted_34386 | Not-determined |
| S13_extracted_34391 | Not-determined |
| S13_extracted_34400 | Not-determined |
| S13_extracted_34407 | Not-determined |
| S13_extracted_34415 | Not-determined |
| S13_extracted_34416 | Not-determined |
| S13_extracted_34427 | Not-determined |
| S13_extracted_34442 | Not-determined |
| S13_extracted_34452 | Not-determined |
| S13_extracted_34454 | Low-quality |
| S13_extracted_34462 | Not-determined |
| S13_extracted_34467 | Low-quality |
| S13_extracted_34479 | Not-determined |
| S13_extracted_34482 | Not-determined |
| S13_extracted_34485 | Not-determined |
| S13_extracted_34501 | Not-determined |
| S13_extracted_34510 | Not-determined |
| S13_extracted_34514 | Not-determined |
| S13_extracted_34515 | Not-determined |
| S13_extracted_34526 | Not-determined |
| S13_extracted_34535 | Not-determined |
| S13_extracted_34541 | Not-determined |
| S13_extracted_34542 | Not-determined |
| S13_extracted_34547 | Not-determined |
| S13_extracted_34556 | Not-determined |
| S13_extracted_34558 | Not-determined |
| S13_extracted_34562 | Not-determined |
| S13_extracted_34586 | Low-quality |
| S13_extracted_34591 | Not-determined |
| S13_extracted_34608 | Not-determined |
| S13_extracted_34612 | Not-determined |
| S13_extracted_34629 | Not-determined |
| S13_extracted_34632 | Not-determined |
| S13_extracted_34636 | Not-determined |
| S13_extracted_34637 | Low-quality |
| S13_extracted_34638 | Not-determined |
| S13_extracted_34649 | Not-determined |
| S13_extracted_34658 | Not-determined |
| S13_extracted_34665 | Low-quality |
| S13_extracted_34671 | Not-determined |
| S13_extracted_34679 | Not-determined |
| S13_extracted_34683 | Not-determined |
| S13_extracted_34684 | Not-determined |
| S13_extracted_34700 | Not-determined |
| S13_extracted_34702 | Not-determined |
| S13_extracted_34712 | Not-determined |
| S13_extracted_34717 | Not-determined |
| S13_extracted_34723 | Not-determined |
| S13_extracted_34735 | Not-determined |
| S13_extracted_34739 | Not-determined |
| S13_extracted_34743 | Not-determined |
| S13_extracted_34745 | Low-quality |
| S13_extracted_34753 | Low-quality |
| S13_extracted_34760 | Not-determined |
| S13_extracted_34773 | Not-determined |
| S13_extracted_34776 | Low-quality |
| S13_extracted_34796 | Not-determined |
| S13_extracted_34797 | Low-quality |
| S13_extracted_34805 | Not-determined |
| S13_extracted_34813 | Not-determined |
| S13_extracted_34831 | Not-determined |
| S13_extracted_34837 | Not-determined |
| S13_extracted_34839 | Not-determined |
| S13_extracted_34848 | Not-determined |
| S13_extracted_34850 | Not-determined |
| S13_extracted_34851 | Not-determined |
| S13_extracted_34863 | Not-determined |
| S13_extracted_34866 | Not-determined |
| S13_extracted_34891 | Not-determined |
| S13_extracted_34898 | Not-determined |
| S13_extracted_34907 | Not-determined |
| S13_extracted_34912 | Not-determined |
| S13_extracted_34917 | Not-determined |
| S13_extracted_34924 | Low-quality |
| S13_extracted_34929 | Not-determined |
| S13_extracted_34966 | Not-determined |
| S13_extracted_34973 | Not-determined |
| S13_extracted_34982 | Not-determined |
| S13_extracted_34985 | Not-determined |
| S13_extracted_34988 | Low-quality |
| S13_extracted_34990 | Not-determined |
| S13_extracted_35002 | Not-determined |
| S13_extracted_35024 | Low-quality |
| S13_extracted_35037 | Not-determined |
| S13_extracted_35054 | Not-determined |
| S13_extracted_35065 | Not-determined |
| S13_extracted_35070 | Low-quality |
| S13_extracted_35074 | Not-determined |
| S13_extracted_35079 | Not-determined |
| S13_extracted_35081 | Not-determined |
| S13_extracted_35083 | Low-quality |
| S13_extracted_35096 | Not-determined |
| S13_extracted_35106 | Not-determined |
| S13_extracted_35107 | Not-determined |
| S13_extracted_35118 | Not-determined |
| S13_extracted_35127 | Not-determined |
| S13_extracted_35129 | Not-determined |
| S13_extracted_35134 | Low-quality |
| S13_extracted_35144 | Not-determined |
| S13_extracted_35145 | Not-determined |
| S13_extracted_35168 | Not-determined |
| S13_extracted_35169 | Not-determined |
| S13_extracted_35183 | Not-determined |
| S13_extracted_35186 | Not-determined |
| S13_extracted_35187 | Not-determined |
| S13_extracted_35192 | Not-determined |
| S13_extracted_35201 | Not-determined |
| S13_extracted_35202 | Not-determined |
| S13_extracted_35216 | Low-quality |
| S13_extracted_35217 | Not-determined |
| S13_extracted_35222 | Not-determined |
| S13_extracted_35227 | Not-determined |
| S13_extracted_35234 | Low-quality |
| S13_extracted_35236 | Not-determined |
| S13_extracted_35246 | Not-determined |
| S13_extracted_35255 | Not-determined |
| S13_extracted_35268 | Low-quality |
| S13_extracted_35278 | Not-determined |
| S13_extracted_35283 | Not-determined |

| | |
|---|---|
| S13_extracted_35295 | Not-determined |
| S13_extracted_35302 | Not-determined |
| S13_extracted_35308 | Not-determined |
| S13_extracted_35312 | Not-determined |
| S13_extracted_35315 | Not-determined |
| S13_extracted_35326 | Not-determined |
| S13_extracted_35335 | Not-determined |
| S13_extracted_35340 | Low-quality |
| S13_extracted_35344 | Not-determined |
| S13_extracted_35358 | Low-quality |
| S13_extracted_35361 | Not-determined |
| S13_extracted_35375 | Not-determined |
| S13_extracted_35377 | Not-determined |
| S13_extracted_35383 | Not-determined |
| S13_extracted_35386 | Not-determined |
| S13_extracted_35387 | Not-determined |
| S13_extracted_35388 | Low-quality |
| S13_extracted_35399 | Not-determined |
| S13_extracted_35402 | Not-determined |
| S13_extracted_35431 | Not-determined |
| S13_extracted_35432 | Not-determined |
| S13_extracted_35436 | Not-determined |
| S13_extracted_35439 | Not-determined |
| S13_extracted_35454 | Not-determined |
| S13_extracted_35458 | Not-determined |
| S13_extracted_35459 | Not-determined |
| S13_extracted_35468 | Not-determined |
| S13_extracted_35472 | Not-determined |
| S13_extracted_35477 | Not-determined |
| S13_extracted_35492 | Not-determined |
| S13_extracted_35510 | Not-determined |
| S13_extracted_35516 | Not-determined |
| S13_extracted_35521 | Not-determined |
| S13_extracted_35530 | Not-determined |
| S13_extracted_35558 | Not-determined |
| S13_extracted_35584 | Not-determined |
| S13_extracted_35589 | Not-determined |
| S13_extracted_35593 | Not-determined |
| S13_extracted_35615 | Not-determined |
| S13_extracted_35621 | Not-determined |
| S13_extracted_35649 | Not-determined |
| S13_extracted_35651 | Not-determined |
| S13_extracted_35661 | Not-determined |
| S13_extracted_35664 | Low-quality |
| S13_extracted_35667 | Not-determined |
| S13_extracted_35671 | Not-determined |
| S13_extracted_35686 | Not-determined |
| S13_extracted_35740 | Not-determined |
| S13_extracted_35744 | Not-determined |
| S13_extracted_35745 | Not-determined |
| S13_extracted_35746 | Not-determined |
| S13_extracted_35763 | Not-determined |
| S13_extracted_35777 | Not-determined |
| S13_extracted_35784 | Not-determined |
| S13_extracted_35789 | Not-determined |
| S13_extracted_35800 | Not-determined |
| S13_extracted_35808 | Low-quality |
| S13_extracted_35809 | Not-determined |
| S13_extracted_35810 | Low-quality |
| S13_extracted_35812 | Not-determined |
| S13_extracted_35813 | Not-determined |
| S13_extracted_35841 | Not-determined |
| S13_extracted_35844 | Not-determined |
| S13_extracted_35847 | Not-determined |
| S13_extracted_35869 | Not-determined |
| S13_extracted_35871 | Not-determined |
| S13_extracted_35878 | Low-quality |
| S13_extracted_35879 | Low-quality |
| S13_extracted_35897 | Low-quality |
| S13_extracted_35899 | Not-determined |
| S13_extracted_35900 | Not-determined |
| S13_extracted_35914 | Not-determined |
| S13_extracted_35922 | Not-determined |
| S13_extracted_35926 | Not-determined |
| S13_extracted_35927 | Low-quality |
| S13_extracted_35935 | Not-determined |
| S13_extracted_35942 | Not-determined |
| S13_extracted_35956 | Not-determined |
| S13_extracted_35961 | Not-determined |
| S13_extracted_35962 | Not-determined |
| S13_extracted_35966 | Not-determined |
| S13_extracted_35971 | Low-quality |
| S13_extracted_35973 | Not-determined |
| S13_extracted_35985 | Not-determined |
| S13_extracted_36000 | Not-determined |
| S13_extracted_36003 | Not-determined |
| S13_extracted_36010 | Low-quality |
| S13_extracted_36013 | Not-determined |
| S13_extracted_36017 | Not-determined |
| S13_extracted_36022 | Not-determined |
| S13_extracted_36060 | Not-determined |
| S13_extracted_36076 | Not-determined |
| S13_extracted_36081 | Not-determined |
| S13_extracted_36096 | Not-determined |
| S13_extracted_36102 | Not-determined |
| S13_extracted_36137 | Not-determined |
| S13_extracted_36138 | Not-determined |
| S13_extracted_36140 | Not-determined |
| S13_extracted_36143 | Not-determined |
| S13_extracted_36151 | Not-determined |
| S13_extracted_36152 | Not-determined |
| S13_extracted_36156 | Not-determined |
| S13_extracted_36157 | Not-determined |
| S13_extracted_36163 | Not-determined |
| S13_extracted_36168 | Not-determined |
| S13_extracted_36175 | Not-determined |
| S13_extracted_36190 | Not-determined |
| S13_extracted_36194 | Not-determined |
| S13_extracted_36196 | Not-determined |
| S15_vs_2032\|\|full | High-quality |
| S13_extracted_36200 | Not-determined |
| S13_extracted_36207 | Not-determined |
| S13_extracted_36214 | Not-determined |
| S13_extracted_36240 | Not-determined |
| S13_extracted_36242 | Not-determined |
| S13_extracted_36249 | Not-determined |
| S13_extracted_36250 | Not-determined |
| S13_extracted_36273 | Not-determined |
| S13_extracted_36282 | Not-determined |
| S13_extracted_36287 | Not-determined |
| S13_extracted_36290 | Not-determined |
| S13_extracted_36298 | Not-determined |
| S13_extracted_36302 | Not-determined |
| S13_extracted_36312 | Not-determined |
| S13_extracted_36330 | Not-determined |
| S13_extracted_36333 | Not-determined |
| S13_extracted_36348 | Not-determined |
| S13_extracted_36351 | Not-determined |
| S13_extracted_36372 | Not-determined |
| S13_extracted_36377 | Not-determined |
| S13_extracted_36390 | Not-determined |
| S13_extracted_36397 | Not-determined |
| S13_extracted_36406 | Not-determined |
| S13_extracted_36417 | Low-quality |
| S13_extracted_36421 | Not-determined |
| S13_extracted_36437 | Not-determined |
| S13_extracted_36442 | Not-determined |
| S13_extracted_36452 | Not-determined |
| S13_extracted_36455 | Low-quality |
| S13_extracted_36465 | Not-determined |
| S13_extracted_36500 | Not-determined |
| S13_extracted_36508 | Not-determined |
| S13_extracted_36518 | Not-determined |
| S13_extracted_36521 | Not-determined |
| S13_extracted_36537 | Not-determined |
| S13_extracted_36548 | Low-quality |
| S13_extracted_36562 | Not-determined |
| S13_extracted_36569 | Not-determined |
| S13_extracted_36576 | Low-quality |
| S13_extracted_36596 | Not-determined |
| S13_extracted_36610 | Low-quality |
| S13_extracted_36620 | Not-determined |
| S13_extracted_36646 | Not-determined |
| S13_extracted_36650 | Not-determined |
| S13_extracted_36656 | Not-determined |
| S13_extracted_36677 | Low-quality |
| S13_extracted_36679 | Not-determined |
| S13_extracted_36689 | Not-determined |
| S13_extracted_36698 | Not-determined |
| S13_extracted_36705 | Not-determined |
| S13_extracted_36707 | Not-determined |
| S13_extracted_36709 | Not-determined |
| S13_extracted_36710 | Not-determined |
| S13_extracted_36714 | Not-determined |
| S13_extracted_36715 | Not-determined |
| S13_extracted_36738 | Not-determined |
| S13_extracted_36767 | Not-determined |
| S13_extracted_36806 | Not-determined |
| S13_extracted_36818 | Not-determined |
| S13_extracted_36836 | Not-determined |
| S13_extracted_36838 | Not-determined |
| S13_extracted_36839 | Not-determined |
| S13_extracted_36851 | Low-quality |
| S13_extracted_36854 | Not-determined |
| S13_extracted_36856 | Not-determined |
| S13_extracted_36861 | Not-determined |
| S13_extracted_36867 | Not-determined |
| S13_extracted_36877 | Not-determined |
| S13_extracted_36885 | Low-quality |
| S13_extracted_36888 | Not-determined |
| S13_extracted_36894 | Not-determined |
| S13_extracted_36899 | Low-quality |
| S13_extracted_36904 | Not-determined |
| S13_extracted_36917 | Not-determined |
| S13_extracted_36920 | Not-determined |
| S13_extracted_36924 | Not-determined |
| S13_extracted_36939 | Not-determined |
| S13_extracted_36940 | Not-determined |
| S13_extracted_36942 | Not-determined |
| S13_extracted_36945 | Not-determined |
| S13_extracted_36949 | Not-determined |
| S13_extracted_36954 | Not-determined |
| S13_extracted_36982 | Not-determined |
| S13_extracted_36986 | Not-determined |
| S13_extracted_36998 | Not-determined |
| S13_extracted_37000 | Not-determined |
| S13_extracted_37014 | Not-determined |
| S13_extracted_37022 | Not-determined |
| S13_extracted_37027 | Not-determined |
| S13_extracted_37035 | Not-determined |
| S13_extracted_37036 | Not-determined |
| S13_extracted_37041 | Not-determined |
| S13_extracted_37042 | Not-determined |
| S13_extracted_37048 | Not-determined |
| S13_extracted_37053 | Not-determined |
| S13_extracted_37054 | Not-determined |
| S13_extracted_37063 | Not-determined |
| S13_extracted_37065 | Not-determined |
| S13_extracted_37066 | Low-quality |
| S13_extracted_37068 | Not-determined |
| S13_extracted_37074 | Not-determined |
| S13_extracted_37079 | Not-determined |
| S13_extracted_37081 | Low-quality |
| S13_extracted_37086 | Not-determined |
| S13_extracted_37100 | Not-determined |
| S13_extracted_37103 | Not-determined |
| S13_extracted_37105 | Low-quality |
| S13_extracted_37109 | Not-determined |
| S13_extracted_37111 | Not-determined |
| S13_extracted_37116 | Not-determined |
| S13_extracted_37135 | Not-determined |
| S13_extracted_37139 | Not-determined |
| S13_extracted_37146 | Not-determined |
| S13_extracted_37150 | Not-determined |
| S13_extracted_37151 | Not-determined |
| S13_extracted_37158 | Not-determined |
| S13_extracted_37164 | Not-determined |
| S13_extracted_37171 | Not-determined |
| S13_extracted_37186 | Not-determined |
| S13_extracted_37205 | Not-determined |
| S13_extracted_37216 | Not-determined |
| S13_extracted_37222 | Not-determined |
| S13_extracted_37223 | Not-determined |
| S13_extracted_37241 | Not-determined |
| S13_extracted_37254 | Not-determined |
| S13_extracted_37269 | Not-determined |
| S13_extracted_37273 | Not-determined |
| S13_extracted_37277 | Not-determined |
| S13_extracted_37282 | Not-determined |
| S13_extracted_37288 | Not-determined |
| S13_extracted_37289 | Not-determined |
| S13_extracted_37290 | Not-determined |
| S13_extracted_37293 | Not-determined |
| S13_extracted_37295 | Not-determined |
| S13_extracted_37302 | Not-determined |
| S13_extracted_37313 | Not-determined |
| S13_extracted_37315 | Not-determined |
| S13_extracted_37325 | Not-determined |
| S13_extracted_37327 | Not-determined |
| S13_extracted_37339 | Not-determined |
| S13_extracted_37345 | Low-quality |
| S13_extracted_37372 | Not-determined |
| S13_extracted_37388 | Not-determined |
| S13_extracted_37403 | Not-determined |
| S13_extracted_37409 | Low-quality |
| S13_extracted_37411 | Not-determined |
| S13_extracted_37437 | Not-determined |
| S13_extracted_37446 | Not-determined |
| S13_extracted_37454 | Not-determined |
| S13_extracted_37456 | Low-quality |
| S13_extracted_37472 | Not-determined |
| S13_extracted_37480 | Not-determined |
| S13_extracted_37486 | Not-determined |
| S13_extracted_37494 | Not-determined |
| S13_extracted_37502 | Not-determined |
| S13_extracted_37506 | Low-quality |
| S13_extracted_37511 | Not-determined |
| S13_extracted_37520 | Not-determined |
| S13_extracted_37522 | Not-determined |
| S13_extracted_37538 | Not-determined |
| S13_extracted_37547 | Not-determined |
| S13_extracted_37557 | Not-determined |
| S13_extracted_37561 | Not-determined |
| S13_extracted_37575 | Not-determined |
| S13_extracted_37576 | Not-determined |
| S13_extracted_37584 | Not-determined |
| S13_extracted_37591 | Not-determined |
| S13_extracted_37596 | Not-determined |
| S13_extracted_37602 | Not-determined |
| S13_extracted_37619 | Not-determined |
| S13_extracted_37621 | Low-quality |
| S13_extracted_37623 | Not-determined |
| S13_extracted_37625 | Not-determined |
| S13_extracted_37632 | Not-determined |
| S13_extracted_37642 | Not-determined |
| S13_extracted_37646 | Not-determined |
| S13_extracted_37649 | Not-determined |
| S13_extracted_37669 | Not-determined |
| S13_extracted_37672 | Not-determined |
| S13_extracted_37677 | Not-determined |
| S13_extracted_37689 | Not-determined |
| S13_extracted_37698 | Not-determined |
| S13_extracted_37706 | Not-determined |
| S13_extracted_37719 | Low-quality |
| S13_extracted_37722 | Not-determined |
| S13_extracted_37726 | Not-determined |
| S13_extracted_37730 | Not-determined |
| S13_extracted_37737 | Not-determined |
| S13_extracted_37739 | Not-determined |
| S13_extracted_37743 | Not-determined |
| S13_extracted_37751 | Not-determined |
| S13_extracted_37755 | Not-determined |
| S13_extracted_37756 | Not-determined |
| S13_extracted_37758 | Not-determined |
| S13_extracted_37762 | Low-quality |
| S13_extracted_37774 | Not-determined |
| S13_extracted_37783 | Not-determined |
| S13_extracted_37787 | Not-determined |
| S13_extracted_37811 | Not-determined |
| S13_extracted_37813 | Low-quality |
| S13_extracted_37814 | Not-determined |
| S13_extracted_37828 | Not-determined |
| S13_extracted_37837 | Low-quality |
| S13_extracted_37870 | Not-determined |
| S13_extracted_37880 | Low-quality |
| S13_extracted_37890 | Low-quality |
| S13_extracted_37894 | Not-determined |
| S13_extracted_37901 | Low-quality |
| S13_extracted_37906 | Not-determined |
| S13_extracted_37912 | Not-determined |
| S13_extracted_37919 | Not-determined |
| S13_extracted_37932 | Not-determined |
| S13_extracted_37934 | Not-determined |
| S13_extracted_37936 | Not-determined |
| S13_extracted_37937 | Low-quality |
| S13_extracted_37943 | Low-quality |
| S13_extracted_37951 | Not-determined |
| S13_extracted_37959 | Not-determined |
| S13_extracted_37960 | Not-determined |
| S13_extracted_37979 | Not-determined |
| S13_extracted_38008 | Not-determined |
| S13_extracted_38012 | Not-determined |
| S13_extracted_38022 | Not-determined |
| S13_extracted_38029 | Not-determined |
| S13_extracted_38039 | Not-determined |
| S13_extracted_38044 | Not-determined |
| S13_extracted_38048 | Not-determined |
| S13_extracted_38049 | Not-determined |
| S13_extracted_38058 | Not-determined |
| S13_extracted_38065 | Low-quality |
| S13_extracted_38076 | Low-quality |
| S13_extracted_38080 | Not-determined |
| S13_extracted_38087 | Low-quality |
| S13_extracted_38106 | Not-determined |
| S13_extracted_38112 | Not-determined |
| S13_extracted_38126 | Low-quality |
| S13_extracted_38141 | Not-determined |
| S13_extracted_38143 | Not-determined |
| S13_extracted_38147 | Low-quality |
| S13_extracted_38156 | Not-determined |
| S13_extracted_38166 | Not-determined |
| S13_extracted_38209 | Not-determined |
| S13_extracted_38210 | Not-determined |
| S13_extracted_38227 | Not-determined |
| S13_extracted_38255 | Low-quality |
| S13_extracted_38259 | Not-determined |
| S13_extracted_38268 | Not-determined |
| S13_extracted_38270 | Not-determined |
| S13_extracted_38279 | Not-determined |
| S13_extracted_38288 | Not-determined |
| S13_extracted_38299 | Not-determined |
| S13_extracted_38304 | Low-quality |
| S13_extracted_38307 | Not-determined |
| S13_extracted_38309 | Not-determined |
| S13_extracted_38310 | Low-quality |
| S13_extracted_38312 | Not-determined |
| S13_extracted_38313 | Not-determined |
| S13_extracted_38317 | Not-determined |
| S13_extracted_38320 | Not-determined |
| S13_extracted_38321 | Not-determined |
| S13_extracted_38329 | Not-determined |
| S13_extracted_38332 | Not-determined |
| S13_extracted_38349 | Low-quality |
| S13_extracted_38373 | Low-quality |
| S13_extracted_38384 | Not-determined |
| S13_extracted_38408 | Not-determined |
| S13_extracted_38430 | Not-determined |
| S13_extracted_38433 | Low-quality |
| S13_extracted_38458 | Low-quality |
| S13_extracted_38462 | Not-determined |
| S13_extracted_38464 | Not-determined |
| S13_extracted_38472 | Not-determined |
| S13_extracted_38481 | Low-quality |
| S13_extracted_38482 | Low-quality |
| S13_extracted_38487 | Not-determined |
| S13_extracted_38488 | Not-determined |
| S13_extracted_38500 | Low-quality |
| S13_extracted_38501 | Low-quality |
| S13_extracted_38508 | Not-determined |
| S13_extracted_38519 | Not-determined |
| S13_extracted_38521 | Not-determined |
| S13_extracted_38524 | Not-determined |
| S13_extracted_38526 | Not-determined |
| S13_extracted_38531 | Not-determined |
| S13_extracted_38534 | Low-quality |
| S13_extracted_38535 | Not-determined |
| S13_extracted_38540 | Not-determined |
| S13_extracted_38549 | Not-determined |
| S13_extracted_38556 | Not-determined |
| S13_extracted_38557 | Not-determined |
| S13_extracted_38560 | Not-determined |
| S13_extracted_38561 | Not-determined |
| S13_extracted_38568 | Not-determined |
| S13_extracted_38572 | Not-determined |
| S13_extracted_38600 | Not-determined |
| S13_extracted_38612 | Low-quality |
| S13_extracted_38618 | Not-determined |
| S13_extracted_38635 | Low-quality |
| S13_extracted_38658 | Not-determined |
| S13_extracted_38660 | Not-determined |
| S13_extracted_38670 | Not-determined |
| S13_extracted_38672 | Low-quality |
| S13_extracted_38694 | Not-determined |
| S13_extracted_38699 | Not-determined |
| S13_extracted_38700 | Not-determined |
| S13_extracted_38706 | Not-determined |
| S12_extracted_k141_188830 | Complete |
| S13_extracted_38709 | Not-determined |
| S13_extracted_38710 | Not-determined |
| S13_extracted_38727 | Not-determined |
| S13_extracted_38740 | Not-determined |
| S13_extracted_38744 | Low-quality |
| S13_extracted_38754 | Not-determined |
| S13_extracted_38763 | Not-determined |
| S13_extracted_38767 | Not-determined |
| S13_extracted_38773 | Not-determined |
| S13_extracted_38774 | Not-determined |
| S13_extracted_38777 | Not-determined |
| S13_extracted_38791 | Not-determined |
| S13_extracted_38811 | Low-quality |
| S13_extracted_38827 | Not-determined |
| S13_extracted_38845 | Not-determined |
| S13_extracted_38848 | Not-determined |
| S13_extracted_38863 | Not-determined |
| S13_extracted_38864 | Not-determined |
| S13_extracted_38869 | Not-determined |
| S13_extracted_38878 | Low-quality |
| S13_extracted_38883 | Not-determined |
| S13_extracted_38899 | Not-determined |
| S13_extracted_38901 | Not-determined |
| S13_extracted_38903 | Not-determined |
| S13_extracted_38908 | Low-quality |
| S13_extracted_38932 | Not-determined |
| S13_extracted_38939 | Not-determined |
| S13_extracted_38951 | Not-determined |
| S13_extracted_38953 | Not-determined |
| S13_extracted_38957 | Not-determined |
| S13_extracted_38958 | Not-determined |
| S13_extracted_38973 | Not-determined |
| S13_extracted_38978 | Not-determined |
| S13_extracted_38984 | Not-determined |
| S13_extracted_38987 | Not-determined |
| S13_extracted_39000 | Not-determined |
| S13_extracted_39006 | Not-determined |
| S13_extracted_39025 | Not-determined |
| S13_extracted_39035 | Not-determined |
| S13_extracted_39039 | Not-determined |
| S13_extracted_39041 | Not-determined |
| S13_extracted_39045 | Not-determined |
| S13_extracted_39055 | Not-determined |
| S13_extracted_39065 | Not-determined |
| S13_extracted_39068 | Not-determined |
| S13_extracted_39069 | Not-determined |
| S13_extracted_39086 | Not-determined |
| S13_extracted_39090 | Low-quality |
| S13_extracted_39091 | Not-determined |
| S13_extracted_39104 | Not-determined |
| S13_extracted_39108 | Not-determined |
| S13_extracted_39113 | Not-determined |
| S13_extracted_39126 | Not-determined |
| S13_extracted_39138 | Low-quality |
| S13_extracted_39150 | Low-quality |
| S13_extracted_39152 | Not-determined |
| S13_extracted_39161 | Low-quality |
| S13_extracted_39174 | Not-determined |
| S13_extracted_39177 | Not-determined |
| S13_extracted_39192 | Not-determined |
| S13_extracted_39196 | Not-determined |
| S13_extracted_39198 | Low-quality |
| S13_extracted_39201 | Not-determined |
| S13_extracted_39203 | Not-determined |
| S13_extracted_39225 | Not-determined |
| S13_extracted_39229 | Not-determined |
| S13_extracted_39249 | Not-determined |
| S13_extracted_39255 | Not-determined |
| S13_extracted_39279 | Not-determined |
| S13_extracted_39290 | Not-determined |
| S13_extracted_39291 | Not-determined |
| S13_extracted_39311 | Not-determined |
| S13_extracted_39313 | Not-determined |
| S13_extracted_39316 | Not-determined |
| S13_extracted_39352 | Not-determined |
| S13_extracted_39366 | Not-determined |
| S13_extracted_39367 | Not-determined |
| S13_extracted_39372 | Not-determined |
| S13_extracted_39420 | Not-determined |
| S13_extracted_39422 | Low-quality |
| S13_extracted_39426 | Low-quality |
| S13_extracted_39428 | Not-determined |
| S13_extracted_39434 | Not-determined |
| S13_extracted_39435 | Not-determined |
| S13_extracted_39440 | Not-determined |
| S13_extracted_39446 | Low-quality |
| S13_extracted_39451 | Not-determined |
| S13_extracted_39463 | Not-determined |
| S13_extracted_39467 | Not-determined |
| S13_extracted_39474 | Low-quality |
| S13_extracted_39475 | Not-determined |
| S13_extracted_39477 | Not-determined |
| S13_extracted_39496 | Not-determined |
| S13_extracted_39506 | Low-quality |
| S13_extracted_39523 | Low-quality |
| S13_extracted_39531 | Not-determined |
| S13_extracted_39533 | Not-determined |
| S13_extracted_39538 | Not-determined |
| S13_extracted_39544 | Not-determined |
| S13_extracted_39579 | Not-determined |
| S13_extracted_39611 | Not-determined |
| S13_extracted_39619 | Low-quality |
| S13_extracted_39622 | Not-determined |
| S13_extracted_39627 | Not-determined |
| S13_extracted_39628 | Low-quality |
| S13_extracted_39634 | Not-determined |
| S13_extracted_39638 | Not-determined |
| S13_extracted_39653 | Not-determined |
| S13_extracted_39654 | Not-determined |
| S13_extracted_39656 | Not-determined |
| S13_extracted_39659 | Not-determined |
| S13_extracted_39662 | Not-determined |
| S13_extracted_39663 | Not-determined |
| S13_extracted_39675 | Not-determined |
| S13_extracted_39686 | Not-determined |
| S13_extracted_39693 | Not-determined |
| S13_extracted_39700 | Not-determined |
| S13_extracted_39741 | Not-determined |
| S13_extracted_39746 | Not-determined |
| S13_extracted_39752 | Not-determined |
| S13_extracted_39753 | Not-determined |
| S13_extracted_39769 | Not-determined |
| S13_extracted_39773 | Not-determined |
| S13_extracted_39790 | Not-determined |
| S13_extracted_39793 | Not-determined |
| S13_extracted_39801 | Low-quality |
| S13_extracted_39803 | Not-determined |
| S13_extracted_39846 | Low-quality |
| S13_extracted_39872 | Not-determined |
| S13_extracted_39873 | Not-determined |
| S13_extracted_39881 | Not-determined |
| S13_extracted_39890 | Not-determined |
| S13_extracted_39903 | Not-determined |
| S13_extracted_39906 | Not-determined |
| S13_extracted_39908 | Not-determined |
| S13_extracted_39909 | Not-determined |
| S13_extracted_39914 | Not-determined |
| S13_extracted_39917 | Not-determined |
| S13_extracted_39924 | Not-determined |
| S13_extracted_39931 | Not-determined |
| S13_extracted_39935 | Not-determined |
| S13_extracted_39936 | Not-determined |
| S13_extracted_39944 | Not-determined |
| S13_extracted_39971 | Not-determined |
| S13_extracted_39972 | Not-determined |
| S13_extracted_39980 | Not-determined |
| S13_extracted_39983 | Not-determined |
| S13_extracted_39987 | Not-determined |
| S13_extracted_39990 | Not-determined |
| S13_extracted_39997 | Not-determined |
| S13_extracted_40002 | Not-determined |
| S13_extracted_40017 | Not-determined |
| S13_extracted_40045 | Not-determined |
| S13_extracted_40051 | Low-quality |
| S13_extracted_40052 | Not-determined |
| S13_extracted_40065 | Not-determined |
| S13_extracted_40081 | Low-quality |
| S13_extracted_40082 | Low-quality |
| S13_extracted_40085 | Not-determined |
| S13_extracted_40106 | Not-determined |
| S13_extracted_40120 | Not-determined |
| S13_extracted_40129 | Not-determined |
| S13_extracted_40139 | Not-determined |
| S13_extracted_40173 | Not-determined |
| S13_extracted_40189 | Not-determined |
| S13_extracted_40192 | Low-quality |
| S13_extracted_40193 | Not-determined |
| S13_extracted_40197 | Not-determined |
| S13_extracted_40202 | Not-determined |
| S13_extracted_40211 | Not-determined |
| S13_extracted_40220 | Not-determined |
| S13_extracted_40232 | Not-determined |

| | |
|---|---|
| S13_extracted_40247 | Low-quality |
| S12_vs_k141_188830||full | High-quality |
| S13_extracted_40276 | Not-determined |
| S13_extracted_40285 | Low-quality |
| S13_extracted_40291 | Not-determined |
| S13_extracted_40297 | Not-determined |
| S13_extracted_40304 | Not-determined |
| S13_extracted_40307 | Not-determined |
| S13_extracted_40353 | Not-determined |
| S13_extracted_40362 | Not-determined |
| S13_extracted_40363 | Not-determined |
| S13_extracted_40372 | Not-determined |
| S13_extracted_40384 | Not-determined |
| S13_extracted_40387 | Not-determined |
| S13_extracted_40393 | Low-quality |
| S13_extracted_40398 | Not-determined |
| S13_extracted_40401 | Low-quality |
| S13_extracted_40421 | Not-determined |
| S13_extracted_40431 | Not-determined |
| S13_extracted_40432 | Not-determined |
| S13_extracted_40438 | Not-determined |
| S13_extracted_40443 | Not-determined |
| S13_extracted_40452 | Not-determined |
| S13_extracted_40463 | Not-determined |
| S13_extracted_40467 | Not-determined |
| S13_extracted_40469 | Not-determined |
| S13_extracted_40476 | Not-determined |
| S13_extracted_40478 | Not-determined |
| S13_extracted_40497 | Not-determined |
| S13_extracted_40504 | Low-quality |
| S13_extracted_40519 | Low-quality |
| S13_extracted_40528 | Not-determined |
| S13_extracted_40541 | Not-determined |
| S13_extracted_40543 | Not-determined |
| S13_extracted_40558 | Low-quality |
| S13_extracted_40562 | Not-determined |
| S13_extracted_40565 | Not-determined |
| S13_extracted_40570 | Not-determined |
| S13_extracted_40574 | Not-determined |
| S13_extracted_40577 | Not-determined |
| S13_extracted_40592 | Not-determined |
| S13_extracted_40595 | Not-determined |
| S13_extracted_40607 | Not-determined |
| S13_extracted_40612 | Not-determined |
| S13_extracted_40628 | Not-determined |
| S13_extracted_40639 | Not-determined |
| S13_extracted_40648 | Not-determined |
| S13_extracted_40661 | Not-determined |
| S13_extracted_40676 | Not-determined |
| S13_extracted_40679 | Not-determined |
| S13_extracted_40680 | Not-determined |
| S13_extracted_40689 | Not-determined |
| S13_extracted_40690 | Not-determined |
| S13_extracted_40706 | Not-determined |
| S13_extracted_40709 | Low-quality |
| S13_extracted_40720 | Not-determined |
| S13_extracted_40723 | Not-determined |
| S13_extracted_40730 | Not-determined |
| S13_extracted_40732 | Low-quality |
| S13_extracted_40738 | Not-determined |
| S13_extracted_40748 | Not-determined |
| S13_extracted_40763 | Not-determined |
| S13_extracted_40766 | Not-determined |
| S13_extracted_40772 | Not-determined |
| S13_extracted_40785 | Not-determined |
| S13_extracted_40791 | Not-determined |
| S13_extracted_40798 | Not-determined |
| S13_extracted_40804 | Not-determined |
| S13_extracted_40823 | Not-determined |
| S13_extracted_40837 | Not-determined |
| S13_extracted_40840 | Not-determined |
| S13_extracted_40846 | Not-determined |
| S13_extracted_40850 | Not-determined |
| S13_extracted_40877 | Not-determined |
| S13_extracted_40879 | Not-determined |
| S13_extracted_40899 | Not-determined |
| S13_extracted_40900 | Not-determined |
| S13_extracted_40901 | Not-determined |
| S13_extracted_40925 | Not-determined |
| S13_extracted_40935 | Not-determined |
| S13_extracted_40937 | Not-determined |
| S13_extracted_40956 | Not-determined |
| S13_extracted_40958 | Not-determined |
| S13_extracted_40977 | Not-determined |
| S13_extracted_40983 | Not-determined |
| S13_extracted_40990 | Not-determined |
| S13_extracted_41000 | Not-determined |
| S13_extracted_41011 | Not-determined |
| S13_extracted_41015 | Not-determined |
| S13_extracted_41016 | Not-determined |
| S13_extracted_41021 | Not-determined |
| S13_extracted_41024 | Not-determined |
| S13_extracted_41028 | Low-quality |
| S13_extracted_41049 | Low-quality |
| S13_extracted_41076 | Not-determined |
| S13_extracted_41081 | Low-quality |
| S13_extracted_41082 | Not-determined |
| S13_extracted_41093 | Not-determined |
| S13_extracted_41104 | Not-determined |
| S13_extracted_41105 | Not-determined |
| S13_extracted_41106 | Not-determined |
| S13_extracted_41108 | Not-determined |
| S13_extracted_41126 | Not-determined |
| S13_extracted_41127 | Not-determined |
| S13_extracted_41138 | Not-determined |
| S13_extracted_41144 | Not-determined |
| S13_extracted_41155 | Low-quality |
| S13_extracted_41158 | Not-determined |
| S13_extracted_41163 | Low-quality |
| S13_extracted_41170 | Not-determined |
| S13_extracted_41174 | Not-determined |
| S13_extracted_41183 | Not-determined |
| S13_extracted_41196 | Not-determined |
| S13_extracted_41207 | Low-quality |
| S13_extracted_41209 | Not-determined |
| S13_extracted_41221 | Not-determined |
| S13_extracted_41226 | Not-determined |
| S13_extracted_41231 | Not-determined |
| S13_extracted_41245 | Not-determined |
| S13_extracted_41260 | Low-quality |
| S13_extracted_41273 | Not-determined |
| S13_extracted_41276 | Not-determined |
| S13_extracted_41290 | Not-determined |
| S13_extracted_41301 | Not-determined |
| S13_extracted_41306 | Not-determined |
| S13_extracted_41308 | Not-determined |
| S13_extracted_41311 | Not-determined |
| S13_extracted_41321 | Not-determined |
| S13_extracted_41326 | Not-determined |
| S13_extracted_41334 | Not-determined |
| S13_extracted_41345 | Not-determined |
| S13_extracted_41371 | Not-determined |
| S13_extracted_41374 | Not-determined |
| S13_extracted_41404 | Not-determined |
| S13_extracted_41413 | Not-determined |
| S13_extracted_41414 | Not-determined |
| S13_extracted_41420 | Not-determined |
| S13_extracted_41422 | Not-determined |
| S13_extracted_41432 | Not-determined |
| S13_extracted_41444 | Not-determined |
| S13_extracted_41449 | Not-determined |
| S13_extracted_41451 | Not-determined |
| S13_extracted_41466 | Not-determined |
| S13_extracted_41469 | Not-determined |
| S13_extracted_41471 | Not-determined |
| S13_extracted_41475 | Not-determined |
| S13_extracted_41480 | Not-determined |
| S13_extracted_41492 | Not-determined |
| S13_extracted_41500 | Not-determined |
| S13_extracted_41501 | Not-determined |
| S13_extracted_41514 | Not-determined |
| S13_extracted_41530 | Not-determined |
| S13_extracted_41539 | Not-determined |
| S13_extracted_41541 | Not-determined |
| S13_extracted_41542 | Not-determined |
| S13_extracted_41547 | Not-determined |
| S13_extracted_41552 | Not-determined |
| S13_extracted_41565 | Low-quality |
| S13_extracted_41566 | Not-determined |
| S13_extracted_41576 | Not-determined |
| S13_extracted_41578 | Not-determined |
| S13_extracted_41582 | Not-determined |
| S13_extracted_41584 | Not-determined |
| S13_extracted_41587 | Low-quality |
| S13_extracted_41595 | Not-determined |
| S13_extracted_41598 | Not-determined |
| S13_extracted_41601 | Not-determined |
| S13_extracted_41610 | Not-determined |
| S13_extracted_41620 | Not-determined |
| S13_extracted_41637 | Not-determined |
| S13_extracted_41641 | Not-determined |
| S13_extracted_41644 | Not-determined |
| S13_extracted_41651 | Not-determined |
| S13_extracted_41655 | Not-determined |
| S13_extracted_41667 | Not-determined |
| S13_extracted_41670 | Not-determined |
| S13_extracted_41673 | Not-determined |
| S13_extracted_41682 | Not-determined |
| S13_extracted_41683 | Not-determined |
| S13_extracted_41685 | Not-determined |
| S13_extracted_41693 | Not-determined |
| S13_extracted_41697 | Not-determined |
| S13_extracted_41705 | Low-quality |
| S13_extracted_41714 | Not-determined |
| S13_extracted_41716 | Low-quality |
| S13_extracted_41760 | Not-determined |
| S13_extracted_41771 | Not-determined |
| S13_extracted_41779 | Not-determined |
| S13_extracted_41783 | Not-determined |
| S13_extracted_41800 | Not-determined |
| S13_extracted_41802 | Not-determined |
| S13_extracted_41804 | Not-determined |
| S13_extracted_41805 | Not-determined |
| S13_extracted_41808 | Not-determined |
| S13_extracted_41839 | Not-determined |
| S13_extracted_41842 | Not-determined |
| S13_extracted_41848 | Low-quality |
| S13_extracted_41852 | Not-determined |
| S13_extracted_41866 | Low-quality |
| S13_extracted_41869 | Not-determined |
| S13_extracted_41890 | Not-determined |
| S13_extracted_41892 | Not-determined |
| S13_extracted_41897 | Low-quality |
| S13_extracted_41905 | Not-determined |
| S13_extracted_41907 | Not-determined |
| S13_extracted_41917 | Not-determined |
| S13_extracted_41921 | Not-determined |
| S13_extracted_41937 | Not-determined |
| S13_extracted_41952 | Not-determined |
| S13_extracted_41961 | Not-determined |
| S13_extracted_41974 | Not-determined |
| S13_extracted_41980 | Not-determined |
| S13_extracted_41989 | Not-determined |
| S13_extracted_41994 | Low-quality |
| S13_extracted_41997 | Not-determined |
| S13_extracted_42018 | Not-determined |
| S13_extracted_42020 | Not-determined |
| S13_extracted_42021 | Low-quality |
| S13_extracted_42022 | Not-determined |
| S13_extracted_42023 | Not-determined |
| S13_extracted_42054 | Low-quality |
| S13_extracted_42066 | Not-determined |
| S13_extracted_42087 | Not-determined |
| S13_extracted_42088 | Not-determined |
| S13_extracted_42098 | Not-determined |
| S13_extracted_42104 | Not-determined |
| S13_extracted_42125 | Not-determined |
| S13_extracted_42127 | Low-quality |
| S13_extracted_42135 | Not-determined |
| S13_extracted_42137 | Not-determined |
| S13_extracted_42148 | Not-determined |
| S13_extracted_42151 | Not-determined |
| S13_extracted_42160 | Not-determined |
| S13_extracted_42232 | Not-determined |
| S13_extracted_42238 | Not-determined |
| S13_extracted_42242 | Not-determined |
| S13_extracted_42248 | Not-determined |
| S13_extracted_42250 | Not-determined |
| S13_extracted_42252 | Not-determined |
| S13_extracted_42269 | Not-determined |
| S13_extracted_42276 | Not-determined |
| S13_extracted_42279 | Low-quality |
| S13_extracted_42285 | Not-determined |
| S13_extracted_42288 | Not-determined |
| S13_extracted_42296 | Low-quality |
| S13_extracted_42301 | Low-quality |
| S13_extracted_42317 | Not-determined |
| S13_extracted_42318 | Not-determined |
| S13_extracted_42321 | Low-quality |
| S13_extracted_42330 | Not-determined |
| S13_extracted_42332 | Not-determined |
| S13_extracted_42337 | Not-determined |
| S13_extracted_42339 | Not-determined |
| S13_extracted_42349 | Not-determined |
| S13_extracted_42355 | Not-determined |
| S13_extracted_42359 | Not-determined |
| S13_extracted_42367 | Not-determined |
| S13_extracted_42368 | Not-determined |
| S13_extracted_42374 | Not-determined |
| S13_extracted_42378 | Low-quality |
| S13_extracted_42382 | Not-determined |
| S13_extracted_42384 | Not-determined |
| S13_extracted_42388 | Not-determined |
| S13_extracted_42394 | Not-determined |
| S13_extracted_42398 | Not-determined |
| S13_extracted_42400 | Not-determined |
| S13_extracted_42416 | Not-determined |
| S13_extracted_42434 | Not-determined |
| S13_extracted_42435 | Not-determined |
| S13_extracted_42450 | Not-determined |
| S13_extracted_42452 | Not-determined |
| S13_extracted_42467 | Not-determined |
| S13_extracted_42486 | Not-determined |
| S13_extracted_42489 | Not-determined |
| S13_extracted_42501 | Not-determined |
| S13_extracted_42513 | Not-determined |
| S13_extracted_42534 | Not-determined |
| S13_extracted_42551 | Not-determined |
| S13_extracted_42552 | Not-determined |
| S13_extracted_42553 | Not-determined |
| S13_extracted_42564 | Not-determined |
| S13_extracted_42571 | Not-determined |
| S13_extracted_42590 | Not-determined |
| S13_extracted_42593 | Not-determined |
| S13_extracted_42607 | Not-determined |
| S13_extracted_42610 | Not-determined |
| S13_extracted_42612 | Not-determined |
| S13_extracted_42649 | Low-quality |
| S13_extracted_42655 | Not-determined |
| S13_extracted_42657 | Not-determined |
| S13_extracted_42672 | Not-determined |
| S13_extracted_42686 | Low-quality |
| S13_extracted_42690 | Not-determined |
| S13_extracted_42691 | Not-determined |
| S13_extracted_42692 | Not-determined |
| S13_extracted_42718 | Not-determined |
| S13_extracted_42722 | Low-quality |
| S13_extracted_42724 | Not-determined |
| S13_extracted_42731 | Not-determined |
| S13_extracted_42735 | Not-determined |
| S13_extracted_42741 | Not-determined |
| S13_extracted_42749 | Not-determined |
| S13_extracted_42750 | Low-quality |
| S13_extracted_42752 | Low-quality |
| S13_extracted_42753 | Not-determined |
| S13_extracted_42754 | Not-determined |
| S13_extracted_42759 | Not-determined |
| S13_extracted_42760 | Low-quality |
| S13_extracted_42776 | Low-quality |
| S13_extracted_42788 | Not-determined |
| S13_extracted_42806 | Not-determined |
| S13_extracted_42818 | Not-determined |
| S13_extracted_42837 | Not-determined |
| S13_extracted_42855 | Not-determined |
| S13_extracted_42859 | Not-determined |
| S13_extracted_42861 | Not-determined |
| S13_extracted_42864 | Not-determined |
| S13_extracted_42884 | Not-determined |
| S13_extracted_42893 | Not-determined |
| S13_extracted_42897 | Not-determined |
| S13_extracted_42918 | Not-determined |
| S13_extracted_42920 | Low-quality |
| S13_extracted_42949 | Not-determined |
| S13_extracted_42956 | Not-determined |
| S13_extracted_42957 | Not-determined |
| S13_extracted_42977 | Not-determined |
| S13_extracted_42981 | Not-determined |
| S13_extracted_42987 | Not-determined |
| S13_extracted_42988 | Not-determined |
| S13_extracted_42991 | Not-determined |
| S13_extracted_43001 | Not-determined |
| S13_extracted_43004 | Not-determined |
| S13_extracted_43056 | Not-determined |
| S13_extracted_43057 | Not-determined |
| S13_extracted_43072 | Not-determined |
| S13_extracted_43087 | Low-quality |
| S13_extracted_43089 | Not-determined |
| S13_extracted_43100 | Low-quality |
| S13_extracted_43112 | Not-determined |
| S13_extracted_43115 | Not-determined |
| S13_extracted_43116 | Not-determined |
| S13_extracted_43118 | Not-determined |
| S13_extracted_43122 | Not-determined |
| S13_extracted_43129 | Not-determined |
| S13_extracted_43131 | Not-determined |
| S13_extracted_43141 | Not-determined |
| S13_extracted_43145 | Not-determined |
| S13_extracted_43146 | Not-determined |
| S13_extracted_43171 | Not-determined |
| S13_extracted_43190 | Not-determined |
| S13_extracted_43193 | Not-determined |
| S13_extracted_43195 | Low-quality |
| S13_extracted_43198 | Not-determined |
| S13_extracted_43213 | Not-determined |
| S13_extracted_43221 | Not-determined |
| S13_extracted_43229 | Not-determined |
| S13_extracted_43263 | Not-determined |
| S13_extracted_43265 | Not-determined |
| S13_extracted_43268 | Not-determined |
| S13_extracted_43272 | Not-determined |
| S13_extracted_43289 | Not-determined |
| S13_extracted_43336 | Not-determined |
| S13_extracted_43363 | Not-determined |
| S13_extracted_43366 | Not-determined |
| S13_extracted_43385 | Not-determined |
| S13_extracted_43391 | Low-quality |
| S13_extracted_43394 | Not-determined |
| S13_extracted_43422 | Not-determined |
| S13_extracted_43447 | Not-determined |
| S13_extracted_43455 | Low-quality |
| S13_extracted_43456 | Not-determined |
| S13_extracted_43457 | Low-quality |
| S13_extracted_43478 | Not-determined |
| S13_extracted_43480 | Low-quality |
| S13_extracted_43492 | Not-determined |
| S13_extracted_43497 | Not-determined |
| S13_extracted_43500 | Not-determined |
| S13_extracted_43506 | Low-quality |
| S13_extracted_43507 | Not-determined |
| S13_extracted_43514 | Not-determined |
| S13_extracted_43532 | Low-quality |
| S13_extracted_43538 | Not-determined |
| S13_extracted_43545 | Not-determined |
| S13_extracted_43549 | Not-determined |
| S13_extracted_43563 | Not-determined |
| S13_extracted_43580 | Not-determined |
| S13_extracted_43586 | Not-determined |
| S13_extracted_43590 | Not-determined |
| S13_extracted_43600 | Not-determined |
| S13_extracted_43601 | Not-determined |
| S13_extracted_43609 | Not-determined |
| S13_extracted_43627 | Not-determined |
| S13_extracted_43629 | Not-determined |
| S13_extracted_43633 | Not-determined |
| S13_extracted_43636 | Not-determined |
| S13_extracted_43637 | Not-determined |
| S13_extracted_43657 | Not-determined |
| S13_extracted_43669 | Not-determined |
| S13_extracted_43691 | Not-determined |
| S13_extracted_43693 | Not-determined |
| S13_extracted_43701 | Not-determined |
| S13_extracted_43713 | Not-determined |
| S13_extracted_43720 | Not-determined |
| S13_extracted_43725 | Not-determined |
| S13_extracted_43749 | Not-determined |
| S13_extracted_43750 | Not-determined |
| S13_extracted_43755 | Not-determined |
| S13_extracted_43770 | Not-determined |
| S13_extracted_43788 | Low-quality |
| S13_extracted_43790 | Not-determined |
| S13_extracted_43803 | Not-determined |
| S13_extracted_43805 | Not-determined |
| S13_extracted_43817 | Not-determined |
| S13_extracted_43829 | Not-determined |
| S13_extracted_43843 | Not-determined |
| S13_extracted_43854 | Not-determined |
| S13_extracted_43858 | Not-determined |
| S13_extracted_43871 | Not-determined |
| S13_extracted_43882 | Not-determined |
| S13_extracted_43884 | Not-determined |
| S13_extracted_43889 | Not-determined |
| S13_extracted_43892 | Not-determined |
| S13_extracted_43910 | Not-determined |
| S13_extracted_43925 | Not-determined |
| S13_extracted_43928 | Not-determined |
| S13_extracted_43952 | Not-determined |
| S13_extracted_43972 | Not-determined |
| S13_extracted_43973 | Not-determined |
| S13_extracted_43983 | Not-determined |
| S13_extracted_43988 | Not-determined |
| S13_extracted_43994 | Not-determined |
| S13_extracted_43995 | Not-determined |
| S13_extracted_44034 | Not-determined |
| S13_extracted_44035 | Not-determined |
| S13_extracted_44040 | Not-determined |
| S13_extracted_44054 | Not-determined |
| S13_extracted_44061 | Not-determined |
| S13_extracted_44078 | Not-determined |
| S13_extracted_44080 | Low-quality |
| S13_extracted_44086 | Low-quality |
| S13_extracted_44108 | Not-determined |
| S13_extracted_44114 | Not-determined |
| S13_extracted_44115 | Not-determined |
| S13_extracted_44135 | Not-determined |
| S13_extracted_44139 | Not-determined |
| S13_extracted_44140 | Not-determined |
| S13_extracted_44142 | Not-determined |
| S13_extracted_44148 | Not-determined |
| S13_extracted_44173 | Not-determined |
| S13_extracted_44176 | Not-determined |
| S13_extracted_44181 | Not-determined |
| S13_extracted_44184 | Not-determined |
| S13_extracted_44214 | Not-determined |
| S13_extracted_44220 | Not-determined |
| S13_extracted_44251 | Not-determined |
| S13_extracted_44254 | Low-quality |
| S13_extracted_44257 | Not-determined |
| S13_extracted_44270 | Not-determined |
| S13_extracted_44274 | Not-determined |
| S13_extracted_44282 | Not-determined |
| S13_extracted_44300 | Not-determined |
| S13_extracted_44301 | Not-determined |
| S13_extracted_44304 | Not-determined |
| S13_extracted_44315 | Not-determined |
| S13_extracted_44327 | Not-determined |
| S13_extracted_44331 | Not-determined |
| S13_extracted_44345 | Not-determined |
| S13_extracted_44349 | Not-determined |
| S13_extracted_44353 | Not-determined |
| S13_extracted_44359 | Not-determined |
| S13_extracted_44367 | Not-determined |
| S13_extracted_44374 | Low-quality |
| S13_extracted_44381 | Not-determined |
| S13_extracted_44394 | Not-determined |
| S13_extracted_44395 | Not-determined |
| S13_extracted_44401 | Not-determined |
| S13_extracted_44402 | Not-determined |
| S13_extracted_44411 | Not-determined |
| S13_extracted_44419 | Not-determined |
| S13_extracted_44421 | Not-determined |
| S13_extracted_44422 | Low-quality |
| S13_extracted_44446 | Not-determined |
| S13_extracted_44452 | Not-determined |
| S13_extracted_44465 | Not-determined |
| S13_extracted_44467 | Not-determined |
| S13_extracted_44508 | Low-quality |
| S13_extracted_44516 | Not-determined |
| S13_extracted_44536 | Not-determined |
| S13_extracted_44546 | Not-determined |
| S13_extracted_44551 | Not-determined |
| S13_extracted_44560 | Not-determined |
| S13_extracted_44570 | Not-determined |
| S13_extracted_44576 | Not-determined |
| S13_extracted_44577 | Low-quality |
| S13_extracted_44585 | Not-determined |
| S13_extracted_44597 | Not-determined |
| S13_extracted_44603 | Not-determined |
| S13_extracted_44606 | Low-quality |
| S13_extracted_44614 | Not-determined |
| S13_extracted_44626 | Not-determined |
| S13_extracted_44639 | Not-determined |
| S13_extracted_44661 | Low-quality |
| S13_extracted_44664 | Not-determined |
| S13_extracted_44678 | Not-determined |
| S13_extracted_44679 | Not-determined |
| S13_extracted_44707 | Not-determined |
| S13_extracted_44708 | Not-determined |
| S13_extracted_44719 | Not-determined |
| S13_extracted_44721 | Not-determined |
| S13_extracted_44725 | Not-determined |
| S13_extracted_44745 | Not-determined |
| S13_extracted_44748 | Not-determined |
| S13_extracted_44753 | Not-determined |
| S13_extracted_44754 | Not-determined |
| S13_extracted_44765 | Not-determined |
| S13_extracted_44768 | Not-determined |
| S13_extracted_44771 | Not-determined |
| S13_extracted_44778 | Not-determined |
| S13_extracted_44786 | Not-determined |
| S13_extracted_44809 | Not-determined |
| S13_extracted_44813 | Not-determined |
| S13_extracted_44822 | Not-determined |
| S13_extracted_44824 | Not-determined |
| S13_extracted_44832 | Not-determined |
| S13_extracted_44836 | Not-determined |
| S13_extracted_44869 | Not-determined |
| S10_vs_2898||full | High-quality |
| S13_extracted_44874 | Low-quality |
| S13_extracted_44879 | Not-determined |
| S13_extracted_44881 | Not-determined |
| S13_extracted_44893 | Not-determined |
| S13_extracted_44907 | Not-determined |
| S13_extracted_44911 | Not-determined |
| S13_extracted_44920 | Not-determined |
| S13_extracted_44922 | Low-quality |
| S13_extracted_44932 | Not-determined |
| S13_extracted_44936 | Not-determined |
| S13_extracted_44940 | Not-determined |
| S13_extracted_44944 | Not-determined |
| S13_extracted_44965 | Not-determined |
| S13_extracted_44968 | Not-determined |
| S13_extracted_44971 | Low-quality |
| S13_extracted_44986 | Not-determined |
| S13_extracted_44996 | Not-determined |
| S13_extracted_45020 | Not-determined |
| S13_extracted_45026 | Not-determined |
| S13_extracted_45027 | Not-determined |
| S13_extracted_45035 | Not-determined |
| S13_extracted_45037 | Not-determined |
| S13_extracted_45039 | Not-determined |
| S13_extracted_45041 | Not-determined |
| S13_extracted_45060 | Low-quality |
| S13_extracted_45069 | Not-determined |
| S13_extracted_45086 | Not-determined |
| S13_extracted_45087 | Not-determined |
| S13_extracted_45094 | Not-determined |
| S13_extracted_45097 | Not-determined |
| S13_extracted_45102 | Not-determined |
| S13_extracted_45106 | Not-determined |
| S13_extracted_45136 | Not-determined |
| S13_extracted_45155 | Low-quality |
| S13_extracted_45164 | Not-determined |
| S13_extracted_45165 | Not-determined |
| S13_extracted_45177 | Not-determined |
| S13_extracted_45180 | Not-determined |
| S13_extracted_45181 | Not-determined |
| S13_extracted_45182 | Not-determined |
| S13_extracted_45186 | Not-determined |
| S13_extracted_45190 | Not-determined |
| S13_extracted_45193 | Not-determined |
| S13_extracted_45216 | Not-determined |
| S13_extracted_45227 | Not-determined |
| S13_extracted_45230 | Not-determined |
| S13_extracted_45236 | Not-determined |
| S13_extracted_45247 | Not-determined |
| S13_extracted_45248 | Not-determined |
| S13_extracted_45259 | Not-determined |
| S13_extracted_45265 | Not-determined |
| S13_extracted_45281 | Not-determined |
| S13_extracted_45283 | Low-quality |
| S13_extracted_45297 | Low-quality |
| S13_extracted_45301 | Not-determined |
| S13_extracted_45319 | Low-quality |
| S13_extracted_45321 | Low-quality |
| S13_extracted_45326 | Not-determined |
| S13_extracted_45330 | Not-determined |
| S13_extracted_45331 | Not-determined |
| S13_extracted_45336 | Not-determined |
| S13_extracted_45346 | Not-determined |

| | |
|---|---|
| S13_extracted_45350 | Not-determined |
| S13_extracted_45357 | Not-determined |
| S13_extracted_45386 | Not-determined |
| S13_extracted_45393 | Not-determined |
| S13_extracted_45405 | Not-determined |
| S13_extracted_45409 | Not-determined |
| S13_extracted_45410 | Low-quality |
| S13_extracted_45424 | Not-determined |
| S13_extracted_45426 | Not-determined |
| S13_extracted_45429 | Not-determined |
| S13_extracted_45435 | Not-determined |
| S13_extracted_45437 | Not-determined |
| S13_extracted_45442 | Not-determined |
| S13_extracted_45462 | Low-quality |
| S13_extracted_45468 | Not-determined |
| S13_extracted_45473 | Not-determined |
| S13_extracted_45477 | Not-determined |
| S13_extracted_45485 | Not-determined |
| S13_extracted_45492 | Low-quality |
| S13_extracted_45494 | Low-quality |
| S13_extracted_45498 | Not-determined |
| S13_extracted_45502 | Low-quality |
| S13_extracted_45512 | Not-determined |
| S13_extracted_45517 | Low-quality |
| S13_extracted_45522 | Not-determined |
| S13_extracted_45524 | Not-determined |
| S13_extracted_45526 | Not-determined |
| S13_extracted_45529 | Not-determined |
| S13_extracted_45535 | Low-quality |
| S13_extracted_45541 | Not-determined |
| S13_extracted_45544 | Not-determined |
| S13_extracted_45567 | Not-determined |
| S13_extracted_45568 | Not-determined |
| S13_extracted_45575 | Low-quality |
| S13_extracted_45582 | Not-determined |
| S13_extracted_45611 | Low-quality |
| S13_extracted_45617 | Not-determined |
| S13_extracted_45646 | Not-determined |
| S13_extracted_45648 | Not-determined |
| S13_extracted_45650 | Not-determined |
| S13_extracted_45654 | Not-determined |
| S13_extracted_45660 | Not-determined |
| S13_extracted_45670 | Not-determined |
| S13_extracted_45685 | Not-determined |
| S13_extracted_45696 | Low-quality |
| S13_extracted_45707 | Not-determined |
| S13_extracted_45713 | Not-determined |
| S13_extracted_45717 | Not-determined |
| S13_extracted_45723 | Not-determined |
| S13_extracted_45734 | Not-determined |
| S13_extracted_45747 | Not-determined |
| S13_extracted_45750 | Not-determined |
| S13_extracted_45763 | Not-determined |
| S13_extracted_45779 | Not-determined |
| S13_extracted_45787 | Low-quality |
| S13_extracted_45790 | Not-determined |
| S13_extracted_45794 | Not-determined |
| S13_extracted_45806 | Not-determined |
| S13_extracted_45807 | Low-quality |
| S13_extracted_45808 | Not-determined |
| S13_extracted_45811 | Not-determined |
| S13_extracted_45814 | Not-determined |
| S13_extracted_45824 | Not-determined |
| S13_extracted_45826 | Not-determined |
| S13_extracted_45828 | Low-quality |
| S13_extracted_45830 | Not-determined |
| S13_extracted_45838 | Not-determined |
| S13_extracted_45841 | Not-determined |
| S13_extracted_45844 | Not-determined |
| S13_extracted_45846 | Low-quality |
| S13_extracted_45860 | Not-determined |
| S13_extracted_45874 | Low-quality |
| S13_extracted_45875 | Not-determined |
| S13_extracted_45896 | Not-determined |
| S13_extracted_45897 | Not-determined |
| S13_extracted_45898 | Not-determined |
| S13_extracted_45906 | Not-determined |
| S13_extracted_45907 | Low-quality |
| S13_extracted_45910 | Low-quality |
| S13_extracted_45920 | Not-determined |
| S13_extracted_45926 | Not-determined |
| S13_extracted_45928 | Not-determined |
| S13_extracted_45941 | Not-determined |
| S13_extracted_45964 | Not-determined |
| S13_extracted_45966 | Not-determined |
| S13_extracted_45986 | Not-determined |
| S13_extracted_45989 | Not-determined |
| S13_extracted_45994 | Low-quality |
| S13_extracted_45999 | Not-determined |
| S13_extracted_46003 | Low-quality |
| S13_extracted_46014 | Not-determined |
| S13_extracted_46028 | Not-determined |
| S13_extracted_46053 | Low-quality |
| S13_extracted_46055 | Not-determined |
| S13_extracted_46061 | Not-determined |
| S13_extracted_46084 | Not-determined |
| S13_extracted_46088 | Not-determined |
| S13_extracted_46116 | Not-determined |
| S13_extracted_46124 | Not-determined |
| S13_extracted_46139 | Not-determined |
| S13_extracted_46140 | Not-determined |
| S13_extracted_46143 | Not-determined |
| S13_extracted_46156 | Not-determined |
| S13_extracted_46158 | Not-determined |
| S13_extracted_46177 | Not-determined |
| S13_extracted_46186 | Not-determined |
| S13_extracted_46190 | Not-determined |
| S13_extracted_46203 | Not-determined |
| S13_extracted_46225 | Not-determined |
| S13_extracted_46231 | Not-determined |
| S13_extracted_46234 | Not-determined |
| S13_extracted_46239 | Low-quality |
| S13_extracted_46245 | Not-determined |
| S13_extracted_46248 | Not-determined |
| S13_extracted_46255 | Not-determined |
| S13_extracted_46260 | Not-determined |
| S13_extracted_46264 | Not-determined |
| S13_extracted_46295 | Not-determined |
| S13_extracted_46326 | Not-determined |
| S13_extracted_46334 | Not-determined |
| S13_extracted_46337 | Not-determined |
| S13_extracted_46343 | Not-determined |
| S13_extracted_46353 | Not-determined |
| S13_extracted_46356 | Not-determined |
| S13_extracted_46358 | Not-determined |
| S13_extracted_46367 | Not-determined |
| S13_extracted_46368 | Not-determined |
| S13_extracted_46397 | Not-determined |
| S13_extracted_46416 | Not-determined |
| S13_extracted_46428 | Not-determined |
| S13_extracted_46434 | Not-determined |
| S13_extracted_46435 | Low-quality |
| S13_extracted_46447 | Low-quality |
| S13_extracted_46473 | Not-determined |
| S13_extracted_46479 | Not-determined |
| S13_extracted_46486 | Not-determined |
| S13_extracted_46525 | Not-determined |
| S13_extracted_46532 | Not-determined |
| S13_extracted_46543 | Not-determined |
| S13_extracted_46561 | Not-determined |
| S13_extracted_46568 | Not-determined |
| S13_extracted_46584 | Not-determined |
| S13_extracted_46586 | Not-determined |
| S13_extracted_46587 | Low-quality |
| S13_extracted_46602 | Not-determined |
| S13_extracted_46617 | Not-determined |
| S13_extracted_46620 | Not-determined |
| S13_extracted_46622 | Not-determined |
| S13_extracted_46626 | Not-determined |
| S13_extracted_46634 | Not-determined |
| S13_extracted_46646 | Low-quality |
| S13_extracted_46648 | Not-determined |
| S13_extracted_46657 | Not-determined |
| S13_extracted_46670 | Not-determined |
| S13_extracted_46696 | Not-determined |
| S13_extracted_46700 | Not-determined |
| S13_extracted_46708 | Low-quality |
| S13_extracted_46709 | Not-determined |
| S13_extracted_46730 | Not-determined |
| S13_extracted_46737 | Not-determined |
| S13_extracted_46738 | Not-determined |
| S13_extracted_46739 | Not-determined |
| S13_extracted_46751 | Not-determined |
| S13_extracted_46753 | Not-determined |
| S13_extracted_46760 | Not-determined |
| S13_extracted_46767 | Not-determined |
| S13_extracted_46772 | Not-determined |
| S13_extracted_46792 | Not-determined |
| S13_extracted_46795 | Not-determined |
| S13_extracted_46813 | Not-determined |
| S13_extracted_46829 | Not-determined |
| S13_extracted_46834 | Low-quality |
| S13_extracted_46838 | Not-determined |
| S13_extracted_46840 | Not-determined |
| S13_extracted_46848 | Not-determined |
| S13_extracted_46850 | Not-determined |
| S13_extracted_46865 | Not-determined |
| S13_extracted_46889 | Not-determined |
| S13_extracted_46897 | Not-determined |
| S13_extracted_46898 | Not-determined |
| S13_extracted_46911 | Not-determined |
| S13_extracted_46924 | Not-determined |
| S13_extracted_46930 | Not-determined |
| S13_extracted_46932 | Not-determined |
| S13_extracted_46935 | Low-quality |
| S13_extracted_46956 | Not-determined |
| S13_extracted_46957 | Not-determined |
| S13_extracted_46962 | Not-determined |
| S13_extracted_46972 | Not-determined |
| S13_extracted_46973 | Not-determined |
| S13_extracted_46985 | Not-determined |
| S13_extracted_46988 | Not-determined |
| S13_extracted_46989 | Low-quality |
| S13_extracted_46990 | Not-determined |
| S13_extracted_46998 | Not-determined |
| S13_extracted_47010 | Low-quality |
| S13_extracted_47014 | Low-quality |
| S13_extracted_47015 | Not-determined |
| S13_extracted_47017 | Not-determined |
| S13_extracted_47025 | Not-determined |
| S13_extracted_47027 | Low-quality |
| S13_extracted_47046 | Low-quality |
| S13_extracted_47053 | Low-quality |
| S13_extracted_47066 | Low-quality |
| S13_extracted_47079 | Not-determined |
| S13_extracted_47090 | Not-determined |
| S13_extracted_47091 | Low-quality |
| S13_extracted_47097 | Not-determined |
| S13_extracted_47098 | Low-quality |
| S13_extracted_47113 | Not-determined |
| S13_extracted_47116 | Not-determined |
| S13_extracted_47120 | Not-determined |
| S13_extracted_47125 | Not-determined |
| S13_extracted_47131 | Low-quality |
| S13_extracted_47173 | Not-determined |
| S13_extracted_47174 | Low-quality |
| S13_extracted_47182 | Not-determined |
| S13_extracted_47184 | Not-determined |
| S13_extracted_47191 | Not-determined |
| S13_extracted_47196 | Not-determined |
| S13_extracted_47197 | Not-determined |
| S13_extracted_47201 | Not-determined |
| S13_extracted_47221 | Not-determined |
| S13_extracted_47222 | Not-determined |
| S13_extracted_47223 | Not-determined |
| S13_extracted_47231 | Not-determined |
| S13_extracted_47251 | Not-determined |
| S13_extracted_47277 | Not-determined |
| S13_extracted_47296 | Not-determined |
| S13_extracted_47308 | Not-determined |
| S13_extracted_47322 | Not-determined |
| S13_extracted_47328 | Not-determined |
| S13_extracted_47332 | Not-determined |
| S13_extracted_47343 | Not-determined |
| S13_extracted_47348 | Not-determined |
| S13_extracted_47350 | Low-quality |
| S13_extracted_47356 | Not-determined |
| S13_extracted_47357 | Low-quality |
| S13_extracted_47369 | Not-determined |
| S13_extracted_47370 | Not-determined |
| S13_extracted_47384 | Low-quality |
| S13_extracted_47405 | Not-determined |
| S13_extracted_47411 | Low-quality |
| S13_extracted_47428 | Not-determined |
| S13_extracted_47437 | Not-determined |
| S13_extracted_47444 | Not-determined |
| S13_extracted_47455 | Not-determined |
| S13_extracted_47456 | Not-determined |
| S13_extracted_47460 | Not-determined |
| S13_extracted_47466 | Not-determined |
| S13_extracted_47469 | Not-determined |
| S13_extracted_47487 | Not-determined |
| S13_extracted_47489 | Not-determined |
| S13_extracted_47491 | Not-determined |
| S13_extracted_47493 | Not-determined |
| S13_extracted_47494 | Low-quality |
| S13_extracted_47497 | Not-determined |
| S13_extracted_47505 | Low-quality |
| S13_extracted_47511 | Not-determined |
| S13_extracted_47516 | Low-quality |
| S13_extracted_47522 | Not-determined |
| S13_extracted_47545 | Low-quality |
| S13_extracted_47553 | Not-determined |
| S13_extracted_47556 | Low-quality |
| S13_extracted_47562 | Not-determined |
| S13_extracted_47569 | Not-determined |
| S13_extracted_47570 | Not-determined |
| S13_extracted_47574 | Not-determined |
| S13_extracted_47576 | Low-quality |
| S13_extracted_47591 | Not-determined |
| S13_extracted_47614 | Not-determined |
| S13_extracted_47626 | Not-determined |
| S13_extracted_47662 | Not-determined |
| S13_extracted_47667 | Not-determined |
| S13_extracted_47668 | Low-quality |
| S13_extracted_47677 | Not-determined |
| S13_extracted_47715 | Not-determined |
| S13_extracted_47720 | Not-determined |
| S13_extracted_47725 | Not-determined |
| S13_extracted_47731 | Not-determined |
| S13_extracted_47737 | Not-determined |
| S13_extracted_47739 | Not-determined |
| S13_extracted_47740 | Not-determined |
| S13_extracted_47747 | Not-determined |
| S13_extracted_47754 | Not-determined |
| S13_extracted_47779 | Not-determined |
| S13_extracted_47791 | Not-determined |
| S13_extracted_47794 | Not-determined |
| S13_extracted_47802 | Not-determined |
| S13_extracted_47868 | Not-determined |
| S13_extracted_47883 | Not-determined |
| S13_extracted_47885 | Not-determined |
| S13_extracted_47907 | Not-determined |
| S13_extracted_47910 | Not-determined |
| S13_extracted_47913 | Not-determined |
| S13_extracted_47937 | Not-determined |
| S13_extracted_47938 | Low-quality |
| S13_extracted_47946 | Not-determined |
| S13_extracted_47960 | Low-quality |
| S13_extracted_47966 | Not-determined |
| S13_extracted_47971 | Low-quality |
| S13_extracted_47994 | Not-determined |
| S13_extracted_47995 | Not-determined |
| S13_extracted_48004 | Not-determined |
| S13_extracted_48006 | Not-determined |
| S13_extracted_48010 | Not-determined |
| S13_extracted_48016 | Not-determined |
| S13_extracted_48032 | Not-determined |
| S13_extracted_48035 | Not-determined |
| S13_extracted_48067 | Not-determined |
| S13_extracted_48078 | Not-determined |
| S13_extracted_48092 | Not-determined |
| S13_extracted_48100 | Not-determined |
| S13_extracted_48101 | Not-determined |
| S13_extracted_48103 | Not-determined |
| S13_extracted_48105 | Not-determined |
| S13_extracted_48108 | Not-determined |
| S13_extracted_48114 | Not-determined |
| S13_extracted_48118 | Not-determined |
| S13_extracted_48120 | Not-determined |
| S13_extracted_48122 | Not-determined |
| S13_extracted_48125 | Not-determined |
| S13_extracted_48144 | Not-determined |
| S13_extracted_48145 | Not-determined |
| S13_extracted_48151 | Not-determined |
| S13_extracted_48152 | Not-determined |
| S13_extracted_48156 | Not-determined |
| S13_extracted_48160 | Not-determined |
| S13_extracted_48168 | Not-determined |
| S13_extracted_48169 | Not-determined |
| S13_extracted_48174 | Not-determined |
| S13_extracted_48175 | Low-quality |
| S13_extracted_48178 | Not-determined |
| S13_extracted_48182 | Not-determined |
| S13_extracted_48192 | Not-determined |
| S13_extracted_48206 | Not-determined |
| S13_extracted_48214 | Not-determined |
| S13_extracted_48227 | Not-determined |
| S13_extracted_48236 | Low-quality |
| S13_extracted_48245 | Not-determined |
| S13_extracted_48248 | Low-quality |
| S13_extracted_48249 | Not-determined |
| S13_extracted_48252 | Not-determined |
| S13_extracted_48256 | Not-determined |
| S13_extracted_48261 | Not-determined |
| S13_extracted_48263 | Not-determined |
| S13_extracted_48267 | Not-determined |
| S13_extracted_48273 | Not-determined |
| S13_extracted_48286 | Low-quality |
| S13_extracted_48291 | Not-determined |
| S13_extracted_48311 | Not-determined |
| S13_extracted_48313 | Not-determined |
| S13_extracted_48325 | Not-determined |
| S13_extracted_48328 | Not-determined |
| S13_extracted_48333 | Not-determined |
| S13_extracted_48334 | Not-determined |
| S13_extracted_48358 | Not-determined |
| S13_extracted_48359 | Not-determined |
| S13_extracted_48379 | Not-determined |
| S13_extracted_48382 | Not-determined |
| S13_extracted_48388 | Not-determined |
| S13_extracted_48396 | Not-determined |
| S13_extracted_48416 | Not-determined |
| S13_extracted_48430 | Not-determined |
| S13_extracted_48434 | Not-determined |
| S13_extracted_48447 | Not-determined |
| S13_extracted_48449 | Low-quality |
| S13_extracted_48451 | Not-determined |
| S13_extracted_48454 | Not-determined |
| S13_extracted_48457 | Not-determined |
| S13_extracted_48463 | Low-quality |
| S13_extracted_48464 | Not-determined |
| S13_extracted_48474 | Not-determined |
| S13_extracted_48479 | Not-determined |
| S13_extracted_48480 | Not-determined |
| S13_extracted_48484 | Not-determined |
| S13_extracted_48488 | Not-determined |
| S13_extracted_48490 | Not-determined |
| S13_extracted_48494 | Not-determined |
| S13_extracted_48498 | Not-determined |
| S13_extracted_48512 | Not-determined |
| S13_extracted_48516 | Not-determined |
| S13_extracted_48532 | Not-determined |
| S13_extracted_48578 | Not-determined |
| S13_extracted_48588 | Low-quality |
| S13_extracted_48592 | Not-determined |
| S13_extracted_48599 | Not-determined |
| S13_extracted_48600 | Not-determined |
| S13_extracted_48610 | Not-determined |
| S13_extracted_48614 | Not-determined |
| S13_extracted_48631 | Not-determined |
| S13_extracted_48635 | Not-determined |
| S13_extracted_48645 | Not-determined |
| S13_extracted_48646 | Not-determined |
| S13_extracted_48652 | Not-determined |
| S13_extracted_48657 | Not-determined |
| S13_extracted_48663 | Not-determined |
| S13_extracted_48666 | Not-determined |
| S13_extracted_48688 | Not-determined |
| S13_extracted_48692 | Not-determined |
| S13_extracted_48695 | Not-determined |
| S13_extracted_48704 | Not-determined |
| S13_extracted_48706 | Not-determined |
| S13_extracted_48715 | Not-determined |
| S13_extracted_48716 | Not-determined |
| S13_extracted_48730 | Not-determined |
| S13_extracted_48738 | Not-determined |
| S13_extracted_48765 | Not-determined |
| S13_extracted_48813 | Not-determined |
| S13_extracted_48825 | Not-determined |
| S13_extracted_48827 | Not-determined |
| S13_extracted_48829 | Not-determined |
| S13_extracted_48845 | Not-determined |
| S13_extracted_48856 | Not-determined |
| S13_extracted_48859 | Not-determined |
| S13_extracted_48867 | Not-determined |
| S13_extracted_48869 | Not-determined |
| S13_extracted_48882 | Not-determined |
| S13_extracted_48884 | Not-determined |
| S13_extracted_48885 | Not-determined |
| S13_extracted_48895 | Not-determined |
| S13_extracted_48898 | Low-quality |
| S13_extracted_48916 | Not-determined |
| S13_extracted_48921 | Not-determined |
| S13_extracted_48924 | Not-determined |
| S13_extracted_48940 | Not-determined |
| S13_extracted_48943 | Not-determined |
| S13_extracted_48946 | Not-determined |
| S13_extracted_48955 | Not-determined |
| S13_extracted_48959 | Not-determined |
| S13_extracted_48967 | Not-determined |
| S13_extracted_48976 | Not-determined |
| S13_extracted_48989 | Not-determined |
| S13_extracted_48997 | Not-determined |
| S13_extracted_49013 | Not-determined |
| S13_extracted_49029 | Not-determined |
| S13_extracted_49041 | Not-determined |
| S13_extracted_49043 | Not-determined |
| S13_extracted_49063 | Not-determined |
| S13_extracted_49073 | Not-determined |
| S13_extracted_49097 | Not-determined |
| S13_extracted_49121 | Not-determined |
| S13_extracted_49123 | Not-determined |
| S13_extracted_49126 | Not-determined |
| S13_extracted_49134 | Not-determined |
| S13_extracted_49138 | Not-determined |
| S13_extracted_49144 | Not-determined |
| S13_extracted_49145 | Not-determined |
| S13_extracted_49153 | Low-quality |
| S13_extracted_49155 | Low-quality |
| S13_extracted_49167 | Not-determined |
| S13_extracted_49178 | Not-determined |
| S13_extracted_49181 | Not-determined |
| S13_extracted_49185 | Not-determined |
| S13_extracted_49186 | Not-determined |
| S13_extracted_49197 | Not-determined |
| S13_extracted_49199 | Not-determined |
| S13_extracted_49209 | Not-determined |
| S13_extracted_49210 | Not-determined |
| S13_extracted_49211 | Not-determined |
| S13_extracted_49213 | Not-determined |
| S13_extracted_49222 | Not-determined |
| S13_extracted_49242 | Not-determined |
| S13_extracted_49253 | Not-determined |
| S13_extracted_49271 | Not-determined |
| S13_extracted_49272 | Not-determined |
| S13_extracted_49281 | Not-determined |
| S13_extracted_49282 | Not-determined |
| S13_extracted_49286 | Low-quality |
| S13_extracted_49292 | Not-determined |
| S13_extracted_49302 | Not-determined |
| S13_extracted_49305 | Not-determined |
| S13_extracted_49309 | Not-determined |
| S13_extracted_49317 | Not-determined |
| S13_extracted_49326 | Not-determined |
| S13_extracted_49328 | Not-determined |
| S13_extracted_49333 | Not-determined |
| S13_extracted_49334 | Not-determined |
| S13_extracted_49345 | Not-determined |
| S13_extracted_49346 | Not-determined |
| S13_extracted_49355 | Low-quality |
| S13_extracted_49357 | Not-determined |
| S13_extracted_49380 | Not-determined |
| S13_extracted_49387 | Not-determined |
| S13_extracted_49391 | Not-determined |
| S13_extracted_49394 | Not-determined |
| S13_extracted_49409 | Not-determined |
| S13_extracted_49437 | Not-determined |
| S13_extracted_49445 | Not-determined |
| S13_extracted_49457 | Not-determined |
| S13_extracted_49472 | Not-determined |
| S13_extracted_49473 | Not-determined |
| S13_extracted_49474 | Not-determined |
| S13_extracted_49484 | Low-quality |
| S13_extracted_49487 | Low-quality |
| S13_extracted_49493 | Not-determined |
| S13_extracted_49494 | Not-determined |
| S13_extracted_49500 | Not-determined |
| S13_extracted_49501 | Not-determined |
| S13_extracted_49505 | Low-quality |
| S13_extracted_49526 | Not-determined |
| S13_extracted_49549 | Not-determined |
| S13_extracted_49571 | Not-determined |
| S13_extracted_49575 | Low-quality |
| S13_extracted_49591 | Not-determined |
| S13_extracted_49598 | Low-quality |
| S13_extracted_49600 | Not-determined |
| S13_extracted_49621 | Not-determined |
| S13_extracted_49624 | Not-determined |
| S13_extracted_49627 | Not-determined |
| S13_extracted_49638 | Not-determined |
| S13_extracted_49639 | Low-quality |
| S13_extracted_49647 | Not-determined |
| S13_extracted_49650 | Not-determined |
| S13_extracted_49665 | Not-determined |
| S13_extracted_49678 | Low-quality |
| S13_extracted_49692 | Not-determined |
| S13_extracted_49699 | Not-determined |
| S13_extracted_49712 | Not-determined |
| S13_extracted_49715 | Not-determined |
| S13_extracted_49716 | Low-quality |
| S13_extracted_49722 | Not-determined |
| S13_extracted_49725 | Not-determined |
| S13_extracted_49728 | Not-determined |
| S13_extracted_49744 | Not-determined |
| S13_extracted_49753 | Not-determined |
| S13_extracted_49762 | Low-quality |
| S13_extracted_49770 | Not-determined |
| S13_extracted_49778 | Not-determined |
| S13_extracted_49807 | Not-determined |
| S13_extracted_49812 | Not-determined |
| S13_extracted_49831 | Not-determined |
| S13_extracted_49859 | Not-determined |
| S13_extracted_49865 | Not-determined |
| S13_extracted_49909 | Not-determined |
| S13_extracted_49916 | Not-determined |
| S13_extracted_49929 | Not-determined |
| S13_extracted_49932 | Not-determined |
| S13_extracted_49941 | Not-determined |
| S13_extracted_49943 | Low-quality |
| S13_extracted_49947 | Not-determined |
| S13_extracted_49963 | Not-determined |
| S13_extracted_49964 | Low-quality |
| S13_extracted_49979 | Not-determined |
| S13_extracted_49994 | Not-determined |
| S13_extracted_49996 | Not-determined |
| S13_extracted_49999 | Not-determined |
| S13_extracted_50003 | Not-determined |
| S13_extracted_50006 | Not-determined |
| S13_extracted_50016 | Not-determined |
| S13_extracted_50030 | Low-quality |
| S13_extracted_50038 | Not-determined |
| S13_extracted_50039 | Not-determined |
| S13_extracted_50041 | Not-determined |
| S13_extracted_50046 | Low-quality |
| S13_extracted_50053 | Not-determined |
| S13_extracted_50062 | Low-quality |
| S13_extracted_50065 | Not-determined |
| S13_extracted_50071 | Low-quality |
| S13_extracted_50081 | Low-quality |
| S13_extracted_50102 | Low-quality |
| S13_extracted_50112 | Low-quality |
| S13_extracted_50123 | Not-determined |
| S13_extracted_50134 | Not-determined |
| S13_extracted_50138 | Low-quality |
| S13_extracted_50140 | Not-determined |
| S13_extracted_50141 | Not-determined |
| S13_extracted_50146 | Not-determined |
| S13_extracted_50151 | Not-determined |
| S13_extracted_50180 | Not-determined |
| S13_extracted_50186 | Not-determined |
| S13_extracted_50202 | Not-determined |
| S13_extracted_50206 | Not-determined |
| S13_extracted_50246 | Not-determined |
| S13_extracted_50248 | Not-determined |
| S13_extracted_50251 | Not-determined |
| S13_extracted_50255 | Not-determined |
| S13_extracted_50266 | Not-determined |
| S13_extracted_50270 | Not-determined |
| S13_extracted_50272 | Not-determined |
| S13_extracted_50285 | Not-determined |
| S13_extracted_50301 | Not-determined |
| S13_extracted_50327 | Not-determined |
| S13_extracted_50392 | Not-determined |
| S13_extracted_50409 | Not-determined |
| S13_extracted_50418 | Low-quality |
| S13_extracted_50452 | Not-determined |
| S13_extracted_50453 | Not-determined |
| S13_extracted_50454 | Not-determined |

| | |
|---|---|
| S13_extracted_S0461 | Not-determined |
| S13_extracted_S0464 | Not-determined |
| S13_extracted_S0472 | Not-determined |
| S13_extracted_S0473 | Not-determined |
| S13_extracted_S0484 | Not-determined |
| S13_extracted_S0505 | Low-quality |
| S13_extracted_S0510 | Not-determined |
| S13_extracted_S0515 | Not-determined |
| S13_extracted_S0518 | Low-quality |
| S13_extracted_S0519 | Not-determined |
| S13_extracted_S0527 | Not-determined |
| S13_extracted_S0531 | Not-determined |
| S13_extracted_S0536 | Not-determined |
| S13_extracted_S0540 | Not-determined |
| S13_extracted_S0543 | Not-determined |
| S13_extracted_S0546 | Not-determined |
| S13_extracted_S0548 | Not-determined |
| S13_extracted_S0564 | Not-determined |
| S13_extracted_S0566 | Low-quality |
| S13_extracted_S0585 | Not-determined |
| S13_extracted_S0591 | Low-quality |
| S13_extracted_S0625 | Not-determined |
| S13_extracted_S0629 | Not-determined |
| S13_extracted_S0633 | Not-determined |
| S13_extracted_S0635 | Not-determined |
| S13_extracted_S0649 | Not-determined |
| S13_extracted_S0650 | Not-determined |
| S13_extracted_S0680 | Low-quality |
| S13_extracted_S0681 | Not-determined |
| S13_extracted_S0695 | Not-determined |
| S13_extracted_S0702 | Not-determined |
| S13_extracted_S0717 | Not-determined |
| S13_extracted_S0722 | Not-determined |
| S13_extracted_S0726 | Not-determined |
| S13_extracted_S0728 | Not-determined |
| S13_extracted_S0733 | Not-determined |
| S13_extracted_S0738 | Not-determined |
| S13_extracted_S0746 | Not-determined |
| S13_extracted_S0753 | Not-determined |
| S13_extracted_S0772 | Not-determined |
| S13_extracted_S0778 | Not-determined |
| S13_extracted_S0793 | Not-determined |
| S13_extracted_S0816 | Not-determined |
| S13_extracted_S0821 | Not-determined |
| S13_extracted_S0829 | Low-quality |
| S13_extracted_S0830 | Not-determined |
| S13_extracted_S0832 | Not-determined |
| S13_extracted_S0836 | Not-determined |
| S13_extracted_S0844 | Not-determined |
| S13_extracted_S0848 | Low-quality |
| S13_extracted_S0858 | Not-determined |
| S13_extracted_S0864 | Not-determined |
| S13_extracted_S0874 | Not-determined |
| S13_extracted_S0888 | Not-determined |
| S13_extracted_S0889 | Not-determined |
| S13_extracted_S0892 | Not-determined |
| S13_extracted_S0903 | Not-determined |
| S13_extracted_S0914 | Not-determined |
| S13_extracted_S0922 | Not-determined |
| S13_extracted_S0925 | Low-quality |
| S13_extracted_S0934 | Not-determined |
| S13_extracted_S0951 | Not-determined |
| S13_extracted_S0959 | Low-quality |
| S13_extracted_S0961 | Not-determined |
| S13_extracted_S0965 | Not-determined |
| S13_extracted_S0967 | Low-quality |
| S13_extracted_S0972 | Not-determined |
| S13_extracted_S0985 | Not-determined |
| S13_extracted_S0986 | Not-determined |
| S13_extracted_S0989 | Not-determined |
| S13_extracted_S1003 | Not-determined |
| S13_extracted_S1010 | Not-determined |
| S13_extracted_S1011 | Not-determined |
| S13_extracted_S1018 | Not-determined |
| S13_extracted_S1022 | Not-determined |
| S13_extracted_S1026 | Not-determined |
| S13_extracted_S1027 | Not-determined |
| S13_extracted_S1032 | Not-determined |
| S13_extracted_S1043 | Not-determined |
| S13_extracted_S1046 | Not-determined |
| S13_extracted_S1058 | Not-determined |
| S13_extracted_S1074 | Not-determined |
| S13_extracted_S1115 | Not-determined |
| S13_extracted_S1118 | Not-determined |
| S13_extracted_S1133 | Not-determined |
| S13_extracted_S1142 | Not-determined |
| S13_extracted_S1149 | Not-determined |
| S13_extracted_S1154 | Not-determined |
| S13_extracted_S1168 | Not-determined |
| S13_extracted_S1169 | Not-determined |
| S13_extracted_S1171 | Not-determined |
| S13_extracted_S1179 | Not-determined |
| S13_extracted_S1218 | Not-determined |
| S13_extracted_S1224 | Not-determined |
| S13_extracted_S1238 | Not-determined |
| S13_extracted_S1239 | Not-determined |
| S13_extracted_S1248 | Not-determined |
| S13_extracted_S1251 | Not-determined |
| S13_extracted_S1252 | Not-determined |
| S13_extracted_S1253 | Not-determined |
| S13_extracted_S1273 | Not-determined |
| S13_extracted_S1289 | Not-determined |
| S13_extracted_S1307 | Not-determined |
| S13_extracted_S1319 | Low-quality |
| S13_extracted_S1322 | Not-determined |
| S13_extracted_S1338 | Not-determined |
| S13_extracted_S1340 | Not-determined |
| S13_extracted_S1344 | Not-determined |
| S13_extracted_S1346 | Not-determined |
| S13_extracted_S1352 | Low-quality |
| S13_extracted_S1353 | Not-determined |
| S13_extracted_S1356 | Not-determined |
| S13_extracted_S1365 | Not-determined |
| S13_extracted_S1371 | Not-determined |
| S13_extracted_S1374 | Not-determined |
| S13_extracted_S1384 | Not-determined |
| S13_extracted_S1388 | Low-quality |
| S13_extracted_S1391 | Not-determined |
| S13_extracted_S1397 | Not-determined |
| S13_extracted_S1403 | Not-determined |
| S13_extracted_S1405 | Low-quality |
| S13_extracted_S1409 | Not-determined |
| S13_extracted_S1417 | Not-determined |
| S13_extracted_S1439 | Not-determined |
| S13_extracted_S1445 | Not-determined |
| S13_extracted_S1456 | Not-determined |
| S13_extracted_S1461 | Not-determined |
| S13_extracted_S1468 | Not-determined |
| S13_extracted_S1470 | Not-determined |
| S13_extracted_S1483 | Not-determined |
| S13_extracted_S1487 | Not-determined |
| S13_extracted_S1509 | Not-determined |
| S13_extracted_S1517 | Not-determined |
| S13_extracted_S1529 | Not-determined |
| S13_extracted_S1535 | Low-quality |
| S13_extracted_S1543 | Not-determined |
| S13_extracted_S1567 | Not-determined |
| S13_extracted_S1574 | Low-quality |
| S13_extracted_S1587 | Not-determined |
| S13_extracted_S1592 | Not-determined |
| S13_extracted_S1596 | Not-determined |
| S13_extracted_S1599 | Not-determined |
| S13_extracted_S1603 | Not-determined |
| S13_extracted_S1605 | Low-quality |
| S13_extracted_S1626 | Not-determined |
| S13_extracted_S1633 | Low-quality |
| S13_extracted_S1661 | Not-determined |
| S13_extracted_S1665 | Not-determined |
| S13_extracted_S1667 | Low-quality |
| S13_extracted_S1668 | Not-determined |
| S13_extracted_S1669 | Not-determined |
| S13_extracted_S1679 | Low-quality |
| S13_extracted_S1686 | Not-determined |
| S13_extracted_S1688 | Not-determined |
| S13_extracted_S1699 | Not-determined |
| S13_extracted_S1702 | Not-determined |
| S13_extracted_S1703 | Not-determined |
| S13_extracted_S1709 | Not-determined |
| S13_extracted_S1730 | Not-determined |
| S13_extracted_S1735 | Not-determined |
| S13_extracted_S1737 | Not-determined |
| S13_extracted_S1739 | Not-determined |
| S13_extracted_S1759 | Not-determined |
| S13_extracted_S1762 | Not-determined |
| S13_extracted_S1778 | Not-determined |
| S13_extracted_S1797 | Not-determined |
| S13_extracted_S1813 | Not-determined |
| S13_extracted_S1829 | Not-determined |
| S13_extracted_S1852 | Not-determined |
| S13_extracted_S1861 | Low-quality |
| S13_extracted_S1868 | Not-determined |
| S13_extracted_S1876 | Low-quality |
| S13_extracted_S1879 | Not-determined |
| S13_extracted_S1900 | Not-determined |
| S13_extracted_S1902 | Not-determined |
| S13_extracted_S1912 | Not-determined |
| S13_extracted_S1919 | Not-determined |
| S13_extracted_S1933 | Not-determined |
| S13_extracted_S1935 | Not-determined |
| S13_extracted_S1950 | Not-determined |
| S13_extracted_S1955 | Low-quality |
| S13_extracted_S1969 | Not-determined |
| S13_extracted_S1974 | Not-determined |
| S13_extracted_S1976 | Not-determined |
| S13_extracted_S1979 | Not-determined |
| S13_extracted_S1993 | Low-quality |
| S13_extracted_S2004 | Not-determined |
| S13_extracted_S2009 | Low-quality |
| S13_extracted_S2015 | Not-determined |
| S13_extracted_S2017 | Not-determined |
| S13_extracted_S2022 | Not-determined |
| S13_extracted_S2039 | Not-determined |
| S13_extracted_S2047 | Not-determined |
| S13_extracted_S2057 | Not-determined |
| S13_extracted_S2074 | Not-determined |
| S13_extracted_S2102 | Not-determined |
| S13_extracted_S2109 | Not-determined |
| S13_extracted_S2123 | Not-determined |
| S13_extracted_S2128 | Not-determined |
| S13_extracted_S2133 | Low-quality |
| S13_extracted_S2137 | Not-determined |
| S13_extracted_S2138 | Not-determined |
| S13_extracted_S2148 | Not-determined |
| S13_extracted_S2153 | Not-determined |
| S13_extracted_S2154 | Not-determined |
| S13_extracted_S2162 | Low-quality |
| S13_extracted_S2165 | Not-determined |
| S13_extracted_S2183 | Not-determined |
| S13_extracted_S2184 | Not-determined |
| S13_extracted_S2197 | Not-determined |
| S13_extracted_S2218 | Not-determined |
| S13_extracted_S2220 | Not-determined |
| S13_extracted_S2231 | Not-determined |
| S13_extracted_S2233 | Not-determined |
| S13_extracted_S2245 | Not-determined |
| S13_extracted_S2252 | Not-determined |
| S13_extracted_S2253 | Not-determined |
| S13_extracted_S2263 | Not-determined |
| S13_extracted_S2268 | Not-determined |
| S13_extracted_S2271 | Not-determined |
| S13_extracted_S2280 | Not-determined |
| S13_extracted_S2281 | Not-determined |
| S13_extracted_S2302 | Not-determined |
| S13_extracted_S2303 | Not-determined |
| S13_extracted_S2312 | Not-determined |
| S13_extracted_S2313 | Not-determined |
| S13_extracted_S2320 | Not-determined |
| S13_extracted_S2353 | Not-determined |
| S13_extracted_S2354 | Not-determined |
| S13_extracted_S2357 | Not-determined |
| S13_extracted_S2361 | Not-determined |
| S13_extracted_S2367 | Not-determined |
| S13_extracted_S2374 | Not-determined |
| S13_extracted_S2383 | Not-determined |
| S13_extracted_S2384 | Not-determined |
| S13_extracted_S2398 | Not-determined |
| S13_extracted_S2410 | Not-determined |
| S13_extracted_S2419 | Not-determined |
| S13_extracted_S2421 | Not-determined |
| S13_extracted_S2424 | Low-quality |
| S13_extracted_S2425 | Not-determined |
| S13_extracted_S2443 | Not-determined |
| S13_extracted_S2444 | Not-determined |
| S13_extracted_S2450 | Not-determined |
| S13_extracted_S2451 | Not-determined |
| S13_extracted_S2471 | Not-determined |
| S13_extracted_S2472 | Not-determined |
| S13_extracted_S2484 | Not-determined |
| S13_extracted_S2491 | Not-determined |
| S13_extracted_S2501 | Not-determined |
| S13_extracted_S2507 | Not-determined |
| S13_extracted_S2522 | Not-determined |
| S13_extracted_S2529 | Not-determined |
| S13_extracted_S2546 | Not-determined |
| S13_extracted_S2560 | Not-determined |
| S13_extracted_S2567 | Not-determined |
| S13_extracted_S2570 | Not-determined |
| S13_extracted_S2576 | Low-quality |
| S13_extracted_S2581 | Not-determined |
| S13_extracted_S2588 | Not-determined |
| S13_extracted_S2594 | Not-determined |
| S13_extracted_S2597 | Not-determined |
| S13_extracted_S2602 | Low-quality |
| S13_extracted_S2612 | Not-determined |
| S13_extracted_S2621 | Not-determined |
| S13_extracted_S2622 | Not-determined |
| S13_extracted_S2628 | Not-determined |
| S13_extracted_S2635 | Not-determined |
| S13_extracted_S2638 | Not-determined |
| S13_extracted_S2644 | Not-determined |
| S13_extracted_S2648 | Not-determined |
| S13_extracted_S2649 | Low-quality |
| S13_extracted_S2672 | Not-determined |
| S13_extracted_S2682 | Not-determined |
| S13_extracted_S2692 | Not-determined |
| S13_extracted_S2694 | Not-determined |
| S13_extracted_S2697 | Not-determined |
| S13_extracted_S2708 | Not-determined |
| S13_extracted_S2722 | Not-determined |
| S13_extracted_S2724 | Not-determined |
| S13_extracted_S2741 | Not-determined |
| S13_extracted_S2744 | Not-determined |
| S13_extracted_S2761 | Not-determined |
| S13_extracted_S2762 | Not-determined |
| S13_extracted_S2767 | Low-quality |
| S13_extracted_S2780 | Not-determined |
| S13_extracted_S2783 | Not-determined |
| S13_extracted_S2788 | Not-determined |
| S13_extracted_S2794 | Not-determined |
| S13_extracted_S2795 | Low-quality |
| S13_extracted_S2798 | Not-determined |
| S13_extracted_S2799 | Not-determined |
| S13_extracted_S2801 | Not-determined |
| S13_extracted_S2803 | Not-determined |
| S13_extracted_S2813 | Not-determined |
| S13_extracted_S2824 | Not-determined |
| S13_extracted_S2826 | Not-determined |
| S13_extracted_S2831 | Not-determined |
| S13_extracted_S2854 | Not-determined |
| S13_extracted_S2855 | Not-determined |
| S13_extracted_S2882 | Not-determined |
| S13_extracted_S2885 | Not-determined |
| S13_extracted_S2918 | Not-determined |
| S13_extracted_S2923 | Not-determined |
| S13_extracted_S2958 | Not-determined |
| S13_extracted_S2966 | Low-quality |
| S13_extracted_S2971 | Not-determined |
| S13_extracted_S2982 | Not-determined |
| S13_extracted_S2993 | Not-determined |
| S13_extracted_S2995 | Not-determined |
| S13_extracted_S3000 | Not-determined |
| S13_extracted_S3001 | Not-determined |
| S13_extracted_S3008 | Not-determined |
| S13_extracted_S3012 | Low-quality |
| S13_extracted_S3013 | Not-determined |
| S13_extracted_S3015 | Not-determined |
| S13_extracted_S3023 | Not-determined |
| S13_extracted_S3025 | Not-determined |
| S13_extracted_S3051 | Not-determined |
| S13_extracted_S3053 | Not-determined |
| S13_extracted_S3054 | Not-determined |
| S13_extracted_S3071 | Not-determined |
| S13_extracted_S3072 | Not-determined |
| S13_extracted_S3073 | Not-determined |
| S13_extracted_S3077 | Not-determined |
| S13_extracted_S3087 | Not-determined |
| S13_extracted_S3088 | Low-quality |
| S13_extracted_S3090 | Not-determined |
| S13_extracted_S3097 | Not-determined |
| S13_extracted_S3120 | Low-quality |
| S13_extracted_S3124 | Not-determined |
| S13_extracted_S3131 | Not-determined |
| S13_extracted_S3148 | Not-determined |
| S13_extracted_S3168 | Not-determined |
| S13_extracted_S3172 | Not-determined |
| S13_extracted_S3173 | Not-determined |
| S13_extracted_S3177 | Not-determined |
| S13_extracted_S3180 | Not-determined |
| S13_extracted_S3185 | Not-determined |
| S13_extracted_S3200 | Not-determined |
| S13_extracted_S3210 | Not-determined |
| S13_extracted_S3231 | Not-determined |
| S13_extracted_S3236 | Not-determined |
| S13_extracted_S3249 | Not-determined |
| S13_extracted_S3257 | Not-determined |
| S13_extracted_S3261 | Not-determined |
| S13_extracted_S3281 | Not-determined |
| S13_extracted_S3283 | Not-determined |
| S13_extracted_S3284 | Not-determined |
| S13_extracted_S3289 | Not-determined |
| S13_extracted_S3312 | Not-determined |
| S13_extracted_S3318 | Not-determined |
| S13_extracted_S3328 | Not-determined |
| S13_extracted_S3330 | Not-determined |
| S13_extracted_S3340 | Not-determined |
| S13_extracted_S3341 | Not-determined |
| S13_extracted_S3354 | Not-determined |
| S13_extracted_S3356 | Not-determined |
| S13_extracted_S3365 | Not-determined |
| S13_extracted_S3367 | Not-determined |
| S13_extracted_S3368 | Not-determined |
| S13_extracted_S3378 | Not-determined |
| S13_extracted_S3379 | Not-determined |
| S13_extracted_S3383 | Not-determined |
| S13_extracted_S3384 | Not-determined |
| S13_extracted_S3392 | Low-quality |
| S13_extracted_S3396 | Not-determined |
| S13_extracted_S3405 | Not-determined |
| S13_extracted_S3407 | Not-determined |
| S13_extracted_S3419 | Not-determined |
| S13_extracted_S3421 | Not-determined |
| S13_extracted_S3430 | Not-determined |
| S13_extracted_S3437 | Not-determined |
| S13_extracted_S3438 | Low-quality |
| S13_extracted_S3442 | Not-determined |
| S13_extracted_S3456 | Not-determined |
| S13_extracted_S3460 | Not-determined |
| S13_extracted_S3527 | Not-determined |
| S13_extracted_S3546 | Not-determined |
| S13_extracted_S3568 | Not-determined |
| S13_extracted_S3602 | Not-determined |
| S13_extracted_S3606 | Not-determined |
| S13_extracted_S3616 | Not-determined |
| S13_extracted_S3620 | Not-determined |
| S13_extracted_S3624 | Not-determined |
| S13_extracted_S3633 | Not-determined |
| S13_extracted_S3636 | Not-determined |
| S13_extracted_S3639 | Low-quality |
| S13_extracted_S3644 | Not-determined |
| S13_extracted_S3649 | Not-determined |
| S13_extracted_S3650 | Not-determined |
| S13_extracted_S3670 | Not-determined |
| S13_extracted_S3701 | Low-quality |
| S13_extracted_S3705 | Not-determined |
| S13_extracted_S3710 | Not-determined |
| S13_extracted_S3713 | Not-determined |
| S13_extracted_S3721 | Not-determined |
| S13_extracted_S3723 | Not-determined |
| S13_extracted_S3724 | Low-quality |
| S13_extracted_S3727 | Not-determined |
| S13_extracted_S3729 | Not-determined |
| S13_extracted_S3734 | Not-determined |
| S13_extracted_S3741 | Not-determined |
| S13_extracted_S3754 | Not-determined |
| S13_extracted_S3755 | Not-determined |
| S13_extracted_S3757 | Not-determined |
| S13_extracted_S3765 | Low-quality |
| S13_extracted_S3769 | Not-determined |
| S13_extracted_S3770 | Not-determined |
| S13_extracted_S3783 | Not-determined |
| S13_extracted_S3791 | Not-determined |
| S13_extracted_S3800 | Not-determined |
| S13_extracted_S3801 | Not-determined |
| S13_extracted_S3806 | Low-quality |
| S13_extracted_S3820 | Low-quality |
| S13_extracted_S3832 | Not-determined |
| S13_extracted_S3835 | Not-determined |
| S13_extracted_S3837 | Low-quality |
| S13_extracted_S3842 | Not-determined |
| S13_extracted_S3843 | Not-determined |
| S13_extracted_S3854 | Not-determined |
| S13_extracted_S3863 | Not-determined |
| S13_extracted_S3869 | Not-determined |
| S13_extracted_S3886 | Not-determined |
| S13_extracted_S3897 | Not-determined |
| S13_extracted_S3904 | Low-quality |
| S13_extracted_S3905 | Not-determined |
| S13_extracted_S3915 | Not-determined |
| S13_extracted_S3916 | Low-quality |
| S13_extracted_S3925 | Not-determined |
| S13_extracted_S3926 | Not-determined |
| S13_extracted_S3931 | Not-determined |
| S13_extracted_S3947 | Not-determined |
| S13_extracted_S3954 | Not-determined |
| S13_extracted_S3955 | Not-determined |
| S13_extracted_S3959 | Not-determined |
| S13_extracted_S3973 | Not-determined |
| S13_extracted_S3980 | Not-determined |
| S13_extracted_S3983 | Not-determined |
| S13_extracted_S4001 | Not-determined |
| S13_extracted_S4003 | Not-determined |
| S13_extracted_S4006 | Not-determined |
| S13_extracted_S4009 | Not-determined |
| S13_extracted_S4012 | Not-determined |
| S13_extracted_S4013 | Not-determined |
| S13_extracted_S4024 | Not-determined |
| S13_extracted_S4058 | Not-determined |
| S13_extracted_S4069 | Not-determined |
| S13_extracted_S4073 | Not-determined |
| S13_extracted_S4074 | Not-determined |
| S13_extracted_S4080 | Not-determined |
| S13_extracted_S4092 | Not-determined |
| S13_extracted_S4100 | Low-quality |
| S13_extracted_S4104 | Low-quality |
| S13_extracted_S4115 | Low-quality |
| S13_extracted_S4124 | Not-determined |
| S13_extracted_S4140 | Not-determined |
| S13_extracted_S4147 | Not-determined |
| S13_extracted_S4153 | Not-determined |
| S13_extracted_S4156 | Not-determined |
| S13_extracted_S4165 | Low-quality |
| S13_extracted_S4168 | Not-determined |
| S13_extracted_S4170 | Not-determined |
| S13_extracted_S4171 | Not-determined |
| S13_extracted_S4184 | Not-determined |
| S13_extracted_S4200 | Low-quality |
| S3_extracted_k141_61860 | Complete |
| S13_extracted_S4202 | Not-determined |
| S13_extracted_S4237 | Not-determined |
| S13_extracted_S4239 | Not-determined |
| S13_extracted_S4243 | Not-determined |
| S13_extracted_S4244 | Not-determined |
| S13_extracted_S4247 | Not-determined |
| S13_extracted_S4257 | Not-determined |
| S13_extracted_S4260 | Not-determined |
| S13_extracted_S4266 | Not-determined |
| S13_extracted_S4273 | Not-determined |
| S13_extracted_S4285 | Not-determined |
| S13_extracted_S4286 | Not-determined |
| S13_extracted_S4304 | Low-quality |
| S13_extracted_S4305 | Not-determined |
| S13_extracted_S4308 | Not-determined |
| S13_extracted_S4312 | Low-quality |
| S13_extracted_S4319 | Not-determined |
| S13_extracted_S4328 | Not-determined |
| S13_extracted_S4356 | Not-determined |
| S13_extracted_S4359 | Not-determined |
| S13_extracted_S4361 | Not-determined |
| S13_extracted_S4371 | Not-determined |
| S13_extracted_S4387 | Low-quality |
| S13_extracted_S4396 | Not-determined |
| S13_extracted_S4400 | Not-determined |
| S13_extracted_S4407 | Not-determined |
| S13_extracted_S4410 | Not-determined |
| S13_extracted_S4438 | Low-quality |
| S13_extracted_S4439 | Not-determined |
| S13_extracted_S4466 | Not-determined |
| S13_extracted_S4467 | Not-determined |
| S13_extracted_S4480 | Not-determined |
| S13_extracted_S4490 | Not-determined |
| S13_extracted_S4496 | Low-quality |
| S13_extracted_S4507 | Not-determined |
| S13_extracted_S4510 | Low-quality |
| S13_extracted_S4514 | Not-determined |
| S13_extracted_S4516 | Low-quality |
| S13_extracted_S4535 | Low-quality |
| S13_extracted_S4541 | Not-determined |
| S13_extracted_S4542 | Not-determined |
| S13_extracted_S4554 | Not-determined |
| S13_extracted_S4575 | Not-determined |
| S13_extracted_S4585 | Not-determined |
| S13_extracted_S4586 | Low-quality |
| S13_extracted_S4608 | Not-determined |
| S13_extracted_S4609 | Not-determined |
| S13_extracted_S4617 | Not-determined |
| S13_extracted_S4622 | Not-determined |
| S13_extracted_S4625 | Low-quality |
| S13_extracted_S4626 | Low-quality |
| S13_extracted_S4635 | Low-quality |
| S13_extracted_S4638 | Low-quality |
| S13_extracted_S4641 | Not-determined |
| S13_extracted_S4653 | Not-determined |
| S13_extracted_S4655 | Not-determined |
| S13_extracted_S4658 | Not-determined |
| S13_extracted_S4664 | Not-determined |
| S13_extracted_S4667 | Not-determined |
| S13_extracted_S4670 | Not-determined |
| S13_extracted_S4672 | Not-determined |
| S13_extracted_S4711 | Not-determined |
| S13_extracted_S4713 | Not-determined |
| S13_extracted_S4751 | Low-quality |
| S13_extracted_S4753 | Not-determined |
| S13_extracted_S4754 | Low-quality |
| S13_extracted_S4755 | Low-quality |
| S13_extracted_S4761 | Not-determined |
| S13_extracted_S4766 | Low-quality |
| S13_extracted_S4787 | Not-determined |
| S13_extracted_S4789 | Not-determined |
| S13_extracted_S4800 | Not-determined |
| S13_extracted_S4807 | Not-determined |
| S13_extracted_S4821 | Not-determined |
| S13_extracted_S4828 | Not-determined |
| S13_extracted_S4837 | Not-determined |
| S13_extracted_S4839 | Not-determined |
| S13_extracted_S4847 | Not-determined |
| S13_extracted_S4850 | Not-determined |
| S13_extracted_S4851 | Low-quality |
| S13_extracted_S4854 | Not-determined |
| S13_extracted_S4860 | Not-determined |
| S13_extracted_S4866 | Not-determined |
| S13_extracted_S4882 | Not-determined |
| S13_extracted_S4892 | Not-determined |
| S13_extracted_S4896 | Not-determined |
| S13_extracted_S4897 | Not-determined |
| S13_extracted_S4899 | Not-determined |
| S13_extracted_S4900 | Not-determined |
| S13_extracted_S4905 | Low-quality |
| S13_extracted_S4906 | Not-determined |
| S13_extracted_S4917 | Not-determined |
| S13_extracted_S4919 | Not-determined |
| S13_extracted_S4921 | Not-determined |
| S13_extracted_S4927 | Low-quality |
| S13_extracted_S4937 | Not-determined |
| S13_extracted_S4951 | Not-determined |
| S13_extracted_S4953 | Not-determined |
| S13_extracted_S4970 | Not-determined |
| S13_extracted_S4972 | Low-quality |
| S13_extracted_S4990 | Low-quality |
| S13_extracted_S4993 | Low-quality |
| S13_extracted_S5003 | Low-quality |
| S13_extracted_S5011 | Low-quality |
| S13_extracted_S5023 | Not-determined |
| S13_extracted_S5037 | Not-determined |
| S13_extracted_S5057 | Not-determined |
| S13_extracted_S5060 | Low-quality |
| S13_extracted_S5064 | Not-determined |
| S13_extracted_S5071 | Not-determined |
| S13_extracted_S5091 | Not-determined |
| S13_extracted_S5093 | Not-determined |
| S13_extracted_S5104 | Low-quality |
| S13_extracted_S5105 | Low-quality |
| S13_extracted_S5114 | Not-determined |
| S13_extracted_S5139 | Not-determined |
| S13_extracted_S5148 | Not-determined |
| S13_extracted_S5159 | Not-determined |
| S13_extracted_S5161 | Low-quality |
| S13_extracted_S5180 | Not-determined |
| S13_extracted_S5183 | Not-determined |
| S13_extracted_S5195 | Not-determined |

| | |
|---|---|
| S13_extracted_55210 | Not-determined |
| S13_extracted_55215 | Not-determined |
| S13_extracted_55228 | Not-determined |
| S13_extracted_55237 | Not-determined |
| S13_extracted_55251 | Low-quality |
| S13_extracted_55253 | Not-determined |
| S13_extracted_55258 | Not-determined |
| S13_extracted_55263 | Not-determined |
| S13_extracted_55265 | Not-determined |
| S13_extracted_55275 | Not-determined |
| S13_extracted_55278 | Not-determined |
| S13_extracted_55280 | Not-determined |
| S13_extracted_55288 | Not-determined |
| S13_extracted_55295 | Not-determined |
| S13_extracted_55313 | Low-quality |
| S13_extracted_55331 | Not-determined |
| S13_extracted_55345 | Not-determined |
| S13_extracted_55351 | Not-determined |
| S13_extracted_55352 | Not-determined |
| S13_extracted_55378 | Low-quality |
| S13_extracted_55393 | Not-determined |
| S13_extracted_55395 | Not-determined |
| S13_extracted_55397 | Low-quality |
| S13_extracted_55398 | Not-determined |
| S13_extracted_55415 | Not-determined |
| S13_extracted_55416 | Not-determined |
| S13_extracted_55421 | Not-determined |
| S13_extracted_55430 | Not-determined |
| S13_extracted_55432 | Not-determined |
| S13_extracted_55440 | Not-determined |
| S13_extracted_55445 | Not-determined |
| S13_extracted_55457 | Not-determined |
| S13_extracted_55461 | Not-determined |
| S13_extracted_55474 | Not-determined |
| S13_extracted_55501 | Not-determined |
| S13_extracted_55502 | Low-quality |
| S13_extracted_55522 | Not-determined |
| S13_extracted_55545 | Not-determined |
| S13_extracted_55547 | Not-determined |
| S13_extracted_55553 | Not-determined |
| S13_extracted_55574 | Not-determined |
| S13_extracted_55582 | Not-determined |
| S13_extracted_55592 | Not-determined |
| S13_extracted_55594 | Not-determined |
| S13_extracted_55603 | Not-determined |
| S13_extracted_55609 | Not-determined |
| S13_extracted_55624 | Not-determined |
| S13_extracted_55632 | Not-determined |
| S13_extracted_55634 | Not-determined |
| S13_extracted_55635 | Not-determined |
| S13_extracted_55637 | Not-determined |
| S13_extracted_55641 | Not-determined |
| S13_extracted_55653 | Not-determined |
| S13_extracted_55680 | Not-determined |
| S13_extracted_55689 | Not-determined |
| S13_extracted_55690 | Low-quality |
| S13_extracted_55703 | Not-determined |
| S13_extracted_55711 | Not-determined |
| S13_extracted_55730 | Not-determined |
| S13_extracted_55745 | Not-determined |
| S13_extracted_55753 | Low-quality |
| S13_extracted_55759 | Not-determined |
| S13_extracted_55778 | Not-determined |
| S13_extracted_55790 | Not-determined |
| S13_extracted_55798 | Not-determined |
| S13_extracted_55802 | Not-determined |
| S13_extracted_55808 | Not-determined |
| S13_extracted_55821 | Not-determined |
| S13_extracted_55823 | Not-determined |
| S13_extracted_55830 | Not-determined |
| S13_extracted_55831 | Not-determined |
| S13_extracted_55835 | Not-determined |
| S13_extracted_55837 | Low-quality |
| S13_extracted_55863 | Not-determined |
| S13_extracted_55864 | Not-determined |
| S13_extracted_55868 | Not-determined |
| S13_extracted_55869 | Not-determined |
| S13_extracted_55879 | Not-determined |
| S13_extracted_55884 | Not-determined |
| S13_extracted_55885 | Not-determined |
| S13_extracted_55903 | Not-determined |
| S13_extracted_55910 | Not-determined |
| S13_extracted_55919 | Not-determined |
| S13_extracted_55921 | Not-determined |
| S13_extracted_55924 | Not-determined |
| S13_extracted_55934 | Not-determined |
| S13_extracted_55939 | Not-determined |
| S13_extracted_55942 | Not-determined |
| S13_extracted_55961 | Not-determined |
| S13_extracted_55962 | Not-determined |
| S13_extracted_55966 | Not-determined |
| S13_extracted_55969 | Not-determined |
| S13_extracted_55971 | Not-determined |
| S13_extracted_55980 | Not-determined |
| S13_extracted_56006 | Not-determined |
| S13_extracted_56015 | Not-determined |
| S13_extracted_56016 | Not-determined |
| S13_extracted_56021 | Not-determined |
| S13_extracted_56023 | Not-determined |
| S13_extracted_56024 | Low-quality |
| S13_extracted_56073 | Not-determined |
| S3_vs_k141_42569\|\|full | High-quality |
| S13_extracted_56082 | Not-determined |
| S13_extracted_56096 | Not-determined |
| S13_extracted_56099 | Not-determined |
| S13_extracted_56100 | Not-determined |
| S13_extracted_56122 | Low-quality |
| S13_extracted_56160 | Not-determined |
| S13_extracted_56170 | Not-determined |
| S13_extracted_56177 | Not-determined |
| S13_extracted_56185 | Not-determined |
| S13_extracted_56198 | Not-determined |
| S13_extracted_56209 | Not-determined |
| S13_extracted_56212 | Not-determined |
| S13_extracted_56216 | Not-determined |
| S13_extracted_56220 | Not-determined |
| S13_extracted_56222 | Not-determined |
| S13_extracted_56229 | Not-determined |
| S13_extracted_56230 | Not-determined |
| S13_extracted_56234 | Low-quality |
| S13_extracted_56244 | Not-determined |
| S13_extracted_56246 | Not-determined |
| S13_extracted_56252 | Low-quality |
| S13_extracted_56260 | Not-determined |
| S13_extracted_56273 | Not-determined |
| S13_extracted_56274 | Not-determined |
| S13_extracted_56282 | Not-determined |
| S13_extracted_56287 | Low-quality |
| S13_extracted_56292 | Not-determined |
| S13_extracted_56313 | Not-determined |
| S13_extracted_56320 | Not-determined |
| S13_extracted_56339 | Not-determined |
| S13_extracted_56346 | Not-determined |
| S13_extracted_56354 | Not-determined |
| S13_extracted_56356 | Not-determined |
| S13_extracted_56372 | Not-determined |
| S13_extracted_56385 | Not-determined |
| S13_extracted_56390 | Not-determined |
| S13_extracted_56408 | Not-determined |
| S13_extracted_56413 | Not-determined |
| S13_extracted_56417 | Not-determined |
| S13_extracted_56426 | Not-determined |
| S13_extracted_56434 | Not-determined |
| S13_extracted_56438 | Not-determined |
| S13_extracted_56449 | Not-determined |
| S13_extracted_56480 | Not-determined |
| S13_extracted_56482 | Not-determined |
| S13_extracted_56485 | Low-quality |
| S13_extracted_56511 | Not-determined |
| S13_extracted_56517 | Not-determined |
| S13_extracted_56526 | Not-determined |
| S13_extracted_56546 | Not-determined |
| S13_extracted_56549 | Not-determined |
| S13_extracted_56557 | Not-determined |
| S13_extracted_56562 | Not-determined |
| S13_extracted_56571 | Not-determined |
| S13_extracted_56572 | Not-determined |
| S13_extracted_56598 | Not-determined |
| S13_extracted_56605 | Not-determined |
| S13_extracted_56614 | Not-determined |
| S13_extracted_56620 | Low-quality |
| S13_extracted_56621 | Not-determined |
| S13_extracted_56625 | Not-determined |
| S13_extracted_56626 | Not-determined |
| S13_extracted_56635 | Not-determined |
| S13_extracted_56638 | Not-determined |
| S13_extracted_56645 | Not-determined |
| S13_extracted_56648 | Not-determined |
| S13_extracted_56683 | Not-determined |
| S13_extracted_56692 | Not-determined |
| S13_extracted_56707 | Not-determined |
| S13_extracted_56708 | Not-determined |
| S13_extracted_56710 | Not-determined |
| S13_extracted_56719 | Not-determined |
| S13_extracted_56725 | Not-determined |
| S13_extracted_56745 | Not-determined |
| S13_extracted_56750 | Not-determined |
| S13_extracted_56764 | Not-determined |
| S13_extracted_56770 | Not-determined |
| S13_extracted_56789 | Not-determined |
| S13_extracted_56790 | Not-determined |
| S13_extracted_56792 | Not-determined |
| S13_extracted_56799 | Not-determined |
| S13_extracted_56805 | Not-determined |
| S13_extracted_56808 | Not-determined |
| S13_extracted_56814 | Not-determined |
| S13_extracted_56826 | Not-determined |
| S13_extracted_56828 | Not-determined |
| S13_extracted_56831 | Not-determined |
| S13_extracted_56834 | Not-determined |
| S13_extracted_56836 | Not-determined |
| S13_extracted_56866 | Not-determined |
| S13_extracted_56873 | Not-determined |
| S13_extracted_56901 | Low-quality |
| S13_extracted_56903 | Not-determined |
| S13_extracted_56924 | Not-determined |
| S13_extracted_56927 | Not-determined |
| S13_extracted_56963 | Not-determined |
| S13_extracted_56965 | Not-determined |
| S13_extracted_56966 | Not-determined |
| S13_extracted_56984 | Not-determined |
| S13_extracted_56987 | Not-determined |
| S13_extracted_57010 | Not-determined |
| S13_extracted_57011 | Not-determined |
| S13_extracted_57014 | Low-quality |
| S13_extracted_57017 | Not-determined |
| S13_extracted_57018 | Low-quality |
| S13_extracted_57021 | Not-determined |
| S13_extracted_57026 | Not-determined |
| S13_extracted_57028 | Not-determined |
| S13_extracted_57033 | Not-determined |
| S13_extracted_57034 | Not-determined |
| S13_extracted_57041 | Not-determined |
| S13_extracted_57044 | Not-determined |
| S13_extracted_57049 | Not-determined |
| S13_extracted_57051 | Not-determined |
| S13_extracted_57070 | Not-determined |
| S13_extracted_57078 | Not-determined |
| S13_extracted_57090 | Not-determined |
| S13_extracted_57097 | Not-determined |
| S13_extracted_57101 | Not-determined |
| S13_extracted_57103 | Not-determined |
| S13_extracted_57107 | Not-determined |
| S13_extracted_57109 | Not-determined |
| S13_extracted_57111 | Not-determined |
| S13_extracted_57117 | Not-determined |
| S13_extracted_57129 | Not-determined |
| S13_extracted_57137 | Not-determined |
| S13_extracted_57150 | Not-determined |
| S13_extracted_57151 | Not-determined |
| S13_extracted_57154 | Low-quality |
| S13_extracted_57158 | Not-determined |
| S13_extracted_57166 | Not-determined |
| S13_extracted_57175 | Not-determined |
| S13_extracted_57185 | Not-determined |
| S13_extracted_57198 | Not-determined |
| S13_extracted_57205 | Not-determined |
| S13_extracted_57216 | Not-determined |
| S13_extracted_57224 | Not-determined |
| S13_extracted_57227 | Not-determined |
| S13_extracted_57228 | Not-determined |
| S13_extracted_57234 | Not-determined |
| S13_extracted_57276 | Not-determined |
| S13_extracted_57285 | Not-determined |
| S13_extracted_57288 | Not-determined |
| S13_extracted_57292 | Not-determined |
| S13_extracted_57321 | Not-determined |
| S13_extracted_57330 | Not-determined |
| S13_extracted_57338 | Not-determined |
| S13_extracted_57346 | Not-determined |
| S13_extracted_57352 | Not-determined |
| S13_extracted_57353 | Not-determined |
| S13_extracted_57358 | Not-determined |
| S13_extracted_57372 | Not-determined |
| S13_extracted_57374 | Not-determined |
| S13_extracted_57386 | Not-determined |
| S13_extracted_57409 | Not-determined |
| S13_extracted_57415 | Not-determined |
| S13_extracted_57421 | Low-quality |
| S13_extracted_57426 | Not-determined |
| S13_extracted_57427 | Not-determined |
| S13_extracted_57432 | Not-determined |
| S13_extracted_57433 | Not-determined |
| S13_extracted_57438 | Not-determined |
| S13_extracted_57444 | Not-determined |
| S13_extracted_57469 | Not-determined |
| S13_extracted_57484 | Not-determined |
| S13_extracted_57528 | Not-determined |
| S13_extracted_57530 | Not-determined |
| S13_extracted_57533 | Not-determined |
| S13_extracted_57542 | Not-determined |
| S13_extracted_57545 | Not-determined |
| S13_extracted_57548 | Not-determined |
| S13_extracted_57551 | Not-determined |
| S13_extracted_57563 | Not-determined |
| S13_extracted_57565 | Not-determined |
| S13_extracted_57570 | Not-determined |
| S13_extracted_57572 | Not-determined |
| S13_extracted_57575 | Not-determined |
| S13_extracted_57583 | Not-determined |
| S13_extracted_57585 | Not-determined |
| S13_extracted_57594 | Not-determined |
| S13_extracted_57602 | Not-determined |
| S13_extracted_57609 | Not-determined |
| S13_extracted_57610 | Not-determined |
| S13_extracted_57634 | Not-determined |
| S13_extracted_57638 | Not-determined |
| S13_extracted_57642 | Low-quality |
| S13_extracted_57649 | Not-determined |
| S13_extracted_57652 | Not-determined |
| S13_extracted_57657 | Not-determined |
| S13_extracted_57662 | Not-determined |
| S13_extracted_57673 | Not-determined |
| S13_extracted_57674 | Not-determined |
| S13_extracted_57680 | Not-determined |
| S13_extracted_57686 | Not-determined |
| S13_extracted_57690 | Not-determined |
| S13_extracted_57699 | Not-determined |
| S13_extracted_57700 | Low-quality |
| S13_extracted_57711 | Not-determined |
| S13_extracted_57722 | Not-determined |
| S13_extracted_57723 | Not-determined |
| S13_extracted_57724 | Low-quality |
| S13_extracted_57727 | Not-determined |
| S13_extracted_57743 | Low-quality |
| S13_extracted_57755 | Not-determined |
| S13_extracted_57764 | Not-determined |
| S13_extracted_57766 | Not-determined |
| S13_extracted_57782 | Not-determined |
| S13_extracted_57786 | Not-determined |
| S13_extracted_57787 | Not-determined |
| S13_extracted_57792 | Not-determined |
| S13_extracted_57798 | Not-determined |
| S13_extracted_57823 | Not-determined |
| S13_extracted_57826 | Not-determined |
| S13_extracted_57831 | Not-determined |
| S13_extracted_57832 | Not-determined |
| S13_extracted_57862 | Not-determined |
| S13_extracted_57865 | Not-determined |
| S13_extracted_57867 | Not-determined |
| S13_extracted_57874 | Not-determined |
| S13_extracted_57881 | Not-determined |
| S13_extracted_57899 | Not-determined |
| S13_extracted_57902 | Not-determined |
| S13_extracted_57910 | Not-determined |
| S13_extracted_57913 | Not-determined |
| S13_extracted_57919 | Not-determined |
| S13_extracted_57924 | Not-determined |
| S13_extracted_57925 | Low-quality |
| S13_extracted_57927 | Not-determined |
| S13_extracted_57934 | Not-determined |
| S13_extracted_57935 | Not-determined |
| S13_extracted_57941 | Low-quality |
| S13_extracted_57957 | Not-determined |
| S13_extracted_57961 | Not-determined |
| S13_extracted_57964 | Not-determined |
| S13_extracted_57965 | Not-determined |
| S13_extracted_57970 | Not-determined |
| S13_extracted_57973 | Not-determined |
| S13_extracted_57975 | Not-determined |
| S13_extracted_57986 | Not-determined |
| S13_extracted_57987 | Low-quality |
| S13_extracted_57992 | Not-determined |
| S13_extracted_57999 | Not-determined |
| S13_extracted_58004 | Not-determined |
| S13_extracted_58016 | Not-determined |
| S13_extracted_58017 | Not-determined |
| S13_extracted_58020 | Low-quality |
| S13_extracted_58022 | Low-quality |
| S13_extracted_58027 | Not-determined |
| S13_extracted_58029 | Low-quality |
| S13_extracted_58030 | Not-determined |
| S13_extracted_58034 | Low-quality |
| S13_extracted_58038 | Not-determined |
| S13_extracted_58063 | Low-quality |
| S13_extracted_58075 | Not-determined |
| S13_extracted_58102 | Not-determined |
| S13_extracted_58111 | Not-determined |
| S13_extracted_58122 | Not-determined |
| S13_extracted_58123 | Not-determined |
| S13_extracted_58124 | Not-determined |
| S13_extracted_58152 | Not-determined |
| S13_extracted_58160 | Not-determined |
| S13_extracted_58164 | Not-determined |
| S13_extracted_58166 | Not-determined |
| S13_extracted_58169 | Not-determined |
| S13_extracted_58191 | Low-quality |
| S13_extracted_58194 | Not-determined |
| S13_extracted_58207 | Not-determined |
| S13_extracted_58265 | Not-determined |
| S13_extracted_58268 | Not-determined |
| S13_extracted_58274 | Not-determined |
| S13_extracted_58280 | Not-determined |
| S13_extracted_58281 | Low-quality |
| S13_extracted_58289 | Not-determined |
| S13_extracted_58295 | Not-determined |
| S13_extracted_58296 | Not-determined |
| S13_extracted_58303 | Not-determined |
| S13_extracted_58306 | Not-determined |
| S13_extracted_58307 | Low-quality |
| S13_extracted_58308 | Not-determined |
| S13_extracted_58322 | Not-determined |
| S13_extracted_58351 | Not-determined |
| S13_extracted_58376 | Not-determined |
| S13_extracted_58386 | Not-determined |
| S13_extracted_58389 | Not-determined |
| S13_extracted_58396 | Not-determined |
| S13_extracted_58407 | Low-quality |
| S13_extracted_58408 | Low-quality |
| S13_extracted_58423 | Not-determined |
| S13_extracted_58436 | Not-determined |
| S13_extracted_58437 | Not-determined |
| S13_extracted_58449 | Not-determined |
| S13_extracted_58466 | Not-determined |
| S13_extracted_58467 | Not-determined |
| S13_extracted_58469 | Not-determined |
| S13_extracted_58470 | Not-determined |
| S13_extracted_58471 | Not-determined |
| S13_extracted_58477 | Not-determined |
| S13_extracted_58486 | Not-determined |
| S13_extracted_58489 | Not-determined |
| S13_extracted_58494 | Not-determined |
| S13_extracted_58495 | Not-determined |
| S13_extracted_58501 | Not-determined |
| S13_extracted_58504 | Not-determined |
| S13_extracted_58511 | Low-quality |
| S13_extracted_58512 | Low-quality |
| S13_extracted_58523 | Not-determined |
| S13_extracted_58527 | Not-determined |
| S13_extracted_58536 | Low-quality |
| S13_extracted_58538 | Not-determined |
| S13_extracted_58540 | Not-determined |
| S13_extracted_58542 | Not-determined |
| S13_extracted_58550 | Not-determined |
| S13_extracted_58557 | Not-determined |
| S13_extracted_58572 | Not-determined |
| S13_extracted_58577 | Not-determined |
| S13_extracted_58578 | Not-determined |
| S13_extracted_58581 | Not-determined |
| S13_extracted_58588 | Not-determined |
| S13_extracted_58595 | Not-determined |
| S13_extracted_58597 | Not-determined |
| S13_extracted_58610 | Not-determined |
| S13_extracted_58621 | Low-quality |
| S13_extracted_58624 | Not-determined |
| S13_extracted_58626 | Not-determined |
| S13_extracted_58631 | Low-quality |
| S13_extracted_58639 | Not-determined |
| S13_extracted_58650 | Not-determined |
| S13_extracted_58671 | Low-quality |
| S13_extracted_58680 | Low-quality |
| S13_extracted_58704 | Not-determined |
| S13_extracted_58719 | Not-determined |
| S13_extracted_58747 | Not-determined |
| S13_extracted_58770 | Low-quality |
| S13_extracted_58771 | Not-determined |
| S13_extracted_58779 | Not-determined |
| S13_extracted_58797 | Not-determined |
| S13_extracted_58810 | Not-determined |
| S13_extracted_58815 | Not-determined |
| S13_extracted_58818 | Not-determined |
| S13_extracted_58824 | Not-determined |
| S13_extracted_58828 | Not-determined |
| S13_extracted_58836 | Not-determined |
| S13_extracted_58841 | Low-quality |
| S13_extracted_58852 | Not-determined |
| S13_extracted_58853 | Low-quality |
| S13_extracted_58859 | Not-determined |
| S13_extracted_58862 | Not-determined |
| S13_extracted_58866 | Not-determined |
| S13_extracted_58874 | Not-determined |
| S13_extracted_58878 | Not-determined |
| S13_extracted_58906 | Low-quality |
| S13_extracted_58918 | Not-determined |
| S13_extracted_58919 | Not-determined |
| S13_extracted_58927 | Not-determined |
| S13_extracted_58941 | Not-determined |
| S13_extracted_58945 | Not-determined |
| S13_extracted_58950 | Not-determined |
| S13_extracted_58953 | Not-determined |
| S13_extracted_58971 | Not-determined |
| S13_extracted_58986 | Not-determined |
| S13_extracted_58987 | Low-quality |
| S13_extracted_59001 | Not-determined |
| S13_extracted_59008 | Not-determined |
| S13_extracted_59017 | Not-determined |
| S13_extracted_59022 | Not-determined |
| S13_extracted_59054 | Not-determined |
| S13_extracted_59062 | Not-determined |
| S13_extracted_59064 | Not-determined |
| S13_extracted_59069 | Not-determined |
| S13_extracted_59075 | Not-determined |
| S13_extracted_59077 | Not-determined |
| S13_extracted_59085 | Not-determined |
| S13_extracted_59088 | Not-determined |
| S13_extracted_59100 | Low-quality |
| S13_extracted_59111 | Not-determined |
| S13_extracted_59118 | Low-quality |
| S13_extracted_59119 | Not-determined |
| S13_extracted_59133 | Not-determined |
| S13_extracted_59151 | Not-determined |
| S13_extracted_59175 | Not-determined |
| S13_extracted_59178 | Not-determined |
| S13_extracted_59182 | Not-determined |
| S13_extracted_59188 | Not-determined |
| S13_extracted_59201 | Low-quality |
| S13_extracted_59209 | Not-determined |
| S13_extracted_59212 | Not-determined |
| S13_extracted_59215 | Low-quality |
| S13_extracted_59244 | Not-determined |
| S13_extracted_59245 | Not-determined |
| S13_extracted_59255 | Not-determined |
| S13_extracted_59257 | Not-determined |
| S13_extracted_59270 | Not-determined |
| S13_extracted_59310 | Not-determined |
| S13_extracted_59312 | Not-determined |
| S13_extracted_59319 | Low-quality |
| S13_extracted_59324 | Not-determined |
| S13_extracted_59326 | Low-quality |
| S13_extracted_59343 | Low-quality |
| S13_extracted_59346 | Not-determined |
| S13_extracted_59349 | Low-quality |
| S13_extracted_59358 | Not-determined |
| S13_extracted_59362 | Not-determined |
| S13_extracted_59363 | Not-determined |
| S13_extracted_59369 | Not-determined |
| S13_extracted_59374 | Not-determined |
| S13_extracted_59375 | Not-determined |
| S13_extracted_59399 | Not-determined |
| S13_extracted_59403 | Low-quality |
| S13_extracted_59408 | Not-determined |
| S13_extracted_59413 | Not-determined |
| S13_extracted_59418 | Not-determined |
| S13_extracted_59419 | Not-determined |
| S13_extracted_59426 | Not-determined |
| S13_extracted_59428 | Not-determined |
| S13_extracted_59434 | Low-quality |
| S13_extracted_59438 | Not-determined |
| S13_extracted_59441 | Not-determined |
| S13_extracted_59443 | Not-determined |
| S13_extracted_59450 | Not-determined |
| S13_extracted_59464 | Not-determined |
| S13_extracted_59471 | Not-determined |
| S13_extracted_59473 | Not-determined |
| S13_extracted_59482 | Not-determined |
| S13_extracted_59483 | Not-determined |
| S13_extracted_59500 | Low-quality |
| S13_extracted_59526 | Not-determined |
| S13_extracted_59530 | Not-determined |
| S13_extracted_59536 | Not-determined |
| S13_extracted_59548 | Not-determined |
| S13_extracted_59560 | Low-quality |
| S13_extracted_59561 | Not-determined |
| S13_extracted_59562 | Not-determined |
| S13_extracted_59565 | Not-determined |
| S13_extracted_59570 | Not-determined |
| S13_extracted_59580 | Not-determined |
| S13_extracted_59597 | Not-determined |
| S13_extracted_59602 | Low-quality |
| S13_extracted_59611 | Low-quality |
| S13_extracted_59615 | Not-determined |
| S13_extracted_59642 | Not-determined |
| S13_extracted_59643 | Not-determined |
| S13_extracted_59644 | Not-determined |
| S13_extracted_59659 | Not-determined |
| S13_extracted_59663 | Low-quality |
| S13_extracted_59683 | Not-determined |
| S13_extracted_59688 | Not-determined |
| S13_extracted_59689 | Low-quality |
| S13_extracted_59712 | Not-determined |
| S13_extracted_59727 | Not-determined |
| S13_extracted_59730 | Not-determined |
| S13_extracted_59732 | Not-determined |
| S13_extracted_59737 | Not-determined |
| S13_extracted_59751 | Low-quality |
| S13_extracted_59769 | Not-determined |
| S13_extracted_59770 | Not-determined |
| S13_extracted_59784 | Not-determined |
| S13_extracted_59790 | Low-quality |
| S13_extracted_59792 | Not-determined |
| S13_extracted_59800 | Not-determined |
| S13_extracted_59823 | Not-determined |
| S13_extracted_59825 | Not-determined |
| S13_extracted_59837 | Not-determined |
| S13_extracted_59855 | Not-determined |
| S13_extracted_59862 | Not-determined |
| S13_extracted_59890 | Not-determined |
| S13_extracted_59894 | Not-determined |
| S13_extracted_59896 | Not-determined |
| S13_extracted_59944 | Not-determined |
| S13_extracted_59946 | Not-determined |
| S13_extracted_59948 | Not-determined |
| S13_extracted_59949 | Not-determined |
| S13_extracted_59973 | Not-determined |
| S13_extracted_59979 | Not-determined |
| S13_extracted_59995 | Not-determined |
| S13_extracted_60004 | Not-determined |
| S13_extracted_60009 | Not-determined |
| S13_extracted_60011 | Low-quality |
| S13_extracted_60013 | Not-determined |
| S13_extracted_60014 | Not-determined |
| S13_extracted_60017 | Not-determined |
| S13_extracted_60039 | Not-determined |
| S13_extracted_60043 | Not-determined |
| S13_extracted_60075 | Not-determined |
| S13_extracted_60080 | Low-quality |
| S13_extracted_60089 | Not-determined |
| S13_extracted_60101 | Not-determined |
| S13_extracted_60105 | Not-determined |

| | |
|---|---|
| S13_extracted_60108 | Not-determined |
| S13_extracted_60111 | Not-determined |
| S13_extracted_60128 | Not-determined |
| S13_extracted_60138 | Not-determined |
| S13_extracted_60155 | Low-quality |
| S13_extracted_60162 | Not-determined |
| S13_extracted_60187 | Not-determined |
| S13_extracted_60194 | Not-determined |
| S13_extracted_60204 | Not-determined |
| S13_extracted_60229 | Low-quality |
| S13_extracted_60240 | Not-determined |
| S13_extracted_60241 | Not-determined |
| S13_extracted_60247 | Not-determined |
| S13_extracted_60250 | Not-determined |
| S13_extracted_60254 | Not-determined |
| S13_extracted_60276 | Low-quality |
| S13_extracted_60282 | Not-determined |
| S13_extracted_60289 | Not-determined |
| S13_extracted_60294 | Not-determined |
| S13_extracted_60305 | Low-quality |
| S13_extracted_60311 | Not-determined |
| S13_extracted_60327 | Not-determined |
| S13_extracted_60341 | Not-determined |
| S13_extracted_60342 | Not-determined |
| S13_extracted_60344 | Not-determined |
| S13_extracted_60351 | Low-quality |
| S13_extracted_60356 | Not-determined |
| S13_extracted_60394 | Not-determined |
| S13_extracted_60395 | Not-determined |
| S13_extracted_60401 | Low-quality |
| S13_extracted_60406 | Not-determined |
| S13_extracted_60419 | Not-determined |
| S13_extracted_60427 | Not-determined |
| S13_extracted_60483 | Low-quality |
| S13_extracted_60485 | Not-determined |
| S13_extracted_60487 | Not-determined |
| S13_extracted_60490 | Not-determined |
| S13_extracted_60496 | Not-determined |
| S13_extracted_60516 | Not-determined |
| S13_extracted_60517 | Not-determined |
| S13_extracted_60530 | Not-determined |
| S13_extracted_60532 | Not-determined |
| S13_extracted_60542 | Not-determined |
| S13_extracted_60546 | Not-determined |
| S13_extracted_60549 | Not-determined |
| S13_extracted_60594 | Not-determined |
| S13_extracted_60600 | Not-determined |
| S13_extracted_60608 | Not-determined |
| S13_extracted_60611 | Not-determined |
| S13_extracted_60619 | Not-determined |
| S13_extracted_60623 | Low-quality |
| S13_extracted_60643 | Not-determined |
| S13_extracted_60655 | Not-determined |
| S13_extracted_60674 | Not-determined |
| S13_extracted_60676 | Not-determined |
| S13_extracted_60677 | Not-determined |
| S13_extracted_60680 | Not-determined |
| S13_extracted_60684 | Not-determined |
| S13_extracted_60690 | Not-determined |
| S13_extracted_60697 | Not-determined |
| S13_extracted_60699 | Not-determined |
| S13_extracted_60700 | Not-determined |
| S13_extracted_60710 | Not-determined |
| S13_extracted_60732 | Not-determined |
| S13_extracted_60764 | Low-quality |
| S13_extracted_60765 | Not-determined |
| S13_extracted_60767 | Not-determined |
| S13_extracted_60769 | Not-determined |
| S13_extracted_60772 | Low-quality |
| S13_extracted_60778 | Not-determined |
| S13_extracted_60797 | Not-determined |
| S13_extracted_60798 | Not-determined |
| S13_extracted_60816 | Not-determined |
| S13_extracted_60817 | Not-determined |
| S13_extracted_60822 | Not-determined |
| S13_extracted_60826 | Not-determined |
| S13_extracted_60827 | Not-determined |
| S13_extracted_60830 | Low-quality |
| S13_extracted_60841 | Not-determined |
| S13_extracted_60843 | Not-determined |
| S13_extracted_60844 | Not-determined |
| S13_extracted_60867 | Not-determined |
| S13_extracted_60872 | Not-determined |
| S13_extracted_60880 | Not-determined |
| S13_extracted_60883 | Not-determined |
| S13_extracted_60888 | Not-determined |
| S13_extracted_60891 | Not-determined |
| S13_extracted_60897 | Not-determined |
| S13_extracted_60920 | Low-quality |
| S13_extracted_60925 | Not-determined |
| S13_extracted_60953 | Not-determined |
| S13_extracted_60970 | Not-determined |
| S13_extracted_60991 | Not-determined |
| S13_extracted_60995 | Low-quality |
| S13_extracted_60999 | Not-determined |
| S13_extracted_61001 | Not-determined |
| S13_extracted_61012 | Not-determined |
| S13_extracted_61014 | Low-quality |
| S13_extracted_61016 | Not-determined |
| S13_extracted_61024 | Low-quality |
| S13_extracted_61034 | Not-determined |
| S13_extracted_61042 | Not-determined |
| S13_extracted_61044 | Low-quality |
| S13_extracted_61045 | Not-determined |
| S13_extracted_61048 | Not-determined |
| S13_extracted_61052 | Low-quality |
| S13_extracted_61057 | Not-determined |
| S13_extracted_61071 | Not-determined |
| S13_extracted_61072 | Not-determined |
| S13_extracted_61081 | Not-determined |
| S13_extracted_61085 | Not-determined |
| S13_extracted_61087 | Not-determined |
| S13_extracted_61094 | Not-determined |
| S13_extracted_61100 | Not-determined |
| S13_extracted_61101 | Not-determined |
| S13_extracted_61108 | Low-quality |
| S13_extracted_61109 | Not-determined |
| S13_extracted_61115 | Not-determined |
| S13_extracted_61117 | Not-determined |
| S13_extracted_61118 | Not-determined |
| S13_extracted_61120 | Not-determined |
| S13_extracted_61134 | Not-determined |
| S13_extracted_61139 | Not-determined |
| S13_extracted_61140 | Not-determined |
| S13_extracted_61144 | Not-determined |
| S13_extracted_61158 | Not-determined |
| S13_extracted_61181 | Not-determined |
| S13_extracted_61216 | Low-quality |
| S13_extracted_61229 | Not-determined |
| S13_extracted_61230 | Not-determined |
| S13_extracted_61231 | Not-determined |
| S13_extracted_61233 | Low-quality |
| S13_extracted_61240 | Not-determined |
| S13_extracted_61256 | Not-determined |
| S13_extracted_61258 | Low-quality |
| S13_extracted_61276 | Not-determined |
| S13_extracted_61288 | Not-determined |
| S13_extracted_61295 | Not-determined |
| S13_extracted_61297 | Not-determined |
| S13_extracted_61313 | Not-determined |
| S13_extracted_61319 | Not-determined |
| S13_extracted_61323 | Not-determined |
| S13_extracted_61328 | Not-determined |
| S13_extracted_61329 | Not-determined |
| S13_extracted_61331 | Not-determined |
| S13_extracted_61333 | Low-quality |
| S13_extracted_61344 | Not-determined |
| S13_extracted_61359 | Not-determined |
| S13_extracted_61362 | Low-quality |
| S13_extracted_61368 | Not-determined |
| S13_extracted_61369 | Not-determined |
| S13_extracted_61370 | Not-determined |
| S13_extracted_61371 | Not-determined |
| S13_extracted_61373 | Not-determined |
| S13_extracted_61378 | Not-determined |
| S13_extracted_61386 | Not-determined |
| S13_extracted_61392 | Low-quality |
| S13_extracted_61405 | Not-determined |
| S13_extracted_61409 | Not-determined |
| S13_extracted_61425 | Not-determined |
| S13_extracted_61429 | Not-determined |
| S13_extracted_61447 | Not-determined |
| S13_extracted_61451 | Not-determined |
| S13_extracted_61452 | Not-determined |
| S13_extracted_61474 | Not-determined |
| S13_extracted_61475 | Not-determined |
| S13_extracted_61479 | Not-determined |
| S13_extracted_61481 | Not-determined |
| S13_extracted_61496 | Not-determined |
| S13_extracted_61501 | Not-determined |
| S13_extracted_61502 | Not-determined |
| S13_extracted_61504 | Not-determined |
| S13_extracted_61509 | Not-determined |
| S13_extracted_61518 | Not-determined |
| S13_extracted_61521 | Not-determined |
| S13_extracted_61524 | Not-determined |
| S13_extracted_61536 | Not-determined |
| S13_extracted_61546 | Not-determined |
| S13_extracted_61563 | Not-determined |
| S13_extracted_61588 | Not-determined |
| S13_extracted_61593 | Not-determined |
| S13_extracted_61612 | Not-determined |
| S13_extracted_61613 | Not-determined |
| S13_extracted_61622 | Not-determined |
| S13_extracted_61628 | Not-determined |
| S13_extracted_61643 | Low-quality |
| S13_extracted_61658 | Not-determined |
| S13_extracted_61660 | Not-determined |
| S13_extracted_61664 | Not-determined |
| S13_extracted_61680 | Low-quality |
| S13_extracted_61682 | Not-determined |
| S13_extracted_61684 | Not-determined |
| S13_extracted_61686 | Not-determined |
| S13_extracted_61690 | Low-quality |
| S13_extracted_61695 | Low-quality |
| S13_extracted_61719 | Not-determined |
| S13_extracted_61725 | Not-determined |
| S13_extracted_61728 | Not-determined |
| S13_extracted_61731 | Not-determined |
| S13_extracted_61733 | Not-determined |
| S13_extracted_61762 | Not-determined |
| S13_extracted_61765 | Not-determined |
| S13_extracted_61789 | Low-quality |
| S13_extracted_61791 | Not-determined |
| S13_extracted_61801 | Not-determined |
| S13_extracted_61814 | Not-determined |
| S13_extracted_61829 | Not-determined |
| S13_extracted_61831 | Not-determined |
| S13_extracted_61840 | Not-determined |
| S13_extracted_61848 | Not-determined |
| S13_extracted_61880 | Low-quality |
| S13_extracted_61886 | Not-determined |
| S13_extracted_61890 | Not-determined |
| S13_extracted_61904 | Not-determined |
| S13_extracted_61910 | Not-determined |
| S13_extracted_61913 | Low-quality |
| S13_extracted_61928 | Not-determined |
| S13_extracted_61931 | Not-determined |
| S13_extracted_61956 | Not-determined |
| S13_extracted_61957 | Not-determined |
| S13_extracted_61961 | Not-determined |
| S13_extracted_61962 | Low-quality |
| S13_extracted_61970 | Not-determined |
| S13_extracted_61972 | Not-determined |
| S13_extracted_61976 | Not-determined |
| S13_extracted_61991 | Not-determined |
| S13_extracted_61993 | Not-determined |
| S13_extracted_62001 | Not-determined |
| S13_extracted_62005 | Not-determined |
| S13_extracted_62006 | Not-determined |
| S13_extracted_62038 | Low-quality |
| S13_extracted_62043 | Not-determined |
| S13_extracted_62045 | Not-determined |
| S13_extracted_62048 | Not-determined |
| S13_extracted_62050 | Not-determined |
| S13_extracted_62056 | Not-determined |
| S13_extracted_62060 | Not-determined |
| S13_extracted_62072 | Not-determined |
| S13_extracted_62075 | Not-determined |
| S13_extracted_62090 | Not-determined |
| S13_extracted_62093 | Not-determined |
| S13_extracted_62095 | Not-determined |
| S13_extracted_62116 | Low-quality |
| S13_extracted_62128 | Not-determined |
| S13_extracted_62135 | Not-determined |
| S13_extracted_62155 | Not-determined |
| S13_extracted_62172 | Not-determined |
| S13_extracted_62176 | Not-determined |
| S13_extracted_62179 | Not-determined |
| S13_extracted_62180 | Not-determined |
| S13_extracted_62189 | Not-determined |
| S13_extracted_62212 | Not-determined |
| S13_extracted_62245 | Not-determined |
| S13_extracted_62255 | Low-quality |
| S13_extracted_62262 | Not-determined |
| S13_extracted_62271 | Not-determined |
| S13_extracted_62307 | Not-determined |
| S13_extracted_62319 | Not-determined |
| S13_extracted_62321 | Not-determined |
| S13_extracted_62325 | Low-quality |
| S13_extracted_62328 | Not-determined |
| S13_extracted_62373 | Not-determined |
| S13_extracted_62391 | Not-determined |
| S13_extracted_62392 | Low-quality |
| S13_extracted_62411 | Not-determined |
| S13_extracted_62413 | Not-determined |
| S13_extracted_62414 | Not-determined |
| S13_extracted_62415 | Not-determined |
| S13_extracted_62424 | Not-determined |
| S13_extracted_62432 | Not-determined |
| S13_extracted_62434 | Not-determined |
| S13_extracted_62445 | Not-determined |
| S13_extracted_62451 | Not-determined |
| S13_extracted_62452 | Not-determined |
| S13_extracted_62462 | Not-determined |
| S13_extracted_62471 | Low-quality |
| S13_extracted_62472 | Not-determined |
| S13_extracted_62482 | Not-determined |
| S13_extracted_62492 | Low-quality |
| S13_extracted_62500 | Not-determined |
| S13_extracted_62528 | Not-determined |
| S13_extracted_62543 | Low-quality |
| S13_extracted_62547 | Not-determined |
| S13_extracted_62560 | Not-determined |
| S13_extracted_62562 | Not-determined |
| S13_extracted_62565 | Not-determined |
| S13_extracted_62581 | Not-determined |
| S13_extracted_62585 | Low-quality |
| S13_extracted_62600 | Low-quality |
| S13_extracted_62608 | Not-determined |
| S13_extracted_62617 | Not-determined |
| S13_extracted_62629 | Not-determined |
| S13_extracted_62631 | Not-determined |
| S13_extracted_62633 | Not-determined |
| S13_extracted_62644 | Not-determined |
| S13_extracted_62649 | Not-determined |
| S13_extracted_62660 | Not-determined |
| S13_extracted_62669 | Not-determined |
| S13_extracted_62675 | Not-determined |
| S13_extracted_62677 | Low-quality |
| S13_extracted_62682 | Not-determined |
| S13_extracted_62685 | Not-determined |
| S13_extracted_62695 | Not-determined |
| S13_extracted_62720 | Not-determined |
| S13_extracted_62731 | Not-determined |
| S13_extracted_62736 | Not-determined |
| S13_extracted_62746 | Not-determined |
| S13_extracted_62752 | Not-determined |
| S13_extracted_62766 | Not-determined |
| S13_extracted_62777 | Not-determined |
| S13_extracted_62783 | Not-determined |
| S13_extracted_62785 | Not-determined |
| S13_extracted_62799 | Not-determined |
| S13_extracted_62802 | Not-determined |
| S13_extracted_62803 | Not-determined |
| S13_extracted_62808 | Low-quality |
| S13_extracted_62820 | Not-determined |
| S13_extracted_62822 | Low-quality |
| S13_extracted_62835 | Not-determined |
| S13_extracted_62843 | Not-determined |
| S13_extracted_62849 | Not-determined |
| S13_extracted_62856 | Not-determined |
| S13_extracted_62858 | Not-determined |
| S13_extracted_62867 | Not-determined |
| S13_extracted_62874 | Not-determined |
| S13_extracted_62882 | Not-determined |
| S13_extracted_62907 | Not-determined |
| S13_extracted_62908 | Not-determined |
| S13_extracted_62911 | Not-determined |
| S13_extracted_62930 | Not-determined |
| S13_extracted_62933 | Not-determined |
| S13_extracted_62960 | Not-determined |
| S13_extracted_62971 | Low-quality |
| S13_extracted_62973 | Low-quality |
| S13_extracted_62975 | Low-quality |
| S13_extracted_62984 | Not-determined |
| S13_extracted_62985 | Not-determined |
| S13_extracted_62995 | Not-determined |
| S13_extracted_63012 | Not-determined |
| S13_extracted_63013 | Not-determined |
| S13_extracted_63015 | Not-determined |
| S13_extracted_63023 | Not-determined |
| S13_extracted_63028 | Not-determined |
| S13_extracted_63044 | Low-quality |
| S13_extracted_63047 | Not-determined |
| S13_extracted_63060 | Not-determined |
| S13_extracted_63062 | Not-determined |
| S13_extracted_63064 | Not-determined |
| S13_extracted_63074 | Not-determined |
| S13_extracted_63075 | Low-quality |
| S13_extracted_63084 | Not-determined |
| S13_extracted_63099 | Not-determined |
| S13_extracted_63104 | Not-determined |
| S13_extracted_63108 | Not-determined |
| S13_extracted_63119 | Not-determined |
| S13_extracted_63154 | Not-determined |
| S13_extracted_63155 | Not-determined |
| S13_extracted_63167 | Not-determined |
| S13_extracted_63194 | Not-determined |
| S13_extracted_63205 | Not-determined |
| S13_extracted_63215 | Not-determined |
| S13_extracted_63258 | Not-determined |
| S13_extracted_63264 | Not-determined |
| S13_extracted_63266 | Not-determined |
| S13_extracted_63268 | Not-determined |
| S13_extracted_63276 | Low-quality |
| S13_extracted_63294 | Low-quality |
| S13_extracted_63297 | Not-determined |
| S13_extracted_63300 | Not-determined |
| S13_extracted_63308 | Not-determined |
| S13_extracted_63309 | Not-determined |
| S13_extracted_63321 | Not-determined |
| S13_extracted_63335 | Low-quality |
| S13_extracted_63338 | Not-determined |
| S13_extracted_63339 | Low-quality |
| S13_extracted_63344 | Not-determined |
| S13_extracted_63347 | Low-quality |
| S13_extracted_63368 | Low-quality |
| S13_extracted_63376 | Not-determined |
| S13_extracted_63393 | Not-determined |
| S13_extracted_63395 | Not-determined |
| S13_extracted_63398 | Not-determined |
| S13_extracted_63402 | Not-determined |
| S13_extracted_63404 | Not-determined |
| S13_extracted_63408 | Not-determined |
| S13_extracted_63429 | Not-determined |
| S13_extracted_63437 | Low-quality |
| S13_extracted_63440 | Not-determined |
| S13_extracted_63452 | Not-determined |
| S13_extracted_63461 | Low-quality |
| S13_extracted_63463 | Not-determined |
| S13_extracted_63472 | Low-quality |
| S13_extracted_63478 | Not-determined |
| S13_extracted_63480 | Not-determined |
| S13_extracted_63495 | Not-determined |
| S13_extracted_63500 | Low-quality |
| S13_extracted_63514 | Not-determined |
| S13_extracted_63519 | Not-determined |
| S13_extracted_63529 | Not-determined |
| S13_extracted_63538 | Low-quality |
| S13_extracted_63546 | Not-determined |
| S13_extracted_63559 | Not-determined |
| S13_extracted_63565 | Not-determined |
| S13_extracted_63573 | Not-determined |
| S13_extracted_63582 | Not-determined |
| S13_extracted_63584 | Not-determined |
| S13_extracted_63587 | Not-determined |
| S13_extracted_63588 | Not-determined |
| S13_extracted_63610 | Not-determined |
| S13_extracted_63612 | Low-quality |
| S13_extracted_63616 | Not-determined |
| S13_extracted_63620 | Not-determined |
| S13_extracted_63634 | Not-determined |
| S13_extracted_63635 | Not-determined |
| S13_extracted_63660 | Not-determined |
| S13_extracted_63666 | Not-determined |
| S13_extracted_63670 | Not-determined |
| S13_extracted_63678 | Not-determined |
| S13_extracted_63692 | Not-determined |
| S13_extracted_63701 | Not-determined |
| S13_extracted_63703 | Low-quality |
| S13_extracted_63712 | Not-determined |
| S13_extracted_63723 | Not-determined |
| S13_extracted_63745 | Not-determined |
| S13_extracted_63750 | Low-quality |
| S13_extracted_63755 | Not-determined |
| S13_extracted_63759 | Not-determined |
| S13_extracted_63774 | Not-determined |
| S13_extracted_63785 | Low-quality |
| S13_extracted_63787 | Not-determined |
| S13_extracted_63789 | Not-determined |
| S13_extracted_63795 | Not-determined |
| S13_extracted_63813 | Not-determined |
| S13_extracted_63819 | Not-determined |
| S13_extracted_63849 | Low-quality |
| S13_extracted_63854 | Not-determined |
| S13_extracted_63856 | Not-determined |
| S13_extracted_63870 | Not-determined |
| S13_extracted_63895 | Not-determined |
| S13_extracted_63910 | Not-determined |
| S13_extracted_63916 | Not-determined |
| S13_extracted_63932 | Not-determined |
| S13_extracted_63937 | Not-determined |
| S13_extracted_63947 | Low-quality |
| S13_extracted_63956 | Not-determined |
| S13_extracted_63960 | Not-determined |
| S13_extracted_63970 | Not-determined |
| S13_extracted_63971 | Not-determined |
| S13_extracted_63976 | Not-determined |
| S13_extracted_63977 | Not-determined |
| S13_extracted_63983 | Not-determined |
| S13_extracted_63984 | Not-determined |
| S13_extracted_63992 | Not-determined |
| S13_extracted_64010 | Not-determined |
| S13_extracted_64018 | Not-determined |
| S13_extracted_64019 | Not-determined |
| S13_extracted_64020 | Not-determined |
| S13_extracted_64022 | Not-determined |
| S13_extracted_64048 | Not-determined |
| S13_extracted_64072 | Not-determined |
| S13_extracted_64084 | Not-determined |
| S13_extracted_64091 | Low-quality |
| S13_extracted_64098 | Not-determined |
| S13_extracted_64109 | Not-determined |
| S13_extracted_64114 | Not-determined |
| S13_extracted_64141 | Not-determined |
| S13_extracted_64148 | Not-determined |
| S13_extracted_64163 | Not-determined |
| S13_extracted_64168 | Not-determined |
| S13_extracted_64170 | Low-quality |
| S13_extracted_64177 | Not-determined |
| S13_extracted_64182 | Not-determined |
| S13_extracted_64188 | Low-quality |
| S13_extracted_64202 | Not-determined |
| S13_extracted_64203 | Not-determined |
| S13_extracted_64221 | Not-determined |
| S13_extracted_64224 | Not-determined |
| S13_extracted_64234 | Not-determined |
| S13_extracted_64236 | Not-determined |
| S13_extracted_64237 | Not-determined |
| S13_extracted_64248 | Not-determined |
| S13_extracted_64257 | Not-determined |
| S13_extracted_64266 | Not-determined |
| S13_extracted_64270 | Low-quality |
| S13_extracted_64273 | Not-determined |
| S13_extracted_64277 | Not-determined |
| S13_extracted_64280 | Not-determined |
| S13_extracted_64282 | Not-determined |
| S13_extracted_64293 | Not-determined |
| S13_extracted_64294 | Not-determined |
| S13_extracted_64298 | Not-determined |
| S13_extracted_64301 | Not-determined |
| S13_extracted_64304 | Not-determined |
| S13_extracted_64310 | Not-determined |
| S13_extracted_64323 | Not-determined |
| S13_extracted_64341 | Not-determined |
| S13_extracted_64344 | Not-determined |
| S13_extracted_64357 | Not-determined |
| S13_extracted_64360 | Not-determined |
| S13_extracted_64382 | Low-quality |
| S13_extracted_64399 | Not-determined |
| S13_extracted_64411 | Low-quality |
| S13_extracted_64453 | Not-determined |
| S13_extracted_64464 | Not-determined |
| S13_extracted_64475 | Not-determined |
| S13_extracted_64478 | Not-determined |
| S13_extracted_64491 | Not-determined |
| S13_extracted_64494 | Not-determined |
| S13_extracted_64497 | Not-determined |
| S13_extracted_64503 | Low-quality |
| S13_extracted_64510 | Not-determined |
| S13_extracted_64519 | Not-determined |
| S13_extracted_64523 | Not-determined |
| S13_extracted_64524 | Not-determined |
| S13_extracted_64533 | Not-determined |
| S13_extracted_64536 | Not-determined |
| S13_extracted_64554 | Not-determined |
| S13_extracted_64561 | Not-determined |
| S13_extracted_64587 | Not-determined |
| S13_extracted_64596 | Not-determined |
| S13_extracted_64604 | Not-determined |
| S13_extracted_64609 | Not-determined |
| S13_extracted_64612 | Not-determined |
| S13_extracted_64624 | Low-quality |
| S13_extracted_64627 | Not-determined |
| S13_extracted_64642 | Not-determined |
| S13_extracted_64656 | Low-quality |
| S13_extracted_64659 | Not-determined |
| S13_extracted_64664 | Not-determined |
| S13_extracted_64668 | Not-determined |
| S13_extracted_64676 | Not-determined |
| S13_extracted_64685 | Not-determined |
| S13_extracted_64688 | Not-determined |
| S13_extracted_64693 | Not-determined |
| S13_extracted_64694 | Not-determined |
| S13_extracted_64702 | Not-determined |
| S13_extracted_64711 | Not-determined |
| S13_extracted_64712 | Not-determined |
| S13_extracted_64714 | Not-determined |
| S13_extracted_64718 | Not-determined |
| S13_extracted_64725 | Low-quality |
| S13_extracted_64728 | Not-determined |
| S13_extracted_64738 | Not-determined |
| S13_extracted_64747 | Not-determined |
| S13_extracted_64749 | Not-determined |
| S13_extracted_64750 | Not-determined |
| S13_extracted_64764 | Not-determined |
| S13_extracted_64765 | Not-determined |
| S13_extracted_64779 | Not-determined |
| S13_extracted_64791 | Not-determined |
| S13_extracted_64795 | Not-determined |
| S13_extracted_64804 | Not-determined |
| S13_extracted_64812 | Not-determined |
| S13_extracted_64836 | Not-determined |
| S13_extracted_64858 | Low-quality |
| S13_extracted_64864 | Not-determined |
| S13_extracted_64865 | Not-determined |
| S13_extracted_64872 | Not-determined |
| S13_extracted_64877 | Not-determined |
| S13_extracted_64886 | Not-determined |
| S13_extracted_64888 | Low-quality |
| S3_vs_k141_61860\|\|full | High-quality |
| S13_extracted_64894 | Low-quality |
| S13_extracted_64901 | Not-determined |
| S13_extracted_64903 | Not-determined |
| S13_extracted_64911 | Not-determined |
| S13_extracted_64916 | Not-determined |
| S13_extracted_64924 | Not-determined |
| S13_extracted_64937 | Not-determined |
| S13_extracted_64941 | Not-determined |
| S13_extracted_64944 | Low-quality |
| S13_extracted_64952 | Not-determined |
| S13_extracted_64954 | Not-determined |
| S13_extracted_64959 | Not-determined |
| S13_extracted_64962 | Not-determined |
| S13_extracted_64965 | Not-determined |
| S13_extracted_64966 | Not-determined |
| S13_extracted_64973 | Not-determined |
| S13_extracted_64977 | Low-quality |
| S13_extracted_64978 | Not-determined |
| S13_extracted_64989 | Low-quality |
| S13_extracted_65007 | Not-determined |
| S13_extracted_65011 | Not-determined |
| S13_extracted_65016 | Not-determined |
| S13_extracted_65044 | Low-quality |
| S13_extracted_65049 | Not-determined |
| S13_extracted_65059 | Not-determined |
| S13_extracted_65067 | Not-determined |
| S13_extracted_65071 | Not-determined |

| | |
|---|---|
| S13_extracted_65084 | Low-quality |
| S13_extracted_65093 | Not-determined |
| S13_extracted_65095 | Low-quality |
| S13_extracted_65096 | Not-determined |
| S13_extracted_65108 | Not-determined |
| S13_extracted_65115 | Not-determined |
| S13_extracted_65120 | Not-determined |
| S13_extracted_65122 | Not-determined |
| S13_extracted_65124 | Not-determined |
| S13_extracted_65138 | Not-determined |
| S13_extracted_65141 | Low-quality |
| S13_extracted_65151 | Not-determined |
| S13_extracted_65152 | Not-determined |
| S13_extracted_65160 | Not-determined |
| S13_extracted_65167 | Not-determined |
| S13_extracted_65175 | Not-determined |
| S13_extracted_65196 | Not-determined |
| S13_extracted_65197 | Not-determined |
| S13_extracted_65198 | Not-determined |
| S13_extracted_65203 | Not-determined |
| S13_extracted_65220 | Low-quality |
| S13_extracted_65231 | Not-determined |
| S13_extracted_65246 | Not-determined |
| S13_extracted_65254 | Low-quality |
| S13_extracted_65268 | Not-determined |
| S13_extracted_65275 | Not-determined |
| S13_extracted_65279 | Not-determined |
| S13_extracted_65291 | Low-quality |
| S13_extracted_65295 | Not-determined |
| S13_extracted_65300 | Not-determined |
| S13_extracted_65319 | Not-determined |
| S13_extracted_65329 | Not-determined |
| S13_extracted_65333 | Not-determined |
| S13_extracted_65335 | Not-determined |
| S13_extracted_65357 | Not-determined |
| S13_extracted_65377 | Low-quality |
| S13_extracted_65392 | Not-determined |
| S13_extracted_65393 | Not-determined |
| S13_extracted_65398 | Not-determined |
| S13_extracted_65400 | Not-determined |
| S13_extracted_65411 | Not-determined |
| S13_extracted_65418 | Not-determined |
| S13_extracted_65427 | Not-determined |
| S13_extracted_65429 | Not-determined |
| S13_extracted_65430 | Not-determined |
| S13_extracted_65440 | Not-determined |
| S13_extracted_65443 | Not-determined |
| S13_extracted_65458 | Not-determined |
| S13_extracted_65465 | Not-determined |
| S13_extracted_65469 | Not-determined |
| S13_extracted_65490 | Low-quality |
| S13_extracted_65494 | Low-quality |
| S13_extracted_65504 | Not-determined |
| S13_extracted_65515 | Not-determined |
| S13_extracted_65518 | Not-determined |
| S13_extracted_65542 | Not-determined |
| S13_extracted_65553 | Low-quality |
| S13_extracted_65561 | Not-determined |
| S13_extracted_65562 | Not-determined |
| S13_extracted_65573 | Not-determined |
| S13_extracted_65574 | Low-quality |
| S13_extracted_65577 | Not-determined |
| S13_extracted_65578 | Not-determined |
| S13_extracted_65589 | Not-determined |
| S13_extracted_65611 | Not-determined |
| S13_extracted_65613 | Not-determined |
| S13_extracted_65619 | Low-quality |
| S13_extracted_65639 | Not-determined |
| S13_extracted_65649 | Not-determined |
| S13_extracted_65652 | Not-determined |
| S13_extracted_65667 | Not-determined |
| S13_extracted_65674 | Not-determined |
| S13_extracted_65700 | Low-quality |
| S13_extracted_65703 | Not-determined |
| S13_extracted_65704 | Low-quality |
| S13_extracted_65708 | Not-determined |
| S13_extracted_65712 | Not-determined |
| S13_extracted_65713 | Not-determined |
| S13_extracted_65722 | Not-determined |
| S13_extracted_65723 | Not-determined |
| S13_extracted_65738 | Not-determined |
| S13_extracted_65742 | Not-determined |
| S13_extracted_65748 | Low-quality |
| S13_extracted_65758 | Low-quality |
| S13_extracted_65759 | Not-determined |
| S13_extracted_65765 | Not-determined |
| S13_extracted_65777 | Not-determined |
| S13_extracted_65797 | Not-determined |
| S13_extracted_65815 | Not-determined |
| S13_extracted_65816 | Not-determined |
| S13_extracted_65819 | Not-determined |
| S13_extracted_65821 | Not-determined |
| S13_extracted_65827 | Not-determined |
| S13_extracted_65831 | Not-determined |
| S13_extracted_65851 | Not-determined |
| S13_extracted_65857 | Low-quality |
| S13_extracted_65875 | Not-determined |
| S13_extracted_65902 | Not-determined |
| S13_extracted_65905 | Not-determined |
| S13_extracted_65914 | Not-determined |
| S13_extracted_65916 | Low-quality |
| S13_extracted_65919 | Not-determined |
| S13_extracted_65927 | Not-determined |
| S13_extracted_65935 | Not-determined |
| S13_extracted_65938 | Not-determined |
| S13_extracted_65948 | Not-determined |
| S13_extracted_65951 | Not-determined |
| S13_extracted_65962 | Not-determined |
| S13_extracted_65964 | Low-quality |
| S13_extracted_65970 | Not-determined |
| S13_extracted_65973 | Not-determined |
| S13_extracted_65975 | Not-determined |
| S13_extracted_66009 | Not-determined |
| S13_extracted_66010 | Not-determined |
| S13_extracted_66017 | Not-determined |
| S13_extracted_66039 | Not-determined |
| S13_extracted_66070 | Not-determined |
| S13_extracted_66073 | Low-quality |
| S13_extracted_66093 | Not-determined |
| S13_extracted_66095 | Low-quality |
| S13_extracted_66111 | Not-determined |
| S13_extracted_66123 | Not-determined |
| S13_extracted_66125 | Not-determined |
| S13_extracted_66131 | Low-quality |
| S13_extracted_66132 | Not-determined |
| S13_extracted_66135 | Not-determined |
| S13_extracted_66142 | Not-determined |
| S13_extracted_66170 | Not-determined |
| S13_extracted_66173 | Not-determined |
| S13_extracted_66190 | Not-determined |
| S13_extracted_66195 | Not-determined |
| S13_extracted_66207 | Not-determined |
| S13_extracted_66214 | Not-determined |
| S13_extracted_66229 | Not-determined |
| S13_extracted_66237 | Not-determined |
| S13_extracted_66239 | Not-determined |
| S13_extracted_66246 | Not-determined |
| S13_extracted_66251 | Not-determined |
| S13_extracted_66264 | Not-determined |
| S13_extracted_66273 | Low-quality |
| S13_extracted_66279 | Not-determined |
| S13_extracted_66283 | Not-determined |
| S13_extracted_66284 | Low-quality |
| S13_extracted_66292 | Not-determined |
| S13_extracted_66301 | Not-determined |
| S13_extracted_66306 | Low-quality |
| S13_extracted_66312 | Not-determined |
| S13_extracted_66319 | Not-determined |
| S13_extracted_66329 | Not-determined |
| S13_extracted_66335 | Not-determined |
| S13_extracted_66339 | Not-determined |
| S13_extracted_66342 | Not-determined |
| S13_extracted_66343 | Not-determined |
| S13_extracted_66348 | Not-determined |
| S13_extracted_66353 | Not-determined |
| S13_extracted_66359 | Not-determined |
| S13_extracted_66365 | Not-determined |
| S13_extracted_66372 | Not-determined |
| S13_extracted_66382 | Not-determined |
| S13_extracted_66393 | Not-determined |
| S13_extracted_66394 | Not-determined |
| S13_extracted_66410 | Not-determined |
| S13_extracted_66411 | Not-determined |
| S13_extracted_66412 | Not-determined |
| S13_extracted_66432 | Not-determined |
| S13_extracted_66443 | Not-determined |
| S13_extracted_66449 | Not-determined |
| S13_extracted_66464 | Not-determined |
| S13_extracted_66465 | Not-determined |
| S13_extracted_66471 | Low-quality |
| S13_extracted_66472 | Not-determined |
| S13_extracted_66484 | Not-determined |
| S13_extracted_66493 | Not-determined |
| S13_extracted_66502 | Not-determined |
| S13_extracted_7670 | High-quality |
| S13_extracted_66510 | Not-determined |
| S13_extracted_66521 | Not-determined |
| S13_extracted_66524 | Not-determined |
| S13_extracted_66528 | Not-determined |
| S13_extracted_66539 | Not-determined |
| S13_extracted_66543 | Low-quality |
| S13_extracted_66546 | Not-determined |
| S13_extracted_66549 | Not-determined |
| S13_extracted_66552 | Not-determined |
| S13_extracted_66555 | Not-determined |
| S13_extracted_66556 | Not-determined |
| S13_extracted_66578 | Not-determined |
| S13_extracted_66597 | Not-determined |
| S13_extracted_66612 | Low-quality |
| S13_extracted_66618 | Not-determined |
| S13_extracted_66623 | Not-determined |
| S13_extracted_66651 | Not-determined |
| S13_extracted_66683 | Not-determined |
| S13_extracted_66690 | Not-determined |
| S13_extracted_66698 | Not-determined |
| S13_extracted_66711 | Not-determined |
| S13_extracted_66727 | Low-quality |
| S13_extracted_66738 | Not-determined |
| S13_extracted_66741 | Not-determined |
| S13_extracted_66747 | Not-determined |
| S13_extracted_66749 | Not-determined |
| S13_extracted_66753 | Not-determined |
| S13_extracted_66771 | Not-determined |
| S13_extracted_66773 | Not-determined |
| S13_extracted_66774 | Not-determined |
| S13_extracted_66791 | Not-determined |
| S13_extracted_66798 | Not-determined |
| S13_extracted_66802 | Not-determined |
| S13_extracted_66808 | Not-determined |
| S13_extracted_66809 | Not-determined |
| S13_extracted_66830 | Low-quality |
| S13_extracted_66833 | Low-quality |
| S13_extracted_66846 | Not-determined |
| S13_extracted_66853 | Not-determined |
| S13_extracted_66861 | Not-determined |
| S13_extracted_66865 | Not-determined |
| S13_extracted_66870 | Not-determined |
| S13_extracted_66874 | Not-determined |
| S13_extracted_66879 | Not-determined |
| S13_extracted_66886 | Not-determined |
| S13_extracted_66888 | Not-determined |
| S13_extracted_66896 | Not-determined |
| S13_extracted_66902 | Not-determined |
| S13_extracted_66929 | Not-determined |
| S13_extracted_66946 | Not-determined |
| S13_extracted_66964 | Not-determined |
| S13_extracted_66968 | Low-quality |
| S13_extracted_66970 | Not-determined |
| S13_extracted_66971 | Not-determined |
| S13_extracted_66997 | Not-determined |
| S13_extracted_67029 | Not-determined |
| S13_extracted_67031 | Not-determined |
| S13_extracted_67052 | Not-determined |
| S13_extracted_67074 | Not-determined |
| S13_extracted_67077 | Not-determined |
| S13_extracted_67086 | Low-quality |
| S13_extracted_67088 | Not-determined |
| S13_extracted_67092 | Not-determined |
| S13_extracted_67095 | Not-determined |
| S13_extracted_67098 | Not-determined |
| S13_extracted_67105 | Not-determined |
| S13_extracted_67127 | Not-determined |
| S13_extracted_67132 | Low-quality |
| S13_extracted_67134 | Not-determined |
| S13_extracted_67138 | Not-determined |
| S13_extracted_67141 | Low-quality |
| S13_extracted_67149 | Low-quality |
| S13_extracted_67155 | Not-determined |
| S13_extracted_67185 | Not-determined |
| S13_extracted_67193 | Not-determined |
| S13_extracted_67203 | Not-determined |
| S13_extracted_67221 | Not-determined |
| S13_extracted_67226 | Not-determined |
| S13_extracted_67244 | Not-determined |
| S13_extracted_67259 | Not-determined |
| S13_extracted_67260 | Not-determined |
| S13_extracted_67263 | Not-determined |
| S13_extracted_67267 | Not-determined |
| S13_extracted_67278 | Not-determined |
| S13_extracted_67280 | Not-determined |
| S13_extracted_67286 | Not-determined |
| S13_extracted_67290 | Not-determined |
| S13_extracted_67296 | Not-determined |
| S13_extracted_67297 | Not-determined |
| S13_extracted_67316 | Not-determined |
| S13_extracted_67323 | Not-determined |
| S13_extracted_67333 | Not-determined |
| S13_extracted_67341 | Not-determined |
| S13_extracted_67372 | Not-determined |
| S13_extracted_67381 | Not-determined |
| S13_extracted_67389 | Not-determined |
| S13_extracted_67399 | Not-determined |
| S13_extracted_67415 | Low-quality |
| S13_extracted_67437 | Not-determined |
| S13_extracted_67449 | Not-determined |
| S13_extracted_67453 | Not-determined |
| S13_extracted_67458 | Not-determined |
| S13_extracted_67465 | Not-determined |
| S13_extracted_67473 | Not-determined |
| S13_extracted_67492 | Low-quality |
| S13_extracted_67493 | Not-determined |
| S13_extracted_67503 | Not-determined |
| S13_extracted_67548 | Not-determined |
| S13_extracted_67555 | Not-determined |
| S13_extracted_67584 | Not-determined |
| S13_extracted_67593 | Not-determined |
| S13_extracted_67594 | Not-determined |
| S13_extracted_67612 | Not-determined |
| S13_extracted_67630 | Not-determined |
| S13_extracted_67641 | Not-determined |
| S13_extracted_67647 | Not-determined |
| S13_extracted_67650 | Low-quality |
| S13_extracted_67654 | Not-determined |
| S13_extracted_67657 | Low-quality |
| S13_extracted_67658 | Not-determined |
| S13_extracted_67664 | Not-determined |
| S13_extracted_67667 | Not-determined |
| S13_extracted_67670 | Not-determined |
| S13_extracted_67671 | Not-determined |
| S13_extracted_67679 | Not-determined |
| S13_extracted_67681 | Low-quality |
| S13_extracted_67682 | Not-determined |
| S13_extracted_67683 | Not-determined |
| S13_extracted_67689 | Not-determined |
| S13_extracted_67701 | Not-determined |
| S13_extracted_67703 | Low-quality |
| S13_extracted_67713 | Not-determined |
| S13_extracted_67719 | Not-determined |
| S13_extracted_67724 | Low-quality |
| S13_extracted_67729 | Not-determined |
| S13_extracted_67730 | Not-determined |
| S13_extracted_67735 | Not-determined |
| S13_extracted_67752 | Not-determined |
| S13_extracted_67766 | Not-determined |
| S13_extracted_67769 | Low-quality |
| S13_extracted_67785 | Not-determined |
| S13_extracted_67795 | Not-determined |
| S13_extracted_67811 | Not-determined |
| S13_extracted_67814 | Not-determined |
| S13_extracted_67819 | Not-determined |
| S13_extracted_67827 | Not-determined |
| S13_extracted_67841 | Not-determined |
| S13_extracted_67844 | Not-determined |
| S13_extracted_67856 | Not-determined |
| S13_extracted_67862 | Not-determined |
| S13_extracted_67867 | Not-determined |
| S13_extracted_67888 | Not-determined |
| S13_extracted_67893 | Low-quality |
| S13_extracted_67896 | Not-determined |
| S13_extracted_67902 | Not-determined |
| S13_extracted_67923 | Not-determined |
| S13_extracted_67924 | Not-determined |
| S13_extracted_67934 | Not-determined |
| S13_extracted_67935 | Not-determined |
| S13_extracted_67939 | Not-determined |
| S13_extracted_67945 | Not-determined |
| S13_extracted_67951 | Not-determined |
| S13_extracted_67960 | Not-determined |
| S13_extracted_67974 | Not-determined |
| S13_extracted_67981 | Not-determined |
| S13_extracted_67987 | Low-quality |
| S13_extracted_67990 | Not-determined |
| S13_extracted_67999 | Not-determined |
| S13_extracted_68001 | Not-determined |
| S13_extracted_68027 | Not-determined |
| S13_extracted_68052 | Not-determined |
| S13_extracted_68055 | Not-determined |
| S13_extracted_68068 | Not-determined |
| S13_extracted_68069 | Not-determined |
| S13_extracted_68070 | Not-determined |
| S13_extracted_68072 | Not-determined |
| S13_extracted_68106 | Not-determined |
| S13_extracted_68117 | Not-determined |
| S13_extracted_68119 | Not-determined |
| S13_extracted_68123 | Low-quality |
| S13_extracted_68129 | Not-determined |
| S13_extracted_68131 | Not-determined |
| S13_extracted_68135 | Not-determined |
| S13_extracted_68139 | Low-quality |
| S13_extracted_68153 | Not-determined |
| S13_extracted_68155 | Not-determined |
| S13_extracted_68163 | Not-determined |
| S13_extracted_68168 | Not-determined |
| S13_extracted_68173 | Not-determined |
| S13_extracted_68183 | Not-determined |
| S13_extracted_68186 | Low-quality |
| S13_extracted_68206 | Not-determined |
| S13_extracted_68215 | Not-determined |
| S13_extracted_68227 | Not-determined |
| S13_extracted_68230 | Low-quality |
| S13_extracted_68234 | Not-determined |
| S13_extracted_68239 | Not-determined |
| S13_extracted_68248 | Not-determined |
| S13_extracted_68260 | Not-determined |
| S13_extracted_68281 | Not-determined |
| S13_extracted_68286 | Low-quality |
| S13_extracted_68291 | Low-quality |
| S13_extracted_68298 | Not-determined |
| S13_extracted_68308 | Not-determined |
| S13_extracted_68313 | Not-determined |
| S13_extracted_68318 | Not-determined |
| S13_extracted_68330 | Not-determined |
| S13_extracted_68334 | Not-determined |
| S13_extracted_68341 | Not-determined |
| S13_extracted_68356 | Low-quality |
| S13_extracted_68358 | Not-determined |
| S13_extracted_68359 | Low-quality |
| S13_extracted_68367 | Not-determined |
| S13_extracted_68375 | Not-determined |
| S13_extracted_68384 | Not-determined |
| S13_extracted_68386 | Not-determined |
| S13_extracted_68388 | Not-determined |
| S13_extracted_68394 | Not-determined |
| S13_extracted_68401 | Not-determined |
| S13_extracted_68403 | Not-determined |
| S13_extracted_68417 | Not-determined |
| S13_extracted_68418 | Not-determined |
| S13_extracted_68429 | Not-determined |
| S13_extracted_68434 | Not-determined |
| S13_extracted_68445 | Low-quality |
| S13_extracted_68452 | Not-determined |
| S13_extracted_68457 | Not-determined |
| S13_extracted_68460 | Not-determined |
| S13_extracted_68470 | Not-determined |
| S13_extracted_68472 | Low-quality |
| S13_extracted_68501 | Not-determined |
| S13_extracted_68520 | Not-determined |
| S13_extracted_68521 | Low-quality |
| S13_extracted_68522 | Low-quality |
| S13_extracted_68527 | Not-determined |
| S13_extracted_68532 | Not-determined |
| S13_extracted_68538 | Low-quality |
| S13_extracted_68544 | Low-quality |
| S13_extracted_68557 | Not-determined |
| S13_extracted_68565 | Not-determined |
| S13_extracted_68573 | Not-determined |
| S13_extracted_68575 | Not-determined |
| S13_extracted_68580 | Not-determined |
| S13_extracted_68585 | Not-determined |
| S13_extracted_68589 | Low-quality |
| S13_extracted_68593 | Not-determined |
| S13_extracted_68595 | Not-determined |
| S13_extracted_68598 | Not-determined |
| S13_extracted_68599 | Low-quality |
| S13_extracted_68606 | Not-determined |
| S13_extracted_68609 | Not-determined |
| S13_extracted_68634 | Not-determined |
| S13_extracted_68636 | Not-determined |
| S13_extracted_68650 | Not-determined |
| S13_extracted_68651 | Not-determined |
| S13_extracted_68653 | Low-quality |
| S13_extracted_68656 | Not-determined |
| S13_extracted_68659 | Low-quality |
| S13_extracted_68663 | Not-determined |
| S13_extracted_68667 | Not-determined |
| S13_extracted_68691 | Not-determined |
| S13_extracted_68707 | Not-determined |
| S13_extracted_68715 | Not-determined |
| S13_extracted_68718 | Not-determined |
| S13_extracted_68719 | Not-determined |
| S13_extracted_68740 | Not-determined |
| S13_extracted_68746 | Not-determined |
| S13_extracted_68751 | Low-quality |
| S13_extracted_68756 | Not-determined |
| S13_extracted_68757 | Not-determined |
| S13_extracted_68758 | Low-quality |
| S13_extracted_68766 | Not-determined |
| S13_extracted_68809 | Not-determined |
| S13_extracted_68810 | Not-determined |
| S13_extracted_68828 | Not-determined |
| S13_extracted_68860 | Not-determined |
| S13_extracted_68865 | Not-determined |
| S13_extracted_68889 | Not-determined |
| S13_extracted_68899 | Not-determined |
| S13_extracted_68925 | Not-determined |
| S13_extracted_68927 | Not-determined |
| S13_extracted_68946 | Not-determined |
| S13_extracted_68950 | Low-quality |
| S13_extracted_68958 | Not-determined |
| S13_extracted_68974 | Not-determined |
| S13_extracted_68977 | Not-determined |
| S13_extracted_68984 | Not-determined |
| S13_extracted_68992 | Not-determined |
| S13_extracted_68993 | Not-determined |
| S13_extracted_68997 | Not-determined |
| S13_extracted_68998 | Not-determined |
| S13_extracted_68999 | Low-quality |
| S13_extracted_69006 | Not-determined |
| S13_extracted_69008 | Not-determined |
| S13_extracted_69024 | Not-determined |
| S13_extracted_69026 | Not-determined |
| S13_extracted_69031 | Not-determined |
| S13_extracted_69047 | Not-determined |
| S13_extracted_69055 | Not-determined |
| S13_extracted_69061 | Not-determined |
| S13_extracted_69062 | Not-determined |
| S13_extracted_69063 | Not-determined |
| S13_extracted_69066 | Not-determined |
| S13_extracted_69092 | Not-determined |
| S13_extracted_69103 | Not-determined |
| S13_extracted_69104 | Not-determined |
| S13_extracted_69118 | Low-quality |
| S13_extracted_69155 | Not-determined |
| S13_extracted_69169 | Not-determined |
| S13_extracted_69179 | Not-determined |
| S13_extracted_69180 | Low-quality |
| S13_extracted_69181 | Not-determined |
| S13_extracted_69201 | Not-determined |
| S13_extracted_69215 | Not-determined |
| S13_extracted_69225 | Not-determined |
| S13_extracted_69238 | Not-determined |
| S13_extracted_69241 | Not-determined |
| S13_extracted_69250 | Not-determined |
| S13_extracted_69254 | Not-determined |
| S13_extracted_69264 | Not-determined |
| S13_extracted_69279 | Not-determined |
| S13_extracted_69283 | Not-determined |
| S13_extracted_69290 | Not-determined |
| S13_extracted_69300 | Not-determined |
| S13_extracted_69311 | Not-determined |
| S13_extracted_69313 | Low-quality |
| S13_extracted_69320 | Low-quality |
| S13_extracted_69350 | Not-determined |
| S13_extracted_69353 | Not-determined |
| S13_extracted_69354 | Not-determined |
| S13_extracted_69373 | Not-determined |
| S13_extracted_69390 | Not-determined |
| S13_extracted_69391 | Not-determined |
| S13_extracted_69412 | Not-determined |
| S13_extracted_69416 | Not-determined |
| S13_extracted_69424 | Not-determined |
| S13_extracted_69430 | Not-determined |
| S13_extracted_69432 | Not-determined |
| S13_extracted_69443 | Not-determined |
| S13_extracted_69458 | Not-determined |
| S13_extracted_69465 | Not-determined |
| S13_extracted_69467 | Not-determined |
| S13_extracted_69478 | Not-determined |
| S13_extracted_69479 | Not-determined |
| S13_extracted_69491 | Low-quality |
| S13_extracted_69503 | Not-determined |
| S13_extracted_69514 | Not-determined |
| S13_extracted_69530 | Not-determined |
| S13_extracted_69532 | Not-determined |
| S13_extracted_69542 | Not-determined |
| S13_extracted_69555 | Not-determined |
| S13_extracted_69558 | Not-determined |
| S13_extracted_69574 | Not-determined |
| S13_extracted_69590 | Not-determined |
| S13_extracted_69606 | Not-determined |
| S13_extracted_69620 | Not-determined |
| S13_extracted_69628 | Not-determined |
| S13_extracted_69630 | Not-determined |
| S13_extracted_69648 | Not-determined |
| S13_extracted_69655 | Not-determined |
| S13_extracted_69686 | Not-determined |
| S13_extracted_69710 | Not-determined |
| S13_extracted_69714 | Not-determined |
| S13_extracted_69723 | Not-determined |
| S13_extracted_69732 | Not-determined |
| S13_extracted_69744 | Not-determined |
| S13_extracted_69746 | Not-determined |
| S13_extracted_69780 | Not-determined |
| S13_extracted_69784 | Not-determined |
| S13_extracted_69789 | Not-determined |
| S13_extracted_69796 | Not-determined |
| S13_extracted_69822 | Not-determined |
| S13_extracted_69832 | Not-determined |
| S13_extracted_69840 | Not-determined |
| S13_extracted_69850 | Not-determined |
| S13_extracted_69851 | Not-determined |
| S13_extracted_69862 | Not-determined |
| S13_extracted_69866 | Not-determined |
| S13_extracted_69886 | Not-determined |
| S13_extracted_69898 | Not-determined |
| S13_extracted_69926 | Not-determined |
| S13_extracted_69929 | Not-determined |
| S13_extracted_69933 | Not-determined |
| S13_extracted_69934 | Not-determined |
| S13_extracted_69943 | Low-quality |
| S13_extracted_69951 | Low-quality |
| S13_extracted_69955 | Not-determined |
| S13_extracted_69978 | Not-determined |
| S13_extracted_70004 | Not-determined |
| S13_extracted_70007 | Not-determined |
| S13_extracted_70008 | Not-determined |
| S13_extracted_70013 | Not-determined |
| S13_extracted_70020 | Not-determined |
| S13_extracted_70023 | Not-determined |
| S13_extracted_70027 | Low-quality |
| S13_extracted_70040 | Not-determined |
| S13_extracted_70041 | Not-determined |
| S13_extracted_70054 | Not-determined |
| S13_extracted_70055 | Not-determined |
| S13_extracted_70058 | Not-determined |
| S13_extracted_70074 | Not-determined |
| S13_extracted_70081 | Not-determined |
| S13_extracted_70085 | Not-determined |
| S13_extracted_70090 | Not-determined |
| S13_extracted_70094 | Not-determined |
| S13_extracted_70099 | Low-quality |
| S13_extracted_70101 | Low-quality |
| S13_extracted_70102 | Not-determined |
| S13_extracted_70103 | Not-determined |
| S13_extracted_70106 | Not-determined |
| S13_extracted_70122 | Not-determined |
| S13_extracted_70126 | Not-determined |
| S13_extracted_70132 | Not-determined |
| S13_extracted_70144 | Not-determined |

| | |
|---|---|
| S13_extracted_70157 | Not-determined |
| S13_extracted_70158 | Low-quality |
| S13_extracted_70162 | Not-determined |
| S13_extracted_70169 | Not-determined |
| S13_extracted_70184 | Not-determined |
| S13_extracted_70186 | Not-determined |
| S13_extracted_70190 | Not-determined |
| S13_extracted_70211 | Not-determined |
| S13_extracted_70223 | Not-determined |
| S13_extracted_70235 | Not-determined |
| S13_extracted_70237 | Not-determined |
| S13_extracted_70267 | Not-determined |
| S13_extracted_70274 | Not-determined |
| S13_extracted_70287 | Not-determined |
| S13_extracted_70293 | Not-determined |
| S13_extracted_70299 | Low-quality |
| S13_extracted_70300 | Not-determined |
| S13_extracted_70311 | Not-determined |
| S13_extracted_70312 | Not-determined |
| S13_extracted_70315 | Not-determined |
| S13_extracted_70334 | Not-determined |
| S13_extracted_70341 | Not-determined |
| S13_extracted_70342 | Not-determined |
| S13_extracted_70344 | Not-determined |
| S13_extracted_70349 | Not-determined |
| S13_extracted_70370 | Not-determined |
| S13_extracted_70381 | Not-determined |
| S13_extracted_70382 | Not-determined |
| S13_extracted_70385 | Not-determined |
| S13_extracted_70388 | Not-determined |
| S13_extracted_70393 | Not-determined |
| S13_extracted_70396 | Not-determined |
| S13_extracted_70399 | Not-determined |
| S13_extracted_70408 | Not-determined |
| S13_extracted_70433 | Not-determined |
| S13_extracted_70443 | Not-determined |
| S13_extracted_70468 | Not-determined |
| S13_extracted_70474 | Not-determined |
| S13_extracted_70477 | Not-determined |
| S13_extracted_70484 | Not-determined |
| S13_extracted_70486 | Not-determined |
| S13_extracted_70499 | Not-determined |
| S13_extracted_70502 | Not-determined |
| S13_extracted_70511 | Not-determined |
| S13_extracted_70512 | Not-determined |
| S13_extracted_70513 | Not-determined |
| S13_extracted_70540 | Not-determined |
| S13_extracted_70542 | Low-quality |
| S13_extracted_70552 | Not-determined |
| S13_extracted_70561 | Not-determined |
| S13_extracted_70569 | Not-determined |
| S13_extracted_70576 | Not-determined |
| S13_extracted_70589 | Not-determined |
| S13_extracted_70598 | Not-determined |
| S13_extracted_70608 | Not-determined |
| S13_extracted_70610 | Low-quality |
| S13_extracted_70617 | Not-determined |
| S13_extracted_70621 | Not-determined |
| S13_extracted_70625 | Not-determined |
| S13_extracted_70630 | Not-determined |
| S13_extracted_70632 | Not-determined |
| S13_extracted_70644 | Not-determined |
| S13_extracted_70645 | Not-determined |
| S13_extracted_70646 | Not-determined |
| S13_extracted_70652 | Not-determined |
| S13_extracted_70656 | Not-determined |
| S13_extracted_70657 | Not-determined |
| S13_extracted_70659 | Not-determined |
| S13_extracted_70668 | Low-quality |
| S13_extracted_70670 | Not-determined |
| S13_extracted_70686 | Not-determined |
| S13_extracted_70692 | Not-determined |
| S13_extracted_70703 | Not-determined |
| S13_extracted_70707 | Not-determined |
| S13_extracted_70709 | Low-quality |
| S13_extracted_70712 | Not-determined |
| S13_extracted_70764 | Not-determined |
| S13_extracted_70769 | Not-determined |
| S13_extracted_70770 | Not-determined |
| S13_extracted_70773 | Not-determined |
| S13_extracted_70784 | Not-determined |
| S13_extracted_70789 | Not-determined |
| S13_extracted_70806 | Not-determined |
| S13_extracted_70815 | Not-determined |
| S13_extracted_70819 | Not-determined |
| S13_extracted_70825 | Not-determined |
| S13_extracted_70839 | Low-quality |
| S13_extracted_70840 | Low-quality |
| S13_extracted_70842 | Not-determined |
| S13_extracted_70856 | Not-determined |
| S13_extracted_70861 | Not-determined |
| S13_extracted_70862 | Not-determined |
| S13_extracted_70874 | Not-determined |
| S13_extracted_70875 | Not-determined |
| S13_extracted_70886 | Not-determined |
| S13_extracted_70895 | Not-determined |
| S13_extracted_70899 | Not-determined |
| S13_extracted_70908 | Not-determined |
| S13_extracted_70915 | Not-determined |
| S13_extracted_70916 | Not-determined |
| S13_extracted_70924 | Not-determined |
| S13_extracted_70932 | Not-determined |
| S13_extracted_70941 | Not-determined |
| S13_extracted_70948 | Not-determined |
| S13_extracted_70952 | Not-determined |
| S13_extracted_70961 | Not-determined |
| S13_extracted_70967 | Low-quality |
| S13_extracted_70970 | Not-determined |
| S13_extracted_70974 | Not-determined |
| S13_extracted_70978 | Not-determined |
| S13_extracted_70979 | Not-determined |
| S13_extracted_70983 | Not-determined |
| S13_extracted_70987 | Low-quality |
| S13_extracted_70994 | Low-quality |
| S13_extracted_71011 | Not-determined |
| S13_extracted_71019 | Not-determined |
| S13_extracted_71034 | Not-determined |
| S13_extracted_71037 | Not-determined |
| S13_extracted_71043 | Not-determined |
| S13_extracted_71053 | Low-quality |
| S13_extracted_71066 | Not-determined |
| S13_extracted_71071 | Not-determined |
| S13_extracted_71079 | Not-determined |
| S13_extracted_71097 | Not-determined |
| S13_extracted_71101 | Not-determined |
| S13_extracted_71121 | Not-determined |
| S13_extracted_71127 | Not-determined |
| S13_extracted_71130 | Not-determined |
| S13_extracted_71142 | Not-determined |
| S13_extracted_71151 | Not-determined |
| S13_extracted_71158 | Not-determined |
| S13_extracted_71170 | Not-determined |
| S13_extracted_71173 | Not-determined |
| S13_extracted_71178 | Not-determined |
| S13_extracted_71179 | Not-determined |
| S13_extracted_71181 | Low-quality |
| S13_extracted_71197 | Low-quality |
| S13_extracted_71221 | Not-determined |
| S13_extracted_71231 | Not-determined |
| S13_extracted_71248 | Not-determined |
| S13_extracted_71254 | Not-determined |
| S13_extracted_71261 | Not-determined |
| S13_extracted_71264 | Not-determined |
| S13_extracted_71269 | Not-determined |
| S13_extracted_71281 | Not-determined |
| S13_extracted_71288 | Not-determined |
| S13_extracted_71299 | Low-quality |
| S13_extracted_71311 | Not-determined |
| S13_extracted_71335 | Not-determined |
| S13_extracted_71342 | Not-determined |
| S13_extracted_71343 | Not-determined |
| S13_extracted_71349 | Not-determined |
| S13_extracted_71372 | Not-determined |
| S13_extracted_71379 | Low-quality |
| S13_extracted_71388 | Not-determined |
| S13_extracted_71393 | Not-determined |
| S13_extracted_71441 | Not-determined |
| S13_extracted_71446 | Not-determined |
| S13_extracted_71447 | Not-determined |
| S13_extracted_71457 | Not-determined |
| S13_extracted_71463 | Low-quality |
| S13_extracted_71484 | Not-determined |
| S13_extracted_71489 | Not-determined |
| S13_extracted_71493 | Not-determined |
| S13_extracted_71496 | Not-determined |
| S13_extracted_71499 | Not-determined |
| S13_extracted_71500 | Not-determined |
| S13_extracted_71510 | Not-determined |
| S13_extracted_71515 | Not-determined |
| S13_extracted_71542 | Not-determined |
| S13_extracted_71544 | Not-determined |
| S13_extracted_71545 | Low-quality |
| S13_extracted_71548 | Not-determined |
| S13_extracted_71560 | Not-determined |
| S13_extracted_71567 | Not-determined |
| S13_extracted_71573 | Not-determined |
| S13_extracted_71579 | Not-determined |
| S13_extracted_71584 | Not-determined |
| S13_extracted_71585 | Not-determined |
| S13_extracted_71596 | Not-determined |
| S13_extracted_71599 | Not-determined |
| S13_extracted_71601 | Not-determined |
| S13_extracted_71605 | Not-determined |
| S13_extracted_71615 | Not-determined |
| S13_extracted_71623 | Not-determined |
| S13_extracted_71636 | Not-determined |
| S13_extracted_71649 | Low-quality |
| S13_extracted_71653 | Not-determined |
| S13_extracted_71660 | Not-determined |
| S13_extracted_71680 | Not-determined |
| S13_extracted_71691 | Not-determined |
| S13_extracted_71695 | Not-determined |
| S13_extracted_71697 | Low-quality |
| S13_extracted_71705 | Not-determined |
| S13_extracted_71707 | Not-determined |
| S13_extracted_71727 | Not-determined |
| S13_extracted_71729 | Not-determined |
| S13_extracted_71732 | Low-quality |
| S13_extracted_71739 | Low-quality |
| S13_extracted_71749 | Not-determined |
| S13_extracted_71753 | Not-determined |
| S13_extracted_71754 | Not-determined |
| S13_extracted_71770 | Not-determined |
| S13_extracted_71776 | Not-determined |
| S13_extracted_71781 | Low-quality |
| S13_extracted_71785 | Not-determined |
| S13_extracted_71789 | Not-determined |
| S13_extracted_71795 | Not-determined |
| S13_extracted_71798 | Not-determined |
| S13_extracted_71800 | Not-determined |
| S13_extracted_71802 | Not-determined |
| S13_extracted_71812 | Not-determined |
| S13_extracted_71819 | Not-determined |
| S13_extracted_71826 | Low-quality |
| S13_extracted_71835 | Not-determined |
| S13_extracted_71848 | Not-determined |
| S13_extracted_71853 | Not-determined |
| S13_extracted_71857 | Not-determined |
| S13_extracted_71859 | Not-determined |
| S13_extracted_71864 | Not-determined |
| S13_extracted_71866 | Not-determined |
| S13_extracted_71889 | Not-determined |
| S13_extracted_71914 | Not-determined |
| S13_extracted_71918 | Not-determined |
| S13_extracted_71960 | Not-determined |
| S13_extracted_71963 | Not-determined |
| S13_extracted_71980 | Not-determined |
| S13_extracted_71990 | Not-determined |
| S13_extracted_72004 | Not-determined |
| S13_extracted_72009 | Low-quality |
| S13_extracted_72010 | Low-quality |
| S13_extracted_72023 | Not-determined |
| S13_extracted_72032 | Not-determined |
| S13_extracted_72033 | Not-determined |
| S13_extracted_72046 | Not-determined |
| S13_extracted_72061 | Not-determined |
| S13_extracted_72063 | Not-determined |
| S13_extracted_72067 | Not-determined |
| S13_extracted_72077 | Not-determined |
| S13_extracted_72079 | Not-determined |
| S13_extracted_72088 | Not-determined |
| S13_extracted_72102 | Not-determined |
| S13_extracted_72106 | Not-determined |
| S13_extracted_72111 | Not-determined |
| S13_extracted_72119 | Not-determined |
| S13_extracted_72131 | Not-determined |
| S13_extracted_72135 | Low-quality |
| S13_extracted_72136 | Not-determined |
| S13_extracted_72149 | Not-determined |
| S13_extracted_72175 | Not-determined |
| S13_extracted_72183 | Low-quality |
| S13_extracted_72193 | Not-determined |
| S13_extracted_72199 | Not-determined |
| S13_extracted_72201 | Not-determined |
| S13_extracted_72207 | Not-determined |
| S13_extracted_72208 | Not-determined |
| S13_extracted_72214 | Not-determined |
| S13_extracted_72233 | Not-determined |
| S13_extracted_72251 | Not-determined |
| S13_extracted_72255 | Not-determined |
| S13_extracted_72275 | Not-determined |
| S13_extracted_72278 | Not-determined |
| S13_extracted_72282 | Not-determined |
| S13_extracted_72283 | Not-determined |
| S13_extracted_72292 | Not-determined |
| S13_extracted_72301 | Low-quality |
| S13_extracted_72307 | Not-determined |
| S13_extracted_72308 | Not-determined |
| S13_extracted_72311 | Not-determined |
| S13_extracted_72316 | Not-determined |
| S13_extracted_72319 | Low-quality |
| S13_extracted_72320 | Low-quality |
| S13_extracted_72325 | Not-determined |
| S13_extracted_72326 | Not-determined |
| S13_extracted_72344 | Not-determined |
| S13_extracted_72347 | Not-determined |
| S13_extracted_72349 | Not-determined |
| S13_extracted_72352 | Not-determined |
| S13_extracted_72374 | Not-determined |
| S13_extracted_72390 | Not-determined |
| S13_extracted_72398 | Not-determined |
| S13_extracted_72402 | Not-determined |
| S13_extracted_72407 | Not-determined |
| S13_extracted_72421 | Not-determined |
| S13_extracted_72439 | Not-determined |
| S13_extracted_72462 | Not-determined |
| S13_extracted_72478 | Not-determined |
| S13_extracted_72508 | Not-determined |
| S13_extracted_72513 | Not-determined |
| S13_extracted_72528 | Not-determined |
| S13_extracted_72548 | Not-determined |
| S13_extracted_72571 | Low-quality |
| S13_extracted_72578 | Not-determined |
| S13_extracted_72600 | Low-quality |
| S13_extracted_72615 | Low-quality |
| S13_extracted_72624 | Not-determined |
| S13_extracted_72629 | Not-determined |
| S13_extracted_72633 | Not-determined |
| S13_extracted_72640 | Not-determined |
| S13_extracted_72659 | Not-determined |
| S13_extracted_72670 | Not-determined |
| S13_extracted_72676 | Not-determined |
| S13_extracted_72687 | Not-determined |
| S13_extracted_72694 | Low-quality |
| S13_extracted_72701 | Not-determined |
| S13_extracted_72702 | Not-determined |
| S13_extracted_72713 | Not-determined |
| S13_extracted_72722 | Not-determined |
| S13_extracted_72746 | Not-determined |
| S13_extracted_72748 | Not-determined |
| S13_extracted_72761 | Not-determined |
| S13_extracted_72771 | Not-determined |
| S13_extracted_72772 | Not-determined |
| S13_extracted_72774 | Not-determined |
| S13_extracted_72776 | Not-determined |
| S13_extracted_72777 | Low-quality |
| S13_extracted_72781 | Not-determined |
| S13_extracted_72793 | Not-determined |
| S13_extracted_72800 | Not-determined |
| S13_extracted_72803 | Not-determined |
| S13_extracted_72809 | Not-determined |
| S13_extracted_72830 | Not-determined |
| S13_extracted_72856 | Not-determined |
| S13_extracted_72858 | Not-determined |
| S13_extracted_72864 | Not-determined |
| S13_extracted_72872 | Not-determined |
| S13_extracted_72884 | Not-determined |
| S13_extracted_72894 | Not-determined |
| S13_extracted_72913 | Not-determined |
| S13_extracted_72917 | Not-determined |
| S13_extracted_72920 | Not-determined |
| S13_extracted_72950 | Not-determined |
| S13_extracted_72968 | Low-quality |
| S13_extracted_72969 | Not-determined |
| S13_extracted_72982 | Not-determined |
| S13_extracted_72989 | Not-determined |
| S13_extracted_72991 | Not-determined |
| S13_extracted_72994 | Not-determined |
| S13_extracted_72997 | Not-determined |
| S13_extracted_73001 | Not-determined |
| S13_extracted_73002 | Not-determined |
| S13_extracted_73010 | Not-determined |
| S13_extracted_73017 | Low-quality |
| S13_extracted_73028 | Not-determined |
| S13_extracted_73029 | Not-determined |
| S13_extracted_73035 | Not-determined |
| S13_extracted_73043 | Not-determined |
| S13_extracted_73052 | Not-determined |
| S13_extracted_73058 | Not-determined |
| S13_extracted_73077 | Not-determined |
| S13_extracted_73079 | Not-determined |
| S13_extracted_73080 | Not-determined |
| S13_extracted_73103 | Not-determined |
| S13_extracted_73106 | Not-determined |
| S13_extracted_73112 | Not-determined |
| S13_extracted_73114 | Low-quality |
| S13_extracted_73117 | Not-determined |
| S13_extracted_73122 | Not-determined |
| S13_extracted_73124 | Not-determined |
| S13_extracted_73128 | Not-determined |
| S13_extracted_73132 | Not-determined |
| S13_extracted_73171 | Not-determined |
| S13_extracted_73172 | Not-determined |
| S13_extracted_73178 | Not-determined |
| S13_extracted_73179 | Not-determined |
| S13_extracted_73182 | Low-quality |
| S13_extracted_73183 | Low-quality |
| S13_extracted_73197 | Not-determined |
| S13_extracted_73209 | Not-determined |
| S13_extracted_73211 | Not-determined |
| S13_extracted_73222 | Not-determined |
| S13_extracted_73235 | Not-determined |
| S13_extracted_73248 | Not-determined |
| S13_extracted_73253 | Not-determined |
| S13_extracted_73271 | Not-determined |
| S13_extracted_73272 | Not-determined |
| S13_extracted_73284 | Not-determined |
| S13_extracted_73287 | Not-determined |
| S13_extracted_73288 | Not-determined |
| S13_extracted_73302 | Not-determined |
| S13_extracted_73320 | Not-determined |
| S13_extracted_73357 | Not-determined |
| S13_extracted_73362 | Not-determined |
| S13_extracted_73370 | Not-determined |
| S13_extracted_73391 | Not-determined |
| S13_extracted_73403 | Low-quality |
| S13_extracted_73409 | Low-quality |
| S13_extracted_73422 | Not-determined |
| S13_extracted_73440 | Not-determined |
| S13_extracted_73442 | Not-determined |
| S13_extracted_73447 | Not-determined |
| S13_extracted_73452 | Low-quality |
| S13_extracted_73457 | Not-determined |
| S13_extracted_73461 | Not-determined |
| S13_extracted_73462 | Not-determined |
| S13_extracted_73468 | Not-determined |
| S13_extracted_73469 | Not-determined |
| S13_extracted_73486 | Not-determined |
| S13_extracted_73500 | Low-quality |
| S13_extracted_73509 | Not-determined |
| S13_extracted_73510 | Not-determined |
| S13_extracted_73522 | Low-quality |
| S13_extracted_73523 | Not-determined |
| S13_extracted_73525 | Not-determined |
| S13_extracted_73528 | Not-determined |
| S13_extracted_73530 | Not-determined |
| S13_extracted_73543 | Not-determined |
| S13_extracted_73559 | Low-quality |
| S13_extracted_73571 | Not-determined |
| S13_extracted_73579 | Not-determined |
| S13_extracted_73585 | Not-determined |
| S13_extracted_73588 | Not-determined |
| S13_extracted_73593 | Not-determined |
| S13_extracted_73595 | Not-determined |
| S13_extracted_73618 | Not-determined |
| S13_extracted_73619 | Low-quality |
| S13_extracted_73631 | Not-determined |
| S13_extracted_73641 | Not-determined |
| S13_extracted_73669 | Not-determined |
| S13_extracted_73682 | Not-determined |
| S13_extracted_73690 | Not-determined |
| S13_extracted_73692 | Not-determined |
| S13_extracted_73702 | Not-determined |
| S13_extracted_73717 | Low-quality |
| S13_extracted_73730 | Low-quality |
| S13_extracted_73732 | Not-determined |
| S13_extracted_73733 | Not-determined |
| S13_extracted_73738 | Not-determined |
| S13_extracted_73742 | Not-determined |
| S13_extracted_73748 | Not-determined |
| S13_extracted_73761 | Not-determined |
| S13_extracted_73768 | Not-determined |
| S13_extracted_73777 | Not-determined |
| S13_extracted_73779 | Not-determined |
| S13_extracted_73788 | Not-determined |
| S13_extracted_73802 | Not-determined |
| S13_extracted_73818 | Not-determined |
| S13_extracted_73835 | Not-determined |
| S13_extracted_73842 | Not-determined |
| S13_extracted_73870 | Low-quality |
| S13_extracted_73885 | Not-determined |
| S13_extracted_73886 | Not-determined |
| S13_extracted_73901 | Not-determined |
| S13_extracted_73913 | Not-determined |
| S13_extracted_73921 | Not-determined |
| S13_extracted_73930 | Not-determined |
| S13_extracted_73938 | Not-determined |
| S13_extracted_73943 | Not-determined |
| S13_extracted_73960 | Not-determined |
| S13_extracted_73968 | Not-determined |
| S13_extracted_73971 | Not-determined |
| S13_extracted_73973 | Not-determined |
| S13_extracted_73992 | Not-determined |
| S13_extracted_73994 | Not-determined |
| S13_extracted_74012 | Not-determined |
| S13_extracted_74017 | Not-determined |
| S13_extracted_74020 | Not-determined |
| S13_extracted_74058 | Low-quality |
| S13_extracted_74063 | Not-determined |
| S13_extracted_74065 | Not-determined |
| S13_extracted_74066 | Not-determined |
| S13_extracted_74067 | Not-determined |
| S13_extracted_74074 | Low-quality |
| S13_extracted_74076 | Not-determined |
| S13_extracted_74089 | Not-determined |
| S13_extracted_74094 | Not-determined |
| S13_extracted_74102 | Not-determined |
| S13_extracted_74108 | Not-determined |
| S13_extracted_74115 | Low-quality |
| S13_extracted_74117 | Not-determined |
| S13_extracted_74128 | Not-determined |
| S13_extracted_74136 | Not-determined |
| S13_extracted_74149 | Not-determined |
| S13_extracted_74182 | Not-determined |
| S13_extracted_74198 | Not-determined |
| S13_extracted_74204 | Not-determined |
| S13_extracted_74224 | Not-determined |
| S13_extracted_74267 | Not-determined |
| S13_extracted_74275 | Not-determined |
| S13_extracted_74278 | Low-quality |
| S13_extracted_74281 | Not-determined |
| S13_extracted_74294 | Not-determined |
| S13_extracted_74301 | Not-determined |
| S13_extracted_74315 | Not-determined |
| S13_extracted_74323 | Not-determined |
| S13_extracted_74326 | Not-determined |
| S13_extracted_74374 | Not-determined |
| S13_extracted_74375 | Not-determined |
| S13_extracted_74383 | Not-determined |
| S13_extracted_74386 | Not-determined |
| S13_extracted_74406 | Not-determined |
| S13_extracted_74411 | Not-determined |
| S13_extracted_74414 | Not-determined |
| S13_extracted_74420 | Not-determined |
| S13_extracted_74427 | Not-determined |
| S13_extracted_74451 | Not-determined |
| S13_extracted_74461 | Not-determined |
| S13_extracted_74471 | Not-determined |
| S13_extracted_74476 | Not-determined |
| S13_extracted_74487 | Not-determined |
| S13_extracted_74499 | Not-determined |
| S13_extracted_74508 | Low-quality |
| S13_extracted_74534 | Not-determined |
| S13_extracted_74546 | Not-determined |
| S13_extracted_74560 | Low-quality |
| S13_extracted_74561 | Not-determined |
| S13_extracted_74562 | Not-determined |
| S13_extracted_74563 | Not-determined |
| S13_extracted_74569 | Not-determined |
| S13_extracted_74574 | Not-determined |
| S13_extracted_74577 | Low-quality |
| S13_extracted_74591 | Low-quality |
| S13_extracted_74598 | Not-determined |
| S13_extracted_74618 | Not-determined |
| S13_extracted_74630 | Not-determined |
| S13_extracted_74638 | Not-determined |
| S13_extracted_74648 | Not-determined |
| S13_extracted_74655 | Not-determined |
| S13_extracted_74666 | Not-determined |
| S13_extracted_74684 | Not-determined |
| S13_extracted_74685 | Not-determined |
| S13_extracted_74686 | Not-determined |
| S13_extracted_74709 | Not-determined |
| S13_extracted_74718 | Not-determined |
| S13_extracted_74722 | Not-determined |
| S13_extracted_74723 | Not-determined |
| S13_extracted_74725 | Not-determined |
| S13_extracted_74736 | Not-determined |
| S13_extracted_74740 | Not-determined |
| S13_extracted_74766 | Not-determined |
| S13_extracted_74769 | Not-determined |
| S13_extracted_74771 | Not-determined |
| S13_extracted_74781 | Not-determined |
| S13_extracted_74784 | Not-determined |
| S13_extracted_74786 | Not-determined |
| S13_extracted_74789 | Low-quality |
| S13_extracted_74807 | Not-determined |
| S13_extracted_74817 | Not-determined |
| S13_extracted_74820 | Not-determined |
| S13_extracted_74821 | Not-determined |
| S13_extracted_74822 | Not-determined |
| S13_extracted_74826 | Not-determined |
| S13_extracted_74830 | Not-determined |
| S13_extracted_74835 | Not-determined |
| S13_extracted_74841 | Not-determined |
| S13_extracted_74847 | Not-determined |
| S13_extracted_74850 | Not-determined |
| S13_extracted_74851 | Not-determined |
| S13_extracted_74869 | Not-determined |
| S13_extracted_74872 | Not-determined |
| S13_extracted_74874 | Not-determined |
| S13_extracted_74882 | Not-determined |
| S13_extracted_74885 | Not-determined |
| S13_extracted_74894 | Not-determined |
| S13_extracted_74915 | Not-determined |
| S13_extracted_74916 | Not-determined |
| S13_extracted_74929 | Not-determined |
| S13_extracted_74931 | Not-determined |
| S13_extracted_74933 | Not-determined |
| S13_extracted_74935 | Not-determined |
| S13_extracted_74940 | Not-determined |
| S13_extracted_74941 | Not-determined |
| S13_extracted_74945 | Not-determined |
| S13_extracted_74950 | Not-determined |
| S13_extracted_74952 | Low-quality |
| S13_extracted_74955 | Not-determined |
| S13_extracted_74962 | Not-determined |
| S13_extracted_74965 | Not-determined |
| S13_extracted_74973 | Low-quality |
| S13_extracted_74977 | Not-determined |
| S13_extracted_74980 | Low-quality |
| S13_extracted_74982 | Not-determined |
| S13_extracted_74985 | Not-determined |
| S13_extracted_74986 | Not-determined |
| S13_extracted_74991 | Not-determined |
| S13_extracted_74995 | Not-determined |
| S13_extracted_74996 | Not-determined |
| S13_extracted_75023 | Not-determined |
| S13_extracted_75027 | Not-determined |
| S13_extracted_75035 | Low-quality |
| S13_extracted_75040 | Not-determined |
| S13_extracted_75050 | Not-determined |
| S13_extracted_75088 | Not-determined |
| S13_extracted_75095 | Not-determined |
| S13_extracted_75097 | Not-determined |
| S13_extracted_75101 | Not-determined |
| S13_extracted_75106 | Not-determined |
| S13_extracted_75117 | Not-determined |
| S13_extracted_75135 | Not-determined |

| | |
|---|---|
| S13_extracted_75139 | Not-determined |
| S13_extracted_75144 | Not-determined |
| S13_extracted_75153 | Not-determined |
| S13_extracted_75156 | Not-determined |
| S13_extracted_75168 | Not-determined |
| S13_extracted_75169 | Not-determined |
| S13_extracted_75203 | Low-quality |
| S13_extracted_75207 | Not-determined |
| S13_extracted_75210 | Not-determined |
| S13_extracted_75215 | Not-determined |
| S13_extracted_75222 | Not-determined |
| S13_extracted_75224 | Low-quality |
| S13_extracted_75229 | Not-determined |
| S13_extracted_75230 | Not-determined |
| S13_extracted_75231 | Not-determined |
| S13_extracted_75256 | Not-determined |
| S13_extracted_75258 | Not-determined |
| S13_extracted_75265 | Not-determined |
| S13_extracted_75268 | Not-determined |
| S13_extracted_75274 | Not-determined |
| S13_extracted_75278 | Not-determined |
| S13_extracted_75304 | Not-determined |
| S13_extracted_75311 | Not-determined |
| S13_extracted_75327 | Not-determined |
| S13_extracted_75328 | Not-determined |
| S13_extracted_75331 | Not-determined |
| S13_extracted_75352 | Not-determined |
| S13_extracted_75372 | Not-determined |
| S13_extracted_75376 | Low-quality |
| S13_extracted_75377 | Low-quality |
| S13_extracted_75400 | Not-determined |
| S13_extracted_75419 | Not-determined |
| S13_extracted_75434 | Not-determined |
| S13_extracted_75464 | Not-determined |
| S13_extracted_75470 | Not-determined |
| S13_extracted_75485 | Not-determined |
| S13_extracted_75489 | Not-determined |
| S13_extracted_75492 | Not-determined |
| S13_extracted_75499 | Not-determined |
| S13_extracted_75512 | Not-determined |
| S13_extracted_75518 | Not-determined |
| S13_extracted_75520 | Not-determined |
| S13_extracted_75530 | Not-determined |
| S13_extracted_75533 | Not-determined |
| S13_extracted_75546 | Low-quality |
| S13_extracted_75573 | Not-determined |
| S13_extracted_75574 | Not-determined |
| S13_extracted_75577 | Not-determined |
| S13_extracted_75593 | Not-determined |
| S13_extracted_75596 | Not-determined |
| S13_extracted_75606 | Not-determined |
| S13_extracted_75610 | Not-determined |
| S13_extracted_75616 | Not-determined |
| S13_extracted_75653 | Not-determined |
| S13_extracted_75656 | Not-determined |
| S13_extracted_75698 | Not-determined |
| S13_extracted_75732 | Not-determined |
| S13_extracted_75763 | Not-determined |
| S13_extracted_75767 | Low-quality |
| S13_extracted_75771 | Not-determined |
| S13_extracted_75778 | Not-determined |
| S13_extracted_75783 | Not-determined |
| S13_extracted_75789 | Not-determined |
| S13_extracted_75794 | Not-determined |
| S13_extracted_75803 | Not-determined |
| S13_extracted_75805 | Not-determined |
| S13_extracted_75816 | Not-determined |
| S13_extracted_75819 | Not-determined |
| S13_extracted_75824 | Not-determined |
| S13_extracted_75827 | Not-determined |
| S13_extracted_75831 | Not-determined |
| S13_extracted_75833 | Not-determined |
| S13_extracted_75856 | Not-determined |
| S13_extracted_75861 | Not-determined |
| S13_extracted_75866 | Not-determined |
| S13_extracted_75879 | Not-determined |
| S13_extracted_75882 | Not-determined |
| S13_extracted_75891 | Not-determined |
| S13_extracted_75895 | Not-determined |
| S13_extracted_75897 | Not-determined |
| S13_extracted_75907 | Low-quality |
| S13_extracted_75910 | Not-determined |
| S13_extracted_75912 | Not-determined |
| S13_extracted_75913 | Not-determined |
| S13_extracted_75921 | Not-determined |
| S13_extracted_75927 | Not-determined |
| S13_extracted_75931 | Not-determined |
| S13_extracted_75942 | Not-determined |
| S13_extracted_75953 | Not-determined |
| S13_extracted_75984 | Not-determined |
| S13_extracted_75990 | Not-determined |
| S13_extracted_76002 | Not-determined |
| S13_extracted_76006 | Not-determined |
| S13_extracted_76018 | Not-determined |
| S13_extracted_76037 | Not-determined |
| S13_extracted_76044 | Not-determined |
| S13_extracted_76057 | Not-determined |
| S13_extracted_76068 | Not-determined |
| S13_extracted_76076 | Not-determined |
| S13_extracted_76081 | Not-determined |
| S13_extracted_76094 | Low-quality |
| S13_extracted_76096 | Not-determined |
| S13_extracted_76103 | Not-determined |
| S13_extracted_76124 | Not-determined |
| S13_extracted_76127 | Not-determined |
| S13_extracted_76131 | Not-determined |
| S13_extracted_76137 | Not-determined |
| S13_extracted_76141 | Not-determined |
| S13_extracted_76153 | Not-determined |
| S13_extracted_76160 | Not-determined |
| S13_extracted_76175 | Not-determined |
| S13_extracted_76178 | Low-quality |
| S13_extracted_76186 | Not-determined |
| S13_extracted_76197 | Not-determined |
| S13_extracted_76203 | Not-determined |
| S13_extracted_76244 | Not-determined |
| S13_extracted_76259 | Low-quality |
| S13_extracted_76265 | Not-determined |
| S13_extracted_76269 | Not-determined |
| S13_extracted_76275 | Not-determined |
| S13_extracted_76276 | Not-determined |
| S13_extracted_76281 | Not-determined |
| S13_extracted_76291 | Not-determined |
| S13_extracted_76296 | Not-determined |
| S13_extracted_76300 | Not-determined |
| S13_extracted_76303 | Not-determined |
| S13_extracted_76306 | Not-determined |
| S13_extracted_76319 | Not-determined |
| S13_extracted_76326 | Low-quality |
| S13_extracted_76329 | Low-quality |
| S13_extracted_76334 | Low-quality |
| S13_extracted_76364 | Not-determined |
| S13_extracted_76366 | Not-determined |
| S13_extracted_76378 | Not-determined |
| S13_extracted_76390 | Low-quality |
| S13_extracted_76392 | Not-determined |
| S13_extracted_76399 | Not-determined |
| S13_extracted_76406 | Not-determined |
| S13_extracted_76418 | Not-determined |
| S13_extracted_76421 | Not-determined |
| S13_extracted_76431 | Not-determined |
| S13_extracted_76436 | Not-determined |
| S13_extracted_76440 | Not-determined |
| S13_extracted_76451 | Not-determined |
| S13_extracted_76470 | Low-quality |
| S13_extracted_76476 | Not-determined |
| S13_extracted_76480 | Not-determined |
| S13_extracted_76481 | Not-determined |
| S13_extracted_76486 | Low-quality |
| S13_extracted_76495 | Not-determined |
| S13_extracted_76496 | Not-determined |
| S13_extracted_76509 | Not-determined |
| S13_extracted_76511 | Not-determined |
| S13_extracted_76524 | Not-determined |
| S13_extracted_76531 | Not-determined |
| S13_extracted_76555 | Not-determined |
| S13_extracted_76559 | Not-determined |
| S13_extracted_76563 | Not-determined |
| S13_extracted_76568 | Not-determined |
| S13_extracted_76571 | Not-determined |
| S13_extracted_76580 | Not-determined |
| S13_extracted_76588 | Not-determined |
| S13_extracted_76592 | Not-determined |
| S13_extracted_76597 | Not-determined |
| S13_extracted_76603 | Not-determined |
| S13_extracted_76606 | Not-determined |
| S13_extracted_76626 | Not-determined |
| S13_extracted_76640 | Not-determined |
| S13_extracted_76641 | Low-quality |
| S13_extracted_76643 | Not-determined |
| S13_extracted_76676 | Not-determined |
| S13_extracted_76680 | Not-determined |
| S13_extracted_76684 | Not-determined |
| S13_extracted_76698 | Low-quality |
| S13_extracted_76713 | Low-quality |
| S13_extracted_76722 | Not-determined |
| S13_extracted_76728 | Not-determined |
| S13_extracted_76733 | Low-quality |
| S13_extracted_76738 | Not-determined |
| S13_extracted_76758 | Not-determined |
| S13_extracted_76768 | Not-determined |
| S13_extracted_76772 | Not-determined |
| S13_extracted_76780 | Low-quality |
| S13_extracted_76781 | Not-determined |
| S13_extracted_76785 | Not-determined |
| S13_extracted_76795 | Not-determined |
| S13_extracted_76802 | Not-determined |
| S13_extracted_76812 | Not-determined |
| S13_extracted_76820 | Low-quality |
| S13_extracted_76822 | Not-determined |
| S13_extracted_76832 | Not-determined |
| S13_extracted_76836 | Not-determined |
| S13_extracted_76840 | Low-quality |
| S13_extracted_76846 | Not-determined |
| S13_extracted_76852 | Not-determined |
| S13_extracted_76867 | Not-determined |
| S13_extracted_76874 | Low-quality |
| S13_extracted_76893 | Not-determined |
| S13_extracted_76894 | Not-determined |
| S13_extracted_76901 | Not-determined |
| S13_extracted_76906 | Not-determined |
| S13_extracted_76916 | Not-determined |
| S13_extracted_76917 | Not-determined |
| S13_extracted_76937 | Not-determined |
| S13_extracted_76941 | Not-determined |
| S13_extracted_76984 | Low-quality |
| S13_extracted_76994 | Not-determined |
| S13_extracted_77004 | Not-determined |
| S13_extracted_77016 | Low-quality |
| S13_extracted_77022 | Not-determined |
| S13_extracted_77027 | Not-determined |
| S13_extracted_77031 | Not-determined |
| S13_extracted_77034 | Not-determined |
| S13_extracted_77042 | Not-determined |
| S13_extracted_77054 | Not-determined |
| S13_extracted_77058 | Not-determined |
| S13_extracted_77064 | Not-determined |
| S13_extracted_77076 | Not-determined |
| S13_extracted_77080 | Low-quality |
| S13_extracted_77087 | Low-quality |
| S13_extracted_77094 | Not-determined |
| S13_extracted_77116 | Not-determined |
| S13_extracted_77125 | Not-determined |
| S13_extracted_77127 | Not-determined |
| S13_extracted_77133 | Not-determined |
| S13_extracted_77137 | Not-determined |
| S13_extracted_77138 | Not-determined |
| S13_extracted_77149 | Low-quality |
| S13_extracted_77155 | Not-determined |
| S13_extracted_77160 | Not-determined |
| S13_extracted_77169 | Low-quality |
| S13_extracted_77172 | Not-determined |
| S13_extracted_77177 | Not-determined |
| S13_extracted_77186 | Not-determined |
| S13_extracted_77194 | Not-determined |
| S13_extracted_77199 | Not-determined |
| S13_extracted_77200 | Low-quality |
| S13_extracted_77201 | Not-determined |
| S13_extracted_77203 | Not-determined |
| S13_extracted_77206 | Not-determined |
| S13_extracted_77214 | Not-determined |
| S13_extracted_77228 | Low-quality |
| S13_extracted_77232 | Not-determined |
| S13_extracted_77240 | Not-determined |
| S13_extracted_77241 | Low-quality |
| S13_extracted_77249 | Not-determined |
| S13_extracted_77263 | Not-determined |
| S13_extracted_77275 | Not-determined |
| S13_extracted_77288 | Not-determined |
| S13_extracted_77309 | Not-determined |
| S13_extracted_77312 | Not-determined |
| S13_extracted_77315 | Not-determined |
| S13_extracted_77325 | Not-determined |
| S13_extracted_77352 | Not-determined |
| S13_extracted_77365 | Not-determined |
| S13_extracted_77375 | Not-determined |
| S13_extracted_77380 | Not-determined |
| S13_extracted_77395 | Not-determined |
| S13_extracted_77406 | Not-determined |
| S13_extracted_77420 | Low-quality |
| S13_extracted_77441 | Not-determined |
| S13_extracted_77445 | Not-determined |
| S13_extracted_77459 | Not-determined |
| S13_extracted_77466 | Not-determined |
| S13_extracted_77471 | Not-determined |
| S13_extracted_77481 | Low-quality |
| S13_extracted_77496 | Not-determined |
| S13_extracted_77499 | Low-quality |
| S13_extracted_77516 | Not-determined |
| S13_extracted_77517 | Not-determined |
| S13_extracted_77528 | Not-determined |
| S13_extracted_77531 | Not-determined |
| S13_extracted_77544 | Not-determined |
| S13_extracted_77557 | Not-determined |
| S13_extracted_77571 | Low-quality |
| S13_extracted_77595 | Not-determined |
| S13_extracted_77597 | Not-determined |
| S13_extracted_77610 | Not-determined |
| S13_extracted_77612 | Low-quality |
| S13_extracted_77618 | Low-quality |
| S13_extracted_77623 | Low-quality |
| S13_extracted_77631 | Not-determined |
| S13_extracted_77653 | Not-determined |
| S13_extracted_77663 | Not-determined |
| S13_extracted_77691 | Not-determined |
| S13_extracted_77695 | Not-determined |
| S13_extracted_77703 | Not-determined |
| S13_extracted_77718 | Not-determined |
| S13_extracted_77733 | Low-quality |
| S13_extracted_77765 | Not-determined |
| S13_extracted_77802 | Not-determined |
| S13_extracted_77811 | Not-determined |
| S13_extracted_77812 | Not-determined |
| S13_extracted_77813 | Not-determined |
| S13_extracted_77819 | Not-determined |
| S13_extracted_77821 | Not-determined |
| S13_extracted_77826 | Not-determined |
| S13_extracted_77828 | Not-determined |
| S13_extracted_77829 | Not-determined |
| S13_extracted_77831 | Not-determined |
| S13_extracted_77846 | Not-determined |
| S13_extracted_77849 | Not-determined |
| S13_extracted_77854 | Not-determined |
| S13_extracted_77867 | Not-determined |
| S13_extracted_77874 | Not-determined |
| S13_extracted_77888 | Not-determined |
| S13_extracted_77894 | Not-determined |
| S13_extracted_77896 | Not-determined |
| S13_extracted_77901 | Not-determined |
| S13_extracted_77906 | Not-determined |
| S13_extracted_77912 | Not-determined |
| S13_extracted_77923 | Not-determined |
| S13_extracted_77924 | Not-determined |
| S13_extracted_77925 | Not-determined |
| S13_extracted_77926 | Not-determined |
| S13_extracted_77937 | Not-determined |
| S13_extracted_77946 | Not-determined |
| S13_extracted_77947 | Not-determined |
| S13_extracted_77951 | Not-determined |
| S13_extracted_77955 | Not-determined |
| S13_extracted_77960 | Not-determined |
| S13_extracted_77973 | Not-determined |
| S13_extracted_77974 | Not-determined |
| S13_extracted_77977 | Not-determined |
| S13_extracted_77979 | Not-determined |
| S13_extracted_78000 | Not-determined |
| S13_extracted_78009 | Not-determined |
| S13_extracted_78010 | Not-determined |
| S13_extracted_78050 | Not-determined |
| S13_extracted_78053 | Not-determined |
| S13_extracted_78062 | Not-determined |
| S13_extracted_78067 | Not-determined |
| S13_extracted_78070 | Not-determined |
| S13_extracted_78079 | Not-determined |
| S13_extracted_78087 | Low-quality |
| S13_extracted_78092 | Not-determined |
| S13_extracted_78098 | Not-determined |
| S13_extracted_78099 | Not-determined |
| S13_extracted_78103 | Not-determined |
| S13_extracted_78104 | Not-determined |
| S13_extracted_78130 | Not-determined |
| S13_extracted_78132 | Not-determined |
| S13_extracted_78146 | Not-determined |
| S13_extracted_78147 | Not-determined |
| S13_extracted_78148 | Not-determined |
| S13_extracted_78151 | Not-determined |
| S13_extracted_78180 | Not-determined |
| S13_extracted_78186 | Not-determined |
| S13_extracted_78189 | Not-determined |
| S13_extracted_78195 | Not-determined |
| S13_extracted_78197 | Not-determined |
| S13_extracted_78207 | Not-determined |
| S13_extracted_78235 | Not-determined |
| S13_extracted_78269 | Low-quality |
| S13_extracted_78273 | Not-determined |
| S13_extracted_78275 | Not-determined |
| S13_extracted_78277 | Not-determined |
| S13_extracted_78304 | Not-determined |
| S13_extracted_78307 | Low-quality |
| S13_extracted_78329 | Not-determined |
| S13_extracted_78347 | Low-quality |
| S13_extracted_78368 | Low-quality |
| S13_extracted_78379 | Not-determined |
| S13_extracted_78386 | Not-determined |
| S13_extracted_78392 | Not-determined |
| S13_extracted_78397 | Low-quality |
| S13_extracted_78412 | Not-determined |
| S13_extracted_78415 | Not-determined |
| S13_extracted_78417 | Not-determined |
| S13_extracted_78424 | Not-determined |
| S13_extracted_78436 | Not-determined |
| S13_extracted_78444 | Not-determined |
| S13_extracted_78448 | Not-determined |
| S13_extracted_78460 | Not-determined |
| S13_extracted_78463 | Not-determined |
| S13_extracted_78466 | Not-determined |
| S13_extracted_78471 | Not-determined |
| S13_extracted_78486 | Not-determined |
| S13_extracted_78489 | Low-quality |
| S13_extracted_78490 | Not-determined |
| S13_extracted_78502 | Low-quality |
| S13_extracted_78509 | Not-determined |
| S13_extracted_78511 | Not-determined |
| S13_extracted_78524 | Low-quality |
| S13_extracted_78533 | Not-determined |
| S13_extracted_78535 | Not-determined |
| S13_extracted_78538 | Not-determined |
| S13_extracted_78541 | Not-determined |
| S13_extracted_78543 | Not-determined |
| S13_extracted_78569 | Not-determined |
| S13_extracted_78573 | Not-determined |
| S13_extracted_78583 | Not-determined |
| S13_extracted_78585 | Not-determined |
| S13_extracted_78586 | Not-determined |
| S13_extracted_78587 | Not-determined |
| S13_extracted_78600 | Not-determined |
| S13_extracted_78603 | Not-determined |
| S13_extracted_78614 | Not-determined |
| S13_extracted_78617 | Not-determined |
| S13_extracted_78618 | Not-determined |
| S13_extracted_78621 | Not-determined |
| S13_extracted_78657 | Not-determined |
| S13_extracted_78667 | Not-determined |
| S13_extracted_78672 | Not-determined |
| S13_extracted_78675 | Not-determined |
| S13_extracted_78676 | Not-determined |
| S13_extracted_78684 | Not-determined |
| S13_extracted_78686 | Not-determined |
| S13_extracted_78689 | Not-determined |
| S13_extracted_78702 | Not-determined |
| S13_extracted_78722 | Not-determined |
| S13_extracted_78726 | Not-determined |
| S13_extracted_78730 | Not-determined |
| S13_extracted_78735 | Not-determined |
| S13_extracted_78747 | Not-determined |
| S13_extracted_78751 | Not-determined |
| S13_extracted_78752 | Not-determined |
| S13_extracted_78760 | Not-determined |
| S13_extracted_78782 | Not-determined |
| S13_extracted_78784 | Low-quality |
| S13_extracted_78786 | Not-determined |
| S13_extracted_78789 | Not-determined |
| S13_extracted_78795 | Not-determined |
| S13_extracted_78806 | Not-determined |
| S13_extracted_78816 | Not-determined |
| S13_extracted_78820 | Not-determined |
| S13_extracted_78824 | Low-quality |
| S13_extracted_78826 | Not-determined |
| S13_extracted_78833 | Not-determined |
| S13_extracted_78845 | Not-determined |
| S13_extracted_78846 | Not-determined |
| S13_extracted_78861 | Not-determined |
| S13_extracted_78871 | Not-determined |
| S13_extracted_78897 | Not-determined |
| S13_extracted_78905 | Not-determined |
| S13_extracted_78915 | Not-determined |
| S13_extracted_78916 | Not-determined |
| S13_extracted_78918 | Not-determined |
| S13_extracted_78920 | Not-determined |
| S13_extracted_78924 | Not-determined |
| S13_extracted_78925 | Not-determined |
| S13_extracted_78931 | Not-determined |
| S13_extracted_78942 | Not-determined |
| S13_extracted_78943 | Low-quality |
| S13_extracted_78950 | Not-determined |
| S13_extracted_78956 | Not-determined |
| S13_extracted_78957 | Not-determined |
| S13_extracted_78975 | Low-quality |
| S13_extracted_78980 | Not-determined |
| S13_extracted_78984 | Not-determined |
| S13_extracted_78992 | Not-determined |
| S13_extracted_78994 | Not-determined |
| S13_extracted_79022 | Not-determined |
| S13_extracted_79023 | Not-determined |
| S13_extracted_79024 | Not-determined |
| S13_extracted_79038 | Not-determined |
| S13_extracted_79041 | Not-determined |
| S13_extracted_79043 | Low-quality |
| S13_extracted_79063 | Not-determined |
| S13_extracted_79070 | Not-determined |
| S13_extracted_79092 | Not-determined |
| S13_extracted_79096 | Low-quality |
| S13_extracted_79097 | Not-determined |
| S13_extracted_79100 | Not-determined |
| S13_extracted_79107 | Not-determined |
| S13_extracted_79114 | Not-determined |
| S13_extracted_79125 | Not-determined |
| S13_extracted_79127 | Not-determined |
| S13_extracted_79131 | Not-determined |
| S13_extracted_79134 | Not-determined |
| S13_extracted_79157 | Not-determined |
| S13_extracted_79172 | Not-determined |
| S13_extracted_79173 | Not-determined |
| S13_extracted_79191 | Not-determined |
| S13_extracted_79196 | Not-determined |
| S13_extracted_79214 | Not-determined |
| S13_extracted_79215 | Not-determined |
| S13_extracted_79216 | Not-determined |
| S13_extracted_79221 | Not-determined |
| S13_extracted_79222 | Not-determined |
| S13_extracted_79226 | Not-determined |
| S13_extracted_79233 | Not-determined |
| S13_extracted_79238 | Not-determined |
| S13_extracted_79239 | Not-determined |
| S13_extracted_79242 | Low-quality |
| S13_extracted_79244 | Not-determined |
| S13_extracted_79252 | Low-quality |
| S13_extracted_79261 | Not-determined |
| S13_extracted_79266 | Not-determined |
| S13_extracted_79268 | Not-determined |
| S13_extracted_79280 | Not-determined |
| S13_extracted_79281 | Not-determined |
| S13_extracted_79294 | Not-determined |
| S13_extracted_79319 | Not-determined |
| S13_extracted_79321 | Low-quality |
| S13_extracted_79328 | Not-determined |
| S13_extracted_79331 | Low-quality |
| S13_extracted_79354 | Not-determined |
| S13_extracted_79374 | Not-determined |
| S13_extracted_79387 | Low-quality |
| S13_extracted_79395 | Not-determined |
| S13_extracted_79403 | Not-determined |
| S13_extracted_79414 | Not-determined |
| S13_extracted_79419 | Not-determined |
| S13_extracted_79442 | Not-determined |
| S13_extracted_79449 | Not-determined |
| S13_extracted_79450 | Not-determined |
| S13_extracted_79458 | Not-determined |
| S13_extracted_79466 | Not-determined |
| S13_extracted_79474 | Not-determined |
| S13_extracted_79483 | Not-determined |
| S13_extracted_79486 | Low-quality |
| S13_extracted_79487 | Not-determined |
| S13_extracted_79494 | Not-determined |
| S13_extracted_79510 | Low-quality |
| S13_extracted_79527 | Not-determined |
| S13_extracted_79537 | Not-determined |
| S13_extracted_79555 | Not-determined |
| S13_extracted_79561 | Not-determined |
| S13_extracted_79563 | Not-determined |
| S13_extracted_79568 | Low-quality |
| S13_extracted_79572 | Not-determined |
| S13_extracted_79573 | Low-quality |
| S13_extracted_79583 | Not-determined |
| S13_extracted_79585 | Not-determined |
| S13_extracted_79592 | Not-determined |
| S13_extracted_79596 | Not-determined |
| S13_extracted_79618 | Not-determined |
| S13_extracted_79619 | Low-quality |
| S13_extracted_79626 | Not-determined |
| S13_extracted_79633 | Not-determined |
| S13_extracted_79644 | Not-determined |
| S13_extracted_79649 | Not-determined |
| S13_extracted_79659 | Low-quality |
| S13_extracted_79661 | Not-determined |
| S13_extracted_79689 | Not-determined |
| S13_extracted_79692 | Low-quality |
| S13_extracted_79704 | Not-determined |
| S13_extracted_79709 | Low-quality |
| S13_extracted_79713 | Not-determined |
| S13_extracted_79715 | Not-determined |
| S13_extracted_79719 | Not-determined |
| S13_extracted_79724 | Not-determined |
| S13_extracted_79735 | Not-determined |
| S13_extracted_79737 | Not-determined |
| S13_extracted_79744 | Not-determined |
| S13_extracted_79754 | Not-determined |
| S13_extracted_79765 | Not-determined |
| S13_extracted_79766 | Not-determined |
| S13_extracted_79778 | Not-determined |
| S13_extracted_79789 | Not-determined |
| S13_extracted_79792 | Not-determined |
| S13_extracted_79824 | Low-quality |
| S13_extracted_79826 | Not-determined |
| S13_extracted_79844 | Not-determined |
| S13_extracted_79845 | Not-determined |
| S13_extracted_79848 | Not-determined |
| S13_extracted_79865 | Not-determined |
| S13_extracted_79874 | Low-quality |
| S13_extracted_79890 | Not-determined |
| S13_extracted_79898 | Not-determined |
| S13_extracted_79902 | Not-determined |
| S13_extracted_79917 | Not-determined |
| S13_extracted_79935 | Not-determined |
| S13_extracted_79943 | Not-determined |
| S13_extracted_79961 | Not-determined |
| S13_extracted_79986 | Not-determined |
| S13_extracted_79994 | Low-quality |
| S13_extracted_80010 | Not-determined |
| S13_extracted_80015 | Not-determined |
| S13_extracted_80021 | Low-quality |
| S13_extracted_80043 | Not-determined |
| S13_extracted_80057 | Not-determined |
| S13_extracted_80062 | Not-determined |
| S13_extracted_80065 | Not-determined |
| S13_extracted_80077 | Not-determined |
| S13_extracted_80087 | Not-determined |
| S13_extracted_80117 | Not-determined |
| S13_extracted_80169 | Not-determined |
| S13_extracted_80179 | Not-determined |
| S13_extracted_80180 | Not-determined |
| S13_extracted_80183 | Not-determined |
| S13_extracted_80185 | Not-determined |
| S13_extracted_80186 | Not-determined |
| S13_extracted_80190 | Low-quality |
| S13_extracted_80195 | Not-determined |
| S13_extracted_80210 | Not-determined |
| S13_extracted_80238 | Not-determined |
| S13_extracted_80241 | Not-determined |

| | |
|---|---|
| S13_extracted_80243 | Not-determined |
| S13_extracted_80245 | Not-determined |
| S13_extracted_80261 | Not-determined |
| S13_extracted_80277 | Not-determined |
| S13_extracted_80285 | Not-determined |
| S13_extracted_80286 | Not-determined |
| S13_extracted_80310 | Not-determined |
| S13_extracted_80326 | Low-quality |
| S13_extracted_80351 | Not-determined |
| S13_extracted_80364 | Low-quality |
| S13_extracted_80374 | Low-quality |
| S13_extracted_80375 | Not-determined |
| S13_extracted_80378 | Not-determined |
| S13_extracted_80383 | Not-determined |
| S13_extracted_80386 | Not-determined |
| S13_extracted_80403 | Not-determined |
| S13_extracted_80415 | Not-determined |
| S13_extracted_80417 | Not-determined |
| S13_extracted_80432 | Not-determined |
| S13_extracted_80450 | Not-determined |
| S13_extracted_80473 | Low-quality |
| S13_extracted_80474 | Not-determined |
| S13_extracted_80483 | Not-determined |
| S13_extracted_80489 | Not-determined |
| S13_extracted_80505 | Not-determined |
| S13_extracted_80514 | Not-determined |
| S13_extracted_80516 | Not-determined |
| S13_extracted_80517 | Not-determined |
| S13_extracted_80539 | Not-determined |
| S13_extracted_80552 | Low-quality |
| S13_extracted_80557 | Not-determined |
| S13_extracted_80563 | Not-determined |
| S13_extracted_80564 | Not-determined |
| S13_extracted_80577 | Not-determined |
| S13_extracted_80587 | Not-determined |
| S13_extracted_80590 | Not-determined |
| S13_extracted_80603 | Not-determined |
| S13_extracted_80606 | Not-determined |
| S13_extracted_80619 | Not-determined |
| S13_extracted_80626 | Not-determined |
| S13_extracted_80634 | Low-quality |
| S13_extracted_80659 | Not-determined |
| S13_extracted_80668 | Not-determined |
| S13_extracted_80669 | Not-determined |
| S13_extracted_80671 | Not-determined |
| S13_extracted_80672 | Not-determined |
| S13_extracted_80678 | Not-determined |
| S13_extracted_80701 | Not-determined |
| S13_extracted_80705 | Not-determined |
| S13_extracted_80710 | Low-quality |
| S13_extracted_80713 | Not-determined |
| S13_extracted_80733 | Not-determined |
| S13_extracted_80734 | Not-determined |
| S13_extracted_80749 | Not-determined |
| S13_extracted_80750 | Low-quality |
| S13_extracted_80759 | Not-determined |
| S13_extracted_80785 | Not-determined |
| S13_extracted_80789 | Not-determined |
| S13_extracted_80791 | Not-determined |
| S13_extracted_80794 | Not-determined |
| S13_extracted_80810 | Not-determined |
| S13_extracted_80819 | Not-determined |
| S13_extracted_80822 | Not-determined |
| S13_extracted_80833 | Not-determined |
| S13_extracted_80851 | Not-determined |
| S13_extracted_80852 | Not-determined |
| S13_extracted_80854 | Not-determined |
| S13_extracted_80865 | Not-determined |
| S13_extracted_80880 | Not-determined |
| S13_extracted_80884 | Not-determined |
| S13_extracted_80901 | Not-determined |
| S13_extracted_80907 | Not-determined |
| S13_extracted_80910 | Low-quality |
| S13_extracted_80917 | Not-determined |
| S13_extracted_80918 | Not-determined |
| S13_extracted_80919 | Low-quality |
| S13_extracted_80923 | Not-determined |
| S13_extracted_80931 | Not-determined |
| S13_extracted_80933 | Not-determined |
| S13_extracted_80937 | Low-quality |
| S13_extracted_80939 | Not-determined |
| S13_extracted_80943 | Not-determined |
| S13_extracted_80948 | Not-determined |
| S13_extracted_80956 | Not-determined |
| S13_extracted_80963 | Not-determined |
| S13_extracted_80969 | Not-determined |
| S13_extracted_80994 | Not-determined |
| S13_extracted_81002 | Not-determined |
| S13_extracted_81014 | Not-determined |
| S13_extracted_81015 | Not-determined |
| S13_extracted_81016 | Not-determined |
| S13_extracted_81045 | Not-determined |
| S13_extracted_81081 | Not-determined |
| S13_extracted_81082 | Not-determined |
| S13_extracted_81084 | Not-determined |
| S13_extracted_81093 | Not-determined |
| S13_extracted_81100 | Not-determined |
| S13_extracted_81107 | Not-determined |
| S13_extracted_81116 | Not-determined |
| S13_extracted_81122 | Low-quality |
| S13_extracted_81123 | Not-determined |
| S13_extracted_81124 | Not-determined |
| S13_extracted_81126 | Not-determined |
| S13_extracted_81128 | Not-determined |
| S13_extracted_81130 | Not-determined |
| S13_extracted_81138 | Not-determined |
| S13_extracted_81144 | Not-determined |
| S13_extracted_81160 | Low-quality |
| S13_extracted_81181 | Not-determined |
| S13_extracted_81185 | Low-quality |
| S13_extracted_81204 | Not-determined |
| S13_extracted_81214 | Not-determined |
| S13_extracted_81246 | Not-determined |
| S13_extracted_81247 | Not-determined |
| S13_extracted_81257 | Not-determined |
| S13_extracted_81269 | Not-determined |
| S13_extracted_81270 | Not-determined |
| S13_extracted_81273 | Not-determined |
| S13_extracted_81278 | Not-determined |
| S13_extracted_81283 | Not-determined |
| S13_extracted_81293 | Not-determined |
| S13_extracted_81334 | Not-determined |
| S13_extracted_81343 | Not-determined |
| S13_extracted_81359 | Not-determined |
| S13_extracted_81370 | Not-determined |
| S13_extracted_81372 | Not-determined |
| S13_extracted_81374 | Not-determined |
| S13_extracted_81375 | Not-determined |
| S13_extracted_81388 | Low-quality |
| S13_extracted_81397 | Low-quality |
| S13_extracted_81402 | Not-determined |
| S13_extracted_81403 | Not-determined |
| S13_extracted_81406 | Not-determined |
| S13_extracted_81425 | Not-determined |
| S13_extracted_81436 | Not-determined |
| S13_extracted_81437 | Not-determined |
| S13_extracted_81445 | Not-determined |
| S13_extracted_81460 | Not-determined |
| S13_extracted_81462 | Not-determined |
| S13_extracted_81463 | Not-determined |
| S13_extracted_81466 | Low-quality |
| S13_extracted_81468 | Not-determined |
| S13_extracted_81471 | Not-determined |
| S13_extracted_81479 | Not-determined |
| S13_extracted_81481 | Not-determined |
| S13_extracted_81486 | Low-quality |
| S13_extracted_81505 | Not-determined |
| S13_extracted_81510 | Low-quality |
| S13_extracted_81511 | Not-determined |
| S13_extracted_81528 | Not-determined |
| S13_extracted_81545 | Not-determined |
| S13_extracted_81553 | Not-determined |
| S13_extracted_81557 | Not-determined |
| S13_extracted_81575 | Not-determined |
| S13_extracted_81591 | Not-determined |
| S13_extracted_81597 | Not-determined |
| S13_extracted_81606 | Not-determined |
| S13_extracted_81611 | Not-determined |
| S13_extracted_81617 | Not-determined |
| S13_extracted_81622 | Not-determined |
| S13_extracted_81641 | Not-determined |
| S13_extracted_81648 | Not-determined |
| S13_extracted_81649 | Not-determined |
| S13_extracted_81656 | Not-determined |
| S13_extracted_81658 | Not-determined |
| S13_extracted_81660 | Not-determined |
| S13_extracted_81669 | Low-quality |
| S13_extracted_81686 | Not-determined |
| S13_extracted_81696 | Low-quality |
| S13_extracted_81703 | Not-determined |
| S13_extracted_81707 | Not-determined |
| S13_extracted_81709 | Not-determined |
| S13_extracted_81711 | Not-determined |
| S13_extracted_81715 | Not-determined |
| S13_extracted_81743 | Not-determined |
| S13_extracted_81763 | Not-determined |
| S13_extracted_81765 | Not-determined |
| S13_extracted_81768 | Not-determined |
| S13_extracted_81769 | Not-determined |
| S13_extracted_81790 | Not-determined |
| S13_extracted_81794 | Not-determined |
| S13_extracted_81811 | Not-determined |
| S13_extracted_81823 | Not-determined |
| S13_extracted_81838 | Not-determined |
| S13_extracted_81840 | Not-determined |
| S13_extracted_81870 | Not-determined |
| S13_extracted_81879 | Not-determined |
| S13_extracted_81891 | Not-determined |
| S13_extracted_81895 | Not-determined |
| S13_extracted_81904 | Low-quality |
| S13_extracted_81906 | Not-determined |
| S13_extracted_81911 | Not-determined |
| S13_extracted_81918 | Not-determined |
| S13_extracted_81924 | Not-determined |
| S13_extracted_81926 | Not-determined |
| S13_extracted_81938 | Not-determined |
| S13_extracted_81942 | Not-determined |
| S13_extracted_81950 | Not-determined |
| S13_extracted_81966 | Not-determined |
| S13_extracted_81971 | Not-determined |
| S13_extracted_81982 | Not-determined |
| S13_extracted_81991 | Not-determined |
| S13_extracted_81999 | Low-quality |
| S13_extracted_82006 | Not-determined |
| S13_extracted_82008 | Not-determined |
| S13_extracted_82011 | Not-determined |
| S13_extracted_82021 | Not-determined |
| S13_extracted_82032 | Not-determined |
| S13_extracted_82060 | Not-determined |
| S13_extracted_82066 | Not-determined |
| S13_extracted_82067 | Low-quality |
| S13_extracted_82079 | Not-determined |
| S13_extracted_82089 | Not-determined |
| S13_extracted_82098 | Low-quality |
| S13_extracted_82099 | Not-determined |
| S13_extracted_82112 | Low-quality |
| S13_extracted_82117 | Not-determined |
| S13_extracted_82121 | Not-determined |
| S13_extracted_82124 | Not-determined |
| S13_extracted_82129 | Not-determined |
| S13_extracted_82131 | Not-determined |
| S13_extracted_82132 | Not-determined |
| S13_extracted_82159 | Not-determined |
| S13_extracted_82183 | Not-determined |
| S13_extracted_82199 | Not-determined |
| S13_extracted_82207 | Not-determined |
| S13_extracted_82208 | Not-determined |
| S13_extracted_82217 | Not-determined |
| S13_extracted_82222 | Not-determined |
| S13_extracted_82241 | Not-determined |
| S13_extracted_82246 | Not-determined |
| S13_extracted_82249 | Low-quality |
| S13_extracted_82254 | Not-determined |
| S13_extracted_82256 | Low-quality |
| S13_extracted_82258 | Not-determined |
| S13_extracted_82267 | Not-determined |
| S13_extracted_82275 | Not-determined |
| S13_extracted_82288 | Not-determined |
| S13_extracted_82296 | Not-determined |
| S13_extracted_82300 | Not-determined |
| S13_extracted_82303 | Not-determined |
| S13_extracted_82309 | Low-quality |
| S13_extracted_82321 | Not-determined |
| S13_extracted_82326 | Not-determined |
| S13_extracted_82334 | Not-determined |
| S13_extracted_82337 | Not-determined |
| S13_extracted_82339 | Not-determined |
| S13_extracted_82346 | Not-determined |
| S13_extracted_82348 | Not-determined |
| S13_extracted_82354 | Not-determined |
| S13_extracted_82355 | Not-determined |
| S13_extracted_82365 | Not-determined |
| S13_extracted_82368 | Not-determined |
| S13_extracted_82372 | Not-determined |
| S13_extracted_82380 | Not-determined |
| S13_extracted_82381 | Not-determined |
| S13_extracted_82397 | Not-determined |
| S13_extracted_82399 | Not-determined |
| S13_extracted_82404 | Not-determined |
| S13_extracted_82435 | Not-determined |
| S13_extracted_82443 | Not-determined |
| S13_extracted_82453 | Not-determined |
| S13_extracted_82454 | Not-determined |
| S13_extracted_82464 | Not-determined |
| S13_extracted_82499 | Not-determined |
| S13_extracted_82508 | Not-determined |
| S13_extracted_82534 | Not-determined |
| S13_extracted_82541 | Not-determined |
| S13_extracted_82571 | Not-determined |
| S13_extracted_82574 | Low-quality |
| S13_extracted_82600 | Low-quality |
| S13_extracted_82608 | Not-determined |
| S13_extracted_82625 | Not-determined |
| S13_extracted_82631 | Not-determined |
| S13_extracted_82632 | Not-determined |
| S13_extracted_82653 | Not-determined |
| S13_extracted_82655 | Not-determined |
| S13_extracted_82665 | Not-determined |
| S13_extracted_82667 | Not-determined |
| S13_extracted_82678 | Not-determined |
| S13_extracted_82695 | Not-determined |
| S13_extracted_82726 | Not-determined |
| S13_extracted_82732 | Not-determined |
| S13_extracted_82740 | Not-determined |
| S13_extracted_82754 | Not-determined |
| S13_extracted_82762 | Not-determined |
| S13_extracted_82770 | Not-determined |
| S13_extracted_82779 | Not-determined |
| S13_extracted_82809 | Not-determined |
| S13_extracted_82820 | Not-determined |
| S13_extracted_82842 | Not-determined |
| S13_extracted_82856 | Low-quality |
| S13_extracted_82859 | Not-determined |
| S13_extracted_82870 | Not-determined |
| S13_extracted_82875 | Not-determined |
| S13_extracted_82881 | Not-determined |
| S13_extracted_82891 | Low-quality |
| S13_extracted_82935 | Not-determined |
| S13_extracted_82936 | Not-determined |
| S13_extracted_82948 | Not-determined |
| S13_extracted_82949 | Not-determined |
| S13_extracted_82973 | Not-determined |
| S13_extracted_82989 | Not-determined |
| S13_extracted_83005 | Not-determined |
| S13_extracted_83016 | Not-determined |
| S13_extracted_83036 | Not-determined |
| S13_extracted_83038 | Not-determined |
| S13_extracted_83040 | Low-quality |
| S13_extracted_83061 | Not-determined |
| S13_extracted_83085 | Not-determined |
| S13_extracted_83098 | Not-determined |
| S13_extracted_83100 | Not-determined |
| S13_extracted_83106 | Low-quality |
| S13_extracted_83113 | Not-determined |
| S13_extracted_83116 | Not-determined |
| S13_extracted_83126 | Not-determined |
| S13_extracted_83134 | Not-determined |
| S13_extracted_83161 | Not-determined |
| S13_extracted_83178 | Not-determined |
| S13_extracted_83187 | Not-determined |
| S13_extracted_83209 | Not-determined |
| S13_extracted_83211 | Low-quality |
| S13_extracted_83217 | Not-determined |
| S13_extracted_83223 | Not-determined |
| S13_extracted_83225 | Not-determined |
| S13_extracted_83230 | Not-determined |
| S13_extracted_83243 | Not-determined |
| S13_extracted_83249 | Not-determined |
| S13_extracted_83250 | Not-determined |
| S13_extracted_83260 | Not-determined |
| S13_extracted_83266 | Not-determined |
| S13_extracted_83287 | Not-determined |
| S13_extracted_83290 | Not-determined |
| S13_extracted_83295 | Not-determined |
| S13_extracted_83296 | Not-determined |
| S13_extracted_83297 | Not-determined |
| S13_extracted_83303 | Not-determined |
| S13_extracted_83307 | Not-determined |
| S13_extracted_83319 | Not-determined |
| S13_extracted_83323 | Not-determined |
| S13_extracted_83343 | Not-determined |
| S13_extracted_83345 | Not-determined |
| S13_extracted_83359 | Low-quality |
| S13_extracted_83400 | Not-determined |
| S13_extracted_83428 | Not-determined |
| S13_extracted_83433 | Not-determined |
| S13_extracted_83434 | Not-determined |
| S13_extracted_83445 | Not-determined |
| S13_extracted_83451 | Not-determined |
| S13_extracted_83457 | Not-determined |
| S13_extracted_83458 | Not-determined |
| S13_extracted_83467 | Not-determined |
| S13_extracted_83480 | Not-determined |
| S13_extracted_83509 | Not-determined |
| S13_extracted_83511 | Not-determined |
| S13_extracted_83517 | Not-determined |
| S13_extracted_83520 | Not-determined |
| S13_extracted_83526 | Not-determined |
| S13_extracted_83529 | Not-determined |
| S13_extracted_83531 | Not-determined |
| S13_extracted_83532 | Not-determined |
| S13_extracted_83540 | Not-determined |
| S13_extracted_83541 | Not-determined |
| S13_extracted_83554 | Not-determined |
| S13_extracted_83570 | Not-determined |
| S13_extracted_83582 | Not-determined |
| S13_extracted_83602 | Low-quality |
| S13_extracted_83614 | Not-determined |
| S13_extracted_83617 | Not-determined |
| S13_extracted_83629 | Not-determined |
| S13_extracted_83631 | Not-determined |
| S13_extracted_83635 | Not-determined |
| S13_extracted_83648 | Not-determined |
| S13_extracted_83656 | Not-determined |
| S13_extracted_83662 | Not-determined |
| S13_extracted_83697 | Not-determined |
| S13_extracted_83700 | Not-determined |
| S13_extracted_83715 | Low-quality |
| S13_extracted_83724 | Not-determined |
| S13_extracted_83747 | Not-determined |
| S13_extracted_83749 | Not-determined |
| S13_extracted_83753 | Not-determined |
| S13_extracted_83768 | Low-quality |
| S13_extracted_83788 | Not-determined |
| S13_extracted_83795 | Not-determined |
| S13_extracted_83803 | Not-determined |
| S13_extracted_83808 | Not-determined |
| S13_extracted_83817 | Not-determined |
| S13_extracted_83831 | Not-determined |
| S13_extracted_83841 | Not-determined |
| S13_extracted_83847 | Not-determined |
| S13_extracted_83852 | Not-determined |
| S13_extracted_83853 | Not-determined |
| S13_extracted_83861 | Not-determined |
| S13_extracted_83872 | Not-determined |
| S13_extracted_83874 | Not-determined |
| S13_extracted_83876 | Not-determined |
| S13_extracted_83879 | Not-determined |
| S13_extracted_83883 | Not-determined |
| S13_extracted_83884 | Not-determined |
| S13_extracted_83893 | Not-determined |
| S13_extracted_83898 | Not-determined |
| S13_extracted_83908 | Not-determined |
| S13_extracted_83923 | Not-determined |
| S13_extracted_83926 | Not-determined |
| S13_extracted_83932 | Not-determined |
| S13_extracted_83933 | Not-determined |
| S13_extracted_83941 | Not-determined |
| S13_extracted_83959 | Not-determined |
| S13_extracted_83964 | Not-determined |
| S13_extracted_83974 | Not-determined |
| S13_extracted_83988 | Not-determined |
| S13_extracted_83998 | Not-determined |
| S13_extracted_84003 | Not-determined |
| S13_extracted_84005 | Low-quality |
| S13_extracted_84012 | Not-determined |
| S13_extracted_84018 | Not-determined |
| S13_extracted_84019 | Not-determined |
| S13_extracted_84021 | Not-determined |
| S13_extracted_84030 | Not-determined |
| S13_extracted_84032 | Not-determined |
| S13_extracted_84033 | Not-determined |
| S13_extracted_84045 | Not-determined |
| S13_extracted_84088 | Not-determined |
| S13_extracted_84092 | Not-determined |
| S13_extracted_84099 | Not-determined |
| S13_extracted_84102 | Not-determined |
| S13_extracted_84108 | Not-determined |
| S13_extracted_84120 | Not-determined |
| S13_extracted_84148 | Low-quality |
| S13_extracted_84158 | Not-determined |
| S13_extracted_84176 | Not-determined |
| S13_extracted_84223 | Not-determined |
| S13_extracted_84225 | Not-determined |
| S13_extracted_84242 | Not-determined |
| S13_extracted_84243 | Not-determined |
| S13_extracted_84265 | Not-determined |
| S13_extracted_84273 | Not-determined |
| S13_extracted_84283 | Not-determined |
| S13_extracted_84288 | Not-determined |
| S13_extracted_84291 | Not-determined |
| S13_extracted_84304 | Not-determined |
| S13_extracted_84305 | Not-determined |
| S13_extracted_84311 | Not-determined |
| S13_extracted_84316 | Not-determined |
| S13_extracted_84321 | Low-quality |
| S13_extracted_84355 | Not-determined |
| S13_extracted_84372 | Not-determined |
| S13_extracted_84378 | Not-determined |
| S13_extracted_84400 | Not-determined |
| S13_extracted_84405 | Not-determined |
| S13_extracted_84409 | Not-determined |
| S13_extracted_84411 | Not-determined |
| S13_extracted_84419 | Not-determined |
| S13_extracted_84420 | Not-determined |
| S13_extracted_84428 | Not-determined |
| S13_extracted_84439 | Not-determined |
| S13_extracted_84459 | Not-determined |
| S13_extracted_84463 | Not-determined |
| S13_extracted_84475 | Not-determined |
| S13_extracted_84478 | Low-quality |
| S13_extracted_84481 | Not-determined |
| S13_extracted_84493 | Not-determined |
| S13_extracted_84499 | Not-determined |
| S13_extracted_84507 | Not-determined |
| S13_extracted_84511 | Not-determined |
| S13_extracted_84519 | Not-determined |
| S13_extracted_84529 | Low-quality |
| S13_extracted_84530 | Not-determined |
| S13_extracted_84537 | Not-determined |
| S13_extracted_84544 | Not-determined |
| S13_extracted_84568 | Not-determined |
| S13_extracted_84573 | Not-determined |
| S13_extracted_84574 | Not-determined |
| S13_extracted_84583 | Not-determined |
| S13_extracted_84590 | Not-determined |
| S13_extracted_84594 | Not-determined |
| S13_extracted_84604 | Not-determined |
| S13_extracted_84607 | Not-determined |
| S13_extracted_84621 | Not-determined |
| S13_extracted_84622 | Not-determined |
| S13_extracted_84623 | Not-determined |
| S13_extracted_84625 | Not-determined |
| S13_extracted_84626 | Not-determined |
| S13_extracted_84630 | Not-determined |
| S13_extracted_84636 | Not-determined |
| S13_extracted_84639 | Not-determined |
| S13_extracted_84647 | Not-determined |
| S13_extracted_84649 | Not-determined |
| S13_extracted_84657 | Not-determined |
| S13_extracted_84659 | Not-determined |
| S13_extracted_84676 | Not-determined |
| S13_extracted_84677 | Not-determined |
| S13_extracted_84678 | Not-determined |
| S13_extracted_84688 | Not-determined |
| S13_extracted_84702 | Not-determined |
| S13_extracted_84721 | Not-determined |
| S13_extracted_84725 | Not-determined |
| S13_extracted_84727 | Not-determined |
| S13_extracted_84728 | Low-quality |
| S13_extracted_84777 | Not-determined |
| S13_extracted_84779 | Not-determined |
| S13_extracted_84780 | Not-determined |
| S13_extracted_84814 | Not-determined |
| S13_extracted_84817 | Not-determined |
| S13_extracted_84823 | Not-determined |
| S13_extracted_84829 | Not-determined |
| S13_extracted_84830 | Low-quality |
| S13_extracted_84840 | Not-determined |
| S13_extracted_84846 | Not-determined |
| S13_extracted_84859 | Not-determined |
| S13_extracted_84861 | Not-determined |
| S13_extracted_84866 | Not-determined |
| S13_extracted_84870 | Not-determined |
| S13_extracted_84901 | Not-determined |
| S13_extracted_84965 | Not-determined |
| S13_extracted_84973 | Not-determined |
| S13_extracted_84980 | Low-quality |
| S13_extracted_84994 | Not-determined |
| S13_extracted_84995 | Not-determined |
| S13_extracted_85005 | Not-determined |
| S13_extracted_85007 | Not-determined |
| S13_extracted_85011 | Not-determined |
| S13_extracted_85014 | Not-determined |
| S13_extracted_85040 | Low-quality |
| S13_extracted_85045 | Low-quality |
| S13_extracted_85048 | Not-determined |
| S13_extracted_85061 | Not-determined |
| S13_extracted_85078 | Not-determined |
| S16_extracted_87190 | Medium-quality |
| S13_extracted_85108 | Not-determined |
| S13_extracted_85121 | Not-determined |
| S13_extracted_85133 | Not-determined |
| S13_extracted_85141 | Not-determined |
| S13_extracted_85146 | Not-determined |
| S13_extracted_85153 | Low-quality |
| S13_extracted_85155 | Not-determined |
| S13_extracted_85166 | Not-determined |
| S13_extracted_85171 | Not-determined |
| S13_extracted_85188 | Not-determined |
| S13_extracted_85192 | Not-determined |
| S13_extracted_85201 | Not-determined |
| S13_extracted_85209 | Low-quality |
| S13_extracted_85211 | Low-quality |
| S13_extracted_85220 | Not-determined |
| S13_extracted_85224 | Not-determined |
| S13_extracted_85236 | Not-determined |
| S13_extracted_85239 | Low-quality |
| S13_extracted_85246 | Not-determined |
| S13_extracted_85261 | Not-determined |
| S13_extracted_85266 | Low-quality |
| S13_extracted_85269 | Not-determined |
| S13_extracted_85272 | Low-quality |
| S13_extracted_85275 | Not-determined |
| S13_extracted_85277 | Not-determined |
| S13_extracted_85281 | Not-determined |
| S13_extracted_85287 | Not-determined |
| S13_extracted_85289 | Not-determined |
| S13_extracted_85307 | Not-determined |
| S13_extracted_85312 | Not-determined |
| S13_extracted_85317 | Not-determined |
| S13_extracted_85318 | Not-determined |
| S13_extracted_85352 | Low-quality |
| S13_extracted_85365 | Not-determined |
| S13_extracted_85368 | Not-determined |
| S13_extracted_85369 | Not-determined |
| S13_extracted_85376 | Not-determined |
| S13_extracted_85378 | Not-determined |
| S13_extracted_85386 | Not-determined |
| S13_extracted_85389 | Not-determined |
| S13_extracted_85399 | Not-determined |
| S13_extracted_85400 | Not-determined |
| S13_extracted_85413 | Low-quality |
| S13_extracted_85414 | Not-determined |
| S13_extracted_85417 | Not-determined |
| S13_extracted_85421 | Not-determined |
| S13_extracted_85466 | Not-determined |
| S13_extracted_85474 | Not-determined |
| S13_extracted_85475 | Not-determined |
| S13_extracted_85480 | Not-determined |
| S13_extracted_85481 | Not-determined |
| S13_extracted_85489 | Not-determined |
| S13_extracted_85501 | Not-determined |
| S13_extracted_85502 | Not-determined |
| S13_extracted_85504 | Not-determined |
| S13_extracted_85505 | Not-determined |
| S13_extracted_85511 | Not-determined |
| S13_extracted_85525 | Not-determined |
| S13_extracted_85555 | Low-quality |

| | |
|---|---|
| S13_extracted_85558 | Not-determined |
| S13_extracted_85563 | Not-determined |
| S13_extracted_85571 | Not-determined |
| S13_extracted_85579 | Not-determined |
| S13_extracted_85585 | Not-determined |
| S13_extracted_85599 | Not-determined |
| S13_extracted_85608 | Not-determined |
| S13_extracted_85618 | Not-determined |
| S13_extracted_85624 | Not-determined |
| S13_extracted_85626 | Not-determined |
| S13_extracted_85629 | Not-determined |
| S13_extracted_85635 | Not-determined |
| S13_extracted_85637 | Not-determined |
| S13_extracted_85638 | Not-determined |
| S13_extracted_85646 | Not-determined |
| S13_extracted_85660 | Not-determined |
| S13_extracted_85661 | Not-determined |
| S13_extracted_85668 | Not-determined |
| S13_extracted_85669 | Low-quality |
| S13_extracted_85707 | Not-determined |
| S13_extracted_85726 | Not-determined |
| S13_extracted_85748 | Not-determined |
| S13_extracted_85749 | Not-determined |
| S13_extracted_85752 | Not-determined |
| S13_extracted_85759 | Low-quality |
| S13_extracted_85801 | Not-determined |
| S13_extracted_85807 | Not-determined |
| S13_extracted_85808 | Low-quality |
| S13_extracted_85837 | Not-determined |
| S13_extracted_85839 | Not-determined |
| S13_extracted_85842 | Not-determined |
| S13_extracted_85846 | Not-determined |
| S13_extracted_85849 | Not-determined |
| S13_extracted_85858 | Not-determined |
| S13_extracted_85860 | Not-determined |
| S13_extracted_85876 | Not-determined |
| S13_extracted_85878 | Not-determined |
| S13_extracted_85886 | Not-determined |
| S13_extracted_85896 | Not-determined |
| S13_extracted_85899 | Not-determined |
| S13_extracted_85905 | Not-determined |
| S13_extracted_85909 | Not-determined |
| S13_extracted_85913 | Not-determined |
| S13_extracted_85925 | Not-determined |
| S13_extracted_85928 | Not-determined |
| S13_extracted_85936 | Not-determined |
| S13_extracted_85946 | Not-determined |
| S13_extracted_85952 | Not-determined |
| S13_extracted_85975 | Not-determined |
| S13_extracted_85980 | Not-determined |
| S13_extracted_85986 | Not-determined |
| S13_extracted_85987 | Not-determined |
| S13_extracted_85991 | Not-determined |
| S13_extracted_85995 | Low-quality |
| S13_extracted_86003 | Not-determined |
| S13_extracted_86005 | Not-determined |
| S13_extracted_86008 | Not-determined |
| S13_extracted_86023 | Not-determined |
| S13_extracted_86024 | Not-determined |
| S13_extracted_86027 | Not-determined |
| S13_extracted_86030 | Not-determined |
| S13_extracted_86039 | Not-determined |
| S13_extracted_86051 | Not-determined |
| S13_extracted_86054 | Not-determined |
| S13_extracted_86057 | Not-determined |
| S13_extracted_86059 | Not-determined |
| S13_extracted_86061 | Not-determined |
| S13_extracted_86064 | Not-determined |
| S13_extracted_86066 | Not-determined |
| S13_extracted_86073 | Not-determined |
| S13_extracted_86077 | Not-determined |
| S13_extracted_86084 | Not-determined |
| S13_extracted_86085 | Not-determined |
| S13_extracted_86088 | Not-determined |
| S13_extracted_86091 | Low-quality |
| S13_extracted_86093 | Not-determined |
| S13_extracted_86103 | Not-determined |
| S13_extracted_86114 | Not-determined |
| S13_extracted_86120 | Not-determined |
| S13_extracted_86128 | Not-determined |
| S13_extracted_86131 | Not-determined |
| S13_extracted_86148 | Not-determined |
| S13_extracted_86154 | Not-determined |
| S13_extracted_86166 | Not-determined |
| S13_extracted_86173 | Not-determined |
| S13_extracted_86177 | Not-determined |
| S13_extracted_86178 | Not-determined |
| S13_extracted_86195 | Not-determined |
| S13_extracted_86208 | Not-determined |
| S13_extracted_86211 | Not-determined |
| S13_extracted_86215 | Not-determined |
| S13_extracted_86221 | Not-determined |
| S13_extracted_86225 | Not-determined |
| S13_extracted_86245 | Not-determined |
| S13_extracted_86252 | Not-determined |
| S13_extracted_86256 | Not-determined |
| S13_extracted_86266 | Not-determined |
| S13_extracted_86275 | Not-determined |
| S13_extracted_86279 | Not-determined |
| S13_extracted_86287 | Not-determined |
| S13_extracted_86304 | Low-quality |
| S13_extracted_86315 | Not-determined |
| S13_extracted_86316 | Not-determined |
| S13_extracted_86323 | Not-determined |
| S13_extracted_86332 | Not-determined |
| S13_extracted_86336 | Not-determined |
| S13_extracted_86350 | Not-determined |
| S13_extracted_86362 | Not-determined |
| S13_extracted_86367 | Not-determined |
| S13_extracted_86371 | Not-determined |
| S13_extracted_86375 | Not-determined |
| S13_extracted_86386 | Not-determined |
| S13_extracted_86392 | Not-determined |
| S13_extracted_86420 | Not-determined |
| S13_extracted_86426 | Not-determined |
| S13_extracted_86436 | Not-determined |
| S13_extracted_86438 | Not-determined |
| S13_extracted_86445 | Not-determined |
| S13_extracted_86448 | Not-determined |
| S13_extracted_86460 | Not-determined |
| S13_extracted_86475 | Not-determined |
| S13_extracted_86527 | Low-quality |
| S13_extracted_86534 | Not-determined |
| S13_extracted_86535 | Not-determined |
| S13_extracted_86546 | Not-determined |
| S13_extracted_86563 | Not-determined |
| S13_extracted_86575 | Low-quality |
| S13_extracted_86581 | Not-determined |
| S13_extracted_86585 | Not-determined |
| S13_extracted_86613 | Not-determined |
| S13_extracted_86616 | Not-determined |
| S13_extracted_86637 | Low-quality |
| S13_extracted_86642 | Not-determined |
| S13_extracted_86643 | Not-determined |
| S13_extracted_86649 | Not-determined |
| S13_extracted_86654 | Not-determined |
| S13_extracted_86667 | Not-determined |
| S13_extracted_86692 | Not-determined |
| S13_extracted_86715 | Not-determined |
| S13_extracted_86741 | Not-determined |
| S13_extracted_86776 | Low-quality |
| S13_extracted_86805 | Not-determined |
| S13_extracted_86814 | Not-determined |
| S13_extracted_86816 | Not-determined |
| S13_extracted_86819 | Not-determined |
| S13_extracted_86826 | Not-determined |
| S13_extracted_86853 | Not-determined |
| S13_extracted_86858 | Not-determined |
| S13_extracted_86874 | Not-determined |
| S13_extracted_86876 | Not-determined |
| S13_extracted_86901 | Low-quality |
| S13_extracted_86908 | Not-determined |
| S13_extracted_86916 | Not-determined |
| S13_extracted_86927 | Not-determined |
| S13_extracted_86955 | Not-determined |
| S13_extracted_86959 | Not-determined |
| S13_extracted_86961 | Not-determined |
| S13_extracted_86965 | Not-determined |
| S13_extracted_86968 | Not-determined |
| S13_extracted_87005 | Not-determined |
| S13_extracted_87017 | Low-quality |
| S13_extracted_87022 | Not-determined |
| S13_extracted_87043 | Not-determined |
| S13_extracted_87079 | Low-quality |
| S13_extracted_87107 | Not-determined |
| S13_extracted_87110 | Not-determined |
| S13_extracted_87132 | Not-determined |
| S13_extracted_87137 | Not-determined |
| S13_extracted_87147 | Not-determined |
| S13_extracted_87149 | Not-determined |
| S13_extracted_87171 | Not-determined |
| S13_extracted_87178 | Not-determined |
| S13_extracted_87186 | Not-determined |
| S13_extracted_87202 | Not-determined |
| S13_extracted_87213 | Not-determined |
| S13_extracted_87235 | Not-determined |
| S13_extracted_87282 | Not-determined |
| S13_extracted_87284 | Not-determined |
| S13_extracted_87298 | Not-determined |
| S13_extracted_87303 | Not-determined |
| S13_extracted_87316 | Not-determined |
| S13_extracted_87325 | Not-determined |
| S13_extracted_87336 | Low-quality |
| S13_extracted_87337 | Not-determined |
| S13_extracted_87343 | Not-determined |
| S13_extracted_87348 | Not-determined |
| S13_extracted_87349 | Not-determined |
| S13_extracted_87357 | Low-quality |
| S13_extracted_87360 | Not-determined |
| S13_extracted_87369 | Not-determined |
| S13_extracted_87370 | Not-determined |
| S13_extracted_87379 | Not-determined |
| S13_extracted_87407 | Not-determined |
| S13_extracted_87421 | Not-determined |
| S13_extracted_87437 | Not-determined |
| S13_extracted_87443 | Low-quality |
| S13_extracted_87444 | Not-determined |
| S13_extracted_87451 | Low-quality |
| S13_extracted_87455 | Low-quality |
| S13_extracted_87459 | Not-determined |
| S13_extracted_87461 | Not-determined |
| S13_extracted_87465 | Not-determined |
| S13_extracted_87475 | Not-determined |
| S13_extracted_87481 | Not-determined |
| S13_extracted_87482 | Not-determined |
| S13_extracted_87494 | Low-quality |
| S13_extracted_87503 | Not-determined |
| S13_extracted_87504 | Not-determined |
| S13_extracted_87505 | Not-determined |
| S13_extracted_87506 | Low-quality |
| S13_extracted_87510 | Not-determined |
| S13_extracted_87517 | Low-quality |
| S13_extracted_87520 | Not-determined |
| S13_extracted_87550 | Not-determined |
| S13_extracted_87561 | Not-determined |
| S13_extracted_87570 | Not-determined |
| S13_extracted_87573 | Low-quality |
| S13_extracted_87590 | Low-quality |
| S13_extracted_87592 | Not-determined |
| S13_extracted_87595 | Not-determined |
| S13_extracted_87598 | Not-determined |
| S13_extracted_87601 | Not-determined |
| S13_extracted_87612 | Low-quality |
| S13_extracted_87619 | Not-determined |
| S13_extracted_87621 | Not-determined |
| S13_extracted_87627 | Not-determined |
| S13_extracted_87632 | Low-quality |
| S13_extracted_87635 | Not-determined |
| S13_extracted_87640 | Not-determined |
| S13_extracted_87641 | Not-determined |
| S13_extracted_87642 | Not-determined |
| S13_extracted_87644 | Not-determined |
| S13_extracted_87677 | Not-determined |
| S13_extracted_87681 | Low-quality |
| S13_extracted_87691 | Low-quality |
| S13_extracted_87696 | Not-determined |
| S13_extracted_87700 | Not-determined |
| S13_extracted_87707 | Not-determined |
| S13_extracted_87709 | Not-determined |
| S13_extracted_87712 | Not-determined |
| S13_extracted_87714 | Not-determined |
| S13_extracted_87726 | Not-determined |
| S13_extracted_87732 | Low-quality |
| S13_extracted_87736 | Not-determined |
| S13_extracted_87759 | Not-determined |
| S13_extracted_87772 | Not-determined |
| S13_extracted_87775 | Not-determined |
| S13_extracted_87807 | Not-determined |
| S13_extracted_87808 | Not-determined |
| S13_extracted_87813 | Not-determined |
| S13_extracted_87815 | Not-determined |
| S13_extracted_87816 | Not-determined |
| S13_extracted_87818 | Not-determined |
| S13_extracted_87820 | Not-determined |
| S13_extracted_87822 | Not-determined |
| S13_extracted_87827 | Not-determined |
| S13_extracted_87838 | Not-determined |
| S13_extracted_87839 | Not-determined |
| S13_extracted_87840 | Not-determined |
| S13_extracted_87852 | Not-determined |
| S13_extracted_87856 | Not-determined |
| S13_extracted_87866 | Not-determined |
| S13_extracted_87869 | Low-quality |
| S16_vs_87190\|\|full | Medium-quality |
| S13_extracted_87897 | Not-determined |
| S13_extracted_87903 | Low-quality |
| S13_extracted_87908 | Not-determined |
| S13_extracted_87920 | Not-determined |
| S13_extracted_87926 | Low-quality |
| S13_extracted_87927 | Not-determined |
| S13_extracted_87942 | Not-determined |
| S13_extracted_87952 | Not-determined |
| S13_extracted_87955 | Not-determined |
| S13_extracted_87958 | Not-determined |
| S13_extracted_87961 | Not-determined |
| S13_extracted_87962 | Low-quality |
| S13_extracted_87966 | Not-determined |
| S13_extracted_87969 | Not-determined |
| S13_extracted_87974 | Not-determined |
| S13_extracted_87990 | Not-determined |
| S13_extracted_87992 | Not-determined |
| S13_extracted_88019 | Not-determined |
| S13_extracted_88021 | Not-determined |
| S13_extracted_88031 | Not-determined |
| S13_extracted_88039 | Not-determined |
| S13_extracted_88041 | Low-quality |
| S13_extracted_88065 | Not-determined |
| S13_extracted_88073 | Not-determined |
| S13_extracted_88076 | Not-determined |
| S13_extracted_88082 | Not-determined |
| S13_extracted_88102 | Not-determined |
| S13_extracted_88105 | Not-determined |
| S13_extracted_88110 | Not-determined |
| S13_extracted_88111 | Not-determined |
| S13_extracted_88119 | Low-quality |
| S13_extracted_88128 | Low-quality |
| S13_extracted_88136 | Low-quality |
| S13_extracted_88145 | Low-quality |
| S13_extracted_88150 | Not-determined |
| S13_extracted_88153 | Not-determined |
| S13_extracted_88154 | Not-determined |
| S13_extracted_88160 | Not-determined |
| S13_extracted_88161 | Not-determined |
| S13_extracted_88163 | Not-determined |
| S13_extracted_88168 | Not-determined |
| S13_extracted_88172 | Not-determined |
| S13_extracted_88173 | Low-quality |
| S13_extracted_88178 | Not-determined |
| S13_extracted_88184 | Not-determined |
| S13_extracted_88189 | Not-determined |
| S13_extracted_88225 | Not-determined |
| S13_extracted_88228 | Not-determined |
| S13_extracted_88233 | Low-quality |
| S13_extracted_88235 | Not-determined |
| S13_extracted_88246 | Not-determined |
| S13_extracted_88274 | Low-quality |
| S13_extracted_88280 | Not-determined |
| S13_extracted_88286 | Not-determined |
| S13_extracted_88303 | Not-determined |
| S13_extracted_88306 | Not-determined |
| S13_extracted_88307 | Low-quality |
| S13_extracted_88309 | Not-determined |
| S13_extracted_88311 | Not-determined |
| S13_extracted_88327 | Not-determined |
| S13_extracted_88341 | Low-quality |
| S13_extracted_88349 | Not-determined |
| S13_extracted_88397 | Not-determined |
| S13_extracted_88401 | Not-determined |
| S13_extracted_88404 | Not-determined |
| S13_extracted_88418 | Not-determined |
| S13_extracted_88426 | Not-determined |
| S13_extracted_88449 | Not-determined |
| S13_extracted_88450 | Not-determined |
| S13_extracted_88451 | Not-determined |
| S13_extracted_88453 | Not-determined |
| S13_extracted_88455 | Not-determined |
| S13_extracted_88462 | Not-determined |
| S13_extracted_88479 | Not-determined |
| S13_extracted_88481 | Not-determined |
| S13_extracted_88492 | Not-determined |
| S13_extracted_88493 | Low-quality |
| S13_extracted_88537 | Low-quality |
| S13_extracted_88559 | Not-determined |
| S13_extracted_88579 | Not-determined |
| S13_extracted_88589 | Not-determined |
| S13_extracted_88592 | Not-determined |
| S13_extracted_88593 | Not-determined |
| S13_extracted_88605 | Not-determined |
| S13_extracted_88613 | Not-determined |
| S13_extracted_88633 | Not-determined |
| S13_extracted_88637 | Not-determined |
| S13_extracted_88639 | Not-determined |
| S13_extracted_88650 | Not-determined |
| S13_extracted_88657 | Not-determined |
| S13_extracted_88658 | Not-determined |
| S13_extracted_88669 | Not-determined |
| S13_extracted_88672 | Not-determined |
| S13_extracted_88675 | Low-quality |
| S13_extracted_88698 | Not-determined |
| S13_extracted_88699 | Not-determined |
| S13_extracted_88701 | Not-determined |
| S13_extracted_88706 | Not-determined |
| S13_extracted_88738 | Not-determined |
| S13_extracted_88752 | Not-determined |
| S13_extracted_88764 | Not-determined |
| S13_extracted_88782 | Not-determined |
| S13_extracted_88791 | Low-quality |
| S13_extracted_88793 | Low-quality |
| S13_extracted_88797 | Low-quality |
| S13_extracted_88818 | Not-determined |
| S13_extracted_88833 | Low-quality |
| S13_extracted_88835 | Not-determined |
| S13_extracted_88839 | Not-determined |
| S13_extracted_88864 | Not-determined |
| S13_extracted_88870 | Not-determined |
| S13_extracted_88871 | Not-determined |
| S13_extracted_88888 | Not-determined |
| S13_extracted_88893 | Not-determined |
| S13_extracted_88900 | Not-determined |
| S13_extracted_88925 | Not-determined |
| S13_extracted_88928 | Not-determined |
| S13_extracted_88932 | Not-determined |
| S13_extracted_88941 | Not-determined |
| S13_extracted_88948 | Not-determined |
| S13_extracted_88964 | Not-determined |
| S13_extracted_88974 | Not-determined |
| S13_extracted_88978 | Not-determined |
| S13_extracted_88985 | Not-determined |
| S13_extracted_89001 | Not-determined |
| S13_extracted_89015 | Not-determined |
| S13_extracted_89028 | Low-quality |
| S13_extracted_89047 | Not-determined |
| S13_extracted_89051 | Not-determined |
| S13_extracted_89084 | Not-determined |
| S13_extracted_89097 | Not-determined |
| S13_extracted_89102 | Not-determined |
| S13_extracted_89119 | Not-determined |
| S13_extracted_89133 | Not-determined |
| S13_extracted_89136 | Not-determined |
| S13_extracted_89158 | Not-determined |
| S13_extracted_89176 | Low-quality |
| S13_extracted_89182 | Not-determined |
| S13_extracted_89185 | Not-determined |
| S13_extracted_89187 | Not-determined |
| S13_extracted_89192 | Not-determined |
| S13_extracted_89194 | Not-determined |
| S13_extracted_89198 | Not-determined |
| S13_extracted_89201 | Low-quality |
| S13_extracted_89210 | Low-quality |
| S13_extracted_89222 | Not-determined |
| S13_extracted_89246 | Not-determined |
| S13_extracted_89247 | Not-determined |
| S13_extracted_89250 | Low-quality |
| S13_extracted_89258 | Not-determined |
| S13_extracted_89262 | Not-determined |
| S13_extracted_89266 | Not-determined |
| S13_extracted_89278 | Not-determined |
| S13_extracted_89297 | Not-determined |
| S13_extracted_89306 | Not-determined |
| S13_extracted_89329 | Not-determined |
| S13_extracted_89333 | Not-determined |
| S13_extracted_89364 | Not-determined |
| S13_extracted_89386 | Not-determined |
| S13_extracted_89392 | Not-determined |
| S13_extracted_89409 | Not-determined |
| S13_extracted_89410 | Not-determined |
| S13_extracted_89412 | Not-determined |
| S13_extracted_89421 | Not-determined |
| S13_extracted_89424 | Not-determined |
| S13_extracted_89429 | Not-determined |
| S13_extracted_89432 | Not-determined |
| S13_extracted_89440 | Not-determined |
| S13_extracted_89444 | Not-determined |
| S13_extracted_89449 | Low-quality |
| S13_extracted_89463 | Not-determined |
| S13_extracted_89476 | Not-determined |
| S13_extracted_89493 | Not-determined |
| S13_extracted_89497 | Not-determined |
| S13_extracted_89505 | Not-determined |
| S13_extracted_89522 | Not-determined |
| S13_extracted_89536 | Not-determined |
| S13_extracted_89540 | Not-determined |
| S13_extracted_89566 | Not-determined |
| S13_extracted_89568 | Not-determined |
| S13_extracted_89569 | Not-determined |
| S13_extracted_89575 | Not-determined |
| S13_extracted_89590 | Low-quality |
| S13_extracted_89597 | Not-determined |
| S13_extracted_89605 | Not-determined |
| S13_extracted_89609 | Not-determined |
| S13_extracted_89618 | Low-quality |
| S13_extracted_89625 | Not-determined |
| S13_extracted_89629 | Not-determined |
| S13_extracted_89631 | Not-determined |
| S13_extracted_89636 | Low-quality |
| S13_extracted_89637 | Low-quality |
| S13_extracted_89658 | Not-determined |
| S13_extracted_89676 | Not-determined |
| S13_extracted_89679 | Not-determined |
| S13_extracted_89680 | Not-determined |
| S13_extracted_89681 | Not-determined |
| S13_extracted_89683 | Not-determined |
| S13_extracted_89691 | Low-quality |
| S13_extracted_89693 | Low-quality |
| S13_extracted_89695 | Not-determined |
| S13_extracted_89696 | Not-determined |
| S13_extracted_89719 | Not-determined |
| S13_extracted_89728 | Not-determined |
| S13_extracted_89739 | Low-quality |
| S13_extracted_89760 | Not-determined |
| S13_extracted_89766 | Not-determined |
| S13_extracted_89770 | Not-determined |
| S13_extracted_89774 | Not-determined |
| S13_extracted_89796 | Not-determined |
| S13_extracted_89799 | Not-determined |
| S13_extracted_89810 | Low-quality |
| S13_extracted_89817 | Not-determined |
| S13_extracted_89861 | Not-determined |
| S13_extracted_89874 | Not-determined |
| S13_extracted_89876 | Not-determined |
| S13_extracted_89877 | Not-determined |
| S13_extracted_89908 | Not-determined |
| S13_extracted_89912 | Not-determined |
| S13_extracted_89913 | Not-determined |
| S13_extracted_89916 | Not-determined |
| S13_extracted_89918 | Not-determined |
| S13_extracted_89936 | Not-determined |
| S13_extracted_89948 | Not-determined |
| S13_extracted_89955 | Not-determined |
| S13_extracted_89965 | Not-determined |
| S13_extracted_89971 | Not-determined |
| S13_extracted_89975 | Not-determined |
| S13_extracted_89976 | Not-determined |
| S13_extracted_89979 | Not-determined |
| S13_extracted_89981 | Not-determined |
| S13_extracted_89982 | Low-quality |
| S13_extracted_89990 | Not-determined |
| S13_extracted_89992 | Not-determined |
| S13_extracted_89997 | Not-determined |
| S13_extracted_89998 | Not-determined |
| S13_extracted_90006 | Not-determined |
| S13_extracted_90022 | Not-determined |
| S13_extracted_90024 | Not-determined |
| S13_extracted_90037 | Not-determined |
| S13_extracted_90041 | Not-determined |
| S13_extracted_90055 | Not-determined |
| S13_extracted_90059 | Not-determined |
| S13_extracted_90071 | Not-determined |
| S13_extracted_90098 | Not-determined |
| S13_extracted_90109 | Not-determined |
| S13_extracted_90116 | Not-determined |
| S13_extracted_90119 | Not-determined |
| S13_extracted_90123 | Not-determined |
| S13_extracted_90129 | Not-determined |
| S13_extracted_90130 | Low-quality |
| S13_extracted_90138 | Low-quality |
| S13_extracted_90144 | Not-determined |
| S13_extracted_90149 | Not-determined |
| S13_extracted_90234 | Not-determined |
| S13_extracted_90239 | Not-determined |
| S13_extracted_90253 | Not-determined |
| S13_extracted_90254 | Not-determined |
| S13_extracted_90271 | Not-determined |
| S13_extracted_90272 | Not-determined |
| S13_extracted_90273 | Not-determined |
| S13_extracted_90277 | Low-quality |
| S13_extracted_90279 | Not-determined |
| S13_extracted_90299 | Not-determined |
| S13_extracted_90308 | Not-determined |
| S13_extracted_90318 | Not-determined |
| S13_extracted_90320 | Not-determined |
| S13_extracted_90323 | Not-determined |
| S13_extracted_90331 | Low-quality |
| S13_extracted_90335 | Not-determined |
| S13_extracted_90337 | Not-determined |
| S13_extracted_90342 | Low-quality |
| S13_extracted_90346 | Low-quality |
| S13_extracted_90353 | Not-determined |
| S13_extracted_90364 | Not-determined |
| S13_extracted_90384 | Not-determined |
| S13_extracted_90396 | Not-determined |
| S13_extracted_90404 | Not-determined |
| S13_extracted_90414 | Not-determined |
| S13_extracted_90435 | Low-quality |
| S13_extracted_90436 | Not-determined |
| S13_extracted_90437 | Not-determined |
| S13_extracted_90449 | Not-determined |
| S13_extracted_90450 | Not-determined |
| S13_extracted_90463 | Low-quality |
| S13_extracted_90475 | Not-determined |
| S13_extracted_90481 | Not-determined |
| S13_extracted_90484 | Not-determined |
| S13_extracted_90488 | Not-determined |
| S13_extracted_90501 | Not-determined |
| S13_extracted_90506 | Not-determined |
| S13_extracted_90515 | Not-determined |
| S13_extracted_90517 | Not-determined |
| S13_extracted_90556 | Not-determined |
| S13_extracted_90577 | Not-determined |
| S13_extracted_90578 | Not-determined |
| S13_extracted_90579 | Not-determined |
| S13_extracted_90580 | Not-determined |
| S13_extracted_90583 | Not-determined |
| S13_extracted_90584 | Low-quality |
| S13_extracted_90595 | Not-determined |
| S13_extracted_90599 | Not-determined |
| S13_extracted_90600 | Not-determined |
| S13_extracted_90622 | Not-determined |
| S13_extracted_90632 | Low-quality |
| S13_extracted_90648 | Low-quality |
| S13_extracted_90660 | Not-determined |
| S13_extracted_90663 | Not-determined |
| S13_extracted_90669 | Not-determined |
| S13_extracted_90680 | Not-determined |
| S13_extracted_90688 | Not-determined |
| S13_extracted_90691 | Low-quality |
| S13_extracted_90692 | Low-quality |
| S13_extracted_90724 | Not-determined |
| S13_extracted_90726 | Low-quality |
| S13_extracted_90740 | Low-quality |
| S13_extracted_90749 | Low-quality |
| S13_extracted_90758 | Not-determined |
| S13_extracted_90782 | Not-determined |
| S13_extracted_90787 | Not-determined |
| S13_extracted_90788 | Low-quality |
| S13_extracted_90794 | Not-determined |
| S13_extracted_90797 | Not-determined |
| S13_extracted_90798 | Not-determined |
| S13_extracted_90802 | Not-determined |

| | |
|---|---|
| S13_extracted_90812 | Not-determined |
| S13_extracted_90836 | Not-determined |
| S13_extracted_90837 | Not-determined |
| S13_extracted_90839 | Not-determined |
| S13_extracted_90850 | Not-determined |
| S13_extracted_90856 | Not-determined |
| S13_extracted_90858 | Not-determined |
| S13_extracted_90865 | Not-determined |
| S13_extracted_90883 | Not-determined |
| S13_extracted_90904 | Not-determined |
| S13_extracted_90905 | Not-determined |
| S13_extracted_90909 | Not-determined |
| S13_extracted_90912 | Not-determined |
| S13_extracted_90918 | Not-determined |
| S13_extracted_90919 | Not-determined |
| S13_extracted_90925 | Not-determined |
| S13_extracted_90926 | Not-determined |
| S13_extracted_90933 | Low-quality |
| S13_extracted_90935 | Not-determined |
| S13_extracted_90951 | Not-determined |
| S13_extracted_90953 | Not-determined |
| S13_extracted_90954 | Not-determined |
| S13_extracted_90958 | Not-determined |
| S13_extracted_90972 | Not-determined |
| S13_extracted_90983 | Not-determined |
| S13_extracted_90986 | Not-determined |
| S13_extracted_90990 | Not-determined |
| S13_extracted_91004 | Not-determined |
| S13_extracted_91009 | Low-quality |
| S13_extracted_91018 | Not-determined |
| S13_extracted_91042 | Low-quality |
| S13_extracted_91056 | Not-determined |
| S13_extracted_91064 | Not-determined |
| S13_extracted_91068 | Not-determined |
| S13_extracted_91092 | Not-determined |
| S13_extracted_91093 | Not-determined |
| S13_extracted_91101 | Not-determined |
| S13_extracted_91107 | Not-determined |
| S13_extracted_91112 | Low-quality |
| S13_extracted_91117 | Not-determined |
| S13_extracted_91119 | Not-determined |
| S13_extracted_91129 | Not-determined |
| S13_extracted_91131 | Not-determined |
| S13_extracted_91132 | Not-determined |
| S13_extracted_91133 | Not-determined |
| S13_extracted_91154 | Not-determined |
| S13_extracted_91156 | Not-determined |
| S13_extracted_91168 | Not-determined |
| S13_extracted_91169 | Not-determined |
| S13_extracted_91180 | Not-determined |
| S13_extracted_91183 | Not-determined |
| S13_extracted_91186 | Not-determined |
| S13_extracted_91187 | Not-determined |
| S14_vs_k141_66208\|\|full | Complete |
| S13_extracted_91200 | Not-determined |
| S13_extracted_91202 | Not-determined |
| S13_extracted_91208 | Not-determined |
| S13_extracted_91215 | Low-quality |
| S13_extracted_91223 | Not-determined |
| S13_extracted_91228 | Not-determined |
| S13_extracted_91229 | Not-determined |
| S13_extracted_91236 | Not-determined |
| S13_extracted_91250 | Not-determined |
| S13_extracted_91252 | Not-determined |
| S13_extracted_91260 | Not-determined |
| S13_extracted_91285 | Not-determined |
| S13_extracted_91286 | Not-determined |
| S13_extracted_91296 | Not-determined |
| S13_extracted_91300 | Not-determined |
| S13_extracted_91301 | Not-determined |
| S13_extracted_91304 | Low-quality |
| S13_extracted_91308 | Not-determined |
| S13_extracted_91320 | Not-determined |
| S13_extracted_91322 | Not-determined |
| S13_extracted_91336 | Not-determined |
| S13_extracted_91337 | Not-determined |
| S13_extracted_91353 | Not-determined |
| S13_extracted_91363 | Not-determined |
| S13_extracted_91366 | Not-determined |
| S13_extracted_91374 | Not-determined |
| S13_extracted_91378 | Not-determined |
| S13_extracted_91383 | Not-determined |
| S13_extracted_91388 | Not-determined |
| S13_extracted_91414 | Not-determined |
| S13_extracted_91415 | Not-determined |
| S13_extracted_91425 | Not-determined |
| S13_extracted_91431 | Not-determined |
| S13_extracted_91438 | Not-determined |
| S13_extracted_91442 | Not-determined |
| S13_extracted_91451 | Not-determined |
| S13_extracted_91455 | Low-quality |
| S13_extracted_91458 | Low-quality |
| S13_extracted_91474 | Not-determined |
| S13_extracted_91485 | Not-determined |
| S13_extracted_91491 | Not-determined |
| S13_extracted_91520 | Not-determined |
| S13_extracted_91525 | Not-determined |
| S13_extracted_91529 | Not-determined |
| S13_extracted_91531 | Not-determined |
| S13_extracted_91539 | Not-determined |
| S13_extracted_91563 | Not-determined |
| S13_extracted_91565 | Not-determined |
| S13_extracted_91570 | Not-determined |
| S13_extracted_91572 | Not-determined |
| S13_extracted_91574 | Not-determined |
| S13_extracted_91576 | Not-determined |
| S13_extracted_91590 | Not-determined |
| S13_extracted_91593 | Not-determined |
| S13_extracted_91621 | Not-determined |
| S13_extracted_91624 | Not-determined |
| S13_extracted_91625 | Not-determined |
| S13_extracted_91653 | Not-determined |
| S13_extracted_91664 | Low-quality |
| S13_extracted_91669 | Not-determined |
| S13_extracted_91679 | Not-determined |
| S13_extracted_91683 | Not-determined |
| S13_extracted_91686 | Not-determined |
| S13_extracted_91703 | Not-determined |
| S13_extracted_91706 | Low-quality |
| S13_extracted_91710 | Not-determined |
| S13_extracted_91712 | Not-determined |
| S13_extracted_91713 | Not-determined |
| S13_extracted_91715 | Not-determined |
| S13_extracted_91722 | Not-determined |
| S13_extracted_91730 | Not-determined |
| S13_extracted_91743 | Not-determined |
| S13_extracted_91746 | Not-determined |
| S13_extracted_91752 | Not-determined |
| S13_extracted_91755 | Not-determined |
| S13_extracted_91757 | Low-quality |
| S13_extracted_91761 | Not-determined |
| S13_extracted_91763 | Not-determined |
| S13_extracted_91766 | Not-determined |
| S13_extracted_91770 | Not-determined |
| S13_extracted_91780 | Not-determined |
| S13_extracted_91787 | Not-determined |
| S13_extracted_91788 | Not-determined |
| S13_extracted_91789 | Not-determined |
| S13_extracted_91794 | Not-determined |
| S13_extracted_91795 | Not-determined |
| S13_extracted_91806 | Not-determined |
| S13_extracted_91811 | Not-determined |
| S13_extracted_91812 | Not-determined |
| S13_extracted_91815 | Not-determined |
| S13_extracted_91828 | Not-determined |
| S13_extracted_91835 | Not-determined |
| S13_extracted_91847 | Low-quality |
| S13_extracted_91854 | Not-determined |
| S13_extracted_91855 | Not-determined |
| S13_extracted_91863 | Not-determined |
| S13_extracted_91881 | Not-determined |
| S13_extracted_91882 | Not-determined |
| S13_extracted_91886 | Not-determined |
| S13_extracted_91889 | Not-determined |
| S13_extracted_91895 | Not-determined |
| S13_extracted_91901 | Not-determined |
| S13_extracted_91904 | Low-quality |
| S13_extracted_91905 | Not-determined |
| S13_extracted_91922 | Not-determined |
| S13_extracted_91935 | Not-determined |
| S13_extracted_91936 | Not-determined |
| S13_extracted_91939 | Not-determined |
| S13_extracted_91940 | Not-determined |
| S13_extracted_91955 | Not-determined |
| S13_extracted_91959 | Low-quality |
| S13_extracted_91973 | Not-determined |
| S13_extracted_91978 | Not-determined |
| S13_extracted_91984 | Low-quality |
| S13_extracted_91986 | Not-determined |
| S13_extracted_91994 | Not-determined |
| S13_extracted_92008 | Not-determined |
| S13_extracted_92020 | Not-determined |
| S13_extracted_92031 | Not-determined |
| S13_extracted_92034 | Not-determined |
| S13_extracted_92048 | Not-determined |
| S13_extracted_92057 | Not-determined |
| S13_extracted_92065 | Not-determined |
| S13_extracted_92102 | Not-determined |
| S13_extracted_92105 | Not-determined |
| S13_extracted_92107 | Not-determined |
| S13_extracted_92118 | Low-quality |
| S13_extracted_92119 | Low-quality |
| S13_extracted_92123 | Not-determined |
| S13_extracted_92126 | Not-determined |
| S13_extracted_92137 | Low-quality |
| S13_extracted_92143 | Not-determined |
| S13_extracted_92152 | Not-determined |
| S13_extracted_92158 | Not-determined |
| S13_extracted_92159 | Not-determined |
| S13_extracted_92165 | Not-determined |
| S13_extracted_92169 | Not-determined |
| S13_extracted_92170 | Not-determined |
| S13_extracted_92177 | Not-determined |
| S13_extracted_92186 | Not-determined |
| S13_extracted_92191 | Not-determined |
| S13_extracted_92205 | Not-determined |
| S13_extracted_92216 | Not-determined |
| S13_extracted_92218 | Not-determined |
| S13_extracted_92226 | Not-determined |
| S13_extracted_92227 | Not-determined |
| S13_extracted_92232 | Not-determined |
| S13_extracted_92234 | Not-determined |
| S13_extracted_92241 | Not-determined |
| S13_extracted_92242 | Not-determined |
| S13_extracted_92256 | Not-determined |
| S13_extracted_92262 | Not-determined |
| S13_extracted_92296 | Not-determined |
| S13_extracted_92304 | Not-determined |
| S13_extracted_92313 | Not-determined |
| S13_extracted_92320 | Not-determined |
| S13_extracted_92327 | Not-determined |
| S13_extracted_92343 | Not-determined |
| S13_extracted_92361 | Not-determined |
| S13_extracted_92382 | Low-quality |
| S13_extracted_92385 | Not-determined |
| S13_extracted_92395 | Not-determined |
| S13_extracted_92401 | Not-determined |
| S13_extracted_92403 | Not-determined |
| S13_extracted_92411 | Not-determined |
| S13_extracted_92429 | Not-determined |
| S13_extracted_92435 | Not-determined |
| S13_extracted_92439 | Not-determined |
| S13_extracted_92444 | Not-determined |
| S13_extracted_92448 | Not-determined |
| S13_extracted_92452 | Not-determined |
| S13_extracted_92458 | Low-quality |
| S13_extracted_92464 | Not-determined |
| S13_extracted_92467 | Not-determined |
| S13_extracted_92472 | Not-determined |
| S13_extracted_92502 | Not-determined |
| S13_extracted_92509 | Not-determined |
| S13_extracted_92516 | Not-determined |
| S13_extracted_92523 | Not-determined |
| S13_extracted_92525 | Not-determined |
| S13_extracted_92530 | Not-determined |
| S13_extracted_92543 | Not-determined |
| S13_extracted_92555 | Not-determined |
| S13_extracted_92556 | Not-determined |
| S13_extracted_92557 | Not-determined |
| S13_extracted_92566 | Not-determined |
| S13_extracted_92568 | Not-determined |
| S13_extracted_92579 | Not-determined |
| S13_extracted_92590 | Low-quality |
| S13_extracted_92602 | Not-determined |
| S13_extracted_92614 | Not-determined |
| S13_extracted_92618 | Not-determined |
| S13_extracted_92625 | Not-determined |
| S13_extracted_92631 | Not-determined |
| S13_extracted_92653 | Not-determined |
| S13_extracted_92658 | Not-determined |
| S13_extracted_92661 | Not-determined |
| S13_extracted_92670 | Not-determined |
| S13_extracted_92671 | Not-determined |
| S13_extracted_92691 | Not-determined |
| S13_extracted_92709 | Not-determined |
| S13_extracted_92721 | Not-determined |
| S13_extracted_92731 | Low-quality |
| S13_extracted_92741 | Not-determined |
| S13_extracted_92776 | Not-determined |
| S13_extracted_92783 | Not-determined |
| S13_extracted_92791 | Not-determined |
| S13_extracted_92796 | Not-determined |
| S13_extracted_92797 | Not-determined |
| S13_extracted_92800 | Not-determined |
| S13_extracted_92806 | Not-determined |
| S13_extracted_92808 | Not-determined |
| S13_extracted_92811 | Low-quality |
| S13_extracted_92813 | Not-determined |
| S13_extracted_92830 | Not-determined |
| S13_extracted_92831 | Not-determined |
| S13_extracted_92852 | Not-determined |
| S13_extracted_92867 | Not-determined |
| S13_extracted_92873 | Not-determined |
| S13_extracted_92874 | Not-determined |
| S13_extracted_92876 | Not-determined |
| S13_extracted_92884 | Not-determined |
| S13_extracted_92918 | Not-determined |
| S13_extracted_92924 | Not-determined |
| S13_extracted_92926 | Not-determined |
| S13_extracted_92927 | Not-determined |
| S13_extracted_92942 | Not-determined |
| S13_extracted_92945 | Not-determined |
| S13_extracted_92946 | Not-determined |
| S13_extracted_92951 | Not-determined |
| S13_extracted_92956 | Not-determined |
| S13_extracted_92968 | Not-determined |
| S13_extracted_92971 | Not-determined |
| S13_extracted_92973 | Low-quality |
| S13_extracted_92982 | Low-quality |
| S13_extracted_92992 | Not-determined |
| S13_extracted_92999 | Not-determined |
| S13_extracted_93000 | Not-determined |
| S13_extracted_93004 | Not-determined |
| S13_extracted_93010 | Not-determined |
| S13_extracted_93021 | Not-determined |
| S13_extracted_93035 | Not-determined |
| S13_extracted_93038 | Not-determined |
| S13_extracted_93042 | Not-determined |
| S13_extracted_93044 | Not-determined |
| S13_extracted_93050 | Not-determined |
| S13_extracted_93057 | Not-determined |
| S13_extracted_93058 | Not-determined |
| S13_extracted_93060 | Not-determined |
| S13_extracted_93066 | Not-determined |
| S13_extracted_93082 | Not-determined |
| S13_extracted_93096 | Not-determined |
| S13_extracted_93109 | Not-determined |
| S13_extracted_93118 | Not-determined |
| S13_extracted_93120 | Not-determined |
| S13_extracted_93131 | Low-quality |
| S13_extracted_93134 | Not-determined |
| S13_extracted_93154 | Not-determined |
| S13_extracted_93164 | Not-determined |
| S13_extracted_93174 | Not-determined |
| S13_extracted_93178 | Not-determined |
| S13_extracted_93185 | Not-determined |
| S13_extracted_93199 | Not-determined |
| S13_extracted_93208 | Not-determined |
| S13_extracted_93218 | Not-determined |
| S13_extracted_93220 | Not-determined |
| S13_extracted_93221 | Not-determined |
| S13_extracted_93222 | Not-determined |
| S13_extracted_93226 | Not-determined |
| S13_extracted_93229 | Not-determined |
| S13_extracted_93230 | Low-quality |
| S13_extracted_93251 | Not-determined |
| S13_extracted_93254 | Not-determined |
| S13_extracted_93258 | Not-determined |
| S13_extracted_93262 | Not-determined |
| S13_extracted_93267 | Not-determined |
| S13_extracted_93293 | Not-determined |
| S13_extracted_93298 | Not-determined |
| S13_extracted_93308 | Not-determined |
| S13_extracted_93324 | Low-quality |
| S13_extracted_93325 | Low-quality |
| S13_extracted_93327 | Not-determined |
| S13_extracted_93329 | Not-determined |
| S13_extracted_93335 | Not-determined |
| S13_extracted_93336 | Not-determined |
| S13_extracted_93364 | Low-quality |
| S13_extracted_93365 | Not-determined |
| S13_extracted_93373 | Not-determined |
| S13_extracted_93381 | Low-quality |
| S13_extracted_93382 | Not-determined |
| S13_extracted_93390 | Low-quality |
| S13_extracted_93400 | Not-determined |
| S13_extracted_93414 | Not-determined |
| S13_extracted_93417 | Not-determined |
| S13_extracted_93427 | Low-quality |
| S13_extracted_93428 | Not-determined |
| S13_extracted_93433 | Low-quality |
| S13_extracted_93434 | Not-determined |
| S13_extracted_93437 | Not-determined |
| S13_extracted_93451 | Low-quality |
| S13_extracted_93453 | Not-determined |
| S13_extracted_93455 | Not-determined |
| S13_extracted_93459 | Not-determined |
| S13_extracted_93460 | Not-determined |
| S13_extracted_93476 | Low-quality |
| S13_extracted_93482 | Low-quality |
| S13_extracted_93483 | Not-determined |
| S13_extracted_93484 | Not-determined |
| S13_extracted_93493 | Not-determined |
| S13_extracted_93496 | Not-determined |
| S13_extracted_93506 | Not-determined |
| S13_extracted_93514 | Not-determined |
| S13_extracted_93524 | Not-determined |
| S13_extracted_93535 | Not-determined |
| S13_extracted_93551 | Not-determined |
| S13_extracted_93579 | Not-determined |
| S13_extracted_93625 | Not-determined |
| S13_extracted_93640 | Not-determined |
| S13_extracted_93641 | Not-determined |
| S13_extracted_93644 | Not-determined |
| S13_extracted_93662 | Low-quality |
| S13_extracted_93681 | Not-determined |
| S13_extracted_93690 | Not-determined |
| S13_extracted_93696 | Not-determined |
| S13_extracted_93704 | Not-determined |
| S13_extracted_93744 | Not-determined |
| S13_extracted_93781 | Not-determined |
| S13_extracted_93799 | Not-determined |
| S13_extracted_93801 | Low-quality |
| S13_extracted_93824 | Not-determined |
| S13_extracted_93831 | Not-determined |
| S13_extracted_93859 | Not-determined |
| S13_extracted_93877 | Not-determined |
| S13_extracted_93881 | Not-determined |
| S13_extracted_93882 | Not-determined |
| S13_extracted_93883 | Not-determined |
| S13_extracted_93890 | Not-determined |
| S13_extracted_93897 | Not-determined |
| S13_extracted_93901 | Low-quality |
| S13_extracted_93908 | Low-quality |
| S13_extracted_93919 | Not-determined |
| S13_extracted_93930 | Not-determined |
| S13_extracted_93939 | Not-determined |
| S13_extracted_93947 | Not-determined |
| S13_extracted_93956 | Not-determined |
| S13_extracted_93961 | Not-determined |
| S13_extracted_93976 | Not-determined |
| S13_extracted_93980 | Not-determined |
| S13_extracted_93990 | Not-determined |
| S13_extracted_93993 | Not-determined |
| S13_extracted_93995 | Not-determined |
| S13_extracted_94000 | Not-determined |
| S13_extracted_94001 | Not-determined |
| S13_extracted_94007 | Not-determined |
| S13_extracted_94028 | Not-determined |
| S13_extracted_94039 | Not-determined |
| S13_extracted_94041 | Not-determined |
| S13_extracted_94050 | Not-determined |
| S13_extracted_94053 | Not-determined |
| S13_extracted_94062 | Not-determined |
| S13_extracted_94065 | Low-quality |
| S13_extracted_94072 | Not-determined |
| S13_extracted_94076 | Low-quality |
| S13_extracted_94085 | Not-determined |
| S13_extracted_94090 | Not-determined |
| S13_extracted_94096 | Not-determined |
| S13_extracted_94099 | Not-determined |
| S13_extracted_94111 | Low-quality |
| S13_extracted_94118 | Low-quality |
| S13_extracted_94129 | Not-determined |
| S13_extracted_94130 | Not-determined |
| S13_extracted_94139 | Not-determined |
| S13_extracted_94144 | Not-determined |
| S13_extracted_94146 | Low-quality |
| S13_extracted_94148 | Not-determined |
| S13_extracted_94149 | Not-determined |
| S13_extracted_94190 | Not-determined |
| S13_extracted_94221 | Not-determined |
| S13_extracted_94256 | Not-determined |
| S13_extracted_94269 | Not-determined |
| S13_extracted_94279 | Not-determined |
| S13_extracted_94285 | Not-determined |
| S13_extracted_94288 | Not-determined |
| S13_extracted_94291 | Not-determined |
| S13_extracted_94293 | Not-determined |
| S13_extracted_94303 | Not-determined |
| S13_extracted_94308 | Not-determined |
| S13_extracted_94338 | Not-determined |
| S13_extracted_94342 | Not-determined |
| S13_extracted_94349 | Not-determined |
| S13_extracted_94350 | Not-determined |
| S13_extracted_94355 | Low-quality |
| S13_extracted_94358 | Not-determined |
| S13_extracted_94367 | Not-determined |
| S13_extracted_94375 | Not-determined |
| S13_extracted_94378 | Not-determined |
| S13_extracted_94386 | Not-determined |
| S13_extracted_94403 | Not-determined |
| S13_extracted_94410 | Not-determined |
| S13_extracted_94411 | Not-determined |
| S13_extracted_94420 | Not-determined |
| S13_extracted_94443 | Not-determined |
| S13_extracted_94454 | Not-determined |
| S13_extracted_94467 | Not-determined |
| S13_extracted_94490 | Not-determined |
| S13_extracted_94492 | Not-determined |
| S13_extracted_94499 | Low-quality |
| S13_extracted_94504 | Not-determined |
| S13_extracted_94509 | Not-determined |
| S13_extracted_94511 | Not-determined |
| S13_extracted_94527 | Not-determined |
| S13_extracted_94529 | Not-determined |
| S13_extracted_94543 | Not-determined |
| S13_extracted_94550 | Low-quality |
| S13_extracted_94556 | Not-determined |
| S13_extracted_94559 | Not-determined |
| S13_extracted_94576 | Not-determined |
| S13_extracted_94577 | Low-quality |
| S13_extracted_94588 | Not-determined |
| S13_extracted_94591 | Not-determined |
| S13_extracted_94600 | Not-determined |
| S13_extracted_94618 | Not-determined |
| S13_extracted_94622 | Not-determined |
| S13_extracted_94623 | Low-quality |
| S13_extracted_94636 | Not-determined |
| S13_extracted_94656 | Low-quality |
| S13_extracted_94659 | Not-determined |
| S13_extracted_94667 | Not-determined |
| S13_extracted_94669 | Low-quality |
| S13_extracted_94671 | Not-determined |
| S13_extracted_94674 | Not-determined |
| S13_extracted_94693 | Not-determined |
| S13_extracted_94698 | Not-determined |
| S13_extracted_94707 | Not-determined |
| S13_extracted_94709 | Low-quality |
| S13_extracted_94729 | Not-determined |
| S13_extracted_94730 | Not-determined |
| S13_extracted_94746 | Not-determined |
| S13_extracted_94751 | Not-determined |
| S13_extracted_94767 | Not-determined |
| S13_extracted_94770 | Not-determined |
| S13_extracted_94783 | Low-quality |
| S13_extracted_94792 | Not-determined |
| S13_extracted_94794 | Not-determined |
| S13_extracted_94797 | Not-determined |
| S13_extracted_94798 | Not-determined |
| S13_extracted_94812 | Not-determined |
| S13_extracted_94827 | Not-determined |
| S13_extracted_94829 | Not-determined |
| S13_extracted_94847 | Not-determined |
| S13_extracted_94853 | Not-determined |
| S13_extracted_94854 | Not-determined |
| S13_extracted_94860 | Low-quality |
| S13_extracted_94861 | Not-determined |
| S13_extracted_94864 | Not-determined |
| S13_extracted_94882 | Not-determined |
| S13_extracted_94911 | Low-quality |
| S13_extracted_94912 | Not-determined |
| S13_extracted_94929 | Not-determined |
| S13_extracted_94931 | Not-determined |
| S13_extracted_94935 | Low-quality |
| S13_extracted_94942 | Not-determined |
| S13_extracted_94945 | Not-determined |
| S13_extracted_94953 | Not-determined |
| S13_extracted_94955 | Low-quality |
| S13_extracted_94957 | Not-determined |
| S13_extracted_94965 | Not-determined |
| S13_extracted_94967 | Not-determined |
| S13_extracted_94968 | Not-determined |
| S13_extracted_94974 | Not-determined |
| S13_extracted_94989 | Not-determined |
| S13_extracted_94997 | Not-determined |
| S13_extracted_95000 | Not-determined |
| S13_extracted_95003 | Not-determined |
| S13_extracted_95005 | Not-determined |
| S13_extracted_95008 | Not-determined |
| S13_extracted_95013 | Not-determined |
| S13_extracted_95021 | Not-determined |
| S13_extracted_95033 | Not-determined |
| S13_extracted_95036 | Not-determined |
| S13_extracted_95041 | Not-determined |
| S13_extracted_95042 | Not-determined |
| S13_extracted_95044 | Not-determined |
| S13_extracted_95046 | Not-determined |
| S13_extracted_95051 | Not-determined |
| S13_extracted_95052 | Not-determined |
| S13_extracted_95092 | Not-determined |
| S13_extracted_95093 | Not-determined |
| S13_extracted_95102 | Low-quality |
| S13_extracted_95107 | Not-determined |
| S13_extracted_95113 | Not-determined |
| S13_extracted_95118 | Not-determined |
| S13_extracted_95139 | Not-determined |
| S13_extracted_95145 | Not-determined |
| S13_extracted_95152 | Not-determined |
| S13_extracted_95160 | Not-determined |
| S13_extracted_95172 | Not-determined |
| S13_extracted_95177 | Not-determined |
| S13_extracted_95178 | Not-determined |
| S13_extracted_95180 | Not-determined |
| S13_extracted_95191 | Not-determined |
| S13_extracted_95212 | Not-determined |
| S13_extracted_95226 | Not-determined |
| S13_extracted_95232 | Low-quality |
| S13_extracted_95242 | Not-determined |
| S13_extracted_95245 | Not-determined |
| S13_extracted_95248 | Not-determined |
| S13_extracted_95257 | Not-determined |
| S13_extracted_95267 | Not-determined |
| S13_extracted_95281 | Not-determined |
| S13_extracted_95282 | Low-quality |
| S13_extracted_95287 | Not-determined |
| S13_extracted_95292 | Not-determined |
| S13_extracted_95319 | Not-determined |
| S13_extracted_95327 | Not-determined |
| S13_extracted_95369 | Not-determined |
| S13_extracted_95375 | Low-quality |
| S13_extracted_95378 | Not-determined |
| S13_extracted_95379 | Low-quality |
| S13_extracted_95411 | Low-quality |
| S13_extracted_95412 | Not-determined |
| S13_extracted_95417 | Low-quality |
| S13_extracted_95448 | Not-determined |
| S13_extracted_95457 | Not-determined |
| S13_extracted_95464 | Not-determined |
| S13_extracted_95471 | Not-determined |
| S13_extracted_95476 | Not-determined |
| S13_extracted_95480 | Not-determined |
| S13_extracted_95496 | Not-determined |
| S13_extracted_95517 | Not-determined |
| S13_extracted_95518 | Low-quality |
| S13_extracted_95528 | Not-determined |
| S13_extracted_95539 | Low-quality |
| S13_extracted_95548 | Not-determined |
| S13_extracted_95552 | Not-determined |

| | |
|---|---|
| S13_extracted_95554 | Low-quality |
| S13_extracted_95555 | Not-determined |
| S13_extracted_95557 | Not-determined |
| S13_extracted_95559 | Not-determined |
| S13_extracted_95570 | Not-determined |
| S13_extracted_95591 | Not-determined |
| S13_extracted_95596 | Not-determined |
| S13_extracted_95607 | Not-determined |
| S13_extracted_95613 | Not-determined |
| S13_extracted_95617 | Not-determined |
| S13_extracted_95621 | Not-determined |
| S13_extracted_95630 | Not-determined |
| S13_extracted_95635 | Not-determined |
| S13_extracted_95646 | Not-determined |
| S13_extracted_95651 | Not-determined |
| S13_extracted_95665 | Not-determined |
| S13_extracted_95667 | Low-quality |
| S13_extracted_95681 | Not-determined |
| S13_extracted_95683 | Not-determined |
| S13_extracted_95687 | Not-determined |
| S13_extracted_95688 | Not-determined |
| S13_extracted_95699 | Not-determined |
| S13_extracted_95701 | Not-determined |
| S13_extracted_95712 | Not-determined |
| S13_extracted_95724 | Not-determined |
| S13_extracted_95727 | Not-determined |
| S13_extracted_95730 | Not-determined |
| S13_extracted_95731 | Not-determined |
| S13_extracted_95735 | Not-determined |
| S13_extracted_95746 | Not-determined |
| S13_extracted_95753 | Not-determined |
| S13_extracted_95770 | Not-determined |
| S13_extracted_95787 | Not-determined |
| S13_extracted_95799 | Not-determined |
| S13_extracted_95802 | Not-determined |
| S13_extracted_95815 | Not-determined |
| S13_extracted_95841 | Not-determined |
| S13_extracted_95863 | Not-determined |
| S13_extracted_95868 | Not-determined |
| S13_extracted_95881 | Not-determined |
| S13_extracted_95888 | Not-determined |
| S13_extracted_95891 | Not-determined |
| S13_extracted_95895 | Not-determined |
| S13_extracted_95907 | Low-quality |
| S13_extracted_95909 | Not-determined |
| S13_extracted_95922 | Not-determined |
| S13_extracted_95924 | Not-determined |
| S13_extracted_95932 | Not-determined |
| S13_extracted_95949 | Low-quality |
| S13_extracted_95951 | Not-determined |
| S13_extracted_95973 | Not-determined |
| S13_extracted_95984 | Not-determined |
| S13_extracted_95989 | Not-determined |
| S13_extracted_95998 | Not-determined |
| S13_extracted_95999 | Not-determined |
| S13_extracted_96000 | Low-quality |
| S13_extracted_96002 | Low-quality |
| S13_extracted_96024 | Not-determined |
| S13_extracted_96029 | Not-determined |
| S13_extracted_96037 | Not-determined |
| S13_extracted_96040 | Not-determined |
| S13_extracted_96050 | Not-determined |
| S13_extracted_96073 | Not-determined |
| S13_extracted_96078 | Not-determined |
| S13_extracted_96086 | Not-determined |
| S13_extracted_96089 | Not-determined |
| S13_extracted_96092 | Not-determined |
| S13_extracted_96095 | Not-determined |
| S13_extracted_96110 | Not-determined |
| S13_extracted_96112 | Not-determined |
| S13_extracted_96131 | Not-determined |
| S13_extracted_96137 | Not-determined |
| S13_extracted_96167 | Not-determined |
| S13_extracted_96169 | Not-determined |
| S13_extracted_96185 | Low-quality |
| S13_extracted_96205 | Not-determined |
| S13_extracted_96247 | Low-quality |
| S13_extracted_96255 | Not-determined |
| S13_extracted_96258 | Low-quality |
| S13_extracted_96261 | Not-determined |
| S13_extracted_96272 | Not-determined |
| S13_extracted_96289 | Not-determined |
| S13_extracted_96292 | Not-determined |
| S13_extracted_96293 | Not-determined |
| S13_extracted_96310 | Not-determined |
| S13_extracted_96318 | Not-determined |
| S13_extracted_96326 | Not-determined |
| S13_extracted_96328 | Low-quality |
| S13_extracted_96346 | Not-determined |
| S13_extracted_96348 | Not-determined |
| S13_extracted_96350 | Not-determined |
| S13_extracted_96351 | Not-determined |
| S13_extracted_96352 | Not-determined |
| S13_extracted_96366 | Not-determined |
| S13_extracted_96383 | Low-quality |
| S13_extracted_96386 | Not-determined |
| S13_extracted_96394 | Not-determined |
| S13_extracted_96404 | Low-quality |
| S13_extracted_96436 | Low-quality |
| S13_extracted_96442 | Not-determined |
| S13_extracted_96452 | Not-determined |
| S13_extracted_96453 | Not-determined |
| S13_extracted_96454 | Not-determined |
| S13_extracted_96456 | Not-determined |
| S13_extracted_96462 | Not-determined |
| S13_extracted_96465 | Not-determined |
| S13_extracted_96467 | Low-quality |
| S13_extracted_96468 | Not-determined |
| S13_extracted_96479 | Not-determined |
| S13_extracted_96481 | Not-determined |
| S13_extracted_96483 | Not-determined |
| S13_extracted_96498 | Not-determined |
| S13_extracted_96505 | Not-determined |
| S13_extracted_96507 | Not-determined |
| S13_extracted_96521 | Not-determined |
| S13_extracted_96526 | Not-determined |
| S13_extracted_96542 | Not-determined |
| S13_extracted_96554 | Not-determined |
| S13_extracted_96556 | Not-determined |
| S13_extracted_96562 | Not-determined |
| S13_extracted_96567 | Low-quality |
| S13_extracted_96574 | Not-determined |
| S13_extracted_96576 | Not-determined |
| S13_extracted_96581 | Not-determined |
| S13_extracted_96587 | Not-determined |
| S13_extracted_96591 | Not-determined |
| S13_extracted_96604 | Not-determined |
| S13_extracted_96611 | Not-determined |
| S13_extracted_96617 | Low-quality |
| S13_extracted_96640 | Not-determined |
| S13_extracted_96644 | Not-determined |
| S13_extracted_96646 | Not-determined |
| S13_extracted_96659 | Not-determined |
| S13_extracted_96664 | Not-determined |
| S13_extracted_96690 | Low-quality |
| S13_extracted_96694 | Not-determined |
| S13_extracted_96696 | Not-determined |
| S13_extracted_96704 | Not-determined |
| S13_extracted_96722 | Low-quality |
| S13_extracted_96725 | Not-determined |
| S13_extracted_96733 | Low-quality |
| S13_extracted_96753 | Not-determined |
| S13_extracted_96762 | Not-determined |
| S13_extracted_96769 | Not-determined |
| S13_extracted_96780 | Not-determined |
| S13_extracted_96788 | Low-quality |
| S13_extracted_96791 | Not-determined |
| S13_extracted_96796 | Not-determined |
| S13_extracted_96800 | Not-determined |
| S13_extracted_96803 | Not-determined |
| S13_extracted_96810 | Not-determined |
| S13_extracted_96820 | Not-determined |
| S13_extracted_96828 | Not-determined |
| S13_extracted_96836 | Not-determined |
| S13_extracted_96841 | Not-determined |
| S13_extracted_96846 | Not-determined |
| S13_extracted_96853 | Low-quality |
| S13_extracted_96858 | Not-determined |
| S13_extracted_96865 | Not-determined |
| S13_extracted_96888 | Not-determined |
| S13_extracted_96889 | Not-determined |
| S13_extracted_96891 | Not-determined |
| S13_extracted_96901 | Low-quality |
| S13_extracted_96904 | Not-determined |
| S13_extracted_96922 | Not-determined |
| S13_extracted_96928 | Low-quality |
| S13_extracted_96935 | Not-determined |
| S13_extracted_96942 | Not-determined |
| S13_extracted_96951 | Not-determined |
| S13_extracted_96953 | Not-determined |
| S13_extracted_96966 | Not-determined |
| S13_extracted_96973 | Not-determined |
| S13_extracted_96975 | Not-determined |
| S13_extracted_96988 | Not-determined |
| S13_extracted_96994 | Not-determined |
| S13_extracted_97001 | Not-determined |
| S13_extracted_97028 | Not-determined |
| S13_extracted_97030 | Not-determined |
| S13_extracted_97031 | Low-quality |
| S13_extracted_97032 | Not-determined |
| S13_extracted_97040 | Not-determined |
| S13_extracted_97044 | Not-determined |
| S13_extracted_97046 | Not-determined |
| S13_extracted_97047 | Not-determined |
| S13_extracted_97055 | Not-determined |
| S13_extracted_97059 | Not-determined |
| S13_extracted_97077 | Not-determined |
| S13_extracted_97078 | Low-quality |
| S13_extracted_97084 | Not-determined |
| S13_extracted_97085 | Not-determined |
| S13_extracted_97087 | Not-determined |
| S13_extracted_97103 | Not-determined |
| S13_extracted_97113 | Low-quality |
| S13_extracted_97118 | Not-determined |
| S13_extracted_97124 | Not-determined |
| S13_extracted_97127 | Not-determined |
| S13_extracted_97143 | Not-determined |
| S13_extracted_97149 | Not-determined |
| S13_extracted_97163 | Not-determined |
| S13_extracted_97166 | Not-determined |
| S13_extracted_97175 | Not-determined |
| S13_extracted_97184 | Not-determined |
| S13_extracted_97188 | Not-determined |
| S13_extracted_97191 | Not-determined |
| S13_extracted_97194 | Low-quality |
| S13_extracted_97200 | Not-determined |
| S13_extracted_97202 | Not-determined |
| S13_extracted_97206 | Not-determined |
| S13_extracted_97208 | Not-determined |
| S13_extracted_97222 | Not-determined |
| S13_extracted_97241 | Not-determined |
| S13_extracted_97268 | Not-determined |
| S13_extracted_97283 | Not-determined |
| S13_extracted_97287 | Not-determined |
| S13_extracted_97291 | Not-determined |
| S13_extracted_97295 | Not-determined |
| S13_extracted_97300 | Not-determined |
| S13_extracted_97303 | Not-determined |
| S13_extracted_97308 | Not-determined |
| S13_extracted_97309 | Not-determined |
| S13_extracted_97311 | Not-determined |
| S13_extracted_97317 | Not-determined |
| S13_extracted_97333 | Not-determined |
| S13_extracted_97340 | Not-determined |
| S13_extracted_97352 | Not-determined |
| S13_extracted_97357 | Not-determined |
| S13_extracted_97359 | Not-determined |
| S13_extracted_97364 | Not-determined |
| S13_extracted_97373 | Not-determined |
| S13_extracted_97383 | Not-determined |
| S13_extracted_97386 | Not-determined |
| S13_extracted_97391 | Not-determined |
| S13_extracted_97395 | Not-determined |
| S13_extracted_97412 | Not-determined |
| S13_extracted_97430 | Not-determined |
| S13_extracted_97432 | Not-determined |
| S13_extracted_97438 | Not-determined |
| S13_extracted_97463 | Not-determined |
| S13_extracted_97475 | Not-determined |
| S13_extracted_97501 | Low-quality |
| S13_extracted_97504 | Low-quality |
| S13_extracted_97518 | Not-determined |
| S13_extracted_97520 | Not-determined |
| S13_extracted_97522 | Not-determined |
| S13_extracted_97523 | Not-determined |
| S13_extracted_97543 | Not-determined |
| S13_extracted_97559 | Not-determined |
| S13_extracted_97571 | Not-determined |
| S13_extracted_97585 | Low-quality |
| S13_extracted_97586 | Not-determined |
| S13_extracted_97590 | Low-quality |
| S13_extracted_97596 | Not-determined |
| S13_extracted_97598 | Not-determined |
| S13_extracted_97600 | Not-determined |
| S13_extracted_97602 | Low-quality |
| S13_extracted_97611 | Not-determined |
| S13_extracted_97612 | Not-determined |
| S13_extracted_97619 | Low-quality |
| S13_extracted_97626 | Not-determined |
| S13_extracted_97629 | Not-determined |
| S13_extracted_97642 | Not-determined |
| S13_extracted_97650 | Not-determined |
| S13_extracted_97678 | Not-determined |
| S13_extracted_97689 | Not-determined |
| S13_extracted_97695 | Not-determined |
| S12_vs_k141_26336\|\|full | Medium-quality |
| S13_extracted_97710 | Not-determined |
| S13_extracted_97743 | Not-determined |
| S13_extracted_97768 | Not-determined |
| S13_extracted_97774 | Not-determined |
| S13_extracted_97780 | Not-determined |
| S13_extracted_97789 | Not-determined |
| S13_extracted_97800 | Not-determined |
| S13_extracted_97809 | Not-determined |
| S13_extracted_97817 | Not-determined |
| S13_extracted_97824 | Not-determined |
| S13_extracted_97830 | Not-determined |
| S13_extracted_97840 | Not-determined |
| S13_extracted_97846 | Not-determined |
| S13_extracted_97863 | Not-determined |
| S13_extracted_97870 | Low-quality |
| S13_extracted_97878 | Not-determined |
| S13_extracted_97879 | Not-determined |
| S13_extracted_97891 | Not-determined |
| S13_extracted_97896 | Not-determined |
| S13_extracted_97916 | Not-determined |
| S13_extracted_97934 | Not-determined |
| S13_extracted_97950 | Not-determined |
| S13_extracted_97954 | Not-determined |
| S13_extracted_97957 | Not-determined |
| S13_extracted_97959 | Not-determined |
| S13_extracted_97962 | Not-determined |
| S13_extracted_97969 | Low-quality |
| S13_extracted_97984 | Not-determined |
| S13_extracted_97985 | Not-determined |
| S13_extracted_97987 | Not-determined |
| S13_extracted_97993 | Not-determined |
| S13_extracted_97995 | Not-determined |
| S13_extracted_98006 | Not-determined |
| S13_extracted_98009 | Not-determined |
| S13_extracted_98012 | Not-determined |
| S13_extracted_98025 | Not-determined |
| S13_extracted_98032 | Not-determined |
| S13_extracted_98035 | Low-quality |
| S13_extracted_98070 | Not-determined |
| S13_extracted_98083 | Not-determined |
| S13_extracted_98085 | Not-determined |
| S13_extracted_98092 | Not-determined |
| S13_extracted_98105 | Not-determined |
| S13_extracted_98124 | Not-determined |
| S13_extracted_98163 | Not-determined |
| S13_extracted_98168 | Not-determined |
| S13_extracted_98169 | Not-determined |
| S13_extracted_98171 | Not-determined |
| S13_extracted_98174 | Not-determined |
| S13_extracted_98186 | Not-determined |
| S13_extracted_98187 | Not-determined |
| S13_extracted_98199 | Not-determined |
| S13_extracted_98207 | Not-determined |
| S13_extracted_98212 | Not-determined |
| S13_extracted_98214 | Not-determined |
| S13_extracted_98215 | Not-determined |
| S13_extracted_98224 | Not-determined |
| S13_extracted_98243 | Not-determined |
| S13_extracted_98247 | Not-determined |
| S13_extracted_98251 | Not-determined |
| S13_extracted_98272 | Not-determined |
| S13_extracted_98284 | Not-determined |
| S13_extracted_98300 | Not-determined |
| S13_extracted_98301 | Not-determined |
| S13_extracted_98304 | Not-determined |
| S13_extracted_98315 | Not-determined |
| S13_extracted_98325 | Not-determined |
| S13_extracted_98331 | Not-determined |
| S13_extracted_98332 | Not-determined |
| S13_extracted_98334 | Not-determined |
| S13_extracted_98335 | Not-determined |
| S13_extracted_98337 | Not-determined |
| S13_extracted_98338 | Not-determined |
| S13_extracted_98340 | Not-determined |
| S13_extracted_98359 | Not-determined |
| S13_extracted_98362 | Not-determined |
| S13_extracted_98368 | Not-determined |
| S13_extracted_98386 | Not-determined |
| S13_extracted_98390 | Not-determined |
| S13_extracted_98391 | Not-determined |
| S13_extracted_98405 | Not-determined |
| S13_extracted_98411 | Not-determined |
| S13_extracted_98427 | Not-determined |
| S13_extracted_98447 | Not-determined |
| S13_extracted_98449 | Not-determined |
| S13_extracted_98453 | Not-determined |
| S13_extracted_98473 | Low-quality |
| S13_extracted_98476 | Not-determined |
| S13_extracted_98477 | Not-determined |
| S13_extracted_98478 | Not-determined |
| S13_extracted_98488 | Not-determined |
| S13_extracted_98491 | Not-determined |
| S13_extracted_98495 | Not-determined |
| S13_extracted_98499 | Not-determined |
| S13_extracted_98502 | Not-determined |
| S13_extracted_98508 | Not-determined |
| S13_extracted_98516 | Low-quality |
| S13_extracted_98519 | Not-determined |
| S13_extracted_98535 | Low-quality |
| S13_extracted_98542 | Not-determined |
| S13_extracted_98545 | Not-determined |
| S13_extracted_98556 | Not-determined |
| S13_extracted_98568 | Low-quality |
| S13_extracted_98571 | Not-determined |
| S13_extracted_98599 | Not-determined |
| S13_extracted_98604 | Not-determined |
| S13_extracted_98610 | Not-determined |
| S13_extracted_98613 | Low-quality |
| S13_extracted_98617 | Not-determined |
| S13_extracted_98637 | Not-determined |
| S13_extracted_98642 | Not-determined |
| S13_extracted_98651 | Low-quality |
| S13_extracted_98653 | Not-determined |
| S13_extracted_98675 | Not-determined |
| S13_extracted_98698 | Not-determined |
| S13_extracted_98701 | Not-determined |
| S13_extracted_98702 | Not-determined |
| S13_extracted_98719 | Not-determined |
| S13_extracted_98720 | Not-determined |
| S13_extracted_98725 | Low-quality |
| S13_extracted_98738 | Not-determined |
| S13_extracted_98742 | Not-determined |
| S13_extracted_98748 | Not-determined |
| S13_extracted_98750 | Not-determined |
| S13_extracted_98754 | Not-determined |
| S13_extracted_98757 | Not-determined |
| S13_extracted_98771 | Not-determined |
| S13_extracted_98773 | Not-determined |
| S13_extracted_98788 | Not-determined |
| S13_extracted_98816 | Not-determined |
| S13_extracted_98825 | Not-determined |
| S13_extracted_98846 | Not-determined |
| S13_extracted_98848 | Not-determined |
| S13_extracted_98849 | Not-determined |
| S13_extracted_98853 | Not-determined |
| S13_extracted_98854 | Not-determined |
| S13_extracted_98870 | Not-determined |
| S13_extracted_98876 | Not-determined |
| S13_extracted_98887 | Not-determined |
| S13_extracted_98890 | Not-determined |
| S13_extracted_98897 | Not-determined |
| S13_extracted_98909 | Not-determined |
| S13_extracted_98910 | Not-determined |
| S13_extracted_98919 | Not-determined |
| S13_extracted_98938 | Not-determined |
| S13_extracted_98942 | Not-determined |
| S13_extracted_98947 | Not-determined |
| S13_extracted_98959 | Not-determined |
| S13_extracted_98964 | Not-determined |
| S13_extracted_98981 | Not-determined |
| S13_extracted_98994 | Not-determined |
| S13_extracted_99007 | Not-determined |
| S13_extracted_99011 | Not-determined |
| S13_extracted_99031 | Not-determined |
| S13_extracted_99032 | Not-determined |
| S13_extracted_99034 | Not-determined |
| S13_extracted_99042 | Not-determined |
| S13_extracted_99043 | Not-determined |
| S13_extracted_99057 | Not-determined |
| S13_extracted_99074 | Not-determined |
| S13_extracted_99100 | Not-determined |
| S13_extracted_99139 | Not-determined |
| S13_extracted_99150 | Not-determined |
| S13_extracted_99172 | Not-determined |
| S13_extracted_99188 | Not-determined |
| S13_extracted_99191 | Not-determined |
| S13_extracted_99203 | Not-determined |
| S13_extracted_99209 | Not-determined |
| S13_extracted_99211 | Not-determined |
| S13_extracted_99221 | Not-determined |
| S13_extracted_99225 | Not-determined |
| S13_extracted_99238 | Not-determined |
| S13_extracted_99241 | Not-determined |
| S13_extracted_99245 | Not-determined |
| S13_extracted_99254 | Not-determined |
| S13_extracted_99258 | Not-determined |
| S13_extracted_99262 | Not-determined |
| S13_extracted_99264 | Not-determined |
| S13_extracted_99267 | Not-determined |
| S13_extracted_99278 | Not-determined |
| S13_extracted_99280 | Not-determined |
| S13_extracted_99283 | Not-determined |
| S13_extracted_99285 | Not-determined |
| S13_extracted_99288 | Not-determined |
| S13_extracted_99289 | Not-determined |
| S13_extracted_99294 | Not-determined |
| S13_extracted_99300 | Not-determined |
| S13_extracted_99311 | Not-determined |
| S13_extracted_99320 | Not-determined |
| S13_extracted_99321 | Not-determined |
| S13_extracted_99359 | Not-determined |
| S13_extracted_99364 | Not-determined |
| S13_extracted_99369 | Not-determined |
| S13_extracted_99372 | Not-determined |
| S13_extracted_99384 | Not-determined |
| S13_extracted_99397 | Not-determined |
| S13_extracted_99409 | Not-determined |
| S13_extracted_99411 | Low-quality |
| S13_extracted_99420 | Not-determined |
| S13_extracted_99436 | Not-determined |
| S13_extracted_99443 | Not-determined |
| S13_extracted_99451 | Not-determined |
| S13_extracted_99453 | Not-determined |
| S13_extracted_99467 | Not-determined |
| S13_extracted_99469 | Not-determined |
| S13_extracted_99473 | Not-determined |
| S13_extracted_99505 | Not-determined |
| S13_extracted_99510 | Not-determined |
| S13_extracted_99524 | Low-quality |
| S13_extracted_99527 | Not-determined |
| S13_extracted_99528 | Not-determined |
| S13_extracted_99531 | Not-determined |
| S13_extracted_99538 | Not-determined |
| S13_extracted_99557 | Low-quality |
| S13_extracted_99562 | Not-determined |
| S13_extracted_99564 | Not-determined |
| S13_extracted_99573 | Not-determined |
| S13_extracted_99574 | Not-determined |
| S13_extracted_99581 | Not-determined |
| S13_extracted_99590 | Not-determined |
| S13_extracted_99619 | Not-determined |
| S13_extracted_99621 | Not-determined |
| S13_extracted_99622 | Low-quality |
| S13_extracted_99634 | Not-determined |
| S13_extracted_99653 | Not-determined |
| S13_extracted_99656 | Low-quality |
| S13_extracted_99663 | Low-quality |
| S13_extracted_99671 | Not-determined |
| S13_extracted_99680 | Not-determined |
| S13_extracted_99682 | Not-determined |
| S13_extracted_99686 | Not-determined |
| S13_extracted_99690 | Not-determined |
| S13_extracted_99693 | Not-determined |
| S13_extracted_99709 | Not-determined |
| S13_extracted_99736 | Not-determined |
| S13_extracted_99744 | Not-determined |
| S13_extracted_99745 | Not-determined |
| S13_extracted_99746 | Not-determined |
| S13_extracted_99752 | Not-determined |
| S13_extracted_99755 | Not-determined |
| S13_extracted_99761 | Not-determined |
| S13_extracted_99798 | Not-determined |
| S13_extracted_99805 | Not-determined |
| S13_extracted_99833 | Not-determined |
| S13_extracted_99834 | Not-determined |
| S13_extracted_99842 | Not-determined |
| S13_extracted_99846 | Not-determined |
| S13_extracted_99855 | Not-determined |
| S13_extracted_99857 | Low-quality |
| S13_extracted_99858 | Not-determined |
| S13_extracted_99877 | Low-quality |
| S13_extracted_99878 | Not-determined |
| S13_extracted_99883 | Not-determined |
| S13_extracted_99889 | Not-determined |
| S13_extracted_99891 | Not-determined |
| S13_extracted_99896 | Not-determined |
| S13_extracted_99902 | Not-determined |
| S13_extracted_99916 | Not-determined |
| S13_extracted_99923 | Not-determined |
| S13_extracted_99930 | Not-determined |
| S13_extracted_99934 | Not-determined |
| S13_extracted_99941 | Not-determined |
| S13_extracted_99950 | Not-determined |
| S13_extracted_99951 | Not-determined |
| S13_extracted_99957 | Not-determined |
| S13_extracted_99965 | Not-determined |
| S13_extracted_99973 | Not-determined |
| S13_extracted_99981 | Not-determined |
| S13_extracted_99985 | Not-determined |
| S13_extracted_99987 | Low-quality |
| S13_extracted_99996 | Not-determined |
| S13_extracted_100012 | Not-determined |
| S13_extracted_100015 | Not-determined |
| S13_extracted_100024 | Not-determined |
| S13_extracted_100027 | Low-quality |
| S13_extracted_100035 | Not-determined |
| S13_extracted_100044 | Low-quality |
| S13_extracted_100046 | Low-quality |
| S13_extracted_100047 | Not-determined |
| S13_extracted_100057 | Not-determined |
| S13_extracted_100059 | Not-determined |
| S13_extracted_100063 | Not-determined |
| S13_extracted_100064 | Not-determined |
| S13_extracted_100066 | Not-determined |
| S13_extracted_100067 | Not-determined |
| S13_extracted_100078 | Not-determined |
| S13_extracted_100079 | Not-determined |
| S13_extracted_100082 | Not-determined |
| S13_extracted_100094 | Not-determined |
| S13_extracted_100107 | Not-determined |
| S13_extracted_100120 | Not-determined |
| S13_extracted_100127 | Not-determined |
| S13_extracted_100153 | Not-determined |
| S13_extracted_100155 | Not-determined |
| S13_extracted_100158 | Not-determined |
| S13_extracted_100161 | Not-determined |
| S13_extracted_100162 | Not-determined |
| S13_extracted_100163 | Not-determined |
| S13_extracted_100168 | Not-determined |
| S13_extracted_100177 | Not-determined |
| S13_extracted_100179 | Not-determined |
| S13_extracted_100198 | Not-determined |
| S13_extracted_100209 | Not-determined |
| S13_extracted_100225 | Not-determined |
| S13_extracted_100227 | Not-determined |
| S13_extracted_100229 | Not-determined |
| S13_extracted_100232 | Not-determined |
| S13_extracted_100245 | Not-determined |
| S13_extracted_100246 | Not-determined |
| S13_extracted_100255 | Low-quality |
| S13_extracted_100258 | Not-determined |
| S13_extracted_100265 | Not-determined |
| S13_extracted_100271 | Not-determined |
| S13_extracted_100272 | Not-determined |
| S13_extracted_100273 | Not-determined |
| S13_extracted_100282 | Not-determined |
| S13_extracted_100318 | Not-determined |
| S13_extracted_100319 | Not-determined |
| S13_extracted_100328 | Not-determined |
| S13_extracted_100331 | Not-determined |
| S13_extracted_100333 | Low-quality |
| S13_extracted_100335 | Not-determined |
| S13_extracted_100343 | Not-determined |

| | |
|---|---|
| S13_extracted_100345 | Not-determined |
| S13_extracted_100346 | Not-determined |
| S13_extracted_100347 | Not-determined |
| S13_extracted_100362 | Not-determined |
| S13_extracted_100363 | Not-determined |
| S13_extracted_100365 | Not-determined |
| S13_extracted_100399 | Not-determined |
| S13_extracted_100411 | Not-determined |
| S13_extracted_100437 | Not-determined |
| S13_extracted_100439 | Not-determined |
| S13_extracted_100442 | Not-determined |
| S13_extracted_100445 | Not-determined |
| S13_extracted_100471 | Not-determined |
| S13_extracted_100483 | Not-determined |
| S13_extracted_100491 | Not-determined |
| S13_extracted_100492 | Not-determined |
| S13_extracted_100503 | Not-determined |
| S13_extracted_100508 | Not-determined |
| S13_extracted_100510 | Not-determined |
| S13_extracted_100515 | Not-determined |
| S13_extracted_100527 | Not-determined |
| S13_extracted_100531 | Not-determined |
| S13_extracted_100535 | Not-determined |
| S13_extracted_100541 | Not-determined |
| S13_extracted_100548 | Not-determined |
| S13_extracted_100553 | Not-determined |
| S13_extracted_100555 | Not-determined |
| S13_extracted_100565 | Not-determined |
| S13_extracted_100566 | Not-determined |
| S13_extracted_100572 | Not-determined |
| S13_extracted_100584 | Not-determined |
| S13_extracted_100605 | Low-quality |
| S13_extracted_100615 | Not-determined |
| S13_extracted_100617 | Not-determined |
| S13_extracted_100618 | Not-determined |
| S13_extracted_100621 | Not-determined |
| S13_extracted_100630 | Low-quality |
| S13_extracted_100640 | Not-determined |
| S13_extracted_100644 | Low-quality |
| S13_extracted_100646 | Low-quality |
| S13_extracted_100663 | Not-determined |
| S13_extracted_100666 | Not-determined |
| S13_extracted_100670 | Not-determined |
| S13_extracted_100674 | Not-determined |
| S13_extracted_100676 | Not-determined |
| S13_extracted_100683 | Not-determined |
| S13_extracted_100687 | Not-determined |
| S13_extracted_100688 | Not-determined |
| S13_extracted_100709 | Not-determined |
| S13_extracted_100710 | Not-determined |
| S13_extracted_100728 | Not-determined |
| S13_extracted_100729 | Not-determined |
| S13_extracted_100738 | Not-determined |
| S13_extracted_100746 | Low-quality |
| S13_extracted_100756 | Not-determined |
| S13_extracted_100782 | Not-determined |
| S13_extracted_100804 | Not-determined |
| S13_extracted_100818 | Not-determined |
| S13_extracted_100826 | Low-quality |
| S13_extracted_100841 | Not-determined |
| S13_extracted_100848 | Not-determined |
| S13_extracted_100855 | Not-determined |
| S13_extracted_100856 | Not-determined |
| S13_extracted_100876 | Low-quality |
| S13_extracted_100885 | Not-determined |
| S13_extracted_100890 | Not-determined |
| S13_extracted_100908 | Not-determined |
| S13_extracted_100925 | Not-determined |
| S13_extracted_100939 | Not-determined |
| S13_extracted_100956 | Low-quality |
| S13_extracted_100973 | Not-determined |
| S13_extracted_100987 | Not-determined |
| S13_extracted_100992 | Not-determined |
| S13_extracted_100995 | Not-determined |
| S13_extracted_101030 | Not-determined |
| S13_extracted_101034 | Not-determined |
| S13_extracted_101036 | Not-determined |
| S13_extracted_101043 | Not-determined |
| S13_extracted_101065 | Not-determined |
| S13_extracted_101071 | Low-quality |
| S13_extracted_101073 | Not-determined |
| S13_extracted_101082 | Not-determined |
| S13_extracted_101085 | Not-determined |
| S13_extracted_101103 | Not-determined |
| S13_extracted_101106 | Low-quality |
| S13_extracted_101108 | Not-determined |
| S13_extracted_101116 | Not-determined |
| S13_extracted_101119 | Not-determined |
| S13_extracted_101125 | Not-determined |
| S13_extracted_101127 | Not-determined |
| S13_extracted_101142 | Low-quality |
| S13_extracted_101151 | Not-determined |
| S13_extracted_101153 | Not-determined |
| S13_extracted_101161 | Not-determined |
| S13_extracted_101174 | Not-determined |
| S13_extracted_101194 | Low-quality |
| S13_extracted_101211 | Not-determined |
| S13_extracted_101214 | Not-determined |
| S13_extracted_101234 | Not-determined |
| S13_extracted_101239 | Low-quality |
| S13_extracted_101248 | Not-determined |
| S13_extracted_101249 | Not-determined |
| S13_extracted_101250 | Low-quality |
| S13_extracted_101252 | Not-determined |
| S13_extracted_101268 | Not-determined |
| S13_extracted_101276 | Not-determined |
| S13_extracted_101278 | Low-quality |
| S13_extracted_101285 | Not-determined |
| S13_extracted_101288 | Low-quality |
| S13_extracted_101293 | Low-quality |
| S13_extracted_101300 | Not-determined |
| S13_extracted_101304 | Not-determined |
| S13_extracted_101306 | Not-determined |
| S13_extracted_101316 | Not-determined |
| S13_extracted_101322 | Not-determined |
| S13_extracted_101325 | Not-determined |
| S13_extracted_101331 | Not-determined |
| S13_extracted_101349 | Not-determined |
| S13_extracted_101361 | Not-determined |
| S13_extracted_101373 | Not-determined |
| S13_extracted_101377 | Not-determined |
| S13_extracted_101378 | Not-determined |
| S13_extracted_101381 | Not-determined |
| S13_extracted_101388 | Not-determined |
| S13_extracted_101401 | Not-determined |
| S13_extracted_101409 | Not-determined |
| S13_extracted_101410 | Not-determined |
| S13_extracted_101411 | Low-quality |
| S13_extracted_101417 | Low-quality |
| S13_extracted_101425 | Not-determined |
| S13_extracted_101434 | Not-determined |
| S13_extracted_101472 | Not-determined |
| S13_extracted_101479 | Not-determined |
| S13_extracted_101482 | Not-determined |
| S13_extracted_101489 | Not-determined |
| S13_extracted_101493 | Not-determined |
| S13_extracted_101499 | Not-determined |
| S13_extracted_101505 | Not-determined |
| S13_extracted_101506 | Not-determined |
| S13_extracted_101512 | Not-determined |
| S13_extracted_101513 | Not-determined |
| S13_extracted_101520 | Not-determined |
| S13_extracted_101532 | Not-determined |
| S13_extracted_101542 | Not-determined |
| S13_extracted_101562 | Not-determined |
| S13_extracted_101568 | Not-determined |
| S13_extracted_101570 | Not-determined |
| S13_extracted_101571 | Not-determined |
| S13_extracted_101575 | Not-determined |
| S13_extracted_101579 | Low-quality |
| S13_extracted_101589 | Not-determined |
| S13_extracted_101593 | Not-determined |
| S13_extracted_101596 | Low-quality |
| S13_extracted_101598 | Not-determined |
| S13_extracted_101602 | Low-quality |
| S13_extracted_101622 | Not-determined |
| S13_extracted_101634 | Not-determined |
| S13_extracted_101652 | Not-determined |
| S13_extracted_101654 | Not-determined |
| S13_extracted_101660 | Not-determined |
| S13_extracted_101667 | Not-determined |
| S13_extracted_101684 | Not-determined |
| S13_extracted_101685 | Not-determined |
| S13_extracted_101710 | Not-determined |
| S13_extracted_101717 | Not-determined |
| S13_extracted_101727 | Not-determined |
| S13_extracted_101739 | Not-determined |
| S13_extracted_101740 | Low-quality |
| S13_extracted_101745 | Not-determined |
| S13_extracted_101746 | Not-determined |
| S13_extracted_101751 | Not-determined |
| S13_extracted_101755 | Not-determined |
| S13_extracted_101765 | Not-determined |
| S13_extracted_101768 | Not-determined |
| S13_extracted_101778 | Not-determined |
| S13_extracted_101792 | Not-determined |
| S13_extracted_101802 | Not-determined |
| S13_vs_7670\|\|full | High-quality |
| S13_extracted_101824 | Not-determined |
| S13_extracted_101837 | Not-determined |
| S13_extracted_101840 | Low-quality |
| S13_extracted_101858 | Low-quality |
| S13_extracted_101862 | Not-determined |
| S13_extracted_101877 | Not-determined |
| S13_extracted_101892 | Not-determined |
| S13_extracted_101895 | Not-determined |
| S13_extracted_101909 | Not-determined |
| S13_extracted_101914 | Not-determined |
| S13_extracted_101918 | Not-determined |
| S13_extracted_101926 | Not-determined |
| S13_extracted_101930 | Not-determined |
| S13_extracted_101933 | Not-determined |
| S13_extracted_101954 | Not-determined |
| S13_extracted_101960 | Not-determined |
| S13_extracted_101961 | Not-determined |
| S13_extracted_101982 | Low-quality |
| S13_extracted_101986 | Not-determined |
| S13_extracted_101997 | Not-determined |
| S13_extracted_102007 | Not-determined |
| S13_extracted_102018 | Low-quality |
| S13_extracted_102023 | Not-determined |
| S13_extracted_102027 | Not-determined |
| S13_extracted_102044 | Not-determined |
| S13_extracted_102047 | Low-quality |
| S13_extracted_102071 | Not-determined |
| S13_extracted_102081 | Not-determined |
| S13_extracted_102083 | Not-determined |
| S13_extracted_102088 | Not-determined |
| S13_extracted_102089 | Not-determined |
| S13_extracted_102091 | Not-determined |
| S13_extracted_102101 | Not-determined |
| S13_extracted_102106 | Not-determined |
| S13_extracted_102120 | Not-determined |
| S13_extracted_102140 | Not-determined |
| S13_extracted_102143 | Not-determined |
| S13_extracted_102157 | Not-determined |
| S13_extracted_102159 | Not-determined |
| S13_extracted_102174 | Not-determined |
| S13_extracted_102175 | Not-determined |
| S13_extracted_102188 | Low-quality |
| S13_extracted_102190 | Not-determined |
| S13_extracted_102199 | Not-determined |
| S13_extracted_102205 | Not-determined |
| S13_extracted_102209 | Low-quality |
| S13_extracted_102212 | Not-determined |
| S13_extracted_102215 | Not-determined |
| S13_extracted_102225 | Not-determined |
| S13_extracted_102228 | Not-determined |
| S13_extracted_102254 | Not-determined |
| S13_extracted_102269 | Not-determined |
| S13_extracted_102270 | Not-determined |
| S13_extracted_102272 | Not-determined |
| S13_extracted_102311 | Not-determined |
| S13_extracted_102321 | Not-determined |
| S13_extracted_102331 | Not-determined |
| S13_extracted_102336 | Not-determined |
| S13_extracted_102339 | Not-determined |
| S13_extracted_102355 | Not-determined |
| S13_extracted_102363 | Not-determined |
| S13_extracted_102364 | Not-determined |
| S13_extracted_102370 | Not-determined |
| S13_extracted_102377 | Not-determined |
| S13_extracted_102380 | Not-determined |
| S13_extracted_102400 | Not-determined |
| S13_extracted_102405 | Not-determined |
| S13_extracted_102407 | Not-determined |
| S13_extracted_102416 | Not-determined |
| S13_extracted_102427 | Not-determined |
| S13_extracted_102430 | Not-determined |
| S13_extracted_102437 | Low-quality |
| S13_extracted_102460 | Not-determined |
| S13_extracted_102475 | Low-quality |
| S13_extracted_102479 | Not-determined |
| S13_extracted_102490 | Not-determined |
| S13_extracted_102507 | Low-quality |
| S13_extracted_102537 | Not-determined |
| S13_extracted_102541 | Low-quality |
| S13_extracted_102543 | Not-determined |
| S13_extracted_102553 | Not-determined |
| S13_extracted_102558 | Low-quality |
| S13_extracted_102561 | Not-determined |
| S13_extracted_102563 | Not-determined |
| S13_extracted_102571 | Not-determined |
| S13_extracted_102587 | Not-determined |
| S13_extracted_102590 | Not-determined |
| S13_extracted_102594 | Not-determined |
| S13_extracted_102597 | Not-determined |
| S13_extracted_102609 | Not-determined |
| S13_extracted_102615 | Not-determined |
| S13_extracted_102647 | Low-quality |
| S13_extracted_102649 | Not-determined |
| S13_extracted_102659 | Not-determined |
| S13_extracted_102684 | Low-quality |
| S13_extracted_102689 | Low-quality |
| S13_extracted_102691 | Not-determined |
| S13_extracted_102703 | Not-determined |
| S13_extracted_102705 | Low-quality |
| S13_extracted_102745 | Not-determined |
| S13_extracted_102752 | Not-determined |
| S13_extracted_102759 | Not-determined |
| S13_extracted_102769 | Not-determined |
| S13_extracted_102796 | Not-determined |
| S13_extracted_102832 | Not-determined |
| S13_extracted_102837 | Not-determined |
| S13_extracted_102840 | Low-quality |
| S13_extracted_102857 | Not-determined |
| S13_extracted_102877 | Not-determined |
| S13_extracted_102910 | Not-determined |
| S13_extracted_102923 | Not-determined |
| S13_extracted_102931 | Not-determined |
| S13_extracted_102934 | Not-determined |
| S13_extracted_102947 | Not-determined |
| S13_extracted_102950 | Low-quality |
| S13_extracted_102953 | Low-quality |
| S13_extracted_102954 | Not-determined |
| S13_extracted_102964 | Not-determined |
| S13_extracted_102978 | Low-quality |
| S13_extracted_102983 | Not-determined |
| S13_extracted_102988 | Not-determined |
| S13_extracted_102996 | Not-determined |
| S13_extracted_103030 | Not-determined |
| S13_extracted_103031 | Not-determined |
| S13_extracted_103032 | Not-determined |
| S13_extracted_103039 | Not-determined |
| S13_extracted_103042 | Not-determined |
| S13_extracted_103053 | Not-determined |
| S13_extracted_103073 | Not-determined |
| S13_extracted_103075 | Not-determined |
| S13_extracted_103094 | Not-determined |
| S13_extracted_103095 | Not-determined |
| S13_extracted_103138 | Not-determined |
| S13_extracted_103150 | Not-determined |
| S13_extracted_103152 | Not-determined |
| S13_extracted_103156 | Not-determined |
| S13_extracted_103166 | Not-determined |
| S13_extracted_103174 | Not-determined |
| S13_extracted_103185 | Not-determined |
| S13_extracted_103192 | Not-determined |
| S13_extracted_103220 | Not-determined |
| S13_extracted_103226 | Not-determined |
| S13_extracted_103237 | Not-determined |
| S13_extracted_103252 | Not-determined |
| S13_extracted_103258 | Not-determined |
| S13_extracted_103267 | Not-determined |
| S13_extracted_103288 | Not-determined |
| S13_extracted_103289 | Not-determined |
| S13_extracted_103299 | Not-determined |
| S13_extracted_103304 | Not-determined |
| S13_extracted_103306 | Not-determined |
| S13_extracted_103319 | Not-determined |
| S13_extracted_103332 | Not-determined |
| S13_extracted_103341 | Not-determined |
| S13_extracted_103346 | Not-determined |
| S13_extracted_103351 | Not-determined |
| S13_extracted_103356 | Not-determined |
| S13_extracted_103379 | Not-determined |
| S13_extracted_103385 | Not-determined |
| S13_extracted_103400 | Low-quality |
| S13_extracted_103405 | Not-determined |
| S13_extracted_103421 | Not-determined |
| S13_extracted_103426 | Not-determined |
| S13_extracted_103435 | Not-determined |
| S13_extracted_103444 | Not-determined |
| S13_extracted_103446 | Not-determined |
| S13_extracted_103458 | Not-determined |
| S13_extracted_103459 | Not-determined |
| S13_extracted_103482 | Low-quality |
| S13_extracted_103501 | Not-determined |
| S13_extracted_103529 | Not-determined |
| S13_extracted_103543 | Not-determined |
| S13_extracted_103544 | Not-determined |
| S13_extracted_103556 | Low-quality |
| S13_extracted_103570 | Not-determined |
| S13_extracted_103590 | Not-determined |
| S13_extracted_103592 | Not-determined |
| S13_extracted_103593 | Not-determined |
| S13_extracted_103594 | Not-determined |
| S13_extracted_103608 | Not-determined |
| S13_extracted_103617 | Not-determined |
| S13_extracted_103620 | Not-determined |
| S13_extracted_103621 | Not-determined |
| S13_extracted_103622 | Not-determined |
| S13_extracted_103632 | Not-determined |
| S13_extracted_103635 | Not-determined |
| S13_extracted_103645 | Not-determined |
| S13_extracted_103646 | Not-determined |
| S13_extracted_103688 | Low-quality |
| S13_extracted_103690 | Not-determined |
| S13_extracted_103692 | Not-determined |
| S13_extracted_103704 | Not-determined |
| S13_extracted_103714 | Not-determined |
| S13_extracted_103728 | Not-determined |
| S13_extracted_103737 | Low-quality |
| S13_extracted_103747 | Low-quality |
| S13_extracted_103756 | Not-determined |
| S13_extracted_103758 | Not-determined |
| S13_extracted_103780 | Not-determined |
| S13_extracted_103788 | Not-determined |
| S13_extracted_103790 | Not-determined |
| S13_extracted_103797 | Not-determined |
| S13_extracted_103798 | Not-determined |
| S13_extracted_103800 | Not-determined |
| S13_extracted_103801 | Not-determined |
| S13_extracted_103804 | Not-determined |
| S13_extracted_103844 | Not-determined |
| S13_extracted_103849 | Not-determined |
| S13_extracted_103867 | Not-determined |
| S13_extracted_103871 | Low-quality |
| S13_extracted_103879 | Not-determined |
| S13_extracted_103892 | Not-determined |
| S13_extracted_103897 | Low-quality |
| S13_extracted_103904 | Not-determined |
| S13_extracted_103906 | Not-determined |
| S13_extracted_103908 | Not-determined |
| S13_extracted_103934 | Not-determined |
| S13_extracted_103953 | Not-determined |
| S13_extracted_103960 | Not-determined |
| S13_extracted_103966 | Not-determined |
| S13_extracted_103969 | Low-quality |
| S13_extracted_103981 | Low-quality |
| S13_extracted_104004 | Not-determined |
| S13_extracted_104007 | Not-determined |
| S13_extracted_104013 | Not-determined |
| S13_extracted_104032 | Not-determined |
| S13_extracted_104034 | Not-determined |
| S13_extracted_104035 | Not-determined |
| S13_extracted_104036 | Not-determined |
| S13_extracted_104052 | Not-determined |
| S13_extracted_104068 | Not-determined |
| S13_extracted_104071 | Not-determined |
| S13_extracted_104076 | Not-determined |
| S13_extracted_104077 | Not-determined |
| S13_extracted_104078 | Not-determined |
| S13_extracted_104098 | Not-determined |
| S13_extracted_104100 | Not-determined |
| S13_extracted_104114 | Not-determined |
| S13_extracted_104123 | Not-determined |
| S13_extracted_104132 | Not-determined |
| S13_extracted_104140 | Not-determined |
| S13_extracted_104142 | Not-determined |
| S13_extracted_104151 | Not-determined |
| S13_extracted_104163 | Not-determined |
| S13_extracted_104168 | Not-determined |
| S13_extracted_104172 | Low-quality |
| S13_extracted_104176 | Not-determined |
| S13_extracted_104178 | Not-determined |
| S13_extracted_104185 | Not-determined |
| S13_extracted_104196 | Not-determined |
| S13_extracted_104210 | Low-quality |
| S13_extracted_104215 | Not-determined |
| S13_extracted_104227 | Not-determined |
| S13_extracted_104229 | Not-determined |
| S13_extracted_104231 | Not-determined |
| S13_extracted_104253 | Not-determined |
| S13_extracted_104255 | Not-determined |
| S13_extracted_104263 | Not-determined |
| S13_extracted_104265 | Low-quality |
| S13_extracted_104277 | Not-determined |
| S13_extracted_104288 | Not-determined |
| S13_extracted_104295 | Not-determined |
| S13_extracted_104299 | Not-determined |
| S13_extracted_104302 | Low-quality |
| S13_extracted_104309 | Not-determined |
| S13_extracted_104318 | Not-determined |
| S13_extracted_104325 | Not-determined |
| S13_extracted_104347 | Not-determined |
| S13_extracted_104352 | Not-determined |
| S13_extracted_104355 | Not-determined |
| S13_extracted_104363 | Low-quality |
| S13_extracted_104366 | Not-determined |
| S13_extracted_104378 | Low-quality |
| S13_extracted_104384 | Not-determined |
| S13_extracted_104392 | Not-determined |
| S13_extracted_104395 | Not-determined |
| S13_extracted_104398 | Not-determined |
| S13_extracted_104411 | Not-determined |
| S13_extracted_104437 | Low-quality |
| S13_extracted_104442 | Not-determined |
| S13_extracted_104454 | Low-quality |
| S13_extracted_104455 | Not-determined |
| S13_extracted_104462 | Not-determined |
| S13_extracted_104476 | Not-determined |
| S13_extracted_104497 | Not-determined |
| S13_extracted_104498 | Not-determined |
| S13_extracted_104511 | Not-determined |
| S13_extracted_104512 | Not-determined |
| S13_extracted_104513 | Low-quality |
| S13_extracted_104525 | Not-determined |
| S13_extracted_104530 | Not-determined |
| S13_extracted_104549 | Low-quality |
| S13_extracted_104554 | Not-determined |
| S13_extracted_104560 | Not-determined |
| S13_extracted_104569 | Not-determined |
| S13_extracted_104575 | Not-determined |
| S13_extracted_104607 | Low-quality |
| S13_extracted_104613 | Not-determined |
| S13_extracted_104621 | Not-determined |
| S13_extracted_104622 | Not-determined |
| S13_extracted_104623 | Not-determined |
| S13_extracted_104626 | Low-quality |
| S13_extracted_104629 | Not-determined |
| S13_extracted_104636 | Not-determined |
| S13_extracted_104659 | Not-determined |
| S13_extracted_104670 | Not-determined |
| S13_extracted_104693 | Not-determined |
| S13_extracted_104698 | Not-determined |
| S13_extracted_104706 | Not-determined |
| S13_extracted_104720 | Not-determined |
| S13_extracted_104727 | Not-determined |
| S13_extracted_104737 | Not-determined |
| S13_extracted_104746 | Not-determined |
| S13_extracted_104758 | Not-determined |
| S13_extracted_104759 | Not-determined |
| S13_extracted_104768 | Not-determined |
| S13_extracted_104772 | Not-determined |
| S13_extracted_104779 | Not-determined |
| S13_extracted_104784 | Not-determined |
| S13_extracted_104785 | Not-determined |
| S13_extracted_104792 | Not-determined |
| S13_extracted_104810 | Not-determined |
| S13_extracted_104816 | Not-determined |
| S13_extracted_104829 | Not-determined |
| S13_extracted_104834 | Not-determined |
| S13_extracted_104860 | Not-determined |
| S13_extracted_104861 | Not-determined |
| S13_extracted_104869 | Not-determined |
| S13_extracted_104882 | Not-determined |
| S13_extracted_104883 | Not-determined |
| S13_extracted_104887 | Not-determined |
| S13_extracted_104906 | Low-quality |
| S13_extracted_104907 | Not-determined |
| S13_extracted_104908 | Not-determined |
| S13_extracted_104911 | Not-determined |
| S13_extracted_104928 | Low-quality |
| S13_extracted_104935 | Low-quality |
| S13_extracted_104967 | Not-determined |
| S13_extracted_104972 | Not-determined |
| S13_extracted_104975 | Not-determined |
| S13_extracted_105013 | Not-determined |
| S13_extracted_105019 | Not-determined |
| S13_extracted_105025 | Not-determined |
| S13_extracted_105038 | Not-determined |
| S13_extracted_105049 | Not-determined |
| S13_extracted_105056 | Not-determined |
| S13_extracted_105058 | Low-quality |
| S13_extracted_105061 | Not-determined |
| S13_extracted_105073 | Low-quality |
| S13_extracted_105081 | Not-determined |
| S13_extracted_105083 | Low-quality |
| S13_extracted_105084 | Not-determined |
| S13_extracted_105092 | Not-determined |
| S13_extracted_105095 | Not-determined |
| S13_extracted_105112 | Not-determined |
| S13_extracted_105123 | Not-determined |
| S13_extracted_105137 | Not-determined |
| S13_extracted_105143 | Not-determined |
| S13_extracted_105146 | Not-determined |
| S13_extracted_105156 | Not-determined |
| S13_extracted_105166 | Not-determined |
| S13_extracted_105193 | Low-quality |
| S13_extracted_105209 | Not-determined |
| S13_extracted_105212 | Not-determined |
| S13_extracted_105213 | Not-determined |
| S13_extracted_105215 | Not-determined |
| S13_extracted_105223 | Not-determined |
| S13_extracted_105225 | Not-determined |
| S13_extracted_105258 | Low-quality |
| S13_extracted_105262 | Low-quality |
| S13_extracted_105265 | Not-determined |
| S13_extracted_105271 | Not-determined |
| S13_extracted_105275 | Not-determined |
| S13_extracted_105284 | Low-quality |
| S13_extracted_105302 | Not-determined |
| S13_extracted_105307 | Not-determined |
| S13_extracted_105311 | Low-quality |
| S13_extracted_105328 | Not-determined |
| S13_extracted_105330 | Not-determined |
| S13_extracted_105336 | Not-determined |
| S13_extracted_105340 | Not-determined |
| S13_extracted_105377 | Not-determined |
| S13_extracted_105378 | Not-determined |
| S13_extracted_105384 | Low-quality |
| S13_extracted_105388 | Not-determined |
| S13_extracted_105390 | Not-determined |
| S13_extracted_105393 | Not-determined |
| S13_extracted_105401 | Not-determined |
| S13_extracted_105423 | Not-determined |
| S13_extracted_105435 | Not-determined |
| S13_extracted_105437 | Not-determined |
| S13_extracted_105439 | Not-determined |
| S13_extracted_105442 | Not-determined |
| S13_extracted_105448 | Not-determined |
| S13_extracted_105449 | Not-determined |
| S13_extracted_105452 | Not-determined |
| S13_extracted_105454 | Low-quality |
| S13_extracted_105469 | Not-determined |
| S13_extracted_105474 | Not-determined |
| S13_extracted_105480 | Not-determined |
| S13_extracted_105484 | Low-quality |
| S13_extracted_105506 | Not-determined |
| S13_extracted_105514 | Not-determined |
| S13_extracted_105515 | Not-determined |
| S13_extracted_105520 | Low-quality |
| S13_extracted_105523 | Not-determined |
| S13_extracted_105539 | Not-determined |
| S13_extracted_105544 | Not-determined |
| S13_extracted_105556 | Not-determined |
| S13_extracted_105578 | Low-quality |
| S13_extracted_105589 | Not-determined |

| | |
|---|---|
| S13_extracted_105611 | Not-determined |
| S13_extracted_105614 | Not-determined |
| S13_extracted_105627 | Not-determined |
| S13_extracted_105638 | Not-determined |
| S13_extracted_105639 | Low-quality |
| S13_extracted_105646 | Not-determined |
| S13_extracted_105648 | Not-determined |
| S13_extracted_105682 | Not-determined |
| S13_extracted_105694 | Not-determined |
| S13_extracted_105696 | Not-determined |
| S13_extracted_105726 | Not-determined |
| S13_extracted_105727 | Not-determined |
| S13_extracted_105733 | Not-determined |
| S13_extracted_105738 | Not-determined |
| S13_extracted_105742 | Not-determined |
| S13_extracted_105750 | Not-determined |
| S13_extracted_105774 | Not-determined |
| S13_extracted_105789 | Low-quality |
| S13_extracted_105790 | Not-determined |
| S13_extracted_105794 | Not-determined |
| S13_extracted_105806 | Not-determined |
| S13_extracted_105818 | Not-determined |
| S13_extracted_105825 | Not-determined |
| S13_extracted_105832 | Not-determined |
| S13_extracted_105838 | Not-determined |
| S13_extracted_105842 | Not-determined |
| S13_extracted_105847 | Not-determined |
| S13_extracted_105859 | Low-quality |
| S13_extracted_105863 | Low-quality |
| S13_extracted_105870 | Not-determined |
| S13_extracted_105878 | Not-determined |
| S13_extracted_105883 | Not-determined |
| S13_extracted_105894 | Not-determined |
| S13_extracted_105916 | Not-determined |
| S13_extracted_105923 | Not-determined |
| S13_extracted_105924 | Low-quality |
| S13_extracted_105930 | Not-determined |
| S13_extracted_105934 | Not-determined |
| S13_extracted_105935 | Not-determined |
| S13_extracted_105937 | Not-determined |
| S13_extracted_105940 | Not-determined |
| S13_extracted_105944 | Not-determined |
| S13_extracted_105953 | Not-determined |
| S13_extracted_105957 | Not-determined |
| S13_extracted_105961 | Not-determined |
| S13_extracted_105981 | Not-determined |
| S13_extracted_105988 | Not-determined |
| S13_extracted_105992 | Not-determined |
| S13_extracted_106031 | Not-determined |
| S13_extracted_106034 | Not-determined |
| S13_extracted_106045 | Not-determined |
| S13_extracted_106048 | Not-determined |
| S13_extracted_106081 | Not-determined |
| S13_extracted_106098 | Not-determined |
| S13_extracted_106100 | Not-determined |
| S13_extracted_106108 | Not-determined |
| S13_extracted_106111 | Not-determined |
| S13_extracted_106115 | Not-determined |
| S13_extracted_106137 | Not-determined |
| S13_extracted_106141 | Not-determined |
| S13_extracted_106142 | Not-determined |
| S13_extracted_106145 | Low-quality |
| S13_extracted_106154 | Not-determined |
| S13_extracted_106155 | Not-determined |
| S13_extracted_106170 | Low-quality |
| S13_extracted_106171 | Not-determined |
| S13_extracted_106179 | Not-determined |
| S13_extracted_106182 | Not-determined |
| S13_extracted_106191 | Not-determined |
| S13_extracted_106192 | Not-determined |
| S13_extracted_106196 | Not-determined |
| S13_extracted_106229 | Not-determined |
| S13_extracted_106239 | Not-determined |
| S13_extracted_106241 | Not-determined |
| S13_extracted_106251 | Not-determined |
| S13_extracted_106252 | Low-quality |
| S13_extracted_106266 | Low-quality |
| S13_extracted_106274 | Not-determined |
| S13_extracted_106276 | Not-determined |
| S13_extracted_106280 | Low-quality |
| S13_extracted_106298 | Not-determined |
| S13_extracted_106317 | Not-determined |
| S13_extracted_106320 | Low-quality |
| S13_extracted_106321 | Not-determined |
| S13_extracted_106336 | Not-determined |
| S13_extracted_106340 | Not-determined |
| S13_extracted_106348 | Not-determined |
| S13_extracted_106356 | Not-determined |
| S13_extracted_106360 | Not-determined |
| S13_extracted_106362 | Not-determined |
| S13_extracted_106380 | Low-quality |
| S13_extracted_106383 | Not-determined |
| S13_extracted_106386 | Not-determined |
| S13_extracted_106391 | Not-determined |
| S13_extracted_106411 | Not-determined |
| S13_extracted_106416 | Low-quality |
| S13_extracted_106421 | Not-determined |
| S13_extracted_106435 | Not-determined |
| S13_extracted_106449 | Not-determined |
| S13_extracted_106452 | Not-determined |
| S13_extracted_106456 | Not-determined |
| S13_extracted_106458 | Not-determined |
| S13_extracted_106464 | Not-determined |
| S13_extracted_106475 | Not-determined |
| S13_extracted_106484 | Not-determined |
| S13_extracted_106496 | Not-determined |
| S13_extracted_106499 | Not-determined |
| S13_extracted_106514 | Not-determined |
| S13_extracted_106518 | Not-determined |
| S13_extracted_106525 | Not-determined |
| S13_extracted_106527 | Not-determined |
| S13_extracted_106529 | Not-determined |
| S13_extracted_106534 | Low-quality |
| S13_extracted_106537 | Not-determined |
| S13_extracted_106553 | Not-determined |
| S13_extracted_106555 | Not-determined |
| S13_extracted_106582 | Not-determined |
| S13_extracted_106586 | Not-determined |
| S13_extracted_106587 | Not-determined |
| S13_extracted_106591 | Not-determined |
| S13_extracted_106593 | Not-determined |
| S13_extracted_106595 | Not-determined |
| S13_extracted_106617 | Not-determined |
| S13_extracted_106623 | Not-determined |
| S13_extracted_106633 | Not-determined |
| S13_extracted_106646 | Not-determined |
| S13_extracted_106654 | Not-determined |
| S13_extracted_106658 | Not-determined |
| S13_extracted_106664 | Not-determined |
| S13_extracted_106680 | Not-determined |
| S13_extracted_106681 | Not-determined |
| S13_extracted_106686 | Not-determined |
| S13_extracted_106693 | Low-quality |
| S13_extracted_106703 | Not-determined |
| S13_extracted_106708 | Not-determined |
| S13_extracted_106720 | Not-determined |
| S13_extracted_106734 | Not-determined |
| S13_extracted_106737 | Not-determined |
| S13_extracted_106738 | Not-determined |
| S13_extracted_106752 | Not-determined |
| S13_extracted_106753 | Low-quality |
| S13_extracted_106780 | Not-determined |
| S13_extracted_106789 | Not-determined |
| S13_extracted_106791 | Not-determined |
| S13_extracted_106795 | Not-determined |
| S13_extracted_106796 | Low-quality |
| S13_extracted_106797 | Not-determined |
| S13_extracted_106814 | Not-determined |
| S13_extracted_106818 | Not-determined |
| S13_extracted_106830 | Low-quality |
| S13_extracted_106835 | Not-determined |
| S13_extracted_106837 | Not-determined |
| S13_extracted_106842 | Not-determined |
| S13_extracted_106852 | Not-determined |
| S13_extracted_106856 | Not-determined |
| S13_extracted_106861 | Low-quality |
| S13_extracted_106871 | Not-determined |
| S13_extracted_106878 | Not-determined |
| S13_extracted_106882 | Not-determined |
| S13_extracted_106891 | Not-determined |
| S13_extracted_106896 | Not-determined |
| S13_extracted_106898 | Not-determined |
| S13_extracted_106900 | Not-determined |
| S13_extracted_106903 | Not-determined |
| S13_extracted_106911 | Low-quality |
| S13_extracted_106917 | Not-determined |
| S13_extracted_106933 | Not-determined |
| S13_extracted_106935 | Not-determined |
| S13_extracted_106938 | Not-determined |
| S13_extracted_106951 | Not-determined |
| S13_extracted_106954 | Not-determined |
| S13_extracted_106962 | Not-determined |
| S13_extracted_106969 | Not-determined |
| S13_extracted_106975 | Not-determined |
| S13_extracted_106979 | Not-determined |
| S13_extracted_106982 | Not-determined |
| S13_extracted_107014 | Not-determined |
| S13_extracted_107015 | Low-quality |
| S13_extracted_107023 | Not-determined |
| S13_extracted_107029 | Not-determined |
| S13_extracted_107031 | Not-determined |
| S13_extracted_107037 | Not-determined |
| S13_extracted_107044 | Not-determined |
| S13_extracted_107055 | Low-quality |
| S13_extracted_107057 | Not-determined |
| S13_extracted_107058 | Not-determined |
| S13_extracted_107060 | Not-determined |
| S13_extracted_107066 | Low-quality |
| S13_extracted_107074 | Not-determined |
| S13_extracted_107080 | Not-determined |
| S13_extracted_107082 | Not-determined |
| S13_extracted_107084 | Not-determined |
| S13_extracted_107092 | Not-determined |
| S13_extracted_107093 | Not-determined |
| S13_extracted_107099 | Not-determined |
| S13_extracted_107101 | Not-determined |
| S13_extracted_107102 | Not-determined |
| S13_extracted_107111 | Low-quality |
| S13_extracted_107130 | Not-determined |
| S13_extracted_107133 | Not-determined |
| S13_extracted_107151 | Not-determined |
| S13_extracted_107165 | Not-determined |
| S13_extracted_107171 | Not-determined |
| S13_extracted_107182 | Not-determined |
| S13_extracted_107183 | Not-determined |
| S13_extracted_107186 | Not-determined |
| S13_extracted_107188 | Not-determined |
| S9_extracted_k141_3124 | Medium-quality |
| S13_extracted_107197 | Not-determined |
| S13_extracted_107201 | Low-quality |
| S13_extracted_107206 | Not-determined |
| S13_extracted_107211 | Low-quality |
| S13_extracted_107228 | Not-determined |
| S13_extracted_107232 | Not-determined |
| S13_extracted_107237 | Not-determined |
| S13_extracted_107262 | Not-determined |
| S13_extracted_107263 | Not-determined |
| S13_extracted_107265 | Not-determined |
| S13_extracted_107267 | Not-determined |
| S13_extracted_107270 | Not-determined |
| S13_extracted_107273 | Not-determined |
| S13_extracted_107278 | Not-determined |
| S13_extracted_107295 | Not-determined |
| S13_extracted_107298 | Not-determined |
| S13_extracted_107299 | Not-determined |
| S13_extracted_107323 | Not-determined |
| S13_extracted_107341 | Not-determined |
| S14_vs_k141_11510\|\|full | Medium-quality |
| S13_extracted_107356 | Not-determined |
| S13_extracted_107367 | Not-determined |
| S13_extracted_107371 | Not-determined |
| S13_extracted_107373 | Not-determined |
| S13_extracted_107383 | Not-determined |
| S13_extracted_107409 | Not-determined |
| S13_extracted_107419 | Not-determined |
| S13_extracted_107420 | Not-determined |
| S13_extracted_107424 | Not-determined |
| S13_extracted_107441 | Not-determined |
| S13_extracted_107451 | Not-determined |
| S13_extracted_107460 | Not-determined |
| S13_extracted_107461 | Not-determined |
| S13_extracted_107463 | Not-determined |
| S13_extracted_107467 | Not-determined |
| S13_extracted_107471 | Not-determined |
| S13_extracted_107488 | Low-quality |
| S13_extracted_107494 | Not-determined |
| S13_extracted_107513 | Not-determined |
| S13_extracted_107515 | Low-quality |
| S13_extracted_107525 | Not-determined |
| S13_extracted_107529 | Not-determined |
| S13_extracted_107556 | Not-determined |
| S13_extracted_107557 | Low-quality |
| S13_extracted_107558 | Not-determined |
| S13_extracted_107575 | Not-determined |
| S13_extracted_107585 | Not-determined |
| S13_extracted_107587 | Not-determined |
| S13_extracted_107591 | Not-determined |
| S13_extracted_107599 | Not-determined |
| S13_extracted_107618 | Not-determined |
| S13_extracted_107636 | Not-determined |
| S13_extracted_107654 | Not-determined |
| S13_extracted_107666 | Not-determined |
| S13_extracted_107694 | Not-determined |
| S13_extracted_107699 | Not-determined |
| S13_extracted_107714 | Not-determined |
| S13_extracted_107721 | Low-quality |
| S13_extracted_107726 | Not-determined |
| S13_extracted_107729 | Not-determined |
| S13_extracted_107734 | Not-determined |
| S13_extracted_107742 | Not-determined |
| S13_extracted_107744 | Low-quality |
| S13_extracted_107756 | Not-determined |
| S13_extracted_107763 | Not-determined |
| S13_extracted_107770 | Not-determined |
| S13_extracted_107771 | Not-determined |
| S13_extracted_107816 | Not-determined |
| S13_extracted_107818 | Not-determined |
| S13_extracted_107853 | Not-determined |
| S13_extracted_107860 | Not-determined |
| S13_extracted_107866 | Not-determined |
| S13_extracted_107867 | Not-determined |
| S13_extracted_107871 | Low-quality |
| S13_extracted_107884 | Low-quality |
| S13_extracted_107912 | Not-determined |
| S13_extracted_107919 | Not-determined |
| S13_extracted_107922 | Not-determined |
| S13_extracted_107928 | Low-quality |
| S13_extracted_107935 | Not-determined |
| S13_extracted_107938 | Not-determined |
| S13_extracted_107941 | Not-determined |
| S13_extracted_107942 | Not-determined |
| S13_extracted_107948 | Low-quality |
| S13_extracted_107980 | Not-determined |
| S13_extracted_107983 | Not-determined |
| S13_extracted_107985 | Low-quality |
| S13_extracted_107988 | Not-determined |
| S13_extracted_107989 | Not-determined |
| S13_extracted_107995 | Not-determined |
| S13_extracted_108021 | Not-determined |
| S13_extracted_108022 | Not-determined |
| S13_extracted_108056 | Not-determined |
| S13_extracted_108078 | Not-determined |
| S13_extracted_108082 | Not-determined |
| S13_extracted_108085 | Not-determined |
| S13_extracted_108093 | Not-determined |
| S13_extracted_108096 | Not-determined |
| S13_extracted_108098 | Not-determined |
| S13_extracted_108110 | Not-determined |
| S13_extracted_108118 | Not-determined |
| S13_extracted_108131 | Not-determined |
| S13_extracted_108133 | Not-determined |
| S13_extracted_108156 | Not-determined |
| S13_extracted_108168 | Not-determined |
| S13_extracted_108173 | Low-quality |
| S13_extracted_108174 | Not-determined |
| S13_extracted_108176 | Not-determined |
| S13_extracted_108177 | Not-determined |
| S13_extracted_108187 | Not-determined |
| S13_extracted_108188 | Low-quality |
| S13_extracted_108189 | Not-determined |
| S13_extracted_108191 | Not-determined |
| S13_extracted_108208 | Not-determined |
| S13_extracted_108217 | Low-quality |
| S13_extracted_108231 | Not-determined |
| S13_extracted_108232 | Not-determined |
| S13_extracted_108233 | Not-determined |
| S13_extracted_108259 | Not-determined |
| S13_extracted_108272 | Not-determined |
| S13_extracted_108281 | Not-determined |
| S13_extracted_108287 | Not-determined |
| S13_extracted_108288 | Not-determined |
| S13_extracted_108291 | Not-determined |
| S13_extracted_108303 | Not-determined |
| S13_extracted_108333 | Not-determined |
| S13_extracted_108337 | Not-determined |
| S13_extracted_108343 | Not-determined |
| S13_extracted_108346 | Not-determined |
| S13_extracted_108348 | Not-determined |
| S13_extracted_108353 | Not-determined |
| S13_extracted_108364 | Low-quality |
| S13_extracted_108374 | Low-quality |
| S13_extracted_108393 | Not-determined |
| S13_extracted_108398 | Not-determined |
| S13_extracted_108407 | Not-determined |
| S13_extracted_108410 | Not-determined |
| S13_extracted_108414 | Not-determined |
| S13_extracted_108423 | Not-determined |
| S13_extracted_108426 | Low-quality |
| S13_extracted_108438 | Not-determined |
| S13_extracted_108447 | Not-determined |
| S13_extracted_108451 | Not-determined |
| S13_extracted_108452 | Low-quality |
| S13_extracted_108456 | Not-determined |
| S13_extracted_108468 | Not-determined |
| S13_extracted_108473 | Not-determined |
| S13_extracted_108507 | Low-quality |
| S13_extracted_108512 | Not-determined |
| S13_extracted_108516 | Not-determined |
| S13_extracted_108527 | Not-determined |
| S13_extracted_108535 | Not-determined |
| S13_extracted_108536 | Not-determined |
| S13_extracted_108538 | Not-determined |
| S13_extracted_108540 | Not-determined |
| S13_extracted_108556 | Not-determined |
| S13_extracted_108559 | Not-determined |
| S13_extracted_108594 | Not-determined |
| S13_extracted_108605 | Not-determined |
| S13_extracted_108606 | Not-determined |
| S13_extracted_108608 | Not-determined |
| S13_extracted_108612 | Low-quality |
| S13_extracted_108617 | Not-determined |
| S13_extracted_108627 | Low-quality |
| S13_extracted_108646 | Not-determined |
| S13_extracted_108652 | Not-determined |
| S13_extracted_108653 | Not-determined |
| S13_extracted_108661 | Not-determined |
| S13_extracted_108670 | Not-determined |
| S13_extracted_108677 | Not-determined |
| S13_extracted_108678 | Not-determined |
| S13_extracted_108679 | Not-determined |
| S13_extracted_108680 | Not-determined |
| S13_extracted_108687 | Not-determined |
| S13_extracted_108697 | Not-determined |
| S13_extracted_108699 | Not-determined |
| S13_extracted_108705 | Not-determined |
| S13_extracted_108712 | Not-determined |
| S13_extracted_108714 | Not-determined |
| S13_extracted_108725 | Not-determined |
| S13_extracted_108727 | Not-determined |
| S13_extracted_108736 | Not-determined |
| S13_extracted_108745 | Not-determined |
| S13_extracted_108751 | Low-quality |
| S13_extracted_108760 | Not-determined |
| S13_extracted_108792 | Not-determined |
| S13_extracted_108795 | Not-determined |
| S13_extracted_108800 | Not-determined |
| S13_extracted_108801 | Low-quality |
| S13_extracted_108810 | Not-determined |
| S13_extracted_108812 | Low-quality |
| S13_extracted_108823 | Not-determined |
| S13_extracted_108825 | Not-determined |
| S13_extracted_108832 | Low-quality |
| S13_extracted_108834 | Not-determined |
| S13_extracted_108838 | Low-quality |
| S13_extracted_108840 | Not-determined |
| S13_extracted_108851 | Not-determined |
| S13_extracted_108852 | Low-quality |
| S13_extracted_108856 | Not-determined |
| S13_extracted_108857 | Not-determined |
| S13_extracted_108862 | Not-determined |
| S13_extracted_108867 | Not-determined |
| S13_extracted_108877 | Not-determined |
| S13_extracted_108878 | Not-determined |
| S13_extracted_108898 | Not-determined |
| S13_extracted_108932 | Not-determined |
| S13_extracted_108937 | Not-determined |
| S13_extracted_108939 | Not-determined |
| S13_extracted_108954 | Low-quality |
| S13_extracted_108965 | Not-determined |
| S13_extracted_108970 | Not-determined |
| S13_extracted_108977 | Not-determined |
| S13_extracted_108980 | Low-quality |
| S13_extracted_108992 | Low-quality |
| S13_extracted_108994 | Not-determined |
| S13_extracted_109000 | Not-determined |
| S13_extracted_109001 | Not-determined |
| S13_extracted_109029 | Not-determined |
| S13_extracted_109037 | Not-determined |
| S13_extracted_109042 | Low-quality |
| S13_extracted_109058 | Not-determined |
| S13_extracted_109062 | Not-determined |
| S13_extracted_109070 | Not-determined |
| S13_extracted_109085 | Not-determined |
| S13_extracted_109092 | Not-determined |
| S13_extracted_109098 | Not-determined |
| S13_extracted_109102 | Not-determined |
| S13_extracted_109121 | Not-determined |
| S13_extracted_109126 | Not-determined |
| S13_extracted_109133 | Not-determined |
| S13_extracted_109142 | Not-determined |
| S13_extracted_109149 | Not-determined |
| S13_extracted_109165 | Not-determined |
| S13_extracted_109169 | Not-determined |
| S13_extracted_109173 | Not-determined |
| S13_extracted_109174 | Low-quality |
| S13_extracted_109185 | Not-determined |
| S13_extracted_109186 | Not-determined |
| S13_extracted_109221 | Not-determined |
| S13_extracted_109232 | Not-determined |
| S12_vs_k141_99219\|\|full | High-quality |
| S13_extracted_109250 | Not-determined |
| S13_extracted_109269 | Not-determined |
| S13_extracted_109282 | Low-quality |
| S13_extracted_109316 | Not-determined |
| S13_extracted_109327 | Not-determined |
| S13_extracted_109353 | Low-quality |
| S13_extracted_109359 | Not-determined |
| S13_extracted_109364 | Low-quality |
| S13_extracted_109373 | Not-determined |
| S13_extracted_109389 | Low-quality |
| S13_extracted_109394 | Low-quality |
| S13_extracted_109406 | Not-determined |
| S13_extracted_109411 | Low-quality |
| S13_extracted_109412 | Not-determined |
| S13_extracted_109431 | Not-determined |
| S13_extracted_109435 | Not-determined |
| S13_extracted_109438 | Low-quality |
| S13_extracted_109439 | Not-determined |
| S13_extracted_109441 | Low-quality |
| S13_extracted_109446 | Not-determined |
| S13_extracted_109448 | Not-determined |
| S13_extracted_109458 | Not-determined |
| S13_extracted_109465 | Not-determined |
| S13_extracted_109483 | Not-determined |
| S13_extracted_109505 | Not-determined |
| S13_extracted_109507 | Not-determined |
| S13_extracted_109514 | Low-quality |
| S13_extracted_109524 | Not-determined |
| S13_extracted_109532 | Not-determined |
| S13_extracted_109545 | Not-determined |
| S13_extracted_109548 | Not-determined |
| S13_extracted_109567 | Low-quality |
| S13_extracted_109576 | Not-determined |
| S13_extracted_109577 | Not-determined |
| S13_extracted_109596 | Not-determined |
| S13_extracted_109603 | Not-determined |
| S13_extracted_109607 | Low-quality |
| S13_extracted_109609 | Not-determined |
| S13_extracted_109611 | Not-determined |
| S13_extracted_109628 | Not-determined |
| S13_extracted_109631 | Not-determined |
| S13_extracted_109636 | Not-determined |
| S13_extracted_109641 | Not-determined |
| S13_extracted_109648 | Not-determined |
| S13_extracted_109659 | Not-determined |
| S13_extracted_109687 | Low-quality |
| S13_extracted_109692 | Not-determined |
| S13_extracted_109697 | Not-determined |
| S13_extracted_109699 | Not-determined |
| S13_extracted_109703 | Low-quality |
| S13_extracted_109716 | Not-determined |
| S13_extracted_109724 | Not-determined |
| S13_extracted_109733 | Not-determined |
| S13_extracted_109738 | Not-determined |
| S13_extracted_109757 | Not-determined |
| S13_extracted_109770 | Not-determined |
| S13_extracted_109774 | Not-determined |
| S13_extracted_109780 | Low-quality |
| S13_extracted_109814 | Not-determined |
| S13_extracted_109815 | Low-quality |
| S13_extracted_109823 | Low-quality |
| S13_extracted_109825 | Not-determined |
| S13_extracted_109828 | Not-determined |
| S13_extracted_109841 | Not-determined |
| S13_extracted_109843 | Not-determined |
| S13_extracted_109855 | Not-determined |
| S13_extracted_109869 | Not-determined |
| S13_extracted_109875 | Not-determined |
| S13_extracted_109879 | Not-determined |
| S13_extracted_109886 | Not-determined |
| S13_extracted_109897 | Not-determined |
| S13_extracted_109905 | Not-determined |
| S13_extracted_109908 | Not-determined |
| S13_extracted_109936 | Not-determined |
| S13_extracted_109951 | Not-determined |
| S13_extracted_109953 | Not-determined |
| S13_extracted_109972 | Not-determined |
| S13_extracted_109976 | Not-determined |
| S13_extracted_109990 | Not-determined |
| S13_extracted_110021 | Not-determined |
| S13_extracted_110023 | Not-determined |
| S13_extracted_110031 | Not-determined |
| S13_extracted_110046 | Not-determined |
| S13_extracted_110048 | Not-determined |
| S13_extracted_110052 | Not-determined |
| S13_extracted_110054 | Not-determined |
| S13_extracted_110068 | Low-quality |
| S13_extracted_110070 | Not-determined |
| S13_extracted_110083 | Not-determined |
| S13_extracted_110087 | Not-determined |
| S13_extracted_110108 | Not-determined |
| S13_extracted_110124 | Not-determined |
| S13_extracted_110133 | Not-determined |
| S13_extracted_110139 | Not-determined |
| S13_extracted_110152 | Not-determined |
| S13_extracted_110159 | Not-determined |
| S13_extracted_110170 | Low-quality |
| S13_extracted_110173 | Low-quality |
| S13_extracted_110178 | Not-determined |
| S13_extracted_110206 | Not-determined |
| S13_extracted_110214 | Low-quality |
| S13_extracted_110217 | Not-determined |
| S13_extracted_110219 | Not-determined |
| S13_extracted_110220 | Not-determined |
| S13_extracted_110221 | Not-determined |
| S13_extracted_110222 | Not-determined |
| S13_extracted_110227 | Not-determined |
| S13_extracted_110235 | Not-determined |
| S13_extracted_110243 | Low-quality |
| S13_extracted_110276 | Not-determined |
| S13_extracted_110280 | Not-determined |
| S13_extracted_110290 | Not-determined |
| S13_extracted_110299 | Not-determined |
| S13_extracted_110303 | Low-quality |
| S13_extracted_110313 | Not-determined |
| S13_extracted_110314 | Low-quality |
| S13_extracted_110316 | Not-determined |
| S13_extracted_110329 | Not-determined |
| S13_extracted_110334 | Not-determined |
| S13_extracted_110336 | Not-determined |
| S13_extracted_110339 | Not-determined |
| S13_extracted_110345 | Not-determined |
| S13_extracted_110361 | Not-determined |
| S13_extracted_110362 | Not-determined |
| S13_extracted_110381 | Not-determined |
| S13_extracted_110396 | Not-determined |
| S13_extracted_110401 | Not-determined |
| S13_extracted_110404 | Not-determined |
| S13_extracted_110408 | Not-determined |
| S13_extracted_110423 | Not-determined |
| S13_extracted_110433 | Not-determined |
| S13_extracted_110442 | Not-determined |
| S13_extracted_110454 | Not-determined |
| S13_extracted_110462 | Not-determined |
| S13_extracted_110466 | Not-determined |
| S13_extracted_110471 | Not-determined |
| S13_extracted_110499 | Not-determined |
| S13_extracted_110501 | Not-determined |
| S13_extracted_110508 | Not-determined |

| | |
|---|---|
| S13_extracted_110513 | Not-determined |
| S13_extracted_110536 | Not-determined |
| S13_extracted_110541 | Low-quality |
| S13_extracted_110555 | Not-determined |
| S13_extracted_110556 | Low-quality |
| S13_extracted_110557 | Not-determined |
| S13_extracted_110565 | Not-determined |
| S13_extracted_110578 | Not-determined |
| S13_extracted_110592 | Not-determined |
| S13_extracted_110593 | Not-determined |
| S13_extracted_110596 | Not-determined |
| S13_extracted_110601 | Not-determined |
| S13_extracted_110625 | Not-determined |
| S13_extracted_110641 | Not-determined |
| S13_extracted_110644 | Not-determined |
| S13_extracted_110662 | Not-determined |
| S13_extracted_110670 | Not-determined |
| S13_extracted_110672 | Not-determined |
| S13_extracted_110682 | Not-determined |
| S13_extracted_110695 | Not-determined |
| S13_extracted_110696 | Not-determined |
| S13_extracted_110701 | Not-determined |
| S13_extracted_110706 | Not-determined |
| S13_extracted_110708 | Not-determined |
| S13_extracted_110710 | Not-determined |
| S13_extracted_110720 | Not-determined |
| S13_extracted_110727 | Low-quality |
| S13_extracted_110728 | Not-determined |
| S13_extracted_110747 | Not-determined |
| S13_extracted_110748 | Low-quality |
| S13_extracted_110752 | Not-determined |
| S13_extracted_110753 | Not-determined |
| S13_extracted_110756 | Low-quality |
| S13_extracted_110763 | Low-quality |
| S13_extracted_110773 | Not-determined |
| S13_extracted_110776 | Not-determined |
| S13_extracted_110795 | Not-determined |
| S13_extracted_110796 | Low-quality |
| S13_extracted_110808 | Not-determined |
| S13_extracted_110812 | Not-determined |
| S13_extracted_110816 | Low-quality |
| S13_extracted_110822 | Not-determined |
| S13_extracted_110833 | Not-determined |
| S13_extracted_110845 | Not-determined |
| S13_extracted_110855 | Not-determined |
| S13_extracted_110860 | Not-determined |
| S13_extracted_110862 | Not-determined |
| S13_extracted_110872 | Not-determined |
| S13_extracted_110874 | Not-determined |
| S13_extracted_110880 | Not-determined |
| S13_extracted_110881 | Not-determined |
| S13_extracted_110882 | Not-determined |
| S13_extracted_110887 | Not-determined |
| S13_extracted_110888 | Not-determined |
| S13_extracted_110893 | Not-determined |
| S13_extracted_110897 | Not-determined |
| S13_extracted_110911 | Not-determined |
| S13_extracted_110920 | Not-determined |
| S13_extracted_110929 | Not-determined |
| S13_extracted_110933 | Not-determined |
| S13_extracted_110938 | Not-determined |
| S13_extracted_110940 | Not-determined |
| S13_extracted_110945 | Low-quality |
| S13_extracted_110955 | Not-determined |
| S13_extracted_110962 | Low-quality |
| S13_extracted_110989 | Low-quality |
| S13_extracted_111007 | Not-determined |
| S13_extracted_111012 | Not-determined |
| S13_extracted_111016 | Not-determined |
| S13_extracted_111024 | Not-determined |
| S13_extracted_111028 | Not-determined |
| S13_extracted_111038 | Not-determined |
| S13_extracted_111042 | Not-determined |
| S13_extracted_111045 | Not-determined |
| S13_extracted_111050 | Not-determined |
| S13_extracted_111056 | Low-quality |
| S13_extracted_111057 | Low-quality |
| S13_extracted_111085 | Not-determined |
| S13_extracted_111086 | Low-quality |
| S13_extracted_111097 | Not-determined |
| S13_extracted_111104 | Not-determined |
| S13_extracted_111107 | Not-determined |
| S13_extracted_111109 | Not-determined |
| S13_extracted_111122 | Not-determined |
| S13_extracted_111125 | Not-determined |
| S13_extracted_111129 | Not-determined |
| S13_extracted_111161 | Low-quality |
| S13_extracted_111170 | Not-determined |
| S13_extracted_111173 | Low-quality |
| S13_extracted_111184 | Not-determined |
| S13_extracted_111189 | Low-quality |
| S13_extracted_111192 | Not-determined |
| S13_extracted_111193 | Low-quality |
| S13_extracted_111197 | Not-determined |
| S13_extracted_111207 | Not-determined |
| S13_extracted_111223 | Not-determined |
| S13_extracted_111227 | Not-determined |
| S13_extracted_111232 | Not-determined |
| S13_extracted_111234 | Not-determined |
| S13_extracted_111247 | Low-quality |
| S13_extracted_111269 | Not-determined |
| S13_extracted_111284 | Low-quality |
| S13_extracted_111287 | Not-determined |
| S13_extracted_111293 | Not-determined |
| S13_extracted_111295 | Not-determined |
| S13_extracted_111296 | Not-determined |
| S13_extracted_111306 | Not-determined |
| S13_extracted_111327 | Not-determined |
| S13_extracted_111334 | Not-determined |
| S13_extracted_111353 | Not-determined |
| S13_extracted_111355 | Low-quality |
| S13_extracted_111364 | Not-determined |
| S13_extracted_111366 | Not-determined |
| S13_extracted_111368 | Not-determined |
| S13_extracted_111390 | Not-determined |
| S13_extracted_111402 | Not-determined |
| S13_extracted_111415 | Not-determined |
| S13_extracted_111419 | Not-determined |
| S13_extracted_111420 | Not-determined |
| S13_extracted_111431 | Not-determined |
| S13_extracted_111432 | Not-determined |
| S13_extracted_111438 | Not-determined |
| S13_extracted_111443 | Not-determined |
| S13_extracted_111480 | Not-determined |
| S13_extracted_111491 | Low-quality |
| S13_extracted_111492 | Low-quality |
| S13_extracted_111501 | Not-determined |
| S13_extracted_111506 | Not-determined |
| S13_extracted_111512 | Not-determined |
| S13_extracted_111515 | Not-determined |
| S13_extracted_111527 | Not-determined |
| S13_extracted_111539 | Not-determined |
| S13_extracted_111549 | Not-determined |
| S13_extracted_111555 | Not-determined |
| S13_extracted_111556 | Not-determined |
| S13_extracted_111557 | Not-determined |
| S13_extracted_111563 | Not-determined |
| S13_extracted_111565 | Not-determined |
| S13_extracted_111566 | Not-determined |
| S13_extracted_111578 | Not-determined |
| S13_extracted_111589 | Not-determined |
| S13_extracted_111593 | Not-determined |
| S13_extracted_111596 | Low-quality |
| S13_extracted_111625 | Not-determined |
| S13_extracted_111626 | Not-determined |
| S13_extracted_111630 | Not-determined |
| S13_extracted_111634 | Not-determined |
| S13_extracted_111671 | Not-determined |
| S13_extracted_111675 | Not-determined |
| S13_extracted_111676 | Not-determined |
| S13_extracted_111683 | Not-determined |
| S13_extracted_111699 | Not-determined |
| S13_extracted_111716 | Not-determined |
| S13_extracted_111774 | Not-determined |
| S13_extracted_111775 | Not-determined |
| S13_extracted_111796 | Not-determined |
| S13_extracted_111802 | Not-determined |
| S13_extracted_111804 | Not-determined |
| S13_extracted_111819 | Not-determined |
| S13_extracted_111828 | Not-determined |
| S13_extracted_111832 | Not-determined |
| S13_extracted_111841 | Not-determined |
| S13_extracted_111846 | Not-determined |
| S13_extracted_111857 | Not-determined |
| S13_extracted_111860 | Not-determined |
| S13_extracted_111865 | Not-determined |
| S13_extracted_111871 | Not-determined |
| S13_extracted_111876 | Not-determined |
| S13_extracted_111903 | Not-determined |
| S13_extracted_111907 | Not-determined |
| S13_extracted_111914 | Not-determined |
| S13_extracted_111927 | Low-quality |
| S13_extracted_111929 | Not-determined |
| S13_extracted_111944 | Not-determined |
| S13_extracted_111960 | Not-determined |
| S13_extracted_112004 | Not-determined |
| S13_extracted_112023 | Not-determined |
| S13_extracted_112028 | Low-quality |
| S13_extracted_112030 | Not-determined |
| S13_extracted_112047 | Not-determined |
| S13_extracted_112061 | Low-quality |
| S13_extracted_112063 | Not-determined |
| S13_extracted_112067 | Not-determined |
| S13_extracted_112073 | Not-determined |
| S13_extracted_112080 | Not-determined |
| S13_extracted_112086 | Not-determined |
| S13_extracted_112096 | Not-determined |
| S13_extracted_112130 | Not-determined |
| S13_extracted_112139 | Not-determined |
| S13_extracted_112141 | Not-determined |
| S13_extracted_112155 | Not-determined |
| S13_extracted_112156 | Not-determined |
| S13_extracted_112160 | Low-quality |
| S13_extracted_112161 | Not-determined |
| S13_extracted_112165 | Not-determined |
| S13_extracted_112172 | Not-determined |
| S13_extracted_112175 | Not-determined |
| S13_extracted_112178 | Not-determined |
| S13_extracted_112186 | Not-determined |
| S13_extracted_112190 | Not-determined |
| S13_extracted_112196 | Not-determined |
| S13_extracted_112203 | Not-determined |
| S13_extracted_112215 | Not-determined |
| S13_extracted_112218 | Not-determined |
| S13_extracted_112222 | Not-determined |
| S13_extracted_112240 | Not-determined |
| S13_extracted_112242 | Not-determined |
| S13_extracted_112252 | Not-determined |
| S13_extracted_112272 | Not-determined |
| S13_extracted_112283 | Not-determined |
| S13_extracted_112286 | Not-determined |
| S13_extracted_112290 | Not-determined |
| S13_extracted_112299 | Low-quality |
| S13_extracted_112305 | Not-determined |
| S13_extracted_112311 | Not-determined |
| S13_extracted_112326 | Not-determined |
| S13_extracted_112347 | Not-determined |
| S13_extracted_112388 | Not-determined |
| S13_extracted_112397 | Not-determined |
| S13_extracted_112411 | Not-determined |
| S13_extracted_112423 | Not-determined |
| S13_extracted_112440 | Not-determined |
| S13_extracted_112446 | Not-determined |
| S13_extracted_112452 | Not-determined |
| S13_extracted_112482 | Not-determined |
| S13_extracted_112484 | Not-determined |
| S13_extracted_112491 | Not-determined |
| S13_extracted_112494 | Not-determined |
| S13_extracted_112499 | Not-determined |
| S13_extracted_112500 | Not-determined |
| S13_extracted_112525 | Not-determined |
| S13_extracted_112539 | Low-quality |
| S13_extracted_112546 | Not-determined |
| S13_extracted_112555 | Not-determined |
| S13_extracted_112560 | Low-quality |
| S13_extracted_112561 | Not-determined |
| S13_extracted_112574 | Not-determined |
| S13_extracted_112587 | Low-quality |
| S13_extracted_112589 | Not-determined |
| S13_extracted_112594 | Not-determined |
| S13_extracted_112600 | Not-determined |
| S13_extracted_112617 | Low-quality |
| S13_extracted_112622 | Low-quality |
| S13_extracted_112625 | Not-determined |
| S13_extracted_112629 | Not-determined |
| S13_extracted_112641 | Not-determined |
| S13_extracted_112642 | Not-determined |
| S13_extracted_112645 | Not-determined |
| S13_extracted_112657 | Not-determined |
| S13_extracted_112661 | Not-determined |
| S13_extracted_112665 | Not-determined |
| S13_extracted_112684 | Not-determined |
| S13_extracted_112696 | Not-determined |
| S13_extracted_112703 | Not-determined |
| S13_extracted_112710 | Low-quality |
| S13_extracted_112715 | Not-determined |
| S13_extracted_112728 | Not-determined |
| S13_extracted_112731 | Not-determined |
| S13_extracted_112736 | Not-determined |
| S13_extracted_112748 | Not-determined |
| S13_extracted_112750 | Not-determined |
| S13_extracted_112757 | Not-determined |
| S13_extracted_112778 | Not-determined |
| S13_extracted_112780 | Not-determined |
| S13_extracted_112790 | Not-determined |
| S13_extracted_112801 | Not-determined |
| S13_extracted_112810 | Not-determined |
| S13_extracted_112829 | Not-determined |
| S13_extracted_112834 | Not-determined |
| S13_extracted_112844 | Not-determined |
| S13_extracted_112847 | Not-determined |
| S13_extracted_112849 | Not-determined |
| S13_extracted_112852 | Not-determined |
| S13_extracted_112854 | Not-determined |
| S13_extracted_112860 | Not-determined |
| S13_extracted_112874 | Not-determined |
| S13_extracted_112883 | Not-determined |
| S13_extracted_112887 | Not-determined |
| S13_extracted_112889 | Not-determined |
| S13_extracted_112892 | Not-determined |
| S13_extracted_112900 | Not-determined |
| S13_extracted_112907 | Not-determined |
| S13_extracted_112908 | Not-determined |
| S13_extracted_112913 | Not-determined |
| S13_extracted_112914 | Not-determined |
| S13_extracted_112935 | Not-determined |
| S13_extracted_112940 | Not-determined |
| S13_extracted_112942 | Low-quality |
| S13_extracted_112953 | Not-determined |
| S13_extracted_112970 | Not-determined |
| S13_extracted_112985 | Not-determined |
| S13_extracted_112994 | Not-determined |
| S13_extracted_112996 | Not-determined |
| S13_extracted_113009 | Not-determined |
| S13_extracted_113011 | Not-determined |
| S13_extracted_113014 | Low-quality |
| S13_extracted_113030 | Not-determined |
| S13_extracted_113034 | Not-determined |
| S13_extracted_113039 | Not-determined |
| S13_extracted_113045 | Not-determined |
| S13_extracted_113062 | Not-determined |
| S13_extracted_113063 | Not-determined |
| S13_extracted_113067 | Not-determined |
| S13_extracted_113072 | Not-determined |
| S13_extracted_113084 | Not-determined |
| S13_extracted_113113 | Not-determined |
| S13_extracted_113142 | Not-determined |
| S13_extracted_113144 | Not-determined |
| S13_extracted_113153 | Not-determined |
| S13_extracted_113208 | Low-quality |
| S13_extracted_113210 | Not-determined |
| S13_extracted_113212 | Low-quality |
| S13_extracted_113221 | Not-determined |
| S13_extracted_113222 | Not-determined |
| S13_extracted_113237 | Not-determined |
| S13_extracted_113245 | Not-determined |
| S13_extracted_113257 | Low-quality |
| S13_extracted_113260 | Not-determined |
| S13_extracted_113263 | Not-determined |
| S13_extracted_113292 | Not-determined |
| S13_extracted_113298 | Low-quality |
| S13_extracted_113302 | Not-determined |
| S13_extracted_113305 | Not-determined |
| S13_extracted_113321 | Not-determined |
| S13_extracted_113347 | Not-determined |
| S13_extracted_113348 | Not-determined |
| S13_extracted_113349 | Not-determined |
| S13_extracted_113406 | Not-determined |
| S13_extracted_113407 | Not-determined |
| S13_extracted_113449 | Not-determined |
| S13_extracted_113450 | Not-determined |
| S13_extracted_113453 | Not-determined |
| S13_extracted_113454 | Not-determined |
| S13_extracted_113472 | Not-determined |
| S13_extracted_113483 | Low-quality |
| S13_extracted_113492 | Not-determined |
| S13_extracted_113513 | Not-determined |
| S13_extracted_113543 | Not-determined |
| S13_extracted_113550 | Not-determined |
| S13_extracted_113567 | Not-determined |
| S13_extracted_113573 | Not-determined |
| S13_extracted_113575 | Not-determined |
| S13_extracted_113577 | Not-determined |
| S13_extracted_113583 | Not-determined |
| S13_extracted_113584 | Low-quality |
| S13_extracted_113606 | Not-determined |
| S13_extracted_113611 | Not-determined |
| S13_extracted_113613 | Low-quality |
| S13_extracted_113631 | Low-quality |
| S13_extracted_113646 | Not-determined |
| S13_extracted_113649 | Low-quality |
| S13_extracted_113664 | Not-determined |
| S13_extracted_113671 | Not-determined |
| S13_extracted_113678 | Not-determined |
| S13_extracted_113683 | Not-determined |
| S13_extracted_113696 | Not-determined |
| S13_extracted_113698 | Not-determined |
| S13_extracted_113699 | Not-determined |
| S13_extracted_113714 | Not-determined |
| S13_extracted_113716 | Not-determined |
| S13_extracted_113722 | Not-determined |
| S13_extracted_113726 | Not-determined |
| S13_extracted_113727 | Not-determined |
| S13_extracted_113747 | Not-determined |
| S13_extracted_113749 | Not-determined |
| S13_extracted_113755 | Not-determined |
| S13_extracted_113759 | Not-determined |
| S13_extracted_113760 | Not-determined |
| S13_extracted_113764 | Not-determined |
| S13_extracted_113768 | Low-quality |
| S13_extracted_113776 | Not-determined |
| S13_extracted_113781 | Not-determined |
| S13_extracted_113794 | Not-determined |
| S13_extracted_113804 | Not-determined |
| S13_extracted_113805 | Not-determined |
| S13_extracted_113807 | Low-quality |
| S13_extracted_113828 | Low-quality |
| S13_extracted_113843 | Not-determined |
| S13_extracted_113847 | Not-determined |
| S13_extracted_113861 | Not-determined |
| S13_extracted_113864 | Not-determined |
| S13_extracted_113871 | Not-determined |
| S13_extracted_113879 | Not-determined |
| S13_extracted_113888 | Low-quality |
| S13_extracted_113890 | Not-determined |
| S13_extracted_113910 | Not-determined |
| S13_extracted_113922 | Not-determined |
| S13_extracted_113926 | Not-determined |
| S13_extracted_113931 | Not-determined |
| S13_extracted_113933 | Low-quality |
| S13_extracted_113944 | Low-quality |
| S13_extracted_113952 | Not-determined |
| S13_extracted_113958 | Not-determined |
| S13_extracted_113965 | Not-determined |
| S13_extracted_113985 | Not-determined |
| S13_extracted_113996 | Not-determined |
| S13_extracted_114007 | Not-determined |
| S13_extracted_114035 | Not-determined |
| S13_extracted_114038 | Not-determined |
| S13_extracted_114063 | Not-determined |
| S13_extracted_114078 | Not-determined |
| S13_extracted_114093 | Not-determined |
| S13_extracted_114097 | Not-determined |
| S13_extracted_114102 | Not-determined |
| S13_extracted_114127 | Not-determined |
| S13_extracted_114137 | Not-determined |
| S13_extracted_114144 | Not-determined |
| S13_extracted_114153 | Not-determined |
| S13_extracted_114161 | Not-determined |
| S13_extracted_114163 | Not-determined |
| S13_extracted_114177 | Not-determined |
| S13_extracted_114178 | Not-determined |
| S13_extracted_114183 | Not-determined |
| S13_extracted_114186 | Not-determined |
| S13_extracted_114193 | Not-determined |
| S13_extracted_114197 | Not-determined |
| S13_extracted_114198 | Not-determined |
| S13_extracted_114203 | Not-determined |
| S13_extracted_114214 | Not-determined |
| S13_extracted_114218 | Not-determined |
| S13_extracted_114221 | Not-determined |
| S13_extracted_114222 | Not-determined |
| S13_extracted_114265 | Not-determined |
| S13_extracted_114266 | Not-determined |
| S13_extracted_114290 | Not-determined |
| S13_extracted_114291 | Not-determined |
| S13_extracted_114294 | Not-determined |
| S13_extracted_114302 | Not-determined |
| S13_extracted_114304 | Not-determined |
| S13_extracted_114324 | Not-determined |
| S13_extracted_114343 | Not-determined |
| S13_extracted_114362 | Not-determined |
| S13_extracted_114365 | Not-determined |
| S13_extracted_114392 | Low-quality |
| S13_extracted_114402 | Not-determined |
| S13_extracted_114421 | Not-determined |
| S13_extracted_114423 | Low-quality |
| S13_extracted_114425 | Not-determined |
| S13_extracted_114444 | Not-determined |
| S13_extracted_114447 | Not-determined |
| S13_extracted_114455 | Not-determined |
| S13_extracted_114459 | Not-determined |
| S13_extracted_114464 | Not-determined |
| S13_extracted_114468 | Not-determined |
| S13_extracted_114474 | Not-determined |
| S13_extracted_114477 | Not-determined |
| S13_extracted_114480 | Not-determined |
| S13_extracted_114483 | Not-determined |
| S13_extracted_114490 | Not-determined |
| S13_extracted_114496 | Not-determined |
| S13_extracted_114513 | Not-determined |
| S13_extracted_114526 | Low-quality |
| SG_vs_k141_54606\|\|full | Medium-quality |
| S13_extracted_114549 | Not-determined |
| S13_extracted_114557 | Not-determined |
| S13_extracted_114558 | Not-determined |
| S13_extracted_114574 | Not-determined |
| S13_extracted_114581 | Not-determined |
| S13_extracted_114588 | Not-determined |
| S13_extracted_114591 | Not-determined |
| S13_extracted_114600 | Not-determined |
| S13_extracted_114613 | Low-quality |
| S13_extracted_114617 | Low-quality |
| S13_extracted_114620 | Low-quality |
| S13_extracted_114634 | Not-determined |
| S13_extracted_114638 | Not-determined |
| S13_extracted_114639 | Not-determined |
| S13_extracted_114641 | Not-determined |
| S13_extracted_114644 | Not-determined |
| S13_extracted_114650 | Not-determined |
| S13_extracted_114660 | Not-determined |
| S13_extracted_114675 | Not-determined |
| S13_extracted_114685 | Not-determined |
| S13_extracted_114694 | Not-determined |
| S13_extracted_114695 | Not-determined |
| S13_extracted_114712 | Not-determined |
| S13_extracted_114720 | Not-determined |
| S13_extracted_114735 | Not-determined |
| S13_extracted_114739 | Not-determined |
| S13_extracted_114745 | Not-determined |
| S13_extracted_114764 | Not-determined |
| S13_extracted_114766 | Not-determined |
| S13_extracted_114768 | Not-determined |
| S13_extracted_114785 | Not-determined |
| S13_extracted_114788 | Not-determined |
| S13_extracted_114796 | Low-quality |
| S13_extracted_114799 | Not-determined |
| S13_extracted_114801 | Not-determined |
| S13_extracted_114821 | Not-determined |
| S13_extracted_114835 | Not-determined |
| S13_extracted_114836 | Not-determined |
| S13_extracted_114839 | Not-determined |
| S13_extracted_114841 | Low-quality |
| S13_extracted_114846 | Not-determined |
| S13_extracted_114856 | Not-determined |
| S13_extracted_114858 | Not-determined |
| S13_extracted_114876 | Not-determined |
| S13_extracted_114894 | Not-determined |
| S13_extracted_114900 | Not-determined |
| S13_extracted_114905 | Not-determined |
| S13_extracted_114911 | Not-determined |
| S13_extracted_114930 | Not-determined |
| S13_extracted_114931 | Not-determined |
| S13_extracted_114934 | Not-determined |
| S13_extracted_114935 | Not-determined |
| S13_extracted_114948 | Not-determined |
| S13_extracted_114953 | Low-quality |
| S13_extracted_114965 | Not-determined |
| S13_extracted_114984 | Not-determined |
| S13_extracted_114991 | Not-determined |
| S13_extracted_115007 | Not-determined |
| S13_extracted_115014 | Not-determined |
| S13_extracted_115016 | Not-determined |
| S13_extracted_115018 | Not-determined |
| S13_extracted_115019 | Not-determined |
| S13_extracted_115021 | Not-determined |
| S13_extracted_115028 | Low-quality |
| S13_extracted_115036 | Not-determined |
| S13_extracted_115044 | Low-quality |
| S13_extracted_115047 | Not-determined |
| S13_extracted_115052 | Not-determined |
| S13_extracted_115064 | Not-determined |
| S13_extracted_115081 | Not-determined |
| S13_extracted_115092 | Not-determined |
| S13_extracted_115106 | Not-determined |
| S13_extracted_115120 | Not-determined |
| S13_extracted_115131 | Not-determined |
| S13_extracted_115144 | Low-quality |
| S13_extracted_115159 | Not-determined |
| S13_extracted_115164 | Not-determined |
| S13_extracted_115167 | Not-determined |
| S13_extracted_115170 | Not-determined |
| S13_extracted_115173 | Not-determined |
| S13_extracted_115174 | Not-determined |
| S13_extracted_115204 | Not-determined |
| S13_extracted_115207 | Low-quality |
| S13_extracted_115208 | Not-determined |
| S13_extracted_115216 | Not-determined |
| S13_extracted_115229 | Not-determined |
| S13_extracted_115238 | Low-quality |
| S13_extracted_115240 | Not-determined |
| S13_extracted_115252 | Not-determined |
| S13_extracted_115272 | Not-determined |
| S13_extracted_115275 | Not-determined |
| S13_extracted_115277 | Not-determined |
| S13_extracted_115293 | Not-determined |
| S13_extracted_115298 | Low-quality |
| S13_extracted_115303 | Not-determined |
| S13_extracted_115307 | Not-determined |
| S13_extracted_115314 | Not-determined |
| S13_extracted_115332 | Not-determined |
| S13_extracted_115342 | Not-determined |
| S13_extracted_115348 | Not-determined |
| S13_extracted_115351 | Not-determined |
| S13_extracted_115352 | Low-quality |
| S13_extracted_115364 | Not-determined |
| S13_extracted_115369 | Not-determined |
| S13_extracted_115373 | Not-determined |
| S13_extracted_115391 | Not-determined |
| S13_extracted_115393 | Not-determined |
| S13_extracted_115395 | Low-quality |
| S13_extracted_115400 | Not-determined |
| S13_extracted_115403 | Not-determined |
| S13_extracted_115405 | Not-determined |
| S13_extracted_115409 | Not-determined |
| S13_extracted_115411 | Not-determined |
| S13_extracted_115423 | Low-quality |
| S13_extracted_115424 | Not-determined |
| S13_extracted_115433 | Not-determined |
| S13_extracted_115440 | Not-determined |
| S13_extracted_115448 | Not-determined |
| S13_extracted_115454 | Low-quality |
| S13_extracted_115461 | Low-quality |
| S13_extracted_115469 | Not-determined |
| S13_extracted_115483 | Not-determined |
| S13_extracted_115501 | Not-determined |
| S13_extracted_115502 | Not-determined |
| S13_extracted_115505 | Not-determined |
| S13_extracted_115520 | Not-determined |
| S13_extracted_115533 | Not-determined |
| S13_extracted_115539 | Not-determined |
| S13_extracted_115560 | Not-determined |
| S13_extracted_115561 | Low-quality |
| S13_extracted_115575 | Not-determined |
| S13_extracted_115577 | Not-determined |
| S13_extracted_115578 | Not-determined |
| S13_extracted_115599 | Low-quality |
| S13_extracted_115600 | Not-determined |
| S13_extracted_115634 | Not-determined |

| | |
|---|---|
| S13_extracted_115656 | Not-determined |
| S13_extracted_115669 | Not-determined |
| S13_extracted_115674 | Not-determined |
| S13_extracted_115692 | Not-determined |
| S13_extracted_115696 | Not-determined |
| S13_extracted_115698 | Not-determined |
| S13_extracted_115707 | Not-determined |
| S13_extracted_115719 | Not-determined |
| S13_extracted_115727 | Not-determined |
| S13_extracted_115728 | Not-determined |
| S13_extracted_115738 | Not-determined |
| S13_extracted_115742 | Not-determined |
| S13_extracted_115743 | Not-determined |
| S13_extracted_115757 | Not-determined |
| S13_extracted_115767 | Not-determined |
| S13_extracted_115776 | Not-determined |
| S13_extracted_115792 | Not-determined |
| S13_extracted_115795 | Not-determined |
| S13_extracted_115804 | Not-determined |
| S13_extracted_115805 | Not-determined |
| S13_extracted_115818 | Not-determined |
| S13_extracted_115828 | Not-determined |
| S13_extracted_115839 | Not-determined |
| S13_extracted_115858 | Low-quality |
| S13_extracted_115865 | Not-determined |
| S13_extracted_115873 | Not-determined |
| S13_extracted_115898 | Not-determined |
| S13_extracted_115915 | Low-quality |
| S13_extracted_115928 | Not-determined |
| S13_extracted_115929 | Low-quality |
| S13_extracted_115964 | Not-determined |
| S13_extracted_115970 | Not-determined |
| S13_extracted_115972 | Not-determined |
| S13_extracted_115991 | Not-determined |
| S13_extracted_115997 | Low-quality |
| S13_extracted_116007 | Not-determined |
| S13_extracted_116013 | Not-determined |
| S13_extracted_116022 | Not-determined |
| S13_extracted_116031 | Not-determined |
| S13_extracted_116038 | Not-determined |
| S13_extracted_116043 | Not-determined |
| S13_extracted_116067 | Not-determined |
| S13_extracted_116071 | Not-determined |
| S13_extracted_116075 | Not-determined |
| S13_extracted_116079 | Not-determined |
| S13_extracted_116084 | Not-determined |
| S13_extracted_116095 | Not-determined |
| S13_extracted_116107 | Low-quality |
| S13_extracted_116109 | Not-determined |
| S13_extracted_116117 | Not-determined |
| S13_extracted_116119 | Not-determined |
| S13_extracted_116129 | Low-quality |
| S13_extracted_116132 | Not-determined |
| S13_extracted_116150 | Not-determined |
| S13_extracted_116155 | Low-quality |
| S13_extracted_116158 | Not-determined |
| S13_extracted_116162 | Not-determined |
| S13_extracted_116171 | Not-determined |
| S13_extracted_116175 | Not-determined |
| S13_extracted_116179 | Not-determined |
| S13_extracted_116180 | Not-determined |
| S13_extracted_116181 | Not-determined |
| S13_extracted_116184 | Low-quality |
| S13_extracted_116190 | Not-determined |
| S13_extracted_116193 | Not-determined |
| S13_extracted_116194 | Not-determined |
| S13_extracted_116196 | Not-determined |
| S13_extracted_116210 | Low-quality |
| S13_extracted_116217 | Not-determined |
| S13_extracted_116221 | Not-determined |
| S13_extracted_116223 | Not-determined |
| S13_extracted_116234 | Not-determined |
| S13_extracted_116260 | Not-determined |
| S13_extracted_116261 | Not-determined |
| S13_extracted_116265 | Not-determined |
| S13_extracted_116267 | Not-determined |
| S13_extracted_116278 | Not-determined |
| S13_extracted_116279 | Not-determined |
| S13_extracted_116289 | Not-determined |
| S13_extracted_116298 | Not-determined |
| S13_extracted_116306 | Not-determined |
| S13_extracted_116309 | Low-quality |
| S13_extracted_116311 | Not-determined |
| S13_extracted_116318 | Not-determined |
| S13_extracted_116327 | Not-determined |
| S13_extracted_116329 | Not-determined |
| S13_extracted_116333 | Not-determined |
| S13_extracted_116344 | Not-determined |
| S13_extracted_116356 | Not-determined |
| S13_extracted_116361 | Not-determined |
| S13_extracted_116374 | Not-determined |
| S13_extracted_116383 | Not-determined |
| S13_extracted_116424 | Not-determined |
| S13_extracted_116425 | Not-determined |
| S13_extracted_116457 | Not-determined |
| S13_extracted_116472 | Not-determined |
| S13_extracted_116490 | Not-determined |
| S13_extracted_116493 | Not-determined |
| S13_extracted_116495 | Not-determined |
| S13_extracted_116496 | Low-quality |
| S13_extracted_116511 | Not-determined |
| S13_extracted_116526 | Not-determined |
| S13_extracted_116533 | Not-determined |
| S13_extracted_116538 | Not-determined |
| S13_extracted_116559 | Not-determined |
| S13_extracted_116570 | Low-quality |
| S13_extracted_116571 | Not-determined |
| S13_extracted_116586 | Not-determined |
| S13_extracted_116593 | Not-determined |
| S13_extracted_116595 | Not-determined |
| S13_extracted_116600 | Not-determined |
| S13_extracted_116622 | Not-determined |
| S13_extracted_116630 | Not-determined |
| S13_extracted_116643 | Not-determined |
| S13_extracted_116649 | Not-determined |
| S13_extracted_116650 | Not-determined |
| S13_extracted_116654 | Not-determined |
| S13_extracted_116655 | Low-quality |
| S13_extracted_116667 | Not-determined |
| S13_extracted_116671 | Not-determined |
| S13_extracted_116681 | Not-determined |
| S13_extracted_116698 | Not-determined |
| S13_extracted_116711 | Not-determined |
| S13_extracted_116722 | Not-determined |
| S13_extracted_116731 | Not-determined |
| S13_extracted_116734 | Not-determined |
| S13_extracted_116740 | Not-determined |
| S13_extracted_116772 | Not-determined |
| S13_extracted_116798 | Not-determined |
| S13_extracted_116799 | Not-determined |
| S13_extracted_116814 | Not-determined |
| S13_extracted_116816 | Not-determined |
| S13_extracted_116817 | Not-determined |
| S13_extracted_116820 | Not-determined |
| S13_extracted_116825 | Not-determined |
| S13_extracted_116828 | Not-determined |
| S13_extracted_116830 | Not-determined |
| S13_extracted_116840 | Not-determined |
| S13_extracted_116851 | Low-quality |
| S13_extracted_116857 | Not-determined |
| S13_extracted_116865 | Not-determined |
| S13_extracted_116872 | Not-determined |
| S13_extracted_116875 | Not-determined |
| S13_extracted_116876 | Not-determined |
| S13_extracted_116890 | Low-quality |
| S13_extracted_116894 | Not-determined |
| S13_extracted_116895 | Not-determined |
| S13_extracted_116901 | Not-determined |
| S13_extracted_116903 | Not-determined |
| S13_extracted_116926 | Not-determined |
| S13_extracted_116935 | Not-determined |
| S13_extracted_116949 | Low-quality |
| S13_extracted_117009 | Not-determined |
| S13_extracted_117016 | Not-determined |
| S13_extracted_117033 | Not-determined |
| S13_extracted_117035 | Not-determined |
| S13_extracted_117058 | Low-quality |
| S13_extracted_117073 | Not-determined |
| S13_extracted_117074 | Not-determined |
| S13_extracted_117088 | Not-determined |
| S13_extracted_117093 | Not-determined |
| S13_extracted_117094 | Not-determined |
| S13_extracted_117097 | Not-determined |
| S13_extracted_117103 | Not-determined |
| S13_extracted_117108 | Not-determined |
| S13_extracted_117109 | Not-determined |
| S13_extracted_117127 | Not-determined |
| S13_extracted_117145 | Not-determined |
| S13_extracted_117146 | Not-determined |
| S13_extracted_117153 | Not-determined |
| S13_extracted_117167 | Not-determined |
| S13_extracted_117175 | Not-determined |
| S13_extracted_117189 | Not-determined |
| S13_extracted_117190 | Not-determined |
| S13_extracted_117197 | Not-determined |
| S13_extracted_117212 | Not-determined |
| S13_extracted_117217 | Not-determined |
| S13_extracted_117218 | Not-determined |
| S13_extracted_117237 | Not-determined |
| S13_extracted_117244 | Not-determined |
| S13_extracted_117246 | Not-determined |
| S13_extracted_117256 | Not-determined |
| S13_extracted_117283 | Not-determined |
| S13_extracted_117292 | Not-determined |
| S13_extracted_117300 | Not-determined |
| S13_extracted_117307 | Low-quality |
| S13_extracted_117310 | Not-determined |
| S13_extracted_117315 | Not-determined |
| S13_extracted_117318 | Not-determined |
| S13_extracted_117327 | Not-determined |
| S13_extracted_117335 | Not-determined |
| S13_extracted_117337 | Not-determined |
| S13_extracted_117346 | Not-determined |
| S13_extracted_117360 | Not-determined |
| S13_extracted_117364 | Not-determined |
| S13_extracted_117367 | Low-quality |
| S13_extracted_117372 | Not-determined |
| S13_extracted_117390 | Not-determined |
| S13_extracted_117392 | Not-determined |
| S13_extracted_117393 | Not-determined |
| S13_extracted_117434 | Not-determined |
| S13_extracted_117452 | Not-determined |
| S13_extracted_117454 | Not-determined |
| S13_extracted_117456 | Not-determined |
| S13_extracted_117463 | Not-determined |
| S13_extracted_117467 | Not-determined |
| S13_extracted_117477 | Low-quality |
| S13_extracted_117501 | Not-determined |
| S13_extracted_117503 | Not-determined |
| S13_extracted_117509 | Not-determined |
| S13_extracted_117530 | Not-determined |
| S13_extracted_117546 | Not-determined |
| S13_extracted_117549 | Not-determined |
| S13_extracted_117561 | Not-determined |
| S13_extracted_117567 | Not-determined |
| S13_extracted_117569 | Not-determined |
| S13_extracted_117580 | Not-determined |
| S13_extracted_117581 | Not-determined |
| S13_extracted_117585 | Low-quality |
| S13_extracted_117586 | Not-determined |
| S13_extracted_117590 | Not-determined |
| S13_extracted_117596 | Not-determined |
| S13_extracted_117597 | Not-determined |
| S13_extracted_117598 | Not-determined |
| S13_extracted_117599 | Low-quality |
| S13_extracted_117610 | Not-determined |
| S13_extracted_117616 | Not-determined |
| S13_extracted_117627 | Not-determined |
| S13_extracted_117643 | Not-determined |
| S13_extracted_117648 | Not-determined |
| S13_extracted_117658 | Not-determined |
| S13_extracted_117670 | Not-determined |
| S13_extracted_117694 | Low-quality |
| S13_extracted_117715 | Not-determined |
| S13_extracted_117719 | Not-determined |
| S13_extracted_117736 | Not-determined |
| S13_extracted_117737 | Not-determined |
| S13_extracted_117742 | Not-determined |
| S13_extracted_117764 | Not-determined |
| S13_extracted_117771 | Not-determined |
| S13_extracted_117772 | Not-determined |
| S13_extracted_117790 | Not-determined |
| S13_extracted_117791 | Not-determined |
| S13_extracted_117801 | Low-quality |
| S13_extracted_117802 | Not-determined |
| S13_extracted_117815 | Not-determined |
| S13_extracted_117841 | Not-determined |
| S13_extracted_117843 | Not-determined |
| S13_extracted_117849 | Not-determined |
| S13_extracted_117865 | Not-determined |
| S13_extracted_117880 | Not-determined |
| S13_extracted_117881 | Not-determined |
| S13_extracted_117887 | Not-determined |
| S13_extracted_117893 | Not-determined |
| S13_extracted_117899 | Not-determined |
| S13_extracted_117930 | Not-determined |
| S13_extracted_117947 | Not-determined |
| S13_extracted_117985 | Not-determined |
| S13_extracted_118001 | Not-determined |
| S13_extracted_118003 | Not-determined |
| S13_extracted_118017 | Not-determined |
| S13_extracted_118037 | Not-determined |
| S13_extracted_118044 | Not-determined |
| S13_extracted_118045 | Not-determined |
| S13_extracted_118047 | Not-determined |
| S13_extracted_118049 | Not-determined |
| S13_extracted_118053 | Low-quality |
| S13_extracted_118075 | Not-determined |
| S13_extracted_118092 | Not-determined |
| S13_extracted_118102 | Not-determined |
| S13_extracted_118104 | Not-determined |
| S13_extracted_118108 | Not-determined |
| S13_extracted_118129 | Not-determined |
| S13_extracted_118147 | Not-determined |
| S13_extracted_118173 | Not-determined |
| S13_extracted_118178 | Not-determined |
| S13_extracted_118184 | Not-determined |
| S13_extracted_118188 | Not-determined |
| S13_extracted_118201 | Not-determined |
| S13_extracted_118204 | Low-quality |
| S13_extracted_118206 | Not-determined |
| S13_extracted_118213 | Not-determined |
| S13_extracted_118219 | Not-determined |
| S13_extracted_118223 | Not-determined |
| S13_extracted_118227 | Not-determined |
| S13_extracted_118269 | Not-determined |
| S13_extracted_118271 | Not-determined |
| S13_extracted_118280 | Not-determined |
| S13_extracted_118290 | Not-determined |
| S13_extracted_118303 | Low-quality |
| S13_extracted_118305 | Not-determined |
| S13_extracted_118326 | Not-determined |
| S13_extracted_118328 | Not-determined |
| S13_extracted_118351 | Not-determined |
| S13_extracted_118352 | Not-determined |
| S13_extracted_118359 | Not-determined |
| S13_extracted_118362 | Not-determined |
| S13_extracted_118366 | Not-determined |
| S13_extracted_118373 | Not-determined |
| S13_extracted_118376 | Not-determined |
| S13_extracted_118398 | Not-determined |
| S13_extracted_118405 | Not-determined |
| S13_extracted_118407 | Not-determined |
| S13_extracted_118412 | Not-determined |
| S13_extracted_118415 | Not-determined |
| S13_extracted_118429 | Not-determined |
| S13_extracted_118436 | Not-determined |
| S13_extracted_118450 | Not-determined |
| S13_extracted_118453 | Not-determined |
| S13_extracted_118458 | Not-determined |
| S13_extracted_118468 | Low-quality |
| S13_extracted_118480 | Not-determined |
| S13_extracted_118481 | Not-determined |
| S13_extracted_118485 | Low-quality |
| S13_extracted_118501 | Not-determined |
| S13_extracted_118516 | Not-determined |
| S13_extracted_118524 | Not-determined |
| S13_extracted_118534 | Not-determined |
| S13_extracted_118536 | Not-determined |
| S13_extracted_118538 | Not-determined |
| S13_extracted_118539 | Not-determined |
| S13_extracted_118543 | Not-determined |
| S13_extracted_118547 | Low-quality |
| S13_extracted_118548 | Low-quality |
| S13_extracted_118560 | Not-determined |
| S13_extracted_118563 | Not-determined |
| S13_extracted_118564 | Not-determined |
| S13_extracted_118568 | Not-determined |
| S13_extracted_118569 | Not-determined |
| S13_extracted_118572 | Not-determined |
| S13_extracted_118583 | Not-determined |
| S13_extracted_118610 | Not-determined |
| S13_extracted_118613 | Not-determined |
| S13_extracted_118626 | Not-determined |
| S13_extracted_118632 | Not-determined |
| S13_extracted_118638 | Not-determined |
| S13_extracted_118640 | Not-determined |
| S13_extracted_118661 | Not-determined |
| S13_extracted_118668 | Not-determined |
| S13_extracted_118672 | Not-determined |
| S13_extracted_118679 | Not-determined |
| S13_extracted_118691 | Not-determined |
| S13_extracted_118707 | Low-quality |
| S13_extracted_118708 | Not-determined |
| S13_extracted_118713 | Not-determined |
| S13_extracted_118753 | Not-determined |
| S13_extracted_118764 | Not-determined |
| S13_extracted_118791 | Not-determined |
| S13_extracted_118798 | Not-determined |
| S13_extracted_118803 | Not-determined |
| S13_extracted_118811 | Not-determined |
| S13_extracted_118813 | Low-quality |
| S13_extracted_118818 | Not-determined |
| S13_extracted_118820 | Not-determined |
| S13_extracted_118824 | Not-determined |
| S13_extracted_118833 | Not-determined |
| S13_extracted_118836 | Not-determined |
| S13_extracted_118864 | Not-determined |
| S13_extracted_118865 | Not-determined |
| S13_extracted_118877 | Not-determined |
| S13_extracted_118882 | Not-determined |
| S13_extracted_118883 | Not-determined |
| S13_extracted_118890 | Not-determined |
| S13_extracted_118894 | Not-determined |
| S13_extracted_118912 | Not-determined |
| S13_extracted_118920 | Not-determined |
| S13_extracted_118921 | Not-determined |
| S13_extracted_118944 | Not-determined |
| S13_extracted_118982 | Not-determined |
| S13_extracted_118989 | Not-determined |
| S13_extracted_118993 | Not-determined |
| S13_extracted_119002 | Not-determined |
| S13_extracted_119003 | Not-determined |
| S13_extracted_119008 | Low-quality |
| S13_extracted_119018 | Not-determined |
| S13_extracted_119045 | Not-determined |
| S13_extracted_119056 | Not-determined |
| S13_extracted_119057 | Not-determined |
| S13_extracted_119061 | Not-determined |
| S13_extracted_119075 | Not-determined |
| S13_extracted_119076 | Not-determined |
| S13_extracted_119082 | Not-determined |
| S13_extracted_119088 | Not-determined |
| S13_extracted_119095 | Not-determined |
| S13_extracted_119099 | Not-determined |
| S13_extracted_119101 | Not-determined |
| S13_extracted_119106 | Not-determined |
| S13_extracted_119107 | Not-determined |
| S13_extracted_119119 | Not-determined |
| S13_extracted_119121 | Not-determined |
| S13_extracted_119137 | Not-determined |
| S13_extracted_119139 | Not-determined |
| S13_extracted_119141 | Low-quality |
| S13_extracted_119146 | Not-determined |
| S13_extracted_119150 | Not-determined |
| S13_extracted_119154 | Not-determined |
| S13_extracted_119155 | Not-determined |
| S13_extracted_119156 | Not-determined |
| S13_extracted_119178 | Low-quality |
| S13_extracted_119183 | Not-determined |
| S13_extracted_119207 | Not-determined |
| S13_extracted_119211 | Not-determined |
| S13_extracted_119214 | Not-determined |
| S13_extracted_119220 | Low-quality |
| S13_extracted_119227 | Not-determined |
| S13_extracted_119230 | Not-determined |
| S13_extracted_119232 | Not-determined |
| S13_extracted_119240 | Not-determined |
| S13_extracted_119241 | Not-determined |
| S13_extracted_119268 | Not-determined |
| S13_extracted_119281 | Not-determined |
| S13_extracted_119294 | Not-determined |
| S13_extracted_119295 | Not-determined |
| S13_extracted_119297 | Not-determined |
| S13_extracted_119300 | Not-determined |
| S13_extracted_119313 | Not-determined |
| S13_extracted_119320 | Not-determined |
| S13_extracted_119321 | Not-determined |
| S13_extracted_119339 | Not-determined |
| S13_extracted_119344 | Not-determined |
| S13_extracted_119350 | Not-determined |
| S13_extracted_119386 | Not-determined |
| S13_extracted_119395 | Not-determined |
| S13_extracted_119397 | Not-determined |
| S13_extracted_119449 | Not-determined |
| S13_extracted_119450 | Not-determined |
| S13_extracted_119453 | Not-determined |
| S13_extracted_119454 | Not-determined |
| S13_extracted_119461 | Not-determined |
| S13_extracted_119470 | Not-determined |
| S13_extracted_119479 | Low-quality |
| S13_extracted_119497 | Not-determined |
| S13_extracted_119520 | Not-determined |
| S13_extracted_119523 | Not-determined |
| S13_extracted_119534 | Not-determined |
| S13_extracted_119573 | Not-determined |
| S13_extracted_119577 | Not-determined |
| S13_extracted_119588 | Not-determined |
| S13_extracted_119589 | Low-quality |
| S13_extracted_119591 | Low-quality |
| S13_extracted_119595 | Not-determined |
| S13_extracted_119615 | Not-determined |
| S13_extracted_119616 | Not-determined |
| S13_extracted_119619 | Not-determined |
| S13_extracted_119628 | Not-determined |
| S13_extracted_119630 | Not-determined |
| S13_extracted_119632 | Not-determined |
| S13_extracted_119644 | Not-determined |
| S13_extracted_119649 | Not-determined |
| S13_extracted_119706 | Not-determined |
| S13_extracted_119709 | Not-determined |
| S13_extracted_119711 | Low-quality |
| S13_extracted_119732 | Not-determined |
| S13_extracted_119737 | Not-determined |
| S13_extracted_119742 | Low-quality |
| S13_extracted_119750 | Not-determined |
| S13_extracted_119752 | Not-determined |
| S13_extracted_119763 | Not-determined |
| S13_extracted_119764 | Low-quality |
| S13_extracted_119796 | Low-quality |
| S13_extracted_119797 | Not-determined |
| S13_extracted_119808 | Not-determined |
| S13_extracted_119818 | Not-determined |
| S13_extracted_119820 | Not-determined |
| S13_extracted_119835 | Not-determined |
| S13_extracted_119838 | Not-determined |
| S13_extracted_119841 | Not-determined |
| S13_extracted_119851 | Not-determined |
| S13_extracted_119853 | Not-determined |
| S13_extracted_119857 | Not-determined |
| S13_extracted_119872 | Not-determined |
| S13_extracted_119884 | Not-determined |
| S13_extracted_119887 | Low-quality |
| S13_extracted_119889 | Low-quality |
| S13_extracted_119890 | Not-determined |
| S13_extracted_119893 | Not-determined |
| S13_extracted_119898 | Not-determined |
| S13_extracted_119907 | Not-determined |
| S13_extracted_119923 | Low-quality |
| S13_extracted_119924 | Low-quality |
| S13_extracted_119925 | Not-determined |
| S13_extracted_119928 | Not-determined |
| S13_extracted_119929 | Not-determined |
| S13_extracted_119936 | Not-determined |
| S13_extracted_119944 | Not-determined |
| S13_extracted_119949 | Not-determined |
| S13_extracted_119969 | Low-quality |
| S13_extracted_119973 | Not-determined |
| S13_extracted_119976 | Not-determined |
| S13_extracted_119978 | Not-determined |
| S13_extracted_119992 | Not-determined |
| S13_extracted_119993 | Not-determined |
| S13_extracted_119994 | Not-determined |
| S13_extracted_120002 | Not-determined |
| S13_extracted_120003 | Not-determined |
| S13_extracted_120013 | Not-determined |
| S13_extracted_120017 | Not-determined |
| S13_extracted_120025 | Not-determined |
| S13_extracted_120027 | Not-determined |
| S13_extracted_120035 | Not-determined |
| S13_extracted_120036 | Not-determined |
| S13_extracted_120051 | Not-determined |
| S13_extracted_120053 | Not-determined |
| S13_extracted_120055 | Not-determined |
| S13_extracted_120056 | Not-determined |
| S13_extracted_120057 | Not-determined |
| S13_extracted_120060 | Not-determined |
| S13_extracted_120064 | Not-determined |
| S13_extracted_120067 | Not-determined |
| S13_extracted_120086 | Not-determined |
| S13_extracted_120099 | Not-determined |
| S13_extracted_120115 | Not-determined |
| S13_extracted_120126 | Low-quality |
| S13_extracted_120136 | Not-determined |
| S13_extracted_120137 | Not-determined |
| S13_extracted_120148 | Not-determined |
| S13_extracted_120155 | Not-determined |
| S13_extracted_120159 | Not-determined |
| S13_extracted_120168 | Not-determined |
| S13_extracted_120180 | Not-determined |
| S13_extracted_120190 | Not-determined |
| S13_extracted_120207 | Not-determined |
| S13_extracted_120211 | Not-determined |
| S13_extracted_120212 | Not-determined |
| S13_extracted_120223 | Not-determined |
| S13_extracted_120234 | Not-determined |
| S13_extracted_120241 | Not-determined |
| S13_extracted_120248 | Not-determined |
| S13_extracted_120257 | Not-determined |
| S13_extracted_120273 | Not-determined |
| S13_extracted_120282 | Not-determined |
| S13_extracted_120295 | Not-determined |
| S13_extracted_120305 | Not-determined |
| S13_extracted_120320 | Not-determined |
| S13_extracted_120321 | Low-quality |
| S13_extracted_120322 | Not-determined |
| S13_extracted_120336 | Not-determined |
| S13_extracted_120345 | Low-quality |
| S13_extracted_120347 | Not-determined |
| S13_extracted_120352 | Not-determined |
| S13_extracted_120376 | Not-determined |
| S13_extracted_120387 | Not-determined |
| S13_extracted_120390 | Not-determined |
| S13_extracted_120391 | Not-determined |
| S13_extracted_120397 | Not-determined |
| S13_extracted_120399 | Not-determined |
| S13_extracted_120424 | Not-determined |
| S13_extracted_120426 | Not-determined |
| S13_extracted_120428 | Not-determined |
| S13_extracted_120429 | Not-determined |
| S13_extracted_120431 | Not-determined |
| S13_extracted_120434 | Not-determined |
| S13_extracted_120440 | Not-determined |
| S13_extracted_120441 | Not-determined |
| S13_extracted_120442 | Not-determined |
| S13_extracted_120452 | Not-determined |
| S13_extracted_120454 | Not-determined |
| S13_extracted_120455 | Not-determined |
| S13_extracted_120456 | Not-determined |
| S13_extracted_120460 | Not-determined |
| S13_extracted_120473 | Low-quality |
| S13_extracted_120474 | Not-determined |
| S13_extracted_120477 | Low-quality |
| S13_extracted_120484 | Not-determined |
| S13_extracted_120485 | Not-determined |
| S13_extracted_120486 | Not-determined |
| S13_extracted_120488 | Not-determined |
| S13_extracted_120501 | Not-determined |
| S13_extracted_120506 | Not-determined |
| S13_extracted_120507 | Not-determined |
| S13_extracted_120532 | Not-determined |
| S13_extracted_120547 | Low-quality |
| S8_extracted_k141_79769 | Complete |
| S13_extracted_120552 | Not-determined |
| S13_extracted_120553 | Low-quality |
| S13_extracted_120554 | Low-quality |
| S13_extracted_120581 | Low-quality |
| S13_extracted_120583 | Not-determined |
| S13_extracted_120593 | Not-determined |
| S13_extracted_120626 | Low-quality |
| S13_extracted_120630 | Not-determined |
| S13_extracted_120635 | Low-quality |
| S13_extracted_120677 | Not-determined |
| S13_extracted_120680 | Not-determined |
| S13_extracted_120685 | Not-determined |
| S13_extracted_120688 | Not-determined |
| S13_extracted_120692 | Low-quality |
| S13_extracted_120698 | Not-determined |
| S13_extracted_120702 | Not-determined |

| | |
|---|---|
| S13_extracted_120703 | Low-quality |
| S13_extracted_120719 | Low-quality |
| S13_extracted_120723 | Not-determined |
| S13_extracted_120724 | Not-determined |
| S13_extracted_120732 | Not-determined |
| S13_extracted_120747 | Not-determined |
| S13_extracted_120754 | Not-determined |
| S13_extracted_120755 | Not-determined |
| S13_extracted_120760 | Not-determined |
| S13_extracted_120766 | Not-determined |
| S13_extracted_120771 | Not-determined |
| S13_extracted_120774 | Not-determined |
| S13_extracted_120777 | Not-determined |
| S13_extracted_120783 | Not-determined |
| S13_extracted_120784 | Not-determined |
| S13_extracted_120790 | Not-determined |
| S13_extracted_120800 | Not-determined |
| S13_extracted_120801 | Not-determined |
| S13_extracted_120804 | Not-determined |
| S13_extracted_120817 | Not-determined |
| S13_extracted_120840 | Not-determined |
| S13_extracted_120852 | Not-determined |
| S13_extracted_120862 | Not-determined |
| S13_extracted_120867 | Not-determined |
| S13_extracted_120877 | Not-determined |
| S13_extracted_120902 | Low-quality |
| S13_extracted_120912 | Not-determined |
| S13_extracted_120916 | Not-determined |
| S13_extracted_120931 | Not-determined |
| S13_extracted_120936 | Not-determined |
| S13_extracted_120941 | Not-determined |
| S13_extracted_120944 | Not-determined |
| S13_extracted_120958 | Not-determined |
| S13_extracted_120984 | Not-determined |
| S13_extracted_120997 | Not-determined |
| S13_extracted_120999 | Not-determined |
| S13_extracted_121003 | Not-determined |
| S13_extracted_121005 | Not-determined |
| S13_extracted_121008 | Not-determined |
| S13_extracted_121018 | Not-determined |
| S13_extracted_121031 | Low-quality |
| S13_extracted_121042 | Low-quality |
| S13_extracted_121050 | Not-determined |
| S13_extracted_121052 | Not-determined |
| S13_extracted_121053 | Not-determined |
| S13_extracted_121057 | Not-determined |
| S13_extracted_121059 | Not-determined |
| S13_extracted_121066 | Low-quality |
| S13_extracted_121075 | Not-determined |
| S13_extracted_121080 | Not-determined |
| S13_extracted_121101 | Not-determined |
| S13_extracted_121108 | Not-determined |
| S13_extracted_121129 | Not-determined |
| S13_extracted_121131 | Not-determined |
| S13_extracted_121156 | Not-determined |
| S13_extracted_121157 | Not-determined |
| S13_extracted_121167 | Not-determined |
| S13_extracted_121170 | Not-determined |
| S13_extracted_121178 | Not-determined |
| S13_extracted_121183 | Not-determined |
| S13_extracted_121208 | Not-determined |
| S13_extracted_121223 | Not-determined |
| S13_extracted_121226 | Not-determined |
| S13_extracted_121227 | Low-quality |
| S13_extracted_121235 | Not-determined |
| S13_extracted_121237 | Not-determined |
| S13_extracted_121255 | Not-determined |
| S13_extracted_121260 | Not-determined |
| S13_extracted_121265 | Not-determined |
| S13_extracted_121267 | Not-determined |
| S13_extracted_121268 | Not-determined |
| S13_extracted_121270 | Not-determined |
| S13_extracted_121273 | Low-quality |
| S13_extracted_121280 | Not-determined |
| S13_extracted_121298 | Not-determined |
| S13_extracted_121321 | Not-determined |
| S13_extracted_121322 | Not-determined |
| S13_extracted_121334 | Not-determined |
| S13_extracted_121336 | Not-determined |
| S13_extracted_121337 | Not-determined |
| S13_extracted_121352 | Not-determined |
| S13_extracted_121376 | Not-determined |
| S13_extracted_121388 | Not-determined |
| S13_extracted_121390 | Low-quality |
| S13_extracted_121399 | Low-quality |
| S13_extracted_121411 | Not-determined |
| S13_extracted_121418 | Not-determined |
| S13_extracted_121419 | Not-determined |
| S13_extracted_121424 | Low-quality |
| S13_extracted_121431 | Not-determined |
| S13_extracted_121438 | Not-determined |
| S13_extracted_121439 | Low-quality |
| S13_extracted_121453 | Not-determined |
| S13_extracted_121462 | Not-determined |
| S13_extracted_121473 | Not-determined |
| S13_extracted_121481 | Not-determined |
| S13_extracted_121485 | Not-determined |
| S13_extracted_121496 | Low-quality |
| S13_extracted_121500 | Not-determined |
| S13_extracted_121505 | Not-determined |
| S13_extracted_121526 | Not-determined |
| S13_extracted_121535 | Not-determined |
| S13_extracted_121540 | Not-determined |
| S13_extracted_121544 | Not-determined |
| S13_extracted_121556 | Not-determined |
| S13_extracted_121567 | Not-determined |
| S13_extracted_121569 | Not-determined |
| S13_extracted_121577 | Not-determined |
| S13_extracted_121580 | Not-determined |
| S13_extracted_121587 | Not-determined |
| S13_extracted_121594 | Not-determined |
| S13_extracted_121606 | Not-determined |
| S13_extracted_121607 | Not-determined |
| S13_extracted_121614 | Not-determined |
| S13_extracted_121621 | Low-quality |
| S13_extracted_121626 | Not-determined |
| S13_extracted_121628 | Not-determined |
| S13_extracted_121633 | Not-determined |
| S13_extracted_121644 | Not-determined |
| S13_extracted_121648 | Not-determined |
| S13_extracted_121653 | Low-quality |
| S13_extracted_121674 | Low-quality |
| S13_extracted_121687 | Not-determined |
| S13_extracted_121691 | Low-quality |
| S13_extracted_121701 | Not-determined |
| S13_extracted_121713 | Not-determined |
| S13_extracted_121730 | Not-determined |
| S13_extracted_121733 | Not-determined |
| S13_extracted_121751 | Not-determined |
| S13_extracted_121758 | Not-determined |
| S13_extracted_121760 | Not-determined |
| S13_extracted_121764 | Not-determined |
| S13_extracted_121780 | Not-determined |
| S13_extracted_121786 | Not-determined |
| S13_extracted_121792 | Not-determined |
| S13_extracted_121795 | Not-determined |
| S13_extracted_121799 | Not-determined |
| S13_extracted_121806 | Not-determined |
| S13_extracted_121814 | Not-determined |
| S13_extracted_121846 | Low-quality |
| S13_extracted_121851 | Not-determined |
| S13_extracted_121857 | Low-quality |
| S13_extracted_121858 | Not-determined |
| S13_extracted_121863 | Low-quality |
| S13_extracted_121879 | Not-determined |
| S13_extracted_121884 | Not-determined |
| S13_extracted_121887 | Not-determined |
| S13_extracted_121900 | Not-determined |
| S13_extracted_121937 | Not-determined |
| S13_extracted_121942 | Not-determined |
| S13_extracted_121945 | Not-determined |
| S13_extracted_121952 | Not-determined |
| S13_extracted_121970 | Not-determined |
| S13_extracted_121973 | Not-determined |
| S13_extracted_121977 | Not-determined |
| S13_extracted_121993 | Not-determined |
| S13_extracted_121994 | Not-determined |
| S13_extracted_122022 | Not-determined |
| S13_extracted_122027 | Not-determined |
| S13_extracted_122031 | Not-determined |
| S13_extracted_122033 | Not-determined |
| S13_extracted_122035 | Low-quality |
| S13_extracted_122041 | Not-determined |
| S13_extracted_122054 | Not-determined |
| S13_extracted_122056 | Not-determined |
| S13_extracted_122063 | Not-determined |
| S13_extracted_122067 | Not-determined |
| S13_extracted_122087 | Low-quality |
| S13_extracted_122090 | Not-determined |
| S13_extracted_122092 | Low-quality |
| S13_extracted_122102 | Not-determined |
| S13_extracted_122117 | Not-determined |
| S13_extracted_122151 | Not-determined |
| S13_extracted_122156 | Not-determined |
| S13_extracted_122158 | Low-quality |
| S13_extracted_122170 | Low-quality |
| S13_extracted_122177 | Low-quality |
| S13_extracted_122193 | Not-determined |
| S13_extracted_122196 | Not-determined |
| S13_extracted_122209 | Not-determined |
| S13_extracted_122210 | Not-determined |
| S13_extracted_122215 | Low-quality |
| S13_extracted_122224 | Low-quality |
| S13_extracted_122227 | Low-quality |
| S13_extracted_122232 | Low-quality |
| S13_extracted_122244 | Not-determined |
| S13_extracted_122252 | Not-determined |
| S13_extracted_122254 | Not-determined |
| S13_extracted_122255 | Not-determined |
| S13_extracted_122259 | Not-determined |
| S13_extracted_122260 | Not-determined |
| S13_extracted_122271 | Not-determined |
| S13_extracted_122277 | Not-determined |
| S13_extracted_122286 | Not-determined |
| S13_extracted_122302 | Not-determined |
| S13_extracted_122310 | Not-determined |
| S13_extracted_122314 | Not-determined |
| S13_extracted_122320 | Not-determined |
| S13_extracted_122362 | Not-determined |
| S13_extracted_122369 | Not-determined |
| S13_extracted_122370 | Not-determined |
| S13_extracted_122372 | Not-determined |
| S13_extracted_122392 | Not-determined |
| S13_extracted_122393 | Not-determined |
| S13_extracted_122396 | Not-determined |
| S13_extracted_122397 | Not-determined |
| S13_extracted_122424 | Not-determined |
| S13_extracted_122426 | Not-determined |
| S13_extracted_122427 | Not-determined |
| S13_extracted_122435 | Not-determined |
| S13_extracted_122436 | Low-quality |
| S13_extracted_122441 | Not-determined |
| S13_extracted_122458 | Not-determined |
| S13_extracted_122459 | Low-quality |
| S13_extracted_122504 | Not-determined |
| S13_extracted_122507 | Not-determined |
| S13_extracted_122512 | Low-quality |
| S13_extracted_122514 | Low-quality |
| S13_extracted_122527 | Not-determined |
| S13_extracted_122530 | Not-determined |
| S13_extracted_122563 | Not-determined |
| S13_extracted_122564 | Not-determined |
| S13_extracted_122567 | Not-determined |
| S13_extracted_122577 | Not-determined |
| S13_extracted_122580 | Not-determined |
| S13_extracted_122585 | Not-determined |
| S13_extracted_122591 | Not-determined |
| S13_extracted_122594 | Not-determined |
| S13_extracted_122609 | Not-determined |
| S13_extracted_122614 | Low-quality |
| S13_extracted_122615 | Low-quality |
| S13_extracted_122617 | Not-determined |
| S13_extracted_122621 | Not-determined |
| S13_extracted_122643 | Not-determined |
| S13_extracted_122653 | Not-determined |
| S13_extracted_122660 | Not-determined |
| S13_extracted_122661 | Not-determined |
| S13_extracted_122663 | Not-determined |
| S13_extracted_122673 | Not-determined |
| S13_extracted_122677 | Not-determined |
| S13_extracted_122682 | Not-determined |
| S13_extracted_122687 | Not-determined |
| S13_extracted_122694 | Not-determined |
| S13_extracted_122698 | Not-determined |
| S13_extracted_122704 | Not-determined |
| S13_extracted_122705 | Not-determined |
| S13_extracted_122719 | Not-determined |
| S13_extracted_122727 | Not-determined |
| S13_extracted_122730 | Not-determined |
| S13_extracted_122746 | Not-determined |
| S13_extracted_122753 | Low-quality |
| S13_extracted_122761 | Not-determined |
| S13_extracted_122762 | Not-determined |
| S13_extracted_122763 | Not-determined |
| S13_extracted_122774 | Not-determined |
| S13_extracted_122785 | Not-determined |
| S13_extracted_122786 | Not-determined |
| S13_extracted_122787 | Not-determined |
| S13_extracted_122794 | Not-determined |
| S13_extracted_122802 | Not-determined |
| S13_extracted_122804 | Not-determined |
| S13_extracted_122810 | Not-determined |
| S13_extracted_122813 | Not-determined |
| S13_extracted_122826 | Not-determined |
| S13_extracted_122831 | Not-determined |
| S13_extracted_122837 | Not-determined |
| S13_extracted_122865 | Not-determined |
| S13_extracted_122888 | Not-determined |
| S13_extracted_122893 | Not-determined |
| S13_extracted_122894 | Not-determined |
| S13_extracted_122898 | Not-determined |
| S13_extracted_122917 | Not-determined |
| S13_extracted_122926 | Low-quality |
| S13_extracted_122934 | Not-determined |
| S13_extracted_122938 | Not-determined |
| S13_extracted_122963 | Not-determined |
| S13_extracted_122990 | Not-determined |
| S13_extracted_122995 | Not-determined |
| S13_extracted_122999 | Not-determined |
| S13_extracted_123009 | Low-quality |
| S13_extracted_123023 | Not-determined |
| S13_extracted_123024 | Not-determined |
| S13_extracted_123025 | Not-determined |
| S13_extracted_123040 | Not-determined |
| S13_extracted_123046 | Not-determined |
| S13_extracted_123059 | Not-determined |
| S13_extracted_123060 | Not-determined |
| S13_extracted_123063 | Not-determined |
| S13_extracted_123070 | Not-determined |
| S13_extracted_123071 | Not-determined |
| S13_extracted_123082 | Low-quality |
| S13_extracted_123084 | Not-determined |
| S13_extracted_123088 | Not-determined |
| S13_extracted_123093 | Not-determined |
| S13_extracted_123101 | Not-determined |
| S13_extracted_123129 | Not-determined |
| S13_extracted_123137 | Not-determined |
| S13_extracted_123140 | Not-determined |
| S13_extracted_123146 | Not-determined |
| S13_extracted_123147 | Not-determined |
| S13_extracted_123151 | Low-quality |
| S13_extracted_123155 | Not-determined |
| S13_extracted_123165 | Not-determined |
| S13_extracted_123191 | Not-determined |
| S13_extracted_123200 | Not-determined |
| S13_extracted_123203 | Not-determined |
| S13_extracted_123204 | Low-quality |
| S13_extracted_123209 | Not-determined |
| S13_extracted_123211 | Not-determined |
| S13_extracted_123218 | Not-determined |
| S13_extracted_123228 | Not-determined |
| S13_extracted_123229 | Not-determined |
| S13_extracted_123236 | Not-determined |
| S13_extracted_123244 | Not-determined |
| S13_extracted_123252 | Not-determined |
| S13_extracted_123264 | Not-determined |
| S13_extracted_123271 | Not-determined |
| S13_extracted_123275 | Not-determined |
| S13_extracted_123282 | Not-determined |
| S13_extracted_123289 | Not-determined |
| S13_extracted_123291 | Not-determined |
| S13_extracted_123294 | Not-determined |
| S13_extracted_123300 | Not-determined |
| S13_extracted_123302 | Not-determined |
| S13_extracted_123307 | Not-determined |
| [illegible] | [illegible] |
| S13_extracted_123365 | Low-quality |
| S13_extracted_123366 | Not-determined |
| S13_extracted_123367 | Not-determined |
| S13_extracted_123370 | Not-determined |
| S13_extracted_123375 | Not-determined |
| S13_extracted_123377 | Not-determined |
| S13_extracted_123382 | Not-determined |
| S13_extracted_123386 | Not-determined |
| S13_extracted_123408 | Not-determined |
| S13_extracted_123413 | Not-determined |
| S13_extracted_123420 | Not-determined |
| S13_extracted_123424 | Not-determined |
| S13_extracted_123427 | Not-determined |
| S13_extracted_123430 | Not-determined |
| S13_extracted_123437 | Not-determined |
| S13_extracted_123441 | Not-determined |
| S13_extracted_123443 | Not-determined |
| S13_extracted_123450 | Not-determined |
| S13_extracted_123462 | Not-determined |
| S13_extracted_123470 | Not-determined |
| S13_extracted_123487 | Not-determined |
| S13_extracted_123489 | Not-determined |
| S13_extracted_123490 | Not-determined |
| S13_extracted_123491 | Not-determined |
| S13_extracted_123496 | Not-determined |
| S13_extracted_123505 | Not-determined |
| S13_extracted_123508 | Not-determined |
| S13_extracted_123509 | Low-quality |
| S13_extracted_123514 | Not-determined |
| S13_extracted_123517 | Not-determined |
| S13_extracted_123537 | Not-determined |
| S13_extracted_123554 | Not-determined |
| S13_extracted_123567 | Not-determined |
| S13_extracted_123571 | Not-determined |
| S13_extracted_123577 | Not-determined |
| S13_extracted_123588 | Not-determined |
| S13_extracted_123595 | Not-determined |
| S13_extracted_123602 | Not-determined |
| S13_extracted_123614 | Low-quality |
| S13_extracted_123632 | Not-determined |
| S13_extracted_123635 | Low-quality |
| S13_extracted_123636 | Not-determined |
| S13_extracted_123646 | Not-determined |
| S13_extracted_123647 | Low-quality |
| S13_extracted_123654 | Not-determined |
| S13_extracted_123658 | Not-determined |
| S13_extracted_123665 | Not-determined |
| S13_extracted_123666 | Not-determined |
| S13_extracted_123668 | Not-determined |
| S13_extracted_123669 | Not-determined |
| S13_extracted_123678 | Not-determined |
| S13_extracted_123699 | Not-determined |
| S13_extracted_123702 | Not-determined |
| S13_extracted_123706 | Not-determined |
| S13_extracted_123707 | Not-determined |
| S13_extracted_123710 | Not-determined |
| S13_extracted_123726 | Not-determined |
| S13_extracted_123728 | Low-quality |
| S13_extracted_123732 | Low-quality |
| S13_extracted_123735 | Not-determined |
| S13_extracted_123762 | Not-determined |
| S13_extracted_123764 | Not-determined |
| S13_extracted_123766 | Not-determined |
| S13_extracted_123775 | Not-determined |
| S13_extracted_123779 | Not-determined |
| S13_extracted_123811 | Not-determined |
| S13_extracted_123818 | Not-determined |
| S13_extracted_123822 | Not-determined |
| S13_extracted_123824 | Not-determined |
| S13_extracted_123825 | Not-determined |
| S13_extracted_123831 | Low-quality |
| S13_extracted_123832 | Low-quality |
| S13_extracted_123833 | Not-determined |
| S13_extracted_123835 | Not-determined |
| S13_extracted_123863 | Low-quality |
| S13_extracted_123865 | Not-determined |
| S13_extracted_123872 | Not-determined |
| S13_extracted_123895 | Not-determined |
| S13_extracted_123897 | Not-determined |
| S13_extracted_123899 | Low-quality |
| S13_extracted_123904 | Not-determined |
| S13_extracted_123908 | Not-determined |
| S13_extracted_123921 | Not-determined |
| S13_extracted_123925 | Not-determined |
| S13_extracted_123939 | Not-determined |
| S13_extracted_123942 | Not-determined |
| S13_extracted_123960 | Not-determined |
| S13_extracted_123963 | Not-determined |
| S13_extracted_123964 | Low-quality |
| S13_extracted_123971 | Low-quality |
| S13_extracted_123989 | Not-determined |
| S13_extracted_124010 | Not-determined |
| S13_extracted_124023 | Not-determined |
| S13_extracted_124024 | Not-determined |
| S13_extracted_124028 | Not-determined |
| S13_extracted_124030 | Low-quality |
| S13_extracted_124048 | Not-determined |
| S13_extracted_124057 | Not-determined |
| S13_extracted_124066 | Not-determined |
| S13_extracted_124068 | Not-determined |
| S13_extracted_124069 | Low-quality |
| S13_extracted_124072 | Not-determined |
| S13_extracted_124085 | Not-determined |
| S13_extracted_124086 | Not-determined |
| S13_extracted_124106 | Not-determined |
| S13_extracted_124109 | Not-determined |
| S13_extracted_124118 | Not-determined |
| S13_extracted_124121 | Not-determined |
| S13_extracted_124122 | Not-determined |
| S13_extracted_124124 | Not-determined |
| S13_extracted_124129 | Not-determined |
| S13_extracted_124130 | Not-determined |
| S13_extracted_124134 | Not-determined |
| S13_extracted_124136 | Not-determined |
| S13_extracted_124138 | Not-determined |
| S13_extracted_124149 | Low-quality |
| S13_extracted_124161 | Not-determined |
| S13_extracted_124182 | Not-determined |
| S13_extracted_124184 | Not-determined |
| S13_extracted_124185 | Low-quality |
| S13_extracted_124187 | Not-determined |
| S13_extracted_124195 | Not-determined |
| S13_extracted_124198 | Not-determined |
| S13_extracted_124199 | Low-quality |
| S13_extracted_124203 | Not-determined |
| S13_extracted_124205 | Not-determined |
| S13_extracted_124208 | Not-determined |
| S13_extracted_124229 | Not-determined |
| S13_extracted_124234 | Not-determined |
| S13_extracted_124255 | Not-determined |
| S13_extracted_124258 | Not-determined |
| S13_extracted_124261 | Not-determined |
| S13_extracted_124263 | Not-determined |
| S13_extracted_124274 | Not-determined |
| S13_extracted_124287 | Not-determined |
| S13_extracted_124302 | Not-determined |
| S13_extracted_124304 | Not-determined |
| S13_extracted_124325 | Not-determined |
| S13_extracted_124327 | Not-determined |
| S13_extracted_124335 | Low-quality |
| S13_extracted_124346 | Not-determined |
| S13_extracted_124352 | Not-determined |
| S13_extracted_124353 | Not-determined |
| S13_extracted_124357 | Not-determined |
| S13_extracted_124368 | Not-determined |
| S13_extracted_124372 | Not-determined |
| S13_extracted_124379 | Not-determined |
| S13_extracted_124422 | Not-determined |
| S13_extracted_124423 | Not-determined |
| S13_extracted_124434 | Not-determined |
| S13_extracted_124443 | Not-determined |
| S13_extracted_124448 | Not-determined |
| S13_extracted_124459 | Low-quality |
| S13_extracted_124465 | Not-determined |
| S13_extracted_124466 | Not-determined |
| S13_extracted_124468 | Not-determined |
| S13_extracted_124497 | Not-determined |
| S13_extracted_124500 | Not-determined |
| S13_extracted_124503 | Not-determined |
| S13_extracted_124505 | Low-quality |
| S13_extracted_124507 | Not-determined |
| S13_extracted_124508 | Not-determined |
| S13_extracted_124516 | Not-determined |
| S13_extracted_124542 | Low-quality |
| S13_extracted_124547 | Low-quality |
| S13_extracted_124548 | Not-determined |
| S13_extracted_124555 | Not-determined |
| S13_extracted_124563 | Low-quality |
| S13_extracted_124572 | Not-determined |
| S13_extracted_124581 | Not-determined |
| S13_extracted_124584 | Low-quality |
| S13_extracted_124590 | Not-determined |
| S13_extracted_124597 | Not-determined |
| S13_extracted_124598 | Not-determined |
| S13_extracted_124608 | Not-determined |
| S13_extracted_124611 | Not-determined |
| S13_extracted_124612 | Not-determined |
| S13_extracted_124616 | Not-determined |
| S13_extracted_124621 | Not-determined |
| S13_extracted_124639 | Not-determined |
| S13_extracted_124655 | Low-quality |
| S13_extracted_124658 | Low-quality |
| S13_extracted_124664 | Not-determined |
| S13_extracted_124671 | Not-determined |
| S13_extracted_124691 | Not-determined |
| S13_extracted_124692 | Not-determined |
| S13_extracted_124696 | Low-quality |
| S13_extracted_124705 | Not-determined |
| S13_extracted_124706 | Not-determined |
| S13_extracted_124713 | Not-determined |
| S13_extracted_124714 | Not-determined |
| S13_extracted_124715 | Not-determined |
| S13_extracted_124718 | Low-quality |
| S13_extracted_124719 | Not-determined |
| S13_extracted_124720 | Low-quality |
| S13_extracted_124732 | Not-determined |
| S13_extracted_124734 | Not-determined |
| S13_extracted_124737 | Low-quality |
| S13_extracted_124741 | Not-determined |
| S13_extracted_124759 | Not-determined |
| S13_extracted_124761 | Not-determined |
| S13_extracted_124764 | Low-quality |
| S13_extracted_124773 | Low-quality |
| S13_extracted_124784 | Low-quality |
| S13_extracted_124791 | Not-determined |
| S13_extracted_124805 | Low-quality |
| S13_extracted_124810 | Not-determined |
| S13_extracted_124812 | Not-determined |
| S13_extracted_124836 | Not-determined |
| S13_extracted_124842 | Not-determined |
| S13_extracted_124847 | Not-determined |
| S13_extracted_124850 | Low-quality |
| S13_extracted_124858 | Not-determined |
| S13_extracted_124861 | Not-determined |
| S13_extracted_124864 | Not-determined |
| S13_extracted_124869 | Not-determined |
| S13_extracted_124877 | Not-determined |
| S13_extracted_124879 | Low-quality |
| S13_extracted_124881 | Not-determined |
| S13_extracted_124882 | Not-determined |
| S13_extracted_124889 | Not-determined |
| S13_extracted_124899 | Not-determined |
| S13_extracted_124904 | Not-determined |
| S13_extracted_124916 | Not-determined |
| S13_extracted_124917 | Not-determined |
| S13_extracted_124924 | Not-determined |
| S13_extracted_124934 | Not-determined |
| S13_extracted_124938 | Not-determined |
| S13_extracted_124939 | Not-determined |
| S13_extracted_124945 | Low-quality |
| S13_extracted_124949 | Not-determined |
| S13_extracted_124960 | Low-quality |
| S13_extracted_124963 | Low-quality |
| S13_extracted_124966 | Not-determined |
| S13_extracted_124978 | Low-quality |
| S13_extracted_124981 | Not-determined |
| S13_extracted_125011 | Not-determined |
| S13_extracted_125012 | Low-quality |
| S13_extracted_125014 | Not-determined |
| S13_extracted_125018 | Not-determined |
| S13_extracted_125031 | Not-determined |
| S13_extracted_125042 | Not-determined |
| S13_extracted_125045 | Not-determined |
| S13_extracted_125046 | Not-determined |
| S13_extracted_125049 | Not-determined |
| S13_extracted_125073 | Not-determined |
| S13_extracted_125077 | Not-determined |
| S13_extracted_125080 | Not-determined |
| S13_extracted_125087 | Low-quality |
| S13_extracted_125092 | Not-determined |
| S13_extracted_125095 | Not-determined |
| S13_extracted_125097 | Not-determined |
| S13_extracted_125105 | Not-determined |
| S13_extracted_125116 | Low-quality |
| S13_extracted_125128 | Not-determined |
| S13_extracted_125140 | Not-determined |
| S13_extracted_125142 | Not-determined |
| S13_extracted_125143 | Not-determined |

| | |
|---|---|
| S13_extracted_125147 | Not-determined |
| S13_extracted_125151 | Not-determined |
| S13_extracted_125159 | Not-determined |
| S13_extracted_125160 | Not-determined |
| S13_extracted_125170 | Not-determined |
| S13_extracted_125188 | Not-determined |
| S13_extracted_125190 | Not-determined |
| S13_extracted_125204 | Not-determined |
| S13_extracted_125217 | Not-determined |
| S13_extracted_125223 | Not-determined |
| S13_extracted_125245 | Not-determined |
| S13_extracted_125246 | Not-determined |
| S13_extracted_125265 | Not-determined |
| S13_extracted_125270 | Not-determined |
| S13_extracted_125288 | Not-determined |
| S13_extracted_125294 | Not-determined |
| S13_extracted_125299 | Not-determined |
| S13_extracted_125304 | Not-determined |
| S13_extracted_125312 | Not-determined |
| S13_extracted_125326 | Not-determined |
| S13_extracted_125332 | Not-determined |
| S13_extracted_125333 | Not-determined |
| S13_extracted_125337 | Not-determined |
| S13_extracted_125352 | Not-determined |
| S13_extracted_125360 | Not-determined |
| S13_extracted_125364 | Not-determined |
| S13_extracted_125366 | Not-determined |
| S13_extracted_125367 | Not-determined |
| S13_extracted_125372 | Not-determined |
| S13_extracted_125374 | Not-determined |
| S13_extracted_125383 | Low-quality |
| S13_extracted_125385 | Not-determined |
| S13_extracted_125387 | Low-quality |
| S13_extracted_125396 | Not-determined |
| S13_extracted_125422 | Not-determined |
| S13_extracted_125424 | Not-determined |
| S13_extracted_125430 | Not-determined |
| S13_extracted_125441 | Not-determined |
| S13_extracted_125449 | Not-determined |
| S13_extracted_125456 | Not-determined |
| S13_extracted_125457 | Not-determined |
| S13_extracted_125461 | Not-determined |
| S13_extracted_125467 | Not-determined |
| S13_extracted_125491 | Not-determined |
| S13_extracted_125492 | Not-determined |
| S13_extracted_125495 | Not-determined |
| S13_extracted_125496 | Not-determined |
| S13_extracted_125497 | Low-quality |
| S13_extracted_125514 | Low-quality |
| S13_extracted_125516 | Not-determined |
| S13_extracted_125519 | Not-determined |
| S13_extracted_125521 | Not-determined |
| S13_extracted_125524 | Low-quality |
| S13_extracted_125527 | Not-determined |
| S13_extracted_125529 | Not-determined |
| S13_extracted_125540 | Low-quality |
| S13_extracted_125542 | Not-determined |
| S13_extracted_125546 | Not-determined |
| S13_extracted_125551 | Not-determined |
| S13_extracted_125553 | Not-determined |
| S13_extracted_125557 | Not-determined |
| S13_extracted_125566 | Not-determined |
| S13_extracted_125568 | Not-determined |
| S13_extracted_125569 | Not-determined |
| S13_extracted_125571 | Not-determined |
| S13_extracted_125632 | Not-determined |
| S13_extracted_125643 | Not-determined |
| S13_extracted_125645 | Not-determined |
| S13_extracted_125648 | Not-determined |
| S13_extracted_125679 | Not-determined |
| S13_extracted_125682 | Not-determined |
| S13_extracted_125691 | Not-determined |
| S13_extracted_125694 | Not-determined |
| S13_extracted_125695 | Not-determined |
| S13_extracted_125711 | Not-determined |
| S13_extracted_125715 | Not-determined |
| S13_extracted_125721 | Not-determined |
| S13_extracted_125726 | Not-determined |
| S13_extracted_125758 | Low-quality |
| S13_extracted_125763 | Not-determined |
| S13_extracted_125778 | Not-determined |
| S13_extracted_125787 | Low-quality |
| S13_extracted_125789 | Not-determined |
| S13_extracted_125798 | Not-determined |
| S13_extracted_125814 | Not-determined |
| S13_extracted_125836 | Low-quality |
| S13_extracted_125844 | Low-quality |
| S13_extracted_125862 | Not-determined |
| S13_extracted_125868 | Not-determined |
| S13_extracted_125875 | Not-determined |
| S13_extracted_125876 | Not-determined |
| S13_extracted_125877 | Low-quality |
| S13_extracted_125901 | Not-determined |
| S13_extracted_125905 | Not-determined |
| S13_extracted_125908 | Not-determined |
| S13_extracted_125917 | Not-determined |
| S13_extracted_125923 | Low-quality |
| S13_extracted_125924 | Not-determined |
| S13_extracted_125928 | Low-quality |
| S13_extracted_125937 | Not-determined |
| S13_extracted_125952 | Not-determined |
| S13_extracted_125953 | Not-determined |
| S13_extracted_125955 | Not-determined |
| S13_extracted_125973 | Not-determined |
| S13_extracted_125979 | Not-determined |
| S13_extracted_125981 | Low-quality |
| S13_extracted_125983 | Not-determined |
| S13_extracted_125989 | Not-determined |
| S13_extracted_125998 | Not-determined |
| S13_extracted_126012 | Not-determined |
| S13_extracted_126013 | Not-determined |
| S13_extracted_126014 | Not-determined |
| S13_extracted_126021 | Not-determined |
| S13_extracted_126032 | Not-determined |
| S13_extracted_126041 | Not-determined |
| S13_extracted_126042 | Not-determined |
| S13_extracted_126044 | Not-determined |
| S13_extracted_126047 | Not-determined |
| S13_extracted_126049 | Not-determined |
| S13_extracted_126065 | Not-determined |
| S13_extracted_126086 | Not-determined |
| S13_extracted_126087 | Not-determined |
| S13_extracted_126091 | Low-quality |
| S13_extracted_126106 | Not-determined |
| S13_extracted_126111 | Not-determined |
| S13_extracted_126125 | Low-quality |
| S13_extracted_126137 | Not-determined |
| S13_extracted_126155 | Not-determined |
| S13_extracted_126160 | Not-determined |
| S13_extracted_126167 | Not-determined |
| S13_extracted_126168 | Not-determined |
| S13_extracted_126184 | Not-determined |
| S13_extracted_126197 | Not-determined |
| S13_extracted_126212 | Not-determined |
| S13_extracted_126226 | Not-determined |
| S13_extracted_126232 | Not-determined |
| S13_extracted_126233 | Not-determined |
| S13_extracted_126237 | Not-determined |
| S13_extracted_126247 | Low-quality |
| S13_extracted_126251 | Low-quality |
| S13_extracted_126259 | Not-determined |
| S13_extracted_126266 | Not-determined |
| S13_extracted_126267 | Not-determined |
| S13_extracted_126274 | Not-determined |
| S13_extracted_126285 | Not-determined |
| S13_extracted_126290 | Low-quality |
| S13_extracted_126293 | Not-determined |
| S13_extracted_126299 | Not-determined |
| S13_extracted_126301 | Not-determined |
| S13_extracted_126311 | Not-determined |
| S13_extracted_126316 | Not-determined |
| S13_extracted_126324 | Not-determined |
| S13_extracted_126327 | Not-determined |
| S13_extracted_126328 | Not-determined |
| S13_extracted_126331 | Low-quality |
| S13_extracted_126333 | Not-determined |
| S13_extracted_126348 | Not-determined |
| S13_extracted_126350 | Not-determined |
| S13_extracted_126351 | Not-determined |
| S13_extracted_126374 | Not-determined |
| S13_extracted_126379 | Not-determined |
| S13_extracted_126381 | Not-determined |
| S13_extracted_126387 | Not-determined |
| S13_extracted_126391 | Low-quality |
| S13_extracted_126397 | Not-determined |
| S13_extracted_126399 | Not-determined |
| S13_extracted_126409 | Low-quality |
| S13_extracted_126421 | Not-determined |
| S13_extracted_126429 | Not-determined |
| S13_extracted_126437 | Not-determined |
| S13_extracted_126438 | Not-determined |
| S13_extracted_126444 | Not-determined |
| S13_extracted_126450 | Not-determined |
| S13_extracted_126451 | Not-determined |
| S13_extracted_126459 | Not-determined |
| S13_extracted_126479 | Not-determined |
| S13_extracted_126480 | Not-determined |
| S13_extracted_126487 | Low-quality |
| S13_extracted_126489 | Not-determined |
| S13_extracted_126493 | Not-determined |
| S13_extracted_126494 | Not-determined |
| S13_extracted_126502 | Low-quality |
| S13_extracted_126505 | Not-determined |
| S13_extracted_126508 | Not-determined |
| S13_extracted_126518 | Low-quality |
| S13_extracted_126524 | Not-determined |
| S13_extracted_126538 | Not-determined |
| S13_extracted_126543 | Not-determined |
| S13_extracted_126552 | Not-determined |
| S13_extracted_126565 | Low-quality |
| S13_extracted_126566 | Not-determined |
| S13_extracted_126573 | Not-determined |
| S13_extracted_126584 | Not-determined |
| S13_extracted_126588 | Not-determined |
| S13_extracted_126591 | Not-determined |
| S13_extracted_126601 | Not-determined |
| S13_extracted_126603 | Not-determined |
| S13_extracted_126609 | Not-determined |
| S13_extracted_126620 | Not-determined |
| S13_extracted_126627 | Not-determined |
| S13_extracted_126647 | Not-determined |
| S13_extracted_126659 | Not-determined |
| S13_extracted_126665 | Not-determined |
| S13_extracted_126684 | Not-determined |
| S13_extracted_126686 | Not-determined |
| S13_extracted_126692 | Not-determined |
| S13_extracted_126693 | Not-determined |
| S13_extracted_126698 | Not-determined |
| S13_extracted_126708 | Low-quality |
| S13_extracted_126717 | Not-determined |
| S13_extracted_126725 | Not-determined |
| S13_extracted_126727 | Not-determined |
| S13_extracted_126744 | Not-determined |
| S13_extracted_126754 | Not-determined |
| S13_extracted_126760 | Not-determined |
| S13_extracted_126763 | Not-determined |
| S13_extracted_126766 | Not-determined |
| S13_extracted_126769 | Not-determined |
| S13_extracted_126774 | Not-determined |
| S13_extracted_126783 | Not-determined |
| S13_extracted_126792 | Not-determined |
| S13_extracted_126801 | Not-determined |
| S13_extracted_126814 | Low-quality |
| S13_extracted_126817 | Not-determined |
| S13_extracted_126825 | Not-determined |
| S13_extracted_126827 | Not-determined |
| S13_extracted_126831 | Not-determined |
| S13_extracted_126833 | Low-quality |
| S13_extracted_126839 | Not-determined |
| S13_extracted_126869 | Not-determined |
| S13_extracted_126872 | Not-determined |
| S13_extracted_126873 | Not-determined |
| S13_extracted_126874 | Not-determined |
| S13_extracted_126879 | Not-determined |
| S13_extracted_126882 | Not-determined |
| S13_extracted_126904 | Not-determined |
| S13_extracted_126919 | Not-determined |
| S13_extracted_126923 | Low-quality |
| S13_extracted_126925 | Not-determined |
| S13_extracted_126928 | Not-determined |
| S13_extracted_126929 | Not-determined |
| S13_extracted_126930 | Not-determined |
| S13_extracted_126931 | Not-determined |
| S13_extracted_126945 | Not-determined |
| S13_extracted_126949 | Not-determined |
| S13_extracted_126965 | Low-quality |
| S13_extracted_126973 | Not-determined |
| S13_extracted_126990 | Low-quality |
| S13_extracted_126993 | Not-determined |
| S13_extracted_126998 | Not-determined |
| S13_extracted_127002 | Not-determined |
| S13_extracted_127020 | Not-determined |
| S13_extracted_127026 | Not-determined |
| S13_extracted_127027 | Not-determined |
| S13_extracted_127028 | Not-determined |
| S13_extracted_127034 | Not-determined |
| S13_extracted_127037 | Not-determined |
| S13_extracted_127039 | Not-determined |
| S13_extracted_127049 | Not-determined |
| S13_extracted_127066 | Not-determined |
| S13_extracted_127074 | Not-determined |
| S13_extracted_127078 | Not-determined |
| S13_extracted_127083 | Not-determined |
| S13_extracted_127092 | Not-determined |
| S13_extracted_127098 | Not-determined |
| S13_extracted_127109 | Not-determined |
| S13_extracted_127114 | Not-determined |
| S13_extracted_127117 | Low-quality |
| S13_extracted_127142 | Not-determined |
| S13_extracted_127150 | Not-determined |
| S13_extracted_127168 | Not-determined |
| S13_extracted_127171 | Not-determined |
| S13_extracted_127185 | Not-determined |
| S13_extracted_127200 | Not-determined |
| S13_extracted_127209 | Not-determined |
| S13_extracted_127212 | Not-determined |
| S13_extracted_127218 | Not-determined |
| S13_extracted_127222 | Not-determined |
| S13_extracted_127232 | Low-quality |
| S13_extracted_127236 | Low-quality |
| S13_extracted_127238 | Not-determined |
| S13_extracted_127258 | Not-determined |
| S13_extracted_127262 | Low-quality |
| S13_extracted_127277 | Not-determined |
| S13_extracted_127286 | Not-determined |
| S13_extracted_127288 | Not-determined |
| S13_extracted_127306 | Not-determined |
| S13_extracted_127316 | Not-determined |
| S13_extracted_127324 | Not-determined |
| S13_extracted_127336 | Not-determined |
| S13_extracted_127342 | Not-determined |
| S13_extracted_127344 | Not-determined |
| S13_extracted_127347 | Low-quality |
| S13_extracted_127353 | Not-determined |
| S13_extracted_127362 | Not-determined |
| S13_extracted_127369 | Not-determined |
| S13_extracted_127370 | Not-determined |
| S13_extracted_127373 | Not-determined |
| S13_extracted_127374 | Not-determined |
| S13_extracted_127376 | Not-determined |
| S13_extracted_127393 | Not-determined |
| S13_extracted_127402 | Not-determined |
| S13_extracted_127419 | Not-determined |
| S13_extracted_127422 | Not-determined |
| S13_extracted_127423 | Low-quality |
| S13_extracted_127435 | Low-quality |
| S13_extracted_127441 | Not-determined |
| S13_extracted_127449 | Not-determined |
| S13_extracted_127458 | Not-determined |
| S13_extracted_127460 | Not-determined |
| S13_extracted_127481 | Low-quality |
| S13_extracted_127487 | Not-determined |
| S13_extracted_127488 | Not-determined |
| S13_extracted_127492 | Not-determined |
| S13_extracted_127493 | Low-quality |
| S13_extracted_127497 | Low-quality |
| S13_extracted_127513 | Not-determined |
| S13_extracted_127519 | Not-determined |
| S13_extracted_127520 | Not-determined |
| S13_extracted_127531 | Not-determined |
| S13_extracted_127532 | Low-quality |
| S13_extracted_127542 | Not-determined |
| S13_extracted_127543 | Not-determined |
| S13_extracted_127556 | Not-determined |
| S13_extracted_127558 | Not-determined |
| S13_extracted_127599 | Not-determined |
| S13_extracted_127616 | Not-determined |
| S13_extracted_127630 | Not-determined |
| S13_extracted_127646 | Not-determined |
| S13_extracted_127665 | Low-quality |
| S13_extracted_127668 | Not-determined |
| S13_extracted_127677 | Not-determined |
| S13_extracted_127707 | Not-determined |
| S13_extracted_127711 | Not-determined |
| S13_extracted_127734 | Not-determined |
| S13_extracted_127744 | Not-determined |
| S13_extracted_127750 | Not-determined |
| S13_extracted_127759 | Low-quality |
| S13_extracted_127788 | Not-determined |
| S13_extracted_127796 | Not-determined |
| S13_extracted_127798 | Low-quality |
| S13_extracted_127804 | Not-determined |
| S13_extracted_127809 | Not-determined |
| S13_extracted_127820 | Not-determined |
| S13_extracted_127840 | Not-determined |
| S13_extracted_127849 | Not-determined |
| S13_extracted_127870 | Not-determined |
| S13_extracted_127875 | Not-determined |
| S13_extracted_127878 | Not-determined |
| S13_extracted_127881 | Not-determined |
| S13_extracted_127884 | Not-determined |
| S13_extracted_127887 | Not-determined |
| S13_extracted_127902 | Not-determined |
| S13_extracted_127907 | Not-determined |
| S13_extracted_127926 | Not-determined |
| S13_extracted_127954 | Low-quality |
| S13_extracted_127956 | Not-determined |
| S13_extracted_127959 | Not-determined |
| S13_extracted_127978 | Not-determined |
| S13_extracted_127989 | Not-determined |
| S13_extracted_127992 | Low-quality |
| S13_extracted_128010 | Not-determined |
| S13_extracted_128011 | Not-determined |
| S13_extracted_128018 | Not-determined |
| S13_extracted_128022 | Not-determined |
| S13_extracted_128030 | Not-determined |
| S13_extracted_128031 | Not-determined |
| S13_extracted_128043 | Low-quality |
| S13_extracted_128045 | Not-determined |
| S13_extracted_128048 | Low-quality |
| S13_extracted_128050 | Low-quality |
| S13_extracted_128086 | Not-determined |
| S13_extracted_128094 | Not-determined |
| S13_extracted_128101 | Low-quality |
| S13_extracted_128104 | Not-determined |
| S13_extracted_128109 | Not-determined |
| S13_extracted_128130 | Low-quality |
| S13_extracted_128131 | Not-determined |
| S13_extracted_128142 | Not-determined |
| S13_extracted_128157 | Not-determined |
| S13_extracted_128163 | Not-determined |
| S13_extracted_128174 | Not-determined |
| S13_extracted_128187 | Not-determined |
| S13_extracted_128194 | Not-determined |
| S13_extracted_128202 | Not-determined |
| S13_extracted_128203 | Not-determined |
| S13_extracted_128211 | Not-determined |
| S13_extracted_128212 | Low-quality |
| S13_extracted_128222 | Not-determined |
| S13_extracted_128234 | Not-determined |
| S13_extracted_128254 | Not-determined |
| S13_extracted_128260 | Not-determined |
| S13_extracted_128262 | Not-determined |
| S13_extracted_128272 | Not-determined |
| S13_extracted_128287 | Not-determined |
| S13_extracted_128298 | Not-determined |
| S13_extracted_128322 | Not-determined |
| S13_extracted_128323 | Not-determined |
| S13_extracted_128328 | Not-determined |
| S13_extracted_128330 | Not-determined |
| S13_extracted_128334 | Not-determined |
| S13_extracted_128337 | Not-determined |
| S13_extracted_128342 | Not-determined |
| S13_extracted_128360 | Low-quality |
| S13_extracted_128368 | Not-determined |
| S13_extracted_128369 | Not-determined |
| S13_extracted_128375 | Not-determined |
| S13_extracted_128383 | Not-determined |
| S13_extracted_128393 | Not-determined |
| S13_extracted_128397 | Not-determined |
| S13_extracted_128402 | Not-determined |
| S13_extracted_128443 | Not-determined |
| S13_extracted_128449 | Not-determined |
| S13_extracted_128450 | Not-determined |
| S13_extracted_128453 | Not-determined |
| S13_extracted_128456 | Not-determined |
| S13_extracted_128465 | Low-quality |
| S13_extracted_128468 | Not-determined |
| S13_extracted_128470 | Not-determined |
| S13_extracted_128475 | Not-determined |
| S13_extracted_128476 | Not-determined |
| S13_extracted_128480 | Not-determined |
| S13_extracted_128507 | Not-determined |
| S13_extracted_128513 | Not-determined |
| S13_extracted_128514 | Not-determined |
| S13_extracted_128526 | Not-determined |
| S13_extracted_128528 | Not-determined |
| S13_extracted_128529 | Not-determined |
| S13_extracted_128539 | Not-determined |
| S13_extracted_128542 | Not-determined |
| S13_extracted_128550 | Not-determined |
| S13_extracted_128569 | Not-determined |
| S13_extracted_128572 | Not-determined |
| S13_extracted_128582 | Not-determined |
| S13_extracted_128593 | Not-determined |
| S13_extracted_128604 | Not-determined |
| S13_extracted_128612 | Not-determined |
| S13_extracted_128617 | Not-determined |
| S13_extracted_128622 | Not-determined |
| S13_extracted_128663 | Not-determined |
| S13_extracted_128677 | Low-quality |
| S13_extracted_128696 | Not-determined |
| S13_extracted_128697 | Not-determined |
| S13_extracted_128711 | Not-determined |
| S13_extracted_128713 | Low-quality |
| S13_extracted_128736 | Not-determined |
| S13_extracted_128758 | Not-determined |
| S13_extracted_128759 | Not-determined |
| S13_extracted_128764 | Not-determined |
| S13_extracted_128765 | Not-determined |
| S13_extracted_128769 | Not-determined |
| S13_extracted_128772 | Low-quality |
| S13_extracted_128783 | Not-determined |
| S13_extracted_128788 | Not-determined |
| S13_extracted_128791 | Not-determined |
| S13_extracted_128799 | Not-determined |
| S13_extracted_128805 | Not-determined |
| S13_extracted_128809 | Not-determined |
| S13_extracted_128812 | Not-determined |
| S13_extracted_128813 | Not-determined |
| S13_extracted_128815 | Not-determined |
| S13_extracted_128831 | Low-quality |
| S13_extracted_128833 | Not-determined |
| S13_extracted_128834 | Not-determined |
| S13_extracted_128835 | Not-determined |
| S13_extracted_128841 | Not-determined |
| S13_extracted_128858 | Not-determined |
| S13_extracted_128867 | Not-determined |
| S13_extracted_128876 | Not-determined |
| S13_extracted_128884 | Not-determined |
| S13_extracted_128889 | Not-determined |
| S13_extracted_128894 | Low-quality |
| S13_extracted_128896 | Low-quality |
| S13_extracted_128919 | Low-quality |
| S13_extracted_128922 | Low-quality |
| S13_extracted_128936 | Not-determined |
| S13_extracted_128950 | Not-determined |
| S13_extracted_128962 | Not-determined |
| S13_extracted_128972 | Not-determined |
| S13_extracted_128983 | Not-determined |
| S13_extracted_129001 | Not-determined |
| S13_extracted_129002 | Not-determined |
| S13_extracted_129005 | Low-quality |
| S13_extracted_129007 | Not-determined |
| S13_extracted_129011 | Not-determined |
| S13_extracted_129042 | Not-determined |
| S13_extracted_129047 | Not-determined |
| S13_extracted_129049 | Low-quality |
| S13_extracted_129053 | Not-determined |
| S13_extracted_129056 | Not-determined |
| S13_extracted_129062 | Not-determined |
| S13_extracted_129070 | Not-determined |
| S13_extracted_129088 | Not-determined |
| S13_extracted_129092 | Not-determined |
| S13_extracted_129108 | Not-determined |
| S13_extracted_129115 | Not-determined |
| S13_extracted_129130 | Low-quality |
| S13_extracted_129154 | Not-determined |
| S13_extracted_129155 | Not-determined |
| S13_extracted_129167 | Not-determined |
| S13_extracted_129187 | Not-determined |
| S13_extracted_129200 | Low-quality |
| S13_extracted_129201 | Low-quality |
| S13_extracted_129213 | Not-determined |
| S13_extracted_129217 | Not-determined |
| S13_extracted_129221 | Not-determined |
| S13_extracted_129237 | Not-determined |
| S13_extracted_129249 | Low-quality |
| S13_extracted_129253 | Not-determined |
| S13_extracted_129255 | Not-determined |
| S13_extracted_129263 | Not-determined |
| S13_extracted_129276 | Low-quality |
| S13_extracted_129284 | Not-determined |
| S13_extracted_129289 | Not-determined |
| S13_extracted_129307 | Not-determined |
| S13_extracted_129308 | Not-determined |
| S13_extracted_129313 | Not-determined |
| S13_extracted_129324 | Not-determined |
| S13_extracted_129328 | Not-determined |
| S13_extracted_129329 | Not-determined |
| S13_extracted_129346 | Not-determined |
| S13_extracted_129351 | Not-determined |
| S13_extracted_129352 | Not-determined |
| S13_extracted_129353 | Not-determined |
| S13_extracted_129359 | Not-determined |
| S13_extracted_129361 | Not-determined |
| S13_extracted_129377 | Not-determined |
| S13_extracted_129403 | Not-determined |
| S13_extracted_129419 | Not-determined |
| S13_extracted_129434 | Low-quality |
| S13_extracted_129441 | Not-determined |
| S13_extracted_129460 | Not-determined |
| S13_extracted_129466 | Not-determined |
| S13_extracted_129480 | Not-determined |
| S13_extracted_129489 | Not-determined |
| S13_extracted_129510 | Not-determined |
| S13_extracted_129523 | Not-determined |
| S13_extracted_129529 | Not-determined |
| S13_extracted_129534 | Not-determined |
| S13_extracted_129544 | Not-determined |
| S13_extracted_129564 | Not-determined |
| S13_extracted_129568 | Not-determined |
| S13_extracted_129587 | Not-determined |
| S13_extracted_129588 | Not-determined |
| S13_extracted_129597 | Not-determined |
| S13_extracted_129644 | Not-determined |
| S13_extracted_129648 | Not-determined |
| S13_extracted_129653 | Low-quality |
| S13_extracted_129655 | Not-determined |
| S13_extracted_129662 | Not-determined |
| S13_extracted_129666 | Not-determined |
| S13_extracted_129682 | Low-quality |
| S13_extracted_129699 | Not-determined |
| S13_extracted_129706 | Not-determined |
| S13_extracted_129711 | Not-determined |
| S13_extracted_129716 | Not-determined |
| S13_extracted_129723 | Not-determined |
| S13_extracted_129729 | Not-determined |
| S13_extracted_129730 | Not-determined |
| S13_extracted_129736 | Not-determined |
| S13_extracted_129741 | Not-determined |
| S13_extracted_129744 | Not-determined |
| S13_extracted_129751 | Not-determined |
| S13_extracted_129752 | Not-determined |
| S13_extracted_129765 | Not-determined |
| S13_extracted_129792 | Not-determined |
| S13_extracted_129799 | Not-determined |
| S13_extracted_129801 | Not-determined |
| S13_extracted_129805 | Low-quality |
| S13_extracted_129830 | Not-determined |
| S13_extracted_129838 | Not-determined |
| S13_extracted_129840 | Not-determined |
| S13_extracted_129850 | Not-determined |
| S13_extracted_129851 | Low-quality |
| S13_extracted_129859 | Not-determined |
| S13_extracted_129860 | Not-determined |
| S13_extracted_129882 | Not-determined |
| S13_extracted_129886 | Not-determined |
| S13_extracted_129889 | Not-determined |
| S13_extracted_129905 | Not-determined |
| S13_extracted_129908 | Not-determined |
| S13_extracted_129935 | Not-determined |
| S13_extracted_129938 | Low-quality |
| S13_extracted_129941 | Not-determined |
| S13_extracted_129946 | Not-determined |
| S13_extracted_129975 | Not-determined |
| S13_extracted_129977 | Not-determined |
| S13_extracted_130004 | Low-quality |
| S13_extracted_130013 | Not-determined |

| | |
|---|---|
| S13_extracted_130032 | Not-determined |
| S13_extracted_130035 | Not-determined |
| S13_extracted_130047 | Not-determined |
| S13_extracted_130049 | Not-determined |
| S13_extracted_130054 | Not-determined |
| S13_extracted_130070 | Not-determined |
| S13_extracted_130075 | Not-determined |
| S13_extracted_130079 | Not-determined |
| S13_extracted_130080 | Not-determined |
| S13_extracted_130082 | Low-quality |
| S13_extracted_130086 | Not-determined |
| S13_extracted_130091 | Not-determined |
| S13_extracted_130095 | Not-determined |
| S13_extracted_130111 | Not-determined |
| S13_extracted_130124 | Not-determined |
| S13_extracted_130135 | Not-determined |
| S13_extracted_130141 | Not-determined |
| S13_extracted_130143 | Not-determined |
| S13_extracted_130145 | Low-quality |
| S13_extracted_130159 | Not-determined |
| S13_extracted_130164 | Not-determined |
| S13_extracted_130168 | Not-determined |
| S13_extracted_130171 | Not-determined |
| S13_extracted_130191 | Not-determined |
| S13_extracted_130198 | Low-quality |
| S13_extracted_130210 | Not-determined |
| S13_extracted_130214 | Not-determined |
| S13_extracted_130221 | Not-determined |
| S13_extracted_130224 | Not-determined |
| S13_extracted_130244 | Not-determined |
| S13_extracted_130246 | Low-quality |
| S13_extracted_130257 | Not-determined |
| S13_extracted_130272 | Not-determined |
| S13_extracted_130278 | Not-determined |
| S13_extracted_130284 | Low-quality |
| S13_extracted_130286 | Low-quality |
| S13_extracted_130287 | Not-determined |
| S13_extracted_130291 | Not-determined |
| S13_extracted_130296 | Not-determined |
| S13_extracted_130299 | Not-determined |
| S13_extracted_130310 | Not-determined |
| S13_extracted_130331 | Not-determined |
| S13_extracted_130357 | Not-determined |
| S13_extracted_130361 | Not-determined |
| S13_extracted_130371 | Not-determined |
| S13_extracted_130373 | Not-determined |
| S13_extracted_130376 | Not-determined |
| S13_extracted_130380 | Not-determined |
| S13_extracted_130384 | Low-quality |
| S13_extracted_130386 | Not-determined |
| S13_extracted_130387 | Not-determined |
| S13_extracted_130427 | Not-determined |
| S13_extracted_130429 | Not-determined |
| S13_extracted_130432 | Not-determined |
| S13_extracted_130437 | Low-quality |
| S13_extracted_130458 | Not-determined |
| S13_extracted_130466 | Not-determined |
| S13_extracted_130471 | Low-quality |
| S13_extracted_130483 | Not-determined |
| S13_extracted_130486 | Not-determined |
| S13_extracted_130495 | Not-determined |
| S13_extracted_130499 | Not-determined |
| S13_extracted_130501 | Low-quality |
| S13_extracted_130505 | Not-determined |
| S13_extracted_130510 | Low-quality |
| S13_extracted_130513 | Not-determined |
| S13_extracted_130545 | Not-determined |
| S13_extracted_130550 | Not-determined |
| S13_extracted_130554 | Not-determined |
| S13_extracted_130582 | Low-quality |
| S13_extracted_130594 | Not-determined |
| S13_extracted_130595 | Not-determined |
| S13_extracted_130597 | Not-determined |
| S13_extracted_130598 | Not-determined |
| S13_extracted_130602 | Not-determined |
| S13_extracted_130609 | Not-determined |
| S13_extracted_130613 | Not-determined |
| S13_extracted_130614 | Not-determined |
| S13_extracted_130625 | Not-determined |
| S13_extracted_130628 | Not-determined |
| S13_extracted_130634 | Not-determined |
| S13_extracted_130640 | Not-determined |
| S13_extracted_130654 | Not-determined |
| S13_extracted_130664 | Not-determined |
| S13_extracted_130670 | Not-determined |
| S13_extracted_130677 | Not-determined |
| S13_extracted_130688 | Not-determined |
| S13_extracted_130692 | Not-determined |
| S13_extracted_130701 | Not-determined |
| S13_extracted_130709 | Not-determined |
| S13_extracted_130724 | Low-quality |
| S13_extracted_130733 | Not-determined |
| S13_extracted_130738 | Low-quality |
| S13_extracted_130747 | Not-determined |
| S13_extracted_130760 | Not-determined |
| S13_extracted_130768 | Not-determined |
| S13_extracted_130769 | Not-determined |
| S13_extracted_130771 | Not-determined |
| S13_extracted_130774 | Low-quality |
| S13_extracted_130796 | Not-determined |
| S13_extracted_130801 | Not-determined |
| S13_extracted_130811 | Low-quality |
| S13_extracted_130818 | Not-determined |
| S13_extracted_130831 | Not-determined |
| S13_extracted_130838 | Not-determined |
| S13_extracted_130849 | Not-determined |
| S13_extracted_130851 | Not-determined |
| S13_extracted_130856 | Not-determined |
| S13_extracted_130860 | Not-determined |
| S13_extracted_130864 | Not-determined |
| S13_extracted_130867 | Not-determined |
| S13_extracted_130891 | Not-determined |
| S13_extracted_130934 | Not-determined |
| S13_extracted_130935 | Not-determined |
| S13_extracted_130937 | Not-determined |
| S13_extracted_130946 | Not-determined |
| S13_extracted_130951 | Not-determined |
| S13_extracted_130965 | Low-quality |
| S13_extracted_130968 | Not-determined |
| S13_extracted_130978 | Not-determined |
| S13_extracted_130983 | Not-determined |
| S13_extracted_130986 | Low-quality |
| S13_extracted_130992 | Not-determined |
| S13_extracted_131012 | Not-determined |
| S13_extracted_131017 | Not-determined |
| S13_extracted_131018 | Not-determined |
| S13_extracted_131020 | Not-determined |
| S13_extracted_131023 | Not-determined |
| S13_extracted_131048 | Not-determined |
| S13_extracted_131053 | Not-determined |
| S13_extracted_131055 | Not-determined |
| S13_extracted_131062 | Not-determined |
| S13_extracted_131066 | Not-determined |
| S13_extracted_131070 | Not-determined |
| S13_extracted_131086 | Not-determined |
| S13_extracted_131094 | Low-quality |
| S13_extracted_131101 | Not-determined |
| S13_extracted_131107 | Not-determined |
| S13_extracted_131108 | Not-determined |
| S13_extracted_131112 | Not-determined |
| S13_extracted_131124 | Not-determined |
| S13_extracted_131131 | Not-determined |
| S13_extracted_131145 | Not-determined |
| S13_extracted_131158 | Not-determined |
| S13_extracted_131163 | Not-determined |
| S13_extracted_131172 | Not-determined |
| S11_vs_k141_48109\|\|full | Medium-quality |
| S13_extracted_131214 | Not-determined |
| S13_extracted_131224 | Low-quality |
| S13_extracted_131280 | Not-determined |
| S13_extracted_131294 | Not-determined |
| S13_extracted_131298 | Low-quality |
| S13_extracted_131300 | Not-determined |
| S13_extracted_131311 | Not-determined |
| S13_extracted_131313 | Not-determined |
| S13_extracted_131317 | Low-quality |
| S13_extracted_131326 | Not-determined |
| S13_extracted_131348 | Low-quality |
| S13_extracted_131380 | Not-determined |
| S13_extracted_131389 | Not-determined |
| S13_extracted_131390 | Low-quality |
| S13_extracted_131399 | Low-quality |
| S13_extracted_131402 | Not-determined |
| S13_extracted_131413 | Low-quality |
| S13_extracted_131416 | Low-quality |
| S13_extracted_131420 | Not-determined |
| S13_extracted_131425 | Low-quality |
| S13_extracted_131447 | Not-determined |
| S13_extracted_131449 | Not-determined |
| S13_extracted_131467 | Not-determined |
| S13_extracted_131479 | Not-determined |
| S13_extracted_131495 | Not-determined |
| S13_extracted_131514 | Low-quality |
| S13_extracted_131516 | Not-determined |
| S13_extracted_131520 | Not-determined |
| S13_extracted_131524 | Not-determined |
| S13_extracted_131566 | Not-determined |
| S13_extracted_131598 | Not-determined |
| S13_extracted_131599 | Low-quality |
| S13_extracted_131610 | Not-determined |
| S13_extracted_131614 | Low-quality |
| S13_extracted_131618 | Not-determined |
| S13_extracted_131628 | Not-determined |
| S13_extracted_131633 | Not-determined |
| S13_extracted_131636 | Low-quality |
| S13_extracted_131650 | Not-determined |
| S13_extracted_131656 | Not-determined |
| S13_extracted_131681 | Not-determined |
| S13_extracted_131682 | Not-determined |
| S13_extracted_131686 | Not-determined |
| S13_extracted_131704 | Not-determined |
| S13_extracted_131706 | Not-determined |
| S13_extracted_131708 | Not-determined |
| S13_extracted_131729 | Not-determined |
| S13_extracted_131732 | Not-determined |
| S13_extracted_131733 | Not-determined |
| S13_extracted_131737 | Low-quality |
| S13_extracted_131738 | Not-determined |
| S13_extracted_131743 | Not-determined |
| S13_extracted_131753 | Not-determined |
| S13_extracted_131763 | Not-determined |
| S13_extracted_131771 | Not-determined |
| S13_extracted_131801 | Not-determined |
| S13_extracted_131803 | Not-determined |
| S13_extracted_131807 | Not-determined |
| S13_extracted_131813 | Not-determined |
| S13_extracted_131819 | Not-determined |
| S13_extracted_131832 | Not-determined |
| S13_extracted_131850 | Not-determined |
| S13_extracted_131854 | Not-determined |
| S13_extracted_131859 | Not-determined |
| S13_extracted_131861 | Not-determined |
| S13_extracted_131862 | Not-determined |
| S13_extracted_131867 | Not-determined |
| S13_extracted_131869 | Not-determined |
| S13_extracted_131871 | Not-determined |
| S13_extracted_131889 | Not-determined |
| S13_extracted_131901 | Not-determined |
| S13_extracted_131924 | Not-determined |
| S13_extracted_131935 | Low-quality |
| S13_extracted_131938 | Not-determined |
| S13_extracted_131939 | Not-determined |
| S13_extracted_131949 | Not-determined |
| S13_extracted_131950 | Not-determined |
| S13_extracted_131957 | Not-determined |
| S13_extracted_131958 | Not-determined |
| S13_extracted_131964 | Not-determined |
| S13_extracted_131965 | Not-determined |
| S13_extracted_131968 | Not-determined |
| S13_extracted_131976 | Not-determined |
| S13_extracted_131988 | Not-determined |
| S13_extracted_132004 | Not-determined |
| S13_extracted_132014 | Not-determined |
| S13_extracted_132020 | Not-determined |
| S13_extracted_132022 | Not-determined |
| S13_extracted_132035 | Not-determined |
| S13_extracted_132040 | Not-determined |
| S13_extracted_132044 | Not-determined |
| S13_extracted_132056 | Low-quality |
| S13_extracted_132058 | Not-determined |
| S13_extracted_132059 | Low-quality |
| S13_extracted_132075 | Not-determined |
| S13_extracted_132085 | Not-determined |
| S13_extracted_132096 | Not-determined |
| S13_extracted_132108 | Low-quality |
| S13_extracted_132111 | Low-quality |
| S13_extracted_132121 | Not-determined |
| S13_extracted_132130 | Not-determined |
| S13_extracted_132140 | Not-determined |
| S13_extracted_132147 | Not-determined |
| S13_extracted_132152 | Not-determined |
| S13_extracted_132153 | Not-determined |
| S13_extracted_132163 | Low-quality |
| S13_extracted_132175 | Not-determined |
| S13_extracted_132184 | Not-determined |
| S13_extracted_132198 | Not-determined |
| S13_extracted_132220 | Not-determined |
| S13_extracted_132224 | Not-determined |
| S13_extracted_132229 | Not-determined |
| S13_extracted_132235 | Not-determined |
| S13_extracted_132245 | Not-determined |
| S13_extracted_132282 | Not-determined |
| S13_extracted_132284 | Low-quality |
| S13_extracted_132287 | Not-determined |
| S13_extracted_132290 | Low-quality |
| S13_extracted_132293 | Not-determined |
| S13_extracted_132298 | Not-determined |
| S13_extracted_132299 | Not-determined |
| S13_extracted_132340 | Not-determined |
| S13_extracted_132343 | Low-quality |
| S13_extracted_132358 | Not-determined |
| S13_extracted_132360 | Not-determined |
| S13_extracted_132368 | Not-determined |
| S13_extracted_132371 | Not-determined |
| S13_extracted_132372 | Not-determined |
| S13_extracted_132407 | Not-determined |
| S13_extracted_132422 | Not-determined |
| S13_extracted_132423 | Not-determined |
| S13_extracted_132431 | Not-determined |
| S13_extracted_132454 | Low-quality |
| S13_extracted_132458 | Not-determined |
| S13_extracted_132474 | Not-determined |
| S13_extracted_132475 | Not-determined |
| S13_extracted_132483 | Not-determined |
| S13_extracted_132490 | Not-determined |
| S13_extracted_132491 | Not-determined |
| S13_extracted_132500 | Not-determined |
| S13_extracted_132521 | Not-determined |
| S13_extracted_132535 | Low-quality |
| S13_extracted_132549 | Not-determined |
| S13_extracted_132558 | Low-quality |
| S13_extracted_132578 | Not-determined |
| S13_extracted_132589 | Not-determined |
| S13_extracted_132598 | Not-determined |
| S13_extracted_132600 | Low-quality |
| S13_extracted_132601 | Not-determined |
| S13_extracted_132612 | Not-determined |
| S13_extracted_132617 | Not-determined |
| S13_extracted_132630 | Not-determined |
| S13_extracted_132632 | Not-determined |
| S13_extracted_132635 | Low-quality |
| S13_extracted_132643 | Not-determined |
| S13_extracted_132656 | Not-determined |
| S13_extracted_132695 | Not-determined |
| S13_extracted_132716 | Not-determined |
| S13_extracted_132719 | Not-determined |
| S13_extracted_132749 | Not-determined |
| S13_extracted_132750 | Low-quality |
| S13_extracted_132755 | Not-determined |
| S13_extracted_132762 | Not-determined |
| S13_extracted_132778 | Low-quality |
| S13_extracted_132789 | Not-determined |
| S13_extracted_132791 | Not-determined |
| S13_extracted_132808 | Not-determined |
| S13_extracted_132818 | Not-determined |
| S13_extracted_132820 | Not-determined |
| S13_extracted_132821 | Not-determined |
| S13_extracted_132831 | Not-determined |
| S13_extracted_132840 | Not-determined |
| S13_extracted_132851 | Not-determined |
| S13_extracted_132853 | Not-determined |
| S13_extracted_132854 | Not-determined |
| S13_extracted_132857 | Not-determined |
| S13_extracted_132858 | Not-determined |
| S13_extracted_132859 | Not-determined |
| S13_extracted_132861 | Not-determined |
| S13_extracted_132876 | Not-determined |
| S13_extracted_132877 | Not-determined |
| S13_extracted_132889 | Not-determined |
| S13_extracted_132893 | Not-determined |
| S13_extracted_132896 | Not-determined |
| S13_extracted_132911 | Not-determined |
| S13_extracted_132915 | Not-determined |
| S13_extracted_132925 | Not-determined |
| S13_extracted_132928 | Not-determined |
| S13_extracted_132942 | Not-determined |
| S13_extracted_132943 | Not-determined |
| S13_extracted_132944 | Not-determined |
| S13_extracted_132962 | Low-quality |
| S13_extracted_132965 | Low-quality |
| S13_extracted_132971 | Not-determined |
| S13_extracted_132973 | Not-determined |
| S13_extracted_132975 | Not-determined |
| S13_extracted_132976 | Not-determined |
| S13_extracted_132977 | Not-determined |
| S13_extracted_132991 | Not-determined |
| S13_extracted_133001 | Not-determined |
| S13_extracted_133025 | Low-quality |
| S13_extracted_133028 | Not-determined |
| S13_extracted_133036 | Not-determined |
| S13_extracted_133052 | Low-quality |
| S13_extracted_133077 | Not-determined |
| S13_extracted_133082 | Not-determined |
| S13_extracted_133094 | Not-determined |
| S13_extracted_133102 | Not-determined |
| S13_extracted_133107 | Not-determined |
| S13_extracted_133115 | Not-determined |
| S13_extracted_133116 | Not-determined |
| S13_extracted_133120 | Low-quality |
| S13_extracted_133128 | Not-determined |
| S13_extracted_133136 | Not-determined |
| S13_extracted_133149 | Not-determined |
| S13_extracted_133161 | Low-quality |
| S13_extracted_133182 | Not-determined |
| S13_extracted_133198 | Not-determined |
| S13_extracted_133202 | Not-determined |
| S13_extracted_133203 | Not-determined |
| S13_extracted_133206 | Not-determined |
| S13_extracted_133210 | Low-quality |
| S13_extracted_133211 | Not-determined |
| S13_extracted_133212 | Not-determined |
| S13_extracted_133220 | Not-determined |
| S13_extracted_133221 | Low-quality |
| S13_extracted_133239 | Not-determined |
| S13_extracted_133240 | Not-determined |
| S13_extracted_133257 | Not-determined |
| S13_extracted_133260 | Low-quality |
| S13_extracted_133261 | Not-determined |
| S13_extracted_133274 | Not-determined |
| S13_extracted_133282 | Not-determined |
| S13_extracted_133287 | Low-quality |
| S13_extracted_133292 | Not-determined |
| S13_extracted_133297 | Not-determined |
| S13_extracted_133311 | Low-quality |
| S13_extracted_133313 | Not-determined |
| S13_extracted_133315 | Not-determined |
| S13_extracted_133319 | Not-determined |
| S13_extracted_133337 | Not-determined |
| S13_extracted_133342 | Not-determined |
| S13_extracted_133344 | Not-determined |
| S13_extracted_133361 | Not-determined |
| S13_extracted_133362 | Not-determined |
| S13_extracted_133384 | Not-determined |
| S13_extracted_133434 | Not-determined |
| S13_extracted_133444 | Not-determined |
| S13_extracted_133452 | Not-determined |
| S13_extracted_133459 | Low-quality |
| S13_extracted_133464 | Not-determined |
| S13_extracted_133473 | Not-determined |
| S13_extracted_133477 | Not-determined |
| S13_extracted_133503 | Not-determined |
| S13_extracted_133509 | Not-determined |
| S13_extracted_133510 | Not-determined |
| S13_extracted_133529 | Low-quality |
| S13_extracted_133535 | Low-quality |
| S13_extracted_133562 | Not-determined |
| S13_extracted_133564 | Not-determined |
| S13_extracted_133576 | Not-determined |
| S13_extracted_133580 | Not-determined |
| S13_extracted_133584 | Not-determined |
| S13_extracted_133608 | Not-determined |
| S13_extracted_133609 | Not-determined |
| S13_extracted_133621 | Not-determined |
| S13_extracted_133625 | Low-quality |
| S13_extracted_133631 | Low-quality |
| S13_extracted_133633 | Low-quality |
| S13_extracted_133635 | Not-determined |
| S13_extracted_133703 | Not-determined |
| S13_extracted_133732 | Not-determined |
| S13_extracted_133737 | Low-quality |
| S13_extracted_133738 | Not-determined |
| S13_extracted_133776 | Not-determined |
| S13_extracted_133777 | Not-determined |
| S13_extracted_133778 | Not-determined |
| S13_extracted_133779 | Not-determined |
| S13_extracted_133785 | Not-determined |
| S13_extracted_133787 | Not-determined |
| S13_extracted_133790 | Not-determined |
| S13_extracted_133800 | Not-determined |
| S13_extracted_133802 | Not-determined |
| S13_extracted_133809 | Not-determined |
| S13_extracted_133814 | Not-determined |
| S13_extracted_133817 | Not-determined |
| S13_extracted_133831 | Not-determined |
| S13_extracted_133835 | Low-quality |
| S13_extracted_133836 | Not-determined |
| S13_extracted_133840 | Not-determined |
| S13_extracted_133847 | Not-determined |
| S13_extracted_133868 | Not-determined |
| S13_extracted_133877 | Not-determined |
| S13_extracted_133886 | Not-determined |
| S13_extracted_133910 | Not-determined |
| S13_extracted_133929 | Not-determined |
| S13_extracted_133943 | Not-determined |
| S13_extracted_133947 | Not-determined |
| S13_extracted_133963 | Low-quality |
| S13_extracted_133965 | Not-determined |
| S13_extracted_133969 | Not-determined |
| S13_extracted_133980 | Not-determined |
| S13_extracted_133987 | Not-determined |
| S13_extracted_133989 | Not-determined |
| S13_extracted_133994 | Not-determined |
| S13_extracted_134001 | Not-determined |
| S13_extracted_134015 | Not-determined |
| S13_extracted_134016 | Not-determined |
| S13_extracted_134021 | Not-determined |
| S13_extracted_134033 | Not-determined |
| S13_extracted_134035 | Not-determined |
| S13_extracted_134036 | Not-determined |
| S13_extracted_134043 | Low-quality |
| S13_extracted_134053 | Low-quality |
| S13_extracted_134065 | Not-determined |
| S13_extracted_134074 | Not-determined |
| S13_extracted_134081 | Not-determined |
| S13_extracted_134083 | Not-determined |
| S13_extracted_134084 | Not-determined |
| S13_extracted_134098 | Not-determined |
| S13_extracted_134104 | Not-determined |
| S13_extracted_134116 | Not-determined |
| S13_extracted_134154 | Low-quality |
| S13_extracted_134156 | Not-determined |
| S13_extracted_134171 | Not-determined |
| S13_extracted_134183 | Not-determined |
| S13_extracted_134193 | Not-determined |
| S13_extracted_134207 | Low-quality |
| S13_extracted_134216 | Low-quality |
| S13_extracted_134227 | Low-quality |
| S13_extracted_134232 | Not-determined |
| S13_extracted_134236 | Not-determined |
| S13_extracted_134245 | Not-determined |
| S13_extracted_134262 | Not-determined |
| S13_extracted_134268 | Not-determined |
| S13_extracted_134287 | Not-determined |
| S13_extracted_134300 | Not-determined |
| S13_extracted_134306 | Not-determined |
| S13_extracted_134321 | Not-determined |
| S13_extracted_134331 | Not-determined |
| S13_extracted_134347 | Not-determined |
| S13_extracted_134395 | Not-determined |
| S13_extracted_134401 | Not-determined |
| S13_extracted_134409 | Low-quality |
| S13_extracted_134411 | Not-determined |
| S13_extracted_134413 | Not-determined |
| S13_extracted_134418 | Low-quality |
| S13_extracted_134431 | Not-determined |
| S13_extracted_134442 | Not-determined |
| S13_extracted_134443 | Not-determined |
| S13_extracted_134444 | Not-determined |
| S13_extracted_134468 | Not-determined |
| S13_extracted_134471 | Not-determined |
| S13_extracted_134489 | Not-determined |
| S13_extracted_134491 | Not-determined |
| S13_extracted_134500 | Not-determined |
| S13_extracted_134510 | Low-quality |
| S13_extracted_134511 | Not-determined |
| S13_extracted_134514 | Not-determined |
| S13_extracted_134516 | Not-determined |
| S13_extracted_134521 | Not-determined |
| S13_extracted_134527 | Not-determined |
| S13_extracted_134541 | Not-determined |
| S13_extracted_134543 | Not-determined |
| S13_extracted_134548 | Low-quality |
| S13_extracted_134552 | Not-determined |
| S13_extracted_134554 | Not-determined |
| S13_extracted_134558 | Low-quality |
| S13_extracted_134560 | Low-quality |
| S13_extracted_134561 | Not-determined |
| S13_extracted_134565 | Not-determined |
| S13_extracted_134569 | Not-determined |
| S13_extracted_134570 | Not-determined |
| S13_extracted_134575 | Not-determined |
| S13_extracted_134591 | Not-determined |
| S13_extracted_134598 | Not-determined |
| S13_extracted_134602 | Not-determined |
| S13_extracted_134617 | Not-determined |
| S13_extracted_134620 | Low-quality |
| S13_extracted_134621 | Not-determined |
| S13_extracted_134624 | Not-determined |
| S13_extracted_134634 | Not-determined |
| S13_extracted_134641 | Not-determined |
| S13_extracted_134642 | Not-determined |
| S13_extracted_134645 | Not-determined |
| S13_extracted_134650 | Not-determined |
| S13_extracted_134664 | Not-determined |
| S13_extracted_134679 | Not-determined |
| S13_extracted_134690 | Not-determined |
| S13_extracted_134691 | Not-determined |
| S13_extracted_134693 | Not-determined |
| S13_extracted_134694 | Not-determined |
| S13_extracted_134722 | Not-determined |
| S13_extracted_134726 | Not-determined |
| S13_extracted_134729 | Not-determined |
| S13_extracted_134753 | Not-determined |
| S13_extracted_134759 | Not-determined |
| S13_extracted_134766 | Not-determined |
| S13_extracted_134781 | Not-determined |
| S13_extracted_134786 | Not-determined |
| S13_extracted_134792 | Not-determined |
| S13_extracted_134794 | Not-determined |
| S13_extracted_134806 | Low-quality |
| S13_extracted_134812 | Low-quality |
| S13_extracted_134815 | Not-determined |
| S13_extracted_134829 | Not-determined |
| S13_extracted_134838 | Not-determined |
| S13_extracted_134841 | Not-determined |
| S13_extracted_134844 | Not-determined |
| S13_extracted_134854 | Not-determined |
| S13_extracted_134861 | Not-determined |
| S13_extracted_134896 | Not-determined |
| S13_extracted_134914 | Not-determined |
| S13_extracted_134916 | Not-determined |
| S13_extracted_134921 | Not-determined |
| S13_extracted_134927 | Low-quality |
| S13_extracted_134932 | Not-determined |
| S13_extracted_134949 | Not-determined |
| S13_extracted_134963 | Not-determined |
| S13_extracted_134971 | Not-determined |
| S13_extracted_134976 | Not-determined |
| S13_extracted_134977 | Not-determined |
| S13_extracted_134998 | Not-determined |
| S13_extracted_135000 | Not-determined |
| S13_extracted_135001 | Low-quality |
| S13_extracted_135009 | Not-determined |
| S13_extracted_135022 | Not-determined |
| S13_extracted_135031 | Not-determined |
| S13_extracted_135036 | Not-determined |
| S13_extracted_135055 | Low-quality |
| S13_extracted_135072 | Not-determined |
| S13_extracted_135075 | Low-quality |
| S13_extracted_135077 | Not-determined |
| S13_extracted_135090 | Not-determined |
| S13_extracted_135092 | Not-determined |
| S13_extracted_135122 | Not-determined |
| S13_extracted_135128 | Not-determined |
| S13_extracted_135147 | Not-determined |
| S13_extracted_135150 | Not-determined |
| S13_extracted_135155 | Not-determined |
| S13_extracted_135171 | Not-determined |
| S13_extracted_135180 | Low-quality |

| | |
|---|---|
| S13_extracted_135194 | Not-determined |
| S13_extracted_135212 | Not-determined |
| S13_extracted_135217 | Not-determined |
| S13_extracted_135220 | Not-determined |
| S13_extracted_135239 | Not-determined |
| S13_extracted_135240 | Not-determined |
| S13_extracted_135244 | Low-quality |
| S13_extracted_135246 | Not-determined |
| S13_extracted_135254 | Not-determined |
| S13_extracted_135256 | Not-determined |
| S13_extracted_135262 | Not-determined |
| S13_extracted_135287 | Not-determined |
| S13_extracted_135311 | Not-determined |
| S13_extracted_135324 | Not-determined |
| S13_extracted_135325 | Not-determined |
| S13_extracted_135334 | Not-determined |
| S13_extracted_135343 | Not-determined |
| S13_extracted_135367 | Not-determined |
| S11_extracted_k141_3790 | Medium-quality |
| S13_extracted_135378 | Not-determined |
| S13_extracted_135384 | Not-determined |
| S13_extracted_135397 | Not-determined |
| S13_extracted_135405 | Not-determined |
| S13_extracted_135426 | Not-determined |
| S13_extracted_135436 | Low-quality |
| S13_extracted_135444 | Not-determined |
| S13_extracted_135455 | Not-determined |
| S13_extracted_135462 | Not-determined |
| S13_extracted_135469 | Low-quality |
| S13_extracted_135473 | Low-quality |
| S13_extracted_135476 | Not-determined |
| S13_extracted_135478 | Not-determined |
| S13_extracted_135497 | Not-determined |
| S13_extracted_135512 | Not-determined |
| S13_extracted_135520 | Low-quality |
| S13_extracted_135540 | Not-determined |
| S13_extracted_135543 | Not-determined |
| S13_extracted_135562 | Not-determined |
| S13_extracted_135575 | Not-determined |
| S13_extracted_135580 | Not-determined |
| S13_extracted_135589 | Not-determined |
| S13_extracted_135604 | Not-determined |
| S13_extracted_135608 | Not-determined |
| S13_extracted_135624 | Not-determined |
| S13_extracted_135627 | Not-determined |
| S13_extracted_135642 | Not-determined |
| S13_extracted_135643 | Not-determined |
| S13_extracted_135650 | Not-determined |
| S13_extracted_135654 | Not-determined |
| S13_extracted_135658 | Not-determined |
| S13_extracted_135669 | Not-determined |
| S13_extracted_135680 | Not-determined |
| S13_extracted_135689 | Not-determined |
| S13_extracted_135691 | Not-determined |
| S13_extracted_135704 | Not-determined |
| S13_extracted_135708 | Not-determined |
| S13_extracted_135710 | Low-quality |
| S13_extracted_135727 | Low-quality |
| S13_extracted_135733 | Not-determined |
| S13_extracted_135735 | Not-determined |
| S13_extracted_135744 | Not-determined |
| S13_extracted_135752 | Not-determined |
| S13_extracted_135757 | Not-determined |
| S13_extracted_135760 | Not-determined |
| S13_extracted_135762 | Not-determined |
| S13_extracted_135772 | Low-quality |
| S13_extracted_135783 | Not-determined |
| S13_extracted_135821 | Not-determined |
| S13_extracted_135823 | Not-determined |
| S13_extracted_135824 | Not-determined |
| S13_extracted_135830 | Not-determined |
| S13_extracted_135838 | Not-determined |
| S13_extracted_135848 | Not-determined |
| S13_extracted_135851 | Not-determined |
| S13_extracted_135864 | Not-determined |
| S13_extracted_135899 | Not-determined |
| S13_extracted_135900 | Not-determined |
| S13_extracted_135901 | Not-determined |
| S13_extracted_135902 | Not-determined |
| S13_extracted_135905 | Not-determined |
| S13_extracted_135921 | Not-determined |
| S13_extracted_135929 | Not-determined |
| S13_extracted_135930 | Not-determined |
| S13_extracted_135935 | Not-determined |
| S13_extracted_135938 | Not-determined |
| S13_extracted_135960 | Not-determined |
| S13_extracted_135962 | Not-determined |
| S13_extracted_135975 | Not-determined |
| S13_extracted_135982 | Not-determined |
| S13_extracted_135987 | Not-determined |
| S13_extracted_135996 | Not-determined |
| S13_extracted_135999 | Not-determined |
| S13_extracted_136015 | Not-determined |
| S13_extracted_136034 | Not-determined |
| S13_extracted_136047 | Not-determined |
| S13_extracted_136057 | Not-determined |
| S13_extracted_136068 | Not-determined |
| S13_extracted_136074 | Not-determined |
| S13_extracted_136077 | Not-determined |
| S13_extracted_136083 | Not-determined |
| S13_extracted_136112 | Not-determined |
| S13_extracted_136113 | Not-determined |
| S13_extracted_136117 | Low-quality |
| S13_extracted_136134 | Not-determined |
| S13_extracted_136142 | Not-determined |
| S13_extracted_136162 | Not-determined |
| S13_extracted_136171 | Not-determined |
| S13_extracted_136180 | Not-determined |
| S13_extracted_136182 | Not-determined |
| S13_extracted_136183 | Not-determined |
| S13_extracted_136186 | Not-determined |
| S13_extracted_136204 | Not-determined |
| S13_extracted_136206 | Not-determined |
| S13_extracted_136211 | Not-determined |
| S13_extracted_136240 | Not-determined |
| S13_extracted_136243 | Not-determined |
| S13_extracted_136258 | Low-quality |
| S13_extracted_136265 | Not-determined |
| S13_extracted_136272 | Not-determined |
| S13_extracted_136288 | Not-determined |
| S13_extracted_136297 | Not-determined |
| S13_extracted_136301 | Not-determined |
| S13_extracted_136320 | Not-determined |
| S13_extracted_136334 | Not-determined |
| S13_extracted_136342 | Not-determined |
| S13_extracted_136351 | Not-determined |
| S13_extracted_136352 | Low-quality |
| S13_extracted_136361 | Not-determined |
| S13_extracted_136369 | Not-determined |
| S13_extracted_136380 | Not-determined |
| S13_extracted_136381 | Not-determined |
| S13_extracted_136388 | Not-determined |
| S13_extracted_136405 | Not-determined |
| S13_extracted_136407 | Not-determined |
| S13_extracted_136419 | Not-determined |
| S13_extracted_136424 | Not-determined |
| S13_extracted_136425 | Not-determined |
| S13_extracted_136427 | Not-determined |
| S13_extracted_136440 | Not-determined |
| S13_extracted_136448 | Low-quality |
| S13_extracted_136460 | Low-quality |
| S13_extracted_136463 | Not-determined |
| S13_extracted_136464 | Not-determined |
| S13_extracted_136470 | Not-determined |
| S13_extracted_136474 | Not-determined |
| S13_extracted_136477 | Not-determined |
| S13_extracted_136487 | Low-quality |
| S13_extracted_136490 | Not-determined |
| S13_extracted_136491 | Not-determined |
| S13_extracted_136506 | Not-determined |
| S13_extracted_136508 | Not-determined |
| S13_extracted_136512 | Not-determined |
| S13_extracted_136521 | Not-determined |
| S13_extracted_136527 | Not-determined |
| S13_extracted_136543 | Not-determined |
| S13_extracted_136556 | Not-determined |
| S13_extracted_136579 | Not-determined |
| S13_extracted_136591 | Not-determined |
| S13_extracted_136592 | Not-determined |
| S13_extracted_136630 | Not-determined |
| S13_extracted_136631 | Not-determined |
| S13_extracted_136632 | Not-determined |
| S13_extracted_136643 | Not-determined |
| S13_extracted_136658 | Not-determined |
| S13_extracted_136663 | Not-determined |
| S13_extracted_136680 | Not-determined |
| S13_extracted_136693 | Not-determined |
| S13_extracted_136699 | Not-determined |
| S13_extracted_136709 | Not-determined |
| S13_extracted_136710 | Not-determined |
| S13_extracted_136716 | Not-determined |
| S13_extracted_136720 | Not-determined |
| S13_extracted_136723 | Not-determined |
| S13_extracted_136724 | Not-determined |
| S13_extracted_136730 | Not-determined |
| S13_extracted_136734 | Not-determined |
| S13_extracted_136736 | Not-determined |
| S13_extracted_136749 | Not-determined |
| S13_extracted_136764 | Low-quality |
| S13_extracted_136766 | Not-determined |
| S13_extracted_136769 | Not-determined |
| S13_extracted_136773 | Low-quality |
| S13_extracted_136776 | Not-determined |
| S13_extracted_136778 | Not-determined |
| S13_extracted_136791 | Not-determined |
| S13_extracted_136806 | Not-determined |
| S13_extracted_136812 | Not-determined |
| S13_extracted_136817 | Not-determined |
| S13_extracted_136819 | Not-determined |
| S13_extracted_136826 | Not-determined |
| S13_extracted_136834 | Not-determined |
| S13_extracted_136835 | Not-determined |
| S13_extracted_136847 | Not-determined |
| S13_extracted_136850 | Not-determined |
| S13_extracted_136853 | Low-quality |
| S13_extracted_136866 | Not-determined |
| S13_extracted_136876 | Not-determined |
| S13_extracted_136877 | Not-determined |
| S13_extracted_136882 | Low-quality |
| S13_extracted_136887 | Not-determined |
| S13_extracted_136902 | Not-determined |
| S13_extracted_136905 | Not-determined |
| S13_extracted_136908 | Not-determined |
| S13_extracted_136947 | Not-determined |
| S13_extracted_136951 | Not-determined |
| S13_extracted_136963 | Not-determined |
| S13_extracted_136965 | Low-quality |
| S13_extracted_136973 | Not-determined |
| S13_extracted_136974 | Not-determined |
| S13_extracted_136980 | Not-determined |
| S13_extracted_136985 | Not-determined |
| S13_extracted_136991 | Not-determined |
| S13_extracted_136995 | Low-quality |
| S13_extracted_136998 | Not-determined |
| S13_extracted_136999 | Not-determined |
| S13_extracted_137000 | Not-determined |
| S13_extracted_137005 | Not-determined |
| S13_extracted_137013 | Not-determined |
| S13_extracted_137015 | Not-determined |
| S13_extracted_137016 | Not-determined |
| S13_extracted_137021 | Not-determined |
| S13_extracted_137023 | Not-determined |
| S13_extracted_137045 | Not-determined |
| S13_extracted_137072 | Not-determined |
| S13_extracted_137091 | Not-determined |
| S13_extracted_137093 | Not-determined |
| S13_extracted_137105 | Low-quality |
| S13_extracted_137106 | Not-determined |
| S13_extracted_137112 | Not-determined |
| S13_extracted_137124 | Not-determined |
| S13_extracted_137130 | Not-determined |
| S13_extracted_137144 | Not-determined |
| S13_extracted_137162 | Low-quality |
| S13_extracted_137168 | Not-determined |
| S13_extracted_137171 | Not-determined |
| S13_extracted_137177 | Not-determined |
| S13_extracted_137182 | Not-determined |
| S13_extracted_137194 | Not-determined |
| S13_extracted_137212 | Not-determined |
| S13_extracted_137215 | Not-determined |
| S13_extracted_137220 | Not-determined |
| S13_extracted_137231 | Not-determined |
| S13_extracted_137241 | Not-determined |
| S13_extracted_137263 | Not-determined |
| S13_extracted_137301 | Low-quality |
| S13_extracted_137302 | Not-determined |
| S13_extracted_137332 | Not-determined |
| S13_extracted_137339 | Not-determined |
| S13_extracted_137357 | Not-determined |
| S13_extracted_137359 | Not-determined |
| S13_extracted_137361 | Not-determined |
| S13_extracted_137383 | Not-determined |
| S13_extracted_137399 | Not-determined |
| S13_extracted_137416 | Not-determined |
| S13_extracted_137426 | Not-determined |
| S13_extracted_137427 | Not-determined |
| S13_extracted_137435 | Not-determined |
| S13_extracted_137444 | Not-determined |
| S13_extracted_137445 | Low-quality |
| S13_extracted_137453 | Not-determined |
| S13_extracted_137462 | Not-determined |
| S13_extracted_137471 | Not-determined |
| S13_extracted_137486 | Not-determined |
| S13_extracted_137492 | Not-determined |
| S13_extracted_137501 | Not-determined |
| S13_extracted_137507 | Not-determined |
| S13_extracted_137524 | Not-determined |
| S13_extracted_137525 | Not-determined |
| S13_extracted_137533 | Not-determined |
| S13_extracted_137538 | Not-determined |
| S13_extracted_137545 | Not-determined |
| S13_extracted_137552 | Not-determined |
| S13_extracted_137560 | Not-determined |
| S13_extracted_137564 | Not-determined |
| S13_extracted_137565 | Not-determined |
| S13_extracted_137576 | Not-determined |
| S13_extracted_137595 | Low-quality |
| S13_extracted_137608 | Low-quality |
| S13_extracted_137634 | Not-determined |
| S13_extracted_137639 | Not-determined |
| S13_extracted_137659 | Not-determined |
| S13_extracted_137667 | Not-determined |
| S13_extracted_137684 | Not-determined |
| S13_extracted_137689 | Not-determined |
| S13_extracted_137699 | Not-determined |
| S13_extracted_137701 | Not-determined |
| S13_extracted_137716 | Not-determined |
| S13_extracted_137725 | Not-determined |
| S13_extracted_137734 | Not-determined |
| S13_extracted_137739 | Not-determined |
| S13_extracted_137743 | Not-determined |
| S13_extracted_137745 | Not-determined |
| S13_extracted_137748 | Low-quality |
| S13_extracted_137750 | Not-determined |
| S13_extracted_137752 | Not-determined |
| S13_extracted_137762 | Not-determined |
| S13_extracted_137763 | Not-determined |
| S13_extracted_137766 | Not-determined |
| S13_extracted_137767 | Not-determined |
| S13_extracted_137768 | Not-determined |
| S13_extracted_137773 | Not-determined |
| S13_extracted_137793 | Not-determined |
| S13_extracted_137795 | Not-determined |
| S13_extracted_137829 | Low-quality |
| S13_extracted_137836 | Not-determined |
| S13_extracted_137843 | Not-determined |
| S13_extracted_137846 | Not-determined |
| S13_extracted_137853 | Not-determined |
| S13_extracted_137856 | Not-determined |
| S13_extracted_137860 | Not-determined |
| S13_extracted_137863 | Not-determined |
| S13_extracted_137865 | Not-determined |
| S13_extracted_137869 | Not-determined |
| S13_extracted_137872 | Not-determined |
| S13_extracted_137877 | Not-determined |
| S13_extracted_137898 | Not-determined |
| S13_extracted_137905 | Not-determined |
| S13_extracted_137906 | Not-determined |
| S13_extracted_137915 | Low-quality |
| S13_extracted_137920 | Not-determined |
| S13_extracted_137921 | Low-quality |
| S13_extracted_137923 | Not-determined |
| S13_extracted_137939 | Not-determined |
| S13_extracted_137940 | Low-quality |
| S13_extracted_137945 | Not-determined |
| S13_extracted_137951 | Not-determined |
| S13_extracted_137967 | Not-determined |
| S13_extracted_137974 | Not-determined |
| S13_extracted_137977 | Not-determined |
| S13_extracted_137981 | Not-determined |
| S13_extracted_137983 | Not-determined |
| S13_extracted_137999 | Not-determined |
| S13_extracted_138006 | Low-quality |
| S13_extracted_138007 | Not-determined |
| S13_extracted_138012 | Not-determined |
| S13_extracted_138015 | Not-determined |
| S13_extracted_138027 | Not-determined |
| S13_extracted_138028 | Not-determined |
| S13_extracted_138031 | Not-determined |
| S13_extracted_138037 | Not-determined |
| S13_extracted_138043 | Low-quality |
| S13_extracted_138051 | Not-determined |
| S13_extracted_138052 | Not-determined |
| S13_extracted_138055 | Not-determined |
| S13_extracted_138057 | Not-determined |
| S13_extracted_138059 | Not-determined |
| S13_extracted_138068 | Not-determined |
| S13_extracted_138094 | Not-determined |
| S13_extracted_138113 | Not-determined |
| S13_extracted_138114 | Not-determined |
| S13_extracted_138116 | Not-determined |
| S13_extracted_138123 | Not-determined |
| S13_extracted_138127 | Not-determined |
| S13_extracted_138128 | Not-determined |
| S13_extracted_138129 | Not-determined |
| S13_extracted_138140 | Not-determined |
| S13_extracted_138144 | Low-quality |
| S13_extracted_138148 | Low-quality |
| S13_extracted_138150 | Not-determined |
| S8_vs_k141_79769\|\|full | High-quality |
| S13_extracted_138157 | Not-determined |
| S13_extracted_138158 | Not-determined |
| S13_extracted_138180 | Not-determined |
| S13_extracted_138181 | Not-determined |
| S13_extracted_138196 | Low-quality |
| S13_extracted_138200 | Not-determined |
| S13_extracted_138202 | Not-determined |
| S13_extracted_138209 | Not-determined |
| S13_extracted_138237 | Low-quality |
| S13_extracted_138238 | Not-determined |
| S13_extracted_138272 | Not-determined |
| S13_extracted_138288 | Not-determined |
| S13_extracted_138300 | Not-determined |
| S13_extracted_138315 | Not-determined |
| S13_extracted_138317 | Not-determined |
| S13_extracted_138331 | Low-quality |
| S13_extracted_138340 | Not-determined |
| S13_extracted_138357 | Not-determined |
| S13_extracted_138366 | Not-determined |
| S13_extracted_138386 | Not-determined |
| S13_extracted_138391 | Not-determined |
| S13_extracted_138415 | Low-quality |
| S13_extracted_138420 | Not-determined |
| S13_extracted_138422 | Not-determined |
| S13_extracted_138435 | Not-determined |
| S13_extracted_138437 | Not-determined |
| S13_extracted_138440 | Not-determined |
| S13_extracted_138443 | Not-determined |
| S13_extracted_138446 | Not-determined |
| S13_extracted_138449 | Not-determined |
| S13_extracted_138457 | Not-determined |
| S13_extracted_138470 | Low-quality |
| S13_extracted_138477 | Not-determined |
| S13_extracted_138478 | Not-determined |
| S13_extracted_138482 | Not-determined |
| S13_extracted_138487 | Not-determined |
| S13_extracted_138497 | Not-determined |
| S13_extracted_138509 | Not-determined |
| S13_extracted_138521 | Not-determined |
| S13_extracted_138546 | Not-determined |
| S13_extracted_138560 | Not-determined |
| S13_extracted_138564 | Not-determined |
| S13_extracted_138573 | Not-determined |
| S13_extracted_138582 | Not-determined |
| S13_extracted_138583 | Not-determined |
| S13_extracted_138597 | Not-determined |
| S13_extracted_138607 | Not-determined |
| S13_extracted_138611 | Not-determined |
| S13_extracted_138613 | Not-determined |
| S13_extracted_138614 | Not-determined |
| S13_extracted_138628 | Not-determined |
| S13_extracted_138629 | Not-determined |
| S13_extracted_138632 | Not-determined |
| S13_extracted_138635 | Not-determined |
| S13_extracted_138638 | Low-quality |
| S13_extracted_138654 | Not-determined |
| S13_extracted_138656 | Not-determined |
| S13_extracted_138660 | Not-determined |
| S13_extracted_138666 | Not-determined |
| S13_extracted_138675 | Not-determined |
| S13_extracted_138687 | Not-determined |
| S13_extracted_138690 | Not-determined |
| S13_extracted_138693 | Low-quality |
| S13_extracted_138701 | Not-determined |
| S13_extracted_138703 | Not-determined |
| S13_extracted_138712 | Not-determined |
| S13_extracted_138730 | Not-determined |
| S13_extracted_138740 | Low-quality |
| S13_extracted_138752 | Not-determined |
| S13_extracted_138760 | Not-determined |
| S13_extracted_138763 | Not-determined |
| S13_extracted_138769 | Low-quality |
| S13_extracted_138778 | Not-determined |
| S13_extracted_138811 | Not-determined |
| S13_extracted_138827 | Not-determined |
| S13_extracted_138834 | Not-determined |
| S13_extracted_138849 | Not-determined |
| S13_extracted_138864 | Not-determined |
| S13_extracted_138868 | Not-determined |
| S13_extracted_138870 | Not-determined |
| S13_extracted_138885 | Low-quality |
| S13_extracted_138888 | Not-determined |
| S13_extracted_138894 | Not-determined |
| S13_extracted_138904 | Not-determined |
| S13_extracted_138914 | Not-determined |
| S13_extracted_138941 | Not-determined |
| S13_extracted_138944 | Not-determined |
| S13_extracted_138952 | Not-determined |
| S13_extracted_138961 | Not-determined |
| S13_extracted_138972 | Not-determined |
| S13_extracted_138973 | Not-determined |
| S13_extracted_138974 | Not-determined |
| S13_extracted_138984 | Not-determined |
| S13_extracted_138992 | Not-determined |
| S13_extracted_138994 | Not-determined |
| S13_extracted_138998 | Not-determined |
| S13_extracted_139013 | Not-determined |
| S13_extracted_139021 | Not-determined |
| S13_extracted_139022 | Not-determined |
| S13_extracted_139030 | Not-determined |
| S13_extracted_139032 | Low-quality |
| S13_extracted_139034 | Not-determined |
| S13_extracted_139048 | Not-determined |
| S13_extracted_139051 | Not-determined |
| S13_extracted_139061 | Not-determined |
| S13_extracted_139073 | Not-determined |
| S13_extracted_139074 | Not-determined |
| S13_extracted_139094 | Not-determined |
| S13_extracted_139103 | Not-determined |
| S13_extracted_139104 | Not-determined |
| S13_extracted_139105 | Not-determined |
| S13_extracted_139123 | Not-determined |
| S13_extracted_139131 | Not-determined |
| S13_extracted_139136 | Not-determined |
| S13_extracted_139151 | Not-determined |
| S13_extracted_139168 | Low-quality |
| S13_extracted_139174 | Not-determined |
| S13_extracted_139176 | Not-determined |
| S13_extracted_139190 | Not-determined |
| S13_extracted_139194 | Not-determined |
| S13_extracted_139214 | Not-determined |
| S13_extracted_139233 | Not-determined |
| S13_extracted_139243 | Not-determined |
| S13_extracted_139267 | Not-determined |
| S13_extracted_139309 | Not-determined |
| S13_extracted_139314 | Not-determined |
| S13_extracted_139332 | Not-determined |
| S13_extracted_139336 | Not-determined |
| S13_extracted_139339 | Not-determined |
| S13_extracted_139345 | Not-determined |
| S13_extracted_139352 | Not-determined |
| S13_extracted_139371 | Not-determined |
| S13_extracted_139391 | Not-determined |
| S13_extracted_139413 | Not-determined |
| S13_extracted_139423 | Not-determined |
| S13_extracted_139450 | Not-determined |
| S13_extracted_139453 | Not-determined |
| S13_extracted_139457 | Not-determined |
| S13_extracted_139460 | Not-determined |
| S13_extracted_139478 | Not-determined |
| S13_extracted_139482 | Not-determined |
| S13_extracted_139491 | Not-determined |
| S13_extracted_139494 | Not-determined |
| S13_extracted_139500 | Not-determined |
| S13_extracted_139508 | Not-determined |
| S13_extracted_139516 | Not-determined |
| S13_extracted_139523 | Not-determined |
| S13_extracted_139531 | Not-determined |
| S13_extracted_139533 | Not-determined |
| S13_extracted_139538 | Not-determined |
| S13_extracted_139548 | Not-determined |
| S13_extracted_139550 | Not-determined |
| S13_extracted_139552 | Not-determined |
| S13_extracted_139555 | Not-determined |
| S13_extracted_139556 | Low-quality |
| S13_extracted_139566 | Not-determined |
| S13_extracted_139567 | Not-determined |
| S13_extracted_139568 | Not-determined |
| S13_extracted_139571 | Not-determined |
| S13_extracted_139576 | Not-determined |
| S13_extracted_139578 | Low-quality |
| S13_extracted_139587 | Not-determined |
| S13_extracted_139633 | Not-determined |
| S13_extracted_139651 | Not-determined |
| S13_extracted_139654 | Not-determined |
| S13_extracted_139655 | Not-determined |
| S13_extracted_139664 | Not-determined |
| S13_extracted_139670 | Not-determined |
| S13_extracted_139681 | Low-quality |
| S13_extracted_139690 | Not-determined |
| S13_extracted_139724 | Not-determined |
| S13_extracted_139728 | Not-determined |
| S13_extracted_139747 | Not-determined |
| S13_extracted_139749 | Not-determined |
| S13_extracted_139752 | Not-determined |
| S13_extracted_139762 | Not-determined |
| S13_extracted_139765 | Not-determined |
| S13_extracted_139768 | Not-determined |
| S13_extracted_139769 | Not-determined |
| S13_extracted_139775 | Not-determined |
| S13_extracted_139780 | Not-determined |
| S13_extracted_139791 | Not-determined |
| S13_extracted_139798 | Not-determined |
| S13_extracted_139800 | Not-determined |
| S13_extracted_139805 | Low-quality |
| S13_extracted_139814 | Not-determined |
| S13_extracted_139821 | Not-determined |
| S13_extracted_139828 | Not-determined |
| S13_extracted_139834 | Not-determined |
| S13_extracted_139848 | Low-quality |
| S13_extracted_139852 | Not-determined |
| S13_extracted_139876 | Not-determined |
| S13_extracted_139886 | Not-determined |
| S13_extracted_139896 | Not-determined |
| S13_extracted_139897 | Not-determined |
| S13_extracted_139898 | Not-determined |
| S13_extracted_139904 | Not-determined |
| S13_extracted_139915 | Not-determined |
| S13_extracted_139921 | Not-determined |
| S13_extracted_139923 | Low-quality |
| S13_extracted_139924 | Not-determined |
| S13_extracted_139942 | Not-determined |
| S13_extracted_139951 | Not-determined |
| S13_extracted_139956 | Not-determined |
| S13_extracted_139962 | Not-determined |
| S13_extracted_139975 | Not-determined |
| S13_extracted_139978 | Not-determined |
| S13_extracted_139980 | Low-quality |
| S13_extracted_139982 | Not-determined |
| S13_extracted_139995 | Not-determined |
| S13_extracted_140002 | Not-determined |
| S13_extracted_140020 | Not-determined |
| S13_extracted_140033 | Low-quality |
| S13_extracted_140050 | Not-determined |
| S13_extracted_140062 | Not-determined |
| S13_extracted_140068 | Not-determined |
| S13_extracted_140072 | Not-determined |
| S13_extracted_140090 | Not-determined |
| S13_extracted_140114 | Low-quality |
| S13_extracted_140115 | Not-determined |
| S13_extracted_140118 | Not-determined |
| S13_extracted_140120 | Not-determined |
| S13_extracted_140125 | Not-determined |
| S13_extracted_140128 | Not-determined |
| S13_extracted_140130 | Not-determined |
| S13_extracted_140136 | Not-determined |
| S13_extracted_140167 | Not-determined |
| S13_extracted_140178 | Not-determined |
| S13_extracted_140187 | Not-determined |

| | |
|---|---|
| S13_extracted_140199 | Not-determined |
| S13_extracted_140203 | Not-determined |
| S13_extracted_140212 | Not-determined |
| S13_extracted_140214 | Not-determined |
| S13_extracted_140216 | Not-determined |
| S13_extracted_140217 | Not-determined |
| S13_extracted_140234 | Not-determined |
| S13_extracted_140244 | Not-determined |
| S13_extracted_140252 | Low-quality |
| S13_extracted_140263 | Not-determined |
| S13_extracted_140267 | Low-quality |
| S13_extracted_140292 | Not-determined |
| S13_extracted_140294 | Not-determined |
| S13_extracted_140295 | Not-determined |
| S13_extracted_140298 | Not-determined |
| S13_extracted_140300 | Not-determined |
| S13_extracted_140313 | Not-determined |
| S13_extracted_140345 | Not-determined |
| S13_extracted_140365 | Not-determined |
| S13_extracted_140376 | Not-determined |
| S13_extracted_140396 | Not-determined |
| S13_extracted_140399 | Not-determined |
| S13_extracted_140401 | Not-determined |
| S13_extracted_140407 | Not-determined |
| S13_extracted_140414 | Not-determined |
| S13_extracted_140425 | Not-determined |
| S13_extracted_140428 | Not-determined |
| S13_extracted_140461 | Not-determined |
| S13_extracted_140462 | Low-quality |
| S13_extracted_140463 | Not-determined |
| S13_extracted_140464 | Not-determined |
| S13_extracted_140481 | Not-determined |
| S13_extracted_140482 | Not-determined |
| S13_extracted_140484 | Not-determined |
| S13_extracted_140493 | Not-determined |
| S13_extracted_140495 | Not-determined |
| S13_extracted_140518 | Not-determined |
| S13_extracted_140522 | Low-quality |
| S13_extracted_140523 | Not-determined |
| S13_extracted_140527 | Not-determined |
| S13_extracted_140541 | Not-determined |
| S13_extracted_140545 | Not-determined |
| S13_extracted_140551 | Not-determined |
| S13_extracted_140571 | Not-determined |
| S13_extracted_140584 | Not-determined |
| S13_extracted_140603 | Not-determined |
| S13_extracted_140611 | Not-determined |
| S13_extracted_140615 | Not-determined |
| S13_extracted_140618 | Not-determined |
| S13_extracted_140626 | Not-determined |
| S13_extracted_140642 | Not-determined |
| S13_extracted_140643 | Not-determined |
| S13_extracted_140647 | Not-determined |
| S13_extracted_140660 | Low-quality |
| S13_extracted_140665 | Not-determined |
| S13_extracted_140669 | Not-determined |
| S13_extracted_140697 | Not-determined |
| S13_extracted_140714 | Not-determined |
| S13_extracted_140724 | Low-quality |
| S13_extracted_140730 | Not-determined |
| S13_extracted_140738 | Not-determined |
| S13_extracted_140740 | Not-determined |
| S13_extracted_140757 | Not-determined |
| S13_extracted_140769 | Not-determined |
| S13_extracted_140772 | Not-determined |
| S13_extracted_140787 | Not-determined |
| S13_extracted_140811 | Not-determined |
| S13_extracted_140814 | Not-determined |
| S13_extracted_140854 | Not-determined |
| S13_extracted_140856 | Not-determined |
| S13_extracted_140858 | Not-determined |
| S13_extracted_140882 | Not-determined |
| S13_extracted_140883 | Not-determined |
| S13_extracted_140891 | Not-determined |
| S13_extracted_140899 | Not-determined |
| S13_extracted_140900 | Not-determined |
| S13_extracted_140901 | Not-determined |
| S13_extracted_140916 | Not-determined |
| S13_extracted_140926 | Low-quality |
| S13_extracted_140928 | Not-determined |
| S13_extracted_140939 | Not-determined |
| S13_extracted_140954 | Not-determined |
| S13_extracted_140958 | Not-determined |
| S13_extracted_140975 | Low-quality |
| S13_extracted_140980 | Not-determined |
| S13_extracted_140996 | Not-determined |
| S13_extracted_141007 | Not-determined |
| S13_extracted_141016 | Not-determined |
| S13_extracted_141018 | Not-determined |
| S13_extracted_141019 | Not-determined |
| S13_extracted_141030 | Not-determined |
| S13_extracted_141046 | Not-determined |
| S13_extracted_141052 | Not-determined |
| S13_extracted_141065 | Not-determined |
| S13_extracted_141069 | Not-determined |
| S13_extracted_141092 | Not-determined |
| S13_extracted_141093 | Low-quality |
| S13_extracted_141094 | Not-determined |
| S13_extracted_141095 | Not-determined |
| S13_extracted_141096 | Not-determined |
| S13_extracted_141098 | Not-determined |
| S13_extracted_141099 | Low-quality |
| S13_extracted_141115 | Not-determined |
| S13_extracted_141120 | Low-quality |
| S13_extracted_141121 | Low-quality |
| S13_extracted_141129 | Low-quality |
| S13_extracted_141133 | Not-determined |
| S13_extracted_141140 | Not-determined |
| S13_extracted_141147 | Not-determined |
| S13_extracted_141151 | Not-determined |
| S13_extracted_141155 | Not-determined |
| S13_extracted_141178 | Not-determined |
| S13_extracted_141184 | Not-determined |
| S13_extracted_141190 | Not-determined |
| S13_extracted_141201 | Not-determined |
| S13_extracted_141203 | Low-quality |
| S13_extracted_141214 | Low-quality |
| S13_extracted_141229 | Not-determined |
| S13_extracted_141233 | Not-determined |
| S13_extracted_141258 | Not-determined |
| S13_extracted_141260 | Not-determined |
| S13_extracted_141262 | Not-determined |
| S13_extracted_141286 | Not-determined |
| S13_extracted_141294 | Low-quality |
| S13_extracted_141296 | Not-determined |
| S13_extracted_141300 | Not-determined |
| S13_extracted_141303 | Not-determined |
| S13_extracted_141305 | Not-determined |
| S13_extracted_141307 | Not-determined |
| S13_extracted_141309 | Not-determined |
| S13_extracted_141322 | Not-determined |
| S13_extracted_141330 | Not-determined |
| S13_extracted_141343 | Not-determined |
| S13_extracted_141350 | Not-determined |
| S13_extracted_141355 | Not-determined |
| S13_extracted_141391 | Not-determined |
| S13_extracted_141393 | Not-determined |
| S13_extracted_141394 | Not-determined |
| S13_extracted_141396 | Not-determined |
| S13_extracted_141397 | Not-determined |
| S13_extracted_141408 | Not-determined |
| S13_extracted_141409 | Not-determined |
| S13_extracted_141433 | Not-determined |
| S13_extracted_141439 | Not-determined |
| S13_extracted_141444 | Not-determined |
| S13_extracted_141453 | Not-determined |
| S13_extracted_141454 | Not-determined |
| S13_extracted_141459 | Low-quality |
| S13_extracted_141462 | Not-determined |
| S13_extracted_141479 | Not-determined |
| S13_extracted_141499 | Not-determined |
| S13_extracted_141507 | Not-determined |
| S13_extracted_141512 | Not-determined |
| S13_extracted_141513 | Not-determined |
| S13_extracted_141514 | Not-determined |
| S13_extracted_141526 | Not-determined |
| S13_extracted_141551 | Not-determined |
| S13_extracted_141566 | Not-determined |
| S13_extracted_141569 | Not-determined |
| S13_extracted_141571 | Not-determined |
| S13_extracted_141572 | Not-determined |
| S13_extracted_141573 | Not-determined |
| S13_extracted_141590 | Not-determined |
| S13_extracted_141597 | Not-determined |
| S13_extracted_141599 | Not-determined |
| S13_extracted_141643 | Not-determined |
| S13_extracted_141654 | Not-determined |
| S13_extracted_141664 | Not-determined |
| S13_extracted_141670 | Not-determined |
| S13_extracted_141680 | Not-determined |
| S13_extracted_141681 | Not-determined |
| S13_extracted_141683 | Not-determined |
| S13_extracted_141686 | Not-determined |
| S13_extracted_141687 | Not-determined |
| S13_extracted_141688 | Not-determined |
| S13_extracted_141699 | Not-determined |
| S13_extracted_141700 | Not-determined |
| S13_extracted_141701 | Not-determined |
| S13_extracted_141715 | Low-quality |
| S13_extracted_141718 | Not-determined |
| S13_extracted_141719 | Not-determined |
| S13_extracted_141723 | Not-determined |
| S13_extracted_141775 | Not-determined |
| S13_extracted_141793 | Not-determined |
| S13_extracted_141804 | Not-determined |
| S13_extracted_141808 | Not-determined |
| S13_extracted_141814 | Not-determined |
| S13_extracted_141835 | Not-determined |
| S13_extracted_141839 | Not-determined |
| S13_extracted_141857 | Not-determined |
| S13_extracted_141859 | Not-determined |
| S13_extracted_141860 | Not-determined |
| S13_extracted_141862 | Not-determined |
| S13_extracted_141871 | Not-determined |
| S13_extracted_141897 | Not-determined |
| S13_extracted_141898 | Not-determined |
| S13_extracted_141900 | Not-determined |
| S13_extracted_141914 | Not-determined |
| S13_extracted_141916 | Not-determined |
| S13_extracted_141917 | Low-quality |
| S13_extracted_141925 | Not-determined |
| S13_extracted_141926 | Not-determined |
| S13_extracted_141932 | Not-determined |
| S13_extracted_141935 | Not-determined |
| S13_extracted_141942 | Not-determined |
| S13_extracted_141948 | Not-determined |
| S13_extracted_141989 | Low-quality |
| S13_extracted_142014 | Not-determined |
| S13_extracted_142042 | Not-determined |
| S13_extracted_142046 | Not-determined |
| S13_extracted_142067 | Not-determined |
| S13_extracted_142079 | Not-determined |
| S13_extracted_142084 | Not-determined |
| S13_extracted_142089 | Not-determined |
| S13_extracted_142095 | Not-determined |
| S13_extracted_142100 | Not-determined |
| S13_extracted_142105 | Not-determined |
| S13_extracted_142117 | Not-determined |
| S13_extracted_142127 | Low-quality |
| S13_extracted_142129 | Not-determined |
| S13_extracted_142134 | Not-determined |
| S13_extracted_142135 | Not-determined |
| S13_extracted_142138 | Not-determined |
| S13_extracted_142144 | Not-determined |
| S13_extracted_142149 | Not-determined |
| S13_extracted_142161 | Not-determined |
| S13_extracted_142162 | Low-quality |
| S13_extracted_142165 | Not-determined |
| S13_extracted_142172 | Low-quality |
| S13_extracted_142176 | Not-determined |
| S13_extracted_142180 | Not-determined |
| S13_extracted_142183 | Not-determined |
| S13_extracted_142191 | Not-determined |
| S13_extracted_142195 | Not-determined |
| S13_extracted_142197 | Not-determined |
| S13_extracted_142210 | Not-determined |
| S13_extracted_142212 | Not-determined |
| S13_extracted_142218 | Not-determined |
| S13_extracted_142228 | Not-determined |
| S13_extracted_142229 | Not-determined |
| S13_extracted_142236 | Not-determined |
| S13_extracted_142265 | Not-determined |
| S13_extracted_142271 | Not-determined |
| S13_extracted_142276 | Not-determined |
| S13_extracted_142281 | Low-quality |
| S13_extracted_142287 | Not-determined |
| S13_extracted_142319 | Not-determined |
| S13_extracted_142338 | Not-determined |
| S13_extracted_142349 | Low-quality |
| S13_extracted_142363 | Not-determined |
| S13_extracted_142396 | Not-determined |
| S13_extracted_142397 | Not-determined |
| S13_extracted_142415 | Not-determined |
| S13_extracted_142421 | Not-determined |
| S13_extracted_142441 | Not-determined |
| S13_extracted_142445 | Not-determined |
| S13_extracted_142469 | Not-determined |
| S13_extracted_142471 | Not-determined |
| S13_extracted_142472 | Not-determined |
| S13_extracted_142476 | Not-determined |
| S13_extracted_142492 | Not-determined |
| S13_extracted_142497 | Not-determined |
| S13_extracted_142504 | Not-determined |
| S13_extracted_142514 | Not-determined |
| S13_extracted_142528 | Not-determined |
| S13_extracted_142537 | Not-determined |
| S13_extracted_142539 | Not-determined |
| S13_extracted_142547 | Not-determined |
| S13_extracted_142548 | Low-quality |
| S13_extracted_142563 | Not-determined |
| S13_extracted_142569 | Not-determined |
| S13_extracted_142573 | Not-determined |
| S13_extracted_142575 | Not-determined |
| S13_extracted_142619 | Not-determined |
| S13_extracted_142621 | Not-determined |
| S13_extracted_142664 | Low-quality |
| S13_extracted_142700 | Not-determined |
| S13_extracted_142706 | Not-determined |
| S13_extracted_142707 | Not-determined |
| S13_extracted_142711 | Not-determined |
| S13_extracted_142714 | Not-determined |
| S13_extracted_142726 | Not-determined |
| S13_extracted_142730 | Not-determined |
| S13_extracted_142750 | Not-determined |
| S13_extracted_142753 | Not-determined |
| S13_extracted_142756 | Not-determined |
| S13_extracted_142761 | Not-determined |
| S13_extracted_142764 | Not-determined |
| S13_extracted_142794 | Not-determined |
| S13_extracted_142805 | Not-determined |
| S13_extracted_142813 | Not-determined |
| S13_extracted_142814 | Not-determined |
| S13_extracted_142817 | Not-determined |
| S13_extracted_142820 | Not-determined |
| S13_extracted_142833 | Not-determined |
| S13_extracted_142835 | Not-determined |
| S13_extracted_142836 | Not-determined |
| S13_extracted_142840 | Not-determined |
| S13_extracted_142848 | Not-determined |
| S13_extracted_142856 | Low-quality |
| S13_extracted_142864 | Not-determined |
| S13_extracted_142887 | Not-determined |
| S13_extracted_142904 | Not-determined |
| S13_extracted_142915 | Low-quality |
| S13_extracted_142922 | Not-determined |
| S13_extracted_142946 | Not-determined |
| S13_extracted_142947 | Not-determined |
| S13_extracted_142949 | Not-determined |
| S13_extracted_142952 | Not-determined |
| S13_extracted_142960 | Not-determined |
| S13_extracted_142971 | Not-determined |
| S13_extracted_142975 | Not-determined |
| S13_extracted_142980 | Not-determined |
| S13_extracted_142986 | Not-determined |
| S13_extracted_142994 | Not-determined |
| S13_extracted_142995 | Not-determined |
| S13_extracted_142996 | Not-determined |
| S13_extracted_142997 | Not-determined |
| S13_extracted_143013 | Not-determined |
| S13_extracted_143016 | Not-determined |
| S13_extracted_143021 | Low-quality |
| S13_extracted_143022 | Not-determined |
| S13_extracted_143029 | Not-determined |
| S13_extracted_143036 | Not-determined |
| S13_extracted_143041 | Not-determined |
| S13_extracted_143052 | Not-determined |
| S13_extracted_143064 | Not-determined |
| S13_extracted_143080 | Not-determined |
| S13_extracted_143097 | Not-determined |
| S13_extracted_143101 | Not-determined |
| S13_extracted_143104 | Not-determined |
| S13_extracted_143116 | Not-determined |
| S13_extracted_143121 | Not-determined |
| S13_extracted_143129 | Not-determined |
| S13_extracted_143130 | Not-determined |
| S13_extracted_143140 | Not-determined |
| S13_extracted_143142 | Not-determined |
| S13_extracted_143155 | Not-determined |
| S13_extracted_143178 | Not-determined |
| S13_extracted_143180 | Not-determined |
| S13_extracted_143195 | Not-determined |
| S13_extracted_143224 | Not-determined |
| S13_extracted_143227 | Not-determined |
| S13_extracted_143240 | Not-determined |
| S13_extracted_143242 | Not-determined |
| S13_extracted_143248 | Not-determined |
| S13_extracted_143282 | Not-determined |
| S13_extracted_143293 | Not-determined |
| S13_extracted_143294 | Not-determined |
| S13_extracted_143308 | Not-determined |
| S13_extracted_143310 | Not-determined |
| S13_extracted_143311 | Not-determined |
| S13_extracted_143312 | Not-determined |
| S13_extracted_143313 | Not-determined |
| S13_extracted_143318 | Not-determined |
| S13_extracted_143321 | Not-determined |
| S13_extracted_143335 | Not-determined |
| S13_extracted_143340 | Low-quality |
| S13_extracted_143357 | Not-determined |
| S13_extracted_143374 | Not-determined |
| S13_extracted_143377 | Not-determined |
| S13_extracted_143380 | Not-determined |
| S13_extracted_143402 | Not-determined |
| S11_vs_k141_3790\|\|full | Medium-quality |
| S13_extracted_143434 | Not-determined |
| S13_extracted_143435 | Not-determined |
| S13_extracted_143437 | Not-determined |
| S13_extracted_143459 | Not-determined |
| S13_extracted_143460 | Not-determined |
| S13_extracted_143461 | Not-determined |
| S13_extracted_143466 | Not-determined |
| S13_extracted_143469 | Not-determined |
| S13_extracted_143486 | Not-determined |
| S13_extracted_143497 | Not-determined |
| S13_extracted_143498 | Not-determined |
| S13_extracted_143499 | Not-determined |
| S13_extracted_143502 | Not-determined |
| S13_extracted_143518 | Not-determined |
| S13_extracted_143519 | Not-determined |
| S13_extracted_143530 | Not-determined |
| S13_extracted_143535 | Not-determined |
| S13_extracted_143536 | Not-determined |
| S13_extracted_143547 | Not-determined |
| S13_extracted_143548 | Not-determined |
| S13_extracted_143552 | Low-quality |
| S13_extracted_143553 | Not-determined |
| S13_extracted_143558 | Not-determined |
| S13_extracted_143567 | Not-determined |
| S13_extracted_143570 | Not-determined |
| S13_extracted_143578 | Not-determined |
| S13_extracted_143599 | Not-determined |
| S13_extracted_143629 | Not-determined |
| S13_extracted_143633 | Not-determined |
| S13_extracted_143643 | Not-determined |
| S13_extracted_143644 | Not-determined |
| S13_extracted_143648 | Not-determined |
| S13_extracted_143649 | Not-determined |
| S13_extracted_143655 | Low-quality |
| S13_extracted_143664 | Not-determined |
| S13_extracted_143672 | Not-determined |
| S13_extracted_143678 | Not-determined |
| S13_extracted_143682 | Not-determined |
| S13_extracted_143691 | Not-determined |
| S13_extracted_143692 | Not-determined |
| S13_extracted_143701 | Not-determined |
| S13_extracted_143702 | Not-determined |
| S13_extracted_143715 | Not-determined |
| S13_extracted_143722 | Not-determined |
| S13_extracted_143724 | Not-determined |
| S13_extracted_143743 | Not-determined |
| S13_extracted_143745 | Low-quality |
| S13_extracted_143772 | Low-quality |
| S13_extracted_143779 | Not-determined |
| S13_extracted_143784 | Low-quality |
| S13_extracted_143793 | Not-determined |
| S13_extracted_143795 | Not-determined |
| S13_extracted_143813 | Not-determined |
| S13_extracted_143817 | Low-quality |
| S13_extracted_143819 | Not-determined |
| S13_extracted_143838 | Not-determined |
| S13_extracted_143853 | Not-determined |
| S13_extracted_143857 | Not-determined |
| S13_extracted_143862 | Not-determined |
| S13_extracted_143892 | Not-determined |
| S13_extracted_143895 | Not-determined |
| S13_extracted_143898 | Not-determined |
| S13_extracted_143899 | Not-determined |
| S13_extracted_143908 | Not-determined |
| S13_extracted_143909 | Not-determined |
| S13_extracted_143912 | Not-determined |
| S13_extracted_143919 | Not-determined |
| S13_extracted_143931 | Not-determined |
| S13_extracted_143941 | Not-determined |
| S13_extracted_143946 | Not-determined |
| S13_extracted_143949 | Not-determined |
| S13_extracted_143951 | Not-determined |
| S13_extracted_143957 | Not-determined |
| S13_extracted_143962 | Not-determined |
| S13_extracted_143964 | Not-determined |
| S13_extracted_143968 | Not-determined |
| S13_extracted_143971 | Not-determined |
| S13_extracted_143973 | Not-determined |
| S13_extracted_143987 | Not-determined |
| S13_extracted_143989 | Not-determined |
| S13_extracted_143992 | Not-determined |
| S13_extracted_143995 | Low-quality |
| S13_extracted_144010 | Low-quality |
| S13_extracted_144011 | Not-determined |
| S13_extracted_144015 | Not-determined |
| S13_extracted_144020 | Not-determined |
| S13_extracted_144038 | Low-quality |
| S13_extracted_144048 | Not-determined |
| S13_extracted_144075 | Not-determined |
| S13_extracted_144081 | Not-determined |
| S13_extracted_144091 | Not-determined |
| S13_extracted_144094 | Not-determined |
| S13_extracted_144096 | Not-determined |
| S13_extracted_144097 | Low-quality |
| S13_extracted_144105 | Not-determined |
| S13_extracted_144109 | Low-quality |
| S13_extracted_144113 | Not-determined |
| S13_extracted_144116 | Not-determined |
| S13_extracted_144121 | Low-quality |
| S13_extracted_144132 | Not-determined |
| S13_extracted_144142 | Low-quality |
| S13_extracted_144147 | Not-determined |
| S13_extracted_144157 | Not-determined |
| S13_extracted_144158 | Not-determined |
| S13_extracted_144162 | Not-determined |
| S13_extracted_144168 | Not-determined |
| S13_extracted_144170 | Low-quality |
| S13_extracted_144195 | Low-quality |
| S13_extracted_144201 | Not-determined |
| S13_extracted_144205 | Not-determined |
| S13_extracted_144206 | Not-determined |
| S13_extracted_144224 | Not-determined |
| S13_extracted_144227 | Not-determined |
| S13_extracted_144235 | Not-determined |
| S13_extracted_144241 | Not-determined |
| S13_extracted_144254 | Not-determined |
| S13_extracted_144259 | Not-determined |
| S13_extracted_144287 | Not-determined |
| S13_extracted_144290 | Not-determined |
| S13_extracted_144292 | Not-determined |
| S13_extracted_144294 | Low-quality |
| S13_extracted_144302 | Not-determined |
| S13_extracted_144315 | Not-determined |
| S13_extracted_144316 | Low-quality |
| S13_extracted_144317 | Not-determined |
| S13_extracted_144323 | Not-determined |
| S13_extracted_144331 | Not-determined |
| S13_extracted_144340 | Not-determined |
| S13_extracted_144346 | Low-quality |
| S13_extracted_144349 | Not-determined |
| S13_extracted_144360 | Not-determined |
| S13_extracted_144361 | Not-determined |
| S13_extracted_144364 | Low-quality |
| S13_extracted_144367 | Not-determined |
| S13_extracted_144372 | Not-determined |
| S13_extracted_144386 | Not-determined |
| S13_extracted_144399 | Not-determined |
| S13_extracted_144401 | Not-determined |
| S13_extracted_144408 | Not-determined |
| S13_extracted_144413 | Not-determined |
| S13_extracted_144419 | Not-determined |
| S13_extracted_144457 | Not-determined |
| S13_extracted_144466 | Not-determined |
| S13_extracted_144474 | Not-determined |
| S13_extracted_144484 | Not-determined |
| S13_extracted_144486 | Low-quality |
| S13_extracted_144504 | Not-determined |
| S13_extracted_144522 | Not-determined |
| S13_extracted_144524 | Low-quality |
| S13_extracted_144538 | Not-determined |
| S13_extracted_144540 | Not-determined |
| S13_extracted_144551 | Not-determined |
| S13_extracted_144554 | Not-determined |
| S13_extracted_144575 | Not-determined |
| S13_extracted_144576 | Not-determined |
| S13_extracted_144577 | Low-quality |
| S13_extracted_144578 | Not-determined |
| S13_extracted_144581 | Not-determined |
| S13_extracted_144598 | Not-determined |
| S13_extracted_144609 | Not-determined |
| S13_extracted_144615 | Not-determined |
| S13_extracted_144617 | Not-determined |
| S13_extracted_144640 | Not-determined |
| S13_extracted_144652 | Not-determined |
| S13_extracted_144674 | Not-determined |
| S13_extracted_144726 | Not-determined |
| S13_extracted_144732 | Not-determined |
| S13_extracted_144738 | Not-determined |
| S13_extracted_144750 | Not-determined |
| S13_extracted_144752 | Not-determined |
| S13_extracted_144758 | Not-determined |
| S13_extracted_144763 | Not-determined |
| S13_extracted_144764 | Not-determined |
| S13_extracted_144767 | Not-determined |
| S13_extracted_144777 | Not-determined |
| S13_extracted_144793 | Not-determined |
| S13_extracted_144795 | Not-determined |
| S13_extracted_144837 | Not-determined |
| S13_extracted_144869 | Not-determined |
| S13_extracted_144874 | Not-determined |
| S13_extracted_144888 | Not-determined |
| S13_extracted_144902 | Not-determined |
| S13_extracted_144905 | Not-determined |
| S13_extracted_144919 | Not-determined |
| S13_extracted_144920 | Not-determined |
| S13_extracted_144921 | Not-determined |
| S13_extracted_144925 | Not-determined |
| S13_extracted_144926 | Not-determined |
| S13_extracted_144928 | Not-determined |
| S13_extracted_144941 | Not-determined |
| S13_extracted_144956 | Not-determined |
| S13_extracted_144957 | Not-determined |
| S13_extracted_144970 | Not-determined |
| S13_extracted_144988 | Low-quality |
| S13_extracted_144989 | Not-determined |
| S13_extracted_144998 | Not-determined |
| S13_extracted_144999 | Not-determined |
| S13_extracted_145005 | Not-determined |
| S13_extracted_145008 | Not-determined |
| S13_extracted_145010 | Not-determined |
| S13_extracted_145017 | Not-determined |
| S13_extracted_145018 | Not-determined |
| S13_extracted_145022 | Not-determined |
| S13_extracted_145031 | Not-determined |
| S13_extracted_145040 | Not-determined |
| S13_extracted_145050 | Low-quality |
| S13_extracted_145061 | Not-determined |
| S13_extracted_145083 | Not-determined |
| S13_extracted_145096 | Not-determined |
| S13_extracted_145104 | Not-determined |
| S13_extracted_145105 | Not-determined |
| S13_extracted_145110 | Not-determined |
| S13_extracted_145115 | Not-determined |
| S13_extracted_145120 | Not-determined |
| S13_extracted_145121 | Not-determined |
| S13_extracted_145133 | Not-determined |
| S13_extracted_145143 | Not-determined |
| S13_extracted_145156 | Not-determined |
| S13_extracted_145158 | Not-determined |
| S13_extracted_145162 | Not-determined |
| S13_extracted_145164 | Not-determined |
| S13_extracted_145170 | Low-quality |
| S13_extracted_145181 | Not-determined |
| S13_extracted_145182 | Not-determined |
| S13_extracted_145192 | Not-determined |

| | |
|---|---|
| S13_extracted_145195 | Not-determined |
| S13_extracted_145196 | Not-determined |
| S13_extracted_145197 | Not-determined |
| S13_extracted_145202 | Not-determined |
| S13_extracted_145214 | Not-determined |
| S13_extracted_145224 | Not-determined |
| S13_extracted_145225 | Not-determined |
| S13_extracted_145230 | Not-determined |
| S13_extracted_145254 | Not-determined |
| S13_extracted_145275 | Not-determined |
| S13_extracted_145300 | Not-determined |
| S13_extracted_145307 | Low-quality |
| S13_extracted_145319 | Not-determined |
| S13_extracted_145334 | Low-quality |
| S13_extracted_145352 | Not-determined |
| S13_extracted_145353 | Not-determined |
| S13_extracted_145363 | Not-determined |
| S13_extracted_145381 | Not-determined |
| S13_extracted_145384 | Not-determined |
| S13_extracted_145389 | Not-determined |
| S13_extracted_145394 | Not-determined |
| S13_extracted_145398 | Not-determined |
| S13_extracted_145400 | Low-quality |
| S13_extracted_145401 | Not-determined |
| S13_extracted_145413 | Not-determined |
| S13_extracted_145420 | Not-determined |
| S13_extracted_145421 | Not-determined |
| S13_extracted_145425 | Not-determined |
| S13_extracted_145445 | Not-determined |
| S13_extracted_145446 | Not-determined |
| S13_extracted_145453 | Not-determined |
| S13_extracted_145459 | Not-determined |
| S13_extracted_145478 | Not-determined |
| S13_extracted_145509 | Not-determined |
| S13_extracted_145533 | Not-determined |
| S13_extracted_145536 | Not-determined |
| S13_extracted_145567 | Not-determined |
| S13_extracted_145571 | Not-determined |
| S13_extracted_145572 | Not-determined |
| S13_extracted_145573 | Not-determined |
| S13_extracted_145588 | Not-determined |
| S13_extracted_145624 | Not-determined |
| S13_extracted_145652 | Not-determined |
| S13_extracted_145668 | Not-determined |
| S13_extracted_145669 | Not-determined |
| S13_extracted_145681 | Not-determined |
| S13_extracted_145685 | Not-determined |
| S13_extracted_145696 | Low-quality |
| S13_extracted_145697 | Not-determined |
| S13_extracted_145706 | Not-determined |
| S13_extracted_145710 | Low-quality |
| S13_extracted_145714 | Not-determined |
| S13_extracted_145717 | Not-determined |
| S13_extracted_145722 | Not-determined |
| S13_extracted_145743 | Low-quality |
| S13_extracted_145746 | Not-determined |
| S13_extracted_145747 | Not-determined |
| S13_extracted_145750 | Not-determined |
| S13_extracted_145752 | Not-determined |
| S13_extracted_145754 | Not-determined |
| S13_extracted_145765 | Not-determined |
| S13_extracted_145767 | Not-determined |
| S13_extracted_145771 | Not-determined |
| S13_extracted_145793 | Not-determined |
| S13_extracted_145805 | Not-determined |
| S13_extracted_145811 | Not-determined |
| S13_extracted_145815 | Not-determined |
| S13_extracted_145820 | Low-quality |
| S13_extracted_145836 | Not-determined |
| S13_extracted_145859 | Not-determined |
| S13_extracted_145870 | Not-determined |
| S13_extracted_145873 | Not-determined |
| S13_extracted_145891 | Not-determined |
| S13_extracted_145894 | Not-determined |
| S13_extracted_145898 | Not-determined |
| S13_extracted_145905 | Not-determined |
| S13_extracted_145906 | Not-determined |
| S13_extracted_145909 | Not-determined |
| S13_extracted_145922 | Not-determined |
| S13_extracted_145923 | Not-determined |
| S13_extracted_145932 | Not-determined |
| S13_extracted_145934 | Not-determined |
| S13_extracted_145940 | Not-determined |
| S13_extracted_145948 | Not-determined |
| S13_extracted_145956 | Not-determined |
| S13_extracted_145962 | Low-quality |
| S13_extracted_145971 | Not-determined |
| S13_extracted_146002 | Not-determined |
| S13_extracted_146024 | Not-determined |
| S13_extracted_146030 | Not-determined |
| S13_extracted_146043 | Not-determined |
| S13_extracted_146045 | Not-determined |
| S13_extracted_146052 | Not-determined |
| S13_extracted_146057 | Not-determined |
| S13_extracted_146058 | Not-determined |
| S13_extracted_146074 | Low-quality |
| S13_extracted_146089 | Not-determined |
| S13_extracted_146093 | Not-determined |
| S13_extracted_146094 | Low-quality |
| S13_extracted_146102 | Not-determined |
| S13_extracted_146107 | Not-determined |
| S13_extracted_146125 | Not-determined |
| S13_extracted_146139 | Not-determined |
| S13_extracted_146145 | Not-determined |
| S13_extracted_146160 | Not-determined |
| S13_extracted_146163 | Not-determined |
| S13_extracted_146200 | Not-determined |
| S13_extracted_146207 | Not-determined |
| S13_extracted_146226 | Low-quality |
| S13_extracted_146227 | Not-determined |
| S13_extracted_146229 | Not-determined |
| S13_extracted_146245 | Not-determined |
| S13_extracted_146260 | Not-determined |
| S13_extracted_146273 | Not-determined |
| S13_extracted_146277 | Not-determined |
| S13_extracted_146295 | Not-determined |
| S13_extracted_146308 | Not-determined |
| S13_extracted_146313 | Not-determined |
| S13_extracted_146316 | Not-determined |
| S13_extracted_146321 | Not-determined |
| S13_extracted_146362 | Not-determined |
| S13_extracted_146364 | Not-determined |
| S13_extracted_146385 | Not-determined |
| S13_extracted_146405 | Not-determined |
| S13_extracted_146409 | Not-determined |
| S13_extracted_146421 | Not-determined |
| S13_extracted_146437 | Not-determined |
| S13_extracted_146444 | Low-quality |
| S13_extracted_146449 | Not-determined |
| S13_extracted_146451 | Not-determined |
| S13_extracted_146453 | Not-determined |
| S13_extracted_146459 | Not-determined |
| S13_extracted_146465 | Not-determined |
| S13_extracted_146467 | Not-determined |
| S13_extracted_146475 | Not-determined |
| S13_extracted_146490 | Not-determined |
| S13_extracted_146496 | Low-quality |
| S13_extracted_146503 | Not-determined |
| S13_extracted_146514 | Low-quality |
| S13_extracted_146520 | Not-determined |
| S13_extracted_146529 | Not-determined |
| S13_extracted_146533 | Low-quality |
| S13_extracted_146540 | Not-determined |
| S13_extracted_146541 | Not-determined |
| S13_extracted_146542 | Low-quality |
| S13_extracted_146550 | Not-determined |
| S13_extracted_146558 | Not-determined |
| S13_extracted_146561 | Not-determined |
| S13_extracted_146563 | Not-determined |
| S13_extracted_146592 | Not-determined |
| S13_extracted_146598 | Not-determined |
| S13_extracted_146627 | Not-determined |
| S13_extracted_146658 | Not-determined |
| S13_extracted_146667 | Low-quality |
| S13_extracted_146674 | Low-quality |
| S13_extracted_146679 | Not-determined |
| S13_extracted_146681 | Not-determined |
| S13_extracted_146697 | Not-determined |
| S13_extracted_146703 | Not-determined |
| S13_extracted_146710 | Not-determined |
| S13_extracted_146731 | Not-determined |
| S13_extracted_146741 | Not-determined |
| S13_extracted_146767 | Not-determined |
| S13_extracted_146786 | Not-determined |
| S13_extracted_146788 | Not-determined |
| S13_extracted_146791 | Not-determined |
| S13_extracted_146803 | Not-determined |
| S13_extracted_146805 | Low-quality |
| S13_extracted_146808 | Not-determined |
| S13_extracted_146817 | Not-determined |
| S13_extracted_146821 | Not-determined |
| S13_extracted_146823 | Not-determined |
| S13_extracted_146827 | Low-quality |
| S13_extracted_146838 | Not-determined |
| S13_extracted_146843 | Not-determined |
| S13_extracted_146857 | Not-determined |
| S13_extracted_146865 | Not-determined |
| S13_extracted_146866 | Low-quality |
| S13_extracted_146876 | Low-quality |
| S13_extracted_146894 | Not-determined |
| S13_extracted_146900 | Not-determined |
| S13_extracted_146930 | Not-determined |
| S13_extracted_146962 | Low-quality |
| S13_extracted_146967 | Not-determined |
| S13_extracted_146971 | Not-determined |
| S13_extracted_147004 | Not-determined |
| S13_extracted_147012 | Not-determined |
| S13_extracted_147020 | Not-determined |
| S13_extracted_147026 | Low-quality |
| S13_extracted_147032 | Not-determined |
| S13_extracted_147035 | Not-determined |
| S13_extracted_147042 | Not-determined |
| S13_extracted_147053 | Low-quality |
| S13_extracted_147056 | Not-determined |
| S13_extracted_147059 | Not-determined |
| S13_extracted_147070 | Not-determined |
| S13_extracted_147074 | Not-determined |
| S13_extracted_147075 | Not-determined |
| S13_extracted_147077 | Not-determined |
| S13_extracted_147086 | Low-quality |
| S13_extracted_147090 | Not-determined |
| S13_extracted_147094 | Not-determined |
| S13_extracted_147098 | Not-determined |
| S13_extracted_147107 | Not-determined |
| S13_extracted_147108 | Not-determined |
| S13_extracted_147112 | Not-determined |
| S13_extracted_147113 | Low-quality |
| S13_extracted_147119 | Not-determined |
| S13_extracted_147120 | Not-determined |
| S13_extracted_147133 | Low-quality |
| S13_extracted_147136 | Low-quality |
| S13_extracted_147153 | Not-determined |
| S13_extracted_147163 | Not-determined |
| S13_extracted_147183 | Not-determined |
| S13_extracted_147185 | Not-determined |
| S13_extracted_147198 | Not-determined |
| S13_extracted_147211 | Not-determined |
| S13_extracted_147233 | Not-determined |
| S13_extracted_147235 | Not-determined |
| S13_extracted_147253 | Not-determined |
| S13_extracted_147259 | Not-determined |
| S13_extracted_147262 | Not-determined |
| S13_extracted_147267 | Not-determined |
| S13_extracted_147285 | Not-determined |
| S13_extracted_147290 | Not-determined |
| S13_extracted_147293 | Not-determined |
| S13_extracted_147296 | Not-determined |
| S13_extracted_147306 | Not-determined |
| S13_extracted_147312 | Not-determined |
| S13_extracted_147330 | Not-determined |
| S13_extracted_147336 | Not-determined |
| S13_extracted_147343 | Not-determined |
| S13_extracted_147349 | Not-determined |
| S13_extracted_147351 | Not-determined |
| S13_extracted_147359 | Not-determined |
| S13_extracted_147360 | Not-determined |
| S13_extracted_147362 | Low-quality |
| S13_extracted_147367 | Not-determined |
| S13_extracted_147369 | Not-determined |
| S13_extracted_147379 | Not-determined |
| S13_extracted_147385 | Not-determined |
| S13_extracted_147386 | Not-determined |
| S13_extracted_147392 | Not-determined |
| S6_vs_k141_34453\|\|full | High-quality |
| S13_extracted_147404 | Not-determined |
| S13_extracted_147450 | Not-determined |
| S13_extracted_147460 | Not-determined |
| S13_extracted_147469 | Not-determined |
| S13_extracted_147485 | Not-determined |
| S13_extracted_147503 | Not-determined |
| S13_extracted_147516 | Low-quality |
| S13_extracted_147524 | Not-determined |
| S13_extracted_147538 | Not-determined |
| S13_extracted_147540 | Not-determined |
| S13_extracted_147543 | Not-determined |
| S13_extracted_147551 | Not-determined |
| S13_extracted_147552 | Not-determined |
| S13_extracted_147554 | Not-determined |
| S13_extracted_147568 | Low-quality |
| S13_extracted_147574 | Not-determined |
| S12_vs_k141_111286\|\|full | High-quality |
| S13_extracted_147592 | Low-quality |
| S13_extracted_147593 | Not-determined |
| S13_extracted_147594 | Not-determined |
| S13_extracted_147627 | Not-determined |
| S13_extracted_147630 | Not-determined |
| S13_extracted_147639 | Not-determined |
| S13_extracted_147641 | Not-determined |
| S13_extracted_147643 | Not-determined |
| S13_extracted_147644 | Not-determined |
| S13_extracted_147649 | Not-determined |
| S13_extracted_147656 | Not-determined |
| S13_extracted_147660 | Not-determined |
| S13_extracted_147664 | Low-quality |
| S13_extracted_147686 | Low-quality |
| S13_extracted_147689 | Not-determined |
| S13_extracted_147702 | Not-determined |
| S13_extracted_147724 | Not-determined |
| S13_extracted_147727 | Not-determined |
| S13_extracted_147736 | Not-determined |
| S13_extracted_147740 | Not-determined |
| S13_extracted_147745 | Not-determined |
| S13_extracted_147755 | Not-determined |
| S13_extracted_147757 | Not-determined |
| S13_extracted_147762 | Low-quality |
| S13_extracted_147768 | Not-determined |
| S13_extracted_147774 | Not-determined |
| S13_extracted_147777 | Not-determined |
| S13_extracted_147792 | Not-determined |
| S13_extracted_147797 | Not-determined |
| S13_extracted_147801 | Not-determined |
| S13_extracted_147810 | Not-determined |
| S13_extracted_147821 | Not-determined |
| S13_extracted_147825 | Not-determined |
| S13_extracted_147828 | Not-determined |
| S13_extracted_147831 | Not-determined |
| S13_extracted_147834 | Not-determined |
| S13_extracted_147835 | Not-determined |
| S13_extracted_147842 | Not-determined |
| S13_extracted_147853 | Not-determined |
| S13_extracted_147864 | Low-quality |
| S13_extracted_147868 | Low-quality |
| S13_extracted_147887 | Not-determined |
| S13_extracted_147900 | Not-determined |
| S13_extracted_147920 | Not-determined |
| S13_extracted_147927 | Not-determined |
| S13_extracted_147932 | Not-determined |
| S13_extracted_147943 | Not-determined |
| S13_extracted_147949 | Not-determined |
| S13_extracted_147957 | Not-determined |
| S13_extracted_147964 | Not-determined |
| S13_extracted_147967 | Not-determined |
| S13_extracted_148007 | Not-determined |
| S13_extracted_148018 | Not-determined |
| S13_extracted_148054 | Not-determined |
| S13_extracted_148066 | Low-quality |
| S13_extracted_148067 | Not-determined |
| S13_extracted_148068 | Not-determined |
| S13_extracted_148093 | Not-determined |
| S13_extracted_148097 | Not-determined |
| S13_extracted_148105 | Not-determined |
| S13_extracted_148107 | Not-determined |
| S13_extracted_148112 | Not-determined |
| S13_extracted_148130 | Not-determined |
| S13_extracted_148135 | Not-determined |
| S13_extracted_148146 | Not-determined |
| S13_extracted_148150 | Not-determined |
| S13_extracted_148151 | Not-determined |
| S13_extracted_148155 | Low-quality |
| S13_extracted_148158 | Not-determined |
| S13_extracted_148159 | Not-determined |
| S13_extracted_148164 | Not-determined |
| S13_extracted_148166 | Not-determined |
| S13_extracted_148187 | Not-determined |
| S13_extracted_148190 | Not-determined |
| S13_extracted_148193 | Not-determined |
| S13_extracted_148205 | Not-determined |
| S13_extracted_148220 | Not-determined |
| S13_extracted_148232 | Not-determined |
| S13_extracted_148234 | Not-determined |
| S13_extracted_148235 | Low-quality |
| S13_extracted_148247 | Not-determined |
| S13_extracted_148259 | Not-determined |
| S13_extracted_148267 | Not-determined |
| S13_extracted_148272 | Not-determined |
| S13_extracted_148282 | Not-determined |
| S13_extracted_148285 | Not-determined |
| S13_extracted_148296 | Not-determined |
| S13_extracted_148298 | Not-determined |
| S13_extracted_148302 | Not-determined |
| S13_extracted_148309 | Not-determined |
| S13_extracted_148313 | Not-determined |
| S13_extracted_148317 | Not-determined |
| S13_extracted_148335 | Not-determined |
| S13_extracted_148338 | Not-determined |
| S13_extracted_148339 | Not-determined |
| S13_extracted_148344 | Not-determined |
| S13_extracted_148353 | Not-determined |
| S13_extracted_148362 | Not-determined |
| S13_extracted_148363 | Not-determined |
| S13_extracted_148382 | Not-determined |
| S13_extracted_148387 | Not-determined |
| S13_extracted_148389 | Low-quality |
| S13_extracted_148394 | Low-quality |
| S13_extracted_148408 | Not-determined |
| S13_extracted_148415 | Not-determined |
| S13_extracted_148427 | Not-determined |
| S13_extracted_148431 | Not-determined |
| S13_extracted_148436 | Not-determined |
| S13_extracted_148438 | Not-determined |
| S13_extracted_148452 | Not-determined |
| S13_extracted_148459 | Not-determined |
| S13_extracted_148473 | Not-determined |
| S13_extracted_148489 | Not-determined |
| S13_extracted_148515 | Low-quality |
| S13_extracted_148520 | Low-quality |
| S13_extracted_148540 | Not-determined |
| S13_extracted_148545 | Not-determined |
| S13_extracted_148552 | Not-determined |
| S13_extracted_148557 | Not-determined |
| S13_extracted_148565 | Not-determined |
| S13_extracted_148568 | Not-determined |
| S13_extracted_148579 | Not-determined |
| S13_extracted_148582 | Not-determined |
| S13_extracted_148587 | Not-determined |
| S13_extracted_148592 | Not-determined |
| S13_extracted_148594 | Not-determined |
| S13_extracted_148601 | Not-determined |
| S13_extracted_148606 | Not-determined |
| S13_extracted_148614 | Low-quality |
| S13_extracted_148631 | Not-determined |
| S13_extracted_148637 | Not-determined |
| S13_extracted_148652 | Not-determined |
| S13_extracted_148653 | Not-determined |
| S13_extracted_148655 | Not-determined |
| S13_extracted_148664 | Not-determined |
| S13_extracted_148681 | Not-determined |
| S13_extracted_148686 | Low-quality |
| S13_extracted_148694 | Not-determined |
| S13_extracted_148701 | Not-determined |
| S13_extracted_148704 | Not-determined |
| S13_extracted_148712 | Not-determined |
| S13_extracted_148723 | Not-determined |
| S13_extracted_148724 | Not-determined |
| S13_extracted_148725 | Low-quality |
| S13_extracted_148736 | Not-determined |
| S13_extracted_148742 | Low-quality |
| S13_extracted_148745 | Not-determined |
| S13_extracted_148753 | Low-quality |
| S13_extracted_148774 | Not-determined |
| S13_extracted_148775 | Not-determined |
| S13_extracted_148778 | Not-determined |
| S13_extracted_148792 | Not-determined |
| S13_extracted_148799 | Not-determined |
| S13_extracted_148808 | Not-determined |
| S13_extracted_148814 | Not-determined |
| S13_extracted_148816 | Low-quality |
| S13_extracted_148824 | Not-determined |
| S13_extracted_148827 | Not-determined |
| S13_extracted_148833 | Not-determined |
| S13_extracted_148840 | Not-determined |
| S13_extracted_148851 | Not-determined |
| S13_extracted_148855 | Not-determined |
| S13_extracted_148894 | Low-quality |
| S13_extracted_148896 | Not-determined |
| S13_extracted_148898 | Not-determined |
| S13_extracted_148906 | Not-determined |
| S13_extracted_148910 | Not-determined |
| S13_extracted_148923 | Not-determined |
| S13_extracted_148924 | Not-determined |
| S13_extracted_148928 | Not-determined |
| S13_extracted_148934 | Not-determined |
| S13_extracted_148936 | Not-determined |
| S13_extracted_148941 | Not-determined |
| S13_extracted_148949 | Not-determined |
| S13_extracted_148954 | Not-determined |
| S13_extracted_148964 | Not-determined |
| S13_extracted_148966 | Not-determined |
| S13_extracted_148978 | Not-determined |
| S13_extracted_148979 | Not-determined |
| S13_extracted_148994 | Not-determined |
| S13_extracted_148997 | Not-determined |
| S13_extracted_149025 | Not-determined |
| S13_extracted_149057 | Not-determined |
| S13_extracted_149060 | Not-determined |
| S13_extracted_149067 | Not-determined |
| S13_extracted_149071 | Not-determined |
| S13_extracted_149086 | Not-determined |
| S13_extracted_149106 | Low-quality |
| S13_extracted_149111 | Not-determined |
| S13_extracted_149119 | Not-determined |
| S13_extracted_149123 | Low-quality |
| S13_extracted_149146 | Not-determined |
| S13_extracted_149158 | Not-determined |
| S13_extracted_149173 | Not-determined |
| S13_extracted_149187 | Not-determined |
| S13_extracted_149195 | Not-determined |
| S13_extracted_149202 | Not-determined |
| S13_extracted_149215 | Not-determined |
| S13_extracted_149226 | Not-determined |
| S13_extracted_149227 | Low-quality |
| S13_extracted_149228 | Low-quality |
| S13_extracted_149231 | Not-determined |
| S13_extracted_149235 | Not-determined |
| S13_extracted_149247 | Not-determined |
| S13_extracted_149248 | Not-determined |
| S13_extracted_149267 | Not-determined |
| S13_extracted_149275 | Not-determined |
| S13_extracted_149279 | Not-determined |
| S13_extracted_149316 | Not-determined |
| S13_extracted_149344 | Low-quality |
| S13_extracted_149360 | Not-determined |
| S13_extracted_149365 | Not-determined |
| S13_extracted_149372 | Not-determined |
| S13_extracted_149379 | Not-determined |
| S13_extracted_149388 | Not-determined |
| S13_extracted_149394 | Not-determined |
| S13_extracted_149395 | Not-determined |
| S13_extracted_149400 | Not-determined |
| S13_extracted_149424 | Not-determined |
| S13_extracted_149425 | Not-determined |
| S13_extracted_149436 | Not-determined |
| S13_extracted_149440 | Not-determined |
| S13_extracted_149466 | Low-quality |
| S13_extracted_149469 | Not-determined |
| S13_extracted_149474 | Not-determined |
| S13_extracted_149475 | Not-determined |
| S13_extracted_149496 | Not-determined |
| S13_extracted_149501 | Not-determined |
| S13_extracted_149502 | Not-determined |
| S13_extracted_149505 | Not-determined |
| S13_extracted_149531 | Low-quality |
| S13_extracted_149539 | Not-determined |
| S13_extracted_149541 | Not-determined |
| S13_extracted_149545 | Not-determined |
| S13_extracted_149551 | Low-quality |
| S13_extracted_149570 | Low-quality |
| S13_extracted_149572 | Low-quality |
| S13_extracted_149574 | Low-quality |
| S13_extracted_149587 | Not-determined |
| S13_extracted_149601 | Not-determined |
| S13_extracted_149603 | Not-determined |
| S13_extracted_149611 | Not-determined |
| S13_extracted_149619 | Not-determined |
| S13_extracted_149631 | Not-determined |
| S13_extracted_149647 | Not-determined |
| S13_extracted_149652 | Not-determined |
| S13_extracted_149654 | Low-quality |
| S13_extracted_149667 | Not-determined |
| S13_extracted_149679 | Not-determined |
| S13_extracted_149680 | Not-determined |
| S13_extracted_149682 | Not-determined |
| S13_extracted_149698 | Not-determined |
| S13_extracted_149706 | Low-quality |
| S13_extracted_149713 | Not-determined |
| S13_extracted_149721 | Not-determined |
| S13_extracted_149727 | Low-quality |
| S13_extracted_149742 | Not-determined |
| S13_extracted_149745 | Not-determined |
| S13_extracted_149759 | Not-determined |
| S13_extracted_149762 | Low-quality |
| S13_extracted_149779 | Low-quality |
| S13_extracted_149780 | Low-quality |
| S13_extracted_149784 | Not-determined |
| S13_extracted_149790 | Low-quality |
| S13_extracted_149796 | Low-quality |
| S13_extracted_149804 | Low-quality |
| S13_extracted_149812 | Not-determined |
| S13_extracted_149817 | Not-determined |
| S13_extracted_149830 | Low-quality |
| S13_extracted_149832 | Not-determined |
| S13_extracted_149840 | Not-determined |
| S13_extracted_149849 | Not-determined |
| S13_extracted_149876 | Not-determined |
| S13_extracted_149879 | Not-determined |
| S13_extracted_149888 | Not-determined |
| S13_extracted_149897 | Not-determined |
| S13_extracted_149899 | Not-determined |
| S13_extracted_149903 | Not-determined |
| S13_extracted_149912 | Not-determined |
| S13_extracted_149917 | Not-determined |
| S13_extracted_149927 | Not-determined |
| S13_extracted_149930 | Low-quality |
| S13_extracted_149942 | Low-quality |
| S13_extracted_149951 | Not-determined |
| S13_extracted_149953 | Not-determined |
| S13_extracted_149956 | Not-determined |
| S13_extracted_149958 | Low-quality |
| S13_extracted_149965 | Not-determined |
| S13_extracted_149977 | Not-determined |
| S13_extracted_149988 | Not-determined |
| S13_extracted_149991 | Not-determined |
| S13_extracted_149992 | Not-determined |
| S13_extracted_149996 | Not-determined |
| S13_extracted_149997 | Not-determined |
| S13_extracted_150005 | Not-determined |
| S13_extracted_150014 | Not-determined |
| S13_extracted_150024 | Not-determined |
| S13_extracted_150027 | Not-determined |
| S13_extracted_150038 | Not-determined |
| S13_extracted_150040 | Not-determined |
| S13_extracted_150059 | Not-determined |
| S13_extracted_150078 | Not-determined |
| S13_extracted_150083 | Not-determined |
| S13_extracted_150096 | Not-determined |
| S13_extracted_150109 | Not-determined |
| S13_extracted_150111 | Not-determined |
| S13_extracted_150130 | Not-determined |
| S13_extracted_150133 | Not-determined |
| S13_extracted_150140 | Low-quality |
| S13_extracted_150143 | Low-quality |
| S13_extracted_150149 | Not-determined |
| S13_extracted_150150 | Not-determined |
| S13_extracted_150153 | Not-determined |
| S13_extracted_150159 | Not-determined |
| S13_extracted_150164 | Not-determined |
| S13_extracted_150168 | Not-determined |
| S13_extracted_150171 | Not-determined |
| S13_extracted_150183 | Not-determined |
| S13_extracted_150185 | Not-determined |
| S13_extracted_150193 | Not-determined |
| S13_extracted_150203 | Not-determined |
| S13_extracted_150205 | Not-determined |
| S13_extracted_150217 | Not-determined |
| S13_extracted_150246 | Not-determined |
| S13_extracted_150247 | Low-quality |
| S13_extracted_150255 | Not-determined |
| S13_extracted_150266 | Not-determined |
| S13_extracted_150272 | Not-determined |

| | |
|---|---|
| S13_extracted_150276 | Not-determined |
| S13_extracted_150294 | Not-determined |
| S13_extracted_150295 | Not-determined |
| S13_extracted_150296 | Not-determined |
| S13_extracted_150311 | Not-determined |
| S13_extracted_150317 | Not-determined |
| S13_extracted_150323 | Not-determined |
| S13_extracted_150363 | Not-determined |
| S12_extracted_k141_10887 | Complete |
| S13_extracted_150384 | Not-determined |
| S13_extracted_150385 | Not-determined |
| S13_extracted_150386 | Not-determined |
| S5_extracted_k141_145585 | Complete |
| S13_extracted_150417 | Low-quality |
| S13_extracted_150435 | Low-quality |
| S13_extracted_150442 | Not-determined |
| S13_extracted_150444 | Not-determined |
| S13_extracted_150454 | Not-determined |
| S13_extracted_150461 | Not-determined |
| S13_extracted_150467 | Not-determined |
| S13_extracted_150473 | Not-determined |
| S13_extracted_150475 | Not-determined |
| S13_extracted_150476 | Not-determined |
| S13_extracted_150481 | Not-determined |
| S13_extracted_150492 | Not-determined |
| S13_extracted_150494 | Not-determined |
| S13_extracted_150502 | Not-determined |
| S13_extracted_150505 | Not-determined |
| S13_extracted_150506 | Not-determined |
| S13_extracted_150521 | Low-quality |
| S13_extracted_150529 | Not-determined |
| S13_extracted_150560 | Not-determined |
| S13_extracted_150562 | Not-determined |
| S13_extracted_150578 | Not-determined |
| S13_extracted_150583 | Not-determined |
| S13_extracted_150594 | Not-determined |
| S13_extracted_150596 | Not-determined |
| S13_extracted_150603 | Not-determined |
| S13_extracted_150609 | Not-determined |
| S13_extracted_150643 | Not-determined |
| S13_extracted_150644 | Not-determined |
| S13_extracted_150661 | Not-determined |
| S13_extracted_150662 | Low-quality |
| S13_extracted_150683 | Low-quality |
| S13_extracted_150691 | Not-determined |
| S13_extracted_150709 | Low-quality |
| S13_extracted_150721 | Not-determined |
| S13_extracted_150725 | Not-determined |
| S13_extracted_150728 | Not-determined |
| S13_extracted_150736 | Not-determined |
| S13_extracted_150737 | Low-quality |
| S13_extracted_150750 | Low-quality |
| S13_extracted_150767 | Not-determined |
| S13_extracted_150777 | Not-determined |
| S13_extracted_150779 | Not-determined |
| S13_extracted_150795 | Not-determined |
| S13_extracted_150804 | Not-determined |
| S13_extracted_150805 | Not-determined |
| S13_extracted_150813 | Not-determined |
| S13_extracted_150825 | Low-quality |
| S13_extracted_150830 | Not-determined |
| S13_extracted_150831 | Low-quality |
| S13_extracted_150838 | Not-determined |
| S13_extracted_150839 | Not-determined |
| S13_extracted_150850 | Not-determined |
| S13_extracted_150856 | Not-determined |
| S13_extracted_150860 | Not-determined |
| S13_extracted_150870 | Not-determined |
| S13_extracted_150874 | Not-determined |
| S13_extracted_150876 | Not-determined |
| S13_extracted_150889 | Not-determined |
| S13_extracted_150896 | Not-determined |
| S13_extracted_150905 | Not-determined |
| S13_extracted_150908 | Not-determined |
| S13_extracted_150917 | Not-determined |
| S13_extracted_150920 | Low-quality |
| S13_extracted_150931 | Not-determined |
| S13_extracted_150937 | Low-quality |
| S13_extracted_150960 | Not-determined |
| S13_extracted_150964 | Not-determined |
| S13_extracted_150965 | Low-quality |
| S13_extracted_150987 | Not-determined |
| S13_extracted_150992 | Not-determined |
| S13_extracted_150999 | Not-determined |
| S13_extracted_151010 | Not-determined |
| S13_extracted_151013 | Not-determined |
| S13_extracted_151019 | Not-determined |
| S13_extracted_151022 | Not-determined |
| S13_extracted_151026 | Not-determined |
| S6_extracted_k141_56487 | Complete |
| S13_extracted_151047 | Not-determined |
| S13_extracted_151070 | Not-determined |
| S13_extracted_151072 | Not-determined |
| S13_extracted_151075 | Not-determined |
| S13_extracted_151086 | Not-determined |
| S13_extracted_151101 | Not-determined |
| S13_extracted_151103 | Not-determined |
| S13_extracted_151107 | Not-determined |
| S13_extracted_151138 | Not-determined |
| S13_extracted_151148 | Not-determined |
| S13_extracted_151151 | Not-determined |
| S13_extracted_151165 | Not-determined |
| S13_extracted_151169 | Not-determined |
| S13_extracted_151172 | Not-determined |
| S13_extracted_151176 | Not-determined |
| S13_extracted_151184 | Not-determined |
| S13_extracted_151186 | Not-determined |
| S13_extracted_151187 | Low-quality |
| S13_extracted_151194 | Not-determined |
| S13_extracted_151212 | Not-determined |
| S13_extracted_151243 | Not-determined |
| S13_extracted_151266 | Not-determined |
| S13_extracted_151274 | Not-determined |
| S13_extracted_151282 | Not-determined |
| S13_extracted_151287 | Not-determined |
| S13_extracted_151299 | Not-determined |
| S13_extracted_151300 | Not-determined |
| S13_extracted_151311 | Not-determined |
| S13_extracted_151312 | Not-determined |
| S13_extracted_151321 | Not-determined |
| S13_extracted_151331 | Not-determined |
| S13_extracted_151344 | Not-determined |
| S13_extracted_151348 | Low-quality |
| S13_extracted_151355 | Not-determined |
| S13_extracted_151384 | Not-determined |
| S13_extracted_151391 | Not-determined |
| S13_extracted_151394 | Not-determined |
| S13_extracted_151407 | Not-determined |
| S13_extracted_151413 | Not-determined |
| S13_extracted_151418 | Not-determined |
| S13_extracted_151422 | Not-determined |
| S13_extracted_151427 | Not-determined |
| S13_extracted_151449 | Not-determined |
| S13_extracted_151469 | Not-determined |
| S13_extracted_151488 | Not-determined |
| S13_extracted_151494 | Not-determined |
| S13_extracted_151496 | Low-quality |
| S13_extracted_151499 | Low-quality |
| S13_extracted_151514 | Not-determined |
| S13_extracted_151522 | Not-determined |
| S13_extracted_151526 | Not-determined |
| S13_extracted_151533 | Not-determined |
| S13_extracted_151560 | Not-determined |
| S13_extracted_151565 | Not-determined |
| S13_extracted_151572 | Not-determined |
| S13_extracted_151573 | Not-determined |
| S13_extracted_151576 | Not-determined |
| S13_extracted_151578 | Low-quality |
| S13_extracted_151586 | Not-determined |
| S13_extracted_151589 | Not-determined |
| S13_extracted_151593 | Not-determined |
| S13_extracted_151642 | Low-quality |
| S13_extracted_151651 | Not-determined |
| S13_extracted_151652 | Not-determined |
| S13_extracted_151662 | Not-determined |
| S13_extracted_151673 | Not-determined |
| S13_extracted_151676 | Not-determined |
| S13_extracted_151683 | Not-determined |
| S13_extracted_151697 | Not-determined |
| S13_extracted_151706 | Low-quality |
| S13_extracted_151716 | Not-determined |
| S13_extracted_151721 | Not-determined |
| S13_extracted_151739 | Not-determined |
| S13_extracted_151744 | Not-determined |
| S13_extracted_151745 | Not-determined |
| S13_extracted_151748 | Not-determined |
| S13_extracted_151754 | Not-determined |
| S12_vs_k141_10887\|\|full | High-quality |
| S13_extracted_151759 | Not-determined |
| S13_extracted_151774 | Not-determined |
| S13_extracted_151784 | Low-quality |
| S13_extracted_151786 | Not-determined |
| S13_extracted_151811 | Not-determined |
| S13_extracted_151813 | Not-determined |
| S13_extracted_151818 | Not-determined |
| S13_extracted_151828 | Not-determined |
| S13_extracted_151833 | Not-determined |
| S13_extracted_151847 | Not-determined |
| S13_extracted_151849 | Not-determined |
| S13_extracted_151854 | Not-determined |
| S13_extracted_151878 | Not-determined |
| S13_extracted_151887 | Not-determined |
| S13_extracted_151901 | Not-determined |
| S13_extracted_151904 | Not-determined |
| S13_extracted_151916 | Not-determined |
| S13_extracted_151920 | Not-determined |
| S13_extracted_151923 | Not-determined |
| S13_extracted_151941 | Not-determined |
| S13_extracted_151964 | Not-determined |
| S13_extracted_151972 | Not-determined |
| S13_extracted_151986 | Not-determined |
| S13_extracted_151989 | Not-determined |
| S13_extracted_151992 | Not-determined |
| S13_vs_27462\|\|full | Medium-quality |
| S13_extracted_152006 | Low-quality |
| S13_extracted_152007 | Low-quality |
| S13_extracted_152022 | Not-determined |
| S13_extracted_152025 | Low-quality |
| S13_extracted_152028 | Not-determined |
| S13_extracted_152031 | Not-determined |
| S13_extracted_152035 | Not-determined |
| S13_extracted_152044 | Low-quality |
| S13_extracted_152047 | Not-determined |
| S13_extracted_152055 | Not-determined |
| S13_extracted_152065 | Not-determined |
| S13_extracted_152066 | Not-determined |
| S13_extracted_152069 | Not-determined |
| S13_extracted_152071 | Not-determined |
| S13_extracted_152077 | Not-determined |
| S13_extracted_152083 | Not-determined |
| S13_extracted_152090 | Not-determined |
| S13_extracted_152095 | Not-determined |
| S13_extracted_152100 | Not-determined |
| S13_extracted_152102 | Not-determined |
| S13_extracted_152104 | Not-determined |
| S13_extracted_152125 | Not-determined |
| S13_extracted_152131 | Low-quality |
| S13_extracted_152151 | Not-determined |
| S13_extracted_152152 | Not-determined |
| S13_extracted_152159 | Not-determined |
| S13_extracted_152160 | Not-determined |
| S13_extracted_152161 | Not-determined |
| S13_extracted_152163 | Not-determined |
| S13_extracted_152164 | Not-determined |
| S13_extracted_152181 | Not-determined |
| S13_extracted_152182 | Not-determined |
| S13_extracted_152183 | Not-determined |
| S13_extracted_152191 | Not-determined |
| S13_extracted_152195 | Low-quality |
| S13_extracted_152200 | Not-determined |
| S13_extracted_152204 | Not-determined |
| S13_extracted_152211 | Not-determined |
| S13_extracted_152213 | Not-determined |
| S13_extracted_152218 | Not-determined |
| S13_extracted_152230 | Not-determined |
| S13_extracted_152242 | Not-determined |
| S13_extracted_152246 | Not-determined |
| S13_extracted_152247 | Not-determined |
| S13_extracted_152250 | Not-determined |
| S13_extracted_152258 | Not-determined |
| S13_extracted_152261 | Not-determined |
| S13_extracted_152266 | Not-determined |
| S13_extracted_152291 | Not-determined |
| S13_extracted_152300 | Not-determined |
| S13_extracted_152305 | Not-determined |
| S13_extracted_152310 | Not-determined |
| S13_extracted_152313 | Not-determined |
| S13_extracted_152318 | Not-determined |
| S13_extracted_152319 | Not-determined |
| S13_extracted_152325 | Not-determined |
| S13_extracted_152330 | Not-determined |
| S13_extracted_152331 | Not-determined |
| S13_extracted_152333 | Not-determined |
| S13_extracted_152337 | Not-determined |
| S13_extracted_152338 | Not-determined |
| S13_extracted_152339 | Low-quality |
| S13_extracted_152340 | Not-determined |
| S13_extracted_152346 | Not-determined |
| S13_extracted_152351 | Not-determined |
| S13_extracted_152352 | Not-determined |
| S13_extracted_152353 | Not-determined |
| S13_extracted_152356 | Not-determined |
| S13_extracted_152369 | Not-determined |
| S13_extracted_152371 | Not-determined |
| S13_extracted_152380 | Not-determined |
| S13_extracted_152391 | Not-determined |
| S13_extracted_152418 | Not-determined |
| S13_extracted_152423 | Not-determined |
| S13_extracted_152427 | Not-determined |
| S13_extracted_152435 | Not-determined |
| S13_extracted_152438 | Low-quality |
| S13_extracted_152447 | Not-determined |
| S13_extracted_152450 | Not-determined |
| S13_extracted_152474 | Not-determined |
| S13_extracted_152480 | Not-determined |
| S13_extracted_152482 | Not-determined |
| S13_extracted_152485 | Not-determined |
| S13_extracted_152511 | Low-quality |
| S13_extracted_152517 | Not-determined |
| S13_extracted_152522 | Not-determined |
| S13_extracted_152524 | Not-determined |
| S13_extracted_152536 | Not-determined |
| S13_extracted_152538 | Not-determined |
| S13_extracted_152568 | Not-determined |
| S13_extracted_152579 | Not-determined |
| S13_extracted_152580 | Not-determined |
| S13_extracted_152582 | Not-determined |
| S13_extracted_152585 | Not-determined |
| S13_extracted_152590 | Not-determined |
| S13_extracted_152594 | Not-determined |
| S13_extracted_152601 | Not-determined |
| S13_extracted_152610 | Not-determined |
| S13_extracted_152616 | Not-determined |
| S13_extracted_152629 | Not-determined |
| S13_extracted_152634 | Not-determined |
| S13_extracted_152651 | Not-determined |
| S13_extracted_152659 | Not-determined |
| S13_extracted_152660 | Not-determined |
| S13_extracted_152669 | Not-determined |
| S13_extracted_152684 | Not-determined |
| S13_extracted_152690 | Not-determined |
| S13_extracted_152699 | Not-determined |
| S13_extracted_152704 | Not-determined |
| S13_extracted_152707 | Low-quality |
| S13_extracted_152710 | Not-determined |
| S13_extracted_152736 | Not-determined |
| S13_extracted_152747 | Not-determined |
| S13_extracted_152755 | Low-quality |
| S13_extracted_152762 | Not-determined |
| S13_extracted_152771 | Not-determined |
| S13_extracted_152781 | Not-determined |
| S13_extracted_152782 | Not-determined |
| S13_extracted_152787 | Not-determined |
| S13_extracted_152793 | Not-determined |
| S13_extracted_152811 | Not-determined |
| S13_extracted_152814 | Not-determined |
| S13_extracted_152824 | Not-determined |
| S13_extracted_152832 | Not-determined |
| S13_extracted_152837 | Not-determined |
| S13_extracted_152844 | Not-determined |
| S13_extracted_152847 | Low-quality |
| S13_extracted_152864 | Not-determined |
| S13_extracted_152869 | Not-determined |
| S13_extracted_152873 | Not-determined |
| S13_extracted_152874 | Not-determined |
| S13_extracted_152902 | Not-determined |
| S13_extracted_152908 | Not-determined |
| S13_extracted_152915 | Not-determined |
| S13_extracted_152918 | Not-determined |
| S13_extracted_152920 | Not-determined |
| S13_extracted_152921 | Not-determined |
| S13_extracted_152923 | Not-determined |
| S13_extracted_152934 | Not-determined |
| S13_extracted_152972 | Not-determined |
| S13_extracted_152981 | Not-determined |
| S13_extracted_152992 | Not-determined |
| S13_extracted_152994 | Not-determined |
| S13_extracted_153001 | Low-quality |
| S13_extracted_153002 | Not-determined |
| S13_extracted_153020 | Not-determined |
| S13_extracted_153021 | Low-quality |
| S13_extracted_153026 | Not-determined |
| S13_extracted_153027 | Not-determined |
| S13_extracted_153036 | Not-determined |
| S13_extracted_153039 | Low-quality |
| S13_extracted_153046 | Low-quality |
| S13_extracted_153051 | Not-determined |
| S13_extracted_153057 | Not-determined |
| S13_extracted_153076 | Not-determined |
| S13_extracted_153078 | Not-determined |
| S13_extracted_153113 | Not-determined |
| S13_extracted_153116 | Not-determined |
| S13_extracted_153125 | Not-determined |
| S13_extracted_153138 | Not-determined |
| S13_extracted_153142 | Not-determined |
| S13_extracted_153145 | Low-quality |
| S13_extracted_153154 | Not-determined |
| S13_extracted_153155 | Not-determined |
| S13_extracted_153156 | Not-determined |
| S13_extracted_153179 | Not-determined |
| S13_extracted_153180 | Not-determined |
| S13_extracted_153182 | Not-determined |
| S13_extracted_153189 | Not-determined |
| S13_extracted_153198 | Low-quality |
| S13_extracted_153199 | Not-determined |
| S13_extracted_153205 | Low-quality |
| S13_extracted_153213 | Not-determined |
| S13_extracted_153218 | Not-determined |
| S13_extracted_153219 | Not-determined |
| S13_extracted_153222 | Not-determined |
| S13_extracted_153230 | Not-determined |
| S13_extracted_153239 | Not-determined |
| S13_extracted_153248 | Not-determined |
| S13_extracted_153267 | Not-determined |
| S13_extracted_153270 | Not-determined |
| S13_extracted_153272 | Not-determined |
| S13_extracted_153278 | Not-determined |
| S13_extracted_153285 | Not-determined |
| S13_extracted_153286 | Not-determined |
| S13_extracted_153288 | Not-determined |
| S13_extracted_153308 | Not-determined |
| S13_extracted_153309 | Not-determined |
| S13_extracted_153316 | Not-determined |
| S13_extracted_153322 | Low-quality |
| S13_extracted_153344 | Not-determined |
| S13_extracted_153346 | Not-determined |
| S13_extracted_153356 | Not-determined |
| S13_extracted_153357 | Not-determined |
| S13_extracted_153367 | Not-determined |
| S13_extracted_153373 | Not-determined |
| S13_extracted_153380 | Not-determined |
| S13_extracted_153385 | Not-determined |
| S13_extracted_153388 | Not-determined |
| S13_extracted_153390 | Not-determined |
| S13_extracted_153403 | Not-determined |
| S13_extracted_153413 | Not-determined |
| S13_extracted_153416 | Not-determined |
| S13_extracted_153441 | Low-quality |
| S13_extracted_153450 | Not-determined |
| S13_extracted_153455 | Low-quality |
| S13_extracted_153470 | Not-determined |
| S13_extracted_153508 | Low-quality |
| S13_extracted_153525 | Not-determined |
| S13_extracted_153526 | Not-determined |
| S13_extracted_153532 | Not-determined |
| S13_extracted_153538 | Not-determined |
| S13_extracted_153553 | Low-quality |
| S13_extracted_153555 | Not-determined |
| S13_extracted_153566 | Not-determined |
| S13_extracted_153575 | Not-determined |
| S13_extracted_153577 | Not-determined |
| S13_extracted_153581 | Not-determined |
| S13_extracted_153586 | Not-determined |
| S13_extracted_153597 | Not-determined |
| S13_extracted_153602 | Not-determined |
| S13_extracted_153609 | Not-determined |
| S13_extracted_153611 | Not-determined |
| S13_extracted_153612 | Not-determined |
| S13_extracted_153622 | Not-determined |
| S13_extracted_153624 | Not-determined |
| S13_extracted_153630 | Not-determined |
| S13_extracted_153634 | Not-determined |
| S13_extracted_153638 | Not-determined |
| S13_extracted_153648 | Not-determined |
| S13_extracted_153652 | Not-determined |
| S13_extracted_153657 | Not-determined |
| S13_extracted_153664 | Not-determined |
| S13_extracted_153667 | Not-determined |
| S13_extracted_153702 | Not-determined |
| S13_extracted_153705 | Low-quality |
| S13_extracted_153719 | Not-determined |
| S13_extracted_153727 | Not-determined |
| S13_extracted_153734 | Low-quality |
| S13_extracted_153735 | Low-quality |
| S13_extracted_153740 | Not-determined |
| S13_extracted_153741 | Not-determined |
| S13_extracted_153743 | Not-determined |
| S13_extracted_153746 | Not-determined |
| S13_extracted_153759 | Not-determined |
| S13_extracted_153803 | Not-determined |
| S13_extracted_153809 | Not-determined |
| S13_extracted_153821 | Not-determined |
| S13_extracted_153822 | Not-determined |
| S13_extracted_153823 | Not-determined |
| S13_extracted_153827 | Not-determined |
| S13_extracted_153838 | Not-determined |
| S13_extracted_153849 | Not-determined |
| S13_extracted_153852 | Not-determined |
| S13_extracted_153855 | Not-determined |
| S13_extracted_153874 | Not-determined |
| S13_extracted_153886 | Not-determined |
| S13_extracted_153932 | Low-quality |
| S13_extracted_153936 | Not-determined |
| S13_extracted_153937 | Not-determined |
| S13_extracted_153939 | Low-quality |
| S13_extracted_153941 | Not-determined |
| S13_extracted_153942 | Not-determined |
| S13_extracted_153965 | Low-quality |
| S13_extracted_153967 | Not-determined |
| S13_extracted_153968 | Not-determined |
| S13_extracted_153983 | Not-determined |
| S13_extracted_153988 | Not-determined |
| S13_extracted_153990 | Low-quality |
| S13_extracted_154011 | Not-determined |
| S13_extracted_154012 | Low-quality |
| S13_extracted_154014 | Low-quality |
| S13_extracted_154015 | Low-quality |
| S13_extracted_154017 | Not-determined |
| S13_extracted_154021 | Low-quality |
| S13_extracted_154023 | Not-determined |
| S13_extracted_154032 | Low-quality |
| S13_extracted_154039 | Not-determined |
| S13_extracted_154047 | Not-determined |
| S13_extracted_154059 | Low-quality |
| S13_extracted_154062 | Not-determined |
| S13_extracted_154063 | Not-determined |
| S13_extracted_154065 | Not-determined |
| S13_extracted_154091 | Not-determined |
| S13_extracted_154092 | Not-determined |
| S13_extracted_154093 | Not-determined |
| S13_extracted_154099 | Low-quality |
| S13_extracted_154105 | Low-quality |
| S13_extracted_154127 | Not-determined |
| S13_extracted_154129 | Not-determined |
| S13_extracted_154130 | Not-determined |
| S13_extracted_154132 | Low-quality |
| S13_extracted_154146 | Not-determined |
| S13_extracted_154156 | Not-determined |
| S13_extracted_154159 | Not-determined |
| S13_extracted_154161 | Not-determined |
| S13_extracted_154164 | Not-determined |
| S13_extracted_154189 | Not-determined |
| S13_extracted_154205 | Not-determined |
| S13_extracted_154218 | Not-determined |
| S13_extracted_154219 | Not-determined |
| S13_extracted_154221 | Not-determined |
| S13_extracted_154231 | Not-determined |
| S13_extracted_154233 | Not-determined |
| S13_extracted_154237 | Not-determined |
| S13_extracted_154244 | Not-determined |
| S13_extracted_154245 | Not-determined |
| S13_extracted_154250 | Low-quality |
| S13_extracted_154264 | Not-determined |
| S13_extracted_154265 | Not-determined |
| S13_extracted_154269 | Not-determined |
| S13_extracted_154281 | Not-determined |
| S13_extracted_154283 | Not-determined |
| S13_extracted_154303 | Not-determined |
| S13_extracted_154315 | Not-determined |
| S13_extracted_154317 | Not-determined |
| S13_extracted_154321 | Not-determined |
| S13_extracted_154325 | Low-quality |
| S13_extracted_154326 | Not-determined |
| S13_extracted_154327 | Not-determined |
| S13_extracted_154342 | Not-determined |
| S13_extracted_154343 | Not-determined |
| S13_extracted_154348 | Not-determined |
| S13_extracted_154368 | Low-quality |
| S13_extracted_154387 | Not-determined |
| S13_extracted_154391 | Not-determined |
| S13_extracted_154394 | Not-determined |
| S13_extracted_154400 | Not-determined |
| S10_vs_40829\|\|full | Medium-quality |
| S13_vs_9029\|\|full | Not-determined |
| S5_vs_k141_145585\|\|full | High-quality |
| S6_vs_k141_56487\|\|full | High-quality |
| S9_vs_k141_2124\|\|full | Medium-quality |
| S13_vs_21329\|\|full | Not-determined |
| S17_extracted_k141_174033 | Medium-quality |
| S16_extracted_9270 | Complete |
| S13_vs_28316\|\|full | Not-determined |
| S11_extracted_k141_42630 | Medium-quality |
| S5_vs_k141_9747\|\|full | High-quality |
| S5_vs_k141_76500\|\|0_partial | Medium-quality |
| S11_vs_k141_42630\|\|full | Medium-quality |
| S13_vs_82403\|\|full | Not-determined |
| S16_vs_9270\|\|full | High-quality |
| S13_vs_132621\|\|full | Low-quality |
| S6_vs_k141_17550\|\|full | High-quality |
| S2_extracted_k141_21511 | High-quality |
| S8_extracted_k141_134943 | High-quality |
| S7_extracted_5801 | Complete |
| S12_vs_k141_151292\|\|full | Medium-quality |
| S2_vs_k141_21511\|\|full | High-quality |
| S7_vs_5801\|\|full | High-quality |
| S13_vs_150477\|\|full | Not-determined |
| S16_vs_127905\|\|full | High-quality |
| S8_extracted_k141_163760 | Complete |
| S8_vs_k141_134943\|\|full | High-quality |
| S12_vs_k141_23796\|\|0_partial | Medium-quality |
| S13_vs_152067\|\|full | Not-determined |
| S13_vs_30\|\|full | Low-quality |
| S13_vs_74\|\|full | Low-quality |
| S13_vs_77\|\|full | Low-quality |
| S13_vs_133\|\|full | Low-quality |
| S13_vs_146\|\|full | Low-quality |
| S13_vs_152\|\|full | Not-determined |
| S13_vs_276\|\|full | Low-quality |
| S13_vs_346\|\|full | Low-quality |
| S13_vs_367\|\|full | Low-quality |
| S13_vs_379\|\|full | Low-quality |
| S13_vs_441\|\|full | Low-quality |
| S13_vs_477\|\|full | Low-quality |
| S13_vs_491\|\|full | Not-determined |
| S13_vs_502\|\|full | Not-determined |
| S13_vs_546\|\|full | Low-quality |
| S13_vs_587\|\|full | Not-determined |
| S13_vs_617\|\|full | Not-determined |
| S13_vs_702\|\|full | Not-determined |
| S13_vs_763\|\|full | Not-determined |
| S13_vs_808\|\|full | Low-quality |
| S13_vs_824\|\|full | Low-quality |
| S13_vs_837\|\|full | Not-determined |
| S13_vs_845\|\|full | Not-determined |
| S13_vs_852\|\|full | Not-determined |
| S13_vs_912\|\|full | Not-determined |
| S13_vs_921\|\|full | Low-quality |
| S13_vs_965\|\|full | Low-quality |
| S13_vs_981\|\|full | Not-determined |
| S13_vs_1000\|\|full | Low-quality |
| S13_vs_1092\|\|full | Not-determined |
| S13_vs_1145\|\|full | Low-quality |
| S13_vs_1200\|\|full | Not-determined |
| S13_vs_1226\|\|full | Low-quality |
| S13_vs_1453\|\|full | Low-quality |
| S13_vs_1612\|\|full | Not-determined |
| S13_vs_1672\|\|full | Low-quality |
| S13_vs_1676\|\|full | Not-determined |
| S13_vs_1736\|\|full | Not-determined |

| | |
|---|---|
| S8_vs_k141_163760\|\|full | High-quality |
| S13_vs_1806\|\|full | Low-quality |
| S13_vs_1851\|\|full | Low-quality |
| S13_vs_1910\|\|full | Low-quality |
| S13_vs_1960\|\|full | Low-quality |
| S17_extracted_k141_68049 | High-quality |
| S13_vs_2066\|\|full | Low-quality |
| S13_vs_2089\|\|full | Low-quality |
| S13_vs_2114\|\|full | Low-quality |
| S13_vs_2124\|\|full | Not-determined |
| S13_vs_2132\|\|full | Low-quality |
| S13_vs_2153\|\|full | Not-determined |
| S13_vs_2215\|\|full | Not-determined |
| S13_vs_2276\|\|full | Not-determined |
| S13_vs_2292\|\|full | Low-quality |
| S13_vs_2380\|\|full | Not-determined |
| S13_vs_2416\|\|full | Not-determined |
| S13_vs_2497\|\|full | Low-quality |
| S13_vs_2543\|\|full | Not-determined |
| S13_vs_2577\|\|full | Low-quality |
| S13_vs_2644\|\|full | Low-quality |
| S13_vs_2690\|\|full | Low-quality |
| S13_vs_2841\|\|full | Not-determined |
| S13_vs_2873\|\|full | Not-determined |
| S13_vs_2894\|\|full | Low-quality |
| S13_vs_2896\|\|full | Low-quality |
| S13_vs_2913\|\|full | Not-determined |
| S13_vs_2939\|\|full | Low-quality |
| S13_vs_3058\|\|full | Low-quality |
| S13_vs_3212\|\|full | Not-determined |
| S13_vs_3242\|\|full | Low-quality |
| S13_vs_3423\|\|full | Not-determined |
| S13_vs_3450\|\|full | Low-quality |
| S13_vs_3468\|\|full | Low-quality |
| S13_vs_3488\|\|full | Not-determined |
| S13_vs_3598\|\|full | Low-quality |
| S13_vs_3630\|\|full | Not-determined |
| S13_vs_3654\|\|full | Not-determined |
| S13_vs_3666\|\|full | Not-determined |
| S13_vs_3708\|\|full | Low-quality |
| S13_vs_3723\|\|full | Low-quality |
| S13_vs_3733\|\|full | Not-determined |
| S13_vs_3823\|\|full | Not-determined |
| S13_vs_3884\|\|full | Not-determined |
| S13_vs_3899\|\|full | Not-determined |
| S13_vs_3910\|\|full | Not-determined |
| S13_vs_3961\|\|full | Low-quality |
| S13_vs_4031\|\|full | Low-quality |
| S13_vs_4092\|\|full | Not-determined |
| S13_vs_4121\|\|full | Low-quality |
| S13_vs_4181\|\|full | Not-determined |
| S13_vs_4234\|\|full | Low-quality |
| S13_vs_4314\|\|full | Not-determined |
| S13_vs_4321\|\|full | Low-quality |
| S13_vs_4364\|\|full | Low-quality |
| S13_vs_4371\|\|full | Low-quality |
| S13_vs_4410\|\|full | Low-quality |
| S13_vs_4474\|\|full | Low-quality |
| S13_vs_4516\|\|full | Not-determined |
| S13_vs_4534\|\|full | Not-determined |
| S13_vs_4539\|\|full | Low-quality |
| S13_vs_4554\|\|full | Low-quality |
| S13_vs_4576\|\|full | Low-quality |
| S13_vs_4591\|\|full | Not-determined |
| S13_vs_4655\|\|full | Low-quality |
| S13_vs_4666\|\|full | Low-quality |
| S13_vs_4695\|\|full | Low-quality |
| S13_vs_4716\|\|full | Not-determined |
| S13_vs_4721\|\|full | Low-quality |
| S13_vs_4729\|\|full | Not-determined |
| S13_vs_4788\|\|full | Low-quality |
| S13_vs_4837\|\|full | Not-determined |
| S15_vs_73469\|\|full | Medium-quality |
| S13_vs_4910\|\|full | Not-determined |
| S13_vs_4947\|\|full | Not-determined |
| S13_vs_4968\|\|full | Not-determined |
| S13_vs_4991\|\|full | Low-quality |
| S13_vs_5047\|\|full | Not-determined |
| S13_vs_5074\|\|full | Low-quality |
| S13_vs_5173\|\|full | Low-quality |
| S13_vs_5181\|\|full | Not-determined |
| S13_vs_5296\|\|full | Low-quality |
| S13_vs_5336\|\|full | Not-determined |
| S13_vs_5348\|\|full | Low-quality |
| S13_vs_5350\|\|full | Not-determined |
| S13_vs_5352\|\|full | Not-determined |
| S13_vs_5394\|\|full | Not-determined |
| S13_vs_5404\|\|full | Not-determined |
| S13_vs_5440\|\|full | Low-quality |
| S13_vs_5559\|\|full | Low-quality |
| S13_vs_5631\|\|full | Not-determined |
| S13_vs_5642\|\|full | Not-determined |
| S13_vs_5788\|\|full | Low-quality |
| S13_vs_5882\|\|full | Low-quality |
| S13_vs_5898\|\|full | Not-determined |
| S13_vs_5959\|\|full | Low-quality |
| S13_vs_5976\|\|full | Not-determined |
| S13_vs_5992\|\|full | Low-quality |
| S13_vs_6013\|\|full | Low-quality |
| S13_vs_6138\|\|full | Low-quality |
| S13_vs_6141\|\|full | Not-determined |
| S13_vs_6144\|\|full | Not-determined |
| S13_vs_6287\|\|full | Not-determined |
| S13_vs_6293\|\|full | Low-quality |
| S13_vs_6313\|\|full | Low-quality |
| S13_vs_6365\|\|full | Low-quality |
| S13_vs_6366\|\|full | Not-determined |
| S13_vs_6385\|\|full | Not-determined |
| S13_vs_6438\|\|full | Low-quality |
| S13_vs_6463\|\|full | Low-quality |
| S13_vs_6499\|\|full | Low-quality |
| S13_vs_6529\|\|full | Low-quality |
| S13_vs_6558\|\|full | Not-determined |
| S13_vs_6641\|\|full | Not-determined |
| S13_vs_6679\|\|full | Low-quality |
| S13_vs_6695\|\|full | Not-determined |
| S13_vs_6698\|\|full | Not-determined |
| S13_vs_6769\|\|full | Not-determined |
| S13_vs_6791\|\|full | Low-quality |
| S13_vs_6794\|\|full | Not-determined |
| S13_vs_6816\|\|full | Not-determined |
| S13_vs_7062\|\|full | Low-quality |
| S13_vs_7140\|\|full | Low-quality |
| S13_vs_7164\|\|full | Not-determined |
| S13_vs_7272\|\|full | Low-quality |
| S13_vs_7293\|\|full | Low-quality |
| S13_vs_7335\|\|full | Low-quality |
| S13_vs_7407\|\|full | Not-determined |
| S13_vs_7415\|\|full | Not-determined |
| S13_vs_7448\|\|full | Low-quality |
| S13_vs_7449\|\|full | Not-determined |
| S13_vs_7485\|\|full | Low-quality |
| S13_vs_7494\|\|full | Low-quality |
| S13_vs_7574\|\|full | Not-determined |
| S13_vs_7581\|\|full | Not-determined |
| S13_vs_7607\|\|full | Low-quality |
| S13_vs_7659\|\|full | Not-determined |
| S4_extracted_20440 | Medium-quality |
| S13_vs_7689\|\|full | Low-quality |
| S13_vs_7762\|\|full | Not-determined |
| S13_vs_7863\|\|full | Not-determined |
| S13_vs_7876\|\|full | Low-quality |
| S13_vs_7888\|\|full | Low-quality |
| S13_vs_7911\|\|full | Not-determined |
| S13_vs_7915\|\|full | Not-determined |
| S13_vs_7941\|\|full | Low-quality |
| S13_vs_7947\|\|full | Low-quality |
| S13_vs_7963\|\|full | Not-determined |
| S13_vs_7970\|\|full | Not-determined |
| S13_vs_8022\|\|full | Not-determined |
| S13_vs_8070\|\|full | Not-determined |
| S13_vs_8166\|\|full | Low-quality |
| S13_vs_8233\|\|full | Low-quality |
| S13_vs_8259\|\|full | Not-determined |
| S13_vs_8262\|\|full | Not-determined |
| S13_vs_8299\|\|full | Not-determined |
| S13_vs_8315\|\|full | Low-quality |
| S13_vs_8351\|\|full | Low-quality |
| S13_vs_8368\|\|full | Low-quality |
| S13_vs_8508\|\|full | Low-quality |
| S13_vs_8520\|\|full | Not-determined |
| S13_vs_8549\|\|full | Low-quality |
| S13_vs_8625\|\|full | Low-quality |
| S13_vs_8638\|\|full | Not-determined |
| S13_vs_8652\|\|full | Not-determined |
| S13_vs_8704\|\|full | Low-quality |
| S13_vs_8785\|\|full | Not-determined |
| S13_vs_8788\|\|full | Not-determined |
| S13_vs_8809\|\|full | Not-determined |
| S13_vs_8817\|\|full | Not-determined |
| S13_vs_8839\|\|full | Not-determined |
| S13_vs_8881\|\|full | Low-quality |
| S13_vs_8915\|\|full | Low-quality |
| S13_vs_8991\|\|full | Low-quality |
| S13_vs_9055\|\|full | Not-determined |
| S13_vs_9066\|\|full | Not-determined |
| S13_vs_9082\|\|full | Not-determined |
| S13_vs_9109\|\|full | Low-quality |
| S13_vs_9225\|\|full | Not-determined |
| S13_vs_9231\|\|full | Low-quality |
| S13_vs_9262\|\|full | Low-quality |
| S13_vs_9301\|\|full | Low-quality |
| S13_vs_9302\|\|full | Low-quality |
| S13_vs_9335\|\|full | Not-determined |
| S13_vs_9341\|\|full | Low-quality |
| S13_vs_9391\|\|full | Low-quality |
| S4_vs_VIRSorter_20440-cat_2 | Medium-quality |
| S13_vs_9474\|\|full | Low-quality |
| S13_vs_9475\|\|full | Not-determined |
| S13_vs_9511\|\|full | Low-quality |
| S13_vs_9536\|\|full | Low-quality |
| S13_vs_9544\|\|full | Low-quality |
| S13_vs_9631\|\|full | Not-determined |
| S13_vs_9638\|\|full | Low-quality |
| S13_vs_9653\|\|full | Not-determined |
| S13_vs_9701\|\|full | Low-quality |
| S13_vs_9751\|\|full | Not-determined |
| S13_vs_9789\|\|full | Low-quality |
| S13_vs_9791\|\|full | Not-determined |
| S13_vs_9802\|\|full | Low-quality |
| S13_vs_9868\|\|full | Low-quality |
| S13_vs_9896\|\|full | Not-determined |
| S13_vs_9908\|\|full | Low-quality |
| S13_vs_10001\|\|full | Not-determined |
| S13_vs_10114\|\|full | Low-quality |
| S13_vs_10116\|\|full | Low-quality |
| S13_vs_10120\|\|full | Low-quality |
| S13_vs_10122\|\|full | Low-quality |
| S13_vs_10181\|\|full | Low-quality |
| S13_vs_10183\|\|full | Low-quality |
| S13_vs_10280\|\|full | Low-quality |
| S13_vs_10330\|\|full | Low-quality |
| S13_vs_10334\|\|full | Not-determined |
| S13_vs_10400\|\|full | Low-quality |
| S13_vs_10498\|\|full | Not-determined |
| S13_vs_10518\|\|full | Not-determined |
| S13_vs_10555\|\|full | Not-determined |
| S13_vs_10602\|\|full | Low-quality |
| S13_vs_10740\|\|full | Low-quality |
| S13_vs_10762\|\|full | Not-determined |
| S13_vs_10823\|\|full | Low-quality |
| S13_vs_10864\|\|full | Low-quality |
| S13_vs_10920\|\|full | Low-quality |
| S13_vs_10995\|\|full | Not-determined |
| S13_vs_11033\|\|full | Low-quality |
| S13_vs_11099\|\|full | Not-determined |
| S13_vs_11107\|\|full | Low-quality |
| S13_vs_11133\|\|full | Low-quality |
| S13_vs_11145\|\|full | Low-quality |
| S13_vs_11274\|\|full | Low-quality |
| S13_vs_11310\|\|full | Not-determined |
| S13_vs_11349\|\|full | Low-quality |
| S13_vs_11402\|\|full | Low-quality |
| S13_vs_11421\|\|full | Not-determined |
| S13_vs_11453\|\|full | Low-quality |
| S13_vs_11491\|\|full | Not-determined |
| S13_vs_11499\|\|full | Low-quality |
| S13_vs_11527\|\|full | Low-quality |
| S13_vs_11592\|\|full | Not-determined |
| S13_vs_11605\|\|full | Low-quality |
| S13_vs_11653\|\|full | Not-determined |
| S13_vs_11655\|\|full | Low-quality |
| S13_vs_11739\|\|full | Low-quality |
| S13_vs_11748\|\|full | Low-quality |
| S13_vs_11753\|\|full | Low-quality |
| S13_vs_11809\|\|full | Not-determined |
| S13_vs_11872\|\|full | Low-quality |
| S13_vs_11917\|\|full | Not-determined |
| S13_vs_11921\|\|full | Not-determined |
| S13_vs_11935\|\|full | Low-quality |
| S13_vs_11943\|\|full | Low-quality |
| S13_vs_11947\|\|full | Not-determined |
| S13_vs_12005\|\|full | Low-quality |
| S13_vs_12015\|\|full | Not-determined |
| S13_vs_12037\|\|full | Not-determined |
| S13_vs_12042\|\|full | Low-quality |
| S13_vs_12119\|\|full | Low-quality |
| S13_vs_12183\|\|full | Low-quality |
| S13_vs_12190\|\|full | Low-quality |
| S13_vs_12198\|\|full | Not-determined |
| S13_vs_12215\|\|full | Not-determined |
| S13_vs_12239\|\|full | Low-quality |
| S13_vs_12323\|\|full | Not-determined |
| S13_vs_12329\|\|full | Low-quality |
| S13_vs_12336\|\|full | Low-quality |
| S13_vs_12359\|\|full | Low-quality |
| S13_vs_12406\|\|full | Not-determined |
| S13_vs_12437\|\|full | Not-determined |
| S13_vs_12474\|\|full | Not-determined |
| S13_vs_12527\|\|full | Not-determined |
| S13_vs_12531\|\|full | Low-quality |
| S13_vs_12547\|\|full | Low-quality |
| S13_vs_12570\|\|full | Not-determined |
| S13_vs_12641\|\|full | Low-quality |
| S13_vs_12642\|\|full | Not-determined |
| S13_vs_12646\|\|full | Low-quality |
| S13_vs_12724\|\|full | Not-determined |
| S13_vs_12784\|\|full | Low-quality |
| S13_vs_12792\|\|full | Low-quality |
| S13_vs_12805\|\|full | Not-determined |
| S13_vs_12830\|\|full | Low-quality |
| S13_vs_12867\|\|full | Not-determined |
| S13_vs_12876\|\|full | Low-quality |
| S13_vs_12889\|\|full | Low-quality |
| S13_vs_12977\|\|full | Not-determined |
| S13_vs_12980\|\|full | Low-quality |
| S13_vs_13070\|\|full | Low-quality |
| S13_vs_13096\|\|full | Low-quality |
| S12_vs_k141_138261\|\|full | High-quality |
| S13_vs_13247\|\|full | Low-quality |
| S13_vs_13289\|\|full | Low-quality |
| S13_vs_13330\|\|full | Not-determined |
| S13_vs_13352\|\|full | Low-quality |
| S13_vs_13453\|\|full | Not-determined |
| S13_vs_13494\|\|full | Not-determined |
| S13_vs_13498\|\|full | Not-determined |
| S13_vs_13514\|\|full | Not-determined |
| S13_vs_13552\|\|full | Low-quality |
| S13_vs_13580\|\|full | Low-quality |
| S13_vs_13609\|\|full | Low-quality |
| S13_vs_13685\|\|full | Low-quality |
| S13_vs_13693\|\|full | Not-determined |
| S13_vs_13710\|\|full | Not-determined |
| S13_vs_13725\|\|full | Not-determined |
| S13_vs_13825\|\|full | Not-determined |
| S13_vs_13838\|\|full | Not-determined |
| S13_vs_13947\|\|full | Low-quality |
| S13_vs_14031\|\|full | Low-quality |
| S13_vs_14086\|\|full | Not-determined |
| S13_vs_14117\|\|full | Not-determined |
| S13_vs_14128\|\|full | Low-quality |
| S13_vs_14131\|\|full | Low-quality |
| S13_vs_14166\|\|full | Low-quality |
| S13_vs_14177\|\|full | Low-quality |
| S13_vs_14207\|\|full | Not-determined |
| S13_vs_14295\|\|full | Low-quality |
| S13_vs_14296\|\|full | Low-quality |
| S13_vs_14325\|\|full | Low-quality |
| S13_vs_14338\|\|full | Low-quality |
| S13_vs_14388\|\|full | Low-quality |
| S13_vs_14391\|\|full | Low-quality |
| S13_vs_14427\|\|full | Low-quality |
| S13_vs_14477\|\|full | Low-quality |
| S13_vs_14493\|\|full | Low-quality |
| S13_vs_14599\|\|full | Low-quality |
| S13_vs_14624\|\|full | Low-quality |
| S13_vs_14701\|\|full | Not-determined |
| S13_vs_14744\|\|full | Low-quality |
| S13_vs_14774\|\|full | Not-determined |
| S10_vs_62776\|\|full | High-quality |
| S13_vs_14800\|\|full | Not-determined |
| S13_vs_14802\|\|full | Low-quality |
| S13_vs_14967\|\|full | Low-quality |
| S13_vs_14975\|\|full | Not-determined |
| S13_vs_14981\|\|full | Not-determined |
| S13_vs_15108\|\|full | Not-determined |
| S13_vs_15121\|\|full | Low-quality |
| S13_vs_15122\|\|full | Low-quality |
| S13_vs_15124\|\|full | Not-determined |
| S13_vs_15135\|\|full | Not-determined |
| S13_vs_15162\|\|full | Not-determined |
| S13_vs_15422\|\|full | Low-quality |
| S13_vs_15438\|\|full | Low-quality |
| S13_vs_15459\|\|full | Not-determined |
| S13_vs_15494\|\|full | Low-quality |
| S13_vs_15502\|\|full | Not-determined |
| S13_vs_15547\|\|full | Low-quality |
| S13_vs_15569\|\|full | Low-quality |
| S13_vs_15612\|\|full | Not-determined |
| S13_vs_15637\|\|full | Low-quality |
| S13_vs_15695\|\|full | Not-determined |
| S13_vs_15712\|\|full | Low-quality |
| S13_vs_15743\|\|full | Low-quality |
| S13_vs_15781\|\|full | Not-determined |
| S13_vs_15802\|\|full | Not-determined |
| S13_vs_15852\|\|full | Not-determined |
| S13_vs_15898\|\|full | Low-quality |
| S13_vs_15938\|\|full | Low-quality |
| S13_vs_16030\|\|full | Not-determined |
| S13_vs_16063\|\|full | Not-determined |
| S13_vs_16148\|\|full | Low-quality |
| S13_vs_16168\|\|full | Not-determined |
| S13_vs_16211\|\|full | Low-quality |
| S13_vs_16269\|\|full | Low-quality |
| S13_vs_16282\|\|full | Not-determined |
| S13_vs_16307\|\|full | Low-quality |
| S13_vs_16317\|\|full | Low-quality |
| S13_vs_16335\|\|full | Low-quality |
| S13_vs_16364\|\|full | Low-quality |
| S13_vs_16423\|\|full | Low-quality |
| S13_vs_16543\|\|full | Low-quality |
| S13_vs_16572\|\|full | Not-determined |
| S13_vs_16626\|\|full | Not-determined |
| S13_vs_16671\|\|full | Low-quality |
| S13_vs_16704\|\|full | Low-quality |
| S13_vs_16726\|\|full | Low-quality |
| S13_vs_16730\|\|full | Low-quality |
| S13_vs_16798\|\|full | Low-quality |
| S13_vs_16814\|\|full | Low-quality |
| S13_vs_16848\|\|full | Low-quality |
| S13_vs_16864\|\|full | Not-determined |
| S13_vs_17024\|\|full | Low-quality |
| S13_vs_17122\|\|full | Not-determined |
| S13_vs_17154\|\|full | Low-quality |
| S13_vs_17205\|\|full | Low-quality |
| S13_vs_17289\|\|full | Not-determined |
| S13_vs_17408\|\|full | Low-quality |
| S13_vs_17422\|\|full | Not-determined |
| S13_vs_17442\|\|full | Low-quality |
| S13_vs_17444\|\|full | Low-quality |
| S13_vs_17497\|\|full | Low-quality |
| S13_vs_17526\|\|full | Low-quality |
| S13_vs_17708\|\|full | Low-quality |
| S13_vs_17713\|\|full | Not-determined |
| S13_vs_17735\|\|full | Low-quality |
| S13_vs_17761\|\|full | Low-quality |
| S13_vs_17846\|\|full | Low-quality |
| S13_vs_17914\|\|full | Not-determined |
| S13_vs_17928\|\|full | Low-quality |
| S13_vs_17956\|\|full | Low-quality |
| S13_vs_18052\|\|full | Low-quality |
| S13_vs_18143\|\|full | Not-determined |
| S13_vs_18161\|\|full | Not-determined |
| S13_vs_18186\|\|full | Low-quality |
| S13_vs_18196\|\|full | Not-determined |
| S13_vs_18211\|\|full | Low-quality |
| S13_vs_18213\|\|full | Low-quality |
| S13_vs_18217\|\|full | Not-determined |
| S13_vs_18318\|\|full | Not-determined |
| S13_vs_18429\|\|full | Not-determined |
| S13_vs_18475\|\|full | Low-quality |
| S13_vs_18500\|\|full | Low-quality |
| S13_vs_18535\|\|full | Not-determined |
| S13_vs_18568\|\|full | Low-quality |
| S13_vs_18580\|\|full | Low-quality |
| S13_vs_18587\|\|full | Low-quality |
| S13_vs_18600\|\|full | Not-determined |
| S13_vs_18607\|\|full | Low-quality |
| S13_vs_18613\|\|full | Not-determined |
| S13_vs_18624\|\|full | Low-quality |
| S13_vs_18650\|\|full | Low-quality |
| S13_vs_18693\|\|full | Not-determined |
| S13_vs_18709\|\|full | Low-quality |
| S13_vs_18715\|\|full | Low-quality |
| S13_vs_18744\|\|full | Not-determined |
| S13_vs_18771\|\|full | Low-quality |
| S13_vs_18776\|\|full | Low-quality |
| S13_vs_18873\|\|full | Low-quality |
| S13_vs_18909\|\|full | Low-quality |
| S13_vs_18931\|\|full | Low-quality |
| S13_vs_18975\|\|full | Not-determined |
| S13_vs_19008\|\|full | Not-determined |
| S13_vs_19051\|\|full | Not-determined |
| S13_vs_19215\|\|full | Low-quality |
| S13_vs_19256\|\|full | Not-determined |
| S13_vs_19288\|\|full | Low-quality |
| S13_vs_19348\|\|full | Low-quality |
| S13_vs_19355\|\|full | Low-quality |
| S13_vs_19376\|\|full | Low-quality |
| S13_vs_19396\|\|full | Low-quality |
| S13_vs_19427\|\|full | Low-quality |
| S13_vs_19428\|\|full | Low-quality |
| S13_vs_19431\|\|full | Not-determined |
| S13_vs_19544\|\|full | Low-quality |
| S13_vs_19565\|\|full | Low-quality |
| S13_vs_19606\|\|full | Low-quality |
| S13_vs_19622\|\|full | Low-quality |
| S13_vs_19646\|\|full | Low-quality |
| S13_vs_19689\|\|full | Low-quality |
| S13_vs_19692\|\|full | Not-determined |
| S13_vs_19701\|\|full | Low-quality |
| S13_vs_19709\|\|full | Not-determined |
| S13_vs_19784\|\|full | Not-determined |
| S13_vs_19808\|\|full | Not-determined |
| S13_vs_19821\|\|full | Low-quality |
| S13_vs_19823\|\|full | Low-quality |
| S13_vs_19886\|\|full | Not-determined |
| S13_vs_19953\|\|full | Not-determined |
| S13_vs_19965\|\|full | Not-determined |
| S13_vs_20045\|\|full | Low-quality |
| S13_vs_20058\|\|full | Low-quality |
| S13_vs_20088\|\|full | Low-quality |
| S13_vs_20101\|\|full | Not-determined |
| S13_vs_20158\|\|full | Low-quality |
| S13_vs_20167\|\|full | Not-determined |
| S13_vs_20171\|\|full | Not-determined |
| S13_vs_20205\|\|full | Low-quality |
| S13_vs_20223\|\|full | Low-quality |
| S13_vs_20247\|\|full | Low-quality |
| S13_vs_20249\|\|full | Low-quality |
| S13_vs_20268\|\|full | Low-quality |
| S13_vs_20301\|\|full | Not-determined |
| S13_vs_20316\|\|full | Low-quality |
| S13_vs_20454\|\|full | Low-quality |
| S13_vs_20602\|\|full | Low-quality |
| S13_vs_20631\|\|full | Low-quality |
| S13_vs_20678\|\|full | Low-quality |
| S13_vs_20689\|\|full | Not-determined |
| S13_vs_20790\|\|full | Not-determined |
| S13_vs_20935\|\|full | Not-determined |
| S13_vs_21015\|\|full | Low-quality |
| S13_vs_21051\|\|full | Low-quality |
| S13_vs_21175\|\|full | Not-determined |
| S13_vs_21179\|\|full | Low-quality |
| S13_vs_21187\|\|full | Low-quality |
| S13_vs_21203\|\|full | Low-quality |
| S13_vs_21226\|\|full | Low-quality |
| S13_vs_21237\|\|full | Not-determined |
| S13_vs_21263\|\|full | Low-quality |
| S13_vs_21287\|\|full | Low-quality |
| S13_vs_21454\|\|full | Not-determined |
| S13_vs_21489\|\|full | Low-quality |
| S13_vs_21560\|\|full | Low-quality |
| S13_vs_21614\|\|full | Not-determined |
| S13_vs_21657\|\|full | Not-determined |
| S13_vs_21737\|\|full | Low-quality |
| S13_vs_21840\|\|full | Low-quality |
| S13_vs_21844\|\|full | Not-determined |
| S13_vs_21924\|\|full | Not-determined |
| S13_vs_21953\|\|full | Not-determined |
| S13_vs_21965\|\|full | Not-determined |
| S13_vs_21999\|\|full | Not-determined |
| S13_vs_22020\|\|full | Not-determined |
| S13_vs_22084\|\|full | Low-quality |
| S17_vs_k141_68049\|\|full | High-quality |
| S13_vs_22352\|\|full | Low-quality |
| S13_vs_22439\|\|full | Low-quality |
| S13_vs_22451\|\|full | Not-determined |
| S13_vs_22455\|\|full | Not-determined |
| S13_vs_22476\|\|full | Not-determined |
| S13_vs_22627\|\|full | Low-quality |
| S13_vs_22629\|\|full | Low-quality |
| S13_vs_22681\|\|full | Low-quality |
| S13_vs_22707\|\|full | Low-quality |
| S13_vs_22748\|\|full | Not-determined |
| S13_vs_22775\|\|full | Low-quality |
| S13_vs_22923\|\|full | Low-quality |
| S13_vs_22954\|\|full | Low-quality |
| S13_vs_22968\|\|full | Low-quality |
| S13_vs_22969\|\|full | Low-quality |
| S13_vs_22974\|\|full | Low-quality |
| S13_vs_22983\|\|full | Low-quality |
| S13_vs_22995\|\|full | Not-determined |
| S13_vs_23010\|\|full | Low-quality |
| S13_vs_23014\|\|full | Not-determined |
| S13_vs_23077\|\|full | Low-quality |
| S13_vs_23086\|\|full | Low-quality |
| S13_vs_23142\|\|full | Low-quality |
| S13_vs_23181\|\|full | Low-quality |
| S13_vs_23195\|\|full | Low-quality |
| S13_vs_23211\|\|full | Not-determined |
| S5_vs_k141_120584\|\|full | High-quality |
| S13_vs_23327\|\|full | Not-determined |
| S13_vs_23340\|\|full | Low-quality |
| S13_vs_23357\|\|full | Not-determined |
| S13_vs_23410\|\|full | Low-quality |
| S13_vs_23423\|\|full | Not-determined |
| S13_vs_23467\|\|full | Not-determined |
| S13_vs_23508\|\|full | Low-quality |
| S13_vs_23669\|\|full | Low-quality |
| S13_vs_23747\|\|full | Low-quality |
| S13_vs_23754\|\|full | Not-determined |
| S13_vs_23862\|\|full | Not-determined |
| S13_vs_23880\|\|full | Low-quality |
| S13_vs_23914\|\|full | Low-quality |
| S13_vs_24002\|\|full | Not-determined |
| S13_vs_24034\|\|full | Low-quality |
| S13_vs_24107\|\|full | Low-quality |
| S13_vs_24110\|\|full | Low-quality |
| S13_vs_24178\|\|full | Low-quality |
| S13_vs_24202\|\|full | Not-determined |
| S13_vs_24229\|\|full | Not-determined |
| S13_vs_24234\|\|full | Low-quality |
| S13_vs_24237\|\|full | Low-quality |
| S13_vs_24324\|\|full | Low-quality |
| S13_vs_24334\|\|full | Low-quality |
| S13_vs_24423\|\|full | Not-determined |
| S13_vs_24549\|\|full | Low-quality |
| S13_vs_24554\|\|full | Low-quality |
| S13_vs_24671\|\|full | Not-determined |
| S13_vs_24733\|\|full | Low-quality |
| S13_vs_24804\|\|full | Low-quality |
| S13_vs_24812\|\|full | Low-quality |
| S13_vs_24914\|\|full | Not-determined |
| S4_vs_VIRSorter_3006-cat_1 | High-quality |
| S13_vs_24934\|\|full | Not-determined |
| S13_vs_25004\|\|full | Not-determined |
| S13_vs_25035\|\|full | Not-determined |
| S13_vs_25076\|\|full | Not-determined |
| S13_vs_25088\|\|full | Low-quality |
| S13_vs_25099\|\|full | Low-quality |
| S13_vs_25129\|\|full | Low-quality |
| S13_vs_25208\|\|full | Low-quality |
| S13_vs_25266\|\|full | Not-determined |
| S13_vs_25271\|\|full | Not-determined |
| S13_vs_25332\|\|full | Not-determined |
| S13_vs_25561\|\|full | Low-quality |
| S13_vs_25581\|\|full | Low-quality |
| S13_vs_25595\|\|full | Not-determined |
| S13_vs_25599\|\|full | Not-determined |
| S13_vs_25617\|\|full | Low-quality |
| S13_vs_25618\|\|full | Low-quality |
| S13_vs_25673\|\|full | Low-quality |
| S13_vs_25675\|\|full | Not-determined |
| S13_vs_25687\|\|full | Not-determined |
| S13_vs_25708\|\|full | Not-determined |
| S13_vs_25752\|\|full | Low-quality |
| S13_vs_25848\|\|full | Low-quality |
| S13_vs_25862\|\|full | Not-determined |

| | |
|---|---|
| S13_vs_25894\|\|full | Not-determined |
| S13_vs_25911\|\|full | Low-quality |
| S13_vs_25915\|\|full | Low-quality |
| S13_vs_25983\|\|full | Low-quality |
| S13_vs_26034\|\|full | Low-quality |
| S13_vs_26039\|\|full | Low-quality |
| S13_vs_26046\|\|full | Low-quality |
| S13_vs_26057\|\|full | Not-determined |
| S13_vs_26067\|\|full | Low-quality |
| S13_vs_26110\|\|full | Low-quality |
| S13_vs_26127\|\|full | Not-determined |
| S13_vs_26216\|\|full | Not-determined |
| S13_vs_26323\|\|full | Low-quality |
| S13_vs_26348\|\|full | Low-quality |
| S13_vs_26488\|\|full | Low-quality |
| S13_vs_26535\|\|full | Low-quality |
| S13_vs_26641\|\|full | Low-quality |
| S13_vs_26734\|\|full | Not-determined |
| S5_vs_k141_120175\|\|full | Medium-quality |
| S13_vs_26796\|\|full | Low-quality |
| S13_vs_26799\|\|full | Low-quality |
| S13_vs_26806\|\|full | Low-quality |
| S13_vs_26900\|\|full | Not-determined |
| S13_vs_26903\|\|full | Low-quality |
| S13_vs_26909\|\|full | Not-determined |
| S13_vs_26957\|\|full | Low-quality |
| S13_vs_26971\|\|full | Low-quality |
| S13_vs_27002\|\|full | Low-quality |
| S13_vs_27017\|\|full | Low-quality |
| S13_vs_27046\|\|full | Low-quality |
| S13_vs_27107\|\|full | Not-determined |
| S13_vs_27115\|\|full | Not-determined |
| S13_vs_27135\|\|full | Low-quality |
| S13_vs_27154\|\|full | Low-quality |
| S13_vs_27160\|\|full | Low-quality |
| S13_vs_27262\|\|full | Low-quality |
| S13_vs_27342\|\|full | Low-quality |
| S13_vs_27357\|\|full | Low-quality |
| S13_vs_27367\|\|full | Not-determined |
| S13_vs_27383\|\|full | Low-quality |
| S13_vs_27385\|\|full | Low-quality |
| S13_vs_27410\|\|full | Low-quality |
| S13_vs_27427\|\|full | Not-determined |
| S13_vs_27433\|\|full | Low-quality |
| S13_vs_27461\|\|full | Low-quality |
| S13_vs_27468\|\|full | Low-quality |
| S13_vs_27511\|\|full | Low-quality |
| S13_vs_27534\|\|full | Not-determined |
| S13_vs_27539\|\|full | Not-determined |
| S13_vs_27659\|\|full | Not-determined |
| S13_vs_27698\|\|full | Not-determined |
| S13_vs_27849\|\|full | Not-determined |
| S13_vs_27991\|\|full | Low-quality |
| S13_vs_28027\|\|full | Low-quality |
| S13_vs_28090\|\|full | Not-determined |
| S13_vs_28171\|\|full | Low-quality |
| S13_vs_28175\|\|full | Not-determined |
| S13_vs_28188\|\|full | Low-quality |
| S13_vs_28267\|\|full | Low-quality |
| S8_extracted_k141_208111 | Complete |
| S13_vs_28361\|\|full | Low-quality |
| S13_vs_28401\|\|full | Low-quality |
| S13_vs_28431\|\|full | Low-quality |
| S13_vs_28449\|\|full | Low-quality |
| S13_vs_28456\|\|full | Low-quality |
| S13_vs_28460\|\|full | Low-quality |
| S13_vs_28539\|\|full | Low-quality |
| S13_vs_28628\|\|full | Not-determined |
| S13_vs_28634\|\|full | Low-quality |
| S13_vs_28682\|\|full | Not-determined |
| S13_vs_28710\|\|full | Not-determined |
| S13_vs_28775\|\|full | Low-quality |
| S13_vs_28815\|\|full | Low-quality |
| S13_vs_28852\|\|full | Low-quality |
| S13_vs_28873\|\|full | Not-determined |
| S13_vs_29021\|\|full | Low-quality |
| S13_vs_29035\|\|full | Low-quality |
| S13_vs_29075\|\|full | Low-quality |
| S13_vs_29089\|\|full | Not-determined |
| S13_vs_29156\|\|full | Low-quality |
| S13_vs_29168\|\|full | Low-quality |
| S13_vs_29196\|\|full | Low-quality |
| S13_vs_29221\|\|full | Low-quality |
| S13_vs_29250\|\|full | Low-quality |
| S13_vs_29425\|\|full | Not-determined |
| S13_vs_29572\|\|full | Low-quality |
| S13_vs_29601\|\|full | Not-determined |
| S13_vs_29604\|\|full | Low-quality |
| S13_vs_29636\|\|full | Not-determined |
| S13_vs_29733\|\|full | Low-quality |
| S13_vs_29874\|\|full | Low-quality |
| S13_vs_29887\|\|full | Low-quality |
| S13_vs_29921\|\|full | Not-determined |
| S13_vs_29974\|\|full | Not-determined |
| S13_vs_29985\|\|full | Not-determined |
| S8_vs_k141_208111\|\|full | High-quality |
| S13_vs_30080\|\|full | Not-determined |
| S5_vs_k141_75530\|\|full | High-quality |
| S13_vs_30136\|\|full | Not-determined |
| S13_vs_30167\|\|full | High-quality |
| S7_vs_6626\|\|full | High-quality |
| S13_vs_30241\|\|full | Not-determined |
| S13_vs_30367\|\|full | Not-determined |
| S13_vs_30417\|\|full | Low-quality |
| S13_vs_30419\|\|full | Low-quality |
| S13_vs_30505\|\|full | Low-quality |
| S13_vs_30515\|\|full | Not-determined |
| S13_vs_30532\|\|full | Low-quality |
| S13_vs_30685\|\|full | Low-quality |
| S13_vs_30709\|\|full | Not-determined |
| S13_vs_30712\|\|full | Low-quality |
| S13_vs_30829\|\|full | Low-quality |
| S13_vs_30893\|\|full | Low-quality |
| S13_vs_31033\|\|full | Not-determined |
| S13_vs_31064\|\|full | Low-quality |
| S13_vs_31065\|\|full | Not-determined |
| S13_vs_31139\|\|full | Low-quality |
| S13_vs_31169\|\|full | Low-quality |
| S13_vs_31218\|\|full | Not-determined |
| S13_vs_31235\|\|full | Low-quality |
| S13_vs_31238\|\|full | Low-quality |
| S13_vs_31241\|\|full | Not-determined |
| S13_vs_31262\|\|full | Low-quality |
| S13_vs_31317\|\|full | Not-determined |
| S13_vs_31328\|\|full | Low-quality |
| S13_vs_31337\|\|full | Low-quality |
| S13_vs_31448\|\|full | Low-quality |
| S13_vs_31463\|\|full | Not-determined |
| S13_vs_31514\|\|full | Low-quality |
| S13_vs_31567\|\|full | Low-quality |
| S13_vs_31656\|\|full | Not-determined |
| S13_vs_31673\|\|full | Low-quality |
| S13_vs_31803\|\|full | Low-quality |
| S13_vs_31804\|\|full | Not-determined |
| S13_vs_31824\|\|full | Not-determined |
| S13_vs_31848\|\|full | Not-determined |
| S13_vs_31967\|\|full | Not-determined |
| S13_vs_31988\|\|full | Low-quality |
| S13_vs_32037\|\|full | Not-determined |
| S13_vs_32081\|\|full | Not-determined |
| S13_vs_32133\|\|full | Low-quality |
| S13_vs_32150\|\|full | Not-determined |
| S13_vs_32207\|\|full | Not-determined |
| S13_vs_32257\|\|full | Low-quality |
| S13_vs_32269\|\|full | Low-quality |
| S13_vs_32408\|\|full | Low-quality |
| S13_vs_32450\|\|full | Low-quality |
| S13_vs_32458\|\|full | Low-quality |
| S13_vs_32488\|\|full | Not-determined |
| S13_vs_32685\|\|full | Low-quality |
| S13_vs_32726\|\|full | Low-quality |
| S13_vs_32747\|\|full | Low-quality |
| S13_vs_32931\|\|full | Low-quality |
| S13_vs_32966\|\|full | Low-quality |
| S13_vs_33032\|\|full | Low-quality |
| S13_vs_33145\|\|full | Not-determined |
| S13_vs_33147\|\|full | Low-quality |
| S13_vs_33241\|\|full | Low-quality |
| S13_vs_33277\|\|full | Low-quality |
| S13_vs_33334\|\|full | Low-quality |
| S13_vs_33367\|\|full | Low-quality |
| S13_vs_33417\|\|full | Not-determined |
| S13_vs_33519\|\|full | Low-quality |
| S13_vs_33522\|\|full | Low-quality |
| S13_vs_33542\|\|full | Not-determined |
| S13_vs_33558\|\|full | Not-determined |
| S13_vs_33611\|\|full | Low-quality |
| S13_vs_33640\|\|full | Not-determined |
| S13_vs_33645\|\|full | Not-determined |
| S13_vs_33650\|\|full | Low-quality |
| S13_vs_33656\|\|full | Low-quality |
| S13_vs_33670\|\|full | Not-determined |
| S13_vs_33677\|\|full | Not-determined |
| S13_vs_33704\|\|full | Not-determined |
| S13_vs_33706\|\|full | Low-quality |
| S13_vs_33785\|\|full | Low-quality |
| S13_vs_33816\|\|full | Not-determined |
| S13_vs_33821\|\|full | Low-quality |
| S13_vs_33845\|\|full | Low-quality |
| S13_vs_33858\|\|full | Low-quality |
| S8_extracted_k141_41376 | High-quality |
| S13_vs_33955\|\|full | Low-quality |
| S13_vs_33957\|\|full | Low-quality |
| S13_vs_33969\|\|full | Not-determined |
| S13_vs_34092\|\|full | Low-quality |
| S13_vs_34106\|\|full | Low-quality |
| S13_vs_34111\|\|full | Not-determined |
| S13_vs_34117\|\|full | Low-quality |
| S13_vs_34284\|\|full | Low-quality |
| S13_vs_34344\|\|full | Not-determined |
| S13_vs_34349\|\|full | Low-quality |
| S13_vs_34421\|\|full | Low-quality |
| S13_vs_34471\|\|full | Low-quality |
| S13_vs_34483\|\|full | Low-quality |
| S13_vs_34512\|\|full | Not-determined |
| S13_vs_34545\|\|full | Low-quality |
| S13_vs_34627\|\|full | Not-determined |
| S13_vs_34701\|\|full | Low-quality |
| S13_vs_34708\|\|full | Low-quality |
| S13_vs_34832\|\|full | Low-quality |
| S13_vs_35024\|\|full | Low-quality |
| S13_vs_35098\|\|full | Not-determined |
| S13_vs_35130\|\|full | Not-determined |
| S13_vs_35134\|\|full | Low-quality |
| S13_vs_35179\|\|full | Low-quality |
| S13_vs_35207\|\|full | Low-quality |
| S13_vs_35216\|\|full | Low-quality |
| S13_vs_35225\|\|full | Not-determined |
| S13_vs_35234\|\|full | Low-quality |
| S13_vs_35282\|\|full | Low-quality |
| S13_vs_35345\|\|full | Low-quality |
| S13_vs_35347\|\|full | Not-determined |
| S13_vs_35464\|\|full | Not-determined |
| S13_vs_35491\|\|full | Not-determined |
| S13_vs_35551\|\|full | Not-determined |
| S13_vs_35590\|\|full | Low-quality |
| S13_vs_35622\|\|full | Low-quality |
| S13_vs_35761\|\|full | Low-quality |
| S16_vs_173079\|\|full | High-quality |
| S13_vs_35831\|\|full | Low-quality |
| S13_vs_35843\|\|full | Not-determined |
| S13_vs_35927\|\|full | Low-quality |
| S13_vs_36005\|\|full | Low-quality |
| S13_vs_36010\|\|full | Low-quality |
| S14_vs_k141_81408\|\|full | High-quality |
| S13_vs_36203\|\|full | Low-quality |
| S13_vs_36251\|\|full | Not-determined |
| S13_vs_36254\|\|full | Low-quality |
| S13_vs_36291\|\|full | Low-quality |
| S13_vs_36297\|\|full | Not-determined |
| S13_vs_36303\|\|full | Not-determined |
| S13_vs_36387\|\|full | Not-determined |
| S13_vs_36395\|\|full | Low-quality |
| S13_vs_36460\|\|full | Not-determined |
| S13_vs_36497\|\|full | Low-quality |
| S13_vs_36548\|\|full | Low-quality |
| S13_vs_36576\|\|full | Low-quality |
| S13_vs_36626\|\|full | Not-determined |
| S13_vs_36630\|\|full | Low-quality |
| S13_vs_36683\|\|full | Low-quality |
| S13_vs_36719\|\|full | Not-determined |
| S13_vs_36816\|\|full | Not-determined |
| S13_vs_36916\|\|full | Low-quality |
| S13_vs_36932\|\|full | Not-determined |
| S13_vs_36972\|\|full | Low-quality |
| S13_vs_37198\|\|full | Low-quality |
| S13_vs_37275\|\|full | Not-determined |
| S13_vs_37340\|\|full | Not-determined |
| S13_vs_37367\|\|full | Low-quality |
| S13_vs_37495\|\|full | Low-quality |
| S13_vs_37534\|\|full | Low-quality |
| S13_vs_37633\|\|full | Low-quality |
| S13_vs_37703\|\|full | Low-quality |
| S13_vs_37759\|\|full | Low-quality |
| S13_vs_37813\|\|full | Low-quality |
| S13_vs_37853\|\|full | Not-determined |
| S13_vs_38057\|\|full | Low-quality |
| S13_vs_38087\|\|full | Low-quality |
| S13_vs_38096\|\|full | Not-determined |
| S13_vs_38125\|\|full | Low-quality |
| S13_vs_38153\|\|full | Not-determined |
| S13_vs_38187\|\|full | Not-determined |
| S13_vs_38192\|\|full | Low-quality |
| S13_vs_38235\|\|full | Low-quality |
| S13_vs_38244\|\|full | Low-quality |
| S13_vs_38255\|\|full | Low-quality |
| S13_vs_38310\|\|full | Low-quality |
| S13_vs_38349\|\|full | Low-quality |
| S13_vs_38356\|\|full | Low-quality |
| S13_vs_38373\|\|full | Low-quality |
| S13_vs_38433\|\|full | Low-quality |
| S13_vs_38485\|\|full | Low-quality |
| S13_vs_38565\|\|full | Not-determined |
| S13_vs_38579\|\|full | Not-determined |
| S13_vs_38603\|\|full | Low-quality |
| S13_vs_38651\|\|full | Low-quality |
| S13_vs_38664\|\|full | Not-determined |
| S13_vs_38672\|\|full | Low-quality |
| S13_vs_38685\|\|full | Low-quality |
| S13_vs_38692\|\|full | Low-quality |
| S3_extracted_k141_108959 | Complete |
| S13_vs_38724\|\|full | Not-determined |
| S13_vs_38790\|\|full | Not-determined |
| S13_vs_38851\|\|full | Low-quality |
| S13_vs_38971\|\|full | Not-determined |
| S13_vs_39145\|\|full | Low-quality |
| S13_vs_39196\|\|full | Not-determined |
| S13_vs_39245\|\|full | Not-determined |
| S13_vs_39399\|\|full | Not-determined |
| S13_vs_39425\|\|full | Low-quality |
| S13_vs_39450\|\|full | Not-determined |
| S13_vs_39455\|\|full | Low-quality |
| S13_vs_39460\|\|full | Not-determined |
| S13_vs_39474\|\|full | Low-quality |
| S13_vs_39506\|\|full | Low-quality |
| S13_vs_39517\|\|full | Low-quality |
| S13_vs_39527\|\|full | Low-quality |
| S13_vs_39531\|\|full | Not-determined |
| S13_vs_39536\|\|full | Not-determined |
| S13_vs_39558\|\|full | Low-quality |
| S13_vs_39603\|\|full | Low-quality |
| S13_vs_39624\|\|full | Low-quality |
| S13_vs_39702\|\|full | Not-determined |
| S13_vs_39800\|\|full | Not-determined |
| S13_vs_39802\|\|full | Low-quality |
| S13_vs_39884\|\|full | Not-determined |
| S13_vs_39940\|\|full | Not-determined |
| S13_vs_39954\|\|full | Not-determined |
| S13_vs_39970\|\|full | Not-determined |
| S13_vs_40006\|\|full | Not-determined |
| S13_vs_40008\|\|full | Not-determined |
| S13_vs_40043\|\|full | Not-determined |
| S13_vs_40142\|\|full | Not-determined |
| S13_vs_40355\|\|full | Low-quality |
| S13_vs_40359\|\|full | Low-quality |
| S13_vs_40364\|\|full | Low-quality |
| S13_vs_40401\|\|full | Low-quality |
| S13_vs_40441\|\|full | Low-quality |
| S13_vs_40443\|\|full | Not-determined |
| S13_vs_40460\|\|full | Not-determined |
| S13_vs_40526\|\|full | Not-determined |
| S13_vs_40564\|\|full | Low-quality |
| S13_vs_40597\|\|full | Not-determined |
| S13_vs_40625\|\|full | Low-quality |
| S13_vs_40645\|\|full | Not-determined |
| S13_vs_40654\|\|full | Low-quality |
| S13_vs_40705\|\|full | Not-determined |
| S13_vs_40709\|\|full | Low-quality |
| S13_vs_40759\|\|full | Low-quality |
| S13_vs_40906\|\|full | Not-determined |
| S13_vs_40998\|\|full | Low-quality |
| S13_vs_41010\|\|full | Low-quality |
| S13_vs_41012\|\|full | Low-quality |
| S13_vs_41049\|\|full | Low-quality |
| S13_vs_41063\|\|full | Low-quality |
| S13_vs_41089\|\|full | Low-quality |
| S13_vs_41103\|\|full | Low-quality |
| S13_vs_41163\|\|full | Low-quality |
| S13_vs_41176\|\|full | Low-quality |
| S13_vs_41177\|\|full | Low-quality |
| S13_vs_41207\|\|full | Low-quality |
| S13_vs_41229\|\|full | Not-determined |
| S13_vs_41325\|\|full | Low-quality |
| S13_vs_41408\|\|full | Low-quality |
| S13_vs_41412\|\|full | Low-quality |
| S13_vs_41418\|\|full | Not-determined |
| S13_vs_41499\|\|full | Low-quality |
| S13_vs_41768\|\|full | Not-determined |
| S13_vs_41825\|\|full | Low-quality |
| S13_vs_41838\|\|full | Low-quality |
| S13_vs_41866\|\|full | Low-quality |
| S13_vs_41926\|\|full | Not-determined |
| S13_vs_42028\|\|full | Not-determined |
| S13_vs_42034\|\|full | Not-determined |
| S13_vs_42165\|\|full | Not-determined |
| S13_vs_42181\|\|full | Low-quality |
| S13_vs_42233\|\|full | Low-quality |
| S13_vs_42284\|\|full | Low-quality |
| S13_vs_42289\|\|full | Not-determined |
| S13_vs_42301\|\|full | Low-quality |
| S13_vs_42345\|\|full | Not-determined |
| S13_vs_42476\|\|full | Low-quality |
| S13_vs_42582\|\|full | Not-determined |
| S13_vs_42617\|\|full | Low-quality |
| S13_vs_42686\|\|full | Low-quality |
| S13_vs_42752\|\|full | Low-quality |
| S13_vs_42983\|\|full | Not-determined |
| S13_vs_43040\|\|full | Low-quality |
| S13_vs_43087\|\|full | Low-quality |
| S13_vs_43152\|\|full | Low-quality |
| S13_vs_43158\|\|full | Low-quality |
| S13_vs_43188\|\|full | Low-quality |
| S13_vs_43194\|\|full | Not-determined |
| S13_vs_43195\|\|full | Low-quality |
| S13_vs_43201\|\|full | Low-quality |
| S13_vs_43315\|\|full | Not-determined |
| S13_vs_43383\|\|full | Low-quality |
| S13_vs_43391\|\|full | Low-quality |
| S13_vs_43414\|\|full | Not-determined |
| S13_vs_43431\|\|full | Low-quality |
| S13_vs_43468\|\|full | Not-determined |
| S13_vs_43480\|\|full | Low-quality |
| S13_vs_43506\|\|full | Low-quality |
| S13_vs_43648\|\|full | Not-determined |
| S13_vs_43768\|\|full | Not-determined |
| S13_vs_43783\|\|full | Not-determined |
| S13_vs_43828\|\|full | Not-determined |
| S13_vs_43911\|\|full | Low-quality |
| S13_vs_43920\|\|full | Not-determined |
| S13_vs_43933\|\|full | Low-quality |
| S13_vs_43981\|\|full | Low-quality |
| S13_vs_44019\|\|full | Not-determined |
| S13_vs_44080\|\|full | Low-quality |
| S13_vs_44084\|\|full | Not-determined |
| S13_vs_44103\|\|full | Low-quality |
| S13_vs_44107\|\|full | Not-determined |
| S13_vs_44154\|\|full | Not-determined |
| S13_vs_44263\|\|full | Low-quality |
| S13_vs_44278\|\|full | Not-determined |
| S13_vs_44325\|\|full | Low-quality |
| S13_vs_44374\|\|full | Low-quality |
| S13_vs_44406\|\|full | Low-quality |
| S13_vs_44532\|\|full | Low-quality |
| S13_vs_44563\|\|full | Low-quality |
| S13_vs_44587\|\|full | Low-quality |
| S13_vs_44593\|\|full | Low-quality |
| S13_vs_44629\|\|full | Not-determined |
| S13_vs_44661\|\|full | Low-quality |
| S13_vs_44739\|\|full | Low-quality |
| S13_vs_44756\|\|full | Low-quality |
| S13_vs_44758\|\|full | Low-quality |
| S13_vs_44843\|\|full | Not-determined |
| S13_vs_44888\|\|full | Low-quality |
| S13_vs_44939\|\|full | Not-determined |
| S13_vs_44971\|\|full | Low-quality |
| S13_vs_45045\|\|full | Low-quality |
| S13_vs_45152\|\|full | Low-quality |
| S13_vs_45155\|\|full | Low-quality |
| S13_vs_45198\|\|full | Low-quality |
| S13_vs_45268\|\|full | Not-determined |
| S13_vs_45282\|\|full | Not-determined |
| S13_vs_45297\|\|full | Low-quality |
| S13_vs_45306\|\|full | Low-quality |
| S13_vs_45319\|\|full | Low-quality |
| S13_vs_45320\|\|full | Not-determined |
| S13_vs_45335\|\|full | Not-determined |
| S13_vs_45363\|\|full | Not-determined |
| S13_vs_45410\|\|full | Low-quality |
| S13_vs_45415\|\|full | Not-determined |
| S13_vs_45460\|\|full | Not-determined |
| S8_vs_k141_41376\|\|full | High-quality |
| S13_vs_45517\|\|full | Low-quality |
| S13_vs_45543\|\|full | Low-quality |
| S13_vs_45544\|\|full | Not-determined |
| S13_vs_45566\|\|full | Low-quality |
| S13_vs_45600\|\|full | Not-determined |
| S13_vs_45609\|\|full | Not-determined |
| S13_vs_45640\|\|full | Low-quality |
| S13_vs_45671\|\|full | Not-determined |
| S13_vs_45678\|\|full | Low-quality |
| S13_vs_45692\|\|full | Low-quality |
| S13_vs_45746\|\|full | Low-quality |
| S13_vs_45778\|\|full | Not-determined |
| S13_vs_45790\|\|full | Not-determined |
| S13_vs_45795\|\|full | Low-quality |
| S13_vs_45868\|\|full | Not-determined |
| S13_vs_45909\|\|full | Low-quality |
| S13_vs_45952\|\|full | Not-determined |
| S13_vs_45953\|\|full | Low-quality |
| S13_vs_45970\|\|full | Low-quality |
| S13_vs_46003\|\|full | Low-quality |
| S13_vs_46023\|\|full | Low-quality |
| S13_vs_46079\|\|full | Not-determined |
| S13_vs_46181\|\|full | Low-quality |
| S13_vs_46239\|\|full | Low-quality |
| S13_vs_46268\|\|full | Low-quality |
| S13_vs_46348\|\|full | Not-determined |
| S13_vs_46489\|\|full | Low-quality |
| S13_vs_46551\|\|full | Not-determined |
| S13_vs_46581\|\|full | Low-quality |
| S13_vs_46603\|\|full | Low-quality |
| S13_vs_46639\|\|full | Not-determined |
| S13_vs_46653\|\|full | Low-quality |
| S13_vs_46672\|\|full | Not-determined |
| S13_vs_46713\|\|full | Low-quality |
| S13_vs_46817\|\|full | Low-quality |
| S13_vs_46866\|\|full | Not-determined |
| S13_vs_46985\|\|full | Not-determined |
| S13_vs_47010\|\|full | Low-quality |
| S13_vs_47014\|\|full | Low-quality |
| S13_vs_47031\|\|full | Low-quality |
| S13_vs_47059\|\|full | Low-quality |
| S13_vs_47062\|\|full | Low-quality |
| S13_vs_47083\|\|full | Low-quality |
| S13_vs_47098\|\|full | Low-quality |
| S13_vs_47161\|\|full | Low-quality |
| S13_vs_47168\|\|full | Low-quality |
| S13_vs_47217\|\|full | Low-quality |
| S13_vs_47262\|\|full | Low-quality |
| S13_vs_47337\|\|full | Not-determined |
| S13_vs_47364\|\|full | Low-quality |
| S13_vs_47384\|\|full | Low-quality |
| S13_vs_47390\|\|full | Not-determined |
| S13_vs_47404\|\|full | Low-quality |
| S13_vs_47494\|\|full | Low-quality |
| S13_vs_47556\|\|full | Low-quality |
| S13_vs_47565\|\|full | Low-quality |
| S13_vs_47576\|\|full | Low-quality |
| S13_vs_47612\|\|full | Not-determined |
| S13_vs_47615\|\|full | Not-determined |
| S13_vs_47622\|\|full | Low-quality |
| S13_vs_47631\|\|full | Not-determined |
| S13_vs_47672\|\|full | Low-quality |
| S13_vs_47762\|\|full | Not-determined |
| S13_vs_47960\|\|full | Low-quality |
| S13_vs_47971\|\|full | Low-quality |
| S13_vs_47999\|\|full | Not-determined |
| S13_vs_48187\|\|full | Not-determined |
| S13_vs_48224\|\|full | Not-determined |
| S13_vs_48230\|\|full | Low-quality |
| S13_vs_48283\|\|full | Low-quality |
| S13_vs_48345\|\|full | Not-determined |
| S13_vs_48449\|\|full | Low-quality |
| S13_vs_48471\|\|full | Not-determined |
| S13_vs_48500\|\|full | Not-determined |
| S13_vs_48542\|\|full | Low-quality |
| S13_vs_48544\|\|full | Not-determined |
| S13_vs_48580\|\|full | Not-determined |
| S13_vs_48588\|\|full | Low-quality |
| S13_vs_48672\|\|full | Low-quality |
| S13_vs_48680\|\|full | Not-determined |
| S13_vs_48754\|\|full | Low-quality |
| S13_vs_48790\|\|full | Not-determined |
| S13_vs_48809\|\|full | Low-quality |
| S13_vs_48816\|\|full | Low-quality |
| S13_vs_48840\|\|full | Low-quality |
| S13_vs_48932\|\|full | Not-determined |
| S13_vs_49000\|\|full | Low-quality |
| S13_vs_49005\|\|full | Not-determined |
| S13_vs_49019\|\|full | Not-determined |
| S13_vs_49053\|\|full | Low-quality |
| S13_vs_49447\|\|full | Low-quality |
| S13_vs_49484\|\|full | Low-quality |
| S13_vs_49486\|\|full | Low-quality |
| S13_vs_49513\|\|full | Not-determined |
| S13_vs_49536\|\|full | Low-quality |
| S13_vs_49675\|\|full | Not-determined |
| S13_vs_49716\|\|full | Low-quality |
| S13_vs_49718\|\|full | Not-determined |
| S13_vs_49760\|\|full | Low-quality |
| S13_vs_49823\|\|full | Not-determined |
| S13_vs_49887\|\|full | Low-quality |
| S4_vs_VIRSorter_4839-cat_2 | High-quality |
| S13_vs_49919\|\|full | Not-determined |
| S13_vs_49943\|\|full | Low-quality |
| S13_vs_49958\|\|full | Low-quality |
| S13_vs_50062\|\|full | Low-quality |
| S13_vs_50071\|\|full | Low-quality |
| S13_vs_50121\|\|full | Low-quality |
| S13_vs_50173\|\|full | Not-determined |
| S13_vs_50405\|\|full | Low-quality |
| S13_vs_50438\|\|full | Low-quality |
| S13_vs_50451\|\|full | Not-determined |
| S13_vs_50517\|\|full | Not-determined |
| S13_vs_50566\|\|full | Low-quality |
| S13_vs_50580\|\|full | Low-quality |
| S13_vs_50591\|\|full | Low-quality |
| S13_vs_50662\|\|full | Not-determined |
| S13_vs_50693\|\|full | Not-determined |
| S13_vs_50746\|\|full | Not-determined |
| S13_vs_50782\|\|full | Not-determined |
| S13_vs_50817\|\|full | Low-quality |
| S13_vs_50829\|\|full | Low-quality |
| S13_vs_50848\|\|full | Low-quality |
| S13_vs_50967\|\|full | Low-quality |
| S13_vs_51105\|\|full | Low-quality |
| S13_vs_51114\|\|full | Low-quality |
| S13_vs_51168\|\|full | Not-determined |
| S13_vs_51241\|\|full | Low-quality |
| S13_vs_51258\|\|full | Low-quality |
| S13_vs_51315\|\|full | Low-quality |
| S13_vs_51320\|\|full | Low-quality |
| S13_vs_51352\|\|full | Low-quality |
| S13_vs_51387\|\|full | Not-determined |
| S13_vs_51389\|\|full | Not-determined |
| S13_vs_51417\|\|full | Not-determined |
| S13_vs_51444\|\|full | Low-quality |
| S13_vs_51473\|\|full | Low-quality |
| S13_vs_51490\|\|full | Not-determined |
| S13_vs_51840\|\|full | Low-quality |
| S13_vs_51869\|\|full | Low-quality |
| S13_vs_51939\|\|full | Not-determined |
| S13_vs_51975\|\|full | Low-quality |
| S13_vs_52002\|\|full | Low-quality |
| S13_vs_52009\|\|full | Low-quality |
| S13_vs_52051\|\|full | Not-determined |
| S13_vs_52088\|\|full | Low-quality |
| S13_vs_52244\|\|full | Low-quality |
| S13_vs_52276\|\|full | Low-quality |
| S13_vs_52326\|\|full | Low-quality |
| S13_vs_52331\|\|full | Not-determined |
| S13_vs_52353\|\|full | Not-determined |
| S13_vs_52439\|\|full | Not-determined |
| S13_vs_52473\|\|full | Low-quality |
| S13_vs_52521\|\|full | Low-quality |
| S13_vs_52526\|\|full | Low-quality |
| S13_vs_52568\|\|full | Low-quality |
| S13_vs_52652\|\|full | Not-determined |
| S13_vs_52666\|\|full | Not-determined |

S13_vs_52708||full Not-determined
S13_vs_52767||full Low-quality
S13_vs_52792||full Not-determined
S13_vs_52806||full Low-quality
S13_vs_52912||full Not-determined
S13_vs_52918||full Not-determined
S13_vs_52924||full Not-determined
S13_vs_52966||full Low-quality
S13_vs_53012||full Low-quality
S13_vs_53120||full Low-quality
S13_vs_53182||full Low-quality
S13_vs_53255||full Low-quality
S13_vs_53271||full Low-quality
S13_vs_53376||full Low-quality
S13_vs_53406||full Low-quality
S13_vs_53434||full Low-quality
S13_vs_53462||full Low-quality
S13_vs_53549||full Low-quality
S13_vs_53639||full Low-quality
S13_vs_53676||full Not-determined
S13_vs_53690||full Low-quality
S13_vs_53701||full Low-quality
S13_vs_53731||full Low-quality
S13_vs_53761||full Low-quality
S13_vs_53808||full Low-quality
S13_vs_53832||full Not-determined
S13_vs_53857||full Not-determined
S13_vs_53892||full Not-determined
S13_vs_53901||full Low-quality
S13_vs_53904||full Low-quality
S13_vs_53916||full Low-quality
S13_vs_53947||full Not-determined
S13_vs_53968||full Not-determined
S13_vs_54076||full Not-determined
S13_vs_54104||full Low-quality
S13_vs_54183||full Not-determined
S11_vs_k141_68151||full High-quality
S13_vs_54208||full Not-determined
S13_vs_54264||full Low-quality
S13_vs_54272||full Low-quality
S13_vs_54466||full Not-determined
S13_vs_54516||full Low-quality
S13_vs_54532||full Not-determined
S13_vs_54535||full Low-quality
S13_vs_54555||full Low-quality
S13_vs_54568||full Low-quality
S13_vs_54602||full Not-determined
S13_vs_54638||full Low-quality
S13_vs_54823||full Not-determined
S13_vs_54826||full Not-determined
S13_vs_54830||full Low-quality
S13_vs_54851||full Low-quality
S13_vs_54919||full Not-determined
S13_vs_54920||full Not-determined
S13_vs_54993||full Low-quality
S13_vs_55029||full Not-determined
S13_vs_55038||full Not-determined
S13_vs_55104||full Low-quality
S13_vs_55232||full Low-quality
S13_vs_55264||full Low-quality
S13_vs_55273||full Not-determined
S13_vs_55283||full Low-quality
S13_vs_55332||full Low-quality
S13_vs_55378||full Low-quality
S13_vs_55392||full Low-quality
S13_vs_55399||full Not-determined
S13_vs_55484||full Not-determined
S13_vs_55502||full Low-quality
S13_vs_55549||full Low-quality
S13_vs_55592||full Not-determined
S13_vs_55658||full Not-determined
S13_vs_55660||full Not-determined
S13_vs_55690||full Low-quality
S13_vs_55753||full Low-quality
S13_vs_55767||full Low-quality
S13_vs_55805||full Low-quality
S13_vs_55811||full Low-quality
S13_vs_55887||full Not-determined
S13_vs_55896||full Low-quality
S13_vs_55906||full Not-determined
S13_vs_55937||full Low-quality
S13_vs_55997||full Not-determined
S13_vs_56005||full Low-quality
S13_vs_56016||full Not-determined
S13_vs_56020||full Not-determined
S13_vs_56095||full Not-determined
S13_vs_56228||full Not-determined
S13_vs_56260||full Not-determined
S13_vs_56265||full Not-determined
S13_vs_56287||full Low-quality
S13_vs_56327||full Not-determined
S13_vs_56415||full Low-quality
S13_vs_56506||full Not-determined
S13_vs_56539||full Low-quality
S13_vs_56549||full Not-determined
S13_vs_56606||full Not-determined
S13_vs_56702||full Low-quality
S13_vs_56727||full Low-quality
S13_vs_56737||full Low-quality
S13_vs_56783||full Low-quality
S13_vs_56792||full Not-determined
S13_vs_56821||full Not-determined
S13_vs_56827||full Low-quality
S13_vs_56845||full Low-quality
S13_vs_56872||full Low-quality
S13_vs_56928||full Not-determined
S13_vs_57007||full Low-quality
S13_vs_57067||full Low-quality
S13_vs_57079||full Not-determined
S13_vs_57090||full Not-determined
S13_vs_57120||full Low-quality
S13_vs_57133||full Not-determined
S13_vs_57144||full Not-determined
S13_vs_57182||full Low-quality
S13_vs_57190||full Low-quality
S13_vs_57251||full Not-determined
S13_vs_57259||full Not-determined
S9_vs_k141_11702||full High-quality
S13_vs_57316||full Not-determined
S13_vs_57367||full Not-determined
S13_vs_57370||full Low-quality
S13_vs_57417||full Not-determined
S13_vs_57421||full Low-quality
S13_vs_57423||full Low-quality
S13_vs_57457||full Low-quality
S13_vs_57700||full Low-quality
S13_vs_57754||full Not-determined
S13_vs_57889||full Not-determined
S13_vs_57941||full Low-quality
S13_vs_57966||full Low-quality
S13_vs_58022||full Low-quality
S13_vs_58034||full Low-quality
S13_vs_58036||full Not-determined
S13_vs_58042||full Low-quality
S13_vs_58051||full Low-quality
S13_vs_58081||full Low-quality
S13_vs_58129||full Not-determined
S13_vs_58173||full Low-quality
S13_vs_58230||full Not-determined
S13_vs_58312||full Low-quality
S13_vs_58462||full Low-quality
S13_vs_58493||full Not-determined
S13_vs_58526||full Not-determined
S13_vs_58547||full Low-quality
S13_vs_58609||full Not-determined
S13_vs_58621||full Low-quality
S13_vs_58631||full Low-quality
S13_vs_58699||full Not-determined
S13_vs_58770||full Low-quality
S13_vs_58807||full Low-quality
S13_vs_58809||full Low-quality
S13_vs_58864||full Not-determined
S13_vs_58884||full Low-quality
S13_vs_58904||full Not-determined
S13_vs_58993||full Low-quality
S13_vs_59025||full Low-quality
S13_vs_59027||full Low-quality
S13_vs_59075||full Not-determined
S13_vs_59102||full Low-quality
S13_vs_59103||full Not-determined
S13_vs_59118||full Low-quality
S13_vs_59201||full Low-quality
S13_vs_59223||full Low-quality
S13_vs_59225||full Not-determined
S13_vs_59266||full Low-quality
S13_vs_59312||full Not-determined
S13_vs_59317||full Low-quality
S13_vs_59388||full Low-quality
S13_vs_59403||full Low-quality
S13_vs_59418||full Not-determined
S13_vs_59429||full Low-quality
S13_vs_59442||full Low-quality
S13_vs_59490||full Not-determined
S13_vs_59547||full Not-determined
S13_vs_59611||full Low-quality
S13_vs_59663||full Low-quality
S13_vs_59664||full Low-quality
S13_vs_59669||full Low-quality
S13_vs_59707||full Low-quality
S13_vs_59794||full Low-quality
S13_vs_59894||full Not-determined
S13_vs_59910||full Low-quality
S13_vs_59961||full Not-determined
S13_vs_59975||full Not-determined
S13_vs_60001||full Not-determined
S13_vs_60011||full Low-quality
S13_vs_60080||full Low-quality
S13_vs_60084||full Low-quality
S13_vs_60093||full Low-quality
S13_vs_60133||full Low-quality
S13_vs_60156||full Low-quality
S13_vs_60281||full Not-determined
S13_vs_60305||full Low-quality
S13_vs_60332||full Low-quality
S13_vs_60368||full Not-determined
S13_vs_60452||full Not-determined
S13_vs_60467||full Not-determined
S13_vs_60521||full Not-determined
S13_vs_60534||full Not-determined
S13_vs_60535||full Not-determined
S13_vs_60600||full Not-determined
S13_vs_60775||full Low-quality
S13_vs_60776||full Low-quality
S13_vs_60808||full Low-quality
S13_vs_60858||full Low-quality
S13_vs_60917||full Low-quality
S13_vs_60997||full Not-determined
S13_vs_61038||full Low-quality
S13_vs_61044||full Low-quality
S13_vs_61110||full Low-quality
S13_vs_61114||full Low-quality
S13_vs_61169||full Low-quality
S13_vs_61225||full Not-determined
S13_vs_61233||full Low-quality
S13_vs_61262||full Not-determined
S13_vs_61271||full Not-determined
S13_vs_61306||full Low-quality
S13_vs_61324||full Low-quality
S13_vs_61342||full Low-quality
S13_vs_61392||full Low-quality
S13_vs_61494||full Low-quality
S13_vs_61531||full Low-quality
S13_vs_61621||full Low-quality
S13_vs_61661||full Not-determined
S13_vs_61677||full Not-determined
S13_vs_61690||full Low-quality
S13_vs_61695||full Low-quality
S13_vs_61730||full Low-quality
S13_vs_61835||full Low-quality
S13_vs_61906||full Low-quality
S13_vs_61913||full Low-quality
S13_vs_62031||full Not-determined
S13_vs_62053||full Low-quality
S13_vs_62142||full Not-determined
S13_vs_62189||full Not-determined
S13_vs_62201||full Low-quality
S13_vs_62243||full Not-determined
S13_vs_62382||full Low-quality
S13_vs_62435||full Low-quality
S13_vs_62445||full Not-determined
S13_vs_62455||full Not-determined
S13_vs_62471||full Low-quality
S13_vs_62489||full Not-determined
S13_vs_62492||full Low-quality
S13_vs_62728||full Not-determined
S13_vs_62806||full Low-quality
S13_vs_62865||full Not-determined
S13_vs_62884||full Not-determined
S13_vs_62912||full Low-quality
S13_vs_62956||full Low-quality
S13_vs_62971||full Low-quality
S13_vs_62973||full Low-quality
S13_vs_62975||full Low-quality
S13_vs_63037||full Low-quality
S13_vs_63076||full Not-determined
S13_vs_63080||full Not-determined
S13_vs_63175||full Low-quality
S13_vs_63248||full Low-quality
S13_vs_63334||full Low-quality
S13_vs_63352||full Low-quality
S13_vs_63425||full Not-determined
S13_vs_63438||full Not-determined
S13_vs_63455||full Not-determined
S13_vs_63457||full Low-quality
S13_vs_63461||full Low-quality
S13_vs_63479||full Low-quality
S13_vs_63553||full Not-determined
S13_vs_63563||full Low-quality
S13_vs_63574||full Low-quality
S13_vs_63620||full Not-determined
S13_vs_63676||full Not-determined
S13_vs_63703||full Low-quality
S13_vs_63739||full Not-determined
S13_vs_63755||full Not-determined
S13_vs_63789||full Not-determined
S13_vs_63792||full Low-quality
S13_vs_63794||full Not-determined
S13_vs_63796||full Low-quality
S13_vs_63810||full Low-quality
S13_vs_63825||full Low-quality
S13_vs_63828||full Low-quality
S13_vs_63833||full Not-determined
S13_vs_63851||full Low-quality
S13_vs_63909||full Low-quality
S13_vs_63949||full Not-determined
S13_vs_63958||full Low-quality
S13_vs_64024||full Not-determined
S13_vs_64070||full Low-quality
S13_vs_64129||full Not-determined
S13_vs_64150||full Low-quality
S13_vs_64160||full Low-quality
S13_vs_64363||full Low-quality
S13_vs_64380||full Not-determined
S13_vs_64381||full Not-determined
S13_vs_64382||full Low-quality
S13_vs_64445||full Low-quality
S13_vs_64477||full Low-quality
S13_vs_64503||full Low-quality
S13_vs_64514||full Not-determined
S13_vs_64676||full Not-determined
S13_vs_64692||full Not-determined
S13_vs_64701||full Not-determined
S13_vs_64745||full Not-determined
S13_vs_64752||full Not-determined
S3_vs_k141_9158||full High-quality
S13_vs_64905||full Low-quality
S13_vs_64926||full Low-quality
S13_vs_64944||full Low-quality
S13_vs_64989||full Low-quality
S13_vs_64993||full Low-quality
S13_vs_64995||full Low-quality
S13_vs_65024||full Low-quality
S13_vs_65044||full Low-quality
S13_vs_65084||full Low-quality
S13_vs_65160||full Not-determined
S13_vs_65215||full Not-determined
S13_vs_65220||full Low-quality
S13_vs_65240||full Low-quality
S13_vs_65333||full Not-determined
S13_vs_65346||full Low-quality
S13_vs_65370||full Low-quality
S13_vs_65421||full Low-quality
S13_vs_65463||full Low-quality
S13_vs_65490||full Low-quality
S13_vs_65500||full Low-quality
S13_vs_65501||full Low-quality
S13_vs_65553||full Low-quality
S13_vs_65574||full Low-quality
S13_vs_65619||full Low-quality
S13_vs_65638||full Low-quality
S13_vs_65681||full Low-quality
S13_vs_65782||full Not-determined
S13_vs_65800||full Not-determined
S13_vs_65904||full Not-determined
S13_vs_65924||full Low-quality
S13_vs_65977||full Low-quality
S13_vs_65990||full Not-determined
S13_vs_66063||full Low-quality
S13_vs_66117||full Low-quality
S13_vs_66174||full Low-quality
S13_vs_66243||full Low-quality
S13_vs_66291||full Low-quality
S13_vs_66424||full Low-quality
S13_vs_66458||full Low-quality
S13_vs_66485||full Not-determined
S3_vs_k141_108959||full High-quality
S13_vs_66544||full Low-quality
S13_vs_66612||full Low-quality
S13_vs_66616||full Low-quality
S13_vs_66647||full Low-quality
S13_vs_66697||full Low-quality
S13_vs_66727||full Low-quality
S13_vs_66735||full Low-quality
S13_vs_66766||full Low-quality
S13_vs_66866||full Low-quality
S13_vs_66919||full Low-quality
S13_vs_66934||full Not-determined
S13_vs_66959||full Low-quality
S13_vs_66994||full Not-determined
S13_vs_67018||full Low-quality
S13_vs_67054||full Not-determined
S13_vs_67086||full Low-quality
S13_vs_67088||full Not-determined
S13_vs_67097||full Not-determined
S13_vs_67112||full Low-quality
S13_vs_67136||full Low-quality
S13_vs_67201||full Not-determined
S13_vs_67222||full Low-quality
S13_vs_67227||full Low-quality
S13_vs_67277||full Not-determined
S13_vs_67350||full Low-quality
S13_vs_67417||full Low-quality
S13_vs_67418||full Low-quality
S12_vs_k141_153780||full High-quality
S13_vs_67436||full Not-determined
S13_vs_67502||full Not-determined
S13_vs_67604||full Not-determined
S13_vs_67610||full Low-quality
S13_vs_67616||full Low-quality
S13_vs_67620||full Low-quality
S13_vs_67681||full Low-quality
S13_vs_67724||full Low-quality
S13_vs_67734||full Low-quality
S13_vs_67772||full Low-quality
S13_vs_67787||full Not-determined
S13_vs_67854||full Not-determined
S13_vs_67875||full Not-determined
S13_vs_67965||full Not-determined
S12_vs_k141_163990||full Medium-quality
S13_vs_68142||full Not-determined
S13_vs_68179||full Not-determined
S13_vs_68184||full Not-determined
S13_vs_68186||full Low-quality
S13_vs_68232||full Not-determined
S13_vs_68257||full Low-quality
S13_vs_68275||full Low-quality
S13_vs_68291||full Low-quality
S13_vs_68356||full Low-quality
S13_vs_68360||full Low-quality
S13_vs_68444||full Low-quality
S13_vs_68492||full Not-determined
S13_vs_68503||full Low-quality
S13_vs_68522||full Low-quality
S13_vs_68599||full Low-quality
S13_vs_68615||full Not-determined
S13_vs_68630||full Not-determined
S13_vs_68653||full Low-quality
S13_vs_68703||full Not-determined
S13_vs_68715||full Not-determined
S13_vs_68718||full Not-determined
S13_vs_68741||full Low-quality
S13_vs_68752||full Not-determined
S13_vs_68772||full Low-quality
S13_vs_68779||full Low-quality
S13_vs_68786||full Low-quality
S13_vs_68813||full Not-determined
S13_vs_68908||full Low-quality
S13_vs_68950||full Low-quality
S13_vs_68986||full Not-determined
S13_vs_68999||full Low-quality
S13_vs_69027||full Not-determined
S13_vs_69031||full Not-determined
S13_vs_69042||full Low-quality
S13_vs_69068||full Low-quality
S13_vs_69086||full Low-quality
S13_vs_69186||full Not-determined
S13_vs_69197||full Not-determined
S13_vs_69218||full Low-quality
S13_vs_69220||full Not-determined
S13_vs_69320||full Low-quality
S13_vs_69345||full Low-quality
S13_vs_69595||full Not-determined
S13_vs_69722||full Not-determined
S13_vs_69731||full Not-determined
S13_vs_69774||full Low-quality
S13_vs_69782||full Low-quality
S13_vs_69818||full Low-quality
S13_vs_69956||full Not-determined
S13_vs_69959||full Not-determined
S13_vs_69998||full Not-determined
S13_vs_70001||full Not-determined
S13_vs_70002||full Low-quality
S13_vs_70036||full Low-quality
S13_vs_70063||full Not-determined
S13_vs_70128||full Low-quality
S13_vs_70158||full Low-quality
S13_vs_70161||full Low-quality
S13_vs_70168||full Low-quality
S13_vs_70303||full Low-quality
S13_vs_70347||full Not-determined
S13_vs_70348||full Low-quality
S13_vs_70386||full Low-quality
S13_vs_70444||full Not-determined
S13_vs_70472||full Low-quality
S13_vs_70539||full Low-quality
S13_vs_70605||full Not-determined
S13_vs_70685||full Not-determined
S13_vs_70694||full Low-quality
S13_vs_70713||full Low-quality
S13_vs_70839||full Low-quality
S13_vs_70840||full Low-quality
S13_vs_70940||full Low-quality
S13_vs_70972||full Low-quality
S13_vs_70994||full Low-quality
S13_vs_71003||full Not-determined
S13_vs_71045||full Low-quality
S13_vs_71060||full Not-determined
S13_vs_71086||full Not-determined
S13_vs_71094||full Low-quality
S13_vs_71246||full Not-determined
S13_vs_71250||full Not-determined
S13_vs_71319||full Not-determined
S13_vs_71379||full Low-quality
S13_vs_71402||full Not-determined
S13_vs_71448||full Not-determined
S13_vs_71450||full Not-determined
S13_vs_71476||full Not-determined
S13_vs_71522||full Not-determined
S13_vs_71528||full Not-determined
S13_vs_71560||full Not-determined
S13_vs_71614||full Low-quality
S13_vs_71641||full Not-determined
S13_vs_71648||full Not-determined
S13_vs_71649||full Low-quality
S13_vs_71739||full Low-quality
S13_vs_71751||full Low-quality
S13_vs_71781||full Low-quality
S13_vs_71826||full Low-quality
S13_vs_71831||full Low-quality
S13_vs_71851||full Not-determined
S13_vs_71870||full Low-quality
S13_vs_71990||full Not-determined
S13_vs_72005||full Low-quality
S13_vs_72010||full Low-quality
S13_vs_72030||full Low-quality
S13_vs_72135||full Low-quality
S13_vs_72212||full Not-determined
S13_vs_72232||full Low-quality
S13_vs_72276||full Not-determined
S13_vs_72315||full Low-quality
S13_vs_72325||full Not-determined
S13_vs_72386||full Not-determined
S13_vs_72530||full Low-quality
S13_vs_72605||full Low-quality
S13_vs_72637||full Not-determined
S13_vs_72654||full Not-determined
S13_vs_72668||full Not-determined
S13_vs_72804||full Low-quality
S13_vs_72848||full Not-determined
S13_vs_72996||full Not-determined
S13_vs_73090||full Low-quality
S13_vs_73114||full Low-quality
S13_vs_73145||full Low-quality
S13_vs_73159||full Not-determined
S13_vs_73183||full Low-quality
S13_vs_73204||full Low-quality
S13_vs_73261||full Low-quality
S13_vs_73280||full Low-quality
S13_vs_73321||full Low-quality
S13_vs_73324||full Not-determined
S13_vs_73333||full Not-determined
S13_vs_73347||full Not-determined
S13_vs_73394||full Low-quality
S13_vs_73409||full Low-quality
S13_vs_73418||full Not-determined
S13_vs_73444||full Low-quality
S13_vs_73497||full Not-determined
S13_vs_73517||full Not-determined
S13_vs_73545||full Not-determined
S13_vs_73559||full Low-quality
S13_vs_73629||full Low-quality
S13_vs_73658||full Not-determined
S13_vs_73717||full Low-quality
S13_vs_73771||full Not-determined
S13_vs_73810||full Low-quality
S13_vs_73837||full Not-determined
S13_vs_73855||full Not-determined
S13_vs_73864||full Not-determined
S13_vs_73870||full Low-quality
S13_vs_73911||full Not-determined
S13_vs_73999||full Not-determined
S13_vs_74007||full Low-quality
S13_vs_74109||full Not-determined
S13_vs_74115||full Low-quality
S13_vs_74376||full Low-quality
S13_vs_74441||full Low-quality
S13_vs_74508||full Low-quality
S13_vs_74518||full Not-determined
S13_vs_74542||full Not-determined
S13_vs_74580||full Not-determined
S13_vs_74584||full Low-quality
S13_vs_74591||full Low-quality
S13_vs_74687||full Not-determined
S13_vs_74707||full Not-determined
S13_vs_74789||full Low-quality
S13_vs_74834||full Low-quality
S13_vs_74849||full Not-determined
S13_vs_74868||full Low-quality
S13_vs_74939||full Not-determined
S13_vs_74971||full Not-determined
S13_vs_75012||full Not-determined
S13_vs_75016||full Low-quality
S13_vs_75044||full Low-quality
S13_vs_75058||full Low-quality
S13_vs_75230||full Not-determined
S13_vs_75306||full Low-quality
S13_vs_75349||full Low-quality
S13_vs_75369||full Not-determined
S13_vs_75376||full Low-quality
S13_vs_75546||full Low-quality
S13_vs_75626||full Low-quality
S13_vs_75629||full Low-quality
S13_vs_75643||full Low-quality
S13_vs_75654||full Not-determined
S13_vs_75659||full Low-quality
S13_vs_75675||full Low-quality
S13_vs_75677||full Low-quality
S13_vs_75681||full Not-determined
S13_vs_75695||full Not-determined
S13_vs_75720||full Low-quality
S13_vs_75728||full Not-determined
S13_vs_75788||full Low-quality
S13_vs_75854||full Not-determined
S13_vs_75867||full Not-determined
S13_vs_75872||full Low-quality
S13_vs_75997||full Low-quality
S13_vs_76004||full Not-determined
S13_vs_76017||full Low-quality
S13_vs_76099||full Low-quality

| | |
|---|---|
| S13_vs_76115\|\|full | Not-determined |
| S13_vs_76133\|\|full | Not-determined |
| S13_vs_76182\|\|full | Low-quality |
| S13_vs_76193\|\|full | Not-determined |
| S13_vs_76200\|\|full | Not-determined |
| S13_vs_76265\|\|full | Not-determined |
| S13_vs_76307\|\|full | Not-determined |
| S13_vs_76342\|\|full | Low-quality |
| S13_vs_76364\|\|full | Not-determined |
| S13_vs_76470\|\|full | Low-quality |
| S13_vs_76515\|\|full | Low-quality |
| S13_vs_76554\|\|full | Low-quality |
| S13_vs_76573\|\|full | Not-determined |
| S13_vs_76595\|\|full | Not-determined |
| S13_vs_76655\|\|full | Not-determined |
| S13_vs_76716\|\|full | Low-quality |
| S13_vs_76759\|\|full | Not-determined |
| S13_vs_76780\|\|full | Low-quality |
| S13_vs_76781\|\|full | Not-determined |
| S13_vs_76800\|\|full | Low-quality |
| S13_vs_76807\|\|full | Low-quality |
| S13_vs_76820\|\|full | Low-quality |
| S13_vs_76912\|\|full | Low-quality |
| S13_vs_76931\|\|full | Not-determined |
| S13_vs_76934\|\|full | Not-determined |
| S13_vs_76950\|\|full | Not-determined |
| S13_vs_76965\|\|full | Low-quality |
| S13_vs_76971\|\|full | Low-quality |
| S13_vs_76980\|\|full | Not-determined |
| S13_vs_77087\|\|full | Low-quality |
| S13_vs_77102\|\|full | Not-determined |
| S13_vs_77128\|\|full | Not-determined |
| S13_vs_77229\|\|full | Low-quality |
| S13_vs_77234\|\|full | Low-quality |
| S13_vs_77247\|\|full | Low-quality |
| S13_vs_77308\|\|full | Low-quality |
| S13_vs_77349\|\|full | Low-quality |
| S13_vs_77355\|\|full | Low-quality |
| S13_vs_77368\|\|full | Low-quality |
| S13_vs_77370\|\|full | Not-determined |
| S13_vs_77420\|\|full | Low-quality |
| S13_vs_77451\|\|full | Not-determined |
| S13_vs_77607\|\|full | Not-determined |
| S13_vs_77619\|\|full | Low-quality |
| S13_vs_77655\|\|full | Low-quality |
| S13_vs_77764\|\|full | Low-quality |
| S13_vs_77843\|\|full | Not-determined |
| S13_vs_77860\|\|full | Low-quality |
| S13_vs_77923\|\|full | Not-determined |
| S13_vs_77981\|\|full | Not-determined |
| S13_vs_78013\|\|full | Low-quality |
| S13_vs_78016\|\|full | Low-quality |
| S13_vs_78017\|\|full | Not-determined |
| S13_vs_78055\|\|full | Not-determined |
| S13_vs_78073\|\|full | Not-determined |
| S13_vs_78177\|\|full | Not-determined |
| S13_vs_78204\|\|full | Low-quality |
| S13_vs_78368\|\|full | Low-quality |
| S13_vs_78397\|\|full | Low-quality |
| S13_vs_78414\|\|full | Not-determined |
| S13_vs_78453\|\|full | Not-determined |
| S13_vs_78560\|\|full | Low-quality |
| S13_vs_78571\|\|full | Low-quality |
| S13_vs_78581\|\|full | Low-quality |
| S13_vs_78620\|\|full | Not-determined |
| S13_vs_78636\|\|full | Not-determined |
| S13_vs_78659\|\|full | Not-determined |
| S13_vs_78675\|\|full | Not-determined |
| S13_vs_78798\|\|full | Not-determined |
| S13_vs_78884\|\|full | Low-quality |
| S13_vs_78895\|\|full | Not-determined |
| S13_vs_78968\|\|full | Not-determined |
| S13_vs_78982\|\|full | Low-quality |
| S13_vs_79043\|\|full | Low-quality |
| S13_vs_79331\|\|full | Low-quality |
| S13_vs_79332\|\|full | Low-quality |
| S13_vs_79347\|\|full | Low-quality |
| S13_vs_79387\|\|full | Low-quality |
| S13_vs_79483\|\|full | Not-determined |
| S13_vs_79594\|\|full | Low-quality |
| S13_vs_79780\|\|full | Low-quality |
| S13_vs_79793\|\|full | Not-determined |
| S13_vs_79864\|\|full | Not-determined |
| S13_vs_79868\|\|full | Low-quality |
| S13_vs_79874\|\|full | Low-quality |
| S13_vs_79966\|\|full | Not-determined |
| S13_vs_80028\|\|full | Low-quality |
| S13_vs_80078\|\|full | Not-determined |
| S13_vs_80140\|\|full | Not-determined |
| S13_vs_80144\|\|full | Not-determined |
| S13_vs_80173\|\|full | Not-determined |
| S13_vs_80238\|\|full | Not-determined |
| S13_vs_80352\|\|full | Low-quality |
| S13_vs_80446\|\|full | Not-determined |
| S13_vs_80456\|\|full | Not-determined |
| S13_vs_80468\|\|full | Low-quality |
| S13_vs_80520\|\|full | Low-quality |
| S13_vs_80537\|\|full | Low-quality |
| S13_vs_80570\|\|full | Not-determined |
| S13_vs_80579\|\|full | Not-determined |
| S13_vs_80626\|\|full | Not-determined |
| S13_vs_80645\|\|full | Not-determined |
| S13_vs_80721\|\|full | Low-quality |
| S13_vs_80727\|\|full | Not-determined |
| S13_vs_80755\|\|full | Low-quality |
| S13_vs_80798\|\|full | Low-quality |
| S13_vs_80807\|\|full | Low-quality |
| S13_vs_80826\|\|full | Low-quality |
| S13_vs_80839\|\|full | Not-determined |
| S13_vs_80910\|\|full | Low-quality |
| S13_vs_80933\|\|full | Not-determined |
| S13_vs_80937\|\|full | Low-quality |
| S13_vs_80984\|\|full | Low-quality |
| S13_vs_81048\|\|full | Low-quality |
| S13_vs_81082\|\|full | Not-determined |
| S13_vs_81140\|\|full | Not-determined |
| S13_vs_81160\|\|full | Low-quality |
| S13_vs_81185\|\|full | Low-quality |
| S13_vs_81194\|\|full | Low-quality |
| S13_vs_81222\|\|full | Not-determined |
| S13_vs_81254\|\|full | Low-quality |
| S13_vs_81258\|\|full | Not-determined |
| S13_vs_81330\|\|full | Low-quality |
| S13_vs_81364\|\|full | Not-determined |
| S13_vs_81388\|\|full | Low-quality |
| S13_vs_81466\|\|full | Low-quality |
| S13_vs_81494\|\|full | Not-determined |
| S13_vs_81510\|\|full | Low-quality |
| S13_vs_81514\|\|full | Low-quality |
| S13_vs_81627\|\|full | Low-quality |
| S13_vs_81665\|\|full | Low-quality |
| S13_vs_81696\|\|full | Low-quality |
| S13_vs_81708\|\|full | Low-quality |
| S13_vs_81716\|\|full | Low-quality |
| S13_vs_81736\|\|full | Low-quality |
| S13_vs_81789\|\|full | Not-determined |
| S13_vs_81796\|\|full | Low-quality |
| S13_vs_81896\|\|full | Low-quality |
| S13_vs_82034\|\|full | Low-quality |
| S13_vs_82039\|\|full | Not-determined |
| S13_vs_82118\|\|full | Low-quality |
| S13_vs_82256\|\|full | Low-quality |
| S13_vs_82259\|\|full | Not-determined |
| S13_vs_82277\|\|full | Not-determined |
| S13_vs_82376\|\|full | Not-determined |
| S13_vs_82498\|\|full | Not-determined |
| S13_vs_82507\|\|full | Low-quality |
| S13_vs_82520\|\|full | Not-determined |
| S13_vs_82574\|\|full | Low-quality |
| S13_vs_82607\|\|full | Low-quality |
| S13_vs_82676\|\|full | Not-determined |
| S13_vs_82891\|\|full | Low-quality |
| S13_vs_83104\|\|full | Not-determined |
| S13_vs_83106\|\|full | Low-quality |
| S13_vs_83211\|\|full | Low-quality |
| S13_vs_83228\|\|full | Not-determined |
| S13_vs_83238\|\|full | Low-quality |
| S13_vs_83263\|\|full | Low-quality |
| S13_vs_83303\|\|full | Not-determined |
| S13_vs_83308\|\|full | Not-determined |
| S13_vs_83326\|\|full | Not-determined |
| S13_vs_83336\|\|full | Low-quality |
| S13_vs_83381\|\|full | Low-quality |
| S13_vs_83404\|\|full | Not-determined |
| S7_vs_79296\|\|full | High-quality |
| S13_vs_83564\|\|full | Not-determined |
| S13_vs_83609\|\|full | Low-quality |
| S13_vs_83622\|\|full | Not-determined |
| S13_vs_83637\|\|full | Low-quality |
| S13_vs_83715\|\|full | Low-quality |
| S13_vs_83736\|\|full | Low-quality |
| S13_vs_83768\|\|full | Low-quality |
| S13_vs_83794\|\|full | Low-quality |
| S13_vs_83809\|\|full | Not-determined |
| S13_vs_83868\|\|full | Low-quality |
| S13_vs_83984\|\|full | Low-quality |
| S13_vs_84082\|\|full | Not-determined |
| S13_vs_84148\|\|full | Low-quality |
| S13_vs_84187\|\|full | Not-determined |
| S13_vs_84321\|\|full | Low-quality |
| S13_vs_84458\|\|full | Not-determined |
| S13_vs_84539\|\|full | Not-determined |
| S13_vs_84613\|\|full | Low-quality |
| S13_vs_84634\|\|full | Not-determined |
| S13_vs_84673\|\|full | Low-quality |
| S13_vs_84712\|\|full | Not-determined |
| S13_vs_84876\|\|full | Low-quality |
| S13_vs_84934\|\|full | Low-quality |
| S13_vs_85028\|\|full | Low-quality |
| S13_vs_85039\|\|full | Low-quality |
| S13_vs_85087\|\|full | Low-quality |
| S9_vs_k141_4558\|\|full | High-quality |
| S13_vs_85204\|\|full | Not-determined |
| S13_vs_85218\|\|full | Not-determined |
| S13_vs_85240\|\|full | Not-determined |
| S13_vs_85252\|\|full | Not-determined |
| S13_vs_85254\|\|full | Not-determined |
| S13_vs_85276\|\|full | Not-determined |
| S13_vs_85351\|\|full | Low-quality |
| S13_vs_85352\|\|full | Low-quality |
| S13_vs_85358\|\|full | Not-determined |
| S13_vs_85392\|\|full | Low-quality |
| S13_vs_85436\|\|full | Low-quality |
| S13_vs_85462\|\|full | Low-quality |
| S13_vs_85489\|\|full | Not-determined |
| S13_vs_85512\|\|full | Not-determined |
| S13_vs_85535\|\|full | Not-determined |
| S13_vs_85566\|\|full | Low-quality |
| S13_vs_85576\|\|full | Low-quality |
| S13_vs_85647\|\|full | Not-determined |
| S13_vs_85777\|\|full | Low-quality |
| S13_vs_85853\|\|full | Not-determined |
| S13_vs_85861\|\|full | Low-quality |
| S13_vs_85869\|\|full | Low-quality |
| S13_vs_85895\|\|full | Not-determined |
| S13_vs_85897\|\|full | Low-quality |
| S13_vs_85916\|\|full | Low-quality |
| S13_vs_86084\|\|full | Not-determined |
| S13_vs_86100\|\|full | Not-determined |
| S13_vs_86129\|\|full | Low-quality |
| S13_vs_86212\|\|full | Not-determined |
| S13_vs_86217\|\|full | Not-determined |
| S13_vs_86222\|\|full | Not-determined |
| S13_vs_86251\|\|full | Not-determined |
| S13_vs_86377\|\|full | Low-quality |
| S13_vs_86462\|\|full | Low-quality |
| S13_vs_86637\|\|full | Low-quality |
| S13_vs_86638\|\|full | Not-determined |
| S13_vs_86639\|\|full | Low-quality |
| S13_vs_86717\|\|full | Low-quality |
| S13_vs_86723\|\|full | Low-quality |
| S13_vs_86747\|\|full | Not-determined |
| S13_vs_86776\|\|full | Low-quality |
| S13_vs_86796\|\|full | Not-determined |
| S13_vs_86823\|\|full | Low-quality |
| S13_vs_86872\|\|full | Not-determined |
| S13_vs_86883\|\|full | Low-quality |
| S13_vs_86954\|\|full | Low-quality |
| S13_vs_86980\|\|full | Low-quality |
| S13_vs_87067\|\|full | Not-determined |
| S13_vs_87084\|\|full | Not-determined |
| S13_vs_87098\|\|full | Low-quality |
| S13_vs_152223\|\|full | High-quality |
| S13_vs_87153\|\|full | Low-quality |
| S13_vs_87197\|\|full | Not-determined |
| S13_vs_87198\|\|full | Low-quality |
| S13_vs_87204\|\|full | Not-determined |
| S13_vs_87321\|\|full | Low-quality |
| S13_vs_87443\|\|full | Low-quality |
| S13_vs_87486\|\|full | Not-determined |
| S13_vs_87555\|\|full | Low-quality |
| S13_vs_87569\|\|full | Not-determined |
| S13_vs_87573\|\|full | Low-quality |
| S13_vs_87578\|\|full | Low-quality |
| S13_vs_87605\|\|full | Low-quality |
| S13_vs_87735\|\|full | Not-determined |
| S13_vs_87782\|\|full | Not-determined |
| S13_vs_87851\|\|full | Low-quality |
| S13_vs_87859\|\|full | Low-quality |
| S12_extracted_k141_188822 | Complete |
| S13_vs_87903\|\|full | Low-quality |
| S13_vs_87923\|\|full | Low-quality |
| S13_vs_87981\|\|full | Not-determined |
| S13_vs_88012\|\|full | Not-determined |
| S13_vs_88025\|\|full | Not-determined |
| S13_vs_88059\|\|full | Not-determined |
| S13_vs_88097\|\|full | Not-determined |
| S13_vs_88101\|\|full | Not-determined |
| S13_vs_88103\|\|full | Not-determined |
| S13_vs_88145\|\|full | Low-quality |
| S13_vs_88164\|\|full | Low-quality |
| S13_vs_88234\|\|full | Not-determined |
| S13_vs_88472\|\|full | Not-determined |
| S13_vs_88501\|\|full | Low-quality |
| S13_vs_88503\|\|full | Low-quality |
| S13_vs_88523\|\|full | Low-quality |
| S13_vs_88541\|\|full | Low-quality |
| S13_vs_88573\|\|full | Not-determined |
| S13_vs_88581\|\|full | Low-quality |
| S13_vs_88638\|\|full | Not-determined |
| S13_vs_88674\|\|full | Not-determined |
| S13_vs_88675\|\|full | Low-quality |
| S13_vs_88741\|\|full | Low-quality |
| S13_vs_88755\|\|full | Not-determined |
| S13_vs_88783\|\|full | Not-determined |
| S13_vs_88794\|\|full | Low-quality |
| S13_vs_88803\|\|full | Low-quality |
| S13_vs_88898\|\|full | Low-quality |
| S13_vs_88907\|\|full | Not-determined |
| S13_vs_88914\|\|full | Low-quality |
| S13_vs_88917\|\|full | Not-determined |
| S13_vs_88952\|\|full | Low-quality |
| S13_vs_88973\|\|full | Low-quality |
| S13_vs_89028\|\|full | Low-quality |
| S13_vs_89088\|\|full | Low-quality |
| S13_vs_89168\|\|full | Low-quality |
| S13_vs_89221\|\|full | Low-quality |
| S13_vs_89247\|\|full | Not-determined |
| S13_vs_89291\|\|full | Low-quality |
| S13_vs_89292\|\|full | Low-quality |
| S13_vs_89299\|\|full | Not-determined |
| S13_vs_89476\|\|full | Not-determined |
| S13_vs_89584\|\|full | Low-quality |
| S13_vs_89590\|\|full | Low-quality |
| S13_vs_89618\|\|full | Low-quality |
| S13_vs_89624\|\|full | Low-quality |
| S13_vs_89636\|\|full | Low-quality |
| S13_vs_89637\|\|full | Low-quality |
| S13_vs_89656\|\|full | Not-determined |
| S13_vs_89721\|\|full | Low-quality |
| S13_vs_89871\|\|full | Not-determined |
| S13_vs_90141\|\|full | Not-determined |
| S13_vs_90146\|\|full | Low-quality |
| S13_vs_90172\|\|full | Not-determined |
| S13_vs_90196\|\|full | Low-quality |
| S13_vs_90223\|\|full | Not-determined |
| S13_vs_90245\|\|full | Low-quality |
| S13_vs_90249\|\|full | Low-quality |
| S13_vs_90280\|\|full | Not-determined |
| S13_vs_90346\|\|full | Low-quality |
| S13_vs_90355\|\|full | Not-determined |
| S13_vs_90430\|\|full | Not-determined |
| S13_vs_90441\|\|full | Not-determined |
| S13_vs_90471\|\|full | Low-quality |
| S13_vs_90489\|\|full | Not-determined |
| S13_vs_90500\|\|full | Not-determined |
| S13_vs_90515\|\|full | Not-determined |
| S13_vs_90596\|\|full | Low-quality |
| S13_vs_90659\|\|full | Low-quality |
| S13_vs_90752\|\|full | Low-quality |
| S13_vs_90762\|\|full | Low-quality |
| S13_vs_90933\|\|full | Low-quality |
| S2_vs_k141_2872\|\|full | High-quality |
| S13_vs_91133\|\|full | Not-determined |
| S13_vs_91158\|\|full | Not-determined |
| S8_vs_k141_126582\|\|full | Medium-quality |
| S13_vs_91288\|\|full | Low-quality |
| S13_vs_91292\|\|full | Low-quality |
| S13_vs_91304\|\|full | Low-quality |
| S13_vs_91316\|\|full | Low-quality |
| S13_vs_91317\|\|full | Low-quality |
| S13_vs_91417\|\|full | Not-determined |
| S13_vs_91428\|\|full | Low-quality |
| S13_vs_91436\|\|full | Not-determined |
| S13_vs_91455\|\|full | Low-quality |
| S13_vs_91521\|\|full | Low-quality |
| S13_vs_91633\|\|full | Not-determined |
| S13_vs_91776\|\|full | Low-quality |
| S13_vs_91924\|\|full | Low-quality |
| S13_vs_91981\|\|full | Not-determined |
| S13_vs_91998\|\|full | Not-determined |
| S13_vs_92065\|\|full | Not-determined |
| S13_vs_92121\|\|full | Low-quality |
| S13_vs_92136\|\|full | Not-determined |
| S13_vs_92209\|\|full | Not-determined |
| S13_vs_92212\|\|full | Low-quality |
| S13_vs_92264\|\|full | Not-determined |
| S13_vs_92270\|\|full | Low-quality |
| S13_vs_92307\|\|full | Not-determined |
| S13_vs_92358\|\|full | Not-determined |
| S13_vs_92484\|\|full | Low-quality |
| S13_vs_92542\|\|full | Low-quality |
| S13_vs_92597\|\|full | Low-quality |
| S13_vs_92621\|\|full | Low-quality |
| S13_vs_92628\|\|full | Low-quality |
| S13_vs_92693\|\|full | Not-determined |
| S13_vs_92807\|\|full | Low-quality |
| S13_vs_92846\|\|full | Not-determined |
| S13_vs_92862\|\|full | Low-quality |
| S13_vs_92942\|\|full | Not-determined |
| S13_vs_93049\|\|full | Low-quality |
| S13_vs_93081\|\|full | Not-determined |
| S13_vs_93156\|\|full | Low-quality |
| S13_vs_93182\|\|full | Not-determined |
| S13_vs_93230\|\|full | Low-quality |
| S13_vs_93249\|\|full | Not-determined |
| S13_vs_93326\|\|full | Not-determined |
| S13_vs_93347\|\|full | Not-determined |
| S13_vs_93369\|\|full | Not-determined |
| S13_vs_93446\|\|full | Low-quality |
| S13_vs_93466\|\|full | Not-determined |
| S13_vs_93476\|\|full | Low-quality |
| S13_vs_93482\|\|full | Low-quality |
| S13_vs_93547\|\|full | Low-quality |
| S13_vs_93602\|\|full | Low-quality |
| S13_vs_93683\|\|full | Low-quality |
| S13_vs_93782\|\|full | Not-determined |
| S13_vs_93818\|\|full | Low-quality |
| S13_vs_93876\|\|full | Not-determined |
| S13_vs_93893\|\|full | Low-quality |
| S13_vs_93924\|\|full | Low-quality |
| S13_vs_94011\|\|full | Not-determined |
| S13_vs_94076\|\|full | Low-quality |
| S13_vs_94146\|\|full | Low-quality |
| S13_vs_94225\|\|full | Low-quality |
| S13_vs_94233\|\|full | Not-determined |
| S13_vs_94238\|\|full | Low-quality |
| S13_vs_94251\|\|full | Low-quality |
| S13_vs_94331\|\|full | Low-quality |
| S13_vs_94416\|\|full | Low-quality |
| S13_vs_94499\|\|full | Low-quality |
| S13_vs_94522\|\|full | Not-determined |
| S13_vs_94563\|\|full | Low-quality |
| S13_vs_94564\|\|full | Low-quality |
| S13_vs_94577\|\|full | Low-quality |
| S13_vs_94656\|\|full | Low-quality |
| S13_vs_94669\|\|full | Low-quality |
| S13_vs_94695\|\|full | Low-quality |
| S13_vs_94709\|\|full | Low-quality |
| S13_vs_94736\|\|full | Low-quality |
| S13_vs_94764\|\|full | Not-determined |
| S13_vs_94817\|\|full | Low-quality |
| S13_vs_94836\|\|full | Low-quality |
| S13_vs_94899\|\|full | Not-determined |
| S13_vs_94981\|\|full | Not-determined |
| S13_vs_95041\|\|full | Not-determined |
| S13_vs_95106\|\|full | Low-quality |
| S13_vs_95131\|\|full | Low-quality |
| S13_vs_95163\|\|full | Low-quality |
| S13_vs_95217\|\|full | Not-determined |
| S13_vs_95221\|\|full | Low-quality |
| S13_vs_95274\|\|full | Not-determined |
| S13_vs_95279\|\|full | Low-quality |
| S13_vs_95282\|\|full | Low-quality |
| S13_vs_95293\|\|full | Not-determined |
| S13_vs_95379\|\|full | Low-quality |
| S13_vs_95411\|\|full | Low-quality |
| S13_vs_95488\|\|full | Low-quality |
| S13_vs_95520\|\|full | Not-determined |
| S13_vs_95570\|\|full | Not-determined |
| S13_vs_95629\|\|full | Low-quality |
| S13_vs_95633\|\|full | Low-quality |
| S13_vs_95638\|\|full | Not-determined |
| S13_vs_95667\|\|full | Low-quality |
| S13_vs_95782\|\|full | Low-quality |
| S13_vs_95923\|\|full | Not-determined |
| S13_vs_96099\|\|full | Low-quality |
| S13_vs_96119\|\|full | Low-quality |
| S13_vs_96206\|\|full | Low-quality |
| S13_vs_96242\|\|full | Not-determined |
| S13_vs_96258\|\|full | Low-quality |
| S13_vs_96317\|\|full | Not-determined |
| S13_vs_96383\|\|full | Low-quality |
| S13_vs_96404\|\|full | Low-quality |
| S13_vs_96436\|\|full | Low-quality |
| S13_vs_96719\|\|full | Low-quality |
| S13_vs_96726\|\|full | Low-quality |
| S13_vs_96730\|\|full | Low-quality |
| S13_vs_96733\|\|full | Low-quality |
| S13_vs_96788\|\|full | Low-quality |
| S13_vs_96853\|\|full | Low-quality |
| S13_vs_96873\|\|full | Low-quality |
| S13_vs_96928\|\|full | Low-quality |
| S13_vs_96972\|\|full | Not-determined |
| S13_vs_96984\|\|full | Low-quality |
| S13_vs_97031\|\|full | Low-quality |
| S13_vs_97066\|\|full | Low-quality |
| S13_vs_97113\|\|full | Low-quality |
| S13_vs_97140\|\|full | Low-quality |
| S13_vs_97142\|\|full | Not-determined |
| S13_vs_97225\|\|full | Low-quality |
| S13_vs_97247\|\|full | Not-determined |
| S13_vs_97313\|\|full | Not-determined |
| S13_vs_97314\|\|full | Low-quality |
| S13_vs_97332\|\|full | Low-quality |
| S13_vs_97359\|\|full | Not-determined |
| S13_vs_97379\|\|full | Low-quality |
| S13_vs_97401\|\|full | Low-quality |
| S13_vs_97403\|\|full | Low-quality |
| S13_vs_97437\|\|full | Low-quality |
| S13_vs_97504\|\|full | Low-quality |
| S13_vs_97532\|\|full | Not-determined |
| S13_vs_97545\|\|full | Low-quality |
| S13_vs_97547\|\|full | Low-quality |
| S13_vs_97612\|\|full | Not-determined |
| S16_vs_6910\|\|full | High-quality |
| S13_vs_97832\|\|full | Not-determined |
| S13_vs_97854\|\|full | Not-determined |
| S13_vs_97943\|\|full | Not-determined |
| S13_vs_97957\|\|full | Not-determined |
| S13_vs_98093\|\|full | Low-quality |
| S13_vs_98144\|\|full | Low-quality |
| S13_vs_98160\|\|full | Not-determined |
| S13_vs_98172\|\|full | Low-quality |
| S13_vs_98299\|\|full | Low-quality |
| S13_vs_98422\|\|full | Low-quality |
| S13_vs_98473\|\|full | Low-quality |
| S13_vs_98568\|\|full | Low-quality |
| S12_vs_k141_188822\|\|full | High-quality |
| S13_vs_98603\|\|full | Low-quality |
| S13_vs_98608\|\|full | Not-determined |
| S13_vs_98613\|\|full | Low-quality |
| S13_vs_98618\|\|full | Low-quality |
| S13_vs_98656\|\|full | Low-quality |
| S13_vs_98720\|\|full | Not-determined |
| S13_vs_98755\|\|full | Low-quality |
| S13_vs_98782\|\|full | Low-quality |
| S13_vs_98832\|\|full | Not-determined |
| S13_vs_98880\|\|full | Not-determined |
| S13_vs_98886\|\|full | Not-determined |
| S13_vs_98911\|\|full | Low-quality |
| S13_vs_98927\|\|full | Low-quality |
| S13_vs_98957\|\|full | Not-determined |
| S13_vs_98986\|\|full | Not-determined |
| S13_vs_99094\|\|full | Low-quality |
| S13_vs_99104\|\|full | Low-quality |
| S13_vs_99125\|\|full | Low-quality |
| S13_vs_99240\|\|full | Not-determined |
| S13_vs_99320\|\|full | Not-determined |
| S13_vs_99394\|\|full | Low-quality |
| S13_vs_99429\|\|full | Low-quality |
| S13_vs_99506\|\|full | Low-quality |
| S13_vs_99539\|\|full | Not-determined |
| S13_vs_99614\|\|full | Low-quality |
| S13_vs_99623\|\|full | Low-quality |
| S13_vs_99650\|\|full | Low-quality |
| S13_vs_99677\|\|full | Low-quality |
| S13_vs_99716\|\|full | Low-quality |
| S13_vs_99735\|\|full | Low-quality |
| S13_vs_99805\|\|full | Not-determined |
| S13_vs_99806\|\|full | Low-quality |
| S13_vs_99845\|\|full | Low-quality |
| S13_vs_99891\|\|full | Not-determined |
| S13_vs_100013\|\|full | Not-determined |
| S13_vs_100015\|\|full | Low-quality |
| S13_vs_100073\|\|full | Low-quality |
| S13_vs_100077\|\|full | Not-determined |
| S13_vs_100196\|\|full | Low-quality |
| S13_vs_100308\|\|full | Low-quality |
| S13_vs_100367\|\|full | Not-determined |
| S13_vs_100371\|\|full | Not-determined |
| S13_vs_100423\|\|full | Not-determined |
| S13_vs_100468\|\|full | Not-determined |
| S13_vs_100559\|\|full | Not-determined |
| S13_vs_100561\|\|full | Not-determined |
| S13_vs_100631\|\|full | Not-determined |
| S13_vs_100690\|\|full | Not-determined |
| S13_vs_100746\|\|full | Low-quality |
| S13_vs_100778\|\|full | Not-determined |
| S13_vs_100883\|\|full | Low-quality |
| S13_vs_100903\|\|full | Low-quality |
| S13_vs_100956\|\|full | Low-quality |
| S13_vs_101039\|\|full | Low-quality |
| S13_vs_101112\|\|full | Low-quality |
| S13_vs_101142\|\|full | Low-quality |
| S13_vs_101194\|\|full | Low-quality |
| S13_vs_101199\|\|full | Low-quality |
| S13_vs_101224\|\|full | Not-determined |
| S13_vs_101241\|\|full | Low-quality |
| S13_vs_101256\|\|full | Not-determined |
| S13_vs_101278\|\|full | Low-quality |
| S13_vs_101282\|\|full | Low-quality |
| S13_vs_101491\|\|full | Not-determined |
| S13_vs_101514\|\|full | Low-quality |
| S13_vs_101541\|\|full | Not-determined |
| S13_vs_101766\|\|full | Low-quality |
| S13_vs_101796\|\|full | Low-quality |
| S5_vs_k141_41663\|\|full | High-quality |
| S13_vs_101820\|\|full | Low-quality |
| S13_vs_101982\|\|full | Low-quality |
| S13_vs_102013\|\|full | Not-determined |
| S13_vs_102021\|\|full | Low-quality |
| S13_vs_102037\|\|full | Not-determined |
| S13_vs_102047\|\|full | Low-quality |
| S13_vs_102072\|\|full | Not-determined |
| S13_vs_102116\|\|full | Low-quality |
| S13_vs_102146\|\|full | Not-determined |
| S13_vs_102231\|\|full | Not-determined |
| S13_vs_102287\|\|full | Not-determined |
| S13_vs_102312\|\|full | Not-determined |
| S13_vs_102356\|\|full | Not-determined |
| S13_vs_102374\|\|full | Low-quality |
| S13_vs_102423\|\|full | Not-determined |
| S13_vs_102500\|\|full | Low-quality |
| S13_vs_102558\|\|full | Low-quality |
| S13_vs_102595\|\|full | Low-quality |
| S13_vs_102629\|\|full | Not-determined |
| S13_vs_102684\|\|full | Low-quality |
| S13_vs_102705\|\|full | Low-quality |
| S13_vs_102797\|\|full | Low-quality |
| S13_vs_102854\|\|full | Low-quality |
| S13_vs_102884\|\|full | Not-determined |
| S13_vs_102899\|\|full | Not-determined |
| S13_vs_102920\|\|full | Low-quality |
| S13_vs_102953\|\|full | Low-quality |
| S13_vs_102955\|\|full | Low-quality |
| S13_vs_103000\|\|full | Not-determined |
| S13_vs_103009\|\|full | Low-quality |
| S13_vs_103022\|\|full | Low-quality |

| | |
|---|---|
| S13_vs_103092\|\|full | Low-quality |
| S13_vs_103161\|\|full | Not-determined |
| S13_vs_103278\|\|full | Not-determined |
| S13_vs_103322\|\|full | Not-determined |
| S13_vs_103400\|\|full | Low-quality |
| S13_vs_103478\|\|full | Not-determined |
| S13_vs_103494\|\|full | Low-quality |
| S13_vs_103538\|\|full | Low-quality |
| S13_vs_103551\|\|full | Low-quality |
| S13_vs_103578\|\|full | Low-quality |
| S13_vs_103595\|\|full | Not-determined |
| S13_vs_103603\|\|full | Low-quality |
| S13_vs_103651\|\|full | Not-determined |
| S13_vs_103715\|\|full | Low-quality |
| S13_vs_103747\|\|full | Low-quality |
| S13_vs_103751\|\|full | Not-determined |
| S13_vs_103800\|\|full | Not-determined |
| S13_vs_103822\|\|full | Low-quality |
| S13_vs_103871\|\|full | Low-quality |
| S13_vs_103896\|\|full | Low-quality |
| S13_vs_103956\|\|full | Low-quality |
| S13_vs_104022\|\|full | Low-quality |
| S13_vs_104023\|\|full | Low-quality |
| S13_vs_104062\|\|full | Low-quality |
| S13_vs_104066\|\|full | Low-quality |
| S13_vs_104198\|\|full | Not-determined |
| S13_vs_104367\|\|full | Not-determined |
| S13_vs_104383\|\|full | Not-determined |
| S13_vs_104389\|\|full | Low-quality |
| S13_vs_104390\|\|full | Low-quality |
| S13_vs_104405\|\|full | Low-quality |
| S13_vs_104418\|\|full | Not-determined |
| S13_vs_104426\|\|full | Low-quality |
| S13_vs_104433\|\|full | Not-determined |
| S13_vs_104483\|\|full | Not-determined |
| S13_vs_104513\|\|full | Low-quality |
| S13_vs_104517\|\|full | Not-determined |
| S13_vs_104546\|\|full | Low-quality |
| S13_vs_104569\|\|full | Not-determined |
| S13_vs_104571\|\|full | Low-quality |
| S13_vs_104606\|\|full | Not-determined |
| S13_vs_104626\|\|full | Low-quality |
| S13_vs_104793\|\|full | Low-quality |
| S13_vs_104869\|\|full | Not-determined |
| S13_vs_104879\|\|full | Low-quality |
| S13_vs_104940\|\|full | Not-determined |
| S13_vs_104946\|\|full | Not-determined |
| S13_vs_104987\|\|full | Low-quality |
| S13_vs_105015\|\|full | Not-determined |
| S13_vs_105097\|\|full | Not-determined |
| S13_vs_105192\|\|full | Not-determined |
| S13_vs_105228\|\|full | Not-determined |
| S13_vs_105235\|\|full | Low-quality |
| S13_vs_105337\|\|full | Not-determined |
| S13_vs_105374\|\|full | Low-quality |
| S13_vs_105387\|\|full | Low-quality |
| S13_vs_105436\|\|full | Low-quality |
| S13_vs_105438\|\|full | Low-quality |
| S13_vs_105454\|\|full | Low-quality |
| S13_vs_105479\|\|full | Low-quality |
| S13_vs_105484\|\|full | Low-quality |
| S13_vs_105491\|\|full | Not-determined |
| S13_vs_105558\|\|full | Low-quality |
| S13_vs_105664\|\|full | Not-determined |
| S13_vs_105671\|\|full | Not-determined |
| S13_vs_105673\|\|full | Low-quality |
| S13_vs_105758\|\|full | Not-determined |
| S13_vs_105770\|\|full | Not-determined |
| S13_vs_105886\|\|full | Not-determined |
| S13_vs_105913\|\|full | Low-quality |
| S13_vs_105920\|\|full | Low-quality |
| S13_vs_105974\|\|full | Not-determined |
| S13_vs_105992\|\|full | Not-determined |
| S13_vs_106016\|\|full | Low-quality |
| S13_vs_106074\|\|full | Low-quality |
| S13_vs_106113\|\|full | Low-quality |
| S13_vs_106129\|\|full | Not-determined |
| S13_vs_106147\|\|full | Low-quality |
| S13_vs_106168\|\|full | Not-determined |
| S13_vs_106222\|\|full | Low-quality |
| S13_vs_106253\|\|full | Low-quality |
| S13_vs_106280\|\|full | Low-quality |
| S13_vs_106307\|\|full | Not-determined |
| S13_vs_106320\|\|full | Low-quality |
| S13_vs_106366\|\|full | Low-quality |
| S13_vs_106474\|\|full | Not-determined |
| S13_vs_106504\|\|full | Not-determined |
| S13_vs_106534\|\|full | Low-quality |
| S13_vs_106548\|\|full | Low-quality |
| S13_vs_106558\|\|full | Low-quality |
| S13_vs_106567\|\|full | Low-quality |
| S13_vs_106667\|\|full | Not-determined |
| S13_vs_106830\|\|full | Low-quality |
| S13_vs_106963\|\|full | Low-quality |
| S13_vs_107103\|\|full | Low-quality |
| S13_vs_107105\|\|full | Not-determined |
| S7_extracted_38697 | Complete |
| S13_vs_107223\|\|full | Not-determined |
| S16_vs_35860\|\|full | High-quality |
| S13_vs_107405\|\|full | Not-determined |
| S13_vs_107419\|\|full | Not-determined |
| S13_vs_107465\|\|full | Not-determined |
| S13_vs_107488\|\|full | Low-quality |
| S13_vs_107490\|\|full | Low-quality |
| S13_vs_107530\|\|full | Low-quality |
| S13_vs_107616\|\|full | Low-quality |
| S13_vs_107635\|\|full | Low-quality |
| S13_vs_107676\|\|full | Low-quality |
| S13_vs_107700\|\|full | Low-quality |
| S13_vs_107719\|\|full | Not-determined |
| S13_vs_107727\|\|full | Not-determined |
| S13_vs_107744\|\|full | Low-quality |
| S13_vs_107799\|\|full | Not-determined |
| S13_vs_107848\|\|full | Low-quality |
| S13_vs_107881\|\|full | Not-determined |
| S13_vs_107948\|\|full | Low-quality |
| S13_vs_107963\|\|full | Not-determined |
| S13_vs_107982\|\|full | Low-quality |
| S13_vs_107990\|\|full | Not-determined |
| S13_vs_108031\|\|full | Not-determined |
| S13_vs_108032\|\|full | Low-quality |
| S13_vs_108036\|\|full | Low-quality |
| S13_vs_108047\|\|full | Not-determined |
| S13_vs_108069\|\|full | Not-determined |
| S13_vs_108120\|\|full | Not-determined |
| S13_vs_108188\|\|full | Low-quality |
| S13_vs_108217\|\|full | Low-quality |
| S13_vs_108252\|\|full | Not-determined |
| S13_vs_108263\|\|full | Low-quality |
| S13_vs_108374\|\|full | Low-quality |
| S13_vs_108376\|\|full | Low-quality |
| S13_vs_108381\|\|full | Not-determined |
| S13_vs_108445\|\|full | Not-determined |
| S13_vs_108463\|\|full | Not-determined |
| S13_vs_108482\|\|full | Low-quality |
| S13_vs_108507\|\|full | Low-quality |
| S13_vs_108577\|\|full | Low-quality |
| S13_vs_108581\|\|full | Low-quality |
| S13_vs_108612\|\|full | Low-quality |
| S13_vs_108733\|\|full | Low-quality |
| S13_vs_108754\|\|full | Not-determined |
| S13_vs_108780\|\|full | Low-quality |
| S13_vs_108832\|\|full | Low-quality |
| S13_vs_108838\|\|full | Low-quality |
| S13_vs_108990\|\|full | Low-quality |
| S13_vs_109093\|\|full | Low-quality |
| S13_vs_109133\|\|full | Not-determined |
| S13_vs_109152\|\|full | Low-quality |
| S13_vs_109160\|\|full | Not-determined |
| S13_vs_109174\|\|full | Low-quality |
| S13_vs_109253\|\|full | Low-quality |
| S13_vs_109289\|\|full | Low-quality |
| S13_vs_109317\|\|full | Low-quality |
| S13_vs_109428\|\|full | Low-quality |
| S13_vs_109514\|\|full | Low-quality |
| S13_vs_109515\|\|full | Not-determined |
| S13_vs_109572\|\|full | Low-quality |
| S13_vs_109620\|\|full | Not-determined |
| S13_vs_109661\|\|full | Not-determined |
| S13_vs_109703\|\|full | Low-quality |
| S13_vs_109780\|\|full | Low-quality |
| S13_vs_109815\|\|full | Low-quality |
| S13_vs_109857\|\|full | Low-quality |
| S13_vs_109880\|\|full | Low-quality |
| S13_vs_109937\|\|full | Low-quality |
| S13_vs_109971\|\|full | Not-determined |
| S13_vs_110059\|\|full | Not-determined |
| S13_vs_110068\|\|full | Low-quality |
| S13_vs_110212\|\|full | Low-quality |
| S13_vs_110261\|\|full | Not-determined |
| S13_vs_110289\|\|full | Low-quality |
| S13_vs_110314\|\|full | Low-quality |
| S13_vs_110358\|\|full | Low-quality |
| S13_vs_110381\|\|full | Not-determined |
| S13_vs_110393\|\|full | Low-quality |
| S13_vs_110470\|\|full | Low-quality |
| S13_vs_110504\|\|full | Low-quality |
| S13_vs_110563\|\|full | Not-determined |
| S13_vs_110572\|\|full | Not-determined |
| S13_vs_110598\|\|full | Low-quality |
| S13_vs_110622\|\|full | Not-determined |
| S13_vs_110673\|\|full | Not-determined |
| S13_vs_110722\|\|full | Low-quality |
| S13_vs_110724\|\|full | Not-determined |
| S13_vs_110741\|\|full | Not-determined |
| S13_vs_110748\|\|full | Low-quality |
| S13_vs_110796\|\|full | Low-quality |
| S13_vs_110818\|\|full | Not-determined |
| S13_vs_110830\|\|full | Low-quality |
| S13_vs_110928\|\|full | Low-quality |
| S13_vs_110945\|\|full | Low-quality |
| S13_vs_110957\|\|full | Low-quality |
| S13_vs_110989\|\|full | Low-quality |
| S13_vs_110993\|\|full | Low-quality |
| S13_vs_111001\|\|full | Low-quality |
| S13_vs_111037\|\|full | Not-determined |
| S13_vs_111078\|\|full | Low-quality |
| S13_vs_111086\|\|full | Low-quality |
| S13_vs_111098\|\|full | Not-determined |
| S13_vs_111128\|\|full | Not-determined |
| S13_vs_111135\|\|full | Low-quality |
| S13_vs_111161\|\|full | Low-quality |
| S13_vs_111173\|\|full | Low-quality |
| S13_vs_111195\|\|full | Low-quality |
| S13_vs_111206\|\|full | Low-quality |
| S13_vs_111336\|\|full | Low-quality |
| S13_vs_111363\|\|full | Low-quality |
| S13_vs_111389\|\|full | Low-quality |
| S13_vs_111398\|\|full | Not-determined |
| S13_vs_111457\|\|full | Low-quality |
| S13_vs_111491\|\|full | Low-quality |
| S13_vs_111523\|\|full | Low-quality |
| S13_vs_111538\|\|full | Not-determined |
| S13_vs_111546\|\|full | Not-determined |
| S13_vs_111830\|\|full | Low-quality |
| S13_vs_111845\|\|full | Low-quality |
| S13_vs_111863\|\|full | Not-determined |
| S13_vs_111997\|\|full | Not-determined |
| S13_vs_112026\|\|full | Low-quality |
| S13_vs_112028\|\|full | Low-quality |
| S13_vs_112032\|\|full | Low-quality |
| S13_vs_112045\|\|full | Not-determined |
| S13_vs_112051\|\|full | Low-quality |
| S13_vs_112061\|\|full | Low-quality |
| S13_vs_112079\|\|full | Low-quality |
| S13_vs_112125\|\|full | Low-quality |
| S13_vs_112313\|\|full | Low-quality |
| S13_vs_112381\|\|full | Low-quality |
| S13_vs_112463\|\|full | Not-determined |
| S13_vs_112470\|\|full | Not-determined |
| S13_vs_112652\|\|full | Not-determined |
| S13_vs_112701\|\|full | Not-determined |
| S13_vs_112817\|\|full | Not-determined |
| S13_vs_112820\|\|full | Not-determined |
| S13_vs_112827\|\|full | Low-quality |
| S13_vs_113003\|\|full | Low-quality |
| S13_vs_113046\|\|full | Low-quality |
| S13_vs_113085\|\|full | Not-determined |
| S13_vs_113157\|\|full | Low-quality |
| S13_vs_113163\|\|full | Low-quality |
| S13_vs_113171\|\|full | Not-determined |
| S13_vs_113185\|\|full | Not-determined |
| S13_vs_113212\|\|full | Low-quality |
| S13_vs_113225\|\|full | Low-quality |
| S13_vs_113228\|\|full | Low-quality |
| S13_vs_113269\|\|full | Low-quality |
| S13_vs_113287\|\|full | Not-determined |
| S13_vs_113324\|\|full | Low-quality |
| S13_vs_113422\|\|full | Low-quality |
| S13_vs_113482\|\|full | Not-determined |
| S13_vs_113516\|\|full | Low-quality |
| S13_vs_113527\|\|full | Low-quality |
| S13_vs_113536\|\|full | Not-determined |
| S9_vs_k141_2799\|\|full | High-quality |
| S13_vs_113628\|\|full | Low-quality |
| S13_vs_113703\|\|full | Low-quality |
| S13_vs_113754\|\|full | Low-quality |
| S13_vs_113768\|\|full | Low-quality |
| S13_vs_113814\|\|full | Low-quality |
| S13_vs_113828\|\|full | Low-quality |
| S13_vs_113933\|\|full | Low-quality |
| S13_vs_114017\|\|full | Low-quality |
| S13_vs_114070\|\|full | Not-determined |
| S13_vs_114076\|\|full | Not-determined |
| S13_vs_114150\|\|full | Low-quality |
| S13_vs_114201\|\|full | Low-quality |
| S13_vs_114202\|\|full | Low-quality |
| S7_vs_38697\|\|full | High-quality |
| S13_vs_114294\|\|full | Not-determined |
| S13_vs_114471\|\|full | Not-determined |
| S5_vs_k141_46394\|\|full | High-quality |
| S13_vs_114567\|\|full | Low-quality |
| S13_vs_114613\|\|full | Low-quality |
| S13_vs_114648\|\|full | Not-determined |
| S13_vs_114670\|\|full | Low-quality |
| S13_vs_114743\|\|full | Low-quality |
| S13_vs_114751\|\|full | Low-quality |
| S13_vs_114766\|\|full | Not-determined |
| S13_vs_114796\|\|full | Low-quality |
| S13_vs_114808\|\|full | Not-determined |
| S13_vs_114874\|\|full | Not-determined |
| S13_vs_114886\|\|full | Low-quality |
| S13_vs_114899\|\|full | Not-determined |
| S13_vs_114912\|\|full | Not-determined |
| S13_vs_114953\|\|full | Low-quality |
| S13_vs_114958\|\|full | Not-determined |
| S13_vs_115006\|\|full | Low-quality |
| S13_vs_115098\|\|full | Low-quality |
| S13_vs_115136\|\|full | Not-determined |
| S13_vs_115230\|\|full | Not-determined |
| S13_vs_115238\|\|full | Low-quality |
| S13_vs_115283\|\|full | Low-quality |
| S13_vs_115285\|\|full | Not-determined |
| S13_vs_115305\|\|full | Low-quality |
| S13_vs_115310\|\|full | Not-determined |
| S13_vs_115373\|\|full | Not-determined |
| S13_vs_115584\|\|full | Not-determined |
| S13_vs_115599\|\|full | Low-quality |
| S13_vs_115635\|\|full | Low-quality |
| S13_vs_115675\|\|full | Not-determined |
| S13_vs_115722\|\|full | Low-quality |
| S13_vs_115730\|\|full | Not-determined |
| S13_vs_115745\|\|full | Low-quality |
| S13_vs_115836\|\|full | Low-quality |
| S13_vs_115915\|\|full | Low-quality |
| S13_vs_115940\|\|full | Low-quality |
| S13_vs_116049\|\|full | Not-determined |
| S13_vs_116102\|\|full | Not-determined |
| S13_vs_116107\|\|full | Low-quality |
| S13_vs_116121\|\|full | Not-determined |
| S13_vs_116184\|\|full | Low-quality |
| S13_vs_116220\|\|full | Low-quality |
| S13_vs_116294\|\|full | Not-determined |
| S13_vs_116300\|\|full | Low-quality |
| S13_vs_116364\|\|full | Not-determined |
| S13_vs_116458\|\|full | Low-quality |
| S13_vs_116535\|\|full | Low-quality |
| S13_vs_116541\|\|full | Low-quality |
| S13_vs_116577\|\|full | Low-quality |
| S13_vs_116609\|\|full | Not-determined |
| S13_vs_116655\|\|full | Not-determined |
| S13_vs_116729\|\|full | Low-quality |
| S13_vs_116745\|\|full | Low-quality |
| S13_vs_116770\|\|full | Not-determined |
| S13_vs_116949\|\|full | Low-quality |
| S13_vs_116962\|\|full | Low-quality |
| S13_vs_116968\|\|full | Not-determined |
| S13_vs_116975\|\|full | Low-quality |
| S13_vs_117035\|\|full | Not-determined |
| S13_vs_117058\|\|full | Low-quality |
| S13_vs_117100\|\|full | Low-quality |
| S13_vs_117109\|\|full | Not-determined |
| S13_vs_117115\|\|full | Low-quality |
| S13_vs_117199\|\|full | Low-quality |
| S13_vs_117291\|\|full | Low-quality |
| S13_vs_117361\|\|full | Not-determined |
| S13_vs_117422\|\|full | Not-determined |
| S13_vs_117528\|\|full | Not-determined |
| S13_vs_117599\|\|full | Low-quality |
| S13_vs_117683\|\|full | Not-determined |
| S13_vs_117733\|\|full | Low-quality |
| S13_vs_117753\|\|full | Low-quality |
| S13_vs_117799\|\|full | Low-quality |
| S13_vs_117826\|\|full | Low-quality |
| S13_vs_117873\|\|full | Not-determined |
| S13_vs_117918\|\|full | Low-quality |
| S13_vs_117964\|\|full | Low-quality |
| S13_vs_118041\|\|full | Low-quality |
| S13_vs_118085\|\|full | Not-determined |
| S13_vs_118087\|\|full | Low-quality |
| S13_vs_118094\|\|full | Low-quality |
| S13_vs_118265\|\|full | Low-quality |
| S13_vs_118292\|\|full | Low-quality |
| S13_vs_118387\|\|full | Low-quality |
| S13_vs_118409\|\|full | Low-quality |
| S13_vs_118442\|\|full | Not-determined |
| S13_vs_118457\|\|full | Low-quality |
| S13_vs_118461\|\|full | Not-determined |
| S13_vs_118462\|\|full | Not-determined |
| S13_vs_118468\|\|full | Not-determined |
| S13_vs_118471\|\|full | Low-quality |
| S13_vs_118548\|\|full | Low-quality |
| S13_vs_118632\|\|full | Not-determined |
| S13_vs_118655\|\|full | Low-quality |
| S13_vs_118721\|\|full | Not-determined |
| S13_vs_118808\|\|full | Low-quality |
| S13_vs_118905\|\|full | Low-quality |
| S13_vs_119008\|\|full | Low-quality |
| S13_vs_119019\|\|full | Low-quality |
| S13_vs_119066\|\|full | Not-determined |
| S13_vs_119087\|\|full | Low-quality |
| S13_vs_119122\|\|full | Not-determined |
| S13_vs_119144\|\|full | Not-determined |
| S13_vs_119182\|\|full | Not-determined |
| S13_vs_119220\|\|full | Low-quality |
| S13_vs_119255\|\|full | Not-determined |
| S13_vs_119401\|\|full | Not-determined |
| S13_vs_119414\|\|full | Low-quality |
| S13_vs_119528\|\|full | Low-quality |
| S13_vs_119560\|\|full | Low-quality |
| S13_vs_119589\|\|full | Low-quality |
| S13_vs_119591\|\|full | Low-quality |
| S13_vs_119672\|\|full | Not-determined |
| S13_vs_119708\|\|full | Low-quality |
| S13_vs_119746\|\|full | Low-quality |
| S13_vs_119774\|\|full | Low-quality |
| S13_vs_119802\|\|full | Low-quality |
| S13_vs_119887\|\|full | Low-quality |
| S13_vs_119905\|\|full | Not-determined |
| S13_vs_119923\|\|full | Low-quality |
| S13_vs_119938\|\|full | Low-quality |
| S13_vs_119990\|\|full | Not-determined |
| S13_vs_120001\|\|full | Not-determined |
| S13_vs_120032\|\|full | Low-quality |
| S13_vs_120100\|\|full | Low-quality |
| S13_vs_120103\|\|full | Low-quality |
| S13_vs_120150\|\|full | Low-quality |
| S13_vs_120202\|\|full | Low-quality |
| S13_vs_120386\|\|full | Not-determined |
| S13_vs_120521\|\|full | Not-determined |
| S13_vs_120547\|\|full | Low-quality |
| S11_extracted_k141_13033 | High-quality |
| S13_vs_120599\|\|full | Low-quality |
| S13_vs_120626\|\|full | Low-quality |
| S13_vs_120684\|\|full | Low-quality |
| S13_vs_120692\|\|full | Low-quality |
| S13_vs_120725\|\|full | Not-determined |
| S13_vs_120787\|\|full | Not-determined |
| S13_vs_120793\|\|full | Not-determined |
| S13_vs_120831\|\|full | Low-quality |
| S13_vs_120915\|\|full | Low-quality |
| S13_vs_120975\|\|full | Not-determined |
| S13_vs_121022\|\|full | Low-quality |
| S13_vs_121028\|\|full | Low-quality |
| S13_vs_121120\|\|full | Low-quality |
| S13_vs_121186\|\|full | Low-quality |
| S13_vs_121227\|\|full | Low-quality |
| S3_vs_k141_127391\|\|full | High-quality |
| S13_vs_121361\|\|full | Low-quality |
| S13_vs_121392\|\|full | Not-determined |
| S13_vs_121399\|\|full | Low-quality |
| S13_vs_121608\|\|full | Not-determined |
| S13_vs_121807\|\|full | Low-quality |
| S13_vs_121819\|\|full | Low-quality |
| S13_vs_121857\|\|full | Low-quality |
| S13_vs_121863\|\|full | Low-quality |
| S13_vs_121954\|\|full | Low-quality |
| S13_vs_121968\|\|full | Low-quality |
| S13_vs_122001\|\|full | Not-determined |
| S13_vs_122036\|\|full | Not-determined |
| S13_vs_122124\|\|full | Not-determined |
| S13_vs_122168\|\|full | Low-quality |
| S13_vs_122177\|\|full | Low-quality |
| S13_vs_122181\|\|full | Low-quality |
| S13_vs_122224\|\|full | Low-quality |
| S13_vs_122276\|\|full | Low-quality |
| S13_vs_122296\|\|full | Low-quality |
| S13_vs_122355\|\|full | Not-determined |
| S13_vs_122387\|\|full | Not-determined |
| S13_vs_122404\|\|full | Low-quality |
| S13_vs_122436\|\|full | Low-quality |
| S13_vs_122514\|\|full | Low-quality |
| S13_vs_122562\|\|full | Low-quality |
| S13_vs_122580\|\|full | Not-determined |
| S13_vs_122581\|\|full | Not-determined |
| S13_vs_122600\|\|full | Not-determined |
| S13_vs_122614\|\|full | Low-quality |
| S13_vs_122615\|\|full | Low-quality |
| S13_vs_122661\|\|full | Not-determined |
| S13_vs_122690\|\|full | Low-quality |
| S13_vs_122777\|\|full | Low-quality |
| S13_vs_122880\|\|full | Low-quality |
| S13_vs_122910\|\|full | Not-determined |
| S13_vs_123006\|\|full | Low-quality |
| S13_vs_123058\|\|full | Not-determined |
| S13_vs_123076\|\|full | Low-quality |
| S13_vs_123173\|\|full | Low-quality |
| S13_vs_123203\|\|full | Not-determined |
| S13_vs_123247\|\|full | Low-quality |
| S13_vs_123251\|\|full | Not-determined |
| S13_vs_123253\|\|full | Not-determined |
| S13_vs_123255\|\|full | Low-quality |
| S13_vs_123300\|\|full | Not-determined |
| S13_vs_123304\|\|full | Not-determined |
| S13_vs_123347\|\|full | Not-determined |
| S13_vs_123365\|\|full | Low-quality |
| S13_vs_123375\|\|full | Not-determined |
| S13_vs_123486\|\|full | Not-determined |
| S13_vs_123495\|\|full | Not-determined |
| S13_vs_123509\|\|full | Low-quality |
| S13_vs_123526\|\|full | Low-quality |
| S13_vs_123594\|\|full | Low-quality |
| S13_vs_123618\|\|full | Low-quality |
| S13_vs_123623\|\|full | Low-quality |
| S13_vs_123651\|\|full | Not-determined |
| S13_vs_123663\|\|full | Low-quality |
| S13_vs_123668\|\|full | Not-determined |
| S13_vs_123863\|\|full | Low-quality |
| S13_vs_123901\|\|full | Low-quality |
| S13_vs_123916\|\|full | Low-quality |
| S13_vs_123945\|\|full | Low-quality |
| S13_vs_123987\|\|full | Not-determined |
| S13_vs_124037\|\|full | Low-quality |
| S13_vs_124069\|\|full | Low-quality |
| S13_vs_124125\|\|full | Low-quality |
| S13_vs_124170\|\|full | Not-determined |
| S13_vs_124252\|\|full | Low-quality |
| S13_vs_124264\|\|full | Not-determined |
| S13_vs_124354\|\|full | Low-quality |
| S13_vs_124380\|\|full | Low-quality |
| S13_vs_124403\|\|full | Low-quality |
| S13_vs_124523\|\|full | Low-quality |
| S13_vs_124527\|\|full | Not-determined |
| S13_vs_124539\|\|full | Low-quality |
| S13_vs_124549\|\|full | Low-quality |
| S13_vs_124553\|\|full | Low-quality |
| S13_vs_124584\|\|full | Low-quality |
| S13_vs_124594\|\|full | Low-quality |
| S13_vs_124618\|\|full | Low-quality |
| S13_vs_124622\|\|full | Not-determined |
| S13_vs_124641\|\|full | Low-quality |
| S13_vs_124655\|\|full | Low-quality |
| S13_vs_124658\|\|full | Low-quality |
| S13_vs_124664\|\|full | Not-determined |
| S13_vs_124674\|\|full | Low-quality |
| S13_vs_124676\|\|full | Low-quality |
| S13_vs_124771\|\|full | Low-quality |
| S13_vs_124830\|\|full | Not-determined |
| S13_vs_124879\|\|full | Low-quality |
| S13_vs_124898\|\|full | Not-determined |
| S13_vs_124929\|\|full | Not-determined |
| S13_vs_124962\|\|full | Low-quality |
| S13_vs_124963\|\|full | Low-quality |
| S13_vs_124974\|\|full | Not-determined |
| S13_vs_125012\|\|full | Low-quality |
| S13_vs_125082\|\|full | Low-quality |
| S13_vs_125093\|\|full | Low-quality |
| S13_vs_125201\|\|full | Low-quality |
| S13_vs_125252\|\|full | Not-determined |
| S13_vs_125459\|\|full | Not-determined |
| S13_vs_125497\|\|full | Low-quality |
| S13_vs_125508\|\|full | Low-quality |
| S13_vs_125524\|\|full | Low-quality |
| S13_vs_125553\|\|full | Not-determined |
| S13_vs_125581\|\|full | Low-quality |
| S13_vs_125836\|\|full | Low-quality |
| S13_vs_125877\|\|full | Low-quality |
| S13_vs_125904\|\|full | Low-quality |
| S13_vs_125928\|\|full | Low-quality |
| S13_vs_125981\|\|full | Low-quality |
| S13_vs_126016\|\|full | Not-determined |
| S13_vs_126059\|\|full | Not-determined |
| S13_vs_126060\|\|full | Low-quality |
| S13_vs_126166\|\|full | Not-determined |
| S13_vs_126245\|\|full | Not-determined |
| S13_vs_126251\|\|full | Low-quality |
| S13_vs_126302\|\|full | Low-quality |
| S13_vs_126331\|\|full | Low-quality |
| S13_vs_126340\|\|full | Not-determined |
| S13_vs_126380\|\|full | Not-determined |
| S13_vs_126464\|\|full | Not-determined |
| S13_vs_126499\|\|full | Low-quality |
| S13_vs_126526\|\|full | Low-quality |
| S13_vs_126565\|\|full | Low-quality |
| S13_vs_126620\|\|full | Not-determined |
| S13_vs_126678\|\|full | Low-quality |
| S13_vs_126828\|\|full | Low-quality |
| S13_vs_126847\|\|full | Low-quality |
| S13_vs_126858\|\|full | Not-determined |
| S13_vs_126870\|\|full | Not-determined |
| S13_vs_126944\|\|full | Low-quality |
| S13_vs_126991\|\|full | Low-quality |
| S13_vs_127009\|\|full | Not-determined |
| S13_vs_127054\|\|full | Not-determined |
| S13_vs_127088\|\|full | Not-determined |
| S13_vs_127170\|\|full | Low-quality |
| S13_vs_127196\|\|full | Not-determined |
| S13_vs_127236\|\|full | Low-quality |
| S13_vs_127352\|\|full | Not-determined |
| S13_vs_127354\|\|full | Not-determined |
| S13_vs_127410\|\|full | Low-quality |
| S13_vs_127445\|\|full | Not-determined |
| S13_vs_127497\|\|full | Low-quality |
| S13_vs_127516\|\|full | Low-quality |
| S13_vs_127569\|\|full | Low-quality |
| S13_vs_127603\|\|full | Not-determined |
| S13_vs_127619\|\|full | Low-quality |
| S13_vs_127622\|\|full | Low-quality |
| S13_vs_127672\|\|full | Low-quality |
| S13_vs_127761\|\|full | Low-quality |
| S13_vs_127817\|\|full | Not-determined |
| S13_vs_127846\|\|full | Low-quality |
| S13_vs_127862\|\|full | Not-determined |
| S13_vs_127871\|\|full | Not-determined |
| S13_vs_127992\|\|full | Low-quality |
| S13_vs_128033\|\|full | Low-quality |
| S13_vs_128039\|\|full | Low-quality |
| S13_vs_128041\|\|full | Not-determined |
| S13_vs_128195\|\|full | Low-quality |
| S13_vs_128212\|\|full | Low-quality |
| S13_vs_128267\|\|full | Low-quality |
| S13_vs_128358\|\|full | Not-determined |
| S13_vs_128421\|\|full | Low-quality |
| S13_vs_128440\|\|full | Low-quality |
| S13_vs_128473\|\|full | Low-quality |
| S13_vs_128552\|\|full | Not-determined |
| S13_vs_128578\|\|full | Low-quality |
| S13_vs_128579\|\|full | Not-determined |
| S13_vs_128590\|\|full | Not-determined |

S13_vs_128598||full Low-quality
S13_vs_128637||full Not-determined
S13_vs_128711||full Not-determined
S13_vs_128713||full Low-quality
S13_vs_128781||full Not-determined
S13_vs_128885||full Low-quality
S13_vs_128922||full Low-quality
S13_vs_129061||full Low-quality
S13_vs_129130||full Low-quality
S13_vs_129132||full Not-determined
S13_vs_129182||full Low-quality
S13_vs_129220||full Low-quality
S13_vs_129291||full Low-quality
S13_vs_129325||full Low-quality
S13_vs_129367||full Not-determined
S13_vs_129410||full Not-determined
S13_vs_129498||full Not-determined
S13_vs_129595||full Low-quality
S13_vs_129602||full Not-determined
S13_vs_129705||full Not-determined
S13_vs_129760||full Low-quality
S13_vs_129805||full Low-quality
S13_vs_129867||full Not-determined
S13_vs_129872||full Not-determined
S13_vs_129953||full Low-quality
S13_vs_129961||full Not-determined
S13_vs_129978||full Not-determined
S13_vs_130008||full Low-quality
S13_vs_130107||full Low-quality
S13_vs_130179||full Not-determined
S13_vs_130246||full Low-quality
S13_vs_130289||full Low-quality
S13_vs_130299||full Not-determined
S13_vs_130403||full Low-quality
S13_vs_130445||full Not-determined
S13_vs_130462||full Not-determined
S13_vs_130471||full Low-quality
S13_vs_130501||full Low-quality
S13_vs_130515||full Low-quality
S13_vs_130582||full Low-quality
S13_vs_130614||full Not-determined
S13_vs_130635||full Low-quality
S13_vs_130669||full Low-quality
S13_vs_130713||full Not-determined
S13_vs_130775||full Low-quality
S13_vs_130808||full Not-determined
S13_vs_130811||full Low-quality
S13_vs_130850||full Not-determined
S13_vs_130917||full Low-quality
S13_vs_130925||full Not-determined
S13_vs_130939||full Not-determined
S13_vs_130962||full Low-quality
S13_vs_130974||full Low-quality
S13_vs_131149||full Low-quality
S13_vs_131191||full Not-determined
S11_extracted_k141_27845 High-quality
S13_vs_131210||full Low-quality
S13_vs_131227||full Not-determined
S13_vs_131391||full Low-quality
S13_vs_131392||full Low-quality
S13_vs_131404||full Low-quality
S13_vs_131409||full Low-quality
S13_vs_131413||full Low-quality
S13_vs_131416||full Low-quality
S13_vs_131447||full Not-determined
S13_vs_131469||full Not-determined
S13_vs_131533||full Low-quality
S13_vs_131548||full Low-quality
S13_vs_131605||full Low-quality
S13_vs_131615||full Not-determined
S13_vs_131633||full Not-determined
S13_vs_131640||full Low-quality
S13_vs_131689||full Not-determined
S13_vs_131708||full Not-determined
S13_vs_131724||full Not-determined
S13_vs_131752||full Low-quality
S13_vs_131795||full Low-quality
S13_vs_131799||full Low-quality
S13_vs_131839||full Not-determined
S13_vs_131999||full Not-determined
S13_vs_132039||full Low-quality
S13_vs_132065||full Low-quality
S13_vs_132092||full Low-quality
S13_vs_132105||full Not-determined
S13_vs_132163||full Low-quality
S13_vs_132182||full Not-determined
S13_vs_132198||full Not-determined
S13_vs_132236||full Low-quality
S13_vs_132460||full Low-quality
S13_vs_132543||full Low-quality
S13_vs_132600||full Low-quality
S13_vs_132616||full Low-quality
S13_vs_132635||full Low-quality
S13_vs_132786||full Low-quality
S13_vs_132799||full Not-determined
S13_vs_132819||full Not-determined
S13_vs_132887||full Low-quality
S13_vs_132964||full Not-determined
S13_vs_132988||full Low-quality
S13_vs_133097||full Low-quality
S13_vs_133105||full Low-quality
S13_vs_133143||full Low-quality
S13_vs_133150||full Low-quality
S13_vs_133210||full Low-quality
S13_vs_133281||full Low-quality
S13_vs_133300||full Not-determined
S13_vs_133375||full Not-determined
S13_vs_133522||full Low-quality
S13_vs_133529||full Low-quality
S13_vs_133535||full Low-quality
S13_vs_133554||full Low-quality
S13_vs_133562||full Not-determined
S13_vs_133614||full Low-quality
S13_vs_133633||full Low-quality
S13_vs_133682||full Not-determined
S13_vs_133788||full Low-quality
S13_vs_133917||full Not-determined
S13_vs_133937||full Not-determined
S13_vs_133980||full Not-determined
S13_vs_134004||full Not-determined
S13_vs_134115||full Not-determined
S13_vs_134138||full Low-quality
S13_vs_134140||full Low-quality
S13_vs_134227||full Low-quality
S13_vs_134311||full Not-determined
S13_vs_134357||full Low-quality
S13_vs_134386||full Not-determined
S13_vs_134499||full Low-quality
S13_vs_134515||full Not-determined
S13_vs_134560||full Low-quality
S13_vs_134734||full Low-quality
S13_vs_134737||full Low-quality
S13_vs_134794||full Not-determined
S13_vs_134827||full Not-determined
S13_vs_134884||full Low-quality
S13_vs_134908||full Low-quality
S13_vs_134948||full Not-determined
S13_vs_135000||full Not-determined
S13_vs_135018||full Not-determined
S13_vs_135042||full Low-quality
S13_vs_135055||full Low-quality
S13_vs_135093||full Low-quality
S13_vs_135119||full Not-determined
S13_vs_135157||full Low-quality
S13_vs_135190||full Not-determined
S13_vs_135200||full Not-determined
S13_vs_135297||full Not-determined
S7_extracted_34520 Complete
S13_vs_135436||full Low-quality
S13_vs_135460||full Low-quality
S13_vs_135484||full Low-quality
S13_vs_135507||full Not-determined
S13_vs_135644||full Low-quality
S13_vs_135703||full Not-determined
S13_vs_135727||full Low-quality
S13_vs_135750||full Not-determined
S13_vs_135772||full Low-quality
S13_vs_135923||full Low-quality
S13_vs_135946||full Low-quality
S13_vs_136154||full Low-quality
S13_vs_136187||full Not-determined
S13_vs_136254||full Not-determined
S13_vs_136271||full Not-determined
S13_vs_136290||full Not-determined
S13_vs_136352||full Low-quality
S13_vs_136377||full Low-quality
S13_vs_136448||full Low-quality
S13_vs_136462||full Not-determined
S13_vs_136487||full Low-quality
S13_vs_136501||full Low-quality
S13_vs_136502||full Not-determined
S13_vs_136558||full Not-determined
S13_vs_136561||full Not-determined
S13_vs_136579||full Not-determined
S13_vs_136583||full Not-determined
S13_vs_136617||full Low-quality
S13_vs_136691||full Low-quality
S13_vs_136824||full Not-determined
S13_vs_136851||full Low-quality
S13_vs_136856||full Low-quality
S13_vs_136888||full Low-quality
S13_vs_136914||full Not-determined
S13_vs_136915||full Low-quality
S13_vs_136965||full Low-quality
S13_vs_136969||full Low-quality
S13_vs_137086||full Not-determined
S13_vs_137105||full Low-quality
S13_vs_137108||full Low-quality
S13_vs_137153||full Low-quality
S13_vs_137178||full Not-determined
S13_vs_137211||full Not-determined
S13_vs_137271||full Not-determined
S13_vs_137288||full Not-determined
S13_vs_137301||full Not-determined
S13_vs_137322||full Low-quality
S13_vs_137323||full Low-quality
S13_vs_137325||full Not-determined
S13_vs_137365||full Not-determined
S13_vs_137372||full Low-quality
S13_vs_137452||full Not-determined
S13_vs_137539||full Not-determined
S13_vs_137587||full Not-determined
S13_vs_137595||full Low-quality
S13_vs_137621||full Low-quality
S13_vs_137777||full Low-quality
S13_vs_137798||full Low-quality
S13_vs_137808||full Low-quality
S13_vs_137865||full Not-determined
S13_vs_137899||full Low-quality
S13_vs_137909||full Low-quality
S13_vs_137913||full Not-determined
S13_vs_138006||full Low-quality
S13_vs_138058||full Not-determined
S13_vs_138128||full Not-determined
S13_vs_138148||full Low-quality
S13_vs_138152||full Not-determined
S13_vs_138161||full Not-determined
S13_vs_138205||full Not-determined
S13_vs_138219||full Low-quality
S13_vs_138235||full Low-quality
S13_vs_138331||full Low-quality
S13_vs_138332||full Low-quality
S13_vs_138356||full Not-determined
S13_vs_138363||full Low-quality
S13_vs_138415||full Low-quality
S13_vs_138536||full Not-determined
S13_vs_138693||full Low-quality
S13_vs_138714||full Low-quality
S13_vs_138740||full Low-quality
S13_vs_138783||full Low-quality
S13_vs_138801||full Not-determined
S13_vs_138810||full Not-determined
S13_vs_138825||full Not-determined
S13_vs_138846||full Low-quality
S13_vs_138862||full Not-determined
S13_vs_138877||full Not-determined
S13_vs_138972||full Not-determined
S13_vs_139016||full Low-quality
S13_vs_139091||full Not-determined
S13_vs_139151||full Not-determined
S13_vs_139181||full Not-determined
S13_vs_139208||full Low-quality
S13_vs_139321||full Low-quality
S13_vs_139337||full Low-quality
S13_vs_139366||full Not-determined
S13_vs_139367||full Low-quality
S13_vs_139536||full Not-determined
S13_vs_139562||full Not-determined
S13_vs_139687||full Not-determined
S13_vs_139693||full Not-determined
S13_vs_139750||full Not-determined
S13_vs_139766||full Not-determined
S13_vs_139807||full Low-quality
S13_vs_139923||full Low-quality
S13_vs_139938||full Low-quality
S13_vs_139976||full Low-quality
S13_vs_140033||full Low-quality
S13_vs_140102||full Low-quality
S13_vs_140131||full Low-quality
S13_vs_140229||full Low-quality
S13_vs_140372||full Low-quality
S13_vs_140381||full Low-quality
S13_vs_140391||full Not-determined
S13_vs_140419||full Low-quality
S13_vs_140451||full Low-quality
S13_vs_140457||full Not-determined
S13_vs_140462||full Low-quality
S13_vs_140564||full Not-determined
S13_vs_140697||full Not-determined
S13_vs_140755||full Low-quality
S13_vs_140806||full Not-determined
S13_vs_140807||full Low-quality
S13_vs_140816||full Low-quality
S13_vs_140844||full Low-quality
S13_vs_140918||full Low-quality
S13_vs_140966||full Low-quality
S13_vs_140998||full Low-quality
S13_vs_141006||full Low-quality
S13_vs_141050||full Not-determined
S13_vs_141081||full Low-quality
S13_vs_141093||full Low-quality
S13_vs_141106||full Not-determined
S13_vs_141153||full Not-determined
S13_vs_141198||full Low-quality
S13_vs_141203||full Low-quality
S13_vs_141214||full Low-quality
S13_vs_141257||full Not-determined
S13_vs_141297||full Low-quality
S13_vs_141329||full Not-determined
S13_vs_141362||full Not-determined
S13_vs_141394||full Not-determined
S13_vs_141459||full Low-quality
S13_vs_141576||full Low-quality
S13_vs_141580||full Not-determined
S13_vs_141631||full Not-determined
S13_vs_141710||full Low-quality
S13_vs_141727||full Low-quality
S13_vs_141772||full Not-determined
S13_vs_141825||full Not-determined
S13_vs_141843||full Low-quality
S13_vs_141861||full Not-determined
S13_vs_141881||full Low-quality
S13_vs_141917||full Low-quality
S13_vs_141933||full Not-determined
S13_vs_141976||full Low-quality
S13_vs_142010||full Low-quality
S13_vs_142028||full Not-determined
S13_vs_142082||full Not-determined
S13_vs_142127||full Low-quality
S13_vs_142214||full Not-determined
S13_vs_142240||full Low-quality
S13_vs_142291||full Not-determined
S13_vs_142306||full Low-quality
S13_vs_142359||full Not-determined
S13_vs_142439||full Not-determined
S13_vs_142485||full Not-determined
S13_vs_142606||full Low-quality
S13_vs_142648||full Low-quality
S13_vs_142655||full Low-quality
S13_vs_142664||full Low-quality
S13_vs_142701||full Low-quality
S13_vs_142740||full Not-determined
S13_vs_142808||full Not-determined
S13_vs_142856||full Low-quality
S13_vs_142865||full Not-determined
S13_vs_142913||full Not-determined
S13_vs_142933||full Low-quality
S13_vs_142962||full Not-determined
S13_vs_142963||full Low-quality
S13_vs_143030||full Not-determined
S13_vs_143068||full Not-determined
S13_vs_143108||full Low-quality
S13_vs_143182||full Low-quality
S13_vs_143229||full Low-quality
S13_vs_143287||full Not-determined
S13_vs_143304||full Not-determined
S13_vs_143385||full Not-determined
S13_vs_143408||full Low-quality
S5_vs_k141_144819||full High-quality
S13_vs_143428||full Low-quality
S13_vs_143552||full Low-quality
S13_vs_143655||full Low-quality
S13_vs_143722||full Not-determined
S13_vs_143739||full Not-determined
S13_vs_143784||full Low-quality
S13_vs_143817||full Low-quality
S13_vs_144010||full Low-quality
S13_vs_144025||full Low-quality
S13_vs_144126||full Low-quality
S13_vs_144195||full Low-quality
S13_vs_144207||full Low-quality
S13_vs_144237||full Low-quality
S13_vs_144282||full Low-quality
S13_vs_144316||full Low-quality
S13_vs_144364||full Low-quality
S13_vs_144378||full Low-quality
S13_vs_144384||full Not-determined
S13_vs_144433||full Low-quality
S13_vs_144482||full Low-quality
S13_vs_144544||full Low-quality
S13_vs_144552||full Not-determined
S13_vs_144572||full Not-determined
S13_vs_144639||full Low-quality
S13_vs_144645||full Not-determined
S13_vs_144747||full Not-determined
S13_vs_144844||full Not-determined
S13_vs_144853||full Not-determined
S13_vs_144896||full Low-quality
S13_vs_144920||full Not-determined
S13_vs_144941||full Not-determined
S13_vs_144988||full Low-quality
S13_vs_144993||full Low-quality
S13_vs_145001||full Not-determined
S13_vs_145047||full Not-determined
S13_vs_145058||full Low-quality
S13_vs_145106||full Not-determined
S13_vs_145170||full Low-quality
S13_vs_145228||full Low-quality
S13_vs_145250||full Not-determined
S13_vs_145278||full Not-determined
S13_vs_145288||full Not-determined
S13_vs_145304||full Low-quality
S13_vs_145447||full Low-quality
S13_vs_145507||full Low-quality
S13_vs_145546||full Not-determined
S13_vs_145575||full Low-quality
S13_vs_145594||full Not-determined
S13_vs_145605||full Low-quality
S13_vs_145743||full Low-quality
S13_vs_145774||full Low-quality
S13_vs_145884||full Low-quality
S13_vs_145888||full Low-quality
S13_vs_145899||full Not-determined
S13_vs_146034||full Low-quality
S13_vs_146182||full Low-quality
S13_vs_146187||full Not-determined
S13_vs_146376||full Low-quality
S13_vs_146472||full Low-quality
S13_vs_146514||full Low-quality
S13_vs_146542||full Low-quality
S13_vs_146543||full Low-quality
S13_vs_146579||full Not-determined
S13_vs_146666||full Not-determined
S13_vs_146667||full Low-quality
S13_vs_146677||full Low-quality
S13_vs_146682||full Low-quality
S13_vs_146761||full Not-determined
S13_vs_146803||full Not-determined
S13_vs_146827||full Low-quality
S13_vs_146829||full Low-quality
S13_vs_146855||full Low-quality
S13_vs_146856||full Not-determined
S13_vs_146915||full Low-quality
S13_vs_146971||full Not-determined
S13_vs_146972||full Low-quality
S13_vs_146995||full Not-determined
S13_vs_147046||full Not-determined
S13_vs_147113||full Low-quality
S13_vs_147131||full Not-determined
S13_vs_147133||full Low-quality
S13_vs_147136||full Low-quality
S13_vs_147159||full Not-determined
S13_vs_147245||full Not-determined
S13_vs_147249||full Low-quality
S13_vs_147392||full Not-determined
S13_vs_147414||full Low-quality
S13_vs_147516||full Low-quality
S13_vs_147686||full Low-quality
S13_vs_147701||full Low-quality
S13_vs_147712||full Not-determined
S13_vs_147838||full Low-quality
S13_vs_147840||full Low-quality
S13_vs_147842||full Not-determined
S13_vs_147945||full Not-determined
S13_vs_148054||full Not-determined
S13_vs_148128||full Low-quality
S13_vs_148155||full Low-quality
S13_vs_148230||full Not-determined
S13_vs_148235||full Low-quality
S13_vs_148258||full Low-quality
S13_vs_148260||full Not-determined
S13_vs_148336||full Low-quality
S13_vs_148352||full Low-quality
S13_vs_148446||full Not-determined
S13_vs_148485||full Not-determined
S13_vs_148495||full Not-determined
S13_vs_148499||full Low-quality
S13_vs_148517||full Low-quality
S13_vs_148539||full Low-quality
S13_vs_148614||full Low-quality
S13_vs_148622||full Not-determined
S13_vs_148648||full Not-determined
S13_vs_148879||full Low-quality
S13_vs_148911||full Not-determined
S13_vs_148952||full Not-determined
S13_vs_148955||full Not-determined
S13_vs_149028||full Low-quality
S13_vs_149054||full Low-quality
S13_vs_149085||full Not-determined
S13_vs_149086||full Not-determined
S13_vs_149123||full Low-quality
S13_vs_149245||full Low-quality
S13_vs_149285||full Low-quality
S13_vs_149345||full Not-determined
S13_vs_149361||full Low-quality
S13_vs_149407||full Low-quality
S13_vs_149550||full Low-quality
S13_vs_149551||full Low-quality
S13_vs_149572||full Low-quality
S13_vs_149596||full Not-determined
S13_vs_149597||full Not-determined
S13_vs_149605||full Low-quality
S13_vs_149691||full Low-quality
S13_vs_149780||full Low-quality
S13_vs_149804||full Low-quality
S13_vs_149822||full Low-quality
S13_vs_149911||full Low-quality
S13_vs_149929||full Low-quality
S13_vs_149930||full Low-quality
S13_vs_149935||full Not-determined
S13_vs_149942||full Low-quality
S13_vs_149986||full Low-quality
S13_vs_149999||full Low-quality
S13_vs_150073||full Not-determined
S13_vs_150082||full Low-quality
S13_vs_150094||full Not-determined
S13_vs_150216||full Not-determined
S13_vs_150247||full Low-quality
S13_vs_150252||full Not-determined
S13_vs_150290||full Low-quality
S13_vs_150317||full Not-determined
S13_vs_150326||full Low-quality
S13_vs_150379||full Not-determined
S13_vs_150479||full Not-determined
S13_vs_150496||full Low-quality
S13_vs_150521||full Low-quality
S13_vs_150577||full Low-quality
S13_vs_150586||full Low-quality
S13_vs_150664||full Low-quality
S13_vs_150686||full Low-quality
S13_vs_150750||full Low-quality
S13_vs_150825||full Low-quality
S13_vs_150916||full Not-determined
S13_vs_150969||full Not-determined
S13_vs_150977||full Not-determined
S13_vs_151011||full Low-quality
S13_vs_151081||full Not-determined
S13_vs_151124||full Low-quality
S13_vs_151150||full Not-determined
S13_vs_151166||full Not-determined
S13_vs_151217||full Not-determined
S13_vs_151308||full Low-quality
S13_vs_151534||full Low-quality
S13_vs_151578||full Low-quality
S13_vs_151601||full Not-determined
S13_vs_151613||full Low-quality
S13_vs_151723||full Low-quality
S13_vs_151937||full Low-quality
S13_vs_152006||full Low-quality
S13_vs_152029||full Low-quality
S13_vs_152044||full Low-quality
S13_vs_152089||full Not-determined
S13_vs_152187||full Low-quality
S13_vs_152195||full Low-quality
S13_vs_152214||full Not-determined
S17_extracted_k141_175097 Medium-quality
S13_vs_152287||full Not-determined
S13_vs_152382||full Not-determined
S13_vs_152414||full Not-determined
S13_vs_152436||full Not-determined
S13_vs_152450||full Not-determined
S13_vs_152574||full Not-determined
S13_vs_152580||full Not-determined
S13_vs_152697||full Not-determined
S13_vs_152700||full Low-quality
S13_vs_152718||full Not-determined
S13_vs_152725||full Not-determined
S13_vs_152750||full Not-determined
S13_vs_152769||full Low-quality
S13_vs_152847||full Low-quality
S13_vs_152855||full Low-quality
S13_vs_152885||full Not-determined
S13_vs_152899||full Low-quality
S13_vs_152907||full Low-quality
S13_vs_152946||full Not-determined
S13_vs_152962||full Low-quality
S13_vs_153039||full Low-quality
S13_vs_153094||full Not-determined
S13_vs_153118||full Low-quality
S13_vs_153121||full Low-quality
S13_vs_153132||full Low-quality
S13_vs_153321||full Not-determined
S13_vs_153332||full Not-determined
S13_vs_153372||full Not-determined
S13_vs_153405||full Not-determined
S13_vs_153429||full Low-quality
S13_vs_153455||full Low-quality
S13_vs_153478||full Low-quality
S13_vs_153488||full Not-determined
S13_vs_153508||full Low-quality
S13_vs_153511||full Low-quality
S13_vs_153560||full Low-quality
S13_vs_153584||full Not-determined
S13_vs_153603||full Not-determined
S13_vs_153722||full Not-determined
S13_vs_153734||full Low-quality
S13_vs_153743||full Not-determined
S13_vs_153766||full Low-quality
S13_vs_153828||full Not-determined
S13_vs_153832||full Low-quality
S13_vs_153906||full Not-determined
S13_vs_153907||full Not-determined
S13_vs_153952||full Not-determined
S13_vs_153990||full Low-quality
S13_vs_154014||full Low-quality
S13_vs_154019||full Not-determined
S13_vs_154021||full Low-quality
S13_vs_154095||full Low-quality
S13_vs_154165||full Low-quality
S13_vs_154194||full Not-determined
S13_vs_154229||full Not-determined
S13_vs_154325||full Low-quality
S13_vs_154341||full Low-quality
S13_vs_4814||0_partial Low-quality
S13_vs_22523||0_partial Not-determined
S13_vs_42736||0_partial Low-quality
S13_vs_43656||0_partial Low-quality
S13_vs_45161||0_partial Low-quality
S13_vs_53416||0_partial Low-quality

| | |
|---|---|
| S13_vs_58880\|\|0_partial | Low-quality |
| S13_vs_66966\|\|0_partial | Not-determined |
| S13_vs_92925\|\|0_partial | Not-determined |
| S13_vs_114932\|\|0_partial | Low-quality |
| S13_vs_123344\|\|0_partial | Not-determined |
| S13_vs_128784\|\|0_partial | Low-quality |
| S13_vs_139426\|\|0_partial | Low-quality |
| S13_vs_148640\|\|0_partial | Low-quality |
| S13_vs_149741\|\|0_partial | Not-determined |
| S13_vs_186\|\|lt2gene | Not-determined |
| S13_vs_274\|\|lt2gene | Not-determined |
| S13_vs_470\|\|lt2gene | Not-determined |
| S13_vs_527\|\|lt2gene | Low-quality |
| S13_vs_531\|\|lt2gene | Low-quality |
| S13_vs_540\|\|lt2gene | Not-determined |
| S13_vs_631\|\|lt2gene | Low-quality |
| S13_vs_962\|\|lt2gene | Not-determined |
| S13_vs_1005\|\|lt2gene | Low-quality |
| S13_vs_1008\|\|lt2gene | Low-quality |
| S13_vs_1066\|\|lt2gene | Not-determined |
| S13_vs_1079\|\|lt2gene | Low-quality |
| S13_vs_1093\|\|lt2gene | Low-quality |
| S13_vs_1117\|\|lt2gene | Low-quality |
| S13_vs_1127\|\|lt2gene | Low-quality |
| S13_vs_1275\|\|lt2gene | Low-quality |
| S13_vs_1475\|\|lt2gene | Low-quality |
| S13_vs_1523\|\|lt2gene | Low-quality |
| S13_vs_1550\|\|lt2gene | Low-quality |
| S13_vs_1568\|\|lt2gene | Low-quality |
| S13_vs_1705\|\|lt2gene | Low-quality |
| S13_vs_1765\|\|lt2gene | Not-determined |
| S13_vs_1908\|\|lt2gene | Low-quality |
| S13_vs_1984\|\|lt2gene | Not-determined |
| S13_vs_1993\|\|lt2gene | Low-quality |
| S13_vs_2362\|\|lt2gene | Low-quality |
| S13_vs_2443\|\|lt2gene | Not-determined |
| S13_vs_2503\|\|lt2gene | Low-quality |
| S13_vs_2619\|\|lt2gene | Low-quality |
| S13_vs_2661\|\|lt2gene | Not-determined |
| S13_vs_2848\|\|lt2gene | Low-quality |
| S13_vs_2925\|\|lt2gene | Low-quality |
| S13_vs_3028\|\|lt2gene | Low-quality |
| S13_vs_3410\|\|lt2gene | Low-quality |
| S13_vs_3640\|\|lt2gene | Low-quality |
| S13_vs_3811\|\|lt2gene | Low-quality |
| S13_vs_3839\|\|lt2gene | Low-quality |
| S13_vs_3876\|\|lt2gene | Low-quality |
| S13_vs_3992\|\|lt2gene | Low-quality |
| S13_vs_4134\|\|lt2gene | Low-quality |
| S13_vs_4211\|\|lt2gene | Low-quality |
| S13_vs_4229\|\|lt2gene | Low-quality |
| S13_vs_4384\|\|lt2gene | Low-quality |
| S13_vs_4528\|\|lt2gene | Not-determined |
| S13_vs_4835\|\|lt2gene | Not-determined |
| S13_vs_5078\|\|lt2gene | Low-quality |
| S13_vs_5099\|\|lt2gene | Low-quality |
| S13_vs_5405\|\|lt2gene | Low-quality |
| S13_vs_5429\|\|lt2gene | Low-quality |
| S13_vs_5529\|\|lt2gene | Low-quality |
| S13_vs_5543\|\|lt2gene | Low-quality |
| S13_vs_5699\|\|lt2gene | Low-quality |
| S13_vs_5762\|\|lt2gene | Low-quality |
| S13_vs_5810\|\|lt2gene | Not-determined |
| S13_vs_6033\|\|lt2gene | Low-quality |
| S13_vs_6614\|\|lt2gene | Low-quality |
| S13_vs_6706\|\|lt2gene | Low-quality |
| S13_vs_6717\|\|lt2gene | Not-determined |
| S13_vs_6817\|\|lt2gene | Low-quality |
| S13_vs_6825\|\|lt2gene | Low-quality |
| S13_vs_7023\|\|lt2gene | Not-determined |
| S13_vs_7232\|\|lt2gene | Not-determined |
| S13_vs_7350\|\|lt2gene | Low-quality |
| S13_vs_7362\|\|lt2gene | Low-quality |
| S13_vs_7411\|\|lt2gene | Low-quality |
| S13_vs_7590\|\|lt2gene | Low-quality |
| S13_vs_7725\|\|lt2gene | Low-quality |
| S13_vs_7789\|\|lt2gene | Not-determined |
| S13_vs_7853\|\|lt2gene | Low-quality |
| S13_vs_8020\|\|lt2gene | Low-quality |
| S13_vs_8093\|\|lt2gene | Low-quality |
| S13_vs_8125\|\|lt2gene | Low-quality |
| S13_vs_8198\|\|lt2gene | Low-quality |
| S13_vs_8381\|\|lt2gene | Low-quality |
| S13_vs_8401\|\|lt2gene | Not-determined |
| S13_vs_8465\|\|lt2gene | Low-quality |
| S13_vs_8539\|\|lt2gene | Not-determined |
| S13_vs_8566\|\|lt2gene | Not-determined |
| S13_vs_8698\|\|lt2gene | Not-determined |
| S13_vs_8797\|\|lt2gene | Not-determined |
| S13_vs_8984\|\|lt2gene | Low-quality |
| S13_vs_9106\|\|lt2gene | Low-quality |
| S13_vs_9114\|\|lt2gene | Low-quality |
| S13_vs_9146\|\|lt2gene | Low-quality |
| S13_vs_9219\|\|lt2gene | Low-quality |
| S13_vs_9245\|\|lt2gene | Low-quality |
| S13_vs_9274\|\|lt2gene | Not-determined |
| S13_vs_9317\|\|lt2gene | Not-determined |
| S13_vs_9319\|\|lt2gene | Low-quality |
| S13_vs_9334\|\|lt2gene | Not-determined |
| S13_vs_9340\|\|lt2gene | Not-determined |
| S13_vs_9369\|\|lt2gene | Not-determined |
| S13_vs_9520\|\|lt2gene | Not-determined |
| S13_vs_9570\|\|lt2gene | Low-quality |
| S13_vs_9579\|\|lt2gene | Low-quality |
| S13_vs_9709\|\|lt2gene | Low-quality |
| S13_vs_9752\|\|lt2gene | Low-quality |
| S13_vs_9827\|\|lt2gene | Low-quality |
| S13_vs_10091\|\|lt2gene | Low-quality |
| S13_vs_10269\|\|lt2gene | Low-quality |
| S13_vs_10309\|\|lt2gene | Low-quality |
| S13_vs_10351\|\|lt2gene | Low-quality |
| S13_vs_10536\|\|lt2gene | Low-quality |
| S13_vs_10803\|\|lt2gene | Low-quality |
| S13_vs_10903\|\|lt2gene | Low-quality |
| S13_vs_11004\|\|lt2gene | Not-determined |
| S13_vs_11106\|\|lt2gene | Low-quality |
| S13_vs_11115\|\|lt2gene | Not-determined |
| S13_vs_11194\|\|lt2gene | Low-quality |
| S13_vs_11294\|\|lt2gene | Not-determined |
| S13_vs_11304\|\|lt2gene | Low-quality |
| S13_vs_11501\|\|lt2gene | Low-quality |
| S13_vs_11642\|\|lt2gene | Low-quality |
| S13_vs_11662\|\|lt2gene | Not-determined |
| S13_vs_11724\|\|lt2gene | Low-quality |
| S13_vs_11743\|\|lt2gene | Low-quality |
| S13_vs_11769\|\|lt2gene | Not-determined |
| S13_vs_11781\|\|lt2gene | Not-determined |
| S13_vs_11794\|\|lt2gene | Not-determined |
| S13_vs_12091\|\|lt2gene | Low-quality |
| S13_vs_12304\|\|lt2gene | Low-quality |
| S13_vs_12353\|\|lt2gene | Low-quality |
| S13_vs_12355\|\|lt2gene | Not-determined |
| S13_vs_12469\|\|lt2gene | Low-quality |
| S13_vs_12504\|\|lt2gene | Low-quality |
| S13_vs_12521\|\|lt2gene | Not-determined |
| S13_vs_12600\|\|lt2gene | Low-quality |
| S13_vs_12610\|\|lt2gene | Not-determined |
| S13_vs_12625\|\|lt2gene | Not-determined |
| S13_vs_12732\|\|lt2gene | Low-quality |
| S13_vs_13024\|\|lt2gene | Low-quality |
| S13_vs_13277\|\|lt2gene | Low-quality |
| S13_vs_13324\|\|lt2gene | Not-determined |
| S13_vs_13484\|\|lt2gene | Low-quality |
| S13_vs_13489\|\|lt2gene | Low-quality |
| S13_vs_13507\|\|lt2gene | Low-quality |
| S13_vs_13606\|\|lt2gene | Not-determined |
| S13_vs_13635\|\|lt2gene | Low-quality |
| S13_vs_13648\|\|lt2gene | Low-quality |
| S13_vs_13729\|\|lt2gene | Not-determined |
| S13_vs_13764\|\|lt2gene | Low-quality |
| S13_vs_13775\|\|lt2gene | Not-determined |
| S13_vs_13945\|\|lt2gene | Not-determined |
| S13_vs_13966\|\|lt2gene | Low-quality |
| S13_vs_13999\|\|lt2gene | Low-quality |
| S13_vs_14008\|\|lt2gene | Low-quality |
| S13_vs_14871\|\|lt2gene | Low-quality |
| S13_vs_15012\|\|lt2gene | Not-determined |
| S13_vs_15212\|\|lt2gene | Low-quality |
| S13_vs_15225\|\|lt2gene | Not-determined |
| S13_vs_15240\|\|lt2gene | Low-quality |
| S13_vs_15283\|\|lt2gene | Low-quality |
| S13_vs_15435\|\|lt2gene | Low-quality |
| S13_vs_15474\|\|lt2gene | Low-quality |
| S13_vs_15599\|\|lt2gene | Low-quality |
| S13_vs_15718\|\|lt2gene | Low-quality |
| S13_vs_15720\|\|lt2gene | Low-quality |
| S13_vs_15899\|\|lt2gene | Not-determined |
| S13_vs_15949\|\|lt2gene | Not-determined |
| S13_vs_16257\|\|lt2gene | Not-determined |
| S13_vs_16274\|\|lt2gene | Low-quality |
| S13_vs_16308\|\|lt2gene | Low-quality |
| S13_vs_16452\|\|lt2gene | Low-quality |
| S13_vs_16602\|\|lt2gene | Not-determined |
| S13_vs_16753\|\|lt2gene | Low-quality |
| S13_vs_16824\|\|lt2gene | Not-determined |
| S13_vs_16858\|\|lt2gene | Low-quality |
| S13_vs_16972\|\|lt2gene | Low-quality |
| S13_vs_17060\|\|lt2gene | Low-quality |
| S13_vs_17103\|\|lt2gene | Not-determined |
| S13_vs_17291\|\|lt2gene | Not-determined |
| S13_vs_17367\|\|lt2gene | Low-quality |
| S13_vs_17413\|\|lt2gene | Low-quality |
| S13_vs_17574\|\|lt2gene | Not-determined |
| S13_vs_17630\|\|lt2gene | Low-quality |
| S13_vs_17727\|\|lt2gene | Low-quality |
| S13_vs_17760\|\|lt2gene | Low-quality |
| S13_vs_17811\|\|lt2gene | Low-quality |
| S13_vs_17841\|\|lt2gene | Low-quality |
| S13_vs_17968\|\|lt2gene | Low-quality |
| S13_vs_18082\|\|lt2gene | Low-quality |
| S13_vs_18152\|\|lt2gene | Low-quality |
| S13_vs_18216\|\|lt2gene | Low-quality |
| S13_vs_18297\|\|lt2gene | Low-quality |
| S13_vs_18357\|\|lt2gene | Low-quality |
| S13_vs_18657\|\|lt2gene | Low-quality |
| S13_vs_18778\|\|lt2gene | Not-determined |
| S13_vs_18957\|\|lt2gene | Low-quality |
| S13_vs_19025\|\|lt2gene | Low-quality |
| S13_vs_19270\|\|lt2gene | Low-quality |
| S13_vs_19302\|\|lt2gene | Low-quality |
| S13_vs_19337\|\|lt2gene | Low-quality |
| S13_vs_19445\|\|lt2gene | Not-determined |
| S13_vs_19634\|\|lt2gene | Low-quality |
| S13_vs_19680\|\|lt2gene | Not-determined |
| S13_vs_19796\|\|lt2gene | Low-quality |
| S13_vs_19804\|\|lt2gene | Not-determined |
| S13_vs_19878\|\|lt2gene | Low-quality |
| S13_vs_19928\|\|lt2gene | Low-quality |
| S13_vs_19939\|\|lt2gene | Low-quality |
| S13_vs_20156\|\|lt2gene | Not-determined |
| S13_vs_20225\|\|lt2gene | Low-quality |
| S13_vs_20308\|\|lt2gene | Low-quality |
| S13_vs_20363\|\|lt2gene | Low-quality |
| S13_vs_20390\|\|lt2gene | Low-quality |
| S13_vs_20440\|\|lt2gene | Low-quality |
| S13_vs_20517\|\|lt2gene | Not-determined |
| S13_vs_20520\|\|lt2gene | Low-quality |
| S13_vs_20533\|\|lt2gene | Low-quality |
| S13_vs_20537\|\|lt2gene | Low-quality |
| S13_vs_20684\|\|lt2gene | Low-quality |
| S13_vs_20897\|\|lt2gene | Low-quality |
| S13_vs_20973\|\|lt2gene | Not-determined |
| S13_vs_21026\|\|lt2gene | Low-quality |
| S13_vs_21212\|\|lt2gene | Low-quality |
| S13_vs_21228\|\|lt2gene | Low-quality |
| S13_vs_21278\|\|lt2gene | Not-determined |
| S13_vs_21432\|\|lt2gene | Not-determined |
| S13_vs_21616\|\|lt2gene | Low-quality |
| S13_vs_21710\|\|lt2gene | Low-quality |
| S13_vs_21939\|\|lt2gene | Low-quality |
| S13_vs_22173\|\|lt2gene | Low-quality |
| S13_vs_22369\|\|lt2gene | Not-determined |
| S13_vs_22405\|\|lt2gene | Not-determined |
| S13_vs_22447\|\|lt2gene | Low-quality |
| S13_vs_22453\|\|lt2gene | Low-quality |
| S13_vs_22470\|\|lt2gene | Low-quality |
| S13_vs_22519\|\|lt2gene | Low-quality |
| S13_vs_22568\|\|lt2gene | Low-quality |
| S13_vs_22672\|\|lt2gene | Not-determined |
| S13_vs_22725\|\|lt2gene | Not-determined |
| S13_vs_22942\|\|lt2gene | Low-quality |
| S13_vs_22957\|\|lt2gene | Not-determined |
| S13_vs_22976\|\|lt2gene | Low-quality |
| S13_vs_23103\|\|lt2gene | Not-determined |
| S13_vs_23158\|\|lt2gene | Low-quality |
| S13_vs_23453\|\|lt2gene | Low-quality |
| S13_vs_23663\|\|lt2gene | Low-quality |
| S13_vs_23758\|\|lt2gene | Low-quality |
| S13_vs_23819\|\|lt2gene | Low-quality |
| S13_vs_23855\|\|lt2gene | Low-quality |
| S13_vs_23875\|\|lt2gene | Low-quality |
| S13_vs_23896\|\|lt2gene | Low-quality |
| S13_vs_24003\|\|lt2gene | Low-quality |
| S13_vs_24041\|\|lt2gene | Low-quality |
| S13_vs_24096\|\|lt2gene | Low-quality |
| S13_vs_24253\|\|lt2gene | Low-quality |
| S13_vs_24269\|\|lt2gene | Low-quality |
| S13_vs_24407\|\|lt2gene | Low-quality |
| S13_vs_24652\|\|lt2gene | Low-quality |
| S13_vs_24743\|\|lt2gene | Not-determined |
| S13_vs_24986\|\|lt2gene | Low-quality |
| S13_vs_25295\|\|lt2gene | Not-determined |
| S13_vs_25368\|\|lt2gene | Low-quality |
| S13_vs_25372\|\|lt2gene | Low-quality |
| S13_vs_25555\|\|lt2gene | Low-quality |
| S13_vs_25583\|\|lt2gene | Not-determined |
| S13_vs_25670\|\|lt2gene | Low-quality |
| S13_vs_25679\|\|lt2gene | Low-quality |
| S13_vs_25700\|\|lt2gene | Low-quality |
| S13_vs_25719\|\|lt2gene | Low-quality |
| S13_vs_25780\|\|lt2gene | Low-quality |
| S13_vs_25783\|\|lt2gene | Low-quality |
| S13_vs_25837\|\|lt2gene | Low-quality |
| S13_vs_25877\|\|lt2gene | Not-determined |
| S13_vs_25993\|\|lt2gene | Low-quality |
| S13_vs_26002\|\|lt2gene | Not-determined |
| S13_vs_26120\|\|lt2gene | Low-quality |
| S13_vs_26130\|\|lt2gene | Low-quality |
| S13_vs_26265\|\|lt2gene | Low-quality |
| S13_vs_26324\|\|lt2gene | Low-quality |
| S13_vs_26385\|\|lt2gene | Low-quality |
| S13_vs_26572\|\|lt2gene | Low-quality |
| S13_vs_26588\|\|lt2gene | Low-quality |
| S13_vs_26667\|\|lt2gene | Not-determined |
| S13_vs_26834\|\|lt2gene | Low-quality |
| S13_vs_27029\|\|lt2gene | Low-quality |
| S13_vs_27109\|\|lt2gene | Not-determined |
| S13_vs_27235\|\|lt2gene | Low-quality |
| S13_vs_27304\|\|lt2gene | Low-quality |
| S13_vs_27341\|\|lt2gene | Low-quality |
| S13_vs_27446\|\|lt2gene | Not-determined |
| S13_vs_27551\|\|lt2gene | Not-determined |
| S13_vs_27729\|\|lt2gene | Low-quality |
| S13_vs_27790\|\|lt2gene | Low-quality |
| S13_vs_27898\|\|lt2gene | Not-determined |
| S13_vs_27937\|\|lt2gene | Low-quality |
| S13_vs_27970\|\|lt2gene | Low-quality |
| S13_vs_28360\|\|lt2gene | Low-quality |
| S13_vs_28471\|\|lt2gene | Low-quality |
| S13_vs_28554\|\|lt2gene | Not-determined |
| S13_vs_28677\|\|lt2gene | Not-determined |
| S13_vs_28727\|\|lt2gene | Not-determined |
| S13_vs_28803\|\|lt2gene | Low-quality |
| S13_vs_29082\|\|lt2gene | Not-determined |
| S13_vs_29090\|\|lt2gene | Low-quality |
| S13_vs_29165\|\|lt2gene | Low-quality |
| S13_vs_29205\|\|lt2gene | Low-quality |
| S13_vs_29260\|\|lt2gene | Low-quality |
| S13_vs_29348\|\|lt2gene | Not-determined |
| S13_vs_29444\|\|lt2gene | Not-determined |
| S13_vs_29488\|\|lt2gene | Low-quality |
| S13_vs_29557\|\|lt2gene | Not-determined |
| S13_vs_29708\|\|lt2gene | Not-determined |
| S13_vs_29809\|\|lt2gene | Not-determined |
| S13_vs_29913\|\|lt2gene | Not-determined |
| S13_vs_29968\|\|lt2gene | Low-quality |
| S13_vs_30027\|\|lt2gene | Low-quality |
| S13_vs_30082\|\|lt2gene | Low-quality |
| S13_vs_30144\|\|lt2gene | Low-quality |
| S13_vs_30329\|\|lt2gene | Low-quality |
| S13_vs_30616\|\|lt2gene | Low-quality |
| S13_vs_30662\|\|lt2gene | Not-determined |
| S13_vs_30670\|\|lt2gene | Low-quality |
| S13_vs_30746\|\|lt2gene | Low-quality |
| S13_vs_30783\|\|lt2gene | Low-quality |
| S13_vs_30832\|\|lt2gene | Low-quality |
| S13_vs_31007\|\|lt2gene | Not-determined |
| S13_vs_31040\|\|lt2gene | Low-quality |
| S13_vs_31347\|\|lt2gene | Low-quality |
| S13_vs_31364\|\|lt2gene | Low-quality |
| S13_vs_31377\|\|lt2gene | Not-determined |
| S13_vs_31411\|\|lt2gene | Not-determined |
| S13_vs_31503\|\|lt2gene | Low-quality |
| S13_vs_31530\|\|lt2gene | Low-quality |
| S13_vs_31672\|\|lt2gene | Low-quality |
| S13_vs_31940\|\|lt2gene | Low-quality |
| S13_vs_32022\|\|lt2gene | Low-quality |
| S13_vs_32104\|\|lt2gene | Low-quality |
| S13_vs_32169\|\|lt2gene | Not-determined |
| S13_vs_32383\|\|lt2gene | Low-quality |
| S13_vs_32388\|\|lt2gene | Low-quality |
| S13_vs_32506\|\|lt2gene | Not-determined |
| S13_vs_32572\|\|lt2gene | Not-determined |
| S13_vs_32573\|\|lt2gene | Low-quality |
| S13_vs_32642\|\|lt2gene | Not-determined |
| S13_vs_32668\|\|lt2gene | Low-quality |
| S13_vs_32721\|\|lt2gene | Low-quality |
| S13_vs_32786\|\|lt2gene | Low-quality |
| S13_vs_32827\|\|lt2gene | Low-quality |
| S13_vs_32976\|\|lt2gene | Low-quality |
| S13_vs_33128\|\|lt2gene | Low-quality |
| S13_vs_33192\|\|lt2gene | Low-quality |
| S13_vs_33335\|\|lt2gene | Low-quality |
| S13_vs_33411\|\|lt2gene | Not-determined |
| S13_vs_33444\|\|lt2gene | Low-quality |
| S13_vs_33499\|\|lt2gene | Low-quality |
| S13_vs_33660\|\|lt2gene | Low-quality |
| S13_vs_33695\|\|lt2gene | Not-determined |
| S13_vs_33778\|\|lt2gene | Not-determined |
| S13_vs_33915\|\|lt2gene | Low-quality |
| S13_vs_33994\|\|lt2gene | Low-quality |
| S13_vs_33999\|\|lt2gene | Low-quality |
| S13_vs_34010\|\|lt2gene | Low-quality |
| S13_vs_34203\|\|lt2gene | Not-determined |
| S13_vs_34208\|\|lt2gene | Low-quality |
| S13_vs_34255\|\|lt2gene | Not-determined |
| S13_vs_34359\|\|lt2gene | Low-quality |
| S13_vs_34368\|\|lt2gene | Not-determined |
| S13_vs_34650\|\|lt2gene | Low-quality |
| S13_vs_34697\|\|lt2gene | Not-determined |
| S13_vs_34797\|\|lt2gene | Low-quality |
| S13_vs_34850\|\|lt2gene | Not-determined |
| S13_vs_35055\|\|lt2gene | Low-quality |
| S13_vs_35160\|\|lt2gene | Low-quality |
| S13_vs_35255\|\|lt2gene | Not-determined |
| S13_vs_35268\|\|lt2gene | Low-quality |
| S13_vs_35374\|\|lt2gene | Not-determined |
| S13_vs_35576\|\|lt2gene | Low-quality |
| S13_vs_35803\|\|lt2gene | Low-quality |
| S13_vs_35879\|\|lt2gene | Low-quality |
| S13_vs_35897\|\|lt2gene | Low-quality |
| S13_vs_36066\|\|lt2gene | Low-quality |
| S13_vs_36094\|\|lt2gene | Not-determined |
| S13_vs_36210\|\|lt2gene | Low-quality |
| S13_vs_36240\|\|lt2gene | Not-determined |
| S13_vs_36246\|\|lt2gene | Not-determined |
| S13_vs_36281\|\|lt2gene | Low-quality |
| S13_vs_36415\|\|lt2gene | Low-quality |
| S13_vs_36417\|\|lt2gene | Low-quality |
| S13_vs_36455\|\|lt2gene | Low-quality |
| S13_vs_36686\|\|lt2gene | Low-quality |
| S13_vs_36690\|\|lt2gene | Low-quality |
| S13_vs_36739\|\|lt2gene | Low-quality |
| S13_vs_36756\|\|lt2gene | Not-determined |
| S13_vs_36899\|\|lt2gene | Low-quality |
| S13_vs_37007\|\|lt2gene | Low-quality |
| S13_vs_37105\|\|lt2gene | Low-quality |
| S13_vs_37256\|\|lt2gene | Low-quality |
| S13_vs_37301\|\|lt2gene | Low-quality |
| S13_vs_37409\|\|lt2gene | Low-quality |
| S13_vs_37452\|\|lt2gene | Low-quality |
| S13_vs_37456\|\|lt2gene | Low-quality |
| S13_vs_37608\|\|lt2gene | Low-quality |
| S13_vs_37615\|\|lt2gene | Not-determined |
| S13_vs_37646\|\|lt2gene | Not-determined |
| S13_vs_37762\|\|lt2gene | Low-quality |
| S13_vs_37775\|\|lt2gene | Low-quality |
| S13_vs_37783\|\|lt2gene | Not-determined |
| S13_vs_37894\|\|lt2gene | Not-determined |
| S13_vs_37937\|\|lt2gene | Low-quality |
| S13_vs_37954\|\|lt2gene | Not-determined |
| S13_vs_37987\|\|lt2gene | Not-determined |
| S13_vs_38015\|\|lt2gene | Not-determined |
| S13_vs_38113\|\|lt2gene | Low-quality |
| S13_vs_38165\|\|lt2gene | Not-determined |
| S13_vs_38344\|\|lt2gene | Not-determined |
| S13_vs_38425\|\|lt2gene | Low-quality |
| S13_vs_38578\|\|lt2gene | Not-determined |
| S13_vs_38801\|\|lt2gene | Low-quality |
| S13_vs_38813\|\|lt2gene | Low-quality |
| S13_vs_38940\|\|lt2gene | Low-quality |
| S13_vs_39079\|\|lt2gene | Low-quality |
| S13_vs_39090\|\|lt2gene | Low-quality |
| S13_vs_39110\|\|lt2gene | Not-determined |
| S13_vs_39319\|\|lt2gene | Not-determined |
| S13_vs_39462\|\|lt2gene | Not-determined |
| S13_vs_39482\|\|lt2gene | Not-determined |
| S13_vs_39515\|\|lt2gene | Low-quality |
| S13_vs_39523\|\|lt2gene | Low-quality |
| S13_vs_39564\|\|lt2gene | Low-quality |
| S13_vs_39625\|\|lt2gene | Low-quality |
| S13_vs_39750\|\|lt2gene | Not-determined |
| S13_vs_39801\|\|lt2gene | Low-quality |
| S13_vs_39883\|\|lt2gene | Low-quality |
| S13_vs_40075\|\|lt2gene | Low-quality |
| S13_vs_40130\|\|lt2gene | Not-determined |
| S13_vs_40182\|\|lt2gene | Not-determined |
| S13_vs_40278\|\|lt2gene | Low-quality |
| S13_vs_40410\|\|lt2gene | Low-quality |
| S13_vs_40463\|\|lt2gene | Not-determined |
| S13_vs_40477\|\|lt2gene | Not-determined |
| S13_vs_40511\|\|lt2gene | Not-determined |
| S13_vs_40836\|\|lt2gene | Not-determined |
| S13_vs_41128\|\|lt2gene | Not-determined |
| S13_vs_41338\|\|lt2gene | Not-determined |
| S13_vs_41343\|\|lt2gene | Low-quality |
| S13_vs_41603\|\|lt2gene | Not-determined |
| S13_vs_42020\|\|lt2gene | Not-determined |
| S13_vs_42127\|\|lt2gene | Low-quality |
| S13_vs_42143\|\|lt2gene | Low-quality |
| S13_vs_42492\|\|lt2gene | Not-determined |
| S13_vs_42496\|\|lt2gene | Not-determined |
| S13_vs_42510\|\|lt2gene | Low-quality |
| S13_vs_42689\|\|lt2gene | Low-quality |
| S13_vs_42975\|\|lt2gene | Low-quality |
| S13_vs_43159\|\|lt2gene | Low-quality |
| S13_vs_43282\|\|lt2gene | Low-quality |
| S13_vs_43284\|\|lt2gene | Low-quality |
| S13_vs_43462\|\|lt2gene | Low-quality |
| S13_vs_43564\|\|lt2gene | Not-determined |
| S13_vs_43900\|\|lt2gene | Low-quality |
| S13_vs_44044\|\|lt2gene | Low-quality |
| S13_vs_44207\|\|lt2gene | Low-quality |
| S13_vs_44214\|\|lt2gene | Not-determined |
| S13_vs_44256\|\|lt2gene | Low-quality |
| S13_vs_44270\|\|lt2gene | Not-determined |
| S13_vs_44307\|\|lt2gene | Low-quality |
| S13_vs_44350\|\|lt2gene | Low-quality |
| S13_vs_44606\|\|lt2gene | Low-quality |
| S13_vs_44927\|\|lt2gene | Low-quality |
| S13_vs_45027\|\|lt2gene | Not-determined |
| S13_vs_45258\|\|lt2gene | Low-quality |
| S13_vs_45312\|\|lt2gene | Low-quality |
| S13_vs_45473\|\|lt2gene | Not-determined |
| S13_vs_45502\|\|lt2gene | Low-quality |
| S13_vs_45608\|\|lt2gene | Low-quality |
| S13_vs_45752\|\|lt2gene | Not-determined |
| S13_vs_46071\|\|lt2gene | Low-quality |
| S13_vs_46407\|\|lt2gene | Low-quality |
| S13_vs_46686\|\|lt2gene | Low-quality |
| S13_vs_46818\|\|lt2gene | Not-determined |
| S13_vs_46883\|\|lt2gene | Low-quality |
| S13_vs_47039\|\|lt2gene | Low-quality |
| S13_vs_47066\|\|lt2gene | Low-quality |
| S13_vs_47245\|\|lt2gene | Not-determined |
| S13_vs_47264\|\|lt2gene | Not-determined |
| S13_vs_47286\|\|lt2gene | Not-determined |
| S13_vs_47350\|\|lt2gene | Low-quality |
| S13_vs_47447\|\|lt2gene | Low-quality |
| S13_vs_47498\|\|lt2gene | Not-determined |
| S13_vs_47678\|\|lt2gene | Not-determined |
| S13_vs_47708\|\|lt2gene | Low-quality |
| S13_vs_47736\|\|lt2gene | Low-quality |
| S13_vs_47766\|\|lt2gene | Low-quality |
| S13_vs_47801\|\|lt2gene | Not-determined |
| S13_vs_47938\|\|lt2gene | Low-quality |
| S13_vs_48017\|\|lt2gene | Not-determined |
| S13_vs_48407\|\|lt2gene | Not-determined |
| S13_vs_48448\|\|lt2gene | Low-quality |
| S13_vs_48480\|\|lt2gene | Not-determined |
| S13_vs_48694\|\|lt2gene | Low-quality |
| S13_vs_48786\|\|lt2gene | Low-quality |
| S13_vs_49338\|\|lt2gene | Low-quality |
| S13_vs_49429\|\|lt2gene | Low-quality |
| S13_vs_49505\|\|lt2gene | Low-quality |
| S13_vs_49602\|\|lt2gene | Low-quality |
| S13_vs_49647\|\|lt2gene | Not-determined |
| S13_vs_49748\|\|lt2gene | Not-determined |
| S13_vs_49985\|\|lt2gene | Not-determined |
| S13_vs_50112\|\|lt2gene | Low-quality |
| S13_vs_50138\|\|lt2gene | Low-quality |
| S13_vs_50376\|\|lt2gene | Low-quality |
| S13_vs_50433\|\|lt2gene | Low-quality |
| S13_vs_50639\|\|lt2gene | Low-quality |
| S13_vs_51203\|\|lt2gene | Not-determined |
| S13_vs_51309\|\|lt2gene | Not-determined |
| S13_vs_51561\|\|lt2gene | Low-quality |
| S13_vs_51574\|\|lt2gene | Low-quality |
| S13_vs_51636\|\|lt2gene | Not-determined |
| S13_vs_51669\|\|lt2gene | Not-determined |
| S13_vs_51955\|\|lt2gene | Low-quality |
| S13_vs_52039\|\|lt2gene | Not-determined |
| S13_vs_52080\|\|lt2gene | Low-quality |
| S13_vs_52226\|\|lt2gene | Not-determined |
| S13_vs_52253\|\|lt2gene | Not-determined |
| S13_vs_52307\|\|lt2gene | Low-quality |
| S13_vs_52468\|\|lt2gene | Not-determined |
| S13_vs_52530\|\|lt2gene | Low-quality |
| S13_vs_52565\|\|lt2gene | Low-quality |
| S13_vs_52602\|\|lt2gene | Low-quality |
| S13_vs_52611\|\|lt2gene | Not-determined |
| S13_vs_52662\|\|lt2gene | Not-determined |
| S13_vs_52769\|\|lt2gene | Low-quality |
| S13_vs_52795\|\|lt2gene | Low-quality |
| S13_vs_53211\|\|lt2gene | Not-determined |
| S13_vs_53444\|\|lt2gene | Low-quality |
| S13_vs_53762\|\|lt2gene | Low-quality |
| S13_vs_53810\|\|lt2gene | Low-quality |
| S13_vs_53818\|\|lt2gene | Low-quality |
| S13_vs_54246\|\|lt2gene | Not-determined |
| S13_vs_54438\|\|lt2gene | Low-quality |
| S13_vs_54605\|\|lt2gene | Low-quality |
| S13_vs_54635\|\|lt2gene | Low-quality |
| S13_vs_54663\|\|lt2gene | Low-quality |
| S13_vs_54787\|\|lt2gene | Not-determined |
| S13_vs_55012\|\|lt2gene | Low-quality |
| S13_vs_55031\|\|lt2gene | Low-quality |
| S13_vs_55244\|\|lt2gene | Not-determined |
| S13_vs_55314\|\|lt2gene | Not-determined |
| S13_vs_55414\|\|lt2gene | Not-determined |
| S13_vs_55505\|\|lt2gene | Not-determined |
| S13_vs_55614\|\|lt2gene | Low-quality |
| S13_vs_55665\|\|lt2gene | Not-determined |
| S13_vs_55820\|\|lt2gene | Low-quality |
| S13_vs_55959\|\|lt2gene | Low-quality |
| S13_vs_56024\|\|lt2gene | Low-quality |
| S13_vs_56585\|\|lt2gene | Not-determined |
| S13_vs_56612\|\|lt2gene | Low-quality |
| S13_vs_56744\|\|lt2gene | Low-quality |
| S13_vs_56807\|\|lt2gene | Low-quality |
| S13_vs_56901\|\|lt2gene | Low-quality |
| S13_vs_57076\|\|lt2gene | Low-quality |
| S13_vs_57105\|\|lt2gene | Low-quality |
| S13_vs_57154\|\|lt2gene | Low-quality |
| S13_vs_57184\|\|lt2gene | Not-determined |
| S13_vs_57245\|\|lt2gene | Not-determined |
| S13_vs_57307\|\|lt2gene | Low-quality |
| S13_vs_57461\|\|lt2gene | Not-determined |
| S13_vs_57487\|\|lt2gene | Not-determined |
| S13_vs_57531\|\|lt2gene | Low-quality |
| S13_vs_57780\|\|lt2gene | Low-quality |
| S13_vs_57888\|\|lt2gene | Low-quality |
| S13_vs_58103\|\|lt2gene | Low-quality |
| S13_vs_58128\|\|lt2gene | Low-quality |
| S13_vs_58503\|\|lt2gene | Low-quality |
| S13_vs_58600\|\|lt2gene | Not-determined |
| S13_vs_58910\|\|lt2gene | Low-quality |
| S13_vs_58917\|\|lt2gene | Low-quality |
| S13_vs_59104\|\|lt2gene | Not-determined |
| S13_vs_59163\|\|lt2gene | Not-determined |
| S13_vs_59361\|\|lt2gene | Low-quality |
| S13_vs_59434\|\|lt2gene | Low-quality |
| S13_vs_59532\|\|lt2gene | Low-quality |
| S13_vs_59560\|\|lt2gene | Low-quality |
| S13_vs_59625\|\|lt2gene | Low-quality |
| S13_vs_60276\|\|lt2gene | Low-quality |
| S13_vs_60311\|\|lt2gene | Not-determined |
| S13_vs_60351\|\|lt2gene | Low-quality |
| S13_vs_60370\|\|lt2gene | Low-quality |

<table>
<tr><td>S13_vs_60689||lt2gene</td><td>Low-quality</td></tr>
<tr><td>S13_vs_60714||lt2gene</td><td>Not-determined</td></tr>
<tr><td>S13_vs_60772||lt2gene</td><td>Low-quality</td></tr>
<tr><td>S13_vs_60797||lt2gene</td><td>Not-determined</td></tr>
<tr><td>S13_vs_60961||lt2gene</td><td>Low-quality</td></tr>
<tr><td>S13_vs_61102||lt2gene</td><td>Low-quality</td></tr>
<tr><td>S13_vs_61200||lt2gene</td><td>Low-quality</td></tr>
<tr><td>S13_vs_61288||lt2gene</td><td>Not-determined</td></tr>
<tr><td>S13_vs_61492||lt2gene</td><td>Not-determined</td></tr>
<tr><td>S13_vs_61585||lt2gene</td><td>Low-quality</td></tr>
<tr><td>S13_vs_61607||lt2gene</td><td>Low-quality</td></tr>
<tr><td>S13_vs_61643||lt2gene</td><td>Low-quality</td></tr>
<tr><td>S13_vs_61659||lt2gene</td><td>Low-quality</td></tr>
<tr><td>S13_vs_61670||lt2gene</td><td>Low-quality</td></tr>
<tr><td>S13_vs_62023||lt2gene</td><td>Low-quality</td></tr>
<tr><td>S13_vs_62038||lt2gene</td><td>Low-quality</td></tr>
<tr><td>S13_vs_62087||lt2gene</td><td>Low-quality</td></tr>
<tr><td>S13_vs_62178||lt2gene</td><td>Low-quality</td></tr>
<tr><td>S13_vs_62255||lt2gene</td><td>Low-quality</td></tr>
<tr><td>S13_vs_62266||lt2gene</td><td>Low-quality</td></tr>
<tr><td>S13_vs_62551||lt2gene</td><td>Not-determined</td></tr>
<tr><td>S13_vs_62585||lt2gene</td><td>Low-quality</td></tr>
<tr><td>S13_vs_62600||lt2gene</td><td>Low-quality</td></tr>
<tr><td>S13_vs_62665||lt2gene</td><td>Low-quality</td></tr>
<tr><td>S13_vs_62766||lt2gene</td><td>Not-determined</td></tr>
<tr><td>S13_vs_62812||lt2gene</td><td>Not-determined</td></tr>
<tr><td>S13_vs_62886||lt2gene</td><td>Low-quality</td></tr>
<tr><td>S13_vs_63042||lt2gene</td><td>Low-quality</td></tr>
<tr><td>S13_vs_63055||lt2gene</td><td>Not-determined</td></tr>
<tr><td>S13_vs_63070||lt2gene</td><td>Low-quality</td></tr>
<tr><td>S13_vs_63073||lt2gene</td><td>Low-quality</td></tr>
<tr><td>S13_vs_63147||lt2gene</td><td>Not-determined</td></tr>
<tr><td>S13_vs_63203||lt2gene</td><td>Low-quality</td></tr>
<tr><td>S13_vs_63245||lt2gene</td><td>Low-quality</td></tr>
<tr><td>S13_vs_63362||lt2gene</td><td>Not-determined</td></tr>
<tr><td>S13_vs_63612||lt2gene</td><td>Low-quality</td></tr>
<tr><td>S13_vs_63785||lt2gene</td><td>Low-quality</td></tr>
<tr><td>S13_vs_64050||lt2gene</td><td>Not-determined</td></tr>
<tr><td>S13_vs_64091||lt2gene</td><td>Low-quality</td></tr>
<tr><td>S13_vs_64153||lt2gene</td><td>Low-quality</td></tr>
<tr><td>S13_vs_64165||lt2gene</td><td>Not-determined</td></tr>
<tr><td>S13_vs_64304||lt2gene</td><td>Not-determined</td></tr>
<tr><td>S13_vs_64315||lt2gene</td><td>Low-quality</td></tr>
<tr><td>S13_vs_64338||lt2gene</td><td>Not-determined</td></tr>
<tr><td>S13_vs_64414||lt2gene</td><td>Low-quality</td></tr>
<tr><td>S13_vs_64417||lt2gene</td><td>Not-determined</td></tr>
<tr><td>S13_vs_64510||lt2gene</td><td>Not-determined</td></tr>
<tr><td>S13_vs_64533||lt2gene</td><td>Not-determined</td></tr>
<tr><td>S13_vs_64624||lt2gene</td><td>Low-quality</td></tr>
<tr><td>S13_vs_64725||lt2gene</td><td>Low-quality</td></tr>
<tr><td>S13_vs_64799||lt2gene</td><td>Not-determined</td></tr>
<tr><td>S13_vs_64828||lt2gene</td><td>Low-quality</td></tr>
<tr><td>S13_vs_64858||lt2gene</td><td>Low-quality</td></tr>
<tr><td>S13_vs_64946||lt2gene</td><td>Not-determined</td></tr>
<tr><td>S13_vs_64977||lt2gene</td><td>Low-quality</td></tr>
<tr><td>S13_vs_65037||lt2gene</td><td>Low-quality</td></tr>
<tr><td>S13_vs_65095||lt2gene</td><td>Low-quality</td></tr>
<tr><td>S13_vs_65145||lt2gene</td><td>Low-quality</td></tr>
<tr><td>S13_vs_65175||lt2gene</td><td>Not-determined</td></tr>
<tr><td>S13_vs_65207||lt2gene</td><td>Not-determined</td></tr>
<tr><td>S13_vs_65226||lt2gene</td><td>Low-quality</td></tr>
<tr><td>S13_vs_65291||lt2gene</td><td>Low-quality</td></tr>
<tr><td>S13_vs_65622||lt2gene</td><td>Low-quality</td></tr>
<tr><td>S13_vs_65795||lt2gene</td><td>Not-determined</td></tr>
<tr><td>S13_vs_65833||lt2gene</td><td>Low-quality</td></tr>
<tr><td>S13_vs_65959||lt2gene</td><td>Not-determined</td></tr>
<tr><td>S13_vs_65964||lt2gene</td><td>Low-quality</td></tr>
<tr><td>S13_vs_66068||lt2gene</td><td>Low-quality</td></tr>
<tr><td>S13_vs_66186||lt2gene</td><td>Low-quality</td></tr>
<tr><td>S13_vs_66255||lt2gene</td><td>Not-determined</td></tr>
<tr><td>S13_vs_66273||lt2gene</td><td>Low-quality</td></tr>
<tr><td>S13_vs_66298||lt2gene</td><td>Not-determined</td></tr>
<tr><td>S13_vs_66306||lt2gene</td><td>Low-quality</td></tr>
<tr><td>S13_vs_66389||lt2gene</td><td>Low-quality</td></tr>
<tr><td>S13_vs_66609||lt2gene</td><td>Not-determined</td></tr>
<tr><td>S13_vs_66670||lt2gene</td><td>Low-quality</td></tr>
<tr><td>S13_vs_66820||lt2gene</td><td>Low-quality</td></tr>
<tr><td>S13_vs_67032||lt2gene</td><td>Low-quality</td></tr>
<tr><td>S13_vs_67204||lt2gene</td><td>Low-quality</td></tr>
<tr><td>S13_vs_67317||lt2gene</td><td>Low-quality</td></tr>
<tr><td>S13_vs_67526||lt2gene</td><td>Not-determined</td></tr>
<tr><td>S13_vs_67657||lt2gene</td><td>Low-quality</td></tr>
<tr><td>S13_vs_67783||lt2gene</td><td>Low-quality</td></tr>
<tr><td>S13_vs_67866||lt2gene</td><td>Not-determined</td></tr>
<tr><td>S13_vs_68080||lt2gene</td><td>Not-determined</td></tr>
<tr><td>S13_vs_68306||lt2gene</td><td>Low-quality</td></tr>
<tr><td>S13_vs_68331||lt2gene</td><td>Low-quality</td></tr>
<tr><td>S13_vs_68445||lt2gene</td><td>Low-quality</td></tr>
<tr><td>S13_vs_68446||lt2gene</td><td>Not-determined</td></tr>
<tr><td>S13_vs_68521||lt2gene</td><td>Low-quality</td></tr>
<tr><td>S13_vs_68533||lt2gene</td><td>Low-quality</td></tr>
<tr><td>S13_vs_68589||lt2gene</td><td>Low-quality</td></tr>
<tr><td>S13_vs_68651||lt2gene</td><td>Not-determined</td></tr>
<tr><td>S13_vs_68736||lt2gene</td><td>Low-quality</td></tr>
<tr><td>S13_vs_68744||lt2gene</td><td>Low-quality</td></tr>
<tr><td>S13_vs_69222||lt2gene</td><td>Low-quality</td></tr>
<tr><td>S13_vs_69344||lt2gene</td><td>Low-quality</td></tr>
<tr><td>S13_vs_69369||lt2gene</td><td>Low-quality</td></tr>
<tr><td>S13_vs_69647||lt2gene</td><td>Low-quality</td></tr>
<tr><td>S13_vs_69911||lt2gene</td><td>Low-quality</td></tr>
<tr><td>S13_vs_70064||lt2gene</td><td>Low-quality</td></tr>
<tr><td>S13_vs_70145||lt2gene</td><td>Low-quality</td></tr>
<tr><td>S13_vs_70261||lt2gene</td><td>Low-quality</td></tr>
<tr><td>S13_vs_70496||lt2gene</td><td>Low-quality</td></tr>
<tr><td>S13_vs_70536||lt2gene</td><td>Low-quality</td></tr>
<tr><td>S13_vs_70542||lt2gene</td><td>Low-quality</td></tr>
<tr><td>S13_vs_70610||lt2gene</td><td>Low-quality</td></tr>
<tr><td>S13_vs_70788||lt2gene</td><td>Low-quality</td></tr>
<tr><td>S13_vs_70888||lt2gene</td><td>Low-quality</td></tr>
<tr><td>S13_vs_70935||lt2gene</td><td>Low-quality</td></tr>
<tr><td>S13_vs_70979||lt2gene</td><td>Not-determined</td></tr>
<tr><td>S13_vs_71256||lt2gene</td><td>Not-determined</td></tr>
<tr><td>S13_vs_71546||lt2gene</td><td>Low-quality</td></tr>
<tr><td>S13_vs_71601||lt2gene</td><td>Not-determined</td></tr>
<tr><td>S13_vs_71714||lt2gene</td><td>Low-quality</td></tr>
<tr><td>S13_vs_72004||lt2gene</td><td>Not-determined</td></tr>
<tr><td>S13_vs_72301||lt2gene</td><td>Low-quality</td></tr>
<tr><td>S13_vs_72323||lt2gene</td><td>Low-quality</td></tr>
<tr><td>S13_vs_72396||lt2gene</td><td>Low-quality</td></tr>
<tr><td>S13_vs_72397||lt2gene</td><td>Low-quality</td></tr>
<tr><td>S13_vs_72481||lt2gene</td><td>Low-quality</td></tr>
<tr><td>S13_vs_72535||lt2gene</td><td>Low-quality</td></tr>
<tr><td>S13_vs_72694||lt2gene</td><td>Low-quality</td></tr>
<tr><td>S13_vs_72695||lt2gene</td><td>Low-quality</td></tr>
<tr><td>S13_vs_72738||lt2gene</td><td>Low-quality</td></tr>
<tr><td>S13_vs_72798||lt2gene</td><td>Low-quality</td></tr>
<tr><td>S13_vs_73527||lt2gene</td><td>Low-quality</td></tr>
<tr><td>S13_vs_73730||lt2gene</td><td>Low-quality</td></tr>
<tr><td>S13_vs_73731||lt2gene</td><td>Low-quality</td></tr>
<tr><td>S13_vs_73882||lt2gene</td><td>Not-determined</td></tr>
<tr><td>S13_vs_73960||lt2gene</td><td>Not-determined</td></tr>
<tr><td>S13_vs_74265||lt2gene</td><td>Not-determined</td></tr>
<tr><td>S13_vs_74338||lt2gene</td><td>Not-determined</td></tr>
<tr><td>S13_vs_74654||lt2gene</td><td>Not-determined</td></tr>
<tr><td>S13_vs_74902||lt2gene</td><td>Low-quality</td></tr>
<tr><td>S13_vs_74973||lt2gene</td><td>Low-quality</td></tr>
<tr><td>S13_vs_75018||lt2gene</td><td>Low-quality</td></tr>
<tr><td>S13_vs_75047||lt2gene</td><td>Low-quality</td></tr>
<tr><td>S13_vs_75075||lt2gene</td><td>Low-quality</td></tr>
<tr><td>S13_vs_75172||lt2gene</td><td>Low-quality</td></tr>
<tr><td>S13_vs_75196||lt2gene</td><td>Not-determined</td></tr>
<tr><td>S13_vs_75341||lt2gene</td><td>Low-quality</td></tr>
<tr><td>S13_vs_75406||lt2gene</td><td>Low-quality</td></tr>
<tr><td>S13_vs_75443||lt2gene</td><td>Low-quality</td></tr>
<tr><td>S13_vs_75767||lt2gene</td><td>Low-quality</td></tr>
<tr><td>S13_vs_75771||lt2gene</td><td>Not-determined</td></tr>
<tr><td>S13_vs_76029||lt2gene</td><td>Low-quality</td></tr>
<tr><td>S13_vs_76157||lt2gene</td><td>Low-quality</td></tr>
<tr><td>S13_vs_76248||lt2gene</td><td>Low-quality</td></tr>
<tr><td>S13_vs_76325||lt2gene</td><td>Low-quality</td></tr>
<tr><td>S13_vs_76334||lt2gene</td><td>Low-quality</td></tr>
<tr><td>S13_vs_76387||lt2gene</td><td>Low-quality</td></tr>
<tr><td>S13_vs_76417||lt2gene</td><td>Low-quality</td></tr>
<tr><td>S13_vs_76500||lt2gene</td><td>Low-quality</td></tr>
<tr><td>S13_vs_76585||lt2gene</td><td>Not-determined</td></tr>
<tr><td>S13_vs_76641||lt2gene</td><td>Low-quality</td></tr>
<tr><td>S13_vs_76673||lt2gene</td><td>Not-determined</td></tr>
<tr><td>S13_vs_76704||lt2gene</td><td>Not-determined</td></tr>
<tr><td>S13_vs_76818||lt2gene</td><td>Low-quality</td></tr>
<tr><td>S13_vs_76850||lt2gene</td><td>Low-quality</td></tr>
<tr><td>S13_vs_76880||lt2gene</td><td>Not-determined</td></tr>
<tr><td>S13_vs_76904||lt2gene</td><td>Low-quality</td></tr>
<tr><td>S13_vs_77038||lt2gene</td><td>Low-quality</td></tr>
<tr><td>S13_vs_77068||lt2gene</td><td>Low-quality</td></tr>
<tr><td>S13_vs_77164||lt2gene</td><td>Low-quality</td></tr>
<tr><td>S13_vs_77280||lt2gene</td><td>Not-determined</td></tr>
<tr><td>S13_vs_77286||lt2gene</td><td>Low-quality</td></tr>
<tr><td>S13_vs_77478||lt2gene</td><td>Low-quality</td></tr>
<tr><td>S13_vs_77499||lt2gene</td><td>Low-quality</td></tr>
<tr><td>S13_vs_77580||lt2gene</td><td>Low-quality</td></tr>
<tr><td>S13_vs_77620||lt2gene</td><td>Not-determined</td></tr>
<tr><td>S13_vs_77650||lt2gene</td><td>Low-quality</td></tr>
<tr><td>S13_vs_77853||lt2gene</td><td>Low-quality</td></tr>
<tr><td>S13_vs_77987||lt2gene</td><td>Low-quality</td></tr>
<tr><td>S13_vs_78110||lt2gene</td><td>Low-quality</td></tr>
<tr><td>S13_vs_78225||lt2gene</td><td>Not-determined</td></tr>
<tr><td>S13_vs_78269||lt2gene</td><td>Low-quality</td></tr>
<tr><td>S13_vs_78313||lt2gene</td><td>Not-determined</td></tr>
<tr><td>S13_vs_78458||lt2gene</td><td>Low-quality</td></tr>
<tr><td>S13_vs_78477||lt2gene</td><td>Low-quality</td></tr>
<tr><td>S13_vs_78524||lt2gene</td><td>Low-quality</td></tr>
<tr><td>S13_vs_78562||lt2gene</td><td>Low-quality</td></tr>
<tr><td>S13_vs_78567||lt2gene</td><td>Low-quality</td></tr>
<tr><td>S13_vs_78943||lt2gene</td><td>Low-quality</td></tr>
<tr><td>S13_vs_78975||lt2gene</td><td>Low-quality</td></tr>
<tr><td>S13_vs_79023||lt2gene</td><td>Not-determined</td></tr>
<tr><td>S13_vs_79207||lt2gene</td><td>Low-quality</td></tr>
<tr><td>S13_vs_79251||lt2gene</td><td>Low-quality</td></tr>
<tr><td>S13_vs_79359||lt2gene</td><td>Low-quality</td></tr>
<tr><td>S13_vs_79522||lt2gene</td><td>Low-quality</td></tr>
<tr><td>S13_vs_79539||lt2gene</td><td>Low-quality</td></tr>
<tr><td>S13_vs_79751||lt2gene</td><td>Low-quality</td></tr>
<tr><td>S13_vs_79765||lt2gene</td><td>Not-determined</td></tr>
<tr><td>S13_vs_79937||lt2gene</td><td>Low-quality</td></tr>
<tr><td>S13_vs_79956||lt2gene</td><td>Not-determined</td></tr>
<tr><td>S13_vs_80005||lt2gene</td><td>Low-quality</td></tr>
<tr><td>S13_vs_80188||lt2gene</td><td>Not-determined</td></tr>
<tr><td>S13_vs_80298||lt2gene</td><td>Low-quality</td></tr>
<tr><td>S13_vs_80360||lt2gene</td><td>Low-quality</td></tr>
<tr><td>S13_vs_80533||lt2gene</td><td>Low-quality</td></tr>
<tr><td>S13_vs_80738||lt2gene</td><td>Low-quality</td></tr>
<tr><td>S13_vs_80750||lt2gene</td><td>Low-quality</td></tr>
<tr><td>S13_vs_80759||lt2gene</td><td>Not-determined</td></tr>
<tr><td>S13_vs_81122||lt2gene</td><td>Low-quality</td></tr>
<tr><td>S13_vs_81353||lt2gene</td><td>Low-quality</td></tr>
<tr><td>S13_vs_81640||lt2gene</td><td>Low-quality</td></tr>
<tr><td>S13_vs_81715||lt2gene</td><td>Not-determined</td></tr>
<tr><td>S13_vs_81753||lt2gene</td><td>Not-determined</td></tr>
<tr><td>S13_vs_81881||lt2gene</td><td>Not-determined</td></tr>
<tr><td>S13_vs_81925||lt2gene</td><td>Not-determined</td></tr>
<tr><td>S13_vs_81929||lt2gene</td><td>Low-quality</td></tr>
<tr><td>S13_vs_81999||lt2gene</td><td>Low-quality</td></tr>
<tr><td>S13_vs_82083||lt2gene</td><td>Low-quality</td></tr>
<tr><td>S13_vs_82098||lt2gene</td><td>Low-quality</td></tr>
<tr><td>S13_vs_82204||lt2gene</td><td>Low-quality</td></tr>
<tr><td>S13_vs_82280||lt2gene</td><td>Low-quality</td></tr>
<tr><td>S13_vs_82300||lt2gene</td><td>Not-determined</td></tr>
<tr><td>S13_vs_82309||lt2gene</td><td>Low-quality</td></tr>
<tr><td>S13_vs_82352||lt2gene</td><td>Not-determined</td></tr>
<tr><td>S13_vs_82377||lt2gene</td><td>Not-determined</td></tr>
<tr><td>S13_vs_82437||lt2gene</td><td>Low-quality</td></tr>
<tr><td>S13_vs_82679||lt2gene</td><td>Low-quality</td></tr>
<tr><td>S13_vs_82820||lt2gene</td><td>Not-determined</td></tr>
<tr><td>S13_vs_82851||lt2gene</td><td>Low-quality</td></tr>
<tr><td>S13_vs_82856||lt2gene</td><td>Low-quality</td></tr>
<tr><td>S13_vs_83020||lt2gene</td><td>Low-quality</td></tr>
<tr><td>S13_vs_83040||lt2gene</td><td>Low-quality</td></tr>
<tr><td>S13_vs_83205||lt2gene</td><td>Not-determined</td></tr>
<tr><td>S13_vs_83359||lt2gene</td><td>Low-quality</td></tr>
<tr><td>S13_vs_83390||lt2gene</td><td>Low-quality</td></tr>
<tr><td>S13_vs_83423||lt2gene</td><td>Low-quality</td></tr>
<tr><td>S13_vs_83969||lt2gene</td><td>Not-determined</td></tr>
<tr><td>S13_vs_84021||lt2gene</td><td>Not-determined</td></tr>
<tr><td>S13_vs_84032||lt2gene</td><td>Not-determined</td></tr>
<tr><td>S13_vs_84057||lt2gene</td><td>Not-determined</td></tr>
<tr><td>S13_vs_84221||lt2gene</td><td>Low-quality</td></tr>
<tr><td>S13_vs_84307||lt2gene</td><td>Low-quality</td></tr>
<tr><td>S13_vs_84313||lt2gene</td><td>Not-determined</td></tr>
<tr><td>S13_vs_84453||lt2gene</td><td>Low-quality</td></tr>
<tr><td>S13_vs_84540||lt2gene</td><td>Low-quality</td></tr>
<tr><td>S13_vs_84561||lt2gene</td><td>Low-quality</td></tr>
<tr><td>S13_vs_84596||lt2gene</td><td>Low-quality</td></tr>
<tr><td>S13_vs_84945||lt2gene</td><td>Low-quality</td></tr>
<tr><td>S13_vs_85223||lt2gene</td><td>Low-quality</td></tr>
<tr><td>S13_vs_85239||lt2gene</td><td>Low-quality</td></tr>
<tr><td>S13_vs_85680||lt2gene</td><td>Not-determined</td></tr>
<tr><td>S13_vs_85806||lt2gene</td><td>Low-quality</td></tr>
<tr><td>S13_vs_85808||lt2gene</td><td>Low-quality</td></tr>
<tr><td>S13_vs_85867||lt2gene</td><td>Low-quality</td></tr>
<tr><td>S13_vs_86014||lt2gene</td><td>Low-quality</td></tr>
<tr><td>S13_vs_86083||lt2gene</td><td>Low-quality</td></tr>
<tr><td>S13_vs_86188||lt2gene</td><td>Low-quality</td></tr>
<tr><td>S13_vs_86246||lt2gene</td><td>Not-determined</td></tr>
<tr><td>S13_vs_86314||lt2gene</td><td>Low-quality</td></tr>
<tr><td>S13_vs_86327||lt2gene</td><td>Low-quality</td></tr>
<tr><td>S13_vs_86494||lt2gene</td><td>Not-determined</td></tr>
<tr><td>S13_vs_86516||lt2gene</td><td>Not-determined</td></tr>
<tr><td>S13_vs_86577||lt2gene</td><td>Low-quality</td></tr>
<tr><td>S13_vs_86929||lt2gene</td><td>Not-determined</td></tr>
<tr><td>S13_vs_86961||lt2gene</td><td>Not-determined</td></tr>
<tr><td>S13_vs_87100||lt2gene</td><td>Not-determined</td></tr>
<tr><td>S13_vs_87184||lt2gene</td><td>Not-determined</td></tr>
<tr><td>S13_vs_87273||lt2gene</td><td>Low-quality</td></tr>
<tr><td>S13_vs_87288||lt2gene</td><td>Not-determined</td></tr>
<tr><td>S13_vs_87304||lt2gene</td><td>Low-quality</td></tr>
<tr><td>S13_vs_87355||lt2gene</td><td>Low-quality</td></tr>
<tr><td>S13_vs_87589||lt2gene</td><td>Low-quality</td></tr>
<tr><td>S13_vs_87691||lt2gene</td><td>Low-quality</td></tr>
<tr><td>S13_vs_87943||lt2gene</td><td>Low-quality</td></tr>
<tr><td>S13_vs_88090||lt2gene</td><td>Low-quality</td></tr>
<tr><td>S13_vs_88128||lt2gene</td><td>Low-quality</td></tr>
<tr><td>S13_vs_88187||lt2gene</td><td>Not-determined</td></tr>
<tr><td>S13_vs_88236||lt2gene</td><td>Low-quality</td></tr>
<tr><td>S13_vs_88274||lt2gene</td><td>Low-quality</td></tr>
<tr><td>S13_vs_88285||lt2gene</td><td>Low-quality</td></tr>
<tr><td>S13_vs_88320||lt2gene</td><td>Not-determined</td></tr>
<tr><td>S13_vs_88372||lt2gene</td><td>Not-determined</td></tr>
<tr><td>S13_vs_88451||lt2gene</td><td>Not-determined</td></tr>
<tr><td>S13_vs_88500||lt2gene</td><td>Not-determined</td></tr>
<tr><td>S13_vs_88505||lt2gene</td><td>Not-determined</td></tr>
<tr><td>S13_vs_88663||lt2gene</td><td>Low-quality</td></tr>
<tr><td>S13_vs_88729||lt2gene</td><td>Low-quality</td></tr>
<tr><td>S13_vs_88793||lt2gene</td><td>Low-quality</td></tr>
<tr><td>S13_vs_88797||lt2gene</td><td>Low-quality</td></tr>
<tr><td>S13_vs_88834||lt2gene</td><td>Low-quality</td></tr>
<tr><td>S13_vs_88949||lt2gene</td><td>Not-determined</td></tr>
<tr><td>S13_vs_89031||lt2gene</td><td>Low-quality</td></tr>
<tr><td>S13_vs_89201||lt2gene</td><td>Low-quality</td></tr>
<tr><td>S13_vs_89253||lt2gene</td><td>Low-quality</td></tr>
<tr><td>S13_vs_89254||lt2gene</td><td>Low-quality</td></tr>
<tr><td>S13_vs_89601||lt2gene</td><td>Low-quality</td></tr>
<tr><td>S13_vs_89642||lt2gene</td><td>Not-determined</td></tr>
<tr><td>S13_vs_89693||lt2gene</td><td>Low-quality</td></tr>
<tr><td>S13_vs_89707||lt2gene</td><td>Low-quality</td></tr>
<tr><td>S13_vs_89725||lt2gene</td><td>Not-determined</td></tr>
<tr><td>S13_vs_89829||lt2gene</td><td>Low-quality</td></tr>
<tr><td>S13_vs_89910||lt2gene</td><td>Low-quality</td></tr>
<tr><td>S13_vs_90128||lt2gene</td><td>Low-quality</td></tr>
<tr><td>S13_vs_90277||lt2gene</td><td>Low-quality</td></tr>
<tr><td>S13_vs_90427||lt2gene</td><td>Low-quality</td></tr>
<tr><td>S13_vs_90463||lt2gene</td><td>Low-quality</td></tr>
<tr><td>S13_vs_90736||lt2gene</td><td>Not-determined</td></tr>
<tr><td>S13_vs_90808||lt2gene</td><td>Not-determined</td></tr>
<tr><td>S13_vs_90949||lt2gene</td><td>Low-quality</td></tr>
<tr><td>S13_vs_91138||lt2gene</td><td>Low-quality</td></tr>
<tr><td>S13_vs_91192||lt2gene</td><td>Low-quality</td></tr>
<tr><td>S13_vs_91453||lt2gene</td><td>Not-determined</td></tr>
<tr><td>S13_vs_91458||lt2gene</td><td>Low-quality</td></tr>
<tr><td>S13_vs_91511||lt2gene</td><td>Low-quality</td></tr>
<tr><td>S13_vs_91571||lt2gene</td><td>Not-determined</td></tr>
<tr><td>S13_vs_91624||lt2gene</td><td>Not-determined</td></tr>
<tr><td>S13_vs_91807||lt2gene</td><td>Low-quality</td></tr>
<tr><td>S13_vs_91896||lt2gene</td><td>Not-determined</td></tr>
<tr><td>S13_vs_91904||lt2gene</td><td>Low-quality</td></tr>
<tr><td>S13_vs_92137||lt2gene</td><td>Low-quality</td></tr>
<tr><td>S13_vs_92224||lt2gene</td><td>Not-determined</td></tr>
<tr><td>S13_vs_92255||lt2gene</td><td>Low-quality</td></tr>
<tr><td>S13_vs_92299||lt2gene</td><td>Low-quality</td></tr>
<tr><td>S13_vs_92369||lt2gene</td><td>Low-quality</td></tr>
<tr><td>S13_vs_92478||lt2gene</td><td>Not-determined</td></tr>
<tr><td>S13_vs_92760||lt2gene</td><td>Low-quality</td></tr>
<tr><td>S13_vs_92784||lt2gene</td><td>Not-determined</td></tr>
<tr><td>S13_vs_92816||lt2gene</td><td>Low-quality</td></tr>
<tr><td>S13_vs_92908||lt2gene</td><td>Low-quality</td></tr>
<tr><td>S13_vs_93063||lt2gene</td><td>Not-determined</td></tr>
<tr><td>S13_vs_93077||lt2gene</td><td>Low-quality</td></tr>
<tr><td>S13_vs_93114||lt2gene</td><td>Low-quality</td></tr>
<tr><td>S13_vs_93361||lt2gene</td><td>Low-quality</td></tr>
<tr><td>S13_vs_93375||lt2gene</td><td>Low-quality</td></tr>
<tr><td>S13_vs_93451||lt2gene</td><td>Low-quality</td></tr>
<tr><td>S13_vs_93542||lt2gene</td><td>Low-quality</td></tr>
<tr><td>S13_vs_94073||lt2gene</td><td>Low-quality</td></tr>
<tr><td>S13_vs_94107||lt2gene</td><td>Low-quality</td></tr>
<tr><td>S13_vs_94355||lt2gene</td><td>Low-quality</td></tr>
<tr><td>S13_vs_94361||lt2gene</td><td>Low-quality</td></tr>
<tr><td>S13_vs_94596||lt2gene</td><td>Not-determined</td></tr>
<tr><td>S13_vs_94678||lt2gene</td><td>Low-quality</td></tr>
<tr><td>S13_vs_94682||lt2gene</td><td>Not-determined</td></tr>
<tr><td>S13_vs_94911||lt2gene</td><td>Low-quality</td></tr>
<tr><td>S13_vs_94928||lt2gene</td><td>Low-quality</td></tr>
<tr><td>S13_vs_95102||lt2gene</td><td>Low-quality</td></tr>
<tr><td>S13_vs_95314||lt2gene</td><td>Low-quality</td></tr>
<tr><td>S13_vs_95320||lt2gene</td><td>Low-quality</td></tr>
<tr><td>S13_vs_95409||lt2gene</td><td>Low-quality</td></tr>
<tr><td>S13_vs_95519||lt2gene</td><td>Low-quality</td></tr>
<tr><td>S13_vs_95554||lt2gene</td><td>Low-quality</td></tr>
<tr><td>S13_vs_95710||lt2gene</td><td>Low-quality</td></tr>
<tr><td>S13_vs_95886||lt2gene</td><td>Low-quality</td></tr>
<tr><td>S13_vs_95907||lt2gene</td><td>Low-quality</td></tr>
<tr><td>S13_vs_96013||lt2gene</td><td>Low-quality</td></tr>
<tr><td>S13_vs_96248||lt2gene</td><td>Low-quality</td></tr>
<tr><td>S13_vs_96309||lt2gene</td><td>Low-quality</td></tr>
<tr><td>S13_vs_96448||lt2gene</td><td>Low-quality</td></tr>
<tr><td>S13_vs_96543||lt2gene</td><td>Low-quality</td></tr>
<tr><td>S13_vs_96627||lt2gene</td><td>Low-quality</td></tr>
<tr><td>S13_vs_96690||lt2gene</td><td>Low-quality</td></tr>
<tr><td>S13_vs_96727||lt2gene</td><td>Low-quality</td></tr>
<tr><td>S13_vs_96839||lt2gene</td><td>Low-quality</td></tr>
<tr><td>S13_vs_96901||lt2gene</td><td>Low-quality</td></tr>
<tr><td>S13_vs_96912||lt2gene</td><td>Low-quality</td></tr>
<tr><td>S13_vs_96929||lt2gene</td><td>Low-quality</td></tr>
<tr><td>S13_vs_97000||lt2gene</td><td>Not-determined</td></tr>
<tr><td>S13_vs_97078||lt2gene</td><td>Low-quality</td></tr>
<tr><td>S13_vs_97151||lt2gene</td><td>Low-quality</td></tr>
<tr><td>S13_vs_97161||lt2gene</td><td>Low-quality</td></tr>
<tr><td>S13_vs_97272||lt2gene</td><td>Low-quality</td></tr>
<tr><td>S13_vs_97602||lt2gene</td><td>Low-quality</td></tr>
<tr><td>S13_vs_97636||lt2gene</td><td>Low-quality</td></tr>
<tr><td>S13_vs_97744||lt2gene</td><td>Low-quality</td></tr>
<tr><td>S13_vs_97754||lt2gene</td><td>Not-determined</td></tr>
<tr><td>S13_vs_98076||lt2gene</td><td>Low-quality</td></tr>
<tr><td>S13_vs_98236||lt2gene</td><td>Low-quality</td></tr>
<tr><td>S13_vs_98251||lt2gene</td><td>Not-determined</td></tr>
<tr><td>S13_vs_98347||lt2gene</td><td>Low-quality</td></tr>
<tr><td>S13_vs_98697||lt2gene</td><td>Low-quality</td></tr>
<tr><td>S13_vs_98732||lt2gene</td><td>Not-determined</td></tr>
<tr><td>S13_vs_98874||lt2gene</td><td>Low-quality</td></tr>
<tr><td>S13_vs_98882||lt2gene</td><td>Low-quality</td></tr>
<tr><td>S13_vs_99159||lt2gene</td><td>Low-quality</td></tr>
<tr><td>S13_vs_99247||lt2gene</td><td>Low-quality</td></tr>
<tr><td>S13_vs_99736||lt2gene</td><td>Not-determined</td></tr>
<tr><td>S13_vs_99840||lt2gene</td><td>Low-quality</td></tr>
<tr><td>S13_vs_99875||lt2gene</td><td>Low-quality</td></tr>
<tr><td>S13_vs_100022||lt2gene</td><td>Not-determined</td></tr>
<tr><td>S13_vs_100333||lt2gene</td><td>Low-quality</td></tr>
<tr><td>S13_vs_100467||lt2gene</td><td>Low-quality</td></tr>
<tr><td>S13_vs_100630||lt2gene</td><td>Low-quality</td></tr>
<tr><td>S13_vs_100759||lt2gene</td><td>Low-quality</td></tr>
<tr><td>S13_vs_100826||lt2gene</td><td>Low-quality</td></tr>
<tr><td>S13_vs_101013||lt2gene</td><td>Not-determined</td></tr>
<tr><td>S13_vs_101071||lt2gene</td><td>Low-quality</td></tr>
<tr><td>S13_vs_101079||lt2gene</td><td>Low-quality</td></tr>
<tr><td>S13_vs_101190||lt2gene</td><td>Not-determined</td></tr>
<tr><td>S13_vs_101313||lt2gene</td><td>Not-determined</td></tr>
<tr><td>S13_vs_101368||lt2gene</td><td>Low-quality</td></tr>
<tr><td>S13_vs_101411||lt2gene</td><td>Low-quality</td></tr>
<tr><td>S13_vs_101740||lt2gene</td><td>Low-quality</td></tr>
<tr><td>S13_vs_102029||lt2gene</td><td>Low-quality</td></tr>
<tr><td>S13_vs_102133||lt2gene</td><td>Not-determined</td></tr>
<tr><td>S13_vs_102254||lt2gene</td><td>Not-determined</td></tr>
<tr><td>S13_vs_102345||lt2gene</td><td>Not-determined</td></tr>
<tr><td>S13_vs_102541||lt2gene</td><td>Low-quality</td></tr>
<tr><td>S13_vs_102542||lt2gene</td><td>Low-quality</td></tr>
<tr><td>S13_vs_102575||lt2gene</td><td>Low-quality</td></tr>
<tr><td>S13_vs_102578||lt2gene</td><td>Low-quality</td></tr>
<tr><td>S13_vs_102945||lt2gene</td><td>Low-quality</td></tr>
<tr><td>S13_vs_103165||lt2gene</td><td>Low-quality</td></tr>
<tr><td>S13_vs_103475||lt2gene</td><td>Not-determined</td></tr>
<tr><td>S13_vs_103570||lt2gene</td><td>Not-determined</td></tr>
<tr><td>S13_vs_103634||lt2gene</td><td>Low-quality</td></tr>
<tr><td>S13_vs_103796||lt2gene</td><td>Not-determined</td></tr>
<tr><td>S13_vs_103819||lt2gene</td><td>Low-quality</td></tr>
<tr><td>S13_vs_103847||lt2gene</td><td>Low-quality</td></tr>
<tr><td>S13_vs_103863||lt2gene</td><td>Not-determined</td></tr>
<tr><td>S13_vs_103996||lt2gene</td><td>Not-determined</td></tr>
<tr><td>S13_vs_104117||lt2gene</td><td>Low-quality</td></tr>
<tr><td>S13_vs_104237||lt2gene</td><td>Low-quality</td></tr>
<tr><td>S13_vs_104437||lt2gene</td><td>Low-quality</td></tr>
<tr><td>S13_vs_104623||lt2gene</td><td>Not-determined</td></tr>
<tr><td>S13_vs_104739||lt2gene</td><td>Not-determined</td></tr>
<tr><td>S13_vs_104767||lt2gene</td><td>Not-determined</td></tr>
<tr><td>S13_vs_104995||lt2gene</td><td>Not-determined</td></tr>
<tr><td>S13_vs_105047||lt2gene</td><td>Low-quality</td></tr>
<tr><td>S13_vs_105073||lt2gene</td><td>Low-quality</td></tr>
<tr><td>S13_vs_105130||lt2gene</td><td>Low-quality</td></tr>
<tr><td>S13_vs_105144||lt2gene</td><td>Not-determined</td></tr>
<tr><td>S13_vs_105430||lt2gene</td><td>Low-quality</td></tr>
<tr><td>S13_vs_105545||lt2gene</td><td>Low-quality</td></tr>
<tr><td>S13_vs_105728||lt2gene</td><td>Low-quality</td></tr>
<tr><td>S13_vs_105764||lt2gene</td><td>Not-determined</td></tr>
<tr><td>S13_vs_105856||lt2gene</td><td>Low-quality</td></tr>
<tr><td>S13_vs_106070||lt2gene</td><td>Not-determined</td></tr>
<tr><td>S13_vs_106170||lt2gene</td><td>Low-quality</td></tr>
<tr><td>S13_vs_106195||lt2gene</td><td>Not-determined</td></tr>
<tr><td>S13_vs_106355||lt2gene</td><td>Low-quality</td></tr>
<tr><td>S13_vs_106576||lt2gene</td><td>Low-quality</td></tr>
<tr><td>S13_vs_107055||lt2gene</td><td>Low-quality</td></tr>
<tr><td>S13_vs_107201||lt2gene</td><td>Low-quality</td></tr>
<tr><td>S13_vs_107364||lt2gene</td><td>Not-determined</td></tr>
<tr><td>S13_vs_107380||lt2gene</td><td>Low-quality</td></tr>
<tr><td>S13_vs_107401||lt2gene</td><td>Not-determined</td></tr>
<tr><td>S13_vs_107610||lt2gene</td><td>Not-determined</td></tr>
<tr><td>S13_vs_107639||lt2gene</td><td>Low-quality</td></tr>
<tr><td>S13_vs_107815||lt2gene</td><td>Not-determined</td></tr>
<tr><td>S13_vs_107834||lt2gene</td><td>Low-quality</td></tr>
<tr><td>S13_vs_107985||lt2gene</td><td>Low-quality</td></tr>
<tr><td>S13_vs_108014||lt2gene</td><td>Low-quality</td></tr>
<tr><td>S13_vs_108159||lt2gene</td><td>Not-determined</td></tr>
<tr><td>S13_vs_108812||lt2gene</td><td>Low-quality</td></tr>
<tr><td>S13_vs_108872||lt2gene</td><td>Not-determined</td></tr>
<tr><td>S13_vs_109024||lt2gene</td><td>Low-quality</td></tr>
<tr><td>S13_vs_109107||lt2gene</td><td>Low-quality</td></tr>
<tr><td>S13_vs_109184||lt2gene</td><td>Not-determined</td></tr>
<tr><td>S13_vs_109282||lt2gene</td><td>Low-quality</td></tr>
<tr><td>S13_vs_109364||lt2gene</td><td>Low-quality</td></tr>
<tr><td>S13_vs_109369||lt2gene</td><td>Low-quality</td></tr>
<tr><td>S13_vs_109615||lt2gene</td><td>Low-quality</td></tr>
<tr><td>S13_vs_109687||lt2gene</td><td>Low-quality</td></tr>
<tr><td>S13_vs_109692||lt2gene</td><td>Not-determined</td></tr>
<tr><td>S13_vs_109856||lt2gene</td><td>Low-quality</td></tr>
<tr><td>S13_vs_110063||lt2gene</td><td>Low-quality</td></tr>
<tr><td>S13_vs_110089||lt2gene</td><td>Not-determined</td></tr>
<tr><td>S13_vs_110172||lt2gene</td><td>Low-quality</td></tr>
<tr><td>S13_vs_110173||lt2gene</td><td>Low-quality</td></tr>
<tr><td>S13_vs_110224||lt2gene</td><td>Low-quality</td></tr>
<tr><td>S13_vs_110373||lt2gene</td><td>Low-quality</td></tr>
<tr><td>S13_vs_110386||lt2gene</td><td>Not-determined</td></tr>
<tr><td>S13_vs_110456||lt2gene</td><td>Low-quality</td></tr>
<tr><td>S13_vs_110649||lt2gene</td><td>Low-quality</td></tr>
<tr><td>S13_vs_110763||lt2gene</td><td>Low-quality</td></tr>
<tr><td>S13_vs_110844||lt2gene</td><td>Not-determined</td></tr>
<tr><td>S13_vs_110911||lt2gene</td><td>Not-determined</td></tr>
<tr><td>S13_vs_111027||lt2gene</td><td>Low-quality</td></tr>
<tr><td>S13_vs_111056||lt2gene</td><td>Low-quality</td></tr>
<tr><td>S13_vs_111121||lt2gene</td><td>Low-quality</td></tr>
<tr><td>S13_vs_111275||lt2gene</td><td>Low-quality</td></tr>
<tr><td>S13_vs_111433||lt2gene</td><td>Not-determined</td></tr>
<tr><td>S13_vs_111635||lt2gene</td><td>Low-quality</td></tr>
<tr><td>S13_vs_111643||lt2gene</td><td>Low-quality</td></tr>
<tr><td>S13_vs_111708||lt2gene</td><td>Low-quality</td></tr>
<tr><td>S13_vs_111857||lt2gene</td><td>Not-determined</td></tr>
<tr><td>S13_vs_111900||lt2gene</td><td>Low-quality</td></tr>
<tr><td>S13_vs_111929||lt2gene</td><td>Not-determined</td></tr>
<tr><td>S13_vs_112131||lt2gene</td><td>Low-quality</td></tr>
<tr><td>S13_vs_112282||lt2gene</td><td>Low-quality</td></tr>
<tr><td>S13_vs_112434||lt2gene</td><td>Not-determined</td></tr>
<tr><td>S13_vs_112552||lt2gene</td><td>Not-determined</td></tr>
<tr><td>S13_vs_112778||lt2gene</td><td>Not-determined</td></tr>
<tr><td>S13_vs_112929||lt2gene</td><td>Not-determined</td></tr>
<tr><td>S13_vs_113041||lt2gene</td><td>Low-quality</td></tr>
<tr><td>S13_vs_113322||lt2gene</td><td>Low-quality</td></tr>
<tr><td>S13_vs_113398||lt2gene</td><td>Not-determined</td></tr>
<tr><td>S13_vs_113483||lt2gene</td><td>Low-quality</td></tr>
<tr><td>S13_vs_113485||lt2gene</td><td>Low-quality</td></tr>
<tr><td>S13_vs_113603||lt2gene</td><td>Low-quality</td></tr>
<tr><td>S13_vs_113613||lt2gene</td><td>Low-quality</td></tr>
<tr><td>S13_vs_113631||lt2gene</td><td>Low-quality</td></tr>
<tr><td>S13_vs_113649||lt2gene</td><td>Low-quality</td></tr>
<tr><td>S13_vs_113673||lt2gene</td><td>Low-quality</td></tr>
<tr><td>S13_vs_113751||lt2gene</td><td>Low-quality</td></tr>
<tr><td>S13_vs_113960||lt2gene</td><td>Low-quality</td></tr>
<tr><td>S13_vs_114052||lt2gene</td><td>Not-determined</td></tr>
<tr><td>S13_vs_114085||lt2gene</td><td>Not-determined</td></tr>
<tr><td>S13_vs_114379||lt2gene</td><td>Low-quality</td></tr>
<tr><td>S13_vs_114423||lt2gene</td><td>Low-quality</td></tr>
<tr><td>S13_vs_114455||lt2gene</td><td>Not-determined</td></tr>
<tr><td>S13_vs_114528||lt2gene</td><td>Not-determined</td></tr>
<tr><td>S13_vs_114595||lt2gene</td><td>Not-determined</td></tr>
<tr><td>S13_vs_114633||lt2gene</td><td>Low-quality</td></tr>
<tr><td>S13_vs_114712||lt2gene</td><td>Not-determined</td></tr>
<tr><td>S13_vs_114837||lt2gene</td><td>Low-quality</td></tr>
<tr><td>S13_vs_115044||lt2gene</td><td>Low-quality</td></tr>
<tr><td>S13_vs_115200||lt2gene</td><td>Low-quality</td></tr>
<tr><td>S13_vs_115271||lt2gene</td><td>Low-quality</td></tr>
<tr><td>S13_vs_115338||lt2gene</td><td>Low-quality</td></tr>
<tr><td>S13_vs_115350||lt2gene</td><td>Not-determined</td></tr>
<tr><td>S13_vs_115395||lt2gene</td><td>Low-quality</td></tr>
<tr><td>S13_vs_115463||lt2gene</td><td>Low-quality</td></tr>
<tr><td>S13_vs_115794||lt2gene</td><td>Not-determined</td></tr>
<tr><td>S13_vs_116037||lt2gene</td><td>Low-quality</td></tr>
<tr><td>S13_vs_116496||lt2gene</td><td>Low-quality</td></tr>
<tr><td>S13_vs_116580||lt2gene</td><td>Low-quality</td></tr>
<tr><td>S13_vs_116664||lt2gene</td><td>Low-quality</td></tr>
<tr><td>S13_vs_116733||lt2gene</td><td>Not-determined</td></tr>
<tr><td>S13_vs_116752||lt2gene</td><td>Not-determined</td></tr>
<tr><td>S13_vs_116818||lt2gene</td><td>Low-quality</td></tr>
<tr><td>S13_vs_116851||lt2gene</td><td>Low-quality</td></tr>
<tr><td>S13_vs_116882||lt2gene</td><td>Low-quality</td></tr>
<tr><td>S13_vs_116907||lt2gene</td><td>Low-quality</td></tr>
<tr><td>S13_vs_116993||lt2gene</td><td>Not-determined</td></tr>
<tr><td>S13_vs_117030||lt2gene</td><td>Low-quality</td></tr>
<tr><td>S13_vs_117064||lt2gene</td><td>Low-quality</td></tr>
<tr><td>S13_vs_117307||lt2gene</td><td>Low-quality</td></tr>
<tr><td>S13_vs_117392||lt2gene</td><td>Not-determined</td></tr>
<tr><td>S13_vs_117643||lt2gene</td><td>Not-determined</td></tr>
<tr><td>S13_vs_117694||lt2gene</td><td>Low-quality</td></tr>
<tr><td>S13_vs_118025||lt2gene</td><td>Low-quality</td></tr>
<tr><td>S13_vs_118169||lt2gene</td><td>Not-determined</td></tr>
<tr><td>S13_vs_118430||lt2gene</td><td>Low-quality</td></tr>
<tr><td>S13_vs_118547||lt2gene</td><td>Low-quality</td></tr>
<tr><td>S13_vs_118888||lt2gene</td><td>Low-quality</td></tr>
<tr><td>S13_vs_118929||lt2gene</td><td>Low-quality</td></tr>
<tr><td>S13_vs_118943||lt2gene</td><td>Low-quality</td></tr>
<tr><td>S13_vs_119139||lt2gene</td><td>Not-determined</td></tr>
<tr><td>S13_vs_119155||lt2gene</td><td>Not-determined</td></tr>
<tr><td>S13_vs_119293||lt2gene</td><td>Low-quality</td></tr>
<tr><td>S13_vs_119410||lt2gene</td><td>Not-determined</td></tr>
<tr><td>S13_vs_119463||lt2gene</td><td>Low-quality</td></tr>
<tr><td>S13_vs_119563||lt2gene</td><td>Low-quality</td></tr>
<tr><td>S13_vs_119703||lt2gene</td><td>Low-quality</td></tr>
<tr><td>S13_vs_119742||lt2gene</td><td>Low-quality</td></tr>
<tr><td>S13_vs_119773||lt2gene</td><td>Low-quality</td></tr>
<tr><td>S13_vs_119776||lt2gene</td><td>Low-quality</td></tr>
<tr><td>S13_vs_119796||lt2gene</td><td>Low-quality</td></tr>
<tr><td>S13_vs_119827||lt2gene</td><td>Low-quality</td></tr>
<tr><td>S13_vs_119832||lt2gene</td><td>Not-determined</td></tr>
<tr><td>S13_vs_120178||lt2gene</td><td>Not-determined</td></tr>
<tr><td>S13_vs_120251||lt2gene</td><td>Low-quality</td></tr>
<tr><td>S13_vs_120480||lt2gene</td><td>Low-quality</td></tr>
<tr><td>S13_vs_120675||lt2gene</td><td>Low-quality</td></tr>
<tr><td>S13_vs_120686||lt2gene</td><td>Low-quality</td></tr>
<tr><td>S13_vs_120695||lt2gene</td><td>Low-quality</td></tr>
<tr><td>S13_vs_120797||lt2gene</td><td>Not-determined</td></tr>
<tr><td>S13_vs_120874||lt2gene</td><td>Low-quality</td></tr>
<tr><td>S13_vs_121268||lt2gene</td><td>Not-determined</td></tr>
<tr><td>S13_vs_121276||lt2gene</td><td>Low-quality</td></tr>
<tr><td>S13_vs_121370||lt2gene</td><td>Low-quality</td></tr>
<tr><td>S13_vs_121443||lt2gene</td><td>Low-quality</td></tr>
<tr><td>S13_vs_121621||lt2gene</td><td>Low-quality</td></tr>
<tr><td>S13_vs_121691||lt2gene</td><td>Low-quality</td></tr>
<tr><td>S13_vs_122097||lt2gene</td><td>Not-determined</td></tr>
<tr><td>S13_vs_122142||lt2gene</td><td>Low-quality</td></tr>
<tr><td>S13_vs_122202||lt2gene</td><td>Low-quality</td></tr>
<tr><td>S13_vs_122215||lt2gene</td><td>Low-quality</td></tr>
<tr><td>S13_vs_122394||lt2gene</td><td>Low-quality</td></tr>
<tr><td>S13_vs_122861||lt2gene</td><td>Low-quality</td></tr>
<tr><td>S13_vs_123065||lt2gene</td><td>Low-quality</td></tr>
<tr><td>S13_vs_123368||lt2gene</td><td>Not-determined</td></tr>
<tr><td>S13_vs_123581||lt2gene</td><td>Low-quality</td></tr>
<tr><td>S13_vs_123635||lt2gene</td><td>Low-quality</td></tr>
<tr><td>S13_vs_123696||lt2gene</td><td>Low-quality</td></tr>
<tr><td>S13_vs_124046||lt2gene</td><td>Low-quality</td></tr>
<tr><td>S13_vs_124194||lt2gene</td><td>Not-determined</td></tr>
<tr><td>S13_vs_124445||lt2gene</td><td>Low-quality</td></tr>
<tr><td>S13_vs_124484||lt2gene</td><td>Low-quality</td></tr>
<tr><td>S13_vs_124491||lt2gene</td><td>Low-quality</td></tr>
<tr><td>S13_vs_124525||lt2gene</td><td>Low-quality</td></tr>
<tr><td>S13_vs_124533||lt2gene</td><td>Low-quality</td></tr>
<tr><td>S13_vs_124546||lt2gene</td><td>Low-quality</td></tr>
</table>

| | |
|---|---|
| S13_vs_124558\|\|lt2gene | Low-quality |
| S13_vs_124659\|\|lt2gene | Not-determined |
| S13_vs_124696\|\|lt2gene | Low-quality |
| S13_vs_124737\|\|lt2gene | Low-quality |
| S13_vs_124809\|\|lt2gene | Low-quality |
| S13_vs_125102\|\|lt2gene | Not-determined |
| S13_vs_125290\|\|lt2gene | Low-quality |
| S13_vs_125514\|\|lt2gene | Low-quality |
| S13_vs_125534\|\|lt2gene | Not-determined |
| S13_vs_125544\|\|lt2gene | Low-quality |
| S13_vs_125660\|\|lt2gene | Low-quality |
| S13_vs_125706\|\|lt2gene | Low-quality |
| S13_vs_125744\|\|lt2gene | Low-quality |
| S13_vs_125978\|\|lt2gene | Not-determined |
| S13_vs_126191\|\|lt2gene | Low-quality |
| S13_vs_126290\|\|lt2gene | Low-quality |
| S13_vs_126341\|\|lt2gene | Low-quality |
| S13_vs_126651\|\|lt2gene | Not-determined |
| S13_vs_126761\|\|lt2gene | Low-quality |
| S13_vs_126961\|\|lt2gene | Not-determined |
| S13_vs_127139\|\|lt2gene | Low-quality |
| S13_vs_127159\|\|lt2gene | Low-quality |
| S13_vs_127266\|\|lt2gene | Low-quality |
| S13_vs_127312\|\|lt2gene | Not-determined |
| S13_vs_127416\|\|lt2gene | Low-quality |
| S13_vs_127418\|\|lt2gene | Low-quality |
| S13_vs_127463\|\|lt2gene | Low-quality |
| S13_vs_127669\|\|lt2gene | Not-determined |
| S13_vs_127759\|\|lt2gene | Low-quality |
| S13_vs_127798\|\|lt2gene | Low-quality |
| S13_vs_127811\|\|lt2gene | Low-quality |
| S13_vs_128130\|\|lt2gene | Low-quality |
| S13_vs_128361\|\|lt2gene | Not-determined |
| S13_vs_128500\|\|lt2gene | Not-determined |
| S13_vs_128677\|\|lt2gene | Low-quality |
| S13_vs_128694\|\|lt2gene | Low-quality |
| S13_vs_129005\|\|lt2gene | Low-quality |
| S13_vs_129138\|\|lt2gene | Low-quality |
| S13_vs_129201\|\|lt2gene | Low-quality |
| S13_vs_129249\|\|lt2gene | Low-quality |
| S13_vs_129464\|\|lt2gene | Low-quality |
| S13_vs_129466\|\|lt2gene | Not-determined |
| S13_vs_129742\|\|lt2gene | Low-quality |
| S13_vs_129758\|\|lt2gene | Low-quality |
| S13_vs_129888\|\|lt2gene | Low-quality |
| S13_vs_130091\|\|lt2gene | Not-determined |
| S13_vs_130106\|\|lt2gene | Low-quality |
| S13_vs_130200\|\|lt2gene | Not-determined |
| S13_vs_130390\|\|lt2gene | Low-quality |
| S13_vs_130671\|\|lt2gene | Not-determined |
| S13_vs_130852\|\|lt2gene | Low-quality |
| S13_vs_131066\|\|lt2gene | Not-determined |
| S13_vs_131203\|\|lt2gene | Low-quality |
| S13_vs_131271\|\|lt2gene | Not-determined |
| S13_vs_131458\|\|lt2gene | Low-quality |
| S13_vs_131513\|\|lt2gene | Low-quality |
| S13_vs_131604\|\|lt2gene | Low-quality |
| S13_vs_131778\|\|lt2gene | Low-quality |
| S13_vs_131826\|\|lt2gene | Low-quality |
| S13_vs_131864\|\|lt2gene | Low-quality |
| S13_vs_131872\|\|lt2gene | Not-determined |
| S13_vs_131990\|\|lt2gene | Not-determined |
| S13_vs_132050\|\|lt2gene | Not-determined |
| S13_vs_132404\|\|lt2gene | Low-quality |
| S13_vs_132572\|\|lt2gene | Low-quality |
| S13_vs_132575\|\|lt2gene | Not-determined |
| S13_vs_132624\|\|lt2gene | Low-quality |
| S13_vs_132658\|\|lt2gene | Not-determined |
| S13_vs_132663\|\|lt2gene | Low-quality |
| S13_vs_132669\|\|lt2gene | Not-determined |
| S13_vs_132937\|\|lt2gene | Not-determined |
| S13_vs_132979\|\|lt2gene | Low-quality |
| S13_vs_133156\|\|lt2gene | Low-quality |
| S13_vs_133178\|\|lt2gene | Not-determined |
| S13_vs_133279\|\|lt2gene | Low-quality |
| S13_vs_133287\|\|lt2gene | Low-quality |
| S13_vs_133394\|\|lt2gene | Low-quality |
| S13_vs_133640\|\|lt2gene | Low-quality |
| S13_vs_133686\|\|lt2gene | Not-determined |
| S13_vs_133845\|\|lt2gene | Not-determined |
| S13_vs_133987\|\|lt2gene | Not-determined |
| S13_vs_134049\|\|lt2gene | Low-quality |
| S13_vs_134129\|\|lt2gene | Not-determined |
| S13_vs_134216\|\|lt2gene | Low-quality |
| S13_vs_134240\|\|lt2gene | Not-determined |
| S13_vs_134493\|\|lt2gene | Not-determined |
| S13_vs_134543\|\|lt2gene | Not-determined |
| S13_vs_134687\|\|lt2gene | Not-determined |
| S13_vs_134855\|\|lt2gene | Low-quality |
| S13_vs_134955\|\|lt2gene | Not-determined |
| S13_vs_135090\|\|lt2gene | Not-determined |
| S13_vs_135244\|\|lt2gene | Low-quality |
| S13_vs_135520\|\|lt2gene | Low-quality |
| S13_vs_135544\|\|lt2gene | Low-quality |
| S13_vs_135599\|\|lt2gene | Not-determined |
| S13_vs_135645\|\|lt2gene | Low-quality |
| S13_vs_135668\|\|lt2gene | Low-quality |
| S13_vs_135680\|\|lt2gene | Not-determined |
| S13_vs_135710\|\|lt2gene | Low-quality |
| S13_vs_135769\|\|lt2gene | Low-quality |
| S13_vs_135842\|\|lt2gene | Low-quality |
| S13_vs_135859\|\|lt2gene | Low-quality |
| S13_vs_135861\|\|lt2gene | Low-quality |
| S13_vs_135889\|\|lt2gene | Low-quality |
| S13_vs_135988\|\|lt2gene | Not-determined |
| S13_vs_136028\|\|lt2gene | Low-quality |
| S13_vs_136117\|\|lt2gene | Low-quality |
| S13_vs_136177\|\|lt2gene | Low-quality |
| S13_vs_136258\|\|lt2gene | Low-quality |
| S13_vs_136463\|\|lt2gene | Not-determined |
| S13_vs_136529\|\|lt2gene | Low-quality |
| S13_vs_136565\|\|lt2gene | Low-quality |
| S13_vs_136596\|\|lt2gene | Low-quality |
| S13_vs_136611\|\|lt2gene | Not-determined |
| S13_vs_136773\|\|lt2gene | Low-quality |
| S13_vs_136790\|\|lt2gene | Low-quality |
| S13_vs_136836\|\|lt2gene | Low-quality |
| S13_vs_137280\|\|lt2gene | Low-quality |
| S13_vs_137512\|\|lt2gene | Low-quality |
| S13_vs_137797\|\|lt2gene | Low-quality |
| S13_vs_137920\|\|lt2gene | Not-determined |
| S13_vs_138033\|\|lt2gene | Low-quality |
| S13_vs_138121\|\|lt2gene | Not-determined |
| S13_vs_138220\|\|lt2gene | Not-determined |
| S13_vs_138264\|\|lt2gene | Low-quality |
| S13_vs_138470\|\|lt2gene | Low-quality |
| S13_vs_138471\|\|lt2gene | Not-determined |
| S13_vs_138503\|\|lt2gene | Not-determined |
| S13_vs_138595\|\|lt2gene | Low-quality |
| S13_vs_138625\|\|lt2gene | Low-quality |
| S13_vs_138702\|\|lt2gene | Low-quality |
| S13_vs_138788\|\|lt2gene | Low-quality |
| S13_vs_139012\|\|lt2gene | Low-quality |
| S13_vs_139092\|\|lt2gene | Low-quality |
| S13_vs_139100\|\|lt2gene | Low-quality |
| S13_vs_139405\|\|lt2gene | Not-determined |
| S13_vs_139565\|\|lt2gene | Low-quality |
| S13_vs_139705\|\|lt2gene | Low-quality |
| S13_vs_139745\|\|lt2gene | Not-determined |
| S13_vs_139925\|\|lt2gene | Not-determined |
| S13_vs_139979\|\|lt2gene | Low-quality |
| S13_vs_140273\|\|lt2gene | Low-quality |
| S13_vs_140287\|\|lt2gene | Low-quality |
| S13_vs_140293\|\|lt2gene | Low-quality |
| S13_vs_140384\|\|lt2gene | Low-quality |
| S13_vs_140433\|\|lt2gene | Not-determined |
| S13_vs_140471\|\|lt2gene | Low-quality |
| S13_vs_140593\|\|lt2gene | Not-determined |
| S13_vs_140669\|\|lt2gene | Not-determined |
| S13_vs_140688\|\|lt2gene | Low-quality |
| S13_vs_141482\|\|lt2gene | Not-determined |
| S13_vs_141680\|\|lt2gene | Not-determined |
| S13_vs_141752\|\|lt2gene | Not-determined |
| S13_vs_141820\|\|lt2gene | Not-determined |
| S13_vs_141884\|\|lt2gene | Low-quality |
| S13_vs_142156\|\|lt2gene | Not-determined |
| S13_vs_142164\|\|lt2gene | Low-quality |
| S13_vs_142304\|\|lt2gene | Low-quality |
| S13_vs_142307\|\|lt2gene | Not-determined |
| S13_vs_142608\|\|lt2gene | Not-determined |
| S13_vs_142649\|\|lt2gene | Low-quality |
| S13_vs_142699\|\|lt2gene | Low-quality |
| S13_vs_142725\|\|lt2gene | Low-quality |
| S13_vs_142841\|\|lt2gene | Not-determined |
| S13_vs_142908\|\|lt2gene | Low-quality |
| S13_vs_143035\|\|lt2gene | Low-quality |
| S13_vs_143351\|\|lt2gene | Low-quality |
| S13_vs_143434\|\|lt2gene | Not-determined |
| S13_vs_143472\|\|lt2gene | Not-determined |
| S13_vs_143500\|\|lt2gene | Low-quality |
| S13_vs_143516\|\|lt2gene | Not-determined |
| S13_vs_143538\|\|lt2gene | Not-determined |
| S13_vs_143584\|\|lt2gene | Low-quality |
| S13_vs_143647\|\|lt2gene | Low-quality |
| S13_vs_143861\|\|lt2gene | Low-quality |
| S13_vs_143917\|\|lt2gene | Low-quality |
| S13_vs_144108\|\|lt2gene | Low-quality |
| S13_vs_144118\|\|lt2gene | Low-quality |
| S13_vs_144121\|\|lt2gene | Low-quality |
| S13_vs_144285\|\|lt2gene | Low-quality |
| S13_vs_144346\|\|lt2gene | Low-quality |
| S13_vs_144372\|\|lt2gene | Not-determined |
| S13_vs_144524\|\|lt2gene | Low-quality |
| S13_vs_144560\|\|lt2gene | Not-determined |
| S13_vs_144577\|\|lt2gene | Low-quality |
| S13_vs_145061\|\|lt2gene | Not-determined |
| S13_vs_145103\|\|lt2gene | Low-quality |
| S13_vs_145187\|\|lt2gene | Low-quality |
| S13_vs_145433\|\|lt2gene | Not-determined |
| S13_vs_145476\|\|lt2gene | Not-determined |
| S13_vs_145647\|\|lt2gene | Low-quality |
| S13_vs_145681\|\|lt2gene | Not-determined |
| S13_vs_145685\|\|lt2gene | Not-determined |
| S13_vs_145696\|\|lt2gene | Low-quality |
| S13_vs_145744\|\|lt2gene | Not-determined |
| S13_vs_146226\|\|lt2gene | Low-quality |
| S13_vs_146279\|\|lt2gene | Low-quality |
| S13_vs_146387\|\|lt2gene | Low-quality |
| S13_vs_146396\|\|lt2gene | Low-quality |
| S13_vs_146444\|\|lt2gene | Low-quality |
| S13_vs_146763\|\|lt2gene | Not-determined |
| S13_vs_146962\|\|lt2gene | Low-quality |
| S13_vs_147255\|\|lt2gene | Low-quality |
| S13_vs_147569\|\|lt2gene | Not-determined |
| S13_vs_147664\|\|lt2gene | Low-quality |
| S13_vs_147762\|\|lt2gene | Low-quality |
| S13_vs_148184\|\|lt2gene | Low-quality |
| S13_vs_148394\|\|lt2gene | Low-quality |
| S13_vs_148407\|\|lt2gene | Low-quality |
| S13_vs_148479\|\|lt2gene | Low-quality |
| S13_vs_148611\|\|lt2gene | Low-quality |
| S13_vs_148649\|\|lt2gene | Low-quality |
| S13_vs_148746\|\|lt2gene | Low-quality |
| S13_vs_149078\|\|lt2gene | Not-determined |
| S13_vs_149134\|\|lt2gene | Low-quality |
| S13_vs_149227\|\|lt2gene | Low-quality |
| S13_vs_149228\|\|lt2gene | Low-quality |
| S13_vs_149366\|\|lt2gene | Low-quality |
| S13_vs_149438\|\|lt2gene | Low-quality |
| S13_vs_149669\|\|lt2gene | Low-quality |
| S13_vs_149779\|\|lt2gene | Low-quality |
| S13_vs_149830\|\|lt2gene | Low-quality |
| S13_vs_149838\|\|lt2gene | Low-quality |
| S13_vs_149883\|\|lt2gene | Not-determined |
| S13_vs_150071\|\|lt2gene | Low-quality |
| S13_vs_150101\|\|lt2gene | Low-quality |
| S13_vs_150145\|\|lt2gene | Not-determined |
| S13_vs_150267\|\|lt2gene | Low-quality |
| S13_vs_150709\|\|lt2gene | Low-quality |
| S13_vs_150817\|\|lt2gene | Low-quality |
| S13_vs_150875\|\|lt2gene | Low-quality |
| S13_vs_151246\|\|lt2gene | Low-quality |
| S13_vs_151314\|\|lt2gene | Not-determined |
| S13_vs_151346\|\|lt2gene | Low-quality |
| S13_vs_151512\|\|lt2gene | Low-quality |
| S13_vs_151545\|\|lt2gene | Not-determined |
| S13_vs_151593\|\|lt2gene | Not-determined |
| S13_vs_151707\|\|lt2gene | Low-quality |
| S13_vs_151769\|\|lt2gene | Low-quality |
| S13_vs_151878\|\|lt2gene | Not-determined |
| S13_vs_151895\|\|lt2gene | Low-quality |
| S13_vs_152017\|\|lt2gene | Low-quality |
| S13_vs_152059\|\|lt2gene | Low-quality |
| S13_vs_152257\|\|lt2gene | Not-determined |
| S13_vs_152371\|\|lt2gene | Not-determined |
| S13_vs_152579\|\|lt2gene | Not-determined |
| S13_vs_152818\|\|lt2gene | Low-quality |
| S13_vs_153006\|\|lt2gene | Low-quality |
| S13_vs_153338\|\|lt2gene | Low-quality |
| S13_vs_153441\|\|lt2gene | Low-quality |
| S13_vs_153443\|\|lt2gene | Not-determined |
| S13_vs_153671\|\|lt2gene | Low-quality |
| S13_vs_153696\|\|lt2gene | Not-determined |
| S13_vs_153699\|\|lt2gene | Low-quality |
| S13_vs_153738\|\|lt2gene | Low-quality |
| S13_vs_153787\|\|lt2gene | Low-quality |
| S13_vs_153829\|\|lt2gene | Low-quality |
| S13_vs_153835\|\|lt2gene | Low-quality |
| S13_vs_153857\|\|lt2gene | Not-determined |
| S13_vs_153951\|\|lt2gene | Not-determined |
| S13_vs_154069\|\|lt2gene | Not-determined |
| S13_vs_154105\|\|lt2gene | Low-quality |
| S13_vs_154272\|\|lt2gene | Not-determined |
| S14_extracted_k141_0 | Not-determined |
| S14_extracted_k141_36637 | Not-determined |
| S14_extracted_k141_65946 | Low-quality |
| S14_extracted_k141_51294 | Low-quality |
| S14_extracted_k141_4 | Not-determined |
| S14_extracted_k141_102582 | Low-quality |
| S14_extracted_k141_5 | Not-determined |
| S14_extracted_k141_7333 | Not-determined |
| S14_extracted_k141_7336 | Low-quality |
| S14_extracted_k141_12 | Not-determined |
| S14_extracted_k141_21989 | Not-determined |
| S14_extracted_k141_73275 | Low-quality |
| S14_extracted_k141_51296 | Low-quality |
| S14_extracted_k141_21991 | Low-quality |
| S14_extracted_k141_18 | Not-determined |
| S14_extracted_k141_36647 | Not-determined |
| S14_extracted_k141_36648 | Not-determined |
| S14_extracted_k141_80610 | Not-determined |
| S14_extracted_k141_43972 | Not-determined |
| S14_extracted_k141_21 | Not-determined |
| S14_extracted_k141_21994 | Low-quality |
| S14_extracted_k141_36656 | Low-quality |
| S14_extracted_k141_27 | Not-determined |
| S14_extracted_k141_36657 | Not-determined |
| S14_extracted_k141_36658 | Not-determined |
| S14_extracted_k141_36660 | Not-determined |
| S14_extracted_k141_109923 | Not-determined |
| S14_extracted_k141_43984 | Low-quality |
| S14_extracted_k141_43985 | Low-quality |
| S14_extracted_k141_43986 | Low-quality |
| S14_extracted_k141_109927 | Not-determined |
| S14_extracted_k141_80615 | Not-determined |
| S14_extracted_k141_43990 | Not-determined |
| S14_extracted_k141_22011 | Low-quality |
| S14_extracted_k141_45 | Not-determined |
| S14_extracted_k141_109932 | Not-determined |
| S14_extracted_k141_87938 | Not-determined |
| S14_extracted_k141_49 | Not-determined |
| S14_extracted_k141_29324 | Not-determined |
| S14_extracted_k141_57 | Not-determined |
| S14_extracted_k141_43997 | Low-quality |
| S14_extracted_k141_43999 | Not-determined |
| S14_extracted_k141_109936 | Low-quality |
| S14_extracted_k141_51314 | Low-quality |
| S14_extracted_k141_80626 | Not-determined |
| S14_extracted_k141_109940 | Not-determined |
| S14_extracted_k141_109941 | Not-determined |
| S14_extracted_k141_36691 | Not-determined |
| S14_extracted_k141_80627 | Low-quality |
| S14_extracted_k141_36694 | Low-quality |
| S14_extracted_k141_14678 | Not-determined |
| S14_extracted_k141_65976 | Low-quality |
| S14_extracted_k141_14681 | Low-quality |
| S14_extracted_k141_95269 | Low-quality |
| S14_extracted_k141_65980 | Low-quality |
| S14_extracted_k141_65981 | Not-determined |
| S14_extracted_k141_51325 | Not-determined |
| S14_extracted_k141_65984 | Not-determined |
| S14_extracted_k141_102595 | Not-determined |
| S14_extracted_k141_80635 | Not-determined |
| S14_extracted_k141_109949 | Not-determined |
| S14_extracted_k141_95274 | Not-determined |
| S14_extracted_k141_109950 | Low-quality |
| S14_extracted_k141_78 | Not-determined |
| S14_extracted_k141_109953 | Not-determined |
| S14_extracted_k141_86 | Not-determined |
| S14_extracted_k141_80642 | Not-determined |
| S14_extracted_k141_14699 | Low-quality |
| S14_extracted_k141_80645 | Not-determined |
| S14_extracted_k141_73282 | Not-determined |
| S14_extracted_k141_66001 | Low-quality |
| S14_extracted_k141_88 | Not-determined |
| S14_extracted_k141_14711 | Not-determined |
| S14_extracted_k141_7376 | Not-determined |
| S14_extracted_k141_73292 | Not-determined |
| S14_extracted_k141_73293 | Not-determined |
| S14_extracted_k141_80658 | Not-determined |
| S14_extracted_k141_36719 | Not-determined |
| S14_extracted_k141_7381 | Not-determined |
| S14_extracted_k141_58645 | Not-determined |
| S14_extracted_k141_58648 | Not-determined |
| S14_extracted_k141_109960 | Low-quality |
| S14_extracted_k141_44047 | Not-determined |
| S14_extracted_k141_109961 | Not-determined |
| S14_extracted_k141_36722 | Low-quality |
| S14_extracted_k141_95307 | Not-determined |
| S14_extracted_k141_80670 | Not-determined |
| S14_extracted_k141_95308 | Low-quality |
| S14_extracted_k141_109962 | Low-quality |
| S14_extracted_k141_36727 | Not-determined |
| S14_extracted_k141_95313 | Not-determined |
| S14_extracted_k141_102606 | Not-determined |
| S14_extracted_k141_66022 | Low-quality |
| S14_extracted_k141_7395 | Not-determined |
| S14_extracted_k141_44049 | Low-quality |
| S14_extracted_k141_22056 | Not-determined |
| S14_extracted_k141_87943 | Low-quality |
| S14_extracted_k141_7398 | Not-determined |
| S14_extracted_k141_80676 | Not-determined |
| S14_extracted_k141_36739 | Not-determined |
| S14_extracted_k141_7399 | Low-quality |
| S14_extracted_k141_36748 | Not-determined |
| S14_extracted_k141_22061 | Not-determined |
| S14_extracted_k141_29366 | Low-quality |
| S14_extracted_k141_22063 | Not-determined |
| S14_extracted_k141_80683 | Not-determined |
| S14_extracted_k141_29371 | Low-quality |
| S14_extracted_k141_80686 | Not-determined |
| S14_extracted_k141_66026 | Low-quality |
| S14_extracted_k141_44052 | Low-quality |
| S14_extracted_k141_58666 | Low-quality |
| S14_extracted_k141_29376 | Not-determined |
| S14_extracted_k141_66031 | Not-determined |
| S14_extracted_k141_44057 | Not-determined |
| S14_extracted_k141_102615 | Not-determined |
| S14_extracted_k141_44059 | Low-quality |
| S14_extracted_k141_87954 | Not-determined |
| S14_extracted_k141_22072 | Not-determined |
| S14_extracted_k141_120 | Not-determined |
| S14_extracted_k141_95333 | Low-quality |
| S14_extracted_k141_122 | Low-quality |
| S14_extracted_k141_29387 | Low-quality |
| S14_extracted_k141_66038 | Not-determined |
| S14_extracted_k141_73303 | Low-quality |
| S14_extracted_k141_51380 | Not-determined |
| S14_extracted_k141_29393 | Not-determined |
| S14_extracted_k141_102632 | Not-determined |
| S14_extracted_k141_80693 | Not-determined |
| S14_extracted_k141_22090 | Not-determined |
| S14_extracted_k141_7432 | Not-determined |
| S14_extracted_k141_58693 | Low-quality |
| S14_extracted_k141_58694 | Low-quality |
| S14_extracted_k141_44082 | Low-quality |
| S14_extracted_k141_87969 | Low-quality |
| S14_extracted_k141_22093 | Not-determined |
| S14_extracted_k141_80694 | Low-quality |
| S14_extracted_k141_87972 | Not-determined |
| S14_extracted_k141_102639 | Not-determined |
| S14_extracted_k141_58698 | Low-quality |
| S14_extracted_k141_51391 | Not-determined |
| S14_extracted_k141_95351 | Not-determined |
| S14_extracted_k141_36776 | Not-determined |
| S14_extracted_k141_51395 | Not-determined |
| S14_extracted_k141_58701 | Not-determined |
| S14_extracted_k141_102642 | Not-determined |
| S14_extracted_k141_7446 | Low-quality |
| S14_extracted_k141_51405 | Not-determined |
| S14_extracted_k141_29409 | Not-determined |
| S14_extracted_k141_102645 | Low-quality |
| S14_extracted_k141_66053 | Low-quality |
| S14_extracted_k141_58707 | Not-determined |
| S14_extracted_k141_29412 | Not-determined |
| S14_extracted_k141_80700 | Not-determined |
| S14_extracted_k141_51413 | Not-determined |
| S14_extracted_k141_7451 | Low-quality |
| S14_extracted_k141_58710 | Low-quality |
| S14_extracted_k141_102653 | Low-quality |
| S14_extracted_k141_29414 | Low-quality |
| S14_extracted_k141_129 | Not-determined |
| S14_extracted_k141_58712 | Not-determined |
| S14_extracted_k141_44092 | Low-quality |
| S14_extracted_k141_80701 | Low-quality |
| S14_extracted_k141_58715 | Low-quality |
| S14_extracted_k141_14748 | Not-determined |
| S14_extracted_k141_66063 | Low-quality |
| S14_extracted_k141_51417 | Low-quality |
| S14_extracted_k141_36795 | Not-determined |
| S14_extracted_k141_139 | Not-determined |
| S14_extracted_k141_51421 | Not-determined |
| S14_extracted_k141_66069 | Not-determined |
| S14_extracted_k141_29423 | Low-quality |
| S14_extracted_k141_140 | Low-quality |
| S14_extracted_k141_66072 | Not-determined |
| S14_extracted_k141_29425 | Low-quality |
| S14_extracted_k141_102662 | Not-determined |
| S14_extracted_k141_95361 | Not-determined |
| S14_extracted_k141_29430 | Low-quality |
| S14_extracted_k141_29432 | Low-quality |
| S14_extracted_k141_80707 | Not-determined |
| S14_extracted_k141_29439 | Not-determined |
| S14_extracted_k141_66086 | Not-determined |
| S14_extracted_k141_44110 | Low-quality |
| S14_extracted_k141_14762 | Not-determined |
| S14_extracted_k141_29442 | Low-quality |
| S14_extracted_k141_58740 | Not-determined |
| S14_extracted_k141_95372 | Not-determined |
| S14_extracted_k141_95373 | Not-determined |
| S14_extracted_k141_14765 | Low-quality |
| S14_extracted_k141_102678 | Low-quality |
| S14_extracted_k141_58743 | Low-quality |
| S14_extracted_k141_58745 | Not-determined |
| S14_extracted_k141_29445 | Low-quality |
| S14_extracted_k141_66092 | Low-quality |
| S14_extracted_k141_95378 | Not-determined |
| S14_extracted_k141_36816 | Not-determined |
| S14_extracted_k141_58748 | Not-determined |
| S14_extracted_k141_149 | Not-determined |
| S14_extracted_k141_102682 | Not-determined |
| S14_extracted_k141_102683 | Not-determined |
| S14_extracted_k141_7470 | Not-determined |
| S14_extracted_k141_7471 | Low-quality |
| S14_extracted_k141_7473 | Not-determined |
| S14_extracted_k141_66097 | Low-quality |
| S14_extracted_k141_44121 | Low-quality |
| S14_extracted_k141_66101 | Low-quality |
| S14_extracted_k141_102690 | Not-determined |
| S14_extracted_k141_162 | Low-quality |
| S14_extracted_k141_58752 | Not-determined |
| S14_extracted_k141_14776 | Not-determined |
| S14_extracted_k141_44124 | Not-determined |
| S14_extracted_k141_14778 | Not-determined |
| S14_extracted_k141_80722 | Low-quality |
| S14_extracted_k141_44130 | Low-quality |
| S14_extracted_k141_44131 | Not-determined |
| S14_extracted_k141_44133 | Not-determined |
| S14_extracted_k141_66112 | Low-quality |
| S14_extracted_k141_73314 | Low-quality |
| S14_extracted_k141_181 | Low-quality |
| S14_extracted_k141_73315 | Not-determined |
| S14_extracted_k141_36833 | Low-quality |
| S14_extracted_k141_95396 | Not-determined |
| S14_extracted_k141_109977 | Not-determined |
| S14_extracted_k141_66121 | Not-determined |
| S14_extracted_k141_95399 | Not-determined |
| S14_extracted_k141_73324 | Not-determined |
| S14_extracted_k141_95402 | Not-determined |
| S14_extracted_k141_36839 | Not-determined |
| S14_extracted_k141_29460 | Low-quality |
| S14_extracted_k141_36841 | Not-determined |
| S14_extracted_k141_51438 | Not-determined |
| S14_extracted_k141_80731 | Low-quality |
| S14_extracted_k141_58778 | Low-quality |
| S14_extracted_k141_29464 | Not-determined |
| S14_extracted_k141_36845 | Not-determined |
| S14_extracted_k141_36846 | Not-determined |
| S14_extracted_k141_73332 | Low-quality |
| S14_extracted_k141_44139 | Not-determined |
| S14_extracted_k141_80739 | Not-determined |
| S14_extracted_k141_95404 | Not-determined |
| S14_extracted_k141_95405 | Low-quality |
| S14_extracted_k141_36854 | Not-determined |
| S14_extracted_k141_44143 | Not-determined |
| S14_extracted_k141_109989 | Not-determined |
| S14_extracted_k141_36856 | Not-determined |
| S14_extracted_k141_109992 | Not-determined |
| S14_extracted_k141_22124 | Not-determined |
| S14_extracted_k141_58797 | Not-determined |
| S14_extracted_k141_185 | Not-determined |
| S14_extracted_k141_58802 | Low-quality |
| S14_extracted_k141_87991 | Low-quality |
| S14_extracted_k141_36863 | Low-quality |
| S14_extracted_k141_29487 | Low-quality |
| S14_extracted_k141_87996 | Low-quality |
| S14_extracted_k141_58810 | Low-quality |
| S14_extracted_k141_22132 | Low-quality |
| S14_extracted_k141_36869 | Not-determined |
| S14_extracted_k141_51440 | Low-quality |
| S14_extracted_k141_51444 | Low-quality |
| S14_extracted_k141_80741 | Not-determined |
| S14_extracted_k141_51446 | Not-determined |
| S14_extracted_k141_22135 | Low-quality |
| S14_extracted_k141_51447 | Low-quality |
| S14_extracted_k141_80744 | Not-determined |
| S14_extracted_k141_88008 | Not-determined |
| S14_extracted_k141_88013 | Low-quality |
| S14_extracted_k141_88014 | Not-determined |
| S14_extracted_k141_51449 | Low-quality |
| S14_extracted_k141_51451 | Low-quality |
| S14_extracted_k141_29500 | Low-quality |
| S14_extracted_k141_51452 | Not-determined |
| S14_extracted_k141_36880 | Not-determined |
| S14_extracted_k141_102709 | Not-determined |
| S14_extracted_k141_73338 | Not-determined |
| S14_extracted_k141_80757 | Not-determined |
| S14_extracted_k141_73341 | Not-determined |
| S14_extracted_k141_22147 | Not-determined |
| S14_extracted_k141_73344 | Low-quality |
| S14_extracted_k141_7500 | Not-determined |
| S14_extracted_k141_73348 | Low-quality |
| S14_extracted_k141_22148 | Not-determined |
| S14_extracted_k141_22149 | Not-determined |
| S14_extracted_k141_29508 | Low-quality |
| S14_extracted_k141_51464 | Low-quality |
| S14_extracted_k141_102721 | Not-determined |
| S14_extracted_k141_110024 | Not-determined |
| S14_extracted_k141_22156 | Not-determined |
| S14_extracted_k141_51465 | Not-determined |
| S14_extracted_k141_22158 | Not-determined |
| S14_extracted_k141_44157 | Low-quality |
| S14_extracted_k141_44160 | Not-determined |
| S14_extracted_k141_51469 | Not-determined |
| S14_extracted_k141_44161 | Not-determined |
| S14_extracted_k141_51471 | Not-determined |
| S14_extracted_k141_51472 | Low-quality |
| S14_extracted_k141_80769 | Not-determined |
| S14_extracted_k141_51478 | Not-determined |
| S14_extracted_k141_102725 | Not-determined |
| S14_extracted_k141_88025 | Not-determined |
| S14_extracted_k141_44167 | Not-determined |
| S14_extracted_k141_88028 | Not-determined |
| S14_extracted_k141_88029 | Low-quality |
| S14_extracted_k141_58819 | Not-determined |
| S14_extracted_k141_51485 | Low-quality |
| S14_extracted_k141_221 | Low-quality |
| S14_extracted_k141_73357 | Not-determined |
| S14_extracted_k141_102728 | Low-quality |
| S14_extracted_k141_223 | Low-quality |
| S14_extracted_k141_224 | Low-quality |
| S14_extracted_k141_58831 | Not-determined |
| S14_extracted_k141_7526 | Not-determined |
| S14_extracted_k141_29526 | Low-quality |
| S14_extracted_k141_95422 | Low-quality |
| S14_extracted_k141_58837 | Not-determined |
| S14_extracted_k141_88044 | Not-determined |
| S14_extracted_k141_88046 | Low-quality |
| S14_extracted_k141_237 | Not-determined |
| S14_extracted_k141_238 | Not-determined |
| S14_extracted_k141_22179 | Not-determined |
| S14_extracted_k141_242 | Not-determined |
| S14_extracted_k141_22187 | Low-quality |
| S14_extracted_k141_80780 | Low-quality |
| S14_extracted_k141_80786 | Not-determined |
| S14_extracted_k141_110032 | Low-quality |
| S14_extracted_k141_80792 | Low-quality |
| S14_extracted_k141_110035 | Low-quality |
| S14_extracted_k141_80794 | Low-quality |

| | |
|---|---|
| S14_extracted_k141_110041 | Low-quality |
| S14_extracted_k141_58853 | Not-determined |
| S14_extracted_k141_14815 | Not-determined |
| S14_extracted_k141_29530 | Not-determined |
| S14_extracted_k141_95430 | Not-determined |
| S14_extracted_k141_22200 | Low-quality |
| S14_extracted_k141_110052 | Low-quality |
| S14_extracted_k141_95436 | Not-determined |
| S14_extracted_k141_36890 | Not-determined |
| S14_extracted_k141_36891 | Not-determined |
| S14_extracted_k141_14828 | Not-determined |
| S14_extracted_k141_58868 | Not-determined |
| S14_extracted_k141_36893 | Not-determined |
| S14_extracted_k141_7566 | Low-quality |
| S14_extracted_k141_44195 | Low-quality |
| S14_extracted_k141_88057 | Low-quality |
| S14_extracted_k141_110058 | Low-quality |
| S14_extracted_k141_88059 | Not-determined |
| S14_extracted_k141_95444 | Not-determined |
| S14_extracted_k141_263 | Not-determined |
| S14_extracted_k141_36902 | Not-determined |
| S14_extracted_k141_88064 | Not-determined |
| S14_extracted_k141_80798 | Low-quality |
| S14_extracted_k141_51508 | Low-quality |
| S14_extracted_k141_7581 | Not-determined |
| S14_extracted_k141_80803 | Not-determined |
| S14_extracted_k141_51512 | Low-quality |
| S14_extracted_k141_36905 | Not-determined |
| S14_extracted_k141_36907 | Not-determined |
| S14_extracted_k141_36910 | Not-determined |
| S14_extracted_k141_51516 | Low-quality |
| S14_extracted_k141_36914 | Not-determined |
| S14_extracted_k141_95455 | Not-determined |
| S14_extracted_k141_278 | Low-quality |
| S14_extracted_k141_88093 | Not-determined |
| S14_extracted_k141_80813 | Not-determined |
| S14_extracted_k141_95462 | Low-quality |
| S14_extracted_k141_58879 | Not-determined |
| S14_extracted_k141_51535 | Low-quality |
| S14_extracted_k141_51536 | Low-quality |
| S14_extracted_k141_7598 | Not-determined |
| S14_extracted_k141_14837 | Low-quality |
| S14_extracted_k141_29537 | Not-determined |
| S14_extracted_k141_51540 | Low-quality |
| S14_extracted_k141_58892 | Not-determined |
| S14_extracted_k141_88114 | Not-determined |
| S14_extracted_k141_36933 | Not-determined |
| S14_extracted_k141_36934 | Not-determined |
| S14_extracted_k141_88120 | Not-determined |
| S14_extracted_k141_36945 | Not-determined |
| S14_extracted_k141_88126 | Low-quality |
| S14_extracted_k141_36946 | Not-determined |
| S14_extracted_k141_88127 | Not-determined |
| S14_extracted_k141_14850 | Not-determined |
| S14_extracted_k141_298 | Low-quality |
| S14_extracted_k141_302 | Low-quality |
| S14_extracted_k141_29541 | Not-determined |
| S14_extracted_k141_58898 | Low-quality |
| S14_extracted_k141_51552 | Not-determined |
| S14_extracted_k141_29544 | Not-determined |
| S14_extracted_k141_22220 | Low-quality |
| S14_extracted_k141_22222 | Low-quality |
| S14_extracted_k141_51556 | Not-determined |
| S14_extracted_k141_36965 | Not-determined |
| S14_extracted_k141_22225 | Not-determined |
| S14_extracted_k141_36969 | Low-quality |
| S14_extracted_k141_14857 | Not-determined |
| S14_extracted_k141_307 | Not-determined |
| S14_extracted_k141_7612 | Not-determined |
| S14_extracted_k141_310 | Not-determined |
| S14_extracted_k141_29557 | Not-determined |
| S14_extracted_k141_14866 | Low-quality |
| S14_extracted_k141_36987 | Low-quality |
| S14_extracted_k141_80829 | Not-determined |
| S14_extracted_k141_110083 | Not-determined |
| S14_extracted_k141_36991 | Low-quality |
| S14_extracted_k141_22248 | Not-determined |
| S14_extracted_k141_36992 | Not-determined |
| S14_extracted_k141_95497 | Low-quality |
| S14_extracted_k141_51561 | Not-determined |
| S14_extracted_k141_88132 | Low-quality |
| S14_extracted_k141_102743 | Low-quality |
| S14_extracted_k141_88134 | Not-determined |
| S14_extracted_k141_317 | Not-determined |
| S14_extracted_k141_73379 | Not-determined |
| S14_extracted_k141_29569 | Not-determined |
| S14_extracted_k141_95503 | Not-determined |
| S14_extracted_k141_102746 | Not-determined |
| S14_extracted_k141_322 | Not-determined |
| S14_extracted_k141_102748 | Not-determined |
| S14_extracted_k141_29572 | Not-determined |
| S14_extracted_k141_95505 | Low-quality |
| S14_extracted_k141_95508 | Not-determined |
| S14_extracted_k141_95510 | Not-determined |
| S14_extracted_k141_29578 | Not-determined |
| S14_extracted_k141_14874 | Not-determined |
| S14_extracted_k141_73386 | Low-quality |
| S14_extracted_k141_14875 | Not-determined |
| S14_extracted_k141_14876 | Low-quality |
| S14_extracted_k141_14877 | Low-quality |
| S14_extracted_k141_37001 | Not-determined |
| S14_extracted_k141_7632 | Not-determined |
| S14_extracted_k141_110100 | Low-quality |
| S14_extracted_k141_37005 | Low-quality |
| S14_extracted_k141_110102 | Low-quality |
| S14_extracted_k141_51564 | Not-determined |
| S14_extracted_k141_341 | Not-determined |
| S14_extracted_k141_80844 | Low-quality |
| S14_extracted_k141_95520 | Not-determined |
| S14_extracted_k141_7634 | Low-quality |
| S14_extracted_k141_80848 | Low-quality |
| S14_extracted_k141_73402 | Low-quality |
| S14_extracted_k141_73404 | Low-quality |
| S14_extracted_k141_80849 | Low-quality |
| S14_extracted_k141_102765 | Low-quality |
| S14_extracted_k141_73409 | Low-quality |
| S14_extracted_k141_37010 | Low-quality |
| S14_extracted_k141_14903 | Low-quality |
| S14_extracted_k141_102768 | Not-determined |
| S14_extracted_k141_29596 | Not-determined |
| S14_extracted_k141_102771 | Not-determined |
| S14_extracted_k141_102772 | Not-determined |
| S14_extracted_k141_14909 | Not-determined |
| S14_extracted_k141_29601 | Low-quality |
| S14_extracted_k141_29602 | Low-quality |
| S14_extracted_k141_88160 | Low-quality |
| S14_extracted_k141_51581 | Not-determined |
| S14_extracted_k141_102781 | Low-quality |
| S17_vs_k141_175097\|\|full | Medium-quality |
| S14_extracted_k141_7644 | Not-determined |
| S14_extracted_k141_14915 | Not-determined |
| S14_extracted_k141_14916 | Not-determined |
| S14_extracted_k141_51594 | Not-determined |
| S14_extracted_k141_37022 | Not-determined |
| S14_extracted_k141_58930 | Not-determined |
| S14_extracted_k141_51598 | Not-determined |
| S14_extracted_k141_58932 | Not-determined |
| S14_extracted_k141_95530 | Not-determined |
| S14_extracted_k141_58935 | Low-quality |
| S14_extracted_k141_58936 | Not-determined |
| S14_extracted_k141_95535 | Not-determined |
| S14_extracted_k141_88175 | Low-quality |
| S14_extracted_k141_88176 | Not-determined |
| S14_extracted_k141_14923 | Not-determined |
| S14_extracted_k141_51616 | Not-determined |
| S14_extracted_k141_37027 | Not-determined |
| S14_extracted_k141_359 | Not-determined |
| S14_extracted_k141_361 | Not-determined |
| S14_extracted_k141_29609 | Low-quality |
| S14_extracted_k141_22259 | Not-determined |
| S14_extracted_k141_95544 | Low-quality |
| S14_extracted_k141_58946 | Low-quality |
| S14_extracted_k141_58947 | Low-quality |
| S14_extracted_k141_51622 | Low-quality |
| S14_extracted_k141_51624 | Not-determined |
| S14_extracted_k141_88184 | Low-quality |
| S14_extracted_k141_37034 | Not-determined |
| S14_extracted_k141_381 | Not-determined |
| S14_extracted_k141_58953 | Low-quality |
| S14_extracted_k141_102820 | Low-quality |
| S14_extracted_k141_73431 | Low-quality |
| S14_extracted_k141_29614 | Not-determined |
| S14_extracted_k141_14940 | Low-quality |
| S14_extracted_k141_88197 | Low-quality |
| S14_extracted_k141_14946 | Not-determined |
| S14_extracted_k141_88199 | Low-quality |
| S14_extracted_k141_7671 | Not-determined |
| S14_extracted_k141_37043 | Low-quality |
| S14_extracted_k141_7673 | Low-quality |
| S14_extracted_k141_88207 | Not-determined |
| S14_extracted_k141_7678 | Low-quality |
| S14_extracted_k141_80872 | Not-determined |
| S14_extracted_k141_102834 | Not-determined |
| S14_extracted_k141_66144 | Not-determined |
| S14_extracted_k141_14953 | Not-determined |
| S14_extracted_k141_102839 | Low-quality |
| S14_extracted_k141_58968 | Low-quality |
| S14_extracted_k141_29623 | Not-determined |
| S14_extracted_k141_80879 | Low-quality |
| S14_extracted_k141_37052 | Low-quality |
| S14_extracted_k141_80883 | Low-quality |
| S14_extracted_k141_7684 | Not-determined |
| S14_extracted_k141_29633 | Low-quality |
| S14_extracted_k141_394 | Low-quality |
| S14_extracted_k141_66150 | Not-determined |
| S14_extracted_k141_110119 | Low-quality |
| S14_extracted_k141_29636 | Not-determined |
| S14_extracted_k141_51640 | Not-determined |
| S14_extracted_k141_102854 | Not-determined |
| S14_extracted_k141_401 | Not-determined |
| S14_extracted_k141_37061 | Low-quality |
| S14_extracted_k141_22280 | Low-quality |
| S14_extracted_k141_58984 | Not-determined |
| S14_extracted_k141_51647 | Not-determined |
| S14_extracted_k141_58987 | Not-determined |
| S14_extracted_k141_95558 | Not-determined |
| S14_extracted_k141_37068 | Low-quality |
| S14_extracted_k141_58991 | Not-determined |
| S14_extracted_k141_95559 | Low-quality |
| S14_extracted_k141_405 | Low-quality |
| S14_extracted_k141_408 | Low-quality |
| S14_extracted_k141_95565 | Not-determined |
| S14_extracted_k141_412 | Not-determined |
| S14_extracted_k141_413 | Low-quality |
| S14_extracted_k141_37071 | Not-determined |
| S14_extracted_k141_44208 | Not-determined |
| S14_extracted_k141_66170 | Not-determined |
| S14_extracted_k141_88237 | Low-quality |
| S14_extracted_k141_88240 | Not-determined |
| S14_extracted_k141_80891 | Low-quality |
| S14_extracted_k141_423 | Not-determined |
| S14_extracted_k141_7692 | Low-quality |
| S14_extracted_k141_424 | Not-determined |
| S14_extracted_k141_80895 | Low-quality |
| S14_extracted_k141_428 | Not-determined |
| S14_extracted_k141_80897 | Not-determined |
| S14_extracted_k141_37077 | Low-quality |
| S14_extracted_k141_51662 | Low-quality |
| S14_extracted_k141_51665 | Low-quality |
| S14_extracted_k141_29662 | Not-determined |
| S14_extracted_k141_66179 | Not-determined |
| S14_extracted_k141_51668 | Not-determined |
| S14_extracted_k141_59002 | Not-determined |
| S14_extracted_k141_110144 | Not-determined |
| S14_extracted_k141_66183 | Low-quality |
| S14_extracted_k141_110148 | Not-determined |
| S14_extracted_k141_95587 | Low-quality |
| S14_extracted_k141_443 | Not-determined |
| S14_extracted_k141_29679 | Not-determined |
| S14_extracted_k141_80904 | Low-quality |
| S14_extracted_k141_452 | Not-determined |
| S14_extracted_k141_66199 | Low-quality |
| S14_extracted_k141_110158 | Not-determined |
| S14_extracted_k141_29683 | Not-determined |
| S14_extracted_k141_29684 | Low-quality |
| S14_extracted_k141_14970 | Low-quality |
| S14_extracted_k141_110166 | Low-quality |
| S14_extracted_k141_29687 | Not-determined |
| S14_extracted_k141_14976 | Low-quality |
| S14_extracted_k141_102870 | Low-quality |
| S14_extracted_k141_14981 | Low-quality |
| S14_extracted_k141_102871 | Low-quality |
| S14_extracted_k141_110170 | Low-quality |
| S14_extracted_k141_95593 | Low-quality |
| S14_extracted_k141_51688 | Low-quality |
| S14_extracted_k141_29699 | Low-quality |
| S14_extracted_k141_110176 | Low-quality |
| S14_extracted_k141_95598 | Not-determined |
| S14_extracted_k141_29703 | Low-quality |
| S14_extracted_k141_29705 | Low-quality |
| S14_extracted_k141_29706 | Low-quality |
| S14_extracted_k141_29707 | Low-quality |
| S14_extracted_k141_59015 | Not-determined |
| S14_extracted_k141_102880 | Not-determined |
| S14_extracted_k141_95604 | Low-quality |
| S14_extracted_k141_88250 | Not-determined |
| S14_extracted_k141_102882 | Not-determined |
| S14_extracted_k141_80924 | Not-determined |
| S14_extracted_k141_110189 | Low-quality |
| S14_extracted_k141_102885 | Low-quality |
| S14_extracted_k141_102886 | Not-determined |
| S14_extracted_k141_59018 | Not-determined |
| S14_extracted_k141_95611 | Not-determined |
| S14_extracted_k141_110192 | Not-determined |
| S14_extracted_k141_14989 | Not-determined |
| S14_extracted_k141_88257 | Not-determined |
| S14_extracted_k141_88258 | Not-determined |
| S14_extracted_k141_88259 | Low-quality |
| S14_extracted_k141_59024 | Not-determined |
| S14_extracted_k141_14998 | Not-determined |
| S14_extracted_k141_73470 | Not-determined |
| S14_extracted_k141_110200 | Not-determined |
| S14_extracted_k141_73473 | Low-quality |
| S14_extracted_k141_486 | Low-quality |
| S14_extracted_k141_488 | Low-quality |
| S14_extracted_k141_88264 | Not-determined |
| S14_extracted_k141_88267 | Not-determined |
| S14_extracted_k141_15005 | Low-quality |
| S14_extracted_k141_95625 | Not-determined |
| S14_extracted_k141_110210 | Not-determined |
| S14_extracted_k141_102902 | Not-determined |
| S14_extracted_k141_102904 | Not-determined |
| S14_extracted_k141_15019 | Low-quality |
| S14_extracted_k141_80935 | Not-determined |
| S14_extracted_k141_102907 | Not-determined |
| S14_extracted_k141_102909 | Low-quality |
| S14_extracted_k141_15024 | Low-quality |
| S14_extracted_k141_95646 | Not-determined |
| S14_extracted_k141_59046 | Not-determined |
| S14_extracted_k141_44216 | Not-determined |
| S14_extracted_k141_80943 | Not-determined |
| S14_extracted_k141_44217 | Low-quality |
| S14_extracted_k141_7703 | Low-quality |
| S14_extracted_k141_496 | Low-quality |
| S14_extracted_k141_80944 | Low-quality |
| S14_extracted_k141_497 | Not-determined |
| S14_extracted_k141_7704 | Not-determined |
| S14_extracted_k141_88269 | Not-determined |
| S14_extracted_k141_59048 | Low-quality |
| S14_extracted_k141_7707 | Low-quality |
| S14_extracted_k141_59049 | Not-determined |
| S14_extracted_k141_7708 | Low-quality |
| S14_extracted_k141_15026 | Not-determined |
| S14_extracted_k141_73488 | Low-quality |
| S14_extracted_k141_506 | Not-determined |
| S14_extracted_k141_88275 | Not-determined |
| S14_extracted_k141_73494 | Not-determined |
| S14_extracted_k141_102918 | Low-quality |
| S14_extracted_k141_510 | Low-quality |
| S14_extracted_k141_73496 | Low-quality |
| S14_extracted_k141_51723 | Not-determined |
| S14_extracted_k141_512 | Not-determined |
| S14_extracted_k141_29736 | Not-determined |
| S14_extracted_k141_73498 | Low-quality |
| S14_extracted_k141_110225 | Not-determined |
| S14_extracted_k141_102921 | Not-determined |
| S14_extracted_k141_110230 | Low-quality |
| S14_extracted_k141_110231 | Not-determined |
| S14_extracted_k141_80962 | Not-determined |
| S14_extracted_k141_110232 | Not-determined |
| S14_extracted_k141_80963 | Low-quality |
| S14_extracted_k141_51729 | Low-quality |
| S14_extracted_k141_523 | Not-determined |
| S14_extracted_k141_526 | Not-determined |
| S14_extracted_k141_88286 | Not-determined |
| S14_extracted_k141_44250 | Low-quality |
| S14_extracted_k141_531 | Low-quality |
| S14_extracted_k141_110243 | Not-determined |
| S14_extracted_k141_88291 | Not-determined |
| S14_extracted_k141_88292 | Not-determined |
| S14_extracted_k141_44255 | Not-determined |
| S14_extracted_k141_80979 | Not-determined |
| S14_extracted_k141_22306 | Not-determined |
| S14_extracted_k141_95676 | Low-quality |
| S14_extracted_k141_95679 | Low-quality |
| S14_extracted_k141_44263 | Not-determined |
| S14_extracted_k141_102932 | Low-quality |
| S14_extracted_k141_51742 | Not-determined |
| S14_extracted_k141_110253 | Low-quality |
| S14_extracted_k141_110255 | Not-determined |
| S14_extracted_k141_59071 | Low-quality |
| S14_extracted_k141_22313 | Low-quality |
| S14_extracted_k141_545 | Not-determined |
| S14_extracted_k141_44275 | Not-determined |
| S14_extracted_k141_95694 | Not-determined |
| S14_extracted_k141_102941 | Not-determined |
| S14_extracted_k141_551 | Not-determined |
| S14_extracted_k141_22319 | Low-quality |
| S14_extracted_k141_44285 | Low-quality |
| S14_extracted_k141_59081 | Not-determined |
| S14_extracted_k141_37090 | Low-quality |
| S14_extracted_k141_95700 | Low-quality |
| S14_extracted_k141_37092 | Not-determined |
| S14_extracted_k141_37095 | Low-quality |
| S14_extracted_k141_15041 | Not-determined |
| S14_extracted_k141_95704 | Not-determined |
| S14_extracted_k141_44290 | Low-quality |
| S14_extracted_k141_95706 | Low-quality |
| S14_extracted_k141_80994 | Low-quality |
| S14_extracted_k141_95708 | Not-determined |
| S14_extracted_k141_80997 | Not-determined |
| S14_extracted_k141_7725 | Low-quality |
| S14_extracted_k141_37102 | Not-determined |
| S14_extracted_k141_59083 | Not-determined |
| S14_extracted_k141_80999 | Low-quality |
| S14_extracted_k141_560 | Low-quality |
| S14_extracted_k141_44295 | Low-quality |
| S14_extracted_k141_102954 | Not-determined |
| S14_extracted_k141_7728 | Not-determined |
| S14_extracted_k141_564 | Not-determined |
| S14_extracted_k141_73503 | Not-determined |
| S14_extracted_k141_102961 | Low-quality |
| S14_extracted_k141_44299 | Not-determined |
| S14_extracted_k141_44300 | Low-quality |
| S14_extracted_k141_29754 | Not-determined |
| S14_extracted_k141_102967 | Not-determined |
| S14_extracted_k141_29755 | Not-determined |
| S14_extracted_k141_73514 | Not-determined |
| S14_extracted_k141_66217 | Low-quality |
| S14_extracted_k141_73516 | Not-determined |
| S14_extracted_k141_66220 | Not-determined |
| S14_extracted_k141_37114 | Not-determined |
| S14_extracted_k141_22336 | Low-quality |
| S14_extracted_k141_37115 | Low-quality |
| S14_extracted_k141_44311 | Low-quality |
| S14_extracted_k141_577 | Low-quality |
| S14_extracted_k141_102975 | Not-determined |
| S14_extracted_k141_73523 | Not-determined |
| S14_extracted_k141_37117 | Not-determined |
| S14_extracted_k141_37118 | Low-quality |
| S14_extracted_k141_95730 | Low-quality |
| S14_extracted_k141_110263 | Low-quality |
| S14_extracted_k141_88311 | Not-determined |
| S14_extracted_k141_88312 | Not-determined |
| S14_extracted_k141_73527 | Low-quality |
| S14_extracted_k141_81007 | Not-determined |
| S14_extracted_k141_95739 | Not-determined |
| S14_extracted_k141_102984 | Not-determined |
| S14_extracted_k141_22345 | Low-quality |
| S14_extracted_k141_88317 | Low-quality |
| S14_extracted_k141_596 | Not-determined |
| S14_extracted_k141_597 | Not-determined |
| S14_extracted_k141_29763 | Not-determined |
| S14_extracted_k141_599 | Not-determined |
| S14_extracted_k141_95745 | Not-determined |
| S14_extracted_k141_29767 | Not-determined |
| S14_extracted_k141_29768 | Not-determined |
| S14_extracted_k141_88321 | Low-quality |
| S14_extracted_k141_102990 | Not-determined |
| S14_extracted_k141_88323 | Not-determined |
| S14_extracted_k141_102992 | Low-quality |
| S14_extracted_k141_88325 | Low-quality |
| S14_extracted_k141_102993 | Low-quality |
| S14_extracted_k141_88327 | Not-determined |
| S14_extracted_k141_73529 | Not-determined |
| S14_extracted_k141_608 | Low-quality |
| S14_extracted_k141_37129 | Not-determined |
| S14_extracted_k141_73531 | Low-quality |
| S14_extracted_k141_95755 | Low-quality |
| S14_extracted_k141_95758 | Not-determined |
| S14_extracted_k141_612 | Not-determined |
| S14_extracted_k141_29775 | Not-determined |
| S14_extracted_k141_88333 | Low-quality |
| S14_extracted_k141_88334 | Not-determined |
| S14_extracted_k141_37141 | Not-determined |
| S14_extracted_k141_29783 | Not-determined |
| S14_extracted_k141_29784 | Not-determined |
| S14_extracted_k141_95772 | Not-determined |
| S14_extracted_k141_37143 | Low-quality |
| S14_extracted_k141_103004 | Not-determined |
| S14_extracted_k141_103011 | Low-quality |
| S14_extracted_k141_88342 | Not-determined |
| S14_extracted_k141_7737 | Not-determined |
| S14_extracted_k141_44336 | Low-quality |
| S14_extracted_k141_95777 | Low-quality |
| S14_extracted_k141_620 | Not-determined |
| S14_extracted_k141_37153 | Low-quality |
| S14_extracted_k141_95781 | Not-determined |
| S14_extracted_k141_88351 | Low-quality |
| S14_extracted_k141_88353 | Not-determined |
| S14_extracted_k141_37159 | Not-determined |
| S14_extracted_k141_73540 | Not-determined |
| S14_extracted_k141_110285 | Not-determined |
| S14_extracted_k141_37162 | Not-determined |
| S14_extracted_k141_95782 | Low-quality |
| S14_extracted_k141_103017 | Not-determined |
| S14_extracted_k141_103019 | Not-determined |
| S14_extracted_k141_44343 | Not-determined |
| S14_extracted_k141_103022 | Not-determined |
| S14_extracted_k141_103024 | Not-determined |
| S14_extracted_k141_73543 | Low-quality |
| S14_extracted_k141_103025 | Low-quality |
| S14_extracted_k141_73544 | Low-quality |
| S14_extracted_k141_73549 | Not-determined |
| S14_extracted_k141_103028 | Not-determined |
| S14_extracted_k141_44357 | Not-determined |
| S14_extracted_k141_103029 | Low-quality |
| S14_extracted_k141_15062 | Not-determined |
| S14_extracted_k141_88360 | Not-determined |
| S14_extracted_k141_628 | Low-quality |
| S14_extracted_k141_73556 | Not-determined |
| S14_extracted_k141_88365 | Not-determined |
| S14_extracted_k141_22355 | Not-determined |
| S14_extracted_k141_88369 | Not-determined |
| S14_extracted_k141_66229 | Not-determined |
| S14_extracted_k141_103042 | Low-quality |
| S14_extracted_k141_22359 | Not-determined |
| S14_extracted_k141_110287 | Not-determined |
| S14_extracted_k141_88372 | Not-determined |
| S14_extracted_k141_103044 | Not-determined |
| S14_extracted_k141_110289 | Not-determined |
| S14_extracted_k141_15074 | Not-determined |
| S14_extracted_k141_81034 | Not-determined |
| S14_extracted_k141_103050 | Not-determined |
| S14_extracted_k141_81036 | Not-determined |
| S14_extracted_k141_103055 | Not-determined |
| S14_extracted_k141_15085 | Not-determined |
| S14_extracted_k141_81041 | Low-quality |
| S14_extracted_k141_103061 | Not-determined |
| S14_extracted_k141_103065 | Low-quality |
| S14_extracted_k141_15094 | Not-determined |
| S14_extracted_k141_95811 | Not-determined |
| S14_extracted_k141_37188 | Low-quality |
| S14_extracted_k141_103075 | Low-quality |
| S14_extracted_k141_7744 | Not-determined |
| S14_extracted_k141_103080 | Not-determined |
| S14_extracted_k141_103083 | Low-quality |
| S14_extracted_k141_73565 | Not-determined |
| S14_extracted_k141_103084 | Not-determined |
| S14_extracted_k141_22369 | Not-determined |
| S14_extracted_k141_37191 | Low-quality |
| S14_extracted_k141_103085 | Not-determined |
| S14_extracted_k141_95824 | Low-quality |
| S14_extracted_k141_59096 | Not-determined |
| S14_extracted_k141_7749 | Low-quality |
| S14_extracted_k141_73573 | Not-determined |
| S14_extracted_k141_29834 | Not-determined |
| S14_extracted_k141_44364 | Not-determined |
| S14_extracted_k141_103092 | Not-determined |
| S14_extracted_k141_22375 | Low-quality |
| S14_extracted_k141_7754 | Low-quality |
| S14_extracted_k141_22377 | Not-determined |
| S14_extracted_k141_81062 | Low-quality |
| S14_extracted_k141_7758 | Low-quality |
| S14_extracted_k141_29840 | Not-determined |
| S14_extracted_k141_81065 | Not-determined |
| S14_extracted_k141_88398 | Low-quality |
| S14_extracted_k141_29848 | Not-determined |
| S14_extracted_k141_81067 | Low-quality |
| S14_extracted_k141_29849 | Not-determined |
| S14_extracted_k141_634 | Not-determined |
| S14_extracted_k141_95831 | Not-determined |
| S14_extracted_k141_15121 | Not-determined |
| S14_extracted_k141_51757 | Not-determined |
| S14_extracted_k141_22379 | Not-determined |
| S14_extracted_k141_110296 | Not-determined |
| S14_extracted_k141_22385 | Not-determined |
| S14_extracted_k141_88403 | Low-quality |
| S14_extracted_k141_59104 | Low-quality |
| S14_extracted_k141_37217 | Low-quality |
| S14_extracted_k141_15132 | Not-determined |
| S14_extracted_k141_51764 | Low-quality |
| S14_extracted_k141_15137 | Not-determined |
| S14_extracted_k141_110306 | Not-determined |
| S14_extracted_k141_81081 | Not-determined |
| S14_extracted_k141_37223 | Not-determined |
| S14_extracted_k141_81082 | Low-quality |
| S14_extracted_k141_81083 | Not-determined |
| S14_extracted_k141_37225 | Not-determined |
| S14_extracted_k141_66251 | Not-determined |
| S14_extracted_k141_81086 | Low-quality |
| S14_extracted_k141_7787 | Not-determined |
| S14_extracted_k141_51774 | Not-determined |
| S14_extracted_k141_59107 | Low-quality |
| S14_extracted_k141_7791 | Not-determined |
| S14_extracted_k141_37228 | Not-determined |
| S14_extracted_k141_59113 | Not-determined |
| S14_extracted_k141_7794 | Not-determined |
| S14_extracted_k141_22398 | Not-determined |
| S14_extracted_k141_7795 | Not-determined |
| S14_extracted_k141_81089 | Not-determined |
| S14_extracted_k141_95844 | Not-determined |
| S14_extracted_k141_22399 | Low-quality |
| S14_extracted_k141_95846 | Low-quality |
| S14_extracted_k141_73587 | Low-quality |
| S14_extracted_k141_110322 | Not-determined |
| S14_extracted_k141_66264 | Not-determined |
| S14_extracted_k141_44381 | Not-determined |
| S14_extracted_k141_44382 | Not-determined |
| S14_extracted_k141_22405 | Not-determined |
| S14_extracted_k141_88419 | Not-determined |
| S14_extracted_k141_37242 | Low-quality |
| S14_extracted_k141_7805 | Not-determined |
| S14_extracted_k141_59120 | Not-determined |
| S14_extracted_k141_73591 | Not-determined |
| S14_extracted_k141_51782 | Low-quality |
| S14_extracted_k141_22407 | Not-determined |
| S14_extracted_k141_81094 | Low-quality |
| S14_extracted_k141_81097 | Not-determined |
| S14_extracted_k141_110330 | Not-determined |
| S14_extracted_k141_22408 | Low-quality |
| S14_extracted_k141_37243 | Not-determined |
| S14_extracted_k141_37244 | Not-determined |
| S14_extracted_k141_22409 | Not-determined |
| S14_extracted_k141_37247 | Low-quality |
| S14_extracted_k141_37248 | Low-quality |
| S14_extracted_k141_37252 | Not-determined |
| S14_extracted_k141_66272 | Not-determined |
| S14_extracted_k141_88429 | Not-determined |
| S14_extracted_k141_37254 | Low-quality |
| S14_extracted_k141_59137 | Low-quality |
| S14_extracted_k141_37260 | Not-determined |
| S14_extracted_k141_81102 | Not-determined |
| S14_extracted_k141_81103 | Not-determined |
| S14_extracted_k141_59145 | Not-determined |
| S14_extracted_k141_44385 | Not-determined |
| S14_extracted_k141_81107 | Not-determined |
| S14_extracted_k141_22421 | Low-quality |
| S14_extracted_k141_37272 | Low-quality |
| S14_extracted_k141_44388 | Not-determined |
| S14_extracted_k141_59152 | Not-determined |
| S14_extracted_k141_110347 | Low-quality |
| S14_extracted_k141_110348 | Low-quality |
| S14_extracted_k141_66289 | Low-quality |
| S14_extracted_k141_59157 | Low-quality |
| S14_extracted_k141_103109 | Not-determined |
| S14_extracted_k141_7824 | Not-determined |
| S14_extracted_k141_110351 | Low-quality |
| S14_extracted_k141_103111 | Not-determined |

| | |
|---|---|
| S14_extracted_k141_37276 | Low-quality |
| S14_extracted_k141_44390 | Not-determined |
| S14_extracted_k141_37277 | Low-quality |
| S14_extracted_k141_44391 | Not-determined |
| S14_extracted_k141_66294 | Not-determined |
| S14_extracted_k141_51803 | Not-determined |
| S14_extracted_k141_95855 | Not-determined |
| S14_extracted_k141_66296 | Low-quality |
| S14_extracted_k141_29876 | Not-determined |
| S14_extracted_k141_66298 | Low-quality |
| S14_extracted_k141_66300 | Low-quality |
| S14_extracted_k141_88447 | Not-determined |
| S14_extracted_k141_88449 | Low-quality |
| S14_extracted_k141_81119 | Low-quality |
| S14_extracted_k141_81120 | Not-determined |
| S14_extracted_k141_81121 | Not-determined |
| S14_extracted_k141_7830 | Low-quality |
| S14_extracted_k141_73604 | Low-quality |
| S14_extracted_k141_95861 | Not-determined |
| S14_extracted_k141_7832 | Low-quality |
| S14_extracted_k141_88454 | Low-quality |
| S14_extracted_k141_44402 | Low-quality |
| S14_extracted_k141_7835 | Not-determined |
| S14_extracted_k141_73611 | Low-quality |
| S14_extracted_k141_88457 | Low-quality |
| S14_extracted_k141_81128 | Not-determined |
| S14_extracted_k141_81129 | Low-quality |
| S14_extracted_k141_95866 | Not-determined |
| S14_extracted_k141_29886 | Not-determined |
| S14_extracted_k141_29887 | Not-determined |
| S14_extracted_k141_44405 | Not-determined |
| S14_extracted_k141_670 | Not-determined |
| S14_extracted_k141_44407 | Not-determined |
| S14_extracted_k141_15167 | Not-determined |
| S14_extracted_k141_95871 | Not-determined |
| S14_extracted_k141_81135 | Low-quality |
| S14_extracted_k141_81143 | Not-determined |
| S14_extracted_k141_680 | Not-determined |
| S14_extracted_k141_59167 | Not-determined |
| S14_extracted_k141_59168 | Not-determined |
| S14_extracted_k141_103131 | Low-quality |
| S14_extracted_k141_73628 | Low-quality |
| S14_extracted_k141_81147 | Not-determined |
| S14_extracted_k141_51810 | Not-determined |
| S14_extracted_k141_7846 | Not-determined |
| S14_extracted_k141_44423 | Not-determined |
| S14_extracted_k141_110354 | Low-quality |
| S14_extracted_k141_51812 | Not-determined |
| S14_extracted_k141_59172 | Low-quality |
| S14_extracted_k141_81153 | Low-quality |
| S14_extracted_k141_44425 | Not-determined |
| S14_extracted_k141_73636 | Low-quality |
| S14_extracted_k141_73638 | Low-quality |
| S14_extracted_k141_15175 | Low-quality |
| S14_extracted_k141_66325 | Not-determined |
| S14_extracted_k141_88473 | Not-determined |
| S14_extracted_k141_103143 | Not-determined |
| S14_extracted_k141_73643 | Not-determined |
| S14_extracted_k141_51830 | Low-quality |
| S14_extracted_k141_103146 | Low-quality |
| S14_extracted_k141_22428 | Not-determined |
| S14_extracted_k141_15178 | Low-quality |
| S14_extracted_k141_15180 | Not-determined |
| S14_extracted_k141_15181 | Not-determined |
| S14_extracted_k141_110368 | Not-determined |
| S14_extracted_k141_88498 | Low-quality |
| S14_extracted_k141_44431 | Not-determined |
| S14_extracted_k141_88499 | Not-determined |
| S14_extracted_k141_88501 | Not-determined |
| S14_extracted_k141_51860 | Not-determined |
| S14_extracted_k141_51861 | Not-determined |
| S14_extracted_k141_73668 | Low-quality |
| S14_extracted_k141_22439 | Not-determined |
| S14_extracted_k141_73669 | Low-quality |
| S14_extracted_k141_81172 | Not-determined |
| S14_extracted_k141_103154 | Not-determined |
| S14_extracted_k141_22445 | Not-determined |
| S14_extracted_k141_22449 | Not-determined |
| S14_extracted_k141_110374 | Low-quality |
| S14_extracted_k141_110375 | Low-quality |
| S14_extracted_k141_73673 | Low-quality |
| S14_extracted_k141_73674 | Low-quality |
| S14_extracted_k141_103162 | Low-quality |
| S14_extracted_k141_73675 | Low-quality |
| S14_extracted_k141_110386 | Not-determined |
| S14_extracted_k141_73676 | Low-quality |
| S14_extracted_k141_59180 | Not-determined |
| S14_extracted_k141_73680 | Not-determined |
| S14_extracted_k141_59182 | Not-determined |
| S14_extracted_k141_59183 | Not-determined |
| S14_extracted_k141_73682 | Low-quality |
| S14_extracted_k141_22460 | Low-quality |
| S14_extracted_k141_59184 | Not-determined |
| S14_extracted_k141_22464 | Not-determined |
| S14_extracted_k141_103169 | Not-determined |
| S14_extracted_k141_51869 | Not-determined |
| S14_extracted_k141_59188 | Not-determined |
| S14_extracted_k141_88506 | Low-quality |
| S14_extracted_k141_73687 | Not-determined |
| S14_extracted_k141_7878 | Not-determined |
| S14_extracted_k141_73689 | Not-determined |
| S14_extracted_k141_88510 | Not-determined |
| S14_extracted_k141_7882 | Not-determined |
| S14_extracted_k141_59197 | Not-determined |
| S14_extracted_k141_51872 | Not-determined |
| S14_extracted_k141_51873 | Not-determined |
| S14_extracted_k141_88517 | Not-determined |
| S14_extracted_k141_15220 | Not-determined |
| S14_extracted_k141_44442 | Not-determined |
| S14_extracted_k141_37290 | Low-quality |
| S14_extracted_k141_37292 | Not-determined |
| S14_extracted_k141_22480 | Not-determined |
| S14_extracted_k141_110404 | Low-quality |
| S14_extracted_k141_51887 | Not-determined |
| S14_extracted_k141_51889 | Not-determined |
| S14_extracted_k141_51890 | Low-quality |
| S14_extracted_k141_37302 | Low-quality |
| S14_extracted_k141_88545 | Not-determined |
| S14_extracted_k141_88549 | Low-quality |
| S14_extracted_k141_59210 | Not-determined |
| S14_extracted_k141_15226 | Not-determined |
| S14_extracted_k141_37309 | Low-quality |
| S14_extracted_k141_59214 | Not-determined |
| S14_extracted_k141_59216 | Not-determined |
| S14_extracted_k141_732 | Not-determined |
| S14_extracted_k141_735 | Low-quality |
| S14_extracted_k141_66343 | Not-determined |
| S14_extracted_k141_29953 | Low-quality |
| S14_extracted_k141_29955 | Not-determined |
| S14_extracted_k141_29959 | Low-quality |
| S14_extracted_k141_81194 | Not-determined |
| S14_extracted_k141_66360 | Low-quality |
| S14_extracted_k141_66362 | Low-quality |
| S14_extracted_k141_66365 | Low-quality |
| S14_extracted_k141_37315 | Not-determined |
| S11_vs_k141_13033\|\|full | High-quality |
| S14_extracted_k141_59235 | Low-quality |
| S14_extracted_k141_44453 | Not-determined |
| S14_extracted_k141_744 | Low-quality |
| S14_extracted_k141_746 | Not-determined |
| S14_extracted_k141_37323 | Not-determined |
| S14_extracted_k141_103190 | Not-determined |
| S14_extracted_k141_88566 | Low-quality |
| S14_extracted_k141_751 | Not-determined |
| S14_extracted_k141_51912 | Low-quality |
| S14_extracted_k141_22499 | Not-determined |
| S14_extracted_k141_752 | Low-quality |
| S14_extracted_k141_88571 | Low-quality |
| S14_extracted_k141_44461 | Not-determined |
| S14_extracted_k141_22501 | Not-determined |
| S14_extracted_k141_44464 | Not-determined |
| S14_extracted_k141_22502 | Low-quality |
| S14_extracted_k141_759 | Low-quality |
| S14_extracted_k141_770 | Not-determined |
| S14_extracted_k141_773 | Not-determined |
| S14_extracted_k141_775 | Not-determined |
| S14_extracted_k141_59255 | Not-determined |
| S14_extracted_k141_51929 | Low-quality |
| S14_extracted_k141_51931 | Not-determined |
| S14_extracted_k141_110422 | Low-quality |
| S14_extracted_k141_110423 | Not-determined |
| S14_extracted_k141_51936 | Not-determined |
| S14_extracted_k141_59260 | Low-quality |
| S14_extracted_k141_88586 | Low-quality |
| S14_extracted_k141_66392 | Not-determined |
| S14_extracted_k141_66396 | Not-determined |
| S14_extracted_k141_792 | Low-quality |
| S14_extracted_k141_110436 | Not-determined |
| S14_extracted_k141_110439 | Not-determined |
| S14_extracted_k141_51962 | Not-determined |
| S14_extracted_k141_51966 | Not-determined |
| S14_extracted_k141_51973 | Low-quality |
| S14_extracted_k141_73730 | Not-determined |
| S14_extracted_k141_66399 | Not-determined |
| S14_extracted_k141_73734 | Not-determined |
| S14_extracted_k141_81205 | Not-determined |
| S14_extracted_k141_73736 | Not-determined |
| S14_extracted_k141_73738 | Not-determined |
| S14_extracted_k141_51977 | Not-determined |
| S14_extracted_k141_59278 | Low-quality |
| S14_extracted_k141_81207 | Low-quality |
| S14_extracted_k141_51981 | Not-determined |
| S14_extracted_k141_37326 | Not-determined |
| S14_extracted_k141_37327 | Low-quality |
| S14_extracted_k141_51983 | Not-determined |
| S14_extracted_k141_66400 | Low-quality |
| S14_extracted_k141_66403 | Low-quality |
| S14_extracted_k141_59287 | Low-quality |
| S14_extracted_k141_110454 | Not-determined |
| S14_extracted_k141_110457 | Not-determined |
| S14_extracted_k141_66405 | Low-quality |
| S14_extracted_k141_37343 | Low-quality |
| S14_extracted_k141_110464 | Not-determined |
| S14_extracted_k141_37344 | Not-determined |
| S14_extracted_k141_110468 | Not-determined |
| S14_extracted_k141_73752 | Low-quality |
| S14_extracted_k141_37353 | Not-determined |
| S14_extracted_k141_66411 | Not-determined |
| S14_extracted_k141_51997 | Not-determined |
| S14_extracted_k141_110476 | Not-determined |
| S14_extracted_k141_66414 | Not-determined |
| S14_extracted_k141_52000 | Not-determined |
| S14_extracted_k141_103202 | Not-determined |
| S14_extracted_k141_110479 | Not-determined |
| S14_extracted_k141_66419 | Low-quality |
| S14_extracted_k141_88605 | Low-quality |
| S14_extracted_k141_52010 | Low-quality |
| S14_extracted_k141_88614 | Not-determined |
| S14_extracted_k141_59294 | Not-determined |
| S14_extracted_k141_59295 | Low-quality |
| S14_extracted_k141_59296 | Not-determined |
| S14_extracted_k141_81222 | Not-determined |
| S14_extracted_k141_81224 | Low-quality |
| S14_extracted_k141_7969 | Not-determined |
| S14_extracted_k141_110487 | Low-quality |
| S14_extracted_k141_88619 | Not-determined |
| S14_extracted_k141_73777 | Not-determined |
| S14_extracted_k141_81229 | Not-determined |
| S14_extracted_k141_22520 | Not-determined |
| S14_extracted_k141_7974 | Not-determined |
| S14_extracted_k141_81231 | Not-determined |
| S14_extracted_k141_816 | Low-quality |
| S14_extracted_k141_7976 | Low-quality |
| S14_extracted_k141_88627 | Not-determined |
| S14_extracted_k141_819 | Low-quality |
| S14_extracted_k141_95885 | Not-determined |
| S14_extracted_k141_95886 | Not-determined |
| S14_extracted_k141_52011 | Not-determined |
| S14_extracted_k141_37387 | Not-determined |
| S14_extracted_k141_30001 | Not-determined |
| S14_extracted_k141_52017 | Low-quality |
| S14_extracted_k141_95890 | Not-determined |
| S14_extracted_k141_44500 | Low-quality |
| S14_extracted_k141_44501 | Not-determined |
| S14_extracted_k141_88636 | Not-determined |
| S14_extracted_k141_52025 | Low-quality |
| S14_extracted_k141_7994 | Not-determined |
| S14_extracted_k141_44506 | Low-quality |
| S14_extracted_k141_110508 | Not-determined |
| S14_extracted_k141_7995 | Not-determined |
| S14_extracted_k141_22530 | Not-determined |
| S14_extracted_k141_95899 | Not-determined |
| S14_extracted_k141_95900 | Not-determined |
| S14_extracted_k141_95903 | Not-determined |
| S14_extracted_k141_30011 | Not-determined |
| S14_extracted_k141_88642 | Low-quality |
| S14_extracted_k141_52026 | Low-quality |
| S14_extracted_k141_37400 | Low-quality |
| S14_extracted_k141_22535 | Not-determined |
| S14_extracted_k141_52029 | Low-quality |
| S14_extracted_k141_37406 | Not-determined |
| S14_extracted_k141_88649 | Low-quality |
| S14_extracted_k141_44513 | Not-determined |
| S14_extracted_k141_44514 | Not-determined |
| S14_extracted_k141_44516 | Not-determined |
| S14_extracted_k141_88654 | Not-determined |
| S14_extracted_k141_44517 | Not-determined |
| S14_extracted_k141_81252 | Low-quality |
| S14_extracted_k141_110512 | Not-determined |
| S14_extracted_k141_22541 | Not-determined |
| S14_extracted_k141_22543 | Not-determined |
| S14_extracted_k141_110513 | Not-determined |
| S14_extracted_k141_8007 | Not-determined |
| S14_extracted_k141_59308 | Low-quality |
| S14_extracted_k141_88663 | Not-determined |
| S14_extracted_k141_81262 | Not-determined |
| S14_extracted_k141_30023 | Not-determined |
| S14_extracted_k141_823 | Not-determined |
| S14_extracted_k141_81274 | Not-determined |
| S14_extracted_k141_30029 | Low-quality |
| S14_extracted_k141_81279 | Low-quality |
| S14_extracted_k141_52052 | Not-determined |
| S14_extracted_k141_81282 | Not-determined |
| S14_extracted_k141_52055 | Low-quality |
| S14_extracted_k141_831 | Not-determined |
| S14_extracted_k141_81287 | Not-determined |
| S14_extracted_k141_30031 | Not-determined |
| S14_extracted_k141_52056 | Not-determined |
| S14_extracted_k141_81293 | Low-quality |
| S14_extracted_k141_37433 | Low-quality |
| S14_extracted_k141_81295 | Low-quality |
| S14_extracted_k141_110540 | Not-determined |
| S14_extracted_k141_37437 | Low-quality |
| S14_extracted_k141_81299 | Low-quality |
| S14_extracted_k141_81304 | Not-determined |
| S14_extracted_k141_59328 | Not-determined |
| S14_extracted_k141_37442 | Low-quality |
| S14_extracted_k141_840 | Not-determined |
| S14_extracted_k141_52071 | Not-determined |
| S14_extracted_k141_103221 | Low-quality |
| S14_extracted_k141_52077 | Low-quality |
| S14_extracted_k141_846 | Low-quality |
| S14_extracted_k141_52081 | Not-determined |
| S14_extracted_k141_37452 | Not-determined |
| S14_extracted_k141_81321 | Not-determined |
| S14_extracted_k141_15251 | Not-determined |
| S14_extracted_k141_103231 | Not-determined |
| S14_extracted_k141_52094 | Not-determined |
| S14_extracted_k141_59341 | Low-quality |
| S14_extracted_k141_8020 | Not-determined |
| S14_extracted_k141_8021 | Low-quality |
| S14_extracted_k141_52105 | Not-determined |
| S14_extracted_k141_73795 | Not-determined |
| S14_extracted_k141_73797 | Not-determined |
| S14_extracted_k141_88670 | Low-quality |
| S14_extracted_k141_30061 | Not-determined |
| S14_extracted_k141_88674 | Not-determined |
| S14_extracted_k141_30064 | Low-quality |
| S14_extracted_k141_37466 | Not-determined |
| S14_extracted_k141_52114 | Not-determined |
| S14_extracted_k141_37467 | Not-determined |
| S14_extracted_k141_73803 | Not-determined |
| S14_extracted_k141_30074 | Not-determined |
| S14_extracted_k141_73805 | Not-determined |
| S14_extracted_k141_30078 | Low-quality |
| S14_extracted_k141_73807 | Low-quality |
| S14_extracted_k141_862 | Not-determined |
| S14_extracted_k141_88686 | Low-quality |
| S14_extracted_k141_110566 | Not-determined |
| S14_extracted_k141_110567 | Not-determined |
| S14_extracted_k141_866 | Not-determined |
| S14_extracted_k141_44554 | Not-determined |
| S14_extracted_k141_73810 | Not-determined |
| S14_extracted_k141_52129 | Not-determined |
| S14_extracted_k141_37483 | Not-determined |
| S14_extracted_k141_73814 | Not-determined |
| S14_extracted_k141_52131 | Low-quality |
| S14_extracted_k141_52132 | Low-quality |
| S14_extracted_k141_37490 | Low-quality |
| S14_extracted_k141_37491 | Not-determined |
| S14_extracted_k141_52139 | Low-quality |
| S14_extracted_k141_8036 | Not-determined |
| S14_extracted_k141_30095 | Not-determined |
| S14_extracted_k141_8043 | Low-quality |
| S14_extracted_k141_73838 | Not-determined |
| S14_extracted_k141_15293 | Not-determined |
| S14_extracted_k141_15294 | Low-quality |
| S14_extracted_k141_15295 | Not-determined |
| S14_extracted_k141_73841 | Not-determined |
| S14_extracted_k141_88696 | Low-quality |
| S14_extracted_k141_8044 | Low-quality |
| S7_vs_34520\|\|full | High-quality |
| S14_extracted_k141_8047 | Low-quality |
| S14_extracted_k141_52156 | Not-determined |
| S14_extracted_k141_88697 | Low-quality |
| S14_extracted_k141_88698 | Low-quality |
| S14_extracted_k141_37504 | Not-determined |
| S14_extracted_k141_869 | Not-determined |
| S14_extracted_k141_8051 | Not-determined |
| S14_extracted_k141_59349 | Not-determined |
| S14_extracted_k141_73848 | Not-determined |
| S14_extracted_k141_88709 | Not-determined |
| S14_extracted_k141_37513 | Not-determined |
| S14_extracted_k141_15316 | Not-determined |
| S14_extracted_k141_882 | Not-determined |
| S14_extracted_k141_8063 | Not-determined |
| S14_extracted_k141_15324 | Not-determined |
| S14_extracted_k141_103234 | Low-quality |
| S14_extracted_k141_59364 | Not-determined |
| S14_extracted_k141_15326 | Low-quality |
| S14_extracted_k141_103239 | Low-quality |
| S14_extracted_k141_15329 | Not-determined |
| S14_extracted_k141_103243 | Low-quality |
| S14_extracted_k141_15332 | Not-determined |
| S14_extracted_k141_103246 | Not-determined |
| S14_extracted_k141_887 | Not-determined |
| S14_extracted_k141_103247 | Not-determined |
| S14_extracted_k141_8074 | Not-determined |
| S14_extracted_k141_15337 | Not-determined |
| S14_extracted_k141_110575 | Not-determined |
| S14_extracted_k141_890 | Not-determined |
| S14_extracted_k141_15344 | Not-determined |
| S14_extracted_k141_73870 | Not-determined |
| S14_extracted_k141_15349 | Low-quality |
| S14_extracted_k141_44563 | Not-determined |
| S14_extracted_k141_15352 | Low-quality |
| S14_extracted_k141_66453 | Low-quality |
| S14_extracted_k141_37534 | Low-quality |
| S14_extracted_k141_37538 | Low-quality |
| S14_extracted_k141_30111 | Not-determined |
| S14_extracted_k141_30112 | Low-quality |
| S14_extracted_k141_66457 | Not-determined |
| S14_extracted_k141_88724 | Not-determined |
| S14_extracted_k141_44574 | Not-determined |
| S14_extracted_k141_103264 | Low-quality |
| S14_extracted_k141_103266 | Not-determined |
| S14_extracted_k141_44576 | Low-quality |
| S14_extracted_k141_110583 | Not-determined |
| S14_extracted_k141_44579 | Low-quality |
| S14_extracted_k141_15362 | Low-quality |
| S14_extracted_k141_88739 | Not-determined |
| S14_extracted_k141_15364 | Not-determined |
| S14_extracted_k141_103273 | Not-determined |
| S14_extracted_k141_52172 | Not-determined |
| S14_extracted_k141_103280 | Low-quality |
| S14_extracted_k141_44587 | Not-determined |
| S14_extracted_k141_44588 | Not-determined |
| S14_extracted_k141_44591 | Not-determined |
| S14_extracted_k141_15370 | Low-quality |
| S14_extracted_k141_30131 | Low-quality |
| S14_extracted_k141_110592 | Low-quality |
| S14_extracted_k141_103285 | Not-determined |
| S14_extracted_k141_73887 | Not-determined |
| S14_extracted_k141_103286 | Not-determined |
| S14_extracted_k141_15380 | Not-determined |
| S14_extracted_k141_52184 | Low-quality |
| S14_extracted_k141_103295 | Not-determined |
| S14_extracted_k141_103296 | Not-determined |
| S14_extracted_k141_73890 | Low-quality |
| S14_extracted_k141_103298 | Not-determined |
| S14_extracted_k141_110605 | Not-determined |
| S14_extracted_k141_37554 | Low-quality |
| S14_extracted_k141_73899 | Not-determined |
| S14_extracted_k141_103308 | Not-determined |
| S14_extracted_k141_103315 | Not-determined |
| S14_extracted_k141_59379 | Low-quality |
| S14_extracted_k141_103317 | Not-determined |
| S14_extracted_k141_103323 | Not-determined |
| S14_extracted_k141_103324 | Not-determined |
| S14_extracted_k141_52190 | Not-determined |
| S14_extracted_k141_81329 | Not-determined |
| S14_extracted_k141_44603 | Low-quality |
| S14_extracted_k141_52194 | Low-quality |
| S14_extracted_k141_52195 | Low-quality |
| S14_extracted_k141_52196 | Not-determined |
| S14_extracted_k141_73916 | Not-determined |
| S14_extracted_k141_95926 | Not-determined |
| S14_extracted_k141_22550 | Low-quality |
| S14_extracted_k141_73926 | Low-quality |
| S14_extracted_k141_8099 | Not-determined |
| S14_extracted_k141_73930 | Not-determined |
| S14_extracted_k141_59389 | Not-determined |
| S14_extracted_k141_88758 | Not-determined |
| S14_extracted_k141_73932 | Low-quality |
| S14_extracted_k141_59391 | Not-determined |
| S14_extracted_k141_920 | Not-determined |
| S14_extracted_k141_44613 | Not-determined |
| S14_extracted_k141_22557 | Not-determined |
| S14_extracted_k141_44617 | Not-determined |
| S14_extracted_k141_44618 | Not-determined |
| S14_extracted_k141_22560 | Low-quality |
| S14_extracted_k141_44620 | Not-determined |
| S14_extracted_k141_22562 | Not-determined |
| S14_extracted_k141_22563 | Not-determined |
| S14_extracted_k141_88760 | Low-quality |
| S14_extracted_k141_30138 | Low-quality |
| S14_extracted_k141_59396 | Low-quality |
| S14_extracted_k141_73944 | Not-determined |
| S14_extracted_k141_30142 | Low-quality |
| S14_extracted_k141_95934 | Low-quality |
| S14_extracted_k141_110628 | Low-quality |
| S14_extracted_k141_73947 | Not-determined |
| S14_extracted_k141_66460 | Low-quality |
| S14_extracted_k141_52217 | Low-quality |
| S14_extracted_k141_95941 | Low-quality |
| S14_extracted_k141_66466 | Low-quality |
| S14_extracted_k141_52224 | Not-determined |
| S14_extracted_k141_81344 | Low-quality |
| S14_extracted_k141_66470 | Not-determined |
| S14_extracted_k141_37574 | Not-determined |
| S14_extracted_k141_8110 | Not-determined |
| S14_extracted_k141_59408 | Low-quality |
| S14_extracted_k141_59410 | Not-determined |
| S14_extracted_k141_110638 | Not-determined |
| S14_extracted_k141_110640 | Not-determined |
| S14_extracted_k141_103349 | Not-determined |
| S14_extracted_k141_15392 | Not-determined |
| S14_extracted_k141_81352 | Not-determined |
| S14_extracted_k141_52237 | Low-quality |
| S14_extracted_k141_103353 | Low-quality |
| S14_extracted_k141_37586 | Not-determined |
| S14_extracted_k141_59419 | Not-determined |
| S14_extracted_k141_88774 | Low-quality |
| S14_extracted_k141_103358 | Not-determined |
| S14_extracted_k141_88775 | Not-determined |
| S14_extracted_k141_103359 | Not-determined |
| S14_extracted_k141_37590 | Not-determined |
| S14_extracted_k141_8130 | Not-determined |
| S14_extracted_k141_88778 | Not-determined |
| S14_extracted_k141_15402 | Not-determined |
| S14_extracted_k141_88782 | Low-quality |
| S14_extracted_k141_8134 | Not-determined |
| S14_extracted_k141_52253 | Not-determined |
| S14_extracted_k141_110656 | Not-determined |
| S14_extracted_k141_59426 | Not-determined |
| S14_extracted_k141_103368 | Not-determined |
| S14_extracted_k141_30157 | Not-determined |
| S14_extracted_k141_37596 | Low-quality |
| S14_extracted_k141_52258 | Low-quality |
| S14_extracted_k141_52260 | Not-determined |
| S14_extracted_k141_81373 | Low-quality |
| S14_extracted_k141_52265 | Not-determined |
| S14_extracted_k141_110667 | Not-determined |
| S14_extracted_k141_95949 | Not-determined |
| S14_extracted_k141_52269 | Not-determined |
| S14_extracted_k141_110678 | Not-determined |
| S14_extracted_k141_22587 | Low-quality |
| S14_extracted_k141_30162 | Not-determined |
| S14_extracted_k141_73980 | Low-quality |
| S14_extracted_k141_44648 | Low-quality |
| S14_extracted_k141_103383 | Low-quality |
| S14_extracted_k141_943 | Not-determined |
| S14_extracted_k141_22617 | Not-determined |
| S14_extracted_k141_52277 | Not-determined |
| S14_extracted_k141_95950 | Low-quality |
| S14_extracted_k141_954 | Low-quality |
| S14_extracted_k141_44655 | Low-quality |
| S14_extracted_k141_73984 | Low-quality |
| S14_extracted_k141_959 | Low-quality |
| S14_extracted_k141_52280 | Low-quality |
| S14_extracted_k141_59468 | Not-determined |
| S14_extracted_k141_52284 | Low-quality |
| S14_extracted_k141_66478 | Low-quality |
| S14_extracted_k141_59472 | Low-quality |
| S14_extracted_k141_52289 | Low-quality |
| S14_extracted_k141_66484 | Not-determined |
| S14_extracted_k141_52297 | Not-determined |
| S14_extracted_k141_66485 | Low-quality |
| S14_extracted_k141_66486 | Not-determined |
| S14_extracted_k141_22628 | Low-quality |
| S14_extracted_k141_22631 | Low-quality |
| S14_extracted_k141_968 | Not-determined |
| S14_extracted_k141_15434 | Not-determined |
| S14_extracted_k141_44662 | Not-determined |
| S14_extracted_k141_15435 | Low-quality |
| S14_extracted_k141_22634 | Not-determined |
| S14_extracted_k141_22635 | Not-determined |
| S14_extracted_k141_103400 | Not-determined |
| S14_extracted_k141_74008 | Not-determined |
| S14_extracted_k141_30175 | Low-quality |
| S14_extracted_k141_88794 | Not-determined |
| S14_extracted_k141_37628 | Not-determined |
| S14_extracted_k141_37632 | Not-determined |
| S14_extracted_k141_81378 | Not-determined |
| S14_extracted_k141_74012 | Not-determined |
| S14_extracted_k141_22646 | Not-determined |
| S14_extracted_k141_74013 | Not-determined |
| S14_extracted_k141_74014 | Low-quality |
| S14_extracted_k141_81381 | Low-quality |
| S14_extracted_k141_30181 | Not-determined |
| S14_extracted_k141_59479 | Low-quality |
| S14_extracted_k141_81386 | Low-quality |
| S14_extracted_k141_81387 | Low-quality |
| S14_extracted_k141_88803 | Not-determined |
| S14_extracted_k141_44684 | Not-determined |
| S14_extracted_k141_88808 | Not-determined |
| S14_extracted_k141_88809 | Not-determined |
| S14_extracted_k141_44687 | Not-determined |
| S14_extracted_k141_88810 | Not-determined |
| S14_extracted_k141_88811 | Low-quality |
| S14_extracted_k141_44691 | Low-quality |
| S14_extracted_k141_15441 | Not-determined |
| S14_extracted_k141_103418 | Low-quality |
| S14_extracted_k141_44692 | Not-determined |
| S14_extracted_k141_44693 | Low-quality |
| S14_extracted_k141_74021 | Low-quality |
| S14_extracted_k141_971 | Not-determined |
| S14_extracted_k141_22665 | Not-determined |
| S14_extracted_k141_103425 | Not-determined |
| S14_extracted_k141_88819 | Low-quality |
| S14_extracted_k141_44699 | Not-determined |
| S14_extracted_k141_66493 | Not-determined |
| S14_extracted_k141_44700 | Not-determined |
| S14_extracted_k141_22672 | Not-determined |
| S14_extracted_k141_15450 | Not-determined |
| S14_extracted_k141_979 | Low-quality |
| S14_extracted_k141_984 | Low-quality |
| S14_extracted_k141_88830 | Low-quality |
| S14_extracted_k141_986 | Low-quality |
| S14_extracted_k141_59496 | Low-quality |
| S14_extracted_k141_59498 | Not-determined |
| S14_extracted_k141_30202 | Not-determined |
| S14_extracted_k141_991 | Not-determined |
| S14_extracted_k141_74046 | Not-determined |
| S14_extracted_k141_30204 | Low-quality |
| S14_extracted_k141_110699 | Not-determined |
| S14_extracted_k141_997 | Low-quality |
| S14_extracted_k141_59503 | Not-determined |
| S14_extracted_k141_15459 | Not-determined |
| S14_extracted_k141_15462 | Not-determined |
| S14_extracted_k141_103431 | Not-determined |
| S14_extracted_k141_110704 | Not-determined |
| S14_extracted_k141_110705 | Not-determined |
| S14_extracted_k141_37668 | Not-determined |
| S14_extracted_k141_15465 | Not-determined |
| S14_extracted_k141_30218 | Low-quality |
| S14_extracted_k141_44725 | Not-determined |
| S14_extracted_k141_81397 | Low-quality |
| S14_extracted_k141_110710 | Low-quality |

| | |
|---|---|
| S14_extracted_k141_88847 | Not-determined |
| S14_extracted_k141_1007 | Not-determined |
| S14_extracted_k141_22685 | Not-determined |
| S14_extracted_k141_81401 | Low-quality |
| S14_extracted_k141_44726 | Not-determined |
| S14_extracted_k141_81402 | Not-determined |
| S14_extracted_k141_59515 | Not-determined |
| S14_extracted_k141_59518 | Not-determined |
| S14_extracted_k141_88849 | Not-determined |
| S14_extracted_k141_74053 | Low-quality |
| S14_extracted_k141_74054 | Low-quality |
| S14_extracted_k141_30229 | Not-determined |
| S14_extracted_k141_88852 | Not-determined |
| S14_extracted_k141_110718 | Not-determined |
| S14_extracted_k141_88861 | Low-quality |
| S14_extracted_k141_88863 | Not-determined |
| S14_extracted_k141_30240 | Low-quality |
| S14_extracted_k141_110722 | Not-determined |
| S14_extracted_k141_88868 | Not-determined |
| S14_extracted_k141_74063 | Not-determined |
| S14_extracted_k141_110727 | Not-determined |
| S14_extracted_k141_30244 | Not-determined |
| S14_extracted_k141_15486 | Low-quality |
| S14_extracted_k141_110731 | Not-determined |
| S14_extracted_k141_103457 | Not-determined |
| S14_extracted_k141_44744 | Low-quality |
| S14_extracted_k141_66513 | Low-quality |
| S14_extracted_k141_44746 | Not-determined |
| S14_extracted_k141_1018 | Low-quality |
| S14_extracted_k141_44748 | Low-quality |
| S14_extracted_k141_44749 | Not-determined |
| S14_extracted_k141_44750 | Low-quality |
| S14_extracted_k141_1021 | Not-determined |
| S14_extracted_k141_95984 | Not-determined |
| S14_extracted_k141_44752 | Low-quality |
| S14_extracted_k141_66521 | Not-determined |
| S14_extracted_k141_88878 | Not-determined |
| S14_extracted_k141_15501 | Low-quality |
| S14_extracted_k141_30245 | Not-determined |
| S14_extracted_k141_15503 | Low-quality |
| S14_extracted_k141_66526 | Not-determined |
| S14_extracted_k141_103473 | Not-determined |
| S14_extracted_k141_95995 | Not-determined |
| S14_extracted_k141_88879 | Not-determined |
| S14_extracted_k141_103475 | Not-determined |
| S14_extracted_k141_59532 | Low-quality |
| S14_extracted_k141_44756 | Not-determined |
| S14_extracted_k141_37688 | Not-determined |
| S14_extracted_k141_44757 | Not-determined |
| S14_extracted_k141_37689 | Not-determined |
| S14_extracted_k141_30256 | Not-determined |
| S14_extracted_k141_30260 | Not-determined |
| S14_extracted_k141_44761 | Not-determined |
| S14_extracted_k141_44762 | Not-determined |
| S14_extracted_k141_44764 | Not-determined |
| S14_extracted_k141_37694 | Not-determined |
| S14_extracted_k141_103485 | Not-determined |
| S14_extracted_k141_103491 | Not-determined |
| S14_extracted_k141_44767 | Low-quality |
| S14_extracted_k141_103493 | Low-quality |
| S14_extracted_k141_66533 | Not-determined |
| S14_extracted_k141_103500 | Low-quality |
| S14_extracted_k141_44775 | Not-determined |
| S14_extracted_k141_30278 | Not-determined |
| S14_extracted_k141_44776 | Not-determined |
| S14_extracted_k141_52329 | Low-quality |
| S14_extracted_k141_44777 | Not-determined |
| S14_extracted_k141_103503 | Not-determined |
| S14_extracted_k141_44779 | Not-determined |
| S14_extracted_k141_1049 | Not-determined |
| S14_extracted_k141_8155 | Low-quality |
| S14_extracted_k141_8156 | Low-quality |
| S14_extracted_k141_103508 | Low-quality |
| S14_extracted_k141_8158 | Not-determined |
| S14_extracted_k141_1052 | Not-determined |
| S14_extracted_k141_59550 | Not-determined |
| S14_extracted_k141_37700 | Low-quality |
| S14_extracted_k141_59552 | Not-determined |
| S14_extracted_k141_1057 | Low-quality |
| S14_extracted_k141_8163 | Not-determined |
| S14_extracted_k141_37703 | Low-quality |
| S14_extracted_k141_110756 | Not-determined |
| S14_extracted_k141_110757 | Not-determined |
| S14_extracted_k141_37705 | Not-determined |
| S14_extracted_k141_103521 | Not-determined |
| S14_extracted_k141_22742 | Low-quality |
| S14_extracted_k141_44786 | Not-determined |
| S14_extracted_k141_103523 | Low-quality |
| S14_extracted_k141_103529 | Not-determined |
| S14_extracted_k141_15526 | Low-quality |
| S14_extracted_k141_15528 | Low-quality |
| S14_extracted_k141_30298 | Not-determined |
| S14_extracted_k141_110782 | Low-quality |
| S14_extracted_k141_30300 | Low-quality |
| S14_extracted_k141_110787 | Not-determined |
| S14_extracted_k141_30302 | Low-quality |
| S14_extracted_k141_110788 | Not-determined |
| S14_extracted_k141_96016 | Not-determined |
| S14_extracted_k141_52343 | Not-determined |
| S14_extracted_k141_15531 | Not-determined |
| S14_extracted_k141_88895 | Not-determined |
| S14_extracted_k141_96025 | Not-determined |
| S14_extracted_k141_88899 | Low-quality |
| S14_extracted_k141_66546 | Low-quality |
| S14_extracted_k141_37715 | Not-determined |
| S14_extracted_k141_15542 | Low-quality |
| S14_extracted_k141_30313 | Low-quality |
| S14_extracted_k141_96035 | Not-determined |
| S14_extracted_k141_52355 | Low-quality |
| S14_extracted_k141_22763 | Not-determined |
| S14_extracted_k141_30316 | Low-quality |
| S14_extracted_k141_15546 | Not-determined |
| S14_extracted_k141_88903 | Not-determined |
| S14_extracted_k141_88906 | Not-determined |
| S14_extracted_k141_30327 | Not-determined |
| S14_extracted_k141_15548 | Low-quality |
| S14_extracted_k141_110808 | Not-determined |
| S14_extracted_k141_81413 | Not-determined |
| S14_extracted_k141_37721 | Low-quality |
| S14_extracted_k141_88914 | Not-determined |
| S14_extracted_k141_15551 | Not-determined |
| S14_extracted_k141_110815 | Not-determined |
| S14_extracted_k141_15553 | Not-determined |
| S14_extracted_k141_110819 | Low-quality |
| S14_extracted_k141_1072 | Not-determined |
| S14_extracted_k141_37727 | Not-determined |
| S14_extracted_k141_44805 | Not-determined |
| S14_extracted_k141_103542 | Not-determined |
| S14_extracted_k141_1079 | Not-determined |
| S14_extracted_k141_1082 | Not-determined |
| S14_extracted_k141_66584 | Not-determined |
| S14_extracted_k141_37734 | Not-determined |
| S14_extracted_k141_44812 | Low-quality |
| S14_extracted_k141_96049 | Not-determined |
| S14_extracted_k141_103559 | Not-determined |
| S14_extracted_k141_81432 | Low-quality |
| S14_extracted_k141_110832 | Not-determined |
| S14_extracted_k141_103572 | Low-quality |
| S14_extracted_k141_103573 | Not-determined |
| S14_extracted_k141_103574 | Low-quality |
| S14_extracted_k141_22769 | Not-determined |
| S14_extracted_k141_110837 | Low-quality |
| S14_extracted_k141_1098 | Not-determined |
| S14_extracted_k141_1099 | Not-determined |
| S14_extracted_k141_96062 | Not-determined |
| S14_extracted_k141_59589 | Not-determined |
| S14_extracted_k141_88928 | Low-quality |
| S14_extracted_k141_44825 | Not-determined |
| S14_extracted_k141_88936 | Not-determined |
| S14_extracted_k141_103584 | Not-determined |
| S14_extracted_k141_44833 | Not-determined |
| S14_extracted_k141_103588 | Not-determined |
| S14_extracted_k141_15575 | Not-determined |
| S14_extracted_k141_1122 | Low-quality |
| S14_extracted_k141_81443 | Low-quality |
| S14_extracted_k141_81445 | Not-determined |
| S14_extracted_k141_103600 | Low-quality |
| S14_extracted_k141_88954 | Not-determined |
| S14_extracted_k141_88955 | Not-determined |
| S14_extracted_k141_88957 | Not-determined |
| S14_extracted_k141_103602 | Not-determined |
| S14_extracted_k141_30351 | Not-determined |
| S14_extracted_k141_30353 | Not-determined |
| S14_extracted_k141_52384 | Low-quality |
| S14_extracted_k141_44846 | Low-quality |
| S14_extracted_k141_110845 | Low-quality |
| S14_extracted_k141_1129 | Not-determined |
| S14_extracted_k141_1132 | Not-determined |
| S14_extracted_k141_103612 | Not-determined |
| S14_extracted_k141_44853 | Not-determined |
| S14_extracted_k141_15592 | Low-quality |
| S14_extracted_k141_15594 | Not-determined |
| S14_extracted_k141_22788 | Not-determined |
| S14_extracted_k141_15595 | Not-determined |
| S14_extracted_k141_88970 | Not-determined |
| S14_extracted_k141_59607 | Not-determined |
| S14_extracted_k141_59608 | Not-determined |
| S14_extracted_k141_88973 | Not-determined |
| S14_extracted_k141_110858 | Low-quality |
| S14_extracted_k141_37765 | Low-quality |
| S14_extracted_k141_59612 | Not-determined |
| S14_extracted_k141_59615 | Not-determined |
| S14_extracted_k141_59618 | Not-determined |
| S14_extracted_k141_103630 | Low-quality |
| S14_extracted_k141_96066 | Not-determined |
| S14_extracted_k141_81451 | Not-determined |
| S14_extracted_k141_8183 | Not-determined |
| S14_extracted_k141_59631 | Not-determined |
| S14_extracted_k141_96081 | Low-quality |
| S14_extracted_k141_52399 | Not-determined |
| S14_extracted_k141_81461 | Low-quality |
| S14_extracted_k141_22800 | Low-quality |
| S14_extracted_k141_8189 | Not-determined |
| S14_extracted_k141_8190 | Not-determined |
| S14_extracted_k141_1142 | Not-determined |
| S14_extracted_k141_110883 | Not-determined |
| S14_extracted_k141_110884 | Not-determined |
| S14_extracted_k141_37778 | Low-quality |
| S14_extracted_k141_15612 | Not-determined |
| S14_extracted_k141_103655 | Not-determined |
| S14_extracted_k141_81469 | Not-determined |
| S14_extracted_k141_103656 | Not-determined |
| S14_extracted_k141_96087 | Not-determined |
| S14_extracted_k141_96088 | Not-determined |
| S14_extracted_k141_22810 | Low-quality |
| S14_extracted_k141_44872 | Low-quality |
| S14_extracted_k141_30365 | Low-quality |
| S14_extracted_k141_96094 | Not-determined |
| S14_extracted_k141_103662 | Low-quality |
| S14_extracted_k141_1147 | Not-determined |
| S14_extracted_k141_88987 | Low-quality |
| S14_extracted_k141_110893 | Low-quality |
| S14_extracted_k141_96101 | Low-quality |
| S14_extracted_k141_52403 | Low-quality |
| S14_extracted_k141_59640 | Low-quality |
| S14_extracted_k141_52405 | Not-determined |
| S14_extracted_k141_110898 | Not-determined |
| S14_extracted_k141_103669 | Not-determined |
| S14_extracted_k141_52409 | Not-determined |
| S14_extracted_k141_103671 | Not-determined |
| S14_extracted_k141_52411 | Not-determined |
| S14_extracted_k141_52415 | Not-determined |
| S14_extracted_k141_96104 | Not-determined |
| S14_extracted_k141_37805 | Not-determined |
| S14_extracted_k141_1174 | Not-determined |
| S14_extracted_k141_8206 | Low-quality |
| S14_extracted_k141_8207 | Not-determined |
| S14_extracted_k141_110910 | Low-quality |
| S14_extracted_k141_59651 | Low-quality |
| S14_extracted_k141_8208 | Low-quality |
| S14_extracted_k141_52423 | Not-determined |
| S14_extracted_k141_30391 | Low-quality |
| S14_extracted_k141_52428 | Low-quality |
| S14_extracted_k141_1180 | Not-determined |
| S14_extracted_k141_103683 | Not-determined |
| S14_extracted_k141_88994 | Not-determined |
| S14_extracted_k141_37814 | Low-quality |
| S14_extracted_k141_88996 | Low-quality |
| S14_extracted_k141_66603 | Not-determined |
| S14_extracted_k141_103694 | Not-determined |
| S14_extracted_k141_66605 | Not-determined |
| S14_extracted_k141_66608 | Not-determined |
| S14_extracted_k141_81481 | Low-quality |
| S14_extracted_k141_52440 | Not-determined |
| S14_extracted_k141_37821 | Not-determined |
| S14_extracted_k141_89003 | Low-quality |
| S14_extracted_k141_81487 | Not-determined |
| S14_extracted_k141_1186 | Low-quality |
| S14_extracted_k141_110916 | Not-determined |
| S14_extracted_k141_110917 | Low-quality |
| S14_extracted_k141_66618 | Low-quality |
| S14_extracted_k141_1191 | Not-determined |
| S14_extracted_k141_89006 | Not-determined |
| S14_extracted_k141_37837 | Low-quality |
| S14_extracted_k141_66622 | Not-determined |
| S14_extracted_k141_66625 | Not-determined |
| S14_extracted_k141_103703 | Not-determined |
| S14_extracted_k141_103704 | Low-quality |
| S14_extracted_k141_30399 | Not-determined |
| S14_extracted_k141_110926 | Low-quality |
| S14_extracted_k141_110930 | Low-quality |
| S14_extracted_k141_103708 | Not-determined |
| S14_extracted_k141_81502 | Not-determined |
| S14_extracted_k141_81503 | Not-determined |
| S14_extracted_k141_1196 | Low-quality |
| S14_extracted_k141_30423 | Low-quality |
| S14_extracted_k141_1197 | Not-determined |
| S14_extracted_k141_8235 | Not-determined |
| S14_extracted_k141_1199 | Not-determined |
| S14_extracted_k141_96112 | Not-determined |
| S14_extracted_k141_1201 | Not-determined |
| S14_extracted_k141_103718 | Low-quality |
| S14_extracted_k141_81510 | Not-determined |
| S14_extracted_k141_22833 | Not-determined |
| S14_extracted_k141_110939 | Not-determined |
| S14_extracted_k141_22834 | Low-quality |
| S14_extracted_k141_30433 | Low-quality |
| S14_extracted_k141_110941 | Not-determined |
| S14_extracted_k141_103727 | Low-quality |
| S14_extracted_k141_110949 | Low-quality |
| S14_extracted_k141_110951 | Not-determined |
| S14_extracted_k141_81517 | Low-quality |
| S14_extracted_k141_110959 | Not-determined |
| S14_extracted_k141_110964 | Not-determined |
| S14_extracted_k141_110974 | Low-quality |
| S14_extracted_k141_59669 | Not-determined |
| S14_extracted_k141_30438 | Low-quality |
| S14_extracted_k141_22844 | Not-determined |
| S14_extracted_k141_59678 | Not-determined |
| S14_extracted_k141_103741 | Low-quality |
| S14_extracted_k141_15657 | Low-quality |
| S14_extracted_k141_59686 | Low-quality |
| S14_extracted_k141_15661 | Low-quality |
| S14_extracted_k141_15662 | Not-determined |
| S14_extracted_k141_44878 | Not-determined |
| S14_extracted_k141_96127 | Low-quality |
| S14_extracted_k141_44890 | Not-determined |
| S14_extracted_k141_22860 | Not-determined |
| S14_extracted_k141_89016 | Low-quality |
| S14_extracted_k141_15673 | Not-determined |
| S14_extracted_k141_103767 | Not-determined |
| S14_extracted_k141_8258 | Low-quality |
| S14_extracted_k141_110987 | Not-determined |
| S14_extracted_k141_15683 | Low-quality |
| S14_extracted_k141_59699 | Not-determined |
| S14_extracted_k141_103771 | Low-quality |
| S14_extracted_k141_103772 | Low-quality |
| S14_extracted_k141_81550 | Not-determined |
| S14_extracted_k141_103777 | Not-determined |
| S14_extracted_k141_111002 | Low-quality |
| S14_extracted_k141_15694 | Low-quality |
| S14_extracted_k141_111007 | Not-determined |
| S14_extracted_k141_15698 | Low-quality |
| S14_extracted_k141_1211 | Low-quality |
| S14_extracted_k141_15702 | Low-quality |
| S14_extracted_k141_59712 | Low-quality |
| S14_extracted_k141_44920 | Not-determined |
| S14_extracted_k141_44921 | Not-determined |
| S14_extracted_k141_44923 | Low-quality |
| S14_extracted_k141_15707 | Not-determined |
| S14_extracted_k141_59720 | Low-quality |
| S14_extracted_k141_8285 | Low-quality |
| S14_extracted_k141_74072 | Low-quality |
| S14_extracted_k141_59726 | Not-determined |
| S14_extracted_k141_8291 | Not-determined |
| S14_extracted_k141_22878 | Low-quality |
| S14_extracted_k141_89053 | Not-determined |
| S14_extracted_k141_1222 | Not-determined |
| S14_extracted_k141_1223 | Not-determined |
| S14_extracted_k141_1225 | Not-determined |
| S14_extracted_k141_89057 | Not-determined |
| S14_extracted_k141_89061 | Low-quality |
| S14_extracted_k141_44932 | Not-determined |
| S14_extracted_k141_66635 | Not-determined |
| S14_extracted_k141_89062 | Not-determined |
| S14_extracted_k141_89063 | Not-determined |
| S14_extracted_k141_44937 | Not-determined |
| S14_extracted_k141_1230 | Low-quality |
| S14_extracted_k141_15726 | Not-determined |
| S14_extracted_k141_15730 | Not-determined |
| S14_extracted_k141_66646 | Low-quality |
| S14_extracted_k141_1242 | Low-quality |
| S14_extracted_k141_37862 | Low-quality |
| S14_extracted_k141_37863 | Not-determined |
| S14_extracted_k141_1245 | Not-determined |
| S14_extracted_k141_103792 | Low-quality |
| S14_extracted_k141_37865 | Not-determined |
| S14_extracted_k141_37867 | Low-quality |
| S14_extracted_k141_59744 | Not-determined |
| S14_extracted_k141_74100 | Low-quality |
| S14_extracted_k141_74101 | Low-quality |
| S14_extracted_k141_22906 | Low-quality |
| S14_extracted_k141_37876 | Not-determined |
| S14_extracted_k141_30480 | Not-determined |
| S14_extracted_k141_66649 | Low-quality |
| S14_extracted_k141_30495 | Not-determined |
| S14_extracted_k141_74116 | Not-determined |
| S14_extracted_k141_111029 | Low-quality |
| S14_extracted_k141_66665 | Not-determined |
| S14_extracted_k141_66667 | Low-quality |
| S14_extracted_k141_74125 | Not-determined |
| S14_extracted_k141_37899 | Not-determined |
| S14_extracted_k141_74127 | Low-quality |
| S14_extracted_k141_59761 | Low-quality |
| S14_extracted_k141_8324 | Not-determined |
| S14_extracted_k141_30502 | Not-determined |
| S14_extracted_k141_103804 | Not-determined |
| S14_extracted_k141_15753 | Not-determined |
| S14_extracted_k141_44959 | Low-quality |
| S14_extracted_k141_44960 | Low-quality |
| S14_extracted_k141_8328 | Low-quality |
| S14_extracted_k141_15758 | Low-quality |
| S14_extracted_k141_66669 | Low-quality |
| S14_extracted_k141_15759 | Not-determined |
| S14_extracted_k141_30507 | Low-quality |
| S14_extracted_k141_74143 | Low-quality |
| S14_extracted_k141_15762 | Not-determined |
| S14_extracted_k141_15763 | Low-quality |
| S14_extracted_k141_37910 | Not-determined |
| S14_extracted_k141_15766 | Not-determined |
| S14_extracted_k141_44968 | Low-quality |
| S14_extracted_k141_44969 | Low-quality |
| S14_extracted_k141_89092 | Low-quality |
| S14_extracted_k141_44971 | Not-determined |
| S14_extracted_k141_96133 | Not-determined |
| S14_extracted_k141_37926 | Not-determined |
| S14_extracted_k141_37927 | Not-determined |
| S14_extracted_k141_44978 | Low-quality |
| S14_extracted_k141_30508 | Low-quality |
| S14_extracted_k141_37928 | Not-determined |
| S14_extracted_k141_44979 | Not-determined |
| S14_extracted_k141_74157 | Not-determined |
| S14_extracted_k141_89095 | Not-determined |
| S14_extracted_k141_74158 | Not-determined |
| S14_extracted_k141_81565 | Not-determined |
| S14_extracted_k141_37932 | Not-determined |
| S14_extracted_k141_15786 | Not-determined |
| S14_extracted_k141_37933 | Not-determined |
| S14_extracted_k141_30512 | Low-quality |
| S14_extracted_k141_81569 | Not-determined |
| S14_extracted_k141_30515 | Not-determined |
| S14_extracted_k141_111045 | Not-determined |
| S14_extracted_k141_89102 | Low-quality |
| S14_extracted_k141_81573 | Not-determined |
| S14_extracted_k141_15793 | Not-determined |
| S14_extracted_k141_66683 | Not-determined |
| S14_extracted_k141_44995 | Low-quality |
| S14_extracted_k141_89107 | Low-quality |
| S14_extracted_k141_44996 | Low-quality |
| S14_extracted_k141_111054 | Not-determined |
| S14_extracted_k141_74168 | Low-quality |
| S14_extracted_k141_89112 | Not-determined |
| S14_extracted_k141_89116 | Not-determined |
| S14_extracted_k141_15803 | Not-determined |
| S14_extracted_k141_89118 | Low-quality |
| S14_extracted_k141_66689 | Not-determined |
| S14_extracted_k141_96147 | Not-determined |
| S14_extracted_k141_74176 | Low-quality |
| S14_extracted_k141_8342 | Low-quality |
| S14_extracted_k141_8344 | Low-quality |
| S14_extracted_k141_66695 | Low-quality |
| S14_extracted_k141_74183 | Not-determined |
| S14_extracted_k141_74186 | Not-determined |
| S14_extracted_k141_81581 | Not-determined |
| S14_extracted_k141_8351 | Not-determined |
| S14_extracted_k141_74189 | Not-determined |
| S14_extracted_k141_81585 | Not-determined |
| S14_extracted_k141_74192 | Not-determined |
| S14_extracted_k141_81587 | Not-determined |
| S14_extracted_k141_89125 | Not-determined |
| S14_extracted_k141_89126 | Not-determined |
| S14_extracted_k141_30535 | Low-quality |
| S14_extracted_k141_37955 | Not-determined |
| S14_extracted_k141_37956 | Not-determined |
| S14_extracted_k141_111080 | Not-determined |
| S14_extracted_k141_111081 | Not-determined |
| S14_extracted_k141_30537 | Not-determined |
| S14_extracted_k141_37958 | Not-determined |
| S14_extracted_k141_74210 | Not-determined |
| S14_extracted_k141_15826 | Low-quality |
| S14_extracted_k141_30542 | Not-determined |
| S14_extracted_k141_74213 | Not-determined |
| S14_extracted_k141_15827 | Not-determined |
| S14_extracted_k141_89129 | Not-determined |
| S14_extracted_k141_15832 | Not-determined |
| S14_extracted_k141_30554 | Not-determined |
| S14_extracted_k141_8374 | Not-determined |
| S14_extracted_k141_8375 | Low-quality |
| S14_extracted_k141_8376 | Low-quality |
| S14_extracted_k141_74221 | Not-determined |
| S14_extracted_k141_8378 | Not-determined |
| S14_extracted_k141_8379 | Low-quality |
| S14_extracted_k141_103831 | Not-determined |
| S14_extracted_k141_103834 | Not-determined |
| S14_extracted_k141_81601 | Low-quality |
| S14_extracted_k141_103839 | Not-determined |
| S14_extracted_k141_81606 | Not-determined |
| S14_extracted_k141_81608 | Not-determined |
| S14_extracted_k141_111095 | Low-quality |
| S14_extracted_k141_22917 | Low-quality |
| S14_extracted_k141_15856 | Not-determined |
| S14_extracted_k141_37969 | Not-determined |
| S14_extracted_k141_103849 | Not-determined |
| S14_extracted_k141_103852 | Low-quality |
| S14_extracted_k141_37977 | Not-determined |
| S14_extracted_k141_111104 | Not-determined |
| S14_extracted_k141_81611 | Not-determined |
| S14_extracted_k141_81612 | Low-quality |
| S14_extracted_k141_37979 | Not-determined |
| S14_extracted_k141_111108 | Not-determined |
| S14_extracted_k141_59780 | Not-determined |
| S14_extracted_k141_22930 | Not-determined |
| S14_extracted_k141_15864 | Low-quality |
| S14_extracted_k141_81618 | Not-determined |
| S14_extracted_k141_111114 | Not-determined |
| S14_extracted_k141_45019 | Low-quality |
| S14_extracted_k141_111120 | Low-quality |
| S14_extracted_k141_22941 | Not-determined |
| S14_extracted_k141_89137 | Not-determined |
| S14_extracted_k141_89139 | Not-determined |
| S14_extracted_k141_8411 | Not-determined |
| S14_extracted_k141_89141 | Not-determined |
| S14_extracted_k141_89142 | Not-determined |
| S14_extracted_k141_8414 | Not-determined |
| S14_extracted_k141_15871 | Not-determined |
| S14_extracted_k141_15872 | Low-quality |
| S14_extracted_k141_81636 | Low-quality |
| S14_extracted_k141_111124 | Not-determined |
| S14_extracted_k141_37981 | Low-quality |
| S14_extracted_k141_15875 | Low-quality |
| S14_extracted_k141_37984 | Low-quality |
| S14_extracted_k141_81639 | Low-quality |
| S14_extracted_k141_15879 | Not-determined |
| S14_extracted_k141_37988 | Low-quality |
| S14_extracted_k141_81646 | Not-determined |
| S14_extracted_k141_66702 | Not-determined |
| S14_extracted_k141_81648 | Low-quality |
| S14_extracted_k141_22950 | Low-quality |
| S14_extracted_k141_66705 | Low-quality |
| S14_extracted_k141_37994 | Not-determined |
| S14_extracted_k141_8425 | Not-determined |
| S14_extracted_k141_15884 | Not-determined |
| S14_extracted_k141_15885 | Low-quality |
| S14_extracted_k141_15886 | Not-determined |
| S14_extracted_k141_66709 | Low-quality |
| S14_extracted_k141_30568 | Not-determined |
| S14_extracted_k141_52491 | Low-quality |
| S14_extracted_k141_66710 | Low-quality |
| S14_extracted_k141_66712 | Not-determined |
| S14_extracted_k141_111138 | Not-determined |
| S14_extracted_k141_66713 | Not-determined |
| S14_extracted_k141_8436 | Not-determined |
| S14_extracted_k141_8437 | Not-determined |
| S14_extracted_k141_30574 | Not-determined |
| S14_extracted_k141_66719 | Low-quality |
| S14_extracted_k141_52502 | Not-determined |
| S14_extracted_k141_52506 | Not-determined |
| S14_extracted_k141_52508 | Not-determined |
| S14_extracted_k141_74225 | Not-determined |
| S14_extracted_k141_103877 | Low-quality |
| S14_extracted_k141_111158 | Not-determined |
| S14_extracted_k141_103879 | Not-determined |
| S14_extracted_k141_74230 | Low-quality |
| S14_extracted_k141_103882 | Not-determined |
| S14_extracted_k141_81678 | Low-quality |
| S14_extracted_k141_103884 | Not-determined |
| S14_extracted_k141_81680 | Not-determined |
| S14_extracted_k141_111165 | Not-determined |
| S14_extracted_k141_38002 | Low-quality |
| S14_extracted_k141_15893 | Low-quality |
| S14_extracted_k141_59821 | Not-determined |
| S14_extracted_k141_81689 | Low-quality |
| S14_extracted_k141_22975 | Not-determined |
| S14_extracted_k141_89157 | Not-determined |
| S14_extracted_k141_111172 | Not-determined |
| S14_extracted_k141_45030 | Not-determined |
| S14_extracted_k141_111174 | Not-determined |
| S14_extracted_k141_111175 | Low-quality |
| S14_extracted_k141_45033 | Not-determined |
| S14_extracted_k141_89163 | Not-determined |
| S14_extracted_k141_15905 | Not-determined |
| S14_extracted_k141_103902 | Not-determined |
| S14_extracted_k141_30589 | Low-quality |
| S14_extracted_k141_59826 | Low-quality |
| S14_extracted_k141_74254 | Low-quality |
| S14_extracted_k141_74256 | Low-quality |
| S14_extracted_k141_74258 | Low-quality |
| S14_extracted_k141_111179 | Low-quality |
| S14_extracted_k141_81701 | Not-determined |
| S14_extracted_k141_22986 | Not-determined |
| S14_extracted_k141_74262 | Low-quality |
| S14_extracted_k141_15914 | Not-determined |
| S14_extracted_k141_89176 | Not-determined |
| S14_extracted_k141_22988 | Not-determined |
| S14_extracted_k141_103913 | Low-quality |
| S14_extracted_k141_103914 | Low-quality |
| S14_extracted_k141_15918 | Low-quality |
| S14_extracted_k141_15919 | Not-determined |
| S14_extracted_k141_74270 | Not-determined |
| S14_extracted_k141_74273 | Not-determined |
| S14_extracted_k141_38016 | Low-quality |
| S14_extracted_k141_30615 | Not-determined |
| S14_extracted_k141_22997 | Low-quality |
| S14_extracted_k141_38019 | Not-determined |
| S14_extracted_k141_23002 | Low-quality |
| S14_extracted_k141_30624 | Not-determined |
| S14_extracted_k141_111193 | Not-determined |
| S14_extracted_k141_81718 | Not-determined |
| S14_extracted_k141_15931 | Low-quality |
| S14_extracted_k141_30627 | Not-determined |
| S14_extracted_k141_23007 | Not-determined |
| S14_extracted_k141_81723 | Not-determined |
| S14_extracted_k141_45053 | Not-determined |
| S14_extracted_k141_111200 | Not-determined |
| S14_extracted_k141_23013 | Not-determined |
| S14_extracted_k141_103919 | Not-determined |
| S14_extracted_k141_15938 | Not-determined |
| S14_extracted_k141_103921 | Not-determined |
| S14_extracted_k141_81727 | Low-quality |
| S14_extracted_k141_15941 | Not-determined |
| S14_extracted_k141_103927 | Not-determined |
| S14_extracted_k141_81729 | Not-determined |
| S14_extracted_k141_15952 | Not-determined |
| S14_extracted_k141_111217 | Not-determined |
| S14_extracted_k141_23021 | Low-quality |
| S14_extracted_k141_30633 | Low-quality |
| S14_extracted_k141_8456 | Not-determined |
| S14_extracted_k141_74284 | Low-quality |
| S14_extracted_k141_111223 | Not-determined |

| | |
|---|---|
| S14_extracted_k141_74286 | Low-quality |
| S14_extracted_k141_45070 | Low-quality |
| S14_extracted_k141_45071 | Low-quality |
| S14_extracted_k141_45072 | Not-determined |
| S14_extracted_k141_89204 | Low-quality |
| S14_extracted_k141_103942 | Low-quality |
| S14_extracted_k141_52516 | Low-quality |
| S14_extracted_k141_74292 | Not-determined |
| S14_extracted_k141_81754 | Low-quality |
| S14_extracted_k141_74294 | Low-quality |
| S14_extracted_k141_111226 | Not-determined |
| S14_extracted_k141_81757 | Not-determined |
| S14_extracted_k141_81759 | Not-determined |
| S14_extracted_k141_81760 | Not-determined |
| S14_extracted_k141_66738 | Not-determined |
| S14_extracted_k141_111232 | Low-quality |
| S14_extracted_k141_8467 | Not-determined |
| S14_extracted_k141_111235 | Not-determined |
| S14_extracted_k141_81768 | Not-determined |
| S14_extracted_k141_89220 | Not-determined |
| S14_extracted_k141_74308 | Low-quality |
| S14_extracted_k141_111238 | Not-determined |
| S14_extracted_k141_66742 | Not-determined |
| S14_extracted_k141_15959 | Low-quality |
| S14_extracted_k141_52523 | Not-determined |
| S14_extracted_k141_52524 | Low-quality |
| S14_extracted_k141_81775 | Not-determined |
| S14_extracted_k141_81776 | Not-determined |
| S14_extracted_k141_66747 | Not-determined |
| S14_extracted_k141_59845 | Not-determined |
| S14_extracted_k141_103971 | Not-determined |
| S14_extracted_k141_52529 | Low-quality |
| S14_extracted_k141_15971 | Not-determined |
| S14_extracted_k141_103974 | Not-determined |
| S14_extracted_k141_89229 | Not-determined |
| S14_extracted_k141_81784 | Not-determined |
| S14_extracted_k141_15973 | Not-determined |
| S14_extracted_k141_81785 | Not-determined |
| S14_extracted_k141_89230 | Not-determined |
| S14_extracted_k141_103977 | Low-quality |
| S14_extracted_k141_59851 | Not-determined |
| S14_extracted_k141_81788 | Not-determined |
| S14_extracted_k141_81790 | Not-determined |
| S14_extracted_k141_15974 | Not-determined |
| S14_extracted_k141_45082 | Low-quality |
| S14_extracted_k141_81793 | Low-quality |
| S14_extracted_k141_15976 | Not-determined |
| S14_extracted_k141_45086 | Low-quality |
| S14_extracted_k141_23057 | Not-determined |
| S14_extracted_k141_96164 | Not-determined |
| S14_extracted_k141_23058 | Not-determined |
| S14_extracted_k141_89239 | Not-determined |
| S14_extracted_k141_81798 | Not-determined |
| S14_extracted_k141_59862 | Not-determined |
| S14_extracted_k141_81801 | Not-determined |
| S14_extracted_k141_96166 | Not-determined |
| S14_extracted_k141_74331 | Not-determined |
| S14_extracted_k141_111253 | Not-determined |
| S14_extracted_k141_15986 | Low-quality |
| S14_extracted_k141_96170 | Low-quality |
| S14_extracted_k141_89244 | Low-quality |
| S14_extracted_k141_81809 | Not-determined |
| S14_extracted_k141_66769 | Not-determined |
| S14_extracted_k141_81811 | Low-quality |
| S14_extracted_k141_74337 | Not-determined |
| S14_extracted_k141_89248 | Low-quality |
| S14_extracted_k141_96175 | Low-quality |
| S14_extracted_k141_89249 | Not-determined |
| S14_extracted_k141_74341 | Not-determined |
| S14_extracted_k141_81814 | Not-determined |
| S14_extracted_k141_96179 | Not-determined |
| S14_extracted_k141_74344 | Not-determined |
| S14_extracted_k141_96188 | Not-determined |
| S14_extracted_k141_111265 | Not-determined |
| S14_extracted_k141_38040 | Low-quality |
| S14_extracted_k141_74346 | Not-determined |
| S14_extracted_k141_59898 | Not-determined |
| S14_extracted_k141_30672 | Not-determined |
| S14_extracted_k141_59908 | Not-determined |
| S14_extracted_k141_74353 | Not-determined |
| S14_extracted_k141_59912 | Not-determined |
| S14_extracted_k141_89274 | Not-determined |
| S14_extracted_k141_38051 | Low-quality |
| S14_extracted_k141_38052 | Not-determined |
| S14_extracted_k141_59918 | Not-determined |
| S14_extracted_k141_59919 | Not-determined |
| S14_extracted_k141_38057 | Not-determined |
| S14_extracted_k141_59922 | Not-determined |
| S14_extracted_k141_59923 | Not-determined |
| S14_extracted_k141_38060 | Not-determined |
| S14_extracted_k141_38063 | Not-determined |
| S14_extracted_k141_52547 | Not-determined |
| S14_extracted_k141_96215 | Not-determined |
| S14_extracted_k141_52548 | Not-determined |
| S14_extracted_k141_23083 | Not-determined |
| S14_extracted_k141_66791 | Not-determined |
| S14_extracted_k141_66794 | Low-quality |
| S14_extracted_k141_23087 | Not-determined |
| S14_extracted_k141_52552 | Not-determined |
| S14_extracted_k141_59936 | Not-determined |
| S14_extracted_k141_74369 | Not-determined |
| S14_extracted_k141_81827 | Low-quality |
| S14_extracted_k141_81828 | Low-quality |
| S14_extracted_k141_52557 | Not-determined |
| S14_extracted_k141_74374 | Low-quality |
| S14_extracted_k141_81836 | Low-quality |
| S14_extracted_k141_38067 | Not-determined |
| S14_extracted_k141_74382 | Not-determined |
| S14_extracted_k141_66796 | Not-determined |
| S14_extracted_k141_38074 | Not-determined |
| S14_extracted_k141_66801 | Low-quality |
| S14_extracted_k141_52562 | Not-determined |
| S14_extracted_k141_111270 | Not-determined |
| S14_extracted_k141_52567 | Not-determined |
| S14_extracted_k141_111271 | Not-determined |
| S14_extracted_k141_81846 | Not-determined |
| S14_extracted_k141_8491 | Low-quality |
| S14_extracted_k141_52572 | Not-determined |
| S14_extracted_k141_74395 | Not-determined |
| S14_extracted_k141_74396 | Not-determined |
| S14_extracted_k141_111277 | Not-determined |
| S14_extracted_k141_52573 | Low-quality |
| S14_extracted_k141_96230 | Not-determined |
| S14_extracted_k141_8498 | Low-quality |
| S14_extracted_k141_96233 | Not-determined |
| S14_extracted_k141_74404 | Low-quality |
| S14_extracted_k141_74405 | Not-determined |
| S14_extracted_k141_81858 | Low-quality |
| S14_extracted_k141_81859 | Low-quality |
| S14_extracted_k141_74409 | Low-quality |
| S14_extracted_k141_23122 | Low-quality |
| S14_extracted_k141_96234 | Low-quality |
| S14_extracted_k141_52577 | Low-quality |
| S14_extracted_k141_23123 | Not-determined |
| S14_extracted_k141_16006 | Not-determined |
| S14_extracted_k141_23128 | Low-quality |
| S14_extracted_k141_111284 | Low-quality |
| S14_extracted_k141_111291 | Not-determined |
| S14_extracted_k141_38098 | Not-determined |
| S14_extracted_k141_16016 | Low-quality |
| S14_extracted_k141_8514 | Low-quality |
| S14_extracted_k141_104011 | Not-determined |
| S14_extracted_k141_104013 | Not-determined |
| S14_extracted_k141_38106 | Not-determined |
| S14_extracted_k141_104017 | Low-quality |
| S14_extracted_k141_104019 | Low-quality |
| S14_extracted_k141_23140 | Not-determined |
| S14_extracted_k141_16029 | Low-quality |
| S14_extracted_k141_66822 | Not-determined |
| S14_extracted_k141_66824 | Low-quality |
| S14_extracted_k141_74431 | Not-determined |
| S14_extracted_k141_89295 | Not-determined |
| S14_extracted_k141_111319 | Not-determined |
| S14_extracted_k141_59991 | Low-quality |
| S14_extracted_k141_23160 | Not-determined |
| S14_extracted_k141_8526 | Not-determined |
| S14_extracted_k141_8527 | Low-quality |
| S14_extracted_k141_111325 | Low-quality |
| S14_extracted_k141_104028 | Low-quality |
| S14_extracted_k141_111328 | Low-quality |
| S14_extracted_k141_74446 | Not-determined |
| S14_extracted_k141_23165 | Not-determined |
| S14_extracted_k141_66839 | Low-quality |
| S14_extracted_k141_8531 | Low-quality |
| S14_extracted_k141_59997 | Not-determined |
| S14_extracted_k141_23168 | Low-quality |
| S14_extracted_k141_74450 | Not-determined |
| S14_extracted_k141_89323 | Not-determined |
| S14_extracted_k141_30680 | Low-quality |
| S14_extracted_k141_96265 | Not-determined |
| S14_extracted_k141_16059 | Low-quality |
| S14_extracted_k141_96273 | Low-quality |
| S14_extracted_k141_111341 | Low-quality |
| S14_extracted_k141_16067 | Not-determined |
| S14_extracted_k141_89336 | Not-determined |
| S14_extracted_k141_30687 | Not-determined |
| S14_extracted_k141_96278 | Not-determined |
| S14_extracted_k141_89339 | Not-determined |
| S14_extracted_k141_111348 | Not-determined |
| S14_extracted_k141_89343 | Not-determined |
| S14_extracted_k141_89344 | Not-determined |
| S14_extracted_k141_96285 | Not-determined |
| S14_extracted_k141_30693 | Not-determined |
| S14_extracted_k141_81886 | Not-determined |
| S14_extracted_k141_30694 | Low-quality |
| S14_extracted_k141_66842 | Low-quality |
| S14_extracted_k141_30696 | Not-determined |
| S14_extracted_k141_52601 | Not-determined |
| S14_extracted_k141_30703 | Not-determined |
| S14_extracted_k141_104059 | Not-determined |
| S14_extracted_k141_81895 | Not-determined |
| S14_extracted_k141_8561 | Not-determined |
| S14_extracted_k141_66847 | Low-quality |
| S14_extracted_k141_30707 | Not-determined |
| S14_extracted_k141_89356 | Low-quality |
| S14_extracted_k141_81899 | Low-quality |
| S14_extracted_k141_66851 | Not-determined |
| S14_extracted_k141_81901 | Not-determined |
| S14_extracted_k141_81905 | Not-determined |
| S14_extracted_k141_111351 | Not-determined |
| S14_extracted_k141_74465 | Low-quality |
| S14_extracted_k141_16078 | Not-determined |
| S14_extracted_k141_16080 | Low-quality |
| S14_extracted_k141_38132 | Low-quality |
| S14_extracted_k141_38134 | Not-determined |
| S14_extracted_k141_111358 | Not-determined |
| S14_extracted_k141_111362 | Not-determined |
| S14_extracted_k141_16085 | Low-quality |
| S14_extracted_k141_81918 | Not-determined |
| S14_extracted_k141_81919 | Not-determined |
| S14_extracted_k141_38136 | Low-quality |
| S14_extracted_k141_111374 | Not-determined |
| S14_extracted_k141_66867 | Low-quality |
| S14_extracted_k141_104079 | Not-determined |
| S14_extracted_k141_104080 | Not-determined |
| S14_extracted_k141_111378 | Low-quality |
| S14_extracted_k141_30721 | Not-determined |
| S14_extracted_k141_104082 | Not-determined |
| S14_extracted_k141_23181 | Not-determined |
| S14_extracted_k141_66872 | Low-quality |
| S14_extracted_k141_81929 | Not-determined |
| S14_extracted_k141_96287 | Not-determined |
| S14_extracted_k141_1265 | Not-determined |
| S14_extracted_k141_111387 | Low-quality |
| S14_extracted_k141_1267 | Not-determined |
| S14_extracted_k141_81931 | Not-determined |
| S14_extracted_k141_8597 | Not-determined |
| S14_extracted_k141_81932 | Not-determined |
| S14_extracted_k141_8598 | Not-determined |
| S14_extracted_k141_96296 | Low-quality |
| S14_extracted_k141_66878 | Not-determined |
| S14_extracted_k141_52615 | Not-determined |
| S14_extracted_k141_16092 | Not-determined |
| S14_extracted_k141_74499 | Not-determined |
| S14_extracted_k141_38142 | Not-determined |
| S14_extracted_k141_8605 | Not-determined |
| S14_extracted_k141_74503 | Low-quality |
| S14_extracted_k141_38146 | Not-determined |
| S14_extracted_k141_16099 | Not-determined |
| S14_extracted_k141_16101 | Not-determined |
| S14_extracted_k141_1274 | Not-determined |
| S14_extracted_k141_38148 | Low-quality |
| S14_extracted_k141_16105 | Not-determined |
| S14_extracted_k141_38151 | Not-determined |
| S14_extracted_k141_8615 | Not-determined |
| S14_extracted_k141_38154 | Not-determined |
| S14_extracted_k141_104103 | Not-determined |
| S14_extracted_k141_8616 | Low-quality |
| S14_extracted_k141_8621 | Not-determined |
| S14_extracted_k141_1283 | Not-determined |
| S14_extracted_k141_38160 | Not-determined |
| S14_extracted_k141_81943 | Not-determined |
| S14_extracted_k141_23202 | Not-determined |
| S14_extracted_k141_38164 | Not-determined |
| S14_extracted_k141_30728 | Low-quality |
| S14_extracted_k141_38165 | Not-determined |
| S14_extracted_k141_104106 | Not-determined |
| S14_extracted_k141_8630 | Not-determined |
| S14_extracted_k141_23210 | Not-determined |
| S14_extracted_k141_74516 | Not-determined |
| S14_extracted_k141_104113 | Low-quality |
| S14_extracted_k141_81959 | Not-determined |
| S14_extracted_k141_81960 | Not-determined |
| S14_extracted_k141_66898 | Low-quality |
| S14_extracted_k141_16123 | Not-determined |
| S14_extracted_k141_8647 | Low-quality |
| S14_extracted_k141_111391 | Not-determined |
| S14_extracted_k141_38175 | Not-determined |
| S14_extracted_k141_66905 | Low-quality |
| S14_extracted_k141_38178 | Not-determined |
| S14_extracted_k141_30752 | Low-quality |
| S14_extracted_k141_96330 | Low-quality |
| S14_extracted_k141_66912 | Not-determined |
| S14_extracted_k141_66913 | Low-quality |
| S14_extracted_k141_96334 | Not-determined |
| S14_extracted_k141_30758 | Not-determined |
| S14_extracted_k141_30759 | Not-determined |
| S14_extracted_k141_30760 | Not-determined |
| S14_extracted_k141_104139 | Not-determined |
| S14_extracted_k141_8650 | Not-determined |
| S14_extracted_k141_38193 | Low-quality |
| S14_extracted_k141_66930 | Low-quality |
| S14_extracted_k141_23218 | Not-determined |
| S14_extracted_k141_66932 | Not-determined |
| S14_extracted_k141_38198 | Low-quality |
| S14_extracted_k141_38199 | Not-determined |
| S14_extracted_k141_81965 | Not-determined |
| S14_extracted_k141_8659 | Not-determined |
| S14_extracted_k141_66940 | Low-quality |
| S14_extracted_k141_16135 | Not-determined |
| S14_extracted_k141_23228 | Not-determined |
| S14_extracted_k141_96349 | Not-determined |
| S14_extracted_k141_104148 | Not-determined |
| S14_extracted_k141_74543 | Not-determined |
| S14_extracted_k141_104153 | Not-determined |
| S14_extracted_k141_8670 | Not-determined |
| S14_extracted_k141_111423 | Not-determined |
| S14_extracted_k141_8677 | Not-determined |
| S14_extracted_k141_1302 | Low-quality |
| S14_extracted_k141_60040 | Low-quality |
| S14_extracted_k141_8685 | Low-quality |
| S14_extracted_k141_23240 | Low-quality |
| S14_extracted_k141_38212 | Not-determined |
| S14_extracted_k141_16144 | Not-determined |
| S14_extracted_k141_104167 | Not-determined |
| S14_extracted_k141_111437 | Not-determined |
| S14_extracted_k141_23250 | Not-determined |
| S14_extracted_k141_52668 | Not-determined |
| S14_extracted_k141_38216 | Low-quality |
| S14_extracted_k141_66950 | Low-quality |
| S14_extracted_k141_23252 | Low-quality |
| S14_extracted_k141_104178 | Not-determined |
| S14_extracted_k141_66953 | Low-quality |
| S14_extracted_k141_1317 | Not-determined |
| S14_extracted_k141_1320 | Not-determined |
| S14_extracted_k141_1321 | Low-quality |
| S14_extracted_k141_1324 | Not-determined |
| S14_extracted_k141_52671 | Not-determined |
| S14_extracted_k141_1325 | Low-quality |
| S14_extracted_k141_60056 | Not-determined |
| S14_extracted_k141_81975 | Not-determined |
| S14_extracted_k141_81976 | Not-determined |
| S14_extracted_k141_52676 | Low-quality |
| S14_extracted_k141_104189 | Not-determined |
| S14_extracted_k141_81984 | Not-determined |
| S14_extracted_k141_104190 | Not-determined |
| S14_extracted_k141_16164 | Not-determined |
| S14_extracted_k141_74564 | Low-quality |
| S14_extracted_k141_52683 | Not-determined |
| S14_extracted_k141_81994 | Low-quality |
| S14_extracted_k141_111458 | Low-quality |
| S14_extracted_k141_52687 | Low-quality |
| S14_extracted_k141_96352 | Low-quality |
| S14_extracted_k141_96355 | Low-quality |
| S14_extracted_k141_66956 | Not-determined |
| S14_extracted_k141_82003 | Not-determined |
| S14_extracted_k141_82007 | Low-quality |
| S14_extracted_k141_104205 | Not-determined |
| S14_extracted_k141_60073 | Not-determined |
| S14_extracted_k141_104211 | Low-quality |
| S14_extracted_k141_104213 | Low-quality |
| S14_extracted_k141_111473 | Low-quality |
| S14_extracted_k141_111474 | Not-determined |
| S14_extracted_k141_111475 | Low-quality |
| S14_extracted_k141_104221 | Not-determined |
| S14_extracted_k141_23268 | Not-determined |
| S14_extracted_k141_23269 | Not-determined |
| S14_extracted_k141_23271 | Not-determined |
| S14_extracted_k141_16175 | Not-determined |
| S14_extracted_k141_60085 | Low-quality |
| S14_extracted_k141_23280 | Not-determined |
| S14_extracted_k141_104226 | Low-quality |
| S14_extracted_k141_111483 | Low-quality |
| S14_extracted_k141_1335 | Not-determined |
| S14_extracted_k141_52700 | Low-quality |
| S14_extracted_k141_52705 | Not-determined |
| S14_extracted_k141_8718 | Low-quality |
| S14_extracted_k141_8721 | Low-quality |
| S14_extracted_k141_52710 | Not-determined |
| S14_extracted_k141_8725 | Low-quality |
| S14_extracted_k141_96384 | Not-determined |
| S14_extracted_k141_66967 | Not-determined |
| S14_extracted_k141_66968 | Low-quality |
| S14_extracted_k141_96387 | Not-determined |
| S14_extracted_k141_16196 | Not-determined |
| S14_extracted_k141_96392 | Not-determined |
| S14_extracted_k141_82021 | Low-quality |
| S14_extracted_k141_82023 | Not-determined |
| S14_extracted_k141_52729 | Not-determined |
| S14_extracted_k141_60111 | Not-determined |
| S14_extracted_k141_52736 | Low-quality |
| S14_extracted_k141_60115 | Low-quality |
| S14_extracted_k141_8751 | Low-quality |
| S14_extracted_k141_16214 | Low-quality |
| S14_extracted_k141_60116 | Not-determined |
| S14_extracted_k141_8753 | Not-determined |
| S14_extracted_k141_111501 | Low-quality |
| S14_extracted_k141_23311 | Not-determined |
| S14_extracted_k141_52748 | Not-determined |
| S14_extracted_k141_23313 | Not-determined |
| S14_extracted_k141_16220 | Not-determined |
| S14_extracted_k141_23314 | Low-quality |
| S14_extracted_k141_82038 | Not-determined |
| S14_extracted_k141_16226 | Low-quality |
| S14_extracted_k141_66977 | Not-determined |
| S14_extracted_k141_96415 | Low-quality |
| S14_extracted_k141_1339 | Not-determined |
| S14_extracted_k141_111509 | Low-quality |
| S14_extracted_k141_52767 | Not-determined |
| S14_extracted_k141_1343 | Not-determined |
| S14_extracted_k141_66988 | Not-determined |
| S14_extracted_k141_8767 | Low-quality |
| S14_extracted_k141_30799 | Low-quality |
| S14_extracted_k141_52778 | Not-determined |
| S14_extracted_k141_1350 | Not-determined |
| S14_extracted_k141_8772 | Not-determined |
| S14_extracted_k141_8773 | Low-quality |
| S14_extracted_k141_30807 | Not-determined |
| S14_extracted_k141_66996 | Not-determined |
| S14_extracted_k141_60131 | Not-determined |
| S14_extracted_k141_1359 | Not-determined |
| S14_extracted_k141_52784 | Not-determined |
| S14_extracted_k141_8782 | Low-quality |
| S14_extracted_k141_67002 | Low-quality |
| S14_extracted_k141_111522 | Low-quality |
| S14_extracted_k141_67004 | Low-quality |
| S14_extracted_k141_16230 | Low-quality |
| S14_extracted_k141_1366 | Not-determined |
| S14_extracted_k141_96425 | Low-quality |
| S14_extracted_k141_111525 | Low-quality |
| S14_extracted_k141_1368 | Low-quality |
| S14_extracted_k141_60136 | Low-quality |
| S14_extracted_k141_16236 | Not-determined |
| S14_extracted_k141_67011 | Low-quality |
| S14_extracted_k141_30816 | Not-determined |
| S14_extracted_k141_67012 | Low-quality |
| S14_extracted_k141_52790 | Not-determined |
| S14_extracted_k141_30821 | Low-quality |
| S14_extracted_k141_52797 | Low-quality |
| S14_extracted_k141_60148 | Low-quality |
| S14_extracted_k141_52799 | Not-determined |
| S14_extracted_k141_60149 | Not-determined |
| S14_extracted_k141_16240 | Not-determined |
| S14_extracted_k141_60153 | Not-determined |
| S14_extracted_k141_111537 | Low-quality |
| S14_extracted_k141_111543 | Not-determined |
| S14_extracted_k141_23333 | Low-quality |
| S14_extracted_k141_52807 | Not-determined |
| S14_extracted_k141_1391 | Not-determined |
| S14_extracted_k141_82060 | Not-determined |
| S14_extracted_k141_30830 | Low-quality |
| S14_extracted_k141_23343 | Not-determined |
| S14_extracted_k141_82062 | Not-determined |
| S14_extracted_k141_111554 | Not-determined |
| S14_extracted_k141_60170 | Low-quality |
| S14_extracted_k141_30831 | Not-determined |
| S14_extracted_k141_104251 | Low-quality |
| S14_extracted_k141_82068 | Not-determined |
| S14_extracted_k141_23349 | Not-determined |
| S14_extracted_k141_30832 | Not-determined |
| S14_extracted_k141_30835 | Not-determined |
| S14_extracted_k141_111560 | Not-determined |
| S14_extracted_k141_30839 | Not-determined |
| S14_extracted_k141_82072 | Not-determined |
| S14_extracted_k141_23356 | Not-determined |
| S14_extracted_k141_23358 | Not-determined |
| S14_extracted_k141_111565 | Low-quality |
| S14_extracted_k141_1398 | Low-quality |
| S14_extracted_k141_104263 | Not-determined |
| S14_extracted_k141_60174 | Low-quality |
| S14_extracted_k141_30850 | Not-determined |
| S14_extracted_k141_111569 | Not-determined |
| S14_extracted_k141_89396 | Low-quality |
| S14_extracted_k141_104268 | Not-determined |
| S14_extracted_k141_89398 | Low-quality |
| S14_extracted_k141_89399 | Low-quality |
| S14_extracted_k141_89400 | Low-quality |
| S14_extracted_k141_89401 | Low-quality |
| S14_extracted_k141_104275 | Not-determined |
| S14_extracted_k141_30862 | Not-determined |
| S14_extracted_k141_89408 | Low-quality |
| S14_extracted_k141_104282 | Low-quality |
| S14_extracted_k141_16254 | Not-determined |
| S14_extracted_k141_38257 | Not-determined |
| S14_extracted_k141_60185 | Low-quality |
| S14_extracted_k141_111578 | Low-quality |
| S14_extracted_k141_60186 | Not-determined |
| S14_extracted_k141_67040 | Not-determined |
| S14_extracted_k141_60189 | Not-determined |
| S14_extracted_k141_104288 | Low-quality |
| S14_extracted_k141_38266 | Not-determined |
| S14_extracted_k141_38267 | Not-determined |
| S14_extracted_k141_104290 | Not-determined |
| S14_extracted_k141_38270 | Not-determined |
| S14_extracted_k141_38271 | Low-quality |
| S14_extracted_k141_30872 | Not-determined |
| S14_extracted_k141_52829 | Low-quality |
| S14_extracted_k141_38273 | Not-determined |
| S14_extracted_k141_104299 | Not-determined |
| S14_extracted_k141_38277 | Not-determined |
| S14_extracted_k141_96446 | Not-determined |
| S14_extracted_k141_52830 | Not-determined |
| S14_extracted_k141_60198 | Low-quality |
| S14_extracted_k141_104301 | Not-determined |
| S14_extracted_k141_104302 | Low-quality |
| S14_extracted_k141_104303 | Not-determined |
| S14_extracted_k141_89427 | Not-determined |
| S14_extracted_k141_52833 | Low-quality |
| S14_extracted_k141_96450 | Low-quality |
| S14_extracted_k141_52838 | Not-determined |
| S14_extracted_k141_30879 | Not-determined |
| S14_extracted_k141_30881 | Not-determined |
| S14_extracted_k141_52843 | Low-quality |
| S14_extracted_k141_104312 | Not-determined |
| S14_extracted_k141_96463 | Low-quality |
| S14_extracted_k141_8817 | Not-determined |
| S14_extracted_k141_96464 | Not-determined |
| S14_extracted_k141_89439 | Low-quality |
| S14_extracted_k141_89440 | Low-quality |
| S14_extracted_k141_67061 | Not-determined |
| S14_extracted_k141_38296 | Not-determined |
| S14_extracted_k141_89444 | Low-quality |
| S14_extracted_k141_74580 | Not-determined |
| S14_extracted_k141_52854 | Not-determined |
| S14_extracted_k141_74581 | Low-quality |
| S14_extracted_k141_45092 | Not-determined |
| S14_extracted_k141_52856 | Not-determined |
| S14_extracted_k141_67063 | Not-determined |
| S14_extracted_k141_67064 | Low-quality |
| S14_extracted_k141_60216 | Low-quality |
| S14_extracted_k141_38309 | Not-determined |
| S14_extracted_k141_30896 | Low-quality |
| S14_extracted_k141_8825 | Low-quality |
| S14_extracted_k141_8827 | Not-determined |
| S14_extracted_k141_67066 | Low-quality |
| S14_extracted_k141_45099 | Not-determined |
| S14_extracted_k141_60221 | Not-determined |
| S14_extracted_k141_74595 | Not-determined |
| S14_extracted_k141_38317 | Not-determined |
| S14_extracted_k141_23392 | Low-quality |
| S14_extracted_k141_74597 | Not-determined |
| S14_extracted_k141_60222 | Not-determined |
| S14_extracted_k141_52865 | Not-determined |
| S14_extracted_k141_8833 | Not-determined |
| S14_extracted_k141_104323 | Not-determined |
| S14_extracted_k141_67070 | Not-determined |
| S14_extracted_k141_67071 | Low-quality |
| S14_extracted_k141_67072 | Not-determined |
| S14_extracted_k141_60224 | Not-determined |
| S14_extracted_k141_67074 | Low-quality |
| S14_extracted_k141_23400 | Low-quality |
| S14_extracted_k141_111595 | Not-determined |
| S14_extracted_k141_23402 | Not-determined |
| S14_extracted_k141_82088 | Not-determined |
| S14_extracted_k141_104330 | Not-determined |
| S14_extracted_k141_104331 | Low-quality |
| S14_extracted_k141_8842 | Not-determined |
| S14_extracted_k141_82093 | Low-quality |
| S14_extracted_k141_67081 | Low-quality |
| S14_extracted_k141_67083 | Low-quality |
| S14_extracted_k141_30909 | Not-determined |
| S14_extracted_k141_1422 | Not-determined |
| S14_extracted_k141_8853 | Not-determined |
| S14_extracted_k141_60241 | Not-determined |
| S14_extracted_k141_60242 | Low-quality |
| S14_extracted_k141_89463 | Low-quality |
| S14_extracted_k141_23420 | Low-quality |
| S14_extracted_k141_52868 | Not-determined |
| S14_extracted_k141_89466 | Low-quality |
| S14_extracted_k141_104349 | Not-determined |
| S14_extracted_k141_96487 | Low-quality |
| S14_extracted_k141_1435 | Not-determined |
| S14_extracted_k141_89472 | Not-determined |
| S14_extracted_k141_23433 | Not-determined |
| S14_extracted_k141_82100 | Low-quality |
| S14_extracted_k141_104353 | Low-quality |
| S14_extracted_k141_52877 | Not-determined |
| S14_extracted_k141_104354 | Not-determined |
| S14_extracted_k141_60253 | Not-determined |
| S14_extracted_k141_23439 | Not-determined |
| S14_extracted_k141_23443 | Not-determined |
| S14_extracted_k141_38320 | Low-quality |
| S14_extracted_k141_89477 | Low-quality |
| S14_extracted_k141_30930 | Not-determined |
| S14_extracted_k141_23450 | Not-determined |
| S14_extracted_k141_52883 | Low-quality |
| S14_extracted_k141_30939 | Low-quality |
| S14_extracted_k141_1439 | Not-determined |
| S14_extracted_k141_104370 | Not-determined |
| S14_extracted_k141_104371 | Not-determined |
| S14_extracted_k141_52892 | Not-determined |
| S14_extracted_k141_96499 | Low-quality |
| S14_extracted_k141_104372 | Not-determined |
| S14_extracted_k141_74615 | Low-quality |
| S14_extracted_k141_96502 | Not-determined |
| S14_extracted_k141_104376 | Not-determined |
| S14_extracted_k141_82111 | Low-quality |
| S14_extracted_k141_45107 | Not-determined |
| S14_extracted_k141_30955 | Low-quality |
| S14_extracted_k141_52902 | Low-quality |
| S14_extracted_k141_45112 | Not-determined |
| S14_extracted_k141_8869 | Not-determined |
| S14_extracted_k141_30964 | Low-quality |
| S14_extracted_k141_52906 | Not-determined |
| S14_extracted_k141_30968 | Not-determined |
| S14_extracted_k141_60273 | Not-determined |
| S14_extracted_k141_8873 | Low-quality |
| S14_extracted_k141_82120 | Not-determined |
| S14_extracted_k141_96517 | Low-quality |

| | |
|---|---|
| S14_extracted_k141_60278 | Not-determined |
| S14_extracted_k141_96518 | Not-determined |
| S14_extracted_k141_16295 | Not-determined |
| S14_extracted_k141_96521 | Not-determined |
| S14_extracted_k141_38344 | Low-quality |
| S14_extracted_k141_82124 | Low-quality |
| S14_extracted_k141_82126 | Not-determined |
| S14_extracted_k141_96525 | Not-determined |
| S14_extracted_k141_96526 | Not-determined |
| S14_extracted_k141_111671 | Not-determined |
| S14_extracted_k141_60279 | Low-quality |
| S14_extracted_k141_104399 | Low-quality |
| S14_extracted_k141_52915 | Not-determined |
| S14_extracted_k141_96530 | Not-determined |
| S14_extracted_k141_16297 | Low-quality |
| S14_extracted_k141_16298 | Not-determined |
| S14_extracted_k141_8889 | Low-quality |
| S14_extracted_k141_8890 | Not-determined |
| S14_extracted_k141_38355 | Low-quality |
| S14_extracted_k141_111677 | Not-determined |
| S14_extracted_k141_60283 | Not-determined |
| S14_extracted_k141_23470 | Not-determined |
| S14_extracted_k141_8895 | Not-determined |
| S14_extracted_k141_8896 | Not-determined |
| S14_extracted_k141_8897 | Low-quality |
| S14_extracted_k141_52926 | Low-quality |
| S14_extracted_k141_60288 | Not-determined |
| S14_extracted_k141_111683 | Low-quality |
| S14_extracted_k141_16304 | Not-determined |
| S14_extracted_k141_111684 | Not-determined |
| S14_extracted_k141_60290 | Not-determined |
| S14_extracted_k141_82141 | Low-quality |
| S14_extracted_k141_8901 | Not-determined |
| S14_extracted_k141_111687 | Not-determined |
| S14_extracted_k141_52932 | Not-determined |
| S14_extracted_k141_8903 | Not-determined |
| S14_extracted_k141_82142 | Not-determined |
| S14_extracted_k141_67111 | Not-determined |
| S14_extracted_k141_96543 | Not-determined |
| S14_extracted_k141_30981 | Low-quality |
| S14_extracted_k141_30982 | Low-quality |
| S14_extracted_k141_16324 | Not-determined |
| S14_extracted_k141_16325 | Not-determined |
| S14_extracted_k141_89493 | Low-quality |
| S14_extracted_k141_111698 | Not-determined |
| S14_extracted_k141_38373 | Not-determined |
| S14_extracted_k141_16331 | Low-quality |
| S14_extracted_k141_89509 | Not-determined |
| S14_extracted_k141_45126 | Low-quality |
| S14_extracted_k141_52945 | Not-determined |
| S14_extracted_k141_45127 | Not-determined |
| S14_extracted_k141_45129 | Not-determined |
| S14_extracted_k141_111704 | Low-quality |
| S14_extracted_k141_45133 | Not-determined |
| S14_extracted_k141_89511 | Not-determined |
| S14_extracted_k141_111708 | Not-determined |
| S14_extracted_k141_52948 | Low-quality |
| S14_extracted_k141_104417 | Not-determined |
| S14_extracted_k141_96561 | Not-determined |
| S14_extracted_k141_16336 | Not-determined |
| S14_extracted_k141_96563 | Not-determined |
| S14_extracted_k141_45144 | Low-quality |
| S14_extracted_k141_89522 | Not-determined |
| S14_extracted_k141_104422 | Low-quality |
| S14_extracted_k141_16349 | Not-determined |
| S14_extracted_k141_45154 | Low-quality |
| S14_extracted_k141_45157 | Low-quality |
| S14_extracted_k141_8906 | Not-determined |
| S14_extracted_k141_74650 | Low-quality |
| S14_extracted_k141_16360 | Low-quality |
| S14_extracted_k141_82144 | Not-determined |
| S14_extracted_k141_74655 | Not-determined |
| S14_extracted_k141_111726 | Low-quality |
| S14_extracted_k141_111729 | Low-quality |
| S14_extracted_k141_45172 | Not-determined |
| S14_extracted_k141_74661 | Not-determined |
| S14_extracted_k141_74662 | Not-determined |
| S14_extracted_k141_74663 | Low-quality |
| S14_extracted_k141_74664 | Low-quality |
| S14_extracted_k141_82151 | Not-determined |
| S14_extracted_k141_82152 | Not-determined |
| S14_extracted_k141_60315 | Not-determined |
| S14_extracted_k141_60316 | Not-determined |
| S14_extracted_k141_52976 | Not-determined |
| S14_extracted_k141_96580 | Low-quality |
| S14_extracted_k141_45181 | Not-determined |
| S14_extracted_k141_96582 | Not-determined |
| S14_extracted_k141_96583 | Not-determined |
| S14_extracted_k141_8922 | Not-determined |
| S14_extracted_k141_96586 | Not-determined |
| S14_extracted_k141_111747 | Not-determined |
| S14_extracted_k141_96588 | Not-determined |
| S14_extracted_k141_96591 | Not-determined |
| S14_extracted_k141_111753 | Low-quality |
| S14_extracted_k141_96594 | Not-determined |
| S14_extracted_k141_96597 | Low-quality |
| S14_extracted_k141_96598 | Low-quality |
| S14_extracted_k141_96606 | Not-determined |
| S14_extracted_k141_89538 | Not-determined |
| S14_extracted_k141_104438 | Low-quality |
| S14_extracted_k141_60322 | Not-determined |
| S14_extracted_k141_52984 | Not-determined |
| S14_extracted_k141_31019 | Not-determined |
| S14_extracted_k141_60325 | Not-determined |
| S14_extracted_k141_74679 | Not-determined |
| S14_extracted_k141_74681 | Not-determined |
| S14_extracted_k141_45193 | Not-determined |
| S14_extracted_k141_52988 | Low-quality |
| S14_extracted_k141_96616 | Not-determined |
| S14_extracted_k141_45196 | Not-determined |
| S14_extracted_k141_52992 | Not-determined |
| S14_extracted_k141_38385 | Not-determined |
| S14_extracted_k141_74685 | Low-quality |
| S14_extracted_k141_74686 | Not-determined |
| S14_extracted_k141_89551 | Not-determined |
| S14_extracted_k141_74687 | Not-determined |
| S14_extracted_k141_74688 | Low-quality |
| S14_extracted_k141_45199 | Not-determined |
| S14_extracted_k141_60328 | Not-determined |
| S14_extracted_k141_60331 | Not-determined |
| S14_extracted_k141_111768 | Not-determined |
| S14_extracted_k141_45203 | Not-determined |
| S14_extracted_k141_16389 | Not-determined |
| S14_extracted_k141_53004 | Not-determined |
| S14_extracted_k141_111775 | Not-determined |
| S14_extracted_k141_16392 | Not-determined |
| S14_extracted_k141_96626 | Not-determined |
| S14_extracted_k141_96629 | Low-quality |
| S14_extracted_k141_96636 | Not-determined |
| S14_extracted_k141_8927 | Not-determined |
| S14_extracted_k141_74703 | Not-determined |
| S14_extracted_k141_38405 | Low-quality |
| S14_extracted_k141_74706 | Not-determined |
| S14_extracted_k141_31043 | Low-quality |
| S14_extracted_k141_82158 | Low-quality |
| S14_extracted_k141_82160 | Low-quality |
| S14_extracted_k141_38411 | Not-determined |
| S14_extracted_k141_67156 | Low-quality |
| S14_extracted_k141_67158 | Not-determined |
| S14_extracted_k141_67159 | Not-determined |
| S14_extracted_k141_8935 | Low-quality |
| S14_extracted_k141_104442 | Low-quality |
| S14_extracted_k141_67162 | Low-quality |
| S14_extracted_k141_67166 | Not-determined |
| S14_extracted_k141_74717 | Low-quality |
| S14_extracted_k141_53009 | Not-determined |
| S14_extracted_k141_89568 | Not-determined |
| S14_extracted_k141_38419 | Low-quality |
| S14_extracted_k141_67170 | Not-determined |
| S14_extracted_k141_60358 | Not-determined |
| S14_extracted_k141_31052 | Low-quality |
| S14_extracted_k141_1456 | Not-determined |
| S14_extracted_k141_89584 | Not-determined |
| S14_extracted_k141_38437 | Not-determined |
| S14_extracted_k141_1472 | Not-determined |
| S14_extracted_k141_111792 | Not-determined |
| S14_extracted_k141_111797 | Low-quality |
| S14_extracted_k141_38447 | Not-determined |
| S14_extracted_k141_89591 | Not-determined |
| S14_extracted_k141_1482 | Low-quality |
| S14_extracted_k141_1483 | Not-determined |
| S14_extracted_k141_31059 | Low-quality |
| S14_extracted_k141_104472 | Not-determined |
| S14_extracted_k141_104473 | Low-quality |
| S14_extracted_k141_104478 | Not-determined |
| S14_extracted_k141_1487 | Low-quality |
| S14_extracted_k141_96657 | Low-quality |
| S14_extracted_k141_74724 | Not-determined |
| S14_extracted_k141_96659 | Not-determined |
| S14_extracted_k141_96662 | Not-determined |
| S14_extracted_k141_23494 | Not-determined |
| S14_extracted_k141_82210 | Low-quality |
| S14_extracted_k141_31065 | Low-quality |
| S14_extracted_k141_31066 | Not-determined |
| S14_extracted_k141_74728 | Low-quality |
| S14_extracted_k141_74730 | Not-determined |
| S14_extracted_k141_104484 | Not-determined |
| S14_extracted_k141_89606 | Low-quality |
| S14_extracted_k141_16437 | Not-determined |
| S14_extracted_k141_74739 | Not-determined |
| S14_extracted_k141_82213 | Low-quality |
| S14_extracted_k141_74742 | Low-quality |
| S14_extracted_k141_104492 | Low-quality |
| S14_extracted_k141_74746 | Not-determined |
| S14_extracted_k141_74750 | Low-quality |
| S14_extracted_k141_104498 | Not-determined |
| S14_extracted_k141_31077 | Low-quality |
| S14_extracted_k141_104502 | Not-determined |
| S14_extracted_k141_74755 | Low-quality |
| S14_extracted_k141_60383 | Not-determined |
| S14_extracted_k141_1515 | Not-determined |
| S14_extracted_k141_60392 | Low-quality |
| S14_extracted_k141_38466 | Not-determined |
| S14_extracted_k141_38467 | Low-quality |
| S14_extracted_k141_104510 | Not-determined |
| S14_extracted_k141_82227 | Low-quality |
| S14_extracted_k141_111814 | Not-determined |
| S14_extracted_k141_104519 | Low-quality |
| S14_extracted_k141_60395 | Low-quality |
| S14_extracted_k141_60396 | Not-determined |
| S14_extracted_k141_67184 | Low-quality |
| S14_extracted_k141_31083 | Not-determined |
| S14_extracted_k141_74765 | Low-quality |
| S14_extracted_k141_38470 | Not-determined |
| S14_extracted_k141_74768 | Not-determined |
| S14_extracted_k141_67186 | Not-determined |
| S14_extracted_k141_82233 | Not-determined |
| S14_extracted_k141_104530 | Not-determined |
| S14_extracted_k141_16474 | Not-determined |
| S14_extracted_k141_31092 | Not-determined |
| S14_extracted_k141_16476 | Not-determined |
| S14_extracted_k141_31094 | Not-determined |
| S14_extracted_k141_31095 | Not-determined |
| S14_extracted_k141_16478 | Low-quality |
| S14_extracted_k141_74780 | Not-determined |
| S14_extracted_k141_82240 | Not-determined |
| S14_extracted_k141_74783 | Not-determined |
| S14_extracted_k141_31099 | Not-determined |
| S14_extracted_k141_82242 | Low-quality |
| S14_extracted_k141_104539 | Low-quality |
| S14_extracted_k141_111817 | Low-quality |
| S14_extracted_k141_67205 | Not-determined |
| S14_extracted_k141_89642 | Not-determined |
| S14_extracted_k141_89643 | Not-determined |
| S14_extracted_k141_89646 | Not-determined |
| S14_extracted_k141_1525 | Not-determined |
| S14_extracted_k141_74791 | Low-quality |
| S14_extracted_k141_104545 | Not-determined |
| S14_extracted_k141_89649 | Low-quality |
| S14_extracted_k141_23508 | Not-determined |
| S14_extracted_k141_67217 | Low-quality |
| S14_extracted_k141_23513 | Not-determined |
| S14_extracted_k141_1533 | Low-quality |
| S14_extracted_k141_1534 | Not-determined |
| S14_extracted_k141_67223 | Not-determined |
| S14_extracted_k141_74805 | Low-quality |
| S14_extracted_k141_74806 | Not-determined |
| S14_extracted_k141_23516 | Low-quality |
| S14_extracted_k141_67225 | Not-determined |
| S14_extracted_k141_89650 | Low-quality |
| S14_extracted_k141_1538 | Low-quality |
| S14_extracted_k141_1539 | Not-determined |
| S14_extracted_k141_1541 | Low-quality |
| S14_extracted_k141_104557 | Not-determined |
| S14_extracted_k141_23523 | Not-determined |
| S14_extracted_k141_111824 | Not-determined |
| S14_extracted_k141_104563 | Not-determined |
| S14_extracted_k141_89657 | Not-determined |
| S14_extracted_k141_16492 | Not-determined |
| S14_extracted_k141_38491 | Low-quality |
| S14_extracted_k141_104567 | Low-quality |
| S14_extracted_k141_38494 | Not-determined |
| S14_extracted_k141_74818 | Low-quality |
| S14_extracted_k141_16497 | Not-determined |
| S14_extracted_k141_89664 | Not-determined |
| S14_extracted_k141_74825 | Low-quality |
| S14_extracted_k141_38501 | Not-determined |
| S14_extracted_k141_111830 | Not-determined |
| S14_extracted_k141_104582 | Not-determined |
| S14_extracted_k141_104584 | Not-determined |
| S14_extracted_k141_111838 | Low-quality |
| S14_extracted_k141_23532 | Low-quality |
| S14_extracted_k141_89680 | Low-quality |
| S14_extracted_k141_67234 | Low-quality |
| S14_extracted_k141_104592 | Not-determined |
| S14_extracted_k141_67238 | Low-quality |
| S14_extracted_k141_82271 | Not-determined |
| S14_extracted_k141_60445 | Low-quality |
| S14_extracted_k141_67246 | Low-quality |
| S14_extracted_k141_82277 | Not-determined |
| S14_extracted_k141_38513 | Low-quality |
| S14_extracted_k141_111847 | Not-determined |
| S14_extracted_k141_38519 | Low-quality |
| S14_extracted_k141_74857 | Low-quality |
| S14_extracted_k141_82285 | Low-quality |
| S14_extracted_k141_82286 | Not-determined |
| S14_extracted_k141_74863 | Not-determined |
| S14_extracted_k141_82287 | Low-quality |
| S14_extracted_k141_82289 | Not-determined |
| S14_extracted_k141_74868 | Low-quality |
| S14_extracted_k141_67262 | Not-determined |
| S14_extracted_k141_1550 | Low-quality |
| S14_extracted_k141_67265 | Low-quality |
| S14_extracted_k141_67266 | Low-quality |
| S14_extracted_k141_111866 | Not-determined |
| S14_extracted_k141_31115 | Low-quality |
| S14_extracted_k141_89688 | Not-determined |
| S14_extracted_k141_74884 | Not-determined |
| S14_extracted_k141_38535 | Not-determined |
| S14_extracted_k141_89698 | Low-quality |
| S14_extracted_k141_111875 | Not-determined |
| S14_extracted_k141_89699 | Not-determined |
| S14_extracted_k141_53017 | Low-quality |
| S14_extracted_k141_74893 | Not-determined |
| S14_extracted_k141_67281 | Not-determined |
| S14_extracted_k141_104623 | Low-quality |
| S14_extracted_k141_111885 | Not-determined |
| S14_extracted_k141_74903 | Not-determined |
| S14_extracted_k141_45231 | Not-determined |
| S14_extracted_k141_60460 | Not-determined |
| S14_extracted_k141_53021 | Not-determined |
| S14_extracted_k141_60462 | Low-quality |
| S14_extracted_k141_74905 | Not-determined |
| S14_extracted_k141_60464 | Low-quality |
| S14_extracted_k141_53022 | Not-determined |
| S14_extracted_k141_16528 | Not-determined |
| S14_extracted_k141_16541 | Low-quality |
| S14_extracted_k141_67300 | Not-determined |
| S14_extracted_k141_67303 | Low-quality |
| S14_extracted_k141_111900 | Not-determined |
| S14_extracted_k141_111903 | Not-determined |
| S14_extracted_k141_89714 | Low-quality |
| S14_extracted_k141_111909 | Low-quality |
| S14_extracted_k141_23564 | Low-quality |
| S14_extracted_k141_111911 | Low-quality |
| S14_extracted_k141_111914 | Low-quality |
| S14_extracted_k141_45246 | Not-determined |
| S14_extracted_k141_45249 | Not-determined |
| S17_vs_k141_52890\|\|full | Medium-quality |
| S14_extracted_k141_1588 | Not-determined |
| S14_extracted_k141_111921 | Not-determined |
| S14_extracted_k141_111922 | Not-determined |
| S14_extracted_k141_67311 | Not-determined |
| S14_extracted_k141_67314 | Low-quality |
| S14_extracted_k141_67315 | Low-quality |
| S14_extracted_k141_89719 | Not-determined |
| S14_extracted_k141_89723 | Low-quality |
| S14_extracted_k141_45255 | Low-quality |
| S14_extracted_k141_45256 | Not-determined |
| S14_extracted_k141_23575 | Low-quality |
| S14_extracted_k141_111929 | Not-determined |
| S14_extracted_k141_60474 | Low-quality |
| S14_extracted_k141_67322 | Low-quality |
| S14_extracted_k141_67323 | Not-determined |
| S14_extracted_k141_53068 | Not-determined |
| S14_extracted_k141_16566 | Not-determined |
| S14_extracted_k141_45261 | Not-determined |
| S14_extracted_k141_111937 | Low-quality |
| S14_extracted_k141_60482 | Low-quality |
| S14_extracted_k141_74923 | Low-quality |
| S14_extracted_k141_96683 | Not-determined |
| S14_extracted_k141_96684 | Not-determined |
| S14_extracted_k141_67329 | Low-quality |
| S14_extracted_k141_67330 | Not-determined |
| S14_extracted_k141_111940 | Low-quality |
| S14_extracted_k141_16575 | Low-quality |
| S14_extracted_k141_60496 | Low-quality |
| S14_extracted_k141_67335 | Low-quality |
| S14_extracted_k141_60498 | Low-quality |
| S14_extracted_k141_60503 | Low-quality |
| S14_extracted_k141_60506 | Low-quality |
| S14_extracted_k141_53080 | Not-determined |
| S14_extracted_k141_31152 | Low-quality |
| S14_extracted_k141_45277 | Not-determined |
| S14_extracted_k141_67343 | Not-determined |
| S14_extracted_k141_45280 | Low-quality |
| S14_extracted_k141_96693 | Not-determined |
| S14_extracted_k141_89748 | Not-determined |
| S14_extracted_k141_31156 | Not-determined |
| S14_extracted_k141_31157 | Not-determined |
| S14_extracted_k141_89750 | Not-determined |
| S14_extracted_k141_38546 | Low-quality |
| S14_extracted_k141_96696 | Low-quality |
| S14_extracted_k141_67351 | Low-quality |
| S14_extracted_k141_96697 | Low-quality |
| S14_extracted_k141_31158 | Not-determined |
| S14_extracted_k141_89755 | Not-determined |
| S14_extracted_k141_23581 | Low-quality |
| S14_extracted_k141_31165 | Not-determined |
| S14_extracted_k141_60522 | Not-determined |
| S14_extracted_k141_60523 | Not-determined |
| S14_extracted_k141_104630 | Not-determined |
| S14_extracted_k141_89758 | Not-determined |
| S14_extracted_k141_96705 | Not-determined |
| S14_extracted_k141_104633 | Not-determined |
| S14_extracted_k141_60528 | Low-quality |
| S14_extracted_k141_104634 | Not-determined |
| S14_extracted_k141_89761 | Not-determined |
| S14_extracted_k141_89763 | Not-determined |
| S14_extracted_k141_60533 | Not-determined |
| S14_extracted_k141_60534 | Not-determined |
| S14_extracted_k141_104638 | Low-quality |
| S14_extracted_k141_60536 | Not-determined |
| S14_extracted_k141_38559 | Not-determined |
| S14_extracted_k141_38566 | Not-determined |
| S14_extracted_k141_96715 | Low-quality |
| S14_extracted_k141_67375 | Not-determined |
| S7_vs_41098\|\|full | High-quality |
| S14_extracted_k141_23594 | Not-determined |
| S14_extracted_k141_104652 | Low-quality |
| S14_extracted_k141_104653 | Not-determined |
| S14_extracted_k141_67382 | Not-determined |
| S14_extracted_k141_45295 | Not-determined |
| S14_extracted_k141_82310 | Not-determined |
| S14_extracted_k141_31168 | Low-quality |
| S14_extracted_k141_82315 | Not-determined |
| S14_extracted_k141_67388 | Not-determined |
| S14_extracted_k141_74961 | Not-determined |
| S14_extracted_k141_82318 | Not-determined |
| S14_extracted_k141_82319 | Low-quality |
| S14_extracted_k141_1622 | Not-determined |
| S14_extracted_k141_53106 | Low-quality |
| S14_extracted_k141_1624 | Not-determined |
| S14_extracted_k141_60551 | Low-quality |
| S14_extracted_k141_1630 | Not-determined |
| S14_extracted_k141_82325 | Not-determined |
| S14_extracted_k141_96724 | Low-quality |
| S14_extracted_k141_96725 | Low-quality |
| S14_extracted_k141_53114 | Not-determined |
| S14_extracted_k141_82327 | Low-quality |
| S14_extracted_k141_53115 | Low-quality |
| S14_extracted_k141_45306 | Low-quality |
| S14_extracted_k141_1637 | Not-determined |
| S14_extracted_k141_53121 | Not-determined |
| S14_extracted_k141_67401 | Low-quality |
| S14_extracted_k141_104679 | Not-determined |
| S14_extracted_k141_53127 | Low-quality |
| S14_extracted_k141_31179 | Low-quality |
| S14_extracted_k141_53129 | Not-determined |
| S14_extracted_k141_111944 | Not-determined |
| S14_extracted_k141_60559 | Low-quality |
| S14_extracted_k141_53131 | Not-determined |
| S14_extracted_k141_45313 | Not-determined |
| S14_extracted_k141_31183 | Low-quality |
| S14_extracted_k141_1648 | Not-determined |
| S14_extracted_k141_60572 | Not-determined |
| S14_extracted_k141_67411 | Low-quality |
| S14_extracted_k141_82345 | Not-determined |
| S14_extracted_k141_31192 | Low-quality |
| S14_extracted_k141_60580 | Not-determined |
| S14_extracted_k141_67416 | Not-determined |
| S14_extracted_k141_1656 | Not-determined |
| S14_extracted_k141_1657 | Not-determined |
| S14_extracted_k141_31197 | Low-quality |
| S14_extracted_k141_1658 | Not-determined |
| S14_extracted_k141_1659 | Low-quality |
| S14_extracted_k141_104685 | Low-quality |
| S14_extracted_k141_104686 | Not-determined |
| S14_extracted_k141_82349 | Not-determined |
| S14_extracted_k141_96747 | Not-determined |
| S14_extracted_k141_82351 | Low-quality |
| S14_extracted_k141_104691 | Not-determined |
| S14_extracted_k141_104693 | Not-determined |
| S14_extracted_k141_60583 | Low-quality |
| S14_extracted_k141_104694 | Not-determined |
| S14_extracted_k141_96752 | Not-determined |
| S14_extracted_k141_53136 | Not-determined |
| S14_extracted_k141_60585 | Low-quality |
| S14_extracted_k141_96755 | Not-determined |
| S14_extracted_k141_111970 | Low-quality |
| S14_extracted_k141_96756 | Low-quality |
| S14_extracted_k141_8939 | Not-determined |
| S14_extracted_k141_74993 | Low-quality |
| S14_extracted_k141_8940 | Not-determined |
| S14_extracted_k141_53142 | Not-determined |
| S14_extracted_k141_96761 | Low-quality |
| S14_extracted_k141_82358 | Low-quality |
| S14_extracted_k141_111974 | Not-determined |
| S14_extracted_k141_8943 | Not-determined |
| S14_extracted_k141_96763 | Not-determined |
| S14_extracted_k141_96764 | Low-quality |
| S14_extracted_k141_75000 | Not-determined |
| S14_extracted_k141_38611 | Not-determined |
| S14_extracted_k141_75005 | Not-determined |
| S14_extracted_k141_8945 | Not-determined |
| S14_extracted_k141_75006 | Not-determined |
| S14_extracted_k141_8946 | Not-determined |
| S14_extracted_k141_67432 | Low-quality |
| S14_extracted_k141_38616 | Low-quality |
| S14_extracted_k141_111998 | Not-determined |
| S14_extracted_k141_53153 | Not-determined |
| S14_extracted_k141_38631 | Not-determined |
| S14_extracted_k141_53154 | Low-quality |
| S14_extracted_k141_112006 | Not-determined |
| S14_extracted_k141_8950 | Not-determined |
| S14_extracted_k141_112009 | Not-determined |
| S14_extracted_k141_45328 | Low-quality |
| S14_extracted_k141_96781 | Not-determined |
| S14_extracted_k141_96783 | Low-quality |
| S14_extracted_k141_1672 | Not-determined |
| S14_extracted_k141_75012 | Low-quality |
| S14_extracted_k141_96786 | Low-quality |
| S14_extracted_k141_112018 | Not-determined |
| S14_extracted_k141_112019 | Low-quality |
| S14_extracted_k141_60596 | Low-quality |
| S14_extracted_k141_82373 | Not-determined |
| S14_extracted_k141_60602 | Not-determined |
| S14_extracted_k141_89825 | Not-determined |
| S14_extracted_k141_89826 | Not-determined |
| S14_extracted_k141_104708 | Not-determined |
| S14_extracted_k141_96793 | Not-determined |
| S14_extracted_k141_31217 | Not-determined |
| S14_extracted_k141_16620 | Not-determined |
| S14_extracted_k141_96802 | Not-determined |
| S14_extracted_k141_82382 | Low-quality |
| S14_extracted_k141_8953 | Not-determined |
| S14_extracted_k141_67450 | Not-determined |
| S14_extracted_k141_67451 | Not-determined |
| S14_extracted_k141_23606 | Not-determined |
| S14_extracted_k141_23607 | Low-quality |
| S14_extracted_k141_82387 | Not-determined |
| S14_extracted_k141_112046 | Not-determined |
| S14_extracted_k141_82388 | Not-determined |
| S14_extracted_k141_23611 | Low-quality |
| S14_extracted_k141_23613 | Low-quality |
| S14_extracted_k141_23615 | Not-determined |
| S14_extracted_k141_16633 | Not-determined |
| S14_extracted_k141_16635 | Not-determined |
| S14_extracted_k141_112050 | Low-quality |
| S14_extracted_k141_96810 | Not-determined |
| S14_extracted_k141_112052 | Not-determined |
| S14_extracted_k141_112053 | Low-quality |
| S14_extracted_k141_67460 | Low-quality |
| S14_extracted_k141_67461 | Low-quality |
| S14_extracted_k141_75015 | Low-quality |
| S14_extracted_k141_67463 | Not-determined |
| S14_extracted_k141_75020 | Not-determined |
| S14_extracted_k141_16645 | Not-determined |
| S14_extracted_k141_45358 | Not-determined |
| S14_extracted_k141_82400 | Not-determined |
| S14_extracted_k141_16647 | Not-determined |
| S14_extracted_k141_96818 | Low-quality |
| S14_extracted_k141_8957 | Not-determined |
| S14_extracted_k141_96819 | Not-determined |
| S14_extracted_k141_60623 | Low-quality |
| S14_extracted_k141_23624 | Low-quality |
| S14_extracted_k141_60624 | Low-quality |
| S14_extracted_k141_31233 | Not-determined |
| S14_extracted_k141_75029 | Low-quality |
| S14_extracted_k141_60632 | Not-determined |
| S14_extracted_k141_75030 | Not-determined |
| S14_extracted_k141_31235 | Low-quality |
| S14_extracted_k141_45366 | Not-determined |
| S14_extracted_k141_23629 | Not-determined |
| S14_extracted_k141_8972 | Not-determined |
| S14_extracted_k141_60634 | Low-quality |
| S14_extracted_k141_16661 | Not-determined |
| S14_extracted_k141_82407 | Low-quality |
| S14_extracted_k141_16665 | Not-determined |
| S14_extracted_k141_38652 | Not-determined |
| S14_extracted_k141_45370 | Low-quality |
| S14_extracted_k141_75043 | Not-determined |
| S14_extracted_k141_75044 | Low-quality |
| S14_extracted_k141_104735 | Not-determined |
| S14_extracted_k141_60639 | Low-quality |
| S14_extracted_k141_45375 | Not-determined |
| S14_extracted_k141_104741 | Not-determined |
| S14_extracted_k141_82411 | Not-determined |
| S14_extracted_k141_31257 | Low-quality |
| S14_extracted_k141_60646 | Not-determined |
| S14_extracted_k141_1716 | Low-quality |
| S14_extracted_k141_23649 | Not-determined |
| S14_extracted_k141_60650 | Not-determined |
| S14_extracted_k141_82418 | Not-determined |
| S14_extracted_k141_82419 | Not-determined |
| S14_extracted_k141_23654 | Not-determined |
| S14_extracted_k141_23655 | Not-determined |
| S14_extracted_k141_53183 | Not-determined |
| S14_extracted_k141_67473 | Low-quality |
| S14_extracted_k141_60661 | Not-determined |
| S14_extracted_k141_45382 | Not-determined |
| S14_extracted_k141_75060 | Low-quality |
| S14_extracted_k141_23659 | Low-quality |
| S14_extracted_k141_67476 | Low-quality |
| S14_extracted_k141_60665 | Not-determined |
| S14_extracted_k141_16691 | Not-determined |
| S14_extracted_k141_82427 | Low-quality |
| S14_extracted_k141_104746 | Low-quality |
| S14_extracted_k141_38668 | Not-determined |
| S14_extracted_k141_9003 | Low-quality |
| S14_extracted_k141_104750 | Low-quality |
| S14_extracted_k141_60673 | Not-determined |
| S14_extracted_k141_53198 | Not-determined |
| S14_extracted_k141_31277 | Low-quality |
| S14_extracted_k141_23670 | Not-determined |
| S14_extracted_k141_23671 | Not-determined |
| S14_extracted_k141_82432 | Low-quality |
| S14_extracted_k141_9010 | Not-determined |
| S14_extracted_k141_16696 | Low-quality |
| S14_extracted_k141_82433 | Not-determined |
| S14_extracted_k141_16700 | Not-determined |
| S14_extracted_k141_31285 | Not-determined |
| S14_extracted_k141_31296 | Not-determined |
| S14_extracted_k141_38681 | Low-quality |
| S14_extracted_k141_75076 | Not-determined |
| S14_extracted_k141_75077 | Low-quality |
| S14_extracted_k141_53213 | Not-determined |
| S14_extracted_k141_31301 | Not-determined |
| S14_extracted_k141_45391 | Not-determined |

| | |
|---|---|
| S14_extracted_k141_38685 | Not-determined |
| S14_extracted_k141_53215 | Not-determined |
| S14_extracted_k141_75081 | Not-determined |
| S14_extracted_k141_23692 | Not-determined |
| S14_extracted_k141_53219 | Not-determined |
| S14_extracted_k141_38693 | Low-quality |
| S14_extracted_k141_53222 | Low-quality |
| S14_extracted_k141_53223 | Low-quality |
| S14_extracted_k141_38694 | Not-determined |
| S14_extracted_k141_31305 | Not-determined |
| S14_extracted_k141_38696 | Low-quality |
| S14_extracted_k141_53226 | Not-determined |
| S14_extracted_k141_38700 | Low-quality |
| S14_extracted_k141_53233 | Not-determined |
| S14_extracted_k141_16719 | Not-determined |
| S14_extracted_k141_60691 | Not-determined |
| S14_extracted_k141_16722 | Low-quality |
| S14_extracted_k141_38706 | Low-quality |
| S14_extracted_k141_82455 | Not-determined |
| S14_extracted_k141_31311 | Not-determined |
| S14_extracted_k141_31313 | Not-determined |
| S14_extracted_k141_31316 | Low-quality |
| S14_extracted_k141_31318 | Not-determined |
| S14_extracted_k141_104765 | Low-quality |
| S14_extracted_k141_53243 | Not-determined |
| S14_extracted_k141_45399 | Low-quality |
| S14_extracted_k141_45400 | Not-determined |
| S14_extracted_k141_53246 | Low-quality |
| S14_extracted_k141_60700 | Not-determined |
| S14_extracted_k141_60701 | Not-determined |
| S14_extracted_k141_82467 | Not-determined |
| S14_extracted_k141_53249 | Not-determined |
| S14_extracted_k141_60703 | Not-determined |
| S14_extracted_k141_82470 | Low-quality |
| S14_extracted_k141_75117 | Not-determined |
| S14_extracted_k141_45408 | Not-determined |
| S14_extracted_k141_31327 | Not-determined |
| S14_extracted_k141_45410 | Not-determined |
| S14_extracted_k141_53262 | Low-quality |
| S14_extracted_k141_38722 | Not-determined |
| S14_extracted_k141_45415 | Not-determined |
| S14_extracted_k141_75128 | Low-quality |
| S14_extracted_k141_45418 | Not-determined |
| S14_extracted_k141_16741 | Not-determined |
| S14_extracted_k141_38726 | Not-determined |
| S14_extracted_k141_60716 | Low-quality |
| S14_extracted_k141_67491 | Not-determined |
| S14_extracted_k141_16745 | Not-determined |
| S14_extracted_k141_82495 | Low-quality |
| S14_extracted_k141_82496 | Low-quality |
| S14_extracted_k141_67501 | Low-quality |
| S14_extracted_k141_82497 | Not-determined |
| S14_extracted_k141_104782 | Low-quality |
| S14_extracted_k141_38743 | Low-quality |
| S14_extracted_k141_16754 | Not-determined |
| S14_extracted_k141_104785 | Not-determined |
| S14_extracted_k141_23735 | Low-quality |
| S14_extracted_k141_23737 | Not-determined |
| S14_extracted_k141_9048 | Not-determined |
| S14_extracted_k141_45427 | Not-determined |
| S14_extracted_k141_67505 | Not-determined |
| S14_extracted_k141_16761 | Not-determined |
| S14_extracted_k141_67509 | Not-determined |
| S14_extracted_k141_67511 | Low-quality |
| S14_extracted_k141_23742 | Low-quality |
| S14_extracted_k141_23743 | Not-determined |
| S14_extracted_k141_82516 | Not-determined |
| S14_extracted_k141_67519 | Not-determined |
| S14_extracted_k141_60738 | Low-quality |
| S14_extracted_k141_60739 | Low-quality |
| S14_extracted_k141_38763 | Not-determined |
| S14_extracted_k141_45440 | Not-determined |
| S14_extracted_k141_82523 | Not-determined |
| S14_extracted_k141_53285 | Not-determined |
| S14_extracted_k141_67522 | Low-quality |
| S14_extracted_k141_38765 | Low-quality |
| S14_extracted_k141_82527 | Not-determined |
| S14_extracted_k141_53287 | Not-determined |
| S14_extracted_k141_38766 | Low-quality |
| S14_extracted_k141_9058 | Not-determined |
| S14_extracted_k141_82529 | Not-determined |
| S14_extracted_k141_67527 | Low-quality |
| S14_extracted_k141_67528 | Low-quality |
| S14_extracted_k141_38769 | Not-determined |
| S14_extracted_k141_60749 | Not-determined |
| S14_extracted_k141_45447 | Not-determined |
| S14_extracted_k141_67533 | Low-quality |
| S14_extracted_k141_31340 | Not-determined |
| S14_extracted_k141_38780 | Not-determined |
| S14_extracted_k141_45449 | Not-determined |
| S14_extracted_k141_45451 | Not-determined |
| S14_extracted_k141_67538 | Low-quality |
| S14_extracted_k141_31343 | Not-determined |
| S14_extracted_k141_31345 | Not-determined |
| S14_extracted_k141_60756 | Not-determined |
| S14_extracted_k141_60757 | Low-quality |
| S14_extracted_k141_60758 | Low-quality |
| S14_extracted_k141_60759 | Low-quality |
| S14_extracted_k141_53299 | Not-determined |
| S14_extracted_k141_38785 | Low-quality |
| S14_extracted_k141_89847 | Not-determined |
| S14_extracted_k141_45456 | Low-quality |
| S14_extracted_k141_45457 | Not-determined |
| S14_extracted_k141_45458 | Not-determined |
| S14_extracted_k141_67542 | Not-determined |
| S14_extracted_k141_16775 | Low-quality |
| S14_extracted_k141_38788 | Not-determined |
| S14_extracted_k141_9080 | Not-determined |
| S14_extracted_k141_82545 | Not-determined |
| S14_extracted_k141_67544 | Not-determined |
| S14_extracted_k141_16776 | Low-quality |
| S14_extracted_k141_104803 | Low-quality |
| S14_extracted_k141_16779 | Not-determined |
| S14_extracted_k141_38791 | Not-determined |
| S14_extracted_k141_9082 | Low-quality |
| S14_extracted_k141_38792 | Not-determined |
| S14_extracted_k141_9083 | Not-determined |
| S14_extracted_k141_31354 | Not-determined |
| S14_extracted_k141_67551 | Low-quality |
| S14_extracted_k141_1761 | Not-determined |
| S14_extracted_k141_1762 | Not-determined |
| S14_extracted_k141_38795 | Not-determined |
| S14_extracted_k141_16785 | Not-determined |
| S14_extracted_k141_60763 | Not-determined |
| S14_extracted_k141_82553 | Not-determined |
| S14_extracted_k141_16787 | Not-determined |
| S14_extracted_k141_104810 | Not-determined |
| S14_extracted_k141_89860 | Low-quality |
| S14_extracted_k141_104813 | Not-determined |
| S14_extracted_k141_89864 | Not-determined |
| S14_extracted_k141_82566 | Not-determined |
| S14_extracted_k141_23748 | Not-determined |
| S14_extracted_k141_31360 | Not-determined |
| S14_extracted_k141_89868 | Not-determined |
| S14_extracted_k141_38805 | Not-determined |
| S14_extracted_k141_67553 | Low-quality |
| S14_extracted_k141_45470 | Not-determined |
| S14_extracted_k141_89876 | Not-determined |
| S14_extracted_k141_23750 | Not-determined |
| S14_extracted_k141_23754 | Not-determined |
| S14_extracted_k141_60772 | Not-determined |
| S14_extracted_k141_104828 | Not-determined |
| S14_extracted_k141_31369 | Not-determined |
| S14_extracted_k141_75157 | Low-quality |
| S14_extracted_k141_75158 | Low-quality |
| S14_extracted_k141_75161 | Low-quality |
| S14_extracted_k141_104834 | Low-quality |
| S14_extracted_k141_75164 | Low-quality |
| S14_extracted_k141_38821 | Not-determined |
| S14_extracted_k141_9092 | Not-determined |
| S14_extracted_k141_38829 | Not-determined |
| S14_extracted_k141_53319 | Not-determined |
| S14_extracted_k141_16792 | Not-determined |
| S14_extracted_k141_82570 | Not-determined |
| S14_extracted_k141_38841 | Not-determined |
| S14_extracted_k141_67560 | Not-determined |
| S14_extracted_k141_104843 | Low-quality |
| S14_extracted_k141_67563 | Not-determined |
| S14_extracted_k141_75174 | Not-determined |
| S14_extracted_k141_38847 | Not-determined |
| S14_extracted_k141_75176 | Low-quality |
| S14_extracted_k141_38849 | Low-quality |
| S14_extracted_k141_31384 | Not-determined |
| S14_extracted_k141_23773 | Not-determined |
| S14_extracted_k141_60790 | Low-quality |
| S14_extracted_k141_31390 | Not-determined |
| S14_extracted_k141_38851 | Not-determined |
| S14_extracted_k141_104859 | Low-quality |
| S14_extracted_k141_16802 | Not-determined |
| S14_extracted_k141_16804 | Low-quality |
| S14_extracted_k141_16805 | Not-determined |
| S14_extracted_k141_38857 | Not-determined |
| S14_extracted_k141_38858 | Not-determined |
| S14_extracted_k141_45512 | Low-quality |
| S14_extracted_k141_53346 | Low-quality |
| S14_extracted_k141_67575 | Low-quality |
| S14_extracted_k141_16810 | Not-determined |
| S14_extracted_k141_38859 | Not-determined |
| S14_extracted_k141_67577 | Low-quality |
| S14_extracted_k141_16813 | Not-determined |
| S14_extracted_k141_53353 | Low-quality |
| S14_extracted_k141_53356 | Low-quality |
| S14_extracted_k141_9103 | Not-determined |
| S14_extracted_k141_75186 | Not-determined |
| S14_extracted_k141_16819 | Not-determined |
| S14_extracted_k141_9107 | Not-determined |
| S14_extracted_k141_9109 | Not-determined |
| S14_extracted_k141_1771 | Not-determined |
| S14_extracted_k141_9111 | Low-quality |
| S14_extracted_k141_67586 | Low-quality |
| S14_extracted_k141_96846 | Not-determined |
| S14_extracted_k141_67593 | Not-determined |
| S14_extracted_k141_67595 | Not-determined |
| S14_extracted_k141_9122 | Not-determined |
| S14_extracted_k141_53368 | Low-quality |
| S14_extracted_k141_67597 | Not-determined |
| S14_extracted_k141_23786 | Not-determined |
| S14_extracted_k141_23787 | Low-quality |
| S14_extracted_k141_9130 | Not-determined |
| S14_extracted_k141_45529 | Not-determined |
| S14_extracted_k141_53379 | Not-determined |
| S14_extracted_k141_53380 | Low-quality |
| S14_extracted_k141_9139 | Not-determined |
| S14_extracted_k141_67609 | Low-quality |
| S14_extracted_k141_53387 | Not-determined |
| S14_extracted_k141_67612 | Not-determined |
| S14_extracted_k141_75222 | Low-quality |
| S14_extracted_k141_67617 | Not-determined |
| S14_extracted_k141_16842 | Low-quality |
| S14_extracted_k141_16844 | Not-determined |
| S14_extracted_k141_9149 | Low-quality |
| S14_extracted_k141_67624 | Low-quality |
| S14_extracted_k141_9152 | Not-determined |
| S14_extracted_k141_89897 | Not-determined |
| S14_extracted_k141_89900 | Low-quality |
| S14_extracted_k141_67643 | Low-quality |
| S14_extracted_k141_67644 | Low-quality |
| S14_extracted_k141_1781 | Not-determined |
| S14_extracted_k141_82595 | Low-quality |
| S14_extracted_k141_104870 | Low-quality |
| S14_extracted_k141_96859 | Not-determined |
| S14_extracted_k141_67652 | Low-quality |
| S14_extracted_k141_1787 | Not-determined |
| S14_extracted_k141_16863 | Not-determined |
| S14_extracted_k141_1788 | Low-quality |
| S14_extracted_k141_82606 | Low-quality |
| S14_extracted_k141_82608 | Not-determined |
| S14_extracted_k141_1793 | Low-quality |
| S14_extracted_k141_1795 | Low-quality |
| S14_extracted_k141_89938 | Not-determined |
| S14_extracted_k141_45542 | Not-determined |
| S14_extracted_k141_31399 | Low-quality |
| S14_extracted_k141_89945 | Not-determined |
| S14_extracted_k141_31405 | Not-determined |
| S14_extracted_k141_75237 | Not-determined |
| S14_extracted_k141_53416 | Not-determined |
| S14_extracted_k141_23798 | Not-determined |
| S14_extracted_k141_75239 | Low-quality |
| S14_extracted_k141_53417 | Not-determined |
| S14_extracted_k141_23800 | Not-determined |
| S14_extracted_k141_38912 | Low-quality |
| S14_extracted_k141_104901 | Not-determined |
| S14_extracted_k141_104904 | Low-quality |
| S14_extracted_k141_104905 | Low-quality |
| S14_extracted_k141_75242 | Low-quality |
| S14_extracted_k141_104908 | Not-determined |
| S14_extracted_k141_45560 | Not-determined |
| S14_extracted_k141_23811 | Not-determined |
| S14_extracted_k141_23813 | Not-determined |
| S14_extracted_k141_104914 | Low-quality |
| S14_extracted_k141_75249 | Low-quality |
| S14_extracted_k141_82631 | Not-determined |
| S14_extracted_k141_89966 | Low-quality |
| S14_extracted_k141_82635 | Not-determined |
| S14_extracted_k141_45572 | Not-determined |
| S14_extracted_k141_23816 | Low-quality |
| S14_extracted_k141_104919 | Not-determined |
| S14_extracted_k141_23819 | Not-determined |
| S14_extracted_k141_104920 | Not-determined |
| S14_extracted_k141_23820 | Not-determined |
| S14_extracted_k141_53439 | Not-determined |
| S14_extracted_k141_104927 | Low-quality |
| S14_extracted_k141_38926 | Not-determined |
| S14_extracted_k141_82646 | Not-determined |
| S14_extracted_k141_89977 | Not-determined |
| S14_extracted_k141_89978 | Not-determined |
| S14_extracted_k141_89979 | Not-determined |
| S14_extracted_k141_82651 | Not-determined |
| S14_extracted_k141_96910 | Not-determined |
| S14_extracted_k141_104931 | Not-determined |
| S14_extracted_k141_45586 | Not-determined |
| S14_extracted_k141_89987 | Low-quality |
| S14_extracted_k141_89988 | Low-quality |
| S14_extracted_k141_45590 | Low-quality |
| S14_extracted_k141_112091 | Low-quality |
| S14_extracted_k141_67663 | Not-determined |
| S14_extracted_k141_45593 | Not-determined |
| S14_extracted_k141_89991 | Low-quality |
| S14_extracted_k141_23832 | Not-determined |
| S14_extracted_k141_23835 | Not-determined |
| S14_extracted_k141_104945 | Not-determined |
| S14_extracted_k141_45598 | Not-determined |
| S14_extracted_k141_45599 | Not-determined |
| S14_extracted_k141_67675 | Low-quality |
| S14_extracted_k141_23839 | Not-determined |
| S14_extracted_k141_67680 | Not-determined |
| S14_extracted_k141_90000 | Not-determined |
| S14_extracted_k141_45604 | Low-quality |
| S14_extracted_k141_67689 | Not-determined |
| S14_extracted_k141_104954 | Not-determined |
| S14_extracted_k141_82672 | Not-determined |
| S14_extracted_k141_67690 | Not-determined |
| S14_extracted_k141_67692 | Not-determined |
| S14_extracted_k141_67694 | Not-determined |
| S14_extracted_k141_53465 | Not-determined |
| S14_extracted_k141_53467 | Low-quality |
| S14_extracted_k141_104960 | Not-determined |
| S14_extracted_k141_90013 | Not-determined |
| S14_extracted_k141_82680 | Not-determined |
| S14_extracted_k141_23853 | Low-quality |
| S14_extracted_k141_23854 | Not-determined |
| S14_extracted_k141_90018 | Low-quality |
| S14_extracted_k141_45642 | Not-determined |
| S14_extracted_k141_45644 | Low-quality |
| S14_extracted_k141_90024 | Not-determined |
| S14_extracted_k141_90025 | Low-quality |
| S14_extracted_k141_112130 | Not-determined |
| S14_extracted_k141_9164 | Not-determined |
| S14_extracted_k141_23865 | Low-quality |
| S14_extracted_k141_90032 | Not-determined |
| S14_extracted_k141_9166 | Not-determined |
| S14_extracted_k141_67720 | Low-quality |
| S14_extracted_k141_38948 | Not-determined |
| S14_extracted_k141_67725 | Low-quality |
| S14_extracted_k141_112131 | Low-quality |
| S14_extracted_k141_112132 | Not-determined |
| S14_extracted_k141_38955 | Not-determined |
| S14_extracted_k141_90042 | Not-determined |
| S14_extracted_k141_38957 | Low-quality |
| S14_extracted_k141_38959 | Not-determined |
| S14_extracted_k141_38960 | Not-determined |
| S14_extracted_k141_90048 | Not-determined |
| S14_extracted_k141_104984 | Low-quality |
| S14_extracted_k141_23882 | Not-determined |
| S14_extracted_k141_67732 | Not-determined |
| S14_extracted_k141_112140 | Not-determined |
| S14_extracted_k141_90054 | Low-quality |
| S14_extracted_k141_67735 | Low-quality |
| S14_extracted_k141_38973 | Not-determined |
| S14_extracted_k141_67738 | Not-determined |
| S14_extracted_k141_82688 | Not-determined |
| S14_extracted_k141_82691 | Not-determined |
| S14_extracted_k141_9171 | Not-determined |
| S14_extracted_k141_112152 | Not-determined |
| S14_extracted_k141_82696 | Not-determined |
| S14_extracted_k141_112155 | Not-determined |
| S14_extracted_k141_38983 | Low-quality |
| S14_extracted_k141_38984 | Low-quality |
| S14_extracted_k141_67742 | Not-determined |
| S14_extracted_k141_82700 | Not-determined |
| S14_extracted_k141_67745 | Not-determined |
| S14_extracted_k141_82705 | Not-determined |
| S14_extracted_k141_82708 | Low-quality |
| S14_extracted_k141_39005 | Not-determined |
| S14_extracted_k141_90073 | Low-quality |
| S14_extracted_k141_90075 | Not-determined |
| S14_extracted_k141_82716 | Not-determined |
| S14_extracted_k141_1831 | Not-determined |
| S14_extracted_k141_1834 | Low-quality |
| S14_extracted_k141_1835 | Low-quality |
| S14_extracted_k141_1836 | Not-determined |
| S14_extracted_k141_112173 | Not-determined |
| S14_extracted_k141_90086 | Low-quality |
| S14_extracted_k141_1842 | Not-determined |
| S14_extracted_k141_112177 | Low-quality |
| S14_extracted_k141_53493 | Not-determined |
| S14_extracted_k141_82726 | Not-determined |
| S14_extracted_k141_1847 | Not-determined |
| S14_extracted_k141_1848 | Not-determined |
| S14_extracted_k141_112182 | Not-determined |
| S14_extracted_k141_112185 | Not-determined |
| S14_extracted_k141_39012 | Low-quality |
| S14_extracted_k141_105002 | Not-determined |
| S14_extracted_k141_39017 | Low-quality |
| S14_extracted_k141_39018 | Not-determined |
| S14_extracted_k141_9203 | Low-quality |
| S14_extracted_k141_23899 | Low-quality |
| S14_extracted_k141_105008 | Low-quality |
| S14_extracted_k141_67779 | Not-determined |
| S14_extracted_k141_39028 | Not-determined |
| S14_extracted_k141_39029 | Low-quality |
| S14_extracted_k141_105015 | Not-determined |
| S14_extracted_k141_39030 | Not-determined |
| S14_extracted_k141_105017 | Not-determined |
| S14_extracted_k141_82736 | Not-determined |
| S14_extracted_k141_105021 | Low-quality |
| S14_extracted_k141_105023 | Not-determined |
| S14_extracted_k141_82741 | Low-quality |
| S14_extracted_k141_39038 | Not-determined |
| S14_extracted_k141_39041 | Low-quality |
| S14_extracted_k141_23917 | Low-quality |
| S14_extracted_k141_39043 | Low-quality |
| S14_extracted_k141_1855 | Not-determined |
| S14_extracted_k141_39046 | Low-quality |
| S14_extracted_k141_39047 | Not-determined |
| S14_extracted_k141_39052 | Not-determined |
| S14_extracted_k141_53506 | Not-determined |
| S14_extracted_k141_9230 | Low-quality |
| S14_extracted_k141_75268 | Not-determined |
| S14_extracted_k141_67790 | Not-determined |
| S14_extracted_k141_75273 | Not-determined |
| S14_extracted_k141_1873 | Not-determined |
| S14_extracted_k141_105047 | Not-determined |
| S14_extracted_k141_75275 | Not-determined |
| S14_extracted_k141_105050 | Low-quality |
| S14_extracted_k141_105051 | Not-determined |
| S14_extracted_k141_1879 | Low-quality |
| S14_extracted_k141_75278 | Not-determined |
| S14_extracted_k141_90102 | Not-determined |
| S14_extracted_k141_96916 | Not-determined |
| S14_extracted_k141_105055 | Not-determined |
| S14_extracted_k141_1881 | Low-quality |
| S14_extracted_k141_96917 | Not-determined |
| S14_extracted_k141_96918 | Not-determined |
| S14_extracted_k141_1886 | Not-determined |
| S14_extracted_k141_96922 | Low-quality |
| S14_extracted_k141_67798 | Low-quality |
| S14_extracted_k141_31422 | Low-quality |
| S14_extracted_k141_105058 | Not-determined |
| S14_extracted_k141_96924 | Low-quality |
| S14_extracted_k141_39063 | Not-determined |
| S14_extracted_k141_1898 | Not-determined |
| S14_extracted_k141_96932 | Not-determined |
| S14_extracted_k141_96934 | Low-quality |
| S14_extracted_k141_23931 | Not-determined |
| S14_extracted_k141_75300 | Not-determined |
| S14_extracted_k141_105076 | Not-determined |
| S14_extracted_k141_31428 | Not-determined |
| S14_extracted_k141_105078 | Not-determined |
| S14_extracted_k141_75303 | Not-determined |
| S14_extracted_k141_112207 | Not-determined |
| S14_extracted_k141_105080 | Not-determined |
| S14_extracted_k141_23942 | Not-determined |
| S14_extracted_k141_53517 | Not-determined |
| S14_extracted_k141_67819 | Not-determined |
| S14_extracted_k141_82759 | Not-determined |
| S14_extracted_k141_31437 | Not-determined |
| S14_extracted_k141_82765 | Not-determined |
| S14_extracted_k141_31439 | Low-quality |
| S14_extracted_k141_23955 | Low-quality |
| S14_extracted_k141_82768 | Not-determined |
| S14_extracted_k141_23957 | Low-quality |
| S14_extracted_k141_82770 | Not-determined |
| S14_extracted_k141_31441 | Not-determined |
| S14_extracted_k141_23960 | Low-quality |
| S14_extracted_k141_67830 | Low-quality |
| S14_extracted_k141_67833 | Not-determined |
| S14_extracted_k141_23963 | Low-quality |
| S14_extracted_k141_53524 | Low-quality |
| S14_extracted_k141_105089 | Not-determined |
| S14_extracted_k141_9242 | Low-quality |
| S14_extracted_k141_53530 | Not-determined |
| S14_extracted_k141_23967 | Not-determined |
| S14_extracted_k141_53533 | Low-quality |
| S14_extracted_k141_1919 | Low-quality |
| S14_extracted_k141_9243 | Low-quality |
| S14_extracted_k141_105098 | Not-determined |
| S14_extracted_k141_82776 | Not-determined |
| S14_extracted_k141_105100 | Not-determined |
| S14_extracted_k141_105108 | Not-determined |
| S14_extracted_k141_9252 | Not-determined |
| S14_extracted_k141_39072 | Not-determined |
| S14_extracted_k141_1927 | Not-determined |
| S14_extracted_k141_96967 | Not-determined |
| S14_extracted_k141_31444 | Not-determined |
| S14_extracted_k141_82791 | Not-determined |
| S14_extracted_k141_96970 | Not-determined |
| S14_extracted_k141_1932 | Low-quality |
| S14_extracted_k141_82797 | Not-determined |
| S14_extracted_k141_82798 | Not-determined |
| S14_extracted_k141_9268 | Not-determined |
| S14_extracted_k141_105117 | Not-determined |
| S14_extracted_k141_9274 | Not-determined |
| S14_extracted_k141_67860 | Low-quality |
| S14_extracted_k141_105122 | Low-quality |
| S14_extracted_k141_53563 | Low-quality |
| S14_extracted_k141_82807 | Not-determined |
| S14_extracted_k141_24010 | Low-quality |
| S14_extracted_k141_39089 | Low-quality |
| S14_extracted_k141_39090 | Not-determined |
| S14_extracted_k141_90110 | Not-determined |
| S14_extracted_k141_82817 | Not-determined |
| S14_extracted_k141_112234 | Not-determined |
| S14_extracted_k141_75344 | Low-quality |
| S14_extracted_k141_67878 | Low-quality |
| S14_extracted_k141_90116 | Low-quality |
| S14_extracted_k141_96999 | Not-determined |
| S14_extracted_k141_97000 | Low-quality |
| S14_extracted_k141_67882 | Not-determined |
| S14_extracted_k141_24026 | Not-determined |
| S14_extracted_k141_31456 | Not-determined |
| S14_extracted_k141_112245 | Low-quality |
| S14_extracted_k141_31459 | Not-determined |
| S14_extracted_k141_53577 | Low-quality |
| S14_extracted_k141_75348 | Not-determined |
| S14_extracted_k141_67892 | Low-quality |
| S14_extracted_k141_90129 | Low-quality |
| S14_extracted_k141_67899 | Not-determined |
| S14_extracted_k141_16901 | Not-determined |
| S14_extracted_k141_82837 | Low-quality |
| S14_extracted_k141_53583 | Not-determined |
| S14_extracted_k141_16908 | Not-determined |
| S14_extracted_k141_75365 | Not-determined |
| S14_extracted_k141_75366 | Low-quality |
| S14_extracted_k141_75368 | Low-quality |
| S14_extracted_k141_75369 | Low-quality |
| S14_extracted_k141_16915 | Low-quality |
| S14_extracted_k141_16919 | Low-quality |
| S14_extracted_k141_16922 | Low-quality |
| S14_extracted_k141_75373 | Low-quality |
| S14_extracted_k141_16924 | Not-determined |
| S14_extracted_k141_90147 | Low-quality |
| S14_extracted_k141_82849 | Not-determined |
| S14_extracted_k141_75376 | Low-quality |
| S14_extracted_k141_82850 | Low-quality |
| S14_extracted_k141_75379 | Low-quality |
| S14_extracted_k141_9298 | Not-determined |
| S14_extracted_k141_9299 | Not-determined |
| S14_extracted_k141_82852 | Not-determined |
| S14_extracted_k141_53595 | Low-quality |
| S14_extracted_k141_82854 | Not-determined |
| S14_extracted_k141_16935 | Not-determined |
| S14_extracted_k141_9305 | Not-determined |
| S14_extracted_k141_16939 | Low-quality |
| S14_extracted_k141_9307 | Not-determined |
| S14_extracted_k141_60849 | Not-determined |
| S14_extracted_k141_82857 | Not-determined |
| S14_extracted_k141_75391 | Low-quality |
| S14_extracted_k141_16945 | Not-determined |
| S14_extracted_k141_82861 | Low-quality |
| S14_extracted_k141_24066 | Low-quality |
| S14_extracted_k141_67912 | Low-quality |
| S14_extracted_k141_60861 | Not-determined |
| S14_extracted_k141_75399 | Not-determined |
| S14_extracted_k141_1955 | Low-quality |
| S14_extracted_k141_53605 | Not-determined |
| S14_extracted_k141_112258 | Not-determined |
| S14_extracted_k141_1957 | Low-quality |
| S14_extracted_k141_82875 | Not-determined |
| S14_extracted_k141_67918 | Not-determined |
| S14_extracted_k141_97023 | Low-quality |
| S14_extracted_k141_9314 | Not-determined |
| S14_extracted_k141_82878 | Not-determined |
| S14_extracted_k141_97028 | Low-quality |
| S14_extracted_k141_112269 | Not-determined |
| S14_extracted_k141_112271 | Low-quality |
| S14_extracted_k141_1967 | Not-determined |
| S14_extracted_k141_97038 | Not-determined |
| S14_extracted_k141_9321 | Not-determined |
| S14_extracted_k141_9323 | Not-determined |
| S14_extracted_k141_31471 | Not-determined |
| S14_extracted_k141_53624 | Not-determined |
| S14_extracted_k141_31473 | Not-determined |
| S14_extracted_k141_97044 | Not-determined |
| S14_extracted_k141_75409 | Low-quality |
| S14_extracted_k141_31476 | Not-determined |
| S14_extracted_k141_105139 | Not-determined |
| S14_extracted_k141_60878 | Low-quality |
| S14_extracted_k141_60882 | Not-determined |
| S14_extracted_k141_60883 | Low-quality |
| S14_extracted_k141_60885 | Low-quality |
| S14_extracted_k141_53632 | Not-determined |
| S14_extracted_k141_9328 | Not-determined |
| S14_extracted_k141_45652 | Low-quality |
| S14_extracted_k141_9329 | Not-determined |
| S14_extracted_k141_9331 | Low-quality |
| S14_extracted_k141_97048 | Not-determined |
| S14_extracted_k141_90166 | Not-determined |
| S14_extracted_k141_67934 | Not-determined |
| S14_extracted_k141_97052 | Not-determined |
| S14_extracted_k141_60891 | Low-quality |
| S14_extracted_k141_9336 | Not-determined |
| S14_extracted_k141_45658 | Not-determined |
| S14_extracted_k141_67939 | Not-determined |
| S14_extracted_k141_53643 | Not-determined |
| S14_extracted_k141_105157 | Not-determined |
| S14_extracted_k141_16954 | Not-determined |
| S14_extracted_k141_97060 | Not-determined |
| S14_extracted_k141_53648 | Not-determined |
| S14_extracted_k141_75425 | Not-determined |
| S14_extracted_k141_97064 | Low-quality |
| S14_extracted_k141_31498 | Low-quality |
| S14_extracted_k141_24079 | Not-determined |
| S14_extracted_k141_16958 | Not-determined |
| S14_extracted_k141_16959 | Not-determined |
| S14_extracted_k141_97067 | Not-determined |
| S14_extracted_k141_75432 | Not-determined |
| S14_extracted_k141_67955 | Low-quality |
| S14_extracted_k141_67957 | Low-quality |
| S14_extracted_k141_97071 | Low-quality |
| S14_extracted_k141_53655 | Not-determined |
| S14_extracted_k141_75438 | Not-determined |
| S14_extracted_k141_31506 | Not-determined |
| S14_extracted_k141_97074 | Low-quality |
| S14_extracted_k141_97075 | Not-determined |
| S14_extracted_k141_60900 | Low-quality |
| S14_extracted_k141_60901 | Not-determined |
| S14_extracted_k141_67962 | Low-quality |
| S14_extracted_k141_75439 | Low-quality |

| | |
|---|---|
| S14_extracted_k141_97077 | Not-determined |
| S14_extracted_k141_9360 | Low-quality |
| S14_extracted_k141_97079 | Not-determined |
| S14_extracted_k141_9361 | Not-determined |
| S14_extracted_k141_97081 | Low-quality |
| S14_extracted_k141_97088 | Not-determined |
| S14_extracted_k141_82883 | Not-determined |
| S14_extracted_k141_97092 | Not-determined |
| S14_extracted_k141_39163 | Low-quality |
| S14_extracted_k141_97097 | Low-quality |
| S14_extracted_k141_75453 | Not-determined |
| S14_extracted_k141_16969 | Not-determined |
| S14_extracted_k141_39169 | Not-determined |
| S14_extracted_k141_53663 | Not-determined |
| S14_extracted_k141_9373 | Low-quality |
| S14_extracted_k141_31512 | Not-determined |
| S14_extracted_k141_31513 | Not-determined |
| S14_extracted_k141_82890 | Low-quality |
| S14_extracted_k141_90190 | Not-determined |
| S14_extracted_k141_31515 | Low-quality |
| S14_extracted_k141_31516 | Not-determined |
| S14_extracted_k141_97109 | Low-quality |
| S14_extracted_k141_53665 | Low-quality |
| S14_extracted_k141_53668 | Low-quality |
| S14_extracted_k141_67977 | Low-quality |
| S14_extracted_k141_97116 | Low-quality |
| S14_extracted_k141_90197 | Not-determined |
| S14_extracted_k141_53672 | Not-determined |
| S14_extracted_k141_75466 | Not-determined |
| S14_extracted_k141_97119 | Not-determined |
| S14_extracted_k141_31526 | Not-determined |
| S14_extracted_k141_9382 | Not-determined |
| S14_extracted_k141_53675 | Low-quality |
| S14_extracted_k141_53678 | Not-determined |
| S14_extracted_k141_16975 | Not-determined |
| S14_extracted_k141_90199 | Not-determined |
| S14_extracted_k141_31538 | Not-determined |
| S14_extracted_k141_9403 | Low-quality |
| S14_extracted_k141_39195 | Low-quality |
| S14_extracted_k141_53687 | Not-determined |
| S14_extracted_k141_9406 | Not-determined |
| S14_extracted_k141_16982 | Not-determined |
| S14_extracted_k141_82897 | Not-determined |
| S14_extracted_k141_9411 | Low-quality |
| S14_extracted_k141_39203 | Low-quality |
| S14_extracted_k141_75485 | Not-determined |
| S14_extracted_k141_39206 | Low-quality |
| S14_extracted_k141_112280 | Not-determined |
| S14_extracted_k141_90209 | Not-determined |
| S14_extracted_k141_31550 | Not-determined |
| S14_extracted_k141_39212 | Low-quality |
| S14_extracted_k141_75496 | Not-determined |
| S14_extracted_k141_39216 | Not-determined |
| S14_extracted_k141_97129 | Not-determined |
| S14_extracted_k141_67988 | Not-determined |
| S14_extracted_k141_97133 | Not-determined |
| S14_extracted_k141_24093 | Low-quality |
| S14_extracted_k141_97142 | Low-quality |
| S14_extracted_k141_1989 | Not-determined |
| S14_extracted_k141_53702 | Not-determined |
| S14_extracted_k141_82904 | Not-determined |
| S14_extracted_k141_82905 | Low-quality |
| S14_extracted_k141_67998 | Low-quality |
| S14_extracted_k141_31563 | Low-quality |
| S14_extracted_k141_24100 | Not-determined |
| S14_extracted_k141_9420 | Low-quality |
| S14_extracted_k141_68006 | Low-quality |
| S14_extracted_k141_68007 | Not-determined |
| S14_extracted_k141_68009 | Low-quality |
| S14_extracted_k141_2004 | Low-quality |
| S14_extracted_k141_17002 | Not-determined |
| S14_extracted_k141_105162 | Low-quality |
| S14_extracted_k141_82917 | Not-determined |
| S14_extracted_k141_82918 | Low-quality |
| S14_extracted_k141_2008 | Low-quality |
| S14_extracted_k141_9426 | Not-determined |
| S14_extracted_k141_31568 | Not-determined |
| S14_extracted_k141_31571 | Low-quality |
| S14_extracted_k141_31573 | Low-quality |
| S14_extracted_k141_97159 | Not-determined |
| S14_extracted_k141_82920 | Low-quality |
| S14_extracted_k141_9430 | Not-determined |
| S14_extracted_k141_17008 | Not-determined |
| S14_extracted_k141_24114 | Not-determined |
| S14_extracted_k141_53717 | Low-quality |
| S14_extracted_k141_45680 | Low-quality |
| S14_extracted_k141_17013 | Not-determined |
| S14_extracted_k141_97166 | Not-determined |
| S14_extracted_k141_75506 | Not-determined |
| S14_extracted_k141_17020 | Low-quality |
| S14_extracted_k141_97172 | Not-determined |
| S14_extracted_k141_75508 | Low-quality |
| S14_extracted_k141_68021 | Not-determined |
| S14_extracted_k141_45693 | Not-determined |
| S14_extracted_k141_82933 | Not-determined |
| S14_extracted_k141_45695 | Not-determined |
| S14_extracted_k141_105168 | Not-determined |
| S14_extracted_k141_75519 | Not-determined |
| S14_extracted_k141_9443 | Low-quality |
| S14_extracted_k141_24124 | Low-quality |
| S14_extracted_k141_39223 | Not-determined |
| S14_extracted_k141_82937 | Not-determined |
| S14_extracted_k141_112314 | Not-determined |
| S14_extracted_k141_75528 | Low-quality |
| S14_extracted_k141_24139 | Not-determined |
| S14_extracted_k141_75532 | Not-determined |
| S14_extracted_k141_105191 | Low-quality |
| S14_extracted_k141_53737 | Not-determined |
| S14_extracted_k141_97184 | Not-determined |
| S14_extracted_k141_60908 | Not-determined |
| S14_extracted_k141_82953 | Not-determined |
| S14_extracted_k141_60910 | Not-determined |
| S14_extracted_k141_60911 | Not-determined |
| S14_extracted_k141_112330 | Low-quality |
| S14_extracted_k141_53738 | Low-quality |
| S14_extracted_k141_82956 | Not-determined |
| S14_extracted_k141_75540 | Not-determined |
| S14_extracted_k141_75543 | Not-determined |
| S14_extracted_k141_24155 | Low-quality |
| S14_extracted_k141_68029 | Low-quality |
| S14_extracted_k141_97191 | Not-determined |
| S14_extracted_k141_112340 | Low-quality |
| S14_extracted_k141_60926 | Not-determined |
| S14_extracted_k141_68031 | Not-determined |
| S14_extracted_k141_97193 | Not-determined |
| S14_extracted_k141_24159 | Low-quality |
| S14_extracted_k141_53748 | Not-determined |
| S14_extracted_k141_90230 | Not-determined |
| S14_extracted_k141_2031 | Not-determined |
| S14_extracted_k141_90241 | Not-determined |
| S14_extracted_k141_105208 | Low-quality |
| S14_extracted_k141_60941 | Low-quality |
| S14_extracted_k141_75564 | Low-quality |
| S14_extracted_k141_9447 | Not-determined |
| S14_extracted_k141_75570 | Not-determined |
| S14_extracted_k141_60955 | Not-determined |
| S14_extracted_k141_105221 | Not-determined |
| S14_extracted_k141_17028 | Not-determined |
| S14_extracted_k141_90255 | Not-determined |
| S14_extracted_k141_75572 | Not-determined |
| S14_extracted_k141_60957 | Not-determined |
| S14_extracted_k141_105225 | Not-determined |
| S14_extracted_k141_53779 | Not-determined |
| S14_extracted_k141_2052 | Not-determined |
| S14_extracted_k141_90260 | Not-determined |
| S14_extracted_k141_90261 | Not-determined |
| S14_extracted_k141_2057 | Not-determined |
| S14_extracted_k141_105243 | Low-quality |
| S14_extracted_k141_97197 | Not-determined |
| S14_extracted_k141_82974 | Not-determined |
| S14_extracted_k141_75584 | Not-determined |
| S14_extracted_k141_105254 | Not-determined |
| S14_extracted_k141_45699 | Low-quality |
| S14_extracted_k141_24179 | Low-quality |
| S14_extracted_k141_45701 | Low-quality |
| S14_extracted_k141_17041 | Not-determined |
| S14_extracted_k141_24183 | Not-determined |
| S14_extracted_k141_97215 | Not-determined |
| S14_extracted_k141_82995 | Low-quality |
| S14_extracted_k141_24192 | Not-determined |
| S14_extracted_k141_82997 | Not-determined |
| S14_extracted_k141_75605 | Not-determined |
| S14_extracted_k141_105274 | Not-determined |
| S14_extracted_k141_45705 | Not-determined |
| S14_extracted_k141_17053 | Low-quality |
| S14_extracted_k141_2089 | Not-determined |
| S14_extracted_k141_105279 | Low-quality |
| S14_extracted_k141_2094 | Not-determined |
| S14_extracted_k141_45713 | Low-quality |
| S14_extracted_k141_17064 | Not-determined |
| S14_extracted_k141_112353 | Low-quality |
| S14_extracted_k141_45715 | Not-determined |
| S14_extracted_k141_105284 | Not-determined |
| S14_extracted_k141_45716 | Not-determined |
| S14_extracted_k141_45717 | Not-determined |
| S14_extracted_k141_112360 | Not-determined |
| S14_extracted_k141_39267 | Not-determined |
| S14_extracted_k141_105292 | Not-determined |
| S14_extracted_k141_45723 | Low-quality |
| S14_extracted_k141_97222 | Not-determined |
| S14_extracted_k141_9468 | Not-determined |
| S14_extracted_k141_83010 | Not-determined |
| S14_extracted_k141_105300 | Not-determined |
| S14_extracted_k141_105301 | Low-quality |
| S14_extracted_k141_83015 | Low-quality |
| S14_extracted_k141_112367 | Not-determined |
| S14_extracted_k141_60976 | Not-determined |
| S14_extracted_k141_17083 | Not-determined |
| S14_extracted_k141_17087 | Low-quality |
| S14_extracted_k141_31581 | Low-quality |
| S14_extracted_k141_60978 | Low-quality |
| S14_extracted_k141_105319 | Low-quality |
| S14_extracted_k141_31592 | Not-determined |
| S14_extracted_k141_68080 | Low-quality |
| S14_extracted_k141_9501 | Low-quality |
| S14_extracted_k141_68082 | Low-quality |
| S14_extracted_k141_83028 | Low-quality |
| S14_extracted_k141_9502 | Not-determined |
| S14_extracted_k141_105332 | Not-determined |
| S14_extracted_k141_45749 | Not-determined |
| S14_extracted_k141_45751 | Not-determined |
| S14_extracted_k141_105338 | Not-determined |
| S14_extracted_k141_105340 | Not-determined |
| S14_extracted_k141_105341 | Not-determined |
| S14_extracted_k141_105343 | Low-quality |
| S14_extracted_k141_105344 | Not-determined |
| S14_extracted_k141_45759 | Not-determined |
| S14_extracted_k141_97232 | Not-determined |
| S14_extracted_k141_105351 | Not-determined |
| S14_extracted_k141_45766 | Not-determined |
| S14_extracted_k141_2137 | Low-quality |
| S14_extracted_k141_112394 | Low-quality |
| S14_extracted_k141_31611 | Low-quality |
| S14_extracted_k141_2143 | Not-determined |
| S14_extracted_k141_112398 | Not-determined |
| S14_extracted_k141_2147 | Low-quality |
| S14_extracted_k141_90267 | Not-determined |
| S14_extracted_k141_9515 | Not-determined |
| S14_extracted_k141_45781 | Not-determined |
| S14_extracted_k141_83046 | Low-quality |
| S14_extracted_k141_31619 | Not-determined |
| S14_extracted_k141_90268 | Not-determined |
| S14_extracted_k141_2151 | Not-determined |
| S14_extracted_k141_97245 | Not-determined |
| S14_extracted_k141_75620 | Low-quality |
| S14_extracted_k141_61002 | Low-quality |
| S14_extracted_k141_90274 | Not-determined |
| S14_extracted_k141_97249 | Not-determined |
| S14_extracted_k141_75628 | Not-determined |
| S14_extracted_k141_90279 | Not-determined |
| S14_extracted_k141_45792 | Not-determined |
| S14_extracted_k141_39305 | Not-determined |
| S14_extracted_k141_39306 | Low-quality |
| S14_extracted_k141_2157 | Not-determined |
| S14_extracted_k141_90283 | Not-determined |
| S14_extracted_k141_112417 | Not-determined |
| S14_extracted_k141_112419 | Low-quality |
| S14_extracted_k141_39314 | Low-quality |
| S14_extracted_k141_45801 | Not-determined |
| S14_extracted_k141_97255 | Low-quality |
| S14_extracted_k141_45803 | Low-quality |
| S14_extracted_k141_68107 | Not-determined |
| S14_extracted_k141_68108 | Not-determined |
| S14_extracted_k141_112429 | Not-determined |
| S14_extracted_k141_90294 | Not-determined |
| S14_extracted_k141_2166 | Not-determined |
| S14_extracted_k141_112436 | Low-quality |
| S14_extracted_k141_90297 | Low-quality |
| S14_extracted_k141_45820 | Low-quality |
| S14_extracted_k141_90300 | Not-determined |
| S14_extracted_k141_2173 | Low-quality |
| S14_extracted_k141_45822 | Not-determined |
| S14_extracted_k141_45823 | Not-determined |
| S14_extracted_k141_75648 | Not-determined |
| S14_extracted_k141_17113 | Low-quality |
| S14_extracted_k141_105369 | Not-determined |
| S14_extracted_k141_61029 | Not-determined |
| S14_extracted_k141_17117 | Not-determined |
| S14_extracted_k141_17120 | Not-determined |
| S14_extracted_k141_75660 | Not-determined |
| S14_extracted_k141_45825 | Not-determined |
| S14_extracted_k141_75662 | Not-determined |
| S14_extracted_k141_9524 | Low-quality |
| S14_extracted_k141_31649 | Not-determined |
| S14_extracted_k141_61041 | Not-determined |
| S14_extracted_k141_31650 | Not-determined |
| S14_extracted_k141_9526 | Not-determined |
| S14_extracted_k141_75668 | Low-quality |
| S14_extracted_k141_75673 | Not-determined |
| S14_extracted_k141_97281 | Not-determined |
| S14_extracted_k141_9535 | Low-quality |
| S14_extracted_k141_17128 | Not-determined |
| S14_extracted_k141_97289 | Not-determined |
| S14_extracted_k141_17131 | Low-quality |
| S14_extracted_k141_9540 | Low-quality |
| S14_extracted_k141_9541 | Low-quality |
| S14_extracted_k141_9543 | Low-quality |
| S14_extracted_k141_17134 | Low-quality |
| S14_extracted_k141_39317 | Not-determined |
| S14_extracted_k141_90305 | Not-determined |
| S14_extracted_k141_83050 | Not-determined |
| S14_extracted_k141_83054 | Not-determined |
| S14_extracted_k141_2189 | Low-quality |
| S14_extracted_k141_105384 | Not-determined |
| S14_extracted_k141_9550 | Low-quality |
| S14_extracted_k141_53788 | Not-determined |
| S14_extracted_k141_9554 | Low-quality |
| S14_extracted_k141_17151 | Not-determined |
| S14_extracted_k141_17152 | Not-determined |
| S14_extracted_k141_9556 | Low-quality |
| S14_extracted_k141_97295 | Not-determined |
| S14_extracted_k141_17155 | Low-quality |
| S14_extracted_k141_9557 | Not-determined |
| S14_extracted_k141_53793 | Not-determined |
| S14_extracted_k141_9559 | Low-quality |
| S14_extracted_k141_2196 | Not-determined |
| S14_extracted_k141_53794 | Low-quality |
| S14_extracted_k141_53795 | Not-determined |
| S14_extracted_k141_53798 | Low-quality |
| S14_extracted_k141_17165 | Low-quality |
| S14_extracted_k141_105385 | Low-quality |
| S14_extracted_k141_45849 | Not-determined |
| S14_extracted_k141_83070 | Low-quality |
| S14_extracted_k141_61048 | Low-quality |
| S14_extracted_k141_17171 | Low-quality |
| S14_extracted_k141_61051 | Not-determined |
| S14_extracted_k141_112454 | Not-determined |
| S14_extracted_k141_53811 | Not-determined |
| S14_extracted_k141_61053 | Not-determined |
| S14_extracted_k141_31672 | Not-determined |
| S14_extracted_k141_31673 | Not-determined |
| S14_extracted_k141_17179 | Not-determined |
| S14_extracted_k141_53814 | Not-determined |
| S14_extracted_k141_53815 | Not-determined |
| S14_extracted_k141_105395 | Not-determined |
| S14_extracted_k141_105400 | Not-determined |
| S14_extracted_k141_61057 | Not-determined |
| S14_extracted_k141_90349 | Low-quality |
| S14_extracted_k141_75688 | Low-quality |
| S14_extracted_k141_9586 | Not-determined |
| S14_extracted_k141_75690 | Not-determined |
| S14_extracted_k141_83084 | Not-determined |
| S14_extracted_k141_83085 | Not-determined |
| S14_extracted_k141_75693 | Low-quality |
| S14_extracted_k141_83087 | Not-determined |
| S14_extracted_k141_9591 | Low-quality |
| S14_extracted_k141_112476 | Not-determined |
| S14_extracted_k141_75696 | Not-determined |
| S14_extracted_k141_112478 | Not-determined |
| S14_extracted_k141_90353 | Not-determined |
| S14_extracted_k141_9598 | Not-determined |
| S14_extracted_k141_90356 | Not-determined |
| S14_extracted_k141_9601 | Low-quality |
| S14_extracted_k141_90361 | Low-quality |
| S14_extracted_k141_90362 | Not-determined |
| S14_extracted_k141_83098 | Not-determined |
| S14_extracted_k141_9605 | Not-determined |
| S14_extracted_k141_61073 | Not-determined |
| S14_extracted_k141_75708 | Not-determined |
| S14_extracted_k141_90368 | Not-determined |
| S14_extracted_k141_39338 | Not-determined |
| S14_extracted_k141_61085 | Not-determined |
| S14_extracted_k141_9614 | Low-quality |
| S14_extracted_k141_90378 | Low-quality |
| S14_extracted_k141_83105 | Low-quality |
| S14_extracted_k141_39341 | Not-determined |
| S14_extracted_k141_105412 | Not-determined |
| S14_extracted_k141_75729 | Low-quality |
| S14_extracted_k141_75732 | Not-determined |
| S14_extracted_k141_9625 | Low-quality |
| S14_extracted_k141_75736 | Low-quality |
| S14_extracted_k141_9627 | Low-quality |
| S14_extracted_k141_9629 | Not-determined |
| S14_extracted_k141_9631 | Low-quality |
| S14_extracted_k141_9632 | Low-quality |
| S14_extracted_k141_45879 | Not-determined |
| S14_extracted_k141_31700 | Not-determined |
| S14_extracted_k141_31702 | Not-determined |
| S14_extracted_k141_45882 | Not-determined |
| S14_extracted_k141_45884 | Not-determined |
| S14_extracted_k141_45886 | Not-determined |
| S14_extracted_k141_61096 | Not-determined |
| S14_extracted_k141_39354 | Not-determined |
| S14_extracted_k141_112499 | Not-determined |
| S14_extracted_k141_97320 | Not-determined |
| S14_extracted_k141_105422 | Not-determined |
| S14_extracted_k141_39357 | Not-determined |
| S14_extracted_k141_112511 | Not-determined |
| S14_extracted_k141_61099 | Low-quality |
| S14_extracted_k141_75763 | Not-determined |
| S14_extracted_k141_75765 | Not-determined |
| S14_extracted_k141_61103 | Not-determined |
| S14_extracted_k141_97330 | Low-quality |
| S14_extracted_k141_97336 | Not-determined |
| S14_extracted_k141_61106 | Not-determined |
| S14_extracted_k141_112523 | Not-determined |
| S14_extracted_k141_112524 | Low-quality |
| S14_extracted_k141_31717 | Not-determined |
| S14_extracted_k141_112527 | Not-determined |
| S14_extracted_k141_17223 | Not-determined |
| S14_extracted_k141_97345 | Not-determined |
| S14_extracted_k141_97346 | Not-determined |
| S14_extracted_k141_68174 | Not-determined |
| S14_extracted_k141_2227 | Low-quality |
| S14_extracted_k141_2229 | Low-quality |
| S14_extracted_k141_2231 | Not-determined |
| S14_extracted_k141_61122 | Not-determined |
| S14_extracted_k141_45899 | Not-determined |
| S14_extracted_k141_97357 | Not-determined |
| S14_extracted_k141_31735 | Not-determined |
| S14_extracted_k141_31736 | Not-determined |
| S14_extracted_k141_31737 | Low-quality |
| S14_extracted_k141_105431 | Low-quality |
| S14_extracted_k141_90432 | Low-quality |
| S14_extracted_k141_39407 | Not-determined |
| S14_extracted_k141_61130 | Low-quality |
| S14_extracted_k141_90433 | Not-determined |
| S14_extracted_k141_61131 | Low-quality |
| S14_extracted_k141_75788 | Not-determined |
| S14_extracted_k141_61134 | Low-quality |
| S14_extracted_k141_24201 | Not-determined |
| S14_extracted_k141_112544 | Not-determined |
| S14_extracted_k141_75793 | Not-determined |
| S14_extracted_k141_2258 | Low-quality |
| S14_extracted_k141_53826 | Not-determined |
| S14_extracted_k141_45916 | Not-determined |
| S14_extracted_k141_9662 | Low-quality |
| S14_extracted_k141_45919 | Low-quality |
| S14_extracted_k141_75799 | Low-quality |
| S14_extracted_k141_61143 | Not-determined |
| S14_extracted_k141_75800 | Not-determined |
| S14_extracted_k141_45923 | Low-quality |
| S14_extracted_k141_61145 | Not-determined |
| S14_extracted_k141_24212 | Not-determined |
| S14_extracted_k141_61147 | Not-determined |
| S14_extracted_k141_68193 | Not-determined |
| S14_extracted_k141_9667 | Low-quality |
| S14_extracted_k141_53831 | Not-determined |
| S14_extracted_k141_17235 | Not-determined |
| S14_extracted_k141_9668 | Not-determined |
| S14_extracted_k141_105445 | Not-determined |
| S14_extracted_k141_75807 | Low-quality |
| S14_extracted_k141_105447 | Not-determined |
| S14_extracted_k141_112554 | Not-determined |
| S14_extracted_k141_105453 | Low-quality |
| S14_extracted_k141_61161 | Low-quality |
| S14_extracted_k141_105458 | Low-quality |
| S14_extracted_k141_112558 | Low-quality |
| S14_extracted_k141_75810 | Not-determined |
| S14_extracted_k141_39414 | Not-determined |
| S14_extracted_k141_61170 | Low-quality |
| S14_extracted_k141_75817 | Not-determined |
| S14_extracted_k141_112560 | Not-determined |
| S14_extracted_k141_75819 | Not-determined |
| S14_extracted_k141_105461 | Not-determined |
| S14_extracted_k141_105463 | Not-determined |
| S14_extracted_k141_53849 | Not-determined |
| S14_extracted_k141_61173 | Low-quality |
| S14_extracted_k141_83126 | Not-determined |
| S14_extracted_k141_39420 | Not-determined |
| S14_extracted_k141_17250 | Not-determined |
| S14_extracted_k141_68215 | Not-determined |
| S14_extracted_k141_53853 | Not-determined |
| S14_extracted_k141_90444 | Not-determined |
| S14_extracted_k141_75831 | Not-determined |
| S14_extracted_k141_105470 | Not-determined |
| S14_extracted_k141_97375 | Low-quality |
| S14_extracted_k141_105473 | Not-determined |
| S14_extracted_k141_39426 | Low-quality |
| S14_extracted_k141_61182 | Not-determined |
| S14_extracted_k141_105479 | Not-determined |
| S14_extracted_k141_68222 | Not-determined |
| S14_extracted_k141_17259 | Not-determined |
| S14_extracted_k141_105485 | Not-determined |
| S14_extracted_k141_90455 | Low-quality |
| S14_extracted_k141_75845 | Not-determined |
| S14_extracted_k141_68229 | Not-determined |
| S14_extracted_k141_45936 | Not-determined |
| S14_extracted_k141_83139 | Low-quality |
| S14_extracted_k141_112565 | Low-quality |
| S14_extracted_k141_112566 | Low-quality |
| S14_extracted_k141_75859 | Not-determined |
| S14_extracted_k141_31764 | Not-determined |
| S14_extracted_k141_24228 | Not-determined |
| S14_extracted_k141_24229 | Low-quality |
| S14_extracted_k141_45948 | Not-determined |
| S14_extracted_k141_53863 | Low-quality |
| S14_extracted_k141_45949 | Low-quality |
| S14_extracted_k141_112572 | Not-determined |
| S14_extracted_k141_90467 | Not-determined |
| S14_extracted_k141_105496 | Not-determined |
| S14_extracted_k141_68259 | Not-determined |
| S14_extracted_k141_112579 | Low-quality |
| S14_extracted_k141_17277 | Not-determined |
| S14_extracted_k141_75862 | Not-determined |
| S14_extracted_k141_90477 | Low-quality |
| S14_extracted_k141_83164 | Not-determined |
| S14_extracted_k141_39431 | Not-determined |
| S14_extracted_k141_31777 | Low-quality |
| S14_extracted_k141_105503 | Not-determined |
| S14_extracted_k141_17288 | Not-determined |
| S14_extracted_k141_105509 | Low-quality |
| S14_extracted_k141_105510 | Low-quality |
| S14_extracted_k141_39437 | Low-quality |
| S14_extracted_k141_90485 | Not-determined |
| S14_extracted_k141_97398 | Not-determined |
| S14_extracted_k141_112586 | Not-determined |
| S14_extracted_k141_83173 | Not-determined |
| S14_extracted_k141_24247 | Not-determined |
| S14_extracted_k141_112590 | Not-determined |
| S14_extracted_k141_17295 | Low-quality |
| S14_extracted_k141_112593 | Low-quality |
| S14_extracted_k141_9679 | Not-determined |
| S14_extracted_k141_75887 | Not-determined |
| S14_extracted_k141_17301 | Not-determined |
| S14_extracted_k141_90498 | Not-determined |
| S14_extracted_k141_68264 | Low-quality |
| S14_extracted_k141_24252 | Not-determined |
| S14_extracted_k141_90504 | Low-quality |
| S14_extracted_k141_68266 | Not-determined |
| S14_extracted_k141_83182 | Low-quality |
| S14_extracted_k141_112598 | Low-quality |
| S14_extracted_k141_112599 | Not-determined |
| S14_extracted_k141_112600 | Not-determined |
| S14_extracted_k141_17310 | Not-determined |
| S14_extracted_k141_53887 | Not-determined |
| S14_extracted_k141_45972 | Not-determined |
| S14_extracted_k141_17311 | Not-determined |
| S14_extracted_k141_53891 | Low-quality |
| S14_extracted_k141_75894 | Not-determined |
| S14_extracted_k141_2273 | Low-quality |
| S14_extracted_k141_2274 | Not-determined |
| S14_extracted_k141_17317 | Low-quality |
| S14_extracted_k141_24254 | Not-determined |
| S14_extracted_k141_68274 | Not-determined |
| S14_extracted_k141_17324 | Low-quality |
| S14_extracted_k141_90510 | Not-determined |
| S14_extracted_k141_53905 | Not-determined |
| S14_extracted_k141_90514 | Not-determined |
| S14_extracted_k141_45989 | Not-determined |
| S14_extracted_k141_31814 | Not-determined |
| S14_extracted_k141_53917 | Not-determined |
| S14_extracted_k141_53921 | Not-determined |
| S14_extracted_k141_2292 | Not-determined |
| S14_extracted_k141_90529 | Not-determined |
| S14_extracted_k141_45998 | Low-quality |
| S14_extracted_k141_31817 | Not-determined |
| S14_extracted_k141_53925 | Not-determined |
| S14_extracted_k141_83209 | Low-quality |
| S14_extracted_k141_112604 | Low-quality |
| S14_extracted_k141_97422 | Low-quality |
| S14_extracted_k141_24278 | Not-determined |
| S14_extracted_k141_9687 | Low-quality |
| S14_extracted_k141_97424 | Low-quality |
| S14_extracted_k141_31824 | Not-determined |
| S14_extracted_k141_31825 | Not-determined |
| S14_extracted_k141_31831 | Not-determined |
| S14_extracted_k141_31832 | Low-quality |
| S14_extracted_k141_31834 | Low-quality |
| S14_extracted_k141_46010 | Low-quality |
| S14_extracted_k141_31839 | Not-determined |
| S14_extracted_k141_68297 | Not-determined |
| S14_extracted_k141_68298 | Not-determined |
| S14_extracted_k141_53929 | Not-determined |
| S14_extracted_k141_31843 | Low-quality |
| S14_extracted_k141_31844 | Not-determined |
| S14_extracted_k141_9699 | Not-determined |
| S14_extracted_k141_9701 | Not-determined |
| S14_extracted_k141_105532 | Low-quality |
| S14_extracted_k141_75932 | Not-determined |
| S14_extracted_k141_97438 | Not-determined |
| S14_extracted_k141_75936 | Not-determined |
| S14_extracted_k141_68311 | Not-determined |
| S14_extracted_k141_24288 | Not-determined |
| S14_extracted_k141_24289 | Not-determined |
| S14_extracted_k141_105537 | Not-determined |
| S14_extracted_k141_75945 | Not-determined |
| S14_extracted_k141_105540 | Low-quality |
| S14_extracted_k141_83212 | Low-quality |
| S14_extracted_k141_83213 | Low-quality |
| S14_extracted_k141_83214 | Not-determined |
| S14_extracted_k141_105548 | Not-determined |
| S14_extracted_k141_97449 | Not-determined |
| S14_extracted_k141_9722 | Low-quality |
| S14_extracted_k141_9725 | Low-quality |
| S14_extracted_k141_90556 | Not-determined |
| S14_extracted_k141_17329 | Low-quality |
| S14_extracted_k141_24312 | Not-determined |
| S14_extracted_k141_105560 | Not-determined |
| S14_extracted_k141_2318 | Not-determined |
| S14_extracted_k141_112615 | Low-quality |
| S14_extracted_k141_24314 | Low-quality |
| S14_extracted_k141_2321 | Not-determined |
| S14_extracted_k141_24315 | Not-determined |
| S14_extracted_k141_75969 | Not-determined |
| S14_extracted_k141_9742 | Not-determined |
| S14_extracted_k141_9745 | Not-determined |
| S14_extracted_k141_39468 | Not-determined |
| S14_extracted_k141_9747 | Low-quality |
| S14_extracted_k141_105570 | Not-determined |
| S14_extracted_k141_9748 | Not-determined |
| S14_extracted_k141_112623 | Not-determined |
| S14_extracted_k141_9749 | Not-determined |
| S14_extracted_k141_97461 | Not-determined |
| S14_extracted_k141_39472 | Not-determined |
| S14_extracted_k141_105574 | Low-quality |
| S14_extracted_k141_75975 | Not-determined |
| S14_extracted_k141_105579 | Not-determined |
| S14_extracted_k141_2334 | Not-determined |

| | |
|---|---|
| S14_extracted_k141_75979 | Low-quality |
| S14_extracted_k141_68344 | Not-determined |
| S14_extracted_k141_105582 | Not-determined |
| S14_extracted_k141_97470 | Not-determined |
| S14_extracted_k141_112631 | Low-quality |
| S14_extracted_k141_90563 | Not-determined |
| S14_extracted_k141_24329 | Not-determined |
| S14_extracted_k141_68353 | Not-determined |
| S14_extracted_k141_75986 | Not-determined |
| S14_extracted_k141_68361 | Low-quality |
| S14_extracted_k141_17337 | Low-quality |
| S14_extracted_k141_2347 | Not-determined |
| S14_extracted_k141_17338 | Not-determined |
| S14_extracted_k141_105585 | Not-determined |
| S14_extracted_k141_90574 | Low-quality |
| S14_extracted_k141_105588 | Not-determined |
| S14_extracted_k141_105589 | Low-quality |
| S14_extracted_k141_24337 | Not-determined |
| S14_extracted_k141_90579 | Low-quality |
| S14_extracted_k141_2358 | Not-determined |
| S14_extracted_k141_53968 | Not-determined |
| S14_extracted_k141_97479 | Low-quality |
| S14_extracted_k141_2365 | Not-determined |
| S14_extracted_k141_24343 | Low-quality |
| S14_extracted_k141_24349 | Not-determined |
| S14_extracted_k141_31851 | Not-determined |
| S14_extracted_k141_31852 | Low-quality |
| S14_extracted_k141_31854 | Not-determined |
| S14_extracted_k141_17360 | Not-determined |
| S14_extracted_k141_31860 | Not-determined |
| S14_extracted_k141_68384 | Low-quality |
| S14_extracted_k141_90598 | Not-determined |
| S14_extracted_k141_9766 | Not-determined |
| S14_extracted_k141_112633 | Not-determined |
| S14_extracted_k141_31868 | Low-quality |
| S14_extracted_k141_53979 | Not-determined |
| S14_extracted_k141_76004 | Not-determined |
| S14_extracted_k141_68400 | Low-quality |
| S14_extracted_k141_97502 | Not-determined |
| S14_extracted_k141_68406 | Not-determined |
| S14_extracted_k141_53989 | Low-quality |
| S14_extracted_k141_53990 | Low-quality |
| S14_extracted_k141_97505 | Not-determined |
| S14_extracted_k141_76011 | Not-determined |
| S14_extracted_k141_112647 | Not-determined |
| S14_extracted_k141_83236 | Low-quality |
| S14_extracted_k141_90614 | Low-quality |
| S14_extracted_k141_76012 | Not-determined |
| S14_extracted_k141_97511 | Low-quality |
| S14_extracted_k141_9779 | Low-quality |
| S14_extracted_k141_68415 | Not-determined |
| S14_extracted_k141_53994 | Not-determined |
| S14_extracted_k141_24357 | Not-determined |
| S14_extracted_k141_112659 | Low-quality |
| S14_extracted_k141_68420 | Low-quality |
| S14_extracted_k141_68423 | Not-determined |
| S14_extracted_k141_112664 | Low-quality |
| S14_extracted_k141_9787 | Not-determined |
| S14_extracted_k141_97516 | Not-determined |
| S14_extracted_k141_17370 | Not-determined |
| S14_extracted_k141_17376 | Not-determined |
| S14_extracted_k141_46046 | Low-quality |
| S14_extracted_k141_9791 | Not-determined |
| S14_extracted_k141_76028 | Not-determined |
| S14_extracted_k141_90638 | Not-determined |
| S14_extracted_k141_24372 | Not-determined |
| S14_extracted_k141_76032 | Low-quality |
| S14_extracted_k141_83252 | Low-quality |
| S14_extracted_k141_90643 | Not-determined |
| S14_extracted_k141_9796 | Not-determined |
| S14_extracted_k141_68441 | Not-determined |
| S14_extracted_k141_9800 | Low-quality |
| S14_extracted_k141_68444 | Not-determined |
| S14_extracted_k141_31906 | Not-determined |
| S14_extracted_k141_24390 | Low-quality |
| S14_extracted_k141_31910 | Not-determined |
| S14_extracted_k141_17386 | Low-quality |
| S14_extracted_k141_68450 | Low-quality |
| S14_extracted_k141_83263 | Not-determined |
| S14_extracted_k141_46053 | Not-determined |
| S14_extracted_k141_46054 | Low-quality |
| S14_extracted_k141_17391 | Not-determined |
| S14_extracted_k141_2386 | Low-quality |
| S14_extracted_k141_2387 | Not-determined |
| S14_extracted_k141_76056 | Not-determined |
| S14_extracted_k141_17394 | Not-determined |
| S14_extracted_k141_68462 | Low-quality |
| S14_extracted_k141_31913 | Not-determined |
| S14_extracted_k141_83274 | Not-determined |
| S14_extracted_k141_31916 | Not-determined |
| S14_extracted_k141_2391 | Not-determined |
| S14_extracted_k141_46063 | Not-determined |
| S14_extracted_k141_2394 | Low-quality |
| S14_extracted_k141_2396 | Not-determined |
| S14_extracted_k141_97532 | Not-determined |
| S14_extracted_k141_76062 | Low-quality |
| S14_extracted_k141_83286 | Not-determined |
| S14_extracted_k141_9822 | Not-determined |
| S14_extracted_k141_83289 | Not-determined |
| S14_extracted_k141_61205 | Not-determined |
| S14_extracted_k141_31918 | Not-determined |
| S14_extracted_k141_97538 | Not-determined |
| S14_extracted_k141_90656 | Not-determined |
| S14_extracted_k141_54040 | Not-determined |
| S14_extracted_k141_46077 | Not-determined |
| S14_extracted_k141_46081 | Low-quality |
| S14_extracted_k141_61212 | Not-determined |
| S14_extracted_k141_97546 | Low-quality |
| S14_extracted_k141_97547 | Low-quality |
| S14_extracted_k141_68479 | Not-determined |
| S14_extracted_k141_68480 | Not-determined |
| S14_extracted_k141_68482 | Not-determined |
| S14_extracted_k141_68483 | Not-determined |
| S14_extracted_k141_54048 | Not-determined |
| S14_extracted_k141_68484 | Not-determined |
| S14_extracted_k141_39516 | Low-quality |
| S14_extracted_k141_90671 | Low-quality |
| S14_extracted_k141_39520 | Low-quality |
| S14_extracted_k141_68491 | Not-determined |
| S14_extracted_k141_83310 | Not-determined |
| S14_extracted_k141_83311 | Not-determined |
| S14_extracted_k141_9838 | Low-quality |
| S14_extracted_k141_76076 | Low-quality |
| S14_extracted_k141_54060 | Low-quality |
| S14_extracted_k141_83313 | Low-quality |
| S14_extracted_k141_105593 | Low-quality |
| S14_extracted_k141_90675 | Not-determined |
| S14_extracted_k141_76078 | Low-quality |
| S14_extracted_k141_46097 | Not-determined |
| S14_extracted_k141_68502 | Not-determined |
| S14_extracted_k141_9841 | Low-quality |
| S14_extracted_k141_9842 | Low-quality |
| S14_extracted_k141_68504 | Not-determined |
| S14_extracted_k141_90682 | Low-quality |
| S14_extracted_k141_83317 | Low-quality |
| S14_extracted_k141_112681 | Not-determined |
| S14_extracted_k141_97572 | Not-determined |
| S14_extracted_k141_97573 | Not-determined |
| S14_extracted_k141_105602 | Not-determined |
| S14_extracted_k141_105603 | Not-determined |
| S14_extracted_k141_2418 | Not-determined |
| S14_extracted_k141_61226 | Not-determined |
| S14_extracted_k141_39524 | Not-determined |
| S14_extracted_k141_2423 | Low-quality |
| S14_extracted_k141_46127 | Not-determined |
| S14_extracted_k141_39528 | Low-quality |
| S14_extracted_k141_2425 | Not-determined |
| S14_extracted_k141_17449 | Not-determined |
| S14_extracted_k141_39530 | Not-determined |
| S14_extracted_k141_17450 | Not-determined |
| S14_extracted_k141_17452 | Low-quality |
| S14_extracted_k141_39534 | Low-quality |
| S14_extracted_k141_97583 | Low-quality |
| S14_extracted_k141_17454 | Not-determined |
| S14_extracted_k141_61231 | Not-determined |
| S14_extracted_k141_112686 | Not-determined |
| S14_extracted_k141_105616 | Not-determined |
| S14_extracted_k141_54080 | Not-determined |
| S14_extracted_k141_17459 | Not-determined |
| S14_extracted_k141_68516 | Low-quality |
| S14_extracted_k141_112691 | Not-determined |
| S14_extracted_k141_2434 | Low-quality |
| S14_extracted_k141_2435 | Not-determined |
| S14_extracted_k141_68523 | Not-determined |
| S14_extracted_k141_68525 | Low-quality |
| S14_extracted_k141_97594 | Not-determined |
| S14_extracted_k141_68527 | Low-quality |
| S14_extracted_k141_90691 | Not-determined |
| S14_extracted_k141_90693 | Not-determined |
| S14_extracted_k141_2447 | Not-determined |
| S14_extracted_k141_46137 | Low-quality |
| S14_extracted_k141_46138 | Not-determined |
| S14_extracted_k141_46139 | Low-quality |
| S14_extracted_k141_90699 | Low-quality |
| S14_extracted_k141_68537 | Low-quality |
| S14_extracted_k141_90702 | Low-quality |
| S14_extracted_k141_68540 | Not-determined |
| S14_extracted_k141_54090 | Not-determined |
| S14_extracted_k141_54091 | Not-determined |
| S14_extracted_k141_31929 | Not-determined |
| S14_extracted_k141_61246 | Not-determined |
| S14_extracted_k141_105635 | Low-quality |
| S14_extracted_k141_76100 | Not-determined |
| S14_extracted_k141_9850 | Not-determined |
| S14_extracted_k141_83329 | Not-determined |
| S14_extracted_k141_2465 | Not-determined |
| S14_extracted_k141_61251 | Not-determined |
| S14_extracted_k141_83335 | Not-determined |
| S14_extracted_k141_83337 | Not-determined |
| S14_extracted_k141_17473 | Low-quality |
| S14_extracted_k141_90709 | Low-quality |
| S14_extracted_k141_112714 | Not-determined |
| S14_extracted_k141_76104 | Not-determined |
| S14_extracted_k141_76105 | Low-quality |
| S14_extracted_k141_112715 | Not-determined |
| S14_extracted_k141_39540 | Not-determined |
| S14_extracted_k141_39542 | Low-quality |
| S14_extracted_k141_54101 | Low-quality |
| S14_extracted_k141_76109 | Not-determined |
| S14_extracted_k141_90720 | Low-quality |
| S14_extracted_k141_83344 | Not-determined |
| S14_extracted_k141_9863 | Not-determined |
| S14_extracted_k141_76114 | Not-determined |
| S14_extracted_k141_17478 | Not-determined |
| S14_extracted_k141_39546 | Low-quality |
| S14_extracted_k141_24402 | Low-quality |
| S14_extracted_k141_2471 | Not-determined |
| S14_extracted_k141_39549 | Low-quality |
| S14_extracted_k141_31947 | Low-quality |
| S14_extracted_k141_46154 | Not-determined |
| S14_extracted_k141_39550 | Not-determined |
| S14_extracted_k141_90725 | Low-quality |
| S14_extracted_k141_39551 | Low-quality |
| S14_extracted_k141_105641 | Low-quality |
| S14_extracted_k141_76127 | Low-quality |
| S14_extracted_k141_76129 | Not-determined |
| S14_extracted_k141_76132 | Not-determined |
| S14_extracted_k141_24418 | Low-quality |
| S14_extracted_k141_24419 | Low-quality |
| S14_extracted_k141_83356 | Low-quality |
| S14_extracted_k141_76136 | Low-quality |
| S14_extracted_k141_2489 | Not-determined |
| S14_extracted_k141_2490 | Not-determined |
| S14_extracted_k141_61268 | Not-determined |
| S14_extracted_k141_31953 | Not-determined |
| S14_extracted_k141_54118 | Not-determined |
| S14_extracted_k141_24429 | Low-quality |
| S14_extracted_k141_24430 | Low-quality |
| S14_extracted_k141_61271 | Not-determined |
| S14_extracted_k141_24431 | Not-determined |
| S14_extracted_k141_83361 | Not-determined |
| S14_extracted_k141_105652 | Not-determined |
| S14_extracted_k141_76143 | Not-determined |
| S14_extracted_k141_24434 | Low-quality |
| S14_extracted_k141_31956 | Not-determined |
| S14_extracted_k141_24436 | Low-quality |
| S14_extracted_k141_2500 | Low-quality |
| S14_extracted_k141_46172 | Low-quality |
| S14_extracted_k141_105660 | Not-determined |
| S14_extracted_k141_68546 | Low-quality |
| S14_extracted_k141_31968 | Low-quality |
| S14_extracted_k141_39570 | Not-determined |
| S14_extracted_k141_54137 | Not-determined |
| S14_extracted_k141_61285 | Not-determined |
| S14_extracted_k141_31971 | Not-determined |
| S14_extracted_k141_76153 | Not-determined |
| S14_extracted_k141_31972 | Not-determined |
| S14_extracted_k141_39573 | Not-determined |
| S14_extracted_k141_105668 | Not-determined |
| S14_extracted_k141_76155 | Not-determined |
| S14_extracted_k141_46187 | Low-quality |
| S14_extracted_k141_112721 | Not-determined |
| S14_extracted_k141_61290 | Not-determined |
| S14_extracted_k141_112723 | Not-determined |
| S14_extracted_k141_61291 | Not-determined |
| S14_extracted_k141_24452 | Not-determined |
| S14_extracted_k141_54145 | Not-determined |
| S14_extracted_k141_97633 | Low-quality |
| S14_extracted_k141_2513 | Not-determined |
| S14_extracted_k141_54147 | Not-determined |
| S14_extracted_k141_54148 | Not-determined |
| S14_extracted_k141_46190 | Not-determined |
| S14_extracted_k141_105670 | Not-determined |
| S14_extracted_k141_105671 | Not-determined |
| S14_extracted_k141_97634 | Low-quality |
| S14_extracted_k141_39583 | Low-quality |
| S14_extracted_k141_112728 | Low-quality |
| S14_extracted_k141_61298 | Not-determined |
| S14_extracted_k141_54153 | Low-quality |
| S14_extracted_k141_68555 | Low-quality |
| S14_extracted_k141_112730 | Not-determined |
| S14_extracted_k141_46194 | Low-quality |
| S14_extracted_k141_97635 | Not-determined |
| S14_extracted_k141_46196 | Low-quality |
| S14_extracted_k141_54159 | Not-determined |
| S14_extracted_k141_2520 | Not-determined |
| S14_extracted_k141_54163 | Not-determined |
| S14_extracted_k141_68558 | Low-quality |
| S14_extracted_k141_54170 | Not-determined |
| S14_extracted_k141_105681 | Not-determined |
| S14_extracted_k141_68563 | Not-determined |
| S14_extracted_k141_68564 | Not-determined |
| S14_extracted_k141_76174 | Not-determined |
| S14_extracted_k141_46206 | Low-quality |
| S14_extracted_k141_76178 | Not-determined |
| S14_extracted_k141_61310 | Not-determined |
| S14_extracted_k141_17489 | Not-determined |
| S14_extracted_k141_97646 | Not-determined |
| S14_extracted_k141_24472 | Not-determined |
| S14_extracted_k141_54171 | Not-determined |
| S14_extracted_k141_97651 | Not-determined |
| S14_extracted_k141_97652 | Not-determined |
| S14_extracted_k141_2532 | Not-determined |
| S14_extracted_k141_54176 | Not-determined |
| S14_extracted_k141_46210 | Low-quality |
| S14_extracted_k141_39597 | Low-quality |
| S14_extracted_k141_46214 | Not-determined |
| S14_extracted_k141_68566 | Not-determined |
| S14_extracted_k141_105690 | Not-determined |
| S14_extracted_k141_112737 | Low-quality |
| S14_extracted_k141_17501 | Not-determined |
| S14_extracted_k141_105691 | Low-quality |
| S14_extracted_k141_97654 | Not-determined |
| S14_extracted_k141_31991 | Not-determined |
| S14_extracted_k141_31993 | Not-determined |
| S14_extracted_k141_83378 | Not-determined |
| S14_extracted_k141_24484 | Not-determined |
| S14_extracted_k141_9887 | Not-determined |
| S14_extracted_k141_46220 | Low-quality |
| S14_extracted_k141_112738 | Low-quality |
| S14_extracted_k141_68573 | Not-determined |
| S14_extracted_k141_31995 | Low-quality |
| S14_extracted_k141_61317 | Not-determined |
| S14_extracted_k141_112741 | Low-quality |
| S14_extracted_k141_112742 | Not-determined |
| S14_extracted_k141_76184 | Not-determined |
| S14_extracted_k141_2547 | Not-determined |
| S14_extracted_k141_9890 | Not-determined |
| S14_extracted_k141_32001 | Not-determined |
| S14_extracted_k141_105699 | Low-quality |
| S14_extracted_k141_32002 | Not-determined |
| S14_extracted_k141_17512 | Not-determined |
| S14_extracted_k141_32006 | Not-determined |
| S14_extracted_k141_61325 | Not-determined |
| S14_extracted_k141_9897 | Not-determined |
| S14_extracted_k141_105703 | Low-quality |
| S14_extracted_k141_17517 | Low-quality |
| S14_extracted_k141_76190 | Not-determined |
| S14_extracted_k141_9902 | Not-determined |
| S14_extracted_k141_9903 | Not-determined |
| S14_extracted_k141_76193 | Low-quality |
| S14_extracted_k141_68576 | Low-quality |
| S14_extracted_k141_17521 | Not-determined |
| S14_extracted_k141_68578 | Low-quality |
| S14_extracted_k141_68579 | Not-determined |
| S14_extracted_k141_83393 | Not-determined |
| S14_extracted_k141_54190 | Not-determined |
| S14_extracted_k141_9908 | Low-quality |
| S14_extracted_k141_61334 | Not-determined |
| S14_extracted_k141_9909 | Low-quality |
| S14_extracted_k141_105711 | Not-determined |
| S14_extracted_k141_68583 | Low-quality |
| S14_extracted_k141_112753 | Not-determined |
| S14_extracted_k141_97673 | Not-determined |
| S14_extracted_k141_17530 | Not-determined |
| S14_extracted_k141_90747 | Low-quality |
| S14_extracted_k141_90749 | Not-determined |
| S14_extracted_k141_105713 | Low-quality |
| S14_extracted_k141_112757 | Not-determined |
| S14_extracted_k141_90752 | Not-determined |
| S14_extracted_k141_76206 | Not-determined |
| S14_extracted_k141_90758 | Not-determined |
| S14_extracted_k141_90760 | Not-determined |
| S14_extracted_k141_54201 | Low-quality |
| S14_extracted_k141_61346 | Not-determined |
| S14_extracted_k141_32023 | Not-determined |
| S14_extracted_k141_61347 | Not-determined |
| S14_extracted_k141_17536 | Not-determined |
| S14_extracted_k141_54204 | Low-quality |
| S14_extracted_k141_17542 | Low-quality |
| S14_extracted_k141_90771 | Low-quality |
| S14_extracted_k141_105726 | Not-determined |
| S14_extracted_k141_39606 | Low-quality |
| S14_extracted_k141_105729 | Not-determined |
| S14_extracted_k141_97685 | Low-quality |
| S14_extracted_k141_105730 | Low-quality |
| S14_extracted_k141_17545 | Not-determined |
| S14_extracted_k141_9928 | Not-determined |
| S14_extracted_k141_9930 | Not-determined |
| S14_extracted_k141_54206 | Not-determined |
| S14_extracted_k141_112771 | Not-determined |
| S14_extracted_k141_17552 | Low-quality |
| S14_extracted_k141_9934 | Low-quality |
| S14_extracted_k141_17555 | Not-determined |
| S14_extracted_k141_97689 | Not-determined |
| S14_extracted_k141_68602 | Not-determined |
| S14_extracted_k141_46237 | Not-determined |
| S14_extracted_k141_97696 | Not-determined |
| S14_extracted_k141_97699 | Low-quality |
| S14_extracted_k141_54213 | Not-determined |
| S14_extracted_k141_54214 | Low-quality |
| S14_extracted_k141_83414 | Not-determined |
| S14_extracted_k141_46244 | Not-determined |
| S14_extracted_k141_61360 | Not-determined |
| S14_extracted_k141_61362 | Low-quality |
| S14_extracted_k141_97702 | Not-determined |
| S14_extracted_k141_112787 | Not-determined |
| S14_extracted_k141_32048 | Low-quality |
| S14_extracted_k141_24513 | Not-determined |
| S14_extracted_k141_90775 | Not-determined |
| S14_extracted_k141_112790 | Low-quality |
| S14_extracted_k141_68611 | Not-determined |
| S14_extracted_k141_2571 | Not-determined |
| S14_extracted_k141_97707 | Low-quality |
| S14_extracted_k141_90779 | Low-quality |
| S14_extracted_k141_2574 | Not-determined |
| S14_extracted_k141_32054 | Not-determined |
| S14_extracted_k141_90781 | Not-determined |
| S14_extracted_k141_54230 | Not-determined |
| S14_extracted_k141_32058 | Not-determined |
| S14_extracted_k141_39617 | Not-determined |
| S14_extracted_k141_90783 | Not-determined |
| S14_extracted_k141_2576 | Not-determined |
| S14_extracted_k141_32061 | Low-quality |
| S14_extracted_k141_39620 | Not-determined |
| S14_extracted_k141_90791 | Low-quality |
| S14_extracted_k141_39622 | Low-quality |
| S14_extracted_k141_9950 | Not-determined |
| S14_extracted_k141_9952 | Not-determined |
| S14_extracted_k141_9956 | Not-determined |
| S14_extracted_k141_97717 | Low-quality |
| S14_extracted_k141_105733 | Not-determined |
| S14_extracted_k141_97718 | Not-determined |
| S14_extracted_k141_24528 | Not-determined |
| S14_extracted_k141_105737 | Not-determined |
| S14_extracted_k141_97723 | Low-quality |
| S14_extracted_k141_17573 | Low-quality |
| S14_extracted_k141_24530 | Low-quality |
| S14_extracted_k141_97729 | Not-determined |
| S14_extracted_k141_24533 | Low-quality |
| S14_extracted_k141_46267 | Low-quality |
| S14_extracted_k141_97733 | Not-determined |
| S14_extracted_k141_24537 | Not-determined |
| S14_extracted_k141_112818 | Low-quality |
| S14_extracted_k141_112819 | Low-quality |
| S14_extracted_k141_76241 | Not-determined |
| S14_extracted_k141_112821 | Low-quality |
| S14_extracted_k141_112823 | Low-quality |
| S14_extracted_k141_9974 | Not-determined |
| S14_extracted_k141_105753 | Not-determined |
| S14_extracted_k141_90806 | Not-determined |
| S14_extracted_k141_105758 | Not-determined |
| S14_extracted_k141_39637 | Low-quality |
| S14_extracted_k141_76248 | Not-determined |
| S14_extracted_k141_9979 | Low-quality |
| S14_extracted_k141_76254 | Not-determined |
| S14_extracted_k141_2591 | Not-determined |
| S14_extracted_k141_97744 | Not-determined |
| S14_extracted_k141_17592 | Low-quality |
| S14_extracted_k141_17594 | Not-determined |
| S14_extracted_k141_105770 | Not-determined |
| S14_extracted_k141_17595 | Low-quality |
| S14_extracted_k141_2594 | Not-determined |
| S14_extracted_k141_46285 | Low-quality |
| S14_extracted_k141_17600 | Not-determined |
| S14_extracted_k141_17602 | Not-determined |
| S14_extracted_k141_2602 | Not-determined |
| S14_extracted_k141_83436 | Not-determined |
| S14_extracted_k141_68615 | Not-determined |
| S14_extracted_k141_46287 | Not-determined |
| S14_extracted_k141_2606 | Not-determined |
| S14_extracted_k141_83438 | Not-determined |
| S14_extracted_k141_24557 | Not-determined |
| S14_extracted_k141_9985 | Low-quality |
| S14_extracted_k141_68622 | Not-determined |
| S14_extracted_k141_90819 | Not-determined |
| S14_extracted_k141_39652 | Low-quality |
| S14_extracted_k141_76273 | Low-quality |
| S14_extracted_k141_83439 | Not-determined |
| S14_extracted_k141_32089 | Not-determined |
| S14_extracted_k141_90829 | Not-determined |
| S14_extracted_k141_90831 | Low-quality |
| S14_extracted_k141_2614 | Not-determined |
| S14_extracted_k141_54280 | Low-quality |
| S14_extracted_k141_54281 | Not-determined |
| S14_extracted_k141_9988 | Not-determined |
| S14_extracted_k141_76278 | Not-determined |
| S14_extracted_k141_112837 | Low-quality |
| S14_extracted_k141_76280 | Not-determined |
| S14_extracted_k141_32094 | Not-determined |
| S14_extracted_k141_112838 | Not-determined |
| S14_extracted_k141_32095 | Low-quality |
| S14_extracted_k141_76282 | Not-determined |
| S14_extracted_k141_32097 | Low-quality |
| S14_extracted_k141_9994 | Not-determined |
| S14_extracted_k141_9995 | Not-determined |
| S14_extracted_k141_32099 | Low-quality |
| S14_extracted_k141_2617 | Not-determined |
| S14_extracted_k141_54289 | Not-determined |
| S14_extracted_k141_83443 | Not-determined |
| S14_extracted_k141_90841 | Not-determined |
| S14_extracted_k141_97763 | Low-quality |
| S14_extracted_k141_10003 | Not-determined |
| S14_extracted_k141_83444 | Low-quality |
| S14_extracted_k141_83445 | Not-determined |
| S14_extracted_k141_76289 | Low-quality |
| S14_extracted_k141_24578 | Not-determined |
| S14_extracted_k141_97764 | Low-quality |
| S14_extracted_k141_39657 | Not-determined |
| S14_extracted_k141_17613 | Not-determined |
| S14_extracted_k141_17614 | Not-determined |
| S14_extracted_k141_32110 | Not-determined |
| S14_extracted_k141_17615 | Not-determined |
| S14_extracted_k141_105792 | Low-quality |
| S14_extracted_k141_17622 | Not-determined |
| S14_extracted_k141_76300 | Not-determined |
| S14_extracted_k141_112856 | Not-determined |
| S14_extracted_k141_112857 | Low-quality |
| S14_extracted_k141_68631 | Low-quality |
| S14_extracted_k141_68632 | Low-quality |
| S14_extracted_k141_83462 | Not-determined |
| S14_extracted_k141_68634 | Not-determined |
| S14_extracted_k141_76309 | Not-determined |
| S14_extracted_k141_83467 | Not-determined |
| S14_extracted_k141_105804 | Low-quality |
| S14_extracted_k141_76313 | Not-determined |
| S14_extracted_k141_68644 | Not-determined |
| S14_extracted_k141_68645 | Low-quality |
| S14_extracted_k141_17634 | Not-determined |
| S14_extracted_k141_105812 | Low-quality |
| S14_extracted_k141_24582 | Not-determined |
| S14_extracted_k141_97769 | Not-determined |
| S14_extracted_k141_39691 | Not-determined |
| S14_extracted_k141_97773 | Not-determined |
| S14_extracted_k141_39694 | Low-quality |
| S14_extracted_k141_90866 | Not-determined |
| S14_extracted_k141_39703 | Not-determined |
| S14_extracted_k141_54336 | Low-quality |
| S14_extracted_k141_97784 | Not-determined |
| S14_extracted_k141_68652 | Not-determined |
| S14_extracted_k141_39706 | Not-determined |
| S14_extracted_k141_68654 | Not-determined |
| S14_extracted_k141_90874 | Low-quality |
| S14_extracted_k141_10015 | Low-quality |
| S14_extracted_k141_83485 | Not-determined |
| S14_extracted_k141_32135 | Not-determined |
| S14_extracted_k141_54344 | Not-determined |
| S14_extracted_k141_10021 | Not-determined |
| S14_extracted_k141_97796 | Not-determined |
| S14_extracted_k141_32142 | Not-determined |
| S14_extracted_k141_61371 | Not-determined |
| S14_extracted_k141_32145 | Not-determined |
| S14_extracted_k141_46311 | Low-quality |
| S14_extracted_k141_46312 | Low-quality |
| S14_extracted_k141_39713 | Not-determined |
| S14_extracted_k141_54357 | Not-determined |
| S14_extracted_k141_97801 | Not-determined |
| S14_extracted_k141_32149 | Not-determined |
| S14_extracted_k141_32150 | Not-determined |
| S14_extracted_k141_32152 | Not-determined |
| S14_extracted_k141_97805 | Not-determined |
| S14_extracted_k141_24595 | Not-determined |
| S14_extracted_k141_39718 | Not-determined |
| S14_extracted_k141_105824 | Low-quality |
| S14_extracted_k141_97807 | Not-determined |
| S14_extracted_k141_112865 | Not-determined |
| S14_extracted_k141_83490 | Not-determined |
| S14_extracted_k141_112867 | Low-quality |
| S14_extracted_k141_46317 | Low-quality |
| S14_extracted_k141_39721 | Not-determined |
| S14_extracted_k141_97811 | Not-determined |
| S14_extracted_k141_97813 | Not-determined |
| S14_extracted_k141_39724 | Not-determined |
| S14_extracted_k141_39728 | Not-determined |
| S14_extracted_k141_24611 | Not-determined |
| S14_extracted_k141_61380 | Not-determined |
| S14_extracted_k141_90888 | Low-quality |
| S14_extracted_k141_90890 | Low-quality |
| S14_extracted_k141_32166 | Not-determined |
| S14_extracted_k141_17649 | Low-quality |
| S14_extracted_k141_68679 | Not-determined |
| S14_extracted_k141_112874 | Low-quality |
| S14_extracted_k141_90897 | Not-determined |
| S14_extracted_k141_68685 | Low-quality |
| S14_extracted_k141_46323 | Low-quality |
| S14_extracted_k141_46324 | Not-determined |
| S14_extracted_k141_68688 | Not-determined |
| S14_extracted_k141_24621 | Not-determined |
| S14_extracted_k141_46333 | Not-determined |
| S14_extracted_k141_54377 | Not-determined |
| S14_extracted_k141_83510 | Not-determined |
| S14_extracted_k141_24631 | Low-quality |
| S14_extracted_k141_68699 | Not-determined |
| S14_extracted_k141_76323 | Not-determined |
| S14_extracted_k141_112881 | Not-determined |
| S14_extracted_k141_24639 | Low-quality |
| S14_extracted_k141_90916 | Not-determined |
| S14_extracted_k141_46339 | Not-determined |
| S14_extracted_k141_83522 | Low-quality |
| S14_extracted_k141_83524 | Low-quality |
| S14_extracted_k141_46344 | Not-determined |
| S14_extracted_k141_32177 | Low-quality |
| S14_extracted_k141_68707 | Low-quality |
| S14_extracted_k141_32178 | Not-determined |
| S14_extracted_k141_17656 | Not-determined |
| S14_extracted_k141_46349 | Not-determined |
| S14_extracted_k141_39753 | Not-determined |
| S14_extracted_k141_90927 | Low-quality |
| S14_extracted_k141_83533 | Low-quality |

| | |
|---|---|
| S14_extracted_k141_61423 | Not-determined |
| S14_extracted_k141_39763 | Low-quality |
| S14_extracted_k141_24659 | Not-determined |
| S14_extracted_k141_46353 | Not-determined |
| S14_extracted_k141_46354 | Not-determined |
| S14_extracted_k141_17667 | Not-determined |
| S14_extracted_k141_32185 | Low-quality |
| S14_extracted_k141_39766 | Not-determined |
| S14_extracted_k141_46356 | Low-quality |
| S14_extracted_k141_76343 | Not-determined |
| S14_extracted_k141_46358 | Low-quality |
| S14_extracted_k141_32188 | Not-determined |
| S14_extracted_k141_68723 | Low-quality |
| S14_extracted_k141_68724 | Low-quality |
| S14_extracted_k141_10028 | Not-determined |
| S14_extracted_k141_97830 | Low-quality |
| S14_extracted_k141_76347 | Low-quality |
| S14_extracted_k141_39777 | Low-quality |
| S14_extracted_k141_61433 | Not-determined |
| S14_extracted_k141_61434 | Not-determined |
| S14_extracted_k141_83554 | Not-determined |
| S14_extracted_k141_10036 | Low-quality |
| S14_extracted_k141_76355 | Low-quality |
| S14_extracted_k141_83562 | Not-determined |
| S14_extracted_k141_105834 | Not-determined |
| S14_extracted_k141_83566 | Not-determined |
| S14_extracted_k141_54410 | Not-determined |
| S14_extracted_k141_61440 | Not-determined |
| S14_extracted_k141_61441 | Not-determined |
| S14_extracted_k141_10048 | Low-quality |
| S14_extracted_k141_39786 | Low-quality |
| S14_extracted_k141_76366 | Not-determined |
| S14_extracted_k141_39788 | Not-determined |
| S14_extracted_k141_46367 | Not-determined |
| S14_extracted_k141_46369 | Not-determined |
| S14_extracted_k141_61448 | Not-determined |
| S14_extracted_k141_112894 | Low-quality |
| S14_extracted_k141_61449 | Low-quality |
| S14_extracted_k141_2636 | Not-determined |
| S14_extracted_k141_61451 | Low-quality |
| S14_extracted_k141_54418 | Not-determined |
| S14_extracted_k141_61452 | Not-determined |
| S14_extracted_k141_46377 | Not-determined |
| S14_extracted_k141_61453 | Low-quality |
| S14_extracted_k141_39796 | Low-quality |
| S14_extracted_k141_112901 | Not-determined |
| S14_extracted_k141_10071 | Not-determined |
| S14_extracted_k141_46380 | Not-determined |
| S14_extracted_k141_2640 | Not-determined |
| S14_extracted_k141_83574 | Not-determined |
| S14_extracted_k141_32232 | Not-determined |
| S14_extracted_k141_10073 | Not-determined |
| S14_extracted_k141_54426 | Low-quality |
| S14_extracted_k141_54427 | Low-quality |
| S14_extracted_k141_112902 | Low-quality |
| S14_extracted_k141_105849 | Not-determined |
| S14_extracted_k141_105851 | Not-determined |
| S14_extracted_k141_2646 | Low-quality |
| S14_extracted_k141_112912 | Not-determined |
| S14_extracted_k141_24677 | Not-determined |
| S14_extracted_k141_2648 | Low-quality |
| S14_extracted_k141_24679 | Not-determined |
| S14_extracted_k141_46397 | Low-quality |
| S14_extracted_k141_112917 | Low-quality |
| S14_extracted_k141_2654 | Not-determined |
| S14_extracted_k141_54440 | Not-determined |
| S14_extracted_k141_10093 | Not-determined |
| S14_extracted_k141_54444 | Not-determined |
| S14_extracted_k141_54447 | Low-quality |
| S14_extracted_k141_32250 | Not-determined |
| S14_extracted_k141_76389 | Not-determined |
| S14_extracted_k141_2658 | Low-quality |
| S14_extracted_k141_10097 | Not-determined |
| S14_extracted_k141_2660 | Not-determined |
| S14_extracted_k141_61458 | Low-quality |
| S14_extracted_k141_83593 | Not-determined |
| S14_extracted_k141_61459 | Not-determined |
| S14_extracted_k141_68737 | Not-determined |
| S14_extracted_k141_54454 | Not-determined |
| S14_extracted_k141_76404 | Low-quality |
| S14_extracted_k141_68742 | Not-determined |
| S14_extracted_k141_61466 | Not-determined |
| S14_extracted_k141_68745 | Not-determined |
| S14_extracted_k141_112945 | Not-determined |
| S14_extracted_k141_10102 | Not-determined |
| S14_extracted_k141_112946 | Not-determined |
| S14_extracted_k141_112948 | Low-quality |
| S14_extracted_k141_10106 | Not-determined |
| S14_extracted_k141_54463 | Not-determined |
| S14_extracted_k141_76413 | Not-determined |
| S14_extracted_k141_68748 | Not-determined |
| S14_extracted_k141_32271 | Not-determined |
| S14_extracted_k141_68753 | Low-quality |
| S14_extracted_k141_90970 | Low-quality |
| S14_extracted_k141_2691 | Not-determined |
| S14_extracted_k141_112956 | Low-quality |
| S14_extracted_k141_46404 | Low-quality |
| S14_extracted_k141_46405 | Not-determined |
| S14_extracted_k141_61480 | Not-determined |
| S14_extracted_k141_2702 | Not-determined |
| S14_extracted_k141_17714 | Low-quality |
| S14_extracted_k141_112961 | Low-quality |
| S14_extracted_k141_17716 | Low-quality |
| S14_extracted_k141_32279 | Low-quality |
| S14_extracted_k141_32283 | Not-determined |
| S14_extracted_k141_112968 | Not-determined |
| S14_extracted_k141_83609 | Not-determined |
| S14_extracted_k141_83610 | Not-determined |
| S14_extracted_k141_17727 | Not-determined |
| S14_extracted_k141_10134 | Not-determined |
| S14_extracted_k141_105895 | Not-determined |
| S14_extracted_k141_39802 | Low-quality |
| S14_extracted_k141_76449 | Low-quality |
| S14_extracted_k141_68776 | Not-determined |
| S14_extracted_k141_10144 | Not-determined |
| S14_extracted_k141_46415 | Low-quality |
| S14_extracted_k141_39811 | Low-quality |
| S14_extracted_k141_39817 | Not-determined |
| S14_extracted_k141_46423 | Not-determined |
| S14_extracted_k141_54485 | Not-determined |
| S14_extracted_k141_2730 | Low-quality |
| S14_extracted_k141_24689 | Not-determined |
| S14_extracted_k141_68787 | Not-determined |
| S14_extracted_k141_24691 | Low-quality |
| S14_extracted_k141_2734 | Not-determined |
| S14_extracted_k141_76464 | Not-determined |
| S14_extracted_k141_2735 | Not-determined |
| S14_extracted_k141_24696 | Not-determined |
| S14_extracted_k141_105904 | Low-quality |
| S14_extracted_k141_46428 | Not-determined |
| S14_extracted_k141_68796 | Not-determined |
| S14_extracted_k141_83629 | Not-determined |
| S14_extracted_k141_39830 | Low-quality |
| S14_extracted_k141_24705 | Not-determined |
| S14_extracted_k141_105911 | Low-quality |
| S14_extracted_k141_105912 | Not-determined |
| S14_extracted_k141_24709 | Low-quality |
| S14_extracted_k141_105917 | Low-quality |
| S14_extracted_k141_76470 | Not-determined |
| S14_extracted_k141_32292 | Not-determined |
| S14_extracted_k141_105919 | Not-determined |
| S14_extracted_k141_32293 | Not-determined |
| S14_extracted_k141_105922 | Not-determined |
| S14_extracted_k141_54507 | Not-determined |
| S14_extracted_k141_105925 | Low-quality |
| S14_extracted_k141_105929 | Not-determined |
| S14_extracted_k141_76478 | Not-determined |
| S14_extracted_k141_32295 | Not-determined |
| S14_extracted_k141_76480 | Not-determined |
| S14_extracted_k141_105937 | Not-determined |
| S14_extracted_k141_54508 | Not-determined |
| S14_extracted_k141_24717 | Not-determined |
| S14_extracted_k141_46445 | Low-quality |
| S14_extracted_k141_24719 | Not-determined |
| S14_extracted_k141_46446 | Low-quality |
| S14_extracted_k141_39844 | Low-quality |
| S14_extracted_k141_83650 | Not-determined |
| S14_extracted_k141_83651 | Not-determined |
| S14_extracted_k141_76485 | Not-determined |
| S14_extracted_k141_76486 | Not-determined |
| S14_extracted_k141_24721 | Not-determined |
| S14_extracted_k141_90973 | Low-quality |
| S14_extracted_k141_90974 | Low-quality |
| S14_extracted_k141_54515 | Low-quality |
| S14_extracted_k141_32302 | Low-quality |
| S14_extracted_k141_39853 | Not-determined |
| S14_extracted_k141_68821 | Not-determined |
| S14_extracted_k141_39854 | Not-determined |
| S14_extracted_k141_39856 | Not-determined |
| S14_extracted_k141_32305 | Low-quality |
| S14_extracted_k141_46456 | Not-determined |
| S14_extracted_k141_83665 | Low-quality |
| S14_extracted_k141_32310 | Low-quality |
| S14_extracted_k141_46461 | Not-determined |
| S14_extracted_k141_46462 | Low-quality |
| S14_extracted_k141_76489 | Not-determined |
| S14_extracted_k141_24723 | Not-determined |
| S14_extracted_k141_24727 | Low-quality |
| S14_extracted_k141_10151 | Low-quality |
| S14_extracted_k141_10152 | Low-quality |
| S14_extracted_k141_76491 | Not-determined |
| S14_extracted_k141_83679 | Not-determined |
| S14_extracted_k141_112993 | Not-determined |
| S14_extracted_k141_68830 | Not-determined |
| S14_extracted_k141_10157 | Low-quality |
| S14_extracted_k141_76495 | Not-determined |
| S14_extracted_k141_10159 | Low-quality |
| S14_extracted_k141_2754 | Not-determined |
| S14_extracted_k141_76497 | Not-determined |
| S14_extracted_k141_54532 | Not-determined |
| S14_extracted_k141_54533 | Not-determined |
| S14_extracted_k141_39860 | Low-quality |
| S14_extracted_k141_68834 | Not-determined |
| S14_extracted_k141_54534 | Not-determined |
| S14_extracted_k141_39865 | Low-quality |
| S14_extracted_k141_68839 | Low-quality |
| S14_extracted_k141_32323 | Low-quality |
| S14_extracted_k141_46476 | Low-quality |
| S14_extracted_k141_39869 | Low-quality |
| S14_extracted_k141_68842 | Low-quality |
| S14_extracted_k141_61493 | Not-determined |
| S14_extracted_k141_54545 | Low-quality |
| S14_extracted_k141_46478 | Not-determined |
| S14_extracted_k141_32331 | Not-determined |
| S14_extracted_k141_54546 | Low-quality |
| S14_extracted_k141_39875 | Not-determined |
| S14_extracted_k141_32332 | Low-quality |
| S14_extracted_k141_54549 | Not-determined |
| S14_extracted_k141_54551 | Not-determined |
| S14_extracted_k141_68846 | Not-determined |
| S14_extracted_k141_54553 | Not-determined |
| S14_extracted_k141_54555 | Not-determined |
| S14_extracted_k141_68855 | Not-determined |
| S14_extracted_k141_68856 | Not-determined |
| S14_extracted_k141_24742 | Not-determined |
| S14_extracted_k141_54562 | Not-determined |
| S14_extracted_k141_32344 | Not-determined |
| S14_extracted_k141_83699 | Not-determined |
| S14_extracted_k141_105970 | Low-quality |
| S14_extracted_k141_32346 | Not-determined |
| S14_extracted_k141_68859 | Not-determined |
| S14_extracted_k141_54568 | Not-determined |
| S14_extracted_k141_39883 | Not-determined |
| S14_extracted_k141_2758 | Not-determined |
| S14_extracted_k141_32352 | Not-determined |
| S14_extracted_k141_68862 | Not-determined |
| S14_extracted_k141_54570 | Not-determined |
| S14_extracted_k141_39887 | Not-determined |
| S14_extracted_k141_39888 | Not-determined |
| S14_extracted_k141_68869 | Low-quality |
| S14_extracted_k141_90999 | Low-quality |
| S14_extracted_k141_68873 | Not-determined |
| S14_extracted_k141_54577 | Not-determined |
| S14_extracted_k141_68874 | Low-quality |
| S14_extracted_k141_46499 | Low-quality |
| S14_extracted_k141_24755 | Low-quality |
| S14_extracted_k141_54579 | Not-determined |
| S14_extracted_k141_54581 | Not-determined |
| S14_extracted_k141_61498 | Low-quality |
| S14_extracted_k141_54586 | Not-determined |
| S14_extracted_k141_61502 | Low-quality |
| S14_extracted_k141_17741 | Low-quality |
| S14_extracted_k141_10186 | Not-determined |
| S14_extracted_k141_10189 | Not-determined |
| S14_extracted_k141_17742 | Not-determined |
| S14_extracted_k141_68882 | Not-determined |
| S14_extracted_k141_68884 | Not-determined |
| S14_extracted_k141_10194 | Not-determined |
| S14_extracted_k141_10195 | Low-quality |
| S14_extracted_k141_24774 | Not-determined |
| S14_extracted_k141_46521 | Not-determined |
| S14_extracted_k141_54590 | Not-determined |
| S14_extracted_k141_39900 | Low-quality |
| S14_extracted_k141_46525 | Low-quality |
| S14_extracted_k141_24777 | Not-determined |
| S14_extracted_k141_46528 | Not-determined |
| S14_extracted_k141_76526 | Not-determined |
| S14_extracted_k141_39905 | Not-determined |
| S14_extracted_k141_46531 | Low-quality |
| S14_extracted_k141_76530 | Low-quality |
| S14_extracted_k141_76538 | Not-determined |
| S14_extracted_k141_68898 | Not-determined |
| S14_extracted_k141_2792 | Not-determined |
| S14_extracted_k141_105990 | Not-determined |
| S14_extracted_k141_68906 | Not-determined |
| S14_extracted_k141_2796 | Low-quality |
| S14_extracted_k141_2798 | Not-determined |
| S14_extracted_k141_105991 | Not-determined |
| S14_extracted_k141_68909 | Not-determined |
| S14_extracted_k141_68910 | Not-determined |
| S14_extracted_k141_68911 | Not-determined |
| S14_extracted_k141_105994 | Low-quality |
| S14_extracted_k141_39919 | Not-determined |
| S14_extracted_k141_105996 | Low-quality |
| S14_extracted_k141_32358 | Not-determined |
| S14_extracted_k141_105999 | Low-quality |
| S14_extracted_k141_83727 | Low-quality |
| S14_extracted_k141_68913 | Not-determined |
| S14_extracted_k141_83729 | Not-determined |
| S14_extracted_k141_76543 | Not-determined |
| S14_extracted_k141_97861 | Not-determined |
| S14_extracted_k141_32365 | Not-determined |
| S14_extracted_k141_76546 | Not-determined |
| S14_extracted_k141_83735 | Not-determined |
| S14_extracted_k141_2810 | Not-determined |
| S14_extracted_k141_2811 | Low-quality |
| S14_extracted_k141_46543 | Not-determined |
| S14_extracted_k141_10205 | Low-quality |
| S14_extracted_k141_68923 | Not-determined |
| S14_extracted_k141_39938 | Not-determined |
| S14_extracted_k141_97869 | Not-determined |
| S14_extracted_k141_24794 | Low-quality |
| S14_extracted_k141_97871 | Not-determined |
| S14_extracted_k141_39940 | Not-determined |
| S14_extracted_k141_32380 | Not-determined |
| S14_extracted_k141_97873 | Not-determined |
| S14_extracted_k141_10209 | Not-determined |
| S14_extracted_k141_10210 | Not-determined |
| S14_extracted_k141_106019 | Low-quality |
| S14_extracted_k141_97877 | Low-quality |
| S14_extracted_k141_46556 | Low-quality |
| S14_extracted_k141_46558 | Not-determined |
| S14_extracted_k141_106023 | Not-determined |
| S14_extracted_k141_46560 | Low-quality |
| S14_extracted_k141_17754 | Not-determined |
| S14_extracted_k141_46562 | Not-determined |
| S14_extracted_k141_97884 | Not-determined |
| S14_extracted_k141_68934 | Not-determined |
| S14_extracted_k141_17756 | Low-quality |
| S14_extracted_k141_10215 | Not-determined |
| S14_extracted_k141_17757 | Not-determined |
| S14_extracted_k141_83749 | Not-determined |
| S14_extracted_k141_97887 | Not-determined |
| S14_extracted_k141_32393 | Low-quality |
| S14_extracted_k141_68941 | Low-quality |
| S14_extracted_k141_2824 | Not-determined |
| S14_extracted_k141_24808 | Low-quality |
| S14_extracted_k141_2826 | Low-quality |
| S14_extracted_k141_97892 | Not-determined |
| S14_extracted_k141_46569 | Not-determined |
| S14_extracted_k141_46571 | Not-determined |
| S14_extracted_k141_68945 | Low-quality |
| S14_extracted_k141_2830 | Low-quality |
| S14_extracted_k141_2831 | Not-determined |
| S14_extracted_k141_83755 | Low-quality |
| S14_extracted_k141_24820 | Low-quality |
| S14_extracted_k141_24823 | Not-determined |
| S14_extracted_k141_10236 | Low-quality |
| S14_extracted_k141_24826 | Not-determined |
| S14_extracted_k141_10238 | Not-determined |
| S14_extracted_k141_10240 | Not-determined |
| S14_extracted_k141_46581 | Not-determined |
| S14_extracted_k141_39954 | Low-quality |
| S14_extracted_k141_76554 | Not-determined |
| S14_extracted_k141_76557 | Not-determined |
| S14_extracted_k141_106034 | Low-quality |
| S14_extracted_k141_106035 | Not-determined |
| S14_extracted_k141_91025 | Low-quality |
| S14_extracted_k141_91027 | Not-determined |
| S14_extracted_k141_91030 | Not-determined |
| S14_extracted_k141_17781 | Not-determined |
| S14_extracted_k141_10246 | Low-quality |
| S14_extracted_k141_39960 | Not-determined |
| S14_extracted_k141_76565 | Not-determined |
| S14_extracted_k141_17785 | Not-determined |
| S14_extracted_k141_91032 | Not-determined |
| S14_extracted_k141_17787 | Not-determined |
| S14_extracted_k141_91040 | Low-quality |
| S14_extracted_k141_83774 | Not-determined |
| S14_extracted_k141_91043 | Not-determined |
| S14_extracted_k141_17803 | Not-determined |
| S14_extracted_k141_106055 | Not-determined |
| S14_extracted_k141_17807 | Not-determined |
| S14_extracted_k141_106057 | Not-determined |
| S14_extracted_k141_68948 | Low-quality |
| S14_extracted_k141_10269 | Not-determined |
| S14_extracted_k141_106062 | Low-quality |
| S14_extracted_k141_106066 | Not-determined |
| S14_extracted_k141_91060 | Not-determined |
| S14_extracted_k141_54636 | Low-quality |
| S14_extracted_k141_17816 | Not-determined |
| S14_extracted_k141_83790 | Not-determined |
| S14_extracted_k141_54638 | Low-quality |
| S14_extracted_k141_46601 | Low-quality |
| S14_extracted_k141_54640 | Not-determined |
| S14_extracted_k141_83796 | Low-quality |
| S14_extracted_k141_76571 | Low-quality |
| S14_extracted_k141_97903 | Not-determined |
| S14_extracted_k141_46606 | Not-determined |
| S14_extracted_k141_17818 | Low-quality |
| S14_extracted_k141_76575 | Not-determined |
| S14_extracted_k141_83806 | Not-determined |
| S14_extracted_k141_2864 | Not-determined |
| S14_extracted_k141_83812 | Not-determined |
| S1_extracted_k141_2995 | High-quality |
| S14_extracted_k141_54654 | Not-determined |
| S14_extracted_k141_10281 | Not-determined |
| S14_extracted_k141_91084 | Low-quality |
| S14_extracted_k141_91085 | Not-determined |
| S14_extracted_k141_32433 | Not-determined |
| S14_extracted_k141_97922 | Not-determined |
| S14_extracted_k141_76581 | Low-quality |
| S14_extracted_k141_10288 | Not-determined |
| S14_extracted_k141_106081 | Not-determined |
| S14_extracted_k141_39992 | Not-determined |
| S14_extracted_k141_54666 | Low-quality |
| S14_extracted_k141_76582 | Low-quality |
| S14_extracted_k141_113015 | Low-quality |
| S14_extracted_k141_40000 | Low-quality |
| S14_extracted_k141_97929 | Low-quality |
| S14_extracted_k141_32446 | Low-quality |
| S14_extracted_k141_97931 | Not-determined |
| S14_extracted_k141_97933 | Low-quality |
| S14_extracted_k141_113022 | Not-determined |
| S14_extracted_k141_91090 | Low-quality |
| S14_extracted_k141_32450 | Not-determined |
| S14_extracted_k141_97935 | Not-determined |
| S14_extracted_k141_97936 | Low-quality |
| S14_extracted_k141_40009 | Low-quality |
| S14_extracted_k141_91093 | Not-determined |
| S14_extracted_k141_24853 | Not-determined |
| S14_extracted_k141_76587 | Not-determined |
| S14_extracted_k141_76590 | Low-quality |
| S14_extracted_k141_24854 | Low-quality |
| S14_extracted_k141_76592 | Not-determined |
| S14_extracted_k141_32456 | Not-determined |
| S14_extracted_k141_40014 | Low-quality |
| S14_extracted_k141_106087 | Low-quality |
| S14_extracted_k141_40019 | Not-determined |
| S14_extracted_k141_40021 | Low-quality |
| S14_extracted_k141_54680 | Not-determined |
| S14_extracted_k141_24864 | Not-determined |
| S14_extracted_k141_24865 | Not-determined |
| S14_extracted_k141_17854 | Low-quality |
| S14_extracted_k141_106090 | Not-determined |
| S14_extracted_k141_17856 | Not-determined |
| S14_extracted_k141_76597 | Low-quality |
| S14_extracted_k141_76599 | Low-quality |
| S14_extracted_k141_76600 | Not-determined |
| S14_extracted_k141_76601 | Not-determined |
| S14_extracted_k141_113036 | Low-quality |
| S14_extracted_k141_24872 | Not-determined |
| S14_extracted_k141_24873 | Not-determined |
| S14_extracted_k141_24877 | Low-quality |
| S14_extracted_k141_76603 | Low-quality |
| S14_extracted_k141_54694 | Not-determined |
| S14_extracted_k141_83842 | Not-determined |
| S14_extracted_k141_54695 | Not-determined |
| S14_extracted_k141_91114 | Not-determined |
| S14_extracted_k141_54701 | Not-determined |
| S14_extracted_k141_91115 | Not-determined |
| S14_extracted_k141_10311 | Not-determined |
| S14_extracted_k141_91121 | Low-quality |
| S14_extracted_k141_83856 | Low-quality |
| S14_extracted_k141_76606 | Not-determined |
| S14_extracted_k141_113047 | Low-quality |
| S14_extracted_k141_32472 | Not-determined |
| S14_extracted_k141_76609 | Low-quality |
| S14_extracted_k141_91126 | Not-determined |
| S14_extracted_k141_76612 | Low-quality |
| S14_extracted_k141_40036 | Not-determined |
| S14_extracted_k141_83859 | Not-determined |
| S14_extracted_k141_97949 | Low-quality |
| S14_extracted_k141_46640 | Not-determined |
| S14_extracted_k141_32484 | Not-determined |
| S14_extracted_k141_32486 | Not-determined |
| S14_extracted_k141_24885 | Not-determined |
| S14_extracted_k141_106099 | Not-determined |
| S14_extracted_k141_83867 | Not-determined |
| S14_extracted_k141_91135 | Low-quality |
| S14_extracted_k141_91136 | Low-quality |
| S14_extracted_k141_40047 | Not-determined |
| S14_extracted_k141_46658 | Not-determined |
| S14_extracted_k141_46671 | Low-quality |
| S14_extracted_k141_113066 | Not-determined |
| S14_extracted_k141_46672 | Low-quality |
| S14_extracted_k141_68962 | Low-quality |
| S14_extracted_k141_113072 | Not-determined |
| S14_extracted_k141_113074 | Not-determined |
| S14_extracted_k141_76629 | Not-determined |
| S14_extracted_k141_113075 | Not-determined |
| S14_extracted_k141_91153 | Not-determined |
| S14_extracted_k141_97975 | Not-determined |
| S14_extracted_k141_24893 | Not-determined |
| S14_extracted_k141_97979 | Low-quality |
| S14_extracted_k141_83885 | Not-determined |
| S14_extracted_k141_76637 | Not-determined |
| S14_extracted_k141_76646 | Not-determined |
| S14_extracted_k141_46690 | Not-determined |
| S14_extracted_k141_68985 | Not-determined |
| S14_extracted_k141_76652 | Not-determined |
| S14_extracted_k141_76653 | Not-determined |
| S14_extracted_k141_40062 | Low-quality |
| S14_extracted_k141_76655 | Not-determined |
| S14_extracted_k141_97995 | Not-determined |
| S14_extracted_k141_68990 | Not-determined |
| S14_extracted_k141_68991 | Not-determined |
| S14_extracted_k141_17891 | Not-determined |
| S14_extracted_k141_113086 | Low-quality |
| S14_extracted_k141_76660 | Not-determined |
| S14_extracted_k141_46695 | Not-determined |
| S14_extracted_k141_17895 | Not-determined |
| S14_extracted_k141_68995 | Not-determined |
| S14_extracted_k141_68996 | Not-determined |
| S14_extracted_k141_17902 | Not-determined |
| S14_extracted_k141_113100 | Not-determined |
| S14_extracted_k141_24914 | Not-determined |
| S14_extracted_k141_91163 | Not-determined |
| S14_extracted_k141_98014 | Not-determined |
| S14_extracted_k141_113103 | Not-determined |
| S14_extracted_k141_69008 | Not-determined |
| S14_extracted_k141_40083 | Not-determined |
| S14_extracted_k141_69011 | Low-quality |
| S14_extracted_k141_91168 | Not-determined |
| S14_extracted_k141_40089 | Not-determined |
| S14_extracted_k141_106133 | Not-determined |
| S14_extracted_k141_106134 | Low-quality |
| S14_extracted_k141_83909 | Not-determined |
| S14_extracted_k141_69021 | Low-quality |
| S14_extracted_k141_113110 | Not-determined |
| S14_extracted_k141_113111 | Not-determined |
| S14_extracted_k141_113112 | Low-quality |
| S14_extracted_k141_113114 | Low-quality |
| S14_extracted_k141_69029 | Not-determined |
| S14_extracted_k141_83918 | Low-quality |
| S14_extracted_k141_69031 | Low-quality |
| S14_extracted_k141_106149 | Not-determined |
| S14_extracted_k141_83922 | Low-quality |
| S14_extracted_k141_40101 | Not-determined |
| S14_extracted_k141_106150 | Low-quality |
| S14_extracted_k141_17909 | Not-determined |
| S14_extracted_k141_40104 | Not-determined |
| S14_extracted_k141_69040 | Not-determined |
| S14_extracted_k141_83925 | Not-determined |
| S14_extracted_k141_76699 | Not-determined |
| S14_extracted_k141_83928 | Not-determined |
| S14_extracted_k141_46698 | Low-quality |
| S14_extracted_k141_113121 | Not-determined |
| S14_extracted_k141_98032 | Not-determined |
| S14_extracted_k141_83931 | Not-determined |
| S14_extracted_k141_69047 | Not-determined |
| S14_extracted_k141_69048 | Low-quality |
| S14_extracted_k141_98036 | Not-determined |
| S14_extracted_k141_24930 | Low-quality |
| S14_extracted_k141_98037 | Not-determined |
| S14_extracted_k141_2876 | Low-quality |
| S14_extracted_k141_98038 | Low-quality |
| S14_extracted_k141_91182 | Not-determined |
| S14_extracted_k141_46702 | Not-determined |
| S14_extracted_k141_69056 | Not-determined |
| S14_extracted_k141_76707 | Not-determined |
| S14_extracted_k141_24935 | Low-quality |
| S14_extracted_k141_17931 | Not-determined |
| S14_extracted_k141_76709 | Not-determined |
| S14_extracted_k141_24937 | Not-determined |
| S14_extracted_k141_76712 | Not-determined |
| S14_extracted_k141_24938 | Not-determined |
| S14_extracted_k141_69057 | Low-quality |
| S14_extracted_k141_69059 | Not-determined |
| S14_extracted_k141_61567 | Not-determined |
| S14_extracted_k141_69061 | Not-determined |
| S14_extracted_k141_76722 | Not-determined |
| S14_extracted_k141_69068 | Low-quality |
| S14_extracted_k141_24946 | Low-quality |
| S14_extracted_k141_76727 | Not-determined |
| S14_extracted_k141_61579 | Low-quality |
| S14_extracted_k141_69073 | Not-determined |
| S14_extracted_k141_98048 | Not-determined |
| S14_extracted_k141_46710 | Not-determined |
| S14_extracted_k141_61582 | Low-quality |
| S14_extracted_k141_98050 | Not-determined |
| S14_extracted_k141_61583 | Low-quality |
| S14_extracted_k141_24951 | Not-determined |
| S14_extracted_k141_61585 | Not-determined |
| S14_extracted_k141_91205 | Not-determined |
| S14_extracted_k141_17953 | Not-determined |
| S14_extracted_k141_2883 | Not-determined |
| S14_extracted_k141_46718 | Low-quality |
| S14_extracted_k141_40131 | Low-quality |
| S14_extracted_k141_24957 | Not-determined |
| S14_extracted_k141_46721 | Not-determined |
| S14_extracted_k141_113135 | Low-quality |
| S14_extracted_k141_32506 | Not-determined |
| S14_extracted_k141_76750 | Not-determined |
| S14_extracted_k141_61599 | Low-quality |
| S14_extracted_k141_76751 | Not-determined |
| S14_extracted_k141_46726 | Low-quality |
| S14_extracted_k141_40143 | Not-determined |
| S14_extracted_k141_113142 | Not-determined |
| S14_extracted_k141_98068 | Low-quality |
| S14_extracted_k141_69082 | Not-determined |
| S14_extracted_k141_69084 | Not-determined |
| S14_extracted_k141_24963 | Low-quality |
| S14_extracted_k141_98070 | Not-determined |
| S14_extracted_k141_91207 | Low-quality |
| S14_extracted_k141_2890 | Not-determined |
| S14_extracted_k141_113149 | Not-determined |
| S14_extracted_k141_69091 | Not-determined |
| S14_extracted_k141_32515 | Low-quality |
| S14_extracted_k141_46727 | Low-quality |
| S14_extracted_k141_76760 | Not-determined |
| S14_extracted_k141_32518 | Not-determined |
| S14_extracted_k141_91216 | Not-determined |
| S14_extracted_k141_46731 | Not-determined |
| S14_extracted_k141_10362 | Not-determined |
| S14_extracted_k141_69100 | Low-quality |
| S14_extracted_k141_61619 | Not-determined |
| S14_extracted_k141_46735 | Not-determined |
| S14_extracted_k141_69101 | Low-quality |

| | |
|---|---|
| S14_extracted_k141_91228 | Not-determined |
| S8_extracted_k141_170878 | High-quality |
| S14_extracted_k141_2894 | Not-determined |
| S14_extracted_k141_2897 | Low-quality |
| S14_extracted_k141_106157 | Not-determined |
| S14_extracted_k141_113158 | Not-determined |
| S14_extracted_k141_98082 | Not-determined |
| S14_extracted_k141_61627 | Low-quality |
| S14_extracted_k141_24975 | Not-determined |
| S14_extracted_k141_24976 | Not-determined |
| S14_extracted_k141_69104 | Not-determined |
| S14_extracted_k141_106161 | Not-determined |
| S14_extracted_k141_32520 | Not-determined |
| S14_extracted_k141_32521 | Not-determined |
| S14_extracted_k141_61630 | Low-quality |
| S14_extracted_k141_113168 | Not-determined |
| S14_extracted_k141_76776 | Not-determined |
| S14_extracted_k141_69108 | Low-quality |
| S14_extracted_k141_32523 | Low-quality |
| S14_extracted_k141_10372 | Low-quality |
| S14_extracted_k141_69112 | Not-determined |
| S14_extracted_k141_91241 | Low-quality |
| S14_extracted_k141_106165 | Low-quality |
| S14_extracted_k141_106166 | Not-determined |
| S14_extracted_k141_106170 | Low-quality |
| S14_extracted_k141_106172 | Not-determined |
| S14_extracted_k141_61638 | Not-determined |
| S14_extracted_k141_54719 | Low-quality |
| S14_extracted_k141_91251 | Not-determined |
| S14_extracted_k141_10381 | Not-determined |
| S14_extracted_k141_54725 | Low-quality |
| S14_extracted_k141_83939 | Low-quality |
| S14_extracted_k141_106179 | Not-determined |
| S14_extracted_k141_10385 | Not-determined |
| S14_extracted_k141_113177 | Not-determined |
| S14_extracted_k141_10386 | Not-determined |
| S14_extracted_k141_106181 | Low-quality |
| S14_extracted_k141_91261 | Not-determined |
| S14_extracted_k141_54728 | Not-determined |
| S14_extracted_k141_54729 | Not-determined |
| S14_extracted_k141_17964 | Not-determined |
| S14_extracted_k141_113181 | Not-determined |
| S14_extracted_k141_83943 | Low-quality |
| S14_extracted_k141_106184 | Not-determined |
| S14_extracted_k141_106186 | Low-quality |
| S14_extracted_k141_54736 | Low-quality |
| S14_extracted_k141_10397 | Low-quality |
| S14_extracted_k141_69129 | Not-determined |
| S14_extracted_k141_91268 | Not-determined |
| S14_extracted_k141_2904 | Low-quality |
| S14_extracted_k141_113191 | Not-determined |
| S14_extracted_k141_2907 | Not-determined |
| S14_extracted_k141_98105 | Not-determined |
| S14_extracted_k141_17979 | Not-determined |
| S14_extracted_k141_40175 | Not-determined |
| S14_extracted_k141_17983 | Not-determined |
| S14_extracted_k141_32550 | Low-quality |
| S14_extracted_k141_76784 | Not-determined |
| S14_extracted_k141_40180 | Not-determined |
| S14_extracted_k141_98109 | Not-determined |
| S14_extracted_k141_113198 | Not-determined |
| S14_extracted_k141_17988 | Not-determined |
| S14_extracted_k141_91271 | Not-determined |
| S14_extracted_k141_98111 | Low-quality |
| S14_extracted_k141_17989 | Not-determined |
| S14_extracted_k141_113204 | Not-determined |
| S14_extracted_k141_40187 | Not-determined |
| S14_extracted_k141_91283 | Not-determined |
| S14_extracted_k141_10407 | Low-quality |
| S14_extracted_k141_76790 | Not-determined |
| S14_extracted_k141_54747 | Not-determined |
| S14_extracted_k141_83947 | Not-determined |
| S14_extracted_k141_91287 | Not-determined |
| S14_extracted_k141_17992 | Not-determined |
| S14_extracted_k141_113207 | Not-determined |
| S14_extracted_k141_113210 | Not-determined |
| S14_extracted_k141_76799 | Not-determined |
| S14_extracted_k141_61679 | Not-determined |
| S14_extracted_k141_18000 | Low-quality |
| S14_extracted_k141_18001 | Low-quality |
| S14_extracted_k141_113212 | Low-quality |
| S14_extracted_k141_18006 | Low-quality |
| S14_extracted_k141_2917 | Low-quality |
| S14_extracted_k141_32566 | Not-determined |
| S14_extracted_k141_76807 | Low-quality |
| S14_extracted_k141_113216 | Not-determined |
| S14_extracted_k141_54754 | Not-determined |
| S14_extracted_k141_32569 | Not-determined |
| S14_extracted_k141_113220 | Low-quality |
| S14_extracted_k141_113221 | Not-determined |
| S14_extracted_k141_76816 | Not-determined |
| S14_extracted_k141_32575 | Not-determined |
| S14_extracted_k141_98125 | Not-determined |
| S14_extracted_k141_76817 | Not-determined |
| S14_extracted_k141_24983 | Not-determined |
| S14_extracted_k141_54767 | Low-quality |
| S14_extracted_k141_69142 | Not-determined |
| S14_extracted_k141_98128 | Low-quality |
| S14_extracted_k141_46750 | Not-determined |
| S14_extracted_k141_40202 | Not-determined |
| S14_extracted_k141_24993 | Not-determined |
| S14_extracted_k141_40203 | Low-quality |
| S14_extracted_k141_61691 | Not-determined |
| S14_extracted_k141_98131 | Not-determined |
| S14_extracted_k141_61695 | Not-determined |
| S14_extracted_k141_106197 | Not-determined |
| S14_extracted_k141_61698 | Not-determined |
| S14_extracted_k141_54778 | Not-determined |
| S14_extracted_k141_61700 | Not-determined |
| S14_extracted_k141_2943 | Low-quality |
| S14_extracted_k141_54782 | Low-quality |
| S14_extracted_k141_40207 | Low-quality |
| S14_extracted_k141_61704 | Not-determined |
| S14_extracted_k141_24998 | Low-quality |
| S14_extracted_k141_69148 | Low-quality |
| S14_extracted_k141_40210 | Not-determined |
| S14_extracted_k141_69149 | Not-determined |
| S14_extracted_k141_61706 | Low-quality |
| S14_extracted_k141_25002 | Not-determined |
| S14_extracted_k141_2949 | Low-quality |
| S14_extracted_k141_2951 | Not-determined |
| S14_extracted_k141_106205 | Not-determined |
| S14_extracted_k141_83963 | Not-determined |
| S14_extracted_k141_54790 | Not-determined |
| S14_extracted_k141_61714 | Low-quality |
| S14_extracted_k141_113238 | Not-determined |
| S14_extracted_k141_69151 | Not-determined |
| S14_extracted_k141_113240 | Not-determined |
| S14_extracted_k141_113242 | Low-quality |
| S14_extracted_k141_10429 | Not-determined |
| S14_extracted_k141_54802 | Not-determined |
| S14_extracted_k141_69156 | Not-determined |
| S14_extracted_k141_25012 | Not-determined |
| S14_extracted_k141_54806 | Not-determined |
| S14_extracted_k141_83974 | Not-determined |
| S14_extracted_k141_61719 | Not-determined |
| S14_extracted_k141_25021 | Not-determined |
| S14_extracted_k141_46785 | Low-quality |
| S14_extracted_k141_83980 | Low-quality |
| S14_extracted_k141_40221 | Not-determined |
| S14_extracted_k141_40228 | Not-determined |
| S14_extracted_k141_61728 | Low-quality |
| S14_extracted_k141_83989 | Not-determined |
| S14_extracted_k141_2968 | Not-determined |
| S14_extracted_k141_91323 | Not-determined |
| S14_extracted_k141_40232 | Not-determined |
| S14_extracted_k141_69169 | Not-determined |
| S14_extracted_k141_69171 | Not-determined |
| S14_extracted_k141_106220 | Not-determined |
| S14_extracted_k141_76835 | Low-quality |
| S14_extracted_k141_91325 | Low-quality |
| S14_extracted_k141_40238 | Not-determined |
| S14_extracted_k141_2972 | Not-determined |
| S14_extracted_k141_98153 | Low-quality |
| S14_extracted_k141_91329 | Low-quality |
| S14_extracted_k141_113273 | Low-quality |
| S14_extracted_k141_2977 | Not-determined |
| S14_extracted_k141_40242 | Not-determined |
| S14_extracted_k141_98157 | Not-determined |
| S14_extracted_k141_83999 | Not-determined |
| S14_extracted_k141_84000 | Not-determined |
| S14_extracted_k141_40243 | Not-determined |
| S14_extracted_k141_84004 | Low-quality |
| S14_extracted_k141_91334 | Not-determined |
| S14_extracted_k141_98159 | Low-quality |
| S14_extracted_k141_10457 | Not-determined |
| S14_extracted_k141_10458 | Low-quality |
| S14_extracted_k141_98166 | Not-determined |
| S14_extracted_k141_98168 | Not-determined |
| S14_extracted_k141_76849 | Low-quality |
| S14_extracted_k141_69188 | Not-determined |
| S14_extracted_k141_69189 | Not-determined |
| S14_extracted_k141_113292 | Not-determined |
| S14_extracted_k141_46798 | Low-quality |
| S14_extracted_k141_98174 | Not-determined |
| S14_extracted_k141_69191 | Not-determined |
| S14_extracted_k141_69192 | Not-determined |
| S14_extracted_k141_106225 | Not-determined |
| S14_extracted_k141_84019 | Not-determined |
| S14_extracted_k141_113301 | Not-determined |
| S14_extracted_k141_69196 | Not-determined |
| S14_extracted_k141_98180 | Not-determined |
| S14_extracted_k141_84023 | Not-determined |
| S14_extracted_k141_98181 | Not-determined |
| S14_extracted_k141_84024 | Not-determined |
| S14_extracted_k141_113307 | Low-quality |
| S14_extracted_k141_106234 | Low-quality |
| S14_extracted_k141_61749 | Low-quality |
| S14_extracted_k141_54816 | Not-determined |
| S14_extracted_k141_54817 | Not-determined |
| S14_extracted_k141_98189 | Not-determined |
| S14_extracted_k141_84032 | Low-quality |
| S14_extracted_k141_32589 | Not-determined |
| S14_extracted_k141_40275 | Low-quality |
| S14_extracted_k141_3019 | Not-determined |
| S14_extracted_k141_69202 | Not-determined |
| S14_extracted_k141_84040 | Not-determined |
| S14_extracted_k141_84041 | Not-determined |
| S14_extracted_k141_40282 | Not-determined |
| S14_extracted_k141_32597 | Not-determined |
| S14_extracted_k141_40284 | Not-determined |
| S14_extracted_k141_84043 | Not-determined |
| S14_extracted_k141_106239 | Not-determined |
| S14_extracted_k141_54825 | Low-quality |
| S14_extracted_k141_32599 | Low-quality |
| S14_extracted_k141_84051 | Not-determined |
| S14_extracted_k141_84052 | Not-determined |
| S14_extracted_k141_40292 | Low-quality |
| S14_extracted_k141_32604 | Not-determined |
| S14_extracted_k141_91383 | Not-determined |
| S14_extracted_k141_91384 | Low-quality |
| S14_extracted_k141_69210 | Not-determined |
| S14_extracted_k141_91386 | Low-quality |
| S14_extracted_k141_40297 | Not-determined |
| S14_extracted_k141_10471 | Not-determined |
| S14_extracted_k141_91389 | Not-determined |
| S14_extracted_k141_84062 | Not-determined |
| S14_extracted_k141_61771 | Low-quality |
| S14_extracted_k141_3048 | Not-determined |
| S14_extracted_k141_106255 | Low-quality |
| S14_extracted_k141_113315 | Not-determined |
| S14_extracted_k141_113318 | Not-determined |
| S14_extracted_k141_46826 | Not-determined |
| S14_extracted_k141_84073 | Not-determined |
| S14_extracted_k141_113324 | Low-quality |
| S14_extracted_k141_18074 | Not-determined |
| S14_extracted_k141_40315 | Low-quality |
| S14_extracted_k141_10487 | Not-determined |
| S14_extracted_k141_69229 | Not-determined |
| S14_extracted_k141_76868 | Not-determined |
| S14_extracted_k141_18079 | Not-determined |
| S14_extracted_k141_40324 | Not-determined |
| S14_extracted_k141_91401 | Low-quality |
| S14_extracted_k141_113325 | Not-determined |
| S14_extracted_k141_113326 | Low-quality |
| S14_extracted_k141_46841 | Low-quality |
| S14_extracted_k141_84089 | Not-determined |
| S14_extracted_k141_46843 | Not-determined |
| S14_extracted_k141_98217 | Not-determined |
| S14_extracted_k141_46844 | Low-quality |
| S14_extracted_k141_18089 | Not-determined |
| S14_extracted_k141_25052 | Not-determined |
| S14_extracted_k141_46849 | Not-determined |
| S14_extracted_k141_25055 | Not-determined |
| S14_extracted_k141_40336 | Not-determined |
| S14_extracted_k141_46853 | Low-quality |
| S14_extracted_k141_46855 | Low-quality |
| S14_extracted_k141_18095 | Not-determined |
| S14_extracted_k141_46859 | Not-determined |
| S14_extracted_k141_84099 | Not-determined |
| S14_extracted_k141_40338 | Low-quality |
| S14_extracted_k141_69245 | Low-quality |
| S14_extracted_k141_84100 | Not-determined |
| S14_extracted_k141_69246 | Not-determined |
| S14_extracted_k141_76885 | Low-quality |
| S14_extracted_k141_91413 | Not-determined |
| S14_extracted_k141_76889 | Low-quality |
| S14_extracted_k141_69250 | Low-quality |
| S14_extracted_k141_40343 | Not-determined |
| S14_extracted_k141_10510 | Not-determined |
| S14_extracted_k141_40344 | Low-quality |
| S14_extracted_k141_32617 | Not-determined |
| S14_extracted_k141_32618 | Low-quality |
| S14_extracted_k141_76892 | Not-determined |
| S14_extracted_k141_76894 | Low-quality |
| S14_extracted_k141_69253 | Not-determined |
| S14_extracted_k141_69254 | Low-quality |
| S14_extracted_k141_10515 | Low-quality |
| S14_extracted_k141_46869 | Low-quality |
| S14_extracted_k141_46870 | Not-determined |
| S14_extracted_k141_76901 | Low-quality |
| S14_extracted_k141_98234 | Not-determined |
| S14_extracted_k141_61779 | Not-determined |
| S14_extracted_k141_113348 | Not-determined |
| S14_extracted_k141_40356 | Low-quality |
| S14_extracted_k141_113350 | Not-determined |
| S14_extracted_k141_46874 | Low-quality |
| S14_extracted_k141_69265 | Low-quality |
| S14_extracted_k141_61783 | Not-determined |
| S14_extracted_k141_69270 | Low-quality |
| S14_extracted_k141_46880 | Not-determined |
| S14_extracted_k141_98245 | Low-quality |
| S14_extracted_k141_69272 | Not-determined |
| S14_extracted_k141_32627 | Not-determined |
| S14_extracted_k141_98247 | Low-quality |
| S14_extracted_k141_69274 | Not-determined |
| S14_extracted_k141_69275 | Low-quality |
| S14_extracted_k141_3071 | Low-quality |
| S14_extracted_k141_18105 | Low-quality |
| S14_extracted_k141_25072 | Low-quality |
| S14_extracted_k141_18106 | Low-quality |
| S14_extracted_k141_25074 | Low-quality |
| S14_extracted_k141_69282 | Low-quality |
| S14_extracted_k141_25075 | Low-quality |
| S14_extracted_k141_18109 | Not-determined |
| S14_extracted_k141_69284 | Low-quality |
| S14_extracted_k141_18110 | Not-determined |
| S14_extracted_k141_25078 | Not-determined |
| S14_extracted_k141_25080 | Not-determined |
| S14_extracted_k141_40370 | Not-determined |
| S14_extracted_k141_46888 | Low-quality |
| S14_extracted_k141_76912 | Low-quality |
| S14_extracted_k141_69288 | Low-quality |
| S14_extracted_k141_32642 | Not-determined |
| S14_extracted_k141_76914 | Not-determined |
| S14_extracted_k141_25095 | Low-quality |
| S14_extracted_k141_61790 | Not-determined |
| S14_extracted_k141_32650 | Not-determined |
| S14_extracted_k141_25101 | Not-determined |
| S14_extracted_k141_46897 | Low-quality |
| S14_extracted_k141_32653 | Not-determined |
| S14_extracted_k141_46899 | Low-quality |
| S14_extracted_k141_32655 | Not-determined |
| S14_extracted_k141_69301 | Low-quality |
| S14_extracted_k141_84105 | Not-determined |
| S14_extracted_k141_76936 | Not-determined |
| S14_extracted_k141_25108 | Not-determined |
| S14_extracted_k141_3078 | Not-determined |
| S14_extracted_k141_84112 | Not-determined |
| S14_extracted_k141_61799 | Low-quality |
| S14_extracted_k141_18130 | Not-determined |
| S14_extracted_k141_54837 | Not-determined |
| S14_extracted_k141_106274 | Not-determined |
| S14_extracted_k141_3087 | Low-quality |
| S14_extracted_k141_54845 | Not-determined |
| S14_extracted_k141_113379 | Not-determined |
| S14_extracted_k141_46907 | Low-quality |
| S14_extracted_k141_46908 | Low-quality |
| S14_extracted_k141_106279 | Not-determined |
| S14_extracted_k141_76947 | Not-determined |
| S14_extracted_k141_91450 | Not-determined |
| S14_extracted_k141_84128 | Not-determined |
| S14_extracted_k141_3101 | Not-determined |
| S14_extracted_k141_91468 | Not-determined |
| S14_extracted_k141_18137 | Low-quality |
| S14_extracted_k141_25112 | Not-determined |
| S14_extracted_k141_69309 | Not-determined |
| S14_extracted_k141_69311 | Not-determined |
| S14_extracted_k141_84135 | Low-quality |
| S14_extracted_k141_18142 | Not-determined |
| S14_extracted_k141_54850 | Not-determined |
| S14_extracted_k141_18143 | Not-determined |
| S14_extracted_k141_18144 | Not-determined |
| S14_extracted_k141_84139 | Low-quality |
| S14_extracted_k141_18146 | Not-determined |
| S14_extracted_k141_91478 | Not-determined |
| S14_extracted_k141_3115 | Low-quality |
| S14_extracted_k141_3118 | Low-quality |
| S14_extracted_k141_10538 | Not-determined |
| S14_extracted_k141_10539 | Not-determined |
| S14_extracted_k141_76975 | Not-determined |
| S14_extracted_k141_46919 | Not-determined |
| S14_extracted_k141_46920 | Not-determined |
| S14_extracted_k141_98267 | Not-determined |
| S14_extracted_k141_69318 | Low-quality |
| S14_extracted_k141_46921 | Not-determined |
| S14_extracted_k141_18162 | Low-quality |
| S14_extracted_k141_54871 | Not-determined |
| S14_extracted_k141_69319 | Not-determined |
| S14_extracted_k141_113397 | Not-determined |
| S14_extracted_k141_113404 | Not-determined |
| S14_extracted_k141_3138 | Low-quality |
| S14_extracted_k141_3145 | Not-determined |
| S14_extracted_k141_91482 | Not-determined |
| S14_extracted_k141_3146 | Not-determined |
| S14_extracted_k141_98276 | Not-determined |
| S14_extracted_k141_46938 | Not-determined |
| S14_extracted_k141_98279 | Not-determined |
| S14_extracted_k141_91483 | Not-determined |
| S14_extracted_k141_61816 | Not-determined |
| S14_extracted_k141_46940 | Not-determined |
| S14_extracted_k141_84150 | Low-quality |
| S14_extracted_k141_69328 | Not-determined |
| S14_extracted_k141_25124 | Not-determined |
| S14_extracted_k141_98285 | Low-quality |
| S14_extracted_k141_98286 | Not-determined |
| S14_extracted_k141_3152 | Not-determined |
| S14_extracted_k141_113420 | Not-determined |
| S14_extracted_k141_69331 | Low-quality |
| S14_extracted_k141_113422 | Not-determined |
| S14_extracted_k141_106290 | Not-determined |
| S14_extracted_k141_106291 | Low-quality |
| S14_extracted_k141_113424 | Not-determined |
| S14_extracted_k141_91489 | Not-determined |
| S14_extracted_k141_84151 | Low-quality |
| S14_extracted_k141_3156 | Low-quality |
| S14_extracted_k141_10561 | Not-determined |
| S14_extracted_k141_91493 | Not-determined |
| S14_extracted_k141_61826 | Not-determined |
| S14_extracted_k141_61827 | Not-determined |
| S14_extracted_k141_106298 | Not-determined |
| S14_extracted_k141_106300 | Not-determined |
| S14_extracted_k141_91495 | Not-determined |
| S14_extracted_k141_18174 | Not-determined |
| S14_extracted_k141_98294 | Not-determined |
| S14_extracted_k141_98295 | Not-determined |
| S14_extracted_k141_98296 | Low-quality |
| S14_extracted_k141_25133 | Low-quality |
| S14_extracted_k141_91506 | Low-quality |
| S14_extracted_k141_3170 | Not-determined |
| S14_extracted_k141_91508 | Low-quality |
| S14_extracted_k141_106303 | Not-determined |
| S14_extracted_k141_40382 | Not-determined |
| S14_extracted_k141_106308 | Low-quality |
| S14_extracted_k141_46944 | Low-quality |
| S14_extracted_k141_10574 | Not-determined |
| S14_extracted_k141_40385 | Not-determined |
| S14_extracted_k141_91517 | Low-quality |
| S14_extracted_k141_40386 | Not-determined |
| S14_extracted_k141_10577 | Not-determined |
| S14_extracted_k141_106313 | Not-determined |
| S14_extracted_k141_46946 | Not-determined |
| S14_extracted_k141_98307 | Not-determined |
| S14_extracted_k141_91522 | Not-determined |
| S14_extracted_k141_98310 | Not-determined |
| S14_extracted_k141_113438 | Low-quality |
| S14_extracted_k141_91523 | Not-determined |
| S14_extracted_k141_69352 | Not-determined |
| S14_extracted_k141_98313 | Not-determined |
| S14_extracted_k141_18183 | Not-determined |
| S14_extracted_k141_10581 | Not-determined |
| S14_extracted_k141_54880 | Low-quality |
| S14_extracted_k141_77007 | Not-determined |
| S14_extracted_k141_106328 | Not-determined |
| S14_extracted_k141_46953 | Not-determined |
| S14_extracted_k141_54890 | Not-determined |
| S14_extracted_k141_77012 | Low-quality |
| S14_extracted_k141_106330 | Not-determined |
| S14_extracted_k141_46956 | Not-determined |
| S14_extracted_k141_46959 | Not-determined |
| S14_extracted_k141_46960 | Not-determined |
| S14_extracted_k141_40400 | Not-determined |
| S14_extracted_k141_98320 | Not-determined |
| S14_extracted_k141_84170 | Low-quality |
| S14_extracted_k141_98322 | Not-determined |
| S14_extracted_k141_84174 | Low-quality |
| S14_extracted_k141_18195 | Low-quality |
| S14_extracted_k141_25152 | Not-determined |
| S14_extracted_k141_98329 | Not-determined |
| S14_extracted_k141_84182 | Not-determined |
| S14_extracted_k141_25156 | Not-determined |
| S14_extracted_k141_91531 | Not-determined |
| S14_extracted_k141_113452 | Not-determined |
| S14_extracted_k141_84184 | Not-determined |
| S14_extracted_k141_98339 | Not-determined |
| S14_extracted_k141_98340 | Not-determined |
| S14_extracted_k141_84192 | Not-determined |
| S14_extracted_k141_25164 | Low-quality |
| S14_extracted_k141_84194 | Low-quality |
| S14_extracted_k141_10601 | Not-determined |
| S14_extracted_k141_10602 | Not-determined |
| S14_extracted_k141_91541 | Not-determined |
| S14_extracted_k141_69388 | Low-quality |
| S14_extracted_k141_98350 | Not-determined |
| S14_extracted_k141_98355 | Not-determined |
| S14_extracted_k141_10611 | Low-quality |
| S14_extracted_k141_77034 | Not-determined |
| S14_extracted_k141_77036 | Low-quality |
| S14_extracted_k141_98357 | Low-quality |
| S14_extracted_k141_46974 | Not-determined |
| S14_extracted_k141_91551 | Not-determined |
| S14_extracted_k141_77042 | Not-determined |
| S14_extracted_k141_10628 | Low-quality |
| S14_extracted_k141_84215 | Low-quality |
| S14_extracted_k141_98359 | Not-determined |
| S14_extracted_k141_84217 | Not-determined |
| S14_extracted_k141_25189 | Not-determined |
| S14_extracted_k141_84220 | Not-determined |
| S14_extracted_k141_54906 | Not-determined |
| S14_extracted_k141_61833 | Not-determined |
| S14_extracted_k141_84222 | Not-determined |
| S14_extracted_k141_69401 | Low-quality |
| S14_extracted_k141_25193 | Low-quality |
| S14_extracted_k141_69402 | Low-quality |
| S14_extracted_k141_84224 | Not-determined |
| S14_extracted_k141_54910 | Low-quality |
| S14_extracted_k141_69405 | Not-determined |
| S14_extracted_k141_10640 | Low-quality |
| S14_extracted_k141_113495 | Not-determined |
| S14_extracted_k141_3205 | Not-determined |
| S14_extracted_k141_40406 | Not-determined |
| S14_extracted_k141_3208 | Not-determined |
| S14_extracted_k141_25202 | Not-determined |
| S14_extracted_k141_113497 | Not-determined |
| S14_extracted_k141_69413 | Not-determined |
| S14_extracted_k141_69415 | Not-determined |
| S14_extracted_k141_46983 | Not-determined |
| S14_extracted_k141_40410 | Not-determined |
| S14_extracted_k141_40412 | Not-determined |
| S14_extracted_k141_46986 | Low-quality |
| S14_extracted_k141_113499 | Low-quality |
| S14_extracted_k141_113502 | Not-determined |
| S14_extracted_k141_3218 | Not-determined |
| S14_extracted_k141_46989 | Low-quality |
| S14_extracted_k141_46994 | Not-determined |
| S14_extracted_k141_106350 | Not-determined |
| S14_extracted_k141_32714 | Not-determined |
| S14_extracted_k141_69419 | Not-determined |
| S14_extracted_k141_18236 | Not-determined |
| S14_extracted_k141_106352 | Not-determined |
| S14_extracted_k141_32718 | Not-determined |
| S14_extracted_k141_77065 | Not-determined |
| S14_extracted_k141_47003 | Not-determined |
| S14_extracted_k141_106355 | Low-quality |
| S14_extracted_k141_10654 | Low-quality |
| S14_extracted_k141_47005 | Not-determined |
| S14_extracted_k141_69420 | Not-determined |
| S14_extracted_k141_18245 | Not-determined |
| S14_extracted_k141_18246 | Low-quality |
| S14_extracted_k141_106362 | Not-determined |
| S14_extracted_k141_32728 | Low-quality |
| S14_extracted_k141_84240 | Not-determined |
| S14_extracted_k141_106365 | Low-quality |
| S14_extracted_k141_98378 | Not-determined |
| S14_extracted_k141_3229 | Not-determined |
| S14_extracted_k141_47006 | Not-determined |
| S14_extracted_k141_47009 | Low-quality |
| S14_extracted_k141_47011 | Not-determined |
| S14_extracted_k141_47012 | Not-determined |
| S14_extracted_k141_113516 | Not-determined |
| S14_extracted_k141_32750 | Low-quality |
| S14_extracted_k141_84247 | Not-determined |
| S14_extracted_k141_84251 | Not-determined |
| S14_extracted_k141_32756 | Not-determined |
| S14_extracted_k141_18278 | Not-determined |
| S14_extracted_k141_32760 | Not-determined |
| S14_extracted_k141_113522 | Not-determined |
| S14_extracted_k141_106385 | Low-quality |
| S14_extracted_k141_47025 | Not-determined |
| S14_extracted_k141_113525 | Low-quality |
| S14_extracted_k141_84255 | Low-quality |
| S14_extracted_k141_47029 | Low-quality |
| S14_extracted_k141_84256 | Not-determined |
| S14_extracted_k141_98410 | Not-determined |
| S14_extracted_k141_47034 | Not-determined |
| S14_extracted_k141_3246 | Not-determined |
| S14_extracted_k141_69445 | Not-determined |
| S14_extracted_k141_10660 | Low-quality |
| S14_extracted_k141_69446 | Not-determined |
| S14_extracted_k141_10661 | Not-determined |
| S14_extracted_k141_84267 | Not-determined |
| S14_extracted_k141_18303 | Not-determined |
| S14_extracted_k141_10662 | Not-determined |
| S14_extracted_k141_98415 | Not-determined |
| S14_extracted_k141_47035 | Not-determined |
| S14_extracted_k141_106393 | Low-quality |
| S14_extracted_k141_10666 | Not-determined |
| S14_extracted_k141_106396 | Low-quality |
| S14_extracted_k141_69453 | Low-quality |
| S14_extracted_k141_25249 | Not-determined |
| S14_extracted_k141_69455 | Low-quality |
| S14_extracted_k141_69457 | Not-determined |
| S14_extracted_k141_18310 | Not-determined |
| S14_extracted_k141_69458 | Low-quality |
| S14_extracted_k141_25250 | Not-determined |
| S14_extracted_k141_18313 | Not-determined |
| S14_extracted_k141_18315 | Not-determined |
| S14_extracted_k141_69462 | Not-determined |
| S14_extracted_k141_61851 | Not-determined |
| S14_extracted_k141_32774 | Not-determined |
| S14_extracted_k141_47042 | Not-determined |
| S14_extracted_k141_18318 | Low-quality |
| S14_extracted_k141_61860 | Low-quality |
| S14_extracted_k141_32781 | Not-determined |
| S14_extracted_k141_54941 | Not-determined |
| S14_extracted_k141_84279 | Low-quality |
| S14_extracted_k141_32783 | Not-determined |
| S14_extracted_k141_47050 | Not-determined |
| S14_extracted_k141_91567 | Low-quality |
| S14_extracted_k141_61864 | Low-quality |
| S14_extracted_k141_61865 | Low-quality |
| S14_extracted_k141_10680 | Not-determined |
| S14_extracted_k141_47058 | Low-quality |
| S14_extracted_k141_18324 | Not-determined |
| S14_extracted_k141_98421 | Not-determined |
| S14_extracted_k141_98422 | Low-quality |
| S14_extracted_k141_106400 | Low-quality |
| S14_extracted_k141_106402 | Low-quality |
| S14_extracted_k141_47059 | Not-determined |
| S14_extracted_k141_47060 | Low-quality |
| S14_extracted_k141_98425 | Not-determined |
| S14_extracted_k141_40421 | Not-determined |
| S14_extracted_k141_47062 | Not-determined |
| S14_extracted_k141_98430 | Not-determined |
| S14_extracted_k141_69469 | Not-determined |
| S14_extracted_k141_91576 | Not-determined |
| S14_extracted_k141_84291 | Not-determined |
| S14_extracted_k141_47067 | Not-determined |
| S14_extracted_k141_47069 | Low-quality |

| | |
|---|---|
| S14_extracted_k141_47071 | Low-quality |
| S14_extracted_k141_18335 | Not-determined |
| S14_extracted_k141_47075 | Low-quality |
| S14_extracted_k141_47076 | Not-determined |
| S14_extracted_k141_18338 | Not-determined |
| S14_extracted_k141_47077 | Not-determined |
| S14_extracted_k141_98438 | Low-quality |
| S14_extracted_k141_25270 | Not-determined |
| S14_extracted_k141_61886 | Not-determined |
| S14_extracted_k141_61888 | Not-determined |
| S14_extracted_k141_98444 | Not-determined |
| S14_extracted_k141_98447 | Not-determined |
| S14_extracted_k141_98448 | Low-quality |
| S14_extracted_k141_32805 | Not-determined |
| S14_extracted_k141_69481 | Low-quality |
| S14_extracted_k141_106411 | Not-determined |
| S14_extracted_k141_69482 | Not-determined |
| S14_extracted_k141_106413 | Low-quality |
| S14_extracted_k141_98452 | Not-determined |
| S14_extracted_k141_84301 | Low-quality |
| S14_extracted_k141_32808 | Not-determined |
| S14_extracted_k141_3252 | Not-determined |
| S14_extracted_k141_69486 | Not-determined |
| S14_extracted_k141_113537 | Not-determined |
| S14_extracted_k141_40424 | Not-determined |
| S14_extracted_k141_113538 | Not-determined |
| S14_extracted_k141_54961 | Low-quality |
| S14_extracted_k141_98457 | Not-determined |
| S14_extracted_k141_3257 | Low-quality |
| S14_extracted_k141_91599 | Not-determined |
| S14_extracted_k141_47085 | Low-quality |
| S14_extracted_k141_106423 | Not-determined |
| S14_extracted_k141_40431 | Not-determined |
| S14_extracted_k141_54966 | Not-determined |
| S14_extracted_k141_47090 | Not-determined |
| S14_extracted_k141_25288 | Not-determined |
| S14_extracted_k141_32820 | Not-determined |
| S14_extracted_k141_106429 | Not-determined |
| S14_extracted_k141_91605 | Not-determined |
| S14_extracted_k141_32824 | Not-determined |
| S14_extracted_k141_25295 | Not-determined |
| S14_extracted_k141_91607 | Not-determined |
| S14_extracted_k141_10711 | Low-quality |
| S14_extracted_k141_91609 | Not-determined |
| S14_extracted_k141_47097 | Low-quality |
| S14_extracted_k141_10716 | Low-quality |
| S14_extracted_k141_91616 | Not-determined |
| S14_extracted_k141_91618 | Low-quality |
| S14_extracted_k141_47102 | Low-quality |
| S14_extracted_k141_40445 | Not-determined |
| S14_extracted_k141_32833 | Not-determined |
| S14_extracted_k141_113556 | Not-determined |
| S14_extracted_k141_106437 | Not-determined |
| S14_extracted_k141_10724 | Not-determined |
| S14_extracted_k141_113558 | Low-quality |
| S14_extracted_k141_40449 | Not-determined |
| S14_extracted_k141_40450 | Low-quality |
| S14_extracted_k141_113560 | Not-determined |
| S14_extracted_k141_91627 | Low-quality |
| S14_extracted_k141_91628 | Not-determined |
| S14_extracted_k141_91630 | Not-determined |
| S14_extracted_k141_25315 | Not-determined |
| S14_extracted_k141_113563 | Not-determined |
| S14_extracted_k141_32844 | Low-quality |
| S14_extracted_k141_47113 | Low-quality |
| S14_extracted_k141_47118 | Not-determined |
| S14_extracted_k141_25323 | Low-quality |
| S14_extracted_k141_47121 | Not-determined |
| S14_extracted_k141_3277 | Not-determined |
| S14_extracted_k141_40462 | Low-quality |
| S14_extracted_k141_32849 | Low-quality |
| S14_extracted_k141_61897 | Not-determined |
| S14_extracted_k141_47129 | Low-quality |
| S14_extracted_k141_106446 | Low-quality |
| S14_extracted_k141_69493 | Not-determined |
| S14_extracted_k141_106450 | Low-quality |
| S14_extracted_k141_47138 | Not-determined |
| S14_extracted_k141_40479 | Not-determined |
| S14_extracted_k141_47141 | Not-determined |
| S14_extracted_k141_84310 | Not-determined |
| S14_extracted_k141_84311 | Low-quality |
| S14_extracted_k141_84312 | Low-quality |
| S14_extracted_k141_113580 | Not-determined |
| S14_extracted_k141_84314 | Not-determined |
| S14_extracted_k141_84315 | Not-determined |
| S14_extracted_k141_3296 | Not-determined |
| S14_extracted_k141_3297 | Not-determined |
| S14_extracted_k141_47151 | Not-determined |
| S14_extracted_k141_106456 | Low-quality |
| S14_extracted_k141_106459 | Not-determined |
| S14_extracted_k141_69503 | Not-determined |
| S14_extracted_k141_69504 | Not-determined |
| S14_extracted_k141_18371 | Not-determined |
| S14_extracted_k141_47166 | Not-determined |
| S14_extracted_k141_91640 | Not-determined |
| S14_extracted_k141_47169 | Low-quality |
| S14_extracted_k141_40491 | Not-determined |
| S14_extracted_k141_47171 | Not-determined |
| S14_extracted_k141_40497 | Not-determined |
| S14_extracted_k141_10746 | Not-determined |
| S14_extracted_k141_10747 | Not-determined |
| S14_extracted_k141_61910 | Low-quality |
| S14_extracted_k141_61913 | Not-determined |
| S14_extracted_k141_98491 | Not-determined |
| S14_extracted_k141_10756 | Low-quality |
| S14_extracted_k141_98494 | Low-quality |
| S14_extracted_k141_40512 | Not-determined |
| S14_extracted_k141_61919 | Low-quality |
| S14_extracted_k141_106472 | Not-determined |
| S14_extracted_k141_113592 | Not-determined |
| S14_extracted_k141_47190 | Not-determined |
| S14_extracted_k141_113596 | Not-determined |
| S14_extracted_k141_106479 | Not-determined |
| S14_extracted_k141_40518 | Low-quality |
| S14_extracted_k141_10759 | Not-determined |
| S14_extracted_k141_47194 | Low-quality |
| S14_extracted_k141_47196 | Not-determined |
| S14_extracted_k141_47197 | Low-quality |
| S14_extracted_k141_47198 | Low-quality |
| S14_extracted_k141_18385 | Not-determined |
| S14_extracted_k141_69527 | Not-determined |
| S14_extracted_k141_91667 | Not-determined |
| S14_extracted_k141_47203 | Low-quality |
| S14_extracted_k141_10769 | Low-quality |
| S14_extracted_k141_18392 | Not-determined |
| S14_extracted_k141_84331 | Not-determined |
| S14_extracted_k141_69530 | Low-quality |
| S14_extracted_k141_18397 | Not-determined |
| S14_extracted_k141_98502 | Not-determined |
| S14_extracted_k141_10776 | Not-determined |
| S14_extracted_k141_10777 | Not-determined |
| S14_extracted_k141_10778 | Not-determined |
| S14_extracted_k141_113610 | Low-quality |
| S14_extracted_k141_84333 | Not-determined |
| S14_extracted_k141_69539 | Not-determined |
| S14_extracted_k141_3325 | Not-determined |
| S14_extracted_k141_84337 | Not-determined |
| S14_extracted_k141_106509 | Not-determined |
| S14_extracted_k141_18404 | Not-determined |
| S14_extracted_k141_84338 | Low-quality |
| S14_extracted_k141_84339 | Not-determined |
| S14_extracted_k141_61939 | Not-determined |
| S14_extracted_k141_69545 | Not-determined |
| S14_extracted_k141_61940 | Low-quality |
| S14_extracted_k141_91688 | Low-quality |
| S14_extracted_k141_18407 | Not-determined |
| S14_extracted_k141_61942 | Not-determined |
| S14_extracted_k141_69554 | Not-determined |
| S14_extracted_k141_61944 | Not-determined |
| S14_extracted_k141_18410 | Low-quality |
| S14_extracted_k141_10792 | Not-determined |
| S14_extracted_k141_54977 | Not-determined |
| S14_extracted_k141_54979 | Low-quality |
| S14_extracted_k141_54980 | Not-determined |
| S14_extracted_k141_113624 | Not-determined |
| S14_extracted_k141_47220 | Low-quality |
| S14_extracted_k141_32903 | Not-determined |
| S14_extracted_k141_91698 | Not-determined |
| S14_extracted_k141_40558 | Not-determined |
| S14_extracted_k141_10796 | Not-determined |
| S14_extracted_k141_3332 | Low-quality |
| S14_extracted_k141_3334 | Not-determined |
| S14_extracted_k141_3335 | Not-determined |
| S14_extracted_k141_18421 | Not-determined |
| S14_extracted_k141_18422 | Not-determined |
| S14_extracted_k141_40562 | Not-determined |
| S14_extracted_k141_3341 | Not-determined |
| S14_extracted_k141_18424 | Not-determined |
| S14_extracted_k141_10803 | Not-determined |
| S14_extracted_k141_18425 | Not-determined |
| S14_extracted_k141_18426 | Not-determined |
| S14_extracted_k141_98521 | Not-determined |
| S14_extracted_k141_106517 | Low-quality |
| S14_extracted_k141_47236 | Not-determined |
| S14_extracted_k141_84342 | Low-quality |
| S14_extracted_k141_91701 | Not-determined |
| S14_extracted_k141_106519 | Low-quality |
| S14_extracted_k141_106520 | Low-quality |
| S14_extracted_k141_113638 | Not-determined |
| S14_extracted_k141_10816 | Not-determined |
| S14_extracted_k141_69577 | Low-quality |
| S14_extracted_k141_113640 | Not-determined |
| S14_extracted_k141_113641 | Not-determined |
| S14_extracted_k141_113642 | Low-quality |
| S14_extracted_k141_98531 | Not-determined |
| S14_extracted_k141_61952 | Not-determined |
| S14_extracted_k141_106523 | Low-quality |
| S14_extracted_k141_84348 | Not-determined |
| S14_extracted_k141_113646 | Not-determined |
| S14_extracted_k141_10819 | Not-determined |
| S14_extracted_k141_10820 | Not-determined |
| S14_extracted_k141_113651 | Low-quality |
| S14_extracted_k141_98540 | Low-quality |
| S14_extracted_k141_61962 | Not-determined |
| S14_extracted_k141_10829 | Not-determined |
| S14_extracted_k141_98544 | Not-determined |
| S14_extracted_k141_3369 | Not-determined |
| S14_extracted_k141_47248 | Not-determined |
| S14_extracted_k141_47249 | Not-determined |
| S14_extracted_k141_10833 | Not-determined |
| S14_extracted_k141_3376 | Not-determined |
| S14_extracted_k141_10840 | Low-quality |
| S14_extracted_k141_91737 | Not-determined |
| S14_extracted_k141_98556 | Low-quality |
| S14_extracted_k141_98558 | Low-quality |
| S14_extracted_k141_10844 | Not-determined |
| S14_extracted_k141_84363 | Low-quality |
| S14_extracted_k141_113664 | Not-determined |
| S14_extracted_k141_18441 | Low-quality |
| S14_extracted_k141_113665 | Not-determined |
| S14_extracted_k141_32918 | Not-determined |
| S14_extracted_k141_98560 | Not-determined |
| S14_extracted_k141_3384 | Low-quality |
| S14_extracted_k141_98561 | Not-determined |
| S14_extracted_k141_98562 | Low-quality |
| S14_extracted_k141_47254 | Not-determined |
| S14_extracted_k141_69583 | Not-determined |
| S14_extracted_k141_98568 | Not-determined |
| S14_extracted_k141_61985 | Low-quality |
| S14_extracted_k141_106538 | Low-quality |
| S14_extracted_k141_10854 | Not-determined |
| S14_extracted_k141_61987 | Low-quality |
| S14_extracted_k141_91761 | Low-quality |
| S14_extracted_k141_32928 | Low-quality |
| S14_extracted_k141_106545 | Low-quality |
| S14_extracted_k141_3394 | Not-determined |
| S14_extracted_k141_61993 | Low-quality |
| S14_extracted_k141_32936 | Low-quality |
| S14_extracted_k141_84373 | Not-determined |
| S14_extracted_k141_98572 | Low-quality |
| S14_extracted_k141_3400 | Not-determined |
| S14_extracted_k141_3406 | Not-determined |
| S14_extracted_k141_10864 | Low-quality |
| S14_extracted_k141_61999 | Not-determined |
| S14_extracted_k141_84380 | Not-determined |
| S14_extracted_k141_91780 | Low-quality |
| S14_extracted_k141_10867 | Not-determined |
| S14_extracted_k141_40571 | Not-determined |
| S14_extracted_k141_18461 | Not-determined |
| S14_extracted_k141_54994 | Not-determined |
| S14_extracted_k141_69596 | Not-determined |
| S14_extracted_k141_91791 | Not-determined |
| S14_extracted_k141_18463 | Low-quality |
| S14_extracted_k141_18465 | Not-determined |
| S14_extracted_k141_18467 | Not-determined |
| S14_extracted_k141_69608 | Low-quality |
| S14_extracted_k141_18468 | Low-quality |
| S14_extracted_k141_18469 | Not-determined |
| S14_extracted_k141_84401 | Low-quality |
| S14_extracted_k141_40576 | Low-quality |
| S14_extracted_k141_113680 | Not-determined |
| S14_extracted_k141_113681 | Not-determined |
| S14_extracted_k141_10878 | Not-determined |
| S14_extracted_k141_55010 | Not-determined |
| S14_extracted_k141_106551 | Not-determined |
| S14_extracted_k141_25347 | Not-determined |
| S14_extracted_k141_113687 | Low-quality |
| S14_extracted_k141_32955 | Low-quality |
| S14_extracted_k141_84408 | Low-quality |
| S14_extracted_k141_62010 | Low-quality |
| S14_extracted_k141_10882 | Not-determined |
| S14_extracted_k141_18481 | Not-determined |
| S14_extracted_k141_55016 | Not-determined |
| S14_extracted_k141_106556 | Not-determined |
| S14_extracted_k141_62013 | Not-determined |
| S14_extracted_k141_18484 | Low-quality |
| S14_extracted_k141_69617 | Low-quality |
| S14_extracted_k141_55018 | Not-determined |
| S14_extracted_k141_47291 | Not-determined |
| S14_extracted_k141_62022 | Not-determined |
| S14_extracted_k141_47293 | Low-quality |
| S14_extracted_k141_25355 | Not-determined |
| S14_extracted_k141_55022 | Not-determined |
| S14_extracted_k141_25356 | Not-determined |
| S14_extracted_k141_47296 | Not-determined |
| S14_extracted_k141_98581 | Not-determined |
| S14_extracted_k141_25361 | Not-determined |
| S14_extracted_k141_40596 | Low-quality |
| S14_extracted_k141_40599 | Not-determined |
| S14_extracted_k141_84413 | Low-quality |
| S14_extracted_k141_47304 | Low-quality |
| S14_extracted_k141_25365 | Not-determined |
| S14_extracted_k141_113703 | Low-quality |
| S14_extracted_k141_113704 | Low-quality |
| S14_extracted_k141_84418 | Not-determined |
| S14_extracted_k141_98585 | Low-quality |
| S14_extracted_k141_10903 | Not-determined |
| S14_extracted_k141_106566 | Not-determined |
| S14_extracted_k141_98589 | Not-determined |
| S14_extracted_k141_84426 | Not-determined |
| S14_extracted_k141_69634 | Not-determined |
| S14_extracted_k141_32979 | Not-determined |
| S14_extracted_k141_25375 | Not-determined |
| S14_extracted_k141_25382 | Not-determined |
| S14_extracted_k141_113716 | Not-determined |
| S14_extracted_k141_32987 | Not-determined |
| S14_extracted_k141_84437 | Not-determined |
| S14_extracted_k141_10917 | Not-determined |
| S14_extracted_k141_69646 | Low-quality |
| S14_extracted_k141_113718 | Not-determined |
| S14_extracted_k141_84439 | Not-determined |
| S14_extracted_k141_10919 | Not-determined |
| S14_extracted_k141_84442 | Not-determined |
| S14_extracted_k141_40622 | Low-quality |
| S14_extracted_k141_84446 | Low-quality |
| S14_extracted_k141_10922 | Low-quality |
| S14_extracted_k141_10923 | Low-quality |
| S14_extracted_k141_25389 | Not-determined |
| S14_extracted_k141_40625 | Low-quality |
| S14_extracted_k141_84450 | Not-determined |
| S14_extracted_k141_84451 | Not-determined |
| S14_extracted_k141_10925 | Not-determined |
| S14_extracted_k141_113723 | Not-determined |
| S14_extracted_k141_69660 | Not-determined |
| S14_extracted_k141_113727 | Not-determined |
| S14_extracted_k141_40633 | Not-determined |
| S14_extracted_k141_113728 | Not-determined |
| S14_extracted_k141_40636 | Not-determined |
| S14_extracted_k141_69667 | Not-determined |
| S14_extracted_k141_113731 | Not-determined |
| S14_extracted_k141_69668 | Not-determined |
| S14_extracted_k141_69669 | Low-quality |
| S14_extracted_k141_62042 | Low-quality |
| S14_extracted_k141_25398 | Low-quality |
| S14_extracted_k141_25403 | Not-determined |
| S14_extracted_k141_40643 | Low-quality |
| S14_extracted_k141_10930 | Low-quality |
| S14_extracted_k141_40644 | Not-determined |
| S14_extracted_k141_40645 | Not-determined |
| S14_extracted_k141_40646 | Low-quality |
| S14_extracted_k141_113743 | Not-determined |
| S14_extracted_k141_98607 | Low-quality |
| S14_extracted_k141_98608 | Low-quality |
| S14_extracted_k141_40650 | Not-determined |
| S14_extracted_k141_98609 | Not-determined |
| S14_extracted_k141_98610 | Low-quality |
| S14_extracted_k141_25409 | Low-quality |
| S14_extracted_k141_55052 | Low-quality |
| S14_extracted_k141_62047 | Not-determined |
| S14_extracted_k141_62048 | Not-determined |
| S14_extracted_k141_55055 | Not-determined |
| S14_extracted_k141_98621 | Low-quality |
| S14_extracted_k141_55057 | Not-determined |
| S14_extracted_k141_55058 | Not-determined |
| S14_extracted_k141_55059 | Not-determined |
| S14_extracted_k141_10943 | Not-determined |
| S14_extracted_k141_55064 | Low-quality |
| S14_extracted_k141_113760 | Not-determined |
| S14_extracted_k141_47311 | Not-determined |
| S14_extracted_k141_47312 | Not-determined |
| S14_extracted_k141_84472 | Not-determined |
| S14_extracted_k141_40659 | Low-quality |
| S14_extracted_k141_10946 | Not-determined |
| S14_extracted_k141_32997 | Low-quality |
| S14_extracted_k141_113768 | Low-quality |
| S14_extracted_k141_84477 | Not-determined |
| S14_extracted_k141_40668 | Not-determined |
| S14_extracted_k141_47326 | Not-determined |
| S14_extracted_k141_10956 | Low-quality |
| S14_extracted_k141_69697 | Not-determined |
| S14_extracted_k141_106584 | Not-determined |
| S14_extracted_k141_113773 | Low-quality |
| S14_extracted_k141_98645 | Not-determined |
| S14_extracted_k141_113774 | Not-determined |
| S14_extracted_k141_106590 | Low-quality |
| S14_extracted_k141_47330 | Not-determined |
| S14_extracted_k141_84500 | Not-determined |
| S14_extracted_k141_25426 | Low-quality |
| S14_extracted_k141_113783 | Not-determined |
| S14_extracted_k141_84505 | Not-determined |
| S14_extracted_k141_98657 | Not-determined |
| S14_extracted_k141_84513 | Low-quality |
| S14_extracted_k141_40686 | Not-determined |
| S14_extracted_k141_106607 | Low-quality |
| S14_extracted_k141_40687 | Not-determined |
| S14_extracted_k141_40688 | Low-quality |
| S14_extracted_k141_55084 | Not-determined |
| S14_extracted_k141_113800 | Low-quality |
| S14_extracted_k141_62070 | Not-determined |
| S14_extracted_k141_113802 | Not-determined |
| S14_extracted_k141_84522 | Low-quality |
| S14_extracted_k141_18516 | Not-determined |
| S14_extracted_k141_55095 | Low-quality |
| S14_extracted_k141_62071 | Low-quality |
| S14_extracted_k141_47352 | Not-determined |
| S14_extracted_k141_62073 | Not-determined |
| S14_extracted_k141_113808 | Not-determined |
| S14_extracted_k141_113809 | Not-determined |
| S14_extracted_k141_62077 | Not-determined |
| S14_extracted_k141_98679 | Not-determined |
| S14_extracted_k141_106620 | Not-determined |
| S14_extracted_k141_47360 | Low-quality |
| S14_extracted_k141_69712 | Not-determined |
| S14_extracted_k141_47361 | Low-quality |
| S14_extracted_k141_69715 | Not-determined |
| S14_extracted_k141_18526 | Not-determined |
| S14_extracted_k141_47366 | Low-quality |
| S14_extracted_k141_33002 | Low-quality |
| S14_extracted_k141_33005 | Not-determined |
| S14_extracted_k141_33006 | Low-quality |
| S14_extracted_k141_113824 | Not-determined |
| S14_extracted_k141_113825 | Not-determined |
| S14_extracted_k141_10963 | Not-determined |
| S14_extracted_k141_18544 | Not-determined |
| S14_extracted_k141_40717 | Not-determined |
| S14_extracted_k141_106634 | Not-determined |
| S14_extracted_k141_40718 | Low-quality |
| S14_extracted_k141_62088 | Low-quality |
| S14_extracted_k141_18548 | Low-quality |
| S14_extracted_k141_18549 | Low-quality |
| S14_extracted_k141_18551 | Not-determined |
| S14_extracted_k141_40724 | Not-determined |
| S14_extracted_k141_33011 | Not-determined |
| S14_extracted_k141_33012 | Low-quality |
| S14_extracted_k141_10973 | Not-determined |
| S14_extracted_k141_33021 | Not-determined |
| S14_extracted_k141_62094 | Low-quality |
| S14_extracted_k141_25442 | Not-determined |
| S14_extracted_k141_62097 | Not-determined |
| S14_extracted_k141_84536 | Not-determined |
| S14_extracted_k141_47387 | Not-determined |
| S14_extracted_k141_98695 | Not-determined |
| S14_extracted_k141_10982 | Low-quality |
| S14_extracted_k141_62099 | Low-quality |
| S14_extracted_k141_10983 | Low-quality |
| S14_extracted_k141_10985 | Not-determined |
| S14_extracted_k141_18568 | Not-determined |
| S14_extracted_k141_91868 | Not-determined |
| S14_extracted_k141_40735 | Not-determined |
| S14_extracted_k141_77080 | Not-determined |
| S14_extracted_k141_10994 | Not-determined |
| S14_extracted_k141_47394 | Not-determined |
| S14_extracted_k141_18577 | Not-determined |
| S14_extracted_k141_106652 | Not-determined |
| S14_extracted_k141_33028 | Not-determined |
| S14_extracted_k141_25460 | Low-quality |
| S14_extracted_k141_18579 | Not-determined |
| S14_extracted_k141_40748 | Not-determined |
| S14_extracted_k141_77085 | Not-determined |
| S14_extracted_k141_77086 | Not-determined |
| S14_extracted_k141_106657 | Not-determined |
| S14_extracted_k141_33032 | Not-determined |
| S14_extracted_k141_18585 | Low-quality |
| S14_extracted_k141_106659 | Not-determined |
| S14_extracted_k141_18586 | Not-determined |
| S14_extracted_k141_69740 | Not-determined |
| S14_extracted_k141_18587 | Not-determined |
| S14_extracted_k141_40755 | Low-quality |
| S14_extracted_k141_40756 | Low-quality |
| S14_extracted_k141_40757 | Not-determined |
| S14_extracted_k141_11009 | Not-determined |
| S14_extracted_k141_25466 | Not-determined |
| S14_extracted_k141_106663 | Not-determined |
| S14_extracted_k141_3424 | Low-quality |
| S14_extracted_k141_98711 | Not-determined |
| S14_extracted_k141_33039 | Not-determined |
| S14_extracted_k141_98712 | Low-quality |
| S14_extracted_k141_18591 | Not-determined |
| S14_extracted_k141_33041 | Not-determined |
| S14_extracted_k141_40760 | Low-quality |
| S14_extracted_k141_47398 | Not-determined |
| S14_extracted_k141_98714 | Not-determined |
| S14_extracted_k141_106666 | Not-determined |
| S14_extracted_k141_106667 | Low-quality |
| S14_extracted_k141_62112 | Low-quality |
| S14_extracted_k141_47403 | Low-quality |
| S14_extracted_k141_33052 | Not-determined |
| S14_extracted_k141_11029 | Not-determined |
| S14_extracted_k141_3441 | Not-determined |
| S14_extracted_k141_11032 | Not-determined |
| S14_extracted_k141_69749 | Not-determined |
| S14_extracted_k141_25468 | Low-quality |
| S14_extracted_k141_3449 | Not-determined |
| S14_extracted_k141_62123 | Low-quality |
| S14_extracted_k141_62124 | Not-determined |
| S14_extracted_k141_40764 | Not-determined |
| S14_extracted_k141_98722 | Not-determined |
| S14_extracted_k141_106679 | Not-determined |
| S14_extracted_k141_25476 | Not-determined |
| S14_extracted_k141_47413 | Not-determined |
| S14_extracted_k141_62128 | Not-determined |
| S14_extracted_k141_40767 | Not-determined |
| S14_extracted_k141_47416 | Not-determined |
| S14_extracted_k141_47417 | Not-determined |
| S14_extracted_k141_47418 | Low-quality |
| S14_extracted_k141_77092 | Low-quality |
| S14_extracted_k141_3457 | Not-determined |
| S14_extracted_k141_40775 | Not-determined |
| S14_extracted_k141_25486 | Not-determined |
| S14_extracted_k141_3462 | Not-determined |
| S14_extracted_k141_25489 | Low-quality |
| S14_extracted_k141_33066 | Not-determined |
| S14_extracted_k141_25495 | Not-determined |
| S14_extracted_k141_98730 | Not-determined |
| S14_extracted_k141_33071 | Low-quality |
| S14_extracted_k141_40788 | Not-determined |
| S14_extracted_k141_69767 | Not-determined |
| S14_extracted_k141_84553 | Low-quality |
| S14_extracted_k141_113828 | Low-quality |
| S14_extracted_k141_40793 | Not-determined |
| S14_extracted_k141_77094 | Low-quality |
| S14_extracted_k141_69774 | Low-quality |
| S14_extracted_k141_33074 | Low-quality |
| S14_extracted_k141_77096 | Low-quality |
| S14_extracted_k141_77097 | Not-determined |
| S14_extracted_k141_98736 | Low-quality |
| S14_extracted_k141_62145 | Not-determined |
| S14_extracted_k141_33077 | Not-determined |
| S14_extracted_k141_18611 | Not-determined |
| S14_extracted_k141_69787 | Low-quality |
| S14_extracted_k141_55115 | Not-determined |
| S14_extracted_k141_25511 | Not-determined |
| S14_extracted_k141_62148 | Low-quality |
| S14_extracted_k141_18613 | Not-determined |
| S14_extracted_k141_3479 | Not-determined |
| S14_extracted_k141_3480 | Not-determined |
| S14_extracted_k141_62154 | Low-quality |
| S14_extracted_k141_84575 | Low-quality |
| S14_extracted_k141_33081 | Not-determined |
| S14_extracted_k141_47440 | Not-determined |
| S14_extracted_k141_84577 | Not-determined |
| S14_extracted_k141_33082 | Not-determined |
| S14_extracted_k141_77106 | Low-quality |
| S14_extracted_k141_84578 | Not-determined |
| S14_extracted_k141_62157 | Not-determined |
| S14_extracted_k141_18619 | Not-determined |
| S14_extracted_k141_33085 | Low-quality |
| S14_extracted_k141_69793 | Not-determined |
| S14_extracted_k141_62159 | Low-quality |
| S14_extracted_k141_62160 | Not-determined |
| S14_extracted_k141_11083 | Not-determined |
| S14_extracted_k141_77109 | Not-determined |
| S14_extracted_k141_11085 | Not-determined |
| S14_extracted_k141_77110 | Not-determined |
| S14_extracted_k141_18629 | Not-determined |
| S14_extracted_k141_113851 | Not-determined |
| S14_extracted_k141_55120 | Low-quality |
| S14_extracted_k141_62166 | Not-determined |
| S14_extracted_k141_106687 | Not-determined |
| S14_extracted_k141_33096 | Not-determined |
| S14_extracted_k141_40815 | Not-determined |
| S14_extracted_k141_113859 | Low-quality |
| S14_extracted_k141_69804 | Not-determined |
| S14_extracted_k141_62169 | Not-determined |
| S14_extracted_k141_40825 | Not-determined |
| S14_extracted_k141_18642 | Not-determined |
| S14_extracted_k141_77117 | Not-determined |
| S14_extracted_k141_77118 | Not-determined |
| S14_extracted_k141_77119 | Not-determined |
| S14_extracted_k141_106694 | Not-determined |
| S14_extracted_k141_40829 | Not-determined |
| S14_extracted_k141_62175 | Low-quality |
| S14_extracted_k141_11092 | Not-determined |
| S14_extracted_k141_77123 | Not-determined |
| S14_extracted_k141_11095 | Not-determined |
| S14_extracted_k141_55139 | Not-determined |
| S14_extracted_k141_3500 | Not-determined |
| S14_extracted_k141_91892 | Not-determined |
| S14_extracted_k141_106699 | Not-determined |
| S14_extracted_k141_40831 | Low-quality |
| S14_extracted_k141_55142 | Not-determined |
| S14_extracted_k141_55144 | Not-determined |
| S14_extracted_k141_3508 | Not-determined |
| S14_extracted_k141_55148 | Not-determined |
| S14_extracted_k141_69811 | Not-determined |
| S14_extracted_k141_113870 | Not-determined |
| S14_extracted_k141_113871 | Low-quality |
| S14_extracted_k141_40837 | Not-determined |
| S14_extracted_k141_40838 | Not-determined |
| S14_extracted_k141_25546 | Not-determined |
| S14_extracted_k141_3513 | Not-determined |
| S14_extracted_k141_77133 | Not-determined |
| S14_extracted_k141_77134 | Not-determined |
| S14_extracted_k141_40843 | Not-determined |
| S14_extracted_k141_3516 | Not-determined |
| S14_extracted_k141_98751 | Not-determined |
| S14_extracted_k141_3518 | Not-determined |
| S14_extracted_k141_25550 | Low-quality |
| S14_extracted_k141_40849 | Not-determined |
| S14_extracted_k141_98757 | Not-determined |
| S14_extracted_k141_98760 | Not-determined |
| S14_extracted_k141_18651 | Not-determined |
| S14_extracted_k141_98761 | Not-determined |
| S14_extracted_k141_84595 | Not-determined |
| S14_extracted_k141_55162 | Not-determined |
| S14_extracted_k141_84599 | Not-determined |
| S14_extracted_k141_40870 | Low-quality |
| S14_extracted_k141_98765 | Not-determined |
| S14_extracted_k141_55177 | Low-quality |

| | |
|---|---|
| S14_extracted_k141_98770 | Not-determined |
| S14_extracted_k141_25568 | Low-quality |
| S14_extracted_k141_98775 | Not-determined |
| S14_extracted_k141_18664 | Not-determined |
| S14_extracted_k141_18665 | Not-determined |
| S14_extracted_k141_47454 | Low-quality |
| S14_extracted_k141_113899 | Low-quality |
| S14_extracted_k141_91912 | Not-determined |
| S14_extracted_k141_91913 | Not-determined |
| S14_extracted_k141_11107 | Not-determined |
| S14_extracted_k141_55182 | Low-quality |
| S14_extracted_k141_91914 | Not-determined |
| S14_extracted_k141_11108 | Not-determined |
| S14_extracted_k141_91915 | Low-quality |
| S14_extracted_k141_18671 | Low-quality |
| S14_extracted_k141_69860 | Not-determined |
| S14_extracted_k141_40879 | Not-determined |
| S14_extracted_k141_84608 | Low-quality |
| S14_extracted_k141_40882 | Not-determined |
| S14_extracted_k141_55189 | Not-determined |
| S14_extracted_k141_40884 | Not-determined |
| S14_extracted_k141_18680 | Not-determined |
| S14_extracted_k141_47465 | Low-quality |
| S14_extracted_k141_40887 | Not-determined |
| S14_extracted_k141_47470 | Not-determined |
| S14_extracted_k141_47471 | Low-quality |
| S14_extracted_k141_77149 | Not-determined |
| S14_extracted_k141_25575 | Not-determined |
| S14_extracted_k141_113907 | Not-determined |
| S14_extracted_k141_40889 | Not-determined |
| S14_extracted_k141_106720 | Not-determined |
| S14_extracted_k141_106721 | Low-quality |
| S14_extracted_k141_55197 | Low-quality |
| S14_extracted_k141_91927 | Low-quality |
| S14_extracted_k141_106725 | Not-determined |
| S14_extracted_k141_40893 | Low-quality |
| S14_extracted_k141_55198 | Not-determined |
| S14_extracted_k141_40896 | Not-determined |
| S14_extracted_k141_11127 | Not-determined |
| S14_extracted_k141_91930 | Low-quality |
| S14_extracted_k141_62181 | Low-quality |
| S14_extracted_k141_25585 | Not-determined |
| S14_extracted_k141_33118 | Not-determined |
| S14_extracted_k141_33119 | Not-determined |
| S14_extracted_k141_62185 | Low-quality |
| S14_extracted_k141_33122 | Not-determined |
| S14_extracted_k141_77158 | Not-determined |
| S14_extracted_k141_69876 | Not-determined |
| S14_extracted_k141_77160 | Not-determined |
| S14_extracted_k141_77161 | Not-determined |
| S14_extracted_k141_77162 | Low-quality |
| S14_extracted_k141_47474 | Not-determined |
| S14_extracted_k141_25595 | Not-determined |
| S14_extracted_k141_62192 | Not-determined |
| S14_extracted_k141_62193 | Not-determined |
| S14_extracted_k141_25597 | Not-determined |
| S14_extracted_k141_47477 | Low-quality |
| S14_extracted_k141_11134 | Not-determined |
| S14_extracted_k141_47479 | Low-quality |
| S14_extracted_k141_77170 | Not-determined |
| S14_extracted_k141_11137 | Not-determined |
| S14_extracted_k141_77173 | Not-determined |
| S14_extracted_k141_25599 | Not-determined |
| S14_extracted_k141_47483 | Low-quality |
| S14_extracted_k141_55205 | Low-quality |
| S14_extracted_k141_91945 | Low-quality |
| S14_extracted_k141_25602 | Not-determined |
| S14_extracted_k141_113922 | Not-determined |
| S14_extracted_k141_84628 | Not-determined |
| S14_extracted_k141_77175 | Low-quality |
| S14_extracted_k141_55211 | Low-quality |
| S14_extracted_k141_3527 | Not-determined |
| S14_extracted_k141_55214 | Not-determined |
| S14_extracted_k141_84634 | Not-determined |
| S14_extracted_k141_91956 | Low-quality |
| S14_extracted_k141_91957 | Low-quality |
| S14_extracted_k141_55217 | Low-quality |
| S14_extracted_k141_113926 | Not-determined |
| S14_extracted_k141_91962 | Low-quality |
| S14_extracted_k141_113929 | Not-determined |
| S14_extracted_k141_3536 | Low-quality |
| S14_extracted_k141_3539 | Low-quality |
| S14_extracted_k141_33144 | Not-determined |
| S14_extracted_k141_91966 | Not-determined |
| S14_extracted_k141_11151 | Low-quality |
| S14_extracted_k141_11153 | Low-quality |
| S14_extracted_k141_33147 | Not-determined |
| S14_extracted_k141_77189 | Not-determined |
| S14_extracted_k141_40922 | Not-determined |
| S14_extracted_k141_55227 | Not-determined |
| S14_extracted_k141_84651 | Low-quality |
| S14_extracted_k141_47504 | Low-quality |
| S14_extracted_k141_55233 | Not-determined |
| S14_extracted_k141_106751 | Not-determined |
| S14_extracted_k141_11165 | Low-quality |
| S14_extracted_k141_47506 | Not-determined |
| S14_extracted_k141_91968 | Low-quality |
| S14_extracted_k141_98783 | Not-determined |
| S14_extracted_k141_3556 | Not-determined |
| S14_extracted_k141_3558 | Low-quality |
| S14_extracted_k141_77202 | Not-determined |
| S14_extracted_k141_33153 | Low-quality |
| S14_extracted_k141_47510 | Not-determined |
| S14_extracted_k141_25609 | Not-determined |
| S14_extracted_k141_113937 | Not-determined |
| S14_extracted_k141_113940 | Low-quality |
| S14_extracted_k141_47519 | Not-determined |
| S14_extracted_k141_47521 | Low-quality |
| S14_extracted_k141_25615 | Not-determined |
| S14_extracted_k141_62219 | Not-determined |
| S14_extracted_k141_91982 | Not-determined |
| S14_extracted_k141_98796 | Low-quality |
| S14_extracted_k141_106760 | Not-determined |
| S14_extracted_k141_47523 | Not-determined |
| S14_extracted_k141_106762 | Not-determined |
| S14_extracted_k141_11179 | Not-determined |
| S14_extracted_k141_98801 | Not-determined |
| S14_extracted_k141_47525 | Not-determined |
| S14_extracted_k141_11180 | Not-determined |
| S14_extracted_k141_33157 | Not-determined |
| S14_extracted_k141_113946 | Not-determined |
| S14_extracted_k141_62221 | Not-determined |
| S14_extracted_k141_113949 | Not-determined |
| S14_extracted_k141_106770 | Low-quality |
| S14_extracted_k141_3572 | Not-determined |
| S14_extracted_k141_84661 | Low-quality |
| S14_extracted_k141_98806 | Not-determined |
| S14_extracted_k141_25623 | Not-determined |
| S14_extracted_k141_47535 | Not-determined |
| S14_extracted_k141_47536 | Not-determined |
| S14_extracted_k141_33161 | Not-determined |
| S14_extracted_k141_3577 | Not-determined |
| S14_extracted_k141_55245 | Not-determined |
| S14_extracted_k141_77215 | Low-quality |
| S14_extracted_k141_55246 | Not-determined |
| S14_extracted_k141_3579 | Low-quality |
| S14_extracted_k141_84669 | Low-quality |
| S14_extracted_k141_25634 | Not-determined |
| S14_extracted_k141_25635 | Not-determined |
| S14_extracted_k141_113956 | Not-determined |
| S14_extracted_k141_98813 | Not-determined |
| S14_extracted_k141_40953 | Low-quality |
| S14_extracted_k141_55259 | Not-determined |
| S14_extracted_k141_11205 | Not-determined |
| S14_extracted_k141_3591 | Not-determined |
| S14_extracted_k141_40959 | Not-determined |
| S14_extracted_k141_62238 | Low-quality |
| S14_extracted_k141_98823 | Not-determined |
| S14_extracted_k141_98824 | Not-determined |
| S14_extracted_k141_62239 | Low-quality |
| S14_extracted_k141_3595 | Not-determined |
| S14_extracted_k141_113960 | Not-determined |
| S14_extracted_k141_62241 | Not-determined |
| S14_extracted_k141_3597 | Not-determined |
| S14_extracted_k141_62242 | Low-quality |
| S14_extracted_k141_62245 | Not-determined |
| S14_extracted_k141_25651 | Low-quality |
| S14_extracted_k141_106797 | Not-determined |
| S14_extracted_k141_98829 | Low-quality |
| S14_extracted_k141_92004 | Not-determined |
| S14_extracted_k141_40969 | Low-quality |
| S14_extracted_k141_11210 | Low-quality |
| S14_extracted_k141_11212 | Not-determined |
| S14_extracted_k141_33187 | Not-determined |
| S14_extracted_k141_33188 | Not-determined |
| S14_extracted_k141_3600 | Not-determined |
| S14_extracted_k141_77228 | Not-determined |
| S14_extracted_k141_84682 | Not-determined |
| S14_extracted_k141_40976 | Low-quality |
| S14_extracted_k141_113975 | Low-quality |
| S14_extracted_k141_62252 | Low-quality |
| S14_extracted_k141_55274 | Not-determined |
| S14_extracted_k141_40983 | Not-determined |
| S14_extracted_k141_11223 | Not-determined |
| S14_extracted_k141_11226 | Low-quality |
| S14_extracted_k141_11229 | Not-determined |
| S14_extracted_k141_98842 | Not-determined |
| S14_extracted_k141_77244 | Low-quality |
| S14_extracted_k141_55283 | Not-determined |
| S14_extracted_k141_25671 | Low-quality |
| S14_extracted_k141_3622 | Not-determined |
| S14_extracted_k141_18738 | Not-determined |
| S14_extracted_k141_77253 | Not-determined |
| S14_extracted_k141_77254 | Low-quality |
| S14_extracted_k141_25675 | Not-determined |
| S14_extracted_k141_25676 | Not-determined |
| S14_extracted_k141_77256 | Not-determined |
| S14_extracted_k141_77257 | Not-determined |
| S14_extracted_k141_18746 | Not-determined |
| S14_extracted_k141_55297 | Not-determined |
| S14_extracted_k141_18752 | Not-determined |
| S14_extracted_k141_25680 | Not-determined |
| S14_extracted_k141_25681 | Not-determined |
| S14_extracted_k141_18754 | Not-determined |
| S14_extracted_k141_40998 | Not-determined |
| S14_extracted_k141_77269 | Low-quality |
| S14_extracted_k141_55304 | Not-determined |
| S14_extracted_k141_77271 | Low-quality |
| S14_extracted_k141_3648 | Not-determined |
| S14_extracted_k141_77274 | Not-determined |
| S14_extracted_k141_106805 | Low-quality |
| S14_extracted_k141_106806 | Low-quality |
| S14_extracted_k141_3652 | Not-determined |
| S14_extracted_k141_18760 | Not-determined |
| S14_extracted_k141_3654 | Low-quality |
| S14_extracted_k141_18761 | Not-determined |
| S14_extracted_k141_106811 | Not-determined |
| S14_extracted_k141_62277 | Not-determined |
| S14_extracted_k141_3658 | Low-quality |
| S14_extracted_k141_55312 | Low-quality |
| S14_extracted_k141_77280 | Low-quality |
| S14_extracted_k141_41014 | Low-quality |
| S14_extracted_k141_113991 | Not-determined |
| S14_extracted_k141_113992 | Not-determined |
| S14_extracted_k141_33192 | Not-determined |
| S14_extracted_k141_41022 | Low-quality |
| S14_extracted_k141_77288 | Low-quality |
| S14_extracted_k141_92026 | Low-quality |
| S14_extracted_k141_62287 | Low-quality |
| S14_extracted_k141_92028 | Not-determined |
| S14_extracted_k141_18769 | Not-determined |
| S14_extracted_k141_18776 | Not-determined |
| S14_extracted_k141_106824 | Low-quality |
| S14_extracted_k141_84702 | Not-determined |
| S14_extracted_k141_11257 | Not-determined |
| S14_extracted_k141_11258 | Not-determined |
| S14_extracted_k141_98873 | Not-determined |
| S14_extracted_k141_92047 | Not-determined |
| S14_extracted_k141_98879 | Not-determined |
| S14_extracted_k141_114009 | Not-determined |
| S14_extracted_k141_33207 | Not-determined |
| S14_extracted_k141_106826 | Not-determined |
| S14_extracted_k141_41038 | Not-determined |
| S14_extracted_k141_106830 | Not-determined |
| S14_extracted_k141_77303 | Not-determined |
| S14_extracted_k141_3687 | Low-quality |
| S14_extracted_k141_33217 | Not-determined |
| S14_extracted_k141_114017 | Not-determined |
| S14_extracted_k141_3690 | Low-quality |
| S14_extracted_k141_106837 | Not-determined |
| S14_extracted_k141_41048 | Not-determined |
| S14_extracted_k141_84719 | Not-determined |
| S14_extracted_k141_33239 | Not-determined |
| S14_extracted_k141_18792 | Not-determined |
| S14_extracted_k141_84721 | Not-determined |
| S14_extracted_k141_18793 | Low-quality |
| S14_extracted_k141_41056 | Not-determined |
| S14_extracted_k141_18794 | Not-determined |
| S14_extracted_k141_92067 | Low-quality |
| S14_extracted_k141_3705 | Not-determined |
| S14_extracted_k141_92068 | Not-determined |
| S14_extracted_k141_114032 | Low-quality |
| S14_extracted_k141_92069 | Low-quality |
| S14_extracted_k141_84722 | Low-quality |
| S14_extracted_k141_25687 | Low-quality |
| S14_extracted_k141_18800 | Low-quality |
| S14_extracted_k141_84723 | Not-determined |
| S14_extracted_k141_92073 | Not-determined |
| S14_extracted_k141_92075 | Not-determined |
| S14_extracted_k141_25693 | Low-quality |
| S14_extracted_k141_3711 | Not-determined |
| S14_extracted_k141_92081 | Not-determined |
| S14_extracted_k141_11279 | Not-determined |
| S14_extracted_k141_25698 | Low-quality |
| S14_extracted_k141_98898 | Low-quality |
| S14_extracted_k141_41066 | Not-determined |
| S14_extracted_k141_18814 | Not-determined |
| S14_extracted_k141_41068 | Low-quality |
| S14_extracted_k141_114044 | Not-determined |
| S14_extracted_k141_84735 | Low-quality |
| S14_extracted_k141_41073 | Not-determined |
| S14_extracted_k141_114047 | Low-quality |
| S14_extracted_k141_18824 | Not-determined |
| S14_extracted_k141_3717 | Not-determined |
| S14_extracted_k141_18828 | Low-quality |
| S14_extracted_k141_92092 | Not-determined |
| S14_extracted_k141_92093 | Not-determined |
| S14_extracted_k141_114052 | Not-determined |
| S14_extracted_k141_106866 | Low-quality |
| S14_extracted_k141_92096 | Not-determined |
| S14_extracted_k141_98914 | Not-determined |
| S14_extracted_k141_98915 | Low-quality |
| S14_extracted_k141_11298 | Not-determined |
| S14_extracted_k141_62309 | Low-quality |
| S14_extracted_k141_11299 | Not-determined |
| S14_extracted_k141_114057 | Not-determined |
| S14_extracted_k141_62312 | Not-determined |
| S14_extracted_k141_98917 | Not-determined |
| S14_extracted_k141_98919 | Not-determined |
| S14_extracted_k141_98920 | Not-determined |
| S14_extracted_k141_106878 | Not-determined |
| S14_extracted_k141_3733 | Not-determined |
| S14_extracted_k141_62320 | Low-quality |
| S14_extracted_k141_92098 | Not-determined |
| S14_extracted_k141_62322 | Low-quality |
| S14_extracted_k141_106882 | Not-determined |
| S14_extracted_k141_114071 | Not-determined |
| S14_extracted_k141_25716 | Not-determined |
| S14_extracted_k141_114072 | Not-determined |
| S14_extracted_k141_106886 | Not-determined |
| S14_extracted_k141_3739 | Not-determined |
| S14_extracted_k141_98929 | Low-quality |
| S14_extracted_k141_98931 | Not-determined |
| S14_extracted_k141_25724 | Low-quality |
| S14_extracted_k141_25728 | Low-quality |
| S14_extracted_k141_25729 | Low-quality |
| S14_extracted_k141_98934 | Not-determined |
| S14_extracted_k141_25733 | Low-quality |
| S14_extracted_k141_25736 | Not-determined |
| S14_extracted_k141_41109 | Not-determined |
| S14_extracted_k141_18865 | Low-quality |
| S14_extracted_k141_18867 | Not-determined |
| S14_extracted_k141_92119 | Low-quality |
| S14_extracted_k141_114096 | Not-determined |
| S14_extracted_k141_77369 | Not-determined |
| S14_extracted_k141_3746 | Not-determined |
| S14_extracted_k141_3749 | Low-quality |
| S14_extracted_k141_92129 | Not-determined |
| S14_extracted_k141_18886 | Not-determined |
| S14_extracted_k141_3752 | Not-determined |
| S14_extracted_k141_18890 | Low-quality |
| S14_extracted_k141_18894 | Not-determined |
| S14_extracted_k141_55330 | Not-determined |
| S14_extracted_k141_55331 | Not-determined |
| S14_extracted_k141_3759 | Not-determined |
| S14_extracted_k141_18898 | Not-determined |
| S14_extracted_k141_92140 | Low-quality |
| S14_extracted_k141_11339 | Not-determined |
| S14_extracted_k141_11345 | Not-determined |
| S14_extracted_k141_92145 | Not-determined |
| S14_extracted_k141_77393 | Low-quality |
| S14_extracted_k141_55338 | Not-determined |
| S14_extracted_k141_114116 | Not-determined |
| S14_extracted_k141_25755 | Not-determined |
| S14_extracted_k141_25756 | Low-quality |
| S14_extracted_k141_18915 | Not-determined |
| S14_extracted_k141_84753 | Not-determined |
| S14_extracted_k141_106891 | Low-quality |
| S14_extracted_k141_18921 | Not-determined |
| S14_extracted_k141_84756 | Not-determined |
| S14_extracted_k141_77400 | Low-quality |
| S14_extracted_k141_18935 | Not-determined |
| S14_extracted_k141_92159 | Not-determined |
| S14_extracted_k141_18936 | Not-determined |
| S14_extracted_k141_106900 | Low-quality |
| S14_extracted_k141_33251 | Low-quality |
| S14_extracted_k141_33252 | Not-determined |
| S14_extracted_k141_92164 | Low-quality |
| S14_extracted_k141_33256 | Not-determined |
| S14_extracted_k141_92166 | Not-determined |
| S14_extracted_k141_98944 | Low-quality |
| S14_extracted_k141_106906 | Low-quality |
| S14_extracted_k141_18951 | Not-determined |
| S14_extracted_k141_92171 | Not-determined |
| S14_extracted_k141_55361 | Not-determined |
| S14_extracted_k141_92172 | Low-quality |
| S14_extracted_k141_25775 | Low-quality |
| S14_extracted_k141_84775 | Low-quality |
| S14_extracted_k141_114149 | Not-determined |
| S14_extracted_k141_55364 | Not-determined |
| S14_extracted_k141_106910 | Not-determined |
| S14_extracted_k141_98948 | Low-quality |
| S14_extracted_k141_25780 | Not-determined |
| S14_extracted_k141_114154 | Not-determined |
| S14_extracted_k141_25783 | Low-quality |
| S14_extracted_k141_114155 | Not-determined |
| S14_extracted_k141_114156 | Not-determined |
| S14_extracted_k141_25785 | Low-quality |
| S14_extracted_k141_33271 | Not-determined |
| S14_extracted_k141_55369 | Not-determined |
| S14_extracted_k141_25789 | Low-quality |
| S14_extracted_k141_33278 | Not-determined |
| S14_extracted_k141_106915 | Low-quality |
| S14_extracted_k141_114162 | Low-quality |
| S14_extracted_k141_25792 | Low-quality |
| S14_extracted_k141_3778 | Not-determined |
| S14_extracted_k141_25793 | Low-quality |
| S14_extracted_k141_92184 | Not-determined |
| S14_extracted_k141_114166 | Not-determined |
| S14_extracted_k141_25796 | Not-determined |
| S14_extracted_k141_3782 | Low-quality |
| S14_extracted_k141_98959 | Not-determined |
| S14_extracted_k141_84785 | Not-determined |
| S14_extracted_k141_18961 | Low-quality |
| S14_extracted_k141_41121 | Low-quality |
| S14_extracted_k141_98962 | Not-determined |
| S14_extracted_k141_98964 | Not-determined |
| S14_extracted_k141_106921 | Not-determined |
| S14_extracted_k141_33288 | Low-quality |
| S14_extracted_k141_41126 | Low-quality |
| S14_extracted_k141_11381 | Low-quality |
| S14_extracted_k141_69901 | Not-determined |
| S14_extracted_k141_84794 | Not-determined |
| S14_extracted_k141_25801 | Not-determined |
| S14_extracted_k141_84800 | Not-determined |
| S14_extracted_k141_106940 | Not-determined |
| S14_extracted_k141_33296 | Not-determined |
| S14_extracted_k141_77432 | Not-determined |
| S14_extracted_k141_33302 | Low-quality |
| S14_extracted_k141_84805 | Low-quality |
| S14_extracted_k141_77436 | Not-determined |
| S14_extracted_k141_77437 | Not-determined |
| S14_extracted_k141_84809 | Low-quality |
| S14_extracted_k141_106948 | Not-determined |
| S14_extracted_k141_106950 | Low-quality |
| S14_extracted_k141_69921 | Low-quality |
| S14_extracted_k141_84812 | Not-determined |
| S14_extracted_k141_69922 | Not-determined |
| S14_extracted_k141_106951 | Low-quality |
| S14_extracted_k141_3785 | Not-determined |
| S14_extracted_k141_77442 | Not-determined |
| S14_extracted_k141_69925 | Low-quality |
| S14_extracted_k141_77447 | Not-determined |
| S14_extracted_k141_77450 | Not-determined |
| S14_extracted_k141_77456 | Not-determined |
| S14_extracted_k141_55409 | Low-quality |
| S14_extracted_k141_3797 | Not-determined |
| S14_extracted_k141_98981 | Not-determined |
| S14_extracted_k141_84824 | Not-determined |
| S14_extracted_k141_92211 | Low-quality |
| S14_extracted_k141_77457 | Low-quality |
| S14_extracted_k141_84828 | Not-determined |
| S14_extracted_k141_98988 | Low-quality |
| S14_extracted_k141_92215 | Not-determined |
| S14_extracted_k141_106956 | Low-quality |
| S14_extracted_k141_18971 | Not-determined |
| S14_extracted_k141_33314 | Not-determined |
| S14_extracted_k141_106957 | Not-determined |
| S14_extracted_k141_55419 | Low-quality |
| S14_extracted_k141_18976 | Not-determined |
| S14_extracted_k141_33317 | Not-determined |
| S14_extracted_k141_33321 | Not-determined |
| S14_extracted_k141_84836 | Low-quality |
| S14_extracted_k141_55427 | Not-determined |
| S14_extracted_k141_3820 | Not-determined |
| S14_extracted_k141_3824 | Not-determined |
| S14_extracted_k141_77476 | Not-determined |
| S14_extracted_k141_3828 | Not-determined |
| S14_extracted_k141_77479 | Not-determined |
| S14_extracted_k141_55437 | Low-quality |
| S14_extracted_k141_3834 | Not-determined |
| S14_extracted_k141_11409 | Not-determined |
| S14_extracted_k141_84849 | Not-determined |
| S14_extracted_k141_11410 | Not-determined |
| S14_extracted_k141_84851 | Low-quality |
| S14_extracted_k141_92232 | Not-determined |
| S14_extracted_k141_99001 | Not-determined |
| S14_extracted_k141_77487 | Not-determined |
| S14_extracted_k141_55451 | Not-determined |
| S14_extracted_k141_77488 | Not-determined |
| S14_extracted_k141_55454 | Not-determined |
| S14_extracted_k141_84862 | Not-determined |
| S14_extracted_k141_84864 | Not-determined |
| S14_extracted_k141_41134 | Low-quality |
| S14_extracted_k141_55458 | Low-quality |
| S14_extracted_k141_3845 | Not-determined |
| S14_extracted_k141_55460 | Not-determined |
| S14_extracted_k141_99006 | Not-determined |
| S14_extracted_k141_55473 | Not-determined |
| S14_extracted_k141_33345 | Not-determined |
| S14_extracted_k141_3856 | Not-determined |
| S14_extracted_k141_19017 | Low-quality |
| S14_extracted_k141_99014 | Low-quality |
| S14_extracted_k141_69945 | Not-determined |
| S14_extracted_k141_77502 | Not-determined |
| S14_extracted_k141_69949 | Not-determined |
| S14_extracted_k141_77503 | Not-determined |
| S14_extracted_k141_77506 | Not-determined |
| S14_extracted_k141_3867 | Not-determined |
| S14_extracted_k141_69954 | Not-determined |
| S14_extracted_k141_25835 | Low-quality |
| S14_extracted_k141_47586 | Not-determined |
| S14_extracted_k141_25840 | Low-quality |
| S14_extracted_k141_62335 | Low-quality |
| S14_extracted_k141_84879 | Low-quality |
| S14_extracted_k141_84880 | Not-determined |
| S14_extracted_k141_33359 | Not-determined |
| S14_extracted_k141_25850 | Low-quality |
| S14_extracted_k141_62340 | Not-determined |
| S14_extracted_k141_84888 | Not-determined |
| S14_extracted_k141_84889 | Low-quality |
| S14_extracted_k141_47592 | Not-determined |
| S14_extracted_k141_84899 | Not-determined |
| S14_extracted_k141_33365 | Low-quality |
| S14_extracted_k141_55494 | Not-determined |
| S14_extracted_k141_84906 | Low-quality |
| S14_extracted_k141_84907 | Not-determined |
| S14_extracted_k141_33373 | Not-determined |
| S3_vs_k141_16825\|\|full | High-quality |
| S14_extracted_k141_92244 | Not-determined |
| S14_extracted_k141_41141 | Not-determined |
| S14_extracted_k141_47609 | Not-determined |
| S14_extracted_k141_11430 | Low-quality |
| S14_extracted_k141_3903 | Not-determined |
| S14_extracted_k141_55499 | Not-determined |
| S14_extracted_k141_55500 | Low-quality |
| S14_extracted_k141_33386 | Not-determined |
| S14_extracted_k141_3905 | Not-determined |
| S14_extracted_k141_3906 | Low-quality |
| S14_extracted_k141_47615 | Low-quality |
| S14_extracted_k141_99041 | Low-quality |
| S14_extracted_k141_77513 | Not-determined |
| S14_extracted_k141_11431 | Not-determined |
| S14_extracted_k141_77515 | Not-determined |
| S14_extracted_k141_69971 | Not-determined |
| S14_extracted_k141_69974 | Not-determined |
| S14_extracted_k141_77520 | Not-determined |
| S14_extracted_k141_55506 | Not-determined |
| S14_extracted_k141_92253 | Not-determined |
| S14_extracted_k141_3916 | Not-determined |
| S14_extracted_k141_3917 | Low-quality |
| S14_extracted_k141_92256 | Low-quality |
| S14_extracted_k141_19065 | Not-determined |
| S14_extracted_k141_99051 | Not-determined |
| S14_extracted_k141_99052 | Not-determined |
| S14_extracted_k141_92263 | Not-determined |
| S14_extracted_k141_47625 | Not-determined |
| S14_extracted_k141_69979 | Not-determined |
| S14_extracted_k141_114210 | Not-determined |
| S14_extracted_k141_11450 | Low-quality |
| S14_extracted_k141_62367 | Not-determined |
| S14_extracted_k141_47629 | Not-determined |
| S14_extracted_k141_114215 | Low-quality |
| S14_extracted_k141_47631 | Not-determined |
| S14_extracted_k141_62369 | Not-determined |
| S14_extracted_k141_19073 | Not-determined |
| S14_extracted_k141_41150 | Low-quality |
| S14_extracted_k141_62370 | Not-determined |
| S14_extracted_k141_99057 | Not-determined |
| S14_extracted_k141_33410 | Not-determined |
| S14_extracted_k141_19075 | Not-determined |
| S14_extracted_k141_62374 | Low-quality |
| S14_extracted_k141_3936 | Not-determined |
| S14_extracted_k141_41156 | Low-quality |
| S14_extracted_k141_77538 | Not-determined |
| S14_extracted_k141_84937 | Not-determined |
| S14_extracted_k141_41163 | Not-determined |
| S14_extracted_k141_62380 | Not-determined |
| S14_extracted_k141_11470 | Not-determined |
| S14_extracted_k141_19087 | Low-quality |
| S14_extracted_k141_77549 | Not-determined |
| S14_extracted_k141_3944 | Not-determined |
| S14_extracted_k141_62384 | Low-quality |
| S14_extracted_k141_114229 | Low-quality |
| S14_extracted_k141_92270 | Low-quality |
| S14_extracted_k141_3947 | Not-determined |
| S14_extracted_k141_92271 | Low-quality |
| S14_extracted_k141_47656 | Low-quality |
| S14_extracted_k141_114233 | Not-determined |
| S14_extracted_k141_114234 | Low-quality |
| S14_extracted_k141_114235 | Low-quality |
| S14_extracted_k141_84945 | Low-quality |
| S14_extracted_k141_114237 | Low-quality |
| S14_extracted_k141_62395 | Not-determined |
| S14_extracted_k141_77561 | Low-quality |
| S14_extracted_k141_114239 | Not-determined |
| S14_extracted_k141_114243 | Not-determined |
| S14_extracted_k141_92280 | Not-determined |
| S14_extracted_k141_77565 | Low-quality |
| S14_extracted_k141_84954 | Not-determined |
| S14_extracted_k141_92288 | Low-quality |
| S14_extracted_k141_92292 | Not-determined |
| S14_extracted_k141_92293 | Low-quality |
| S14_extracted_k141_19110 | Not-determined |
| S14_extracted_k141_3961 | Low-quality |
| S14_extracted_k141_62405 | Not-determined |
| S14_extracted_k141_62406 | Not-determined |
| S14_extracted_k141_41182 | Not-determined |
| S14_extracted_k141_92298 | Not-determined |
| S14_extracted_k141_41184 | Low-quality |
| S14_extracted_k141_84974 | Not-determined |
| S14_extracted_k141_19125 | Not-determined |
| S14_extracted_k141_84975 | Low-quality |
| S14_extracted_k141_33430 | Not-determined |
| S14_extracted_k141_3966 | Not-determined |
| S14_extracted_k141_41188 | Not-determined |
| S14_extracted_k141_77577 | Not-determined |
| S14_extracted_k141_77578 | Low-quality |
| S14_extracted_k141_62410 | Not-determined |
| S14_extracted_k141_3972 | Not-determined |
| S14_extracted_k141_11491 | Not-determined |
| S14_extracted_k141_41194 | Low-quality |
| S14_extracted_k141_41195 | Not-determined |
| S14_extracted_k141_99089 | Not-determined |
| S14_extracted_k141_84993 | Not-determined |
| S14_extracted_k141_3981 | Low-quality |
| S14_extracted_k141_11495 | Not-determined |
| S14_extracted_k141_3984 | Not-determined |
| S14_extracted_k141_3985 | Low-quality |

| | |
|---|---|
| S14_extracted_k141_70004 | Not-determined |
| S14_extracted_k141_11496 | Not-determined |
| S14_extracted_k141_33433 | Not-determined |
| S14_extracted_k141_70008 | Not-determined |
| S14_extracted_k141_3991 | Not-determined |
| S14_extracted_k141_33436 | Not-determined |
| S14_extracted_k141_70012 | Not-determined |
| S14_extracted_k141_19137 | Not-determined |
| S14_extracted_k141_55525 | Not-determined |
| S14_extracted_k141_3997 | Not-determined |
| S14_extracted_k141_19141 | Low-quality |
| S14_extracted_k141_99104 | Low-quality |
| S14_extracted_k141_70020 | Not-determined |
| S14_extracted_k141_19142 | Low-quality |
| S14_extracted_k141_70021 | Not-determined |
| S14_extracted_k141_41203 | Not-determined |
| S14_extracted_k141_41206 | Not-determined |
| S14_extracted_k141_47678 | Not-determined |
| S14_extracted_k141_41209 | Not-determined |
| S14_extracted_k141_62440 | Low-quality |
| S14_extracted_k141_33443 | Low-quality |
| S14_extracted_k141_99108 | Low-quality |
| S14_extracted_k141_70032 | Not-determined |
| S14_extracted_k141_19148 | Not-determined |
| S14_extracted_k141_92322 | Not-determined |
| S14_extracted_k141_41212 | Not-determined |
| S14_extracted_k141_70036 | Not-determined |
| S14_extracted_k141_55537 | Not-determined |
| S14_extracted_k141_92326 | Not-determined |
| S14_extracted_k141_33447 | Not-determined |
| S14_extracted_k141_47685 | Not-determined |
| S14_extracted_k141_99109 | Not-determined |
| S14_extracted_k141_19161 | Not-determined |
| S14_extracted_k141_55546 | Not-determined |
| S14_extracted_k141_33456 | Not-determined |
| S14_extracted_k141_106994 | Not-determined |
| S14_extracted_k141_106997 | Not-determined |
| S14_extracted_k141_62448 | Not-determined |
| S14_extracted_k141_114276 | Not-determined |
| S14_extracted_k141_114278 | Not-determined |
| S14_extracted_k141_41222 | Low-quality |
| S14_extracted_k141_47693 | Low-quality |
| S14_extracted_k141_47694 | Not-determined |
| S14_extracted_k141_19170 | Not-determined |
| S14_extracted_k141_70045 | Low-quality |
| S14_extracted_k141_41225 | Not-determined |
| S14_extracted_k141_107006 | Not-determined |
| S14_extracted_k141_4002 | Low-quality |
| S14_extracted_k141_85011 | Not-determined |
| S14_extracted_k141_70052 | Not-determined |
| S14_extracted_k141_19181 | Not-determined |
| S14_extracted_k141_47710 | Low-quality |
| S14_extracted_k141_55554 | Not-determined |
| S14_extracted_k141_107011 | Not-determined |
| S14_extracted_k141_55555 | Low-quality |
| S14_extracted_k141_85019 | Not-determined |
| S14_extracted_k141_70066 | Not-determined |
| S14_extracted_k141_70068 | Not-determined |
| S14_extracted_k141_107016 | Not-determined |
| S14_extracted_k141_107026 | Low-quality |
| S14_extracted_k141_62472 | Low-quality |
| S14_extracted_k141_47717 | Not-determined |
| S14_extracted_k141_47718 | Not-determined |
| S14_extracted_k141_47719 | Not-determined |
| S14_extracted_k141_62475 | Not-determined |
| S14_extracted_k141_41238 | Not-determined |
| S14_extracted_k141_85020 | Not-determined |
| S14_extracted_k141_85027 | Not-determined |
| S14_extracted_k141_55570 | Low-quality |
| S14_extracted_k141_19202 | Not-determined |
| S14_extracted_k141_85033 | Not-determined |
| S14_extracted_k141_70082 | Not-determined |
| S14_extracted_k141_85034 | Low-quality |
| S14_extracted_k141_33479 | Low-quality |
| S14_extracted_k141_99117 | Not-determined |
| S14_extracted_k141_4019 | Low-quality |
| S14_extracted_k141_4020 | Not-determined |
| S14_extracted_k141_70085 | Low-quality |
| S14_extracted_k141_55575 | Low-quality |
| S14_extracted_k141_4021 | Low-quality |
| S14_extracted_k141_25872 | Not-determined |
| S14_extracted_k141_70088 | Low-quality |
| S14_extracted_k141_25874 | Not-determined |
| S14_extracted_k141_99123 | Not-determined |
| S14_extracted_k141_25875 | Not-determined |
| S14_extracted_k141_70090 | Not-determined |
| S14_extracted_k141_85048 | Not-determined |
| S14_extracted_k141_62496 | Low-quality |
| S14_extracted_k141_77594 | Not-determined |
| S14_extracted_k141_107042 | Not-determined |
| S14_extracted_k141_92346 | Low-quality |
| S14_extracted_k141_70098 | Not-determined |
| S14_extracted_k141_114309 | Not-determined |
| S14_extracted_k141_33491 | Low-quality |
| S14_extracted_k141_70101 | Low-quality |
| S14_extracted_k141_25883 | Not-determined |
| S14_extracted_k141_114312 | Not-determined |
| S14_extracted_k141_99127 | Not-determined |
| S14_extracted_k141_4029 | Not-determined |
| S14_extracted_k141_41253 | Low-quality |
| S14_extracted_k141_114314 | Low-quality |
| S14_extracted_k141_62508 | Not-determined |
| S14_extracted_k141_114317 | Not-determined |
| S14_extracted_k141_114318 | Low-quality |
| S14_extracted_k141_4031 | Not-determined |
| S14_extracted_k141_85050 | Low-quality |
| S14_extracted_k141_62512 | Not-determined |
| S14_extracted_k141_55584 | Not-determined |
| S14_extracted_k141_77604 | Not-determined |
| S14_extracted_k141_33501 | Not-determined |
| S14_extracted_k141_99133 | Low-quality |
| S14_extracted_k141_55587 | Low-quality |
| S14_extracted_k141_85053 | Low-quality |
| S14_extracted_k141_55589 | Not-determined |
| S14_extracted_k141_77606 | Low-quality |
| S14_extracted_k141_99138 | Not-determined |
| S14_extracted_k141_33506 | Not-determined |
| S14_extracted_k141_114321 | Not-determined |
| S14_extracted_k141_77610 | Not-determined |
| S14_extracted_k141_77612 | Not-determined |
| S14_extracted_k141_19234 | Not-determined |
| S14_extracted_k141_11514 | Not-determined |
| S14_extracted_k141_107044 | Low-quality |
| S14_extracted_k141_62524 | Not-determined |
| S14_extracted_k141_62525 | Not-determined |
| S14_extracted_k141_62526 | Low-quality |
| S14_extracted_k141_11517 | Not-determined |
| S14_extracted_k141_70115 | Not-determined |
| S14_extracted_k141_33513 | Not-determined |
| S14_extracted_k141_11518 | Not-determined |
| S14_extracted_k141_114327 | Not-determined |
| S14_extracted_k141_107046 | Low-quality |
| S14_extracted_k141_99145 | Low-quality |
| S14_extracted_k141_70116 | Not-determined |
| S14_extracted_k141_11520 | Not-determined |
| S14_extracted_k141_70118 | Low-quality |
| S14_extracted_k141_62532 | Low-quality |
| S14_extracted_k141_77618 | Low-quality |
| S14_extracted_k141_41281 | Not-determined |
| S14_extracted_k141_70125 | Low-quality |
| S14_extracted_k141_77621 | Not-determined |
| S14_extracted_k141_77622 | Low-quality |
| S14_extracted_k141_55595 | Not-determined |
| S14_extracted_k141_114328 | Not-determined |
| S14_extracted_k141_62537 | Not-determined |
| S14_extracted_k141_85063 | Not-determined |
| S14_extracted_k141_11527 | Low-quality |
| S14_extracted_k141_114334 | Not-determined |
| S14_extracted_k141_4043 | Not-determined |
| S14_extracted_k141_11532 | Not-determined |
| S14_extracted_k141_77630 | Not-determined |
| S14_extracted_k141_85066 | Not-determined |
| S14_extracted_k141_41297 | Not-determined |
| S14_extracted_k141_4049 | Not-determined |
| S14_extracted_k141_41298 | Not-determined |
| S14_extracted_k141_85070 | Not-determined |
| S14_extracted_k141_33527 | Not-determined |
| S14_extracted_k141_92354 | Not-determined |
| S14_extracted_k141_19248 | Not-determined |
| S14_extracted_k141_4057 | Not-determined |
| S14_extracted_k141_47748 | Not-determined |
| S14_extracted_k141_41307 | Low-quality |
| S14_extracted_k141_19251 | Not-determined |
| S14_extracted_k141_114346 | Low-quality |
| S14_extracted_k141_114347 | Not-determined |
| S14_extracted_k141_19255 | Not-determined |
| S14_extracted_k141_92362 | Not-determined |
| S14_extracted_k141_55614 | Not-determined |
| S14_extracted_k141_41317 | Not-determined |
| S14_extracted_k141_55618 | Not-determined |
| S14_extracted_k141_41321 | Not-determined |
| S14_extracted_k141_19265 | Not-determined |
| S14_extracted_k141_25889 | Low-quality |
| S14_extracted_k141_19266 | Not-determined |
| S14_extracted_k141_19267 | Not-determined |
| S14_extracted_k141_107068 | Not-determined |
| S14_extracted_k141_114360 | Not-determined |
| S14_extracted_k141_55623 | Low-quality |
| S14_extracted_k141_55624 | Not-determined |
| S14_extracted_k141_55626 | Not-determined |
| S14_extracted_k141_70136 | Not-determined |
| S14_extracted_k141_55628 | Not-determined |
| S14_extracted_k141_99169 | Not-determined |
| S14_extracted_k141_4076 | Not-determined |
| S14_extracted_k141_70140 | Not-determined |
| S14_extracted_k141_55634 | Low-quality |
| S14_extracted_k141_114362 | Low-quality |
| S14_extracted_k141_107071 | Low-quality |
| S14_extracted_k141_99174 | Not-determined |
| S14_extracted_k141_41335 | Not-determined |
| S14_extracted_k141_107073 | Not-determined |
| S14_extracted_k141_70148 | Not-determined |
| S14_extracted_k141_99179 | Low-quality |
| S14_extracted_k141_33539 | Not-determined |
| S14_extracted_k141_99180 | Low-quality |
| S14_extracted_k141_99181 | Not-determined |
| S14_extracted_k141_77655 | Low-quality |
| S14_extracted_k141_70153 | Not-determined |
| S14_extracted_k141_99183 | Low-quality |
| S14_extracted_k141_99184 | Not-determined |
| S14_extracted_k141_25899 | Not-determined |
| S14_extracted_k141_47760 | Not-determined |
| S14_extracted_k141_77663 | Low-quality |
| S14_extracted_k141_4087 | Not-determined |
| S14_extracted_k141_19290 | Not-determined |
| S14_extracted_k141_25909 | Not-determined |
| S14_extracted_k141_33542 | Not-determined |
| S14_extracted_k141_25910 | Not-determined |
| S14_extracted_k141_55645 | Not-determined |
| S14_extracted_k141_77665 | Not-determined |
| S14_extracted_k141_11557 | Low-quality |
| S14_extracted_k141_11559 | Not-determined |
| S14_extracted_k141_4092 | Low-quality |
| S14_extracted_k141_107096 | Low-quality |
| S14_extracted_k141_11562 | Not-determined |
| S14_extracted_k141_107099 | Low-quality |
| S14_extracted_k141_77669 | Not-determined |
| S14_extracted_k141_107101 | Not-determined |
| S14_extracted_k141_70170 | Low-quality |
| S14_extracted_k141_11567 | Low-quality |
| S14_extracted_k141_85095 | Not-determined |
| S14_extracted_k141_70174 | Low-quality |
| S14_extracted_k141_19299 | Low-quality |
| S14_extracted_k141_70177 | Low-quality |
| S14_extracted_k141_70180 | Low-quality |
| S14_extracted_k141_70181 | Low-quality |
| S14_extracted_k141_47781 | Not-determined |
| S14_extracted_k141_85101 | Not-determined |
| S14_extracted_k141_85102 | Not-determined |
| S14_extracted_k141_11574 | Low-quality |
| S14_extracted_k141_55652 | Not-determined |
| S14_extracted_k141_19309 | Not-determined |
| S14_extracted_k141_19311 | Not-determined |
| S14_extracted_k141_99201 | Not-determined |
| S14_extracted_k141_4104 | Low-quality |
| S14_extracted_k141_107109 | Low-quality |
| S14_extracted_k141_19315 | Not-determined |
| S14_extracted_k141_33556 | Not-determined |
| S14_extracted_k141_77679 | Not-determined |
| S14_extracted_k141_25913 | Not-determined |
| S14_extracted_k141_33559 | Not-determined |
| S14_extracted_k141_107112 | Not-determined |
| S14_extracted_k141_19320 | Not-determined |
| S14_extracted_k141_107113 | Low-quality |
| S14_extracted_k141_33563 | Low-quality |
| S14_extracted_k141_62570 | Low-quality |
| S14_extracted_k141_85112 | Low-quality |
| S14_extracted_k141_107119 | Not-determined |
| S14_extracted_k141_11594 | Low-quality |
| S14_extracted_k141_33570 | Not-determined |
| S14_extracted_k141_107120 | Not-determined |
| S14_extracted_k141_55662 | Low-quality |
| S14_extracted_k141_11596 | Low-quality |
| S14_extracted_k141_99204 | Low-quality |
| S14_extracted_k141_55663 | Not-determined |
| S14_extracted_k141_77688 | Not-determined |
| S14_extracted_k141_25923 | Not-determined |
| S14_extracted_k141_70199 | Low-quality |
| S14_extracted_k141_77693 | Low-quality |
| S14_extracted_k141_11600 | Not-determined |
| S14_extracted_k141_55668 | Not-determined |
| S14_extracted_k141_11602 | Not-determined |
| S14_extracted_k141_70201 | Not-determined |
| S14_extracted_k141_99208 | Not-determined |
| S14_extracted_k141_107130 | Not-determined |
| S14_extracted_k141_11611 | Low-quality |
| S14_extracted_k141_107136 | Low-quality |
| S14_extracted_k141_62589 | Not-determined |
| S14_extracted_k141_19341 | Low-quality |
| S14_extracted_k141_47784 | Low-quality |
| S14_extracted_k141_19342 | Low-quality |
| S14_extracted_k141_41355 | Not-determined |
| S14_extracted_k141_92417 | Not-determined |
| S14_extracted_k141_19344 | Not-determined |
| S14_extracted_k141_70209 | Not-determined |
| S14_extracted_k141_47787 | Not-determined |
| S14_extracted_k141_19349 | Not-determined |
| S14_extracted_k141_70216 | Not-determined |
| S14_extracted_k141_114373 | Not-determined |
| S14_extracted_k141_55684 | Not-determined |
| S14_extracted_k141_62599 | Not-determined |
| S14_extracted_k141_19354 | Low-quality |
| S14_extracted_k141_62600 | Not-determined |
| S14_extracted_k141_77711 | Not-determined |
| S14_extracted_k141_41365 | Low-quality |
| S14_extracted_k141_70225 | Not-determined |
| S14_extracted_k141_11618 | Low-quality |
| S14_extracted_k141_70226 | Not-determined |
| S14_extracted_k141_92434 | Not-determined |
| S14_extracted_k141_11625 | Low-quality |
| S14_extracted_k141_19364 | Not-determined |
| S14_extracted_k141_107159 | Low-quality |
| S14_extracted_k141_62614 | Not-determined |
| S14_extracted_k141_41367 | Low-quality |
| S14_extracted_k141_55690 | Not-determined |
| S14_extracted_k141_55692 | Not-determined |
| S14_extracted_k141_25939 | Not-determined |
| S14_extracted_k141_19374 | Low-quality |
| S14_extracted_k141_114387 | Not-determined |
| S14_extracted_k141_11631 | Not-determined |
| S14_extracted_k141_41379 | Not-determined |
| S14_extracted_k141_19378 | Low-quality |
| S14_extracted_k141_114393 | Not-determined |
| S14_extracted_k141_19381 | Low-quality |
| S14_extracted_k141_25949 | Low-quality |
| S14_extracted_k141_92446 | Low-quality |
| S14_extracted_k141_99251 | Not-determined |
| S14_extracted_k141_99255 | Not-determined |
| S14_extracted_k141_19383 | Not-determined |
| S14_extracted_k141_62630 | Low-quality |
| S14_extracted_k141_99257 | Low-quality |
| S14_extracted_k141_19388 | Not-determined |
| S14_extracted_k141_19389 | Low-quality |
| S14_extracted_k141_25961 | Low-quality |
| S14_extracted_k141_62634 | Not-determined |
| S14_extracted_k141_117157 | Complete |
| S14_extracted_k141_11633 | Low-quality |
| S14_extracted_k141_19395 | Not-determined |
| S14_extracted_k141_11634 | Not-determined |
| S14_extracted_k141_41391 | Not-determined |
| S14_extracted_k141_11638 | Not-determined |
| S14_extracted_k141_62647 | Not-determined |
| S14_extracted_k141_62648 | Low-quality |
| S14_extracted_k141_33594 | Not-determined |
| S14_extracted_k141_77729 | Low-quality |
| S14_extracted_k141_107177 | Not-determined |
| S14_extracted_k141_85136 | Not-determined |
| S14_extracted_k141_41394 | Not-determined |
| S14_extracted_k141_25977 | Low-quality |
| S14_extracted_k141_55716 | Not-determined |
| S14_extracted_k141_55717 | Not-determined |
| S14_extracted_k141_62657 | Low-quality |
| S14_extracted_k141_19415 | Not-determined |
| S14_extracted_k141_55719 | Not-determined |
| S14_extracted_k141_41404 | Not-determined |
| S14_extracted_k141_62666 | Not-determined |
| S14_extracted_k141_92471 | Not-determined |
| S14_extracted_k141_77746 | Low-quality |
| S14_extracted_k141_92474 | Low-quality |
| S14_extracted_k141_62668 | Not-determined |
| S14_extracted_k141_62671 | Not-determined |
| S14_extracted_k141_99297 | Not-determined |
| S14_extracted_k141_114417 | Low-quality |
| S14_extracted_k141_77751 | Not-determined |
| S14_extracted_k141_41409 | Not-determined |
| S14_extracted_k141_41410 | Not-determined |
| S14_extracted_k141_41411 | Not-determined |
| S14_extracted_k141_62679 | Not-determined |
| S14_extracted_k141_62680 | Not-determined |
| S14_extracted_k141_41415 | Not-determined |
| S14_extracted_k141_77756 | Not-determined |
| S14_extracted_k141_41418 | Not-determined |
| S14_extracted_k141_92491 | Not-determined |
| S14_extracted_k141_92493 | Not-determined |
| S14_extracted_k141_77761 | Not-determined |
| S14_extracted_k141_62687 | Not-determined |
| S14_extracted_k141_62688 | Not-determined |
| S14_extracted_k141_107199 | Low-quality |
| S14_extracted_k141_62692 | Low-quality |
| S14_extracted_k141_107203 | Not-determined |
| S14_extracted_k141_92497 | Not-determined |
| S14_extracted_k141_62696 | Low-quality |
| S14_extracted_k141_92498 | Not-determined |
| S14_extracted_k141_92500 | Not-determined |
| S14_extracted_k141_41423 | Not-determined |
| S14_extracted_k141_41424 | Not-determined |
| S14_extracted_k141_41425 | Not-determined |
| S14_extracted_k141_107211 | Not-determined |
| S14_extracted_k141_47823 | Not-determined |
| S14_extracted_k141_99311 | Not-determined |
| S14_extracted_k141_33615 | Not-determined |
| S14_extracted_k141_47826 | Low-quality |
| S14_extracted_k141_55738 | Low-quality |
| S14_extracted_k141_55739 | Low-quality |
| S14_extracted_k141_77771 | Not-determined |
| S14_extracted_k141_25994 | Not-determined |
| S14_extracted_k141_114436 | Low-quality |
| S14_extracted_k141_114437 | Not-determined |
| S14_extracted_k141_41427 | Not-determined |
| S14_extracted_k141_107213 | Low-quality |
| S14_extracted_k141_19433 | Not-determined |
| S14_extracted_k141_99322 | Low-quality |
| S14_extracted_k141_85164 | Not-determined |
| S14_extracted_k141_99324 | Low-quality |
| S14_extracted_k141_92508 | Low-quality |
| S14_extracted_k141_41435 | Low-quality |
| S14_extracted_k141_99329 | Low-quality |
| S14_extracted_k141_19444 | Not-determined |
| S14_extracted_k141_47842 | Not-determined |
| S14_extracted_k141_19449 | Not-determined |
| S14_extracted_k141_114445 | Low-quality |
| S14_extracted_k141_19457 | Not-determined |
| S14_extracted_k141_114449 | Not-determined |
| S14_extracted_k141_62705 | Not-determined |
| S14_extracted_k141_107232 | Not-determined |
| S14_extracted_k141_19467 | Low-quality |
| S14_extracted_k141_41445 | Not-determined |
| S14_extracted_k141_33631 | Low-quality |
| S14_extracted_k141_33633 | Low-quality |
| S14_extracted_k141_107241 | Not-determined |
| S14_extracted_k141_77814 | Low-quality |
| S14_extracted_k141_41455 | Not-determined |
| S14_extracted_k141_70237 | Low-quality |
| S14_extracted_k141_77817 | Not-determined |
| S14_extracted_k141_77818 | Low-quality |
| S14_extracted_k141_19481 | Low-quality |
| S14_extracted_k141_33639 | Not-determined |
| S14_extracted_k141_33640 | Low-quality |
| S14_extracted_k141_41461 | Low-quality |
| S14_extracted_k141_114459 | Not-determined |
| S14_extracted_k141_77825 | Not-determined |
| S14_extracted_k141_85172 | Low-quality |
| S14_extracted_k141_26000 | Not-determined |
| S14_extracted_k141_85175 | Not-determined |
| S14_extracted_k141_33647 | Not-determined |
| S14_extracted_k141_41464 | Not-determined |
| S14_extracted_k141_114465 | Not-determined |
| S14_extracted_k141_41468 | Low-quality |
| S14_extracted_k141_92524 | Not-determined |
| S14_extracted_k141_26004 | Not-determined |
| S14_extracted_k141_85179 | Low-quality |
| S14_extracted_k141_114467 | Not-determined |
| S14_extracted_k141_62723 | Not-determined |
| S14_extracted_k141_92527 | Low-quality |
| S14_extracted_k141_107257 | Low-quality |
| S14_extracted_k141_77831 | Not-determined |
| S14_extracted_k141_77832 | Not-determined |
| S14_extracted_k141_92532 | Low-quality |
| S14_extracted_k141_114476 | Not-determined |
| S14_extracted_k141_26014 | Not-determined |
| S14_extracted_k141_41472 | Low-quality |
| S14_extracted_k141_19498 | Not-determined |
| S14_extracted_k141_70248 | Not-determined |
| S11_extracted_k141_43997 | Medium-quality |
| S14_extracted_k141_26016 | Not-determined |
| S14_extracted_k141_114481 | Low-quality |
| S14_extracted_k141_19500 | Not-determined |
| S14_extracted_k141_62732 | Low-quality |
| S14_extracted_k141_62733 | Not-determined |
| S14_extracted_k141_19502 | Not-determined |
| S14_extracted_k141_99353 | Not-determined |
| S14_extracted_k141_26027 | Not-determined |
| S14_extracted_k141_55766 | Not-determined |
| S14_extracted_k141_107260 | Not-determined |
| S14_extracted_k141_19509 | Not-determined |
| S14_extracted_k141_70253 | Not-determined |
| S14_extracted_k141_70254 | Low-quality |
| S14_extracted_k141_99361 | Not-determined |
| S14_extracted_k141_4152 | Not-determined |
| S14_extracted_k141_107263 | Low-quality |
| S14_extracted_k141_55770 | Not-determined |
| S14_extracted_k141_55772 | Low-quality |
| S14_extracted_k141_4157 | Low-quality |
| S14_extracted_k141_70261 | Not-determined |
| S14_extracted_k141_85184 | Not-determined |
| S14_extracted_k141_70263 | Not-determined |
| S14_extracted_k141_41489 | Not-determined |
| S14_extracted_k141_26042 | Not-determined |
| S14_extracted_k141_85189 | Not-determined |
| S14_extracted_k141_55786 | Low-quality |
| S14_extracted_k141_77853 | Low-quality |
| S14_extracted_k141_99366 | Not-determined |
| S14_extracted_k141_41494 | Not-determined |
| S14_extracted_k141_4168 | Not-determined |
| S14_extracted_k141_62737 | Not-determined |
| S14_extracted_k141_77859 | Not-determined |
| S14_extracted_k141_26050 | Low-quality |
| S14_extracted_k141_19526 | Not-determined |
| S14_extracted_k141_26052 | Low-quality |
| S14_extracted_k141_62742 | Not-determined |
| S14_extracted_k141_41504 | Not-determined |
| S14_extracted_k141_55796 | Low-quality |
| S14_extracted_k141_41506 | Not-determined |
| S14_extracted_k141_77866 | Low-quality |
| S14_extracted_k141_4180 | Low-quality |
| S14_extracted_k141_26056 | Not-determined |
| S14_extracted_k141_41510 | Not-determined |
| S14_extracted_k141_92551 | Not-determined |
| S14_extracted_k141_92552 | Low-quality |
| S14_extracted_k141_70279 | Low-quality |
| S14_extracted_k141_41512 | Not-determined |
| S14_extracted_k141_92553 | Low-quality |
| S14_extracted_k141_26063 | Not-determined |
| S14_extracted_k141_4186 | Low-quality |
| S14_extracted_k141_55807 | Not-determined |
| S14_extracted_k141_92555 | Low-quality |
| S14_extracted_k141_62746 | Not-determined |
| S14_extracted_k141_70282 | Low-quality |
| S14_extracted_k141_77874 | Not-determined |
| S14_extracted_k141_4188 | Not-determined |
| S14_extracted_k141_77875 | Not-determined |
| S11_vs_k141_43997\|\|full | Medium-quality |
| S14_extracted_k141_107280 | Not-determined |
| S14_extracted_k141_107281 | Not-determined |
| S14_extracted_k141_62750 | Low-quality |
| S14_extracted_k141_55813 | Low-quality |
| S14_extracted_k141_99394 | Not-determined |
| S14_extracted_k141_85206 | Low-quality |
| S14_extracted_k141_62754 | Low-quality |
| S14_extracted_k141_107286 | Not-determined |
| S14_extracted_k141_62756 | Low-quality |
| S14_extracted_k141_41517 | Not-determined |
| S14_extracted_k141_62758 | Low-quality |
| S14_extracted_k141_77884 | Not-determined |
| S14_extracted_k141_55819 | Not-determined |
| S14_extracted_k141_41520 | Not-determined |
| S14_extracted_k141_41521 | Not-determined |
| S14_extracted_k141_19547 | Low-quality |
| S14_extracted_k141_55822 | Not-determined |
| S14_extracted_k141_55823 | Not-determined |
| S14_extracted_k141_107294 | Low-quality |
| S14_extracted_k141_85213 | Not-determined |
| S14_extracted_k141_85215 | Not-determined |
| S14_extracted_k141_114498 | Low-quality |
| S14_extracted_k141_62767 | Low-quality |
| S14_extracted_k141_99403 | Not-determined |
| S14_extracted_k141_70313 | Low-quality |
| S14_extracted_k141_99406 | Low-quality |
| S14_extracted_k141_70315 | Not-determined |
| S14_extracted_k141_19557 | Not-determined |
| S14_extracted_k141_4196 | Not-determined |
| S14_extracted_k141_4197 | Not-determined |
| S14_extracted_k141_92574 | Not-determined |
| S14_extracted_k141_26072 | Not-determined |
| S14_extracted_k141_26074 | Not-determined |
| S14_extracted_k141_4202 | Not-determined |
| S14_extracted_k141_26077 | Low-quality |
| S14_extracted_k141_85225 | Not-determined |
| S14_extracted_k141_70320 | Not-determined |
| S14_extracted_k141_99411 | Not-determined |
| S14_extracted_k141_114516 | Not-determined |
| S14_extracted_k141_107310 | Not-determined |
| S14_extracted_k141_85233 | Not-determined |
| S14_extracted_k141_70328 | Low-quality |
| S14_extracted_k141_85242 | Not-determined |
| S14_extracted_k141_26085 | Not-determined |
| S14_extracted_k141_4220 | Not-determined |
| S14_extracted_k141_55853 | Not-determined |
| S14_extracted_k141_99421 | Low-quality |
| S14_extracted_k141_92591 | Low-quality |
| S14_extracted_k141_85246 | Not-determined |
| S14_extracted_k141_99424 | Not-determined |
| S14_extracted_k141_4224 | Not-determined |
| S14_extracted_k141_114524 | Not-determined |
| S14_extracted_k141_55859 | Not-determined |
| S14_extracted_k141_26100 | Not-determined |
| S14_extracted_k141_92593 | Not-determined |
| S14_extracted_k141_114530 | Low-quality |
| S14_extracted_k141_4234 | Not-determined |
| S14_extracted_k141_99426 | Low-quality |
| S14_extracted_k141_4235 | Not-determined |
| S14_extracted_k141_114535 | Not-determined |
| S14_extracted_k141_85256 | Not-determined |
| S14_extracted_k141_85257 | Low-quality |
| S14_extracted_k141_114537 | Low-quality |
| S14_extracted_k141_26110 | Not-determined |
| S14_extracted_k141_114538 | Not-determined |
| S14_extracted_k141_70345 | Not-determined |
| S14_extracted_k141_4237 | Not-determined |
| S14_extracted_k141_85261 | Low-quality |
| S14_extracted_k141_70348 | Low-quality |
| S14_extracted_k141_26116 | Not-determined |
| S14_extracted_k141_99431 | Not-determined |
| S14_extracted_k141_77896 | Low-quality |
| S14_extracted_k141_70352 | Not-determined |
| S14_extracted_k141_77898 | Not-determined |
| S14_extracted_k141_4242 | Not-determined |
| S14_extracted_k141_99435 | Low-quality |
| S14_extracted_k141_62784 | Low-quality |
| S14_extracted_k141_99438 | Not-determined |
| S14_extracted_k141_62787 | Not-determined |
| S14_extracted_k141_99439 | Not-determined |
| S14_extracted_k141_107330 | Not-determined |
| S14_extracted_k141_99442 | Not-determined |
| S14_extracted_k141_99444 | Not-determined |
| S14_extracted_k141_47856 | Not-determined |
| S14_extracted_k141_4255 | Not-determined |
| S14_extracted_k141_4259 | Low-quality |
| S14_extracted_k141_92622 | Not-determined |
| S14_extracted_k141_47860 | Low-quality |
| S14_extracted_k141_47861 | Low-quality |
| S14_extracted_k141_92633 | Not-determined |
| S14_extracted_k141_70368 | Not-determined |
| S14_extracted_k141_70369 | Low-quality |
| S14_extracted_k141_70373 | Low-quality |
| S14_extracted_k141_114547 | Not-determined |
| S14_extracted_k141_114548 | Not-determined |
| S14_extracted_k141_47872 | Not-determined |
| S14_extracted_k141_114549 | Low-quality |

| | |
|---|---|
| S14_extracted_k141_92644 | Not-determined |
| S14_extracted_k141_107348 | Low-quality |
| S14_extracted_k141_114552 | Not-determined |
| S14_extracted_k141_55876 | Not-determined |
| S14_extracted_k141_19580 | Not-determined |
| S14_extracted_k141_77937 | Low-quality |
| S14_extracted_k141_33661 | Not-determined |
| S14_extracted_k141_55883 | Not-determined |
| S14_extracted_k141_114563 | Not-determined |
| S14_extracted_k141_62810 | Low-quality |
| S14_extracted_k141_77940 | Not-determined |
| S14_extracted_k141_70379 | Low-quality |
| S14_extracted_k141_77942 | Not-determined |
| S14_extracted_k141_77943 | Low-quality |
| S14_extracted_k141_92660 | Not-determined |
| S14_extracted_k141_62815 | Not-determined |
| S14_extracted_k141_19582 | Not-determined |
| S14_extracted_k141_92661 | Low-quality |
| S14_extracted_k141_70387 | Not-determined |
| S14_extracted_k141_55885 | Not-determined |
| S14_extracted_k141_62820 | Low-quality |
| S14_extracted_k141_92662 | Low-quality |
| S14_extracted_k141_62823 | Not-determined |
| S14_extracted_k141_92663 | Low-quality |
| S14_extracted_k141_26121 | Not-determined |
| S14_extracted_k141_85299 | Not-determined |
| S14_extracted_k141_70399 | Not-determined |
| S14_extracted_k141_26123 | Not-determined |
| S14_extracted_k141_4266 | Not-determined |
| S14_extracted_k141_107359 | Not-determined |
| S14_extracted_k141_114581 | Not-determined |
| S14_extracted_k141_26125 | Low-quality |
| S14_extracted_k141_62830 | Low-quality |
| S14_extracted_k141_4269 | Not-determined |
| S14_extracted_k141_99460 | Not-determined |
| S14_extracted_k141_99462 | Not-determined |
| S14_extracted_k141_114583 | Not-determined |
| S14_extracted_k141_62833 | Low-quality |
| S14_extracted_k141_99463 | Low-quality |
| S14_extracted_k141_107369 | Not-determined |
| S14_extracted_k141_99465 | Low-quality |
| S14_extracted_k141_114586 | Not-determined |
| S14_extracted_k141_26132 | Not-determined |
| S14_extracted_k141_99467 | Low-quality |
| S14_extracted_k141_55895 | Low-quality |
| S14_extracted_k141_77965 | Low-quality |
| S14_extracted_k141_107381 | Not-determined |
| S14_extracted_k141_47883 | Not-determined |
| S14_extracted_k141_85310 | Not-determined |
| S14_extracted_k141_47890 | Low-quality |
| S14_extracted_k141_114594 | Not-determined |
| S14_extracted_k141_114595 | Not-determined |
| S14_extracted_k141_114598 | Low-quality |
| S14_extracted_k141_19599 | Low-quality |
| S14_extracted_k141_4280 | Not-determined |
| S14_extracted_k141_19602 | Not-determined |
| S14_extracted_k141_19605 | Not-determined |
| S14_extracted_k141_47898 | Low-quality |
| S14_extracted_k141_85322 | Not-determined |
| S14_extracted_k141_26137 | Not-determined |
| S14_extracted_k141_85325 | Low-quality |
| S14_extracted_k141_92670 | Low-quality |
| S14_extracted_k141_70435 | Not-determined |
| S14_extracted_k141_107398 | Not-determined |
| S14_extracted_k141_47902 | Low-quality |
| S14_extracted_k141_114616 | Not-determined |
| S14_extracted_k141_4297 | Low-quality |
| S14_extracted_k141_99472 | Not-determined |
| S14_extracted_k141_107410 | Not-determined |
| S14_extracted_k141_99476 | Low-quality |
| S14_extracted_k141_55902 | Not-determined |
| S14_extracted_k141_47918 | Not-determined |
| S14_extracted_k141_62874 | Low-quality |
| S14_extracted_k141_99485 | Not-determined |
| S14_extracted_k141_92692 | Not-determined |
| S14_extracted_k141_11647 | Not-determined |
| S14_extracted_k141_85335 | Low-quality |
| S14_extracted_k141_92693 | Not-determined |
| S14_extracted_k141_11650 | Low-quality |
| S14_extracted_k141_107413 | Low-quality |
| S14_extracted_k141_11651 | Low-quality |
| S14_extracted_k141_33677 | Not-determined |
| S14_extracted_k141_85338 | Not-determined |
| S14_extracted_k141_62880 | Not-determined |
| S14_extracted_k141_85340 | Not-determined |
| S14_extracted_k141_26148 | Not-determined |
| S14_extracted_k141_26152 | Low-quality |
| S14_extracted_k141_107419 | Not-determined |
| S14_extracted_k141_33685 | Not-determined |
| S14_extracted_k141_85346 | Not-determined |
| S14_extracted_k141_26154 | Not-determined |
| S14_extracted_k141_70454 | Not-determined |
| S14_extracted_k141_19611 | Not-determined |
| S14_extracted_k141_85349 | Low-quality |
| S14_extracted_k141_85350 | Low-quality |
| S14_extracted_k141_114634 | Not-determined |
| S14_extracted_k141_114636 | Not-determined |
| S14_extracted_k141_33689 | Not-determined |
| S14_extracted_k141_70457 | Low-quality |
| S14_extracted_k141_70458 | Not-determined |
| S14_extracted_k141_70460 | Not-determined |
| S14_extracted_k141_11668 | Not-determined |
| S14_extracted_k141_55925 | Low-quality |
| S14_extracted_k141_41592 | Not-determined |
| S14_extracted_k141_47942 | Not-determined |
| S14_extracted_k141_92711 | Not-determined |
| S14_extracted_k141_4300 | Not-determined |
| S14_extracted_k141_70466 | Low-quality |
| S14_extracted_k141_77989 | Not-determined |
| S14_extracted_k141_92714 | Not-determined |
| S14_extracted_k141_70470 | Low-quality |
| S14_extracted_k141_70472 | Not-determined |
| S14_extracted_k141_11675 | Not-determined |
| S14_extracted_k141_4306 | Not-determined |
| S14_extracted_k141_77996 | Not-determined |
| S14_extracted_k141_4309 | Low-quality |
| S14_extracted_k141_4311 | Low-quality |
| S14_extracted_k141_55939 | Not-determined |
| S14_extracted_k141_55940 | Low-quality |
| S14_extracted_k141_92731 | Not-determined |
| S8_vs_k141_170878\|\|full | High-quality |
| S14_extracted_k141_11685 | Not-determined |
| S14_extracted_k141_4320 | Not-determined |
| S14_extracted_k141_70489 | Not-determined |
| S14_extracted_k141_70490 | Not-determined |
| S14_extracted_k141_4326 | Low-quality |
| S14_extracted_k141_114651 | Not-determined |
| S14_extracted_k141_55947 | Low-quality |
| S14_extracted_k141_4328 | Not-determined |
| S14_extracted_k141_33698 | Not-determined |
| S14_extracted_k141_19620 | Not-determined |
| S14_extracted_k141_41607 | Not-determined |
| S14_extracted_k141_99523 | Not-determined |
| S14_extracted_k141_47948 | Low-quality |
| S14_extracted_k141_33706 | Not-determined |
| S14_extracted_k141_78021 | Not-determined |
| S14_extracted_k141_33708 | Low-quality |
| S14_extracted_k141_92751 | Not-determined |
| S14_extracted_k141_70506 | Not-determined |
| S14_extracted_k141_47955 | Not-determined |
| S14_extracted_k141_4344 | Low-quality |
| S14_extracted_k141_11693 | Not-determined |
| S14_extracted_k141_41625 | Not-determined |
| S14_extracted_k141_107449 | Not-determined |
| S14_extracted_k141_4348 | Not-determined |
| S14_extracted_k141_11695 | Low-quality |
| S14_extracted_k141_107452 | Low-quality |
| S14_extracted_k141_4350 | Low-quality |
| S14_extracted_k141_107454 | Not-determined |
| S14_extracted_k141_70514 | Low-quality |
| S14_extracted_k141_4353 | Low-quality |
| S14_extracted_k141_4357 | Low-quality |
| S14_extracted_k141_114659 | Not-determined |
| S14_extracted_k141_41639 | Not-determined |
| S14_extracted_k141_33714 | Low-quality |
| S14_extracted_k141_4369 | Low-quality |
| S14_extracted_k141_85361 | Not-determined |
| S14_extracted_k141_85363 | Not-determined |
| S14_extracted_k141_47957 | Low-quality |
| S14_extracted_k141_4375 | Not-determined |
| S14_extracted_k141_4378 | Not-determined |
| S14_extracted_k141_4379 | Low-quality |
| S14_extracted_k141_19635 | Not-determined |
| S14_extracted_k141_4382 | Not-determined |
| S14_extracted_k141_19637 | Not-determined |
| S14_extracted_k141_47961 | Not-determined |
| S14_extracted_k141_4387 | Low-quality |
| S14_extracted_k141_85368 | Low-quality |
| S14_extracted_k141_85369 | Not-determined |
| S14_extracted_k141_4390 | Not-determined |
| S14_extracted_k141_33724 | Not-determined |
| S14_extracted_k141_4391 | Not-determined |
| S14_extracted_k141_4393 | Low-quality |
| S14_extracted_k141_114679 | Low-quality |
| S14_extracted_k141_4396 | Not-determined |
| S14_extracted_k141_114681 | Not-determined |
| S14_extracted_k141_33731 | Not-determined |
| S14_extracted_k141_19642 | Not-determined |
| S14_extracted_k141_114686 | Not-determined |
| S14_extracted_k141_33740 | Not-determined |
| S14_extracted_k141_33741 | Low-quality |
| S14_extracted_k141_92790 | Not-determined |
| S14_extracted_k141_47970 | Not-determined |
| S14_extracted_k141_114697 | Not-determined |
| S14_extracted_k141_47974 | Not-determined |
| S14_extracted_k141_26166 | Not-determined |
| S14_extracted_k141_78036 | Low-quality |
| S14_extracted_k141_85380 | Not-determined |
| S14_extracted_k141_41663 | Low-quality |
| S14_extracted_k141_78039 | Not-determined |
| S14_extracted_k141_70547 | Not-determined |
| S14_extracted_k141_92803 | Low-quality |
| S14_extracted_k141_85384 | Not-determined |
| S14_extracted_k141_85387 | Low-quality |
| S14_extracted_k141_62947 | Not-determined |
| S14_extracted_k141_4418 | Not-determined |
| S14_extracted_k141_92815 | Not-determined |
| S14_extracted_k141_62957 | Not-determined |
| S14_extracted_k141_99561 | Not-determined |
| S14_extracted_k141_19668 | Not-determined |
| S14_extracted_k141_99565 | Not-determined |
| S14_extracted_k141_107479 | Low-quality |
| S14_extracted_k141_107480 | Not-determined |
| S14_extracted_k141_62963 | Not-determined |
| S14_extracted_k141_19677 | Not-determined |
| S14_extracted_k141_41689 | Low-quality |
| S14_extracted_k141_107484 | Not-determined |
| S14_extracted_k141_70559 | Low-quality |
| S14_extracted_k141_70560 | Not-determined |
| S14_extracted_k141_19679 | Not-determined |
| S14_extracted_k141_70561 | Not-determined |
| S14_extracted_k141_4443 | Not-determined |
| S14_extracted_k141_19683 | Not-determined |
| S14_extracted_k141_19684 | Low-quality |
| S14_extracted_k141_70566 | Low-quality |
| S14_extracted_k141_85404 | Not-determined |
| S14_extracted_k141_55973 | Low-quality |
| S14_extracted_k141_55974 | Not-determined |
| S14_extracted_k141_19688 | Low-quality |
| S14_extracted_k141_48005 | Low-quality |
| S14_extracted_k141_107490 | Not-determined |
| S14_extracted_k141_48006 | Not-determined |
| S14_extracted_k141_4453 | Not-determined |
| S14_extracted_k141_48009 | Low-quality |
| S14_extracted_k141_107493 | Low-quality |
| S14_extracted_k141_107494 | Not-determined |
| S14_extracted_k141_107495 | Not-determined |
| S14_extracted_k141_26187 | Low-quality |
| S14_extracted_k141_99576 | Not-determined |
| S14_extracted_k141_26193 | Low-quality |
| S14_extracted_k141_70576 | Low-quality |
| S14_extracted_k141_19702 | Not-determined |
| S14_extracted_k141_78076 | Not-determined |
| S14_extracted_k141_33782 | Not-determined |
| S14_extracted_k141_114715 | Low-quality |
| S14_extracted_k141_114722 | Not-determined |
| S14_extracted_k141_19712 | Not-determined |
| S14_extracted_k141_114726 | Not-determined |
| S14_extracted_k141_62978 | Low-quality |
| S14_extracted_k141_62980 | Low-quality |
| S14_extracted_k141_114728 | Not-determined |
| S14_extracted_k141_26196 | Low-quality |
| S14_extracted_k141_4476 | Low-quality |
| S14_extracted_k141_78084 | Not-determined |
| S14_extracted_k141_33785 | Not-determined |
| S14_extracted_k141_70597 | Low-quality |
| S14_extracted_k141_4480 | Low-quality |
| S14_extracted_k141_4483 | Low-quality |
| S14_extracted_k141_26202 | Not-determined |
| S14_extracted_k141_26203 | Low-quality |
| S14_extracted_k141_41699 | Low-quality |
| S14_extracted_k141_99595 | Not-determined |
| S14_extracted_k141_41703 | Not-determined |
| S14_extracted_k141_107501 | Not-determined |
| S14_extracted_k141_41705 | Low-quality |
| S14_extracted_k141_107502 | Low-quality |
| S14_extracted_k141_33792 | Low-quality |
| S14_extracted_k141_70604 | Low-quality |
| S14_extracted_k141_19723 | Low-quality |
| S14_extracted_k141_33794 | Not-determined |
| S14_extracted_k141_33796 | Not-determined |
| S14_extracted_k141_70608 | Low-quality |
| S14_extracted_k141_85414 | Not-determined |
| S14_extracted_k141_62986 | Not-determined |
| S14_extracted_k141_26210 | Not-determined |
| S14_extracted_k141_92868 | Low-quality |
| S14_extracted_k141_26213 | Not-determined |
| S14_extracted_k141_114742 | Low-quality |
| S14_extracted_k141_114743 | Not-determined |
| S14_extracted_k141_114744 | Not-determined |
| S14_extracted_k141_4492 | Not-determined |
| S14_extracted_k141_99606 | Low-quality |
| S14_extracted_k141_11711 | Low-quality |
| S14_extracted_k141_19736 | Low-quality |
| S14_extracted_k141_19737 | Not-determined |
| S14_extracted_k141_11715 | Not-determined |
| S14_extracted_k141_114749 | Low-quality |
| S14_extracted_k141_4494 | Not-determined |
| S14_extracted_k141_41719 | Not-determined |
| S14_extracted_k141_26227 | Not-determined |
| S14_extracted_k141_41724 | Not-determined |
| S14_extracted_k141_33808 | Low-quality |
| S14_extracted_k141_70635 | Not-determined |
| S14_extracted_k141_70637 | Low-quality |
| S14_extracted_k141_114759 | Low-quality |
| S14_extracted_k141_78114 | Not-determined |
| S14_extracted_k141_41727 | Low-quality |
| S14_extracted_k141_11727 | Not-determined |
| S14_extracted_k141_107519 | Not-determined |
| S14_extracted_k141_26233 | Not-determined |
| S14_extracted_k141_11728 | Low-quality |
| S14_extracted_k141_11729 | Low-quality |
| S14_extracted_k141_114762 | Not-determined |
| S14_extracted_k141_70640 | Not-determined |
| S14_extracted_k141_85432 | Low-quality |
| S14_extracted_k141_4514 | Not-determined |
| S14_extracted_k141_107523 | Not-determined |
| S14_extracted_k141_26236 | Low-quality |
| S14_extracted_k141_33819 | Low-quality |
| S14_extracted_k141_70643 | Not-determined |
| S14_extracted_k141_4518 | Not-determined |
| S14_extracted_k141_107527 | Not-determined |
| S14_extracted_k141_26238 | Low-quality |
| S14_extracted_k141_55996 | Low-quality |
| S14_extracted_k141_26240 | Low-quality |
| S14_extracted_k141_55997 | Not-determined |
| S14_extracted_k141_70649 | Not-determined |
| S14_extracted_k141_107531 | Not-determined |
| S14_extracted_k141_4522 | Low-quality |
| S14_extracted_k141_56001 | Not-determined |
| S14_extracted_k141_70650 | Not-determined |
| S14_extracted_k141_70651 | Not-determined |
| S14_extracted_k141_56003 | Low-quality |
| S14_extracted_k141_33827 | Not-determined |
| S14_extracted_k141_26248 | Low-quality |
| S14_extracted_k141_107538 | Not-determined |
| S14_extracted_k141_107539 | Low-quality |
| S14_extracted_k141_70656 | Not-determined |
| S14_extracted_k141_62995 | Low-quality |
| S14_extracted_k141_78124 | Not-determined |
| S14_extracted_k141_62996 | Low-quality |
| S14_extracted_k141_41746 | Not-determined |
| S14_extracted_k141_33834 | Low-quality |
| S14_extracted_k141_33836 | Not-determined |
| S14_extracted_k141_70664 | Not-determined |
| S14_extracted_k141_62999 | Low-quality |
| S14_extracted_k141_19753 | Not-determined |
| S14_extracted_k141_41752 | Not-determined |
| S14_extracted_k141_19755 | Not-determined |
| S14_extracted_k141_41754 | Not-determined |
| S14_extracted_k141_41755 | Not-determined |
| S14_extracted_k141_63002 | Not-determined |
| S14_extracted_k141_19757 | Not-determined |
| S14_extracted_k141_33844 | Low-quality |
| S14_extracted_k141_63009 | Not-determined |
| S14_extracted_k141_107543 | Low-quality |
| S14_extracted_k141_56020 | Low-quality |
| S14_extracted_k141_33851 | Low-quality |
| S14_extracted_k141_11733 | Low-quality |
| S14_extracted_k141_56024 | Not-determined |
| S14_extracted_k141_63017 | Not-determined |
| S14_extracted_k141_26258 | Low-quality |
| S14_extracted_k141_56028 | Not-determined |
| S14_extracted_k141_56031 | Low-quality |
| S14_extracted_k141_19777 | Low-quality |
| S14_extracted_k141_41769 | Low-quality |
| S14_extracted_k141_11738 | Low-quality |
| S14_extracted_k141_56035 | Low-quality |
| S14_extracted_k141_41772 | Not-determined |
| S14_extracted_k141_63025 | Not-determined |
| S14_extracted_k141_56037 | Low-quality |
| S14_extracted_k141_114770 | Not-determined |
| S14_extracted_k141_56038 | Not-determined |
| S14_extracted_k141_78130 | Not-determined |
| S14_extracted_k141_48025 | Not-determined |
| S14_extracted_k141_26273 | Not-determined |
| S14_extracted_k141_78131 | Not-determined |
| S14_extracted_k141_63028 | Not-determined |
| S14_extracted_k141_99628 | Low-quality |
| S14_extracted_k141_11744 | Low-quality |
| S14_extracted_k141_26279 | Low-quality |
| S14_extracted_k141_19786 | Not-determined |
| S14_extracted_k141_11746 | Not-determined |
| S14_extracted_k141_107562 | Low-quality |
| S14_extracted_k141_11753 | Not-determined |
| S14_extracted_k141_99636 | Not-determined |
| S14_extracted_k141_19790 | Low-quality |
| S14_extracted_k141_56050 | Not-determined |
| S14_extracted_k141_33873 | Low-quality |
| S14_extracted_k141_107567 | Low-quality |
| S14_extracted_k141_48033 | Not-determined |
| S14_extracted_k141_56055 | Not-determined |
| S14_extracted_k141_48034 | Not-determined |
| S14_extracted_k141_11760 | Not-determined |
| S14_extracted_k141_48039 | Not-determined |
| S14_extracted_k141_56060 | Not-determined |
| S14_extracted_k141_114783 | Low-quality |
| S14_extracted_k141_48043 | Low-quality |
| S14_extracted_k141_78138 | Not-determined |
| S14_extracted_k141_70695 | Not-determined |
| S14_extracted_k141_48046 | Low-quality |
| S14_extracted_k141_99647 | Not-determined |
| S14_extracted_k141_85437 | Not-determined |
| S14_extracted_k141_26296 | Low-quality |
| S14_extracted_k141_11762 | Not-determined |
| S14_extracted_k141_48050 | Not-determined |
| S14_extracted_k141_41781 | Not-determined |
| S14_extracted_k141_33882 | Not-determined |
| S14_extracted_k141_11765 | Low-quality |
| S14_extracted_k141_70703 | Not-determined |
| S14_extracted_k141_70704 | Not-determined |
| S14_extracted_k141_70708 | Low-quality |
| S14_extracted_k141_56076 | Not-determined |
| S14_extracted_k141_11770 | Not-determined |
| S14_extracted_k141_99654 | Not-determined |
| S14_extracted_k141_26308 | Not-determined |
| S14_extracted_k141_48060 | Low-quality |
| S14_extracted_k141_48061 | Not-determined |
| S14_extracted_k141_70712 | Low-quality |
| S14_extracted_k141_48062 | Not-determined |
| S14_extracted_k141_107568 | Low-quality |
| S14_extracted_k141_70715 | Not-determined |
| S14_extracted_k141_41794 | Low-quality |
| S14_extracted_k141_85442 | Not-determined |
| S14_extracted_k141_99660 | Not-determined |
| S14_extracted_k141_107577 | Not-determined |
| S14_extracted_k141_107578 | Not-determined |
| S14_extracted_k141_85446 | Not-determined |
| S14_extracted_k141_63063 | Not-determined |
| S14_extracted_k141_85447 | Not-determined |
| S14_extracted_k141_41800 | Not-determined |
| S14_extracted_k141_11783 | Not-determined |
| S14_extracted_k141_63067 | Low-quality |
| S14_extracted_k141_107583 | Not-determined |
| S14_extracted_k141_63069 | Low-quality |
| S14_extracted_k141_99674 | Low-quality |
| S14_extracted_k141_99675 | Low-quality |
| S14_extracted_k141_114789 | Low-quality |
| S14_extracted_k141_99678 | Low-quality |
| S14_extracted_k141_11791 | Not-determined |
| S14_extracted_k141_70741 | Not-determined |
| S14_extracted_k141_33902 | Not-determined |
| S14_extracted_k141_70743 | Not-determined |
| S14_extracted_k141_11799 | Not-determined |
| S14_extracted_k141_107595 | Not-determined |
| S14_extracted_k141_33907 | Not-determined |
| S14_extracted_k141_114801 | Not-determined |
| S14_extracted_k141_33908 | Low-quality |
| S14_extracted_k141_70755 | Not-determined |
| S14_extracted_k141_19801 | Not-determined |
| S14_extracted_k141_48082 | Not-determined |
| S14_extracted_k141_48084 | Not-determined |
| S14_extracted_k141_33916 | Low-quality |
| S14_extracted_k141_41809 | Low-quality |
| S14_extracted_k141_63085 | Not-determined |
| S14_extracted_k141_114813 | Not-determined |
| S14_extracted_k141_48093 | Not-determined |
| S14_extracted_k141_48096 | Low-quality |
| S14_extracted_k141_48097 | Not-determined |
| S14_extracted_k141_63090 | Not-determined |
| S14_extracted_k141_41815 | Not-determined |
| S14_extracted_k141_19822 | Not-determined |
| S14_extracted_k141_107610 | Not-determined |
| S14_extracted_k141_114818 | Not-determined |
| S14_extracted_k141_33929 | Not-determined |
| S14_extracted_k141_48101 | Not-determined |
| S14_extracted_k141_19827 | Not-determined |
| S14_extracted_k141_33930 | Not-determined |
| S14_extracted_k141_48102 | Not-determined |
| S14_extracted_k141_19828 | Not-determined |
| S14_extracted_k141_41821 | Not-determined |
| S14_extracted_k141_63103 | Low-quality |
| S14_extracted_k141_33937 | Not-determined |
| S14_extracted_k141_41826 | Not-determined |
| S14_extracted_k141_107617 | Not-determined |
| S14_extracted_k141_107620 | Low-quality |
| S14_extracted_k141_63106 | Low-quality |
| S14_extracted_k141_85486 | Not-determined |
| S14_extracted_k141_107628 | Low-quality |
| S14_extracted_k141_107629 | Low-quality |
| S14_extracted_k141_107630 | Low-quality |
| S6_vs_k141_9784\|\|full | Complete |
| S14_extracted_k141_41839 | Not-determined |
| S14_extracted_k141_85493 | Low-quality |
| S14_extracted_k141_41843 | Low-quality |
| S14_extracted_k141_41845 | Not-determined |
| S14_extracted_k141_85501 | Low-quality |
| S14_extracted_k141_85504 | Low-quality |
| S14_extracted_k141_85505 | Low-quality |
| S14_extracted_k141_41850 | Not-determined |
| S14_extracted_k141_85506 | Low-quality |
| S14_extracted_k141_4544 | Not-determined |
| S14_extracted_k141_70778 | Not-determined |
| S14_extracted_k141_41854 | Not-determined |
| S14_extracted_k141_107644 | Not-determined |
| S14_extracted_k141_19845 | Not-determined |
| S14_extracted_k141_19848 | Not-determined |
| S14_extracted_k141_107646 | Not-determined |
| S14_extracted_k141_70783 | Low-quality |
| S14_extracted_k141_4549 | Not-determined |
| S14_extracted_k141_19856 | Low-quality |
| S14_extracted_k141_56084 | Not-determined |
| S14_extracted_k141_114843 | Low-quality |
| S14_extracted_k141_85516 | Not-determined |
| S14_extracted_k141_78145 | Low-quality |
| S14_extracted_k141_107652 | Low-quality |
| S14_extracted_k141_4559 | Low-quality |
| S14_extracted_k141_63123 | Not-determined |
| S14_extracted_k141_33974 | Not-determined |
| S14_extracted_k141_70790 | Not-determined |
| S14_extracted_k141_11806 | Low-quality |
| S14_extracted_k141_99699 | Not-determined |
| S14_extracted_k141_85521 | Not-determined |
| S14_extracted_k141_48110 | Low-quality |
| S14_extracted_k141_48111 | Not-determined |
| S14_extracted_k141_114860 | Not-determined |
| S14_extracted_k141_78153 | Not-determined |
| S14_extracted_k141_92884 | Not-determined |
| S14_extracted_k141_99704 | Not-determined |
| S14_extracted_k141_107662 | Not-determined |
| S14_extracted_k141_92887 | Not-determined |
| S14_extracted_k141_114865 | Not-determined |
| S14_extracted_k141_48117 | Low-quality |
| S14_extracted_k141_48120 | Not-determined |
| S14_extracted_k141_92893 | Not-determined |
| S14_extracted_k141_48122 | Low-quality |
| S14_extracted_k141_92901 | Not-determined |
| S14_extracted_k141_19870 | Not-determined |
| S14_extracted_k141_92903 | Not-determined |
| S14_extracted_k141_114872 | Not-determined |
| S14_extracted_k141_63143 | Low-quality |
| S14_extracted_k141_70813 | Not-determined |
| S14_extracted_k141_92916 | Not-determined |
| S14_extracted_k141_70820 | Not-determined |
| S14_extracted_k141_114887 | Low-quality |
| S14_extracted_k141_114888 | Low-quality |
| S14_extracted_k141_63150 | Low-quality |
| S14_extracted_k141_70825 | Not-determined |
| S14_extracted_k141_11832 | Not-determined |
| S14_extracted_k141_33985 | Low-quality |
| S14_extracted_k141_92923 | Low-quality |
| S14_extracted_k141_85543 | Not-determined |
| S14_extracted_k141_11834 | Not-determined |
| S14_extracted_k141_33986 | Not-determined |
| S14_extracted_k141_114893 | Low-quality |
| S14_extracted_k141_11840 | Not-determined |
| S14_extracted_k141_33992 | Not-determined |
| S14_extracted_k141_33997 | Not-determined |
| S14_extracted_k141_107688 | Not-determined |
| S14_extracted_k141_107690 | Not-determined |
| S14_extracted_k141_114902 | Not-determined |
| S14_extracted_k141_92929 | Not-determined |
| S14_extracted_k141_85556 | Not-determined |
| S14_extracted_k141_70844 | Low-quality |
| S14_extracted_k141_19895 | Not-determined |
| S14_extracted_k141_11852 | Low-quality |
| S14_extracted_k141_4570 | Low-quality |
| S14_extracted_k141_11853 | Not-determined |
| S14_extracted_k141_11857 | Low-quality |
| S14_extracted_k141_11858 | Not-determined |
| S14_extracted_k141_4576 | Low-quality |
| S14_extracted_k141_92937 | Not-determined |
| S14_extracted_k141_11863 | Low-quality |
| S14_extracted_k141_92938 | Not-determined |
| S14_extracted_k141_107699 | Low-quality |
| S14_extracted_k141_107702 | Not-determined |
| S14_extracted_k141_19905 | Not-determined |
| S14_extracted_k141_56125 | Low-quality |
| S14_extracted_k141_19910 | Not-determined |
| S14_extracted_k141_63168 | Low-quality |
| S14_extracted_k141_92953 | Not-determined |
| S14_extracted_k141_107708 | Low-quality |
| S14_extracted_k141_19914 | Not-determined |
| S14_extracted_k141_11872 | Not-determined |
| S14_extracted_k141_85579 | Low-quality |
| S14_extracted_k141_92958 | Low-quality |
| S14_extracted_k141_85585 | Low-quality |
| S14_extracted_k141_41884 | Low-quality |
| S14_extracted_k141_107712 | Not-determined |
| S14_extracted_k141_92967 | Low-quality |
| S14_extracted_k141_92968 | Not-determined |
| S14_extracted_k141_41886 | Not-determined |
| S14_extracted_k141_85590 | Not-determined |
| S14_extracted_k141_48145 | Not-determined |
| S14_extracted_k141_11880 | Low-quality |
| S14_extracted_k141_107714 | Not-determined |
| S14_extracted_k141_107718 | Not-determined |
| S14_extracted_k141_63187 | Not-determined |
| S14_extracted_k141_92983 | Low-quality |
| S14_extracted_k141_19928 | Not-determined |
| S14_extracted_k141_85602 | Not-determined |
| S14_extracted_k141_92987 | Not-determined |
| S14_extracted_k141_19933 | Not-determined |
| S14_extracted_k141_107723 | Not-determined |
| S14_extracted_k141_63195 | Not-determined |
| S14_extracted_k141_92994 | Not-determined |
| S14_extracted_k141_107725 | Not-determined |
| S14_extracted_k141_70886 | Not-determined |
| S14_extracted_k141_85606 | Not-determined |
| S14_extracted_k141_85607 | Not-determined |
| S14_extracted_k141_107731 | Not-determined |
| S14_extracted_k141_107732 | Low-quality |
| S14_extracted_k141_107733 | Not-determined |
| S14_extracted_k141_63198 | Low-quality |
| S14_extracted_k141_70888 | Not-determined |
| S14_extracted_k141_93000 | Not-determined |
| S14_extracted_k141_99717 | Not-determined |
| S14_extracted_k141_99721 | Not-determined |
| S14_extracted_k141_93006 | Not-determined |

| | |
|---|---|
| S14_extracted_k141_99725 | Low-quality |
| S14_extracted_k141_85615 | Not-determined |
| S14_extracted_k141_99727 | Low-quality |
| S14_extracted_k141_70898 | Not-determined |
| S14_extracted_k141_19939 | Low-quality |
| S14_extracted_k141_99731 | Not-determined |
| S14_extracted_k141_99733 | Not-determined |
| S14_extracted_k141_70907 | Low-quality |
| S14_extracted_k141_11891 | Not-determined |
| S14_extracted_k141_24013 | Not-determined |
| S14_extracted_k141_99739 | Not-determined |
| S14_extracted_k141_70912 | Low-quality |
| S14_extracted_k141_41896 | Low-quality |
| S14_extracted_k141_70921 | Low-quality |
| S14_extracted_k141_48156 | Low-quality |
| S14_extracted_k141_70922 | Not-determined |
| S14_extracted_k141_85625 | Not-determined |
| S14_extracted_k141_85626 | Not-determined |
| S14_extracted_k141_48159 | Low-quality |
| S14_extracted_k141_4629 | Low-quality |
| S14_extracted_k141_85633 | Low-quality |
| S14_extracted_k141_11902 | Not-determined |
| S14_extracted_k141_24022 | Not-determined |
| S14_extracted_k141_93045 | Low-quality |
| S14_extracted_k141_85636 | Low-quality |
| S14_extracted_k141_93053 | Not-determined |
| S14_extracted_k141_48175 | Not-determined |
| S14_extracted_k141_107743 | Low-quality |
| S14_vs_k141_117157\|\|full | High-quality |
| S14_extracted_k141_107746 | Low-quality |
| S14_extracted_k141_11906 | Not-determined |
| S14_extracted_k141_70940 | Low-quality |
| S14_extracted_k141_41913 | Not-determined |
| S14_extracted_k141_4639 | Not-determined |
| S14_extracted_k141_41915 | Not-determined |
| S14_extracted_k141_4643 | Not-determined |
| S14_extracted_k141_93057 | Low-quality |
| S14_extracted_k141_48189 | Not-determined |
| S14_extracted_k141_93063 | Not-determined |
| S14_extracted_k141_93065 | Not-determined |
| S14_extracted_k141_93066 | Not-determined |
| S14_extracted_k141_4655 | Not-determined |
| S14_extracted_k141_63207 | Low-quality |
| S14_extracted_k141_93070 | Not-determined |
| S14_extracted_k141_11919 | Not-determined |
| S14_extracted_k141_93073 | Low-quality |
| S14_extracted_k141_93074 | Low-quality |
| S14_extracted_k141_114920 | Low-quality |
| S14_extracted_k141_114921 | Low-quality |
| S14_extracted_k141_70959 | Not-determined |
| S14_extracted_k141_19979 | Not-determined |
| S14_extracted_k141_24025 | Not-determined |
| S14_extracted_k141_41930 | Not-determined |
| S14_extracted_k141_48204 | Low-quality |
| S14_extracted_k141_4674 | Low-quality |
| S14_extracted_k141_48205 | Not-determined |
| S14_extracted_k141_78177 | Not-determined |
| S14_extracted_k141_11941 | Low-quality |
| S14_extracted_k141_93078 | Not-determined |
| S14_extracted_k141_4683 | Not-determined |
| S14_extracted_k141_41941 | Not-determined |
| S14_extracted_k141_4686 | Low-quality |
| S14_extracted_k141_93082 | Not-determined |
| S14_extracted_k141_24036 | Low-quality |
| S14_extracted_k141_78183 | Not-determined |
| S14_extracted_k141_63228 | Low-quality |
| S14_extracted_k141_26322 | Not-determined |
| S14_extracted_k141_11950 | Low-quality |
| S14_extracted_k141_26324 | Not-determined |
| S14_extracted_k141_78190 | Low-quality |
| S14_extracted_k141_41948 | Not-determined |
| S14_extracted_k141_26325 | Low-quality |
| S14_extracted_k141_41954 | Not-determined |
| S14_extracted_k141_26328 | Not-determined |
| S14_extracted_k141_78195 | Not-determined |
| S14_extracted_k141_26330 | Not-determined |
| S14_extracted_k141_93095 | Not-determined |
| S14_extracted_k141_70984 | Low-quality |
| S14_extracted_k141_93097 | Not-determined |
| S14_extracted_k141_4710 | Not-determined |
| S14_extracted_k141_4711 | Low-quality |
| S14_extracted_k141_4713 | Not-determined |
| S14_extracted_k141_48227 | Not-determined |
| S14_extracted_k141_78212 | Not-determined |
| S14_extracted_k141_48230 | Not-determined |
| S14_extracted_k141_26336 | Not-determined |
| S14_extracted_k141_93119 | Not-determined |
| S14_extracted_k141_48234 | Not-determined |
| S14_extracted_k141_93120 | Low-quality |
| S14_extracted_k141_26338 | Not-determined |
| S14_extracted_k141_4717 | Not-determined |
| S14_extracted_k141_107748 | Low-quality |
| S14_extracted_k141_107753 | Not-determined |
| S14_extracted_k141_26341 | Low-quality |
| S14_extracted_k141_78217 | Not-determined |
| S14_extracted_k141_26342 | Not-determined |
| S14_extracted_k141_26344 | Not-determined |
| S14_extracted_k141_114953 | Not-determined |
| S14_extracted_k141_93130 | Not-determined |
| S14_extracted_k141_85664 | Low-quality |
| S14_extracted_k141_41968 | Low-quality |
| S14_extracted_k141_107761 | Low-quality |
| S14_extracted_k141_85666 | Low-quality |
| S14_extracted_k141_48239 | Not-determined |
| S14_extracted_k141_56135 | Not-determined |
| S14_extracted_k141_99793 | Not-determined |
| S14_extracted_k141_11956 | Low-quality |
| S14_extracted_k141_11961 | Low-quality |
| S14_extracted_k141_26358 | Not-determined |
| S14_extracted_k141_41980 | Low-quality |
| S14_extracted_k141_41982 | Low-quality |
| S14_extracted_k141_4750 | Not-determined |
| S14_extracted_k141_107774 | Low-quality |
| S14_extracted_k141_78230 | Not-determined |
| S14_extracted_k141_85677 | Not-determined |
| S14_extracted_k141_56155 | Not-determined |
| S14_extracted_k141_107776 | Low-quality |
| S14_extracted_k141_107778 | Low-quality |
| S14_extracted_k141_41984 | Not-determined |
| S12_extracted_k141_16374 | High-quality |
| S14_extracted_k141_56159 | Not-determined |
| S14_extracted_k141_24057 | Not-determined |
| S14_extracted_k141_24059 | Not-determined |
| S14_extracted_k141_85683 | Low-quality |
| S14_extracted_k141_85685 | Not-determined |
| S14_extracted_k141_41986 | Not-determined |
| S14_extracted_k141_24066 | Low-quality |
| S14_extracted_k141_107789 | Not-determined |
| S14_extracted_k141_93138 | Not-determined |
| S14_extracted_k141_11983 | Low-quality |
| S14_extracted_k141_48257 | Not-determined |
| S14_extracted_k141_26375 | Not-determined |
| S14_extracted_k141_48258 | Low-quality |
| S14_extracted_k141_26376 | Low-quality |
| S14_extracted_k141_24073 | Not-determined |
| S14_extracted_k141_56170 | Not-determined |
| S14_extracted_k141_85695 | Not-determined |
| S14_extracted_k141_56172 | Not-determined |
| S14_extracted_k141_26382 | Not-determined |
| S14_extracted_k141_93146 | Low-quality |
| S14_extracted_k141_48265 | Not-determined |
| S14_extracted_k141_24081 | Not-determined |
| S14_extracted_k141_85699 | Not-determined |
| S14_extracted_k141_114964 | Not-determined |
| S14_extracted_k141_93148 | Low-quality |
| S14_extracted_k141_4768 | Not-determined |
| S14_extracted_k141_24084 | Low-quality |
| S14_extracted_k141_42000 | Not-determined |
| S14_extracted_k141_99815 | Not-determined |
| S14_extracted_k141_42001 | Not-determined |
| S14_extracted_k141_85702 | Not-determined |
| S14_extracted_k141_48271 | Not-determined |
| S14_extracted_k141_85704 | Not-determined |
| S14_extracted_k141_4769 | Low-quality |
| S14_extracted_k141_24091 | Low-quality |
| S14_extracted_k141_99819 | Not-determined |
| S14_extracted_k141_85707 | Not-determined |
| S14_extracted_k141_42006 | Not-determined |
| S14_extracted_k141_78259 | Not-determined |
| S14_extracted_k141_114972 | Not-determined |
| S14_extracted_k141_26388 | Low-quality |
| S14_extracted_k141_114974 | Not-determined |
| S14_extracted_k141_48278 | Low-quality |
| S14_extracted_k141_63243 | Not-determined |
| S14_extracted_k141_85713 | Low-quality |
| S14_extracted_k141_85715 | Low-quality |
| S14_extracted_k141_99833 | Not-determined |
| S14_extracted_k141_63248 | Not-determined |
| S14_extracted_k141_26393 | Low-quality |
| S14_extracted_k141_85720 | Not-determined |
| S14_extracted_k141_11997 | Not-determined |
| S14_extracted_k141_114981 | Not-determined |
| S14_extracted_k141_63249 | Low-quality |
| S14_extracted_k141_99840 | Low-quality |
| S14_extracted_k141_63254 | Not-determined |
| S14_extracted_k141_63255 | Not-determined |
| S14_extracted_k141_24100 | Low-quality |
| S14_extracted_k141_99846 | Not-determined |
| S14_extracted_k141_42025 | Not-determined |
| S14_extracted_k141_114987 | Not-determined |
| S14_extracted_k141_85726 | Low-quality |
| S14_extracted_k141_24104 | Low-quality |
| S14_extracted_k141_114992 | Not-determined |
| S14_extracted_k141_24108 | Not-determined |
| S14_extracted_k141_99851 | Low-quality |
| S14_extracted_k141_85735 | Not-determined |
| S14_extracted_k141_114994 | Not-determined |
| S14_extracted_k141_26421 | Low-quality |
| S14_extracted_k141_99855 | Not-determined |
| S14_extracted_k141_26426 | Not-determined |
| S14_extracted_k141_93166 | Low-quality |
| S14_extracted_k141_93168 | Not-determined |
| S14_extracted_k141_4787 | Not-determined |
| S14_extracted_k141_56202 | Not-determined |
| S14_extracted_k141_93171 | Low-quality |
| S14_extracted_k141_26433 | Not-determined |
| S14_extracted_k141_4795 | Not-determined |
| S14_extracted_k141_63274 | Low-quality |
| S14_extracted_k141_56206 | Not-determined |
| S14_extracted_k141_20009 | Low-quality |
| S14_extracted_k141_56207 | Low-quality |
| S14_extracted_k141_115000 | Not-determined |
| S14_extracted_k141_85746 | Low-quality |
| S14_extracted_k141_26436 | Low-quality |
| S14_extracted_k141_26439 | Not-determined |
| S14_extracted_k141_56212 | Low-quality |
| S14_extracted_k141_12016 | Low-quality |
| S14_extracted_k141_107804 | Low-quality |
| S14_extracted_k141_24132 | Not-determined |
| S14_extracted_k141_12018 | Not-determined |
| S14_extracted_k141_24135 | Not-determined |
| S14_extracted_k141_107809 | Low-quality |
| S14_extracted_k141_107810 | Low-quality |
| S14_extracted_k141_107811 | Not-determined |
| S14_extracted_k141_115009 | Not-determined |
| S14_extracted_k141_24138 | Low-quality |
| S14_extracted_k141_4803 | Not-determined |
| S14_extracted_k141_99869 | Not-determined |
| S14_extracted_k141_56218 | Not-determined |
| S14_extracted_k141_4809 | Not-determined |
| S14_extracted_k141_115016 | Low-quality |
| S14_extracted_k141_99877 | Not-determined |
| S14_extracted_k141_85758 | Low-quality |
| S14_extracted_k141_63284 | Not-determined |
| S14_extracted_k141_42063 | Not-determined |
| S14_extracted_k141_56225 | Not-determined |
| S14_extracted_k141_4818 | Low-quality |
| S14_extracted_k141_12024 | Low-quality |
| S14_extracted_k141_42068 | Not-determined |
| S14_extracted_k141_42073 | Low-quality |
| S14_extracted_k141_99886 | Not-determined |
| S14_extracted_k141_48325 | Not-determined |
| S14_extracted_k141_63293 | Not-determined |
| S14_extracted_k141_93196 | Not-determined |
| S14_extracted_k141_85761 | Not-determined |
| S14_extracted_k141_99894 | Not-determined |
| S14_extracted_k141_93199 | Not-determined |
| S14_extracted_k141_99897 | Low-quality |
| S14_extracted_k141_93202 | Not-determined |
| S14_extracted_k141_12038 | Not-determined |
| S14_extracted_k141_4834 | Not-determined |
| S14_extracted_k141_107824 | Not-determined |
| S14_extracted_k141_63303 | Low-quality |
| S14_extracted_k141_93208 | Not-determined |
| S14_extracted_k141_93209 | Not-determined |
| S14_extracted_k141_24150 | Not-determined |
| S14_extracted_k141_56233 | Not-determined |
| S14_extracted_k141_12044 | Not-determined |
| S14_extracted_k141_93212 | Low-quality |
| S14_extracted_k141_107833 | Not-determined |
| S14_extracted_k141_24161 | Not-determined |
| S14_extracted_k141_107835 | Not-determined |
| S14_extracted_k141_24167 | Low-quality |
| S14_extracted_k141_12045 | Low-quality |
| S14_extracted_k141_107840 | Not-determined |
| S14_extracted_k141_78285 | Not-determined |
| S14_extracted_k141_107841 | Low-quality |
| S14_extracted_k141_93220 | Not-determined |
| S14_extracted_k141_99917 | Not-determined |
| S14_extracted_k141_99920 | Not-determined |
| S14_extracted_k141_12053 | Not-determined |
| S14_extracted_k141_63311 | Low-quality |
| S14_extracted_k141_4858 | Not-determined |
| S14_extracted_k141_48337 | Not-determined |
| S14_extracted_k141_4862 | Not-determined |
| S14_extracted_k141_12060 | Low-quality |
| S14_extracted_k141_56246 | Low-quality |
| S14_extracted_k141_56247 | Not-determined |
| S14_extracted_k141_63315 | Not-determined |
| S14_extracted_k141_12066 | Low-quality |
| S14_extracted_k141_42098 | Low-quality |
| S14_extracted_k141_93230 | Not-determined |
| S14_extracted_k141_42099 | Not-determined |
| S14_extracted_k141_63319 | Low-quality |
| S14_extracted_k141_4869 | Not-determined |
| S14_extracted_k141_63323 | Not-determined |
| S14_extracted_k141_115022 | Low-quality |
| S14_extracted_k141_115026 | Not-determined |
| S14_extracted_k141_4873 | Low-quality |
| S14_extracted_k141_93236 | Low-quality |
| S14_extracted_k141_4880 | Not-determined |
| S14_extracted_k141_78302 | Not-determined |
| S14_extracted_k141_107860 | Not-determined |
| S14_extracted_k141_115032 | Low-quality |
| S14_extracted_k141_115034 | Low-quality |
| S14_extracted_k141_93246 | Not-determined |
| S14_extracted_k141_4881 | Low-quality |
| S14_extracted_k141_115038 | Low-quality |
| S14_extracted_k141_26450 | Not-determined |
| S14_extracted_k141_26451 | Not-determined |
| S14_extracted_k141_12085 | Not-determined |
| S14_extracted_k141_26454 | Low-quality |
| S14_extracted_k141_99933 | Low-quality |
| S14_extracted_k141_48347 | Not-determined |
| S14_extracted_k141_107872 | Low-quality |
| S14_extracted_k141_78319 | Not-determined |
| S14_extracted_k141_26456 | Low-quality |
| S14_extracted_k141_78325 | Not-determined |
| S14_extracted_k141_99939 | Low-quality |
| S14_extracted_k141_115045 | Not-determined |
| S14_extracted_k141_93254 | Low-quality |
| S14_extracted_k141_93255 | Low-quality |
| S14_extracted_k141_93256 | Low-quality |
| S14_extracted_k141_93257 | Not-determined |
| S14_extracted_k141_93259 | Low-quality |
| S14_extracted_k141_107875 | Not-determined |
| S14_extracted_k141_93264 | Not-determined |
| S14_extracted_k141_12092 | Not-determined |
| S14_extracted_k141_107877 | Not-determined |
| S14_extracted_k141_99952 | Low-quality |
| S14_extracted_k141_115052 | Not-determined |
| S14_extracted_k141_12097 | Not-determined |
| S14_extracted_k141_63353 | Not-determined |
| S14_extracted_k141_63355 | Low-quality |
| S14_extracted_k141_63357 | Not-determined |
| S14_extracted_k141_99956 | Not-determined |
| S14_extracted_k141_115058 | Not-determined |
| S14_extracted_k141_107886 | Low-quality |
| S14_extracted_k141_63361 | Low-quality |
| S14_extracted_k141_56252 | Not-determined |
| S14_extracted_k141_99958 | Not-determined |
| S14_extracted_k141_78352 | Not-determined |
| S14_extracted_k141_63371 | Not-determined |
| S14_extracted_k141_107893 | Not-determined |
| S14_extracted_k141_107894 | Not-determined |
| S14_extracted_k141_12104 | Not-determined |
| S14_extracted_k141_115070 | Not-determined |
| S14_extracted_k141_12106 | Not-determined |
| S14_extracted_k141_78357 | Low-quality |
| S14_extracted_k141_115071 | Low-quality |
| S14_extracted_k141_78361 | Not-determined |
| S14_extracted_k141_56263 | Not-determined |
| S14_extracted_k141_24185 | Low-quality |
| S14_extracted_k141_78364 | Not-determined |
| S14_extracted_k141_107904 | Not-determined |
| S14_extracted_k141_12119 | Not-determined |
| S14_extracted_k141_24194 | Not-determined |
| S14_extracted_k141_24195 | Low-quality |
| S14_extracted_k141_12122 | Not-determined |
| S14_extracted_k141_56280 | Not-determined |
| S14_extracted_k141_115083 | Not-determined |
| S14_extracted_k141_70994 | Not-determined |
| S14_extracted_k141_93279 | Not-determined |
| S14_extracted_k141_70997 | Not-determined |
| S14_extracted_k141_70998 | Low-quality |
| S14_extracted_k141_63393 | Not-determined |
| S14_extracted_k141_24208 | Low-quality |
| S14_extracted_k141_115093 | Not-determined |
| S14_extracted_k141_56288 | Not-determined |
| S14_extracted_k141_42139 | Not-determined |
| S14_extracted_k141_48357 | Low-quality |
| S14_extracted_k141_93288 | Not-determined |
| S14_extracted_k141_107910 | Not-determined |
| S14_extracted_k141_24214 | Not-determined |
| S14_extracted_k141_93294 | Low-quality |
| S14_extracted_k141_48367 | Not-determined |
| S14_extracted_k141_42149 | Not-determined |
| S14_extracted_k141_99993 | Not-determined |
| S14_extracted_k141_93298 | Low-quality |
| S14_extracted_k141_78378 | Not-determined |
| S14_extracted_k141_42151 | Not-determined |
| S14_extracted_k141_24217 | Not-determined |
| S14_extracted_k141_107919 | Not-determined |
| S14_extracted_k141_107921 | Not-determined |
| S14_extracted_k141_107922 | Not-determined |
| S14_extracted_k141_63412 | Not-determined |
| S14_extracted_k141_93308 | Low-quality |
| S14_extracted_k141_48381 | Low-quality |
| S14_extracted_k141_56317 | Low-quality |
| S14_extracted_k141_56318 | Not-determined |
| S14_extracted_k141_115118 | Not-determined |
| S14_extracted_k141_56320 | Not-determined |
| S14_extracted_k141_107933 | Low-quality |
| S14_extracted_k141_78390 | Low-quality |
| S14_extracted_k141_100006 | Low-quality |
| S14_extracted_k141_71002 | Not-determined |
| S14_extracted_k141_71003 | Low-quality |
| S14_extracted_k141_107934 | Not-determined |
| S14_extracted_k141_100007 | Not-determined |
| S14_extracted_k141_4911 | Low-quality |
| S14_extracted_k141_100008 | Not-determined |
| S14_extracted_k141_107937 | Low-quality |
| S14_extracted_k141_56327 | Not-determined |
| S14_extracted_k141_48392 | Not-determined |
| S14_extracted_k141_63419 | Not-determined |
| S14_extracted_k141_78398 | Low-quality |
| S14_extracted_k141_42169 | Not-determined |
| S14_extracted_k141_48395 | Not-determined |
| S14_extracted_k141_48396 | Low-quality |
| S14_extracted_k141_93322 | Not-determined |
| S14_extracted_k141_100015 | Not-determined |
| S14_extracted_k141_107943 | Not-determined |
| S14_extracted_k141_42172 | Not-determined |
| S14_extracted_k141_42173 | Low-quality |
| S14_extracted_k141_115126 | Not-determined |
| S14_extracted_k141_78402 | Not-determined |
| S14_extracted_k141_48401 | Low-quality |
| S14_extracted_k141_115127 | Not-determined |
| S14_extracted_k141_115128 | Not-determined |
| S14_extracted_k141_56334 | Not-determined |
| S14_extracted_k141_100018 | Low-quality |
| S14_extracted_k141_115130 | Low-quality |
| S14_extracted_k141_115131 | Low-quality |
| S14_extracted_k141_100020 | Low-quality |
| S14_extracted_k141_71016 | Not-determined |
| S14_extracted_k141_63422 | Low-quality |
| S14_extracted_k141_115132 | Low-quality |
| S14_extracted_k141_115133 | Not-determined |
| S14_extracted_k141_71018 | Low-quality |
| S14_extracted_k141_48406 | Not-determined |
| S14_extracted_k141_100022 | Low-quality |
| S14_extracted_k141_48408 | Not-determined |
| S14_extracted_k141_71021 | Not-determined |
| S14_extracted_k141_78409 | Not-determined |
| S14_extracted_k141_71022 | Not-determined |
| S14_extracted_k141_71023 | Not-determined |
| S14_extracted_k141_93329 | Not-determined |
| S14_extracted_k141_100026 | Not-determined |
| S14_extracted_k141_24251 | Not-determined |
| S14_extracted_k141_56347 | Not-determined |
| S14_extracted_k141_78413 | Not-determined |
| S14_extracted_k141_78414 | Not-determined |
| S14_extracted_k141_63435 | Not-determined |
| S14_extracted_k141_4917 | Not-determined |
| S14_extracted_k141_63437 | Not-determined |
| S14_extracted_k141_93334 | Low-quality |
| S14_extracted_k141_78420 | Not-determined |
| S14_extracted_k141_56351 | Not-determined |
| S14_extracted_k141_24257 | Low-quality |
| S14_extracted_k141_78427 | Not-determined |
| S14_extracted_k141_63447 | Not-determined |
| S14_extracted_k141_63451 | Low-quality |
| S14_extracted_k141_71034 | Low-quality |
| S14_extracted_k141_42192 | Not-determined |
| S14_extracted_k141_56364 | Not-determined |
| S14_extracted_k141_78432 | Low-quality |
| S14_extracted_k141_26466 | Not-determined |
| S14_extracted_k141_115145 | Not-determined |
| S14_extracted_k141_26469 | Low-quality |
| S14_extracted_k141_63459 | Not-determined |
| S14_extracted_k141_26471 | Low-quality |
| S14_extracted_k141_24267 | Low-quality |
| S14_extracted_k141_4934 | Not-determined |
| S14_extracted_k141_100042 | Low-quality |
| S14_extracted_k141_24269 | Not-determined |
| S14_extracted_k141_26473 | Low-quality |
| S14_extracted_k141_4939 | Not-determined |
| S14_extracted_k141_26475 | Low-quality |
| S14_extracted_k141_24272 | Not-determined |
| S14_extracted_k141_115149 | Not-determined |
| S14_extracted_k141_78441 | Not-determined |
| S14_extracted_k141_4942 | Not-determined |
| S14_extracted_k141_115158 | Not-determined |
| S14_extracted_k141_115159 | Not-determined |
| S14_extracted_k141_42210 | Low-quality |
| S14_extracted_k141_48420 | Low-quality |
| S14_extracted_k141_115162 | Not-determined |
| S14_extracted_k141_4947 | Low-quality |
| S14_extracted_k141_4950 | Low-quality |
| S14_extracted_k141_100061 | Not-determined |
| S14_extracted_k141_56373 | Low-quality |
| S14_extracted_k141_78447 | Not-determined |
| S14_extracted_k141_63485 | Low-quality |
| S14_extracted_k141_115171 | Not-determined |
| S14_extracted_k141_24298 | Low-quality |
| S14_extracted_k141_4960 | Not-determined |
| S14_extracted_k141_100075 | Not-determined |
| S14_extracted_k141_26490 | Low-quality |
| S14_extracted_k141_56386 | Not-determined |
| S14_extracted_k141_115179 | Not-determined |
| S14_extracted_k141_78455 | Low-quality |
| S14_extracted_k141_42240 | Not-determined |
| S14_extracted_k141_24301 | Not-determined |
| S14_extracted_k141_12175 | Low-quality |
| S14_extracted_k141_63495 | Not-determined |
| S14_extracted_k141_78459 | Low-quality |
| S14_extracted_k141_100082 | Low-quality |
| S14_extracted_k141_115185 | Not-determined |
| S14_extracted_k141_107973 | Not-determined |
| S14_extracted_k141_24309 | Not-determined |
| S14_extracted_k141_71063 | Not-determined |
| S14_extracted_k141_107976 | Low-quality |
| S14_extracted_k141_107977 | Low-quality |
| S14_extracted_k141_12178 | Not-determined |
| S14_extracted_k141_26501 | Low-quality |
| S14_extracted_k141_42250 | Low-quality |
| S14_extracted_k141_42251 | Not-determined |
| S14_extracted_k141_24318 | Not-determined |
| S14_extracted_k141_42255 | Not-determined |
| S14_extracted_k141_4977 | Not-determined |
| S14_extracted_k141_4978 | Not-determined |
| S14_extracted_k141_107981 | Not-determined |
| S14_extracted_k141_26510 | Not-determined |
| S14_extracted_k141_42261 | Low-quality |
| S14_extracted_k141_42263 | Not-determined |
| S14_extracted_k141_4981 | Not-determined |
| S14_extracted_k141_48440 | Low-quality |
| S14_extracted_k141_107989 | Not-determined |
| S14_extracted_k141_48446 | Not-determined |
| S14_extracted_k141_26518 | Low-quality |
| S14_extracted_k141_78476 | Not-determined |
| S14_extracted_k141_93358 | Low-quality |
| S14_extracted_k141_93361 | Low-quality |
| S14_extracted_k141_115199 | Low-quality |
| S14_extracted_k141_107994 | Not-determined |
| S14_extracted_k141_42278 | Not-determined |
| S14_extracted_k141_42280 | Not-determined |
| S14_extracted_k141_56404 | Not-determined |
| S14_extracted_k141_107999 | Low-quality |
| S14_extracted_k141_24352 | Not-determined |
| S14_extracted_k141_26520 | Low-quality |
| S14_extracted_k141_24356 | Not-determined |
| S14_extracted_k141_56409 | Low-quality |
| S14_extracted_k141_42286 | Low-quality |
| S14_extracted_k141_78486 | Not-determined |
| S14_extracted_k141_42289 | Low-quality |
| S14_extracted_k141_24367 | Not-determined |
| S14_extracted_k141_24368 | Not-determined |
| S14_extracted_k141_78489 | Not-determined |
| S14_extracted_k141_100094 | Low-quality |
| S14_extracted_k141_26523 | Low-quality |
| S14_extracted_k141_24372 | Low-quality |
| S14_extracted_k141_71075 | Not-determined |
| S14_extracted_k141_48453 | Not-determined |
| S14_extracted_k141_48454 | Not-determined |
| S14_extracted_k141_48456 | Not-determined |
| S14_extracted_k141_4999 | Low-quality |
| S14_extracted_k141_5005 | Not-determined |
| S14_extracted_k141_48467 | Not-determined |
| S14_extracted_k141_48477 | Low-quality |
| S14_extracted_k141_24378 | Not-determined |
| S14_extracted_k141_48481 | Not-determined |
| S14_extracted_k141_71085 | Not-determined |
| S14_extracted_k141_100105 | Not-determined |
| S14_extracted_k141_24381 | Not-determined |
| S14_extracted_k141_63531 | Not-determined |
| S14_extracted_k141_63533 | Not-determined |
| S14_extracted_k141_48482 | Not-determined |
| S14_extracted_k141_42297 | Low-quality |
| S14_extracted_k141_56414 | Not-determined |
| S14_extracted_k141_12207 | Not-determined |
| S14_extracted_k141_42301 | Low-quality |
| S14_extracted_k141_48487 | Not-determined |
| S14_extracted_k141_48489 | Low-quality |
| S14_extracted_k141_12210 | Low-quality |
| S14_extracted_k141_48492 | Not-determined |
| S14_extracted_k141_100119 | Not-determined |
| S14_extracted_k141_78505 | Not-determined |
| S14_extracted_k141_42315 | Not-determined |
| S14_extracted_k141_78510 | Low-quality |
| S14_extracted_k141_78512 | Not-determined |
| S14_extracted_k141_42318 | Low-quality |
| S14_extracted_k141_78515 | Low-quality |
| S14_extracted_k141_78517 | Not-determined |
| S14_extracted_k141_42322 | Low-quality |
| S14_extracted_k141_78519 | Low-quality |
| S14_extracted_k141_24406 | Not-determined |
| S14_extracted_k141_108026 | Not-determined |
| S14_extracted_k141_78524 | Not-determined |
| S14_extracted_k141_24412 | Not-determined |
| S14_extracted_k141_115222 | Not-determined |
| S14_extracted_k141_24413 | Low-quality |
| S14_extracted_k141_100136 | Low-quality |
| S14_extracted_k141_24417 | Low-quality |
| S14_extracted_k141_100139 | Not-determined |
| S14_extracted_k141_63550 | Not-determined |
| S14_extracted_k141_63551 | Not-determined |
| S14_extracted_k141_56427 | Low-quality |
| S14_extracted_k141_85797 | Not-determined |
| S14_extracted_k141_56428 | Not-determined |
| S14_extracted_k141_85799 | Not-determined |
| S14_extracted_k141_48518 | Not-determined |
| S14_extracted_k141_115231 | Not-determined |
| S14_extracted_k141_85801 | Not-determined |
| S14_extracted_k141_115234 | Not-determined |
| S14_extracted_k141_115236 | Not-determined |
| S14_extracted_k141_100145 | Not-determined |
| S14_extracted_k141_26553 | Not-determined |
| S14_extracted_k141_108036 | Not-determined |
| S14_extracted_k141_5033 | Low-quality |
| S14_extracted_k141_26554 | Not-determined |
| S14_extracted_k141_24432 | Not-determined |
| S14_extracted_k141_24433 | Low-quality |
| S14_extracted_k141_12218 | Low-quality |
| S14_extracted_k141_100150 | Not-determined |
| S14_extracted_k141_48525 | Low-quality |
| S14_extracted_k141_56433 | Not-determined |
| S14_extracted_k141_108044 | Not-determined |
| S14_extracted_k141_78529 | Not-determined |

| | |
|---|---|
| S14_extracted_k141_5042 | Low-quality |
| S14_extracted_k141_78532 | Low-quality |
| S14_extracted_k141_5044 | Not-determined |
| S14_extracted_k141_71091 | Not-determined |
| S14_extracted_k141_5045 | Not-determined |
| S14_extracted_k141_42338 | Low-quality |
| S14_extracted_k141_108048 | Low-quality |
| S14_extracted_k141_93399 | Not-determined |
| S14_extracted_k141_42343 | Low-quality |
| S14_extracted_k141_100162 | Not-determined |
| S14_extracted_k141_100163 | Not-determined |
| S14_extracted_k141_100165 | Not-determined |
| S14_extracted_k141_78541 | Not-determined |
| S14_extracted_k141_78542 | Low-quality |
| S14_extracted_k141_100168 | Not-determined |
| S14_extracted_k141_56439 | Not-determined |
| S14_extracted_k141_100170 | Not-determined |
| S14_extracted_k141_93407 | Low-quality |
| S14_extracted_k141_115239 | Not-determined |
| S14_extracted_k141_42358 | Not-determined |
| S14_extracted_k141_63573 | Not-determined |
| S14_extracted_k141_56446 | Not-determined |
| S14_extracted_k141_63575 | Low-quality |
| S14_extracted_k141_56447 | Low-quality |
| S14_extracted_k141_63576 | Low-quality |
| S14_extracted_k141_71097 | Not-determined |
| S14_extracted_k141_115242 | Low-quality |
| S14_extracted_k141_108056 | Not-determined |
| S14_extracted_k141_115243 | Not-determined |
| S14_extracted_k141_85825 | Low-quality |
| S14_extracted_k141_71099 | Low-quality |
| S14_extracted_k141_85827 | Not-determined |
| S14_extracted_k141_63579 | Not-determined |
| S14_extracted_k141_34444 | Low-quality |
| S14_extracted_k141_34446 | Not-determined |
| S14_extracted_k141_42370 | Not-determined |
| S14_extracted_k141_78555 | Low-quality |
| S14_extracted_k141_71106 | Not-determined |
| S14_extracted_k141_115250 | Not-determined |
| S14_extracted_k141_71109 | Not-determined |
| S14_extracted_k141_12244 | Low-quality |
| S14_extracted_k141_71114 | Low-quality |
| S14_extracted_k141_115258 | Low-quality |
| S14_extracted_k141_56454 | Not-determined |
| S14_extracted_k141_63590 | Not-determined |
| [illegible] | [illegible] |

| | |
|---|---|
| S14_extracted_k141_93710 | Not-determined |
| S14_extracted_k141_12634 | Low-quality |
| S14_extracted_k141_93712 | Not-determined |
| S14_extracted_k141_63891 | Not-determined |
| S14_extracted_k141_115483 | Not-determined |
| S14_extracted_k141_42765 | Not-determined |
| S14_extracted_k141_86228 | Not-determined |
| S14_extracted_k141_48796 | Not-determined |
| S14_extracted_k141_63896 | Not-determined |
| S14_extracted_k141_48799 | Not-determined |
| S14_extracted_k141_100530 | Low-quality |
| S14_extracted_k141_34821 | Low-quality |
| S14_extracted_k141_20337 | Not-determined |
| S14_extracted_k141_115491 | Not-determined |
| S14_extracted_k141_108331 | Low-quality |
| S14_extracted_k141_48810 | Not-determined |
| S14_extracted_k141_12647 | Not-determined |
| S14_extracted_k141_56746 | Not-determined |
| S14_extracted_k141_108336 | Not-determined |
| S14_extracted_k141_42780 | Not-determined |
| S14_extracted_k141_100539 | Low-quality |
| S14_extracted_k141_12649 | Not-determined |
| S14_extracted_k141_20340 | Low-quality |
| S14_extracted_k141_115496 | Not-determined |
| S14_extracted_k141_108340 | Not-determined |
| S14_extracted_k141_12653 | Not-determined |
| S14_extracted_k141_42783 | Not-determined |
| S14_extracted_k141_115500 | Low-quality |
| S14_extracted_k141_20345 | Not-determined |
| S14_extracted_k141_20346 | Not-determined |
| S14_extracted_k141_12656 | Low-quality |
| S14_extracted_k141_48819 | Not-determined |
| S14_extracted_k141_20348 | Not-determined |
| S14_extracted_k141_12660 | Low-quality |
| S14_extracted_k141_48825 | Not-determined |
| S14_extracted_k141_93721 | Low-quality |
| S14_extracted_k141_115512 | Not-determined |
| S14_extracted_k141_78717 | Low-quality |
| S14_extracted_k141_56755 | Low-quality |
| S14_extracted_k141_27004 | Not-determined |
| S14_extracted_k141_78720 | Not-determined |
| S14_extracted_k141_71462 | Not-determined |
| S14_extracted_k141_20361 | Not-determined |
| S14_extracted_k141_34833 | Not-determined |
| S14_extracted_k141_115516 | Not-determined |
| S14_extracted_k141_115517 | Not-determined |
| S14_extracted_k141_71465 | Not-determined |
| S14_extracted_k141_100558 | Not-determined |
| S14_extracted_k141_27007 | Not-determined |
| S14_extracted_k141_27009 | Not-determined |
| S14_extracted_k141_5359 | Not-determined |
| S14_extracted_k141_5361 | Low-quality |
| S14_extracted_k141_34845 | Not-determined |
| S14_extracted_k141_27021 | Not-determined |
| S14_extracted_k141_100562 | Not-determined |
| S14_extracted_k141_78729 | Low-quality |
| S14_extracted_k141_63916 | Not-determined |
| S14_extracted_k141_100567 | Not-determined |
| S14_extracted_k141_27024 | Not-determined |
| S14_extracted_k141_56765 | Not-determined |
| S14_extracted_k141_34850 | Not-determined |
| S14_extracted_k141_56766 | Not-determined |
| S14_extracted_k141_108352 | Not-determined |
| S14_extracted_k141_34852 | Not-determined |
| S14_extracted_k141_5373 | Low-quality |
| S14_extracted_k141_5374 | Not-determined |
| S14_extracted_k141_71490 | Not-determined |
| S14_extracted_k141_56768 | Low-quality |
| S14_extracted_k141_5376 | Not-determined |
| S14_extracted_k141_42804 | Low-quality |
| S14_extracted_k141_100569 | Not-determined |
| S14_extracted_k141_100571 | Not-determined |
| S14_extracted_k141_5386 | Not-determined |
| S14_extracted_k141_5387 | Low-quality |
| S14_extracted_k141_93739 | Not-determined |
| S14_extracted_k141_20375 | Low-quality |
| S14_extracted_k141_5389 | Low-quality |
| S14_extracted_k141_71496 | Low-quality |
| S14_extracted_k141_115541 | Not-determined |
| S14_extracted_k141_115543 | Not-determined |
| S14_extracted_k141_34867 | Not-determined |
| S14_extracted_k141_108366 | Not-determined |
| S14_extracted_k141_48838 | Low-quality |
| S14_extracted_k141_48839 | Not-determined |
| S14_extracted_k141_48841 | Not-determined |
| S14_extracted_k141_115552 | Not-determined |
| S14_extracted_k141_100589 | Not-determined |
| S14_extracted_k141_34883 | Low-quality |
| S14_extracted_k141_93753 | Low-quality |
| S14_extracted_k141_93755 | Not-determined |
| S14_extracted_k141_5409 | Low-quality |
| S14_extracted_k141_63926 | Low-quality |
| S14_extracted_k141_42823 | Not-determined |
| S14_extracted_k141_93759 | Low-quality |
| S14_extracted_k141_63929 | Low-quality |
| S14_extracted_k141_63930 | Not-determined |
| S14_extracted_k141_5413 | Not-determined |
| S14_extracted_k141_93760 | Low-quality |
| S14_extracted_k141_115560 | Not-determined |
| S14_extracted_k141_56793 | Low-quality |
| S14_extracted_k141_71514 | Low-quality |
| S14_extracted_k141_86255 | Not-determined |
| S14_extracted_k141_71518 | Not-determined |
| S14_extracted_k141_86260 | Not-determined |
| S14_extracted_k141_34901 | Not-determined |
| S14_extracted_k141_34903 | Not-determined |
| S14_extracted_k141_86265 | Not-determined |
| S14_extracted_k141_48853 | Not-determined |
| S14_extracted_k141_48854 | Not-determined |
| S14_extracted_k141_71527 | Not-determined |
| S14_extracted_k141_63941 | Low-quality |
| S14_extracted_k141_12710 | Not-determined |
| S14_extracted_k141_115573 | Low-quality |
| S14_extracted_k141_115574 | Not-determined |
| S14_extracted_k141_63947 | Not-determined |
| S14_extracted_k141_34906 | Not-determined |
| S14_extracted_k141_34908 | Not-determined |
| S14_extracted_k141_115585 | Not-determined |
| S14_extracted_k141_34910 | Low-quality |
| S14_extracted_k141_48867 | Low-quality |
| S14_extracted_k141_56803 | Not-determined |
| S14_extracted_k141_115587 | Not-determined |
| S14_extracted_k141_115588 | Low-quality |
| S14_extracted_k141_42846 | Not-determined |
| S14_extracted_k141_34913 | Low-quality |
| S14_extracted_k141_12722 | Not-determined |
| S14_extracted_k141_115591 | Low-quality |
| S14_extracted_k141_48872 | Not-determined |
| S14_extracted_k141_115594 | Not-determined |
| S14_extracted_k141_71541 | Not-determined |
| S14_extracted_k141_71543 | Not-determined |
| S14_extracted_k141_48877 | Low-quality |
| S14_extracted_k141_71545 | Not-determined |
| S14_extracted_k141_115602 | Low-quality |
| S14_extracted_k141_71550 | Not-determined |
| S14_extracted_k141_48884 | Low-quality |
| S14_extracted_k141_48886 | Low-quality |
| S14_extracted_k141_42857 | Not-determined |
| S14_extracted_k141_48890 | Not-determined |
| S14_extracted_k141_56808 | Low-quality |
| S14_extracted_k141_86279 | Not-determined |
| S14_extracted_k141_27064 | Low-quality |
| S14_extracted_k141_108397 | Not-determined |
| S14_extracted_k141_48898 | Not-determined |
| S14_extracted_k141_27072 | Not-determined |
| S14_extracted_k141_34936 | Not-determined |
| S14_extracted_k141_12734 | Low-quality |
| S14_extracted_k141_34937 | Not-determined |
| S14_extracted_k141_48905 | Not-determined |
| S5_extracted_k141_145587 | Complete |
| S14_extracted_k141_100634 | Low-quality |
| S14_extracted_k141_20384 | Not-determined |
| S14_extracted_k141_12737 | Not-determined |
| S14_extracted_k141_115618 | Low-quality |
| S14_extracted_k141_108408 | Not-determined |
| S14_extracted_k141_108410 | Not-determined |
| S14_extracted_k141_5415 | Low-quality |
| S14_extracted_k141_86293 | Not-determined |
| S14_extracted_k141_34945 | Low-quality |
| S14_extracted_k141_78761 | Not-determined |
| S14_extracted_k141_108416 | Not-determined |
| S14_extracted_k141_100643 | Not-determined |
| S14_extracted_k141_42870 | Low-quality |
| S14_extracted_k141_108417 | Low-quality |
| S14_extracted_k141_115628 | Not-determined |
| S14_extracted_k141_108419 | Low-quality |
| S14_extracted_k141_108420 | Not-determined |
| S14_extracted_k141_42875 | Not-determined |
| S14_extracted_k141_5417 | Not-determined |
| S14_extracted_k141_115632 | Not-determined |
| S14_extracted_k141_12749 | Not-determined |
| S14_extracted_k141_108424 | Not-determined |
| S14_extracted_k141_71575 | Not-determined |
| S14_extracted_k141_86298 | Not-determined |
| S14_extracted_k141_108426 | Not-determined |
| S14_extracted_k141_56820 | Low-quality |
| S14_extracted_k141_78768 | Not-determined |
| S14_extracted_k141_86303 | Low-quality |
| S14_extracted_k141_12756 | Not-determined |
| S14_extracted_k141_78770 | Low-quality |
| S14_extracted_k141_56822 | Low-quality |
| S14_extracted_k141_93782 | Low-quality |
| S14_extracted_k141_78771 | Low-quality |
| S14_extracted_k141_56825 | Not-determined |
| S14_extracted_k141_56826 | Low-quality |
| S14_extracted_k141_78772 | Low-quality |
| S14_extracted_k141_78775 | Low-quality |
| S14_extracted_k141_71584 | Not-determined |
| S14_extracted_k141_12762 | Low-quality |
| S14_extracted_k141_115642 | Low-quality |
| S14_extracted_k141_115644 | Not-determined |
| S14_extracted_k141_108430 | Low-quality |
| S14_extracted_k141_86313 | Not-determined |
| S14_extracted_k141_27105 | Not-determined |
| S14_extracted_k141_78783 | Not-determined |
| S14_extracted_k141_93787 | Not-determined |
| S14_extracted_k141_86315 | Low-quality |
| S14_extracted_k141_34967 | Not-determined |
| S14_extracted_k141_12771 | Low-quality |
| S14_extracted_k141_93791 | Not-determined |
| S14_extracted_k141_115646 | Not-determined |
| S14_extracted_k141_12774 | Not-determined |
| S14_extracted_k141_93798 | Not-determined |
| S14_extracted_k141_20408 | Not-determined |
| S14_extracted_k141_27120 | Not-determined |
| S14_extracted_k141_86324 | Not-determined |
| S14_extracted_k141_12787 | Not-determined |
| S14_extracted_k141_78792 | Not-determined |
| S14_extracted_k141_20421 | Not-determined |
| S14_extracted_k141_12791 | Low-quality |
| S14_extracted_k141_48910 | Not-determined |
| S14_extracted_k141_48911 | Not-determined |
| S14_extracted_k141_93812 | Not-determined |
| S14_extracted_k141_27131 | Low-quality |
| S14_extracted_k141_48918 | Not-determined |
| S14_extracted_k141_93818 | Not-determined |
| S14_extracted_k141_48923 | Low-quality |
| S14_extracted_k141_48930 | Not-determined |
| S14_extracted_k141_20434 | Not-determined |
| S14_extracted_k141_48932 | Not-determined |
| S14_extracted_k141_71601 | Low-quality |
| S14_extracted_k141_108450 | Low-quality |
| S14_extracted_k141_20439 | Not-determined |
| S14_extracted_k141_5452 | Low-quality |
| S14_extracted_k141_48944 | Low-quality |
| S14_extracted_k141_42914 | Not-determined |
| S14_extracted_k141_20443 | Not-determined |
| S14_extracted_k141_42920 | Low-quality |
| S14_extracted_k141_71607 | Low-quality |
| S14_extracted_k141_78801 | Not-determined |
| S14_extracted_k141_20447 | Not-determined |
| S14_extracted_k141_78804 | Not-determined |
| S14_extracted_k141_78805 | Not-determined |
| S14_extracted_k141_86345 | Not-determined |
| S14_extracted_k141_115653 | Not-determined |
| S14_extracted_k141_86353 | Not-determined |
| S14_extracted_k141_108458 | Not-determined |
| S14_extracted_k141_86355 | Not-determined |
| S14_extracted_k141_108461 | Low-quality |
| S14_extracted_k141_56841 | Low-quality |
| S14_extracted_k141_56842 | Low-quality |
| S14_extracted_k141_48965 | Low-quality |
| S14_extracted_k141_100671 | Low-quality |
| S14_extracted_k141_100672 | Low-quality |
| S14_extracted_k141_48967 | Low-quality |
| S14_extracted_k141_100675 | Low-quality |
| S14_extracted_k141_48970 | Low-quality |
| S14_extracted_k141_56846 | Low-quality |
| S14_extracted_k141_34991 | Not-determined |
| S14_extracted_k141_12818 | Low-quality |
| S14_extracted_k141_115670 | Not-determined |
| S14_extracted_k141_93843 | Not-determined |
| S14_extracted_k141_115673 | Not-determined |
| S14_extracted_k141_42938 | Low-quality |
| S14_extracted_k141_108472 | Low-quality |
| S14_extracted_k141_108473 | Not-determined |
| S14_extracted_k141_56854 | Low-quality |
| S14_extracted_k141_64005 | Low-quality |
| S14_extracted_k141_86360 | Not-determined |
| S14_extracted_k141_93845 | Low-quality |
| S14_extracted_k141_108478 | Not-determined |
| S14_extracted_k141_20470 | Low-quality |
| S14_extracted_k141_86367 | Not-determined |
| S14_extracted_k141_93850 | Low-quality |
| S14_extracted_k141_20478 | Not-determined |
| S14_extracted_k141_115685 | Not-determined |
| S14_extracted_k141_48993 | Not-determined |
| S14_extracted_k141_64028 | Not-determined |
| S14_extracted_k141_20484 | Not-determined |
| S14_extracted_k141_64030 | Not-determined |
| S14_extracted_k141_48997 | Not-determined |
| S14_extracted_k141_48999 | Low-quality |
| S14_extracted_k141_71630 | Low-quality |
| S14_extracted_k141_42950 | Low-quality |
| S14_extracted_k141_100685 | Low-quality |
| S14_extracted_k141_93857 | Not-determined |
| S14_extracted_k141_115695 | Not-determined |
| S14_extracted_k141_115697 | Not-determined |
| S14_extracted_k141_108490 | Low-quality |
| S14_extracted_k141_93860 | Not-determined |
| S14_extracted_k141_115698 | Not-determined |
| S14_extracted_k141_100688 | Not-determined |
| S14_extracted_k141_93862 | Low-quality |
| S14_extracted_k141_42956 | Not-determined |
| S14_extracted_k141_71637 | Not-determined |
| S14_extracted_k141_93865 | Not-determined |
| S14_extracted_k141_71638 | Low-quality |
| S14_extracted_k141_108494 | Not-determined |
| S14_extracted_k141_93866 | Not-determined |
| S14_extracted_k141_71641 | Not-determined |
| S14_extracted_k141_20494 | Low-quality |
| S14_extracted_k141_20496 | Not-determined |
| S14_extracted_k141_56866 | Low-quality |
| S14_extracted_k141_93873 | Not-determined |
| S14_extracted_k141_64051 | Not-determined |
| S14_extracted_k141_56873 | Low-quality |
| S14_extracted_k141_12847 | Not-determined |
| S14_extracted_k141_115709 | Low-quality |
| S14_extracted_k141_115711 | Not-determined |
| S14_extracted_k141_20505 | Not-determined |
| S14_extracted_k141_27157 | Not-determined |
| S14_extracted_k141_35006 | Not-determined |
| S14_extracted_k141_35007 | Low-quality |
| S14_extracted_k141_71645 | Low-quality |
| S14_extracted_k141_42972 | Low-quality |
| S14_extracted_k141_56882 | Not-determined |
| S14_extracted_k141_12861 | Not-determined |
| S14_extracted_k141_20514 | Not-determined |
| S14_extracted_k141_71649 | Low-quality |
| S14_extracted_k141_93886 | Not-determined |
| S14_extracted_k141_12866 | Low-quality |
| S14_extracted_k141_42984 | Not-determined |
| S14_extracted_k141_42985 | Low-quality |
| S14_extracted_k141_56896 | Not-determined |
| S14_extracted_k141_12876 | Low-quality |
| S14_extracted_k141_27168 | Not-determined |
| S14_extracted_k141_35030 | Low-quality |
| S14_extracted_k141_12877 | Low-quality |
| S14_extracted_k141_27170 | Low-quality |
| S14_extracted_k141_35031 | Not-determined |
| S14_extracted_k141_78836 | Not-determined |
| S14_extracted_k141_35033 | Not-determined |
| S14_extracted_k141_12882 | Not-determined |
| S14_extracted_k141_64068 | Low-quality |
| S14_extracted_k141_71664 | Not-determined |
| S14_extracted_k141_35039 | Not-determined |
| S14_extracted_k141_108523 | Not-determined |
| S14_extracted_k141_71665 | Low-quality |
| S14_extracted_k141_108524 | Not-determined |
| S14_extracted_k141_64071 | Low-quality |
| S14_extracted_k141_108528 | Low-quality |
| S14_extracted_k141_49029 | Not-determined |
| S14_extracted_k141_86419 | Not-determined |
| S14_extracted_k141_20528 | Not-determined |
| S14_extracted_k141_42993 | Not-determined |
| S14_extracted_k141_108530 | Low-quality |
| S14_extracted_k141_78847 | Not-determined |
| S14_extracted_k141_42996 | Not-determined |
| S14_extracted_k141_108534 | Low-quality |
| S14_extracted_k141_20536 | Not-determined |
| S14_extracted_k141_108537 | Low-quality |
| S14_extracted_k141_20538 | Not-determined |
| S14_extracted_k141_35059 | Not-determined |
| S14_extracted_k141_20540 | Not-determined |
| S14_extracted_k141_86423 | Not-determined |
| S14_extracted_k141_20541 | Not-determined |
| S14_extracted_k141_93901 | Low-quality |
| S14_extracted_k141_49045 | Not-determined |
| S14_extracted_k141_43005 | Not-determined |
| S14_extracted_k141_71669 | Not-determined |
| S14_extracted_k141_71671 | Not-determined |
| S14_extracted_k141_43012 | Not-determined |
| S14_extracted_k141_12895 | Not-determined |
| S14_extracted_k141_93918 | Low-quality |
| S14_extracted_k141_5488 | Not-determined |
| S14_extracted_k141_5492 | Not-determined |
| S14_extracted_k141_78867 | Low-quality |
| S14_extracted_k141_71680 | Not-determined |
| S14_extracted_k141_5493 | Low-quality |
| S14_extracted_k141_35080 | Not-determined |
| S14_extracted_k141_5494 | Not-determined |
| S14_extracted_k141_35082 | Low-quality |
| S14_extracted_k141_27189 | Not-determined |
| S14_extracted_k141_27190 | Low-quality |
| S14_extracted_k141_12902 | Not-determined |
| S14_extracted_k141_49049 | Not-determined |
| S14_extracted_k141_71686 | Not-determined |
| S14_extracted_k141_43028 | Not-determined |
| S14_extracted_k141_27199 | Not-determined |
| S14_extracted_k141_71687 | Low-quality |
| S14_extracted_k141_43029 | Not-determined |
| S14_extracted_k141_64081 | Not-determined |
| S14_extracted_k141_78877 | Not-determined |
| S14_extracted_k141_93934 | Not-determined |
| S14_extracted_k141_78882 | Not-determined |
| S14_extracted_k141_12913 | Not-determined |
| S14_extracted_k141_43038 | Not-determined |
| S14_extracted_k141_12916 | Not-determined |
| S14_extracted_k141_64090 | Not-determined |
| S14_extracted_k141_43040 | Not-determined |
| S14_extracted_k141_20558 | Low-quality |
| S14_extracted_k141_115725 | Low-quality |
| S14_extracted_k141_43041 | Not-determined |
| S14_extracted_k141_115728 | Not-determined |
| S14_extracted_k141_12921 | Not-determined |
| S14_extracted_k141_12923 | Not-determined |
| S14_extracted_k141_5506 | Low-quality |
| S14_extracted_k141_64096 | Not-determined |
| S14_extracted_k141_49064 | Not-determined |
| S14_extracted_k141_115730 | Low-quality |
| S14_extracted_k141_12927 | Low-quality |
| S14_extracted_k141_49067 | Not-determined |
| S14_extracted_k141_93944 | Not-determined |
| S14_extracted_k141_49070 | Low-quality |
| S14_extracted_k141_71701 | Not-determined |
| S14_extracted_k141_71705 | Not-determined |
| S14_extracted_k141_64101 | Low-quality |
| S14_extracted_k141_93946 | Not-determined |
| S14_extracted_k141_43044 | Not-determined |
| S14_extracted_k141_27218 | Low-quality |
| S14_extracted_k141_27220 | Low-quality |
| S14_extracted_k141_49081 | Low-quality |
| S14_extracted_k141_93952 | Low-quality |
| S14_extracted_k141_100738 | Not-determined |
| S14_extracted_k141_43048 | Not-determined |
| S14_extracted_k141_93957 | Not-determined |
| S14_extracted_k141_12943 | Not-determined |
| S14_extracted_k141_93961 | Not-determined |
| S14_extracted_k141_35100 | Not-determined |
| S14_extracted_k141_35104 | Low-quality |
| S14_extracted_k141_86456 | Low-quality |
| S14_extracted_k141_35106 | Not-determined |
| S14_extracted_k141_56925 | Low-quality |
| S14_extracted_k141_20573 | Not-determined |
| S14_extracted_k141_100753 | Not-determined |
| S14_extracted_k141_43069 | Not-determined |
| S14_extracted_k141_100756 | Not-determined |
| S14_extracted_k141_56931 | Not-determined |
| S14_extracted_k141_100757 | Not-determined |
| S14_extracted_k141_56932 | Not-determined |
| S14_extracted_k141_20579 | Low-quality |
| S14_extracted_k141_93973 | Low-quality |
| S14_extracted_k141_12959 | Low-quality |
| S14_extracted_k141_12961 | Not-determined |
| S14_extracted_k141_12966 | Not-determined |
| S14_extracted_k141_27237 | Not-determined |
| S14_extracted_k141_100763 | Not-determined |
| S14_extracted_k141_5522 | Low-quality |
| S14_extracted_k141_86460 | Not-determined |
| S14_extracted_k141_100766 | Low-quality |
| S14_extracted_k141_86467 | Low-quality |
| S14_extracted_k141_64118 | Not-determined |
| S14_extracted_k141_12982 | Not-determined |
| S14_extracted_k141_43094 | Low-quality |
| S14_extracted_k141_93991 | Not-determined |
| S14_extracted_k141_43096 | Low-quality |
| S14_extracted_k141_5527 | Not-determined |
| S14_extracted_k141_20588 | Low-quality |
| S14_extracted_k141_20589 | Not-determined |
| S14_extracted_k141_5532 | Not-determined |
| S14_extracted_k141_43103 | Not-determined |
| S14_extracted_k141_108555 | Not-determined |
| S14_extracted_k141_27258 | Not-determined |
| S14_extracted_k141_93992 | Low-quality |
| S14_extracted_k141_100775 | Low-quality |
| S14_extracted_k141_12988 | Not-determined |
| S14_extracted_k141_100776 | Not-determined |
| S14_extracted_k141_108556 | Not-determined |
| S14_extracted_k141_94000 | Not-determined |
| S14_extracted_k141_12989 | Not-determined |
| S14_extracted_k141_56939 | Low-quality |
| S14_extracted_k141_20590 | Low-quality |
| S14_extracted_k141_71746 | Not-determined |
| S14_extracted_k141_86483 | Not-determined |
| S14_extracted_k141_20593 | Not-determined |
| S14_extracted_k141_27273 | Low-quality |
| S14_extracted_k141_71752 | Low-quality |
| S14_extracted_k141_71753 | Low-quality |
| S14_extracted_k141_71757 | Not-determined |
| S14_extracted_k141_86488 | Not-determined |
| S14_extracted_k141_94020 | Low-quality |
| S14_extracted_k141_100796 | Not-determined |
| S6_extracted_k141_56483 | Complete |
| S14_extracted_k141_94023 | Not-determined |
| S14_extracted_k141_115737 | Not-determined |
| S14_extracted_k141_71760 | Not-determined |
| S14_extracted_k141_27277 | Not-determined |
| S14_extracted_k141_100798 | Not-determined |
| S14_extracted_k141_71762 | Not-determined |
| S14_extracted_k141_94028 | Not-determined |
| S14_extracted_k141_71763 | Not-determined |
| S14_extracted_k141_115742 | Low-quality |
| S14_extracted_k141_100811 | Not-determined |
| S14_extracted_k141_43114 | Not-determined |
| S14_extracted_k141_43120 | Not-determined |
| S14_extracted_k141_86500 | Not-determined |
| S14_extracted_k141_71777 | Not-determined |
| S14_extracted_k141_86501 | Not-determined |
| S14_extracted_k141_86504 | Not-determined |
| S14_extracted_k141_56952 | Low-quality |
| S14_extracted_k141_56953 | Low-quality |
| S14_extracted_k141_94041 | Not-determined |
| S14_extracted_k141_108586 | Low-quality |
| S14_extracted_k141_43134 | Low-quality |
| S14_extracted_k141_100814 | Low-quality |
| S14_extracted_k141_94042 | Low-quality |
| S14_extracted_k141_94044 | Not-determined |
| S14_extracted_k141_56958 | Not-determined |
| S14_extracted_k141_35128 | Low-quality |
| S14_extracted_k141_35130 | Low-quality |
| S14_extracted_k141_100817 | Not-determined |
| S14_extracted_k141_100818 | Low-quality |
| S14_extracted_k141_56963 | Not-determined |
| S14_extracted_k141_56965 | Not-determined |
| S14_extracted_k141_115760 | Not-determined |
| S14_extracted_k141_56969 | Low-quality |
| S14_extracted_k141_78905 | Low-quality |
| S14_extracted_k141_78906 | Not-determined |
| S14_extracted_k141_94054 | Not-determined |
| S14_extracted_k141_43148 | Not-determined |
| S14_extracted_k141_115766 | Low-quality |
| S14_extracted_k141_20626 | Low-quality |
| S14_extracted_k141_71782 | Not-determined |
| S14_extracted_k141_115769 | Not-determined |
| S14_extracted_k141_71785 | Not-determined |
| S14_extracted_k141_43156 | Not-determined |
| S14_extracted_k141_5563 | Not-determined |
| S14_extracted_k141_5566 | Not-determined |
| S14_extracted_k141_43158 | Low-quality |
| S14_extracted_k141_71793 | Not-determined |
| S14_extracted_k141_20632 | Not-determined |
| S14_extracted_k141_27309 | Not-determined |
| S14_extracted_k141_108606 | Low-quality |
| S14_extracted_k141_56973 | Not-determined |
| S14_extracted_k141_20637 | Low-quality |
| S14_extracted_k141_108607 | Not-determined |
| S14_extracted_k141_5568 | Not-determined |
| S14_extracted_k141_5570 | Not-determined |
| S14_extracted_k141_13035 | Not-determined |
| S14_extracted_k141_27314 | Not-determined |
| S14_extracted_k141_35163 | Not-determined |
| S14_extracted_k141_78923 | Not-determined |
| S14_extracted_k141_20640 | Not-determined |
| S14_extracted_k141_56984 | Not-determined |
| S14_extracted_k141_78925 | Not-determined |
| S14_extracted_k141_35166 | Low-quality |
| S14_extracted_k141_20644 | Not-determined |
| S14_extracted_k141_35168 | Not-determined |
| S14_extracted_k141_5581 | Not-determined |
| S14_extracted_k141_27324 | Low-quality |
| S14_extracted_k141_71809 | Low-quality |
| S14_extracted_k141_56987 | Not-determined |
| S14_extracted_k141_13048 | Low-quality |
| S14_extracted_k141_20648 | Low-quality |
| S14_extracted_k141_27333 | Not-determined |
| S14_extracted_k141_94089 | Not-determined |
| S14_extracted_k141_56996 | Not-determined |
| S14_extracted_k141_94094 | Not-determined |
| S14_extracted_k141_100866 | Not-determined |
| S14_extracted_k141_78934 | Not-determined |
| S14_extracted_k141_108624 | Not-determined |
| S14_extracted_k141_100874 | Not-determined |
| S14_extracted_k141_94106 | Not-determined |
| S14_extracted_k141_71825 | Not-determined |
| S14_extracted_k141_27346 | Low-quality |
| S14_extracted_k141_27348 | Low-quality |
| S14_extracted_k141_64138 | Not-determined |
| S14_extracted_k141_20657 | Not-determined |
| S14_extracted_k141_20660 | Low-quality |
| S14_extracted_k141_94114 | Low-quality |
| S14_extracted_k141_20661 | Low-quality |
| S14_extracted_k141_100883 | Not-determined |
| S14_extracted_k141_35182 | Not-determined |
| S14_extracted_k141_64146 | Not-determined |
| S14_extracted_k141_115791 | Not-determined |
| S14_extracted_k141_86530 | Not-determined |
| S14_extracted_k141_5593 | Low-quality |
| S14_extracted_k141_43179 | Not-determined |
| S14_extracted_k141_115800 | Not-determined |
| S14_extracted_k141_94122 | Not-determined |
| S14_extracted_k141_5597 | Low-quality |
| S14_extracted_k141_100889 | Not-determined |
| S14_extracted_k141_71832 | Low-quality |
| S14_extracted_k141_71833 | Low-quality |
| S14_extracted_k141_100892 | Not-determined |
| S14_extracted_k141_20669 | Not-determined |
| S14_extracted_k141_115806 | Low-quality |
| S14_extracted_k141_43182 | Not-determined |
| S14_extracted_k141_108632 | Not-determined |
| S14_extracted_k141_86540 | Low-quality |
| S14_extracted_k141_94130 | Not-determined |
| S14_extracted_k141_43184 | Not-determined |
| S14_extracted_k141_5600 | Low-quality |
| S14_extracted_k141_108637 | Low-quality |
| S14_extracted_k141_86544 | Not-determined |
| S14_extracted_k141_27356 | Not-determined |
| S14_extracted_k141_13063 | Not-determined |
| S14_extracted_k141_115811 | Not-determined |
| S14_extracted_k141_115814 | Not-determined |
| S14_extracted_k141_108646 | Low-quality |
| S14_extracted_k141_13068 | Not-determined |
| S14_extracted_k141_5611 | Not-determined |
| S14_extracted_k141_108648 | Not-determined |
| S14_extracted_k141_20683 | Not-determined |
| S14_extracted_k141_78957 | Not-determined |
| S14_extracted_k141_35206 | Not-determined |
| S14_extracted_k141_115819 | Not-determined |
| S14_extracted_k141_78958 | Not-determined |
| S14_extracted_k141_43191 | Not-determined |

| | |
|---|---|
| S14_extracted_k141_115821 | Not-determined |
| S14_extracted_k141_20684 | Not-determined |
| S14_extracted_k141_108661 | Not-determined |
| S14_extracted_k141_108663 | Low-quality |
| S14_extracted_k141_108665 | Not-determined |
| S14_extracted_k141_86555 | Low-quality |
| S14_extracted_k141_43199 | Not-determined |
| S14_extracted_k141_71857 | Not-determined |
| S14_extracted_k141_108669 | Not-determined |
| S14_extracted_k141_5626 | Not-determined |
| S14_extracted_k141_108671 | Not-determined |
| S14_extracted_k141_35218 | Low-quality |
| S14_extracted_k141_35219 | Low-quality |
| S14_extracted_k141_115828 | Not-determined |
| S14_extracted_k141_43207 | Not-determined |
| S14_extracted_k141_71861 | Not-determined |
| S14_extracted_k141_108676 | Low-quality |
| S14_extracted_k141_20694 | Not-determined |
| S14_extracted_k141_71866 | Low-quality |
| S14_extracted_k141_115833 | Not-determined |
| S14_extracted_k141_71868 | Low-quality |
| S14_extracted_k141_86570 | Not-determined |
| S14_extracted_k141_43212 | Not-determined |
| S14_extracted_k141_13076 | Not-determined |
| S14_extracted_k141_86573 | Not-determined |
| S14_extracted_k141_108683 | Not-determined |
| S14_extracted_k141_86577 | Not-determined |
| S14_extracted_k141_13079 | Low-quality |
| S14_extracted_k141_13084 | Not-determined |
| S14_extracted_k141_13085 | Not-determined |
| S14_extracted_k141_71874 | Low-quality |
| S14_extracted_k141_100918 | Low-quality |
| S14_extracted_k141_108686 | Not-determined |
| S14_extracted_k141_35232 | Not-determined |
| S14_extracted_k141_71886 | Not-determined |
| S14_extracted_k141_35243 | Not-determined |
| S14_extracted_k141_86586 | Not-determined |
| S14_extracted_k141_100931 | Low-quality |
| S14_extracted_k141_71887 | Not-determined |
| S14_extracted_k141_13095 | Low-quality |
| S14_extracted_k141_115854 | Not-determined |
| S14_extracted_k141_71889 | Low-quality |
| S14_extracted_k141_35253 | Not-determined |
| S14_extracted_k141_78974 | Low-quality |
| S14_extracted_k141_43220 | Not-determined |
| S14_extracted_k141_115855 | Low-quality |
| S14_extracted_k141_78975 | Not-determined |
| S14_extracted_k141_78977 | Low-quality |
| S14_extracted_k141_43223 | Low-quality |
| S14_extracted_k141_71893 | Not-determined |
| S14_extracted_k141_86598 | Low-quality |
| S14_extracted_k141_115860 | Not-determined |
| S14_extracted_k141_5651 | Low-quality |
| S14_extracted_k141_20723 | Not-determined |
| S14_extracted_k141_115862 | Low-quality |
| S14_extracted_k141_35261 | Not-determined |
| S14_extracted_k141_78980 | Low-quality |
| S14_extracted_k141_71901 | Not-determined |
| S14_extracted_k141_71904 | Not-determined |
| S14_extracted_k141_20726 | Low-quality |
| S14_extracted_k141_71910 | Not-determined |
| S14_extracted_k141_108697 | Not-determined |
| S14_extracted_k141_71913 | Low-quality |
| S14_extracted_k141_43235 | Not-determined |
| S14_extracted_k141_71914 | Low-quality |
| S14_extracted_k141_100947 | Not-determined |
| S14_extracted_k141_35274 | Not-determined |
| S14_extracted_k141_49103 | Not-determined |
| S14_extracted_k141_35276 | Low-quality |
| S14_extracted_k141_49104 | Not-determined |
| S14_extracted_k141_43238 | Not-determined |
| S14_extracted_k141_71924 | Not-determined |
| S14_extracted_k141_20740 | Not-determined |
| S14_extracted_k141_27393 | Not-determined |
| S14_extracted_k141_94175 | Not-determined |
| S14_extracted_k141_86613 | Low-quality |
| S14_extracted_k141_108716 | Not-determined |
| S14_extracted_k141_100961 | Not-determined |
| S14_extracted_k141_86616 | Not-determined |
| S14_extracted_k141_13099 | Not-determined |
| S14_extracted_k141_13100 | Not-determined |
| S14_extracted_k141_49111 | Not-determined |
| S14_extracted_k141_5654 | Not-determined |
| S14_extracted_k141_13102 | Not-determined |
| S14_extracted_k141_20755 | Low-quality |
| S14_extracted_k141_5659 | Low-quality |
| S14_extracted_k141_94189 | Low-quality |
| S14_extracted_k141_78989 | Not-determined |
| S14_extracted_k141_94190 | Not-determined |
| S14_extracted_k141_78991 | Not-determined |
| S14_extracted_k141_5664 | Low-quality |
| S14_extracted_k141_43253 | Not-determined |
| S14_extracted_k141_5666 | Low-quality |
| S14_extracted_k141_49121 | Not-determined |
| S14_extracted_k141_108733 | Not-determined |
| S14_extracted_k141_49122 | Low-quality |
| S14_extracted_k141_94197 | Low-quality |
| S14_extracted_k141_5673 | Low-quality |
| S14_extracted_k141_94200 | Low-quality |
| S14_extracted_k141_94203 | Not-determined |
| S14_extracted_k141_79005 | Not-determined |
| S14_extracted_k141_94204 | Low-quality |
| S14_extracted_k141_43263 | Low-quality |
| S14_extracted_k141_43265 | Not-determined |
| S14_extracted_k141_94209 | Not-determined |
| S14_extracted_k141_64181 | Not-determined |
| S14_extracted_k141_94213 | Not-determined |
| S14_extracted_k141_100992 | Not-determined |
| S14_extracted_k141_5688 | Not-determined |
| S14_extracted_k141_57011 | Not-determined |
| S14_extracted_k141_79017 | Not-determined |
| S14_extracted_k141_27422 | Low-quality |
| S14_extracted_k141_20778 | Not-determined |
| S14_extracted_k141_94216 | Low-quality |
| S14_extracted_k141_27427 | Low-quality |
| S14_extracted_k141_64195 | Not-determined |
| S14_extracted_k141_64196 | Not-determined |
| S14_extracted_k141_71935 | Low-quality |
| S14_extracted_k141_27430 | Low-quality |
| S14_extracted_k141_57021 | Not-determined |
| S14_extracted_k141_108740 | Not-determined |
| S14_extracted_k141_27433 | Not-determined |
| S14_extracted_k141_64202 | Not-determined |
| S14_extracted_k141_86650 | Not-determined |
| S14_extracted_k141_49158 | Low-quality |
| S14_extracted_k141_108747 | Not-determined |
| S14_extracted_k141_35287 | Not-determined |
| S14_extracted_k141_86655 | Low-quality |
| S14_extracted_k141_64207 | Not-determined |
| S14_extracted_k141_35288 | Not-determined |
| S14_extracted_k141_71947 | Not-determined |
| S14_extracted_k141_108755 | Low-quality |
| S14_extracted_k141_86659 | Not-determined |
| S14_extracted_k141_27447 | Not-determined |
| S14_extracted_k141_115914 | Low-quality |
| S14_extracted_k141_64217 | Low-quality |
| S14_extracted_k141_101026 | Not-determined |
| S14_extracted_k141_27456 | Not-determined |
| S14_extracted_k141_86665 | Not-determined |
| S14_extracted_k141_49167 | Low-quality |
| S14_extracted_k141_43306 | Low-quality |
| S14_extracted_k141_108764 | Not-determined |
| S14_extracted_k141_35306 | Low-quality |
| S14_extracted_k141_43308 | Low-quality |
| S14_extracted_k141_43315 | Low-quality |
| S14_extracted_k141_43317 | Not-determined |
| S14_extracted_k141_20794 | Not-determined |
| S14_extracted_k141_43319 | Not-determined |
| S14_extracted_k141_43321 | Low-quality |
| S14_extracted_k141_49184 | Not-determined |
| S14_extracted_k141_43322 | Not-determined |
| S14_extracted_k141_27469 | Low-quality |
| S14_extracted_k141_115927 | Not-determined |
| S14_extracted_k141_115929 | Not-determined |
| S14_extracted_k141_86682 | Not-determined |
| S14_extracted_k141_86683 | Not-determined |
| S14_extracted_k141_64230 | Not-determined |
| S14_extracted_k141_115932 | Not-determined |
| S14_extracted_k141_108787 | Not-determined |
| S14_extracted_k141_64232 | Low-quality |
| S14_extracted_k141_71964 | Low-quality |
| S14_extracted_k141_64233 | Not-determined |
| S14_extracted_k141_64237 | Not-determined |
| S14_extracted_k141_108790 | Low-quality |
| S14_extracted_k141_94227 | Not-determined |
| S14_extracted_k141_71967 | Low-quality |
| S14_extracted_k141_115936 | Not-determined |
| S14_extracted_k141_71970 | Low-quality |
| S14_extracted_k141_115937 | Not-determined |
| S14_extracted_k141_57044 | Not-determined |
| S14_extracted_k141_79062 | Not-determined |
| S14_extracted_k141_108796 | Not-determined |
| S14_extracted_k141_27479 | Not-determined |
| S14_extracted_k141_79064 | Not-determined |
| S14_extracted_k141_57046 | Not-determined |
| S14_extracted_k141_20800 | Not-determined |
| S14_extracted_k141_49195 | Not-determined |
| S14_extracted_k141_115940 | Not-determined |
| S14_extracted_k141_79069 | Not-determined |
| S14_extracted_k141_108803 | Low-quality |
| S14_extracted_k141_71976 | Not-determined |
| S14_extracted_k141_115945 | Low-quality |
| S14_extracted_k141_71978 | Not-determined |
| S14_extracted_k141_86701 | Not-determined |
| S14_extracted_k141_94246 | Not-determined |
| S14_extracted_k141_49205 | Not-determined |
| S14_extracted_k141_49207 | Not-determined |
| S14_extracted_k141_49208 | Not-determined |
| S14_extracted_k141_5750 | Not-determined |
| S14_extracted_k141_49210 | Low-quality |
| S14_extracted_k141_79078 | Not-determined |
| S14_extracted_k141_20818 | Low-quality |
| S14_extracted_k141_108809 | Not-determined |
| S14_extracted_k141_57056 | Not-determined |
| S14_extracted_k141_49212 | Low-quality |
| S14_extracted_k141_57058 | Not-determined |
| S14_extracted_k141_86711 | Not-determined |
| S14_extracted_k141_5754 | Not-determined |
| S14_extracted_k141_27488 | Low-quality |
| S14_extracted_k141_108814 | Not-determined |
| S14_extracted_k141_5758 | Not-determined |
| S14_extracted_k141_20828 | Not-determined |
| S14_extracted_k141_57064 | Not-determined |
| S4_vs_VIRSorter_31825-circular_ge | Medium-quality |
| S14_extracted_k141_71981 | Not-determined |
| S14_extracted_k141_108818 | Low-quality |
| S14_extracted_k141_94259 | Low-quality |
| S14_extracted_k141_108819 | Low-quality |
| S14_extracted_k141_108820 | Low-quality |
| S14_extracted_k141_94260 | Not-determined |
| S14_extracted_k141_108821 | Not-determined |
| S14_extracted_k141_35314 | Low-quality |
| S14_extracted_k141_94261 | Low-quality |
| S14_extracted_k141_64246 | Low-quality |
| S14_extracted_k141_108825 | Not-determined |
| S14_extracted_k141_94268 | Low-quality |
| S14_extracted_k141_27502 | Not-determined |
| S14_extracted_k141_35318 | Not-determined |
| S14_extracted_k141_13131 | Not-determined |
| S14_extracted_k141_86723 | Not-determined |
| S14_extracted_k141_71992 | Low-quality |
| S14_extracted_k141_35323 | Low-quality |
| S14_extracted_k141_57076 | Not-determined |
| S14_extracted_k141_57077 | Not-determined |
| S14_extracted_k141_108837 | Not-determined |
| S14_extracted_k141_57080 | Not-determined |
| S14_extracted_k141_57081 | Not-determined |
| S14_extracted_k141_20857 | Not-determined |
| S14_extracted_k141_27509 | Not-determined |
| S14_extracted_k141_57085 | Low-quality |
| S14_extracted_k141_5774 | Not-determined |
| S14_extracted_k141_108845 | Not-determined |
| S14_extracted_k141_57088 | Not-determined |
| S14_extracted_k141_27513 | Not-determined |
| S14_extracted_k141_86735 | Not-determined |
| S14_extracted_k141_115980 | Not-determined |
| S14_extracted_k141_57095 | Not-determined |
| S14_extracted_k141_27519 | Not-determined |
| S14_extracted_k141_108851 | Not-determined |
| S14_extracted_k141_35337 | Not-determined |
| S14_extracted_k141_5779 | Low-quality |
| S14_extracted_k141_108853 | Not-determined |
| S14_extracted_k141_94286 | Not-determined |
| S14_extracted_k141_13142 | Not-determined |
| S14_extracted_k141_101045 | Not-determined |
| S14_extracted_k141_72014 | Not-determined |
| S14_extracted_k141_86741 | Low-quality |
| S14_extracted_k141_5786 | Low-quality |
| S14_extracted_k141_72015 | Not-determined |
| S14_extracted_k141_101050 | Not-determined |
| S14_extracted_k141_86743 | Low-quality |
| S14_extracted_k141_13148 | Low-quality |
| S14_extracted_k141_57109 | Not-determined |
| S14_extracted_k141_5788 | Not-determined |
| S14_extracted_k141_108869 | Not-determined |
| S14_extracted_k141_64265 | Not-determined |
| S14_extracted_k141_57111 | Not-determined |
| S14_extracted_k141_115988 | Low-quality |
| S14_extracted_k141_72017 | Low-quality |
| S14_extracted_k141_72018 | Not-determined |
| S14_extracted_k141_64268 | Not-determined |
| S14_extracted_k141_64269 | Not-determined |
| S14_extracted_k141_13156 | Low-quality |
| S14_extracted_k141_20884 | Not-determined |
| S14_extracted_k141_27539 | Not-determined |
| S14_extracted_k141_27545 | Not-determined |
| S14_extracted_k141_13161 | Low-quality |
| S14_extracted_k141_13162 | Low-quality |
| S14_extracted_k141_108883 | Not-determined |
| S14_extracted_k141_86755 | Not-determined |
| S14_extracted_k141_64278 | Low-quality |
| S14_extracted_k141_64279 | Low-quality |
| S14_extracted_k141_86759 | Not-determined |
| S14_extracted_k141_13172 | Not-determined |
| S14_extracted_k141_20888 | Not-determined |
| S14_extracted_k141_13175 | Not-determined |
| S14_extracted_k141_79091 | Low-quality |
| S8_extracted_k141_132697 | Complete |
| S14_extracted_k141_20892 | Not-determined |
| S14_extracted_k141_94304 | Not-determined |
| S14_extracted_k141_43344 | Not-determined |
| S14_extracted_k141_27557 | Not-determined |
| S14_extracted_k141_13183 | Not-determined |
| S14_extracted_k141_64282 | Not-determined |
| S14_extracted_k141_101065 | Not-determined |
| S14_extracted_k141_86769 | Low-quality |
| S14_extracted_k141_101066 | Not-determined |
| S14_extracted_k141_101067 | Not-determined |
| S14_extracted_k141_116008 | Not-determined |
| S14_extracted_k141_116009 | Not-determined |
| S14_extracted_k141_64288 | Not-determined |
| S14_extracted_k141_27566 | Low-quality |
| S14_extracted_k141_5811 | Not-determined |
| S14_extracted_k141_116011 | Not-determined |
| S14_extracted_k141_79098 | Not-determined |
| S14_extracted_k141_27571 | Not-determined |
| S14_extracted_k141_5815 | Not-determined |
| S14_extracted_k141_5816 | Low-quality |
| S14_extracted_k141_79103 | Not-determined |
| S14_extracted_k141_43355 | Not-determined |
| S14_extracted_k141_57129 | Low-quality |
| S14_extracted_k141_27579 | Not-determined |
| S14_extracted_k141_43357 | Not-determined |
| S14_extracted_k141_43358 | Not-determined |
| S14_extracted_k141_5821 | Low-quality |
| S14_extracted_k141_43361 | Not-determined |
| S14_extracted_k141_79107 | Not-determined |
| S14_extracted_k141_43362 | Not-determined |
| S14_extracted_k141_94318 | Not-determined |
| S14_extracted_k141_86786 | Low-quality |
| S14_extracted_k141_108900 | Not-determined |
| S14_extracted_k141_79112 | Low-quality |
| S14_extracted_k141_57136 | Not-determined |
| S14_extracted_k141_108902 | Low-quality |
| S14_extracted_k141_101081 | Low-quality |
| S14_extracted_k141_43371 | Not-determined |
| S14_extracted_k141_108906 | Low-quality |
| S14_extracted_k141_101084 | Low-quality |
| S14_extracted_k141_27591 | Low-quality |
| S14_extracted_k141_86795 | Not-determined |
| S14_extracted_k141_27596 | Not-determined |
| S14_extracted_k141_20921 | Low-quality |
| S14_extracted_k141_5830 | Not-determined |
| S14_extracted_k141_20924 | Not-determined |
| S14_extracted_k141_35347 | Not-determined |
| S14_extracted_k141_108915 | Not-determined |
| S14_extracted_k141_57142 | Not-determined |
| S14_extracted_k141_43388 | Low-quality |
| S14_extracted_k141_79124 | Not-determined |
| S14_extracted_k141_43390 | Not-determined |
| S14_extracted_k141_79126 | Not-determined |
| S14_extracted_k141_27605 | Not-determined |
| S14_extracted_k141_86805 | Low-quality |
| S14_extracted_k141_43391 | Low-quality |
| S14_extracted_k141_79127 | Not-determined |
| S14_extracted_k141_108921 | Not-determined |
| S14_extracted_k141_116030 | Not-determined |
| S14_extracted_k141_13197 | Not-determined |
| S14_extracted_k141_13198 | Low-quality |
| S14_extracted_k141_79131 | Not-determined |
| S14_extracted_k141_86811 | Not-determined |
| S14_extracted_k141_20931 | Low-quality |
| S14_extracted_k141_20938 | Low-quality |
| S14_extracted_k141_5843 | Not-determined |
| S14_extracted_k141_43398 | Not-determined |
| S14_extracted_k141_94351 | Not-determined |
| S14_extracted_k141_108934 | Not-determined |
| S14_extracted_k141_57154 | Not-determined |
| S14_extracted_k141_108936 | Not-determined |
| S14_extracted_k141_13212 | Not-determined |
| S14_extracted_k141_86823 | Not-determined |
| S14_extracted_k141_35363 | Not-determined |
| S14_extracted_k141_57163 | Not-determined |
| S14_extracted_k141_35366 | Not-determined |
| S14_extracted_k141_43408 | Not-determined |
| S14_extracted_k141_79135 | Low-quality |
| S14_extracted_k141_86827 | Not-determined |
| S14_extracted_k141_35370 | Low-quality |
| S14_extracted_k141_64327 | Low-quality |
| S14_extracted_k141_57170 | Low-quality |
| S14_extracted_k141_57171 | Not-determined |
| S14_extracted_k141_86830 | Low-quality |
| S14_extracted_k141_101108 | Not-determined |
| S14_extracted_k141_86832 | Not-determined |
| S14_extracted_k141_43413 | Not-determined |
| S14_extracted_k141_49253 | Not-determined |
| S14_extracted_k141_43414 | Low-quality |
| S14_extracted_k141_49254 | Not-determined |
| S14_extracted_k141_57176 | Not-determined |
| S14_extracted_k141_108954 | Low-quality |
| S14_extracted_k141_101116 | Low-quality |
| S14_extracted_k141_43415 | Low-quality |
| S14_extracted_k141_20959 | Not-determined |
| S14_extracted_k141_101122 | Not-determined |
| S14_extracted_k141_35382 | Not-determined |
| S14_extracted_k141_5853 | Not-determined |
| S14_extracted_k141_79139 | Not-determined |
| S14_extracted_k141_86843 | Low-quality |
| S14_extracted_k141_108966 | Not-determined |
| S14_extracted_k141_101127 | Not-determined |
| S14_extracted_k141_43426 | Not-determined |
| S14_extracted_k141_108969 | Not-determined |
| S14_extracted_k141_20966 | Not-determined |
| S14_extracted_k141_35393 | Not-determined |
| S14_extracted_k141_35394 | Not-determined |
| S14_extracted_k141_5864 | Not-determined |
| S14_extracted_k141_5865 | Not-determined |
| S14_extracted_k141_35400 | Not-determined |
| S14_extracted_k141_49265 | Not-determined |
| S14_extracted_k141_108973 | Not-determined |
| S14_extracted_k141_5869 | Not-determined |
| S14_extracted_k141_5870 | Not-determined |
| S14_extracted_k141_79160 | Low-quality |
| S14_extracted_k141_64350 | Not-determined |
| S14_extracted_k141_108980 | Not-determined |
| S14_extracted_k141_57204 | Not-determined |
| S14_extracted_k141_35414 | Not-determined |
| S14_extracted_k141_27638 | Not-determined |
| S14_extracted_k141_35419 | Not-determined |
| S14_extracted_k141_20979 | Not-determined |
| S14_extracted_k141_35420 | Not-determined |
| S14_extracted_k141_79174 | Not-determined |
| S14_extracted_k141_43438 | Low-quality |
| S14_extracted_k141_101153 | Not-determined |
| S14_extracted_k141_79177 | Not-determined |
| S14_extracted_k141_49275 | Low-quality |
| S14_extracted_k141_35427 | Not-determined |
| S14_extracted_k141_57213 | Not-determined |
| S14_extracted_k141_94370 | Not-determined |
| S14_extracted_k141_57216 | Not-determined |
| S14_extracted_k141_116058 | Not-determined |
| S14_extracted_k141_49291 | Not-determined |
| S14_extracted_k141_27652 | Not-determined |
| S14_extracted_k141_79188 | Not-determined |
| S14_extracted_k141_27656 | Not-determined |
| S14_extracted_k141_49292 | Low-quality |
| S14_extracted_k141_5888 | Not-determined |
| S14_extracted_k141_116060 | Low-quality |
| S14_extracted_k141_5890 | Not-determined |
| S14_extracted_k141_79193 | Not-determined |
| S14_extracted_k141_35442 | Low-quality |
| S14_extracted_k141_116062 | Not-determined |
| S14_extracted_k141_57225 | Low-quality |
| S14_extracted_k141_116063 | Not-determined |
| S14_extracted_k141_5891 | Not-determined |
| S14_extracted_k141_101161 | Low-quality |
| S14_extracted_k141_57232 | Not-determined |
| S14_extracted_k141_27660 | Low-quality |
| S14_extracted_k141_57236 | Not-determined |
| S14_extracted_k141_27661 | Not-determined |
| S14_extracted_k141_57237 | Not-determined |
| S14_extracted_k141_94371 | Low-quality |
| S14_extracted_k141_27663 | Low-quality |
| S14_extracted_k141_79199 | Not-determined |
| S14_extracted_k141_79202 | Not-determined |
| S14_extracted_k141_79205 | Not-determined |
| S14_extracted_k141_86870 | Not-determined |
| S14_extracted_k141_43464 | Not-determined |
| S14_extracted_k141_86874 | Not-determined |
| S14_extracted_k141_13242 | Low-quality |
| S14_extracted_k141_116076 | Low-quality |
| S14_extracted_k141_94388 | Not-determined |
| S14_extracted_k141_49310 | Not-determined |
| S14_extracted_k141_116082 | Not-determined |
| S14_extracted_k141_116085 | Not-determined |
| S14_extracted_k141_57240 | Not-determined |
| S14_extracted_k141_49316 | Not-determined |
| S14_extracted_k141_57243 | Not-determined |
| S14_extracted_k141_49319 | Not-determined |
| S14_extracted_k141_49321 | Not-determined |
| S14_extracted_k141_86895 | Low-quality |
| S14_extracted_k141_57250 | Not-determined |
| S14_extracted_k141_86899 | Low-quality |
| S14_extracted_k141_43481 | Not-determined |
| S14_extracted_k141_13261 | Not-determined |
| S14_extracted_k141_35469 | Low-quality |
| S14_extracted_k141_35470 | Not-determined |
| S14_extracted_k141_13264 | Low-quality |
| S14_extracted_k141_35473 | Not-determined |
| S14_extracted_k141_13268 | Low-quality |
| S14_extracted_k141_49330 | Low-quality |
| S14_extracted_k141_43485 | Not-determined |
| S14_extracted_k141_13273 | Low-quality |
| S14_extracted_k141_43488 | Not-determined |
| S14_extracted_k141_43489 | Not-determined |
| S14_extracted_k141_35475 | Not-determined |
| S14_extracted_k141_13277 | Not-determined |
| S14_extracted_k141_49342 | Not-determined |
| S14_extracted_k141_101173 | Low-quality |
| S14_extracted_k141_49344 | Not-determined |
| S14_extracted_k141_86915 | Low-quality |
| S14_extracted_k141_43496 | Low-quality |
| S14_extracted_k141_49346 | Not-determined |
| S14_extracted_k141_5922 | Low-quality |
| S14_extracted_k141_43498 | Low-quality |
| S14_extracted_k141_101179 | Low-quality |
| S14_extracted_k141_49350 | Low-quality |
| S14_extracted_k141_5928 | Not-determined |
| S14_extracted_k141_116105 | Not-determined |
| S14_extracted_k141_13289 | Low-quality |
| S14_extracted_k141_5934 | Low-quality |
| S14_extracted_k141_86926 | Not-determined |
| S14_extracted_k141_49355 | Not-determined |
| S14_extracted_k141_27673 | Not-determined |
| S14_extracted_k141_57276 | Not-determined |
| S14_extracted_k141_20998 | Not-determined |
| S14_extracted_k141_86932 | Not-determined |
| S14_extracted_k141_57278 | Not-determined |
| S14_extracted_k141_86935 | Low-quality |
| S14_extracted_k141_27683 | Not-determined |
| S14_extracted_k141_49359 | Not-determined |
| S14_extracted_k141_86939 | Not-determined |
| S14_extracted_k141_49360 | Not-determined |
| S14_extracted_k141_116118 | Low-quality |
| S14_extracted_k141_21000 | Not-determined |
| S14_extracted_k141_5943 | Not-determined |
| S14_extracted_k141_43507 | Not-determined |
| S14_extracted_k141_86942 | Not-determined |
| S14_extracted_k141_79246 | Low-quality |
| S14_extracted_k141_79247 | Low-quality |
| S14_extracted_k141_86944 | Low-quality |
| S14_extracted_k141_21002 | Not-determined |
| S14_extracted_k141_86946 | Not-determined |
| S14_extracted_k141_116125 | Not-determined |
| S14_extracted_k141_101183 | Low-quality |
| S14_extracted_k141_72035 | Low-quality |
| S14_extracted_k141_86950 | Not-determined |
| S14_extracted_k141_57286 | Low-quality |
| S14_extracted_k141_13307 | Not-determined |
| S14_extracted_k141_13308 | Not-determined |
| S14_extracted_k141_94416 | Not-determined |
| S14_extracted_k141_109007 | Not-determined |
| S14_extracted_k141_72043 | Not-determined |
| S14_extracted_k141_5954 | Not-determined |
| S14_extracted_k141_94420 | Low-quality |
| S14_extracted_k141_21012 | Not-determined |
| S14_extracted_k141_79255 | Low-quality |
| S14_extracted_k141_79257 | Not-determined |
| S14_extracted_k141_101192 | Not-determined |
| S14_extracted_k141_101194 | Not-determined |
| S14_extracted_k141_94427 | Low-quality |
| S14_extracted_k141_57297 | Low-quality |
| S14_extracted_k141_86972 | Not-determined |
| S14_extracted_k141_79260 | Not-determined |
| S14_extracted_k141_86975 | Not-determined |
| S14_extracted_k141_79261 | Low-quality |
| S14_extracted_k141_101201 | Not-determined |
| S14_extracted_k141_5964 | Not-determined |
| S14_extracted_k141_64353 | Not-determined |
| S14_extracted_k141_64354 | Not-determined |
| S14_extracted_k141_35486 | Not-determined |
| S14_extracted_k141_49379 | Low-quality |
| S14_extracted_k141_13330 | Not-determined |
| S14_extracted_k141_64359 | Low-quality |
| S14_extracted_k141_109017 | Not-determined |
| S14_extracted_k141_35492 | Not-determined |
| S14_extracted_k141_116140 | Not-determined |
| S14_extracted_k141_79271 | Low-quality |
| S14_extracted_k141_94441 | Low-quality |
| S14_extracted_k141_116143 | Not-determined |
| S14_extracted_k141_101212 | Not-determined |
| S14_extracted_k141_86984 | Not-determined |
| S14_extracted_k141_94446 | Not-determined |
| S14_extracted_k141_57313 | Low-quality |
| S14_extracted_k141_64364 | Not-determined |
| S14_extracted_k141_27703 | Low-quality |
| S14_extracted_k141_13337 | Not-determined |
| S14_extracted_k141_57316 | Not-determined |
| S14_extracted_k141_79275 | Not-determined |
| S14_extracted_k141_27707 | Not-determined |
| S14_extracted_k141_13340 | Not-determined |
| S14_extracted_k141_94450 | Low-quality |
| S14_extracted_k141_79277 | Not-determined |
| S14_extracted_k141_116149 | Not-determined |
| S14_extracted_k141_64368 | Low-quality |
| S14_extracted_k141_116150 | Not-determined |
| S14_extracted_k141_94454 | Low-quality |
| S14_extracted_k141_79278 | Not-determined |
| S14_extracted_k141_94456 | Not-determined |
| S14_extracted_k141_101216 | Not-determined |
| S14_extracted_k141_79280 | Low-quality |
| S14_extracted_k141_94457 | Low-quality |
| S14_extracted_k141_116155 | Not-determined |
| S14_extracted_k141_86990 | Not-determined |
| S14_extracted_k141_27715 | Not-determined |
| S14_extracted_k141_5978 | Not-determined |
| S14_extracted_k141_57321 | Not-determined |
| S14_extracted_k141_57322 | Not-determined |
| S14_extracted_k141_79285 | Not-determined |
| S14_extracted_k141_13349 | Not-determined |
| S14_extracted_k141_94462 | Not-determined |
| S14_extracted_k141_109034 | Not-determined |
| S14_extracted_k141_35509 | Low-quality |
| S14_extracted_k141_101220 | Not-determined |
| S14_extracted_k141_13354 | Not-determined |
| S14_extracted_k141_101221 | Low-quality |
| S14_extracted_k141_27720 | Not-determined |
| S14_extracted_k141_27722 | Not-determined |
| S14_extracted_k141_79294 | Low-quality |
| S14_extracted_k141_87000 | Not-determined |
| S14_extracted_k141_57336 | Not-determined |
| S14_extracted_k141_27730 | Not-determined |
| S14_extracted_k141_101223 | Not-determined |
| S14_extracted_k141_101226 | Not-determined |
| S14_extracted_k141_87009 | Low-quality |
| S14_extracted_k141_101231 | Low-quality |
| S14_extracted_k141_35519 | Not-determined |
| S14_extracted_k141_35520 | Not-determined |
| S14_extracted_k141_27733 | Low-quality |

| | |
|---|---|
| S14_extracted_k141_87012 | Not-determined |
| S14_extracted_k141_27735 | Not-determined |
| S14_extracted_k141_101235 | Low-quality |
| S14_extracted_k141_109044 | Not-determined |
| S14_extracted_k141_35522 | Not-determined |
| S14_extracted_k141_109046 | Low-quality |
| S14_extracted_k141_35524 | Low-quality |
| S14_extracted_k141_94476 | Not-determined |
| S14_extracted_k141_116173 | Low-quality |
| S14_extracted_k141_116174 | Not-determined |
| S14_extracted_k141_49428 | Low-quality |
| S14_extracted_k141_6007 | Low-quality |
| S14_extracted_k141_27740 | Not-determined |
| S14_extracted_k141_79305 | Not-determined |
| S14_extracted_k141_21037 | Not-determined |
| S14_extracted_k141_49435 | Not-determined |
| S14_extracted_k141_109058 | Not-determined |
| S14_extracted_k141_64372 | Not-determined |
| S14_extracted_k141_109060 | Not-determined |
| S14_extracted_k141_94489 | Low-quality |
| S14_extracted_k141_6014 | Low-quality |
| S14_extracted_k141_21044 | Not-determined |
| S14_extracted_k141_64375 | Not-determined |
| S14_extracted_k141_27745 | Low-quality |
| S14_extracted_k141_6021 | Low-quality |
| S14_extracted_k141_49442 | Not-determined |
| S14_extracted_k141_35537 | Low-quality |
| S14_extracted_k141_101258 | Low-quality |
| S14_extracted_k141_64382 | Not-determined |
| S14_extracted_k141_101266 | Low-quality |
| S14_extracted_k141_57369 | Low-quality |
| S14_extracted_k141_109070 | Not-determined |
| S14_extracted_k141_101267 | Low-quality |
| S14_extracted_k141_109072 | Low-quality |
| S14_extracted_k141_43511 | Not-determined |
| S14_extracted_k141_21061 | Not-determined |
| S14_extracted_k141_101269 | Low-quality |
| S14_extracted_k141_109076 | Low-quality |
| S14_extracted_k141_101271 | Not-determined |
| S14_extracted_k141_43515 | Not-determined |
| S14_extracted_k141_27756 | Not-determined |
| S14_extracted_k141_49455 | Low-quality |
| S14_extracted_k141_101275 | Low-quality |
| S14_extracted_k141_87036 | Not-determined |
| S14_extracted_k141_27759 | Not-determined |
| S14_extracted_k141_109085 | Low-quality |
| S14_extracted_k141_35556 | Not-determined |
| S14_extracted_k141_43519 | Not-determined |
| S14_extracted_k141_43521 | Not-determined |
| S14_extracted_k141_109090 | Not-determined |
| S14_extracted_k141_94506 | Low-quality |
| S14_extracted_k141_87042 | Low-quality |
| S14_extracted_k141_72074 | Not-determined |
| S14_extracted_k141_57381 | Not-determined |
| S14_extracted_k141_6028 | Not-determined |
| S14_extracted_k141_87043 | Not-determined |
| S14_extracted_k141_27773 | Not-determined |
| S14_extracted_k141_87046 | Not-determined |
| S14_extracted_k141_49467 | Not-determined |
| S14_extracted_k141_49469 | Low-quality |
| S14_extracted_k141_27776 | Not-determined |
| S14_extracted_k141_79335 | Not-determined |
| S14_extracted_k141_43531 | Not-determined |
| S14_extracted_k141_64390 | Not-determined |
| S14_extracted_k141_43534 | Not-determined |
| S14_extracted_k141_35569 | Low-quality |
| S14_extracted_k141_87052 | Not-determined |
| S14_extracted_k141_116196 | Not-determined |
| S14_extracted_k141_116199 | Low-quality |
| S14_extracted_k141_21091 | Low-quality |
| S14_extracted_k141_116201 | Low-quality |
| S14_extracted_k141_116202 | Not-determined |
| S14_extracted_k141_35576 | Not-determined |
| S14_extracted_k141_21094 | Low-quality |
| S14_extracted_k141_57400 | Not-determined |
| S14_extracted_k141_116205 | Low-quality |
| S14_extracted_k141_101294 | Low-quality |
| S14_extracted_k141_49487 | Not-determined |
| S14_extracted_k141_49494 | Not-determined |
| S14_extracted_k141_43549 | Not-determined |
| S14_extracted_k141_109123 | Not-determined |
| S14_extracted_k141_6056 | Not-determined |
| S14_extracted_k141_72092 | Not-determined |
| S14_extracted_k141_116211 | Not-determined |
| S14_extracted_k141_35583 | Not-determined |
| S14_extracted_k141_87066 | Not-determined |
| S14_extracted_k141_94524 | Not-determined |
| S14_extracted_k141_35587 | Not-determined |
| S14_extracted_k141_109127 | Not-determined |
| S14_extracted_k141_6060 | Not-determined |
| S14_extracted_k141_57406 | Not-determined |
| S14_extracted_k141_27801 | Not-determined |
| S14_extracted_k141_49503 | Not-determined |
| S14_extracted_k141_116221 | Not-determined |
| S14_extracted_k141_79367 | Low-quality |
| S14_extracted_k141_116223 | Not-determined |
| S14_extracted_k141_79370 | Not-determined |
| S14_extracted_k141_87083 | Not-determined |
| S14_extracted_k141_49506 | Not-determined |
| S14_extracted_k141_109133 | Not-determined |
| S14_extracted_k141_94534 | Not-determined |
| S14_extracted_k141_49508 | Not-determined |
| S14_extracted_k141_72101 | Not-determined |
| S14_extracted_k141_109137 | Not-determined |
| S14_extracted_k141_35605 | Low-quality |
| S14_extracted_k141_72104 | Not-determined |
| S14_extracted_k141_94537 | Not-determined |
| S14_extracted_k141_94543 | Not-determined |
| S14_extracted_k141_116240 | Not-determined |
| S14_extracted_k141_87096 | Not-determined |
| S14_extracted_k141_79384 | Low-quality |
| S14_extracted_k141_79386 | Low-quality |
| S14_extracted_k141_116244 | Low-quality |
| S14_extracted_k141_116245 | Low-quality |
| S14_extracted_k141_49519 | Low-quality |
| S14_extracted_k141_94547 | Not-determined |
| S14_extracted_k141_94548 | Not-determined |
| S14_extracted_k141_49523 | Not-determined |
| S14_extracted_k141_94551 | Not-determined |
| S14_extracted_k141_94552 | Not-determined |
| S14_extracted_k141_79394 | Not-determined |
| S14_extracted_k141_79396 | Not-determined |
| S14_extracted_k141_116252 | Not-determined |
| S14_extracted_k141_35619 | Not-determined |
| S14_extracted_k141_116253 | Low-quality |
| S14_extracted_k141_35623 | Low-quality |
| S14_extracted_k141_64401 | Not-determined |
| S14_extracted_k141_79398 | Not-determined |
| S14_extracted_k141_64403 | Low-quality |
| S14_extracted_k141_94568 | Not-determined |
| S14_extracted_k141_49537 | Not-determined |
| S14_extracted_k141_57411 | Low-quality |
| S14_extracted_k141_101311 | Not-determined |
| S14_extracted_k141_6081 | Low-quality |
| S14_extracted_k141_43572 | Low-quality |
| S14_extracted_k141_116264 | Not-determined |
| S14_extracted_k141_35633 | Not-determined |
| S14_extracted_k141_64411 | Low-quality |
| S14_extracted_k141_87117 | Low-quality |
| S14_extracted_k141_49555 | Low-quality |
| S14_extracted_k141_79415 | Low-quality |
| S14_extracted_k141_49556 | Not-determined |
| S14_extracted_k141_35644 | Not-determined |
| S14_extracted_k141_35646 | Not-determined |
| S14_extracted_k141_64416 | Not-determined |
| S14_extracted_k141_27831 | Low-quality |
| S14_extracted_k141_35649 | Not-determined |
| S14_extracted_k141_27833 | Not-determined |
| S14_extracted_k141_35650 | Not-determined |
| S14_extracted_k141_116276 | Not-determined |
| S14_extracted_k141_109157 | Low-quality |
| S14_extracted_k141_72115 | Not-determined |
| S14_extracted_k141_64419 | Not-determined |
| S14_extracted_k141_21126 | Low-quality |
| S14_extracted_k141_43585 | Not-determined |
| S14_extracted_k141_57423 | Not-determined |
| S14_extracted_k141_109163 | Low-quality |
| S14_extracted_k141_72122 | Not-determined |
| S14_extracted_k141_57425 | Low-quality |
| S14_extracted_k141_49575 | Not-determined |
| S14_extracted_k141_72124 | Low-quality |
| S14_extracted_k141_109167 | Not-determined |
| S14_extracted_k141_72126 | Low-quality |
| S14_extracted_k141_101340 | Low-quality |
| S14_extracted_k141_87136 | Not-determined |
| S14_extracted_k141_79434 | Low-quality |
| S14_extracted_k141_94580 | Low-quality |
| S14_extracted_k141_79436 | Low-quality |
| S14_extracted_k141_101342 | Low-quality |
| S14_extracted_k141_94584 | Not-determined |
| S14_extracted_k141_64425 | Not-determined |
| S14_extracted_k141_6101 | Not-determined |
| S14_extracted_k141_27850 | Not-determined |
| S14_extracted_k141_64426 | Not-determined |
| S14_extracted_k141_87140 | Low-quality |
| S14_extracted_k141_21131 | Not-determined |
| S14_extracted_k141_27858 | Not-determined |
| S14_extracted_k141_6106 | Low-quality |
| S14_extracted_k141_109170 | Low-quality |
| S14_extracted_k141_101352 | Low-quality |
| S14_extracted_k141_87145 | Not-determined |
| S14_extracted_k141_6112 | Not-determined |
| S14_extracted_k141_101354 | Low-quality |
| S14_extracted_k141_116292 | Not-determined |
| S14_extracted_k141_27866 | Not-determined |
| S14_extracted_k141_116293 | Not-determined |
| S14_extracted_k141_6117 | Not-determined |
| S14_extracted_k141_101360 | Not-determined |
| S14_extracted_k141_6122 | Low-quality |
| S14_extracted_k141_94599 | Not-determined |
| S14_extracted_k141_72134 | Not-determined |
| S14_extracted_k141_87156 | Low-quality |
| S14_extracted_k141_35659 | Not-determined |
| S14_extracted_k141_57446 | Low-quality |
| S14_extracted_k141_72139 | Not-determined |
| S14_extracted_k141_116303 | Not-determined |
| S14_extracted_k141_49606 | Low-quality |
| S14_extracted_k141_49608 | Low-quality |
| S14_extracted_k141_94602 | Not-determined |
| S14_extracted_k141_109179 | Not-determined |
| S14_extracted_k141_27884 | Not-determined |
| S14_extracted_k141_6137 | Not-determined |
| S14_extracted_k141_6138 | Not-determined |
| S14_extracted_k141_79468 | Not-determined |
| S14_extracted_k141_35675 | Low-quality |
| S14_extracted_k141_79469 | Not-determined |
| S14_extracted_k141_6141 | Not-determined |
| S14_extracted_k141_94605 | Not-determined |
| S14_extracted_k141_87162 | Not-determined |
| S14_extracted_k141_87163 | Not-determined |
| S14_extracted_k141_79477 | Not-determined |
| S14_extracted_k141_87167 | Not-determined |
| S14_extracted_k141_35680 | Not-determined |
| S14_extracted_k141_72158 | Not-determined |
| S14_extracted_k141_72159 | Low-quality |
| S14_extracted_k141_79482 | Not-determined |
| S14_extracted_k141_6150 | Not-determined |
| S14_extracted_k141_116320 | Low-quality |
| S14_extracted_k141_101369 | Not-determined |
| S14_extracted_k141_35689 | Not-determined |
| S14_extracted_k141_109190 | Not-determined |
| S14_extracted_k141_109191 | Low-quality |
| S14_extracted_k141_94611 | Low-quality |
| S14_extracted_k141_94612 | Not-determined |
| S14_extracted_k141_109193 | Not-determined |
| S14_extracted_k141_21156 | Not-determined |
| S14_extracted_k141_109197 | Not-determined |
| S14_extracted_k141_101376 | Not-determined |
| S14_extracted_k141_27891 | Not-determined |
| S14_extracted_k141_35696 | Low-quality |
| S14_extracted_k141_79493 | Not-determined |
| S14_extracted_k141_79499 | Not-determined |
| S14_extracted_k141_43643 | Not-determined |
| S14_extracted_k141_79500 | Not-determined |
| S14_extracted_k141_6164 | Not-determined |
| S14_extracted_k141_64431 | Not-determined |
| S14_extracted_k141_43650 | Low-quality |
| S14_extracted_k141_43652 | Low-quality |
| S14_extracted_k141_94624 | Low-quality |
| S14_extracted_k141_35704 | Not-determined |
| S14_extracted_k141_57452 | Not-determined |
| S14_extracted_k141_116341 | Not-determined |
| S14_extracted_k141_94627 | Not-determined |
| S14_extracted_k141_87190 | Not-determined |
| S14_extracted_k141_43656 | Low-quality |
| S14_extracted_k141_109207 | Not-determined |
| S14_extracted_k141_6175 | Low-quality |
| S14_extracted_k141_35707 | Not-determined |
| S14_extracted_k141_64436 | Not-determined |
| S14_extracted_k141_35709 | Not-determined |
| S14_extracted_k141_35710 | Not-determined |
| S14_extracted_k141_72186 | Not-determined |
| S14_extracted_k141_116354 | Not-determined |
| S14_extracted_k141_116356 | Not-determined |
| S14_extracted_k141_49656 | Not-determined |
| S14_extracted_k141_13405 | Low-quality |
| S14_extracted_k141_101394 | Not-determined |
| S14_extracted_k141_87206 | Not-determined |
| S14_extracted_k141_57462 | Not-determined |
| S14_extracted_k141_43669 | Not-determined |
| S14_extracted_k141_101399 | Low-quality |
| S14_extracted_k141_21188 | Not-determined |
| S14_extracted_k141_64453 | Not-determined |
| S14_extracted_k141_13415 | Low-quality |
| S14_extracted_k141_109218 | Not-determined |
| S14_extracted_k141_27910 | Low-quality |
| S14_extracted_k141_101405 | Low-quality |
| S14_extracted_k141_94651 | Not-determined |
| S14_extracted_k141_35729 | Low-quality |
| S14_extracted_k141_43679 | Not-determined |
| S14_extracted_k141_49666 | Not-determined |
| S14_extracted_k141_109231 | Low-quality |
| S14_extracted_k141_21195 | Not-determined |
| S14_extracted_k141_109233 | Not-determined |
| S14_extracted_k141_94658 | Low-quality |
| S14_extracted_k141_87217 | Not-determined |
| S14_extracted_k141_109234 | Not-determined |
| S14_extracted_k141_116372 | Not-determined |
| S14_extracted_k141_109238 | Low-quality |
| S14_extracted_k141_109240 | Not-determined |
| S14_extracted_k141_35739 | Low-quality |
| S14_extracted_k141_94661 | Not-determined |
| S14_extracted_k141_6184 | Not-determined |
| S14_extracted_k141_57481 | Not-determined |
| S14_extracted_k141_27923 | Not-determined |
| S14_extracted_k141_109243 | Not-determined |
| S14_extracted_k141_94667 | Not-determined |
| S14_extracted_k141_43687 | Low-quality |
| S14_extracted_k141_6193 | Low-quality |
| S14_extracted_k141_109247 | Not-determined |
| S14_extracted_k141_101406 | Low-quality |
| S14_extracted_k141_101408 | Low-quality |
| S14_extracted_k141_87226 | Low-quality |
| S14_extracted_k141_35754 | Not-determined |
| S14_extracted_k141_49676 | Not-determined |
| S14_extracted_k141_49678 | Not-determined |
| S14_extracted_k141_49682 | Low-quality |
| S14_extracted_k141_87229 | Not-determined |
| S14_extracted_k141_109258 | Not-determined |
| S14_extracted_k141_87230 | Not-determined |
| S14_extracted_k141_27933 | Not-determined |
| S14_extracted_k141_57493 | Not-determined |
| S14_extracted_k141_87232 | Not-determined |
| S14_extracted_k141_57497 | Not-determined |
| S14_extracted_k141_57498 | Low-quality |
| S14_extracted_k141_94676 | Not-determined |
| S14_extracted_k141_49689 | Not-determined |
| S14_extracted_k141_6201 | Low-quality |
| S14_extracted_k141_116395 | Not-determined |
| S14_extracted_k141_43709 | Not-determined |
| S14_extracted_k141_109263 | Not-determined |
| S14_extracted_k141_6204 | Not-determined |
| S14_extracted_k141_6205 | Low-quality |
| S14_extracted_k141_64481 | Low-quality |
| S14_extracted_k141_27936 | Low-quality |
| S14_extracted_k141_87241 | Low-quality |
| S14_extracted_k141_109265 | Low-quality |
| S14_extracted_k141_6208 | Low-quality |
| S14_extracted_k141_13447 | Low-quality |
| S14_extracted_k141_13449 | Not-determined |
| S14_extracted_k141_35776 | Not-determined |
| S14_extracted_k141_87244 | Not-determined |
| S14_extracted_k141_6210 | Not-determined |
| S14_extracted_k141_64485 | Not-determined |
| S14_extracted_k141_64486 | Not-determined |
| S14_extracted_k141_64490 | Low-quality |
| S14_extracted_k141_64491 | Not-determined |
| S14_extracted_k141_94689 | Low-quality |
| S14_extracted_k141_72215 | Low-quality |
| S14_extracted_k141_64493 | Not-determined |
| S14_extracted_k141_43712 | Not-determined |
| S14_extracted_k141_64496 | Low-quality |
| S14_extracted_k141_49695 | Low-quality |
| S14_extracted_k141_64497 | Not-determined |
| S14_extracted_k141_72221 | Not-determined |
| S14_extracted_k141_43715 | Not-determined |
| S14_extracted_k141_43717 | Low-quality |
| S14_extracted_k141_49699 | Not-determined |
| S14_extracted_k141_6217 | Low-quality |
| S14_extracted_k141_43721 | Not-determined |
| S14_extracted_k141_21206 | Not-determined |
| S14_extracted_k141_109278 | Not-determined |
| S14_extracted_k141_6219 | Not-determined |
| S14_extracted_k141_72228 | Low-quality |
| S14_extracted_k141_21212 | Not-determined |
| S14_extracted_k141_21213 | Not-determined |
| S14_extracted_k141_116414 | Not-determined |
| S14_extracted_k141_21216 | Not-determined |
| S14_extracted_k141_94704 | Not-determined |
| S14_extracted_k141_94706 | Low-quality |
| S14_extracted_k141_116416 | Not-determined |
| S14_extracted_k141_6231 | Not-determined |
| S14_extracted_k141_64501 | Not-determined |
| S14_extracted_k141_6232 | Not-determined |
| S14_extracted_k141_13467 | Not-determined |
| S14_extracted_k141_49714 | Low-quality |
| S14_extracted_k141_13470 | Not-determined |
| S14_extracted_k141_64504 | Low-quality |
| S14_extracted_k141_35787 | Low-quality |
| S14_extracted_k141_116425 | Not-determined |
| S14_extracted_k141_35789 | Not-determined |
| S14_extracted_k141_109292 | Low-quality |
| S14_extracted_k141_6240 | Not-determined |
| S14_extracted_k141_49725 | Low-quality |
| S14_extracted_k141_13475 | Low-quality |
| S14_extracted_k141_49728 | Not-determined |
| S14_extracted_k141_6247 | Not-determined |
| S14_extracted_k141_6255 | Not-determined |
| S14_extracted_k141_49734 | Low-quality |
| S14_extracted_k141_21238 | Not-determined |
| S14_extracted_k141_21240 | Not-determined |
| S14_extracted_k141_21242 | Not-determined |
| S14_extracted_k141_116441 | Not-determined |
| S14_extracted_k141_64519 | Not-determined |
| S14_extracted_k141_13485 | Low-quality |
| S14_extracted_k141_13488 | Not-determined |
| S14_extracted_k141_57543 | Low-quality |
| S14_extracted_k141_116445 | Low-quality |
| S14_extracted_k141_21250 | Not-determined |
| S14_extracted_k141_87266 | Not-determined |
| S14_extracted_k141_101442 | Not-determined |
| S14_extracted_k141_94719 | Not-determined |
| S14_extracted_k141_57549 | Not-determined |
| S14_extracted_k141_94722 | Not-determined |
| S14_extracted_k141_57552 | Not-determined |
| S14_extracted_k141_13495 | Not-determined |
| S14_extracted_k141_64531 | Not-determined |
| S14_extracted_k141_64533 | Low-quality |
| S14_extracted_k141_35803 | Not-determined |
| S14_extracted_k141_72251 | Low-quality |
| S14_extracted_k141_94729 | Not-determined |
| S14_extracted_k141_72252 | Low-quality |
| S14_extracted_k141_64542 | Low-quality |
| S14_extracted_k141_79527 | Not-determined |
| S14_extracted_k141_94734 | Not-determined |
| S14_extracted_k141_101456 | Not-determined |
| S14_extracted_k141_101458 | Low-quality |
| S14_extracted_k141_72253 | Not-determined |
| S14_extracted_k141_43770 | Low-quality |
| S14_extracted_k141_87276 | Not-determined |
| S14_extracted_k141_57571 | Not-determined |
| S14_extracted_k141_43774 | Not-determined |
| S14_extracted_k141_57573 | Not-determined |
| S14_extracted_k141_27974 | Low-quality |
| S14_extracted_k141_21255 | Not-determined |
| S14_extracted_k141_72255 | Not-determined |
| S14_extracted_k141_72257 | Low-quality |
| S14_extracted_k141_101474 | Not-determined |
| S14_extracted_k141_35825 | Not-determined |
| S14_extracted_k141_27979 | Not-determined |
| S14_extracted_k141_94750 | Not-determined |
| S14_extracted_k141_43787 | Not-determined |
| S14_extracted_k141_27981 | Not-determined |
| S14_extracted_k141_72260 | Not-determined |
| S14_extracted_k141_57591 | Not-determined |
| S14_extracted_k141_57592 | Low-quality |
| S14_extracted_k141_27983 | Not-determined |
| S14_extracted_k141_21263 | Not-determined |
| S14_extracted_k141_27985 | Not-determined |
| S14_extracted_k141_57594 | Low-quality |
| S14_extracted_k141_49760 | Low-quality |
| S14_extracted_k141_21265 | Not-determined |
| S14_extracted_k141_79534 | Not-determined |
| S14_extracted_k141_13526 | Not-determined |
| S14_extracted_k141_13527 | Low-quality |
| S14_extracted_k141_57603 | Not-determined |
| S14_extracted_k141_21271 | Not-determined |
| S14_extracted_k141_57605 | Not-determined |
| S14_extracted_k141_79538 | Low-quality |
| S14_extracted_k141_57606 | Low-quality |
| S14_extracted_k141_116458 | Low-quality |
| S14_extracted_k141_79545 | Not-determined |
| S14_extracted_k141_116463 | Low-quality |
| S14_extracted_k141_109325 | Low-quality |
| S14_extracted_k141_87300 | Low-quality |
| S14_extracted_k141_87301 | Not-determined |
| S14_extracted_k141_94760 | Not-determined |
| S14_extracted_k141_116469 | Low-quality |
| S14_extracted_k141_87306 | Not-determined |
| S14_extracted_k141_57617 | Not-determined |
| S14_extracted_k141_79557 | Low-quality |
| S14_extracted_k141_21279 | Not-determined |
| S14_extracted_k141_79559 | Not-determined |
| S14_extracted_k141_79562 | Not-determined |
| S14_extracted_k141_13535 | Not-determined |
| S14_extracted_k141_35829 | Not-determined |
| S14_extracted_k141_35830 | Low-quality |
| S14_extracted_k141_43818 | Low-quality |
| S14_extracted_k141_35831 | Low-quality |
| S14_extracted_k141_64576 | Not-determined |
| S14_extracted_k141_57624 | Not-determined |
| S14_extracted_k141_64577 | Low-quality |
| S14_extracted_k141_35833 | Low-quality |
| S14_extracted_k141_13539 | Not-determined |
| S14_extracted_k141_64579 | Low-quality |
| S14_extracted_k141_21281 | Low-quality |
| S14_extracted_k141_64582 | Not-determined |
| S14_extracted_k141_6316 | Low-quality |
| S14_extracted_k141_57629 | Not-determined |
| S14_extracted_k141_13546 | Not-determined |
| S14_extracted_k141_21287 | Not-determined |
| S14_extracted_k141_79571 | Not-determined |
| S14_extracted_k141_43829 | Low-quality |
| S14_extracted_k141_21288 | Not-determined |
| S14_extracted_k141_87312 | Not-determined |
| S14_extracted_k141_64586 | Low-quality |
| S14_extracted_k141_57631 | Low-quality |
| S14_extracted_k141_64589 | Not-determined |
| S14_extracted_k141_49798 | Not-determined |
| S14_extracted_k141_101485 | Not-determined |
| S14_extracted_k141_64597 | Not-determined |
| S14_extracted_k141_49806 | Not-determined |
| S14_extracted_k141_21300 | Not-determined |
| S14_extracted_k141_21301 | Low-quality |
| S14_extracted_k141_79576 | Not-determined |
| S14_extracted_k141_79578 | Not-determined |
| S14_extracted_k141_13557 | Not-determined |
| S14_extracted_k141_79582 | Not-determined |
| S14_extracted_k141_21307 | Not-determined |
| S14_extracted_k141_21310 | Not-determined |
| S14_extracted_k141_87325 | Not-determined |
| S14_extracted_k141_49812 | Low-quality |
| S14_extracted_k141_87327 | Low-quality |
| S14_extracted_k141_87329 | Not-determined |
| S14_extracted_k141_79588 | Low-quality |
| S14_extracted_k141_79590 | Not-determined |
| S14_extracted_k141_87330 | Not-determined |
| S14_extracted_k141_21320 | Low-quality |
| S14_extracted_k141_116486 | Low-quality |
| S14_extracted_k141_87332 | Not-determined |
| S14_extracted_k141_79594 | Low-quality |
| S14_extracted_k141_94800 | Not-determined |
| S14_extracted_k141_43838 | Not-determined |
| S14_extracted_k141_87339 | Not-determined |
| S14_extracted_k141_43842 | Not-determined |
| S14_extracted_k141_87340 | Low-quality |
| S14_extracted_k141_43843 | Low-quality |
| S14_extracted_k141_27997 | Not-determined |
| S14_extracted_k141_87343 | Not-determined |
| S14_extracted_k141_94815 | Not-determined |
| S14_extracted_k141_6342 | Not-determined |
| S14_extracted_k141_109332 | Low-quality |
| S14_extracted_k141_94818 | Low-quality |
| S14_extracted_k141_116501 | Not-determined |
| S14_extracted_k141_43854 | Not-determined |
| S14_extracted_k141_64603 | Not-determined |
| S14_extracted_k141_116503 | Low-quality |
| S14_extracted_k141_109338 | Low-quality |
| S14_extracted_k141_79600 | Not-determined |
| S14_extracted_k141_79603 | Low-quality |
| S14_extracted_k141_6352 | Low-quality |
| S14_extracted_k141_6354 | Not-determined |
| S14_extracted_k141_6355 | Not-determined |
| S14_extracted_k141_6356 | Low-quality |
| S14_extracted_k141_116510 | Not-determined |
| S14_extracted_k141_6357 | Not-determined |
| S14_extracted_k141_87365 | Not-determined |
| S14_extracted_k141_35857 | Low-quality |
| S14_extracted_k141_101496 | Not-determined |
| S14_extracted_k141_64614 | Low-quality |
| S14_extracted_k141_109344 | Low-quality |
| S14_extracted_k141_49827 | Not-determined |
| S14_extracted_k141_64621 | Not-determined |
| S14_extracted_k141_49831 | Not-determined |
| S14_extracted_k141_109349 | Not-determined |
| S14_extracted_k141_49832 | Not-determined |
| S14_extracted_k141_109353 | Not-determined |
| S14_extracted_k141_13563 | Not-determined |
| S14_extracted_k141_72268 | Not-determined |
| S14_extracted_k141_101501 | Not-determined |
| S14_extracted_k141_101502 | Not-determined |
| S14_extracted_k141_87371 | Not-determined |
| S14_extracted_k141_13570 | Not-determined |
| S14_extracted_k141_79625 | Not-determined |
| S14_extracted_k141_87375 | Not-determined |
| S14_extracted_k141_28018 | Low-quality |
| S14_extracted_k141_35881 | Not-determined |
| S5_vs_k141_145587\|\|full | Medium-quality |
| S14_extracted_k141_101517 | Not-determined |
| S14_extracted_k141_79630 | Low-quality |
| S14_extracted_k141_101518 | Not-determined |
| S14_extracted_k141_35884 | Not-determined |
| S14_extracted_k141_49851 | Low-quality |
| S14_extracted_k141_109363 | Not-determined |
| S14_extracted_k141_79636 | Not-determined |
| S14_extracted_k141_116530 | Not-determined |
| S14_extracted_k141_28029 | Not-determined |
| S14_extracted_k141_57649 | Low-quality |
| S14_extracted_k141_57650 | Not-determined |
| S14_extracted_k141_72306 | Low-quality |
| S14_extracted_k141_79640 | Not-determined |
| S14_extracted_k141_72308 | Not-determined |
| S14_extracted_k141_116537 | Not-determined |
| S14_extracted_k141_116538 | Not-determined |
| S14_extracted_k141_35900 | Low-quality |
| S14_extracted_k141_72309 | Not-determined |
| S14_extracted_k141_57658 | Not-determined |
| S14_extracted_k141_6382 | Not-determined |
| S14_extracted_k141_116541 | Not-determined |
| S14_extracted_k141_21358 | Not-determined |
| S14_extracted_k141_6384 | Low-quality |
| S14_extracted_k141_116542 | Not-determined |
| S14_extracted_k141_21361 | Low-quality |
| S14_extracted_k141_79650 | Not-determined |
| S14_extracted_k141_87385 | Not-determined |
| S14_extracted_k141_101531 | Not-determined |
| S14_extracted_k141_116553 | Not-determined |
| S14_extracted_k141_21364 | Not-determined |
| S14_extracted_k141_21365 | Not-determined |
| S14_extracted_k141_43891 | Not-determined |
| S14_extracted_k141_6396 | Not-determined |
| S14_extracted_k141_101535 | Not-determined |
| S14_extracted_k141_6398 | Not-determined |
| S14_extracted_k141_79660 | Low-quality |
| S14_extracted_k141_21377 | Low-quality |
| S14_extracted_k141_64639 | Not-determined |
| S14_extracted_k141_101537 | Not-determined |
| S14_extracted_k141_79662 | Not-determined |
| S14_extracted_k141_64640 | Low-quality |
| S14_extracted_k141_28060 | Not-determined |
| S14_extracted_k141_43901 | Not-determined |
| S14_extracted_k141_109382 | Not-determined |
| S14_extracted_k141_13604 | Not-determined |
| S14_extracted_k141_57683 | Not-determined |
| S14_extracted_k141_109385 | Not-determined |
| S14_extracted_k141_87403 | Low-quality |
| S14_extracted_k141_109390 | Not-determined |
| S14_extracted_k141_49874 | Not-determined |
| S14_extracted_k141_109397 | Not-determined |

| | |
|---|---|
| S14_extracted_k141_28067 | Not-determined |
| S14_extracted_k141_43907 | Not-determined |
| S14_extracted_k141_87418 | Low-quality |
| S14_extracted_k141_109405 | Not-determined |
| S14_extracted_k141_79677 | Not-determined |
| S14_extracted_k141_21398 | Not-determined |
| S14_extracted_k141_109410 | Not-determined |
| S14_extracted_k141_64663 | Not-determined |
| S14_extracted_k141_87421 | Low-quality |
| S14_extracted_k141_21402 | Not-determined |
| S14_extracted_k141_79682 | Not-determined |
| S14_extracted_k141_109416 | Low-quality |
| S14_extracted_k141_21403 | Low-quality |
| S14_extracted_k141_79685 | Low-quality |
| S14_extracted_k141_43915 | Low-quality |
| S14_extracted_k141_101547 | Not-determined |
| S14_extracted_k141_28075 | Not-determined |
| S14_extracted_k141_6429 | Not-determined |
| S14_extracted_k141_43918 | Low-quality |
| S14_extracted_k141_21405 | Low-quality |
| S14_extracted_k141_79689 | Not-determined |
| S14_extracted_k141_43919 | Low-quality |
| S14_extracted_k141_109425 | Not-determined |
| S14_extracted_k141_101556 | Not-determined |
| S14_extracted_k141_28090 | Not-determined |
| S14_extracted_k141_109431 | Not-determined |
| S14_extracted_k141_87439 | Not-determined |
| S14_extracted_k141_35908 | Not-determined |
| S14_extracted_k141_87444 | Low-quality |
| S14_extracted_k141_109436 | Not-determined |
| S14_extracted_k141_79691 | Low-quality |
| S14_extracted_k141_87446 | Not-determined |
| S14_extracted_k141_79693 | Low-quality |
| S14_extracted_k141_28111 | Not-determined |
| S14_extracted_k141_28116 | Low-quality |
| S14_extracted_k141_35919 | Not-determined |
| S14_extracted_k141_116585 | Not-determined |
| S14_extracted_k141_6449 | Not-determined |
| S14_extracted_k141_64673 | Not-determined |
| S14_extracted_k141_6450 | Not-determined |
| S14_extracted_k141_64674 | Low-quality |
| S14_extracted_k141_35925 | Not-determined |
| S6_vs_k141_56483\|\|full | Medium-quality |
| S14_extracted_k141_35926 | Not-determined |
| S14_extracted_k141_64676 | Not-determined |
| S14_extracted_k141_13630 | Not-determined |
| S14_extracted_k141_28119 | Not-determined |
| S14_extracted_k141_28120 | Not-determined |
| S14_extracted_k141_57693 | Not-determined |
| S14_extracted_k141_94873 | Low-quality |
| S14_extracted_k141_64682 | Low-quality |
| S14_extracted_k141_79706 | Not-determined |
| S14_extracted_k141_21432 | Not-determined |
| S14_extracted_k141_43935 | Low-quality |
| S14_extracted_k141_21435 | Low-quality |
| S14_extracted_k141_109445 | Not-determined |
| S14_extracted_k141_94882 | Low-quality |
| S14_extracted_k141_94883 | Low-quality |
| S14_extracted_k141_6462 | Low-quality |
| S14_extracted_k141_64687 | Not-determined |
| S14_extracted_k141_109447 | Not-determined |
| S14_extracted_k141_109452 | Not-determined |
| S14_extracted_k141_21451 | Not-determined |
| S14_extracted_k141_35944 | Not-determined |
| S14_extracted_k141_6472 | Not-determined |
| S14_extracted_k141_35945 | Low-quality |
| S14_extracted_k141_13638 | Low-quality |
| S14_extracted_k141_101573 | Not-determined |
| S14_extracted_k141_79726 | Low-quality |
| S14_extracted_k141_116619 | Not-determined |
| S14_extracted_k141_13641 | Not-determined |
| S14_extracted_k141_109461 | Low-quality |
| S14_extracted_k141_116624 | Not-determined |
| S14_extracted_k141_116625 | Not-determined |
| S14_extracted_k141_101577 | Not-determined |
| S14_extracted_k141_116627 | Low-quality |
| S14_extracted_k141_109467 | Not-determined |
| S14_extracted_k141_64691 | Not-determined |
| S14_extracted_k141_109470 | Not-determined |
| S14_extracted_k141_13644 | Not-determined |
| S14_extracted_k141_64697 | Not-determined |
| S14_extracted_k141_116633 | Not-determined |
| S14_extracted_k141_101584 | Not-determined |
| S14_extracted_k141_101587 | Not-determined |
| S14_extracted_k141_6485 | Not-determined |
| S14_extracted_k141_101592 | Not-determined |
| S14_extracted_k141_64707 | Not-determined |
| S14_extracted_k141_43949 | Not-determined |
| S14_extracted_k141_21473 | Not-determined |
| S14_extracted_k141_43951 | Not-determined |
| S14_extracted_k141_79750 | Low-quality |
| S14_extracted_k141_79751 | Not-determined |
| S14_extracted_k141_79754 | Low-quality |
| S14_extracted_k141_43955 | Not-determined |
| S14_extracted_k141_13667 | Not-determined |
| S14_extracted_k141_109480 | Not-determined |
| S14_extracted_k141_6493 | Not-determined |
| S14_extracted_k141_87469 | Not-determined |
| S14_extracted_k141_35971 | Not-determined |
| S14_extracted_k141_35973 | Not-determined |
| S14_extracted_k141_116641 | Low-quality |
| S14_extracted_k141_94912 | Not-determined |
| S14_extracted_k141_109491 | Not-determined |
| S14_extracted_k141_109492 | Low-quality |
| S14_extracted_k141_49889 | Low-quality |
| S14_extracted_k141_28138 | Low-quality |
| S14_extracted_k141_57746 | Not-determined |
| S14_extracted_k141_94924 | Not-determined |
| S14_extracted_k141_79781 | Low-quality |
| S14_extracted_k141_57749 | Not-determined |
| S14_extracted_k141_109504 | Not-determined |
| S14_extracted_k141_21492 | Not-determined |
| S14_extracted_k141_79786 | Low-quality |
| S14_extracted_k141_13689 | Not-determined |
| S14_extracted_k141_79796 | Not-determined |
| S14_extracted_k141_57764 | Not-determined |
| S14_extracted_k141_57765 | Not-determined |
| S14_extracted_k141_116650 | Low-quality |
| S14_extracted_k141_13702 | Low-quality |
| S14_extracted_k141_101605 | Not-determined |
| S14_extracted_k141_109516 | Not-determined |
| S14_extracted_k141_109517 | Low-quality |
| S14_extracted_k141_101611 | Low-quality |
| S14_extracted_k141_101612 | Low-quality |
| S14_extracted_k141_21517 | Not-determined |
| S14_extracted_k141_79806 | Low-quality |
| S14_extracted_k141_35991 | Not-determined |
| S14_extracted_k141_116657 | Not-determined |
| S14_extracted_k141_79812 | Low-quality |
| S14_extracted_k141_57782 | Not-determined |
| S14_extracted_k141_109523 | Low-quality |
| S14_extracted_k141_109524 | Low-quality |
| S14_extracted_k141_101621 | Low-quality |
| S14_extracted_k141_49907 | Not-determined |
| S14_extracted_k141_49914 | Not-determined |
| S14_extracted_k141_13713 | Not-determined |
| S14_extracted_k141_28151 | Not-determined |
| S14_extracted_k141_109534 | Not-determined |
| S14_extracted_k141_109541 | Low-quality |
| S14_extracted_k141_94956 | Not-determined |
| S14_extracted_k141_94958 | Low-quality |
| S14_extracted_k141_94959 | Low-quality |
| S14_extracted_k141_21555 | Low-quality |
| S14_extracted_k141_101644 | Not-determined |
| S14_extracted_k141_94967 | Not-determined |
| S14_extracted_k141_94968 | Not-determined |
| S14_extracted_k141_21559 | Not-determined |
| S14_extracted_k141_36016 | Not-determined |
| S14_extracted_k141_94973 | Low-quality |
| S14_extracted_k141_79844 | Not-determined |
| S14_extracted_k141_28158 | Low-quality |
| S14_extracted_k141_94977 | Low-quality |
| S14_extracted_k141_101654 | Not-determined |
| S14_extracted_k141_21568 | Not-determined |
| S14_extracted_k141_101660 | Low-quality |
| S14_extracted_k141_49928 | Not-determined |
| S14_extracted_k141_6532 | Not-determined |
| S14_extracted_k141_101661 | Low-quality |
| S14_extracted_k141_101662 | Low-quality |
| S14_extracted_k141_79847 | Not-determined |
| S14_extracted_k141_101663 | Not-determined |
| S14_extracted_k141_28166 | Not-determined |
| S14_extracted_k141_6539 | Low-quality |
| S14_extracted_k141_13740 | Not-determined |
| S14_extracted_k141_13741 | Not-determined |
| S14_extracted_k141_13745 | Not-determined |
| S14_extracted_k141_21572 | Not-determined |
| S14_extracted_k141_13748 | Not-determined |
| S14_extracted_k141_101681 | Not-determined |
| S14_extracted_k141_57830 | Not-determined |
| S14_extracted_k141_116680 | Not-determined |
| S14_extracted_k141_116683 | Not-determined |
| S14_extracted_k141_49936 | Not-determined |
| S14_extracted_k141_21579 | Low-quality |
| S8_extracted_k141_126240 | High-quality |
| S14_extracted_k141_109547 | Not-determined |
| S14_extracted_k141_79854 | Not-determined |
| S14_extracted_k141_109548 | Not-determined |
| S14_extracted_k141_79856 | Low-quality |
| S14_extracted_k141_13758 | Not-determined |
| S14_extracted_k141_49941 | Low-quality |
| S14_extracted_k141_79858 | Low-quality |
| S14_extracted_k141_13760 | Not-determined |
| S14_extracted_k141_57840 | Not-determined |
| S14_extracted_k141_49943 | Low-quality |
| S14_extracted_k141_109552 | Not-determined |
| S14_extracted_k141_13762 | Not-determined |
| S14_extracted_k141_36051 | Low-quality |
| S14_extracted_k141_57845 | Not-determined |
| S14_extracted_k141_49950 | Not-determined |
| S14_extracted_k141_28194 | Not-determined |
| S14_extracted_k141_116691 | Not-determined |
| S14_extracted_k141_13764 | Not-determined |
| S14_extracted_k141_36055 | Low-quality |
| S14_extracted_k141_21585 | Not-determined |
| S14_extracted_k141_95007 | Not-determined |
| S14_extracted_k141_64733 | Low-quality |
| S14_extracted_k141_36060 | Not-determined |
| S14_extracted_k141_28207 | Low-quality |
| S14_extracted_k141_36066 | Low-quality |
| S14_extracted_k141_64738 | Not-determined |
| S14_extracted_k141_21592 | Not-determined |
| S14_extracted_k141_57857 | Not-determined |
| S14_extracted_k141_57858 | Not-determined |
| S14_extracted_k141_79883 | Low-quality |
| S14_extracted_k141_64744 | Not-determined |
| S14_extracted_k141_95015 | Not-determined |
| S14_extracted_k141_116701 | Not-determined |
| S14_extracted_k141_64750 | Not-determined |
| S14_extracted_k141_49972 | Low-quality |
| S14_extracted_k141_95021 | Not-determined |
| S14_extracted_k141_95024 | Not-determined |
| S14_extracted_k141_116707 | Low-quality |
| S14_extracted_k141_95027 | Not-determined |
| S14_extracted_k141_49976 | Not-determined |
| S14_extracted_k141_95031 | Not-determined |
| S14_extracted_k141_79898 | Not-determined |
| S14_extracted_k141_49981 | Not-determined |
| S14_extracted_k141_101686 | Not-determined |
| S14_extracted_k141_49982 | Not-determined |
| S14_extracted_k141_95034 | Not-determined |
| S14_extracted_k141_95035 | Low-quality |
| S14_extracted_k141_79901 | Not-determined |
| S14_extracted_k141_49984 | Low-quality |
| S14_extracted_k141_57874 | Low-quality |
| S14_extracted_k141_64765 | Not-determined |
| S14_extracted_k141_116714 | Low-quality |
| S14_extracted_k141_49987 | Not-determined |
| S14_extracted_k141_64770 | Not-determined |
| S14_extracted_k141_6543 | Not-determined |
| S14_extracted_k141_109581 | Not-determined |
| S14_extracted_k141_57882 | Not-determined |
| S14_extracted_k141_6546 | Low-quality |
| S14_extracted_k141_36090 | Low-quality |
| S14_extracted_k141_36091 | Not-determined |
| S14_extracted_k141_109586 | Not-determined |
| S14_extracted_k141_109587 | Low-quality |
| S14_extracted_k141_6552 | Not-determined |
| S14_extracted_k141_36098 | Low-quality |
| S14_extracted_k141_13793 | Low-quality |
| S14_extracted_k141_6561 | Low-quality |
| S14_extracted_k141_50004 | Not-determined |
| S14_extracted_k141_101705 | Low-quality |
| S14_extracted_k141_50007 | Not-determined |
| S14_extracted_k141_79913 | Not-determined |
| S14_extracted_k141_36108 | Low-quality |
| S14_extracted_k141_101714 | Not-determined |
| S14_extracted_k141_101715 | Not-determined |
| S14_extracted_k141_64798 | Not-determined |
| S14_extracted_k141_109607 | Not-determined |
| S8_vs_k141_132697\|\|full | High-quality |
| S14_extracted_k141_64802 | Not-determined |
| S14_extracted_k141_79927 | Low-quality |
| S14_extracted_k141_64805 | Not-determined |
| S14_extracted_k141_79933 | Low-quality |
| S14_extracted_k141_79934 | Not-determined |
| S14_extracted_k141_13804 | Low-quality |
| S14_extracted_k141_13805 | Not-determined |
| S14_extracted_k141_6587 | Not-determined |
| S14_extracted_k141_28238 | Low-quality |
| S14_extracted_k141_36118 | Not-determined |
| S14_extracted_k141_64818 | Low-quality |
| S14_extracted_k141_72345 | Low-quality |
| S14_extracted_k141_116735 | Not-determined |
| S14_extracted_k141_72346 | Not-determined |
| S14_extracted_k141_57910 | Not-determined |
| S14_extracted_k141_116739 | Low-quality |
| S14_extracted_k141_57912 | Low-quality |
| S14_extracted_k141_64821 | Not-determined |
| S14_extracted_k141_64822 | Not-determined |
| S14_extracted_k141_36129 | Not-determined |
| S14_extracted_k141_116744 | Not-determined |
| S14_extracted_k141_13813 | Low-quality |
| S14_extracted_k141_64827 | Low-quality |
| S14_extracted_k141_13817 | Low-quality |
| S14_extracted_k141_13821 | Low-quality |
| S14_extracted_k141_116747 | Not-determined |
| S14_extracted_k141_64833 | Not-determined |
| S14_extracted_k141_28248 | Not-determined |
| S14_extracted_k141_79948 | Low-quality |
| S14_extracted_k141_101745 | Not-determined |
| S14_extracted_k141_6604 | Low-quality |
| S14_extracted_k141_36131 | Not-determined |
| S14_extracted_k141_101751 | Low-quality |
| S14_extracted_k141_116759 | Not-determined |
| S14_extracted_k141_13825 | Not-determined |
| S14_extracted_k141_64839 | Not-determined |
| S14_extracted_k141_36140 | Not-determined |
| S14_extracted_k141_28261 | Not-determined |
| S14_extracted_k141_95056 | Not-determined |
| S14_extracted_k141_116763 | Not-determined |
| S14_extracted_k141_72362 | Not-determined |
| S14_extracted_k141_6612 | Not-determined |
| S14_extracted_k141_50022 | Not-determined |
| S14_extracted_k141_72364 | Low-quality |
| S14_extracted_k141_6615 | Low-quality |
| S14_extracted_k141_101767 | Not-determined |
| S14_extracted_k141_116766 | Not-determined |
| S14_extracted_k141_101770 | Not-determined |
| S14_extracted_k141_64854 | Not-determined |
| S14_extracted_k141_72372 | Not-determined |
| S14_extracted_k141_50036 | Not-determined |
| S14_extracted_k141_116773 | Not-determined |
| S14_extracted_k141_13838 | Not-determined |
| S14_extracted_k141_109659 | Not-determined |
| S14_extracted_k141_116778 | Not-determined |
| S14_extracted_k141_95079 | Not-determined |
| S14_extracted_k141_64862 | Low-quality |
| S14_extracted_k141_109667 | Not-determined |
| S14_extracted_k141_109668 | Not-determined |
| S14_extracted_k141_109670 | Not-determined |
| S14_extracted_k141_28279 | Low-quality |
| S14_extracted_k141_6633 | Low-quality |
| S14_extracted_k141_57942 | Not-determined |
| S14_extracted_k141_50040 | Low-quality |
| S14_extracted_k141_57946 | Not-determined |
| S14_extracted_k141_57947 | Low-quality |
| S14_extracted_k141_64879 | Not-determined |
| S14_extracted_k141_21632 | Not-determined |
| S14_extracted_k141_72404 | Not-determined |
| S14_extracted_k141_64884 | Not-determined |
| S14_extracted_k141_50053 | Low-quality |
| S14_extracted_k141_50055 | Low-quality |
| S14_extracted_k141_6638 | Not-determined |
| S14_extracted_k141_64888 | Not-determined |
| S14_extracted_k141_116783 | Not-determined |
| S14_extracted_k141_21640 | Not-determined |
| S14_extracted_k141_116786 | Low-quality |
| S14_extracted_k141_79987 | Not-determined |
| S14_extracted_k141_95082 | Not-determined |
| S14_extracted_k141_116788 | Low-quality |
| S14_extracted_k141_95083 | Not-determined |
| S14_extracted_k141_28300 | Low-quality |
| S14_extracted_k141_95084 | Not-determined |
| S14_extracted_k141_116792 | Low-quality |
| S14_extracted_k141_28302 | Not-determined |
| S14_extracted_k141_64897 | Not-determined |
| S14_extracted_k141_13850 | Not-determined |
| S14_extracted_k141_50063 | Low-quality |
| S14_extracted_k141_64898 | Not-determined |
| S14_extracted_k141_28309 | Not-determined |
| S14_extracted_k141_116797 | Not-determined |
| S14_extracted_k141_72426 | Not-determined |
| S14_extracted_k141_72429 | Not-determined |
| S14_extracted_k141_116801 | Not-determined |
| S14_extracted_k141_13855 | Low-quality |
| S14_extracted_k141_6653 | Not-determined |
| S14_extracted_k141_109715 | Not-determined |
| S14_extracted_k141_95095 | Low-quality |
| S14_extracted_k141_101802 | Low-quality |
| S14_extracted_k141_28318 | Low-quality |
| S14_extracted_k141_21651 | Not-determined |
| S14_extracted_k141_64916 | Low-quality |
| S14_extracted_k141_6661 | Not-determined |
| S14_extracted_k141_95096 | Not-determined |
| S14_extracted_k141_95097 | Not-determined |
| S14_extracted_k141_116807 | Low-quality |
| S14_extracted_k141_116808 | Not-determined |
| S14_extracted_k141_64919 | Not-determined |
| S14_extracted_k141_101807 | Not-determined |
| S14_extracted_k141_116813 | Not-determined |
| S14_extracted_k141_28321 | Not-determined |
| S14_extracted_k141_95104 | Not-determined |
| S14_extracted_k141_36162 | Low-quality |
| S14_extracted_k141_36163 | Not-determined |
| S14_extracted_k141_50069 | Not-determined |
| S14_extracted_k141_109721 | Not-determined |
| S14_extracted_k141_36172 | Not-determined |
| S14_extracted_k141_36173 | Not-determined |
| S14_extracted_k141_36174 | Not-determined |
| S14_extracted_k141_109727 | Low-quality |
| S14_extracted_k141_57970 | Low-quality |
| S14_extracted_k141_109732 | Not-determined |
| S14_extracted_k141_72458 | Low-quality |
| S14_extracted_k141_64938 | Low-quality |
| S14_extracted_k141_36183 | Not-determined |
| S14_extracted_k141_80000 | Not-determined |
| S14_extracted_k141_80002 | Not-determined |
| S14_extracted_k141_95120 | Low-quality |
| S14_extracted_k141_116826 | Not-determined |
| S14_extracted_k141_28329 | Low-quality |
| S14_extracted_k141_109741 | Not-determined |
| S14_extracted_k141_116832 | Not-determined |
| S14_extracted_k141_116833 | Not-determined |
| S14_extracted_k141_80010 | Not-determined |
| S14_extracted_k141_6676 | Not-determined |
| S14_extracted_k141_95126 | Low-quality |
| S14_extracted_k141_36196 | Not-determined |
| S14_extracted_k141_116835 | Not-determined |
| S14_extracted_k141_50085 | Not-determined |
| S14_extracted_k141_101833 | Low-quality |
| S14_extracted_k141_13880 | Not-determined |
| S14_extracted_k141_50093 | Low-quality |
| S14_extracted_k141_116840 | Not-determined |
| S14_extracted_k141_13884 | Low-quality |
| S14_extracted_k141_57987 | Not-determined |
| S14_extracted_k141_6679 | Low-quality |
| S14_extracted_k141_72471 | Low-quality |
| S14_extracted_k141_13887 | Low-quality |
| S14_extracted_k141_13889 | Low-quality |
| S14_extracted_k141_80022 | Not-determined |
| S14_extracted_k141_6682 | Not-determined |
| S14_extracted_k141_21679 | Low-quality |
| S14_extracted_k141_80024 | Not-determined |
| S14_extracted_k141_80025 | Low-quality |
| S14_extracted_k141_109753 | Not-determined |
| S14_extracted_k141_109756 | Low-quality |
| S14_extracted_k141_21684 | Not-determined |
| S14_extracted_k141_6687 | Not-determined |
| S14_extracted_k141_116845 | Low-quality |
| S14_extracted_k141_21687 | Not-determined |
| S14_extracted_k141_6695 | Low-quality |
| S14_extracted_k141_80028 | Not-determined |
| S14_extracted_k141_57993 | Not-determined |
| S14_extracted_k141_13892 | Low-quality |
| S14_extracted_k141_13895 | Not-determined |
| S14_extracted_k141_21700 | Not-determined |
| S14_extracted_k141_50104 | Not-determined |
| S14_extracted_k141_109767 | Not-determined |
| S14_extracted_k141_101845 | Low-quality |
| S14_extracted_k141_87490 | Low-quality |
| S14_extracted_k141_6710 | Not-determined |
| S14_extracted_k141_80039 | Not-determined |
| S14_extracted_k141_87492 | Not-determined |
| S14_extracted_k141_21705 | Low-quality |
| S14_extracted_k141_116855 | Not-determined |
| S14_extracted_k141_116856 | Not-determined |
| S14_extracted_k141_36214 | Not-determined |
| S14_extracted_k141_58008 | Low-quality |
| S14_extracted_k141_80046 | Not-determined |
| S14_extracted_k141_95152 | Not-determined |
| S14_extracted_k141_80048 | Low-quality |
| S14_extracted_k141_109777 | Not-determined |
| S14_extracted_k141_28343 | Not-determined |
| S14_extracted_k141_87502 | Not-determined |
| S14_extracted_k141_36220 | Not-determined |
| S14_extracted_k141_58012 | Low-quality |
| S14_extracted_k141_58013 | Low-quality |
| S14_extracted_k141_36224 | Low-quality |
| S14_extracted_k141_87506 | Not-determined |
| S14_extracted_k141_64961 | Not-determined |
| S14_extracted_k141_28349 | Not-determined |
| S14_extracted_k141_21717 | Not-determined |
| S14_extracted_k141_58015 | Not-determined |
| S14_extracted_k141_64963 | Not-determined |
| S14_extracted_k141_95162 | Not-determined |
| S14_extracted_k141_80055 | Not-determined |
| S14_extracted_k141_101850 | Low-quality |
| S14_extracted_k141_87513 | Not-determined |
| S14_extracted_k141_58027 | Not-determined |
| S14_extracted_k141_72485 | Low-quality |
| S14_extracted_k141_116865 | Not-determined |
| S14_extracted_k141_50117 | Not-determined |
| S14_extracted_k141_36243 | Not-determined |
| S14_extracted_k141_80065 | Low-quality |
| S14_extracted_k141_50124 | Not-determined |
| S14_extracted_k141_6727 | Not-determined |
| S14_extracted_k141_95173 | Not-determined |
| S14_extracted_k141_6730 | Not-determined |
| S14_extracted_k141_58041 | Low-quality |
| S14_extracted_k141_101862 | Not-determined |
| S14_extracted_k141_72506 | Not-determined |
| S14_extracted_k141_58043 | Not-determined |
| S14_extracted_k141_58044 | Not-determined |
| S14_extracted_k141_50131 | Low-quality |
| S14_extracted_k141_101865 | Not-determined |
| S14_extracted_k141_58049 | Not-determined |
| S14_extracted_k141_109795 | Not-determined |
| S14_extracted_k141_95175 | Not-determined |
| S14_extracted_k141_87530 | Not-determined |
| S14_extracted_k141_95176 | Not-determined |
| S14_extracted_k141_58052 | Not-determined |
| S14_extracted_k141_109802 | Low-quality |
| S14_extracted_k141_36252 | Not-determined |
| S14_extracted_k141_87535 | Low-quality |
| S14_extracted_k141_87536 | Low-quality |
| S14_extracted_k141_6741 | Not-determined |
| S14_extracted_k141_101874 | Low-quality |
| S14_extracted_k141_6742 | Not-determined |
| S14_extracted_k141_80080 | Low-quality |
| S14_extracted_k141_116867 | Not-determined |
| S14_extracted_k141_109813 | Low-quality |
| S14_extracted_k141_109814 | Not-determined |
| S14_extracted_k141_80085 | Not-determined |
| S14_extracted_k141_109817 | Not-determined |
| S14_extracted_k141_36258 | Not-determined |
| S14_extracted_k141_116873 | Low-quality |
| S14_extracted_k141_50150 | Not-determined |
| S14_extracted_k141_80088 | Not-determined |
| S14_extracted_k141_28367 | Low-quality |
| S14_extracted_k141_58064 | Not-determined |
| S14_extracted_k141_36273 | Not-determined |
| S14_extracted_k141_6759 | Low-quality |
| S14_extracted_k141_58073 | Low-quality |
| S14_extracted_k141_72534 | Low-quality |
| S14_extracted_k141_36281 | Not-determined |
| S14_extracted_k141_95192 | Not-determined |
| S14_extracted_k141_95193 | Not-determined |
| S14_extracted_k141_21751 | Not-determined |
| S14_extracted_k141_109827 | Low-quality |
| S14_extracted_k141_21753 | Not-determined |
| S14_extracted_k141_36285 | Not-determined |
| S14_extracted_k141_58076 | Not-determined |
| S14_extracted_k141_65010 | Not-determined |
| S14_extracted_k141_36293 | Not-determined |
| S14_extracted_k141_87565 | Not-determined |
| S14_extracted_k141_36297 | Low-quality |
| S14_extracted_k141_28373 | Not-determined |
| S14_extracted_k141_28374 | Low-quality |
| S14_extracted_k141_109834 | Not-determined |
| S14_extracted_k141_101880 | Not-determined |
| S14_extracted_k141_58088 | Low-quality |
| S14_extracted_k141_50175 | Low-quality |
| S14_extracted_k141_50176 | Not-determined |
| S14_extracted_k141_109837 | Not-determined |
| S14_extracted_k141_58090 | Low-quality |
| S14_extracted_k141_109839 | Not-determined |
| S14_extracted_k141_109840 | Not-determined |
| S14_extracted_k141_28384 | Low-quality |
| S14_extracted_k141_13920 | Low-quality |
| S14_extracted_k141_50180 | Not-determined |
| S14_extracted_k141_21771 | Low-quality |
| S14_extracted_k141_13926 | Low-quality |
| S14_extracted_k141_87578 | Not-determined |
| S14_extracted_k141_28391 | Not-determined |
| S14_extracted_k141_87582 | Not-determined |
| S14_extracted_k141_95211 | Not-determined |
| S14_extracted_k141_80102 | Not-determined |
| S14_extracted_k141_109854 | Not-determined |
| S14_extracted_k141_87586 | Not-determined |
| S14_extracted_k141_101890 | Not-determined |
| S14_extracted_k141_80109 | Not-determined |
| S14_extracted_k141_87591 | Not-determined |
| S14_extracted_k141_28396 | Not-determined |
| S14_extracted_k141_87594 | Low-quality |
| S14_extracted_k141_28398 | Low-quality |
| S14_extracted_k141_109864 | Not-determined |
| S14_extracted_k141_101899 | Low-quality |
| S14_extracted_k141_109867 | Low-quality |
| S14_extracted_k141_101901 | Not-determined |
| S14_extracted_k141_21780 | Low-quality |
| S14_extracted_k141_65015 | Not-determined |
| S14_extracted_k141_101903 | Not-determined |
| S14_extracted_k141_116890 | Not-determined |
| S14_extracted_k141_72552 | Low-quality |
| S14_extracted_k141_95232 | Not-determined |
| S14_extracted_k141_116897 | Low-quality |
| S14_extracted_k141_116899 | Not-determined |
| S14_extracted_k141_13937 | Low-quality |
| S14_extracted_k141_116900 | Not-determined |
| S14_extracted_k141_36312 | Not-determined |
| S14_extracted_k141_50205 | Not-determined |
| S14_extracted_k141_50206 | Not-determined |
| S14_extracted_k141_36315 | Not-determined |
| S14_extracted_k141_65031 | Not-determined |
| S14_extracted_k141_13941 | Not-determined |
| S14_extracted_k141_95240 | Not-determined |
| S14_extracted_k141_36321 | Not-determined |
| S14_extracted_k141_36322 | Not-determined |
| S14_extracted_k141_109889 | Low-quality |
| S14_extracted_k141_87609 | Low-quality |
| S14_extracted_k141_50217 | Not-determined |
| S14_extracted_k141_65039 | Low-quality |
| S14_extracted_k141_6783 | Low-quality |
| S14_extracted_k141_87616 | Low-quality |
| S14_extracted_k141_13954 | Not-determined |
| S14_extracted_k141_65043 | Not-determined |
| S14_extracted_k141_109896 | Low-quality |
| S14_extracted_k141_87619 | Low-quality |
| S14_extracted_k141_6784 | Low-quality |
| S14_extracted_k141_116920 | Not-determined |
| S14_extracted_k141_65050 | Low-quality |
| S14_extracted_k141_65053 | Not-determined |
| S14_extracted_k141_6790 | Not-determined |
| S14_extracted_k141_21802 | Not-determined |
| S14_extracted_k141_6793 | Not-determined |
| S14_extracted_k141_28421 | Not-determined |
| S14_extracted_k141_65060 | Not-determined |
| S14_extracted_k141_87625 | Not-determined |
| S14_extracted_k141_21808 | Low-quality |
| S14_extracted_k141_72580 | Not-determined |
| S14_extracted_k141_6800 | Low-quality |
| S14_extracted_k141_50222 | Low-quality |
| S14_extracted_k141_80131 | Not-determined |
| S14_extracted_k141_13974 | Not-determined |
| S14_extracted_k141_65069 | Low-quality |
| S14_extracted_k141_116948 | Not-determined |
| S14_extracted_k141_80142 | Not-determined |
| S14_extracted_k141_50235 | Not-determined |

| | |
|---|---|
| S14_extracted_k141_28435 | Not-determined |
| S14_extracted_k141_116951 | Not-determined |
| S14_extracted_k141_50237 | Not-determined |
| S14_extracted_k141_58122 | Not-determined |
| S14_extracted_k141_6817 | Not-determined |
| S14_extracted_k141_58128 | Not-determined |
| S14_extracted_k141_116954 | Not-determined |
| S14_extracted_k141_13981 | Not-determined |
| S14_extracted_k141_13982 | Not-determined |
| S14_extracted_k141_28440 | Not-determined |
| S14_extracted_k141_28441 | Low-quality |
| S14_extracted_k141_36327 | Not-determined |
| S14_extracted_k141_58136 | Low-quality |
| S14_extracted_k141_116963 | Not-determined |
| S14_extracted_k141_13987 | Low-quality |
| S14_extracted_k141_28444 | Not-determined |
| S14_extracted_k141_58138 | Not-determined |
| S14_extracted_k141_116966 | Not-determined |
| S14_extracted_k141_13992 | Not-determined |
| S14_extracted_k141_13993 | Not-determined |
| S14_extracted_k141_116969 | Low-quality |
| S14_extracted_k141_58143 | Not-determined |
| S14_extracted_k141_21832 | Not-determined |
| S14_extracted_k141_58144 | Not-determined |
| S14_extracted_k141_36332 | Not-determined |
| S14_extracted_k141_21833 | Not-determined |
| S14_extracted_k141_50245 | Low-quality |
| S14_extracted_k141_116973 | Not-determined |
| S14_extracted_k141_6833 | Not-determined |
| S14_extracted_k141_21839 | Low-quality |
| S14_extracted_k141_116976 | Not-determined |
| S14_extracted_k141_6838 | Not-determined |
| S14_extracted_k141_50251 | Not-determined |
| S14_extracted_k141_58153 | Low-quality |
| S14_extracted_k141_80155 | Not-determined |
| S14_extracted_k141_65076 | Not-determined |
| S14_extracted_k141_65077 | Low-quality |
| S14_extracted_k141_6845 | Not-determined |
| S14_extracted_k141_6846 | Not-determined |
| S14_extracted_k141_36338 | Low-quality |
| S14_extracted_k141_14000 | Low-quality |
| S14_extracted_k141_14004 | Low-quality |
| S14_extracted_k141_6851 | Not-determined |
| S14_extracted_k141_14005 | Not-determined |
| S14_extracted_k141_80159 | Not-determined |
| S14_extracted_k141_21847 | Not-determined |
| S14_extracted_k141_21850 | Low-quality |
| S14_extracted_k141_80166 | Not-determined |
| S14_extracted_k141_14010 | Low-quality |
| S14_extracted_k141_80169 | Low-quality |
| S14_extracted_k141_72607 | Low-quality |
| S14_extracted_k141_72610 | Not-determined |
| S14_extracted_k141_14014 | Not-determined |
| S14_extracted_k141_58176 | Not-determined |
| S14_extracted_k141_58178 | Not-determined |
| S14_extracted_k141_80173 | Low-quality |
| S14_extracted_k141_50256 | Not-determined |
| S14_extracted_k141_65099 | Not-determined |
| S14_extracted_k141_50259 | Not-determined |
| S14_extracted_k141_21853 | Not-determined |
| S14_extracted_k141_21856 | Low-quality |
| S14_extracted_k141_36347 | Low-quality |
| S14_extracted_k141_65108 | Low-quality |
| S14_extracted_k141_50264 | Not-determined |
| S14_extracted_k141_65109 | Not-determined |
| S14_extracted_k141_50265 | Not-determined |
| S14_extracted_k141_80181 | Not-determined |
| S14_extracted_k141_58190 | Not-determined |
| S14_extracted_k141_65115 | Low-quality |
| S14_extracted_k141_21866 | Low-quality |
| S14_extracted_k141_65118 | Low-quality |
| S14_extracted_k141_72632 | Not-determined |
| S14_extracted_k141_65121 | Not-determined |
| S14_extracted_k141_21871 | Not-determined |
| S14_extracted_k141_87664 | Low-quality |
| S14_extracted_k141_101930 | Not-determined |
| S14_extracted_k141_116994 | Low-quality |
| S14_extracted_k141_101931 | Not-determined |
| S14_extracted_k141_36364 | Not-determined |
| S14_extracted_k141_36366 | Low-quality |
| S14_extracted_k141_36367 | Low-quality |
| S14_extracted_k141_87670 | Not-determined |
| S14_extracted_k141_87672 | Not-determined |
| S14_extracted_k141_36372 | Not-determined |
| S14_extracted_k141_72644 | Not-determined |
| S14_extracted_k141_36373 | Not-determined |
| S14_extracted_k141_101943 | Low-quality |
| S14_extracted_k141_72646 | Low-quality |
| S14_extracted_k141_50271 | Low-quality |
| S14_extracted_k141_101951 | Not-determined |
| S14_extracted_k141_117010 | Not-determined |
| S14_extracted_k141_117012 | Low-quality |
| S14_extracted_k141_101954 | Low-quality |
| S14_extracted_k141_72653 | Low-quality |
| S14_extracted_k141_21894 | Not-determined |
| S14_extracted_k141_36383 | Low-quality |
| S14_extracted_k141_117016 | Not-determined |
| S14_extracted_k141_14052 | Not-determined |
| S14_extracted_k141_101958 | Not-determined |
| S14_extracted_k141_117017 | Not-determined |
| S14_extracted_k141_117020 | Low-quality |
| S14_extracted_k141_21897 | Not-determined |
| S14_extracted_k141_101962 | Not-determined |
| S14_extracted_k141_21900 | Not-determined |
| S14_extracted_k141_6875 | Not-determined |
| S14_extracted_k141_101968 | Not-determined |
| S14_extracted_k141_14060 | Not-determined |
| S14_extracted_k141_14063 | Low-quality |
| S14_extracted_k141_58223 | Low-quality |
| S14_extracted_k141_14066 | Not-determined |
| S14_extracted_k141_117027 | Not-determined |
| S14_extracted_k141_72675 | Not-determined |
| S14_extracted_k141_117032 | Not-determined |
| S14_extracted_k141_36402 | Not-determined |
| S14_extracted_k141_50292 | Not-determined |
| S14_extracted_k141_50294 | Low-quality |
| S14_extracted_k141_21908 | Not-determined |
| S14_extracted_k141_101977 | Not-determined |
| S14_extracted_k141_21915 | Not-determined |
| S14_extracted_k141_50299 | Not-determined |
| S14_extracted_k141_58241 | Not-determined |
| S14_extracted_k141_72685 | Not-determined |
| S14_extracted_k141_58243 | Not-determined |
| S14_extracted_k141_36418 | Not-determined |
| S14_extracted_k141_58245 | Not-determined |
| S14_extracted_k141_14097 | Low-quality |
| S14_extracted_k141_21928 | Not-determined |
| S14_extracted_k141_14099 | Low-quality |
| S14_extracted_k141_58246 | Not-determined |
| S14_extracted_k141_58247 | Not-determined |
| S14_extracted_k141_58248 | Low-quality |
| S14_extracted_k141_117046 | Not-determined |
| S14_extracted_k141_14103 | Not-determined |
| S14_extracted_k141_6879 | Not-determined |
| S14_extracted_k141_80193 | Low-quality |
| S14_extracted_k141_21932 | Not-determined |
| S14_extracted_k141_87718 | Not-determined |
| S14_extracted_k141_58253 | Not-determined |
| S14_extracted_k141_36426 | Low-quality |
| S14_extracted_k141_36429 | Not-determined |
| S14_extracted_k141_58259 | Not-determined |
| S14_extracted_k141_6895 | Low-quality |
| S14_extracted_k141_87724 | Low-quality |
| S14_extracted_k141_21941 | Low-quality |
| S14_extracted_k141_14108 | Low-quality |
| S14_extracted_k141_80210 | Not-determined |
| S14_extracted_k141_21944 | Low-quality |
| S14_extracted_k141_87731 | Not-determined |
| S14_extracted_k141_101990 | Not-determined |
| S14_extracted_k141_87732 | Not-determined |
| S14_extracted_k141_50327 | Not-determined |
| S14_extracted_k141_117058 | Not-determined |
| S14_extracted_k141_101992 | Not-determined |
| S14_extracted_k141_65138 | Not-determined |
| S14_extracted_k141_101995 | Not-determined |
| S14_extracted_k141_50330 | Not-determined |
| S14_extracted_k141_50331 | Not-determined |
| S14_extracted_k141_50334 | Not-determined |
| S14_extracted_k141_50337 | Not-determined |
| S14_extracted_k141_80214 | Not-determined |
| S14_extracted_k141_50339 | Not-determined |
| S14_extracted_k141_65141 | Low-quality |
| S14_extracted_k141_117062 | Not-determined |
| S14_extracted_k141_72729 | Not-determined |
| S14_extracted_k141_80216 | Not-determined |
| S14_extracted_k141_36446 | Low-quality |
| S14_extracted_k141_36447 | Low-quality |
| S14_extracted_k141_36450 | Low-quality |
| S14_extracted_k141_102002 | Not-determined |
| S14_extracted_k141_117068 | Low-quality |
| S14_extracted_k141_36456 | Low-quality |
| S14_extracted_k141_50347 | Not-determined |
| S14_extracted_k141_102008 | Low-quality |
| S14_extracted_k141_36459 | Not-determined |
| S14_extracted_k141_117072 | Not-determined |
| S14_extracted_k141_72736 | Not-determined |
| S14_extracted_k141_117076 | Low-quality |
| S14_extracted_k141_117077 | Not-determined |
| S14_extracted_k141_6903 | Low-quality |
| S14_extracted_k141_117079 | Not-determined |
| S14_extracted_k141_65153 | Not-determined |
| S14_extracted_k141_87757 | Not-determined |
| S14_extracted_k141_36463 | Not-determined |
| S14_extracted_k141_50353 | Not-determined |
| S14_extracted_k141_72743 | Low-quality |
| S14_extracted_k141_14122 | Not-determined |
| S14_extracted_k141_65157 | Low-quality |
| S14_extracted_k141_65159 | Not-determined |
| S14_extracted_k141_58273 | Low-quality |
| S14_extracted_k141_72753 | Not-determined |
| S14_extracted_k141_72754 | Not-determined |
| S14_extracted_k141_72756 | Not-determined |
| S14_extracted_k141_65163 | Low-quality |
| S14_extracted_k141_87765 | Not-determined |
| S14_extracted_k141_21962 | Low-quality |
| S14_extracted_k141_87770 | Low-quality |
| S14_extracted_k141_87772 | Not-determined |
| S14_extracted_k141_14136 | Not-determined |
| S14_extracted_k141_58281 | Low-quality |
| S14_extracted_k141_72766 | Not-determined |
| S14_extracted_k141_50364 | Not-determined |
| S14_extracted_k141_87774 | Not-determined |
| S14_extracted_k141_87776 | Not-determined |
| S14_extracted_k141_72770 | Low-quality |
| S14_extracted_k141_117083 | Not-determined |
| S14_extracted_k141_36484 | Not-determined |
| S14_extracted_k141_65170 | Low-quality |
| S14_extracted_k141_6923 | Not-determined |
| S14_extracted_k141_58290 | Not-determined |
| S14_extracted_k141_65172 | Not-determined |
| S14_extracted_k141_65173 | Not-determined |
| S14_extracted_k141_58293 | Not-determined |
| S14_extracted_k141_102020 | Low-quality |
| S14_extracted_k141_36490 | Not-determined |
| S14_extracted_k141_102021 | Not-determined |
| S14_extracted_k141_80243 | Not-determined |
| S14_extracted_k141_102025 | Low-quality |
| S14_extracted_k141_117095 | Low-quality |
| S14_extracted_k141_58300 | Not-determined |
| S14_extracted_k141_72785 | Not-determined |
| S14_extracted_k141_102028 | Not-determined |
| S14_extracted_k141_14147 | Low-quality |
| S14_extracted_k141_72795 | Not-determined |
| S14_extracted_k141_65179 | Not-determined |
| S14_extracted_k141_36500 | Low-quality |
| S14_extracted_k141_36507 | Not-determined |
| S14_extracted_k141_117111 | Not-determined |
| S14_extracted_k141_6951 | Not-determined |
| S14_extracted_k141_28460 | Low-quality |
| S14_extracted_k141_80249 | Not-determined |
| S14_extracted_k141_58307 | Not-determined |
| S14_extracted_k141_14160 | Not-determined |
| S14_extracted_k141_36512 | Not-determined |
| S14_extracted_k141_50380 | Low-quality |
| S14_extracted_k141_72810 | Not-determined |
| S14_extracted_k141_6964 | Not-determined |
| S14_extracted_k141_65202 | Not-determined |
| S14_extracted_k141_28467 | Not-determined |
| S14_extracted_k141_14171 | Not-determined |
| S14_extracted_k141_28470 | Not-determined |
| S14_extracted_k141_80262 | Not-determined |
| S14_extracted_k141_117128 | Not-determined |
| S14_extracted_k141_28471 | Not-determined |
| S14_extracted_k141_6970 | Low-quality |
| S14_extracted_k141_117130 | Not-determined |
| S14_extracted_k141_117132 | Low-quality |
| S14_extracted_k141_117133 | Not-determined |
| S14_extracted_k141_36524 | Low-quality |
| S14_extracted_k141_117135 | Not-determined |
| S14_extracted_k141_28474 | Low-quality |
| S14_extracted_k141_80263 | Not-determined |
| S14_extracted_k141_50392 | Low-quality |
| S14_extracted_k141_102060 | Not-determined |
| S14_extracted_k141_80271 | Not-determined |
| S14_extracted_k141_50398 | Low-quality |
| S14_extracted_k141_80276 | Low-quality |
| S14_extracted_k141_28486 | Not-determined |
| S14_extracted_k141_102066 | Low-quality |
| S14_extracted_k141_50407 | Not-determined |
| S14_extracted_k141_6976 | Not-determined |
| S14_extracted_k141_87782 | Not-determined |
| S14_extracted_k141_102076 | Low-quality |
| S14_extracted_k141_6980 | Low-quality |
| S14_extracted_k141_117141 | Low-quality |
| S14_extracted_k141_28494 | Not-determined |
| S14_extracted_k141_102081 | Low-quality |
| S14_extracted_k141_102082 | Not-determined |
| S14_extracted_k141_28499 | Not-determined |
| S14_extracted_k141_36546 | Low-quality |
| S14_extracted_k141_80295 | Not-determined |
| S14_extracted_k141_72812 | Low-quality |
| S14_extracted_k141_58331 | Low-quality |
| S14_extracted_k141_102089 | Not-determined |
| S14_extracted_k141_14179 | Not-determined |
| S14_extracted_k141_87793 | Low-quality |
| S14_extracted_k141_58336 | Not-determined |
| S14_extracted_k141_36555 | Not-determined |
| S14_extracted_k141_87796 | Low-quality |
| S14_extracted_k141_58339 | Not-determined |
| S14_extracted_k141_6982 | Low-quality |
| S14_extracted_k141_58341 | Not-determined |
| S14_extracted_k141_58344 | Not-determined |
| S14_extracted_k141_80305 | Not-determined |
| S14_extracted_k141_58347 | Not-determined |
| S14_extracted_k141_87808 | Not-determined |
| S14_extracted_k141_14191 | Not-determined |
| S14_extracted_k141_6994 | Not-determined |
| S14_extracted_k141_50431 | Not-determined |
| S14_extracted_k141_50434 | Not-determined |
| S14_extracted_k141_50435 | Not-determined |
| S14_extracted_k141_58353 | Not-determined |
| S14_extracted_k141_6998 | Low-quality |
| S14_extracted_k141_6999 | Low-quality |
| S14_extracted_k141_7000 | Low-quality |
| S14_extracted_k141_7002 | Not-determined |
| S14_extracted_k141_50439 | Not-determined |
| S14_extracted_k141_80312 | Low-quality |
| S14_extracted_k141_14196 | Low-quality |
| S14_extracted_k141_50443 | Not-determined |
| S14_extracted_k141_7006 | Not-determined |
| S14_extracted_k141_65225 | Low-quality |
| S14_extracted_k141_14200 | Not-determined |
| S14_extracted_k141_14201 | Low-quality |
| S14_extracted_k141_65228 | Not-determined |
| S14_extracted_k141_7011 | Low-quality |
| S14_extracted_k141_72834 | Not-determined |
| S14_extracted_k141_58365 | Low-quality |
| S14_extracted_k141_28518 | Not-determined |
| S14_extracted_k141_14209 | Low-quality |
| S14_extracted_k141_14210 | Low-quality |
| S14_extracted_k141_58369 | Low-quality |
| S14_extracted_k141_14213 | Not-determined |
| S14_extracted_k141_87818 | Low-quality |
| S14_extracted_k141_14214 | Low-quality |
| S14_extracted_k141_50458 | Low-quality |
| S14_extracted_k141_50459 | Low-quality |
| S14_extracted_k141_65246 | Not-determined |
| S14_extracted_k141_50464 | Not-determined |
| S14_extracted_k141_36582 | Low-quality |
| S14_extracted_k141_50466 | Not-determined |
| S14_extracted_k141_80332 | Low-quality |
| S14_extracted_k141_14225 | Not-determined |
| S14_extracted_k141_65249 | Not-determined |
| S14_extracted_k141_65252 | Low-quality |
| S14_extracted_k141_80340 | Low-quality |
| S14_extracted_k141_58377 | Low-quality |
| S14_extracted_k141_58379 | Not-determined |
| S14_extracted_k141_50473 | Not-determined |
| S14_extracted_k141_58385 | Not-determined |
| S14_extracted_k141_80347 | Low-quality |
| S14_extracted_k141_80351 | Low-quality |
| S14_extracted_k141_72866 | Low-quality |
| S14_extracted_k141_87836 | Not-determined |
| S14_extracted_k141_50487 | Not-determined |
| S14_extracted_k141_28524 | Not-determined |
| S7_extracted_138069 | Complete |
| S14_extracted_k141_80360 | Low-quality |
| S14_extracted_k141_14245 | Low-quality |
| S14_extracted_k141_72874 | Not-determined |
| S14_extracted_k141_7016 | Low-quality |
| S14_extracted_k141_50498 | Not-determined |
| S14_extracted_k141_28529 | Not-determined |
| S14_extracted_k141_65272 | Not-determined |
| S14_extracted_k141_7019 | Low-quality |
| S14_extracted_k141_50508 | Low-quality |
| S14_extracted_k141_102120 | Not-determined |
| S14_extracted_k141_80366 | Low-quality |
| S14_extracted_k141_102122 | Not-determined |
| S14_extracted_k141_80370 | Not-determined |
| S14_extracted_k141_80372 | Low-quality |
| S14_extracted_k141_36609 | Not-determined |
| S14_extracted_k141_80374 | Low-quality |
| S14_extracted_k141_36612 | Not-determined |
| S14_extracted_k141_65281 | Low-quality |
| S14_extracted_k141_65283 | Not-determined |
| S14_extracted_k141_72899 | Not-determined |
| S14_extracted_k141_65284 | Not-determined |
| S14_extracted_k141_36617 | Low-quality |
| S14_extracted_k141_28539 | Not-determined |
| S14_extracted_k141_28540 | Not-determined |
| S14_extracted_k141_87848 | Not-determined |
| S14_extracted_k141_87849 | Not-determined |
| S14_extracted_k141_72906 | Not-determined |
| S14_extracted_k141_7024 | Low-quality |
| S14_extracted_k141_65296 | Low-quality |
| S14_extracted_k141_28552 | Not-determined |
| S14_extracted_k141_28554 | Not-determined |
| S14_extracted_k141_102141 | Not-determined |
| S14_extracted_k141_102145 | Not-determined |
| S14_extracted_k141_36625 | Low-quality |
| S14_extracted_k141_87854 | Low-quality |
| S14_extracted_k141_87856 | Low-quality |
| S14_extracted_k141_80395 | Not-determined |
| S14_extracted_k141_28566 | Low-quality |
| S14_extracted_k141_14299 | Not-determined |
| S14_extracted_k141_102151 | Not-determined |
| S14_extracted_k141_28575 | Low-quality |
| S14_extracted_k141_72909 | Not-determined |
| S14_extracted_k141_65315 | Low-quality |
| S14_extracted_k141_7047 | Not-determined |
| S14_extracted_k141_65318 | Not-determined |
| S14_extracted_k141_50530 | Low-quality |
| S14_extracted_k141_72918 | Not-determined |
| S14_extracted_k141_72919 | Low-quality |
| S14_extracted_k141_72921 | Not-determined |
| S14_extracted_k141_72924 | Low-quality |
| S14_extracted_k141_7064 | Low-quality |
| S14_extracted_k141_102160 | Not-determined |
| S14_extracted_k141_72929 | Low-quality |
| S14_extracted_k141_87872 | Low-quality |
| S14_extracted_k141_7073 | Low-quality |
| S14_extracted_k141_72939 | Not-determined |
| S14_extracted_k141_14328 | Low-quality |
| S14_extracted_k141_28591 | Not-determined |
| S14_extracted_k141_28596 | Low-quality |
| S14_extracted_k141_72947 | Low-quality |
| S14_extracted_k141_102178 | Not-determined |
| S14_extracted_k141_102180 | Not-determined |
| S14_extracted_k141_72951 | Not-determined |
| S14_extracted_k141_50538 | Not-determined |
| S14_extracted_k141_28599 | Low-quality |
| S14_extracted_k141_28602 | Not-determined |
| S14_extracted_k141_87883 | Low-quality |
| S14_extracted_k141_87885 | Low-quality |
| S14_extracted_k141_87886 | Not-determined |
| S14_extracted_k141_102190 | Not-determined |
| S14_extracted_k141_7087 | Low-quality |
| S14_extracted_k141_58435 | Not-determined |
| S14_extracted_k141_50545 | Not-determined |
| S14_extracted_k141_7093 | Low-quality |
| S14_extracted_k141_87894 | Not-determined |
| S14_extracted_k141_50554 | Not-determined |
| S14_extracted_k141_58440 | Not-determined |
| S14_extracted_k141_7100 | Not-determined |
| S14_extracted_k141_14356 | Not-determined |
| S14_extracted_k141_50558 | Low-quality |
| S1_vs_k141_2995\|\|full | High-quality |
| S14_extracted_k141_58444 | Not-determined |
| S14_extracted_k141_14362 | Not-determined |
| S14_extracted_k141_50565 | Not-determined |
| S14_extracted_k141_28606 | Low-quality |
| S14_extracted_k141_14364 | Low-quality |
| S14_extracted_k141_14365 | Not-determined |
| S14_extracted_k141_28610 | Low-quality |
| S14_extracted_k141_7113 | Not-determined |
| S14_extracted_k141_102205 | Not-determined |
| S14_extracted_k141_7116 | Not-determined |
| S14_extracted_k141_14374 | Low-quality |
| S14_extracted_k141_7118 | Not-determined |
| S14_extracted_k141_28619 | Not-determined |
| S14_extracted_k141_7124 | Not-determined |
| S14_extracted_k141_14385 | Not-determined |
| S14_extracted_k141_14386 | Low-quality |
| S14_extracted_k141_14387 | Not-determined |
| S14_extracted_k141_102216 | Not-determined |
| S14_extracted_k141_7130 | Not-determined |
| S14_extracted_k141_102221 | Not-determined |
| S14_extracted_k141_50581 | Low-quality |
| S14_extracted_k141_28625 | Low-quality |
| S14_extracted_k141_7141 | Not-determined |
| S14_extracted_k141_80413 | Not-determined |
| S14_extracted_k141_50585 | Not-determined |
| S14_extracted_k141_7144 | Low-quality |
| S8_vs_k141_126240\|\|full | High-quality |
| S14_extracted_k141_72954 | Not-determined |
| S14_extracted_k141_7152 | Not-determined |
| S14_extracted_k141_65361 | Low-quality |
| S14_extracted_k141_7156 | Low-quality |
| S14_extracted_k141_50595 | Not-determined |
| S14_extracted_k141_7159 | Low-quality |
| S14_extracted_k141_50598 | Not-determined |
| S14_extracted_k141_7161 | Low-quality |
| S14_extracted_k141_65373 | Low-quality |
| S14_extracted_k141_14390 | Not-determined |
| S14_extracted_k141_50606 | Not-determined |
| S14_extracted_k141_14395 | Low-quality |
| S14_extracted_k141_50609 | Not-determined |
| S14_extracted_k141_7170 | Low-quality |
| S14_extracted_k141_65379 | Not-determined |
| S14_extracted_k141_7173 | Not-determined |
| S14_extracted_k141_7178 | Not-determined |
| S14_extracted_k141_50616 | Not-determined |
| S14_extracted_k141_58464 | Not-determined |
| S14_extracted_k141_65384 | Not-determined |
| S14_extracted_k141_28657 | Not-determined |
| S14_extracted_k141_7188 | Not-determined |
| S14_extracted_k141_28659 | Low-quality |
| S14_extracted_k141_28660 | Not-determined |
| S14_extracted_k141_58472 | Not-determined |
| S14_extracted_k141_58479 | Not-determined |
| S14_extracted_k141_50628 | Not-determined |
| S14_extracted_k141_102258 | Not-determined |
| S14_extracted_k141_72990 | Not-determined |
| S14_extracted_k141_102266 | Not-determined |
| S14_extracted_k141_28662 | Low-quality |
| S14_extracted_k141_72995 | Low-quality |
| S14_extracted_k141_7194 | Low-quality |
| S14_extracted_k141_58495 | Not-determined |
| S14_extracted_k141_80448 | Not-determined |
| S14_extracted_k141_65398 | Not-determined |
| S14_extracted_k141_80450 | Low-quality |
| S14_extracted_k141_102281 | Not-determined |
| S14_extracted_k141_58501 | Not-determined |
| S14_extracted_k141_58506 | Low-quality |
| S14_extracted_k141_58509 | Low-quality |
| S14_extracted_k141_7205 | Low-quality |
| S14_extracted_k141_50634 | Low-quality |
| S14_extracted_k141_14425 | Low-quality |
| S14_extracted_k141_50635 | Not-determined |
| S14_extracted_k141_73010 | Low-quality |
| S14_extracted_k141_58515 | Low-quality |
| S14_extracted_k141_73011 | Not-determined |
| S14_extracted_k141_65405 | Not-determined |
| S14_extracted_k141_58516 | Low-quality |
| S14_extracted_k141_58518 | Not-determined |
| S14_extracted_k141_58520 | Not-determined |
| S14_extracted_k141_102303 | Not-determined |
| S14_extracted_k141_50640 | Low-quality |
| S14_extracted_k141_50643 | Not-determined |
| S14_extracted_k141_65424 | Not-determined |
| S14_extracted_k141_7221 | Not-determined |
| S14_extracted_k141_80463 | Not-determined |
| S14_extracted_k141_50649 | Not-determined |
| S14_extracted_k141_73028 | Not-determined |
| S14_extracted_k141_73029 | Not-determined |
| S14_extracted_k141_58534 | Not-determined |
| S14_extracted_k141_73033 | Not-determined |
| S14_extracted_k141_58537 | Not-determined |
| S14_extracted_k141_7228 | Low-quality |
| S14_extracted_k141_73034 | Low-quality |
| S14_extracted_k141_65431 | Not-determined |
| S14_extracted_k141_50657 | Low-quality |
| S14_extracted_k141_58541 | Low-quality |
| S14_extracted_k141_73043 | Low-quality |
| S14_extracted_k141_58543 | Low-quality |
| S14_extracted_k141_102319 | Not-determined |
| S14_extracted_k141_28683 | Not-determined |
| S14_extracted_k141_28685 | Not-determined |
| S14_extracted_k141_28687 | Not-determined |
| S14_extracted_k141_65435 | Not-determined |
| S14_extracted_k141_65436 | Not-determined |
| S14_extracted_k141_7240 | Not-determined |
| S14_extracted_k141_73050 | Not-determined |
| S14_extracted_k141_7243 | Not-determined |
| S14_extracted_k141_7245 | Low-quality |
| S14_extracted_k141_28708 | Not-determined |
| S14_extracted_k141_80473 | Not-determined |
| S14_extracted_k141_73053 | Low-quality |
| S14_extracted_k141_7248 | Low-quality |
| S14_extracted_k141_50684 | Not-determined |
| S14_extracted_k141_50685 | Not-determined |
| S14_extracted_k141_102335 | Not-determined |
| S14_extracted_k141_102336 | Not-determined |
| S14_extracted_k141_80485 | Not-determined |
| S14_extracted_k141_80487 | Low-quality |
| S14_extracted_k141_7262 | Not-determined |
| S14_extracted_k141_102340 | Not-determined |
| S14_extracted_k141_7264 | Not-determined |
| S14_extracted_k141_80488 | Not-determined |
| S14_extracted_k141_102345 | Low-quality |
| S14_extracted_k141_7265 | Not-determined |
| S14_extracted_k141_50696 | Low-quality |
| S14_extracted_k141_102347 | Low-quality |
| S14_extracted_k141_50698 | Not-determined |
| S14_extracted_k141_7267 | Not-determined |
| S14_extracted_k141_102348 | Not-determined |
| S14_extracted_k141_50704 | Low-quality |
| S14_extracted_k141_80489 | Not-determined |
| S14_extracted_k141_73088 | Low-quality |
| S14_extracted_k141_73091 | Low-quality |
| S14_extracted_k141_14448 | Not-determined |
| S14_extracted_k141_28740 | Not-determined |
| S14_extracted_k141_50713 | Not-determined |
| S14_extracted_k141_73094 | Low-quality |
| S14_extracted_k141_7281 | Not-determined |
| S14_extracted_k141_7283 | Not-determined |
| S14_extracted_k141_7284 | Low-quality |
| S14_extracted_k141_14452 | Not-determined |
| S14_extracted_k141_7287 | Not-determined |
| S14_extracted_k141_14455 | Not-determined |
| S14_extracted_k141_102362 | Not-determined |
| S14_extracted_k141_7296 | Low-quality |
| S14_extracted_k141_50728 | Low-quality |
| S14_extracted_k141_73114 | Not-determined |
| S14_extracted_k141_80514 | Not-determined |
| S14_extracted_k141_65486 | Not-determined |
| S14_extracted_k141_28751 | Low-quality |
| S14_extracted_k141_73119 | Low-quality |
| S14_extracted_k141_7312 | Low-quality |
| S14_extracted_k141_80526 | Not-determined |
| S14_extracted_k141_14465 | Not-determined |
| S14_extracted_k141_14466 | Not-determined |
| S14_extracted_k141_65495 | Not-determined |
| S14_extracted_k141_14473 | Low-quality |
| S14_extracted_k141_80530 | Low-quality |
| S14_extracted_k141_65501 | Low-quality |
| S14_extracted_k141_14476 | Low-quality |
| S14_extracted_k141_102381 | Not-determined |
| S14_extracted_k141_80532 | Not-determined |
| S14_extracted_k141_58591 | Not-determined |
| S14_extracted_k141_28783 | Low-quality |
| S14_extracted_k141_58598 | Low-quality |
| S14_extracted_k141_28786 | Not-determined |
| S14_extracted_k141_58603 | Not-determined |
| S14_extracted_k141_50741 | Low-quality |
| S14_extracted_k141_50744 | Low-quality |
| S14_extracted_k141_80554 | Low-quality |
| S14_extracted_k141_50746 | Not-determined |
| S14_extracted_k141_28792 | Low-quality |

| | |
|---|---|
| S14_extracted_k141_73137 | Not-determined |
| S14_extracted_k141_102391 | Low-quality |
| S14_extracted_k141_28797 | Not-determined |
| S14_extracted_k141_7323 | Low-quality |
| S14_extracted_k141_14492 | Low-quality |
| S14_extracted_k141_102402 | Not-determined |
| S14_extracted_k141_50755 | Low-quality |
| S14_extracted_k141_14499 | Low-quality |
| S14_extracted_k141_73143 | Not-determined |
| S14_extracted_k141_28801 | Not-determined |
| S14_extracted_k141_28805 | Low-quality |
| S14_extracted_k141_102416 | Not-determined |
| S14_extracted_k141_80570 | Not-determined |
| S14_extracted_k141_80571 | Not-determined |
| S14_extracted_k141_73155 | Low-quality |
| S14_extracted_k141_80573 | Not-determined |
| S14_extracted_k141_102419 | Not-determined |
| S14_extracted_k141_73158 | Not-determined |
| S14_extracted_k141_80575 | Low-quality |
| S14_extracted_k141_102426 | Low-quality |
| S14_extracted_k141_14503 | Not-determined |
| S14_extracted_k141_102427 | Not-determined |
| S14_extracted_k141_14506 | Not-determined |
| S14_extracted_k141_80582 | Not-determined |
| S14_extracted_k141_80584 | Not-determined |
| S14_extracted_k141_28820 | Not-determined |
| S14_extracted_k141_80590 | Not-determined |
| S12_extracted_k141_108501 | Complete |
| S14_extracted_k141_50768 | Not-determined |
| S14_extracted_k141_14517 | Not-determined |
| S14_extracted_k141_50778 | Not-determined |
| S14_extracted_k141_28830 | Not-determined |
| S14_extracted_k141_14523 | Not-determined |
| S14_extracted_k141_73176 | Not-determined |
| S14_extracted_k141_28833 | Not-determined |
| S14_extracted_k141_102452 | Not-determined |
| S14_extracted_k141_102453 | Not-determined |
| S14_extracted_k141_28840 | Low-quality |
| S14_extracted_k141_28841 | Not-determined |
| S14_extracted_k141_14537 | Low-quality |
| S14_extracted_k141_28843 | Not-determined |
| S14_extracted_k141_73191 | Low-quality |
| S14_extracted_k141_102460 | Not-determined |
| S14_extracted_k141_102461 | Low-quality |
| S14_extracted_k141_73196 | Not-determined |
| S14_extracted_k141_73201 | Low-quality |
| S14_extracted_k141_28848 | Low-quality |
| S14_extracted_k141_102487 | Not-determined |
| S14_extracted_k141_102488 | Not-determined |
| S14_extracted_k141_28855 | Not-determined |
| S14_extracted_k141_102492 | Low-quality |
| S14_extracted_k141_50818 | Low-quality |
| S14_extracted_k141_28870 | Not-determined |
| S14_extracted_k141_28872 | Low-quality |
| S14_extracted_k141_50823 | Low-quality |
| S14_extracted_k141_14563 | Not-determined |
| S14_extracted_k141_73218 | Low-quality |
| S14_extracted_k141_50828 | Low-quality |
| S14_extracted_k141_50829 | Not-determined |
| S14_extracted_k141_14572 | Not-determined |
| S14_extracted_k141_28883 | Not-determined |
| S14_extracted_k141_73224 | Not-determined |
| S14_extracted_k141_50835 | Not-determined |
| S14_extracted_k141_50839 | Low-quality |
| S14_extracted_k141_102505 | Low-quality |
| S14_extracted_k141_50843 | Not-determined |
| S14_extracted_k141_65510 | Low-quality |
| S14_extracted_k141_65511 | Low-quality |
| S14_extracted_k141_28893 | Not-determined |
| S14_extracted_k141_14587 | Not-determined |
| S14_extracted_k141_50852 | Not-determined |
| S14_extracted_k141_65526 | Not-determined |
| S14_extracted_k141_28910 | Not-determined |
| S14_extracted_k141_73245 | Low-quality |
| S14_extracted_k141_73247 | Low-quality |
| S14_extracted_k141_14593 | Not-determined |
| S14_extracted_k141_28915 | Low-quality |
| S14_extracted_k141_14597 | Not-determined |
| S14_extracted_k141_65532 | Low-quality |
| S14_extracted_k141_28921 | Not-determined |
| S14_extracted_k141_14609 | Not-determined |
| S14_extracted_k141_28933 | Low-quality |
| S14_extracted_k141_65542 | Not-determined |
| S3_extracted_k141_35073 | High-quality |
| S14_extracted_k141_65543 | Low-quality |
| S14_extracted_k141_50863 | Low-quality |
| S14_extracted_k141_65549 | Not-determined |
| S14_extracted_k141_50866 | Not-determined |
| S14_extracted_k141_65557 | Low-quality |
| S14_extracted_k141_28938 | Low-quality |
| S14_extracted_k141_50872 | Not-determined |
| S14_extracted_k141_73262 | Not-determined |
| S14_extracted_k141_28941 | Low-quality |
| S14_extracted_k141_102539 | Low-quality |
| S14_extracted_k141_102541 | Not-determined |
| S14_extracted_k141_102543 | Low-quality |
| S14_extracted_k141_28949 | Not-determined |
| S14_extracted_k141_65565 | Not-determined |
| S14_extracted_k141_65567 | Not-determined |
| S14_extracted_k141_102555 | Not-determined |
| S14_extracted_k141_14625 | Not-determined |
| S14_extracted_k141_50888 | Low-quality |
| S14_extracted_k141_50890 | Low-quality |
| S14_extracted_k141_65576 | Low-quality |
| S14_extracted_k141_65579 | Low-quality |
| S14_extracted_k141_65582 | Low-quality |
| S14_extracted_k141_65585 | Not-determined |
| S14_extracted_k141_28958 | Low-quality |
| S14_extracted_k141_28960 | Low-quality |
| S14_extracted_k141_28966 | Not-determined |
| S14_extracted_k141_28974 | Not-determined |
| S14_extracted_k141_28977 | Low-quality |
| S14_extracted_k141_50913 | Not-determined |
| S14_extracted_k141_50917 | Low-quality |
| S14_extracted_k141_50918 | Low-quality |
| S14_extracted_k141_28990 | Not-determined |
| S14_extracted_k141_65597 | Not-determined |
| S14_extracted_k141_65598 | Low-quality |
| S14_extracted_k141_28997 | Not-determined |
| S14_extracted_k141_50926 | Low-quality |
| S14_extracted_k141_65617 | Not-determined |
| S14_extracted_k141_50932 | Not-determined |
| S14_extracted_k141_50938 | Not-determined |
| S14_extracted_k141_50943 | Not-determined |
| S14_extracted_k141_50945 | Not-determined |
| S14_extracted_k141_50950 | Low-quality |
| S14_extracted_k141_50953 | Low-quality |
| S14_extracted_k141_50955 | Low-quality |
| S14_extracted_k141_102567 | Not-determined |
| S14_extracted_k141_29028 | Not-determined |
| S14_extracted_k141_29029 | Not-determined |
| S14_extracted_k141_50962 | Low-quality |
| S14_extracted_k141_65636 | Not-determined |
| S14_extracted_k141_65637 | Not-determined |
| S14_extracted_k141_65639 | Low-quality |
| S14_extracted_k141_65645 | Not-determined |
| S14_extracted_k141_65649 | Not-determined |
| S14_extracted_k141_65656 | Low-quality |
| S14_extracted_k141_50985 | Not-determined |
| S14_extracted_k141_29049 | Low-quality |
| S14_extracted_k141_50991 | Not-determined |
| S14_extracted_k141_65674 | Not-determined |
| S14_extracted_k141_65676 | Not-determined |
| S14_extracted_k141_65680 | Low-quality |
| S14_extracted_k141_29060 | Low-quality |
| S14_extracted_k141_65685 | Not-determined |
| S14_extracted_k141_51007 | Not-determined |
| S14_extracted_k141_65690 | Not-determined |
| S14_extracted_k141_65693 | Not-determined |
| S14_extracted_k141_51013 | Not-determined |
| S14_extracted_k141_51026 | Low-quality |
| S14_extracted_k141_29081 | Not-determined |
| S14_extracted_k141_65705 | Not-determined |
| S14_extracted_k141_51035 | Not-determined |
| S14_extracted_k141_65711 | Not-determined |
| S14_extracted_k141_65712 | Low-quality |
| S14_extracted_k141_65715 | Not-determined |
| S14_extracted_k141_51060 | Low-quality |
| S14_extracted_k141_51067 | Not-determined |
| S14_extracted_k141_51070 | Not-determined |
| S14_extracted_k141_29120 | Low-quality |
| S14_extracted_k141_65722 | Low-quality |
| S14_extracted_k141_51076 | Low-quality |
| S14_extracted_k141_29126 | Not-determined |
| S14_extracted_k141_65729 | Not-determined |
| S14_extracted_k141_51079 | Not-determined |
| S14_extracted_k141_29127 | Not-determined |
| S14_extracted_k141_65739 | Not-determined |
| S14_extracted_k141_51089 | Not-determined |
| S14_extracted_k141_51092 | Low-quality |
| S14_extracted_k141_29142 | Not-determined |
| S14_extracted_k141_29145 | Low-quality |
| S14_extracted_k141_51102 | Not-determined |
| S14_extracted_k141_51104 | Not-determined |
| S14_extracted_k141_51105 | Not-determined |
| S14_extracted_k141_51108 | Not-determined |
| S14_extracted_k141_29153 | Not-determined |
| S12_vs_k141_188821\|\|full | High-quality |
| S14_extracted_k141_51121 | Low-quality |
| S14_extracted_k141_51122 | Low-quality |
| S14_extracted_k141_29162 | Low-quality |
| S14_extracted_k141_29164 | Not-determined |
| S14_extracted_k141_65757 | Not-determined |
| S14_extracted_k141_29182 | Low-quality |
| S14_extracted_k141_29184 | Low-quality |
| S14_extracted_k141_29193 | Not-determined |
| S7_vs_138069\|\|full | High-quality |
| S14_extracted_k141_29200 | Low-quality |
| S14_extracted_k141_29206 | Not-determined |
| S14_extracted_k141_29210 | Not-determined |
| S12_vs_k141_108501\|\|full | High-quality |
| S14_extracted_k141_65781 | Low-quality |
| S14_extracted_k141_51145 | Not-determined |
| S14_extracted_k141_65787 | Not-determined |
| S14_extracted_k141_65788 | Low-quality |
| S14_extracted_k141_29229 | Not-determined |
| S14_extracted_k141_65795 | Low-quality |
| S14_extracted_k141_65799 | Low-quality |
| S14_extracted_k141_65801 | Not-determined |
| S14_extracted_k141_29234 | Low-quality |
| S14_extracted_k141_29236 | Low-quality |
| S14_extracted_k141_65809 | Low-quality |
| S14_extracted_k141_65812 | Not-determined |
| S14_extracted_k141_65819 | Not-determined |
| S14_extracted_k141_51175 | Not-determined |
| S14_extracted_k141_51176 | Not-determined |
| S14_extracted_k141_65847 | Not-determined |
| S14_extracted_k141_65848 | Not-determined |
| S14_extracted_k141_65851 | Not-determined |
| S14_extracted_k141_65860 | Low-quality |
| S14_extracted_k141_51196 | Low-quality |
| S14_extracted_k141_65861 | Not-determined |
| S14_extracted_k141_51199 | Low-quality |
| S14_extracted_k141_51200 | Low-quality |
| S14_extracted_k141_65869 | Low-quality |
| S14_extracted_k141_29300 | Not-determined |
| S14_extracted_k141_65882 | Low-quality |
| S14_extracted_k141_29305 | Not-determined |
| S14_extracted_k141_51249 | Not-determined |
| S14_extracted_k141_65904 | Not-determined |
| S14_extracted_k141_65909 | Low-quality |
| S14_extracted_k141_117194 | Not-determined |
| S14_extracted_k141_117197 | Not-determined |
| S14_extracted_k141_117205 | Not-determined |
| S14_extracted_k141_117207 | Not-determined |
| S14_extracted_k141_65924 | Not-determined |
| S14_extracted_k141_51271 | Not-determined |
| S14_extracted_k141_117213 | Not-determined |
| S14_extracted_k141_117216 | Not-determined |
| S14_extracted_k141_117220 | Not-determined |
| S14_extracted_k141_51281 | Not-determined |
| S14_extracted_k141_51282 | Not-determined |
| S14_extracted_k141_117226 | Not-determined |
| S14_extracted_k141_65925 | Low-quality |
| S14_extracted_k141_65926 | Not-determined |
| S14_extracted_k141_117230 | Not-determined |
| S14_extracted_k141_51287 | Not-determined |
| S14_extracted_k141_117232 | Not-determined |
| S14_extracted_k141_65938 | Not-determined |
| S10_extracted_64371 | Complete |
| S3_vs_k141_35073\|\|full | High-quality |
| S14_vs_k141_1013\|\|full | Not-determined |
| S9_extracted_k141_45558 | Complete |
| S14_vs_k141_59845\|\|full | Not-determined |
| S10_vs_64371\|\|full | High-quality |
| S14_vs_k141_96640\|\|full | Not-determined |
| S9_vs_k141_45558\|\|full | High-quality |
| S14_vs_k141_17386\|\|full | Low-quality |
| S16_extracted_1037 | Complete |
| S14_vs_k141_69166\|\|full | Not-determined |
| S16_vs_1037\|\|full | High-quality |
| S14_vs_k141_40939\|\|full | Not-determined |
| S4_vs_VIRSorter_31976-circular-cat | High-quality |
| S10_vs_26365\|\|full | High-quality |
| S8_vs_k141_205257\|\|full | High-quality |
| S14_vs_k141_42401\|\|full | Not-determined |
| S8_vs_k141_9987\|\|full | High-quality |
| S14_vs_k141_94397\|\|full | Not-determined |
| S3_vs_k141_98472\|\|full | High-quality |
| S14_vs_k141_117148\|\|full | Low-quality |
| S14_vs_k141_117149\|\|full | Low-quality |
| S11_vs_k141_104064\|\|full | High-quality |
| S4_vs_VIRSorter_2441-circular_gen | Medium-quality |
| S16_vs_16840\|\|full | High-quality |
| S3_vs_k141_2156\|\|full | High-quality |
| S14_vs_k141_117156\|\|full | Low-quality |
| S11_extracted_k141_48781 | High-quality |
| S11_vs_k141_27845\|\|full | High-quality |
| S5_vs_k141_145582\|\|full | High-quality |
| S7_vs_11081\|\|full | High-quality |
| S5_vs_k141_85065\|\|full | High-quality |
| S5_extracted_k141_69162 | Medium-quality |
| S14_vs_k141_117163\|\|full | Not-determined |
| S5_vs_k141_69162\|\|full | Medium-quality |
| S15_vs_3868\|\|full | High-quality |
| S3_extracted_k141_54353 | High-quality |
| S7_vs_49426\|\|full | High-quality |
| S14_vs_k141_117171\|\|full | Not-determined |
| S14_vs_k141_117194\|\|full | Not-determined |
| S14_vs_k141_95256\|\|full | Low-quality |
| S14_vs_k141_7331\|\|full | Not-determined |
| S14_vs_k141_13\|\|full | Low-quality |
| S14_vs_k141_80607\|\|full | Low-quality |
| S14_vs_k141_73275\|\|full | Low-quality |
| S14_vs_k141_58618\|\|full | Not-determined |
| S14_vs_k141_65962\|\|full | Low-quality |
| S14_vs_k141_73280\|\|full | Low-quality |
| S14_vs_k141_21994\|\|full | Low-quality |
| S14_vs_k141_27\|\|full | Low-quality |
| S14_vs_k141_36663\|\|full | Not-determined |
| S14_vs_k141_29315\|\|full | Not-determined |
| S14_vs_k141_22006\|\|full | Not-determined |
| S14_vs_k141_49\|\|full | Not-determined |
| S14_vs_k141_7346\|\|full | Not-determined |
| S14_vs_k141_43992\|\|full | Not-determined |
| S14_vs_k141_51314\|\|full | Low-quality |
| S14_vs_k141_14675\|\|full | Low-quality |
| S14_vs_k141_80627\|\|full | Low-quality |
| S14_vs_k141_66\|\|full | Not-determined |
| S14_vs_k141_95269\|\|full | Low-quality |
| S14_vs_k141_65985\|\|full | Not-determined |
| S14_vs_k141_44020\|\|full | Not-determined |
| S14_vs_k141_95278\|\|full | Not-determined |
| S14_vs_k141_58629\|\|full | Low-quality |
| S14_vs_k141_14699\|\|full | Low-quality |
| S14_vs_k141_58631\|\|full | Low-quality |
| S14_vs_k141_91\|\|full | Low-quality |
| S14_vs_k141_58635\|\|full | Low-quality |
| S14_vs_k141_36709\|\|full | Low-quality |
| S14_vs_k141_73290\|\|full | Low-quality |
| S14_vs_k141_109960\|\|full | Low-quality |
| S14_vs_k141_36722\|\|full | Low-quality |
| S14_vs_k141_22051\|\|full | Low-quality |
| S14_vs_k141_66022\|\|full | Low-quality |
| S14_vs_k141_29356\|\|full | Not-determined |
| S14_vs_k141_44049\|\|full | Low-quality |
| S14_vs_k141_29358\|\|full | Low-quality |
| S14_vs_k141_7398\|\|full | Not-determined |
| S14_vs_k141_80680\|\|full | Not-determined |
| S14_vs_k141_36748\|\|full | Not-determined |
| S14_vs_k141_29366\|\|full | Low-quality |
| S14_vs_k141_7403\|\|full | Not-determined |
| S14_vs_k141_29371\|\|full | Low-quality |
| S14_vs_k141_44052\|\|full | Low-quality |
| S14_vs_k141_58666\|\|full | Low-quality |
| S14_vs_k141_7410\|\|full | Low-quality |
| S14_vs_k141_58668\|\|full | Not-determined |
| S14_vs_k141_87956\|\|full | Not-determined |
| S3_vs_k141_54353\|\|full | High-quality |
| S14_vs_k141_36754\|\|full | Not-determined |
| S14_vs_k141_73303\|\|full | Low-quality |
| S14_vs_k141_29392\|\|full | Low-quality |
| S14_vs_k141_102630\|\|full | Low-quality |
| S14_vs_k141_58690\|\|full | Not-determined |
| S14_vs_k141_29398\|\|full | Not-determined |
| S14_vs_k141_22091\|\|full | Low-quality |
| S14_vs_k141_102637\|\|full | Low-quality |
| S14_vs_k141_44084\|\|full | Low-quality |
| S14_vs_k141_36772\|\|full | Low-quality |
| S14_vs_k141_80694\|\|full | Low-quality |
| S4_vs_VIRSorter_6698-cat_2 | Medium-quality |
| S14_vs_k141_36785\|\|full | Low-quality |
| S14_vs_k141_7451\|\|full | Low-quality |
| S14_vs_k141_29414\|\|full | Low-quality |
| S14_vs_k141_44092\|\|full | Low-quality |
| S14_vs_k141_80701\|\|full | Low-quality |
| S14_vs_k141_36795\|\|full | Not-determined |
| S14_vs_k141_51418\|\|full | Not-determined |
| S14_vs_k141_58723\|\|full | Low-quality |
| S14_vs_k141_29430\|\|full | Low-quality |
| S14_vs_k141_80705\|\|full | Low-quality |
| S14_vs_k141_29431\|\|full | Low-quality |
| S14_vs_k141_29434\|\|full | Low-quality |
| S14_vs_k141_102673\|\|full | Low-quality |
| S14_vs_k141_36807\|\|full | Not-determined |
| S14_vs_k141_44110\|\|full | Low-quality |
| S14_vs_k141_29442\|\|full | Low-quality |
| S14_vs_k141_44112\|\|full | Low-quality |
| S14_vs_k141_7465\|\|full | Not-determined |
| S14_vs_k141_29445\|\|full | Low-quality |
| S14_vs_k141_66092\|\|full | Low-quality |
| S14_vs_k141_14768\|\|full | Low-quality |
| S14_vs_k141_102681\|\|full | Low-quality |
| S14_vs_k141_102687\|\|full | Not-determined |
| S14_vs_k141_66097\|\|full | Low-quality |
| S14_vs_k141_36822\|\|full | Not-determined |
| S14_vs_k141_66107\|\|full | Low-quality |
| S14_vs_k141_44130\|\|full | Low-quality |
| S14_vs_k141_66112\|\|full | Low-quality |
| S14_vs_k141_73314\|\|full | Low-quality |
| S14_vs_k141_181\|\|full | Low-quality |
| S14_vs_k141_102697\|\|full | Low-quality |
| S14_vs_k141_22112\|\|full | Low-quality |
| S14_vs_k141_73332\|\|full | Low-quality |
| S14_vs_k141_66125\|\|full | Low-quality |
| S14_vs_k141_183\|\|full | Not-determined |
| S14_vs_k141_95415\|\|full | Not-determined |
| S14_vs_k141_87991\|\|full | Low-quality |
| S14_vs_k141_36863\|\|full | Low-quality |
| S14_vs_k141_29487\|\|full | Low-quality |
| S14_vs_k141_22132\|\|full | Low-quality |
| S14_vs_k141_51440\|\|full | Low-quality |
| S15_vs_27510\|\|full | High-quality |
| S14_vs_k141_36876\|\|full | Low-quality |
| S14_vs_k141_51454\|\|full | Low-quality |
| S14_vs_k141_51463\|\|full | Not-determined |
| S14_vs_k141_44155\|\|full | Low-quality |
| S14_vs_k141_44157\|\|full | Low-quality |
| S14_vs_k141_51472\|\|full | Low-quality |
| S14_vs_k141_212\|\|full | Not-determined |
| S14_vs_k141_58815\|\|full | Not-determined |
| S14_vs_k141_44164\|\|full | Low-quality |
| S14_vs_k141_88029\|\|full | Low-quality |
| S14_vs_k141_88034\|\|full | Not-determined |
| S14_vs_k141_58821\|\|full | Low-quality |
| S14_vs_k141_102726\|\|full | Not-determined |
| S14_vs_k141_102728\|\|full | Low-quality |
| S14_vs_k141_22168\|\|full | Low-quality |
| S6_vs_k141_39025\|\|full | Medium-quality |
| S14_vs_k141_7526\|\|full | Not-determined |
| S14_vs_k141_7527\|\|full | Low-quality |
| S14_vs_k141_88044\|\|full | Not-determined |
| S14_vs_k141_246\|\|full | Not-determined |
| S14_vs_k141_44175\|\|full | Low-quality |
| S14_vs_k141_110034\|\|full | Low-quality |
| S14_vs_k141_253\|\|full | Low-quality |
| S14_vs_k141_110049\|\|full | Low-quality |
| S14_vs_k141_22200\|\|full | Low-quality |
| S14_vs_k141_58867\|\|full | Not-determined |
| S14_vs_k141_110058\|\|full | Low-quality |
| S14_vs_k141_7571\|\|full | Not-determined |
| S14_vs_k141_58870\|\|full | Not-determined |
| S14_vs_k141_7574\|\|full | Low-quality |
| S14_vs_k141_51510\|\|full | Low-quality |
| S14_vs_k141_80799\|\|full | Not-determined |
| S14_vs_k141_80801\|\|full | Low-quality |
| S14_vs_k141_51515\|\|full | Not-determined |
| S14_vs_k141_88079\|\|full | Low-quality |
| S14_vs_k141_51519\|\|full | Low-quality |
| S14_vs_k141_22206\|\|full | Low-quality |
| S14_vs_k141_80815\|\|full | Not-determined |
| S14_vs_k141_51535\|\|full | Low-quality |
| S14_vs_k141_95466\|\|full | Not-determined |
| S14_vs_k141_14840\|\|full | Not-determined |
| S14_vs_k141_7605\|\|full | Not-determined |
| S14_vs_k141_302\|\|full | Low-quality |
| S14_vs_k141_14853\|\|full | Not-determined |
| S14_vs_k141_305\|\|full | Low-quality |
| S14_vs_k141_22222\|\|full | Low-quality |
| S14_vs_k141_58913\|\|full | Low-quality |
| S14_vs_k141_58917\|\|full | Low-quality |
| S14_vs_k141_22245\|\|full | Not-determined |
| S14_vs_k141_14866\|\|full | Low-quality |
| S14_vs_k141_88130\|\|full | Low-quality |
| S14_vs_k141_73372\|\|full | Low-quality |
| S14_vs_k141_95500\|\|full | Low-quality |
| S14_vs_k141_73380\|\|full | Low-quality |
| S14_vs_k141_102744\|\|full | Not-determined |
| S14_vs_k141_102751\|\|full | Low-quality |
| S14_vs_k141_29575\|\|full | Not-determined |
| S14_vs_k141_73382\|\|full | Low-quality |
| S14_vs_k141_80831\|\|full | Not-determined |
| S14_vs_k141_332\|\|full | Not-determined |
| S14_vs_k141_73386\|\|full | Low-quality |
| S14_vs_k141_36999\|\|full | Low-quality |
| S14_vs_k141_80838\|\|full | Low-quality |
| S14_vs_k141_110103\|\|full | Low-quality |
| S14_vs_k141_7634\|\|full | Low-quality |
| S14_vs_k141_14894\|\|full | Low-quality |
| S14_vs_k141_37006\|\|full | Low-quality |
| S14_vs_k141_80849\|\|full | Low-quality |
| S14_vs_k141_14898\|\|full | Low-quality |
| S14_vs_k141_88153\|\|full | Low-quality |
| S14_vs_k141_88156\|\|full | Not-determined |
| S14_vs_k141_29595\|\|full | Not-determined |
| S14_vs_k141_7635\|\|full | Not-determined |
| S14_vs_k141_7641\|\|full | Not-determined |
| S8_extracted_k141_131009 | Complete |
| S14_vs_k141_29605\|\|full | Not-determined |
| S14_vs_k141_58925\|\|full | Low-quality |
| S14_vs_k141_102790\|\|full | Low-quality |
| S14_vs_k141_51602\|\|full | Not-determined |
| S14_vs_k141_88171\|\|full | Not-determined |
| S14_vs_k141_51606\|\|full | Low-quality |
| S14_vs_k141_102804\|\|full | Not-determined |
| S14_vs_k141_7656\|\|full | Low-quality |
| S14_vs_k141_29609\|\|full | Low-quality |
| S14_vs_k141_22259\|\|full | Not-determined |
| S14_vs_k141_58947\|\|full | Low-quality |
| S14_vs_k141_22263\|\|full | Low-quality |
| S14_vs_k141_14937\|\|full | Low-quality |
| S14_vs_k141_372\|\|full | Low-quality |
| S14_vs_k141_88184\|\|full | Low-quality |
| S14_vs_k141_73426\|\|full | Low-quality |
| S14_vs_k141_80857\|\|full | Low-quality |
| S14_vs_k141_58953\|\|full | Low-quality |
| S14_vs_k141_102820\|\|full | Low-quality |
| S14_vs_k141_37038\|\|full | Not-determined |
| S14_vs_k141_88197\|\|full | Low-quality |
| S14_vs_k141_37040\|\|full | Low-quality |
| S14_vs_k141_37043\|\|full | Low-quality |
| S14_vs_k141_88202\|\|full | Not-determined |
| S11_vs_k141_48781\|\|full | High-quality |
| S14_vs_k141_22271\|\|full | Not-determined |
| S14_vs_k141_58958\|\|full | Not-determined |
| S14_vs_k141_7678\|\|full | Low-quality |
| S14_vs_k141_102833\|\|full | Low-quality |
| S14_vs_k141_37052\|\|full | Low-quality |
| S14_vs_k141_58976\|\|full | Not-determined |
| S14_vs_k141_73446\|\|full | Low-quality |
| S14_vs_k141_58977\|\|full | Low-quality |
| S14_vs_k141_7683\|\|full | Not-determined |
| S14_vs_k141_29633\|\|full | Low-quality |
| S14_vs_k141_394\|\|full | Low-quality |
| S14_vs_k141_66152\|\|full | Not-determined |
| S14_vs_k141_51641\|\|full | Low-quality |
| S14_vs_k141_58983\|\|full | Low-quality |
| S14_vs_k141_29640\|\|full | Low-quality |
| S14_vs_k141_95559\|\|full | Low-quality |
| S14_vs_k141_405\|\|full | Low-quality |
| S14_vs_k141_88222\|\|full | Low-quality |
| S14_vs_k141_419\|\|full | Low-quality |
| S14_vs_k141_29655\|\|full | Low-quality |
| S14_vs_k141_110139\|\|full | Not-determined |
| S14_vs_k141_44211\|\|full | Low-quality |
| S14_vs_k141_51657\|\|full | Not-determined |
| S14_vs_k141_80896\|\|full | Not-determined |
| S14_vs_k141_37077\|\|full | Low-quality |
| S14_vs_k141_434\|\|full | Low-quality |
| S14_vs_k141_37079\|\|full | Low-quality |
| S14_vs_k141_95589\|\|full | Not-determined |
| S14_vs_k141_450\|\|full | Low-quality |
| S14_vs_k141_80905\|\|full | Not-determined |
| S14_vs_k141_102869\|\|full | Not-determined |
| S14_vs_k141_51676\|\|full | Not-determined |
| S14_vs_k141_102870\|\|full | Low-quality |
| S14_vs_k141_80917\|\|full | Not-determined |
| S14_vs_k141_102871\|\|full | Low-quality |
| S14_vs_k141_102872\|\|full | Not-determined |
| S14_vs_k141_14985\|\|full | Not-determined |
| S14_vs_k141_95608\|\|full | Low-quality |
| S14_vs_k141_110189\|\|full | Low-quality |
| S14_vs_k141_73466\|\|full | Not-determined |
| S14_vs_k141_486\|\|full | Low-quality |
| S14_vs_k141_88263\|\|full | Low-quality |
| S14_vs_k141_59029\|\|full | Not-determined |
| S14_vs_k141_102899\|\|full | Not-determined |
| S14_vs_k141_15019\|\|full | Low-quality |
| S14_vs_k141_491\|\|full | Not-determined |
| S14_vs_k141_80937\|\|full | Low-quality |
| S14_vs_k141_59041\|\|full | Not-determined |
| S14_vs_k141_102909\|\|full | Low-quality |
| S14_vs_k141_95648\|\|full | Low-quality |
| S14_vs_k141_44218\|\|full | Not-determined |
| S14_vs_k141_51718\|\|full | Not-determined |
| S14_vs_k141_102917\|\|full | Not-determined |
| S14_vs_k141_110231\|\|full | Not-determined |
| S14_vs_k141_80963\|\|full | Low-quality |
| S14_vs_k141_520\|\|full | Low-quality |
| S14_vs_k141_95659\|\|full | Low-quality |
| S14_vs_k141_51729\|\|full | Low-quality |
| S14_vs_k141_88282\|\|full | Not-determined |
| S14_vs_k141_525\|\|full | Low-quality |
| S14_vs_k141_51741\|\|full | Not-determined |
| S14_vs_k141_102932\|\|full | Low-quality |
| S14_vs_k141_22313\|\|full | Low-quality |
| S14_vs_k141_545\|\|full | Not-determined |
| S14_vs_k141_95694\|\|full | Not-determined |
| S14_vs_k141_547\|\|full | Not-determined |
| S15_extracted_2513 | High-quality |
| S14_vs_k141_44282\|\|full | Low-quality |
| S14_vs_k141_102943\|\|full | Low-quality |
| S14_vs_k141_51749\|\|full | Low-quality |
| S14_vs_k141_7724\|\|full | Low-quality |
| S14_vs_k141_80999\|\|full | Low-quality |
| S14_vs_k141_561\|\|full | Low-quality |
| S14_vs_k141_102954\|\|full | Not-determined |
| S14_vs_k141_29751\|\|full | Not-determined |
| S14_vs_k141_565\|\|full | Not-determined |
| S14_vs_k141_66216\|\|full | Low-quality |
| S14_vs_k141_44305\|\|full | Low-quality |
| S14_vs_k141_66217\|\|full | Low-quality |
| S14_vs_k141_22336\|\|full | Low-quality |
| S14_vs_k141_577\|\|full | Low-quality |
| S14_vs_k141_7734\|\|full | Low-quality |
| S14_vs_k141_95730\|\|full | Low-quality |
| S14_vs_k141_583\|\|full | Low-quality |
| S14_vs_k141_102981\|\|full | Low-quality |
| S14_vs_k141_110263\|\|full | Low-quality |
| S14_vs_k141_44321\|\|full | Not-determined |
| S14_vs_k141_44322\|\|full | Low-quality |
| S14_vs_k141_88321\|\|full | Low-quality |
| S14_vs_k141_44323\|\|full | Not-determined |
| S14_vs_k141_110264\|\|full | Low-quality |
| S14_vs_k141_95751\|\|full | Low-quality |
| S14_vs_k141_29772\|\|full | Not-determined |
| S14_vs_k141_73534\|\|full | Low-quality |
| S14_vs_k141_81016\|\|full | Low-quality |
| S14_vs_k141_88333\|\|full | Low-quality |
| S14_vs_k141_103003\|\|full | Not-determined |
| S14_vs_k141_37145\|\|full | Low-quality |
| S14_vs_k141_110277\|\|full | Low-quality |
| S14_vs_k141_44336\|\|full | Low-quality |
| S14_vs_k141_88351\|\|full | Low-quality |
| S14_vs_k141_95782\|\|full | Low-quality |
| S14_vs_k141_73543\|\|full | Low-quality |
| S14_vs_k141_73544\|\|full | Low-quality |
| S14_vs_k141_626\|\|full | Low-quality |
| S14_vs_k141_103026\|\|full | Low-quality |

S14_vs_k141_29805||full Not-determined
S14_vs_k141_15067||full Not-determined
S14_vs_k141_15071||full Low-quality
S14_vs_k141_15072||full Low-quality
S14_vs_k141_29817||full Low-quality
S14_vs_k141_15080||full Low-quality
S14_vs_k141_81047||full Not-determined
S14_vs_k141_22370||full Low-quality
S14_vs_k141_37191||full Low-quality
S14_vs_k141_29833||full Not-determined
S14_vs_k141_7754||full Low-quality
S14_vs_k141_88387||full Not-determined
S14_vs_k141_37199||full Not-determined
S14_vs_k141_103102||full Low-quality
S14_vs_k141_29841||full Low-quality
S14_vs_k141_88393||full Low-quality
S14_vs_k141_88394||full Low-quality
S14_vs_k141_95827||full Low-quality
S14_vs_k141_37207||full Low-quality
S4_extracted_9444 Complete
S15_vs_2513||full High-quality
S14_vs_k141_7771||full Not-determined
S14_vs_k141_37214||full Not-determined
S14_vs_k141_103105||full Low-quality
S14_vs_k141_88403||full Low-quality
S14_vs_k141_37217||full Low-quality
S14_vs_k141_95835||full Not-determined
S14_vs_k141_81076||full Low-quality
S14_vs_k141_29854||full Low-quality
S14_vs_k141_95839||full Low-quality
S14_vs_k141_59107||full Low-quality
S14_vs_k141_88409||full Not-determined
S14_vs_k141_7796||full Low-quality
S14_vs_k141_59115||full Low-quality
S14_vs_k141_22406||full Low-quality
S14_vs_k141_51782||full Low-quality
S14_vs_k141_7808||full Not-determined
S14_vs_k141_22408||full Low-quality
S14_vs_k141_110331||full Low-quality
S14_vs_k141_110338||full Low-quality
S14_vs_k141_73593||full Low-quality
S14_vs_k141_29861||full Low-quality
S14_vs_k141_44383||full Not-determined
S14_vs_k141_88437||full Low-quality
S14_vs_k141_66286||full Low-quality
S14_vs_k141_110346||full Low-quality
S14_vs_k141_88444||full Low-quality
S14_vs_k141_81118||full Not-determined
S14_vs_k141_81119||full Low-quality
S14_vs_k141_81122||full Not-determined
S14_vs_k141_95859||full Low-quality
S14_vs_k141_7830||full Low-quality
S14_vs_k141_88452||full Low-quality
S14_vs_k141_7832||full Low-quality
S14_vs_k141_88454||full Low-quality
S14_vs_k141_7836||full Low-quality
S14_vs_k141_103121||full Low-quality
S14_vs_k141_37283||full Low-quality
S14_vs_k141_103131||full Low-quality
S14_vs_k141_15168||full Low-quality
S14_vs_k141_73628||full Low-quality
S14_vs_k141_687||full Low-quality
S14_vs_k141_110354||full Low-quality
S14_vs_k141_44424||full Not-determined
S14_vs_k141_73636||full Low-quality
S14_vs_k141_692||full Low-quality
S14_vs_k141_51823||full Low-quality
S14_vs_k141_88471||full Low-quality
S14_vs_k141_51827||full Not-determined
S14_vs_k141_51830||full Low-quality
S14_vs_k141_88484||full Not-determined
S14_vs_k141_51836||full Not-determined
S14_vs_k141_51852||full Low-quality
S14_vs_k141_73669||full Low-quality
S14_vs_k141_103154||full Not-determined
S14_vs_k141_73673||full Low-quality
S14_vs_k141_103162||full Low-quality
S14_vs_k141_59178||full Not-determined
S14_vs_k141_22460||full Low-quality
S14_vs_k141_59185||full Not-determined
S14_vs_k141_81188||full Not-determined
S14_vs_k141_59196||full Low-quality
S14_vs_k141_110390||full Low-quality
S14_vs_k141_88513||full Low-quality
S14_vs_k141_15215||full Low-quality
S14_vs_k141_29918||full Low-quality
S14_vs_k141_110394||full Low-quality
S14_vs_k141_37290||full Low-quality
S14_vs_k141_51883||full Not-determined
S14_vs_k141_88538||full Low-quality
S14_vs_k141_37302||full Low-quality
S14_vs_k141_88549||full Low-quality
S14_vs_k141_29937||full Not-determined
S14_vs_k141_29959||full Low-quality
S14_vs_k141_66365||full Low-quality
S14_vs_k141_29966||full Low-quality
S8_vs_k141_131009||full High-quality
S14_vs_k141_37317||full Not-determined
S14_vs_k141_743||full Low-quality
S14_vs_k141_744||full Low-quality
S14_vs_k141_22491||full Low-quality
S14_vs_k141_73711||full Low-quality
S14_vs_k141_7910||full Low-quality
S14_vs_k141_752||full Low-quality
S14_vs_k141_110410||full Not-determined
S14_vs_k141_759||full Low-quality
S14_vs_k141_44472||full Low-quality
S14_vs_k141_22511||full Low-quality
S14_vs_k141_59243||full Low-quality
S14_vs_k141_66379||full Not-determined
S14_vs_k141_51926||full Low-quality
S14_vs_k141_110422||full Low-quality
S14_vs_k141_88586||full Low-quality
S14_vs_k141_51939||full Low-quality
S14_vs_k141_782||full Low-quality
S14_vs_k141_110424||full Not-determined
S14_vs_k141_795||full Low-quality
S14_vs_k141_51952||full Not-determined
S14_vs_k141_59267||full Low-quality
S14_vs_k141_51964||full Low-quality
S14_vs_k141_73731||full Low-quality
S14_vs_k141_66400||full Low-quality
S14_vs_k141_7943||full Low-quality
S14_vs_k141_66403||full Low-quality
S14_vs_k141_59292||full Not-determined
S14_vs_k141_66405||full Low-quality
S14_vs_k141_37334||full Not-determined
S14_vs_k141_103199||full Not-determined
S14_vs_k141_73757||full Low-quality
S14_vs_k141_66412||full Not-determined
S14_vs_k141_7962||full Low-quality
S14_vs_k141_88604||full Low-quality
S14_vs_k141_66419||full Low-quality
S14_vs_k141_88605||full Low-quality
S14_vs_k141_7966||full Low-quality
S14_vs_k141_37365||full Low-quality
S14_vs_k141_59300||full Not-determined
S14_vs_k141_81224||full Low-quality
S14_vs_k141_110487||full Low-quality
S14_vs_k141_816||full Low-quality
S14_vs_k141_818||full Not-determined
S14_vs_k141_110493||full Not-determined
S14_vs_k141_819||full Low-quality
S14_vs_k141_37385||full Not-determined
S14_vs_k141_7991||full Low-quality
S14_vs_k141_88631||full Not-determined
S14_vs_k141_44500||full Low-quality
S14_vs_k141_22525||full Low-quality
S14_vs_k141_110509||full Low-quality
S14_vs_k141_88639||full Not-determined
S14_vs_k141_52026||full Low-quality
S14_vs_k141_110510||full Low-quality
S14_vs_k141_81252||full Low-quality
S14_vs_k141_88655||full Low-quality
S14_vs_k141_103213||full Low-quality
S14_vs_k141_44526||full Low-quality
S14_vs_k141_110514||full Not-determined
S14_vs_k141_110520||full Low-quality
S14_vs_k141_73784||full Not-determined
S14_vs_k141_59318||full Low-quality
S14_vs_k141_110522||full Not-determined
S14_vs_k141_110525||full Low-quality
S14_vs_k141_30032||full Not-determined
S14_vs_k141_52058||full Not-determined
S14_vs_k141_59324||full Low-quality
S14_vs_k141_15237||full Not-determined
S14_vs_k141_81299||full Low-quality
S14_vs_k141_44531||full Not-determined
S14_vs_k141_59327||full Not-determined
S14_vs_k141_37442||full Low-quality
S14_vs_k141_37445||full Low-quality
S14_vs_k141_30041||full Low-quality
S14_vs_k141_30044||full Not-determined
S14_vs_k141_8018||full Not-determined
S14_vs_k141_30049||full Not-determined
S14_vs_k141_30059||full Low-quality
S14_vs_k141_73802||full Low-quality
S14_vs_k141_52114||full Not-determined
S14_vs_k141_852||full Low-quality
S14_vs_k141_30074||full Not-determined
S14_vs_k141_855||full Low-quality
S14_vs_k141_52127||full Low-quality
S14_vs_k141_8024||full Not-determined
S14_vs_k141_52131||full Low-quality
S14_vs_k141_73821||full Not-determined
S14_vs_k141_52133||full Not-determined
S14_vs_k141_52136||full Low-quality
S14_vs_k141_37495||full Low-quality
S14_vs_k141_88695||full Not-determined
S14_vs_k141_8044||full Low-quality
S13_vs_35816||full Complete
S14_vs_k141_52155||full Low-quality
S14_vs_k141_88697||full Low-quality
S3_vs_k141_70508||full High-quality
S14_vs_k141_15307||full Low-quality
S14_vs_k141_88706||full Low-quality
S14_vs_k141_8055||full Low-quality
S14_vs_k141_59358||full Low-quality
S14_vs_k141_73857||full Not-determined
S14_vs_k141_883||full Not-determined
S14_vs_k141_103234||full Low-quality
S14_vs_k141_73862||full Not-determined
S14_vs_k141_15326||full Low-quality
S14_vs_k141_37522||full Not-determined
S14_vs_k141_893||full Low-quality
S4_vs_VIRSorter_9444-circular-cat_ High-quality
S14_vs_k141_88714||full Low-quality
S14_vs_k141_66453||full Low-quality
S14_vs_k141_66454||full Low-quality
S14_vs_k141_44564||full Low-quality
S14_vs_k141_15354||full Not-determined
S14_vs_k141_52160||full Not-determined
S14_vs_k141_30114||full Low-quality
S14_vs_k141_103264||full Low-quality
S14_vs_k141_110584||full Low-quality
S14_vs_k141_103280||full Low-quality
S14_vs_k141_15370||full Low-quality
S14_vs_k141_52177||full Not-determined
S14_vs_k141_73888||full Low-quality
S14_vs_k141_73896||full Low-quality
S14_vs_k141_73903||full Low-quality
S14_vs_k141_59379||full Low-quality
S14_vs_k141_59380||full Low-quality
S14_vs_k141_95916||full Low-quality
S14_vs_k141_901||full Low-quality
S14_vs_k141_95921||full Low-quality
S14_vs_k141_22550||full Low-quality
S14_vs_k141_909||full Low-quality
S14_vs_k141_59383||full Low-quality
S14_vs_k141_88753||full Low-quality
S14_vs_k141_44606||full Low-quality
S14_vs_k141_95932||full Not-determined
S14_vs_k141_59387||full Low-quality
S14_vs_k141_88756||full Low-quality
S14_vs_k141_8104||full Not-determined
S14_vs_k141_59396||full Low-quality
S14_vs_k141_30146||full Not-determined
S14_vs_k141_110634||full Not-determined
S14_vs_k141_73958||full Low-quality
S14_vs_k141_88767||full Not-determined
S14_vs_k141_8111||full Not-determined
S14_vs_k141_934||full Low-quality
S14_vs_k141_59414||full Low-quality
S14_vs_k141_30152||full Not-determined
S14_vs_k141_15408||full Low-quality
S14_vs_k141_73973||full Low-quality
S14_vs_k141_73974||full Low-quality
S14_vs_k141_22587||full Low-quality
S14_vs_k141_30159||full Low-quality
S14_vs_k141_15417||full Low-quality
S14_vs_k141_44640||full Not-determined
S14_vs_k141_59445||full Low-quality
S14_vs_k141_59448||full Not-determined
S14_vs_k141_103381||full Not-determined
S14_vs_k141_95950||full Low-quality
S14_vs_k141_44655||full Low-quality
S14_vs_k141_73984||full Low-quality
S14_vs_k141_52280||full Low-quality
S14_vs_k141_961||full Low-quality
S14_vs_k141_37607||full Not-determined
S14_vs_k141_52288||full Low-quality
S14_vs_k141_52289||full Low-quality
S14_vs_k141_15427||full Not-determined
S14_vs_k141_66485||full Low-quality
S14_vs_k141_22628||full Low-quality
S14_vs_k141_37619||full Low-quality
S14_vs_k141_44661||full Not-determined
S14_vs_k141_37627||full Not-determined
S14_vs_k141_74009||full Low-quality
S14_vs_k141_44677||full Low-quality
S14_vs_k141_81387||full Low-quality
S14_vs_k141_22667||full Low-quality
S14_vs_k141_59486||full Low-quality
S14_vs_k141_88819||full Low-quality
S14_vs_k141_88830||full Low-quality
S14_vs_k141_74041||full Low-quality
S14_vs_k141_74042||full Low-quality
S14_vs_k141_44718||full Not-determined
S14_vs_k141_997||full Low-quality
S14_vs_k141_15457||full Not-determined
S14_vs_k141_998||full Low-quality
S14_vs_k141_59503||full Not-determined
S14_vs_k141_44722||full Not-determined
S14_vs_k141_1001||full Not-determined
S14_vs_k141_44724||full Not-determined
S14_vs_k141_1002||full Low-quality
S14_vs_k141_81401||full Low-quality
S14_vs_k141_44726||full Not-determined
S14_vs_k141_88848||full Low-quality
S14_vs_k141_110714||full Not-determined
S14_vs_k141_74053||full Low-quality
S14_vs_k141_30230||full Low-quality
S14_vs_k141_103439||full Not-determined
S14_vs_k141_88862||full Low-quality
S14_vs_k141_74061||full Low-quality
S14_vs_k141_88870||full Not-determined
S14_vs_k141_103450||full Not-determined
S14_vs_k141_15486||full Low-quality
S14_vs_k141_66510||full Not-determined
S14_vs_k141_103462||full Low-quality
S14_vs_k141_66513||full Low-quality
S14_vs_k141_44750||full Low-quality
S14_vs_k141_52308||full Low-quality
S14_vs_k141_15501||full Low-quality
S14_vs_k141_30245||full Not-determined
S14_vs_k141_15503||full Low-quality
S14_vs_k141_103474||full Low-quality
S14_vs_k141_52312||full Low-quality
S14_vs_k141_15505||full Low-quality
S14_vs_k141_59531||full Low-quality
S14_vs_k141_1031||full Not-determined
S14_vs_k141_59532||full Low-quality
S14_vs_k141_44755||full Low-quality
S14_vs_k141_52314||full Not-determined
S14_vs_k141_44760||full Not-determined
S14_vs_k141_1037||full Not-determined
S14_vs_k141_95998||full Not-determined
S14_vs_k141_52317||full Not-determined
S14_vs_k141_44767||full Low-quality
S14_vs_k141_110744||full Low-quality
S14_vs_k141_1043||full Low-quality
S14_vs_k141_44771||full Not-determined
S14_vs_k141_1046||full Low-quality
S14_vs_k141_52329||full Low-quality
S14_vs_k141_8155||full Low-quality
S14_vs_k141_59549||full Not-determined
S14_vs_k141_37703||full Low-quality
S14_vs_k141_59556||full Not-determined
S14_vs_k141_1060||full Low-quality
S14_vs_k141_59557||full Not-determined
S14_vs_k141_30297||full Low-quality
S14_vs_k141_110781||full Low-quality
S14_vs_k141_59561||full Not-determined
S14_vs_k141_30300||full Low-quality
S14_vs_k141_30302||full Low-quality
S14_vs_k141_88892||full Low-quality
S14_vs_k141_15536||full Low-quality
S16_extracted_108554 High-quality
S14_vs_k141_22755||full Low-quality
S14_vs_k141_52352||full Not-determined
S14_vs_k141_15540||full Low-quality
S14_vs_k141_66546||full Low-quality
S14_vs_k141_22761||full Low-quality
S14_vs_k141_15541||full Not-determined
S14_vs_k141_30313||full Low-quality
S14_vs_k141_52357||full Low-quality
S14_vs_k141_52360||full Not-determined
S14_vs_k141_15548||full Low-quality
S14_vs_k141_30332||full Low-quality
S14_vs_k141_44798||full Not-determined
S14_vs_k141_66572||full Low-quality
S14_vs_k141_88918||full Low-quality
S14_vs_k141_110819||full Low-quality
S14_vs_k141_1071||full Not-determined
S14_vs_k141_110825||full Not-determined
S14_vs_k141_37733||full Not-determined
S14_vs_k141_81430||full Not-determined
S14_vs_k141_103549||full Not-determined
S14_vs_k141_1083||full Not-determined
S14_vs_k141_66590||full Not-determined
S14_vs_k141_30336||full Not-determined
S14_vs_k141_1089||full Low-quality
S14_vs_k141_1091||full Low-quality
S14_vs_k141_103571||full Low-quality
S14_vs_k141_44819||full Low-quality
S14_vs_k141_81436||full Low-quality
S14_vs_k141_44823||full Low-quality
S14_vs_k141_22776||full Not-determined
S14_vs_k141_81441||full Low-quality
S14_vs_k141_1122||full Low-quality
S14_vs_k141_59605||full Not-determined
S14_vs_k141_110845||full Low-quality
S14_vs_k141_1133||full Low-quality
S14_vs_k141_15590||full Low-quality
S14_vs_k141_37764||full Not-determined
S14_vs_k141_103625||full Not-determined
S14_vs_k141_8169||full Low-quality
S14_vs_k141_59619||full Low-quality
S14_vs_k141_15599||full Not-determined
S14_vs_k141_30360||full Low-quality
S14_vs_k141_59630||full Low-quality
S14_vs_k141_81461||full Low-quality
S14_vs_k141_22800||full Low-quality
S14_vs_k141_44863||full Not-determined
S14_vs_k141_8197||full Not-determined
S14_vs_k141_37778||full Low-quality
S14_vs_k141_1145||full Low-quality
S14_vs_k141_44871||full Low-quality
S14_vs_k141_44872||full Low-quality
S14_vs_k141_30364||full Low-quality
S14_vs_k141_103660||full Low-quality
S14_vs_k141_96095||full Low-quality
S14_vs_k141_1158||full Low-quality
S14_vs_k141_103672||full Not-determined
S14_vs_k141_15620||full Not-determined
S14_vs_k141_15631||full Low-quality
S14_vs_k141_52421||full Low-quality
S14_vs_k141_8212||full Not-determined
S14_vs_k141_1183||full Not-determined
S14_vs_k141_52429||full Not-determined
S14_vs_k141_88996||full Low-quality
S14_vs_k141_52435||full Low-quality
S14_vs_k141_81481||full Low-quality
S14_vs_k141_89000||full Low-quality
S14_vs_k141_110917||full Low-quality
S14_vs_k141_66618||full Low-quality
S14_vs_k141_52451||full Not-determined
S14_vs_k141_103700||full Not-determined
S14_vs_k141_8228||full Low-quality
S14_vs_k141_1196||full Low-quality
S16_vs_108554||full High-quality
S14_vs_k141_22824||full Low-quality
S14_vs_k141_103718||full Low-quality
S14_vs_k141_30431||full Low-quality
S14_vs_k141_30433||full Low-quality
S14_vs_k141_59667||full Not-determined
S14_vs_k141_30438||full Low-quality
S14_vs_k141_15657||full Low-quality
S14_vs_k141_103750||full Not-determined
S14_vs_k141_8250||full Low-quality
S14_vs_k141_89014||full Low-quality
S14_vs_k141_96127||full Low-quality
S14_vs_k141_89015||full Low-quality
S14_vs_k141_89016||full Low-quality
S14_vs_k141_59695||full Not-determined
S14_vs_k141_89021||full Low-quality
S14_vs_k141_89023||full Not-determined
S14_vs_k141_44907||full Not-determined
S14_vs_k141_59707||full Low-quality
S14_vs_k141_103781||full Low-quality
S14_vs_k141_8270||full Not-determined
S14_vs_k141_111011||full Low-quality
S14_vs_k141_1212||full Low-quality
S3_extracted_k141_73597 Medium-quality
S14_vs_k141_111017||full Not-determined
S14_vs_k141_74069||full Not-determined
S14_vs_k141_22878||full Low-quality
S14_vs_k141_59730||full Low-quality
S14_vs_k141_1221||full Not-determined
S14_vs_k141_89055||full Low-quality
S14_vs_k141_74092||full Low-quality
S14_vs_k141_103786||full Low-quality
S14_vs_k141_1231||full Not-determined
S14_vs_k141_22893||full Low-quality
S14_vs_k141_37862||full Low-quality
S14_vs_k141_103792||full Low-quality
S14_vs_k141_74100||full Low-quality
S14_vs_k141_59747||full Low-quality
S14_vs_k141_37881||full Low-quality
S14_vs_k141_111020||full Low-quality
S14_vs_k141_111025||full Low-quality
S14_vs_k141_66662||full Not-determined
S14_vs_k141_111029||full Low-quality
S14_vs_k141_15745||full Not-determined
S14_vs_k141_37896||full Not-determined
S14_vs_k141_59756||full Not-determined
S14_vs_k141_1261||full Not-determined
S14_vs_k141_111033||full Low-quality
S14_vs_k141_103805||full Low-quality
S14_vs_k141_15757||full Not-determined
S14_vs_k141_103822||full Low-quality
S14_vs_k141_30508||full Low-quality
S14_vs_k141_89093||full Low-quality
S14_vs_k141_111042||full Low-quality
S14_vs_k141_66679||full Low-quality
S14_vs_k141_111050||full Not-determined
S14_vs_k141_30519||full Low-quality
S14_vs_k141_37935||full Not-determined
S14_vs_k141_74168||full Low-quality
S14_vs_k141_30523||full Not-determined
S14_vs_k141_8342||full Low-quality
S14_vs_k141_8344||full Low-quality
S14_vs_k141_111060||full Low-quality
S14_vs_k141_8348||full Low-quality
S14_vs_k141_45000||full Low-quality
S14_vs_k141_45002||full Low-quality
S14_vs_k141_89123||full Low-quality
S14_vs_k141_74195||full Low-quality
S14_vs_k141_111074||full Low-quality
S14_vs_k141_111075||full Not-determined
S14_vs_k141_52461||full Not-determined
S14_vs_k141_30546||full Low-quality
S14_vs_k141_8365||full Low-quality
S14_vs_k141_30559||full Low-quality
S14_vs_k141_81601||full Low-quality
S14_vs_k141_81605||full Low-quality
S14_vs_k141_111095||full Low-quality
S14_vs_k141_103848||full Low-quality
S14_vs_k141_81610||full Low-quality
S14_vs_k141_45012||full Low-quality
S14_vs_k141_59793||full Low-quality
S14_vs_k141_37981||full Low-quality
S14_vs_k141_15875||full Low-quality
S14_vs_k141_66702||full Not-determined
S14_vs_k141_66704||full Low-quality
S14_vs_k141_22950||full Low-quality
S14_vs_k141_66705||full Low-quality
S14_vs_k141_37993||full Not-determined
S14_vs_k141_15882||full Not-determined
S11_extracted_k141_66160 High-quality
S14_vs_k141_59799||full Low-quality
S14_vs_k141_8436||full Not-determined
S14_vs_k141_59802||full Not-determined
S14_vs_k141_22963||full Not-determined
S14_vs_k141_30579||full Not-determined
S14_vs_k141_103877||full Low-quality
S14_vs_k141_22974||full Low-quality
S14_vs_k141_103883||full Low-quality
S14_vs_k141_38002||full Low-quality
S14_vs_k141_81689||full Low-quality
S14_vs_k141_15896||full Not-determined
S14_vs_k141_111171||full Low-quality
S14_vs_k141_59822||full Low-quality
S14_vs_k141_103904||full Low-quality
S14_vs_k141_15907||full Low-quality
S14_vs_k141_30598||full Not-determined
S14_vs_k141_81700||full Low-quality
S14_vs_k141_30601||full Not-determined
S14_vs_k141_22988||full Not-determined
S14_vs_k141_74269||full Low-quality
S14_vs_k141_45045||full Not-determined
S14_vs_k141_111189||full Low-quality
S14_vs_k141_111196||full Low-quality
S14_vs_k141_81721||full Low-quality
S14_vs_k141_81725||full Low-quality
S14_vs_k141_103922||full Not-determined
S14_vs_k141_45054||full Not-determined
S14_vs_k141_15945||full Low-quality
S14_vs_k141_30632||full Low-quality
S14_vs_k141_30633||full Low-quality
S14_vs_k141_30634||full Not-determined
S14_vs_k141_74284||full Low-quality
S14_vs_k141_8459||full Not-determined
S14_vs_k141_30639||full Low-quality
S14_vs_k141_89202||full Low-quality
S14_vs_k141_81747||full Low-quality
S14_vs_k141_45073||full Not-determined
S14_vs_k141_8466||full Low-quality
S14_vs_k141_74294||full Low-quality
S14_vs_k141_89208||full Not-determined
S14_vs_k141_74308||full Low-quality
S14_vs_k141_52524||full Low-quality
S14_vs_k141_15965||full Not-determined
S14_vs_k141_103976||full Low-quality
S14_vs_k141_66757||full Not-determined
S14_vs_k141_89235||full Low-quality
S14_vs_k141_74328||full Not-determined
S14_vs_k141_66768||full Not-determined
S14_vs_k141_59883||full Low-quality
S14_vs_k141_89250||full Low-quality
S14_vs_k141_89252||full Low-quality
S14_vs_k141_52540||full Low-quality
S14_vs_k141_59896||full Low-quality
S14_vs_k141_96192||full Low-quality
S14_vs_k141_74347||full Not-determined
S14_vs_k141_96194||full Not-determined
S14_vs_k141_89270||full Low-quality
S14_vs_k141_89271||full Not-determined
S14_vs_k141_96198||full Low-quality
S14_vs_k141_89278||full Low-quality
S14_vs_k141_52550||full Low-quality
S14_vs_k141_66794||full Low-quality
S14_vs_k141_23105||full Not-determined
S14_vs_k141_38072||full Not-determined
S14_vs_k141_23110||full Low-quality
S14_vs_k141_111271||full Not-determined
S14_vs_k141_38076||full Low-quality
S14_vs_k141_111272||full Low-quality
S14_vs_k141_38083||full Not-determined
S14_vs_k141_111276||full Low-quality
S14_vs_k141_52573||full Low-quality
S14_vs_k141_23115||full Not-determined
S14_vs_k141_8498||full Low-quality
S14_vs_k141_52574||full Low-quality
S14_vs_k141_81857||full Low-quality
S14_vs_k141_81858||full Low-quality
S14_vs_k141_96234||full Low-quality
S14_vs_k141_52577||full Low-quality
S14_vs_k141_8502||full Low-quality
S14_vs_k141_23128||full Low-quality
S14_vs_k141_23133||full Low-quality
S14_vs_k141_38094||full Low-quality
S14_vs_k141_104005||full Low-quality
S14_vs_k141_59977||full Not-determined
S14_vs_k141_8513||full Low-quality
S14_vs_k141_96238||full Low-quality
S14_vs_k141_23148||full Low-quality
S14_vs_k141_59982||full Low-quality
S14_vs_k141_8525||full Low-quality

S14_vs_k141_89302||full Not-determined
S14_vs_k141_74444||full Low-quality
S14_vs_k141_66839||full Low-quality
S14_vs_k141_104029||full Low-quality
S14_vs_k141_8531||full Low-quality
S14_vs_k141_74447||full Low-quality
S14_vs_k141_23166||full Low-quality
S14_vs_k141_96265||full Not-determined
S14_vs_k141_16059||full Low-quality
S14_vs_k141_104053||full Not-determined
S11_vs_k141_66160||full High-quality
S14_vs_k141_104059||full Not-determined
S14_vs_k141_30708||full Low-quality
S14_vs_k141_81907||full Low-quality
S14_vs_k141_38132||full Low-quality
S14_vs_k141_38133||full Low-quality
S14_vs_k141_16085||full Low-quality
S14_vs_k141_8572||full Low-quality
S14_vs_k141_81919||full Not-determined
S14_vs_k141_104078||full Low-quality
S14_vs_k141_52610||full Low-quality
S14_vs_k141_23188||full Not-determined
S14_vs_k141_66880||full Low-quality
S14_vs_k141_74498||full Low-quality
S14_vs_k141_104092||full Not-determined
S14_vs_k141_81942||full Low-quality
S14_vs_k141_23195||full Not-determined
S14_vs_k141_8616||full Low-quality
S14_vs_k141_38162||full Not-determined
S14_vs_k141_23208||full Not-determined
S14_vs_k141_23213||full Low-quality
S14_vs_k141_96317||full Not-determined
S14_vs_k141_16122||full Not-determined
S14_vs_k141_104120||full Low-quality
S14_vs_k141_16127||full Not-determined
S14_vs_k141_38180||full Low-quality
S14_vs_k141_96330||full Low-quality
S14_vs_k141_38182||full Not-determined
S14_vs_k141_111399||full Not-determined
S14_vs_k141_8657||full Low-quality
S14_vs_k141_23222||full Not-determined
S14_vs_k141_104144||full Low-quality
S14_vs_k141_23227||full Not-determined
S14_vs_k141_81967||full Low-quality
S14_vs_k141_30764||full Low-quality
S14_vs_k141_60036||full Not-determined
S14_vs_k141_38206||full Low-quality
S14_vs_k141_38207||full Low-quality
S14_vs_k141_74550||full Low-quality
S14_vs_k141_23240||full Low-quality
S14_vs_k141_1313||full Low-quality
S14_vs_k141_74556||full Low-quality
S14_vs_k141_38216||full Low-quality
S14_vs_k141_66950||full Low-quality
S14_vs_k141_23252||full Low-quality
S14_vs_k141_66951||full Not-determined
S14_vs_k141_104179||full Low-quality
S14_vs_k141_104180||full Low-quality
S14_vs_k141_111448||full Not-determined
S14_vs_k141_52676||full Low-quality
S14_vs_k141_52679||full Not-determined
S14_vs_k141_104192||full Low-quality
S14_vs_k141_74563||full Low-quality
S14_vs_k141_60066||full Not-determined
S14_vs_k141_60068||full Not-determined
S14_vs_k141_60069||full Low-quality
S14_vs_k141_82007||full Low-quality
S14_vs_k141_96358||full Not-determined
S14_vs_k141_8705||full Low-quality
S14_vs_k141_96364||full Not-determined
S14_vs_k141_104222||full Low-quality
S14_vs_k141_8711||full Low-quality
S14_vs_k141_104226||full Low-quality
S14_vs_k141_111483||full Low-quality
S14_vs_k141_66964||full Not-determined
S14_vs_k141_16193||full Low-quality
S14_vs_k141_66968||full Low-quality
S14_vs_k141_82021||full Low-quality
S14_vs_k141_23295||full Low-quality
S14_vs_k141_111497||full Not-determined
S14_vs_k141_111498||full Low-quality
S14_vs_k141_60115||full Low-quality
S14_vs_k141_52739||full Low-quality
S14_vs_k141_8752||full Low-quality
S14_vs_k141_16219||full Low-quality
S14_vs_k141_23314||full Low-quality
S14_vs_k141_96407||full Low-quality
S14_vs_k141_111506||full Low-quality
S14_vs_k141_111509||full Low-quality
S14_vs_k141_8767||full Low-quality
S14_vs_k141_8777||full Low-quality
S14_vs_k141_8782||full Low-quality
S14_vs_k141_8783||full Not-determined
S14_vs_k141_111525||full Low-quality
S14_vs_k141_60136||full Low-quality
S14_vs_k141_60146||full Low-quality
S14_vs_k141_60148||full Low-quality
S14_vs_k141_52800||full Not-determined
S14_vs_k141_82046||full Low-quality
S14_vs_k141_23330||full Low-quality
S14_vs_k141_23333||full Low-quality
S14_vs_k141_111545||full Low-quality
S14_vs_k141_16241||full Not-determined
S14_vs_k141_104246||full Low-quality
S14_vs_k141_16244||full Low-quality
S14_vs_k141_60170||full Low-quality
S14_vs_k141_104258||full Not-determined
S14_vs_k141_82071||full Not-determined
S14_vs_k141_38235||full Low-quality
S14_vs_k141_52814||full Not-determined
S14_vs_k141_111565||full Low-quality
S14_vs_k141_104264||full Not-determined
S14_vs_k141_60174||full Low-quality
S14_vs_k141_89399||full Low-quality
S14_vs_k141_30852||full Not-determined
S14_vs_k141_8785||full Low-quality
S14_vs_k141_67031||full Low-quality
S14_vs_k141_89408||full Low-quality
S14_vs_k141_60185||full Low-quality
S14_vs_k141_16256||full Low-quality
S14_vs_k141_89414||full Low-quality
S14_vs_k141_52829||full Low-quality
S14_vs_k141_89421||full Low-quality
S14_vs_k141_89424||full Low-quality
S14_vs_k141_16267||full Low-quality
S14_vs_k141_111589||full Low-quality
S14_vs_k141_111590||full Low-quality
S14_vs_k141_8818||full Not-determined
S14_vs_k141_30891||full Low-quality
S14_vs_k141_89450||full Low-quality
S14_vs_k141_8821||full Low-quality
S14_vs_k141_30896||full Low-quality
S14_vs_k141_82079||full Low-quality
S14_vs_k141_60220||full Low-quality
S14_vs_k141_74599||full Not-determined
S14_vs_k141_1419||full Not-determined
S14_vs_k141_82087||full Low-quality
S14_vs_k141_30904||full Not-determined
S14_vs_k141_8844||full Low-quality
S14_vs_k141_23411||full Low-quality
S14_vs_k141_74610||full Low-quality
S14_vs_k141_111617||full Not-determined
S14_vs_k141_104343||full Not-determined
S14_vs_k141_96481||full Not-determined
S14_vs_k141_1429||full Not-determined
S14_vs_k141_104348||full Low-quality
S14_vs_k141_111621||full Not-determined
S14_vs_k141_82100||full Low-quality
S14_vs_k141_23436||full Low-quality
S14_vs_k141_111631||full Low-quality
S14_vs_k141_60257||full Low-quality
S14_vs_k141_60258||full Low-quality
S14_vs_k141_96492||full Not-determined
S14_vs_k141_30933||full Not-determined
S14_vs_k141_38328||full Not-determined
S14_vs_k141_82107||full Low-quality
S14_vs_k141_74615||full Low-quality
S14_vs_k141_111651||full Low-quality
S14_vs_k141_1441||full Not-determined
S14_vs_k141_111657||full Low-quality
S14_vs_k141_52902||full Low-quality
S14_vs_k141_96509||full Low-quality
S14_vs_k141_82117||full Low-quality
S14_vs_k141_82119||full Low-quality
S14_vs_k141_23457||full Not-determined
S14_vs_k141_8877||full Low-quality
S14_vs_k141_82124||full Low-quality
S14_vs_k141_60279||full Low-quality
S14_vs_k141_38352||full Low-quality
S14_vs_k141_104402||full Not-determined
S14_vs_k141_67100||full Not-determined
S14_vs_k141_82142||full Not-determined
S14_vs_k141_96545||full Low-quality
S14_vs_k141_89493||full Low-quality
S14_vs_k141_60301||full Not-determined
S14_vs_k141_45125||full Low-quality
S14_vs_k141_45126||full Low-quality
S14_vs_k141_111704||full Low-quality
S14_vs_k141_96557||full Not-determined
S14_vs_k141_52947||full Not-determined
S14_vs_k141_52948||full Low-quality
S14_vs_k141_16335||full Low-quality
S14_vs_k141_30990||full Low-quality
S14_vs_k141_45144||full Low-quality
S14_vs_k141_74643||full Not-determined
S14_vs_k141_104419||full Low-quality
S14_vs_k141_96565||full Low-quality
S14_vs_k141_45151||full Low-quality
S14_vs_k141_45154||full Low-quality
S14_vs_k141_16354||full Low-quality
S14_vs_k141_67134||full Not-determined
S14_vs_k141_16360||full Low-quality
S14_vs_k141_82148||full Not-determined
S14_vs_k141_16364||full Low-quality
S14_vs_k141_60318||full Low-quality
S14_vs_k141_52977||full Low-quality
S14_vs_k141_67148||full Low-quality
S14_vs_k141_74671||full Not-determined
S14_vs_k141_111756||full Low-quality
S14_vs_k141_89537||full Low-quality
S14_vs_k141_52988||full Low-quality
S14_vs_k141_89552||full Not-determined
S14_vs_k141_96618||full Low-quality
S14_vs_k141_111766||full Not-determined
S14_vs_k141_45207||full Not-determined
S14_vs_k141_31036||full Low-quality
S14_vs_k141_96625||full Low-quality
S14_vs_k141_89556||full Not-determined
S14_vs_k141_38397||full Not-determined
S14_vs_k141_31040||full Low-quality
S14_vs_k141_8929||full Not-determined
S14_vs_k141_31043||full Low-quality
S14_vs_k141_82161||full Not-determined
S14_vs_k141_31048||full Low-quality
S14_vs_k141_60345||full Low-quality
S14_vs_k141_82170||full Low-quality
S14_vs_k141_111784||full Low-quality
S14_vs_k141_111785||full Low-quality
S14_vs_k141_67169||full Low-quality
S14_vs_k141_38422||full Low-quality
S14_vs_k141_60360||full Not-determined
S14_vs_k141_82183||full Low-quality
S14_vs_k141_82187||full Not-determined
S14_vs_k141_16412||full Low-quality
S14_vs_k141_111800||full Low-quality
S14_vs_k141_38447||full Not-determined
S14_vs_k141_82207||full Low-quality
S14_vs_k141_82210||full Low-quality
S14_vs_k141_96666||full Low-quality
S14_vs_k141_74728||full Low-quality
S14_vs_k141_89604||full Low-quality
S14_vs_k141_82211||full Not-determined
S14_vs_k141_16434||full Low-quality
S14_vs_k141_104486||full Not-determined
S14_vs_k141_23497||full Not-determined
S14_vs_k141_16439||full Not-determined
S14_vs_k141_1497||full Not-determined
S14_vs_k141_82214||full Low-quality
S14_vs_k141_1502||full Low-quality
S14_vs_k141_74750||full Low-quality
S14_vs_k141_82222||full Not-determined
S14_vs_k141_96674||full Not-determined
S14_vs_k141_31077||full Low-quality
S14_vs_k141_74755||full Low-quality
S14_vs_k141_16453||full Low-quality
S14_vs_k141_60390||full Not-determined
S14_vs_k141_1519||full Low-quality
S14_vs_k141_38464||full Low-quality
S14_vs_k141_82227||full Low-quality
S14_vs_k141_74763||full Low-quality
S14_vs_k141_60395||full Low-quality
S14_vs_k141_74765||full Low-quality
S14_vs_k141_31086||full Low-quality
S14_vs_k141_31088||full Low-quality
S14_vs_k141_74770||full Low-quality
S14_vs_k141_89620||full Not-determined
S14_vs_k141_67200||full Not-determined
S14_vs_k141_60407||full Low-quality
S14_vs_k141_89631||full Not-determined
S14_vs_k141_89632||full Not-determined
S14_vs_k141_104540||full Low-quality
S14_vs_k141_104542||full Low-quality
S14_vs_k141_38480||full Not-determined
S14_vs_k141_31104||full Not-determined
S14_vs_k141_67214||full Low-quality
S14_vs_k141_23516||full Low-quality
S14_vs_k141_74807||full Low-quality
S14_vs_k141_89650||full Low-quality
S14_vs_k141_74814||full Low-quality
S14_vs_k141_16496||full Not-determined
S14_vs_k141_45217||full Low-quality
S14_vs_k141_111831||full Low-quality
S14_vs_k141_82265||full Low-quality
S14_vs_k141_82267||full Low-quality
S14_vs_k141_74839||full Not-determined
S14_vs_k141_67244||full Low-quality
S14_vs_k141_38519||full Low-quality
S14_vs_k141_74857||full Low-quality
S14_vs_k141_74868||full Low-quality
S14_vs_k141_16523||full Not-determined
S14_vs_k141_111863||full Low-quality
S14_vs_k141_74875||full Low-quality
S14_vs_k141_53013||full Low-quality
S14_vs_k141_89698||full Low-quality
S14_vs_k141_53017||full Low-quality
S14_vs_k141_111883||full Not-determined
S14_vs_k141_111884||full Low-quality
S14_vs_k141_53020||full Low-quality
S14_vs_k141_111887||full Low-quality
S14_vs_k141_111897||full Low-quality
S7_vs_7112||full High-quality
S14_vs_k141_89714||full Low-quality
S14_vs_k141_53034||full Not-determined
S14_vs_k141_111915||full Low-quality
S5_vs_k141_1338||full Medium-quality
S14_vs_k141_67318||full Low-quality
S14_vs_k141_89723||full Low-quality
S14_vs_k141_111927||full Not-determined
S14_vs_k141_16562||full Not-determined
S14_vs_k141_45255||full Low-quality
S14_vs_k141_23575||full Low-quality
S14_vs_k141_67324||full Not-determined
S14_vs_k141_60482||full Low-quality
S14_vs_k141_16569||full Low-quality
S14_vs_k141_31134||full Not-determined
S14_vs_k141_111940||full Low-quality
S14_vs_k141_16570||full Low-quality
S14_vs_k141_74925||full Not-determined
S14_vs_k141_67332||full Low-quality
S14_vs_k141_67335||full Low-quality
S14_vs_k141_60498||full Low-quality
S14_vs_k141_60501||full Not-determined
S14_vs_k141_31145||full Not-determined
S14_vs_k141_53080||full Not-determined
S14_vs_k141_60507||full Not-determined
S14_vs_k141_53087||full Low-quality
S14_vs_k141_96695||full Low-quality
S14_vs_k141_38546||full Low-quality
S14_vs_k141_16585||full Low-quality
S14_vs_k141_60518||full Low-quality
S14_vs_k141_89759||full Low-quality
S14_vs_k141_16596||full Low-quality
S14_vs_k141_96712||full Low-quality
S14_vs_k141_104649||full Low-quality
S14_vs_k141_96715||full Low-quality
S3_vs_k141_73597||full Medium-quality
S14_vs_k141_53096||full Not-determined
S14_vs_k141_82321||full Not-determined
S14_vs_k141_60551||full Low-quality
S14_vs_k141_104664||full Low-quality
S14_vs_k141_45304||full Low-quality
S14_vs_k141_38581||full Not-determined
S14_vs_k141_45306||full Low-quality
S14_vs_k141_104682||full Low-quality
S14_vs_k141_60559||full Low-quality
S14_vs_k141_67402||full Not-determined
S14_vs_k141_111946||full Low-quality
S14_vs_k141_31183||full Low-quality
S14_vs_k141_111951||full Low-quality
S14_vs_k141_1651||full Low-quality
S14_vs_k141_96740||full Low-quality
S14_vs_k141_60581||full Low-quality
S14_vs_k141_1660||full Low-quality
S14_vs_k141_104685||full Low-quality
S14_vs_k141_38607||full Not-determined
S14_vs_k141_53134||full Low-quality
S14_vs_k141_104696||full Low-quality
S14_vs_k141_31199||full Not-determined
S14_vs_k141_111970||full Low-quality
S14_vs_k141_89794||full Not-determined
S14_vs_k141_31202||full Low-quality
S14_vs_k141_38611||full Low-quality
S14_vs_k141_67429||full Low-quality
S14_vs_k141_75004||full Not-determined
S14_vs_k141_45320||full Low-quality
S14_vs_k141_67439||full Not-determined
S14_vs_k141_53154||full Low-quality
S14_vs_k141_1668||full Low-quality
S14_vs_k141_45329||full Not-determined
S14_vs_k141_53160||full Not-determined
S14_vs_k141_75012||full Low-quality
S14_vs_k141_31208||full Not-determined
S14_vs_k141_60593||full Low-quality
S14_vs_k141_104703||full Not-determined
S14_vs_k141_16610||full Low-quality
S14_vs_k141_82374||full Not-determined
S14_vs_k141_104713||full Low-quality
S14_vs_k141_8951||full Low-quality
S14_vs_k141_82380||full Low-quality
S14_vs_k141_31221||full Not-determined
S14_vs_k141_104719||full Low-quality
S14_vs_k141_75014||full Not-determined
S14_vs_k141_75015||full Low-quality
S14_vs_k141_112054||full Low-quality
S14_vs_k141_82401||full Not-determined
S14_vs_k141_60623||full Low-quality
S14_vs_k141_23624||full Low-quality
S14_vs_k141_31231||full Not-determined
S14_vs_k141_75029||full Low-quality
S14_vs_k141_104728||full Low-quality
S14_vs_k141_8983||full Not-determined
S14_vs_k141_104732||full Not-determined
S14_vs_k141_75043||full Low-quality
S14_vs_k141_60639||full Low-quality
S14_vs_k141_23641||full Low-quality
S14_vs_k141_60642||full Low-quality
S14_vs_k141_1716||full Low-quality
S14_vs_k141_75052||full Low-quality
S14_vs_k141_67473||full Low-quality
S14_vs_k141_23659||full Low-quality
S14_vs_k141_67476||full Low-quality
S14_vs_k141_82427||full Low-quality
S14_vs_k141_45387||full Not-determined
S14_vs_k141_104746||full Low-quality
S14_vs_k141_9003||full Low-quality
S14_vs_k141_9004||full Low-quality
S14_vs_k141_38675||full Low-quality
S14_vs_k141_60676||full Not-determined
S14_vs_k141_82432||full Low-quality
S14_vs_k141_16694||full Not-determined
S14_vs_k141_38681||full Low-quality
S14_vs_k141_38691||full Low-quality
S14_vs_k141_9012||full Low-quality
S14_vs_k141_16724||full Low-quality
S14_vs_k141_16732||full Low-quality
S14_vs_k141_31314||full Not-determined
S14_vs_k141_[illegible]||full Low-quality
S14_vs_k141_104770||full Low-quality
S14_vs_k141_23710||full Not-determined
S7_vs_54322||full High-quality
S14_vs_k141_82496||full Low-quality
S14_vs_k141_45422||full Low-quality
S14_vs_k141_82506||full Low-quality
S14_vs_k141_16759||full Not-determined
S14_vs_k141_16763||full Not-determined
S14_vs_k141_23742||full Low-quality
S14_vs_k141_82514||full Low-quality
S14_vs_k141_67520||full Low-quality
S14_vs_k141_38764||full Low-quality
S14_vs_k141_31343||full Not-determined
S14_vs_k141_53298||full Not-determined
S14_vs_k141_45455||full Low-quality
S14_vs_k141_38785||full Low-quality
S14_vs_k141_67541||full Not-determined
S14_vs_k141_16776||full Low-quality
S14_vs_k141_67545||full Low-quality
S14_vs_k141_31350||full Low-quality
S14_vs_k141_9082||full Low-quality
S14_vs_k141_89856||full Low-quality
S14_vs_k141_45459||full Low-quality
S14_vs_k141_1768||full Not-determined
S14_vs_k141_75151||full Low-quality
S14_vs_k141_31367||full Low-quality
S14_vs_k141_75157||full Low-quality
S14_vs_k141_45479||full Low-quality
S14_vs_k141_75158||full Low-quality
S14_vs_k141_60776||full Not-determined
S14_vs_k141_89885||full Low-quality
S14_vs_k141_104835||full Not-determined
S14_vs_k141_53311||full Not-determined
S14_vs_k141_104841||full Not-determined
S14_vs_k141_23767||full Low-quality
S14_vs_k141_60783||full Low-quality
S14_vs_k141_82569||full Low-quality
S14_vs_k141_75166||full Low-quality
S14_vs_k141_104843||full Low-quality
S14_vs_k141_75176||full Low-quality
S14_vs_k141_38849||full Low-quality
S14_vs_k141_45506||full Low-quality
S14_vs_k141_75179||full Low-quality
S14_vs_k141_67575||full Low-quality
S14_vs_k141_53349||full Not-determined
S14_vs_k141_75183||full Not-determined
S14_vs_k141_53353||full Low-quality
S14_vs_k141_53365||full Not-determined
S14_vs_k141_53370||full Low-quality
S14_vs_k141_82571||full Low-quality
S14_vs_k141_45526||full Low-quality
S14_vs_k141_82577||full Low-quality
S14_vs_k141_67601||full Not-determined
S14_vs_k141_53380||full Low-quality
S14_vs_k141_9143||full Low-quality
S14_vs_k141_16842||full Low-quality
S14_vs_k141_89896||full Low-quality
S14_vs_k141_75227||full Low-quality
S14_vs_k141_16856||full Low-quality
S14_vs_k141_16859||full Not-determined
S14_vs_k141_67657||full Low-quality
S14_vs_k141_16871||full Low-quality
S14_vs_k141_75233||full Low-quality
S14_vs_k141_82611||full Low-quality
S14_vs_k141_104878||full Low-quality
S14_vs_k141_82612||full Low-quality
S14_vs_k141_45534||full Not-determined
S14_vs_k141_82615||full Low-quality
S14_vs_k141_1795||full Low-quality
S14_vs_k141_45539||full Low-quality
S14_vs_k141_45543||full Low-quality
S14_vs_k141_104884||full Low-quality
S3_vs_k141_80228||full High-quality
S14_vs_k141_104887||full Low-quality
S14_vs_k141_1804||full Low-quality
S14_vs_k141_23805||full Not-determined
S14_vs_k141_104904||full Low-quality
S14_vs_k141_53419||full Not-determined
S14_vs_k141_38917||full Low-quality
S14_vs_k141_53424||full Low-quality
S14_vs_k141_1828||full Not-determined
S14_vs_k141_75254||full Low-quality
S14_vs_k141_23823||full Low-quality
S14_vs_k141_82652||full Low-quality
S14_vs_k141_53444||full Low-quality
S14_vs_k141_67670||full Not-determined
S14_vs_k141_112103||full Low-quality
S14_vs_k141_90006||full Not-determined
S14_vs_k141_112106||full Low-quality
S14_vs_k141_45610||full Not-determined
S14_vs_k141_67690||full Not-determined
S14_vs_k141_38934||full Low-quality
S14_vs_k141_82684||full Not-determined
S14_vs_k141_45640||full Low-quality
S14_vs_k141_53480||full Low-quality
S14_vs_k141_112128||full Low-quality
S3_extracted_k141_67333 Complete
S14_vs_k141_38946||full Not-determined
S14_vs_k141_112131||full Low-quality
S14_vs_k141_23870||full Low-quality
S14_vs_k141_67727||full Not-determined
S14_vs_k141_90043||full Low-quality
S14_vs_k141_104984||full Low-quality
S14_vs_k141_67734||full Low-quality
S13_vs_83447||full High-quality
S14_vs_k141_9168||full Low-quality
S14_vs_k141_23890||full Low-quality
S14_vs_k141_82708||full Low-quality
S14_vs_k141_82713||full Low-quality
S14_vs_k141_90084||full Low-quality
S14_vs_k141_112176||full Not-determined
S14_vs_k141_90086||full Low-quality
S14_vs_k141_53490||full Not-determined
S14_vs_k141_112179||full Low-quality
S14_vs_k141_1849||full Low-quality
S14_vs_k141_105001||full Low-quality
S14_vs_k141_82735||full Not-determined
S14_vs_k141_67777||full Low-quality
S14_vs_k141_105012||full Low-quality
S14_vs_k141_67780||full Low-quality
S14_vs_k141_82736||full Not-determined
S14_vs_k141_67781||full Low-quality
S14_vs_k141_9212||full Low-quality
S14_vs_k141_82741||full Low-quality
S14_vs_k141_105024||full Low-quality
S14_vs_k141_23917||full Low-quality
S14_vs_k141_39046||full Low-quality
S14_vs_k141_105033||full Low-quality
S14_vs_k141_9229||full Low-quality
S14_vs_k141_1862||full Low-quality
S14_vs_k141_75272||full Low-quality
S14_vs_k141_105050||full Low-quality
S14_vs_k141_96922||full Low-quality
S14_vs_k141_31422||full Low-quality
S14_vs_k141_96924||full Low-quality
S14_vs_k141_105061||full Low-quality
S14_vs_k141_31426||full Low-quality
S14_vs_k141_1900||full Low-quality
S14_vs_k141_67804||full Low-quality
S14_vs_k141_96933||full Low-quality
S14_vs_k141_96934||full Low-quality
S14_vs_k141_96942||full Low-quality
S14_vs_k141_105079||full Low-quality
S14_vs_k141_31431||full Not-determined
S14_vs_k141_67815||full Low-quality
S14_vs_k141_105086||full Low-quality
S14_vs_k141_67830||full Low-quality
S14_vs_k141_53524||full Low-quality
S14_vs_k141_9242||full Low-quality
S14_vs_k141_96957||full Low-quality
S14_vs_k141_9243||full Low-quality
S14_vs_k141_67843||full Low-quality
S14_vs_k141_53538||full Not-determined
S14_vs_k141_96959||full Not-determined
S14_vs_k141_9255||full Not-determined
S14_vs_k141_105116||full Not-determined
S14_vs_k141_105120||full Not-determined
S14_vs_k141_105122||full Low-quality
S14_vs_k141_53559||full Low-quality
S14_vs_k141_24010||full Low-quality
S14_vs_k141_90109||full Not-determined
S14_vs_k141_96997||full Not-determined
S14_vs_k141_53567||full Not-determined
S14_vs_k141_75344||full Low-quality
S14_vs_k141_112242||full Not-determined
S14_vs_k141_53569||full Low-quality
S14_vs_k141_53575||full Not-determined
S14_vs_k141_97003||full Low-quality
S14_vs_k141_24032||full Low-quality
S14_vs_k141_67892||full Low-quality
S14_vs_k141_90123||full Not-determined
S14_vs_k141_24043||full Not-determined
S14_vs_k141_82837||full Low-quality
S14_vs_k141_53580||full Not-determined
S14_vs_k141_90131||full Low-quality
S14_vs_k141_16913||full Low-quality
S14_vs_k141_9286||full Not-determined
S14_vs_k141_82850||full Low-quality
S14_vs_k141_97007||full Low-quality
S14_vs_k141_97009||full Not-determined
S14_vs_k141_67905||full Not-determined
S13_extracted_114546 High-quality

S14_vs_k141_112253||full Low-quality
S14_vs_k141_1947||full Low-quality
S14_vs_k141_90151||full Low-quality
S14_vs_k141_75392||full Low-quality
S14_vs_k141_60851||full Low-quality
S14_vs_k141_67908||full Low-quality
S14_vs_k141_97012||full Low-quality
S14_vs_k141_60862||full Not-determined
S14_vs_k141_1955||full Low-quality
S14_vs_k141_1957||full Low-quality
S14_vs_k141_112259||full Low-quality
S14_vs_k141_39113||full Low-quality
S14_vs_k141_97021||full Not-determined
S14_vs_k141_31464||full Not-determined
S14_vs_k141_97028||full Low-quality
S14_vs_k141_53611||full Not-determined
S14_vs_k141_60867||full Low-quality
S14_vs_k141_105134||full Low-quality
S14_vs_k141_1974||full Low-quality
S14_vs_k141_45652||full Low-quality
S14_vs_k141_75419||full Not-determined
S14_vs_k141_39132||full Not-determined
S14_vs_k141_39138||full Not-determined
S14_vs_k141_45664||full Low-quality
S14_vs_k141_53651||full Low-quality
S14_vs_k141_75439||full Low-quality
S14_vs_k141_9360||full Low-quality
S14_vs_k141_60904||full Not-determined
S14_vs_k141_67965||full Low-quality
S14_vs_k141_53659||full Low-quality
S14_vs_k141_97102||full Not-determined
S14_vs_k141_9372||full Low-quality
S14_vs_k141_31515||full Low-quality
S14_vs_k141_39177||full Low-quality
S14_vs_k141_75475||full Not-determined
S14_vs_k141_9411||full Low-quality
S14_vs_k141_9413||full Not-determined
S14_vs_k141_75490||full Not-determined
S14_vs_k141_39212||full Low-quality
S14_vs_k141_112281||full Low-quality
S14_vs_k141_53688||full Not-determined
S14_vs_k141_31563||full Low-quality
S14_vs_k141_1996||full Low-quality
S14_vs_k141_75499||full Low-quality
S14_vs_k141_97153||full Low-quality
S14_vs_k141_31567||full Low-quality
S14_vs_k141_68014||full Low-quality
S14_vs_k141_2009||full Low-quality
S14_vs_k141_53726||full Low-quality
S14_vs_k141_45687||full Low-quality
S14_vs_k141_75514||full Low-quality
S14_vs_k141_97176||full Low-quality
S14_vs_k141_24123||full Not-determined
S14_vs_k141_53731||full Not-determined
S14_vs_k141_112319||full Low-quality
S14_vs_k141_97182||full Not-determined
S14_vs_k141_75528||full Low-quality
S14_vs_k141_2016||full Not-determined
S14_vs_k141_82953||full Not-determined
S14_vs_k141_112330||full Low-quality
S14_vs_k141_53738||full Low-quality
S14_vs_k141_75539||full Low-quality
S14_vs_k141_24159||full Low-quality
S14_vs_k141_60930||full Not-determined
S14_vs_k141_68035||full Low-quality
S14_vs_k141_105208||full Low-quality
S14_vs_k141_24171||full Low-quality
S14_vs_k141_2037||full Not-determined
S14_vs_k141_39260||full Not-determined
S14_vs_k141_68041||full Low-quality
S14_vs_k141_2043||full Not-determined
S14_vs_k141_17026||full Low-quality
S14_vs_k141_68050||full Low-quality
S14_vs_k141_68052||full Low-quality
S14_vs_k141_90264||full Not-determined
S14_vs_k141_2065||full Low-quality
S14_vs_k141_68060||full Low-quality
S14_vs_k141_2069||full Not-determined
S14_vs_k141_24179||full Low-quality
S14_vs_k141_45701||full Low-quality
S14_vs_k141_9453||full Not-determined
S14_vs_k141_24184||full Low-quality
S14_vs_k141_60961||full Not-determined
S14_vs_k141_75597||full Low-quality
S14_vs_k141_97217||full Low-quality
S14_vs_k141_24191||full Not-determined
S14_vs_k141_24193||full Low-quality
S14_vs_k141_17049||full Not-determined
S13_vs_114546||full High-quality
S14_vs_k141_45713||full Low-quality
S14_vs_k141_105287||full Not-determined
S3_vs_k141_117894||0_partial Medium-quality
S14_vs_k141_9464||full Not-determined
S14_vs_k141_9466||full Not-determined
S14_vs_k141_75607||full Low-quality
S14_vs_k141_31579||full Low-quality
S14_vs_k141_60983||full Low-quality
S14_vs_k141_2115||full Not-determined
S14_vs_k141_97223||full Low-quality
S14_vs_k141_39293||full Low-quality
S14_vs_k141_31608||full Low-quality
S14_vs_k141_9507||full Low-quality
S14_vs_k141_83036||full Not-determined
S14_vs_k141_9511||full Low-quality
S14_vs_k141_112401||full Low-quality
S14_vs_k141_83046||full Low-quality
S14_vs_k141_2151||full Not-determined
S14_vs_k141_9518||full Low-quality
S14_vs_k141_31623||full Not-determined
S14_vs_k141_9521||full Low-quality
S14_vs_k141_97254||full Not-determined
S14_vs_k141_45791||full Low-quality
S14_vs_k141_112419||full Low-quality
S14_vs_k141_97255||full Low-quality
S14_vs_k141_97256||full Not-determined
S14_vs_k141_45812||full Low-quality
S14_vs_k141_105358||full Low-quality
S14_vs_k141_90297||full Low-quality
S14_vs_k141_112441||full Low-quality
S14_vs_k141_105362||full Low-quality
S14_vs_k141_97257||full Low-quality
S14_vs_k141_2173||full Low-quality
S14_vs_k141_17113||full Low-quality
S14_vs_k141_17121||full Not-determined
S14_vs_k141_2179||full Low-quality
S14_vs_k141_9524||full Low-quality
S14_vs_k141_75667||full Low-quality
S14_vs_k141_90303||full Low-quality
S14_vs_k141_2191||full Not-determined
S14_vs_k141_45844||full Low-quality
S14_vs_k141_53794||full Low-quality
S14_vs_k141_53798||full Low-quality
S14_vs_k141_105385||full Low-quality
S14_vs_k141_90326||full Low-quality
S14_vs_k141_9573||full Low-quality
S14_vs_k141_61048||full Low-quality
S14_vs_k141_105387||full Low-quality
S14_vs_k141_31666||full Not-determined
S14_vs_k141_75686||full Not-determined
S14_vs_k141_9574||full Not-determined
S14_vs_k141_83076||full Low-quality
S14_vs_k141_90340||full Low-quality
S14_vs_k141_9582||full Low-quality
S14_vs_k141_75688||full Low-quality
S14_vs_k141_9585||full Not-determined
S14_vs_k141_105404||full Low-quality
S14_vs_k141_68148||full Low-quality
S14_vs_k141_90361||full Low-quality
S14_vs_k141_83103||full Low-quality
S14_vs_k141_61083||full Low-quality
S14_vs_k141_75716||full Low-quality
S14_vs_k141_9613||full Low-quality
S14_vs_k141_83105||full Low-quality
S14_vs_k141_61088||full Not-determined
S14_vs_k141_39342||full Not-determined
S14_vs_k141_9622||full Low-quality
S14_vs_k141_31688||full Not-determined
S14_vs_k141_9626||full Low-quality
S14_vs_k141_97316||full Low-quality
S14_vs_k141_83117||full Not-determined
S14_vs_k141_75742||full Low-quality
S14_vs_k141_31706||full Low-quality
S14_vs_k141_97324||full Not-determined
S14_vs_k141_97326||full Low-quality
S14_vs_k141_68168||full Low-quality
S14_vs_k141_112522||full Not-determined
S14_vs_k141_17221||full Low-quality
S14_vs_k141_75768||full Low-quality
S14_vs_k141_90404||full Low-quality
S14_vs_k141_61114||full Not-determined
S14_vs_k141_2227||full Low-quality
S14_vs_k141_31725||full Not-determined
S14_vs_k141_90414||full Low-quality
S14_vs_k141_97349||full Not-determined
S14_vs_k141_90418||full Low-quality
S14_vs_k141_61124||full Low-quality
S14_vs_k141_97352||full Not-determined
S14_vs_k141_2247||full Low-quality
S14_vs_k141_31732||full Not-determined
S14_vs_k141_90429||full Low-quality
S14_vs_k141_45908||full Low-quality
S14_vs_k141_53823||full Low-quality
S14_vs_k141_53828||full Low-quality
S14_vs_k141_75799||full Low-quality
S14_vs_k141_39409||full Low-quality
S14_vs_k141_75803||full Not-determined
S14_vs_k141_45925||full Low-quality
S14_vs_k141_17239||full Low-quality
S14_vs_k141_53833||full Low-quality
S14_vs_k141_9671||full Low-quality
S14_vs_k141_97360||full Not-determined
S14_vs_k141_75808||full Not-determined
S14_vs_k141_112557||full Low-quality
S14_vs_k141_68202||full Low-quality
S14_vs_k141_24213||full Low-quality
S14_vs_k141_97367||full Low-quality
S14_vs_k141_61173||full Low-quality
S14_vs_k141_97371||full Not-determined
S14_vs_k141_61176||full Low-quality
S14_vs_k141_75830||full Low-quality
S14_vs_k141_39426||full Low-quality
S14_vs_k141_53854||full Not-determined
S14_vs_k141_83138||full Not-determined
S14_vs_k141_83139||full Low-quality
S14_vs_k141_53857||full Not-determined
S14_vs_k141_31768||full Not-determined
S14_vs_k141_17266||full Low-quality
S14_vs_k141_53863||full Low-quality
S14_vs_k141_68255||full Low-quality
S14_vs_k141_90477||full Low-quality
S14_vs_k141_83171||full Not-determined
S14_vs_k141_17287||full Not-determined
S14_vs_k141_31778||full Low-quality
S14_vs_k141_112586||full Not-determined
S14_vs_k141_31786||full Low-quality
S14_vs_k141_112594||full Not-determined
S14_vs_k141_90494||full Low-quality
S14_vs_k141_83178||full Low-quality
S14_vs_k141_17307||full Not-determined
S14_vs_k141_17308||full Low-quality
S14_vs_k141_83189||full Low-quality
S14_vs_k141_90506||full Low-quality
S14_vs_k141_75891||full Low-quality
S14_vs_k141_45981||full Low-quality
S14_vs_k141_24254||full Not-determined
S14_vs_k141_90513||full Low-quality
S14_vs_k141_31811||full Not-determined
S14_vs_k141_24261||full Low-quality
S14_vs_k141_24264||full Low-quality
S14_vs_k141_97415||full Low-quality
S14_vs_k141_2294||full Not-determined
S14_vs_k141_90534||full Low-quality
S14_vs_k141_9685||full Low-quality
S14_vs_k141_9687||full Low-quality
S14_vs_k141_9688||full Low-quality
S14_vs_k141_90546||full Low-quality
S14_vs_k141_31827||full Low-quality
S14_vs_k141_31832||full Low-quality
S14_vs_k141_97429||full Not-determined
S14_vs_k141_31835||full Low-quality
S14_vs_k141_31836||full Low-quality
S14_vs_k141_97439||full Low-quality
S14_vs_k141_9702||full Not-determined
S14_vs_k141_9704||full Not-determined
S14_vs_k141_105536||full Not-determined
S14_vs_k141_9716||full Low-quality
S14_vs_k141_68318||full Not-determined
S14_vs_k141_9720||full Low-quality
S14_vs_k141_105549||full Not-determined
S14_vs_k141_9725||full Low-quality
S14_vs_k141_17325||full Low-quality
S14_vs_k141_90559||full Low-quality
S14_vs_k141_75965||full Not-determined
S14_vs_k141_97459||full Low-quality
S14_vs_k141_75967||full Not-determined
S14_vs_k141_24314||full Low-quality
S14_vs_k141_68324||full Low-quality
S14_vs_k141_31846||full Not-determined
S14_vs_k141_53953||full Not-determined
S14_vs_k141_17330||full Low-quality
S14_vs_k141_31848||full Not-determined
S14_vs_k141_105587||full Low-quality
S14_vs_k141_90574||full Low-quality
S14_vs_k141_53966||full Low-quality
S14_vs_k141_24343||full Low-quality
S14_vs_k141_53969||full Low-quality
S14_vs_k141_31857||full Low-quality
S14_vs_k141_31861||full Low-quality
S14_vs_k141_53975||full Low-quality
S14_vs_k141_31865||full Low-quality
S14_vs_k141_53977||full Low-quality
S14_vs_k141_76001||full Low-quality
S14_vs_k141_112635||full Low-quality
S14_vs_k141_68397||full Low-quality
S14_vs_k141_68407||full Not-determined
S14_vs_k141_9772||full Low-quality
S14_vs_k141_68414||full Low-quality
S14_vs_k141_24356||full Not-determined
S14_vs_k141_90623||full Low-quality
S14_vs_k141_17365||full Low-quality
S14_vs_k141_54000||full Not-determined
S14_vs_k141_46046||full Low-quality
S14_vs_k141_24370||full Low-quality
S14_vs_k141_46048||full Low-quality
S14_vs_k141_54006||full Low-quality
S14_vs_k141_24380||full Not-determined
S14_vs_k141_31905||full Low-quality
S14_vs_k141_2383||full Not-determined
S14_vs_k141_68453||full Not-determined
S14_vs_k141_83276||full Not-determined
S14_vs_k141_9817||full Low-quality
S14_vs_k141_68467||full Low-quality
S14_vs_k141_76062||full Low-quality
S14_vs_k141_31917||full Not-determined
S14_vs_k141_31919||full Low-quality
S14_vs_k141_39502||full Low-quality
S14_vs_k141_46077||full Not-determined
S14_vs_k141_68470||full Not-determined
S14_vs_k141_46088||full Low-quality
S14_vs_k141_17414||full Low-quality
S14_vs_k141_39516||full Low-quality
S14_vs_k141_76068||full Not-determined
S14_vs_k141_68490||full Low-quality
S14_vs_k141_54057||full Not-determined
S14_vs_k141_54058||full Low-quality
S14_vs_k141_46095||full Low-quality
S14_vs_k141_83313||full Low-quality
S14_vs_k141_83317||full Low-quality
S14_vs_k141_39523||full Low-quality
S14_vs_k141_17440||full Low-quality
S14_vs_k141_61225||full Low-quality
S14_vs_k141_39524||full Not-determined
S14_vs_k141_68515||full Low-quality
S14_vs_k141_76091||full Not-determined
S14_vs_k141_76092||full Not-determined
S14_vs_k141_61238||full Low-quality
S14_vs_k141_112700||full Low-quality
S14_vs_k141_2451||full Not-determined
S14_vs_k141_105634||full Not-determined
S14_vs_k141_54093||full Not-determined
S14_vs_k141_39537||full Low-quality
S14_vs_k141_90709||full Low-quality
S14_vs_k141_76106||full Low-quality
S14_vs_k141_9859||full Low-quality
S14_vs_k141_31947||full Low-quality
S14_vs_k141_24406||full Low-quality
S14_vs_k141_39551||full Low-quality
S14_vs_k141_83347||full Not-determined
S14_vs_k141_105641||full Low-quality
S14_vs_k141_46163||full Not-determined
S14_vs_k141_24416||full Low-quality
S14_vs_k141_76136||full Low-quality
S14_vs_k141_54113||full Low-quality
S14_vs_k141_83360||full Low-quality
S14_vs_k141_76141||full Low-quality
S14_vs_k141_31959||full Not-determined
S14_vs_k141_2500||full Low-quality
S14_vs_k141_31968||full Low-quality
S14_vs_k141_83369||full Low-quality
S14_vs_k141_76156||full Low-quality
S14_vs_k141_54144||full Low-quality
S14_vs_k141_90733||full Low-quality
S14_vs_k141_54153||full Low-quality
S14_vs_k141_105672||full Not-determined
S14_vs_k141_97635||full Not-determined
S14_vs_k141_17484||full Low-quality
S14_vs_k141_97636||full Low-quality
S14_vs_k141_54160||full Low-quality
S14_vs_k141_61302||full Not-determined
S14_vs_k141_68558||full Low-quality
S14_vs_k141_2523||full Not-determined
S14_vs_k141_39594||full Low-quality
S14_vs_k141_83374||full Not-determined
S14_vs_k141_46220||full Low-quality
S14_vs_k141_112738||full Low-quality
S14_vs_k141_112741||full Low-quality
S14_vs_k141_31999||full Low-quality
S14_vs_k141_105701||full Not-determined
S14_vs_k141_97664||full Not-determined
S14_vs_k141_17517||full Low-quality
S14_vs_k141_68574||full Low-quality
S14_vs_k141_39602||full Not-determined
S14_vs_k141_24499||full Low-quality
S14_vs_k141_68576||full Low-quality
S14_vs_k141_9908||full Low-quality
S14_vs_k141_83396||full Low-quality
S14_vs_k141_105714||full Low-quality
S14_vs_k141_61337||full Low-quality
S14_vs_k141_83398||full Low-quality
S14_vs_k141_54197||full Low-quality
S14_vs_k141_76214||full Low-quality
S14_vs_k141_9916||full Not-determined
S14_vs_k141_32020||full Low-quality
S14_vs_k141_90766||full Low-quality
S14_vs_k141_17545||full Not-determined
S14_vs_k141_54205||full Low-quality
S14_vs_k141_46234||full Low-quality
S14_vs_k141_112778||full Not-determined
S14_vs_k141_97699||full Low-quality
S14_vs_k141_68605||full Low-quality
S14_vs_k141_24512||full Low-quality
S14_vs_k141_90774||full Low-quality
S14_vs_k141_61363||full Not-determined
S14_vs_k141_32051||full Not-determined
S14_vs_k141_39615||full Low-quality
S14_vs_k141_97710||full Low-quality
S14_vs_k141_112796||full Low-quality
S14_vs_k141_112800||full Not-determined
S14_vs_k141_17573||full Low-quality
S14_vs_k141_105739||full Not-determined
S14_vs_k141_54246||full Low-quality
S14_vs_k141_17574||full Not-determined
S14_vs_k141_24533||full Low-quality
S14_vs_k141_46267||full Low-quality
S14_vs_k141_76234||full Not-determined
S14_vs_k141_39635||full Low-quality
S14_vs_k141_105749||full Low-quality
S14_vs_k141_105750||full Low-quality
S14_vs_k141_76250||full Low-quality
S14_vs_k141_112830||full Low-quality
S14_vs_k141_17592||full Low-quality
S14_vs_k141_54262||full Low-quality
S14_vs_k141_46285||full Low-quality
S14_vs_k141_24554||full Low-quality
S14_vs_k141_39652||full Low-quality
S14_vs_k141_90827||full Low-quality
S14_vs_k141_46292||full Low-quality
S14_vs_k141_112837||full Low-quality
S14_vs_k141_112845||full Low-quality
S14_vs_k141_2621||full Not-determined
S14_vs_k141_32105||full Low-quality
S14_vs_k141_97764||full Low-quality
S14_vs_k141_83449||full Low-quality
S14_vs_k141_90842||full Low-quality
S14_vs_k141_76294||full Low-quality
S14_vs_k141_32110||full Not-determined
S14_vs_k141_112857||full Low-quality
S14_vs_k141_90845||full Low-quality
S14_vs_k141_68631||full Low-quality
S14_vs_k141_90846||full Low-quality
S14_vs_k141_97766||full Low-quality
S14_vs_k141_39694||full Low-quality
S14_vs_k141_90872||full Low-quality
S14_vs_k141_54340||full Low-quality
S14_vs_k141_32134||full Not-determined
S14_vs_k141_90878||full Not-determined
S14_vs_k141_83488||full Not-determined
S14_vs_k141_46311||full Low-quality
S14_vs_k141_24592||full Low-quality
S14_vs_k141_39718||full Not-determined
S14_vs_k141_68668||full Low-quality
S14_vs_k141_24602||full Low-quality
S14_vs_k141_68670||full Not-determined
S14_vs_k141_54361||full Not-determined
S14_vs_k141_46319||full Low-quality
S14_vs_k141_83502||full Low-quality
S14_vs_k141_112874||full Low-quality
S14_vs_k141_17652||full Not-determined
S14_vs_k141_90898||full Low-quality
S14_vs_k141_24620||full Not-determined
S14_vs_k141_83504||full Low-quality
S14_vs_k141_76319||full Low-quality
S14_vs_k141_24629||full Not-determined
S14_vs_k141_90910||full Not-determined
S14_vs_k141_39743||full Low-quality
S14_vs_k141_61400||full Not-determined
S14_vs_k141_24642||full Not-determined
S14_vs_k141_17657||full Low-quality
S14_vs_k141_90927||full Low-quality
S14_vs_k141_61420||full Not-determined
S14_vs_k141_76342||full Not-determined
S14_vs_k141_17668||full Not-determined
S14_vs_k141_39777||full Low-quality
S14_vs_k141_39784||full Not-determined
S14_vs_k141_39786||full Low-quality
S14_vs_k141_32220||full Not-determined
S14_vs_k141_61445||full Not-determined
S14_vs_k141_17694||full Low-quality
S14_vs_k141_32225||full Low-quality
S14_vs_k141_17695||full Not-determined
S14_vs_k141_2639||full Low-quality
S14_vs_k141_61453||full Low-quality
S14_vs_k141_83574||full Not-determined
S14_vs_k141_10072||full Low-quality
S14_vs_k141_54425||full Low-quality
S14_vs_k141_54426||full Low-quality
S14_vs_k141_112902||full Low-quality
S14_vs_k141_76374||full Low-quality
S4_extracted_31978 Complete
S14_vs_k141_54431||full Low-quality
S14_vs_k141_10091||full Low-quality
S14_vs_k141_2658||full Low-quality
S14_vs_k141_105866||full Not-determined
S14_vs_k141_83593||full Not-determined
S14_vs_k141_2668||full Low-quality
S14_vs_k141_61463||full Low-quality
S14_vs_k141_83599||full Not-determined
S14_vs_k141_2676||full Not-determined
S14_vs_k141_32266||full Low-quality
S14_vs_k141_2682||full Low-quality
S14_vs_k141_32269||full Not-determined
S14_vs_k141_10110||full Low-quality
S14_vs_k141_68753||full Low-quality
S14_vs_k141_105880||full Low-quality
S14_vs_k141_54467||full Low-quality
S14_vs_k141_112956||full Low-quality
S14_vs_k141_10121||full Not-determined
S14_vs_k141_17713||full Low-quality
S14_vs_k141_2703||full Low-quality
S14_vs_k141_83601||full Low-quality
S14_vs_k141_112964||full Not-determined
S14_vs_k141_61487||full Not-determined
S5_vs_k141_30490||full Medium-quality
S14_vs_k141_76444||full Low-quality
S14_vs_k141_105892||full Low-quality
S14_vs_k141_68771||full Low-quality
S14_vs_k141_10132||full Low-quality
S14_vs_k141_2720||full Low-quality
S14_vs_k141_68777||full Not-determined
S14_vs_k141_105897||full Low-quality
S14_vs_k141_46413||full Low-quality
S14_vs_k141_2723||full Low-quality
S14_vs_k141_2726||full Low-quality
S14_vs_k141_68779||full Low-quality
S14_vs_k141_2730||full Low-quality
S14_vs_k141_68794||full Low-quality
S14_vs_k141_112976||full Not-determined
S14_vs_k141_39828||full Low-quality
S14_vs_k141_39830||full Low-quality
S14_vs_k141_105918||full Not-determined
S14_vs_k141_68802||full Low-quality
S14_vs_k141_83645||full Low-quality
S14_vs_k141_105938||full Low-quality
S14_vs_k141_46448||full Low-quality
S14_vs_k141_76483||full Low-quality
S14_vs_k141_68812||full Low-quality
S14_vs_k141_32301||full Low-quality
S14_vs_k141_68824||full Not-determined
S14_vs_k141_83676||full Not-determined
S14_vs_k141_10151||full Low-quality
S14_vs_k141_54526||full Not-determined
S14_vs_k141_68832||full Low-quality
S14_vs_k141_39857||full Not-determined
S14_vs_k141_39869||full Not-determined
S12_extracted_k141_187941 Medium-quality
S14_vs_k141_83685||full Low-quality
S14_vs_k141_39877||full Low-quality
S14_vs_k141_105967||full Low-quality
S14_vs_k141_24744||full Not-determined
S14_vs_k141_54565||full Low-quality
S14_vs_k141_105970||full Low-quality
S14_vs_k141_32345||full Low-quality
S14_vs_k141_68858||full Not-determined
S14_vs_k141_46494||full Low-quality
S14_vs_k141_54569||full Not-determined
S14_vs_k141_90998||full Low-quality
S14_vs_k141_91001||full Not-determined
S14_vs_k141_39891||full Low-quality
S14_vs_k141_2765||full Low-quality
S8_extracted_k141_122206 High-quality
S14_vs_k141_39894||full Low-quality
S14_vs_k141_10194||full Not-determined
S14_vs_k141_10195||full Low-quality
S14_vs_k141_97841||full Low-quality
S14_vs_k141_24772||full Not-determined
S14_vs_k141_17749||full Low-quality
S14_vs_k141_97846||full Low-quality
S14_vs_k141_46531||full Low-quality
S14_vs_k141_76530||full Low-quality
S14_vs_k141_2790||full Not-determined
S14_vs_k141_46532||full Low-quality
S14_vs_k141_46534||full Low-quality
S14_vs_k141_83720||full Low-quality
S14_vs_k141_105992||full Not-determined
S14_vs_k141_24785||full Not-determined
S14_vs_k141_76543||full Not-determined
S14_vs_k141_32370||full Not-determined
S14_vs_k141_68920||full Low-quality
S14_vs_k141_68922||full Low-quality
S14_vs_k141_10205||full Low-quality
S14_vs_k141_97866||full Low-quality
S14_vs_k141_10207||full Low-quality
S14_vs_k141_68930||full Low-quality
S14_vs_k141_24799||full Low-quality
S14_vs_k141_24800||full Not-determined
S14_vs_k141_2822||full Not-determined
S14_vs_k141_2830||full Low-quality
S14_vs_k141_83755||full Low-quality
S14_vs_k141_106025||full Low-quality
S14_vs_k141_46577||full Not-determined
S14_vs_k141_106031||full Low-quality
S14_vs_k141_54607||full Low-quality
S14_vs_k141_39954||full Low-quality
S14_vs_k141_39957||full Low-quality
S14_vs_k141_39961||full Low-quality
S14_vs_k141_10248||full Low-quality
S14_vs_k141_32417||full Low-quality
S14_vs_k141_46599||full Not-determined
S14_vs_k141_10276||full Low-quality
S14_vs_k141_76570||full Low-quality
S14_vs_k141_83797||full Low-quality
S14_vs_k141_76573||full Low-quality
S14_vs_k141_2859||full Low-quality
S14_vs_k141_97915||full Low-quality
S14_vs_k141_91083||full Low-quality
S14_vs_k141_76580||full Low-quality
S14_vs_k141_83819||full Low-quality
S14_vs_k141_83822||full Low-quality
S14_vs_k141_76582||full Low-quality

| | |
|---|---|
| S14_vs_k141_113015||full | Low-quality |
| S14_vs_k141_97928||full | Low-quality |
| S14_vs_k141_113016||full | Low-quality |
| S14_vs_k141_17836||full | Not-determined |
| S14_vs_k141_32449||full | Low-quality |
| S14_vs_k141_97938||full | Low-quality |
| S14_vs_k141_76590||full | Low-quality |
| S14_vs_k141_54672||full | Low-quality |
| S14_vs_k141_40014||full | Low-quality |
| S14_vs_k141_106087||full | Low-quality |
| S14_vs_k141_54673||full | Low-quality |
| S14_vs_k141_40021||full | Low-quality |
| S14_vs_k141_76599||full | Not-determined |
| S14_vs_k141_10308||full | Low-quality |
| S14_vs_k141_91116||full | Not-determined |
| S14_vs_k141_83852||full | Low-quality |
| S14_vs_k141_10330||full | Not-determined |
| S14_vs_k141_32496||full | Low-quality |
| S14_vs_k141_83863||full | Not-determined |
| S14_vs_k141_32497||full | Low-quality |
| S14_vs_k141_61529||full | Not-determined |
| S14_vs_k141_113058||full | Low-quality |
| S14_vs_k141_46672||full | Low-quality |
| S14_vs_k141_17877||full | Not-determined |
| S14_vs_k141_83875||full | Not-determined |
| S14_vs_k141_91154||full | Low-quality |
| S14_vs_k141_97975||full | Not-determined |
| S14_vs_k141_106122||full | Not-determined |
| S14_vs_k141_46681||full | Not-determined |
| S14_vs_k141_76638||full | Low-quality |
| S14_vs_k141_68974||full | Not-determined |
| S14_vs_k141_83895||full | Not-determined |
| S14_vs_k141_97991||full | Low-quality |
| S14_vs_k141_83896||full | Low-quality |
| S14_vs_k141_113085||full | Not-determined |
| S14_vs_k141_46693||full | Not-determined |
| S16_vs_107235||full | Medium-quality |
| S14_vs_k141_113090||full | Low-quality |
| S4_vs_VIRSorter_31978-circular-cat | High-quality |
| S14_vs_k141_76672||full | Low-quality |
| S14_vs_k141_113098||full | Low-quality |
| S14_vs_k141_24922||full | Not-determined |
| S14_vs_k141_83909||full | Low-quality |
| S14_vs_k141_69021||full | Low-quality |
| S14_vs_k141_106136||full | Not-determined |
| S14_vs_k141_113112||full | Low-quality |
| S14_vs_k141_69027||full | Low-quality |
| S14_vs_k141_17906||full | Not-determined |
| S14_vs_k141_106150||full | Low-quality |
| S14_vs_k141_91179||full | Not-determined |
| S14_vs_k141_113117||full | Low-quality |
| S14_vs_k141_98032||full | Not-determined |
| S14_vs_k141_106153||full | Low-quality |
| S14_vs_k141_40109||full | Not-determined |
| S14_vs_k141_24930||full | Low-quality |
| S14_vs_k141_76700||full | Not-determined |
| S14_vs_k141_10335||full | Low-quality |
| S14_vs_k141_69051||full | Low-quality |
| S14_vs_k141_98038||full | Low-quality |
| S14_vs_k141_69057||full | Low-quality |
| S14_vs_k141_17936||full | Low-quality |
| S14_vs_k141_91189||full | Low-quality |
| S14_vs_k141_61576||full | Low-quality |
| S14_vs_k141_69068||full | Low-quality |
| S14_vs_k141_24946||full | Low-quality |
| S14_vs_k141_61579||full | Low-quality |
| S14_vs_k141_40128||full | Low-quality |
| S14_vs_k141_24947||full | Low-quality |
| S14_vs_k141_61583||full | Low-quality |
| S14_vs_k141_61585||full | Not-determined |
| S14_vs_k141_17954||full | Low-quality |
| S14_vs_k141_40131||full | Low-quality |
| S3_vs_k141_119605||full | High-quality |
| S14_vs_k141_98056||full | Low-quality |
| S14_vs_k141_61595||full | Low-quality |
| S14_vs_k141_98068||full | Low-quality |
| S14_vs_k141_24963||full | Low-quality |
| S14_vs_k141_32509||full | Low-quality |
| S14_vs_k141_69087||full | Low-quality |
| S14_vs_k141_40152||full | Low-quality |
| S14_vs_k141_46727||full | Low-quality |
| S14_vs_k141_24969||full | Not-determined |
| S14_vs_k141_10364||full | Low-quality |
| S14_vs_k141_69101||full | Low-quality |
| S3_vs_k141_67333||full | High-quality |
| S14_vs_k141_24974||full | Low-quality |
| S14_vs_k141_61629||full | Not-determined |
| S14_vs_k141_91236||full | Low-quality |
| S14_vs_k141_32522||full | Not-determined |
| S14_vs_k141_32523||full | Low-quality |
| S14_vs_k141_98091||full | Not-determined |
| S14_vs_k141_69109||full | Low-quality |
| S14_vs_k141_113169||full | Not-determined |
| S14_vs_k141_98092||full | Not-determined |
| S14_vs_k141_98097||full | Low-quality |
| S14_vs_k141_54721||full | Low-quality |
| S14_vs_k141_10390||full | Low-quality |
| S14_vs_k141_69127||full | Low-quality |
| S14_vs_k141_61662||full | Not-determined |
| S14_vs_k141_113192||full | Low-quality |
| S14_vs_k141_61668||full | Not-determined |
| S14_vs_k141_91274||full | Not-determined |
| S14_vs_k141_76788||full | Low-quality |
| S14_vs_k141_76792||full | Low-quality |
| S14_vs_k141_69139||full | Low-quality |
| S14_vs_k141_98119||full | Low-quality |
| S14_vs_k141_113212||full | Low-quality |
| S14_vs_k141_76806||full | Low-quality |
| S14_vs_k141_113216||full | Not-determined |
| S14_vs_k141_113219||full | Low-quality |
| S14_vs_k141_40194||full | Low-quality |
| S14_vs_k141_76814||full | Low-quality |
| S14_vs_k141_18007||full | Low-quality |
| S14_vs_k141_54767||full | Low-quality |
| S14_vs_k141_98128||full | Low-quality |
| S14_vs_k141_113226||full | Not-determined |
| S14_vs_k141_113227||full | Not-determined |
| S14_vs_k141_83953||full | Low-quality |
| S14_vs_k141_18015||full | Low-quality |
| S14_vs_k141_10416||full | Low-quality |
| S14_vs_k141_24998||full | Low-quality |
| S14_vs_k141_61714||full | Low-quality |
| S14_vs_k141_46771||full | Low-quality |
| S14_vs_k141_91308||full | Not-determined |
| S14_vs_k141_40216||full | Low-quality |
| S14_vs_k141_83976||full | Not-determined |
| S14_vs_k141_61720||full | Low-quality |
| S14_vs_k141_98150||full | Not-determined |
| S14_vs_k141_113258||full | Not-determined |
| S14_vs_k141_76835||full | Low-quality |
| S14_vs_k141_83995||full | Low-quality |
| S14_vs_k141_91325||full | Low-quality |
| S14_vs_k141_91327||full | Low-quality |
| S14_vs_k141_2971||full | Not-determined |
| S14_vs_k141_98153||full | Low-quality |
| S14_vs_k141_83996||full | Not-determined |
| S14_vs_k141_69182||full | Low-quality |
| S14_vs_k141_84002||full | Low-quality |
| S14_vs_k141_18056||full | Low-quality |
| S14_vs_k141_91334||full | Low-quality |
| S14_vs_k141_98159||full | Low-quality |
| S14_vs_k141_40250||full | Low-quality |
| S14_vs_k141_69183||full | Not-determined |
| S14_vs_k141_18059||full | Low-quality |
| S14_vs_k141_18063||full | Low-quality |
| S14_vs_k141_69184||full | Not-determined |
| S14_vs_k141_98169||full | Not-determined |
| S14_vs_k141_46798||full | Low-quality |
| S14_vs_k141_76854||full | Low-quality |
| S14_vs_k141_91350||full | Not-determined |
| S14_vs_k141_106224||full | Low-quality |
| S14_vs_k141_18066||full | Not-determined |
| S14_vs_k141_54820||full | Low-quality |
| S14_vs_k141_40277||full | Low-quality |
| S14_vs_k141_32599||full | Low-quality |
| S14_vs_k141_32600||full | Not-determined |
| S14_vs_k141_69206||full | Low-quality |
| S14_vs_k141_106240||full | Not-determined |
| S14_vs_k141_106243||full | Low-quality |
| S14_vs_k141_46812||full | Not-determined |
| S14_vs_k141_3041||full | Low-quality |
| S14_vs_k141_46813||full | Low-quality |
| S14_vs_k141_69209||full | Low-quality |
| S14_vs_k141_106248||full | Not-determined |
| S14_vs_k141_46816||full | Not-determined |
| S14_vs_k141_3046||full | Low-quality |
| S14_vs_k141_106255||full | Low-quality |
| S14_vs_k141_84071||full | Low-quality |
| S14_vs_k141_40315||full | Low-quality |
| S14_vs_k141_84080||full | Low-quality |
| S14_vs_k141_3049||full | Low-quality |
| S14_vs_k141_91401||full | Low-quality |
| S14_vs_k141_98213||full | Low-quality |
| S14_vs_k141_113326||full | Low-quality |
| S14_vs_k141_91404||full | Not-determined |
| S14_vs_k141_76874||full | Not-determined |
| S14_vs_k141_46842||full | Low-quality |
| S14_vs_k141_98222||full | Low-quality |
| S14_vs_k141_25062||full | Low-quality |
| S14_vs_k141_40338||full | Low-quality |
| S14_vs_k141_91412||full | Low-quality |
| S14_vs_k141_76889||full | Low-quality |
| S14_vs_k141_69250||full | Low-quality |
| S14_vs_k141_10514||full | Low-quality |
| S14_vs_k141_91424||full | Low-quality |
| S14_vs_k141_69262||full | Not-determined |
| S14_vs_k141_98241||full | Low-quality |
| S14_vs_k141_32621||full | Not-determined |
| S14_vs_k141_98245||full | Low-quality |
| S14_vs_k141_3071||full | Low-quality |
| S14_vs_k141_40365||full | Low-quality |
| S14_vs_k141_18106||full | Low-quality |
| S14_vs_k141_40368||full | Low-quality |
| S14_vs_k141_18114||full | Low-quality |
| S14_vs_k141_25088||full | Not-determined |
| S14_vs_k141_25089||full | Low-quality |
| S14_vs_k141_69299||full | Low-quality |
| S14_vs_k141_113368||full | Not-determined |
| S14_vs_k141_10527||full | Not-determined |
| S14_vs_k141_18128||full | Not-determined |
| S14_vs_k141_76938||full | Not-determined |
| S14_vs_k141_3087||full | Low-quality |
| S14_vs_k141_46907||full | Low-quality |
| S14_vs_k141_84126||full | Not-determined |
| S14_vs_k141_113384||full | Not-determined |
| S14_vs_k141_54846||full | Not-determined |
| S14_vs_k141_18137||full | Low-quality |
| S14_vs_k141_18139||full | Low-quality |
| S14_vs_k141_98261||full | Not-determined |
| S14_vs_k141_91476||full | Not-determined |
| S14_vs_k141_3115||full | Low-quality |
| S14_vs_k141_69318||full | Low-quality |
| S14_vs_k141_3122||full | Not-determined |
| S14_vs_k141_18162||full | Low-quality |
| S14_vs_k141_76981||full | Not-determined |
| S14_vs_k141_46926||full | Low-quality |
| S14_vs_k141_76994||full | Not-determined |
| S14_vs_k141_69331||full | Low-quality |
| S14_vs_k141_46941||full | Not-determined |
| S14_vs_k141_84151||full | Low-quality |
| S14_vs_k141_91495||full | Not-determined |
| S14_vs_k141_113430||full | Low-quality |
| S14_vs_k141_25133||full | Low-quality |
| S14_vs_k141_54872||full | Low-quality |
| S14_vs_k141_46944||full | Low-quality |
| S14_vs_k141_10572||full | Low-quality |
| S14_vs_k141_91517||full | Low-quality |
| S14_vs_k141_40389||full | Not-determined |
| S14_vs_k141_113438||full | Low-quality |
| S14_vs_k141_98312||full | Low-quality |
| S14_vs_k141_54880||full | Low-quality |
| S14_vs_k141_10582||full | Not-determined |
| S14_vs_k141_84162||full | Low-quality |
| S14_vs_k141_25139||full | Low-quality |
| S14_vs_k141_98316||full | Not-determined |
| S14_vs_k141_84166||full | Low-quality |
| S14_vs_k141_54891||full | Low-quality |
| S14_vs_k141_98324||full | Low-quality |
| S14_vs_k141_113447||full | Low-quality |
| S14_vs_k141_3196||full | Low-quality |
| S14_vs_k141_18199||full | Not-determined |
| S14_vs_k141_91536||full | Low-quality |
| S14_vs_k141_113456||full | Not-determined |
| S14_vs_k141_18212||full | Not-determined |
| S14_vs_k141_69388||full | Low-quality |
| S14_vs_k141_25168||full | Low-quality |
| S14_vs_k141_91543||full | Not-determined |
| S14_vs_k141_69395||full | Low-quality |
| S14_vs_k141_113479||full | Low-quality |
| S14_vs_k141_84218||full | Low-quality |
| S14_vs_k141_10637||full | Low-quality |
| S14_vs_k141_10640||full | Low-quality |
| S14_vs_k141_61836||full | Low-quality |
| S14_vs_k141_46982||full | Low-quality |
| S14_vs_k141_3209||full | Not-determined |
| S14_vs_k141_25213||full | Not-determined |
| S14_vs_k141_77063||full | Not-determined |
| S14_vs_k141_106355||full | Low-quality |
| S14_vs_k141_18245||full | Not-determined |
| S14_vs_k141_18249||full | Not-determined |
| S14_vs_k141_106363||full | Low-quality |
| S14_vs_k141_32748||full | Not-determined |
| S14_vs_k141_106384||full | Low-quality |
| S9_vs_k141_50439||full | High-quality |
| S14_vs_k141_47026||full | Not-determined |
| S14_vs_k141_84255||full | Low-quality |
| S14_vs_k141_98404||full | Low-quality |
| S8_vs_k141_122206||full | High-quality |
| S14_vs_k141_25239||full | Not-determined |
| S14_vs_k141_10657||full | Low-quality |
| S14_vs_k141_10660||full | Low-quality |
| S14_vs_k141_25243||full | Low-quality |
| S14_vs_k141_106393||full | Low-quality |
| S14_vs_k141_69453||full | Low-quality |
| S14_vs_k141_54938||full | Low-quality |
| S14_vs_k141_25251||full | Low-quality |
| S14_vs_k141_18318||full | Low-quality |
| S14_vs_k141_84279||full | Low-quality |
| S14_vs_k141_47052||full | Low-quality |
| S14_vs_k141_18319||full | Low-quality |
| S14_vs_k141_47057||full | Not-determined |
| S14_vs_k141_32795||full | Not-determined |
| S14_vs_k141_25262||full | Low-quality |
| S14_vs_k141_69466||full | Not-determined |
| S14_vs_k141_84289||full | Not-determined |
| S14_vs_k141_84295||full | Low-quality |
| S14_vs_k141_18333||full | Not-determined |
| S14_vs_k141_98431||full | Low-quality |
| S14_vs_k141_32799||full | Not-determined |
| S14_vs_k141_25269||full | Not-determined |
| S14_vs_k141_54951||full | Not-determined |
| S14_vs_k141_47079||full | Low-quality |
| S14_vs_k141_54953||full | Low-quality |
| S14_vs_k141_84297||full | Low-quality |
| S14_vs_k141_10693||full | Not-determined |
| S14_vs_k141_91585||full | Not-determined |
| S14_vs_k141_69483||full | Not-determined |
| S14_vs_k141_54958||full | Low-quality |
| S14_vs_k141_10700||full | Not-determined |
| S14_vs_k141_113539||full | Low-quality |
| S14_vs_k141_47085||full | Low-quality |
| S14_vs_k141_113541||full | Not-determined |
| S14_vs_k141_40431||full | Not-determined |
| S14_vs_k141_10711||full | Low-quality |
| S14_vs_k141_113546||full | Low-quality |
| S14_vs_k141_10717||full | Not-determined |
| S14_vs_k141_91620||full | Low-quality |
| S14_vs_k141_32840||full | Low-quality |
| S14_vs_k141_40452||full | Low-quality |
| S14_vs_k141_98469||full | Low-quality |
| S14_vs_k141_32844||full | Low-quality |
| S14_vs_k141_10729||full | Not-determined |
| S14_vs_k141_98473||full | Low-quality |
| S14_vs_k141_32851||full | Not-determined |
| S14_vs_k141_40466||full | Low-quality |
| S14_vs_k141_40471||full | Not-determined |
| S14_vs_k141_84319||full | Low-quality |
| S14_vs_k141_69510||full | Low-quality |
| S14_vs_k141_47168||full | Not-determined |
| S14_vs_k141_61906||full | Low-quality |
| S14_vs_k141_47169||full | Low-quality |
| S14_vs_k141_106466||full | Not-determined |
| S14_vs_k141_113588||full | Not-determined |
| S14_vs_k141_69519||full | Not-determined |
| S14_vs_k141_47190||full | Not-determined |
| S14_vs_k141_47194||full | Low-quality |
| S14_vs_k141_47197||full | Low-quality |
| S14_vs_k141_91663||full | Low-quality |
| S14_vs_k141_47199||full | Low-quality |
| S14_vs_k141_106490||full | Low-quality |
| S14_vs_k141_84329||full | Low-quality |
| S14_vs_k141_98500||full | Not-determined |
| S14_vs_k141_69530||full | Low-quality |
| S14_vs_k141_3318||full | Not-determined |
| S14_vs_k141_113610||full | Low-quality |
| S14_vs_k141_84332||full | Not-determined |
| S14_vs_k141_69536||full | Low-quality |
| S14_vs_k141_91678||full | Low-quality |
| S14_vs_k141_84338||full | Low-quality |
| S14_vs_k141_54974||full | Low-quality |
| S14_vs_k141_32896||full | Low-quality |
| S14_vs_k141_40553||full | Low-quality |
| S14_vs_k141_69557||full | Low-quality |
| S14_vs_k141_47220||full | Low-quality |
| S14_vs_k141_3332||full | Low-quality |
| S14_vs_k141_106512||full | Low-quality |
| S14_vs_k141_84342||full | Low-quality |
| S14_vs_k141_10810||full | Low-quality |
| S14_vs_k141_98524||full | Low-quality |
| S14_vs_k141_98540||full | Low-quality |
| S14_vs_k141_40567||full | Low-quality |
| S14_vs_k141_10823||full | Low-quality |
| S14_vs_k141_47242||full | Low-quality |
| S14_vs_k141_32911||full | Low-quality |
| S14_vs_k141_84360||full | Low-quality |
| S14_vs_k141_47250||full | Not-determined |
| S14_vs_k141_18441||full | Low-quality |
| S14_vs_k141_47251||full | Low-quality |
| S14_vs_k141_98559||full | Not-determined |
| S14_vs_k141_3384||full | Low-quality |
| S14_vs_k141_98562||full | Low-quality |
| S14_vs_k141_106533||full | Low-quality |
| S14_vs_k141_32920||full | Low-quality |
| S14_vs_k141_18451||full | Not-determined |
| S14_vs_k141_113670||full | Low-quality |
| S14_vs_k141_47256||full | Low-quality |
| S14_vs_k141_10853||full | Low-quality |
| S14_vs_k141_106538||full | Low-quality |
| S14_vs_k141_32922||full | Not-determined |
| S14_vs_k141_47259||full | Not-determined |
| S14_vs_k141_84366||full | Low-quality |
| S14_vs_k141_113672||full | Low-quality |
| S14_vs_k141_47260||full | Not-determined |
| S14_vs_k141_61993||full | Low-quality |
| S14_vs_k141_32936||full | Low-quality |
| S14_vs_k141_98571||full | Low-quality |
| S14_vs_k141_98572||full | Low-quality |
| S14_vs_k141_18459||full | Low-quality |
| S14_vs_k141_98573||full | Low-quality |
| S14_vs_k141_47266||full | Low-quality |
| S14_vs_k141_10866||full | Not-determined |
| S14_vs_k141_54998||full | Low-quality |
| S14_vs_k141_55003||full | Low-quality |
| S14_vs_k141_69605||full | Low-quality |
| S14_vs_k141_47282||full | Low-quality |
| S14_vs_k141_40576||full | Low-quality |
| S14_vs_k141_113680||full | Not-determined |
| S14_vs_k141_113681||full | Not-determined |
| S14_vs_k141_91799||full | Not-determined |
| S14_vs_k141_10887||full | Low-quality |
| S14_vs_k141_40586||full | Low-quality |
| S14_vs_k141_106563||full | Low-quality |
| S14_vs_k141_10901||full | Low-quality |
| S14_vs_k141_32970||full | Not-determined |
| S14_vs_k141_25370||full | Low-quality |
| S14_vs_k141_84432||full | Not-determined |
| S14_vs_k141_62034||full | Low-quality |
| S14_vs_k141_84436||full | Low-quality |
| S14_vs_k141_69644||full | Low-quality |
| S14_vs_k141_91818||full | Low-quality |
| S14_vs_k141_25384||full | Low-quality |
| S14_vs_k141_98593||full | Low-quality |
| S14_vs_k141_25385||full | Low-quality |
| S14_vs_k141_98596||full | Low-quality |
| S14_vs_k141_40625||full | Low-quality |
| S14_vs_k141_98597||full | Not-determined |
| S14_vs_k141_69659||full | Not-determined |
| S14_vs_k141_98601||full | Low-quality |
| S14_vs_k141_113733||full | Not-determined |
| S14_vs_k141_62042||full | Low-quality |
| S14_vs_k141_62043||full | Low-quality |
| S14_vs_k141_40643||full | Low-quality |
| S14_vs_k141_10931||full | Low-quality |
| S14_vs_k141_55049||full | Not-determined |
| S14_vs_k141_25409||full | Low-quality |
| S14_vs_k141_98616||full | Low-quality |
| S14_vs_k141_40653||full | Low-quality |
| S14_vs_k141_91847||full | Low-quality |
| S14_vs_k141_106574||full | Low-quality |
| S14_vs_k141_10942||full | Not-determined |
| S14_vs_k141_69695||full | Not-determined |
| S14_vs_k141_40659||full | Low-quality |
| S14_vs_k141_106576||full | Not-determined |
| S14_vs_k141_32997||full | Low-quality |
| S14_vs_k141_98640||full | Not-determined |
| S14_vs_k141_10958||full | Low-quality |
| S14_vs_k141_69698||full | Low-quality |
| S14_vs_k141_113773||full | Low-quality |
| S14_vs_k141_113779||full | Low-quality |
| S14_vs_k141_84504||full | Low-quality |
| S14_vs_k141_25432||full | Low-quality |
| S14_vs_k141_106607||full | Low-quality |
| S14_vs_k141_98666||full | Not-determined |
| S14_vs_k141_62059||full | Low-quality |
| S14_vs_k141_55083||full | Low-quality |
| S14_vs_k141_62066||full | Low-quality |
| S14_vs_k141_84519||full | Not-determined |
| S14_vs_k141_84522||full | Low-quality |
| S14_vs_k141_98677||full | Low-quality |
| S14_vs_k141_62071||full | Low-quality |
| S14_vs_k141_47349||full | Low-quality |
| S14_vs_k141_55098||full | Low-quality |
| S14_vs_k141_18520||full | Low-quality |
| S14_vs_k141_40706||full | Low-quality |
| S17_vs_k141_126212||full | High-quality |
| S14_vs_k141_106626||full | Low-quality |
| S14_vs_k141_47366||full | Low-quality |
| S14_vs_k141_98685||full | Not-determined |
| S14_vs_k141_106635||full | Not-determined |
| S14_vs_k141_33012||full | Low-quality |
| S14_vs_k141_77071||full | Low-quality |
| S14_vs_k141_91858||full | Not-determined |
| S14_vs_k141_10971||full | Low-quality |
| S14_vs_k141_98691||full | Low-quality |
| S14_vs_k141_47388||full | Low-quality |
| S14_vs_k141_40734||full | Low-quality |
| S14_vs_k141_33023||full | Low-quality |
| S14_vs_k141_10983||full | Low-quality |
| S14_vs_k141_47392||full | Not-determined |
| S14_vs_k141_91874||full | Low-quality |
| S14_vs_k141_98701||full | Low-quality |
| S14_vs_k141_18576||full | Not-determined |
| S14_vs_k141_98712||full | Low-quality |
| S14_vs_k141_33050||full | Low-quality |
| S14_vs_k141_47402||full | Not-determined |
| S14_vs_k141_62112||full | Low-quality |
| S14_vs_k141_47403||full | Low-quality |
| S14_vs_k141_98715||full | Low-quality |
| S14_vs_k141_62123||full | Low-quality |
| S14_vs_k141_69755||full | Not-determined |
| S14_vs_k141_3453||full | Not-determined |
| S14_vs_k141_25481||full | Low-quality |
| S14_vs_k141_69758||full | Not-determined |
| S14_vs_k141_98727||full | Not-determined |
| S14_vs_k141_62137||full | Low-quality |
| S14_vs_k141_33071||full | Low-quality |
| S14_vs_k141_25499||full | Low-quality |
| S14_vs_k141_69772||full | Low-quality |
| S14_vs_k141_25505||full | Low-quality |
| S14_vs_k141_33079||full | Not-determined |
| S14_vs_k141_55119||full | Not-determined |
| S14_vs_k141_77105||full | Not-determined |
| S14_vs_k141_69791||full | Low-quality |
| S14_vs_k141_11081||full | Low-quality |
| S14_vs_k141_62159||full | Low-quality |
| S14_vs_k141_40812||full | Not-determined |
| S14_vs_k141_55120||full | Low-quality |
| S14_vs_k141_18634||full | Low-quality |
| S14_vs_k141_18636||full | Not-determined |
| S14_vs_k141_77114||full | Low-quality |
| S14_vs_k141_106691||full | Low-quality |
| S14_vs_k141_62171||full | Low-quality |
| S14_vs_k141_106697||full | Not-determined |
| S14_vs_k141_113872||full | Low-quality |
| S14_vs_k141_106703||full | Low-quality |
| S14_vs_k141_25550||full | Low-quality |
| S14_vs_k141_3519||full | Low-quality |
| S14_vs_k141_84590||full | Low-quality |
| S14_vs_k141_69834||full | Low-quality |
| S14_vs_k141_40870||full | Low-quality |
| S14_vs_k141_106711||full | Not-determined |
| S14_vs_k141_69847||full | Not-determined |
| S14_vs_k141_91904||full | Low-quality |
| S14_vs_k141_113899||full | Low-quality |
| S14_vs_k141_55180||full | Low-quality |
| S14_vs_k141_91917||full | Low-quality |
| S14_vs_k141_84608||full | Low-quality |
| S14_vs_k141_77141||full | Low-quality |
| S14_vs_k141_47461||full | Low-quality |
| S14_vs_k141_69864||full | Not-determined |
| S14_vs_k141_47465||full | Low-quality |
| S14_vs_k141_47466||full | Not-determined |
| S14_vs_k141_113904||full | Low-quality |
| S14_vs_k141_77148||full | Not-determined |
| S11_vs_k141_42927||full | Medium-quality |
| S14_vs_k141_33111||full | Low-quality |
| S14_vs_k141_55197||full | Low-quality |
| S14_vs_k141_69870||full | Not-determined |
| S14_vs_k141_91928||full | Not-determined |
| S14_vs_k141_62181||full | Low-quality |
| S14_vs_k141_55199||full | Not-determined |
| S14_vs_k141_11132||full | Low-quality |
| S14_vs_k141_25600||full | Low-quality |
| S14_vs_k141_55205||full | Low-quality |
| S14_vs_k141_47488||full | Not-determined |
| S14_vs_k141_55210||full | Low-quality |
| S14_vs_k141_33133||full | Not-determined |
| S14_vs_k141_47489||full | Low-quality |
| S14_vs_k141_40918||full | Not-determined |
| S14_vs_k141_113931||full | Low-quality |
| S14_vs_k141_40926||full | Low-quality |
| S14_vs_k141_40927||full | Low-quality |
| S14_vs_k141_11173||full | Low-quality |
| S14_vs_k141_25606||full | Low-quality |
| S14_vs_k141_106752||full | Low-quality |
| S14_vs_k141_91978||full | Low-quality |
| S14_vs_k141_84654||full | Not-determined |
| S14_vs_k141_84655||full | Low-quality |
| S14_vs_k141_62220||full | Low-quality |
| S14_vs_k141_106770||full | Low-quality |
| S14_vs_k141_91990||full | Low-quality |
| S14_vs_k141_84671||full | Low-quality |
| S14_vs_k141_3585||full | Low-quality |
| S14_vs_k141_47549||full | Low-quality |
| S14_vs_k141_113960||full | Not-determined |
| S14_vs_k141_47560||full | Low-quality |
| S14_vs_k141_25651||full | Low-quality |
| S14_vs_k141_113963||full | Low-quality |
| S14_vs_k141_11210||full | Low-quality |
| S14_vs_k141_3598||full | Low-quality |
| S14_vs_k141_40970||full | Low-quality |
| S14_vs_k141_55267||full | Not-determined |
| S14_vs_k141_3604||full | Low-quality |
| S14_vs_k141_62252||full | Low-quality |
| S14_vs_k141_62253||full | Not-determined |
| S14_vs_k141_55275||full | Low-quality |
| S14_vs_k141_18734||full | Not-determined |
| S14_vs_k141_3620||full | Not-determined |
| S14_vs_k141_77247||full | Low-quality |
| S14_vs_k141_18735||full | Low-quality |
| S14_vs_k141_62264||full | Not-determined |
| S14_vs_k141_98849||full | Low-quality |
| S14_vs_k141_77263||full | Not-determined |
| S14_vs_k141_113984||full | Not-determined |
| S14_vs_k141_62268||full | Low-quality |
| S14_vs_k141_11234||full | Low-quality |
| S14_vs_k141_113987||full | Low-quality |
| S14_vs_k141_41014||full | Low-quality |
| S14_vs_k141_41025||full | Low-quality |
| S14_vs_k141_11248||full | Not-determined |
| S14_vs_k141_106819||full | Low-quality |
| S14_vs_k141_62295||full | Low-quality |
| S9_extracted_k141_57277 | Complete |
| S14_vs_k141_106824||full | Low-quality |
| S14_vs_k141_77304||full | Not-determined |
| S14_vs_k141_77305||full | Low-quality |
| S14_vs_k141_114022||full | Not-determined |
| S14_vs_k141_92059||full | Not-determined |
| S14_vs_k141_3698||full | Low-quality |
| S14_vs_k141_18789||full | Not-determined |
| S14_vs_k141_41053||full | Low-quality |
| S14_vs_k141_18793||full | Low-quality |
| S14_vs_k141_77321||full | Low-quality |
| S14_vs_k141_92069||full | Low-quality |
| S14_vs_k141_84722||full | Low-quality |
| S14_vs_k141_3707||full | Low-quality |
| S14_vs_k141_92076||full | Not-determined |
| S14_vs_k141_3713||full | Low-quality |
| S14_vs_k141_77345||full | Low-quality |

| | |
|---|---|
| S14_vs_k141_92091\|\|full | Not-determined |
| S14_vs_k141_98906\|\|full | Low-quality |
| S14_vs_k141_98909\|\|full | Not-determined |
| S14_vs_k141_62305\|\|full | Low-quality |
| S14_vs_k141_106866\|\|full | Low-quality |
| S14_vs_k141_98915\|\|full | Low-quality |
| S14_vs_k141_18833\|\|full | Low-quality |
| S14_vs_k141_25710\|\|full | Low-quality |
| S14_vs_k141_114061\|\|full | Low-quality |
| S14_vs_k141_62314\|\|full | Not-determined |
| S14_vs_k141_25712\|\|full | Low-quality |
| S14_vs_k141_3732\|\|full | Not-determined |
| S14_vs_k141_25713\|\|full | Low-quality |
| S14_vs_k141_92100\|\|full | Low-quality |
| S14_vs_k141_18845\|\|full | Not-determined |
| S14_vs_k141_98929\|\|full | Low-quality |
| S14_vs_k141_98930\|\|full | Low-quality |
| S14_vs_k141_18853\|\|full | Low-quality |
| S14_vs_k141_25724\|\|full | Low-quality |
| S14_vs_k141_98933\|\|full | Not-determined |
| S14_vs_k141_41099\|\|full | Low-quality |
| S14_vs_k141_11324\|\|full | Low-quality |
| S14_vs_k141_25741\|\|full | Not-determined |
| S14_vs_k141_18890\|\|full | Low-quality |
| S14_vs_k141_77384\|\|full | Low-quality |
| S14_vs_k141_55333\|\|full | Low-quality |
| S14_vs_k141_92145\|\|full | Not-determined |
| S14_vs_k141_11365\|\|full | Not-determined |
| S14_vs_k141_55353\|\|full | Low-quality |
| S14_vs_k141_92164\|\|full | Low-quality |
| S14_vs_k141_98944\|\|full | Low-quality |
| S14_vs_k141_106906\|\|full | Low-quality |
| S14_vs_k141_25773\|\|full | Low-quality |
| S14_vs_k141_25775\|\|full | Low-quality |
| S14_vs_k141_106907\|\|full | Low-quality |
| S14_vs_k141_98948\|\|full | Low-quality |
| S14_vs_k141_3776\|\|full | Not-determined |
| S14_vs_k141_33280\|\|full | Not-determined |
| S14_vs_k141_84784\|\|full | Low-quality |
| S14_vs_k141_55376\|\|full | Not-determined |
| S14_vs_k141_18962\|\|full | Not-determined |
| S14_vs_k141_41126\|\|full | Low-quality |
| S14_vs_k141_98965\|\|full | Not-determined |
| S14_vs_k141_92186\|\|full | Low-quality |
| S14_vs_k141_69914\|\|full | Not-determined |
| S14_vs_k141_77439\|\|full | Not-determined |
| S14_vs_k141_69921\|\|full | Low-quality |
| S14_vs_k141_55407\|\|full | Not-determined |
| S14_vs_k141_55410\|\|full | Low-quality |
| S14_vs_k141_77457\|\|full | Low-quality |
| S14_vs_k141_92218\|\|full | Not-determined |
| S14_vs_k141_92222\|\|full | Not-determined |
| S14_vs_k141_11400\|\|full | Low-quality |
| S14_vs_k141_84846\|\|full | Not-determined |
| S14_vs_k141_106971\|\|full | Not-determined |
| S14_vs_k141_98995\|\|full | Low-quality |
| S14_vs_k141_18986\|\|full | Low-quality |
| S14_vs_k141_41134\|\|full | Low-quality |
| S14_vs_k141_3849\|\|full | Low-quality |
| S14_vs_k141_99003\|\|full | Not-determined |
| S14_vs_k141_55472\|\|full | Low-quality |
| S14_vs_k141_77496\|\|full | Low-quality |
| S14_vs_k141_11417\|\|full | Low-quality |
| S14_vs_k141_99022\|\|full | Not-determined |
| S14_vs_k141_62335\|\|full | Low-quality |
| S14_vs_k141_84879\|\|full | Low-quality |
| S14_vs_k141_84883\|\|full | Low-quality |
| S14_vs_k141_77511\|\|full | Low-quality |
| S14_vs_k141_84906\|\|full | Low-quality |
| S14_vs_k141_19041\|\|full | Low-quality |
| S14_vs_k141_99033\|\|full | Low-quality |
| S9_vs_k141_57277\|\|full | High-quality |
| S14_vs_k141_69959\|\|full | Not-determined |
| S14_vs_k141_84912\|\|full | Low-quality |
| S14_vs_k141_55498\|\|full | Not-determined |
| S14_vs_k141_99040\|\|full | Not-determined |
| S14_vs_k141_47615\|\|full | Low-quality |
| S14_vs_k141_41144\|\|full | Low-quality |
| S14_vs_k141_55502\|\|full | Not-determined |
| S14_vs_k141_55503\|\|full | Not-determined |
| S14_vs_k141_92253\|\|full | Not-determined |
| S14_vs_k141_114204\|\|full | Low-quality |
| S14_vs_k141_19064\|\|full | Low-quality |
| S14_vs_k141_99048\|\|full | Not-determined |
| S14_vs_k141_55512\|\|full | Low-quality |
| S14_vs_k141_99050\|\|full | Low-quality |
| S14_vs_k141_69976\|\|full | Low-quality |
| S14_vs_k141_3928\|\|full | Not-determined |
| S14_vs_k141_11455\|\|full | Low-quality |
| S14_vs_k141_11469\|\|full | Low-quality |
| S14_vs_k141_99067\|\|full | Low-quality |
| S14_vs_k141_92270\|\|full | Low-quality |
| S14_vs_k141_92271\|\|full | Low-quality |
| S14_vs_k141_114234\|\|full | Low-quality |
| S14_vs_k141_114235\|\|full | Low-quality |
| S14_vs_k141_84945\|\|full | Low-quality |
| S14_vs_k141_114239\|\|full | Not-determined |
| S14_vs_k141_3952\|\|full | Low-quality |
| S14_vs_k141_11476\|\|full | Low-quality |
| S14_vs_k141_41178\|\|full | Not-determined |
| S14_vs_k141_77569\|\|full | Low-quality |
| S14_vs_k141_19126\|\|full | Low-quality |
| S14_vs_k141_11489\|\|full | Not-determined |
| S14_vs_k141_3971\|\|full | Not-determined |
| S14_vs_k141_11491\|\|full | Not-determined |
| S14_vs_k141_3973\|\|full | Not-determined |
| S14_vs_k141_84998\|\|full | Not-determined |
| S14_vs_k141_99105\|\|full | Low-quality |
| S14_vs_k141_62437\|\|full | Not-determined |
| S14_vs_k141_99108\|\|full | Low-quality |
| S14_vs_k141_70031\|\|full | Not-determined |
| S14_vs_k141_70033\|\|full | Not-determined |
| S14_vs_k141_85002\|\|full | Low-quality |
| S14_vs_k141_92323\|\|full | Low-quality |
| S14_vs_k141_19163\|\|full | Not-determined |
| S14_vs_k141_70044\|\|full | Low-quality |
| S14_vs_k141_85005\|\|full | Not-determined |
| S14_vs_k141_62457\|\|full | Low-quality |
| S14_vs_k141_19175\|\|full | Not-determined |
| S14_vs_k141_70056\|\|full | Low-quality |
| S14_vs_k141_55555\|\|full | Low-quality |
| S14_vs_k141_33472\|\|full | Low-quality |
| S14_vs_k141_62472\|\|full | Low-quality |
| S16_extracted_5270 | Medium-quality |
| S14_vs_k141_62476\|\|full | Low-quality |
| S14_vs_k141_99110\|\|full | Low-quality |
| S14_vs_k141_55569\|\|full | Low-quality |
| S14_vs_k141_41248\|\|full | Low-quality |
| S14_vs_k141_25869\|\|full | Not-determined |
| S14_vs_k141_92342\|\|full | Not-determined |
| S14_vs_k141_4019\|\|full | Low-quality |
| S14_vs_k141_55575\|\|full | Low-quality |
| S14_vs_k141_70088\|\|full | Low-quality |
| S14_vs_k141_19209\|\|full | Low-quality |
| S14_vs_k141_107041\|\|full | Not-determined |
| S14_vs_k141_25877\|\|full | Not-determined |
| S14_vs_k141_92346\|\|full | Low-quality |
| S14_vs_k141_85050\|\|full | Low-quality |
| S14_vs_k141_19221\|\|full | Not-determined |
| S14_vs_k141_55587\|\|full | Low-quality |
| S14_vs_k141_19223\|\|full | Not-determined |
| S14_vs_k141_85054\|\|full | Low-quality |
| S13_vs_113593\|\|full | High-quality |
| S14_vs_k141_70109\|\|full | Low-quality |
| S14_vs_k141_107044\|\|full | Low-quality |
| S14_vs_k141_33510\|\|full | Low-quality |
| S14_vs_k141_41276\|\|full | Low-quality |
| S14_vs_k141_99146\|\|full | Low-quality |
| S14_vs_k141_70118\|\|full | Low-quality |
| S14_vs_k141_77618\|\|full | Low-quality |
| S14_vs_k141_4037\|\|full | Not-determined |
| S14_vs_k141_11522\|\|full | Not-determined |
| S14_vs_k141_4043\|\|full | Not-determined |
| S14_vs_k141_107051\|\|full | Not-determined |
| S14_vs_k141_47745\|\|full | Not-determined |
| S14_vs_k141_4056\|\|full | Low-quality |
| S14_vs_k141_85078\|\|full | Not-determined |
| S14_vs_k141_4065\|\|full | Low-quality |
| S14_vs_k141_107063\|\|full | Low-quality |
| S14_vs_k141_25889\|\|full | Low-quality |
| S14_vs_k141_85083\|\|full | Not-determined |
| S14_vs_k141_77646\|\|full | Low-quality |
| S14_vs_k141_41331\|\|full | Low-quality |
| S14_vs_k141_99173\|\|full | Low-quality |
| S14_vs_k141_70147\|\|full | Low-quality |
| S14_vs_k141_77655\|\|full | Low-quality |
| S14_vs_k141_55636\|\|full | Not-determined |
| S14_vs_k141_85090\|\|full | Low-quality |
| S14_vs_k141_70159\|\|full | Not-determined |
| S14_vs_k141_25903\|\|full | Low-quality |
| S14_vs_k141_47764\|\|full | Low-quality |
| S14_vs_k141_33541\|\|full | Not-determined |
| S14_vs_k141_4090\|\|full | Not-determined |
| S14_vs_k141_77667\|\|full | Low-quality |
| S14_vs_k141_107098\|\|full | Not-determined |
| S14_vs_k141_107107\|\|full | Low-quality |
| S14_vs_k141_85100\|\|full | Low-quality |
| S14_vs_k141_11574\|\|full | Low-quality |
| S14_vs_k141_4097\|\|full | Not-determined |
| S14_vs_k141_33555\|\|full | Low-quality |
| S14_vs_k141_11581\|\|full | Not-determined |
| S14_vs_k141_11583\|\|full | Low-quality |
| S14_vs_k141_107109\|\|full | Low-quality |
| S14_vs_k141_25912\|\|full | Low-quality |
| S14_vs_k141_62570\|\|full | Low-quality |
| S14_vs_k141_4118\|\|full | Low-quality |
| S14_vs_k141_62577\|\|full | Not-determined |
| S14_vs_k141_99205\|\|full | Low-quality |
| S14_vs_k141_25920\|\|full | Low-quality |
| S14_vs_k141_19329\|\|full | Not-determined |
| S14_vs_k141_62586\|\|full | Low-quality |
| S14_vs_k141_41354\|\|full | Not-determined |
| S14_vs_k141_19345\|\|full | Low-quality |
| S14_vs_k141_41356\|\|full | Low-quality |
| S14_vs_k141_11614\|\|full | Low-quality |
| S14_vs_k141_41358\|\|full | Low-quality |
| S14_vs_k141_11616\|\|full | Low-quality |
| S14_vs_k141_99228\|\|full | Low-quality |
| S14_vs_k141_25933\|\|full | Not-determined |
| S14_vs_k141_41367\|\|full | Low-quality |
| S14_vs_k141_47799\|\|full | Low-quality |
| S14_vs_k141_47803\|\|full | Low-quality |
| S14_vs_k141_47808\|\|full | Low-quality |
| S14_vs_k141_70233\|\|full | Low-quality |
| S14_vs_k141_25949\|\|full | Low-quality |
| S14_vs_k141_62625\|\|full | Low-quality |
| S14_vs_k141_41386\|\|full | Low-quality |
| S14_vs_k141_92450\|\|full | Low-quality |
| S16_vs_5270\|\|full | Medium-quality |
| S14_vs_k141_107175\|\|full | Not-determined |
| S14_vs_k141_92455\|\|full | Low-quality |
| S7_vs_135322\|\|full | High-quality |
| S14_vs_k141_19400\|\|full | Low-quality |
| S14_vs_k141_55710\|\|full | Low-quality |
| S14_vs_k141_25974\|\|full | Low-quality |
| S14_vs_k141_92465\|\|full | Not-determined |
| S14_vs_k141_62657\|\|full | Low-quality |
| S14_vs_k141_33606\|\|full | Low-quality |
| S14_vs_k141_92469\|\|full | Low-quality |
| S14_vs_k141_55727\|\|full | Low-quality |
| S14_vs_k141_19424\|\|full | Not-determined |
| S14_vs_k141_55728\|\|full | Low-quality |
| S14_vs_k141_114417\|\|full | Low-quality |
| S14_vs_k141_62676\|\|full | Low-quality |
| S14_vs_k141_99299\|\|full | Low-quality |
| S14_vs_k141_41420\|\|full | Not-determined |
| S14_vs_k141_114422\|\|full | Not-determined |
| S14_vs_k141_25985\|\|full | Not-determined |
| S14_vs_k141_41422\|\|full | Not-determined |
| S14_vs_k141_77769\|\|full | Low-quality |
| S14_vs_k141_33617\|\|full | Not-determined |
| S14_vs_k141_92502\|\|full | Low-quality |
| S14_vs_k141_99322\|\|full | Low-quality |
| S14_vs_k141_85162\|\|full | Low-quality |
| S14_vs_k141_55748\|\|full | Low-quality |
| S14_vs_k141_99328\|\|full | Low-quality |
| S14_vs_k141_19465\|\|full | Not-determined |
| S14_vs_k141_114445\|\|full | Low-quality |
| S14_vs_k141_41437\|\|full | Not-determined |
| S14_vs_k141_77798\|\|full | Low-quality |
| S14_vs_k141_41439\|\|full | Low-quality |
| S14_vs_k141_92517\|\|full | Not-determined |
| S14_vs_k141_77800\|\|full | Low-quality |
| S14_vs_k141_77812\|\|full | Low-quality |
| S14_vs_k141_62709\|\|full | Low-quality |
| S14_vs_k141_33640\|\|full | Low-quality |
| S14_vs_k141_107244\|\|full | Low-quality |
| S14_vs_k141_85172\|\|full | Low-quality |
| S14_vs_k141_107252\|\|full | Low-quality |
| S14_vs_k141_70246\|\|full | Low-quality |
| S3_extracted_k141_2186 | Medium-quality |
| S14_vs_k141_77836\|\|full | Low-quality |
| S14_vs_k141_4143\|\|full | Low-quality |
| S14_vs_k141_62734\|\|full | Not-determined |
| S14_vs_k141_107261\|\|full | Low-quality |
| S14_vs_k141_41484\|\|full | Low-quality |
| S14_vs_k141_26041\|\|full | Low-quality |
| S14_vs_k141_41488\|\|full | Low-quality |
| S14_vs_k141_19517\|\|full | Not-determined |
| S14_vs_k141_70272\|\|full | Not-determined |
| S14_vs_k141_26050\|\|full | Low-quality |
| S14_vs_k141_41499\|\|full | Low-quality |
| S14_vs_k141_55796\|\|full | Low-quality |
| S14_vs_k141_107274\|\|full | Low-quality |
| S14_vs_k141_41509\|\|full | Low-quality |
| S14_vs_k141_70279\|\|full | Low-quality |
| S14_vs_k141_62743\|\|full | Not-determined |
| S7_vs_21521\|\|full | Medium-quality |
| S14_vs_k141_77877\|\|full | Low-quality |
| S14_vs_k141_62755\|\|full | Low-quality |
| S14_vs_k141_77883\|\|full | Low-quality |
| S14_vs_k141_41522\|\|full | Low-quality |
| S14_vs_k141_114498\|\|full | Low-quality |
| S14_vs_k141_114499\|\|full | Not-determined |
| S14_vs_k141_19552\|\|full | Not-determined |
| S14_vs_k141_4191\|\|full | Not-determined |
| S14_vs_k141_107305\|\|full | Low-quality |
| S14_vs_k141_62771\|\|full | Not-determined |
| S14_vs_k141_55833\|\|full | Low-quality |
| S14_vs_k141_99411\|\|full | Not-determined |
| S14_vs_k141_92583\|\|full | Low-quality |
| S14_vs_k141_55848\|\|full | Low-quality |
| S14_vs_k141_92591\|\|full | Low-quality |
| S14_vs_k141_26099\|\|full | Low-quality |
| S14_vs_k141_92593\|\|full | Not-determined |
| S14_vs_k141_114537\|\|full | Low-quality |
| S14_vs_k141_4237\|\|full | Not-determined |
| S14_vs_k141_99430\|\|full | Low-quality |
| S14_vs_k141_70354\|\|full | Not-determined |
| S14_vs_k141_85269\|\|full | Not-determined |
| S14_vs_k141_4253\|\|full | Not-determined |
| S14_vs_k141_85272\|\|full | Low-quality |
| S14_vs_k141_114544\|\|full | Low-quality |
| S14_vs_k141_114549\|\|full | Low-quality |
| S14_vs_k141_70375\|\|full | Low-quality |
| S14_vs_k141_77935\|\|full | Low-quality |
| S14_vs_k141_114560\|\|full | Not-determined |
| S14_vs_k141_55884\|\|full | Not-determined |
| S14_vs_k141_62811\|\|full | Low-quality |
| S14_vs_k141_114568\|\|full | Low-quality |
| S14_vs_k141_92662\|\|full | Low-quality |
| S14_vs_k141_4266\|\|full | Not-determined |
| S14_vs_k141_114582\|\|full | Low-quality |
| S14_vs_k141_99467\|\|full | Low-quality |
| S14_vs_k141_77965\|\|full | Low-quality |
| S14_vs_k141_62840\|\|full | Not-determined |
| S14_vs_k141_19598\|\|full | Not-determined |
| S14_vs_k141_19599\|\|full | Low-quality |
| S14_vs_k141_114602\|\|full | Low-quality |
| S14_vs_k141_107398\|\|full | Not-determined |
| S14_vs_k141_47902\|\|full | Low-quality |
| S14_vs_k141_4291\|\|full | Not-determined |
| S14_vs_k141_55896\|\|full | Low-quality |
| S14_vs_k141_62866\|\|full | Low-quality |
| S14_vs_k141_99476\|\|full | Low-quality |
| S14_vs_k141_62872\|\|full | Low-quality |
| S14_vs_k141_99477\|\|full | Low-quality |
| S14_vs_k141_99489\|\|full | Not-determined |
| S14_vs_k141_41576\|\|full | Low-quality |
| S14_vs_k141_107416\|\|full | Low-quality |
| S14_vs_k141_77985\|\|full | Not-determined |
| S14_vs_k141_33682\|\|full | Not-determined |
| S14_vs_k141_11660\|\|full | Not-determined |
| S14_vs_k141_47931\|\|full | Low-quality |
| S14_vs_k141_11662\|\|full | Low-quality |
| S14_vs_k141_70455\|\|full | Not-determined |
| S14_vs_k141_99507\|\|full | Low-quality |
| S14_vs_k141_4310\|\|full | Low-quality |
| S14_vs_k141_55935\|\|full | Not-determined |
| S12_vs_k141_182611\|\|full | High-quality |
| S14_vs_k141_99519\|\|full | Low-quality |
| S14_vs_k141_70484\|\|full | Low-quality |
| S14_vs_k141_41599\|\|full | Low-quality |
| S14_vs_k141_114648\|\|full | Not-determined |
| S14_vs_k141_4332\|\|full | Low-quality |
| S14_vs_k141_70495\|\|full | Low-quality |
| S14_vs_k141_55950\|\|full | Not-determined |
| S14_vs_k141_33707\|\|full | Not-determined |
| S14_vs_k141_41620\|\|full | Low-quality |
| S14_vs_k141_33709\|\|full | Low-quality |
| S14_vs_k141_47950\|\|full | Not-determined |
| S14_vs_k141_92752\|\|full | Low-quality |
| S14_vs_k141_47951\|\|full | Low-quality |
| S14_vs_k141_4342\|\|full | Not-determined |
| S14_vs_k141_33712\|\|full | Low-quality |
| S14_vs_k141_62901\|\|full | Low-quality |
| S14_vs_k141_55952\|\|full | Low-quality |
| S14_vs_k141_4344\|\|full | Low-quality |
| S14_vs_k141_41626\|\|full | Low-quality |
| S14_vs_k141_114657\|\|full | Not-determined |
| S14_vs_k141_114660\|\|full | Low-quality |
| S14_vs_k141_33714\|\|full | Low-quality |
| S14_vs_k141_4365\|\|full | Low-quality |
| S14_vs_k141_92769\|\|full | Low-quality |
| S14_vs_k141_33717\|\|full | Low-quality |
| S14_vs_k141_4368\|\|full | Low-quality |
| S14_vs_k141_33719\|\|full | Not-determined |
| S14_vs_k141_47957\|\|full | Low-quality |
| S14_vs_k141_33720\|\|full | Low-quality |
| S14_vs_k141_92775\|\|full | Low-quality |
| S14_vs_k141_70518\|\|full | Not-determined |
| S14_vs_k141_107471\|\|full | Low-quality |
| S14_vs_k141_107472\|\|full | Not-determined |
| S14_vs_k141_33741\|\|full | Low-quality |
| S14_vs_k141_85373\|\|full | Not-determined |
| S14_vs_k141_92796\|\|full | Not-determined |
| S14_vs_k141_92797\|\|full | Not-determined |
| S14_vs_k141_78036\|\|full | Low-quality |
| S14_vs_k141_41663\|\|full | Low-quality |
| S14_vs_k141_62948\|\|full | Low-quality |
| S14_vs_k141_85392\|\|full | Low-quality |
| S14_vs_k141_47990\|\|full | Low-quality |
| S14_vs_k141_47996\|\|full | Low-quality |
| S14_vs_k141_41689\|\|full | Low-quality |
| S14_vs_k141_70559\|\|full | Low-quality |
| S14_vs_k141_70566\|\|full | Low-quality |
| S14_vs_k141_55971\|\|full | Low-quality |
| S14_vs_k141_92821\|\|full | Not-determined |
| S14_vs_k141_26180\|\|full | Not-determined |
| S14_vs_k141_19688\|\|full | Low-quality |
| S14_vs_k141_48009\|\|full | Low-quality |
| S14_vs_k141_107493\|\|full | Low-quality |
| S14_vs_k141_19691\|\|full | Low-quality |
| S14_vs_k141_70579\|\|full | Not-determined |
| S14_vs_k141_26194\|\|full | Low-quality |
| S14_vs_k141_78078\|\|full | Not-determined |
| S14_vs_k141_62978\|\|full | Low-quality |
| S14_vs_k141_4476\|\|full | Low-quality |
| S14_vs_k141_26199\|\|full | Low-quality |
| S14_vs_k141_99593\|\|full | Low-quality |
| S14_vs_k141_107498\|\|full | Low-quality |
| S14_vs_k141_41705\|\|full | Low-quality |
| S14_vs_k141_99599\|\|full | Low-quality |
| S14_vs_k141_107503\|\|full | Low-quality |
| S14_vs_k141_26208\|\|full | Not-determined |
| S14_vs_k141_33797\|\|full | Low-quality |
| S14_vs_k141_19724\|\|full | Not-determined |
| S14_vs_k141_78100\|\|full | Low-quality |
| S14_vs_k141_11711\|\|full | Low-quality |
| S14_vs_k141_19736\|\|full | Low-quality |
| S14_vs_k141_85422\|\|full | Low-quality |
| S14_vs_k141_114748\|\|full | Low-quality |
| S14_vs_k141_78107\|\|full | Low-quality |
| S14_vs_k141_85424\|\|full | Not-determined |
| S14_vs_k141_85425\|\|full | Low-quality |
| S14_vs_k141_70632\|\|full | Low-quality |
| S14_vs_k141_4504\|\|full | Low-quality |
| S14_vs_k141_99614\|\|full | Low-quality |
| S14_vs_k141_26228\|\|full | Low-quality |
| S14_vs_k141_11724\|\|full | Low-quality |
| S14_vs_k141_41727\|\|full | Low-quality |
| S14_vs_k141_55994\|\|full | Not-determined |
| S14_vs_k141_41729\|\|full | Not-determined |
| S14_vs_k141_26238\|\|full | Low-quality |
| S14_vs_k141_55996\|\|full | Low-quality |
| S14_vs_k141_26249\|\|full | Low-quality |
| S14_vs_k141_107540\|\|full | Not-determined |
| S14_vs_k141_41743\|\|full | Not-determined |
| S14_vs_k141_62996\|\|full | Low-quality |
| S14_vs_k141_56015\|\|full | Low-quality |
| S14_vs_k141_48012\|\|full | Low-quality |
| S14_vs_k141_33851\|\|full | Low-quality |
| S14_vs_k141_48013\|\|full | Not-determined |
| S14_vs_k141_26258\|\|full | Low-quality |
| S14_vs_k141_107544\|\|full | Not-determined |
| S14_vs_k141_107552\|\|full | Not-determined |
| S14_vs_k141_19784\|\|full | Low-quality |
| S14_vs_k141_56044\|\|full | Low-quality |
| S14_vs_k141_107562\|\|full | Low-quality |
| S14_vs_k141_19787\|\|full | Not-determined |
| S14_vs_k141_99632\|\|full | Low-quality |
| S14_vs_k141_26283\|\|full | Not-determined |
| S14_vs_k141_107564\|\|full | Not-determined |
| S14_vs_k141_107565\|\|full | Low-quality |
| S14_vs_k141_107567\|\|full | Low-quality |
| S14_vs_k141_70684\|\|full | Low-quality |
| S14_vs_k141_114781\|\|full | Low-quality |
| S14_vs_k141_114783\|\|full | Low-quality |
| S14_vs_k141_11773\|\|full | Low-quality |
| S14_vs_k141_107568\|\|full | Low-quality |
| S14_vs_k141_107573\|\|full | Not-determined |
| S14_vs_k141_63061\|\|full | Not-determined |
| S14_vs_k141_11784\|\|full | Low-quality |
| S14_vs_k141_70742\|\|full | Not-determined |
| S14_vs_k141_11792\|\|full | Low-quality |
| S14_vs_k141_107595\|\|full | Not-determined |
| S14_vs_k141_114806\|\|full | Low-quality |
| S14_vs_k141_70763\|\|full | Low-quality |
| S14_vs_k141_107607\|\|full | Low-quality |
| S14_vs_k141_107611\|\|full | Not-determined |
| S14_vs_k141_41819\|\|full | Not-determined |
| S14_vs_k141_107617\|\|full | Not-determined |
| S14_vs_k141_41831\|\|full | Not-determined |
| S14_vs_k141_41833\|\|full | Not-determined |
| S12_vs_k141_187941\|\|full | Medium-quality |
| S14_vs_k141_85493\|\|full | Low-quality |
| S14_vs_k141_85495\|\|full | Low-quality |
| S14_vs_k141_41842\|\|full | Low-quality |
| S14_vs_k141_4545\|\|full | Low-quality |
| S14_vs_k141_85509\|\|full | Not-determined |
| S14_vs_k141_70787\|\|full | Low-quality |
| S14_vs_k141_48104\|\|full | Low-quality |
| S14_vs_k141_11809\|\|full | Low-quality |
| S14_vs_k141_48117\|\|full | Low-quality |
| S14_vs_k141_70801\|\|full | Not-determined |
| S14_vs_k141_11819\|\|full | Low-quality |
| S14_vs_k141_107669\|\|full | Not-determined |
| S14_vs_k141_70810\|\|full | Not-determined |
| S14_vs_k141_11828\|\|full | Low-quality |
| S14_vs_k141_41869\|\|full | Not-determined |
| S14_vs_k141_114886\|\|full | Not-determined |
| S14_vs_k141_63150\|\|full | Low-quality |
| S14_vs_k141_92918\|\|full | Low-quality |
| S14_vs_k141_19879\|\|full | Low-quality |
| S14_vs_k141_33985\|\|full | Low-quality |
| S14_vs_k141_41877\|\|full | Low-quality |
| S14_vs_k141_114893\|\|full | Low-quality |
| S14_vs_k141_33991\|\|full | Low-quality |
| S14_vs_k141_19884\|\|full | Low-quality |
| S14_vs_k141_78159\|\|full | Not-determined |
| S14_vs_k141_19888\|\|full | Low-quality |
| S14_vs_k141_56112\|\|full | Low-quality |
| S14_vs_k141_85558\|\|full | Not-determined |
| S14_vs_k141_34001\|\|full | Not-determined |
| S14_vs_k141_56125\|\|full | Low-quality |
| S14_vs_k141_70859\|\|full | Low-quality |
| S14_vs_k141_63168\|\|full | Low-quality |
| S14_vs_k141_63172\|\|full | Low-quality |
| S14_vs_k141_92971\|\|full | Not-determined |
| S14_vs_k141_41888\|\|full | Not-determined |
| S14_vs_k141_70870\|\|full | Low-quality |
| S14_vs_k141_85595\|\|full | Low-quality |
| S14_vs_k141_4603\|\|full | Not-determined |
| S14_vs_k141_4604\|\|full | Not-determined |
| S14_vs_k141_4607\|\|full | Not-determined |
| S14_vs_k141_4608\|\|full | Low-quality |
| S14_vs_k141_63198\|\|full | Low-quality |
| S14_vs_k141_99725\|\|full | Low-quality |
| S3_vs_k141_2186\|\|full | Medium-quality |
| S14_vs_k141_85617\|\|full | Not-determined |
| S14_vs_k141_48149\|\|full | Low-quality |
| S16_vs_32764\|\|full | Medium-quality |
| S14_vs_k141_34015\|\|full | Not-determined |
| S14_vs_k141_19952\|\|full | Not-determined |
| S14_vs_k141_11900\|\|full | Low-quality |
| S14_vs_k141_4629\|\|full | Low-quality |
| S14_vs_k141_11901\|\|full | Low-quality |
| S14_vs_k141_19955\|\|full | Low-quality |
| S14_vs_k141_93053\|\|full | Not-determined |
| S8_vs_k141_147697\|\|full | Medium-quality |
| S14_vs_k141_41910\|\|full | Not-determined |
| S14_vs_k141_19962\|\|full | Low-quality |
| S14_vs_k141_93057\|\|full | Low-quality |
| S14_vs_k141_99756\|\|full | Low-quality |
| S14_vs_k141_19974\|\|full | Not-determined |
| S14_vs_k141_114920\|\|full | Low-quality |
| S14_vs_k141_41916\|\|full | Low-quality |
| S14_vs_k141_48196\|\|full | Not-determined |
| S14_vs_k141_4674\|\|full | Low-quality |
| S14_vs_k141_78178\|\|full | Low-quality |
| S14_vs_k141_34036\|\|full | Low-quality |
| S14_vs_k141_70975\|\|full | Low-quality |
| S14_vs_k141_11947\|\|full | Not-determined |
| S14_vs_k141_34039\|\|full | Low-quality |
| S14_vs_k141_41951\|\|full | Low-quality |
| S14_vs_k141_4706\|\|full | Low-quality |
| S14_vs_k141_85640\|\|full | Low-quality |
| S14_vs_k141_93104\|\|full | Low-quality |
| S14_vs_k141_85648\|\|full | Low-quality |
| S14_vs_k141_48232\|\|full | Low-quality |
| S14_vs_k141_78215\|\|full | Not-determined |
| S14_vs_k141_114951\|\|full | Not-determined |
| S14_vs_k141_26346\|\|full | Low-quality |
| S14_vs_k141_78218\|\|full | Not-determined |
| S14_vs_k141_26348\|\|full | Not-determined |
| S14_vs_k141_56138\|\|full | Low-quality |
| S14_vs_k141_26351\|\|full | Not-determined |
| S14_vs_k141_4745\|\|full | Not-determined |
| S14_vs_k141_41984\|\|full | Not-determined |
| S11_extracted_k141_26512 | Medium-quality |
| S14_vs_k141_11983\|\|full | Low-quality |
| S14_vs_k141_41990\|\|full | Low-quality |
| S14_vs_k141_34073\|\|full | Not-determined |
| S14_vs_k141_99812\|\|full | Low-quality |
| S14_vs_k141_26380\|\|full | Low-quality |
| S14_vs_k141_85694\|\|full | Low-quality |
| S14_vs_k141_34078\|\|full | Not-determined |
| S14_vs_k141_56176\|\|full | Not-determined |
| S14_vs_k141_4769\|\|full | Low-quality |
| S14_vs_k141_42008\|\|full | Not-determined |
| S14_vs_k141_48278\|\|full | Low-quality |
| S14_vs_k141_48283\|\|full | Low-quality |
| S14_vs_k141_4775\|\|full | Low-quality |
| S14_vs_k141_85726\|\|full | Low-quality |
| S14_vs_k141_56191\|\|full | Not-determined |
| S14_vs_k141_85727\|\|full | Low-quality |
| S14_vs_k141_99849\|\|full | Not-determined |
| S14_vs_k141_34111\|\|full | Low-quality |
| S14_vs_k141_12004\|\|full | Not-determined |
| S14_vs_k141_85740\|\|full | Low-quality |
| S14_vs_k141_56199\|\|full | Low-quality |
| S14_vs_k141_34117\|\|full | Not-determined |
| S14_vs_k141_85743\|\|full | Low-quality |
| S14_vs_k141_56203\|\|full | Low-quality |
| S14_vs_k141_26432\|\|full | Low-quality |
| S14_vs_k141_93170\|\|full | Not-determined |
| S14_vs_k141_56206\|\|full | Not-determined |
| S14_vs_k141_85746\|\|full | Low-quality |
| S14_vs_k141_4798\|\|full | Low-quality |
| S14_vs_k141_26438\|\|full | Low-quality |
| S14_vs_k141_56212\|\|full | Low-quality |
| S14_vs_k141_42052\|\|full | Not-determined |
| S14_vs_k141_4799\|\|full | Not-determined |
| S14_vs_k141_42054\|\|full | Low-quality |
| S14_vs_k141_63281\|\|full | Low-quality |
| S14_vs_k141_4814\|\|full | Low-quality |
| S14_vs_k141_63283\|\|full | Low-quality |
| S14_vs_k141_56223\|\|full | Not-determined |
| S14_vs_k141_42064\|\|full | Low-quality |
| S14_vs_k141_4818\|\|full | Low-quality |
| S14_vs_k141_85759\|\|full | Not-determined |
| S14_vs_k141_63297\|\|full | Low-quality |
| S14_vs_k141_93204\|\|full | Not-determined |
| S14_vs_k141_93211\|\|full | Not-determined |
| S14_vs_k141_99908\|\|full | Not-determined |
| S14_vs_k141_107829\|\|full | Not-determined |
| S14_vs_k141_107832\|\|full | Low-quality |
| S14_vs_k141_4847\|\|full | Not-determined |
| S14_vs_k141_78282\|\|full | Not-determined |

| | |
|---|---|
| S14_vs_k141_34167\|\|full | Low-quality |
| S14_vs_k141_12045\|\|full | Low-quality |
| S14_vs_k141_63310\|\|full | Low-quality |
| S14_vs_k141_78291\|\|full | Low-quality |
| S14_vs_k141_63311\|\|full | Low-quality |
| S14_vs_k141_78296\|\|full | Not-determined |
| S14_vs_k141_4873\|\|full | Low-quality |
| S14_vs_k141_93236\|\|full | Low-quality |
| S14_vs_k141_93237\|\|full | Not-determined |
| S14_vs_k141_115030\|\|full | Low-quality |
| S14_vs_k141_4881\|\|full | Low-quality |
| S14_vs_k141_93248\|\|full | Low-quality |
| S14_vs_k141_48346\|\|full | Low-quality |
| S14_vs_k141_99933\|\|full | Low-quality |
| S14_vs_k141_12087\|\|full | Not-determined |
| S14_vs_k141_63340\|\|full | Low-quality |
| S14_vs_k141_12090\|\|full | Not-determined |
| S14_vs_k141_107874\|\|full | Low-quality |
| S14_vs_k141_115049\|\|full | Low-quality |
| S14_vs_k141_99951\|\|full | Not-determined |
| S14_vs_k141_115050\|\|full | Low-quality |
| S14_vs_k141_78337\|\|full | Low-quality |
| S14_vs_k141_107878\|\|full | Not-determined |
| S14_vs_k141_99954\|\|full | Low-quality |
| S14_vs_k141_93273\|\|full | Low-quality |
| S14_vs_k141_63361\|\|full | Low-quality |
| S11_vs_k141_26512\|\|full | Medium-quality |
| S14_vs_k141_115062\|\|full | Not-determined |
| S14_vs_k141_78356\|\|full | Low-quality |
| S14_vs_k141_115071\|\|full | Low-quality |
| S14_vs_k141_56261\|\|full | Not-determined |
| S14_vs_k141_99966\|\|full | Not-determined |
| S14_vs_k141_99968\|\|full | Not-determined |
| S14_vs_k141_12113\|\|full | Low-quality |
| S14_vs_k141_115075\|\|full | Not-determined |
| S14_vs_k141_42127\|\|full | Low-quality |
| S3_extracted_k141_127392 | Complete |
| S14_vs_k141_48351\|\|full | Low-quality |
| S14_vs_k141_78370\|\|full | Low-quality |
| S14_vs_k141_34208\|\|full | Low-quality |
| S14_vs_k141_93284\|\|full | Not-determined |
| S14_vs_k141_115093\|\|full | Not-determined |
| S14_vs_k141_56287\|\|full | Low-quality |
| S14_vs_k141_63402\|\|full | Not-determined |
| S14_vs_k141_4897\|\|full | Low-quality |
| S9_vs_k141_48424\|\|full | High-quality |
| S14_vs_k141_78386\|\|full | Low-quality |
| S14_vs_k141_71000\|\|full | Not-determined |
| S14_vs_k141_78390\|\|full | Low-quality |
| S14_vs_k141_115120\|\|full | Not-determined |
| S14_vs_k141_34235\|\|full | Low-quality |
| S14_vs_k141_48389\|\|full | Low-quality |
| S14_vs_k141_115122\|\|full | Not-determined |
| S14_vs_k141_4912\|\|full | Low-quality |
| S14_vs_k141_12147\|\|full | Low-quality |
| S14_vs_k141_93323\|\|full | Low-quality |
| S14_vs_k141_100017\|\|full | Low-quality |
| S14_vs_k141_115129\|\|full | Low-quality |
| S14_vs_k141_48402\|\|full | Low-quality |
| S14_vs_k141_34243\|\|full | Low-quality |
| S14_vs_k141_34248\|\|full | Low-quality |
| S14_vs_k141_78410\|\|full | Low-quality |
| S14_vs_k141_56348\|\|full | Low-quality |
| S14_vs_k141_93334\|\|full | Low-quality |
| S14_vs_k141_63439\|\|full | Low-quality |
| S14_vs_k141_56362\|\|full | Low-quality |
| S14_vs_k141_42195\|\|full | Not-determined |
| S14_vs_k141_26471\|\|full | Low-quality |
| S14_vs_k141_100042\|\|full | Low-quality |
| S14_vs_k141_63462\|\|full | Low-quality |
| S14_vs_k141_78435\|\|full | Low-quality |
| S14_vs_k141_93354\|\|full | Not-determined |
| S14_vs_k141_48416\|\|full | Not-determined |
| S14_vs_k141_71043\|\|full | Low-quality |
| S14_vs_k141_63482\|\|full | Low-quality |
| S14_vs_k141_100065\|\|full | Low-quality |
| S14_vs_k141_12167\|\|full | Not-determined |
| S14_vs_k141_71051\|\|full | Not-determined |
| S14_vs_k141_26487\|\|full | Low-quality |
| S14_vs_k141_78451\|\|full | Not-determined |
| S14_vs_k141_56385\|\|full | Not-determined |
| S14_vs_k141_34300\|\|full | Not-determined |
| S14_vs_k141_100079\|\|full | Low-quality |
| S14_vs_k141_26497\|\|full | Low-quality |
| S14_vs_k141_26507\|\|full | Low-quality |
| S14_vs_k141_78467\|\|full | Low-quality |
| S14_vs_k141_12187\|\|full | Low-quality |
| S14_vs_k141_48440\|\|full | Low-quality |
| S14_vs_k141_34333\|\|full | Not-determined |
| S14_vs_k141_115195\|\|full | Low-quality |
| S14_vs_k141_56398\|\|full | Low-quality |
| S14_vs_k141_26518\|\|full | Low-quality |
| S14_vs_k141_63500\|\|full | Low-quality |
| S14_vs_k141_107992\|\|full | Low-quality |
| S14_vs_k141_48447\|\|full | Low-quality |
| S14_vs_k141_56407\|\|full | Not-determined |
| S14_vs_k141_108002\|\|full | Low-quality |
| S14_vs_k141_100085\|\|full | Low-quality |
| S14_vs_k141_115202\|\|full | Not-determined |
| S14_vs_k141_42285\|\|full | Not-determined |
| S14_vs_k141_42286\|\|full | Low-quality |
| S14_vs_k141_78486\|\|full | Not-determined |
| S14_vs_k141_115203\|\|full | Low-quality |
| S14_vs_k141_26522\|\|full | Not-determined |
| S14_vs_k141_78492\|\|full | Low-quality |
| S14_vs_k141_100095\|\|full | Low-quality |
| S14_vs_k141_63514\|\|full | Not-determined |
| S14_vs_k141_26537\|\|full | Low-quality |
| S14_vs_k141_63521\|\|full | Low-quality |
| S3_vs_k141_127392\|\|full | High-quality |
| S14_vs_k141_34382\|\|full | Low-quality |
| S14_vs_k141_78499\|\|full | Low-quality |
| S14_vs_k141_78500\|\|full | Not-determined |
| S14_vs_k141_42301\|\|full | Low-quality |
| S14_vs_k141_12208\|\|full | Low-quality |
| S14_vs_k141_78501\|\|full | Low-quality |
| S14_vs_k141_85778\|\|full | Low-quality |
| S14_vs_k141_100121\|\|full | Low-quality |
| S14_vs_k141_78515\|\|full | Low-quality |
| S14_vs_k141_12212\|\|full | Low-quality |
| S14_vs_k141_71087\|\|full | Low-quality |
| S14_vs_k141_34417\|\|full | Low-quality |
| S14_vs_k141_26547\|\|full | Not-determined |
| S14_vs_k141_56427\|\|full | Low-quality |
| S14_vs_k141_34423\|\|full | Not-determined |
| S14_vs_k141_108029\|\|full | Not-determined |
| S14_vs_k141_5031\|\|full | Low-quality |
| S14_vs_k141_12216\|\|full | Not-determined |
| S14_vs_k141_12218\|\|full | Low-quality |
| S14_vs_k141_100154\|\|full | Low-quality |
| S14_vs_k141_108048\|\|full | Low-quality |
| S14_vs_k141_100157\|\|full | Low-quality |
| S14_vs_k141_12224\|\|full | Low-quality |
| S14_vs_k141_42353\|\|full | Low-quality |
| S14_vs_k141_115240\|\|full | Not-determined |
| S14_vs_k141_108054\|\|full | Not-determined |
| S14_vs_k141_12236\|\|full | Low-quality |
| S14_vs_k141_71098\|\|full | Not-determined |
| S14_vs_k141_63578\|\|full | Not-determined |
| S14_vs_k141_108062\|\|full | Low-quality |
| S14_vs_k141_63588\|\|full | Low-quality |
| S14_vs_k141_85835\|\|full | Not-determined |
| S14_vs_k141_5076\|\|full | Not-determined |
| S14_vs_k141_42386\|\|full | Not-determined |
| S14_vs_k141_5085\|\|full | Not-determined |
| S14_vs_k141_85840\|\|full | Low-quality |
| S14_vs_k141_12261\|\|full | Not-determined |
| S14_vs_k141_12266\|\|full | Low-quality |
| S14_vs_k141_12273\|\|full | Low-quality |
| S14_vs_k141_78588\|\|full | Low-quality |
| S14_vs_k141_78589\|\|full | Not-determined |
| S14_vs_k141_85849\|\|full | Low-quality |
| S14_vs_k141_108072\|\|full | Low-quality |
| S14_vs_k141_26585\|\|full | Not-determined |
| S14_vs_k141_85852\|\|full | Low-quality |
| S8_extracted_k141_87755 | Medium-quality |
| S14_vs_k141_100221\|\|full | Low-quality |
| S14_vs_k141_26609\|\|full | Not-determined |
| S14_vs_k141_115289\|\|full | Low-quality |
| S14_vs_k141_42414\|\|full | Low-quality |
| S14_vs_k141_108080\|\|full | Not-determined |
| S14_vs_k141_85876\|\|full | Low-quality |
| S14_vs_k141_26617\|\|full | Low-quality |
| S14_vs_k141_42435\|\|full | Low-quality |
| S14_vs_k141_26626\|\|full | Low-quality |
| S14_vs_k141_56474\|\|full | Low-quality |
| S14_vs_k141_85880\|\|full | Low-quality |
| S14_vs_k141_48559\|\|full | Low-quality |
| S14_vs_k141_63637\|\|full | Low-quality |
| S14_vs_k141_48569\|\|full | Low-quality |
| S8_vs_k141_87755\|\|full | Medium-quality |
| S14_vs_k141_108098\|\|full | Low-quality |
| S14_vs_k141_48578\|\|full | Low-quality |
| S14_vs_k141_115329\|\|full | Low-quality |
| S14_vs_k141_85902\|\|full | Not-determined |
| S14_vs_k141_100233\|\|full | Low-quality |
| S14_vs_k141_85911\|\|full | Low-quality |
| S14_vs_k141_42462\|\|full | Low-quality |
| S14_vs_k141_26673\|\|full | Low-quality |
| S14_vs_k141_20066\|\|full | Low-quality |
| S14_vs_k141_26679\|\|full | Low-quality |
| S14_vs_k141_26680\|\|full | Low-quality |
| S14_vs_k141_115363\|\|full | Not-determined |
| S14_vs_k141_108135\|\|full | Low-quality |
| S14_vs_k141_115370\|\|full | Not-determined |
| S14_vs_k141_42466\|\|full | Not-determined |
| S14_vs_k141_85942\|\|full | Low-quality |
| S14_vs_k141_26706\|\|full | Low-quality |
| S14_vs_k141_71229\|\|full | Low-quality |
| S14_vs_k141_85948\|\|full | Not-determined |
| S14_vs_k141_85950\|\|full | Not-determined |
| S14_vs_k141_71242\|\|full | Low-quality |
| S14_vs_k141_100254\|\|full | Low-quality |
| S14_vs_k141_108142\|\|full | Not-determined |
| S14_vs_k141_42489\|\|full | Not-determined |
| S14_vs_k141_48634\|\|full | Low-quality |
| S14_vs_k141_108145\|\|full | Not-determined |
| S14_vs_k141_71252\|\|full | Low-quality |
| S14_vs_k141_100260\|\|full | Low-quality |
| S14_vs_k141_20092\|\|full | Low-quality |
| S14_vs_k141_56520\|\|full | Low-quality |
| S14_vs_k141_20097\|\|full | Low-quality |
| S14_vs_k141_5091\|\|full | Low-quality |
| S14_vs_k141_93490\|\|full | Low-quality |
| S14_vs_k141_12383\|\|full | Low-quality |
| S14_vs_k141_63726\|\|full | Not-determined |
| S14_vs_k141_108159\|\|full | Not-determined |
| S5_vs_k141_138980\|\|full | High-quality |
| S14_vs_k141_48657\|\|full | Low-quality |
| S14_vs_k141_85977\|\|full | Low-quality |
| S14_vs_k141_12387\|\|full | Low-quality |
| S14_vs_k141_26734\|\|full | Low-quality |
| S14_vs_k141_63729\|\|full | Low-quality |
| S14_vs_k141_34548\|\|full | Not-determined |
| S14_vs_k141_20113\|\|full | Low-quality |
| S14_vs_k141_20114\|\|full | Low-quality |
| S14_vs_k141_71287\|\|full | Low-quality |
| S14_vs_k141_12397\|\|full | Low-quality |
| S14_vs_k141_115387\|\|full | Low-quality |
| S14_vs_k141_63741\|\|full | Not-determined |
| S14_vs_k141_48688\|\|full | Low-quality |
| S14_vs_k141_100305\|\|full | Low-quality |
| S14_vs_k141_5101\|\|full | Low-quality |
| S14_vs_k141_34568\|\|full | Not-determined |
| S14_vs_k141_34578\|\|full | Low-quality |
| S14_vs_k141_42558\|\|full | Low-quality |
| S14_vs_k141_5116\|\|full | Low-quality |
| S14_vs_k141_42566\|\|full | Low-quality |
| S14_vs_k141_34583\|\|full | Not-determined |
| S14_vs_k141_26768\|\|full | Not-determined |
| S14_vs_k141_26777\|\|full | Not-determined |
| S14_vs_k141_108193\|\|full | Not-determined |
| S14_vs_k141_26782\|\|full | Low-quality |
| S14_vs_k141_12464\|\|full | Low-quality |
| S14_vs_k141_63776\|\|full | Not-determined |
| S14_vs_k141_12475\|\|full | Low-quality |
| S14_vs_k141_42577\|\|full | Not-determined |
| S14_vs_k141_12481\|\|full | Low-quality |
| S14_vs_k141_42579\|\|full | Low-quality |
| S14_vs_k141_100342\|\|full | Low-quality |
| S14_vs_k141_71332\|\|full | Low-quality |
| S14_vs_k141_108218\|\|full | Not-determined |
| S14_vs_k141_93558\|\|full | Low-quality |
| S14_vs_k141_20184\|\|full | Low-quality |
| S14_vs_k141_12506\|\|full | Low-quality |
| S14_vs_k141_42627\|\|full | Low-quality |
| S14_vs_k141_71377\|\|full | Not-determined |
| S14_vs_k141_86076\|\|full | Low-quality |
| S14_vs_k141_56602\|\|full | Low-quality |
| S14_vs_k141_42641\|\|full | Low-quality |
| S16_vs_76823\|\|full | High-quality |
| S14_vs_k141_71388\|\|full | Low-quality |
| S14_vs_k141_42646\|\|full | Not-determined |
| S14_vs_k141_86098\|\|full | Low-quality |
| S14_vs_k141_93571\|\|full | Not-determined |
| S14_vs_k141_34675\|\|full | Low-quality |
| S14_vs_k141_71397\|\|full | Not-determined |
| S14_vs_k141_56629\|\|full | Not-determined |
| S14_vs_k141_34676\|\|full | Low-quality |
| S14_vs_k141_71398\|\|full | Low-quality |
| S14_vs_k141_56642\|\|full | Not-determined |
| S14_vs_k141_71407\|\|full | Not-determined |
| S14_vs_k141_12538\|\|full | Low-quality |
| S14_vs_k141_56643\|\|full | Not-determined |
| S14_vs_k141_100401\|\|full | Low-quality |
| S14_vs_k141_12542\|\|full | Not-determined |
| S12_extracted_k141_60224 | Complete |
| S14_vs_k141_42696\|\|full | Low-quality |
| S14_vs_k141_26872\|\|full | Not-determined |
| S14_vs_k141_93590\|\|full | Low-quality |
| S14_vs_k141_12553\|\|full | Not-determined |
| S14_vs_k141_93593\|\|full | Low-quality |
| S14_vs_k141_42703\|\|full | Low-quality |
| S14_vs_k141_100437\|\|full | Not-determined |
| S14_vs_k141_34721\|\|full | Low-quality |
| S14_vs_k141_34725\|\|full | Not-determined |
| S14_vs_k141_42705\|\|full | Not-determined |
| S14_vs_k141_93616\|\|full | Low-quality |
| S14_vs_k141_5278\|\|full | Low-quality |
| S14_vs_k141_20226\|\|full | Low-quality |
| S14_vs_k141_86151\|\|full | Not-determined |
| S14_vs_k141_115454\|\|full | Low-quality |
| S14_vs_k141_20254\|\|full | Low-quality |
| S14_vs_k141_26901\|\|full | Low-quality |
| S14_vs_k141_108259\|\|full | Not-determined |
| S14_vs_k141_26906\|\|full | Low-quality |
| S14_vs_k141_12587\|\|full | Low-quality |
| S14_vs_k141_42718\|\|full | Not-determined |
| S14_vs_k141_20278\|\|full | Not-determined |
| S14_vs_k141_100470\|\|full | Not-determined |
| S14_vs_k141_100477\|\|full | Not-determined |
| S14_vs_k141_34768\|\|full | Not-determined |
| S14_vs_k141_56684\|\|full | Low-quality |
| S14_vs_k141_86188\|\|full | Low-quality |
| S14_vs_k141_93661\|\|full | Not-determined |
| S14_vs_k141_48775\|\|full | Low-quality |
| S14_vs_k141_5307\|\|full | Low-quality |
| S14_vs_k141_93675\|\|full | Not-determined |
| S14_vs_k141_108288\|\|full | Low-quality |
| S14_vs_k141_48780\|\|full | Not-determined |
| S14_vs_k141_12616\|\|full | Not-determined |
| S14_vs_k141_86202\|\|full | Not-determined |
| S14_vs_k141_26957\|\|full | Low-quality |
| S14_vs_k141_5315\|\|full | Low-quality |
| S14_vs_k141_5321\|\|full | Low-quality |
| S12_vs_k141_60224\|\|full | Complete |
| S14_vs_k141_71425\|\|full | Low-quality |
| S14_vs_k141_100488\|\|full | Not-determined |
| S14_vs_k141_56710\|\|full | Low-quality |
| S14_vs_k141_86210\|\|full | Low-quality |
| S14_vs_k141_5326\|\|full | Low-quality |
| S14_vs_k141_108309\|\|full | Low-quality |
| S14_vs_k141_71437\|\|full | Low-quality |
| S14_vs_k141_20330\|\|full | Low-quality |
| S14_vs_k141_71438\|\|full | Not-determined |
| S14_vs_k141_5339\|\|full | Not-determined |
| S14_vs_k141_5340\|\|full | Low-quality |
| S14_vs_k141_26993\|\|full | Not-determined |
| S14_vs_k141_56733\|\|full | Not-determined |
| S14_vs_k141_93711\|\|full | Low-quality |
| S14_vs_k141_115481\|\|full | Low-quality |
| S14_vs_k141_12639\|\|full | Not-determined |
| S14_vs_k141_12640\|\|full | Low-quality |
| S14_vs_k141_12643\|\|full | Not-determined |
| S14_vs_k141_108331\|\|full | Low-quality |
| S14_vs_k141_12644\|\|full | Low-quality |
| S14_vs_k141_115493\|\|full | Low-quality |
| S14_vs_k141_108337\|\|full | Not-determined |
| S14_vs_k141_20340\|\|full | Low-quality |
| S14_vs_k141_115500\|\|full | Low-quality |
| S14_vs_k141_78705\|\|full | Not-determined |
| S14_vs_k141_71453\|\|full | Low-quality |
| S14_vs_k141_56751\|\|full | Not-determined |
| S14_vs_k141_12662\|\|full | Not-determined |
| S14_vs_k141_42784\|\|full | Low-quality |
| S14_vs_k141_100556\|\|full | Low-quality |
| S14_vs_k141_5359\|\|full | Not-determined |
| S14_vs_k141_34848\|\|full | Low-quality |
| S14_vs_k141_48826\|\|full | Not-determined |
| S14_vs_k141_100565\|\|full | Low-quality |
| S14_vs_k141_56768\|\|full | Low-quality |
| S14_vs_k141_5376\|\|full | Not-determined |
| S14_vs_k141_42804\|\|full | Low-quality |
| S14_vs_k141_20374\|\|full | Low-quality |
| S14_vs_k141_20375\|\|full | Low-quality |
| S14_vs_k141_48829\|\|full | Low-quality |
| S14_vs_k141_71499\|\|full | Low-quality |
| S14_vs_k141_27035\|\|full | Low-quality |
| S14_vs_k141_100590\|\|full | Low-quality |
| S14_vs_k141_5409\|\|full | Low-quality |
| S14_vs_k141_63926\|\|full | Low-quality |
| S14_vs_k141_71509\|\|full | Low-quality |
| S14_vs_k141_93760\|\|full | Low-quality |
| S14_vs_k141_56793\|\|full | Low-quality |
| S14_vs_k141_27046\|\|full | Low-quality |
| S14_vs_k141_34899\|\|full | Not-determined |
| S14_vs_k141_12708\|\|full | Low-quality |
| S14_vs_k141_27049\|\|full | Not-determined |
| S14_vs_k141_63941\|\|full | Low-quality |
| S11_vs_k141_11994\|\|full | High-quality |
| S14_vs_k141_63952\|\|full | Low-quality |
| S14_vs_k141_115588\|\|full | Low-quality |
| S14_vs_k141_27056\|\|full | Low-quality |
| S14_vs_k141_34914\|\|full | Low-quality |
| S14_vs_k141_56805\|\|full | Low-quality |
| S14_vs_k141_27058\|\|full | Not-determined |
| S14_vs_k141_63957\|\|full | Low-quality |
| S14_vs_k141_71537\|\|full | Not-determined |
| S14_vs_k141_78735\|\|full | Not-determined |
| S14_vs_k141_27062\|\|full | Not-determined |
| S14_vs_k141_63959\|\|full | Low-quality |
| S14_vs_k141_78739\|\|full | Not-determined |
| S14_vs_k141_34922\|\|full | Low-quality |
| S14_vs_k141_108396\|\|full | Not-determined |
| S14_vs_k141_56808\|\|full | Low-quality |
| S14_vs_k141_27064\|\|full | Low-quality |
| S14_vs_k141_48894\|\|full | Not-determined |
| S14_vs_k141_12733\|\|full | Low-quality |
| S14_vs_k141_34934\|\|full | Not-determined |
| S14_vs_k141_86284\|\|full | Low-quality |
| S14_vs_k141_12734\|\|full | Low-quality |
| S16_vs_19968\|\|full | Medium-quality |
| S14_vs_k141_108412\|\|full | Low-quality |
| S13_extracted_143422 | Medium-quality |
| S14_vs_k141_5415\|\|full | Low-quality |
| S14_vs_k141_56811\|\|full | Low-quality |
| S14_vs_k141_108417\|\|full | Low-quality |
| S14_vs_k141_100644\|\|full | Low-quality |
| S14_vs_k141_108421\|\|full | Low-quality |
| S14_vs_k141_5418\|\|full | Not-determined |
| S14_vs_k141_63981\|\|full | Not-determined |
| S14_vs_k141_93779\|\|full | Not-determined |
| S14_vs_k141_56816\|\|full | Not-determined |
| S14_vs_k141_42881\|\|full | Not-determined |
| S14_vs_k141_93780\|\|full | Low-quality |
| S14_vs_k141_78771\|\|full | Low-quality |
| S14_vs_k141_63984\|\|full | Low-quality |
| S14_vs_k141_78777\|\|full | Low-quality |
| S14_vs_k141_20402\|\|full | Not-determined |
| S14_vs_k141_12764\|\|full | Low-quality |
| S14_vs_k141_108427\|\|full | Low-quality |
| S14_vs_k141_56832\|\|full | Low-quality |
| S14_vs_k141_56834\|\|full | Not-determined |
| S14_vs_k141_12775\|\|full | Low-quality |
| S14_vs_k141_12782\|\|full | Low-quality |
| S14_vs_k141_78789\|\|full | Not-determined |
| S14_vs_k141_93806\|\|full | Low-quality |
| S14_vs_k141_12788\|\|full | Not-determined |
| S14_vs_k141_48917\|\|full | Low-quality |
| S14_vs_k141_71592\|\|full | Not-determined |
| S14_vs_k141_71597\|\|full | Not-determined |
| S14_vs_k141_93820\|\|full | Low-quality |
| S14_vs_k141_108450\|\|full | Low-quality |
| S14_vs_k141_20437\|\|full | Not-determined |
| S14_vs_k141_86335\|\|full | Not-determined |
| S14_vs_k141_86336\|\|full | Not-determined |
| S14_vs_k141_27153\|\|full | Not-determined |
| S14_vs_k141_100667\|\|full | Low-quality |
| S14_vs_k141_71621\|\|full | Low-quality |
| S14_vs_k141_20458\|\|full | Low-quality |
| S14_vs_k141_93840\|\|full | Low-quality |
| S14_vs_k141_48977\|\|full | Low-quality |
| S14_vs_k141_12821\|\|full | Low-quality |
| S14_vs_k141_86359\|\|full | Low-quality |
| S14_vs_k141_42940\|\|full | Not-determined |
| S14_vs_k141_86365\|\|full | Low-quality |
| S14_vs_k141_42941\|\|full | Low-quality |
| S14_vs_k141_78822\|\|full | Not-determined |
| S14_vs_k141_42945\|\|full | Low-quality |
| S14_vs_k141_71630\|\|full | Low-quality |
| S14_vs_k141_71631\|\|full | Not-determined |
| S14_vs_k141_93862\|\|full | Low-quality |
| S14_vs_k141_20494\|\|full | Low-quality |
| S14_vs_k141_49004\|\|full | Not-determined |
| S14_vs_k141_115703\|\|full | Low-quality |
| S14_vs_k141_56865\|\|full | Low-quality |
| S14_vs_k141_100699\|\|full | Low-quality |
| S14_vs_k141_56878\|\|full | Low-quality |
| S14_vs_k141_42971\|\|full | Low-quality |
| S14_vs_k141_71645\|\|full | Low-quality |
| S14_vs_k141_86399\|\|full | Not-determined |
| S14_vs_k141_42972\|\|full | Low-quality |
| S14_vs_k141_12855\|\|full | Not-determined |
| S14_vs_k141_93880\|\|full | Low-quality |
| S14_vs_k141_71649\|\|full | Low-quality |
| S14_vs_k141_49018\|\|full | Low-quality |
| S14_vs_k141_12862\|\|full | Not-determined |
| S14_vs_k141_56892\|\|full | Low-quality |
| S14_vs_k141_12868\|\|full | Not-determined |
| S14_vs_k141_93890\|\|full | Not-determined |
| S14_vs_k141_78833\|\|full | Not-determined |
| S14_vs_k141_71653\|\|full | Not-determined |
| S14_vs_k141_35029\|\|full | Low-quality |
| S14_vs_k141_12877\|\|full | Low-quality |
| S14_vs_k141_64067\|\|full | Low-quality |
| S14_vs_k141_20523\|\|full | Low-quality |
| S14_vs_k141_71662\|\|full | Low-quality |
| S14_vs_k141_49023\|\|full | Not-determined |
| S14_vs_k141_20536\|\|full | Not-determined |
| S14_vs_k141_86421\|\|full | Not-determined |
| S14_vs_k141_93901\|\|full | Low-quality |
| S14_vs_k141_27178\|\|full | Not-determined |
| S14_vs_k141_20544\|\|full | Low-quality |
| S14_vs_k141_35065\|\|full | Low-quality |
| S14_vs_k141_78858\|\|full | Low-quality |
| S14_vs_k141_27179\|\|full | Not-determined |
| S14_vs_k141_12891\|\|full | Not-determined |
| S14_vs_k141_100709\|\|full | Low-quality |
| S14_vs_k141_71678\|\|full | Not-determined |
| S14_vs_k141_93920\|\|full | Low-quality |
| S14_vs_k141_12899\|\|full | Low-quality |
| S14_vs_k141_5497\|\|full | Not-determined |
| S14_vs_k141_27195\|\|full | Not-determined |
| S14_vs_k141_100715\|\|full | Not-determined |
| S14_vs_k141_71687\|\|full | Low-quality |
| S14_vs_k141_20553\|\|full | Low-quality |
| S14_vs_k141_43035\|\|full | Low-quality |
| S14_vs_k141_71691\|\|full | Low-quality |
| S14_vs_k141_12914\|\|full | Not-determined |
| S14_vs_k141_100722\|\|full | Low-quality |
| S14_vs_k141_93937\|\|full | Not-determined |
| S14_vs_k141_20558\|\|full | Low-quality |
| S14_vs_k141_78884\|\|full | Not-determined |
| S14_vs_k141_78885\|\|full | Low-quality |
| S14_vs_k141_49072\|\|full | Low-quality |
| S14_vs_k141_43042\|\|full | Low-quality |
| S14_vs_k141_49075\|\|full | Low-quality |
| S14_vs_k141_56906\|\|full | Not-determined |
| S14_vs_k141_20563\|\|full | Not-determined |
| S14_vs_k141_93952\|\|full | Low-quality |
| S14_vs_k141_100752\|\|full | Low-quality |
| S14_vs_k141_20579\|\|full | Low-quality |
| S14_vs_k141_93973\|\|full | Low-quality |
| S14_vs_k141_12959\|\|full | Low-quality |
| S14_vs_k141_64107\|\|full | Low-quality |
| S14_vs_k141_27241\|\|full | Not-determined |
| S14_vs_k141_43092\|\|full | Low-quality |
| S14_vs_k141_71729\|\|full | Not-determined |
| S14_vs_k141_108553\|\|full | Not-determined |
| S14_vs_k141_86476\|\|full | Low-quality |
| S14_vs_k141_94003\|\|full | Low-quality |
| S14_vs_k141_56938\|\|full | Low-quality |
| S14_vs_k141_94007\|\|full | Low-quality |
| S14_vs_k141_94020\|\|full | Low-quality |
| S14_vs_k141_100797\|\|full | Low-quality |
| S14_vs_k141_43111\|\|full | Not-determined |
| S14_vs_k141_43112\|\|full | Not-determined |
| S14_vs_k141_108570\|\|full | Not-determined |
| S14_vs_k141_13006\|\|full | Low-quality |
| S14_vs_k141_56946\|\|full | Not-determined |
| S14_vs_k141_94036\|\|full | Low-quality |
| S14_vs_k141_27279\|\|full | Not-determined |
| S14_vs_k141_94040\|\|full | Not-determined |
| S14_vs_k141_43130\|\|full | Low-quality |
| S14_vs_k141_100812\|\|full | Low-quality |
| S14_vs_k141_100814\|\|full | Low-quality |
| S14_vs_k141_35127\|\|full | Low-quality |
| S14_vs_k141_115757\|\|full | Low-quality |
| S14_vs_k141_78897\|\|full | Low-quality |
| S14_vs_k141_94056\|\|full | Low-quality |
| S14_vs_k141_35144\|\|full | Not-determined |
| S14_vs_k141_13027\|\|full | Low-quality |
| S14_vs_k141_35152\|\|full | Low-quality |
| S14_vs_k141_27304\|\|full | Low-quality |
| S14_vs_k141_78914\|\|full | Not-determined |
| S14_vs_k141_27312\|\|full | Low-quality |
| S14_vs_k141_20639\|\|full | Low-quality |
| S14_vs_k141_56985\|\|full | Low-quality |
| S14_vs_k141_27324\|\|full | Low-quality |
| S14_vs_k141_78927\|\|full | Low-quality |
| S14_vs_k141_56989\|\|full | Not-determined |
| S14_vs_k141_100858\|\|full | Low-quality |
| S14_vs_k141_108620\|\|full | Low-quality |
| S14_vs_k141_86523\|\|full | Low-quality |
| S14_vs_k141_35177\|\|full | Low-quality |
| S14_vs_k141_94114\|\|full | Low-quality |
| S14_vs_k141_35185\|\|full | Low-quality |
| S14_vs_k141_94118\|\|full | Low-quality |
| S14_vs_k141_5597\|\|full | Low-quality |
| S14_vs_k141_86540\|\|full | Low-quality |
| S14_vs_k141_78951\|\|full | Low-quality |
| S14_vs_k141_5599\|\|full | Low-quality |
| S14_vs_k141_86543\|\|full | Not-determined |
| S14_vs_k141_20673\|\|full | Not-determined |
| S14_vs_k141_100897\|\|full | Low-quality |
| S14_vs_k141_43189\|\|full | Not-determined |
| S14_vs_k141_35204\|\|full | Not-determined |
| S14_vs_k141_115815\|\|full | Not-determined |
| S14_vs_k141_27363\|\|full | Not-determined |
| S14_vs_k141_27364\|\|full | Low-quality |
| S14_vs_k141_115822\|\|full | Low-quality |
| S14_vs_k141_5616\|\|full | Not-determined |
| S14_vs_k141_78961\|\|full | Low-quality |
| S14_vs_k141_35208\|\|full | Low-quality |
| S14_vs_k141_71855\|\|full | Low-quality |
| S14_vs_k141_78967\|\|full | Low-quality |
| S14_vs_k141_71859\|\|full | Low-quality |
| S14_vs_k141_108676\|\|full | Low-quality |
| S14_vs_k141_64161\|\|full | Not-determined |
| S14_vs_k141_20695\|\|full | Low-quality |
| S14_vs_k141_108680\|\|full | Low-quality |
| S14_vs_k141_13079\|\|full | Low-quality |
| S14_vs_k141_100911\|\|full | Not-determined |
| S14_vs_k141_20707\|\|full | Low-quality |
| S14_vs_k141_13086\|\|full | Low-quality |
| S14_vs_k141_100925\|\|full | Low-quality |
| S14_vs_k141_43219\|\|full | Low-quality |
| S14_vs_k141_13095\|\|full | Low-quality |
| S14_vs_k141_43220\|\|full | Not-determined |
| S14_vs_k141_78977\|\|full | Low-quality |
| S14_vs_k141_71897\|\|full | Low-quality |
| S14_vs_k141_43225\|\|full | Not-determined |
| S14_vs_k141_115862\|\|full | Low-quality |
| S14_vs_k141_78979\|\|full | Not-determined |
| S14_vs_k141_43229\|\|full | Low-quality |
| S14_vs_k141_27377\|\|full | Low-quality |
| S14_vs_k141_86599\|\|full | Low-quality |
| S14_vs_k141_71914\|\|full | Low-quality |
| S14_vs_k141_20733\|\|full | Not-determined |
| S14_vs_k141_20745\|\|full | Low-quality |
| S14_vs_k141_78984\|\|full | Not-determined |
| S14_vs_k141_100965\|\|full | Not-determined |
| S14_vs_k141_86624\|\|full | Low-quality |
| S14_vs_k141_78994\|\|full | Low-quality |
| S14_vs_k141_5679\|\|full | Low-quality |
| S14_vs_k141_27412\|\|full | Low-quality |
| S14_vs_k141_115885\|\|full | Low-quality |
| S14_vs_k141_57011\|\|full | Not-determined |
| S14_vs_k141_43269\|\|full | Not-determined |

| | |
|---|---|
| S14_vs_k141_71932\|\|full | Low-quality |
| S14_vs_k141_64199\|\|full | Not-determined |
| S14_vs_k141_86648\|\|full | Not-determined |
| S14_vs_k141_49158\|\|full | Low-quality |
| S14_vs_k141_108745\|\|full | Low-quality |
| S14_vs_k141_115910\|\|full | Low-quality |
| S14_vs_k141_27446\|\|full | Not-determined |
| S14_vs_k141_115917\|\|full | Low-quality |
| S14_vs_k141_43299\|\|full | Not-determined |
| S14_vs_k141_43300\|\|full | Not-determined |
| S14_vs_k141_49167\|\|full | Low-quality |
| S14_vs_k141_108763\|\|full | Not-determined |
| S14_vs_k141_20793\|\|full | Low-quality |
| S14_vs_k141_49179\|\|full | Low-quality |
| S14_vs_k141_49182\|\|full | Low-quality |
| S14_vs_k141_27469\|\|full | Low-quality |
| S14_vs_k141_43328\|\|full | Low-quality |
| S14_vs_k141_86708\|\|full | Low-quality |
| S14_vs_k141_79082\|\|full | Low-quality |
| S13_vs_143422\|\|full | Medium-quality |
| S14_vs_k141_20836\|\|full | Not-determined |
| S14_vs_k141_94258\|\|full | Low-quality |
| S14_vs_k141_108818\|\|full | Low-quality |
| S14_vs_k141_27505\|\|full | Low-quality |
| S14_vs_k141_57074\|\|full | Not-determined |
| S14_vs_k141_20847\|\|full | Low-quality |
| S14_vs_k141_115968\|\|full | Low-quality |
| S14_vs_k141_94273\|\|full | Not-determined |
| S14_vs_k141_5769\|\|full | Low-quality |
| S14_vs_k141_71998\|\|full | Low-quality |
| S14_vs_k141_20849\|\|full | Not-determined |
| S14_vs_k141_20854\|\|full | Low-quality |
| S14_vs_k141_94277\|\|full | Low-quality |
| S14_vs_k141_86730\|\|full | Low-quality |
| S14_vs_k141_94281\|\|full | Low-quality |
| S14_vs_k141_57091\|\|full | Low-quality |
| S14_vs_k141_35330\|\|full | Low-quality |
| S14_vs_k141_5777\|\|full | Low-quality |
| S14_vs_k141_115982\|\|full | Low-quality |
| S14_vs_k141_86740\|\|full | Not-determined |
| S14_vs_k141_94290\|\|full | Low-quality |
| S14_vs_k141_72017\|\|full | Low-quality |
| S14_vs_k141_64272\|\|full | Low-quality |
| S14_vs_k141_43337\|\|full | Low-quality |
| S14_vs_k141_72019\|\|full | Low-quality |
| S14_vs_k141_64276\|\|full | Low-quality |
| S14_vs_k141_79091\|\|full | Low-quality |
| S16_extracted_84462 | Complete |
| S14_vs_k141_101062\|\|full | Low-quality |
| S14_vs_k141_57126\|\|full | Low-quality |
| S14_vs_k141_86777\|\|full | Not-determined |
| S14_vs_k141_20901\|\|full | Not-determined |
| S14_vs_k141_57129\|\|full | Low-quality |
| S14_vs_k141_108893\|\|full | Not-determined |
| S14_vs_k141_116019\|\|full | Not-determined |
| S14_vs_k141_27580\|\|full | Low-quality |
| S14_vs_k141_27583\|\|full | Low-quality |
| S14_vs_k141_94320\|\|full | Not-determined |
| S14_vs_k141_86797\|\|full | Not-determined |
| S14_vs_k141_35348\|\|full | Not-determined |
| S14_vs_k141_5836\|\|full | Low-quality |
| S14_vs_k141_86814\|\|full | Low-quality |
| S14_vs_k141_20938\|\|full | Low-quality |
| S14_vs_k141_13208\|\|full | Low-quality |
| S14_vs_k141_108934\|\|full | Not-determined |
| S14_vs_k141_5846\|\|full | Low-quality |
| S14_vs_k141_57157\|\|full | Low-quality |
| S14_vs_k141_64314\|\|full | Low-quality |
| S14_vs_k141_20942\|\|full | Not-determined |
| S14_vs_k141_101103\|\|full | Not-determined |
| S14_vs_k141_57165\|\|full | Not-determined |
| S14_vs_k141_64319\|\|full | Low-quality |
| S14_vs_k141_79135\|\|full | Low-quality |
| S14_vs_k141_94361\|\|full | Low-quality |
| S14_vs_k141_101105\|\|full | Not-determined |
| S14_vs_k141_86830\|\|full | Low-quality |
| S14_vs_k141_64335\|\|full | Low-quality |
| S14_vs_k141_35378\|\|full | Low-quality |
| S14_vs_k141_43415\|\|full | Low-quality |
| S14_vs_k141_13227\|\|full | Low-quality |
| S14_vs_k141_79138\|\|full | Low-quality |
| S14_vs_k141_86850\|\|full | Low-quality |
| S14_vs_k141_27630\|\|full | Low-quality |
| S14_vs_k141_57197\|\|full | Low-quality |
| S14_vs_k141_64348\|\|full | Low-quality |
| S14_vs_k141_5869\|\|full | Not-determined |
| S14_vs_k141_35410\|\|full | Not-determined |
| S14_vs_k141_43438\|\|full | Low-quality |
| S14_vs_k141_35431\|\|full | Low-quality |
| S14_vs_k141_101155\|\|full | Low-quality |
| S14_vs_k141_5887\|\|full | Not-determined |
| S14_vs_k141_35441\|\|full | Low-quality |
| S14_vs_k141_101161\|\|full | Low-quality |
| S14_vs_k141_116066\|\|full | Not-determined |
| S14_vs_k141_101163\|\|full | Low-quality |
| S14_vs_k141_94371\|\|full | Low-quality |
| S14_vs_k141_94376\|\|full | Not-determined |
| S14_vs_k141_94380\|\|full | Low-quality |
| S14_vs_k141_116076\|\|full | Low-quality |
| S14_vs_k141_79213\|\|full | Not-determined |
| S14_vs_k141_49309\|\|full | Not-determined |
| S14_vs_k141_35460\|\|full | Low-quality |
| S14_vs_k141_5902\|\|full | Low-quality |
| S14_vs_k141_86882\|\|full | Low-quality |
| S14_vs_k141_13248\|\|full | Not-determined |
| S14_vs_k141_94393\|\|full | Not-determined |
| S14_vs_k141_79221\|\|full | Low-quality |
| S14_vs_k141_116091\|\|full | Low-quality |
| S14_vs_k141_57247\|\|full | Low-quality |
| S14_vs_k141_35463\|\|full | Not-determined |
| S14_vs_k141_86898\|\|full | Not-determined |
| S14_vs_k141_13263\|\|full | Low-quality |
| S14_vs_k141_35469\|\|full | Low-quality |
| S14_vs_k141_49330\|\|full | Low-quality |
| S14_vs_k141_5915\|\|full | Not-determined |
| S14_vs_k141_5916\|\|full | Low-quality |
| S14_vs_k141_35474\|\|full | Low-quality |
| S14_vs_k141_101172\|\|full | Not-determined |
| S14_vs_k141_94409\|\|full | Not-determined |
| S14_vs_k141_5919\|\|full | Low-quality |
| S14_vs_k141_101178\|\|full | Low-quality |
| S14_vs_k141_57266\|\|full | Not-determined |
| S14_vs_k141_49350\|\|full | Low-quality |
| S14_vs_k141_13289\|\|full | Low-quality |
| S14_vs_k141_57270\|\|full | Low-quality |
| S14_vs_k141_86944\|\|full | Low-quality |
| S14_vs_k141_72035\|\|full | Low-quality |
| S14_vs_k141_57286\|\|full | Low-quality |
| S14_vs_k141_13306\|\|full | Not-determined |
| S14_vs_k141_86955\|\|full | Low-quality |
| S14_vs_k141_13310\|\|full | Not-determined |
| S14_vs_k141_94423\|\|full | Low-quality |
| S14_vs_k141_21021\|\|full | Not-determined |
| S14_vs_k141_35485\|\|full | Low-quality |
| S14_vs_k141_116136\|\|full | Not-determined |
| S14_vs_k141_13332\|\|full | Low-quality |
| S14_vs_k141_94441\|\|full | Low-quality |
| S14_vs_k141_35495\|\|full | Not-determined |
| S14_vs_k141_101211\|\|full | Not-determined |
| S14_vs_k141_5965\|\|full | Low-quality |
| S14_vs_k141_13339\|\|full | Low-quality |
| S14_vs_k141_94450\|\|full | Low-quality |
| S14_vs_k141_64368\|\|full | Low-quality |
| S14_vs_k141_79280\|\|full | Low-quality |
| S14_vs_k141_49403\|\|full | Low-quality |
| S14_vs_k141_94458\|\|full | Not-determined |
| S14_vs_k141_27716\|\|full | Low-quality |
| S14_vs_k141_57325\|\|full | Not-determined |
| S14_vs_k141_101221\|\|full | Low-quality |
| S14_vs_k141_94471\|\|full | Not-determined |
| S14_vs_k141_5988\|\|full | Low-quality |
| S14_vs_k141_57341\|\|full | Not-determined |
| S14_vs_k141_57342\|\|full | Not-determined |
| S14_vs_k141_13370\|\|full | Low-quality |
| S14_vs_k141_5990\|\|full | Not-determined |
| S14_vs_k141_27733\|\|full | Low-quality |
| S14_vs_k141_94475\|\|full | Not-determined |
| S14_vs_k141_116173\|\|full | Low-quality |
| S14_vs_k141_27742\|\|full | Low-quality |
| S14_vs_k141_57351\|\|full | Low-quality |
| S14_vs_k141_79306\|\|full | Low-quality |
| S14_vs_k141_6013\|\|full | Low-quality |
| S14_vs_k141_21040\|\|full | Not-determined |
| S14_vs_k141_13382\|\|full | Low-quality |
| S14_vs_k141_101256\|\|full | Not-determined |
| S14_vs_k141_21058\|\|full | Not-determined |
| S14_vs_k141_116187\|\|full | Low-quality |
| S14_vs_k141_109076\|\|full | Low-quality |
| S14_vs_k141_72063\|\|full | Low-quality |
| S14_vs_k141_35551\|\|full | Low-quality |
| S14_vs_k141_21067\|\|full | Not-determined |
| S14_vs_k141_94505\|\|full | Low-quality |
| S14_vs_k141_72072\|\|full | Not-determined |
| S14_vs_k141_79331\|\|full | Not-determined |
| S14_vs_k141_49465\|\|full | Low-quality |
| S14_vs_k141_94513\|\|full | Low-quality |
| S14_vs_k141_64389\|\|full | Not-determined |
| S14_vs_k141_87047\|\|full | Low-quality |
| S14_vs_k141_94518\|\|full | Low-quality |
| S14_vs_k141_94519\|\|full | Not-determined |
| S14_vs_k141_101282\|\|full | Low-quality |
| S14_vs_k141_35569\|\|full | Low-quality |
| S14_vs_k141_57396\|\|full | Not-determined |
| S14_vs_k141_87049\|\|full | Low-quality |
| S14_vs_k141_27790\|\|full | Not-determined |
| S14_vs_k141_87059\|\|full | Low-quality |
| S14_vs_k141_79362\|\|full | Low-quality |
| S14_vs_k141_35593\|\|full | Low-quality |
| S14_vs_k141_116218\|\|full | Low-quality |
| S14_vs_k141_109138\|\|full | Low-quality |
| S14_vs_k141_6071\|\|full | Low-quality |
| S14_vs_k141_87100\|\|full | Not-determined |
| S14_vs_k141_49524\|\|full | Low-quality |
| S14_vs_k141_94553\|\|full | Low-quality |
| S14_vs_k141_116253\|\|full | Low-quality |
| S14_vs_k141_87103\|\|full | Low-quality |
| S14_vs_k141_79397\|\|full | Not-determined |
| S14_vs_k141_64403\|\|full | Low-quality |
| S14_vs_k141_79401\|\|full | Low-quality |
| S14_vs_k141_116265\|\|full | Not-determined |
| S14_vs_k141_27825\|\|full | Low-quality |
| S14_vs_k141_87114\|\|full | Low-quality |
| S14_vs_k141_101324\|\|full | Not-determined |
| S14_vs_k141_43577\|\|full | Low-quality |
| S14_vs_k141_64414\|\|full | Low-quality |
| S14_vs_k141_49559\|\|full | Low-quality |
| S14_vs_k141_72113\|\|full | Not-determined |
| S14_vs_k141_27837\|\|full | Not-determined |
| S14_vs_k141_35653\|\|full | Low-quality |
| S14_vs_k141_94577\|\|full | Not-determined |
| S14_vs_k141_6095\|\|full | Not-determined |
| S14_vs_k141_109163\|\|full | Low-quality |
| S14_vs_k141_72121\|\|full | Low-quality |
| S14_vs_k141_101338\|\|full | Low-quality |
| S14_vs_k141_79433\|\|full | Low-quality |
| S14_vs_k141_43590\|\|full | Not-determined |
| S14_vs_k141_101341\|\|full | Low-quality |
| S14_vs_k141_21128\|\|full | Low-quality |
| S14_vs_k141_87142\|\|full | Not-determined |
| S14_vs_k141_6106\|\|full | Low-quality |
| S14_vs_k141_49588\|\|full | Low-quality |
| S14_vs_k141_27862\|\|full | Not-determined |
| S14_vs_k141_101352\|\|full | Low-quality |
| S14_vs_k141_21138\|\|full | Low-quality |
| S14_vs_k141_27867\|\|full | Not-determined |
| S14_vs_k141_49597\|\|full | Low-quality |
| S14_vs_k141_57446\|\|full | Low-quality |
| S14_vs_k141_6134\|\|full | Low-quality |
| S14_vs_k141_49608\|\|full | Low-quality |
| S14_vs_k141_35669\|\|full | Not-determined |
| S14_vs_k141_6135\|\|full | Low-quality |
| S14_vs_k141_109181\|\|full | Not-determined |
| S14_vs_k141_94606\|\|full | Low-quality |
| S14_vs_k141_72154\|\|full | Low-quality |
| S14_vs_k141_116315\|\|full | Low-quality |
| S14_vs_k141_116320\|\|full | Low-quality |
| S14_vs_k141_94608\|\|full | Low-quality |
| S14_vs_k141_101376\|\|full | Not-determined |
| S14_vs_k141_109198\|\|full | Not-determined |
| S14_vs_k141_116327\|\|full | Not-determined |
| S14_vs_k141_87175\|\|full | Low-quality |
| S14_vs_k141_49637\|\|full | Not-determined |
| S14_vs_k141_116336\|\|full | Not-determined |
| S14_vs_k141_57452\|\|full | Not-determined |
| S14_vs_k141_6173\|\|full | Low-quality |
| S14_vs_k141_57454\|\|full | Not-determined |
| S14_vs_k141_64438\|\|full | Not-determined |
| S14_vs_k141_13399\|\|full | Low-quality |
| S14_vs_k141_57458\|\|full | Not-determined |
| S14_vs_k141_27910\|\|full | Low-quality |
| S14_vs_k141_49662\|\|full | Not-determined |
| S14_vs_k141_72204\|\|full | Not-determined |
| S14_vs_k141_87214\|\|full | Low-quality |
| S14_vs_k141_27917\|\|full | Low-quality |
| S14_vs_k141_94660\|\|full | Not-determined |
| S14_vs_k141_116376\|\|full | Not-determined |
| S14_vs_k141_13430\|\|full | Not-determined |
| S14_vs_k141_64465\|\|full | Low-quality |
| S14_vs_k141_94663\|\|full | Low-quality |
| S14_vs_k141_27920\|\|full | Not-determined |
| S14_vs_k141_87221\|\|full | Not-determined |
| S14_vs_k141_35746\|\|full | Low-quality |
| S14_vs_k141_101406\|\|full | Low-quality |
| S14_vs_k141_27925\|\|full | Not-determined |
| S14_vs_k141_49674\|\|full | Low-quality |
| S14_vs_k141_64472\|\|full | Not-determined |
| S14_vs_k141_6194\|\|full | Not-determined |
| S14_vs_k141_27933\|\|full | Not-determined |
| S14_vs_k141_13436\|\|full | Not-determined |
| S14_vs_k141_94673\|\|full | Not-determined |
| S11_vs_k141_27176\|\|full | High-quality |
| S14_vs_k141_27936\|\|full | Low-quality |
| S14_vs_k141_35775\|\|full | Low-quality |
| S14_vs_k141_116397\|\|full | Low-quality |
| S14_vs_k141_13451\|\|full | Not-determined |
| S14_vs_k141_35780\|\|full | Not-determined |
| S14_vs_k141_87250\|\|full | Not-determined |
| S14_vs_k141_57506\|\|full | Low-quality |
| S14_vs_k141_72219\|\|full | Not-determined |
| S14_vs_k141_43720\|\|full | Low-quality |
| S14_vs_k141_6217\|\|full | Low-quality |
| S14_vs_k141_49704\|\|full | Not-determined |
| S14_vs_k141_94709\|\|full | Low-quality |
| S14_vs_k141_49714\|\|full | Low-quality |
| S14_vs_k141_116423\|\|full | Low-quality |
| S14_vs_k141_64505\|\|full | Not-determined |
| S14_vs_k141_6236\|\|full | Low-quality |
| S14_vs_k141_21224\|\|full | Not-determined |
| S14_vs_k141_57530\|\|full | Low-quality |
| S14_vs_k141_13479\|\|full | Low-quality |
| S14_vs_k141_94713\|\|full | Not-determined |
| S14_vs_k141_116439\|\|full | Low-quality |
| S14_vs_k141_57540\|\|full | Not-determined |
| S14_vs_k141_64518\|\|full | Low-quality |
| S14_vs_k141_27951\|\|full | Not-determined |
| S14_vs_k141_116445\|\|full | Low-quality |
| S14_vs_k141_64535\|\|full | Not-determined |
| S14_vs_k141_72252\|\|full | Low-quality |
| S14_vs_k141_43763\|\|full | Low-quality |
| S14_vs_k141_64541\|\|full | Low-quality |
| S14_vs_k141_79524\|\|full | Not-determined |
| S14_vs_k141_27964\|\|full | Low-quality |
| S14_vs_k141_64542\|\|full | Low-quality |
| S14_vs_k141_35809\|\|full | Low-quality |
| S14_vs_k141_57566\|\|full | Low-quality |
| S14_vs_k141_21252\|\|full | Not-determined |
| S14_vs_k141_101465\|\|full | Not-determined |
| S14_vs_k141_87280\|\|full | Not-determined |
| S14_vs_k141_35822\|\|full | Not-determined |
| S14_vs_k141_64558\|\|full | Low-quality |
| S14_vs_k141_43797\|\|full | Low-quality |
| S14_vs_k141_13520\|\|full | Low-quality |
| S14_vs_k141_116453\|\|full | Low-quality |
| S14_vs_k141_109316\|\|full | Not-determined |
| S14_vs_k141_21278\|\|full | Not-determined |
| S14_vs_k141_13533\|\|full | Not-determined |
| S14_vs_k141_79558\|\|full | Not-determined |
| S14_vs_k141_49781\|\|full | Low-quality |
| S14_vs_k141_64579\|\|full | Low-quality |
| S14_vs_k141_6313\|\|full | Low-quality |
| S14_vs_k141_43825\|\|full | Low-quality |
| S14_vs_k141_21286\|\|full | Low-quality |
| S11_vs_k141_104037\|\|full | High-quality |
| S14_vs_k141_49808\|\|full | Low-quality |
| S14_vs_k141_79585\|\|full | Low-quality |
| S14_vs_k141_94798\|\|full | Not-determined |
| S14_vs_k141_94804\|\|full | Not-determined |
| S14_vs_k141_87340\|\|full | Low-quality |
| S14_vs_k141_49820\|\|full | Low-quality |
| S14_vs_k141_6352\|\|full | Low-quality |
| S14_vs_k141_101488\|\|full | Not-determined |
| S14_vs_k141_109344\|\|full | Low-quality |
| S14_vs_k141_6363\|\|full | Low-quality |
| S14_vs_k141_43874\|\|full | Low-quality |
| S14_vs_k141_43876\|\|full | Not-determined |
| S14_vs_k141_28016\|\|full | Not-determined |
| S14_vs_k141_13579\|\|full | Not-determined |
| S14_vs_k141_35874\|\|full | Low-quality |
| S14_vs_k141_72286\|\|full | Low-quality |
| S9_vs_k141_32588\|\|full | Medium-quality |
| S14_vs_k141_72292\|\|full | Not-determined |
| S14_vs_k141_49849\|\|full | Low-quality |
| S14_vs_k141_101522\|\|full | Not-determined |
| S14_vs_k141_116526\|\|full | Not-determined |
| S14_vs_k141_79637\|\|full | Not-determined |
| S14_vs_k141_101524\|\|full | Low-quality |
| S14_vs_k141_79638\|\|full | Not-determined |
| S14_vs_k141_72309\|\|full | Not-determined |
| S14_vs_k141_87380\|\|full | Low-quality |
| S14_vs_k141_28037\|\|full | Low-quality |
| S14_vs_k141_21363\|\|full | Low-quality |
| S14_vs_k141_43890\|\|full | Not-determined |
| S14_vs_k141_43894\|\|full | Low-quality |
| S14_vs_k141_43898\|\|full | Low-quality |
| S14_vs_k141_94829\|\|full | Not-determined |
| S14_vs_k141_6417\|\|full | Not-determined |
| S14_vs_k141_13609\|\|full | Low-quality |
| S14_vs_k141_72330\|\|full | Not-determined |
| S14_vs_k141_43908\|\|full | Not-determined |
| S14_vs_k141_94848\|\|full | Low-quality |
| S14_vs_k141_21403\|\|full | Low-quality |
| S14_vs_k141_13616\|\|full | Low-quality |
| S14_vs_k141_79686\|\|full | Low-quality |
| S14_vs_k141_21405\|\|full | Low-quality |
| S14_vs_k141_43921\|\|full | Low-quality |
| S14_vs_k141_109431\|\|full | Not-determined |
| S14_vs_k141_6433\|\|full | Low-quality |
| S14_vs_k141_87442\|\|full | Low-quality |
| S14_vs_k141_21408\|\|full | Low-quality |
| S14_vs_k141_94866\|\|full | Low-quality |
| S14_vs_k141_28112\|\|full | Low-quality |
| S14_vs_k141_35917\|\|full | Low-quality |
| S14_vs_k141_13626\|\|full | Low-quality |
| S16_vs_120622\|\|full | High-quality |
| S14_vs_k141_87448\|\|full | Not-determined |
| S14_vs_k141_28118\|\|full | Not-determined |
| S14_vs_k141_101570\|\|full | Low-quality |
| S14_vs_k141_94873\|\|full | Low-quality |
| S14_vs_k141_35932\|\|full | Low-quality |
| S14_vs_k141_43935\|\|full | Low-quality |
| S14_vs_k141_94884\|\|full | Low-quality |
| S14_vs_k141_35945\|\|full | Low-quality |
| S14_vs_k141_43936\|\|full | Low-quality |
| S14_vs_k141_101576\|\|full | Low-quality |
| S14_vs_k141_57712\|\|full | Low-quality |
| S14_vs_k141_116627\|\|full | Not-determined |
| S14_vs_k141_13642\|\|full | Low-quality |
| S14_vs_k141_6476\|\|full | Low-quality |
| S14_vs_k141_109471\|\|full | Low-quality |
| S14_vs_k141_13648\|\|full | Low-quality |
| S14_vs_k141_101593\|\|full | Low-quality |
| S14_vs_k141_13664\|\|full | Low-quality |
| S14_vs_k141_21476\|\|full | Not-determined |
| S14_vs_k141_21477\|\|full | Low-quality |
| S14_vs_k141_21478\|\|full | Low-quality |
| S14_vs_k141_35972\|\|full | Low-quality |
| S14_vs_k141_57724\|\|full | Low-quality |
| S14_vs_k141_21484\|\|full | Not-determined |
| S14_vs_k141_35979\|\|full | Low-quality |
| S14_vs_k141_109503\|\|full | Not-determined |
| S14_vs_k141_13689\|\|full | Not-determined |
| S14_vs_k141_57767\|\|full | Not-determined |
| S14_vs_k141_28144\|\|full | Low-quality |
| S14_vs_k141_49899\|\|full | Low-quality |
| S14_vs_k141_28145\|\|full | Not-determined |
| S14_vs_k141_116655\|\|full | Low-quality |
| S14_vs_k141_6520\|\|full | Low-quality |
| S14_vs_k141_6521\|\|full | Low-quality |
| S14_vs_k141_101621\|\|full | Low-quality |
| S14_vs_k141_13706\|\|full | Not-determined |
| S14_vs_k141_13707\|\|full | Low-quality |
| S14_vs_k141_79824\|\|full | Low-quality |
| S14_vs_k141_21552\|\|full | Low-quality |
| S14_vs_k141_36007\|\|full | Low-quality |
| S14_vs_k141_94958\|\|full | Low-quality |
| S14_vs_k141_21555\|\|full | Low-quality |
| S14_vs_k141_28158\|\|full | Low-quality |
| S14_vs_k141_49927\|\|full | Not-determined |
| S14_vs_k141_6532\|\|full | Not-determined |
| S14_vs_k141_13737\|\|full | Not-determined |
| S5_vs_k141_79992\|\|full | High-quality |
| S14_vs_k141_36046\|\|full | Low-quality |
| S14_vs_k141_79868\|\|full | Low-quality |
| S14_vs_k141_116693\|\|full | Not-determined |
| S14_vs_k141_49952\|\|full | Low-quality |
| S14_vs_k141_21588\|\|full | Low-quality |
| S14_vs_k141_57853\|\|full | Not-determined |
| S14_vs_k141_79875\|\|full | Not-determined |
| S14_vs_k141_49953\|\|full | Not-determined |
| S14_vs_k141_95019\|\|full | Low-quality |
| S14_vs_k141_49977\|\|full | Not-determined |
| S14_vs_k141_109568\|\|full | Not-determined |
| S14_vs_k141_109571\|\|full | Not-determined |
| S14_vs_k141_116714\|\|full | Low-quality |
| S14_vs_k141_36087\|\|full | Not-determined |
| S14_vs_k141_101696\|\|full | Low-quality |
| S14_vs_k141_109592\|\|full | Not-determined |
| S14_vs_k141_13793\|\|full | Low-quality |
| S14_vs_k141_36104\|\|full | Low-quality |
| S14_vs_k141_101705\|\|full | Low-quality |
| S14_vs_k141_101711\|\|full | Low-quality |
| S14_vs_k141_6568\|\|full | Not-determined |
| S14_vs_k141_36109\|\|full | Low-quality |
| S14_vs_k141_116718\|\|full | Not-determined |
| S14_vs_k141_6576\|\|full | Low-quality |
| S7_vs_87279\|\|full | Medium-quality |
| S14_vs_k141_64806\|\|full | Not-determined |
| S14_vs_k141_64807\|\|full | Low-quality |
| S14_vs_k141_109617\|\|full | Low-quality |
| S14_vs_k141_57905\|\|full | Not-determined |
| S14_vs_k141_72345\|\|full | Low-quality |
| S14_vs_k141_101733\|\|full | Low-quality |
| S14_vs_k141_13811\|\|full | Low-quality |
| S14_vs_k141_64836\|\|full | Not-determined |
| S14_vs_k141_57926\|\|full | Not-determined |
| S13_extracted_120549 | High-quality |
| S14_vs_k141_57927\|\|full | Low-quality |
| S14_vs_k141_101768\|\|full | Not-determined |
| S14_vs_k141_79966\|\|full | Not-determined |
| S14_vs_k141_109674\|\|full | Not-determined |
| S14_vs_k141_64885\|\|full | Low-quality |
| S14_vs_k141_64889\|\|full | Low-quality |
| S14_vs_k141_6647\|\|full | Low-quality |
| S14_vs_k141_72418\|\|full | Low-quality |
| S14_vs_k141_116792\|\|full | Low-quality |
| S14_vs_k141_72419\|\|full | Low-quality |
| S14_vs_k141_13852\|\|full | Not-determined |
| S14_vs_k141_6652\|\|full | Not-determined |
| S14_vs_k141_72436\|\|full | Low-quality |
| S14_vs_k141_21651\|\|full | Not-determined |
| S14_vs_k141_116805\|\|full | Not-determined |
| S14_vs_k141_13863\|\|full | Not-determined |
| S14_vs_k141_101810\|\|full | Low-quality |
| S14_vs_k141_36162\|\|full | Low-quality |
| S14_vs_k141_57970\|\|full | Low-quality |
| S14_vs_k141_95115\|\|full | Low-quality |
| S14_vs_k141_36187\|\|full | Low-quality |
| S14_vs_k141_101829\|\|full | Not-determined |
| S14_vs_k141_13877\|\|full | Not-determined |
| S14_vs_k141_87479\|\|full | Low-quality |
| S14_vs_k141_21677\|\|full | Not-determined |
| S14_vs_k141_13884\|\|full | Low-quality |
| S14_vs_k141_6679\|\|full | Low-quality |
| S14_vs_k141_13889\|\|full | Low-quality |
| S14_vs_k141_116845\|\|full | Low-quality |
| S14_vs_k141_87481\|\|full | Low-quality |
| S14_vs_k141_80027\|\|full | Not-determined |
| S14_vs_k141_101843\|\|full | Not-determined |
| S14_vs_k141_80033\|\|full | Not-determined |
| S14_vs_k141_80035\|\|full | Low-quality |
| S14_vs_k141_109770\|\|full | Not-determined |
| S14_vs_k141_6713\|\|full | Low-quality |
| S14_vs_k141_95159\|\|full | Not-determined |
| S14_vs_k141_21718\|\|full | Low-quality |
| S14_vs_k141_80056\|\|full | Low-quality |
| S14_vs_k141_21722\|\|full | Low-quality |
| S14_vs_k141_58026\|\|full | Not-determined |
| S14_vs_k141_95164\|\|full | Low-quality |
| S14_vs_k141_87517\|\|full | Not-determined |
| S14_vs_k141_21728\|\|full | Low-quality |
| S14_vs_k141_21729\|\|full | Not-determined |
| S14_vs_k141_80073\|\|full | Low-quality |
| S14_vs_k141_109793\|\|full | Low-quality |
| S14_vs_k141_64981\|\|full | Not-determined |
| S14_vs_k141_72511\|\|full | Not-determined |
| S14_vs_k141_87535\|\|full | Low-quality |
| S14_vs_k141_50141\|\|full | Low-quality |
| S14_vs_k141_80080\|\|full | Low-quality |
| S14_vs_k141_28350\|\|full | Low-quality |
| S14_vs_k141_36259\|\|full | Low-quality |
| S14_vs_k141_116873\|\|full | Low-quality |
| S14_vs_k141_87550\|\|full | Low-quality |
| S14_vs_k141_109827\|\|full | Low-quality |
| S14_vs_k141_6761\|\|full | Low-quality |
| S14_vs_k141_6762\|\|full | Not-determined |
| S14_vs_k141_109830\|\|full | Not-determined |
| S14_vs_k141_109833\|\|full | Low-quality |
| S14_vs_k141_36298\|\|full | Low-quality |
| S14_vs_k141_13927\|\|full | Not-determined |
| S14_vs_k141_21774\|\|full | Not-determined |
| S14_vs_k141_109867\|\|full | Low-quality |
| S14_vs_k141_72553\|\|full | Low-quality |
| S14_vs_k141_6767\|\|full | Not-determined |
| S14_vs_k141_116897\|\|full | Low-quality |
| S14_vs_k141_13937\|\|full | Low-quality |
| S14_vs_k141_21796\|\|full | Not-determined |
| S14_vs_k141_65030\|\|full | Not-determined |
| S14_vs_k141_109883\|\|full | Low-quality |
| S14_vs_k141_116910\|\|full | Not-determined |
| S14_vs_k141_95245\|\|full | Low-quality |
| S14_vs_k141_109894\|\|full | Low-quality |
| S14_vs_k141_109896\|\|full | Low-quality |
| S14_vs_k141_13958\|\|full | Low-quality |
| S14_vs_k141_95249\|\|full | Low-quality |
| S14_vs_k141_28423\|\|full | Low-quality |
| S14_vs_k141_58114\|\|full | Not-determined |
| S14_vs_k141_116943\|\|full | Not-determined |
| S14_vs_k141_28426\|\|full | Low-quality |
| S14_vs_k141_21811\|\|full | Not-determined |
| S14_vs_k141_87627\|\|full | Low-quality |
| S14_vs_k141_6816\|\|full | Low-quality |
| S14_vs_k141_21824\|\|full | Not-determined |
| S14_vs_k141_116961\|\|full | Low-quality |
| S14_vs_k141_28442\|\|full | Not-determined |
| S14_vs_k141_50245\|\|full | Low-quality |
| S14_vs_k141_6834\|\|full | Low-quality |
| S14_vs_k141_50246\|\|full | Low-quality |
| S14_vs_k141_65072\|\|full | Not-determined |
| S14_vs_k141_72600\|\|full | Low-quality |
| S14_vs_k141_36336\|\|full | Not-determined |
| S14_vs_k141_36338\|\|full | Low-quality |
| S14_vs_k141_21843\|\|full | Not-determined |
| S14_vs_k141_116977\|\|full | Low-quality |
| S14_vs_k141_80165\|\|full | Not-determined |
| S14_vs_k141_72607\|\|full | Low-quality |
| S14_vs_k141_72612\|\|full | Low-quality |
| S14_vs_k141_87657\|\|full | Not-determined |
| S14_vs_k141_58189\|\|full | Low-quality |
| S14_vs_k141_58192\|\|full | Low-quality |
| S14_vs_k141_21866\|\|full | Low-quality |
| S14_vs_k141_65118\|\|full | Low-quality |
| S14_vs_k141_87661\|\|full | Not-determined |
| S13_vs_120549\|\|full | High-quality |
| S14_vs_k141_116993\|\|full | Low-quality |
| S14_vs_k141_72636\|\|full | Low-quality |
| S14_vs_k141_21874\|\|full | Low-quality |
| S14_vs_k141_36366\|\|full | Low-quality |
| S14_vs_k141_58206\|\|full | Not-determined |
| S14_vs_k141_36380\|\|full | Not-determined |
| S14_vs_k141_58212\|\|full | Low-quality |
| S14_vs_k141_101954\|\|full | Low-quality |
| S14_vs_k141_21892\|\|full | Not-determined |
| S14_vs_k141_36382\|\|full | Not-determined |
| S14_vs_k141_36383\|\|full | Low-quality |
| S14_vs_k141_14053\|\|full | Low-quality |
| S14_vs_k141_72665\|\|full | Not-determined |
| S14_vs_k141_65128\|\|full | Low-quality |
| S14_vs_k141_14072\|\|full | Low-quality |
| S14_vs_k141_14074\|\|full | Not-determined |
| S14_vs_k141_14077\|\|full | Low-quality |
| S14_vs_k141_87711\|\|full | Not-determined |
| S14_vs_k141_50307\|\|full | Not-determined |
| S14_vs_k141_6877\|\|full | Low-quality |
| S14_vs_k141_6879\|\|full | Not-determined |
| S14_vs_k141_87715\|\|full | Low-quality |
| S14_vs_k141_72707\|\|full | Not-determined |
| S14_vs_k141_21935\|\|full | Not-determined |
| S14_vs_k141_36442\|\|full | Low-quality |
| S14_vs_k141_6903\|\|full | Low-quality |

| | |
|---|---|
| S14_vs_k141_58273\|\|full | Low-quality |
| S14_vs_k141_50360\|\|full | Low-quality |
| S14_vs_k141_87773\|\|full | Not-determined |
| S14_vs_k141_58285\|\|full | Low-quality |
| S14_vs_k141_36482\|\|full | Not-determined |
| S14_vs_k141_72769\|\|full | Low-quality |
| S14_vs_k141_21978\|\|full | Not-determined |
| S14_vs_k141_72776\|\|full | Not-determined |
| S14_vs_k141_21980\|\|full | Low-quality |
| S14_vs_k141_102020\|\|full | Low-quality |
| S14_vs_k141_117092\|\|full | Low-quality |
| S14_vs_k141_36496\|\|full | Low-quality |
| S14_vs_k141_14147\|\|full | Low-quality |
| S14_vs_k141_117106\|\|full | Not-determined |
| S14_vs_k141_58305\|\|full | Not-determined |
| S14_vs_k141_28460\|\|full | Low-quality |
| S14_vs_k141_50373\|\|full | Low-quality |
| S14_vs_k141_6960\|\|full | Low-quality |
| S14_vs_k141_65198\|\|full | Low-quality |
| S14_vs_k141_50384\|\|full | Low-quality |
| S14_vs_k141_14169\|\|full | Low-quality |
| S14_vs_k141_14170\|\|full | Not-determined |
| S14_vs_k141_102047\|\|full | Low-quality |
| S14_vs_k141_50390\|\|full | Low-quality |
| S14_vs_k141_28474\|\|full | Low-quality |
| S14_vs_k141_28485\|\|full | Low-quality |
| S14_vs_k141_80278\|\|full | Low-quality |
| S14_vs_k141_6971\|\|full | Low-quality |
| S14_vs_k141_102080\|\|full | Low-quality |
| S14_vs_k141_102088\|\|full | Low-quality |
| S14_vs_k141_87796\|\|full | Low-quality |
| S14_vs_k141_80302\|\|full | Low-quality |
| S14_vs_k141_6981\|\|full | Not-determined |
| S14_vs_k141_50423\|\|full | Low-quality |
| S14_vs_k141_65219\|\|full | Low-quality |
| S14_vs_k141_65229\|\|full | Low-quality |
| S14_vs_k141_65232\|\|full | Not-determined |
| S14_vs_k141_58369\|\|full | Low-quality |
| S14_vs_k141_80332\|\|full | Low-quality |
| S14_vs_k141_65248\|\|full | Not-determined |
| S14_vs_k141_58375\|\|full | Low-quality |
| S14_vs_k141_58376\|\|full | Not-determined |
| S14_vs_k141_80340\|\|full | Low-quality |
| S14_vs_k141_87827\|\|full | Not-determined |
| S14_vs_k141_80342\|\|full | Not-determined |
| S14_vs_k141_14231\|\|full | Not-determined |
| S14_vs_k141_65255\|\|full | Low-quality |
| S14_vs_k141_80362\|\|full | Not-determined |
| S14_vs_k141_14249\|\|full | Low-quality |
| S14_vs_k141_7016\|\|full | Low-quality |
| S14_vs_k141_58392\|\|full | Not-determined |
| S14_vs_k141_28528\|\|full | Low-quality |
| S14_vs_k141_72886\|\|full | Low-quality |
| S16_vs_84462\|\|full | High-quality |
| S14_vs_k141_7019\|\|full | Low-quality |
| S14_vs_k141_80366\|\|full | Low-quality |
| S14_vs_k141_80373\|\|full | Low-quality |
| S14_vs_k141_65281\|\|full | Low-quality |
| S14_vs_k141_28536\|\|full | Low-quality |
| S14_vs_k141_36614\|\|full | Low-quality |
| S14_vs_k141_14271\|\|full | Low-quality |
| S14_vs_k141_87847\|\|full | Low-quality |
| S14_vs_k141_65290\|\|full | Not-determined |
| S14_vs_k141_87850\|\|full | Low-quality |
| S14_vs_k141_28548\|\|full | Not-determined |
| S14_vs_k141_36625\|\|full | Low-quality |
| S14_vs_k141_28561\|\|full | Low-quality |
| S14_vs_k141_28566\|\|full | Low-quality |
| S14_vs_k141_72909\|\|full | Not-determined |
| S14_vs_k141_7046\|\|full | Low-quality |
| S14_vs_k141_65315\|\|full | Low-quality |
| S14_vs_k141_36632\|\|full | Not-determined |
| S14_vs_k141_87859\|\|full | Low-quality |
| S14_vs_k141_50530\|\|full | Low-quality |
| S14_vs_k141_50531\|\|full | Not-determined |
| S14_vs_k141_102181\|\|full | Not-determined |
| S14_vs_k141_28599\|\|full | Low-quality |
| S14_vs_k141_7083\|\|full | Low-quality |
| S14_vs_k141_14355\|\|full | Not-determined |
| S14_vs_k141_50557\|\|full | Low-quality |
| S14_vs_k141_28606\|\|full | Low-quality |
| S14_vs_k141_7109\|\|full | Not-determined |
| S14_vs_k141_28610\|\|full | Low-quality |
| S14_vs_k141_65348\|\|full | Low-quality |
| S14_vs_k141_28629\|\|full | Low-quality |
| S14_vs_k141_50587\|\|full | Not-determined |
| S16_vs_80244\|\|full | High-quality |
| S14_vs_k141_80415\|\|full | Not-determined |
| S14_vs_k141_28635\|\|full | Not-determined |
| S14_vs_k141_102235\|\|full | Low-quality |
| S14_vs_k141_102246\|\|full | Not-determined |
| S14_vs_k141_14394\|\|full | Low-quality |
| S14_vs_k141_58459\|\|full | Not-determined |
| S14_vs_k141_80431\|\|full | Not-determined |
| S14_vs_k141_72990\|\|full | Not-determined |
| S14_vs_k141_7192\|\|full | Not-determined |
| S14_vs_k141_72995\|\|full | Low-quality |
| S14_vs_k141_14403\|\|full | Not-determined |
| S14_vs_k141_65390\|\|full | Low-quality |
| S14_vs_k141_58499\|\|full | Not-determined |
| S14_vs_k141_7200\|\|full | Not-determined |
| S14_vs_k141_14419\|\|full | Low-quality |
| S14_vs_k141_58507\|\|full | Low-quality |
| S14_vs_k141_58509\|\|full | Low-quality |
| S14_vs_k141_14422\|\|full | Low-quality |
| S14_vs_k141_14425\|\|full | Low-quality |
| S14_vs_k141_102300\|\|full | Not-determined |
| S14_vs_k141_73010\|\|full | Low-quality |
| S14_vs_k141_65411\|\|full | Not-determined |
| S14_vs_k141_50639\|\|full | Low-quality |
| S14_vs_k141_73014\|\|full | Not-determined |
| S14_vs_k141_7223\|\|full | Low-quality |
| S14_vs_k141_80468\|\|full | Not-determined |
| S14_vs_k141_73040\|\|full | Not-determined |
| S14_vs_k141_58541\|\|full | Low-quality |
| S14_vs_k141_7229\|\|full | Not-determined |
| S14_vs_k141_50677\|\|full | Not-determined |
| S14_vs_k141_7242\|\|full | Not-determined |
| S14_vs_k141_73053\|\|full | Low-quality |
| S14_vs_k141_102322\|\|full | Low-quality |
| S14_vs_k141_80478\|\|full | Not-determined |
| S14_vs_k141_65452\|\|full | Not-determined |
| S14_vs_k141_73061\|\|full | Low-quality |
| S14_vs_k141_102337\|\|full | Low-quality |
| S14_vs_k141_73063\|\|full | Low-quality |
| S14_vs_k141_28727\|\|full | Not-determined |
| S14_vs_k141_73106\|\|full | Not-determined |
| S14_vs_k141_65483\|\|full | Not-determined |
| S14_vs_k141_102373\|\|full | Low-quality |
| S14_vs_k141_14468\|\|full | Not-determined |
| S14_vs_k141_65501\|\|full | Low-quality |
| S14_vs_k141_102379\|\|full | Low-quality |
| S14_vs_k141_73126\|\|full | Not-determined |
| S14_vs_k141_7323\|\|full | Low-quality |
| S14_vs_k141_7325\|\|full | Not-determined |
| S14_vs_k141_28805\|\|full | Low-quality |
| S14_vs_k141_80575\|\|full | Low-quality |
| S14_vs_k141_80580\|\|full | Not-determined |
| S14_vs_k141_80588\|\|full | Not-determined |
| S14_vs_k141_80594\|\|full | Not-determined |
| S14_vs_k141_102450\|\|full | Low-quality |
| S14_vs_k141_50787\|\|full | Not-determined |
| S14_vs_k141_28839\|\|full | Not-determined |
| S14_vs_k141_102458\|\|full | Low-quality |
| S14_vs_k141_73201\|\|full | Low-quality |
| S14_vs_k141_28848\|\|full | Low-quality |
| S14_vs_k141_73202\|\|full | Low-quality |
| S14_vs_k141_73214\|\|full | Not-determined |
| S14_vs_k141_50811\|\|full | Low-quality |
| S14_vs_k141_28871\|\|full | Low-quality |
| S14_vs_k141_50825\|\|full | Not-determined |
| S14_vs_k141_50833\|\|full | Not-determined |
| S14_vs_k141_50834\|\|full | Low-quality |
| S14_vs_k141_28891\|\|full | Low-quality |
| S14_vs_k141_14585\|\|full | Low-quality |
| S14_vs_k141_28905\|\|full | Low-quality |
| S14_vs_k141_28909\|\|full | Low-quality |
| S14_vs_k141_28911\|\|full | Not-determined |
| S14_vs_k141_14607\|\|full | Low-quality |
| S14_vs_k141_65541\|\|full | Not-determined |
| S14_vs_k141_50862\|\|full | Low-quality |
| S14_vs_k141_73261\|\|full | Not-determined |
| S14_vs_k141_28937\|\|full | Low-quality |
| S14_vs_k141_73265\|\|full | Low-quality |
| S14_vs_k141_65561\|\|full | Not-determined |
| S14_vs_k141_50896\|\|full | Not-determined |
| S14_vs_k141_28960\|\|full | Low-quality |
| S14_vs_k141_50900\|\|full | Not-determined |
| S14_vs_k141_14643\|\|full | Low-quality |
| S14_vs_k141_28977\|\|full | Low-quality |
| S14_vs_k141_28989\|\|full | Not-determined |
| S14_vs_k141_50926\|\|full | Low-quality |
| S14_vs_k141_50931\|\|full | Low-quality |
| S14_vs_k141_65616\|\|full | Not-determined |
| S14_vs_k141_29030\|\|full | Low-quality |
| S14_vs_k141_65639\|\|full | Low-quality |
| S14_vs_k141_65641\|\|full | Low-quality |
| S14_vs_k141_29064\|\|full | Not-determined |
| S14_vs_k141_65694\|\|full | Not-determined |
| S14_vs_k141_51027\|\|full | Not-determined |
| S14_vs_k141_29083\|\|full | Low-quality |
| S14_vs_k141_51052\|\|full | Not-determined |
| S14_vs_k141_51066\|\|full | Not-determined |
| S14_vs_k141_29118\|\|full | Low-quality |
| S14_vs_k141_29120\|\|full | Low-quality |
| S14_vs_k141_65723\|\|full | Not-determined |
| S14_vs_k141_65725\|\|full | Not-determined |
| S14_vs_k141_51076\|\|full | Low-quality |
| S14_vs_k141_51085\|\|full | Low-quality |
| S14_vs_k141_29137\|\|full | Low-quality |
| S14_vs_k141_51092\|\|full | Low-quality |
| S14_vs_k141_51100\|\|full | Low-quality |
| S14_vs_k141_51106\|\|full | Low-quality |
| S14_vs_k141_29162\|\|full | Low-quality |
| S14_vs_k141_65751\|\|full | Not-determined |
| S14_vs_k141_65754\|\|full | Low-quality |
| S14_vs_k141_65759\|\|full | Low-quality |
| S14_vs_k141_65781\|\|full | Low-quality |
| S14_vs_k141_51154\|\|full | Low-quality |
| S14_vs_k141_65785\|\|full | Low-quality |
| S14_vs_k141_65795\|\|full | Low-quality |
| S14_vs_k141_29244\|\|full | Low-quality |
| S14_vs_k141_65816\|\|full | Not-determined |
| S14_vs_k141_65827\|\|full | Low-quality |
| S14_vs_k141_65839\|\|full | Low-quality |
| S14_vs_k141_65842\|\|full | Low-quality |
| S14_vs_k141_29270\|\|full | Low-quality |
| S14_vs_k141_51199\|\|full | Low-quality |
| S14_vs_k141_51201\|\|full | Low-quality |
| S14_vs_k141_65866\|\|full | Not-determined |
| S14_vs_k141_51211\|\|full | Not-determined |
| S14_vs_k141_65882\|\|full | Low-quality |
| S14_vs_k141_65925\|\|full | Low-quality |
| S14_vs_k141_65928\|\|full | Low-quality |
| S14_vs_k141_117151\|\|0_partial | Not-determined |
| S14_vs_k141_44198\|\|0_partial | Low-quality |
| S14_vs_k141_95555\|\|0_partial | Low-quality |
| S14_vs_k141_22544\|\|0_partial | Low-quality |
| S14_vs_k141_38028\|\|0_partial | Not-determined |
| S14_vs_k141_30677\|\|0_partial | Not-determined |
| S14_vs_k141_16287\|\|0_partial | Not-determined |
| S11_vs_k141_62573\|\|full | High-quality |
| S14_vs_k141_45208\|\|0_partial | Not-determined |
| S14_vs_k141_45208\|\|1_partial | Low-quality |
| S14_vs_k141_82295\|\|0_partial | Low-quality |
| S14_vs_k141_82295\|\|1_partial | Low-quality |
| S14_vs_k141_89890\|\|0_partial | Low-quality |
| S14_vs_k141_60822\|\|0_partial | Not-determined |
| S14_vs_k141_112443\|\|0_partial | Low-quality |
| S14_vs_k141_61186\|\|0_partial | Low-quality |
| S14_vs_k141_54597\|\|0_partial | Not-determined |
| S14_vs_k141_106399\|\|0_partial | Not-determined |
| S14_vs_k141_47308\|\|0_partial | Low-quality |
| S14_vs_k141_4124\|\|0_partial | Low-quality |
| S14_vs_k141_26120\|\|0_partial | Not-determined |
| S14_vs_k141_56082\|\|0_partial | Low-quality |
| S14_vs_k141_35119\|\|0_partial | Low-quality |
| S14_vs_k141_72262\|\|0_partial | Low-quality |
| S14_vs_k141_101601\|\|0_partial | Low-quality |
| S14_vs_k141_28459\|\|0_partial | Low-quality |
| S14_vs_k141_0\|\|lt2gene | Not-determined |
| S14_vs_k141_80599\|\|lt2gene | Low-quality |
| S14_vs_k141_14658\|\|lt2gene | Low-quality |
| S14_vs_k141_80605\|\|lt2gene | Not-determined |
| S14_vs_k141_7333\|\|lt2gene | Not-determined |
| S14_vs_k141_36642\|\|lt2gene | Low-quality |
| S14_vs_k141_58621\|\|lt2gene | Low-quality |
| S14_vs_k141_58622\|\|lt2gene | Low-quality |
| S14_vs_k141_80611\|\|lt2gene | Low-quality |
| S14_vs_k141_35\|\|lt2gene | Low-quality |
| S14_vs_k141_109923\|\|lt2gene | Not-determined |
| S14_vs_k141_22005\|\|lt2gene | Low-quality |
| S14_vs_k141_43984\|\|lt2gene | Low-quality |
| S14_vs_k141_43985\|\|lt2gene | Low-quality |
| S14_vs_k141_109926\|\|lt2gene | Not-determined |
| S14_vs_k141_43986\|\|lt2gene | Low-quality |
| S14_vs_k141_42\|\|lt2gene | Low-quality |
| S14_vs_k141_43990\|\|lt2gene | Not-determined |
| S14_vs_k141_55\|\|lt2gene | Low-quality |
| S14_vs_k141_56\|\|lt2gene | Low-quality |
| S14_vs_k141_43997\|\|lt2gene | Low-quality |
| S14_vs_k141_109936\|\|lt2gene | Low-quality |
| S14_vs_k141_36684\|\|lt2gene | Low-quality |
| S14_vs_k141_44001\|\|lt2gene | Low-quality |
| S14_vs_k141_36694\|\|lt2gene | Low-quality |
| S14_vs_k141_64\|\|lt2gene | Low-quality |
| S14_vs_k141_80635\|\|lt2gene | Not-determined |
| S14_vs_k141_95274\|\|lt2gene | Not-determined |
| S14_vs_k141_65997\|\|lt2gene | Low-quality |
| S14_vs_k141_66001\|\|lt2gene | Low-quality |
| S14_vs_k141_73292\|\|lt2gene | Not-determined |
| S14_vs_k141_95300\|\|lt2gene | Low-quality |
| S14_vs_k141_66018\|\|lt2gene | Low-quality |
| S14_vs_k141_22045\|\|lt2gene | Low-quality |
| S14_vs_k141_58649\|\|lt2gene | Low-quality |
| S14_vs_k141_7395\|\|lt2gene | Not-determined |
| S14_vs_k141_95317\|\|lt2gene | Low-quality |
| S14_vs_k141_87943\|\|lt2gene | Low-quality |
| S14_vs_k141_36739\|\|lt2gene | Not-determined |
| S14_vs_k141_22059\|\|lt2gene | Low-quality |
| S14_vs_k141_58657\|\|lt2gene | Not-determined |
| S14_vs_k141_58660\|\|lt2gene | Not-determined |
| S14_vs_k141_51358\|\|lt2gene | Low-quality |
| S14_vs_k141_66027\|\|lt2gene | Low-quality |
| S14_vs_k141_51367\|\|lt2gene | Not-determined |
| S14_vs_k141_58672\|\|lt2gene | Low-quality |
| S14_vs_k141_102632\|\|lt2gene | Not-determined |
| S14_vs_k141_44082\|\|lt2gene | Low-quality |
| S14_vs_k141_95351\|\|lt2gene | Not-determined |
| S14_vs_k141_51394\|\|lt2gene | Low-quality |
| S14_vs_k141_44088\|\|lt2gene | Low-quality |
| S14_vs_k141_51400\|\|lt2gene | Low-quality |
| S14_vs_k141_66053\|\|lt2gene | Low-quality |
| S14_vs_k141_58707\|\|lt2gene | Not-determined |
| S14_vs_k141_58708\|\|lt2gene | Low-quality |
| S14_vs_k141_58710\|\|lt2gene | Low-quality |
| S14_vs_k141_102653\|\|lt2gene | Low-quality |
| S14_vs_k141_130\|\|lt2gene | Low-quality |
| S14_vs_k141_87977\|\|lt2gene | Low-quality |
| S14_vs_k141_58715\|\|lt2gene | Low-quality |
| S14_vs_k141_66063\|\|lt2gene | Low-quality |
| S14_vs_k141_140\|\|lt2gene | Low-quality |
| S14_vs_k141_66075\|\|lt2gene | Low-quality |
| S14_vs_k141_14759\|\|lt2gene | Low-quality |
| S14_vs_k141_66079\|\|lt2gene | Low-quality |
| S14_vs_k141_58743\|\|lt2gene | Low-quality |
| S14_vs_k141_102682\|\|lt2gene | Not-determined |
| S14_vs_k141_44124\|\|lt2gene | Not-determined |
| S14_vs_k141_7485\|\|lt2gene | Low-quality |
| S14_vs_k141_14793\|\|lt2gene | Low-quality |
| S14_vs_k141_51436\|\|lt2gene | Low-quality |
| S14_vs_k141_109982\|\|lt2gene | Low-quality |
| S14_vs_k141_58784\|\|lt2gene | Low-quality |
| S14_vs_k141_66135\|\|lt2gene | Not-determined |
| S14_vs_k141_87993\|\|lt2gene | Low-quality |
| S14_vs_k141_51444\|\|lt2gene | Low-quality |
| S14_vs_k141_88013\|\|lt2gene | Low-quality |
| S14_vs_k141_51461\|\|lt2gene | Low-quality |
| S14_vs_k141_73344\|\|lt2gene | Low-quality |
| S14_vs_k141_110016\|\|lt2gene | Low-quality |
| S14_vs_k141_200\|\|lt2gene | Low-quality |
| S14_vs_k141_51464\|\|lt2gene | Low-quality |
| S14_vs_k141_51486\|\|lt2gene | Low-quality |
| S14_vs_k141_73354\|\|lt2gene | Low-quality |
| S14_vs_k141_73357\|\|lt2gene | Not-determined |
| S14_vs_k141_58825\|\|lt2gene | Low-quality |
| S14_vs_k141_223\|\|lt2gene | Low-quality |
| S14_vs_k141_224\|\|lt2gene | Low-quality |
| S14_vs_k141_88039\|\|lt2gene | Low-quality |
| S14_vs_k141_95422\|\|lt2gene | Low-quality |
| S14_vs_k141_88046\|\|lt2gene | Low-quality |
| S14_vs_k141_22183\|\|lt2gene | Low-quality |
| S14_vs_k141_80780\|\|lt2gene | Low-quality |
| S14_vs_k141_110030\|\|lt2gene | Not-determined |
| S14_vs_k141_110032\|\|lt2gene | Low-quality |
| S14_vs_k141_80790\|\|lt2gene | Low-quality |
| S14_vs_k141_80792\|\|lt2gene | Low-quality |
| S14_vs_k141_110035\|\|lt2gene | Low-quality |
| S14_vs_k141_22197\|\|lt2gene | Low-quality |
| S14_vs_k141_88048\|\|lt2gene | Not-determined |
| S14_vs_k141_110052\|\|lt2gene | Low-quality |
| S14_vs_k141_36893\|\|lt2gene | Not-determined |
| S14_vs_k141_36898\|\|lt2gene | Low-quality |
| S14_vs_k141_266\|\|lt2gene | Low-quality |
| S14_vs_k141_95448\|\|lt2gene | Low-quality |
| S14_vs_k141_51512\|\|lt2gene | Low-quality |
| S14_vs_k141_80806\|\|lt2gene | Low-quality |
| S14_vs_k141_51516\|\|lt2gene | Low-quality |
| S14_vs_k141_36914\|\|lt2gene | Not-determined |
| S14_vs_k141_7592\|\|lt2gene | Low-quality |
| S14_vs_k141_278\|\|lt2gene | Low-quality |
| S14_vs_k141_88091\|\|lt2gene | Low-quality |
| S14_vs_k141_51526\|\|lt2gene | Low-quality |
| S14_vs_k141_51528\|\|lt2gene | Low-quality |
| S14_vs_k141_95463\|\|lt2gene | Low-quality |
| S14_vs_k141_51536\|\|lt2gene | Low-quality |
| S14_vs_k141_88126\|\|lt2gene | Low-quality |
| S14_vs_k141_36946\|\|lt2gene | Not-determined |
| S14_vs_k141_14851\|\|lt2gene | Not-determined |
| S14_vs_k141_36959\|\|lt2gene | Low-quality |
| S14_vs_k141_95476\|\|lt2gene | Low-quality |
| S14_vs_k141_36960\|\|lt2gene | Low-quality |
| S14_vs_k141_22225\|\|lt2gene | Not-determined |
| S14_vs_k141_36978\|\|lt2gene | Low-quality |
| S14_vs_k141_95497\|\|lt2gene | Low-quality |
| S14_vs_k141_102743\|\|lt2gene | Low-quality |
| S14_vs_k141_22253\|\|lt2gene | Low-quality |
| S14_vs_k141_321\|\|lt2gene | Low-quality |
| S14_vs_k141_327\|\|lt2gene | Not-determined |
| S14_vs_k141_95505\|\|lt2gene | Low-quality |
| S14_vs_k141_328\|\|lt2gene | Low-quality |
| S14_vs_k141_95513\|\|lt2gene | Low-quality |
| S14_vs_k141_29586\|\|lt2gene | Low-quality |
| S14_vs_k141_102765\|\|lt2gene | Low-quality |
| S14_vs_k141_7648\|\|lt2gene | Not-determined |
| S14_vs_k141_73421\|\|lt2gene | Low-quality |
| S14_vs_k141_102794\|\|lt2gene | Low-quality |
| S14_vs_k141_95530\|\|lt2gene | Not-determined |
| S14_vs_k141_88175\|\|lt2gene | Low-quality |
| S14_vs_k141_37027\|\|lt2gene | Not-determined |
| S14_vs_k141_95538\|\|lt2gene | Low-quality |
| S14_vs_k141_22260\|\|lt2gene | Low-quality |
| S14_vs_k141_58946\|\|lt2gene | Low-quality |
| S14_vs_k141_73431\|\|lt2gene | Low-quality |
| S14_vs_k141_88199\|\|lt2gene | Low-quality |
| S14_vs_k141_37044\|\|lt2gene | Low-quality |
| S14_vs_k141_110119\|\|lt2gene | Low-quality |
| S14_vs_k141_37061\|\|lt2gene | Low-quality |
| S14_vs_k141_37068\|\|lt2gene | Low-quality |
| S14_vs_k141_110133\|\|lt2gene | Low-quality |
| S14_vs_k141_95563\|\|lt2gene | Low-quality |
| S14_vs_k141_80891\|\|lt2gene | Low-quality |
| S14_vs_k141_80892\|\|lt2gene | Low-quality |
| S14_vs_k141_80895\|\|lt2gene | Low-quality |
| S14_vs_k141_66183\|\|lt2gene | Low-quality |
| S14_vs_k141_73456\|\|lt2gene | Not-determined |
| S14_vs_k141_95587\|\|lt2gene | Low-quality |
| S14_vs_k141_29679\|\|lt2gene | Not-determined |
| S14_vs_k141_80904\|\|lt2gene | Low-quality |
| S14_vs_k141_66199\|\|lt2gene | Low-quality |
| S14_vs_k141_14970\|\|lt2gene | Low-quality |
| S14_vs_k141_110166\|\|lt2gene | Low-quality |
| S14_vs_k141_95593\|\|lt2gene | Low-quality |
| S14_vs_k141_95594\|\|lt2gene | Low-quality |
| S14_vs_k141_95604\|\|lt2gene | Low-quality |
| S14_vs_k141_95606\|\|lt2gene | Low-quality |
| S14_vs_k141_102885\|\|lt2gene | Low-quality |
| S14_vs_k141_484\|\|lt2gene | Low-quality |
| S14_vs_k141_95613\|\|lt2gene | Low-quality |
| S14_vs_k141_73473\|\|lt2gene | Low-quality |
| S14_vs_k141_488\|\|lt2gene | Low-quality |
| S14_vs_k141_15005\|\|lt2gene | Low-quality |
| S14_vs_k141_59037\|\|lt2gene | Low-quality |
| S14_vs_k141_44217\|\|lt2gene | Low-quality |
| S14_vs_k141_80944\|\|lt2gene | Low-quality |
| S14_vs_k141_102912\|\|lt2gene | Low-quality |
| S14_vs_k141_29728\|\|lt2gene | Not-determined |
| S14_vs_k141_102918\|\|lt2gene | Low-quality |
| S14_vs_k141_510\|\|lt2gene | Low-quality |
| S14_vs_k141_51724\|\|lt2gene | Not-determined |
| S14_vs_k141_516\|\|lt2gene | Low-quality |
| S14_vs_k141_102923\|\|lt2gene | Low-quality |
| S14_vs_k141_29741\|\|lt2gene | Low-quality |
| S14_vs_k141_88285\|\|lt2gene | Low-quality |
| S14_vs_k141_51733\|\|lt2gene | Low-quality |
| S14_vs_k141_110237\|\|lt2gene | Low-quality |
| S14_vs_k141_95674\|\|lt2gene | Low-quality |
| S14_vs_k141_110253\|\|lt2gene | Low-quality |
| S14_vs_k141_51744\|\|lt2gene | Low-quality |
| S14_vs_k141_59071\|\|lt2gene | Low-quality |
| S14_vs_k141_102939\|\|lt2gene | Not-determined |
| S14_vs_k141_548\|\|lt2gene | Low-quality |
| S14_vs_k141_37095\|\|lt2gene | Low-quality |
| S14_vs_k141_80991\|\|lt2gene | Not-determined |
| S14_vs_k141_44290\|\|lt2gene | Low-quality |
| S14_vs_k141_95708\|\|lt2gene | Not-determined |
| S14_vs_k141_95714\|\|lt2gene | Low-quality |
| S14_vs_k141_560\|\|lt2gene | Low-quality |
| S14_vs_k141_73505\|\|lt2gene | Low-quality |
| S14_vs_k141_73507\|\|lt2gene | Not-determined |
| S14_vs_k141_37115\|\|lt2gene | Low-quality |
| S14_vs_k141_73517\|\|lt2gene | Low-quality |
| S14_vs_k141_66223\|\|lt2gene | Low-quality |
| S14_vs_k141_66226\|\|lt2gene | Low-quality |
| S14_vs_k141_37118\|\|lt2gene | Low-quality |
| S14_vs_k141_95733\|\|lt2gene | Low-quality |
| S14_vs_k141_73527\|\|lt2gene | Low-quality |
| S14_vs_k141_95737\|\|lt2gene | Low-quality |
| S14_vs_k141_95739\|\|lt2gene | Not-determined |
| S14_vs_k141_591\|\|lt2gene | Not-determined |
| S14_vs_k141_22345\|\|lt2gene | Low-quality |
| S14_vs_k141_102985\|\|lt2gene | Not-determined |
| S14_vs_k141_593\|\|lt2gene | Not-determined |
| S14_vs_k141_88320\|\|lt2gene | Not-determined |
| S14_vs_k141_596\|\|lt2gene | Not-determined |
| S14_vs_k141_597\|\|lt2gene | Not-determined |
| S14_vs_k141_599\|\|lt2gene | Not-determined |
| S14_vs_k141_29768\|\|lt2gene | Not-determined |
| S14_vs_k141_608\|\|lt2gene | Low-quality |
| S14_vs_k141_37139\|\|lt2gene | Not-determined |
| S14_vs_k141_37141\|\|lt2gene | Not-determined |
| S14_vs_k141_81026\|\|lt2gene | Low-quality |
| S14_vs_k141_37143\|\|lt2gene | Low-quality |
| S14_vs_k141_110274\|\|lt2gene | Not-determined |
| S14_vs_k141_103011\|\|lt2gene | Low-quality |
| S14_vs_k141_95777\|\|lt2gene | Low-quality |
| S14_vs_k141_37150\|\|lt2gene | Low-quality |
| S14_vs_k141_37153\|\|lt2gene | Low-quality |
| S14_vs_k141_103025\|\|lt2gene | Low-quality |
| S14_vs_k141_103029\|\|lt2gene | Low-quality |
| S14_vs_k141_29810\|\|lt2gene | Not-determined |
| S14_vs_k141_66230\|\|lt2gene | Low-quality |
| S14_vs_k141_22360\|\|lt2gene | Low-quality |
| S14_vs_k141_81041\|\|lt2gene | Low-quality |
| S14_vs_k141_103065\|\|lt2gene | Low-quality |
| S14_vs_k141_73558\|\|lt2gene | Low-quality |
| S14_vs_k141_103075\|\|lt2gene | Low-quality |
| S14_vs_k141_103086\|\|lt2gene | Low-quality |
| S14_vs_k141_44363\|\|lt2gene | Low-quality |
| S14_vs_k141_22375\|\|lt2gene | Low-quality |
| S14_vs_k141_103100\|\|lt2gene | Not-determined |
| S14_vs_k141_81062\|\|lt2gene | Low-quality |
| S14_vs_k141_7758\|\|lt2gene | Low-quality |
| S14_vs_k141_88398\|\|lt2gene | Low-quality |
| S14_vs_k141_81067\|\|lt2gene | Low-quality |
| S14_vs_k141_637\|\|lt2gene | Low-quality |
| S14_vs_k141_51757\|\|lt2gene | Not-determined |
| S14_vs_k141_37210\|\|lt2gene | Low-quality |
| S14_vs_k141_59102\|\|lt2gene | Low-quality |
| S14_vs_k141_59104\|\|lt2gene | Low-quality |
| S14_vs_k141_81082\|\|lt2gene | Low-quality |
| S14_vs_k141_51767\|\|lt2gene | Low-quality |
| S14_vs_k141_44374\|\|lt2gene | Low-quality |
| S14_vs_k141_110311\|\|lt2gene | Low-quality |
| S14_vs_k141_81086\|\|lt2gene | Low-quality |
| S14_vs_k141_59114\|\|lt2gene | Low-quality |
| S14_vs_k141_22399\|\|lt2gene | Low-quality |
| S14_vs_k141_95846\|\|lt2gene | Low-quality |
| S14_vs_k141_73587\|\|lt2gene | Low-quality |
| S14_vs_k141_37242\|\|lt2gene | Low-quality |
| S14_vs_k141_37244\|\|lt2gene | Not-determined |
| S14_vs_k141_37248\|\|lt2gene | Low-quality |
| S14_vs_k141_37254\|\|lt2gene | Low-quality |
| S14_vs_k141_73597\|\|lt2gene | Low-quality |
| S14_vs_k141_110340\|\|lt2gene | Low-quality |
| S14_vs_k141_59146\|\|lt2gene | Not-determined |
| S14_vs_k141_7819\|\|lt2gene | Not-determined |
| S14_vs_k141_37265\|\|lt2gene | Low-quality |
| S14_vs_k141_37272\|\|lt2gene | Low-quality |
| S14_vs_k141_59150\|\|lt2gene | Low-quality |
| S14_vs_k141_110347\|\|lt2gene | Low-quality |
| S14_vs_k141_59157\|\|lt2gene | Low-quality |
| S14_vs_k141_110351\|\|lt2gene | Low-quality |
| S14_vs_k141_37276\|\|lt2gene | Low-quality |
| S14_vs_k141_7827\|\|lt2gene | Low-quality |
| S14_vs_k141_37277\|\|lt2gene | Low-quality |
| S14_vs_k141_66296\|\|lt2gene | Low-quality |
| S14_vs_k141_37279\|\|lt2gene | Low-quality |
| S14_vs_k141_44393\|\|lt2gene | Low-quality |
| S14_vs_k141_66300\|\|lt2gene | Low-quality |
| S14_vs_k141_88449\|\|lt2gene | Low-quality |
| S14_vs_k141_88457\|\|lt2gene | Low-quality |
| S14_vs_k141_95864\|\|lt2gene | Low-quality |
| S14_vs_k141_81134\|\|lt2gene | Low-quality |
| S14_vs_k141_29894\|\|lt2gene | Low-quality |
| S14_vs_k141_81150\|\|lt2gene | Not-determined |
| S14_vs_k141_51814\|\|lt2gene | Low-quality |
| S14_vs_k141_73638\|\|lt2gene | Low-quality |
| S14_vs_k141_15175\|\|lt2gene | Low-quality |
| S14_vs_k141_88474\|\|lt2gene | Low-quality |
| S14_vs_k141_88477\|\|lt2gene | Low-quality |
| S14_vs_k141_73644\|\|lt2gene | Low-quality |
| S14_vs_k141_22424\|\|lt2gene | Low-quality |
| S14_vs_k141_51837\|\|lt2gene | Low-quality |
| S14_vs_k141_44429\|\|lt2gene | Low-quality |
| S14_vs_k141_73654\|\|lt2gene | Low-quality |
| S14_vs_k141_29896\|\|lt2gene | Low-quality |
| S14_vs_k141_51858\|\|lt2gene | Low-quality |
| S14_vs_k141_15197\|\|lt2gene | Low-quality |
| S14_vs_k141_715\|\|lt2gene | Low-quality |
| S14_vs_k141_81181\|\|lt2gene | Low-quality |
| S14_vs_k141_73674\|\|lt2gene | Low-quality |
| S14_vs_k141_73675\|\|lt2gene | Low-quality |
| S14_vs_k141_59180\|\|lt2gene | Not-determined |
| S14_vs_k141_59181\|\|lt2gene | Low-quality |
| S14_vs_k141_59182\|\|lt2gene | Not-determined |
| S14_vs_k141_7869\|\|lt2gene | Not-determined |
| S14_vs_k141_15214\|\|lt2gene | Low-quality |
| S14_vs_k141_59193\|\|lt2gene | Low-quality |
| S14_vs_k141_59194\|\|lt2gene | Low-quality |
| S14_vs_k141_73687\|\|lt2gene | Not-determined |
| S14_vs_k141_73689\|\|lt2gene | Not-determined |
| S14_vs_k141_51890\|\|lt2gene | Low-quality |
| S14_vs_k141_51894\|\|lt2gene | Low-quality |
| S14_vs_k141_88551\|\|lt2gene | Low-quality |
| S14_vs_k141_727\|\|lt2gene | Low-quality |
| S14_vs_k141_742\|\|lt2gene | Not-determined |
| S14_vs_k141_59235\|\|lt2gene | Low-quality |
| S14_vs_k141_51912\|\|lt2gene | Low-quality |
| S14_vs_k141_22499\|\|lt2gene | Not-determined |
| S14_vs_k141_66384\|\|lt2gene | Low-quality |
| S14_vs_k141_110430\|\|lt2gene | Low-quality |
| S14_vs_k141_66398\|\|lt2gene | Low-quality |
| S14_vs_k141_110442\|\|lt2gene | Not-determined |
| S14_vs_k141_51959\|\|lt2gene | Low-quality |
| S14_vs_k141_804\|\|lt2gene | Low-quality |
| S14_vs_k141_59270\|\|lt2gene | Not-determined |
| S14_vs_k141_51973\|\|lt2gene | Low-quality |
| S14_vs_k141_81205\|\|lt2gene | Not-determined |
| S14_vs_k141_51977\|\|lt2gene | Not-determined |
| S14_vs_k141_81207\|\|lt2gene | Low-quality |
| S14_vs_k141_110448\|\|lt2gene | Not-determined |
| S14_vs_k141_37344\|\|lt2gene | Not-determined |
| S14_vs_k141_51993\|\|lt2gene | Low-quality |
| S14_vs_k141_7970\|\|lt2gene | Low-quality |
| S14_vs_k141_7973\|\|lt2gene | Low-quality |
| S14_vs_k141_7976\|\|lt2gene | Low-quality |
| S14_vs_k141_7977\|\|lt2gene | Low-quality |
| S14_vs_k141_7978\|\|lt2gene | Low-quality |
| S14_vs_k141_37393\|\|lt2gene | Not-determined |
| S14_vs_k141_52025\|\|lt2gene | Low-quality |
| S14_vs_k141_7996\|\|lt2gene | Low-quality |
| S14_vs_k141_95907\|\|lt2gene | Low-quality |
| S14_vs_k141_88649\|\|lt2gene | Low-quality |
| S14_vs_k141_37413\|\|lt2gene | Low-quality |
| S14_vs_k141_30017\|\|lt2gene | Low-quality |
| S14_vs_k141_37422\|\|lt2gene | Low-quality |
| S14_vs_k141_37426\|\|lt2gene | Not-determined |
| S14_vs_k141_110530\|\|lt2gene | Not-determined |

| | |
|---|---|
| S14_vs_k141_52055\|\|lt2gene | Low-quality |
| S14_vs_k141_81293\|\|lt2gene | Low-quality |
| S14_vs_k141_81295\|\|lt2gene | Low-quality |
| S14_vs_k141_37437\|\|lt2gene | Low-quality |
| S14_vs_k141_44532\|\|lt2gene | Low-quality |
| S14_vs_k141_103221\|\|lt2gene | Low-quality |
| S14_vs_k141_52077\|\|lt2gene | Low-quality |
| S14_vs_k141_52078\|\|lt2gene | Not-determined |
| S14_vs_k141_845\|\|lt2gene | Low-quality |
| S14_vs_k141_37451\|\|lt2gene | Low-quality |
| S14_vs_k141_15251\|\|lt2gene | Not-determined |
| S14_vs_k141_44535\|\|lt2gene | Low-quality |
| S14_vs_k141_59339\|\|lt2gene | Not-determined |
| S14_vs_k141_15254\|\|lt2gene | Low-quality |
| S14_vs_k141_15260\|\|lt2gene | Low-quality |
| S14_vs_k141_44542\|\|lt2gene | Low-quality |
| S14_vs_k141_88670\|\|lt2gene | Low-quality |
| S14_vs_k141_15268\|\|lt2gene | Low-quality |
| S14_vs_k141_15272\|\|lt2gene | Low-quality |
| S14_vs_k141_73805\|\|lt2gene | Not-determined |
| S14_vs_k141_73807\|\|lt2gene | Low-quality |
| S14_vs_k141_52126\|\|lt2gene | Low-quality |
| S14_vs_k141_88686\|\|lt2gene | Low-quality |
| S14_vs_k141_73809\|\|lt2gene | Low-quality |
| S14_vs_k141_88694\|\|lt2gene | Low-quality |
| S14_vs_k141_8050\|\|lt2gene | Low-quality |
| S14_vs_k141_59353\|\|lt2gene | Low-quality |
| S14_vs_k141_875\|\|lt2gene | Not-determined |
| S14_vs_k141_8065\|\|lt2gene | Low-quality |
| S14_vs_k141_15332\|\|lt2gene | Not-determined |
| S14_vs_k141_15333\|\|lt2gene | Low-quality |
| S14_vs_k141_15349\|\|lt2gene | Low-quality |
| S14_vs_k141_15352\|\|lt2gene | Low-quality |
| S14_vs_k141_37538\|\|lt2gene | Low-quality |
| S14_vs_k141_30112\|\|lt2gene | Low-quality |
| S14_vs_k141_88737\|\|lt2gene | Low-quality |
| S14_vs_k141_15362\|\|lt2gene | Low-quality |
| S14_vs_k141_30125\|\|lt2gene | Low-quality |
| S14_vs_k141_88749\|\|lt2gene | Low-quality |
| S14_vs_k141_110592\|\|lt2gene | Low-quality |
| S14_vs_k141_59368\|\|lt2gene | Low-quality |
| S14_vs_k141_59372\|\|lt2gene | Low-quality |
| S14_vs_k141_52184\|\|lt2gene | Low-quality |
| S14_vs_k141_73890\|\|lt2gene | Low-quality |
| S14_vs_k141_103317\|\|lt2gene | Not-determined |
| S14_vs_k141_52190\|\|lt2gene | Not-determined |
| S14_vs_k141_103327\|\|lt2gene | Low-quality |
| S14_vs_k141_44603\|\|lt2gene | Low-quality |
| S14_vs_k141_52194\|\|lt2gene | Low-quality |
| S14_vs_k141_52195\|\|lt2gene | Low-quality |
| S14_vs_k141_918\|\|lt2gene | Low-quality |
| S14_vs_k141_73929\|\|lt2gene | Not-determined |
| S14_vs_k141_73932\|\|lt2gene | Low-quality |
| S14_vs_k141_73933\|\|lt2gene | Not-determined |
| S14_vs_k141_22554\|\|lt2gene | Low-quality |
| S14_vs_k141_110622\|\|lt2gene | Low-quality |
| S14_vs_k141_22560\|\|lt2gene | Low-quality |
| S14_vs_k141_73940\|\|lt2gene | Low-quality |
| S14_vs_k141_30138\|\|lt2gene | Low-quality |
| S14_vs_k141_44622\|\|lt2gene | Low-quality |
| S14_vs_k141_95935\|\|lt2gene | Low-quality |
| S14_vs_k141_95936\|\|lt2gene | Low-quality |
| S14_vs_k141_88764\|\|lt2gene | Low-quality |
| S14_vs_k141_66460\|\|lt2gene | Low-quality |
| S14_vs_k141_37561\|\|lt2gene | Low-quality |
| S14_vs_k141_95941\|\|lt2gene | Low-quality |
| S14_vs_k141_30151\|\|lt2gene | Not-determined |
| S14_vs_k141_81346\|\|lt2gene | Low-quality |
| S14_vs_k141_59408\|\|lt2gene | Low-quality |
| S14_vs_k141_103346\|\|lt2gene | Low-quality |
| S14_vs_k141_103347\|\|lt2gene | Low-quality |
| S14_vs_k141_81351\|\|lt2gene | Low-quality |
| S14_vs_k141_59419\|\|lt2gene | Not-determined |
| S14_vs_k141_15410\|\|lt2gene | Low-quality |
| S14_vs_k141_22600\|\|lt2gene | Not-determined |
| S14_vs_k141_44642\|\|lt2gene | Low-quality |
| S14_vs_k141_949\|\|lt2gene | Low-quality |
| S14_vs_k141_959\|\|lt2gene | Low-quality |
| S14_vs_k141_73989\|\|lt2gene | Low-quality |
| S14_vs_k141_59472\|\|lt2gene | Low-quality |
| S14_vs_k141_22631\|\|lt2gene | Low-quality |
| S14_vs_k141_15433\|\|lt2gene | Low-quality |
| S14_vs_k141_15435\|\|lt2gene | Low-quality |
| S14_vs_k141_81381\|\|lt2gene | Low-quality |
| S14_vs_k141_81385\|\|lt2gene | Low-quality |
| S14_vs_k141_88811\|\|lt2gene | Low-quality |
| S14_vs_k141_88813\|\|lt2gene | Low-quality |
| S14_vs_k141_44691\|\|lt2gene | Low-quality |
| S14_vs_k141_103418\|\|lt2gene | Low-quality |
| S14_vs_k141_44693\|\|lt2gene | Low-quality |
| S14_vs_k141_979\|\|lt2gene | Low-quality |
| S14_vs_k141_15451\|\|lt2gene | Low-quality |
| S14_vs_k141_15455\|\|lt2gene | Low-quality |
| S14_vs_k141_984\|\|lt2gene | Low-quality |
| S14_vs_k141_986\|\|lt2gene | Low-quality |
| S14_vs_k141_987\|\|lt2gene | Low-quality |
| S14_vs_k141_990\|\|lt2gene | Low-quality |
| S14_vs_k141_30204\|\|lt2gene | Low-quality |
| S14_vs_k141_110713\|\|lt2gene | Low-quality |
| S14_vs_k141_59518\|\|lt2gene | Not-determined |
| S14_vs_k141_15476\|\|lt2gene | Low-quality |
| S14_vs_k141_74062\|\|lt2gene | Low-quality |
| S14_vs_k141_1011\|\|lt2gene | Low-quality |
| S14_vs_k141_59523\|\|lt2gene | Low-quality |
| S14_vs_k141_1018\|\|lt2gene | Low-quality |
| S14_vs_k141_22700\|\|lt2gene | Not-determined |
| S14_vs_k141_1021\|\|lt2gene | Not-determined |
| S14_vs_k141_66521\|\|lt2gene | Not-determined |
| S14_vs_k141_1029\|\|lt2gene | Not-determined |
| S14_vs_k141_52313\|\|lt2gene | Low-quality |
| S14_vs_k141_37698\|\|lt2gene | Low-quality |
| S14_vs_k141_52320\|\|lt2gene | Not-determined |
| S14_vs_k141_52321\|\|lt2gene | Not-determined |
| S14_vs_k141_52323\|\|lt2gene | Not-determined |
| S14_vs_k141_110747\|\|lt2gene | Low-quality |
| S14_vs_k141_30276\|\|lt2gene | Low-quality |
| S14_vs_k141_30283\|\|lt2gene | Not-determined |
| S14_vs_k141_103508\|\|lt2gene | Low-quality |
| S14_vs_k141_59550\|\|lt2gene | Not-determined |
| S14_vs_k141_103512\|\|lt2gene | Low-quality |
| S14_vs_k141_59552\|\|lt2gene | Not-determined |
| S14_vs_k141_1057\|\|lt2gene | Low-quality |
| S14_vs_k141_66537\|\|lt2gene | Low-quality |
| S14_vs_k141_103523\|\|lt2gene | Low-quality |
| S14_vs_k141_30291\|\|lt2gene | Low-quality |
| S14_vs_k141_15528\|\|lt2gene | Low-quality |
| S14_vs_k141_110798\|\|lt2gene | Not-determined |
| S14_vs_k141_15551\|\|lt2gene | Not-determined |
| S14_vs_k141_37730\|\|lt2gene | Low-quality |
| S14_vs_k141_96046\|\|lt2gene | Low-quality |
| S14_vs_k141_81432\|\|lt2gene | Low-quality |
| S14_vs_k141_1092\|\|lt2gene | Low-quality |
| S14_vs_k141_15559\|\|lt2gene | Low-quality |
| S14_vs_k141_88928\|\|lt2gene | Low-quality |
| S14_vs_k141_1115\|\|lt2gene | Low-quality |
| S14_vs_k141_103590\|\|lt2gene | Not-determined |
| S14_vs_k141_88951\|\|lt2gene | Low-quality |
| S14_vs_k141_88954\|\|lt2gene | Not-determined |
| S14_vs_k141_88955\|\|lt2gene | Not-determined |
| S14_vs_k141_59604\|\|lt2gene | Low-quality |
| S14_vs_k141_44846\|\|lt2gene | Low-quality |
| S14_vs_k141_15592\|\|lt2gene | Low-quality |
| S14_vs_k141_103624\|\|lt2gene | Low-quality |
| S14_vs_k141_110858\|\|lt2gene | Low-quality |
| S14_vs_k141_59615\|\|lt2gene | Not-determined |
| S14_vs_k141_52388\|\|lt2gene | Low-quality |
| S14_vs_k141_8171\|\|lt2gene | Low-quality |
| S14_vs_k141_59626\|\|lt2gene | Low-quality |
| S14_vs_k141_88978\|\|lt2gene | Low-quality |
| S14_vs_k141_110883\|\|lt2gene | Not-determined |
| S14_vs_k141_110884\|\|lt2gene | Not-determined |
| S14_vs_k141_22810\|\|lt2gene | Low-quality |
| S14_vs_k141_103662\|\|lt2gene | Low-quality |
| S14_vs_k141_110893\|\|lt2gene | Low-quality |
| S14_vs_k141_1148\|\|lt2gene | Not-determined |
| S14_vs_k141_96101\|\|lt2gene | Low-quality |
| S14_vs_k141_59640\|\|lt2gene | Low-quality |
| S14_vs_k141_52410\|\|lt2gene | Low-quality |
| S14_vs_k141_15636\|\|lt2gene | Low-quality |
| S14_vs_k141_89003\|\|lt2gene | Low-quality |
| S14_vs_k141_37837\|\|lt2gene | Low-quality |
| S14_vs_k141_81494\|\|lt2gene | Not-determined |
| S14_vs_k141_103704\|\|lt2gene | Low-quality |
| S14_vs_k141_81502\|\|lt2gene | Not-determined |
| S14_vs_k141_1197\|\|lt2gene | Not-determined |
| S14_vs_k141_1199\|\|lt2gene | Not-determined |
| S14_vs_k141_8241\|\|lt2gene | Low-quality |
| S14_vs_k141_103721\|\|lt2gene | Low-quality |
| S14_vs_k141_110954\|\|lt2gene | Low-quality |
| S14_vs_k141_110960\|\|lt2gene | Not-determined |
| S14_vs_k141_15647\|\|lt2gene | Not-determined |
| S14_vs_k141_110964\|\|lt2gene | Not-determined |
| S14_vs_k141_110974\|\|lt2gene | Low-quality |
| S14_vs_k141_103735\|\|lt2gene | Low-quality |
| S14_vs_k141_59683\|\|lt2gene | Not-determined |
| S14_vs_k141_44878\|\|lt2gene | Not-determined |
| S14_vs_k141_15680\|\|lt2gene | Low-quality |
| S14_vs_k141_110986\|\|lt2gene | Low-quality |
| S14_vs_k141_103778\|\|lt2gene | Not-determined |
| S14_vs_k141_103780\|\|lt2gene | Not-determined |
| S14_vs_k141_15698\|\|lt2gene | Low-quality |
| S14_vs_k141_59712\|\|lt2gene | Low-quality |
| S14_vs_k141_74095\|\|lt2gene | Not-determined |
| S14_vs_k141_22889\|\|lt2gene | Low-quality |
| S14_vs_k141_66646\|\|lt2gene | Low-quality |
| S14_vs_k141_1242\|\|lt2gene | Low-quality |
| S14_vs_k141_44948\|\|lt2gene | Low-quality |
| S14_vs_k141_37867\|\|lt2gene | Low-quality |
| S14_vs_k141_66649\|\|lt2gene | Low-quality |
| S14_vs_k141_30498\|\|lt2gene | Low-quality |
| S14_vs_k141_66667\|\|lt2gene | Low-quality |
| S14_vs_k141_59757\|\|lt2gene | Low-quality |
| S14_vs_k141_74125\|\|lt2gene | Not-determined |
| S14_vs_k141_74127\|\|lt2gene | Low-quality |
| S14_vs_k141_74139\|\|lt2gene | Not-determined |
| S14_vs_k141_15758\|\|lt2gene | Low-quality |
| S14_vs_k141_66669\|\|lt2gene | Low-quality |
| S14_vs_k141_30507\|\|lt2gene | Low-quality |
| S14_vs_k141_8331\|\|lt2gene | Not-determined |
| S14_vs_k141_15763\|\|lt2gene | Low-quality |
| S14_vs_k141_8336\|\|lt2gene | Not-determined |
| S14_vs_k141_89091\|\|lt2gene | Low-quality |
| S14_vs_k141_89092\|\|lt2gene | Low-quality |
| S14_vs_k141_74149\|\|lt2gene | Low-quality |
| S14_vs_k141_81557\|\|lt2gene | Low-quality |
| S14_vs_k141_44973\|\|lt2gene | Low-quality |
| S14_vs_k141_44976\|\|lt2gene | Low-quality |
| S14_vs_k141_44978\|\|lt2gene | Low-quality |
| S14_vs_k141_15786\|\|lt2gene | Not-determined |
| S14_vs_k141_37964\|\|lt2gene | Low-quality |
| S14_vs_k141_66694\|\|lt2gene | Low-quality |
| S14_vs_k141_74182\|\|lt2gene | Low-quality |
| S14_vs_k141_74183\|\|lt2gene | Not-determined |
| S14_vs_k141_74186\|\|lt2gene | Not-determined |
| S14_vs_k141_74189\|\|lt2gene | Not-determined |
| S14_vs_k141_111068\|\|lt2gene | Low-quality |
| S14_vs_k141_111069\|\|lt2gene | Low-quality |
| S14_vs_k141_89126\|\|lt2gene | Not-determined |
| S14_vs_k141_30535\|\|lt2gene | Low-quality |
| S14_vs_k141_111078\|\|lt2gene | Low-quality |
| S14_vs_k141_74206\|\|lt2gene | Not-determined |
| S14_vs_k141_15826\|\|lt2gene | Low-quality |
| S14_vs_k141_52468\|\|lt2gene | Low-quality |
| S14_vs_k141_8376\|\|lt2gene | Low-quality |
| S14_vs_k141_52480\|\|lt2gene | Low-quality |
| S14_vs_k141_103840\|\|lt2gene | Low-quality |
| S14_vs_k141_22917\|\|lt2gene | Low-quality |
| S14_vs_k141_8398\|\|lt2gene | Not-determined |
| S14_vs_k141_103852\|\|lt2gene | Low-quality |
| S14_vs_k141_111104\|\|lt2gene | Not-determined |
| S14_vs_k141_8401\|\|lt2gene | Not-determined |
| S14_vs_k141_22929\|\|lt2gene | Low-quality |
| S14_vs_k141_111130\|\|lt2gene | Low-quality |
| S14_vs_k141_15883\|\|lt2gene | Low-quality |
| S14_vs_k141_66718\|\|lt2gene | Not-determined |
| S14_vs_k141_30574\|\|lt2gene | Not-determined |
| S14_vs_k141_59805\|\|lt2gene | Low-quality |
| S14_vs_k141_52511\|\|lt2gene | Not-determined |
| S14_vs_k141_59810\|\|lt2gene | Low-quality |
| S14_vs_k141_74230\|\|lt2gene | Low-quality |
| S14_vs_k141_81678\|\|lt2gene | Low-quality |
| S14_vs_k141_81684\|\|lt2gene | Low-quality |
| S14_vs_k141_103888\|\|lt2gene | Low-quality |
| S14_vs_k141_111175\|\|lt2gene | Low-quality |
| S14_vs_k141_59826\|\|lt2gene | Low-quality |
| S14_vs_k141_74254\|\|lt2gene | Low-quality |
| S14_vs_k141_74256\|\|lt2gene | Low-quality |
| S14_vs_k141_74258\|\|lt2gene | Low-quality |
| S14_vs_k141_111179\|\|lt2gene | Low-quality |
| S14_vs_k141_74262\|\|lt2gene | Low-quality |
| S14_vs_k141_30606\|\|lt2gene | Not-determined |
| S14_vs_k141_15918\|\|lt2gene | Low-quality |
| S14_vs_k141_38016\|\|lt2gene | Low-quality |
| S14_vs_k141_81713\|\|lt2gene | Low-quality |
| S14_vs_k141_23002\|\|lt2gene | Low-quality |
| S14_vs_k141_15928\|\|lt2gene | Low-quality |
| S14_vs_k141_89184\|\|lt2gene | Low-quality |
| S14_vs_k141_81727\|\|lt2gene | Low-quality |
| S14_vs_k141_23021\|\|lt2gene | Low-quality |
| S14_vs_k141_111218\|\|lt2gene | Not-determined |
| S14_vs_k141_81745\|\|lt2gene | Low-quality |
| S14_vs_k141_8464\|\|lt2gene | Not-determined |
| S14_vs_k141_30643\|\|lt2gene | Low-quality |
| S14_vs_k141_81751\|\|lt2gene | Low-quality |
| S14_vs_k141_89211\|\|lt2gene | Low-quality |
| S14_vs_k141_8477\|\|lt2gene | Low-quality |
| S14_vs_k141_74311\|\|lt2gene | Low-quality |
| S14_vs_k141_15959\|\|lt2gene | Low-quality |
| S14_vs_k141_15962\|\|lt2gene | Low-quality |
| S14_vs_k141_23049\|\|lt2gene | Low-quality |
| S14_vs_k141_103968\|\|lt2gene | Low-quality |
| S14_vs_k141_103975\|\|lt2gene | Low-quality |
| S14_vs_k141_74331\|\|lt2gene | Not-determined |
| S14_vs_k141_96170\|\|lt2gene | Low-quality |
| S14_vs_k141_96175\|\|lt2gene | Low-quality |
| S14_vs_k141_74339\|\|lt2gene | Low-quality |
| S14_vs_k141_96182\|\|lt2gene | Low-quality |
| S14_vs_k141_30664\|\|lt2gene | Low-quality |
| S14_vs_k141_103986\|\|lt2gene | Low-quality |
| S14_vs_k141_103998\|\|lt2gene | Low-quality |
| S14_vs_k141_59918\|\|lt2gene | Not-determined |
| S14_vs_k141_59923\|\|lt2gene | Not-determined |
| S14_vs_k141_38060\|\|lt2gene | Not-determined |
| S14_vs_k141_104003\|\|lt2gene | Low-quality |
| S14_vs_k141_23090\|\|lt2gene | Low-quality |
| S14_vs_k141_81827\|\|lt2gene | Low-quality |
| S14_vs_k141_81828\|\|lt2gene | Low-quality |
| S14_vs_k141_23096\|\|lt2gene | Not-determined |
| S14_vs_k141_23097\|\|lt2gene | Low-quality |
| S14_vs_k141_74374\|\|lt2gene | Low-quality |
| S14_vs_k141_81836\|\|lt2gene | Low-quality |
| S14_vs_k141_52563\|\|lt2gene | Low-quality |
| S14_vs_k141_81854\|\|lt2gene | Low-quality |
| S14_vs_k141_16003\|\|lt2gene | Low-quality |
| S14_vs_k141_111284\|\|lt2gene | Low-quality |
| S14_vs_k141_38100\|\|lt2gene | Low-quality |
| S14_vs_k141_16016\|\|lt2gene | Low-quality |
| S14_vs_k141_38103\|\|lt2gene | Low-quality |
| S14_vs_k141_59980\|\|lt2gene | Low-quality |
| S14_vs_k141_104017\|\|lt2gene | Low-quality |
| S14_vs_k141_16029\|\|lt2gene | Low-quality |
| S14_vs_k141_66836\|\|lt2gene | Low-quality |
| S14_vs_k141_16046\|\|lt2gene | Low-quality |
| S14_vs_k141_59991\|\|lt2gene | Low-quality |
| S14_vs_k141_23167\|\|lt2gene | Low-quality |
| S14_vs_k141_89326\|\|lt2gene | Low-quality |
| S14_vs_k141_89333\|\|lt2gene | Low-quality |
| S14_vs_k141_111340\|\|lt2gene | Low-quality |
| S14_vs_k141_30694\|\|lt2gene | Low-quality |
| S14_vs_k141_81887\|\|lt2gene | Low-quality |
| S14_vs_k141_66842\|\|lt2gene | Low-quality |
| S14_vs_k141_30703\|\|lt2gene | Not-determined |
| S14_vs_k141_66847\|\|lt2gene | Low-quality |
| S14_vs_k141_81899\|\|lt2gene | Low-quality |
| S14_vs_k141_16076\|\|lt2gene | Low-quality |
| S14_vs_k141_16078\|\|lt2gene | Not-determined |
| S14_vs_k141_74467\|\|lt2gene | Low-quality |
| S14_vs_k141_89362\|\|lt2gene | Low-quality |
| S14_vs_k141_74473\|\|lt2gene | Low-quality |
| S14_vs_k141_66867\|\|lt2gene | Low-quality |
| S14_vs_k141_104079\|\|lt2gene | Not-determined |
| S14_vs_k141_111378\|\|lt2gene | Low-quality |
| S14_vs_k141_111387\|\|lt2gene | Low-quality |
| S14_vs_k141_52613\|\|lt2gene | Low-quality |
| S14_vs_k141_16099\|\|lt2gene | Not-determined |
| S14_vs_k141_1274\|\|lt2gene | Not-determined |
| S14_vs_k141_89388\|\|lt2gene | Low-quality |
| S14_vs_k141_104100\|\|lt2gene | Low-quality |
| S14_vs_k141_38156\|\|lt2gene | Not-determined |
| S14_vs_k141_104108\|\|lt2gene | Low-quality |
| S14_vs_k141_104109\|\|lt2gene | Low-quality |
| S14_vs_k141_30734\|\|lt2gene | Not-determined |
| S14_vs_k141_30735\|\|lt2gene | Low-quality |
| S14_vs_k141_104114\|\|lt2gene | Not-determined |
| S14_vs_k141_81957\|\|lt2gene | Low-quality |
| S14_vs_k141_104122\|\|lt2gene | Low-quality |
| S14_vs_k141_66905\|\|lt2gene | Low-quality |
| S14_vs_k141_66909\|\|lt2gene | Low-quality |
| S14_vs_k141_66913\|\|lt2gene | Low-quality |
| S14_vs_k141_66915\|\|lt2gene | Low-quality |
| S14_vs_k141_111407\|\|lt2gene | Not-determined |
| S14_vs_k141_111409\|\|lt2gene | Low-quality |
| S14_vs_k141_38201\|\|lt2gene | Low-quality |
| S14_vs_k141_8685\|\|lt2gene | Low-quality |
| S14_vs_k141_16144\|\|lt2gene | Not-determined |
| S14_vs_k141_23249\|\|lt2gene | Not-determined |
| S14_vs_k141_60054\|\|lt2gene | Low-quality |
| S14_vs_k141_1323\|\|lt2gene | Low-quality |
| S14_vs_k141_1325\|\|lt2gene | Low-quality |
| S14_vs_k141_74564\|\|lt2gene | Low-quality |
| S14_vs_k141_52685\|\|lt2gene | Low-quality |
| S14_vs_k141_52688\|\|lt2gene | Low-quality |
| S14_vs_k141_30789\|\|lt2gene | Not-determined |
| S14_vs_k141_111482\|\|lt2gene | Low-quality |
| S14_vs_k141_52709\|\|lt2gene | Not-determined |
| S14_vs_k141_16190\|\|lt2gene | Not-determined |
| S14_vs_k141_52729\|\|lt2gene | Not-determined |
| S14_vs_k141_60113\|\|lt2gene | Low-quality |
| S14_vs_k141_52736\|\|lt2gene | Low-quality |
| S14_vs_k141_38233\|\|lt2gene | Low-quality |
| S14_vs_k141_111501\|\|lt2gene | Low-quality |
| S14_vs_k141_23313\|\|lt2gene | Not-determined |
| S14_vs_k141_82038\|\|lt2gene | Not-determined |
| S14_vs_k141_52756\|\|lt2gene | Low-quality |
| S14_vs_k141_66982\|\|lt2gene | Low-quality |
| S14_vs_k141_8766\|\|lt2gene | Low-quality |
| S14_vs_k141_1368\|\|lt2gene | Low-quality |
| S14_vs_k141_67011\|\|lt2gene | Low-quality |
| S14_vs_k141_52792\|\|lt2gene | Low-quality |
| S14_vs_k141_52794\|\|lt2gene | Low-quality |
| S14_vs_k141_111537\|\|lt2gene | Low-quality |
| S14_vs_k141_1384\|\|lt2gene | Low-quality |
| S14_vs_k141_60164\|\|lt2gene | Low-quality |
| S14_vs_k141_82063\|\|lt2gene | Low-quality |
| S14_vs_k141_60173\|\|lt2gene | Low-quality |
| S14_vs_k141_30837\|\|lt2gene | Low-quality |
| S14_vs_k141_89396\|\|lt2gene | Low-quality |
| S14_vs_k141_89398\|\|lt2gene | Low-quality |
| S14_vs_k141_89400\|\|lt2gene | Low-quality |
| S14_vs_k141_89401\|\|lt2gene | Low-quality |
| S14_vs_k141_96434\|\|lt2gene | Low-quality |
| S14_vs_k141_111578\|\|lt2gene | Low-quality |
| S14_vs_k141_67041\|\|lt2gene | Low-quality |
| S14_vs_k141_104288\|\|lt2gene | Low-quality |
| S14_vs_k141_60198\|\|lt2gene | Low-quality |
| S14_vs_k141_52835\|\|lt2gene | Low-quality |
| S14_vs_k141_104305\|\|lt2gene | Low-quality |
| S14_vs_k141_96463\|\|lt2gene | Low-quality |
| S14_vs_k141_89444\|\|lt2gene | Low-quality |
| S14_vs_k141_74581\|\|lt2gene | Low-quality |
| S14_vs_k141_60212\|\|lt2gene | Low-quality |
| S14_vs_k141_67064\|\|lt2gene | Low-quality |
| S14_vs_k141_60216\|\|lt2gene | Low-quality |
| S14_vs_k141_82080\|\|lt2gene | Low-quality |
| S14_vs_k141_23392\|\|lt2gene | Low-quality |
| S14_vs_k141_23400\|\|lt2gene | Low-quality |
| S14_vs_k141_60226\|\|lt2gene | Low-quality |
| S14_vs_k141_104331\|\|lt2gene | Low-quality |
| S14_vs_k141_67081\|\|lt2gene | Low-quality |
| S14_vs_k141_67083\|\|lt2gene | Low-quality |
| S14_vs_k141_1422\|\|lt2gene | Not-determined |
| S14_vs_k141_96489\|\|lt2gene | Not-determined |
| S14_vs_k141_60251\|\|lt2gene | Low-quality |
| S14_vs_k141_60253\|\|lt2gene | Not-determined |
| S14_vs_k141_38320\|\|lt2gene | Low-quality |
| S14_vs_k141_111645\|\|lt2gene | Low-quality |
| S14_vs_k141_82111\|\|lt2gene | Low-quality |
| S14_vs_k141_67092\|\|lt2gene | Not-determined |
| S14_vs_k141_96510\|\|lt2gene | Not-determined |
| S14_vs_k141_30965\|\|lt2gene | Low-quality |
| S14_vs_k141_111659\|\|lt2gene | Low-quality |
| S14_vs_k141_96514\|\|lt2gene | Low-quality |
| S14_vs_k141_16290\|\|lt2gene | Not-determined |
| S14_vs_k141_96517\|\|lt2gene | Low-quality |
| S14_vs_k141_89485\|\|lt2gene | Low-quality |
| S14_vs_k141_111679\|\|lt2gene | Not-determined |
| S14_vs_k141_8897\|\|lt2gene | Low-quality |
| S14_vs_k141_111683\|\|lt2gene | Low-quality |
| S14_vs_k141_38358\|\|lt2gene | Low-quality |
| S14_vs_k141_38359\|\|lt2gene | Low-quality |
| S14_vs_k141_30982\|\|lt2gene | Low-quality |
| S14_vs_k141_45131\|\|lt2gene | Not-determined |
| S14_vs_k141_89521\|\|lt2gene | Low-quality |
| S14_vs_k141_16343\|\|lt2gene | Low-quality |
| S14_vs_k141_104422\|\|lt2gene | Low-quality |
| S14_vs_k141_67132\|\|lt2gene | Low-quality |
| S14_vs_k141_104424\|\|lt2gene | Low-quality |
| S14_vs_k141_74650\|\|lt2gene | Low-quality |
| S14_vs_k141_82144\|\|lt2gene | Not-determined |
| S14_vs_k141_111726\|\|lt2gene | Low-quality |
| S14_vs_k141_16362\|\|lt2gene | Low-quality |
| S14_vs_k141_104430\|\|lt2gene | Low-quality |
| S14_vs_k141_74664\|\|lt2gene | Low-quality |
| S14_vs_k141_96579\|\|lt2gene | Low-quality |
| S14_vs_k141_89527\|\|lt2gene | Low-quality |
| S14_vs_k141_104438\|\|lt2gene | Low-quality |
| S14_vs_k141_74703\|\|lt2gene | Not-determined |
| S14_vs_k141_74706\|\|lt2gene | Not-determined |
| S14_vs_k141_8931\|\|lt2gene | Not-determined |
| S14_vs_k141_8932\|\|lt2gene | Not-determined |
| S14_vs_k141_82160\|\|lt2gene | Low-quality |
| S14_vs_k141_67156\|\|lt2gene | Low-quality |
| S14_vs_k141_104442\|\|lt2gene | Low-quality |
| S14_vs_k141_74717\|\|lt2gene | Low-quality |
| S14_vs_k141_38419\|\|lt2gene | Low-quality |
| S14_vs_k141_31052\|\|lt2gene | Low-quality |
| S14_vs_k141_89576\|\|lt2gene | Not-determined |
| S14_vs_k141_38439\|\|lt2gene | Low-quality |
| S14_vs_k141_111797\|\|lt2gene | Low-quality |
| S14_vs_k141_1482\|\|lt2gene | Low-quality |
| S14_vs_k141_104473\|\|lt2gene | Low-quality |
| S14_vs_k141_31063\|\|lt2gene | Low-quality |
| S14_vs_k141_96656\|\|lt2gene | Low-quality |
| S14_vs_k141_96657\|\|lt2gene | Low-quality |
| S14_vs_k141_31069\|\|lt2gene | Not-determined |
| S14_vs_k141_89606\|\|lt2gene | Low-quality |
| S14_vs_k141_16445\|\|lt2gene | Low-quality |
| S14_vs_k141_31081\|\|lt2gene | Low-quality |
| S14_vs_k141_60392\|\|lt2gene | Low-quality |
| S14_vs_k141_74761\|\|lt2gene | Not-determined |
| S14_vs_k141_16462\|\|lt2gene | Low-quality |
| S14_vs_k141_16466\|\|lt2gene | Low-quality |
| S14_vs_k141_82233\|\|lt2gene | Not-determined |
| S14_vs_k141_16474\|\|lt2gene | Not-determined |
| S14_vs_k141_74785\|\|lt2gene | Low-quality |
| S14_vs_k141_82245\|\|lt2gene | Low-quality |
| S14_vs_k141_67210\|\|lt2gene | Not-determined |
| S14_vs_k141_74791\|\|lt2gene | Low-quality |
| S14_vs_k141_74801\|\|lt2gene | Not-determined |
| S14_vs_k141_23525\|\|lt2gene | Not-determined |
| S14_vs_k141_82253\|\|lt2gene | Not-determined |
| S14_vs_k141_60426\|\|lt2gene | Low-quality |
| S14_vs_k141_111838\|\|lt2gene | Low-quality |
| S14_vs_k141_82269\|\|lt2gene | Low-quality |
| S14_vs_k141_60445\|\|lt2gene | Low-quality |
| S14_vs_k141_38513\|\|lt2gene | Low-quality |
| S14_vs_k141_82285\|\|lt2gene | Low-quality |
| S14_vs_k141_104604\|\|lt2gene | Low-quality |
| S14_vs_k141_23548\|\|lt2gene | Low-quality |
| S14_vs_k141_111852\|\|lt2gene | Low-quality |
| S14_vs_k141_82287\|\|lt2gene | Low-quality |
| S14_vs_k141_16522\|\|lt2gene | Low-quality |
| S14_vs_k141_67266\|\|lt2gene | Low-quality |
| S14_vs_k141_1561\|\|lt2gene | Low-quality |
| S14_vs_k141_67277\|\|lt2gene | Not-determined |
| S14_vs_k141_74893\|\|lt2gene | Not-determined |
| S14_vs_k141_60462\|\|lt2gene | Low-quality |
| S14_vs_k141_45236\|\|lt2gene | Low-quality |
| S14_vs_k141_60471\|\|lt2gene | Not-determined |
| S14_vs_k141_16541\|\|lt2gene | Low-quality |
| S14_vs_k141_67302\|\|lt2gene | Low-quality |
| S14_vs_k141_111906\|\|lt2gene | Low-quality |
| S14_vs_k141_23564\|\|lt2gene | Low-quality |
| S14_vs_k141_111911\|\|lt2gene | Low-quality |
| S14_vs_k141_111914\|\|lt2gene | Low-quality |
| S14_vs_k141_53051\|\|lt2gene | Low-quality |
| S14_vs_k141_1589\|\|lt2gene | Low-quality |
| S14_vs_k141_53061\|\|lt2gene | Low-quality |
| S14_vs_k141_89719\|\|lt2gene | Not-determined |
| S14_vs_k141_89720\|\|lt2gene | Low-quality |
| S14_vs_k141_89724\|\|lt2gene | Low-quality |
| S14_vs_k141_60474\|\|lt2gene | Low-quality |
| S14_vs_k141_67322\|\|lt2gene | Low-quality |
| S14_vs_k141_1598\|\|lt2gene | Low-quality |
| S14_vs_k141_96683\|\|lt2gene | Not-determined |
| S14_vs_k141_96684\|\|lt2gene | Not-determined |
| S14_vs_k141_1603\|\|lt2gene | Low-quality |
| S14_vs_k141_16575\|\|lt2gene | Low-quality |
| S14_vs_k141_31139\|\|lt2gene | Low-quality |
| S14_vs_k141_31140\|\|lt2gene | Low-quality |
| S14_vs_k141_60496\|\|lt2gene | Low-quality |
| S14_vs_k141_60503\|\|lt2gene | Low-quality |
| S14_vs_k141_31149\|\|lt2gene | Not-determined |
| S14_vs_k141_31152\|\|lt2gene | Low-quality |
| S14_vs_k141_31153\|\|lt2gene | Low-quality |
| S14_vs_k141_89748\|\|lt2gene | Not-determined |
| S14_vs_k141_82296\|\|lt2gene | Low-quality |
| S14_vs_k141_74935\|\|lt2gene | Low-quality |
| S14_vs_k141_96707\|\|lt2gene | Low-quality |
| S14_vs_k141_60535\|\|lt2gene | Low-quality |
| S14_vs_k141_104638\|\|lt2gene | Low-quality |
| S14_vs_k141_67369\|\|lt2gene | Low-quality |
| S14_vs_k141_67380\|\|lt2gene | Low-quality |
| S14_vs_k141_31168\|\|lt2gene | Low-quality |
| S14_vs_k141_74958\|\|lt2gene | Low-quality |
| S14_vs_k141_74959\|\|lt2gene | Not-determined |
| S14_vs_k141_74963\|\|lt2gene | Not-determined |
| S14_vs_k141_53106\|\|lt2gene | Low-quality |
| S14_vs_k141_74971\|\|lt2gene | Low-quality |
| S14_vs_k141_96724\|\|lt2gene | Low-quality |
| S14_vs_k141_96725\|\|lt2gene | Low-quality |
| S14_vs_k141_53120\|\|lt2gene | Low-quality |
| S14_vs_k141_96731\|\|lt2gene | Not-determined |
| S14_vs_k141_53124\|\|lt2gene | Low-quality |
| S14_vs_k141_31179\|\|lt2gene | Low-quality |
| S14_vs_k141_60571\|\|lt2gene | Low-quality |
| S14_vs_k141_31192\|\|lt2gene | Low-quality |
| S14_vs_k141_31194\|\|lt2gene | Low-quality |
| S14_vs_k141_82351\|\|lt2gene | Low-quality |
| S14_vs_k141_104693\|\|lt2gene | Not-determined |
| S14_vs_k141_96761\|\|lt2gene | Low-quality |
| S14_vs_k141_96764\|\|lt2gene | Low-quality |
| S14_vs_k141_67430\|\|lt2gene | Low-quality |
| S14_vs_k141_38616\|\|lt2gene | Low-quality |
| S14_vs_k141_38630\|\|lt2gene | Low-quality |
| S14_vs_k141_67440\|\|lt2gene | Low-quality |
| S14_vs_k141_89812\|\|lt2gene | Low-quality |
| S14_vs_k141_96783\|\|lt2gene | Low-quality |
| S14_vs_k141_96786\|\|lt2gene | Low-quality |
| S14_vs_k141_45331\|\|lt2gene | Low-quality |
| S14_vs_k141_112019\|\|lt2gene | Low-quality |
| S14_vs_k141_60596\|\|lt2gene | Low-quality |
| S14_vs_k141_38639\|\|lt2gene | Not-determined |
| S14_vs_k141_96800\|\|lt2gene | Low-quality |
| S14_vs_k141_31217\|\|lt2gene | Not-determined |
| S14_vs_k141_23607\|\|lt2gene | Low-quality |
| S14_vs_k141_82388\|\|lt2gene | Not-determined |
| S14_vs_k141_23611\|\|lt2gene | Low-quality |
| S14_vs_k141_23613\|\|lt2gene | Low-quality |
| S14_vs_k141_82394\|\|lt2gene | Low-quality |
| S14_vs_k141_67460\|\|lt2gene | Low-quality |
| S14_vs_k141_67461\|\|lt2gene | Low-quality |
| S14_vs_k141_112058\|\|lt2gene | Low-quality |
| S14_vs_k141_8979\|\|lt2gene | Not-determined |
| S14_vs_k141_23634\|\|lt2gene | Low-quality |
| S14_vs_k141_8987\|\|lt2gene | Low-quality |
| S14_vs_k141_38652\|\|lt2gene | Not-determined |
| S14_vs_k141_75044\|\|lt2gene | Low-quality |
| S14_vs_k141_8991\|\|lt2gene | Low-quality |
| S14_vs_k141_60643\|\|lt2gene | Low-quality |
| S14_vs_k141_23652\|\|lt2gene | Low-quality |
| S14_vs_k141_82418\|\|lt2gene | Not-determined |
| S14_vs_k141_82419\|\|lt2gene | Not-determined |
| S14_vs_k141_16691\|\|lt2gene | Not-determined |
| S14_vs_k141_53194\|\|lt2gene | Not-determined |
| S14_vs_k141_31272\|\|lt2gene | Low-quality |
| S14_vs_k141_31277\|\|lt2gene | Low-quality |
| S14_vs_k141_31298\|\|lt2gene | Low-quality |
| S14_vs_k141_23695\|\|lt2gene | Low-quality |
| S14_vs_k141_38693\|\|lt2gene | Low-quality |
| S14_vs_k141_38697\|\|lt2gene | Low-quality |
| S14_vs_k141_38699\|\|lt2gene | Low-quality |
| S14_vs_k141_53234\|\|lt2gene | Low-quality |
| S14_vs_k141_16722\|\|lt2gene | Low-quality |
| S14_vs_k141_16729\|\|lt2gene | Low-quality |
| S14_vs_k141_104765\|\|lt2gene | Low-quality |
| S14_vs_k141_53249\|\|lt2gene | Not-determined |
| S14_vs_k141_45419\|\|lt2gene | Low-quality |
| S14_vs_k141_16740\|\|lt2gene | Low-quality |

| | |
|---|---|
| S14_vs_k141_60716||lt2gene | Low-quality |
| S14_vs_k141_112077||lt2gene | Low-quality |
| S14_vs_k141_9030||lt2gene | Not-determined |
| S14_vs_k141_38733||lt2gene | Low-quality |
| S14_vs_k141_82495||lt2gene | Low-quality |
| S14_vs_k141_67501||lt2gene | Low-quality |
| S14_vs_k141_104777||lt2gene | Low-quality |
| S14_vs_k141_16752||lt2gene | Not-determined |
| S14_vs_k141_104782||lt2gene | Low-quality |
| S14_vs_k141_38743||lt2gene | Low-quality |
| S14_vs_k141_45423||lt2gene | Low-quality |
| S14_vs_k141_67511||lt2gene | Low-quality |
| S14_vs_k141_9050||lt2gene | Not-determined |
| S14_vs_k141_16764||lt2gene | Low-quality |
| S14_vs_k141_38757||lt2gene | Low-quality |
| S14_vs_k141_60738||lt2gene | Low-quality |
| S14_vs_k141_82518||lt2gene | Low-quality |
| S14_vs_k141_60739||lt2gene | Low-quality |
| S14_vs_k141_38765||lt2gene | Low-quality |
| S14_vs_k141_67527||lt2gene | Low-quality |
| S14_vs_k141_38775||lt2gene | Low-quality |
| S14_vs_k141_31340||lt2gene | Not-determined |
| S14_vs_k141_9070||lt2gene | Low-quality |
| S14_vs_k141_67538||lt2gene | Low-quality |
| S14_vs_k141_60758||lt2gene | Low-quality |
| S14_vs_k141_60759||lt2gene | Low-quality |
| S14_vs_k141_45456||lt2gene | Low-quality |
| S14_vs_k141_104797||lt2gene | Not-determined |
| S14_vs_k141_16775||lt2gene | Low-quality |
| S14_vs_k141_9081||lt2gene | Not-determined |
| S14_vs_k141_89868||lt2gene | Not-determined |
| S14_vs_k141_67553||lt2gene | Low-quality |
| S14_vs_k141_23751||lt2gene | Not-determined |
| S14_vs_k141_104834||lt2gene | Low-quality |
| S14_vs_k141_89887||lt2gene | Low-quality |
| S14_vs_k141_38823||lt2gene | Low-quality |
| S14_vs_k141_9093||lt2gene | Not-determined |
| S14_vs_k141_38837||lt2gene | Not-determined |
| S14_vs_k141_16797||lt2gene | Low-quality |
| S14_vs_k141_31384||lt2gene | Not-determined |
| S14_vs_k141_60790||lt2gene | Low-quality |
| S14_vs_k141_16801||lt2gene | Low-quality |
| S14_vs_k141_104859||lt2gene | Low-quality |
| S14_vs_k141_16802||lt2gene | Not-determined |
| S14_vs_k141_16804||lt2gene | Low-quality |
| S14_vs_k141_45512||lt2gene | Low-quality |
| S14_vs_k141_53346||lt2gene | Low-quality |
| S14_vs_k141_67578||lt2gene | Not-determined |
| S14_vs_k141_53356||lt2gene | Low-quality |
| S14_vs_k141_67584||lt2gene | Not-determined |
| S14_vs_k141_9111||lt2gene | Low-quality |
| S14_vs_k141_67586||lt2gene | Low-quality |
| S14_vs_k141_60804||lt2gene | Low-quality |
| S14_vs_k141_67595||lt2gene | Not-determined |
| S14_vs_k141_75200||lt2gene | Low-quality |
| S14_vs_k141_53368||lt2gene | Low-quality |
| S14_vs_k141_23787||lt2gene | Low-quality |
| S14_vs_k141_38874||lt2gene | Low-quality |
| S14_vs_k141_75208||lt2gene | Not-determined |
| S14_vs_k141_82579||lt2gene | Low-quality |
| S14_vs_k141_38889||lt2gene | Low-quality |
| S14_vs_k141_67609||lt2gene | Low-quality |
| S14_vs_k141_38893||lt2gene | Low-quality |
| S14_vs_k141_75222||lt2gene | Low-quality |
| S14_vs_k141_16844||lt2gene | Not-determined |
| S14_vs_k141_16851||lt2gene | Low-quality |
| S14_vs_k141_82595||lt2gene | Low-quality |
| S14_vs_k141_96857||lt2gene | Low-quality |
| S14_vs_k141_67652||lt2gene | Low-quality |
| S14_vs_k141_1789||lt2gene | Not-determined |
| S14_vs_k141_82599||lt2gene | Not-determined |
| S14_vs_k141_82622||lt2gene | Low-quality |
| S14_vs_k141_89942||lt2gene | Low-quality |
| S14_vs_k141_75239||lt2gene | Low-quality |
| S14_vs_k141_104905||lt2gene | Low-quality |
| S14_vs_k141_96888||lt2gene | Low-quality |
| S14_vs_k141_104914||lt2gene | Low-quality |
| S14_vs_k141_89966||lt2gene | Low-quality |
| S14_vs_k141_82638||lt2gene | Not-determined |
| S14_vs_k141_89973||lt2gene | Low-quality |
| S14_vs_k141_104926||lt2gene | Low-quality |
| S14_vs_k141_104933||lt2gene | Low-quality |
| S14_vs_k141_112091||lt2gene | Low-quality |
| S14_vs_k141_89994||lt2gene | Low-quality |
| S14_vs_k141_89995||lt2gene | Not-determined |
| S14_vs_k141_45604||lt2gene | Low-quality |
| S14_vs_k141_53467||lt2gene | Low-quality |
| S14_vs_k141_90013||lt2gene | Not-determined |
| S14_vs_k141_23857||lt2gene | Not-determined |
| S14_vs_k141_67707||lt2gene | Not-determined |
| S14_vs_k141_112127||lt2gene | Low-quality |
| S14_vs_k141_23862||lt2gene | Low-quality |
| S14_vs_k141_9164||lt2gene | Not-determined |
| S14_vs_k141_90033||lt2gene | Low-quality |
| S14_vs_k141_104980||lt2gene | Low-quality |
| S14_vs_k141_104982||lt2gene | Low-quality |
| S14_vs_k141_90054||lt2gene | Low-quality |
| S14_vs_k141_23886||lt2gene | Low-quality |
| S14_vs_k141_82700||lt2gene | Not-determined |
| S14_vs_k141_90066||lt2gene | Low-quality |
| S14_vs_k141_1834||lt2gene | Low-quality |
| S14_vs_k141_1835||lt2gene | Low-quality |
| S14_vs_k141_9181||lt2gene | Not-determined |
| S14_vs_k141_9188||lt2gene | Low-quality |
| S14_vs_k141_1847||lt2gene | Not-determined |
| S14_vs_k141_105008||lt2gene | Low-quality |
| S14_vs_k141_23905||lt2gene | Low-quality |
| S14_vs_k141_39029||lt2gene | Low-quality |
| S14_vs_k141_39032||lt2gene | Low-quality |
| S14_vs_k141_82738||lt2gene | Low-quality |
| S14_vs_k141_105021||lt2gene | Low-quality |
| S14_vs_k141_39040||lt2gene | Low-quality |
| S14_vs_k141_39043||lt2gene | Low-quality |
| S14_vs_k141_1855||lt2gene | Not-determined |
| S14_vs_k141_53505||lt2gene | Low-quality |
| S14_vs_k141_9230||lt2gene | Low-quality |
| S14_vs_k141_96915||lt2gene | Low-quality |
| S14_vs_k141_1881||lt2gene | Low-quality |
| S14_vs_k141_31418||lt2gene | Low-quality |
| S14_vs_k141_67798||lt2gene | Low-quality |
| S14_vs_k141_39063||lt2gene | Not-determined |
| S14_vs_k141_1913||lt2gene | Low-quality |
| S14_vs_k141_23939||lt2gene | Not-determined |
| S14_vs_k141_23955||lt2gene | Low-quality |
| S14_vs_k141_82767||lt2gene | Not-determined |
| S14_vs_k141_23957||lt2gene | Low-quality |
| S14_vs_k141_23963||lt2gene | Low-quality |
| S14_vs_k141_23964||lt2gene | Low-quality |
| S14_vs_k141_82775||lt2gene | Not-determined |
| S14_vs_k141_23979||lt2gene | Low-quality |
| S14_vs_k141_1935||lt2gene | Low-quality |
| S14_vs_k141_1939||lt2gene | Not-determined |
| S14_vs_k141_53555||lt2gene | Low-quality |
| S14_vs_k141_9278||lt2gene | Low-quality |
| S14_vs_k141_90111||lt2gene | Low-quality |
| S14_vs_k141_60829||lt2gene | Low-quality |
| S14_vs_k141_90120||lt2gene | Low-quality |
| S14_vs_k141_53577||lt2gene | Low-quality |
| S14_vs_k141_16889||lt2gene | Low-quality |
| S14_vs_k141_24034||lt2gene | Not-determined |
| S14_vs_k141_90129||lt2gene | Low-quality |
| S14_vs_k141_75362||lt2gene | Low-quality |
| S14_vs_k141_9285||lt2gene | Low-quality |
| S14_vs_k141_75366||lt2gene | Low-quality |
| S14_vs_k141_75368||lt2gene | Low-quality |
| S14_vs_k141_75369||lt2gene | Low-quality |
| S14_vs_k141_75373||lt2gene | Low-quality |
| S14_vs_k141_75376||lt2gene | Low-quality |
| S14_vs_k141_75379||lt2gene | Low-quality |
| S14_vs_k141_16932||lt2gene | Low-quality |
| S14_vs_k141_16934||lt2gene | Not-determined |
| S14_vs_k141_16939||lt2gene | Low-quality |
| S14_vs_k141_75391||lt2gene | Low-quality |
| S14_vs_k141_24066||lt2gene | Low-quality |
| S14_vs_k141_67916||lt2gene | Not-determined |
| S14_vs_k141_97020||lt2gene | Low-quality |
| S14_vs_k141_112271||lt2gene | Low-quality |
| S14_vs_k141_39115||lt2gene | Low-quality |
| S14_vs_k141_67926||lt2gene | Not-determined |
| S14_vs_k141_39120||lt2gene | Low-quality |
| S14_vs_k141_75409||lt2gene | Low-quality |
| S14_vs_k141_31478||lt2gene | Low-quality |
| S14_vs_k141_60878||lt2gene | Low-quality |
| S14_vs_k141_60883||lt2gene | Low-quality |
| S14_vs_k141_60885||lt2gene | Low-quality |
| S14_vs_k141_31483||lt2gene | Low-quality |
| S14_vs_k141_9331||lt2gene | Low-quality |
| S14_vs_k141_39131||lt2gene | Low-quality |
| S14_vs_k141_16949||lt2gene | Not-determined |
| S14_vs_k141_16951||lt2gene | Low-quality |
| S14_vs_k141_9341||lt2gene | Low-quality |
| S14_vs_k141_9346||lt2gene | Not-determined |
| S14_vs_k141_97060||lt2gene | Not-determined |
| S14_vs_k141_45665||lt2gene | Low-quality |
| S14_vs_k141_67955||lt2gene | Low-quality |
| S14_vs_k141_67957||lt2gene | Low-quality |
| S14_vs_k141_97071||lt2gene | Low-quality |
| S14_vs_k141_97074||lt2gene | Low-quality |
| S14_vs_k141_67962||lt2gene | Low-quality |
| S14_vs_k141_82883||lt2gene | Not-determined |
| S14_vs_k141_97094||lt2gene | Not-determined |
| S14_vs_k141_53664||lt2gene | Low-quality |
| S14_vs_k141_75455||lt2gene | Not-determined |
| S14_vs_k141_9377||lt2gene | Low-quality |
| S14_vs_k141_9382||lt2gene | Not-determined |
| S14_vs_k141_53675||lt2gene | Low-quality |
| S14_vs_k141_9400||lt2gene | Low-quality |
| S14_vs_k141_9403||lt2gene | Low-quality |
| S14_vs_k141_9404||lt2gene | Not-determined |
| S14_vs_k141_39203||lt2gene | Low-quality |
| S14_vs_k141_39205||lt2gene | Low-quality |
| S14_vs_k141_90209||lt2gene | Not-determined |
| S14_vs_k141_75495||lt2gene | Low-quality |
| S14_vs_k141_67993||lt2gene | Low-quality |
| S14_vs_k141_112291||lt2gene | Low-quality |
| S14_vs_k141_67995||lt2gene | Low-quality |
| S14_vs_k141_67998||lt2gene | Low-quality |
| S14_vs_k141_9420||lt2gene | Low-quality |
| S14_vs_k141_68006||lt2gene | Low-quality |
| S14_vs_k141_68008||lt2gene | Low-quality |
| S14_vs_k141_68009||lt2gene | Low-quality |
| S14_vs_k141_82918||lt2gene | Low-quality |
| S14_vs_k141_24110||lt2gene | Not-determined |
| S14_vs_k141_2008||lt2gene | Low-quality |
| S14_vs_k141_31571||lt2gene | Low-quality |
| S14_vs_k141_31573||lt2gene | Low-quality |
| S14_vs_k141_97163||lt2gene | Not-determined |
| S14_vs_k141_53718||lt2gene | Low-quality |
| S14_vs_k141_9435||lt2gene | Low-quality |
| S14_vs_k141_45684||lt2gene | Low-quality |
| S14_vs_k141_17020||lt2gene | Low-quality |
| S14_vs_k141_75508||lt2gene | Low-quality |
| S14_vs_k141_24124||lt2gene | Low-quality |
| S14_vs_k141_105179||lt2gene | Low-quality |
| S14_vs_k141_39240||lt2gene | Low-quality |
| S14_vs_k141_24146||lt2gene | Low-quality |
| S14_vs_k141_112331||lt2gene | Not-determined |
| S14_vs_k141_75540||lt2gene | Not-determined |
| S14_vs_k141_75543||lt2gene | Not-determined |
| S14_vs_k141_68029||lt2gene | Low-quality |
| S14_vs_k141_112340||lt2gene | Low-quality |
| S14_vs_k141_24169||lt2gene | Low-quality |
| S14_vs_k141_90230||lt2gene | Not-determined |
| S14_vs_k141_68036||lt2gene | Low-quality |
| S14_vs_k141_60932||lt2gene | Low-quality |
| S14_vs_k141_24172||lt2gene | Low-quality |
| S14_vs_k141_90255||lt2gene | Not-determined |
| S14_vs_k141_17030||lt2gene | Low-quality |
| S14_vs_k141_105232||lt2gene | Low-quality |
| S14_vs_k141_97197||lt2gene | Not-determined |
| S14_vs_k141_68058||lt2gene | Low-quality |
| S14_vs_k141_17045||lt2gene | Low-quality |
| S14_vs_k141_105279||lt2gene | Low-quality |
| S14_vs_k141_2101||lt2gene | Low-quality |
| S14_vs_k141_105284||lt2gene | Not-determined |
| S14_vs_k141_83002||lt2gene | Not-determined |
| S14_vs_k141_105293||lt2gene | Low-quality |
| S14_vs_k141_83015||lt2gene | Low-quality |
| S14_vs_k141_105303||lt2gene | Low-quality |
| S14_vs_k141_53783||lt2gene | Low-quality |
| S14_vs_k141_83018||lt2gene | Low-quality |
| S14_vs_k141_31589||lt2gene | Low-quality |
| S14_vs_k141_105319||lt2gene | Low-quality |
| S14_vs_k141_83021||lt2gene | Low-quality |
| S14_vs_k141_83024||lt2gene | Low-quality |
| S14_vs_k141_112373||lt2gene | Low-quality |
| S14_vs_k141_9501||lt2gene | Low-quality |
| S14_vs_k141_68082||lt2gene | Low-quality |
| S14_vs_k141_83028||lt2gene | Low-quality |
| S14_vs_k141_39284||lt2gene | Low-quality |
| S14_vs_k141_31600||lt2gene | Low-quality |
| S14_vs_k141_105344||lt2gene | Not-determined |
| S14_vs_k141_45759||lt2gene | Not-determined |
| S14_vs_k141_112387||lt2gene | Low-quality |
| S14_vs_k141_68092||lt2gene | Not-determined |
| S14_vs_k141_45766||lt2gene | Not-determined |
| S14_vs_k141_2143||lt2gene | Not-determined |
| S14_vs_k141_2147||lt2gene | Low-quality |
| S14_vs_k141_17093||lt2gene | Not-determined |
| S14_vs_k141_31617||lt2gene | Low-quality |
| S14_vs_k141_9515||lt2gene | Not-determined |
| S14_vs_k141_61002||lt2gene | Low-quality |
| S14_vs_k141_90269||lt2gene | Low-quality |
| S14_vs_k141_90270||lt2gene | Low-quality |
| S14_vs_k141_75623||lt2gene | Low-quality |
| S14_vs_k141_75628||lt2gene | Not-determined |
| S14_vs_k141_75629||lt2gene | Low-quality |
| S14_vs_k141_39305||lt2gene | Not-determined |
| S14_vs_k141_39306||lt2gene | Low-quality |
| S14_vs_k141_112417||lt2gene | Not-determined |
| S14_vs_k141_39309||lt2gene | Low-quality |
| S14_vs_k141_45810||lt2gene | Low-quality |
| S14_vs_k141_112436||lt2gene | Low-quality |
| S14_vs_k141_97268||lt2gene | Low-quality |
| S14_vs_k141_17131||lt2gene | Low-quality |
| S14_vs_k141_17132||lt2gene | Not-determined |
| S14_vs_k141_9540||lt2gene | Low-quality |
| S14_vs_k141_9543||lt2gene | Low-quality |
| S14_vs_k141_17134||lt2gene | Low-quality |
| S14_vs_k141_9548||lt2gene | Low-quality |
| S14_vs_k141_45840||lt2gene | Not-determined |
| S14_vs_k141_9554||lt2gene | Low-quality |
| S14_vs_k141_9555||lt2gene | Low-quality |
| S14_vs_k141_9556||lt2gene | Low-quality |
| S14_vs_k141_90314||lt2gene | Low-quality |
| S14_vs_k141_17155||lt2gene | Low-quality |
| S14_vs_k141_9559||lt2gene | Low-quality |
| S14_vs_k141_61043||lt2gene | Low-quality |
| S14_vs_k141_53800||lt2gene | Low-quality |
| S14_vs_k141_53804||lt2gene | Not-determined |
| S14_vs_k141_53808||lt2gene | Not-determined |
| S14_vs_k141_53810||lt2gene | Not-determined |
| S14_vs_k141_53815||lt2gene | Not-determined |
| S14_vs_k141_112470||lt2gene | Low-quality |
| S14_vs_k141_112471||lt2gene | Low-quality |
| S14_vs_k141_75693||lt2gene | Low-quality |
| S14_vs_k141_75702||lt2gene | Low-quality |
| S14_vs_k141_61069||lt2gene | Low-quality |
| S14_vs_k141_75715||lt2gene | Low-quality |
| S14_vs_k141_75721||lt2gene | Low-quality |
| S14_vs_k141_90384||lt2gene | Low-quality |
| S14_vs_k141_68162||lt2gene | Low-quality |
| S14_vs_k141_75729||lt2gene | Low-quality |
| S14_vs_k141_75731||lt2gene | Not-determined |
| S14_vs_k141_9625||lt2gene | Low-quality |
| S14_vs_k141_9627||lt2gene | Low-quality |
| S14_vs_k141_31697||lt2gene | Not-determined |
| S14_vs_k141_9631||lt2gene | Low-quality |
| S14_vs_k141_9632||lt2gene | Low-quality |
| S14_vs_k141_45881||lt2gene | Low-quality |
| S14_vs_k141_45885||lt2gene | Low-quality |
| S14_vs_k141_112503||lt2gene | Not-determined |
| S14_vs_k141_97336||lt2gene | Not-determined |
| S14_vs_k141_39391||lt2gene | Low-quality |
| S14_vs_k141_90410||lt2gene | Low-quality |
| S14_vs_k141_2233||lt2gene | Low-quality |
| S14_vs_k141_2241||lt2gene | Not-determined |
| S14_vs_k141_31735||lt2gene | Not-determined |
| S14_vs_k141_31736||lt2gene | Not-determined |
| S14_vs_k141_90432||lt2gene | Low-quality |
| S14_vs_k141_75788||lt2gene | Not-determined |
| S14_vs_k141_75793||lt2gene | Not-determined |
| S14_vs_k141_24208||lt2gene | Low-quality |
| S14_vs_k141_45916||lt2gene | Not-determined |
| S14_vs_k141_61142||lt2gene | Low-quality |
| S14_vs_k141_68195||lt2gene | Low-quality |
| S14_vs_k141_9667||lt2gene | Low-quality |
| S14_vs_k141_105445||lt2gene | Not-determined |
| S14_vs_k141_105447||lt2gene | Not-determined |
| S14_vs_k141_61154||lt2gene | Low-quality |
| S14_vs_k141_105453||lt2gene | Low-quality |
| S14_vs_k141_105458||lt2gene | Low-quality |
| S14_vs_k141_75809||lt2gene | Low-quality |
| S14_vs_k141_68216||lt2gene | Not-determined |
| S14_vs_k141_105474||lt2gene | Low-quality |
| S14_vs_k141_45933||lt2gene | Low-quality |
| S14_vs_k141_105485||lt2gene | Not-determined |
| S14_vs_k141_31758||lt2gene | Low-quality |
| S14_vs_k141_45941||lt2gene | Low-quality |
| S14_vs_k141_31763||lt2gene | Low-quality |
| S14_vs_k141_83149||lt2gene | Low-quality |
| S14_vs_k141_17275||lt2gene | Not-determined |
| S14_vs_k141_105497||lt2gene | Low-quality |
| S14_vs_k141_105509||lt2gene | Low-quality |
| S14_vs_k141_24235||lt2gene | Low-quality |
| S14_vs_k141_105510||lt2gene | Low-quality |
| S14_vs_k141_31780||lt2gene | Not-determined |
| S14_vs_k141_112593||lt2gene | Low-quality |
| S14_vs_k141_68264||lt2gene | Low-quality |
| S14_vs_k141_90504||lt2gene | Low-quality |
| S14_vs_k141_112598||lt2gene | Low-quality |
| S14_vs_k141_105521||lt2gene | Low-quality |
| S14_vs_k141_45975||lt2gene | Not-determined |
| S14_vs_k141_53891||lt2gene | Low-quality |
| S14_vs_k141_2270||lt2gene | Low-quality |
| S14_vs_k141_83196||lt2gene | Low-quality |
| S14_vs_k141_75899||lt2gene | Low-quality |
| S14_vs_k141_2283||lt2gene | Low-quality |
| S14_vs_k141_17324||lt2gene | Low-quality |
| S14_vs_k141_90511||lt2gene | Low-quality |
| S14_vs_k141_90518||lt2gene | Not-determined |
| S14_vs_k141_75913||lt2gene | Low-quality |
| S14_vs_k141_97422||lt2gene | Low-quality |
| S14_vs_k141_53930||lt2gene | Low-quality |
| S14_vs_k141_105532||lt2gene | Low-quality |
| S14_vs_k141_68303||lt2gene | Low-quality |
| S14_vs_k141_39459||lt2gene | Low-quality |
| S14_vs_k141_83212||lt2gene | Low-quality |
| S14_vs_k141_105547||lt2gene | Low-quality |
| S14_vs_k141_9722||lt2gene | Low-quality |
| S14_vs_k141_2318||lt2gene | Not-determined |
| S14_vs_k141_112615||lt2gene | Low-quality |
| S14_vs_k141_2321||lt2gene | Not-determined |
| S14_vs_k141_53950||lt2gene | Low-quality |
| S14_vs_k141_112621||lt2gene | Low-quality |
| S14_vs_k141_9747||lt2gene | Low-quality |
| S14_vs_k141_105574||lt2gene | Low-quality |
| S14_vs_k141_68340||lt2gene | Not-determined |
| S14_vs_k141_2332||lt2gene | Not-determined |
| S14_vs_k141_53954||lt2gene | Low-quality |
| S14_vs_k141_68361||lt2gene | Low-quality |
| S14_vs_k141_90579||lt2gene | Low-quality |
| S14_vs_k141_53968||lt2gene | Not-determined |
| S14_vs_k141_90582||lt2gene | Low-quality |
| S14_vs_k141_97484||lt2gene | Low-quality |
| S14_vs_k141_68377||lt2gene | Low-quality |
| S14_vs_k141_9766||lt2gene | Not-determined |
| S14_vs_k141_75999||lt2gene | Low-quality |
| S14_vs_k141_83229||lt2gene | Not-determined |
| S14_vs_k141_112644||lt2gene | Low-quality |
| S14_vs_k141_76011||lt2gene | Not-determined |
| S14_vs_k141_112648||lt2gene | Low-quality |
| S14_vs_k141_24354||lt2gene | Not-determined |
| S14_vs_k141_83240||lt2gene | Low-quality |
| S14_vs_k141_9779||lt2gene | Low-quality |
| S14_vs_k141_76021||lt2gene | Not-determined |
| S14_vs_k141_112665||lt2gene | Low-quality |
| S14_vs_k141_68426||lt2gene | Low-quality |
| S14_vs_k141_46040||lt2gene | Low-quality |
| S14_vs_k141_17376||lt2gene | Not-determined |
| S14_vs_k141_76029||lt2gene | Low-quality |
| S14_vs_k141_83256||lt2gene | Low-quality |
| S14_vs_k141_17383||lt2gene | Low-quality |
| S14_vs_k141_54010||lt2gene | Low-quality |
| S14_vs_k141_24390||lt2gene | Low-quality |
| S14_vs_k141_68450||lt2gene | Low-quality |
| S14_vs_k141_39491||lt2gene | Low-quality |
| S14_vs_k141_68462||lt2gene | Low-quality |
| S14_vs_k141_2394||lt2gene | Low-quality |
| S14_vs_k141_83286||lt2gene | Not-determined |
| S14_vs_k141_46075||lt2gene | Low-quality |
| S14_vs_k141_31920||lt2gene | Low-quality |
| S14_vs_k141_9831||lt2gene | Low-quality |
| S14_vs_k141_90659||lt2gene | Low-quality |
| S14_vs_k141_97546||lt2gene | Low-quality |
| S14_vs_k141_68480||lt2gene | Not-determined |
| S14_vs_k141_68482||lt2gene | Not-determined |
| S14_vs_k141_68484||lt2gene | Not-determined |
| S14_vs_k141_68493||lt2gene | Low-quality |
| S14_vs_k141_9838||lt2gene | Low-quality |
| S14_vs_k141_105593||lt2gene | Low-quality |
| S14_vs_k141_68500||lt2gene | Not-determined |
| S14_vs_k141_76078||lt2gene | Low-quality |
| S14_vs_k141_46097||lt2gene | Not-determined |
| S14_vs_k141_9841||lt2gene | Low-quality |
| S14_vs_k141_9842||lt2gene | Low-quality |
| S14_vs_k141_46101||lt2gene | Not-determined |
| S14_vs_k141_97572||lt2gene | Not-determined |
| S14_vs_k141_97579||lt2gene | Low-quality |
| S14_vs_k141_17447||lt2gene | Low-quality |
| S14_vs_k141_17452||lt2gene | Low-quality |
| S14_vs_k141_17462||lt2gene | Low-quality |
| S14_vs_k141_68525||lt2gene | Low-quality |
| S14_vs_k141_68527||lt2gene | Low-quality |
| S14_vs_k141_90691||lt2gene | Not-determined |
| S14_vs_k141_46140||lt2gene | Low-quality |
| S14_vs_k141_112706||lt2gene | Not-determined |
| S14_vs_k141_61248||lt2gene | Low-quality |
| S14_vs_k141_2465||lt2gene | Not-determined |
| S14_vs_k141_17473||lt2gene | Low-quality |
| S14_vs_k141_112714||lt2gene | Not-determined |
| S14_vs_k141_76105||lt2gene | Low-quality |
| S14_vs_k141_39540||lt2gene | Not-determined |
| S14_vs_k141_54101||lt2gene | Low-quality |
| S14_vs_k141_17478||lt2gene | Not-determined |
| S14_vs_k141_24405||lt2gene | Low-quality |
| S14_vs_k141_90725||lt2gene | Low-quality |
| S14_vs_k141_83348||lt2gene | Low-quality |
| S14_vs_k141_61268||lt2gene | Not-determined |
| S14_vs_k141_97620||lt2gene | Low-quality |
| S14_vs_k141_31953||lt2gene | Not-determined |
| S14_vs_k141_61271||lt2gene | Not-determined |
| S14_vs_k141_24431||lt2gene | Not-determined |
| S14_vs_k141_31956||lt2gene | Not-determined |
| S14_vs_k141_61279||lt2gene | Low-quality |
| S14_vs_k141_90729||lt2gene | Low-quality |
| S14_vs_k141_90732||lt2gene | Low-quality |
| S14_vs_k141_2512||lt2gene | Not-determined |
| S14_vs_k141_39583||lt2gene | Low-quality |
| S14_vs_k141_68555||lt2gene | Low-quality |
| S14_vs_k141_54169||lt2gene | Low-quality |
| S14_vs_k141_2528||lt2gene | Low-quality |
| S14_vs_k141_2531||lt2gene | Low-quality |
| S14_vs_k141_39597||lt2gene | Low-quality |
| S14_vs_k141_105690||lt2gene | Not-determined |
| S14_vs_k141_112737||lt2gene | Low-quality |
| S14_vs_k141_105692||lt2gene | Low-quality |
| S14_vs_k141_54187||lt2gene | Not-determined |
| S14_vs_k141_105699||lt2gene | Low-quality |
| S14_vs_k141_90737||lt2gene | Low-quality |
| S14_vs_k141_17514||lt2gene | Low-quality |
| S14_vs_k141_105703||lt2gene | Low-quality |
| S14_vs_k141_76190||lt2gene | Not-determined |
| S14_vs_k141_105706||lt2gene | Low-quality |
| S14_vs_k141_68578||lt2gene | Low-quality |
| S14_vs_k141_76198||lt2gene | Low-quality |
| S14_vs_k141_9909||lt2gene | Low-quality |
| S14_vs_k141_105717||lt2gene | Low-quality |
| S14_vs_k141_97679||lt2gene | Low-quality |
| S14_vs_k141_90761||lt2gene | Low-quality |
| S14_vs_k141_54201||lt2gene | Low-quality |
| S14_vs_k141_54204||lt2gene | Low-quality |
| S14_vs_k141_105730||lt2gene | Low-quality |
| S14_vs_k141_32035||lt2gene | Low-quality |
| S14_vs_k141_17552||lt2gene | Low-quality |
| S14_vs_k141_9934||lt2gene | Low-quality |
| S14_vs_k141_17555||lt2gene | Not-determined |
| S14_vs_k141_97697||lt2gene | Low-quality |
| S14_vs_k141_2566||lt2gene | Low-quality |
| S14_vs_k141_46248||lt2gene | Low-quality |
| S14_vs_k141_97707||lt2gene | Low-quality |
| S14_vs_k141_90779||lt2gene | Low-quality |
| S14_vs_k141_32061||lt2gene | Low-quality |
| S14_vs_k141_39624||lt2gene | Low-quality |
| S14_vs_k141_112802||lt2gene | Low-quality |
| S14_vs_k141_97717||lt2gene | Low-quality |
| S14_vs_k141_112805||lt2gene | Low-quality |
| S14_vs_k141_76225||lt2gene | Not-determined |
| S14_vs_k141_9959||lt2gene | Low-quality |
| S14_vs_k141_97723||lt2gene | Low-quality |
| S14_vs_k141_112807||lt2gene | Low-quality |
| S14_vs_k141_24530||lt2gene | Low-quality |
| S14_vs_k141_97725||lt2gene | Low-quality |
| S14_vs_k141_76230||lt2gene | Not-determined |
| S14_vs_k141_46265||lt2gene | Not-determined |
| S14_vs_k141_76235||lt2gene | Not-determined |
| S14_vs_k141_112818||lt2gene | Low-quality |
| S14_vs_k141_112819||lt2gene | Low-quality |
| S14_vs_k141_9970||lt2gene | Low-quality |
| S14_vs_k141_105757||lt2gene | Low-quality |
| S14_vs_k141_46281||lt2gene | Low-quality |
| S14_vs_k141_54258||lt2gene | Low-quality |
| S14_vs_k141_68613||lt2gene | Low-quality |
| S14_vs_k141_24553||lt2gene | Low-quality |
| S14_vs_k141_9985||lt2gene | Low-quality |
| S14_vs_k141_24561||lt2gene | Not-determined |
| S14_vs_k141_76273||lt2gene | Low-quality |
| S14_vs_k141_32089||lt2gene | Not-determined |
| S14_vs_k141_2613||lt2gene | Low-quality |
| S14_vs_k141_32095||lt2gene | Low-quality |
| S14_vs_k141_32097||lt2gene | Low-quality |
| S14_vs_k141_32099||lt2gene | Low-quality |
| S14_vs_k141_9996||lt2gene | Not-determined |
| S14_vs_k141_24568||lt2gene | Low-quality |
| S14_vs_k141_54293||lt2gene | Low-quality |
| S14_vs_k141_112851||lt2gene | Low-quality |
| S14_vs_k141_105792||lt2gene | Low-quality |
| S14_vs_k141_39663||lt2gene | Low-quality |
| S14_vs_k141_105801||lt2gene | Not-determined |
| S14_vs_k141_105804||lt2gene | Low-quality |
| S14_vs_k141_76317||lt2gene | Not-determined |
| S14_vs_k141_54327||lt2gene | Low-quality |
| S14_vs_k141_46297||lt2gene | Low-quality |
| S14_vs_k141_90867||lt2gene | Low-quality |
| S14_vs_k141_32131||lt2gene | Not-determined |
| S14_vs_k141_10014||lt2gene | Low-quality |
| S14_vs_k141_32135||lt2gene | Not-determined |
| S14_vs_k141_61367||lt2gene | Low-quality |
| S14_vs_k141_46312||lt2gene | Low-quality |
| S14_vs_k141_97801||lt2gene | Not-determined |
| S14_vs_k141_105820||lt2gene | Low-quality |
| S14_vs_k141_46314||lt2gene | Low-quality |
| S14_vs_k141_32150||lt2gene | Not-determined |
| S14_vs_k141_32153||lt2gene | Low-quality |
| S14_vs_k141_112867||lt2gene | Low-quality |
| S14_vs_k141_46317||lt2gene | Low-quality |
| S14_vs_k141_39723||lt2gene | Low-quality |
| S14_vs_k141_17640||lt2gene | Low-quality |
| S14_vs_k141_17642||lt2gene | Low-quality |
| S14_vs_k141_90890||lt2gene | Low-quality |
| S14_vs_k141_17649||lt2gene | Low-quality |
| S14_vs_k141_54368||lt2gene | Not-determined |
| S14_vs_k141_68685||lt2gene | Low-quality |
| S14_vs_k141_54369||lt2gene | Low-quality |
| S14_vs_k141_61396||lt2gene | Low-quality |
| S14_vs_k141_24631||lt2gene | Low-quality |
| S14_vs_k141_39745||lt2gene | Not-determined |
| S14_vs_k141_46338||lt2gene | Not-determined |
| S14_vs_k141_32177||lt2gene | Low-quality |
| S14_vs_k141_83530||lt2gene | Not-determined |
| S14_vs_k141_83533||lt2gene | Low-quality |
| S14_vs_k141_17664||lt2gene | Low-quality |
| S14_vs_k141_61426||lt2gene | Low-quality |
| S14_vs_k141_32184||lt2gene | Low-quality |
| S14_vs_k141_83545||lt2gene | Low-quality |
| S14_vs_k141_10036||lt2gene | Low-quality |
| S14_vs_k141_32215||lt2gene | Low-quality |
| S14_vs_k141_61449||lt2gene | Low-quality |
| S14_vs_k141_61451||lt2gene | Low-quality |
| S14_vs_k141_10062||lt2gene | Low-quality |
| S14_vs_k141_39796||lt2gene | Low-quality |
| S14_vs_k141_112903||lt2gene | Low-quality |
| S14_vs_k141_2646||lt2gene | Low-quality |
| S14_vs_k141_32240||lt2gene | Low-quality |
| S14_vs_k141_24681||lt2gene | Low-quality |
| S14_vs_k141_46397||lt2gene | Low-quality |
| S14_vs_k141_112917||lt2gene | Low-quality |
| S14_vs_k141_76389||lt2gene | Not-determined |
| S14_vs_k141_68732||lt2gene | Not-determined |
| S14_vs_k141_76404||lt2gene | Low-quality |
| S14_vs_k141_68745||lt2gene | Not-determined |
| S14_vs_k141_112948||lt2gene | Low-quality |
| S14_vs_k141_54462||lt2gene | Low-quality |
| S14_vs_k141_112951||lt2gene | Low-quality |
| S14_vs_k141_76417||lt2gene | Low-quality |
| S14_vs_k141_10114||lt2gene | Low-quality |
| S14_vs_k141_32273||lt2gene | Low-quality |
| S14_vs_k141_76424||lt2gene | Low-quality |
| S14_vs_k141_2699||lt2gene | Low-quality |

| | |
|---|---|
| S14_vs_k141_76431\|\|lt2gene | Not-determined |
| S14_vs_k141_76432\|\|lt2gene | Low-quality |
| S14_vs_k141_17714\|\|lt2gene | Low-quality |
| S14_vs_k141_105888\|\|lt2gene | Low-quality |
| S14_vs_k141_32279\|\|lt2gene | Low-quality |
| S14_vs_k141_112965\|\|lt2gene | Low-quality |
| S14_vs_k141_68768\|\|lt2gene | Not-determined |
| S14_vs_k141_83613\|\|lt2gene | Low-quality |
| S14_vs_k141_17728\|\|lt2gene | Low-quality |
| S14_vs_k141_68778\|\|lt2gene | Low-quality |
| S14_vs_k141_83625\|\|lt2gene | Low-quality |
| S14_vs_k141_112978\|\|lt2gene | Low-quality |
| S14_vs_k141_105911\|\|lt2gene | Low-quality |
| S14_vs_k141_24709\|\|lt2gene | Low-quality |
| S14_vs_k141_105919\|\|lt2gene | Not-determined |
| S14_vs_k141_105922\|\|lt2gene | Not-determined |
| S14_vs_k141_105925\|\|lt2gene | Low-quality |
| S14_vs_k141_105928\|\|lt2gene | Low-quality |
| S14_vs_k141_76474\|\|lt2gene | Low-quality |
| S14_vs_k141_105929\|\|lt2gene | Not-determined |
| S14_vs_k141_68804\|\|lt2gene | Not-determined |
| S14_vs_k141_46446\|\|lt2gene | Low-quality |
| S14_vs_k141_39844\|\|lt2gene | Low-quality |
| S14_vs_k141_54515\|\|lt2gene | Low-quality |
| S14_vs_k141_83656\|\|lt2gene | Low-quality |
| S14_vs_k141_68819\|\|lt2gene | Low-quality |
| S14_vs_k141_32302\|\|lt2gene | Low-quality |
| S14_vs_k141_32311\|\|lt2gene | Low-quality |
| S14_vs_k141_32314\|\|lt2gene | Low-quality |
| S14_vs_k141_10152\|\|lt2gene | Low-quality |
| S14_vs_k141_112992\|\|lt2gene | Not-determined |
| S14_vs_k141_2750\|\|lt2gene | Low-quality |
| S14_vs_k141_10159\|\|lt2gene | Low-quality |
| S14_vs_k141_39860\|\|lt2gene | Low-quality |
| S14_vs_k141_32323\|\|lt2gene | Low-quality |
| S14_vs_k141_54537\|\|lt2gene | Low-quality |
| S14_vs_k141_54543\|\|lt2gene | Low-quality |
| S14_vs_k141_54549\|\|lt2gene | Not-determined |
| S14_vs_k141_54551\|\|lt2gene | Not-determined |
| S14_vs_k141_54553\|\|lt2gene | Not-determined |
| S14_vs_k141_54555\|\|lt2gene | Not-determined |
| S14_vs_k141_10176\|\|lt2gene | Low-quality |
| S14_vs_k141_105969\|\|lt2gene | Low-quality |
| S14_vs_k141_54558\|\|lt2gene | Low-quality |
| S14_vs_k141_113003\|\|lt2gene | Not-determined |
| S14_vs_k141_68869\|\|lt2gene | Low-quality |
| S14_vs_k141_90999\|\|lt2gene | Low-quality |
| S14_vs_k141_39892\|\|lt2gene | Low-quality |
| S14_vs_k141_2766\|\|lt2gene | Low-quality |
| S14_vs_k141_61498\|\|lt2gene | Low-quality |
| S14_vs_k141_76514\|\|lt2gene | Low-quality |
| S14_vs_k141_61502\|\|lt2gene | Low-quality |
| S14_vs_k141_10185\|\|lt2gene | Low-quality |
| S14_vs_k141_68890\|\|lt2gene | Low-quality |
| S14_vs_k141_24776\|\|lt2gene | Low-quality |
| S14_vs_k141_76531\|\|lt2gene | Low-quality |
| S14_vs_k141_68893\|\|lt2gene | Not-determined |
| S14_vs_k141_76535\|\|lt2gene | Low-quality |
| S14_vs_k141_46535\|\|lt2gene | Low-quality |
| S14_vs_k141_2797\|\|lt2gene | Low-quality |
| S14_vs_k141_105995\|\|lt2gene | Low-quality |
| S14_vs_k141_105996\|\|lt2gene | Low-quality |
| S14_vs_k141_76542\|\|lt2gene | Low-quality |
| S14_vs_k141_76546\|\|lt2gene | Not-determined |
| S14_vs_k141_39933\|\|lt2gene | Low-quality |
| S14_vs_k141_24794\|\|lt2gene | Low-quality |
| S14_vs_k141_97871\|\|lt2gene | Not-determined |
| S14_vs_k141_17751\|\|lt2gene | Low-quality |
| S14_vs_k141_97877\|\|lt2gene | Low-quality |
| S14_vs_k141_39944\|\|lt2gene | Not-determined |
| S14_vs_k141_46557\|\|lt2gene | Low-quality |
| S14_vs_k141_32386\|\|lt2gene | Low-quality |
| S14_vs_k141_97884\|\|lt2gene | Not-determined |
| S14_vs_k141_91009\|\|lt2gene | Low-quality |
| S14_vs_k141_68941\|\|lt2gene | Low-quality |
| S14_vs_k141_2826\|\|lt2gene | Low-quality |
| S14_vs_k141_54600\|\|lt2gene | Low-quality |
| S14_vs_k141_24817\|\|lt2gene | Low-quality |
| S14_vs_k141_32404\|\|lt2gene | Low-quality |
| S14_vs_k141_24820\|\|lt2gene | Low-quality |
| S14_vs_k141_106027\|\|lt2gene | Not-determined |
| S14_vs_k141_106033\|\|lt2gene | Low-quality |
| S14_vs_k141_91025\|\|lt2gene | Low-quality |
| S14_vs_k141_10246\|\|lt2gene | Low-quality |
| S14_vs_k141_39962\|\|lt2gene | Low-quality |
| S14_vs_k141_106046\|\|lt2gene | Low-quality |
| S14_vs_k141_39972\|\|lt2gene | Low-quality |
| S14_vs_k141_91039\|\|lt2gene | Low-quality |
| S14_vs_k141_54620\|\|lt2gene | Low-quality |
| S14_vs_k141_10262\|\|lt2gene | Low-quality |
| S14_vs_k141_46592\|\|lt2gene | Low-quality |
| S14_vs_k141_106066\|\|lt2gene | Not-determined |
| S14_vs_k141_68955\|\|lt2gene | Low-quality |
| S14_vs_k141_106069\|\|lt2gene | Not-determined |
| S14_vs_k141_46601\|\|lt2gene | Low-quality |
| S14_vs_k141_46611\|\|lt2gene | Low-quality |
| S14_vs_k141_32426\|\|lt2gene | Low-quality |
| S14_vs_k141_17818\|\|lt2gene | Low-quality |
| S14_vs_k141_91085\|\|lt2gene | Not-determined |
| S14_vs_k141_32432\|\|lt2gene | Low-quality |
| S14_vs_k141_76581\|\|lt2gene | Low-quality |
| S14_vs_k141_54666\|\|lt2gene | Low-quality |
| S14_vs_k141_32446\|\|lt2gene | Low-quality |
| S14_vs_k141_106086\|\|lt2gene | Low-quality |
| S14_vs_k141_97933\|\|lt2gene | Low-quality |
| S14_vs_k141_32450\|\|lt2gene | Not-determined |
| S14_vs_k141_113024\|\|lt2gene | Low-quality |
| S14_vs_k141_24854\|\|lt2gene | Low-quality |
| S14_vs_k141_54674\|\|lt2gene | Low-quality |
| S14_vs_k141_83827\|\|lt2gene | Low-quality |
| S14_vs_k141_46621\|\|lt2gene | Not-determined |
| S14_vs_k141_24865\|\|lt2gene | Not-determined |
| S14_vs_k141_17854\|\|lt2gene | Low-quality |
| S14_vs_k141_76603\|\|lt2gene | Low-quality |
| S14_vs_k141_54694\|\|lt2gene | Not-determined |
| S14_vs_k141_10313\|\|lt2gene | Low-quality |
| S14_vs_k141_91121\|\|lt2gene | Low-quality |
| S14_vs_k141_97949\|\|lt2gene | Low-quality |
| S14_vs_k141_32483\|\|lt2gene | Low-quality |
| S14_vs_k141_61521\|\|lt2gene | Low-quality |
| S14_vs_k141_61525\|\|lt2gene | Low-quality |
| S14_vs_k141_83864\|\|lt2gene | Low-quality |
| S14_vs_k141_91135\|\|lt2gene | Low-quality |
| S14_vs_k141_91136\|\|lt2gene | Low-quality |
| S14_vs_k141_83873\|\|lt2gene | Low-quality |
| S14_vs_k141_46661\|\|lt2gene | Low-quality |
| S14_vs_k141_46667\|\|lt2gene | Low-quality |
| S14_vs_k141_68962\|\|lt2gene | Low-quality |
| S14_vs_k141_91149\|\|lt2gene | Not-determined |
| S14_vs_k141_97979\|\|lt2gene | Low-quality |
| S14_vs_k141_76641\|\|lt2gene | Not-determined |
| S14_vs_k141_76652\|\|lt2gene | Not-determined |
| S14_vs_k141_68986\|\|lt2gene | Low-quality |
| S14_vs_k141_113086\|\|lt2gene | Low-quality |
| S14_vs_k141_98001\|\|lt2gene | Not-determined |
| S14_vs_k141_113104\|\|lt2gene | Low-quality |
| S14_vs_k141_69011\|\|lt2gene | Low-quality |
| S14_vs_k141_69032\|\|lt2gene | Low-quality |
| S14_vs_k141_83922\|\|lt2gene | Low-quality |
| S14_vs_k141_61553\|\|lt2gene | Low-quality |
| S14_vs_k141_2876\|\|lt2gene | Low-quality |
| S14_vs_k141_61571\|\|lt2gene | Not-determined |
| S14_vs_k141_2878\|\|lt2gene | Low-quality |
| S14_vs_k141_46710\|\|lt2gene | Not-determined |
| S14_vs_k141_61582\|\|lt2gene | Low-quality |
| S14_vs_k141_46713\|\|lt2gene | Low-quality |
| S14_vs_k141_46718\|\|lt2gene | Low-quality |
| S14_vs_k141_61590\|\|lt2gene | Low-quality |
| S14_vs_k141_113135\|\|lt2gene | Low-quality |
| S14_vs_k141_46726\|\|lt2gene | Low-quality |
| S14_vs_k141_24961\|\|lt2gene | Low-quality |
| S14_vs_k141_24962\|\|lt2gene | Not-determined |
| S14_vs_k141_69084\|\|lt2gene | Not-determined |
| S14_vs_k141_2887\|\|lt2gene | Low-quality |
| S14_vs_k141_113148\|\|lt2gene | Low-quality |
| S14_vs_k141_69090\|\|lt2gene | Low-quality |
| S14_vs_k141_91212\|\|lt2gene | Low-quality |
| S14_vs_k141_69094\|\|lt2gene | Low-quality |
| S14_vs_k141_69097\|\|lt2gene | Low-quality |
| S14_vs_k141_24971\|\|lt2gene | Low-quality |
| S14_vs_k141_61627\|\|lt2gene | Low-quality |
| S14_vs_k141_113159\|\|lt2gene | Low-quality |
| S14_vs_k141_113162\|\|lt2gene | Not-determined |
| S14_vs_k141_113164\|\|lt2gene | Low-quality |
| S14_vs_k141_61630\|\|lt2gene | Low-quality |
| S14_vs_k141_113168\|\|lt2gene | Not-determined |
| S14_vs_k141_10372\|\|lt2gene | Low-quality |
| S14_vs_k141_69112\|\|lt2gene | Not-determined |
| S14_vs_k141_10373\|\|lt2gene | Low-quality |
| S14_vs_k141_91245\|\|lt2gene | Low-quality |
| S14_vs_k141_106165\|\|lt2gene | Low-quality |
| S14_vs_k141_91247\|\|lt2gene | Low-quality |
| S14_vs_k141_106170\|\|lt2gene | Low-quality |
| S14_vs_k141_54725\|\|lt2gene | Low-quality |
| S14_vs_k141_113178\|\|lt2gene | Not-determined |
| S14_vs_k141_10397\|\|lt2gene | Low-quality |
| S14_vs_k141_2904\|\|lt2gene | Low-quality |
| S14_vs_k141_32550\|\|lt2gene | Low-quality |
| S14_vs_k141_10403\|\|lt2gene | Not-determined |
| S14_vs_k141_98111\|\|lt2gene | Low-quality |
| S14_vs_k141_76787\|\|lt2gene | Low-quality |
| S14_vs_k141_113205\|\|lt2gene | Low-quality |
| S14_vs_k141_10407\|\|lt2gene | Low-quality |
| S14_vs_k141_17993\|\|lt2gene | Not-determined |
| S14_vs_k141_18000\|\|lt2gene | Low-quality |
| S14_vs_k141_18001\|\|lt2gene | Low-quality |
| S14_vs_k141_18006\|\|lt2gene | Low-quality |
| S14_vs_k141_76807\|\|lt2gene | Low-quality |
| S14_vs_k141_2922\|\|lt2gene | Low-quality |
| S14_vs_k141_54772\|\|lt2gene | Low-quality |
| S14_vs_k141_46750\|\|lt2gene | Not-determined |
| S14_vs_k141_24992\|\|lt2gene | Low-quality |
| S14_vs_k141_40203\|\|lt2gene | Low-quality |
| S14_vs_k141_98131\|\|lt2gene | Not-determined |
| S14_vs_k141_61695\|\|lt2gene | Not-determined |
| S14_vs_k141_2940\|\|lt2gene | Not-determined |
| S14_vs_k141_18020\|\|lt2gene | Low-quality |
| S14_vs_k141_2943\|\|lt2gene | Low-quality |
| S14_vs_k141_40207\|\|lt2gene | Low-quality |
| S14_vs_k141_18024\|\|lt2gene | Low-quality |
| S14_vs_k141_46763\|\|lt2gene | Low-quality |
| S14_vs_k141_69148\|\|lt2gene | Low-quality |
| S14_vs_k141_91301\|\|lt2gene | Not-determined |
| S14_vs_k141_91302\|\|lt2gene | Not-determined |
| S14_vs_k141_83964\|\|lt2gene | Low-quality |
| S14_vs_k141_40211\|\|lt2gene | Low-quality |
| S14_vs_k141_106211\|\|lt2gene | Low-quality |
| S14_vs_k141_69153\|\|lt2gene | Low-quality |
| S14_vs_k141_25019\|\|lt2gene | Low-quality |
| S14_vs_k141_83977\|\|lt2gene | Not-determined |
| S14_vs_k141_25022\|\|lt2gene | Low-quality |
| S14_vs_k141_83989\|\|lt2gene | Not-determined |
| S14_vs_k141_91329\|\|lt2gene | Low-quality |
| S14_vs_k141_113279\|\|lt2gene | Low-quality |
| S14_vs_k141_84004\|\|lt2gene | Low-quality |
| S14_vs_k141_18058\|\|lt2gene | Low-quality |
| S14_vs_k141_10458\|\|lt2gene | Low-quality |
| S14_vs_k141_76849\|\|lt2gene | Low-quality |
| S14_vs_k141_46794\|\|lt2gene | Not-determined |
| S14_vs_k141_69188\|\|lt2gene | Not-determined |
| S14_vs_k141_91348\|\|lt2gene | Low-quality |
| S14_vs_k141_106234\|\|lt2gene | Low-quality |
| S14_vs_k141_54815\|\|lt2gene | Low-quality |
| S14_vs_k141_91368\|\|lt2gene | Not-determined |
| S14_vs_k141_40275\|\|lt2gene | Low-quality |
| S14_vs_k141_84040\|\|lt2gene | Not-determined |
| S14_vs_k141_84043\|\|lt2gene | Not-determined |
| S14_vs_k141_54825\|\|lt2gene | Low-quality |
| S14_vs_k141_84051\|\|lt2gene | Not-determined |
| S14_vs_k141_40292\|\|lt2gene | Low-quality |
| S14_vs_k141_3038\|\|lt2gene | Low-quality |
| S14_vs_k141_46814\|\|lt2gene | Not-determined |
| S14_vs_k141_40304\|\|lt2gene | Low-quality |
| S14_vs_k141_10480\|\|lt2gene | Low-quality |
| S14_vs_k141_10481\|\|lt2gene | Not-determined |
| S14_vs_k141_18079\|\|lt2gene | Not-determined |
| S14_vs_k141_46841\|\|lt2gene | Low-quality |
| S14_vs_k141_46844\|\|lt2gene | Low-quality |
| S14_vs_k141_84090\|\|lt2gene | Low-quality |
| S14_vs_k141_106262\|\|lt2gene | Low-quality |
| S14_vs_k141_46853\|\|lt2gene | Low-quality |
| S14_vs_k141_46855\|\|lt2gene | Low-quality |
| S14_vs_k141_76885\|\|lt2gene | Low-quality |
| S14_vs_k141_32618\|\|lt2gene | Low-quality |
| S14_vs_k141_69254\|\|lt2gene | Low-quality |
| S14_vs_k141_46869\|\|lt2gene | Low-quality |
| S14_vs_k141_98249\|\|lt2gene | Not-determined |
| S14_vs_k141_25072\|\|lt2gene | Low-quality |
| S14_vs_k141_113358\|\|lt2gene | Low-quality |
| S14_vs_k141_25074\|\|lt2gene | Low-quality |
| S14_vs_k141_69282\|\|lt2gene | Low-quality |
| S14_vs_k141_25078\|\|lt2gene | Not-determined |
| S14_vs_k141_40375\|\|lt2gene | Low-quality |
| S14_vs_k141_25090\|\|lt2gene | Low-quality |
| S14_vs_k141_32642\|\|lt2gene | Not-determined |
| S14_vs_k141_76922\|\|lt2gene | Not-determined |
| S14_vs_k141_32650\|\|lt2gene | Not-determined |
| S14_vs_k141_32653\|\|lt2gene | Not-determined |
| S14_vs_k141_32655\|\|lt2gene | Not-determined |
| S14_vs_k141_61807\|\|lt2gene | Low-quality |
| S14_vs_k141_76947\|\|lt2gene | Not-determined |
| S14_vs_k141_91456\|\|lt2gene | Low-quality |
| S14_vs_k141_91460\|\|lt2gene | Low-quality |
| S14_vs_k141_18147\|\|lt2gene | Not-determined |
| S14_vs_k141_46915\|\|lt2gene | Low-quality |
| S14_vs_k141_76978\|\|lt2gene | Low-quality |
| S14_vs_k141_76980\|\|lt2gene | Low-quality |
| S14_vs_k141_18163\|\|lt2gene | Low-quality |
| S14_vs_k141_3134\|\|lt2gene | Low-quality |
| S14_vs_k141_3137\|\|lt2gene | Low-quality |
| S14_vs_k141_98276\|\|lt2gene | Not-determined |
| S14_vs_k141_10550\|\|lt2gene | Not-determined |
| S14_vs_k141_10552\|\|lt2gene | Low-quality |
| S14_vs_k141_84150\|\|lt2gene | Low-quality |
| S14_vs_k141_98285\|\|lt2gene | Low-quality |
| S14_vs_k141_98286\|\|lt2gene | Not-determined |
| S14_vs_k141_113419\|\|lt2gene | Low-quality |
| S14_vs_k141_3153\|\|lt2gene | Low-quality |
| S14_vs_k141_61824\|\|lt2gene | Low-quality |
| S14_vs_k141_25128\|\|lt2gene | Low-quality |
| S14_vs_k141_25130\|\|lt2gene | Not-determined |
| S14_vs_k141_10570\|\|lt2gene | Not-determined |
| S14_vs_k141_91506\|\|lt2gene | Low-quality |
| S14_vs_k141_91508\|\|lt2gene | Low-quality |
| S14_vs_k141_106303\|\|lt2gene | Not-determined |
| S14_vs_k141_40391\|\|lt2gene | Not-determined |
| S14_vs_k141_106322\|\|lt2gene | Low-quality |
| S14_vs_k141_106327\|\|lt2gene | Not-determined |
| S14_vs_k141_77010\|\|lt2gene | Low-quality |
| S14_vs_k141_77012\|\|lt2gene | Low-quality |
| S14_vs_k141_84170\|\|lt2gene | Low-quality |
| S14_vs_k141_54894\|\|lt2gene | Low-quality |
| S14_vs_k141_18191\|\|lt2gene | Low-quality |
| S14_vs_k141_69368\|\|lt2gene | Low-quality |
| S14_vs_k141_84174\|\|lt2gene | Low-quality |
| S14_vs_k141_18193\|\|lt2gene | Low-quality |
| S14_vs_k141_18195\|\|lt2gene | Low-quality |
| S14_vs_k141_10614\|\|lt2gene | Low-quality |
| S14_vs_k141_98357\|\|lt2gene | Low-quality |
| S14_vs_k141_113473\|\|lt2gene | Low-quality |
| S14_vs_k141_10630\|\|lt2gene | Not-determined |
| S14_vs_k141_18223\|\|lt2gene | Low-quality |
| S14_vs_k141_113489\|\|lt2gene | Low-quality |
| S14_vs_k141_25193\|\|lt2gene | Low-quality |
| S14_vs_k141_54908\|\|lt2gene | Low-quality |
| S14_vs_k141_10639\|\|lt2gene | Low-quality |
| S14_vs_k141_32698\|\|lt2gene | Low-quality |
| S14_vs_k141_113499\|\|lt2gene | Low-quality |
| S14_vs_k141_46989\|\|lt2gene | Low-quality |
| S14_vs_k141_46994\|\|lt2gene | Not-determined |
| S14_vs_k141_32722\|\|lt2gene | Low-quality |
| S14_vs_k141_18246\|\|lt2gene | Low-quality |
| S14_vs_k141_3224\|\|lt2gene | Low-quality |
| S14_vs_k141_32728\|\|lt2gene | Low-quality |
| S14_vs_k141_3225\|\|lt2gene | Low-quality |
| S14_vs_k141_113512\|\|lt2gene | Low-quality |
| S14_vs_k141_47010\|\|lt2gene | Low-quality |
| S14_vs_k141_32750\|\|lt2gene | Low-quality |
| S14_vs_k141_98392\|\|lt2gene | Low-quality |
| S14_vs_k141_47028\|\|lt2gene | Low-quality |
| S14_vs_k141_84263\|\|lt2gene | Low-quality |
| S14_vs_k141_10658\|\|lt2gene | Not-determined |
| S14_vs_k141_18294\|\|lt2gene | Low-quality |
| S14_vs_k141_18297\|\|lt2gene | Low-quality |
| S14_vs_k141_84265\|\|lt2gene | Not-determined |
| S14_vs_k141_18300\|\|lt2gene | Low-quality |
| S14_vs_k141_84268\|\|lt2gene | Low-quality |
| S14_vs_k141_69458\|\|lt2gene | Low-quality |
| S14_vs_k141_32772\|\|lt2gene | Low-quality |
| S14_vs_k141_61865\|\|lt2gene | Low-quality |
| S14_vs_k141_91573\|\|lt2gene | Low-quality |
| S14_vs_k141_25266\|\|lt2gene | Low-quality |
| S14_vs_k141_61880\|\|lt2gene | Low-quality |
| S14_vs_k141_47071\|\|lt2gene | Low-quality |
| S14_vs_k141_47075\|\|lt2gene | Low-quality |
| S14_vs_k141_98448\|\|lt2gene | Low-quality |
| S14_vs_k141_18352\|\|lt2gene | Low-quality |
| S14_vs_k141_113535\|\|lt2gene | Not-determined |
| S14_vs_k141_54961\|\|lt2gene | Low-quality |
| S14_vs_k141_18360\|\|lt2gene | Low-quality |
| S14_vs_k141_47097\|\|lt2gene | Low-quality |
| S14_vs_k141_113549\|\|lt2gene | Low-quality |
| S14_vs_k141_25308\|\|lt2gene | Not-determined |
| S14_vs_k141_47115\|\|lt2gene | Low-quality |
| S14_vs_k141_3272\|\|lt2gene | Low-quality |
| S14_vs_k141_47121\|\|lt2gene | Not-determined |
| S14_vs_k141_32849\|\|lt2gene | Low-quality |
| S14_vs_k141_47129\|\|lt2gene | Low-quality |
| S14_vs_k141_106446\|\|lt2gene | Low-quality |
| S14_vs_k141_113569\|\|lt2gene | Low-quality |
| S14_vs_k141_84312\|\|lt2gene | Low-quality |
| S14_vs_k141_106459\|\|lt2gene | Not-determined |
| S14_vs_k141_106460\|\|lt2gene | Low-quality |
| S14_vs_k141_47162\|\|lt2gene | Low-quality |
| S14_vs_k141_25338\|\|lt2gene | Low-quality |
| S14_vs_k141_40489\|\|lt2gene | Low-quality |
| S14_vs_k141_40503\|\|lt2gene | Low-quality |
| S14_vs_k141_10756\|\|lt2gene | Low-quality |
| S14_vs_k141_61919\|\|lt2gene | Low-quality |
| S14_vs_k141_61921\|\|lt2gene | Not-determined |
| S14_vs_k141_47203\|\|lt2gene | Low-quality |
| S14_vs_k141_32880\|\|lt2gene | Low-quality |
| S14_vs_k141_61939\|\|lt2gene | Not-determined |
| S14_vs_k141_69547\|\|lt2gene | Not-determined |
| S14_vs_k141_91695\|\|lt2gene | Low-quality |
| S14_vs_k141_32903\|\|lt2gene | Not-determined |
| S14_vs_k141_18420\|\|lt2gene | Low-quality |
| S14_vs_k141_18421\|\|lt2gene | Not-determined |
| S14_vs_k141_18422\|\|lt2gene | Not-determined |
| S14_vs_k141_18424\|\|lt2gene | Not-determined |
| S14_vs_k141_18425\|\|lt2gene | Not-determined |
| S14_vs_k141_106519\|\|lt2gene | Low-quality |
| S14_vs_k141_69577\|\|lt2gene | Low-quality |
| S14_vs_k141_98528\|\|lt2gene | Low-quality |
| S14_vs_k141_113641\|\|lt2gene | Not-determined |
| S14_vs_k141_98532\|\|lt2gene | Low-quality |
| S14_vs_k141_3363\|\|lt2gene | Low-quality |
| S14_vs_k141_10822\|\|lt2gene | Not-determined |
| S14_vs_k141_98545\|\|lt2gene | Low-quality |
| S14_vs_k141_61978\|\|lt2gene | Low-quality |
| S14_vs_k141_10849\|\|lt2gene | Low-quality |
| S14_vs_k141_113664\|\|lt2gene | Not-determined |
| S14_vs_k141_3400\|\|lt2gene | Not-determined |
| S14_vs_k141_3401\|\|lt2gene | Low-quality |
| S14_vs_k141_18467\|\|lt2gene | Not-determined |
| S14_vs_k141_18469\|\|lt2gene | Not-determined |
| S14_vs_k141_62003\|\|lt2gene | Low-quality |
| S14_vs_k141_62007\|\|lt2gene | Not-determined |
| S14_vs_k141_32955\|\|lt2gene | Low-quality |
| S14_vs_k141_62016\|\|lt2gene | Low-quality |
| S14_vs_k141_10886\|\|lt2gene | Low-quality |
| S14_vs_k141_18485\|\|lt2gene | Low-quality |
| S14_vs_k141_47293\|\|lt2gene | Low-quality |
| S14_vs_k141_47297\|\|lt2gene | Low-quality |
| S14_vs_k141_40598\|\|lt2gene | Not-determined |
| S14_vs_k141_40600\|\|lt2gene | Low-quality |
| S14_vs_k141_47304\|\|lt2gene | Low-quality |
| S14_vs_k141_113702\|\|lt2gene | Low-quality |
| S14_vs_k141_113704\|\|lt2gene | Low-quality |
| S14_vs_k141_98583\|\|lt2gene | Low-quality |
| S14_vs_k141_98585\|\|lt2gene | Low-quality |
| S14_vs_k141_113709\|\|lt2gene | Low-quality |
| S14_vs_k141_69646\|\|lt2gene | Low-quality |
| S14_vs_k141_55037\|\|lt2gene | Low-quality |
| S14_vs_k141_91819\|\|lt2gene | Not-determined |
| S14_vs_k141_98594\|\|lt2gene | Not-determined |
| S14_vs_k141_84446\|\|lt2gene | Low-quality |
| S14_vs_k141_10922\|\|lt2gene | Low-quality |
| S14_vs_k141_10923\|\|lt2gene | Low-quality |
| S14_vs_k141_113728\|\|lt2gene | Not-determined |
| S14_vs_k141_69669\|\|lt2gene | Low-quality |
| S14_vs_k141_91835\|\|lt2gene | Low-quality |
| S14_vs_k141_69678\|\|lt2gene | Not-determined |
| S14_vs_k141_40646\|\|lt2gene | Low-quality |
| S14_vs_k141_98610\|\|lt2gene | Low-quality |
| S14_vs_k141_18509\|\|lt2gene | Low-quality |
| S14_vs_k141_84465\|\|lt2gene | Low-quality |
| S14_vs_k141_55064\|\|lt2gene | Low-quality |
| S14_vs_k141_55065\|\|lt2gene | Low-quality |
| S14_vs_k141_98646\|\|lt2gene | Low-quality |
| S14_vs_k141_106590\|\|lt2gene | Low-quality |
| S14_vs_k141_98657\|\|lt2gene | Not-determined |
| S14_vs_k141_62069\|\|lt2gene | Not-determined |
| S14_vs_k141_47360\|\|lt2gene | Low-quality |
| S14_vs_k141_47361\|\|lt2gene | Low-quality |
| S14_vs_k141_33001\|\|lt2gene | Not-determined |
| S14_vs_k141_33002\|\|lt2gene | Low-quality |
| S14_vs_k141_33006\|\|lt2gene | Low-quality |
| S14_vs_k141_18532\|\|lt2gene | Not-determined |
| S14_vs_k141_47370\|\|lt2gene | Low-quality |
| S14_vs_k141_18534\|\|lt2gene | Not-determined |
| S14_vs_k141_47371\|\|lt2gene | Not-determined |
| S14_vs_k141_18546\|\|lt2gene | Not-determined |
| S14_vs_k141_18553\|\|lt2gene | Low-quality |
| S14_vs_k141_91860\|\|lt2gene | Low-quality |
| S14_vs_k141_62094\|\|lt2gene | Low-quality |
| S14_vs_k141_84536\|\|lt2gene | Not-determined |
| S14_vs_k141_77080\|\|lt2gene | Not-determined |
| S14_vs_k141_69730\|\|lt2gene | Low-quality |
| S14_vs_k141_25457\|\|lt2gene | Low-quality |
| S14_vs_k141_62103\|\|lt2gene | Low-quality |
| S14_vs_k141_33044\|\|lt2gene | Low-quality |
| S14_vs_k141_106667\|\|lt2gene | Low-quality |
| S14_vs_k141_106672\|\|lt2gene | Low-quality |
| S14_vs_k141_11029\|\|lt2gene | Not-determined |
| S14_vs_k141_3449\|\|lt2gene | Not-determined |
| S14_vs_k141_40767\|\|lt2gene | Not-determined |
| S14_vs_k141_33058\|\|lt2gene | Low-quality |
| S14_vs_k141_3457\|\|lt2gene | Not-determined |
| S14_vs_k141_62135\|\|lt2gene | Low-quality |
| S14_vs_k141_33065\|\|lt2gene | Low-quality |
| S14_vs_k141_98733\|\|lt2gene | Low-quality |
| S14_vs_k141_113828\|\|lt2gene | Low-quality |
| S14_vs_k141_113829\|\|lt2gene | Low-quality |
| S14_vs_k141_33074\|\|lt2gene | Low-quality |
| S14_vs_k141_77096\|\|lt2gene | Low-quality |
| S14_vs_k141_98736\|\|lt2gene | Low-quality |
| S14_vs_k141_84565\|\|lt2gene | Low-quality |
| S14_vs_k141_3476\|\|lt2gene | Low-quality |
| S14_vs_k141_47437\|\|lt2gene | Low-quality |
| S14_vs_k141_55117\|\|lt2gene | Low-quality |
| S14_vs_k141_40811\|\|lt2gene | Not-determined |
| S14_vs_k141_84579\|\|lt2gene | Low-quality |
| S14_vs_k141_113859\|\|lt2gene | Low-quality |
| S14_vs_k141_62168\|\|lt2gene | Not-determined |
| S14_vs_k141_62170\|\|lt2gene | Low-quality |
| S14_vs_k141_62175\|\|lt2gene | Low-quality |
| S14_vs_k141_113871\|\|lt2gene | Low-quality |
| S14_vs_k141_25551\|\|lt2gene | Not-determined |
| S14_vs_k141_84592\|\|lt2gene | Low-quality |
| S14_vs_k141_113887\|\|lt2gene | Low-quality |
| S14_vs_k141_91901\|\|lt2gene | Not-determined |
| S14_vs_k141_55177\|\|lt2gene | Low-quality |
| S14_vs_k141_47454\|\|lt2gene | Low-quality |
| S14_vs_k141_55182\|\|lt2gene | Low-quality |
| S14_vs_k141_106721\|\|lt2gene | Low-quality |
| S14_vs_k141_91927\|\|lt2gene | Low-quality |
| S14_vs_k141_106730\|\|lt2gene | Low-quality |
| S14_vs_k141_91930\|\|lt2gene | Low-quality |
| S14_vs_k141_33127\|\|lt2gene | Low-quality |
| S14_vs_k141_55203\|\|lt2gene | Low-quality |
| S14_vs_k141_77173\|\|lt2gene | Not-determined |
| S14_vs_k141_47483\|\|lt2gene | Low-quality |
| S14_vs_k141_91945\|\|lt2gene | Low-quality |
| S14_vs_k141_40908\|\|lt2gene | Low-quality |
| S14_vs_k141_55211\|\|lt2gene | Low-quality |
| S14_vs_k141_55213\|\|lt2gene | Low-quality |
| S14_vs_k141_91956\|\|lt2gene | Low-quality |
| S14_vs_k141_91957\|\|lt2gene | Low-quality |
| S14_vs_k141_55217\|\|lt2gene | Low-quality |
| S14_vs_k141_55219\|\|lt2gene | Low-quality |
| S14_vs_k141_84638\|\|lt2gene | Low-quality |
| S14_vs_k141_3536\|\|lt2gene | Low-quality |
| S14_vs_k141_18709\|\|lt2gene | Low-quality |
| S14_vs_k141_55227\|\|lt2gene | Not-determined |
| S14_vs_k141_84651\|\|lt2gene | Low-quality |
| S14_vs_k141_47504\|\|lt2gene | Low-quality |
| S14_vs_k141_11164\|\|lt2gene | Low-quality |
| S14_vs_k141_47507\|\|lt2gene | Low-quality |
| S14_vs_k141_3557\|\|lt2gene | Low-quality |
| S14_vs_k141_33153\|\|lt2gene | Low-quality |
| S14_vs_k141_91973\|\|lt2gene | Low-quality |
| S14_vs_k141_40935\|\|lt2gene | Low-quality |
| S14_vs_k141_47518\|\|lt2gene | Not-determined |
| S14_vs_k141_47521\|\|lt2gene | Low-quality |
| S14_vs_k141_98796\|\|lt2gene | Low-quality |
| S14_vs_k141_77206\|\|lt2gene | Not-determined |
| S14_vs_k141_55241\|\|lt2gene | Low-quality |
| S14_vs_k141_33161\|\|lt2gene | Not-determined |
| S14_vs_k141_25632\|\|lt2gene | Low-quality |
| S14_vs_k141_84669\|\|lt2gene | Low-quality |
| S14_vs_k141_25635\|\|lt2gene | Not-determined |
| S14_vs_k141_47545\|\|lt2gene | Not-determined |
| S14_vs_k141_47546\|\|lt2gene | Low-quality |
| S14_vs_k141_47547\|\|lt2gene | Not-determined |
| S14_vs_k141_47550\|\|lt2gene | Low-quality |
| S14_vs_k141_40960\|\|lt2gene | Not-determined |
| S14_vs_k141_3595\|\|lt2gene | Not-determined |
| S14_vs_k141_3597\|\|lt2gene | Not-determined |
| S14_vs_k141_62242\|\|lt2gene | Low-quality |
| S14_vs_k141_40967\|\|lt2gene | Not-determined |
| S14_vs_k141_92006\|\|lt2gene | Low-quality |
| S14_vs_k141_92013\|\|lt2gene | Not-determined |
| S14_vs_k141_3601\|\|lt2gene | Low-quality |
| S14_vs_k141_18724\|\|lt2gene | Low-quality |
| S14_vs_k141_18730\|\|lt2gene | Not-determined |
| S14_vs_k141_3612\|\|lt2gene | Not-determined |
| S14_vs_k141_98842\|\|lt2gene | Not-determined |
| S14_vs_k141_25666\|\|lt2gene | Low-quality |
| S14_vs_k141_77244\|\|lt2gene | Low-quality |
| S14_vs_k141_55286\|\|lt2gene | Low-quality |
| S14_vs_k141_55293\|\|lt2gene | Low-quality |
| S14_vs_k141_77265\|\|lt2gene | Low-quality |
| S14_vs_k141_77266\|\|lt2gene | Low-quality |
| S14_vs_k141_77273\|\|lt2gene | Not-determined |
| S14_vs_k141_3658\|\|lt2gene | Low-quality |
| S14_vs_k141_62284\|\|lt2gene | Low-quality |
| S14_vs_k141_18765\|\|lt2gene | Not-determined |
| S14_vs_k141_41030\|\|lt2gene | Low-quality |
| S14_vs_k141_84704\|\|lt2gene | Low-quality |
| S14_vs_k141_3680\|\|lt2gene | Low-quality |
| S14_vs_k141_84705\|\|lt2gene | Low-quality |
| S14_vs_k141_98880\|\|lt2gene | Low-quality |
| S14_vs_k141_33213\|\|lt2gene | Low-quality |
| S14_vs_k141_3690\|\|lt2gene | Low-quality |
| S14_vs_k141_18788\|\|lt2gene | Not-determined |
| S14_vs_k141_41042\|\|lt2gene | Low-quality |
| S14_vs_k141_18790\|\|lt2gene | Low-quality |
| S14_vs_k141_33239\|\|lt2gene | Not-determined |
| S14_vs_k141_92067\|\|lt2gene | Low-quality |
| S14_vs_k141_25685\|\|lt2gene | Low-quality |
| S14_vs_k141_25687\|\|lt2gene | Low-quality |
| S14_vs_k141_18800\|\|lt2gene | Low-quality |
| S14_vs_k141_84724\|\|lt2gene | Low-quality |
| S14_vs_k141_114040\|\|lt2gene | Low-quality |
| S14_vs_k141_25698\|\|lt2gene | Low-quality |
| S14_vs_k141_84737\|\|lt2gene | Not-determined |
| S14_vs_k141_25703\|\|lt2gene | Low-quality |
| S14_vs_k141_25709\|\|lt2gene | Low-quality |
| S14_vs_k141_25718\|\|lt2gene | Low-quality |
| S14_vs_k141_92117\|\|lt2gene | Low-quality |
| S14_vs_k141_18868\|\|lt2gene | Low-quality |
| S14_vs_k141_92119\|\|lt2gene | Low-quality |
| S14_vs_k141_11348\|\|lt2gene | Not-determined |
| S14_vs_k141_11351\|\|lt2gene | Not-determined |
| S14_vs_k141_11373\|\|lt2gene | Low-quality |
| S14_vs_k141_106891\|\|lt2gene | Low-quality |
| S14_vs_k141_18923\|\|lt2gene | Low-quality |
| S14_vs_k141_77400\|\|lt2gene | Low-quality |
| S14_vs_k141_18944\|\|lt2gene | Low-quality |
| S14_vs_k141_55361\|\|lt2gene | Not-determined |
| S14_vs_k141_92172\|\|lt2gene | Low-quality |
| S14_vs_k141_84775\|\|lt2gene | Low-quality |
| S14_vs_k141_55364\|\|lt2gene | Not-determined |
| S14_vs_k141_25783\|\|lt2gene | Low-quality |
| S14_vs_k141_25785\|\|lt2gene | Low-quality |
| S14_vs_k141_25789\|\|lt2gene | Low-quality |
| S14_vs_k141_114164\|\|lt2gene | Low-quality |
| S14_vs_k141_25793\|\|lt2gene | Low-quality |
| S14_vs_k141_98959\|\|lt2gene | Not-determined |
| S14_vs_k141_18961\|\|lt2gene | Low-quality |
| S14_vs_k141_114174\|\|lt2gene | Low-quality |
| S14_vs_k141_55385\|\|lt2gene | Low-quality |
| S14_vs_k141_55393\|\|lt2gene | Not-determined |
| S14_vs_k141_33302\|\|lt2gene | Low-quality |
| S14_vs_k141_84805\|\|lt2gene | Low-quality |
| S14_vs_k141_77436\|\|lt2gene | Not-determined |
| S14_vs_k141_84809\|\|lt2gene | Low-quality |
| S14_vs_k141_106950\|\|lt2gene | Low-quality |

S14_vs_k141_106952||lt2gene Low-quality
S14_vs_k141_77442||lt2gene Not-determined
S14_vs_k141_77447||lt2gene Not-determined
S14_vs_k141_77450||lt2gene Not-determined
S14_vs_k141_55408||lt2gene Low-quality
S14_vs_k141_84820||lt2gene Low-quality
S14_vs_k141_55409||lt2gene Low-quality
S14_vs_k141_3797||lt2gene Not-determined
S14_vs_k141_41132||lt2gene Low-quality
S14_vs_k141_92211||lt2gene Low-quality
S14_vs_k141_98988||lt2gene Low-quality
S14_vs_k141_92215||lt2gene Not-determined
S14_vs_k141_18968||lt2gene Low-quality
S14_vs_k141_33310||lt2gene Low-quality
S14_vs_k141_3810||lt2gene Low-quality
S14_vs_k141_55419||lt2gene Low-quality
S14_vs_k141_84836||lt2gene Low-quality
S14_vs_k141_55437||lt2gene Low-quality
S14_vs_k141_77482||lt2gene Not-determined
S14_vs_k141_77484||lt2gene Low-quality
S14_vs_k141_55454||lt2gene Not-determined
S14_vs_k141_19005||lt2gene Low-quality
S14_vs_k141_3850||lt2gene Low-quality
S14_vs_k141_99006||lt2gene Not-determined
S14_vs_k141_99007||lt2gene Low-quality
S14_vs_k141_19017||lt2gene Low-quality
S14_vs_k141_19021||lt2gene Low-quality
S14_vs_k141_69953||lt2gene Low-quality
S14_vs_k141_25840||lt2gene Low-quality
S14_vs_k141_84884||lt2gene Low-quality
S14_vs_k141_47605||lt2gene Low-quality
S14_vs_k141_11430||lt2gene Low-quality
S14_vs_k141_84913||lt2gene Low-quality
S14_vs_k141_99041||lt2gene Low-quality
S14_vs_k141_55506||lt2gene Not-determined
S14_vs_k141_92256||lt2gene Low-quality
S14_vs_k141_33399||lt2gene Low-quality
S14_vs_k141_11450||lt2gene Low-quality
S14_vs_k141_19074||lt2gene Not-determined
S14_vs_k141_41156||lt2gene Low-quality
S14_vs_k141_77538||lt2gene Not-determined
S14_vs_k141_84937||lt2gene Not-determined
S14_vs_k141_41165||lt2gene Low-quality
S14_vs_k141_62384||lt2gene Low-quality
S14_vs_k141_77560||lt2gene Not-determined
S14_vs_k141_77562||lt2gene Not-determined
S14_vs_k141_92295||lt2gene Low-quality
S14_vs_k141_3961||lt2gene Low-quality
S14_vs_k141_84974||lt2gene Not-determined
S14_vs_k141_84979||lt2gene Not-determined
S14_vs_k141_77578||lt2gene Low-quality
S14_vs_k141_84981||lt2gene Low-quality
S14_vs_k141_11497||lt2gene Low-quality
S14_vs_k141_99104||lt2gene Low-quality
S14_vs_k141_62440||lt2gene Low-quality
S14_vs_k141_85003||lt2gene Not-determined
S14_vs_k141_41214||lt2gene Low-quality
S14_vs_k141_107001||lt2gene Low-quality
S14_vs_k141_114278||lt2gene Not-determined
S14_vs_k141_70045||lt2gene Low-quality
S14_vs_k141_47702||lt2gene Low-quality
S14_vs_k141_47706||lt2gene Low-quality
S14_vs_k141_4014||lt2gene Low-quality
S14_vs_k141_107019||lt2gene Low-quality
S14_vs_k141_107024||lt2gene Not-determined
S14_vs_k141_41238||lt2gene Not-determined
S14_vs_k141_70079||lt2gene Low-quality
S14_vs_k141_55570||lt2gene Low-quality
S14_vs_k141_85032||lt2gene Low-quality
S14_vs_k141_114303||lt2gene Low-quality
S14_vs_k141_33491||lt2gene Low-quality
S14_vs_k141_25882||lt2gene Low-quality
S14_vs_k141_114314||lt2gene Low-quality
S14_vs_k141_99130||lt2gene Not-determined
S14_vs_k141_77606||lt2gene Low-quality
S14_vs_k141_114322||lt2gene Low-quality
S14_vs_k141_41272||lt2gene Low-quality
S14_vs_k141_62526||lt2gene Low-quality
S14_vs_k141_99145||lt2gene Low-quality
S14_vs_k141_62532||lt2gene Low-quality
S14_vs_k141_70125||lt2gene Low-quality
S14_vs_k141_77622||lt2gene Low-quality
S14_vs_k141_62540||lt2gene Low-quality
S14_vs_k141_41291||lt2gene Not-determined
S14_vs_k141_107054||lt2gene Low-quality
S14_vs_k141_62547||lt2gene Low-quality
S14_vs_k141_4060||lt2gene Low-quality
S14_vs_k141_114346||lt2gene Low-quality
S14_vs_k141_107057||lt2gene Not-determined
S14_vs_k141_55608||lt2gene Low-quality
S14_vs_k141_99162||lt2gene Not-determined
S14_vs_k141_55618||lt2gene Not-determined
S14_vs_k141_55626||lt2gene Not-determined
S14_vs_k141_55628||lt2gene Not-determined
S14_vs_k141_55634||lt2gene Low-quality
S14_vs_k141_114362||lt2gene Low-quality
S14_vs_k141_85084||lt2gene Low-quality
S14_vs_k141_70148||lt2gene Not-determined
S14_vs_k141_99179||lt2gene Low-quality
S14_vs_k141_55638||lt2gene Low-quality
S14_vs_k141_11557||lt2gene Low-quality
S14_vs_k141_107099||lt2gene Low-quality
S14_vs_k141_70174||lt2gene Low-quality
S14_vs_k141_11570||lt2gene Low-quality
S14_vs_k141_47778||lt2gene Not-determined
S14_vs_k141_70177||lt2gene Low-quality
S14_vs_k141_70180||lt2gene Low-quality
S14_vs_k141_70181||lt2gene Low-quality
S14_vs_k141_19309||lt2gene Not-determined
S14_vs_k141_77680||lt2gene Not-determined
S14_vs_k141_41343||lt2gene Low-quality
S14_vs_k141_33559||lt2gene Not-determined
S14_vs_k141_11594||lt2gene Low-quality
S14_vs_k141_11596||lt2gene Low-quality
S14_vs_k141_77693||lt2gene Low-quality
S14_vs_k141_107127||lt2gene Low-quality
S14_vs_k141_25929||lt2gene Low-quality
S14_vs_k141_70202||lt2gene Not-determined
S14_vs_k141_11611||lt2gene Low-quality
S14_vs_k141_19349||lt2gene Not-determined
S14_vs_k141_41365||lt2gene Low-quality
S14_vs_k141_107147||lt2gene Not-determined
S14_vs_k141_11618||lt2gene Low-quality
S14_vs_k141_11623||lt2gene Low-quality
S14_vs_k141_70231||lt2gene Low-quality
S14_vs_k141_19378||lt2gene Low-quality
S14_vs_k141_19379||lt2gene Low-quality
S14_vs_k141_114392||lt2gene Low-quality
S14_vs_k141_33584||lt2gene Not-determined
S14_vs_k141_19381||lt2gene Low-quality
S14_vs_k141_92446||lt2gene Low-quality
S14_vs_k141_62630||lt2gene Low-quality
S14_vs_k141_19384||lt2gene Low-quality
S14_vs_k141_99257||lt2gene Low-quality
S14_vs_k141_19389||lt2gene Low-quality
S14_vs_k141_25959||lt2gene Low-quality
S14_vs_k141_62638||lt2gene Low-quality
S14_vs_k141_25968||lt2gene Not-determined
S14_vs_k141_62648||lt2gene Low-quality
S14_vs_k141_77729||lt2gene Low-quality
S14_vs_k141_85133||lt2gene Not-determined
S14_vs_k141_19411||lt2gene Not-determined
S14_vs_k141_62655||lt2gene Low-quality
S14_vs_k141_25977||lt2gene Low-quality
S14_vs_k141_25978||lt2gene Low-quality
S14_vs_k141_33607||lt2gene Not-determined
S14_vs_k141_62660||lt2gene Low-quality
S14_vs_k141_85149||lt2gene Low-quality
S14_vs_k141_62692||lt2gene Low-quality
S14_vs_k141_107202||lt2gene Low-quality
S14_vs_k141_62696||lt2gene Low-quality
S14_vs_k141_114429||lt2gene Low-quality
S14_vs_k141_99310||lt2gene Low-quality
S14_vs_k141_55739||lt2gene Low-quality
S14_vs_k141_77775||lt2gene Low-quality
S14_vs_k141_77778||lt2gene Low-quality
S14_vs_k141_107227||lt2gene Low-quality
S14_vs_k141_62705||lt2gene Not-determined
S14_vs_k141_19467||lt2gene Low-quality
S14_vs_k141_33630||lt2gene Low-quality
S14_vs_k141_77814||lt2gene Low-quality
S14_vs_k141_70237||lt2gene Low-quality
S14_vs_k141_114455||lt2gene Not-determined
S14_vs_k141_41461||lt2gene Low-quality
S14_vs_k141_114459||lt2gene Not-determined
S14_vs_k141_85179||lt2gene Low-quality
S14_vs_k141_107257||lt2gene Low-quality
S14_vs_k141_92534||lt2gene Low-quality
S14_vs_k141_41472||lt2gene Low-quality
S14_vs_k141_70254||lt2gene Low-quality
S14_vs_k141_55770||lt2gene Not-determined
S14_vs_k141_55772||lt2gene Low-quality
S14_vs_k141_85181||lt2gene Low-quality
S14_vs_k141_4157||lt2gene Low-quality
S14_vs_k141_41492||lt2gene Not-determined
S14_vs_k141_19519||lt2gene Low-quality
S14_vs_k141_99371||lt2gene Low-quality
S14_vs_k141_26049||lt2gene Low-quality
S14_vs_k141_26052||lt2gene Low-quality
S14_vs_k141_4172||lt2gene Not-determined
S14_vs_k141_19527||lt2gene Low-quality
S14_vs_k141_70282||lt2gene Low-quality
S14_vs_k141_85206||lt2gene Low-quality
S14_vs_k141_62758||lt2gene Low-quality
S14_vs_k141_19547||lt2gene Low-quality
S14_vs_k141_70298||lt2gene Low-quality
S14_vs_k141_55823||lt2gene Not-determined
S14_vs_k141_70301||lt2gene Low-quality
S14_vs_k141_107295||lt2gene Low-quality
S14_vs_k141_62767||lt2gene Low-quality
S14_vs_k141_41528||lt2gene Low-quality
S14_vs_k141_26071||lt2gene Low-quality
S14_vs_k141_114506||lt2gene Low-quality
S14_vs_k141_26077||lt2gene Low-quality
S14_vs_k141_4207||lt2gene Not-determined
S14_vs_k141_107311||lt2gene Low-quality
S14_vs_k141_70325||lt2gene Low-quality
S14_vs_k141_55844||lt2gene Not-determined
S14_vs_k141_85235||lt2gene Low-quality
S14_vs_k141_26106||lt2gene Low-quality
S14_vs_k141_85257||lt2gene Low-quality
S14_vs_k141_26111||lt2gene Not-determined
S14_vs_k141_70348||lt2gene Low-quality
S14_vs_k141_41555||lt2gene Not-determined
S14_vs_k141_92604||lt2gene Low-quality
S14_vs_k141_47845||lt2gene Low-quality
S14_vs_k141_62784||lt2gene Low-quality
S14_vs_k141_47853||lt2gene Low-quality
S14_vs_k141_99448||lt2gene Low-quality
S14_vs_k141_47860||lt2gene Low-quality
S14_vs_k141_47861||lt2gene Low-quality
S14_vs_k141_107348||lt2gene Low-quality
S14_vs_k141_107353||lt2gene Low-quality
S14_vs_k141_55880||lt2gene Low-quality
S14_vs_k141_77937||lt2gene Low-quality
S14_vs_k141_70379||lt2gene Low-quality
S14_vs_k141_77943||lt2gene Low-quality
S14_vs_k141_92661||lt2gene Low-quality
S14_vs_k141_4265||lt2gene Low-quality
S14_vs_k141_92663||lt2gene Low-quality
S14_vs_k141_26121||lt2gene Not-determined
S14_vs_k141_62833||lt2gene Low-quality
S14_vs_k141_4277||lt2gene Not-determined
S14_vs_k141_114598||lt2gene Low-quality
S14_vs_k141_70420||lt2gene Low-quality
S14_vs_k141_47898||lt2gene Low-quality
S14_vs_k141_107393||lt2gene Not-determined
S14_vs_k141_92670||lt2gene Low-quality
S14_vs_k141_99475||lt2gene Low-quality
S14_vs_k141_107410||lt2gene Not-determined
S14_vs_k141_62874||lt2gene Low-quality
S14_vs_k141_47920||lt2gene Low-quality
S14_vs_k141_92692||lt2gene Not-determined
S14_vs_k141_92694||lt2gene Low-quality
S14_vs_k141_107413||lt2gene Low-quality
S14_vs_k141_11651||lt2gene Low-quality
S14_vs_k141_26142||lt2gene Low-quality
S14_vs_k141_26148||lt2gene Not-determined
S14_vs_k141_26149||lt2gene Not-determined
S14_vs_k141_26152||lt2gene Low-quality
S14_vs_k141_85348||lt2gene Low-quality
S14_vs_k141_92710||lt2gene Low-quality
S14_vs_k141_77989||lt2gene Not-determined
S14_vs_k141_55947||lt2gene Low-quality
S14_vs_k141_4330||lt2gene Low-quality
S14_vs_k141_19623||lt2gene Not-determined
S14_vs_k141_92756||lt2gene Low-quality
S14_vs_k141_19631||lt2gene Not-determined
S14_vs_k141_4350||lt2gene Low-quality
S14_vs_k141_4351||lt2gene Low-quality
S14_vs_k141_92765||lt2gene Low-quality
S14_vs_k141_70514||lt2gene Low-quality
S14_vs_k141_4353||lt2gene Low-quality
S14_vs_k141_41634||lt2gene Low-quality
S14_vs_k141_4357||lt2gene Low-quality
S14_vs_k141_4358||lt2gene Low-quality
S14_vs_k141_92766||lt2gene Low-quality
S14_vs_k141_85357||lt2gene Low-quality
S14_vs_k141_41647||lt2gene Low-quality
S14_vs_k141_85363||lt2gene Not-determined
S14_vs_k141_4379||lt2gene Low-quality
S14_vs_k141_4393||lt2gene Low-quality
S14_vs_k141_114681||lt2gene Not-determined
S14_vs_k141_47965||lt2gene Low-quality
S14_vs_k141_62926||lt2gene Low-quality
S14_vs_k141_70525||lt2gene Low-quality
S14_vs_k141_114688||lt2gene Not-determined
S14_vs_k141_19649||lt2gene Low-quality
S14_vs_k141_114690||lt2gene Low-quality
S14_vs_k141_62935||lt2gene Low-quality
S14_vs_k141_62937||lt2gene Not-determined
S14_vs_k141_47975||lt2gene Low-quality
S14_vs_k141_114702||lt2gene Not-determined
S14_vs_k141_47982||lt2gene Not-determined
S14_vs_k141_92803||lt2gene Low-quality
S14_vs_k141_78044||lt2gene Not-determined
S14_vs_k141_85385||lt2gene Low-quality
S14_vs_k141_4413||lt2gene Low-quality
S14_vs_k141_41669||lt2gene Low-quality
S14_vs_k141_62947||lt2gene Not-determined
S14_vs_k141_85389||lt2gene Not-determined
S14_vs_k141_4429||lt2gene Not-determined
S14_vs_k141_19674||lt2gene Low-quality
S14_vs_k141_55972||lt2gene Low-quality
S14_vs_k141_62971||lt2gene Low-quality
S14_vs_k141_26185||lt2gene Low-quality
S14_vs_k141_4456||lt2gene Low-quality
S14_vs_k141_26187||lt2gene Low-quality
S14_vs_k141_78067||lt2gene Low-quality
S14_vs_k141_26193||lt2gene Low-quality
S14_vs_k141_70576||lt2gene Low-quality
S14_vs_k141_62975||lt2gene Low-quality
S14_vs_k141_114715||lt2gene Low-quality
S14_vs_k141_85408||lt2gene Low-quality
S14_vs_k141_62979||lt2gene Not-determined
S14_vs_k141_62980||lt2gene Low-quality
S14_vs_k141_26196||lt2gene Low-quality
S14_vs_k141_92856||lt2gene Low-quality
S14_vs_k141_26203||lt2gene Low-quality
S14_vs_k141_99595||lt2gene Not-determined
S14_vs_k141_41701||lt2gene Low-quality
S14_vs_k141_19718||lt2gene Low-quality
S14_vs_k141_33792||lt2gene Low-quality
S14_vs_k141_70608||lt2gene Low-quality
S14_vs_k141_33798||lt2gene Not-determined
S14_vs_k141_92868||lt2gene Low-quality
S14_vs_k141_70615||lt2gene Low-quality
S14_vs_k141_41710||lt2gene Low-quality
S14_vs_k141_114749||lt2gene Low-quality
S14_vs_k141_4499||lt2gene Low-quality
S14_vs_k141_11718||lt2gene Low-quality
S14_vs_k141_33808||lt2gene Low-quality
S14_vs_k141_70637||lt2gene Low-quality
S14_vs_k141_107514||lt2gene Low-quality
S14_vs_k141_4514||lt2gene Not-determined
S14_vs_k141_4518||lt2gene Not-determined
S14_vs_k141_55997||lt2gene Not-determined
S14_vs_k141_4522||lt2gene Low-quality
S14_vs_k141_107539||lt2gene Low-quality
S14_vs_k141_26251||lt2gene Not-determined
S14_vs_k141_62995||lt2gene Low-quality
S14_vs_k141_33834||lt2gene Low-quality
S14_vs_k141_41750||lt2gene Low-quality
S14_vs_k141_41752||lt2gene Not-determined
S14_vs_k141_41754||lt2gene Not-determined
S14_vs_k141_41755||lt2gene Not-determined
S14_vs_k141_33844||lt2gene Low-quality
S14_vs_k141_56020||lt2gene Low-quality
S14_vs_k141_19773||lt2gene Low-quality
S14_vs_k141_19777||lt2gene Low-quality
S14_vs_k141_41769||lt2gene Low-quality
S14_vs_k141_33858||lt2gene Low-quality
S14_vs_k141_56037||lt2gene Low-quality
S14_vs_k141_26279||lt2gene Low-quality
S14_vs_k141_99634||lt2gene Low-quality
S14_vs_k141_19790||lt2gene Low-quality
S14_vs_k141_70691||lt2gene Not-determined
S14_vs_k141_70695||lt2gene Not-determined
S14_vs_k141_26294||lt2gene Low-quality
S14_vs_k141_48046||lt2gene Low-quality
S14_vs_k141_26300||lt2gene Low-quality
S14_vs_k141_48056||lt2gene Low-quality
S14_vs_k141_48060||lt2gene Low-quality
S14_vs_k141_26314||lt2gene Not-determined
S14_vs_k141_70718||lt2gene Low-quality
S14_vs_k141_85446||lt2gene Not-determined
S14_vs_k141_63067||lt2gene Low-quality
S14_vs_k141_11785||lt2gene Low-quality
S14_vs_k141_70735||lt2gene Low-quality
S14_vs_k141_48081||lt2gene Low-quality
S14_vs_k141_19805||lt2gene Low-quality
S14_vs_k141_33916||lt2gene Low-quality
S14_vs_k141_48092||lt2gene Low-quality
S14_vs_k141_19820||lt2gene Low-quality
S14_vs_k141_63103||lt2gene Low-quality
S14_vs_k141_33939||lt2gene Low-quality
S14_vs_k141_107620||lt2gene Low-quality
S14_vs_k141_107625||lt2gene Low-quality
S14_vs_k141_107626||lt2gene Low-quality
S14_vs_k141_107629||lt2gene Low-quality
S14_vs_k141_107630||lt2gene Low-quality
S14_vs_k141_107632||lt2gene Low-quality
S14_vs_k141_41843||lt2gene Low-quality
S14_vs_k141_85506||lt2gene Low-quality
S14_vs_k141_19856||lt2gene Low-quality
S14_vs_k141_70786||lt2gene Low-quality
S14_vs_k141_114843||lt2gene Low-quality
S14_vs_k141_48105||lt2gene Low-quality
S14_vs_k141_78145||lt2gene Low-quality
S14_vs_k141_107652||lt2gene Low-quality
S14_vs_k141_4559||lt2gene Low-quality
S14_vs_k141_99700||lt2gene Not-determined
S14_vs_k141_63126||lt2gene Not-determined
S14_vs_k141_92887||lt2gene Not-determined
S14_vs_k141_70799||lt2gene Low-quality
S14_vs_k141_92892||lt2gene Low-quality
S14_vs_k141_92897||lt2gene Not-determined
S14_vs_k141_19873||lt2gene Low-quality
S14_vs_k141_114876||lt2gene Not-determined
S14_vs_k141_63143||lt2gene Low-quality
S14_vs_k141_70813||lt2gene Not-determined
S14_vs_k141_114885||lt2gene Not-determined
S14_vs_k141_114888||lt2gene Low-quality
S14_vs_k141_107676||lt2gene Low-quality
S14_vs_k141_92923||lt2gene Low-quality
S14_vs_k141_33986||lt2gene Not-determined
S14_vs_k141_85553||lt2gene Low-quality
S14_vs_k141_70841||lt2gene Low-quality
S14_vs_k141_85556||lt2gene Not-determined
S14_vs_k141_70844||lt2gene Low-quality
S14_vs_k141_11856||lt2gene Low-quality
S14_vs_k141_11857||lt2gene Low-quality
S14_vs_k141_92939||lt2gene Low-quality
S14_vs_k141_107702||lt2gene Not-determined
S14_vs_k141_4584||lt2gene Low-quality
S14_vs_k141_70861||lt2gene Low-quality
S14_vs_k141_11872||lt2gene Not-determined
S14_vs_k141_85579||lt2gene Low-quality
S14_vs_k141_85585||lt2gene Low-quality
S14_vs_k141_63173||lt2gene Low-quality
S14_vs_k141_92966||lt2gene Low-quality
S14_vs_k141_92970||lt2gene Not-determined
S14_vs_k141_63177||lt2gene Low-quality
S14_vs_k141_85592||lt2gene Low-quality
S14_vs_k141_11880||lt2gene Low-quality
S14_vs_k141_41893||lt2gene Not-determined
S14_vs_k141_92976||lt2gene Low-quality
S14_vs_k141_92983||lt2gene Low-quality
S14_vs_k141_70888||lt2gene Not-determined
S14_vs_k141_93000||lt2gene Not-determined
S14_vs_k141_4616||lt2gene Low-quality
S14_vs_k141_19949||lt2gene Low-quality
S14_vs_k141_70919||lt2gene Low-quality
S14_vs_k141_99744||lt2gene Low-quality
S14_vs_k141_70921||lt2gene Low-quality
S14_vs_k141_85628||lt2gene Low-quality
S14_vs_k141_34020||lt2gene Not-determined
S14_vs_k141_93045||lt2gene Low-quality
S14_vs_k141_41909||lt2gene Low-quality
S14_vs_k141_4634||lt2gene Not-determined
S14_vs_k141_70940||lt2gene Low-quality
S14_vs_k141_41914||lt2gene Not-determined
S14_vs_k141_4645||lt2gene Low-quality
S14_vs_k141_93066||lt2gene Not-determined
S14_vs_k141_93073||lt2gene Low-quality
S14_vs_k141_41917||lt2gene Low-quality
S14_vs_k141_11926||lt2gene Low-quality
S14_vs_k141_48204||lt2gene Low-quality
S14_vs_k141_4678||lt2gene Low-quality
S14_vs_k141_70971||lt2gene Low-quality
S14_vs_k141_34033||lt2gene Low-quality
S14_vs_k141_4686||lt2gene Low-quality
S14_vs_k141_26322||lt2gene Not-determined
S14_vs_k141_11950||lt2gene Low-quality
S14_vs_k141_26324||lt2gene Not-determined
S14_vs_k141_99770||lt2gene Low-quality
S14_vs_k141_93096||lt2gene Low-quality
S14_vs_k141_70984||lt2gene Low-quality
S14_vs_k141_93099||lt2gene Low-quality
S14_vs_k141_93102||lt2gene Low-quality
S14_vs_k141_4710||lt2gene Not-determined
S14_vs_k141_78204||lt2gene Low-quality
S14_vs_k141_93120||lt2gene Low-quality
S14_vs_k141_107748||lt2gene Low-quality
S14_vs_k141_99791||lt2gene Low-quality
S14_vs_k141_11956||lt2gene Low-quality
S14_vs_k141_11964||lt2gene Low-quality
S14_vs_k141_41980||lt2gene Low-quality
S14_vs_k141_107770||lt2gene Low-quality
S14_vs_k141_85677||lt2gene Not-determined
S14_vs_k141_4755||lt2gene Low-quality
S14_vs_k141_107778||lt2gene Low-quality
S14_vs_k141_4758||lt2gene Low-quality
S14_vs_k141_34066||lt2gene Low-quality
S14_vs_k141_34080||lt2gene Low-quality
S14_vs_k141_93146||lt2gene Low-quality
S14_vs_k141_41997||lt2gene Low-quality
S14_vs_k141_93148||lt2gene Low-quality
S14_vs_k141_4768||lt2gene Not-determined
S14_vs_k141_34084||lt2gene Low-quality
S14_vs_k141_93160||lt2gene Not-determined
S14_vs_k141_85713||lt2gene Low-quality
S14_vs_k141_85715||lt2gene Low-quality
S14_vs_k141_63254||lt2gene Not-determined
S14_vs_k141_26403||lt2gene Low-quality
S14_vs_k141_93166||lt2gene Low-quality
S14_vs_k141_12016||lt2gene Low-quality
S14_vs_k141_115004||lt2gene Not-determined
S14_vs_k141_115007||lt2gene Low-quality
S14_vs_k141_34138||lt2gene Low-quality
S14_vs_k141_93179||lt2gene Low-quality
S14_vs_k141_4808||lt2gene Low-quality
S14_vs_k141_42061||lt2gene Low-quality
S14_vs_k141_56224||lt2gene Low-quality
S14_vs_k141_42068||lt2gene Not-determined
S14_vs_k141_99895||lt2gene Not-determined
S14_vs_k141_99897||lt2gene Low-quality
S14_vs_k141_63296||lt2gene Not-determined
S14_vs_k141_93206||lt2gene Low-quality
S14_vs_k141_48327||lt2gene Low-quality
S14_vs_k141_107835||lt2gene Not-determined
S14_vs_k141_78294||lt2gene Low-quality
S14_vs_k141_56243||lt2gene Low-quality
S14_vs_k141_42098||lt2gene Low-quality
S14_vs_k141_63321||lt2gene Low-quality
S14_vs_k141_63323||lt2gene Not-determined
S14_vs_k141_115032||lt2gene Low-quality
S14_vs_k141_115034||lt2gene Low-quality
S14_vs_k141_26448||lt2gene Low-quality
S14_vs_k141_93245||lt2gene Low-quality
S14_vs_k141_42106||lt2gene Low-quality
S14_vs_k141_26454||lt2gene Low-quality
S14_vs_k141_107872||lt2gene Low-quality
S14_vs_k141_26456||lt2gene Low-quality
S14_vs_k141_78322||lt2gene Low-quality
S14_vs_k141_63333||lt2gene Not-determined
S14_vs_k141_26461||lt2gene Not-determined
S14_vs_k141_93254||lt2gene Low-quality
S14_vs_k141_26463||lt2gene Low-quality
S14_vs_k141_93259||lt2gene Low-quality
S14_vs_k141_78336||lt2gene Low-quality
S14_vs_k141_107886||lt2gene Low-quality
S14_vs_k141_78350||lt2gene Low-quality
S14_vs_k141_93281||lt2gene Low-quality
S14_vs_k141_115090||lt2gene Low-quality
S14_vs_k141_12133||lt2gene Low-quality
S14_vs_k141_107911||lt2gene Low-quality
S14_vs_k141_78375||lt2gene Low-quality
S14_vs_k141_93298||lt2gene Low-quality
S14_vs_k141_34227||lt2gene Low-quality
S14_vs_k141_48381||lt2gene Low-quality
S14_vs_k141_56317||lt2gene Low-quality
S14_vs_k141_56320||lt2gene Not-determined
S14_vs_k141_107933||lt2gene Low-quality
S14_vs_k141_12141||lt2gene Low-quality
S14_vs_k141_100007||lt2gene Not-determined
S14_vs_k141_78398||lt2gene Low-quality
S14_vs_k141_100015||lt2gene Not-determined
S14_vs_k141_42173||lt2gene Low-quality
S14_vs_k141_115130||lt2gene Low-quality
S14_vs_k141_115131||lt2gene Low-quality
S14_vs_k141_63422||lt2gene Low-quality
S14_vs_k141_115132||lt2gene Low-quality
S14_vs_k141_107948||lt2gene Low-quality
S14_vs_k141_71018||lt2gene Low-quality
S14_vs_k141_48406||lt2gene Not-determined
S14_vs_k141_93335||lt2gene Low-quality
S14_vs_k141_4924||lt2gene Low-quality
S14_vs_k141_78424||lt2gene Low-quality
S14_vs_k141_63443||lt2gene Low-quality
S14_vs_k141_42186||lt2gene Low-quality
S14_vs_k141_34258||lt2gene Low-quality
S14_vs_k141_63451||lt2gene Low-quality
S14_vs_k141_78430||lt2gene Not-determined
S14_vs_k141_42199||lt2gene Low-quality
S14_vs_k141_34276||lt2gene Low-quality
S14_vs_k141_42210||lt2gene Low-quality
S14_vs_k141_48420||lt2gene Low-quality
S14_vs_k141_4947||lt2gene Low-quality
S14_vs_k141_4950||lt2gene Low-quality
S14_vs_k141_56373||lt2gene Low-quality
S14_vs_k141_100064||lt2gene Low-quality
S14_vs_k141_42224||lt2gene Low-quality
S14_vs_k141_56376||lt2gene Low-quality
S14_vs_k141_34298||lt2gene Low-quality
S14_vs_k141_115175||lt2gene Low-quality
S14_vs_k141_26491||lt2gene Low-quality
S14_vs_k141_78455||lt2gene Low-quality
S14_vs_k141_78459||lt2gene Low-quality
S14_vs_k141_100082||lt2gene Low-quality
S14_vs_k141_107977||lt2gene Low-quality
S14_vs_k141_34320||lt2gene Low-quality
S14_vs_k141_107983||lt2gene Low-quality
S14_vs_k141_93358||lt2gene Low-quality
S14_vs_k141_115199||lt2gene Low-quality
S14_vs_k141_93363||lt2gene Low-quality
S14_vs_k141_56402||lt2gene Low-quality
S14_vs_k141_107999||lt2gene Low-quality
S14_vs_k141_108003||lt2gene Low-quality
S14_vs_k141_42289||lt2gene Low-quality
S14_vs_k141_71076||lt2gene Low-quality
S14_vs_k141_26527||lt2gene Not-determined
S14_vs_k141_5025||lt2gene Low-quality
S14_vs_k141_63534||lt2gene Not-determined
S14_vs_k141_48489||lt2gene Low-quality
S14_vs_k141_48491||lt2gene Low-quality
S14_vs_k141_78505||lt2gene Not-determined
S14_vs_k141_34397||lt2gene Low-quality
S14_vs_k141_78510||lt2gene Low-quality
S14_vs_k141_78512||lt2gene Not-determined
S14_vs_k141_100136||lt2gene Low-quality
S14_vs_k141_48511||lt2gene Low-quality
S14_vs_k141_115234||lt2gene Not-determined
S14_vs_k141_115236||lt2gene Not-determined
S14_vs_k141_5033||lt2gene Low-quality
S14_vs_k141_34433||lt2gene Low-quality
S14_vs_k141_48526||lt2gene Low-quality
S14_vs_k141_5042||lt2gene Low-quality
S14_vs_k141_78534||lt2gene Low-quality
S14_vs_k141_42338||lt2gene Low-quality
S14_vs_k141_78536||lt2gene Low-quality
S14_vs_k141_42343||lt2gene Low-quality
S14_vs_k141_78542||lt2gene Low-quality
S14_vs_k141_115239||lt2gene Not-determined
S14_vs_k141_56443||lt2gene Not-determined
S14_vs_k141_56446||lt2gene Not-determined
S14_vs_k141_56447||lt2gene Low-quality
S14_vs_k141_34444||lt2gene Low-quality
S14_vs_k141_12239||lt2gene Low-quality

S14_vs_k141_108069||lt2gene Low-quality
S14_vs_k141_71114||lt2gene Low-quality
S14_vs_k141_5081||lt2gene Not-determined
S14_vs_k141_12256||lt2gene Low-quality
S14_vs_k141_5084||lt2gene Low-quality
S14_vs_k141_85841||lt2gene Low-quality
S14_vs_k141_71145||lt2gene Low-quality
S14_vs_k141_26568||lt2gene Low-quality
S14_vs_k141_12269||lt2gene Not-determined
S14_vs_k141_12279||lt2gene Low-quality
S14_vs_k141_85850||lt2gene Low-quality
S14_vs_k141_26588||lt2gene Low-quality
S14_vs_k141_108077||lt2gene Low-quality
S14_vs_k141_12284||lt2gene Low-quality
S14_vs_k141_71175||lt2gene Low-quality
S14_vs_k141_26605||lt2gene Low-quality
S14_vs_k141_26606||lt2gene Low-quality
S14_vs_k141_71182||lt2gene Low-quality
S14_vs_k141_42416||lt2gene Low-quality
S14_vs_k141_71189||lt2gene Low-quality
S14_vs_k141_71191||lt2gene Low-quality
S14_vs_k141_12302||lt2gene Not-determined
S14_vs_k141_34461||lt2gene Low-quality
S14_vs_k141_63623||lt2gene Low-quality
S14_vs_k141_34475||lt2gene Low-quality
S14_vs_k141_20021||lt2gene Low-quality
S14_vs_k141_20024||lt2gene Not-determined
S14_vs_k141_12316||lt2gene Low-quality
S14_vs_k141_78630||lt2gene Low-quality
S14_vs_k141_34491||lt2gene Low-quality
S14_vs_k141_34494||lt2gene Low-quality
S14_vs_k141_93434||lt2gene Not-determined
S14_vs_k141_12331||lt2gene Low-quality
S14_vs_k141_26631||lt2gene Low-quality
S14_vs_k141_26634||lt2gene Low-quality
S14_vs_k141_26636||lt2gene Low-quality
S14_vs_k141_26642||lt2gene Low-quality
S14_vs_k141_100228||lt2gene Low-quality
S14_vs_k141_115328||lt2gene Low-quality
S14_vs_k141_26651||lt2gene Low-quality
S14_vs_k141_93450||lt2gene Low-quality
S14_vs_k141_93451||lt2gene Low-quality
S14_vs_k141_63660||lt2gene Low-quality
S14_vs_k141_93461||lt2gene Low-quality
S14_vs_k141_26658||lt2gene Not-determined
S14_vs_k141_115345||lt2gene Low-quality
S14_vs_k141_63670||lt2gene Not-determined
S14_vs_k141_115353||lt2gene Not-determined
S14_vs_k141_115356||lt2gene Low-quality
S14_vs_k141_85920||lt2gene Low-quality
S14_vs_k141_26670||lt2gene Low-quality
S14_vs_k141_115358||lt2gene Low-quality
S14_vs_k141_71213||lt2gene Low-quality
S14_vs_k141_115364||lt2gene Low-quality
S14_vs_k141_20072||lt2gene Low-quality
S14_vs_k141_85934||lt2gene Low-quality
S14_vs_k141_20076||lt2gene Low-quality
S14_vs_k141_26700||lt2gene Low-quality
S14_vs_k141_48626||lt2gene Low-quality
S14_vs_k141_42479||lt2gene Not-determined
S14_vs_k141_26708||lt2gene Low-quality
S14_vs_k141_48632||lt2gene Low-quality
S14_vs_k141_71248||lt2gene Not-determined
S14_vs_k141_93470||lt2gene Low-quality
S14_vs_k141_42495||lt2gene Low-quality
S14_vs_k141_34526||lt2gene Low-quality
S14_vs_k141_71263||lt2gene Low-quality
S14_vs_k141_63718||lt2gene Not-determined
S14_vs_k141_71265||lt2gene Low-quality
S14_vs_k141_100277||lt2gene Low-quality
S14_vs_k141_5092||lt2gene Low-quality
S14_vs_k141_34541||lt2gene Low-quality
S14_vs_k141_71271||lt2gene Low-quality
S14_vs_k141_85975||lt2gene Low-quality
S14_vs_k141_108162||lt2gene Low-quality
S14_vs_k141_108166||lt2gene Low-quality
S14_vs_k141_108167||lt2gene Low-quality
S14_vs_k141_26738||lt2gene Low-quality
S14_vs_k141_42520||lt2gene Low-quality
S14_vs_k141_63732||lt2gene Low-quality
S14_vs_k141_56549||lt2gene Low-quality
S14_vs_k141_42522||lt2gene Low-quality
S14_vs_k141_48667||lt2gene Low-quality
S14_vs_k141_100290||lt2gene Low-quality
S14_vs_k141_85981||lt2gene Low-quality
S14_vs_k141_100300||lt2gene Low-quality
S14_vs_k141_48681||lt2gene Low-quality
S14_vs_k141_115391||lt2gene Low-quality
S14_vs_k141_100304||lt2gene Low-quality
S14_vs_k141_34565||lt2gene Low-quality
S14_vs_k141_5102||lt2gene Low-quality
S14_vs_k141_5105||lt2gene Low-quality
S14_vs_k141_26765||lt2gene Low-quality
S14_vs_k141_42549||lt2gene Low-quality
S14_vs_k141_42555||lt2gene Low-quality
S14_vs_k141_42560||lt2gene Low-quality
S14_vs_k141_93524||lt2gene Low-quality
S14_vs_k141_93525||lt2gene Low-quality
S14_vs_k141_86036||lt2gene Not-determined
S14_vs_k141_108198||lt2gene Low-quality
S14_vs_k141_34617||lt2gene Low-quality
S14_vs_k141_34618||lt2gene Low-quality
S14_vs_k141_56577||lt2gene Low-quality
S14_vs_k141_5142||lt2gene Low-quality
S14_vs_k141_93545||lt2gene Not-determined
S14_vs_k141_5162||lt2gene Low-quality
S14_vs_k141_5164||lt2gene Low-quality
S14_vs_k141_20167||lt2gene Low-quality
S14_vs_k141_5179||lt2gene Low-quality
S14_vs_k141_71358||lt2gene Not-determined
S14_vs_k141_63795||lt2gene Low-quality
S14_vs_k141_100375||lt2gene Not-determined
S14_vs_k141_26797||lt2gene Not-determined
S14_vs_k141_34670||lt2gene Low-quality
S14_vs_k141_5191||lt2gene Not-determined
S14_vs_k141_5194||lt2gene Low-quality
S14_vs_k141_26807||lt2gene Not-determined
S14_vs_k141_86082||lt2gene Low-quality
S14_vs_k141_71386||lt2gene Low-quality
S14_vs_k141_56616||lt2gene Low-quality
S14_vs_k141_63821||lt2gene Low-quality
S14_vs_k141_34679||lt2gene Not-determined
S14_vs_k141_71399||lt2gene Low-quality
S14_vs_k141_26842||lt2gene Low-quality
S14_vs_k141_5233||lt2gene Not-determined
S14_vs_k141_26852||lt2gene Low-quality
S14_vs_k141_56651||lt2gene Low-quality
S14_vs_k141_12543||lt2gene Low-quality
S14_vs_k141_26861||lt2gene Not-determined
S14_vs_k141_100410||lt2gene Low-quality
S14_vs_k141_5245||lt2gene Low-quality
S14_vs_k141_56656||lt2gene Low-quality
S14_vs_k141_86113||lt2gene Low-quality
S14_vs_k141_93589||lt2gene Low-quality
S14_vs_k141_115409||lt2gene Low-quality
S14_vs_k141_48764||lt2gene Not-determined
S14_vs_k141_93591||lt2gene Not-determined
S14_vs_k141_48766||lt2gene Not-determined
S14_vs_k141_48767||lt2gene Not-determined
S14_vs_k141_5257||lt2gene Not-determined
S14_vs_k141_26877||lt2gene Low-quality
S14_vs_k141_26878||lt2gene Not-determined
S14_vs_k141_86119||lt2gene Low-quality
S14_vs_k141_12559||lt2gene Not-determined
S14_vs_k141_63840||lt2gene Low-quality
S14_vs_k141_86131||lt2gene Not-determined
S14_vs_k141_63844||lt2gene Low-quality
S14_vs_k141_86134||lt2gene Low-quality
S14_vs_k141_86135||lt2gene Low-quality
S14_vs_k141_42709||lt2gene Low-quality
S14_vs_k141_115435||lt2gene Low-quality
S14_vs_k141_26891||lt2gene Low-quality
S14_vs_k141_12578||lt2gene Low-quality
S14_vs_k141_26896||lt2gene Low-quality
S14_vs_k141_26897||lt2gene Low-quality
S14_vs_k141_42716||lt2gene Low-quality
S14_vs_k141_108276||lt2gene Low-quality
S14_vs_k141_56680||lt2gene Low-quality
S14_vs_k141_20289||lt2gene Not-determined
S14_vs_k141_20303||lt2gene Not-determined
S14_vs_k141_93674||lt2gene Not-determined
S14_vs_k141_93676||lt2gene Low-quality
S14_vs_k141_78664||lt2gene Low-quality
S14_vs_k141_42743||lt2gene Low-quality
S14_vs_k141_78667||lt2gene Low-quality
S14_vs_k141_12618||lt2gene Low-quality
S14_vs_k141_56708||lt2gene Low-quality
S14_vs_k141_12623||lt2gene Low-quality
S14_vs_k141_12625||lt2gene Low-quality
S14_vs_k141_86206||lt2gene Low-quality
S14_vs_k141_48788||lt2gene Low-quality
S14_vs_k141_12630||lt2gene Low-quality
S14_vs_k141_34798||lt2gene Not-determined
S14_vs_k141_42754||lt2gene Low-quality
S14_vs_k141_78677||lt2gene Low-quality
S14_vs_k141_108313||lt2gene Not-determined
S14_vs_k141_100500||lt2gene Low-quality
S14_vs_k141_100507||lt2gene Low-quality
S14_vs_k141_78688||lt2gene Low-quality
S14_vs_k141_86232||lt2gene Low-quality
S14_vs_k141_42776||lt2gene Low-quality
S14_vs_k141_100539||lt2gene Low-quality
S14_vs_k141_48811||lt2gene Low-quality
S14_vs_k141_42783||lt2gene Not-determined
S14_vs_k141_86245||lt2gene Low-quality
S14_vs_k141_12656||lt2gene Low-quality
S14_vs_k141_34827||lt2gene Low-quality
S14_vs_k141_12660||lt2gene Low-quality
S14_vs_k141_78717||lt2gene Low-quality
S14_vs_k141_56755||lt2gene Low-quality
S14_vs_k141_27004||lt2gene Not-determined
S14_vs_k141_20363||lt2gene Low-quality
S14_vs_k141_12675||lt2gene Low-quality
S14_vs_k141_5354||lt2gene Low-quality
S14_vs_k141_27014||lt2gene Low-quality
S14_vs_k141_5361||lt2gene Low-quality
S14_vs_k141_5373||lt2gene Low-quality
S14_vs_k141_34855||lt2gene Low-quality
S14_vs_k141_5387||lt2gene Low-quality
S14_vs_k141_5388||lt2gene Low-quality
S14_vs_k141_5389||lt2gene Low-quality
S14_vs_k141_108359||lt2gene Low-quality
S14_vs_k141_108361||lt2gene Not-determined
S14_vs_k141_71501||lt2gene Low-quality
S14_vs_k141_100582||lt2gene Low-quality
S14_vs_k141_100586||lt2gene Low-quality
S14_vs_k141_34883||lt2gene Low-quality
S14_vs_k141_63924||lt2gene Low-quality
S14_vs_k141_93759||lt2gene Low-quality
S14_vs_k141_63929||lt2gene Low-quality
S14_vs_k141_100600||lt2gene Low-quality
S14_vs_k141_42831||lt2gene Low-quality
S14_vs_k141_42833||lt2gene Not-determined
S14_vs_k141_86259||lt2gene Low-quality
S14_vs_k141_100611||lt2gene Low-quality
S14_vs_k141_86269||lt2gene Low-quality
S14_vs_k141_12711||lt2gene Low-quality
S14_vs_k141_12712||lt2gene Low-quality
S14_vs_k141_115574||lt2gene Not-determined
S14_vs_k141_12719||lt2gene Low-quality
S14_vs_k141_34913||lt2gene Low-quality
S14_vs_k141_12722||lt2gene Not-determined
S14_vs_k141_115594||lt2gene Not-determined
S14_vs_k141_115602||lt2gene Low-quality
S14_vs_k141_71555||lt2gene Low-quality
S14_vs_k141_100634||lt2gene Low-quality
S14_vs_k141_108408||lt2gene Not-determined
S14_vs_k141_12740||lt2gene Low-quality
S14_vs_k141_63976||lt2gene Low-quality
S14_vs_k141_108419||lt2gene Low-quality
S14_vs_k141_56812||lt2gene Low-quality
S14_vs_k141_108420||lt2gene Not-determined
S14_vs_k141_115636||lt2gene Low-quality
S14_vs_k141_42878||lt2gene Low-quality
S14_vs_k141_34953||lt2gene Not-determined
S14_vs_k141_71575||lt2gene Not-determined
S14_vs_k141_86301||lt2gene Not-determined
S14_vs_k141_63987||lt2gene Not-determined
S14_vs_k141_56827||lt2gene Low-quality
S14_vs_k141_86307||lt2gene Low-quality
S14_vs_k141_12762||lt2gene Low-quality
S14_vs_k141_115642||lt2gene Low-quality
S14_vs_k141_63989||lt2gene Low-quality
S14_vs_k141_86315||lt2gene Low-quality
S14_vs_k141_108434||lt2gene Low-quality
S14_vs_k141_93795||lt2gene Low-quality
S14_vs_k141_93801||lt2gene Low-quality
S14_vs_k141_12791||lt2gene Low-quality
S14_vs_k141_71591||lt2gene Low-quality
S14_vs_k141_93814||lt2gene Low-quality
S14_vs_k141_108445||lt2gene Low-quality
S14_vs_k141_71599||lt2gene Low-quality
S14_vs_k141_71601||lt2gene Low-quality
S14_vs_k141_5443||lt2gene Low-quality
S14_vs_k141_20441||lt2gene Not-determined
S14_vs_k141_20444||lt2gene Low-quality
S14_vs_k141_27154||lt2gene Not-determined
S14_vs_k141_71615||lt2gene Low-quality
S14_vs_k141_56841||lt2gene Low-quality
S14_vs_k141_56842||lt2gene Low-quality
S14_vs_k141_48968||lt2gene Not-determined
S14_vs_k141_115663||lt2gene Low-quality
S14_vs_k141_56846||lt2gene Low-quality
S14_vs_k141_12818||lt2gene Low-quality
S14_vs_k141_108470||lt2gene Low-quality
S14_vs_k141_42938||lt2gene Low-quality
S14_vs_k141_20466||lt2gene Low-quality
S14_vs_k141_93845||lt2gene Low-quality
S14_vs_k141_86363||lt2gene Low-quality
S14_vs_k141_20470||lt2gene Low-quality
S14_vs_k141_71629||lt2gene Not-determined
S14_vs_k141_48999||lt2gene Low-quality
S14_vs_k141_115698||lt2gene Not-determined
S14_vs_k141_42955||lt2gene Low-quality
S14_vs_k141_71632||lt2gene Not-determined
S14_vs_k141_100691||lt2gene Low-quality
S14_vs_k141_71638||lt2gene Low-quality
S14_vs_k141_93869||lt2gene Low-quality
S14_vs_k141_56866||lt2gene Low-quality
S14_vs_k141_12843||lt2gene Low-quality
S14_vs_k141_56873||lt2gene Low-quality
S14_vs_k141_86393||lt2gene Low-quality
S14_vs_k141_35007||lt2gene Low-quality
S14_vs_k141_35023||lt2gene Low-quality
S14_vs_k141_27170||lt2gene Low-quality
S14_vs_k141_35033||lt2gene Not-determined
S14_vs_k141_108522||lt2gene Low-quality
S14_vs_k141_35039||lt2gene Not-determined
S14_vs_k141_64071||lt2gene Low-quality
S14_vs_k141_108528||lt2gene Low-quality
S14_vs_k141_108530||lt2gene Low-quality
S14_vs_k141_108534||lt2gene Low-quality
S14_vs_k141_49037||lt2gene Low-quality
S14_vs_k141_108537||lt2gene Low-quality
S14_vs_k141_49047||lt2gene Low-quality
S14_vs_k141_5483||lt2gene Low-quality
S14_vs_k141_93918||lt2gene Low-quality
S14_vs_k141_5491||lt2gene Low-quality
S14_vs_k141_5494||lt2gene Not-determined
S14_vs_k141_35084||lt2gene Low-quality
S14_vs_k141_43030||lt2gene Low-quality
S14_vs_k141_115725||lt2gene Low-quality
S14_vs_k141_12927||lt2gene Low-quality
S14_vs_k141_49070||lt2gene Low-quality
S14_vs_k141_5513||lt2gene Low-quality
S14_vs_k141_5514||lt2gene Low-quality
S14_vs_k141_49081||lt2gene Low-quality
S14_vs_k141_35092||lt2gene Low-quality
S14_vs_k141_12945||lt2gene Low-quality
S14_vs_k141_35102||lt2gene Low-quality
S14_vs_k141_35104||lt2gene Low-quality
S14_vs_k141_100751||lt2gene Low-quality
S14_vs_k141_35110||lt2gene Low-quality
S14_vs_k141_100753||lt2gene Not-determined
S14_vs_k141_100756||lt2gene Not-determined
S14_vs_k141_27240||lt2gene Not-determined
S14_vs_k141_100766||lt2gene Low-quality
S14_vs_k141_12977||lt2gene Low-quality
S14_vs_k141_86467||lt2gene Low-quality
S14_vs_k141_27248||lt2gene Low-quality
S14_vs_k141_86471||lt2gene Low-quality
S14_vs_k141_43098||lt2gene Low-quality
S14_vs_k141_5532||lt2gene Not-determined
S14_vs_k141_100775||lt2gene Low-quality
S14_vs_k141_12987||lt2gene Low-quality
S14_vs_k141_5535||lt2gene Not-determined
S14_vs_k141_5536||lt2gene Not-determined
S14_vs_k141_100780||lt2gene Low-quality
S14_vs_k141_5539||lt2gene Not-determined
S14_vs_k141_20595||lt2gene Low-quality
S14_vs_k141_71752||lt2gene Low-quality
S14_vs_k141_71753||lt2gene Low-quality
S14_vs_k141_115739||lt2gene Low-quality
S14_vs_k141_71765||lt2gene Low-quality
S14_vs_k141_115742||lt2gene Low-quality
S14_vs_k141_56947||lt2gene Not-determined
S14_vs_k141_71777||lt2gene Not-determined
S14_vs_k141_86503||lt2gene Low-quality
S14_vs_k141_56952||lt2gene Low-quality
S14_vs_k141_20616||lt2gene Low-quality
S14_vs_k141_35128||lt2gene Low-quality
S14_vs_k141_100818||lt2gene Low-quality
S14_vs_k141_115761||lt2gene Low-quality
S14_vs_k141_71782||lt2gene Not-determined
S14_vs_k141_100831||lt2gene Not-determined
S14_vs_k141_94067||lt2gene Not-determined
S14_vs_k141_35163||lt2gene Not-determined
S14_vs_k141_115776||lt2gene Low-quality
S14_vs_k141_78930||lt2gene Not-determined
S14_vs_k141_71811||lt2gene Not-determined
S14_vs_k141_94085||lt2gene Not-determined
S14_vs_k141_27333||lt2gene Not-determined
S14_vs_k141_35172||lt2gene Low-quality
S14_vs_k141_20660||lt2gene Low-quality
S14_vs_k141_115793||lt2gene Not-determined
S14_vs_k141_43176||lt2gene Low-quality
S14_vs_k141_94122||lt2gene Not-determined
S14_vs_k141_108635||lt2gene Low-quality
S14_vs_k141_108638||lt2gene Low-quality
S14_vs_k141_5611||lt2gene Not-determined
S14_vs_k141_35206||lt2gene Not-determined
S14_vs_k141_94148||lt2gene Low-quality
S14_vs_k141_5620||lt2gene Low-quality
S14_vs_k141_94154||lt2gene Low-quality
S14_vs_k141_86555||lt2gene Low-quality
S14_vs_k141_71857||lt2gene Not-determined
S14_vs_k141_86558||lt2gene Not-determined
S14_vs_k141_86559||lt2gene Low-quality
S14_vs_k141_5626||lt2gene Not-determined
S14_vs_k141_35221||lt2gene Not-determined
S14_vs_k141_71861||lt2gene Not-determined
S14_vs_k141_13072||lt2gene Low-quality
S14_vs_k141_108683||lt2gene Not-determined
S14_vs_k141_86577||lt2gene Not-determined
S14_vs_k141_71870||lt2gene Low-quality
S14_vs_k141_20705||lt2gene Low-quality
S14_vs_k141_71874||lt2gene Low-quality
S14_vs_k141_20710||lt2gene Low-quality
S14_vs_k141_35247||lt2gene Low-quality
S14_vs_k141_100931||lt2gene Low-quality
S14_vs_k141_115855||lt2gene Low-quality
S14_vs_k141_43223||lt2gene Low-quality
S14_vs_k141_78980||lt2gene Low-quality
S14_vs_k141_35266||lt2gene Not-determined
S14_vs_k141_94167||lt2gene Low-quality
S14_vs_k141_43242||lt2gene Low-quality
S14_vs_k141_43246||lt2gene Low-quality
S14_vs_k141_94189||lt2gene Low-quality
S14_vs_k141_5664||lt2gene Low-quality
S14_vs_k141_5666||lt2gene Low-quality
S14_vs_k141_94200||lt2gene Low-quality
S14_vs_k141_86629||lt2gene Low-quality
S14_vs_k141_115880||lt2gene Low-quality
S14_vs_k141_94204||lt2gene Low-quality
S14_vs_k141_100981||lt2gene Low-quality
S14_vs_k141_79015||lt2gene Low-quality
S14_vs_k141_64191||lt2gene Not-determined
S14_vs_k141_94216||lt2gene Low-quality
S14_vs_k141_115890||lt2gene Low-quality
S14_vs_k141_71935||lt2gene Low-quality
S14_vs_k141_115894||lt2gene Low-quality
S14_vs_k141_86642||lt2gene Not-determined
S14_vs_k141_43283||lt2gene Low-quality
S14_vs_k141_86655||lt2gene Low-quality
S14_vs_k141_108755||lt2gene Low-quality
S14_vs_k141_64217||lt2gene Low-quality
S14_vs_k141_49166||lt2gene Not-determined
S14_vs_k141_43306||lt2gene Low-quality
S14_vs_k141_35306||lt2gene Low-quality
S14_vs_k141_43308||lt2gene Low-quality
S14_vs_k141_43314||lt2gene Low-quality
S14_vs_k141_5720||lt2gene Low-quality
S14_vs_k141_43316||lt2gene Low-quality
S14_vs_k141_43322||lt2gene Not-determined
S14_vs_k141_71962||lt2gene Low-quality
S14_vs_k141_64232||lt2gene Low-quality
S14_vs_k141_71964||lt2gene Low-quality
S14_vs_k141_108790||lt2gene Low-quality
S14_vs_k141_13107||lt2gene Low-quality
S14_vs_k141_71967||lt2gene Low-quality
S14_vs_k141_71970||lt2gene Low-quality
S14_vs_k141_94232||lt2gene Not-determined
S14_vs_k141_79070||lt2gene Not-determined
S14_vs_k141_108803||lt2gene Low-quality
S14_vs_k141_5750||lt2gene Not-determined
S14_vs_k141_49210||lt2gene Low-quality
S14_vs_k141_79078||lt2gene Not-determined
S14_vs_k141_20818||lt2gene Low-quality
S14_vs_k141_49212||lt2gene Low-quality
S14_vs_k141_20827||lt2gene Low-quality
S14_vs_k141_20830||lt2gene Low-quality
S14_vs_k141_64246||lt2gene Low-quality
S14_vs_k141_94266||lt2gene Low-quality
S14_vs_k141_71992||lt2gene Low-quality
S14_vs_k141_35323||lt2gene Low-quality
S14_vs_k141_13135||lt2gene Low-quality
S14_vs_k141_57080||lt2gene Not-determined
S14_vs_k141_108843||lt2gene Low-quality
S14_vs_k141_57085||lt2gene Low-quality
S14_vs_k141_27518||lt2gene Low-quality
S14_vs_k141_72009||lt2gene Not-determined
S14_vs_k141_35337||lt2gene Not-determined
S14_vs_k141_49237||lt2gene Low-quality
S14_vs_k141_86741||lt2gene Low-quality
S14_vs_k141_86743||lt2gene Low-quality
S14_vs_k141_115986||lt2gene Low-quality
S14_vs_k141_108867||lt2gene Not-determined
S14_vs_k141_108869||lt2gene Not-determined
S14_vs_k141_57110||lt2gene Low-quality
S14_vs_k141_86746||lt2gene Low-quality
S14_vs_k141_5793||lt2gene Low-quality
S14_vs_k141_27547||lt2gene Low-quality
S14_vs_k141_13164||lt2gene Low-quality
S14_vs_k141_43341||lt2gene Low-quality
S14_vs_k141_43344||lt2gene Not-determined
S14_vs_k141_43349||lt2gene Not-determined
S14_vs_k141_79097||lt2gene Low-quality
S14_vs_k141_27571||lt2gene Not-determined
S14_vs_k141_5816||lt2gene Low-quality
S14_vs_k141_43355||lt2gene Not-determined
S14_vs_k141_27577||lt2gene Not-determined
S14_vs_k141_43358||lt2gene Not-determined
S14_vs_k141_5821||lt2gene Low-quality
S14_vs_k141_94316||lt2gene Low-quality
S14_vs_k141_43361||lt2gene Not-determined
S14_vs_k141_94317||lt2gene Not-determined
S14_vs_k141_43362||lt2gene Not-determined
S14_vs_k141_101081||lt2gene Low-quality
S14_vs_k141_108906||lt2gene Low-quality
S14_vs_k141_116025||lt2gene Low-quality
S14_vs_k141_20922||lt2gene Not-determined
S14_vs_k141_20923||lt2gene Low-quality
S14_vs_k141_108918||lt2gene Not-determined
S14_vs_k141_43388||lt2gene Low-quality
S14_vs_k141_79125||lt2gene Low-quality
S14_vs_k141_13198||lt2gene Low-quality
S14_vs_k141_116034||lt2gene Low-quality
S14_vs_k141_116036||lt2gene Low-quality
S14_vs_k141_108927||lt2gene Low-quality
S14_vs_k141_35352||lt2gene Low-quality
S14_vs_k141_5845||lt2gene Low-quality
S14_vs_k141_57155||lt2gene Low-quality
S14_vs_k141_35357||lt2gene Low-quality
S14_vs_k141_86823||lt2gene Not-determined
S14_vs_k141_35363||lt2gene Not-determined
S14_vs_k141_108940||lt2gene Low-quality
S14_vs_k141_64327||lt2gene Low-quality
S14_vs_k141_57170||lt2gene Low-quality
S14_vs_k141_108951||lt2gene Not-determined
S14_vs_k141_101116||lt2gene Low-quality
S14_vs_k141_35393||lt2gene Not-determined
S14_vs_k141_35394||lt2gene Not-determined
S14_vs_k141_5864||lt2gene Not-determined
S14_vs_k141_5865||lt2gene Not-determined
S14_vs_k141_5870||lt2gene Not-determined
S14_vs_k141_108980||lt2gene Not-determined
S14_vs_k141_35420||lt2gene Not-determined
S14_vs_k141_5876||lt2gene Not-determined
S14_vs_k141_116056||lt2gene Low-quality
S14_vs_k141_5885||lt2gene Low-quality
S14_vs_k141_94370||lt2gene Not-determined
S14_vs_k141_49291||lt2gene Not-determined
S14_vs_k141_101156||lt2gene Not-determined
S14_vs_k141_86869||lt2gene Low-quality
S14_vs_k141_57225||lt2gene Low-quality
S14_vs_k141_57235||lt2gene Low-quality
S14_vs_k141_27660||lt2gene Low-quality
S14_vs_k141_13233||lt2gene Low-quality
S14_vs_k141_116073||lt2gene Low-quality
S14_vs_k141_35459||lt2gene Low-quality
S14_vs_k141_49310||lt2gene Not-determined
S14_vs_k141_43476||lt2gene Not-determined
S14_vs_k141_49323||lt2gene Low-quality
S14_vs_k141_57256||lt2gene Low-quality
S14_vs_k141_49332||lt2gene Not-determined
S14_vs_k141_13273||lt2gene Low-quality
S14_vs_k141_86915||lt2gene Low-quality
S14_vs_k141_86916||lt2gene Low-quality
S14_vs_k141_5927||lt2gene Low-quality
S14_vs_k141_5931||lt2gene Low-quality
S14_vs_k141_49356||lt2gene Low-quality
S14_vs_k141_57276||lt2gene Not-determined
S14_vs_k141_86932||lt2gene Not-determined
S14_vs_k141_86935||lt2gene Low-quality
S14_vs_k141_27683||lt2gene Not-determined
S14_vs_k141_79240||lt2gene Low-quality
S14_vs_k141_116121||lt2gene Low-quality
S14_vs_k141_5945||lt2gene Not-determined
S14_vs_k141_86945||lt2gene Not-determined
S14_vs_k141_94417||lt2gene Low-quality
S14_vs_k141_109007||lt2gene Not-determined
S14_vs_k141_21013||lt2gene Low-quality
S14_vs_k141_86964||lt2gene Low-quality
S14_vs_k141_101194||lt2gene Not-determined
S14_vs_k141_86973||lt2gene Low-quality
S14_vs_k141_79261||lt2gene Low-quality
S14_vs_k141_94438||lt2gene Low-quality
S14_vs_k141_64356||lt2gene Low-quality
S14_vs_k141_49379||lt2gene Low-quality
S14_vs_k141_64359||lt2gene Low-quality
S14_vs_k141_79271||lt2gene Low-quality
S14_vs_k141_64362||lt2gene Low-quality
S14_vs_k141_27703||lt2gene Low-quality
S14_vs_k141_27722||lt2gene Not-determined
S14_vs_k141_79293||lt2gene Low-quality
S14_vs_k141_27729||lt2gene Not-determined
S14_vs_k141_101237||lt2gene Low-quality
S14_vs_k141_109046||lt2gene Low-quality
S14_vs_k141_49428||lt2gene Low-quality
S14_vs_k141_6007||lt2gene Low-quality
S14_vs_k141_21036||lt2gene Low-quality
S14_vs_k141_49431||lt2gene Low-quality
S14_vs_k141_6014||lt2gene Low-quality
S14_vs_k141_27745||lt2gene Low-quality
S14_vs_k141_49450||lt2gene Low-quality
S14_vs_k141_13391||lt2gene Low-quality
S14_vs_k141_94498||lt2gene Low-quality
S14_vs_k141_21064||lt2gene Low-quality
S14_vs_k141_109078||lt2gene Low-quality
S14_vs_k141_49455||lt2gene Low-quality
S14_vs_k141_109085||lt2gene Low-quality
S14_vs_k141_87042||lt2gene Low-quality
S14_vs_k141_109095||lt2gene Not-determined
S14_vs_k141_49469||lt2gene Low-quality
S14_vs_k141_49471||lt2gene Low-quality
S14_vs_k141_21086||lt2gene Low-quality
S14_vs_k141_116196||lt2gene Not-determined
S14_vs_k141_49488||lt2gene Not-determined
S14_vs_k141_49495||lt2gene Low-quality
S14_vs_k141_72092||lt2gene Not-determined
S14_vs_k141_72095||lt2gene Low-quality
S14_vs_k141_57403||lt2gene Low-quality
S14_vs_k141_116221||lt2gene Not-determined
S14_vs_k141_79367||lt2gene Low-quality
S14_vs_k141_87082||lt2gene Low-quality
S14_vs_k141_87089||lt2gene Low-quality
S14_vs_k141_94539||lt2gene Not-determined
S14_vs_k141_116240||lt2gene Not-determined
S14_vs_k141_79384||lt2gene Low-quality
S14_vs_k141_94546||lt2gene Low-quality
S14_vs_k141_79394||lt2gene Not-determined
S14_vs_k141_79396||lt2gene Not-determined
S14_vs_k141_49528||lt2gene Low-quality
S14_vs_k141_35623||lt2gene Low-quality
S14_vs_k141_49535||lt2gene Low-quality
S14_vs_k141_57411||lt2gene Low-quality
S14_vs_k141_116261||lt2gene Low-quality
S14_vs_k141_6081||lt2gene Low-quality
S14_vs_k141_49550||lt2gene Low-quality

| | |
|---|---|
| S14_vs_k141_43579||lt2gene | Not-determined |
| S14_vs_k141_27843||lt2gene | Low-quality |
| S14_vs_k141_79434||lt2gene | Low-quality |
| S14_vs_k141_94580||lt2gene | Low-quality |
| S14_vs_k141_79436||lt2gene | Low-quality |
| S14_vs_k141_101342||lt2gene | Low-quality |
| S14_vs_k141_27848||lt2gene | Low-quality |
| S14_vs_k141_6100||lt2gene | Low-quality |
| S14_vs_k141_87140||lt2gene | Low-quality |
| S14_vs_k141_87141||lt2gene | Low-quality |
| S14_vs_k141_109170||lt2gene | Low-quality |
| S14_vs_k141_43598||lt2gene | Low-quality |
| S14_vs_k141_57436||lt2gene | Low-quality |
| S14_vs_k141_6117||lt2gene | Not-determined |
| S14_vs_k141_87148||lt2gene | Low-quality |
| S14_vs_k141_116303||lt2gene | Not-determined |
| S14_vs_k141_116305||lt2gene | Not-determined |
| S14_vs_k141_49612||lt2gene | Low-quality |
| S14_vs_k141_109183||lt2gene | Low-quality |
| S14_vs_k141_6138||lt2gene | Not-determined |
| S14_vs_k141_79478||lt2gene | Not-determined |
| S14_vs_k141_109189||lt2gene | Not-determined |
| S14_vs_k141_94611||lt2gene | Low-quality |
| S14_vs_k141_109197||lt2gene | Not-determined |
| S14_vs_k141_43642||lt2gene | Low-quality |
| S14_vs_k141_79499||lt2gene | Not-determined |
| S14_vs_k141_79502||lt2gene | Low-quality |
| S14_vs_k141_6164||lt2gene | Not-determined |
| S14_vs_k141_27899||lt2gene | Not-determined |
| S14_vs_k141_6167||lt2gene | Low-quality |
| S14_vs_k141_43649||lt2gene | Not-determined |
| S14_vs_k141_6172||lt2gene | Low-quality |
| S14_vs_k141_43652||lt2gene | Low-quality |
| S14_vs_k141_72181||lt2gene | Low-quality |
| S14_vs_k141_43658||lt2gene | Low-quality |
| S14_vs_k141_109209||lt2gene | Low-quality |
| S14_vs_k141_21179||lt2gene | Low-quality |
| S14_vs_k141_13405||lt2gene | Low-quality |
| S14_vs_k141_35723||lt2gene | Low-quality |
| S14_vs_k141_109220||lt2gene | Low-quality |
| S14_vs_k141_35729||lt2gene | Low-quality |
| S14_vs_k141_109225||lt2gene | Low-quality |
| S14_vs_k141_49668||lt2gene | Low-quality |
| S14_vs_k141_27916||lt2gene | Low-quality |
| S14_vs_k141_94658||lt2gene | Low-quality |
| S14_vs_k141_109238||lt2gene | Low-quality |
| S14_vs_k141_35739||lt2gene | Low-quality |
| S14_vs_k141_35747||lt2gene | Not-determined |
| S14_vs_k141_6193||lt2gene | Low-quality |
| S14_vs_k141_43692||lt2gene | Low-quality |
| S14_vs_k141_35769||lt2gene | Low-quality |
| S14_vs_k141_101421||lt2gene | Low-quality |
| S14_vs_k141_27935||lt2gene | Low-quality |
| S14_vs_k141_6201||lt2gene | Low-quality |
| S14_vs_k141_6203||lt2gene | Low-quality |
| S14_vs_k141_27937||lt2gene | Low-quality |
| S14_vs_k141_94689||lt2gene | Low-quality |
| S14_vs_k141_13456||lt2gene | Low-quality |
| S14_vs_k141_109272||lt2gene | Low-quality |
| S14_vs_k141_43717||lt2gene | Low-quality |
| S14_vs_k141_72228||lt2gene | Low-quality |
| S14_vs_k141_57516||lt2gene | Low-quality |
| S14_vs_k141_13472||lt2gene | Low-quality |
| S14_vs_k141_35787||lt2gene | Low-quality |
| S14_vs_k141_13474||lt2gene | Low-quality |
| S14_vs_k141_109292||lt2gene | Low-quality |
| S14_vs_k141_57526||lt2gene | Low-quality |
| S14_vs_k141_116430||lt2gene | Low-quality |
| S14_vs_k141_64511||lt2gene | Low-quality |
| S14_vs_k141_6255||lt2gene | Not-determined |
| S14_vs_k141_116435||lt2gene | Low-quality |
| S14_vs_k141_21238||lt2gene | Not-determined |
| S14_vs_k141_49736||lt2gene | Low-quality |
| S14_vs_k141_13485||lt2gene | Low-quality |
| S14_vs_k141_13489||lt2gene | Low-quality |
| S14_vs_k141_43756||lt2gene | Low-quality |
| S14_vs_k141_64536||lt2gene | Not-determined |
| S14_vs_k141_94732||lt2gene | Low-quality |
| S14_vs_k141_79527||lt2gene | Not-determined |
| S14_vs_k141_57563||lt2gene | Not-determined |
| S14_vs_k141_6285||lt2gene | Not-determined |
| S14_vs_k141_35823||lt2gene | Not-determined |
| S14_vs_k141_27980||lt2gene | Not-determined |
| S14_vs_k141_43792||lt2gene | Low-quality |
| S14_vs_k141_49760||lt2gene | Low-quality |
| S14_vs_k141_64560||lt2gene | Low-quality |
| S14_vs_k141_6294||lt2gene | Low-quality |
| S14_vs_k141_57606||lt2gene | Low-quality |
| S14_vs_k141_116458||lt2gene | Low-quality |
| S14_vs_k141_109324||lt2gene | Low-quality |
| S14_vs_k141_87300||lt2gene | Low-quality |
| S14_vs_k141_116469||lt2gene | Low-quality |
| S14_vs_k141_57615||lt2gene | Low-quality |
| S14_vs_k141_79557||lt2gene | Low-quality |
| S14_vs_k141_64569||lt2gene | Not-determined |
| S14_vs_k141_64570||lt2gene | Low-quality |
| S14_vs_k141_13538||lt2gene | Low-quality |
| S14_vs_k141_43829||lt2gene | Low-quality |
| S14_vs_k141_94778||lt2gene | Low-quality |
| S14_vs_k141_64586||lt2gene | Low-quality |
| S14_vs_k141_57631||lt2gene | Low-quality |
| S14_vs_k141_13548||lt2gene | Low-quality |
| S14_vs_k141_64597||lt2gene | Not-determined |
| S14_vs_k141_21300||lt2gene | Not-determined |
| S14_vs_k141_49810||lt2gene | Low-quality |
| S14_vs_k141_49812||lt2gene | Low-quality |
| S14_vs_k141_87327||lt2gene | Low-quality |
| S14_vs_k141_79587||lt2gene | Not-determined |
| S14_vs_k141_87329||lt2gene | Not-determined |
| S14_vs_k141_87332||lt2gene | Not-determined |
| S14_vs_k141_43842||lt2gene | Not-determined |
| S14_vs_k141_109332||lt2gene | Low-quality |
| S14_vs_k141_116501||lt2gene | Not-determined |
| S14_vs_k141_43852||lt2gene | Low-quality |
| S14_vs_k141_116503||lt2gene | Low-quality |
| S14_vs_k141_109338||lt2gene | Low-quality |
| S14_vs_k141_64614||lt2gene | Low-quality |
| S14_vs_k141_64615||lt2gene | Low-quality |
| S14_vs_k141_116514||lt2gene | Low-quality |
| S14_vs_k141_64619||lt2gene | Low-quality |
| S14_vs_k141_109346||lt2gene | Not-determined |
| S14_vs_k141_101501||lt2gene | Not-determined |
| S14_vs_k141_72273||lt2gene | Low-quality |
| S14_vs_k141_13572||lt2gene | Low-quality |
| S14_vs_k141_35870||lt2gene | Not-determined |
| S14_vs_k141_13585||lt2gene | Not-determined |
| S14_vs_k141_79634||lt2gene | Low-quality |
| S14_vs_k141_101523||lt2gene | Low-quality |
| S14_vs_k141_49851||lt2gene | Low-quality |
| S14_vs_k141_72306||lt2gene | Low-quality |
| S14_vs_k141_35899||lt2gene | Not-determined |
| S14_vs_k141_116539||lt2gene | Not-determined |
| S14_vs_k141_116542||lt2gene | Not-determined |
| S14_vs_k141_116545||lt2gene | Not-determined |
| S14_vs_k141_28039||lt2gene | Low-quality |
| S14_vs_k141_87385||lt2gene | Not-determined |
| S14_vs_k141_43889||lt2gene | Low-quality |
| S14_vs_k141_116560||lt2gene | Not-determined |
| S14_vs_k141_116565||lt2gene | Low-quality |
| S14_vs_k141_21377||lt2gene | Low-quality |
| S14_vs_k141_79664||lt2gene | Low-quality |
| S14_vs_k141_13599||lt2gene | Not-determined |
| S14_vs_k141_72322||lt2gene | Low-quality |
| S14_vs_k141_87403||lt2gene | Low-quality |
| S14_vs_k141_21392||lt2gene | Low-quality |
| S14_vs_k141_87411||lt2gene | Low-quality |
| S14_vs_k141_21399||lt2gene | Low-quality |
| S14_vs_k141_87427||lt2gene | Low-quality |
| S14_vs_k141_101545||lt2gene | Not-determined |
| S14_vs_k141_43915||lt2gene | Low-quality |
| S14_vs_k141_101547||lt2gene | Not-determined |
| S14_vs_k141_43919||lt2gene | Low-quality |
| S14_vs_k141_101553||lt2gene | Low-quality |
| S14_vs_k141_101558||lt2gene | Low-quality |
| S14_vs_k141_101561||lt2gene | Not-determined |
| S14_vs_k141_87444||lt2gene | Low-quality |
| S14_vs_k141_6437||lt2gene | Low-quality |
| S14_vs_k141_79691||lt2gene | Low-quality |
| S14_vs_k141_79699||lt2gene | Low-quality |
| S14_vs_k141_64674||lt2gene | Low-quality |
| S14_vs_k141_13631||lt2gene | Low-quality |
| S14_vs_k141_35933||lt2gene | Low-quality |
| S14_vs_k141_94882||lt2gene | Low-quality |
| S14_vs_k141_94883||lt2gene | Low-quality |
| S14_vs_k141_21437||lt2gene | Low-quality |
| S14_vs_k141_6462||lt2gene | Low-quality |
| S14_vs_k141_21438||lt2gene | Low-quality |
| S14_vs_k141_21441||lt2gene | Low-quality |
| S14_vs_k141_6467||lt2gene | Low-quality |
| S14_vs_k141_116614||lt2gene | Low-quality |
| S14_vs_k141_13644||lt2gene | Not-determined |
| S14_vs_k141_6485||lt2gene | Not-determined |
| S14_vs_k141_13660||lt2gene | Low-quality |
| S14_vs_k141_79754||lt2gene | Low-quality |
| S14_vs_k141_35965||lt2gene | Low-quality |
| S14_vs_k141_6493||lt2gene | Not-determined |
| S14_vs_k141_87473||lt2gene | Low-quality |
| S14_vs_k141_35971||lt2gene | Not-determined |
| S14_vs_k141_35973||lt2gene | Not-determined |
| S14_vs_k141_79769||lt2gene | Low-quality |
| S14_vs_k141_49889||lt2gene | Low-quality |
| S14_vs_k141_57755||lt2gene | Low-quality |
| S14_vs_k141_57759||lt2gene | Not-determined |
| S14_vs_k141_79795||lt2gene | Low-quality |
| S14_vs_k141_57764||lt2gene | Not-determined |
| S14_vs_k141_57765||lt2gene | Not-determined |
| S14_vs_k141_21506||lt2gene | Low-quality |
| S14_vs_k141_13702||lt2gene | Low-quality |
| S14_vs_k141_109517||lt2gene | Low-quality |
| S14_vs_k141_79806||lt2gene | Low-quality |
| S14_vs_k141_21522||lt2gene | Low-quality |
| S14_vs_k141_57778||lt2gene | Not-determined |
| S14_vs_k141_79812||lt2gene | Low-quality |
| S14_vs_k141_57782||lt2gene | Not-determined |
| S14_vs_k141_109526||lt2gene | Low-quality |
| S14_vs_k141_79822||lt2gene | Not-determined |
| S14_vs_k141_49914||lt2gene | Not-determined |
| S14_vs_k141_6529||lt2gene | Low-quality |
| S14_vs_k141_36000||lt2gene | Not-determined |
| S14_vs_k141_109535||lt2gene | Low-quality |
| S14_vs_k141_57798||lt2gene | Low-quality |
| S14_vs_k141_109541||lt2gene | Low-quality |
| S14_vs_k141_79833||lt2gene | Low-quality |
| S14_vs_k141_94959||lt2gene | Low-quality |
| S14_vs_k141_21554||lt2gene | Low-quality |
| S14_vs_k141_101644||lt2gene | Not-determined |
| S14_vs_k141_49925||lt2gene | Low-quality |
| S14_vs_k141_94977||lt2gene | Low-quality |
| S14_vs_k141_101662||lt2gene | Low-quality |
| S14_vs_k141_28167||lt2gene | Low-quality |
| S14_vs_k141_57829||lt2gene | Low-quality |
| S14_vs_k141_6536||lt2gene | Not-determined |
| S14_vs_k141_94987||lt2gene | Low-quality |
| S14_vs_k141_49936||lt2gene | Not-determined |
| S14_vs_k141_21579||lt2gene | Low-quality |
| S14_vs_k141_49941||lt2gene | Low-quality |
| S14_vs_k141_49943||lt2gene | Low-quality |
| S14_vs_k141_94997||lt2gene | Low-quality |
| S14_vs_k141_49947||lt2gene | Low-quality |
| S14_vs_k141_28196||lt2gene | Low-quality |
| S14_vs_k141_95001||lt2gene | Not-determined |
| S14_vs_k141_36055||lt2gene | Low-quality |
| S14_vs_k141_64733||lt2gene | Low-quality |
| S14_vs_k141_13779||lt2gene | Low-quality |
| S14_vs_k141_49972||lt2gene | Low-quality |
| S14_vs_k141_116707||lt2gene | Low-quality |
| S14_vs_k141_36080||lt2gene | Low-quality |
| S14_vs_k141_57874||lt2gene | Low-quality |
| S14_vs_k141_6542||lt2gene | Low-quality |
| S14_vs_k141_6550||lt2gene | Low-quality |
| S14_vs_k141_109587||lt2gene | Low-quality |
| S14_vs_k141_36098||lt2gene | Low-quality |
| S14_vs_k141_36101||lt2gene | Low-quality |
| S14_vs_k141_95039||lt2gene | Low-quality |
| S14_vs_k141_36108||lt2gene | Low-quality |
| S14_vs_k141_6572||lt2gene | Low-quality |
| S14_vs_k141_64798||lt2gene | Not-determined |
| S14_vs_k141_64802||lt2gene | Not-determined |
| S14_vs_k141_79933||lt2gene | Low-quality |
| S14_vs_k141_72340||lt2gene | Not-determined |
| S14_vs_k141_64809||lt2gene | Low-quality |
| S14_vs_k141_13803||lt2gene | Low-quality |
| S14_vs_k141_72343||lt2gene | Low-quality |
| S14_vs_k141_64818||lt2gene | Low-quality |
| S14_vs_k141_57912||lt2gene | Low-quality |
| S14_vs_k141_36129||lt2gene | Not-determined |
| S14_vs_k141_13813||lt2gene | Low-quality |
| S14_vs_k141_64827||lt2gene | Low-quality |
| S14_vs_k141_13817||lt2gene | Low-quality |
| S14_vs_k141_13821||lt2gene | Low-quality |
| S14_vs_k141_64833||lt2gene | Not-determined |
| S14_vs_k141_57921||lt2gene | Low-quality |
| S14_vs_k141_36135||lt2gene | Low-quality |
| S14_vs_k141_101758||lt2gene | Low-quality |
| S14_vs_k141_50022||lt2gene | Not-determined |
| S14_vs_k141_50024||lt2gene | Not-determined |
| S14_vs_k141_101778||lt2gene | Not-determined |
| S14_vs_k141_64862||lt2gene | Low-quality |
| S14_vs_k141_109667||lt2gene | Not-determined |
| S14_vs_k141_109670||lt2gene | Not-determined |
| S14_vs_k141_28279||lt2gene | Low-quality |
| S14_vs_k141_57941||lt2gene | Low-quality |
| S14_vs_k141_109679||lt2gene | Not-determined |
| S14_vs_k141_57947||lt2gene | Low-quality |
| S14_vs_k141_64880||lt2gene | Low-quality |
| S14_vs_k141_28292||lt2gene | Low-quality |
| S14_vs_k141_72402||lt2gene | Low-quality |
| S14_vs_k141_101786||lt2gene | Low-quality |
| S14_vs_k141_6642||lt2gene | Low-quality |
| S14_vs_k141_109692||lt2gene | Low-quality |
| S14_vs_k141_28298||lt2gene | Low-quality |
| S14_vs_k141_28302||lt2gene | Not-determined |
| S14_vs_k141_28305||lt2gene | Not-determined |
| S14_vs_k141_109711||lt2gene | Low-quality |
| S14_vs_k141_64908||lt2gene | Low-quality |
| S14_vs_k141_64910||lt2gene | Low-quality |
| S14_vs_k141_109715||lt2gene | Not-determined |
| S14_vs_k141_28318||lt2gene | Low-quality |
| S14_vs_k141_95097||lt2gene | Not-determined |
| S14_vs_k141_6663||lt2gene | Low-quality |
| S14_vs_k141_116807||lt2gene | Low-quality |
| S14_vs_k141_36175||lt2gene | Low-quality |
| S14_vs_k141_36183||lt2gene | Not-determined |
| S14_vs_k141_80001||lt2gene | Low-quality |
| S14_vs_k141_80002||lt2gene | Not-determined |
| S14_vs_k141_95120||lt2gene | Low-quality |
| S14_vs_k141_95126||lt2gene | Low-quality |
| S14_vs_k141_57979||lt2gene | Low-quality |
| S14_vs_k141_50089||lt2gene | Not-determined |
| S14_vs_k141_13882||lt2gene | Low-quality |
| S14_vs_k141_50093||lt2gene | Low-quality |
| S14_vs_k141_13887||lt2gene | Low-quality |
| S14_vs_k141_80025||lt2gene | Low-quality |
| S14_vs_k141_72474||lt2gene | Not-determined |
| S14_vs_k141_101836||lt2gene | Low-quality |
| S14_vs_k141_109756||lt2gene | Low-quality |
| S14_vs_k141_6695||lt2gene | Low-quality |
| S14_vs_k141_6701||lt2gene | Low-quality |
| S14_vs_k141_13892||lt2gene | Low-quality |
| S14_vs_k141_6707||lt2gene | Not-determined |
| S14_vs_k141_87490||lt2gene | Low-quality |
| S14_vs_k141_21709||lt2gene | Not-determined |
| S14_vs_k141_21715||lt2gene | Low-quality |
| S14_vs_k141_36224||lt2gene | Low-quality |
| S14_vs_k141_87507||lt2gene | Low-quality |
| S14_vs_k141_21717||lt2gene | Not-determined |
| S14_vs_k141_50111||lt2gene | Low-quality |
| S14_vs_k141_80062||lt2gene | Low-quality |
| S14_vs_k141_58032||lt2gene | Low-quality |
| S14_vs_k141_36244||lt2gene | Not-determined |
| S14_vs_k141_95167||lt2gene | Low-quality |
| S14_vs_k141_64984||lt2gene | Not-determined |
| S14_vs_k141_36252||lt2gene | Not-determined |
| S14_vs_k141_87536||lt2gene | Low-quality |
| S14_vs_k141_87538||lt2gene | Low-quality |
| S14_vs_k141_28367||lt2gene | Low-quality |
| S14_vs_k141_72529||lt2gene | Low-quality |
| S14_vs_k141_95192||lt2gene | Not-determined |
| S14_vs_k141_95193||lt2gene | Not-determined |
| S14_vs_k141_36297||lt2gene | Low-quality |
| S14_vs_k141_50175||lt2gene | Low-quality |
| S14_vs_k141_13926||lt2gene | Low-quality |
| S14_vs_k141_87582||lt2gene | Not-determined |
| S14_vs_k141_50185||lt2gene | Low-quality |
| S14_vs_k141_109859||lt2gene | Low-quality |
| S14_vs_k141_21780||lt2gene | Low-quality |
| S14_vs_k141_21781||lt2gene | Not-determined |
| S14_vs_k141_116890||lt2gene | Not-determined |
| S14_vs_k141_65027||lt2gene | Low-quality |
| S14_vs_k141_65029||lt2gene | Not-determined |
| S14_vs_k141_36310||lt2gene | Low-quality |
| S14_vs_k141_116903||lt2gene | Low-quality |
| S14_vs_k141_36312||lt2gene | Not-determined |
| S14_vs_k141_36314||lt2gene | Not-determined |
| S14_vs_k141_109889||lt2gene | Low-quality |
| S14_vs_k141_6783||lt2gene | Low-quality |
| S14_vs_k141_87616||lt2gene | Low-quality |
| S14_vs_k141_87619||lt2gene | Low-quality |
| S14_vs_k141_65048||lt2gene | Low-quality |
| S14_vs_k141_116926||lt2gene | Low-quality |
| S14_vs_k141_65050||lt2gene | Low-quality |
| S14_vs_k141_65053||lt2gene | Not-determined |
| S14_vs_k141_72578||lt2gene | Low-quality |
| S14_vs_k141_65060||lt2gene | Not-determined |
| S14_vs_k141_13970||lt2gene | Low-quality |
| S14_vs_k141_80124||lt2gene | Low-quality |
| S14_vs_k141_21810||lt2gene | Low-quality |
| S14_vs_k141_58116||lt2gene | Not-determined |
| S14_vs_k141_21812||lt2gene | Low-quality |
| S14_vs_k141_65069||lt2gene | Low-quality |
| S14_vs_k141_116954||lt2gene | Not-determined |
| S14_vs_k141_36327||lt2gene | Not-determined |
| S14_vs_k141_58136||lt2gene | Low-quality |
| S14_vs_k141_58139||lt2gene | Low-quality |
| S14_vs_k141_21839||lt2gene | Low-quality |
| S14_vs_k141_58153||lt2gene | Low-quality |
| S14_vs_k141_65077||lt2gene | Low-quality |
| S14_vs_k141_14002||lt2gene | Not-determined |
| S14_vs_k141_14004||lt2gene | Low-quality |
| S14_vs_k141_21850||lt2gene | Low-quality |
| S14_vs_k141_80169||lt2gene | Low-quality |
| S14_vs_k141_65092||lt2gene | Low-quality |
| S14_vs_k141_72614||lt2gene | Low-quality |
| S14_vs_k141_65107||lt2gene | Low-quality |
| S14_vs_k141_36347||lt2gene | Low-quality |
| S14_vs_k141_50265||lt2gene | Not-determined |
| S14_vs_k141_50266||lt2gene | Not-determined |
| S14_vs_k141_87664||lt2gene | Low-quality |
| S14_vs_k141_116994||lt2gene | Low-quality |
| S14_vs_k141_101931||lt2gene | Not-determined |
| S14_vs_k141_116995||lt2gene | Low-quality |
| S14_vs_k141_87671||lt2gene | Low-quality |
| S14_vs_k141_21881||lt2gene | Low-quality |
| S14_vs_k141_36373||lt2gene | Not-determined |
| S14_vs_k141_50279||lt2gene | Low-quality |
| S14_vs_k141_117015||lt2gene | Low-quality |
| S14_vs_k141_36386||lt2gene | Low-quality |
| S14_vs_k141_87700||lt2gene | Low-quality |
| S14_vs_k141_14063||lt2gene | Low-quality |
| S14_vs_k141_14068||lt2gene | Low-quality |
| S14_vs_k141_14071||lt2gene | Low-quality |
| S14_vs_k141_21920||lt2gene | Low-quality |
| S14_vs_k141_21924||lt2gene | Low-quality |
| S14_vs_k141_14093||lt2gene | Low-quality |
| S14_vs_k141_21927||lt2gene | Low-quality |
| S14_vs_k141_14097||lt2gene | Low-quality |
| S14_vs_k141_21928||lt2gene | Not-determined |
| S14_vs_k141_14100||lt2gene | Low-quality |
| S14_vs_k141_14103||lt2gene | Not-determined |
| S14_vs_k141_50316||lt2gene | Low-quality |
| S14_vs_k141_36426||lt2gene | Low-quality |
| S14_vs_k141_6895||lt2gene | Low-quality |
| S14_vs_k141_87724||lt2gene | Low-quality |
| S14_vs_k141_21941||lt2gene | Low-quality |
| S14_vs_k141_50325||lt2gene | Low-quality |
| S14_vs_k141_50329||lt2gene | Low-quality |
| S14_vs_k141_36447||lt2gene | Low-quality |
| S14_vs_k141_80222||lt2gene | Low-quality |
| S14_vs_k141_65148||lt2gene | Low-quality |
| S14_vs_k141_80227||lt2gene | Low-quality |
| S14_vs_k141_36460||lt2gene | Low-quality |
| S14_vs_k141_14117||lt2gene | Low-quality |
| S14_vs_k141_72743||lt2gene | Low-quality |
| S14_vs_k141_50357||lt2gene | Low-quality |
| S14_vs_k141_72746||lt2gene | Low-quality |
| S14_vs_k141_65158||lt2gene | Low-quality |
| S14_vs_k141_72754||lt2gene | Not-determined |
| S14_vs_k141_6913||lt2gene | Low-quality |
| S14_vs_k141_21961||lt2gene | Low-quality |
| S14_vs_k141_65170||lt2gene | Low-quality |
| S14_vs_k141_102025||lt2gene | Low-quality |
| S14_vs_k141_117095||lt2gene | Low-quality |
| S14_vs_k141_102029||lt2gene | Low-quality |
| S14_vs_k141_117104||lt2gene | Low-quality |
| S14_vs_k141_58309||lt2gene | Low-quality |
| S14_vs_k141_50380||lt2gene | Low-quality |
| S14_vs_k141_72810||lt2gene | Not-determined |
| S14_vs_k141_6963||lt2gene | Low-quality |
| S14_vs_k141_58319||lt2gene | Low-quality |
| S14_vs_k141_28472||lt2gene | Low-quality |
| S14_vs_k141_36523||lt2gene | Low-quality |
| S14_vs_k141_50392||lt2gene | Low-quality |
| S14_vs_k141_117137||lt2gene | Low-quality |
| S14_vs_k141_102058||lt2gene | Low-quality |
| S14_vs_k141_50398||lt2gene | Low-quality |
| S14_vs_k141_80279||lt2gene | Low-quality |
| S14_vs_k141_14173||lt2gene | Low-quality |
| S14_vs_k141_36536||lt2gene | Low-quality |
| S14_vs_k141_102076||lt2gene | Low-quality |
| S14_vs_k141_102077||lt2gene | Low-quality |
| S14_vs_k141_102081||lt2gene | Low-quality |
| S14_vs_k141_87794||lt2gene | Low-quality |
| S14_vs_k141_6982||lt2gene | Low-quality |
| S14_vs_k141_6985||lt2gene | Low-quality |
| S14_vs_k141_58348||lt2gene | Low-quality |
| S14_vs_k141_102105||lt2gene | Low-quality |
| S14_vs_k141_50434||lt2gene | Not-determined |
| S14_vs_k141_50435||lt2gene | Not-determined |
| S14_vs_k141_58354||lt2gene | Low-quality |
| S14_vs_k141_14196||lt2gene | Low-quality |
| S14_vs_k141_65225||lt2gene | Low-quality |
| S14_vs_k141_7011||lt2gene | Low-quality |
| S14_vs_k141_72839||lt2gene | Low-quality |
| S14_vs_k141_14210||lt2gene | Low-quality |
| S14_vs_k141_14214||lt2gene | Low-quality |
| S14_vs_k141_36582||lt2gene | Low-quality |
| S14_vs_k141_50466||lt2gene | Not-determined |
| S14_vs_k141_58377||lt2gene | Low-quality |
| S14_vs_k141_72864||lt2gene | Low-quality |
| S14_vs_k141_87833||lt2gene | Low-quality |
| S14_vs_k141_50486||lt2gene | Not-determined |
| S14_vs_k141_65264||lt2gene | Low-quality |
| S14_vs_k141_50491||lt2gene | Low-quality |
| S14_vs_k141_50501||lt2gene | Not-determined |
| S14_vs_k141_65276||lt2gene | Not-determined |
| S14_vs_k141_102121||lt2gene | Not-determined |
| S14_vs_k141_80372||lt2gene | Low-quality |
| S14_vs_k141_87848||lt2gene | Not-determined |
| S14_vs_k141_87849||lt2gene | Not-determined |
| S14_vs_k141_28545||lt2gene | Not-determined |
| S14_vs_k141_7024||lt2gene | Low-quality |
| S14_vs_k141_87854||lt2gene | Low-quality |
| S14_vs_k141_50514||lt2gene | Low-quality |
| S14_vs_k141_87856||lt2gene | Low-quality |
| S14_vs_k141_28570||lt2gene | Low-quality |
| S14_vs_k141_72919||lt2gene | Low-quality |
| S14_vs_k141_87862||lt2gene | Not-determined |
| S14_vs_k141_72924||lt2gene | Low-quality |
| S14_vs_k141_28594||lt2gene | Low-quality |
| S14_vs_k141_87883||lt2gene | Low-quality |
| S14_vs_k141_87885||lt2gene | Low-quality |
| S14_vs_k141_7087||lt2gene | Low-quality |
| S14_vs_k141_7097||lt2gene | Low-quality |
| S14_vs_k141_7103||lt2gene | Not-determined |
| S14_vs_k141_50558||lt2gene | Low-quality |
| S14_vs_k141_87902||lt2gene | Low-quality |
| S14_vs_k141_14369||lt2gene | Low-quality |
| S14_vs_k141_65350||lt2gene | Low-quality |
| S14_vs_k141_102211||lt2gene | Low-quality |
| S14_vs_k141_14385||lt2gene | Not-determined |
| S14_vs_k141_14386||lt2gene | Low-quality |
| S14_vs_k141_14387||lt2gene | Not-determined |
| S14_vs_k141_87908||lt2gene | Low-quality |
| S14_vs_k141_7148||lt2gene | Low-quality |
| S14_vs_k141_7156||lt2gene | Low-quality |
| S14_vs_k141_7161||lt2gene | Low-quality |
| S14_vs_k141_102243||lt2gene | Not-determined |
| S14_vs_k141_72978||lt2gene | Low-quality |
| S14_vs_k141_14395||lt2gene | Low-quality |
| S14_vs_k141_50609||lt2gene | Not-determined |
| S14_vs_k141_7170||lt2gene | Low-quality |
| S14_vs_k141_7194||lt2gene | Low-quality |
| S14_vs_k141_58493||lt2gene | Not-determined |
| S14_vs_k141_58495||lt2gene | Not-determined |
| S14_vs_k141_58497||lt2gene | Low-quality |
| S14_vs_k141_14411||lt2gene | Low-quality |
| S14_vs_k141_7205||lt2gene | Low-quality |
| S14_vs_k141_50634||lt2gene | Low-quality |
| S14_vs_k141_50636||lt2gene | Low-quality |
| S14_vs_k141_58515||lt2gene | Low-quality |
| S14_vs_k141_50640||lt2gene | Low-quality |
| S14_vs_k141_7226||lt2gene | Low-quality |
| S14_vs_k141_28672||lt2gene | Low-quality |
| S14_vs_k141_102312||lt2gene | Low-quality |
| S14_vs_k141_50667||lt2gene | Low-quality |
| S14_vs_k141_28687||lt2gene | Not-determined |
| S14_vs_k141_28693||lt2gene | Not-determined |
| S14_vs_k141_80472||lt2gene | Not-determined |
| S14_vs_k141_28708||lt2gene | Not-determined |
| S14_vs_k141_28726||lt2gene | Low-quality |
| S14_vs_k141_50696||lt2gene | Low-quality |
| S14_vs_k141_102347||lt2gene | Low-quality |
| S14_vs_k141_7269||lt2gene | Low-quality |
| S14_vs_k141_65467||lt2gene | Low-quality |
| S14_vs_k141_73094||lt2gene | Low-quality |
| S14_vs_k141_58565||lt2gene | Not-determined |
| S14_vs_k141_73097||lt2gene | Low-quality |
| S14_vs_k141_7283||lt2gene | Not-determined |
| S14_vs_k141_14458||lt2gene | Not-determined |
| S14_vs_k141_80506||lt2gene | Low-quality |
| S14_vs_k141_7296||lt2gene | Low-quality |
| S14_vs_k141_50728||lt2gene | Low-quality |
| S14_vs_k141_73113||lt2gene | Low-quality |
| S14_vs_k141_80514||lt2gene | Not-determined |
| S14_vs_k141_73119||lt2gene | Low-quality |
| S14_vs_k141_28762||lt2gene | Not-determined |
| S14_vs_k141_80543||lt2gene | Not-determined |
| S14_vs_k141_58611||lt2gene | Low-quality |
| S14_vs_k141_80554||lt2gene | Low-quality |
| S14_vs_k141_102391||lt2gene | Low-quality |
| S14_vs_k141_102405||lt2gene | Low-quality |
| S14_vs_k141_102412||lt2gene | Low-quality |
| S14_vs_k141_102413||lt2gene | Low-quality |
| S14_vs_k141_80596||lt2gene | Low-quality |
| S14_vs_k141_28830||lt2gene | Not-determined |
| S14_vs_k141_14523||lt2gene | Not-determined |
| S14_vs_k141_50784||lt2gene | Not-determined |
| S14_vs_k141_73184||lt2gene | Low-quality |
| S14_vs_k141_28833||lt2gene | Not-determined |
| S14_vs_k141_28841||lt2gene | Not-determined |
| S14_vs_k141_102456||lt2gene | Low-quality |
| S14_vs_k141_102459||lt2gene | Low-quality |
| S14_vs_k141_28852||lt2gene | Low-quality |
| S14_vs_k141_50804||lt2gene | Low-quality |
| S14_vs_k141_50814||lt2gene | Not-determined |
| S14_vs_k141_50815||lt2gene | Low-quality |
| S14_vs_k141_28872||lt2gene | Low-quality |
| S14_vs_k141_50823||lt2gene | Low-quality |
| S14_vs_k141_50824||lt2gene | Not-determined |
| S14_vs_k141_73218||lt2gene | Low-quality |
| S14_vs_k141_50828||lt2gene | Low-quality |
| S14_vs_k141_73221||lt2gene | Low-quality |
| S14_vs_k141_102505||lt2gene | Low-quality |
| S14_vs_k141_102509||lt2gene | Low-quality |
| S14_vs_k141_73245||lt2gene | Low-quality |
| S14_vs_k141_73247||lt2gene | Low-quality |
| S14_vs_k141_65532||lt2gene | Low-quality |
| S14_vs_k141_28933||lt2gene | Low-quality |
| S14_vs_k141_65552||lt2gene | Low-quality |
| S14_vs_k141_102539||lt2gene | Low-quality |
| S14_vs_k141_73268||lt2gene | Not-determined |
| S14_vs_k141_102555||lt2gene | Not-determined |
| S14_vs_k141_65579||lt2gene | Low-quality |
| S14_vs_k141_65582||lt2gene | Low-quality |
| S14_vs_k141_50899||lt2gene | Low-quality |
| S14_vs_k141_65593||lt2gene | Low-quality |
| S14_vs_k141_28972||lt2gene | Low-quality |
| S14_vs_k141_28974||lt2gene | Not-determined |
| S14_vs_k141_50917||lt2gene | Low-quality |
| S14_vs_k141_50944||lt2gene | Low-quality |
| S14_vs_k141_50953||lt2gene | Low-quality |
| S14_vs_k141_50955||lt2gene | Low-quality |
| S14_vs_k141_50959||lt2gene | Low-quality |
| S14_vs_k141_50962||lt2gene | Low-quality |
| S14_vs_k141_50966||lt2gene | Not-determined |
| S14_vs_k141_50975||lt2gene | Not-determined |
| S14_vs_k141_65656||lt2gene | Low-quality |
| S14_vs_k141_29043||lt2gene | Low-quality |
| S14_vs_k141_50995||lt2gene | Low-quality |
| S14_vs_k141_29063||lt2gene | Low-quality |
| S14_vs_k141_65707||lt2gene | Low-quality |
| S14_vs_k141_65712||lt2gene | Low-quality |
| S14_vs_k141_51070||lt2gene | Not-determined |
| S14_vs_k141_51102||lt2gene | Not-determined |
| S14_vs_k141_51104||lt2gene | Not-determined |
| S14_vs_k141_51107||lt2gene | Low-quality |
| S14_vs_k141_51122||lt2gene | Low-quality |
| S14_vs_k141_29200||lt2gene | Low-quality |
| S14_vs_k141_29234||lt2gene | Low-quality |

| | |
|---|---|
| S14_vs_k141_29235\|\|lt2gene | Low-quality |
| S14_vs_k141_29237\|\|lt2gene | Not-determined |
| S14_vs_k141_65812\|\|lt2gene | Not-determined |
| S14_vs_k141_65813\|\|lt2gene | Low-quality |
| S14_vs_k141_65877\|\|lt2gene | Low-quality |
| S14_vs_k141_51230\|\|lt2gene | Low-quality |
| S14_vs_k141_51235\|\|lt2gene | Low-quality |
| S14_vs_k141_51239\|\|lt2gene | Low-quality |
| S14_vs_k141_51249\|\|lt2gene | Not-determined |
| S14_vs_k141_65898\|\|lt2gene | Low-quality |
| S14_vs_k141_65909\|\|lt2gene | Low-quality |
| S14_vs_k141_65912\|\|lt2gene | Not-determined |
| S14_vs_k141_65924\|\|lt2gene | Not-determined |
| S1_extracted_k141_0 | Not-determined |
| S1_extracted_k141_113475 | Not-determined |
| S1_extracted_k141_33375 | Not-determined |
| S1_extracted_k141_153521 | Not-determined |
| S1_extracted_k141_126825 | Not-determined |
| S1_extracted_k141_120151 | Not-determined |
| S1_extracted_k141_66750 | Low-quality |
| S1_extracted_k141_106801 | Not-determined |
| S1_extracted_k141_46726 | Not-determined |
| S1_extracted_k141_153522 | Not-determined |
| S1_extracted_k141_20028 | Not-determined |
| S1_extracted_k141_60075 | Not-determined |
| S1_extracted_k141_100130 | Not-determined |
| S1_extracted_k141_60076 | Not-determined |
| S1_extracted_k141_106803 | Not-determined |
| S1_extracted_k141_80106 | Not-determined |
| S1_extracted_k141_26703 | Not-determined |
| S1_extracted_k141_153523 | Not-determined |
| S1_extracted_k141_100133 | Not-determined |
| S1_extracted_k141_6680 | Not-determined |
| S1_extracted_k141_53406 | Not-determined |
| S1_extracted_k141_46732 | Not-determined |
| S1_extracted_k141_93453 | Not-determined |
| S1_extracted_k141_113482 | Not-determined |
| S1_extracted_k141_80110 | Low-quality |
| S1_extracted_k141_6682 | Not-determined |
| S1_extracted_k141_153527 | Low-quality |
| S1_extracted_k141_46736 | Not-determined |
| S1_extracted_k141_15 | Not-determined |
| S1_extracted_k141_153532 | Not-determined |
| S1_extracted_k141_93458 | Not-determined |
| S1_extracted_k141_53414 | Not-determined |
| S1_extracted_k141_20041 | Low-quality |
| S1_extracted_k141_46740 | Not-determined |
| S1_extracted_k141_80115 | Not-determined |
| S1_extracted_k141_120162 | Low-quality |
| S1_extracted_k141_93463 | Not-determined |
| S1_extracted_k141_13368 | Not-determined |
| S1_extracted_k141_20045 | Not-determined |
| S1_extracted_k141_133507 | Low-quality |
| S1_extracted_k141_126830 | Not-determined |
| S1_extracted_k141_33390 | Not-determined |
| S1_extracted_k141_126831 | Not-determined |
| S1_extracted_k141_140191 | Not-determined |
| S1_extracted_k141_93467 | Not-determined |
| S1_extracted_k141_106814 | Low-quality |
| S1_extracted_k141_53419 | Not-determined |
| S1_extracted_k141_153535 | Low-quality |
| S1_extracted_k141_26712 | Not-determined |
| S1_extracted_k141_153537 | Low-quality |
| S1_extracted_k141_80121 | Low-quality |
| S1_extracted_k141_133514 | Not-determined |
| S1_extracted_k141_32 | Not-determined |
| S1_extracted_k141_33395 | Not-determined |
| S1_extracted_k141_146860 | Not-determined |
| S1_extracted_k141_93471 | Not-determined |
| S1_extracted_k141_106818 | Low-quality |
| S1_extracted_k141_46748 | Low-quality |
| S1_extracted_k141_106819 | Not-determined |
| S1_extracted_k141_26715 | Not-determined |
| S1_extracted_k141_6699 | Not-determined |
| S1_extracted_k141_106822 | Not-determined |
| S1_extracted_k141_146864 | Not-determined |
| S1_extracted_k141_86793 | Not-determined |
| S1_extracted_k141_146865 | Not-determined |
| S1_extracted_k141_6701 | Low-quality |
| S1_extracted_k141_46755 | Not-determined |
| S1_extracted_k141_133522 | Not-determined |
| S1_extracted_k141_33400 | Not-determined |
| S1_extracted_k141_93480 | Not-determined |
| S1_extracted_k141_126848 | Not-determined |
| S1_extracted_k141_140205 | Not-determined |
| S1_extracted_k141_133526 | Not-determined |
| S1_extracted_k141_113502 | Not-determined |
| S1_extracted_k141_133527 | Not-determined |
| S1_extracted_k141_53424 | Not-determined |
| S1_extracted_k141_60087 | Not-determined |
| S1_extracted_k141_26720 | Not-determined |
| S1_extracted_k141_126854 | Low-quality |
| S1_extracted_k141_46 | Low-quality |
| S1_extracted_k141_66766 | Not-determined |
| S1_extracted_k141_48 | Not-determined |
| S1_extracted_k141_53434 | Not-determined |
| S1_extracted_k141_66767 | Not-determined |
| S1_extracted_k141_33406 | Not-determined |
| S1_extracted_k141_106838 | Not-determined |
| S1_extracted_k141_113512 | Low-quality |
| S1_extracted_k141_80139 | Not-determined |
| S1_extracted_k141_153552 | Not-determined |
| S1_extracted_k141_40058 | Low-quality |
| S1_extracted_k141_80141 | Not-determined |
| S1_extracted_k141_140219 | Low-quality |
| S1_extracted_k141_133546 | Low-quality |
| S1_extracted_k141_46772 | Not-determined |
| S1_extracted_k141_33413 | Low-quality |
| S1_extracted_k141_46776 | Not-determined |
| S1_extracted_k141_113517 | Not-determined |
| S1_extracted_k141_40067 | Not-determined |
| S1_extracted_k141_20067 | Not-determined |
| S1_extracted_k141_60 | Low-quality |
| S1_extracted_k141_66776 | Not-determined |
| S1_extracted_k141_113518 | Low-quality |
| S1_extracted_k141_86812 | Low-quality |
| S1_extracted_k141_140229 | Not-determined |
| S1_extracted_k141_93497 | Not-determined |
| S1_extracted_k141_86813 | Not-determined |
| S1_extracted_k141_13370 | Not-determined |
| S1_extracted_k141_93500 | Not-determined |
| S1_extracted_k141_106849 | Not-determined |
| S1_extracted_k141_40072 | Not-determined |
| S1_extracted_k141_66780 | Not-determined |
| S1_extracted_k141_133555 | Not-determined |
| S1_extracted_k141_46786 | Not-determined |
| S1_extracted_k141_93505 | Not-determined |
| S1_extracted_k141_20073 | Not-determined |
| S1_extracted_k141_126887 | Low-quality |
| S1_extracted_k141_140234 | Low-quality |
| S1_extracted_k141_106855 | Not-determined |
| S1_extracted_k141_86825 | Not-determined |
| S1_extracted_k141_13376 | Not-determined |
| S1_extracted_k141_46790 | Not-determined |
| S1_extracted_k141_66791 | Not-determined |
| S1_extracted_k141_86827 | Not-determined |
| S1_extracted_k141_140237 | Low-quality |
| S1_extracted_k141_153574 | Low-quality |
| S1_extracted_k141_6749 | Not-determined |
| S1_extracted_k141_140239 | Not-determined |
| S1_extracted_k141_73464 | Not-determined |
| S1_extracted_k141_66795 | Not-determined |
| S1_extracted_k141_73467 | Not-determined |
| S1_extracted_k141_86833 | Not-determined |
| S1_extracted_k141_66796 | Low-quality |
| S1_extracted_k141_80 | Not-determined |
| S8_vs_k141_179529\|\|full | High-quality |
| S1_extracted_k141_13382 | Not-determined |
| S1_extracted_k141_81 | Low-quality |
| S1_extracted_k141_73469 | Not-determined |
| S1_extracted_k141_73470 | Not-determined |
| S1_extracted_k141_83 | Not-determined |
| S1_extracted_k141_133563 | Not-determined |
| S1_extracted_k141_13385 | Not-determined |
| S1_extracted_k141_33430 | Not-determined |
| S1_extracted_k141_13387 | Not-determined |
| S1_extracted_k141_86838 | Not-determined |
| S1_extracted_k141_106866 | Not-determined |
| S1_extracted_k141_80177 | Not-determined |
| S1_extracted_k141_153577 | Low-quality |
| S1_extracted_k141_87 | Not-determined |
| S1_extracted_k141_26748 | Not-determined |
| S1_extracted_k141_66804 | Not-determined |
| S1_extracted_k141_53472 | Not-determined |
| S1_extracted_k141_120210 | Not-determined |
| S1_extracted_k141_60095 | Low-quality |
| S1_extracted_k141_80182 | Low-quality |
| S1_extracted_k141_100161 | Not-determined |
| S1_extracted_k141_153581 | Low-quality |
| S1_extracted_k141_126904 | Not-determined |
| S1_extracted_k141_13396 | Not-determined |
| S1_extracted_k141_96 | Not-determined |
| S1_extracted_k141_53480 | Not-determined |
| S1_extracted_k141_20089 | Not-determined |
| S1_extracted_k141_93530 | Not-determined |
| S1_extracted_k141_106871 | Not-determined |
| S1_extracted_k141_33436 | Not-determined |
| S1_extracted_k141_153586 | Low-quality |
| S1_extracted_k141_100166 | Low-quality |
| S1_extracted_k141_106874 | Low-quality |
| S1_extracted_k141_33438 | Not-determined |
| S1_extracted_k141_86848 | Not-determined |
| S1_extracted_k141_100 | Low-quality |
| S1_extracted_k141_13401 | Not-determined |
| S1_extracted_k141_140255 | Low-quality |
| S1_extracted_k141_113540 | Not-determined |
| S1_extracted_k141_146896 | Low-quality |
| S1_extracted_k141_60097 | Low-quality |
| S1_extracted_k141_120221 | Not-determined |
| S1_extracted_k141_100168 | Not-determined |
| S1_extracted_k141_66810 | Low-quality |
| S1_extracted_k141_100172 | Not-determined |
| S1_extracted_k141_80198 | Low-quality |
| S1_extracted_k141_66811 | Not-determined |
| S1_extracted_k141_46800 | Low-quality |
| S9_extracted_k141_74235 | Medium-quality |
| S1_extracted_k141_93538 | Not-determined |
| S1_extracted_k141_106885 | Not-determined |
| S1_extracted_k141_26760 | Not-determined |
| S1_extracted_k141_13418 | Not-determined |
| S1_extracted_k141_146906 | Not-determined |
| S1_extracted_k141_93542 | Not-determined |
| S1_extracted_k141_13420 | Not-determined |
| S1_extracted_k141_66816 | Not-determined |
| S1_extracted_k141_106887 | Not-determined |
| S1_extracted_k141_106889 | Low-quality |
| S1_extracted_k141_153592 | Low-quality |
| S1_extracted_k141_33448 | Not-determined |
| S1_extracted_k141_120235 | Not-determined |
| S1_extracted_k141_113556 | Not-determined |
| S1_extracted_k141_86854 | Not-determined |
| S1_extracted_k141_66821 | Low-quality |
| S1_extracted_k141_53506 | Low-quality |
| S1_extracted_k141_86857 | Low-quality |
| S1_extracted_k141_6791 | Not-determined |
| S1_extracted_k141_33452 | Not-determined |
| S1_extracted_k141_120240 | Not-determined |
| S1_extracted_k141_80212 | Not-determined |
| S1_extracted_k141_113 | Not-determined |
| S1_extracted_k141_120243 | Not-determined |
| S1_extracted_k141_40107 | Not-determined |
| S1_extracted_k141_40108 | Not-determined |
| S1_extracted_k141_53513 | Not-determined |
| S1_extracted_k141_13429 | Not-determined |
| S1_extracted_k141_120245 | Not-determined |
| S1_extracted_k141_33457 | Not-determined |
| S1_extracted_k141_26767 | Not-determined |
| S1_extracted_k141_86860 | Not-determined |
| S1_extracted_k141_120246 | Low-quality |
| S1_extracted_k141_146919 | Not-determined |
| S1_extracted_k141_33458 | Not-determined |
| S1_extracted_k141_66834 | Not-determined |
| S1_extracted_k141_100188 | Low-quality |
| S1_extracted_k141_100189 | Not-determined |
| S1_extracted_k141_113564 | Not-determined |
| S1_extracted_k141_13435 | Not-determined |
| S1_extracted_k141_80219 | Not-determined |
| S1_extracted_k141_46820 | Low-quality |
| S1_extracted_k141_6800 | Not-determined |
| S1_extracted_k141_53519 | Not-determined |
| S1_extracted_k141_40115 | Not-determined |
| S1_extracted_k141_86871 | Not-determined |
| S1_extracted_k141_106895 | Not-determined |
| S1_extracted_k141_46821 | Not-determined |
| S1_extracted_k141_118 | Not-determined |
| S1_extracted_k141_113574 | Not-determined |
| S1_extracted_k141_93565 | Not-determined |
| S1_extracted_k141_53522 | Not-determined |
| S1_extracted_k141_6806 | Low-quality |
| S1_extracted_k141_13447 | Not-determined |
| S1_extracted_k141_120256 | Not-determined |
| S1_extracted_k141_46833 | Not-determined |
| S1_extracted_k141_86880 | Not-determined |
| S1_extracted_k141_100203 | Not-determined |
| S1_extracted_k141_86883 | Not-determined |
| S1_extracted_k141_93575 | Not-determined |
| S1_extracted_k141_120258 | Not-determined |
| S1_extracted_k141_120259 | Not-determined |
| S1_extracted_k141_113587 | Not-determined |
| S1_extracted_k141_93577 | Not-determined |
| S1_extracted_k141_53530 | Not-determined |
| S1_extracted_k141_40126 | Not-determined |
| S1_extracted_k141_13456 | Not-determined |
| S1_extracted_k141_124 | Low-quality |
| S1_extracted_k141_33475 | Not-determined |
| S1_extracted_k141_60104 | Low-quality |
| S1_extracted_k141_73519 | Not-determined |
| S1_extracted_k141_33476 | Not-determined |
| S1_extracted_k141_153595 | Not-determined |
| S1_extracted_k141_40129 | Not-determined |
| S1_extracted_k141_73520 | Not-determined |
| S1_extracted_k141_153596 | Not-determined |
| S1_extracted_k141_140295 | Not-determined |
| S1_extracted_k141_66848 | Not-determined |
| S1_extracted_k141_33479 | Not-determined |
| S1_extracted_k141_126958 | Not-determined |
| S1_extracted_k141_26777 | Not-determined |
| S1_extracted_k141_106910 | Not-determined |
| S1_extracted_k141_93586 | Not-determined |
| S1_extracted_k141_26779 | Not-determined |
| S1_extracted_k141_140300 | Not-determined |
| S1_extracted_k141_40134 | Not-determined |
| S1_extracted_k141_100211 | Not-determined |
| S1_extracted_k141_113596 | Not-determined |
| S1_extracted_k141_120265 | Not-determined |
| S1_extracted_k141_40135 | Not-determined |
| S1_extracted_k141_26781 | Not-determined |
| S1_extracted_k141_126961 | Not-determined |
| S1_extracted_k141_80236 | Low-quality |
| S1_extracted_k141_130 | Not-determined |
| S1_extracted_k141_13465 | Not-determined |
| S1_extracted_k141_26783 | Not-determined |
| S1_extracted_k141_100219 | Low-quality |
| S1_extracted_k141_120266 | Low-quality |
| S1_extracted_k141_6822 | Low-quality |
| S1_extracted_k141_66853 | Low-quality |
| S1_extracted_k141_66855 | Low-quality |
| S1_extracted_k141_136 | Low-quality |
| S1_extracted_k141_46849 | Not-determined |
| S1_extracted_k141_106914 | Not-determined |
| S1_extracted_k141_46850 | Not-determined |
| S1_extracted_k141_138 | Not-determined |
| S1_extracted_k141_53549 | Not-determined |
| S1_extracted_k141_20127 | Low-quality |
| S1_extracted_k141_113601 | Not-determined |
| S1_extracted_k141_120274 | Not-determined |
| S1_extracted_k141_73532 | Not-determined |
| S1_extracted_k141_26788 | Low-quality |
| S1_extracted_k141_120277 | Not-determined |
| S1_extracted_k141_26790 | Not-determined |
| S1_extracted_k141_40151 | Not-determined |
| S1_extracted_k141_33488 | Not-determined |
| S1_extracted_k141_73539 | Not-determined |
| S1_extracted_k141_120282 | Not-determined |
| S1_extracted_k141_93611 | Not-determined |
| S1_extracted_k141_146944 | Not-determined |
| S1_extracted_k141_46856 | Low-quality |
| S1_extracted_k141_126974 | Not-determined |
| S1_extracted_k141_140313 | Not-determined |
| S1_extracted_k141_100226 | Not-determined |
| S1_extracted_k141_133579 | Not-determined |
| S1_extracted_k141_40160 | Not-determined |
| S1_extracted_k141_46864 | Not-determined |
| S1_extracted_k141_33498 | Low-quality |
| S1_extracted_k141_40161 | Not-determined |
| S1_extracted_k141_6843 | Not-determined |
| S1_extracted_k141_80261 | Not-determined |
| S1_extracted_k141_26798 | Not-determined |
| S1_extracted_k141_33499 | Not-determined |
| S1_extracted_k141_13488 | Not-determined |
| S1_extracted_k141_151 | Not-determined |
| S1_extracted_k141_126984 | Not-determined |
| S1_extracted_k141_40164 | Not-determined |
| S1_extracted_k141_146947 | Not-determined |
| S1_extracted_k141_106933 | Not-determined |
| S1_extracted_k141_80265 | Not-determined |
| S1_extracted_k141_120293 | Not-determined |
| S1_extracted_k141_80266 | Not-determined |
| S1_extracted_k141_73548 | Low-quality |
| S1_extracted_k141_120295 | Not-determined |
| S1_extracted_k141_140323 | Not-determined |
| S1_extracted_k141_6851 | Not-determined |
| S1_extracted_k141_120296 | Low-quality |
| S1_extracted_k141_73551 | Not-determined |
| S1_extracted_k141_140325 | Not-determined |
| S1_extracted_k141_33503 | Not-determined |
| S1_extracted_k141_53562 | Not-determined |
| S1_extracted_k141_13500 | Not-determined |
| S1_extracted_k141_140328 | Not-determined |
| S1_extracted_k141_100237 | Low-quality |
| S1_extracted_k141_6854 | Not-determined |
| S1_extracted_k141_158 | Not-determined |
| S1_extracted_k141_159 | Not-determined |
| S1_extracted_k141_106935 | Not-determined |
| S1_extracted_k141_160 | Not-determined |
| S1_extracted_k141_140330 | Not-determined |
| S1_extracted_k141_126994 | Not-determined |
| S1_extracted_k141_20146 | Not-determined |
| S1_extracted_k141_80274 | Not-determined |
| S1_extracted_k141_73557 | Not-determined |
| S1_extracted_k141_113624 | Low-quality |
| S1_extracted_k141_13507 | Not-determined |
| S1_extracted_k141_120302 | Not-determined |
| S1_extracted_k141_162 | Not-determined |
| S1_extracted_k141_106936 | Not-determined |
| S1_extracted_k141_126998 | Not-determined |
| S1_extracted_k141_153612 | Not-determined |
| S1_extracted_k141_86931 | Not-determined |
| S1_extracted_k141_93628 | Not-determined |
| S1_extracted_k141_80280 | Not-determined |
| S1_extracted_k141_93629 | Not-determined |
| S1_extracted_k141_120310 | Not-determined |
| S1_extracted_k141_86933 | Not-determined |
| S1_extracted_k141_66876 | Not-determined |
| S1_extracted_k141_73566 | Not-determined |
| S1_extracted_k141_80283 | Not-determined |
| S1_extracted_k141_133591 | Not-determined |
| S1_extracted_k141_106943 | Not-determined |
| S1_extracted_k141_100253 | Low-quality |
| S1_extracted_k141_46882 | Not-determined |
| S1_extracted_k141_33515 | Not-determined |
| S1_extracted_k141_146959 | Not-determined |
| S1_extracted_k141_120315 | Not-determined |
| S1_extracted_k141_33520 | Not-determined |
| S1_extracted_k141_46887 | Not-determined |
| S1_extracted_k141_146964 | Low-quality |
| S1_extracted_k141_120318 | Not-determined |
| S1_extracted_k141_66884 | Not-determined |
| S1_extracted_k141_20165 | Not-determined |
| S1_extracted_k141_73570 | Not-determined |
| S1_extracted_k141_93648 | Not-determined |
| S1_extracted_k141_20167 | Not-determined |
| S1_extracted_k141_33524 | Not-determined |
| S1_extracted_k141_20169 | Not-determined |
| S1_extracted_k141_140340 | Low-quality |
| S1_extracted_k141_53578 | Low-quality |
| S1_extracted_k141_53579 | Not-determined |
| S1_extracted_k141_179 | Not-determined |
| S1_extracted_k141_80295 | Low-quality |
| S1_extracted_k141_73578 | Not-determined |
| S1_extracted_k141_80298 | Not-determined |
| S1_extracted_k141_26825 | Low-quality |
| S1_extracted_k141_93653 | Not-determined |
| S1_extracted_k141_86954 | Low-quality |
| S1_extracted_k141_40197 | Not-determined |
| S1_extracted_k141_93657 | Not-determined |
| S1_extracted_k141_127021 | Not-determined |
| S1_extracted_k141_188 | Not-determined |
| S1_extracted_k141_127023 | Not-determined |
| S1_extracted_k141_93660 | Not-determined |
| S1_extracted_k141_73579 | Not-determined |
| S1_extracted_k141_66896 | Not-determined |
| S1_extracted_k141_20182 | Not-determined |
| S1_extracted_k141_93663 | Not-determined |
| S1_extracted_k141_113643 | Not-determined |
| S1_extracted_k141_86958 | Low-quality |
| S1_extracted_k141_140352 | Not-determined |
| S1_extracted_k141_13525 | Not-determined |
| S1_extracted_k141_120342 | Not-determined |
| S1_extracted_k141_113645 | Not-determined |
| S1_extracted_k141_106964 | Not-determined |
| S1_extracted_k141_127028 | Not-determined |
| S1_extracted_k141_86961 | Not-determined |
| S1_extracted_k141_46907 | Not-determined |
| S1_extracted_k141_153623 | Not-determined |
| S1_extracted_k141_20189 | Not-determined |
| S1_extracted_k141_60144 | Not-determined |
| S1_extracted_k141_53593 | Not-determined |
| S1_extracted_k141_106967 | Not-determined |
| S1_extracted_k141_46912 | Not-determined |
| S1_extracted_k141_201 | Not-determined |
| S1_extracted_k141_120347 | Not-determined |
| S1_extracted_k141_13534 | Not-determined |
| S1_extracted_k141_106968 | Not-determined |
| S1_extracted_k141_93668 | Not-determined |
| S1_extracted_k141_127032 | Not-determined |
| S1_extracted_k141_73593 | Not-determined |
| S1_extracted_k141_100291 | Not-determined |
| S1_extracted_k141_20195 | Not-determined |
| S1_extracted_k141_60149 | Not-determined |
| S1_extracted_k141_133612 | Not-determined |
| S1_extracted_k141_20196 | Low-quality |
| S1_extracted_k141_13539 | Not-determined |
| S1_extracted_k141_66907 | Not-determined |
| S1_extracted_k141_40224 | Not-determined |
| S1_extracted_k141_46914 | Not-determined |
| S1_extracted_k141_140358 | Low-quality |
| S1_extracted_k141_127036 | Not-determined |
| S1_extracted_k141_40226 | Not-determined |
| S1_extracted_k141_20199 | Not-determined |
| S1_extracted_k141_207 | Not-determined |
| S1_extracted_k141_100299 | Not-determined |
| S1_extracted_k141_133618 | Not-determined |
| S1_extracted_k141_146974 | Not-determined |
| S1_extracted_k141_106977 | Not-determined |
| S1_extracted_k141_13542 | Not-determined |
| S1_extracted_k141_40231 | Not-determined |
| S1_extracted_k141_13545 | Not-determined |
| S1_extracted_k141_133623 | Not-determined |
| S1_extracted_k141_127042 | Not-determined |
| S1_extracted_k141_73603 | Not-determined |
| S1_extracted_k141_140363 | Not-determined |
| S1_extracted_k141_73604 | Not-determined |
| S1_extracted_k141_133625 | Not-determined |
| S1_extracted_k141_140365 | Not-determined |
| S1_extracted_k141_40235 | Low-quality |
| S1_extracted_k141_86980 | Not-determined |
| S1_extracted_k141_40236 | Not-determined |
| S1_extracted_k141_6918 | Not-determined |
| S1_extracted_k141_40237 | Not-determined |
| S1_extracted_k141_120359 | Not-determined |
| S1_extracted_k141_66921 | Not-determined |
| S1_extracted_k141_100306 | Not-determined |
| S1_extracted_k141_60161 | Not-determined |
| S1_extracted_k141_100307 | Not-determined |
| S1_extracted_k141_40240 | Not-determined |
| S1_extracted_k141_146979 | Not-determined |
| S1_extracted_k141_100309 | Not-determined |
| S1_extracted_k141_66925 | Not-determined |
| S1_extracted_k141_127049 | Not-determined |
| S1_extracted_k141_60162 | Low-quality |
| S1_extracted_k141_86986 | Low-quality |
| S1_extracted_k141_93687 | Not-determined |
| S1_extracted_k141_120364 | Not-determined |
| S1_extracted_k141_153626 | Not-determined |
| S1_extracted_k141_127054 | Not-determined |
| S1_extracted_k141_133632 | Not-determined |
| S1_extracted_k141_127055 | Not-determined |
| S1_extracted_k141_93690 | Low-quality |
| S1_extracted_k141_20215 | Low-quality |
| S1_extracted_k141_133634 | Not-determined |
| S1_extracted_k141_6925 | Not-determined |
| S1_extracted_k141_73614 | Not-determined |
| S1_extracted_k141_40246 | Low-quality |
| S1_extracted_k141_53622 | Not-determined |
| S1_extracted_k141_40248 | Not-determined |
| S1_extracted_k141_13565 | Not-determined |
| S1_extracted_k141_33555 | Not-determined |
| S1_extracted_k141_100315 | Not-determined |
| S1_extracted_k141_6930 | Not-determined |
| S1_extracted_k141_66938 | Not-determined |
| S1_extracted_k141_127063 | Not-determined |
| S1_extracted_k141_80337 | Not-determined |
| S1_extracted_k141_93695 | Not-determined |
| S1_extracted_k141_80339 | Not-determined |
| S1_extracted_k141_93696 | Not-determined |
| S1_extracted_k141_140389 | Not-determined |
| S1_extracted_k141_93698 | Not-determined |
| S1_extracted_k141_146993 | Not-determined |
| S1_extracted_k141_127068 | Not-determined |
| S1_extracted_k141_20225 | Not-determined |
| S1_extracted_k141_6942 | Not-determined |
| S1_extracted_k141_113658 | Not-determined |
| S1_extracted_k141_33559 | Low-quality |
| S1_extracted_k141_87001 | Not-determined |
| S1_extracted_k141_233 | Not-determined |
| S1_extracted_k141_66945 | Not-determined |
| S1_extracted_k141_113661 | Not-determined |
| S1_extracted_k141_107002 | Not-determined |
| S1_extracted_k141_33561 | Low-quality |
| S1_extracted_k141_113662 | Not-determined |
| S1_extracted_k141_100323 | Not-determined |
| S1_extracted_k141_238 | Not-determined |
| S1_extracted_k141_107005 | Low-quality |
| S1_extracted_k141_146998 | Low-quality |
| S1_extracted_k141_26847 | Not-determined |
| S1_extracted_k141_93705 | Not-determined |
| S1_extracted_k141_6950 | Not-determined |
| S1_extracted_k141_93710 | Not-determined |
| S1_extracted_k141_107014 | Not-determined |
| S1_extracted_k141_100328 | Not-determined |
| S1_extracted_k141_100329 | Not-determined |
| S1_extracted_k141_127078 | Not-determined |
| S1_extracted_k141_153634 | Low-quality |
| S1_extracted_k141_113674 | Not-determined |
| S1_extracted_k141_140411 | Low-quality |
| S1_extracted_k141_107018 | Not-determined |
| S1_extracted_k141_249 | Not-determined |
| S1_extracted_k141_20241 | Not-determined |
| S1_extracted_k141_140413 | Not-determined |
| S1_extracted_k141_100334 | Not-determined |
| S1_extracted_k141_73636 | Not-determined |
| S1_extracted_k141_20248 | Not-determined |
| S1_extracted_k141_80363 | Not-determined |
| S1_extracted_k141_26850 | Not-determined |
| S1_extracted_k141_46931 | Not-determined |
| S1_extracted_k141_107023 | Not-determined |
| S1_extracted_k141_66969 | Low-quality |
| S1_extracted_k141_26853 | Not-determined |
| S1_extracted_k141_257 | Not-determined |
| S1_extracted_k141_46938 | Not-determined |
| S1_extracted_k141_133646 | Not-determined |
| S1_extracted_k141_46940 | Not-determined |
| S1_extracted_k141_147006 | Not-determined |
| S1_extracted_k141_73651 | Not-determined |
| S1_extracted_k141_127092 | Not-determined |
| S1_extracted_k141_133651 | Not-determined |
| S1_extracted_k141_120417 | Not-determined |
| S1_extracted_k141_53656 | Low-quality |
| S1_extracted_k141_140423 | Not-determined |
| S1_extracted_k141_133652 | Not-determined |
| S1_extracted_k141_33580 | Not-determined |
| S1_extracted_k141_40277 | Low-quality |
| S1_extracted_k141_113694 | Not-determined |
| S1_extracted_k141_120420 | Not-determined |
| S1_extracted_k141_40278 | Not-determined |
| S1_extracted_k141_87033 | Not-determined |
| S1_extracted_k141_267 | Not-determined |
| S1_extracted_k141_147011 | Not-determined |
| S1_extracted_k141_20259 | Not-determined |
| S1_extracted_k141_100348 | Not-determined |
| S1_extracted_k141_100350 | Not-determined |
| S1_extracted_k141_140427 | Not-determined |
| S1_extracted_k141_87040 | Not-determined |
| S1_extracted_k141_140428 | Not-determined |
| S1_extracted_k141_26863 | Low-quality |
| S1_extracted_k141_66977 | Low-quality |
| S1_extracted_k141_93728 | Not-determined |
| S1_extracted_k141_100352 | Low-quality |
| S1_extracted_k141_107030 | Low-quality |
| S1_extracted_k141_140430 | Not-determined |
| S1_extracted_k141_107032 | Not-determined |
| S1_extracted_k141_113701 | Not-determined |
| S1_extracted_k141_20266 | Not-determined |
| S1_extracted_k141_40290 | Low-quality |
| S1_extracted_k141_20267 | Not-determined |
| S1_extracted_k141_40291 | Not-determined |
| S1_extracted_k141_53671 | Not-determined |
| S1_extracted_k141_87047 | Not-determined |

| | |
|---|---|
| S1_extracted_k141_127116 | Not-determined |
| S1_extracted_k141_87048 | Not-determined |
| S1_extracted_k141_13605 | Not-determined |
| S1_extracted_k141_113708 | Not-determined |
| S1_extracted_k141_87049 | Low-quality |
| S1_extracted_k141_147019 | Not-determined |
| S1_extracted_k141_53674 | Not-determined |
| S1_extracted_k141_140439 | Not-determined |
| S1_extracted_k141_120433 | Not-determined |
| S1_extracted_k141_147024 | Not-determined |
| S1_extracted_k141_153648 | Not-determined |
| S1_extracted_k141_133667 | Low-quality |
| S1_extracted_k141_107043 | Not-determined |
| S1_extracted_k141_274 | Not-determined |
| S1_extracted_k141_133669 | Not-determined |
| S1_extracted_k141_147026 | Not-determined |
| S1_extracted_k141_275 | Low-quality |
| S1_extracted_k141_80382 | Low-quality |
| S1_extracted_k141_127126 | Not-determined |
| S1_extracted_k141_87055 | Not-determined |
| S1_extracted_k141_120440 | Not-determined |
| S1_extracted_k141_133673 | Not-determined |
| S1_extracted_k141_73668 | Not-determined |
| S1_extracted_k141_73669 | Not-determined |
| S1_extracted_k141_133675 | Low-quality |
| S1_extracted_k141_153654 | Low-quality |
| S1_extracted_k141_40302 | Not-determined |
| S1_extracted_k141_20275 | Low-quality |
| S1_extracted_k141_53682 | Not-determined |
| S1_extracted_k141_107051 | Not-determined |
| S1_extracted_k141_93745 | Not-determined |
| S1_extracted_k141_13615 | Not-determined |
| S1_extracted_k141_113720 | Not-determined |
| S1_extracted_k141_133686 | Not-determined |
| S1_extracted_k141_100378 | Not-determined |
| S1_extracted_k141_40308 | Not-determined |
| S1_extracted_k141_73680 | Not-determined |
| S1_extracted_k141_80395 | Not-determined |
| S1_extracted_k141_127133 | Not-determined |
| S1_extracted_k141_153656 | Not-determined |
| S1_extracted_k141_73683 | Not-determined |
| S1_extracted_k141_46965 | Not-determined |
| S1_extracted_k141_107059 | Not-determined |
| S1_extracted_k141_73686 | Not-determined |
| S1_extracted_k141_113725 | Not-determined |
| S1_extracted_k141_20287 | Not-determined |
| S1_extracted_k141_40319 | Not-determined |
| S1_extracted_k141_73688 | Not-determined |
| S1_extracted_k141_80398 | Not-determined |
| S1_extracted_k141_13628 | Not-determined |
| S1_extracted_k141_100387 | Not-determined |
| S1_extracted_k141_140460 | Not-determined |
| S1_extracted_k141_13632 | Not-determined |
| S1_extracted_k141_73694 | Not-determined |
| S1_extracted_k141_127144 | Not-determined |
| S1_extracted_k141_46971 | Not-determined |
| S1_extracted_k141_20292 | Not-determined |
| S1_extracted_k141_33619 | Not-determined |
| S1_extracted_k141_46974 | Not-determined |
| S1_extracted_k141_33621 | Not-determined |
| S1_extracted_k141_73701 | Not-determined |
| S1_extracted_k141_13640 | Not-determined |
| S1_extracted_k141_140466 | Not-determined |
| S1_extracted_k141_140467 | Not-determined |
| S1_extracted_k141_147034 | Low-quality |
| S1_extracted_k141_113735 | Not-determined |
| S1_extracted_k141_60190 | Not-determined |
| S1_extracted_k141_60192 | Not-determined |
| S1_extracted_k141_20301 | Low-quality |
| S1_extracted_k141_26891 | Not-determined |
| S1_extracted_k141_80411 | Not-determined |
| S1_extracted_k141_40328 | Not-determined |
| S1_extracted_k141_133705 | Not-determined |
| S1_extracted_k141_53709 | Not-determined |
| S1_extracted_k141_87075 | Not-determined |
| S1_extracted_k141_93768 | Not-determined |
| S1_extracted_k141_13655 | Not-determined |
| S1_extracted_k141_53714 | Not-determined |
| S1_extracted_k141_133710 | Not-determined |
| S1_extracted_k141_73708 | Not-determined |
| S1_extracted_k141_46989 | Not-determined |
| S1_extracted_k141_20304 | Low-quality |
| S14_extracted_k141_78139 | High-quality |
| S1_extracted_k141_46991 | Low-quality |
| S1_extracted_k141_20306 | Not-determined |
| S1_extracted_k141_53718 | Not-determined |
| S1_extracted_k141_80421 | Not-determined |
| S1_extracted_k141_113748 | Low-quality |
| S1_extracted_k141_33636 | Low-quality |
| S1_extracted_k141_93777 | Not-determined |
| S1_extracted_k141_87085 | Not-determined |
| S1_extracted_k141_60197 | Not-determined |
| S1_extracted_k141_53724 | Low-quality |
| S1_extracted_k141_20311 | Not-determined |
| S1_extracted_k141_127165 | Not-determined |
| S1_extracted_k141_120451 | Low-quality |
| S1_extracted_k141_20315 | Not-determined |
| S1_extracted_k141_80428 | Not-determined |
| S1_extracted_k141_26906 | Low-quality |
| S1_extracted_k141_120453 | Low-quality |
| S1_extracted_k141_13666 | Not-determined |
| S1_extracted_k141_26907 | Not-determined |
| S1_extracted_k141_20318 | Not-determined |
| S1_extracted_k141_60199 | Low-quality |
| S1_extracted_k141_127171 | Not-determined |
| S1_extracted_k141_60200 | Not-determined |
| S1_extracted_k141_107089 | Not-determined |
| S1_extracted_k141_140469 | Low-quality |
| S1_extracted_k141_93783 | Not-determined |
| S1_extracted_k141_87092 | Not-determined |
| S1_extracted_k141_147046 | Not-determined |
| S1_extracted_k141_127175 | Not-determined |
| S1_extracted_k141_26912 | Not-determined |
| S1_extracted_k141_40350 | Not-determined |
| S1_extracted_k141_312 | Not-determined |
| S1_extracted_k141_67010 | Not-determined |
| S1_extracted_k141_67011 | Low-quality |
| S1_extracted_k141_93787 | Not-determined |
| S1_extracted_k141_26915 | Not-determined |
| S1_extracted_k141_93789 | Not-determined |
| S1_extracted_k141_107095 | Not-determined |
| S1_extracted_k141_315 | Not-determined |
| S1_extracted_k141_147051 | Not-determined |
| S1_extracted_k141_140473 | Low-quality |
| S1_extracted_k141_33647 | Not-determined |
| S1_extracted_k141_13673 | Not-determined |
| S1_extracted_k141_33649 | Not-determined |
| S1_extracted_k141_100413 | Not-determined |
| S1_extracted_k141_6975 | Not-determined |
| S1_extracted_k141_6976 | Not-determined |
| S1_extracted_k141_13677 | Not-determined |
| S1_extracted_k141_120468 | Not-determined |
| S1_extracted_k141_100417 | Not-determined |
| S1_extracted_k141_20331 | Not-determined |
| S1_extracted_k141_73726 | Not-determined |
| S1_extracted_k141_53734 | Not-determined |
| S1_extracted_k141_100420 | Not-determined |
| S1_extracted_k141_127183 | Low-quality |
| S1_extracted_k141_133723 | Not-determined |
| S1_extracted_k141_322 | Not-determined |
| S1_extracted_k141_47008 | Not-determined |
| S1_extracted_k141_153680 | Not-determined |
| S1_extracted_k141_140480 | Not-determined |
| S1_extracted_k141_153682 | Not-determined |
| S1_extracted_k141_147060 | Not-determined |
| S1_extracted_k141_53740 | Not-determined |
| S1_extracted_k141_6982 | Not-determined |
| S1_extracted_k141_100425 | Not-determined |
| S1_extracted_k141_53742 | Not-determined |
| S1_extracted_k141_113762 | Not-determined |
| S1_extracted_k141_113763 | Not-determined |
| S1_extracted_k141_153684 | Not-determined |
| S1_extracted_k141_53747 | Not-determined |
| S1_extracted_k141_26930 | Not-determined |
| S1_extracted_k141_53748 | Not-determined |
| S1_extracted_k141_113765 | Not-determined |
| S1_extracted_k141_13687 | Not-determined |
| S1_extracted_k141_20339 | Not-determined |
| S1_extracted_k141_33662 | Not-determined |
| S1_extracted_k141_87114 | Not-determined |
| S1_extracted_k141_6993 | Not-determined |
| S1_extracted_k141_53754 | Not-determined |
| S1_extracted_k141_80467 | Not-determined |
| S1_extracted_k141_147069 | Not-determined |
| S1_extracted_k141_73737 | Not-determined |
| S1_extracted_k141_147070 | Not-determined |
| S1_extracted_k141_33663 | Not-determined |
| S1_extracted_k141_87117 | Not-determined |
| S1_extracted_k141_87118 | Not-determined |
| S1_extracted_k141_127198 | Not-determined |
| S1_extracted_k141_60209 | Not-determined |
| S1_extracted_k141_140482 | Not-determined |
| S1_extracted_k141_73741 | Not-determined |
| S1_extracted_k141_53762 | Not-determined |
| S1_extracted_k141_26942 | Not-determined |
| S1_extracted_k141_120487 | Not-determined |
| S1_extracted_k141_87125 | Not-determined |
| S1_extracted_k141_73745 | Not-determined |
| S1_extracted_k141_140487 | Not-determined |
| S1_extracted_k141_13703 | Not-determined |
| S1_extracted_k141_127206 | Not-determined |
| S1_extracted_k141_147078 | Not-determined |
| S1_extracted_k141_107122 | Not-determined |
| S1_extracted_k141_87127 | Not-determined |
| S1_extracted_k141_113775 | Not-determined |
| S1_extracted_k141_53767 | Not-determined |
| S1_extracted_k141_7001 | Not-determined |
| S1_extracted_k141_93825 | Not-determined |
| S1_extracted_k141_13705 | Low-quality |
| S1_extracted_k141_140493 | Not-determined |
| S1_extracted_k141_53772 | Not-determined |
| S1_extracted_k141_73750 | Not-determined |
| S1_extracted_k141_20355 | Not-determined |
| S1_extracted_k141_100449 | Not-determined |
| S1_extracted_k141_40386 | Not-determined |
| S1_extracted_k141_53776 | Not-determined |
| S1_extracted_k141_80486 | Not-determined |
| S1_extracted_k141_26954 | Not-determined |
| S1_extracted_k141_40389 | Not-determined |
| S1_extracted_k141_133735 | Low-quality |
| S1_extracted_k141_153691 | Low-quality |
| S1_extracted_k141_87138 | Low-quality |
| S1_extracted_k141_133738 | Low-quality |
| S1_extracted_k141_53784 | Low-quality |
| S1_extracted_k141_13717 | Not-determined |
| S1_extracted_k141_140496 | Not-determined |
| S1_extracted_k141_93839 | Not-determined |
| S1_extracted_k141_40393 | Not-determined |
| S1_extracted_k141_87144 | Not-determined |
| S1_extracted_k141_13722 | Not-determined |
| S1_extracted_k141_113787 | Not-determined |
| S1_extracted_k141_53790 | Not-determined |
| S1_extracted_k141_26965 | Not-determined |
| S1_extracted_k141_60218 | Low-quality |
| S1_extracted_k141_33678 | Low-quality |
| S1_extracted_k141_87148 | Not-determined |
| S1_extracted_k141_60219 | Not-determined |
| S1_extracted_k141_133746 | Not-determined |
| S1_extracted_k141_26967 | Not-determined |
| S1_extracted_k141_93845 | Not-determined |
| S1_extracted_k141_113791 | Not-determined |
| S1_extracted_k141_80494 | Not-determined |
| S1_extracted_k141_351 | Not-determined |
| S1_extracted_k141_73759 | Not-determined |
| S1_extracted_k141_26971 | Not-determined |
| S1_extracted_k141_100465 | Not-determined |
| S1_extracted_k141_107138 | Low-quality |
| S1_extracted_k141_80498 | Not-determined |
| S1_extracted_k141_40407 | Not-determined |
| S1_extracted_k141_127224 | Not-determined |
| S1_extracted_k141_100469 | Not-determined |
| S1_extracted_k141_120514 | Not-determined |
| S1_extracted_k141_127226 | Not-determined |
| S1_extracted_k141_13737 | Not-determined |
| S1_extracted_k141_93849 | Not-determined |
| S1_extracted_k141_120516 | Low-quality |
| S1_extracted_k141_127227 | Not-determined |
| S1_extracted_k141_33689 | Not-determined |
| S1_extracted_k141_133755 | Low-quality |
| S1_extracted_k141_133756 | Not-determined |
| S1_extracted_k141_33690 | Not-determined |
| S1_extracted_k141_47060 | Not-determined |
| S1_extracted_k141_133757 | Not-determined |
| S1_extracted_k141_40413 | Not-determined |
| S1_extracted_k141_26984 | Low-quality |
| S1_extracted_k141_133758 | Low-quality |
| S1_extracted_k141_133760 | Not-determined |
| S1_extracted_k141_53800 | Not-determined |
| S1_extracted_k141_26988 | Not-determined |
| S1_extracted_k141_67057 | Low-quality |
| S1_extracted_k141_80503 | Low-quality |
| S1_extracted_k141_47066 | Not-determined |
| S1_extracted_k141_26990 | Not-determined |
| S1_extracted_k141_107145 | Low-quality |
| S1_extracted_k141_67058 | Not-determined |
| S1_extracted_k141_60241 | Not-determined |
| S1_extracted_k141_33695 | Not-determined |
| S1_extracted_k141_26995 | Not-determined |
| S1_extracted_k141_93856 | Not-determined |
| S1_extracted_k141_120527 | Not-determined |
| S1_extracted_k141_100482 | Low-quality |
| S1_extracted_k141_53811 | Not-determined |
| S1_extracted_k141_80512 | Not-determined |
| S1_extracted_k141_27001 | Not-determined |
| S1_extracted_k141_53814 | Not-determined |
| S1_extracted_k141_67065 | Not-determined |
| S1_extracted_k141_107148 | Not-determined |
| S1_extracted_k141_7044 | Not-determined |
| S1_extracted_k141_100486 | Not-determined |
| S1_extracted_k141_147095 | Low-quality |
| S1_extracted_k141_93866 | Not-determined |
| S1_extracted_k141_147096 | Not-determined |
| S1_extracted_k141_40430 | Not-determined |
| S1_extracted_k141_100488 | Not-determined |
| S1_extracted_k141_40431 | Not-determined |
| S1_extracted_k141_27004 | Low-quality |
| S1_extracted_k141_47079 | Not-determined |
| S1_extracted_k141_67069 | Not-determined |
| S1_extracted_k141_113810 | Not-determined |
| S1_extracted_k141_47081 | Not-determined |
| S1_extracted_k141_80522 | Not-determined |
| S1_extracted_k141_140516 | Low-quality |
| S1_extracted_k141_100492 | Not-determined |
| S1_extracted_k141_107149 | Low-quality |
| S1_extracted_k141_53825 | Low-quality |
| S1_extracted_k141_93870 | Not-determined |
| S1_extracted_k141_60258 | Not-determined |
| S1_extracted_k141_100495 | Not-determined |
| S1_extracted_k141_80524 | Not-determined |
| S1_extracted_k141_133768 | Low-quality |
| S1_extracted_k141_73791 | Not-determined |
| S1_extracted_k141_382 | Not-determined |
| S1_extracted_k141_127242 | Not-determined |
| S1_extracted_k141_73793 | Low-quality |
| S1_extracted_k141_383 | Not-determined |
| S1_extracted_k141_127243 | Not-determined |
| S1_extracted_k141_140522 | Low-quality |
| S1_extracted_k141_60262 | Low-quality |
| S1_extracted_k141_80527 | Not-determined |
| S1_extracted_k141_140523 | Not-determined |
| S1_extracted_k141_140525 | Not-determined |
| S1_extracted_k141_93876 | Not-determined |
| S1_extracted_k141_67072 | Not-determined |
| S1_extracted_k141_67073 | Not-determined |
| S1_extracted_k141_133771 | Not-determined |
| S1_extracted_k141_67074 | Not-determined |
| S1_extracted_k141_87172 | Not-determined |
| S1_extracted_k141_388 | Not-determined |
| S1_extracted_k141_93879 | Not-determined |
| S1_extracted_k141_53838 | Not-determined |
| S1_extracted_k141_107159 | Not-determined |
| S1_extracted_k141_113817 | Not-determined |
| S1_extracted_k141_27013 | Not-determined |
| S1_extracted_k141_100506 | Not-determined |
| S1_extracted_k141_53841 | Low-quality |
| S1_extracted_k141_67081 | Not-determined |
| S1_extracted_k141_80538 | Low-quality |
| S1_extracted_k141_33707 | Not-determined |
| S1_extracted_k141_73807 | Not-determined |
| S1_extracted_k141_140535 | Not-determined |
| S1_extracted_k141_127255 | Not-determined |
| S1_extracted_k141_13769 | Low-quality |
| S1_extracted_k141_60268 | Not-determined |
| S1_extracted_k141_73811 | Not-determined |
| S1_extracted_k141_113822 | Low-quality |
| S1_extracted_k141_60269 | Not-determined |
| S1_extracted_k141_33710 | Not-determined |
| S1_extracted_k141_73815 | Not-determined |
| S1_extracted_k141_13775 | Low-quality |
| S1_extracted_k141_133779 | Not-determined |
| S1_extracted_k141_53848 | Not-determined |
| S1_extracted_k141_20403 | Low-quality |
| S1_extracted_k141_13778 | Not-determined |
| S1_extracted_k141_140536 | Low-quality |
| S1_extracted_k141_47093 | Low-quality |
| S1_extracted_k141_73819 | Not-determined |
| S1_extracted_k141_40459 | Not-determined |
| S1_extracted_k141_107171 | Not-determined |
| S1_extracted_k141_7070 | Not-determined |
| S1_extracted_k141_133785 | Not-determined |
| S1_extracted_k141_93898 | Not-determined |
| S1_extracted_k141_153723 | Not-determined |
| S1_extracted_k141_93899 | Low-quality |
| S1_extracted_k141_113830 | Low-quality |
| S1_extracted_k141_47096 | Not-determined |
| S1_extracted_k141_153726 | Low-quality |
| S1_extracted_k141_113832 | Low-quality |
| S1_extracted_k141_80551 | Low-quality |
| S1_extracted_k141_113834 | Low-quality |
| S1_extracted_k141_127269 | Not-determined |
| S1_extracted_k141_13788 | Not-determined |
| S1_extracted_k141_40466 | Low-quality |
| S1_extracted_k141_40467 | Not-determined |
| S1_extracted_k141_133793 | Low-quality |
| S1_extracted_k141_113836 | Low-quality |
| S1_extracted_k141_93900 | Low-quality |
| S1_extracted_k141_392 | Not-determined |
| S1_extracted_k141_73827 | Not-determined |
| S1_extracted_k141_40468 | Low-quality |
| S1_extracted_k141_20416 | Not-determined |
| S1_extracted_k141_73830 | Not-determined |
| S1_extracted_k141_40471 | Not-determined |
| S1_extracted_k141_133798 | Not-determined |
| S1_extracted_k141_53859 | Low-quality |
| S1_extracted_k141_33716 | Not-determined |
| S1_extracted_k141_7083 | Not-determined |
| S1_extracted_k141_27034 | Not-determined |
| S1_extracted_k141_397 | Not-determined |
| S1_extracted_k141_398 | Not-determined |
| S1_extracted_k141_93904 | Not-determined |
| S1_extracted_k141_67100 | Not-determined |
| S1_extracted_k141_33719 | Not-determined |
| S1_extracted_k141_133802 | Not-determined |
| S1_extracted_k141_93906 | Not-determined |
| S1_extracted_k141_20423 | Not-determined |
| S1_extracted_k141_153729 | Not-determined |
| S1_extracted_k141_20424 | Not-determined |
| S1_extracted_k141_13797 | Not-determined |
| S1_extracted_k141_87192 | Not-determined |
| S1_extracted_k141_140542 | Not-determined |
| S1_extracted_k141_40474 | Not-determined |
| S1_extracted_k141_33725 | Not-determined |
| S1_extracted_k141_20428 | Not-determined |
| S1_extracted_k141_27047 | Low-quality |
| S1_extracted_k141_120559 | Not-determined |
| S1_extracted_k141_20430 | Not-determined |
| S1_extracted_k141_107195 | Not-determined |
| S1_extracted_k141_133809 | Not-determined |
| S1_extracted_k141_87202 | Not-determined |
| S1_extracted_k141_87205 | Not-determined |
| S1_extracted_k141_47113 | Low-quality |
| S1_extracted_k141_140552 | Not-determined |
| S1_extracted_k141_40487 | Not-determined |
| S1_extracted_k141_153742 | Not-determined |
| S1_extracted_k141_40488 | Low-quality |
| S1_extracted_k141_113854 | Low-quality |
| S1_extracted_k141_7097 | Not-determined |
| S1_extracted_k141_113857 | Not-determined |
| S1_extracted_k141_93915 | Low-quality |
| S1_extracted_k141_53871 | Not-determined |
| S1_extracted_k141_60293 | Not-determined |
| S1_extracted_k141_133822 | Not-determined |
| S1_extracted_k141_67122 | Low-quality |
| S1_extracted_k141_53877 | Not-determined |
| S1_extracted_k141_40495 | Not-determined |
| S1_extracted_k141_80578 | Low-quality |
| S1_extracted_k141_27055 | Not-determined |
| S1_extracted_k141_20439 | Not-determined |
| S1_extracted_k141_120575 | Low-quality |
| S1_extracted_k141_120578 | Not-determined |
| S1_extracted_k141_67128 | Not-determined |
| S1_extracted_k141_113869 | Not-determined |
| S1_extracted_k141_100550 | Not-determined |
| S1_extracted_k141_40502 | Not-determined |
| S1_extracted_k141_60304 | Not-determined |
| S1_extracted_k141_13829 | Not-determined |
| S1_extracted_k141_27060 | Low-quality |
| S1_extracted_k141_20443 | Not-determined |
| S1_extracted_k141_13832 | Not-determined |
| S1_extracted_k141_20446 | Not-determined |
| S1_extracted_k141_13835 | Low-quality |
| S1_extracted_k141_120583 | Not-determined |
| S1_extracted_k141_53891 | Low-quality |
| S1_extracted_k141_147112 | Not-determined |
| S1_extracted_k141_133836 | Low-quality |
| S1_extracted_k141_93934 | Not-determined |
| S1_extracted_k141_13836 | Low-quality |
| S1_extracted_k141_27065 | Low-quality |
| S1_extracted_k141_40511 | Low-quality |
| S1_extracted_k141_107213 | Not-determined |
| S1_extracted_k141_93935 | Not-determined |
| S1_extracted_k141_27066 | Not-determined |
| S1_extracted_k141_13837 | Low-quality |
| S1_extracted_k141_20449 | Not-determined |
| S1_extracted_k141_87219 | Not-determined |
| S1_extracted_k141_67138 | Not-determined |
| S1_extracted_k141_27067 | Low-quality |
| S1_extracted_k141_127286 | Not-determined |
| S1_extracted_k141_93937 | Not-determined |
| S1_extracted_k141_53894 | Not-determined |
| S1_extracted_k141_40512 | Not-determined |
| S1_extracted_k141_13840 | Not-determined |
| S1_extracted_k141_93938 | Low-quality |
| S1_extracted_k141_93940 | Not-determined |
| S1_extracted_k141_7128 | Low-quality |
| S1_extracted_k141_33747 | Low-quality |
| S1_extracted_k141_107219 | Not-determined |
| S1_extracted_k141_33748 | Not-determined |
| S1_extracted_k141_107220 | Not-determined |
| S1_extracted_k141_40517 | Not-determined |
| S1_extracted_k141_100568 | Not-determined |
| S1_extracted_k141_140573 | Low-quality |
| S1_extracted_k141_113882 | Not-determined |
| S1_extracted_k141_87225 | Low-quality |
| S1_extracted_k141_107226 | Not-determined |
| S1_extracted_k141_73852 | Not-determined |
| S1_extracted_k141_100570 | Not-determined |
| S1_extracted_k141_147120 | Low-quality |
| S1_extracted_k141_100572 | Low-quality |
| S1_extracted_k141_147121 | Not-determined |
| S1_extracted_k141_33753 | Not-determined |
| S1_extracted_k141_107230 | Low-quality |
| S1_extracted_k141_20459 | Not-determined |
| S1_extracted_k141_153743 | Not-determined |
| S1_extracted_k141_60321 | Not-determined |
| S1_extracted_k141_53904 | Not-determined |
| S1_extracted_k141_113884 | Low-quality |
| S1_extracted_k141_153745 | Not-determined |
| S1_extracted_k141_47143 | Not-determined |
| S1_extracted_k141_60324 | Low-quality |
| S1_extracted_k141_113888 | Not-determined |
| S1_extracted_k141_120591 | Low-quality |
| S1_extracted_k141_7134 | Not-determined |
| S1_extracted_k141_60328 | Not-determined |
| S1_extracted_k141_27085 | Not-determined |
| S1_extracted_k141_13860 | Not-determined |
| S1_extracted_k141_27086 | Not-determined |
| S1_extracted_k141_100575 | Not-determined |
| S1_extracted_k141_53911 | Not-determined |
| S1_extracted_k141_7137 | Not-determined |
| S1_extracted_k141_100576 | Not-determined |
| S1_extracted_k141_438 | Not-determined |
| S1_extracted_k141_60330 | Not-determined |
| S1_extracted_k141_87236 | Not-determined |
| S1_extracted_k141_53914 | Not-determined |
| S1_extracted_k141_133856 | Low-quality |
| S1_extracted_k141_80608 | Not-determined |
| S1_extracted_k141_100581 | Not-determined |
| S1_extracted_k141_80609 | Not-determined |
| S1_extracted_k141_107240 | Not-determined |
| S1_extracted_k141_80610 | Not-determined |
| S1_extracted_k141_33757 | Not-determined |
| S1_extracted_k141_73868 | Not-determined |
| S1_extracted_k141_107241 | Not-determined |
| S1_extracted_k141_40537 | Not-determined |
| S1_extracted_k141_73870 | Not-determined |
| S1_extracted_k141_33760 | Not-determined |
| S1_extracted_k141_93952 | Not-determined |
| S1_extracted_k141_13862 | Low-quality |
| S1_extracted_k141_53922 | Not-determined |
| S1_extracted_k141_33762 | Not-determined |
| S1_extracted_k141_7143 | Low-quality |
| S1_extracted_k141_120600 | Not-determined |
| S1_extracted_k141_444 | Not-determined |
| S1_extracted_k141_73873 | Low-quality |
| S1_extracted_k141_87246 | Not-determined |
| S1_extracted_k141_127312 | Low-quality |
| S1_extracted_k141_73874 | Not-determined |
| S1_extracted_k141_27096 | Not-determined |
| S1_extracted_k141_87248 | Not-determined |
| S1_extracted_k141_100589 | Not-determined |
| S1_extracted_k141_47162 | Not-determined |
| S1_extracted_k141_113891 | Not-determined |
| S1_extracted_k141_33767 | Not-determined |
| S1_extracted_k141_127316 | Not-determined |
| S1_extracted_k141_127319 | Not-determined |
| S1_extracted_k141_7151 | Low-quality |
| S1_extracted_k141_33771 | Not-determined |
| S1_extracted_k141_100596 | Not-determined |
| S1_extracted_k141_73881 | Not-determined |
| S1_extracted_k141_33772 | Not-determined |
| S1_extracted_k141_140595 | Not-determined |
| S1_extracted_k141_100599 | Not-determined |
| S1_extracted_k141_93960 | Not-determined |
| S1_extracted_k141_113898 | Not-determined |
| S1_extracted_k141_107256 | Not-determined |
| S1_extracted_k141_7155 | Not-determined |
| S1_extracted_k141_20469 | Low-quality |
| S1_extracted_k141_53938 | Not-determined |
| S1_extracted_k141_127332 | Not-determined |
| S1_extracted_k141_27105 | Low-quality |
| S1_extracted_k141_133873 | Not-determined |
| S1_extracted_k141_107261 | Not-determined |
| S1_extracted_k141_27106 | Not-determined |
| S1_extracted_k141_127335 | Not-determined |
| S1_extracted_k141_452 | Not-determined |
| S1_extracted_k141_107262 | Not-determined |
| S1_extracted_k141_93964 | Not-determined |
| S1_extracted_k141_33780 | Not-determined |
| S1_extracted_k141_27112 | Not-determined |
| S1_extracted_k141_13880 | Not-determined |
| S1_extracted_k141_40563 | Low-quality |
| S1_extracted_k141_120609 | Low-quality |
| S1_extracted_k141_73885 | Not-determined |
| S1_extracted_k141_80630 | Not-determined |
| S1_extracted_k141_27118 | Not-determined |
| S1_extracted_k141_33785 | Not-determined |
| S1_extracted_k141_87267 | Not-determined |
| S1_extracted_k141_73888 | Not-determined |
| S1_extracted_k141_53946 | Not-determined |
| S1_extracted_k141_120612 | Not-determined |
| S1_extracted_k141_13885 | Low-quality |
| S1_extracted_k141_127339 | Low-quality |
| S1_extracted_k141_107266 | Not-determined |
| S1_extracted_k141_7166 | Low-quality |
| S1_extracted_k141_33788 | Low-quality |
| S1_extracted_k141_100616 | Low-quality |
| S1_extracted_k141_133889 | Not-determined |
| S1_extracted_k141_140621 | Not-determined |
| S1_extracted_k141_100617 | Not-determined |
| S1_extracted_k141_133890 | Not-determined |
| S1_extracted_k141_73894 | Not-determined |
| S1_extracted_k141_133891 | Not-determined |
| S1_extracted_k141_120617 | Not-determined |
| S1_extracted_k141_7172 | Low-quality |
| S1_extracted_k141_87276 | Not-determined |
| S1_extracted_k141_80639 | Not-determined |
| S1_extracted_k141_13895 | Low-quality |
| S1_extracted_k141_120620 | Not-determined |
| S1_extracted_k141_80642 | Not-determined |
| S1_extracted_k141_53965 | Not-determined |
| S1_extracted_k141_60344 | Not-determined |
| S1_extracted_k141_27129 | Not-determined |
| S1_extracted_k141_53967 | Low-quality |
| S1_extracted_k141_140633 | Not-determined |
| S1_extracted_k141_113928 | Not-determined |
| S1_extracted_k141_33803 | Not-determined |
| S1_extracted_k141_133906 | Not-determined |
| S1_extracted_k141_80647 | Not-determined |
| S1_extracted_k141_100633 | Not-determined |
| S1_extracted_k141_80652 | Not-determined |
| S1_extracted_k141_120624 | Not-determined |
| S1_extracted_k141_100634 | Not-determined |
| S1_extracted_k141_67173 | Not-determined |
| S1_extracted_k141_153755 | Low-quality |
| S1_extracted_k141_113934 | Not-determined |
| S1_extracted_k141_93977 | Low-quality |
| S1_extracted_k141_80655 | Not-determined |
| S1_extracted_k141_140644 | Not-determined |

| | |
|---|---|
| S1_extracted_k141_93979 | Not-determined |
| S1_extracted_k141_100640 | Low-quality |
| S1_extracted_k141_60350 | Not-determined |
| S1_extracted_k141_153759 | Not-determined |
| S1_extracted_k141_40587 | Low-quality |
| S1_extracted_k141_100642 | Not-determined |
| S1_extracted_k141_73914 | Not-determined |
| S1_extracted_k141_153761 | Not-determined |
| S1_extracted_k141_33815 | Not-determined |
| S1_extracted_k141_73915 | Not-determined |
| S1_extracted_k141_20513 | Not-determined |
| S1_extracted_k141_80660 | Not-determined |
| S1_extracted_k141_53978 | Not-determined |
| S1_extracted_k141_107267 | Low-quality |
| S1_extracted_k141_107268 | Not-determined |
| S1_extracted_k141_40592 | Not-determined |
| S1_extracted_k141_93982 | Not-determined |
| S1_extracted_k141_40594 | Not-determined |
| S1_extracted_k141_40595 | Low-quality |
| S1_extracted_k141_60355 | Low-quality |
| S1_extracted_k141_140652 | Low-quality |
| S1_extracted_k141_40597 | Not-determined |
| S1_extracted_k141_478 | Not-determined |
| S1_extracted_k141_140654 | Not-determined |
| S1_extracted_k141_140655 | Not-determined |
| S1_extracted_k141_67185 | Not-determined |
| S1_extracted_k141_93990 | Not-determined |
| S1_extracted_k141_140656 | Low-quality |
| S1_extracted_k141_40600 | Not-determined |
| S1_extracted_k141_93991 | Not-determined |
| S1_extracted_k141_40601 | Low-quality |
| S1_extracted_k141_60359 | Not-determined |
| S1_extracted_k141_153766 | Not-determined |
| S1_extracted_k141_33823 | Not-determined |
| S1_extracted_k141_113950 | Not-determined |
| S1_extracted_k141_100654 | Not-determined |
| S1_extracted_k141_87309 | Low-quality |
| S1_extracted_k141_73922 | Not-determined |
| S1_extracted_k141_13916 | Not-determined |
| S1_extracted_k141_40610 | Not-determined |
| S1_extracted_k141_120640 | Low-quality |
| S1_extracted_k141_7202 | Low-quality |
| S1_extracted_k141_93996 | Not-determined |
| S1_extracted_k141_140664 | Not-determined |
| S1_extracted_k141_13919 | Not-determined |
| S1_extracted_k141_120645 | Not-determined |
| S1_extracted_k141_87316 | Low-quality |
| S1_extracted_k141_47209 | Not-determined |
| S1_extracted_k141_40622 | Not-determined |
| S1_extracted_k141_140669 | Not-determined |
| S1_extracted_k141_80669 | Not-determined |
| S1_extracted_k141_140670 | Not-determined |
| S1_extracted_k141_127368 | Not-determined |
| S1_extracted_k141_140671 | Not-determined |
| S1_extracted_k141_153779 | Not-determined |
| S1_extracted_k141_13931 | Not-determined |
| S1_extracted_k141_153780 | Low-quality |
| S1_extracted_k141_140675 | Low-quality |
| S1_extracted_k141_60372 | Not-determined |
| S1_extracted_k141_47213 | Not-determined |
| S1_extracted_k141_13933 | Not-determined |
| S1_extracted_k141_20525 | Not-determined |
| S1_extracted_k141_153782 | Not-determined |
| S1_extracted_k141_94008 | Not-determined |
| S1_extracted_k141_7216 | Low-quality |
| S1_extracted_k141_60373 | Low-quality |
| S1_extracted_k141_60374 | Not-determined |
| S1_extracted_k141_87322 | Not-determined |
| S1_extracted_k141_67209 | Not-determined |
| S1_extracted_k141_94012 | Not-determined |
| S1_extracted_k141_100662 | Not-determined |
| S1_extracted_k141_73943 | Low-quality |
| S1_extracted_k141_100663 | Not-determined |
| S1_extracted_k141_100664 | Not-determined |
| S1_extracted_k141_7219 | Not-determined |
| S1_extracted_k141_140685 | Not-determined |
| S1_extracted_k141_60378 | Not-determined |
| S1_extracted_k141_127374 | Low-quality |
| S1_extracted_k141_7221 | Not-determined |
| S1_extracted_k141_113957 | Not-determined |
| S1_extracted_k141_140687 | Not-determined |
| S1_extracted_k141_147137 | Not-determined |
| S1_extracted_k141_133938 | Not-determined |
| S1_extracted_k141_133940 | Not-determined |
| S1_extracted_k141_100666 | Not-determined |
| S1_extracted_k141_113959 | Low-quality |
| S1_extracted_k141_503 | Not-determined |
| S1_extracted_k141_113961 | Not-determined |
| S1_extracted_k141_133942 | Not-determined |
| S1_extracted_k141_140695 | Not-determined |
| S1_extracted_k141_60380 | Low-quality |
| S1_extracted_k141_13959 | Not-determined |
| S1_extracted_k141_100673 | Low-quality |
| S1_extracted_k141_140698 | Not-determined |
| S1_extracted_k141_127382 | Not-determined |
| S1_extracted_k141_113971 | Not-determined |
| S1_extracted_k141_87334 | Not-determined |
| S1_extracted_k141_27179 | Not-determined |
| S1_extracted_k141_127384 | Not-determined |
| S1_extracted_k141_133951 | Low-quality |
| S1_extracted_k141_87336 | Not-determined |
| S1_extracted_k141_120660 | Low-quality |
| S1_extracted_k141_33850 | Not-determined |
| S1_extracted_k141_7243 | Not-determined |
| S1_extracted_k141_508 | Not-determined |
| S1_extracted_k141_153805 | Not-determined |
| S1_extracted_k141_7246 | Not-determined |
| S1_extracted_k141_27184 | Not-determined |
| S1_extracted_k141_87341 | Not-determined |
| S1_extracted_k141_47235 | Not-determined |
| S1_extracted_k141_147143 | Low-quality |
| S1_extracted_k141_147144 | Not-determined |
| S1_extracted_k141_33852 | Not-determined |
| S1_extracted_k141_27190 | Not-determined |
| S1_extracted_k141_40643 | Not-determined |
| S1_extracted_k141_33853 | Not-determined |
| S1_extracted_k141_80691 | Low-quality |
| S1_extracted_k141_67231 | Not-determined |
| S1_extracted_k141_27195 | Low-quality |
| S1_extracted_k141_94036 | Low-quality |
| S1_extracted_k141_27196 | Not-determined |
| S1_extracted_k141_127395 | Not-determined |
| S1_extracted_k141_127396 | Not-determined |
| S1_extracted_k141_515 | Not-determined |
| S1_extracted_k141_67236 | Not-determined |
| S1_extracted_k141_516 | Low-quality |
| S1_extracted_k141_113974 | Low-quality |
| S1_extracted_k141_140707 | Not-determined |
| S1_extracted_k141_67237 | Not-determined |
| S1_extracted_k141_67239 | Low-quality |
| S1_extracted_k141_80699 | Low-quality |
| S1_extracted_k141_127402 | Not-determined |
| S1_extracted_k141_7264 | Low-quality |
| S1_extracted_k141_67243 | Not-determined |
| S1_extracted_k141_94048 | Not-determined |
| S1_extracted_k141_120678 | Not-determined |
| S1_extracted_k141_113979 | Not-determined |
| S1_extracted_k141_153811 | Low-quality |
| S1_extracted_k141_47245 | Not-determined |
| S1_extracted_k141_153812 | Low-quality |
| S1_extracted_k141_7268 | Not-determined |
| S1_extracted_k141_33866 | Not-determined |
| S1_extracted_k141_113983 | Not-determined |
| S1_extracted_k141_80708 | Not-determined |
| S1_extracted_k141_73969 | Not-determined |
| S1_extracted_k141_80709 | Not-determined |
| S1_extracted_k141_133960 | Not-determined |
| S1_extracted_k141_87353 | Low-quality |
| S1_extracted_k141_54014 | Not-determined |
| S1_extracted_k141_113985 | Not-determined |
| S1_extracted_k141_80713 | Not-determined |
| S1_extracted_k141_153816 | Not-determined |
| S1_extracted_k141_54016 | Not-determined |
| S1_extracted_k141_47253 | Not-determined |
| S1_extracted_k141_120683 | Not-determined |
| S1_extracted_k141_113993 | Not-determined |
| S1_extracted_k141_107305 | Not-determined |
| S1_extracted_k141_40670 | Not-determined |
| S1_extracted_k141_14001 | Not-determined |
| S1_extracted_k141_7279 | Not-determined |
| S1_extracted_k141_113998 | Not-determined |
| S1_extracted_k141_133971 | Not-determined |
| S1_extracted_k141_7280 | Not-determined |
| S1_extracted_k141_54020 | Low-quality |
| S1_extracted_k141_127424 | Low-quality |
| S1_extracted_k141_67256 | Low-quality |
| S1_extracted_k141_120685 | Low-quality |
| S1_extracted_k141_127425 | Not-determined |
| S1_extracted_k141_120686 | Not-determined |
| S1_extracted_k141_114002 | Not-determined |
| S1_extracted_k141_539 | Not-determined |
| S1_extracted_k141_114005 | Not-determined |
| S1_extracted_k141_67257 | Low-quality |
| S1_extracted_k141_27218 | Low-quality |
| S1_extracted_k141_114007 | Not-determined |
| S1_extracted_k141_120695 | Low-quality |
| S1_extracted_k141_120698 | Not-determined |
| S1_extracted_k141_60386 | Not-determined |
| S1_extracted_k141_27223 | Low-quality |
| S1_extracted_k141_140730 | Not-determined |
| S1_extracted_k141_27224 | Not-determined |
| S1_extracted_k141_127438 | Not-determined |
| S1_extracted_k141_140732 | Not-determined |
| S1_extracted_k141_40684 | Low-quality |
| S1_extracted_k141_40685 | Low-quality |
| S1_extracted_k141_7293 | Not-determined |
| S1_extracted_k141_153834 | Not-determined |
| S1_extracted_k141_14014 | Low-quality |
| S1_extracted_k141_47274 | Not-determined |
| S1_extracted_k141_73999 | Not-determined |
| S1_extracted_k141_20552 | Not-determined |
| S1_extracted_k141_120703 | Not-determined |
| S1_extracted_k141_100717 | Not-determined |
| S1_extracted_k141_100719 | Not-determined |
| S1_extracted_k141_133986 | Not-determined |
| S1_extracted_k141_100720 | Low-quality |
| S1_extracted_k141_100722 | Not-determined |
| S1_extracted_k141_27233 | Not-determined |
| S1_extracted_k141_67265 | Not-determined |
| S1_extracted_k141_100723 | Not-determined |
| S1_extracted_k141_27234 | Not-determined |
| S1_extracted_k141_80739 | Not-determined |
| S1_extracted_k141_54039 | Low-quality |
| S1_extracted_k141_127449 | Not-determined |
| S1_extracted_k141_33883 | Not-determined |
| S1_extracted_k141_100727 | Not-determined |
| S1_extracted_k141_74008 | Not-determined |
| S1_extracted_k141_127450 | Low-quality |
| S1_extracted_k141_67268 | Not-determined |
| S1_extracted_k141_74010 | Not-determined |
| S1_extracted_k141_33887 | Not-determined |
| S1_extracted_k141_67270 | Not-determined |
| S1_extracted_k141_54044 | Not-determined |
| S1_extracted_k141_27241 | Not-determined |
| S1_extracted_k141_100732 | Not-determined |
| S1_extracted_k141_47287 | Not-determined |
| S1_extracted_k141_133997 | Not-determined |
| S1_extracted_k141_47288 | Not-determined |
| S1_extracted_k141_120712 | Not-determined |
| S1_extracted_k141_140748 | Not-determined |
| S1_extracted_k141_14033 | Not-determined |
| S1_extracted_k141_47292 | Not-determined |
| S1_extracted_k141_7313 | Not-determined |
| S1_extracted_k141_114028 | Not-determined |
| S1_extracted_k141_127465 | Low-quality |
| S1_extracted_k141_120721 | Not-determined |
| S1_extracted_k141_127467 | Not-determined |
| S1_extracted_k141_100741 | Low-quality |
| S1_extracted_k141_7318 | Not-determined |
| S1_extracted_k141_100742 | Not-determined |
| S1_extracted_k141_7319 | Not-determined |
| S1_extracted_k141_74028 | Not-determined |
| S1_extracted_k141_100746 | Not-determined |
| S1_extracted_k141_100747 | Not-determined |
| S1_extracted_k141_87394 | Not-determined |
| S1_extracted_k141_153849 | Not-determined |
| S1_extracted_k141_40709 | Not-determined |
| S1_extracted_k141_127475 | Not-determined |
| S1_extracted_k141_134012 | Low-quality |
| S1_extracted_k141_80761 | Not-determined |
| S1_extracted_k141_127477 | Not-determined |
| S1_extracted_k141_7328 | Not-determined |
| S1_extracted_k141_134015 | Not-determined |
| S1_extracted_k141_127481 | Not-determined |
| S1_extracted_k141_563 | Not-determined |
| S1_extracted_k141_67285 | Not-determined |
| S1_extracted_k141_134018 | Not-determined |
| S1_extracted_k141_67287 | Not-determined |
| S1_extracted_k141_100761 | Low-quality |
| S1_extracted_k141_67288 | Not-determined |
| S1_extracted_k141_127487 | Not-determined |
| S1_extracted_k141_74039 | Not-determined |
| S1_extracted_k141_27269 | Not-determined |
| S1_extracted_k141_14056 | Not-determined |
| S1_extracted_k141_114042 | Low-quality |
| S1_extracted_k141_20574 | Not-determined |
| S1_extracted_k141_14058 | Not-determined |
| S1_extracted_k141_54075 | Not-determined |
| S1_extracted_k141_87415 | Not-determined |
| S1_extracted_k141_14059 | Not-determined |
| S1_extracted_k141_140767 | Low-quality |
| S1_extracted_k141_80776 | Not-determined |
| S1_extracted_k141_7341 | Not-determined |
| S1_extracted_k141_14065 | Low-quality |
| S1_extracted_k141_120737 | Low-quality |
| S1_extracted_k141_120741 | Not-determined |
| S1_extracted_k141_114048 | Not-determined |
| S1_extracted_k141_568 | Not-determined |
| S1_extracted_k141_140772 | Not-determined |
| S1_extracted_k141_20576 | Low-quality |
| S1_extracted_k141_7346 | Not-determined |
| S1_extracted_k141_27283 | Low-quality |
| S1_extracted_k141_100769 | Low-quality |
| S1_extracted_k141_87417 | Not-determined |
| S1_extracted_k141_120745 | Not-determined |
| S1_extracted_k141_7351 | Low-quality |
| S1_extracted_k141_87418 | Low-quality |
| S1_extracted_k141_54088 | Not-determined |
| S1_extracted_k141_114051 | Not-determined |
| S1_extracted_k141_60393 | Not-determined |
| S1_extracted_k141_120747 | Not-determined |
| S1_extracted_k141_80788 | Not-determined |
| S1_extracted_k141_127509 | Not-determined |
| S1_extracted_k141_54094 | Not-determined |
| S1_extracted_k141_27286 | Not-determined |
| S1_extracted_k141_54097 | Not-determined |
| S1_extracted_k141_87426 | Not-determined |
| S1_extracted_k141_120753 | Not-determined |
| S1_extracted_k141_74059 | Not-determined |
| S1_extracted_k141_20585 | Not-determined |
| S1_extracted_k141_7358 | Not-determined |
| S1_extracted_k141_40735 | Low-quality |
| S1_extracted_k141_127518 | Not-determined |
| S1_extracted_k141_80796 | Not-determined |
| S1_extracted_k141_47332 | Not-determined |
| S1_extracted_k141_114070 | Not-determined |
| S1_extracted_k141_67303 | Low-quality |
| S1_extracted_k141_114071 | Not-determined |
| S1_extracted_k141_40737 | Not-determined |
| S1_extracted_k141_127520 | Low-quality |
| S1_extracted_k141_67307 | Not-determined |
| S1_extracted_k141_87434 | Not-determined |
| S1_extracted_k141_60406 | Not-determined |
| S1_extracted_k141_100782 | Not-determined |
| S1_extracted_k141_140791 | Not-determined |
| S1_extracted_k141_578 | Low-quality |
| S1_extracted_k141_140792 | Not-determined |
| S1_extracted_k141_100783 | Not-determined |
| S1_extracted_k141_33917 | Not-determined |
| S1_extracted_k141_33918 | Not-determined |
| S1_extracted_k141_47340 | Not-determined |
| S1_extracted_k141_74073 | Not-determined |
| S1_extracted_k141_120766 | Low-quality |
| S1_extracted_k141_134030 | Not-determined |
| S1_extracted_k141_14092 | Not-determined |
| S1_extracted_k141_27303 | Not-determined |
| S12_extracted_k141_79003 | Medium-quality |
| S1_extracted_k141_134032 | Not-determined |
| S1_extracted_k141_140797 | Low-quality |
| S1_extracted_k141_114080 | Not-determined |
| S1_extracted_k141_40748 | Not-determined |
| S1_extracted_k141_127528 | Not-determined |
| S1_extracted_k141_40749 | Not-determined |
| S1_extracted_k141_127529 | Not-determined |
| S1_extracted_k141_27307 | Not-determined |
| S1_extracted_k141_140801 | Not-determined |
| S1_extracted_k141_127531 | Not-determined |
| S1_extracted_k141_100791 | Not-determined |
| S1_extracted_k141_74084 | Not-determined |
| S1_extracted_k141_27309 | Not-determined |
| S1_extracted_k141_147171 | Not-determined |
| S1_extracted_k141_120777 | Not-determined |
| S1_extracted_k141_147172 | Not-determined |
| S1_extracted_k141_87442 | Not-determined |
| S1_extracted_k141_127533 | Not-determined |
| S1_extracted_k141_40756 | Low-quality |
| S1_extracted_k141_47352 | Low-quality |
| S1_extracted_k141_100796 | Not-determined |
| S1_extracted_k141_60415 | Not-determined |
| S1_extracted_k141_67325 | Low-quality |
| S1_extracted_k141_33930 | Low-quality |
| S1_extracted_k141_87446 | Low-quality |
| S1_extracted_k141_60417 | Not-determined |
| S1_extracted_k141_7384 | Not-determined |
| S1_extracted_k141_100799 | Not-determined |
| S1_extracted_k141_94127 | Not-determined |
| S1_extracted_k141_33935 | Not-determined |
| S1_extracted_k141_47361 | Low-quality |
| S1_extracted_k141_67330 | Low-quality |
| S1_extracted_k141_14114 | Not-determined |
| S1_extracted_k141_153886 | Low-quality |
| S1_extracted_k141_107334 | Not-determined |
| S1_extracted_k141_120788 | Not-determined |
| S1_extracted_k141_33939 | Not-determined |
| S1_extracted_k141_27315 | Not-determined |
| S1_extracted_k141_33941 | Not-determined |
| S1_extracted_k141_100810 | Not-determined |
| S1_extracted_k141_592 | Not-determined |
| S1_extracted_k141_134057 | Not-determined |
| S1_extracted_k141_67338 | Low-quality |
| S1_extracted_k141_127548 | Not-determined |
| S1_extracted_k141_54133 | Not-determined |
| S1_extracted_k141_153893 | Low-quality |
| S1_extracted_k141_120793 | Not-determined |
| S1_extracted_k141_153894 | Not-determined |
| S1_extracted_k141_14126 | Not-determined |
| S1_extracted_k141_40765 | Low-quality |
| S1_extracted_k141_74107 | Not-determined |
| S1_extracted_k141_40766 | Not-determined |
| S1_extracted_k141_47375 | Not-determined |
| S1_extracted_k141_87453 | Not-determined |
| S1_extracted_k141_100816 | Not-determined |
| S1_extracted_k141_100817 | Not-determined |
| S1_extracted_k141_127555 | Not-determined |
| S1_extracted_k141_14133 | Low-quality |
| S1_extracted_k141_20632 | Not-determined |
| S1_extracted_k141_27321 | Low-quality |
| S1_extracted_k141_20633 | Low-quality |
| S1_extracted_k141_134069 | Not-determined |
| S1_extracted_k141_47380 | Not-determined |
| S1_extracted_k141_127560 | Not-determined |
| S1_extracted_k141_40771 | Not-determined |
| S1_extracted_k141_47381 | Not-determined |
| S1_extracted_k141_33950 | Low-quality |
| S1_extracted_k141_47382 | Not-determined |
| S1_extracted_k141_47383 | Not-determined |
| S1_extracted_k141_140825 | Not-determined |
| S1_extracted_k141_100820 | Low-quality |
| S1_extracted_k141_134075 | Not-determined |
| S1_extracted_k141_87463 | Not-determined |
| S1_extracted_k141_47386 | Not-determined |
| S1_extracted_k141_74113 | Low-quality |
| S1_extracted_k141_134078 | Not-determined |
| S1_extracted_k141_33955 | Not-determined |
| S1_extracted_k141_100823 | Low-quality |
| S1_extracted_k141_40776 | Not-determined |
| S1_extracted_k141_107358 | Not-determined |
| S1_extracted_k141_94153 | Low-quality |
| S1_extracted_k141_7407 | Not-determined |
| S1_extracted_k141_100825 | Not-determined |
| S1_extracted_k141_100826 | Not-determined |
| S1_extracted_k141_94156 | Not-determined |
| S1_extracted_k141_114108 | Not-determined |
| S1_extracted_k141_47396 | Not-determined |
| S1_extracted_k141_134084 | Not-determined |
| S1_extracted_k141_54161 | Not-determined |
| S1_extracted_k141_134086 | Not-determined |
| S1_extracted_k141_134088 | Not-determined |
| S1_extracted_k141_611 | Not-determined |
| S1_extracted_k141_127577 | Not-determined |
| S1_extracted_k141_94166 | Not-determined |
| S1_extracted_k141_14152 | Not-determined |
| S1_extracted_k141_87483 | Not-determined |
| S1_extracted_k141_74126 | Not-determined |
| S1_extracted_k141_87486 | Not-determined |
| S1_extracted_k141_47402 | Low-quality |
| S1_extracted_k141_134094 | Not-determined |
| S1_extracted_k141_20655 | Not-determined |
| S1_extracted_k141_140847 | Not-determined |
| S1_extracted_k141_40789 | Low-quality |
| S1_extracted_k141_140852 | Not-determined |
| S1_extracted_k141_153899 | Not-determined |
| S1_extracted_k141_134097 | Not-determined |
| S1_extracted_k141_67361 | Low-quality |
| S1_extracted_k141_47406 | Not-determined |
| S1_extracted_k141_107376 | Not-determined |
| S1_extracted_k141_100841 | Not-determined |
| S1_extracted_k141_14164 | Not-determined |
| S1_extracted_k141_27348 | Low-quality |
| S1_extracted_k141_54169 | Not-determined |
| S1_extracted_k141_67364 | Not-determined |
| S1_extracted_k141_107380 | Not-determined |
| S1_extracted_k141_87493 | Low-quality |
| S1_extracted_k141_14170 | Not-determined |
| S1_extracted_k141_60439 | Low-quality |
| S1_extracted_k141_54176 | Not-determined |
| S1_extracted_k141_87495 | Not-determined |
| S1_extracted_k141_74136 | Low-quality |
| S1_extracted_k141_107384 | Not-determined |
| S1_extracted_k141_100844 | Low-quality |
| S1_extracted_k141_134104 | Not-determined |
| S1_extracted_k141_67367 | Not-determined |
| S1_extracted_k141_40799 | Not-determined |
| S1_extracted_k141_60441 | Low-quality |
| S1_extracted_k141_74140 | Not-determined |
| S1_extracted_k141_100848 | Not-determined |
| S1_extracted_k141_33984 | Not-determined |
| S1_extracted_k141_120818 | Not-determined |
| S1_extracted_k141_54182 | Low-quality |
| S1_extracted_k141_74145 | Low-quality |
| S1_extracted_k141_107387 | Low-quality |
| S1_extracted_k141_20670 | Not-determined |
| S1_extracted_k141_120823 | Not-determined |
| S1_extracted_k141_114116 | Low-quality |
| S1_extracted_k141_633 | Not-determined |
| S1_extracted_k141_67370 | Not-determined |
| S1_extracted_k141_634 | Not-determined |
| S1_extracted_k141_100854 | Not-determined |
| S1_extracted_k141_140865 | Not-determined |
| S1_extracted_k141_20674 | Not-determined |
| S1_extracted_k141_114119 | Not-determined |
| S1_extracted_k141_114120 | Not-determined |
| S1_extracted_k141_67372 | Low-quality |
| S1_extracted_k141_140868 | Low-quality |
| S1_extracted_k141_67373 | Not-determined |
| S1_extracted_k141_94185 | Not-determined |
| S1_extracted_k141_153901 | Not-determined |
| S1_extracted_k141_114122 | Not-determined |
| S1_extracted_k141_80836 | Not-determined |
| S1_extracted_k141_87510 | Not-determined |
| S1_extracted_k141_14182 | Not-determined |
| S1_extracted_k141_140870 | Not-determined |
| S1_extracted_k141_80838 | Not-determined |
| S1_extracted_k141_67377 | Not-determined |
| S1_extracted_k141_120826 | Not-determined |
| S1_extracted_k141_14183 | Not-determined |
| S1_extracted_k141_54190 | Not-determined |
| S1_extracted_k141_33997 | Low-quality |
| S1_extracted_k141_100856 | Not-determined |
| S1_extracted_k141_114129 | Not-determined |
| S1_extracted_k141_114130 | Not-determined |
| S1_extracted_k141_74154 | Not-determined |
| S1_extracted_k141_47439 | Not-determined |
| S1_extracted_k141_14187 | Not-determined |
| S1_extracted_k141_80844 | Low-quality |
| S1_extracted_k141_54194 | Not-determined |
| S1_extracted_k141_47442 | Not-determined |
| S1_extracted_k141_67385 | Not-determined |
| S1_extracted_k141_100860 | Low-quality |
| S1_extracted_k141_134120 | Not-determined |
| S1_extracted_k141_54197 | Not-determined |
| S1_extracted_k141_60454 | Not-determined |
| S1_extracted_k141_100863 | Not-determined |
| S1_extracted_k141_60455 | Low-quality |
| S1_extracted_k141_40821 | Not-determined |
| S1_extracted_k141_643 | Not-determined |
| S1_extracted_k141_60456 | Not-determined |
| S1_extracted_k141_80854 | Not-determined |
| S1_extracted_k141_140879 | Low-quality |
| S1_extracted_k141_645 | Not-determined |
| S1_extracted_k141_14201 | Low-quality |
| S1_extracted_k141_80859 | Not-determined |
| S1_extracted_k141_60460 | Not-determined |
| S1_extracted_k141_140884 | Not-determined |
| S1_extracted_k141_649 | Not-determined |
| S1_extracted_k141_54204 | Not-determined |
| S1_extracted_k141_40824 | Low-quality |
| S1_extracted_k141_74171 | Not-determined |
| S1_extracted_k141_134128 | Low-quality |
| S1_extracted_k141_54206 | Not-determined |
| S1_extracted_k141_60462 | Not-determined |
| S1_extracted_k141_34012 | Not-determined |
| S1_extracted_k141_27381 | Not-determined |
| S1_extracted_k141_147202 | Low-quality |
| S1_extracted_k141_47450 | Not-determined |
| S1_extracted_k141_100878 | Not-determined |
| S1_extracted_k141_60466 | Not-determined |
| S1_extracted_k141_47453 | Not-determined |
| S1_extracted_k141_147205 | Not-determined |
| S1_extracted_k141_657 | Low-quality |
| S1_extracted_k141_20705 | Not-determined |
| S1_extracted_k141_100887 | Not-determined |
| S1_extracted_k141_34021 | Not-determined |
| S1_extracted_k141_20708 | Not-determined |
| S1_extracted_k141_20709 | Not-determined |
| S1_extracted_k141_94215 | Not-determined |
| S1_extracted_k141_20711 | Low-quality |
| S1_extracted_k141_134137 | Not-determined |
| S1_extracted_k141_140900 | Low-quality |
| S1_extracted_k141_87542 | Not-determined |
| S1_extracted_k141_14216 | Not-determined |
| S1_extracted_k141_140903 | Not-determined |
| S1_extracted_k141_94219 | Not-determined |
| S1_extracted_k141_54221 | Not-determined |
| S1_extracted_k141_87545 | Low-quality |
| S1_extracted_k141_94220 | Not-determined |
| S1_extracted_k141_20716 | Not-determined |
| S1_extracted_k141_20718 | Not-determined |
| S1_extracted_k141_67405 | Not-determined |
| S1_extracted_k141_140907 | Not-determined |
| S1_extracted_k141_34024 | Low-quality |
| S1_extracted_k141_7440 | Not-determined |
| S1_extracted_k141_134146 | Not-determined |
| S1_extracted_k141_87550 | Not-determined |
| S1_extracted_k141_127617 | Not-determined |
| S1_extracted_k141_94224 | Low-quality |
| S1_extracted_k141_40842 | Not-determined |
| S1_extracted_k141_20722 | Low-quality |
| S1_extracted_k141_120828 | Low-quality |
| S1_extracted_k141_47464 | Not-determined |
| S1_extracted_k141_34030 | Not-determined |
| S1_extracted_k141_664 | Low-quality |
| S1_extracted_k141_120831 | Not-determined |
| S1_extracted_k141_134149 | Not-determined |
| S1_extracted_k141_153912 | Not-determined |
| S1_extracted_k141_40849 | Low-quality |
| S1_extracted_k141_7445 | Not-determined |
| S1_extracted_k141_27396 | Not-determined |
| S1_extracted_k141_80881 | Not-determined |
| S1_extracted_k141_87557 | Not-determined |
| S1_extracted_k141_34031 | Low-quality |
| S1_extracted_k141_54235 | Low-quality |
| S1_extracted_k141_140921 | Not-determined |
| S1_extracted_k141_80884 | Not-determined |
| S1_extracted_k141_107422 | Not-determined |
| S1_extracted_k141_153916 | Not-determined |
| S1_extracted_k141_67413 | Not-determined |
| S1_extracted_k141_14232 | Not-determined |
| S1_extracted_k141_107424 | Not-determined |
| S1_extracted_k141_27402 | Low-quality |
| S1_extracted_k141_127628 | Not-determined |
| S1_extracted_k141_114143 | Not-determined |
| S1_extracted_k141_27404 | Not-determined |
| S1_extracted_k141_80888 | Not-determined |
| S1_extracted_k141_14236 | Not-determined |
| S1_extracted_k141_100913 | Not-determined |
| S1_extracted_k141_100915 | Not-determined |
| S1_extracted_k141_54242 | Not-determined |
| S1_extracted_k141_94238 | Low-quality |
| S1_extracted_k141_100916 | Not-determined |
| S1_extracted_k141_127631 | Not-determined |
| S1_extracted_k141_54243 | Not-determined |
| S1_extracted_k141_80892 | Not-determined |
| S1_extracted_k141_54245 | Not-determined |
| S1_extracted_k141_14238 | Not-determined |
| S1_extracted_k141_114149 | Not-determined |
| S1_extracted_k141_27407 | Not-determined |
| S1_extracted_k141_107429 | Not-determined |
| S1_extracted_k141_94242 | Low-quality |

| | |
|---|---|
| S1_extracted_k141_100922 | Not-determined |
| S1_extracted_k141_94243 | Not-determined |
| S1_extracted_k141_94245 | Not-determined |
| S1_extracted_k141_14240 | Not-determined |
| S1_extracted_k141_94246 | Not-determined |
| S1_extracted_k141_14241 | Not-determined |
| S1_extracted_k141_94250 | Not-determined |
| S1_extracted_k141_20742 | Not-determined |
| S1_extracted_k141_7460 | Not-determined |
| S1_extracted_k141_134165 | Low-quality |
| S1_extracted_k141_67428 | Not-determined |
| S1_extracted_k141_27415 | Not-determined |
| S1_extracted_k141_87569 | Low-quality |
| S1_extracted_k141_147224 | Low-quality |
| S1_extracted_k141_114160 | Not-determined |
| S1_extracted_k141_114161 | Low-quality |
| S1_extracted_k141_7471 | Not-determined |
| S1_extracted_k141_100935 | Not-determined |
| S1_extracted_k141_67433 | Low-quality |
| S1_extracted_k141_67434 | Not-determined |
| S1_extracted_k141_100938 | Not-determined |
| S1_extracted_k141_67436 | Not-determined |
| S1_extracted_k141_147228 | Not-determined |
| S1_extracted_k141_100942 | Not-determined |
| S1_extracted_k141_140935 | Not-determined |
| S1_extracted_k141_47494 | Not-determined |
| S1_extracted_k141_14262 | Not-determined |
| S1_extracted_k141_80919 | Not-determined |
| S1_extracted_k141_20758 | Not-determined |
| S1_extracted_k141_54263 | Not-determined |
| S1_extracted_k141_80921 | Not-determined |
| S1_extracted_k141_14266 | Not-determined |
| S1_extracted_k141_134174 | Not-determined |
| S1_extracted_k141_60513 | Not-determined |
| S1_extracted_k141_140945 | Not-determined |
| S1_extracted_k141_54265 | Not-determined |
| S1_extracted_k141_114176 | Not-determined |
| S1_extracted_k141_54267 | Not-determined |
| S1_extracted_k141_60514 | Low-quality |
| S1_extracted_k141_27433 | Not-determined |
| S1_extracted_k141_153927 | Not-determined |
| S1_extracted_k141_27434 | Not-determined |
| S1_extracted_k141_60515 | Not-determined |
| S1_extracted_k141_94272 | Not-determined |
| S1_extracted_k141_120844 | Not-determined |
| S1_extracted_k141_20768 | Not-determined |
| S1_extracted_k141_54270 | Low-quality |
| S1_extracted_k141_47509 | Not-determined |
| S1_extracted_k141_67450 | Not-determined |
| S1_extracted_k141_688 | Low-quality |
| S1_extracted_k141_153930 | Not-determined |
| S1_extracted_k141_94275 | Not-determined |
| S1_extracted_k141_40880 | Not-determined |
| S1_extracted_k141_67454 | Low-quality |
| S1_extracted_k141_14277 | Not-determined |
| S1_extracted_k141_14278 | Not-determined |
| S1_extracted_k141_67457 | Not-determined |
| S1_extracted_k141_80925 | Low-quality |
| S1_extracted_k141_153933 | Not-determined |
| S1_extracted_k141_80927 | Low-quality |
| S1_extracted_k141_80928 | Not-determined |
| S1_extracted_k141_34054 | Not-determined |
| S1_extracted_k141_27440 | Not-determined |
| S1_extracted_k141_20780 | Not-determined |
| S1_extracted_k141_100960 | Low-quality |
| S1_extracted_k141_54279 | Not-determined |
| S1_extracted_k141_94283 | Not-determined |
| S1_extracted_k141_100963 | Not-determined |
| S1_extracted_k141_127660 | Not-determined |
| S1_extracted_k141_14291 | Not-determined |
| S1_extracted_k141_127663 | Not-determined |
| S1_extracted_k141_54283 | Low-quality |
| S1_extracted_k141_54285 | Not-determined |
| S1_extracted_k141_127668 | Not-determined |
| S1_extracted_k141_87580 | Not-determined |
| S1_extracted_k141_114187 | Not-determined |
| S1_extracted_k141_27446 | Not-determined |
| S1_extracted_k141_94294 | Not-determined |
| S1_extracted_k141_127674 | Not-determined |
| S1_extracted_k141_20795 | Not-determined |
| S1_extracted_k141_60524 | Not-determined |
| S1_extracted_k141_107456 | Not-determined |
| S1_extracted_k141_127680 | Not-determined |
| S1_extracted_k141_27450 | Not-determined |
| S1_extracted_k141_120854 | Not-determined |
| S1_extracted_k141_67477 | Low-quality |
| S1_extracted_k141_47526 | Not-determined |
| S1_extracted_k141_20801 | Not-determined |
| S1_extracted_k141_27453 | Not-determined |
| S1_extracted_k141_153944 | Low-quality |
| S1_extracted_k141_107461 | Not-determined |
| S1_extracted_k141_134199 | Not-determined |
| S1_extracted_k141_127684 | Low-quality |
| S1_extracted_k141_114200 | Low-quality |
| S1_extracted_k141_134201 | Not-determined |
| S1_extracted_k141_20805 | Low-quality |
| S1_extracted_k141_107463 | Low-quality |
| S1_extracted_k141_100980 | Not-determined |
| S1_extracted_k141_80946 | Not-determined |
| S1_extracted_k141_47532 | Not-determined |
| S1_extracted_k141_134204 | Not-determined |
| S1_extracted_k141_134205 | Not-determined |
| S1_extracted_k141_718 | Not-determined |
| S1_extracted_k141_127690 | Not-determined |
| S1_extracted_k141_720 | Not-determined |
| S1_extracted_k141_147235 | Not-determined |
| S1_extracted_k141_87591 | Not-determined |
| S1_extracted_k141_127691 | Not-determined |
| S1_extracted_k141_120874 | Not-determined |
| S1_extracted_k141_80952 | Not-determined |
| S1_extracted_k141_140964 | Not-determined |
| S1_extracted_k141_94305 | Low-quality |
| S1_extracted_k141_120876 | Not-determined |
| S1_extracted_k141_47536 | Low-quality |
| S1_extracted_k141_140967 | Not-determined |
| S1_extracted_k141_14325 | Not-determined |
| S1_extracted_k141_7508 | Not-determined |
| S1_extracted_k141_34081 | Not-determined |
| S1_extracted_k141_34082 | Not-determined |
| S1_extracted_k141_114207 | Not-determined |
| S1_extracted_k141_87599 | Low-quality |
| S1_extracted_k141_114208 | Not-determined |
| S1_extracted_k141_7511 | Not-determined |
| S1_extracted_k141_94312 | Not-determined |
| S1_extracted_k141_100995 | Not-determined |
| S1_extracted_k141_7514 | Not-determined |
| S1_extracted_k141_87603 | Low-quality |
| S1_extracted_k141_134214 | Not-determined |
| S1_extracted_k141_47543 | Low-quality |
| S1_extracted_k141_7517 | Low-quality |
| S1_extracted_k141_87605 | Not-determined |
| S1_extracted_k141_74239 | Not-determined |
| S1_extracted_k141_60542 | Not-determined |
| S1_extracted_k141_14333 | Not-determined |
| S1_extracted_k141_14335 | Low-quality |
| S1_extracted_k141_147242 | Low-quality |
| S1_extracted_k141_80972 | Not-determined |
| S1_extracted_k141_140979 | Not-determined |
| S1_extracted_k141_47550 | Not-determined |
| S1_extracted_k141_54308 | Not-determined |
| S1_extracted_k141_87612 | Not-determined |
| S1_extracted_k141_67493 | Not-determined |
| S1_extracted_k141_140982 | Not-determined |
| S1_extracted_k141_34097 | Not-determined |
| S1_extracted_k141_107484 | Low-quality |
| S1_extracted_k141_153949 | Not-determined |
| S1_extracted_k141_60544 | Not-determined |
| S1_extracted_k141_742 | Low-quality |
| S1_extracted_k141_60545 | Not-determined |
| S1_extracted_k141_101010 | Not-determined |
| S1_extracted_k141_101011 | Low-quality |
| S1_extracted_k141_153953 | Not-determined |
| S1_extracted_k141_14349 | Not-determined |
| S1_extracted_k141_80982 | Low-quality |
| S1_extracted_k141_748 | Low-quality |
| S1_extracted_k141_40915 | Not-determined |
| S1_extracted_k141_54318 | Not-determined |
| S1_extracted_k141_7530 | Not-determined |
| S1_extracted_k141_101016 | Low-quality |
| S1_extracted_k141_7533 | Not-determined |
| S1_extracted_k141_60549 | Low-quality |
| S1_extracted_k141_60550 | Not-determined |
| S1_extracted_k141_27487 | Not-determined |
| S1_extracted_k141_34103 | Low-quality |
| S1_extracted_k141_140992 | Low-quality |
| S1_extracted_k141_114226 | Not-determined |
| S1_extracted_k141_153958 | Low-quality |
| S1_extracted_k141_153959 | Not-determined |
| S1_extracted_k141_755 | Not-determined |
| S1_extracted_k141_20844 | Low-quality |
| S1_extracted_k141_34104 | Low-quality |
| S1_extracted_k141_7540 | Not-determined |
| S1_extracted_k141_7541 | Not-determined |
| S1_extracted_k141_153961 | Not-determined |
| S1_extracted_k141_7542 | Low-quality |
| S1_extracted_k141_87634 | Not-determined |
| S1_extracted_k141_7543 | Not-determined |
| S1_extracted_k141_114228 | Not-determined |
| S1_extracted_k141_153962 | Not-determined |
| S1_extracted_k141_60553 | Not-determined |
| S1_extracted_k141_40925 | Not-determined |
| S1_extracted_k141_153964 | Not-determined |
| S1_extracted_k141_40928 | Low-quality |
| S1_extracted_k141_763 | Not-determined |
| S1_extracted_k141_60556 | Not-determined |
| S1_extracted_k141_40931 | Not-determined |
| S1_extracted_k141_7552 | Not-determined |
| S1_extracted_k141_134231 | Not-determined |
| S1_extracted_k141_74253 | Not-determined |
| S1_extracted_k141_114237 | Not-determined |
| S1_extracted_k141_7554 | Not-determined |
| S1_extracted_k141_147244 | Not-determined |
| S1_extracted_k141_114238 | Not-determined |
| S1_extracted_k141_27498 | Not-determined |
| S1_extracted_k141_54331 | Not-determined |
| S1_extracted_k141_114239 | Not-determined |
| S1_extracted_k141_74257 | Low-quality |
| S1_extracted_k141_47574 | Not-determined |
| S1_extracted_k141_54333 | Not-determined |
| S1_extracted_k141_60561 | Not-determined |
| S1_extracted_k141_147247 | Not-determined |
| S1_extracted_k141_60562 | Not-determined |
| S1_extracted_k141_27506 | Not-determined |
| S1_extracted_k141_7566 | Not-determined |
| S1_extracted_k141_101038 | Not-determined |
| S1_extracted_k141_40943 | Not-determined |
| S1_extracted_k141_127721 | Not-determined |
| S1_extracted_k141_7569 | Not-determined |
| S1_extracted_k141_27509 | Not-determined |
| S1_extracted_k141_101045 | Not-determined |
| S1_extracted_k141_107512 | Not-determined |
| S1_extracted_k141_54343 | Low-quality |
| S1_extracted_k141_67525 | Not-determined |
| S1_extracted_k141_94345 | Not-determined |
| S1_extracted_k141_7576 | Not-determined |
| S1_extracted_k141_94347 | Low-quality |
| S1_extracted_k141_7577 | Not-determined |
| S1_extracted_k141_120877 | Low-quality |
| S1_extracted_k141_40957 | Not-determined |
| S1_extracted_k141_94348 | Not-determined |
| S1_extracted_k141_94349 | Not-determined |
| S1_extracted_k141_114251 | Not-determined |
| S1_extracted_k141_127729 | Low-quality |
| S1_extracted_k141_34118 | Low-quality |
| S1_extracted_k141_134241 | Not-determined |
| S1_extracted_k141_7581 | Not-determined |
| S1_extracted_k141_786 | Low-quality |
| S1_extracted_k141_74274 | Low-quality |
| S1_extracted_k141_20871 | Not-determined |
| S1_extracted_k141_141017 | Not-determined |
| S1_extracted_k141_787 | Not-determined |
| S1_extracted_k141_87662 | Not-determined |
| S1_extracted_k141_60570 | Not-determined |
| S1_extracted_k141_94352 | Low-quality |
| S1_extracted_k141_107518 | Not-determined |
| S1_extracted_k141_54348 | Not-determined |
| S1_extracted_k141_14394 | Not-determined |
| S1_extracted_k141_7584 | Not-determined |
| S1_extracted_k141_141019 | Not-determined |
| S1_extracted_k141_27518 | Not-determined |
| S1_extracted_k141_81023 | Not-determined |
| S1_extracted_k141_87667 | Not-determined |
| S1_extracted_k141_87668 | Low-quality |
| S1_extracted_k141_87670 | Low-quality |
| S1_extracted_k141_107525 | Not-determined |
| S1_extracted_k141_34123 | Not-determined |
| S1_extracted_k141_54350 | Not-determined |
| S1_extracted_k141_134248 | Not-determined |
| S1_extracted_k141_120882 | Low-quality |
| S1_extracted_k141_114255 | Not-determined |
| S1_extracted_k141_134250 | Not-determined |
| S1_extracted_k141_127734 | Not-determined |
| S1_extracted_k141_795 | Not-determined |
| S1_extracted_k141_134252 | Not-determined |
| S1_extracted_k141_7598 | Not-determined |
| S1_extracted_k141_114258 | Low-quality |
| S1_extracted_k141_94360 | Not-determined |
| S1_extracted_k141_141027 | Not-determined |
| S1_extracted_k141_81028 | Not-determined |
| S1_extracted_k141_87678 | Not-determined |
| S1_extracted_k141_800 | Low-quality |
| S1_extracted_k141_107529 | Not-determined |
| S1_extracted_k141_114264 | Not-determined |
| S1_extracted_k141_107530 | Not-determined |
| S1_extracted_k141_34129 | Not-determined |
| S1_extracted_k141_74293 | Not-determined |
| S1_extracted_k141_20889 | Not-determined |
| S1_extracted_k141_127739 | Not-determined |
| S1_extracted_k141_127740 | Not-determined |
| S1_extracted_k141_120891 | Not-determined |
| S1_extracted_k141_87681 | Not-determined |
| S1_extracted_k141_120893 | Not-determined |
| S1_extracted_k141_81042 | Not-determined |
| S1_extracted_k141_27525 | Low-quality |
| S1_extracted_k141_81044 | Low-quality |
| S1_extracted_k141_60574 | Low-quality |
| S1_extracted_k141_34135 | Not-determined |
| S1_extracted_k141_803 | Low-quality |
| S1_extracted_k141_101065 | Not-determined |
| S1_extracted_k141_87688 | Not-determined |
| S1_extracted_k141_74303 | Not-determined |
| S1_extracted_k141_141040 | Not-determined |
| S1_extracted_k141_127747 | Not-determined |
| S1_extracted_k141_120900 | Not-determined |
| S1_extracted_k141_60580 | Not-determined |
| S1_extracted_k141_114275 | Not-determined |
| S1_extracted_k141_141044 | Low-quality |
| S1_extracted_k141_107539 | Not-determined |
| S1_extracted_k141_14420 | Not-determined |
| S1_extracted_k141_101070 | Not-determined |
| S1_extracted_k141_114278 | Not-determined |
| S1_extracted_k141_94376 | Not-determined |
| S1_extracted_k141_7610 | Not-determined |
| S1_extracted_k141_81054 | Not-determined |
| S1_extracted_k141_7611 | Not-determined |
| S1_extracted_k141_67559 | Not-determined |
| S1_extracted_k141_87694 | Not-determined |
| S1_extracted_k141_147260 | Low-quality |
| S1_extracted_k141_94379 | Not-determined |
| S1_extracted_k141_27536 | Not-determined |
| S1_extracted_k141_20902 | Not-determined |
| S1_extracted_k141_94382 | Not-determined |
| S1_extracted_k141_47599 | Not-determined |
| S1_extracted_k141_134278 | Not-determined |
| S1_extracted_k141_34144 | Not-determined |
| S1_extracted_k141_120908 | Not-determined |
| S1_extracted_k141_127756 | Not-determined |
| S1_extracted_k141_40978 | Not-determined |
| S1_extracted_k141_20908 | Not-determined |
| S1_extracted_k141_14431 | Not-determined |
| S1_extracted_k141_127757 | Not-determined |
| S1_extracted_k141_120909 | Low-quality |
| S1_extracted_k141_120910 | Not-determined |
| S1_extracted_k141_147268 | Not-determined |
| S1_extracted_k141_14433 | Not-determined |
| S1_extracted_k141_34149 | Not-determined |
| S1_extracted_k141_107546 | Low-quality |
| S1_extracted_k141_27548 | Not-determined |
| S1_extracted_k141_7620 | Not-determined |
| S1_extracted_k141_816 | Not-determined |
| S1_extracted_k141_127765 | Not-determined |
| S1_extracted_k141_81062 | Not-determined |
| S1_extracted_k141_14439 | Not-determined |
| S1_extracted_k141_40985 | Not-determined |
| S1_extracted_k141_74310 | Not-determined |
| S1_extracted_k141_147271 | Not-determined |
| S1_extracted_k141_134288 | Not-determined |
| S1_extracted_k141_7624 | Not-determined |
| S1_extracted_k141_47611 | Not-determined |
| S1_extracted_k141_114295 | Not-determined |
| S1_extracted_k141_101077 | Not-determined |
| S1_extracted_k141_101078 | Not-determined |
| S1_extracted_k141_107556 | Not-determined |
| S1_extracted_k141_101079 | Low-quality |
| S1_extracted_k141_94397 | Low-quality |
| S1_extracted_k141_134292 | Low-quality |
| S1_extracted_k141_67580 | Not-determined |
| S1_extracted_k141_81071 | Not-determined |
| S1_extracted_k141_114301 | Low-quality |
| S1_extracted_k141_40994 | Not-determined |
| S1_extracted_k141_81074 | Not-determined |
| S1_extracted_k141_147275 | Low-quality |
| S1_extracted_k141_14451 | Not-determined |
| S1_extracted_k141_87710 | Not-determined |
| S1_extracted_k141_147276 | Low-quality |
| S1_extracted_k141_40997 | Not-determined |
| S1_extracted_k141_47616 | Not-determined |
| S1_extracted_k141_822 | Not-determined |
| S1_extracted_k141_94403 | Not-determined |
| S1_extracted_k141_120919 | Low-quality |
| S1_extracted_k141_134301 | Low-quality |
| S1_extracted_k141_141064 | Not-determined |
| S1_extracted_k141_107563 | Not-determined |
| S1_extracted_k141_14455 | Not-determined |
| S1_extracted_k141_120921 | Low-quality |
| S1_extracted_k141_147277 | Not-determined |
| S1_extracted_k141_94411 | Not-determined |
| S1_extracted_k141_141067 | Not-determined |
| S1_extracted_k141_14459 | Not-determined |
| S1_extracted_k141_141068 | Not-determined |
| S1_extracted_k141_87717 | Not-determined |
| S1_extracted_k141_60597 | Low-quality |
| S1_extracted_k141_41007 | Not-determined |
| S1_extracted_k141_60598 | Not-determined |
| S1_extracted_k141_54397 | Not-determined |
| S1_extracted_k141_826 | Not-determined |
| S1_extracted_k141_60600 | Not-determined |
| S1_extracted_k141_47627 | Not-determined |
| S1_extracted_k141_134309 | Not-determined |
| S1_extracted_k141_60601 | Not-determined |
| S1_extracted_k141_114310 | Not-determined |
| S1_extracted_k141_7634 | Not-determined |
| S1_extracted_k141_54402 | Low-quality |
| S1_extracted_k141_127786 | Low-quality |
| S1_extracted_k141_134312 | Not-determined |
| S1_extracted_k141_41011 | Low-quality |
| S1_extracted_k141_120937 | Low-quality |
| S1_extracted_k141_87724 | Not-determined |
| S1_extracted_k141_141072 | Not-determined |
| S1_extracted_k141_34168 | Not-determined |
| S1_extracted_k141_67593 | Not-determined |
| S1_extracted_k141_114315 | Not-determined |
| S1_extracted_k141_14473 | Not-determined |
| S1_extracted_k141_7638 | Not-determined |
| S1_extracted_k141_127793 | Not-determined |
| S1_extracted_k141_127794 | Not-determined |
| S1_extracted_k141_74340 | Not-determined |
| S1_extracted_k141_14478 | Not-determined |
| S1_extracted_k141_20929 | Not-determined |
| S1_extracted_k141_20931 | Not-determined |
| S1_extracted_k141_27563 | Low-quality |
| S1_extracted_k141_47636 | Not-determined |
| S1_extracted_k141_20932 | Not-determined |
| S1_extracted_k141_94426 | Not-determined |
| S1_extracted_k141_141080 | Not-determined |
| S1_extracted_k141_81100 | Not-determined |
| S1_extracted_k141_134322 | Not-determined |
| S1_extracted_k141_120951 | Not-determined |
| S1_extracted_k141_153992 | Not-determined |
| S1_extracted_k141_134324 | Not-determined |
| S1_extracted_k141_94433 | Not-determined |
| S1_extracted_k141_141086 | Not-determined |
| S1_extracted_k141_127803 | Not-determined |
| S1_extracted_k141_147291 | Not-determined |
| S1_extracted_k141_107576 | Low-quality |
| S1_extracted_k141_81104 | Not-determined |
| S1_extracted_k141_14488 | Not-determined |
| S1_extracted_k141_141090 | Not-determined |
| S1_extracted_k141_81106 | Not-determined |
| S1_extracted_k141_67601 | Not-determined |
| S1_extracted_k141_120954 | Not-determined |
| S1_extracted_k141_47648 | Low-quality |
| S1_extracted_k141_41022 | Not-determined |
| S1_extracted_k141_114325 | Not-determined |
| S1_extracted_k141_67603 | Not-determined |
| S1_extracted_k141_41024 | Not-determined |
| S1_extracted_k141_94437 | Low-quality |
| S1_extracted_k141_134334 | Low-quality |
| S1_extracted_k141_107579 | Low-quality |
| S1_extracted_k141_20937 | Not-determined |
| S1_extracted_k141_94438 | Not-determined |
| S1_extracted_k141_134335 | Not-determined |
| S1_extracted_k141_114327 | Not-determined |
| S1_extracted_k141_54417 | Low-quality |
| S1_extracted_k141_141094 | Not-determined |
| S1_extracted_k141_101114 | Not-determined |
| S1_extracted_k141_107583 | Low-quality |
| S1_extracted_k141_60617 | Not-determined |
| S1_extracted_k141_854 | Not-determined |
| S1_extracted_k141_855 | Not-determined |
| S1_extracted_k141_107587 | Not-determined |
| S1_extracted_k141_94447 | Not-determined |
| S1_extracted_k141_67610 | Not-determined |
| S1_extracted_k141_857 | Not-determined |
| S1_extracted_k141_60622 | Not-determined |
| S1_extracted_k141_154004 | Low-quality |
| S1_extracted_k141_94451 | Not-determined |
| S1_extracted_k141_60625 | Not-determined |
| S1_extracted_k141_127817 | Not-determined |
| S1_extracted_k141_34185 | Low-quality |
| S1_extracted_k141_20954 | Not-determined |
| S1_extracted_k141_20957 | Not-determined |
| S1_extracted_k141_34186 | Not-determined |
| S1_extracted_k141_7674 | Low-quality |
| S1_extracted_k141_94458 | Not-determined |
| S1_extracted_k141_107600 | Not-determined |
| S1_extracted_k141_7678 | Not-determined |
| S1_extracted_k141_14520 | Not-determined |
| S1_extracted_k141_81129 | Low-quality |
| S1_extracted_k141_94461 | Not-determined |
| S1_extracted_k141_81130 | Not-determined |
| S1_extracted_k141_7684 | Not-determined |
| S1_extracted_k141_67628 | Not-determined |
| S1_extracted_k141_67629 | Not-determined |
| S1_extracted_k141_67631 | Not-determined |
| S1_extracted_k141_94464 | Not-determined |
| S1_extracted_k141_134358 | Not-determined |
| S1_extracted_k141_120968 | Not-determined |
| S1_extracted_k141_154009 | Not-determined |
| S1_extracted_k141_60631 | Not-determined |
| S1_extracted_k141_54429 | Not-determined |
| S1_extracted_k141_41042 | Not-determined |
| S1_extracted_k141_870 | Not-determined |
| S1_extracted_k141_20965 | Not-determined |
| S1_extracted_k141_120972 | Not-determined |
| S1_extracted_k141_127825 | Not-determined |
| S1_extracted_k141_14531 | Not-determined |
| S1_extracted_k141_81140 | Not-determined |
| S1_extracted_k141_41045 | Not-determined |
| S1_extracted_k141_14532 | Not-determined |
| S1_extracted_k141_94470 | Low-quality |
| S1_extracted_k141_20966 | Low-quality |
| S1_extracted_k141_74358 | Not-determined |
| S1_extracted_k141_94471 | Not-determined |
| S1_extracted_k141_87739 | Not-determined |
| S1_extracted_k141_67640 | Not-determined |
| S1_extracted_k141_47666 | Not-determined |
| S1_extracted_k141_41048 | Low-quality |
| S1_extracted_k141_127828 | Low-quality |
| S1_extracted_k141_41049 | Low-quality |
| S1_extracted_k141_87742 | Low-quality |
| S1_extracted_k141_74366 | Not-determined |
| S1_extracted_k141_20971 | Low-quality |
| S1_extracted_k141_74368 | Not-determined |
| S1_extracted_k141_41058 | Low-quality |
| S1_extracted_k141_134362 | Low-quality |
| S1_extracted_k141_127841 | Not-determined |
| S1_extracted_k141_107620 | Not-determined |
| S1_extracted_k141_81157 | Not-determined |
| S1_extracted_k141_94482 | Not-determined |
| S1_extracted_k141_60640 | Not-determined |
| S1_extracted_k141_101144 | Not-determined |
| S1_extracted_k141_14546 | Low-quality |
| S1_extracted_k141_120978 | Low-quality |
| S1_extracted_k141_107623 | Low-quality |
| S1_extracted_k141_81162 | Not-determined |
| S1_extracted_k141_27594 | Not-determined |
| S1_extracted_k141_101145 | Not-determined |
| S1_extracted_k141_34207 | Not-determined |
| S1_extracted_k141_34208 | Not-determined |
| S1_extracted_k141_67655 | Not-determined |
| S1_extracted_k141_141145 | Not-determined |
| S1_extracted_k141_14549 | Not-determined |
| S1_extracted_k141_27601 | Not-determined |
| S1_extracted_k141_20980 | Not-determined |
| S1_extracted_k141_107633 | Not-determined |
| S1_extracted_k141_34212 | Not-determined |
| S1_extracted_k141_81168 | Not-determined |
| S1_extracted_k141_41074 | Not-determined |
| S1_extracted_k141_67666 | Low-quality |
| S1_extracted_k141_94498 | Low-quality |
| S1_extracted_k141_47671 | Not-determined |
| S1_extracted_k141_147306 | Not-determined |
| S1_extracted_k141_27605 | Not-determined |
| S1_extracted_k141_107642 | Not-determined |
| S1_extracted_k141_34219 | Low-quality |
| S1_extracted_k141_47674 | Not-determined |
| S1_extracted_k141_34220 | Not-determined |
| S1_extracted_k141_47675 | Not-determined |
| S1_extracted_k141_120990 | Low-quality |
| S1_extracted_k141_47676 | Not-determined |
| S1_extracted_k141_114360 | Not-determined |
| S1_extracted_k141_7721 | Low-quality |
| S1_extracted_k141_67669 | Low-quality |
| S1_extracted_k141_94507 | Not-determined |
| S1_extracted_k141_120995 | Low-quality |
| S1_extracted_k141_60652 | Not-determined |
| S1_extracted_k141_47680 | Not-determined |
| S1_extracted_k141_94508 | Not-determined |
| S1_extracted_k141_141154 | Not-determined |
| S1_extracted_k141_20992 | Not-determined |
| S1_extracted_k141_47683 | Not-determined |
| S1_extracted_k141_27617 | Low-quality |
| S1_extracted_k141_101160 | Not-determined |
| S1_extracted_k141_87774 | Not-determined |
| S1_extracted_k141_60656 | Not-determined |
| S1_extracted_k141_7724 | Not-determined |
| S1_extracted_k141_81178 | Not-determined |
| S1_extracted_k141_41097 | Not-determined |
| S1_extracted_k141_60662 | Not-determined |
| S1_extracted_k141_27623 | Not-determined |
| S1_extracted_k141_120999 | Not-determined |
| S1_extracted_k141_27625 | Not-determined |
| S1_extracted_k141_101166 | Low-quality |
| S1_extracted_k141_27627 | Not-determined |
| S1_extracted_k141_87779 | Not-determined |
| S1_extracted_k141_20994 | Not-determined |
| S1_extracted_k141_41104 | Not-determined |
| S1_extracted_k141_114367 | Not-determined |
| S1_extracted_k141_27633 | Not-determined |
| S1_extracted_k141_67670 | Low-quality |
| S1_extracted_k141_67671 | Not-determined |
| S1_extracted_k141_114369 | Not-determined |
| S1_extracted_k141_141169 | Not-determined |
| S1_extracted_k141_81191 | Not-determined |
| S1_extracted_k141_74399 | Not-determined |
| S1_extracted_k141_141172 | Not-determined |
| S1_extracted_k141_87781 | Not-determined |
| S1_extracted_k141_67673 | Low-quality |
| S1_extracted_k141_21000 | Not-determined |
| S1_extracted_k141_134370 | Not-determined |
| S1_extracted_k141_141174 | Not-determined |
| S1_extracted_k141_94521 | Low-quality |
| S1_extracted_k141_7737 | Not-determined |
| S1_extracted_k141_101174 | Not-determined |
| S1_extracted_k141_60673 | Not-determined |
| S1_extracted_k141_101176 | Not-determined |
| S1_extracted_k141_41121 | Not-determined |
| S1_extracted_k141_121018 | Not-determined |
| S1_extracted_k141_41124 | Not-determined |
| S1_extracted_k141_107672 | Not-determined |
| S1_extracted_k141_60677 | Not-determined |
| S1_extracted_k141_60678 | Not-determined |
| S1_extracted_k141_154035 | Not-determined |
| S1_extracted_k141_14599 | Not-determined |
| S1_extracted_k141_74413 | Not-determined |
| S1_extracted_k141_14601 | Low-quality |
| S1_extracted_k141_134378 | Not-determined |
| S1_extracted_k141_7746 | Not-determined |
| S1_extracted_k141_101186 | Not-determined |
| S1_extracted_k141_7749 | Not-determined |
| S1_extracted_k141_101187 | Low-quality |
| S1_extracted_k141_134383 | Not-determined |
| S1_extracted_k141_67692 | Not-determined |
| S1_extracted_k141_107677 | Not-determined |
| S1_extracted_k141_47711 | Not-determined |
| S1_extracted_k141_21014 | Not-determined |
| S1_extracted_k141_41134 | Not-determined |
| S1_extracted_k141_7756 | Not-determined |

| | |
|---|---|
| S1_extracted_k141_7757 | Low-quality |
| S1_extracted_k141_74420 | Not-determined |
| S1_extracted_k141_134389 | Not-determined |
| S1_extracted_k141_34246 | Not-determined |
| S1_extracted_k141_21020 | Not-determined |
| S1_extracted_k141_141194 | Not-determined |
| S1_extracted_k141_141195 | Not-determined |
| S1_extracted_k141_134391 | Not-determined |
| S1_extracted_k141_14615 | Not-determined |
| S1_extracted_k141_27657 | Not-determined |
| S1_extracted_k141_81205 | Low-quality |
| S1_extracted_k141_147317 | Not-determined |
| S1_extracted_k141_34248 | Not-determined |
| S1_extracted_k141_134396 | Low-quality |
| S1_extracted_k141_141201 | Not-determined |
| S1_extracted_k141_34251 | Not-determined |
| S1_extracted_k141_147318 | Not-determined |
| S1_extracted_k141_107693 | Not-determined |
| S1_extracted_k141_14623 | Not-determined |
| S1_extracted_k141_101189 | Low-quality |
| S1_extracted_k141_87814 | Not-determined |
| S1_extracted_k141_127856 | Not-determined |
| S1_extracted_k141_127857 | Not-determined |
| S1_extracted_k141_905 | Not-determined |
| S1_extracted_k141_47727 | Not-determined |
| S1_extracted_k141_41142 | Not-determined |
| S1_extracted_k141_127858 | Not-determined |
| S1_extracted_k141_94552 | Not-determined |
| S1_extracted_k141_107700 | Not-determined |
| S1_extracted_k141_94553 | Low-quality |
| S1_extracted_k141_907 | Low-quality |
| S1_extracted_k141_47728 | Low-quality |
| S1_extracted_k141_141211 | Not-determined |
| S1_extracted_k141_134403 | Not-determined |
| S1_extracted_k141_141212 | Not-determined |
| S1_extracted_k141_81208 | Not-determined |
| S1_extracted_k141_134404 | Not-determined |
| S1_extracted_k141_27670 | Not-determined |
| S1_extracted_k141_154057 | Not-determined |
| S1_extracted_k141_154059 | Not-determined |
| S1_extracted_k141_121025 | Not-determined |
| S1_extracted_k141_47732 | Not-determined |
| S1_extracted_k141_107705 | Not-determined |
| S1_extracted_k141_94561 | Not-determined |
| S1_extracted_k141_14632 | Not-determined |
| S1_extracted_k141_81211 | Not-determined |
| S1_extracted_k141_81212 | Not-determined |
| S1_extracted_k141_107707 | Not-determined |
| S1_extracted_k141_47736 | Not-determined |
| S1_extracted_k141_141220 | Not-determined |
| S1_extracted_k141_41151 | Not-determined |
| S1_extracted_k141_67709 | Not-determined |
| S1_extracted_k141_87822 | Not-determined |
| S1_extracted_k141_81215 | Not-determined |
| S1_extracted_k141_14637 | Not-determined |
| S1_extracted_k141_134413 | Not-determined |
| S1_extracted_k141_7787 | Not-determined |
| S1_extracted_k141_87824 | Not-determined |
| S1_extracted_k141_27677 | Not-determined |
| S1_extracted_k141_81219 | Not-determined |
| S1_extracted_k141_67716 | Not-determined |
| S1_extracted_k141_74440 | Not-determined |
| S1_extracted_k141_154070 | Not-determined |
| S1_extracted_k141_54470 | Not-determined |
| S1_extracted_k141_107716 | Not-determined |
| S1_extracted_k141_134417 | Not-determined |
| S1_extracted_k141_54472 | Not-determined |
| S1_extracted_k141_34271 | Low-quality |
| S1_extracted_k141_101208 | Not-determined |
| S1_extracted_k141_7796 | Not-determined |
| S1_extracted_k141_34273 | Not-determined |
| S1_extracted_k141_121040 | Not-determined |
| S1_extracted_k141_7798 | Not-determined |
| S1_extracted_k141_94573 | Not-determined |
| S1_extracted_k141_41165 | Not-determined |
| S1_extracted_k141_94574 | Not-determined |
| S1_extracted_k141_154077 | Not-determined |
| S1_extracted_k141_101212 | Not-determined |
| S1_extracted_k141_47748 | Low-quality |
| S1_extracted_k141_41168 | Not-determined |
| S1_extracted_k141_101213 | Low-quality |
| S1_extracted_k141_94577 | Low-quality |
| S1_extracted_k141_114419 | Not-determined |
| S1_extracted_k141_7805 | Low-quality |
| S1_extracted_k141_14655 | Low-quality |
| S1_extracted_k141_134426 | Not-determined |
| S1_extracted_k141_114423 | Not-determined |
| S1_extracted_k141_41176 | Not-determined |
| S1_extracted_k141_94582 | Low-quality |
| S1_extracted_k141_27691 | Low-quality |
| S1_extracted_k141_154084 | Low-quality |
| S1_extracted_k141_34278 | Not-determined |
| S1_extracted_k141_154085 | Not-determined |
| S1_extracted_k141_114426 | Not-determined |
| S1_extracted_k141_101217 | Low-quality |
| S1_extracted_k141_27695 | Not-determined |
| S1_extracted_k141_74461 | Not-determined |
| S1_extracted_k141_101220 | Not-determined |
| S1_extracted_k141_94591 | Not-determined |
| S1_extracted_k141_74463 | Not-determined |
| S1_extracted_k141_107728 | Not-determined |
| S1_extracted_k141_94594 | Not-determined |
| S1_extracted_k141_917 | Not-determined |
| S1_extracted_k141_121052 | Not-determined |
| S1_extracted_k141_134434 | Not-determined |
| S1_extracted_k141_107731 | Not-determined |
| S1_extracted_k141_154092 | Not-determined |
| S1_extracted_k141_60695 | Low-quality |
| S1_extracted_k141_121055 | Not-determined |
| S1_extracted_k141_81230 | Not-determined |
| S1_extracted_k141_121056 | Not-determined |
| S1_extracted_k141_54490 | Not-determined |
| S1_extracted_k141_134437 | Not-determined |
| S1_extracted_k141_134438 | Not-determined |
| S1_extracted_k141_121058 | Not-determined |
| S1_extracted_k141_134439 | Low-quality |
| S1_extracted_k141_94600 | Not-determined |
| S1_extracted_k141_141241 | Not-determined |
| S1_extracted_k141_14678 | Not-determined |
| S1_extracted_k141_101231 | Not-determined |
| S1_extracted_k141_34279 | Low-quality |
| S1_extracted_k141_41199 | Low-quality |
| S1_extracted_k141_81236 | Low-quality |
| S1_extracted_k141_54495 | Not-determined |
| S1_extracted_k141_74468 | Not-determined |
| S1_extracted_k141_41201 | Not-determined |
| S1_extracted_k141_7823 | Not-determined |
| S1_extracted_k141_154099 | Not-determined |
| S1_extracted_k141_67743 | Not-determined |
| S1_extracted_k141_114447 | Not-determined |
| S1_extracted_k141_107741 | Not-determined |
| S1_extracted_k141_926 | Not-determined |
| S1_extracted_k141_47773 | Not-determined |
| S1_extracted_k141_47774 | Not-determined |
| S1_extracted_k141_101237 | Low-quality |
| S1_extracted_k141_107745 | Not-determined |
| S1_extracted_k141_154105 | Not-determined |
| S1_extracted_k141_87856 | Not-determined |
| S1_extracted_k141_41210 | Not-determined |
| S1_extracted_k141_81245 | Not-determined |
| S1_extracted_k141_21087 | Not-determined |
| S1_extracted_k141_127906 | Not-determined |
| S1_extracted_k141_154107 | Low-quality |
| S1_extracted_k141_81247 | Low-quality |
| S1_extracted_k141_74481 | Not-determined |
| S1_extracted_k141_107752 | Not-determined |
| S1_extracted_k141_134447 | Not-determined |
| S1_extracted_k141_147344 | Not-determined |
| S1_extracted_k141_7832 | Not-determined |
| S1_extracted_k141_141249 | Not-determined |
| S1_extracted_k141_101246 | Not-determined |
| S1_extracted_k141_7833 | Not-determined |
| S1_extracted_k141_141251 | Not-determined |
| S1_extracted_k141_114458 | Not-determined |
| S1_extracted_k141_7834 | Not-determined |
| S1_extracted_k141_81250 | Low-quality |
| S1_extracted_k141_14700 | Not-determined |
| S1_extracted_k141_154111 | Not-determined |
| S1_extracted_k141_67753 | Not-determined |
| S1_extracted_k141_41219 | Low-quality |
| S1_extracted_k141_47787 | Low-quality |
| S1_extracted_k141_74486 | Not-determined |
| S1_extracted_k141_127919 | Not-determined |
| S1_extracted_k141_87872 | Not-determined |
| S1_extracted_k141_141260 | Not-determined |
| S1_extracted_k141_101252 | Low-quality |
| S1_extracted_k141_21101 | Not-determined |
| S1_extracted_k141_41226 | Not-determined |
| S1_extracted_k141_114465 | Not-determined |
| S1_extracted_k141_41227 | Not-determined |
| S1_extracted_k141_94616 | Not-determined |
| S1_extracted_k141_27718 | Not-determined |
| S1_extracted_k141_134456 | Not-determined |
| S1_extracted_k141_939 | Low-quality |
| S1_extracted_k141_47795 | Not-determined |
| S1_extracted_k141_41232 | Not-determined |
| S1_extracted_k141_127929 | Not-determined |
| S1_extracted_k141_943 | Not-determined |
| S1_extracted_k141_67762 | Not-determined |
| S1_extracted_k141_101260 | Low-quality |
| S1_extracted_k141_47800 | Not-determined |
| S1_extracted_k141_41238 | Not-determined |
| S1_extracted_k141_114476 | Not-determined |
| S1_extracted_k141_27726 | Not-determined |
| S1_extracted_k141_60706 | Not-determined |
| S1_extracted_k141_21113 | Not-determined |
| S1_extracted_k141_47808 | Not-determined |
| S1_extracted_k141_7853 | Low-quality |
| S1_extracted_k141_47809 | Not-determined |
| S1_extracted_k141_47810 | Not-determined |
| S1_extracted_k141_41244 | Not-determined |
| S1_extracted_k141_948 | Low-quality |
| S1_extracted_k141_121087 | Low-quality |
| S1_extracted_k141_21120 | Low-quality |
| S1_extracted_k141_41247 | Not-determined |
| S1_extracted_k141_114484 | Not-determined |
| S1_extracted_k141_34303 | Not-determined |
| S1_extracted_k141_7860 | Not-determined |
| S1_extracted_k141_54530 | Not-determined |
| S1_extracted_k141_7862 | Not-determined |
| S1_extracted_k141_121095 | Low-quality |
| S1_extracted_k141_154123 | Not-determined |
| S1_extracted_k141_47814 | Low-quality |
| S1_extracted_k141_114491 | Not-determined |
| S1_extracted_k141_101266 | Low-quality |
| S1_extracted_k141_101267 | Not-determined |
| S1_extracted_k141_141275 | Low-quality |
| S1_extracted_k141_7869 | Low-quality |
| S1_extracted_k141_54537 | Low-quality |
| S1_extracted_k141_134471 | Low-quality |
| S1_extracted_k141_94636 | Not-determined |
| S1_extracted_k141_147347 | Not-determined |
| S1_extracted_k141_101268 | Not-determined |
| S1_extracted_k141_107785 | Not-determined |
| S1_extracted_k141_27742 | Not-determined |
| S1_extracted_k141_953 | Not-determined |
| S1_extracted_k141_147348 | Not-determined |
| S1_extracted_k141_107788 | Not-determined |
| S1_extracted_k141_21132 | Not-determined |
| S1_extracted_k141_21135 | Not-determined |
| S1_extracted_k141_7879 | Not-determined |
| S1_extracted_k141_14729 | Not-determined |
| S1_extracted_k141_121109 | Not-determined |
| S1_extracted_k141_34313 | Not-determined |
| S1_extracted_k141_14730 | Not-determined |
| S1_extracted_k141_21139 | Low-quality |
| S1_extracted_k141_107797 | Not-determined |
| S1_extracted_k141_87899 | Low-quality |
| S1_extracted_k141_7881 | Not-determined |
| S1_extracted_k141_121112 | Not-determined |
| S1_extracted_k141_141288 | Not-determined |
| S1_extracted_k141_87901 | Low-quality |
| S1_extracted_k141_101278 | Low-quality |
| S1_extracted_k141_154139 | Not-determined |
| S1_extracted_k141_74510 | Not-determined |
| S1_extracted_k141_81290 | Not-determined |
| S1_extracted_k141_107802 | Low-quality |
| S1_extracted_k141_94648 | Not-determined |
| S1_extracted_k141_67804 | Low-quality |
| S1_extracted_k141_54550 | Low-quality |
| S1_extracted_k141_41260 | Not-determined |
| S1_extracted_k141_60719 | Low-quality |
| S1_extracted_k141_127940 | Not-determined |
| S1_extracted_k141_965 | Low-quality |
| S1_extracted_k141_27748 | Not-determined |
| S1_extracted_k141_147352 | Not-determined |
| S1_extracted_k141_54553 | Not-determined |
| S1_extracted_k141_966 | Not-determined |
| S1_extracted_k141_81299 | Not-determined |
| S1_extracted_k141_141293 | Not-determined |
| S1_extracted_k141_94656 | Not-determined |
| S1_extracted_k141_970 | Low-quality |
| S1_extracted_k141_94657 | Not-determined |
| S1_extracted_k141_147355 | Low-quality |
| S1_extracted_k141_94661 | Not-determined |
| S1_extracted_k141_87910 | Low-quality |
| S1_extracted_k141_47838 | Low-quality |
| S1_extracted_k141_127950 | Low-quality |
| S1_extracted_k141_67814 | Low-quality |
| S1_extracted_k141_81310 | Not-determined |
| S1_extracted_k141_87916 | Low-quality |
| S1_extracted_k141_60721 | Not-determined |
| S1_extracted_k141_27761 | Not-determined |
| S1_extracted_k141_54570 | Not-determined |
| S1_extracted_k141_134499 | Not-determined |
| S1_extracted_k141_94667 | Low-quality |
| S1_extracted_k141_107825 | Not-determined |
| S1_extracted_k141_47846 | Not-determined |
| S1_extracted_k141_101300 | Low-quality |
| S1_extracted_k141_134504 | Not-determined |
| S1_extracted_k141_127961 | Low-quality |
| S1_extracted_k141_154153 | Not-determined |
| S1_extracted_k141_81317 | Low-quality |
| S1_extracted_k141_101302 | Low-quality |
| S1_extracted_k141_94674 | Not-determined |
| S1_extracted_k141_147360 | Low-quality |
| S1_extracted_k141_54574 | Not-determined |
| S1_extracted_k141_47852 | Not-determined |
| S1_extracted_k141_134509 | Not-determined |
| S1_extracted_k141_27768 | Not-determined |
| S1_extracted_k141_67819 | Low-quality |
| S1_extracted_k141_27770 | Low-quality |
| S1_extracted_k141_67820 | Not-determined |
| S1_extracted_k141_7913 | Not-determined |
| S1_extracted_k141_134511 | Not-determined |
| S1_extracted_k141_87924 | Low-quality |
| S1_extracted_k141_54579 | Not-determined |
| S1_extracted_k141_147364 | Not-determined |
| S1_extracted_k141_107837 | Not-determined |
| S1_extracted_k141_54581 | Not-determined |
| S1_extracted_k141_81320 | Not-determined |
| S1_extracted_k141_67826 | Not-determined |
| S1_extracted_k141_34345 | Not-determined |
| S1_extracted_k141_67828 | Not-determined |
| S1_extracted_k141_87927 | Low-quality |
| S1_extracted_k141_127978 | Not-determined |
| S1_extracted_k141_81323 | Not-determined |
| S1_extracted_k141_21165 | Not-determined |
| S1_extracted_k141_41294 | Not-determined |
| S1_extracted_k141_34348 | Not-determined |
| S1_extracted_k141_74528 | Not-determined |
| S1_extracted_k141_67831 | Not-determined |
| S1_extracted_k141_41298 | Not-determined |
| S1_extracted_k141_14778 | Not-determined |
| S1_extracted_k141_60727 | Not-determined |
| S1_extracted_k141_101318 | Not-determined |
| S1_extracted_k141_21170 | Not-determined |
| S1_extracted_k141_54584 | Not-determined |
| S1_extracted_k141_114518 | Not-determined |
| S1_extracted_k141_21172 | Not-determined |
| S1_extracted_k141_14781 | Not-determined |
| S1_extracted_k141_27782 | Not-determined |
| S1_extracted_k141_94684 | Not-determined |
| S1_extracted_k141_14782 | Not-determined |
| S1_extracted_k141_154162 | Not-determined |
| S1_extracted_k141_67836 | Not-determined |
| S1_extracted_k141_134515 | Not-determined |
| S1_extracted_k141_27785 | Not-determined |
| S1_extracted_k141_54588 | Not-determined |
| S1_extracted_k141_81329 | Not-determined |
| S1_extracted_k141_101320 | Low-quality |
| S1_extracted_k141_14785 | Low-quality |
| S1_extracted_k141_107839 | Low-quality |
| S1_extracted_k141_134517 | Low-quality |
| S1_extracted_k141_107840 | Not-determined |
| S1_extracted_k141_14789 | Not-determined |
| S1_extracted_k141_21175 | Not-determined |
| S1_extracted_k141_7928 | Not-determined |
| S1_extracted_k141_101325 | Not-determined |
| S1_extracted_k141_21177 | Not-determined |
| S1_extracted_k141_107843 | Not-determined |
| S1_extracted_k141_134518 | Low-quality |
| S1_extracted_k141_47865 | Not-determined |
| S1_extracted_k141_114527 | Not-determined |
| S1_extracted_k141_54592 | Not-determined |
| S1_extracted_k141_141315 | Not-determined |
| S1_extracted_k141_74533 | Not-determined |
| S1_extracted_k141_60734 | Not-determined |
| S1_extracted_k141_47869 | Not-determined |
| S1_extracted_k141_27790 | Not-determined |
| S1_extracted_k141_134522 | Not-determined |
| S1_extracted_k141_60736 | Not-determined |
| S1_extracted_k141_974 | Not-determined |
| S1_extracted_k141_7937 | Not-determined |
| S1_extracted_k141_7938 | Not-determined |
| S1_extracted_k141_154174 | Not-determined |
| S1_extracted_k141_7939 | Not-determined |
| S1_extracted_k141_47873 | Not-determined |
| S1_extracted_k141_27796 | Not-determined |
| S1_extracted_k141_94695 | Low-quality |
| S1_extracted_k141_134524 | Not-determined |
| S1_extracted_k141_47875 | Not-determined |
| S1_extracted_k141_7943 | Low-quality |
| S1_extracted_k141_60738 | Not-determined |
| S1_extracted_k141_7945 | Not-determined |
| S1_extracted_k141_41313 | Not-determined |
| S1_extracted_k141_74537 | Not-determined |
| S1_extracted_k141_7946 | Not-determined |
| S1_extracted_k141_60740 | Not-determined |
| S1_extracted_k141_74540 | Not-determined |
| S1_extracted_k141_982 | Not-determined |
| S1_extracted_k141_47879 | Low-quality |
| S1_extracted_k141_21185 | Low-quality |
| S1_extracted_k141_74542 | Low-quality |
| S1_extracted_k141_101343 | Not-determined |
| S1_extracted_k141_94697 | Not-determined |
| S1_extracted_k141_94698 | Not-determined |
| S1_extracted_k141_21189 | Not-determined |
| S1_extracted_k141_21193 | Not-determined |
| S1_extracted_k141_107853 | Not-determined |
| S1_extracted_k141_134532 | Not-determined |
| S1_extracted_k141_114539 | Low-quality |
| S1_extracted_k141_67854 | Not-determined |
| S1_extracted_k141_87957 | Not-determined |
| S1_extracted_k141_21196 | Not-determined |
| S1_extracted_k141_134538 | Not-determined |
| S1_extracted_k141_41326 | Not-determined |
| S1_extracted_k141_147377 | Low-quality |
| S1_extracted_k141_34360 | Not-determined |
| S1_extracted_k141_993 | Not-determined |
| S1_extracted_k141_101357 | Not-determined |
| S1_extracted_k141_994 | Low-quality |
| S1_extracted_k141_154193 | Not-determined |
| S1_extracted_k141_14828 | Low-quality |
| S1_extracted_k141_74560 | Not-determined |
| S1_extracted_k141_47901 | Not-determined |
| S1_extracted_k141_67864 | Not-determined |
| S1_extracted_k141_114548 | Not-determined |
| S1_extracted_k141_21204 | Not-determined |
| S1_extracted_k141_74561 | Not-determined |
| S1_extracted_k141_7967 | Not-determined |
| S1_extracted_k141_81357 | Not-determined |
| S1_extracted_k141_134547 | Not-determined |
| S1_extracted_k141_67866 | Not-determined |
| S1_extracted_k141_128005 | Not-determined |
| S1_extracted_k141_21208 | Low-quality |
| S1_extracted_k141_81362 | Not-determined |
| S1_extracted_k141_101362 | Not-determined |
| S1_extracted_k141_154201 | Not-determined |
| S1_extracted_k141_41337 | Not-determined |
| S1_extracted_k141_107866 | Not-determined |
| S1_extracted_k141_134552 | Not-determined |
| S1_extracted_k141_107868 | Not-determined |
| S1_extracted_k141_87970 | Not-determined |
| S1_extracted_k141_47912 | Low-quality |
| S1_extracted_k141_114557 | Low-quality |
| S1_extracted_k141_94712 | Not-determined |
| S1_extracted_k141_41341 | Not-determined |
| S1_extracted_k141_27819 | Not-determined |
| S1_extracted_k141_94714 | Not-determined |
| S1_extracted_k141_101368 | Not-determined |
| S1_extracted_k141_141331 | Low-quality |
| S1_extracted_k141_128013 | Not-determined |
| S1_extracted_k141_154202 | Not-determined |
| S1_extracted_k141_34369 | Not-determined |
| S1_extracted_k141_41346 | Not-determined |
| S1_extracted_k141_101369 | Not-determined |
| S1_extracted_k141_81367 | Not-determined |
| S1_extracted_k141_27823 | Low-quality |
| S1_extracted_k141_1011 | Not-determined |
| S1_extracted_k141_114562 | Not-determined |
| S1_extracted_k141_14841 | Not-determined |
| S1_extracted_k141_54616 | Not-determined |
| S1_extracted_k141_94715 | Not-determined |
| S1_extracted_k141_7982 | Not-determined |
| S1_extracted_k141_1013 | Not-determined |
| S1_extracted_k141_74578 | Not-determined |
| S1_extracted_k141_74580 | Not-determined |
| S1_extracted_k141_81373 | Not-determined |
| S1_extracted_k141_14844 | Not-determined |
| S1_extracted_k141_60747 | Not-determined |
| S1_extracted_k141_81376 | Not-determined |
| S1_extracted_k141_7986 | Not-determined |
| S1_extracted_k141_94718 | Not-determined |
| S1_extracted_k141_128028 | Not-determined |
| S1_extracted_k141_14850 | Low-quality |
| S1_extracted_k141_34375 | Low-quality |
| S1_extracted_k141_81380 | Not-determined |
| S1_extracted_k141_7988 | Not-determined |
| S1_extracted_k141_7989 | Not-determined |
| S1_extracted_k141_101376 | Not-determined |
| S1_extracted_k141_34377 | Not-determined |
| S1_extracted_k141_154205 | Low-quality |
| S1_extracted_k141_27842 | Not-determined |
| S1_extracted_k141_47929 | Low-quality |
| S1_extracted_k141_47930 | Not-determined |
| S1_extracted_k141_21219 | Low-quality |
| S1_extracted_k141_41358 | Low-quality |
| S1_extracted_k141_7993 | Low-quality |
| S1_extracted_k141_128035 | Not-determined |
| S1_extracted_k141_141332 | Low-quality |
| S1_extracted_k141_94726 | Not-determined |
| S1_extracted_k141_101380 | Not-determined |
| S1_extracted_k141_81386 | Not-determined |
| S1_extracted_k141_7998 | Not-determined |
| S1_extracted_k141_67892 | Not-determined |
| S1_extracted_k141_14864 | Not-determined |
| S1_extracted_k141_154208 | Not-determined |
| S1_extracted_k141_114583 | Low-quality |
| S1_extracted_k141_121138 | Not-determined |
| S1_extracted_k141_14867 | Not-determined |
| S1_extracted_k141_21222 | Not-determined |
| S1_extracted_k141_8004 | Not-determined |
| S1_extracted_k141_134566 | Not-determined |
| S1_extracted_k141_121142 | Not-determined |
| S1_extracted_k141_67897 | Low-quality |
| S1_extracted_k141_101387 | Not-determined |
| S1_extracted_k141_94729 | Not-determined |
| S1_extracted_k141_134569 | Not-determined |
| S1_extracted_k141_154213 | Low-quality |
| S1_extracted_k141_60768 | Not-determined |
| S1_extracted_k141_27852 | Not-determined |
| S1_extracted_k141_94731 | Not-determined |
| S1_extracted_k141_27853 | Low-quality |
| S1_extracted_k141_54635 | Not-determined |
| S1_extracted_k141_87998 | Not-determined |
| S1_extracted_k141_54636 | Not-determined |
| S1_extracted_k141_107912 | Not-determined |
| S1_extracted_k141_121145 | Not-determined |
| S1_extracted_k141_60770 | Not-determined |
| S1_extracted_k141_1036 | Not-determined |
| S1_extracted_k141_101391 | Not-determined |
| S1_extracted_k141_74605 | Not-determined |
| S1_extracted_k141_88002 | Not-determined |
| S1_extracted_k141_94735 | Not-determined |
| S1_extracted_k141_101394 | Not-determined |
| S1_extracted_k141_67907 | Not-determined |
| S1_extracted_k141_34392 | Not-determined |
| S1_extracted_k141_1041 | Not-determined |
| S1_extracted_k141_101396 | Not-determined |
| S1_extracted_k141_14879 | Not-determined |
| S1_extracted_k141_101399 | Not-determined |
| S1_extracted_k141_21232 | Not-determined |
| S1_extracted_k141_14881 | Not-determined |
| S1_extracted_k141_41366 | Not-determined |
| S1_extracted_k141_74611 | Not-determined |
| S1_extracted_k141_41368 | Not-determined |
| S1_extracted_k141_67911 | Low-quality |
| S1_extracted_k141_81399 | Low-quality |
| S1_extracted_k141_101401 | Not-determined |
| S1_extracted_k141_60778 | Not-determined |
| S1_extracted_k141_27863 | Low-quality |
| S1_extracted_k141_107915 | Not-determined |
| S1_extracted_k141_41372 | Not-determined |
| S1_extracted_k141_128049 | Not-determined |
| S1_extracted_k141_41373 | Not-determined |
| S1_extracted_k141_107918 | Not-determined |
| S1_extracted_k141_121158 | Not-determined |
| S1_extracted_k141_128053 | Not-determined |
| S1_extracted_k141_54651 | Not-determined |
| S1_extracted_k141_27869 | Low-quality |
| S1_extracted_k141_47960 | Low-quality |
| S1_extracted_k141_88015 | Not-determined |
| S1_extracted_k141_88016 | Not-determined |
| S1_extracted_k141_88017 | Not-determined |
| S1_extracted_k141_88018 | Not-determined |
| S1_extracted_k141_8021 | Not-determined |
| S1_extracted_k141_107922 | Low-quality |
| S1_extracted_k141_81409 | Low-quality |
| S1_extracted_k141_134587 | Low-quality |
| S1_extracted_k141_114601 | Not-determined |
| S1_extracted_k141_107923 | Not-determined |
| S1_extracted_k141_101418 | Not-determined |
| S1_extracted_k141_94751 | Not-determined |
| S1_extracted_k141_47965 | Low-quality |
| S1_extracted_k141_67925 | Not-determined |
| S1_extracted_k141_94752 | Not-determined |
| S1_extracted_k141_107924 | Not-determined |
| S1_extracted_k141_134591 | Not-determined |
| S1_extracted_k141_47968 | Not-determined |
| S1_extracted_k141_114602 | Not-determined |
| S1_extracted_k141_88024 | Not-determined |
| S1_extracted_k141_14888 | Low-quality |
| S1_extracted_k141_21250 | Not-determined |
| S1_extracted_k141_141349 | Not-determined |
| S1_extracted_k141_60792 | Not-determined |
| S1_extracted_k141_128062 | Not-determined |
| S1_extracted_k141_121170 | Low-quality |
| S1_extracted_k141_128063 | Low-quality |
| S1_extracted_k141_134594 | Not-determined |
| S1_extracted_k141_47973 | Not-determined |
| S1_extracted_k141_114608 | Not-determined |
| S1_extracted_k141_101426 | Not-determined |
| S1_extracted_k141_47977 | Not-determined |
| S1_extracted_k141_67935 | Not-determined |
| S1_extracted_k141_88025 | Low-quality |
| S1_extracted_k141_67937 | Not-determined |
| S1_extracted_k141_47980 | Not-determined |
| S1_extracted_k141_14901 | Not-determined |
| S1_extracted_k141_14902 | Not-determined |
| S1_extracted_k141_27891 | Not-determined |
| S1_extracted_k141_114620 | Not-determined |
| S1_extracted_k141_47985 | Low-quality |
| S1_extracted_k141_107928 | Not-determined |
| S1_extracted_k141_41381 | Low-quality |
| S1_extracted_k141_81415 | Low-quality |
| S1_extracted_k141_107931 | Low-quality |
| S1_extracted_k141_27897 | Not-determined |
| S1_extracted_k141_67943 | Not-determined |
| S1_extracted_k141_54653 | Low-quality |
| S1_extracted_k141_74642 | Not-determined |
| S1_extracted_k141_8040 | Low-quality |
| S1_extracted_k141_128078 | Not-determined |
| S1_extracted_k141_34414 | Low-quality |
| S1_extracted_k141_134597 | Not-determined |
| S1_extracted_k141_41387 | Not-determined |
| S1_extracted_k141_34415 | Not-determined |
| S1_extracted_k141_54655 | Not-determined |
| S1_extracted_k141_134598 | Not-determined |
| S1_extracted_k141_121192 | Not-determined |
| S1_extracted_k141_107938 | Not-determined |
| S1_extracted_k141_74646 | Not-determined |
| S1_extracted_k141_60806 | Not-determined |
| S1_extracted_k141_74649 | Not-determined |
| S1_extracted_k141_141358 | Low-quality |
| S1_extracted_k141_60807 | Not-determined |
| S1_extracted_k141_67948 | Not-determined |
| S1_extracted_k141_107943 | Not-determined |
| S1_extracted_k141_27903 | Low-quality |
| S1_extracted_k141_41391 | Low-quality |
| S1_extracted_k141_54662 | Not-determined |
| S1_extracted_k141_14914 | Not-determined |
| S1_extracted_k141_34423 | Not-determined |
| S1_extracted_k141_154229 | Not-determined |
| S1_extracted_k141_114626 | Not-determined |
| S1_extracted_k141_114627 | Not-determined |
| S1_extracted_k141_81426 | Not-determined |
| S1_extracted_k141_67954 | Not-determined |
| S1_extracted_k141_8054 | Not-determined |

| | |
|---|---|
| S1_extracted_k141_88042 | Not-determined |
| S1_extracted_k141_34428 | Not-determined |
| S1_extracted_k141_81428 | Not-determined |
| S1_extracted_k141_114630 | Not-determined |
| S1_extracted_k141_34429 | Not-determined |
| S1_extracted_k141_141366 | Not-determined |
| S1_extracted_k141_60811 | Low-quality |
| S1_extracted_k141_114632 | Low-quality |
| S1_extracted_k141_81429 | Not-determined |
| S1_extracted_k141_154232 | Low-quality |
| S1_extracted_k141_141370 | Low-quality |
| S1_extracted_k141_60815 | Low-quality |
| S1_extracted_k141_34433 | Not-determined |
| S1_extracted_k141_121193 | Low-quality |
| S1_extracted_k141_41411 | Low-quality |
| S1_extracted_k141_107958 | Not-determined |
| S1_extracted_k141_81431 | Not-determined |
| S1_extracted_k141_21267 | Not-determined |
| S1_extracted_k141_21268 | Not-determined |
| S1_extracted_k141_121196 | Low-quality |
| S1_extracted_k141_114645 | Low-quality |
| S1_extracted_k141_1060 | Not-determined |
| S1_extracted_k141_67966 | Not-determined |
| S1_extracted_k141_88051 | Not-determined |
| S1_extracted_k141_81435 | Not-determined |
| S1_extracted_k141_107963 | Not-determined |
| S1_extracted_k141_88053 | Low-quality |
| S1_extracted_k141_21272 | Not-determined |
| S1_extracted_k141_41417 | Not-determined |
| S1_extracted_k141_88054 | Not-determined |
| S1_extracted_k141_134625 | Not-determined |
| S1_extracted_k141_134626 | Not-determined |
| S1_extracted_k141_154241 | Not-determined |
| S1_extracted_k141_27924 | Not-determined |
| S1_extracted_k141_88057 | Not-determined |
| S1_extracted_k141_54679 | Not-determined |
| S1_extracted_k141_114653 | Not-determined |
| S1_extracted_k141_74681 | Not-determined |
| S1_extracted_k141_81437 | Low-quality |
| S1_extracted_k141_14940 | Not-determined |
| S1_extracted_k141_121209 | Not-determined |
| S1_extracted_k141_128107 | Not-determined |
| S1_extracted_k141_134631 | Not-determined |
| S1_extracted_k141_74683 | Not-determined |
| S1_extracted_k141_128109 | Low-quality |
| S1_extracted_k141_67980 | Not-determined |
| S1_extracted_k141_34448 | Not-determined |
| S1_extracted_k141_81444 | Not-determined |
| S1_extracted_k141_34450 | Not-determined |
| S1_extracted_k141_121212 | Not-determined |
| S1_extracted_k141_107972 | Low-quality |
| S1_extracted_k141_128114 | Not-determined |
| S1_extracted_k141_27931 | Not-determined |
| S1_extracted_k141_21283 | Not-determined |
| S1_extracted_k141_27932 | Not-determined |
| S1_extracted_k141_134640 | Not-determined |
| S1_extracted_k141_94798 | Not-determined |
| S1_extracted_k141_21285 | Not-determined |
| S1_extracted_k141_60825 | Not-determined |
| S1_extracted_k141_1081 | Not-determined |
| S1_extracted_k141_114670 | Not-determined |
| S1_extracted_k141_1082 | Not-determined |
| S1_extracted_k141_60826 | Not-determined |
| S1_extracted_k141_88066 | Not-determined |
| S1_extracted_k141_14958 | Not-determined |
| S1_extracted_k141_141402 | Not-determined |
| S1_extracted_k141_34458 | Not-determined |
| S1_extracted_k141_101494 | Not-determined |
| S1_extracted_k141_141403 | Low-quality |
| S1_extracted_k141_107982 | Not-determined |
| S1_extracted_k141_48030 | Not-determined |
| S1_extracted_k141_141405 | Not-determined |
| S1_extracted_k141_67989 | Not-determined |
| S1_extracted_k141_147430 | Not-determined |
| S1_extracted_k141_88071 | Not-determined |
| S1_extracted_k141_34461 | Not-determined |
| S1_extracted_k141_54701 | Not-determined |
| S1_extracted_k141_1089 | Not-determined |
| S1_extracted_k141_67993 | Not-determined |
| S1_extracted_k141_21297 | Not-determined |
| S1_extracted_k141_41449 | Low-quality |
| S1_extracted_k141_107992 | Not-determined |
| S1_extracted_k141_1092 | Not-determined |
| S1_extracted_k141_107995 | Not-determined |
| S1_extracted_k141_81460 | Not-determined |
| S1_extracted_k141_114684 | Not-determined |
| S1_extracted_k141_114685 | Not-determined |
| S1_extracted_k141_107997 | Not-determined |
| S1_extracted_k141_54707 | Not-determined |
| S1_extracted_k141_114687 | Not-determined |
| S1_extracted_k141_60828 | Not-determined |
| S1_extracted_k141_81465 | Not-determined |
| S1_extracted_k141_121224 | Not-determined |
| S1_extracted_k141_121225 | Not-determined |
| S1_extracted_k141_14972 | Not-determined |
| S1_extracted_k141_81467 | Not-determined |
| S1_extracted_k141_27944 | Not-determined |
| S1_extracted_k141_81469 | Low-quality |
| S1_extracted_k141_114691 | Not-determined |
| S1_extracted_k141_141416 | Not-determined |
| S1_extracted_k141_34469 | Not-determined |
| S1_extracted_k141_141418 | Not-determined |
| S1_extracted_k141_141419 | Not-determined |
| S1_extracted_k141_121230 | Not-determined |
| S1_extracted_k141_101517 | Low-quality |
| S1_extracted_k141_14978 | Not-determined |
| S1_extracted_k141_108006 | Not-determined |
| S1_extracted_k141_147432 | Low-quality |
| S1_extracted_k141_114701 | Not-determined |
| S1_extracted_k141_88092 | Not-determined |
| S1_extracted_k141_88093 | Not-determined |
| S1_extracted_k141_114704 | Not-determined |
| S1_extracted_k141_114705 | Not-determined |
| S1_extracted_k141_88096 | Not-determined |
| S1_extracted_k141_114707 | Low-quality |
| S1_extracted_k141_121238 | Not-determined |
| S1_extracted_k141_41461 | Not-determined |
| S1_extracted_k141_88098 | Low-quality |
| S1_extracted_k141_14982 | Low-quality |
| S1_extracted_k141_134671 | Not-determined |
| S1_extracted_k141_60840 | Not-determined |
| S1_extracted_k141_27962 | Not-determined |
| S1_extracted_k141_14984 | Not-determined |
| S1_extracted_k141_8100 | Not-determined |
| S1_extracted_k141_54718 | Not-determined |
| S1_extracted_k141_81475 | Not-determined |
| S1_extracted_k141_147438 | Low-quality |
| S1_extracted_k141_41463 | Not-determined |
| S1_extracted_k141_121242 | Not-determined |
| S1_extracted_k141_108007 | Low-quality |
| S1_extracted_k141_8103 | Not-determined |
| S1_extracted_k141_154262 | Not-determined |
| S1_extracted_k141_114711 | Not-determined |
| S1_extracted_k141_94826 | Not-determined |
| S1_extracted_k141_21328 | Not-determined |
| S1_extracted_k141_27970 | Not-determined |
| S1_extracted_k141_128140 | Not-determined |
| S1_extracted_k141_14992 | Not-determined |
| S1_extracted_k141_134679 | Not-determined |
| S1_extracted_k141_147441 | Not-determined |
| S1_extracted_k141_8109 | Not-determined |
| S1_extracted_k141_27974 | Not-determined |
| S1_extracted_k141_34484 | Not-determined |
| S1_extracted_k141_154263 | Not-determined |
| S1_extracted_k141_121243 | Low-quality |
| S1_extracted_k141_94833 | Not-determined |
| S1_extracted_k141_114718 | Not-determined |
| S1_extracted_k141_94835 | Not-determined |
| S1_extracted_k141_14998 | Not-determined |
| S1_extracted_k141_81483 | Low-quality |
| S1_extracted_k141_81484 | Low-quality |
| S1_extracted_k141_88111 | Not-determined |
| S1_extracted_k141_21343 | Not-determined |
| S1_extracted_k141_54727 | Not-determined |
| S1_extracted_k141_141438 | Not-determined |
| S1_extracted_k141_68027 | Low-quality |
| S1_extracted_k141_68028 | Low-quality |
| S1_extracted_k141_21350 | Not-determined |
| S1_extracted_k141_141440 | Not-determined |
| S1_extracted_k141_27988 | Not-determined |
| S1_extracted_k141_41474 | Not-determined |
| S1_extracted_k141_21354 | Not-determined |
| S1_extracted_k141_68032 | Not-determined |
| S1_extracted_k141_141441 | Low-quality |
| S1_extracted_k141_154275 | Not-determined |
| S1_extracted_k141_34496 | Not-determined |
| S1_extracted_k141_114732 | Low-quality |
| S1_extracted_k141_74742 | Not-determined |
| S1_extracted_k141_114733 | Not-determined |
| S1_extracted_k141_27994 | Low-quality |
| S1_extracted_k141_134699 | Low-quality |
| S1_extracted_k141_101538 | Not-determined |
| S1_extracted_k141_121251 | Not-determined |
| S1_extracted_k141_27996 | Not-determined |
| S1_extracted_k141_121252 | Not-determined |
| S1_extracted_k141_48070 | Not-determined |
| S1_extracted_k141_54737 | Not-determined |
| S1_extracted_k141_88119 | Not-determined |
| S1_extracted_k141_154280 | Not-determined |
| S1_extracted_k141_48073 | Not-determined |
| S1_extracted_k141_141451 | Not-determined |
| S1_extracted_k141_141452 | Not-determined |
| S1_extracted_k141_101545 | Low-quality |
| S1_extracted_k141_134706 | Not-determined |
| S1_extracted_k141_81497 | Not-determined |
| S1_extracted_k141_68050 | Not-determined |
| S1_extracted_k141_48078 | Not-determined |
| S1_extracted_k141_74756 | Not-determined |
| S1_extracted_k141_101548 | Low-quality |
| S1_extracted_k141_154283 | Not-determined |
| S1_extracted_k141_1132 | Not-determined |
| S1_extracted_k141_15025 | Not-determined |
| S1_extracted_k141_108033 | Not-determined |
| S1_extracted_k141_94861 | Not-determined |
| S1_extracted_k141_101552 | Not-determined |
| S1_extracted_k141_41497 | Not-determined |
| S1_extracted_k141_68053 | Not-determined |
| S1_extracted_k141_121264 | Not-determined |
| S1_extracted_k141_128150 | Not-determined |
| S1_extracted_k141_94865 | Not-determined |
| S1_extracted_k141_114747 | Not-determined |
| S1_extracted_k141_54753 | Not-determined |
| S1_extracted_k141_94867 | Not-determined |
| S1_extracted_k141_54754 | Not-determined |
| S1_extracted_k141_134715 | Low-quality |
| S1_extracted_k141_101557 | Not-determined |
| S1_extracted_k141_41502 | Not-determined |
| S1_extracted_k141_114753 | Low-quality |
| S1_extracted_k141_68058 | Not-determined |
| S1_extracted_k141_81507 | Not-determined |
| S1_extracted_k141_21377 | Not-determined |
| S1_extracted_k141_121269 | Not-determined |
| S1_extracted_k141_81508 | Low-quality |
| S1_extracted_k141_114756 | Not-determined |
| S1_extracted_k141_141465 | Not-determined |
| S1_extracted_k141_141466 | Not-determined |
| S1_extracted_k141_121270 | Not-determined |
| S1_extracted_k141_101562 | Low-quality |
| S1_extracted_k141_28011 | Not-determined |
| S1_extracted_k141_8142 | Not-determined |
| S1_extracted_k141_28012 | Not-determined |
| S1_extracted_k141_54766 | Not-determined |
| S1_extracted_k141_28014 | Not-determined |
| S1_extracted_k141_121273 | Not-determined |
| S1_extracted_k141_128160 | Not-determined |
| S1_extracted_k141_8143 | Low-quality |
| S1_extracted_k141_121275 | Not-determined |
| S1_extracted_k141_15037 | Low-quality |
| S1_extracted_k141_68068 | Not-determined |
| S1_extracted_k141_8147 | Not-determined |
| S1_extracted_k141_121278 | Not-determined |
| S1_extracted_k141_1142 | Not-determined |
| S1_extracted_k141_34509 | Low-quality |
| S1_extracted_k141_154297 | Not-determined |
| S1_extracted_k141_121280 | Not-determined |
| S1_extracted_k141_34511 | Low-quality |
| S1_extracted_k141_154299 | Not-determined |
| S1_extracted_k141_48091 | Not-determined |
| S1_extracted_k141_88130 | Low-quality |
| S1_extracted_k141_94880 | Not-determined |
| S1_extracted_k141_101572 | Not-determined |
| S1_extracted_k141_74771 | Low-quality |
| S1_extracted_k141_81523 | Not-determined |
| S1_extracted_k141_141477 | Not-determined |
| S1_extracted_k141_134731 | Low-quality |
| S1_extracted_k141_81524 | Not-determined |
| S1_extracted_k141_81525 | Not-determined |
| S1_extracted_k141_108051 | Low-quality |
| S1_extracted_k141_134732 | Not-determined |
| S1_extracted_k141_94883 | Not-determined |
| S1_extracted_k141_88134 | Not-determined |
| S1_extracted_k141_94884 | Not-determined |
| S1_extracted_k141_74773 | Not-determined |
| S1_extracted_k141_121286 | Not-determined |
| S1_extracted_k141_68082 | Not-determined |
| S1_extracted_k141_134734 | Low-quality |
| S1_extracted_k141_101579 | Not-determined |
| S1_extracted_k141_81527 | Not-determined |
| S1_extracted_k141_101582 | Not-determined |
| S1_extracted_k141_68084 | Not-determined |
| S1_extracted_k141_154303 | Low-quality |
| S1_extracted_k141_94892 | Not-determined |
| S1_extracted_k141_28025 | Not-determined |
| S1_extracted_k141_88138 | Not-determined |
| S1_extracted_k141_154304 | Not-determined |
| S1_extracted_k141_28026 | Not-determined |
| S1_extracted_k141_154305 | Not-determined |
| S1_extracted_k141_21404 | Low-quality |
| S1_extracted_k141_8160 | Not-determined |
| S1_extracted_k141_141487 | Not-determined |
| S1_extracted_k141_128168 | Not-determined |
| S1_extracted_k141_54785 | Low-quality |
| S1_extracted_k141_54786 | Not-determined |
| S1_extracted_k141_34524 | Low-quality |
| S1_extracted_k141_68085 | Low-quality |
| S1_extracted_k141_15049 | Not-determined |
| S1_extracted_k141_54788 | Not-determined |
| S1_extracted_k141_41535 | Not-determined |
| S1_extracted_k141_48102 | Not-determined |
| S1_extracted_k141_28032 | Low-quality |
| S1_extracted_k141_15052 | Not-determined |
| S1_extracted_k141_1149 | Not-determined |
| S1_extracted_k141_94900 | Not-determined |
| S1_extracted_k141_101585 | Not-determined |
| S1_extracted_k141_81534 | Not-determined |
| S1_extracted_k141_108063 | Not-determined |
| S1_extracted_k141_108064 | Not-determined |
| S1_extracted_k141_34530 | Not-determined |
| S1_extracted_k141_121299 | Not-determined |
| S1_extracted_k141_60858 | Not-determined |
| S1_extracted_k141_128172 | Low-quality |
| S1_extracted_k141_134750 | Not-determined |
| S1_extracted_k141_128176 | Not-determined |
| S1_extracted_k141_134753 | Not-determined |
| S1_extracted_k141_48114 | Not-determined |
| S1_extracted_k141_114792 | Not-determined |
| S1_extracted_k141_141498 | Not-determined |
| S1_extracted_k141_94911 | Not-determined |
| S1_extracted_k141_154312 | Not-determined |
| S1_extracted_k141_1162 | Not-determined |
| S1_extracted_k141_114797 | Not-determined |
| S1_extracted_k141_141501 | Not-determined |
| S1_extracted_k141_68097 | Not-determined |
| S1_extracted_k141_101595 | Not-determined |
| S1_extracted_k141_54798 | Not-determined |
| S1_extracted_k141_68099 | Low-quality |
| S1_extracted_k141_121312 | Not-determined |
| S1_extracted_k141_54801 | Not-determined |
| S1_extracted_k141_48119 | Low-quality |
| S1_extracted_k141_28049 | Not-determined |
| S1_extracted_k141_88158 | Not-determined |
| S1_extracted_k141_81554 | Not-determined |
| S1_extracted_k141_34538 | Low-quality |
| S1_extracted_k141_21410 | Not-determined |
| S1_extracted_k141_15061 | Low-quality |
| S1_extracted_k141_128184 | Low-quality |
| S1_extracted_k141_101599 | Low-quality |
| S1_extracted_k141_28053 | Not-determined |
| S1_extracted_k141_154319 | Low-quality |
| S1_extracted_k141_28054 | Low-quality |
| S1_extracted_k141_121315 | Not-determined |
| S1_extracted_k141_81559 | Not-determined |
| S1_extracted_k141_88164 | Not-determined |
| S1_extracted_k141_88165 | Not-determined |
| S1_extracted_k141_147470 | Not-determined |
| S1_extracted_k141_121317 | Not-determined |
| S1_extracted_k141_147471 | Not-determined |
| S1_extracted_k141_94922 | Not-determined |
| S1_extracted_k141_147472 | Not-determined |
| S1_extracted_k141_34543 | Not-determined |
| S1_extracted_k141_28058 | Not-determined |
| S1_extracted_k141_1170 | Low-quality |
| S1_extracted_k141_60866 | Not-determined |
| S1_extracted_k141_88169 | Not-determined |
| S1_extracted_k141_1172 | Not-determined |
| S1_extracted_k141_34547 | Not-determined |
| S1_extracted_k141_101605 | Not-determined |
| S1_extracted_k141_154321 | Not-determined |
| S1_extracted_k141_48129 | Not-determined |
| S1_extracted_k141_88176 | Low-quality |
| S1_extracted_k141_94935 | Low-quality |
| S1_extracted_k141_41562 | Not-determined |
| S1_extracted_k141_41564 | Low-quality |
| S1_extracted_k141_147478 | Not-determined |
| S1_extracted_k141_28070 | Not-determined |
| S1_extracted_k141_15077 | Not-determined |
| S1_extracted_k141_8196 | Not-determined |
| S1_extracted_k141_15079 | Not-determined |
| S1_extracted_k141_41569 | Not-determined |
| S1_extracted_k141_41570 | Not-determined |
| S1_extracted_k141_81570 | Low-quality |
| S1_extracted_k141_28075 | Not-determined |
| S1_extracted_k141_101618 | Not-determined |
| S1_extracted_k141_121337 | Low-quality |
| S1_extracted_k141_108090 | Not-determined |
| S1_extracted_k141_147481 | Low-quality |
| S1_extracted_k141_114818 | Not-determined |
| S1_extracted_k141_147482 | Low-quality |
| S1_extracted_k141_41577 | Not-determined |
| S1_extracted_k141_15085 | Not-determined |
| S1_extracted_k141_141510 | Not-determined |
| S1_extracted_k141_108092 | Not-determined |
| S1_extracted_k141_28079 | Not-determined |
| S1_extracted_k141_101622 | Not-determined |
| S1_extracted_k141_28080 | Not-determined |
| S1_extracted_k141_114822 | Not-determined |
| S1_extracted_k141_15087 | Not-determined |
| S1_extracted_k141_48144 | Not-determined |
| S1_extracted_k141_74815 | Not-determined |
| S1_extracted_k141_41581 | Low-quality |
| S1_extracted_k141_134782 | Not-determined |
| S1_extracted_k141_101626 | Not-determined |
| S1_extracted_k141_154338 | Low-quality |
| S1_extracted_k141_74821 | Low-quality |
| S1_extracted_k141_94939 | Not-determined |
| S1_extracted_k141_74823 | Not-determined |
| S1_extracted_k141_28088 | Not-determined |
| S1_extracted_k141_1174 | Low-quality |
| S1_extracted_k141_48152 | Not-determined |
| S1_extracted_k141_48153 | Not-determined |
| S1_extracted_k141_121349 | Not-determined |
| S1_extracted_k141_48155 | Not-determined |
| S1_extracted_k141_94946 | Not-determined |
| S1_extracted_k141_128226 | Not-determined |
| S1_extracted_k141_88183 | Not-determined |
| S1_extracted_k141_128227 | Not-determined |
| S1_extracted_k141_134790 | Not-determined |
| S1_extracted_k141_15097 | Low-quality |
| S1_extracted_k141_154347 | Not-determined |
| S1_extracted_k141_141535 | Not-determined |
| S1_extracted_k141_68132 | Low-quality |
| S1_extracted_k141_114828 | Not-determined |
| S1_extracted_k141_101637 | Not-determined |
| S1_extracted_k141_74835 | Not-determined |
| S1_extracted_k141_41588 | Low-quality |
| S1_extracted_k141_88190 | Not-determined |
| S1_extracted_k141_154351 | Not-determined |
| S1_extracted_k141_101639 | Low-quality |
| S1_extracted_k141_74838 | Not-determined |
| S1_extracted_k141_101641 | Not-determined |
| S1_extracted_k141_128238 | Not-determined |
| S1_extracted_k141_147489 | Not-determined |
| S1_extracted_k141_128240 | Not-determined |
| S1_extracted_k141_141540 | Not-determined |
| S1_extracted_k141_15112 | Not-determined |
| S1_extracted_k141_147492 | Not-determined |
| S1_extracted_k141_1188 | Not-determined |
| S1_extracted_k141_28100 | Not-determined |
| S1_extracted_k141_128241 | Low-quality |
| S1_extracted_k141_101645 | Not-determined |
| S1_extracted_k141_88198 | Not-determined |
| S1_extracted_k141_134809 | Not-determined |
| S1_extracted_k141_48178 | Low-quality |
| S1_extracted_k141_74844 | Not-determined |
| S1_extracted_k141_1193 | Not-determined |
| S1_extracted_k141_28107 | Not-determined |
| S1_extracted_k141_94964 | Low-quality |
| S1_extracted_k141_34588 | Not-determined |
| S1_extracted_k141_101652 | Not-determined |
| S1_extracted_k141_54858 | Low-quality |
| S1_extracted_k141_54859 | Low-quality |
| S1_extracted_k141_141549 | Low-quality |
| S1_extracted_k141_54862 | Not-determined |
| S1_extracted_k141_134815 | Not-determined |
| S1_extracted_k141_1202 | Not-determined |
| S1_extracted_k141_154362 | Not-determined |
| S1_extracted_k141_134817 | Not-determined |
| S1_extracted_k141_114851 | Not-determined |
| S1_extracted_k141_154363 | Not-determined |
| S1_extracted_k141_147499 | Not-determined |
| S1_extracted_k141_108126 | Not-determined |
| S1_extracted_k141_108131 | Not-determined |
| S1_extracted_k141_101660 | Low-quality |
| S1_extracted_k141_48194 | Not-determined |
| S1_extracted_k141_15138 | Low-quality |
| S1_extracted_k141_134820 | Low-quality |
| S1_extracted_k141_94979 | Not-determined |
| S1_extracted_k141_1212 | Not-determined |
| S1_extracted_k141_134821 | Not-determined |
| S1_extracted_k141_134822 | Not-determined |
| S1_extracted_k141_147505 | Not-determined |
| S1_extracted_k141_128248 | Not-determined |
| S1_extracted_k141_8251 | Not-determined |
| S1_extracted_k141_74858 | Not-determined |
| S1_extracted_k141_60888 | Not-determined |
| S1_extracted_k141_88218 | Not-determined |
| S1_extracted_k141_15148 | Not-determined |
| S1_extracted_k141_1219 | Not-determined |
| S1_extracted_k141_48203 | Not-determined |
| S1_extracted_k141_15149 | Not-determined |
| S1_extracted_k141_141563 | Not-determined |
| S1_extracted_k141_15150 | Low-quality |
| S1_extracted_k141_101673 | Not-determined |
| S1_extracted_k141_154367 | Not-determined |
| S1_extracted_k141_15154 | Not-determined |
| S1_extracted_k141_88224 | Not-determined |
| S1_extracted_k141_88226 | Not-determined |
| S1_extracted_k141_8264 | Low-quality |
| S1_extracted_k141_101677 | Not-determined |
| S1_extracted_k141_74867 | Low-quality |
| S1_extracted_k141_74868 | Not-determined |
| S1_extracted_k141_34611 | Not-determined |
| S1_extracted_k141_1228 | Not-determined |
| S1_extracted_k141_88231 | Not-determined |
| S1_extracted_k141_88232 | Low-quality |
| S1_extracted_k141_34613 | Not-determined |
| S1_extracted_k141_60897 | Low-quality |
| S1_extracted_k141_15161 | Not-determined |
| S1_extracted_k141_74870 | Not-determined |
| S1_extracted_k141_41602 | Not-determined |
| S1_extracted_k141_74872 | Not-determined |
| S1_extracted_k141_41603 | Not-determined |
| S1_extracted_k141_28134 | Low-quality |
| S1_extracted_k141_74873 | Not-determined |
| S1_extracted_k141_88237 | Not-determined |
| S1_extracted_k141_114871 | Not-determined |
| S1_extracted_k141_134843 | Not-determined |
| S1_extracted_k141_95003 | Not-determined |
| S1_extracted_k141_108142 | Not-determined |
| S1_extracted_k141_114873 | Not-determined |
| S1_extracted_k141_108143 | Low-quality |
| S1_extracted_k141_48220 | Not-determined |
| S1_extracted_k141_41609 | Not-determined |
| S1_extracted_k141_34616 | Not-determined |
| S1_extracted_k141_74878 | Not-determined |
| S1_extracted_k141_1235 | Not-determined |
| S1_extracted_k141_41611 | Low-quality |
| S1_extracted_k141_134846 | Not-determined |
| S1_extracted_k141_128266 | Low-quality |
| S1_extracted_k141_128267 | Not-determined |
| S1_extracted_k141_34620 | Not-determined |
| S1_extracted_k141_8275 | Low-quality |
| S1_extracted_k141_108152 | Not-determined |
| S1_extracted_k141_48225 | Not-determined |
| S1_extracted_k141_68172 | Not-determined |
| S1_extracted_k141_21445 | Not-determined |
| S1_extracted_k141_114883 | Not-determined |
| S1_extracted_k141_108154 | Low-quality |
| S1_extracted_k141_68177 | Not-determined |
| S1_extracted_k141_108155 | Not-determined |
| S1_extracted_k141_54870 | Not-determined |
| S1_extracted_k141_8280 | Not-determined |
| S1_extracted_k141_28149 | Not-determined |
| S1_extracted_k141_68179 | Not-determined |
| S1_extracted_k141_88241 | Not-determined |
| S1_extracted_k141_48236 | Not-determined |
| S1_extracted_k141_101696 | Not-determined |
| S1_extracted_k141_41624 | Not-determined |
| S1_extracted_k141_15178 | Not-determined |
| S1_extracted_k141_95013 | Not-determined |
| S1_extracted_k141_101699 | Not-determined |
| S1_extracted_k141_88245 | Not-determined |
| S1_extracted_k141_28157 | Not-determined |
| S1_extracted_k141_101703 | Not-determined |
| S1_extracted_k141_74894 | Not-determined |
| S1_extracted_k141_41626 | Not-determined |
| S1_extracted_k141_8285 | Not-determined |
| S1_extracted_k141_60914 | Not-determined |
| S1_extracted_k141_21455 | Not-determined |
| S1_extracted_k141_108168 | Not-determined |
| S1_extracted_k141_95017 | Not-determined |
| S1_extracted_k141_8288 | Not-determined |
| S1_extracted_k141_68182 | Not-determined |
| S1_extracted_k141_108172 | Not-determined |
| S1_extracted_k141_21463 | Not-determined |
| S1_extracted_k141_81600 | Low-quality |
| S1_extracted_k141_88251 | Not-determined |
| S1_extracted_k141_121408 | Not-determined |
| S1_extracted_k141_21466 | Not-determined |
| S1_extracted_k141_41631 | Not-determined |
| S1_extracted_k141_134869 | Low-quality |
| S1_extracted_k141_48251 | Not-determined |
| S1_extracted_k141_81604 | Not-determined |
| S1_extracted_k141_147516 | Not-determined |
| S1_extracted_k141_54883 | Not-determined |
| S1_extracted_k141_101713 | Not-determined |
| S1_extracted_k141_1258 | Not-determined |
| S1_extracted_k141_121412 | Not-determined |
| S1_extracted_k141_74904 | Not-determined |
| S1_extracted_k141_54886 | Not-determined |
| S1_extracted_k141_81611 | Not-determined |
| S1_extracted_k141_81612 | Not-determined |
| S1_extracted_k141_34642 | Not-determined |
| S1_extracted_k141_128297 | Low-quality |
| S1_extracted_k141_81615 | Low-quality |
| S1_extracted_k141_28164 | Low-quality |
| S1_extracted_k141_21472 | Not-determined |
| S1_extracted_k141_81617 | Low-quality |
| S1_extracted_k141_8301 | Not-determined |
| S1_extracted_k141_154392 | Not-determined |
| S1_extracted_k141_134874 | Not-determined |
| S1_extracted_k141_54892 | Not-determined |
| S1_extracted_k141_68197 | Low-quality |
| S1_extracted_k141_74913 | Not-determined |
| S1_extracted_k141_1259 | Not-determined |
| S1_extracted_k141_95036 | Not-determined |
| S1_extracted_k141_101719 | Not-determined |
| S1_extracted_k141_95038 | Not-determined |
| S1_extracted_k141_108181 | Low-quality |
| S1_extracted_k141_48266 | Not-determined |
| S1_extracted_k141_128306 | Low-quality |
| S1_extracted_k141_128307 | Not-determined |
| S1_extracted_k141_15198 | Not-determined |
| S1_extracted_k141_60919 | Not-determined |
| S1_extracted_k141_81628 | Not-determined |
| S1_extracted_k141_41641 | Low-quality |
| S1_extracted_k141_121420 | Not-determined |
| S1_extracted_k141_141612 | Not-determined |
| S1_extracted_k141_68204 | Not-determined |
| S1_extracted_k141_21484 | Not-determined |
| S1_extracted_k141_81630 | Not-determined |
| S1_extracted_k141_28176 | Low-quality |
| S1_extracted_k141_88262 | Low-quality |
| S1_extracted_k141_147526 | Not-determined |
| S1_extracted_k141_41648 | Not-determined |
| S1_extracted_k141_95045 | Low-quality |
| S1_extracted_k141_8309 | Not-determined |
| S1_extracted_k141_74927 | Not-determined |
| S1_extracted_k141_154399 | Not-determined |
| S1_extracted_k141_114922 | Not-determined |
| S1_extracted_k141_81637 | Not-determined |
| S1_extracted_k141_154401 | Not-determined |
| S1_extracted_k141_48273 | Low-quality |
| S1_extracted_k141_108187 | Not-determined |
| S1_extracted_k141_147530 | Not-determined |
| S1_extracted_k141_8314 | Not-determined |
| S1_extracted_k141_108188 | Not-determined |
| S1_extracted_k141_48278 | Low-quality |
| S1_extracted_k141_121431 | Not-determined |

| | |
|---|---|
| S1_extracted_k141_8319 | Not-determined |
| S1_extracted_k141_101736 | Not-determined |
| S1_extracted_k141_8322 | Not-determined |
| S1_extracted_k141_54914 | Not-determined |
| S1_extracted_k141_74939 | Not-determined |
| S1_extracted_k141_147536 | Not-determined |
| S1_extracted_k141_28198 | Not-determined |
| S1_extracted_k141_147537 | Not-determined |
| S1_extracted_k141_88278 | Not-determined |
| S1_extracted_k141_114926 | Low-quality |
| S1_extracted_k141_28200 | Not-determined |
| S1_extracted_k141_121438 | Low-quality |
| S1_extracted_k141_74943 | Not-determined |
| S1_extracted_k141_54921 | Not-determined |
| S1_extracted_k141_74944 | Not-determined |
| S1_extracted_k141_81649 | Not-determined |
| S1_extracted_k141_34663 | Not-determined |
| S1_extracted_k141_34664 | Not-determined |
| S1_extracted_k141_41654 | Not-determined |
| S1_extracted_k141_128330 | Not-determined |
| S1_extracted_k141_88283 | Not-determined |
| S1_extracted_k141_28206 | Not-determined |
| S1_extracted_k141_108203 | Not-determined |
| S1_extracted_k141_108204 | Not-determined |
| S1_extracted_k141_88285 | Not-determined |
| S1_extracted_k141_154411 | Not-determined |
| S1_extracted_k141_41661 | Not-determined |
| S1_extracted_k141_101740 | Not-determined |
| S1_extracted_k141_128338 | Not-determined |
| S1_extracted_k141_8333 | Not-determined |
| S1_extracted_k141_21499 | Not-determined |
| S1_extracted_k141_128340 | Not-determined |
| S1_extracted_k141_134891 | Not-determined |
| S1_extracted_k141_41667 | Low-quality |
| S1_extracted_k141_21502 | Not-determined |
| S1_extracted_k141_114945 | Not-determined |
| S1_extracted_k141_134895 | Not-determined |
| S1_extracted_k141_121448 | Not-determined |
| S1_extracted_k141_108210 | Low-quality |
| S1_extracted_k141_74959 | Low-quality |
| S1_extracted_k141_8338 | Not-determined |
| S1_extracted_k141_15210 | Low-quality |
| S1_extracted_k141_1290 | Not-determined |
| S1_extracted_k141_34671 | Not-determined |
| S1_extracted_k141_8341 | Not-determined |
| S1_extracted_k141_21506 | Not-determined |
| S1_extracted_k141_95075 | Not-determined |
| S1_extracted_k141_1293 | Not-determined |
| S1_extracted_k141_81650 | Low-quality |
| S1_extracted_k141_108220 | Not-determined |
| S1_extracted_k141_108221 | Not-determined |
| S1_extracted_k141_41672 | Not-determined |
| S1_extracted_k141_21511 | Not-determined |
| S1_extracted_k141_60934 | Not-determined |
| S1_extracted_k141_48301 | Not-determined |
| S1_extracted_k141_101745 | Not-determined |
| S1_extracted_k141_28219 | Low-quality |
| S1_extracted_k141_141647 | Not-determined |
| S1_extracted_k141_128342 | Not-determined |
| S1_extracted_k141_134908 | Not-determined |
| S1_extracted_k141_128345 | Not-determined |
| S1_extracted_k141_108227 | Low-quality |
| S1_extracted_k141_28222 | Not-determined |
| S1_extracted_k141_128347 | Not-determined |
| S1_extracted_k141_8358 | Not-determined |
| S1_extracted_k141_121465 | Low-quality |
| S1_extracted_k141_41677 | Not-determined |
| S1_extracted_k141_41678 | Not-determined |
| S1_extracted_k141_88308 | Not-determined |
| S1_extracted_k141_114965 | Not-determined |
| S1_extracted_k141_74970 | Not-determined |
| S1_extracted_k141_81670 | Not-determined |
| S1_extracted_k141_8370 | Not-determined |
| S1_extracted_k141_88314 | Not-determined |
| S1_extracted_k141_74975 | Low-quality |
| S1_extracted_k141_88316 | Low-quality |
| S1_extracted_k141_54954 | Low-quality |
| S1_extracted_k141_74978 | Low-quality |
| S1_extracted_k141_95084 | Low-quality |
| S1_extracted_k141_74980 | Not-determined |
| S1_extracted_k141_101755 | Not-determined |
| S1_extracted_k141_101757 | Not-determined |
| S1_extracted_k141_8374 | Low-quality |
| S1_extracted_k141_128354 | Low-quality |
| S1_extracted_k141_34696 | Not-determined |
| S1_extracted_k141_41696 | Not-determined |
| S1_extracted_k141_95092 | Not-determined |
| S1_extracted_k141_74990 | Not-determined |
| S1_extracted_k141_68240 | Not-determined |
| S1_extracted_k141_21532 | Not-determined |
| S1_extracted_k141_128359 | Not-determined |
| S1_extracted_k141_74992 | Not-determined |
| S1_extracted_k141_68245 | Not-determined |
| S1_extracted_k141_121485 | Low-quality |
| S1_extracted_k141_121487 | Not-determined |
| S1_extracted_k141_81692 | Not-determined |
| S1_extracted_k141_141662 | Low-quality |
| S1_extracted_k141_134919 | Not-determined |
| S1_extracted_k141_88319 | Not-determined |
| S1_extracted_k141_81693 | Not-determined |
| S1_extracted_k141_8386 | Not-determined |
| S1_extracted_k141_21534 | Low-quality |
| S1_extracted_k141_74999 | Not-determined |
| S1_extracted_k141_28245 | Not-determined |
| S1_extracted_k141_68251 | Low-quality |
| S1_extracted_k141_95100 | Low-quality |
| S1_extracted_k141_95102 | Low-quality |
| S1_extracted_k141_54976 | Not-determined |
| S1_extracted_k141_141670 | Not-determined |
| S1_extracted_k141_68254 | Not-determined |
| S1_extracted_k141_114985 | Not-determined |
| S1_extracted_k141_95105 | Not-determined |
| S1_extracted_k141_21544 | Not-determined |
| S1_extracted_k141_95107 | Not-determined |
| S1_extracted_k141_68257 | Not-determined |
| S1_extracted_k141_154439 | Not-determined |
| S1_extracted_k141_28255 | Not-determined |
| S1_extracted_k141_128374 | Not-determined |
| S1_extracted_k141_81705 | Not-determined |
| S1_extracted_k141_28256 | Low-quality |
| S1_extracted_k141_68260 | Not-determined |
| S1_extracted_k141_8400 | Not-determined |
| S1_extracted_k141_75017 | Not-determined |
| S1_extracted_k141_60955 | Not-determined |
| S1_extracted_k141_81710 | Not-determined |
| S1_extracted_k141_114994 | Not-determined |
| S1_extracted_k141_75021 | Not-determined |
| S1_extracted_k141_81711 | Not-determined |
| S1_extracted_k141_68264 | Low-quality |
| S1_extracted_k141_114998 | Not-determined |
| S1_extracted_k141_147543 | Low-quality |
| S1_extracted_k141_68267 | Not-determined |
| S1_extracted_k141_21552 | Not-determined |
| S1_extracted_k141_60959 | Not-determined |
| S1_extracted_k141_115002 | Not-determined |
| S1_extracted_k141_28266 | Not-determined |
| S1_extracted_k141_34708 | Not-determined |
| S1_extracted_k141_95118 | Not-determined |
| S1_extracted_k141_54989 | Not-determined |
| S1_extracted_k141_115007 | Not-determined |
| S1_extracted_k141_128384 | Not-determined |
| S1_extracted_k141_15271 | Not-determined |
| S1_extracted_k141_95120 | Not-determined |
| S1_extracted_k141_21554 | Low-quality |
| S1_extracted_k141_21555 | Not-determined |
| S1_extracted_k141_115010 | Not-determined |
| S1_extracted_k141_15273 | Not-determined |
| S1_extracted_k141_8416 | Not-determined |
| S1_extracted_k141_15274 | Low-quality |
| S1_extracted_k141_1344 | Low-quality |
| S1_extracted_k141_1345 | Not-determined |
| S1_extracted_k141_60964 | Low-quality |
| S1_extracted_k141_108263 | Low-quality |
| S1_extracted_k141_128392 | Not-determined |
| S1_extracted_k141_15280 | Not-determined |
| S1_extracted_k141_15281 | Not-determined |
| S1_extracted_k141_141690 | Not-determined |
| S1_extracted_k141_75038 | Not-determined |
| S1_extracted_k141_21562 | Not-determined |
| S1_extracted_k141_8424 | Not-determined |
| S1_extracted_k141_115017 | Not-determined |
| S1_extracted_k141_48345 | Low-quality |
| S1_extracted_k141_154441 | Not-determined |
| S1_extracted_k141_115020 | Low-quality |
| S1_extracted_k141_147551 | Low-quality |
| S1_extracted_k141_81739 | Not-determined |
| S1_extracted_k141_141698 | Not-determined |
| S1_extracted_k141_81740 | Low-quality |
| S1_extracted_k141_115022 | Not-determined |
| S1_extracted_k141_108269 | Not-determined |
| S1_extracted_k141_81743 | Not-determined |
| S1_extracted_k141_8428 | Not-determined |
| S1_extracted_k141_15293 | Not-determined |
| S1_extracted_k141_108272 | Not-determined |
| S1_extracted_k141_68280 | Not-determined |
| S1_extracted_k141_21572 | Not-determined |
| S1_extracted_k141_154450 | Not-determined |
| S1_extracted_k141_8434 | Not-determined |
| S1_extracted_k141_108276 | Low-quality |
| S1_extracted_k141_41724 | Not-determined |
| S1_extracted_k141_121505 | Not-determined |
| S1_extracted_k141_88333 | Low-quality |
| S1_extracted_k141_121508 | Not-determined |
| S1_extracted_k141_55012 | Not-determined |
| S1_extracted_k141_41727 | Low-quality |
| S1_extracted_k141_8436 | Not-determined |
| S1_extracted_k141_41731 | Not-determined |
| S1_extracted_k141_15307 | Not-determined |
| S1_extracted_k141_108286 | Not-determined |
| S1_extracted_k141_48366 | Not-determined |
| S1_extracted_k141_68291 | Not-determined |
| S1_extracted_k141_154460 | Low-quality |
| S1_extracted_k141_101779 | Not-determined |
| S1_extracted_k141_68293 | Not-determined |
| S1_extracted_k141_75063 | Not-determined |
| S1_extracted_k141_48371 | Not-determined |
| S1_extracted_k141_88339 | Not-determined |
| S1_extracted_k141_141716 | Not-determined |
| S1_extracted_k141_15314 | Not-determined |
| S1_extracted_k141_108293 | Not-determined |
| S1_extracted_k141_21584 | Not-determined |
| S1_extracted_k141_34727 | Low-quality |
| S1_extracted_k141_81765 | Low-quality |
| S1_extracted_k141_128414 | Not-determined |
| S1_extracted_k141_55027 | Low-quality |
| S1_extracted_k141_115045 | Low-quality |
| S1_extracted_k141_81768 | Not-determined |
| S1_extracted_k141_48380 | Not-determined |
| S1_extracted_k141_154469 | Not-determined |
| S1_extracted_k141_95139 | Low-quality |
| S1_extracted_k141_154470 | Not-determined |
| S1_extracted_k141_154471 | Low-quality |
| S1_extracted_k141_75074 | Not-determined |
| S1_extracted_k141_68301 | Not-determined |
| S1_extracted_k141_81770 | Not-determined |
| S1_extracted_k141_128420 | Not-determined |
| S1_extracted_k141_108302 | Not-determined |
| S1_extracted_k141_147557 | Low-quality |
| S1_extracted_k141_128422 | Low-quality |
| S1_extracted_k141_134982 | Not-determined |
| S1_extracted_k141_95141 | Low-quality |
| S1_extracted_k141_48387 | Not-determined |
| S1_extracted_k141_48388 | Not-determined |
| S1_extracted_k141_68313 | Not-determined |
| S1_extracted_k141_28301 | Not-determined |
| S1_extracted_k141_95144 | Not-determined |
| S1_extracted_k141_101790 | Not-determined |
| S1_extracted_k141_21593 | Not-determined |
| S1_extracted_k141_68316 | Not-determined |
| S1_extracted_k141_1361 | Low-quality |
| S1_extracted_k141_154482 | Not-determined |
| S1_extracted_k141_41748 | Not-determined |
| S1_extracted_k141_115058 | Not-determined |
| S1_extracted_k141_134993 | Not-determined |
| S1_extracted_k141_101797 | Not-determined |
| S1_extracted_k141_41752 | Not-determined |
| S1_extracted_k141_28311 | Not-determined |
| S1_extracted_k141_41753 | Not-determined |
| S1_extracted_k141_81783 | Not-determined |
| S1_extracted_k141_34740 | Not-determined |
| S1_extracted_k141_101798 | Low-quality |
| S1_extracted_k141_88352 | Not-determined |
| S1_extracted_k141_81786 | Not-determined |
| S1_extracted_k141_60986 | Not-determined |
| S1_extracted_k141_15336 | Not-determined |
| S1_extracted_k141_88356 | Low-quality |
| S1_extracted_k141_21601 | Not-determined |
| S1_extracted_k141_128443 | Not-determined |
| S1_extracted_k141_141748 | Low-quality |
| S1_extracted_k141_108321 | Low-quality |
| S1_extracted_k141_81790 | Not-determined |
| S1_extracted_k141_55049 | Not-determined |
| S1_extracted_k141_135005 | Not-determined |
| S1_extracted_k141_147568 | Not-determined |
| S1_extracted_k141_128448 | Not-determined |
| S1_extracted_k141_1367 | Not-determined |
| S1_extracted_k141_55053 | Not-determined |
| S1_extracted_k141_121542 | Not-determined |
| S1_extracted_k141_68331 | Not-determined |
| S1_extracted_k141_147571 | Not-determined |
| S1_extracted_k141_75091 | Low-quality |
| S1_extracted_k141_75092 | Low-quality |
| S1_extracted_k141_21607 | Not-determined |
| S1_extracted_k141_81795 | Not-determined |
| S1_extracted_k141_154497 | Not-determined |
| S1_extracted_k141_141757 | Not-determined |
| S1_extracted_k141_41756 | Not-determined |
| S1_extracted_k141_55057 | Low-quality |
| S1_extracted_k141_108329 | Not-determined |
| S1_extracted_k141_81799 | Low-quality |
| S1_extracted_k141_147574 | Not-determined |
| S1_extracted_k141_28324 | Low-quality |
| S1_extracted_k141_128456 | Not-determined |
| S1_extracted_k141_154500 | Not-determined |
| S1_extracted_k141_55060 | Not-determined |
| S1_extracted_k141_135007 | Low-quality |
| S1_extracted_k141_68339 | Not-determined |
| S1_extracted_k141_88363 | Low-quality |
| S1_extracted_k141_34757 | Not-determined |
| S1_extracted_k141_135009 | Not-determined |
| S1_extracted_k141_15345 | Low-quality |
| S1_extracted_k141_28329 | Not-determined |
| S1_extracted_k141_21620 | Low-quality |
| S1_extracted_k141_15346 | Low-quality |
| S1_extracted_k141_115081 | Not-determined |
| S1_extracted_k141_115082 | Not-determined |
| S1_extracted_k141_75103 | Not-determined |
| S1_extracted_k141_121561 | Not-determined |
| S1_extracted_k141_141772 | Not-determined |
| S1_extracted_k141_68343 | Not-determined |
| S1_extracted_k141_88366 | Low-quality |
| S1_extracted_k141_81807 | Not-determined |
| S1_extracted_k141_108335 | Not-determined |
| S1_extracted_k141_128460 | Not-determined |
| S1_extracted_k141_108337 | Not-determined |
| S1_extracted_k141_21628 | Not-determined |
| S1_extracted_k141_21629 | Not-determined |
| S1_extracted_k141_128462 | Not-determined |
| S1_extracted_k141_1383 | Not-determined |
| S1_extracted_k141_34762 | Not-determined |
| S1_extracted_k141_115089 | Low-quality |
| S1_extracted_k141_121568 | Not-determined |
| S1_extracted_k141_95159 | Not-determined |
| S1_extracted_k141_154507 | Low-quality |
| S1_extracted_k141_81814 | Not-determined |
| S1_extracted_k141_101821 | Not-determined |
| S1_extracted_k141_15361 | Not-determined |
| S1_extracted_k141_95163 | Low-quality |
| S1_extracted_k141_28344 | Low-quality |
| S1_extracted_k141_147578 | Not-determined |
| S1_extracted_k141_108347 | Not-determined |
| S1_extracted_k141_68358 | Not-determined |
| S1_extracted_k141_21632 | Low-quality |
| S1_extracted_k141_95167 | Not-determined |
| S1_extracted_k141_101824 | Not-determined |
| S1_extracted_k141_8444 | Not-determined |
| S1_extracted_k141_95168 | Not-determined |
| S1_extracted_k141_88376 | Low-quality |
| S1_extracted_k141_21636 | Low-quality |
| S1_extracted_k141_141784 | Not-determined |
| S1_extracted_k141_88378 | Not-determined |
| S1_extracted_k141_88379 | Not-determined |
| S1_extracted_k141_28355 | Not-determined |
| S1_extracted_k141_154514 | Not-determined |
| S1_extracted_k141_128481 | Not-determined |
| S1_extracted_k141_101829 | Not-determined |
| S1_extracted_k141_101830 | Not-determined |
| S1_extracted_k141_81828 | Low-quality |
| S1_extracted_k141_115110 | Not-determined |
| S1_extracted_k141_135042 | Not-determined |
| S1_extracted_k141_108357 | Not-determined |
| S1_extracted_k141_101836 | Not-determined |
| S1_extracted_k141_108358 | Low-quality |
| S1_extracted_k141_41790 | Not-determined |
| S1_extracted_k141_41791 | Not-determined |
| S1_extracted_k141_135044 | Not-determined |
| S1_extracted_k141_15376 | Not-determined |
| S1_extracted_k141_108360 | Not-determined |
| S1_extracted_k141_128488 | Not-determined |
| S1_extracted_k141_34779 | Not-determined |
| S1_extracted_k141_101840 | Not-determined |
| S1_extracted_k141_135050 | Not-determined |
| S1_extracted_k141_101841 | Not-determined |
| S1_extracted_k141_55089 | Not-determined |
| S1_extracted_k141_128491 | Low-quality |
| S1_extracted_k141_135053 | Not-determined |
| S1_extracted_k141_108367 | Not-determined |
| S1_extracted_k141_55091 | Not-determined |
| S1_extracted_k141_81832 | Not-determined |
| S1_extracted_k141_75117 | Not-determined |
| S1_extracted_k141_128495 | Not-determined |
| S1_extracted_k141_28375 | Not-determined |
| S1_extracted_k141_154530 | Not-determined |
| S1_extracted_k141_101847 | Low-quality |
| S1_extracted_k141_135056 | Not-determined |
| S1_extracted_k141_21652 | Not-determined |
| S1_extracted_k141_121599 | Not-determined |
| S1_extracted_k141_88393 | Not-determined |
| S1_extracted_k141_115114 | Not-determined |
| S1_extracted_k141_81837 | Not-determined |
| S1_extracted_k141_121602 | Not-determined |
| S1_extracted_k141_88396 | Low-quality |
| S1_extracted_k141_34785 | Not-determined |
| S1_extracted_k141_8466 | Not-determined |
| S1_extracted_k141_128502 | Not-determined |
| S1_extracted_k141_15392 | Not-determined |
| S1_extracted_k141_95194 | Not-determined |
| S1_extracted_k141_41809 | Not-determined |
| S1_extracted_k141_101855 | Not-determined |
| S1_extracted_k141_154545 | Low-quality |
| S1_extracted_k141_95197 | Not-determined |
| S1_extracted_k141_115121 | Low-quality |
| S1_extracted_k141_154547 | Not-determined |
| S1_extracted_k141_141807 | Not-determined |
| S1_extracted_k141_154548 | Not-determined |
| S1_extracted_k141_28388 | Not-determined |
| S1_extracted_k141_75133 | Not-determined |
| S1_extracted_k141_55103 | Not-determined |
| S1_extracted_k141_154549 | Low-quality |
| S1_extracted_k141_141810 | Low-quality |
| S1_extracted_k141_101862 | Not-determined |
| S1_extracted_k141_135063 | Not-determined |
| S1_extracted_k141_61006 | Not-determined |
| S1_extracted_k141_108381 | Not-determined |
| S1_extracted_k141_8476 | Not-determined |
| S1_extracted_k141_68395 | Low-quality |
| S1_extracted_k141_95205 | Not-determined |
| S1_extracted_k141_75141 | Low-quality |
| S1_extracted_k141_108385 | Not-determined |
| S1_extracted_k141_154556 | Not-determined |
| S1_extracted_k141_135067 | Not-determined |
| S1_extracted_k141_8484 | Not-determined |
| S1_extracted_k141_135069 | Low-quality |
| S1_extracted_k141_75144 | Not-determined |
| S1_extracted_k141_128514 | Not-determined |
| S1_extracted_k141_48473 | Not-determined |
| S1_extracted_k141_34792 | Not-determined |
| S1_extracted_k141_68401 | Low-quality |
| S1_extracted_k141_88413 | Not-determined |
| S1_extracted_k141_154562 | Low-quality |
| S1_extracted_k141_1404 | Low-quality |
| S1_extracted_k141_81857 | Not-determined |
| S1_extracted_k141_121621 | Not-determined |
| S1_extracted_k141_55112 | Not-determined |
| S1_extracted_k141_15404 | Not-determined |
| S1_extracted_k141_21677 | Not-determined |
| S1_extracted_k141_61009 | Low-quality |
| S1_extracted_k141_61010 | Not-determined |
| S1_extracted_k141_48477 | Not-determined |
| S1_extracted_k141_55114 | Not-determined |
| S1_extracted_k141_55115 | Not-determined |
| S1_extracted_k141_108396 | Not-determined |
| S1_extracted_k141_1412 | Not-determined |
| S1_extracted_k141_121629 | Not-determined |
| S1_extracted_k141_101877 | Not-determined |
| S1_extracted_k141_128526 | Not-determined |
| S1_extracted_k141_128527 | Not-determined |
| S1_extracted_k141_141822 | Not-determined |
| S1_extracted_k141_141823 | Not-determined |
| S1_extracted_k141_81865 | Not-determined |
| S1_extracted_k141_21686 | Not-determined |
| S1_extracted_k141_8494 | Not-determined |
| S1_extracted_k141_34801 | Not-determined |
| S1_extracted_k141_68411 | Not-determined |
| S1_extracted_k141_135078 | Not-determined |
| S1_extracted_k141_135079 | Not-determined |
| S1_extracted_k141_108399 | Not-determined |
| S1_extracted_k141_88420 | Not-determined |
| S1_extracted_k141_21687 | Not-determined |
| S1_extracted_k141_68417 | Not-determined |
| S1_extracted_k141_41831 | Not-determined |
| S1_extracted_k141_21688 | Low-quality |
| S1_extracted_k141_8502 | Not-determined |
| S1_extracted_k141_147598 | Not-determined |
| S1_extracted_k141_1421 | Not-determined |
| S1_extracted_k141_135081 | Not-determined |
| S1_extracted_k141_21692 | Not-determined |
| S1_extracted_k141_135083 | Low-quality |
| S1_extracted_k141_147600 | Not-determined |
| S1_extracted_k141_21693 | Not-determined |
| S1_extracted_k141_101890 | Not-determined |
| S1_extracted_k141_95214 | Not-determined |
| S1_extracted_k141_1424 | Not-determined |
| S1_extracted_k141_121638 | Not-determined |
| S1_extracted_k141_34812 | Low-quality |
| S1_extracted_k141_115150 | Not-determined |
| S1_extracted_k141_1429 | Not-determined |
| S1_extracted_k141_21695 | Not-determined |
| S1_extracted_k141_154570 | Not-determined |
| S1_extracted_k141_75157 | Not-determined |
| S1_extracted_k141_88432 | Not-determined |
| S1_extracted_k141_121644 | Not-determined |
| S1_extracted_k141_147606 | Not-determined |
| S1_extracted_k141_141836 | Low-quality |
| S1_extracted_k141_75161 | Not-determined |
| S1_extracted_k141_128543 | Not-determined |
| S1_extracted_k141_8514 | Not-determined |
| S1_extracted_k141_75164 | Not-determined |
| S1_extracted_k141_48505 | Not-determined |
| S1_extracted_k141_135088 | Not-determined |
| S1_extracted_k141_88441 | Not-determined |
| S1_extracted_k141_101897 | Not-determined |
| S1_extracted_k141_21699 | Not-determined |
| S1_extracted_k141_108416 | Not-determined |
| S1_extracted_k141_75166 | Not-determined |
| S1_extracted_k141_135089 | Not-determined |
| S1_extracted_k141_75167 | Not-determined |
| S1_extracted_k141_128549 | Not-determined |
| S1_extracted_k141_95220 | Not-determined |
| S1_extracted_k141_41842 | Low-quality |
| S1_extracted_k141_108417 | Not-determined |
| S1_extracted_k141_115159 | Not-determined |
| S1_extracted_k141_55143 | Not-determined |
| S1_extracted_k141_128551 | Not-determined |
| S1_extracted_k141_147612 | Not-determined |
| S1_extracted_k141_15430 | Not-determined |
| S1_extracted_k141_115162 | Not-determined |
| S1_extracted_k141_88445 | Low-quality |
| S1_extracted_k141_101903 | Not-determined |
| S1_extracted_k141_81888 | Low-quality |
| S1_extracted_k141_55148 | Not-determined |
| S1_extracted_k141_8521 | Not-determined |
| S1_extracted_k141_135098 | Low-quality |
| S1_extracted_k141_41853 | Not-determined |
| S1_extracted_k141_55152 | Not-determined |
| S1_extracted_k141_8525 | Not-determined |
| S1_extracted_k141_81894 | Not-determined |
| S1_extracted_k141_41857 | Not-determined |
| S1_extracted_k141_81896 | Not-determined |
| S1_extracted_k141_95225 | Not-determined |
| S1_extracted_k141_68443 | Not-determined |
| S1_extracted_k141_141848 | Not-determined |
| S1_extracted_k141_21711 | Not-determined |
| S1_extracted_k141_154578 | Not-determined |
| S1_extracted_k141_68445 | Not-determined |
| S1_extracted_k141_115168 | Not-determined |
| S1_extracted_k141_21712 | Not-determined |
| S1_extracted_k141_61024 | Not-determined |
| S1_extracted_k141_154579 | Low-quality |
| S1_extracted_k141_41862 | Not-determined |
| S1_extracted_k141_1444 | Not-determined |
| S1_extracted_k141_75174 | Not-determined |
| S1_extracted_k141_1446 | Not-determined |
| S1_extracted_k141_147616 | Not-determined |
| S1_extracted_k141_115175 | Not-determined |
| S1_extracted_k141_128559 | Not-determined |
| S1_extracted_k141_141857 | Not-determined |
| S1_extracted_k141_48527 | Not-determined |
| S1_extracted_k141_147618 | Not-determined |
| S12_vs_k141_79003\|\|full | Medium-quality |
| S1_extracted_k141_28400 | Not-determined |
| S1_extracted_k141_48531 | Not-determined |
| S1_extracted_k141_101922 | Low-quality |
| S1_extracted_k141_95237 | Not-determined |
| S1_extracted_k141_48532 | Not-determined |
| S1_extracted_k141_55161 | Low-quality |
| S1_extracted_k141_88455 | Not-determined |
| S1_extracted_k141_41865 | Low-quality |
| S1_extracted_k141_41866 | Not-determined |
| S1_extracted_k141_128571 | Not-determined |
| S1_extracted_k141_154582 | Not-determined |
| S1_extracted_k141_135120 | Low-quality |
| S1_extracted_k141_75185 | Not-determined |
| S1_extracted_k141_95241 | Low-quality |
| S1_extracted_k141_101931 | Not-determined |
| S1_extracted_k141_75186 | Not-determined |
| S1_extracted_k141_101932 | Low-quality |
| S1_extracted_k141_115194 | Not-determined |
| S1_extracted_k141_8547 | Low-quality |
| S1_extracted_k141_108433 | Not-determined |
| S1_extracted_k141_154590 | Not-determined |
| S1_extracted_k141_135126 | Not-determined |
| S1_extracted_k141_28410 | Low-quality |
| S1_extracted_k141_101936 | Not-determined |
| S1_extracted_k141_68459 | Not-determined |
| S1_extracted_k141_21719 | Not-determined |
| S1_extracted_k141_75191 | Low-quality |
| S1_extracted_k141_48542 | Not-determined |
| S1_extracted_k141_101938 | Not-determined |
| S1_extracted_k141_34845 | Not-determined |
| S1_extracted_k141_95245 | Not-determined |
| S1_extracted_k141_68461 | Not-determined |
| S1_extracted_k141_8553 | Not-determined |
| S1_extracted_k141_108438 | Not-determined |
| S1_extracted_k141_55171 | Not-determined |
| S1_extracted_k141_34846 | Not-determined |
| S1_extracted_k141_88464 | Not-determined |
| S1_extracted_k141_128586 | Not-determined |
| S1_extracted_k141_55173 | Not-determined |
| S1_extracted_k141_28412 | Not-determined |
| S1_extracted_k141_128587 | Low-quality |
| S1_extracted_k141_15457 | Low-quality |
| S1_extracted_k141_8558 | Not-determined |
| S1_extracted_k141_147629 | Not-determined |
| S1_extracted_k141_8559 | Not-determined |
| S1_extracted_k141_81926 | Not-determined |
| S1_extracted_k141_61034 | Not-determined |
| S1_extracted_k141_115203 | Not-determined |
| S1_extracted_k141_8562 | Not-determined |
| S1_extracted_k141_28417 | Not-determined |
| S1_extracted_k141_55185 | Not-determined |
| S1_extracted_k141_15465 | Not-determined |
| S1_extracted_k141_75198 | Low-quality |
| S1_extracted_k141_108445 | Not-determined |
| S1_extracted_k141_108446 | Not-determined |
| S1_extracted_k141_141891 | Low-quality |
| S1_extracted_k141_121660 | Not-determined |
| S1_extracted_k141_101948 | Not-determined |
| S1_extracted_k141_121661 | Not-determined |
| S1_extracted_k141_88477 | Not-determined |
| S1_extracted_k141_154601 | Low-quality |
| S1_extracted_k141_8572 | Not-determined |
| S1_extracted_k141_128592 | Not-determined |
| S1_extracted_k141_75207 | Low-quality |
| S1_extracted_k141_68482 | Not-determined |
| S1_extracted_k141_108452 | Not-determined |
| S1_extracted_k141_21734 | Not-determined |
| S1_extracted_k141_108454 | Low-quality |
| S1_extracted_k141_108455 | Not-determined |
| S1_extracted_k141_48554 | Not-determined |
| S1_extracted_k141_81934 | Not-determined |
| S1_extracted_k141_154606 | Not-determined |
| S1_extracted_k141_48556 | Not-determined |
| S1_extracted_k141_135144 | Low-quality |
| S1_extracted_k141_128600 | Not-determined |
| S1_extracted_k141_75211 | Not-determined |
| S1_extracted_k141_55196 | Not-determined |
| S1_extracted_k141_95267 | Not-determined |
| S1_extracted_k141_75214 | Not-determined |

| | |
|---|---|
| S1_extracted_k141_1478 | Not-determined |
| S1_extracted_k141_88490 | Not-determined |
| S1_extracted_k141_68488 | Not-determined |
| S1_extracted_k141_81940 | Not-determined |
| S1_extracted_k141_135149 | Not-determined |
| S1_extracted_k141_21744 | Not-determined |
| S1_extracted_k141_121674 | Not-determined |
| S1_extracted_k141_21748 | Not-determined |
| S1_extracted_k141_48565 | Not-determined |
| S1_extracted_k141_34870 | Low-quality |
| S1_extracted_k141_88500 | Not-determined |
| S1_extracted_k141_68492 | Low-quality |
| S1_extracted_k141_34871 | Not-determined |
| S1_extracted_k141_21754 | Low-quality |
| S1_extracted_k141_147632 | Not-determined |
| S1_extracted_k141_108473 | Not-determined |
| S1_extracted_k141_68496 | Not-determined |
| S1_extracted_k141_48569 | Not-determined |
| S1_extracted_k141_108475 | Not-determined |
| S1_extracted_k141_28451 | Low-quality |
| S1_extracted_k141_81953 | Not-determined |
| S1_extracted_k141_68499 | Not-determined |
| S1_extracted_k141_61043 | Not-determined |
| S1_extracted_k141_48573 | Low-quality |
| S1_extracted_k141_28454 | Not-determined |
| S1_extracted_k141_95285 | Not-determined |
| S1_extracted_k141_8577 | Not-determined |
| S1_extracted_k141_141906 | Not-determined |
| S1_extracted_k141_8581 | Not-determined |
| S1_extracted_k141_81960 | Not-determined |
| S1_extracted_k141_41912 | Not-determined |
| S1_extracted_k141_128622 | Not-determined |
| S1_extracted_k141_81962 | Not-determined |
| S1_extracted_k141_48579 | Not-determined |
| S1_extracted_k141_141911 | Not-determined |
| S1_extracted_k141_75227 | Not-determined |
| S1_extracted_k141_8586 | Not-determined |
| S1_extracted_k141_88510 | Not-determined |
| S1_extracted_k141_95294 | Not-determined |
| S1_extracted_k141_147637 | Not-determined |
| S1_extracted_k141_55223 | Not-determined |
| S1_extracted_k141_141917 | Not-determined |
| S1_extracted_k141_95297 | Not-determined |
| S1_extracted_k141_34877 | Not-determined |
| S1_extracted_k141_141918 | Not-determined |
| S1_extracted_k141_68514 | Not-determined |
| S1_extracted_k141_135176 | Low-quality |
| S1_extracted_k141_48588 | Not-determined |
| S1_extracted_k141_141921 | Not-determined |
| S1_extracted_k141_1496 | Not-determined |
| S1_extracted_k141_15508 | Not-determined |
| S1_extracted_k141_141922 | Not-determined |
| S1_extracted_k141_88517 | Not-determined |
| S1_extracted_k141_121700 | Not-determined |
| S1_extracted_k141_55230 | Not-determined |
| S1_extracted_k141_88519 | Not-determined |
| S1_extracted_k141_101985 | Low-quality |
| S1_extracted_k141_101987 | Not-determined |
| S1_extracted_k141_41932 | Not-determined |
| S1_extracted_k141_154622 | Not-determined |
| S1_extracted_k141_101989 | Not-determined |
| S1_extracted_k141_15516 | Not-determined |
| S1_extracted_k141_108493 | Low-quality |
| S1_extracted_k141_115225 | Not-determined |
| S1_extracted_k141_21777 | Not-determined |
| S1_extracted_k141_88521 | Not-determined |
| S1_extracted_k141_135191 | Not-determined |
| S1_extracted_k141_41934 | Low-quality |
| S1_extracted_k141_68520 | Low-quality |
| S1_extracted_k141_135192 | Not-determined |
| S1_extracted_k141_147641 | Not-determined |
| S1_extracted_k141_1501 | Low-quality |
| S1_extracted_k141_41935 | Not-determined |
| S1_extracted_k141_128648 | Not-determined |
| S1_extracted_k141_28479 | Not-determined |
| S1_extracted_k141_68523 | Low-quality |
| S1_extracted_k141_95311 | Not-determined |
| S1_extracted_k141_68528 | Not-determined |
| S1_extracted_k141_8602 | Not-determined |
| S1_extracted_k141_95316 | Not-determined |
| S1_extracted_k141_154623 | Not-determined |
| S1_extracted_k141_8603 | Not-determined |
| S1_extracted_k141_108504 | Not-determined |
| S1_extracted_k141_21785 | Not-determined |
| S1_extracted_k141_48598 | Low-quality |
| S1_extracted_k141_55243 | Not-determined |
| S1_extracted_k141_81983 | Not-determined |
| S1_extracted_k141_141938 | Not-determined |
| S1_extracted_k141_34884 | Low-quality |
| S1_extracted_k141_28489 | Not-determined |
| S1_extracted_k141_1513 | Not-determined |
| S1_extracted_k141_88535 | Not-determined |
| S1_extracted_k141_147646 | Not-determined |
| S1_extracted_k141_128656 | Not-determined |
| S1_extracted_k141_28491 | Not-determined |
| S1_extracted_k141_128658 | Not-determined |
| S1_extracted_k141_95323 | Not-determined |
| S1_extracted_k141_28494 | Not-determined |
| S1_extracted_k141_21790 | Not-determined |
| S1_extracted_k141_75237 | Not-determined |
| S1_extracted_k141_88537 | Low-quality |
| S1_extracted_k141_48604 | Not-determined |
| S1_extracted_k141_21795 | Not-determined |
| S1_extracted_k141_88538 | Low-quality |
| S1_extracted_k141_68537 | Not-determined |
| S1_extracted_k141_68538 | Not-determined |
| S1_extracted_k141_102013 | Not-determined |
| S1_extracted_k141_121728 | Not-determined |
| S1_extracted_k141_41958 | Not-determined |
| S1_extracted_k141_15547 | Not-determined |
| S1_extracted_k141_34892 | Not-determined |
| S1_extracted_k141_95332 | Low-quality |
| S1_extracted_k141_141947 | Low-quality |
| S1_extracted_k141_81992 | Not-determined |
| S1_extracted_k141_102019 | Not-determined |
| S1_extracted_k141_141948 | Not-determined |
| S1_extracted_k141_81993 | Not-determined |
| S1_extracted_k141_88548 | Not-determined |
| S1_extracted_k141_115254 | Not-determined |
| S1_extracted_k141_95339 | Not-determined |
| S1_extracted_k141_135210 | Not-determined |
| S1_extracted_k141_115255 | Not-determined |
| S1_extracted_k141_81995 | Not-determined |
| S1_extracted_k141_135213 | Not-determined |
| S1_extracted_k141_88554 | Not-determined |
| S1_extracted_k141_34899 | Low-quality |
| S1_extracted_k141_21805 | Low-quality |
| S1_extracted_k141_48618 | Not-determined |
| S1_extracted_k141_108526 | Not-determined |
| S1_extracted_k141_128670 | Not-determined |
| S1_extracted_k141_141955 | Not-determined |
| S1_extracted_k141_68554 | Not-determined |
| S1_extracted_k141_15561 | Not-determined |
| S1_extracted_k141_21811 | Not-determined |
| S1_extracted_k141_88560 | Low-quality |
| S1_extracted_k141_102022 | Not-determined |
| S1_extracted_k141_55260 | Low-quality |
| S1_extracted_k141_8615 | Not-determined |
| S1_extracted_k141_128676 | Not-determined |
| S1_extracted_k141_21815 | Not-determined |
| S1_extracted_k141_154645 | Not-determined |
| S1_extracted_k141_102027 | Not-determined |
| S1_extracted_k141_128678 | Not-determined |
| S1_extracted_k141_68564 | Not-determined |
| S1_extracted_k141_95351 | Low-quality |
| S1_extracted_k141_128684 | Not-determined |
| S1_extracted_k141_135224 | Not-determined |
| S1_extracted_k141_34910 | Not-determined |
| S1_extracted_k141_115273 | Not-determined |
| S1_extracted_k141_75264 | Not-determined |
| S1_extracted_k141_154648 | Low-quality |
| S1_extracted_k141_115275 | Not-determined |
| S1_extracted_k141_55270 | Not-determined |
| S1_extracted_k141_88580 | Not-determined |
| S1_extracted_k141_147651 | Low-quality |
| S1_extracted_k141_95355 | Not-determined |
| S1_extracted_k141_8626 | Not-determined |
| S1_extracted_k141_8627 | Not-determined |
| S1_extracted_k141_75268 | Not-determined |
| S1_extracted_k141_128691 | Low-quality |
| S1_extracted_k141_95360 | Not-determined |
| S1_extracted_k141_55271 | Low-quality |
| S1_extracted_k141_128692 | Not-determined |
| S1_extracted_k141_95361 | Not-determined |
| S1_extracted_k141_115281 | Not-determined |
| S1_extracted_k141_75273 | Not-determined |
| S1_extracted_k141_34916 | Not-determined |
| S1_extracted_k141_115284 | Not-determined |
| S1_extracted_k141_141969 | Not-determined |
| S1_extracted_k141_95364 | Low-quality |
| S1_extracted_k141_21823 | Not-determined |
| S1_extracted_k141_141970 | Not-determined |
| S1_extracted_k141_108540 | Low-quality |
| S1_extracted_k141_28511 | Not-determined |
| S1_extracted_k141_95366 | Low-quality |
| S1_extracted_k141_8638 | Not-determined |
| S1_extracted_k141_28512 | Not-determined |
| S1_extracted_k141_102040 | Not-determined |
| S1_extracted_k141_15595 | Low-quality |
| S1_extracted_k141_115287 | Low-quality |
| S1_extracted_k141_21828 | Not-determined |
| S1_extracted_k141_115288 | Low-quality |
| S1_extracted_k141_128703 | Not-determined |
| S1_extracted_k141_154658 | Not-determined |
| S1_extracted_k141_48641 | Not-determined |
| S1_extracted_k141_141976 | Not-determined |
| S1_extracted_k141_102045 | Not-determined |
| S1_extracted_k141_15599 | Not-determined |
| S1_extracted_k141_28521 | Not-determined |
| S1_extracted_k141_55277 | Not-determined |
| S1_extracted_k141_115296 | Not-determined |
| S1_extracted_k141_102049 | Not-determined |
| S1_extracted_k141_41974 | Not-determined |
| S1_extracted_k141_115297 | Low-quality |
| S1_extracted_k141_88598 | Not-determined |
| S1_extracted_k141_48647 | Not-determined |
| S1_extracted_k141_108554 | Not-determined |
| S1_extracted_k141_154670 | Not-determined |
| S1_extracted_k141_41981 | Not-determined |
| S1_extracted_k141_121768 | Not-determined |
| S1_extracted_k141_115306 | Low-quality |
| S1_extracted_k141_128711 | Not-determined |
| S1_extracted_k141_121771 | Not-determined |
| S1_extracted_k141_75292 | Low-quality |
| S1_extracted_k141_135252 | Not-determined |
| S1_extracted_k141_141985 | Not-determined |
| S1_extracted_k141_8650 | Not-determined |
| S1_extracted_k141_82031 | Not-determined |
| S1_extracted_k141_15611 | Not-determined |
| S1_extracted_k141_121773 | Not-determined |
| S1_extracted_k141_102065 | Low-quality |
| S1_extracted_k141_135256 | Not-determined |
| S1_extracted_k141_15612 | Not-determined |
| S1_extracted_k141_121774 | Not-determined |
| S1_extracted_k141_95380 | Not-determined |
| S1_extracted_k141_95381 | Not-determined |
| S1_extracted_k141_82032 | Not-determined |
| S1_extracted_k141_95383 | Not-determined |
| S1_extracted_k141_28540 | Not-determined |
| S1_extracted_k141_75302 | Not-determined |
| S1_extracted_k141_128717 | Not-determined |
| S1_extracted_k141_75303 | Not-determined |
| S1_extracted_k141_28543 | Low-quality |
| S1_extracted_k141_1564 | Not-determined |
| S1_extracted_k141_28546 | Not-determined |
| S1_extracted_k141_34932 | Not-determined |
| S1_extracted_k141_88617 | Not-determined |
| S1_extracted_k141_82040 | Not-determined |
| S1_extracted_k141_41993 | Not-determined |
| S1_extracted_k141_48667 | Low-quality |
| S1_extracted_k141_48668 | Low-quality |
| S1_extracted_k141_1566 | Not-determined |
| S1_extracted_k141_21844 | Not-determined |
| S1_extracted_k141_68592 | Not-determined |
| S1_extracted_k141_135269 | Not-determined |
| S1_extracted_k141_68593 | Low-quality |
| S1_extracted_k141_147672 | Not-determined |
| S1_extracted_k141_88622 | Not-determined |
| S1_extracted_k141_28552 | Not-determined |
| S1_extracted_k141_141998 | Low-quality |
| S1_extracted_k141_68595 | Not-determined |
| S1_extracted_k141_1568 | Low-quality |
| S1_extracted_k141_128730 | Not-determined |
| S1_extracted_k141_75315 | Low-quality |
| S1_extracted_k141_135275 | Low-quality |
| S1_extracted_k141_95393 | Not-determined |
| S1_extracted_k141_102083 | Not-determined |
| S1_extracted_k141_1572 | Not-determined |
| S1_extracted_k141_95395 | Low-quality |
| S1_extracted_k141_121787 | Not-determined |
| S1_extracted_k141_28562 | Not-determined |
| S1_extracted_k141_75319 | Low-quality |
| S1_extracted_k141_28563 | Not-determined |
| S1_extracted_k141_154683 | Not-determined |
| S1_extracted_k141_142007 | Not-determined |
| S1_extracted_k141_21852 | Not-determined |
| S1_extracted_k141_95400 | Low-quality |
| S1_extracted_k141_154685 | Not-determined |
| S1_extracted_k141_102087 | Low-quality |
| S1_extracted_k141_128737 | Not-determined |
| S1_extracted_k141_75323 | Low-quality |
| S1_extracted_k141_121789 | Not-determined |
| S1_extracted_k141_42004 | Not-determined |
| S1_extracted_k141_42005 | Low-quality |
| S1_extracted_k141_108580 | Not-determined |
| S1_extracted_k141_8670 | Low-quality |
| S1_extracted_k141_55311 | Not-determined |
| S1_extracted_k141_142013 | Not-determined |
| S1_extracted_k141_95404 | Not-determined |
| S1_extracted_k141_34946 | Low-quality |
| S1_extracted_k141_88637 | Not-determined |
| S1_extracted_k141_135283 | Not-determined |
| S1_extracted_k141_154688 | Not-determined |
| S1_extracted_k141_21857 | Low-quality |
| S1_extracted_k141_154690 | Low-quality |
| S1_extracted_k141_121794 | Low-quality |
| S1_extracted_k141_95411 | Not-determined |
| S1_extracted_k141_121798 | Not-determined |
| S1_extracted_k141_34954 | Not-determined |
| S1_extracted_k141_95416 | Not-determined |
| S1_extracted_k141_28578 | Not-determined |
| S1_extracted_k141_75337 | Not-determined |
| S1_extracted_k141_8684 | Not-determined |
| S1_extracted_k141_128755 | Low-quality |
| S1_extracted_k141_135299 | Not-determined |
| S1_extracted_k141_128757 | Not-determined |
| S1_extracted_k141_15655 | Not-determined |
| S1_extracted_k141_28584 | Not-determined |
| S1_extracted_k141_88645 | Not-determined |
| S1_extracted_k141_108595 | Not-determined |
| S1_extracted_k141_42009 | Not-determined |
| S1_extracted_k141_61093 | Not-determined |
| S1_extracted_k141_1582 | Not-determined |
| S1_extracted_k141_142029 | Not-determined |
| S1_extracted_k141_88647 | Not-determined |
| S1_extracted_k141_15659 | Not-determined |
| S1_extracted_k141_128759 | Low-quality |
| S1_extracted_k141_147676 | Not-determined |
| S1_extracted_k141_15663 | Not-determined |
| S1_extracted_k141_128762 | Not-determined |
| S1_extracted_k141_15666 | Not-determined |
| S1_extracted_k141_42017 | Not-determined |
| S1_extracted_k141_154706 | Not-determined |
| S1_extracted_k141_95423 | Not-determined |
| S1_extracted_k141_75346 | Not-determined |
| S1_extracted_k141_1587 | Not-determined |
| S1_extracted_k141_128765 | Not-determined |
| S1_extracted_k141_42020 | Low-quality |
| S1_extracted_k141_121805 | Not-determined |
| S1_extracted_k141_75349 | Not-determined |
| S1_extracted_k141_147681 | Not-determined |
| S1_extracted_k141_121808 | Not-determined |
| S1_extracted_k141_48683 | Not-determined |
| S1_extracted_k141_88657 | Not-determined |
| S1_extracted_k141_82073 | Not-determined |
| S1_extracted_k141_42025 | Not-determined |
| S1_extracted_k141_154711 | Not-determined |
| S1_extracted_k141_42027 | Not-determined |
| S1_extracted_k141_108610 | Not-determined |
| S1_extracted_k141_34965 | Not-determined |
| S1_extracted_k141_142040 | Not-determined |
| S1_extracted_k141_61094 | Not-determined |
| S1_extracted_k141_95441 | Not-determined |
| S1_extracted_k141_15679 | Not-determined |
| S1_extracted_k141_34968 | Not-determined |
| S1_extracted_k141_135311 | Not-determined |
| S1_extracted_k141_61097 | Not-determined |
| S1_extracted_k141_82081 | Not-determined |
| S1_extracted_k141_75356 | Low-quality |
| S1_extracted_k141_115335 | Not-determined |
| S1_extracted_k141_61099 | Not-determined |
| S1_extracted_k141_34969 | Not-determined |
| S1_extracted_k141_135318 | Not-determined |
| S1_extracted_k141_135319 | Low-quality |
| S1_extracted_k141_48692 | Low-quality |
| S1_extracted_k141_61101 | Low-quality |
| S1_extracted_k141_55350 | Not-determined |
| S1_extracted_k141_121817 | Low-quality |
| S1_extracted_k141_21885 | Not-determined |
| S1_extracted_k141_82091 | Not-determined |
| S1_extracted_k141_55354 | Not-determined |
| S1_extracted_k141_21887 | Not-determined |
| S1_extracted_k141_135326 | Not-determined |
| S1_extracted_k141_75360 | Not-determined |
| S1_extracted_k141_154728 | Not-determined |
| S1_extracted_k141_102130 | Not-determined |
| S1_extracted_k141_121822 | Not-determined |
| S1_extracted_k141_15693 | Not-determined |
| S1_extracted_k141_154733 | Not-determined |
| S1_extracted_k141_102132 | Not-determined |
| S1_extracted_k141_115349 | Not-determined |
| S1_extracted_k141_21896 | Low-quality |
| S1_extracted_k141_108623 | Not-determined |
| S1_extracted_k141_21897 | Not-determined |
| S1_extracted_k141_142055 | Not-determined |
| S1_extracted_k141_102138 | Not-determined |
| S1_extracted_k141_75370 | Not-determined |
| S1_extracted_k141_154737 | Not-determined |
| S1_extracted_k141_82098 | Not-determined |
| S1_extracted_k141_115355 | Not-determined |
| S1_extracted_k141_82099 | Not-determined |
| S1_extracted_k141_21901 | Not-determined |
| S1_extracted_k141_21902 | Not-determined |
| S1_extracted_k141_75373 | Low-quality |
| S1_extracted_k141_108631 | Not-determined |
| S1_extracted_k141_128782 | Not-determined |
| S1_extracted_k141_28623 | Low-quality |
| S1_extracted_k141_128784 | Low-quality |
| S1_extracted_k141_121833 | Not-determined |
| S1_extracted_k141_15706 | Not-determined |
| S1_extracted_k141_154738 | Not-determined |
| S1_extracted_k141_115364 | Not-determined |
| S1_extracted_k141_115366 | Not-determined |
| S1_extracted_k141_34978 | Low-quality |
| S1_extracted_k141_15708 | Low-quality |
| S1_extracted_k141_135340 | Not-determined |
| S1_extracted_k141_8708 | Low-quality |
| S1_extracted_k141_154742 | Not-determined |
| S1_extracted_k141_55378 | Not-determined |
| S1_extracted_k141_102152 | Low-quality |
| S1_extracted_k141_82110 | Not-determined |
| S1_extracted_k141_75390 | Not-determined |
| S1_extracted_k141_21911 | Not-determined |
| S1_extracted_k141_28631 | Not-determined |
| S1_extracted_k141_135345 | Not-determined |
| S1_extracted_k141_8714 | Not-determined |
| S1_extracted_k141_102156 | Low-quality |
| S1_extracted_k141_82114 | Low-quality |
| S1_extracted_k141_68651 | Not-determined |
| S1_extracted_k141_82115 | Not-determined |
| S1_extracted_k141_15716 | Not-determined |
| S1_extracted_k141_102158 | Low-quality |
| S1_extracted_k141_28634 | Not-determined |
| S1_extracted_k141_48720 | Not-determined |
| S1_extracted_k141_142073 | Not-determined |
| S1_extracted_k141_142075 | Not-determined |
| S1_extracted_k141_42062 | Not-determined |
| S1_extracted_k141_142077 | Not-determined |
| S1_extracted_k141_88690 | Not-determined |
| S1_extracted_k141_82119 | Low-quality |
| S1_extracted_k141_55392 | Not-determined |
| S1_extracted_k141_128805 | Not-determined |
| S1_extracted_k141_88692 | Not-determined |
| S1_extracted_k141_154748 | Not-determined |
| S1_extracted_k141_42065 | Not-determined |
| S1_extracted_k141_147712 | Not-determined |
| S1_extracted_k141_8723 | Low-quality |
| S1_extracted_k141_88696 | Not-determined |
| S1_extracted_k141_42067 | Not-determined |
| S1_extracted_k141_142084 | Not-determined |
| S1_extracted_k141_121862 | Not-determined |
| S1_extracted_k141_15731 | Not-determined |
| S1_extracted_k141_154751 | Not-determined |
| S1_extracted_k141_128808 | Low-quality |
| S1_extracted_k141_121865 | Not-determined |
| S1_extracted_k141_142089 | Low-quality |
| S1_extracted_k141_147721 | Low-quality |
| S1_extracted_k141_128811 | Not-determined |
| S1_extracted_k141_28650 | Low-quality |
| S1_extracted_k141_108651 | Not-determined |
| S1_extracted_k141_8735 | Not-determined |
| S1_extracted_k141_135369 | Not-determined |
| S1_extracted_k141_128814 | Not-determined |
| S1_extracted_k141_48737 | Not-determined |
| S1_extracted_k141_147724 | Not-determined |
| S1_extracted_k141_55412 | Not-determined |
| S1_extracted_k141_88704 | Not-determined |
| S1_extracted_k141_82135 | Not-determined |
| S1_extracted_k141_142097 | Not-determined |
| S1_extracted_k141_15738 | Not-determined |
| S1_extracted_k141_88707 | Not-determined |
| S1_extracted_k141_108657 | Not-determined |
| S1_extracted_k141_88708 | Low-quality |
| S1_extracted_k141_142101 | Not-determined |
| S1_extracted_k141_61128 | Not-determined |
| S1_extracted_k141_48742 | Not-determined |
| S1_extracted_k141_8742 | Low-quality |
| S1_extracted_k141_115405 | Low-quality |
| S1_extracted_k141_95461 | Not-determined |
| S1_extracted_k141_55423 | Not-determined |
| S1_extracted_k141_68680 | Not-determined |
| S1_extracted_k141_48747 | Not-determined |
| S1_extracted_k141_1633 | Low-quality |
| S1_extracted_k141_95464 | Not-determined |
| S1_extracted_k141_142106 | Not-determined |
| S1_extracted_k141_142107 | Not-determined |
| S1_extracted_k141_15754 | Not-determined |
| S1_extracted_k141_128826 | Not-determined |
| S1_extracted_k141_135380 | Low-quality |
| S1_extracted_k141_142111 | Not-determined |
| S1_extracted_k141_108662 | Not-determined |
| S1_extracted_k141_61136 | Not-determined |
| S1_extracted_k141_8753 | Not-determined |
| S1_extracted_k141_95468 | Not-determined |
| S1_extracted_k141_48754 | Not-determined |
| S1_extracted_k141_135386 | Not-determined |
| S1_extracted_k141_82148 | Not-determined |
| S1_extracted_k141_121875 | Not-determined |
| S1_extracted_k141_21924 | Not-determined |
| S1_extracted_k141_8756 | Not-determined |
| S1_extracted_k141_75408 | Not-determined |
| S1_extracted_k141_21928 | Not-determined |
| S1_extracted_k141_82151 | Not-determined |
| S1_extracted_k141_55438 | Not-determined |
| S1_extracted_k141_48760 | Low-quality |
| S1_extracted_k141_82153 | Low-quality |
| S1_extracted_k141_68694 | Not-determined |
| S1_extracted_k141_15775 | Low-quality |
| S1_extracted_k141_121885 | Not-determined |
| S1_extracted_k141_42095 | Not-determined |
| S1_extracted_k141_55442 | Not-determined |
| S1_extracted_k141_75415 | Not-determined |
| S1_extracted_k141_82159 | Not-determined |
| S1_extracted_k141_15779 | Not-determined |
| S1_extracted_k141_28668 | Low-quality |
| S1_extracted_k141_61146 | Not-determined |
| S1_extracted_k141_42100 | Low-quality |
| S1_extracted_k141_154769 | Not-determined |
| S1_extracted_k141_95484 | Not-determined |
| S1_extracted_k141_108674 | Not-determined |
| S1_extracted_k141_75423 | Not-determined |
| S1_extracted_k141_68699 | Not-determined |
| S1_extracted_k141_8767 | Low-quality |
| S1_extracted_k141_1656 | Not-determined |
| S1_extracted_k141_154772 | Not-determined |
| S1_extracted_k141_68701 | Low-quality |
| S1_extracted_k141_135391 | Not-determined |
| S1_extracted_k141_108678 | Low-quality |
| S1_extracted_k141_147740 | Not-determined |
| S1_extracted_k141_1658 | Not-determined |
| S1_extracted_k141_108679 | Not-determined |
| S1_extracted_k141_102179 | Low-quality |
| S1_extracted_k141_108682 | Not-determined |
| S1_extracted_k141_102181 | Not-determined |
| S1_extracted_k141_21932 | Not-determined |
| S1_extracted_k141_42110 | Not-determined |
| S1_extracted_k141_82169 | Not-determined |
| S1_extracted_k141_108688 | Not-determined |
| S1_extracted_k141_15795 | Not-determined |
| S1_extracted_k141_108693 | Low-quality |
| S1_extracted_k141_75435 | Low-quality |
| S1_extracted_k141_142140 | Not-determined |
| S1_extracted_k141_75436 | Not-determined |
| S1_extracted_k141_42119 | Not-determined |
| S1_extracted_k141_15801 | Not-determined |
| S1_extracted_k141_21942 | Not-determined |
| S1_extracted_k141_82180 | Not-determined |
| S1_extracted_k141_135407 | Not-determined |
| S1_extracted_k141_115439 | Not-determined |
| S1_extracted_k141_28693 | Not-determined |
| S1_extracted_k141_108705 | Not-determined |
| S1_extracted_k141_68717 | Not-determined |
| S1_extracted_k141_102194 | Not-determined |
| S1_extracted_k141_95507 | Not-determined |
| S1_extracted_k141_102195 | Not-determined |
| S1_extracted_k141_95508 | Not-determined |
| S1_extracted_k141_28694 | Low-quality |
| S1_extracted_k141_128857 | Not-determined |
| S1_extracted_k141_1678 | Not-determined |
| S1_extracted_k141_48791 | Not-determined |
| S1_extracted_k141_82189 | Not-determined |
| S1_extracted_k141_115451 | Not-determined |
| S1_extracted_k141_135415 | Not-determined |
| S1_extracted_k141_75451 | Not-determined |
| S1_extracted_k141_48797 | Not-determined |
| S1_extracted_k141_108711 | Not-determined |
| S1_extracted_k141_135419 | Not-determined |
| S1_extracted_k141_121918 | Low-quality |
| S1_extracted_k141_61167 | Low-quality |
| S1_extracted_k141_1688 | Not-determined |
| S1_extracted_k141_128863 | Low-quality |
| S1_extracted_k141_82191 | Not-determined |
| S1_extracted_k141_135425 | Not-determined |
| S1_extracted_k141_21959 | Not-determined |
| S1_extracted_k141_1692 | Not-determined |
| S1_extracted_k141_21960 | Not-determined |
| S1_extracted_k141_35018 | Not-determined |
| S1_extracted_k141_128864 | Not-determined |
| S1_extracted_k141_142153 | Not-determined |
| S1_extracted_k141_135427 | Not-determined |
| S1_extracted_k141_82194 | Low-quality |
| S1_extracted_k141_1694 | Not-determined |
| S1_extracted_k141_75460 | Not-determined |
| S1_extracted_k141_95536 | Not-determined |
| S1_extracted_k141_154792 | Not-determined |
| S1_extracted_k141_55473 | Not-determined |
| S1_extracted_k141_15833 | Low-quality |
| S1_extracted_k141_142163 | Not-determined |
| S1_extracted_k141_8793 | Low-quality |
| S1_extracted_k141_154794 | Not-determined |
| S1_extracted_k141_102211 | Low-quality |
| S1_extracted_k141_102212 | Low-quality |
| S1_extracted_k141_102214 | Not-determined |
| S1_extracted_k141_115468 | Not-determined |
| S1_extracted_k141_108722 | Not-determined |
| S1_extracted_k141_82198 | Not-determined |
| S1_extracted_k141_108723 | Not-determined |
| S1_extracted_k141_55475 | Not-determined |
| S1_extracted_k141_1700 | Not-determined |
| S1_extracted_k141_35032 | Not-determined |
| S1_extracted_k141_75475 | Not-determined |
| S1_extracted_k141_42152 | Not-determined |
| S1_extracted_k141_48811 | Low-quality |
| S1_extracted_k141_61174 | Not-determined |
| S1_extracted_k141_28721 | Not-determined |
| S1_extracted_k141_21978 | Not-determined |
| S1_extracted_k141_48815 | Not-determined |
| S1_extracted_k141_15843 | Not-determined |
| S1_extracted_k141_88757 | Not-determined |
| S1_extracted_k141_154800 | Not-determined |
| S1_extracted_k141_88758 | Not-determined |
| S1_extracted_k141_115482 | Not-determined |
| S1_extracted_k141_128876 | Not-determined |
| S1_extracted_k141_115484 | Not-determined |
| S1_extracted_k141_95554 | Not-determined |
| S1_extracted_k141_82202 | Low-quality |
| S1_extracted_k141_108727 | Low-quality |
| S1_extracted_k141_154802 | Not-determined |
| S1_extracted_k141_61180 | Low-quality |
| S1_extracted_k141_128878 | Low-quality |
| S1_extracted_k141_102225 | Not-determined |
| S1_extracted_k141_48819 | Low-quality |
| S1_extracted_k141_21985 | Low-quality |
| S1_extracted_k141_8807 | Not-determined |
| S1_extracted_k141_48821 | Not-determined |
| S1_extracted_k141_68727 | Not-determined |
| S1_extracted_k141_15851 | Not-determined |
| S1_extracted_k141_154807 | Not-determined |
| S1_extracted_k141_48823 | Low-quality |

| | |
|---|---|
| S1_extracted_k141_8809 | Not-determined |
| S1_extracted_k141_142174 | Not-determined |
| S1_extracted_k141_102232 | Not-determined |
| S1_extracted_k141_8814 | Not-determined |
| S1_extracted_k141_68733 | Not-determined |
| S1_extracted_k141_82207 | Not-determined |
| S1_extracted_k141_8816 | Not-determined |
| S1_extracted_k141_8817 | Not-determined |
| S1_extracted_k141_121962 | Low-quality |
| S1_extracted_k141_15860 | Low-quality |
| S1_extracted_k141_1711 | Not-determined |
| S1_extracted_k141_42171 | Not-determined |
| S1_extracted_k141_75491 | Low-quality |
| S1_extracted_k141_108742 | Not-determined |
| S1_extracted_k141_102242 | Not-determined |
| S1_extracted_k141_48833 | Not-determined |
| S1_extracted_k141_108743 | Low-quality |
| S1_extracted_k141_1716 | Not-determined |
| S1_extracted_k141_8819 | Not-determined |
| S1_extracted_k141_95578 | Not-determined |
| S1_extracted_k141_35049 | Not-determined |
| S1_extracted_k141_15872 | Not-determined |
| S1_extracted_k141_108749 | Not-determined |
| S1_extracted_k141_8825 | Not-determined |
| S1_extracted_k141_15874 | Not-determined |
| S1_extracted_k141_8828 | Not-determined |
| S1_extracted_k141_82227 | Not-determined |
| S1_extracted_k141_55503 | Low-quality |
| S1_extracted_k141_102252 | Not-determined |
| S1_extracted_k141_142195 | Not-determined |
| S1_extracted_k141_82235 | Not-determined |
| S1_extracted_k141_88784 | Not-determined |
| S1_extracted_k141_1725 | Not-determined |
| S1_extracted_k141_8836 | Not-determined |
| S1_extracted_k141_108763 | Not-determined |
| S1_extracted_k141_135462 | Not-determined |
| S1_extracted_k141_102257 | Not-determined |
| S1_extracted_k141_128907 | Low-quality |
| S1_extracted_k141_55511 | Not-determined |
| S1_extracted_k141_75508 | Not-determined |
| S1_extracted_k141_135466 | Not-determined |
| S1_extracted_k141_102263 | Not-determined |
| S1_extracted_k141_8846 | Not-determined |
| S1_extracted_k141_48855 | Not-determined |
| S1_extracted_k141_115497 | Not-determined |
| S1_extracted_k141_121983 | Not-determined |
| S1_extracted_k141_68759 | Not-determined |
| S1_extracted_k141_102267 | Not-determined |
| S1_extracted_k141_88792 | Low-quality |
| S1_extracted_k141_82244 | Not-determined |
| S1_extracted_k141_1735 | Low-quality |
| S1_extracted_k141_42184 | Not-determined |
| S1_extracted_k141_95599 | Not-determined |
| S1_extracted_k141_121989 | Not-determined |
| S1_extracted_k141_55521 | Not-determined |
| S1_extracted_k141_154817 | Low-quality |
| S1_extracted_k141_15891 | Not-determined |
| S1_extracted_k141_22032 | Not-determined |
| S1_extracted_k141_55523 | Not-determined |
| S1_extracted_k141_88801 | Not-determined |
| S1_extracted_k141_28746 | Not-determined |
| S1_extracted_k141_88802 | Not-determined |
| S1_extracted_k141_68767 | Not-determined |
| S1_extracted_k141_121992 | Not-determined |
| S1_extracted_k141_68770 | Not-determined |
| S1_extracted_k141_154820 | Not-determined |
| S1_extracted_k141_88804 | Not-determined |
| S1_extracted_k141_128913 | Not-determined |
| S1_extracted_k141_28752 | Not-determined |
| S1_extracted_k141_108782 | Not-determined |
| S1_extracted_k141_15896 | Not-determined |
| S1_extracted_k141_1745 | Low-quality |
| S1_extracted_k141_154823 | Not-determined |
| S1_extracted_k141_35071 | Not-determined |
| S1_extracted_k141_82249 | Not-determined |
| S1_extracted_k141_48872 | Not-determined |
| S1_extracted_k141_95616 | Low-quality |
| S1_extracted_k141_135473 | Not-determined |
| S1_extracted_k141_95617 | Low-quality |
| S1_extracted_k141_1750 | Not-determined |
| S1_extracted_k141_135478 | Not-determined |
| S1_extracted_k141_142209 | Not-determined |
| S1_extracted_k141_142211 | Low-quality |
| S1_extracted_k141_42196 | Not-determined |
| S1_extracted_k141_128921 | Not-determined |
| S1_extracted_k141_22039 | Not-determined |
| S1_extracted_k141_108791 | Not-determined |
| S1_extracted_k141_88816 | Low-quality |
| S1_extracted_k141_128927 | Not-determined |
| S1_extracted_k141_15917 | Not-determined |
| S1_extracted_k141_115527 | Not-determined |
| S1_extracted_k141_128928 | Not-determined |
| S1_extracted_k141_108794 | Low-quality |
| S1_extracted_k141_35088 | Low-quality |
| S1_extracted_k141_22047 | Not-determined |
| S1_extracted_k141_1767 | Not-determined |
| S1_extracted_k141_22048 | Not-determined |
| S1_extracted_k141_1768 | Not-determined |
| S1_extracted_k141_88820 | Low-quality |
| S1_extracted_k141_128933 | Not-determined |
| S1_extracted_k141_15926 | Low-quality |
| S1_extracted_k141_147777 | Not-determined |
| S1_extracted_k141_15927 | Not-determined |
| S1_extracted_k141_42210 | Not-determined |
| S1_extracted_k141_82268 | Not-determined |
| S1_extracted_k141_68795 | Not-determined |
| S1_extracted_k141_115538 | Not-determined |
| S1_extracted_k141_35091 | Not-determined |
| S1_extracted_k141_68796 | Not-determined |
| S1_extracted_k141_68797 | Not-determined |
| S1_extracted_k141_122011 | Not-determined |
| S1_extracted_k141_61245 | Not-determined |
| S1_extracted_k141_75534 | Not-determined |
| S1_extracted_k141_68798 | Not-determined |
| S1_extracted_k141_122012 | Not-determined |
| S1_extracted_k141_154829 | Not-determined |
| S1_extracted_k141_82272 | Not-determined |
| S1_extracted_k141_82274 | Low-quality |
| S1_extracted_k141_42214 | Not-determined |
| S1_extracted_k141_48901 | Not-determined |
| S1_extracted_k141_35096 | Not-determined |
| S1_extracted_k141_154833 | Not-determined |
| S1_extracted_k141_95643 | Not-determined |
| S1_extracted_k141_75543 | Not-determined |
| S1_extracted_k141_82281 | Not-determined |
| S1_extracted_k141_28779 | Low-quality |
| S1_extracted_k141_82283 | Not-determined |
| S1_extracted_k141_95649 | Low-quality |
| S1_extracted_k141_115550 | Low-quality |
| S1_extracted_k141_48906 | Not-determined |
| S1_extracted_k141_102298 | Not-determined |
| S1_extracted_k141_154837 | Low-quality |
| S1_extracted_k141_61257 | Not-determined |
| S1_extracted_k141_48913 | Not-determined |
| S1_extracted_k141_22067 | Not-determined |
| S1_extracted_k141_154839 | Not-determined |
| S1_extracted_k141_135499 | Not-determined |
| S1_extracted_k141_135500 | Low-quality |
| S1_extracted_k141_154840 | Low-quality |
| S1_extracted_k141_8881 | Not-determined |
| S1_extracted_k141_88845 | Not-determined |
| S1_extracted_k141_122031 | Not-determined |
| S1_extracted_k141_135504 | Not-determined |
| S1_extracted_k141_48919 | Not-determined |
| S1_extracted_k141_22072 | Low-quality |
| S1_extracted_k141_1775 | Not-determined |
| S1_extracted_k141_128954 | Not-determined |
| S1_extracted_k141_142251 | Not-determined |
| S1_extracted_k141_68811 | Not-determined |
| S1_extracted_k141_28795 | Not-determined |
| S1_extracted_k141_128956 | Not-determined |
| S1_extracted_k141_8884 | Not-determined |
| S1_extracted_k141_68813 | Not-determined |
| S1_extracted_k141_15938 | Not-determined |
| S1_extracted_k141_8886 | Not-determined |
| S1_extracted_k141_122037 | Not-determined |
| S1_extracted_k141_48925 | Not-determined |
| S1_extracted_k141_102301 | Low-quality |
| S1_extracted_k141_68815 | Not-determined |
| S1_extracted_k141_102303 | Not-determined |
| S1_extracted_k141_55535 | Not-determined |
| S1_extracted_k141_154849 | Not-determined |
| S1_extracted_k141_108820 | Not-determined |
| S1_extracted_k141_88852 | Low-quality |
| S1_extracted_k141_28803 | Not-determined |
| S1_extracted_k141_48927 | Not-determined |
| S1_extracted_k141_147787 | Not-determined |
| S1_extracted_k141_115564 | Not-determined |
| S1_extracted_k141_115565 | Not-determined |
| S1_extracted_k141_147789 | Low-quality |
| S1_extracted_k141_108825 | Low-quality |
| S1_extracted_k141_48932 | Not-determined |
| S1_extracted_k141_68821 | Not-determined |
| S1_extracted_k141_28805 | Low-quality |
| S1_extracted_k141_22085 | Not-determined |
| S1_extracted_k141_75562 | Not-determined |
| S1_extracted_k141_28806 | Not-determined |
| S1_extracted_k141_95659 | Not-determined |
| S1_extracted_k141_135515 | Not-determined |
| S1_extracted_k141_115572 | Not-determined |
| S1_extracted_k141_48937 | Not-determined |
| S1_extracted_k141_48938 | Not-determined |
| S1_extracted_k141_22092 | Not-determined |
| S1_extracted_k141_15947 | Not-determined |
| S1_extracted_k141_95663 | Not-determined |
| S1_extracted_k141_75573 | Not-determined |
| S1_extracted_k141_82306 | Not-determined |
| S1_extracted_k141_48941 | Not-determined |
| S1_extracted_k141_55542 | Not-determined |
| S1_extracted_k141_95665 | Not-determined |
| S1_extracted_k141_68831 | Not-determined |
| S1_extracted_k141_8899 | Not-determined |
| S1_extracted_k141_147796 | Not-determined |
| S1_extracted_k141_102321 | Not-determined |
| S1_extracted_k141_15959 | Not-determined |
| S1_extracted_k141_147798 | Not-determined |
| S1_extracted_k141_28818 | Not-determined |
| S1_extracted_k141_8903 | Not-determined |
| S1_extracted_k141_35115 | Low-quality |
| S1_extracted_k141_15963 | Not-determined |
| S1_extracted_k141_122056 | Low-quality |
| S1_extracted_k141_95674 | Not-determined |
| S1_extracted_k141_102327 | Not-determined |
| S1_extracted_k141_122059 | Not-determined |
| S1_extracted_k141_122060 | Not-determined |
| S1_extracted_k141_68847 | Not-determined |
| S1_extracted_k141_122062 | Not-determined |
| S1_extracted_k141_115583 | Not-determined |
| S1_extracted_k141_142264 | Not-determined |
| S1_extracted_k141_102331 | Low-quality |
| S1_extracted_k141_75587 | Not-determined |
| S1_extracted_k141_75588 | Not-determined |
| S1_extracted_k141_102334 | Not-determined |
| S1_extracted_k141_48959 | Not-determined |
| S1_extracted_k141_75592 | Not-determined |
| S1_extracted_k141_15973 | Not-determined |
| S1_extracted_k141_75593 | Not-determined |
| S1_extracted_k141_15975 | Not-determined |
| S1_extracted_k141_102337 | Low-quality |
| S1_extracted_k141_55564 | Low-quality |
| S1_extracted_k141_154872 | Not-determined |
| S1_extracted_k141_102338 | Not-determined |
| S1_extracted_k141_42233 | Not-determined |
| S1_extracted_k141_42239 | Not-determined |
| S1_extracted_k141_95684 | Not-determined |
| S1_extracted_k141_22110 | Low-quality |
| S1_extracted_k141_102343 | Low-quality |
| S1_extracted_k141_35132 | Low-quality |
| S1_extracted_k141_55569 | Not-determined |
| S1_extracted_k141_147801 | Not-determined |
| S1_extracted_k141_154873 | Not-determined |
| S1_extracted_k141_22113 | Not-determined |
| S1_extracted_k141_22114 | Not-determined |
| S1_extracted_k141_75601 | Low-quality |
| S1_extracted_k141_42246 | Not-determined |
| S1_extracted_k141_128998 | Not-determined |
| S1_extracted_k141_102347 | Low-quality |
| S1_extracted_k141_75603 | Not-determined |
| S1_extracted_k141_42248 | Not-determined |
| S1_extracted_k141_122073 | Low-quality |
| S1_extracted_k141_61275 | Not-determined |
| S1_extracted_k141_95695 | Not-determined |
| S1_extracted_k141_28832 | Not-determined |
| S1_extracted_k141_82321 | Not-determined |
| S1_extracted_k141_1788 | Not-determined |
| S1_extracted_k141_22119 | Not-determined |
| S1_extracted_k141_88892 | Not-determined |
| S1_extracted_k141_147807 | Not-determined |
| S1_extracted_k141_102354 | Not-determined |
| S1_extracted_k141_154878 | Not-determined |
| S1_extracted_k141_8929 | Not-determined |
| S1_extracted_k141_122083 | Not-determined |
| S1_extracted_k141_48966 | Low-quality |
| S1_extracted_k141_8932 | Not-determined |
| S1_extracted_k141_42257 | Not-determined |
| S1_extracted_k141_15991 | Not-determined |
| S1_extracted_k141_8937 | Not-determined |
| S1_extracted_k141_15992 | Low-quality |
| S1_extracted_k141_28844 | Not-determined |
| S1_extracted_k141_22132 | Not-determined |
| S1_extracted_k141_115610 | Not-determined |
| S1_extracted_k141_68857 | Not-determined |
| S1_extracted_k141_122092 | Not-determined |
| S1_extracted_k141_28846 | Not-determined |
| S1_extracted_k141_108861 | Not-determined |
| S1_extracted_k141_129010 | Not-determined |
| S1_extracted_k141_42259 | Not-determined |
| S1_extracted_k141_22136 | Not-determined |
| S1_extracted_k141_42260 | Low-quality |
| S1_extracted_k141_15998 | Not-determined |
| S1_extracted_k141_102363 | Low-quality |
| S1_extracted_k141_68859 | Not-determined |
| S1_extracted_k141_95707 | Not-determined |
| S1_extracted_k141_15999 | Low-quality |
| S1_extracted_k141_102364 | Not-determined |
| S1_extracted_k141_142282 | Not-determined |
| S1_extracted_k141_75608 | Not-determined |
| S1_extracted_k141_35144 | Not-determined |
| S1_extracted_k141_16003 | Not-determined |
| S1_extracted_k141_22138 | Low-quality |
| S1_extracted_k141_55593 | Low-quality |
| S1_extracted_k141_135557 | Not-determined |
| S1_extracted_k141_122102 | Not-determined |
| S1_extracted_k141_35146 | Not-determined |
| S1_extracted_k141_108872 | Low-quality |
| S1_extracted_k141_28861 | Low-quality |
| S1_extracted_k141_16009 | Not-determined |
| S1_extracted_k141_42268 | Not-determined |
| S1_extracted_k141_35150 | Not-determined |
| S1_extracted_k141_48979 | Not-determined |
| S1_extracted_k141_35152 | Not-determined |
| S1_extracted_k141_61284 | Not-determined |
| S1_extracted_k141_42272 | Not-determined |
| S1_extracted_k141_135566 | Low-quality |
| S1_extracted_k141_115629 | Low-quality |
| S1_extracted_k141_122112 | Not-determined |
| S1_extracted_k141_115630 | Not-determined |
| S1_extracted_k141_115631 | Not-determined |
| S1_extracted_k141_122114 | Not-determined |
| S1_extracted_k141_129022 | Not-determined |
| S1_extracted_k141_42280 | Not-determined |
| S1_extracted_k141_1808 | Low-quality |
| S1_extracted_k141_88899 | Not-determined |
| S1_extracted_k141_108882 | Low-quality |
| S1_extracted_k141_68875 | Low-quality |
| S1_extracted_k141_115639 | Not-determined |
| S1_extracted_k141_122124 | Not-determined |
| S1_extracted_k141_75630 | Not-determined |
| S1_extracted_k141_122125 | Not-determined |
| S1_extracted_k141_135573 | Not-determined |
| S1_extracted_k141_28875 | Not-determined |
| S1_extracted_k141_48989 | Not-determined |
| S1_extracted_k141_108884 | Not-determined |
| S1_extracted_k141_135576 | Not-determined |
| S1_extracted_k141_108885 | Low-quality |
| S1_extracted_k141_135577 | Not-determined |
| S1_extracted_k141_147824 | Not-determined |
| S1_extracted_k141_154897 | Low-quality |
| S1_extracted_k141_35164 | Not-determined |
| S1_extracted_k141_108889 | Not-determined |
| S1_extracted_k141_122131 | Not-determined |
| S1_extracted_k141_129033 | Not-determined |
| S1_extracted_k141_122134 | Low-quality |
| S1_extracted_k141_1816 | Not-determined |
| S1_extracted_k141_55607 | Low-quality |
| S1_extracted_k141_35169 | Not-determined |
| S1_extracted_k141_42297 | Low-quality |
| S1_extracted_k141_55612 | Not-determined |
| S1_extracted_k141_95735 | Not-determined |
| S1_extracted_k141_147829 | Not-determined |
| S1_extracted_k141_122146 | Not-determined |
| S1_extracted_k141_61292 | Low-quality |
| S1_extracted_k141_16038 | Not-determined |
| S1_extracted_k141_82370 | Not-determined |
| S1_extracted_k141_122149 | Low-quality |
| S1_extracted_k141_122150 | Not-determined |
| S1_extracted_k141_142294 | Not-determined |
| S1_extracted_k141_61294 | Low-quality |
| S1_extracted_k141_35179 | Not-determined |
| S1_extracted_k141_122152 | Not-determined |
| S1_extracted_k141_49002 | Not-determined |
| S1_extracted_k141_95741 | Not-determined |
| S1_extracted_k141_75653 | Not-determined |
| S1_extracted_k141_49003 | Not-determined |
| S1_extracted_k141_61296 | Not-determined |
| S1_extracted_k141_115648 | Not-determined |
| S1_extracted_k141_122156 | Not-determined |
| S1_extracted_k141_35181 | Low-quality |
| S1_extracted_k141_122160 | Low-quality |
| S1_extracted_k141_28887 | Low-quality |
| S1_extracted_k141_142301 | Not-determined |
| S1_extracted_k141_55625 | Not-determined |
| S1_extracted_k141_95751 | Not-determined |
| S1_extracted_k141_42311 | Not-determined |
| S1_extracted_k141_122165 | Not-determined |
| S1_extracted_k141_135601 | Not-determined |
| S1_extracted_k141_49010 | Low-quality |
| S1_extracted_k141_154909 | Not-determined |
| S1_extracted_k141_88925 | Low-quality |
| S1_extracted_k141_8971 | Not-determined |
| S1_extracted_k141_28900 | Not-determined |
| S1_extracted_k141_16044 | Not-determined |
| S1_extracted_k141_49015 | Not-determined |
| S1_extracted_k141_68890 | Not-determined |
| S1_extracted_k141_61309 | Not-determined |
| S1_extracted_k141_75660 | Low-quality |
| S1_extracted_k141_16047 | Not-determined |
| S1_extracted_k141_28903 | Not-determined |
| S1_extracted_k141_142309 | Not-determined |
| S1_extracted_k141_82391 | Not-determined |
| S1_extracted_k141_115673 | Low-quality |
| S1_extracted_k141_95759 | Not-determined |
| S1_extracted_k141_68898 | Not-determined |
| S1_extracted_k141_68899 | Not-determined |
| S1_extracted_k141_122178 | Not-determined |
| S1_extracted_k141_16052 | Not-determined |
| S1_extracted_k141_102422 | Not-determined |
| S1_extracted_k141_82400 | Not-determined |
| S1_extracted_k141_129066 | Not-determined |
| S1_extracted_k141_8985 | Low-quality |
| S1_extracted_k141_142315 | Not-determined |
| S1_extracted_k141_95763 | Not-determined |
| S1_extracted_k141_49037 | Low-quality |
| S1_extracted_k141_16056 | Not-determined |
| S1_extracted_k141_88940 | Low-quality |
| S1_extracted_k141_122182 | Low-quality |
| S1_extracted_k141_122184 | Low-quality |
| S1_extracted_k141_55644 | Not-determined |
| S1_extracted_k141_8997 | Low-quality |
| S1_extracted_k141_8998 | Not-determined |
| S1_extracted_k141_88944 | Not-determined |
| S1_extracted_k141_42337 | Not-determined |
| S1_extracted_k141_95774 | Low-quality |
| S1_extracted_k141_88947 | Not-determined |
| S1_extracted_k141_49049 | Not-determined |
| S1_extracted_k141_68906 | Not-determined |
| S1_extracted_k141_16062 | Low-quality |
| S1_extracted_k141_42341 | Not-determined |
| S1_extracted_k141_88949 | Low-quality |
| S1_extracted_k141_55652 | Not-determined |
| S1_extracted_k141_42344 | Low-quality |
| S1_extracted_k141_55654 | Low-quality |
| S1_extracted_k141_135631 | Not-determined |
| S1_extracted_k141_16067 | Not-determined |
| S1_extracted_k141_68912 | Not-determined |
| S1_extracted_k141_42349 | Not-determined |
| S1_extracted_k141_142327 | Not-determined |
| S1_extracted_k141_49060 | Not-determined |
| S1_extracted_k141_135633 | Low-quality |
| S1_extracted_k141_55657 | Not-determined |
| S1_extracted_k141_88958 | Low-quality |
| S1_extracted_k141_115694 | Not-determined |
| S1_extracted_k141_108930 | Not-determined |
| S1_extracted_k141_82412 | Not-determined |
| S1_extracted_k141_135635 | Not-determined |
| S1_extracted_k141_102441 | Low-quality |
| S1_extracted_k141_68920 | Not-determined |
| S1_extracted_k141_129083 | Not-determined |
| S1_extracted_k141_16072 | Not-determined |
| S1_extracted_k141_82416 | Low-quality |
| S1_extracted_k141_28923 | Low-quality |
| S1_extracted_k141_88965 | Low-quality |
| S1_extracted_k141_154936 | Low-quality |
| S1_extracted_k141_75689 | Not-determined |
| S1_extracted_k141_122203 | Not-determined |
| S11_extracted_k141_70015 | High-quality |
| S1_extracted_k141_95786 | Not-determined |
| S1_extracted_k141_129090 | Low-quality |
| S1_extracted_k141_75692 | Not-determined |
| S1_extracted_k141_95787 | Low-quality |
| S1_extracted_k141_129091 | Not-determined |
| S1_extracted_k141_22150 | Not-determined |
| S1_extracted_k141_88971 | Not-determined |
| S1_extracted_k141_61332 | Not-determined |
| S1_extracted_k141_102451 | Not-determined |
| S1_extracted_k141_147837 | Not-determined |
| S1_extracted_k141_75696 | Low-quality |
| S1_extracted_k141_129093 | Not-determined |
| S1_extracted_k141_68927 | Not-determined |
| S1_extracted_k141_135651 | Not-determined |
| S1_extracted_k141_88975 | Not-determined |
| S1_extracted_k141_49072 | Not-determined |
| S1_extracted_k141_115712 | Not-determined |
| S1_extracted_k141_68929 | Not-determined |
| S1_extracted_k141_22153 | Not-determined |
| S1_extracted_k141_102452 | Low-quality |
| S1_extracted_k141_75700 | Not-determined |
| S1_extracted_k141_108934 | Not-determined |
| S1_extracted_k141_42358 | Low-quality |
| S1_extracted_k141_1866 | Not-determined |
| S1_extracted_k141_35190 | Low-quality |
| S1_extracted_k141_16073 | Low-quality |
| S1_extracted_k141_68931 | Not-determined |
| S1_extracted_k141_55672 | Not-determined |
| S1_extracted_k141_55673 | Not-determined |
| S1_extracted_k141_75703 | Low-quality |
| S1_extracted_k141_122207 | Not-determined |
| S1_extracted_k141_75705 | Not-determined |
| S1_extracted_k141_154947 | Not-determined |
| S1_extracted_k141_55674 | Not-determined |
| S1_extracted_k141_108939 | Not-determined |
| S1_extracted_k141_28938 | Low-quality |
| S1_extracted_k141_122210 | Low-quality |
| S1_extracted_k141_102460 | Not-determined |
| S1_extracted_k141_95802 | Not-determined |
| S1_extracted_k141_135663 | Low-quality |
| S1_extracted_k141_115719 | Low-quality |
| S1_extracted_k141_115720 | Not-determined |
| S1_extracted_k141_129108 | Not-determined |
| S1_extracted_k141_142343 | Not-determined |
| S1_extracted_k141_82428 | Not-determined |
| S1_extracted_k141_154951 | Not-determined |
| S1_extracted_k141_35204 | Not-determined |
| S1_extracted_k141_122220 | Not-determined |
| S1_extracted_k141_55684 | Not-determined |
| S1_extracted_k141_135672 | Not-determined |
| S1_extracted_k141_82434 | Not-determined |
| S1_extracted_k141_88985 | Low-quality |
| S1_extracted_k141_122223 | Not-determined |
| S1_extracted_k141_95805 | Not-determined |
| S1_extracted_k141_75722 | Not-determined |
| S1_extracted_k141_61335 | Not-determined |
| S1_extracted_k141_129115 | Not-determined |
| S1_extracted_k141_35207 | Not-determined |
| S1_extracted_k141_129117 | Not-determined |
| S1_extracted_k141_122227 | Not-determined |
| S1_extracted_k141_82438 | Not-determined |
| S1_extracted_k141_22163 | Low-quality |
| S1_extracted_k141_108949 | Not-determined |
| S1_extracted_k141_115726 | Not-determined |
| S1_extracted_k141_135680 | Low-quality |
| S1_extracted_k141_61339 | Not-determined |
| S1_extracted_k141_16093 | Not-determined |
| S1_extracted_k141_135682 | Low-quality |
| S1_extracted_k141_108953 | Not-determined |
| S1_extracted_k141_102484 | Not-determined |
| S1_extracted_k141_42377 | Low-quality |
| S1_extracted_k141_75731 | Not-determined |
| S1_extracted_k141_28961 | Not-determined |
| S1_extracted_k141_108959 | Not-determined |
| S1_extracted_k141_82446 | Not-determined |
| S1_extracted_k141_16101 | Not-determined |
| S1_extracted_k141_49081 | Not-determined |
| S1_extracted_k141_9037 | Not-determined |
| S1_extracted_k141_142356 | Low-quality |
| S1_extracted_k141_95821 | Not-determined |
| S1_extracted_k141_154954 | Not-determined |
| S1_extracted_k141_68949 | Not-determined |
| S1_extracted_k141_16105 | Not-determined |
| S1_extracted_k141_147855 | Not-determined |
| S1_extracted_k141_49085 | Not-determined |
| S1_extracted_k141_82451 | Not-determined |
| S1_extracted_k141_102489 | Low-quality |
| S1_extracted_k141_95827 | Not-determined |
| S1_extracted_k141_42393 | Not-determined |
| S1_extracted_k141_28972 | Not-determined |
| S1_extracted_k141_42396 | Not-determined |
| S1_extracted_k141_16116 | Not-determined |
| S1_extracted_k141_82457 | Not-determined |
| S1_extracted_k141_1897 | Not-determined |
| S1_extracted_k141_75735 | Not-determined |
| S1_extracted_k141_49094 | Not-determined |
| S1_extracted_k141_1899 | Not-determined |
| S1_extracted_k141_75738 | Not-determined |
| S1_extracted_k141_154958 | Not-determined |
| S1_extracted_k141_16122 | Not-determined |
| S1_extracted_k141_102490 | Low-quality |
| S1_extracted_k141_1903 | Not-determined |
| S1_extracted_k141_28978 | Not-determined |
| S1_extracted_k141_49102 | Not-determined |
| S1_extracted_k141_75744 | Not-determined |
| S1_extracted_k141_42408 | Not-determined |
| S1_extracted_k141_102494 | Not-determined |
| S1_extracted_k141_122247 | Not-determined |
| S1_extracted_k141_135698 | Not-determined |
| S1_extracted_k141_35228 | Not-determined |
| S1_extracted_k141_61353 | Low-quality |
| S1_extracted_k141_1907 | Low-quality |
| S1_extracted_k141_49105 | Not-determined |
| S1_extracted_k141_42412 | Not-determined |
| S1_extracted_k141_28983 | Not-determined |
| S1_extracted_k141_75746 | Not-determined |
| S1_extracted_k141_129158 | Not-determined |
| S1_extracted_k141_129159 | Not-determined |
| S1_extracted_k141_122250 | Not-determined |
| S1_extracted_k141_68969 | Not-determined |
| S1_extracted_k141_129165 | Not-determined |
| S1_extracted_k141_122256 | Not-determined |
| S1_extracted_k141_9053 | Not-determined |
| S1_extracted_k141_49109 | Not-determined |
| S1_extracted_k141_16132 | Not-determined |
| S1_extracted_k141_1915 | Not-determined |
| S1_extracted_k141_115759 | Not-determined |
| S1_extracted_k141_115760 | Not-determined |
| S1_extracted_k141_35233 | Not-determined |
| S1_extracted_k141_115763 | Not-determined |
| S1_extracted_k141_42422 | Not-determined |
| S1_extracted_k141_95850 | Low-quality |
| S1_extracted_k141_9063 | Not-determined |
| S1_extracted_k141_115767 | Low-quality |
| S1_extracted_k141_122265 | Not-determined |
| S1_extracted_k141_89011 | Low-quality |
| S1_extracted_k141_1922 | Low-quality |
| S1_extracted_k141_35239 | Not-determined |
| S1_extracted_k141_9068 | Not-determined |
| S1_extracted_k141_135711 | Not-determined |
| S1_extracted_k141_16137 | Not-determined |
| S1_extracted_k141_61368 | Not-determined |
| S1_extracted_k141_102508 | Not-determined |
| S1_extracted_k141_82477 | Low-quality |
| S1_extracted_k141_35245 | Not-determined |
| S1_extracted_k141_9076 | Low-quality |
| S1_extracted_k141_55731 | Not-determined |
| S1_extracted_k141_82480 | Not-determined |
| S1_extracted_k141_135717 | Not-determined |
| S1_extracted_k141_42426 | Not-determined |
| S1_extracted_k141_16145 | Not-determined |
| S1_extracted_k141_75766 | Not-determined |
| S1_extracted_k141_42428 | Not-determined |
| S1_extracted_k141_55736 | Not-determined |
| S1_extracted_k141_147875 | Low-quality |
| S1_extracted_k141_122270 | Not-determined |
| S1_extracted_k141_147876 | Not-determined |
| S1_extracted_k141_22200 | Not-determined |
| S1_extracted_k141_95864 | Not-determined |
| S1_extracted_k141_89027 | Not-determined |
| S1_extracted_k141_95867 | Low-quality |
| S1_extracted_k141_68992 | Not-determined |
| S1_extracted_k141_89029 | Not-determined |

| | |
|---|---|
| S1_extracted_k141_16153 | Low-quality |
| S1_extracted_k141_95868 | Not-determined |
| S1_extracted_k141_95869 | Low-quality |
| S1_extracted_k141_61377 | Low-quality |
| S1_extracted_k141_22205 | Low-quality |
| S1_extracted_k141_95870 | Not-determined |
| S1_extracted_k141_35253 | Not-determined |
| S1_extracted_k141_154971 | Not-determined |
| S1_extracted_k141_129190 | Not-determined |
| S1_extracted_k141_22206 | Low-quality |
| S1_extracted_k141_55745 | Not-determined |
| S1_extracted_k141_82495 | Not-determined |
| S1_extracted_k141_135728 | Not-determined |
| S1_extracted_k141_135729 | Not-determined |
| S1_extracted_k141_28997 | Not-determined |
| S1_extracted_k141_61382 | Not-determined |
| S1_extracted_k141_22208 | Not-determined |
| S1_extracted_k141_95874 | Not-determined |
| S1_extracted_k141_95876 | Low-quality |
| S1_extracted_k141_55752 | Not-determined |
| S1_extracted_k141_89033 | Low-quality |
| S1_extracted_k141_55753 | Not-determined |
| S1_extracted_k141_129198 | Not-determined |
| S1_extracted_k141_122279 | Not-determined |
| S1_extracted_k141_9096 | Not-determined |
| S1_extracted_k141_61388 | Not-determined |
| S1_extracted_k141_102529 | Not-determined |
| S1_extracted_k141_75773 | Not-determined |
| S1_extracted_k141_69000 | Not-determined |
| S1_extracted_k141_122282 | Not-determined |
| S1_extracted_k141_42445 | Not-determined |
| S1_extracted_k141_122285 | Low-quality |
| S1_extracted_k141_16165 | Low-quality |
| S1_extracted_k141_82505 | Not-determined |
| S1_extracted_k141_29007 | Not-determined |
| S1_extracted_k141_82507 | Not-determined |
| S1_extracted_k141_122287 | Not-determined |
| S1_extracted_k141_9109 | Not-determined |
| S1_extracted_k141_147884 | Not-determined |
| S1_extracted_k141_147885 | Not-determined |
| S1_extracted_k141_102536 | Low-quality |
| S1_extracted_k141_49129 | Not-determined |
| S1_extracted_k141_49130 | Not-determined |
| S1_extracted_k141_115787 | Low-quality |
| S1_extracted_k141_82512 | Low-quality |
| S1_extracted_k141_102539 | Not-determined |
| S1_extracted_k141_69007 | Not-determined |
| S1_extracted_k141_147890 | Not-determined |
| S1_extracted_k141_42448 | Low-quality |
| S1_extracted_k141_9115 | Not-determined |
| S1_extracted_k141_75784 | Not-determined |
| S1_extracted_k141_102543 | Not-determined |
| S1_extracted_k141_69010 | Low-quality |
| S1_extracted_k141_61398 | Not-determined |
| S1_extracted_k141_89043 | Not-determined |
| S1_extracted_k141_16176 | Not-determined |
| S1_extracted_k141_122294 | Not-determined |
| S1_extracted_k141_55775 | Low-quality |
| S1_extracted_k141_102545 | Not-determined |
| S1_extracted_k141_69012 | Low-quality |
| S1_extracted_k141_16180 | Not-determined |
| S1_extracted_k141_55779 | Low-quality |
| S1_extracted_k141_16181 | Low-quality |
| S1_extracted_k141_61402 | Low-quality |
| S1_extracted_k141_129212 | Not-determined |
| S1_extracted_k141_129213 | Not-determined |
| S1_extracted_k141_75794 | Not-determined |
| S1_extracted_k141_147902 | Not-determined |
| S1_extracted_k141_1956 | Not-determined |
| S1_extracted_k141_89049 | Not-determined |
| S1_extracted_k141_147903 | Not-determined |
| S1_extracted_k141_9128 | Not-determined |
| S1_extracted_k141_89051 | Not-determined |
| S1_extracted_k141_16184 | Not-determined |
| S1_extracted_k141_1960 | Not-determined |
| S1_extracted_k141_75800 | Not-determined |
| S1_extracted_k141_115806 | Not-determined |
| S1_extracted_k141_55788 | Not-determined |
| S1_extracted_k141_55789 | Not-determined |
| S1_extracted_k141_129221 | Not-determined |
| S1_extracted_k141_49149 | Not-determined |
| S1_extracted_k141_42460 | Not-determined |
| S1_extracted_k141_29026 | Low-quality |
| S1_extracted_k141_22221 | Not-determined |
| S1_extracted_k141_89058 | Low-quality |
| S1_extracted_k141_102564 | Low-quality |
| S1_extracted_k141_22223 | Low-quality |
| S1_extracted_k141_102565 | Not-determined |
| S1_extracted_k141_122311 | Not-determined |
| S1_extracted_k141_95914 | Not-determined |
| S1_extracted_k141_1972 | Not-determined |
| S1_extracted_k141_49158 | Not-determined |
| S1_extracted_k141_16193 | Not-determined |
| S1_extracted_k141_95918 | Not-determined |
| S1_extracted_k141_122315 | Not-determined |
| S1_extracted_k141_142438 | Not-determined |
| S1_extracted_k141_1974 | Not-determined |
| S1_extracted_k141_129234 | Low-quality |
| S1_extracted_k141_82528 | Not-determined |
| S1_extracted_k141_16195 | Not-determined |
| S1_extracted_k141_129237 | Not-determined |
| S1_extracted_k141_82531 | Not-determined |
| S1_extracted_k141_147906 | Low-quality |
| S1_extracted_k141_89071 | Not-determined |
| S1_extracted_k141_102575 | Not-determined |
| S1_extracted_k141_102577 | Not-determined |
| S1_extracted_k141_147908 | Not-determined |
| S1_extracted_k141_16200 | Low-quality |
| S1_extracted_k141_22238 | Not-determined |
| S1_extracted_k141_1978 | Not-determined |
| S1_extracted_k141_89072 | Low-quality |
| S1_extracted_k141_109048 | Low-quality |
| S1_extracted_k141_95927 | Not-determined |
| S1_extracted_k141_109049 | Not-determined |
| S1_extracted_k141_55807 | Not-determined |
| S1_extracted_k141_61418 | Not-determined |
| S1_extracted_k141_82541 | Not-determined |
| S1_extracted_k141_154997 | Not-determined |
| S1_extracted_k141_89078 | Not-determined |
| S1_extracted_k141_42485 | Not-determined |
| S1_extracted_k141_109052 | Not-determined |
| S1_extracted_k141_55812 | Not-determined |
| S1_extracted_k141_16205 | Low-quality |
| S1_extracted_k141_142450 | Not-determined |
| S1_extracted_k141_9147 | Not-determined |
| S1_extracted_k141_135748 | Not-determined |
| S1_extracted_k141_129243 | Not-determined |
| S1_extracted_k141_1988 | Not-determined |
| S1_extracted_k141_129244 | Low-quality |
| S1_extracted_k141_89086 | Not-determined |
| S1_extracted_k141_155003 | Not-determined |
| S1_extracted_k141_129246 | Not-determined |
| S1_extracted_k141_69032 | Not-determined |
| S1_extracted_k141_135751 | Not-determined |
| S1_extracted_k141_49172 | Not-determined |
| S1_extracted_k141_29055 | Not-determined |
| S1_extracted_k141_22244 | Low-quality |
| S1_extracted_k141_49175 | Not-determined |
| S1_extracted_k141_1992 | Not-determined |
| S1_extracted_k141_115834 | Not-determined |
| S1_extracted_k141_135756 | Not-determined |
| S1_extracted_k141_75820 | Not-determined |
| S1_extracted_k141_55820 | Not-determined |
| S1_extracted_k141_95940 | Not-determined |
| S1_extracted_k141_9161 | Not-determined |
| S1_extracted_k141_142458 | Low-quality |
| S1_extracted_k141_142459 | Not-determined |
| S1_extracted_k141_9163 | Not-determined |
| S1_extracted_k141_42504 | Not-determined |
| S1_extracted_k141_69040 | Not-determined |
| S1_extracted_k141_55824 | Not-determined |
| S1_extracted_k141_42505 | Not-determined |
| S1_extracted_k141_122325 | Not-determined |
| S1_extracted_k141_35301 | Not-determined |
| S1_extracted_k141_29060 | Not-determined |
| S1_extracted_k141_115845 | Not-determined |
| S1_extracted_k141_49182 | Not-determined |
| S1_extracted_k141_42511 | Low-quality |
| S1_extracted_k141_9172 | Low-quality |
| S1_extracted_k141_49186 | Not-determined |
| S1_extracted_k141_147920 | Not-determined |
| S1_extracted_k141_16221 | Not-determined |
| S1_extracted_k141_55827 | Not-determined |
| S1_extracted_k141_29064 | Not-determined |
| S1_extracted_k141_95953 | Not-determined |
| S1_extracted_k141_122334 | Not-determined |
| S1_extracted_k141_42513 | Not-determined |
| S1_extracted_k141_49192 | Not-determined |
| S1_extracted_k141_122336 | Not-determined |
| S1_extracted_k141_69056 | Low-quality |
| S1_extracted_k141_75843 | Not-determined |
| S1_extracted_k141_89103 | Low-quality |
| S1_extracted_k141_129272 | Not-determined |
| S1_extracted_k141_75845 | Not-determined |
| S1_extracted_k141_22262 | Low-quality |
| S1_extracted_k141_2008 | Not-determined |
| S1_extracted_k141_102591 | Not-determined |
| S1_extracted_k141_2009 | Low-quality |
| S1_extracted_k141_89106 | Not-determined |
| S1_extracted_k141_135783 | Not-determined |
| S1_extracted_k141_16230 | Not-determined |
| S1_extracted_k141_95960 | Not-determined |
| S1_extracted_k141_61445 | Not-determined |
| S1_extracted_k141_142478 | Not-determined |
| S1_extracted_k141_42519 | Not-determined |
| S1_extracted_k141_102597 | Not-determined |
| S1_extracted_k141_55839 | Not-determined |
| S1_extracted_k141_142481 | Not-determined |
| S1_extracted_k141_109098 | Not-determined |
| S1_extracted_k141_142482 | Not-determined |
| S1_extracted_k141_109099 | Not-determined |
| S1_extracted_k141_61448 | Not-determined |
| S1_extracted_k141_89111 | Low-quality |
| S1_extracted_k141_135790 | Not-determined |
| S1_extracted_k141_122346 | Not-determined |
| S1_extracted_k141_142484 | Not-determined |
| S1_extracted_k141_82553 | Not-determined |
| S1_extracted_k141_61450 | Low-quality |
| S1_extracted_k141_2017 | Not-determined |
| S1_extracted_k141_135793 | Not-determined |
| S1_extracted_k141_147926 | Not-determined |
| S1_extracted_k141_61453 | Not-determined |
| S1_extracted_k141_89117 | Low-quality |
| S1_extracted_k141_102606 | Not-determined |
| S1_extracted_k141_95967 | Low-quality |
| S1_extracted_k141_29085 | Not-determined |
| S1_extracted_k141_89118 | Not-determined |
| S1_extracted_k141_115855 | Not-determined |
| S1_extracted_k141_42533 | Not-determined |
| S1_extracted_k141_89120 | Low-quality |
| S1_extracted_k141_129286 | Low-quality |
| S1_extracted_k141_42534 | Not-determined |
| S1_extracted_k141_115859 | Not-determined |
| S1_extracted_k141_109109 | Low-quality |
| S1_extracted_k141_89122 | Not-determined |
| S1_extracted_k141_22280 | Not-determined |
| S1_extracted_k141_69064 | Not-determined |
| S1_extracted_k141_9195 | Not-determined |
| S1_extracted_k141_16245 | Not-determined |
| S1_extracted_k141_142498 | Not-determined |
| S1_extracted_k141_89124 | Low-quality |
| S1_extracted_k141_29095 | Not-determined |
| S1_extracted_k141_129291 | Low-quality |
| S1_extracted_k141_109115 | Low-quality |
| S1_extracted_k141_142501 | Not-determined |
| S1_extracted_k141_135807 | Not-determined |
| S1_extracted_k141_55862 | Not-determined |
| S1_extracted_k141_9208 | Low-quality |
| S1_extracted_k141_142503 | Low-quality |
| S1_extracted_k141_22283 | Not-determined |
| S1_extracted_k141_142505 | Low-quality |
| S1_extracted_k141_135811 | Not-determined |
| S1_extracted_k141_55866 | Not-determined |
| S1_extracted_k141_109122 | Not-determined |
| S1_extracted_k141_115870 | Not-determined |
| S1_extracted_k141_29103 | Not-determined |
| S1_extracted_k141_155037 | Not-determined |
| S1_extracted_k141_35321 | Not-determined |
| S1_extracted_k141_102620 | Not-determined |
| S1_extracted_k141_42553 | Not-determined |
| S1_extracted_k141_135816 | Low-quality |
| S1_extracted_k141_35323 | Not-determined |
| S1_extracted_k141_115875 | Not-determined |
| S1_extracted_k141_129295 | Not-determined |
| S1_extracted_k141_29108 | Not-determined |
| S1_extracted_k141_22291 | Not-determined |
| S1_extracted_k141_16255 | Not-determined |
| S1_extracted_k141_35324 | Not-determined |
| S1_extracted_k141_61463 | Not-determined |
| S1_extracted_k141_95990 | Not-determined |
| S1_extracted_k141_29111 | Not-determined |
| S1_extracted_k141_82575 | Not-determined |
| S1_extracted_k141_22293 | Low-quality |
| S1_extracted_k141_82576 | Low-quality |
| S1_extracted_k141_35326 | Not-determined |
| S1_extracted_k141_95993 | Not-determined |
| S1_extracted_k141_89140 | Not-determined |
| S1_extracted_k141_147930 | Not-determined |
| S1_extracted_k141_147931 | Not-determined |
| S1_extracted_k141_89142 | Not-determined |
| S1_extracted_k141_69080 | Not-determined |
| S1_extracted_k141_69081 | Not-determined |
| S1_extracted_k141_95998 | Not-determined |
| S1_extracted_k141_129301 | Low-quality |
| S1_extracted_k141_89146 | Not-determined |
| S1_extracted_k141_102629 | Low-quality |
| S1_extracted_k141_42555 | Not-determined |
| S1_extracted_k141_9215 | Not-determined |
| S1_extracted_k141_2036 | Not-determined |
| S1_extracted_k141_35333 | Not-determined |
| S1_extracted_k141_16265 | Not-determined |
| S1_extracted_k141_35334 | Low-quality |
| S1_extracted_k141_9216 | Low-quality |
| S1_extracted_k141_69085 | Not-determined |
| S1_extracted_k141_61473 | Not-determined |
| S1_extracted_k141_89149 | Not-determined |
| S1_extracted_k141_82588 | Not-determined |
| S1_extracted_k141_29122 | Low-quality |
| S1_extracted_k141_109138 | Not-determined |
| S1_extracted_k141_35337 | Not-determined |
| S1_extracted_k141_82592 | Not-determined |
| S1_extracted_k141_115887 | Low-quality |
| S1_extracted_k141_155043 | Not-determined |
| S1_extracted_k141_42561 | Low-quality |
| S1_extracted_k141_42563 | Low-quality |
| S1_extracted_k141_115891 | Not-determined |
| S1_extracted_k141_89157 | Not-determined |
| S1_extracted_k141_142521 | Not-determined |
| S1_extracted_k141_42568 | Not-determined |
| S1_extracted_k141_82601 | Not-determined |
| S1_extracted_k141_115897 | Not-determined |
| S1_extracted_k141_69093 | Not-determined |
| S1_extracted_k141_89162 | Not-determined |
| S1_extracted_k141_115898 | Not-determined |
| S1_extracted_k141_22305 | Not-determined |
| S1_extracted_k141_2047 | Low-quality |
| S1_extracted_k141_109150 | Not-determined |
| S1_extracted_k141_29136 | Not-determined |
| S1_extracted_k141_102650 | Not-determined |
| S1_extracted_k141_42571 | Low-quality |
| S1_extracted_k141_69098 | Not-determined |
| S1_extracted_k141_96010 | Not-determined |
| S1_extracted_k141_9224 | Not-determined |
| S1_extracted_k141_135844 | Not-determined |
| S1_extracted_k141_115903 | Not-determined |
| S1_extracted_k141_115904 | Low-quality |
| S1_extracted_k141_135846 | Not-determined |
| S1_extracted_k141_16286 | Not-determined |
| S1_extracted_k141_147938 | Not-determined |
| S1_extracted_k141_115906 | Not-determined |
| S1_extracted_k141_96015 | Low-quality |
| S1_extracted_k141_55894 | Not-determined |
| S1_extracted_k141_2052 | Not-determined |
| S1_extracted_k141_16289 | Not-determined |
| S1_extracted_k141_142529 | Not-determined |
| S1_extracted_k141_49238 | Not-determined |
| S1_extracted_k141_96017 | Not-determined |
| S1_extracted_k141_129314 | Not-determined |
| S1_extracted_k141_9229 | Not-determined |
| S1_extracted_k141_42576 | Not-determined |
| S1_extracted_k141_29144 | Not-determined |
| S1_extracted_k141_49240 | Low-quality |
| S1_extracted_k141_61491 | Not-determined |
| S1_extracted_k141_49241 | Not-determined |
| S1_extracted_k141_2060 | Not-determined |
| S1_extracted_k141_29150 | Not-determined |
| S1_extracted_k141_29151 | Not-determined |
| S1_extracted_k141_115913 | Not-determined |
| S1_extracted_k141_2062 | Not-determined |
| S1_extracted_k141_155049 | Not-determined |
| S1_extracted_k141_22317 | Not-determined |
| S1_extracted_k141_49250 | Not-determined |
| S1_extracted_k141_82615 | Not-determined |
| S1_extracted_k141_69111 | Not-determined |
| S1_extracted_k141_49252 | Not-determined |
| S1_extracted_k141_102659 | Not-determined |
| S1_extracted_k141_82618 | Low-quality |
| S1_extracted_k141_115920 | Not-determined |
| S1_extracted_k141_69114 | Not-determined |
| S1_extracted_k141_122402 | Not-determined |
| S1_extracted_k141_142540 | Not-determined |
| S1_extracted_k141_122403 | Low-quality |
| S1_extracted_k141_96028 | Not-determined |
| S1_extracted_k141_29155 | Not-determined |
| S1_extracted_k141_16303 | Not-determined |
| S1_extracted_k141_9239 | Not-determined |
| S1_extracted_k141_142545 | Not-determined |
| S1_extracted_k141_122408 | Not-determined |
| S1_extracted_k141_42595 | Not-determined |
| S1_extracted_k141_22325 | Not-determined |
| S1_extracted_k141_55914 | Not-determined |
| S1_extracted_k141_42598 | Not-determined |
| S1_extracted_k141_115932 | Not-determined |
| S1_extracted_k141_42601 | Not-determined |
| S1_extracted_k141_96039 | Not-determined |
| S1_extracted_k141_96040 | Not-determined |
| S1_extracted_k141_55920 | Not-determined |
| S1_extracted_k141_16315 | Not-determined |
| S1_extracted_k141_49264 | Not-determined |
| S1_extracted_k141_61507 | Low-quality |
| S1_extracted_k141_42604 | Not-determined |
| S1_extracted_k141_49265 | Not-determined |
| S1_extracted_k141_115935 | Not-determined |
| S1_extracted_k141_135870 | Not-determined |
| S1_extracted_k141_16321 | Not-determined |
| S1_extracted_k141_16322 | Low-quality |
| S1_extracted_k141_109169 | Low-quality |
| S1_extracted_k141_29160 | Low-quality |
| S1_extracted_k141_29161 | Not-determined |
| S1_extracted_k141_55929 | Not-determined |
| S1_extracted_k141_29163 | Not-determined |
| S1_extracted_k141_16326 | Low-quality |
| S1_extracted_k141_147950 | Not-determined |
| S1_extracted_k141_61517 | Not-determined |
| S1_extracted_k141_75887 | Low-quality |
| S1_extracted_k141_22332 | Not-determined |
| S1_extracted_k141_22333 | Not-determined |
| S1_extracted_k141_102669 | Not-determined |
| S1_extracted_k141_75888 | Not-determined |
| S1_extracted_k141_82647 | Not-determined |
| S1_extracted_k141_22334 | Not-determined |
| S1_extracted_k141_142558 | Not-determined |
| S1_extracted_k141_115953 | Low-quality |
| S1_extracted_k141_29169 | Not-determined |
| S1_extracted_k141_35381 | Low-quality |
| S1_extracted_k141_129333 | Not-determined |
| S1_extracted_k141_29170 | Not-determined |
| S1_extracted_k141_16335 | Not-determined |
| S1_extracted_k141_155062 | Low-quality |
| S1_extracted_k141_42622 | Low-quality |
| S1_extracted_k141_115961 | Not-determined |
| S1_extracted_k141_122416 | Not-determined |
| S1_extracted_k141_16340 | Not-determined |
| S1_extracted_k141_49289 | Low-quality |
| S1_extracted_k141_122418 | Low-quality |
| S1_extracted_k141_82656 | Low-quality |
| S1_extracted_k141_49291 | Not-determined |
| S1_extracted_k141_109178 | Not-determined |
| S1_extracted_k141_142566 | Not-determined |
| S1_extracted_k141_129340 | Low-quality |
| S1_extracted_k141_35394 | Low-quality |
| S1_extracted_k141_82659 | Low-quality |
| S1_extracted_k141_135891 | Low-quality |
| S1_extracted_k141_16351 | Not-determined |
| S1_extracted_k141_9270 | Not-determined |
| S1_extracted_k141_42632 | Low-quality |
| S1_extracted_k141_102681 | Not-determined |
| S1_extracted_k141_22347 | Not-determined |
| S1_extracted_k141_122426 | Not-determined |
| S1_extracted_k141_49302 | Not-determined |
| S1_extracted_k141_16353 | Not-determined |
| S1_extracted_k141_42636 | Not-determined |
| S1_extracted_k141_109189 | Not-determined |
| S1_extracted_k141_49305 | Low-quality |
| S1_extracted_k141_115974 | Not-determined |
| S1_extracted_k141_16357 | Not-determined |
| S1_extracted_k141_69154 | Not-determined |
| S1_extracted_k141_96064 | Not-determined |
| S1_extracted_k141_42639 | Not-determined |
| S1_extracted_k141_22352 | Not-determined |
| S1_extracted_k141_96066 | Not-determined |
| S1_extracted_k141_109193 | Not-determined |
| S1_extracted_k141_135904 | Not-determined |
| S1_extracted_k141_122435 | Not-determined |
| S1_extracted_k141_42643 | Low-quality |
| S1_extracted_k141_69159 | Not-determined |
| S1_extracted_k141_29194 | Not-determined |
| S1_extracted_k141_82665 | Not-determined |
| S1_extracted_k141_29195 | Not-determined |
| S1_extracted_k141_22360 | Not-determined |
| S1_extracted_k141_9276 | Low-quality |
| S1_extracted_k141_115981 | Not-determined |
| S1_extracted_k141_42645 | Not-determined |
| S1_extracted_k141_96071 | Not-determined |
| S1_extracted_k141_75925 | Not-determined |
| S1_extracted_k141_82668 | Not-determined |
| S1_extracted_k141_115983 | Not-determined |
| S1_extracted_k141_89180 | Low-quality |
| S1_extracted_k141_96074 | Not-determined |
| S1_extracted_k141_122445 | Not-determined |
| S1_extracted_k141_96075 | Not-determined |
| S1_extracted_k141_147973 | Not-determined |
| S1_extracted_k141_122446 | Low-quality |
| S1_extracted_k141_115988 | Not-determined |
| S1_extracted_k141_89185 | Not-determined |
| S1_extracted_k141_129356 | Not-determined |
| S1_extracted_k141_142583 | Low-quality |
| S1_extracted_k141_9284 | Not-determined |
| S1_extracted_k141_89186 | Low-quality |
| S1_extracted_k141_102693 | Not-determined |
| S1_extracted_k141_82674 | Low-quality |
| S1_extracted_k141_69167 | Not-determined |
| S1_extracted_k141_102695 | Not-determined |
| S1_extracted_k141_9286 | Not-determined |
| S1_extracted_k141_16371 | Not-determined |
| S1_extracted_k141_115990 | Not-determined |
| S1_extracted_k141_29210 | Not-determined |
| S1_extracted_k141_129361 | Not-determined |
| S1_extracted_k141_16373 | Low-quality |
| S1_extracted_k141_16374 | Not-determined |
| S1_extracted_k141_75930 | Low-quality |
| S1_extracted_k141_2099 | Not-determined |
| S1_extracted_k141_109203 | Not-determined |
| S1_extracted_k141_109205 | Not-determined |
| S1_extracted_k141_49323 | Not-determined |
| S1_extracted_k141_82684 | Not-determined |
| S1_extracted_k141_16382 | Not-determined |
| S1_extracted_k141_115991 | Low-quality |
| S1_extracted_k141_135922 | Low-quality |
| S1_extracted_k141_49327 | Not-determined |
| S1_extracted_k141_69176 | Not-determined |
| S1_extracted_k141_49328 | Not-determined |
| S1_extracted_k141_9297 | Low-quality |
| S1_extracted_k141_102704 | Not-determined |
| S1_extracted_k141_29220 | Not-determined |
| S1_extracted_k141_22373 | Not-determined |
| S1_extracted_k141_142595 | Low-quality |
| S1_extracted_k141_55961 | Low-quality |
| S1_extracted_k141_9301 | Not-determined |
| S1_extracted_k141_29223 | Not-determined |
| S1_extracted_k141_142596 | Not-determined |
| S1_extracted_k141_69179 | Not-determined |
| S1_extracted_k141_75942 | Low-quality |
| S1_extracted_k141_16389 | Not-determined |
| S1_extracted_k141_16390 | Not-determined |
| S1_extracted_k141_29227 | Not-determined |
| S1_extracted_k141_16391 | Not-determined |
| S1_extracted_k141_155093 | Not-determined |
| S1_extracted_k141_75947 | Low-quality |
| S1_extracted_k141_16393 | Low-quality |
| S1_extracted_k141_2103 | Low-quality |
| S1_extracted_k141_82692 | Not-determined |
| S1_extracted_k141_55963 | Low-quality |
| S1_extracted_k141_142604 | Low-quality |
| S1_extracted_k141_9313 | Not-determined |
| S1_extracted_k141_102710 | Low-quality |
| S1_extracted_k141_82694 | Low-quality |
| S1_extracted_k141_22384 | Not-determined |
| S1_extracted_k141_109214 | Not-determined |
| S1_extracted_k141_75952 | Not-determined |
| S1_extracted_k141_129372 | Not-determined |
| S1_extracted_k141_96093 | Not-determined |
| S1_extracted_k141_2105 | Low-quality |
| S1_extracted_k141_75956 | Not-determined |
| S1_extracted_k141_35409 | Not-determined |
| S1_extracted_k141_42674 | Low-quality |
| S1_extracted_k141_142610 | Not-determined |
| S1_extracted_k141_82700 | Not-determined |
| S1_extracted_k141_49343 | Not-determined |
| S1_extracted_k141_29237 | Not-determined |
| S1_extracted_k141_55971 | Not-determined |
| S1_extracted_k141_22386 | Not-determined |
| S1_extracted_k141_29240 | Not-determined |
| S1_extracted_k141_55974 | Not-determined |
| S1_extracted_k141_109225 | Not-determined |
| S1_extracted_k141_55977 | Not-determined |
| S1_extracted_k141_129382 | Not-determined |
| S1_extracted_k141_82709 | Not-determined |
| S1_extracted_k141_142618 | Not-determined |
| S1_extracted_k141_129385 | Not-determined |
| S1_extracted_k141_49352 | Not-determined |
| S1_extracted_k141_55986 | Low-quality |
| S1_extracted_k141_109233 | Not-determined |
| S1_extracted_k141_116015 | Low-quality |
| S1_extracted_k141_75973 | Not-determined |
| S1_extracted_k141_129387 | Not-determined |
| S1_extracted_k141_55990 | Not-determined |
| S1_extracted_k141_102720 | Not-determined |
| S1_extracted_k141_16412 | Not-determined |
| S1_extracted_k141_142626 | Not-determined |
| S1_extracted_k141_135927 | Not-determined |
| S1_extracted_k141_75976 | Not-determined |
| S1_extracted_k141_116020 | Not-determined |
| S1_extracted_k141_75979 | Not-determined |
| S1_extracted_k141_96105 | Low-quality |
| S1_extracted_k141_135928 | Low-quality |
| S1_extracted_k141_22395 | Not-determined |
| S1_extracted_k141_69189 | Not-determined |
| S1_extracted_k141_155118 | Low-quality |
| S1_extracted_k141_116023 | Not-determined |
| S1_extracted_k141_142633 | Not-determined |
| S1_extracted_k141_42690 | Low-quality |
| S1_extracted_k141_55995 | Not-determined |
| S1_extracted_k141_35419 | Not-determined |
| S1_extracted_k141_82718 | Not-determined |
| S1_extracted_k141_102728 | Not-determined |
| S1_extracted_k141_16415 | Low-quality |
| S1_extracted_k141_102729 | Not-determined |
| S1_extracted_k141_135935 | Not-determined |
| S1_extracted_k141_116025 | Low-quality |
| S1_extracted_k141_96109 | Not-determined |
| S1_extracted_k141_22408 | Not-determined |
| S1_extracted_k141_142638 | Not-determined |
| S1_extracted_k141_129394 | Low-quality |
| S1_extracted_k141_135944 | Not-determined |
| S1_extracted_k141_82730 | Low-quality |
| S1_extracted_k141_89230 | Not-determined |
| S1_extracted_k141_75993 | Not-determined |
| S1_extracted_k141_82731 | Not-determined |
| S1_extracted_k141_29258 | Low-quality |
| S1_extracted_k141_116031 | Not-determined |
| S1_extracted_k141_96118 | Not-determined |
| S1_extracted_k141_116034 | Not-determined |
| S1_extracted_k141_102733 | Low-quality |
| S1_extracted_k141_42698 | Not-determined |
| S1_extracted_k141_89234 | Not-determined |
| S1_extracted_k141_89236 | Not-determined |
| S1_extracted_k141_129401 | Not-determined |
| S1_extracted_k141_102736 | Not-determined |
| S1_extracted_k141_61560 | Not-determined |
| S1_extracted_k141_89238 | Not-determined |
| S1_extracted_k141_96123 | Not-determined |
| S1_extracted_k141_82745 | Not-determined |
| S1_extracted_k141_49385 | Not-determined |
| S1_extracted_k141_49386 | Not-determined |
| S1_extracted_k141_22424 | Not-determined |
| S1_extracted_k141_61563 | Not-determined |
| S1_extracted_k141_35438 | Not-determined |
| S1_extracted_k141_142647 | Not-determined |
| S1_extracted_k141_96125 | Not-determined |
| S1_extracted_k141_96126 | Low-quality |
| S1_extracted_k141_142651 | Not-determined |
| S1_extracted_k141_102741 | Not-determined |
| S1_extracted_k141_56017 | Not-determined |
| S1_extracted_k141_29282 | Not-determined |
| S1_extracted_k141_49391 | Not-determined |
| S1_extracted_k141_35445 | Not-determined |
| S1_extracted_k141_142654 | Low-quality |
| S1_extracted_k141_35446 | Not-determined |

| | |
|---|---|
| S1_extracted_k141_49393 | Not-determined |
| S1_extracted_k141_35449 | Not-determined |
| S1_extracted_k141_129415 | Not-determined |
| S1_extracted_k141_82759 | Not-determined |
| S1_extracted_k141_29289 | Not-determined |
| S1_extracted_k141_82761 | Not-determined |
| S1_extracted_k141_102754 | Not-determined |
| S1_extracted_k141_56025 | Not-determined |
| S1_extracted_k141_82765 | Not-determined |
| S1_extracted_k141_135963 | Not-determined |
| S1_extracted_k141_35458 | Not-determined |
| S1_extracted_k141_116050 | Not-determined |
| S1_extracted_k141_89257 | Not-determined |
| S1_extracted_k141_42721 | Not-determined |
| S1_extracted_k141_76011 | Not-determined |
| S1_extracted_k141_96141 | Not-determined |
| S1_extracted_k141_9362 | Low-quality |
| S1_extracted_k141_102761 | Not-determined |
| S1_extracted_k141_42724 | Not-determined |
| S1_extracted_k141_116055 | Not-determined |
| S1_extracted_k141_49409 | Not-determined |
| S1_extracted_k141_76014 | Not-determined |
| S1_extracted_k141_35462 | Not-determined |
| S1_extracted_k141_22441 | Not-determined |
| S1_extracted_k141_22442 | Not-determined |
| S1_extracted_k141_135972 | Not-determined |
| S1_extracted_k141_76017 | Not-determined |
| S1_extracted_k141_109273 | Not-determined |
| S1_extracted_k141_69228 | Not-determined |
| S1_extracted_k141_9364 | Low-quality |
| S1_extracted_k141_22444 | Not-determined |
| S1_extracted_k141_109275 | Not-determined |
| S1_extracted_k141_89262 | Not-determined |
| S1_extracted_k141_56034 | Not-determined |
| S1_extracted_k141_155146 | Not-determined |
| S1_extracted_k141_109279 | Not-determined |
| S1_extracted_k141_82774 | Not-determined |
| S1_extracted_k141_135974 | Low-quality |
| S1_extracted_k141_129429 | Not-determined |
| S1_extracted_k141_135975 | Not-determined |
| S1_extracted_k141_76022 | Not-determined |
| S1_extracted_k141_42733 | Not-determined |
| S1_extracted_k141_102774 | Not-determined |
| S1_extracted_k141_142675 | Not-determined |
| S1_extracted_k141_102775 | Not-determined |
| S1_extracted_k141_129431 | Not-determined |
| S1_extracted_k141_129432 | Not-determined |
| S1_extracted_k141_148000 | Not-determined |
| S1_extracted_k141_122518 | Low-quality |
| S1_extracted_k141_148001 | Not-determined |
| S1_extracted_k141_2148 | Not-determined |
| S1_extracted_k141_49424 | Not-determined |
| S1_extracted_k141_9372 | Low-quality |
| S1_extracted_k141_42742 | Low-quality |
| S1_extracted_k141_129436 | Low-quality |
| S1_extracted_k141_129437 | Not-determined |
| S1_extracted_k141_116076 | Low-quality |
| S1_extracted_k141_129441 | Low-quality |
| S1_extracted_k141_29319 | Not-determined |
| S1_extracted_k141_29322 | Not-determined |
| S1_extracted_k141_16464 | Not-determined |
| S1_extracted_k141_42747 | Low-quality |
| S1_extracted_k141_135985 | Not-determined |
| S1_extracted_k141_69238 | Not-determined |
| S1_extracted_k141_35473 | Low-quality |
| S1_extracted_k141_69239 | Not-determined |
| S1_extracted_k141_102787 | Not-determined |
| S1_extracted_k141_35475 | Low-quality |
| S1_extracted_k141_2153 | Not-determined |
| S1_extracted_k141_35476 | Not-determined |
| S1_extracted_k141_2154 | Not-determined |
| S1_extracted_k141_102791 | Not-determined |
| S1_extracted_k141_89272 | Not-determined |
| S1_extracted_k141_82790 | Not-determined |
| S1_extracted_k141_2157 | Not-determined |
| S1_extracted_k141_96149 | Low-quality |
| S1_extracted_k141_2158 | Not-determined |
| S1_extracted_k141_49437 | Not-determined |
| S1_extracted_k141_35483 | Low-quality |
| S1_extracted_k141_102797 | Not-determined |
| S1_extracted_k141_142687 | Not-determined |
| S1_extracted_k141_135999 | Not-determined |
| S1_extracted_k141_76041 | Not-determined |
| S1_extracted_k141_16479 | Not-determined |
| S1_extracted_k141_89275 | Not-determined |
| S1_extracted_k141_136003 | Not-determined |
| S1_extracted_k141_16482 | Not-determined |
| S1_extracted_k141_102802 | Not-determined |
| S1_extracted_k141_96157 | Not-determined |
| S1_extracted_k141_116094 | Low-quality |
| S1_extracted_k141_16483 | Not-determined |
| S1_extracted_k141_9378 | Not-determined |
| S1_extracted_k141_102804 | Low-quality |
| S1_extracted_k141_22471 | Not-determined |
| S1_extracted_k141_16485 | Not-determined |
| S1_extracted_k141_56063 | Not-determined |
| S1_extracted_k141_9384 | Not-determined |
| S1_extracted_k141_69256 | Low-quality |
| S1_extracted_k141_142692 | Not-determined |
| S1_extracted_k141_116098 | Not-determined |
| S1_extracted_k141_49449 | Not-determined |
| S1_extracted_k141_116099 | Not-determined |
| S1_extracted_k141_136008 | Not-determined |
| S1_extracted_k141_82808 | Not-determined |
| S1_extracted_k141_29348 | Not-determined |
| S1_extracted_k141_122531 | Not-determined |
| S1_extracted_k141_29349 | Not-determined |
| S1_extracted_k141_76047 | Low-quality |
| S1_extracted_k141_136013 | Not-determined |
| S1_extracted_k141_29351 | Not-determined |
| S1_extracted_k141_96166 | Low-quality |
| S1_extracted_k141_61591 | Not-determined |
| S1_extracted_k141_82811 | Not-determined |
| S1_extracted_k141_9389 | Not-determined |
| S1_extracted_k141_49458 | Low-quality |
| S1_extracted_k141_136021 | Not-determined |
| S1_extracted_k141_155162 | Not-determined |
| S1_extracted_k141_129462 | Not-determined |
| S1_extracted_k141_136024 | Not-determined |
| S1_extracted_k141_129463 | Not-determined |
| S1_extracted_k141_89282 | Low-quality |
| S1_extracted_k141_22483 | Not-determined |
| S1_extracted_k141_29362 | Not-determined |
| S1_extracted_k141_69267 | Low-quality |
| S1_extracted_k141_96171 | Not-determined |
| S1_extracted_k141_49462 | Not-determined |
| S1_extracted_k141_136029 | Not-determined |
| S1_extracted_k141_69269 | Not-determined |
| S1_extracted_k141_76062 | Not-determined |
| S1_extracted_k141_2179 | Low-quality |
| S1_extracted_k141_42763 | Low-quality |
| S1_extracted_k141_116113 | Not-determined |
| S1_extracted_k141_61603 | Not-determined |
| S1_extracted_k141_102823 | Low-quality |
| S1_extracted_k141_155168 | Not-determined |
| S1_extracted_k141_136038 | Not-determined |
| S1_extracted_k141_136039 | Not-determined |
| S1_extracted_k141_109314 | Not-determined |
| S1_extracted_k141_69278 | Not-determined |
| S1_extracted_k141_89286 | Low-quality |
| S1_extracted_k141_116117 | Low-quality |
| S1_extracted_k141_16495 | Not-determined |
| S1_extracted_k141_2184 | Low-quality |
| S1_extracted_k141_136042 | Low-quality |
| S1_extracted_k141_2185 | Not-determined |
| S1_extracted_k141_69281 | Low-quality |
| S1_extracted_k141_96178 | Not-determined |
| S1_extracted_k141_69284 | Not-determined |
| S1_extracted_k141_49476 | Not-determined |
| S1_extracted_k141_129479 | Not-determined |
| S1_extracted_k141_96179 | Low-quality |
| S1_extracted_k141_76076 | Low-quality |
| S1_extracted_k141_56078 | Not-determined |
| S1_extracted_k141_109320 | Not-determined |
| S1_extracted_k141_56079 | Not-determined |
| S1_extracted_k141_16502 | Not-determined |
| S1_extracted_k141_35495 | Not-determined |
| S1_extracted_k141_129483 | Not-determined |
| S1_extracted_k141_22491 | Not-determined |
| S1_extracted_k141_9414 | Not-determined |
| S1_extracted_k141_56083 | Not-determined |
| S1_extracted_k141_129485 | Not-determined |
| S1_extracted_k141_155179 | Not-determined |
| S1_extracted_k141_49481 | Low-quality |
| S1_extracted_k141_61606 | Low-quality |
| S1_extracted_k141_155181 | Not-determined |
| S1_extracted_k141_116127 | Not-determined |
| S1_extracted_k141_142724 | Not-determined |
| S1_extracted_k141_22496 | Not-determined |
| S1_extracted_k141_109329 | Not-determined |
| S1_extracted_k141_22497 | Low-quality |
| S1_extracted_k141_155182 | Not-determined |
| S1_extracted_k141_102838 | Low-quality |
| S1_extracted_k141_122563 | Not-determined |
| S1_extracted_k141_42775 | Not-determined |
| S1_extracted_k141_2197 | Not-determined |
| S1_extracted_k141_82843 | Low-quality |
| S1_extracted_k141_56092 | Not-determined |
| S1_extracted_k141_2198 | Not-determined |
| S1_extracted_k141_96191 | Low-quality |
| S1_extracted_k141_16510 | Low-quality |
| S1_extracted_k141_82844 | Not-determined |
| S1_extracted_k141_109333 | Not-determined |
| S1_extracted_k141_129495 | Not-determined |
| S1_extracted_k141_2202 | Not-determined |
| S1_extracted_k141_122571 | Not-determined |
| S1_extracted_k141_109337 | Not-determined |
| S1_extracted_k141_22509 | Low-quality |
| S1_extracted_k141_9431 | Low-quality |
| S1_extracted_k141_22510 | Low-quality |
| S1_extracted_k141_82850 | Not-determined |
| S1_extracted_k141_22511 | Not-determined |
| S1_extracted_k141_69301 | Not-determined |
| S1_extracted_k141_89300 | Not-determined |
| S1_extracted_k141_122576 | Not-determined |
| S1_extracted_k141_109347 | Not-determined |
| S1_extracted_k141_122578 | Not-determined |
| S1_extracted_k141_69310 | Not-determined |
| S1_extracted_k141_35518 | Not-determined |
| S1_extracted_k141_29397 | Not-determined |
| S1_extracted_k141_56103 | Not-determined |
| S1_extracted_k141_22521 | Low-quality |
| S1_extracted_k141_122582 | Not-determined |
| S1_extracted_k141_136053 | Not-determined |
| S1_extracted_k141_16529 | Not-determined |
| S1_extracted_k141_16530 | Not-determined |
| S1_extracted_k141_9440 | Low-quality |
| S1_extracted_k141_22522 | Low-quality |
| S1_extracted_k141_82852 | Not-determined |
| S1_extracted_k141_96200 | Not-determined |
| S1_extracted_k141_89308 | Not-determined |
| S1_extracted_k141_136057 | Not-determined |
| S1_extracted_k141_61615 | Not-determined |
| S1_extracted_k141_122587 | Not-determined |
| S1_extracted_k141_49512 | Not-determined |
| S1_extracted_k141_102856 | Not-determined |
| S1_extracted_k141_122589 | Not-determined |
| S1_extracted_k141_96203 | Not-determined |
| S1_extracted_k141_142753 | Low-quality |
| S1_extracted_k141_56107 | Low-quality |
| S1_extracted_k141_16536 | Not-determined |
| S1_extracted_k141_29407 | Not-determined |
| S1_extracted_k141_142756 | Not-determined |
| S1_extracted_k141_96207 | Low-quality |
| S1_extracted_k141_2227 | Not-determined |
| S1_extracted_k141_16541 | Not-determined |
| S1_extracted_k141_96208 | Not-determined |
| S1_extracted_k141_42806 | Not-determined |
| S1_extracted_k141_22538 | Not-determined |
| S1_extracted_k141_76099 | Low-quality |
| S1_extracted_k141_136074 | Not-determined |
| S1_extracted_k141_142764 | Not-determined |
| S1_extracted_k141_29415 | Not-determined |
| S1_extracted_k141_122595 | Low-quality |
| S1_extracted_k141_96212 | Low-quality |
| S1_extracted_k141_9453 | Not-determined |
| S1_extracted_k141_22541 | Low-quality |
| S1_extracted_k141_142771 | Not-determined |
| S1_extracted_k141_9454 | Not-determined |
| S1_extracted_k141_109354 | Not-determined |
| S1_extracted_k141_82869 | Not-determined |
| S1_extracted_k141_116170 | Not-determined |
| S1_extracted_k141_42819 | Not-determined |
| S1_extracted_k141_142777 | Not-determined |
| S1_extracted_k141_109358 | Low-quality |
| S1_extracted_k141_96222 | Not-determined |
| S1_extracted_k141_76107 | Not-determined |
| S1_extracted_k141_136083 | Not-determined |
| S1_extracted_k141_109362 | Not-determined |
| S1_extracted_k141_96227 | Not-determined |
| S1_extracted_k141_76114 | Not-determined |
| S1_extracted_k141_35535 | Not-determined |
| S1_extracted_k141_42829 | Not-determined |
| S1_extracted_k141_102870 | Not-determined |
| S1_extracted_k141_69343 | Not-determined |
| S1_extracted_k141_16557 | Low-quality |
| S1_extracted_k141_136090 | Not-determined |
| S1_extracted_k141_29432 | Not-determined |
| S1_extracted_k141_49533 | Not-determined |
| S1_extracted_k141_56119 | Not-determined |
| S1_extracted_k141_9467 | Low-quality |
| S1_extracted_k141_29434 | Not-determined |
| S1_extracted_k141_61626 | Not-determined |
| S1_extracted_k141_109370 | Not-determined |
| S1_extracted_k141_89336 | Not-determined |
| S1_extracted_k141_136093 | Not-determined |
| S1_extracted_k141_76122 | Not-determined |
| S1_extracted_k141_16566 | Not-determined |
| S1_extracted_k141_89339 | Not-determined |
| S1_extracted_k141_116186 | Not-determined |
| S1_extracted_k141_129531 | Not-determined |
| S1_extracted_k141_116187 | Not-determined |
| S1_extracted_k141_9472 | Not-determined |
| S1_extracted_k141_42835 | Not-determined |
| S1_extracted_k141_82898 | Not-determined |
| S1_extracted_k141_109376 | Not-determined |
| S1_extracted_k141_56124 | Not-determined |
| S1_extracted_k141_96239 | Low-quality |
| S1_extracted_k141_142798 | Not-determined |
| S1_extracted_k141_142800 | Not-determined |
| S1_extracted_k141_89351 | Not-determined |
| S1_extracted_k141_35553 | Not-determined |
| S1_extracted_k141_56129 | Not-determined |
| S1_extracted_k141_22563 | Not-determined |
| S1_extracted_k141_116191 | Not-determined |
| S1_extracted_k141_61635 | Not-determined |
| S1_extracted_k141_9485 | Not-determined |
| S1_extracted_k141_96240 | Not-determined |
| S1_extracted_k141_16567 | Not-determined |
| S1_extracted_k141_29446 | Not-determined |
| S1_extracted_k141_22567 | Low-quality |
| S1_extracted_k141_122622 | Not-determined |
| S1_extracted_k141_96249 | Not-determined |
| S1_extracted_k141_155185 | Not-determined |
| S1_extracted_k141_69368 | Not-determined |
| S1_extracted_k141_9494 | Low-quality |
| S1_extracted_k141_129547 | Not-determined |
| S1_extracted_k141_2254 | Not-determined |
| S1_extracted_k141_9496 | Not-determined |
| S1_extracted_k141_129548 | Not-determined |
| S1_extracted_k141_148035 | Not-determined |
| S1_extracted_k141_35565 | Not-determined |
| S1_extracted_k141_42855 | Not-determined |
| S1_extracted_k141_129550 | Not-determined |
| S1_extracted_k141_42856 | Low-quality |
| S1_extracted_k141_56142 | Not-determined |
| S1_extracted_k141_69374 | Not-determined |
| S1_extracted_k141_96256 | Not-determined |
| S1_extracted_k141_89361 | Not-determined |
| S1_extracted_k141_122632 | Not-determined |
| S1_extracted_k141_129555 | Low-quality |
| S1_extracted_k141_122633 | Not-determined |
| S1_extracted_k141_116205 | Not-determined |
| S1_extracted_k141_96258 | Not-determined |
| S1_extracted_k141_16589 | Not-determined |
| S1_extracted_k141_42859 | Not-determined |
| S1_extracted_k141_16590 | Not-determined |
| S1_extracted_k141_102902 | Not-determined |
| S1_extracted_k141_35570 | Not-determined |
| S1_extracted_k141_116206 | Not-determined |
| S1_extracted_k141_82921 | Not-determined |
| S1_extracted_k141_2266 | Not-determined |
| S1_extracted_k141_148040 | Not-determined |
| S1_extracted_k141_29459 | Not-determined |
| S1_extracted_k141_109390 | Not-determined |
| S1_extracted_k141_89369 | Not-determined |
| S1_extracted_k141_116216 | Not-determined |
| S1_extracted_k141_22584 | Not-determined |
| S1_extracted_k141_148044 | Not-determined |
| S1_extracted_k141_89372 | Not-determined |
| S1_extracted_k141_102914 | Not-determined |
| S1_extracted_k141_56148 | Not-determined |
| S1_extracted_k141_82927 | Low-quality |
| S1_extracted_k141_22588 | Not-determined |
| S1_extracted_k141_116222 | Not-determined |
| S1_extracted_k141_35579 | Not-determined |
| S1_extracted_k141_109394 | Low-quality |
| S1_extracted_k141_148047 | Not-determined |
| S1_extracted_k141_76153 | Not-determined |
| S1_extracted_k141_22597 | Low-quality |
| S1_extracted_k141_56153 | Not-determined |
| S1_extracted_k141_109399 | Not-determined |
| S1_extracted_k141_22600 | Not-determined |
| S1_extracted_k141_56156 | Not-determined |
| S1_extracted_k141_89384 | Not-determined |
| S1_extracted_k141_129574 | Not-determined |
| S1_extracted_k141_2279 | Low-quality |
| S1_extracted_k141_2280 | Not-determined |
| S1_extracted_k141_136113 | Not-determined |
| S1_extracted_k141_9526 | Not-determined |
| S1_extracted_k141_29479 | Not-determined |
| S1_extracted_k141_122644 | Not-determined |
| S1_extracted_k141_82944 | Low-quality |
| S1_extracted_k141_16613 | Not-determined |
| S1_extracted_k141_35593 | Not-determined |
| S1_extracted_k141_76164 | Not-determined |
| S1_extracted_k141_9531 | Not-determined |
| S1_extracted_k141_16615 | Not-determined |
| S1_extracted_k141_49584 | Not-determined |
| S1_extracted_k141_42882 | Not-determined |
| S1_extracted_k141_22608 | Not-determined |
| S1_extracted_k141_129585 | Not-determined |
| S1_extracted_k141_42885 | Not-determined |
| S1_extracted_k141_49589 | Not-determined |
| S1_extracted_k141_136122 | Not-determined |
| S1_extracted_k141_129588 | Not-determined |
| S1_extracted_k141_116250 | Not-determined |
| S1_extracted_k141_109409 | Low-quality |
| S1_extracted_k141_96286 | Not-determined |
| S1_extracted_k141_89396 | Not-determined |
| S1_extracted_k141_155197 | Low-quality |
| S1_extracted_k141_96287 | Not-determined |
| S1_extracted_k141_122652 | Low-quality |
| S1_extracted_k141_76166 | Low-quality |
| S1_extracted_k141_89398 | Low-quality |
| S1_extracted_k141_16622 | Not-determined |
| S1_extracted_k141_136126 | Not-determined |
| S1_extracted_k141_89399 | Not-determined |
| S1_extracted_k141_16623 | Not-determined |
| S1_extracted_k141_35603 | Not-determined |
| S1_extracted_k141_136127 | Low-quality |
| S1_extracted_k141_122657 | Not-determined |
| S1_extracted_k141_49591 | Not-determined |
| S1_extracted_k141_56168 | Low-quality |
| S1_extracted_k141_9542 | Not-determined |
| S1_extracted_k141_22614 | Low-quality |
| S1_extracted_k141_129595 | Low-quality |
| S1_extracted_k141_29487 | Low-quality |
| S1_extracted_k141_42892 | Not-determined |
| S1_extracted_k141_76174 | Not-determined |
| S1_extracted_k141_116259 | Not-determined |
| S1_extracted_k141_96293 | Low-quality |
| S1_extracted_k141_102941 | Not-determined |
| S1_extracted_k141_155209 | Low-quality |
| S1_extracted_k141_102944 | Not-determined |
| S1_extracted_k141_35612 | Low-quality |
| S1_extracted_k141_142833 | Not-determined |
| S1_extracted_k141_122665 | Low-quality |
| S1_extracted_k141_96299 | Not-determined |
| S1_extracted_k141_16636 | Not-determined |
| S1_extracted_k141_2308 | Not-determined |
| S1_extracted_k141_56183 | Not-determined |
| S1_extracted_k141_9558 | Not-determined |
| S1_extracted_k141_116269 | Not-determined |
| S1_extracted_k141_96300 | Not-determined |
| S1_extracted_k141_69393 | Not-determined |
| S1_extracted_k141_49608 | Not-determined |
| S1_extracted_k141_116271 | Not-determined |
| S1_extracted_k141_9562 | Not-determined |
| S1_extracted_k141_35620 | Not-determined |
| S1_extracted_k141_2314 | Not-determined |
| S1_extracted_k141_129612 | Not-determined |
| S1_extracted_k141_129613 | Not-determined |
| S1_extracted_k141_129614 | Not-determined |
| S1_extracted_k141_49611 | Not-determined |
| S1_extracted_k141_136135 | Low-quality |
| S1_extracted_k141_76192 | Low-quality |
| S1_extracted_k141_102953 | Not-determined |
| S1_extracted_k141_155212 | Low-quality |
| S1_extracted_k141_96305 | Not-determined |
| S1_extracted_k141_102956 | Not-determined |
| S1_extracted_k141_2318 | Not-determined |
| S1_extracted_k141_42909 | Low-quality |
| S1_extracted_k141_122673 | Not-determined |
| S1_extracted_k141_61655 | Low-quality |
| S1_extracted_k141_116279 | Not-determined |
| S1_extracted_k141_69397 | Not-determined |
| S1_extracted_k141_56194 | Not-determined |
| S1_extracted_k141_96313 | Not-determined |
| S1_extracted_k141_89421 | Not-determined |
| S1_extracted_k141_102962 | Not-determined |
| S1_extracted_k141_116286 | Low-quality |
| S1_extracted_k141_109428 | Not-determined |
| S1_extracted_k141_155217 | Low-quality |
| S1_extracted_k141_129627 | Not-determined |
| S1_extracted_k141_89428 | Not-determined |
| S1_extracted_k141_116289 | Not-determined |
| S1_extracted_k141_155218 | Not-determined |
| S1_extracted_k141_76203 | Not-determined |
| S1_extracted_k141_122679 | Not-determined |
| S1_extracted_k141_29499 | Not-determined |
| S1_extracted_k141_102971 | Not-determined |
| S1_extracted_k141_69400 | Not-determined |
| S1_extracted_k141_22646 | Not-determined |
| S1_extracted_k141_16660 | Not-determined |
| S1_extracted_k141_76209 | Not-determined |
| S1_extracted_k141_102974 | Low-quality |
| S1_extracted_k141_129632 | Not-determined |
| S1_extracted_k141_89433 | Not-determined |
| S1_extracted_k141_102976 | Not-determined |
| S1_extracted_k141_102977 | Not-determined |
| S1_extracted_k141_116297 | Not-determined |
| S1_extracted_k141_89434 | Not-determined |
| S1_extracted_k141_129634 | Low-quality |
| S1_extracted_k141_122688 | Not-determined |
| S1_extracted_k141_16666 | Not-determined |
| S1_extracted_k141_122689 | Not-determined |
| S1_extracted_k141_9583 | Not-determined |
| S1_extracted_k141_69412 | Not-determined |
| S1_extracted_k141_136159 | Not-determined |
| S1_extracted_k141_82994 | Not-determined |
| S1_extracted_k141_76222 | Not-determined |
| S1_extracted_k141_49641 | Low-quality |
| S1_extracted_k141_96317 | Not-determined |
| S1_extracted_k141_136165 | Low-quality |
| S1_extracted_k141_155231 | Not-determined |
| S1_extracted_k141_136166 | Not-determined |
| S1_extracted_k141_96319 | Not-determined |
| S1_extracted_k141_136168 | Not-determined |
| S1_extracted_k141_56213 | Not-determined |
| S1_extracted_k141_69421 | Not-determined |
| S1_extracted_k141_42929 | Not-determined |
| S1_extracted_k141_102992 | Not-determined |
| S1_extracted_k141_76225 | Not-determined |
| S1_extracted_k141_69423 | Not-determined |
| S1_extracted_k141_49645 | Not-determined |
| S1_extracted_k141_49646 | Not-determined |
| S1_extracted_k141_136173 | Low-quality |
| S1_extracted_k141_69426 | Not-determined |
| S1_extracted_k141_16688 | Not-determined |
| S1_extracted_k141_122700 | Not-determined |
| S1_extracted_k141_102997 | Not-determined |
| S1_extracted_k141_61670 | Not-determined |
| S1_extracted_k141_69434 | Not-determined |
| S1_extracted_k141_103002 | Low-quality |
| S1_extracted_k141_69436 | Low-quality |
| S1_extracted_k141_29526 | Not-determined |
| S1_extracted_k141_16693 | Low-quality |
| S1_extracted_k141_16694 | Not-determined |
| S1_extracted_k141_136176 | Not-determined |
| S1_extracted_k141_89448 | Low-quality |
| S1_extracted_k141_116313 | Not-determined |
| S1_extracted_k141_136179 | Not-determined |
| S1_extracted_k141_49651 | Not-determined |
| S1_extracted_k141_109447 | Not-determined |
| S1_extracted_k141_103010 | Not-determined |
| S1_extracted_k141_61674 | Low-quality |
| S1_extracted_k141_109448 | Not-determined |
| S1_extracted_k141_69442 | Not-determined |
| S1_extracted_k141_109449 | Not-determined |
| S1_extracted_k141_2348 | Low-quality |
| S1_extracted_k141_2349 | Not-determined |
| S1_extracted_k141_142844 | Not-determined |
| S1_extracted_k141_109452 | Not-determined |
| S1_extracted_k141_16706 | Low-quality |
| S1_extracted_k141_136187 | Not-determined |
| S1_extracted_k141_49654 | Not-determined |
| S1_extracted_k141_2351 | Not-determined |
| S1_extracted_k141_16708 | Not-determined |
| S1_extracted_k141_9612 | Not-determined |
| S1_extracted_k141_56234 | Not-determined |
| S1_extracted_k141_49659 | Not-determined |
| S1_extracted_k141_29533 | Low-quality |
| S1_extracted_k141_76241 | Low-quality |
| S1_extracted_k141_56238 | Not-determined |
| S1_extracted_k141_22669 | Low-quality |
| S1_extracted_k141_122718 | Not-determined |
| S1_extracted_k141_2359 | Not-determined |
| S1_extracted_k141_89460 | Not-determined |
| S1_extracted_k141_103024 | Not-determined |
| S1_extracted_k141_155233 | Low-quality |
| S1_extracted_k141_9625 | Not-determined |
| S1_extracted_k141_116324 | Not-determined |
| S1_extracted_k141_109462 | Low-quality |
| S1_extracted_k141_142856 | Not-determined |
| S1_extracted_k141_29540 | Not-determined |
| S1_extracted_k141_148059 | Not-determined |
| S1_extracted_k141_83013 | Not-determined |
| S1_extracted_k141_49671 | Low-quality |
| S1_extracted_k141_103028 | Not-determined |
| S1_extracted_k141_96350 | Not-determined |
| S1_extracted_k141_2362 | Not-determined |
| S1_extracted_k141_29544 | Not-determined |
| S1_extracted_k141_122724 | Not-determined |
| S1_extracted_k141_56248 | Not-determined |
| S1_extracted_k141_129663 | Not-determined |
| S1_extracted_k141_42950 | Not-determined |
| S1_extracted_k141_96354 | Not-determined |
| S1_extracted_k141_109466 | Not-determined |
| S1_extracted_k141_56251 | Not-determined |
| S1_extracted_k141_96356 | Not-determined |
| S1_extracted_k141_109467 | Not-determined |
| S11_vs_k141_70015\|\|full | High-quality |
| S1_extracted_k141_129666 | Not-determined |
| S1_extracted_k141_2366 | Low-quality |
| S1_extracted_k141_56252 | Not-determined |
| S1_extracted_k141_2368 | Not-determined |
| S1_extracted_k141_109470 | Not-determined |
| S1_extracted_k141_116336 | Low-quality |
| S1_extracted_k141_89472 | Not-determined |
| S1_extracted_k141_122728 | Low-quality |
| S1_extracted_k141_122729 | Not-determined |
| S1_extracted_k141_155237 | Not-determined |
| S1_extracted_k141_29552 | Not-determined |
| S1_extracted_k141_42956 | Not-determined |
| S1_extracted_k141_136217 | Low-quality |
| S1_extracted_k141_16724 | Not-determined |
| S1_extracted_k141_116340 | Not-determined |
| S1_extracted_k141_109471 | Not-determined |
| S1_extracted_k141_116341 | Not-determined |
| S1_extracted_k141_22689 | Not-determined |
| S1_extracted_k141_2369 | Not-determined |
| S1_extracted_k141_103042 | Not-determined |
| S1_extracted_k141_22692 | Not-determined |
| S1_extracted_k141_155241 | Not-determined |
| S1_extracted_k141_56258 | Not-determined |
| S1_extracted_k141_155242 | Not-determined |
| S1_extracted_k141_9641 | Not-determined |
| S1_extracted_k141_103047 | Low-quality |
| S1_extracted_k141_96363 | Not-determined |
| S1_extracted_k141_148069 | Not-determined |
| S1_extracted_k141_103049 | Not-determined |
| S1_extracted_k141_42970 | Not-determined |
| S1_extracted_k141_89486 | Not-determined |
| S1_extracted_k141_122739 | Not-determined |
| S1_extracted_k141_29554 | Not-determined |
| S1_extracted_k141_103052 | Not-determined |
| S1_extracted_k141_56261 | Low-quality |
| S1_extracted_k141_109482 | Low-quality |
| S1_extracted_k141_49696 | Not-determined |
| S1_extracted_k141_89487 | Low-quality |
| S1_extracted_k141_49697 | Not-determined |
| S1_extracted_k141_103056 | Low-quality |
| S1_extracted_k141_116350 | Not-determined |
| S1_extracted_k141_116351 | Not-determined |
| S1_extracted_k141_89489 | Not-determined |
| S1_extracted_k141_35648 | Not-determined |
| S1_extracted_k141_89491 | Low-quality |
| S1_extracted_k141_155256 | Not-determined |

| | |
|---|---|
| S1_extracted_k141_76280 | Not-determined |
| S1_extracted_k141_109490 | Not-determined |
| S1_extracted_k141_2382 | Not-determined |
| S1_extracted_k141_61694 | Not-determined |
| S1_extracted_k141_49711 | Not-determined |
| S1_extracted_k141_109492 | Not-determined |
| S1_extracted_k141_122749 | Low-quality |
| S1_extracted_k141_56276 | Not-determined |
| S1_extracted_k141_69481 | Not-determined |
| S1_extracted_k141_49714 | Not-determined |
| S1_extracted_k141_69483 | Not-determined |
| S1_extracted_k141_116357 | Not-determined |
| S1_extracted_k141_109497 | Not-determined |
| S1_extracted_k141_9657 | Not-determined |
| S1_extracted_k141_103066 | Not-determined |
| S1_extracted_k141_76287 | Not-determined |
| S1_extracted_k141_116358 | Not-determined |
| S1_extracted_k141_2389 | Not-determined |
| S1_extracted_k141_129700 | Not-determined |
| S1_extracted_k141_69485 | Not-determined |
| S1_extracted_k141_61699 | Not-determined |
| S1_extracted_k141_42988 | Not-determined |
| S1_extracted_k141_122757 | Not-determined |
| S1_extracted_k141_2392 | Not-determined |
| S1_extracted_k141_96392 | Not-determined |
| S1_extracted_k141_155266 | Not-determined |
| S1_extracted_k141_16756 | Low-quality |
| S1_extracted_k141_56288 | Low-quality |
| S1_extracted_k141_142862 | Low-quality |
| S1_extracted_k141_116364 | Not-determined |
| S1_extracted_k141_29579 | Not-determined |
| S1_extracted_k141_49724 | Not-determined |
| S1_extracted_k141_89514 | Not-determined |
| S1_extracted_k141_89515 | Not-determined |
| S1_extracted_k141_49725 | Not-determined |
| S1_extracted_k141_29582 | Not-determined |
| S1_extracted_k141_155268 | Not-determined |
| S1_extracted_k141_35659 | Not-determined |
| S1_extracted_k141_16762 | Not-determined |
| S1_extracted_k141_136236 | Not-determined |
| S1_extracted_k141_35662 | Not-determined |
| S1_extracted_k141_22702 | Not-determined |
| S1_extracted_k141_9675 | Not-determined |
| S1_extracted_k141_89523 | Not-determined |
| S1_extracted_k141_103083 | Not-determined |
| S1_extracted_k141_103086 | Not-determined |
| S1_extracted_k141_109516 | Not-determined |
| S1_extracted_k141_49736 | Not-determined |
| S1_extracted_k141_136238 | Not-determined |
| S1_extracted_k141_56293 | Not-determined |
| S1_extracted_k141_76295 | Not-determined |
| S1_extracted_k141_35670 | Not-determined |
| S1_extracted_k141_49738 | Not-determined |
| S1_extracted_k141_83059 | Not-determined |
| S1_extracted_k141_69505 | Not-determined |
| S1_extracted_k141_61711 | Not-determined |
| S1_extracted_k141_49739 | Not-determined |
| S1_extracted_k141_155271 | Low-quality |
| S1_extracted_k141_96408 | Not-determined |
| S1_extracted_k141_155272 | Not-determined |
| S1_extracted_k141_142880 | Not-determined |
| S1_extracted_k141_142882 | Not-determined |
| S1_extracted_k141_148075 | Not-determined |
| S1_extracted_k141_103094 | Not-determined |
| S1_extracted_k141_43011 | Not-determined |
| S1_extracted_k141_49745 | Not-determined |
| S1_extracted_k141_148076 | Not-determined |
| S1_extracted_k141_9687 | Not-determined |
| S1_extracted_k141_129709 | Low-quality |
| S1_extracted_k141_116380 | Not-determined |
| S1_extracted_k141_142885 | Not-determined |
| S1_extracted_k141_49749 | Not-determined |
| S1_extracted_k141_148079 | Not-determined |
| S1_extracted_k141_61720 | Not-determined |
| S1_extracted_k141_61721 | Not-determined |
| S1_extracted_k141_76312 | Not-determined |
| S1_extracted_k141_142891 | Not-determined |
| S1_extracted_k141_116388 | Not-determined |
| S1_extracted_k141_142892 | Not-determined |
| S1_extracted_k141_69514 | Low-quality |
| S1_extracted_k141_29599 | Not-determined |
| S1_extracted_k141_109528 | Not-determined |
| S1_extracted_k141_116389 | Not-determined |
| S1_extracted_k141_136261 | Not-determined |
| S1_extracted_k141_2421 | Low-quality |
| S1_extracted_k141_76317 | Not-determined |
| S1_extracted_k141_103103 | Low-quality |
| S1_extracted_k141_129717 | Low-quality |
| S1_extracted_k141_103104 | Not-determined |
| S1_extracted_k141_136263 | Not-determined |
| S1_extracted_k141_16785 | Not-determined |
| S1_extracted_k141_109534 | Not-determined |
| S1_extracted_k141_89544 | Not-determined |
| S1_extracted_k141_22717 | Not-determined |
| S1_extracted_k141_89545 | Low-quality |
| S1_extracted_k141_76324 | Not-determined |
| S1_extracted_k141_116396 | Not-determined |
| S1_extracted_k141_83067 | Not-determined |
| S1_extracted_k141_109538 | Not-determined |
| S1_extracted_k141_29608 | Not-determined |
| S1_extracted_k141_61735 | Low-quality |
| S1_extracted_k141_35683 | Not-determined |
| S1_extracted_k141_83069 | Not-determined |
| S1_extracted_k141_69518 | Low-quality |
| S1_extracted_k141_136274 | Not-determined |
| S1_extracted_k141_103108 | Not-determined |
| S1_extracted_k141_103111 | Not-determined |
| S1_extracted_k141_69520 | Not-determined |
| S1_extracted_k141_29611 | Low-quality |
| S1_extracted_k141_2431 | Not-determined |
| S1_extracted_k141_22728 | Not-determined |
| S1_extracted_k141_89557 | Not-determined |
| S1_extracted_k141_89560 | Not-determined |
| S1_extracted_k141_103114 | Not-determined |
| S1_extracted_k141_35688 | Not-determined |
| S1_extracted_k141_96429 | Not-determined |
| S1_extracted_k141_22729 | Not-determined |
| S1_extracted_k141_16789 | Not-determined |
| S1_extracted_k141_76333 | Low-quality |
| S1_extracted_k141_2434 | Not-determined |
| S1_extracted_k141_89561 | Low-quality |
| S1_extracted_k141_136283 | Low-quality |
| S1_extracted_k141_69527 | Low-quality |
| S1_extracted_k141_61747 | Not-determined |
| S1_extracted_k141_142898 | Not-determined |
| S1_extracted_k141_9697 | Low-quality |
| S1_extracted_k141_103120 | Not-determined |
| S1_extracted_k141_9700 | Not-determined |
| S1_extracted_k141_116407 | Not-determined |
| S1_extracted_k141_9701 | Low-quality |
| S1_extracted_k141_83078 | Not-determined |
| S1_extracted_k141_136289 | Not-determined |
| S1_extracted_k141_109556 | Not-determined |
| S1_extracted_k141_116410 | Not-determined |
| S1_extracted_k141_16800 | Not-determined |
| S1_extracted_k141_116412 | Not-determined |
| S1_extracted_k141_9706 | Not-determined |
| S1_extracted_k141_136293 | Not-determined |
| S1_extracted_k141_96440 | Not-determined |
| S1_extracted_k141_69534 | Low-quality |
| S1_extracted_k141_136294 | Not-determined |
| S1_extracted_k141_29622 | Not-determined |
| S1_extracted_k141_49778 | Low-quality |
| S1_extracted_k141_122808 | Not-determined |
| S1_extracted_k141_43040 | Not-determined |
| S1_extracted_k141_116415 | Not-determined |
| S1_extracted_k141_2448 | Not-determined |
| S1_extracted_k141_116416 | Not-determined |
| S1_extracted_k141_69536 | Not-determined |
| S1_extracted_k141_116417 | Not-determined |
| S1_extracted_k141_9710 | Not-determined |
| S1_extracted_k141_69537 | Not-determined |
| S1_extracted_k141_116418 | Not-determined |
| S1_extracted_k141_49783 | Not-determined |
| S1_extracted_k141_29627 | Not-determined |
| S1_extracted_k141_61757 | Not-determined |
| S1_extracted_k141_16804 | Not-determined |
| S1_extracted_k141_89576 | Not-determined |
| S1_extracted_k141_109565 | Not-determined |
| S1_extracted_k141_43047 | Low-quality |
| S1_extracted_k141_22746 | Not-determined |
| S1_extracted_k141_61760 | Not-determined |
| S1_extracted_k141_69544 | Not-determined |
| S1_extracted_k141_76342 | Not-determined |
| S1_extracted_k141_43051 | Not-determined |
| S1_extracted_k141_109567 | Not-determined |
| S1_extracted_k141_116425 | Not-determined |
| S1_extracted_k141_142917 | Not-determined |
| S1_extracted_k141_35703 | Not-determined |
| S1_extracted_k141_155287 | Not-determined |
| S1_extracted_k141_142919 | Low-quality |
| S1_extracted_k141_22752 | Low-quality |
| S1_extracted_k141_2460 | Not-determined |
| S1_extracted_k141_109574 | Not-determined |
| S1_extracted_k141_61765 | Not-determined |
| S1_extracted_k141_83099 | Not-determined |
| S1_extracted_k141_16815 | Not-determined |
| S1_extracted_k141_29640 | Low-quality |
| S1_extracted_k141_43064 | Not-determined |
| S1_extracted_k141_2462 | Not-determined |
| S1_extracted_k141_49799 | Low-quality |
| S1_extracted_k141_89588 | Not-determined |
| S1_extracted_k141_49801 | Not-determined |
| S1_extracted_k141_43067 | Not-determined |
| S1_extracted_k141_43068 | Not-determined |
| S1_extracted_k141_22753 | Not-determined |
| S1_extracted_k141_89591 | Not-determined |
| S1_extracted_k141_96449 | Low-quality |
| S1_extracted_k141_35711 | Low-quality |
| S1_extracted_k141_142927 | Not-determined |
| S1_extracted_k141_89593 | Not-determined |
| S1_extracted_k141_122829 | Not-determined |
| S1_extracted_k141_109585 | Not-determined |
| S1_extracted_k141_89595 | Not-determined |
| S1_extracted_k141_69558 | Not-determined |
| S1_extracted_k141_83107 | Not-determined |
| S1_extracted_k141_129752 | Not-determined |
| S1_extracted_k141_61778 | Not-determined |
| S1_extracted_k141_83109 | Not-determined |
| S1_extracted_k141_122832 | Not-determined |
| S1_extracted_k141_89597 | Not-determined |
| S1_extracted_k141_103145 | Not-determined |
| S1_extracted_k141_49813 | Low-quality |
| S1_extracted_k141_136312 | Low-quality |
| S1_extracted_k141_43075 | Low-quality |
| S1_extracted_k141_29653 | Not-determined |
| S1_extracted_k141_122834 | Low-quality |
| S1_extracted_k141_29654 | Low-quality |
| S1_extracted_k141_35720 | Not-determined |
| S1_extracted_k141_136314 | Not-determined |
| S1_extracted_k141_35721 | Not-determined |
| S1_extracted_k141_9742 | Not-determined |
| S1_extracted_k141_83112 | Not-determined |
| S1_extracted_k141_69562 | Not-determined |
| S1_extracted_k141_49819 | Not-determined |
| S1_extracted_k141_22763 | Not-determined |
| S1_extracted_k141_116447 | Not-determined |
| S1_extracted_k141_89603 | Not-determined |
| S1_extracted_k141_136318 | Low-quality |
| S1_extracted_k141_69564 | Not-determined |
| S1_extracted_k141_22767 | Not-determined |
| S1_extracted_k141_49823 | Low-quality |
| S1_extracted_k141_129762 | Low-quality |
| S1_extracted_k141_9745 | Low-quality |
| S1_extracted_k141_69569 | Not-determined |
| S1_extracted_k141_49827 | Not-determined |
| S1_extracted_k141_16834 | Not-determined |
| S1_extracted_k141_29663 | Not-determined |
| S1_extracted_k141_136324 | Not-determined |
| S1_extracted_k141_61782 | Not-determined |
| S1_extracted_k141_103156 | Not-determined |
| S1_extracted_k141_69570 | Not-determined |
| S1_extracted_k141_22774 | Not-determined |
| S1_extracted_k141_29665 | Not-determined |
| S1_extracted_k141_122846 | Not-determined |
| S1_extracted_k141_56337 | Not-determined |
| S1_extracted_k141_9747 | Not-determined |
| S1_extracted_k141_43087 | Low-quality |
| S1_extracted_k141_96474 | Not-determined |
| S1_extracted_k141_43089 | Not-determined |
| S1_extracted_k141_16839 | Not-determined |
| S1_extracted_k141_116455 | Low-quality |
| S1_extracted_k141_16841 | Not-determined |
| S1_extracted_k141_16842 | Not-determined |
| S1_extracted_k141_103164 | Not-determined |
| S1_extracted_k141_29669 | Low-quality |
| S1_extracted_k141_76375 | Not-determined |
| S1_extracted_k141_103165 | Low-quality |
| S1_extracted_k141_103166 | Not-determined |
| S1_extracted_k141_109607 | Not-determined |
| S1_extracted_k141_103169 | Not-determined |
| S1_extracted_k141_142947 | Not-determined |
| S1_extracted_k141_129772 | Not-determined |
| S1_extracted_k141_129773 | Not-determined |
| S1_extracted_k141_103172 | Not-determined |
| S1_extracted_k141_69587 | Not-determined |
| S1_extracted_k141_69588 | Not-determined |
| S1_extracted_k141_103175 | Low-quality |
| S1_extracted_k141_76381 | Not-determined |
| S1_extracted_k141_69591 | Not-determined |
| S1_extracted_k141_136336 | Not-determined |
| S1_extracted_k141_83113 | Low-quality |
| S1_extracted_k141_136337 | Not-determined |
| S1_extracted_k141_116465 | Not-determined |
| S1_extracted_k141_2474 | Not-determined |
| S1_extracted_k141_49842 | Low-quality |
| S1_extracted_k141_129781 | Not-determined |
| S1_extracted_k141_43105 | Not-determined |
| S1_extracted_k141_83118 | Not-determined |
| S1_extracted_k141_96490 | Not-determined |
| S1_extracted_k141_56344 | Not-determined |
| S1_extracted_k141_109619 | Not-determined |
| S1_extracted_k141_96493 | Not-determined |
| S1_extracted_k141_103181 | Not-determined |
| S1_extracted_k141_43109 | Not-determined |
| S1_extracted_k141_129786 | Not-determined |
| S1_extracted_k141_76392 | Not-determined |
| S1_extracted_k141_103184 | Not-determined |
| S1_extracted_k141_109621 | Low-quality |
| S1_extracted_k141_9763 | Not-determined |
| S1_extracted_k141_22797 | Low-quality |
| S1_extracted_k141_122864 | Low-quality |
| S1_extracted_k141_136344 | Not-determined |
| S1_extracted_k141_109625 | Not-determined |
| S1_extracted_k141_22800 | Not-determined |
| S1_extracted_k141_83128 | Not-determined |
| S1_extracted_k141_109628 | Low-quality |
| S1_extracted_k141_2487 | Not-determined |
| S1_extracted_k141_83131 | Not-determined |
| S1_extracted_k141_142967 | Not-determined |
| S1_extracted_k141_136348 | Not-determined |
| S1_extracted_k141_56354 | Not-determined |
| S1_extracted_k141_136350 | Not-determined |
| S1_extracted_k141_29699 | Not-determined |
| S1_extracted_k141_61797 | Not-determined |
| S1_extracted_k141_129795 | Not-determined |
| S1_extracted_k141_49860 | Low-quality |
| S1_extracted_k141_83136 | Not-determined |
| S1_extracted_k141_116482 | Not-determined |
| S1_extracted_k141_96501 | Not-determined |
| S1_extracted_k141_103192 | Not-determined |
| S1_extracted_k141_109640 | Not-determined |
| S1_extracted_k141_43121 | Not-determined |
| S1_extracted_k141_49864 | Not-determined |
| S1_extracted_k141_116484 | Not-determined |
| S1_extracted_k141_148124 | Low-quality |
| S1_extracted_k141_103198 | Not-determined |
| S1_extracted_k141_116485 | Not-determined |
| S1_extracted_k141_22821 | Low-quality |
| S1_extracted_k141_96506 | Not-determined |
| S1_extracted_k141_69605 | Not-determined |
| S1_extracted_k141_129807 | Not-determined |
| S1_extracted_k141_122872 | Not-determined |
| S1_extracted_k141_2500 | Not-determined |
| S1_extracted_k141_103206 | Not-determined |
| S1_extracted_k141_122874 | Low-quality |
| S1_extracted_k141_49872 | Not-determined |
| S1_extracted_k141_43128 | Low-quality |
| S1_extracted_k141_103208 | Not-determined |
| S1_extracted_k141_83153 | Not-determined |
| S1_extracted_k141_89645 | Low-quality |
| S1_extracted_k141_35761 | Low-quality |
| S1_extracted_k141_142974 | Not-determined |
| S1_extracted_k141_56371 | Not-determined |
| S1_extracted_k141_83155 | Low-quality |
| S1_extracted_k141_56374 | Low-quality |
| S1_extracted_k141_61819 | Not-determined |
| S1_extracted_k141_89646 | Not-determined |
| S1_extracted_k141_129821 | Not-determined |
| S1_extracted_k141_103219 | Not-determined |
| S1_extracted_k141_22829 | Not-determined |
| S1_extracted_k141_22830 | Not-determined |
| S1_extracted_k141_83159 | Low-quality |
| S1_extracted_k141_136360 | Not-determined |
| S1_extracted_k141_16888 | Not-determined |
| S1_extracted_k141_35763 | Low-quality |
| S1_extracted_k141_56378 | Not-determined |
| S1_extracted_k141_103224 | Not-determined |
| S1_extracted_k141_43140 | Not-determined |
| S1_extracted_k141_69622 | Not-determined |
| S1_extracted_k141_116504 | Low-quality |
| S1_extracted_k141_116505 | Not-determined |
| S1_extracted_k141_129828 | Not-determined |
| S1_extracted_k141_76427 | Not-determined |
| S1_extracted_k141_103225 | Not-determined |
| S1_extracted_k141_136368 | Not-determined |
| S1_extracted_k141_16893 | Low-quality |
| S1_extracted_k141_22837 | Not-determined |
| S1_extracted_k141_49889 | Not-determined |
| S1_extracted_k141_22838 | Not-determined |
| S1_extracted_k141_122885 | Not-determined |
| S1_extracted_k141_116511 | Not-determined |
| S1_extracted_k141_116512 | Not-determined |
| S1_extracted_k141_43143 | Not-determined |
| S1_extracted_k141_56380 | Not-determined |
| S1_extracted_k141_43144 | Low-quality |
| S1_extracted_k141_9800 | Not-determined |
| S1_extracted_k141_83169 | Not-determined |
| S1_extracted_k141_22847 | Low-quality |
| S1_extracted_k141_89662 | Not-determined |
| S1_extracted_k141_56389 | Not-determined |
| S1_extracted_k141_56392 | Not-determined |
| S1_extracted_k141_122892 | Low-quality |
| S1_extracted_k141_109658 | Not-determined |
| S1_extracted_k141_69625 | Not-determined |
| S1_extracted_k141_109661 | Not-determined |
| S1_extracted_k141_116521 | Low-quality |
| S1_extracted_k141_76437 | Not-determined |
| S1_extracted_k141_136377 | Not-determined |
| S1_extracted_k141_22854 | Not-determined |
| S1_extracted_k141_9807 | Not-determined |
| S1_extracted_k141_116523 | Low-quality |
| S1_extracted_k141_69626 | Not-determined |
| S1_extracted_k141_109668 | Not-determined |
| S1_extracted_k141_89674 | Not-determined |
| S1_extracted_k141_103246 | Not-determined |
| S1_extracted_k141_69630 | Not-determined |
| S1_extracted_k141_2532 | Not-determined |
| S1_extracted_k141_76445 | Not-determined |
| S1_extracted_k141_2533 | Not-determined |
| S1_extracted_k141_103247 | Not-determined |
| S1_extracted_k141_76446 | Not-determined |
| S1_extracted_k141_22856 | Low-quality |
| S1_extracted_k141_142987 | Not-determined |
| S1_extracted_k141_122905 | Not-determined |
| S1_extracted_k141_89678 | Low-quality |
| S1_extracted_k141_9816 | Low-quality |
| S1_extracted_k141_129841 | Low-quality |
| S1_extracted_k141_35784 | Low-quality |
| S1_extracted_k141_142990 | Not-determined |
| S1_extracted_k141_22859 | Not-determined |
| S1_extracted_k141_2540 | Low-quality |
| S1_extracted_k141_148150 | Not-determined |
| S1_extracted_k141_122909 | Not-determined |
| S1_extracted_k141_109673 | Not-determined |
| S1_extracted_k141_61842 | Not-determined |
| S1_extracted_k141_16914 | Not-determined |
| S1_extracted_k141_35787 | Not-determined |
| S1_extracted_k141_142995 | Not-determined |
| S1_extracted_k141_122911 | Not-determined |
| S1_extracted_k141_22865 | Not-determined |
| S1_extracted_k141_35790 | Not-determined |
| S1_extracted_k141_96560 | Not-determined |
| S1_extracted_k141_142999 | Not-determined |
| S1_extracted_k141_103260 | Not-determined |
| S1_extracted_k141_22867 | Low-quality |
| S1_extracted_k141_103261 | Not-determined |
| S1_extracted_k141_56411 | Not-determined |
| S1_extracted_k141_43171 | Not-determined |
| S1_extracted_k141_69643 | Low-quality |
| S1_extracted_k141_22869 | Not-determined |
| S1_extracted_k141_103265 | Low-quality |
| S1_extracted_k141_109680 | Not-determined |
| S1_extracted_k141_136406 | Not-determined |
| S1_extracted_k141_16927 | Low-quality |
| S1_extracted_k141_148159 | Not-determined |
| S1_extracted_k141_109683 | Not-determined |
| S1_extracted_k141_9828 | Not-determined |
| S1_extracted_k141_143009 | Not-determined |
| S1_extracted_k141_143010 | Not-determined |
| S1_extracted_k141_89686 | Not-determined |
| S1_extracted_k141_16933 | Not-determined |
| S1_extracted_k141_69649 | Not-determined |
| S1_extracted_k141_122922 | Not-determined |
| S1_extracted_k141_22876 | Not-determined |
| S1_extracted_k141_96567 | Low-quality |
| S1_extracted_k141_35800 | Not-determined |
| S1_extracted_k141_69654 | Not-determined |
| S1_extracted_k141_29725 | Not-determined |
| S1_extracted_k141_61858 | Low-quality |
| S1_extracted_k141_56418 | Not-determined |
| S1_extracted_k141_136413 | Not-determined |
| S1_extracted_k141_49920 | Not-determined |
| S1_extracted_k141_116546 | Not-determined |
| S1_extracted_k141_2560 | Low-quality |
| S1_extracted_k141_56420 | Not-determined |
| S1_extracted_k141_116547 | Not-determined |
| S1_extracted_k141_69658 | Not-determined |
| S1_extracted_k141_22884 | Not-determined |
| S1_extracted_k141_103281 | Not-determined |
| S1_extracted_k141_29730 | Not-determined |
| S1_extracted_k141_129872 | Not-determined |
| S1_extracted_k141_109688 | Low-quality |
| S1_extracted_k141_116549 | Not-determined |
| S1_extracted_k141_22887 | Not-determined |
| S1_extracted_k141_56425 | Not-determined |
| S1_extracted_k141_103284 | Not-determined |
| S1_extracted_k141_103285 | Not-determined |
| S1_extracted_k141_129877 | Not-determined |
| S1_extracted_k141_22890 | Not-determined |
| S1_extracted_k141_76457 | Not-determined |
| S1_extracted_k141_129878 | Not-determined |
| S1_extracted_k141_9848 | Not-determined |
| S1_extracted_k141_56427 | Not-determined |
| S1_extracted_k141_148169 | Not-determined |
| S1_extracted_k141_49928 | Not-determined |
| S1_extracted_k141_56428 | Not-determined |
| S1_extracted_k141_61865 | Not-determined |
| S1_extracted_k141_143023 | Not-determined |
| S1_extracted_k141_43185 | Not-determined |
| S1_extracted_k141_122935 | Not-determined |
| S1_extracted_k141_148170 | Not-determined |
| S1_extracted_k141_61870 | Not-determined |
| S1_extracted_k141_136427 | Not-determined |
| S1_extracted_k141_116559 | Not-determined |
| S1_extracted_k141_103299 | Not-determined |
| S1_extracted_k141_69670 | Low-quality |
| S1_extracted_k141_22893 | Not-determined |
| S1_extracted_k141_22894 | Not-determined |
| S1_extracted_k141_109704 | Not-determined |
| S1_extracted_k141_143032 | Not-determined |
| S1_extracted_k141_103303 | Not-determined |
| S1_extracted_k141_43193 | Low-quality |
| S1_extracted_k141_49939 | Not-determined |
| S1_extracted_k141_103305 | Not-determined |
| S1_extracted_k141_83219 | Not-determined |
| S1_extracted_k141_136429 | Low-quality |
| S1_extracted_k141_129891 | Not-determined |
| S1_extracted_k141_143038 | Low-quality |
| S1_extracted_k141_2573 | Not-determined |
| S1_extracted_k141_143041 | Not-determined |
| S1_extracted_k141_61889 | Not-determined |
| S1_extracted_k141_69678 | Not-determined |
| S1_extracted_k141_89720 | Low-quality |
| S1_extracted_k141_35822 | Not-determined |
| S1_extracted_k141_109713 | Not-determined |
| S1_extracted_k141_16969 | Low-quality |
| S1_extracted_k141_76479 | Not-determined |
| S1_extracted_k141_56446 | Not-determined |
| S1_extracted_k141_129902 | Not-determined |
| S1_extracted_k141_16971 | Not-determined |
| S1_extracted_k141_129903 | Low-quality |
| S1_extracted_k141_116578 | Low-quality |
| S1_extracted_k141_49941 | Low-quality |
| S1_extracted_k141_143047 | Not-determined |
| S1_extracted_k141_143048 | Not-determined |
| S1_extracted_k141_122953 | Low-quality |
| S1_extracted_k141_69684 | Not-determined |
| S1_extracted_k141_83232 | Not-determined |
| S1_extracted_k141_89728 | Not-determined |
| S1_extracted_k141_76484 | Not-determined |
| S1_extracted_k141_29759 | Not-determined |
| S1_extracted_k141_83234 | Low-quality |
| S1_extracted_k141_129911 | Not-determined |
| S1_extracted_k141_109720 | Not-determined |
| S1_extracted_k141_143049 | Not-determined |
| S1_extracted_k141_35832 | Not-determined |
| S1_extracted_k141_69687 | Not-determined |
| S1_extracted_k141_35833 | Not-determined |
| S1_extracted_k141_83237 | Not-determined |
| S1_extracted_k141_143051 | Low-quality |
| S1_extracted_k141_109722 | Low-quality |
| S1_extracted_k141_96602 | Not-determined |
| S1_extracted_k141_89734 | Not-determined |
| S1_extracted_k141_155360 | Not-determined |
| S1_extracted_k141_109725 | Low-quality |
| S1_extracted_k141_116592 | Low-quality |
| S1_extracted_k141_103311 | Low-quality |
| S1_extracted_k141_16988 | Not-determined |
| S1_extracted_k141_83247 | Not-determined |
| S1_extracted_k141_96607 | Low-quality |
| S1_extracted_k141_61907 | Not-determined |
| S1_extracted_k141_29766 | Not-determined |
| S1_extracted_k141_49952 | Not-determined |
| S1_extracted_k141_116595 | Low-quality |
| S1_extracted_k141_56461 | Not-determined |
| S1_extracted_k141_136455 | Not-determined |
| S1_extracted_k141_56462 | Not-determined |
| S1_extracted_k141_35843 | Low-quality |
| S1_extracted_k141_69699 | Not-determined |
| S1_extracted_k141_83249 | Not-determined |
| S1_extracted_k141_76504 | Not-determined |
| S1_extracted_k141_76505 | Low-quality |
| S1_extracted_k141_143065 | Not-determined |
| S1_extracted_k141_2592 | Not-determined |
| S1_extracted_k141_116601 | Not-determined |
| S1_extracted_k141_17001 | Not-determined |
| S1_extracted_k141_56469 | Low-quality |
| S1_extracted_k141_129928 | Not-determined |
| S1_extracted_k141_148186 | Not-determined |
| S1_extracted_k141_17002 | Not-determined |
| S1_extracted_k141_49960 | Not-determined |
| S1_extracted_k141_49961 | Not-determined |
| S1_extracted_k141_49962 | Not-determined |
| S1_extracted_k141_43205 | Low-quality |
| S1_extracted_k141_96615 | Not-determined |
| S1_extracted_k141_49964 | Not-determined |
| S1_extracted_k141_122963 | Not-determined |
| S1_extracted_k141_43207 | Not-determined |
| S1_extracted_k141_116605 | Not-determined |
| S1_extracted_k141_43209 | Not-determined |
| S1_extracted_k141_2598 | Low-quality |
| S1_extracted_k141_83254 | Not-determined |
| S1_extracted_k141_96619 | Not-determined |
| S1_extracted_k141_109747 | Not-determined |
| S1_extracted_k141_96623 | Not-determined |
| S1_extracted_k141_43214 | Not-determined |
| S1_extracted_k141_22901 | Not-determined |
| S1_extracted_k141_49973 | Not-determined |
| S1_extracted_k141_17015 | Low-quality |
| S1_extracted_k141_148194 | Low-quality |
| S1_extracted_k141_129935 | Low-quality |
| S1_extracted_k141_122980 | Not-determined |
| S1_extracted_k141_49977 | Low-quality |
| S1_extracted_k141_129936 | Low-quality |
| S1_extracted_k141_2608 | Not-determined |
| S1_extracted_k141_9880 | Not-determined |
| S1_extracted_k141_122984 | Not-determined |
| S1_extracted_k141_22907 | Not-determined |
| S1_extracted_k141_29791 | Not-determined |
| S1_extracted_k141_76526 | Not-determined |
| S1_extracted_k141_43216 | Low-quality |
| S1_extracted_k141_29793 | Not-determined |
| S1_extracted_k141_83272 | Low-quality |
| S1_extracted_k141_17024 | Not-determined |
| S1_extracted_k141_122988 | Not-determined |
| S1_extracted_k141_129944 | Not-determined |
| S1_extracted_k141_76531 | Low-quality |
| S1_extracted_k141_116620 | Not-determined |
| S1_extracted_k141_61932 | Not-determined |
| S1_extracted_k141_76533 | Low-quality |
| S1_extracted_k141_116622 | Not-determined |
| S1_extracted_k141_143089 | Low-quality |
| S1_extracted_k141_49985 | Not-determined |
| S1_extracted_k141_69708 | Not-determined |
| S1_extracted_k141_76538 | Not-determined |
| S1_extracted_k141_43224 | Not-determined |
| S1_extracted_k141_148205 | Not-determined |
| S1_extracted_k141_89743 | Low-quality |

| | |
|---|---|
| S1_extracted_k141_89744 | Not-determined |
| S1_extracted_k141_49994 | Not-determined |
| S1_extracted_k141_129955 | Not-determined |
| S1_extracted_k141_43231 | Not-determined |
| S1_extracted_k141_122994 | Not-determined |
| S1_extracted_k141_49999 | Low-quality |
| S1_extracted_k141_50000 | Not-determined |
| S1_extracted_k141_43236 | Not-determined |
| S1_extracted_k141_61946 | Not-determined |
| S1_extracted_k141_116636 | Not-determined |
| S1_extracted_k141_43238 | Not-determined |
| S1_extracted_k141_22925 | Not-determined |
| S1_extracted_k141_136491 | Not-determined |
| S1_extracted_k141_109781 | Low-quality |
| S1_extracted_k141_76550 | Not-determined |
| S1_extracted_k141_43241 | Not-determined |
| S1_extracted_k141_43242 | Not-determined |
| S1_extracted_k141_9904 | Low-quality |
| S1_extracted_k141_136493 | Not-determined |
| S1_extracted_k141_29819 | Not-determined |
| S1_extracted_k141_50006 | Low-quality |
| S1_extracted_k141_9906 | Not-determined |
| S1_extracted_k141_123001 | Not-determined |
| S1_extracted_k141_83291 | Not-determined |
| S1_extracted_k141_9909 | Not-determined |
| S1_extracted_k141_103338 | Not-determined |
| S1_extracted_k141_76554 | Not-determined |
| S1_extracted_k141_9910 | Not-determined |
| S1_extracted_k141_89761 | Not-determined |
| S1_extracted_k141_103341 | Low-quality |
| S1_extracted_k141_143100 | Not-determined |
| S1_extracted_k141_89762 | Not-determined |
| S1_extracted_k141_103342 | Not-determined |
| S1_extracted_k141_35886 | Not-determined |
| S1_extracted_k141_9914 | Not-determined |
| S1_extracted_k141_103346 | Not-determined |
| S1_extracted_k141_22931 | Not-determined |
| S1_extracted_k141_96656 | Not-determined |
| S1_extracted_k141_22933 | Low-quality |
| S1_extracted_k141_109791 | Not-determined |
| S1_extracted_k141_89768 | Not-determined |
| S1_extracted_k141_96659 | Low-quality |
| S1_extracted_k141_17052 | Not-determined |
| S1_extracted_k141_83302 | Not-determined |
| S1_extracted_k141_143105 | Not-determined |
| S1_extracted_k141_89770 | Low-quality |
| S1_extracted_k141_61949 | Low-quality |
| S1_extracted_k141_123009 | Not-determined |
| S1_extracted_k141_22936 | Low-quality |
| S1_extracted_k141_76562 | Not-determined |
| S1_extracted_k141_155378 | Not-determined |
| S1_extracted_k141_109797 | Not-determined |
| S1_extracted_k141_56492 | Not-determined |
| S1_extracted_k141_123011 | Low-quality |
| S1_extracted_k141_83307 | Not-determined |
| S1_extracted_k141_96666 | Low-quality |
| S1_extracted_k141_22940 | Low-quality |
| S1_extracted_k141_56493 | Low-quality |
| S1_extracted_k141_56494 | Not-determined |
| S1_extracted_k141_61955 | Not-determined |
| S1_extracted_k141_61956 | Not-determined |
| S1_extracted_k141_76570 | Not-determined |
| S1_extracted_k141_50021 | Low-quality |
| S1_extracted_k141_76572 | Low-quality |
| S1_extracted_k141_22944 | Not-determined |
| S1_extracted_k141_116655 | Low-quality |
| S1_extracted_k141_76573 | Not-determined |
| S1_extracted_k141_109805 | Not-determined |
| S1_extracted_k141_116656 | Not-determined |
| S1_extracted_k141_109806 | Not-determined |
| S1_extracted_k141_143113 | Not-determined |
| S1_extracted_k141_22946 | Not-determined |
| S1_extracted_k141_116658 | Low-quality |
| S1_extracted_k141_143114 | Not-determined |
| S1_extracted_k141_123018 | Not-determined |
| S1_extracted_k141_9928 | Low-quality |
| S1_extracted_k141_22949 | Not-determined |
| S1_extracted_k141_43255 | Not-determined |
| S1_extracted_k141_136505 | Not-determined |
| S1_extracted_k141_17062 | Not-determined |
| S1_extracted_k141_17063 | Low-quality |
| S1_extracted_k141_136507 | Not-determined |
| S1_extracted_k141_148212 | Not-determined |
| S1_extracted_k141_148213 | Not-determined |
| S1_extracted_k141_109809 | Not-determined |
| S1_extracted_k141_43259 | Not-determined |
| S1_extracted_k141_109811 | Not-determined |
| S1_extracted_k141_116670 | Low-quality |
| S1_extracted_k141_17073 | Not-determined |
| S1_extracted_k141_116671 | Low-quality |
| S1_extracted_k141_109813 | Not-determined |
| S1_extracted_k141_17074 | Low-quality |
| S1_extracted_k141_103361 | Not-determined |
| S1_extracted_k141_116672 | Not-determined |
| S1_extracted_k141_43265 | Low-quality |
| S1_extracted_k141_9938 | Not-determined |
| S1_extracted_k141_43266 | Not-determined |
| S1_extracted_k141_50030 | Not-determined |
| S1_extracted_k141_103363 | Low-quality |
| S1_extracted_k141_96688 | Not-determined |
| S1_extracted_k141_103364 | Not-determined |
| S1_extracted_k141_129991 | Not-determined |
| S1_extracted_k141_136520 | Not-determined |
| S1_extracted_k141_35904 | Not-determined |
| S1_extracted_k141_148219 | Not-determined |
| S1_extracted_k141_143122 | Low-quality |
| S1_extracted_k141_43272 | Low-quality |
| S1_extracted_k141_129993 | Not-determined |
| S1_extracted_k141_56518 | Low-quality |
| S1_extracted_k141_103369 | Not-determined |
| S1_extracted_k141_56521 | Not-determined |
| S1_extracted_k141_56522 | Not-determined |
| S1_extracted_k141_123024 | Not-determined |
| S1_extracted_k141_143129 | Low-quality |
| S1_extracted_k141_123025 | Not-determined |
| S1_extracted_k141_148226 | Not-determined |
| S1_extracted_k141_9947 | Not-determined |
| S1_extracted_k141_2631 | Low-quality |
| S1_extracted_k141_148228 | Not-determined |
| S1_extracted_k141_130004 | Not-determined |
| S1_extracted_k141_123032 | Not-determined |
| S1_extracted_k141_2634 | Not-determined |
| S1_extracted_k141_123033 | Not-determined |
| S1_extracted_k141_17080 | Low-quality |
| S1_extracted_k141_2635 | Not-determined |
| S1_extracted_k141_35913 | Not-determined |
| S1_extracted_k141_155388 | Low-quality |
| S1_extracted_k141_2637 | Not-determined |
| S1_extracted_k141_17082 | Not-determined |
| S1_extracted_k141_43289 | Low-quality |
| S1_extracted_k141_109831 | Not-determined |
| S1_extracted_k141_109832 | Not-determined |
| S1_extracted_k141_43291 | Not-determined |
| S1_extracted_k141_130012 | Not-determined |
| S1_extracted_k141_35917 | Not-determined |
| S1_extracted_k141_116698 | Not-determined |
| S1_extracted_k141_89788 | Not-determined |
| S1_extracted_k141_50058 | Not-determined |
| S1_extracted_k141_69738 | Not-determined |
| S1_extracted_k141_109837 | Not-determined |
| S1_extracted_k141_123036 | Not-determined |
| S1_extracted_k141_56544 | Not-determined |
| S1_extracted_k141_76590 | Not-determined |
| S1_extracted_k141_43303 | Not-determined |
| S1_extracted_k141_148240 | Low-quality |
| S1_extracted_k141_43305 | Not-determined |
| S1_extracted_k141_116706 | Not-determined |
| S1_extracted_k141_96722 | Not-determined |
| S1_extracted_k141_148245 | Not-determined |
| S1_extracted_k141_29856 | Not-determined |
| S1_extracted_k141_123042 | Not-determined |
| S1_extracted_k141_29858 | Not-determined |
| S1_extracted_k141_123043 | Not-determined |
| S1_extracted_k141_29859 | Not-determined |
| S1_extracted_k141_9966 | Not-determined |
| S1_extracted_k141_69750 | Not-determined |
| S1_extracted_k141_56550 | Low-quality |
| S1_extracted_k141_29861 | Not-determined |
| S1_extracted_k141_35924 | Not-determined |
| S1_extracted_k141_96732 | Not-determined |
| S1_extracted_k141_76605 | Not-determined |
| S1_extracted_k141_96733 | Not-determined |
| S1_extracted_k141_56556 | Not-determined |
| S1_extracted_k141_148251 | Not-determined |
| S1_extracted_k141_109856 | Not-determined |
| S1_extracted_k141_56558 | Not-determined |
| S1_extracted_k141_109857 | Not-determined |
| S1_extracted_k141_35928 | Low-quality |
| S1_extracted_k141_62018 | Not-determined |
| S1_extracted_k141_89808 | Not-determined |
| S1_extracted_k141_116717 | Not-determined |
| S1_extracted_k141_155402 | Not-determined |
| S1_extracted_k141_89812 | Not-determined |
| S1_extracted_k141_2657 | Not-determined |
| S1_extracted_k141_89814 | Not-determined |
| S1_extracted_k141_116718 | Low-quality |
| S1_extracted_k141_96738 | Not-determined |
| S1_extracted_k141_35933 | Not-determined |
| S1_extracted_k141_155405 | Not-determined |
| S1_extracted_k141_116719 | Not-determined |
| S1_extracted_k141_103399 | Not-determined |
| S1_extracted_k141_116722 | Not-determined |
| S1_extracted_k141_76618 | Low-quality |
| S1_extracted_k141_76619 | Not-determined |
| S1_extracted_k141_123062 | Not-determined |
| S1_extracted_k141_148261 | Not-determined |
| S1_extracted_k141_103403 | Not-determined |
| S1_extracted_k141_96744 | Low-quality |
| S1_extracted_k141_76622 | Not-determined |
| S1_extracted_k141_130035 | Not-determined |
| S1_extracted_k141_103405 | Not-determined |
| S1_extracted_k141_130036 | Low-quality |
| S1_extracted_k141_123066 | Not-determined |
| S1_extracted_k141_130038 | Not-determined |
| S1_extracted_k141_103407 | Not-determined |
| S1_extracted_k141_62034 | Not-determined |
| S1_extracted_k141_76626 | Not-determined |
| S1_extracted_k141_29874 | Low-quality |
| S1_extracted_k141_116726 | Not-determined |
| S1_extracted_k141_155415 | Low-quality |
| S1_extracted_k141_62036 | Not-determined |
| S1_extracted_k141_143151 | Low-quality |
| S1_extracted_k141_89818 | Not-determined |
| S1_extracted_k141_148262 | Not-determined |
| S1_extracted_k141_76630 | Not-determined |
| S1_extracted_k141_103415 | Low-quality |
| S1_extracted_k141_35946 | Not-determined |
| S1_extracted_k141_43333 | Not-determined |
| S1_extracted_k141_130045 | Not-determined |
| S1_extracted_k141_50100 | Not-determined |
| S1_extracted_k141_56577 | Not-determined |
| S1_extracted_k141_29877 | Not-determined |
| S1_extracted_k141_109871 | Not-determined |
| S1_extracted_k141_130047 | Not-determined |
| S1_extracted_k141_29878 | Not-determined |
| S1_extracted_k141_103420 | Not-determined |
| S1_extracted_k141_50103 | Not-determined |
| S1_extracted_k141_29880 | Not-determined |
| S1_extracted_k141_103421 | Not-determined |
| S1_extracted_k141_109875 | Not-determined |
| S1_extracted_k141_69755 | Not-determined |
| S1_extracted_k141_9985 | Not-determined |
| S1_extracted_k141_130055 | Not-determined |
| S1_extracted_k141_116737 | Not-determined |
| S1_extracted_k141_69757 | Not-determined |
| S1_extracted_k141_2677 | Not-determined |
| S1_extracted_k141_9989 | Not-determined |
| S1_extracted_k141_89827 | Low-quality |
| S1_extracted_k141_29885 | Not-determined |
| S1_extracted_k141_109879 | Not-determined |
| S1_extracted_k141_29887 | Not-determined |
| S1_extracted_k141_109881 | Not-determined |
| S1_extracted_k141_83385 | Low-quality |
| S1_extracted_k141_83386 | Not-determined |
| S6_vs_k141_1146\|\|full | High-quality |
| S1_extracted_k141_76646 | Not-determined |
| S1_extracted_k141_130063 | Not-determined |
| S1_extracted_k141_155421 | Not-determined |
| S1_extracted_k141_83387 | Not-determined |
| S1_extracted_k141_22967 | Not-determined |
| S1_extracted_k141_35961 | Not-determined |
| S1_extracted_k141_89835 | Not-determined |
| S1_extracted_k141_116752 | Not-determined |
| S1_extracted_k141_96770 | Not-determined |
| S1_extracted_k141_76653 | Not-determined |
| S1_extracted_k141_89836 | Not-determined |
| S1_extracted_k141_29896 | Not-determined |
| S1_extracted_k141_143166 | Low-quality |
| S1_extracted_k141_123084 | Not-determined |
| S1_extracted_k141_89838 | Not-determined |
| S1_extracted_k141_123085 | Not-determined |
| S1_extracted_k141_96774 | Not-determined |
| S1_extracted_k141_155426 | Not-determined |
| S1_extracted_k141_143168 | Not-determined |
| S1_extracted_k141_103436 | Not-determined |
| S8_extracted_k141_54514 | Complete |
| S1_extracted_k141_35970 | Not-determined |
| S1_extracted_k141_22977 | Not-determined |
| S1_extracted_k141_29900 | Not-determined |
| S1_extracted_k141_103445 | Low-quality |
| S1_extracted_k141_96787 | Not-determined |
| S1_extracted_k141_155431 | Low-quality |
| S1_extracted_k141_155433 | Low-quality |
| S1_extracted_k141_130074 | Not-determined |
| S1_extracted_k141_10010 | Not-determined |
| S1_extracted_k141_103451 | Not-determined |
| S1_extracted_k141_2688 | Not-determined |
| S1_extracted_k141_89857 | Low-quality |
| S1_extracted_k141_89858 | Not-determined |
| S1_extracted_k141_109904 | Not-determined |
| S1_extracted_k141_43367 | Not-determined |
| S1_extracted_k141_103453 | Low-quality |
| S1_extracted_k141_29909 | Not-determined |
| S1_extracted_k141_148280 | Not-determined |
| S1_extracted_k141_56593 | Not-determined |
| S1_extracted_k141_89863 | Not-determined |
| S1_extracted_k141_130079 | Not-determined |
| S1_extracted_k141_155435 | Not-determined |
| S1_extracted_k141_89864 | Not-determined |
| S1_extracted_k141_29913 | Not-determined |
| S1_extracted_k141_130081 | Low-quality |
| S1_extracted_k141_83404 | Low-quality |
| S1_extracted_k141_109911 | Not-determined |
| S1_extracted_k141_43375 | Not-determined |
| S1_extracted_k141_155440 | Not-determined |
| S1_extracted_k141_148282 | Not-determined |
| S1_extracted_k141_116757 | Not-determined |
| S1_extracted_k141_29920 | Not-determined |
| S1_extracted_k141_83406 | Not-determined |
| S1_extracted_k141_22986 | Low-quality |
| S1_extracted_k141_62056 | Not-determined |
| S1_extracted_k141_43376 | Not-determined |
| S1_extracted_k141_10020 | Not-determined |
| S1_extracted_k141_22988 | Low-quality |
| S1_extracted_k141_103465 | Not-determined |
| S1_extracted_k141_83407 | Low-quality |
| S1_extracted_k141_10024 | Not-determined |
| S1_extracted_k141_103466 | Low-quality |
| S1_extracted_k141_83409 | Not-determined |
| S1_extracted_k141_96814 | Not-determined |
| S1_extracted_k141_43385 | Not-determined |
| S1_extracted_k141_10028 | Not-determined |
| S1_extracted_k141_76670 | Low-quality |
| S1_extracted_k141_155451 | Low-quality |
| S1_extracted_k141_130091 | Not-determined |
| S1_extracted_k141_89878 | Not-determined |
| S1_extracted_k141_143198 | Not-determined |
| S1_extracted_k141_143199 | Not-determined |
| S1_extracted_k141_10034 | Not-determined |
| S1_extracted_k141_155453 | Not-determined |
| S1_extracted_k141_89881 | Not-determined |
| S1_extracted_k141_103470 | Not-determined |
| S1_extracted_k141_76673 | Not-determined |
| S1_extracted_k141_62075 | Not-determined |
| S13_extracted_64889 | High-quality |
| S1_extracted_k141_96819 | Low-quality |
| S1_extracted_k141_23002 | Not-determined |
| S1_extracted_k141_56618 | Not-determined |
| S1_extracted_k141_143203 | Low-quality |
| S1_extracted_k141_123117 | Low-quality |
| S1_extracted_k141_35984 | Not-determined |
| S1_extracted_k141_109931 | Not-determined |
| S1_extracted_k141_83421 | Not-determined |
| S1_extracted_k141_123119 | Not-determined |
| S1_extracted_k141_56622 | Not-determined |
| S1_extracted_k141_10048 | Not-determined |
| S1_extracted_k141_143206 | Not-determined |
| S1_extracted_k141_96822 | Not-determined |
| S1_extracted_k141_2709 | Low-quality |
| S1_extracted_k141_109935 | Low-quality |
| S1_extracted_k141_29942 | Not-determined |
| S1_extracted_k141_50143 | Low-quality |
| S1_extracted_k141_35987 | Not-determined |
| S1_extracted_k141_29943 | Not-determined |
| S1_extracted_k141_23008 | Not-determined |
| S1_extracted_k141_10051 | Not-determined |
| S1_extracted_k141_10054 | Not-determined |
| S1_extracted_k141_69783 | Not-determined |
| S1_extracted_k141_69784 | Low-quality |
| S1_extracted_k141_136538 | Not-determined |
| S1_extracted_k141_29950 | Not-determined |
| S1_extracted_k141_143214 | Low-quality |
| S1_extracted_k141_10059 | Not-determined |
| S1_extracted_k141_143215 | Not-determined |
| S1_extracted_k141_109942 | Not-determined |
| S1_extracted_k141_17133 | Not-determined |
| S1_extracted_k141_76695 | Not-determined |
| S1_extracted_k141_109947 | Not-determined |
| S1_extracted_k141_23016 | Not-determined |
| S1_extracted_k141_56630 | Low-quality |
| S1_extracted_k141_29954 | Not-determined |
| S1_extracted_k141_83433 | Not-determined |
| S1_extracted_k141_69790 | Not-determined |
| S1_extracted_k141_23021 | Not-determined |
| S1_extracted_k141_96829 | Not-determined |
| S1_extracted_k141_56635 | Not-determined |
| S1_extracted_k141_17137 | Not-determined |
| S1_extracted_k141_83437 | Not-determined |
| S1_extracted_k141_89904 | Not-determined |
| S1_extracted_k141_143225 | Not-determined |
| S1_extracted_k141_35993 | Not-determined |
| S1_extracted_k141_23026 | Not-determined |
| S1_extracted_k141_17140 | Low-quality |
| S1_extracted_k141_83442 | Not-determined |
| S1_extracted_k141_136550 | Not-determined |
| S1_extracted_k141_43420 | Not-determined |
| S1_extracted_k141_23029 | Not-determined |
| S1_extracted_k141_143228 | Not-determined |
| S1_extracted_k141_130111 | Not-determined |
| S1_extracted_k141_69796 | Not-determined |
| S1_extracted_k141_148299 | Not-determined |
| S1_extracted_k141_96842 | Not-determined |
| S1_extracted_k141_130117 | Not-determined |
| S1_extracted_k141_143233 | Low-quality |
| S1_extracted_k141_83448 | Not-determined |
| S1_extracted_k141_35998 | Not-determined |
| S1_extracted_k141_96845 | Low-quality |
| S1_extracted_k141_35999 | Not-determined |
| S1_extracted_k141_43428 | Not-determined |
| S1_extracted_k141_76711 | Not-determined |
| S1_extracted_k141_89921 | Not-determined |
| S1_extracted_k141_103481 | Not-determined |
| S1_extracted_k141_136571 | Not-determined |
| S1_extracted_k141_23038 | Not-determined |
| S1_extracted_k141_23039 | Not-determined |
| S1_extracted_k141_96853 | Not-determined |
| S1_extracted_k141_116815 | Not-determined |
| S1_extracted_k141_50167 | Not-determined |
| S1_extracted_k141_50169 | Not-determined |
| S1_extracted_k141_17152 | Not-determined |
| S1_extracted_k141_89924 | Not-determined |
| S1_extracted_k141_109970 | Not-determined |
| S1_extracted_k141_10087 | Not-determined |
| S1_extracted_k141_56661 | Not-determined |
| S1_extracted_k141_36008 | Not-determined |
| S1_extracted_k141_83460 | Not-determined |
| S1_extracted_k141_2724 | Low-quality |
| S1_extracted_k141_50178 | Not-determined |
| S1_extracted_k141_76723 | Not-determined |
| S1_extracted_k141_17160 | Not-determined |
| S1_extracted_k141_56667 | Not-determined |
| S1_extracted_k141_69812 | Not-determined |
| S1_extracted_k141_36013 | Low-quality |
| S1_extracted_k141_29979 | Not-determined |
| S1_extracted_k141_143242 | Not-determined |
| S1_extracted_k141_123169 | Low-quality |
| S1_extracted_k141_56675 | Not-determined |
| S1_extracted_k141_23044 | Not-determined |
| S1_extracted_k141_136582 | Not-determined |
| S1_extracted_k141_109983 | Not-determined |
| S1_extracted_k141_56678 | Not-determined |
| S1_extracted_k141_116829 | Not-determined |
| S1_extracted_k141_23046 | Low-quality |
| S1_extracted_k141_56680 | Low-quality |
| S1_extracted_k141_96863 | Not-determined |
| S1_extracted_k141_56681 | Not-determined |
| S1_extracted_k141_96864 | Not-determined |
| S1_extracted_k141_43437 | Low-quality |
| S1_extracted_k141_62116 | Not-determined |
| S1_extracted_k141_143253 | Not-determined |
| S1_extracted_k141_76730 | Low-quality |
| S1_extracted_k141_62117 | Not-determined |
| S1_extracted_k141_10098 | Low-quality |
| S1_extracted_k141_123177 | Not-determined |
| S1_extracted_k141_123178 | Not-determined |
| S1_extracted_k141_130142 | Not-determined |
| S1_extracted_k141_123179 | Not-determined |
| S1_extracted_k141_56686 | Not-determined |
| S1_extracted_k141_17172 | Low-quality |
| S1_extracted_k141_23059 | Not-determined |
| S1_extracted_k141_96872 | Not-determined |
| S1_extracted_k141_23060 | Not-determined |
| S1_extracted_k141_116836 | Not-determined |
| S1_extracted_k141_148325 | Not-determined |
| S1_extracted_k141_23064 | Not-determined |
| S1_extracted_k141_50205 | Low-quality |
| S1_extracted_k141_10108 | Not-determined |
| S1_extracted_k141_36033 | Not-determined |
| S1_extracted_k141_155465 | Not-determined |
| S1_extracted_k141_17180 | Not-determined |
| S1_extracted_k141_10112 | Low-quality |
| S1_extracted_k141_30000 | Not-determined |
| S1_extracted_k141_43448 | Low-quality |
| S1_extracted_k141_96882 | Not-determined |
| S1_extracted_k141_110002 | Not-determined |
| S1_extracted_k141_130146 | Not-determined |
| S1_extracted_k141_23071 | Not-determined |
| S1_extracted_k141_36038 | Low-quality |
| S1_extracted_k141_10117 | Not-determined |
| S1_extracted_k141_148329 | Low-quality |
| S1_extracted_k141_130149 | Low-quality |
| S1_extracted_k141_30006 | Not-determined |
| S1_extracted_k141_76739 | Not-determined |
| S1_extracted_k141_110006 | Not-determined |
| S1_extracted_k141_143267 | Not-determined |
| S1_extracted_k141_2746 | Not-determined |
| S1_extracted_k141_136603 | Not-determined |
| S1_extracted_k141_110008 | Not-determined |
| S1_extracted_k141_30008 | Low-quality |
| S1_extracted_k141_76744 | Not-determined |
| S1_extracted_k141_96888 | Not-determined |
| S1_extracted_k141_143269 | Not-determined |
| S1_extracted_k141_148333 | Not-determined |
| S1_extracted_k141_50221 | Not-determined |
| S1_extracted_k141_155472 | Not-determined |
| S1_extracted_k141_123196 | Not-determined |
| S1_extracted_k141_96891 | Not-determined |
| S1_extracted_k141_148337 | Not-determined |
| S1_extracted_k141_96895 | Not-determined |
| S1_extracted_k141_136610 | Not-determined |
| S1_extracted_k141_50227 | Not-determined |
| S1_extracted_k141_148339 | Not-determined |
| S1_extracted_k141_89949 | Low-quality |
| S1_extracted_k141_2755 | Not-determined |
| S1_extracted_k141_30022 | Not-determined |
| S1_extracted_k141_155479 | Not-determined |
| S1_extracted_k141_2756 | Not-determined |
| S1_extracted_k141_43468 | Not-determined |
| S1_extracted_k141_83498 | Not-determined |
| S1_extracted_k141_110021 | Not-determined |
| S1_extracted_k141_56705 | Low-quality |
| S1_extracted_k141_36045 | Low-quality |
| S1_extracted_k141_76749 | Low-quality |
| S1_extracted_k141_89958 | Not-determined |
| S1_extracted_k141_10138 | Not-determined |
| S1_extracted_k141_130163 | Not-determined |
| S1_extracted_k141_36046 | Low-quality |
| S1_extracted_k141_155483 | Not-determined |
| S1_extracted_k141_17208 | Not-determined |
| S1_extracted_k141_62130 | Not-determined |
| S1_extracted_k141_36048 | Low-quality |
| S1_extracted_k141_23085 | Low-quality |
| S1_extracted_k141_10145 | Low-quality |
| S1_extracted_k141_43475 | Not-determined |
| S1_extracted_k141_116858 | Not-determined |
| S1_extracted_k141_89966 | Not-determined |
| S1_extracted_k141_123221 | Not-determined |
| S1_extracted_k141_56707 | Not-determined |
| S1_extracted_k141_62137 | Not-determined |
| S1_extracted_k141_23092 | Not-determined |
| S1_extracted_k141_69827 | Not-determined |
| S1_extracted_k141_89969 | Not-determined |
| S1_extracted_k141_10154 | Not-determined |
| S1_extracted_k141_62139 | Not-determined |
| S1_extracted_k141_148349 | Not-determined |
| S1_extracted_k141_123228 | Not-determined |
| S1_extracted_k141_110028 | Not-determined |
| S1_extracted_k141_155495 | Not-determined |
| S1_extracted_k141_89976 | Not-determined |
| S1_extracted_k141_148354 | Low-quality |
| S1_extracted_k141_50246 | Not-determined |
| S1_extracted_k141_96911 | Not-determined |
| S1_extracted_k141_110033 | Low-quality |
| S1_extracted_k141_83526 | Not-determined |
| S1_extracted_k141_103546 | Not-determined |
| S1_extracted_k141_30048 | Not-determined |
| S1_extracted_k141_50252 | Not-determined |
| S1_extracted_k141_56718 | Not-determined |
| S1_extracted_k141_103548 | Not-determined |
| S1_extracted_k141_89981 | Low-quality |
| S1_extracted_k141_89984 | Not-determined |
| S1_extracted_k141_36065 | Not-determined |
| S1_extracted_k141_56723 | Not-determined |
| S1_extracted_k141_96920 | Low-quality |
| S1_extracted_k141_30055 | Not-determined |
| S1_extracted_k141_10170 | Not-determined |
| S1_extracted_k141_56725 | Not-determined |
| S1_extracted_k141_43499 | Not-determined |
| S1_extracted_k141_62152 | Not-determined |
| S1_extracted_k141_62153 | Not-determined |
| S1_extracted_k141_17220 | Low-quality |
| S1_extracted_k141_30059 | Not-determined |
| S1_extracted_k141_143293 | Not-determined |
| S1_extracted_k141_110049 | Not-determined |
| S1_extracted_k141_30062 | Not-determined |
| S1_extracted_k141_43510 | Not-determined |
| S1_extracted_k141_110052 | Not-determined |
| S1_extracted_k141_43513 | Not-determined |
| S1_extracted_k141_89997 | Not-determined |
| S1_extracted_k141_143301 | Not-determined |
| S1_extracted_k141_43515 | Not-determined |
| S1_extracted_k141_30065 | Low-quality |
| S1_extracted_k141_17232 | Not-determined |
| S1_extracted_k141_30067 | Not-determined |
| S1_extracted_k141_155507 | Not-determined |
| S1_extracted_k141_83550 | Not-determined |
| S1_extracted_k141_62164 | Low-quality |
| S1_extracted_k141_96942 | Not-determined |
| S1_extracted_k141_30071 | Not-determined |
| S1_extracted_k141_36079 | Not-determined |
| S1_extracted_k141_69838 | Not-determined |
| S1_extracted_k141_17235 | Not-determined |
| S1_extracted_k141_123240 | Not-determined |
| S1_extracted_k141_62167 | Not-determined |
| S1_extracted_k141_130182 | Low-quality |
| S1_extracted_k141_56737 | Not-determined |
| S1_extracted_k141_123241 | Not-determined |
| S1_extracted_k141_69839 | Not-determined |
| S1_extracted_k141_62168 | Not-determined |
| S1_extracted_k141_43524 | Not-determined |
| S1_extracted_k141_110061 | Low-quality |
| S1_extracted_k141_36084 | Low-quality |
| S1_extracted_k141_110064 | Low-quality |
| S1_extracted_k141_116890 | Not-determined |
| S1_extracted_k141_148361 | Not-determined |
| S1_extracted_k141_30075 | Low-quality |
| S1_extracted_k141_30077 | Not-determined |
| S1_extracted_k141_30079 | Not-determined |
| S1_extracted_k141_123249 | Low-quality |
| S1_extracted_k141_143309 | Not-determined |
| S1_extracted_k141_30082 | Not-determined |
| S1_extracted_k141_23119 | Low-quality |
| S1_extracted_k141_155517 | Not-determined |
| S1_extracted_k141_69844 | Not-determined |
| S1_extracted_k141_30084 | Not-determined |
| S1_extracted_k141_148363 | Not-determined |
| S1_extracted_k141_17244 | Not-determined |
| S1_extracted_k141_50276 | Not-determined |
| S1_extracted_k141_136655 | Not-determined |
| S1_extracted_k141_90008 | Not-determined |
| S1_extracted_k141_155521 | Low-quality |
| S1_extracted_k141_56753 | Not-determined |
| S1_extracted_k141_17248 | Not-determined |
| S1_extracted_k141_103586 | Low-quality |
| S1_extracted_k141_50278 | Not-determined |
| S1_extracted_k141_155525 | Low-quality |
| S1_extracted_k141_110077 | Not-determined |
| S1_extracted_k141_116905 | Not-determined |
| S1_extracted_k141_62189 | Not-determined |

| | |
|---|---|
| S1_extracted_k141_36095 | Low-quality |
| S1_extracted_k141_62190 | Not-determined |
| S1_extracted_k141_56760 | Low-quality |
| S1_extracted_k141_76802 | Not-determined |
| S1_extracted_k141_143319 | Not-determined |
| S1_extracted_k141_23127 | Not-determined |
| S1_extracted_k141_50287 | Not-determined |
| S1_extracted_k141_110083 | Low-quality |
| S1_extracted_k141_69853 | Not-determined |
| S1_extracted_k141_36100 | Not-determined |
| S1_extracted_k141_50288 | Not-determined |
| S1_extracted_k141_23133 | Not-determined |
| S1_extracted_k141_69856 | Not-determined |
| S1_extracted_k141_110085 | Low-quality |
| S1_extracted_k141_143328 | Not-determined |
| S1_extracted_k141_123265 | Not-determined |
| S1_extracted_k141_96970 | Not-determined |
| S1_extracted_k141_50292 | Not-determined |
| S1_extracted_k141_96971 | Not-determined |
| S1_extracted_k141_50295 | Not-determined |
| S1_extracted_k141_76816 | Not-determined |
| S1_extracted_k141_56776 | Not-determined |
| S1_extracted_k141_2809 | Not-determined |
| S1_extracted_k141_130193 | Low-quality |
| S1_extracted_k141_136682 | Not-determined |
| S1_extracted_k141_17249 | Low-quality |
| S1_extracted_k141_10199 | Low-quality |
| S1_extracted_k141_148372 | Low-quality |
| S1_extracted_k141_50299 | Not-determined |
| S1_extracted_k141_148373 | Low-quality |
| S1_extracted_k141_90031 | Not-determined |
| S1_extracted_k141_90032 | Low-quality |
| S1_extracted_k141_17254 | Not-determined |
| S1_extracted_k141_96976 | Not-determined |
| S1_extracted_k141_143339 | Not-determined |
| S1_extracted_k141_83586 | Not-determined |
| S1_extracted_k141_83588 | Not-determined |
| S1_extracted_k141_155533 | Not-determined |
| S1_extracted_k141_123274 | Not-determined |
| S1_extracted_k141_56784 | Not-determined |
| S1_extracted_k141_17258 | Not-determined |
| S1_extracted_k141_103612 | Not-determined |
| S1_extracted_k141_76829 | Not-determined |
| S1_extracted_k141_10210 | Not-determined |
| S1_extracted_k141_83591 | Not-determined |
| S1_extracted_k141_103613 | Not-determined |
| S1_extracted_k141_17261 | Not-determined |
| S1_extracted_k141_148378 | Low-quality |
| S1_extracted_k141_136696 | Not-determined |
| S1_extracted_k141_36116 | Not-determined |
| S1_extracted_k141_103614 | Low-quality |
| S1_extracted_k141_155535 | Not-determined |
| S1_extracted_k141_76835 | Not-determined |
| S1_extracted_k141_90042 | Not-determined |
| S1_extracted_k141_56791 | Not-determined |
| S1_extracted_k141_96987 | Not-determined |
| S1_extracted_k141_69865 | Low-quality |
| S1_extracted_k141_23157 | Not-determined |
| S1_extracted_k141_103619 | Not-determined |
| S1_extracted_k141_123283 | Not-determined |
| S1_extracted_k141_136706 | Not-determined |
| S1_extracted_k141_136707 | Not-determined |
| S1_extracted_k141_76841 | Not-determined |
| S1_extracted_k141_23160 | Low-quality |
| S1_extracted_k141_155542 | Low-quality |
| S1_extracted_k141_69867 | Low-quality |
| S1_extracted_k141_136709 | Not-determined |
| S1_extracted_k141_103623 | Not-determined |
| S1_extracted_k141_83601 | Not-determined |
| S1_extracted_k141_130206 | Not-determined |
| S1_extracted_k141_130208 | Not-determined |
| S1_extracted_k141_83603 | Low-quality |
| S1_extracted_k141_50306 | Low-quality |
| S1_extracted_k141_155545 | Not-determined |
| S1_extracted_k141_23163 | Low-quality |
| S1_extracted_k141_90043 | Low-quality |
| S1_extracted_k141_123287 | Not-determined |
| S1_extracted_k141_96994 | Not-determined |
| S1_extracted_k141_69873 | Low-quality |
| S1_extracted_k141_136717 | Not-determined |
| S1_extracted_k141_116941 | Not-determined |
| S1_extracted_k141_43546 | Low-quality |
| S1_extracted_k141_96995 | Not-determined |
| S1_extracted_k141_17279 | Not-determined |
| S1_extracted_k141_123291 | Low-quality |
| S1_extracted_k141_83610 | Not-determined |
| S1_extracted_k141_23171 | Not-determined |
| S1_extracted_k141_123294 | Not-determined |
| S1_extracted_k141_43548 | Not-determined |
| S1_extracted_k141_116946 | Low-quality |
| S1_extracted_k141_50319 | Not-determined |
| S1_extracted_k141_90052 | Low-quality |
| S1_extracted_k141_83615 | Not-determined |
| S1_extracted_k141_30126 | Low-quality |
| S1_extracted_k141_130219 | Not-determined |
| S1_extracted_k141_69884 | Not-determined |
| S1_extracted_k141_155550 | Not-determined |
| S1_extracted_k141_155551 | Not-determined |
| S1_extracted_k141_30130 | Not-determined |
| S1_extracted_k141_123302 | Not-determined |
| S1_extracted_k141_69887 | Not-determined |
| S1_extracted_k141_76856 | Low-quality |
| S1_extracted_k141_43554 | Low-quality |
| S1_extracted_k141_116957 | Not-determined |
| S1_extracted_k141_69889 | Not-determined |
| S1_extracted_k141_30134 | Not-determined |
| S1_extracted_k141_123312 | Not-determined |
| S1_extracted_k141_2830 | Not-determined |
| S1_extracted_k141_123314 | Low-quality |
| S1_extracted_k141_56818 | Low-quality |
| S1_extracted_k141_130230 | Not-determined |
| S1_extracted_k141_62232 | Not-determined |
| S1_extracted_k141_103645 | Not-determined |
| S1_extracted_k141_123318 | Not-determined |
| S1_extracted_k141_143359 | Low-quality |
| S1_extracted_k141_2836 | Not-determined |
| S1_extracted_k141_56825 | Not-determined |
| S1_extracted_k141_10235 | Not-determined |
| S1_extracted_k141_103648 | Not-determined |
| S1_extracted_k141_148390 | Not-determined |
| S1_extracted_k141_123323 | Not-determined |
| S1_extracted_k141_97017 | Low-quality |
| S1_extracted_k141_123325 | Not-determined |
| S1_extracted_k141_62237 | Not-determined |
| S1_extracted_k141_17302 | Not-determined |
| S1_extracted_k141_2838 | Not-determined |
| S1_extracted_k141_17304 | Not-determined |
| S1_extracted_k141_143367 | Not-determined |
| S1_extracted_k141_2841 | Not-determined |
| S1_extracted_k141_76868 | Not-determined |
| S1_extracted_k141_43567 | Not-determined |
| S1_extracted_k141_90059 | Not-determined |
| S1_extracted_k141_62245 | Not-determined |
| S1_extracted_k141_116976 | Not-determined |
| S1_extracted_k141_116977 | Not-determined |
| S1_extracted_k141_123326 | Not-determined |
| S1_extracted_k141_97027 | Not-determined |
| S1_extracted_k141_90062 | Not-determined |
| S1_extracted_k141_97028 | Not-determined |
| S1_extracted_k141_97029 | Low-quality |
| S1_extracted_k141_136751 | Not-determined |
| S1_extracted_k141_23213 | Not-determined |
| S1_extracted_k141_116985 | Not-determined |
| S1_extracted_k141_143380 | Not-determined |
| S1_extracted_k141_76880 | Not-determined |
| S1_extracted_k141_69895 | Not-determined |
| S1_extracted_k141_10253 | Not-determined |
| S1_extracted_k141_30163 | Not-determined |
| S1_extracted_k141_136760 | Low-quality |
| S1_extracted_k141_116988 | Low-quality |
| S1_extracted_k141_62262 | Not-determined |
| S1_extracted_k141_62263 | Not-determined |
| S1_extracted_k141_143388 | Not-determined |
| S1_extracted_k141_43582 | Not-determined |
| S1_extracted_k141_62267 | Low-quality |
| S1_extracted_k141_76886 | Not-determined |
| S1_extracted_k141_116992 | Not-determined |
| S1_extracted_k141_62269 | Not-determined |
| S1_extracted_k141_123340 | Not-determined |
| S1_extracted_k141_2855 | Not-determined |
| S1_extracted_k141_123341 | Low-quality |
| S1_extracted_k141_110138 | Not-determined |
| S1_extracted_k141_43587 | Not-determined |
| S1_extracted_k141_116997 | Not-determined |
| S1_extracted_k141_56848 | Not-determined |
| S1_extracted_k141_90076 | Not-determined |
| S1_extracted_k141_136764 | Not-determined |
| S1_extracted_k141_155582 | Not-determined |
| S1_extracted_k141_90077 | Not-determined |
| S1_extracted_k141_123347 | Not-determined |
| S1_extracted_k141_23234 | Low-quality |
| S1_extracted_k141_36130 | Not-determined |
| S1_extracted_k141_103657 | Low-quality |
| S1_extracted_k141_117001 | Low-quality |
| S1_extracted_k141_30179 | Not-determined |
| S1_extracted_k141_90082 | Not-determined |
| S1_extracted_k141_2862 | Not-determined |
| S1_extracted_k141_123350 | Not-determined |
| S1_extracted_k141_97045 | Not-determined |
| S1_extracted_k141_110145 | Low-quality |
| S1_extracted_k141_97046 | Not-determined |
| S1_extracted_k141_143399 | Not-determined |
| S1_extracted_k141_117008 | Not-determined |
| S1_extracted_k141_56859 | Not-determined |
| S1_extracted_k141_97049 | Not-determined |
| S1_extracted_k141_90090 | Not-determined |
| S1_extracted_k141_143400 | Not-determined |
| S1_extracted_k141_103662 | Low-quality |
| S1_extracted_k141_90091 | Not-determined |
| S1_extracted_k141_83676 | Low-quality |
| S1_extracted_k141_76899 | Not-determined |
| S1_extracted_k141_10278 | Not-determined |
| S1_extracted_k141_103665 | Not-determined |
| S1_extracted_k141_10279 | Low-quality |
| S1_extracted_k141_30189 | Not-determined |
| S1_extracted_k141_56866 | Low-quality |
| S1_extracted_k141_10281 | Not-determined |
| S1_extracted_k141_117015 | Not-determined |
| S1_extracted_k141_43599 | Low-quality |
| S1_extracted_k141_143408 | Not-determined |
| S1_extracted_k141_117017 | Not-determined |
| S1_extracted_k141_155588 | Not-determined |
| S1_extracted_k141_62289 | Not-determined |
| S1_extracted_k141_117020 | Not-determined |
| S1_extracted_k141_90094 | Not-determined |
| S1_extracted_k141_10285 | Low-quality |
| S1_extracted_k141_62290 | Not-determined |
| S1_extracted_k141_83680 | Not-determined |
| S1_extracted_k141_123356 | Not-determined |
| S1_extracted_k141_23248 | Not-determined |
| S1_extracted_k141_17326 | Not-determined |
| S1_extracted_k141_148411 | Low-quality |
| S1_extracted_k141_143415 | Low-quality |
| S1_extracted_k141_130252 | Not-determined |
| S1_extracted_k141_56874 | Not-determined |
| S1_extracted_k141_76915 | Not-determined |
| S1_extracted_k141_90103 | Low-quality |
| S1_extracted_k141_130255 | Not-determined |
| S1_extracted_k141_50335 | Low-quality |
| S1_extracted_k141_56879 | Low-quality |
| S1_extracted_k141_17330 | Low-quality |
| S1_extracted_k141_23257 | Not-determined |
| S1_extracted_k141_83686 | Low-quality |
| S1_extracted_k141_83689 | Low-quality |
| S1_extracted_k141_56885 | Not-determined |
| S1_extracted_k141_17334 | Not-determined |
| S1_extracted_k141_76923 | Not-determined |
| S1_extracted_k141_123368 | Not-determined |
| S1_extracted_k141_155603 | Not-determined |
| S1_extracted_k141_23259 | Low-quality |
| S1_extracted_k141_10297 | Not-determined |
| S1_extracted_k141_123369 | Not-determined |
| S1_extracted_k141_143425 | Not-determined |
| S1_extracted_k141_50342 | Not-determined |
| S1_extracted_k141_76925 | Not-determined |
| S1_extracted_k141_143427 | Low-quality |
| S1_extracted_k141_17339 | Not-determined |
| S1_extracted_k141_23263 | Not-determined |
| S1_extracted_k141_117029 | Not-determined |
| S1_extracted_k141_148417 | Not-determined |
| S1_extracted_k141_155609 | Not-determined |
| S1_extracted_k141_36152 | Not-determined |
| S1_extracted_k141_17342 | Not-determined |
| S1_extracted_k141_2888 | Low-quality |
| S1_extracted_k141_43612 | Low-quality |
| S1_extracted_k141_2890 | Not-determined |
| S1_extracted_k141_155612 | Not-determined |
| S1_extracted_k141_110177 | Not-determined |
| S1_extracted_k141_43616 | Not-determined |
| S1_extracted_k141_136793 | Not-determined |
| S1_extracted_k141_56898 | Not-determined |
| S1_extracted_k141_103694 | Not-determined |
| S1_extracted_k141_43618 | Not-determined |
| S1_extracted_k141_36159 | Not-determined |
| S1_extracted_k141_103696 | Not-determined |
| S1_extracted_k141_136796 | Not-determined |
| S1_extracted_k141_117040 | Not-determined |
| S1_extracted_k141_2896 | Not-determined |
| S1_extracted_k141_117042 | Not-determined |
| S1_extracted_k141_43622 | Not-determined |
| S1_extracted_k141_56901 | Low-quality |
| S1_extracted_k141_155619 | Low-quality |
| S1_extracted_k141_10313 | Not-determined |
| S1_extracted_k141_155621 | Not-determined |
| S1_extracted_k141_56905 | Not-determined |
| S1_extracted_k141_56906 | Not-determined |
| S1_extracted_k141_10316 | Not-determined |
| S1_extracted_k141_23268 | Low-quality |
| S1_extracted_k141_69916 | Not-determined |
| S1_extracted_k141_117048 | Not-determined |
| S1_extracted_k141_123387 | Not-determined |
| S1_extracted_k141_23271 | Not-determined |
| S1_extracted_k141_123388 | Not-determined |
| S1_extracted_k141_123389 | Not-determined |
| S1_extracted_k141_69917 | Not-determined |
| S1_extracted_k141_117052 | Not-determined |
| S1_extracted_k141_62311 | Not-determined |
| S1_extracted_k141_50366 | Not-determined |
| S1_extracted_k141_148421 | Low-quality |
| S1_extracted_k141_136811 | Not-determined |
| S1_extracted_k141_103709 | Low-quality |
| S1_extracted_k141_123395 | Not-determined |
| S1_extracted_k141_83707 | Not-determined |
| S1_extracted_k141_30215 | Not-determined |
| S1_extracted_k141_110191 | Not-determined |
| S1_extracted_k141_17360 | Not-determined |
| S1_extracted_k141_23277 | Low-quality |
| S1_extracted_k141_36161 | Not-determined |
| S1_extracted_k141_23278 | Not-determined |
| S1_extracted_k141_23280 | Not-determined |
| S1_extracted_k141_110192 | Not-determined |
| S1_extracted_k141_43633 | Low-quality |
| S1_extracted_k141_56911 | Not-determined |
| S1_extracted_k141_148424 | Not-determined |
| S1_extracted_k141_117057 | Not-determined |
| S1_extracted_k141_103713 | Not-determined |
| S1_extracted_k141_83713 | Not-determined |
| S1_extracted_k141_117064 | Not-determined |
| S1_extracted_k141_43640 | Not-determined |
| S1_extracted_k141_83718 | Not-determined |
| S1_extracted_k141_23287 | Not-determined |
| S1_extracted_k141_143443 | Not-determined |
| S1_extracted_k141_90137 | Not-determined |
| S1_extracted_k141_2906 | Not-determined |
| S1_extracted_k141_123403 | Low-quality |
| S1_extracted_k141_76969 | Not-determined |
| S1_extracted_k141_90140 | Not-determined |
| S1_extracted_k141_50380 | Not-determined |
| S1_extracted_k141_36172 | Not-determined |
| S1_extracted_k141_2910 | Not-determined |
| S1_extracted_k141_30234 | Not-determined |
| S1_extracted_k141_83723 | Low-quality |
| S1_extracted_k141_117072 | Not-determined |
| S1_extracted_k141_62329 | Not-determined |
| S1_extracted_k141_130307 | Low-quality |
| S1_extracted_k141_117074 | Not-determined |
| S1_extracted_k141_10339 | Not-determined |
| S1_extracted_k141_117075 | Not-determined |
| S1_extracted_k141_43652 | Not-determined |
| S1_extracted_k141_110205 | Low-quality |
| S1_extracted_k141_2913 | Not-determined |
| S1_extracted_k141_76974 | Low-quality |
| S1_extracted_k141_36175 | Low-quality |
| S1_extracted_k141_83728 | Low-quality |
| S1_extracted_k141_90144 | Not-determined |
| S1_extracted_k141_2915 | Not-determined |
| S1_extracted_k141_148435 | Not-determined |
| S1_extracted_k141_83730 | Not-determined |
| S1_extracted_k141_130315 | Not-determined |
| S1_extracted_k141_56930 | Not-determined |
| S1_extracted_k141_36178 | Not-determined |
| S1_extracted_k141_43655 | Not-determined |
| S1_extracted_k141_136825 | Not-determined |
| S1_extracted_k141_123417 | Not-determined |
| S1_extracted_k141_155625 | Not-determined |
| S1_extracted_k141_62340 | Not-determined |
| S1_extracted_k141_10346 | Not-determined |
| S1_extracted_k141_76981 | Not-determined |
| S1_extracted_k141_97101 | Not-determined |
| S1_extracted_k141_10347 | Not-determined |
| S1_extracted_k141_50392 | Not-determined |
| S1_extracted_k141_76983 | Not-determined |
| S1_extracted_k141_36182 | Not-determined |
| S1_extracted_k141_76984 | Not-determined |
| S1_extracted_k141_10350 | Not-determined |
| S1_extracted_k141_2924 | Low-quality |
| S1_extracted_k141_10354 | Low-quality |
| S1_extracted_k141_148441 | Not-determined |
| S1_extracted_k141_110221 | Not-determined |
| S1_extracted_k141_43660 | Not-determined |
| S1_extracted_k141_23310 | Not-determined |
| S1_extracted_k141_90155 | Not-determined |
| S1_extracted_k141_50403 | Low-quality |
| S1_extracted_k141_76993 | Not-determined |
| S1_extracted_k141_83743 | Not-determined |
| S1_extracted_k141_103740 | Not-determined |
| S1_extracted_k141_130328 | Not-determined |
| S1_extracted_k141_148446 | Not-determined |
| S1_extracted_k141_43665 | Not-determined |
| S1_extracted_k141_36189 | Not-determined |
| S1_extracted_k141_50409 | Not-determined |
| S1_extracted_k141_136847 | Low-quality |
| S1_extracted_k141_56943 | Low-quality |
| S1_extracted_k141_136848 | Not-determined |
| S1_extracted_k141_97109 | Low-quality |
| S1_extracted_k141_83750 | Low-quality |
| S1_extracted_k141_30264 | Not-determined |
| S1_extracted_k141_123428 | Low-quality |
| S1_extracted_k141_97114 | Not-determined |
| S1_extracted_k141_17400 | Not-determined |
| S1_extracted_k141_10369 | Not-determined |
| S1_extracted_k141_90164 | Not-determined |
| S1_extracted_k141_83756 | Not-determined |
| S1_extracted_k141_17403 | Not-determined |
| S1_extracted_k141_110240 | Not-determined |
| S1_extracted_k141_117093 | Low-quality |
| S1_extracted_k141_50417 | Low-quality |
| S1_extracted_k141_117094 | Not-determined |
| S1_extracted_k141_2937 | Not-determined |
| S1_extracted_k141_50420 | Not-determined |
| S1_extracted_k141_90172 | Not-determined |
| S1_extracted_k141_17409 | Not-determined |
| S1_extracted_k141_10376 | Not-determined |
| S1_extracted_k141_83763 | Not-determined |
| S1_extracted_k141_56956 | Not-determined |
| S1_extracted_k141_90174 | Not-determined |
| S1_extracted_k141_30281 | Low-quality |
| S1_extracted_k141_90175 | Not-determined |
| S1_extracted_k141_36200 | Low-quality |
| S1_extracted_k141_90178 | Not-determined |
| S1_extracted_k141_97120 | Low-quality |
| S1_extracted_k141_2944 | Low-quality |
| S1_extracted_k141_23335 | Not-determined |
| S1_extracted_k141_30284 | Not-determined |
| S1_extracted_k141_110247 | Not-determined |
| S1_extracted_k141_23338 | Not-determined |
| S1_extracted_k141_17415 | Not-determined |
| S1_extracted_k141_123439 | Not-determined |
| S1_extracted_k141_10385 | Not-determined |
| S1_extracted_k141_123440 | Not-determined |
| S1_extracted_k141_36204 | Not-determined |
| S1_extracted_k141_97127 | Not-determined |
| S1_extracted_k141_136867 | Not-determined |
| S1_extracted_k141_155651 | Not-determined |
| S1_extracted_k141_136868 | Not-determined |
| S1_extracted_k141_136869 | Not-determined |
| S1_extracted_k141_30287 | Not-determined |
| S1_extracted_k141_30288 | Not-determined |
| S1_extracted_k141_56972 | Not-determined |
| S1_extracted_k141_123445 | Low-quality |
| S1_extracted_k141_50434 | Not-determined |
| S1_extracted_k141_117108 | Low-quality |
| S1_extracted_k141_23344 | Not-determined |
| S1_extracted_k141_23345 | Not-determined |
| S1_extracted_k141_56974 | Not-determined |
| S1_extracted_k141_10393 | Not-determined |
| S1_extracted_k141_136872 | Not-determined |
| S1_extracted_k141_10395 | Not-determined |
| S1_extracted_k141_155656 | Not-determined |
| S1_extracted_k141_77012 | Not-determined |
| S1_extracted_k141_2954 | Not-determined |
| S1_extracted_k141_62357 | Not-determined |
| S1_extracted_k141_97131 | Not-determined |
| S1_extracted_k141_130355 | Not-determined |
| S1_extracted_k141_123449 | Not-determined |
| S1_extracted_k141_50439 | Not-determined |
| S1_extracted_k141_148466 | Not-determined |
| S1_extracted_k141_77015 | Not-determined |
| S1_extracted_k141_62359 | Not-determined |
| S1_extracted_k141_123451 | Not-determined |
| S1_extracted_k141_143461 | Low-quality |
| S1_extracted_k141_17426 | Not-determined |
| S1_extracted_k141_43698 | Low-quality |
| S1_extracted_k141_77018 | Not-determined |
| S1_extracted_k141_62364 | Not-determined |
| S1_extracted_k141_110253 | Not-determined |
| S1_extracted_k141_136883 | Not-determined |
| S1_extracted_k141_36216 | Not-determined |
| S1_extracted_k141_56988 | Low-quality |
| S1_extracted_k141_110258 | Not-determined |
| S1_extracted_k141_136887 | Not-determined |
| S1_extracted_k141_43703 | Not-determined |
| S1_extracted_k141_123457 | Not-determined |
| S1_extracted_k141_17435 | Low-quality |
| S1_extracted_k141_43705 | Low-quality |
| S1_extracted_k141_155663 | Not-determined |
| S1_extracted_k141_36218 | Not-determined |
| S1_extracted_k141_143474 | Not-determined |
| S1_extracted_k141_83786 | Low-quality |
| S1_extracted_k141_143475 | Not-determined |
| S1_extracted_k141_77022 | Not-determined |
| S1_extracted_k141_23355 | Not-determined |
| S1_extracted_k141_43709 | Not-determined |
| S1_extracted_k141_123460 | Low-quality |
| S1_extracted_k141_77024 | Not-determined |
| S1_extracted_k141_23358 | Not-determined |
| S1_extracted_k141_123462 | Not-determined |
| S1_extracted_k141_23359 | Not-determined |
| S1_extracted_k141_148472 | Low-quality |
| S1_extracted_k141_2969 | Low-quality |
| S1_extracted_k141_30305 | Not-determined |
| S1_extracted_k141_2970 | Low-quality |
| S1_extracted_k141_10397 | Not-determined |
| S1_extracted_k141_69934 | Not-determined |
| S1_extracted_k141_62383 | Not-determined |
| S1_extracted_k141_117123 | Not-determined |
| S1_extracted_k141_136900 | Not-determined |
| S1_extracted_k141_69936 | Not-determined |
| S1_extracted_k141_97149 | Not-determined |
| S1_extracted_k141_148481 | Not-determined |
| S1_extracted_k141_10402 | Not-determined |
| S1_extracted_k141_69939 | Not-determined |
| S1_extracted_k141_148482 | Not-determined |
| S1_extracted_k141_36226 | Not-determined |
| S1_extracted_k141_77042 | Low-quality |
| S1_extracted_k141_136901 | Low-quality |
| S1_extracted_k141_123481 | Not-determined |
| S1_extracted_k141_10408 | Not-determined |
| S1_extracted_k141_136902 | Not-determined |
| S1_extracted_k141_123482 | Not-determined |
| S1_extracted_k141_110280 | Low-quality |
| S1_extracted_k141_117126 | Low-quality |
| S1_extracted_k141_23374 | Not-determined |
| S1_extracted_k141_23376 | Not-determined |
| S1_extracted_k141_50468 | Not-determined |
| S1_extracted_k141_57011 | Low-quality |
| S1_extracted_k141_117130 | Low-quality |
| S1_extracted_k141_123485 | Not-determined |
| S1_extracted_k141_148488 | Not-determined |
| S1_extracted_k141_57012 | Not-determined |
| S1_extracted_k141_2973 | Low-quality |
| S1_extracted_k141_23377 | Not-determined |
| S1_extracted_k141_10412 | Not-determined |
| S1_extracted_k141_2974 | Not-determined |
| S1_extracted_k141_23378 | Not-determined |
| S1_extracted_k141_83787 | Low-quality |
| S1_extracted_k141_117132 | Not-determined |
| S1_extracted_k141_103761 | Not-determined |
| S1_extracted_k141_57015 | Low-quality |
| S1_extracted_k141_143496 | Not-determined |
| S1_extracted_k141_130373 | Low-quality |
| S1_extracted_k141_148492 | Low-quality |
| S1_extracted_k141_123490 | Low-quality |
| S1_extracted_k141_110290 | Low-quality |
| S1_extracted_k141_136910 | Not-determined |
| S1_extracted_k141_117137 | Not-determined |
| S1_extracted_k141_130375 | Not-determined |
| S1_extracted_k141_97168 | Not-determined |
| S1_extracted_k141_17454 | Not-determined |
| S1_extracted_k141_155673 | Not-determined |
| S1_extracted_k141_148495 | Not-determined |
| S1_extracted_k141_136914 | Not-determined |
| S1_extracted_k141_148497 | Not-determined |
| S1_extracted_k141_83792 | Not-determined |
| S1_extracted_k141_43731 | Not-determined |
| S1_extracted_k141_136918 | Not-determined |
| S1_extracted_k141_136919 | Not-determined |
| S1_extracted_k141_10420 | Low-quality |
| S1_extracted_k141_83797 | Not-determined |
| S1_extracted_k141_43732 | Not-determined |
| S1_extracted_k141_97169 | Not-determined |
| S1_extracted_k141_117142 | Low-quality |
| S1_extracted_k141_17465 | Not-determined |
| S1_extracted_k141_83802 | Not-determined |
| S1_extracted_k141_57031 | Not-determined |
| S1_extracted_k141_136926 | Not-determined |
| S1_extracted_k141_155679 | Low-quality |
| S1_extracted_k141_10426 | Low-quality |
| S1_extracted_k141_57034 | Not-determined |
| S1_extracted_k141_123498 | Not-determined |
| S1_extracted_k141_110301 | Not-determined |
| S1_extracted_k141_136932 | Not-determined |
| S1_extracted_k141_90230 | Not-determined |
| S1_extracted_k141_103781 | Not-determined |
| S1_extracted_k141_103782 | Not-determined |
| S1_extracted_k141_57038 | Not-determined |
| S1_extracted_k141_23392 | Not-determined |
| S1_extracted_k141_43740 | Low-quality |
| S1_extracted_k141_30329 | Not-determined |
| S1_extracted_k141_136938 | Not-determined |
| S1_extracted_k141_30334 | Not-determined |
| S1_extracted_k141_83810 | Not-determined |
| S1_extracted_k141_83811 | Not-determined |
| S1_extracted_k141_30336 | Not-determined |
| S1_extracted_k141_110312 | Not-determined |
| S1_extracted_k141_123504 | Not-determined |
| S1_extracted_k141_30339 | Not-determined |
| S1_extracted_k141_43749 | Not-determined |
| S1_extracted_k141_136943 | Not-determined |
| S1_extracted_k141_43750 | Not-determined |
| S1_extracted_k141_123508 | Low-quality |
| S1_extracted_k141_17476 | Not-determined |
| S1_extracted_k141_136946 | Low-quality |
| S1_extracted_k141_110316 | Low-quality |
| S1_extracted_k141_103795 | Not-determined |
| S1_extracted_k141_97195 | Not-determined |
| S1_extracted_k141_43755 | Not-determined |
| S1_extracted_k141_130399 | Low-quality |
| S1_extracted_k141_43757 | Not-determined |
| S1_extracted_k141_23406 | Not-determined |
| S1_extracted_k141_23407 | Not-determined |
| S1_extracted_k141_50514 | Not-determined |
| S1_extracted_k141_90242 | Not-determined |
| S1_extracted_k141_36252 | Not-determined |
| S1_extracted_k141_103804 | Not-determined |
| S1_extracted_k141_36253 | Not-determined |
| S1_extracted_k141_30351 | Not-determined |
| S1_extracted_k141_130402 | Not-determined |
| S1_extracted_k141_97204 | Not-determined |
| S1_extracted_k141_23410 | Low-quality |
| S1_extracted_k141_97205 | Not-determined |
| S1_extracted_k141_77093 | Not-determined |
| S1_extracted_k141_30353 | Not-determined |
| S1_extracted_k141_155688 | Not-determined |
| S1_extracted_k141_50517 | Not-determined |
| S1_extracted_k141_97207 | Low-quality |
| S1_extracted_k141_136952 | Not-determined |
| S1_extracted_k141_117173 | Not-determined |
| S1_extracted_k141_90248 | Not-determined |
| S1_extracted_k141_97208 | Not-determined |
| S1_extracted_k141_123515 | Not-determined |
| S1_extracted_k141_136955 | Low-quality |
| S1_extracted_k141_123516 | Not-determined |
| S1_extracted_k141_30359 | Low-quality |
| S1_extracted_k141_110324 | Not-determined |
| S1_extracted_k141_97212 | Not-determined |
| S1_extracted_k141_69964 | Not-determined |
| S1_extracted_k141_90251 | Not-determined |
| S1_extracted_k141_97214 | Not-determined |

| | |
|---|---|
| S1_extracted_k141_69966 | Not-determined |
| S1_extracted_k141_123518 | Not-determined |
| S1_extracted_k141_97217 | Not-determined |
| S1_extracted_k141_130411 | Low-quality |
| S1_extracted_k141_136961 | Not-determined |
| S1_extracted_k141_50526 | Low-quality |
| S1_extracted_k141_103814 | Not-determined |
| S1_extracted_k141_130412 | Not-determined |
| S1_extracted_k141_123524 | Not-determined |
| S1_extracted_k141_23424 | Low-quality |
| S1_extracted_k141_90256 | Low-quality |
| S1_extracted_k141_103817 | Not-determined |
| S1_extracted_k141_17495 | Not-determined |
| S1_extracted_k141_110331 | Not-determined |
| S1_extracted_k141_110333 | Not-determined |
| S1_extracted_k141_117183 | Not-determined |
| S1_extracted_k141_17500 | Not-determined |
| S1_extracted_k141_117186 | Not-determined |
| S1_extracted_k141_23432 | Not-determined |
| S1_extracted_k141_90264 | Not-determined |
| S1_extracted_k141_97229 | Not-determined |
| S1_extracted_k141_50528 | Not-determined |
| S1_extracted_k141_69978 | Not-determined |
| S1_extracted_k141_123527 | Not-determined |
| S1_extracted_k141_123528 | Not-determined |
| S1_extracted_k141_143515 | Low-quality |
| S1_extracted_k141_17509 | Not-determined |
| S1_extracted_k141_130426 | Low-quality |
| S1_extracted_k141_10478 | Not-determined |
| S1_extracted_k141_50535 | Low-quality |
| S1_extracted_k141_30384 | Not-determined |
| S1_extracted_k141_110341 | Not-determined |
| S1_extracted_k141_62432 | Not-determined |
| S1_extracted_k141_117192 | Not-determined |
| S1_extracted_k141_97233 | Not-determined |
| S1_extracted_k141_50540 | Not-determined |
| S1_extracted_k141_143518 | Low-quality |
| S1_extracted_k141_97235 | Low-quality |
| S1_extracted_k141_130432 | Low-quality |
| S1_extracted_k141_30387 | Not-determined |
| S1_extracted_k141_143520 | Not-determined |
| S1_extracted_k141_97239 | Not-determined |
| S1_extracted_k141_90272 | Not-determined |
| S1_extracted_k141_23442 | Not-determined |
| S1_extracted_k141_50543 | Not-determined |
| S1_extracted_k141_23443 | Not-determined |
| S1_extracted_k141_123538 | Not-determined |
| S1_extracted_k141_103835 | Low-quality |
| S1_extracted_k141_90276 | Not-determined |
| S1_extracted_k141_62440 | Not-determined |
| S1_extracted_k141_117198 | Low-quality |
| S1_extracted_k141_97241 | Not-determined |
| S1_extracted_k141_30391 | Not-determined |
| S1_extracted_k141_148528 | Not-determined |
| S1_extracted_k141_123542 | Not-determined |
| S1_extracted_k141_117200 | Not-determined |
| S1_extracted_k141_136975 | Low-quality |
| S1_extracted_k141_123546 | Not-determined |
| S1_extracted_k141_143530 | Not-determined |
| S1_extracted_k141_90283 | Not-determined |
| S1_extracted_k141_136976 | Not-determined |
| S1_extracted_k141_62446 | Not-determined |
| S1_extracted_k141_97243 | Not-determined |
| S1_extracted_k141_123551 | Not-determined |
| S1_extracted_k141_90288 | Not-determined |
| S1_extracted_k141_117209 | Not-determined |
| S1_extracted_k141_110361 | Not-determined |
| S1_extracted_k141_17524 | Not-determined |
| S1_extracted_k141_36278 | Low-quality |
| S1_extracted_k141_23455 | Not-determined |
| S1_extracted_k141_62449 | Low-quality |
| S1_extracted_k141_130452 | Not-determined |
| S1_extracted_k141_155699 | Not-determined |
| S1_extracted_k141_117213 | Low-quality |
| S1_extracted_k141_155700 | Not-determined |
| S1_extracted_k141_103845 | Not-determined |
| S1_extracted_k141_117214 | Not-determined |
| S1_extracted_k141_36281 | Not-determined |
| S1_extracted_k141_155701 | Not-determined |
| S1_extracted_k141_23461 | Not-determined |
| S1_extracted_k141_23462 | Not-determined |
| S1_extracted_k141_43767 | Not-determined |
| S1_extracted_k141_136984 | Not-determined |
| S1_extracted_k141_143539 | Low-quality |
| S1_extracted_k141_155703 | Not-determined |
| S1_extracted_k141_50555 | Low-quality |
| S1_extracted_k141_143541 | Not-determined |
| S1_extracted_k141_143542 | Not-determined |
| S1_extracted_k141_23465 | Low-quality |
| S1_extracted_k141_50557 | Not-determined |
| S1_extracted_k141_17538 | Not-determined |
| S1_extracted_k141_110365 | Low-quality |
| S1_extracted_k141_97255 | Not-determined |
| S1_extracted_k141_97256 | Low-quality |
| S1_extracted_k141_83853 | Low-quality |
| S1_extracted_k141_23468 | Not-determined |
| S1_extracted_k141_23469 | Not-determined |
| S1_extracted_k141_70006 | Low-quality |
| S1_extracted_k141_148541 | Not-determined |
| S1_extracted_k141_110370 | Not-determined |
| S1_extracted_k141_30402 | Not-determined |
| S1_extracted_k141_90302 | Low-quality |
| S1_extracted_k141_123560 | Not-determined |
| S1_extracted_k141_90303 | Not-determined |
| S1_extracted_k141_70008 | Not-determined |
| S1_extracted_k141_103851 | Not-determined |
| S1_extracted_k141_30405 | Not-determined |
| S1_extracted_k141_62457 | Not-determined |
| S1_extracted_k141_50566 | Not-determined |
| S1_extracted_k141_103853 | Not-determined |
| S1_extracted_k141_50568 | Not-determined |
| S1_extracted_k141_148542 | Not-determined |
| S1_extracted_k141_83860 | Not-determined |
| S1_extracted_k141_143548 | Not-determined |
| S1_extracted_k141_62461 | Not-determined |
| S1_extracted_k141_97266 | Not-determined |
| S1_extracted_k141_103857 | Not-determined |
| S1_extracted_k141_83865 | Not-determined |
| S1_extracted_k141_17551 | Low-quality |
| S1_extracted_k141_57090 | Not-determined |
| S1_extracted_k141_97271 | Not-determined |
| S1_extracted_k141_62469 | Not-determined |
| S1_extracted_k141_43786 | Low-quality |
| S1_extracted_k141_83869 | Not-determined |
| S1_extracted_k141_83870 | Not-determined |
| S1_extracted_k141_57094 | Not-determined |
| S1_extracted_k141_50580 | Not-determined |
| S1_extracted_k141_148545 | Not-determined |
| S1_extracted_k141_117227 | Not-determined |
| S1_extracted_k141_103860 | Not-determined |
| S1_extracted_k141_10528 | Not-determined |
| S1_extracted_k141_123574 | Not-determined |
| S1_extracted_k141_43793 | Not-determined |
| S1_extracted_k141_97277 | Low-quality |
| S1_extracted_k141_97278 | Low-quality |
| S1_extracted_k141_30412 | Not-determined |
| S1_extracted_k141_43795 | Not-determined |
| S1_extracted_k141_110384 | Not-determined |
| S1_extracted_k141_103864 | Not-determined |
| S1_extracted_k141_97285 | Not-determined |
| S1_extracted_k141_117238 | Not-determined |
| S1_extracted_k141_77132 | Not-determined |
| S1_extracted_k141_30418 | Not-determined |
| S1_extracted_k141_155713 | Not-determined |
| S1_extracted_k141_103867 | Low-quality |
| S1_extracted_k141_117240 | Not-determined |
| S1_extracted_k141_110388 | Low-quality |
| S1_extracted_k141_30421 | Not-determined |
| S1_extracted_k141_103871 | Low-quality |
| S1_extracted_k141_70029 | Low-quality |
| S1_extracted_k141_30422 | Not-determined |
| S1_extracted_k141_90322 | Not-determined |
| S1_extracted_k141_130490 | Not-determined |
| S1_extracted_k141_103872 | Not-determined |
| S1_extracted_k141_57115 | Low-quality |
| S1_extracted_k141_117246 | Not-determined |
| S1_extracted_k141_103877 | Not-determined |
| S1_extracted_k141_30427 | Not-determined |
| S1_extracted_k141_17562 | Low-quality |
| S1_extracted_k141_155721 | Not-determined |
| S1_extracted_k141_57116 | Not-determined |
| S1_extracted_k141_43809 | Not-determined |
| S1_extracted_k141_77142 | Not-determined |
| S1_extracted_k141_10536 | Not-determined |
| S1_extracted_k141_103879 | Not-determined |
| S1_extracted_k141_123589 | Not-determined |
| S1_extracted_k141_117253 | Not-determined |
| S1_extracted_k141_117254 | Not-determined |
| S1_extracted_k141_83888 | Low-quality |
| S1_extracted_k141_77150 | Not-determined |
| S1_extracted_k141_77152 | Not-determined |
| S1_extracted_k141_103886 | Not-determined |
| S1_extracted_k141_77154 | Not-determined |
| S1_extracted_k141_43820 | Not-determined |
| S1_extracted_k141_103888 | Not-determined |
| S1_extracted_k141_123597 | Not-determined |
| S1_extracted_k141_103891 | Not-determined |
| S1_extracted_k141_77158 | Not-determined |
| S1_extracted_k141_155722 | Not-determined |
| S1_extracted_k141_117259 | Not-determined |
| S1_extracted_k141_103892 | Not-determined |
| S1_extracted_k141_143576 | Not-determined |
| S1_extracted_k141_103893 | Not-determined |
| S1_extracted_k141_43825 | Not-determined |
| S1_extracted_k141_43826 | Not-determined |
| S1_extracted_k141_77161 | Not-determined |
| S1_extracted_k141_30432 | Not-determined |
| S1_extracted_k141_137026 | Not-determined |
| S1_extracted_k141_103895 | Not-determined |
| S1_extracted_k141_97303 | Not-determined |
| S1_extracted_k141_17576 | Low-quality |
| S1_extracted_k141_23525 | Not-determined |
| S1_extracted_k141_70037 | Not-determined |
| S1_extracted_k141_90341 | Not-determined |
| S1_extracted_k141_103900 | Not-determined |
| S1_extracted_k141_30441 | Not-determined |
| S1_extracted_k141_10553 | Not-determined |
| S1_extracted_k141_155727 | Not-determined |
| S1_extracted_k141_62501 | Not-determined |
| S1_extracted_k141_17581 | Not-determined |
| S1_extracted_k141_43835 | Not-determined |
| S1_extracted_k141_70043 | Not-determined |
| S1_extracted_k141_137034 | Not-determined |
| S1_extracted_k141_70046 | Not-determined |
| S1_extracted_k141_90352 | Not-determined |
| S1_extracted_k141_90354 | Not-determined |
| S1_extracted_k141_17585 | Not-determined |
| S1_extracted_k141_17587 | Not-determined |
| S1_extracted_k141_97313 | Not-determined |
| S1_extracted_k141_23533 | Not-determined |
| S1_extracted_k141_103906 | Low-quality |
| S1_extracted_k141_97315 | Not-determined |
| S1_extracted_k141_43844 | Not-determined |
| S1_extracted_k141_117274 | Not-determined |
| S1_extracted_k141_83901 | Low-quality |
| S1_extracted_k141_77174 | Low-quality |
| S1_extracted_k141_43846 | Not-determined |
| S1_extracted_k141_23539 | Not-determined |
| S1_extracted_k141_117276 | Not-determined |
| S1_extracted_k141_83905 | Not-determined |
| S1_extracted_k141_97321 | Low-quality |
| S1_extracted_k141_10564 | Not-determined |
| S1_extracted_k141_83907 | Not-determined |
| S1_extracted_k141_43852 | Not-determined |
| S1_extracted_k141_90364 | Not-determined |
| S1_extracted_k141_30452 | Not-determined |
| S1_extracted_k141_10566 | Not-determined |
| S5_vs_k141_72532\|\|full | Medium-quality |
| S1_extracted_k141_83910 | Not-determined |
| S1_extracted_k141_123605 | Low-quality |
| S1_extracted_k141_43855 | Not-determined |
| S1_extracted_k141_90367 | Not-determined |
| S1_extracted_k141_23541 | Not-determined |
| S1_extracted_k141_70054 | Low-quality |
| S1_extracted_k141_90369 | Not-determined |
| S1_extracted_k141_137046 | Not-determined |
| S1_extracted_k141_36311 | Not-determined |
| S1_extracted_k141_57135 | Not-determined |
| S1_extracted_k141_143598 | Not-determined |
| S1_extracted_k141_110433 | Not-determined |
| S1_extracted_k141_123608 | Not-determined |
| S1_extracted_k141_103916 | Low-quality |
| S1_extracted_k141_36314 | Not-determined |
| S1_extracted_k141_143600 | Not-determined |
| S1_extracted_k141_155732 | Not-determined |
| S1_extracted_k141_36315 | Not-determined |
| S1_extracted_k141_143603 | Not-determined |
| S1_extracted_k141_57141 | Not-determined |
| S1_extracted_k141_143604 | Not-determined |
| S13_vs_64889\|\|full | High-quality |
| S1_extracted_k141_23550 | Not-determined |
| S1_extracted_k141_97325 | Not-determined |
| S1_extracted_k141_10572 | Not-determined |
| S1_extracted_k141_83917 | Not-determined |
| S1_extracted_k141_97326 | Not-determined |
| S1_extracted_k141_23553 | Not-determined |
| S1_extracted_k141_137059 | Not-determined |
| S1_extracted_k141_50605 | Not-determined |
| S1_extracted_k141_97330 | Low-quality |
| S1_extracted_k141_57143 | Not-determined |
| S1_extracted_k141_90375 | Not-determined |
| S1_extracted_k141_10578 | Not-determined |
| S1_extracted_k141_130534 | Not-determined |
| S1_extracted_k141_70060 | Not-determined |
| S1_extracted_k141_123617 | Low-quality |
| S1_extracted_k141_83923 | Not-determined |
| S1_extracted_k141_137068 | Not-determined |
| S1_extracted_k141_3004 | Not-determined |
| S1_extracted_k141_130538 | Not-determined |
| S1_extracted_k141_83926 | Not-determined |
| S1_extracted_k141_62529 | Low-quality |
| S1_extracted_k141_50616 | Not-determined |
| S1_extracted_k141_83927 | Not-determined |
| S1_extracted_k141_43873 | Not-determined |
| S1_extracted_k141_57157 | Not-determined |
| S1_extracted_k141_36324 | Low-quality |
| S1_extracted_k141_50620 | Low-quality |
| S1_extracted_k141_90385 | Not-determined |
| S1_extracted_k141_23563 | Low-quality |
| S1_extracted_k141_50622 | Not-determined |
| S1_extracted_k141_110458 | Not-determined |
| S1_extracted_k141_143614 | Low-quality |
| S1_extracted_k141_30484 | Low-quality |
| S1_extracted_k141_143615 | Not-determined |
| S1_extracted_k141_43878 | Not-determined |
| S1_extracted_k141_83932 | Not-determined |
| S1_extracted_k141_50625 | Not-determined |
| S1_extracted_k141_143617 | Not-determined |
| S1_extracted_k141_10592 | Not-determined |
| S1_extracted_k141_143618 | Not-determined |
| S1_extracted_k141_137080 | Not-determined |
| S1_extracted_k141_43881 | Not-determined |
| S1_extracted_k141_143619 | Not-determined |
| S1_extracted_k141_103931 | Not-determined |
| S1_extracted_k141_148579 | Not-determined |
| S1_extracted_k141_77199 | Not-determined |
| S1_extracted_k141_90397 | Not-determined |
| S1_extracted_k141_143623 | Not-determined |
| S1_extracted_k141_137084 | Not-determined |
| S1_extracted_k141_30492 | Low-quality |
| S1_extracted_k141_137088 | Not-determined |
| S1_extracted_k141_3007 | Not-determined |
| S1_extracted_k141_148581 | Not-determined |
| S1_extracted_k141_143627 | Low-quality |
| S1_extracted_k141_3009 | Not-determined |
| S1_extracted_k141_117310 | Low-quality |
| S1_extracted_k141_117311 | Not-determined |
| S1_extracted_k141_97349 | Not-determined |
| S1_extracted_k141_148583 | Not-determined |
| S1_extracted_k141_23573 | Not-determined |
| S1_extracted_k141_3014 | Not-determined |
| S1_extracted_k141_90405 | Not-determined |
| S1_extracted_k141_30501 | Not-determined |
| S1_extracted_k141_143632 | Not-determined |
| S1_extracted_k141_90407 | Low-quality |
| S1_extracted_k141_50648 | Not-determined |
| S1_extracted_k141_90410 | Not-determined |
| S1_extracted_k141_137106 | Not-determined |
| S1_extracted_k141_57173 | Not-determined |
| S1_extracted_k141_90413 | Not-determined |
| S1_extracted_k141_148585 | Not-determined |
| S1_extracted_k141_3021 | Not-determined |
| S1_extracted_k141_137110 | Not-determined |
| S1_extracted_k141_148586 | Not-determined |
| S1_extracted_k141_50651 | Not-determined |
| S1_extracted_k141_103940 | Not-determined |
| S1_extracted_k141_110485 | Not-determined |
| S1_extracted_k141_50655 | Not-determined |
| S1_extracted_k141_90420 | Not-determined |
| S1_extracted_k141_50659 | Not-determined |
| S1_extracted_k141_137116 | Not-determined |
| S1_extracted_k141_103946 | Not-determined |
| S1_extracted_k141_143642 | Not-determined |
| S1_extracted_k141_62552 | Not-determined |
| S1_extracted_k141_30514 | Low-quality |
| S1_extracted_k141_43909 | Not-determined |
| S1_extracted_k141_143647 | Not-determined |
| S1_extracted_k141_43910 | Not-determined |
| S1_extracted_k141_83966 | Not-determined |
| S1_extracted_k141_50663 | Not-determined |
| S1_extracted_k141_62556 | Not-determined |
| S1_extracted_k141_30518 | Low-quality |
| S1_extracted_k141_43913 | Not-determined |
| S1_extracted_k141_57187 | Not-determined |
| S1_extracted_k141_97371 | Low-quality |
| S1_extracted_k141_62558 | Not-determined |
| S1_extracted_k141_36344 | Not-determined |
| S1_extracted_k141_77210 | Not-determined |
| S1_extracted_k141_43917 | Not-determined |
| S1_extracted_k141_110503 | Not-determined |
| S1_extracted_k141_143657 | Not-determined |
| S1_extracted_k141_30523 | Not-determined |
| S1_extracted_k141_110505 | Not-determined |
| S1_extracted_k141_70073 | Not-determined |
| S1_extracted_k141_77213 | Not-determined |
| S1_extracted_k141_155753 | Not-determined |
| S1_extracted_k141_57196 | Not-determined |
| S1_extracted_k141_110510 | Not-determined |
| S1_extracted_k141_103964 | Not-determined |
| S1_extracted_k141_30533 | Low-quality |
| S1_extracted_k141_143666 | Not-determined |
| S1_extracted_k141_70077 | Not-determined |
| S1_extracted_k141_43928 | Not-determined |
| S1_extracted_k141_90436 | Not-determined |
| S1_extracted_k141_57205 | Not-determined |
| S1_extracted_k141_10634 | Not-determined |
| S1_extracted_k141_57207 | Not-determined |
| S1_extracted_k141_90440 | Low-quality |
| S1_extracted_k141_103970 | Not-determined |
| S1_extracted_k141_130593 | Not-determined |
| S1_extracted_k141_62576 | Not-determined |
| S1_extracted_k141_57208 | Not-determined |
| S1_extracted_k141_17608 | Not-determined |
| S1_extracted_k141_17609 | Low-quality |
| S1_extracted_k141_43941 | Not-determined |
| S1_extracted_k141_23606 | Low-quality |
| S1_extracted_k141_43943 | Not-determined |
| S1_extracted_k141_137145 | Not-determined |
| S1_extracted_k141_30542 | Not-determined |
| S1_extracted_k141_57212 | Not-determined |
| S1_extracted_k141_97392 | Not-determined |
| S1_extracted_k141_97393 | Not-determined |
| S1_extracted_k141_10646 | Low-quality |
| S1_extracted_k141_137149 | Not-determined |
| S1_extracted_k141_43945 | Low-quality |
| S1_extracted_k141_50689 | Not-determined |
| S1_extracted_k141_77214 | Low-quality |
| S1_extracted_k141_50691 | Not-determined |
| S1_extracted_k141_36361 | Not-determined |
| S1_extracted_k141_97399 | Not-determined |
| S1_extracted_k141_30549 | Low-quality |
| S1_extracted_k141_3053 | Low-quality |
| S1_extracted_k141_36362 | Not-determined |
| S1_extracted_k141_90455 | Not-determined |
| S1_extracted_k141_17630 | Low-quality |
| S1_extracted_k141_17632 | Not-determined |
| S1_extracted_k141_43958 | Not-determined |
| S1_extracted_k141_50693 | Low-quality |
| S1_extracted_k141_10647 | Low-quality |
| S1_extracted_k141_23625 | Not-determined |
| S1_extracted_k141_77227 | Not-determined |
| S1_extracted_k141_137168 | Not-determined |
| S1_extracted_k141_43966 | Not-determined |
| S1_extracted_k141_123651 | Not-determined |
| S1_extracted_k141_123652 | Not-determined |
| S1_extracted_k141_70099 | Low-quality |
| S1_extracted_k141_143698 | Not-determined |
| S1_extracted_k141_43969 | Low-quality |
| S1_extracted_k141_90463 | Low-quality |
| S1_extracted_k141_43971 | Not-determined |
| S1_extracted_k141_10657 | Not-determined |
| S1_extracted_k141_50698 | Not-determined |
| S1_extracted_k141_43973 | Not-determined |
| S1_extracted_k141_130615 | Not-determined |
| S1_extracted_k141_50699 | Not-determined |
| S1_extracted_k141_17640 | Not-determined |
| S1_extracted_k141_123660 | Low-quality |
| S1_extracted_k141_110542 | Low-quality |
| S1_extracted_k141_110543 | Not-determined |
| S1_extracted_k141_17645 | Low-quality |
| S1_extracted_k141_117320 | Low-quality |
| S1_extracted_k141_90473 | Not-determined |
| S1_extracted_k141_97412 | Low-quality |
| S1_extracted_k141_36387 | Not-determined |
| S1_extracted_k141_50706 | Not-determined |
| S1_extracted_k141_117322 | Not-determined |
| S1_extracted_k141_130626 | Not-determined |
| S1_extracted_k141_30569 | Not-determined |
| S1_extracted_k141_17654 | Not-determined |
| S1_extracted_k141_17657 | Not-determined |
| S1_extracted_k141_17658 | Not-determined |
| S1_extracted_k141_148594 | Not-determined |
| S1_extracted_k141_10682 | Low-quality |
| S1_extracted_k141_50712 | Not-determined |
| S1_extracted_k141_17664 | Not-determined |
| S1_extracted_k141_130635 | Not-determined |
| S1_extracted_k141_143706 | Not-determined |
| S1_extracted_k141_77256 | Not-determined |
| S1_extracted_k141_30575 | Low-quality |
| S1_extracted_k141_30576 | Not-determined |
| S1_extracted_k141_84012 | Not-determined |
| S1_extracted_k141_90488 | Low-quality |
| S1_extracted_k141_130641 | Not-determined |
| S1_extracted_k141_30578 | Not-determined |
| S1_extracted_k141_97429 | Not-determined |
| S1_extracted_k141_130642 | Not-determined |
| S1_extracted_k141_148596 | Not-determined |
| S1_extracted_k141_30580 | Not-determined |
| S1_extracted_k141_130643 | Not-determined |
| S1_extracted_k141_43988 | Not-determined |
| S1_extracted_k141_43989 | Not-determined |
| S1_extracted_k141_50719 | Not-determined |
| S1_extracted_k141_43990 | Not-determined |
| S1_extracted_k141_84018 | Not-determined |
| S1_extracted_k141_84019 | Not-determined |
| S1_extracted_k141_17679 | Low-quality |
| S1_extracted_k141_148598 | Not-determined |
| S1_extracted_k141_137192 | Not-determined |
| S1_extracted_k141_57257 | Not-determined |
| S1_extracted_k141_148599 | Low-quality |
| S1_extracted_k141_104016 | Not-determined |
| S1_extracted_k141_130651 | Low-quality |
| S1_extracted_k141_148601 | Not-determined |
| S1_extracted_k141_36402 | Low-quality |
| S1_extracted_k141_104018 | Not-determined |
| S1_extracted_k141_70117 | Not-determined |
| S1_extracted_k141_117337 | Low-quality |
| S1_extracted_k141_50726 | Not-determined |
| S1_extracted_k141_17686 | Not-determined |
| S1_extracted_k141_137196 | Not-determined |
| S1_extracted_k141_155783 | Not-determined |
| S1_extracted_k141_110559 | Not-determined |
| S1_extracted_k141_77267 | Not-determined |
| S1_extracted_k141_104023 | Not-determined |
| S1_extracted_k141_117342 | Low-quality |
| S1_extracted_k141_90498 | Not-determined |
| S1_extracted_k141_36406 | Not-determined |
| S1_extracted_k141_70123 | Not-determined |
| S1_extracted_k141_97443 | Not-determined |
| S1_extracted_k141_148604 | Not-determined |
| S1_extracted_k141_84030 | Not-determined |
| S1_extracted_k141_23665 | Not-determined |
| S1_extracted_k141_30601 | Not-determined |
| S1_extracted_k141_130662 | Not-determined |
| S1_extracted_k141_77275 | Low-quality |
| S1_extracted_k141_130664 | Not-determined |
| S1_extracted_k141_123685 | Low-quality |
| S1_extracted_k141_44000 | Low-quality |
| S1_extracted_k141_70126 | Not-determined |
| S1_extracted_k141_110569 | Low-quality |
| S1_extracted_k141_155785 | Not-determined |
| S1_extracted_k141_30609 | Low-quality |
| S1_extracted_k141_90505 | Not-determined |
| S1_extracted_k141_70128 | Not-determined |
| S1_extracted_k141_90506 | Not-determined |
| S1_extracted_k141_110573 | Not-determined |
| S1_extracted_k141_84042 | Not-determined |
| S1_extracted_k141_84043 | Not-determined |
| S1_extracted_k141_155788 | Not-determined |
| S1_extracted_k141_155789 | Not-determined |
| S1_extracted_k141_30620 | Not-determined |
| S1_extracted_k141_123695 | Not-determined |
| S1_extracted_k141_117364 | Not-determined |
| S1_extracted_k141_97457 | Not-determined |
| S1_extracted_k141_117365 | Not-determined |
| S1_extracted_k141_143731 | Not-determined |
| S1_extracted_k141_143732 | Not-determined |
| S1_extracted_k141_44012 | Low-quality |
| S1_extracted_k141_17701 | Low-quality |
| S1_extracted_k141_143734 | Not-determined |
| S1_extracted_k141_62630 | Not-determined |
| S1_extracted_k141_143735 | Not-determined |
| S1_extracted_k141_137212 | Not-determined |
| S1_extracted_k141_44017 | Not-determined |
| S1_extracted_k141_117371 | Not-determined |
| S1_extracted_k141_70137 | Low-quality |
| S1_extracted_k141_137213 | Not-determined |
| S1_extracted_k141_70138 | Not-determined |
| S1_extracted_k141_50739 | Not-determined |
| S1_extracted_k141_70139 | Not-determined |
| S1_extracted_k141_97468 | Low-quality |
| S1_extracted_k141_137215 | Low-quality |
| S1_extracted_k141_137216 | Not-determined |
| S1_extracted_k141_62636 | Not-determined |
| S1_extracted_k141_70143 | Not-determined |
| S1_extracted_k141_57280 | Low-quality |
| S1_extracted_k141_84061 | Low-quality |
| S1_extracted_k141_44023 | Not-determined |
| S1_extracted_k141_50748 | Not-determined |
| S1_extracted_k141_130676 | Low-quality |
| S1_extracted_k141_17713 | Not-determined |
| S1_extracted_k141_117381 | Not-determined |
| S1_extracted_k141_110598 | Low-quality |
| S1_extracted_k141_110599 | Not-determined |
| S1_extracted_k141_70150 | Not-determined |
| S1_extracted_k141_30636 | Not-determined |
| S1_extracted_k141_90521 | Not-determined |
| S1_extracted_k141_155791 | Not-determined |
| S1_extracted_k141_97484 | Low-quality |
| S1_extracted_k141_70156 | Not-determined |
| S1_extracted_k141_84068 | Not-determined |
| S1_extracted_k141_23678 | Low-quality |
| S1_extracted_k141_90523 | Not-determined |
| S1_extracted_k141_77294 | Low-quality |
| S1_extracted_k141_110604 | Low-quality |
| S1_extracted_k141_130685 | Not-determined |
| S1_extracted_k141_143741 | Not-determined |
| S1_extracted_k141_110606 | Not-determined |
| S1_extracted_k141_10733 | Low-quality |
| S1_extracted_k141_62652 | Not-determined |
| S1_extracted_k141_90529 | Not-determined |
| S1_extracted_k141_62654 | Not-determined |
| S1_extracted_k141_44037 | Low-quality |
| S1_extracted_k141_130691 | Not-determined |
| S1_extracted_k141_155795 | Not-determined |
| S1_extracted_k141_117392 | Not-determined |
| S1_extracted_k141_36423 | Low-quality |
| S1_extracted_k141_10742 | Not-determined |
| S1_extracted_k141_57296 | Not-determined |
| S1_extracted_k141_30650 | Not-determined |
| S1_extracted_k141_84077 | Not-determined |
| S1_extracted_k141_17730 | Not-determined |
| S1_extracted_k141_50758 | Not-determined |
| S1_extracted_k141_137236 | Not-determined |
| S1_extracted_k141_30655 | Not-determined |
| S1_extracted_k141_143754 | Not-determined |
| S1_extracted_k141_50762 | Not-determined |
| S1_extracted_k141_97501 | Not-determined |
| S1_extracted_k141_17738 | Not-determined |
| S1_extracted_k141_137239 | Not-determined |
| S1_extracted_k141_17739 | Not-determined |
| S1_extracted_k141_90544 | Not-determined |
| S1_extracted_k141_143762 | Not-determined |
| S1_extracted_k141_17740 | Not-determined |
| S1_extracted_k141_44051 | Not-determined |
| S1_extracted_k141_90546 | Not-determined |
| S1_extracted_k141_70168 | Low-quality |
| S1_extracted_k141_57308 | Not-determined |
| S1_extracted_k141_123727 | Not-determined |
| S1_extracted_k141_123728 | Not-determined |
| S1_extracted_k141_148620 | Not-determined |
| S1_extracted_k141_148621 | Not-determined |
| S1_extracted_k141_57310 | Not-determined |
| S1_extracted_k141_44057 | Not-determined |
| S1_extracted_k141_57311 | Not-determined |
| S1_extracted_k141_3093 | Low-quality |
| S1_extracted_k141_110626 | Not-determined |
| S1_extracted_k141_70177 | Not-determined |
| S1_extracted_k141_148627 | Not-determined |

| | |
|---|---|
| S1_extracted_k141_77321 | Not-determined |
| S1_extracted_k141_97512 | Not-determined |
| S1_extracted_k141_44064 | Not-determined |
| S1_extracted_k141_44065 | Low-quality |
| S1_extracted_k141_137248 | Not-determined |
| S1_extracted_k141_110629 | Not-determined |
| S1_extracted_k141_130707 | Low-quality |
| S1_extracted_k141_84086 | Not-determined |
| S1_extracted_k141_62681 | Not-determined |
| S1_extracted_k141_77325 | Not-determined |
| S1_extracted_k141_117414 | Low-quality |
| S1_extracted_k141_84090 | Low-quality |
| S1_extracted_k141_117418 | Low-quality |
| S1_extracted_k141_30679 | Low-quality |
| S1_extracted_k141_70185 | Not-determined |
| S1_extracted_k141_123744 | Not-determined |
| S1_extracted_k141_143783 | Not-determined |
| S1_extracted_k141_57322 | Not-determined |
| S1_extracted_k141_30682 | Not-determined |
| S1_extracted_k141_10769 | Not-determined |
| S1_extracted_k141_143788 | Not-determined |
| S1_extracted_k141_44082 | Not-determined |
| S1_extracted_k141_3103 | Low-quality |
| S1_extracted_k141_143789 | Not-determined |
| S1_extracted_k141_84095 | Not-determined |
| S1_extracted_k141_62689 | Not-determined |
| S1_extracted_k141_77337 | Not-determined |
| S1_extracted_k141_36436 | Not-determined |
| S1_extracted_k141_110643 | Not-determined |
| S1_extracted_k141_137262 | Not-determined |
| S1_extracted_k141_77339 | Low-quality |
| S1_extracted_k141_155812 | Not-determined |
| S1_extracted_k141_17766 | Not-determined |
| S1_extracted_k141_90568 | Not-determined |
| S1_extracted_k141_97528 | Not-determined |
| S1_extracted_k141_70197 | Not-determined |
| S1_extracted_k141_30689 | Not-determined |
| S1_extracted_k141_104061 | Low-quality |
| S1_extracted_k141_117427 | Not-determined |
| S1_extracted_k141_130713 | Low-quality |
| S1_extracted_k141_36441 | Not-determined |
| S1_extracted_k141_30690 | Not-determined |
| S1_extracted_k141_77349 | Not-determined |
| S1_extracted_k141_70200 | Low-quality |
| S1_extracted_k141_50789 | Low-quality |
| S1_extracted_k141_50790 | Not-determined |
| S1_extracted_k141_97534 | Not-determined |
| S1_extracted_k141_50791 | Not-determined |
| S1_extracted_k141_70202 | Not-determined |
| S1_extracted_k141_57329 | Low-quality |
| S1_extracted_k141_30696 | Not-determined |
| S1_extracted_k141_17769 | Not-determined |
| S1_extracted_k141_44092 | Not-determined |
| S1_extracted_k141_10782 | Not-determined |
| S1_extracted_k141_36448 | Not-determined |
| S1_extracted_k141_137267 | Not-determined |
| S1_extracted_k141_97538 | Low-quality |
| S1_extracted_k141_10789 | Not-determined |
| S1_extracted_k141_110651 | Not-determined |
| S1_extracted_k141_50800 | Not-determined |
| S1_extracted_k141_97539 | Not-determined |
| S1_extracted_k141_62706 | Not-determined |
| S1_extracted_k141_17777 | Not-determined |
| S1_extracted_k141_110652 | Not-determined |
| S1_extracted_k141_130726 | Not-determined |
| S1_extracted_k141_44103 | Not-determined |
| S1_extracted_k141_137269 | Not-determined |
| S1_extracted_k141_17786 | Low-quality |
| S1_extracted_k141_62708 | Not-determined |
| S1_extracted_k141_137271 | Not-determined |
| S1_extracted_k141_148639 | Not-determined |
| S1_extracted_k141_84117 | Low-quality |
| S1_extracted_k141_110654 | Not-determined |
| S1_extracted_k141_44105 | Low-quality |
| S1_extracted_k141_17789 | Not-determined |
| S1_extracted_k141_10800 | Not-determined |
| S1_extracted_k141_30705 | Low-quality |
| S1_extracted_k141_50812 | Not-determined |
| S1_extracted_k141_10801 | Not-determined |
| S1_extracted_k141_50814 | Not-determined |
| S1_extracted_k141_50816 | Not-determined |
| S1_extracted_k141_84121 | Not-determined |
| S1_extracted_k141_130735 | Not-determined |
| S1_extracted_k141_77366 | Not-determined |
| S1_extracted_k141_84124 | Low-quality |
| S1_extracted_k141_104079 | Not-determined |
| S1_extracted_k141_97549 | Not-determined |
| S1_extracted_k141_104080 | Not-determined |
| S1_extracted_k141_110664 | Low-quality |
| S1_extracted_k141_110666 | Low-quality |
| S1_extracted_k141_57339 | Not-determined |
| S1_extracted_k141_44115 | Low-quality |
| S1_extracted_k141_30712 | Not-determined |
| S1_extracted_k141_70216 | Not-determined |
| S1_extracted_k141_30714 | Not-determined |
| S1_extracted_k141_44121 | Not-determined |
| S1_extracted_k141_57344 | Not-determined |
| S1_extracted_k141_70221 | Low-quality |
| S1_extracted_k141_62717 | Not-determined |
| S1_extracted_k141_30720 | Low-quality |
| S1_extracted_k141_90594 | Low-quality |
| S1_extracted_k141_3130 | Low-quality |
| S1_extracted_k141_117448 | Low-quality |
| S1_extracted_k141_44125 | Not-determined |
| S1_extracted_k141_36467 | Low-quality |
| S1_extracted_k141_110676 | Not-determined |
| S1_extracted_k141_70227 | Not-determined |
| S1_extracted_k141_117451 | Not-determined |
| S1_extracted_k141_137291 | Not-determined |
| S1_extracted_k141_57353 | Not-determined |
| S1_extracted_k141_143810 | Not-determined |
| S1_extracted_k141_57355 | Not-determined |
| S1_extracted_k141_50835 | Not-determined |
| S1_extracted_k141_57358 | Not-determined |
| S1_extracted_k141_97558 | Not-determined |
| S1_extracted_k141_143815 | Not-determined |
| S1_extracted_k141_23684 | Not-determined |
| S1_extracted_k141_17808 | Not-determined |
| S1_extracted_k141_137297 | Not-determined |
| S1_extracted_k141_23688 | Not-determined |
| S1_extracted_k141_50838 | Low-quality |
| S1_extracted_k141_44134 | Not-determined |
| S1_extracted_k141_137299 | Not-determined |
| S1_extracted_k141_130744 | Not-determined |
| S1_extracted_k141_90612 | Not-determined |
| S1_extracted_k141_57367 | Not-determined |
| S1_extracted_k141_23697 | Not-determined |
| S1_extracted_k141_70234 | Not-determined |
| S1_extracted_k141_148654 | Not-determined |
| S1_extracted_k141_148657 | Not-determined |
| S1_extracted_k141_123774 | Not-determined |
| S1_extracted_k141_117475 | Not-determined |
| S1_extracted_k141_90621 | Not-determined |
| S1_extracted_k141_50854 | Not-determined |
| S1_extracted_k141_50856 | Not-determined |
| S1_extracted_k141_23711 | Not-determined |
| S1_extracted_k141_84153 | Not-determined |
| S1_extracted_k141_97572 | Not-determined |
| S1_extracted_k141_84155 | Not-determined |
| S1_extracted_k141_30749 | Not-determined |
| S1_extracted_k141_44151 | Not-determined |
| S1_extracted_k141_57383 | Not-determined |
| S1_extracted_k141_44154 | Not-determined |
| S1_extracted_k141_130752 | Not-determined |
| S1_extracted_k141_123786 | Not-determined |
| S1_extracted_k141_30753 | Low-quality |
| S1_extracted_k141_123788 | Not-determined |
| S1_extracted_k141_123789 | Not-determined |
| S1_extracted_k141_50866 | Low-quality |
| S1_extracted_k141_17829 | Not-determined |
| S1_extracted_k141_30755 | Not-determined |
| S1_extracted_k141_143834 | Not-determined |
| S1_extracted_k141_62742 | Not-determined |
| S1_extracted_k141_143836 | Not-determined |
| S1_extracted_k141_84166 | Not-determined |
| S1_extracted_k141_62745 | Not-determined |
| S1_extracted_k141_143838 | Not-determined |
| S1_extracted_k141_30761 | Low-quality |
| S1_extracted_k141_44162 | Not-determined |
| S1_extracted_k141_77403 | Not-determined |
| S1_extracted_k141_77404 | Not-determined |
| S1_extracted_k141_110707 | Not-determined |
| S1_extracted_k141_84173 | Not-determined |
| S1_extracted_k141_130761 | Not-determined |
| S1_extracted_k141_130763 | Not-determined |
| S1_extracted_k141_117491 | Not-determined |
| S1_extracted_k141_30767 | Not-determined |
| S1_extracted_k141_130764 | Not-determined |
| S1_extracted_k141_70252 | Not-determined |
| S1_extracted_k141_137327 | Low-quality |
| S1_extracted_k141_17837 | Not-determined |
| S1_extracted_k141_70255 | Not-determined |
| S1_extracted_k141_143850 | Not-determined |
| S1_extracted_k141_62752 | Not-determined |
| S1_extracted_k141_117495 | Low-quality |
| S1_extracted_k141_36507 | Low-quality |
| S1_extracted_k141_117497 | Not-determined |
| S1_extracted_k141_10844 | Low-quality |
| S1_extracted_k141_50883 | Not-determined |
| S1_extracted_k141_50884 | Not-determined |
| S1_extracted_k141_117499 | Not-determined |
| S1_extracted_k141_143853 | Not-determined |
| S1_extracted_k141_70256 | Low-quality |
| S1_extracted_k141_130770 | Not-determined |
| S1_extracted_k141_123800 | Not-determined |
| S1_extracted_k141_3175 | Not-determined |
| S1_extracted_k141_77422 | Not-determined |
| S1_extracted_k141_77423 | Not-determined |
| S1_extracted_k141_104140 | Not-determined |
| S1_extracted_k141_130775 | Not-determined |
| S1_extracted_k141_62759 | Not-determined |
| S1_extracted_k141_148667 | Not-determined |
| S1_extracted_k141_3178 | Not-determined |
| S1_extracted_k141_143857 | Not-determined |
| S1_extracted_k141_70261 | Not-determined |
| S1_extracted_k141_50894 | Not-determined |
| S1_extracted_k141_143858 | Not-determined |
| S1_extracted_k141_17846 | Not-determined |
| S1_extracted_k141_104143 | Not-determined |
| S1_extracted_k141_97607 | Not-determined |
| S1_extracted_k141_70265 | Not-determined |
| S1_extracted_k141_123806 | Low-quality |
| S1_extracted_k141_104145 | Not-determined |
| S1_extracted_k141_57417 | Not-determined |
| S1_extracted_k141_110729 | Low-quality |
| S1_extracted_k141_123808 | Not-determined |
| S1_extracted_k141_110730 | Not-determined |
| S1_extracted_k141_3186 | Not-determined |
| S1_extracted_k141_10856 | Not-determined |
| S1_extracted_k141_137334 | Not-determined |
| S1_extracted_k141_148677 | Not-determined |
| S1_extracted_k141_90649 | Not-determined |
| S1_extracted_k141_130785 | Not-determined |
| S1_extracted_k141_104156 | Not-determined |
| S1_extracted_k141_62774 | Low-quality |
| S1_extracted_k141_23732 | Not-determined |
| S1_extracted_k141_17854 | Low-quality |
| S1_extracted_k141_130791 | Not-determined |
| S1_extracted_k141_130792 | Not-determined |
| S1_extracted_k141_30784 | Not-determined |
| S1_extracted_k141_143862 | Not-determined |
| S1_extracted_k141_30787 | Not-determined |
| S1_extracted_k141_90657 | Low-quality |
| S1_extracted_k141_117506 | Not-determined |
| S1_extracted_k141_97621 | Not-determined |
| S1_extracted_k141_50915 | Not-determined |
| S1_extracted_k141_90660 | Not-determined |
| S1_extracted_k141_117508 | Not-determined |
| S1_extracted_k141_17866 | Not-determined |
| S1_extracted_k141_117510 | Low-quality |
| S1_extracted_k141_30794 | Not-determined |
| S1_extracted_k141_117512 | Low-quality |
| S1_extracted_k141_57436 | Low-quality |
| S1_extracted_k141_62784 | Not-determined |
| S1_extracted_k141_3204 | Not-determined |
| S1_extracted_k141_36535 | Low-quality |
| S1_extracted_k141_57440 | Low-quality |
| S1_extracted_k141_84188 | Low-quality |
| S1_extracted_k141_10874 | Not-determined |
| S1_extracted_k141_30799 | Not-determined |
| S1_extracted_k141_143875 | Not-determined |
| S1_extracted_k141_104169 | Not-determined |
| S1_extracted_k141_90663 | Low-quality |
| S1_extracted_k141_143879 | Not-determined |
| S1_extracted_k141_17871 | Not-determined |
| S1_extracted_k141_70274 | Low-quality |
| S1_extracted_k141_90664 | Not-determined |
| S1_extracted_k141_70275 | Not-determined |
| S1_extracted_k141_62791 | Low-quality |
| S1_extracted_k141_137366 | Not-determined |
| S1_extracted_k141_77439 | Not-determined |
| S1_extracted_k141_123823 | Not-determined |
| S1_extracted_k141_123824 | Not-determined |
| S1_extracted_k141_110745 | Not-determined |
| S1_extracted_k141_143881 | Not-determined |
| S1_extracted_k141_10879 | Not-determined |
| S1_extracted_k141_17876 | Not-determined |
| S1_extracted_k141_84197 | Not-determined |
| S1_extracted_k141_50936 | Not-determined |
| S1_extracted_k141_36539 | Not-determined |
| S1_extracted_k141_143885 | Low-quality |
| S1_extracted_k141_17880 | Not-determined |
| S1_extracted_k141_23766 | Not-determined |
| S1_extracted_k141_70279 | Low-quality |
| S1_extracted_k141_44176 | Not-determined |
| S1_extracted_k141_84203 | Not-determined |
| S1_extracted_k141_148682 | Not-determined |
| S1_extracted_k141_10885 | Not-determined |
| S1_extracted_k141_137373 | Not-determined |
| S1_extracted_k141_36543 | Not-determined |
| S1_extracted_k141_62799 | Not-determined |
| S1_extracted_k141_137375 | Not-determined |
| S1_extracted_k141_110753 | Low-quality |
| S1_extracted_k141_137376 | Not-determined |
| S1_extracted_k141_17890 | Not-determined |
| S1_extracted_k141_77457 | Not-determined |
| S1_extracted_k141_36545 | Not-determined |
| S1_extracted_k141_97641 | Not-determined |
| S1_extracted_k141_77460 | Low-quality |
| S1_extracted_k141_77461 | Low-quality |
| S1_extracted_k141_90680 | Not-determined |
| S1_extracted_k141_57459 | Not-determined |
| S1_extracted_k141_44184 | Not-determined |
| S1_extracted_k141_123831 | Low-quality |
| S1_extracted_k141_97644 | Not-determined |
| S1_extracted_k141_148694 | Not-determined |
| S1_extracted_k141_110765 | Not-determined |
| S1_extracted_k141_70291 | Not-determined |
| S1_extracted_k141_77470 | Low-quality |
| S1_extracted_k141_23776 | Not-determined |
| S1_extracted_k141_90690 | Low-quality |
| S1_extracted_k141_84213 | Not-determined |
| S1_extracted_k141_110776 | Not-determined |
| S1_extracted_k141_77480 | Not-determined |
| S1_extracted_k141_130826 | Not-determined |
| S1_extracted_k141_44198 | Not-determined |
| S1_extracted_k141_123847 | Not-determined |
| S1_extracted_k141_97661 | Not-determined |
| S1_extracted_k141_110781 | Not-determined |
| S1_extracted_k141_23785 | Not-determined |
| S1_extracted_k141_117564 | Not-determined |
| S1_extracted_k141_44201 | Low-quality |
| S1_extracted_k141_104198 | Not-determined |
| S1_extracted_k141_30830 | Not-determined |
| S1_extracted_k141_50966 | Not-determined |
| S1_extracted_k141_77487 | Not-determined |
| S1_extracted_k141_137404 | Not-determined |
| S1_extracted_k141_117568 | Not-determined |
| S1_extracted_k141_17900 | Not-determined |
| S1_extracted_k141_143909 | Not-determined |
| S1_extracted_k141_130836 | Not-determined |
| S1_extracted_k141_17902 | Not-determined |
| S1_extracted_k141_23793 | Not-determined |
| S1_extracted_k141_50971 | Not-determined |
| S1_extracted_k141_110786 | Not-determined |
| S1_extracted_k141_30841 | Not-determined |
| S1_extracted_k141_90699 | Not-determined |
| S1_extracted_k141_97673 | Not-determined |
| S1_extracted_k141_70312 | Not-determined |
| S1_extracted_k141_77494 | Not-determined |
| S1_extracted_k141_143914 | Low-quality |
| S1_extracted_k141_70314 | Not-determined |
| S1_extracted_k141_155869 | Not-determined |
| S1_extracted_k141_130845 | Low-quality |
| S1_extracted_k141_17911 | Not-determined |
| S1_extracted_k141_77496 | Not-determined |
| S1_extracted_k141_137412 | Not-determined |
| S1_extracted_k141_97676 | Low-quality |
| S1_extracted_k141_148701 | Not-determined |
| S1_extracted_k141_57488 | Not-determined |
| S1_extracted_k141_23806 | Not-determined |
| S1_extracted_k141_77499 | Not-determined |
| S1_extracted_k141_123866 | Not-determined |
| S1_extracted_k141_77500 | Not-determined |
| S1_extracted_k141_130847 | Not-determined |
| S1_extracted_k141_30848 | Not-determined |
| S1_extracted_k141_10929 | Not-determined |
| S1_extracted_k141_70316 | Not-determined |
| S1_extracted_k141_3236 | Not-determined |
| S1_extracted_k141_3237 | Not-determined |
| S1_extracted_k141_143924 | Not-determined |
| S1_extracted_k141_90715 | Not-determined |
| S1_extracted_k141_130851 | Not-determined |
| S1_extracted_k141_3240 | Not-determined |
| S1_extracted_k141_84230 | Not-determined |
| S1_extracted_k141_110801 | Not-determined |
| S1_extracted_k141_104208 | Not-determined |
| S1_extracted_k141_90719 | Not-determined |
| S1_extracted_k141_155872 | Not-determined |
| S1_extracted_k141_62807 | Not-determined |
| S1_extracted_k141_90720 | Not-determined |
| S1_extracted_k141_110803 | Low-quality |
| S1_extracted_k141_30851 | Low-quality |
| S1_extracted_k141_104211 | Not-determined |
| S1_extracted_k141_62808 | Not-determined |
| S1_extracted_k141_62809 | Low-quality |
| S1_extracted_k141_143927 | Not-determined |
| S1_extracted_k141_30857 | Low-quality |
| S1_extracted_k141_36579 | Not-determined |
| S1_extracted_k141_10942 | Not-determined |
| S1_extracted_k141_23820 | Not-determined |
| S1_extracted_k141_84236 | Low-quality |
| S1_extracted_k141_62812 | Not-determined |
| S1_extracted_k141_84238 | Not-determined |
| S1_extracted_k141_50994 | Low-quality |
| S1_extracted_k141_70319 | Not-determined |
| S1_extracted_k141_36583 | Not-determined |
| S1_extracted_k141_77511 | Not-determined |
| S1_extracted_k141_123875 | Not-determined |
| S1_extracted_k141_104220 | Low-quality |
| S1_extracted_k141_117595 | Not-determined |
| S1_extracted_k141_84242 | Low-quality |
| S1_extracted_k141_117597 | Not-determined |
| S1_extracted_k141_137433 | Low-quality |
| S1_extracted_k141_10951 | Not-determined |
| S1_extracted_k141_10952 | Not-determined |
| S1_extracted_k141_77521 | Low-quality |
| S1_extracted_k141_44227 | Not-determined |
| S1_extracted_k141_110814 | Not-determined |
| S1_extracted_k141_70324 | Not-determined |
| S1_extracted_k141_44228 | Not-determined |
| S1_extracted_k141_23837 | Not-determined |
| S1_extracted_k141_17936 | Not-determined |
| S1_extracted_k141_36591 | Not-determined |
| S1_extracted_k141_97690 | Not-determined |
| S1_extracted_k141_44234 | Not-determined |
| S1_extracted_k141_57513 | Not-determined |
| S1_extracted_k141_84253 | Not-determined |
| S1_extracted_k141_36593 | Not-determined |
| S1_extracted_k141_84254 | Low-quality |
| S1_extracted_k141_104233 | Low-quality |
| S1_extracted_k141_155877 | Low-quality |
| S1_extracted_k141_117606 | Not-determined |
| S1_extracted_k141_10965 | Low-quality |
| S1_extracted_k141_97694 | Not-determined |
| S1_extracted_k141_104236 | Not-determined |
| S1_extracted_k141_3255 | Not-determined |
| S1_extracted_k141_123892 | Not-determined |
| S1_extracted_k141_130873 | Low-quality |
| S1_extracted_k141_44243 | Not-determined |
| S1_extracted_k141_57529 | Not-determined |
| S1_extracted_k141_51021 | Low-quality |
| S1_extracted_k141_3259 | Not-determined |
| S1_extracted_k141_97703 | Not-determined |
| S1_extracted_k141_117617 | Not-determined |
| S1_extracted_k141_51024 | Not-determined |
| S1_extracted_k141_17942 | Not-determined |
| S1_extracted_k141_90747 | Not-determined |
| S1_extracted_k141_57536 | Not-determined |
| S1_extracted_k141_130876 | Not-determined |
| S1_extracted_k141_84266 | Low-quality |
| S1_extracted_k141_77534 | Low-quality |
| S1_extracted_k141_104249 | Low-quality |
| S1_extracted_k141_104250 | Low-quality |
| S1_extracted_k141_110826 | Low-quality |
| S1_extracted_k141_51032 | Not-determined |
| S1_extracted_k141_23856 | Not-determined |
| S1_extracted_k141_23857 | Not-determined |
| S1_extracted_k141_123898 | Not-determined |
| S1_extracted_k141_77539 | Not-determined |
| S1_extracted_k141_77541 | Not-determined |
| S1_extracted_k141_36608 | Low-quality |
| S1_extracted_k141_90754 | Low-quality |
| S1_extracted_k141_62845 | Low-quality |
| S1_extracted_k141_84272 | Not-determined |
| S1_extracted_k141_10970 | Low-quality |
| S1_extracted_k141_44257 | Not-determined |
| S1_extracted_k141_17951 | Not-determined |
| S1_extracted_k141_117630 | Not-determined |
| S1_extracted_k141_137462 | Not-determined |
| S1_extracted_k141_84276 | Not-determined |
| S1_extracted_k141_77548 | Not-determined |
| S1_extracted_k141_104261 | Not-determined |
| S1_extracted_k141_77550 | Not-determined |
| S1_extracted_k141_110833 | Not-determined |
| S1_extracted_k141_17958 | Not-determined |
| S1_extracted_k141_117635 | Not-determined |
| S1_extracted_k141_44262 | Not-determined |
| S1_extracted_k141_104263 | Not-determined |
| S1_extracted_k141_84285 | Low-quality |
| S1_extracted_k141_23879 | Not-determined |
| S1_extracted_k141_130890 | Not-determined |
| S1_extracted_k141_3275 | Not-determined |
| S1_extracted_k141_110841 | Not-determined |
| S1_extracted_k141_77557 | Not-determined |
| S1_extracted_k141_77558 | Not-determined |
| S1_extracted_k141_104268 | Not-determined |
| S1_extracted_k141_17965 | Not-determined |
| S1_extracted_k141_110848 | Not-determined |
| S1_extracted_k141_90764 | Not-determined |
| S1_extracted_k141_10990 | Low-quality |
| S1_extracted_k141_77564 | Low-quality |
| S1_extracted_k141_130896 | Not-determined |
| S1_extracted_k141_30885 | Not-determined |
| S1_extracted_k141_17973 | Not-determined |
| S1_extracted_k141_148729 | Not-determined |
| S1_extracted_k141_70353 | Low-quality |
| S1_extracted_k141_123912 | Not-determined |
| S1_extracted_k141_90767 | Not-determined |
| S1_extracted_k141_30887 | Not-determined |
| S1_extracted_k141_23892 | Not-determined |
| S1_extracted_k141_97716 | Low-quality |
| S1_extracted_k141_90770 | Not-determined |
| S1_extracted_k141_123917 | Not-determined |
| S1_extracted_k141_104279 | Low-quality |
| S1_extracted_k141_137471 | Not-determined |
| S1_extracted_k141_23896 | Not-determined |
| S1_extracted_k141_117647 | Not-determined |
| S1_extracted_k141_104280 | Not-determined |
| S1_extracted_k141_30893 | Low-quality |
| S1_extracted_k141_110849 | Not-determined |
| S1_extracted_k141_10992 | Not-determined |
| S1_extracted_k141_110850 | Not-determined |
| S1_extracted_k141_3283 | Low-quality |
| S1_extracted_k141_104281 | Low-quality |
| S1_extracted_k141_90777 | Not-determined |
| S1_extracted_k141_51056 | Not-determined |
| S1_extracted_k141_97723 | Not-determined |
| S1_extracted_k141_90780 | Not-determined |
| S1_extracted_k141_117652 | Low-quality |
| S1_extracted_k141_90783 | Low-quality |
| S1_extracted_k141_137482 | Not-determined |
| S1_extracted_k141_110855 | Not-determined |
| S1_extracted_k141_110857 | Not-determined |
| S1_extracted_k141_51061 | Not-determined |
| S1_extracted_k141_51062 | Not-determined |
| S1_extracted_k141_30901 | Not-determined |
| S1_extracted_k141_97726 | Not-determined |
| S1_extracted_k141_117660 | Not-determined |
| S1_extracted_k141_143932 | Low-quality |
| S1_extracted_k141_137488 | Low-quality |
| S1_extracted_k141_130918 | Not-determined |
| S1_extracted_k141_23910 | Not-determined |
| S1_extracted_k141_148738 | Not-determined |
| S1_extracted_k141_90788 | Not-determined |
| S1_extracted_k141_117663 | Low-quality |
| S1_extracted_k141_110867 | Not-determined |
| S1_extracted_k141_110868 | Not-determined |
| S1_extracted_k141_17987 | Not-determined |
| S1_extracted_k141_148743 | Not-determined |
| S1_extracted_k141_51066 | Low-quality |
| S1_extracted_k141_117665 | Not-determined |
| S1_extracted_k141_36641 | Low-quality |
| S1_extracted_k141_30907 | Not-determined |
| S1_extracted_k141_36642 | Not-determined |
| S1_extracted_k141_130921 | Not-determined |
| S1_extracted_k141_23915 | Not-determined |
| S1_extracted_k141_97733 | Not-determined |
| S1_extracted_k141_30909 | Not-determined |
| S1_extracted_k141_23918 | Low-quality |
| S1_extracted_k141_51074 | Not-determined |
| S1_extracted_k141_137500 | Not-determined |
| S1_extracted_k141_110876 | Low-quality |
| S1_extracted_k141_137505 | Not-determined |
| S1_extracted_k141_117677 | Not-determined |
| S1_extracted_k141_36652 | Not-determined |
| S1_extracted_k141_11024 | Not-determined |
| S1_extracted_k141_51081 | Not-determined |
| S1_extracted_k141_148750 | Low-quality |
| S1_extracted_k141_70363 | Not-determined |
| S1_extracted_k141_123930 | Low-quality |
| S8_vs_k141_54514\|\|full | High-quality |
| S1_extracted_k141_137511 | Not-determined |
| S1_extracted_k141_3305 | Not-determined |
| S1_extracted_k141_51084 | Not-determined |
| S1_extracted_k141_36658 | Not-determined |
| S1_extracted_k141_57545 | Low-quality |
| S1_extracted_k141_148758 | Not-determined |
| S1_extracted_k141_130935 | Low-quality |
| S1_extracted_k141_70370 | Not-determined |
| S1_extracted_k141_148759 | Not-determined |
| S1_extracted_k141_155900 | Not-determined |
| S1_extracted_k141_11039 | Not-determined |
| S1_extracted_k141_137517 | Not-determined |
| S1_extracted_k141_84331 | Not-determined |
| S1_extracted_k141_36666 | Not-determined |
| S1_extracted_k141_44309 | Not-determined |
| S1_extracted_k141_123938 | Not-determined |
| S1_extracted_k141_84333 | Not-determined |
| S1_extracted_k141_23934 | Not-determined |
| S1_extracted_k141_104298 | Low-quality |
| S1_extracted_k141_117689 | Not-determined |
| S1_extracted_k141_23936 | Not-determined |
| S1_extracted_k141_44315 | Low-quality |
| S1_extracted_k141_70375 | Not-determined |
| S1_extracted_k141_23941 | Low-quality |
| S1_extracted_k141_104299 | Not-determined |
| S1_extracted_k141_110894 | Not-determined |
| S1_extracted_k141_18006 | Not-determined |
| S1_extracted_k141_30936 | Not-determined |
| S1_extracted_k141_90817 | Not-determined |
| S1_extracted_k141_11053 | Low-quality |
| S1_extracted_k141_18008 | Not-determined |
| S1_extracted_k141_77607 | Not-determined |
| S1_extracted_k141_148762 | Low-quality |
| S1_extracted_k141_143953 | Not-determined |
| S1_extracted_k141_62913 | Not-determined |
| S1_extracted_k141_90819 | Not-determined |
| S1_extracted_k141_104302 | Not-determined |
| S1_extracted_k141_84337 | Not-determined |
| S1_extracted_k141_90821 | Not-determined |
| S1_extracted_k141_97748 | Low-quality |
| S1_extracted_k141_23951 | Not-determined |
| S1_extracted_k141_90823 | Not-determined |
| S1_extracted_k141_23953 | Not-determined |
| S1_extracted_k141_36678 | Not-determined |
| S1_extracted_k141_110900 | Low-quality |
| S1_extracted_k141_30947 | Not-determined |
| S1_extracted_k141_3327 | Low-quality |
| S1_extracted_k141_18022 | Not-determined |
| S1_extracted_k141_23956 | Low-quality |
| S1_extracted_k141_77617 | Not-determined |
| S1_extracted_k141_97759 | Not-determined |
| S1_extracted_k141_51110 | Not-determined |
| S1_extracted_k141_36685 | Not-determined |
| S1_extracted_k141_97762 | Not-determined |
| S1_extracted_k141_62923 | Not-determined |
| S1_extracted_k141_77620 | Not-determined |
| S1_extracted_k141_104313 | Not-determined |
| S1_extracted_k141_90830 | Low-quality |
| S1_extracted_k141_36688 | Not-determined |
| S1_extracted_k141_130960 | Low-quality |
| S1_extracted_k141_3330 | Not-determined |
| S1_extracted_k141_30955 | Not-determined |
| S1_extracted_k141_104318 | Not-determined |
| S1_extracted_k141_77622 | Low-quality |
| S1_extracted_k141_104321 | Low-quality |
| S1_extracted_k141_155913 | Not-determined |
| S1_extracted_k141_36699 | Low-quality |

| | |
|---|---|
| S1_extracted_k141_104323 | Low-quality |
| S1_extracted_k141_130963 | Low-quality |
| S1_extracted_k141_30962 | Low-quality |
| S1_extracted_k141_30963 | Not-determined |
| S1_extracted_k141_123957 | Low-quality |
| S1_extracted_k141_117720 | Not-determined |
| S1_extracted_k141_110920 | Not-determined |
| S1_extracted_k141_77631 | Not-determined |
| S1_extracted_k141_30965 | Not-determined |
| S1_extracted_k141_51119 | Not-determined |
| S1_extracted_k141_143959 | Not-determined |
| S1_extracted_k141_30968 | Not-determined |
| S1_extracted_k141_104329 | Not-determined |
| S1_extracted_k141_90845 | Not-determined |
| S1_extracted_k141_97775 | Not-determined |
| S1_extracted_k141_104331 | Not-determined |
| S1_extracted_k141_11079 | Not-determined |
| S1_extracted_k141_51122 | Not-determined |
| S1_extracted_k141_84365 | Not-determined |
| S1_extracted_k141_18041 | Not-determined |
| S1_extracted_k141_130965 | Not-determined |
| S1_extracted_k141_77636 | Not-determined |
| S1_extracted_k141_30971 | Not-determined |
| S1_extracted_k141_84367 | Not-determined |
| S1_extracted_k141_97779 | Not-determined |
| S1_extracted_k141_90849 | Low-quality |
| S1_extracted_k141_84370 | Not-determined |
| S1_extracted_k141_3338 | Not-determined |
| S1_extracted_k141_84372 | Not-determined |
| S1_extracted_k141_36711 | Not-determined |
| S1_extracted_k141_77648 | Not-determined |
| S1_extracted_k141_90852 | Not-determined |
| S1_extracted_k141_18051 | Not-determined |
| S1_extracted_k141_137553 | Not-determined |
| S1_extracted_k141_143973 | Not-determined |
| S1_extracted_k141_23987 | Not-determined |
| S1_extracted_k141_23988 | Not-determined |
| S1_extracted_k141_117728 | Not-determined |
| S1_extracted_k141_44355 | Not-determined |
| S1_extracted_k141_148767 | Not-determined |
| S1_extracted_k141_90856 | Not-determined |
| S1_extracted_k141_97787 | Not-determined |
| S1_extracted_k141_11087 | Not-determined |
| S1_extracted_k141_84379 | Not-determined |
| S1_extracted_k141_90857 | Not-determined |
| S1_extracted_k141_18056 | Not-determined |
| S1_extracted_k141_23991 | Not-determined |
| S1_extracted_k141_70401 | Not-determined |
| S1_extracted_k141_155923 | Low-quality |
| S1_extracted_k141_90858 | Not-determined |
| S1_extracted_k141_130980 | Not-determined |
| S1_extracted_k141_137565 | Not-determined |
| S17_extracted_k141_177197 | Complete |
| S1_extracted_k141_30987 | Not-determined |
| S1_extracted_k141_51138 | Not-determined |
| S1_extracted_k141_130984 | Low-quality |
| S1_extracted_k141_137569 | Low-quality |
| S1_extracted_k141_130985 | Not-determined |
| S1_extracted_k141_110932 | Not-determined |
| S1_extracted_k141_57553 | Low-quality |
| S1_extracted_k141_30995 | Not-determined |
| S1_extracted_k141_36727 | Not-determined |
| S1_extracted_k141_90867 | Not-determined |
| S1_extracted_k141_97797 | Low-quality |
| S1_extracted_k141_143988 | Not-determined |
| S1_extracted_k141_24000 | Not-determined |
| S1_extracted_k141_62954 | Not-determined |
| S1_extracted_k141_30999 | Not-determined |
| S1_extracted_k141_62955 | Not-determined |
| S1_extracted_k141_148771 | Low-quality |
| S1_extracted_k141_24003 | Not-determined |
| S1_extracted_k141_130991 | Not-determined |
| S1_extracted_k141_57559 | Not-determined |
| S1_extracted_k141_51145 | Not-determined |
| S1_extracted_k141_143994 | Not-determined |
| S1_extracted_k141_130995 | Not-determined |
| S1_extracted_k141_137581 | Not-determined |
| S1_extracted_k141_62961 | Not-determined |
| S1_extracted_k141_31008 | Not-determined |
| S1_extracted_k141_24009 | Not-determined |
| S1_extracted_k141_51149 | Not-determined |
| S1_extracted_k141_130997 | Low-quality |
| S1_extracted_k141_57560 | Low-quality |
| S1_extracted_k141_137584 | Not-determined |
| S1_extracted_k141_51151 | Not-determined |
| S1_extracted_k141_90881 | Not-determined |
| S1_extracted_k141_44362 | Not-determined |
| S1_extracted_k141_137586 | Not-determined |
| S1_extracted_k141_144005 | Not-determined |
| S1_extracted_k141_62969 | Not-determined |
| S1_extracted_k141_104361 | Not-determined |
| S1_extracted_k141_24011 | Low-quality |
| S1_extracted_k141_137588 | Not-determined |
| S1_extracted_k141_137589 | Low-quality |
| S1_extracted_k141_3360 | Not-determined |
| S1_extracted_k141_44369 | Not-determined |
| S1_extracted_k141_84413 | Not-determined |
| S1_extracted_k141_44370 | Not-determined |
| S1_extracted_k141_90888 | Low-quality |
| S1_extracted_k141_51160 | Not-determined |
| S1_extracted_k141_31021 | Not-determined |
| S13_extracted_151995 | Complete |
| S1_extracted_k141_44376 | Low-quality |
| S1_extracted_k141_110950 | Not-determined |
| S1_extracted_k141_110951 | Not-determined |
| S1_extracted_k141_104375 | Not-determined |
| S1_extracted_k141_57568 | Not-determined |
| S1_extracted_k141_131012 | Not-determined |
| S1_extracted_k141_137601 | Low-quality |
| S1_extracted_k141_24018 | Not-determined |
| S1_extracted_k141_31026 | Not-determined |
| S1_extracted_k141_137602 | Not-determined |
| S1_extracted_k141_3369 | Not-determined |
| S1_extracted_k141_110958 | Not-determined |
| S1_extracted_k141_84423 | Low-quality |
| S1_extracted_k141_70428 | Not-determined |
| S1_extracted_k141_155941 | Not-determined |
| S1_extracted_k141_31029 | Not-determined |
| S1_extracted_k141_31030 | Not-determined |
| S1_extracted_k141_70430 | Not-determined |
| S1_extracted_k141_104385 | Not-determined |
| S1_extracted_k141_77665 | Not-determined |
| S1_extracted_k141_24023 | Not-determined |
| S1_extracted_k141_104387 | Not-determined |
| S1_extracted_k141_57578 | Not-determined |
| S1_extracted_k141_110965 | Low-quality |
| S1_extracted_k141_77667 | Not-determined |
| S1_extracted_k141_97804 | Low-quality |
| S1_extracted_k141_104389 | Not-determined |
| S1_extracted_k141_117769 | Not-determined |
| S1_extracted_k141_110970 | Not-determined |
| S1_extracted_k141_104391 | Not-determined |
| S1_extracted_k141_11130 | Low-quality |
| S1_extracted_k141_144028 | Not-determined |
| S1_extracted_k141_84429 | Low-quality |
| S1_extracted_k141_3375 | Low-quality |
| S1_extracted_k141_44390 | Low-quality |
| S1_extracted_k141_131033 | Not-determined |
| S1_extracted_k141_144033 | Not-determined |
| S1_extracted_k141_62992 | Not-determined |
| S1_extracted_k141_123970 | Not-determined |
| S1_extracted_k141_144035 | Not-determined |
| S1_extracted_k141_77680 | Low-quality |
| S1_extracted_k141_3383 | Not-determined |
| S1_extracted_k141_131039 | Not-determined |
| S1_extracted_k141_104403 | Not-determined |
| S1_extracted_k141_44403 | Not-determined |
| S1_extracted_k141_70440 | Not-determined |
| S1_extracted_k141_51179 | Not-determined |
| S1_extracted_k141_3388 | Not-determined |
| S1_extracted_k141_97823 | Not-determined |
| S1_extracted_k141_90904 | Not-determined |
| S1_extracted_k141_84436 | Low-quality |
| S1_extracted_k141_90906 | Not-determined |
| S1_extracted_k141_51182 | Not-determined |
| S1_extracted_k141_70444 | Not-determined |
| S1_extracted_k141_117780 | Not-determined |
| S1_extracted_k141_70446 | Not-determined |
| S1_extracted_k141_51186 | Not-determined |
| S1_extracted_k141_117782 | Not-determined |
| S1_extracted_k141_77692 | Low-quality |
| S1_extracted_k141_117785 | Low-quality |
| S1_extracted_k141_123988 | Not-determined |
| S1_extracted_k141_51188 | Not-determined |
| S1_extracted_k141_90914 | Not-determined |
| S1_extracted_k141_44413 | Not-determined |
| S1_extracted_k141_90917 | Low-quality |
| S1_extracted_k141_51191 | Not-determined |
| S1_extracted_k141_63009 | Not-determined |
| S1_extracted_k141_123996 | Not-determined |
| S1_extracted_k141_11155 | Not-determined |
| S1_extracted_k141_84441 | Not-determined |
| S1_extracted_k141_117794 | Not-determined |
| S1_extracted_k141_123997 | Not-determined |
| S1_extracted_k141_44418 | Not-determined |
| S1_extracted_k141_144064 | Not-determined |
| S1_extracted_k141_90921 | Low-quality |
| S1_extracted_k141_144066 | Not-determined |
| S1_extracted_k141_51197 | Not-determined |
| S1_extracted_k141_90923 | Not-determined |
| S1_extracted_k141_131058 | Low-quality |
| S1_extracted_k141_90924 | Not-determined |
| S1_extracted_k141_24044 | Not-determined |
| S1_extracted_k141_131059 | Not-determined |
| S1_extracted_k141_144068 | Not-determined |
| S1_extracted_k141_18116 | Not-determined |
| S1_extracted_k141_131061 | Not-determined |
| S1_extracted_k141_137619 | Not-determined |
| S1_extracted_k141_11160 | Not-determined |
| S1_extracted_k141_24048 | Not-determined |
| S1_extracted_k141_144069 | Low-quality |
| S1_extracted_k141_36752 | Low-quality |
| S1_extracted_k141_11162 | Not-determined |
| S1_extracted_k141_57591 | Not-determined |
| S1_extracted_k141_11163 | Not-determined |
| S1_extracted_k141_31058 | Not-determined |
| S1_extracted_k141_11165 | Not-determined |
| S1_extracted_k141_63026 | Not-determined |
| S1_extracted_k141_36757 | Not-determined |
| S1_extracted_k141_63027 | Not-determined |
| S1_extracted_k141_124007 | Low-quality |
| S1_extracted_k141_90933 | Not-determined |
| S1_extracted_k141_111001 | Not-determined |
| S1_extracted_k141_90936 | Not-determined |
| S1_extracted_k141_131067 | Not-determined |
| S1_extracted_k141_57597 | Low-quality |
| S1_extracted_k141_131068 | Not-determined |
| S1_extracted_k141_70458 | Low-quality |
| S1_extracted_k141_18130 | Not-determined |
| S1_extracted_k141_97856 | Not-determined |
| S1_extracted_k141_3406 | Not-determined |
| S1_extracted_k141_11173 | Not-determined |
| S1_extracted_k141_144077 | Not-determined |
| S1_extracted_k141_77707 | Not-determined |
| S1_extracted_k141_36768 | Low-quality |
| S1_extracted_k141_117795 | Low-quality |
| S1_extracted_k141_131078 | Not-determined |
| S1_extracted_k141_51208 | Not-determined |
| S1_extracted_k141_104407 | Low-quality |
| S1_extracted_k141_84460 | Low-quality |
| S1_extracted_k141_36770 | Low-quality |
| S1_extracted_k141_144085 | Not-determined |
| S1_extracted_k141_36771 | Low-quality |
| S1_extracted_k141_97869 | Not-determined |
| S1_extracted_k141_148802 | Not-determined |
| S1_extracted_k141_18134 | Low-quality |
| S1_extracted_k141_137644 | Not-determined |
| S1_extracted_k141_144089 | Not-determined |
| S1_extracted_k141_111018 | Not-determined |
| S1_extracted_k141_111020 | Not-determined |
| S1_extracted_k141_51218 | Not-determined |
| S1_extracted_k141_63047 | Not-determined |
| S1_extracted_k141_90952 | Not-determined |
| S1_extracted_k141_44445 | Not-determined |
| S1_extracted_k141_111022 | Not-determined |
| S1_extracted_k141_36772 | Not-determined |
| S1_extracted_k141_84468 | Not-determined |
| S1_extracted_k141_144095 | Not-determined |
| S1_extracted_k141_57616 | Not-determined |
| S1_extracted_k141_31071 | Not-determined |
| S1_extracted_k141_3411 | Low-quality |
| S1_extracted_k141_137648 | Not-determined |
| S1_extracted_k141_131089 | Not-determined |
| S1_extracted_k141_70476 | Low-quality |
| S1_extracted_k141_124031 | Low-quality |
| S1_extracted_k141_57622 | Not-determined |
| S1_extracted_k141_51222 | Low-quality |
| S1_extracted_k141_18140 | Low-quality |
| S1_extracted_k141_124034 | Not-determined |
| S1_extracted_k141_131095 | Not-determined |
| S1_extracted_k141_57627 | Low-quality |
| S1_extracted_k141_97880 | Not-determined |
| S1_extracted_k141_124038 | Low-quality |
| S1_extracted_k141_44454 | Not-determined |
| S1_extracted_k141_104424 | Low-quality |
| S1_extracted_k141_137661 | Not-determined |
| S1_extracted_k141_90967 | Not-determined |
| S1_extracted_k141_90968 | Not-determined |
| S1_extracted_k141_97881 | Low-quality |
| S1_extracted_k141_90969 | Not-determined |
| S1_extracted_k141_117807 | Not-determined |
| S1_extracted_k141_124043 | Low-quality |
| S1_extracted_k141_18152 | Not-determined |
| S1_extracted_k141_155959 | Not-determined |
| S1_extracted_k141_117811 | Not-determined |
| S1_extracted_k141_24063 | Low-quality |
| S1_extracted_k141_111043 | Not-determined |
| S1_extracted_k141_70488 | Low-quality |
| S1_extracted_k141_11206 | Not-determined |
| S1_extracted_k141_3427 | Not-determined |
| S1_extracted_k141_148812 | Not-determined |
| S1_extracted_k141_11209 | Not-determined |
| S1_extracted_k141_31093 | Not-determined |
| S1_extracted_k141_11212 | Not-determined |
| S1_extracted_k141_90975 | Not-determined |
| S1_extracted_k141_51242 | Not-determined |
| S1_extracted_k141_44459 | Not-determined |
| S1_extracted_k141_137669 | Not-determined |
| S1_extracted_k141_148818 | Not-determined |
| S1_extracted_k141_36794 | Not-determined |
| S1_extracted_k141_104436 | Not-determined |
| S1_extracted_k141_104438 | Not-determined |
| S1_extracted_k141_44463 | Not-determined |
| S1_extracted_k141_148825 | Not-determined |
| S1_extracted_k141_124059 | Not-determined |
| S1_extracted_k141_111055 | Not-determined |
| S1_extracted_k141_57630 | Not-determined |
| S1_extracted_k141_70496 | Not-determined |
| S1_extracted_k141_104443 | Not-determined |
| S1_extracted_k141_90984 | Low-quality |
| S1_extracted_k141_84486 | Not-determined |
| S1_extracted_k141_117822 | Not-determined |
| S1_extracted_k141_18166 | Low-quality |
| S1_extracted_k141_18167 | Not-determined |
| S1_extracted_k141_44467 | Low-quality |
| S1_extracted_k141_84490 | Not-determined |
| S1_extracted_k141_144125 | Not-determined |
| S1_extracted_k141_77752 | Low-quality |
| S1_extracted_k141_131111 | Not-determined |
| S1_extracted_k141_90994 | Not-determined |
| S1_extracted_k141_77761 | Not-determined |
| S1_extracted_k141_90995 | Low-quality |
| S1_extracted_k141_77762 | Not-determined |
| S1_extracted_k141_148834 | Not-determined |
| S1_extracted_k141_24083 | Not-determined |
| S1_extracted_k141_111066 | Not-determined |
| S1_extracted_k141_36805 | Not-determined |
| S1_extracted_k141_144134 | Not-determined |
| S1_extracted_k141_77766 | Not-determined |
| S1_extracted_k141_155967 | Not-determined |
| S1_extracted_k141_104449 | Not-determined |
| S1_extracted_k141_18168 | Low-quality |
| S1_extracted_k141_51259 | Not-determined |
| S1_extracted_k141_124070 | Low-quality |
| S1_extracted_k141_3441 | Not-determined |
| S1_extracted_k141_104451 | Not-determined |
| S1_extracted_k141_124073 | Not-determined |
| S1_extracted_k141_11242 | Not-determined |
| S1_extracted_k141_18174 | Not-determined |
| S1_extracted_k141_137686 | Not-determined |
| S1_extracted_k141_77774 | Not-determined |
| S1_extracted_k141_104457 | Not-determined |
| S1_extracted_k141_117839 | Not-determined |
| S1_extracted_k141_44479 | Not-determined |
| S1_extracted_k141_91001 | Low-quality |
| S1_extracted_k141_104460 | Low-quality |
| S1_extracted_k141_131125 | Low-quality |
| S1_extracted_k141_104461 | Not-determined |
| S1_extracted_k141_51274 | Not-determined |
| S1_extracted_k141_44482 | Not-determined |
| S1_extracted_k141_91008 | Not-determined |
| S1_extracted_k141_63087 | Not-determined |
| S1_extracted_k141_77784 | Low-quality |
| S1_extracted_k141_97910 | Not-determined |
| S1_extracted_k141_24093 | Low-quality |
| S1_extracted_k141_3450 | Not-determined |
| S1_extracted_k141_18181 | Low-quality |
| S1_extracted_k141_11256 | Not-determined |
| S1_extracted_k141_51281 | Not-determined |
| S1_extracted_k141_91015 | Low-quality |
| S1_extracted_k141_70517 | Not-determined |
| S1_extracted_k141_57655 | Low-quality |
| S1_extracted_k141_57656 | Not-determined |
| S1_extracted_k141_24101 | Not-determined |
| S1_extracted_k141_24102 | Low-quality |
| S1_extracted_k141_18184 | Not-determined |
| S1_extracted_k141_24104 | Low-quality |
| S1_extracted_k141_144152 | Not-determined |
| S1_extracted_k141_144153 | Not-determined |
| S1_extracted_k141_44493 | Low-quality |
| S1_extracted_k141_111091 | Not-determined |
| S1_extracted_k141_51285 | Not-determined |
| S1_extracted_k141_104476 | Not-determined |
| S1_extracted_k141_51286 | Low-quality |
| S1_extracted_k141_44496 | Not-determined |
| S1_extracted_k141_36839 | Not-determined |
| S1_extracted_k141_77796 | Not-determined |
| S1_extracted_k141_36840 | Not-determined |
| S1_extracted_k141_3457 | Low-quality |
| S1_extracted_k141_70525 | Not-determined |
| S1_extracted_k141_31118 | Not-determined |
| S1_extracted_k141_97923 | Not-determined |
| S1_extracted_k141_36843 | Low-quality |
| S1_extracted_k141_124113 | Not-determined |
| S1_extracted_k141_3462 | Not-determined |
| S1_extracted_k141_44505 | Low-quality |
| S1_extracted_k141_91020 | Not-determined |
| S1_extracted_k141_91021 | Not-determined |
| S1_extracted_k141_70529 | Not-determined |
| S1_extracted_k141_148844 | Not-determined |
| S1_extracted_k141_31123 | Low-quality |
| S1_extracted_k141_91023 | Not-determined |
| S1_extracted_k141_57667 | Not-determined |
| S1_extracted_k141_91024 | Not-determined |
| S1_extracted_k141_137696 | Not-determined |
| S1_extracted_k141_144172 | Not-determined |
| S1_extracted_k141_63101 | Not-determined |
| S1_extracted_k141_31125 | Not-determined |
| S1_extracted_k141_144174 | Not-determined |
| S1_extracted_k141_91028 | Not-determined |
| S1_extracted_k141_63103 | Not-determined |
| S1_extracted_k141_24120 | Not-determined |
| S1_extracted_k141_31129 | Low-quality |
| S1_extracted_k141_97929 | Low-quality |
| S1_extracted_k141_11279 | Not-determined |
| S1_extracted_k141_18202 | Not-determined |
| S1_extracted_k141_84504 | Low-quality |
| S1_extracted_k141_44512 | Not-determined |
| S1_extracted_k141_57674 | Not-determined |
| S1_extracted_k141_24123 | Not-determined |
| S1_extracted_k141_3470 | Not-determined |
| S1_extracted_k141_104491 | Not-determined |
| S1_extracted_k141_97934 | Low-quality |
| S1_extracted_k141_124122 | Not-determined |
| S1_extracted_k141_144182 | Not-determined |
| S1_extracted_k141_124125 | Not-determined |
| S1_extracted_k141_91034 | Not-determined |
| S1_extracted_k141_51298 | Not-determined |
| S1_extracted_k141_124127 | Low-quality |
| S1_extracted_k141_91035 | Not-determined |
| S1_extracted_k141_144187 | Not-determined |
| S1_extracted_k141_124131 | Not-determined |
| S1_extracted_k141_11289 | Not-determined |
| S1_extracted_k141_131160 | Not-determined |
| S1_extracted_k141_57683 | Not-determined |
| S1_extracted_k141_148855 | Not-determined |
| S1_extracted_k141_31141 | Low-quality |
| S1_extracted_k141_84511 | Not-determined |
| S1_extracted_k141_91039 | Not-determined |
| S1_extracted_k141_97941 | Low-quality |
| S1_extracted_k141_144191 | Not-determined |
| S1_extracted_k141_137703 | Not-determined |
| S1_extracted_k141_3485 | Not-determined |
| S1_extracted_k141_36854 | Low-quality |
| S1_extracted_k141_51310 | Not-determined |
| S1_extracted_k141_24137 | Not-determined |
| S1_extracted_k141_124140 | Not-determined |
| S1_extracted_k141_70546 | Not-determined |
| S1_extracted_k141_11295 | Not-determined |
| S1_extracted_k141_31142 | Not-determined |
| S1_extracted_k141_3489 | Not-determined |
| S1_extracted_k141_51316 | Not-determined |
| S1_extracted_k141_77822 | Not-determined |
| S1_extracted_k141_24146 | Not-determined |
| S1_extracted_k141_117878 | Not-determined |
| S1_extracted_k141_77825 | Not-determined |
| S1_extracted_k141_3492 | Not-determined |
| S1_extracted_k141_91049 | Not-determined |
| S1_extracted_k141_3493 | Not-determined |
| S1_extracted_k141_144194 | Not-determined |
| S1_extracted_k141_44525 | Not-determined |
| S1_extracted_k141_18224 | Not-determined |
| S1_extracted_k141_84516 | Not-determined |
| S1_extracted_k141_104511 | Not-determined |
| S1_extracted_k141_36861 | Not-determined |
| S1_extracted_k141_111119 | Not-determined |
| S1_extracted_k141_18226 | Not-determined |
| S1_extracted_k141_57686 | Not-determined |
| S1_extracted_k141_3498 | Not-determined |
| S1_extracted_k141_111122 | Not-determined |
| S1_extracted_k141_155983 | Not-determined |
| S1_extracted_k141_63132 | Not-determined |
| S1_extracted_k141_63135 | Not-determined |
| S1_extracted_k141_51325 | Not-determined |
| S1_extracted_k141_111127 | Not-determined |
| S1_extracted_k141_155985 | Not-determined |
| S1_extracted_k141_91053 | Not-determined |
| S1_extracted_k141_51328 | Not-determined |
| S1_extracted_k141_104519 | Not-determined |
| S1_extracted_k141_104520 | Not-determined |
| S1_extracted_k141_117889 | Not-determined |
| S1_extracted_k141_11312 | Not-determined |
| S1_extracted_k141_91056 | Not-determined |
| S1_extracted_k141_36867 | Not-determined |
| S1_extracted_k141_51330 | Not-determined |
| S1_extracted_k141_104524 | Not-determined |
| S1_extracted_k141_131172 | Not-determined |
| S1_extracted_k141_91059 | Not-determined |
| S1_extracted_k141_117897 | Not-determined |
| S1_extracted_k141_91060 | Not-determined |
| S1_extracted_k141_111135 | Not-determined |
| S1_extracted_k141_97964 | Low-quality |
| S1_extracted_k141_137724 | Not-determined |
| S1_extracted_k141_44532 | Not-determined |
| S1_extracted_k141_57693 | Not-determined |
| S1_extracted_k141_11317 | Not-determined |
| S1_extracted_k141_131178 | Low-quality |
| S1_extracted_k141_91064 | Not-determined |
| S1_extracted_k141_104531 | Not-determined |
| S1_extracted_k141_117905 | Not-determined |
| S1_extracted_k141_77841 | Not-determined |
| S1_extracted_k141_91070 | Not-determined |
| S1_extracted_k141_155994 | Not-determined |
| S1_extracted_k141_91072 | Not-determined |
| S1_extracted_k141_36880 | Not-determined |
| S1_extracted_k141_51348 | Not-determined |
| S1_extracted_k141_24187 | Not-determined |
| S1_extracted_k141_3523 | Not-determined |
| S1_extracted_k141_51356 | Not-determined |
| S1_extracted_k141_124155 | Not-determined |
| S1_extracted_k141_51357 | Low-quality |
| S1_extracted_k141_91079 | Low-quality |
| S1_extracted_k141_57714 | Not-determined |
| S1_extracted_k141_11337 | Not-determined |
| S1_extracted_k141_77859 | Not-determined |
| S1_extracted_k141_57717 | Not-determined |
| S1_extracted_k141_156007 | Low-quality |
| S1_extracted_k141_57718 | Not-determined |
| S1_extracted_k141_84540 | Not-determined |
| S1_extracted_k141_31177 | Not-determined |
| S1_extracted_k141_18266 | Not-determined |
| S1_extracted_k141_57721 | Not-determined |
| S1_extracted_k141_124165 | Not-determined |
| S1_extracted_k141_148860 | Not-determined |
| S1_extracted_k141_31178 | Not-determined |
| S1_extracted_k141_144200 | Not-determined |
| S1_extracted_k141_31180 | Not-determined |
| S1_extracted_k141_111141 | Not-determined |
| S1_extracted_k141_104551 | Low-quality |
| S1_extracted_k141_57727 | Not-determined |
| S1_extracted_k141_124169 | Not-determined |
| S1_extracted_k141_57728 | Low-quality |
| S1_extracted_k141_18267 | Low-quality |
| S1_extracted_k141_84548 | Not-determined |
| S1_extracted_k141_44563 | Not-determined |
| S1_extracted_k141_91091 | Not-determined |
| S1_extracted_k141_63169 | Not-determined |
| S1_extracted_k141_57730 | Not-determined |
| S1_extracted_k141_137748 | Not-determined |
| S1_extracted_k141_137749 | Not-determined |
| S1_extracted_k141_51368 | Not-determined |
| S1_extracted_k141_51369 | Not-determined |
| S1_extracted_k141_111147 | Not-determined |
| S1_extracted_k141_144209 | Not-determined |
| S1_extracted_k141_44568 | Not-determined |
| S1_extracted_k141_148868 | Not-determined |
| S1_extracted_k141_137751 | Low-quality |
| S1_extracted_k141_84551 | Not-determined |
| S1_extracted_k141_131203 | Not-determined |
| S1_extracted_k141_137755 | Not-determined |
| S1_extracted_k141_51373 | Not-determined |
| S1_extracted_k141_24213 | Not-determined |
| S1_extracted_k141_148871 | Not-determined |
| S1_extracted_k141_91099 | Not-determined |
| S1_extracted_k141_131207 | Low-quality |
| S1_extracted_k141_144216 | Not-determined |
| S1_extracted_k141_24218 | Not-determined |
| S1_extracted_k141_51379 | Not-determined |
| S1_extracted_k141_124177 | Low-quality |
| S1_extracted_k141_91102 | Not-determined |
| S1_extracted_k141_104568 | Not-determined |
| S1_extracted_k141_144220 | Not-determined |
| S1_extracted_k141_11360 | Low-quality |
| S1_extracted_k141_84557 | Not-determined |
| S1_extracted_k141_91104 | Low-quality |
| S1_extracted_k141_104570 | Not-determined |
| S1_extracted_k141_3544 | Not-determined |
| S1_extracted_k141_97985 | Not-determined |
| S1_extracted_k141_137767 | Not-determined |
| S1_extracted_k141_36910 | Not-determined |
| S1_extracted_k141_24228 | Not-determined |
| S1_extracted_k141_36911 | Not-determined |
| S1_extracted_k141_31204 | Not-determined |
| S1_extracted_k141_131218 | Not-determined |
| S1_extracted_k141_131219 | Not-determined |
| S1_extracted_k141_84561 | Not-determined |
| S1_extracted_k141_18293 | Low-quality |
| S1_extracted_k141_36913 | Low-quality |
| S1_extracted_k141_131220 | Not-determined |
| S1_extracted_k141_57740 | Low-quality |
| S1_extracted_k141_131222 | Not-determined |
| S1_extracted_k141_18297 | Not-determined |
| S1_extracted_k141_77903 | Low-quality |
| S1_extracted_k141_97989 | Low-quality |
| S1_extracted_k141_18299 | Not-determined |
| S1_extracted_k141_144230 | Not-determined |
| S1_extracted_k141_148881 | Low-quality |
| S1_extracted_k141_44575 | Low-quality |
| S1_extracted_k141_77905 | Not-determined |
| S1_extracted_k141_44576 | Not-determined |
| S1_extracted_k141_24237 | Not-determined |
| S1_extracted_k141_97993 | Not-determined |
| S1_extracted_k141_84571 | Not-determined |
| S1_extracted_k141_84573 | Not-determined |
| S1_extracted_k141_77911 | Not-determined |
| S1_extracted_k141_57752 | Not-determined |
| S1_extracted_k141_97996 | Not-determined |
| S1_extracted_k141_104579 | Not-determined |
| S1_extracted_k141_3561 | Not-determined |
| S1_extracted_k141_77915 | Not-determined |
| S1_extracted_k141_131228 | Not-determined |
| S1_extracted_k141_3564 | Not-determined |
| S1_extracted_k141_117938 | Not-determined |
| S1_extracted_k141_70551 | Not-determined |
| S1_extracted_k141_11379 | Low-quality |
| S1_extracted_k141_111181 | Not-determined |
| S1_extracted_k141_18308 | Not-determined |
| S1_extracted_k141_117941 | Not-determined |
| S1_extracted_k141_70553 | Not-determined |
| S1_extracted_k141_57760 | Not-determined |
| S1_extracted_k141_3569 | Not-determined |
| S1_extracted_k141_98004 | Not-determined |
| S1_extracted_k141_98006 | Not-determined |
| S1_extracted_k141_98007 | Not-determined |
| S1_extracted_k141_70556 | Not-determined |
| S1_extracted_k141_91137 | Low-quality |
| S1_extracted_k141_98008 | Not-determined |
| S1_extracted_k141_148897 | Not-determined |
| S1_extracted_k141_148899 | Low-quality |

| | |
|---|---|
| S1_extracted_k141_18319 | Not-determined |
| S1_extracted_k141_63210 | Not-determined |
| S1_extracted_k141_36944 | Not-determined |
| S1_extracted_k141_18322 | Not-determined |
| S1_extracted_k141_31232 | Low-quality |
| S1_extracted_k141_137805 | Low-quality |
| S1_extracted_k141_36945 | Not-determined |
| S1_extracted_k141_104593 | Not-determined |
| S1_extracted_k141_144260 | Not-determined |
| S1_extracted_k141_124217 | Low-quality |
| S1_extracted_k141_104594 | Not-determined |
| S1_extracted_k141_36946 | Not-determined |
| S1_extracted_k141_144262 | Not-determined |
| S1_extracted_k141_31236 | Not-determined |
| S1_extracted_k141_51424 | Not-determined |
| S1_extracted_k141_24273 | Not-determined |
| S1_extracted_k141_44596 | Low-quality |
| S1_extracted_k141_144265 | Not-determined |
| S1_extracted_k141_137808 | Not-determined |
| S1_extracted_k141_91149 | Not-determined |
| S1_extracted_k141_137809 | Low-quality |
| S1_extracted_k141_156022 | Not-determined |
| S1_extracted_k141_91151 | Not-determined |
| S1_extracted_k141_131253 | Not-determined |
| S1_extracted_k141_131254 | Not-determined |
| S1_extracted_k141_91153 | Not-determined |
| S1_extracted_k141_36955 | Not-determined |
| S1_extracted_k141_98019 | Low-quality |
| S1_extracted_k141_36958 | Not-determined |
| S1_extracted_k141_70575 | Not-determined |
| S1_extracted_k141_137811 | Not-determined |
| S1_extracted_k141_137812 | Not-determined |
| S1_extracted_k141_36960 | Not-determined |
| S1_extracted_k141_104604 | Not-determined |
| S1_extracted_k141_131258 | Not-determined |
| S1_extracted_k141_36964 | Not-determined |
| S1_extracted_k141_57774 | Not-determined |
| S1_extracted_k141_24285 | Not-determined |
| S1_extracted_k141_137818 | Not-determined |
| S1_extracted_k141_57776 | Not-determined |
| S1_extracted_k141_44607 | Not-determined |
| S1_extracted_k141_111200 | Not-determined |
| S1_extracted_k141_70577 | Not-determined |
| S1_extracted_k141_131262 | Not-determined |
| S1_extracted_k141_91167 | Not-determined |
| S1_extracted_k141_131263 | Low-quality |
| S1_extracted_k141_31257 | Not-determined |
| S1_extracted_k141_137822 | Not-determined |
| S1_extracted_k141_24294 | Not-determined |
| S1_extracted_k141_36971 | Not-determined |
| S1_extracted_k141_31262 | Not-determined |
| S1_extracted_k141_156032 | Not-determined |
| S1_extracted_k141_36973 | Not-determined |
| S1_extracted_k141_124243 | Not-determined |
| S1_extracted_k141_31265 | Not-determined |
| S1_extracted_k141_24297 | Not-determined |
| S1_extracted_k141_18346 | Not-determined |
| S1_extracted_k141_51448 | Not-determined |
| S1_extracted_k141_131273 | Not-determined |
| S1_extracted_k141_124247 | Not-determined |
| S1_extracted_k141_137830 | Not-determined |
| S1_extracted_k141_36975 | Not-determined |
| S1_extracted_k141_77962 | Not-determined |
| S1_extracted_k141_31268 | Not-determined |
| S1_extracted_k141_36976 | Low-quality |
| S1_extracted_k141_77964 | Not-determined |
| S1_extracted_k141_124251 | Not-determined |
| S1_extracted_k141_57791 | Not-determined |
| S1_extracted_k141_51450 | Low-quality |
| S1_extracted_k141_44615 | Not-determined |
| S1_extracted_k141_63229 | Not-determined |
| S1_extracted_k141_131278 | Not-determined |
| S1_extracted_k141_84613 | Not-determined |
| S1_extracted_k141_111204 | Not-determined |
| S1_extracted_k141_31273 | Not-determined |
| S1_extracted_k141_111205 | Not-determined |
| S1_extracted_k141_51453 | Not-determined |
| S1_extracted_k141_70590 | Not-determined |
| S1_extracted_k141_11400 | Not-determined |
| S1_extracted_k141_63232 | Not-determined |
| S1_extracted_k141_11402 | Not-determined |
| S1_extracted_k141_51457 | Not-determined |
| S1_extracted_k141_117980 | Not-determined |
| S1_extracted_k141_91188 | Not-determined |
| S1_extracted_k141_84618 | Not-determined |
| S1_extracted_k141_117982 | Not-determined |
| S1_extracted_k141_31277 | Not-determined |
| S1_extracted_k141_44625 | Not-determined |
| S1_extracted_k141_57798 | Not-determined |
| S1_extracted_k141_11408 | Not-determined |
| S1_extracted_k141_24310 | Low-quality |
| S1_extracted_k141_57800 | Not-determined |
| S1_extracted_k141_144295 | Not-determined |
| S1_extracted_k141_77976 | Low-quality |
| S1_extracted_k141_57803 | Not-determined |
| S1_extracted_k141_51461 | Not-determined |
| S1_extracted_k141_131290 | Not-determined |
| S1_extracted_k141_18364 | Not-determined |
| S1_extracted_k141_11413 | Not-determined |
| S1_extracted_k141_117987 | Not-determined |
| S1_extracted_k141_137842 | Not-determined |
| S1_extracted_k141_11414 | Not-determined |
| S1_extracted_k141_31284 | Low-quality |
| S1_extracted_k141_31285 | Not-determined |
| S1_extracted_k141_137845 | Not-determined |
| S1_extracted_k141_117990 | Not-determined |
| S1_extracted_k141_24319 | Not-determined |
| S1_extracted_k141_137847 | Not-determined |
| S1_extracted_k141_84625 | Low-quality |
| S1_extracted_k141_84628 | Low-quality |
| S1_extracted_k141_11421 | Not-determined |
| S1_extracted_k141_18371 | Not-determined |
| S1_extracted_k141_124270 | Not-determined |
| S1_extracted_k141_24324 | Low-quality |
| S1_extracted_k141_11423 | Not-determined |
| S1_extracted_k141_70604 | Not-determined |
| S1_extracted_k141_111224 | Low-quality |
| S1_extracted_k141_11425 | Not-determined |
| S1_extracted_k141_57812 | Not-determined |
| S1_extracted_k141_117995 | Not-determined |
| S1_extracted_k141_84632 | Not-determined |
| S1_extracted_k141_18375 | Not-determined |
| S1_extracted_k141_117996 | Not-determined |
| S1_extracted_k141_144302 | Low-quality |
| S1_extracted_k141_131300 | Not-determined |
| S1_extracted_k141_57816 | Not-determined |
| S1_extracted_k141_31293 | Not-determined |
| S1_extracted_k141_111227 | Not-determined |
| S1_extracted_k141_70611 | Not-determined |
| S1_extracted_k141_70612 | Low-quality |
| S1_extracted_k141_3584 | Not-determined |
| S1_extracted_k141_11431 | Not-determined |
| S1_extracted_k141_57821 | Not-determined |
| S1_extracted_k141_24335 | Not-determined |
| S1_extracted_k141_57823 | Not-determined |
| S1_extracted_k141_63240 | Not-determined |
| S1_extracted_k141_63243 | Not-determined |
| S1_extracted_k141_77998 | Not-determined |
| S1_extracted_k141_131316 | Not-determined |
| S1_extracted_k141_131317 | Not-determined |
| S1_extracted_k141_124281 | Not-determined |
| S1_extracted_k141_70620 | Not-determined |
| S1_extracted_k141_144314 | Not-determined |
| S1_extracted_k141_51498 | Not-determined |
| S1_extracted_k141_91226 | Not-determined |
| S1_extracted_k141_111240 | Low-quality |
| S1_extracted_k141_36994 | Not-determined |
| S1_extracted_k141_131324 | Not-determined |
| S1_extracted_k141_18393 | Not-determined |
| S1_extracted_k141_11442 | Not-determined |
| S1_extracted_k141_91230 | Not-determined |
| S1_extracted_k141_24348 | Not-determined |
| S1_extracted_k141_137865 | Low-quality |
| S1_extracted_k141_148912 | Not-determined |
| S1_extracted_k141_144317 | Low-quality |
| S1_extracted_k141_24351 | Not-determined |
| S1_extracted_k141_98063 | Not-determined |
| S1_extracted_k141_31311 | Low-quality |
| S1_extracted_k141_11446 | Low-quality |
| S1_extracted_k141_44647 | Not-determined |
| S1_extracted_k141_18397 | Not-determined |
| S1_extracted_k141_137867 | Not-determined |
| S1_extracted_k141_70627 | Not-determined |
| S1_extracted_k141_31318 | Not-determined |
| S1_extracted_k141_31321 | Not-determined |
| S1_extracted_k141_144322 | Low-quality |
| S1_extracted_k141_37008 | Not-determined |
| S1_extracted_k141_24359 | Not-determined |
| S1_extracted_k141_31326 | Not-determined |
| S1_extracted_k141_44654 | Not-determined |
| S1_extracted_k141_44655 | Not-determined |
| S1_extracted_k141_131340 | Not-determined |
| S1_extracted_k141_104651 | Low-quality |
| S1_extracted_k141_144329 | Not-determined |
| S1_extracted_k141_51514 | Not-determined |
| S1_extracted_k141_98078 | Not-determined |
| S1_extracted_k141_98079 | Not-determined |
| S1_extracted_k141_118022 | Not-determined |
| S1_extracted_k141_78009 | Not-determined |
| S1_extracted_k141_70638 | Not-determined |
| S1_extracted_k141_98080 | Low-quality |
| S1_extracted_k141_51518 | Not-determined |
| S1_extracted_k141_24366 | Not-determined |
| S1_extracted_k141_111258 | Low-quality |
| S1_extracted_k141_78011 | Not-determined |
| S1_extracted_k141_91235 | Not-determined |
| S1_extracted_k141_44663 | Not-determined |
| S1_extracted_k141_137884 | Not-determined |
| S1_extracted_k141_51521 | Low-quality |
| S1_extracted_k141_98089 | Not-determined |
| S1_extracted_k141_137886 | Not-determined |
| S1_extracted_k141_24373 | Not-determined |
| S1_extracted_k141_31334 | Not-determined |
| S1_extracted_k141_84663 | Not-determined |
| S1_extracted_k141_18420 | Not-determined |
| S1_extracted_k141_44668 | Low-quality |
| S1_extracted_k141_44669 | Not-determined |
| S1_extracted_k141_70642 | Not-determined |
| S1_extracted_k141_37016 | Not-determined |
| S1_extracted_k141_98095 | Low-quality |
| S1_extracted_k141_31340 | Not-determined |
| S1_extracted_k141_63260 | Not-determined |
| S1_extracted_k141_31342 | Not-determined |
| S1_extracted_k141_104669 | Not-determined |
| S1_extracted_k141_24377 | Not-determined |
| S1_extracted_k141_63262 | Low-quality |
| S1_extracted_k141_137895 | Not-determined |
| S1_extracted_k141_18432 | Not-determined |
| S1_extracted_k141_31344 | Not-determined |
| S1_extracted_k141_11469 | Not-determined |
| S1_extracted_k141_156038 | Not-determined |
| S1_extracted_k141_148919 | Not-determined |
| S1_extracted_k141_84670 | Not-determined |
| S1_extracted_k141_11470 | Low-quality |
| S1_extracted_k141_98101 | Low-quality |
| S1_extracted_k141_137900 | Not-determined |
| S1_extracted_k141_24383 | Not-determined |
| S1_extracted_k141_84675 | Low-quality |
| S1_extracted_k141_104674 | Not-determined |
| S1_extracted_k141_11471 | Low-quality |
| S1_extracted_k141_144348 | Not-determined |
| S1_extracted_k141_11472 | Not-determined |
| S1_extracted_k141_63264 | Not-determined |
| S1_extracted_k141_104676 | Not-determined |
| S1_extracted_k141_156045 | Not-determined |
| S1_extracted_k141_124301 | Not-determined |
| S1_extracted_k141_124302 | Not-determined |
| S1_extracted_k141_144350 | Not-determined |
| S1_extracted_k141_63266 | Not-determined |
| S1_extracted_k141_131374 | Low-quality |
| S1_extracted_k141_156046 | Low-quality |
| S1_extracted_k141_51531 | Not-determined |
| S1_extracted_k141_137904 | Not-determined |
| S1_extracted_k141_118036 | Not-determined |
| S1_extracted_k141_44689 | Not-determined |
| S1_extracted_k141_44690 | Not-determined |
| S1_extracted_k141_24386 | Not-determined |
| S1_extracted_k141_131377 | Not-determined |
| S1_extracted_k141_44691 | Not-determined |
| S1_extracted_k141_131378 | Not-determined |
| S1_extracted_k141_91250 | Not-determined |
| S1_extracted_k141_18447 | Not-determined |
| S1_extracted_k141_124307 | Not-determined |
| S1_extracted_k141_84681 | Not-determined |
| S1_extracted_k141_37028 | Low-quality |
| S1_extracted_k141_37029 | Not-determined |
| S1_extracted_k141_24388 | Not-determined |
| S1_extracted_k141_44697 | Not-determined |
| S1_extracted_k141_91255 | Not-determined |
| S1_extracted_k141_18458 | Not-determined |
| S1_extracted_k141_111294 | Not-determined |
| S1_extracted_k141_57895 | Not-determined |
| S1_extracted_k141_18461 | Not-determined |
| S1_extracted_k141_31358 | Not-determined |
| S1_extracted_k141_11490 | Not-determined |
| S1_extracted_k141_98121 | Not-determined |
| S1_extracted_k141_51544 | Not-determined |
| S1_extracted_k141_70651 | Not-determined |
| S1_extracted_k141_156060 | Not-determined |
| S1_extracted_k141_118045 | Not-determined |
| S1_extracted_k141_118046 | Not-determined |
| S1_extracted_k141_44709 | Not-determined |
| S1_extracted_k141_137923 | Not-determined |
| S1_extracted_k141_63283 | Not-determined |
| S1_extracted_k141_98127 | Not-determined |
| S1_extracted_k141_124321 | Low-quality |
| S1_extracted_k141_18470 | Not-determined |
| S1_extracted_k141_84698 | Not-determined |
| S1_extracted_k141_18471 | Not-determined |
| S1_extracted_k141_24400 | Not-determined |
| S1_extracted_k141_124324 | Not-determined |
| S1_extracted_k141_104695 | Not-determined |
| S1_extracted_k141_51551 | Not-determined |
| S1_extracted_k141_57907 | Not-determined |
| S1_extracted_k141_124328 | Not-determined |
| S1_extracted_k141_137933 | Not-determined |
| S1_extracted_k141_137934 | Not-determined |
| S1_extracted_k141_63289 | Not-determined |
| S1_extracted_k141_3644 | Not-determined |
| S1_extracted_k141_148937 | Not-determined |
| S1_extracted_k141_24406 | Not-determined |
| S1_extracted_k141_148938 | Low-quality |
| S1_extracted_k141_11510 | Not-determined |
| S1_extracted_k141_104701 | Not-determined |
| S1_extracted_k141_144373 | Not-determined |
| S1_extracted_k141_63296 | Not-determined |
| S1_extracted_k141_118062 | Not-determined |
| S1_extracted_k141_44718 | Not-determined |
| S1_extracted_k141_51556 | Low-quality |
| S1_extracted_k141_98135 | Low-quality |
| S1_extracted_k141_144375 | Not-determined |
| S1_extracted_k141_78055 | Not-determined |
| S1_extracted_k141_3648 | Not-determined |
| S1_extracted_k141_144376 | Low-quality |
| S1_extracted_k141_44720 | Low-quality |
| S1_extracted_k141_137940 | Not-determined |
| S1_extracted_k141_18474 | Not-determined |
| S1_extracted_k141_156075 | Not-determined |
| S1_extracted_k141_18477 | Not-determined |
| S1_extracted_k141_78059 | Not-determined |
| S1_extracted_k141_37056 | Low-quality |
| S1_extracted_k141_18480 | Not-determined |
| S1_extracted_k141_78061 | Low-quality |
| S1_extracted_k141_144381 | Low-quality |
| S1_extracted_k141_57930 | Not-determined |
| S1_extracted_k141_11523 | Not-determined |
| S1_extracted_k141_44734 | Not-determined |
| S1_extracted_k141_51574 | Low-quality |
| S1_extracted_k141_144383 | Low-quality |
| S1_extracted_k141_84723 | Low-quality |
| S1_extracted_k141_144386 | Not-determined |
| S1_extracted_k141_37060 | Low-quality |
| S1_extracted_k141_18482 | Not-determined |
| S1_extracted_k141_137954 | Low-quality |
| S1_extracted_k141_98145 | Not-determined |
| S1_extracted_k141_44743 | Not-determined |
| S1_extracted_k141_137958 | Not-determined |
| S1_extracted_k141_44744 | Not-determined |
| S1_extracted_k141_144392 | Not-determined |
| S1_extracted_k141_57939 | Not-determined |
| S1_extracted_k141_144393 | Not-determined |
| S1_extracted_k141_91272 | Not-determined |
| S1_extracted_k141_124351 | Not-determined |
| S1_extracted_k141_84729 | Not-determined |
| S1_extracted_k141_3664 | Not-determined |
| S1_extracted_k141_98153 | Low-quality |
| S1_extracted_k141_111325 | Not-determined |
| S1_extracted_k141_63321 | Not-determined |
| S1_extracted_k141_111328 | Not-determined |
| S1_extracted_k141_51582 | Not-determined |
| S1_extracted_k141_63325 | Not-determined |
| S1_extracted_k141_51583 | Not-determined |
| S1_extracted_k141_111330 | Low-quality |
| S1_extracted_k141_24430 | Not-determined |
| S1_extracted_k141_124358 | Not-determined |
| S1_extracted_k141_131407 | Low-quality |
| S1_extracted_k141_24431 | Not-determined |
| S1_extracted_k141_111331 | Not-determined |
| S1_extracted_k141_144406 | Not-determined |
| S1_extracted_k141_18506 | Not-determined |
| S1_extracted_k141_148949 | Not-determined |
| S1_extracted_k141_51586 | Not-determined |
| S1_extracted_k141_63327 | Not-determined |
| S1_extracted_k141_131411 | Not-determined |
| S1_extracted_k141_148951 | Not-determined |
| S1_extracted_k141_104735 | Not-determined |
| S1_extracted_k141_156098 | Not-determined |
| S1_extracted_k141_24433 | Low-quality |
| S1_extracted_k141_18511 | Not-determined |
| S1_extracted_k141_3678 | Not-determined |
| S1_extracted_k141_11546 | Not-determined |
| S1_extracted_k141_124367 | Not-determined |
| S1_extracted_k141_144411 | Not-determined |
| S1_extracted_k141_91283 | Low-quality |
| S1_extracted_k141_137978 | Not-determined |
| S1_extracted_k141_24442 | Not-determined |
| S1_extracted_k141_131418 | Not-determined |
| S1_extracted_k141_37066 | Not-determined |
| S1_extracted_k141_104746 | Not-determined |
| S1_extracted_k141_144416 | Not-determined |
| S1_extracted_k141_124377 | Not-determined |
| S1_extracted_k141_84747 | Low-quality |
| S1_extracted_k141_18522 | Not-determined |
| S1_extracted_k141_137981 | Not-determined |
| S1_extracted_k141_124379 | Low-quality |
| S1_extracted_k141_104749 | Not-determined |
| S1_extracted_k141_118099 | Not-determined |
| S1_extracted_k141_104753 | Low-quality |
| S1_extracted_k141_24451 | Not-determined |
| S1_extracted_k141_111344 | Not-determined |
| S1_extracted_k141_104755 | Not-determined |
| S1_extracted_k141_137983 | Not-determined |
| S1_extracted_k141_24456 | Low-quality |
| S1_extracted_k141_111348 | Low-quality |
| S1_extracted_k141_111349 | Not-determined |
| S1_extracted_k141_91290 | Low-quality |
| S1_extracted_k141_31372 | Low-quality |
| S1_extracted_k141_44765 | Not-determined |
| S1_extracted_k141_111353 | Not-determined |
| S1_extracted_k141_3689 | Not-determined |
| S1_extracted_k141_124386 | Not-determined |
| S1_extracted_k141_84756 | Not-determined |
| S1_extracted_k141_111356 | Not-determined |
| S1_extracted_k141_137991 | Not-determined |
| S1_extracted_k141_144430 | Not-determined |
| S1_extracted_k141_51608 | Not-determined |
| S1_extracted_k141_84758 | Not-determined |
| S1_extracted_k141_24461 | Not-determined |
| S1_extracted_k141_63346 | Not-determined |
| S1_extracted_k141_124391 | Not-determined |
| S1_extracted_k141_18532 | Not-determined |
| S1_extracted_k141_84762 | Low-quality |
| S1_extracted_k141_37083 | Not-determined |
| S1_extracted_k141_84763 | Not-determined |
| S1_extracted_k141_51611 | Not-determined |
| S1_extracted_k141_44772 | Not-determined |
| S1_extracted_k141_111360 | Not-determined |
| S1_extracted_k141_57977 | Not-determined |
| S1_extracted_k141_111361 | Not-determined |
| S1_extracted_k141_57978 | Not-determined |
| S1_extracted_k141_37086 | Not-determined |
| S1_extracted_k141_24468 | Not-determined |
| S1_extracted_k141_44775 | Not-determined |
| S1_extracted_k141_63353 | Low-quality |
| S1_extracted_k141_137997 | Not-determined |
| S1_extracted_k141_111365 | Not-determined |
| S1_extracted_k141_98167 | Not-determined |
| S1_extracted_k141_78085 | Not-determined |
| S1_extracted_k141_78086 | Low-quality |
| S1_extracted_k141_148966 | Low-quality |
| S1_extracted_k141_84774 | Not-determined |
| S1_extracted_k141_111367 | Not-determined |
| S1_extracted_k141_57985 | Not-determined |
| S1_extracted_k141_84775 | Low-quality |
| S1_extracted_k141_91304 | Not-determined |
| S1_extracted_k141_57986 | Not-determined |
| S1_extracted_k141_138005 | Not-determined |
| S1_extracted_k141_144441 | Not-determined |
| S1_extracted_k141_111371 | Not-determined |
| S1_extracted_k141_98170 | Not-determined |
| S1_extracted_k141_70678 | Not-determined |
| S1_extracted_k141_104778 | Not-determined |
| S1_extracted_k141_70680 | Not-determined |
| S1_extracted_k141_63367 | Not-determined |
| S1_extracted_k141_44784 | Low-quality |
| S1_extracted_k141_37092 | Not-determined |
| S1_extracted_k141_131438 | Low-quality |
| S1_extracted_k141_148967 | Not-determined |
| S1_extracted_k141_31383 | Low-quality |
| S1_extracted_k141_98177 | Not-determined |
| S1_extracted_k141_124408 | Not-determined |
| S1_extracted_k141_98181 | Not-determined |
| S1_extracted_k141_24475 | Not-determined |
| S1_extracted_k141_31387 | Not-determined |
| S1_extracted_k141_98182 | Not-determined |
| S1_extracted_k141_24477 | Low-quality |
| S1_extracted_k141_138015 | Not-determined |
| S1_extracted_k141_31389 | Not-determined |
| S1_extracted_k141_124412 | Low-quality |
| S1_extracted_k141_31391 | Not-determined |
| S1_extracted_k141_31392 | Not-determined |
| S1_extracted_k141_58000 | Not-determined |
| S1_extracted_k141_58001 | Not-determined |
| S1_extracted_k141_148974 | Not-determined |
| S1_extracted_k141_84785 | Not-determined |
| S1_extracted_k141_78107 | Not-determined |
| S1_extracted_k141_131447 | Not-determined |
| S1_extracted_k141_58002 | Not-determined |
| S1_extracted_k141_98189 | Not-determined |
| S1_extracted_k141_131448 | Not-determined |
| S1_extracted_k141_144458 | Not-determined |
| S1_extracted_k141_24483 | Not-determined |
| S1_extracted_k141_44794 | Not-determined |
| S1_extracted_k141_11591 | Not-determined |
| S1_extracted_k141_24485 | Not-determined |
| S1_extracted_k141_118132 | Not-determined |
| S1_extracted_k141_131455 | Low-quality |
| S1_extracted_k141_63384 | Not-determined |
| S1_extracted_k141_118138 | Not-determined |
| S1_extracted_k141_51633 | Not-determined |
| S1_extracted_k141_138032 | Not-determined |
| S1_extracted_k141_24493 | Not-determined |
| S1_extracted_k141_144462 | Not-determined |
| S1_extracted_k141_138033 | Not-determined |
| S1_extracted_k141_118142 | Low-quality |
| S1_extracted_k141_24496 | Low-quality |
| S1_extracted_k141_3717 | Low-quality |
| S1_extracted_k141_3719 | Low-quality |
| S1_extracted_k141_70684 | Low-quality |
| S1_extracted_k141_91333 | Not-determined |
| S1_extracted_k141_111393 | Not-determined |
| S1_extracted_k141_63397 | Low-quality |
| S1_extracted_k141_84795 | Low-quality |
| S1_extracted_k141_18562 | Not-determined |
| S1_extracted_k141_111395 | Not-determined |
| S1_extracted_k141_148981 | Not-determined |
| S1_extracted_k141_51640 | Not-determined |
| S1_extracted_k141_111397 | Not-determined |
| S1_extracted_k141_18566 | Not-determined |
| S1_extracted_k141_78137 | Not-determined |
| S1_extracted_k141_118149 | Low-quality |
| S1_extracted_k141_58012 | Low-quality |
| S1_extracted_k141_63406 | Not-determined |
| S1_extracted_k141_124432 | Not-determined |
| S1_extracted_k141_63407 | Not-determined |
| S1_extracted_k141_63408 | Not-determined |
| S1_extracted_k141_144469 | Low-quality |
| S1_extracted_k141_138042 | Not-determined |
| S1_extracted_k141_24522 | Not-determined |
| S1_extracted_k141_124437 | Not-determined |
| S1_extracted_k141_111408 | Not-determined |
| S1_extracted_k141_37117 | Not-determined |
| S1_extracted_k141_37118 | Not-determined |
| S1_extracted_k141_91352 | Not-determined |
| S1_extracted_k141_51649 | Low-quality |
| S1_extracted_k141_131480 | Not-determined |
| S1_extracted_k141_124442 | Not-determined |
| S1_extracted_k141_58025 | Not-determined |
| S1_extracted_k141_58027 | Low-quality |
| S1_extracted_k141_91354 | Not-determined |
| S1_extracted_k141_118160 | Not-determined |
| S1_extracted_k141_131485 | Low-quality |
| S1_extracted_k141_63418 | Low-quality |
| S1_extracted_k141_58030 | Not-determined |
| S1_extracted_k141_63419 | Not-determined |
| S1_extracted_k141_156141 | Not-determined |
| S1_extracted_k141_24537 | Not-determined |
| S1_extracted_k141_3743 | Not-determined |
| S1_extracted_k141_70695 | Not-determined |
| S1_extracted_k141_156142 | Low-quality |
| S1_extracted_k141_31409 | Not-determined |
| S1_extracted_k141_91357 | Not-determined |
| S1_extracted_k141_24539 | Not-determined |
| S1_extracted_k141_31412 | Not-determined |
| S1_extracted_k141_37123 | Not-determined |
| S1_extracted_k141_138057 | Not-determined |
| S1_extracted_k141_111422 | Low-quality |
| S1_extracted_k141_91361 | Not-determined |
| S1_extracted_k141_58036 | Not-determined |
| S1_extracted_k141_70700 | Not-determined |
| S1_extracted_k141_18576 | Not-determined |
| S1_extracted_k141_118171 | Not-determined |
| S1_extracted_k141_24543 | Not-determined |
| S1_extracted_k141_124449 | Not-determined |
| S1_extracted_k141_51663 | Not-determined |
| S1_extracted_k141_118173 | Low-quality |
| S1_extracted_k141_124451 | Not-determined |
| S1_extracted_k141_118174 | Not-determined |
| S1_extracted_k141_148991 | Not-determined |
| S1_extracted_k141_44822 | Not-determined |
| S1_extracted_k141_156151 | Not-determined |
| S1_extracted_k141_131499 | Low-quality |
| S1_extracted_k141_84821 | Not-determined |
| S1_extracted_k141_63432 | Low-quality |
| S1_extracted_k141_131500 | Not-determined |
| S1_extracted_k141_124455 | Not-determined |
| S1_extracted_k141_58044 | Not-determined |
| S1_extracted_k141_98231 | Not-determined |
| S1_extracted_k141_44826 | Not-determined |
| S1_extracted_k141_51668 | Not-determined |
| S1_extracted_k141_44828 | Not-determined |
| S1_extracted_k141_124457 | Low-quality |
| S1_extracted_k141_118179 | Not-determined |
| S1_extracted_k141_70704 | Not-determined |
| S1_extracted_k141_51669 | Not-determined |
| S1_extracted_k141_156153 | Low-quality |
| S1_extracted_k141_104815 | Not-determined |
| S1_extracted_k141_131505 | Low-quality |
| S1_extracted_k141_24554 | Not-determined |
| S1_extracted_k141_131507 | Low-quality |
| S1_extracted_k141_63439 | Not-determined |
| S1_extracted_k141_58051 | Low-quality |
| S1_extracted_k141_63440 | Not-determined |
| S1_extracted_k141_84825 | Not-determined |
| S1_extracted_k141_138067 | Not-determined |
| S1_extracted_k141_104821 | Low-quality |
| S1_extracted_k141_118183 | Not-determined |
| S1_extracted_k141_111430 | Not-determined |
| S1_extracted_k141_78154 | Low-quality |
| S1_extracted_k141_104825 | Not-determined |
| S1_extracted_k141_144505 | Not-determined |
| S1_extracted_k141_51670 | Low-quality |
| S1_extracted_k141_58053 | Not-determined |
| S1_extracted_k141_63443 | Not-determined |
| S1_extracted_k141_3754 | Low-quality |
| S1_extracted_k141_44841 | Not-determined |
| S1_extracted_k141_124465 | Not-determined |
| S1_extracted_k141_118189 | Not-determined |
| S1_extracted_k141_104828 | Not-determined |
| S1_extracted_k141_51674 | Not-determined |
| S1_extracted_k141_31427 | Not-determined |
| S1_extracted_k141_51675 | Not-determined |
| S1_extracted_k141_78165 | Not-determined |
| S1_extracted_k141_63446 | Not-determined |
| S1_extracted_k141_78168 | Not-determined |
| S1_extracted_k141_78169 | Not-determined |
| S1_extracted_k141_131519 | Not-determined |
| S1_extracted_k141_51679 | Low-quality |
| S1_extracted_k141_31432 | Not-determined |
| S1_extracted_k141_31435 | Not-determined |
| S1_extracted_k141_11641 | Not-determined |
| S1_extracted_k141_111439 | Not-determined |
| S1_extracted_k141_24575 | Not-determined |
| S1_extracted_k141_98253 | Not-determined |
| S1_extracted_k141_37145 | Not-determined |
| S1_extracted_k141_118198 | Low-quality |
| S1_extracted_k141_18592 | Not-determined |
| S1_extracted_k141_58063 | Not-determined |
| S1_extracted_k141_91384 | Not-determined |

| | |
|---|---|
| S1_extracted_k141_11643 | Not-determined |
| S1_extracted_k141_18597 | Not-determined |
| S1_extracted_k141_37149 | Not-determined |
| S1_extracted_k141_63456 | Not-determined |
| S1_extracted_k141_58066 | Low-quality |
| S1_extracted_k141_31443 | Not-determined |
| S1_extracted_k141_24581 | Low-quality |
| S1_extracted_k141_124468 | Low-quality |
| S1_extracted_k141_156170 | Not-determined |
| S1_extracted_k141_63461 | Low-quality |
| S1_extracted_k141_131528 | Not-determined |
| S1_extracted_k141_3770 | Not-determined |
| S1_extracted_k141_51683 | Not-determined |
| S1_extracted_k141_118205 | Not-determined |
| S1_extracted_k141_63464 | Not-determined |
| S1_extracted_k141_111447 | Not-determined |
| S1_extracted_k141_118206 | Not-determined |
| S1_extracted_k141_78185 | Not-determined |
| S1_extracted_k141_18605 | Low-quality |
| S1_extracted_k141_111448 | Not-determined |
| S1_extracted_k141_31448 | Not-determined |
| S1_extracted_k141_131531 | Low-quality |
| S1_extracted_k141_63467 | Not-determined |
| S1_extracted_k141_70710 | Not-determined |
| S1_extracted_k141_18609 | Not-determined |
| S1_extracted_k141_98267 | Not-determined |
| S1_extracted_k141_148996 | Not-determined |
| S1_extracted_k141_131537 | Not-determined |
| S1_extracted_k141_84840 | Not-determined |
| S1_extracted_k141_104841 | Not-determined |
| S1_extracted_k141_124478 | Low-quality |
| S1_extracted_k141_91394 | Not-determined |
| S1_extracted_k141_138081 | Not-determined |
| S1_extracted_k141_44857 | Not-determined |
| S1_extracted_k141_18616 | Low-quality |
| S1_extracted_k141_24596 | Not-determined |
| S1_extracted_k141_131545 | Not-determined |
| S1_extracted_k141_91398 | Not-determined |
| S1_extracted_k141_3781 | Not-determined |
| S1_extracted_k141_63481 | Low-quality |
| S1_extracted_k141_111461 | Not-determined |
| S1_extracted_k141_44865 | Not-determined |
| S1_extracted_k141_37163 | Not-determined |
| S1_extracted_k141_84853 | Low-quality |
| S1_extracted_k141_44869 | Not-determined |
| S1_extracted_k141_51698 | Not-determined |
| S1_extracted_k141_84854 | Low-quality |
| S1_extracted_k141_156189 | Not-determined |
| S1_extracted_k141_98272 | Not-determined |
| S1_extracted_k141_138093 | Not-determined |
| S1_extracted_k141_78201 | Not-determined |
| S1_extracted_k141_104853 | Not-determined |
| S1_extracted_k141_70715 | Low-quality |
| S1_extracted_k141_44874 | Low-quality |
| S1_extracted_k141_31461 | Not-determined |
| S1_extracted_k141_104860 | Not-determined |
| S1_extracted_k141_11662 | Not-determined |
| S1_extracted_k141_18633 | Not-determined |
| S1_extracted_k141_63489 | Not-determined |
| S1_extracted_k141_144518 | Low-quality |
| S1_extracted_k141_111473 | Not-determined |
| S1_extracted_k141_104863 | Not-determined |
| S1_extracted_k141_44880 | Not-determined |
| S1_extracted_k141_124489 | Not-determined |
| S1_extracted_k141_44881 | Not-determined |
| S1_extracted_k141_11669 | Low-quality |
| S1_extracted_k141_149005 | Low-quality |
| S1_extracted_k141_58070 | Low-quality |
| S1_extracted_k141_124493 | Not-determined |
| S1_extracted_k141_98286 | Not-determined |
| S1_extracted_k141_104869 | Low-quality |
| S1_extracted_k141_24615 | Not-determined |
| S1_extracted_k141_149006 | Not-determined |
| S1_extracted_k141_44883 | Low-quality |
| S1_extracted_k141_138105 | Not-determined |
| S1_extracted_k141_111479 | Low-quality |
| S1_extracted_k141_98291 | Not-determined |
| S1_extracted_k141_58074 | Not-determined |
| S1_extracted_k141_37173 | Not-determined |
| S1_extracted_k141_11675 | Not-determined |
| S1_extracted_k141_84865 | Not-determined |
| S1_extracted_k141_149012 | Not-determined |
| S1_extracted_k141_24626 | Not-determined |
| S1_extracted_k141_44889 | Not-determined |
| S1_extracted_k141_138112 | Not-determined |
| S1_extracted_k141_138114 | Not-determined |
| S1_extracted_k141_124504 | Not-determined |
| S1_extracted_k141_149014 | Low-quality |
| S1_extracted_k141_138118 | Not-determined |
| S1_extracted_k141_111488 | Not-determined |
| S1_extracted_k141_24632 | Low-quality |
| S1_extracted_k141_44898 | Not-determined |
| S1_extracted_k141_24635 | Not-determined |
| S1_extracted_k141_63507 | Low-quality |
| S1_extracted_k141_24636 | Not-determined |
| S1_extracted_k141_111491 | Not-determined |
| S1_extracted_k141_37185 | Not-determined |
| S1_extracted_k141_98296 | Not-determined |
| S1_extracted_k141_111494 | Not-determined |
| S1_extracted_k141_58088 | Not-determined |
| S1_extracted_k141_111497 | Not-determined |
| S1_extracted_k141_118242 | Not-determined |
| S1_extracted_k141_70718 | Not-determined |
| S1_extracted_k141_44905 | Low-quality |
| S1_extracted_k141_118245 | Not-determined |
| S1_extracted_k141_149026 | Not-determined |
| S1_extracted_k141_131566 | Low-quality |
| S1_extracted_k141_144546 | Not-determined |
| S1_extracted_k141_104894 | Not-determined |
| S1_extracted_k141_24649 | Not-determined |
| S1_extracted_k141_24650 | Not-determined |
| S1_extracted_k141_63514 | Not-determined |
| S1_extracted_k141_91451 | Not-determined |
| S1_extracted_k141_98306 | Not-determined |
| S1_extracted_k141_11694 | Not-determined |
| S1_extracted_k141_70721 | Not-determined |
| S1_extracted_k141_78239 | Not-determined |
| S1_extracted_k141_98311 | Not-determined |
| S1_extracted_k141_70724 | Not-determined |
| S1_extracted_k141_37186 | Low-quality |
| S1_extracted_k141_24657 | Not-determined |
| S1_extracted_k141_31491 | Not-determined |
| S1_extracted_k141_111515 | Low-quality |
| S1_extracted_k141_37189 | Not-determined |
| S1_extracted_k141_118258 | Not-determined |
| S1_extracted_k141_98313 | Low-quality |
| S1_extracted_k141_31497 | Not-determined |
| S1_extracted_k141_149033 | Not-determined |
| S1_extracted_k141_31499 | Not-determined |
| S1_extracted_k141_131578 | Not-determined |
| S1_extracted_k141_104904 | Not-determined |
| S1_extracted_k141_118261 | Not-determined |
| S1_extracted_k141_118262 | Not-determined |
| S1_extracted_k141_78250 | Low-quality |
| S1_extracted_k141_51715 | Not-determined |
| S1_extracted_k141_104905 | Low-quality |
| S1_extracted_k141_58108 | Not-determined |
| S1_extracted_k141_11706 | Not-determined |
| S1_extracted_k141_84882 | Not-determined |
| S1_extracted_k141_63536 | Not-determined |
| S1_extracted_k141_149037 | Not-determined |
| S1_extracted_k141_37195 | Not-determined |
| S1_extracted_k141_104908 | Not-determined |
| S1_extracted_k141_31510 | Not-determined |
| S1_extracted_k141_51717 | Low-quality |
| S1_extracted_k141_149040 | Not-determined |
| S1_extracted_k141_144560 | Not-determined |
| S1_extracted_k141_131589 | Low-quality |
| S1_extracted_k141_63544 | Not-determined |
| S1_extracted_k141_63545 | Not-determined |
| S1_extracted_k141_91470 | Low-quality |
| S1_extracted_k141_91471 | Low-quality |
| S1_extracted_k141_104913 | Not-determined |
| S1_extracted_k141_44919 | Not-determined |
| S1_extracted_k141_111534 | Low-quality |
| S1_extracted_k141_44920 | Not-determined |
| S1_extracted_k141_144565 | Not-determined |
| S1_extracted_k141_11717 | Not-determined |
| S1_extracted_k141_78262 | Not-determined |
| S1_extracted_k141_11720 | Not-determined |
| S1_extracted_k141_24677 | Not-determined |
| S1_extracted_k141_24678 | Low-quality |
| S1_extracted_k141_98330 | Not-determined |
| S1_extracted_k141_51726 | Not-determined |
| S1_extracted_k141_3820 | Not-determined |
| S1_extracted_k141_124529 | Not-determined |
| S1_extracted_k141_104922 | Not-determined |
| S1_extracted_k141_124530 | Not-determined |
| S1_extracted_k141_144575 | Not-determined |
| S1_extracted_k141_91479 | Low-quality |
| S1_extracted_k141_156201 | Not-determined |
| S1_extracted_k141_104924 | Not-determined |
| S1_extracted_k141_24687 | Low-quality |
| S1_extracted_k141_131600 | Not-determined |
| S1_extracted_k141_144582 | Not-determined |
| S1_extracted_k141_124534 | Not-determined |
| S1_extracted_k141_91486 | Not-determined |
| S1_extracted_k141_98342 | Low-quality |
| S1_extracted_k141_51729 | Not-determined |
| S1_extracted_k141_98344 | Not-determined |
| S1_extracted_k141_91489 | Not-determined |
| S1_extracted_k141_51730 | Not-determined |
| S1_extracted_k141_11741 | Not-determined |
| S1_extracted_k141_51732 | Not-determined |
| S1_extracted_k141_118287 | Not-determined |
| S1_extracted_k141_78265 | Not-determined |
| S1_extracted_k141_131611 | Low-quality |
| S1_extracted_k141_131612 | Not-determined |
| S1_extracted_k141_124539 | Low-quality |
| S1_extracted_k141_44935 | Low-quality |
| S1_extracted_k141_37212 | Not-determined |
| S1_extracted_k141_104932 | Not-determined |
| S1_extracted_k141_11745 | Not-determined |
| S1_extracted_k141_149051 | Low-quality |
| S1_extracted_k141_118291 | Not-determined |
| S1_extracted_k141_58130 | Not-determined |
| S1_extracted_k141_37215 | Not-determined |
| S1_extracted_k141_3833 | Not-determined |
| S1_extracted_k141_51739 | Not-determined |
| S1_extracted_k141_63551 | Not-determined |
| S1_extracted_k141_63552 | Not-determined |
| S1_extracted_k141_111558 | Low-quality |
| S1_extracted_k141_149053 | Low-quality |
| S1_extracted_k141_11749 | Not-determined |
| S1_extracted_k141_131619 | Not-determined |
| S1_extracted_k141_11750 | Not-determined |
| S1_extracted_k141_3836 | Not-determined |
| S1_extracted_k141_104937 | Not-determined |
| S1_extracted_k141_91502 | Not-determined |
| S1_extracted_k141_124546 | Not-determined |
| S1_extracted_k141_91504 | Not-determined |
| S1_extracted_k141_44940 | Not-determined |
| S1_extracted_k141_24694 | Not-determined |
| S1_extracted_k141_58138 | Not-determined |
| S1_extracted_k141_63557 | Not-determined |
| S1_extracted_k141_144599 | Not-determined |
| S1_extracted_k141_51747 | Not-determined |
| S1_extracted_k141_11756 | Not-determined |
| S1_extracted_k141_44941 | Low-quality |
| S1_extracted_k141_91507 | Not-determined |
| S1_extracted_k141_91508 | Low-quality |
| S1_extracted_k141_104944 | Not-determined |
| S1_extracted_k141_11757 | Not-determined |
| S1_extracted_k141_31527 | Not-determined |
| S1_extracted_k141_11758 | Not-determined |
| S1_extracted_k141_111566 | Not-determined |
| S1_extracted_k141_18705 | Not-determined |
| S1_extracted_k141_111570 | Not-determined |
| S1_extracted_k141_44948 | Not-determined |
| S1_extracted_k141_18708 | Not-determined |
| S1_extracted_k141_63568 | Not-determined |
| S1_extracted_k141_44950 | Low-quality |
| S1_extracted_k141_98360 | Not-determined |
| S1_extracted_k141_111572 | Not-determined |
| S1_extracted_k141_149054 | Low-quality |
| S1_extracted_k141_63569 | Low-quality |
| S1_extracted_k141_124559 | Low-quality |
| S1_extracted_k141_131637 | Not-determined |
| S1_extracted_k141_44955 | Low-quality |
| S1_extracted_k141_78278 | Not-determined |
| S1_extracted_k141_144608 | Not-determined |
| S1_extracted_k141_11769 | Not-determined |
| S1_extracted_k141_63573 | Not-determined |
| S1_extracted_k141_44959 | Not-determined |
| S1_extracted_k141_24709 | Not-determined |
| S1_extracted_k141_58152 | Not-determined |
| S1_extracted_k141_98371 | Not-determined |
| S1_extracted_k141_91524 | Not-determined |
| S1_extracted_k141_44965 | Not-determined |
| S1_extracted_k141_156224 | Not-determined |
| S1_extracted_k141_131643 | Low-quality |
| S1_extracted_k141_98373 | Not-determined |
| S1_extracted_k141_131645 | Not-determined |
| S1_extracted_k141_111574 | Not-determined |
| S1_extracted_k141_104957 | Not-determined |
| S1_extracted_k141_111575 | Not-determined |
| S1_extracted_k141_63583 | Low-quality |
| S1_extracted_k141_118310 | Not-determined |
| S1_extracted_k141_91532 | Not-determined |
| S1_extracted_k141_104964 | Not-determined |
| S1_extracted_k141_156225 | Not-determined |
| S1_extracted_k141_44976 | Low-quality |
| S1_extracted_k141_111582 | Not-determined |
| S1_extracted_k141_44977 | Not-determined |
| S1_extracted_k141_118314 | Not-determined |
| S1_extracted_k141_91535 | Not-determined |
| S1_extracted_k141_3845 | Low-quality |
| S1_extracted_k141_111583 | Not-determined |
| S1_extracted_k141_58162 | Low-quality |
| S1_extracted_k141_124573 | Not-determined |
| S1_extracted_k141_131653 | Not-determined |
| S1_extracted_k141_84914 | Not-determined |
| S1_extracted_k141_156230 | Not-determined |
| S1_extracted_k141_91538 | Not-determined |
| S1_extracted_k141_104968 | Not-determined |
| S1_extracted_k141_131658 | Not-determined |
| S1_extracted_k141_3849 | Not-determined |
| S1_extracted_k141_131659 | Not-determined |
| S1_extracted_k141_58166 | Not-determined |
| S1_extracted_k141_156231 | Not-determined |
| S1_extracted_k141_78301 | Low-quality |
| S1_extracted_k141_124578 | Low-quality |
| S1_extracted_k141_58167 | Not-determined |
| S1_extracted_k141_31554 | Low-quality |
| S1_extracted_k141_84922 | Not-determined |
| S1_extracted_k141_111588 | Not-determined |
| S1_extracted_k141_131662 | Not-determined |
| S1_extracted_k141_111589 | Not-determined |
| S1_extracted_k141_44983 | Not-determined |
| S1_extracted_k141_111590 | Not-determined |
| S1_extracted_k141_37249 | Not-determined |
| S1_extracted_k141_104975 | Not-determined |
| S8_extracted_k141_143411 | Complete |
| S1_extracted_k141_84923 | Not-determined |
| S1_extracted_k141_144617 | Low-quality |
| S1_extracted_k141_91549 | Not-determined |
| S1_extracted_k141_98387 | Not-determined |
| S1_extracted_k141_44986 | Not-determined |
| S1_extracted_k141_104977 | Not-determined |
| S1_extracted_k141_124587 | Low-quality |
| S1_extracted_k141_131669 | Not-determined |
| S1_extracted_k141_11788 | Not-determined |
| S1_extracted_k141_18718 | Not-determined |
| S1_extracted_k141_11789 | Low-quality |
| S1_extracted_k141_58171 | Not-determined |
| S1_extracted_k141_18719 | Not-determined |
| S1_extracted_k141_37250 | Not-determined |
| S1_extracted_k141_104981 | Low-quality |
| S1_extracted_k141_138163 | Not-determined |
| S1_extracted_k141_156240 | Not-determined |
| S1_extracted_k141_104983 | Not-determined |
| S1_extracted_k141_98393 | Low-quality |
| S1_extracted_k141_44990 | Not-determined |
| S1_extracted_k141_144623 | Low-quality |
| S1_extracted_k141_124594 | Not-determined |
| S1_extracted_k141_118327 | Not-determined |
| S1_extracted_k141_63599 | Not-determined |
| S1_extracted_k141_118328 | Not-determined |
| S1_extracted_k141_31564 | Low-quality |
| S1_extracted_k141_149086 | Not-determined |
| S1_extracted_k141_84936 | Not-determined |
| S1_extracted_k141_131682 | Not-determined |
| S1_extracted_k141_3864 | Not-determined |
| S1_extracted_k141_118331 | Not-determined |
| S1_extracted_k141_24732 | Low-quality |
| S1_extracted_k141_124602 | Not-determined |
| S1_extracted_k141_44999 | Not-determined |
| S1_extracted_k141_45000 | Not-determined |
| S1_extracted_k141_118334 | Not-determined |
| S1_extracted_k141_84942 | Not-determined |
| S1_extracted_k141_37264 | Not-determined |
| S1_extracted_k141_51789 | Not-determined |
| S1_extracted_k141_118335 | Not-determined |
| S1_extracted_k141_18742 | Not-determined |
| S1_extracted_k141_24737 | Not-determined |
| S1_extracted_k141_18744 | Not-determined |
| S1_extracted_k141_124608 | Not-determined |
| S1_extracted_k141_18745 | Not-determined |
| S1_extracted_k141_91569 | Low-quality |
| S1_extracted_k141_3870 | Not-determined |
| S1_extracted_k141_24741 | Low-quality |
| S1_extracted_k141_78317 | Not-determined |
| S1_extracted_k141_111617 | Low-quality |
| S1_extracted_k141_51797 | Not-determined |
| S1_extracted_k141_91574 | Not-determined |
| S1_extracted_k141_149095 | Low-quality |
| S1_extracted_k141_45012 | Not-determined |
| S1_extracted_k141_58179 | Not-determined |
| S1_extracted_k141_31580 | Low-quality |
| S1_extracted_k141_51798 | Not-determined |
| S1_extracted_k141_58181 | Not-determined |
| S1_extracted_k141_78322 | Not-determined |
| S1_extracted_k141_18756 | Not-determined |
| S1_extracted_k141_51802 | Not-determined |
| S1_extracted_k141_138178 | Not-determined |
| S1_extracted_k141_3877 | Not-determined |
| S1_extracted_k141_149098 | Not-determined |
| S1_extracted_k141_78327 | Not-determined |
| S1_extracted_k141_118342 | Low-quality |
| S1_extracted_k141_144631 | Low-quality |
| S1_extracted_k141_84948 | Not-determined |
| S1_extracted_k141_98417 | Not-determined |
| S1_extracted_k141_45021 | Not-determined |
| S1_extracted_k141_131691 | Not-determined |
| S1_extracted_k141_63619 | Not-determined |
| S1_extracted_k141_144633 | Not-determined |
| S1_extracted_k141_18764 | Not-determined |
| S1_extracted_k141_131693 | Not-determined |
| S1_extracted_k141_18766 | Not-determined |
| S1_extracted_k141_24754 | Not-determined |
| S1_extracted_k141_124627 | Not-determined |
| S1_extracted_k141_70753 | Not-determined |
| S1_extracted_k141_104997 | Not-determined |
| S1_extracted_k141_98423 | Not-determined |
| S1_extracted_k141_111634 | Not-determined |
| S1_extracted_k141_98424 | Not-determined |
| S1_extracted_k141_24756 | Low-quality |
| S1_extracted_k141_11825 | Not-determined |
| S1_extracted_k141_124635 | Not-determined |
| S1_extracted_k141_58197 | Not-determined |
| S1_extracted_k141_51810 | Not-determined |
| S1_extracted_k141_91592 | Not-determined |
| S1_extracted_k141_18775 | Not-determined |
| S1_extracted_k141_24762 | Not-determined |
| S1_extracted_k141_58198 | Low-quality |
| S1_extracted_k141_84956 | Not-determined |
| S1_extracted_k141_144643 | Not-determined |
| S1_extracted_k141_131701 | Not-determined |
| S1_extracted_k141_24764 | Not-determined |
| S1_extracted_k141_156251 | Not-determined |
| S1_extracted_k141_84957 | Low-quality |
| S1_extracted_k141_98429 | Not-determined |
| S1_extracted_k141_144645 | Not-determined |
| S1_extracted_k141_51815 | Not-determined |
| S1_extracted_k141_3884 | Not-determined |
| S1_extracted_k141_131703 | Not-determined |
| S1_extracted_k141_138183 | Not-determined |
| S1_extracted_k141_105003 | Not-determined |
| S1_extracted_k141_91596 | Not-determined |
| S1_extracted_k141_24769 | Not-determined |
| S1_extracted_k141_58202 | Not-determined |
| S1_extracted_k141_91598 | Not-determined |
| S1_extracted_k141_24770 | Low-quality |
| S1_extracted_k141_118348 | Low-quality |
| S1_extracted_k141_105008 | Not-determined |
| S1_extracted_k141_149109 | Not-determined |
| S1_extracted_k141_118349 | Low-quality |
| S1_extracted_k141_144654 | Not-determined |
| S1_extracted_k141_91601 | Not-determined |
| S1_extracted_k141_124653 | Not-determined |
| S1_extracted_k141_51822 | Not-determined |
| S1_extracted_k141_24772 | Not-determined |
| S1_extracted_k141_3889 | Not-determined |
| S1_extracted_k141_105014 | Not-determined |
| S1_extracted_k141_58210 | Not-determined |
| S1_extracted_k141_98437 | Not-determined |
| S1_extracted_k141_131715 | Not-determined |
| S1_extracted_k141_11831 | Low-quality |
| S1_extracted_k141_84970 | Not-determined |
| S1_extracted_k141_144660 | Not-determined |
| S1_extracted_k141_111653 | Not-determined |
| S1_extracted_k141_78362 | Not-determined |
| S1_extracted_k141_138196 | Not-determined |
| S1_extracted_k141_24780 | Not-determined |
| S1_extracted_k141_78367 | Not-determined |
| S1_extracted_k141_24782 | Not-determined |
| S1_extracted_k141_118359 | Not-determined |
| S1_extracted_k141_138205 | Not-determined |
| S1_extracted_k141_24783 | Low-quality |
| S1_extracted_k141_98448 | Not-determined |
| S1_extracted_k141_105029 | Not-determined |
| S1_extracted_k141_51838 | Low-quality |
| S1_extracted_k141_124670 | Not-determined |
| S1_extracted_k141_105031 | Not-determined |
| S1_extracted_k141_124673 | Not-determined |
| S1_extracted_k141_118362 | Not-determined |
| S1_extracted_k141_84984 | Low-quality |
| S1_extracted_k141_37295 | Not-determined |
| S1_extracted_k141_124676 | Not-determined |
| S1_extracted_k141_131718 | Low-quality |
| S1_extracted_k141_84986 | Not-determined |
| S1_extracted_k141_131719 | Not-determined |
| S1_extracted_k141_149125 | Not-determined |
| S1_extracted_k141_31610 | Not-determined |
| S1_extracted_k141_18797 | Not-determined |
| S1_extracted_k141_118368 | Not-determined |
| S1_extracted_k141_138213 | Not-determined |
| S1_extracted_k141_58231 | Not-determined |
| S1_extracted_k141_18799 | Not-determined |
| S1_extracted_k141_3907 | Not-determined |
| S1_extracted_k141_118371 | Low-quality |
| S1_extracted_k141_131725 | Not-determined |
| S1_extracted_k141_45053 | Not-determined |
| S1_extracted_k141_11854 | Not-determined |
| S1_extracted_k141_84991 | Low-quality |
| S1_extracted_k141_138216 | Not-determined |
| S1_extracted_k141_11855 | Not-determined |
| S1_extracted_k141_18808 | Not-determined |
| S1_extracted_k141_105045 | Not-determined |
| S1_extracted_k141_11857 | Not-determined |
| S1_extracted_k141_78373 | Not-determined |
| S1_extracted_k141_18813 | Not-determined |
| S1_extracted_k141_11860 | Not-determined |
| S1_extracted_k141_63659 | Low-quality |
| S1_extracted_k141_131735 | Not-determined |
| S1_extracted_k141_3915 | Not-determined |
| S1_extracted_k141_11865 | Not-determined |
| S1_extracted_k141_3916 | Not-determined |
| S1_extracted_k141_11866 | Not-determined |
| S1_extracted_k141_91631 | Not-determined |
| S1_extracted_k141_138225 | Not-determined |
| S1_extracted_k141_144669 | Low-quality |
| S1_extracted_k141_124692 | Low-quality |
| S1_extracted_k141_11870 | Not-determined |
| S1_extracted_k141_11871 | Not-determined |
| S1_extracted_k141_124695 | Not-determined |
| S1_extracted_k141_85006 | Not-determined |
| S1_extracted_k141_111680 | Low-quality |
| S1_extracted_k141_124696 | Not-determined |
| S1_extracted_k141_31625 | Low-quality |
| S1_extracted_k141_58233 | Not-determined |
| S1_extracted_k141_91641 | Not-determined |
| S1_extracted_k141_149141 | Low-quality |
| S1_extracted_k141_124703 | Not-determined |
| S1_extracted_k141_45072 | Low-quality |
| S1_extracted_k141_31631 | Not-determined |
| S1_extracted_k141_78377 | Low-quality |
| S1_extracted_k141_18828 | Low-quality |
| S1_extracted_k141_24803 | Not-determined |
| S1_extracted_k141_156268 | Not-determined |
| S1_extracted_k141_11879 | Low-quality |
| S1_extracted_k141_37313 | Not-determined |
| S1_extracted_k141_156270 | Not-determined |
| S1_extracted_k141_144680 | Not-determined |
| S1_extracted_k141_111693 | Not-determined |
| S1_extracted_k141_31635 | Not-determined |
| S1_extracted_k141_85018 | Not-determined |
| S1_extracted_k141_58240 | Not-determined |
| S1_extracted_k141_63680 | Not-determined |
| S1_extracted_k141_131756 | Not-determined |
| S1_extracted_k141_85021 | Not-determined |
| S1_extracted_k141_24807 | Not-determined |
| S1_extracted_k141_63682 | Not-determined |
| S1_extracted_k141_58244 | Not-determined |
| S1_extracted_k141_85022 | Not-determined |
| S1_extracted_k141_138240 | Not-determined |
| S1_extracted_k141_144687 | Low-quality |
| S1_extracted_k141_91658 | Not-determined |
| S1_extracted_k141_11883 | Not-determined |
| S1_extracted_k141_78386 | Not-determined |
| S1_extracted_k141_124720 | Not-determined |
| S1_extracted_k141_144689 | Not-determined |
| S1_extracted_k141_98475 | Not-determined |
| S1_extracted_k141_131761 | Not-determined |
| S1_extracted_k141_51894 | Not-determined |
| S1_extracted_k141_124723 | Not-determined |
| S1_extracted_k141_149155 | Low-quality |
| S1_extracted_k141_78388 | Not-determined |
| S1_extracted_k141_58250 | Not-determined |
| S1_extracted_k141_3936 | Low-quality |
| S1_extracted_k141_111701 | Not-determined |
| S1_extracted_k141_70772 | Low-quality |
| S1_extracted_k141_58253 | Not-determined |
| S1_extracted_k141_138244 | Not-determined |
| S1_extracted_k141_70775 | Not-determined |
| S1_extracted_k141_11898 | Not-determined |
| S1_extracted_k141_63692 | Not-determined |
| S1_extracted_k141_98486 | Not-determined |
| S1_extracted_k141_91667 | Not-determined |
| S1_extracted_k141_31650 | Not-determined |
| S1_extracted_k141_98488 | Low-quality |
| S1_extracted_k141_149159 | Not-determined |
| S1_extracted_k141_111709 | Not-determined |
| S1_extracted_k141_63694 | Low-quality |
| S1_extracted_k141_63695 | Not-determined |
| S1_extracted_k141_98491 | Not-determined |
| S1_extracted_k141_24816 | Not-determined |
| S1_extracted_k141_144704 | Not-determined |
| S1_extracted_k141_63699 | Low-quality |
| S1_extracted_k141_45095 | Not-determined |
| S1_extracted_k141_98493 | Low-quality |
| S1_extracted_k141_144708 | Not-determined |
| S1_extracted_k141_51916 | Not-determined |
| S1_extracted_k141_98494 | Low-quality |
| S1_extracted_k141_45098 | Not-determined |
| S1_extracted_k141_3952 | Not-determined |
| S1_extracted_k141_78395 | Low-quality |
| S1_extracted_k141_58259 | Not-determined |
| S1_extracted_k141_78397 | Not-determined |
| S1_extracted_k141_45102 | Not-determined |
| S1_extracted_k141_85036 | Low-quality |
| S1_extracted_k141_58262 | Not-determined |
| S1_extracted_k141_18843 | Not-determined |
| S1_extracted_k141_37327 | Not-determined |
| S1_extracted_k141_85041 | Low-quality |
| S1_extracted_k141_58266 | Not-determined |
| S1_extracted_k141_24825 | Low-quality |
| S1_extracted_k141_18851 | Not-determined |
| S1_extracted_k141_63702 | Not-determined |
| S1_extracted_k141_58271 | Not-determined |
| S1_extracted_k141_31666 | Not-determined |
| S1_extracted_k141_24829 | Not-determined |
| S1_extracted_k141_138247 | Low-quality |
| S1_extracted_k141_138248 | Not-determined |
| S1_extracted_k141_91687 | Not-determined |
| S1_extracted_k141_70796 | Not-determined |
| S1_extracted_k141_78406 | Not-determined |
| S1_extracted_k141_131788 | Not-determined |
| S1_extracted_k141_3956 | Low-quality |
| S1_extracted_k141_70798 | Not-determined |
| S1_extracted_k141_118399 | Not-determined |
| S1_extracted_k141_58277 | Not-determined |
| S1_extracted_k141_45109 | Low-quality |
| S1_extracted_k141_131793 | Not-determined |
| S1_extracted_k141_11932 | Low-quality |
| S1_extracted_k141_131794 | Not-determined |
| S1_extracted_k141_11934 | Not-determined |
| S1_extracted_k141_118408 | Low-quality |
| S1_extracted_k141_70806 | Not-determined |
| S1_extracted_k141_98511 | Not-determined |
| S1_extracted_k141_131800 | Not-determined |
| S1_extracted_k141_111718 | Not-determined |
| S1_extracted_k141_63720 | Not-determined |
| S1_extracted_k141_118412 | Not-determined |
| S1_extracted_k141_98515 | Not-determined |
| S1_extracted_k141_51930 | Not-determined |
| S1_extracted_k141_78417 | Not-determined |
| S1_extracted_k141_45121 | Not-determined |
| S1_extracted_k141_51931 | Not-determined |

| | |
|---|---|
| S1_extracted_k141_63723 | Not-determined |
| S1_extracted_k141_105092 | Not-determined |
| S1_extracted_k141_131805 | Not-determined |
| S1_extracted_k141_124753 | Not-determined |
| S1_extracted_k141_149176 | Not-determined |
| S1_extracted_k141_124755 | Not-determined |
| S1_extracted_k141_51934 | Not-determined |
| S1_extracted_k141_63728 | Not-determined |
| S1_extracted_k141_58291 | Not-determined |
| S1_extracted_k141_78423 | Not-determined |
| S1_extracted_k141_91706 | Not-determined |
| S1_extracted_k141_124759 | Low-quality |
| S1_extracted_k141_18883 | Not-determined |
| S1_extracted_k141_31695 | Low-quality |
| S1_extracted_k141_58296 | Not-determined |
| S1_extracted_k141_31696 | Low-quality |
| S1_extracted_k141_144738 | Not-determined |
| S1_extracted_k141_18885 | Not-determined |
| S1_extracted_k141_3972 | Not-determined |
| S1_extracted_k141_58300 | Low-quality |
| S1_extracted_k141_45131 | Not-determined |
| S1_extracted_k141_118425 | Not-determined |
| S1_extracted_k141_118428 | Low-quality |
| S1_extracted_k141_105109 | Not-determined |
| S1_extracted_k141_31698 | Not-determined |
| S1_extracted_k141_78432 | Not-determined |
| S1_extracted_k141_31699 | Low-quality |
| S1_extracted_k141_105111 | Not-determined |
| S1_extracted_k141_124766 | Low-quality |
| S1_extracted_k141_138254 | Not-determined |
| S1_extracted_k141_105113 | Low-quality |
| S1_extracted_k141_149184 | Not-determined |
| S1_extracted_k141_51946 | Not-determined |
| S1_extracted_k141_98538 | Not-determined |
| S1_extracted_k141_37340 | Not-determined |
| S1_extracted_k141_18892 | Not-determined |
| S1_extracted_k141_118435 | Not-determined |
| S1_extracted_k141_24862 | Not-determined |
| S1_extracted_k141_124770 | Not-determined |
| S1_extracted_k141_51950 | Not-determined |
| S1_extracted_k141_51952 | Not-determined |
| S1_extracted_k141_63751 | Not-determined |
| S1_extracted_k141_58313 | Not-determined |
| S1_extracted_k141_11964 | Low-quality |
| S1_extracted_k141_98544 | Not-determined |
| S1_extracted_k141_111739 | Not-determined |
| S1_extracted_k141_45148 | Not-determined |
| S1_extracted_k141_118440 | Low-quality |
| S1_extracted_k141_51954 | Not-determined |
| S1_extracted_k141_11966 | Not-determined |
| S1_extracted_k141_105121 | Not-determined |
| S1_extracted_k141_58317 | Not-determined |
| S1_extracted_k141_63755 | Not-determined |
| S1_extracted_k141_98546 | Not-determined |
| S1_extracted_k141_78445 | Not-determined |
| S1_extracted_k141_118444 | Not-determined |
| S1_extracted_k141_3982 | Not-determined |
| S1_extracted_k141_138266 | Low-quality |
| S1_extracted_k141_118446 | Not-determined |
| S1_extracted_k141_98548 | Not-determined |
| S1_extracted_k141_24869 | Low-quality |
| S1_extracted_k141_3985 | Not-determined |
| S1_extracted_k141_138269 | Not-determined |
| S1_extracted_k141_111747 | Not-determined |
| S1_extracted_k141_124778 | Not-determined |
| S1_extracted_k141_149188 | Not-determined |
| S1_extracted_k141_37350 | Not-determined |
| S1_extracted_k141_37351 | Not-determined |
| S1_extracted_k141_3989 | Low-quality |
| S1_extracted_k141_78453 | Not-determined |
| S1_extracted_k141_131823 | Not-determined |
| S1_extracted_k141_149190 | Low-quality |
| S1_extracted_k141_111751 | Low-quality |
| S1_extracted_k141_91728 | Not-determined |
| S1_extracted_k141_78455 | Not-determined |
| S1_extracted_k141_111753 | Not-determined |
| S1_extracted_k141_63760 | Not-determined |
| S1_extracted_k141_63761 | Not-determined |
| S1_extracted_k141_58329 | Not-determined |
| S1_extracted_k141_58330 | Not-determined |
| S1_extracted_k141_51964 | Not-determined |
| S1_extracted_k141_118451 | Not-determined |
| S1_extracted_k141_45156 | Not-determined |
| S1_extracted_k141_144768 | Not-determined |
| S1_extracted_k141_37358 | Low-quality |
| S1_extracted_k141_98560 | Not-determined |
| S1_extracted_k141_144770 | Not-determined |
| S1_extracted_k141_78462 | Not-determined |
| S1_extracted_k141_85090 | Low-quality |
| S1_extracted_k141_18907 | Not-determined |
| S1_extracted_k141_63765 | Low-quality |
| S1_extracted_k141_131837 | Not-determined |
| S1_extracted_k141_45163 | Not-determined |
| S1_extracted_k141_18909 | Low-quality |
| S1_extracted_k141_131839 | Not-determined |
| S1_extracted_k141_37363 | Not-determined |
| S1_extracted_k141_11974 | Not-determined |
| S1_extracted_k141_144776 | Not-determined |
| S1_extracted_k141_78473 | Not-determined |
| S1_extracted_k141_91743 | Not-determined |
| S1_extracted_k141_58337 | Low-quality |
| S1_extracted_k141_11979 | Not-determined |
| S1_extracted_k141_118467 | Not-determined |
| S1_extracted_k141_63775 | Not-determined |
| S1_extracted_k141_31728 | Not-determined |
| S1_extracted_k141_98572 | Not-determined |
| S1_extracted_k141_51981 | Not-determined |
| S1_extracted_k141_58341 | Not-determined |
| S1_extracted_k141_144783 | Low-quality |
| S1_extracted_k141_37367 | Not-determined |
| S1_extracted_k141_124788 | Not-determined |
| S1_extracted_k141_149195 | Not-determined |
| S1_extracted_k141_78482 | Not-determined |
| S1_extracted_k141_91750 | Not-determined |
| S1_extracted_k141_58344 | Not-determined |
| S1_extracted_k141_98577 | Not-determined |
| S1_extracted_k141_124791 | Not-determined |
| S1_extracted_k141_85104 | Not-determined |
| S1_extracted_k141_124792 | Not-determined |
| S1_extracted_k141_85106 | Not-determined |
| S1_extracted_k141_124794 | Not-determined |
| S1_extracted_k141_111773 | Low-quality |
| S1_extracted_k141_156293 | Not-determined |
| S1_extracted_k141_138285 | Not-determined |
| S1_extracted_k141_105147 | Low-quality |
| S1_extracted_k141_31737 | Not-determined |
| S1_extracted_k141_118477 | Not-determined |
| S1_extracted_k141_91757 | Not-determined |
| S1_extracted_k141_118478 | Not-determined |
| S1_extracted_k141_111776 | Not-determined |
| S1_extracted_k141_4018 | Not-determined |
| S1_extracted_k141_138290 | Low-quality |
| S1_extracted_k141_45180 | Not-determined |
| S1_extracted_k141_105150 | Not-determined |
| S1_extracted_k141_138293 | Not-determined |
| S1_extracted_k141_124805 | Not-determined |
| S1_extracted_k141_70852 | Not-determined |
| S1_extracted_k141_51991 | Not-determined |
| S1_extracted_k141_78492 | Not-determined |
| S1_extracted_k141_51992 | Not-determined |
| S1_extracted_k141_98585 | Not-determined |
| S1_extracted_k141_58357 | Not-determined |
| S1_extracted_k141_131846 | Low-quality |
| S1_extracted_k141_45182 | Low-quality |
| S1_extracted_k141_24902 | Not-determined |
| S1_extracted_k141_63789 | Low-quality |
| S1_extracted_k141_156302 | Not-determined |
| S1_extracted_k141_31744 | Not-determined |
| S1_extracted_k141_37372 | Not-determined |
| S1_extracted_k141_144794 | Low-quality |
| S1_extracted_k141_85114 | Not-determined |
| S1_extracted_k141_4025 | Low-quality |
| S1_extracted_k141_131851 | Not-determined |
| S1_extracted_k141_98593 | Not-determined |
| S1_extracted_k141_18938 | Not-determined |
| S1_extracted_k141_85117 | Not-determined |
| S1_extracted_k141_31751 | Not-determined |
| S1_extracted_k141_70857 | Not-determined |
| S1_extracted_k141_149203 | Not-determined |
| S1_extracted_k141_4032 | Not-determined |
| S1_extracted_k141_131858 | Not-determined |
| S1_extracted_k141_52002 | Low-quality |
| S1_extracted_k141_37385 | Not-determined |
| S1_extracted_k141_124824 | Low-quality |
| S1_extracted_k141_124825 | Not-determined |
| S1_extracted_k141_98601 | Not-determined |
| S1_extracted_k141_144804 | Not-determined |
| S1_extracted_k141_24915 | Not-determined |
| S1_extracted_k141_70860 | Low-quality |
| S1_extracted_k141_111805 | Not-determined |
| S1_extracted_k141_131861 | Not-determined |
| S1_extracted_k141_118488 | Not-determined |
| S1_extracted_k141_31765 | Not-determined |
| S1_extracted_k141_131863 | Not-determined |
| S1_extracted_k141_85126 | Not-determined |
| S1_extracted_k141_144809 | Not-determined |
| S1_extracted_k141_45212 | Not-determined |
| S1_extracted_k141_58368 | Not-determined |
| S1_extracted_k141_45213 | Not-determined |
| S1_extracted_k141_131865 | Not-determined |
| S1_extracted_k141_131866 | Low-quality |
| S1_extracted_k141_118491 | Low-quality |
| S1_extracted_k141_85129 | Not-determined |
| S1_extracted_k141_85130 | Not-determined |
| S1_extracted_k141_124837 | Not-determined |
| S1_extracted_k141_124839 | Not-determined |
| S1_extracted_k141_105182 | Not-determined |
| S1_extracted_k141_63804 | Not-determined |
| S1_extracted_k141_91776 | Low-quality |
| S1_extracted_k141_131875 | Not-determined |
| S1_extracted_k141_63805 | Not-determined |
| S1_extracted_k141_58376 | Not-determined |
| S1_extracted_k141_70872 | Not-determined |
| S1_extracted_k141_24933 | Not-determined |
| S1_extracted_k141_70874 | Not-determined |
| S1_extracted_k141_118501 | Not-determined |
| S1_extracted_k141_118504 | Not-determined |
| S1_extracted_k141_24939 | Not-determined |
| S1_extracted_k141_24941 | Not-determined |
| S1_extracted_k141_124852 | Not-determined |
| S1_extracted_k141_131882 | Not-determined |
| S1_extracted_k141_52024 | Not-determined |
| S1_extracted_k141_138296 | Low-quality |
| S1_extracted_k141_45231 | Not-determined |
| S1_extracted_k141_131884 | Not-determined |
| S1_extracted_k141_18952 | Not-determined |
| S1_extracted_k141_4046 | Not-determined |
| S1_extracted_k141_31792 | Not-determined |
| S1_extracted_k141_24947 | Not-determined |
| S1_extracted_k141_149209 | Low-quality |
| S1_extracted_k141_149210 | Not-determined |
| S1_extracted_k141_105191 | Not-determined |
| S1_extracted_k141_85146 | Not-determined |
| S1_extracted_k141_63817 | Not-determined |
| S1_extracted_k141_70881 | Not-determined |
| S1_extracted_k141_111812 | Not-determined |
| S1_extracted_k141_124859 | Low-quality |
| S1_extracted_k141_18962 | Not-determined |
| S1_extracted_k141_118522 | Not-determined |
| S1_extracted_k141_98637 | Not-determined |
| S1_extracted_k141_91798 | Not-determined |
| S1_extracted_k141_111816 | Low-quality |
| S1_extracted_k141_111817 | Not-determined |
| S1_extracted_k141_52033 | Not-determined |
| S1_extracted_k141_91801 | Not-determined |
| S1_extracted_k141_85159 | Not-determined |
| S1_extracted_k141_70889 | Not-determined |
| S1_extracted_k141_124870 | Not-determined |
| S1_extracted_k141_63827 | Not-determined |
| S1_extracted_k141_131894 | Not-determined |
| S1_extracted_k141_78538 | Not-determined |
| S1_extracted_k141_124873 | Not-determined |
| S1_extracted_k141_118533 | Not-determined |
| S1_extracted_k141_4060 | Not-determined |
| S1_extracted_k141_91806 | Not-determined |
| S1_extracted_k141_131897 | Not-determined |
| S1_extracted_k141_18971 | Not-determined |
| S1_extracted_k141_52038 | Not-determined |
| S1_extracted_k141_85166 | Low-quality |
| S1_extracted_k141_118536 | Not-determined |
| S1_extracted_k141_4061 | Low-quality |
| S1_extracted_k141_37394 | Not-determined |
| S1_extracted_k141_37395 | Not-determined |
| S1_extracted_k141_78543 | Not-determined |
| S1_extracted_k141_98646 | Not-determined |
| S1_extracted_k141_63832 | Not-determined |
| S1_extracted_k141_91809 | Low-quality |
| S1_extracted_k141_105211 | Low-quality |
| S1_extracted_k141_156331 | Low-quality |
| S1_extracted_k141_124879 | Not-determined |
| S1_extracted_k141_4067 | Not-determined |
| S1_extracted_k141_24963 | Not-determined |
| S1_extracted_k141_45243 | Not-determined |
| S1_extracted_k141_52047 | Not-determined |
| S1_extracted_k141_124881 | Not-determined |
| S1_extracted_k141_63834 | Low-quality |
| S1_extracted_k141_45246 | Not-determined |
| S1_extracted_k141_78549 | Not-determined |
| S1_extracted_k141_156334 | Not-determined |
| S1_extracted_k141_118542 | Low-quality |
| S1_extracted_k141_52049 | Not-determined |
| S1_extracted_k141_4075 | Low-quality |
| S1_extracted_k141_70899 | Not-determined |
| S1_extracted_k141_124885 | Not-determined |
| S1_extracted_k141_118546 | Not-determined |
| S1_extracted_k141_78555 | Not-determined |
| S1_extracted_k141_91816 | Not-determined |
| S1_extracted_k141_105217 | Not-determined |
| S1_extracted_k141_124888 | Not-determined |
| S1_extracted_k141_98660 | Not-determined |
| S1_extracted_k141_12010 | Not-determined |
| S1_extracted_k141_31798 | Low-quality |
| S1_extracted_k141_4080 | Not-determined |
| S1_extracted_k141_98662 | Not-determined |
| S1_extracted_k141_58418 | Not-determined |
| S1_extracted_k141_52053 | Low-quality |
| S1_extracted_k141_58419 | Not-determined |
| S1_extracted_k141_124898 | Not-determined |
| S1_extracted_k141_58422 | Not-determined |
| S1_extracted_k141_63851 | Not-determined |
| S1_extracted_k141_4083 | Not-determined |
| S1_extracted_k141_58424 | Not-determined |
| S1_extracted_k141_149230 | Not-determined |
| S1_extracted_k141_24978 | Not-determined |
| S1_extracted_k141_118560 | Not-determined |
| S1_extracted_k141_24980 | Not-determined |
| S1_extracted_k141_91826 | Low-quality |
| S1_extracted_k141_70908 | Low-quality |
| S1_extracted_k141_131926 | Low-quality |
| S1_extracted_k141_124901 | Not-determined |
| S1_extracted_k141_131927 | Not-determined |
| S1_extracted_k141_111848 | Not-determined |
| S1_extracted_k141_52067 | Not-determined |
| S1_extracted_k141_63856 | Not-determined |
| S1_extracted_k141_52069 | Not-determined |
| S1_extracted_k141_91830 | Not-determined |
| S1_extracted_k141_78566 | Not-determined |
| S1_extracted_k141_156338 | Low-quality |
| S1_extracted_k141_131930 | Not-determined |
| S1_extracted_k141_31816 | Not-determined |
| S1_extracted_k141_63862 | Not-determined |
| S1_extracted_k141_118563 | Low-quality |
| S1_extracted_k141_124911 | Not-determined |
| S1_extracted_k141_63867 | Not-determined |
| S1_extracted_k141_131936 | Not-determined |
| S1_extracted_k141_124914 | Not-determined |
| S1_extracted_k141_4091 | Not-determined |
| S1_extracted_k141_63869 | Not-determined |
| S1_extracted_k141_131937 | Low-quality |
| S1_extracted_k141_52075 | Not-determined |
| S1_extracted_k141_18983 | Not-determined |
| S1_extracted_k141_98682 | Not-determined |
| S1_extracted_k141_18984 | Not-determined |
| S1_extracted_k141_105244 | Not-determined |
| S1_extracted_k141_111857 | Not-determined |
| S1_extracted_k141_31822 | Not-determined |
| S1_extracted_k141_24995 | Not-determined |
| S1_extracted_k141_131939 | Not-determined |
| S1_extracted_k141_78570 | Low-quality |
| S1_extracted_k141_144868 | Not-determined |
| S1_extracted_k141_45255 | Not-determined |
| S1_extracted_k141_144869 | Not-determined |
| S1_extracted_k141_18988 | Not-determined |
| S1_extracted_k141_131942 | Not-determined |
| S1_extracted_k141_111859 | Low-quality |
| S1_extracted_k141_78575 | Not-determined |
| S1_extracted_k141_37411 | Not-determined |
| S1_extracted_k141_78576 | Not-determined |
| S1_extracted_k141_58440 | Not-determined |
| S1_extracted_k141_52080 | Not-determined |
| S1_extracted_k141_138338 | Not-determined |
| S1_extracted_k141_138340 | Not-determined |
| S1_extracted_k141_91847 | Low-quality |
| S1_extracted_k141_91848 | Not-determined |
| S1_extracted_k141_12053 | Not-determined |
| S1_extracted_k141_91849 | Not-determined |
| S1_extracted_k141_4102 | Not-determined |
| S1_extracted_k141_25007 | Not-determined |
| S1_extracted_k141_124936 | Not-determined |
| S1_extracted_k141_63890 | Not-determined |
| S1_extracted_k141_85210 | Not-determined |
| S1_extracted_k141_4103 | Not-determined |
| S1_extracted_k141_52089 | Not-determined |
| S1_extracted_k141_138347 | Not-determined |
| S1_extracted_k141_4104 | Not-determined |
| S1_extracted_k141_105258 | Not-determined |
| S1_extracted_k141_78586 | Not-determined |
| S1_extracted_k141_52092 | Not-determined |
| S1_extracted_k141_4107 | Not-determined |
| S1_extracted_k141_131955 | Not-determined |
| S1_extracted_k141_144886 | Not-determined |
| S1_extracted_k141_85214 | Not-determined |
| S1_extracted_k141_25014 | Not-determined |
| S1_extracted_k141_131956 | Not-determined |
| S1_extracted_k141_91857 | Not-determined |
| S1_extracted_k141_131957 | Not-determined |
| S1_extracted_k141_31831 | Not-determined |
| S1_extracted_k141_98701 | Not-determined |
| S1_extracted_k141_118576 | Not-determined |
| S1_extracted_k141_144892 | Not-determined |
| S1_extracted_k141_118578 | Not-determined |
| S1_extracted_k141_45270 | Not-determined |
| S1_extracted_k141_58455 | Low-quality |
| S1_extracted_k141_52100 | Not-determined |
| S1_extracted_k141_37431 | Not-determined |
| S1_extracted_k141_31835 | Low-quality |
| S1_extracted_k141_45273 | Not-determined |
| S1_extracted_k141_98708 | Not-determined |
| S1_extracted_k141_98709 | Not-determined |
| S1_extracted_k141_12072 | Not-determined |
| S1_extracted_k141_118585 | Not-determined |
| S1_extracted_k141_12074 | Not-determined |
| S1_extracted_k141_78601 | Low-quality |
| S1_extracted_k141_37435 | Not-determined |
| S1_extracted_k141_19011 | Low-quality |
| S1_extracted_k141_138365 | Not-determined |
| S1_extracted_k141_118586 | Low-quality |
| S1_extracted_k141_58466 | Not-determined |
| S1_extracted_k141_45279 | Low-quality |
| S1_extracted_k141_37439 | Not-determined |
| S1_extracted_k141_58467 | Low-quality |
| S1_extracted_k141_52109 | Low-quality |
| S1_extracted_k141_63917 | Not-determined |
| S1_extracted_k141_70937 | Not-determined |
| S1_extracted_k141_25024 | Not-determined |
| S1_extracted_k141_138376 | Low-quality |
| S1_extracted_k141_124954 | Not-determined |
| S1_extracted_k141_70939 | Not-determined |
| S1_extracted_k141_58476 | Not-determined |
| S1_extracted_k141_37449 | Not-determined |
| S1_extracted_k141_45290 | Not-determined |
| S1_extracted_k141_85219 | Low-quality |
| S1_extracted_k141_131981 | Not-determined |
| S1_extracted_k141_138384 | Not-determined |
| S1_extracted_k141_12095 | Not-determined |
| S1_extracted_k141_144915 | Not-determined |
| S1_extracted_k141_12097 | Not-determined |
| S1_extracted_k141_37454 | Not-determined |
| S1_extracted_k141_85223 | Not-determined |
| S1_extracted_k141_111876 | Low-quality |
| S1_extracted_k141_85224 | Not-determined |
| S1_extracted_k141_138388 | Not-determined |
| S1_extracted_k141_144919 | Not-determined |
| S1_extracted_k141_58485 | Not-determined |
| S1_extracted_k141_19029 | Not-determined |
| S1_extracted_k141_70942 | Not-determined |
| S1_extracted_k141_131989 | Not-determined |
| S1_extracted_k141_91870 | Not-determined |
| S1_extracted_k141_124970 | Not-determined |
| S1_extracted_k141_144926 | Not-determined |
| S1_extracted_k141_131990 | Not-determined |
| S1_extracted_k141_19032 | Not-determined |
| S1_extracted_k141_78621 | Not-determined |
| S1_extracted_k141_58489 | Not-determined |
| S1_extracted_k141_91874 | Not-determined |
| S1_extracted_k141_156356 | Not-determined |
| S1_extracted_k141_131995 | Low-quality |
| S1_extracted_k141_91878 | Not-determined |
| S1_extracted_k141_52117 | Low-quality |
| S1_extracted_k141_131996 | Not-determined |
| S1_extracted_k141_118597 | Not-determined |
| S1_extracted_k141_118598 | Not-determined |
| S1_extracted_k141_91881 | Not-determined |
| S1_extracted_k141_85231 | Not-determined |
| S1_extracted_k141_111887 | Not-determined |
| S1_extracted_k141_12102 | Low-quality |
| S1_extracted_k141_105298 | Not-determined |
| S1_extracted_k141_45304 | Not-determined |
| S1_extracted_k141_58495 | Not-determined |
| S1_extracted_k141_85235 | Not-determined |
| S1_extracted_k141_111889 | Low-quality |
| S1_extracted_k141_98721 | Low-quality |
| S1_extracted_k141_138406 | Not-determined |
| S1_extracted_k141_45307 | Not-determined |
| S1_extracted_k141_132004 | Not-determined |
| S1_extracted_k141_63942 | Low-quality |
| S1_extracted_k141_85239 | Not-determined |
| S1_extracted_k141_25053 | Not-determined |
| S1_extracted_k141_4140 | Not-determined |
| S1_extracted_k141_63943 | Low-quality |
| S1_extracted_k141_45309 | Low-quality |
| S1_extracted_k141_124985 | Not-determined |
| S1_extracted_k141_31864 | Not-determined |
| S1_extracted_k141_91890 | Not-determined |
| S1_extracted_k141_132007 | Low-quality |
| S1_extracted_k141_91892 | Not-determined |
| S1_extracted_k141_4143 | Not-determined |
| S1_extracted_k141_124989 | Not-determined |
| S1_extracted_k141_78642 | Not-determined |
| S1_extracted_k141_58502 | Low-quality |
| S1_extracted_k141_70958 | Low-quality |
| S1_extracted_k141_31869 | Not-determined |
| S1_extracted_k141_138414 | Not-determined |
| S1_extracted_k141_63950 | Not-determined |
| S1_extracted_k141_31871 | Not-determined |
| S1_extracted_k141_70960 | Not-determined |
| S1_extracted_k141_138416 | Not-determined |
| S1_extracted_k141_37481 | Low-quality |
| S1_extracted_k141_138417 | Low-quality |
| S1_extracted_k141_52132 | Not-determined |
| S1_extracted_k141_91899 | Not-determined |
| S1_extracted_k141_111903 | Not-determined |
| S1_extracted_k141_4151 | Not-determined |
| S1_extracted_k141_25064 | Not-determined |
| S1_extracted_k141_4152 | Not-determined |
| S1_extracted_k141_58508 | Low-quality |
| S1_extracted_k141_149274 | Not-determined |
| S1_extracted_k141_52133 | Not-determined |
| S1_extracted_k141_58509 | Not-determined |
| S1_extracted_k141_45324 | Not-determined |
| S1_extracted_k141_58510 | Not-determined |
| S1_extracted_k141_111906 | Not-determined |
| S1_extracted_k141_31875 | Low-quality |
| S1_extracted_k141_124996 | Not-determined |
| S1_extracted_k141_91904 | Not-determined |
| S1_extracted_k141_98739 | Low-quality |
| S1_extracted_k141_85245 | Not-determined |
| S1_extracted_k141_98741 | Not-determined |
| S1_extracted_k141_138423 | Not-determined |
| S1_extracted_k141_98742 | Not-determined |
| S1_extracted_k141_25071 | Not-determined |
| S1_extracted_k141_132020 | Not-determined |
| S1_extracted_k141_52139 | Not-determined |
| S1_extracted_k141_19062 | Not-determined |
| S1_extracted_k141_132022 | Not-determined |
| S1_extracted_k141_19064 | Not-determined |
| S1_extracted_k141_37489 | Not-determined |
| S1_extracted_k141_52143 | Not-determined |
| S1_extracted_k141_45331 | Not-determined |
| S1_extracted_k141_70969 | Not-determined |
| S1_extracted_k141_31888 | Not-determined |
| S1_extracted_k141_125006 | Not-determined |
| S1_extracted_k141_118625 | Not-determined |
| S1_extracted_k141_98744 | Not-determined |
| S1_extracted_k141_125009 | Not-determined |
| S1_extracted_k141_58519 | Low-quality |
| S1_extracted_k141_105325 | Low-quality |
| S1_extracted_k141_144942 | Not-determined |
| S1_extracted_k141_105326 | Not-determined |
| S1_extracted_k141_138436 | Not-determined |
| S1_extracted_k141_78670 | Low-quality |
| S1_extracted_k141_118634 | Not-determined |
| S1_extracted_k141_125016 | Not-determined |
| S1_extracted_k141_138438 | Not-determined |
| S1_extracted_k141_138439 | Not-determined |
| S1_extracted_k141_31902 | Low-quality |
| S1_extracted_k141_45335 | Not-determined |
| S1_extracted_k141_144946 | Not-determined |
| S1_extracted_k141_144947 | Not-determined |
| S1_extracted_k141_91920 | Not-determined |
| S1_extracted_k141_156374 | Low-quality |
| S1_extracted_k141_85261 | Not-determined |
| S1_extracted_k141_91924 | Low-quality |
| S1_extracted_k141_63976 | Not-determined |
| S1_extracted_k141_91925 | Low-quality |
| S1_extracted_k141_70988 | Low-quality |
| S1_extracted_k141_111915 | Not-determined |
| S1_extracted_k141_105338 | Low-quality |
| S1_extracted_k141_63978 | Not-determined |
| S1_extracted_k141_111916 | Low-quality |
| S1_extracted_k141_19077 | Not-determined |
| S1_extracted_k141_12139 | Low-quality |
| S1_extracted_k141_98763 | Not-determined |
| S1_extracted_k141_118647 | Not-determined |
| S1_extracted_k141_25089 | Not-determined |
| S1_extracted_k141_105342 | Not-determined |
| S1_extracted_k141_144953 | Not-determined |
| S1_extracted_k141_118649 | Not-determined |
| S1_extracted_k141_144954 | Low-quality |
| S1_extracted_k141_78687 | Not-determined |
| S1_extracted_k141_45348 | Not-determined |
| S1_extracted_k141_63987 | Not-determined |
| S1_extracted_k141_98767 | Not-determined |
| S1_extracted_k141_52170 | Not-determined |
| S1_extracted_k141_63989 | Not-determined |
| S1_extracted_k141_12147 | Low-quality |
| S1_extracted_k141_111921 | Not-determined |
| S1_extracted_k141_12149 | Not-determined |
| S1_extracted_k141_63992 | Not-determined |
| S1_extracted_k141_85269 | Not-determined |
| S1_extracted_k141_37502 | Not-determined |
| S1_extracted_k141_118655 | Low-quality |
| S1_extracted_k141_98771 | Not-determined |
| S1_extracted_k141_138455 | Not-determined |
| S1_extracted_k141_138456 | Not-determined |
| S1_extracted_k141_12153 | Not-determined |
| S1_extracted_k141_149287 | Not-determined |
| S1_extracted_k141_132051 | Not-determined |
| S1_extracted_k141_98773 | Low-quality |
| S1_extracted_k141_125041 | Not-determined |
| S1_extracted_k141_125042 | Not-determined |
| S1_extracted_k141_91941 | Not-determined |
| S1_extracted_k141_91944 | Not-determined |
| S1_extracted_k141_118663 | Not-determined |
| S1_extracted_k141_25101 | Low-quality |
| S1_extracted_k141_25102 | Not-determined |
| S1_extracted_k141_58544 | Not-determined |
| S1_extracted_k141_85281 | Not-determined |
| S1_extracted_k141_31914 | Low-quality |
| S1_extracted_k141_12163 | Not-determined |
| S1_extracted_k141_85284 | Not-determined |
| S1_extracted_k141_111932 | Not-determined |
| S1_extracted_k141_78697 | Not-determined |
| S1_extracted_k141_64001 | Low-quality |
| S1_extracted_k141_85286 | Not-determined |
| S1_extracted_k141_138467 | Not-determined |
| S1_extracted_k141_111933 | Not-determined |
| S1_extracted_k141_91953 | Not-determined |
| S1_extracted_k141_85288 | Low-quality |
| S1_extracted_k141_138468 | Low-quality |
| S1_extracted_k141_19103 | Not-determined |
| S1_extracted_k141_91956 | Not-determined |
| S1_extracted_k141_71003 | Not-determined |
| S1_extracted_k141_91957 | Low-quality |
| S1_extracted_k141_118666 | Not-determined |
| S1_extracted_k141_25110 | Not-determined |
| S1_extracted_k141_78703 | Not-determined |
| S1_extracted_k141_105361 | Not-determined |
| S1_extracted_k141_91963 | Low-quality |
| S1_extracted_k141_105363 | Not-determined |
| S1_extracted_k141_144972 | Low-quality |
| S1_extracted_k141_91965 | Not-determined |
| S1_extracted_k141_125055 | Not-determined |
| S1_extracted_k141_12173 | Not-determined |
| S1_extracted_k141_25115 | Low-quality |
| S1_extracted_k141_58547 | Low-quality |
| S1_extracted_k141_144976 | Not-determined |
| S1_extracted_k141_98781 | Not-determined |
| S1_extracted_k141_111936 | Low-quality |

| | |
|---|---|
| S1_extracted_k141_105370 | Not-determined |
| S1_extracted_k141_52183 | Not-determined |
| S1_extracted_k141_31929 | Low-quality |
| S1_extracted_k141_138483 | Low-quality |
| S1_extracted_k141_58550 | Low-quality |
| S1_extracted_k141_64007 | Not-determined |
| S1_extracted_k141_111943 | Not-determined |
| S1_extracted_k141_105375 | Not-determined |
| S1_extracted_k141_132069 | Low-quality |
| S1_extracted_k141_105377 | Not-determined |
| S1_extracted_k141_64010 | Not-determined |
| S1_extracted_k141_132071 | Not-determined |
| S1_extracted_k141_98788 | Not-determined |
| S1_extracted_k141_105379 | Low-quality |
| S1_extracted_k141_45375 | Not-determined |
| S1_extracted_k141_19111 | Not-determined |
| S1_extracted_k141_64013 | Not-determined |
| S1_extracted_k141_31939 | Not-determined |
| S1_extracted_k141_25122 | Not-determined |
| S1_extracted_k141_71017 | Not-determined |
| S1_extracted_k141_132075 | Not-determined |
| S1_extracted_k141_31941 | Not-determined |
| S1_extracted_k141_12182 | Not-determined |
| S1_extracted_k141_85300 | Not-determined |
| S1_extracted_k141_4183 | Not-determined |
| S1_extracted_k141_19114 | Not-determined |
| S1_extracted_k141_91969 | Not-determined |
| S1_extracted_k141_118684 | Low-quality |
| S1_extracted_k141_144985 | Not-determined |
| S1_extracted_k141_125071 | Not-determined |
| S1_extracted_k141_138490 | Not-determined |
| S1_extracted_k141_19118 | Not-determined |
| S1_extracted_k141_111955 | Not-determined |
| S1_extracted_k141_31945 | Not-determined |
| S1_extracted_k141_45387 | Not-determined |
| S1_extracted_k141_64016 | Not-determined |
| S1_extracted_k141_12191 | Not-determined |
| S1_extracted_k141_52200 | Not-determined |
| S1_extracted_k141_19120 | Not-determined |
| S1_extracted_k141_98800 | Not-determined |
| S1_extracted_k141_19121 | Not-determined |
| S1_extracted_k141_118690 | Not-determined |
| S1_extracted_k141_4188 | Not-determined |
| S1_extracted_k141_118691 | Low-quality |
| S1_extracted_k141_25135 | Not-determined |
| S1_extracted_k141_85310 | Not-determined |
| S1_extracted_k141_138495 | Not-determined |
| S1_extracted_k141_71021 | Not-determined |
| S1_extracted_k141_58568 | Not-determined |
| S1_extracted_k141_138496 | Not-determined |
| S1_extracted_k141_19126 | Not-determined |
| S1_extracted_k141_91971 | Not-determined |
| S1_extracted_k141_58570 | Low-quality |
| S1_extracted_k141_64020 | Not-determined |
| S1_extracted_k141_78716 | Not-determined |
| S1_extracted_k141_58575 | Not-determined |
| S1_extracted_k141_98803 | Low-quality |
| S1_extracted_k141_78718 | Not-determined |
| S1_extracted_k141_105397 | Not-determined |
| S1_extracted_k141_64023 | Not-determined |
| S1_extracted_k141_105398 | Not-determined |
| S1_extracted_k141_132082 | Low-quality |
| S1_extracted_k141_98804 | Not-determined |
| S1_extracted_k141_31959 | Low-quality |
| S1_extracted_k141_132084 | Not-determined |
| S1_extracted_k141_31961 | Not-determined |
| S1_extracted_k141_25142 | Not-determined |
| S1_extracted_k141_25143 | Not-determined |
| S1_extracted_k141_71033 | Low-quality |
| S1_extracted_k141_138510 | Not-determined |
| S1_extracted_k141_71035 | Not-determined |
| S1_extracted_k141_85324 | Not-determined |
| S1_extracted_k141_37541 | Not-determined |
| S1_extracted_k141_98807 | Low-quality |
| S1_extracted_k141_4204 | Not-determined |
| S1_extracted_k141_98813 | Low-quality |
| S1_extracted_k141_91989 | Low-quality |
| S1_extracted_k141_58592 | Not-determined |
| S1_extracted_k141_85330 | Low-quality |
| S1_extracted_k141_25145 | Not-determined |
| S1_extracted_k141_31969 | Not-determined |
| S1_extracted_k141_91993 | Not-determined |
| S1_extracted_k141_12218 | Not-determined |
| S1_extracted_k141_64037 | Not-determined |
| S1_extracted_k141_25150 | Not-determined |
| S1_extracted_k141_85332 | Low-quality |
| S1_extracted_k141_58597 | Not-determined |
| S1_extracted_k141_118706 | Not-determined |
| S1_extracted_k141_12221 | Not-determined |
| S1_extracted_k141_98819 | Not-determined |
| S1_extracted_k141_37551 | Low-quality |
| S1_extracted_k141_58601 | Not-determined |
| S1_extracted_k141_25158 | Not-determined |
| S1_extracted_k141_98823 | Not-determined |
| S1_extracted_k141_85340 | Not-determined |
| S1_extracted_k141_145009 | Not-determined |
| S1_extracted_k141_31977 | Not-determined |
| S1_extracted_k141_64045 | Low-quality |
| S1_extracted_k141_64046 | Not-determined |
| S1_extracted_k141_118711 | Not-determined |
| S1_extracted_k141_52216 | Not-determined |
| S1_extracted_k141_45400 | Not-determined |
| S1_extracted_k141_52218 | Low-quality |
| S1_extracted_k141_78747 | Low-quality |
| S1_extracted_k141_118712 | Not-determined |
| S1_extracted_k141_64048 | Not-determined |
| S1_extracted_k141_45402 | Not-determined |
| S1_extracted_k141_125099 | Not-determined |
| S1_extracted_k141_78751 | Not-determined |
| S1_extracted_k141_85349 | Not-determined |
| S1_extracted_k141_125103 | Not-determined |
| S1_extracted_k141_78753 | Not-determined |
| S1_extracted_k141_149306 | Not-determined |
| S1_extracted_k141_19168 | Low-quality |
| S1_extracted_k141_31990 | Not-determined |
| S1_extracted_k141_4207 | Not-determined |
| S1_extracted_k141_58616 | Low-quality |
| S1_extracted_k141_19170 | Low-quality |
| S1_extracted_k141_71050 | Not-determined |
| S1_extracted_k141_118718 | Not-determined |
| S1_extracted_k141_45410 | Not-determined |
| S1_extracted_k141_78760 | Not-determined |
| S1_extracted_k141_105413 | Not-determined |
| S1_extracted_k141_149312 | Not-determined |
| S1_extracted_k141_118720 | Not-determined |
| S1_extracted_k141_78761 | Not-determined |
| S1_extracted_k141_92004 | Not-determined |
| S1_extracted_k141_19178 | Low-quality |
| S1_extracted_k141_19180 | Not-determined |
| S1_extracted_k141_45414 | Not-determined |
| S1_extracted_k141_78762 | Low-quality |
| S1_extracted_k141_118723 | Low-quality |
| S1_extracted_k141_78763 | Not-determined |
| S1_extracted_k141_138527 | Not-determined |
| S1_extracted_k141_105418 | Not-determined |
| S1_extracted_k141_105419 | Not-determined |
| S1_extracted_k141_78765 | Low-quality |
| S1_extracted_k141_125115 | Not-determined |
| S1_extracted_k141_125116 | Not-determined |
| S1_extracted_k141_71056 | Not-determined |
| S1_extracted_k141_64061 | Low-quality |
| S1_extracted_k141_132120 | Not-determined |
| S1_extracted_k141_71057 | Low-quality |
| S1_extracted_k141_85360 | Low-quality |
| S1_extracted_k141_32009 | Not-determined |
| S1_extracted_k141_19184 | Low-quality |
| S1_extracted_k141_112009 | Not-determined |
| S1_extracted_k141_118726 | Not-determined |
| S1_extracted_k141_145030 | Not-determined |
| S1_extracted_k141_105423 | Not-determined |
| S1_extracted_k141_19187 | Not-determined |
| S1_extracted_k141_12246 | Not-determined |
| S1_extracted_k141_64065 | Not-determined |
| S1_extracted_k141_149321 | Not-determined |
| S1_extracted_k141_98839 | Low-quality |
| S1_extracted_k141_25188 | Not-determined |
| S1_extracted_k141_105427 | Not-determined |
| S1_extracted_k141_12249 | Not-determined |
| S1_extracted_k141_64069 | Not-determined |
| S1_extracted_k141_138536 | Not-determined |
| S1_extracted_k141_138537 | Not-determined |
| S1_extracted_k141_45421 | Not-determined |
| S1_extracted_k141_132133 | Not-determined |
| S1_extracted_k141_58633 | Not-determined |
| S1_extracted_k141_52244 | Not-determined |
| S1_extracted_k141_78782 | Not-determined |
| S1_extracted_k141_58636 | Not-determined |
| S1_extracted_k141_32015 | Not-determined |
| S1_extracted_k141_138546 | Not-determined |
| S1_extracted_k141_145039 | Not-determined |
| S1_extracted_k141_132140 | Low-quality |
| S1_extracted_k141_132141 | Not-determined |
| S1_extracted_k141_105437 | Low-quality |
| S1_extracted_k141_12256 | Not-determined |
| S1_extracted_k141_4225 | Not-determined |
| S1_extracted_k141_118736 | Low-quality |
| S1_extracted_k141_125119 | Not-determined |
| S1_extracted_k141_92034 | Not-determined |
| S1_extracted_k141_58642 | Not-determined |
| S1_extracted_k141_132144 | Not-determined |
| S1_extracted_k141_32024 | Low-quality |
| S1_extracted_k141_112024 | Not-determined |
| S1_extracted_k141_45424 | Low-quality |
| S1_extracted_k141_92037 | Not-determined |
| S1_extracted_k141_156390 | Not-determined |
| S1_extracted_k141_45425 | Not-determined |
| S1_extracted_k141_149322 | Low-quality |
| S1_extracted_k141_64080 | Not-determined |
| S1_extracted_k141_64081 | Not-determined |
| S1_extracted_k141_125126 | Not-determined |
| S1_extracted_k141_58649 | Not-determined |
| S1_extracted_k141_118742 | Not-determined |
| S1_extracted_k141_45431 | Not-determined |
| S1_extracted_k141_92043 | Not-determined |
| S1_extracted_k141_45433 | Not-determined |
| S1_extracted_k141_45434 | Low-quality |
| S1_extracted_k141_45435 | Not-determined |
| S1_extracted_k141_4230 | Not-determined |
| S1_extracted_k141_12272 | Not-determined |
| S1_extracted_k141_64089 | Not-determined |
| S1_extracted_k141_19211 | Not-determined |
| S1_extracted_k141_112038 | Not-determined |
| S1_extracted_k141_32032 | Not-determined |
| S1_extracted_k141_45440 | Not-determined |
| S1_extracted_k141_138558 | Not-determined |
| S1_extracted_k141_85386 | Not-determined |
| S1_extracted_k141_71065 | Not-determined |
| S1_extracted_k141_12278 | Not-determined |
| S1_extracted_k141_112043 | Low-quality |
| S1_extracted_k141_71068 | Not-determined |
| S1_extracted_k141_105452 | Low-quality |
| S1_extracted_k141_132161 | Low-quality |
| S1_extracted_k141_118756 | Not-determined |
| S1_extracted_k141_58659 | Not-determined |
| S1_extracted_k141_85391 | Not-determined |
| S1_extracted_k141_52271 | Not-determined |
| S1_extracted_k141_118757 | Not-determined |
| S1_extracted_k141_132164 | Not-determined |
| S1_extracted_k141_12285 | Not-determined |
| S1_extracted_k141_138568 | Not-determined |
| S1_extracted_k141_12287 | Low-quality |
| S1_extracted_k141_78807 | Not-determined |
| S1_extracted_k141_85397 | Not-determined |
| S1_extracted_k141_118764 | Not-determined |
| S1_extracted_k141_132168 | Low-quality |
| S1_extracted_k141_4241 | Low-quality |
| S1_extracted_k141_138570 | Not-determined |
| S1_extracted_k141_52277 | Low-quality |
| S1_extracted_k141_12291 | Not-determined |
| S1_extracted_k141_4243 | Not-determined |
| S1_extracted_k141_138573 | Not-determined |
| S1_extracted_k141_118768 | Not-determined |
| S1_extracted_k141_78811 | Low-quality |
| S1_extracted_k141_105460 | Not-determined |
| S1_extracted_k141_12294 | Not-determined |
| S1_extracted_k141_45457 | Not-determined |
| S1_extracted_k141_138575 | Not-determined |
| S1_extracted_k141_12297 | Low-quality |
| S1_extracted_k141_32055 | Not-determined |
| S1_extracted_k141_37561 | Not-determined |
| S1_extracted_k141_105464 | Low-quality |
| S1_extracted_k141_92065 | Low-quality |
| S1_extracted_k141_92067 | Not-determined |
| S1_extracted_k141_125136 | Low-quality |
| S1_extracted_k141_138579 | Not-determined |
| S1_extracted_k141_105467 | Low-quality |
| S1_extracted_k141_64111 | Not-determined |
| S1_extracted_k141_32057 | Not-determined |
| S1_extracted_k141_138580 | Not-determined |
| S1_extracted_k141_52284 | Not-determined |
| S1_extracted_k141_58671 | Not-determined |
| S1_extracted_k141_112060 | Not-determined |
| S1_extracted_k141_4248 | Not-determined |
| S1_extracted_k141_85412 | Not-determined |
| S1_extracted_k141_78815 | Low-quality |
| S1_extracted_k141_32059 | Not-determined |
| S1_extracted_k141_145061 | Low-quality |
| S1_extracted_k141_19219 | Not-determined |
| S1_extracted_k141_78817 | Not-determined |
| S1_extracted_k141_58673 | Not-determined |
| S1_extracted_k141_149345 | Not-determined |
| S1_extracted_k141_112064 | Not-determined |
| S1_extracted_k141_92077 | Not-determined |
| S1_extracted_k141_105474 | Not-determined |
| S1_extracted_k141_32063 | Not-determined |
| S1_extracted_k141_92080 | Not-determined |
| S1_extracted_k141_118773 | Not-determined |
| S1_extracted_k141_32065 | Not-determined |
| S1_extracted_k141_19224 | Low-quality |
| S1_extracted_k141_25217 | Not-determined |
| S1_extracted_k141_58677 | Not-determined |
| S1_extracted_k141_52293 | Low-quality |
| S1_extracted_k141_78824 | Not-determined |
| S1_extracted_k141_138586 | Not-determined |
| S1_extracted_k141_92082 | Not-determined |
| S1_extracted_k141_78827 | Not-determined |
| S1_extracted_k141_92084 | Not-determined |
| S1_extracted_k141_71081 | Not-determined |
| S1_extracted_k141_156400 | Not-determined |
| S1_extracted_k141_45466 | Low-quality |
| S1_extracted_k141_58684 | Not-determined |
| S1_extracted_k141_92087 | Low-quality |
| S1_extracted_k141_19234 | Low-quality |
| S1_extracted_k141_78832 | Low-quality |
| S1_extracted_k141_64119 | Not-determined |
| S1_extracted_k141_92089 | Not-determined |
| S1_extracted_k141_125152 | Not-determined |
| S1_extracted_k141_37567 | Not-determined |
| S1_extracted_k141_132186 | Not-determined |
| S1_extracted_k141_45472 | Not-determined |
| S1_extracted_k141_37569 | Not-determined |
| S1_extracted_k141_19241 | Not-determined |
| S1_extracted_k141_132188 | Not-determined |
| S1_extracted_k141_156406 | Not-determined |
| S1_extracted_k141_125157 | Not-determined |
| S1_extracted_k141_145077 | Low-quality |
| S1_extracted_k141_138591 | Not-determined |
| S1_extracted_k141_105490 | Not-determined |
| S1_extracted_k141_19246 | Not-determined |
| S1_extracted_k141_98853 | Not-determined |
| S1_extracted_k141_45478 | Low-quality |
| S1_extracted_k141_138592 | Not-determined |
| S1_extracted_k141_19247 | Not-determined |
| S1_extracted_k141_156410 | Not-determined |
| S1_extracted_k141_64124 | Not-determined |
| S1_extracted_k141_25232 | Not-determined |
| S1_extracted_k141_58698 | Not-determined |
| S1_extracted_k141_58699 | Not-determined |
| S1_extracted_k141_52303 | Low-quality |
| S1_extracted_k141_25234 | Not-determined |
| S1_extracted_k141_145079 | Not-determined |
| S1_extracted_k141_25235 | Not-determined |
| S1_extracted_k141_58703 | Not-determined |
| S1_extracted_k141_52306 | Not-determined |
| S1_extracted_k141_25236 | Not-determined |
| S1_extracted_k141_145082 | Not-determined |
| S1_extracted_k141_19253 | Not-determined |
| S1_extracted_k141_45483 | Not-determined |
| S1_extracted_k141_145085 | Not-determined |
| S1_extracted_k141_64131 | Not-determined |
| S1_extracted_k141_19256 | Not-determined |
| S1_extracted_k141_4269 | Not-determined |
| S1_extracted_k141_37580 | Not-determined |
| S1_extracted_k141_156413 | Not-determined |
| S1_extracted_k141_32097 | Not-determined |
| S1_extracted_k141_45490 | Not-determined |
| S1_extracted_k141_52319 | Not-determined |
| S1_extracted_k141_71087 | Not-determined |
| S1_extracted_k141_45494 | Not-determined |
| S1_extracted_k141_19268 | Not-determined |
| S1_extracted_k141_52322 | Not-determined |
| S1_extracted_k141_19269 | Not-determined |
| S1_extracted_k141_132204 | Not-determined |
| S1_extracted_k141_19270 | Not-determined |
| S1_extracted_k141_25253 | Not-determined |
| S1_extracted_k141_32103 | Low-quality |
| S1_extracted_k141_32104 | Not-determined |
| S1_extracted_k141_138608 | Not-determined |
| S1_extracted_k141_78854 | Low-quality |
| S1_extracted_k141_156420 | Not-determined |
| S1_extracted_k141_85443 | Low-quality |
| S1_extracted_k141_125168 | Not-determined |
| S1_extracted_k141_132207 | Low-quality |
| S1_extracted_k141_145101 | Not-determined |
| S1_extracted_k141_52333 | Not-determined |
| S1_extracted_k141_98878 | Not-determined |
| S1_extracted_k141_125170 | Not-determined |
| S1_extracted_k141_118800 | Low-quality |
| S1_extracted_k141_12309 | Low-quality |
| S1_extracted_k141_145106 | Not-determined |
| S1_extracted_k141_71091 | Not-determined |
| S1_extracted_k141_85449 | Not-determined |
| S1_extracted_k141_12313 | Not-determined |
| S1_extracted_k141_98884 | Not-determined |
| S1_extracted_k141_52337 | Not-determined |
| S1_extracted_k141_85452 | Low-quality |
| S1_extracted_k141_32120 | Not-determined |
| S1_extracted_k141_64163 | Not-determined |
| S1_extracted_k141_58730 | Not-determined |
| S1_extracted_k141_32122 | Not-determined |
| S1_extracted_k141_112094 | Not-determined |
| S1_extracted_k141_118810 | Not-determined |
| S1_extracted_k141_71097 | Not-determined |
| S1_extracted_k141_64165 | Low-quality |
| S1_extracted_k141_105512 | Low-quality |
| S1_extracted_k141_98887 | Not-determined |
| S1_extracted_k141_32126 | Low-quality |
| S1_extracted_k141_64170 | Not-determined |
| S1_extracted_k141_58738 | Not-determined |
| S1_extracted_k141_112099 | Not-determined |
| S1_extracted_k141_71106 | Low-quality |
| S1_extracted_k141_78858 | Not-determined |
| S1_extracted_k141_85456 | Low-quality |
| S1_extracted_k141_138632 | Not-determined |
| S1_extracted_k141_145113 | Not-determined |
| S1_extracted_k141_132223 | Not-determined |
| S1_extracted_k141_125191 | Not-determined |
| S1_extracted_k141_64176 | Not-determined |
| S1_extracted_k141_37612 | Not-determined |
| S1_extracted_k141_156431 | Not-determined |
| S1_extracted_k141_156432 | Not-determined |
| S1_extracted_k141_105519 | Low-quality |
| S1_extracted_k141_105520 | Not-determined |
| S1_extracted_k141_85464 | Not-determined |
| S1_extracted_k141_52348 | Not-determined |
| S1_extracted_k141_32135 | Not-determined |
| S1_extracted_k141_19303 | Not-determined |
| S1_extracted_k141_12327 | Low-quality |
| S1_extracted_k141_125194 | Not-determined |
| S1_extracted_k141_125195 | Not-determined |
| S1_extracted_k141_92115 | Not-determined |
| S1_extracted_k141_12330 | Low-quality |
| S1_extracted_k141_98898 | Not-determined |
| S1_extracted_k141_25269 | Not-determined |
| S1_extracted_k141_105524 | Low-quality |
| S1_extracted_k141_32140 | Low-quality |
| S1_extracted_k141_125203 | Not-determined |
| S1_extracted_k141_58754 | Not-determined |
| S1_extracted_k141_92122 | Not-determined |
| S1_extracted_k141_145122 | Not-determined |
| S1_extracted_k141_118825 | Low-quality |
| S1_extracted_k141_118826 | Not-determined |
| S1_extracted_k141_112110 | Not-determined |
| S1_extracted_k141_32146 | Not-determined |
| S1_extracted_k141_58758 | Not-determined |
| S1_extracted_k141_145127 | Not-determined |
| S1_extracted_k141_45524 | Low-quality |
| S1_extracted_k141_125209 | Not-determined |
| S1_extracted_k141_78878 | Low-quality |
| S1_extracted_k141_92129 | Not-determined |
| S1_extracted_k141_52359 | Not-determined |
| S1_extracted_k141_45531 | Not-determined |
| S1_extracted_k141_37622 | Low-quality |
| S1_extracted_k141_92133 | Low-quality |
| S1_extracted_k141_45533 | Not-determined |
| S1_extracted_k141_138638 | Low-quality |
| S1_extracted_k141_85479 | Not-determined |
| S1_extracted_k141_37624 | Not-determined |
| S1_extracted_k141_132235 | Not-determined |
| S1_extracted_k141_132236 | Not-determined |
| S1_extracted_k141_19328 | Not-determined |
| S1_extracted_k141_118832 | Low-quality |
| S1_extracted_k141_85482 | Not-determined |
| S1_extracted_k141_58771 | Not-determined |
| S1_extracted_k141_145136 | Low-quality |
| S1_extracted_k141_45535 | Not-determined |
| S1_extracted_k141_145139 | Not-determined |
| S1_extracted_k141_132241 | Not-determined |
| S1_extracted_k141_125223 | Not-determined |
| S1_extracted_k141_64200 | Not-determined |
| S1_extracted_k141_37627 | Not-determined |
| S1_extracted_k141_105542 | Low-quality |
| S1_extracted_k141_37628 | Not-determined |
| S1_extracted_k141_12353 | Not-determined |
| S1_extracted_k141_149396 | Not-determined |
| S1_extracted_k141_125226 | Not-determined |
| S1_extracted_k141_32157 | Not-determined |
| S1_extracted_k141_12355 | Low-quality |
| S1_extracted_k141_64205 | Low-quality |
| S1_extracted_k141_145143 | Not-determined |
| S1_extracted_k141_98923 | Not-determined |
| S1_extracted_k141_85490 | Not-determined |
| S1_extracted_k141_98925 | Not-determined |
| S1_extracted_k141_85491 | Not-determined |
| S1_extracted_k141_52368 | Not-determined |
| S1_extracted_k141_105546 | Not-determined |
| S1_extracted_k141_19339 | Not-determined |
| S1_extracted_k141_149402 | Not-determined |
| S1_extracted_k141_58784 | Not-determined |
| S1_extracted_k141_45547 | Not-determined |
| S1_extracted_k141_138647 | Not-determined |
| S1_extracted_k141_92144 | Not-determined |
| S1_extracted_k141_145146 | Low-quality |
| S1_extracted_k141_118845 | Not-determined |
| S1_extracted_k141_71111 | Not-determined |
| S1_extracted_k141_78900 | Not-determined |
| S1_extracted_k141_19346 | Not-determined |
| S1_extracted_k141_58788 | Not-determined |
| S1_extracted_k141_52373 | Not-determined |
| S1_extracted_k141_71115 | Not-determined |
| S1_extracted_k141_45558 | Not-determined |
| S1_extracted_k141_52379 | Not-determined |
| S1_extracted_k141_58794 | Low-quality |
| S1_extracted_k141_78910 | Not-determined |
| S1_extracted_k141_12366 | Not-determined |
| S1_extracted_k141_149415 | Not-determined |
| S1_extracted_k141_52381 | Not-determined |
| S1_extracted_k141_19352 | Low-quality |
| S1_extracted_k141_71119 | Low-quality |
| S1_extracted_k141_58798 | Not-determined |
| S1_extracted_k141_19353 | Not-determined |
| S1_extracted_k141_45559 | Not-determined |
| S1_extracted_k141_98938 | Not-determined |
| S1_extracted_k141_25310 | Not-determined |
| S1_extracted_k141_118859 | Not-determined |
| S1_extracted_k141_125239 | Not-determined |
| S1_extracted_k141_19355 | Not-determined |
| S1_extracted_k141_52390 | Low-quality |
| S1_extracted_k141_92158 | Low-quality |
| S1_extracted_k141_85507 | Not-determined |
| S1_extracted_k141_58806 | Not-determined |
| S1_extracted_k141_64223 | Not-determined |
| S1_extracted_k141_37636 | Low-quality |
| S1_extracted_k141_12378 | Not-determined |
| S1_extracted_k141_125243 | Not-determined |
| S1_extracted_k141_58807 | Not-determined |
| S1_extracted_k141_12383 | Not-determined |
| S1_extracted_k141_19364 | Not-determined |
| S1_extracted_k141_145165 | Not-determined |
| S1_extracted_k141_98946 | Not-determined |
| S1_extracted_k141_45571 | Not-determined |
| S1_extracted_k141_85512 | Not-determined |
| S1_extracted_k141_98947 | Not-determined |
| S1_extracted_k141_64228 | Low-quality |
| S1_extracted_k141_12386 | Not-determined |
| S1_extracted_k141_71122 | Not-determined |
| S1_extracted_k141_132265 | Not-determined |
| S1_extracted_k141_92164 | Low-quality |
| S1_extracted_k141_37642 | Not-determined |
| S1_extracted_k141_58816 | Not-determined |
| S1_extracted_k141_85514 | Not-determined |
| S1_extracted_k141_45579 | Not-determined |
| S1_extracted_k141_19370 | Low-quality |
| S1_extracted_k141_85517 | Not-determined |
| S1_extracted_k141_105562 | Not-determined |
| S1_extracted_k141_64231 | Not-determined |
| S1_extracted_k141_149432 | Not-determined |
| S1_extracted_k141_58822 | Not-determined |
| S1_extracted_k141_52407 | Not-determined |
| S1_extracted_k141_25325 | Not-determined |
| S1_extracted_k141_25326 | Not-determined |
| S1_extracted_k141_149435 | Not-determined |
| S1_extracted_k141_156462 | Not-determined |
| S1_extracted_k141_156463 | Low-quality |
| S1_extracted_k141_98967 | Not-determined |
| S1_extracted_k141_156464 | Not-determined |
| S1_extracted_k141_112128 | Not-determined |
| S1_extracted_k141_12403 | Not-determined |
| S1_extracted_k141_112129 | Not-determined |
| S1_extracted_k141_105573 | Not-determined |
| S1_extracted_k141_25329 | Low-quality |
| S1_extracted_k141_85528 | Not-determined |
| S1_extracted_k141_58828 | Low-quality |
| S1_extracted_k141_32196 | Low-quality |
| S1_extracted_k141_112131 | Not-determined |
| S1_extracted_k141_12406 | Not-determined |
| S1_extracted_k141_12407 | Not-determined |
| S1_extracted_k141_112136 | Not-determined |
| S1_extracted_k141_78943 | Not-determined |
| S1_extracted_k141_85534 | Not-determined |
| S1_extracted_k141_19379 | Low-quality |
| S1_extracted_k141_92188 | Not-determined |
| S1_extracted_k141_92189 | Not-determined |
| S1_extracted_k141_64242 | Not-determined |
| S1_extracted_k141_98979 | Not-determined |
| S1_extracted_k141_118893 | Not-determined |
| S1_extracted_k141_105578 | Not-determined |
| S1_extracted_k141_25338 | Not-determined |
| S1_extracted_k141_64246 | Not-determined |
| S1_extracted_k141_125262 | Not-determined |
| S1_extracted_k141_32200 | Low-quality |
| S1_extracted_k141_85544 | Not-determined |
| S1_extracted_k141_125264 | Not-determined |
| S1_extracted_k141_12421 | Not-determined |
| S1_extracted_k141_58840 | Not-determined |
| S1_extracted_k141_125265 | Not-determined |
| S1_extracted_k141_64255 | Not-determined |
| S1_extracted_k141_45603 | Not-determined |
| S1_extracted_k141_4307 | Not-determined |
| S1_extracted_k141_25341 | Not-determined |
| S1_extracted_k141_98994 | Not-determined |
| S1_extracted_k141_58845 | Not-determined |
| S1_extracted_k141_145196 | Not-determined |
| S1_extracted_k141_149454 | Not-determined |
| S1_extracted_k141_19393 | Low-quality |
| S1_extracted_k141_25347 | Low-quality |
| S1_extracted_k141_78957 | Not-determined |
| S1_extracted_k141_85553 | Not-determined |
| S1_extracted_k141_125274 | Not-determined |
| S1_extracted_k141_64264 | Not-determined |
| S1_extracted_k141_112147 | Not-determined |
| S1_extracted_k141_32209 | Low-quality |
| S1_extracted_k141_58849 | Not-determined |
| S1_extracted_k141_92204 | Low-quality |
| S1_extracted_k141_78966 | Not-determined |
| S1_extracted_k141_25354 | Not-determined |
| S1_extracted_k141_4315 | Not-determined |
| S1_extracted_k141_118907 | Not-determined |
| S1_extracted_k141_145201 | Not-determined |
| S1_extracted_k141_19400 | Not-determined |
| S1_extracted_k141_32211 | Not-determined |
| S1_extracted_k141_58853 | Not-determined |
| S1_extracted_k141_145203 | Not-determined |
| S1_extracted_k141_105595 | Not-determined |
| S1_extracted_k141_99009 | Not-determined |
| S1_extracted_k141_112157 | Not-determined |
| S1_extracted_k141_52441 | Not-determined |
| S1_extracted_k141_4322 | Not-determined |
| S1_extracted_k141_37651 | Not-determined |
| S1_extracted_k141_149464 | Not-determined |
| S1_extracted_k141_125284 | Not-determined |
| S1_extracted_k141_52448 | Low-quality |
| S1_extracted_k141_92214 | Low-quality |
| S1_extracted_k141_52449 | Not-determined |
| S1_extracted_k141_19406 | Low-quality |
| S1_extracted_k141_145209 | Low-quality |
| S1_extracted_k141_64272 | Low-quality |
| S1_extracted_k141_52451 | Not-determined |
| S1_extracted_k141_145210 | Low-quality |

| | |
|---|---|
| S1_extracted_k141_112168 | Not-determined |
| S1_extracted_k141_125289 | Not-determined |
| S1_extracted_k141_25372 | Not-determined |
| S1_extracted_k141_85570 | Not-determined |
| S1_extracted_k141_64276 | Not-determined |
| S1_extracted_k141_138686 | Not-determined |
| S1_extracted_k141_32229 | Not-determined |
| S1_extracted_k141_92218 | Not-determined |
| S1_extracted_k141_132296 | Low-quality |
| S1_extracted_k141_37654 | Not-determined |
| S1_extracted_k141_156475 | Not-determined |
| S1_extracted_k141_138689 | Not-determined |
| S1_extracted_k141_99019 | Not-determined |
| S1_extracted_k141_105605 | Not-determined |
| S1_extracted_k141_25375 | Not-determined |
| S1_extracted_k141_85579 | Not-determined |
| S1_extracted_k141_78981 | Not-determined |
| S1_extracted_k141_112173 | Not-determined |
| S1_extracted_k141_85580 | Not-determined |
| S1_extracted_k141_99022 | Not-determined |
| S1_extracted_k141_145215 | Not-determined |
| S1_extracted_k141_12441 | Not-determined |
| S1_extracted_k141_25379 | Not-determined |
| S1_extracted_k141_149478 | Not-determined |
| S1_extracted_k141_156480 | Not-determined |
| S1_extracted_k141_85584 | Not-determined |
| S1_extracted_k141_125299 | Low-quality |
| S1_extracted_k141_145217 | Low-quality |
| S1_extracted_k141_125302 | Low-quality |
| S1_extracted_k141_156483 | Not-determined |
| S1_extracted_k141_125303 | Low-quality |
| S1_extracted_k141_92223 | Not-determined |
| S1_extracted_k141_85591 | Not-determined |
| S1_extracted_k141_132306 | Not-determined |
| S1_extracted_k141_156487 | Not-determined |
| S1_extracted_k141_99028 | Not-determined |
| S1_extracted_k141_12453 | Low-quality |
| S1_extracted_k141_132311 | Not-determined |
| S1_extracted_k141_32250 | Not-determined |
| S1_extracted_k141_78987 | Low-quality |
| S1_extracted_k141_58890 | Not-determined |
| S1_extracted_k141_32252 | Not-determined |
| S1_extracted_k141_78989 | Not-determined |
| S1_extracted_k141_64295 | Not-determined |
| S1_extracted_k141_125308 | Not-determined |
| S1_extracted_k141_156493 | Not-determined |
| S1_extracted_k141_145227 | Not-determined |
| S1_extracted_k141_105626 | Not-determined |
| S1_extracted_k141_105628 | Not-determined |
| S1_extracted_k141_85604 | Not-determined |
| S1_extracted_k141_52477 | Not-determined |
| S1_extracted_k141_118932 | Not-determined |
| S1_extracted_k141_92234 | Not-determined |
| S1_extracted_k141_85608 | Not-determined |
| S1_extracted_k141_125313 | Not-determined |
| S1_extracted_k141_145230 | Not-determined |
| S1_extracted_k141_25397 | Not-determined |
| S1_extracted_k141_92236 | Not-determined |
| S1_extracted_k141_4362 | Not-determined |
| S1_extracted_k141_132326 | Not-determined |
| S1_extracted_k141_125320 | Not-determined |
| S1_extracted_k141_4366 | Not-determined |
| S1_extracted_k141_25399 | Low-quality |
| S1_extracted_k141_58900 | Not-determined |
| S1_extracted_k141_112199 | Not-determined |
| S1_extracted_k141_92241 | Low-quality |
| S1_extracted_k141_125321 | Not-determined |
| S1_extracted_k141_112202 | Not-determined |
| S1_extracted_k141_105641 | Not-determined |
| S1_extracted_k141_78997 | Low-quality |
| S1_extracted_k141_45660 | Not-determined |
| S1_extracted_k141_32272 | Not-determined |
| S1_extracted_k141_105642 | Not-determined |
| S1_extracted_k141_32273 | Not-determined |
| S1_extracted_k141_45662 | Not-determined |
| S1_extracted_k141_4368 | Not-determined |
| S1_extracted_k141_132332 | Low-quality |
| S1_extracted_k141_112207 | Not-determined |
| S1_extracted_k141_92247 | Not-determined |
| S1_extracted_k141_12467 | Not-determined |
| S1_extracted_k141_132336 | Not-determined |
| S1_extracted_k141_92249 | Not-determined |
| S1_extracted_k141_85625 | Low-quality |
| S1_extracted_k141_25406 | Not-determined |
| S1_extracted_k141_37672 | Not-determined |
| S1_extracted_k141_45665 | Not-determined |
| S1_extracted_k141_145244 | Not-determined |
| S1_extracted_k141_138720 | Not-determined |
| S1_extracted_k141_138722 | Not-determined |
| S1_extracted_k141_12475 | Not-determined |
| S1_extracted_k141_99051 | Not-determined |
| S1_extracted_k141_118959 | Not-determined |
| S1_extracted_k141_138725 | Not-determined |
| S1_extracted_k141_37677 | Not-determined |
| S1_extracted_k141_45672 | Not-determined |
| S1_extracted_k141_99055 | Not-determined |
| S1_extracted_k141_138728 | Not-determined |
| S1_extracted_k141_52510 | Not-determined |
| S1_extracted_k141_71166 | Not-determined |
| S1_extracted_k141_138729 | Not-determined |
| S1_extracted_k141_156516 | Not-determined |
| S1_extracted_k141_99060 | Not-determined |
| S1_extracted_k141_138732 | Not-determined |
| S1_extracted_k141_32290 | Not-determined |
| S1_extracted_k141_64325 | Not-determined |
| S1_extracted_k141_92265 | Not-determined |
| S1_extracted_k141_138735 | Not-determined |
| S1_extracted_k141_112231 | Not-determined |
| S1_extracted_k141_138737 | Not-determined |
| S1_extracted_k141_92271 | Not-determined |
| S1_extracted_k141_125341 | Not-determined |
| S1_extracted_k141_25419 | Not-determined |
| S1_extracted_k141_92272 | Not-determined |
| S1_extracted_k141_149486 | Not-determined |
| S1_extracted_k141_138739 | Not-determined |
| S1_extracted_k141_105670 | Not-determined |
| S1_extracted_k141_52520 | Low-quality |
| S1_extracted_k141_118972 | Not-determined |
| S1_extracted_k141_85640 | Not-determined |
| S1_extracted_k141_105672 | Low-quality |
| S1_extracted_k141_37690 | Not-determined |
| S1_extracted_k141_99066 | Not-determined |
| S1_extracted_k141_64335 | Not-determined |
| S1_extracted_k141_52525 | Not-determined |
| S1_extracted_k141_92277 | Not-determined |
| S1_extracted_k141_138743 | Not-determined |
| S1_extracted_k141_118974 | Low-quality |
| S1_extracted_k141_99067 | Low-quality |
| S1_extracted_k141_32304 | Not-determined |
| S1_extracted_k141_105677 | Low-quality |
| S1_extracted_k141_85645 | Low-quality |
| S1_extracted_k141_138748 | Not-determined |
| S1_extracted_k141_45692 | Not-determined |
| S1_extracted_k141_45693 | Low-quality |
| S1_extracted_k141_145268 | Not-determined |
| S1_extracted_k141_99073 | Not-determined |
| S1_extracted_k141_79032 | Not-determined |
| S1_extracted_k141_45696 | Not-determined |
| S1_extracted_k141_64343 | Not-determined |
| S1_extracted_k141_58915 | Low-quality |
| S1_extracted_k141_32313 | Low-quality |
| S1_extracted_k141_118985 | Not-determined |
| S1_extracted_k141_132353 | Not-determined |
| S1_extracted_k141_79039 | Low-quality |
| S1_extracted_k141_12500 | Not-determined |
| S1_extracted_k141_138756 | Not-determined |
| S1_extracted_k141_37696 | Low-quality |
| S1_extracted_k141_32316 | Not-determined |
| S1_extracted_k141_79042 | Not-determined |
| S1_extracted_k141_125353 | Not-determined |
| S1_extracted_k141_85658 | Not-determined |
| S1_extracted_k141_32317 | Not-determined |
| S1_extracted_k141_92285 | Low-quality |
| S1_extracted_k141_132356 | Low-quality |
| S1_extracted_k141_125356 | Not-determined |
| S1_extracted_k141_45706 | Not-determined |
| S1_extracted_k141_45707 | Not-determined |
| S1_extracted_k141_71173 | Not-determined |
| S1_extracted_k141_19460 | Not-determined |
| S1_extracted_k141_71175 | Low-quality |
| S1_extracted_k141_125360 | Not-determined |
| S1_extracted_k141_105688 | Low-quality |
| S1_extracted_k141_58923 | Not-determined |
| S1_extracted_k141_132357 | Not-determined |
| S1_extracted_k141_112247 | Low-quality |
| S1_extracted_k141_85668 | Not-determined |
| S1_extracted_k141_19467 | Not-determined |
| S1_extracted_k141_85669 | Not-determined |
| S1_extracted_k141_112250 | Not-determined |
| S1_extracted_k141_145281 | Not-determined |
| S1_extracted_k141_79049 | Not-determined |
| S1_extracted_k141_125362 | Low-quality |
| S1_extracted_k141_32330 | Low-quality |
| S1_extracted_k141_37705 | Low-quality |
| S1_extracted_k141_118994 | Not-determined |
| S1_extracted_k141_92296 | Not-determined |
| S1_extracted_k141_85673 | Not-determined |
| S1_extracted_k141_12511 | Not-determined |
| S1_extracted_k141_118995 | Not-determined |
| S1_extracted_k141_92299 | Not-determined |
| S1_extracted_k141_58927 | Not-determined |
| S1_extracted_k141_132363 | Not-determined |
| S1_extracted_k141_99087 | Not-determined |
| S1_extracted_k141_118996 | Low-quality |
| S1_extracted_k141_145284 | Not-determined |
| S1_extracted_k141_79054 | Low-quality |
| S1_extracted_k141_79055 | Not-determined |
| S1_extracted_k141_92303 | Low-quality |
| S1_extracted_k141_12515 | Low-quality |
| S1_extracted_k141_99093 | Not-determined |
| S1_extracted_k141_132371 | Not-determined |
| S1_extracted_k141_99095 | Not-determined |
| S1_extracted_k141_99096 | Not-determined |
| S1_extracted_k141_132375 | Not-determined |
| S1_extracted_k141_64357 | Not-determined |
| S1_extracted_k141_92308 | Low-quality |
| S1_extracted_k141_112259 | Not-determined |
| S1_extracted_k141_99100 | Not-determined |
| S1_extracted_k141_156546 | Not-determined |
| S1_extracted_k141_112260 | Not-determined |
| S1_extracted_k141_156547 | Not-determined |
| S1_extracted_k141_138779 | Not-determined |
| S1_extracted_k141_92311 | Not-determined |
| S1_extracted_k141_99104 | Not-determined |
| S1_extracted_k141_64361 | Not-determined |
| S1_extracted_k141_25452 | Not-determined |
| S1_extracted_k141_4401 | Low-quality |
| S1_extracted_k141_58935 | Not-determined |
| S1_extracted_k141_112269 | Not-determined |
| S1_extracted_k141_119008 | Not-determined |
| S1_extracted_k141_58937 | Not-determined |
| S1_extracted_k141_58938 | Not-determined |
| S1_extracted_k141_119010 | Not-determined |
| S1_extracted_k141_156550 | Not-determined |
| S1_extracted_k141_12536 | Not-determined |
| S1_extracted_k141_119011 | Not-determined |
| S1_extracted_k141_125383 | Low-quality |
| S1_extracted_k141_132388 | Not-determined |
| S1_extracted_k141_125384 | Not-determined |
| S1_extracted_k141_112276 | Not-determined |
| S1_extracted_k141_64371 | Not-determined |
| S1_extracted_k141_25460 | Not-determined |
| S1_extracted_k141_19489 | Not-determined |
| S1_extracted_k141_132391 | Not-determined |
| S1_extracted_k141_92325 | Not-determined |
| S1_extracted_k141_99107 | Not-determined |
| S1_extracted_k141_4409 | Not-determined |
| S1_extracted_k141_132392 | Not-determined |
| S1_extracted_k141_85699 | Not-determined |
| S1_extracted_k141_112280 | Not-determined |
| S1_extracted_k141_58946 | Not-determined |
| S1_extracted_k141_79075 | Not-determined |
| S1_extracted_k141_25466 | Not-determined |
| S1_extracted_k141_4415 | Not-determined |
| S1_extracted_k141_25467 | Not-determined |
| S1_extracted_k141_71184 | Low-quality |
| S1_extracted_k141_99111 | Low-quality |
| S1_extracted_k141_85706 | Not-determined |
| S1_extracted_k141_32343 | Not-determined |
| S1_extracted_k141_12550 | Low-quality |
| S1_extracted_k141_92335 | Not-determined |
| S1_extracted_k141_145312 | Not-determined |
| S1_extracted_k141_125396 | Not-determined |
| S1_extracted_k141_45745 | Not-determined |
| S1_extracted_k141_58951 | Low-quality |
| S1_extracted_k141_25475 | Not-determined |
| S1_extracted_k141_125398 | Not-determined |
| S1_extracted_k141_32351 | Low-quality |
| S1_extracted_k141_25477 | Not-determined |
| S1_extracted_k141_85714 | Not-determined |
| S1_extracted_k141_92338 | Not-determined |
| S1_extracted_k141_92339 | Not-determined |
| S1_extracted_k141_45748 | Not-determined |
| S1_extracted_k141_52570 | Not-determined |
| S1_extracted_k141_58957 | Not-determined |
| S1_extracted_k141_92341 | Not-determined |
| S1_extracted_k141_132400 | Not-determined |
| S1_extracted_k141_112288 | Not-determined |
| S1_extracted_k141_132401 | Not-determined |
| S1_extracted_k141_112291 | Low-quality |
| S1_extracted_k141_85716 | Not-determined |
| S1_extracted_k141_105710 | Not-determined |
| S1_extracted_k141_12563 | Not-determined |
| S1_extracted_k141_112294 | Not-determined |
| S1_extracted_k141_132404 | Not-determined |
| S1_extracted_k141_99124 | Not-determined |
| S1_extracted_k141_58958 | Not-determined |
| S1_extracted_k141_138807 | Not-determined |
| S1_extracted_k141_25488 | Not-determined |
| S1_extracted_k141_37741 | Low-quality |
| S1_extracted_k141_52583 | Low-quality |
| S1_extracted_k141_119034 | Not-determined |
| S1_extracted_k141_145326 | Not-determined |
| S1_extracted_k141_19508 | Not-determined |
| S1_extracted_k141_112296 | Not-determined |
| S1_extracted_k141_25490 | Not-determined |
| S1_extracted_k141_32359 | Not-determined |
| S1_extracted_k141_132416 | Not-determined |
| S1_extracted_k141_25492 | Not-determined |
| S1_extracted_k141_119037 | Not-determined |
| S1_extracted_k141_119039 | Not-determined |
| S1_extracted_k141_99135 | Not-determined |
| S1_extracted_k141_112301 | Not-determined |
| S1_extracted_k141_32361 | Not-determined |
| S1_extracted_k141_25495 | Not-determined |
| S1_extracted_k141_105718 | Low-quality |
| S1_extracted_k141_45762 | Low-quality |
| S1_extracted_k141_12579 | Not-determined |
| S1_extracted_k141_138815 | Low-quality |
| S1_extracted_k141_145333 | Not-determined |
| S1_extracted_k141_99139 | Not-determined |
| S1_extracted_k141_99140 | Not-determined |
| S1_extracted_k141_145334 | Not-determined |
| S1_extracted_k141_25501 | Not-determined |
| S1_extracted_k141_45764 | Not-determined |
| S1_extracted_k141_92344 | Not-determined |
| S1_extracted_k141_19519 | Not-determined |
| S1_extracted_k141_112313 | Not-determined |
| S1_extracted_k141_156569 | Not-determined |
| S1_extracted_k141_92347 | Not-determined |
| S1_extracted_k141_112315 | Not-determined |
| S1_extracted_k141_25505 | Not-determined |
| S1_extracted_k141_45766 | Not-determined |
| S1_extracted_k141_25506 | Not-determined |
| S1_extracted_k141_19520 | Not-determined |
| S1_extracted_k141_125421 | Not-determined |
| S1_extracted_k141_105721 | Not-determined |
| S1_extracted_k141_79112 | Not-determined |
| S1_extracted_k141_71201 | Not-determined |
| S1_extracted_k141_125423 | Not-determined |
| S1_extracted_k141_105723 | Not-determined |
| S1_extracted_k141_119050 | Not-determined |
| S1_extracted_k141_58975 | Low-quality |
| S1_extracted_k141_99149 | Not-determined |
| S1_extracted_k141_119051 | Not-determined |
| S1_extracted_k141_125425 | Not-determined |
| S1_extracted_k141_99150 | Not-determined |
| S1_extracted_k141_25510 | Not-determined |
| S1_extracted_k141_25512 | Not-determined |
| S1_extracted_k141_58977 | Not-determined |
| S1_extracted_k141_138823 | Not-determined |
| S1_extracted_k141_25513 | Not-determined |
| S1_extracted_k141_99152 | Not-determined |
| S1_extracted_k141_85744 | Not-determined |
| S1_extracted_k141_79116 | Not-determined |
| S1_extracted_k141_4443 | Low-quality |
| S1_extracted_k141_85747 | Low-quality |
| S1_extracted_k141_64393 | Not-determined |
| S1_extracted_k141_71210 | Not-determined |
| S1_extracted_k141_45777 | Not-determined |
| S1_extracted_k141_25517 | Low-quality |
| S1_extracted_k141_58983 | Not-determined |
| S1_extracted_k141_71213 | Not-determined |
| S1_extracted_k141_71214 | Not-determined |
| S1_extracted_k141_19528 | Not-determined |
| S1_extracted_k141_92360 | Not-determined |
| S1_extracted_k141_4451 | Not-determined |
| S1_extracted_k141_132448 | Not-determined |
| S1_extracted_k141_52602 | Low-quality |
| S1_extracted_k141_105737 | Not-determined |
| S1_extracted_k141_99161 | Low-quality |
| S1_extracted_k141_105739 | Not-determined |
| S1_extracted_k141_119061 | Low-quality |
| S1_extracted_k141_64402 | Low-quality |
| S1_extracted_k141_156576 | Not-determined |
| S1_extracted_k141_32393 | Not-determined |
| S1_extracted_k141_92361 | Low-quality |
| S1_extracted_k141_85752 | Not-determined |
| S1_extracted_k141_138836 | Not-determined |
| S1_extracted_k141_79130 | Not-determined |
| S1_extracted_k141_92364 | Not-determined |
| S1_extracted_k141_85754 | Not-determined |
| S1_extracted_k141_32401 | Not-determined |
| S1_extracted_k141_149555 | Not-determined |
| S1_extracted_k141_85757 | Not-determined |
| S1_extracted_k141_19545 | Not-determined |
| S1_extracted_k141_71221 | Not-determined |
| S1_extracted_k141_85759 | Not-determined |
| S1_extracted_k141_58992 | Not-determined |
| S1_extracted_k141_12595 | Not-determined |
| S1_extracted_k141_19552 | Not-determined |
| S1_extracted_k141_71224 | Low-quality |
| S1_extracted_k141_138849 | Not-determined |
| S1_extracted_k141_52624 | Low-quality |
| S1_extracted_k141_132463 | Not-determined |
| S1_extracted_k141_71226 | Not-determined |
| S1_extracted_k141_52625 | Not-determined |
| S1_extracted_k141_12599 | Not-determined |
| S1_extracted_k141_79145 | Low-quality |
| S1_extracted_k141_99176 | Low-quality |
| S1_extracted_k141_85776 | Not-determined |
| S1_extracted_k141_112351 | Not-determined |
| S1_extracted_k141_156581 | Not-determined |
| S1_extracted_k141_52629 | Not-determined |
| S1_extracted_k141_149570 | Not-determined |
| S1_extracted_k141_112357 | Not-determined |
| S1_extracted_k141_119090 | Not-determined |
| S1_extracted_k141_19561 | Not-determined |
| S1_extracted_k141_79157 | Not-determined |
| S1_extracted_k141_105759 | Low-quality |
| S1_extracted_k141_138851 | Low-quality |
| S1_extracted_k141_125462 | Low-quality |
| S1_extracted_k141_32414 | Not-determined |
| S1_extracted_k141_25552 | Not-determined |
| S1_extracted_k141_149578 | Low-quality |
| S1_extracted_k141_79166 | Not-determined |
| S1_extracted_k141_25554 | Low-quality |
| S1_extracted_k141_125466 | Not-determined |
| S1_extracted_k141_138855 | Low-quality |
| S1_extracted_k141_79168 | Not-determined |
| S1_extracted_k141_125469 | Not-determined |
| S1_extracted_k141_99185 | Low-quality |
| S1_extracted_k141_52646 | Not-determined |
| S1_extracted_k141_125472 | Not-determined |
| S1_extracted_k141_37783 | Not-determined |
| S1_extracted_k141_138858 | Not-determined |
| S1_extracted_k141_45802 | Not-determined |
| S1_extracted_k141_79179 | Not-determined |
| S1_extracted_k141_92378 | Not-determined |
| S1_extracted_k141_64428 | Not-determined |
| S1_extracted_k141_79180 | Low-quality |
| S1_extracted_k141_79183 | Not-determined |
| S1_extracted_k141_112374 | Not-determined |
| S1_extracted_k141_99188 | Low-quality |
| S1_extracted_k141_119097 | Not-determined |
| S1_extracted_k141_119098 | Not-determined |
| S1_extracted_k141_99190 | Not-determined |
| S1_extracted_k141_125480 | Not-determined |
| S1_extracted_k141_52651 | Not-determined |
| S1_extracted_k141_45810 | Not-determined |
| S1_extracted_k141_99192 | Not-determined |
| S1_extracted_k141_132495 | Not-determined |
| S1_extracted_k141_19591 | Not-determined |
| S1_extracted_k141_112381 | Not-determined |
| S1_extracted_k141_59029 | Not-determined |
| S1_extracted_k141_149581 | Not-determined |
| S1_extracted_k141_138870 | Low-quality |
| S1_extracted_k141_45816 | Not-determined |
| S1_extracted_k141_19595 | Not-determined |
| S1_extracted_k141_45817 | Not-determined |
| S1_extracted_k141_145386 | Low-quality |
| S1_extracted_k141_138873 | Not-determined |
| S1_extracted_k141_119108 | Not-determined |
| S1_extracted_k141_145387 | Not-determined |
| S1_extracted_k141_59034 | Not-determined |
| S1_extracted_k141_32442 | Not-determined |
| S1_extracted_k141_105779 | Not-determined |
| S1_extracted_k141_132506 | Not-determined |
| S1_extracted_k141_52661 | Not-determined |
| S1_extracted_k141_112383 | Not-determined |
| S1_extracted_k141_52662 | Not-determined |
| S1_extracted_k141_105780 | Not-determined |
| S1_extracted_k141_32446 | Not-determined |
| S1_extracted_k141_99198 | Not-determined |
| S1_extracted_k141_12606 | Low-quality |
| S1_extracted_k141_52666 | Not-determined |
| S1_extracted_k141_19604 | Not-determined |
| S1_extracted_k141_92401 | Not-determined |
| S1_extracted_k141_105785 | Not-determined |
| S1_extracted_k141_32448 | Not-determined |
| S1_extracted_k141_52670 | Not-determined |
| S1_extracted_k141_119111 | Not-determined |
| S1_extracted_k141_112384 | Not-determined |
| S1_extracted_k141_125490 | Low-quality |
| S1_extracted_k141_37805 | Not-determined |
| S1_extracted_k141_71242 | Not-determined |
| S1_extracted_k141_105788 | Not-determined |
| S1_extracted_k141_92405 | Not-determined |
| S1_extracted_k141_85833 | Not-determined |
| S1_extracted_k141_105790 | Not-determined |
| S1_extracted_k141_59040 | Low-quality |
| S1_extracted_k141_25573 | Low-quality |
| S1_extracted_k141_125497 | Not-determined |
| S1_extracted_k141_12614 | Not-determined |
| S1_extracted_k141_125498 | Not-determined |
| S1_extracted_k141_85836 | Not-determined |
| S1_extracted_k141_145395 | Low-quality |
| S1_extracted_k141_12620 | Not-determined |
| S1_extracted_k141_25581 | Not-determined |
| S1_extracted_k141_59051 | Not-determined |
| S1_extracted_k141_132526 | Not-determined |
| S1_extracted_k141_132527 | Not-determined |
| S1_extracted_k141_45824 | Not-determined |
| S1_extracted_k141_64462 | Low-quality |
| S1_extracted_k141_105804 | Not-determined |
| S1_extracted_k141_32458 | Low-quality |
| S1_extracted_k141_25583 | Not-determined |
| S1_extracted_k141_45827 | Not-determined |
| S1_extracted_k141_92407 | Low-quality |
| S1_extracted_k141_32463 | Not-determined |
| S1_extracted_k141_37817 | Not-determined |
| S1_extracted_k141_105807 | Low-quality |
| S1_extracted_k141_85846 | Not-determined |
| S1_extracted_k141_112397 | Not-determined |
| S1_extracted_k141_37818 | Not-determined |
| S1_extracted_k141_105809 | Not-determined |
| S1_extracted_k141_45832 | Not-determined |
| S1_extracted_k141_37819 | Not-determined |
| S1_extracted_k141_19627 | Not-determined |
| S1_extracted_k141_105811 | Not-determined |
| S1_extracted_k141_45835 | Not-determined |
| S1_extracted_k141_85852 | Not-determined |
| S1_extracted_k141_45836 | Low-quality |
| S1_extracted_k141_132544 | Not-determined |
| S1_extracted_k141_132545 | Not-determined |
| S1_extracted_k141_149587 | Not-determined |
| S1_extracted_k141_119127 | Not-determined |
| S1_extracted_k141_92414 | Not-determined |
| S1_extracted_k141_85859 | Not-determined |
| S1_extracted_k141_138907 | Not-determined |
| S1_extracted_k141_37822 | Low-quality |
| S1_extracted_k141_45845 | Not-determined |
| S1_extracted_k141_119131 | Not-determined |
| S1_extracted_k141_145408 | Not-determined |
| S1_extracted_k141_4489 | Not-determined |
| S1_extracted_k141_25593 | Not-determined |
| S1_extracted_k141_19636 | Not-determined |
| S1_extracted_k141_85862 | Low-quality |
| S1_extracted_k141_64474 | Not-determined |
| S1_extracted_k141_25594 | Not-determined |
| S1_extracted_k141_79231 | Low-quality |
| S1_extracted_k141_156605 | Not-determined |
| S1_extracted_k141_79232 | Not-determined |
| S1_extracted_k141_99235 | Low-quality |
| S1_extracted_k141_119133 | Low-quality |
| S1_extracted_k141_156606 | Low-quality |
| S1_extracted_k141_119134 | Low-quality |
| S1_extracted_k141_59073 | Low-quality |
| S1_extracted_k141_12642 | Not-determined |
| S1_extracted_k141_12643 | Not-determined |
| S1_extracted_k141_112408 | Low-quality |
| S1_extracted_k141_132548 | Low-quality |
| S1_extracted_k141_32485 | Not-determined |
| S1_extracted_k141_112411 | Low-quality |
| S1_extracted_k141_12648 | Low-quality |
| S1_extracted_k141_4499 | Not-determined |
| S1_extracted_k141_92431 | Not-determined |
| S1_extracted_k141_132553 | Not-determined |
| S1_extracted_k141_79240 | Not-determined |
| S1_extracted_k141_52701 | Not-determined |
| S1_extracted_k141_112416 | Not-determined |
| S1_extracted_k141_85871 | Low-quality |
| S1_extracted_k141_99246 | Not-determined |
| S1_extracted_k141_25605 | Not-determined |
| S1_extracted_k141_52707 | Not-determined |
| S1_extracted_k141_25606 | Not-determined |
| S1_extracted_k141_52710 | Not-determined |
| S1_extracted_k141_32498 | Not-determined |
| S1_extracted_k141_25608 | Low-quality |
| S1_extracted_k141_132557 | Not-determined |
| S1_extracted_k141_25609 | Not-determined |
| S1_extracted_k141_79250 | Not-determined |
| S1_extracted_k141_85880 | Not-determined |
| S1_extracted_k141_92437 | Not-determined |
| S1_extracted_k141_25610 | Not-determined |
| S1_extracted_k141_79253 | Not-determined |
| S1_extracted_k141_119153 | Not-determined |
| S1_extracted_k141_132559 | Not-determined |
| S1_extracted_k141_92439 | Low-quality |
| S1_extracted_k141_19655 | Not-determined |
| S1_extracted_k141_25611 | Not-determined |
| S1_extracted_k141_156614 | Not-determined |
| S1_extracted_k141_112422 | Not-determined |
| S1_extracted_k141_156615 | Low-quality |
| S1_extracted_k141_19658 | Not-determined |
| S1_extracted_k141_132565 | Not-determined |
| S1_extracted_k141_19663 | Not-determined |
| S1_extracted_k141_12672 | Low-quality |
| S1_extracted_k141_125541 | Not-determined |
| S1_extracted_k141_64499 | Not-determined |
| S1_extracted_k141_92447 | Not-determined |
| S1_extracted_k141_19666 | Not-determined |
| S1_extracted_k141_85887 | Not-determined |
| S1_extracted_k141_112429 | Not-determined |
| S1_extracted_k141_25615 | Not-determined |
| S1_extracted_k141_79258 | Not-determined |
| S1_extracted_k141_25616 | Not-determined |
| S1_extracted_k141_19669 | Not-determined |
| S1_extracted_k141_59092 | Not-determined |
| S1_extracted_k141_4512 | Not-determined |
| S1_extracted_k141_59094 | Low-quality |
| S1_extracted_k141_59095 | Not-determined |
| S1_extracted_k141_149611 | Not-determined |
| S1_extracted_k141_85890 | Low-quality |
| S1_extracted_k141_59100 | Not-determined |
| S1_extracted_k141_79269 | Not-determined |
| S1_extracted_k141_52726 | Not-determined |
| S1_extracted_k141_119171 | Not-determined |
| S1_extracted_k141_25626 | Low-quality |
| S1_extracted_k141_105852 | Low-quality |
| S1_extracted_k141_12690 | Not-determined |
| S1_extracted_k141_105853 | Not-determined |
| S1_extracted_k141_59103 | Not-determined |
| S1_extracted_k141_79274 | Not-determined |
| S1_extracted_k141_125556 | Not-determined |
| S1_extracted_k141_132575 | Not-determined |
| S1_extracted_k141_25630 | Not-determined |
| S1_extracted_k141_138953 | Not-determined |
| S1_extracted_k141_71277 | Not-determined |
| S1_extracted_k141_59107 | Low-quality |
| S1_extracted_k141_156623 | Not-determined |
| S1_extracted_k141_59108 | Not-determined |
| S1_extracted_k141_105860 | Not-determined |
| S1_extracted_k141_19676 | Not-determined |
| S1_extracted_k141_132582 | Not-determined |
| S1_extracted_k141_125558 | Low-quality |
| S1_extracted_k141_4530 | Not-determined |
| S1_extracted_k141_45875 | Not-determined |
| S1_extracted_k141_19679 | Not-determined |

| | |
|---|---|
| S1_extracted_k141_125559 | Not-determined |
| S1_extracted_k141_25637 | Not-determined |
| S1_extracted_k141_92469 | Not-determined |
| S1_extracted_k141_85903 | Low-quality |
| S1_extracted_k141_125560 | Not-determined |
| S1_extracted_k141_138955 | Not-determined |
| S1_extracted_k141_132587 | Not-determined |
| S1_extracted_k141_52735 | Not-determined |
| S1_extracted_k141_52737 | Not-determined |
| S1_extracted_k141_119189 | Not-determined |
| S1_extracted_k141_45878 | Not-determined |
| S1_extracted_k141_92477 | Not-determined |
| S1_extracted_k141_105870 | Not-determined |
| S1_extracted_k141_25642 | Not-determined |
| S1_extracted_k141_125565 | Not-determined |
| S1_extracted_k141_71288 | Not-determined |
| S1_extracted_k141_19689 | Not-determined |
| S1_extracted_k141_132596 | Not-determined |
| S1_extracted_k141_64525 | Not-determined |
| S1_extracted_k141_12705 | Not-determined |
| S1_extracted_k141_52748 | Not-determined |
| S1_extracted_k141_59120 | Not-determined |
| S1_extracted_k141_59121 | Not-determined |
| S1_extracted_k141_19693 | Not-determined |
| S1_extracted_k141_37866 | Not-determined |
| S1_extracted_k141_19694 | Not-determined |
| S1_extracted_k141_19696 | Not-determined |
| S1_extracted_k141_99264 | Not-determined |
| S1_extracted_k141_85917 | Not-determined |
| S1_extracted_k141_45885 | Not-determined |
| S1_extracted_k141_12713 | Not-determined |
| S1_extracted_k141_45887 | Low-quality |
| S1_extracted_k141_92482 | Not-determined |
| S1_extracted_k141_138965 | Not-determined |
| S1_extracted_k141_25650 | Not-determined |
| S1_extracted_k141_119205 | Not-determined |
| S1_extracted_k141_37874 | Not-determined |
| S1_extracted_k141_119206 | Not-determined |
| S1_extracted_k141_71296 | Not-determined |
| S1_extracted_k141_12722 | Not-determined |
| S1_extracted_k141_85919 | Not-determined |
| S1_extracted_k141_125585 | Not-determined |
| S1_extracted_k141_71301 | Not-determined |
| S1_extracted_k141_32536 | Not-determined |
| S1_extracted_k141_4558 | Not-determined |
| S1_extracted_k141_119212 | Low-quality |
| S1_extracted_k141_59130 | Not-determined |
| S1_extracted_k141_79298 | Low-quality |
| S1_extracted_k141_12725 | Not-determined |
| S1_extracted_k141_59131 | Not-determined |
| S1_extracted_k141_25655 | Not-determined |
| S1_extracted_k141_112457 | Not-determined |
| S1_extracted_k141_156649 | Not-determined |
| S1_extracted_k141_37877 | Not-determined |
| S1_extracted_k141_132615 | Not-determined |
| S1_extracted_k141_4564 | Not-determined |
| S1_extracted_k141_12728 | Low-quality |
| S1_extracted_k141_45898 | Low-quality |
| S1_extracted_k141_132618 | Not-determined |
| S1_extracted_k141_119216 | Not-determined |
| S1_extracted_k141_37880 | Not-determined |
| S1_extracted_k141_132623 | Not-determined |
| S1_extracted_k141_99270 | Low-quality |
| S1_extracted_k141_149619 | Not-determined |
| S1_extracted_k141_79314 | Not-determined |
| S1_extracted_k141_119219 | Not-determined |
| S1_extracted_k141_156658 | Not-determined |
| S1_extracted_k141_25665 | Not-determined |
| S1_extracted_k141_85927 | Not-determined |
| S1_extracted_k141_132629 | Not-determined |
| S1_extracted_k141_37888 | Not-determined |
| S1_extracted_k141_92501 | Not-determined |
| S1_extracted_k141_99272 | Not-determined |
| S1_extracted_k141_79320 | Low-quality |
| S1_extracted_k141_59141 | Not-determined |
| S1_extracted_k141_19735 | Not-determined |
| S1_extracted_k141_25667 | Low-quality |
| S1_extracted_k141_45903 | Not-determined |
| S1_extracted_k141_99275 | Not-determined |
| S1_extracted_k141_19736 | Not-determined |
| S1_extracted_k141_12730 | Low-quality |
| S1_extracted_k141_105905 | Not-determined |
| S1_extracted_k141_19739 | Not-determined |
| S1_extracted_k141_79325 | Not-determined |
| S1_extracted_k141_32565 | Not-determined |
| S1_extracted_k141_37892 | Not-determined |
| S1_extracted_k141_25672 | Not-determined |
| S1_extracted_k141_25673 | Low-quality |
| S1_extracted_k141_32567 | Not-determined |
| S1_extracted_k141_37895 | Not-determined |
| S1_extracted_k141_105912 | Not-determined |
| S1_extracted_k141_85939 | Not-determined |
| S1_extracted_k141_85940 | Low-quality |
| S1_extracted_k141_85941 | Not-determined |
| S1_extracted_k141_112469 | Not-determined |
| S1_extracted_k141_145474 | Not-determined |
| S1_extracted_k141_85943 | Not-determined |
| S1_extracted_k141_4586 | Not-determined |
| S1_extracted_k141_105917 | Not-determined |
| S1_extracted_k141_138975 | Not-determined |
| S1_extracted_k141_85947 | Low-quality |
| S1_extracted_k141_99281 | Not-determined |
| S1_extracted_k141_145476 | Not-determined |
| S1_extracted_k141_25678 | Not-determined |
| S1_extracted_k141_59159 | Not-determined |
| S1_extracted_k141_37905 | Not-determined |
| S1_extracted_k141_32580 | Not-determined |
| S1_extracted_k141_105923 | Not-determined |
| S1_extracted_k141_59161 | Not-determined |
| S1_extracted_k141_125601 | Low-quality |
| S1_extracted_k141_64557 | Not-determined |
| S1_extracted_k141_12748 | Not-determined |
| S1_extracted_k141_138982 | Not-determined |
| S1_extracted_k141_19751 | Not-determined |
| S1_extracted_k141_12749 | Not-determined |
| S1_extracted_k141_119251 | Low-quality |
| S1_extracted_k141_79345 | Not-determined |
| S1_extracted_k141_32585 | Not-determined |
| S1_extracted_k141_52802 | Not-determined |
| S1_extracted_k141_59168 | Not-determined |
| S1_extracted_k141_19756 | Not-determined |
| S1_extracted_k141_92528 | Not-determined |
| S1_extracted_k141_52804 | Not-determined |
| S1_extracted_k141_85957 | Not-determined |
| S1_extracted_k141_125607 | Low-quality |
| S1_extracted_k141_79349 | Low-quality |
| S1_extracted_k141_37910 | Not-determined |
| S1_extracted_k141_25686 | Not-determined |
| S1_extracted_k141_145485 | Not-determined |
| S1_extracted_k141_156684 | Not-determined |
| S1_extracted_k141_37912 | Low-quality |
| S1_extracted_k141_125612 | Not-determined |
| S1_extracted_k141_32592 | Low-quality |
| S1_extracted_k141_25694 | Low-quality |
| S1_extracted_k141_125617 | Not-determined |
| S1_extracted_k141_119261 | Not-determined |
| S1_extracted_k141_45926 | Not-determined |
| S1_extracted_k141_125619 | Not-determined |
| S1_extracted_k141_99305 | Not-determined |
| S1_extracted_k141_99306 | Not-determined |
| S1_extracted_k141_32596 | Not-determined |
| S1_extracted_k141_156686 | Not-determined |
| S1_extracted_k141_45929 | Not-determined |
| S1_extracted_k141_64565 | Low-quality |
| S1_extracted_k141_105934 | Not-determined |
| S1_extracted_k141_25698 | Not-determined |
| S1_extracted_k141_156687 | Low-quality |
| S1_extracted_k141_4601 | Not-determined |
| S1_extracted_k141_64569 | Not-determined |
| S1_extracted_k141_85965 | Low-quality |
| S1_extracted_k141_37921 | Low-quality |
| S1_extracted_k141_85967 | Not-determined |
| S1_extracted_k141_32601 | Not-determined |
| S1_extracted_k141_92551 | Not-determined |
| S1_extracted_k141_32604 | Not-determined |
| S1_extracted_k141_156694 | Not-determined |
| S1_extracted_k141_85971 | Not-determined |
| S1_extracted_k141_59182 | Not-determined |
| S1_extracted_k141_105939 | Low-quality |
| S1_extracted_k141_105942 | Not-determined |
| S1_extracted_k141_64582 | Not-determined |
| S1_extracted_k141_105943 | Not-determined |
| S1_extracted_k141_92560 | Not-determined |
| S1_extracted_k141_37927 | Not-determined |
| S1_extracted_k141_4614 | Not-determined |
| S1_extracted_k141_132673 | Not-determined |
| S1_extracted_k141_37929 | Not-determined |
| S1_extracted_k141_139006 | Not-determined |
| S1_extracted_k141_32615 | Not-determined |
| S1_extracted_k141_79371 | Low-quality |
| S1_extracted_k141_19782 | Not-determined |
| S1_extracted_k141_112486 | Not-determined |
| S1_extracted_k141_52822 | Low-quality |
| S1_extracted_k141_112487 | Not-determined |
| S1_extracted_k141_105945 | Not-determined |
| S1_extracted_k141_19785 | Not-determined |
| S1_extracted_k141_45939 | Not-determined |
| S1_extracted_k141_85977 | Low-quality |
| S1_extracted_k141_52824 | Not-determined |
| S1_extracted_k141_156707 | Not-determined |
| S1_extracted_k141_52826 | Not-determined |
| S1_extracted_k141_145514 | Not-determined |
| S1_extracted_k141_85980 | Not-determined |
| S1_extracted_k141_85981 | Not-determined |
| S1_extracted_k141_52827 | Low-quality |
| S1_extracted_k141_132683 | Not-determined |
| S1_extracted_k141_119286 | Not-determined |
| S1_extracted_k141_59201 | Not-determined |
| S1_extracted_k141_45946 | Not-determined |
| S1_extracted_k141_59202 | Not-determined |
| S1_extracted_k141_64599 | Not-determined |
| S1_extracted_k141_145520 | Not-determined |
| S1_extracted_k141_37944 | Not-determined |
| S1_extracted_k141_59204 | Not-determined |
| S1_extracted_k141_112496 | Not-determined |
| S1_extracted_k141_145521 | Low-quality |
| S1_extracted_k141_79381 | Low-quality |
| S1_extracted_k141_25717 | Low-quality |
| S1_extracted_k141_105952 | Low-quality |
| S1_extracted_k141_156714 | Not-determined |
| S1_extracted_k141_52834 | Not-determined |
| S1_extracted_k141_64604 | Not-determined |
| S1_extracted_k141_59207 | Not-determined |
| S1_extracted_k141_105955 | Not-determined |
| S1_extracted_k141_119293 | Low-quality |
| S1_extracted_k141_19797 | Not-determined |
| S1_extracted_k141_119295 | Not-determined |
| S1_extracted_k141_112509 | Not-determined |
| S1_extracted_k141_99338 | Low-quality |
| S1_extracted_k141_79391 | Not-determined |
| S1_extracted_k141_4634 | Not-determined |
| S1_extracted_k141_156715 | Low-quality |
| S1_extracted_k141_37951 | Not-determined |
| S1_extracted_k141_112512 | Not-determined |
| S1_extracted_k141_99344 | Not-determined |
| S1_extracted_k141_145531 | Not-determined |
| S1_extracted_k141_99348 | Not-determined |
| S1_extracted_k141_105960 | Not-determined |
| S1_extracted_k141_25735 | Not-determined |
| S1_extracted_k141_52848 | Low-quality |
| S1_extracted_k141_112517 | Not-determined |
| S1_extracted_k141_19808 | Not-determined |
| S1_extracted_k141_52850 | Low-quality |
| S1_extracted_k141_32630 | Low-quality |
| S1_extracted_k141_52851 | Not-determined |
| S1_extracted_k141_59218 | Low-quality |
| S1_extracted_k141_4641 | Not-determined |
| S1_extracted_k141_25738 | Not-determined |
| S1_extracted_k141_25739 | Not-determined |
| S1_extracted_k141_139044 | Not-determined |
| S1_extracted_k141_79398 | Not-determined |
| S1_extracted_k141_32634 | Not-determined |
| S1_extracted_k141_139046 | Low-quality |
| S1_extracted_k141_52854 | Not-determined |
| S1_extracted_k141_25743 | Not-determined |
| S1_extracted_k141_19817 | Not-determined |
| S1_extracted_k141_52856 | Low-quality |
| S1_extracted_k141_64620 | Not-determined |
| S1_extracted_k141_52857 | Not-determined |
| S1_extracted_k141_12788 | Not-determined |
| S1_extracted_k141_99355 | Not-determined |
| S1_extracted_k141_32637 | Not-determined |
| S1_extracted_k141_12790 | Low-quality |
| S1_extracted_k141_125643 | Not-determined |
| S1_extracted_k141_64624 | Low-quality |
| S1_extracted_k141_86006 | Low-quality |
| S1_extracted_k141_139054 | Not-determined |
| S1_extracted_k141_59225 | Low-quality |
| S1_extracted_k141_45978 | Not-determined |
| S1_extracted_k141_37963 | Low-quality |
| S1_extracted_k141_86009 | Not-determined |
| S1_extracted_k141_105971 | Not-determined |
| S1_extracted_k141_12798 | Not-determined |
| S1_extracted_k141_92581 | Not-determined |
| S1_extracted_k141_132700 | Low-quality |
| S1_extracted_k141_32646 | Low-quality |
| S1_extracted_k141_149651 | Not-determined |
| S1_extracted_k141_132702 | Not-determined |
| S1_extracted_k141_4653 | Not-determined |
| S1_extracted_k141_45980 | Low-quality |
| S1_extracted_k141_79415 | Low-quality |
| S1_extracted_k141_45981 | Low-quality |
| S1_extracted_k141_105975 | Not-determined |
| S1_extracted_k141_132706 | Not-determined |
| S1_extracted_k141_4655 | Not-determined |
| S1_extracted_k141_132708 | Not-determined |
| S1_extracted_k141_105978 | Not-determined |
| S1_extracted_k141_119319 | Not-determined |
| S1_extracted_k141_4658 | Not-determined |
| S1_extracted_k141_52873 | Not-determined |
| S1_extracted_k141_112538 | Low-quality |
| S1_extracted_k141_149661 | Not-determined |
| S12_vs_k141_111815\|\|full | High-quality |
| S1_extracted_k141_112539 | Not-determined |
| S1_extracted_k141_125651 | Low-quality |
| S1_extracted_k141_112541 | Not-determined |
| S1_extracted_k141_25764 | Low-quality |
| S1_extracted_k141_45982 | Not-determined |
| S1_extracted_k141_12810 | Low-quality |
| S1_extracted_k141_4664 | Not-determined |
| S1_extracted_k141_32663 | Not-determined |
| S1_extracted_k141_59236 | Low-quality |
| S1_extracted_k141_86020 | Not-determined |
| S1_extracted_k141_12812 | Low-quality |
| S1_extracted_k141_86022 | Not-determined |
| S1_extracted_k141_45987 | Not-determined |
| S1_extracted_k141_105985 | Not-determined |
| S1_extracted_k141_59239 | Not-determined |
| S1_extracted_k141_132722 | Not-determined |
| S1_extracted_k141_99374 | Not-determined |
| S1_extracted_k141_125657 | Not-determined |
| S1_extracted_k141_145540 | Not-determined |
| S1_extracted_k141_156726 | Not-determined |
| S1_extracted_k141_52888 | Not-determined |
| S1_extracted_k141_132726 | Not-determined |
| S1_extracted_k141_52891 | Not-determined |
| S1_extracted_k141_119320 | Low-quality |
| S1_extracted_k141_45989 | Low-quality |
| S1_extracted_k141_132732 | Not-determined |
| S1_extracted_k141_59247 | Not-determined |
| S1_extracted_k141_25782 | Not-determined |
| S1_extracted_k141_79441 | Not-determined |
| S1_extracted_k141_79442 | Low-quality |
| S1_extracted_k141_112554 | Not-determined |
| S1_extracted_k141_99390 | Not-determined |
| S1_extracted_k141_86036 | Not-determined |
| S1_extracted_k141_145556 | Not-determined |
| S1_extracted_k141_125670 | Low-quality |
| S1_extracted_k141_12830 | Low-quality |
| S1_extracted_k141_37990 | Not-determined |
| S1_extracted_k141_37991 | Not-determined |
| S1_extracted_k141_45998 | Low-quality |
| S1_extracted_k141_132743 | Not-determined |
| S1_extracted_k141_125673 | Not-determined |
| S1_extracted_k141_132746 | Not-determined |
| S1_extracted_k141_19846 | Not-determined |
| S1_extracted_k141_106007 | Not-determined |
| S1_extracted_k141_92606 | Not-determined |
| S1_extracted_k141_25789 | Not-determined |
| S1_extracted_k141_32687 | Not-determined |
| S1_extracted_k141_25790 | Not-determined |
| S1_extracted_k141_145574 | Not-determined |
| S1_extracted_k141_79459 | Not-determined |
| S1_extracted_k141_25791 | Not-determined |
| S1_extracted_k141_145576 | Not-determined |
| S1_extracted_k141_4686 | Low-quality |
| S1_extracted_k141_79462 | Not-determined |
| S1_extracted_k141_86050 | Low-quality |
| S1_extracted_k141_19851 | Low-quality |
| S1_extracted_k141_12845 | Low-quality |
| S1_extracted_k141_52914 | Low-quality |
| S1_extracted_k141_132756 | Low-quality |
| S1_extracted_k141_59268 | Not-determined |
| S1_extracted_k141_12850 | Not-determined |
| S1_extracted_k141_149679 | Not-determined |
| S1_extracted_k141_37998 | Low-quality |
| S1_extracted_k141_59271 | Not-determined |
| S1_extracted_k141_37999 | Not-determined |
| S1_extracted_k141_64676 | Low-quality |
| S1_extracted_k141_32696 | Low-quality |
| S1_extracted_k141_145589 | Not-determined |
| S1_extracted_k141_12854 | Not-determined |
| S1_extracted_k141_132762 | Not-determined |
| S1_extracted_k141_145590 | Not-determined |
| S1_extracted_k141_99410 | Not-determined |
| S1_extracted_k141_125689 | Not-determined |
| S1_extracted_k141_132763 | Low-quality |
| S1_extracted_k141_25800 | Low-quality |
| S1_extracted_k141_38003 | Not-determined |
| S1_extracted_k141_125691 | Not-determined |
| S1_extracted_k141_106021 | Not-determined |
| S1_extracted_k141_92622 | Not-determined |
| S1_extracted_k141_139095 | Not-determined |
| S1_extracted_k141_145595 | Not-determined |
| S1_extracted_k141_52926 | Not-determined |
| S1_extracted_k141_79482 | Not-determined |
| S1_extracted_k141_79483 | Not-determined |
| S1_extracted_k141_12860 | Not-determined |
| S1_extracted_k141_12862 | Not-determined |
| S1_extracted_k141_19872 | Not-determined |
| S1_extracted_k141_149689 | Not-determined |
| S1_extracted_k141_156747 | Not-determined |
| S1_extracted_k141_59285 | Not-determined |
| S1_extracted_k141_125704 | Not-determined |
| S1_extracted_k141_12869 | Not-determined |
| S1_extracted_k141_32709 | Not-determined |
| S1_extracted_k141_112568 | Low-quality |
| S1_extracted_k141_19877 | Not-determined |
| S1_extracted_k141_112569 | Not-determined |
| S1_extracted_k141_46023 | Not-determined |
| S1_extracted_k141_32711 | Not-determined |
| S1_extracted_k141_125707 | Low-quality |
| S1_extracted_k141_52939 | Not-determined |
| S1_extracted_k141_32714 | Not-determined |
| S1_extracted_k141_125709 | Not-determined |
| S1_extracted_k141_132786 | Not-determined |
| S1_extracted_k141_71372 | Not-determined |
| S1_extracted_k141_99419 | Not-determined |
| S1_extracted_k141_71373 | Not-determined |
| S1_extracted_k141_64696 | Not-determined |
| S1_extracted_k141_79490 | Not-determined |
| S1_extracted_k141_12878 | Not-determined |
| S1_extracted_k141_99422 | Not-determined |
| S1_extracted_k141_139112 | Not-determined |
| S1_extracted_k141_79492 | Not-determined |
| S1_extracted_k141_79494 | Not-determined |
| S1_extracted_k141_106038 | Not-determined |
| S1_extracted_k141_106039 | Not-determined |
| S1_extracted_k141_139115 | Not-determined |
| S1_extracted_k141_59293 | Not-determined |
| S1_extracted_k141_139116 | Not-determined |
| S1_extracted_k141_79497 | Not-determined |
| S1_extracted_k141_71375 | Not-determined |
| S1_extracted_k141_139118 | Not-determined |
| S1_extracted_k141_12885 | Not-determined |
| S1_extracted_k141_132794 | Not-determined |
| S1_extracted_k141_119375 | Not-determined |
| S1_extracted_k141_25824 | Not-determined |
| S1_extracted_k141_25827 | Not-determined |
| S1_extracted_k141_25828 | Not-determined |
| S1_extracted_k141_99426 | Low-quality |
| S1_extracted_k141_52948 | Not-determined |
| S1_extracted_k141_52949 | Not-determined |
| S1_extracted_k141_149704 | Not-determined |
| S1_extracted_k141_112583 | Not-determined |
| S1_extracted_k141_79502 | Low-quality |
| S1_extracted_k141_71379 | Not-determined |
| S1_extracted_k141_46044 | Low-quality |
| S1_extracted_k141_106045 | Not-determined |
| S1_extracted_k141_99430 | Not-determined |
| S1_extracted_k141_92636 | Low-quality |
| S1_extracted_k141_92637 | Not-determined |
| S1_extracted_k141_12895 | Not-determined |
| S1_extracted_k141_86071 | Not-determined |
| S1_extracted_k141_106048 | Not-determined |
| S1_extracted_k141_139127 | Not-determined |
| S1_extracted_k141_19900 | Not-determined |
| S1_extracted_k141_125725 | Not-determined |
| S1_extracted_k141_25834 | Not-determined |
| S1_extracted_k141_52957 | Not-determined |
| S1_extracted_k141_32733 | Not-determined |
| S1_extracted_k141_4698 | Low-quality |
| S1_extracted_k141_149707 | Not-determined |
| S1_extracted_k141_139133 | Not-determined |
| S1_extracted_k141_86073 | Not-determined |
| S1_extracted_k141_149708 | Not-determined |
| S1_extracted_k141_99439 | Not-determined |
| S1_extracted_k141_145629 | Not-determined |
| S1_extracted_k141_139136 | Not-determined |
| S1_extracted_k141_139138 | Not-determined |
| S1_extracted_k141_52962 | Not-determined |
| S1_extracted_k141_132812 | Not-determined |
| S1_extracted_k141_19904 | Not-determined |
| S1_extracted_k141_139143 | Not-determined |
| S1_extracted_k141_99447 | Not-determined |
| S1_extracted_k141_12899 | Not-determined |
| S1_extracted_k141_132820 | Not-determined |
| S1_extracted_k141_99450 | Not-determined |
| S1_extracted_k141_52966 | Not-determined |
| S1_extracted_k141_4707 | Not-determined |
| S1_extracted_k141_119397 | Not-determined |
| S1_extracted_k141_38041 | Not-determined |
| S1_extracted_k141_86080 | Not-determined |
| S1_extracted_k141_132823 | Not-determined |
| S1_extracted_k141_125738 | Not-determined |
| S1_extracted_k141_38044 | Low-quality |
| S1_extracted_k141_19910 | Low-quality |
| S1_extracted_k141_125739 | Low-quality |
| S1_extracted_k141_119401 | Not-determined |
| S1_extracted_k141_46057 | Not-determined |
| S1_extracted_k141_25839 | Not-determined |
| S1_extracted_k141_145639 | Not-determined |
| S1_extracted_k141_59316 | Not-determined |
| S1_extracted_k141_86085 | Low-quality |
| S1_extracted_k141_25841 | Not-determined |
| S1_extracted_k141_112604 | Low-quality |
| S1_extracted_k141_46062 | Not-determined |
| S1_extracted_k141_149722 | Not-determined |
| S1_extracted_k141_19918 | Low-quality |
| S1_extracted_k141_86088 | Not-determined |
| S1_extracted_k141_38053 | Not-determined |
| S1_extracted_k141_38054 | Not-determined |
| S1_extracted_k141_52981 | Not-determined |
| S1_extracted_k141_132837 | Not-determined |
| S1_extracted_k141_149726 | Not-determined |
| S1_extracted_k141_25844 | Not-determined |
| S1_extracted_k141_59323 | Low-quality |
| S1_extracted_k141_79527 | Not-determined |
| S1_extracted_k141_99466 | Not-determined |
| S1_extracted_k141_106078 | Not-determined |
| S1_extracted_k141_112612 | Not-determined |
| S1_extracted_k141_92667 | Not-determined |
| S1_extracted_k141_12914 | Not-determined |
| S1_extracted_k141_125754 | Low-quality |
| S1_extracted_k141_4718 | Not-determined |
| S1_extracted_k141_149734 | Not-determined |
| S1_extracted_k141_12919 | Not-determined |
| S1_extracted_k141_25852 | Not-determined |
| S1_extracted_k141_132849 | Not-determined |
| S1_extracted_k141_59334 | Not-determined |
| S1_extracted_k141_145652 | Low-quality |
| S1_extracted_k141_12924 | Not-determined |
| S1_extracted_k141_52992 | Not-determined |
| S1_extracted_k141_12925 | Not-determined |
| S1_extracted_k141_64712 | Low-quality |
| S1_extracted_k141_125762 | Low-quality |
| S1_extracted_k141_12926 | Not-determined |
| S1_extracted_k141_86102 | Not-determined |
| S1_extracted_k141_32758 | Not-determined |
| S1_extracted_k141_46088 | Not-determined |
| S1_extracted_k141_132853 | Not-determined |
| S1_extracted_k141_4730 | Not-determined |
| S1_extracted_k141_139157 | Not-determined |
| S1_extracted_k141_125766 | Not-determined |
| S1_extracted_k141_86106 | Not-determined |
| S1_extracted_k141_25861 | Not-determined |
| S1_extracted_k141_4732 | Not-determined |
| S1_extracted_k141_38067 | Not-determined |
| S1_extracted_k141_125769 | Not-determined |
| S1_extracted_k141_156769 | Not-determined |
| S1_extracted_k141_106095 | Not-determined |
| S1_extracted_k141_106097 | Not-determined |
| S1_extracted_k141_64724 | Not-determined |
| S1_extracted_k141_79544 | Not-determined |
| S1_extracted_k141_64725 | Not-determined |
| S1_extracted_k141_64727 | Not-determined |
| S1_extracted_k141_4739 | Not-determined |
| S1_extracted_k141_79547 | Not-determined |
| S1_extracted_k141_79551 | Not-determined |
| S1_extracted_k141_99485 | Low-quality |
| S1_extracted_k141_25866 | Not-determined |
| S1_extracted_k141_99486 | Not-determined |
| S1_extracted_k141_64732 | Not-determined |
| S1_extracted_k141_99488 | Not-determined |
| S1_extracted_k141_79555 | Not-determined |
| S1_extracted_k141_132858 | Not-determined |
| S1_extracted_k141_53015 | Not-determined |
| S1_extracted_k141_25868 | Not-determined |
| S1_extracted_k141_4746 | Not-determined |
| S1_extracted_k141_139167 | Not-determined |
| S1_extracted_k141_92696 | Not-determined |
| S1_extracted_k141_32766 | Low-quality |
| S1_extracted_k141_156773 | Not-determined |
| S1_extracted_k141_19950 | Not-determined |
| S1_extracted_k141_53018 | Not-determined |
| S1_extracted_k141_112623 | Not-determined |
| S1_extracted_k141_132859 | Low-quality |
| S1_extracted_k141_38081 | Not-determined |
| S1_extracted_k141_19952 | Not-determined |
| S1_extracted_k141_156775 | Not-determined |
| S1_extracted_k141_99498 | Not-determined |
| S1_extracted_k141_139173 | Not-determined |
| S1_extracted_k141_25879 | Not-determined |
| S1_extracted_k141_99500 | Low-quality |
| S1_extracted_k141_32770 | Not-determined |
| S1_extracted_k141_38086 | Not-determined |
| S1_extracted_k141_4747 | Not-determined |
| S1_extracted_k141_92705 | Not-determined |
| S1_extracted_k141_12956 | Not-determined |
| S1_extracted_k141_149751 | Not-determined |
| S1_extracted_k141_139180 | Not-determined |
| S1_extracted_k141_145677 | Not-determined |
| S1_extracted_k141_46098 | Low-quality |
| S1_extracted_k141_139184 | Not-determined |
| S1_extracted_k141_86129 | Low-quality |
| S1_extracted_k141_4753 | Low-quality |
| S1_extracted_k141_125780 | Not-determined |
| S1_extracted_k141_53032 | Not-determined |
| S1_extracted_k141_12961 | Not-determined |
| S1_extracted_k141_32779 | Not-determined |
| S1_extracted_k141_99511 | Not-determined |
| S1_extracted_k141_125782 | Not-determined |
| S1_extracted_k141_99512 | Not-determined |
| S1_extracted_k141_59346 | Not-determined |
| S1_extracted_k141_46103 | Low-quality |
| S1_extracted_k141_139189 | Not-determined |
| S1_extracted_k141_12963 | Not-determined |
| S1_extracted_k141_92716 | Not-determined |
| S1_extracted_k141_53037 | Not-determined |
| S1_extracted_k141_19962 | Not-determined |
| S1_extracted_k141_12965 | Not-determined |
| S1_extracted_k141_99517 | Not-determined |
| S1_extracted_k141_12966 | Not-determined |
| S1_extracted_k141_125792 | Not-determined |
| S1_extracted_k141_64747 | Not-determined |
| S1_extracted_k141_32784 | Low-quality |
| S1_extracted_k141_59355 | Not-determined |
| S1_extracted_k141_86143 | Not-determined |
| S1_extracted_k141_112638 | Not-determined |
| S1_extracted_k141_92725 | Not-determined |
| S1_extracted_k141_53044 | Not-determined |
| S1_extracted_k141_86149 | Not-determined |
| S1_extracted_k141_38097 | Low-quality |
| S1_extracted_k141_92728 | Not-determined |
| S1_extracted_k141_19980 | Not-determined |
| S1_extracted_k141_145696 | Not-determined |
| S1_extracted_k141_86150 | Low-quality |
| S1_extracted_k141_149767 | Not-determined |
| S1_extracted_k141_38099 | Not-determined |
| S1_extracted_k141_59361 | Not-determined |
| S1_extracted_k141_125806 | Not-determined |
| S1_extracted_k141_125807 | Not-determined |
| S1_extracted_k141_149769 | Not-determined |
| S1_extracted_k141_125808 | Not-determined |
| S1_extracted_k141_32787 | Not-determined |
| S1_extracted_k141_86153 | Low-quality |
| S1_extracted_k141_53048 | Low-quality |
| S1_extracted_k141_99531 | Not-determined |
| S1_extracted_k141_92733 | Low-quality |

| | |
|---|---|
| S1_extracted_k141_53050 | Low-quality |
| S1_extracted_k141_149773 | Not-determined |
| S1_extracted_k141_53053 | Not-determined |
| S1_extracted_k141_119446 | Not-determined |
| S1_extracted_k141_149776 | Low-quality |
| S1_extracted_k141_149777 | Not-determined |
| S1_extracted_k141_86165 | Not-determined |
| S1_extracted_k141_106142 | Low-quality |
| S1_extracted_k141_106143 | Not-determined |
| S1_extracted_k141_53057 | Not-determined |
| S1_extracted_k141_19992 | Not-determined |
| S1_extracted_k141_92742 | Low-quality |
| S1_extracted_k141_19993 | Not-determined |
| S1_extracted_k141_4778 | Low-quality |
| S1_extracted_k141_64763 | Not-determined |
| S1_extracted_k141_86170 | Not-determined |
| S1_extracted_k141_119451 | Not-determined |
| S1_extracted_k141_125817 | Not-determined |
| S1_extracted_k141_64765 | Not-determined |
| S1_extracted_k141_92746 | Not-determined |
| S1_extracted_k141_125820 | Not-determined |
| S1_extracted_k141_112668 | Not-determined |
| S1_extracted_k141_149783 | Not-determined |
| S1_extracted_k141_92749 | Low-quality |
| S1_extracted_k141_132905 | Not-determined |
| S1_extracted_k141_149785 | Not-determined |
| S1_extracted_k141_79564 | Low-quality |
| S1_extracted_k141_32802 | Low-quality |
| S1_extracted_k141_79569 | Not-determined |
| S1_extracted_k141_59380 | Not-determined |
| S1_extracted_k141_79573 | Low-quality |
| S1_extracted_k141_32806 | Not-determined |
| S1_extracted_k141_46137 | Not-determined |
| S1_extracted_k141_59382 | Not-determined |
| S1_extracted_k141_38111 | Not-determined |
| S1_extracted_k141_13000 | Not-determined |
| S1_extracted_k141_46138 | Low-quality |
| S1_extracted_k141_92760 | Not-determined |
| S1_extracted_k141_32810 | Not-determined |
| S1_extracted_k141_132919 | Low-quality |
| S1_extracted_k141_119459 | Not-determined |
| S1_extracted_k141_139226 | Not-determined |
| S1_extracted_k141_86184 | Low-quality |
| S1_extracted_k141_59385 | Not-determined |
| S1_extracted_k141_149797 | Not-determined |
| S1_extracted_k141_86186 | Low-quality |
| S1_extracted_k141_38114 | Not-determined |
| S1_extracted_k141_92769 | Not-determined |
| S1_extracted_k141_71417 | Not-determined |
| S1_extracted_k141_92770 | Not-determined |
| S1_extracted_k141_53078 | Not-determined |
| S1_extracted_k141_71419 | Not-determined |
| S1_extracted_k141_32816 | Not-determined |
| S1_extracted_k141_38120 | Not-determined |
| S1_extracted_k141_53082 | Not-determined |
| S1_extracted_k141_13017 | Not-determined |
| S1_extracted_k141_32820 | Not-determined |
| S1_extracted_k141_32823 | Not-determined |
| S1_extracted_k141_32824 | Not-determined |
| S1_extracted_k141_139236 | Not-determined |
| S1_extracted_k141_46156 | Not-determined |
| S1_extracted_k141_20022 | Not-determined |
| S1_extracted_k141_156811 | Low-quality |
| S1_extracted_k141_149809 | Not-determined |
| S1_extracted_k141_32828 | Low-quality |
| S1_extracted_k141_125844 | Low-quality |
| S1_extracted_k141_92783 | Low-quality |
| S1_extracted_k141_46158 | Not-determined |
| S1_extracted_k141_125847 | Low-quality |
| S1_extracted_k141_112692 | Not-determined |
| S1_extracted_k141_145723 | Not-determined |
| S1_extracted_k141_32831 | Not-determined |
| S1_extracted_k141_38135 | Not-determined |
| S1_extracted_k141_38137 | Not-determined |
| S1_extracted_k141_4811 | Not-determined |
| S1_extracted_k141_92790 | Not-determined |
| S1_extracted_k141_145727 | Not-determined |
| S1_extracted_k141_139242 | Not-determined |
| S1_extracted_k141_25945 | Not-determined |
| S1_extracted_k141_145728 | Not-determined |
| S1_extracted_k141_139243 | Not-determined |
| S1_extracted_k141_92796 | Not-determined |
| S1_extracted_k141_25948 | Not-determined |
| S1_extracted_k141_112698 | Not-determined |
| S1_extracted_k141_25949 | Not-determined |
| S1_extracted_k141_139249 | Not-determined |
| S1_extracted_k141_106184 | Not-determined |
| S1_extracted_k141_149815 | Not-determined |
| S1_extracted_k141_112699 | Not-determined |
| S1_extracted_k141_125860 | Not-determined |
| S1_extracted_k141_132943 | Low-quality |
| S1_extracted_k141_106187 | Not-determined |
| S1_extracted_k141_92801 | Not-determined |
| S1_extracted_k141_125864 | Low-quality |
| S1_extracted_k141_13029 | Low-quality |
| S1_extracted_k141_99559 | Not-determined |
| S1_extracted_k141_13030 | Not-determined |
| S1_extracted_k141_25953 | Not-determined |
| S1_extracted_k141_86214 | Not-determined |
| S1_extracted_k141_25954 | Not-determined |
| S1_extracted_k141_149820 | Not-determined |
| S1_extracted_k141_32849 | Not-determined |
| S1_extracted_k141_132952 | Not-determined |
| S1_extracted_k141_119486 | Not-determined |
| S1_extracted_k141_132953 | Low-quality |
| S1_extracted_k141_156825 | Not-determined |
| S1_extracted_k141_46180 | Not-determined |
| S1_extracted_k141_112709 | Low-quality |
| S1_extracted_k141_86223 | Not-determined |
| S1_extracted_k141_38150 | Not-determined |
| S1_extracted_k141_86224 | Not-determined |
| S1_extracted_k141_125882 | Not-determined |
| S1_extracted_k141_64810 | Not-determined |
| S1_extracted_k141_106199 | Not-determined |
| S1_extracted_k141_132957 | Low-quality |
| S1_extracted_k141_106200 | Not-determined |
| S1_extracted_k141_132958 | Not-determined |
| S1_extracted_k141_64813 | Not-determined |
| S1_extracted_k141_25963 | Not-determined |
| S1_extracted_k141_32859 | Not-determined |
| S1_extracted_k141_79580 | Not-determined |
| S1_extracted_k141_32863 | Not-determined |
| S1_extracted_k141_139269 | Low-quality |
| S1_extracted_k141_38157 | Not-determined |
| S1_extracted_k141_86229 | Not-determined |
| S1_extracted_k141_149832 | Low-quality |
| S1_extracted_k141_132967 | Not-determined |
| S1_extracted_k141_156831 | Not-determined |
| S1_extracted_k141_145746 | Not-determined |
| S1_extracted_k141_92816 | Not-determined |
| S1_extracted_k141_59422 | Low-quality |
| S1_extracted_k141_92817 | Not-determined |
| S1_extracted_k141_99577 | Not-determined |
| S1_extracted_k141_145748 | Low-quality |
| S1_extracted_k141_99580 | Not-determined |
| S1_extracted_k141_4814 | Low-quality |
| S1_extracted_k141_59427 | Not-determined |
| S1_extracted_k141_149840 | Not-determined |
| S1_extracted_k141_132980 | Not-determined |
| S1_extracted_k141_106220 | Not-determined |
| S1_extracted_k141_79590 | Not-determined |
| S1_extracted_k141_86240 | Not-determined |
| S1_extracted_k141_99584 | Not-determined |
| S1_extracted_k141_86242 | Not-determined |
| S1_extracted_k141_79592 | Not-determined |
| S1_extracted_k141_86243 | Not-determined |
| S1_extracted_k141_112724 | Not-determined |
| S1_extracted_k141_145753 | Low-quality |
| S1_extracted_k141_46184 | Low-quality |
| S1_extracted_k141_32876 | Not-determined |
| S1_extracted_k141_125901 | Not-determined |
| S1_extracted_k141_139278 | Not-determined |
| S1_extracted_k141_149846 | Low-quality |
| S1_extracted_k141_92827 | Not-determined |
| S1_extracted_k141_32879 | Not-determined |
| S1_extracted_k141_145758 | Not-determined |
| S1_extracted_k141_59430 | Low-quality |
| S1_extracted_k141_32881 | Not-determined |
| S1_extracted_k141_79599 | Low-quality |
| S1_extracted_k141_149849 | Low-quality |
| S1_extracted_k141_79600 | Not-determined |
| S1_extracted_k141_106232 | Not-determined |
| S1_extracted_k141_32884 | Low-quality |
| S1_extracted_k141_79603 | Not-determined |
| S1_extracted_k141_53114 | Not-determined |
| S1_extracted_k141_119524 | Low-quality |
| S1_extracted_k141_46194 | Not-determined |
| S1_extracted_k141_99593 | Low-quality |
| S1_extracted_k141_79606 | Low-quality |
| S1_extracted_k141_13063 | Not-determined |
| S1_extracted_k141_132993 | Not-determined |
| S1_extracted_k141_112735 | Low-quality |
| S1_extracted_k141_112737 | Not-determined |
| S1_extracted_k141_112739 | Not-determined |
| S1_extracted_k141_125912 | Not-determined |
| S1_extracted_k141_59434 | Not-determined |
| S1_extracted_k141_125913 | Low-quality |
| S1_extracted_k141_119533 | Not-determined |
| S1_extracted_k141_125915 | Not-determined |
| S1_extracted_k141_119534 | Not-determined |
| S1_extracted_k141_106234 | Low-quality |
| S1_extracted_k141_86258 | Not-determined |
| S1_extracted_k141_112741 | Not-determined |
| S1_extracted_k141_149859 | Not-determined |
| S1_extracted_k141_38170 | Low-quality |
| S1_extracted_k141_46209 | Not-determined |
| S1_extracted_k141_106238 | Not-determined |
| S1_extracted_k141_25988 | Not-determined |
| S1_extracted_k141_64844 | Not-determined |
| S1_extracted_k141_25989 | Not-determined |
| S1_extracted_k141_64846 | Not-determined |
| S1_extracted_k141_133000 | Not-determined |
| S1_extracted_k141_92850 | Not-determined |
| S1_extracted_k141_112747 | Not-determined |
| S1_extracted_k141_13079 | Not-determined |
| S1_extracted_k141_79620 | Low-quality |
| S1_extracted_k141_112748 | Not-determined |
| S1_extracted_k141_92852 | Low-quality |
| S1_extracted_k141_139301 | Not-determined |
| S1_extracted_k141_59437 | Not-determined |
| S1_extracted_k141_86266 | Not-determined |
| S1_extracted_k141_139303 | Not-determined |
| S1_extracted_k141_64848 | Not-determined |
| S1_extracted_k141_25998 | Not-determined |
| S1_extracted_k141_32909 | Not-determined |
| S1_extracted_k141_26000 | Not-determined |
| S1_extracted_k141_119547 | Not-determined |
| S1_extracted_k141_133006 | Low-quality |
| S1_extracted_k141_125931 | Not-determined |
| S1_extracted_k141_125932 | Not-determined |
| S1_extracted_k141_4826 | Not-determined |
| S1_extracted_k141_133009 | Not-determined |
| S1_extracted_k141_26002 | Low-quality |
| S1_extracted_k141_46223 | Not-determined |
| S1_extracted_k141_71450 | Low-quality |
| S1_extracted_k141_106251 | Not-determined |
| S1_extracted_k141_112757 | Not-determined |
| S1_extracted_k141_64855 | Not-determined |
| S1_extracted_k141_139315 | Not-determined |
| S1_extracted_k141_26006 | Not-determined |
| S1_extracted_k141_99614 | Not-determined |
| S1_extracted_k141_79630 | Not-determined |
| S1_extracted_k141_133015 | Not-determined |
| S1_extracted_k141_32918 | Not-determined |
| S1_extracted_k141_26010 | Not-determined |
| S1_extracted_k141_32920 | Not-determined |
| S1_extracted_k141_99619 | Not-determined |
| S1_extracted_k141_46228 | Not-determined |
| S1_extracted_k141_59445 | Not-determined |
| S1_extracted_k141_32923 | Not-determined |
| S1_extracted_k141_59447 | Not-determined |
| S1_extracted_k141_71455 | Not-determined |
| S1_extracted_k141_32925 | Not-determined |
| S1_extracted_k141_133019 | Not-determined |
| S1_extracted_k141_64865 | Not-determined |
| S1_extracted_k141_92868 | Not-determined |
| S1_extracted_k141_106261 | Not-determined |
| S1_extracted_k141_86285 | Not-determined |
| S1_extracted_k141_71460 | Low-quality |
| S1_extracted_k141_59457 | Low-quality |
| S1_extracted_k141_38186 | Not-determined |
| S1_extracted_k141_106268 | Low-quality |
| S1_extracted_k141_38188 | Not-determined |
| S1_extracted_k141_26026 | Not-determined |
| S1_extracted_k141_156844 | Not-determined |
| S1_extracted_k141_79643 | Not-determined |
| S1_extracted_k141_86289 | Not-determined |
| S1_extracted_k141_156847 | Not-determined |
| S1_extracted_k141_4839 | Not-determined |
| S1_extracted_k141_125950 | Low-quality |
| S1_extracted_k141_119562 | Not-determined |
| S1_extracted_k141_139339 | Not-determined |
| S1_extracted_k141_26035 | Not-determined |
| S1_extracted_k141_79650 | Not-determined |
| S1_extracted_k141_156851 | Not-determined |
| S1_extracted_k141_79653 | Not-determined |
| S1_extracted_k141_13123 | Not-determined |
| S1_extracted_k141_4844 | Not-determined |
| S1_extracted_k141_32937 | Low-quality |
| S1_extracted_k141_53158 | Not-determined |
| S1_extracted_k141_46235 | Low-quality |
| S1_extracted_k141_53160 | Not-determined |
| S1_extracted_k141_106276 | Not-determined |
| S1_extracted_k141_59475 | Low-quality |
| S1_extracted_k141_38198 | Low-quality |
| S1_extracted_k141_145768 | Low-quality |
| S1_extracted_k141_99655 | Not-determined |
| S1_extracted_k141_119571 | Not-determined |
| S1_extracted_k141_86304 | Not-determined |
| S1_extracted_k141_46239 | Not-determined |
| S1_extracted_k141_106281 | Not-determined |
| S1_extracted_k141_99657 | Not-determined |
| S1_extracted_k141_38202 | Not-determined |
| S1_extracted_k141_4857 | Not-determined |
| S1_extracted_k141_46242 | Low-quality |
| S1_extracted_k141_106282 | Not-determined |
| S1_extracted_k141_71478 | Not-determined |
| S1_extracted_k141_32943 | Not-determined |
| S1_extracted_k141_99661 | Not-determined |
| S1_extracted_k141_112790 | Not-determined |
| S1_extracted_k141_64891 | Not-determined |
| S1_extracted_k141_112792 | Not-determined |
| S1_extracted_k141_119578 | Not-determined |
| S1_extracted_k141_119579 | Not-determined |
| S1_extracted_k141_133045 | Not-determined |
| S1_extracted_k141_125970 | Not-determined |
| S1_extracted_k141_139357 | Not-determined |
| S1_extracted_k141_4862 | Not-determined |
| S1_extracted_k141_92880 | Not-determined |
| S1_extracted_k141_64894 | Low-quality |
| S1_extracted_k141_99662 | Low-quality |
| S1_extracted_k141_106290 | Not-determined |
| S1_extracted_k141_64900 | Not-determined |
| S1_extracted_k141_145782 | Not-determined |
| S1_extracted_k141_119589 | Not-determined |
| S1_extracted_k141_125978 | Not-determined |
| S1_extracted_k141_13149 | Not-determined |
| S1_extracted_k141_125980 | Not-determined |
| S1_extracted_k141_79667 | Low-quality |
| S1_extracted_k141_64903 | Not-determined |
| S1_extracted_k141_106293 | Not-determined |
| S1_extracted_k141_112800 | Not-determined |
| S1_extracted_k141_99665 | Not-determined |
| S1_extracted_k141_32956 | Not-determined |
| S1_extracted_k141_64907 | Not-determined |
| S1_extracted_k141_64909 | Not-determined |
| S1_extracted_k141_4863 | Low-quality |
| S1_extracted_k141_64910 | Not-determined |
| S1_extracted_k141_119594 | Not-determined |
| S1_extracted_k141_112808 | Not-determined |
| S1_extracted_k141_38206 | Not-determined |
| S1_extracted_k141_71488 | Not-determined |
| S1_extracted_k141_112810 | Not-determined |
| S1_extracted_k141_4867 | Not-determined |
| S1_extracted_k141_13162 | Not-determined |
| S1_extracted_k141_71490 | Low-quality |
| S1_extracted_k141_106304 | Not-determined |
| S1_extracted_k141_26056 | Low-quality |
| S1_extracted_k141_79678 | Not-determined |
| S1_extracted_k141_139380 | Not-determined |
| S1_extracted_k141_46267 | Low-quality |
| S1_extracted_k141_71491 | Low-quality |
| S1_extracted_k141_59500 | Not-determined |
| S1_extracted_k141_106307 | Not-determined |
| S1_extracted_k141_64920 | Low-quality |
| S1_extracted_k141_139383 | Low-quality |
| S1_extracted_k141_32963 | Not-determined |
| S1_extracted_k141_38216 | Low-quality |
| S1_extracted_k141_112818 | Not-determined |
| S1_extracted_k141_119607 | Low-quality |
| S1_extracted_k141_139390 | Low-quality |
| S1_extracted_k141_79684 | Not-determined |
| S1_extracted_k141_133082 | Not-determined |
| S1_extracted_k141_119608 | Low-quality |
| S1_extracted_k141_125988 | Not-determined |
| S1_extracted_k141_53184 | Not-determined |
| S1_extracted_k141_79686 | Not-determined |
| S1_extracted_k141_71500 | Not-determined |
| S1_extracted_k141_92898 | Not-determined |
| S1_extracted_k141_99693 | Low-quality |
| S1_extracted_k141_99694 | Not-determined |
| S1_extracted_k141_64935 | Not-determined |
| S1_extracted_k141_13179 | Not-determined |
| S1_extracted_k141_99697 | Not-determined |
| S1_extracted_k141_92906 | Not-determined |
| S1_extracted_k141_86342 | Low-quality |
| S1_extracted_k141_4889 | Not-determined |
| S1_extracted_k141_26069 | Not-determined |
| S1_extracted_k141_46288 | Not-determined |
| S1_extracted_k141_126001 | Not-determined |
| S1_extracted_k141_26070 | Not-determined |
| S1_extracted_k141_112824 | Low-quality |
| S1_extracted_k141_71510 | Not-determined |
| S1_extracted_k141_92910 | Not-determined |
| S1_extracted_k141_59523 | Not-determined |
| S1_extracted_k141_46295 | Not-determined |
| S1_extracted_k141_26073 | Not-determined |
| S1_extracted_k141_53198 | Not-determined |
| S1_extracted_k141_4897 | Not-determined |
| S1_extracted_k141_156868 | Not-determined |
| S1_extracted_k141_59527 | Not-determined |
| S1_extracted_k141_149894 | Not-determined |
| S1_extracted_k141_112834 | Not-determined |
| S1_extracted_k141_133094 | Not-determined |
| S1_extracted_k141_156870 | Not-determined |
| S1_extracted_k141_119619 | Low-quality |
| S1_extracted_k141_99703 | Not-determined |
| S1_extracted_k141_71520 | Not-determined |
| S1_extracted_k141_112842 | Not-determined |
| S1_extracted_k141_126022 | Not-determined |
| S1_extracted_k141_99705 | Not-determined |
| S1_extracted_k141_149896 | Not-determined |
| S1_extracted_k141_32996 | Not-determined |
| S1_extracted_k141_92925 | Low-quality |
| S1_extracted_k141_4907 | Not-determined |
| S1_extracted_k141_99710 | Not-determined |
| S1_extracted_k141_64952 | Not-determined |
| S1_extracted_k141_26089 | Not-determined |
| S1_extracted_k141_133104 | Not-determined |
| S1_extracted_k141_64953 | Not-determined |
| S1_extracted_k141_26091 | Not-determined |
| S1_extracted_k141_71527 | Not-determined |
| S1_extracted_k141_4912 | Not-determined |
| S1_extracted_k141_71528 | Not-determined |
| S1_extracted_k141_53214 | Not-determined |
| S1_extracted_k141_64955 | Low-quality |
| S1_extracted_k141_59539 | Low-quality |
| S1_extracted_k141_99716 | Not-determined |
| S1_extracted_k141_139421 | Not-determined |
| S1_extracted_k141_79725 | Not-determined |
| S1_extracted_k141_71532 | Not-determined |
| S1_extracted_k141_119634 | Not-determined |
| S1_extracted_k141_139424 | Not-determined |
| S1_extracted_k141_33007 | Not-determined |
| S1_extracted_k141_53217 | Not-determined |
| S1_extracted_k141_46307 | Not-determined |
| S1_extracted_k141_133107 | Low-quality |
| S1_extracted_k141_112858 | Low-quality |
| S1_extracted_k141_139426 | Low-quality |
| S1_extracted_k141_46310 | Not-determined |
| S1_extracted_k141_86365 | Not-determined |
| S1_extracted_k141_38244 | Not-determined |
| S1_extracted_k141_71539 | Not-determined |
| S1_extracted_k141_92936 | Not-determined |
| S1_extracted_k141_46313 | Not-determined |
| S1_extracted_k141_86367 | Low-quality |
| S1_extracted_k141_64965 | Not-determined |
| S1_extracted_k141_64967 | Low-quality |
| S1_extracted_k141_79735 | Not-determined |
| S1_extracted_k141_26096 | Low-quality |
| S1_extracted_k141_106340 | Not-determined |
| S1_extracted_k141_46316 | Not-determined |
| S1_extracted_k141_38247 | Not-determined |
| S1_extracted_k141_99728 | Low-quality |
| S1_extracted_k141_79737 | Low-quality |
| S1_extracted_k141_13214 | Not-determined |
| S1_extracted_k141_119646 | Low-quality |
| S1_extracted_k141_106342 | Not-determined |
| S1_extracted_k141_145795 | Low-quality |
| S1_extracted_k141_106343 | Not-determined |
| S1_extracted_k141_139436 | Not-determined |
| S1_extracted_k141_86371 | Low-quality |
| S1_extracted_k141_26098 | Low-quality |
| S1_extracted_k141_4932 | Not-determined |
| S1_extracted_k141_92946 | Not-determined |
| S1_extracted_k141_26099 | Low-quality |
| S1_extracted_k141_38254 | Not-determined |
| S1_extracted_k141_106346 | Not-determined |
| S1_extracted_k141_145801 | Not-determined |
| S1_extracted_k141_26105 | Not-determined |
| S1_extracted_k141_106349 | Not-determined |
| S1_extracted_k141_33027 | Not-determined |
| S1_extracted_k141_46325 | Not-determined |
| S1_extracted_k141_119652 | Not-determined |
| S1_extracted_k141_92951 | Not-determined |
| S1_extracted_k141_53240 | Not-determined |
| S1_extracted_k141_106355 | Not-determined |
| S1_extracted_k141_71556 | Not-determined |
| S1_extracted_k141_46334 | Low-quality |
| S1_extracted_k141_149899 | Not-determined |
| S1_extracted_k141_59564 | Low-quality |
| S1_extracted_k141_38260 | Low-quality |
| S1_extracted_k141_86381 | Low-quality |
| S1_extracted_k141_13229 | Not-determined |
| S1_extracted_k141_53249 | Not-determined |
| S1_extracted_k141_149902 | Not-determined |
| S1_extracted_k141_38264 | Not-determined |
| S1_extracted_k141_13233 | Low-quality |
| S1_extracted_k141_106362 | Not-determined |
| S1_extracted_k141_126053 | Not-determined |
| S1_extracted_k141_71565 | Low-quality |
| S1_extracted_k141_59567 | Not-determined |
| S1_extracted_k141_46340 | Not-determined |
| S1_extracted_k141_59568 | Not-determined |
| S1_extracted_k141_64992 | Not-determined |
| S1_extracted_k141_99755 | Not-determined |
| S1_extracted_k141_33041 | Not-determined |
| S1_extracted_k141_145818 | Not-determined |
| S1_extracted_k141_33044 | Not-determined |
| S1_extracted_k141_133121 | Not-determined |
| S1_extracted_k141_133122 | Low-quality |
| S1_extracted_k141_38274 | Low-quality |
| S1_extracted_k141_13244 | Not-determined |
| S1_extracted_k141_86388 | Not-determined |
| S1_extracted_k141_13247 | Not-determined |
| S1_extracted_k141_46353 | Low-quality |
| S1_extracted_k141_65004 | Not-determined |
| S1_extracted_k141_139470 | Not-determined |
| S1_extracted_k141_26130 | Not-determined |
| S1_extracted_k141_65006 | Not-determined |
| S1_extracted_k141_86391 | Not-determined |
| S1_extracted_k141_53271 | Not-determined |
| S1_extracted_k141_149910 | Not-determined |
| S1_extracted_k141_71572 | Not-determined |
| S1_extracted_k141_126059 | Not-determined |
| S1_extracted_k141_86393 | Low-quality |
| S1_extracted_k141_119672 | Not-determined |
| S1_extracted_k141_112905 | Not-determined |
| S1_extracted_k141_53277 | Not-determined |
| S1_extracted_k141_59586 | Not-determined |
| S1_extracted_k141_99762 | Not-determined |
| S1_extracted_k141_145827 | Not-determined |
| S1_extracted_k141_53280 | Not-determined |
| S1_extracted_k141_149914 | Low-quality |
| S1_extracted_k141_46366 | Not-determined |
| S1_extracted_k141_86402 | Low-quality |
| S1_extracted_k141_65013 | Not-determined |
| S1_extracted_k141_112913 | Not-determined |
| S1_extracted_k141_38278 | Not-determined |
| S1_extracted_k141_26139 | Not-determined |
| S1_extracted_k141_79750 | Low-quality |
| S1_extracted_k141_71585 | Not-determined |
| S1_extracted_k141_126077 | Not-determined |
| S1_extracted_k141_86407 | Not-determined |
| S1_extracted_k141_4941 | Not-determined |
| S1_extracted_k141_99770 | Not-determined |
| S1_extracted_k141_13260 | Low-quality |
| S1_extracted_k141_26146 | Not-determined |
| S1_extracted_k141_112926 | Not-determined |
| S1_extracted_k141_13263 | Not-determined |
| S1_extracted_k141_149918 | Not-determined |
| S1_extracted_k141_119685 | Not-determined |
| S1_extracted_k141_106397 | Not-determined |
| S1_extracted_k141_71589 | Low-quality |
| S1_extracted_k141_33059 | Not-determined |
| S1_extracted_k141_46384 | Not-determined |
| S1_extracted_k141_59601 | Not-determined |
| S1_extracted_k141_86416 | Not-determined |
| S1_extracted_k141_13268 | Not-determined |
| S1_extracted_k141_106399 | Not-determined |
| S1_extracted_k141_112931 | Not-determined |
| S1_extracted_k141_133153 | Low-quality |
| S1_extracted_k141_156878 | Not-determined |
| S1_extracted_k141_86418 | Low-quality |
| S1_extracted_k141_59604 | Low-quality |
| S1_extracted_k141_46389 | Low-quality |
| S1_extracted_k141_38292 | Not-determined |
| S1_extracted_k141_46390 | Not-determined |
| S1_extracted_k141_106403 | Not-determined |
| S1_extracted_k141_149924 | Not-determined |
| S1_extracted_k141_26153 | Not-determined |
| S1_extracted_k141_149925 | Low-quality |
| S1_extracted_k141_106404 | Not-determined |
| S1_extracted_k141_145845 | Not-determined |
| S1_extracted_k141_59608 | Low-quality |
| S1_extracted_k141_106405 | Low-quality |
| S1_extracted_k141_4948 | Not-determined |
| S1_extracted_k141_145848 | Not-determined |
| S1_extracted_k141_26157 | Low-quality |
| S1_extracted_k141_86426 | Not-determined |
| S1_extracted_k141_53299 | Not-determined |
| S1_extracted_k141_119689 | Not-determined |
| S1_extracted_k141_99785 | Not-determined |
| S1_extracted_k141_133161 | Low-quality |
| S1_extracted_k141_133162 | Not-determined |
| S1_extracted_k141_71599 | Not-determined |
| S1_extracted_k141_119691 | Not-determined |
| S1_extracted_k141_65025 | Low-quality |
| S1_extracted_k141_133164 | Not-determined |
| S1_extracted_k141_99788 | Not-determined |
| S1_extracted_k141_4953 | Not-determined |
| S1_extracted_k141_106409 | Not-determined |
| S1_extracted_k141_99790 | Not-determined |
| S1_extracted_k141_46397 | Not-determined |
| S1_extracted_k141_65027 | Not-determined |
| S1_extracted_k141_86434 | Not-determined |
| S1_extracted_k141_26165 | Not-determined |
| S1_extracted_k141_59618 | Not-determined |
| S1_extracted_k141_145854 | Low-quality |
| S1_extracted_k141_106412 | Not-determined |
| S1_extracted_k141_53307 | Not-determined |
| S1_extracted_k141_126098 | Not-determined |
| S1_extracted_k141_33076 | Not-determined |
| S1_extracted_k141_46401 | Not-determined |
| S1_extracted_k141_13283 | Not-determined |
| S1_extracted_k141_86436 | Not-determined |
| S1_extracted_k141_53312 | Not-determined |
| S1_extracted_k141_149926 | Not-determined |
| S1_extracted_k141_133175 | Not-determined |
| S1_extracted_k141_92991 | Low-quality |
| S1_extracted_k141_79777 | Not-determined |
| S1_extracted_k141_13294 | Not-determined |
| S1_extracted_k141_92992 | Not-determined |
| S1_extracted_k141_145860 | Not-determined |
| S1_extracted_k141_33086 | Low-quality |
| S1_extracted_k141_145861 | Not-determined |
| S1_extracted_k141_4964 | Not-determined |
| S1_extracted_k141_139506 | Low-quality |
| S1_extracted_k141_149930 | Not-determined |
| S1_extracted_k141_106424 | Not-determined |
| S1_extracted_k141_106425 | Not-determined |
| S1_extracted_k141_53318 | Not-determined |
| S1_extracted_k141_71613 | Not-determined |
| S1_extracted_k141_149932 | Low-quality |
| S1_extracted_k141_133182 | Not-determined |
| S1_extracted_k141_4966 | Not-determined |
| S1_extracted_k141_112954 | Not-determined |
| S1_extracted_k141_53320 | Not-determined |
| S1_extracted_k141_33091 | Not-determined |
| S1_extracted_k141_13297 | Not-determined |
| S1_extracted_k141_4967 | Not-determined |
| S1_extracted_k141_92997 | Low-quality |
| S1_extracted_k141_119698 | Low-quality |
| S1_extracted_k141_145868 | Not-determined |
| S1_extracted_k141_149935 | Not-determined |
| S1_extracted_k141_99809 | Not-determined |
| S1_extracted_k141_119702 | Not-determined |
| S1_extracted_k141_59624 | Low-quality |
| S1_extracted_k141_126110 | Not-determined |

| | |
|---|---|
| S1_extracted_k141_71619 | Not-determined |
| S1_extracted_k141_53324 | Low-quality |
| S1_extracted_k141_99815 | Low-quality |
| S1_extracted_k141_93004 | Not-determined |
| S1_extracted_k141_33098 | Not-determined |
| S1_extracted_k141_65045 | Not-determined |
| S1_extracted_k141_79788 | Low-quality |
| S1_extracted_k141_65046 | Not-determined |
| S1_extracted_k141_59629 | Not-determined |
| S1_extracted_k141_99819 | Not-determined |
| S1_extracted_k141_71624 | Not-determined |
| S1_extracted_k141_133197 | Not-determined |
| S1_extracted_k141_13313 | Low-quality |
| S1_extracted_k141_139521 | Not-determined |
| S1_extracted_k141_46420 | Not-determined |
| S1_extracted_k141_71626 | Not-determined |
| S1_extracted_k141_86456 | Not-determined |
| S1_extracted_k141_106439 | Low-quality |
| S1_extracted_k141_133201 | Not-determined |
| S1_extracted_k141_149939 | Not-determined |
| S1_extracted_k141_71627 | Not-determined |
| S1_extracted_k141_112972 | Not-determined |
| S1_extracted_k141_38301 | Low-quality |
| S1_extracted_k141_106442 | Not-determined |
| S1_extracted_k141_4973 | Low-quality |
| S1_extracted_k141_53331 | Low-quality |
| S1_extracted_k141_93012 | Not-determined |
| S1_extracted_k141_86459 | Not-determined |
| S1_extracted_k141_139527 | Not-determined |
| S1_extracted_k141_65053 | Low-quality |
| S1_extracted_k141_79794 | Not-determined |
| S1_extracted_k141_33106 | Not-determined |
| S1_extracted_k141_86463 | Not-determined |
| S1_extracted_k141_106446 | Not-determined |
| S1_extracted_k141_99834 | Not-determined |
| S1_extracted_k141_93018 | Not-determined |
| S1_extracted_k141_139530 | Not-determined |
| S1_extracted_k141_149945 | Low-quality |
| S1_extracted_k141_65057 | Not-determined |
| S1_extracted_k141_145886 | Not-determined |
| S1_extracted_k141_46426 | Not-determined |
| S1_extracted_k141_139536 | Not-determined |
| S1_extracted_k141_59642 | Not-determined |
| S1_extracted_k141_133214 | Not-determined |
| S1_extracted_k141_106454 | Not-determined |
| S1_extracted_k141_126125 | Not-determined |
| S1_extracted_k141_133216 | Not-determined |
| S1_extracted_k141_86471 | Not-determined |
| S1_extracted_k141_93026 | Not-determined |
| S1_extracted_k141_126127 | Not-determined |
| S1_extracted_k141_13329 | Not-determined |
| S1_extracted_k141_26209 | Not-determined |
| S1_extracted_k141_38306 | Low-quality |
| S1_extracted_k141_139543 | Low-quality |
| S1_extracted_k141_26211 | Low-quality |
| S1_extracted_k141_93031 | Not-determined |
| S1_extracted_k141_53345 | Not-determined |
| S1_extracted_k141_106464 | Low-quality |
| S1_extracted_k141_71642 | Not-determined |
| S1_extracted_k141_99847 | Not-determined |
| S1_extracted_k141_86477 | Not-determined |
| S1_extracted_k141_65069 | Not-determined |
| S1_extracted_k141_38313 | Low-quality |
| S1_extracted_k141_46436 | Low-quality |
| S1_extracted_k141_145896 | Low-quality |
| S1_extracted_k141_145897 | Not-determined |
| S1_extracted_k141_65072 | Not-determined |
| S1_extracted_k141_93039 | Not-determined |
| S1_extracted_k141_119727 | Not-determined |
| S1_extracted_k141_38317 | Not-determined |
| S1_extracted_k141_119730 | Low-quality |
| S1_extracted_k141_59656 | Not-determined |
| S1_extracted_k141_133228 | Not-determined |
| S1_extracted_k141_86484 | Not-determined |
| S1_extracted_k141_33130 | Not-determined |
| S1_extracted_k141_133233 | Not-determined |
| S1_extracted_k141_79817 | Not-determined |
| S1_extracted_k141_86487 | Not-determined |
| S1_extracted_k141_133234 | Not-determined |
| S1_extracted_k141_93043 | Low-quality |
| S1_extracted_k141_46440 | Not-determined |
| S1_extracted_k141_38323 | Not-determined |
| S1_extracted_k141_139557 | Not-determined |
| S1_extracted_k141_79821 | Low-quality |
| S1_extracted_k141_86492 | Low-quality |
| S1_extracted_k141_26217 | Not-determined |
| S1_extracted_k141_119741 | Not-determined |
| S1_extracted_k141_133235 | Low-quality |
| S1_extracted_k141_26221 | Not-determined |
| S1_extracted_k141_86498 | Not-determined |
| S1_extracted_k141_112997 | Not-determined |
| S1_extracted_k141_149956 | Not-determined |
| S1_extracted_k141_86499 | Low-quality |
| S1_extracted_k141_38329 | Not-determined |
| S1_extracted_k141_33137 | Not-determined |
| S1_extracted_k141_13349 | Not-determined |
| S1_extracted_k141_112999 | Not-determined |
| S1_extracted_k141_149958 | Low-quality |
| S1_extracted_k141_126150 | Not-determined |
| S1_extracted_k141_93050 | Not-determined |
| S1_extracted_k141_113002 | Not-determined |
| S1_extracted_k141_126154 | Not-determined |
| S1_extracted_k141_33141 | Not-determined |
| S1_extracted_k141_38332 | Not-determined |
| S1_extracted_k141_33144 | Not-determined |
| S1_extracted_k141_5007 | Not-determined |
| S1_extracted_k141_99860 | Not-determined |
| S1_extracted_k141_71661 | Not-determined |
| S1_extracted_k141_79831 | Low-quality |
| S1_extracted_k141_139561 | Low-quality |
| S1_extracted_k141_133248 | Not-determined |
| S1_extracted_k141_79834 | Not-determined |
| S1_extracted_k141_145907 | Not-determined |
| S1_extracted_k141_93058 | Not-determined |
| S1_extracted_k141_33147 | Low-quality |
| S1_extracted_k141_133250 | Low-quality |
| S1_extracted_k141_126162 | Low-quality |
| S1_extracted_k141_86513 | Not-determined |
| S1_extracted_k141_145910 | Low-quality |
| S1_extracted_k141_59667 | Not-determined |
| S1_extracted_k141_53380 | Low-quality |
| S1_extracted_k141_113013 | Low-quality |
| S1_extracted_k141_145912 | Not-determined |
| S1_extracted_k141_59669 | Low-quality |
| S1_extracted_k141_26240 | Not-determined |
| S1_extracted_k141_126171 | Not-determined |
| S1_extracted_k141_86524 | Not-determined |
| S1_extracted_k141_93072 | Low-quality |
| S1_extracted_k141_93073 | Not-determined |
| S1_extracted_k141_79845 | Not-determined |
| S1_extracted_k141_59671 | Not-determined |
| S1_extracted_k141_65087 | Not-determined |
| S1_extracted_k141_65088 | Not-determined |
| S1_extracted_k141_79848 | Not-determined |
| S1_extracted_k141_53392 | Not-determined |
| S1_extracted_k141_106496 | Not-determined |
| S1_extracted_k141_86532 | Not-determined |
| S1_extracted_k141_126179 | Not-determined |
| S1_extracted_k141_53394 | Low-quality |
| S1_extracted_k141_86534 | Not-determined |
| S1_extracted_k141_86535 | Not-determined |
| S1_extracted_k141_46455 | Low-quality |
| S1_extracted_k141_145925 | Not-determined |
| S1_extracted_k141_65098 | Not-determined |
| S1_extracted_k141_38354 | Not-determined |
| S1_extracted_k141_145926 | Low-quality |
| S1_extracted_k141_106500 | Not-determined |
| S1_extracted_k141_113035 | Not-determined |
| S1_extracted_k141_133271 | Not-determined |
| S1_extracted_k141_99884 | Not-determined |
| S1_extracted_k141_113037 | Not-determined |
| S1_extracted_k141_65103 | Not-determined |
| S1_extracted_k141_79859 | Not-determined |
| S1_extracted_k141_65107 | Not-determined |
| S1_extracted_k141_79861 | Not-determined |
| S1_extracted_k141_106507 | Not-determined |
| S1_extracted_k141_79864 | Not-determined |
| S1_extracted_k141_38362 | Low-quality |
| S1_extracted_k141_145934 | Not-determined |
| S1_extracted_k141_33161 | Not-determined |
| S1_extracted_k141_26258 | Not-determined |
| S1_extracted_k141_119773 | Low-quality |
| S1_extracted_k141_99889 | Not-determined |
| S1_extracted_k141_26259 | Not-determined |
| S1_extracted_k141_133279 | Not-determined |
| S1_extracted_k141_145937 | Not-determined |
| S1_extracted_k141_65114 | Low-quality |
| S1_extracted_k141_113044 | Low-quality |
| S1_extracted_k141_133281 | Not-determined |
| S1_extracted_k141_86549 | Not-determined |
| S1_extracted_k141_93096 | Not-determined |
| S1_extracted_k141_126200 | Not-determined |
| S1_extracted_k141_86551 | Low-quality |
| S1_extracted_k141_33164 | Low-quality |
| S1_extracted_k141_113055 | Low-quality |
| S1_extracted_k141_149968 | Not-determined |
| S1_extracted_k141_119778 | Not-determined |
| S1_extracted_k141_139597 | Not-determined |
| S1_extracted_k141_133295 | Not-determined |
| S1_extracted_k141_33173 | Not-determined |
| S1_extracted_k141_113056 | Low-quality |
| S1_extracted_k141_139600 | Not-determined |
| S1_extracted_k141_38376 | Low-quality |
| S1_extracted_k141_65126 | Not-determined |
| S1_extracted_k141_139606 | Low-quality |
| S1_extracted_k141_65128 | Not-determined |
| S1_extracted_k141_33174 | Low-quality |
| S1_extracted_k141_149978 | Not-determined |
| S1_extracted_k141_145959 | Not-determined |
| S1_extracted_k141_86568 | Low-quality |
| S1_extracted_k141_59699 | Low-quality |
| S1_extracted_k141_133305 | Not-determined |
| S1_extracted_k141_33176 | Not-determined |
| S1_extracted_k141_38380 | Not-determined |
| S1_extracted_k141_26282 | Not-determined |
| S1_extracted_k141_99921 | Low-quality |
| S1_extracted_k141_139613 | Not-determined |
| S1_extracted_k141_38382 | Not-determined |
| S1_extracted_k141_113070 | Not-determined |
| S1_extracted_k141_38384 | Low-quality |
| S1_extracted_k141_71701 | Not-determined |
| S1_extracted_k141_106529 | Not-determined |
| S1_extracted_k141_59709 | Not-determined |
| S1_extracted_k141_33184 | Low-quality |
| S1_extracted_k141_79899 | Not-determined |
| S1_extracted_k141_106533 | Not-determined |
| S1_extracted_k141_5065 | Not-determined |
| S1_extracted_k141_119796 | Low-quality |
| S1_extracted_k141_46477 | Low-quality |
| S1_extracted_k141_133311 | Low-quality |
| S1_extracted_k141_59714 | Not-determined |
| S1_extracted_k141_139619 | Not-determined |
| S1_extracted_k141_119800 | Not-determined |
| S1_extracted_k141_93118 | Not-determined |
| S1_extracted_k141_93119 | Not-determined |
| S1_extracted_k141_113073 | Not-determined |
| S1_extracted_k141_65146 | Low-quality |
| S1_extracted_k141_106538 | Low-quality |
| S1_extracted_k141_33197 | Not-determined |
| S1_extracted_k141_33199 | Not-determined |
| S1_extracted_k141_113075 | Not-determined |
| S1_extracted_k141_33201 | Low-quality |
| S1_extracted_k141_5073 | Not-determined |
| S1_extracted_k141_65152 | Low-quality |
| S1_extracted_k141_65154 | Not-determined |
| S1_extracted_k141_86583 | Not-determined |
| S1_extracted_k141_38397 | Not-determined |
| S1_extracted_k141_106542 | Not-determined |
| S1_extracted_k141_33209 | Not-determined |
| S1_extracted_k141_59731 | Low-quality |
| S1_extracted_k141_33212 | Not-determined |
| S1_extracted_k141_86585 | Not-determined |
| S1_extracted_k141_139634 | Not-determined |
| S1_extracted_k141_38402 | Not-determined |
| S1_extracted_k141_93127 | Low-quality |
| S1_extracted_k141_145973 | Not-determined |
| S1_extracted_k141_126210 | Not-determined |
| S1_extracted_k141_139639 | Not-determined |
| S1_extracted_k141_38406 | Not-determined |
| S1_extracted_k141_149991 | Low-quality |
| S1_extracted_k141_59737 | Not-determined |
| S1_extracted_k141_65165 | Not-determined |
| S1_extracted_k141_26305 | Not-determined |
| S1_extracted_k141_65172 | Low-quality |
| S1_extracted_k141_65173 | Not-determined |
| S1_extracted_k141_133332 | Not-determined |
| S1_extracted_k141_38411 | Not-determined |
| S1_extracted_k141_93135 | Not-determined |
| S1_extracted_k141_145991 | Low-quality |
| S1_extracted_k141_99947 | Not-determined |
| S1_extracted_k141_26312 | Not-determined |
| S1_extracted_k141_93138 | Low-quality |
| S1_extracted_k141_46497 | Not-determined |
| S1_extracted_k141_79924 | Not-determined |
| S1_extracted_k141_33230 | Not-determined |
| S1_extracted_k141_133339 | Not-determined |
| S1_extracted_k141_119823 | Not-determined |
| S1_extracted_k141_99954 | Low-quality |
| S1_extracted_k141_46502 | Not-determined |
| S1_extracted_k141_113105 | Not-determined |
| S1_extracted_k141_106563 | Not-determined |
| S1_extracted_k141_139654 | Not-determined |
| S1_extracted_k141_38425 | Not-determined |
| S1_extracted_k141_46506 | Not-determined |
| S1_extracted_k141_86600 | Not-determined |
| S1_extracted_k141_26318 | Not-determined |
| S1_extracted_k141_26319 | Not-determined |
| S1_extracted_k141_146001 | Not-determined |
| S1_extracted_k141_59747 | Not-determined |
| S1_extracted_k141_149997 | Not-determined |
| S1_extracted_k141_149998 | Not-determined |
| S1_extracted_k141_86607 | Not-determined |
| S1_extracted_k141_139663 | Not-determined |
| S1_extracted_k141_106571 | Not-determined |
| S1_extracted_k141_133352 | Not-determined |
| S1_extracted_k141_126233 | Not-determined |
| S1_extracted_k141_126234 | Not-determined |
| S1_extracted_k141_46516 | Not-determined |
| S1_extracted_k141_65191 | Low-quality |
| S1_extracted_k141_126235 | Not-determined |
| S1_extracted_k141_146010 | Not-determined |
| S1_extracted_k141_146011 | Not-determined |
| S1_extracted_k141_126238 | Not-determined |
| S1_extracted_k141_126239 | Not-determined |
| S1_extracted_k141_59760 | Not-determined |
| S1_extracted_k141_146015 | Not-determined |
| S1_extracted_k141_133361 | Not-determined |
| S1_extracted_k141_46522 | Not-determined |
| S1_extracted_k141_71724 | Not-determined |
| S1_extracted_k141_93164 | Low-quality |
| S1_extracted_k141_33262 | Not-determined |
| S1_extracted_k141_93165 | Low-quality |
| S1_extracted_k141_146017 | Not-determined |
| S1_extracted_k141_133362 | Not-determined |
| S1_extracted_k141_5110 | Not-determined |
| S1_extracted_k141_65201 | Not-determined |
| S1_extracted_k141_46524 | Not-determined |
| S1_extracted_k141_113130 | Not-determined |
| S1_extracted_k141_5111 | Not-determined |
| S1_extracted_k141_139675 | Not-determined |
| S1_extracted_k141_38437 | Not-determined |
| S1_extracted_k141_106579 | Not-determined |
| S1_extracted_k141_99971 | Low-quality |
| S1_extracted_k141_46532 | Not-determined |
| S1_extracted_k141_33266 | Low-quality |
| S1_extracted_k141_99972 | Not-determined |
| S1_extracted_k141_146023 | Not-determined |
| S1_extracted_k141_86622 | Low-quality |
| S1_extracted_k141_106581 | Not-determined |
| S1_extracted_k141_99973 | Low-quality |
| S1_extracted_k141_79958 | Not-determined |
| S1_extracted_k141_99975 | Not-determined |
| S1_extracted_k141_139681 | Not-determined |
| S1_extracted_k141_113141 | Not-determined |
| S1_extracted_k141_26340 | Not-determined |
| S1_extracted_k141_133375 | Not-determined |
| S1_extracted_k141_126249 | Not-determined |
| S1_extracted_k141_133376 | Not-determined |
| S1_extracted_k141_38448 | Not-determined |
| S1_extracted_k141_86625 | Not-determined |
| S1_extracted_k141_133378 | Not-determined |
| S1_extracted_k141_146030 | Not-determined |
| S1_extracted_k141_59767 | Low-quality |
| S1_extracted_k141_86628 | Not-determined |
| S1_extracted_k141_93182 | Not-determined |
| S1_extracted_k141_86629 | Not-determined |
| S1_extracted_k141_113148 | Low-quality |
| S1_extracted_k141_119863 | Not-determined |
| S1_extracted_k141_156938 | Not-determined |
| S1_extracted_k141_26349 | Not-determined |
| S1_extracted_k141_93186 | Low-quality |
| S1_extracted_k141_65211 | Low-quality |
| S1_extracted_k141_33284 | Not-determined |
| S1_extracted_k141_113152 | Not-determined |
| S1_extracted_k141_99982 | Not-determined |
| S1_extracted_k141_26358 | Not-determined |
| S1_extracted_k141_99984 | Not-determined |
| S1_extracted_k141_93193 | Not-determined |
| S1_extracted_k141_65219 | Not-determined |
| S1_extracted_k141_59778 | Low-quality |
| S1_extracted_k141_146044 | Low-quality |
| S1_extracted_k141_126255 | Not-determined |
| S1_extracted_k141_59779 | Not-determined |
| S1_extracted_k141_33287 | Not-determined |
| S1_extracted_k141_113157 | Not-determined |
| S1_extracted_k141_139701 | Not-determined |
| S1_extracted_k141_33289 | Not-determined |
| S1_extracted_k141_59786 | Not-determined |
| S1_extracted_k141_86650 | Low-quality |
| S1_extracted_k141_33293 | Not-determined |
| S1_extracted_k141_99997 | Not-determined |
| S1_extracted_k141_33294 | Not-determined |
| S1_extracted_k141_126262 | Not-determined |
| S1_extracted_k141_79974 | Low-quality |
| S1_extracted_k141_93201 | Not-determined |
| S1_extracted_k141_133393 | Not-determined |
| S1_extracted_k141_5132 | Not-determined |
| S1_extracted_k141_119878 | Not-determined |
| S1_extracted_k141_5134 | Not-determined |
| S1_extracted_k141_156950 | Not-determined |
| S1_extracted_k141_65236 | Low-quality |
| S1_extracted_k141_156952 | Not-determined |
| S1_extracted_k141_71734 | Low-quality |
| S1_extracted_k141_38457 | Not-determined |
| S1_extracted_k141_100010 | Not-determined |
| S1_extracted_k141_5141 | Not-determined |
| S1_extracted_k141_139714 | Not-determined |
| S1_extracted_k141_26370 | Low-quality |
| S1_extracted_k141_100013 | Not-determined |
| S1_extracted_k141_126272 | Not-determined |
| S1_extracted_k141_100014 | Not-determined |
| S1_extracted_k141_5143 | Not-determined |
| S1_extracted_k141_100015 | Not-determined |
| S1_extracted_k141_38461 | Low-quality |
| S1_extracted_k141_46566 | Low-quality |
| S1_extracted_k141_100016 | Not-determined |
| S1_extracted_k141_139716 | Not-determined |
| S1_extracted_k141_79989 | Not-determined |
| S1_extracted_k141_38463 | Not-determined |
| S1_extracted_k141_46569 | Low-quality |
| S1_extracted_k141_65246 | Not-determined |
| S1_extracted_k141_133400 | Not-determined |
| S1_extracted_k141_126277 | Not-determined |
| S1_extracted_k141_146067 | Not-determined |
| S1_extracted_k141_33314 | Not-determined |
| S1_extracted_k141_126280 | Not-determined |
| S1_extracted_k141_93219 | Not-determined |
| S1_extracted_k141_26382 | Not-determined |
| S1_extracted_k141_71736 | Low-quality |
| S1_extracted_k141_79994 | Not-determined |
| S1_extracted_k141_46580 | Not-determined |
| S1_extracted_k141_38470 | Not-determined |
| S1_extracted_k141_59805 | Not-determined |
| S1_extracted_k141_150016 | Not-determined |
| S1_extracted_k141_133407 | Not-determined |
| S1_extracted_k141_86679 | Low-quality |
| S1_extracted_k141_150017 | Not-determined |
| S1_extracted_k141_156964 | Low-quality |
| S1_extracted_k141_139727 | Not-determined |
| S1_extracted_k141_93228 | Not-determined |
| S1_extracted_k141_5157 | Not-determined |
| S1_extracted_k141_156966 | Not-determined |
| S1_extracted_k141_150021 | Not-determined |
| S1_extracted_k141_126288 | Not-determined |
| S1_extracted_k141_113180 | Not-determined |
| S1_extracted_k141_26395 | Not-determined |
| S1_extracted_k141_139732 | Not-determined |
| S1_extracted_k141_38483 | Not-determined |
| S1_extracted_k141_113184 | Not-determined |
| S1_extracted_k141_133417 | Not-determined |
| S1_extracted_k141_46596 | Not-determined |
| S1_extracted_k141_126291 | Not-determined |
| S1_extracted_k141_46598 | Not-determined |
| S1_extracted_k141_80007 | Not-determined |
| S1_extracted_k141_93241 | Not-determined |
| S1_extracted_k141_46601 | Not-determined |
| S1_extracted_k141_139735 | Not-determined |
| S1_extracted_k141_156975 | Not-determined |
| S1_extracted_k141_5172 | Not-determined |
| S1_extracted_k141_80010 | Low-quality |
| S1_extracted_k141_113195 | Not-determined |
| S1_extracted_k141_113196 | Not-determined |
| S1_extracted_k141_139739 | Low-quality |
| S1_extracted_k141_113197 | Not-determined |
| S1_extracted_k141_93250 | Not-determined |
| S1_extracted_k141_133423 | Not-determined |
| S1_extracted_k141_106633 | Not-determined |
| S1_extracted_k141_113198 | Low-quality |
| S1_extracted_k141_133426 | Not-determined |
| S1_extracted_k141_106639 | Not-determined |
| S1_extracted_k141_46607 | Low-quality |
| S1_extracted_k141_106641 | Not-determined |
| S1_extracted_k141_146090 | Not-determined |
| S1_extracted_k141_33341 | Not-determined |
| S1_extracted_k141_33342 | Not-determined |
| S1_extracted_k141_80013 | Not-determined |
| S1_extracted_k141_150030 | Not-determined |
| S1_extracted_k141_71738 | Not-determined |
| S1_extracted_k141_46616 | Not-determined |
| S1_extracted_k141_133437 | Not-determined |
| S1_extracted_k141_93264 | Low-quality |
| S1_extracted_k141_156983 | Not-determined |
| S1_extracted_k141_71739 | Not-determined |
| S1_extracted_k141_156984 | Low-quality |
| S1_extracted_k141_113204 | Not-determined |
| S1_extracted_k141_106649 | Not-determined |
| S1_extracted_k141_93268 | Not-determined |
| S1_extracted_k141_38494 | Low-quality |
| S1_extracted_k141_80020 | Not-determined |
| S1_extracted_k141_113208 | Not-determined |
| S1_extracted_k141_126315 | Not-determined |
| S1_extracted_k141_33350 | Not-determined |
| S1_extracted_k141_100066 | Not-determined |
| S1_extracted_k141_126317 | Not-determined |
| S1_extracted_k141_93270 | Not-determined |
| S1_extracted_k141_80023 | Not-determined |
| S1_extracted_k141_126319 | Not-determined |
| S1_extracted_k141_38498 | Not-determined |
| S1_extracted_k141_86681 | Not-determined |
| S1_extracted_k141_65269 | Not-determined |
| S1_extracted_k141_59834 | Low-quality |
| S1_extracted_k141_80027 | Not-determined |
| S1_extracted_k141_106651 | Not-determined |
| S1_extracted_k141_38500 | Not-determined |
| S1_extracted_k141_156988 | Not-determined |
| S1_extracted_k141_38502 | Not-determined |
| S1_extracted_k141_106657 | Not-determined |
| S1_extracted_k141_80031 | Not-determined |
| S1_extracted_k141_86684 | Not-determined |
| S1_extracted_k141_100077 | Not-determined |
| S1_extracted_k141_33361 | Not-determined |
| S1_extracted_k141_38506 | Not-determined |
| S1_extracted_k141_80036 | Low-quality |
| S1_extracted_k141_100082 | Not-determined |
| S1_extracted_k141_86688 | Not-determined |
| S1_extracted_k141_156998 | Not-determined |
| S1_extracted_k141_100084 | Low-quality |
| S1_extracted_k141_93282 | Not-determined |
| S1_extracted_k141_146097 | Low-quality |
| S1_extracted_k141_146098 | Not-determined |
| S1_extracted_k141_93284 | Not-determined |
| S1_extracted_k141_65280 | Not-determined |
| S1_extracted_k141_80042 | Low-quality |
| S1_extracted_k141_38511 | Low-quality |
| S1_extracted_k141_157000 | Not-determined |
| S1_extracted_k141_71761 | Not-determined |
| S1_extracted_k141_80044 | Not-determined |
| S1_extracted_k141_100086 | Low-quality |
| S1_extracted_k141_65286 | Low-quality |
| S1_extracted_k141_93288 | Not-determined |
| S1_extracted_k141_86700 | Low-quality |
| S1_extracted_k141_46639 | Not-determined |
| S1_extracted_k141_106666 | Low-quality |
| S1_extracted_k141_113213 | Not-determined |
| S1_extracted_k141_133468 | Not-determined |
| S1_extracted_k141_139765 | Low-quality |
| S1_extracted_k141_113216 | Not-determined |
| S1_extracted_k141_139766 | Not-determined |
| S1_extracted_k141_80051 | Low-quality |
| S1_extracted_k141_59845 | Low-quality |
| S1_extracted_k141_93296 | Not-determined |
| S1_extracted_k141_46644 | Not-determined |
| S1_extracted_k141_86706 | Not-determined |
| S1_extracted_k141_100097 | Not-determined |
| S1_extracted_k141_93299 | Not-determined |
| S1_extracted_k141_65299 | Not-determined |
| S1_extracted_k141_80053 | Not-determined |
| S1_extracted_k141_59847 | Not-determined |
| S1_extracted_k141_113220 | Low-quality |
| S1_extracted_k141_126338 | Not-determined |
| S1_extracted_k141_59848 | Not-determined |
| S1_extracted_k141_86708 | Not-determined |
| S1_extracted_k141_5208 | Not-determined |
| S1_extracted_k141_86711 | Not-determined |
| S1_extracted_k141_126341 | Not-determined |
| S1_extracted_k141_5210 | Not-determined |
| S1_extracted_k141_26432 | Not-determined |
| S1_extracted_k141_38525 | Not-determined |
| S1_extracted_k141_139771 | Not-determined |
| S1_extracted_k141_93304 | Not-determined |
| S1_extracted_k141_113227 | Low-quality |
| S1_extracted_k141_139774 | Not-determined |
| S1_extracted_k141_119927 | Not-determined |
| S1_extracted_k141_139778 | Not-determined |
| S1_extracted_k141_113232 | Not-determined |
| S1_extracted_k141_71775 | Not-determined |
| S1_extracted_k141_113235 | Not-determined |
| S1_extracted_k141_119930 | Not-determined |
| S1_extracted_k141_80063 | Not-determined |
| S1_extracted_k141_93314 | Not-determined |
| S1_extracted_k141_93315 | Low-quality |
| S1_extracted_k141_119932 | Not-determined |
| S1_extracted_k141_106695 | Not-determined |
| S1_extracted_k141_146126 | Not-determined |
| S1_extracted_k141_93316 | Not-determined |
| S1_extracted_k141_46669 | Not-determined |
| S1_extracted_k141_80068 | Not-determined |
| S1_extracted_k141_119936 | Not-determined |
| S1_extracted_k141_100110 | Low-quality |
| S1_extracted_k141_113245 | Not-determined |
| S1_extracted_k141_65312 | Not-determined |
| S1_extracted_k141_133491 | Not-determined |
| S1_extracted_k141_5213 | Not-determined |
| S1_extracted_k141_100115 | Not-determined |
| S1_extracted_k141_93326 | Not-determined |
| S1_extracted_k141_126359 | Not-determined |
| S1_extracted_k141_157032 | Not-determined |
| S1_extracted_k141_113258 | Not-determined |
| S1_extracted_k141_113260 | Not-determined |
| S1_extracted_k141_65323 | Not-determined |
| S1_extracted_k141_113262 | Low-quality |
| S1_extracted_k141_80085 | Not-determined |
| S1_extracted_k141_86739 | Low-quality |
| S1_extracted_k141_113265 | Low-quality |
| S1_extracted_k141_86743 | Not-determined |
| S1_extracted_k141_38541 | Not-determined |
| S1_extracted_k141_5223 | Not-determined |
| S1_extracted_k141_119954 | Not-determined |
| S1_extracted_k141_119955 | Not-determined |
| S1_extracted_k141_113271 | Low-quality |
| S1_extracted_k141_139809 | Not-determined |
| S1_extracted_k141_65337 | Not-determined |
| S1_extracted_k141_119961 | Low-quality |
| S1_extracted_k141_46692 | Not-determined |
| S1_extracted_k141_71805 | Not-determined |
| S1_extracted_k141_46696 | Low-quality |
| S1_extracted_k141_46697 | Not-determined |
| S1_extracted_k141_93362 | Not-determined |
| S1_extracted_k141_65342 | Not-determined |
| S1_extracted_k141_146146 | Not-determined |
| S1_extracted_k141_86758 | Low-quality |
| S1_extracted_k141_80093 | Not-determined |
| S1_extracted_k141_38556 | Not-determined |
| S1_extracted_k141_126373 | Not-determined |
| S1_extracted_k141_65346 | Low-quality |
| S1_extracted_k141_65347 | Not-determined |
| S1_extracted_k141_139821 | Not-determined |
| S1_extracted_k141_113291 | Not-determined |
| S1_extracted_k141_65350 | Not-determined |
| S1_extracted_k141_38561 | Not-determined |
| S1_extracted_k141_93373 | Not-determined |
| S1_extracted_k141_65352 | Not-determined |
| S1_extracted_k141_150049 | Not-determined |
| S1_extracted_k141_26439 | Not-determined |
| S1_extracted_k141_93375 | Not-determined |
| S1_extracted_k141_65356 | Not-determined |
| S1_extracted_k141_106739 | Not-determined |
| S1_extracted_k141_93378 | Not-determined |
| S1_extracted_k141_65360 | Not-determined |

| | |
|---|---|
| S1_extracted_k141_59911 | Low-quality |
| S1_extracted_k141_139832 | Not-determined |
| S1_extracted_k141_71827 | Not-determined |
| S1_extracted_k141_59913 | Low-quality |
| S1_extracted_k141_113302 | Not-determined |
| S1_extracted_k141_86768 | Low-quality |
| S1_extracted_k141_93383 | Not-determined |
| S1_extracted_k141_113305 | Low-quality |
| S1_extracted_k141_26443 | Not-determined |
| S1_extracted_k141_86773 | Not-determined |
| S1_extracted_k141_113307 | Not-determined |
| S1_extracted_k141_46722 | Not-determined |
| S1_extracted_k141_150061 | Not-determined |
| S1_extracted_k141_59918 | Not-determined |
| S1_extracted_k141_139844 | Not-determined |
| S1_extracted_k141_71832 | Not-determined |
| S1_extracted_k141_150062 | Not-determined |
| S1_extracted_k141_71833 | Not-determined |
| S1_extracted_k141_113310 | Not-determined |
| S1_extracted_k141_146167 | Not-determined |
| S1_extracted_k141_59922 | Not-determined |
| S1_extracted_k141_139848 | Low-quality |
| S1_extracted_k141_106755 | Low-quality |
| S1_extracted_k141_146170 | Not-determined |
| S1_extracted_k141_26448 | Not-determined |
| S1_extracted_k141_26454 | Not-determined |
| S1_extracted_k141_146180 | Low-quality |
| S1_extracted_k141_38577 | Not-determined |
| S1_extracted_k141_65377 | Not-determined |
| S1_extracted_k141_93406 | Not-determined |
| S1_extracted_k141_59933 | Low-quality |
| S1_extracted_k141_139851 | Low-quality |
| S1_extracted_k141_38579 | Not-determined |
| S1_extracted_k141_146186 | Not-determined |
| S1_extracted_k141_146188 | Low-quality |
| S1_extracted_k141_113321 | Not-determined |
| S1_extracted_k141_106771 | Low-quality |
| S1_extracted_k141_146192 | Not-determined |
| S1_extracted_k141_26460 | Low-quality |
| S1_extracted_k141_65389 | Not-determined |
| S1_extracted_k141_59944 | Not-determined |
| S1_extracted_k141_93416 | Not-determined |
| S1_extracted_k141_71853 | Not-determined |
| S1_extracted_k141_59952 | Not-determined |
| S1_extracted_k141_71854 | Not-determined |
| S1_extracted_k141_65395 | Not-determined |
| S1_extracted_k141_106779 | Not-determined |
| S1_extracted_k141_26466 | Low-quality |
| S1_extracted_k141_120000 | Low-quality |
| S1_extracted_k141_146202 | Not-determined |
| S1_extracted_k141_113334 | Not-determined |
| S1_extracted_k141_146205 | Not-determined |
| S1_extracted_k141_59957 | Not-determined |
| S1_extracted_k141_26471 | Not-determined |
| S1_extracted_k141_120003 | Low-quality |
| S1_extracted_k141_65402 | Not-determined |
| S1_extracted_k141_93422 | Not-determined |
| S1_extracted_k141_59959 | Not-determined |
| S1_extracted_k141_26475 | Not-determined |
| S1_extracted_k141_59961 | Not-determined |
| S1_extracted_k141_71859 | Not-determined |
| S1_extracted_k141_120007 | Low-quality |
| S1_extracted_k141_120009 | Not-determined |
| S1_extracted_k141_59966 | Not-determined |
| S1_extracted_k141_106792 | Not-determined |
| S1_extracted_k141_71863 | Not-determined |
| S1_extracted_k141_106795 | Not-determined |
| S1_extracted_k141_59970 | Not-determined |
| S1_extracted_k141_113350 | Low-quality |
| S1_extracted_k141_126380 | Not-determined |
| S1_extracted_k141_113354 | Not-determined |
| S1_extracted_k141_59975 | Not-determined |
| S1_extracted_k141_59977 | Low-quality |
| S1_extracted_k141_59979 | Not-determined |
| S1_extracted_k141_120019 | Not-determined |
| S1_extracted_k141_126385 | Not-determined |
| S1_extracted_k141_93438 | Not-determined |
| S1_extracted_k141_120021 | Not-determined |
| S1_extracted_k141_5274 | Low-quality |
| S1_extracted_k141_120022 | Not-determined |
| S1_extracted_k141_120023 | Not-determined |
| S1_extracted_k141_157056 | Not-determined |
| S1_extracted_k141_150069 | Not-determined |
| S1_extracted_k141_71873 | Not-determined |
| S1_extracted_k141_93441 | Not-determined |
| S1_extracted_k141_150074 | Not-determined |
| S1_extracted_k141_113368 | Not-determined |
| S1_extracted_k141_59987 | Not-determined |
| S1_extracted_k141_65437 | Low-quality |
| S1_extracted_k141_59988 | Not-determined |
| S1_extracted_k141_65439 | Not-determined |
| S1_extracted_k141_38609 | Not-determined |
| S1_extracted_k141_150079 | Low-quality |
| S1_extracted_k141_113372 | Not-determined |
| S1_extracted_k141_93445 | Not-determined |
| S1_extracted_k141_120035 | Not-determined |
| S1_extracted_k141_93446 | Not-determined |
| S1_extracted_k141_120038 | Not-determined |
| S1_extracted_k141_150085 | Not-determined |
| S1_extracted_k141_146239 | Not-determined |
| S1_extracted_k141_146240 | Not-determined |
| S1_extracted_k141_150087 | Not-determined |
| S1_extracted_k141_113380 | Not-determined |
| S1_extracted_k141_120044 | Low-quality |
| S1_extracted_k141_65458 | Not-determined |
| S1_extracted_k141_150091 | Not-determined |
| S1_extracted_k141_26496 | Not-determined |
| S1_extracted_k141_65468 | Not-determined |
| S1_extracted_k141_65469 | Not-determined |
| S1_extracted_k141_139886 | Low-quality |
| S1_extracted_k141_126418 | Not-determined |
| S1_extracted_k141_139890 | Not-determined |
| S1_extracted_k141_126422 | Not-determined |
| S1_extracted_k141_120061 | Not-determined |
| S1_extracted_k141_71897 | Not-determined |
| S1_extracted_k141_120064 | Not-determined |
| S1_extracted_k141_65477 | Low-quality |
| S1_extracted_k141_146259 | Not-determined |
| S1_extracted_k141_120070 | Not-determined |
| S1_extracted_k141_71901 | Low-quality |
| S1_extracted_k141_26511 | Not-determined |
| S1_extracted_k141_126434 | Not-determined |
| S1_extracted_k141_5300 | Low-quality |
| S1_extracted_k141_65491 | Not-determined |
| S1_extracted_k141_71906 | Not-determined |
| S1_extracted_k141_71907 | Not-determined |
| S1_extracted_k141_146269 | Low-quality |
| S1_extracted_k141_126444 | Low-quality |
| S1_extracted_k141_26519 | Not-determined |
| S1_extracted_k141_71911 | Not-determined |
| S1_extracted_k141_5308 | Not-determined |
| S1_extracted_k141_5309 | Not-determined |
| S1_extracted_k141_71912 | Not-determined |
| S1_extracted_k141_113397 | Low-quality |
| S1_extracted_k141_60020 | Not-determined |
| S1_extracted_k141_126450 | Not-determined |
| S1_extracted_k141_120088 | Not-determined |
| S1_extracted_k141_60023 | Not-determined |
| S1_extracted_k141_26525 | Low-quality |
| S1_extracted_k141_60028 | Not-determined |
| S1_extracted_k141_120095 | Not-determined |
| S1_extracted_k141_65505 | Not-determined |
| S1_extracted_k141_71919 | Not-determined |
| S1_extracted_k141_146278 | Low-quality |
| S1_extracted_k141_60033 | Not-determined |
| S1_extracted_k141_5321 | Not-determined |
| S1_extracted_k141_26530 | Not-determined |
| S1_extracted_k141_65514 | Not-determined |
| S1_extracted_k141_113411 | Not-determined |
| S1_extracted_k141_26533 | Not-determined |
| S1_extracted_k141_150116 | Not-determined |
| S1_extracted_k141_113415 | Not-determined |
| S1_extracted_k141_146281 | Low-quality |
| S1_extracted_k141_71922 | Low-quality |
| S1_extracted_k141_71926 | Low-quality |
| S1_extracted_k141_120103 | Not-determined |
| S1_extracted_k141_120105 | Not-determined |
| S1_extracted_k141_146286 | Not-determined |
| S1_extracted_k141_26545 | Not-determined |
| S1_extracted_k141_150124 | Not-determined |
| S1_extracted_k141_60049 | Not-determined |
| S1_extracted_k141_146292 | Low-quality |
| S1_extracted_k141_65529 | Not-determined |
| S1_extracted_k141_120109 | Low-quality |
| S1_extracted_k141_146296 | Not-determined |
| S1_extracted_k141_139944 | Not-determined |
| S1_extracted_k141_60060 | Low-quality |
| S1_extracted_k141_139947 | Low-quality |
| S1_extracted_k141_139948 | Not-determined |
| S1_extracted_k141_60061 | Low-quality |
| S1_extracted_k141_139950 | Low-quality |
| S1_extracted_k141_71934 | Low-quality |
| S1_extracted_k141_150136 | Not-determined |
| S1_extracted_k141_5340 | Not-determined |
| S1_extracted_k141_157063 | Not-determined |
| S1_extracted_k141_5341 | Not-determined |
| S1_extracted_k141_5342 | Not-determined |
| S1_extracted_k141_157065 | Not-determined |
| S1_extracted_k141_146310 | Low-quality |
| S1_extracted_k141_146312 | Not-determined |
| S1_extracted_k141_146315 | Not-determined |
| S1_extracted_k141_26560 | Not-determined |
| S1_extracted_k141_65547 | Not-determined |
| S1_extracted_k141_5346 | Not-determined |
| S1_extracted_k141_71943 | Not-determined |
| S1_extracted_k141_71945 | Not-determined |
| S1_extracted_k141_65550 | Not-determined |
| S1_extracted_k141_65551 | Not-determined |
| S1_extracted_k141_146320 | Not-determined |
| S1_extracted_k141_26567 | Not-determined |
| S1_extracted_k141_65552 | Not-determined |
| S1_extracted_k141_5349 | Not-determined |
| S1_extracted_k141_5352 | Not-determined |
| S1_extracted_k141_113431 | Low-quality |
| S1_extracted_k141_5356 | Not-determined |
| S1_extracted_k141_150137 | Not-determined |
| S1_extracted_k141_38634 | Not-determined |
| S1_extracted_k141_120125 | Low-quality |
| S1_extracted_k141_139971 | Not-determined |
| S1_extracted_k141_120131 | Low-quality |
| S1_extracted_k141_139976 | Not-determined |
| S1_extracted_k141_5370 | Not-determined |
| S1_extracted_k141_71964 | Low-quality |
| S1_extracted_k141_146341 | Low-quality |
| S1_extracted_k141_26585 | Low-quality |
| S1_extracted_k141_146345 | Not-determined |
| S1_extracted_k141_65571 | Not-determined |
| S1_extracted_k141_120135 | Not-determined |
| S1_extracted_k141_38646 | Not-determined |
| S1_extracted_k141_71972 | Low-quality |
| S1_extracted_k141_126494 | Low-quality |
| S1_extracted_k141_113453 | Not-determined |
| S1_extracted_k141_139995 | Not-determined |
| S1_extracted_k141_113455 | Not-determined |
| S1_extracted_k141_139999 | Not-determined |
| S1_extracted_k141_26597 | Not-determined |
| S1_extracted_k141_157079 | Not-determined |
| S1_extracted_k141_5387 | Not-determined |
| S1_extracted_k141_65579 | Not-determined |
| S1_extracted_k141_38659 | Not-determined |
| S1_extracted_k141_146369 | Not-determined |
| S1_extracted_k141_113465 | Not-determined |
| S1_extracted_k141_126506 | Not-determined |
| S1_extracted_k141_146379 | Not-determined |
| S1_extracted_k141_150147 | Not-determined |
| S1_extracted_k141_71988 | Not-determined |
| S1_extracted_k141_113470 | Low-quality |
| S1_extracted_k141_146386 | Not-determined |
| S1_extracted_k141_38665 | Not-determined |
| S1_extracted_k141_26614 | Not-determined |
| S1_extracted_k141_126531 | Not-determined |
| S1_extracted_k141_26623 | Low-quality |
| S1_extracted_k141_150151 | Low-quality |
| S1_extracted_k141_38672 | Not-determined |
| S1_extracted_k141_146392 | Not-determined |
| S1_extracted_k141_38673 | Not-determined |
| S1_extracted_k141_126539 | Not-determined |
| S1_extracted_k141_126540 | Not-determined |
| S1_extracted_k141_65618 | Low-quality |
| S1_extracted_k141_126544 | Not-determined |
| S1_extracted_k141_146405 | Low-quality |
| S1_extracted_k141_26632 | Not-determined |
| S1_extracted_k141_38685 | Not-determined |
| S1_extracted_k141_38687 | Not-determined |
| S1_extracted_k141_146409 | Not-determined |
| S1_extracted_k141_38690 | Low-quality |
| S1_extracted_k141_65626 | Low-quality |
| S1_extracted_k141_72016 | Low-quality |
| S1_extracted_k141_26639 | Not-determined |
| S1_extracted_k141_146418 | Not-determined |
| S1_extracted_k141_140043 | Low-quality |
| S1_extracted_k141_5415 | Not-determined |
| S1_extracted_k141_146421 | Low-quality |
| S1_extracted_k141_150162 | Not-determined |
| S1_extracted_k141_5419 | Not-determined |
| S1_extracted_k141_38699 | Not-determined |
| S1_extracted_k141_150164 | Low-quality |
| S1_extracted_k141_150166 | Not-determined |
| S1_extracted_k141_72028 | Not-determined |
| S1_extracted_k141_38705 | Not-determined |
| S1_extracted_k141_38707 | Not-determined |
| S1_extracted_k141_72033 | Not-determined |
| S1_extracted_k141_65647 | Not-determined |
| S1_extracted_k141_38711 | Not-determined |
| S1_extracted_k141_38712 | Not-determined |
| S1_extracted_k141_140058 | Low-quality |
| S1_extracted_k141_146442 | Not-determined |
| S1_extracted_k141_140061 | Not-determined |
| S1_extracted_k141_65654 | Not-determined |
| S1_extracted_k141_26661 | Not-determined |
| S1_extracted_k141_72047 | Not-determined |
| S1_extracted_k141_5440 | Not-determined |
| S1_extracted_k141_126587 | Not-determined |
| S1_extracted_k141_65660 | Not-determined |
| S1_extracted_k141_26668 | Not-determined |
| S1_extracted_k141_38718 | Not-determined |
| S1_extracted_k141_146459 | Not-determined |
| S1_extracted_k141_126601 | Not-determined |
| S1_extracted_k141_126602 | Not-determined |
| S1_extracted_k141_150175 | Not-determined |
| S1_extracted_k141_146468 | Not-determined |
| S1_extracted_k141_150176 | Not-determined |
| S1_extracted_k141_146469 | Not-determined |
| S1_extracted_k141_140079 | Not-determined |
| S1_extracted_k141_150180 | Not-determined |
| S1_extracted_k141_38730 | Low-quality |
| S1_extracted_k141_140085 | Not-determined |
| S1_extracted_k141_5462 | Not-determined |
| S1_extracted_k141_26681 | Not-determined |
| S1_extracted_k141_38736 | Not-determined |
| S1_extracted_k141_5466 | Not-determined |
| S1_extracted_k141_126620 | Low-quality |
| S1_extracted_k141_38741 | Not-determined |
| S1_extracted_k141_26687 | Not-determined |
| S1_extracted_k141_146485 | Not-determined |
| S1_extracted_k141_157104 | Not-determined |
| S1_extracted_k141_26691 | Not-determined |
| S1_extracted_k141_72054 | Not-determined |
| S1_extracted_k141_26695 | Not-determined |
| S1_extracted_k141_126628 | Low-quality |
| S1_extracted_k141_72056 | Not-determined |
| S1_extracted_k141_140100 | Low-quality |
| S1_extracted_k141_140104 | Not-determined |
| S1_extracted_k141_72061 | Low-quality |
| S1_extracted_k141_72062 | Not-determined |
| S1_extracted_k141_140110 | Not-determined |
| S1_extracted_k141_38754 | Not-determined |
| S1_extracted_k141_126633 | Low-quality |
| S1_extracted_k141_38755 | Not-determined |
| S1_extracted_k141_140114 | Not-determined |
| S1_extracted_k141_72069 | Not-determined |
| S1_extracted_k141_126639 | Not-determined |
| S1_extracted_k141_65705 | Low-quality |
| S1_extracted_k141_126641 | Not-determined |
| S1_extracted_k141_150190 | Low-quality |
| S1_extracted_k141_146513 | Not-determined |
| S1_extracted_k141_140123 | Not-determined |
| S1_extracted_k141_126647 | Not-determined |
| S1_extracted_k141_140125 | Not-determined |
| S1_extracted_k141_65711 | Not-determined |
| S1_extracted_k141_38761 | Not-determined |
| S1_extracted_k141_126650 | Not-determined |
| S1_extracted_k141_146518 | Not-determined |
| S1_extracted_k141_72080 | Low-quality |
| S1_extracted_k141_5488 | Not-determined |
| S1_extracted_k141_65713 | Not-determined |
| S1_extracted_k141_65714 | Not-determined |
| S1_extracted_k141_140128 | Low-quality |
| S1_extracted_k141_65717 | Not-determined |
| S1_extracted_k141_65719 | Not-determined |
| S1_extracted_k141_5495 | Not-determined |
| S1_extracted_k141_157133 | Low-quality |
| S1_extracted_k141_5501 | Not-determined |
| S1_extracted_k141_140144 | Not-determined |
| S1_extracted_k141_150191 | Not-determined |
| S1_extracted_k141_65732 | Not-determined |
| S1_extracted_k141_65735 | Not-determined |
| S1_extracted_k141_65736 | Not-determined |
| S1_extracted_k141_140151 | Not-determined |
| S1_extracted_k141_126677 | Not-determined |
| S1_extracted_k141_72108 | Not-determined |
| S1_extracted_k141_140159 | Not-determined |
| S1_extracted_k141_157150 | Low-quality |
| S1_extracted_k141_146555 | Not-determined |
| S1_extracted_k141_5514 | Not-determined |
| S1_extracted_k141_72112 | Not-determined |
| S1_extracted_k141_5516 | Not-determined |
| S1_extracted_k141_126683 | Not-determined |
| S1_extracted_k141_65749 | Not-determined |
| S1_extracted_k141_157163 | Not-determined |
| S1_extracted_k141_65758 | Not-determined |
| S1_extracted_k141_146577 | Not-determined |
| S1_extracted_k141_38800 | Not-determined |
| S1_extracted_k141_126701 | Not-determined |
| S1_extracted_k141_146585 | Not-determined |
| S1_extracted_k141_146588 | Not-determined |
| S1_extracted_k141_5536 | Not-determined |
| S1_extracted_k141_157174 | Not-determined |
| S1_extracted_k141_65767 | Not-determined |
| S1_extracted_k141_38811 | Not-determined |
| S1_extracted_k141_126714 | Not-determined |
| S1_extracted_k141_146599 | Not-determined |
| S1_extracted_k141_146600 | Not-determined |
| S1_extracted_k141_146607 | Low-quality |
| S1_extracted_k141_126725 | Not-determined |
| S1_extracted_k141_150207 | Not-determined |
| S1_extracted_k141_157185 | Low-quality |
| S1_extracted_k141_5542 | Not-determined |
| S1_extracted_k141_150208 | Not-determined |
| S1_extracted_k141_5546 | Not-determined |
| S1_extracted_k141_150216 | Not-determined |
| S1_extracted_k141_38826 | Not-determined |
| S1_extracted_k141_150220 | Not-determined |
| S1_extracted_k141_126734 | Not-determined |
| S1_extracted_k141_65798 | Low-quality |
| S1_extracted_k141_157194 | Low-quality |
| S1_extracted_k141_126737 | Not-determined |
| S1_extracted_k141_38834 | Not-determined |
| S1_extracted_k141_65805 | Not-determined |
| S1_extracted_k141_157197 | Not-determined |
| S1_extracted_k141_65808 | Not-determined |
| S1_extracted_k141_126743 | Not-determined |
| S1_extracted_k141_146623 | Not-determined |
| S1_extracted_k141_146625 | Not-determined |
| S1_extracted_k141_65816 | Not-determined |
| S1_extracted_k141_157206 | Not-determined |
| S1_extracted_k141_5564 | Not-determined |
| S1_extracted_k141_150230 | Not-determined |
| S1_extracted_k141_157211 | Low-quality |
| S1_extracted_k141_150236 | Not-determined |
| S1_extracted_k141_150237 | Not-determined |
| S1_extracted_k141_65828 | Not-determined |
| S1_extracted_k141_126748 | Not-determined |
| S1_extracted_k141_126751 | Not-determined |
| S1_extracted_k141_146630 | Not-determined |
| S1_extracted_k141_65832 | Not-determined |
| S1_extracted_k141_157223 | Not-determined |
| S1_extracted_k141_126757 | Not-determined |
| S1_extracted_k141_65838 | Not-determined |
| S1_extracted_k141_38860 | Low-quality |
| S1_extracted_k141_38863 | Not-determined |
| S1_extracted_k141_157233 | Low-quality |
| S1_extracted_k141_5588 | Not-determined |
| S1_extracted_k141_65851 | Not-determined |
| S1_extracted_k141_126771 | Not-determined |
| S1_extracted_k141_65856 | Not-determined |
| S1_extracted_k141_157237 | Not-determined |
| S1_extracted_k141_65859 | Not-determined |
| S1_extracted_k141_150258 | Low-quality |
| S1_extracted_k141_126780 | Not-determined |
| S1_extracted_k141_146655 | Not-determined |
| S1_extracted_k141_146663 | Not-determined |
| S1_extracted_k141_157246 | Not-determined |
| S1_extracted_k141_146667 | Low-quality |
| S1_extracted_k141_72149 | Not-determined |
| S1_extracted_k141_146674 | Not-determined |
| S1_extracted_k141_126805 | Not-determined |
| S1_extracted_k141_72159 | Not-determined |
| S1_extracted_k141_72161 | Not-determined |
| S1_extracted_k141_146677 | Not-determined |
| S1_extracted_k141_72162 | Not-determined |
| S1_extracted_k141_5619 | Not-determined |
| S1_extracted_k141_126806 | Low-quality |
| S1_extracted_k141_126808 | Not-determined |
| S1_extracted_k141_38903 | Not-determined |
| S1_extracted_k141_65897 | Not-determined |
| S1_extracted_k141_157262 | Low-quality |
| S1_extracted_k141_5625 | Not-determined |
| S1_extracted_k141_5626 | Not-determined |
| S1_extracted_k141_146696 | Low-quality |
| S1_extracted_k141_65912 | Not-determined |
| S1_extracted_k141_72164 | Low-quality |
| S1_extracted_k141_5647 | Not-determined |
| S1_extracted_k141_38913 | Not-determined |
| S1_extracted_k141_150268 | Not-determined |
| S1_extracted_k141_65921 | Not-determined |
| S1_extracted_k141_38921 | Low-quality |
| S1_extracted_k141_65932 | Not-determined |
| S1_extracted_k141_150281 | Not-determined |
| S1_extracted_k141_38931 | Not-determined |
| S1_extracted_k141_146733 | Not-determined |
| S1_extracted_k141_65945 | Not-determined |
| S1_extracted_k141_150289 | Not-determined |
| S1_extracted_k141_146745 | Low-quality |
| S1_extracted_k141_146750 | Not-determined |
| S1_extracted_k141_5682 | Not-determined |
| S1_extracted_k141_146754 | Not-determined |
| S1_extracted_k141_38943 | Not-determined |
| S1_extracted_k141_146762 | Not-determined |
| S1_extracted_k141_157276 | Not-determined |
| S1_extracted_k141_65968 | Not-determined |
| S1_extracted_k141_65970 | Not-determined |
| S1_extracted_k141_5697 | Low-quality |
| S1_extracted_k141_146771 | Low-quality |
| S1_extracted_k141_146773 | Low-quality |
| S1_extracted_k141_5707 | Low-quality |
| S1_extracted_k141_157298 | Not-determined |
| S1_extracted_k141_72186 | Low-quality |
| S1_extracted_k141_157302 | Not-determined |
| S1_extracted_k141_72187 | Not-determined |
| S1_extracted_k141_157306 | Not-determined |
| S1_extracted_k141_5712 | Not-determined |
| S1_extracted_k141_5714 | Not-determined |
| S1_extracted_k141_157313 | Not-determined |
| S1_extracted_k141_157315 | Not-determined |
| S1_extracted_k141_65982 | Not-determined |
| S1_extracted_k141_38962 | Not-determined |
| S1_extracted_k141_38963 | Not-determined |
| S1_extracted_k141_150303 | Low-quality |
| S1_extracted_k141_38966 | Not-determined |
| S1_extracted_k141_146797 | Not-determined |
| S1_extracted_k141_38969 | Not-determined |
| S1_extracted_k141_157328 | Not-determined |
| S1_extracted_k141_146805 | Not-determined |
| S1_extracted_k141_5732 | Low-quality |
| S1_extracted_k141_146819 | Not-determined |
| S1_extracted_k141_66008 | Low-quality |
| S1_extracted_k141_38980 | Low-quality |
| S1_extracted_k141_66029 | Not-determined |
| S1_extracted_k141_66046 | Not-determined |
| S1_extracted_k141_66048 | Not-determined |
| S1_extracted_k141_72212 | Not-determined |
| S1_extracted_k141_146841 | Not-determined |
| S1_extracted_k141_72221 | Not-determined |
| S1_extracted_k141_66069 | Low-quality |
| S1_extracted_k141_66071 | Not-determined |
| S1_extracted_k141_39007 | Not-determined |
| S1_extracted_k141_5759 | Not-determined |
| S1_extracted_k141_157372 | Not-determined |
| S1_extracted_k141_66078 | Not-determined |
| S1_extracted_k141_157379 | Not-determined |
| S1_extracted_k141_157384 | Not-determined |
| S1_extracted_k141_66093 | Not-determined |
| S1_extracted_k141_157393 | Not-determined |
| S1_extracted_k141_157394 | Not-determined |
| S1_extracted_k141_157396 | Not-determined |
| S1_extracted_k141_66098 | Low-quality |
| S1_extracted_k141_39020 | Low-quality |
| S1_extracted_k141_150338 | Not-determined |
| S1_extracted_k141_39028 | Not-determined |
| S1_extracted_k141_5806 | Not-determined |
| S1_extracted_k141_72235 | Not-determined |
| S1_extracted_k141_72240 | Not-determined |
| S1_extracted_k141_5813 | Not-determined |
| S1_extracted_k141_5814 | Low-quality |
| S1_extracted_k141_72245 | Not-determined |
| S1_extracted_k141_72247 | Low-quality |
| S1_extracted_k141_66129 | Not-determined |
| S1_extracted_k141_39045 | Not-determined |
| S1_extracted_k141_66135 | Not-determined |
| S1_extracted_k141_157408 | Not-determined |
| S1_extracted_k141_72255 | Not-determined |
| S1_extracted_k141_66147 | Not-determined |
| S1_extracted_k141_5836 | Not-determined |
| S1_extracted_k141_72267 | Not-determined |
| S1_extracted_k141_5840 | Not-determined |
| S1_extracted_k141_157421 | Not-determined |
| S1_extracted_k141_157422 | Not-determined |
| S1_extracted_k141_39063 | Not-determined |
| S1_extracted_k141_5846 | Not-determined |
| S1_extracted_k141_66157 | Not-determined |
| S1_extracted_k141_39066 | Not-determined |
| S1_extracted_k141_150347 | Not-determined |
| S1_extracted_k141_39068 | Not-determined |
| S1_extracted_k141_39070 | Low-quality |
| S1_extracted_k141_39071 | Not-determined |
| S1_extracted_k141_66166 | Not-determined |
| S1_extracted_k141_39073 | Not-determined |
| S1_extracted_k141_5858 | Not-determined |
| S1_extracted_k141_5859 | Not-determined |
| S1_extracted_k141_66176 | Low-quality |
| S1_extracted_k141_5861 | Not-determined |
| S1_extracted_k141_39083 | Not-determined |
| S1_extracted_k141_150358 | Not-determined |
| S1_extracted_k141_39085 | Not-determined |
| S1_extracted_k141_66184 | Not-determined |
| S1_extracted_k141_66185 | Low-quality |
| S1_extracted_k141_66186 | Low-quality |
| S1_extracted_k141_157436 | Not-determined |
| S1_extracted_k141_39091 | Not-determined |
| S1_extracted_k141_5870 | Not-determined |
| S1_extracted_k141_66193 | Not-determined |
| S1_extracted_k141_66194 | Low-quality |
| S1_extracted_k141_66195 | Not-determined |
| S1_extracted_k141_150376 | Not-determined |
| S1_extracted_k141_66204 | Not-determined |
| S1_extracted_k141_66208 | Not-determined |
| S1_extracted_k141_157455 | Not-determined |
| S1_extracted_k141_66217 | Low-quality |
| S1_extracted_k141_157460 | Not-determined |
| S1_extracted_k141_66220 | Not-determined |
| S1_extracted_k141_66224 | Not-determined |
| S1_extracted_k141_5881 | Low-quality |
| S1_extracted_k141_39116 | Not-determined |
| S1_extracted_k141_39117 | Not-determined |
| S1_extracted_k141_157468 | Not-determined |
| S1_extracted_k141_72281 | Not-determined |
| S1_extracted_k141_72284 | Not-determined |
| S1_extracted_k141_72285 | Not-determined |
| S1_extracted_k141_72286 | Not-determined |
| S1_extracted_k141_39124 | Not-determined |
| S1_extracted_k141_5889 | Low-quality |
| S1_extracted_k141_72291 | Not-determined |
| S1_extracted_k141_39130 | Not-determined |
| S1_extracted_k141_5896 | Not-determined |
| S1_extracted_k141_66247 | Not-determined |
| S1_extracted_k141_5902 | Not-determined |
| S1_extracted_k141_66253 | Not-determined |
| S1_extracted_k141_39141 | Not-determined |
| S1_extracted_k141_5909 | Not-determined |
| S1_extracted_k141_66254 | Not-determined |
| S1_extracted_k141_66258 | Not-determined |
| S1_extracted_k141_157487 | Not-determined |
| S1_extracted_k141_39149 | Not-determined |
| S1_extracted_k141_39151 | Not-determined |
| S1_extracted_k141_39153 | Not-determined |
| S1_extracted_k141_157491 | Not-determined |
| S1_extracted_k141_66259 | Low-quality |
| S1_extracted_k141_150393 | Low-quality |
| S1_extracted_k141_66279 | Low-quality |
| S1_extracted_k141_72309 | Not-determined |
| S1_extracted_k141_39166 | Not-determined |
| S1_extracted_k141_150408 | Not-determined |
| S1_extracted_k141_5940 | Not-determined |
| S1_extracted_k141_150410 | Not-determined |
| S1_extracted_k141_5942 | Not-determined |
| S1_extracted_k141_39177 | Not-determined |
| S1_extracted_k141_72312 | Low-quality |
| S1_extracted_k141_66296 | Not-determined |

| | |
|---|---|
| S1_extracted_k141_72315 | Not-determined |
| S1_extracted_k141_5947 | Not-determined |
| S1_extracted_k141_39185 | Not-determined |
| S1_extracted_k141_39188 | Not-determined |
| S1_extracted_k141_39191 | Low-quality |
| S1_extracted_k141_72340 | Not-determined |
| S1_extracted_k141_5968 | Low-quality |
| S1_extracted_k141_66336 | Not-determined |
| S1_extracted_k141_157510 | Not-determined |
| S1_extracted_k141_39201 | Not-determined |
| S1_extracted_k141_39204 | Not-determined |
| S1_extracted_k141_5974 | Not-determined |
| S1_extracted_k141_5975 | Not-determined |
| S1_extracted_k141_72347 | Not-determined |
| S1_extracted_k141_66353 | Not-determined |
| S1_extracted_k141_5985 | Not-determined |
| S1_extracted_k141_150418 | Not-determined |
| S1_extracted_k141_72355 | Low-quality |
| S1_extracted_k141_66362 | Not-determined |
| S1_extracted_k141_66364 | Not-determined |
| S1_extracted_k141_39222 | Not-determined |
| S1_extracted_k141_72361 | Not-determined |
| S1_extracted_k141_39224 | Low-quality |
| S1_extracted_k141_39225 | Not-determined |
| S1_extracted_k141_66368 | Not-determined |
| S1_extracted_k141_66370 | Not-determined |
| S1_extracted_k141_66372 | Not-determined |
| S1_extracted_k141_66373 | Not-determined |
| S1_extracted_k141_150428 | Not-determined |
| S1_extracted_k141_66383 | Not-determined |
| S1_extracted_k141_150432 | Not-determined |
| S1_extracted_k141_66389 | Not-determined |
| S1_extracted_k141_39238 | Not-determined |
| S1_extracted_k141_6001 | Not-determined |
| S1_extracted_k141_39241 | Not-determined |
| S1_extracted_k141_66394 | Low-quality |
| S1_extracted_k141_72377 | Not-determined |
| S1_extracted_k141_150440 | Not-determined |
| S1_extracted_k141_6005 | Low-quality |
| S1_extracted_k141_66399 | Not-determined |
| S1_extracted_k141_157515 | Not-determined |
| S1_extracted_k141_66411 | Not-determined |
| S1_extracted_k141_157521 | Not-determined |
| S1_extracted_k141_72386 | Not-determined |
| S1_extracted_k141_6022 | Low-quality |
| S1_extracted_k141_66427 | Not-determined |
| S1_extracted_k141_6026 | Not-determined |
| S1_extracted_k141_66429 | Not-determined |
| S1_extracted_k141_66431 | Not-determined |
| S1_extracted_k141_157537 | Not-determined |
| S1_extracted_k141_72407 | Not-determined |
| S1_extracted_k141_66437 | Low-quality |
| S1_extracted_k141_157540 | Not-determined |
| S1_extracted_k141_39249 | Not-determined |
| S1_extracted_k141_66451 | Low-quality |
| S1_extracted_k141_6043 | Not-determined |
| S1_extracted_k141_39266 | Not-determined |
| S1_extracted_k141_66466 | Not-determined |
| S1_extracted_k141_6058 | Not-determined |
| S1_extracted_k141_72433 | Not-determined |
| S1_extracted_k141_72434 | Not-determined |
| S1_extracted_k141_66476 | Not-determined |
| S1_extracted_k141_72436 | Not-determined |
| S1_extracted_k141_66477 | Not-determined |
| S1_extracted_k141_72438 | Low-quality |
| S1_extracted_k141_66481 | Not-determined |
| S1_extracted_k141_66488 | Low-quality |
| S1_extracted_k141_66490 | Not-determined |
| S1_extracted_k141_6076 | Not-determined |
| S1_extracted_k141_66496 | Not-determined |
| S1_extracted_k141_157557 | Not-determined |
| S1_extracted_k141_157559 | Low-quality |
| S1_extracted_k141_6088 | Not-determined |
| S1_extracted_k141_66512 | Not-determined |
| S1_extracted_k141_6091 | Low-quality |
| S1_extracted_k141_157568 | Not-determined |
| S1_extracted_k141_6094 | Not-determined |
| S1_extracted_k141_39271 | Low-quality |
| S1_extracted_k141_39280 | Low-quality |
| S1_extracted_k141_72463 | Not-determined |
| S1_extracted_k141_39282 | Not-determined |
| S1_extracted_k141_72466 | Not-determined |
| S1_extracted_k141_6102 | Not-determined |
| S1_extracted_k141_39287 | Low-quality |
| S1_extracted_k141_39291 | Not-determined |
| S1_extracted_k141_6123 | Not-determined |
| S1_extracted_k141_6124 | Not-determined |
| S1_extracted_k141_39305 | Not-determined |
| S1_extracted_k141_72487 | Not-determined |
| S1_extracted_k141_66548 | Low-quality |
| S1_extracted_k141_150477 | Low-quality |
| S1_extracted_k141_66554 | Not-determined |
| S1_extracted_k141_72494 | Not-determined |
| S1_extracted_k141_66563 | Not-determined |
| S1_extracted_k141_39335 | Not-determined |
| S1_extracted_k141_39340 | Low-quality |
| S1_extracted_k141_66573 | Low-quality |
| S1_extracted_k141_72515 | Not-determined |
| S1_extracted_k141_39346 | Not-determined |
| S1_extracted_k141_66577 | Not-determined |
| S1_extracted_k141_39354 | Not-determined |
| S1_extracted_k141_39356 | Low-quality |
| S1_extracted_k141_6171 | Not-determined |
| S1_extracted_k141_39361 | Low-quality |
| S1_extracted_k141_72521 | Not-determined |
| S1_extracted_k141_72523 | Not-determined |
| S1_extracted_k141_72524 | Not-determined |
| S1_extracted_k141_66599 | Not-determined |
| S1_extracted_k141_6190 | Low-quality |
| S1_extracted_k141_66601 | Not-determined |
| S1_extracted_k141_6193 | Not-determined |
| S1_extracted_k141_39370 | Not-determined |
| S1_extracted_k141_39373 | Not-determined |
| S1_extracted_k141_157580 | Not-determined |
| S1_extracted_k141_39375 | Not-determined |
| S1_extracted_k141_72542 | Low-quality |
| S1_extracted_k141_6211 | Not-determined |
| S1_extracted_k141_39378 | Not-determined |
| S1_extracted_k141_39379 | Not-determined |
| S1_extracted_k141_72547 | Not-determined |
| S1_extracted_k141_6216 | Not-determined |
| S1_extracted_k141_6219 | Not-determined |
| S1_extracted_k141_66631 | Not-determined |
| S1_extracted_k141_66633 | Not-determined |
| S1_extracted_k141_6223 | Not-determined |
| S1_extracted_k141_39393 | Not-determined |
| S1_extracted_k141_66639 | Not-determined |
| S1_extracted_k141_157599 | Not-determined |
| S1_extracted_k141_6231 | Not-determined |
| S1_extracted_k141_6236 | Low-quality |
| S1_extracted_k141_39404 | Low-quality |
| S1_extracted_k141_72578 | Not-determined |
| S1_extracted_k141_6239 | Not-determined |
| S1_extracted_k141_72588 | Not-determined |
| S1_extracted_k141_39421 | Not-determined |
| S1_extracted_k141_72601 | Low-quality |
| S1_extracted_k141_66673 | Not-determined |
| S1_extracted_k141_72612 | Low-quality |
| S1_extracted_k141_72616 | Low-quality |
| S1_extracted_k141_72618 | Low-quality |
| S1_extracted_k141_66686 | Not-determined |
| S1_extracted_k141_66687 | Not-determined |
| S1_extracted_k141_66689 | Low-quality |
| S1_extracted_k141_157620 | Low-quality |
| S1_extracted_k141_72638 | Not-determined |
| S1_extracted_k141_39447 | Not-determined |
| S1_extracted_k141_72641 | Not-determined |
| S1_extracted_k141_6269 | Not-determined |
| S1_extracted_k141_157637 | Not-determined |
| S1_extracted_k141_157643 | Not-determined |
| S1_extracted_k141_150514 | Low-quality |
| S1_extracted_k141_39460 | Not-determined |
| S1_extracted_k141_66731 | Low-quality |
| S1_extracted_k141_72676 | Not-determined |
| S1_extracted_k141_66744 | Low-quality |
| S1_extracted_k141_66747 | Not-determined |
| S1_extracted_k141_39487 | Low-quality |
| S1_extracted_k141_39496 | Not-determined |
| S1_extracted_k141_39500 | Low-quality |
| S1_extracted_k141_157661 | Low-quality |
| S1_extracted_k141_39506 | Low-quality |
| S1_extracted_k141_39509 | Low-quality |
| S1_extracted_k141_72706 | Not-determined |
| S1_extracted_k141_39512 | Not-determined |
| S1_extracted_k141_72721 | Not-determined |
| S17_vs_k141_177197\|\|full | High-quality |
| S1_extracted_k141_39522 | Not-determined |
| S1_extracted_k141_72736 | Not-determined |
| S1_extracted_k141_72741 | Not-determined |
| S1_extracted_k141_6292 | Not-determined |
| S1_extracted_k141_39538 | Not-determined |
| S1_extracted_k141_6297 | Not-determined |
| S1_extracted_k141_157677 | Not-determined |
| S1_extracted_k141_39550 | Not-determined |
| S1_extracted_k141_6300 | Not-determined |
| S1_extracted_k141_39557 | Not-determined |
| S1_extracted_k141_72757 | Not-determined |
| S1_extracted_k141_157687 | Not-determined |
| S1_extracted_k141_39568 | Not-determined |
| S1_extracted_k141_157693 | Not-determined |
| S1_extracted_k141_157697 | Not-determined |
| S1_extracted_k141_39578 | Not-determined |
| S1_extracted_k141_39579 | Low-quality |
| S1_extracted_k141_72769 | Low-quality |
| S1_extracted_k141_6323 | Not-determined |
| S1_extracted_k141_150557 | Not-determined |
| S1_extracted_k141_6327 | Not-determined |
| S1_extracted_k141_150565 | Low-quality |
| S1_extracted_k141_72774 | Low-quality |
| S1_extracted_k141_6346 | Low-quality |
| S1_extracted_k141_157705 | Not-determined |
| S1_extracted_k141_6348 | Low-quality |
| S1_extracted_k141_157713 | Not-determined |
| S1_extracted_k141_157715 | Not-determined |
| S1_extracted_k141_157723 | Low-quality |
| S1_extracted_k141_150580 | Not-determined |
| S1_extracted_k141_157727 | Low-quality |
| S1_extracted_k141_157728 | Not-determined |
| S1_extracted_k141_157729 | Not-determined |
| S1_extracted_k141_39626 | Not-determined |
| S1_extracted_k141_157733 | Low-quality |
| S1_extracted_k141_157734 | Not-determined |
| S1_extracted_k141_6374 | Not-determined |
| S1_extracted_k141_157736 | Not-determined |
| S1_extracted_k141_6379 | Not-determined |
| S1_extracted_k141_72781 | Low-quality |
| S1_extracted_k141_72784 | Not-determined |
| S1_extracted_k141_72785 | Not-determined |
| S1_extracted_k141_72789 | Not-determined |
| S1_extracted_k141_39644 | Not-determined |
| S1_extracted_k141_157755 | Not-determined |
| S1_extracted_k141_6408 | Low-quality |
| S1_extracted_k141_6412 | Low-quality |
| S1_extracted_k141_39658 | Not-determined |
| S1_extracted_k141_39660 | Not-determined |
| S1_extracted_k141_39661 | Not-determined |
| S1_extracted_k141_39662 | Not-determined |
| S1_extracted_k141_72822 | Not-determined |
| S1_extracted_k141_6429 | Not-determined |
| S1_extracted_k141_39691 | Not-determined |
| S1_extracted_k141_72825 | Not-determined |
| S1_extracted_k141_6432 | Not-determined |
| S1_extracted_k141_39694 | Not-determined |
| S1_extracted_k141_150590 | Not-determined |
| S1_extracted_k141_150592 | Not-determined |
| S1_extracted_k141_39696 | Not-determined |
| S1_extracted_k141_157770 | Low-quality |
| S1_extracted_k141_157776 | Not-determined |
| S1_extracted_k141_157779 | Not-determined |
| S1_extracted_k141_150603 | Not-determined |
| S1_extracted_k141_157784 | Not-determined |
| S1_extracted_k141_150604 | Not-determined |
| S1_extracted_k141_6437 | Not-determined |
| S1_extracted_k141_39708 | Low-quality |
| S1_extracted_k141_6444 | Low-quality |
| S1_extracted_k141_6450 | Not-determined |
| S1_extracted_k141_6451 | Not-determined |
| S1_extracted_k141_157802 | Not-determined |
| S1_extracted_k141_157805 | Low-quality |
| S1_extracted_k141_6457 | Not-determined |
| S1_extracted_k141_157807 | Not-determined |
| S1_extracted_k141_157810 | Not-determined |
| S1_extracted_k141_157816 | Not-determined |
| S1_extracted_k141_6470 | Not-determined |
| S1_extracted_k141_6472 | Not-determined |
| S1_extracted_k141_6473 | Not-determined |
| S1_extracted_k141_6480 | Not-determined |
| S1_extracted_k141_6483 | Low-quality |
| S1_extracted_k141_157820 | Not-determined |
| S1_extracted_k141_157822 | Low-quality |
| S1_extracted_k141_39736 | Not-determined |
| S1_extracted_k141_157825 | Not-determined |
| S1_extracted_k141_39739 | Not-determined |
| S1_extracted_k141_150645 | Low-quality |
| S1_extracted_k141_39742 | Low-quality |
| S1_extracted_k141_39743 | Not-determined |
| S1_extracted_k141_150647 | Not-determined |
| S1_extracted_k141_6519 | Not-determined |
| S1_extracted_k141_150655 | Low-quality |
| S1_extracted_k141_39745 | Low-quality |
| S1_extracted_k141_39750 | Low-quality |
| S1_extracted_k141_6534 | Not-determined |
| S1_extracted_k141_6535 | Not-determined |
| S1_extracted_k141_39755 | Not-determined |
| S1_extracted_k141_6541 | Not-determined |
| S1_extracted_k141_157851 | Not-determined |
| S1_extracted_k141_157852 | Low-quality |
| S1_extracted_k141_157853 | Not-determined |
| S1_extracted_k141_39765 | Not-determined |
| S1_extracted_k141_72875 | Not-determined |
| S1_extracted_k141_157863 | Not-determined |
| S1_extracted_k141_39777 | Low-quality |
| S1_extracted_k141_150666 | Not-determined |
| S1_extracted_k141_157881 | Not-determined |
| S1_extracted_k141_6562 | Not-determined |
| S1_extracted_k141_150673 | Not-determined |
| S1_extracted_k141_6565 | Not-determined |
| S1_extracted_k141_6578 | Not-determined |
| S1_extracted_k141_39804 | Not-determined |
| S1_extracted_k141_157902 | Not-determined |
| S1_extracted_k141_39810 | Not-determined |
| S1_extracted_k141_6597 | Not-determined |
| S1_extracted_k141_157905 | Not-determined |
| S1_extracted_k141_72890 | Not-determined |
| S1_extracted_k141_157913 | Not-determined |
| S1_extracted_k141_157916 | Not-determined |
| S1_extracted_k141_72894 | Not-determined |
| S1_extracted_k141_72902 | Not-determined |
| S1_extracted_k141_39830 | Not-determined |
| S1_extracted_k141_6619 | Low-quality |
| S1_extracted_k141_157920 | Low-quality |
| S1_extracted_k141_6630 | Not-determined |
| S1_extracted_k141_150697 | Not-determined |
| S1_extracted_k141_6636 | Not-determined |
| S1_extracted_k141_157934 | Not-determined |
| S1_extracted_k141_6640 | Not-determined |
| S1_extracted_k141_39853 | Not-determined |
| S1_extracted_k141_6652 | Not-determined |
| S1_extracted_k141_39854 | Not-determined |
| S1_extracted_k141_39855 | Not-determined |
| S1_extracted_k141_157948 | Not-determined |
| S1_extracted_k141_6654 | Low-quality |
| S1_extracted_k141_157955 | Not-determined |
| S1_extracted_k141_39864 | Low-quality |
| S1_extracted_k141_150727 | Low-quality |
| S1_extracted_k141_6668 | Not-determined |
| S1_extracted_k141_157963 | Not-determined |
| S1_extracted_k141_157967 | Not-determined |
| S1_extracted_k141_39879 | Low-quality |
| S1_extracted_k141_39888 | Not-determined |
| S1_extracted_k141_157975 | Not-determined |
| S1_extracted_k141_157977 | Not-determined |
| S1_extracted_k141_150732 | Low-quality |
| S1_extracted_k141_157989 | Not-determined |
| S1_extracted_k141_157993 | Not-determined |
| S1_extracted_k141_157994 | Not-determined |
| S1_extracted_k141_150739 | Not-determined |
| S1_extracted_k141_157995 | Not-determined |
| S1_extracted_k141_157997 | Not-determined |
| S1_extracted_k141_150743 | Not-determined |
| S1_extracted_k141_158001 | Low-quality |
| S1_extracted_k141_39915 | Low-quality |
| S1_extracted_k141_150750 | Not-determined |
| S1_extracted_k141_158009 | Not-determined |
| S1_extracted_k141_39921 | Not-determined |
| S1_extracted_k141_158013 | Not-determined |
| S1_extracted_k141_158014 | Not-determined |
| S1_extracted_k141_150760 | Not-determined |
| S1_extracted_k141_158015 | Not-determined |
| S1_extracted_k141_72932 | Not-determined |
| S1_extracted_k141_158018 | Low-quality |
| S1_extracted_k141_158019 | Not-determined |
| S1_extracted_k141_39933 | Not-determined |
| S1_extracted_k141_158026 | Not-determined |
| S1_extracted_k141_158032 | Not-determined |
| S1_extracted_k141_158033 | Not-determined |
| S1_extracted_k141_39959 | Not-determined |
| S1_extracted_k141_150774 | Not-determined |
| S1_extracted_k141_39970 | Not-determined |
| S1_extracted_k141_158053 | Not-determined |
| S1_extracted_k141_158056 | Not-determined |
| S1_extracted_k141_39986 | Not-determined |
| S1_extracted_k141_39989 | Not-determined |
| S1_extracted_k141_39993 | Not-determined |
| S1_extracted_k141_39994 | Not-determined |
| S1_extracted_k141_72955 | Low-quality |
| S1_extracted_k141_39996 | Low-quality |
| S1_extracted_k141_158071 | Not-determined |
| S1_extracted_k141_158075 | Not-determined |
| S1_extracted_k141_40018 | Low-quality |
| S1_extracted_k141_40019 | Low-quality |
| S1_extracted_k141_158079 | Not-determined |
| S1_extracted_k141_40024 | Low-quality |
| S1_extracted_k141_158087 | Not-determined |
| S1_extracted_k141_40035 | Not-determined |
| S1_extracted_k141_158089 | Not-determined |
| S1_extracted_k141_150796 | Not-determined |
| S1_extracted_k141_158091 | Not-determined |
| S1_extracted_k141_40039 | Not-determined |
| S1_extracted_k141_150799 | Not-determined |
| S1_extracted_k141_150808 | Low-quality |
| S1_extracted_k141_158109 | Not-determined |
| S1_extracted_k141_150812 | Not-determined |
| S1_extracted_k141_150827 | Low-quality |
| S1_extracted_k141_150828 | Not-determined |
| S1_extracted_k141_158122 | Low-quality |
| S1_extracted_k141_150837 | Not-determined |
| S1_extracted_k141_158144 | Low-quality |
| S1_extracted_k141_158147 | Not-determined |
| S1_extracted_k141_150846 | Not-determined |
| S1_extracted_k141_73018 | Not-determined |
| S1_extracted_k141_73020 | Not-determined |
| S1_extracted_k141_158150 | Low-quality |
| S1_extracted_k141_73021 | Low-quality |
| S1_extracted_k141_73025 | Not-determined |
| S1_extracted_k141_158155 | Not-determined |
| S1_extracted_k141_73026 | Low-quality |
| S1_extracted_k141_150859 | Not-determined |
| S1_extracted_k141_158161 | Low-quality |
| S1_extracted_k141_150863 | Not-determined |
| S1_extracted_k141_150864 | Not-determined |
| S1_extracted_k141_150880 | Not-determined |
| S1_extracted_k141_73041 | Not-determined |
| S1_extracted_k141_73051 | Not-determined |
| S1_extracted_k141_73052 | Low-quality |
| S1_extracted_k141_158181 | Low-quality |
| S1_extracted_k141_158182 | Not-determined |
| S1_extracted_k141_73060 | Low-quality |
| S1_extracted_k141_73063 | Low-quality |
| S1_extracted_k141_158193 | Not-determined |
| S1_extracted_k141_150897 | Not-determined |
| S1_extracted_k141_158201 | Not-determined |
| S1_extracted_k141_150898 | Low-quality |
| S1_extracted_k141_150900 | Not-determined |
| S1_extracted_k141_150901 | Not-determined |
| S1_extracted_k141_73071 | Low-quality |
| S1_extracted_k141_73073 | Not-determined |
| S1_extracted_k141_150903 | Not-determined |
| S1_extracted_k141_158208 | Not-determined |
| S1_extracted_k141_73078 | Not-determined |
| S1_extracted_k141_158210 | Not-determined |
| S1_extracted_k141_158212 | Not-determined |
| S1_extracted_k141_150908 | Not-determined |
| S1_extracted_k141_73086 | Low-quality |
| S1_extracted_k141_150914 | Not-determined |
| S1_extracted_k141_73093 | Low-quality |
| S1_extracted_k141_73094 | Not-determined |
| S1_extracted_k141_73095 | Not-determined |
| S1_extracted_k141_73097 | Not-determined |
| S1_extracted_k141_150921 | Not-determined |
| S1_extracted_k141_150922 | Not-determined |
| S1_extracted_k141_158228 | Not-determined |
| S1_extracted_k141_158229 | Low-quality |
| S1_extracted_k141_150928 | Low-quality |
| S1_extracted_k141_150934 | Not-determined |
| S1_extracted_k141_158236 | Low-quality |
| S1_extracted_k141_150937 | Low-quality |
| S1_extracted_k141_150939 | Not-determined |
| S1_extracted_k141_150943 | Not-determined |
| S1_extracted_k141_150947 | Low-quality |
| S1_extracted_k141_150948 | Not-determined |
| S1_extracted_k141_73110 | Low-quality |
| S1_extracted_k141_150950 | Not-determined |
| S1_extracted_k141_158250 | Low-quality |
| S1_extracted_k141_150959 | Low-quality |
| S1_extracted_k141_158265 | Low-quality |
| S1_extracted_k141_158267 | Not-determined |
| S1_extracted_k141_158268 | Low-quality |
| S1_extracted_k141_158271 | Not-determined |
| S1_extracted_k141_73157 | Low-quality |
| S1_extracted_k141_150982 | Not-determined |
| S1_extracted_k141_158279 | Not-determined |
| S1_extracted_k141_73182 | Not-determined |
| S1_extracted_k141_158285 | Low-quality |
| S1_extracted_k141_73186 | Not-determined |
| S1_extracted_k141_73188 | Not-determined |
| S1_extracted_k141_158295 | Not-determined |
| S1_extracted_k141_73194 | Not-determined |
| S1_extracted_k141_151012 | Not-determined |
| S1_extracted_k141_151018 | Not-determined |
| S1_extracted_k141_73202 | Not-determined |
| S1_extracted_k141_73206 | Not-determined |
| S1_extracted_k141_151027 | Not-determined |
| S1_extracted_k141_73216 | Not-determined |
| S1_extracted_k141_151039 | Not-determined |
| S1_extracted_k141_151045 | Low-quality |
| S1_extracted_k141_158345 | Not-determined |
| S1_extracted_k141_151051 | Low-quality |
| S1_extracted_k141_158356 | Low-quality |
| S1_extracted_k141_151057 | Not-determined |
| S1_extracted_k141_158369 | Not-determined |
| S1_extracted_k141_158374 | Not-determined |
| S1_extracted_k141_158383 | Not-determined |
| S1_extracted_k141_158394 | Not-determined |
| S1_extracted_k141_73278 | Not-determined |
| S1_extracted_k141_158403 | Low-quality |
| S1_extracted_k141_73284 | Not-determined |
| S1_extracted_k141_73286 | Not-determined |
| S1_extracted_k141_73294 | Not-determined |
| S1_extracted_k141_73308 | Not-determined |
| S1_extracted_k141_158420 | Not-determined |
| S1_extracted_k141_158421 | Low-quality |
| S1_extracted_k141_158424 | Not-determined |
| S1_extracted_k141_158428 | Not-determined |
| S1_extracted_k141_158429 | Not-determined |
| S1_extracted_k141_151076 | Not-determined |
| S1_extracted_k141_158431 | Not-determined |
| S1_extracted_k141_73314 | Not-determined |
| S1_extracted_k141_151090 | Not-determined |
| S1_extracted_k141_151096 | Not-determined |
| S1_extracted_k141_151102 | Not-determined |
| S1_extracted_k141_151106 | Not-determined |
| S1_extracted_k141_151108 | Not-determined |
| S1_extracted_k141_158454 | Not-determined |
| S1_extracted_k141_158456 | Not-determined |
| S1_extracted_k141_158458 | Low-quality |
| S1_extracted_k141_151119 | Not-determined |
| S1_extracted_k141_158462 | Not-determined |
| S1_extracted_k141_158463 | Not-determined |
| S1_extracted_k141_158466 | Not-determined |
| S1_extracted_k141_151127 | Low-quality |
| S1_extracted_k141_151130 | Not-determined |
| S1_extracted_k141_151134 | Low-quality |
| S1_extracted_k141_151140 | Not-determined |
| S1_extracted_k141_158474 | Not-determined |
| S1_extracted_k141_158477 | Not-determined |
| S1_extracted_k141_151161 | Low-quality |
| S1_extracted_k141_151163 | Not-determined |
| S1_extracted_k141_73345 | Not-determined |
| S1_extracted_k141_73348 | Not-determined |
| S1_extracted_k141_158494 | Not-determined |
| S1_extracted_k141_158496 | Not-determined |
| S1_extracted_k141_73362 | Not-determined |
| S1_extracted_k141_73365 | Low-quality |
| S1_extracted_k141_151183 | Not-determined |
| S1_extracted_k141_73376 | Not-determined |
| S1_extracted_k141_73384 | Not-determined |
| S1_extracted_k141_73396 | Not-determined |
| S1_extracted_k141_73397 | Not-determined |
| S1_extracted_k141_158506 | Not-determined |
| S1_extracted_k141_151197 | Not-determined |
| S1_extracted_k141_151202 | Not-determined |
| S1_extracted_k141_151206 | Not-determined |
| S1_extracted_k141_151207 | Not-determined |
| S1_extracted_k141_151209 | Low-quality |
| S1_extracted_k141_158520 | Low-quality |
| S1_extracted_k141_73416 | Not-determined |
| S1_extracted_k141_151237 | Not-determined |
| S1_extracted_k141_151238 | Low-quality |
| S1_extracted_k141_158531 | Not-determined |
| S1_extracted_k141_151250 | Not-determined |
| S1_extracted_k141_151253 | Not-determined |
| S1_extracted_k141_151257 | Not-determined |
| S1_extracted_k141_158542 | Low-quality |
| S1_extracted_k141_158544 | Not-determined |
| S1_extracted_k141_151271 | Not-determined |
| S1_extracted_k141_158564 | Not-determined |
| S1_extracted_k141_158565 | Not-determined |
| S1_extracted_k141_158566 | Not-determined |
| S1_extracted_k141_158569 | Low-quality |
| S1_extracted_k141_158575 | Low-quality |
| S1_extracted_k141_158583 | Low-quality |
| S1_extracted_k141_151294 | Not-determined |
| S1_extracted_k141_151298 | Not-determined |
| S1_extracted_k141_151301 | Not-determined |
| S1_extracted_k141_158587 | Not-determined |
| S1_extracted_k141_158594 | Not-determined |
| S1_extracted_k141_158598 | Not-determined |
| S1_extracted_k141_158602 | Low-quality |
| S1_extracted_k141_158603 | Not-determined |
| S1_extracted_k141_158612 | Not-determined |
| S1_extracted_k141_158613 | Low-quality |
| S1_extracted_k141_158617 | Low-quality |
| S1_extracted_k141_158619 | Not-determined |
| S1_extracted_k141_158628 | Low-quality |
| S1_extracted_k141_158632 | Low-quality |
| S1_extracted_k141_151321 | Not-determined |
| S1_extracted_k141_158636 | Not-determined |
| S1_extracted_k141_151339 | Not-determined |
| S1_extracted_k141_151342 | Low-quality |
| S1_extracted_k141_151348 | Not-determined |
| S1_extracted_k141_151370 | Not-determined |
| S1_extracted_k141_151375 | Low-quality |
| S1_extracted_k141_158666 | Not-determined |
| S1_extracted_k141_158669 | Not-determined |
| S1_extracted_k141_151391 | Not-determined |
| S1_extracted_k141_158687 | Not-determined |
| S1_extracted_k141_158690 | Not-determined |
| S1_extracted_k141_158693 | Not-determined |
| S1_extracted_k141_158695 | Low-quality |
| S1_extracted_k141_158701 | Not-determined |
| S1_extracted_k141_151416 | Not-determined |
| S1_extracted_k141_151423 | Not-determined |
| S1_extracted_k141_151426 | Not-determined |
| S1_extracted_k141_151438 | Not-determined |
| S1_extracted_k141_158721 | Low-quality |
| S1_extracted_k141_151478 | Not-determined |
| S1_extracted_k141_151485 | Not-determined |
| S1_extracted_k141_151492 | Not-determined |
| S1_extracted_k141_151494 | Not-determined |
| S1_extracted_k141_158769 | Low-quality |
| S1_extracted_k141_158780 | Not-determined |
| S1_extracted_k141_158782 | Not-determined |
| S1_extracted_k141_158785 | Not-determined |
| S1_extracted_k141_151514 | Not-determined |
| S1_extracted_k141_158798 | Not-determined |
| S1_extracted_k141_151548 | Low-quality |
| S1_extracted_k141_151559 | Not-determined |
| S1_extracted_k141_151564 | Not-determined |
| S1_extracted_k141_158809 | Not-determined |
| S1_extracted_k141_158812 | Low-quality |
| S1_extracted_k141_151569 | Low-quality |
| S1_extracted_k141_151585 | Not-determined |
| S1_extracted_k141_151587 | Not-determined |
| S1_extracted_k141_151604 | Low-quality |
| S1_extracted_k141_151606 | Not-determined |
| S1_extracted_k141_151613 | Not-determined |
| S1_extracted_k141_151614 | Not-determined |
| S1_extracted_k141_158843 | Not-determined |
| S1_extracted_k141_151624 | Low-quality |
| S1_extracted_k141_158860 | Low-quality |
| S1_extracted_k141_158871 | Low-quality |
| S1_extracted_k141_158873 | Low-quality |
| S1_extracted_k141_158874 | Not-determined |
| S1_extracted_k141_151639 | Not-determined |
| S1_extracted_k141_158880 | Low-quality |
| S1_extracted_k141_158882 | Low-quality |
| S1_extracted_k141_158890 | Not-determined |
| S1_extracted_k141_151672 | Not-determined |

| | |
|---|---|
| S1_extracted_k141_151678 | Not-determined |
| S1_extracted_k141_158902 | Low-quality |
| S1_extracted_k141_151687 | Not-determined |
| S1_extracted_k141_151688 | Low-quality |
| S1_extracted_k141_158914 | Low-quality |
| S1_extracted_k141_158915 | Not-determined |
| S1_extracted_k141_151696 | Not-determined |
| S1_extracted_k141_151702 | Low-quality |
| S1_extracted_k141_151703 | Not-determined |
| S1_extracted_k141_158926 | Low-quality |
| S1_extracted_k141_158929 | Not-determined |
| S1_extracted_k141_158931 | Not-determined |
| S1_extracted_k141_158935 | Low-quality |
| S1_extracted_k141_158941 | Not-determined |
| S1_extracted_k141_158945 | Low-quality |
| S1_extracted_k141_158946 | Not-determined |
| S1_extracted_k141_151708 | Not-determined |
| S1_extracted_k141_151724 | Not-determined |
| S1_extracted_k141_151725 | Not-determined |
| S1_extracted_k141_151745 | Not-determined |
| S1_extracted_k141_151748 | Not-determined |
| S1_extracted_k141_151760 | Low-quality |
| S1_extracted_k141_151765 | Not-determined |
| S1_extracted_k141_151774 | Not-determined |
| S1_extracted_k141_151780 | Not-determined |
| S1_extracted_k141_151785 | Low-quality |
| S13_vs_151995\|\|full | High-quality |
| S1_extracted_k141_158977 | Not-determined |
| S1_extracted_k141_158985 | Not-determined |
| S1_extracted_k141_151812 | Not-determined |
| S1_extracted_k141_151818 | Not-determined |
| S1_extracted_k141_151828 | Not-determined |
| S1_extracted_k141_159001 | Low-quality |
| S1_extracted_k141_151846 | Not-determined |
| S1_extracted_k141_151854 | Not-determined |
| S1_extracted_k141_159037 | Not-determined |
| S1_extracted_k141_159047 | Not-determined |
| S1_extracted_k141_151875 | Not-determined |
| S1_extracted_k141_159058 | Not-determined |
| S1_extracted_k141_151886 | Not-determined |
| S1_extracted_k141_151893 | Not-determined |
| S1_extracted_k141_159069 | Not-determined |
| S1_extracted_k141_151898 | Not-determined |
| S1_extracted_k141_159075 | Low-quality |
| S1_extracted_k141_159085 | Not-determined |
| S1_extracted_k141_159088 | Not-determined |
| S1_extracted_k141_159097 | Not-determined |
| S1_extracted_k141_159100 | Low-quality |
| S1_extracted_k141_151931 | Not-determined |
| S1_extracted_k141_151941 | Not-determined |
| S1_extracted_k141_159111 | Not-determined |
| S1_extracted_k141_151951 | Low-quality |
| S1_extracted_k141_159113 | Low-quality |
| S1_extracted_k141_159114 | Low-quality |
| S1_extracted_k141_151968 | Not-determined |
| S1_extracted_k141_151969 | Not-determined |
| S1_extracted_k141_151973 | Low-quality |
| S1_extracted_k141_151979 | Not-determined |
| S1_extracted_k141_159144 | Not-determined |
| S1_extracted_k141_159145 | Not-determined |
| S1_extracted_k141_159152 | Not-determined |
| S1_extracted_k141_151985 | Not-determined |
| S1_extracted_k141_151987 | Not-determined |
| S1_extracted_k141_151993 | Low-quality |
| S1_extracted_k141_151995 | Not-determined |
| S1_extracted_k141_159177 | Low-quality |
| S1_extracted_k141_152012 | Not-determined |
| S1_extracted_k141_159185 | Not-determined |
| S1_extracted_k141_152033 | Not-determined |
| S1_extracted_k141_152037 | Not-determined |
| S1_extracted_k141_159194 | Not-determined |
| S1_extracted_k141_152041 | Not-determined |
| S1_extracted_k141_159201 | Not-determined |
| S1_extracted_k141_152046 | Not-determined |
| S1_extracted_k141_159202 | Not-determined |
| S1_extracted_k141_152061 | Low-quality |
| S1_extracted_k141_152064 | Not-determined |
| S1_extracted_k141_152069 | Not-determined |
| S1_extracted_k141_152071 | Not-determined |
| S1_extracted_k141_152086 | Low-quality |
| S1_extracted_k141_159223 | Low-quality |
| S1_extracted_k141_152094 | Not-determined |
| S1_extracted_k141_152102 | Not-determined |
| S1_extracted_k141_159234 | Not-determined |
| S1_extracted_k141_159235 | Low-quality |
| S1_extracted_k141_159237 | Low-quality |
| S1_extracted_k141_152109 | Low-quality |
| S1_extracted_k141_152112 | Low-quality |
| S1_extracted_k141_159248 | Low-quality |
| S1_extracted_k141_159259 | Not-determined |
| S1_extracted_k141_159261 | Not-determined |
| S1_extracted_k141_152146 | Low-quality |
| S1_extracted_k141_152171 | Not-determined |
| S1_extracted_k141_159279 | Not-determined |
| S1_extracted_k141_152176 | Not-determined |
| S1_extracted_k141_152178 | Low-quality |
| S1_extracted_k141_152182 | Not-determined |
| S1_extracted_k141_159292 | Not-determined |
| S1_extracted_k141_152189 | Not-determined |
| S1_extracted_k141_152193 | Not-determined |
| S1_extracted_k141_159308 | Not-determined |
| S1_extracted_k141_159309 | Low-quality |
| S1_extracted_k141_159311 | Not-determined |
| S1_extracted_k141_159313 | Not-determined |
| S1_extracted_k141_159316 | Low-quality |
| S1_extracted_k141_159322 | Not-determined |
| S1_extracted_k141_152213 | Not-determined |
| S1_extracted_k141_152215 | Not-determined |
| S1_extracted_k141_159329 | Not-determined |
| S1_extracted_k141_152224 | Not-determined |
| S1_extracted_k141_152230 | Low-quality |
| S1_extracted_k141_159333 | Not-determined |
| S1_extracted_k141_152235 | Low-quality |
| S1_extracted_k141_152236 | Not-determined |
| S1_extracted_k141_152237 | Low-quality |
| S1_extracted_k141_159340 | Low-quality |
| S1_extracted_k141_159342 | Not-determined |
| S1_extracted_k141_152254 | Low-quality |
| S1_extracted_k141_152267 | Not-determined |
| S1_extracted_k141_152268 | Not-determined |
| S1_extracted_k141_159376 | Low-quality |
| S1_extracted_k141_152275 | Not-determined |
| S1_extracted_k141_152291 | Not-determined |
| S1_extracted_k141_152294 | Low-quality |
| S1_extracted_k141_152296 | Low-quality |
| S1_extracted_k141_152298 | Not-determined |
| S1_extracted_k141_152301 | Low-quality |
| S1_extracted_k141_159418 | Not-determined |
| S1_extracted_k141_152308 | Not-determined |
| S1_extracted_k141_159422 | Not-determined |
| S1_extracted_k141_159423 | Low-quality |
| S1_extracted_k141_159425 | Low-quality |
| S1_extracted_k141_159435 | Low-quality |
| S1_extracted_k141_152328 | Not-determined |
| S1_extracted_k141_152329 | Not-determined |
| S1_extracted_k141_159451 | Not-determined |
| S1_extracted_k141_152340 | Low-quality |
| S1_extracted_k141_159458 | Not-determined |
| S1_extracted_k141_159464 | Not-determined |
| S1_extracted_k141_152351 | Not-determined |
| S1_extracted_k141_152357 | Not-determined |
| S1_extracted_k141_159479 | Not-determined |
| S1_extracted_k141_159483 | Not-determined |
| S1_extracted_k141_159486 | Low-quality |
| S1_extracted_k141_152364 | Not-determined |
| S1_extracted_k141_159491 | Low-quality |
| S1_extracted_k141_152377 | Low-quality |
| S1_extracted_k141_152378 | Not-determined |
| S1_extracted_k141_159496 | Not-determined |
| S1_extracted_k141_152382 | Not-determined |
| S1_extracted_k141_159520 | Not-determined |
| S1_extracted_k141_159523 | Not-determined |
| S1_extracted_k141_159524 | Not-determined |
| S1_extracted_k141_159525 | Not-determined |
| S1_extracted_k141_159530 | Not-determined |
| S1_extracted_k141_159543 | Not-determined |
| S1_extracted_k141_159545 | Not-determined |
| S1_extracted_k141_159546 | Low-quality |
| S1_extracted_k141_159548 | Not-determined |
| S1_extracted_k141_152446 | Not-determined |
| S1_extracted_k141_152448 | Not-determined |
| S1_extracted_k141_152452 | Not-determined |
| S1_extracted_k141_152454 | Not-determined |
| S1_extracted_k141_152458 | Not-determined |
| S1_extracted_k141_152461 | Not-determined |
| S1_extracted_k141_152485 | Not-determined |
| S1_extracted_k141_152496 | Not-determined |
| S8_vs_k141_143411\|\|full | High-quality |
| S1_extracted_k141_152522 | Not-determined |
| S1_extracted_k141_159578 | Not-determined |
| S1_extracted_k141_159579 | Not-determined |
| S1_extracted_k141_159585 | Not-determined |
| S1_extracted_k141_152564 | Not-determined |
| S1_extracted_k141_152565 | Not-determined |
| S1_extracted_k141_152568 | Not-determined |
| S1_extracted_k141_159589 | Not-determined |
| S1_extracted_k141_152570 | Not-determined |
| S1_extracted_k141_152571 | Not-determined |
| S1_extracted_k141_152577 | Not-determined |
| S1_extracted_k141_152585 | Not-determined |
| S1_extracted_k141_152594 | Low-quality |
| S1_extracted_k141_159613 | Not-determined |
| S1_extracted_k141_152603 | Low-quality |
| S1_extracted_k141_152613 | Low-quality |
| S1_extracted_k141_152615 | Not-determined |
| S1_extracted_k141_152616 | Not-determined |
| S1_extracted_k141_152618 | Not-determined |
| S1_extracted_k141_159626 | Not-determined |
| S1_extracted_k141_159629 | Not-determined |
| S1_extracted_k141_152626 | Not-determined |
| S1_extracted_k141_152657 | Not-determined |
| S1_extracted_k141_152662 | Low-quality |
| S1_extracted_k141_152663 | Not-determined |
| S1_extracted_k141_159650 | Low-quality |
| S1_extracted_k141_159651 | Not-determined |
| S1_extracted_k141_159657 | Not-determined |
| S1_extracted_k141_159659 | Not-determined |
| S1_extracted_k141_159660 | Not-determined |
| S1_extracted_k141_159667 | Not-determined |
| S1_extracted_k141_152680 | Low-quality |
| S1_extracted_k141_152682 | Not-determined |
| S1_extracted_k141_159668 | Not-determined |
| S1_extracted_k141_152684 | Not-determined |
| S1_extracted_k141_159671 | Not-determined |
| S1_extracted_k141_152687 | Not-determined |
| S1_extracted_k141_152689 | Not-determined |
| S1_extracted_k141_152694 | Not-determined |
| S1_extracted_k141_159673 | Not-determined |
| S1_extracted_k141_152699 | Not-determined |
| S1_extracted_k141_159678 | Low-quality |
| S1_extracted_k141_159680 | Not-determined |
| S1_extracted_k141_152707 | Low-quality |
| S1_extracted_k141_152713 | Not-determined |
| S1_extracted_k141_152714 | Not-determined |
| S1_extracted_k141_152716 | Low-quality |
| S1_extracted_k141_152717 | Not-determined |
| S1_extracted_k141_159689 | Not-determined |
| S1_extracted_k141_152731 | Not-determined |
| S1_extracted_k141_152733 | Not-determined |
| S1_extracted_k141_159694 | Not-determined |
| S1_extracted_k141_159704 | Not-determined |
| S1_extracted_k141_159706 | Not-determined |
| S1_extracted_k141_152750 | Low-quality |
| S1_extracted_k141_159709 | Not-determined |
| S1_extracted_k141_159720 | Not-determined |
| S1_extracted_k141_159731 | Not-determined |
| S1_extracted_k141_152777 | Not-determined |
| S1_extracted_k141_152783 | Not-determined |
| S1_extracted_k141_152786 | Not-determined |
| S1_extracted_k141_152790 | Low-quality |
| S1_extracted_k141_159746 | Low-quality |
| S1_extracted_k141_152792 | Low-quality |
| S1_extracted_k141_152797 | Not-determined |
| S1_extracted_k141_159749 | Not-determined |
| S1_extracted_k141_152806 | Not-determined |
| S1_extracted_k141_152807 | Low-quality |
| S1_extracted_k141_159759 | Not-determined |
| S1_extracted_k141_159766 | Not-determined |
| S1_extracted_k141_152821 | Not-determined |
| S1_extracted_k141_159772 | Not-determined |
| S1_extracted_k141_159782 | Not-determined |
| S1_extracted_k141_152834 | Not-determined |
| S1_extracted_k141_159785 | Not-determined |
| S1_extracted_k141_159788 | Not-determined |
| S1_extracted_k141_159789 | Low-quality |
| S1_extracted_k141_159791 | Not-determined |
| S1_extracted_k141_152840 | Not-determined |
| S1_extracted_k141_159795 | Not-determined |
| S1_extracted_k141_152851 | Not-determined |
| S1_extracted_k141_159802 | Not-determined |
| S1_extracted_k141_159810 | Not-determined |
| S1_extracted_k141_152867 | Not-determined |
| S1_extracted_k141_159821 | Low-quality |
| S1_extracted_k141_159823 | Not-determined |
| S1_extracted_k141_152878 | Not-determined |
| S1_extracted_k141_159824 | Low-quality |
| S1_extracted_k141_159825 | Not-determined |
| S1_extracted_k141_159832 | Not-determined |
| S1_extracted_k141_159839 | Not-determined |
| S1_extracted_k141_152882 | Not-determined |
| S1_extracted_k141_159853 | Not-determined |
| S1_extracted_k141_152902 | Not-determined |
| S1_extracted_k141_152910 | Not-determined |
| S1_extracted_k141_159874 | Low-quality |
| S1_extracted_k141_159875 | Not-determined |
| S1_extracted_k141_159877 | Low-quality |
| S1_extracted_k141_159880 | Not-determined |
| S1_extracted_k141_152922 | Not-determined |
| S1_extracted_k141_159895 | Not-determined |
| S1_extracted_k141_159901 | Not-determined |
| S1_extracted_k141_152948 | Not-determined |
| S1_extracted_k141_159917 | Not-determined |
| S1_extracted_k141_159921 | Low-quality |
| S1_extracted_k141_159925 | Low-quality |
| S1_extracted_k141_159947 | Not-determined |
| S1_extracted_k141_159948 | Not-determined |
| S1_extracted_k141_153020 | Not-determined |
| S1_extracted_k141_153033 | Not-determined |
| S1_extracted_k141_153034 | Low-quality |
| S1_extracted_k141_153052 | Not-determined |
| S1_extracted_k141_153077 | Low-quality |
| S1_extracted_k141_153088 | Not-determined |
| S1_extracted_k141_153109 | Not-determined |
| S1_extracted_k141_153121 | Not-determined |
| S1_extracted_k141_153122 | Low-quality |
| S1_extracted_k141_153133 | Not-determined |
| S1_extracted_k141_153137 | Not-determined |
| S7_vs_103530\|\|full | High-quality |
| S1_extracted_k141_159952 | Not-determined |
| S1_extracted_k141_159957 | Not-determined |
| S1_extracted_k141_153170 | Not-determined |
| S1_extracted_k141_153184 | Not-determined |
| S1_extracted_k141_153189 | Not-determined |
| S1_extracted_k141_153198 | Low-quality |
| S1_extracted_k141_153200 | Not-determined |
| S1_extracted_k141_153228 | Not-determined |
| S1_extracted_k141_153230 | Not-determined |
| S1_extracted_k141_153237 | Not-determined |
| S1_extracted_k141_159959 | Not-determined |
| S1_extracted_k141_159962 | Not-determined |
| S1_extracted_k141_159964 | Not-determined |
| S1_extracted_k141_153245 | Not-determined |
| S1_extracted_k141_159966 | Not-determined |
| S1_extracted_k141_159969 | Not-determined |
| S1_extracted_k141_159971 | Not-determined |
| S1_extracted_k141_159975 | Not-determined |
| S1_extracted_k141_159985 | Not-determined |
| S1_extracted_k141_159986 | Not-determined |
| S1_extracted_k141_153252 | Not-determined |
| S1_extracted_k141_153257 | Not-determined |
| S1_extracted_k141_159996 | Low-quality |
| S1_extracted_k141_160000 | Not-determined |
| S1_extracted_k141_160002 | Not-determined |
| S1_extracted_k141_153268 | Not-determined |
| S1_extracted_k141_153274 | Not-determined |
| S1_extracted_k141_153279 | Not-determined |
| S1_extracted_k141_153282 | Not-determined |
| S1_extracted_k141_153296 | Low-quality |
| S1_extracted_k141_160049 | Not-determined |
| S1_extracted_k141_160053 | Not-determined |
| S1_extracted_k141_160054 | Not-determined |
| S1_extracted_k141_153299 | Not-determined |
| S1_extracted_k141_160061 | Not-determined |
| S1_extracted_k141_160068 | Not-determined |
| S1_extracted_k141_160070 | Not-determined |
| S1_extracted_k141_160082 | Not-determined |
| S1_extracted_k141_153309 | Not-determined |
| S1_extracted_k141_160083 | Not-determined |
| S1_extracted_k141_160089 | Not-determined |
| S1_extracted_k141_160090 | Not-determined |
| S1_extracted_k141_160091 | Low-quality |
| S1_extracted_k141_153318 | Not-determined |
| S1_extracted_k141_160095 | Not-determined |
| S1_extracted_k141_160105 | Not-determined |
| S1_extracted_k141_160106 | Not-determined |
| S1_extracted_k141_160109 | Not-determined |
| S1_extracted_k141_160115 | Not-determined |
| S1_extracted_k141_160116 | Not-determined |
| S1_extracted_k141_160118 | Not-determined |
| S1_extracted_k141_160123 | Not-determined |
| S1_extracted_k141_160125 | Not-determined |
| S1_extracted_k141_160135 | Not-determined |
| S1_extracted_k141_160137 | Not-determined |
| S1_extracted_k141_160139 | Not-determined |
| S1_extracted_k141_153338 | Not-determined |
| S1_extracted_k141_160151 | Not-determined |
| S1_extracted_k141_160152 | Not-determined |
| S1_extracted_k141_160154 | Not-determined |
| S1_extracted_k141_160158 | Not-determined |
| S1_extracted_k141_160160 | Not-determined |
| S1_extracted_k141_160165 | Not-determined |
| S1_extracted_k141_160166 | Not-determined |
| S1_extracted_k141_160168 | Not-determined |
| S1_extracted_k141_160173 | Not-determined |
| S1_extracted_k141_160176 | Not-determined |
| S1_extracted_k141_160180 | Not-determined |
| S1_extracted_k141_160192 | Not-determined |
| S1_extracted_k141_160193 | Not-determined |
| S1_extracted_k141_153373 | Not-determined |
| S1_extracted_k141_153378 | Not-determined |
| S1_extracted_k141_153385 | Low-quality |
| S1_extracted_k141_153386 | Not-determined |
| S1_extracted_k141_153387 | Not-determined |
| S1_extracted_k141_153388 | Not-determined |
| S1_extracted_k141_153389 | Not-determined |
| S1_extracted_k141_153391 | Not-determined |
| S1_extracted_k141_153397 | Not-determined |
| S1_extracted_k141_153398 | Not-determined |
| S1_extracted_k141_153419 | Not-determined |
| S1_extracted_k141_153423 | Not-determined |
| S1_extracted_k141_153425 | Not-determined |
| S1_extracted_k141_153456 | Not-determined |
| S1_extracted_k141_153464 | Not-determined |
| S1_extracted_k141_153465 | Low-quality |
| S1_extracted_k141_153493 | Not-determined |
| S1_extracted_k141_153500 | Not-determined |
| S1_extracted_k141_153508 | Not-determined |
| S5_extracted_k141_122568 | Complete |
| S9_vs_k141_74235\|\|full | Medium-quality |
| S1_vs_k141_143203\|\|full | Low-quality |
| S7_extracted_37293 | High-quality |
| S14_vs_k141_78139\|\|full | High-quality |
| S5_extracted_k141_80727 | Medium-quality |
| S7_vs_37293\|\|full | High-quality |
| S1_vs_k141_159991\|\|full | Not-determined |
| S1_vs_k141_66750\|\|full | Low-quality |
| S1_vs_k141_66751\|\|full | Not-determined |
| S1_vs_k141_100131\|\|full | Not-determined |
| S1_vs_k141_140178\|\|full | Low-quality |
| S1_vs_k141_100132\|\|full | Not-determined |
| S1_vs_k141_153523\|\|full | Not-determined |
| S1_vs_k141_26704\|\|full | Not-determined |
| S1_vs_k141_93456\|\|full | Not-determined |
| S1_vs_k141_17\|\|full | Low-quality |
| S1_vs_k141_80114\|\|full | Not-determined |
| S1_vs_k141_120162\|\|full | Low-quality |
| S1_vs_k141_133507\|\|full | Low-quality |
| S1_vs_k141_126830\|\|full | Not-determined |
| S1_vs_k141_53416\|\|full | Low-quality |
| S1_vs_k141_93468\|\|full | Low-quality |
| S1_vs_k141_153535\|\|full | Low-quality |
| S1_vs_k141_100148\|\|full | Low-quality |
| S1_vs_k141_80121\|\|full | Low-quality |
| S1_vs_k141_146858\|\|full | Not-determined |
| S1_vs_k141_126840\|\|full | Low-quality |
| S1_vs_k141_146867\|\|full | Low-quality |
| S1_vs_k141_133524\|\|full | Low-quality |
| S1_vs_k141_66760\|\|full | Low-quality |
| S1_vs_k141_33401\|\|full | Low-quality |
| S1_vs_k141_33403\|\|full | Not-determined |
| S1_vs_k141_113503\|\|full | Low-quality |
| S1_vs_k141_113507\|\|full | Low-quality |
| S1_vs_k141_93486\|\|full | Low-quality |
| S1_vs_k141_126860\|\|full | Low-quality |
| S1_vs_k141_86808\|\|full | Not-determined |
| S1_vs_k141_26723\|\|full | Not-determined |
| S1_vs_k141_153556\|\|full | Not-determined |
| S1_vs_k141_26726\|\|full | Low-quality |
| S1_vs_k141_133544\|\|full | Not-determined |
| S1_vs_k141_73449\|\|full | Not-determined |
| S1_vs_k141_20064\|\|full | Not-determined |
| S1_vs_k141_140219\|\|full | Low-quality |
| S1_vs_k141_93492\|\|full | Low-quality |
| S1_vs_k141_113515\|\|full | Not-determined |
| S1_vs_k141_146876\|\|full | Not-determined |
| S1_vs_k141_20066\|\|full | Not-determined |
| S1_vs_k141_66774\|\|full | Low-quality |
| S1_vs_k141_153570\|\|full | Not-determined |
| S1_vs_k141_20077\|\|full | Low-quality |
| S1_vs_k141_46789\|\|full | Low-quality |
| S1_vs_k141_153575\|\|full | Not-determined |
| S1_vs_k141_120200\|\|full | Not-determined |
| S1_vs_k141_146884\|\|full | Low-quality |
| S5_vs_k141_122568\|\|full | High-quality |
| S1_vs_k141_106864\|\|full | Low-quality |
| S1_vs_k141_20078\|\|full | Low-quality |
| S1_vs_k141_126895\|\|full | Low-quality |
| S1_vs_k141_6753\|\|full | Low-quality |
| S1_vs_k141_133562\|\|full | Not-determined |
| S1_vs_k141_60093\|\|full | Low-quality |
| S1_vs_k141_93521\|\|full | Low-quality |
| S1_vs_k141_13388\|\|full | Low-quality |
| S1_vs_k141_53478\|\|full | Low-quality |
| S1_vs_k141_153586\|\|full | Low-quality |
| S1_vs_k141_80190\|\|full | Low-quality |
| S1_vs_k141_140255\|\|full | Low-quality |
| S1_vs_k141_26758\|\|full | Low-quality |
| S1_vs_k141_66810\|\|full | Low-quality |
| S12_vs_k141_88085\|\|full | High-quality |
| S1_vs_k141_126918\|\|full | Low-quality |
| S1_vs_k141_20102\|\|full | Low-quality |
| S1_vs_k141_113552\|\|full | Not-determined |
| S1_vs_k141_40097\|\|full | Low-quality |
| S1_vs_k141_73491\|\|full | Not-determined |
| S1_vs_k141_110\|\|full | Not-determined |
| S1_vs_k141_46816\|\|full | Low-quality |
| S1_vs_k141_86859\|\|full | Not-determined |
| S1_vs_k141_66832\|\|full | Not-determined |
| S1_vs_k141_33461\|\|full | Low-quality |
| S1_vs_k141_73502\|\|full | Not-determined |
| S1_vs_k141_93560\|\|full | Not-determined |
| S1_vs_k141_146922\|\|full | Low-quality |
| S1_vs_k141_46821\|\|full | Not-determined |
| S1_vs_k141_6806\|\|full | Low-quality |
| S1_vs_k141_66837\|\|full | Not-determined |
| S1_vs_k141_126944\|\|full | Low-quality |
| S1_vs_k141_100202\|\|full | Low-quality |
| S1_vs_k141_73513\|\|full | Not-determined |
| S1_vs_k141_6812\|\|full | Low-quality |
| S1_vs_k141_153593\|\|full | Low-quality |
| S1_vs_k141_113585\|\|full | Not-determined |
| S1_vs_k141_124\|\|full | Low-quality |
| S1_vs_k141_46837\|\|full | Not-determined |
| S1_vs_k141_53540\|\|full | Low-quality |
| S1_vs_k141_126961\|\|full | Not-determined |
| S1_vs_k141_120266\|\|full | Low-quality |
| S1_vs_k141_66853\|\|full | Low-quality |
| S1_vs_k141_66855\|\|full | Low-quality |
| S1_vs_k141_106912\|\|full | Not-determined |
| S1_vs_k141_140306\|\|full | Not-determined |
| S1_vs_k141_20127\|\|full | Low-quality |
| S1_vs_k141_80245\|\|full | Not-determined |
| S1_vs_k141_26788\|\|full | Low-quality |
| S1_vs_k141_126967\|\|full | Not-determined |
| S1_vs_k141_46856\|\|full | Low-quality |
| S13_vs_23314\|\|full | High-quality |
| S1_vs_k141_73542\|\|full | Not-determined |
| S1_vs_k141_13488\|\|full | Not-determined |
| S1_vs_k141_133581\|\|full | Low-quality |
| S1_vs_k141_120299\|\|full | Low-quality |
| S1_vs_k141_20142\|\|full | Low-quality |
| S1_vs_k141_80273\|\|full | Not-determined |
| S1_vs_k141_33507\|\|full | Low-quality |
| S1_vs_k141_53569\|\|full | Low-quality |
| S1_vs_k141_113634\|\|full | Not-determined |
| S1_vs_k141_26819\|\|full | Not-determined |
| S1_vs_k141_66882\|\|full | Low-quality |
| S1_vs_k141_80290\|\|full | Not-determined |
| S1_vs_k141_146964\|\|full | Low-quality |
| S1_vs_k141_53574\|\|full | Low-quality |
| S1_vs_k141_120320\|\|full | Low-quality |
| S1_vs_k141_140340\|\|full | Low-quality |
| S1_vs_k141_86950\|\|full | Low-quality |
| S1_vs_k141_100277\|\|full | Low-quality |
| S1_vs_k141_66889\|\|full | Not-determined |
| S1_vs_k141_40202\|\|full | Low-quality |
| S1_vs_k141_189\|\|full | Low-quality |
| S1_vs_k141_73579\|\|full | Not-determined |
| S1_vs_k141_86958\|\|full | Low-quality |
| S1_vs_k141_106971\|\|full | Low-quality |
| S1_vs_k141_140356\|\|full | Low-quality |
| S1_vs_k141_204\|\|full | Not-determined |
| S1_vs_k141_140357\|\|full | Low-quality |
| S1_vs_k141_146971\|\|full | Low-quality |
| S1_vs_k141_100297\|\|full | Low-quality |
| S1_vs_k141_140360\|\|full | Low-quality |
| S1_vs_k141_127040\|\|full | Low-quality |
| S1_vs_k141_93690\|\|full | Low-quality |
| S1_vs_k141_20215\|\|full | Low-quality |
| S1_vs_k141_6928\|\|full | Low-quality |
| S1_vs_k141_86995\|\|full | Low-quality |
| S1_vs_k141_33559\|\|full | Low-quality |
| S1_vs_k141_66944\|\|full | Low-quality |
| S1_vs_k141_6945\|\|full | Low-quality |
| S1_vs_k141_33561\|\|full | Low-quality |
| S1_vs_k141_146998\|\|full | Low-quality |
| S1_vs_k141_147001\|\|full | Low-quality |
| S1_vs_k141_33571\|\|full | Low-quality |
| S1_vs_k141_33578\|\|full | Low-quality |
| S1_vs_k141_53655\|\|full | Not-determined |
| S1_vs_k141_40277\|\|full | Low-quality |
| S1_vs_k141_20257\|\|full | Not-determined |
| S1_vs_k141_40284\|\|full | Not-determined |
| S1_vs_k141_120423\|\|full | Low-quality |
| S1_vs_k141_26863\|\|full | Low-quality |
| S1_vs_k141_93728\|\|full | Not-determined |
| S1_vs_k141_100352\|\|full | Low-quality |
| S1_vs_k141_133659\|\|full | Low-quality |
| S1_vs_k141_107030\|\|full | Low-quality |
| S1_vs_k141_153644\|\|full | Low-quality |
| S1_vs_k141_20265\|\|full | Not-determined |
| S1_vs_k141_40290\|\|full | Low-quality |
| S1_vs_k141_87042\|\|full | Not-determined |
| S1_vs_k141_53670\|\|full | Low-quality |
| S1_vs_k141_80379\|\|full | Low-quality |
| S1_vs_k141_273\|\|full | Low-quality |
| S1_vs_k141_73663\|\|full | Not-determined |
| S1_vs_k141_275\|\|full | Low-quality |
| S1_vs_k141_93741\|\|full | Low-quality |
| S1_vs_k141_153654\|\|full | Low-quality |
| S1_vs_k141_100373\|\|full | Low-quality |
| S1_vs_k141_20275\|\|full | Low-quality |
| S1_vs_k141_113719\|\|full | Not-determined |
| S1_vs_k141_127131\|\|full | Not-determined |
| S1_vs_k141_13634\|\|full | Not-determined |
| S1_vs_k141_20289\|\|full | Not-determined |
| S1_vs_k141_33617\|\|full | Not-determined |
| S1_vs_k141_87062\|\|full | Low-quality |
| S1_vs_k141_66989\|\|full | Not-determined |
| S1_vs_k141_107065\|\|full | Not-determined |
| S1_vs_k141_147034\|\|full | Low-quality |
| S1_vs_k141_46978\|\|full | Not-determined |
| S1_vs_k141_26890\|\|full | Not-determined |
| S1_vs_k141_93765\|\|full | Low-quality |
| S1_vs_k141_133706\|\|full | Low-quality |
| S1_vs_k141_40329\|\|full | Not-determined |
| S1_vs_k141_53713\|\|full | Low-quality |
| S1_vs_k141_46988\|\|full | Low-quality |
| S1_vs_k141_66995\|\|full | Not-determined |
| S1_vs_k141_20304\|\|full | Low-quality |
| S1_vs_k141_60195\|\|full | Not-determined |
| S1_vs_k141_13656\|\|full | Low-quality |
| S8_vs_k141_125717\|\|full | Medium-quality |
| S1_vs_k141_46991\|\|full | Low-quality |
| S1_vs_k141_73709\|\|full | Not-determined |
| S1_vs_k141_46993\|\|full | Low-quality |
| S1_vs_k141_113749\|\|full | Not-determined |
| S1_vs_k141_120451\|\|full | Low-quality |
| S1_vs_k141_120452\|\|full | Low-quality |
| S1_vs_k141_87087\|\|full | Low-quality |
| S1_vs_k141_140469\|\|full | Low-quality |
| S1_vs_k141_127174\|\|full | Low-quality |
| S1_vs_k141_133716\|\|full | Not-determined |
| S1_vs_k141_113756\|\|full | Low-quality |
| S1_vs_k141_80450\|\|full | Not-determined |
| S1_vs_k141_127183\|\|full | Low-quality |
| S1_vs_k141_107099\|\|full | Low-quality |
| S1_vs_k141_153684\|\|full | Not-determined |
| S1_vs_k141_100429\|\|full | Low-quality |

| | |
|---|---|
| S1_vs_k141_133731\|\|full | Not-determined |
| S1_vs_k141_127197\|\|full | Not-determined |
| S1_vs_k141_26937\|\|full | Not-determined |
| S1_vs_k141_13693\|\|full | Low-quality |
| S1_vs_k141_60210\|\|full | Low-quality |
| S1_vs_k141_341\|\|full | Low-quality |
| S1_vs_k141_140490\|\|full | Not-determined |
| S1_vs_k141_120494\|\|full | Low-quality |
| S1_vs_k141_120496\|\|full | Not-determined |
| S1_vs_k141_140495\|\|full | Not-determined |
| S1_vs_k141_7015\|\|full | Low-quality |
| S1_vs_k141_350\|\|full | Not-determined |
| S1_vs_k141_73761\|\|full | Not-determined |
| S1_vs_k141_87155\|\|full | Low-quality |
| S1_vs_k141_13735\|\|full | Not-determined |
| S1_vs_k141_93849\|\|full | Not-determined |
| S1_vs_k141_67057\|\|full | Low-quality |
| S1_vs_k141_80503\|\|full | Low-quality |
| S1_vs_k141_33692\|\|full | Not-determined |
| S1_vs_k141_120522\|\|full | Low-quality |
| S1_vs_k141_80507\|\|full | Not-determined |
| S1_vs_k141_100482\|\|full | Low-quality |
| S1_vs_k141_133765\|\|full | Low-quality |
| S1_vs_k141_20380\|\|full | Not-determined |
| S1_vs_k141_107147\|\|full | Low-quality |
| S1_vs_k141_120534\|\|full | Low-quality |
| S1_vs_k141_370\|\|full | Low-quality |
| S1_vs_k141_93868\|\|full | Low-quality |
| S1_vs_k141_27004\|\|full | Low-quality |
| S1_vs_k141_107149\|\|full | Low-quality |
| S1_vs_k141_140517\|\|full | Not-determined |
| S1_vs_k141_80523\|\|full | Low-quality |
| S1_vs_k141_13755\|\|full | Low-quality |
| S1_vs_k141_133768\|\|full | Low-quality |
| S1_vs_k141_87165\|\|full | Low-quality |
| S1_vs_k141_47082\|\|full | Low-quality |
| S1_vs_k141_13759\|\|full | Not-determined |
| S1_vs_k141_113817\|\|full | Not-determined |
| S1_vs_k141_140529\|\|full | Low-quality |
| S1_vs_k141_33705\|\|full | Low-quality |
| S1_vs_k141_113822\|\|full | Low-quality |
| S1_vs_k141_140536\|\|full | Low-quality |
| S1_vs_k141_80546\|\|full | Low-quality |
| S1_vs_k141_27021\|\|full | Not-determined |
| S1_vs_k141_93900\|\|full | Low-quality |
| S1_vs_k141_100517\|\|full | Low-quality |
| S1_vs_k141_133797\|\|full | Low-quality |
| S1_vs_k141_53859\|\|full | Low-quality |
| S1_vs_k141_107192\|\|full | Low-quality |
| S1_vs_k141_53866\|\|full | Not-determined |
| S1_vs_k141_7092\|\|full | Low-quality |
| S1_vs_k141_7093\|\|full | Low-quality |
| S1_vs_k141_113854\|\|full | Low-quality |
| S1_vs_k141_93915\|\|full | Low-quality |
| S1_vs_k141_60292\|\|full | Not-determined |
| S1_vs_k141_60295\|\|full | Not-determined |
| S1_vs_k141_80578\|\|full | Low-quality |
| S1_vs_k141_13820\|\|full | Not-determined |
| S1_vs_k141_33742\|\|full | Low-quality |
| S1_vs_k141_120581\|\|full | Low-quality |
| S1_vs_k141_93932\|\|full | Low-quality |
| S1_vs_k141_13836\|\|full | Low-quality |
| S1_vs_k141_40511\|\|full | Low-quality |
| S1_vs_k141_13837\|\|full | Not-determined |
| S1_vs_k141_20451\|\|full | Not-determined |
| S1_vs_k141_53895\|\|full | Low-quality |
| S1_vs_k141_40521\|\|full | Not-determined |
| S1_vs_k141_113884\|\|full | Low-quality |
| S1_vs_k141_20463\|\|full | Low-quality |
| S1_vs_k141_20466\|\|full | Low-quality |
| S1_vs_k141_73865\|\|full | Not-determined |
| S1_vs_k141_140583\|\|full | Low-quality |
| S1_vs_k141_40539\|\|full | Low-quality |
| S1_vs_k141_120598\|\|full | Low-quality |
| S1_vs_k141_13862\|\|full | Low-quality |
| S1_vs_k141_87249\|\|full | Low-quality |
| S1_vs_k141_53928\|\|full | Low-quality |
| S1_vs_k141_13867\|\|full | Low-quality |
| S1_vs_k141_147125\|\|full | Low-quality |
| S1_vs_k141_47170\|\|full | Not-determined |
| S1_vs_k141_20469\|\|full | Low-quality |
| S1_vs_k141_27105\|\|full | Low-quality |
| S1_vs_k141_100609\|\|full | Not-determined |
| S1_vs_k141_133878\|\|full | Not-determined |
| S1_vs_k141_107263\|\|full | Low-quality |
| S1_vs_k141_133881\|\|full | Low-quality |
| S1_vs_k141_80630\|\|full | Not-determined |
| S1_vs_k141_100614\|\|full | Low-quality |
| S1_vs_k141_7166\|\|full | Low-quality |
| S1_vs_k141_93967\|\|full | Not-determined |
| S1_vs_k141_133894\|\|full | Low-quality |
| S1_vs_k141_73900\|\|full | Low-quality |
| S1_vs_k141_7186\|\|full | Low-quality |
| S1_vs_k141_27137\|\|full | Low-quality |
| S1_vs_k141_93977\|\|full | Low-quality |
| S1_vs_k141_80655\|\|full | Not-determined |
| S1_vs_k141_100640\|\|full | Low-quality |
| S1_vs_k141_113935\|\|full | Not-determined |
| S1_vs_k141_107267\|\|full | Low-quality |
| S1_vs_k141_73917\|\|full | Not-determined |
| S1_vs_k141_120637\|\|full | Not-determined |
| S1_vs_k141_27150\|\|full | Low-quality |
| S1_vs_k141_140658\|\|full | Low-quality |
| S1_vs_k141_107273\|\|full | Low-quality |
| S1_vs_k141_113952\|\|full | Low-quality |
| S1_vs_k141_94003\|\|full | Not-determined |
| S1_vs_k141_100656\|\|full | Low-quality |
| S1_vs_k141_60373\|\|full | Low-quality |
| S1_vs_k141_13939\|\|full | Not-determined |
| S1_vs_k141_127374\|\|full | Low-quality |
| S1_vs_k141_40630\|\|full | Low-quality |
| S1_vs_k141_47221\|\|full | Not-determined |
| S1_vs_k141_40631\|\|full | Low-quality |
| S1_vs_k141_107275\|\|full | Low-quality |
| S1_vs_k141_120654\|\|full | Low-quality |
| S1_vs_k141_53993\|\|full | Low-quality |
| S1_vs_k141_47231\|\|full | Low-quality |
| S1_vs_k141_67224\|\|full | Low-quality |
| S1_vs_k141_100678\|\|full | Low-quality |
| S1_vs_k141_140702\|\|full | Not-determined |
| S1_vs_k141_67229\|\|full | Not-determined |
| S1_vs_k141_100683\|\|full | Not-determined |
| S1_vs_k141_47236\|\|full | Not-determined |
| S1_vs_k141_113974\|\|full | Low-quality |
| S1_vs_k141_94039\|\|full | Not-determined |
| S1_vs_k141_54003\|\|full | Low-quality |
| S1_vs_k141_127397\|\|full | Low-quality |
| S1_vs_k141_27200\|\|full | Not-determined |
| S1_vs_k141_13985\|\|full | Not-determined |
| S1_vs_k141_113975\|\|full | Not-determined |
| S1_vs_k141_94046\|\|full | Not-determined |
| S1_vs_k141_153811\|\|full | Low-quality |
| S1_vs_k141_133958\|\|full | Low-quality |
| S1_vs_k141_87353\|\|full | Low-quality |
| S1_vs_k141_40663\|\|full | Low-quality |
| S1_vs_k141_20537\|\|full | Not-determined |
| S1_vs_k141_54020\|\|full | Low-quality |
| S1_vs_k141_40671\|\|full | Low-quality |
| S1_vs_k141_67256\|\|full | Low-quality |
| S1_vs_k141_127429\|\|full | Low-quality |
| S1_vs_k141_67257\|\|full | Low-quality |
| S1_vs_k141_540\|\|full | Not-determined |
| S1_vs_k141_127436\|\|full | Low-quality |
| S1_vs_k141_7288\|\|full | Not-determined |
| S1_vs_k141_120704\|\|full | Low-quality |
| S1_vs_k141_74005\|\|full | Low-quality |
| S1_vs_k141_27237\|\|full | Not-determined |
| S1_vs_k141_100733\|\|full | Low-quality |
| S1_vs_k141_80751\|\|full | Low-quality |
| S1_vs_k141_133999\|\|full | Low-quality |
| S1_vs_k141_27251\|\|full | Not-determined |
| S1_vs_k141_87391\|\|full | Not-determined |
| S1_vs_k141_54061\|\|full | Low-quality |
| S1_vs_k141_147167\|\|full | Low-quality |
| S1_vs_k141_153850\|\|full | Not-determined |
| S1_vs_k141_114040\|\|full | Not-determined |
| S1_vs_k141_67281\|\|full | Low-quality |
| S1_vs_k141_47305\|\|full | Low-quality |
| S1_vs_k141_14049\|\|full | Low-quality |
| S1_vs_k141_120728\|\|full | Low-quality |
| S1_vs_k141_20571\|\|full | Not-determined |
| S1_vs_k141_120731\|\|full | Not-determined |
| S1_vs_k141_94089\|\|full | Low-quality |
| S1_vs_k141_27281\|\|full | Low-quality |
| S1_vs_k141_20576\|\|full | Low-quality |
| S1_vs_k141_100769\|\|full | Low-quality |
| S1_vs_k141_140774\|\|full | Low-quality |
| S1_vs_k141_114050\|\|full | Low-quality |
| S1_vs_k141_80787\|\|full | Not-determined |
| S1_vs_k141_7354\|\|full | Low-quality |
| S1_vs_k141_33910\|\|full | Low-quality |
| S1_vs_k141_140782\|\|full | Low-quality |
| S1_vs_k141_127516\|\|full | Not-determined |
| S1_vs_k141_153867\|\|full | Low-quality |
| S1_vs_k141_94096\|\|full | Low-quality |
| S1_vs_k141_578\|\|full | Low-quality |
| S1_vs_k141_80806\|\|full | Not-determined |
| S1_vs_k141_14102\|\|full | Low-quality |
| S1_vs_k141_153882\|\|full | Not-determined |
| S1_vs_k141_40755\|\|full | Not-determined |
| S1_vs_k141_134038\|\|full | Low-quality |
| S1_vs_k141_147174\|\|full | Low-quality |
| S1_vs_k141_87447\|\|full | Not-determined |
| S1_vs_k141_67330\|\|full | Low-quality |
| S1_vs_k141_40759\|\|full | Low-quality |
| S1_vs_k141_114095\|\|full | Low-quality |
| S1_vs_k141_20619\|\|full | Low-quality |
| S1_vs_k141_94132\|\|full | Low-quality |
| S1_vs_k141_114101\|\|full | Low-quality |
| S1_vs_k141_114104\|\|full | Low-quality |
| S1_vs_k141_40764\|\|full | Not-determined |
| S1_vs_k141_20633\|\|full | Low-quality |
| S1_vs_k141_67348\|\|full | Low-quality |
| S1_vs_k141_100820\|\|full | Low-quality |
| S1_vs_k141_74113\|\|full | Low-quality |
| S1_vs_k141_127564\|\|full | Low-quality |
| S1_vs_k141_120804\|\|full | Low-quality |
| S1_vs_k141_602\|\|full | Low-quality |
| S1_vs_k141_14149\|\|full | Not-determined |
| S1_vs_k141_100828\|\|full | Low-quality |
| S1_vs_k141_114115\|\|full | Low-quality |
| S1_vs_k141_40790\|\|full | Not-determined |
| S1_vs_k141_67361\|\|full | Low-quality |
| S1_vs_k141_94172\|\|full | Not-determined |
| S1_vs_k141_87493\|\|full | Low-quality |
| S1_vs_k141_74136\|\|full | Low-quality |
| S1_vs_k141_100844\|\|full | Low-quality |
| S1_vs_k141_74138\|\|full | Not-determined |
| S1_vs_k141_20667\|\|full | Low-quality |
| S1_vs_k141_54182\|\|full | Low-quality |
| S1_vs_k141_114116\|\|full | Low-quality |
| S1_vs_k141_67370\|\|full | Not-determined |
| S1_vs_k141_27365\|\|full | Not-determined |
| S1_vs_k141_120825\|\|full | Low-quality |
| S1_vs_k141_74151\|\|full | Low-quality |
| S1_vs_k141_147201\|\|full | Not-determined |
| S1_vs_k141_140883\|\|full | Low-quality |
| S1_vs_k141_147202\|\|full | Low-quality |
| S3_vs_k141_90331\|\|full | High-quality |
| S1_vs_k141_140890\|\|full | Low-quality |
| S1_vs_k141_100881\|\|full | Low-quality |
| S1_vs_k141_74179\|\|full | Not-determined |
| S1_vs_k141_7424\|\|full | Low-quality |
| S1_vs_k141_147210\|\|full | Low-quality |
| S1_vs_k141_140900\|\|full | Low-quality |
| S1_vs_k141_27384\|\|full | Low-quality |
| S1_vs_k141_127610\|\|full | Low-quality |
| S1_vs_k141_67403\|\|full | Not-determined |
| S1_vs_k141_120828\|\|full | Low-quality |
| S1_vs_k141_60479\|\|full | Not-determined |
| S1_vs_k141_120830\|\|full | Low-quality |
| S1_vs_k141_127634\|\|full | Low-quality |
| S1_vs_k141_114154\|\|full | Not-determined |
| S1_vs_k141_54249\|\|full | Low-quality |
| S1_vs_k141_47487\|\|full | Low-quality |
| S1_vs_k141_27413\|\|full | Low-quality |
| S1_vs_k141_67428\|\|full | Not-determined |
| S1_vs_k141_127642\|\|full | Low-quality |
| S1_vs_k141_87569\|\|full | Low-quality |
| S1_vs_k141_140932\|\|full | Not-determined |
| S1_vs_k141_67432\|\|full | Low-quality |
| S1_vs_k141_140933\|\|full | Not-determined |
| S1_vs_k141_114162\|\|full | Not-determined |
| S1_vs_k141_27422\|\|full | Not-determined |
| S1_vs_k141_153918\|\|full | Low-quality |
| S1_vs_k141_27423\|\|full | Low-quality |
| S1_vs_k141_14263\|\|full | Low-quality |
| S1_vs_k141_120838\|\|full | Low-quality |
| S1_vs_k141_688\|\|full | Low-quality |
| S1_vs_k141_120845\|\|full | Low-quality |
| S1_vs_k141_67454\|\|full | Low-quality |
| S1_vs_k141_127657\|\|full | Not-determined |
| S1_vs_k141_80925\|\|full | Low-quality |
| S1_vs_k141_134180\|\|full | Low-quality |
| S1_vs_k141_94280\|\|full | Not-determined |
| S1_vs_k141_54278\|\|full | Low-quality |
| S1_vs_k141_20783\|\|full | Low-quality |
| S1_vs_k141_47516\|\|full | Low-quality |
| S1_vs_k141_107452\|\|full | Not-determined |
| S1_vs_k141_80932\|\|full | Low-quality |
| S1_vs_k141_100973\|\|full | Low-quality |
| S1_vs_k141_27448\|\|full | Low-quality |
| S1_vs_k141_107455\|\|full | Low-quality |
| S1_vs_k141_134196\|\|full | Low-quality |
| S1_vs_k141_127681\|\|full | Not-determined |
| S1_vs_k141_54295\|\|full | Low-quality |
| S1_vs_k141_127682\|\|full | Low-quality |
| S1_vs_k141_107463\|\|full | Low-quality |
| S1_vs_k141_94301\|\|full | Not-determined |
| S1_vs_k141_20806\|\|full | Low-quality |
| S1_vs_k141_87588\|\|full | Low-quality |
| S1_vs_k141_94303\|\|full | Low-quality |
| S1_vs_k141_107471\|\|full | Low-quality |
| S1_vs_k141_54301\|\|full | Low-quality |
| S1_vs_k141_47536\|\|full | Low-quality |
| S1_vs_k141_100990\|\|full | Not-determined |
| S1_vs_k141_80961\|\|full | Low-quality |
| S1_vs_k141_114212\|\|full | Low-quality |
| S1_vs_k141_67485\|\|full | Low-quality |
| S1_vs_k141_80966\|\|full | Low-quality |
| S1_vs_k141_94319\|\|full | Not-determined |
| S1_vs_k141_147242\|\|full | Low-quality |
| S1_vs_k141_7522\|\|full | Not-determined |
| S1_vs_k141_94323\|\|full | Low-quality |
| S1_vs_k141_74241\|\|full | Low-quality |
| S1_vs_k141_107484\|\|full | Low-quality |
| S1_vs_k141_101007\|\|full | Low-quality |
| S1_vs_k141_7525\|\|full | Low-quality |
| S1_vs_k141_34098\|\|full | Low-quality |
| S1_vs_k141_47553\|\|full | Not-determined |
| S1_vs_k141_74243\|\|full | Low-quality |
| S1_vs_k141_60549\|\|full | Low-quality |
| S1_vs_k141_87632\|\|full | Low-quality |
| S1_vs_k141_140995\|\|full | Not-determined |
| S1_vs_k141_101031\|\|full | Low-quality |
| S1_vs_k141_40934\|\|full | Low-quality |
| S1_vs_k141_127715\|\|full | Low-quality |
| S1_vs_k141_769\|\|full | Low-quality |
| S1_vs_k141_7568\|\|full | Not-determined |
| S1_vs_k141_81008\|\|full | Low-quality |
| S1_vs_k141_134240\|\|full | Not-determined |
| S1_vs_k141_134241\|\|full | Not-determined |
| S1_vs_k141_120880\|\|full | Not-determined |
| S1_vs_k141_141016\|\|full | Not-determined |
| S1_vs_k141_67536\|\|full | Low-quality |
| S1_vs_k141_40963\|\|full | Not-determined |
| S1_vs_k141_120882\|\|full | Low-quality |
| S1_vs_k141_127732\|\|full | Not-determined |
| S1_vs_k141_141024\|\|full | Low-quality |
| S1_vs_k141_67543\|\|full | Not-determined |
| S1_vs_k141_27524\|\|full | Not-determined |
| S1_vs_k141_54351\|\|full | Low-quality |
| S1_vs_k141_801\|\|full | Not-determined |
| S1_vs_k141_67547\|\|full | Low-quality |
| S1_vs_k141_27525\|\|full | Low-quality |
| S1_vs_k141_60574\|\|full | Low-quality |
| S1_vs_k141_81047\|\|full | Low-quality |
| S1_vs_k141_27527\|\|full | Low-quality |
| S1_vs_k141_7603\|\|full | Low-quality |
| S1_vs_k141_806\|\|full | Low-quality |
| S1_vs_k141_147260\|\|full | Low-quality |
| S1_vs_k141_114279\|\|full | Low-quality |
| S1_vs_k141_120906\|\|full | Low-quality |
| S1_vs_k141_81058\|\|full | Low-quality |
| S1_vs_k141_20907\|\|full | Low-quality |
| S1_vs_k141_87697\|\|full | Low-quality |
| S1_vs_k141_107546\|\|full | Low-quality |
| S1_vs_k141_20910\|\|full | Not-determined |
| S1_vs_k141_54376\|\|full | Low-quality |
| S1_vs_k141_134292\|\|full | Low-quality |
| S1_vs_k141_120915\|\|full | Low-quality |
| S1_vs_k141_141061\|\|full | Low-quality |
| S1_vs_k141_74319\|\|full | Low-quality |
| S1_vs_k141_87711\|\|full | Not-determined |
| S1_vs_k141_147279\|\|full | Low-quality |
| S1_vs_k141_127786\|\|full | Low-quality |
| S1_vs_k141_120937\|\|full | Low-quality |
| S1_vs_k141_94417\|\|full | Not-determined |
| S1_vs_k141_94420\|\|full | Low-quality |
| S1_vs_k141_27563\|\|full | Low-quality |
| S1_vs_k141_87728\|\|full | Low-quality |
| S1_vs_k141_134315\|\|full | Low-quality |
| S1_vs_k141_87731\|\|full | Low-quality |
| S1_vs_k141_14485\|\|full | Low-quality |
| S1_vs_k141_41019\|\|full | Low-quality |
| S1_vs_k141_120953\|\|full | Low-quality |
| S1_vs_k141_114323\|\|full | Low-quality |
| S1_vs_k141_60613\|\|full | Not-determined |
| S1_vs_k141_34183\|\|full | Low-quality |
| S1_vs_k141_120958\|\|full | Not-determined |
| S1_vs_k141_114332\|\|full | Not-determined |
| S1_vs_k141_81118\|\|full | Low-quality |
| S1_vs_k141_154004\|\|full | Low-quality |
| S1_vs_k141_14513\|\|full | Low-quality |
| S1_vs_k141_94452\|\|full | Not-determined |
| S1_vs_k141_74343\|\|full | Low-quality |
| S1_vs_k141_154006\|\|full | Low-quality |
| S1_vs_k141_34185\|\|full | Low-quality |
| S1_vs_k141_107598\|\|full | Low-quality |
| S1_vs_k141_127818\|\|full | Low-quality |
| S1_vs_k141_74346\|\|full | Low-quality |
| S1_vs_k141_147292\|\|full | Not-determined |
| S1_vs_k141_120965\|\|full | Low-quality |
| S1_vs_k141_94469\|\|full | Not-determined |
| S1_vs_k141_20966\|\|full | Low-quality |
| S1_vs_k141_87738\|\|full | Low-quality |
| S1_vs_k141_27585\|\|full | Low-quality |
| S1_vs_k141_41048\|\|full | Low-quality |
| S1_vs_k141_41049\|\|full | Low-quality |
| S1_vs_k141_107613\|\|full | Not-determined |
| S1_vs_k141_47667\|\|full | Low-quality |
| S1_vs_k141_134362\|\|full | Low-quality |
| S1_vs_k141_120978\|\|full | Low-quality |
| S1_vs_k141_114341\|\|full | Low-quality |
| S1_vs_k141_81166\|\|full | Not-determined |
| S1_vs_k141_47669\|\|full | Low-quality |
| S1_vs_k141_94498\|\|full | Low-quality |
| S1_vs_k141_120989\|\|full | Low-quality |
| S1_vs_k141_141151\|\|full | Not-determined |
| S1_vs_k141_7721\|\|full | Low-quality |
| S1_vs_k141_107647\|\|full | Low-quality |
| S1_vs_k141_101166\|\|full | Low-quality |
| S1_vs_k141_7725\|\|full | Low-quality |
| S1_vs_k141_134365\|\|full | Low-quality |
| S1_vs_k141_67670\|\|full | Low-quality |
| S1_vs_k141_20997\|\|full | Low-quality |
| S1_vs_k141_141177\|\|full | Low-quality |
| S1_vs_k141_14598\|\|full | Low-quality |
| S1_vs_k141_7744\|\|full | Not-determined |
| S1_vs_k141_34233\|\|full | Low-quality |
| S1_vs_k141_60681\|\|full | Low-quality |
| S1_vs_k141_101187\|\|full | Low-quality |
| S1_vs_k141_896\|\|full | Not-determined |
| S1_vs_k141_7760\|\|full | Not-determined |
| S1_vs_k141_21026\|\|full | Not-determined |
| S1_vs_k141_81205\|\|full | Low-quality |
| S1_vs_k141_141202\|\|full | Not-determined |
| S1_vs_k141_101189\|\|full | Low-quality |
| S1_vs_k141_21038\|\|full | Low-quality |
| S1_vs_k141_81207\|\|full | Not-determined |
| S1_vs_k141_74437\|\|full | Low-quality |
| S1_vs_k141_121033\|\|full | Not-determined |
| S1_vs_k141_74438\|\|full | Low-quality |
| S1_vs_k141_21045\|\|full | Low-quality |
| S1_vs_k141_107719\|\|full | Low-quality |
| S1_vs_k141_47748\|\|full | Low-quality |
| S1_vs_k141_54476\|\|full | Low-quality |
| S1_vs_k141_912\|\|full | Not-determined |
| S1_vs_k141_101217\|\|full | Low-quality |
| S1_vs_k141_87833\|\|full | Low-quality |
| S1_vs_k141_81226\|\|full | Not-determined |
| S1_vs_k141_94593\|\|full | Not-determined |
| S1_vs_k141_47761\|\|full | Low-quality |
| S1_vs_k141_134439\|\|full | Low-quality |
| S1_vs_k141_54491\|\|full | Not-determined |
| S1_vs_k141_87839\|\|full | Low-quality |
| S1_vs_k141_67737\|\|full | Low-quality |
| S1_vs_k141_34279\|\|full | Low-quality |
| S1_vs_k141_121066\|\|full | Low-quality |
| S1_vs_k141_67743\|\|full | Not-determined |
| S1_vs_k141_927\|\|full | Low-quality |
| S1_vs_k141_154105\|\|full | Not-determined |
| S1_vs_k141_81243\|\|full | Low-quality |
| S1_vs_k141_7830\|\|full | Not-determined |
| S1_vs_k141_21090\|\|full | Not-determined |
| S1_vs_k141_121073\|\|full | Low-quality |
| S1_vs_k141_101248\|\|full | Not-determined |
| S1_vs_k141_81250\|\|full | Low-quality |
| S1_vs_k141_14700\|\|full | Not-determined |
| S1_vs_k141_87868\|\|full | Not-determined |
| S1_vs_k141_41219\|\|full | Low-quality |
| S1_vs_k141_21099\|\|full | Not-determined |
| S1_vs_k141_54513\|\|full | Low-quality |
| S1_vs_k141_21102\|\|full | Not-determined |
| S1_vs_k141_60701\|\|full | Low-quality |
| S1_vs_k141_114469\|\|full | Low-quality |
| S1_vs_k141_94617\|\|full | Low-quality |
| S1_vs_k141_154117\|\|full | Low-quality |
| S1_vs_k141_101260\|\|full | Low-quality |
| S1_vs_k141_60702\|\|full | Low-quality |
| S1_vs_k141_107771\|\|full | Low-quality |
| S1_vs_k141_7853\|\|full | Low-quality |
| S1_vs_k141_121087\|\|full | Low-quality |
| S1_vs_k141_47813\|\|full | Not-determined |
| S1_vs_k141_101263\|\|full | Low-quality |
| S1_vs_k141_950\|\|full | Not-determined |
| S1_vs_k141_134471\|\|full | Low-quality |
| S1_vs_k141_47819\|\|full | Low-quality |
| S1_vs_k141_94638\|\|full | Not-determined |
| S1_vs_k141_141278\|\|full | Low-quality |
| S1_vs_k141_101269\|\|full | Not-determined |
| S1_vs_k141_957\|\|full | Low-quality |
| S1_vs_k141_14732\|\|full | Low-quality |
| S1_vs_k141_41259\|\|full | Low-quality |
| S1_vs_k141_107800\|\|full | Low-quality |
| S1_vs_k141_54550\|\|full | Low-quality |
| S1_vs_k141_134480\|\|full | Not-determined |
| S1_vs_k141_60719\|\|full | Low-quality |
| S1_vs_k141_965\|\|full | Low-quality |
| S1_vs_k141_47832\|\|full | Not-determined |
| S1_vs_k141_7891\|\|full | Low-quality |
| S1_vs_k141_41263\|\|full | Not-determined |
| S1_vs_k141_107811\|\|full | Low-quality |
| S1_vs_k141_114502\|\|full | Low-quality |
| S1_vs_k141_81303\|\|full | Low-quality |
| S1_vs_k141_14750\|\|full | Low-quality |
| S1_vs_k141_87910\|\|full | Low-quality |
| S1_vs_k141_67814\|\|full | Low-quality |
| S1_vs_k141_87916\|\|full | Low-quality |
| S1_vs_k141_81314\|\|full | Not-determined |
| S1_vs_k141_47849\|\|full | Low-quality |
| S1_vs_k141_147361\|\|full | Low-quality |
| S1_vs_k141_87924\|\|full | Low-quality |
| S1_vs_k141_114509\|\|full | Low-quality |
| S1_vs_k141_41290\|\|full | Not-determined |
| S1_vs_k141_21164\|\|full | Low-quality |
| S1_vs_k141_21166\|\|full | Low-quality |
| S1_vs_k141_141309\|\|full | Not-determined |
| S1_vs_k141_107839\|\|full | Low-quality |
| S1_vs_k141_47862\|\|full | Not-determined |
| S1_vs_k141_21174\|\|full | Low-quality |
| S1_vs_k141_7927\|\|full | Low-quality |
| S1_vs_k141_134518\|\|full | Low-quality |
| S1_vs_k141_134521\|\|full | Not-determined |
| S1_vs_k141_27788\|\|full | Not-determined |
| S1_vs_k141_60734\|\|full | Not-determined |
| S1_vs_k141_154178\|\|full | Low-quality |
| S1_vs_k141_127996\|\|full | Not-determined |
| S1_vs_k141_107851\|\|full | Low-quality |
| S1_vs_k141_27803\|\|full | Low-quality |
| S1_vs_k141_990\|\|full | Low-quality |
| S1_vs_k141_41325\|\|full | Not-determined |
| S1_vs_k141_147377\|\|full | Low-quality |
| S1_vs_k141_47905\|\|full | Low-quality |
| S1_vs_k141_121124\|\|full | Low-quality |
| S1_vs_k141_21208\|\|full | Low-quality |
| S1_vs_k141_128007\|\|full | Low-quality |
| S1_vs_k141_27816\|\|full | Low-quality |
| S1_vs_k141_141331\|\|full | Low-quality |
| S1_vs_k141_27823\|\|full | Low-quality |
| S1_vs_k141_14853\|\|full | Not-determined |
| S1_vs_k141_154205\|\|full | Low-quality |
| S1_vs_k141_60757\|\|full | Low-quality |
| S1_vs_k141_141332\|\|full | Low-quality |
| S1_vs_k141_107898\|\|full | Low-quality |
| S1_vs_k141_87993\|\|full | Low-quality |
| S1_vs_k141_67896\|\|full | Not-determined |
| S1_vs_k141_1033\|\|full | Not-determined |
| S1_vs_k141_34387\|\|full | Low-quality |
| S1_vs_k141_54637\|\|full | Not-determined |
| S1_vs_k141_101400\|\|full | Low-quality |
| S1_vs_k141_128046\|\|full | Not-determined |
| S1_vs_k141_101403\|\|full | Low-quality |
| S1_vs_k141_27866\|\|full | Not-determined |
| S1_vs_k141_47960\|\|full | Low-quality |
| S1_vs_k141_107922\|\|full | Low-quality |
| S1_vs_k141_27872\|\|full | Not-determined |
| S1_vs_k141_141347\|\|full | Low-quality |
| S1_vs_k141_81409\|\|full | Low-quality |
| S1_vs_k141_147408\|\|full | Not-determined |
| S1_vs_k141_121166\|\|full | Low-quality |
| S1_vs_k141_60789\|\|full | Not-determined |
| S1_vs_k141_14888\|\|full | Low-quality |
| S1_vs_k141_121170\|\|full | Low-quality |
| S1_vs_k141_88025\|\|full | Low-quality |
| S1_vs_k141_81415\|\|full | Low-quality |
| S1_vs_k141_54653\|\|full | Low-quality |
| S1_vs_k141_67945\|\|full | Not-determined |
| S1_vs_k141_54657\|\|full | Low-quality |
| S1_vs_k141_141358\|\|full | Low-quality |
| S1_vs_k141_27903\|\|full | Low-quality |
| S1_vs_k141_107944\|\|full | Low-quality |
| S1_vs_k141_154231\|\|full | Not-determined |
| S1_vs_k141_60810\|\|full | Low-quality |
| S1_vs_k141_107954\|\|full | Low-quality |
| S1_vs_k141_54670\|\|full | Low-quality |
| S1_vs_k141_134615\|\|full | Low-quality |
| S1_vs_k141_34430\|\|full | Low-quality |
| S1_vs_k141_21263\|\|full | Not-determined |
| S1_vs_k141_121193\|\|full | Low-quality |
| S1_vs_k141_54672\|\|full | Low-quality |
| S1_vs_k141_60819\|\|full | Not-determined |
| S1_vs_k141_94775\|\|full | Low-quality |
| S1_vs_k141_134623\|\|full | Low-quality |
| S1_vs_k141_8071\|\|full | Not-determined |
| S1_vs_k141_107967\|\|full | Not-determined |
| S1_vs_k141_48018\|\|full | Not-determined |
| S1_vs_k141_147424\|\|full | Low-quality |
| S1_vs_k141_128109\|\|full | Low-quality |
| S1_vs_k141_114659\|\|full | Not-determined |
| S1_vs_k141_107970\|\|full | Low-quality |
| S1_vs_k141_121212\|\|full | Not-determined |
| S1_vs_k141_88063\|\|full | Not-determined |
| S1_vs_k141_128118\|\|full | Not-determined |
| S1_vs_k141_154248\|\|full | Not-determined |
| S1_vs_k141_8082\|\|full | Low-quality |
| S1_vs_k141_88069\|\|full | Low-quality |
| S1_vs_k141_48031\|\|full | Not-determined |
| S1_vs_k141_41443\|\|full | Low-quality |
| S1_vs_k141_74697\|\|full | Low-quality |
| S1_vs_k141_54705\|\|full | Not-determined |
| S1_vs_k141_27940\|\|full | Low-quality |
| S1_vs_k141_88079\|\|full | Low-quality |
| S1_vs_k141_81466\|\|full | Not-determined |
| S1_vs_k141_114691\|\|full | Not-determined |
| S1_vs_k141_54708\|\|full | Not-determined |
| S1_vs_k141_68006\|\|full | Not-determined |
| S1_vs_k141_121232\|\|full | Not-determined |
| S1_vs_k141_48040\|\|full | Low-quality |
| S1_vs_k141_14977\|\|full | Not-determined |
| S1_vs_k141_60838\|\|full | Not-determined |
| S1_vs_k141_21321\|\|full | Low-quality |
| S1_vs_k141_81472\|\|full | Low-quality |
| S1_vs_k141_54717\|\|full | Not-determined |
| S1_vs_k141_34480\|\|full | Low-quality |
| S1_vs_k141_108007\|\|full | Low-quality |
| S1_vs_k141_74720\|\|full | Low-quality |
| S1_vs_k141_41464\|\|full | Low-quality |
| S1_vs_k141_128139\|\|full | Low-quality |
| S1_vs_k141_121243\|\|full | Low-quality |
| S1_vs_k141_141428\|\|full | Not-determined |
| S1_vs_k141_27982\|\|full | Low-quality |
| S1_vs_k141_154269\|\|full | Low-quality |
| S1_vs_k141_88109\|\|full | Low-quality |
| S1_vs_k141_1116\|\|full | Low-quality |
| S1_vs_k141_41473\|\|full | Not-determined |
| S1_vs_k141_101536\|\|full | Low-quality |
| S1_vs_k141_134699\|\|full | Low-quality |
| S1_vs_k141_27995\|\|full | Low-quality |

| | |
|---|---|
| S1_vs_k141_54735\|\|full | Low-quality |
| S1_vs_k141_15011\|\|full | Low-quality |
| S1_vs_k141_94857\|\|full | Low-quality |
| S1_vs_k141_108029\|\|full | Low-quality |
| S1_vs_k141_48078\|\|full | Not-determined |
| S1_vs_k141_128148\|\|full | Low-quality |
| S1_vs_k141_15026\|\|full | Low-quality |
| S1_vs_k141_21373\|\|full | Low-quality |
| S1_vs_k141_41498\|\|full | Low-quality |
| S1_vs_k141_34501\|\|full | Not-determined |
| S1_vs_k141_81508\|\|full | Low-quality |
| S1_vs_k141_54761\|\|full | Low-quality |
| S1_vs_k141_1139\|\|full | Not-determined |
| S1_vs_k141_8143\|\|full | Low-quality |
| S1_vs_k141_74768\|\|full | Not-determined |
| S1_vs_k141_101566\|\|full | Not-determined |
| S1_vs_k141_21387\|\|full | Low-quality |
| S1_vs_k141_34509\|\|full | Low-quality |
| S1_vs_k141_88130\|\|full | Low-quality |
| S1_vs_k141_1144\|\|full | Low-quality |
| S1_vs_k141_34513\|\|full | Low-quality |
| S1_vs_k141_74771\|\|full | Low-quality |
| S1_vs_k141_108051\|\|full | Low-quality |
| S1_vs_k141_154303\|\|full | Low-quality |
| S1_vs_k141_15041\|\|full | Low-quality |
| S1_vs_k141_21404\|\|full | Low-quality |
| S1_vs_k141_101583\|\|full | Not-determined |
| S1_vs_k141_81531\|\|full | Low-quality |
| S1_vs_k141_54785\|\|full | Low-quality |
| S1_vs_k141_68085\|\|full | Low-quality |
| S1_vs_k141_8161\|\|full | Low-quality |
| S1_vs_k141_114773\|\|full | Low-quality |
| S1_vs_k141_81533\|\|full | Not-determined |
| S1_vs_k141_101585\|\|full | Not-determined |
| S1_vs_k141_128170\|\|full | Low-quality |
| S1_vs_k141_141492\|\|full | Not-determined |
| S1_vs_k141_88156\|\|full | Not-determined |
| S1_vs_k141_15061\|\|full | Low-quality |
| S1_vs_k141_128184\|\|full | Low-quality |
| S1_vs_k141_101599\|\|full | Low-quality |
| S1_vs_k141_154319\|\|full | Low-quality |
| S1_vs_k141_41551\|\|full | Low-quality |
| S1_vs_k141_101602\|\|full | Not-determined |
| S1_vs_k141_114804\|\|full | Low-quality |
| S1_vs_k141_21420\|\|full | Low-quality |
| S1_vs_k141_15072\|\|full | Not-determined |
| S1_vs_k141_154323\|\|full | Not-determined |
| S1_vs_k141_134770\|\|full | Low-quality |
| S1_vs_k141_28075\|\|full | Not-determined |
| S1_vs_k141_15081\|\|full | Low-quality |
| S1_vs_k141_48140\|\|full | Not-determined |
| S1_vs_k141_121342\|\|full | Not-determined |
| S1_vs_k141_1174\|\|full | Low-quality |
| S1_vs_k141_88178\|\|full | Low-quality |
| S1_vs_k141_147483\|\|full | Low-quality |
| S1_vs_k141_154342\|\|full | Low-quality |
| S1_vs_k141_81575\|\|full | Not-determined |
| S1_vs_k141_15097\|\|full | Low-quality |
| S1_vs_k141_28093\|\|full | Low-quality |
| S1_vs_k141_60884\|\|full | Not-determined |
| S1_vs_k141_88187\|\|full | Not-determined |
| S1_vs_k141_134800\|\|full | Low-quality |
| S1_vs_k141_1183\|\|full | Low-quality |
| S1_vs_k141_101639\|\|full | Low-quality |
| S1_vs_k141_154353\|\|full | Low-quality |
| S1_vs_k141_128241\|\|full | Low-quality |
| S1_vs_k141_1190\|\|full | Not-determined |
| S1_vs_k141_94964\|\|full | Low-quality |
| S1_vs_k141_94965\|\|full | Low-quality |
| S1_vs_k141_54858\|\|full | Low-quality |
| S1_vs_k141_54859\|\|full | Low-quality |
| S1_vs_k141_121371\|\|full | Low-quality |
| S1_vs_k141_48187\|\|full | Low-quality |
| S1_vs_k141_108123\|\|full | Low-quality |
| S1_vs_k141_134818\|\|full | Low-quality |
| S1_vs_k141_34597\|\|full | Not-determined |
| S1_vs_k141_28120\|\|full | Low-quality |
| S1_vs_k141_94974\|\|full | Low-quality |
| S1_vs_k141_15133\|\|full | Not-determined |
| S1_vs_k141_74849\|\|full | Low-quality |
| S1_vs_k141_68137\|\|full | Not-determined |
| S1_vs_k141_134820\|\|full | Low-quality |
| S1_vs_k141_108132\|\|full | Not-determined |
| S1_vs_k141_114857\|\|full | Not-determined |
| S1_vs_k141_101665\|\|full | Low-quality |
| S1_vs_k141_74865\|\|full | Low-quality |
| S1_vs_k141_154366\|\|full | Not-determined |
| S1_vs_k141_41598\|\|full | Low-quality |
| S1_vs_k141_68149\|\|full | Low-quality |
| S5_vs_k141_80727\|\|full | Medium-quality |
| S1_vs_k141_28134\|\|full | Low-quality |
| S1_vs_k141_54863\|\|full | Low-quality |
| S1_vs_k141_8275\|\|full | Low-quality |
| S1_vs_k141_121394\|\|full | Low-quality |
| S1_vs_k141_108154\|\|full | Low-quality |
| S1_vs_k141_128277\|\|full | Not-determined |
| S1_vs_k141_88245\|\|full | Not-determined |
| S1_vs_k141_101707\|\|full | Low-quality |
| S1_vs_k141_41628\|\|full | Low-quality |
| S1_vs_k141_54880\|\|full | Not-determined |
| S1_vs_k141_81600\|\|full | Low-quality |
| S1_vs_k141_88250\|\|full | Not-determined |
| S1_vs_k141_48250\|\|full | Not-determined |
| S1_vs_k141_134869\|\|full | Low-quality |
| S1_vs_k141_101712\|\|full | Low-quality |
| S1_vs_k141_41639\|\|full | Low-quality |
| S1_vs_k141_15193\|\|full | Not-determined |
| S1_vs_k141_128302\|\|full | Low-quality |
| S1_vs_k141_128304\|\|full | Low-quality |
| S1_vs_k141_108181\|\|full | Low-quality |
| S1_vs_k141_54893\|\|full | Not-determined |
| S1_vs_k141_141610\|\|full | Low-quality |
| S1_vs_k141_41641\|\|full | Low-quality |
| S1_vs_k141_21482\|\|full | Low-quality |
| S1_vs_k141_54900\|\|full | Low-quality |
| S1_vs_k141_121423\|\|full | Not-determined |
| S1_vs_k141_81637\|\|full | Not-determined |
| S1_vs_k141_128316\|\|full | Low-quality |
| S1_vs_k141_147532\|\|full | Not-determined |
| S1_vs_k141_154403\|\|full | Not-determined |
| S1_vs_k141_134887\|\|full | Not-determined |
| S1_vs_k141_41653\|\|full | Low-quality |
| S1_vs_k141_128329\|\|full | Low-quality |
| S1_vs_k141_28205\|\|full | Low-quality |
| S1_vs_k141_8332\|\|full | Low-quality |
| S1_vs_k141_88286\|\|full | Low-quality |
| S1_vs_k141_74957\|\|full | Low-quality |
| S1_vs_k141_95065\|\|full | Not-determined |
| S1_vs_k141_68220\|\|full | Low-quality |
| S1_vs_k141_141635\|\|full | Not-determined |
| S1_vs_k141_15210\|\|full | Low-quality |
| S1_vs_k141_81650\|\|full | Low-quality |
| S1_vs_k141_68222\|\|full | Low-quality |
| S1_vs_k141_154416\|\|full | Low-quality |
| S1_vs_k141_88298\|\|full | Low-quality |
| S1_vs_k141_101747\|\|full | Not-determined |
| S1_vs_k141_95079\|\|full | Low-quality |
| S1_vs_k141_54954\|\|full | Low-quality |
| S1_vs_k141_95084\|\|full | Low-quality |
| S1_vs_k141_128354\|\|full | Low-quality |
| S1_vs_k141_48313\|\|full | Low-quality |
| S1_vs_k141_68239\|\|full | Not-determined |
| S1_vs_k141_15245\|\|full | Low-quality |
| S1_vs_k141_128358\|\|full | Low-quality |
| S1_vs_k141_88317\|\|full | Low-quality |
| S1_vs_k141_68252\|\|full | Low-quality |
| S1_vs_k141_28256\|\|full | Low-quality |
| S1_vs_k141_60954\|\|full | Low-quality |
| S1_vs_k141_41700\|\|full | Low-quality |
| S1_vs_k141_95109\|\|full | Low-quality |
| S1_vs_k141_41701\|\|full | Low-quality |
| S1_vs_k141_28261\|\|full | Low-quality |
| S1_vs_k141_141682\|\|full | Not-determined |
| S1_vs_k141_34707\|\|full | Low-quality |
| S1_vs_k141_134935\|\|full | Not-determined |
| S1_vs_k141_75028\|\|full | Low-quality |
| S1_vs_k141_28271\|\|full | Low-quality |
| S1_vs_k141_21554\|\|full | Low-quality |
| S1_vs_k141_81723\|\|full | Low-quality |
| S1_vs_k141_60964\|\|full | Low-quality |
| S1_vs_k141_54992\|\|full | Low-quality |
| S1_vs_k141_15282\|\|full | Low-quality |
| S1_vs_k141_81740\|\|full | Low-quality |
| S1_vs_k141_101776\|\|full | Low-quality |
| S1_vs_k141_108276\|\|full | Low-quality |
| S1_vs_k141_55011\|\|full | Not-determined |
| S1_vs_k141_60974\|\|full | Low-quality |
| S1_vs_k141_88333\|\|full | Low-quality |
| S1_vs_k141_21578\|\|full | Not-determined |
| S1_vs_k141_121514\|\|full | Low-quality |
| S1_vs_k141_68293\|\|full | Not-determined |
| S1_vs_k141_88337\|\|full | Low-quality |
| S1_vs_k141_108290\|\|full | Low-quality |
| S1_vs_k141_55025\|\|full | Not-determined |
| S1_vs_k141_95141\|\|full | Low-quality |
| S1_vs_k141_75076\|\|full | Low-quality |
| S1_vs_k141_28299\|\|full | Low-quality |
| S1_vs_k141_55032\|\|full | Not-determined |
| S1_vs_k141_1361\|\|full | Low-quality |
| S1_vs_k141_108318\|\|full | Not-determined |
| S1_vs_k141_128440\|\|full | Not-determined |
| S1_vs_k141_88356\|\|full | Low-quality |
| S1_vs_k141_154487\|\|full | Low-quality |
| S1_vs_k141_101802\|\|full | Low-quality |
| S1_vs_k141_68328\|\|full | Low-quality |
| S1_vs_k141_81793\|\|full | Low-quality |
| S1_vs_k141_41754\|\|full | Not-determined |
| S1_vs_k141_1371\|\|full | Low-quality |
| S1_vs_k141_15337\|\|full | Not-determined |
| S1_vs_k141_34749\|\|full | Not-determined |
| S1_vs_k141_121548\|\|full | Low-quality |
| S1_vs_k141_115078\|\|full | Low-quality |
| S1_vs_k141_28324\|\|full | Low-quality |
| S1_vs_k141_68338\|\|full | Low-quality |
| S1_vs_k141_135007\|\|full | Low-quality |
| S1_vs_k141_88363\|\|full | Low-quality |
| S1_vs_k141_1379\|\|full | Not-determined |
| S1_vs_k141_88366\|\|full | Low-quality |
| S1_vs_k141_121565\|\|full | Low-quality |
| S1_vs_k141_81813\|\|full | Low-quality |
| S1_vs_k141_128466\|\|full | Low-quality |
| S1_vs_k141_128469\|\|full | Low-quality |
| S1_vs_k141_121573\|\|full | Not-determined |
| S1_vs_k141_34767\|\|full | Low-quality |
| S1_vs_k141_8443\|\|full | Low-quality |
| S1_vs_k141_121574\|\|full | Low-quality |
| S1_vs_k141_95168\|\|full | Not-determined |
| S1_vs_k141_48428\|\|full | Not-determined |
| S1_vs_k141_15363\|\|full | Low-quality |
| S1_vs_k141_108350\|\|full | Low-quality |
| S1_vs_k141_81828\|\|full | Low-quality |
| S1_vs_k141_15372\|\|full | Low-quality |
| S1_vs_k141_101835\|\|full | Not-determined |
| S1_vs_k141_28362\|\|full | Low-quality |
| S1_vs_k141_60999\|\|full | Low-quality |
| S1_vs_k141_128489\|\|full | Not-determined |
| S1_vs_k141_41796\|\|full | Low-quality |
| S1_vs_k141_8460\|\|full | Low-quality |
| S1_vs_k141_48454\|\|full | Low-quality |
| S1_vs_k141_8461\|\|full | Not-determined |
| S1_vs_k141_48459\|\|full | Low-quality |
| S1_vs_k141_115119\|\|full | Low-quality |
| S1_vs_k141_21661\|\|full | Low-quality |
| S1_vs_k141_1401\|\|full | Low-quality |
| S1_vs_k141_141810\|\|full | Low-quality |
| S1_vs_k141_15399\|\|full | Not-determined |
| S1_vs_k141_88406\|\|full | Low-quality |
| S1_vs_k141_21666\|\|full | Low-quality |
| S1_vs_k141_1404\|\|full | Low-quality |
| S1_vs_k141_48476\|\|full | Low-quality |
| S1_vs_k141_8486\|\|full | Low-quality |
| S1_vs_k141_21678\|\|full | Low-quality |
| S1_vs_k141_101872\|\|full | Low-quality |
| S1_vs_k141_81863\|\|full | Low-quality |
| S1_vs_k141_88417\|\|full | Low-quality |
| S1_vs_k141_135076\|\|full | Low-quality |
| S1_vs_k141_115140\|\|full | Low-quality |
| S1_vs_k141_15413\|\|full | Not-determined |
| S1_vs_k141_75149\|\|full | Not-determined |
| S1_vs_k141_121634\|\|full | Low-quality |
| S1_vs_k141_108406\|\|full | Low-quality |
| S1_vs_k141_68424\|\|full | Low-quality |
| S1_vs_k141_75163\|\|full | Low-quality |
| S1_vs_k141_135087\|\|full | Low-quality |
| S1_vs_k141_88442\|\|full | Not-determined |
| S1_vs_k141_101898\|\|full | Low-quality |
| S1_vs_k141_8517\|\|full | Not-determined |
| S1_vs_k141_95222\|\|full | Not-determined |
| S1_vs_k141_115162\|\|full | Low-quality |
| S1_vs_k141_88445\|\|full | Low-quality |
| S1_vs_k141_75170\|\|full | Low-quality |
| S1_vs_k141_141852\|\|full | Low-quality |
| S1_vs_k141_8534\|\|full | Low-quality |
| S1_vs_k141_128559\|\|full | Not-determined |
| S1_vs_k141_15439\|\|full | Low-quality |
| S16_vs_15093\|\|full | High-quality |
| S1_vs_k141_141861\|\|full | Low-quality |
| S1_vs_k141_55161\|\|full | Low-quality |
| S1_vs_k141_147619\|\|full | Low-quality |
| S1_vs_k141_135117\|\|full | Low-quality |
| S1_vs_k141_135120\|\|full | Low-quality |
| S1_vs_k141_15446\|\|full | Not-determined |
| S1_vs_k141_108432\|\|full | Low-quality |
| S1_vs_k141_68458\|\|full | Low-quality |
| S1_vs_k141_101937\|\|full | Not-determined |
| S1_vs_k141_34845\|\|full | Not-determined |
| S1_vs_k141_108436\|\|full | Low-quality |
| S1_vs_k141_68461\|\|full | Not-determined |
| S1_vs_k141_75193\|\|full | Not-determined |
| S1_vs_k141_15457\|\|full | Low-quality |
| S1_vs_k141_108439\|\|full | Low-quality |
| S1_vs_k141_21723\|\|full | Not-determined |
| S1_vs_k141_88467\|\|full | Not-determined |
| S1_vs_k141_95249\|\|full | Low-quality |
| S1_vs_k141_154596\|\|full | Not-determined |
| S1_vs_k141_88472\|\|full | Low-quality |
| S1_vs_k141_101947\|\|full | Not-determined |
| S1_vs_k141_108448\|\|full | Low-quality |
| S1_vs_k141_34853\|\|full | Low-quality |
| S1_vs_k141_15469\|\|full | Low-quality |
| S1_vs_k141_135150\|\|full | Low-quality |
| S1_vs_k141_95280\|\|full | Low-quality |
| S1_vs_k141_154608\|\|full | Not-determined |
| S1_vs_k141_41902\|\|full | Low-quality |
| S1_vs_k141_115208\|\|full | Low-quality |
| S1_vs_k141_88509\|\|full | Low-quality |
| S1_vs_k141_81961\|\|full | Low-quality |
| S1_vs_k141_108481\|\|full | Not-determined |
| S1_vs_k141_21761\|\|full | Low-quality |
| S1_vs_k141_55221\|\|full | Not-determined |
| S1_vs_k141_88512\|\|full | Not-determined |
| S1_vs_k141_34878\|\|full | Low-quality |
| S1_vs_k141_121696\|\|full | Low-quality |
| S1_vs_k141_28468\|\|full | Low-quality |
| S1_vs_k141_34880\|\|full | Not-determined |
| S1_vs_k141_61059\|\|full | Not-determined |
| S1_vs_k141_48592\|\|full | Low-quality |
| S1_vs_k141_108493\|\|full | Low-quality |
| S1_vs_k141_41933\|\|full | Not-determined |
| S1_vs_k141_8591\|\|full | Not-determined |
| S1_vs_k141_68518\|\|full | Not-determined |
| S1_vs_k141_21781\|\|full | Low-quality |
| S1_vs_k141_135196\|\|full | Low-quality |
| S1_vs_k141_115238\|\|full | Not-determined |
| S1_vs_k141_108500\|\|full | Low-quality |
| S1_vs_k141_48597\|\|full | Not-determined |
| S1_vs_k141_55242\|\|full | Low-quality |
| S1_vs_k141_34884\|\|full | Low-quality |
| S1_vs_k141_28489\|\|full | Not-determined |
| S1_vs_k141_102005\|\|full | Low-quality |
| S1_vs_k141_15540\|\|full | Not-determined |
| S1_vs_k141_48609\|\|full | Low-quality |
| S1_vs_k141_15554\|\|full | Low-quality |
| S1_vs_k141_41966\|\|full | Not-determined |
| S1_vs_k141_41967\|\|full | Low-quality |
| S1_vs_k141_115256\|\|full | Not-determined |
| S1_vs_k141_102022\|\|full | Not-determined |
| S1_vs_k141_34904\|\|full | Low-quality |
| S1_vs_k141_121749\|\|full | Low-quality |
| S1_vs_k141_154648\|\|full | Low-quality |
| S1_vs_k141_95356\|\|full | Low-quality |
| S1_vs_k141_55271\|\|full | Low-quality |
| S1_vs_k141_141964\|\|full | Not-determined |
| S1_vs_k141_28509\|\|full | Low-quality |
| S1_vs_k141_115285\|\|full | Not-determined |
| S1_vs_k141_15592\|\|full | Not-determined |
| S1_vs_k141_135240\|\|full | Low-quality |
| S1_vs_k141_121762\|\|full | Low-quality |
| S1_vs_k141_88599\|\|full | Not-determined |
| S1_vs_k141_21832\|\|full | Not-determined |
| S1_vs_k141_68587\|\|full | Not-determined |
| S1_vs_k141_121782\|\|full | Not-determined |
| S1_vs_k141_1568\|\|full | Low-quality |
| S1_vs_k141_115327\|\|full | Low-quality |
| S1_vs_k141_142003\|\|full | Low-quality |
| S1_vs_k141_42004\|\|full | Not-determined |
| S1_vs_k141_34946\|\|full | Low-quality |
| S1_vs_k141_21855\|\|full | Low-quality |
| S1_vs_k141_135284\|\|full | Low-quality |
| S1_vs_k141_154690\|\|full | Low-quality |
| S1_vs_k141_88639\|\|full | Low-quality |
| S1_vs_k141_154695\|\|full | Low-quality |
| S1_vs_k141_21865\|\|full | Low-quality |
| S1_vs_k141_142023\|\|full | Low-quality |
| S1_vs_k141_128753\|\|full | Low-quality |
| S1_vs_k141_61092\|\|full | Not-determined |
| S1_vs_k141_8688\|\|full | Low-quality |
| S1_vs_k141_128759\|\|full | Low-quality |
| S1_vs_k141_55338\|\|full | Not-determined |
| S1_vs_k141_42027\|\|full | Not-determined |
| S1_vs_k141_55346\|\|full | Low-quality |
| S1_vs_k141_75354\|\|full | Not-determined |
| S1_vs_k141_75356\|\|full | Low-quality |
| S1_vs_k141_28600\|\|full | Not-determined |
| S1_vs_k141_115341\|\|full | Not-determined |
| S1_vs_k141_42042\|\|full | Low-quality |
| S1_vs_k141_68632\|\|full | Low-quality |
| S1_vs_k141_68634\|\|full | Low-quality |
| S1_vs_k141_61111\|\|full | Low-quality |
| S1_vs_k141_8708\|\|full | Low-quality |
| S1_vs_k141_102152\|\|full | Low-quality |
| S1_vs_k141_115369\|\|full | Not-determined |
| S1_vs_k141_8711\|\|full | Low-quality |
| S1_vs_k141_108640\|\|full | Not-determined |
| S1_vs_k141_88678\|\|full | Low-quality |
| S1_vs_k141_102158\|\|full | Low-quality |
| S1_vs_k141_8717\|\|full | Low-quality |
| S1_vs_k141_135351\|\|full | Low-quality |
| S1_vs_k141_115378\|\|full | Low-quality |
| S1_vs_k141_142081\|\|full | Low-quality |
| S1_vs_k141_8723\|\|full | Low-quality |
| S1_vs_k141_75397\|\|full | Low-quality |
| S1_vs_k141_128808\|\|full | Low-quality |
| S1_vs_k141_102159\|\|full | Low-quality |
| S1_vs_k141_55406\|\|full | Low-quality |
| S1_vs_k141_15733\|\|full | Low-quality |
| S1_vs_k141_82130\|\|full | Low-quality |
| S1_vs_k141_95454\|\|full | Low-quality |
| S1_vs_k141_108658\|\|full | Low-quality |
| S1_vs_k141_88718\|\|full | Low-quality |
| S1_vs_k141_75403\|\|full | Not-determined |
| S1_vs_k141_135378\|\|full | Low-quality |
| S1_vs_k141_95466\|\|full | Low-quality |
| S1_vs_k141_68686\|\|full | Not-determined |
| S1_vs_k141_95472\|\|full | Low-quality |
| S1_vs_k141_68690\|\|full | Not-determined |
| S1_vs_k141_75408\|\|full | Not-determined |
| S1_vs_k141_142118\|\|full | Low-quality |
| S1_vs_k141_55437\|\|full | Low-quality |
| S1_vs_k141_8760\|\|full | Not-determined |
| S1_vs_k141_42097\|\|full | Low-quality |
| S1_vs_k141_102170\|\|full | Low-quality |
| S1_vs_k141_108673\|\|full | Low-quality |
| S1_vs_k141_1654\|\|full | Not-determined |
| S1_vs_k141_95489\|\|full | Low-quality |
| S1_vs_k141_34995\|\|full | Low-quality |
| S1_vs_k141_75433\|\|full | Not-determined |
| S1_vs_k141_42112\|\|full | Low-quality |
| S1_vs_k141_135400\|\|full | Low-quality |
| S1_vs_k141_48783\|\|full | Not-determined |
| S1_vs_k141_21936\|\|full | Not-determined |
| S1_vs_k141_61157\|\|full | Low-quality |
| S1_vs_k141_15803\|\|full | Low-quality |
| S1_vs_k141_15814\|\|full | Not-determined |
| S1_vs_k141_102196\|\|full | Not-determined |
| S1_vs_k141_48796\|\|full | Not-determined |
| S1_vs_k141_82190\|\|full | Not-determined |
| S1_vs_k141_142145\|\|full | Low-quality |
| S1_vs_k141_61167\|\|full | Low-quality |
| S1_vs_k141_88745\|\|full | Low-quality |
| S1_vs_k141_115460\|\|full | Low-quality |
| S1_vs_k141_28706\|\|full | Low-quality |
| S1_vs_k141_95537\|\|full | Low-quality |
| S1_vs_k141_1696\|\|full | Not-determined |
| S1_vs_k141_42148\|\|full | Low-quality |
| S1_vs_k141_154793\|\|full | Low-quality |
| S1_vs_k141_102211\|\|full | Low-quality |
| S1_vs_k141_82197\|\|full | Low-quality |
| S1_vs_k141_128868\|\|full | Low-quality |
| S1_vs_k141_21973\|\|full | Low-quality |
| S1_vs_k141_82202\|\|full | Low-quality |
| S1_vs_k141_108727\|\|full | Low-quality |
| S1_vs_k141_28724\|\|full | Low-quality |
| S1_vs_k141_128878\|\|full | Low-quality |
| S1_vs_k141_68723\|\|full | Not-determined |
| S1_vs_k141_135444\|\|full | Not-determined |
| S1_vs_k141_75480\|\|full | Not-determined |
| S1_vs_k141_68724\|\|full | Low-quality |
| S1_vs_k141_95563\|\|full | Not-determined |
| S1_vs_k141_75488\|\|full | Low-quality |
| S1_vs_k141_88772\|\|full | Not-determined |
| S1_vs_k141_121962\|\|full | Low-quality |
| S1_vs_k141_48832\|\|full | Low-quality |
| S1_vs_k141_88777\|\|full | Not-determined |
| S1_vs_k141_95584\|\|full | Low-quality |
| S1_vs_k141_102255\|\|full | Low-quality |
| S1_vs_k141_35057\|\|full | Not-determined |
| S1_vs_k141_75503\|\|full | Low-quality |
| S1_vs_k141_95592\|\|full | Not-determined |
| S1_vs_k141_82240\|\|full | Low-quality |
| S1_vs_k141_61209\|\|full | Low-quality |
| S1_vs_k141_42180\|\|full | Low-quality |
| S1_vs_k141_121990\|\|full | Not-determined |
| S1_vs_k141_15893\|\|full | Not-determined |
| S1_vs_k141_42188\|\|full | Low-quality |
| S1_vs_k141_128919\|\|full | Not-determined |
| S1_vs_k141_42196\|\|full | Not-determined |
| S1_vs_k141_75515\|\|full | Not-determined |
| S1_vs_k141_55527\|\|full | Low-quality |
| S1_vs_k141_82263\|\|full | Not-determined |
| S1_vs_k141_15926\|\|full | Low-quality |
| S1_vs_k141_42214\|\|full | Not-determined |
| S1_vs_k141_82275\|\|full | Not-determined |
| S1_vs_k141_102291\|\|full | Low-quality |
| S1_vs_k141_122022\|\|full | Not-determined |
| S1_vs_k141_68800\|\|full | Low-quality |
| S1_vs_k141_142243\|\|full | Low-quality |
| S1_vs_k141_154837\|\|full | Low-quality |
| S1_vs_k141_128952\|\|full | Low-quality |
| S1_vs_k141_122035\|\|full | Not-determined |
| S1_vs_k141_135509\|\|full | Low-quality |
| S1_vs_k141_95653\|\|full | Low-quality |
| S1_vs_k141_28804\|\|full | Low-quality |
| S1_vs_k141_154851\|\|full | Not-determined |
| S1_vs_k141_22084\|\|full | Not-determined |
| S1_vs_k141_55538\|\|full | Not-determined |
| S1_vs_k141_95661\|\|full | Not-determined |
| S1_vs_k141_22102\|\|full | Low-quality |
| S1_vs_k141_122056\|\|full | Low-quality |
| S1_vs_k141_55552\|\|full | Not-determined |
| S1_vs_k141_8905\|\|full | Low-quality |
| S1_vs_k141_82307\|\|full | Not-determined |
| S1_vs_k141_75582\|\|full | Low-quality |
| S1_vs_k141_28822\|\|full | Not-determined |
| S1_vs_k141_108844\|\|full | Low-quality |
| S1_vs_k141_22105\|\|full | Not-determined |
| S1_vs_k141_42230\|\|full | Low-quality |
| S1_vs_k141_22110\|\|full | Low-quality |
| S1_vs_k141_102343\|\|full | Low-quality |
| S1_vs_k141_55569\|\|full | Not-determined |
| S1_vs_k141_75601\|\|full | Low-quality |
| S1_vs_k141_35134\|\|full | Not-determined |
| S1_vs_k141_68850\|\|full | Low-quality |
| S1_vs_k141_75604\|\|full | Not-determined |
| S1_vs_k141_108858\|\|full | Low-quality |
| S1_vs_k141_147813\|\|full | Low-quality |
| S1_vs_k141_122093\|\|full | Low-quality |
| S1_vs_k141_102370\|\|full | Low-quality |
| S1_vs_k141_22138\|\|full | Low-quality |
| S1_vs_k141_75609\|\|full | Not-determined |
| S1_vs_k141_55594\|\|full | Low-quality |
| S1_vs_k141_22143\|\|full | Low-quality |
| S1_vs_k141_75630\|\|full | Not-determined |
| S1_vs_k141_55601\|\|full | Not-determined |
| S1_vs_k141_82356\|\|full | Low-quality |
| S1_vs_k141_1815\|\|full | Not-determined |
| S1_vs_k141_115641\|\|full | Low-quality |
| S1_vs_k141_8955\|\|full | Not-determined |
| S1_vs_k141_115646\|\|full | Low-quality |
| S1_vs_k141_75646\|\|full | Low-quality |
| S1_vs_k141_102398\|\|full | Low-quality |
| S1_vs_k141_28886\|\|full | Low-quality |
| S1_vs_k141_1819\|\|full | Low-quality |
| S1_vs_k141_135590\|\|full | Low-quality |
| S1_vs_k141_42301\|\|full | Not-determined |
| S1_vs_k141_102403\|\|full | Low-quality |
| S1_vs_k141_28887\|\|full | Low-quality |
| S1_vs_k141_35182\|\|full | Not-determined |
| S1_vs_k141_16041\|\|full | Not-determined |
| S1_vs_k141_142302\|\|full | Not-determined |
| S1_vs_k141_8966\|\|full | Not-determined |
| S1_vs_k141_88925\|\|full | Low-quality |
| S1_vs_k141_142303\|\|full | Low-quality |
| S1_vs_k141_16044\|\|full | Not-determined |
| S1_vs_k141_68889\|\|full | Low-quality |
| S1_vs_k141_82389\|\|full | Not-determined |
| S1_vs_k141_75659\|\|full | Not-determined |
| S1_vs_k141_28902\|\|full | Low-quality |
| S1_vs_k141_55634\|\|full | Low-quality |
| S1_vs_k141_129066\|\|full | Not-determined |
| S1_vs_k141_135623\|\|full | Not-determined |
| S1_vs_k141_88940\|\|full | Low-quality |
| S1_vs_k141_122182\|\|full | Low-quality |
| S1_vs_k141_115682\|\|full | Low-quality |
| S1_vs_k141_102432\|\|full | Low-quality |
| S1_vs_k141_42336\|\|full | Not-determined |
| S1_vs_k141_49044\|\|full | Low-quality |
| S1_vs_k141_8999\|\|full | Not-determined |
| S1_vs_k141_16062\|\|full | Low-quality |
| S1_vs_k141_9002\|\|full | Low-quality |
| S1_vs_k141_82405\|\|full | Not-determined |
| S1_vs_k141_55654\|\|full | Low-quality |
| S1_vs_k141_82407\|\|full | Not-determined |
| S1_vs_k141_122195\|\|full | Not-determined |
| S1_vs_k141_135633\|\|full | Low-quality |
| S1_vs_k141_154933\|\|full | Low-quality |
| S1_vs_k141_102446\|\|full | Low-quality |
| S11_extracted_k141_33117 | Complete |
| S1_vs_k141_102452\|\|full | Low-quality |
| S1_vs_k141_42358\|\|full | Low-quality |
| S1_vs_k141_16073\|\|full | Low-quality |
| S1_vs_k141_122206\|\|full | Not-determined |
| S1_vs_k141_42364\|\|full | Low-quality |
| S1_vs_k141_35193\|\|full | Not-determined |
| S1_vs_k141_82421\|\|full | Low-quality |
| S1_vs_k141_22155\|\|full | Low-quality |
| S1_vs_k141_9021\|\|full | Low-quality |
| S1_vs_k141_68937\|\|full | Low-quality |
| S1_vs_k141_115719\|\|full | Low-quality |
| S1_vs_k141_115721\|\|full | Low-quality |
| S1_vs_k141_22157\|\|full | Low-quality |
| S1_vs_k141_88985\|\|full | Low-quality |
| S1_vs_k141_102470\|\|full | Low-quality |
| S1_vs_k141_142356\|\|full | Low-quality |
| S1_vs_k141_102489\|\|full | Low-quality |
| S1_vs_k141_16117\|\|full | Not-determined |
| S1_vs_k141_102490\|\|full | Low-quality |
| S1_vs_k141_9043\|\|full | Low-quality |
| S1_vs_k141_154964\|\|full | Low-quality |
| S1_vs_k141_68966\|\|full | Low-quality |
| S1_vs_k141_102496\|\|full | Low-quality |
| S1_vs_k141_95842\|\|full | Low-quality |
| S1_vs_k141_42416\|\|full | Low-quality |
| S1_vs_k141_122257\|\|full | Not-determined |
| S1_vs_k141_9062\|\|full | Not-determined |
| S1_vs_k141_55727\|\|full | Not-determined |
| S1_vs_k141_89011\|\|full | Low-quality |
| S1_vs_k141_68977\|\|full | Not-determined |
| S1_vs_k141_115769\|\|full | Low-quality |
| S1_vs_k141_82477\|\|full | Low-quality |
| S1_vs_k141_16140\|\|full | Low-quality |
| S1_vs_k141_1928\|\|full | Low-quality |
| S1_vs_k141_142395\|\|full | Low-quality |
| S1_vs_k141_28994\|\|full | Not-determined |
| S1_vs_k141_61379\|\|full | Low-quality |
| S1_vs_k141_42434\|\|full | Low-quality |
| S1_vs_k141_49114\|\|full | Low-quality |
| S1_vs_k141_102522\|\|full | Not-determined |
| S1_vs_k141_142408\|\|full | Not-determined |
| S1_vs_k141_115785\|\|full | Low-quality |
| S1_vs_k141_42448\|\|full | Low-quality |
| S1_vs_k141_55775\|\|full | Low-quality |
| S1_vs_k141_16183\|\|full | Not-determined |
| S1_vs_k141_35276\|\|full | Low-quality |
| S1_vs_k141_22216\|\|full | Not-determined |
| S1_vs_k141_29023\|\|full | Low-quality |
| S1_vs_k141_75809\|\|full | Not-determined |
| S1_vs_k141_1969\|\|full | Low-quality |
| S1_vs_k141_82519\|\|full | Low-quality |

| | |
|---|---|
| S1_vs_k141_129233\|\|full | Low-quality |
| S1_vs_k141_109039\|\|full | Not-determined |
| S1_vs_k141_42475\|\|full | Low-quality |
| S1_vs_k141_89072\|\|full | Low-quality |
| S1_vs_k141_16203\|\|full | Not-determined |
| S1_vs_k141_16205\|\|full | Low-quality |
| S1_vs_k141_1990\|\|full | Low-quality |
| S1_vs_k141_55819\|\|full | Low-quality |
| S1_vs_k141_22244\|\|full | Low-quality |
| S1_vs_k141_35293\|\|full | Not-determined |
| S1_vs_k141_35294\|\|full | Not-determined |
| S1_vs_k141_142458\|\|full | Low-quality |
| S1_vs_k141_35296\|\|full | Low-quality |
| S1_vs_k141_155010\|\|full | Not-determined |
| S1_vs_k141_16216\|\|full | Not-determined |
| S1_vs_k141_75836\|\|full | Not-determined |
| S1_vs_k141_135772\|\|full | Low-quality |
| S1_vs_k141_69053\|\|full | Not-determined |
| S1_vs_k141_135774\|\|full | Low-quality |
| S1_vs_k141_9177\|\|full | Low-quality |
| S1_vs_k141_129272\|\|full | Not-determined |
| S1_vs_k141_109088\|\|full | Not-determined |
| S1_vs_k141_82546\|\|full | Low-quality |
| S1_vs_k141_42521\|\|full | Low-quality |
| S1_vs_k141_122344\|\|full | Low-quality |
| S1_vs_k141_89111\|\|full | Low-quality |
| S1_vs_k141_61450\|\|full | Low-quality |
| S1_vs_k141_122349\|\|full | Low-quality |
| S1_vs_k141_16236\|\|full | Not-determined |
| S1_vs_k141_122356\|\|full | Low-quality |
| S1_vs_k141_129286\|\|full | Low-quality |
| S1_vs_k141_95975\|\|full | Low-quality |
| S1_vs_k141_55858\|\|full | Not-determined |
| S1_vs_k141_42541\|\|full | Low-quality |
| S1_vs_k141_55861\|\|full | Low-quality |
| S1_vs_k141_42546\|\|full | Not-determined |
| S1_vs_k141_22283\|\|full | Not-determined |
| S1_vs_k141_42547\|\|full | Low-quality |
| S1_vs_k141_61460\|\|full | Low-quality |
| S1_vs_k141_82572\|\|full | Low-quality |
| S1_vs_k141_69079\|\|full | Not-determined |
| S1_vs_k141_2028\|\|full | Low-quality |
| S1_vs_k141_109128\|\|full | Not-determined |
| S1_vs_k141_69082\|\|full | Low-quality |
| S1_vs_k141_22297\|\|full | Low-quality |
| S1_vs_k141_155039\|\|full | Not-determined |
| S1_vs_k141_115882\|\|full | Low-quality |
| S1_vs_k141_9216\|\|full | Low-quality |
| S1_vs_k141_29121\|\|full | Not-determined |
| S1_vs_k141_115887\|\|full | Low-quality |
| S1_vs_k141_96002\|\|full | Not-determined |
| S1_vs_k141_115896\|\|full | Low-quality |
| S1_vs_k141_16284\|\|full | Low-quality |
| S1_vs_k141_82604\|\|full | Not-determined |
| S1_vs_k141_22313\|\|full | Low-quality |
| S1_vs_k141_49240\|\|full | Low-quality |
| S1_vs_k141_55898\|\|full | Low-quality |
| S1_vs_k141_135847\|\|full | Not-determined |
| S1_vs_k141_69108\|\|full | Low-quality |
| S1_vs_k141_122399\|\|full | Low-quality |
| S1_vs_k141_69113\|\|full | Low-quality |
| S1_vs_k141_16300\|\|full | Not-determined |
| S1_vs_k141_122402\|\|full | Not-determined |
| S1_vs_k141_147943\|\|full | Not-determined |
| S1_vs_k141_129321\|\|full | Not-determined |
| S1_vs_k141_82629\|\|full | Low-quality |
| S1_vs_k141_109169\|\|full | Low-quality |
| S1_vs_k141_155058\|\|full | Not-determined |
| S1_vs_k141_61514\|\|full | Not-determined |
| S1_vs_k141_9256\|\|full | Low-quality |
| S1_vs_k141_16329\|\|full | Low-quality |
| S1_vs_k141_2072\|\|full | Not-determined |
| S1_vs_k141_22341\|\|full | Not-determined |
| S1_vs_k141_82656\|\|full | Low-quality |
| S1_vs_k141_129339\|\|full | Not-determined |
| S1_vs_k141_135889\|\|full | Low-quality |
| S1_vs_k141_82659\|\|full | Low-quality |
| S1_vs_k141_135891\|\|full | Low-quality |
| S1_vs_k141_49297\|\|full | Low-quality |
| S1_vs_k141_42632\|\|full | Low-quality |
| S1_vs_k141_96063\|\|full | Low-quality |
| S1_vs_k141_82662\|\|full | Not-determined |
| S1_vs_k141_42640\|\|full | Low-quality |
| S1_vs_k141_55948\|\|full | Not-determined |
| S1_vs_k141_102687\|\|full | Not-determined |
| S1_vs_k141_135905\|\|full | Low-quality |
| S1_vs_k141_42645\|\|full | Not-determined |
| S1_vs_k141_22361\|\|full | Not-determined |
| S1_vs_k141_49310\|\|full | Not-determined |
| S1_vs_k141_155079\|\|full | Low-quality |
| S1_vs_k141_142583\|\|full | Low-quality |
| S1_vs_k141_16373\|\|full | Low-quality |
| S1_vs_k141_75930\|\|full | Low-quality |
| S1_vs_k141_89189\|\|full | Not-determined |
| S1_vs_k141_22371\|\|full | Not-determined |
| S1_vs_k141_115991\|\|full | Low-quality |
| S1_vs_k141_16383\|\|full | Low-quality |
| S1_vs_k141_155087\|\|full | Not-determined |
| S1_vs_k141_82686\|\|full | Low-quality |
| S1_vs_k141_82691\|\|full | Not-determined |
| S1_vs_k141_55963\|\|full | Low-quality |
| S1_vs_k141_142604\|\|full | Low-quality |
| S1_vs_k141_22385\|\|full | Low-quality |
| S1_vs_k141_2107\|\|full | Low-quality |
| S1_vs_k141_109232\|\|full | Not-determined |
| S1_vs_k141_42685\|\|full | Low-quality |
| S1_vs_k141_135923\|\|full | Low-quality |
| S1_vs_k141_116015\|\|full | Low-quality |
| S1_vs_k141_35417\|\|full | Low-quality |
| S1_vs_k141_89215\|\|full | Not-determined |
| S1_vs_k141_135928\|\|full | Low-quality |
| S1_vs_k141_89218\|\|full | Low-quality |
| S1_vs_k141_42690\|\|full | Low-quality |
| S1_vs_k141_55996\|\|full | Not-determined |
| S1_vs_k141_16415\|\|full | Low-quality |
| S1_vs_k141_82720\|\|full | Low-quality |
| S1_vs_k141_49363\|\|full | Low-quality |
| S1_vs_k141_116025\|\|full | Low-quality |
| S1_vs_k141_102733\|\|full | Low-quality |
| S1_vs_k141_61559\|\|full | Low-quality |
| S1_vs_k141_129399\|\|full | Low-quality |
| S1_vs_k141_29268\|\|full | Low-quality |
| S1_vs_k141_129403\|\|full | Low-quality |
| S1_vs_k141_61563\|\|full | Not-determined |
| S1_vs_k141_142647\|\|full | Not-determined |
| S1_vs_k141_9343\|\|full | Not-determined |
| S1_vs_k141_142654\|\|full | Low-quality |
| S1_vs_k141_42713\|\|full | Not-determined |
| S1_vs_k141_135958\|\|full | Low-quality |
| S1_vs_k141_56020\|\|full | Not-determined |
| S1_vs_k141_76006\|\|full | Low-quality |
| S1_vs_k141_142657\|\|full | Low-quality |
| S1_vs_k141_155140\|\|full | Low-quality |
| S1_vs_k141_96139\|\|full | Not-determined |
| S1_vs_k141_69224\|\|full | Low-quality |
| S1_vs_k141_9362\|\|full | Low-quality |
| S1_vs_k141_49407\|\|full | Low-quality |
| S1_vs_k141_9363\|\|full | Low-quality |
| S1_vs_k141_29298\|\|full | Low-quality |
| S1_vs_k141_135971\|\|full | Not-determined |
| S1_vs_k141_109282\|\|full | Not-determined |
| S1_vs_k141_9365\|\|full | Low-quality |
| S1_vs_k141_56041\|\|full | Low-quality |
| S1_vs_k141_69231\|\|full | Not-determined |
| S1_vs_k141_129436\|\|full | Low-quality |
| S1_vs_k141_109289\|\|full | Not-determined |
| S1_vs_k141_22460\|\|full | Low-quality |
| S1_vs_k141_42747\|\|full | Low-quality |
| S1_vs_k141_16466\|\|full | Not-determined |
| S1_vs_k141_116090\|\|full | Low-quality |
| S1_vs_k141_29331\|\|full | Low-quality |
| S1_vs_k141_22467\|\|full | Low-quality |
| S1_vs_k141_56060\|\|full | Not-determined |
| S1_vs_k141_116093\|\|full | Low-quality |
| S1_vs_k141_122526\|\|full | Low-quality |
| S1_vs_k141_136005\|\|full | Low-quality |
| S1_vs_k141_76047\|\|full | Low-quality |
| S1_vs_k141_9386\|\|full | Low-quality |
| S1_vs_k141_155160\|\|full | Low-quality |
| S1_vs_k141_102810\|\|full | Low-quality |
| S1_vs_k141_116107\|\|full | Low-quality |
| S1_vs_k141_49459\|\|full | Low-quality |
| S1_vs_k141_116110\|\|full | Low-quality |
| S1_vs_k141_89282\|\|full | Low-quality |
| S1_vs_k141_82817\|\|full | Low-quality |
| S1_vs_k141_142716\|\|full | Not-determined |
| S1_vs_k141_96175\|\|full | Low-quality |
| S1_vs_k141_9403\|\|full | Not-determined |
| S1_vs_k141_89286\|\|full | Low-quality |
| S1_vs_k141_116117\|\|full | Low-quality |
| S1_vs_k141_56075\|\|full | Low-quality |
| S1_vs_k141_35492\|\|full | Not-determined |
| S1_vs_k141_56077\|\|full | Low-quality |
| S1_vs_k141_102829\|\|full | Low-quality |
| S1_vs_k141_142719\|\|full | Low-quality |
| S1_vs_k141_61606\|\|full | Low-quality |
| S1_vs_k141_96189\|\|full | Low-quality |
| S1_vs_k141_22506\|\|full | Low-quality |
| S1_vs_k141_136043\|\|full | Low-quality |
| S1_vs_k141_69299\|\|full | Low-quality |
| S1_vs_k141_49494\|\|full | Not-determined |
| S1_vs_k141_96192\|\|full | Not-determined |
| S1_vs_k141_148031\|\|full | Low-quality |
| S1_vs_k141_22522\|\|full | Low-quality |
| S1_vs_k141_22531\|\|full | Low-quality |
| S1_vs_k141_116160\|\|full | Low-quality |
| S1_vs_k141_56108\|\|full | Low-quality |
| S1_vs_k141_76099\|\|full | Low-quality |
| S1_vs_k141_122595\|\|full | Low-quality |
| S1_vs_k141_102860\|\|full | Not-determined |
| S1_vs_k141_76106\|\|full | Not-determined |
| S1_vs_k141_56110\|\|full | Not-determined |
| S1_vs_k141_136082\|\|full | Low-quality |
| S1_vs_k141_2233\|\|full | Low-quality |
| S1_vs_k141_61618\|\|full | Low-quality |
| S1_vs_k141_69340\|\|full | Low-quality |
| S1_vs_k141_16557\|\|full | Low-quality |
| S1_vs_k141_96231\|\|full | Not-determined |
| S1_vs_k141_96234\|\|full | Low-quality |
| S1_vs_k141_122603\|\|full | Not-determined |
| S1_vs_k141_61629\|\|full | Low-quality |
| S1_vs_k141_42835\|\|full | Not-determined |
| S1_vs_k141_102879\|\|full | Not-determined |
| S1_vs_k141_142793\|\|full | Not-determined |
| S1_vs_k141_49539\|\|full | Not-determined |
| S1_vs_k141_136102\|\|full | Low-quality |
| S1_vs_k141_69364\|\|full | Low-quality |
| S1_vs_k141_2251\|\|full | Not-determined |
| S12_vs_k141_188820\|\|full | High-quality |
| S1_vs_k141_69367\|\|full | Low-quality |
| S1_vs_k141_122623\|\|full | Not-determined |
| S1_vs_k141_142807\|\|full | Low-quality |
| S1_vs_k141_96250\|\|full | Low-quality |
| S1_vs_k141_16585\|\|full | Not-determined |
| S1_vs_k141_76130\|\|full | Low-quality |
| S1_vs_k141_42858\|\|full | Low-quality |
| S1_vs_k141_56145\|\|full | Not-determined |
| S1_vs_k141_116206\|\|full | Not-determined |
| S1_vs_k141_49565\|\|full | Not-determined |
| S1_vs_k141_16591\|\|full | Low-quality |
| S1_vs_k141_109387\|\|full | Low-quality |
| S1_vs_k141_29459\|\|full | Not-determined |
| S1_vs_k141_82925\|\|full | Low-quality |
| S1_vs_k141_122637\|\|full | Not-determined |
| S1_vs_k141_102915\|\|full | Not-determined |
| S1_vs_k141_109394\|\|full | Low-quality |
| S1_vs_k141_49576\|\|full | Not-determined |
| S1_vs_k141_122638\|\|full | Low-quality |
| S1_vs_k141_16608\|\|full | Low-quality |
| S1_vs_k141_9523\|\|full | Low-quality |
| S1_vs_k141_129574\|\|full | Not-determined |
| S1_vs_k141_22603\|\|full | Low-quality |
| S1_vs_k141_56159\|\|full | Low-quality |
| S1_vs_k141_42880\|\|full | Low-quality |
| S1_vs_k141_49583\|\|full | Low-quality |
| S1_vs_k141_22608\|\|full | Not-determined |
| S1_vs_k141_42883\|\|full | Not-determined |
| S1_vs_k141_109409\|\|full | Low-quality |
| S1_vs_k141_56165\|\|full | Low-quality |
| S1_vs_k141_116252\|\|full | Not-determined |
| S1_vs_k141_76166\|\|full | Low-quality |
| S1_vs_k141_136127\|\|full | Low-quality |
| S1_vs_k141_42888\|\|full | Low-quality |
| S1_vs_k141_109411\|\|full | Low-quality |
| S1_vs_k141_56168\|\|full | Low-quality |
| S1_vs_k141_89400\|\|full | Not-determined |
| S1_vs_k141_29487\|\|full | Low-quality |
| S1_vs_k141_136135\|\|full | Low-quality |
| S1_vs_k141_155212\|\|full | Low-quality |
| S1_vs_k141_89418\|\|full | Low-quality |
| S1_vs_k141_102957\|\|full | Not-determined |
| S1_vs_k141_56194\|\|full | Not-determined |
| S1_vs_k141_155213\|\|full | Not-determined |
| S1_vs_k141_109428\|\|full | Not-determined |
| S1_vs_k141_49621\|\|full | Not-determined |
| S1_vs_k141_9574\|\|full | Not-determined |
| S1_vs_k141_155216\|\|full | Low-quality |
| S1_vs_k141_129626\|\|full | Low-quality |
| S1_vs_k141_61660\|\|full | Low-quality |
| S1_vs_k141_9575\|\|full | Not-determined |
| S1_vs_k141_116292\|\|full | Low-quality |
| S1_vs_k141_129634\|\|full | Low-quality |
| S1_vs_k141_9579\|\|full | Not-determined |
| S1_vs_k141_142839\|\|full | Not-determined |
| S1_vs_k141_89436\|\|full | Low-quality |
| S1_vs_k141_102982\|\|full | Low-quality |
| S1_vs_k141_76225\|\|full | Not-determined |
| S1_vs_k141_56214\|\|full | Low-quality |
| S1_vs_k141_76226\|\|full | Low-quality |
| S1_vs_k141_2345\|\|full | Not-determined |
| S1_vs_k141_129646\|\|full | Not-determined |
| S1_vs_k141_109437\|\|full | Low-quality |
| S1_vs_k141_89447\|\|full | Low-quality |
| S1_vs_k141_16700\|\|full | Low-quality |
| S1_vs_k141_136180\|\|full | Low-quality |
| S1_vs_k141_76233\|\|full | Not-determined |
| S1_vs_k141_9610\|\|full | Not-determined |
| S1_vs_k141_103022\|\|full | Not-determined |
| S1_vs_k141_155233\|\|full | Low-quality |
| S1_vs_k141_42944\|\|full | Low-quality |
| S1_vs_k141_109462\|\|full | Low-quality |
| S1_vs_k141_2362\|\|full | Not-determined |
| S1_vs_k141_103029\|\|full | Not-determined |
| S9_extracted_k141_68272 | Medium-quality |
| S1_vs_k141_42951\|\|full | Low-quality |
| S1_vs_k141_83024\|\|full | Not-determined |
| S1_vs_k141_22684\|\|full | Low-quality |
| S1_vs_k141_9630\|\|full | Low-quality |
| S1_vs_k141_148065\|\|full | Low-quality |
| S1_vs_k141_96357\|\|full | Low-quality |
| S1_vs_k141_9634\|\|full | Low-quality |
| S1_vs_k141_22687\|\|full | Not-determined |
| S1_vs_k141_96360\|\|full | Low-quality |
| S1_vs_k141_83035\|\|full | Not-determined |
| S1_vs_k141_56259\|\|full | Low-quality |
| S1_vs_k141_56261\|\|full | Low-quality |
| S1_vs_k141_89487\|\|full | Low-quality |
| S1_vs_k141_103056\|\|full | Low-quality |
| S1_vs_k141_89491\|\|full | Low-quality |
| S1_vs_k141_76279\|\|full | Low-quality |
| S1_vs_k141_9651\|\|full | Not-determined |
| S1_vs_k141_69477\|\|full | Low-quality |
| S1_vs_k141_122748\|\|full | Not-determined |
| S1_vs_k141_155257\|\|full | Low-quality |
| S1_vs_k141_42979\|\|full | Low-quality |
| S1_vs_k141_116361\|\|full | Not-determined |
| S1_vs_k141_96390\|\|full | Low-quality |
| S1_vs_k141_61700\|\|full | Low-quality |
| S1_vs_k141_69490\|\|full | Not-determined |
| S1_vs_k141_35658\|\|full | Low-quality |
| S1_vs_k141_9668\|\|full | Low-quality |
| S1_vs_k141_42994\|\|full | Low-quality |
| S1_vs_k141_9669\|\|full | Low-quality |
| S1_vs_k141_16762\|\|full | Not-determined |
| S1_vs_k141_76291\|\|full | Low-quality |
| S1_vs_k141_103087\|\|full | Low-quality |
| S1_vs_k141_129704\|\|full | Low-quality |
| S1_vs_k141_29589\|\|full | Low-quality |
| S1_vs_k141_155271\|\|full | Low-quality |
| S1_vs_k141_22711\|\|full | Not-determined |
| S1_vs_k141_69514\|\|full | Low-quality |
| S1_vs_k141_129717\|\|full | Low-quality |
| S1_vs_k141_22716\|\|full | Not-determined |
| S1_vs_k141_142896\|\|full | Low-quality |
| S1_vs_k141_69518\|\|full | Low-quality |
| S1_vs_k141_22725\|\|full | Low-quality |
| S1_vs_k141_9690\|\|full | Low-quality |
| S1_vs_k141_29610\|\|full | Low-quality |
| S1_vs_k141_116400\|\|full | Not-determined |
| S1_vs_k141_96426\|\|full | Low-quality |
| S1_vs_k141_43034\|\|full | Not-determined |
| S1_vs_k141_49763\|\|full | Not-determined |
| S1_vs_k141_76333\|\|full | Low-quality |
| S1_vs_k141_89561\|\|full | Low-quality |
| S1_vs_k141_9696\|\|full | Not-determined |
| S1_vs_k141_22733\|\|full | Low-quality |
| S1_vs_k141_56304\|\|full | Low-quality |
| S1_vs_k141_29621\|\|full | Low-quality |
| S1_vs_k141_61752\|\|full | Low-quality |
| S1_vs_k141_122805\|\|full | Not-determined |
| S1_vs_k141_69532\|\|full | Low-quality |
| S1_vs_k141_83079\|\|full | Not-determined |
| S1_vs_k141_155286\|\|full | Low-quality |
| S1_vs_k141_49795\|\|full | Low-quality |
| S1_vs_k141_69546\|\|full | Low-quality |
| S1_vs_k141_122821\|\|full | Low-quality |
| S1_vs_k141_49799\|\|full | Low-quality |
| S1_vs_k141_96449\|\|full | Low-quality |
| S1_vs_k141_56323\|\|full | Low-quality |
| S1_vs_k141_16820\|\|full | Low-quality |
| S1_vs_k141_148101\|\|full | Not-determined |
| S1_vs_k141_16823\|\|full | Low-quality |
| S1_vs_k141_49823\|\|full | Low-quality |
| S1_vs_k141_148103\|\|full | Not-determined |
| S1_vs_k141_16832\|\|full | Not-determined |
| S1_vs_k141_9745\|\|full | Low-quality |
| S1_vs_k141_116449\|\|full | Low-quality |
| S1_vs_k141_9746\|\|full | Low-quality |
| S1_vs_k141_136327\|\|full | Low-quality |
| S1_vs_k141_76369\|\|full | Not-determined |
| S1_vs_k141_89615\|\|full | Not-determined |
| S1_vs_k141_56338\|\|full | Low-quality |
| S1_vs_k141_96483\|\|full | Low-quality |
| S1_vs_k141_142950\|\|full | Low-quality |
| S1_vs_k141_83113\|\|full | Low-quality |
| S1_vs_k141_35740\|\|full | Low-quality |
| S1_vs_k141_148113\|\|full | Low-quality |
| S1_vs_k141_9762\|\|full | Low-quality |
| S1_vs_k141_83124\|\|full | Not-determined |
| S1_vs_k141_122864\|\|full | Low-quality |
| S1_vs_k141_2485\|\|full | Low-quality |
| S1_vs_k141_136345\|\|full | Not-determined |
| S1_vs_k141_142963\|\|full | Low-quality |
| S1_vs_k141_109628\|\|full | Low-quality |
| S1_vs_k141_61792\|\|full | Not-determined |
| S1_vs_k141_22809\|\|full | Low-quality |
| S1_vs_k141_61799\|\|full | Low-quality |
| S1_vs_k141_89635\|\|full | Low-quality |
| S1_vs_k141_35758\|\|full | Not-determined |
| S1_vs_k141_136354\|\|full | Low-quality |
| S1_vs_k141_96505\|\|full | Low-quality |
| S1_vs_k141_22821\|\|full | Low-quality |
| S1_vs_k141_56364\|\|full | Low-quality |
| S1_vs_k141_122871\|\|full | Not-determined |
| S1_vs_k141_9775\|\|full | Not-determined |
| S1_vs_k141_89645\|\|full | Low-quality |
| S1_vs_k141_35761\|\|full | Low-quality |
| S1_vs_k141_136355\|\|full | Not-determined |
| S1_vs_k141_56370\|\|full | Not-determined |
| S1_vs_k141_16880\|\|full | Not-determined |
| S1_vs_k141_83155\|\|full | Low-quality |
| S1_vs_k141_49882\|\|full | Low-quality |
| S1_vs_k141_116503\|\|full | Low-quality |
| S1_vs_k141_35763\|\|full | Low-quality |
| S1_vs_k141_122876\|\|full | Low-quality |
| S1_vs_k141_109646\|\|full | Low-quality |
| S1_vs_k141_16890\|\|full | Low-quality |
| S1_vs_k141_136365\|\|full | Not-determined |
| S1_vs_k141_76430\|\|full | Not-determined |
| S1_vs_k141_43143\|\|full | Not-determined |
| S1_vs_k141_61824\|\|full | Not-determined |
| S1_vs_k141_122886\|\|full | Low-quality |
| S1_vs_k141_9800\|\|full | Not-determined |
| S1_vs_k141_35773\|\|full | Not-determined |
| S1_vs_k141_69623\|\|full | Low-quality |
| S1_vs_k141_116521\|\|full | Low-quality |
| S1_vs_k141_83173\|\|full | Low-quality |
| S1_vs_k141_122895\|\|full | Low-quality |
| S1_vs_k141_129840\|\|full | Low-quality |
| S1_vs_k141_89678\|\|full | Low-quality |
| S1_vs_k141_9816\|\|full | Low-quality |
| S1_vs_k141_16910\|\|full | Not-determined |
| S1_vs_k141_103255\|\|full | Low-quality |
| S1_vs_k141_136395\|\|full | Low-quality |
| S1_vs_k141_61841\|\|full | Not-determined |
| S1_vs_k141_103265\|\|full | Low-quality |
| S1_vs_k141_129860\|\|full | Not-determined |
| S1_vs_k141_9828\|\|full | Not-determined |
| S1_vs_k141_103272\|\|full | Not-determined |
| S1_vs_k141_109688\|\|full | Low-quality |
| S1_vs_k141_9843\|\|full | Not-determined |
| S1_vs_k141_69662\|\|full | Low-quality |
| S1_vs_k141_2565\|\|full | Not-determined |
| S1_vs_k141_148167\|\|full | Low-quality |
| S1_vs_k141_96578\|\|full | Low-quality |
| S1_vs_k141_69668\|\|full | Not-determined |
| S1_vs_k141_96582\|\|full | Low-quality |
| S1_vs_k141_89710\|\|full | Not-determined |
| S1_vs_k141_49938\|\|full | Not-determined |
| S1_vs_k141_16965\|\|full | Low-quality |
| S1_vs_k141_29746\|\|full | Low-quality |
| S1_vs_k141_129901\|\|full | Low-quality |
| S1_vs_k141_49941\|\|full | Low-quality |
| S1_vs_k141_61897\|\|full | Not-determined |
| S1_vs_k141_89731\|\|full | Not-determined |
| S1_vs_k141_116585\|\|full | Not-determined |
| S1_vs_k141_103310\|\|full | Low-quality |
| S1_vs_k141_116592\|\|full | Low-quality |
| S1_vs_k141_49948\|\|full | Low-quality |
| S1_vs_k141_136449\|\|full | Low-quality |
| S1_vs_k141_116595\|\|full | Low-quality |
| S1_vs_k141_136454\|\|full | Not-determined |
| S1_vs_k141_148182\|\|full | Not-determined |
| S1_vs_k141_83249\|\|full | Not-determined |
| S1_vs_k141_129927\|\|full | Not-determined |
| S1_vs_k141_143066\|\|full | Low-quality |
| S1_vs_k141_103317\|\|full | Low-quality |
| S1_vs_k141_9870\|\|full | Not-determined |
| S1_vs_k141_17015\|\|full | Low-quality |
| S1_vs_k141_129935\|\|full | Low-quality |
| S1_vs_k141_103321\|\|full | Not-determined |
| S1_vs_k141_129936\|\|full | Low-quality |
| S1_vs_k141_148197\|\|full | Low-quality |
| S1_vs_k141_22905\|\|full | Not-determined |
| S1_vs_k141_69704\|\|full | Low-quality |
| S1_vs_k141_116621\|\|full | Low-quality |
| S1_vs_k141_29800\|\|full | Not-determined |
| S1_vs_k141_43222\|\|full | Low-quality |
| S1_vs_k141_83277\|\|full | Not-determined |
| S1_vs_k141_89743\|\|full | Low-quality |
| S1_vs_k141_49999\|\|full | Low-quality |
| S1_vs_k141_122997\|\|full | Low-quality |
| S1_vs_k141_9902\|\|full | Low-quality |
| S1_vs_k141_76551\|\|full | Low-quality |
| S1_vs_k141_50006\|\|full | Low-quality |
| S1_vs_k141_122999\|\|full | Not-determined |
| S1_vs_k141_89759\|\|full | Low-quality |
| S1_vs_k141_22927\|\|full | Low-quality |
| S1_vs_k141_61947\|\|full | Not-determined |
| S3_vs_k141_56292\|\|full | High-quality |
| S1_vs_k141_56493\|\|full | Low-quality |
| S1_vs_k141_129976\|\|full | Low-quality |
| S1_vs_k141_50021\|\|full | Low-quality |
| S1_vs_k141_29833\|\|full | Not-determined |
| S1_vs_k141_116658\|\|full | Low-quality |
| S1_vs_k141_89772\|\|full | Low-quality |
| S1_vs_k141_9928\|\|full | Low-quality |
| S1_vs_k141_129979\|\|full | Not-determined |
| S1_vs_k141_109808\|\|full | Not-determined |
| S1_vs_k141_50028\|\|full | Low-quality |
| S1_vs_k141_50029\|\|full | Low-quality |
| S1_vs_k141_143122\|\|full | Low-quality |
| S1_vs_k141_143129\|\|full | Low-quality |
| S1_vs_k141_103370\|\|full | Low-quality |
| S1_vs_k141_103372\|\|full | Not-determined |
| S1_vs_k141_43284\|\|full | Low-quality |
| S1_vs_k141_17080\|\|full | Low-quality |
| S1_vs_k141_155388\|\|full | Low-quality |
| S1_vs_k141_123034\|\|full | Not-determined |
| S1_vs_k141_109834\|\|full | Low-quality |
| S1_vs_k141_116699\|\|full | Not-determined |
| S1_vs_k141_43297\|\|full | Low-quality |
| S1_vs_k141_76590\|\|full | Not-determined |
| S1_vs_k141_109841\|\|full | Not-determined |
| S1_vs_k141_130024\|\|full | Low-quality |
| S1_vs_k141_43311\|\|full | Low-quality |
| S1_vs_k141_9965\|\|full | Low-quality |
| S1_vs_k141_89806\|\|full | Low-quality |
| S1_vs_k141_35931\|\|full | Not-determined |
| S1_vs_k141_116718\|\|full | Low-quality |
| S1_vs_k141_96739\|\|full | Not-determined |
| S1_vs_k141_76618\|\|full | Low-quality |
| S1_vs_k141_89819\|\|full | Low-quality |
| S1_vs_k141_9981\|\|full | Not-determined |
| S1_vs_k141_143157\|\|full | Not-determined |
| S1_vs_k141_62040\|\|full | Not-determined |
| S1_vs_k141_96766\|\|full | Low-quality |
| S1_vs_k141_143165\|\|full | Low-quality |
| S1_vs_k141_123082\|\|full | Not-determined |
| S1_vs_k141_43354\|\|full | Low-quality |
| S1_vs_k141_148274\|\|full | Low-quality |
| S1_vs_k141_50117\|\|full | Low-quality |
| S11_vs_k141_33117\|\|full | High-quality |
| S1_vs_k141_103445\|\|full | Low-quality |
| S1_vs_k141_103446\|\|full | Low-quality |
| S1_vs_k141_62045\|\|full | Not-determined |
| S1_vs_k141_56589\|\|full | Low-quality |
| S1_vs_k141_17087\|\|full | Low-quality |
| S1_vs_k141_123094\|\|full | Not-determined |
| S1_vs_k141_83404\|\|full | Low-quality |
| S1_vs_k141_22986\|\|full | Low-quality |
| S1_vs_k141_155442\|\|full | Not-determined |
| S1_vs_k141_35975\|\|full | Low-quality |
| S1_vs_k141_123100\|\|full | Not-determined |
| S1_vs_k141_10023\|\|full | Not-determined |
| S1_vs_k141_56611\|\|full | Not-determined |
| S1_vs_k141_69771\|\|full | Not-determined |
| S1_vs_k141_123112\|\|full | Low-quality |
| S5_vs_k141_24951\|\|full | Complete |
| S1_vs_k141_96818\|\|full | Low-quality |
| S1_vs_k141_123115\|\|full | Low-quality |
| S1_vs_k141_109932\|\|full | Not-determined |
| S1_vs_k141_43402\|\|full | Low-quality |
| S1_vs_k141_29942\|\|full | Not-determined |
| S1_vs_k141_50143\|\|full | Low-quality |
| S1_vs_k141_50145\|\|full | Not-determined |
| S1_vs_k141_83426\|\|full | Not-determined |
| S1_vs_k141_29953\|\|full | Low-quality |
| S1_vs_k141_35991\|\|full | Not-determined |
| S1_vs_k141_43414\|\|full | Not-determined |
| S1_vs_k141_130107\|\|full | Low-quality |
| S1_vs_k141_116803\|\|full | Low-quality |
| S1_vs_k141_10083\|\|full | Not-determined |
| S1_vs_k141_116811\|\|full | Low-quality |
| S1_vs_k141_143236\|\|full | Not-determined |
| S1_vs_k141_136576\|\|full | Not-determined |
| S1_vs_k141_148305\|\|full | Low-quality |
| S1_vs_k141_96857\|\|full | Low-quality |
| S1_vs_k141_2724\|\|full | Low-quality |
| S1_vs_k141_83462\|\|full | Low-quality |
| S1_vs_k141_76725\|\|full | Low-quality |
| S1_vs_k141_69816\|\|full | Low-quality |
| S1_vs_k141_2729\|\|full | Not-determined |
| S1_vs_k141_43437\|\|full | Low-quality |
| S1_vs_k141_10098\|\|full | Low-quality |
| S1_vs_k141_96866\|\|full | Low-quality |
| S1_vs_k141_109996\|\|full | Low-quality |
| S1_vs_k141_29992\|\|full | Not-determined |
| S1_vs_k141_155463\|\|full | Not-determined |
| S1_vs_k141_2738\|\|full | Low-quality |
| S1_vs_k141_56688\|\|full | Low-quality |
| S1_vs_k141_76731\|\|full | Low-quality |
| S1_vs_k141_43448\|\|full | Low-quality |
| S1_vs_k141_69819\|\|full | Low-quality |
| S1_vs_k141_143264\|\|full | Low-quality |
| S1_vs_k141_56700\|\|full | Not-determined |
| S1_vs_k141_43453\|\|full | Low-quality |
| S1_vs_k141_103529\|\|full | Low-quality |
| S1_vs_k141_2747\|\|full | Not-determined |
| S1_vs_k141_155473\|\|full | Not-determined |
| S1_vs_k141_116837\|\|full | Low-quality |
| S1_vs_k141_89949\|\|full | Low-quality |
| S1_vs_k141_50229\|\|full | Low-quality |
| S1_vs_k141_17207\|\|full | Low-quality |
| S1_vs_k141_83504\|\|full | Not-determined |
| S1_vs_k141_130164\|\|full | Low-quality |
| S1_vs_k141_96904\|\|full | Not-determined |
| S1_vs_k141_136616\|\|full | Low-quality |
| S1_vs_k141_62139\|\|full | Not-determined |
| S1_vs_k141_123222\|\|full | Low-quality |
| S1_vs_k141_50243\|\|full | Not-determined |
| S1_vs_k141_2771\|\|full | Low-quality |
| S1_vs_k141_143281\|\|full | Low-quality |
| S1_vs_k141_143282\|\|full | Not-determined |
| S1_vs_k141_103550\|\|full | Not-determined |
| S1_vs_k141_69834\|\|full | Low-quality |
| S1_vs_k141_23101\|\|full | Low-quality |
| S1_vs_k141_56728\|\|full | Low-quality |
| S1_vs_k141_62162\|\|full | Low-quality |

S1_vs_k141_56733||full Low-quality
S1_vs_k141_155508||full Not-determined
S1_vs_k141_116884||full Low-quality
S1_vs_k141_96945||full Low-quality
S1_vs_k141_36084||full Low-quality
S1_vs_k141_2797||full Low-quality
S1_vs_k141_143302||full Not-determined
S1_vs_k141_23119||full Low-quality
S1_vs_k141_123250||full Not-determined
S1_vs_k141_96955||full Low-quality
S1_vs_k141_103587||full Not-determined
S1_vs_k141_62187||full Low-quality
S1_vs_k141_23124||full Not-determined
S1_vs_k141_83575||full Not-determined
S1_vs_k141_123264||full Low-quality
S1_vs_k141_50291||full Low-quality
S1_vs_k141_62196||full Low-quality
S1_vs_k141_96972||full Not-determined
S1_vs_k141_130193||full Low-quality
S1_vs_k141_17249||full Low-quality
S1_vs_k141_10199||full Low-quality
S1_vs_k141_36109||full Low-quality
S1_vs_k141_30104||full Low-quality
S1_vs_k141_69860||full Not-determined
S1_vs_k141_130198||full Not-determined
S1_vs_k141_155534||full Not-determined
S1_vs_k141_83594||full Not-determined
S1_vs_k141_103617||full Not-determined
S1_vs_k141_36123||full Low-quality
S1_vs_k141_36124||full Low-quality
S1_vs_k141_43546||full Low-quality
S1_vs_k141_90045||full Low-quality
S1_vs_k141_143352||full Low-quality
S1_vs_k141_136718||full Not-determined
S1_vs_k141_116946||full Low-quality
S1_vs_k141_130216||full Low-quality
S1_vs_k141_76856||full Low-quality
S1_vs_k141_130223||full Not-determined
S1_vs_k141_30131||full Not-determined
S1_vs_k141_110116||full Low-quality
S1_vs_k141_43557||full Low-quality
S1_vs_k141_83632||full Low-quality
S1_vs_k141_17301||full Low-quality
S1_vs_k141_97017||full Low-quality
S1_vs_k141_2838||full Not-determined
S1_vs_k141_76867||full Low-quality
S1_vs_k141_36127||full Not-determined
S1_vs_k141_23215||full Low-quality
S1_vs_k141_23218||full Low-quality
S1_vs_k141_56839||full Low-quality
S1_vs_k141_17313||full Low-quality
S1_vs_k141_62267||full Low-quality
S1_vs_k141_83664||full Low-quality
S1_vs_k141_103657||full Low-quality
S1_vs_k141_17319||full Not-determined
S1_vs_k141_110145||full Low-quality
S1_vs_k141_23244||full Low-quality
S1_vs_k141_123352||full Low-quality
S1_vs_k141_56866||full Low-quality
S1_vs_k141_76904||full Not-determined
S1_vs_k141_136772||full Low-quality
S1_vs_k141_43599||full Low-quality
S1_vs_k141_83677||full Not-determined
S1_vs_k141_97051||full Low-quality
S1_vs_k141_136775||full Not-determined
S1_vs_k141_76911||full Low-quality
S1_vs_k141_148411||full Low-quality
S1_vs_k141_123358||full Low-quality
S1_vs_k141_130252||full Low-quality
S1_vs_k141_10291||full Low-quality
S1_vs_k141_136781||full Not-determined
S1_vs_k141_83685||full Low-quality
S1_vs_k141_110167||full Low-quality
S1_vs_k141_2878||full Low-quality
S1_vs_k141_50339||full Low-quality
S1_vs_k141_23259||full Low-quality
S1_vs_k141_143426||full Not-determined
S1_vs_k141_90107||full Not-determined
S1_vs_k141_143427||full Low-quality
S1_vs_k141_23261||full Low-quality
S1_vs_k141_50343||full Low-quality
S1_vs_k141_123375||full Not-determined
S1_vs_k141_56893||full Not-determined
S1_vs_k141_17342||full Not-determined
S1_vs_k141_2888||full Low-quality
S1_vs_k141_155611||full Not-determined
S1_vs_k141_50351||full Not-determined
S1_vs_k141_50356||full Not-determined
S1_vs_k141_56901||full Low-quality
S1_vs_k141_83704||full Not-determined
S1_vs_k141_43629||full Low-quality
S1_vs_k141_50369||full Low-quality
S1_vs_k141_43632||full Low-quality
S1_vs_k141_148424||full Not-determined
S1_vs_k141_103715||full Low-quality
S1_vs_k141_2905||full Low-quality
S1_vs_k141_50377||full Low-quality
S1_vs_k141_56918||full Low-quality
S1_vs_k141_97088||full Low-quality
S1_vs_k141_90140||full Not-determined
S1_vs_k141_69924||full Low-quality
S1_vs_k141_76974||full Low-quality
S1_vs_k141_136821||full Not-determined
S1_vs_k141_136824||full Low-quality
S1_vs_k141_103731||full Not-determined
S1_vs_k141_117076||full Low-quality
S1_vs_k141_2919||full Not-determined
S1_vs_k141_155626||full Low-quality
S1_vs_k141_10354||full Low-quality
S1_vs_k141_90151||full Not-determined
S1_vs_k141_30257||full Low-quality
S1_vs_k141_97109||full Low-quality
S1_vs_k141_155635||full Low-quality
S1_vs_k141_30265||full Low-quality
S1_vs_k141_50417||full Low-quality
S1_vs_k141_130337||full Low-quality
S1_vs_k141_43668||full Low-quality
S1_vs_k141_30282||full Not-determined
S1_vs_k141_123437||full Low-quality
S1_vs_k141_62356||full Low-quality
S1_vs_k141_10388||full Not-determined
S1_vs_k141_50446||full Not-determined
S1_vs_k141_143473||full Low-quality
S1_vs_k141_83785||full Low-quality
S1_vs_k141_77021||full Not-determined
S1_vs_k141_123460||full Low-quality
S1_vs_k141_103749||full Low-quality
S1_vs_k141_50458||full Not-determined
S1_vs_k141_2970||full Low-quality
S1_vs_k141_136900||full Not-determined
S1_vs_k141_136901||full Low-quality
S1_vs_k141_117126||full Low-quality
S1_vs_k141_136903||full Not-determined
S1_vs_k141_136904||full Not-determined
S1_vs_k141_83787||full Low-quality
S1_vs_k141_117132||full Not-determined
S1_vs_k141_17449||full Low-quality
S1_vs_k141_110286||full Low-quality
S1_vs_k141_123489||full Not-determined
S1_vs_k141_130373||full Low-quality
S1_vs_k141_103765||full Not-determined
S1_vs_k141_10420||full Low-quality
S1_vs_k141_97169||full Not-determined
S1_vs_k141_123498||full Not-determined
S1_vs_k141_103778||full Not-determined
S1_vs_k141_117157||full Low-quality
S1_vs_k141_17466||full Not-determined
S1_vs_k141_123503||full Low-quality
S1_vs_k141_123508||full Low-quality
S1_vs_k141_110316||full Low-quality
S1_vs_k141_130398||full Low-quality
S1_vs_k141_77087||full Low-quality
S1_vs_k141_130399||full Low-quality
S1_vs_k141_123509||full Low-quality
S1_vs_k141_23408||full Not-determined
S16_extracted_96483 Medium-quality
S1_vs_k141_148520||full Low-quality
S1_vs_k141_69970||full Low-quality
S1_vs_k141_50526||full Low-quality
S1_vs_k141_90265||full Not-determined
S1_vs_k141_103827||full Low-quality
S1_vs_k141_117190||full Low-quality
S1_vs_k141_136968||full Not-determined
S1_vs_k141_143517||full Low-quality
S1_vs_k141_30385||full Low-quality
S1_vs_k141_23438||full Low-quality
S1_vs_k141_117197||full Low-quality
S1_vs_k141_130440||full Not-determined
S1_vs_k141_136975||full Low-quality
S1_vs_k141_57065||full Low-quality
S1_vs_k141_103839||full Not-determined
S1_vs_k141_43763||full Low-quality
S1_vs_k141_62449||full Low-quality
S1_vs_k141_130452||full Not-determined
S1_vs_k141_136983||full Not-determined
S1_vs_k141_70002||full Low-quality
S1_vs_k141_50555||full Low-quality
S1_vs_k141_57082||full Not-determined
S1_vs_k141_90305||full Not-determined
S1_vs_k141_136992||full Low-quality
S1_vs_k141_117218||full Low-quality
S1_vs_k141_10517||full Low-quality
S1_vs_k141_30411||full Low-quality
S1_vs_k141_57103||full Low-quality
S1_vs_k141_110388||full Low-quality
S1_vs_k141_43802||full Low-quality
S1_vs_k141_50589||full Not-determined
S1_vs_k141_155719||full Low-quality
S1_vs_k141_17562||full Low-quality
S1_vs_k141_97292||full Low-quality
S1_vs_k141_130497||full Not-determined
S1_vs_k141_57126||full Not-determined
S1_vs_k141_117263||full Low-quality
S1_vs_k141_77166||full Not-determined
S1_vs_k141_23526||full Low-quality
S1_vs_k141_148556||full Low-quality
S1_vs_k141_23532||full Low-quality
S1_vs_k141_83898||full Not-determined
S1_vs_k141_10556||full Not-determined
S1_vs_k141_130521||full Not-determined
S8_vs_k141_201752||full Medium-quality
S1_vs_k141_23540||full Not-determined
S1_vs_k141_77181||full Low-quality
S1_vs_k141_83913||full Low-quality
S1_vs_k141_117288||full Low-quality
S8_extracted_k141_208120 Complete
S1_vs_k141_97324||full Not-determined
S1_vs_k141_143608||full Not-determined
S1_vs_k141_117297||full Not-determined
S1_vs_k141_36321||full Low-quality
S1_vs_k141_36324||full Low-quality
S1_vs_k141_23563||full Low-quality
S1_vs_k141_117303||full Not-determined
S1_vs_k141_30484||full Low-quality
S1_vs_k141_57165||full Low-quality
S1_vs_k141_10607||full Low-quality
S1_vs_k141_50650||full Not-determined
S1_vs_k141_90414||full Not-determined
S1_vs_k141_143634||full Low-quality
S1_vs_k141_103940||full Not-determined
S1_vs_k141_57180||full Not-determined
S1_vs_k141_123626||full Low-quality
S1_vs_k141_30516||full Not-determined
S1_vs_k141_97370||full Not-determined
S1_vs_k141_3034||full Low-quality
S1_vs_k141_17596||full Low-quality
S1_vs_k141_36351||full Low-quality
S1_vs_k141_17597||full Low-quality
S1_vs_k141_143667||full Low-quality
S1_vs_k141_23606||full Low-quality
S1_vs_k141_43945||full Low-quality
S1_vs_k141_77214||full Low-quality
S1_vs_k141_70084||full Not-determined
S1_vs_k141_50693||full Low-quality
S1_vs_k141_10647||full Low-quality
S1_vs_k141_103990||full Low-quality
S1_vs_k141_130612||full Not-determined
S1_vs_k141_90463||full Low-quality
S16_vs_96483||full Medium-quality
S1_vs_k141_110542||full Low-quality
S1_vs_k141_84004||full Not-determined
S1_vs_k141_117320||full Low-quality
S1_vs_k141_10682||full Low-quality
S1_vs_k141_50715||full Low-quality
S1_vs_k141_84010||full Low-quality
S1_vs_k141_84019||full Not-determined
S1_vs_k141_17679||full Low-quality
S1_vs_k141_36402||full Low-quality
S1_vs_k141_117337||full Low-quality
S1_vs_k141_77264||full Low-quality
S1_vs_k141_123683||full Low-quality
S1_vs_k141_143727||full Not-determined
S1_vs_k141_143729||full Not-determined
S1_vs_k141_137204||full Not-determined
S1_vs_k141_44012||full Low-quality
S1_vs_k141_130675||full Low-quality
S1_vs_k141_77278||full Low-quality
S1_vs_k141_17706||full Low-quality
S1_vs_k141_117378||full Low-quality
S1_vs_k141_130676||full Low-quality
S1_vs_k141_110601||full Not-determined
S1_vs_k141_130679||full Low-quality
S1_vs_k141_17717||full Not-determined
S1_vs_k141_110602||full Low-quality
S1_vs_k141_23678||full Low-quality
S1_vs_k141_77294||full Low-quality
S1_vs_k141_23680||full Low-quality
S1_vs_k141_10733||full Low-quality
S1_vs_k141_62653||full Low-quality
S1_vs_k141_50760||full Low-quality
S1_vs_k141_70168||full Low-quality
S1_vs_k141_57307||full Low-quality
S1_vs_k141_84082||full Low-quality
S1_vs_k141_62672||full Not-determined
S1_vs_k141_36427||full Low-quality
S1_vs_k141_3093||full Low-quality
S1_vs_k141_155806||full Low-quality
S1_vs_k141_117415||full Low-quality
S1_vs_k141_117418||full Low-quality
S1_vs_k141_30679||full Low-quality
S1_vs_k141_3103||full Low-quality
S1_vs_k141_104061||full Low-quality
S1_vs_k141_117427||full Not-determined
S1_vs_k141_130713||full Low-quality
S1_vs_k141_137264||full Low-quality
S1_vs_k141_50789||full Low-quality
S1_vs_k141_57328||full Low-quality
S1_vs_k141_110645||full Low-quality
S1_vs_k141_155816||full Low-quality
S1_vs_k141_70208||full Not-determined
S1_vs_k141_97540||full Low-quality
S1_vs_k141_123748||full Low-quality
S1_vs_k141_84117||full Low-quality
S1_vs_k141_30705||full Low-quality
S1_vs_k141_130734||full Not-determined
S1_vs_k141_143797||full Low-quality
S1_vs_k141_143798||full Low-quality
S1_vs_k141_70221||full Low-quality
S1_vs_k141_30720||full Low-quality
S1_vs_k141_110672||full Low-quality
S1_vs_k141_117448||full Low-quality
S1_vs_k141_17797||full Low-quality
S1_vs_k141_155819||full Not-determined
S1_vs_k141_57350||full Low-quality
S1_vs_k141_70228||full Not-determined
S8_extracted_k141_208107 Complete
S1_vs_k141_137293||full Low-quality
S1_vs_k141_44126||full Not-determined
S1_vs_k141_30728||full Not-determined
S1_vs_k141_50838||full Low-quality
S1_vs_k141_84141||full Not-determined
S1_vs_k141_104103||full Not-determined
S1_vs_k141_117462||full Not-determined
S1_vs_k141_44140||full Not-determined
S1_vs_k141_90613||full Low-quality
S1_vs_k141_117465||full Low-quality
S1_vs_k141_148660||full Not-determined
S1_vs_k141_57379||full Low-quality
S1_vs_k141_148662||full Low-quality
S1_vs_k141_30753||full Low-quality
S1_vs_k141_117481||full Low-quality
S1_vs_k141_3159||full Not-determined
S1_vs_k141_130754||full Low-quality
S1_vs_k141_110704||full Not-determined
S1_vs_k141_143847||full Low-quality
S1_vs_k141_77415||full Low-quality
S1_vs_k141_90633||full Low-quality
S1_vs_k141_104135||full Low-quality
S1_vs_k141_36507||full Low-quality
S1_vs_k141_10844||full Low-quality
S1_vs_k141_17838||full Low-quality
S1_vs_k141_3173||full Not-determined
S1_vs_k141_70256||full Low-quality
S1_vs_k141_104138||full Not-determined
S1_vs_k141_10846||full Not-determined
S1_vs_k141_57414||full Low-quality
S1_vs_k141_123806||full Low-quality
S1_vs_k141_97612||full Low-quality
S1_vs_k141_70269||full Low-quality
S1_vs_k141_143859||full Low-quality
S1_vs_k141_36520||full Not-determined
S1_vs_k141_62774||full Low-quality
S1_vs_k141_143860||full Not-determined
S1_vs_k141_90655||full Not-determined
S1_vs_k141_3197||full Low-quality
S1_vs_k141_10865||full Low-quality
S1_vs_k141_84188||full Low-quality
S1_vs_k141_137364||full Low-quality
S1_vs_k141_17870||full Not-determined
S1_vs_k141_90663||full Low-quality
S1_vs_k141_62791||full Low-quality
S1_vs_k141_57448||full Low-quality
S1_vs_k141_104171||full Low-quality
S1_vs_k141_62792||full Low-quality
S1_vs_k141_30800||full Low-quality
S1_vs_k141_143883||full Low-quality
S1_vs_k141_36539||full Not-determined
S1_vs_k141_50942||full Low-quality
S1_vs_k141_143887||full Not-determined
S1_vs_k141_97641||full Not-determined
S1_vs_k141_23772||full Low-quality
S1_vs_k141_57461||full Low-quality
S1_vs_k141_90689||full Low-quality
S1_vs_k141_123842||full Low-quality
S1_vs_k141_117557||full Low-quality
S1_vs_k141_44202||full Low-quality
S1_vs_k141_155866||full Low-quality
S1_vs_k141_90691||full Low-quality
S1_vs_k141_30833||full Low-quality
S1_vs_k141_110783||full Not-determined
S1_vs_k141_137406||full Low-quality
S1_vs_k141_50975||full Low-quality
S1_vs_k141_104200||full Not-determined
S1_vs_k141_84225||full Low-quality
S1_vs_k141_104201||full Low-quality
S1_vs_k141_137420||full Not-determined
S1_vs_k141_30851||full Low-quality
S1_vs_k141_77508||full Low-quality
S1_vs_k141_97681||full Not-determined
S1_vs_k141_62810||full Not-determined
S1_vs_k141_77509||full Not-determined
S1_vs_k141_57499||full Not-determined
S1_vs_k141_36584||full Not-determined
S1_vs_k141_84245||full Low-quality
S1_vs_k141_77523||full Low-quality
S1_vs_k141_84254||full Low-quality
S1_vs_k141_30875||full Low-quality
S1_vs_k141_130873||full Low-quality
S1_vs_k141_90740||full Low-quality
S1_vs_k141_84262||full Low-quality
S1_vs_k141_70338||full Low-quality
S1_vs_k141_10967||full Not-determined
S1_vs_k141_104250||full Low-quality
S1_vs_k141_123896||full Not-determined
S1_vs_k141_36607||full Low-quality
S1_vs_k141_137457||full Low-quality
S1_vs_k141_117629||full Not-determined
S1_vs_k141_110833||full Not-determined
S8_vs_k141_208120||full High-quality
S1_vs_k141_130882||full Not-determined
S1_vs_k141_51040||full Low-quality
S1_vs_k141_10979||full Not-determined
S1_vs_k141_77554||full Low-quality
S1_vs_k141_70348||full Low-quality
S1_vs_k141_90765||full Low-quality
S1_vs_k141_62867||full Low-quality
S1_vs_k141_97713||full Low-quality
S1_vs_k141_110849||full Not-determined
S1_vs_k141_3283||full Low-quality
S1_vs_k141_17979||full Low-quality
S1_vs_k141_117652||full Low-quality
S1_vs_k141_117655||full Low-quality
S1_vs_k141_97725||full Low-quality
S1_vs_k141_130912||full Not-determined
S1_vs_k141_84304||full Low-quality
S1_vs_k141_90786||full Not-determined
S1_vs_k141_90788||full Not-determined
S1_vs_k141_90793||full Low-quality
S1_vs_k141_97740||full Not-determined
S17_extracted_k141_35138 Medium-quality
S1_vs_k141_11025||full Low-quality
S1_vs_k141_123934||full Not-determined
S1_vs_k141_17997||full Not-determined
S1_vs_k141_70371||full Low-quality
S1_vs_k141_36667||full Low-quality
S1_vs_k141_84335||full Low-quality
S1_vs_k141_117694||full Not-determined
S1_vs_k141_11053||full Low-quality
S1_vs_k141_148762||full Low-quality
S1_vs_k141_110900||full Low-quality
S1_vs_k141_3327||full Low-quality
S1_vs_k141_123951||full Not-determined
S1_vs_k141_97758||full Low-quality
S1_vs_k141_130959||full Low-quality
S1_vs_k141_23962||full Not-determined
S1_vs_k141_84346||full Not-determined
S1_vs_k141_51115||full Not-determined
S1_vs_k141_77622||full Low-quality
S1_vs_k141_44334||full Low-quality
S1_vs_k141_123953||full Not-determined
S1_vs_k141_62936||full Low-quality
S1_vs_k141_97769||full Low-quality
S1_vs_k141_104322||full Not-determined
S1_vs_k141_130963||full Low-quality
S1_vs_k141_143956||full Low-quality
S1_vs_k141_123957||full Low-quality
S1_vs_k141_23978||full Low-quality
S1_vs_k141_70393||full Not-determined
S1_vs_k141_117721||full Not-determined
S1_vs_k141_137550||full Low-quality
S1_vs_k141_110922||full Low-quality
S1_vs_k141_51125||full Not-determined
S1_vs_k141_11084||full Not-determined
S1_vs_k141_51133||full Not-determined
S9_vs_k141_68272||full Medium-quality
S1_vs_k141_117734||full Low-quality
S1_vs_k141_23996||full Not-determined
S1_vs_k141_104340||full Low-quality
S1_vs_k141_130984||full Low-quality
S1_vs_k141_155928||full Low-quality
S1_vs_k141_57553||full Low-quality
S1_vs_k141_84393||full Low-quality
S1_vs_k141_97796||full Not-determined
S1_vs_k141_97797||full Low-quality
S1_vs_k141_30996||full Low-quality
S1_vs_k141_148771||full Low-quality
S1_vs_k141_44356||full Low-quality
S1_vs_k141_104349||full Not-determined
S1_vs_k141_104350||full Low-quality
S1_vs_k141_148772||full Low-quality
S1_vs_k141_44358||full Not-determined
S1_vs_k141_57560||full Low-quality
S1_vs_k141_3354||full Low-quality
S1_vs_k141_44364||full Low-quality
S1_vs_k141_84410||full Low-quality
S1_vs_k141_137594||full Low-quality
S8_vs_k141_208107||full High-quality
S1_vs_k141_84417||full Low-quality
S1_vs_k141_57568||full Not-determined
S1_vs_k141_70423||full Low-quality
S1_vs_k141_123963||full Not-determined
S1_vs_k141_77660||full Low-quality
S1_vs_k141_11122||full Low-quality
S1_vs_k141_24019||full Low-quality
S1_vs_k141_97804||full Low-quality
S1_vs_k141_84429||full Low-quality
S1_vs_k141_137611||full Not-determined
S1_vs_k141_57583||full Low-quality
S1_vs_k141_62994||full Not-determined
S1_vs_k141_104399||full Not-determined
S1_vs_k141_90898||full Low-quality
S1_vs_k141_131038||full Not-determined
S1_vs_k141_84436||full Low-quality
S1_vs_k141_144045||full Low-quality
S1_vs_k141_44404||full Not-determined
S1_vs_k141_18097||full Low-quality
S1_vs_k141_117785||full Low-quality
S1_vs_k141_24037||full Low-quality
S1_vs_k141_11152||full Not-determined
S1_vs_k141_155942||full Low-quality
S1_vs_k141_36749||full Low-quality
S1_vs_k141_11159||full Low-quality
S1_vs_k141_57595||full Not-determined
S1_vs_k141_144075||full Low-quality
S1_vs_k141_77700||full Low-quality
S1_vs_k141_70458||full Low-quality
S1_vs_k141_44436||full Not-determined
S1_vs_k141_111010||full Low-quality
S1_vs_k141_117795||full Low-quality
S1_vs_k141_104407||full Low-quality
S1_vs_k141_36770||full Low-quality
S1_vs_k141_84462||full Low-quality
S1_vs_k141_36771||full Low-quality
S1_vs_k141_111016||full Low-quality
S1_vs_k141_90949||full Low-quality
S1_vs_k141_137646||full Not-determined
S1_vs_k141_3411||full Low-quality
S1_vs_k141_70476||full Low-quality
S1_vs_k141_57621||full Low-quality
S1_vs_k141_131093||full Low-quality
S1_vs_k141_57627||full Low-quality
S1_vs_k141_70479||full Not-determined
S1_vs_k141_24058||full Not-determined
S1_vs_k141_31080||full Low-quality
S1_vs_k141_131096||full Not-determined
S1_vs_k141_97881||full Low-quality
S1_vs_k141_124043||full Low-quality
S1_vs_k141_97887||full Low-quality
S1_vs_k141_24066||full Low-quality
S1_vs_k141_124046||full Low-quality
S1_vs_k141_124052||full Low-quality
S1_vs_k141_24074||full Not-determined
S1_vs_k141_131103||full Not-determined
S1_vs_k141_57631||full Low-quality
S1_vs_k141_11224||full Not-determined
S1_vs_k141_18166||full Low-quality
S1_vs_k141_51252||full Low-quality
S1_vs_k141_36800||full Not-determined
S17_vs_k141_35138||full Medium-quality
S1_vs_k141_90995||full Low-quality
S1_vs_k141_117829||full Low-quality
S1_vs_k141_18168||full Low-quality
S1_vs_k141_11239||full Not-determined
S1_vs_k141_124070||full Low-quality
S1_vs_k141_36811||full Low-quality
S1_vs_k141_137685||full Not-determined
S1_vs_k141_91001||full Low-quality
S1_vs_k141_31097||full Low-quality
S1_vs_k141_104460||full Low-quality
S1_vs_k141_155973||full Low-quality
S1_vs_k141_51284||full Low-quality
S1_vs_k141_117848||full Low-quality
S1_vs_k141_111093||full Not-determined
S1_vs_k141_124107||full Low-quality
S1_vs_k141_44502||full Low-quality
S1_vs_k141_91019||full Not-determined
S1_vs_k141_148847||full Low-quality
S1_vs_k141_18200||full Not-determined
S1_vs_k141_77803||full Low-quality
S1_vs_k141_104488||full Low-quality
S1_vs_k141_84504||full Low-quality
S1_vs_k141_36848||full Not-determined
S1_vs_k141_117861||full Not-determined
S1_vs_k141_31136||full Low-quality
S1_vs_k141_97937||full Low-quality
S1_vs_k141_77817||full Not-determined
S1_vs_k141_97941||full Low-quality
S1_vs_k141_11293||full Low-quality
S1_vs_k141_36854||full Low-quality
S1_vs_k141_77820||full Low-quality
S1_vs_k141_18222||full Low-quality
S1_vs_k141_97950||full Low-quality
S1_vs_k141_104511||full Not-determined
S1_vs_k141_137707||full Not-determined
S1_vs_k141_131168||full Low-quality
S1_vs_k141_84524||full Not-determined
S1_vs_k141_44529||full Not-determined
S1_vs_k141_97964||full Low-quality
S1_vs_k141_84529||full Not-determined
S1_vs_k141_131186||full Low-quality
S1_vs_k141_124148||full Low-quality
S1_vs_k141_84535||full Not-determined
S1_vs_k141_131188||full Low-quality
S1_vs_k141_63147||full Low-quality
S1_vs_k141_51347||full Low-quality
S1_vs_k141_91074||full Not-determined
S1_vs_k141_44548||full Not-determined
S1_vs_k141_18262||full Low-quality
S1_vs_k141_36893||full Low-quality

| | |
|---|---|
| S1_vs_k141_3531\|\|full | Not-determined |
| S1_vs_k141_97972\|\|full | Low-quality |
| S1_vs_k141_44555\|\|full | Low-quality |
| S1_vs_k141_124163\|\|full | Not-determined |
| S1_vs_k141_44556\|\|full | Low-quality |
| S1_vs_k141_91087\|\|full | Low-quality |
| S1_vs_k141_36898\|\|full | Not-determined |
| S1_vs_k141_24203\|\|full | Not-determined |
| S1_vs_k141_18267\|\|full | Low-quality |
| S1_vs_k141_11354\|\|full | Low-quality |
| S1_vs_k141_131207\|\|full | Low-quality |
| S1_vs_k141_31201\|\|full | Low-quality |
| S1_vs_k141_148878\|\|full | Not-determined |
| S1_vs_k141_57740\|\|full | Low-quality |
| S1_vs_k141_97989\|\|full | Low-quality |
| S1_vs_k141_137782\|\|full | Not-determined |
| S1_vs_k141_57750\|\|full | Not-determined |
| S1_vs_k141_117929\|\|full | Low-quality |
| S1_vs_k141_91121\|\|full | Low-quality |
| S1_vs_k141_148884\|\|full | Low-quality |
| S1_vs_k141_131225\|\|full | Not-determined |
| S1_vs_k141_24246\|\|full | Low-quality |
| S1_vs_k141_51402\|\|full | Low-quality |
| S1_vs_k141_137793\|\|full | Low-quality |
| S1_vs_k141_44585\|\|full | Not-determined |
| S1_vs_k141_18310\|\|full | Low-quality |
| S1_vs_k141_148892\|\|full | Not-determined |
| S1_vs_k141_111186\|\|full | Not-determined |
| S1_vs_k141_70560\|\|full | Not-determined |
| S1_vs_k141_104590\|\|full | Low-quality |
| S1_vs_k141_137804\|\|full | Low-quality |
| S1_vs_k141_51421\|\|full | Not-determined |
| S1_vs_k141_44596\|\|full | Low-quality |
| S1_vs_k141_18329\|\|full | Not-determined |
| S1_vs_k141_137809\|\|full | Low-quality |
| S1_vs_k141_24279\|\|full | Not-determined |
| S1_vs_k141_124223\|\|full | Not-determined |
| S1_vs_k141_104603\|\|full | Not-determined |
| S1_vs_k141_24289\|\|full | Not-determined |
| S1_vs_k141_51445\|\|full | Low-quality |
| S1_vs_k141_144286\|\|full | Low-quality |
| S1_vs_k141_98028\|\|full | Low-quality |
| S1_vs_k141_44611\|\|full | Low-quality |
| S1_vs_k141_18352\|\|full | Low-quality |
| S1_vs_k141_98032\|\|full | Low-quality |
| S1_vs_k141_91187\|\|full | Not-determined |
| S1_vs_k141_18356\|\|full | Low-quality |
| S1_vs_k141_144293\|\|full | Low-quality |
| S1_vs_k141_51460\|\|full | Low-quality |
| S1_vs_k141_70597\|\|full | Low-quality |
| S1_vs_k141_144296\|\|full | Not-determined |
| S1_vs_k141_131298\|\|full | Not-determined |
| S1_vs_k141_117994\|\|full | Low-quality |
| S1_vs_k141_91202\|\|full | Low-quality |
| S1_vs_k141_98050\|\|full | Low-quality |
| S1_vs_k141_57817\|\|full | Low-quality |
| S1_vs_k141_111229\|\|full | Low-quality |
| S1_vs_k141_144304\|\|full | Low-quality |
| S1_vs_k141_31294\|\|full | Low-quality |
| S1_vs_k141_104627\|\|full | Not-determined |
| S1_vs_k141_18382\|\|full | Not-determined |
| S1_vs_k141_144309\|\|full | Not-determined |
| S1_vs_k141_31305\|\|full | Low-quality |
| S1_vs_k141_144315\|\|full | Low-quality |
| S1_vs_k141_137865\|\|full | Low-quality |
| S1_vs_k141_144317\|\|full | Low-quality |
| S1_vs_k141_78005\|\|full | Not-determined |
| S1_vs_k141_24355\|\|full | Low-quality |
| S1_vs_k141_144322\|\|full | Low-quality |
| S1_vs_k141_104651\|\|full | Low-quality |
| S1_vs_k141_18411\|\|full | Low-quality |
| S1_vs_k141_111258\|\|full | Low-quality |
| S1_vs_k141_98084\|\|full | Not-determined |
| S1_vs_k141_84660\|\|full | Low-quality |
| S1_vs_k141_18416\|\|full | Not-determined |
| S1_vs_k141_144333\|\|full | Not-determined |
| S1_vs_k141_104662\|\|full | Not-determined |
| S1_vs_k141_137892\|\|full | Low-quality |
| S1_vs_k141_144339\|\|full | Low-quality |
| S1_vs_k141_118032\|\|full | Not-determined |
| S1_vs_k141_11470\|\|full | Low-quality |
| S1_vs_k141_137897\|\|full | Not-determined |
| S1_vs_k141_124297\|\|full | Not-determined |
| S1_vs_k141_111277\|\|full | Low-quality |
| S1_vs_k141_18438\|\|full | Low-quality |
| S1_vs_k141_137903\|\|full | Low-quality |
| S1_vs_k141_118035\|\|full | Low-quality |
| S1_vs_k141_131374\|\|full | Low-quality |
| S1_vs_k141_3623\|\|full | Not-determined |
| S1_vs_k141_124306\|\|full | Not-determined |
| S1_vs_k141_84680\|\|full | Not-determined |
| S1_vs_k141_98109\|\|full | Low-quality |
| S1_vs_k141_137908\|\|full | Low-quality |
| S1_vs_k141_24387\|\|full | Not-determined |
| S1_vs_k141_148928\|\|full | Low-quality |
| S1_vs_k141_63271\|\|full | Low-quality |
| S1_vs_k141_78038\|\|full | Low-quality |
| S1_vs_k141_137919\|\|full | Low-quality |
| S1_vs_k141_51544\|\|full | Not-determined |
| S1_vs_k141_98130\|\|full | Low-quality |
| S1_vs_k141_37038\|\|full | Low-quality |
| S1_vs_k141_131379\|\|full | Low-quality |
| S1_vs_k141_131380\|\|full | Not-determined |
| S1_vs_k141_3643\|\|full | Low-quality |
| S1_vs_k141_11508\|\|full | Low-quality |
| S1_vs_k141_124331\|\|full | Low-quality |
| S1_vs_k141_98135\|\|full | Low-quality |
| S1_vs_k141_111313\|\|full | Low-quality |
| S1_vs_k141_144378\|\|full | Low-quality |
| S1_vs_k141_31365\|\|full | Not-determined |
| S1_vs_k141_137956\|\|full | Low-quality |
| S1_vs_k141_84724\|\|full | Low-quality |
| S8_extracted_k141_151384 | High-quality |
| S1_vs_k141_11529\|\|full | Low-quality |
| S1_vs_k141_118080\|\|full | Not-determined |
| S1_vs_k141_37065\|\|full | Not-determined |
| S1_vs_k141_78072\|\|full | Not-determined |
| S1_vs_k141_11539\|\|full | Not-determined |
| S1_vs_k141_156096\|\|full | Low-quality |
| S1_vs_k141_24433\|\|full | Low-quality |
| S1_vs_k141_84741\|\|full | Low-quality |
| S1_vs_k141_63334\|\|full | Not-determined |
| S1_vs_k141_144415\|\|full | Low-quality |
| S1_vs_k141_37066\|\|full | Not-determined |
| S1_vs_k141_3684\|\|full | Not-determined |
| S1_vs_k141_11559\|\|full | Not-determined |
| S1_vs_k141_18524\|\|full | Low-quality |
| S1_vs_k141_57961\|\|full | Low-quality |
| S1_vs_k141_111348\|\|full | Low-quality |
| S1_vs_k141_63345\|\|full | Not-determined |
| S1_vs_k141_57969\|\|full | Low-quality |
| S1_vs_k141_91296\|\|full | Not-determined |
| S1_vs_k141_44768\|\|full | Low-quality |
| S1_vs_k141_156106\|\|full | Not-determined |
| S1_vs_k141_118112\|\|full | Not-determined |
| S1_vs_k141_63353\|\|full | Low-quality |
| S1_vs_k141_18541\|\|full | Low-quality |
| S1_vs_k141_57990\|\|full | Low-quality |
| S1_vs_k141_104779\|\|full | Low-quality |
| S1_vs_k141_44788\|\|full | Low-quality |
| S1_vs_k141_138013\|\|full | Low-quality |
| S1_vs_k141_24480\|\|full | Low-quality |
| S1_vs_k141_24481\|\|full | Low-quality |
| S1_vs_k141_98193\|\|full | Not-determined |
| S1_vs_k141_78122\|\|full | Low-quality |
| S1_vs_k141_118141\|\|full | Low-quality |
| S1_vs_k141_3717\|\|full | Low-quality |
| S1_vs_k141_70684\|\|full | Low-quality |
| S1_vs_k141_138039\|\|full | Low-quality |
| S1_vs_k141_84795\|\|full | Low-quality |
| S1_vs_k141_118147\|\|full | Low-quality |
| S1_vs_k141_156127\|\|full | Low-quality |
| S1_vs_k141_156131\|\|full | Not-determined |
| S1_vs_k141_111398\|\|full | Low-quality |
| S1_vs_k141_58012\|\|full | Low-quality |
| S1_vs_k141_148987\|\|full | Not-determined |
| S1_vs_k141_44805\|\|full | Not-determined |
| S1_vs_k141_98210\|\|full | Not-determined |
| S1_vs_k141_70687\|\|full | Low-quality |
| S1_vs_k141_51649\|\|full | Low-quality |
| S1_vs_k141_51650\|\|full | Low-quality |
| S1_vs_k141_3742\|\|full | Not-determined |
| S1_vs_k141_111419\|\|full | Not-determined |
| S1_vs_k141_104803\|\|full | Low-quality |
| S1_vs_k141_144496\|\|full | Low-quality |
| S1_vs_k141_124457\|\|full | Low-quality |
| S1_vs_k141_156153\|\|full | Low-quality |
| S1_vs_k141_104821\|\|full | Low-quality |
| S1_vs_k141_98239\|\|full | Not-determined |
| S1_vs_k141_3754\|\|full | Low-quality |
| S1_vs_k141_144510\|\|full | Low-quality |
| S1_vs_k141_118193\|\|full | Low-quality |
| S1_vs_k141_104833\|\|full | Low-quality |
| S1_vs_k141_91388\|\|full | Not-determined |
| S1_vs_k141_24585\|\|full | Low-quality |
| S1_vs_k141_18605\|\|full | Low-quality |
| S1_vs_k141_156172\|\|full | Low-quality |
| S1_vs_k141_3777\|\|full | Low-quality |
| S1_vs_k141_104839\|\|full | Low-quality |
| S1_vs_k141_51684\|\|full | Low-quality |
| S1_vs_k141_37157\|\|full | Not-determined |
| S1_vs_k141_18613\|\|full | Not-determined |
| S1_vs_k141_3778\|\|full | Low-quality |
| S1_vs_k141_3779\|\|full | Not-determined |
| S1_vs_k141_84846\|\|full | Low-quality |
| S1_vs_k141_98270\|\|full | Low-quality |
| S1_vs_k141_24601\|\|full | Low-quality |
| S1_vs_k141_18625\|\|full | Low-quality |
| S1_vs_k141_91406\|\|full | Low-quality |
| S1_vs_k141_124481\|\|full | Low-quality |
| S1_vs_k141_63486\|\|full | Low-quality |
| S1_vs_k141_51703\|\|full | Low-quality |
| S1_vs_k141_144515\|\|full | Low-quality |
| S1_vs_k141_91415\|\|full | Not-determined |
| S1_vs_k141_58070\|\|full | Low-quality |
| S1_vs_k141_156191\|\|full | Low-quality |
| S1_vs_k141_44883\|\|full | Low-quality |
| S1_vs_k141_118229\|\|full | Not-determined |
| S1_vs_k141_98292\|\|full | Not-determined |
| S1_vs_k141_144537\|\|full | Not-determined |
| S1_vs_k141_58084\|\|full | Low-quality |
| S1_vs_k141_18654\|\|full | Low-quality |
| S1_vs_k141_131566\|\|full | Low-quality |
| S1_vs_k141_18657\|\|full | Not-determined |
| S1_vs_k141_131570\|\|full | Not-determined |
| S1_vs_k141_37186\|\|full | Low-quality |
| S1_vs_k141_138124\|\|full | Low-quality |
| S1_vs_k141_51713\|\|full | Low-quality |
| S1_vs_k141_18669\|\|full | Low-quality |
| S1_vs_k141_51717\|\|full | Low-quality |
| S1_vs_k141_131589\|\|full | Low-quality |
| S1_vs_k141_58112\|\|full | Low-quality |
| S1_vs_k141_44917\|\|full | Not-determined |
| S1_vs_k141_31515\|\|full | Low-quality |
| S1_vs_k141_18677\|\|full | Low-quality |
| S1_vs_k141_37202\|\|full | Low-quality |
| S1_vs_k141_91479\|\|full | Low-quality |
| S1_vs_k141_18687\|\|full | Not-determined |
| S1_vs_k141_18692\|\|full | Low-quality |
| S1_vs_k141_44929\|\|full | Low-quality |
| S1_vs_k141_149045\|\|full | Not-determined |
| S1_vs_k141_118283\|\|full | Low-quality |
| S1_vs_k141_78263\|\|full | Low-quality |
| S1_vs_k141_149048\|\|full | Low-quality |
| S1_vs_k141_111552\|\|full | Not-determined |
| S1_vs_k141_144589\|\|full | Low-quality |
| S1_vs_k141_63550\|\|full | Low-quality |
| S1_vs_k141_44939\|\|full | Low-quality |
| S1_vs_k141_104936\|\|full | Low-quality |
| S1_vs_k141_18701\|\|full | Not-determined |
| S1_vs_k141_91501\|\|full | Not-determined |
| S1_vs_k141_98354\|\|full | Not-determined |
| S1_vs_k141_124553\|\|full | Low-quality |
| S1_vs_k141_31530\|\|full | Not-determined |
| S1_vs_k141_58139\|\|full | Not-determined |
| S1_vs_k141_149054\|\|full | Low-quality |
| S1_vs_k141_124559\|\|full | Low-quality |
| S1_vs_k141_131638\|\|full | Low-quality |
| S1_vs_k141_104951\|\|full | Not-determined |
| S1_vs_k141_156225\|\|full | Not-determined |
| S1_vs_k141_91539\|\|full | Low-quality |
| S1_vs_k141_118317\|\|full | Low-quality |
| S8_extracted_k141_208102 | Complete |
| S1_vs_k141_118326\|\|full | Not-determined |
| S1_vs_k141_24729\|\|full | Low-quality |
| S1_vs_k141_31562\|\|full | Low-quality |
| S1_vs_k141_138167\|\|full | Not-determined |
| S1_vs_k141_124606\|\|full | Not-determined |
| S1_vs_k141_63605\|\|full | Low-quality |
| S1_vs_k141_3869\|\|full | Not-determined |
| S1_vs_k141_24741\|\|full | Low-quality |
| S1_vs_k141_84944\|\|full | Low-quality |
| S1_vs_k141_78316\|\|full | Low-quality |
| S1_vs_k141_131684\|\|full | Not-determined |
| S1_vs_k141_149095\|\|full | Low-quality |
| S1_vs_k141_84946\|\|full | Low-quality |
| S1_vs_k141_131690\|\|full | Low-quality |
| S1_vs_k141_31587\|\|full | Low-quality |
| S1_vs_k141_11821\|\|full | Not-determined |
| S1_vs_k141_111633\|\|full | Low-quality |
| S1_vs_k141_45023\|\|full | Low-quality |
| S1_vs_k141_144640\|\|full | Low-quality |
| S1_vs_k141_138181\|\|full | Low-quality |
| S1_vs_k141_84957\|\|full | Low-quality |
| S1_vs_k141_45026\|\|full | Not-determined |
| S1_vs_k141_144647\|\|full | Not-determined |
| S1_vs_k141_31597\|\|full | Not-determined |
| S1_vs_k141_84960\|\|full | Low-quality |
| S1_vs_k141_118348\|\|full | Low-quality |
| S1_vs_k141_11831\|\|full | Low-quality |
| S1_vs_k141_31601\|\|full | Low-quality |
| S1_vs_k141_45044\|\|full | Low-quality |
| S1_vs_k141_91608\|\|full | Not-determined |
| S1_vs_k141_70769\|\|full | Low-quality |
| S1_vs_k141_91617\|\|full | Low-quality |
| S1_vs_k141_156258\|\|full | Low-quality |
| S1_vs_k141_144661\|\|full | Low-quality |
| S1_vs_k141_24784\|\|full | Low-quality |
| S1_vs_k141_111676\|\|full | Low-quality |
| S1_vs_k141_111677\|\|full | Low-quality |
| S1_vs_k141_105049\|\|full | Low-quality |
| S1_vs_k141_31620\|\|full | Low-quality |
| S1_vs_k141_124692\|\|full | Low-quality |
| S1_vs_k141_111680\|\|full | Low-quality |
| S1_vs_k141_105057\|\|full | Low-quality |
| S1_vs_k141_18824\|\|full | Not-determined |
| S1_vs_k141_31625\|\|full | Low-quality |
| S1_vs_k141_45072\|\|full | Low-quality |
| S1_vs_k141_156267\|\|full | Low-quality |
| S1_vs_k141_18828\|\|full | Low-quality |
| S1_vs_k141_105063\|\|full | Low-quality |
| S1_vs_k141_91651\|\|full | Low-quality |
| S1_vs_k141_18832\|\|full | Low-quality |
| S1_vs_k141_78382\|\|full | Not-determined |
| S1_vs_k141_111695\|\|full | Low-quality |
| S1_vs_k141_144687\|\|full | Low-quality |
| S1_vs_k141_63685\|\|full | Low-quality |
| S1_vs_k141_98478\|\|full | Not-determined |
| S1_vs_k141_91663\|\|full | Not-determined |
| S1_vs_k141_111706\|\|full | Low-quality |
| S1_vs_k141_149159\|\|full | Not-determined |
| S1_vs_k141_105073\|\|full | Low-quality |
| S1_vs_k141_98494\|\|full | Low-quality |
| S1_vs_k141_105077\|\|full | Low-quality |
| S1_vs_k141_131775\|\|full | Low-quality |
| S1_vs_k141_144711\|\|full | Not-determined |
| S1_vs_k141_31655\|\|full | Low-quality |
| S1_vs_k141_37328\|\|full | Low-quality |
| S1_vs_k141_138247\|\|full | Low-quality |
| S1_vs_k141_58272\|\|full | Not-determined |
| S1_vs_k141_144725\|\|full | Not-determined |
| S1_vs_k141_85054\|\|full | Low-quality |
| S1_vs_k141_144727\|\|full | Low-quality |
| S1_vs_k141_149173\|\|full | Low-quality |
| S1_vs_k141_45122\|\|full | Not-determined |
| S1_vs_k141_58289\|\|full | Low-quality |
| S1_vs_k141_24850\|\|full | Low-quality |
| S1_vs_k141_124759\|\|full | Low-quality |
| S1_vs_k141_31696\|\|full | Low-quality |
| S1_vs_k141_124761\|\|full | Not-determined |
| S1_vs_k141_85065\|\|full | Not-determined |
| S1_vs_k141_138250\|\|full | Not-determined |
| S1_vs_k141_31698\|\|full | Not-determined |
| S1_vs_k141_124765\|\|full | Low-quality |
| S1_vs_k141_58305\|\|full | Low-quality |
| S1_vs_k141_105115\|\|full | Low-quality |
| S1_vs_k141_118436\|\|full | Low-quality |
| S1_vs_k141_63748\|\|full | Not-determined |
| S1_vs_k141_31703\|\|full | Low-quality |
| S1_vs_k141_11965\|\|full | Not-determined |
| S1_vs_k141_78444\|\|full | Low-quality |
| S1_vs_k141_78445\|\|full | Not-determined |
| S1_vs_k141_58318\|\|full | Not-determined |
| S1_vs_k141_118447\|\|full | Not-determined |
| S1_vs_k141_131826\|\|full | Low-quality |
| S1_vs_k141_78458\|\|full | Low-quality |
| S1_vs_k141_98559\|\|full | Low-quality |
| S1_vs_k141_63765\|\|full | Low-quality |
| S1_vs_k141_45162\|\|full | Not-determined |
| S1_vs_k141_18909\|\|full | Low-quality |
| S1_vs_k141_85098\|\|full | Not-determined |
| S1_vs_k141_11977\|\|full | Low-quality |
| S1_vs_k141_31727\|\|full | Low-quality |
| S1_vs_k141_58337\|\|full | Low-quality |
| S1_vs_k141_144783\|\|full | Low-quality |
| S1_vs_k141_63779\|\|full | Low-quality |
| S1_vs_k141_105145\|\|full | Low-quality |
| S1_vs_k141_105147\|\|full | Low-quality |
| S1_vs_k141_144791\|\|full | Not-determined |
| S1_vs_k141_91759\|\|full | Low-quality |
| S1_vs_k141_131846\|\|full | Low-quality |
| S1_vs_k141_37371\|\|full | Low-quality |
| S1_vs_k141_91764\|\|full | Low-quality |
| S1_vs_k141_31749\|\|full | Not-determined |
| S1_vs_k141_18939\|\|full | Not-determined |
| S1_vs_k141_118479\|\|full | Low-quality |
| S1_vs_k141_105170\|\|full | Not-determined |
| S1_vs_k141_58365\|\|full | Not-determined |
| S1_vs_k141_45208\|\|full | Low-quality |
| S1_vs_k141_4033\|\|full | Not-determined |
| S1_vs_k141_70860\|\|full | Low-quality |
| S1_vs_k141_63802\|\|full | Low-quality |
| S1_vs_k141_4040\|\|full | Not-determined |
| S1_vs_k141_63807\|\|full | Low-quality |
| S1_vs_k141_105184\|\|full | Low-quality |
| S1_vs_k141_4042\|\|full | Not-determined |
| S1_vs_k141_131881\|\|full | Low-quality |
| S1_vs_k141_18950\|\|full | Low-quality |
| S1_vs_k141_45230\|\|full | Not-determined |
| S1_vs_k141_52024\|\|full | Not-determined |
| S1_vs_k141_37386\|\|full | Low-quality |
| S1_vs_k141_24958\|\|full | Low-quality |
| S1_vs_k141_45241\|\|full | Low-quality |
| S1_vs_k141_124872\|\|full | Not-determined |
| S1_vs_k141_118532\|\|full | Not-determined |
| S1_vs_k141_111824\|\|full | Low-quality |
| S1_vs_k141_118538\|\|full | Not-determined |
| S1_vs_k141_156331\|\|full | Low-quality |
| S1_vs_k141_63833\|\|full | Low-quality |
| S1_vs_k141_70895\|\|full | Not-determined |
| S1_vs_k141_58404\|\|full | Low-quality |
| S1_vs_k141_98657\|\|full | Low-quality |
| S1_vs_k141_131912\|\|full | Low-quality |
| S1_vs_k141_63844\|\|full | Low-quality |
| S1_vs_k141_52053\|\|full | Low-quality |
| S1_vs_k141_85180\|\|full | Not-determined |
| S1_vs_k141_111842\|\|full | Low-quality |
| S1_vs_k141_98665\|\|full | Not-determined |
| S1_vs_k141_144853\|\|full | Low-quality |
| S1_vs_k141_12021\|\|full | Low-quality |
| S1_vs_k141_91826\|\|full | Low-quality |
| S1_vs_k141_70908\|\|full | Low-quality |
| S1_vs_k141_78563\|\|full | Not-determined |
| S1_vs_k141_111850\|\|full | Low-quality |
| S1_vs_k141_131929\|\|full | Low-quality |
| S1_vs_k141_118563\|\|full | Low-quality |
| S1_vs_k141_78568\|\|full | Low-quality |
| S1_vs_k141_58437\|\|full | Not-determined |
| S1_vs_k141_24993\|\|full | Low-quality |
| S1_vs_k141_91841\|\|full | Low-quality |
| S1_vs_k141_58440\|\|full | Not-determined |
| S1_vs_k141_111861\|\|full | Low-quality |
| S1_vs_k141_25000\|\|full | Not-determined |
| S1_vs_k141_98692\|\|full | Low-quality |
| S1_vs_k141_25005\|\|full | Not-determined |
| S1_vs_k141_78582\|\|full | Not-determined |
| S1_vs_k141_149244\|\|full | Low-quality |
| S1_vs_k141_45260\|\|full | Low-quality |
| S1_vs_k141_124937\|\|full | Not-determined |
| S1_vs_k141_111865\|\|full | Low-quality |
| S1_vs_k141_124942\|\|full | Not-determined |
| S1_vs_k141_111868\|\|full | Not-determined |
| S1_vs_k141_91864\|\|full | Not-determined |
| S1_vs_k141_31837\|\|full | Low-quality |
| S1_vs_k141_111870\|\|full | Low-quality |
| S1_vs_k141_105277\|\|full | Low-quality |
| S1_vs_k141_85219\|\|full | Low-quality |
| S1_vs_k141_37457\|\|full | Not-determined |
| S1_vs_k141_111882\|\|full | Low-quality |
| S1_vs_k141_19037\|\|full | Not-determined |
| S1_vs_k141_52117\|\|full | Low-quality |
| S1_vs_k141_131996\|\|full | Not-determined |
| S1_vs_k141_19045\|\|full | Not-determined |
| S1_vs_k141_111889\|\|full | Low-quality |
| S1_vs_k141_156362\|\|full | Not-determined |
| S1_vs_k141_98721\|\|full | Low-quality |
| S1_vs_k141_118601\|\|full | Low-quality |
| S1_vs_k141_78635\|\|full | Not-determined |
| S1_vs_k141_105302\|\|full | Low-quality |
| S1_vs_k141_138411\|\|full | Low-quality |
| S1_vs_k141_70958\|\|full | Low-quality |
| S1_vs_k141_37481\|\|full | Low-quality |
| S1_vs_k141_58506\|\|full | Low-quality |
| S1_vs_k141_19055\|\|full | Low-quality |
| S1_vs_k141_12121\|\|full | Low-quality |
| S1_vs_k141_31875\|\|full | Low-quality |
| S1_vs_k141_45325\|\|full | Not-determined |
| S1_vs_k141_78661\|\|full | Low-quality |
| S1_vs_k141_91914\|\|full | Low-quality |
| S1_vs_k141_19068\|\|full | Low-quality |
| S1_vs_k141_156374\|\|full | Low-quality |
| S1_vs_k141_91924\|\|full | Low-quality |
| S1_vs_k141_144948\|\|full | Not-determined |
| S1_vs_k141_19082\|\|full | Not-determined |
| S1_vs_k141_25088\|\|full | Low-quality |
| S1_vs_k141_19083\|\|full | Low-quality |
| S1_vs_k141_132049\|\|full | Not-determined |
| S1_vs_k141_118655\|\|full | Low-quality |
| S1_vs_k141_98773\|\|full | Low-quality |
| S1_vs_k141_85275\|\|full | Not-determined |
| S1_vs_k141_25101\|\|full | Low-quality |
| S1_vs_k141_31914\|\|full | Low-quality |
| S1_vs_k141_132055\|\|full | Low-quality |
| S1_vs_k141_85288\|\|full | Low-quality |
| S1_vs_k141_19104\|\|full | Low-quality |
| S1_vs_k141_25115\|\|full | Low-quality |
| S1_vs_k141_58547\|\|full | Low-quality |
| S1_vs_k141_45372\|\|full | Low-quality |
| S1_vs_k141_12180\|\|full | Low-quality |
| S1_vs_k141_85294\|\|full | Low-quality |
| S1_vs_k141_64012\|\|full | Low-quality |
| S1_vs_k141_37522\|\|full | Low-quality |
| S1_vs_k141_85298\|\|full | Low-quality |
| S1_vs_k141_19116\|\|full | Low-quality |
| S1_vs_k141_58565\|\|full | Not-determined |
| S1_vs_k141_78714\|\|full | Low-quality |
| S1_vs_k141_98801\|\|full | Low-quality |
| S1_vs_k141_19128\|\|full | Low-quality |
| S1_vs_k141_138498\|\|full | Low-quality |
| S1_vs_k141_52209\|\|full | Not-determined |
| S1_vs_k141_91989\|\|full | Low-quality |
| S1_vs_k141_156383\|\|full | Low-quality |
| S1_vs_k141_85330\|\|full | Low-quality |
| S1_vs_k141_132095\|\|full | Low-quality |
| S1_vs_k141_19150\|\|full | Low-quality |
| S1_vs_k141_111993\|\|full | Low-quality |
| S1_vs_k141_85332\|\|full | Low-quality |
| S1_vs_k141_45396\|\|full | Not-determined |
| S1_vs_k141_132102\|\|full | Low-quality |
| S1_vs_k141_125096\|\|full | Not-determined |
| S1_vs_k141_78745\|\|full | Not-determined |
| S1_vs_k141_145012\|\|full | Low-quality |
| S1_vs_k141_64047\|\|full | Low-quality |
| S1_vs_k141_52222\|\|full | Not-determined |
| S1_vs_k141_12242\|\|full | Low-quality |
| S1_vs_k141_118722\|\|full | Low-quality |
| S1_vs_k141_64061\|\|full | Low-quality |
| S1_vs_k141_132134\|\|full | Low-quality |
| S1_vs_k141_32013\|\|full | Not-determined |
| S1_vs_k141_145035\|\|full | Not-determined |
| S1_vs_k141_58635\|\|full | Low-quality |
| S1_vs_k141_19194\|\|full | Low-quality |
| S1_vs_k141_45422\|\|full | Not-determined |
| S1_vs_k141_145042\|\|full | Not-determined |
| S1_vs_k141_118736\|\|full | Low-quality |
| S1_vs_k141_132142\|\|full | Low-quality |
| S1_vs_k141_45424\|\|full | Low-quality |
| S1_vs_k141_149322\|\|full | Low-quality |
| S1_vs_k141_125124\|\|full | Low-quality |
| S1_vs_k141_92041\|\|full | Not-determined |
| S1_vs_k141_19209\|\|full | Low-quality |
| S1_vs_k141_19210\|\|full | Not-determined |
| S1_vs_k141_64092\|\|full | Low-quality |
| S1_vs_k141_78800\|\|full | Not-determined |
| S1_vs_k141_105452\|\|full | Low-quality |
| S1_vs_k141_132168\|\|full | Low-quality |
| S1_vs_k141_125129\|\|full | Low-quality |
| S1_vs_k141_4241\|\|full | Low-quality |
| S1_vs_k141_105456\|\|full | Not-determined |
| S1_vs_k141_58665\|\|full | Low-quality |
| S1_vs_k141_92065\|\|full | Low-quality |
| S1_vs_k141_19214\|\|full | Low-quality |
| S1_vs_k141_125136\|\|full | Low-quality |
| S1_vs_k141_85411\|\|full | Low-quality |
| S1_vs_k141_78815\|\|full | Low-quality |
| S1_vs_k141_98843\|\|full | Low-quality |
| S1_vs_k141_64117\|\|full | Not-determined |
| S1_vs_k141_45466\|\|full | Low-quality |
| S1_vs_k141_125150\|\|full | Low-quality |
| S1_vs_k141_132183\|\|full | Low-quality |
| S1_vs_k141_138589\|\|full | Low-quality |
| S1_vs_k141_92090\|\|full | Not-determined |
| S1_vs_k141_98850\|\|full | Not-determined |
| S1_vs_k141_37576\|\|full | Not-determined |
| S1_vs_k141_118787\|\|full | Not-determined |
| S1_vs_k141_4265\|\|full | Low-quality |
| S1_vs_k141_98859\|\|full | Low-quality |
| S1_vs_k141_98863\|\|full | Not-determined |
| S1_vs_k141_4270\|\|full | Low-quality |
| S1_vs_k141_98867\|\|full | Low-quality |
| S1_vs_k141_145089\|\|full | Low-quality |
| S1_vs_k141_118795\|\|full | Not-determined |
| S1_vs_k141_98869\|\|full | Low-quality |
| S1_vs_k141_112086\|\|full | Low-quality |
| S1_vs_k141_58721\|\|full | Low-quality |
| S1_vs_k141_118800\|\|full | Low-quality |
| S1_vs_k141_12309\|\|full | Low-quality |
| S1_vs_k141_92098\|\|full | Low-quality |
| S1_vs_k141_19278\|\|full | Not-determined |
| S1_vs_k141_45502\|\|full | Low-quality |
| S1_vs_k141_149373\|\|full | Low-quality |
| S1_vs_k141_12319\|\|full | Low-quality |
| S1_vs_k141_85456\|\|full | Low-quality |
| S1_vs_k141_45512\|\|full | Low-quality |
| S1_vs_k141_37608\|\|full | Low-quality |
| S1_vs_k141_149378\|\|full | Low-quality |
| S1_vs_k141_125188\|\|full | Low-quality |
| S1_vs_k141_156428\|\|full | Low-quality |
| S1_vs_k141_132221\|\|full | Low-quality |
| S1_vs_k141_138633\|\|full | Low-quality |
| S1_vs_k141_105519\|\|full | Low-quality |
| S1_vs_k141_19302\|\|full | Low-quality |
| S1_vs_k141_25266\|\|full | Low-quality |
| S1_vs_k141_145118\|\|full | Low-quality |
| S1_vs_k141_12330\|\|full | Low-quality |
| S1_vs_k141_45517\|\|full | Low-quality |
| S1_vs_k141_112104\|\|full | Not-determined |
| S1_vs_k141_45520\|\|full | Low-quality |
| S1_vs_k141_32140\|\|full | Low-quality |
| S1_vs_k141_25272\|\|full | Low-quality |
| S1_vs_k141_118825\|\|full | Low-quality |
| S1_vs_k141_92126\|\|full | Low-quality |
| S1_vs_k141_37617\|\|full | Not-determined |
| S1_vs_k141_78878\|\|full | Low-quality |
| S1_vs_k141_25279\|\|full | Not-determined |
| S1_vs_k141_145131\|\|full | Not-determined |
| S1_vs_k141_138637\|\|full | Not-determined |
| S1_vs_k141_118832\|\|full | Low-quality |
| S1_vs_k141_145136\|\|full | Low-quality |
| S1_vs_k141_52363\|\|full | Not-determined |
| S1_vs_k141_58780\|\|full | Not-determined |
| S1_vs_k141_85489\|\|full | Not-determined |
| S1_vs_k141_37629\|\|full | Low-quality |
| S1_vs_k141_132249\|\|full | Low-quality |
| S1_vs_k141_52370\|\|full | Not-determined |
| S1_vs_k141_58791\|\|full | Low-quality |
| S1_vs_k141_78908\|\|full | Not-determined |
| S1_vs_k141_85501\|\|full | Not-determined |
| S1_vs_k141_85503\|\|full | Low-quality |
| S1_vs_k141_125235\|\|full | Low-quality |
| S1_vs_k141_19352\|\|full | Low-quality |
| S1_vs_k141_78912\|\|full | Low-quality |
| S1_vs_k141_98940\|\|full | Low-quality |
| S1_vs_k141_92158\|\|full | Low-quality |
| S5_extracted_k141_60597 | Complete |
| S1_vs_k141_25317\|\|full | Low-quality |
| S1_vs_k141_32183\|\|full | Low-quality |
| S1_vs_k141_145174\|\|full | Low-quality |
| S1_vs_k141_64238\|\|full | Low-quality |

| | |
|---|---|
| S1_vs_k141_85530\|\|full | Low-quality |
| S1_vs_k141_92182\|\|full | Low-quality |
| S1_vs_k141_19378\|\|full | Not-determined |
| S1_vs_k141_138674\|\|full | Low-quality |
| S1_vs_k141_64244\|\|full | Low-quality |
| S1_vs_k141_125258\|\|full | Low-quality |
| S1_vs_k141_92194\|\|full | Not-determined |
| S1_vs_k141_45606\|\|full | Not-determined |
| S1_vs_k141_45610\|\|full | Low-quality |
| S6_extracted_k141_29282 | Complete |
| S1_vs_k141_92204\|\|full | Low-quality |
| S1_vs_k141_132280\|\|full | Low-quality |
| S1_vs_k141_4318\|\|full | Not-determined |
| S1_vs_k141_4320\|\|full | Low-quality |
| S1_vs_k141_99010\|\|full | Low-quality |
| S1_vs_k141_25360\|\|full | Low-quality |
| S1_vs_k141_145209\|\|full | Low-quality |
| S1_vs_k141_64272\|\|full | Low-quality |
| S1_vs_k141_85567\|\|full | Low-quality |
| S1_vs_k141_92215\|\|full | Low-quality |
| S1_vs_k141_138683\|\|full | Low-quality |
| S1_vs_k141_37652\|\|full | Low-quality |
| S1_vs_k141_78976\|\|full | Not-determined |
| S1_vs_k141_37656\|\|full | Low-quality |
| S1_vs_k141_64277\|\|full | Low-quality |
| S1_vs_k141_19419\|\|full | Low-quality |
| S1_vs_k141_85583\|\|full | Not-determined |
| S1_vs_k141_58878\|\|full | Low-quality |
| S1_vs_k141_145217\|\|full | Low-quality |
| S1_vs_k141_78984\|\|full | Low-quality |
| S1_vs_k141_92221\|\|full | Low-quality |
| S1_vs_k141_12453\|\|full | Low-quality |
| S1_vs_k141_45637\|\|full | Not-determined |
| S1_vs_k141_52469\|\|full | Low-quality |
| S1_vs_k141_25391\|\|full | Low-quality |
| S1_vs_k141_78987\|\|full | Low-quality |
| S1_vs_k141_138709\|\|full | Not-determined |
| S1_vs_k141_118929\|\|full | Low-quality |
| S1_vs_k141_58896\|\|full | Not-determined |
| S1_vs_k141_45645\|\|full | Low-quality |
| S1_vs_k141_32263\|\|full | Low-quality |
| S1_vs_k141_37666\|\|full | Not-determined |
| S1_vs_k141_92241\|\|full | Low-quality |
| S1_vs_k141_132329\|\|full | Not-determined |
| S1_vs_k141_52486\|\|full | Low-quality |
| S1_vs_k141_132332\|\|full | Low-quality |
| S1_vs_k141_25402\|\|full | Not-determined |
| S1_vs_k141_37669\|\|full | Low-quality |
| S1_vs_k141_85624\|\|full | Low-quality |
| S1_vs_k141_125329\|\|full | Low-quality |
| S1_vs_k141_12470\|\|full | Not-determined |
| S1_vs_k141_92250\|\|full | Low-quality |
| S1_vs_k141_118957\|\|full | Low-quality |
| S1_vs_k141_145253\|\|full | Low-quality |
| S1_vs_k141_58904\|\|full | Low-quality |
| S1_vs_k141_32291\|\|full | Low-quality |
| S1_vs_k141_99061\|\|full | Not-determined |
| S1_vs_k141_79019\|\|full | Not-determined |
| S1_vs_k141_45677\|\|full | Low-quality |
| S1_vs_k141_105669\|\|full | Low-quality |
| S1_vs_k141_118974\|\|full | Low-quality |
| S1_vs_k141_132344\|\|full | Low-quality |
| S1_vs_k141_92278\|\|full | Low-quality |
| S1_vs_k141_79039\|\|full | Low-quality |
| S1_vs_k141_105688\|\|full | Low-quality |
| S1_vs_k141_132357\|\|full | Not-determined |
| S1_vs_k141_25432\|\|full | Not-determined |
| S1_vs_k141_37703\|\|full | Low-quality |
| S1_vs_k141_145279\|\|full | Not-determined |
| S1_vs_k141_125362\|\|full | Low-quality |
| S1_vs_k141_132361\|\|full | Not-determined |
| S1_vs_k141_125365\|\|full | Low-quality |
| S1_vs_k141_112255\|\|full | Not-determined |
| S1_vs_k141_52546\|\|full | Not-determined |
| S1_vs_k141_85675\|\|full | Not-determined |
| S1_vs_k141_156541\|\|full | Low-quality |
| [illegible] | [illegible] |
| S1_vs_k141_112490\|\|full | Low-quality |
| S1_vs_k141_85986\|\|full | Low-quality |
| S1_vs_k141_92569\|\|full | Low-quality |
| S1_vs_k141_32624\|\|full | Low-quality |
| S1_vs_k141_4629\|\|full | Not-determined |
| S1_vs_k141_59212\|\|full | Low-quality |
| S1_vs_k141_145528\|\|full | Low-quality |
| S1_vs_k141_45960\|\|full | Not-determined |
| S1_vs_k141_85990\|\|full | Not-determined |
| S1_vs_k141_156715\|\|full | Low-quality |
| S1_vs_k141_85992\|\|full | Low-quality |
| S1_vs_k141_149643\|\|full | Low-quality |
| S1_vs_k141_132688\|\|full | Low-quality |
| S1_vs_k141_132690\|\|full | Not-determined |
| S1_vs_k141_92575\|\|full | Not-determined |
| S1_vs_k141_25748\|\|full | Not-determined |
| S1_vs_k141_132692\|\|full | Low-quality |
| S1_vs_k141_19827\|\|full | Low-quality |
| S1_vs_k141_92582\|\|full | Not-determined |
| S1_vs_k141_45980\|\|full | Low-quality |
| S1_vs_k141_112538\|\|full | Low-quality |
| S11_extracted_k141_104045 | Complete |
| S1_vs_k141_25764\|\|full | Low-quality |
| S1_vs_k141_99365\|\|full | Low-quality |
| S1_vs_k141_32665\|\|full | Not-determined |
| S1_vs_k141_37979\|\|full | Not-determined |
| S1_vs_k141_156725\|\|full | Low-quality |
| S1_vs_k141_59242\|\|full | Not-determined |
| S1_vs_k141_112549\|\|full | Not-determined |
| S1_vs_k141_79433\|\|full | Low-quality |
| S1_vs_k141_119320\|\|full | Low-quality |
| S1_vs_k141_45989\|\|full | Low-quality |
| S1_vs_k141_99383\|\|full | Low-quality |
| S1_vs_k141_32675\|\|full | Low-quality |
| S1_vs_k141_37987\|\|full | Low-quality |
| S1_vs_k141_86035\|\|full | Not-determined |
| S1_vs_k141_125670\|\|full | Low-quality |
| S1_vs_k141_25785\|\|full | Low-quality |
| S1_vs_k141_59252\|\|full | Not-determined |
| S1_vs_k141_132753\|\|full | Low-quality |
| S1_vs_k141_12843\|\|full | Low-quality |
| S1_vs_k141_19851\|\|full | Low-quality |
| S1_vs_k141_99402\|\|full | Low-quality |
| S1_vs_k141_119346\|\|full | Low-quality |
| S1_vs_k141_4688\|\|full | Low-quality |
| S1_vs_k141_37998\|\|full | Low-quality |
| S1_vs_k141_139084\|\|full | Low-quality |
| S1_vs_k141_92618\|\|full | Low-quality |
| S1_vs_k141_125688\|\|full | Not-determined |
| S1_vs_k141_32701\|\|full | Not-determined |
| S1_vs_k141_12856\|\|full | Not-determined |
| S1_vs_k141_86061\|\|full | Low-quality |
| S1_vs_k141_92624\|\|full | Low-quality |
| S1_vs_k141_132775\|\|full | Not-determined |
| S1_vs_k141_25808\|\|full | Low-quality |
| S1_vs_k141_156748\|\|full | Low-quality |
| S1_vs_k141_139099\|\|full | Not-determined |
| S1_vs_k141_112568\|\|full | Low-quality |
| S1_vs_k141_38014\|\|full | Low-quality |
| S1_vs_k141_139108\|\|full | Low-quality |
| S1_vs_k141_86066\|\|full | Low-quality |
| S1_vs_k141_64699\|\|full | Low-quality |
| S1_vs_k141_132795\|\|full | Low-quality |
| S1_vs_k141_112580\|\|full | Low-quality |
| S1_vs_k141_99426\|\|full | Low-quality |
| S1_vs_k141_46041\|\|full | Not-determined |
| S1_vs_k141_71377\|\|full | Low-quality |
| S1_vs_k141_79502\|\|full | Low-quality |
| S1_vs_k141_119377\|\|full | Not-determined |
| S1_vs_k141_156755\|\|full | Low-quality |
| S1_vs_k141_92636\|\|full | Low-quality |
| S1_vs_k141_106049\|\|full | Low-quality |
| S1_vs_k141_4698\|\|full | Low-quality |
| S1_vs_k141_46049\|\|full | Low-quality |
| S1_vs_k141_25837\|\|full | Low-quality |
| S1_vs_k141_59312\|\|full | Low-quality |
| S1_vs_k141_25839\|\|full | Not-determined |
| S1_vs_k141_156761\|\|full | Not-determined |
| S1_vs_k141_125744\|\|full | Not-determined |
| S1_vs_k141_112604\|\|full | Low-quality |
| S1_vs_k141_79522\|\|full | Low-quality |
| S1_vs_k141_86086\|\|full | Low-quality |
| S1_vs_k141_92661\|\|full | Low-quality |
| S1_vs_k141_19918\|\|full | Low-quality |
| S1_vs_k141_52979\|\|full | Low-quality |
| S1_vs_k141_92665\|\|full | Low-quality |
| [illegible] | [illegible] |

| | |
|---|---|
| S5_vs_k141_60597\|\|full | High-quality |
| S1_vs_k141_38478\|\|full | Low-quality |
| S1_vs_k141_65248\|\|full | Low-quality |
| S1_vs_k141_126290\|\|full | Low-quality |
| S1_vs_k141_106624\|\|full | Low-quality |
| S1_vs_k141_119898\|\|full | Low-quality |
| S1_vs_k141_38485\|\|full | Not-determined |
| S1_vs_k141_119901\|\|full | Low-quality |
| S1_vs_k141_133422\|\|full | Not-determined |
| S1_vs_k141_126303\|\|full | Low-quality |
| S1_vs_k141_46607\|\|full | Low-quality |
| S1_vs_k141_106644\|\|full | Low-quality |
| S1_vs_k141_106646\|\|full | Not-determined |
| S1_vs_k141_33349\|\|full | Low-quality |
| S1_vs_k141_26418\|\|full | Not-determined |
| S1_vs_k141_100073\|\|full | Low-quality |
| S1_vs_k141_119905\|\|full | Not-determined |
| S1_vs_k141_71747\|\|full | Low-quality |
| S1_vs_k141_26419\|\|full | Not-determined |
| S1_vs_k141_59837\|\|full | Low-quality |
| S1_vs_k141_146096\|\|full | Not-determined |
| S1_vs_k141_100088\|\|full | Low-quality |
| S1_vs_k141_139760\|\|full | Low-quality |
| S1_vs_k141_106666\|\|full | Low-quality |
| S1_vs_k141_46641\|\|full | Low-quality |
| S1_vs_k141_100100\|\|full | Not-determined |
| S1_vs_k141_86712\|\|full | Low-quality |
| S1_vs_k141_65303\|\|full | Low-quality |
| S1_vs_k141_46658\|\|full | Low-quality |
| S1_vs_k141_126342\|\|full | Low-quality |
| S1_vs_k141_126343\|\|full | Low-quality |
| S1_vs_k141_113238\|\|full | Not-determined |
| S1_vs_k141_86719\|\|full | Low-quality |
| S1_vs_k141_80070\|\|full | Low-quality |
| S1_vs_k141_65312\|\|full | Not-determined |
| S1_vs_k141_46676\|\|full | Not-determined |
| S1_vs_k141_106704\|\|full | Low-quality |
| S1_vs_k141_71787\|\|full | Not-determined |
| S1_vs_k141_86729\|\|full | Not-determined |
| S1_vs_k141_93331\|\|full | Not-determined |
| S1_vs_k141_86736\|\|full | Not-determined |
| S1_vs_k141_119949\|\|full | Not-determined |
| S1_vs_k141_86739\|\|full | Low-quality |
| S1_vs_k141_93347\|\|full | Low-quality |
| S1_vs_k141_71799\|\|full | Not-determined |
| S1_vs_k141_38545\|\|full | Low-quality |
| S1_vs_k141_5227\|\|full | Low-quality |
| S1_vs_k141_146146\|\|full | Not-determined |
| S1_vs_k141_86758\|\|full | Low-quality |
| S1_vs_k141_139820\|\|full | Low-quality |
| S1_vs_k141_93374\|\|full | Not-determined |
| S1_vs_k141_150050\|\|full | Low-quality |
| S1_vs_k141_86766\|\|full | Low-quality |
| S1_vs_k141_65361\|\|full | Not-determined |
| S1_vs_k141_46716\|\|full | Not-determined |
| S1_vs_k141_71830\|\|full | Not-determined |
| S1_vs_k141_106747\|\|full | Low-quality |
| S1_vs_k141_119974\|\|full | Not-determined |
| S1_vs_k141_59917\|\|full | Low-quality |
| S1_vs_k141_113310\|\|full | Not-determined |
| S1_vs_k141_65375\|\|full | Low-quality |
| S1_vs_k141_26455\|\|full | Not-determined |
| S1_vs_k141_38578\|\|full | Not-determined |
| S1_vs_k141_71855\|\|full | Low-quality |
| S1_vs_k141_126375\|\|full | Low-quality |
| S1_vs_k141_93429\|\|full | Low-quality |
| S1_vs_k141_59980\|\|full | Low-quality |
| S1_vs_k141_5274\|\|full | Low-quality |
| S1_vs_k141_93439\|\|full | Low-quality |
| S1_vs_k141_65435\|\|full | Not-determined |
| S1_vs_k141_59989\|\|full | Low-quality |
| S1_vs_k141_126407\|\|full | Low-quality |
| S1_vs_k141_139876\|\|full | Not-determined |
| S1_vs_k141_126411\|\|full | Not-determined |
| S1_vs_k141_139884\|\|full | Low-quality |
| S1_vs_k141_150108\|\|full | Low-quality |
| S1_vs_k141_60016\|\|full | Low-quality |
| S1_vs_k141_113397\|\|full | Low-quality |
| S1_vs_k141_146273\|\|full | Low-quality |
| S1_vs_k141_26525\|\|full | Low-quality |
| S1_vs_k141_65506\|\|full | Low-quality |
| S1_vs_k141_146281\|\|full | Low-quality |
| S1_vs_k141_65527\|\|full | Low-quality |
| S1_vs_k141_26552\|\|full | Low-quality |
| S1_vs_k141_120115\|\|full | Low-quality |
| S1_vs_k141_71934\|\|full | Low-quality |
| S1_vs_k141_146318\|\|full | Not-determined |
| S1_vs_k141_126475\|\|full | Not-determined |
| S1_vs_k141_38622\|\|full | Low-quality |
| S1_vs_k141_5355\|\|full | Low-quality |
| S1_vs_k141_113442\|\|full | Low-quality |
| S1_vs_k141_120125\|\|full | Low-quality |
| S1_vs_k141_150143\|\|full | Low-quality |
| S1_vs_k141_157076\|\|full | Not-determined |
| S1_vs_k141_26586\|\|full | Not-determined |
| S1_vs_k141_71972\|\|full | Low-quality |
| S1_vs_k141_146354\|\|full | Not-determined |
| S1_vs_k141_150150\|\|full | Low-quality |
| S1_vs_k141_5403\|\|full | Low-quality |
| S1_vs_k141_26638\|\|full | Not-determined |
| S1_vs_k141_140040\|\|full | Not-determined |
| S1_vs_k141_65636\|\|full | Low-quality |
| S1_vs_k141_26650\|\|full | Not-determined |
| S1_vs_k141_157090\|\|full | Not-determined |
| S1_vs_k141_26652\|\|full | Low-quality |
| S1_vs_k141_5432\|\|full | Not-determined |
| S1_vs_k141_140062\|\|full | Low-quality |
| S1_vs_k141_146444\|\|full | Not-determined |
| S1_vs_k141_65661\|\|full | Low-quality |
| S1_vs_k141_5446\|\|full | Low-quality |
| S1_vs_k141_146475\|\|full | Low-quality |
| S1_vs_k141_126617\|\|full | Not-determined |
| S1_vs_k141_126620\|\|full | Low-quality |
| S1_vs_k141_146491\|\|full | Low-quality |
| S1_vs_k141_126633\|\|full | Low-quality |
| S1_vs_k141_72075\|\|full | Low-quality |
| S1_vs_k141_157124\|\|full | Not-determined |
| S1_vs_k141_157131\|\|full | Not-determined |
| S1_vs_k141_5497\|\|full | Not-determined |
| S1_vs_k141_126659\|\|full | Low-quality |
| S1_vs_k141_140138\|\|full | Low-quality |
| S1_vs_k141_65725\|\|full | Not-determined |
| S1_vs_k141_146547\|\|full | Not-determined |
| S1_vs_k141_140147\|\|full | Low-quality |
| S1_vs_k141_140161\|\|full | Low-quality |
| S1_vs_k141_5512\|\|full | Not-determined |
| S1_vs_k141_140167\|\|full | Not-determined |
| S1_vs_k141_140172\|\|full | Not-determined |
| S1_vs_k141_5524\|\|full | Not-determined |
| S1_vs_k141_38795\|\|full | Not-determined |
| S1_vs_k141_126705\|\|full | Not-determined |
| S1_vs_k141_65763\|\|full | Low-quality |
| S1_vs_k141_126706\|\|full | Low-quality |
| S1_vs_k141_157181\|\|full | Low-quality |
| S1_vs_k141_157192\|\|full | Not-determined |
| S1_vs_k141_126732\|\|full | Low-quality |
| S1_vs_k141_157194\|\|full | Low-quality |
| S1_vs_k141_126746\|\|full | Low-quality |
| S1_vs_k141_65833\|\|full | Low-quality |
| S1_vs_k141_126755\|\|full | Not-determined |
| S1_vs_k141_157233\|\|full | Low-quality |
| S1_vs_k141_38864\|\|full | Low-quality |
| S1_vs_k141_150258\|\|full | Low-quality |
| S1_vs_k141_146644\|\|full | Not-determined |
| S1_vs_k141_126779\|\|full | Low-quality |
| S1_vs_k141_146659\|\|full | Low-quality |
| S1_vs_k141_157242\|\|full | Low-quality |
| S1_vs_k141_146673\|\|full | Low-quality |
| S1_vs_k141_72154\|\|full | Low-quality |
| S1_vs_k141_146677\|\|full | Low-quality |
| S1_vs_k141_126806\|\|full | Low-quality |
| S1_vs_k141_38902\|\|full | Low-quality |
| S1_vs_k141_65899\|\|full | Not-determined |
| S1_vs_k141_65906\|\|full | Low-quality |
| S1_vs_k141_126823\|\|full | Low-quality |
| S1_vs_k141_5646\|\|full | Not-determined |
| S1_vs_k141_72164\|\|full | Low-quality |
| S1_vs_k141_150269\|\|full | Not-determined |
| S1_vs_k141_65923\|\|full | Low-quality |
| S1_vs_k141_65930\|\|full | Low-quality |
| S1_vs_k141_65938\|\|full | Not-determined |
| S1_vs_k141_5665\|\|full | Not-determined |
| S1_vs_k141_65941\|\|full | Low-quality |
| S1_vs_k141_157266\|\|full | Low-quality |
| S1_vs_k141_38938\|\|full | Low-quality |
| S1_vs_k141_38942\|\|full | Low-quality |
| S1_vs_k141_38946\|\|full | Low-quality |
| S1_vs_k141_72169\|\|full | Low-quality |
| S1_vs_k141_5688\|\|full | Low-quality |
| S1_vs_k141_65959\|\|full | Low-quality |
| S1_vs_k141_146764\|\|full | Not-determined |
| S1_vs_k141_157283\|\|full | Not-determined |
| S1_vs_k141_146773\|\|full | Low-quality |
| S1_vs_k141_72186\|\|full | Low-quality |
| S1_vs_k141_72188\|\|full | Low-quality |
| S1_vs_k141_65978\|\|full | Low-quality |
| S1_vs_k141_65984\|\|full | Not-determined |
| S1_vs_k141_65992\|\|full | Low-quality |
| S1_vs_k141_150312\|\|full | Low-quality |
| S1_vs_k141_150314\|\|full | Low-quality |
| S1_vs_k141_66008\|\|full | Low-quality |
| S1_vs_k141_66033\|\|full | Low-quality |
| S1_vs_k141_5757\|\|full | Low-quality |
| S1_vs_k141_66089\|\|full | Not-determined |
| S1_vs_k141_66100\|\|full | Low-quality |
| S1_vs_k141_150338\|\|full | Not-determined |
| S1_vs_k141_5797\|\|full | Low-quality |
| S1_vs_k141_157398\|\|full | Not-determined |
| S1_vs_k141_157401\|\|full | Low-quality |
| S1_vs_k141_39049\|\|full | Low-quality |
| S1_vs_k141_157411\|\|full | Not-determined |
| S1_vs_k141_39057\|\|full | Not-determined |
| S1_vs_k141_72267\|\|full | Not-determined |
| S1_vs_k141_157419\|\|full | Low-quality |
| S1_vs_k141_157429\|\|full | Low-quality |
| S1_vs_k141_66182\|\|full | Not-determined |
| S1_vs_k141_157442\|\|full | Low-quality |
| S1_vs_k141_39095\|\|full | Not-determined |
| S1_vs_k141_66217\|\|full | Low-quality |
| S1_vs_k141_72274\|\|full | Low-quality |
| S1_vs_k141_39109\|\|full | Low-quality |
| S1_vs_k141_72278\|\|full | Low-quality |
| S1_vs_k141_66259\|\|full | Low-quality |
| S1_vs_k141_66279\|\|full | Low-quality |
| S1_vs_k141_72311\|\|full | Low-quality |
| S1_vs_k141_72319\|\|full | Low-quality |
| S1_vs_k141_66316\|\|full | Not-determined |
| S1_vs_k141_66324\|\|full | Low-quality |
| S1_vs_k141_39196\|\|full | Low-quality |
| S1_vs_k141_66340\|\|full | Not-determined |
| S1_vs_k141_72359\|\|full | Low-quality |
| S1_vs_k141_5998\|\|full | Low-quality |
| S1_vs_k141_6002\|\|full | Low-quality |
| S1_vs_k141_6004\|\|full | Low-quality |
| S1_vs_k141_6006\|\|full | Not-determined |
| S1_vs_k141_39244\|\|full | Not-determined |
| S1_vs_k141_6016\|\|full | Low-quality |
| S1_vs_k141_72386\|\|full | Low-quality |
| S1_vs_k141_6019\|\|full | Low-quality |
| S1_vs_k141_66421\|\|full | Not-determined |
| S1_vs_k141_157534\|\|full | Low-quality |
| S1_vs_k141_6030\|\|full | Low-quality |
| S1_vs_k141_6033\|\|full | Not-determined |
| S1_vs_k141_6037\|\|full | Not-determined |
| S1_vs_k141_150458\|\|full | Not-determined |
| S1_vs_k141_66475\|\|full | Not-determined |
| S1_vs_k141_6070\|\|full | Low-quality |
| S1_vs_k141_157548\|\|full | Not-determined |
| S1_vs_k141_39271\|\|full | Low-quality |
| S1_vs_k141_150463\|\|full | Not-determined |
| S1_vs_k141_72470\|\|full | Low-quality |
| S1_vs_k141_72493\|\|full | Low-quality |
| S1_vs_k141_66573\|\|full | Low-quality |
| S1_vs_k141_150499\|\|full | Low-quality |
| S1_vs_k141_72540\|\|full | Not-determined |
| S1_vs_k141_72545\|\|full | Low-quality |
| S1_vs_k141_157591\|\|full | Low-quality |
| S1_vs_k141_39404\|\|full | Low-quality |
| S1_vs_k141_66658\|\|full | Low-quality |
| S1_vs_k141_6259\|\|full | Not-determined |
| S1_vs_k141_72612\|\|full | Low-quality |
| S1_vs_k141_66685\|\|full | Not-determined |
| S1_vs_k141_66689\|\|full | Low-quality |
| S1_vs_k141_39452\|\|full | Low-quality |
| S1_vs_k141_72656\|\|full | Not-determined |
| S1_vs_k141_150530\|\|full | Not-determined |
| S1_vs_k141_72705\|\|full | Low-quality |
| S1_vs_k141_39513\|\|full | Low-quality |
| S1_vs_k141_6281\|\|full | Not-determined |
| S1_vs_k141_39537\|\|full | Not-determined |
| S1_vs_k141_150556\|\|full | Low-quality |
| S1_vs_k141_72762\|\|full | Not-determined |
| S1_vs_k141_72774\|\|full | Low-quality |
| S1_vs_k141_39604\|\|full | Not-determined |
| S1_vs_k141_150572\|\|full | Low-quality |
| S1_vs_k141_6348\|\|full | Low-quality |
| S1_vs_k141_150573\|\|full | Low-quality |
| S1_vs_k141_6356\|\|full | Low-quality |
| S1_vs_k141_72781\|\|full | Low-quality |
| S1_vs_k141_157757\|\|full | Low-quality |
| S1_vs_k141_157764\|\|full | Not-determined |
| S1_vs_k141_150598\|\|full | Low-quality |
| S1_vs_k141_150606\|\|full | Not-determined |
| S1_vs_k141_39708\|\|full | Low-quality |
| S1_vs_k141_157795\|\|full | Not-determined |
| S1_vs_k141_157810\|\|full | Not-determined |
| S1_vs_k141_6465\|\|full | Low-quality |
| S1_vs_k141_157819\|\|full | Low-quality |
| S1_vs_k141_39723\|\|full | Low-quality |
| S1_vs_k141_39730\|\|full | Not-determined |
| S1_vs_k141_72847\|\|full | Low-quality |
| S1_vs_k141_72853\|\|full | Low-quality |
| S1_vs_k141_157821\|\|full | Low-quality |
| S1_vs_k141_150645\|\|full | Low-quality |
| S1_vs_k141_157839\|\|full | Low-quality |
| S1_vs_k141_150655\|\|full | Low-quality |
| S1_vs_k141_157867\|\|full | Low-quality |
| S1_vs_k141_157877\|\|full | Not-determined |
| S1_vs_k141_157901\|\|full | Not-determined |
| S1_vs_k141_39809\|\|full | Not-determined |
| S1_vs_k141_6619\|\|full | Low-quality |
| S1_vs_k141_6632\|\|full | Low-quality |
| S1_vs_k141_150700\|\|full | Low-quality |
| S1_vs_k141_150704\|\|full | Not-determined |
| S1_vs_k141_39865\|\|full | Low-quality |
| S1_vs_k141_6666\|\|full | Not-determined |
| S1_vs_k141_157970\|\|full | Not-determined |
| S1_vs_k141_39887\|\|full | Low-quality |
| S1_vs_k141_72928\|\|full | Low-quality |
| S1_vs_k141_150732\|\|full | Low-quality |
| S1_vs_k141_39913\|\|full | Not-determined |
| S1_vs_k141_72955\|\|full | Low-quality |
| S1_vs_k141_40018\|\|full | Low-quality |
| S1_vs_k141_40019\|\|full | Low-quality |
| S1_vs_k141_150798\|\|full | Low-quality |
| S1_vs_k141_158122\|\|full | Low-quality |
| S1_vs_k141_150835\|\|full | Not-determined |
| S1_vs_k141_158144\|\|full | Low-quality |
| S1_vs_k141_158150\|\|full | Low-quality |
| S1_vs_k141_73026\|\|full | Low-quality |
| S1_vs_k141_150865\|\|full | Not-determined |
| S1_vs_k141_73039\|\|full | Low-quality |
| S1_vs_k141_158178\|\|full | Not-determined |
| S1_vs_k141_73060\|\|full | Low-quality |
| S1_vs_k141_158190\|\|full | Not-determined |
| S1_vs_k141_73071\|\|full | Low-quality |
| S1_vs_k141_150907\|\|full | Low-quality |
| S1_vs_k141_150919\|\|full | Not-determined |
| S1_vs_k141_73100\|\|full | Not-determined |
| S1_vs_k141_158229\|\|full | Low-quality |
| S1_vs_k141_150937\|\|full | Low-quality |
| S1_vs_k141_158241\|\|full | Not-determined |
| S1_vs_k141_73123\|\|full | Low-quality |
| S1_vs_k141_150959\|\|full | Low-quality |
| S1_vs_k141_158314\|\|full | Low-quality |
| S1_vs_k141_73219\|\|full | Low-quality |
| S1_vs_k141_151051\|\|full | Low-quality |
| S1_vs_k141_158356\|\|full | Low-quality |
| S1_vs_k141_158362\|\|full | Not-determined |
| S1_vs_k141_158416\|\|full | Low-quality |
| S1_vs_k141_73309\|\|full | Low-quality |
| S1_vs_k141_158458\|\|full | Low-quality |
| S1_vs_k141_158460\|\|full | Low-quality |
| S1_vs_k141_151127\|\|full | Low-quality |
| S6_vs_k141_29282\|\|full | Medium-quality |
| S1_vs_k141_158492\|\|full | Not-determined |
| S1_vs_k141_158493\|\|full | Not-determined |
| S1_vs_k141_158498\|\|full | Low-quality |
| S1_vs_k141_158500\|\|full | Low-quality |
| S1_vs_k141_73400\|\|full | Not-determined |
| S1_vs_k141_73404\|\|full | Low-quality |
| S1_vs_k141_158520\|\|full | Low-quality |
| S1_vs_k141_151248\|\|full | Low-quality |
| S1_vs_k141_158536\|\|full | Low-quality |
| S1_vs_k141_151255\|\|full | Low-quality |
| S1_vs_k141_158545\|\|full | Low-quality |
| S1_vs_k141_151281\|\|full | Low-quality |
| S1_vs_k141_158583\|\|full | Low-quality |
| S1_vs_k141_151311\|\|full | Low-quality |
| S1_vs_k141_158613\|\|full | Low-quality |
| S1_vs_k141_151354\|\|full | Low-quality |
| S1_vs_k141_158647\|\|full | Low-quality |
| S1_vs_k141_158659\|\|full | Low-quality |
| S1_vs_k141_151396\|\|full | Low-quality |
| S1_vs_k141_158693\|\|full | Not-determined |
| S1_vs_k141_151403\|\|full | Not-determined |
| S1_vs_k141_158736\|\|full | Not-determined |
| S1_vs_k141_151488\|\|full | Low-quality |
| S1_vs_k141_158763\|\|full | Not-determined |
| S1_vs_k141_151566\|\|full | Low-quality |
| S1_vs_k141_151567\|\|full | Low-quality |
| S1_vs_k141_151571\|\|full | Low-quality |
| S1_vs_k141_151609\|\|full | Low-quality |
| S1_vs_k141_158860\|\|full | Low-quality |
| S1_vs_k141_158885\|\|full | Low-quality |
| S1_vs_k141_151674\|\|full | Low-quality |
| S1_vs_k141_158902\|\|full | Low-quality |
| S1_vs_k141_151688\|\|full | Low-quality |
| S1_vs_k141_151692\|\|full | Not-determined |
| S1_vs_k141_158926\|\|full | Low-quality |
| S1_vs_k141_158944\|\|full | Low-quality |
| S1_vs_k141_158945\|\|full | Low-quality |
| S1_vs_k141_151707\|\|full | Low-quality |
| S16_extracted_6102 | High-quality |
| S1_vs_k141_159001\|\|full | Low-quality |
| S1_vs_k141_159004\|\|full | Not-determined |
| S1_vs_k141_151840\|\|full | Not-determined |
| S1_vs_k141_151880\|\|full | Low-quality |
| S1_vs_k141_151883\|\|full | Not-determined |
| S1_vs_k141_159069\|\|full | Not-determined |
| S1_vs_k141_151926\|\|full | Not-determined |
| S1_vs_k141_151943\|\|full | Low-quality |
| S1_vs_k141_151956\|\|full | Low-quality |
| S1_vs_k141_159134\|\|full | Not-determined |
| S1_vs_k141_159171\|\|full | Not-determined |
| S1_vs_k141_159179\|\|full | Low-quality |
| S1_vs_k141_159210\|\|full | Low-quality |
| S1_vs_k141_159211\|\|full | Low-quality |
| S1_vs_k141_159220\|\|full | Not-determined |
| S1_vs_k141_159223\|\|full | Low-quality |
| S1_vs_k141_152103\|\|full | Low-quality |
| S1_vs_k141_159240\|\|full | Low-quality |
| S1_vs_k141_152110\|\|full | Low-quality |
| S1_vs_k141_159244\|\|full | Not-determined |
| S1_vs_k141_152128\|\|full | Not-determined |
| S1_vs_k141_159249\|\|full | Not-determined |
| S1_vs_k141_152138\|\|full | Low-quality |
| S1_vs_k141_152148\|\|full | Low-quality |
| S1_vs_k141_159281\|\|full | Not-determined |
| S1_vs_k141_159305\|\|full | Low-quality |
| S1_vs_k141_159309\|\|full | Low-quality |
| S1_vs_k141_152196\|\|full | Low-quality |
| S1_vs_k141_152211\|\|full | Not-determined |
| S1_vs_k141_152220\|\|full | Low-quality |
| S1_vs_k141_152230\|\|full | Low-quality |
| S1_vs_k141_152244\|\|full | Low-quality |
| S1_vs_k141_152282\|\|full | Low-quality |
| S1_vs_k141_152288\|\|full | Not-determined |
| S1_vs_k141_152320\|\|full | Not-determined |
| S1_vs_k141_159500\|\|full | Not-determined |
| S1_vs_k141_159505\|\|full | Low-quality |
| S1_vs_k141_152399\|\|full | Not-determined |
| S1_vs_k141_159536\|\|full | Low-quality |
| S1_vs_k141_152437\|\|full | Low-quality |
| S1_vs_k141_152456\|\|full | Low-quality |
| S11_vs_k141_104045\|\|full | High-quality |
| S1_vs_k141_159562\|\|full | Low-quality |
| S1_vs_k141_152539\|\|full | Low-quality |
| S1_vs_k141_159593\|\|full | Not-determined |
| S1_vs_k141_152594\|\|full | Low-quality |
| S1_vs_k141_152603\|\|full | Low-quality |
| S1_vs_k141_159643\|\|full | Low-quality |
| S1_vs_k141_159646\|\|full | Low-quality |
| S1_vs_k141_159654\|\|full | Not-determined |
| S1_vs_k141_152678\|\|full | Low-quality |
| S1_vs_k141_152707\|\|full | Low-quality |
| S1_vs_k141_159698\|\|full | Low-quality |
| S1_vs_k141_159702\|\|full | Low-quality |
| S1_vs_k141_152750\|\|full | Low-quality |
| S1_vs_k141_159736\|\|full | Not-determined |
| S1_vs_k141_152792\|\|full | Low-quality |
| S1_vs_k141_159757\|\|full | Low-quality |
| S1_vs_k141_152852\|\|full | Low-quality |
| S1_vs_k141_159807\|\|full | Not-determined |
| S1_vs_k141_152864\|\|full | Not-determined |
| S1_vs_k141_159821\|\|full | Not-determined |
| S1_vs_k141_159824\|\|full | Low-quality |
| S1_vs_k141_159838\|\|full | Low-quality |
| S1_vs_k141_152892\|\|full | Low-quality |
| S1_vs_k141_159877\|\|full | Low-quality |
| S1_vs_k141_159882\|\|full | Not-determined |
| S1_vs_k141_152934\|\|full | Low-quality |
| S1_vs_k141_159912\|\|full | Not-determined |
| S1_vs_k141_159927\|\|full | Not-determined |
| S1_vs_k141_153012\|\|full | Not-determined |
| S1_vs_k141_153070\|\|full | Low-quality |
| S1_vs_k141_153122\|\|full | Low-quality |
| S1_vs_k141_153330\|\|full | Low-quality |
| S1_vs_k141_153339\|\|full | Low-quality |
| S1_vs_k141_153340\|\|full | Not-determined |
| S1_vs_k141_153377\|\|full | Low-quality |
| S1_vs_k141_153385\|\|full | Low-quality |
| S1_vs_k141_153396\|\|full | Low-quality |
| S1_vs_k141_9858\|\|0_partial | Low-quality |
| S1_vs_k141_80103\|\|lt2gene | Low-quality |
| S1_vs_k141_80110\|\|lt2gene | Low-quality |
| S1_vs_k141_93455\|\|lt2gene | Low-quality |
| S1_vs_k141_6683\|\|lt2gene | Low-quality |
| S1_vs_k141_20041\|\|lt2gene | Low-quality |
| S1_vs_k141_26706\|\|lt2gene | Low-quality |
| S1_vs_k141_140188\|\|lt2gene | Low-quality |
| S1_vs_k141_106814\|\|lt2gene | Low-quality |
| S1_vs_k141_140195\|\|lt2gene | Not-determined |
| S1_vs_k141_20053\|\|lt2gene | Low-quality |
| S1_vs_k141_93471\|\|lt2gene | Not-determined |
| S1_vs_k141_46753\|\|lt2gene | Not-determined |
| S1_vs_k141_6703\|\|lt2gene | Low-quality |
| S1_vs_k141_106825\|\|lt2gene | Low-quality |
| S1_vs_k141_153555\|\|lt2gene | Not-determined |
| S1_vs_k141_106845\|\|lt2gene | Low-quality |
| S1_vs_k141_86812\|\|lt2gene | Low-quality |
| S1_vs_k141_140232\|\|lt2gene | Not-determined |
| S1_vs_k141_120198\|\|lt2gene | Low-quality |
| S1_vs_k141_140237\|\|lt2gene | Low-quality |
| S1_vs_k141_93514\|\|lt2gene | Low-quality |
| S1_vs_k141_66796\|\|lt2gene | Low-quality |
| S1_vs_k141_53468\|\|lt2gene | Not-determined |
| S1_vs_k141_113536\|\|lt2gene | Low-quality |
| S1_vs_k141_100166\|\|lt2gene | Low-quality |
| S1_vs_k141_80199\|\|lt2gene | Not-determined |
| S1_vs_k141_53498\|\|lt2gene | Not-determined |
| S1_vs_k141_106889\|\|lt2gene | Low-quality |
| S1_vs_k141_153592\|\|lt2gene | Low-quality |
| S1_vs_k141_66821\|\|lt2gene | Low-quality |
| S1_vs_k141_40103\|\|lt2gene | Low-quality |
| S1_vs_k141_100181\|\|lt2gene | Not-determined |
| S1_vs_k141_93553\|\|lt2gene | Low-quality |
| S1_vs_k141_100188\|\|lt2gene | Low-quality |
| S1_vs_k141_146920\|\|lt2gene | Low-quality |
| S1_vs_k141_116\|\|lt2gene | Not-determined |
| S1_vs_k141_100194\|\|lt2gene | Low-quality |
| S1_vs_k141_73512\|\|lt2gene | Low-quality |
| S1_vs_k141_93574\|\|lt2gene | Not-determined |
| S1_vs_k141_53528\|\|lt2gene | Not-determined |
| S1_vs_k141_53543\|\|lt2gene | Low-quality |
| S1_vs_k141_60105\|\|lt2gene | Not-determined |
| S1_vs_k141_73547\|\|lt2gene | Not-determined |
| S1_vs_k141_26805\|\|lt2gene | Low-quality |
| S1_vs_k141_73553\|\|lt2gene | Low-quality |
| S1_vs_k141_153611\|\|lt2gene | Not-determined |
| S1_vs_k141_53571\|\|lt2gene | Low-quality |
| S1_vs_k141_86941\|\|lt2gene | Low-quality |
| S1_vs_k141_86949\|\|lt2gene | Low-quality |
| S1_vs_k141_26825\|\|lt2gene | Low-quality |
| S1_vs_k141_106959\|\|lt2gene | Low-quality |
| S1_vs_k141_140349\|\|lt2gene | Low-quality |
| S1_vs_k141_93663\|\|lt2gene | Not-determined |
| S1_vs_k141_194\|\|lt2gene | Not-determined |
| S1_vs_k141_133605\|\|lt2gene | Not-determined |
| S1_vs_k141_20190\|\|lt2gene | Not-determined |
| S1_vs_k141_13531\|\|lt2gene | Low-quality |
| S1_vs_k141_73590\|\|lt2gene | Low-quality |
| S1_vs_k141_80313\|\|lt2gene | Not-determined |
| S1_vs_k141_93672\|\|lt2gene | Not-determined |
| S1_vs_k141_60162\|\|lt2gene | Low-quality |
| S1_vs_k141_86986\|\|lt2gene | Low-quality |
| S1_vs_k141_40244\|\|lt2gene | Low-quality |
| S1_vs_k141_66931\|\|lt2gene | Low-quality |
| S1_vs_k141_127060\|\|lt2gene | Low-quality |
| S1_vs_k141_86992\|\|lt2gene | Low-quality |
| S1_vs_k141_87000\|\|lt2gene | Not-determined |
| S1_vs_k141_113660\|\|lt2gene | Low-quality |
| S1_vs_k141_107011\|\|lt2gene | Low-quality |
| S1_vs_k141_127081\|\|lt2gene | Not-determined |
| S1_vs_k141_26851\|\|lt2gene | Not-determined |
| S1_vs_k141_53656\|\|lt2gene | Low-quality |
| S1_vs_k141_100358\|\|lt2gene | Low-quality |
| S1_vs_k141_53671\|\|lt2gene | Not-determined |
| S1_vs_k141_140437\|\|lt2gene | Low-quality |
| S1_vs_k141_53676\|\|lt2gene | Low-quality |
| S1_vs_k141_87055\|\|lt2gene | Not-determined |
| S1_vs_k141_120440\|\|lt2gene | Not-determined |
| S1_vs_k141_127135\|\|lt2gene | Low-quality |
| S1_vs_k141_113726\|\|lt2gene | Low-quality |
| S1_vs_k141_53701\|\|lt2gene | Not-determined |
| S1_vs_k141_87069\|\|lt2gene | Low-quality |
| S1_vs_k141_100400\|\|lt2gene | Not-determined |
| S1_vs_k141_147045\|\|lt2gene | Low-quality |
| S1_vs_k141_20317\|\|lt2gene | Not-determined |
| S1_vs_k141_20319\|\|lt2gene | Not-determined |
| S1_vs_k141_13667\|\|lt2gene | Low-quality |
| S1_vs_k141_67019\|\|lt2gene | Low-quality |
| S1_vs_k141_140477\|\|lt2gene | Low-quality |
| S1_vs_k141_40357\|\|lt2gene | Low-quality |
| S1_vs_k141_40360\|\|lt2gene | Not-determined |
| S1_vs_k141_53742\|\|lt2gene | Not-determined |
| S1_vs_k141_133724\|\|lt2gene | Low-quality |
| S1_vs_k141_13683\|\|lt2gene | Low-quality |
| S1_vs_k141_107103\|\|lt2gene | Low-quality |
| S1_vs_k141_26930\|\|lt2gene | Not-determined |
| S1_vs_k141_100431\|\|lt2gene | Not-determined |
| S1_vs_k141_127208\|\|lt2gene | Low-quality |
| S1_vs_k141_13705\|\|lt2gene | Low-quality |
| S1_vs_k141_60218\|\|lt2gene | Low-quality |
| S1_vs_k141_33678\|\|lt2gene | Low-quality |
| S1_vs_k141_133746\|\|lt2gene | Not-determined |
| S1_vs_k141_351\|\|lt2gene | Not-determined |
| S1_vs_k141_80495\|\|lt2gene | Low-quality |
| S1_vs_k141_13736\|\|lt2gene | Low-quality |
| S1_vs_k141_13741\|\|lt2gene | Low-quality |
| S1_vs_k141_60239\|\|lt2gene | Low-quality |
| S1_vs_k141_53811\|\|lt2gene | Not-determined |
| S1_vs_k141_53814\|\|lt2gene | Not-determined |
| S1_vs_k141_147096\|\|lt2gene | Not-determined |
| S1_vs_k141_80520\|\|lt2gene | Low-quality |
| S1_vs_k141_120541\|\|lt2gene | Low-quality |
| S1_vs_k141_87166\|\|lt2gene | Low-quality |
| S1_vs_k141_93878\|\|lt2gene | Not-determined |
| S1_vs_k141_20403\|\|lt2gene | Low-quality |
| S1_vs_k141_47093\|\|lt2gene | Low-quality |
| S1_vs_k141_67089\|\|lt2gene | Low-quality |
| S1_vs_k141_153723\|\|lt2gene | Not-determined |
| S1_vs_k141_73825\|\|lt2gene | Low-quality |
| S1_vs_k141_40468\|\|lt2gene | Low-quality |
| S1_vs_k141_133799\|\|lt2gene | Not-determined |
| S1_vs_k141_53861\|\|lt2gene | Not-determined |
| S1_vs_k141_140546\|\|lt2gene | Low-quality |
| S1_vs_k141_47113\|\|lt2gene | Low-quality |
| S1_vs_k141_47115\|\|lt2gene | Low-quality |
| S1_vs_k141_27065\|\|lt2gene | Low-quality |
| S1_vs_k141_73850\|\|lt2gene | Low-quality |
| S1_vs_k141_33747\|\|lt2gene | Low-quality |
| S1_vs_k141_140573\|\|lt2gene | Low-quality |
| S1_vs_k141_73853\|\|lt2gene | Low-quality |
| S1_vs_k141_73854\|\|lt2gene | Low-quality |
| S1_vs_k141_107230\|\|lt2gene | Low-quality |
| S1_vs_k141_127319\|\|lt2gene | Not-determined |
| S1_vs_k141_53932\|\|lt2gene | Not-determined |
| S1_vs_k141_127323\|\|lt2gene | Low-quality |
| S1_vs_k141_100600\|\|lt2gene | Low-quality |
| S1_vs_k141_120609\|\|lt2gene | Low-quality |
| S1_vs_k141_133889\|\|lt2gene | Not-determined |
| S1_vs_k141_113924\|\|lt2gene | Low-quality |
| S1_vs_k141_13906\|\|lt2gene | Low-quality |
| S1_vs_k141_20500\|\|lt2gene | Low-quality |
| S1_vs_k141_40587\|\|lt2gene | Low-quality |
| S1_vs_k141_53978\|\|lt2gene | Not-determined |
| S1_vs_k141_33820\|\|lt2gene | Low-quality |
| S1_vs_k141_80665\|\|lt2gene | Low-quality |

S1_vs_k141_93986||lt2gene Low-quality
S1_vs_k141_7201||lt2gene Low-quality
S1_vs_k141_47209||lt2gene Not-determined
S1_vs_k141_113953||lt2gene Low-quality
S1_vs_k141_140675||lt2gene Low-quality
S1_vs_k141_7223||lt2gene Low-quality
S1_vs_k141_13950||lt2gene Low-quality
S1_vs_k141_33847||lt2gene Low-quality
S1_vs_k141_120660||lt2gene Low-quality
S1_vs_k141_140701||lt2gene Low-quality
S1_vs_k141_94031||lt2gene Low-quality
S1_vs_k141_33856||lt2gene Low-quality
S1_vs_k141_94053||lt2gene Low-quality
S1_vs_k141_47250||lt2gene Low-quality
S1_vs_k141_153818||lt2gene Low-quality
S1_vs_k141_94060||lt2gene Low-quality
S1_vs_k141_73975||lt2gene Not-determined
S1_vs_k141_127424||lt2gene Low-quality
S1_vs_k141_80730||lt2gene Low-quality
S1_vs_k141_40685||lt2gene Low-quality
S1_vs_k141_114012||lt2gene Not-determined
S1_vs_k141_74006||lt2gene Not-determined
S1_vs_k141_74011||lt2gene Low-quality
S1_vs_k141_54047||lt2gene Low-quality
S1_vs_k141_54055||lt2gene Low-quality
S1_vs_k141_127465||lt2gene Low-quality
S1_vs_k141_100741||lt2gene Low-quality
S1_vs_k141_54064||lt2gene Not-determined
S1_vs_k141_87399||lt2gene Low-quality
S1_vs_k141_54072||lt2gene Low-quality
S1_vs_k141_134022||lt2gene Low-quality
S1_vs_k141_87424||lt2gene Low-quality
S1_vs_k141_100776||lt2gene Low-quality
S1_vs_k141_47333||lt2gene Not-determined
S1_vs_k141_20599||lt2gene Low-quality
S1_vs_k141_153879||lt2gene Low-quality
S1_vs_k141_60411||lt2gene Low-quality
S1_vs_k141_67325||lt2gene Low-quality
S1_vs_k141_134045||lt2gene Low-quality
S1_vs_k141_47361||lt2gene Low-quality
S1_vs_k141_67347||lt2gene Not-determined
S1_vs_k141_7405||lt2gene Low-quality
S1_vs_k141_40784||lt2gene Low-quality
S1_vs_k141_74126||lt2gene Not-determined
S1_vs_k141_615||lt2gene Low-quality
S1_vs_k141_14156||lt2gene Low-quality
S1_vs_k141_100836||lt2gene Not-determined
S1_vs_k141_27343||lt2gene Low-quality
S1_vs_k141_27344||lt2gene Not-determined
S1_vs_k141_20659||lt2gene Not-determined
S1_vs_k141_67362||lt2gene Low-quality
S1_vs_k141_87496||lt2gene Not-determined
S1_vs_k141_33984||lt2gene Not-determined
S1_vs_k141_114117||lt2gene Low-quality
S1_vs_k141_634||lt2gene Not-determined
S1_vs_k141_67373||lt2gene Not-determined
S1_vs_k141_33997||lt2gene Low-quality
S1_vs_k141_100856||lt2gene Not-determined
S1_vs_k141_60455||lt2gene Low-quality
S1_vs_k141_34018||lt2gene Low-quality
S1_vs_k141_54213||lt2gene Low-quality
S1_vs_k141_7427||lt2gene Low-quality
S1_vs_k141_153906||lt2gene Not-determined
S1_vs_k141_94219||lt2gene Not-determined
S1_vs_k141_87546||lt2gene Not-determined
S1_vs_k141_27390||lt2gene Not-determined
S1_vs_k141_47460||lt2gene Low-quality
S1_vs_k141_40842||lt2gene Not-determined
S1_vs_k141_664||lt2gene Low-quality
S1_vs_k141_100905||lt2gene Not-determined
S1_vs_k141_40849||lt2gene Low-quality
S1_vs_k141_153913||lt2gene Low-quality
S1_vs_k141_27402||lt2gene Low-quality
S1_vs_k141_94241||lt2gene Low-quality
S1_vs_k141_7458||lt2gene Low-quality
S1_vs_k141_87575||lt2gene Low-quality
S1_vs_k141_27432||lt2gene Not-determined
S1_vs_k141_692||lt2gene Low-quality
S1_vs_k141_34055||lt2gene Low-quality
S1_vs_k141_67463||lt2gene Low-quality
S1_vs_k141_114182||lt2gene Not-determined
S1_vs_k141_153940||lt2gene Low-quality
S1_vs_k141_153944||lt2gene Low-quality
S1_vs_k141_87599||lt2gene Low-quality
S1_vs_k141_140977||lt2gene Low-quality
S1_vs_k141_107483||lt2gene Not-determined
S1_vs_k141_153947||lt2gene Low-quality
S1_vs_k141_748||lt2gene Low-quality
S1_vs_k141_134223||lt2gene Low-quality
S1_vs_k141_140992||lt2gene Low-quality
S1_vs_k141_107496||lt2gene Low-quality
S1_vs_k141_101030||lt2gene Low-quality
S1_vs_k141_94347||lt2gene Low-quality
S1_vs_k141_127730||lt2gene Not-determined
S1_vs_k141_789||lt2gene Low-quality
S1_vs_k141_7586||lt2gene Low-quality
S1_vs_k141_20876||lt2gene Low-quality
S1_vs_k141_134246||lt2gene Low-quality
S1_vs_k141_7597||lt2gene Not-determined
S1_vs_k141_87678||lt2gene Not-determined
S1_vs_k141_94363||lt2gene Low-quality
S1_vs_k141_14410||lt2gene Not-determined
S1_vs_k141_14420||lt2gene Not-determined
S1_vs_k141_14431||lt2gene Not-determined
S1_vs_k141_14433||lt2gene Not-determined
S1_vs_k141_127765||lt2gene Not-determined
S1_vs_k141_141059||lt2gene Low-quality
S1_vs_k141_114301||lt2gene Low-quality
S1_vs_k141_60600||lt2gene Not-determined
S1_vs_k141_81090||lt2gene Low-quality
S1_vs_k141_7655||lt2gene Low-quality
S1_vs_k141_54417||lt2gene Low-quality
S1_vs_k141_20950||lt2gene Low-quality
S1_vs_k141_67617||lt2gene Low-quality
S1_vs_k141_41037||lt2gene Low-quality
S1_vs_k141_94459||lt2gene Low-quality
S1_vs_k141_54433||lt2gene Not-determined
S1_vs_k141_127828||lt2gene Low-quality
S1_vs_k141_41058||lt2gene Low-quality
S1_vs_k141_34204||lt2gene Low-quality
S1_vs_k141_87750||lt2gene Not-determined
S1_vs_k141_81155||lt2gene Low-quality
S1_vs_k141_87751||lt2gene Not-determined
S1_vs_k141_27595||lt2gene Low-quality
S1_vs_k141_107632||lt2gene Not-determined
S1_vs_k141_7718||lt2gene Not-determined
S1_vs_k141_94500||lt2gene Low-quality
S1_vs_k141_27610||lt2gene Low-quality
S1_vs_k141_74377||lt2gene Not-determined
S1_vs_k141_101161||lt2gene Low-quality
S1_vs_k141_14570||lt2gene Not-determined
S1_vs_k141_81180||lt2gene Low-quality
S1_vs_k141_60665||lt2gene Low-quality
S1_vs_k141_67673||lt2gene Low-quality
S1_vs_k141_134370||lt2gene Not-determined
S1_vs_k141_107670||lt2gene Low-quality
S1_vs_k141_7765||lt2gene Low-quality
S1_vs_k141_904||lt2gene Not-determined
S1_vs_k141_907||lt2gene Low-quality
S1_vs_k141_127879||lt2gene Low-quality
S1_vs_k141_67738||lt2gene Low-quality
S1_vs_k141_14685||lt2gene Not-determined
S1_vs_k141_47774||lt2gene Not-determined
S1_vs_k141_121070||lt2gene Low-quality
S1_vs_k141_154107||lt2gene Low-quality
S1_vs_k141_21089||lt2gene Not-determined
S1_vs_k141_114456||lt2gene Low-quality
S1_vs_k141_934||lt2gene Low-quality
S1_vs_k141_81251||lt2gene Low-quality
S1_vs_k141_101252||lt2gene Low-quality
S1_vs_k141_939||lt2gene Low-quality
S1_vs_k141_67764||lt2gene Low-quality
S1_vs_k141_67769||lt2gene Low-quality
S1_vs_k141_21120||lt2gene Low-quality
S1_vs_k141_7858||lt2gene Low-quality
S1_vs_k141_41251||lt2gene Low-quality
S1_vs_k141_54537||lt2gene Low-quality
S1_vs_k141_81277||lt2gene Low-quality
S1_vs_k141_81283||lt2gene Low-quality
S1_vs_k141_121114||lt2gene Low-quality
S1_vs_k141_94659||lt2gene Low-quality
S1_vs_k141_47838||lt2gene Low-quality
S1_vs_k141_87911||lt2gene Low-quality
S1_vs_k141_127950||lt2gene Low-quality
S1_vs_k141_134493||lt2gene Low-quality
S1_vs_k141_54566||lt2gene Not-determined
S1_vs_k141_127954||lt2gene Low-quality
S1_vs_k141_107823||lt2gene Not-determined
S1_vs_k141_60723||lt2gene Low-quality
S1_vs_k141_127960||lt2gene Low-quality
S1_vs_k141_147360||lt2gene Low-quality
S1_vs_k141_27770||lt2gene Low-quality
S1_vs_k141_34340||lt2gene Low-quality
S1_vs_k141_41293||lt2gene Low-quality
S1_vs_k141_74528||lt2gene Not-determined
S1_vs_k141_114521||lt2gene Low-quality
S1_vs_k141_134517||lt2gene Low-quality
S1_vs_k141_87943||lt2gene Low-quality
S1_vs_k141_980||lt2gene Low-quality
S1_vs_k141_41320||lt2gene Not-determined
S1_vs_k141_114539||lt2gene Low-quality
S1_vs_k141_21210||lt2gene Low-quality
S1_vs_k141_54610||lt2gene Low-quality
S1_vs_k141_21216||lt2gene Low-quality
S1_vs_k141_7977||lt2gene Low-quality
S1_vs_k141_128025||lt2gene Low-quality
S1_vs_k141_47929||lt2gene Low-quality
S1_vs_k141_41360||lt2gene Low-quality
S1_vs_k141_114576||lt2gene Not-determined
S1_vs_k141_87992||lt2gene Low-quality
S1_vs_k141_54629||lt2gene Not-determined
S1_vs_k141_54630||lt2gene Not-determined
S1_vs_k141_114589||lt2gene Low-quality
S1_vs_k141_27855||lt2gene Low-quality
S1_vs_k141_74604||lt2gene Low-quality
S1_vs_k141_34393||lt2gene Low-quality
S1_vs_k141_21236||lt2gene Not-determined
S1_vs_k141_101409||lt2gene Low-quality
S1_vs_k141_54652||lt2gene Low-quality
S1_vs_k141_47963||lt2gene Low-quality
S1_vs_k141_60788||lt2gene Low-quality
S1_vs_k141_141350||lt2gene Low-quality
S1_vs_k141_67932||lt2gene Not-determined
S1_vs_k141_128068||lt2gene Not-determined
S1_vs_k141_67941||lt2gene Low-quality
S1_vs_k141_8037||lt2gene Low-quality
S1_vs_k141_60808||lt2gene Low-quality
S1_vs_k141_114627||lt2gene Not-determined
S1_vs_k141_94772||lt2gene Low-quality
S1_vs_k141_60815||lt2gene Low-quality
S1_vs_k141_134617||lt2gene Low-quality
S1_vs_k141_101470||lt2gene Not-determined
S1_vs_k141_1063||lt2gene Low-quality
S1_vs_k141_48015||lt2gene Not-determined
S1_vs_k141_141395||lt2gene Low-quality
S1_vs_k141_21280||lt2gene Not-determined
S1_vs_k141_34457||lt2gene Low-quality
S1_vs_k141_114676||lt2gene Not-determined
S1_vs_k141_60832||lt2gene Low-quality
S1_vs_k141_141413||lt2gene Low-quality
S1_vs_k141_88106||lt2gene Low-quality
S1_vs_k141_8110||lt2gene Not-determined
S1_vs_k141_154267||lt2gene Not-determined
S1_vs_k141_81484||lt2gene Low-quality
S1_vs_k141_94839||lt2gene Low-quality
S1_vs_k141_54727||lt2gene Not-determined
S1_vs_k141_74743||lt2gene Low-quality
S1_vs_k141_121251||lt2gene Not-determined
S1_vs_k141_121252||lt2gene Not-determined
S1_vs_k141_101545||lt2gene Low-quality
S1_vs_k141_141458||lt2gene Low-quality
S1_vs_k141_134708||lt2gene Not-determined
S1_vs_k141_128154||lt2gene Not-determined
S1_vs_k141_8139||lt2gene Low-quality
S1_vs_k141_15037||lt2gene Low-quality
S1_vs_k141_34511||lt2gene Low-quality
S1_vs_k141_134739||lt2gene Low-quality
S1_vs_k141_34524||lt2gene Low-quality
S1_vs_k141_88151||lt2gene Low-quality
S1_vs_k141_81545||lt2gene Not-determined
S1_vs_k141_81547||lt2gene Low-quality
S1_vs_k141_88155||lt2gene Low-quality
S1_vs_k141_94918||lt2gene Low-quality
S1_vs_k141_48119||lt2gene Low-quality
S1_vs_k141_28061||lt2gene Low-quality
S1_vs_k141_15070||lt2gene Not-determined
S1_vs_k141_15077||lt2gene Not-determined
S1_vs_k141_121328||lt2gene Low-quality
S1_vs_k141_154328||lt2gene Not-determined
S1_vs_k141_28076||lt2gene Not-determined
S1_vs_k141_147482||lt2gene Low-quality
S1_vs_k141_114821||lt2gene Not-determined
S1_vs_k141_48146||lt2gene Low-quality
S1_vs_k141_74821||lt2gene Low-quality
S1_vs_k141_34571||lt2gene Low-quality
S1_vs_k141_141524||lt2gene Not-determined
S1_vs_k141_15096||lt2gene Low-quality
S1_vs_k141_141531||lt2gene Low-quality
S1_vs_k141_81586||lt2gene Low-quality
S1_vs_k141_121376||lt2gene Low-quality
S1_vs_k141_134828||lt2gene Low-quality
S1_vs_k141_60897||lt2gene Low-quality
S1_vs_k141_108143||lt2gene Low-quality
S1_vs_k141_114877||lt2gene Not-determined
S1_vs_k141_54865||lt2gene Low-quality
S1_vs_k141_114882||lt2gene Not-determined
S1_vs_k141_28145||lt2gene Low-quality
S1_vs_k141_108166||lt2gene Low-quality
S1_vs_k141_101706||lt2gene Low-quality
S1_vs_k141_8290||lt2gene Low-quality
S1_vs_k141_121407||lt2gene Not-determined
S1_vs_k141_121413||lt2gene Low-quality
S1_vs_k141_95029||lt2gene Low-quality
S1_vs_k141_81617||lt2gene Low-quality
S1_vs_k141_74910||lt2gene Not-determined
S1_vs_k141_154393||lt2gene Low-quality
S1_vs_k141_48264||lt2gene Low-quality
S1_vs_k141_41642||lt2gene Not-determined
S1_vs_k141_81631||lt2gene Low-quality
S1_vs_k141_41649||lt2gene Low-quality
S1_vs_k141_74928||lt2gene Low-quality
S1_vs_k141_48273||lt2gene Low-quality
S1_vs_k141_48278||lt2gene Low-quality
S1_vs_k141_95054||lt2gene Not-determined
S1_vs_k141_95055||lt2gene Low-quality
S1_vs_k141_15208||lt2gene Low-quality
S1_vs_k141_95062||lt2gene Low-quality
S1_vs_k141_60924||lt2gene Low-quality
S1_vs_k141_1279||lt2gene Low-quality
S1_vs_k141_28207||lt2gene Low-quality
S1_vs_k141_68216||lt2gene Low-quality
S1_vs_k141_1282||lt2gene Low-quality
S1_vs_k141_74959||lt2gene Low-quality
S1_vs_k141_15212||lt2gene Low-quality
S1_vs_k141_154428||lt2gene Low-quality
S1_vs_k141_95081||lt2gene Not-determined
S1_vs_k141_81682||lt2gene Not-determined
S1_vs_k141_141653||lt2gene Low-quality
S1_vs_k141_128357||lt2gene Low-quality
S1_vs_k141_141659||lt2gene Not-determined
S1_vs_k141_48322||lt2gene Not-determined
S1_vs_k141_134921||lt2gene Low-quality
S1_vs_k141_95100||lt2gene Low-quality
S1_vs_k141_95101||lt2gene Low-quality
S1_vs_k141_121490||lt2gene Not-determined
S1_vs_k141_95103||lt2gene Low-quality
S1_vs_k141_114983||lt2gene Not-determined
S1_vs_k141_128370||lt2gene Low-quality
S1_vs_k141_68258||lt2gene Low-quality
S1_vs_k141_81706||lt2gene Not-determined
S1_vs_k141_81731||lt2gene Low-quality
S1_vs_k141_21563||lt2gene Low-quality
S1_vs_k141_134951||lt2gene Low-quality
S1_vs_k141_115020||lt2gene Low-quality
S1_vs_k141_68279||lt2gene Low-quality
S1_vs_k141_41727||lt2gene Low-quality
S1_vs_k141_75056||lt2gene Not-determined
S1_vs_k141_81765||lt2gene Low-quality
S1_vs_k141_55027||lt2gene Low-quality
S1_vs_k141_88341||lt2gene Low-quality
S1_vs_k141_75070||lt2gene Low-quality
S1_vs_k141_48384||lt2gene Not-determined
S1_vs_k141_128422||lt2gene Low-quality
S1_vs_k141_141734||lt2gene Not-determined
S1_vs_k141_21593||lt2gene Not-determined
S1_vs_k141_15329||lt2gene Not-determined
S1_vs_k141_141736||lt2gene Low-quality
S1_vs_k141_34741||lt2gene Not-determined
S1_vs_k141_34744||lt2gene Low-quality
S1_vs_k141_108320||lt2gene Low-quality
S1_vs_k141_81791||lt2gene Low-quality
S1_vs_k141_15346||lt2gene Low-quality
S1_vs_k141_154503||lt2gene Not-determined
S1_vs_k141_68342||lt2gene Not-determined
S1_vs_k141_141772||lt2gene Not-determined
S1_vs_k141_15357||lt2gene Low-quality
S1_vs_k141_128464||lt2gene Not-determined
S1_vs_k141_135022||lt2gene Low-quality
S1_vs_k141_101819||lt2gene Low-quality
S1_vs_k141_48426||lt2gene Not-determined
S1_vs_k141_88374||lt2gene Not-determined
S1_vs_k141_135034||lt2gene Low-quality
S1_vs_k141_154517||lt2gene Low-quality
S1_vs_k141_48439||lt2gene Not-determined
S1_vs_k141_121590||lt2gene Low-quality
S1_vs_k141_68380||lt2gene Low-quality
S1_vs_k141_48456||lt2gene Low-quality
S1_vs_k141_141803||lt2gene Low-quality
S1_vs_k141_75134||lt2gene Low-quality
S1_vs_k141_154555||lt2gene Not-determined
S1_vs_k141_101867||lt2gene Not-determined
S1_vs_k141_135069||lt2gene Low-quality
S1_vs_k141_1415||lt2gene Low-quality
S1_vs_k141_141830||lt2gene Low-quality
S1_vs_k141_88427||lt2gene Not-determined
S1_vs_k141_115142||lt2gene Low-quality
S1_vs_k141_68419||lt2gene Low-quality
S1_vs_k141_121635||lt2gene Low-quality
S1_vs_k141_141841||lt2gene Not-determined
S1_vs_k141_15431||lt2gene Low-quality
S1_vs_k141_41845||lt2gene Low-quality
S1_vs_k141_81888||lt2gene Low-quality
S1_vs_k141_95232||lt2gene Low-quality
S1_vs_k141_41871||lt2gene Not-determined
S1_vs_k141_75191||lt2gene Low-quality
S1_vs_k141_68465||lt2gene Not-determined
S1_vs_k141_28412||lt2gene Not-determined
S1_vs_k141_41883||lt2gene Low-quality
S1_vs_k141_8565||lt2gene Not-determined
S1_vs_k141_141891||lt2gene Low-quality
S1_vs_k141_95256||lt2gene Low-quality
S1_vs_k141_75207||lt2gene Low-quality
S1_vs_k141_135142||lt2gene Low-quality
S1_vs_k141_88483||lt2gene Not-determined
S1_vs_k141_121666||lt2gene Low-quality
S1_vs_k141_81950||lt2gene Low-quality
S1_vs_k141_41905||lt2gene Not-determined
S1_vs_k141_81958||lt2gene Low-quality
S1_vs_k141_154612||lt2gene Not-determined
S1_vs_k141_21765||lt2gene Low-quality
S1_vs_k141_55224||lt2gene Low-quality
S1_vs_k141_101984||lt2gene Low-quality
S1_vs_k141_101987||lt2gene Not-determined
S1_vs_k141_141929||lt2gene Not-determined
S1_vs_k141_115225||lt2gene Not-determined
S1_vs_k141_41934||lt2gene Low-quality
S1_vs_k141_41935||lt2gene Not-determined
S1_vs_k141_68523||lt2gene Low-quality
S1_vs_k141_81978||lt2gene Low-quality
S1_vs_k141_81980||lt2gene Low-quality
S1_vs_k141_88538||lt2gene Low-quality
S1_vs_k141_68546||lt2gene Low-quality
S1_vs_k141_15558||lt2gene Low-quality
S1_vs_k141_82006||lt2gene Low-quality
S1_vs_k141_88569||lt2gene Not-determined
S1_vs_k141_55265||lt2gene Low-quality
S1_vs_k141_88573||lt2gene Low-quality
S1_vs_k141_95354||lt2gene Low-quality
S1_vs_k141_108532||lt2gene Low-quality
S1_vs_k141_8631||lt2gene Not-determined
S1_vs_k141_154651||lt2gene Low-quality
S1_vs_k141_68575||lt2gene Low-quality
S1_vs_k141_108547||lt2gene Low-quality
S1_vs_k141_95372||lt2gene Low-quality
S1_vs_k141_108548||lt2gene Low-quality
S1_vs_k141_102062||lt2gene Not-determined
S1_vs_k141_147665||lt2gene Low-quality
S1_vs_k141_95381||lt2gene Not-determined
S1_vs_k141_48655||lt2gene Low-quality
S1_vs_k141_115314||lt2gene Low-quality
S1_vs_k141_141998||lt2gene Low-quality
S1_vs_k141_135275||lt2gene Low-quality
S1_vs_k141_15638||lt2gene Low-quality
S1_vs_k141_102087||lt2gene Low-quality
S1_vs_k141_21866||lt2gene Not-determined
S1_vs_k141_128755||lt2gene Low-quality
S1_vs_k141_135300||lt2gene Low-quality
S1_vs_k141_8689||lt2gene Not-determined
S1_vs_k141_42020||lt2gene Low-quality
S1_vs_k141_121805||lt2gene Not-determined
S1_vs_k141_95428||lt2gene Low-quality
S1_vs_k141_154710||lt2gene Not-determined
S1_vs_k141_61094||lt2gene Not-determined
S1_vs_k141_15686||lt2gene Not-determined
S1_vs_k141_61102||lt2gene Low-quality
S1_vs_k141_15697||lt2gene Low-quality
S1_vs_k141_75373||lt2gene Low-quality
S1_vs_k141_28623||lt2gene Low-quality
S1_vs_k141_48705||lt2gene Low-quality
S1_vs_k141_108635||lt2gene Low-quality
S1_vs_k141_115371||lt2gene Low-quality
S1_vs_k141_21914||lt2gene Low-quality
S1_vs_k141_48718||lt2gene Low-quality
S1_vs_k141_8718||lt2gene Not-determined
S1_vs_k141_142077||lt2gene Not-determined
S1_vs_k141_135352||lt2gene Not-determined
S1_vs_k141_82122||lt2gene Not-determined
S1_vs_k141_75401||lt2gene Low-quality
S1_vs_k141_115402||lt2gene Low-quality
S1_vs_k141_15744||lt2gene Low-quality
S1_vs_k141_95461||lt2gene Not-determined
S1_vs_k141_95467||lt2gene Low-quality
S1_vs_k141_121883||lt2gene Low-quality
S1_vs_k141_15774||lt2gene Not-determined
S1_vs_k141_42096||lt2gene Low-quality
S1_vs_k141_61144||lt2gene Not-determined
S1_vs_k141_75414||lt2gene Not-determined
S1_vs_k141_15778||lt2gene Low-quality
S1_vs_k141_75415||lt2gene Not-determined
S1_vs_k141_48768||lt2gene Low-quality
S1_vs_k141_102179||lt2gene Low-quality
S1_vs_k141_75435||lt2gene Low-quality
S1_vs_k141_35000||lt2gene Low-quality
S1_vs_k141_8782||lt2gene Low-quality
S1_vs_k141_128856||lt2gene Low-quality
S1_vs_k141_15825||lt2gene Low-quality
S1_vs_k141_75462||lt2gene Low-quality
S1_vs_k141_8793||lt2gene Low-quality
S1_vs_k141_75474||lt2gene Low-quality
S1_vs_k141_15840||lt2gene Low-quality
S1_vs_k141_128880||lt2gene Low-quality
S1_vs_k141_28742||lt2gene Low-quality
S1_vs_k141_108743||lt2gene Low-quality
S1_vs_k141_128890||lt2gene Low-quality
S1_vs_k141_82218||lt2gene Low-quality
S1_vs_k141_102248||lt2gene Low-quality
S1_vs_k141_55503||lt2gene Low-quality
S1_vs_k141_55504||lt2gene Not-determined
S1_vs_k141_48856||lt2gene Low-quality
S1_vs_k141_82243||lt2gene Not-determined
S1_vs_k141_55519||lt2gene Low-quality
S1_vs_k141_115502||lt2gene Not-determined
S1_vs_k141_88816||lt2gene Low-quality
S1_vs_k141_108797||lt2gene Not-determined
S1_vs_k141_142229||lt2gene Not-determined
S1_vs_k141_82274||lt2gene Low-quality
S1_vs_k141_28774||lt2gene Not-determined
S1_vs_k141_28776||lt2gene Low-quality
S1_vs_k141_48905||lt2gene Low-quality
S1_vs_k141_48908||lt2gene Low-quality
S1_vs_k141_22072||lt2gene Low-quality
S1_vs_k141_8889||lt2gene Low-quality
S1_vs_k141_122058||lt2gene Not-determined
S1_vs_k141_68845||lt2gene Low-quality
S1_vs_k141_102331||lt2gene Low-quality
S1_vs_k141_115594||lt2gene Not-determined
S1_vs_k141_128997||lt2gene Low-quality
S1_vs_k141_122073||lt2gene Low-quality
S1_vs_k141_15979||lt2gene Not-determined
S1_vs_k141_95704||lt2gene Not-determined
S1_vs_k141_28843||lt2gene Not-determined
S1_vs_k141_15999||lt2gene Low-quality
S1_vs_k141_28864||lt2gene Low-quality
S1_vs_k141_135566||lt2gene Low-quality
S1_vs_k141_68871||lt2gene Low-quality
S1_vs_k141_1808||lt2gene Low-quality
S1_vs_k141_108888||lt2gene Low-quality
S1_vs_k141_16026||lt2gene Low-quality
S1_vs_k141_129034||lt2gene Low-quality
S1_vs_k141_55607||lt2gene Low-quality
S1_vs_k141_88913||lt2gene Not-determined
S1_vs_k141_108901||lt2gene Low-quality
S1_vs_k141_122157||lt2gene Low-quality
S1_vs_k141_88924||lt2gene Low-quality
S1_vs_k141_122166||lt2gene Low-quality
S1_vs_k141_49010||lt2gene Low-quality
S1_vs_k141_42313||lt2gene Low-quality
S1_vs_k141_154910||lt2gene Low-quality
S1_vs_k141_42319||lt2gene Low-quality
S1_vs_k141_1832||lt2gene Low-quality
S1_vs_k141_115664||lt2gene Low-quality
S1_vs_k141_122173||lt2gene Not-determined
S1_vs_k141_68895||lt2gene Low-quality
S1_vs_k141_147836||lt2gene Low-quality
S1_vs_k141_1837||lt2gene Low-quality
S1_vs_k141_75675||lt2gene Low-quality
S1_vs_k141_154923||lt2gene Low-quality
S1_vs_k141_35189||lt2gene Not-determined
S1_vs_k141_135628||lt2gene Low-quality
S1_vs_k141_42350||lt2gene Low-quality
S1_vs_k141_88958||lt2gene Low-quality
S1_vs_k141_102440||lt2gene Low-quality
S1_vs_k141_75687||lt2gene Low-quality
S1_vs_k141_42353||lt2gene Not-determined
S1_vs_k141_82415||lt2gene Low-quality
S1_vs_k141_28923||lt2gene Low-quality
S1_vs_k141_9015||lt2gene Low-quality
S1_vs_k141_75696||lt2gene Low-quality
S1_vs_k141_115716||lt2gene Low-quality
S1_vs_k141_122210||lt2gene Low-quality
S1_vs_k141_28939||lt2gene Low-quality
S1_vs_k141_129122||lt2gene Not-determined
S1_vs_k141_135682||lt2gene Low-quality
S1_vs_k141_75733||lt2gene Low-quality
S1_vs_k141_115739||lt2gene Low-quality
S1_vs_k141_9039||lt2gene Low-quality
S1_vs_k141_55698||lt2gene Low-quality
S1_vs_k141_49088||lt2gene Low-quality
S1_vs_k141_9056||lt2gene Not-determined
S1_vs_k141_55725||lt2gene Not-determined
S1_vs_k141_122264||lt2gene Low-quality
S1_vs_k141_102508||lt2gene Not-determined
S1_vs_k141_9075||lt2gene Not-determined
S1_vs_k141_9076||lt2gene Low-quality
S1_vs_k141_55758||lt2gene Not-determined
S1_vs_k141_16166||lt2gene Low-quality
S1_vs_k141_55769||lt2gene Low-quality
S1_vs_k141_9112||lt2gene Low-quality
S1_vs_k141_89041||lt2gene Low-quality
S1_vs_k141_89042||lt2gene Not-determined
S1_vs_k141_61402||lt2gene Low-quality
S1_vs_k141_95900||lt2gene Low-quality
S1_vs_k141_75793||lt2gene Low-quality
S1_vs_k141_75794||lt2gene Not-determined
S1_vs_k141_89058||lt2gene Low-quality
S1_vs_k141_16200||lt2gene Low-quality
S1_vs_k141_129244||lt2gene Low-quality
S1_vs_k141_9154||lt2gene Low-quality
S1_vs_k141_9168||lt2gene Low-quality
S1_vs_k141_22262||lt2gene Low-quality
S1_vs_k141_69064||lt2gene Not-determined
S1_vs_k141_155029||lt2gene Low-quality
S1_vs_k141_129291||lt2gene Low-quality
S1_vs_k141_9208||lt2gene Low-quality
S1_vs_k141_82570||lt2gene Low-quality
S1_vs_k141_115875||lt2gene Not-determined
S1_vs_k141_89139||lt2gene Low-quality
S1_vs_k141_82576||lt2gene Low-quality
S1_vs_k141_82583||lt2gene Not-determined
S1_vs_k141_129301||lt2gene Low-quality
S1_vs_k141_147934||lt2gene Low-quality
S1_vs_k141_102629||lt2gene Low-quality
S1_vs_k141_122376||lt2gene Not-determined
S1_vs_k141_155040||lt2gene Low-quality
S1_vs_k141_122379||lt2gene Low-quality
S1_vs_k141_29123||lt2gene Not-determined
S1_vs_k141_129303||lt2gene Low-quality
S1_vs_k141_42563||lt2gene Low-quality
S1_vs_k141_69091||lt2gene Low-quality
S1_vs_k141_2047||lt2gene Low-quality
S1_vs_k141_55888||lt2gene Not-determined
S1_vs_k141_42571||lt2gene Low-quality
S1_vs_k141_96013||lt2gene Low-quality
S1_vs_k141_82618||lt2gene Low-quality
S1_vs_k141_155053||lt2gene Low-quality
S1_vs_k141_55913||lt2gene Low-quality
S1_vs_k141_61507||lt2gene Low-quality
S1_vs_k141_35376||lt2gene Low-quality
S1_vs_k141_115953||lt2gene Low-quality

S1_vs_k141_69139||lt2gene Not-determined
S1_vs_k141_49289||lt2gene Low-quality
S1_vs_k141_96058||lt2gene Low-quality
S1_vs_k141_49302||lt2gene Not-determined
S1_vs_k141_42641||lt2gene Low-quality
S1_vs_k141_29196||lt2gene Low-quality
S1_vs_k141_2094||lt2gene Low-quality
S1_vs_k141_129351||lt2gene Low-quality
S1_vs_k141_122443||lt2gene Low-quality
S1_vs_k141_89186||lt2gene Low-quality
S1_vs_k141_102696||lt2gene Low-quality
S1_vs_k141_9288||lt2gene Not-determined
S1_vs_k141_102698||lt2gene Not-determined
S1_vs_k141_75932||lt2gene Low-quality
S1_vs_k141_82681||lt2gene Not-determined
S1_vs_k141_102710||lt2gene Low-quality
S1_vs_k141_2105||lt2gene Low-quality
S1_vs_k141_129386||lt2gene Not-determined
S1_vs_k141_82717||lt2gene Not-determined
S1_vs_k141_155123||lt2gene Low-quality
S1_vs_k141_82732||lt2gene Low-quality
S1_vs_k141_129401||lt2gene Not-determined
S1_vs_k141_142648||lt2gene Low-quality
S1_vs_k141_135959||lt2gene Low-quality
S1_vs_k141_129417||lt2gene Low-quality
S1_vs_k141_89257||lt2gene Not-determined
S1_vs_k141_142673||lt2gene Low-quality
S1_vs_k141_135978||lt2gene Low-quality
S1_vs_k141_96146||lt2gene Not-determined
S1_vs_k141_135994||lt2gene Low-quality
S1_vs_k141_2160||lt2gene Not-determined
S1_vs_k141_9374||lt2gene Low-quality
S1_vs_k141_16479||lt2gene Not-determined
S1_vs_k141_49440||lt2gene Low-quality
S1_vs_k141_2162||lt2gene Not-determined
S1_vs_k141_2167||lt2gene Low-quality
S1_vs_k141_96166||lt2gene Low-quality
S1_vs_k141_122536||lt2gene Not-determined
S1_vs_k141_9393||lt2gene Not-determined
S1_vs_k141_61597||lt2gene Low-quality
S1_vs_k141_49464||lt2gene Not-determined
S1_vs_k141_2179||lt2gene Low-quality
S1_vs_k141_136042||lt2gene Low-quality
S1_vs_k141_16499||lt2gene Low-quality
S1_vs_k141_16510||lt2gene Low-quality
S1_vs_k141_82849||lt2gene Low-quality
S1_vs_k141_22510||lt2gene Low-quality
S1_vs_k141_49495||lt2gene Low-quality
S1_vs_k141_69303||lt2gene Low-quality
S1_vs_k141_102851||lt2gene Low-quality
S1_vs_k141_96209||lt2gene Low-quality
S1_vs_k141_35524||lt2gene Not-determined
S1_vs_k141_9451||lt2gene Low-quality
S1_vs_k141_136081||lt2gene Low-quality
S1_vs_k141_42820||lt2gene Not-determined
S1_vs_k141_35533||lt2gene Low-quality
S1_vs_k141_16554||lt2gene Low-quality
S1_vs_k141_82895||lt2gene Low-quality
S1_vs_k141_116189||lt2gene Low-quality
S1_vs_k141_142804||lt2gene Low-quality
S1_vs_k141_69370||lt2gene Not-determined
S1_vs_k141_89358||lt2gene Not-determined
S1_vs_k141_49567||lt2gene Not-determined
S1_vs_k141_9511||lt2gene Low-quality
S1_vs_k141_56150||lt2gene Low-quality
S1_vs_k141_35583||lt2gene Low-quality
S1_vs_k141_42870||lt2gene Low-quality
S1_vs_k141_116230||lt2gene Low-quality
S1_vs_k141_136114||lt2gene Not-determined
S1_vs_k141_35595||lt2gene Not-determined
S1_vs_k141_136119||lt2gene Low-quality
S1_vs_k141_69386||lt2gene Not-determined
S1_vs_k141_35599||lt2gene Low-quality
S1_vs_k141_122652||lt2gene Low-quality
S1_vs_k141_22614||lt2gene Low-quality
S1_vs_k141_42894||lt2gene Low-quality
S1_vs_k141_35621||lt2gene Low-quality
S1_vs_k141_122670||lt2gene Low-quality
S1_vs_k141_49614||lt2gene Low-quality
S1_vs_k141_22630||lt2gene Low-quality
S1_vs_k141_69410||lt2gene Low-quality
S1_vs_k141_49641||lt2gene Low-quality
S1_vs_k141_96318||lt2gene Low-quality
S1_vs_k141_16683||lt2gene Low-quality
S1_vs_k141_29524||lt2gene Low-quality
S1_vs_k141_83001||lt2gene Not-determined
S1_vs_k141_89450||lt2gene Low-quality
S1_vs_k141_69442||lt2gene Not-determined
S1_vs_k141_2348||lt2gene Low-quality
S1_vs_k141_129651||lt2gene Low-quality
S1_vs_k141_16703||lt2gene Low-quality
S1_vs_k141_76241||lt2gene Low-quality
S1_vs_k141_103027||lt2gene Low-quality
S1_vs_k141_49671||lt2gene Low-quality
S1_vs_k141_116334||lt2gene Low-quality
S1_vs_k141_129670||lt2gene Not-determined
S1_vs_k141_83027||lt2gene Low-quality
S1_vs_k141_103047||lt2gene Low-quality
S1_vs_k141_29557||lt2gene Not-determined
S1_vs_k141_42976||lt2gene Low-quality
S1_vs_k141_109486||lt2gene Low-quality
S1_vs_k141_16748||lt2gene Low-quality
S1_vs_k141_42988||lt2gene Not-determined
S1_vs_k141_129701||lt2gene Low-quality
S1_vs_k141_155265||lt2gene Not-determined
S1_vs_k141_16756||lt2gene Low-quality
S1_vs_k141_122761||lt2gene Low-quality
S1_vs_k141_103079||lt2gene Low-quality
S1_vs_k141_16776||lt2gene Low-quality
S1_vs_k141_103103||lt2gene Low-quality
S1_vs_k141_116394||lt2gene Low-quality
S1_vs_k141_116396||lt2gene Not-determined
S1_vs_k141_29608||lt2gene Not-determined
S1_vs_k141_142900||lt2gene Low-quality
S1_vs_k141_122806||lt2gene Low-quality
S1_vs_k141_56313||lt2gene Low-quality
S1_vs_k141_43055||lt2gene Low-quality
S1_vs_k141_56321||lt2gene Low-quality
S1_vs_k141_61770||lt2gene Not-determined
S1_vs_k141_122826||lt2gene Not-determined
S1_vs_k141_136315||lt2gene Low-quality
S1_vs_k141_136325||lt2gene Not-determined
S1_vs_k141_122846||lt2gene Not-determined
S1_vs_k141_35733||lt2gene Low-quality
S1_vs_k141_96477||lt2gene Not-determined
S1_vs_k141_43094||lt2gene Not-determined
S1_vs_k141_129786||lt2gene Not-determined
S1_vs_k141_109621||lt2gene Low-quality
S1_vs_k141_103206||lt2gene Not-determined
S1_vs_k141_16873||lt2gene Low-quality
S1_vs_k141_155317||lt2gene Not-determined
S1_vs_k141_69615||lt2gene Low-quality
S1_vs_k141_2515||lt2gene Low-quality
S1_vs_k141_116517||lt2gene Low-quality
S1_vs_k141_155327||lt2gene Low-quality
S1_vs_k141_122892||lt2gene Low-quality
S1_vs_k141_61830||lt2gene Not-determined
S1_vs_k141_56395||lt2gene Not-determined
S1_vs_k141_109668||lt2gene Not-determined
S1_vs_k141_129841||lt2gene Low-quality
S1_vs_k141_129844||lt2gene Low-quality
S1_vs_k141_56405||lt2gene Not-determined
S1_vs_k141_142993||lt2gene Low-quality
S1_vs_k141_22867||lt2gene Low-quality
S1_vs_k141_129855||lt2gene Not-determined
S1_vs_k141_109681||lt2gene Low-quality
S1_vs_k141_83187||lt2gene Not-determined
S1_vs_k141_83188||lt2gene Low-quality
S1_vs_k141_9834||lt2gene Low-quality
S1_vs_k141_16942||lt2gene Not-determined
S1_vs_k141_69666||lt2gene Low-quality
S1_vs_k141_136423||lt2gene Low-quality
S1_vs_k141_129889||lt2gene Not-determined
S1_vs_k141_61881||lt2gene Low-quality
S1_vs_k141_35818||lt2gene Not-determined
S1_vs_k141_2575||lt2gene Low-quality
S1_vs_k141_116578||lt2gene Low-quality
S1_vs_k141_83233||lt2gene Not-determined
S1_vs_k141_29757||lt2gene Low-quality
S1_vs_k141_29760||lt2gene Low-quality
S1_vs_k141_43195||lt2gene Low-quality
S1_vs_k141_109722||lt2gene Low-quality
S1_vs_k141_89734||lt2gene Not-determined
S1_vs_k141_89735||lt2gene Not-determined
S1_vs_k141_43205||lt2gene Low-quality
S1_vs_k141_103318||lt2gene Low-quality
S1_vs_k141_2598||lt2gene Low-quality
S1_vs_k141_35852||lt2gene Not-determined
S1_vs_k141_49970||lt2gene Not-determined
S1_vs_k141_61923||lt2gene Low-quality
S1_vs_k141_116619||lt2gene Low-quality
S1_vs_k141_2617||lt2gene Low-quality
S1_vs_k141_143089||lt2gene Low-quality
S1_vs_k141_49989||lt2gene Not-determined
S1_vs_k141_2623||lt2gene Low-quality
S1_vs_k141_89753||lt2gene Low-quality
S1_vs_k141_9904||lt2gene Low-quality
S1_vs_k141_143107||lt2gene Low-quality
S1_vs_k141_123012||lt2gene Not-determined
S1_vs_k141_96666||lt2gene Low-quality
S1_vs_k141_22943||lt2gene Not-determined
S1_vs_k141_76570||lt2gene Not-determined
S1_vs_k141_22948||lt2gene Low-quality
S1_vs_k141_17074||lt2gene Low-quality
S1_vs_k141_43273||lt2gene Low-quality
S1_vs_k141_56519||lt2gene Not-determined
S1_vs_k141_35906||lt2gene Not-determined
S1_vs_k141_50041||lt2gene Low-quality
S1_vs_k141_2632||lt2gene Not-determined
S1_vs_k141_83346||lt2gene Low-quality
S1_vs_k141_9959||lt2gene Not-determined
S1_vs_k141_103381||lt2gene Low-quality
S1_vs_k141_116703||lt2gene Low-quality
S1_vs_k141_76597||lt2gene Not-determined
S1_vs_k141_29856||lt2gene Not-determined
S1_vs_k141_123042||lt2gene Not-determined
S1_vs_k141_155398||lt2gene Low-quality
S1_vs_k141_103390||lt2gene Low-quality
S1_vs_k141_96742||lt2gene Low-quality
S1_vs_k141_155411||lt2gene Low-quality
S1_vs_k141_76632||lt2gene Low-quality
S1_vs_k141_96781||lt2gene Low-quality
S1_vs_k141_109896||lt2gene Not-determined
S1_vs_k141_35972||lt2gene Not-determined
S1_vs_k141_43362||lt2gene Low-quality
S1_vs_k141_155433||lt2gene Low-quality
S1_vs_k141_130081||lt2gene Low-quality
S1_vs_k141_29914||lt2gene Not-determined
S1_vs_k141_155443||lt2gene Low-quality
S1_vs_k141_148283||lt2gene Low-quality
S1_vs_k141_10024||lt2gene Not-determined
S1_vs_k141_103466||lt2gene Low-quality
S1_vs_k141_62066||lt2gene Not-determined
S1_vs_k141_76670||lt2gene Low-quality
S1_vs_k141_29936||lt2gene Low-quality
S1_vs_k141_96819||lt2gene Low-quality
S1_vs_k141_62079||lt2gene Not-determined
S1_vs_k141_2709||lt2gene Low-quality
S1_vs_k141_155456||lt2gene Low-quality
S1_vs_k141_29949||lt2gene Not-determined
S1_vs_k141_83431||lt2gene Not-determined
S1_vs_k141_10066||lt2gene Low-quality
S1_vs_k141_123139||lt2gene Not-determined
S1_vs_k141_69796||lt2gene Not-determined
S1_vs_k141_116819||lt2gene Low-quality
S1_vs_k141_83469||lt2gene Not-determined
S1_vs_k141_29984||lt2gene Low-quality
S1_vs_k141_23046||lt2gene Low-quality
S1_vs_k141_56680||lt2gene Low-quality
S1_vs_k141_109987||lt2gene Low-quality
S1_vs_k141_123175||lt2gene Low-quality
S1_vs_k141_36025||lt2gene Not-determined
S1_vs_k141_36028||lt2gene Low-quality
S1_vs_k141_17183||lt2gene Low-quality
S1_vs_k141_10123||lt2gene Not-determined
S1_vs_k141_43455||lt2gene Low-quality
S1_vs_k141_110013||lt2gene Not-determined
S1_vs_k141_76749||lt2gene Low-quality
S1_vs_k141_36048||lt2gene Low-quality
S1_vs_k141_23085||lt2gene Low-quality
S1_vs_k141_89962||lt2gene Not-determined
S1_vs_k141_83516||lt2gene Not-determined
S1_vs_k141_83526||lt2gene Not-determined
S1_vs_k141_89981||lt2gene Low-quality
S1_vs_k141_103556||lt2gene Low-quality
S1_vs_k141_103558||lt2gene Low-quality
S1_vs_k141_136636||lt2gene Not-determined
S1_vs_k141_2796||lt2gene Not-determined
S1_vs_k141_69840||lt2gene Low-quality
S1_vs_k141_83555||lt2gene Low-quality
S1_vs_k141_50269||lt2gene Low-quality
S1_vs_k141_110073||lt2gene Low-quality
S1_vs_k141_23126||lt2gene Low-quality
S1_vs_k141_96963||lt2gene Not-determined
S1_vs_k141_90036||lt2gene Not-determined
S1_vs_k141_148378||lt2gene Low-quality
S1_vs_k141_36115||lt2gene Low-quality
S1_vs_k141_136709||lt2gene Not-determined
S1_vs_k141_90043||lt2gene Low-quality
S1_vs_k141_23166||lt2gene Low-quality
S1_vs_k141_69874||lt2gene Not-determined
S1_vs_k141_62217||lt2gene Not-determined
S1_vs_k141_17283||lt2gene Not-determined
S1_vs_k141_2826||lt2gene Not-determined
S1_vs_k141_43554||lt2gene Low-quality
S1_vs_k141_83629||lt2gene Low-quality
S1_vs_k141_130237||lt2gene Low-quality
S1_vs_k141_110128||lt2gene Low-quality
S1_vs_k141_130245||lt2gene Low-quality
S1_vs_k141_117001||lt2gene Low-quality
S1_vs_k141_36132||lt2gene Not-determined
S1_vs_k141_143404||lt2gene Low-quality
S1_vs_k141_10285||lt2gene Low-quality
S1_vs_k141_23247||lt2gene Not-determined
S1_vs_k141_50330||lt2gene Low-quality
S1_vs_k141_155595||lt2gene Low-quality
S1_vs_k141_110162||lt2gene Low-quality
S1_vs_k141_17330||lt2gene Low-quality
S1_vs_k141_83689||lt2gene Low-quality
S1_vs_k141_56887||lt2gene Low-quality
S1_vs_k141_123370||lt2gene Low-quality
S1_vs_k141_23260||lt2gene Low-quality
S1_vs_k141_17336||lt2gene Low-quality
S1_vs_k141_90110||lt2gene Low-quality
S1_vs_k141_50345||lt2gene Low-quality
S1_vs_k141_76933||lt2gene Low-quality
S1_vs_k141_76938||lt2gene Low-quality
S1_vs_k141_69911||lt2gene Low-quality
S1_vs_k141_76945||lt2gene Not-determined
S1_vs_k141_23268||lt2gene Low-quality
S1_vs_k141_148421||lt2gene Low-quality
S1_vs_k141_90132||lt2gene Low-quality
S1_vs_k141_2906||lt2gene Not-determined
S1_vs_k141_62323||lt2gene Low-quality
S1_vs_k141_30244||lt2gene Not-determined
S1_vs_k141_56936||lt2gene Low-quality
S1_vs_k141_2924||lt2gene Low-quality
S1_vs_k141_76987||lt2gene Low-quality
S1_vs_k141_76999||lt2gene Low-quality
S1_vs_k141_123428||lt2gene Low-quality
S1_vs_k141_30281||lt2gene Low-quality
S1_vs_k141_36200||lt2gene Low-quality
S1_vs_k141_130346||lt2gene Low-quality
S1_vs_k141_30290||lt2gene Low-quality
S1_vs_k141_117108||lt2gene Low-quality
S1_vs_k141_17420||lt2gene Low-quality
S1_vs_k141_136874||lt2gene Not-determined
S1_vs_k141_123453||lt2gene Not-determined
S1_vs_k141_155664||lt2gene Low-quality
S1_vs_k141_30300||lt2gene Low-quality
S1_vs_k141_143486||lt2gene Not-determined
S1_vs_k141_155679||lt2gene Low-quality
S1_vs_k141_57032||lt2gene Low-quality
S1_vs_k141_57036||lt2gene Low-quality
S1_vs_k141_117155||lt2gene Low-quality
S1_vs_k141_57038||lt2gene Not-determined
S1_vs_k141_43740||lt2gene Low-quality
S1_vs_k141_50497||lt2gene Low-quality
S1_vs_k141_83814||lt2gene Not-determined
S1_vs_k141_36247||lt2gene Low-quality
S1_vs_k141_17481||lt2gene Low-quality
S1_vs_k141_10455||lt2gene Low-quality
S1_vs_k141_23433||lt2gene Low-quality
S1_vs_k141_130426||lt2gene Low-quality
S1_vs_k141_97235||lt2gene Low-quality
S1_vs_k141_130432||lt2gene Low-quality
S1_vs_k141_103835||lt2gene Low-quality
S1_vs_k141_148530||lt2gene Low-quality
S1_vs_k141_103841||lt2gene Low-quality
S1_vs_k141_36278||lt2gene Low-quality
S1_vs_k141_83853||lt2gene Low-quality
S1_vs_k141_83864||lt2gene Low-quality
S1_vs_k141_57091||lt2gene Low-quality
S1_vs_k141_97278||lt2gene Low-quality
S1_vs_k141_23491||lt2gene Low-quality
S1_vs_k141_83881||lt2gene Low-quality
S1_vs_k141_62489||lt2gene Low-quality
S1_vs_k141_43806||lt2gene Low-quality
S1_vs_k141_57115||lt2gene Low-quality
S1_vs_k141_77151||lt2gene Low-quality
S1_vs_k141_83890||lt2gene Not-determined
S1_vs_k141_77161||lt2gene Not-determined
S1_vs_k141_30432||lt2gene Not-determined
S1_vs_k141_110422||lt2gene Low-quality
S1_vs_k141_30448||lt2gene Not-determined
S1_vs_k141_90357||lt2gene Low-quality
S1_vs_k141_77183||lt2gene Not-determined
S1_vs_k141_117292||lt2gene Not-determined
S1_vs_k141_110452||lt2gene Not-determined
S1_vs_k141_50634||lt2gene Low-quality
S1_vs_k141_117315||lt2gene Low-quality
S1_vs_k141_50649||lt2gene Low-quality
S1_vs_k141_3020||lt2gene Low-quality
S1_vs_k141_83960||lt2gene Low-quality
S1_vs_k141_103954||lt2gene Not-determined
S1_vs_k141_43909||lt2gene Not-determined
S1_vs_k141_30518||lt2gene Low-quality
S1_vs_k141_23593||lt2gene Not-determined
S1_vs_k141_17609||lt2gene Low-quality
S1_vs_k141_130599||lt2gene Not-determined
S1_vs_k141_3053||lt2gene Low-quality
S1_vs_k141_23621||lt2gene Low-quality
S1_vs_k141_123653||lt2gene Low-quality
S1_vs_k141_3066||lt2gene Low-quality
S1_vs_k141_3067||lt2gene Low-quality
S1_vs_k141_97412||lt2gene Low-quality
S1_vs_k141_77250||lt2gene Low-quality
S1_vs_k141_30574||lt2gene Not-determined
S1_vs_k141_148599||lt2gene Low-quality
S1_vs_k141_50725||lt2gene Low-quality
S1_vs_k141_97439||lt2gene Not-determined
S1_vs_k141_30590||lt2gene Low-quality
S1_vs_k141_117342||lt2gene Low-quality
S1_vs_k141_30597||lt2gene Low-quality
S1_vs_k141_30602||lt2gene Low-quality
S1_vs_k141_77275||lt2gene Low-quality
S1_vs_k141_110569||lt2gene Low-quality
S1_vs_k141_30609||lt2gene Low-quality
S1_vs_k141_30621||lt2gene Low-quality
S1_vs_k141_104036||lt2gene Not-determined
S1_vs_k141_97465||lt2gene Low-quality
S1_vs_k141_97468||lt2gene Low-quality
S1_vs_k141_57280||lt2gene Low-quality
S1_vs_k141_97474||lt2gene Low-quality
S1_vs_k141_97477||lt2gene Low-quality
S1_vs_k141_84064||lt2gene Low-quality
S1_vs_k141_10730||lt2gene Low-quality
S1_vs_k141_17718||lt2gene Low-quality
S1_vs_k141_30641||lt2gene Low-quality
S1_vs_k141_110604||lt2gene Low-quality
S1_vs_k141_30646||lt2gene Not-determined
S1_vs_k141_36423||lt2gene Low-quality
S1_vs_k141_44042||lt2gene Low-quality
S1_vs_k141_130694||lt2gene Low-quality
S1_vs_k141_17735||lt2gene Low-quality
S1_vs_k141_97501||lt2gene Not-determined
S1_vs_k141_77313||lt2gene Not-determined
S1_vs_k141_77315||lt2gene Low-quality
S1_vs_k141_57313||lt2gene Low-quality
S1_vs_k141_3096||lt2gene Low-quality
S1_vs_k141_97521||lt2gene Low-quality
S1_vs_k141_36435||lt2gene Low-quality
S1_vs_k141_36439||lt2gene Low-quality
S1_vs_k141_104062||lt2gene Low-quality
S1_vs_k141_44088||lt2gene Low-quality
S1_vs_k141_70200||lt2gene Low-quality
S1_vs_k141_57329||lt2gene Low-quality
S1_vs_k141_70211||lt2gene Low-quality
S1_vs_k141_137271||lt2gene Not-determined
S1_vs_k141_148639||lt2gene Not-determined
S1_vs_k141_130730||lt2gene Low-quality
S1_vs_k141_3120||lt2gene Low-quality
S1_vs_k141_10803||lt2gene Not-determined
S1_vs_k141_30714||lt2gene Not-determined
S1_vs_k141_3130||lt2gene Low-quality
S1_vs_k141_3133||lt2gene Low-quality
S1_vs_k141_117476||lt2gene Low-quality
S1_vs_k141_97574||lt2gene Low-quality
S1_vs_k141_62738||lt2gene Low-quality
S1_vs_k141_23720||lt2gene Low-quality
S1_vs_k141_77405||lt2gene Low-quality
S1_vs_k141_137327||lt2gene Low-quality
S1_vs_k141_17837||lt2gene Not-determined
S1_vs_k141_90636||lt2gene Low-quality
S1_vs_k141_143852||lt2gene Low-quality
S1_vs_k141_36518||lt2gene Not-determined
S1_vs_k141_17854||lt2gene Low-quality
S1_vs_k141_57429||lt2gene Low-quality
S1_vs_k141_3200||lt2gene Not-determined
S1_vs_k141_117512||lt2gene Low-quality
S1_vs_k141_57440||lt2gene Low-quality
S1_vs_k141_143879||lt2gene Not-determined
S1_vs_k141_110749||lt2gene Low-quality
S1_vs_k141_70279||lt2gene Low-quality
S1_vs_k141_110753||lt2gene Low-quality
S1_vs_k141_104186||lt2gene Not-determined
S1_vs_k141_155860||lt2gene Not-determined
S1_vs_k141_3220||lt2gene Low-quality
S1_vs_k141_57467||lt2gene Low-quality
S1_vs_k141_57475||lt2gene Low-quality
S1_vs_k141_84217||lt2gene Low-quality
S1_vs_k141_36561||lt2gene Low-quality
S1_vs_k141_130845||lt2gene Low-quality
S1_vs_k141_23826||lt2gene Not-determined
S1_vs_k141_84240||lt2gene Not-determined
S1_vs_k141_23831||lt2gene Low-quality
S1_vs_k141_137433||lt2gene Low-quality
S1_vs_k141_90733||lt2gene Low-quality
S1_vs_k141_155877||lt2gene Low-quality
S1_vs_k141_23844||lt2gene Low-quality
S1_vs_k141_130878||lt2gene Low-quality
S1_vs_k141_110826||lt2gene Low-quality
S1_vs_k141_10983||lt2gene Low-quality
S1_vs_k141_117644||lt2gene Low-quality
S1_vs_k141_23885||lt2gene Not-determined
S1_vs_k141_104281||lt2gene Low-quality
S1_vs_k141_90783||lt2gene Low-quality
S1_vs_k141_155894||lt2gene Not-determined
S1_vs_k141_30907||lt2gene Not-determined
S1_vs_k141_23918||lt2gene Low-quality
S1_vs_k141_84319||lt2gene Low-quality
S1_vs_k141_110876||lt2gene Low-quality
S1_vs_k141_44297||lt2gene Not-determined
S1_vs_k141_130939||lt2gene Not-determined
S1_vs_k141_137516||lt2gene Not-determined
S1_vs_k141_51095||lt2gene Low-quality
S1_vs_k141_97748||lt2gene Low-quality
S1_vs_k141_51105||lt2gene Not-determined
S1_vs_k141_51107||lt2gene Low-quality
S1_vs_k141_36680||lt2gene Not-determined
S1_vs_k141_130960||lt2gene Low-quality
S1_vs_k141_90833||lt2gene Low-quality
S1_vs_k141_30958||lt2gene Low-quality
S1_vs_k141_155912||lt2gene Not-determined
S1_vs_k141_117714||lt2gene Not-determined
S1_vs_k141_117715||lt2gene Low-quality
S1_vs_k141_23976||lt2gene Not-determined
S1_vs_k141_30974||lt2gene Not-determined
S1_vs_k141_90849||lt2gene Low-quality
S1_vs_k141_23986||lt2gene Low-quality
S1_vs_k141_51137||lt2gene Low-quality
S1_vs_k141_24007||lt2gene Not-determined
S1_vs_k141_117751||lt2gene Low-quality
S1_vs_k141_90886||lt2gene Low-quality
S1_vs_k141_44375||lt2gene Low-quality
S1_vs_k141_44376||lt2gene Low-quality
S1_vs_k141_31027||lt2gene Low-quality
S1_vs_k141_70426||lt2gene Low-quality
S1_vs_k141_137603||lt2gene Low-quality
S1_vs_k141_131025||lt2gene Low-quality
S1_vs_k141_44390||lt2gene Low-quality
S1_vs_k141_18084||lt2gene Not-determined
S1_vs_k141_90921||lt2gene Low-quality
S1_vs_k141_70450||lt2gene Not-determined
S1_vs_k141_18119||lt2gene Low-quality
S1_vs_k141_124007||lt2gene Low-quality
S1_vs_k141_111011||lt2gene Low-quality
S1_vs_k141_51219||lt2gene Low-quality
S1_vs_k141_31081||lt2gene Low-quality
S1_vs_k141_11201||lt2gene Not-determined
S1_vs_k141_104446||lt2gene Low-quality
S1_vs_k141_57635||lt2gene Not-determined
S1_vs_k141_144137||lt2gene Low-quality
S1_vs_k141_44478||lt2gene Not-determined
S1_vs_k141_97929||lt2gene Low-quality
S1_vs_k141_3470||lt2gene Not-determined
S1_vs_k141_44513||lt2gene Low-quality
S1_vs_k141_3473||lt2gene Low-quality
S1_vs_k141_124127||lt2gene Low-quality
S1_vs_k141_91040||lt2gene Low-quality
S1_vs_k141_117871||lt2gene Low-quality
S1_vs_k141_137708||lt2gene Low-quality
S1_vs_k141_36871||lt2gene Low-quality
S1_vs_k141_111136||lt2gene Not-determined
S1_vs_k141_131178||lt2gene Low-quality
S1_vs_k141_131179||lt2gene Low-quality
S1_vs_k141_63142||lt2gene Low-quality
S1_vs_k141_3514||lt2gene Not-determined
S1_vs_k141_77857||lt2gene Not-determined
S1_vs_k141_156007||lt2gene Low-quality
S1_vs_k141_3538||lt2gene Low-quality
S1_vs_k141_18270||lt2gene Low-quality
S1_vs_k141_77874||lt2gene Not-determined
S1_vs_k141_124173||lt2gene Low-quality
S1_vs_k141_97982||lt2gene Low-quality
S1_vs_k141_31189||lt2gene Not-determined
S1_vs_k141_91100||lt2gene Low-quality
S1_vs_k141_104565||lt2gene Low-quality
S1_vs_k141_18280||lt2gene Low-quality
S1_vs_k141_18285||lt2gene Low-quality
S1_vs_k141_144237||lt2gene Low-quality
S1_vs_k141_3562||lt2gene Not-determined
S1_vs_k141_24247||lt2gene Low-quality
S1_vs_k141_91126||lt2gene Low-quality
S1_vs_k141_11379||lt2gene Low-quality
S1_vs_k141_117944||lt2gene Low-quality
S1_vs_k141_148891||lt2gene Low-quality
S1_vs_k141_98006||lt2gene Not-determined
S1_vs_k141_24256||lt2gene Low-quality
S1_vs_k141_11387||lt2gene Low-quality
S1_vs_k141_24258||lt2gene Low-quality
S1_vs_k141_31231||lt2gene Low-quality
S1_vs_k141_63213||lt2gene Low-quality
S1_vs_k141_104591||lt2gene Low-quality
S1_vs_k141_111187||lt2gene Low-quality
S1_vs_k141_84599||lt2gene Low-quality
S1_vs_k141_51427||lt2gene Not-determined
S1_vs_k141_31243||lt2gene Not-determined
S1_vs_k141_77944||lt2gene Low-quality
S1_vs_k141_36976||lt2gene Low-quality
S1_vs_k141_84612||lt2gene Not-determined
S1_vs_k141_77967||lt2gene Low-quality
S1_vs_k141_111205||lt2gene Not-determined
S1_vs_k141_117982||lt2gene Not-determined
S1_vs_k141_44626||lt2gene Low-quality
S1_vs_k141_3581||lt2gene Low-quality
S1_vs_k141_124273||lt2gene Low-quality
S1_vs_k141_77994||lt2gene Low-quality
S1_vs_k141_57823||lt2gene Not-determined
S1_vs_k141_51495||lt2gene Low-quality
S1_vs_k141_131318||lt2gene Low-quality
S1_vs_k141_144319||lt2gene Low-quality
S1_vs_k141_57843||lt2gene Low-quality
S1_vs_k141_137873||lt2gene Low-quality
S1_vs_k141_98080||lt2gene Low-quality
S1_vs_k141_98085||lt2gene Low-quality
S1_vs_k141_18418||lt2gene Low-quality
S1_vs_k141_44666||lt2gene Low-quality
S1_vs_k141_44668||lt2gene Low-quality
S1_vs_k141_11466||lt2gene Not-determined
S1_vs_k141_131373||lt2gene Not-determined
S1_vs_k141_137904||lt2gene Not-determined
S1_vs_k141_44690||lt2gene Not-determined
S1_vs_k141_137907||lt2gene Not-determined
S1_vs_k141_63269||lt2gene Low-quality
S1_vs_k141_18451||lt2gene Low-quality
S1_vs_k141_137911||lt2gene Low-quality
S1_vs_k141_3627||lt2gene Not-determined
S1_vs_k141_98115||lt2gene Not-determined
S1_vs_k141_31359||lt2gene Low-quality
S1_vs_k141_3635||lt2gene Low-quality
S1_vs_k141_137927||lt2gene Not-determined
S1_vs_k141_24403||lt2gene Low-quality
S1_vs_k141_124333||lt2gene Low-quality
S1_vs_k141_131391||lt2gene Not-determined
S1_vs_k141_18489||lt2gene Low-quality
S1_vs_k141_98153||lt2gene Low-quality
S1_vs_k141_111342||lt2gene Low-quality
S1_vs_k141_118098||lt2gene Not-determined

S1_vs_k141_104753||lt2gene Low-quality
S1_vs_k141_91290||lt2gene Low-quality
S1_vs_k141_31372||lt2gene Low-quality
S1_vs_k141_84759||lt2gene Low-quality
S1_vs_k141_144434||lt2gene Low-quality
S1_vs_k141_51610||lt2gene Low-quality
S1_vs_k141_24465||lt2gene Not-determined
S1_vs_k141_78097||lt2gene Low-quality
S1_vs_k141_131439||lt2gene Low-quality
S1_vs_k141_91316||lt2gene Low-quality
S1_vs_k141_63378||lt2gene Not-determined
S1_vs_k141_131443||lt2gene Low-quality
S1_vs_k141_144455||lt2gene Low-quality
S1_vs_k141_63380||lt2gene Low-quality
S1_vs_k141_131455||lt2gene Low-quality
S1_vs_k141_131460||lt2gene Low-quality
S1_vs_k141_63395||lt2gene Low-quality
S1_vs_k141_91334||lt2gene Low-quality
S1_vs_k141_24507||lt2gene Low-quality
S1_vs_k141_156138||lt2gene Low-quality
S1_vs_k141_24531||lt2gene Low-quality
S1_vs_k141_44812||lt2gene Low-quality
S1_vs_k141_31415||lt2gene Low-quality
S1_vs_k141_144487||lt2gene Low-quality
S1_vs_k141_44821||lt2gene Not-determined
S1_vs_k141_11615||lt2gene Low-quality
S1_vs_k141_144495||lt2gene Low-quality
S1_vs_k141_131507||lt2gene Low-quality
S1_vs_k141_58051||lt2gene Low-quality
S1_vs_k141_118184||lt2gene Low-quality
S1_vs_k141_78156||lt2gene Low-quality
S1_vs_k141_11627||lt2gene Low-quality
S1_vs_k141_63443||lt2gene Not-determined
S1_vs_k141_98253||lt2gene Not-determined
S1_vs_k141_118198||lt2gene Low-quality
S1_vs_k141_78186||lt2gene Low-quality
S1_vs_k141_24597||lt2gene Low-quality
S1_vs_k141_51693||lt2gene Low-quality
S1_vs_k141_91410||lt2gene Low-quality
S1_vs_k141_118219||lt2gene Low-quality
S1_vs_k141_104869||lt2gene Low-quality
S1_vs_k141_3792||lt2gene Low-quality
S1_vs_k141_111479||lt2gene Low-quality
S1_vs_k141_31475||lt2gene Low-quality
S1_vs_k141_124503||lt2gene Low-quality
S1_vs_k141_149014||lt2gene Low-quality
S1_vs_k141_124510||lt2gene Low-quality
S1_vs_k141_124517||lt2gene Low-quality
S1_vs_k141_24648||lt2gene Low-quality
S1_vs_k141_11691||lt2gene Low-quality
S1_vs_k141_24653||lt2gene Low-quality
S1_vs_k141_111518||lt2gene Low-quality
S1_vs_k141_118259||lt2gene Not-determined
S1_vs_k141_37190||lt2gene Not-determined
S1_vs_k141_104905||lt2gene Low-quality
S1_vs_k141_18675||lt2gene Low-quality
S1_vs_k141_44921||lt2gene Low-quality
S1_vs_k141_3815||lt2gene Low-quality
S1_vs_k141_131595||lt2gene Low-quality
S1_vs_k141_37207||lt2gene Low-quality
S1_vs_k141_144586||lt2gene Low-quality
S1_vs_k141_131611||lt2gene Low-quality
S1_vs_k141_124539||lt2gene Low-quality
S1_vs_k141_118292||lt2gene Low-quality
S1_vs_k141_3835||lt2gene Low-quality
S1_vs_k141_63557||lt2gene Not-determined
S1_vs_k141_63569||lt2gene Low-quality
S1_vs_k141_44955||lt2gene Low-quality
S1_vs_k141_91534||lt2gene Not-determined
S1_vs_k141_3845||lt2gene Low-quality
S1_vs_k141_31554||lt2gene Low-quality
S1_vs_k141_11789||lt2gene Low-quality
S1_vs_k141_104981||lt2gene Low-quality
S1_vs_k141_18726||lt2gene Low-quality
S1_vs_k141_84941||lt2gene Not-determined
S1_vs_k141_63602||lt2gene Not-determined
S1_vs_k141_91569||lt2gene Low-quality
S1_vs_k141_58183||lt2gene Low-quality
S1_vs_k141_63624||lt2gene Low-quality
S1_vs_k141_131701||lt2gene Not-determined
S1_vs_k141_58213||lt2gene Low-quality
S1_vs_k141_98446||lt2gene Low-quality
S1_vs_k141_24783||lt2gene Low-quality
S1_vs_k141_63641||lt2gene Low-quality
S1_vs_k141_138211||lt2gene Not-determined
S1_vs_k141_45053||lt2gene Not-determined
S1_vs_k141_31622||lt2gene Low-quality
S1_vs_k141_18817||lt2gene Low-quality
S1_vs_k141_51874||lt2gene Low-quality
S1_vs_k141_149141||lt2gene Low-quality
S1_vs_k141_131761||lt2gene Not-determined
S1_vs_k141_91665||lt2gene Not-determined
S1_vs_k141_98488||lt2gene Low-quality
S1_vs_k141_3946||lt2gene Low-quality
S1_vs_k141_78395||lt2gene Low-quality
S1_vs_k141_85036||lt2gene Low-quality
S1_vs_k141_70792||lt2gene Not-determined
S1_vs_k141_131786||lt2gene Not-determined
S1_vs_k141_85048||lt2gene Not-determined
S1_vs_k141_11932||lt2gene Low-quality
S1_vs_k141_63715||lt2gene Not-determined
S1_vs_k141_98517||lt2gene Low-quality
S1_vs_k141_58291||lt2gene Not-determined
S1_vs_k141_124758||lt2gene Not-determined
S1_vs_k141_91710||lt2gene Not-determined
S1_vs_k141_51944||lt2gene Low-quality
S1_vs_k141_31699||lt2gene Low-quality
S1_vs_k141_11964||lt2gene Low-quality
S1_vs_k141_91715||lt2gene Not-determined
S1_vs_k141_3979||lt2gene Low-quality
S1_vs_k141_111741||lt2gene Low-quality
S1_vs_k141_111744||lt2gene Low-quality
S1_vs_k141_156289||lt2gene Low-quality
S1_vs_k141_131830||lt2gene Low-quality
S1_vs_k141_51967||lt2gene Not-determined
S1_vs_k141_131831||lt2gene Low-quality
S1_vs_k141_85090||lt2gene Low-quality
S1_vs_k141_18920||lt2gene Low-quality
S1_vs_k141_111779||lt2gene Low-quality
S1_vs_k141_144792||lt2gene Low-quality
S1_vs_k141_78493||lt2gene Not-determined
S1_vs_k141_58358||lt2gene Low-quality
S1_vs_k141_63789||lt2gene Low-quality
S1_vs_k141_63791||lt2gene Low-quality
S1_vs_k141_18935||lt2gene Low-quality
S1_vs_k141_52002||lt2gene Low-quality
S1_vs_k141_131859||lt2gene Low-quality
S1_vs_k141_131866||lt2gene Low-quality
S1_vs_k141_70865||lt2gene Low-quality
S1_vs_k141_118496||lt2gene Low-quality
S1_vs_k141_31776||lt2gene Low-quality
S1_vs_k141_98625||lt2gene Low-quality
S1_vs_k141_91787||lt2gene Low-quality
S1_vs_k141_149211||lt2gene Low-quality
S1_vs_k141_37387||lt2gene Low-quality
S1_vs_k141_124866||lt2gene Low-quality
S1_vs_k141_78536||lt2gene Low-quality
S1_vs_k141_78540||lt2gene Low-quality
S1_vs_k141_91809||lt2gene Low-quality
S1_vs_k141_144838||lt2gene Not-determined
S1_vs_k141_118542||lt2gene Low-quality
S1_vs_k141_105229||lt2gene Low-quality
S1_vs_k141_131926||lt2gene Low-quality
S1_vs_k141_12045||lt2gene Not-determined
S1_vs_k141_91847||lt2gene Low-quality
S1_vs_k141_25006||lt2gene Not-determined
S1_vs_k141_91861||lt2gene Low-quality
S1_vs_k141_58455||lt2gene Low-quality
S1_vs_k141_31835||lt2gene Low-quality
S1_vs_k141_124950||lt2gene Low-quality
S1_vs_k141_118586||lt2gene Low-quality
S1_vs_k141_52109||lt2gene Low-quality
S1_vs_k141_19017||lt2gene Low-quality
S1_vs_k141_58474||lt2gene Low-quality
S1_vs_k141_105290||lt2gene Not-determined
S1_vs_k141_156357||lt2gene Not-determined
S1_vs_k141_4138||lt2gene Low-quality
S1_vs_k141_105299||lt2gene Low-quality
S1_vs_k141_63943||lt2gene Low-quality
S1_vs_k141_70951||lt2gene Low-quality
S1_vs_k141_31864||lt2gene Not-determined
S1_vs_k141_132007||lt2gene Low-quality
S1_vs_k141_98732||lt2gene Low-quality
S1_vs_k141_52133||lt2gene Not-determined
S1_vs_k141_52135||lt2gene Low-quality
S1_vs_k141_98739||lt2gene Low-quality
S1_vs_k141_31883||lt2gene Not-determined
S1_vs_k141_45330||lt2gene Not-determined
S1_vs_k141_63960||lt2gene Not-determined
S1_vs_k141_85253||lt2gene Low-quality
S1_vs_k141_125012||lt2gene Not-determined
S1_vs_k141_105325||lt2gene Low-quality
S1_vs_k141_125014||lt2gene Not-determined
S1_vs_k141_78670||lt2gene Low-quality
S1_vs_k141_31902||lt2gene Low-quality
S1_vs_k141_138444||lt2gene Low-quality
S1_vs_k141_118642||lt2gene Low-quality
S1_vs_k141_31911||lt2gene Low-quality
S1_vs_k141_105338||lt2gene Low-quality
S1_vs_k141_98761||lt2gene Low-quality
S1_vs_k141_70990||lt2gene Low-quality
S1_vs_k141_71003||lt2gene Not-determined
S1_vs_k141_125055||lt2gene Not-determined
S1_vs_k141_45368||lt2gene Low-quality
S1_vs_k141_132069||lt2gene Low-quality
S1_vs_k141_105379||lt2gene Low-quality
S1_vs_k141_118684||lt2gene Low-quality
S1_vs_k141_52202||lt2gene Low-quality
S1_vs_k141_4197||lt2gene Low-quality
S1_vs_k141_71031||lt2gene Not-determined
S1_vs_k141_111991||lt2gene Low-quality
S1_vs_k141_19151||lt2gene Low-quality
S1_vs_k141_37551||lt2gene Low-quality
S1_vs_k141_52218||lt2gene Low-quality
S1_vs_k141_52221||lt2gene Not-determined
S1_vs_k141_31986||lt2gene Low-quality
S1_vs_k141_58616||lt2gene Low-quality
S1_vs_k141_19170||lt2gene Low-quality
S1_vs_k141_145021||lt2gene Not-determined
S1_vs_k141_52225||lt2gene Low-quality
S1_vs_k141_19178||lt2gene Low-quality
S1_vs_k141_85353||lt2gene Low-quality
S1_vs_k141_125111||lt2gene Low-quality
S1_vs_k141_138526||lt2gene Low-quality
S1_vs_k141_19184||lt2gene Low-quality
S1_vs_k141_98839||lt2gene Low-quality
S1_vs_k141_12266||lt2gene Low-quality
S1_vs_k141_45434||lt2gene Low-quality
S1_vs_k141_112032||lt2gene Low-quality
S1_vs_k141_112035||lt2gene Low-quality
S1_vs_k141_12277||lt2gene Not-determined
S1_vs_k141_112043||lt2gene Low-quality
S1_vs_k141_132161||lt2gene Low-quality
S1_vs_k141_78811||lt2gene Low-quality
S1_vs_k141_12297||lt2gene Low-quality
S1_vs_k141_85409||lt2gene Not-determined
S1_vs_k141_58670||lt2gene Not-determined
S1_vs_k141_32072||lt2gene Low-quality
S1_vs_k141_37566||lt2gene Low-quality
S1_vs_k141_145076||lt2gene Low-quality
S1_vs_k141_45478||lt2gene Low-quality
S1_vs_k141_145087||lt2gene Not-determined
S1_vs_k141_112078||lt2gene Low-quality
S1_vs_k141_85443||lt2gene Low-quality
S1_vs_k141_98874||lt2gene Low-quality
S1_vs_k141_132210||lt2gene Low-quality
S1_vs_k141_125175||lt2gene Not-determined
S1_vs_k141_32126||lt2gene Low-quality
S1_vs_k141_32129||lt2gene Not-determined
S1_vs_k141_52350||lt2gene Low-quality
S1_vs_k141_45522||lt2gene Low-quality
S1_vs_k141_125216||lt2gene Low-quality
S1_vs_k141_85481||lt2gene Not-determined
S1_vs_k141_92135||lt2gene Not-determined
S1_vs_k141_125222||lt2gene Low-quality
S1_vs_k141_19332||lt2gene Not-determined
S1_vs_k141_85486||lt2gene Low-quality
S1_vs_k141_25287||lt2gene Not-determined
S1_vs_k141_132248||lt2gene Low-quality
S1_vs_k141_92145||lt2gene Not-determined
S1_vs_k141_118853||lt2gene Low-quality
S1_vs_k141_149414||lt2gene Low-quality
S1_vs_k141_12371||lt2gene Low-quality
S1_vs_k141_92162||lt2gene Low-quality
S1_vs_k141_19365||lt2gene Low-quality
S1_vs_k141_58813||lt2gene Low-quality
S1_vs_k141_45578||lt2gene Low-quality
S1_vs_k141_105569||lt2gene Low-quality
S1_vs_k141_98967||lt2gene Not-determined
S1_vs_k141_118886||lt2gene Low-quality
S1_vs_k141_118893||lt2gene Not-determined
S1_vs_k141_32200||lt2gene Low-quality
S1_vs_k141_149452||lt2gene Low-quality
S1_vs_k141_105584||lt2gene Not-determined
S1_vs_k141_92208||lt2gene Not-determined
S1_vs_k141_112167||lt2gene Low-quality
S1_vs_k141_92217||lt2gene Low-quality
S1_vs_k141_156473||lt2gene Low-quality
S1_vs_k141_37654||lt2gene Not-determined
S1_vs_k141_37659||lt2gene Low-quality
S1_vs_k141_125302||lt2gene Low-quality
S1_vs_k141_12449||lt2gene Low-quality
S1_vs_k141_149482||lt2gene Low-quality
S1_vs_k141_105620||lt2gene Not-determined
S1_vs_k141_19432||lt2gene Not-determined
S1_vs_k141_138712||lt2gene Low-quality
S1_vs_k141_45650||lt2gene Not-determined
S1_vs_k141_118939||lt2gene Not-determined
S1_vs_k141_112201||lt2gene Low-quality
S1_vs_k141_78997||lt2gene Low-quality
S1_vs_k141_32275||lt2gene Low-quality
S1_vs_k141_85625||lt2gene Low-quality
S1_vs_k141_52509||lt2gene Low-quality
S1_vs_k141_138729||lt2gene Not-determined
S1_vs_k141_156518||lt2gene Low-quality
S1_vs_k141_52519||lt2gene Low-quality
S1_vs_k141_52520||lt2gene Low-quality
S1_vs_k141_132348||lt2gene Low-quality
S1_vs_k141_45696||lt2gene Not-determined
S1_vs_k141_45700||lt2gene Low-quality
S1_vs_k141_37696||lt2gene Low-quality
S1_vs_k141_92288||lt2gene Not-determined
S1_vs_k141_92289||lt2gene Not-determined
S1_vs_k141_99081||lt2gene Low-quality
S1_vs_k141_92292||lt2gene Not-determined
S1_vs_k141_12513||lt2gene Low-quality
S1_vs_k141_92303||lt2gene Low-quality
S1_vs_k141_132370||lt2gene Not-determined
S1_vs_k141_99097||lt2gene Low-quality
S1_vs_k141_37714||lt2gene Not-determined
S1_vs_k141_156545||lt2gene Not-determined
S1_vs_k141_92308||lt2gene Low-quality
S1_vs_k141_19484||lt2gene Low-quality
S1_vs_k141_112275||lt2gene Not-determined
S1_vs_k141_125385||lt2gene Low-quality
S1_vs_k141_92323||lt2gene Not-determined
S1_vs_k141_37729||lt2gene Low-quality
S1_vs_k141_71186||lt2gene Low-quality
S1_vs_k141_32346||lt2gene Low-quality
S1_vs_k141_149511||lt2gene Not-determined
S1_vs_k141_145315||lt2gene Low-quality
S1_vs_k141_112291||lt2gene Low-quality
S1_vs_k141_64381||lt2gene Low-quality
S1_vs_k141_4431||lt2gene Low-quality
S1_vs_k141_58966||lt2gene Not-determined
S1_vs_k141_99134||lt2gene Low-quality
S1_vs_k141_132434||lt2gene Low-quality
S1_vs_k141_64407||lt2gene Low-quality
S1_vs_k141_132457||lt2gene Low-quality
S1_vs_k141_58993||lt2gene Low-quality
S1_vs_k141_45783||lt2gene Low-quality
S1_vs_k141_32410||lt2gene Low-quality
S1_vs_k141_52643||lt2gene Low-quality
S1_vs_k141_156591||lt2gene Not-determined
S1_vs_k141_156592||lt2gene Low-quality
S1_vs_k141_45805||lt2gene Low-quality
S1_vs_k141_112375||lt2gene Low-quality
S1_vs_k141_99191||lt2gene Low-quality
S1_vs_k141_52651||lt2gene Not-determined
S1_vs_k141_145386||lt2gene Low-quality
S1_vs_k141_105790||lt2gene Not-determined
S1_vs_k141_105802||lt2gene Low-quality
S1_vs_k141_64462||lt2gene Low-quality
S1_vs_k141_19623||lt2gene Low-quality
S1_vs_k141_132540||lt2gene Not-determined
S1_vs_k141_105818||lt2gene Low-quality
S1_vs_k141_138909||lt2gene Low-quality
S1_vs_k141_99232||lt2gene Low-quality
S1_vs_k141_4489||lt2gene Not-determined
S1_vs_k141_85862||lt2gene Low-quality
S1_vs_k141_37832||lt2gene Not-determined
S1_vs_k141_85869||lt2gene Low-quality
S1_vs_k141_105829||lt2gene Low-quality
S1_vs_k141_79242||lt2gene Low-quality
S1_vs_k141_105831||lt2gene Low-quality
S1_vs_k141_85879||lt2gene Low-quality
S1_vs_k141_105849||lt2gene Not-determined
S1_vs_k141_12688||lt2gene Not-determined
S1_vs_k141_105854||lt2gene Low-quality
S1_vs_k141_4536||lt2gene Low-quality
S1_vs_k141_37864||lt2gene Not-determined
S1_vs_k141_52749||lt2gene Low-quality
S1_vs_k141_125569||lt2gene Not-determined
S1_vs_k141_138964||lt2gene Low-quality
S1_vs_k141_59129||lt2gene Not-determined
S1_vs_k141_12728||lt2gene Low-quality
S1_vs_k141_149618||lt2gene Not-determined
S1_vs_k141_19729||lt2gene Low-quality
S1_vs_k141_156661||lt2gene Low-quality
S1_vs_k141_25669||lt2gene Not-determined
S1_vs_k141_32572||lt2gene Not-determined
S1_vs_k141_25685||lt2gene Low-quality
S1_vs_k141_99310||lt2gene Not-determined
S1_vs_k141_37921||lt2gene Low-quality
S1_vs_k141_25700||lt2gene Low-quality
S1_vs_k141_105939||lt2gene Low-quality
S1_vs_k141_71320||lt2gene Not-determined
S1_vs_k141_145510||lt2gene Low-quality
S1_vs_k141_79379||lt2gene Low-quality
S1_vs_k141_52829||lt2gene Not-determined
S1_vs_k141_25717||lt2gene Low-quality
S1_vs_k141_156717||lt2gene Low-quality
S1_vs_k141_119301||lt2gene Low-quality
S1_vs_k141_4641||lt2gene Not-determined
S1_vs_k141_119309||lt2gene Low-quality
S1_vs_k141_99357||lt2gene Low-quality
S1_vs_k141_86006||lt2gene Low-quality
S1_vs_k141_79412||lt2gene Not-determined
S1_vs_k141_79415||lt2gene Low-quality
S1_vs_k141_92586||lt2gene Not-determined
S1_vs_k141_145552||lt2gene Low-quality
S1_vs_k141_19838||lt2gene Low-quality
S1_vs_k141_45998||lt2gene Low-quality
S1_vs_k141_119335||lt2gene Low-quality
S1_vs_k141_32685||lt2gene Not-determined
S1_vs_k141_92612||lt2gene Low-quality
S1_vs_k141_32693||lt2gene Low-quality
S1_vs_k141_99405||lt2gene Low-quality
S1_vs_k141_32696||lt2gene Low-quality
S1_vs_k141_156744||lt2gene Low-quality
S1_vs_k141_145601||lt2gene Low-quality
S1_vs_k141_139107||lt2gene Low-quality
S1_vs_k141_139109||lt2gene Low-quality
S1_vs_k141_59290||lt2gene Low-quality
S1_vs_k141_119372||lt2gene Low-quality
S1_vs_k141_139121||lt2gene Low-quality
S1_vs_k141_52948||lt2gene Not-determined
S1_vs_k141_92645||lt2gene Not-determined
S1_vs_k141_125730||lt2gene Low-quality
S1_vs_k141_86083||lt2gene Not-determined
S1_vs_k141_46058||lt2gene Low-quality
S1_vs_k141_86085||lt2gene Low-quality
S1_vs_k141_156763||lt2gene Low-quality
S1_vs_k141_125749||lt2gene Not-determined
S1_vs_k141_149724||lt2gene Low-quality
S1_vs_k141_32746||lt2gene Low-quality
S1_vs_k141_38059||lt2gene Low-quality
S1_vs_k141_132849||lt2gene Not-determined
S1_vs_k141_145652||lt2gene Low-quality
S1_vs_k141_71403||lt2gene Low-quality
S1_vs_k141_125764||lt2gene Not-determined
S1_vs_k141_79544||lt2gene Not-determined
S1_vs_k141_86118||lt2gene Not-determined
S1_vs_k141_92691||lt2gene Low-quality
S1_vs_k141_38084||lt2gene Low-quality
S1_vs_k141_64734||lt2gene Low-quality
S1_vs_k141_119433||lt2gene Not-determined
S1_vs_k141_4756||lt2gene Low-quality
S1_vs_k141_92713||lt2gene Low-quality
S1_vs_k141_125793||lt2gene Low-quality
S1_vs_k141_59353||lt2gene Low-quality
S1_vs_k141_149766||lt2gene Low-quality
S1_vs_k141_53048||lt2gene Low-quality
S1_vs_k141_119446||lt2gene Not-determined
S1_vs_k141_112661||lt2gene Low-quality
S1_vs_k141_79564||lt2gene Low-quality
S1_vs_k141_25917||lt2gene Not-determined
S1_vs_k141_132915||lt2gene Low-quality
S1_vs_k141_79575||lt2gene Low-quality
S1_vs_k141_46140||lt2gene Not-determined
S1_vs_k141_32812||lt2gene Low-quality
S1_vs_k141_99545||lt2gene Low-quality
S1_vs_k141_139238||lt2gene Low-quality
S1_vs_k141_125847||lt2gene Low-quality
S1_vs_k141_92786||lt2gene Not-determined
S1_vs_k141_156815||lt2gene Not-determined
S1_vs_k141_132937||lt2gene Low-quality
S1_vs_k141_25949||lt2gene Not-determined
S1_vs_k141_125859||lt2gene Low-quality
S1_vs_k141_149817||lt2gene Not-determined
S1_vs_k141_64802||lt2gene Low-quality
S1_vs_k141_132953||lt2gene Low-quality
S1_vs_k141_145737||lt2gene Low-quality
S1_vs_k141_38150||lt2gene Not-determined
S1_vs_k141_149830||lt2gene Low-quality
S1_vs_k141_145752||lt2gene Low-quality
S1_vs_k141_13051||lt2gene Low-quality
S1_vs_k141_32884||lt2gene Low-quality
S1_vs_k141_71438||lt2gene Low-quality
S1_vs_k141_46194||lt2gene Not-determined
S1_vs_k141_92844||lt2gene Low-quality
S1_vs_k141_25981||lt2gene Not-determined
S1_vs_k141_13081||lt2gene Not-determined
S1_vs_k141_13084||lt2gene Low-quality
S1_vs_k141_64856||lt2gene Not-determined
S1_vs_k141_38181||lt2gene Low-quality
S1_vs_k141_38183||lt2gene Low-quality
S1_vs_k141_79641||lt2gene Low-quality
S1_vs_k141_106270||lt2gene Low-quality
S1_vs_k141_149871||lt2gene Low-quality
S1_vs_k141_112776||lt2gene Low-quality
S1_vs_k141_119566||lt2gene Low-quality
S1_vs_k141_59474||lt2gene Not-determined
S1_vs_k141_99654||lt2gene Not-determined
S1_vs_k141_139360||lt2gene Low-quality
S1_vs_k141_79667||lt2gene Low-quality
S1_vs_k141_99673||lt2gene Low-quality
S1_vs_k141_71491||lt2gene Low-quality
S1_vs_k141_86323||lt2gene Not-determined
S1_vs_k141_32971||lt2gene Not-determined
S1_vs_k141_139392||lt2gene Low-quality
S1_vs_k141_92905||lt2gene Low-quality
S1_vs_k141_53195||lt2gene Not-determined
S1_vs_k141_59526||lt2gene Low-quality
S1_vs_k141_112840||lt2gene Low-quality
S1_vs_k141_4906||lt2gene Low-quality
S1_vs_k141_112846||lt2gene Not-determined
S1_vs_k141_112858||lt2gene Low-quality
S1_vs_k141_139426||lt2gene Low-quality
S1_vs_k141_53245||lt2gene Low-quality
S1_vs_k141_106365||lt2gene Low-quality
S1_vs_k141_46352||lt2gene Low-quality
S1_vs_k141_71570||lt2gene Low-quality
S1_vs_k141_65014||lt2gene Low-quality
S1_vs_k141_106393||lt2gene Low-quality
S1_vs_k141_133143||lt2gene Low-quality
S1_vs_k141_13260||lt2gene Low-quality
S1_vs_k141_99773||lt2gene Low-quality
S1_vs_k141_4944||lt2gene Low-quality
S1_vs_k141_26151||lt2gene Low-quality
S1_vs_k141_133153||lt2gene Low-quality
S1_vs_k141_86418||lt2gene Low-quality
S1_vs_k141_92980||lt2gene Not-determined
S1_vs_k141_99796||lt2gene Low-quality
S1_vs_k141_145857||lt2gene Not-determined
S1_vs_k141_133176||lt2gene Low-quality
S1_vs_k141_106432||lt2gene Not-determined
S1_vs_k141_139517||lt2gene Not-determined
S1_vs_k141_86451||lt2gene Low-quality
S1_vs_k141_99817||lt2gene Low-quality
S1_vs_k141_33099||lt2gene Low-quality
S1_vs_k141_46418||lt2gene Low-quality
S1_vs_k141_59631||lt2gene Not-determined
S1_vs_k141_53331||lt2gene Low-quality
S1_vs_k141_149944||lt2gene Low-quality
S1_vs_k141_53344||lt2gene Low-quality
S1_vs_k141_38310||lt2gene Low-quality
S1_vs_k141_79809||lt2gene Low-quality
S1_vs_k141_4990||lt2gene Low-quality
S1_vs_k141_71647||lt2gene Low-quality
S1_vs_k141_13339||lt2gene Low-quality
S1_vs_k141_93043||lt2gene Low-quality
S1_vs_k141_86490||lt2gene Low-quality
S1_vs_k141_59663||lt2gene Low-quality
S1_vs_k141_79825||lt2gene Not-determined
S1_vs_k141_133240||lt2gene Low-quality
S1_vs_k141_86499||lt2gene Low-quality
S1_vs_k141_86506||lt2gene Low-quality
S1_vs_k141_133250||lt2gene Low-quality
S1_vs_k141_156894||lt2gene Low-quality
S1_vs_k141_53380||lt2gene Low-quality
S1_vs_k141_93072||lt2gene Low-quality
S1_vs_k141_106499||lt2gene Low-quality
S1_vs_k141_33169||lt2gene Low-quality
S1_vs_k141_139606||lt2gene Low-quality
S1_vs_k141_59699||lt2gene Low-quality
S1_vs_k141_113067||lt2gene Low-quality
S1_vs_k141_5060||lt2gene Low-quality
S1_vs_k141_106529||lt2gene Not-determined
S1_vs_k141_119799||lt2gene Low-quality
S1_vs_k141_119806||lt2gene Low-quality
S1_vs_k141_65152||lt2gene Low-quality
S1_vs_k141_149991||lt2gene Low-quality
S1_vs_k141_126215||lt2gene Low-quality
S1_vs_k141_5090||lt2gene Low-quality
S1_vs_k141_133337||lt2gene Low-quality
S1_vs_k141_133341||lt2gene Not-determined
S1_vs_k141_33242||lt2gene Low-quality
S1_vs_k141_146010||lt2gene Not-determined
S1_vs_k141_133356||lt2gene Low-quality
S1_vs_k141_5108||lt2gene Not-determined
S1_vs_k141_86613||lt2gene Low-quality
S1_vs_k141_33269||lt2gene Not-determined
S1_vs_k141_156929||lt2gene Not-determined
S1_vs_k141_146026||lt2gene Low-quality
S1_vs_k141_26344||lt2gene Low-quality
S1_vs_k141_113155||lt2gene Low-quality
S1_vs_k141_126256||lt2gene Low-quality
S1_vs_k141_156946||lt2gene Low-quality
S1_vs_k141_156952||lt2gene Not-determined
S1_vs_k141_71734||lt2gene Low-quality
S1_vs_k141_126269||lt2gene Low-quality
S1_vs_k141_38459||lt2gene Low-quality
S1_vs_k141_46566||lt2gene Low-quality
S1_vs_k141_119886||lt2gene Low-quality
S1_vs_k141_133405||lt2gene Low-quality
S1_vs_k141_156964||lt2gene Low-quality
S1_vs_k141_106621||lt2gene Low-quality
S1_vs_k141_93259||lt2gene Low-quality
S1_vs_k141_59825||lt2gene Low-quality
S1_vs_k141_156984||lt2gene Low-quality
S1_vs_k141_100066||lt2gene Not-determined
S1_vs_k141_93272||lt2gene Not-determined
S1_vs_k141_100072||lt2gene Low-quality
S1_vs_k141_93277||lt2gene Low-quality
S1_vs_k141_86685||lt2gene Not-determined
S1_vs_k141_71751||lt2gene Low-quality
S1_vs_k141_80038||lt2gene Low-quality
S1_vs_k141_100084||lt2gene Low-quality
S1_vs_k141_5194||lt2gene Low-quality
S1_vs_k141_38511||lt2gene Low-quality
S1_vs_k141_71760||lt2gene Low-quality
S1_vs_k141_65286||lt2gene Low-quality
S1_vs_k141_86700||lt2gene Low-quality
S1_vs_k141_46640||lt2gene Not-determined
S1_vs_k141_100090||lt2gene Not-determined
S1_vs_k141_146105||lt2gene Not-determined
S1_vs_k141_65306||lt2gene Not-determined
S1_vs_k141_146123||lt2gene Not-determined
S1_vs_k141_93315||lt2gene Low-quality
S1_vs_k141_59872||lt2gene Low-quality
S1_vs_k141_86730||lt2gene Low-quality
S1_vs_k141_146135||lt2gene Low-quality
S1_vs_k141_106710||lt2gene Low-quality
S1_vs_k141_119961||lt2gene Low-quality
S1_vs_k141_113279||lt2gene Low-quality
S1_vs_k141_86761||lt2gene Low-quality
S1_vs_k141_65353||lt2gene Low-quality
S1_vs_k141_38562||lt2gene Low-quality
S1_vs_k141_5240||lt2gene Low-quality
S1_vs_k141_93385||lt2gene Low-quality
S1_vs_k141_93392||lt2gene Not-determined
S1_vs_k141_139848||lt2gene Low-quality
S1_vs_k141_119981||lt2gene Low-quality
S1_vs_k141_38575||lt2gene Low-quality
S1_vs_k141_59928||lt2gene Low-quality
S1_vs_k141_65403||lt2gene Low-quality
S1_vs_k141_126378||lt2gene Low-quality
S1_vs_k141_120013||lt2gene Low-quality

<table>
<tr><td>S1_vs_k141_120023||lt2gene</td><td>Not-determined</td></tr>
<tr><td>S1_vs_k141_150069||lt2gene</td><td>Not-determined</td></tr>
<tr><td>S1_vs_k141_65432||lt2gene</td><td>Low-quality</td></tr>
<tr><td>S1_vs_k141_120025||lt2gene</td><td>Low-quality</td></tr>
<tr><td>S1_vs_k141_71895||lt2gene</td><td>Low-quality</td></tr>
<tr><td>S1_vs_k141_65477||lt2gene</td><td>Low-quality</td></tr>
<tr><td>S1_vs_k141_113395||lt2gene</td><td>Low-quality</td></tr>
<tr><td>S1_vs_k141_5302||lt2gene</td><td>Not-determined</td></tr>
<tr><td>S1_vs_k141_113404||lt2gene</td><td>Not-determined</td></tr>
<tr><td>S1_vs_k141_113413||lt2gene</td><td>Low-quality</td></tr>
<tr><td>S1_vs_k141_146291||lt2gene</td><td>Not-determined</td></tr>
<tr><td>S1_vs_k141_146292||lt2gene</td><td>Low-quality</td></tr>
<tr><td>S1_vs_k141_139941||lt2gene</td><td>Low-quality</td></tr>
<tr><td>S1_vs_k141_5367||lt2gene</td><td>Low-quality</td></tr>
<tr><td>S1_vs_k141_38649||lt2gene</td><td>Low-quality</td></tr>
<tr><td>S1_vs_k141_71976||lt2gene</td><td>Low-quality</td></tr>
<tr><td>S1_vs_k141_146363||lt2gene</td><td>Not-determined</td></tr>
<tr><td>S1_vs_k141_5398||lt2gene</td><td>Low-quality</td></tr>
<tr><td>S1_vs_k141_113470||lt2gene</td><td>Low-quality</td></tr>
<tr><td>S1_vs_k141_146405||lt2gene</td><td>Low-quality</td></tr>
<tr><td>S1_vs_k141_72016||lt2gene</td><td>Low-quality</td></tr>
<tr><td>S1_vs_k141_140058||lt2gene</td><td>Low-quality</td></tr>
<tr><td>S1_vs_k141_26669||lt2gene</td><td>Low-quality</td></tr>
<tr><td>S1_vs_k141_38726||lt2gene</td><td>Not-determined</td></tr>
<tr><td>S1_vs_k141_65674||lt2gene</td><td>Low-quality</td></tr>
<tr><td>S1_vs_k141_65679||lt2gene</td><td>Low-quality</td></tr>
<tr><td>S1_vs_k141_140098||lt2gene</td><td>Low-quality</td></tr>
<tr><td>S1_vs_k141_72061||lt2gene</td><td>Low-quality</td></tr>
<tr><td>S1_vs_k141_65694||lt2gene</td><td>Low-quality</td></tr>
<tr><td>S1_vs_k141_72084||lt2gene</td><td>Low-quality</td></tr>
<tr><td>S1_vs_k141_157168||lt2gene</td><td>Low-quality</td></tr>
<tr><td>S1_vs_k141_72124||lt2gene</td><td>Not-determined</td></tr>
<tr><td>S1_vs_k141_157174||lt2gene</td><td>Not-determined</td></tr>
<tr><td>S1_vs_k141_146595||lt2gene</td><td>Low-quality</td></tr>
<tr><td>S1_vs_k141_126723||lt2gene</td><td>Low-quality</td></tr>
<tr><td>S1_vs_k141_126744||lt2gene</td><td>Low-quality</td></tr>
<tr><td>S1_vs_k141_150229||lt2gene</td><td>Low-quality</td></tr>
<tr><td>S1_vs_k141_157207||lt2gene</td><td>Low-quality</td></tr>
<tr><td>S1_vs_k141_5572||lt2gene</td><td>Low-quality</td></tr>
<tr><td>S1_vs_k141_38860||lt2gene</td><td>Low-quality</td></tr>
<tr><td>S1_vs_k141_150249||lt2gene</td><td>Low-quality</td></tr>
<tr><td>S1_vs_k141_146648||lt2gene</td><td>Low-quality</td></tr>
<tr><td>S1_vs_k141_38882||lt2gene</td><td>Low-quality</td></tr>
<tr><td>S1_vs_k141_72145||lt2gene</td><td>Not-determined</td></tr>
<tr><td>S1_vs_k141_126815||lt2gene</td><td>Low-quality</td></tr>
<tr><td>S1_vs_k141_65911||lt2gene</td><td>Not-determined</td></tr>
<tr><td>S1_vs_k141_150294||lt2gene</td><td>Low-quality</td></tr>
<tr><td>S1_vs_k141_157286||lt2gene</td><td>Not-determined</td></tr>
<tr><td>S1_vs_k141_157295||lt2gene</td><td>Low-quality</td></tr>
<tr><td>S1_vs_k141_157311||lt2gene</td><td>Low-quality</td></tr>
<tr><td>S1_vs_k141_157324||lt2gene</td><td>Low-quality</td></tr>
<tr><td>S1_vs_k141_157339||lt2gene</td><td>Low-quality</td></tr>
<tr><td>S1_vs_k141_72202||lt2gene</td><td>Low-quality</td></tr>
<tr><td>S1_vs_k141_39058||lt2gene</td><td>Not-determined</td></tr>
<tr><td>S1_vs_k141_5854||lt2gene</td><td>Low-quality</td></tr>
<tr><td>S1_vs_k141_150350||lt2gene</td><td>Low-quality</td></tr>
<tr><td>S1_vs_k141_66186||lt2gene</td><td>Low-quality</td></tr>
<tr><td>S1_vs_k141_150375||lt2gene</td><td>Low-quality</td></tr>
<tr><td>S1_vs_k141_150412||lt2gene</td><td>Low-quality</td></tr>
<tr><td>S1_vs_k141_39184||lt2gene</td><td>Low-quality</td></tr>
<tr><td>S1_vs_k141_66315||lt2gene</td><td>Low-quality</td></tr>
<tr><td>S1_vs_k141_5968||lt2gene</td><td>Low-quality</td></tr>
<tr><td>S1_vs_k141_72362||lt2gene</td><td>Low-quality</td></tr>
<tr><td>S1_vs_k141_157521||lt2gene</td><td>Not-determined</td></tr>
<tr><td>S1_vs_k141_66438||lt2gene</td><td>Low-quality</td></tr>
<tr><td>S1_vs_k141_6036||lt2gene</td><td>Low-quality</td></tr>
<tr><td>S1_vs_k141_72438||lt2gene</td><td>Low-quality</td></tr>
<tr><td>S1_vs_k141_66517||lt2gene</td><td>Low-quality</td></tr>
<tr><td>S1_vs_k141_6134||lt2gene</td><td>Not-determined</td></tr>
<tr><td>S1_vs_k141_72496||lt2gene</td><td>Low-quality</td></tr>
<tr><td>S1_vs_k141_39361||lt2gene</td><td>Low-quality</td></tr>
<tr><td>S1_vs_k141_150503||lt2gene</td><td>Low-quality</td></tr>
<tr><td>S1_vs_k141_6220||lt2gene</td><td>Not-determined</td></tr>
<tr><td>S1_vs_k141_6236||lt2gene</td><td>Low-quality</td></tr>
<tr><td>S1_vs_k141_6250||lt2gene</td><td>Low-quality</td></tr>
<tr><td>S1_vs_k141_39423||lt2gene</td><td>Not-determined</td></tr>
<tr><td>S1_vs_k141_72601||lt2gene</td><td>Low-quality</td></tr>
<tr><td>S1_vs_k141_66677||lt2gene</td><td>Low-quality</td></tr>
<tr><td>S1_vs_k141_72628||lt2gene</td><td>Low-quality</td></tr>
<tr><td>S1_vs_k141_39464||lt2gene</td><td>Not-determined</td></tr>
<tr><td>S1_vs_k141_157655||lt2gene</td><td>Low-quality</td></tr>
<tr><td>S1_vs_k141_157661||lt2gene</td><td>Low-quality</td></tr>
<tr><td>S1_vs_k141_157672||lt2gene</td><td>Low-quality</td></tr>
<tr><td>S1_vs_k141_72755||lt2gene</td><td>Low-quality</td></tr>
<tr><td>S1_vs_k141_39579||lt2gene</td><td>Low-quality</td></tr>
<tr><td>S1_vs_k141_39590||lt2gene</td><td>Low-quality</td></tr>
<tr><td>S1_vs_k141_39623||lt2gene</td><td>Low-quality</td></tr>
<tr><td>S1_vs_k141_72822||lt2gene</td><td>Not-determined</td></tr>
<tr><td>S1_vs_k141_157791||lt2gene</td><td>Not-determined</td></tr>
<tr><td>S1_vs_k141_39709||lt2gene</td><td>Low-quality</td></tr>
<tr><td>S1_vs_k141_6483||lt2gene</td><td>Low-quality</td></tr>
<tr><td>S1_vs_k141_39734||lt2gene</td><td>Low-quality</td></tr>
<tr><td>S1_vs_k141_39745||lt2gene</td><td>Low-quality</td></tr>
<tr><td>S1_vs_k141_150670||lt2gene</td><td>Not-determined</td></tr>
<tr><td>S1_vs_k141_39785||lt2gene</td><td>Low-quality</td></tr>
<tr><td>S1_vs_k141_72909||lt2gene</td><td>Not-determined</td></tr>
<tr><td>S1_vs_k141_150696||lt2gene</td><td>Not-determined</td></tr>
<tr><td>S1_vs_k141_157935||lt2gene</td><td>Low-quality</td></tr>
<tr><td>S1_vs_k141_150714||lt2gene</td><td>Low-quality</td></tr>
<tr><td>S1_vs_k141_157983||lt2gene</td><td>Low-quality</td></tr>
<tr><td>S1_vs_k141_150737||lt2gene</td><td>Not-determined</td></tr>
<tr><td>S1_vs_k141_158013||lt2gene</td><td>Not-determined</td></tr>
<tr><td>S1_vs_k141_39949||lt2gene</td><td>Low-quality</td></tr>
<tr><td>S1_vs_k141_39979||lt2gene</td><td>Not-determined</td></tr>
<tr><td>S1_vs_k141_39998||lt2gene</td><td>Low-quality</td></tr>
<tr><td>S1_vs_k141_150801||lt2gene</td><td>Low-quality</td></tr>
<tr><td>S1_vs_k141_158108||lt2gene</td><td>Low-quality</td></tr>
<tr><td>S1_vs_k141_150825||lt2gene</td><td>Low-quality</td></tr>
<tr><td>S1_vs_k141_150828||lt2gene</td><td>Not-determined</td></tr>
<tr><td>S1_vs_k141_73021||lt2gene</td><td>Low-quality</td></tr>
<tr><td>S1_vs_k141_158179||lt2gene</td><td>Low-quality</td></tr>
<tr><td>S1_vs_k141_158192||lt2gene</td><td>Low-quality</td></tr>
<tr><td>S1_vs_k141_150928||lt2gene</td><td>Low-quality</td></tr>
<tr><td>S1_vs_k141_150947||lt2gene</td><td>Low-quality</td></tr>
<tr><td>S1_vs_k141_158265||lt2gene</td><td>Low-quality</td></tr>
<tr><td>S1_vs_k141_158268||lt2gene</td><td>Low-quality</td></tr>
<tr><td>S1_vs_k141_150975||lt2gene</td><td>Low-quality</td></tr>
<tr><td>S1_vs_k141_73160||lt2gene</td><td>Low-quality</td></tr>
<tr><td>S1_vs_k141_158282||lt2gene</td><td>Not-determined</td></tr>
<tr><td>S1_vs_k141_151003||lt2gene</td><td>Low-quality</td></tr>
<tr><td>S1_vs_k141_151048||lt2gene</td><td>Low-quality</td></tr>
<tr><td>S1_vs_k141_73241||lt2gene</td><td>Low-quality</td></tr>
<tr><td>S1_vs_k141_73300||lt2gene</td><td>Low-quality</td></tr>
<tr><td>S1_vs_k141_151155||lt2gene</td><td>Not-determined</td></tr>
<tr><td>S1_vs_k141_158481||lt2gene</td><td>Low-quality</td></tr>
<tr><td>S1_vs_k141_73365||lt2gene</td><td>Low-quality</td></tr>
<tr><td>S1_vs_k141_158569||lt2gene</td><td>Low-quality</td></tr>
<tr><td>S1_vs_k141_151349||lt2gene</td><td>Low-quality</td></tr>
<tr><td>S1_vs_k141_158698||lt2gene</td><td>Low-quality</td></tr>
<tr><td>S1_vs_k141_158718||lt2gene</td><td>Low-quality</td></tr>
<tr><td>S1_vs_k141_158756||lt2gene</td><td>Low-quality</td></tr>
<tr><td>S1_vs_k141_158784||lt2gene</td><td>Low-quality</td></tr>
<tr><td>S1_vs_k141_151531||lt2gene</td><td>Not-determined</td></tr>
<tr><td>S1_vs_k141_158828||lt2gene</td><td>Low-quality</td></tr>
<tr><td>S1_vs_k141_158871||lt2gene</td><td>Low-quality</td></tr>
<tr><td>S1_vs_k141_158880||lt2gene</td><td>Low-quality</td></tr>
<tr><td>S1_vs_k141_158888||lt2gene</td><td>Low-quality</td></tr>
<tr><td>S1_vs_k141_151694||lt2gene</td><td>Not-determined</td></tr>
<tr><td>S1_vs_k141_158938||lt2gene</td><td>Not-determined</td></tr>
<tr><td>S1_vs_k141_158942||lt2gene</td><td>Low-quality</td></tr>
<tr><td>S1_vs_k141_151855||lt2gene</td><td>Low-quality</td></tr>
<tr><td>S1_vs_k141_151863||lt2gene</td><td>Low-quality</td></tr>
<tr><td>S1_vs_k141_151889||lt2gene</td><td>Not-determined</td></tr>
<tr><td>S1_vs_k141_159075||lt2gene</td><td>Low-quality</td></tr>
<tr><td>S1_vs_k141_159173||lt2gene</td><td>Not-determined</td></tr>
<tr><td>S1_vs_k141_151997||lt2gene</td><td>Not-determined</td></tr>
<tr><td>S1_vs_k141_159177||lt2gene</td><td>Low-quality</td></tr>
<tr><td>S1_vs_k141_152021||lt2gene</td><td>Low-quality</td></tr>
<tr><td>S1_vs_k141_152045||lt2gene</td><td>Not-determined</td></tr>
<tr><td>S1_vs_k141_152046||lt2gene</td><td>Not-determined</td></tr>
<tr><td>S1_vs_k141_152061||lt2gene</td><td>Low-quality</td></tr>
<tr><td>S1_vs_k141_152109||lt2gene</td><td>Low-quality</td></tr>
<tr><td>S1_vs_k141_159263||lt2gene</td><td>Not-determined</td></tr>
<tr><td>S1_vs_k141_152231||lt2gene</td><td>Low-quality</td></tr>
<tr><td>S1_vs_k141_152237||lt2gene</td><td>Low-quality</td></tr>
<tr><td>S1_vs_k141_159344||lt2gene</td><td>Not-determined</td></tr>
<tr><td>S1_vs_k141_159377||lt2gene</td><td>Not-determined</td></tr>
<tr><td>S1_vs_k141_152294||lt2gene</td><td>Low-quality</td></tr>
<tr><td>S1_vs_k141_159423||lt2gene</td><td>Low-quality</td></tr>
<tr><td>S1_vs_k141_159435||lt2gene</td><td>Low-quality</td></tr>
<tr><td>S1_vs_k141_152360||lt2gene</td><td>Low-quality</td></tr>
<tr><td>S1_vs_k141_159477||lt2gene</td><td>Low-quality</td></tr>
<tr><td>S1_vs_k141_152377||lt2gene</td><td>Low-quality</td></tr>
<tr><td>S1_vs_k141_159501||lt2gene</td><td>Low-quality</td></tr>
<tr><td>S1_vs_k141_159612||lt2gene</td><td>Not-determined</td></tr>
<tr><td>S1_vs_k141_152680||lt2gene</td><td>Low-quality</td></tr>
<tr><td>S1_vs_k141_152683||lt2gene</td><td>Low-quality</td></tr>
<tr><td>S1_vs_k141_152699||lt2gene</td><td>Not-determined</td></tr>
<tr><td>S1_vs_k141_152709||lt2gene</td><td>Not-determined</td></tr>
<tr><td>S1_vs_k141_159706||lt2gene</td><td>Not-determined</td></tr>
<tr><td>S1_vs_k141_152875||lt2gene</td><td>Low-quality</td></tr>
<tr><td>S1_vs_k141_159840||lt2gene</td><td>Low-quality</td></tr>
<tr><td>S1_vs_k141_152896||lt2gene</td><td>Low-quality</td></tr>
<tr><td>S1_vs_k141_152961||lt2gene</td><td>Low-quality</td></tr>
<tr><td>S1_vs_k141_159925||lt2gene</td><td>Low-quality</td></tr>
<tr><td>S1_vs_k141_159933||lt2gene</td><td>Not-determined</td></tr>
<tr><td>S1_vs_k141_152988||lt2gene</td><td>Low-quality</td></tr>
<tr><td>S1_vs_k141_153008||lt2gene</td><td>Low-quality</td></tr>
<tr><td>S1_vs_k141_153148||lt2gene</td><td>Low-quality</td></tr>
<tr><td>S1_vs_k141_153198||lt2gene</td><td>Low-quality</td></tr>
<tr><td>S1_vs_k141_153213||lt2gene</td><td>Low-quality</td></tr>
<tr><td>S1_vs_k141_153275||lt2gene</td><td>Low-quality</td></tr>
<tr><td>S1_vs_k141_153321||lt2gene</td><td>Low-quality</td></tr>
<tr><td>S1_vs_k141_153515||lt2gene</td><td>Low-quality</td></tr>
<tr><td>S3_extracted_k141_55790</td><td>Not-determined</td></tr>
<tr><td>S3_extracted_k141_23912</td><td>Not-determined</td></tr>
<tr><td>S3_extracted_k141_23913</td><td>Not-determined</td></tr>
<tr><td>S3_extracted_k141_103611</td><td>Low-quality</td></tr>
<tr><td>S3_extracted_k141_55793</td><td>Not-determined</td></tr>
<tr><td>S3_extracted_k141_103613</td><td>Low-quality</td></tr>
<tr><td>S3_extracted_k141_71731</td><td>Not-determined</td></tr>
<tr><td>S3_extracted_k141_7973</td><td>Not-determined</td></tr>
<tr><td>S3_extracted_k141_15942</td><td>Not-determined</td></tr>
<tr><td>S3_extracted_k141_87676</td><td>Not-determined</td></tr>
<tr><td>S3_extracted_k141_7979</td><td>Low-quality</td></tr>
<tr><td>S3_extracted_k141_2</td><td>Not-determined</td></tr>
<tr><td>S3_extracted_k141_3</td><td>Not-determined</td></tr>
<tr><td>S3_extracted_k141_87683</td><td>Not-determined</td></tr>
<tr><td>S3_extracted_k141_23923</td><td>Not-determined</td></tr>
<tr><td>S3_extracted_k141_39859</td><td>Not-determined</td></tr>
<tr><td>S3_extracted_k141_63761</td><td>Not-determined</td></tr>
<tr><td>S3_extracted_k141_63763</td><td>Not-determined</td></tr>
<tr><td>S3_extracted_k141_55804</td><td>Not-determined</td></tr>
<tr><td>S3_extracted_k141_63765</td><td>Low-quality</td></tr>
<tr><td>S3_extracted_k141_103619</td><td>Low-quality</td></tr>
<tr><td>S3_extracted_k141_16</td><td>Not-determined</td></tr>
<tr><td>S3_extracted_k141_19</td><td>Low-quality</td></tr>
<tr><td>S3_extracted_k141_47829</td><td>Not-determined</td></tr>
<tr><td>S3_extracted_k141_47830</td><td>Not-determined</td></tr>
<tr><td>S3_extracted_k141_23933</td><td>Not-determined</td></tr>
<tr><td>S3_extracted_k141_119567</td><td>Not-determined</td></tr>
<tr><td>S3_extracted_k141_111587</td><td>Not-determined</td></tr>
<tr><td>S3_extracted_k141_7996</td><td>Not-determined</td></tr>
<tr><td>S3_extracted_k141_8000</td><td>Not-determined</td></tr>
<tr><td>S3_extracted_k141_8001</td><td>Not-determined</td></tr>
<tr><td>S3_extracted_k141_15957</td><td>Not-determined</td></tr>
<tr><td>S3_extracted_k141_23947</td><td>Not-determined</td></tr>
<tr><td>S3_extracted_k141_8004</td><td>Low-quality</td></tr>
<tr><td>S3_extracted_k141_95664</td><td>Not-determined</td></tr>
<tr><td>S3_extracted_k141_23951</td><td>Not-determined</td></tr>
<tr><td>S3_extracted_k141_23956</td><td>Not-determined</td></tr>
<tr><td>S3_extracted_k141_23958</td><td>Not-determined</td></tr>
<tr><td>S3_extracted_k141_63783</td><td>Not-determined</td></tr>
<tr><td>S3_extracted_k141_23961</td><td>Not-determined</td></tr>
<tr><td>S3_extracted_k141_119577</td><td>Not-determined</td></tr>
<tr><td>S3_extracted_k141_23964</td><td>Not-determined</td></tr>
<tr><td>S3_extracted_k141_23965</td><td>Not-determined</td></tr>
<tr><td>S3_extracted_k141_28</td><td>Low-quality</td></tr>
<tr><td>S3_extracted_k141_15964</td><td>Not-determined</td></tr>
<tr><td>S3_extracted_k141_39862</td><td>Not-determined</td></tr>
<tr><td>S3_extracted_k141_15968</td><td>Not-determined</td></tr>
<tr><td>S3_extracted_k141_71748</td><td>Not-determined</td></tr>
<tr><td>S3_extracted_k141_47854</td><td>Not-determined</td></tr>
<tr><td>S3_extracted_k141_71750</td><td>Not-determined</td></tr>
<tr><td>S3_extracted_k141_95676</td><td>Not-determined</td></tr>
<tr><td>S3_extracted_k141_31912</td><td>Not-determined</td></tr>
<tr><td>S3_extracted_k141_119589</td><td>Not-determined</td></tr>
<tr><td>S3_extracted_k141_23977</td><td>Not-determined</td></tr>
<tr><td>S3_extracted_k141_111597</td><td>Not-determined</td></tr>
<tr><td>S3_extracted_k141_23984</td><td>Not-determined</td></tr>
<tr><td>S3_extracted_k141_55824</td><td>Not-determined</td></tr>
<tr><td>S3_extracted_k141_79714</td><td>Not-determined</td></tr>
<tr><td>S3_extracted_k141_55828</td><td>Not-determined</td></tr>
<tr><td>S3_extracted_k141_55831</td><td>Not-determined</td></tr>
<tr><td>S3_extracted_k141_79722</td><td>Not-determined</td></tr>
<tr><td>S3_extracted_k141_47864</td><td>Not-determined</td></tr>
<tr><td>S3_extracted_k141_111607</td><td>Low-quality</td></tr>
<tr><td>S3_extracted_k141_79725</td><td>Not-determined</td></tr>
<tr><td>S3_extracted_k141_47867</td><td>Not-determined</td></tr>
<tr><td>S3_extracted_k141_47871</td><td>Not-determined</td></tr>
<tr><td>S3_extracted_k141_37</td><td>Low-quality</td></tr>
<tr><td>S3_extracted_k141_38</td><td>Not-determined</td></tr>
<tr><td>S3_extracted_k141_47872</td><td>Low-quality</td></tr>
<tr><td>S3_extracted_k141_55839</td><td>Not-determined</td></tr>
<tr><td>S3_extracted_k141_42</td><td>Not-determined</td></tr>
<tr><td>S3_extracted_k141_71776</td><td>Not-determined</td></tr>
<tr><td>S3_extracted_k141_71777</td><td>Not-determined</td></tr>
<tr><td>S3_extracted_k141_95688</td><td>Not-determined</td></tr>
<tr><td>S3_extracted_k141_103621</td><td>Not-determined</td></tr>
<tr><td>S3_extracted_k141_23994</td><td>Not-determined</td></tr>
<tr><td>S3_extracted_k141_103624</td><td>Low-quality</td></tr>
<tr><td>S3_extracted_k141_111612</td><td>Not-determined</td></tr>
<tr><td>S3_extracted_k141_103625</td><td>Not-determined</td></tr>
<tr><td>S3_extracted_k141_39889</td><td>Not-determined</td></tr>
<tr><td>S3_extracted_k141_111613</td><td>Low-quality</td></tr>
<tr><td>S3_extracted_k141_47887</td><td>Not-determined</td></tr>
<tr><td>S3_extracted_k141_63798</td><td>Not-determined</td></tr>
<tr><td>S3_extracted_k141_23999</td><td>Not-determined</td></tr>
<tr><td>S3_extracted_k141_95703</td><td>Not-determined</td></tr>
<tr><td>S3_extracted_k141_95707</td><td>Not-determined</td></tr>
<tr><td>S3_extracted_k141_95709</td><td>Not-determined</td></tr>
<tr><td>S3_extracted_k141_95711</td><td>Not-determined</td></tr>
<tr><td>S3_extracted_k141_79745</td><td>Low-quality</td></tr>
<tr><td>S3_extracted_k141_79747</td><td>Not-determined</td></tr>
<tr><td>S3_extracted_k141_79750</td><td>Not-determined</td></tr>
<tr><td>S3_extracted_k141_63800</td><td>Not-determined</td></tr>
<tr><td>S3_extracted_k141_24004</td><td>Low-quality</td></tr>
<tr><td>S3_extracted_k141_63802</td><td>Not-determined</td></tr>
<tr><td>S3_extracted_k141_95713</td><td>Not-determined</td></tr>
<tr><td>S3_extracted_k141_71798</td><td>Not-determined</td></tr>
<tr><td>S3_extracted_k141_71807</td><td>Not-determined</td></tr>
<tr><td>S3_extracted_k141_111618</td><td>Low-quality</td></tr>
<tr><td>S3_extracted_k141_111622</td><td>Not-determined</td></tr>
<tr><td>S3_extracted_k141_103632</td><td>Low-quality</td></tr>
<tr><td>S3_extracted_k141_55847</td><td>Low-quality</td></tr>
<tr><td>S3_extracted_k141_111628</td><td>Not-determined</td></tr>
<tr><td>S3_extracted_k141_111631</td><td>Not-determined</td></tr>
<tr><td>S3_extracted_k141_55855</td><td>Not-determined</td></tr>
<tr><td>S3_extracted_k141_24018</td><td>Low-quality</td></tr>
<tr><td>S3_extracted_k141_55861</td><td>Not-determined</td></tr>
<tr><td>S3_extracted_k141_111632</td><td>Not-determined</td></tr>
<tr><td>S3_extracted_k141_47919</td><td>Not-determined</td></tr>
<tr><td>S3_extracted_k141_47924</td><td>Not-determined</td></tr>
<tr><td>S3_extracted_k141_63814</td><td>Not-determined</td></tr>
<tr><td>S3_extracted_k141_24040</td><td>Not-determined</td></tr>
<tr><td>S3_extracted_k141_24041</td><td>Not-determined</td></tr>
<tr><td>S3_extracted_k141_47927</td><td>Not-determined</td></tr>
<tr><td>S3_extracted_k141_71813</td><td>Low-quality</td></tr>
<tr><td>S3_extracted_k141_24046</td><td>Not-determined</td></tr>
<tr><td>S3_extracted_k141_8049</td><td>Not-determined</td></tr>
<tr><td>S3_extracted_k141_63825</td><td>Not-determined</td></tr>
<tr><td>S3_extracted_k141_111640</td><td>Not-determined</td></tr>
<tr><td>S3_extracted_k141_111644</td><td>Low-quality</td></tr>
<tr><td>S3_extracted_k141_24052</td><td>Low-quality</td></tr>
<tr><td>S3_extracted_k141_8056</td><td>Not-determined</td></tr>
<tr><td>S3_extracted_k141_111650</td><td>Not-determined</td></tr>
<tr><td>S3_extracted_k141_77</td><td>Not-determined</td></tr>
<tr><td>S3_extracted_k141_78</td><td>Low-quality</td></tr>
<tr><td>S3_extracted_k141_47948</td><td>Not-determined</td></tr>
<tr><td>S3_extracted_k141_71819</td><td>Not-determined</td></tr>
<tr><td>S3_extracted_k141_24055</td><td>Not-determined</td></tr>
<tr><td>S3_extracted_k141_55879</td><td>Low-quality</td></tr>
<tr><td>S3_extracted_k141_63830</td><td>Not-determined</td></tr>
<tr><td>S3_extracted_k141_15985</td><td>Not-determined</td></tr>
<tr><td>S3_extracted_k141_47967</td><td>Not-determined</td></tr>
<tr><td>S3_extracted_k141_87690</td><td>Not-determined</td></tr>
<tr><td>S3_extracted_k141_79767</td><td>Not-determined</td></tr>
<tr><td>S3_extracted_k141_95729</td><td>Low-quality</td></tr>
<tr><td>S3_extracted_k141_95731</td><td>Not-determined</td></tr>
<tr><td>S3_extracted_k141_47975</td><td>Not-determined</td></tr>
<tr><td>S3_extracted_k141_47980</td><td>Not-determined</td></tr>
<tr><td>S3_extracted_k141_71839</td><td>Not-determined</td></tr>
<tr><td>S3_extracted_k141_111664</td><td>Not-determined</td></tr>
<tr><td>S3_extracted_k141_87692</td><td>Not-determined</td></tr>
<tr><td>S3_extracted_k141_87693</td><td>Not-determined</td></tr>
<tr><td>S3_extracted_k141_79771</td><td>Not-determined</td></tr>
<tr><td>S3_extracted_k141_24071</td><td>Not-determined</td></tr>
<tr><td>S3_extracted_k141_31936</td><td>Not-determined</td></tr>
<tr><td>S3_extracted_k141_79783</td><td>Not-determined</td></tr>
<tr><td>S3_extracted_k141_24076</td><td>Low-quality</td></tr>
<tr><td>S3_extracted_k141_87705</td><td>Low-quality</td></tr>
<tr><td>S3_extracted_k141_24079</td><td>Not-determined</td></tr>
<tr><td>S3_extracted_k141_24080</td><td>Not-determined</td></tr>
<tr><td>S3_extracted_k141_71851</td><td>Not-determined</td></tr>
<tr><td>S3_extracted_k141_119611</td><td>Low-quality</td></tr>
<tr><td>S3_extracted_k141_71854</td><td>Not-determined</td></tr>
<tr><td>S3_extracted_k141_87710</td><td>Low-quality</td></tr>
<tr><td>S3_extracted_k141_47988</td><td>Not-determined</td></tr>
<tr><td>S3_extracted_k141_24090</td><td>Not-determined</td></tr>
<tr><td>S15_extracted_17857</td><td>Medium-quality</td></tr>
<tr><td>S3_extracted_k141_47989</td><td>Not-determined</td></tr>
<tr><td>S3_extracted_k141_47990</td><td>Not-determined</td></tr>
<tr><td>S3_extracted_k141_24093</td><td>Not-determined</td></tr>
<tr><td>S3_extracted_k141_24097</td><td>Not-determined</td></tr>
<tr><td>S3_extracted_k141_47992</td><td>Not-determined</td></tr>
<tr><td>S3_extracted_k141_103646</td><td>Low-quality</td></tr>
<tr><td>S3_extracted_k141_47994</td><td>Low-quality</td></tr>
<tr><td>S3_extracted_k141_103650</td><td>Not-determined</td></tr>
<tr><td>S3_extracted_k141_55913</td><td>Low-quality</td></tr>
<tr><td>S3_extracted_k141_48001</td><td>Not-determined</td></tr>
<tr><td>S3_extracted_k141_39891</td><td>Not-determined</td></tr>
<tr><td>S3_extracted_k141_48004</td><td>Low-quality</td></tr>
<tr><td>S3_extracted_k141_39895</td><td>Not-determined</td></tr>
<tr><td>S3_extracted_k141_87719</td><td>Not-determined</td></tr>
<tr><td>S3_extracted_k141_119629</td><td>Not-determined</td></tr>
<tr><td>S3_extracted_k141_39898</td><td>Not-determined</td></tr>
<tr><td>S3_extracted_k141_8077</td><td>Not-determined</td></tr>
<tr><td>S3_extracted_k141_8078</td><td>Not-determined</td></tr>
<tr><td>S3_extracted_k141_119636</td><td>Not-determined</td></tr>
<tr><td>S3_extracted_k141_63852</td><td>Not-determined</td></tr>
<tr><td>S3_extracted_k141_55917</td><td>Not-determined</td></tr>
<tr><td>S3_extracted_k141_119639</td><td>Not-determined</td></tr>
<tr><td>S3_extracted_k141_71876</td><td>Low-quality</td></tr>
<tr><td>S3_extracted_k141_71877</td><td>Not-determined</td></tr>
<tr><td>S3_extracted_k141_71878</td><td>Not-determined</td></tr>
<tr><td>S3_extracted_k141_55923</td><td>Not-determined</td></tr>
<tr><td>S3_extracted_k141_71881</td><td>Not-determined</td></tr>
<tr><td>S3_extracted_k141_24110</td><td>Not-determined</td></tr>
<tr><td>S3_extracted_k141_103653</td><td>Low-quality</td></tr>
<tr><td>S3_extracted_k141_24115</td><td>Not-determined</td></tr>
<tr><td>S3_extracted_k141_31949</td><td>Not-determined</td></tr>
<tr><td>S3_extracted_k141_103657</td><td>Low-quality</td></tr>
<tr><td>S3_extracted_k141_24130</td><td>Not-determined</td></tr>
<tr><td>S3_extracted_k141_111678</td><td>Not-determined</td></tr>
<tr><td>S3_extracted_k141_8093</td><td>Low-quality</td></tr>
<tr><td>S3_extracted_k141_48032</td><td>Low-quality</td></tr>
<tr><td>S3_extracted_k141_111680</td><td>Not-determined</td></tr>
<tr><td>S3_extracted_k141_8094</td><td>Low-quality</td></tr>
<tr><td>S3_extracted_k141_115</td><td>Not-determined</td></tr>
<tr><td>S3_extracted_k141_71885</td><td>Not-determined</td></tr>
<tr><td>S3_extracted_k141_71886</td><td>Not-determined</td></tr>
<tr><td>S3_extracted_k141_111685</td><td>Not-determined</td></tr>
<tr><td>S3_extracted_k141_24132</td><td>Not-determined</td></tr>
<tr><td>S3_extracted_k141_111686</td><td>Not-determined</td></tr>
<tr><td>S3_extracted_k141_48040</td><td>Not-determined</td></tr>
<tr><td>S3_extracted_k141_8102</td><td>Low-quality</td></tr>
<tr><td>S3_extracted_k141_111691</td><td>Low-quality</td></tr>
<tr><td>S3_extracted_k141_111692</td><td>Not-determined</td></tr>
<tr><td>S3_extracted_k141_55950</td><td>Not-determined</td></tr>
<tr><td>S3_extracted_k141_24135</td><td>Low-quality</td></tr>
<tr><td>S3_extracted_k141_71891</td><td>Not-determined</td></tr>
<tr><td>S3_extracted_k141_71892</td><td>Not-determined</td></tr>
<tr><td>S3_extracted_k141_24136</td><td>Not-determined</td></tr>
<tr><td>S3_extracted_k141_111694</td><td>Not-determined</td></tr>
<tr><td>S3_extracted_k141_71894</td><td>Not-determined</td></tr>
<tr><td>S3_extracted_k141_24139</td><td>Not-determined</td></tr>
<tr><td>S3_extracted_k141_8113</td><td>Not-determined</td></tr>
<tr><td>S3_extracted_k141_71895</td><td>Low-quality</td></tr>
<tr><td>S3_extracted_k141_111696</td><td>Not-determined</td></tr>
<tr><td>S3_extracted_k141_95739</td><td>Low-quality</td></tr>
<tr><td>S3_extracted_k141_63887</td><td>Not-determined</td></tr>
<tr><td>S3_extracted_k141_71900</td><td>Not-determined</td></tr>
<tr><td>S3_extracted_k141_111699</td><td>Low-quality</td></tr>
<tr><td>S3_extracted_k141_119656</td><td>Not-determined</td></tr>
<tr><td>S3_extracted_k141_136</td><td>Not-determined</td></tr>
<tr><td>S3_extracted_k141_137</td><td>Not-determined</td></tr>
<tr><td>S3_extracted_k141_138</td><td>Not-determined</td></tr>
<tr><td>S3_extracted_k141_8118</td><td>Not-determined</td></tr>
<tr><td>S3_extracted_k141_87730</td><td>Not-determined</td></tr>
<tr><td>S3_extracted_k141_111708</td><td>Not-determined</td></tr>
<tr><td>S3_extracted_k141_8122</td><td>Not-determined</td></tr>
<tr><td>S3_extracted_k141_55953</td><td>Not-determined</td></tr>
<tr><td>S3_extracted_k141_8126</td><td>Not-determined</td></tr>
<tr><td>S3_extracted_k141_87738</td><td>Not-determined</td></tr>
<tr><td>S3_extracted_k141_39937</td><td>Not-determined</td></tr>
<tr><td>S3_extracted_k141_87739</td><td>Not-determined</td></tr>
<tr><td>S3_extracted_k141_8132</td><td>Not-determined</td></tr>
<tr><td>S3_extracted_k141_55955</td><td>Not-determined</td></tr>
<tr><td>S3_extracted_k141_8139</td><td>Low-quality</td></tr>
<tr><td>S3_extracted_k141_8141</td><td>Low-quality</td></tr>
<tr><td>S3_extracted_k141_55962</td><td>Not-determined</td></tr>
<tr><td>S3_extracted_k141_79795</td><td>Not-determined</td></tr>
<tr><td>S3_extracted_k141_71911</td><td>Not-determined</td></tr>
<tr><td>S3_extracted_k141_71915</td><td>Not-determined</td></tr>
<tr><td>S3_extracted_k141_103673</td><td>Low-quality</td></tr>
<tr><td>S3_extracted_k141_55976</td><td>Not-determined</td></tr>
<tr><td>S3_extracted_k141_71920</td><td>Low-quality</td></tr>
<tr><td>S3_extracted_k141_71922</td><td>Not-determined</td></tr>
<tr><td>S3_extracted_k141_79804</td><td>Not-determined</td></tr>
<tr><td>S3_extracted_k141_79806</td><td>Not-determined</td></tr>
<tr><td>S3_extracted_k141_63898</td><td>Not-determined</td></tr>
<tr><td>S3_extracted_k141_55987</td><td>Low-quality</td></tr>
<tr><td>S3_extracted_k141_55989</td><td>Not-determined</td></tr>
<tr><td>S3_extracted_k141_63905</td><td>Not-determined</td></tr>
<tr><td>S3_extracted_k141_71930</td><td>Not-determined</td></tr>
<tr><td>S3_extracted_k141_95758</td><td>Not-determined</td></tr>
<tr><td>S3_extracted_k141_95759</td><td>Not-determined</td></tr>
<tr><td>S3_extracted_k141_63911</td><td>Not-determined</td></tr>
<tr><td>S3_extracted_k141_87768</td><td>Low-quality</td></tr>
<tr><td>S3_extracted_k141_55992</td><td>Not-determined</td></tr>
<tr><td>S3_extracted_k141_79811</td><td>Not-determined</td></tr>
<tr><td>S3_extracted_k141_79812</td><td>Not-determined</td></tr>
<tr><td>S3_extracted_k141_55997</td><td>Not-determined</td></tr>
<tr><td>S3_extracted_k141_143</td><td>Not-determined</td></tr>
<tr><td>S3_extracted_k141_87776</td><td>Not-determined</td></tr>
<tr><td>S3_extracted_k141_87781</td><td>Not-determined</td></tr>
<tr><td>S3_extracted_k141_87784</td><td>Low-quality</td></tr>
<tr><td>S3_extracted_k141_111724</td><td>Not-determined</td></tr>
<tr><td>S3_extracted_k141_154</td><td>Not-determined</td></tr>
<tr><td>S3_extracted_k141_48097</td><td>Not-determined</td></tr>
<tr><td>S3_extracted_k141_8157</td><td>Not-determined</td></tr>
<tr><td>S3_extracted_k141_48099</td><td>Low-quality</td></tr>
<tr><td>S3_extracted_k141_95762</td><td>Not-determined</td></tr>
<tr><td>S3_extracted_k141_31976</td><td>Not-determined</td></tr>
<tr><td>S3_extracted_k141_31977</td><td>Not-determined</td></tr>
<tr><td>S3_extracted_k141_119661</td><td>Not-determined</td></tr>
<tr><td>S3_extracted_k141_79820</td><td>Not-determined</td></tr>
<tr><td>S3_extracted_k141_119667</td><td>Not-determined</td></tr>
<tr><td>S3_extracted_k141_48110</td><td>Low-quality</td></tr>
<tr><td>S3_extracted_k141_63931</td><td>Not-determined</td></tr>
<tr><td>S3_extracted_k141_31986</td><td>Not-determined</td></tr>
<tr><td>S3_extracted_k141_63933</td><td>Not-determined</td></tr>
<tr><td>S3_extracted_k141_119678</td><td>Not-determined</td></tr>
<tr><td>S3_extracted_k141_63939</td><td>Not-determined</td></tr>
<tr><td>S3_extracted_k141_31988</td><td>Not-determined</td></tr>
<tr><td>S3_extracted_k141_31991</td><td>Not-determined</td></tr>
<tr><td>S3_extracted_k141_87790</td><td>Low-quality</td></tr>
<tr><td>S3_extracted_k141_63951</td><td>Not-determined</td></tr>
<tr><td>S3_extracted_k141_31999</td><td>Not-determined</td></tr>
<tr><td>S3_extracted_k141_103695</td><td>Not-determined</td></tr>
<tr><td>S3_extracted_k141_39948</td><td>Low-quality</td></tr>
<tr><td>S3_extracted_k141_32008</td><td>Low-quality</td></tr>
<tr><td>S3_extracted_k141_79837</td><td>Not-determined</td></tr>
<tr><td>S3_extracted_k141_63962</td><td>Not-determined</td></tr>
<tr><td>S3_extracted_k141_119684</td><td>Not-determined</td></tr>
<tr><td>S3_extracted_k141_176</td><td>Not-determined</td></tr>
<tr><td>S3_extracted_k141_111729</td><td>Low-quality</td></tr>
<tr><td>S3_extracted_k141_111730</td><td>Low-quality</td></tr>
<tr><td>S3_extracted_k141_56007</td><td>Not-determined</td></tr>
<tr><td>S3_extracted_k141_56008</td><td>Not-determined</td></tr>
<tr><td>S3_extracted_k141_103708</td><td>Not-determined</td></tr>
<tr><td>S3_extracted_k141_111735</td><td>Low-quality</td></tr>
<tr><td>S3_extracted_k141_79846</td><td>Not-determined</td></tr>
<tr><td>S3_extracted_k141_32012</td><td>Low-quality</td></tr>
<tr><td>S3_extracted_k141_103715</td><td>Low-quality</td></tr>
<tr><td>S3_extracted_k141_111741</td><td>Not-determined</td></tr>
<tr><td>S3_extracted_k141_103716</td><td>Not-determined</td></tr>
<tr><td>S3_extracted_k141_32017</td><td>Not-determined</td></tr>
<tr><td>S3_extracted_k141_182</td><td>Not-determined</td></tr>
<tr><td>S3_extracted_k141_103722</td><td>Not-determined</td></tr>
<tr><td>S3_extracted_k141_39966</td><td>Not-determined</td></tr>
<tr><td>S3_extracted_k141_39967</td><td>Not-determined</td></tr>
<tr><td>S3_extracted_k141_39969</td><td>Not-determined</td></tr>
<tr><td>S3_extracted_k141_32032</td><td>Not-determined</td></tr>
<tr><td>S3_extracted_k141_87800</td><td>Not-determined</td></tr>
<tr><td>S3_extracted_k141_103736</td><td>Not-determined</td></tr>
<tr><td>S3_extracted_k141_32039</td><td>Not-determined</td></tr>
<tr><td>S3_extracted_k141_71941</td><td>Low-quality</td></tr>
<tr><td>S3_extracted_k141_79869</td><td>Not-determined</td></tr>
<tr><td>S3_extracted_k141_48138</td><td>Not-determined</td></tr>
<tr><td>S15_vs_17857||full</td><td>Medium-quality</td></tr>
<tr><td>S3_extracted_k141_8169</td><td>Not-determined</td></tr>
<tr><td>S3_extracted_k141_39977</td><td>Not-determined</td></tr>
<tr><td>S3_extracted_k141_79878</td><td>Low-quality</td></tr>
<tr><td>S3_extracted_k141_39979</td><td>Not-determined</td></tr>
<tr><td>S3_extracted_k141_95782</td><td>Not-determined</td></tr>
<tr><td>S3_extracted_k141_39981</td><td>Low-quality</td></tr>
<tr><td>S3_extracted_k141_111762</td><td>Not-determined</td></tr>
<tr><td>S3_extracted_k141_63981</td><td>Low-quality</td></tr>
<tr><td>S3_extracted_k141_103758</td><td>Not-determined</td></tr>
<tr><td>S3_extracted_k141_103759</td><td>Not-determined</td></tr>
<tr><td>S3_extracted_k141_48153</td><td>Not-determined</td></tr>
<tr><td>S3_extracted_k141_203</td><td>Not-determined</td></tr>
<tr><td>S3_extracted_k141_79889</td><td>Not-determined</td></tr>
<tr><td>S3_extracted_k141_119702</td><td>Not-determined</td></tr>
<tr><td>S3_extracted_k141_79891</td><td>Not-determined</td></tr>
<tr><td>S3_extracted_k141_111764</td><td>Not-determined</td></tr>
<tr><td>S3_extracted_k141_48159</td><td>Not-determined</td></tr>
<tr><td>S3_extracted_k141_87806</td><td>Low-quality</td></tr>
<tr><td>S3_extracted_k141_208</td><td>Not-determined</td></tr>
<tr><td>S3_extracted_k141_56017</td><td>Not-determined</td></tr>
<tr><td>S3_extracted_k141_32051</td><td>Not-determined</td></tr>
<tr><td>S3_extracted_k141_32053</td><td>Low-quality</td></tr>
<tr><td>S3_extracted_k141_214</td><td>Not-determined</td></tr>
<tr><td>S3_extracted_k141_79901</td><td>Not-determined</td></tr>
<tr><td>S3_extracted_k141_215</td><td>Not-determined</td></tr>
<tr><td>S3_extracted_k141_39992</td><td>Not-determined</td></tr>
<tr><td>S3_extracted_k141_63997</td><td>Not-determined</td></tr>
<tr><td>S3_extracted_k141_39996</td><td>Not-determined</td></tr>
<tr><td>S3_extracted_k141_24182</td><td>Low-quality</td></tr>
<tr><td>S3_extracted_k141_8177</td><td>Not-determined</td></tr>
<tr><td>S3_extracted_k141_8183</td><td>Not-determined</td></tr>
<tr><td>S3_extracted_k141_56031</td><td>Not-determined</td></tr>
<tr><td>S3_extracted_k141_95809</td><td>Not-determined</td></tr>
<tr><td>S3_extracted_k141_95810</td><td>Not-determined</td></tr>
<tr><td>S3_extracted_k141_56034</td><td>Not-determined</td></tr>
<tr><td>S3_extracted_k141_87808</td><td>Low-quality</td></tr>
<tr><td>S3_extracted_k141_87811</td><td>Not-determined</td></tr>
<tr><td>S3_extracted_k141_87813</td><td>Low-quality</td></tr>
<tr><td>S3_extracted_k141_8194</td><td>Not-determined</td></tr>
<tr><td>S3_extracted_k141_220</td><td>Not-determined</td></tr>
<tr><td>S3_extracted_k141_79918</td><td>Low-quality</td></tr>
<tr><td>S3_extracted_k141_79919</td><td>Not-determined</td></tr>
<tr><td>S3_extracted_k141_79920</td><td>Low-quality</td></tr>
<tr><td>S3_extracted_k141_48189</td><td>Not-determined</td></tr>
<tr><td>S3_extracted_k141_226</td><td>Not-determined</td></tr>
<tr><td>S3_extracted_k141_111769</td><td>Not-determined</td></tr>
<tr><td>S3_extracted_k141_79922</td><td>Low-quality</td></tr>
<tr><td>S3_extracted_k141_71952</td><td>Not-determined</td></tr>
<tr><td>S3_extracted_k141_95844</td><td>Not-determined</td></tr>
<tr><td>S3_extracted_k141_8208</td><td>Not-determined</td></tr>
<tr><td>S3_extracted_k141_64011</td><td>Not-determined</td></tr>
<tr><td>S3_extracted_k141_64013</td><td>Not-determined</td></tr>
<tr><td>S3_extracted_k141_40009</td><td>Not-determined</td></tr>
<tr><td>S3_extracted_k141_24190</td><td>Not-determined</td></tr>
<tr><td>S3_extracted_k141_71961</td><td>Not-determined</td></tr>
<tr><td>S3_extracted_k141_71968</td><td>Not-determined</td></tr>
<tr><td>S3_extracted_k141_71969</td><td>Not-determined</td></tr>
<tr><td>S3_extracted_k141_32076</td><td>Not-determined</td></tr>
<tr><td>S3_extracted_k141_71974</td><td>Not-determined</td></tr>
<tr><td>S3_extracted_k141_79939</td><td>Not-determined</td></tr>
<tr><td>S3_extracted_k141_56041</td><td>Not-determined</td></tr>
<tr><td>S3_extracted_k141_234</td><td>Not-determined</td></tr>
<tr><td>S3_extracted_k141_32085</td><td>Low-quality</td></tr>
<tr><td>S3_extracted_k141_56043</td><td>Low-quality</td></tr>
<tr><td>S3_extracted_k141_24205</td><td>Not-determined</td></tr>
<tr><td>S3_extracted_k141_64018</td><td>Not-determined</td></tr>
<tr><td>S3_extracted_k141_56047</td><td>Not-determined</td></tr>
<tr><td>S3_extracted_k141_243</td><td>Not-determined</td></tr>
<tr><td>S3_extracted_k141_247</td><td>Not-determined</td></tr>
<tr><td>S3_extracted_k141_79942</td><td>Not-determined</td></tr>
<tr><td>S3_extracted_k141_95866</td><td>Not-determined</td></tr>
<tr><td>S3_extracted_k141_79945</td><td>Not-determined</td></tr>
<tr><td>S3_extracted_k141_24213</td><td>Not-determined</td></tr>
<tr><td>S3_extracted_k141_8233</td><td>Not-determined</td></tr>
<tr><td>S3_extracted_k141_40018</td><td>Not-determined</td></tr>
<tr><td>S3_extracted_k141_40021</td><td>Not-determined</td></tr>
<tr><td>S3_extracted_k141_40023</td><td>Not-determined</td></tr>
<tr><td>S3_extracted_k141_259</td><td>Low-quality</td></tr>
<tr><td>S3_extracted_k141_71998</td><td>Low-quality</td></tr>
<tr><td>S3_extracted_k141_95885</td><td>Not-determined</td></tr>
<tr><td>S3_extracted_k141_40028</td><td>Low-quality</td></tr>
<tr><td>S3_extracted_k141_32100</td><td>Not-determined</td></tr>
<tr><td>S3_extracted_k141_64036</td><td>Not-determined</td></tr>
<tr><td>S3_extracted_k141_64037</td><td>Not-determined</td></tr>
<tr><td>S3_extracted_k141_8251</td><td>Not-determined</td></tr>
<tr><td>S3_extracted_k141_32103</td><td>Not-determined</td></tr>
<tr><td>S3_extracted_k141_72003</td><td>Not-determined</td></tr>
<tr><td>S3_extracted_k141_40039</td><td>Not-determined</td></tr>
</table>

| | |
|---|---|
| S3_extracted_k141_40040 | Low-quality |
| S3_extracted_k141_40041 | Low-quality |
| S3_extracted_k141_8258 | Not-determined |
| S3_extracted_k141_40043 | Low-quality |
| S3_extracted_k141_265 | Not-determined |
| S3_extracted_k141_32114 | Not-determined |
| S3_extracted_k141_32115 | Not-determined |
| S3_extracted_k141_268 | Not-determined |
| S3_extracted_k141_40051 | Low-quality |
| S3_extracted_k141_24228 | Not-determined |
| S3_extracted_k141_95914 | Not-determined |
| S3_extracted_k141_64060 | Not-determined |
| S3_extracted_k141_40058 | Low-quality |
| S3_extracted_k141_87840 | Low-quality |
| S3_extracted_k141_40061 | Not-determined |
| S3_extracted_k141_40064 | Not-determined |
| S3_extracted_k141_87845 | Not-determined |
| S3_extracted_k141_64063 | Not-determined |
| S3_extracted_k141_95923 | Low-quality |
| S3_extracted_k141_95926 | Not-determined |
| S3_extracted_k141_8268 | Not-determined |
| S3_extracted_k141_79962 | Not-determined |
| S3_extracted_k141_79966 | Not-determined |
| S3_extracted_k141_72017 | Not-determined |
| S3_extracted_k141_72021 | Not-determined |
| S3_extracted_k141_8280 | Not-determined |
| S3_extracted_k141_103776 | Not-determined |
| S3_extracted_k141_8283 | Not-determined |
| S3_extracted_k141_16009 | Not-determined |
| S3_extracted_k141_8284 | Low-quality |
| S3_extracted_k141_8292 | Not-determined |
| S3_extracted_k141_72023 | Not-determined |
| S3_extracted_k141_72024 | Not-determined |
| S3_extracted_k141_24250 | Not-determined |
| S3_extracted_k141_56078 | Not-determined |
| S3_extracted_k141_95950 | Low-quality |
| S3_extracted_k141_79967 | Not-determined |
| S3_extracted_k141_24255 | Not-determined |
| S3_extracted_k141_95954 | Not-determined |
| S3_extracted_k141_95960 | Not-determined |
| S3_extracted_k141_8299 | Not-determined |
| S3_extracted_k141_95961 | Not-determined |
| S3_extracted_k141_56087 | Not-determined |
| S3_extracted_k141_56089 | Not-determined |
| S3_extracted_k141_56090 | Not-determined |
| S3_extracted_k141_56092 | Not-determined |
| S3_extracted_k141_8307 | Not-determined |
| S3_extracted_k141_87851 | Not-determined |
| S3_extracted_k141_8314 | Not-determined |
| S3_extracted_k141_48260 | Not-determined |
| S3_extracted_k141_8316 | Not-determined |
| S3_extracted_k141_8317 | Not-determined |
| S3_extracted_k141_16035 | Not-determined |
| S3_extracted_k141_72028 | Not-determined |
| S3_extracted_k141_48266 | Not-determined |
| S3_extracted_k141_103806 | Not-determined |
| S3_extracted_k141_16045 | Low-quality |
| S3_extracted_k141_48272 | Not-determined |
| S3_extracted_k141_103811 | Not-determined |
| S3_extracted_k141_48276 | Not-determined |
| S3_extracted_k141_64076 | Not-determined |
| S3_extracted_k141_40090 | Not-determined |
| S3_extracted_k141_103819 | Not-determined |
| S3_extracted_k141_8330 | Not-determined |
| S3_extracted_k141_32132 | Not-determined |
| S3_extracted_k141_103824 | Not-determined |
| S8_vs_k141_151384\|\|full | High-quality |
| S3_extracted_k141_24261 | Not-determined |
| S3_extracted_k141_24268 | Low-quality |
| S3_extracted_k141_24270 | Not-determined |
| S3_extracted_k141_72039 | Not-determined |
| S3_extracted_k141_24274 | Not-determined |
| S3_extracted_k141_72041 | Not-determined |
| S3_extracted_k141_72042 | Not-determined |
| S3_extracted_k141_40109 | Not-determined |
| S3_extracted_k141_119727 | Not-determined |
| S3_extracted_k141_119728 | Not-determined |
| S3_extracted_k141_72046 | Not-determined |
| S3_extracted_k141_16054 | Low-quality |
| S3_extracted_k141_40115 | Not-determined |
| S3_extracted_k141_24277 | Low-quality |
| S3_extracted_k141_119734 | Low-quality |
| S3_extracted_k141_95975 | Low-quality |
| S3_extracted_k141_8347 | Not-determined |
| S3_extracted_k141_48294 | Not-determined |
| S3_extracted_k141_111786 | Low-quality |
| S3_extracted_k141_56107 | Not-determined |
| S3_extracted_k141_8355 | Not-determined |
| S3_extracted_k141_79992 | Not-determined |
| S3_extracted_k141_103848 | Not-determined |
| S3_extracted_k141_72054 | Not-determined |
| S3_extracted_k141_72055 | Not-determined |
| S3_extracted_k141_119735 | Not-determined |
| S3_extracted_k141_95986 | Low-quality |
| S3_extracted_k141_95987 | Not-determined |
| S3_extracted_k141_16070 | Not-determined |
| S3_extracted_k141_48304 | Not-determined |
| S3_extracted_k141_16074 | Not-determined |
| S3_extracted_k141_326 | Not-determined |
| S3_extracted_k141_64106 | Not-determined |
| S3_extracted_k141_16076 | Low-quality |
| S3_extracted_k141_64110 | Not-determined |
| S3_extracted_k141_16078 | Not-determined |
| S3_extracted_k141_48313 | Not-determined |
| S3_extracted_k141_8358 | Low-quality |
| S3_extracted_k141_111799 | Not-determined |
| S3_extracted_k141_48316 | Not-determined |
| S3_extracted_k141_119751 | Not-determined |
| S3_extracted_k141_16089 | Not-determined |
| S3_extracted_k141_345 | Not-determined |
| S3_extracted_k141_119754 | Not-determined |
| S3_extracted_k141_16094 | Not-determined |
| S3_extracted_k141_350 | Not-determined |
| S3_extracted_k141_8371 | Not-determined |
| S3_extracted_k141_48324 | Not-determined |
| S3_extracted_k141_355 | Not-determined |
| S3_extracted_k141_87878 | Not-determined |
| S3_extracted_k141_119766 | Not-determined |
| S3_extracted_k141_358 | Not-determined |
| S3_extracted_k141_56127 | Not-determined |
| S3_extracted_k141_111808 | Not-determined |
| S3_extracted_k141_360 | Not-determined |
| S3_extracted_k141_8381 | Not-determined |
| S3_extracted_k141_96001 | Low-quality |
| S3_extracted_k141_40140 | Not-determined |
| S3_extracted_k141_103862 | Low-quality |
| S3_extracted_k141_103864 | Not-determined |
| S3_extracted_k141_111818 | Not-determined |
| S3_extracted_k141_96025 | Not-determined |
| S3_extracted_k141_103877 | Not-determined |
| S3_extracted_k141_96026 | Low-quality |
| S3_extracted_k141_96027 | Low-quality |
| S3_extracted_k141_56135 | Not-determined |
| S3_extracted_k141_96029 | Not-determined |
| S3_extracted_k141_24282 | Not-determined |
| S3_extracted_k141_24285 | Not-determined |
| S3_extracted_k141_8403 | Not-determined |
| S3_extracted_k141_80006 | Not-determined |
| S3_extracted_k141_24290 | Low-quality |
| S3_extracted_k141_56149 | Not-determined |
| S3_extracted_k141_8409 | Not-determined |
| S3_extracted_k141_80012 | Not-determined |
| S3_extracted_k141_96035 | Not-determined |
| S3_extracted_k141_111824 | Not-determined |
| S3_extracted_k141_72066 | Not-determined |
| S3_extracted_k141_56154 | Not-determined |
| S3_extracted_k141_24295 | Not-determined |
| S3_extracted_k141_16100 | Not-determined |
| S3_extracted_k141_32172 | Not-determined |
| S3_extracted_k141_16102 | Not-determined |
| S3_extracted_k141_16103 | Not-determined |
| S3_extracted_k141_119774 | Not-determined |
| S3_extracted_k141_24301 | Not-determined |
| S3_extracted_k141_32176 | Low-quality |
| S3_extracted_k141_56162 | Low-quality |
| S3_extracted_k141_40151 | Low-quality |
| S3_extracted_k141_32177 | Not-determined |
| S3_extracted_k141_40152 | Not-determined |
| S3_extracted_k141_56164 | Low-quality |
| S3_extracted_k141_72076 | Not-determined |
| S3_extracted_k141_72077 | Low-quality |
| S3_extracted_k141_24303 | Not-determined |
| S3_extracted_k141_24304 | Not-determined |
| S3_extracted_k141_8419 | Not-determined |
| S3_extracted_k141_48366 | Low-quality |
| S3_extracted_k141_103897 | Not-determined |
| S3_extracted_k141_111833 | Not-determined |
| S3_extracted_k141_64126 | Low-quality |
| S3_extracted_k141_8428 | Not-determined |
| S3_extracted_k141_64129 | Not-determined |
| S3_extracted_k141_64130 | Low-quality |
| S3_extracted_k141_32194 | Not-determined |
| S3_extracted_k141_32195 | Not-determined |
| S3_extracted_k141_119787 | Not-determined |
| S3_extracted_k141_119791 | Not-determined |
| S10_extracted_20810 | Complete |
| S3_extracted_k141_40169 | Not-determined |
| S3_extracted_k141_24317 | Not-determined |
| S3_extracted_k141_80029 | Not-determined |
| S3_extracted_k141_24319 | Not-determined |
| S3_extracted_k141_8430 | Low-quality |
| S3_extracted_k141_24321 | Not-determined |
| S3_extracted_k141_8432 | Not-determined |
| S3_extracted_k141_24325 | Not-determined |
| S3_extracted_k141_404 | Low-quality |
| S3_extracted_k141_8440 | Not-determined |
| S3_extracted_k141_8447 | Low-quality |
| S3_extracted_k141_56198 | Not-determined |
| S3_extracted_k141_111839 | Low-quality |
| S3_extracted_k141_96057 | Not-determined |
| S3_extracted_k141_96059 | Not-determined |
| S3_extracted_k141_80037 | Low-quality |
| S3_extracted_k141_103911 | Not-determined |
| S3_extracted_k141_103915 | Not-determined |
| S3_extracted_k141_111845 | Not-determined |
| S3_extracted_k141_111847 | Not-determined |
| S3_extracted_k141_103919 | Not-determined |
| S3_extracted_k141_40174 | Not-determined |
| S3_extracted_k141_111850 | Not-determined |
| S3_extracted_k141_56207 | Not-determined |
| S3_extracted_k141_111851 | Not-determined |
| S3_extracted_k141_56209 | Not-determined |
| S3_extracted_k141_111857 | Low-quality |
| S3_extracted_k141_24338 | Not-determined |
| S3_extracted_k141_111859 | Not-determined |
| S3_extracted_k141_56214 | Not-determined |
| S3_extracted_k141_24348 | Low-quality |
| S3_extracted_k141_96075 | Not-determined |
| S3_extracted_k141_24350 | Low-quality |
| S3_extracted_k141_103929 | Not-determined |
| S3_extracted_k141_32214 | Low-quality |
| S3_extracted_k141_24354 | Not-determined |
| S3_extracted_k141_32222 | Not-determined |
| S3_extracted_k141_111862 | Not-determined |
| S3_extracted_k141_111865 | Not-determined |
| S3_extracted_k141_111866 | Not-determined |
| S3_extracted_k141_40194 | Not-determined |
| S3_extracted_k141_111870 | Not-determined |
| S3_extracted_k141_32233 | Not-determined |
| S3_extracted_k141_40196 | Not-determined |
| S3_extracted_k141_111872 | Not-determined |
| S3_extracted_k141_32237 | Not-determined |
| S3_extracted_k141_119815 | Not-determined |
| S3_extracted_k141_40200 | Low-quality |
| S3_extracted_k141_119819 | Not-determined |
| S3_extracted_k141_96097 | Not-determined |
| S3_extracted_k141_87900 | Not-determined |
| S3_extracted_k141_119821 | Not-determined |
| S3_extracted_k141_87903 | Not-determined |
| S3_extracted_k141_417 | Not-determined |
| S3_extracted_k141_8477 | Not-determined |
| S3_extracted_k141_111887 | Not-determined |
| S3_extracted_k141_111889 | Not-determined |
| S3_extracted_k141_111890 | Not-determined |
| S3_extracted_k141_72096 | Not-determined |
| S3_extracted_k141_40217 | Not-determined |
| S3_extracted_k141_80062 | Not-determined |
| S3_extracted_k141_32252 | Not-determined |
| S3_extracted_k141_32255 | Not-determined |
| S3_extracted_k141_40224 | Not-determined |
| S3_extracted_k141_8487 | Not-determined |
| S3_extracted_k141_40226 | Not-determined |
| S3_extracted_k141_87921 | Not-determined |
| S3_extracted_k141_119826 | Not-determined |
| S3_extracted_k141_24364 | Not-determined |
| S3_extracted_k141_32262 | Not-determined |
| S3_extracted_k141_80076 | Not-determined |
| S3_extracted_k141_16139 | Not-determined |
| S3_extracted_k141_32264 | Not-determined |
| S3_extracted_k141_103951 | Not-determined |
| S3_extracted_k141_96116 | Not-determined |
| S3_extracted_k141_87927 | Not-determined |
| S3_extracted_k141_32272 | Not-determined |
| S3_extracted_k141_24368 | Low-quality |
| S3_extracted_k141_87932 | Low-quality |
| S3_extracted_k141_80094 | Low-quality |
| S3_extracted_k141_103964 | Not-determined |
| S3_extracted_k141_32285 | Not-determined |
| S3_extracted_k141_40249 | Not-determined |
| S3_extracted_k141_80098 | Not-determined |
| S3_extracted_k141_48385 | Not-determined |
| S3_extracted_k141_72104 | Not-determined |
| S3_extracted_k141_119837 | Not-determined |
| S3_extracted_k141_8506 | Not-determined |
| S3_extracted_k141_48390 | Low-quality |
| S3_extracted_k141_24373 | Not-determined |
| S3_extracted_k141_40258 | Not-determined |
| S3_extracted_k141_103981 | Low-quality |
| S3_extracted_k141_40266 | Not-determined |
| S3_extracted_k141_24379 | Not-determined |
| S3_extracted_k141_40269 | Not-determined |
| S3_extracted_k141_24382 | Not-determined |
| S3_extracted_k141_40271 | Not-determined |
| S3_extracted_k141_32288 | Not-determined |
| S3_extracted_k141_48399 | Not-determined |
| S3_extracted_k141_439 | Not-determined |
| S3_extracted_k141_8511 | Low-quality |
| S3_extracted_k141_440 | Not-determined |
| S3_extracted_k141_80111 | Not-determined |
| S3_extracted_k141_87946 | Low-quality |
| S3_extracted_k141_119852 | Low-quality |
| S3_extracted_k141_119855 | Not-determined |
| S3_extracted_k141_80118 | Not-determined |
| S3_extracted_k141_8519 | Low-quality |
| S3_extracted_k141_87953 | Not-determined |
| S3_extracted_k141_445 | Not-determined |
| S3_extracted_k141_87957 | Not-determined |
| S3_extracted_k141_24400 | Low-quality |
| S3_extracted_k141_119864 | Not-determined |
| S3_extracted_k141_87961 | Not-determined |
| S3_extracted_k141_119866 | Not-determined |
| S3_extracted_k141_32312 | Not-determined |
| S3_extracted_k141_103987 | Not-determined |
| S3_extracted_k141_24406 | Not-determined |
| S3_extracted_k141_119875 | Low-quality |
| S3_extracted_k141_32316 | Not-determined |
| S3_extracted_k141_16146 | Not-determined |
| S3_extracted_k141_24413 | Not-determined |
| S3_extracted_k141_24414 | Low-quality |
| S3_extracted_k141_24415 | Not-determined |
| S3_extracted_k141_56234 | Not-determined |
| S3_extracted_k141_87969 | Not-determined |
| S3_extracted_k141_103992 | Not-determined |
| S3_extracted_k141_8531 | Low-quality |
| S3_extracted_k141_87970 | Not-determined |
| S3_extracted_k141_87971 | Not-determined |
| S3_extracted_k141_96123 | Not-determined |
| S3_extracted_k141_87973 | Not-determined |
| S3_extracted_k141_87974 | Not-determined |
| S3_extracted_k141_40280 | Low-quality |
| S3_extracted_k141_8535 | Low-quality |
| S3_extracted_k141_40285 | Not-determined |
| S7_vs_119679\|\|full | High-quality |
| S3_extracted_k141_8539 | Not-determined |
| S3_extracted_k141_56240 | Not-determined |
| S3_extracted_k141_111898 | Not-determined |
| S3_extracted_k141_119881 | Not-determined |
| S3_extracted_k141_111901 | Not-determined |
| S3_extracted_k141_24424 | Low-quality |
| S3_extracted_k141_96131 | Not-determined |
| S3_extracted_k141_40291 | Not-determined |
| S3_extracted_k141_40293 | Not-determined |
| S3_extracted_k141_96132 | Not-determined |
| S3_extracted_k141_64144 | Low-quality |
| S3_extracted_k141_64146 | Low-quality |
| S3_extracted_k141_32344 | Not-determined |
| S3_extracted_k141_64150 | Not-determined |
| S3_extracted_k141_111905 | Not-determined |
| S3_extracted_k141_104010 | Not-determined |
| S3_extracted_k141_24437 | Not-determined |
| S3_extracted_k141_56251 | Not-determined |
| S3_extracted_k141_32352 | Not-determined |
| S3_extracted_k141_8557 | Not-determined |
| S3_extracted_k141_64160 | Not-determined |
| S3_extracted_k141_8559 | Not-determined |
| S3_extracted_k141_40297 | Low-quality |
| S3_extracted_k141_64169 | Not-determined |
| S3_extracted_k141_87977 | Low-quality |
| S3_extracted_k141_87979 | Not-determined |
| S3_extracted_k141_87982 | Not-determined |
| S3_extracted_k141_40303 | Not-determined |
| S3_extracted_k141_87989 | Not-determined |
| S3_extracted_k141_119909 | Not-determined |
| S3_extracted_k141_119910 | Not-determined |
| S3_extracted_k141_473 | Not-determined |
| S3_extracted_k141_87993 | Not-determined |
| S3_extracted_k141_40312 | Not-determined |
| S3_extracted_k141_119917 | Not-determined |
| S3_extracted_k141_96141 | Not-determined |
| S3_extracted_k141_88001 | Not-determined |
| S3_extracted_k141_64175 | Not-determined |
| S3_extracted_k141_32389 | Not-determined |
| S3_extracted_k141_104028 | Not-determined |
| S3_extracted_k141_119931 | Low-quality |
| S3_extracted_k141_8594 | Not-determined |
| S3_extracted_k141_8596 | Not-determined |
| S3_extracted_k141_32397 | Not-determined |
| S3_extracted_k141_119937 | Not-determined |
| S3_extracted_k141_119940 | Not-determined |
| S3_extracted_k141_88004 | Low-quality |
| S3_extracted_k141_88006 | Not-determined |
| S3_extracted_k141_8608 | Not-determined |
| S3_extracted_k141_88009 | Not-determined |
| S3_extracted_k141_32413 | Low-quality |
| S3_extracted_k141_88011 | Not-determined |
| S3_extracted_k141_24456 | Not-determined |
| S3_extracted_k141_40336 | Low-quality |
| S3_extracted_k141_32421 | Not-determined |
| S3_extracted_k141_40342 | Not-determined |
| S3_extracted_k141_111933 | Not-determined |
| S3_extracted_k141_32423 | Not-determined |
| S3_extracted_k141_32427 | Not-determined |
| S3_extracted_k141_8623 | Not-determined |
| S3_extracted_k141_40345 | Low-quality |
| S3_extracted_k141_24463 | Not-determined |
| S3_extracted_k141_88015 | Not-determined |
| S3_extracted_k141_32432 | Not-determined |
| S3_extracted_k141_111941 | Not-determined |
| S3_extracted_k141_96156 | Not-determined |
| S3_extracted_k141_88025 | Not-determined |
| S3_extracted_k141_88026 | Not-determined |
| S3_extracted_k141_508 | Low-quality |
| S3_extracted_k141_119960 | Not-determined |
| S3_extracted_k141_513 | Low-quality |
| S3_extracted_k141_104041 | Not-determined |
| S3_extracted_k141_56253 | Not-determined |
| S3_extracted_k141_56255 | Not-determined |
| S3_extracted_k141_56256 | Not-determined |
| S3_extracted_k141_119965 | Not-determined |
| S3_extracted_k141_40369 | Not-determined |
| S3_extracted_k141_119967 | Not-determined |
| S3_extracted_k141_119968 | Not-determined |
| S3_extracted_k141_104048 | Not-determined |
| S3_extracted_k141_104049 | Not-determined |
| S3_extracted_k141_111944 | Not-determined |
| S3_extracted_k141_56261 | Not-determined |
| S3_extracted_k141_56262 | Not-determined |
| S3_extracted_k141_32439 | Not-determined |
| S3_extracted_k141_111948 | Not-determined |
| S3_extracted_k141_56268 | Not-determined |
| S3_extracted_k141_119976 | Not-determined |
| S3_extracted_k141_32444 | Not-determined |
| S3_extracted_k141_111959 | Not-determined |
| S3_extracted_k141_16166 | Not-determined |
| S3_extracted_k141_32450 | Not-determined |
| S3_extracted_k141_119981 | Low-quality |
| S3_extracted_k141_104056 | Not-determined |
| S3_extracted_k141_119986 | Not-determined |
| S3_extracted_k141_24498 | Low-quality |
| S3_extracted_k141_48414 | Not-determined |
| S3_extracted_k141_64216 | Not-determined |
| S3_extracted_k141_119991 | Not-determined |
| S3_extracted_k141_119992 | Not-determined |
| S3_extracted_k141_104061 | Not-determined |
| S3_extracted_k141_40381 | Low-quality |
| S3_extracted_k141_48431 | Not-determined |
| S3_extracted_k141_32452 | Not-determined |
| S3_extracted_k141_64228 | Not-determined |
| S3_extracted_k141_88032 | Not-determined |
| S3_extracted_k141_88035 | Not-determined |
| S3_extracted_k141_539 | Not-determined |
| S3_extracted_k141_16170 | Not-determined |
| S3_extracted_k141_64230 | Not-determined |
| S3_extracted_k141_547 | Not-determined |
| S3_extracted_k141_553 | Not-determined |
| S3_extracted_k141_72130 | Not-determined |
| S3_extracted_k141_72133 | Not-determined |
| S3_extracted_k141_558 | Not-determined |
| S3_extracted_k141_32469 | Not-determined |
| S3_extracted_k141_24517 | Not-determined |
| S3_extracted_k141_72134 | Not-determined |
| S3_extracted_k141_64245 | Low-quality |
| S3_extracted_k141_8660 | Low-quality |
| S3_extracted_k141_8664 | Not-determined |
| S3_extracted_k141_72137 | Not-determined |
| S3_extracted_k141_8665 | Not-determined |
| S3_extracted_k141_32475 | Not-determined |
| S3_extracted_k141_104069 | Not-determined |
| S3_extracted_k141_64248 | Not-determined |
| S3_extracted_k141_32480 | Not-determined |
| S3_extracted_k141_8669 | Not-determined |
| S3_extracted_k141_32482 | Not-determined |
| S3_extracted_k141_24527 | Not-determined |
| S3_extracted_k141_72146 | Not-determined |
| S3_extracted_k141_72150 | Not-determined |
| S3_extracted_k141_104075 | Not-determined |
| S3_extracted_k141_72155 | Not-determined |
| S3_extracted_k141_40391 | Not-determined |
| S3_extracted_k141_104084 | Not-determined |
| S3_extracted_k141_72159 | Not-determined |
| S3_extracted_k141_569 | Not-determined |
| S3_extracted_k141_88055 | Not-determined |
| S3_extracted_k141_72168 | Not-determined |
| S3_extracted_k141_575 | Not-determined |
| S3_extracted_k141_16193 | Not-determined |
| S3_extracted_k141_48433 | Not-determined |
| S3_extracted_k141_586 | Not-determined |
| S3_extracted_k141_587 | Not-determined |
| S3_extracted_k141_80152 | Not-determined |
| S3_extracted_k141_80159 | Not-determined |
| S3_extracted_k141_96188 | Not-determined |
| S3_extracted_k141_48449 | Not-determined |
| S3_extracted_k141_88065 | Not-determined |
| S3_extracted_k141_80171 | Not-determined |
| S3_extracted_k141_72194 | Not-determined |
| S3_extracted_k141_8703 | Not-determined |
| S3_extracted_k141_8704 | Not-determined |
| S3_extracted_k141_8706 | Not-determined |
| S3_extracted_k141_8708 | Not-determined |
| S3_extracted_k141_88082 | Not-determined |
| S3_extracted_k141_8709 | Not-determined |
| S3_extracted_k141_96194 | Not-determined |
| S3_extracted_k141_8713 | Not-determined |
| S3_extracted_k141_48468 | Not-determined |
| S3_extracted_k141_48471 | Not-determined |
| S3_extracted_k141_32494 | Not-determined |
| S3_extracted_k141_104092 | Not-determined |
| S3_extracted_k141_96202 | Not-determined |
| S3_extracted_k141_104099 | Not-determined |
| S3_extracted_k141_8720 | Low-quality |
| S3_extracted_k141_88099 | Low-quality |
| S3_extracted_k141_80181 | Low-quality |
| S3_extracted_k141_598 | Not-determined |
| S3_extracted_k141_104103 | Not-determined |
| S3_extracted_k141_8728 | Not-determined |
| S3_extracted_k141_104108 | Not-determined |
| S3_extracted_k141_8732 | Not-determined |
| S3_extracted_k141_88101 | Low-quality |
| S3_extracted_k141_72226 | Low-quality |
| S3_extracted_k141_602 | Not-determined |
| S3_extracted_k141_603 | Not-determined |
| S3_extracted_k141_8742 | Not-determined |
| S3_extracted_k141_620 | Not-determined |
| S3_extracted_k141_623 | Not-determined |
| S3_extracted_k141_627 | Not-determined |
| S3_extracted_k141_32512 | Not-determined |
| S3_extracted_k141_8751 | Not-determined |
| S3_extracted_k141_32520 | Not-determined |
| S3_extracted_k141_32523 | Low-quality |
| S3_extracted_k141_56277 | Not-determined |
| S3_extracted_k141_72249 | Low-quality |
| S3_extracted_k141_650 | Not-determined |
| S3_extracted_k141_659 | Not-determined |
| S3_extracted_k141_80186 | Low-quality |
| S3_extracted_k141_111989 | Not-determined |
| S3_extracted_k141_80188 | Low-quality |
| S3_extracted_k141_111991 | Not-determined |
| S3_extracted_k141_80190 | Not-determined |
| S3_extracted_k141_8770 | Not-determined |
| S3_extracted_k141_40452 | Not-determined |
| S3_extracted_k141_80200 | Low-quality |
| S3_extracted_k141_111997 | Low-quality |
| S3_extracted_k141_664 | Not-determined |
| S3_extracted_k141_40458 | Not-determined |
| S3_extracted_k141_72258 | Low-quality |
| S3_extracted_k141_104127 | Not-determined |
| S3_extracted_k141_40463 | Not-determined |
| S3_extracted_k141_104128 | Not-determined |
| S3_extracted_k141_40465 | Not-determined |
| S3_extracted_k141_8789 | Not-determined |
| S3_extracted_k141_72277 | Not-determined |
| S3_extracted_k141_112004 | Not-determined |
| S3_extracted_k141_80208 | Not-determined |
| S3_extracted_k141_104139 | Low-quality |
| S3_extracted_k141_104142 | Not-determined |
| S3_extracted_k141_104143 | Not-determined |
| S3_extracted_k141_64270 | Low-quality |
| S3_extracted_k141_112011 | Not-determined |
| S3_extracted_k141_80215 | Not-determined |
| S3_extracted_k141_8816 | Not-determined |
| S3_extracted_k141_8818 | Not-determined |
| S3_extracted_k141_8819 | Not-determined |
| S3_extracted_k141_112024 | Not-determined |
| S3_extracted_k141_104161 | Low-quality |
| S3_extracted_k141_104164 | Not-determined |
| S3_extracted_k141_8832 | Not-determined |
| S3_extracted_k141_104171 | Low-quality |
| S3_extracted_k141_104174 | Not-determined |
| S3_extracted_k141_112037 | Not-determined |
| S3_extracted_k141_8836 | Not-determined |
| S10_vs_35854\|\|full | Medium-quality |
| S3_extracted_k141_104177 | Low-quality |
| S3_extracted_k141_104179 | Not-determined |
| S3_extracted_k141_96223 | Not-determined |
| S3_extracted_k141_32533 | Not-determined |
| S3_extracted_k141_32538 | Not-determined |
| S3_extracted_k141_16198 | Not-determined |
| S3_extracted_k141_112044 | Not-determined |
| S3_extracted_k141_32547 | Not-determined |
| S3_extracted_k141_32552 | Not-determined |
| S3_extracted_k141_104190 | Low-quality |
| S3_extracted_k141_104194 | Not-determined |
| S3_extracted_k141_8848 | Low-quality |
| S3_extracted_k141_96243 | Low-quality |
| S3_extracted_k141_104198 | Not-determined |
| S3_extracted_k141_72301 | Not-determined |
| S3_extracted_k141_112047 | Low-quality |
| S3_extracted_k141_72303 | Not-determined |
| S3_extracted_k141_88111 | Not-determined |
| S3_extracted_k141_72304 | Not-determined |
| S3_extracted_k141_8858 | Not-determined |
| S3_extracted_k141_72305 | Not-determined |
| S3_extracted_k141_112049 | Low-quality |
| S3_extracted_k141_48526 | Not-determined |
| S3_extracted_k141_32570 | Low-quality |
| S3_extracted_k141_88116 | Not-determined |
| S3_extracted_k141_96252 | Not-determined |
| S3_extracted_k141_88118 | Not-determined |
| S3_extracted_k141_32573 | Low-quality |
| S3_extracted_k141_88120 | Not-determined |
| S3_extracted_k141_88122 | Not-determined |
| S3_extracted_k141_104211 | Not-determined |
| S3_extracted_k141_48539 | Low-quality |
| S3_extracted_k141_705 | Not-determined |
| S3_extracted_k141_24547 | Low-quality |
| S3_extracted_k141_16223 | Not-determined |
| S3_extracted_k141_709 | Not-determined |
| S3_extracted_k141_32580 | Not-determined |
| S3_extracted_k141_8864 | Not-determined |
| S3_extracted_k141_48547 | Not-determined |
| S3_extracted_k141_32589 | Low-quality |
| S3_extracted_k141_88126 | Not-determined |
| S3_extracted_k141_8870 | Not-determined |
| S3_extracted_k141_56297 | Low-quality |
| S3_extracted_k141_72336 | Not-determined |
| S3_extracted_k141_56302 | Not-determined |
| S3_extracted_k141_72343 | Not-determined |
| S3_extracted_k141_56307 | Not-determined |
| S3_extracted_k141_56308 | Not-determined |

| | |
|---|---|
| S3_extracted_k141_48555 | Not-determined |
| S3_extracted_k141_32594 | Low-quality |
| S3_extracted_k141_96278 | Not-determined |
| S3_extracted_k141_72348 | Low-quality |
| S3_extracted_k141_104225 | Not-determined |
| S3_extracted_k141_104228 | Not-determined |
| S3_extracted_k141_48561 | Low-quality |
| S3_extracted_k141_72366 | Not-determined |
| S3_extracted_k141_80245 | Not-determined |
| S3_extracted_k141_72370 | Not-determined |
| S3_extracted_k141_72371 | Low-quality |
| S3_extracted_k141_72377 | Not-determined |
| S3_extracted_k141_88131 | Not-determined |
| S3_extracted_k141_32609 | Low-quality |
| S3_extracted_k141_32610 | Not-determined |
| S3_extracted_k141_56320 | Not-determined |
| S3_extracted_k141_72391 | Low-quality |
| S3_extracted_k141_32617 | Not-determined |
| S3_extracted_k141_721 | Not-determined |
| S3_extracted_k141_48576 | Not-determined |
| S3_extracted_k141_48578 | Not-determined |
| S3_extracted_k141_56324 | Low-quality |
| S3_extracted_k141_56326 | Not-determined |
| S3_extracted_k141_48580 | Not-determined |
| S3_extracted_k141_40503 | Not-determined |
| S3_extracted_k141_48581 | Low-quality |
| S3_extracted_k141_48582 | Not-determined |
| S3_extracted_k141_32622 | Low-quality |
| S3_extracted_k141_728 | Not-determined |
| S3_extracted_k141_56331 | Not-determined |
| S3_extracted_k141_40504 | Not-determined |
| S3_extracted_k141_80273 | Not-determined |
| S3_extracted_k141_72396 | Not-determined |
| S3_extracted_k141_48589 | Not-determined |
| S3_extracted_k141_104238 | Low-quality |
| S3_extracted_k141_104242 | Not-determined |
| S3_extracted_k141_32628 | Not-determined |
| S3_extracted_k141_104244 | Not-determined |
| S3_extracted_k141_56341 | Not-determined |
| S3_extracted_k141_32634 | Not-determined |
| S3_extracted_k141_80286 | Not-determined |
| S3_extracted_k141_24552 | Low-quality |
| S3_extracted_k141_72415 | Not-determined |
| S3_extracted_k141_72417 | Not-determined |
| S3_extracted_k141_40524 | Not-determined |
| S3_extracted_k141_56362 | Not-determined |
| S3_extracted_k141_32655 | Not-determined |
| S3_extracted_k141_120021 | Not-determined |
| S3_extracted_k141_40532 | Not-determined |
| S3_extracted_k141_64281 | Low-quality |
| S3_extracted_k141_88142 | Low-quality |
| S3_extracted_k141_64284 | Low-quality |
| S3_extracted_k141_48616 | Not-determined |
| S3_extracted_k141_72435 | Not-determined |
| S3_extracted_k141_40542 | Not-determined |
| S3_extracted_k141_64287 | Not-determined |
| S3_extracted_k141_64289 | Not-determined |
| S3_extracted_k141_48623 | Not-determined |
| S3_extracted_k141_24561 | Not-determined |
| S3_extracted_k141_48626 | Low-quality |
| S3_extracted_k141_32665 | Low-quality |
| S3_extracted_k141_64295 | Not-determined |
| S3_extracted_k141_40550 | Not-determined |
| S3_extracted_k141_96314 | Low-quality |
| S3_extracted_k141_96317 | Low-quality |
| S3_extracted_k141_8875 | Not-determined |
| S3_extracted_k141_120045 | Low-quality |
| S3_extracted_k141_96320 | Low-quality |
| S3_extracted_k141_8879 | Not-determined |
| S3_extracted_k141_48636 | Low-quality |
| S3_extracted_k141_8881 | Not-determined |
| S3_extracted_k141_32677 | Not-determined |
| S3_extracted_k141_104250 | Not-determined |
| S3_extracted_k141_48639 | Not-determined |
| S3_extracted_k141_96322 | Low-quality |
| S3_extracted_k141_48644 | Not-determined |
| S3_extracted_k141_48645 | Not-determined |
| S3_extracted_k141_96323 | Not-determined |
| S3_extracted_k141_741 | Not-determined |
| S3_extracted_k141_48648 | Not-determined |
| S3_extracted_k141_72454 | Not-determined |
| S3_extracted_k141_64298 | Not-determined |
| S10_vs_20810\|\|full | High-quality |
| S3_extracted_k141_80289 | Not-determined |
| S3_extracted_k141_104259 | Not-determined |
| S3_extracted_k141_80292 | Not-determined |
| S3_extracted_k141_72457 | Not-determined |
| S3_extracted_k141_40567 | Low-quality |
| S3_extracted_k141_80296 | Low-quality |
| S3_extracted_k141_72458 | Not-determined |
| S3_extracted_k141_80298 | Not-determined |
| S3_extracted_k141_72460 | Not-determined |
| S3_extracted_k141_80300 | Not-determined |
| S7_extracted_5439 | Complete |
| S3_extracted_k141_40571 | Not-determined |
| S7_vs_5439\|\|full | High-quality |
| S3_extracted_k141_8907 | Not-determined |
| S3_extracted_k141_80303 | Not-determined |
| S3_extracted_k141_96333 | Not-determined |
| S3_extracted_k141_112070 | Not-determined |
| S3_extracted_k141_751 | Not-determined |
| S3_extracted_k141_56367 | Not-determined |
| S3_extracted_k141_112072 | Low-quality |
| S3_extracted_k141_56368 | Not-determined |
| S3_extracted_k141_8918 | Not-determined |
| S3_extracted_k141_8920 | Not-determined |
| S3_extracted_k141_88161 | Not-determined |
| S3_extracted_k141_56376 | Not-determined |
| S3_extracted_k141_758 | Not-determined |
| S3_extracted_k141_8923 | Not-determined |
| S3_extracted_k141_56380 | Not-determined |
| S3_extracted_k141_40589 | Not-determined |
| S3_extracted_k141_8926 | Low-quality |
| S3_extracted_k141_96344 | Not-determined |
| S3_extracted_k141_40594 | Not-determined |
| S3_extracted_k141_8930 | Not-determined |
| S3_extracted_k141_96347 | Not-determined |
| S3_extracted_k141_48668 | Low-quality |
| S3_extracted_k141_88174 | Not-determined |
| S3_extracted_k141_88175 | Not-determined |
| S3_extracted_k141_80323 | Low-quality |
| S3_extracted_k141_32705 | Not-determined |
| S3_extracted_k141_776 | Not-determined |
| S3_extracted_k141_96353 | Low-quality |
| S3_extracted_k141_8940 | Not-determined |
| S3_extracted_k141_104269 | Not-determined |
| S3_extracted_k141_48670 | Not-determined |
| S3_extracted_k141_72484 | Low-quality |
| S3_extracted_k141_8945 | Not-determined |
| S3_extracted_k141_56392 | Not-determined |
| S3_extracted_k141_104274 | Low-quality |
| S3_extracted_k141_104277 | Not-determined |
| S3_extracted_k141_788 | Not-determined |
| S3_extracted_k141_64326 | Not-determined |
| S3_extracted_k141_88180 | Not-determined |
| S3_extracted_k141_112098 | Not-determined |
| S3_extracted_k141_112100 | Not-determined |
| S3_extracted_k141_96364 | Not-determined |
| S3_extracted_k141_56401 | Not-determined |
| S3_extracted_k141_112108 | Low-quality |
| S3_extracted_k141_793 | Low-quality |
| S3_extracted_k141_24608 | Not-determined |
| S3_extracted_k141_112113 | Not-determined |
| S3_extracted_k141_48684 | Not-determined |
| S3_extracted_k141_88191 | Not-determined |
| S3_extracted_k141_16238 | Not-determined |
| S3_extracted_k141_48688 | Not-determined |
| S3_extracted_k141_120056 | Not-determined |
| S3_extracted_k141_56410 | Low-quality |
| S3_extracted_k141_104292 | Not-determined |
| S3_extracted_k141_40604 | Not-determined |
| S3_extracted_k141_88204 | Not-determined |
| S3_extracted_k141_40605 | Not-determined |
| S3_extracted_k141_798 | Not-determined |
| S3_extracted_k141_800 | Not-determined |
| S3_extracted_k141_32715 | Not-determined |
| S3_extracted_k141_32716 | Low-quality |
| S3_extracted_k141_88213 | Not-determined |
| S3_extracted_k141_48700 | Not-determined |
| S3_extracted_k141_16245 | Not-determined |
| S3_extracted_k141_88215 | Low-quality |
| S3_extracted_k141_40614 | Low-quality |
| S3_extracted_k141_112134 | Not-determined |
| S3_extracted_k141_56421 | Not-determined |
| S3_extracted_k141_40617 | Not-determined |
| S3_extracted_k141_24623 | Low-quality |
| S3_extracted_k141_8950 | Not-determined |
| S3_extracted_k141_32717 | Low-quality |
| S3_extracted_k141_814 | Not-determined |
| S3_extracted_k141_56432 | Low-quality |
| S3_extracted_k141_32723 | Not-determined |
| S3_extracted_k141_8959 | Not-determined |
| S3_extracted_k141_56436 | Not-determined |
| S3_extracted_k141_112145 | Not-determined |
| S3_extracted_k141_32726 | Not-determined |
| S3_extracted_k141_96386 | Not-determined |
| S3_extracted_k141_8968 | Not-determined |
| S3_extracted_k141_112154 | Not-determined |
| S3_extracted_k141_24632 | Not-determined |
| S3_extracted_k141_16250 | Not-determined |
| S3_extracted_k141_56449 | Not-determined |
| S3_extracted_k141_16251 | Not-determined |
| S3_extracted_k141_72510 | Low-quality |
| S3_extracted_k141_8978 | Not-determined |
| S3_extracted_k141_72513 | Not-determined |
| S3_extracted_k141_8979 | Not-determined |
| S3_extracted_k141_8982 | Not-determined |
| S3_extracted_k141_104302 | Low-quality |
| S3_extracted_k141_56457 | Not-determined |
| S3_extracted_k141_72517 | Not-determined |
| S3_extracted_k141_40630 | Low-quality |
| S3_extracted_k141_32729 | Not-determined |
| S3_extracted_k141_48718 | Low-quality |
| S3_extracted_k141_32730 | Not-determined |
| S3_extracted_k141_104303 | Low-quality |
| S3_extracted_k141_88226 | Not-determined |
| S3_extracted_k141_9000 | Not-determined |
| S3_extracted_k141_32740 | Low-quality |
| S3_extracted_k141_72522 | Low-quality |
| S3_extracted_k141_56475 | Not-determined |
| S3_extracted_k141_32750 | Not-determined |
| S3_extracted_k141_16264 | Not-determined |
| S3_extracted_k141_56477 | Low-quality |
| S3_extracted_k141_9006 | Not-determined |
| S3_extracted_k141_96404 | Not-determined |
| S3_extracted_k141_9009 | Not-determined |
| S3_extracted_k141_32757 | Not-determined |
| S3_extracted_k141_32762 | Not-determined |
| S3_extracted_k141_40638 | Not-determined |
| S3_extracted_k141_32764 | Not-determined |
| S3_extracted_k141_112186 | Low-quality |
| S3_extracted_k141_72547 | Low-quality |
| S3_extracted_k141_96420 | Not-determined |
| S3_extracted_k141_40643 | Low-quality |
| S3_extracted_k141_104317 | Not-determined |
| S3_extracted_k141_96426 | Not-determined |
| S3_extracted_k141_96427 | Not-determined |
| S3_extracted_k141_56484 | Not-determined |
| S3_extracted_k141_120099 | Not-determined |
| S3_extracted_k141_72561 | Not-determined |
| S3_extracted_k141_96430 | Not-determined |
| S3_extracted_k141_120103 | Not-determined |
| S3_extracted_k141_9030 | Not-determined |
| S3_extracted_k141_96442 | Not-determined |
| S3_extracted_k141_56503 | Not-determined |
| S3_extracted_k141_96448 | Not-determined |
| S3_extracted_k141_88248 | Not-determined |
| S3_extracted_k141_823 | Not-determined |
| S3_extracted_k141_56505 | Not-determined |
| S3_extracted_k141_72562 | Low-quality |
| S3_extracted_k141_112206 | Not-determined |
| S3_extracted_k141_16269 | Not-determined |
| S3_extracted_k141_56510 | Not-determined |
| S3_extracted_k141_72569 | Not-determined |
| S3_extracted_k141_56512 | Not-determined |
| S3_extracted_k141_80350 | Not-determined |
| S3_extracted_k141_80355 | Not-determined |
| S3_extracted_k141_826 | Low-quality |
| S3_extracted_k141_827 | Not-determined |
| S3_extracted_k141_9046 | Not-determined |
| S3_extracted_k141_16285 | Not-determined |
| S3_extracted_k141_72587 | Low-quality |
| S3_extracted_k141_32803 | Not-determined |
| S3_extracted_k141_32804 | Not-determined |
| S3_extracted_k141_32806 | Not-determined |
| S3_extracted_k141_32807 | Not-determined |
| S3_extracted_k141_32810 | Not-determined |
| S3_extracted_k141_9056 | Not-determined |
| S3_extracted_k141_32814 | Not-determined |
| S3_extracted_k141_40658 | Not-determined |
| S3_extracted_k141_80367 | Low-quality |
| S3_extracted_k141_112213 | Not-determined |
| S3_extracted_k141_32819 | Not-determined |
| S3_extracted_k141_9066 | Not-determined |
| S3_extracted_k141_104341 | Low-quality |
| S3_extracted_k141_9070 | Not-determined |
| S3_extracted_k141_56549 | Not-determined |
| S3_extracted_k141_16302 | Not-determined |
| S3_extracted_k141_56554 | Not-determined |
| S3_extracted_k141_72614 | Not-determined |
| S3_extracted_k141_16306 | Not-determined |
| S3_extracted_k141_834 | Not-determined |
| S3_extracted_k141_72617 | Low-quality |
| S3_extracted_k141_9074 | Not-determined |
| S3_extracted_k141_9076 | Not-determined |
| S3_extracted_k141_839 | Not-determined |
| S3_extracted_k141_72620 | Not-determined |
| S3_extracted_k141_16310 | Not-determined |
| S3_extracted_k141_40672 | Not-determined |
| S3_extracted_k141_32824 | Not-determined |
| S3_extracted_k141_32827 | Not-determined |
| S3_extracted_k141_80377 | Low-quality |
| S3_extracted_k141_120118 | Not-determined |
| S3_extracted_k141_64331 | Not-determined |
| S3_extracted_k141_48756 | Low-quality |
| S3_extracted_k141_96466 | Not-determined |
| S3_extracted_k141_48761 | Not-determined |
| S3_extracted_k141_88259 | Low-quality |
| S3_extracted_k141_80383 | Not-determined |
| S3_extracted_k141_32833 | Not-determined |
| S3_extracted_k141_32834 | Low-quality |
| S3_extracted_k141_72631 | Not-determined |
| S3_extracted_k141_9098 | Not-determined |
| S3_extracted_k141_16328 | Not-determined |
| S3_extracted_k141_72637 | Not-determined |
| S3_extracted_k141_9107 | Not-determined |
| S3_extracted_k141_72640 | Not-determined |
| S3_extracted_k141_16330 | Low-quality |
| S3_extracted_k141_40703 | Not-determined |
| S3_extracted_k141_120124 | Not-determined |
| S3_extracted_k141_96496 | Not-determined |
| S3_extracted_k141_16339 | Not-determined |
| S3_extracted_k141_32859 | Not-determined |
| S3_extracted_k141_80392 | Not-determined |
| S3_extracted_k141_80395 | Low-quality |
| S3_extracted_k141_72650 | Low-quality |
| S3_extracted_k141_64353 | Low-quality |
| S3_extracted_k141_16341 | Not-determined |
| S3_extracted_k141_112240 | Low-quality |
| S3_extracted_k141_80400 | Not-determined |
| S3_extracted_k141_40716 | Not-determined |
| S3_extracted_k141_112247 | Not-determined |
| S3_extracted_k141_16349 | Not-determined |
| S3_extracted_k141_846 | Not-determined |
| S3_extracted_k141_9129 | Not-determined |
| S3_extracted_k141_9132 | Low-quality |
| S3_extracted_k141_40725 | Not-determined |
| S3_extracted_k141_96516 | Low-quality |
| S3_extracted_k141_56571 | Low-quality |
| S3_extracted_k141_96520 | Not-determined |
| S3_extracted_k141_96523 | Not-determined |
| S3_extracted_k141_112249 | Not-determined |
| S3_extracted_k141_16362 | Low-quality |
| S3_extracted_k141_72689 | Not-determined |
| S3_extracted_k141_112254 | Not-determined |
| S3_extracted_k141_112255 | Not-determined |
| S3_extracted_k141_112259 | Not-determined |
| S3_extracted_k141_56579 | Not-determined |
| S3_extracted_k141_56580 | Not-determined |
| S3_extracted_k141_72700 | Not-determined |
| S3_extracted_k141_96531 | Not-determined |
| S3_extracted_k141_16365 | Low-quality |
| S3_extracted_k141_56587 | Not-determined |
| S3_extracted_k141_72708 | Not-determined |
| S3_extracted_k141_32880 | Not-determined |
| S3_extracted_k141_56590 | Low-quality |
| S3_extracted_k141_16368 | Not-determined |
| S3_extracted_k141_112264 | Not-determined |
| S3_extracted_k141_96533 | Low-quality |
| S3_extracted_k141_9147 | Not-determined |
| S3_extracted_k141_112267 | Not-determined |
| S3_extracted_k141_32888 | Not-determined |
| S3_extracted_k141_16373 | Low-quality |
| S3_extracted_k141_88284 | Not-determined |
| S3_extracted_k141_88285 | Not-determined |
| S3_extracted_k141_104356 | Not-determined |
| S3_extracted_k141_9154 | Not-determined |
| S3_extracted_k141_72723 | Low-quality |
| S3_extracted_k141_48769 | Low-quality |
| S3_extracted_k141_48771 | Not-determined |
| S3_extracted_k141_16388 | Not-determined |
| S3_extracted_k141_16390 | Low-quality |
| S3_extracted_k141_56606 | Not-determined |
| S3_extracted_k141_16395 | Not-determined |
| S3_extracted_k141_64379 | Not-determined |
| S3_extracted_k141_24675 | Not-determined |
| S3_extracted_k141_64380 | Not-determined |
| S3_extracted_k141_24676 | Not-determined |
| S3_extracted_k141_88299 | Not-determined |
| S3_extracted_k141_16398 | Not-determined |
| S3_extracted_k141_88301 | Not-determined |
| S3_extracted_k141_88302 | Not-determined |
| S3_extracted_k141_24678 | Not-determined |
| S3_extracted_k141_88303 | Not-determined |
| S3_extracted_k141_24682 | Low-quality |
| S3_extracted_k141_80446 | Not-determined |
| S3_extracted_k141_24687 | Not-determined |
| S3_extracted_k141_56613 | Not-determined |
| S3_extracted_k141_64389 | Not-determined |
| S3_extracted_k141_64390 | Not-determined |
| S3_extracted_k141_88306 | Not-determined |
| S3_extracted_k141_24690 | Not-determined |
| S3_extracted_k141_64391 | Not-determined |
| S3_extracted_k141_80449 | Not-determined |
| S3_extracted_k141_24701 | Not-determined |
| S3_extracted_k141_112279 | Low-quality |
| S3_extracted_k141_16412 | Not-determined |
| S3_extracted_k141_16413 | Not-determined |
| S3_extracted_k141_80459 | Not-determined |
| S3_extracted_k141_40772 | Not-determined |
| S3_extracted_k141_104366 | Not-determined |
| S3_extracted_k141_80463 | Not-determined |
| S3_extracted_k141_112283 | Not-determined |
| S3_extracted_k141_96541 | Low-quality |
| S3_extracted_k141_104375 | Not-determined |
| S3_extracted_k141_48808 | Not-determined |
| S3_extracted_k141_88316 | Not-determined |
| S3_extracted_k141_56639 | Not-determined |
| S3_extracted_k141_120130 | Not-determined |
| S3_extracted_k141_80468 | Not-determined |
| S3_extracted_k141_56650 | Not-determined |
| S3_extracted_k141_56655 | Not-determined |
| S3_extracted_k141_40786 | Not-determined |
| S3_extracted_k141_120134 | Not-determined |
| S3_extracted_k141_48816 | Not-determined |
| S3_extracted_k141_120136 | Low-quality |
| S3_extracted_k141_96559 | Low-quality |
| S3_extracted_k141_32919 | Low-quality |
| S3_extracted_k141_48825 | Not-determined |
| S3_extracted_k141_48826 | Not-determined |
| S3_extracted_k141_48827 | Not-determined |
| S3_extracted_k141_72759 | Not-determined |
| S3_extracted_k141_40788 | Low-quality |
| S3_extracted_k141_72765 | Not-determined |
| S3_extracted_k141_96567 | Low-quality |
| S3_extracted_k141_80472 | Not-determined |
| S3_extracted_k141_32941 | Not-determined |
| S3_extracted_k141_16435 | Not-determined |
| S3_extracted_k141_16440 | Not-determined |
| S3_extracted_k141_40793 | Not-determined |
| S3_extracted_k141_896 | Not-determined |
| S3_extracted_k141_32947 | Not-determined |
| S3_extracted_k141_104383 | Not-determined |
| S3_extracted_k141_56660 | Not-determined |
| S3_extracted_k141_104391 | Low-quality |
| S3_extracted_k141_104396 | Not-determined |
| S3_extracted_k141_72769 | Not-determined |
| S3_extracted_k141_120142 | Low-quality |
| S3_extracted_k141_80485 | Not-determined |
| S3_extracted_k141_48854 | Not-determined |
| S3_extracted_k141_88347 | Not-determined |
| S3_extracted_k141_48855 | Not-determined |
| S3_extracted_k141_88348 | Not-determined |
| S3_extracted_k141_48856 | Not-determined |
| S3_extracted_k141_120151 | Not-determined |
| S3_extracted_k141_56669 | Not-determined |
| S3_extracted_k141_16442 | Not-determined |
| S3_extracted_k141_88357 | Not-determined |
| S3_extracted_k141_56673 | Not-determined |
| S3_extracted_k141_16449 | Not-determined |
| S3_extracted_k141_88359 | Low-quality |
| S3_extracted_k141_898 | Not-determined |
| S3_extracted_k141_72782 | Not-determined |
| S3_extracted_k141_72785 | Not-determined |
| S3_extracted_k141_96573 | Not-determined |
| S3_extracted_k141_120163 | Not-determined |
| S3_extracted_k141_96578 | Not-determined |
| S3_extracted_k141_80503 | Not-determined |
| S3_extracted_k141_48864 | Low-quality |
| S3_extracted_k141_96579 | Not-determined |
| S3_extracted_k141_96580 | Not-determined |
| S3_extracted_k141_80505 | Not-determined |
| S3_extracted_k141_72793 | Not-determined |
| S3_extracted_k141_912 | Low-quality |
| S3_extracted_k141_96585 | Not-determined |
| S3_extracted_k141_80512 | Not-determined |
| S3_extracted_k141_32972 | Not-determined |
| S3_extracted_k141_40823 | Not-determined |
| S3_extracted_k141_40825 | Not-determined |
| S3_extracted_k141_80522 | Not-determined |
| S3_extracted_k141_48869 | Not-determined |
| S3_extracted_k141_48873 | Low-quality |
| S3_extracted_k141_72818 | Not-determined |
| S3_extracted_k141_24722 | Not-determined |
| S3_extracted_k141_104423 | Not-determined |
| S3_extracted_k141_104424 | Not-determined |
| S3_extracted_k141_24725 | Low-quality |
| S3_extracted_k141_56684 | Not-determined |
| S3_extracted_k141_48881 | Not-determined |
| S3_extracted_k141_24730 | Not-determined |
| S3_extracted_k141_72825 | Low-quality |
| S3_extracted_k141_96592 | Not-determined |
| S3_extracted_k141_72828 | Not-determined |
| S3_extracted_k141_96594 | Not-determined |
| S3_extracted_k141_96596 | Not-determined |
| S3_extracted_k141_48892 | Not-determined |
| S3_extracted_k141_96597 | Low-quality |
| S3_extracted_k141_96599 | Low-quality |
| S3_extracted_k141_88373 | Not-determined |
| S3_extracted_k141_48901 | Low-quality |
| S3_extracted_k141_56701 | Not-determined |
| S3_extracted_k141_56702 | Not-determined |
| S3_extracted_k141_24740 | Not-determined |
| S3_extracted_k141_112296 | Not-determined |
| S3_extracted_k141_96606 | Not-determined |
| S3_extracted_k141_112299 | Not-determined |
| S3_extracted_k141_56709 | Not-determined |
| S3_extracted_k141_112301 | Low-quality |
| S3_extracted_k141_56710 | Not-determined |
| S3_extracted_k141_48906 | Not-determined |
| S3_extracted_k141_48907 | Low-quality |
| S3_extracted_k141_56713 | Not-determined |
| S3_extracted_k141_56715 | Not-determined |
| S3_extracted_k141_112312 | Not-determined |
| S3_extracted_k141_56723 | Not-determined |
| S3_extracted_k141_72839 | Not-determined |
| S3_extracted_k141_72845 | Not-determined |
| S3_extracted_k141_112319 | Low-quality |
| S3_extracted_k141_112321 | Not-determined |
| S3_extracted_k141_32997 | Not-determined |
| S3_extracted_k141_96623 | Not-determined |
| S3_extracted_k141_112325 | Not-determined |
| S3_extracted_k141_24755 | Not-determined |
| S3_extracted_k141_24756 | Not-determined |
| S3_extracted_k141_80559 | Low-quality |
| S3_extracted_k141_96634 | Not-determined |
| S3_extracted_k141_112338 | Not-determined |
| S3_extracted_k141_72853 | Not-determined |
| S3_extracted_k141_88388 | Low-quality |
| S3_extracted_k141_56726 | Not-determined |
| S3_extracted_k141_40850 | Not-determined |
| S3_extracted_k141_112340 | Not-determined |
| S3_extracted_k141_72855 | Not-determined |
| S3_extracted_k141_40851 | Not-determined |
| S3_extracted_k141_40858 | Not-determined |
| S3_extracted_k141_96650 | Not-determined |
| S3_extracted_k141_40861 | Not-determined |
| S3_extracted_k141_48928 | Not-determined |
| S3_extracted_k141_40868 | Low-quality |
| S3_extracted_k141_40869 | Low-quality |
| S3_extracted_k141_104447 | Not-determined |
| S3_extracted_k141_104450 | Not-determined |
| S3_extracted_k141_72866 | Not-determined |
| S3_extracted_k141_72867 | Not-determined |
| S3_extracted_k141_9204 | Not-determined |
| S3_extracted_k141_48947 | Not-determined |
| S3_extracted_k141_9211 | Not-determined |
| S3_extracted_k141_33002 | Not-determined |
| S3_extracted_k141_9212 | Not-determined |
| S3_extracted_k141_48950 | Low-quality |
| S3_extracted_k141_88400 | Low-quality |
| S3_extracted_k141_33004 | Not-determined |
| S3_extracted_k141_96657 | Not-determined |
| S3_extracted_k141_112353 | Low-quality |
| S3_extracted_k141_33007 | Low-quality |
| S3_extracted_k141_9222 | Not-determined |
| S3_extracted_k141_72883 | Not-determined |
| S3_extracted_k141_112356 | Not-determined |
| S3_extracted_k141_48960 | Not-determined |
| S3_extracted_k141_40883 | Low-quality |
| S3_extracted_k141_9229 | Not-determined |
| S3_extracted_k141_88408 | Not-determined |
| S3_extracted_k141_48963 | Not-determined |
| S3_extracted_k141_72885 | Not-determined |
| S3_extracted_k141_56733 | Not-determined |
| S3_extracted_k141_96678 | Low-quality |
| S3_extracted_k141_96681 | Not-determined |
| S3_extracted_k141_56735 | Low-quality |
| S3_extracted_k141_9243 | Low-quality |
| S3_extracted_k141_96685 | Not-determined |
| S3_extracted_k141_64431 | Not-determined |
| S3_extracted_k141_72907 | Not-determined |
| S3_extracted_k141_9256 | Not-determined |
| S3_extracted_k141_88421 | Not-determined |
| S3_extracted_k141_72910 | Not-determined |
| S3_extracted_k141_72913 | Not-determined |
| S3_extracted_k141_9261 | Not-determined |
| S3_extracted_k141_9264 | Not-determined |
| S3_extracted_k141_112364 | Not-determined |
| S3_extracted_k141_112367 | Not-determined |
| S3_extracted_k141_56746 | Not-determined |
| S3_extracted_k141_80586 | Not-determined |
| S3_extracted_k141_9265 | Not-determined |
| S3_extracted_k141_48982 | Not-determined |
| S3_extracted_k141_33025 | Not-determined |
| S3_extracted_k141_112369 | Not-determined |
| S3_extracted_k141_48987 | Low-quality |
| S3_extracted_k141_56753 | Low-quality |
| S3_extracted_k141_72923 | Not-determined |
| S3_extracted_k141_80596 | Not-determined |
| S3_extracted_k141_112371 | Low-quality |
| S3_extracted_k141_88430 | Low-quality |
| S3_extracted_k141_33032 | Not-determined |
| S3_extracted_k141_33035 | Not-determined |
| S3_extracted_k141_48992 | Low-quality |
| S3_extracted_k141_56766 | Low-quality |
| S3_extracted_k141_88436 | Not-determined |
| S3_extracted_k141_932 | Not-determined |
| S3_extracted_k141_72933 | Not-determined |
| S3_extracted_k141_40893 | Not-determined |
| S3_extracted_k141_72935 | Not-determined |
| S3_extracted_k141_40897 | Not-determined |
| S3_extracted_k141_49002 | Not-determined |
| S3_extracted_k141_49005 | Low-quality |
| S3_extracted_k141_80608 | Low-quality |
| S3_extracted_k141_933 | Not-determined |
| S3_extracted_k141_24780 | Not-determined |
| S3_extracted_k141_64462 | Not-determined |
| S3_extracted_k141_9282 | Not-determined |
| S3_extracted_k141_72942 | Low-quality |
| S3_extracted_k141_80615 | Low-quality |
| S3_extracted_k141_49019 | Not-determined |
| S3_extracted_k141_49020 | Not-determined |
| S3_extracted_k141_49022 | Not-determined |
| S3_extracted_k141_24787 | Not-determined |
| S3_extracted_k141_9287 | Not-determined |
| S3_extracted_k141_49024 | Not-determined |
| S3_extracted_k141_24788 | Not-determined |
| S3_extracted_k141_24792 | Not-determined |
| S3_extracted_k141_33055 | Low-quality |
| S3_extracted_k141_56784 | Not-determined |
| S3_extracted_k141_40913 | Not-determined |
| S3_extracted_k141_64480 | Not-determined |
| S3_extracted_k141_49031 | Not-determined |
| S3_extracted_k141_120178 | Low-quality |
| S3_extracted_k141_72952 | Not-determined |
| S3_extracted_k141_56789 | Not-determined |
| S3_extracted_k141_120181 | Not-determined |
| S3_extracted_k141_80632 | Not-determined |

| | |
|---|---|
| S3_extracted_k141_40925 | Low-quality |
| S3_extracted_k141_120184 | Low-quality |
| S3_extracted_k141_40932 | Not-determined |
| S3_extracted_k141_9289 | Not-determined |
| S3_extracted_k141_9291 | Not-determined |
| S3_extracted_k141_64493 | Low-quality |
| S3_extracted_k141_88471 | Low-quality |
| S3_extracted_k141_72970 | Not-determined |
| S3_extracted_k141_96709 | Not-determined |
| S3_extracted_k141_88473 | Not-determined |
| S3_extracted_k141_88479 | Low-quality |
| S3_extracted_k141_88480 | Not-determined |
| S3_extracted_k141_96714 | Not-determined |
| S3_extracted_k141_24799 | Low-quality |
| S3_extracted_k141_88484 | Not-determined |
| S3_extracted_k141_955 | Not-determined |
| S3_extracted_k141_64497 | Not-determined |
| S3_extracted_k141_112377 | Low-quality |
| S3_extracted_k141_33072 | Not-determined |
| S3_extracted_k141_958 | Low-quality |
| S3_extracted_k141_56797 | Not-determined |
| S3_extracted_k141_9299 | Not-determined |
| S3_extracted_k141_9301 | Not-determined |
| S3_extracted_k141_24809 | Not-determined |
| S3_extracted_k141_33082 | Low-quality |
| S3_extracted_k141_33083 | Not-determined |
| S3_extracted_k141_64507 | Not-determined |
| S3_extracted_k141_64508 | Not-determined |
| S3_extracted_k141_24817 | Not-determined |
| S3_extracted_k141_104464 | Not-determined |
| S3_extracted_k141_104465 | Not-determined |
| S3_extracted_k141_104467 | Not-determined |
| S3_extracted_k141_9304 | Low-quality |
| S3_extracted_k141_9306 | Not-determined |
| S3_extracted_k141_24830 | Not-determined |
| S3_extracted_k141_16482 | Not-determined |
| S3_extracted_k141_16484 | Not-determined |
| S3_extracted_k141_33095 | Not-determined |
| S3_extracted_k141_104470 | Not-determined |
| S3_extracted_k141_9318 | Low-quality |
| S3_extracted_k141_120200 | Low-quality |
| S3_extracted_k141_112393 | Not-determined |
| S3_extracted_k141_64522 | Not-determined |
| S3_extracted_k141_112395 | Not-determined |
| S3_extracted_k141_72988 | Not-determined |
| S3_extracted_k141_976 | Low-quality |
| S3_extracted_k141_56811 | Low-quality |
| S3_extracted_k141_64530 | Not-determined |
| S3_extracted_k141_64531 | Not-determined |
| S3_extracted_k141_49045 | Not-determined |
| S3_extracted_k141_56814 | Low-quality |
| S3_extracted_k141_112398 | Not-determined |
| S3_extracted_k141_979 | Not-determined |
| S3_extracted_k141_80649 | Low-quality |
| S3_extracted_k141_96725 | Not-determined |
| S3_extracted_k141_104492 | Not-determined |
| S3_extracted_k141_49050 | Not-determined |
| S3_extracted_k141_80653 | Low-quality |
| S3_extracted_k141_72998 | Not-determined |
| S3_extracted_k141_989 | Not-determined |
| S3_extracted_k141_73010 | Not-determined |
| S3_extracted_k141_73012 | Not-determined |
| S3_extracted_k141_991 | Low-quality |
| S3_extracted_k141_24861 | Not-determined |
| S3_extracted_k141_73016 | Not-determined |
| S3_extracted_k141_49062 | Not-determined |
| S3_extracted_k141_56817 | Not-determined |
| S3_extracted_k141_56818 | Not-determined |
| S3_extracted_k141_88502 | Not-determined |
| S3_extracted_k141_88507 | Not-determined |
| S3_extracted_k141_49068 | Not-determined |
| S3_extracted_k141_104505 | Not-determined |
| S3_extracted_k141_49070 | Low-quality |
| S3_extracted_k141_56827 | Not-determined |
| S3_extracted_k141_88517 | Not-determined |
| S3_extracted_k141_9333 | Not-determined |
| S3_extracted_k141_80658 | Not-determined |
| S3_extracted_k141_1009 | Not-determined |
| S3_extracted_k141_88523 | Not-determined |
| S3_extracted_k141_80665 | Not-determined |
| S3_extracted_k141_49077 | Not-determined |
| S3_extracted_k141_56840 | Low-quality |
| S3_extracted_k141_49080 | Not-determined |
| S3_extracted_k141_1026 | Not-determined |
| S3_extracted_k141_9339 | Low-quality |
| S3_extracted_k141_73039 | Not-determined |
| S3_extracted_k141_56843 | Not-determined |
| S3_extracted_k141_9341 | Low-quality |
| S3_extracted_k141_9345 | Not-determined |
| S3_extracted_k141_73046 | Not-determined |
| S3_extracted_k141_73049 | Not-determined |
| S3_extracted_k141_49089 | Not-determined |
| S3_extracted_k141_112406 | Not-determined |
| S3_extracted_k141_73059 | Not-determined |
| S3_extracted_k141_9352 | Not-determined |
| S3_extracted_k141_49093 | Low-quality |
| S3_extracted_k141_56853 | Not-determined |
| S3_extracted_k141_80682 | Low-quality |
| S3_extracted_k141_80683 | Not-determined |
| S3_extracted_k141_49096 | Not-determined |
| S3_extracted_k141_80685 | Not-determined |
| S3_extracted_k141_80686 | Not-determined |
| S3_extracted_k141_104511 | Not-determined |
| S3_extracted_k141_112413 | Not-determined |
| S3_extracted_k141_24889 | Not-determined |
| S3_extracted_k141_9358 | Low-quality |
| S3_extracted_k141_80693 | Not-determined |
| S3_extracted_k141_49105 | Not-determined |
| S3_extracted_k141_80697 | Not-determined |
| S3_extracted_k141_9369 | Not-determined |
| S3_extracted_k141_24896 | Not-determined |
| S3_extracted_k141_49108 | Not-determined |
| S3_extracted_k141_56863 | Not-determined |
| S3_extracted_k141_9375 | Low-quality |
| S3_extracted_k141_104522 | Low-quality |
| S3_extracted_k141_80715 | Not-determined |
| S3_extracted_k141_24901 | Not-determined |
| S3_extracted_k141_73078 | Not-determined |
| S3_extracted_k141_24904 | Not-determined |
| S3_extracted_k141_112417 | Not-determined |
| S3_extracted_k141_80719 | Not-determined |
| S3_extracted_k141_16527 | Not-determined |
| S3_extracted_k141_49130 | Not-determined |
| S3_extracted_k141_9394 | Not-determined |
| S3_extracted_k141_24908 | Low-quality |
| S3_extracted_k141_24909 | Not-determined |
| S3_extracted_k141_56868 | Not-determined |
| S3_extracted_k141_24912 | Not-determined |
| S3_extracted_k141_9410 | Not-determined |
| S3_extracted_k141_49134 | Not-determined |
| S3_extracted_k141_49136 | Not-determined |
| S3_extracted_k141_56876 | Not-determined |
| S3_extracted_k141_73093 | Not-determined |
| S3_extracted_k141_24920 | Not-determined |
| S3_extracted_k141_73099 | Not-determined |
| S3_extracted_k141_64542 | Low-quality |
| S3_extracted_k141_49157 | Not-determined |
| S3_extracted_k141_56896 | Not-determined |
| S3_extracted_k141_73106 | Low-quality |
| S3_extracted_k141_49162 | Not-determined |
| S3_extracted_k141_64548 | Not-determined |
| S3_extracted_k141_73110 | Not-determined |
| S3_extracted_k141_16549 | Low-quality |
| S3_extracted_k141_9431 | Not-determined |
| S3_extracted_k141_64554 | Not-determined |
| S3_extracted_k141_104533 | Low-quality |
| S3_extracted_k141_73121 | Low-quality |
| S3_extracted_k141_104536 | Not-determined |
| S3_extracted_k141_104541 | Not-determined |
| S3_extracted_k141_9434 | Not-determined |
| S3_extracted_k141_56911 | Not-determined |
| S3_extracted_k141_33127 | Not-determined |
| S3_extracted_k141_104547 | Low-quality |
| S3_extracted_k141_80735 | Not-determined |
| S3_extracted_k141_73139 | Low-quality |
| S3_extracted_k141_104551 | Not-determined |
| S3_extracted_k141_9443 | Not-determined |
| S3_extracted_k141_49181 | Not-determined |
| S3_extracted_k141_16572 | Not-determined |
| S3_extracted_k141_9450 | Not-determined |
| S3_extracted_k141_9451 | Not-determined |
| S3_extracted_k141_80745 | Not-determined |
| S3_extracted_k141_96771 | Not-determined |
| S3_extracted_k141_49185 | Not-determined |
| S3_extracted_k141_49186 | Not-determined |
| S3_extracted_k141_80750 | Low-quality |
| S3_extracted_k141_49192 | Not-determined |
| S3_extracted_k141_49193 | Not-determined |
| S3_extracted_k141_104574 | Not-determined |
| S3_extracted_k141_80761 | Not-determined |
| S3_extracted_k141_49195 | Not-determined |
| S3_extracted_k141_56937 | Low-quality |
| S3_extracted_k141_104575 | Not-determined |
| S3_extracted_k141_56939 | Not-determined |
| S3_extracted_k141_80768 | Not-determined |
| S3_extracted_k141_80769 | Not-determined |
| S3_extracted_k141_9462 | Not-determined |
| S3_extracted_k141_56949 | Not-determined |
| S3_extracted_k141_96784 | Not-determined |
| S3_extracted_k141_9468 | Not-determined |
| S3_extracted_k141_96792 | Not-determined |
| S3_extracted_k141_56955 | Not-determined |
| S3_extracted_k141_80784 | Not-determined |
| S3_extracted_k141_33146 | Not-determined |
| S3_extracted_k141_33149 | Not-determined |
| S3_extracted_k141_56962 | Not-determined |
| S3_extracted_k141_33152 | Not-determined |
| S3_extracted_k141_9476 | Not-determined |
| S3_extracted_k141_9480 | Not-determined |
| S3_extracted_k141_9481 | Not-determined |
| S3_extracted_k141_9483 | Not-determined |
| S3_extracted_k141_49203 | Low-quality |
| S3_extracted_k141_16596 | Not-determined |
| S3_extracted_k141_49204 | Low-quality |
| S3_extracted_k141_104584 | Not-determined |
| S3_extracted_k141_80796 | Low-quality |
| S3_extracted_k141_104586 | Not-determined |
| S3_extracted_k141_9489 | Not-determined |
| S3_extracted_k141_96815 | Not-determined |
| S3_extracted_k141_49209 | Not-determined |
| S3_extracted_k141_49210 | Not-determined |
| S3_extracted_k141_49211 | Not-determined |
| S3_extracted_k141_73180 | Not-determined |
| S3_extracted_k141_96825 | Not-determined |
| S3_extracted_k141_49221 | Low-quality |
| S3_extracted_k141_16602 | Not-determined |
| S3_extracted_k141_16610 | Not-determined |
| S3_extracted_k141_96842 | Low-quality |
| S3_extracted_k141_16617 | Not-determined |
| S3_extracted_k141_96849 | Low-quality |
| S3_extracted_k141_9513 | Not-determined |
| S3_extracted_k141_56978 | Not-determined |
| S3_extracted_k141_73204 | Low-quality |
| S3_extracted_k141_96853 | Not-determined |
| S3_extracted_k141_73207 | Low-quality |
| S3_extracted_k141_96855 | Not-determined |
| S3_extracted_k141_9523 | Not-determined |
| S3_extracted_k141_96859 | Not-determined |
| S3_extracted_k141_9526 | Not-determined |
| S3_extracted_k141_9528 | Not-determined |
| S3_extracted_k141_16625 | Not-determined |
| S3_extracted_k141_9532 | Not-determined |
| S3_extracted_k141_120264 | Not-determined |
| S3_extracted_k141_96870 | Not-determined |
| S3_extracted_k141_96871 | Not-determined |
| S3_extracted_k141_49239 | Not-determined |
| S3_extracted_k141_73208 | Not-determined |
| S3_extracted_k141_73210 | Low-quality |
| S3_extracted_k141_96879 | Not-determined |
| S3_extracted_k141_73217 | Not-determined |
| S3_extracted_k141_73221 | Not-determined |
| S3_extracted_k141_73224 | Not-determined |
| S3_extracted_k141_56998 | Not-determined |
| S3_extracted_k141_96883 | Not-determined |
| S3_extracted_k141_9542 | Not-determined |
| S3_extracted_k141_9545 | Low-quality |
| S3_extracted_k141_16647 | Not-determined |
| S3_extracted_k141_16650 | Not-determined |
| S3_extracted_k141_9553 | Low-quality |
| S3_extracted_k141_9555 | Not-determined |
| S3_extracted_k141_16656 | Low-quality |
| S3_extracted_k141_57015 | Not-determined |
| S3_extracted_k141_16664 | Not-determined |
| S3_extracted_k141_16665 | Not-determined |
| S3_extracted_k141_120266 | Not-determined |
| S3_extracted_k141_73245 | Not-determined |
| S3_extracted_k141_80816 | Not-determined |
| S3_extracted_k141_16669 | Not-determined |
| S3_extracted_k141_73259 | Not-determined |
| S3_extracted_k141_33186 | Not-determined |
| S3_extracted_k141_64593 | Low-quality |
| S3_extracted_k141_16670 | Low-quality |
| S3_extracted_k141_73265 | Low-quality |
| S3_extracted_k141_73266 | Low-quality |
| S3_extracted_k141_33192 | Not-determined |
| S3_extracted_k141_49267 | Not-determined |
| S3_extracted_k141_33194 | Not-determined |
| S3_extracted_k141_49270 | Not-determined |
| S3_extracted_k141_80829 | Low-quality |
| S3_extracted_k141_33206 | Not-determined |
| S3_extracted_k141_73278 | Low-quality |
| S3_extracted_k141_57031 | Not-determined |
| S3_extracted_k141_73281 | Low-quality |
| S3_extracted_k141_49279 | Not-determined |
| S3_extracted_k141_104630 | Not-determined |
| S3_extracted_k141_73283 | Not-determined |
| S3_extracted_k141_104632 | Not-determined |
| S3_extracted_k141_104634 | Not-determined |
| S3_extracted_k141_80844 | Not-determined |
| S3_extracted_k141_80848 | Not-determined |
| S3_extracted_k141_73289 | Not-determined |
| S3_extracted_k141_96931 | Not-determined |
| S3_extracted_k141_49292 | Not-determined |
| S3_extracted_k141_120283 | Not-determined |
| S3_extracted_k141_96938 | Not-determined |
| S3_extracted_k141_9580 | Not-determined |
| S3_extracted_k141_1041 | Not-determined |
| S3_extracted_k141_1042 | Not-determined |
| S3_extracted_k141_49302 | Not-determined |
| S3_extracted_k141_120289 | Not-determined |
| S3_extracted_k141_1048 | Low-quality |
| S3_extracted_k141_9585 | Not-determined |
| S3_extracted_k141_64598 | Not-determined |
| S3_extracted_k141_49308 | Not-determined |
| S3_extracted_k141_120291 | Low-quality |
| S3_extracted_k141_9588 | Not-determined |
| S3_extracted_k141_73300 | Not-determined |
| S3_extracted_k141_80855 | Not-determined |
| S3_extracted_k141_64602 | Not-determined |
| S3_extracted_k141_49311 | Not-determined |
| S3_extracted_k141_1051 | Not-determined |
| S3_extracted_k141_96955 | Low-quality |
| S3_extracted_k141_96958 | Not-determined |
| S3_extracted_k141_9601 | Not-determined |
| S3_extracted_k141_80870 | Not-determined |
| S3_extracted_k141_104646 | Low-quality |
| S3_extracted_k141_104648 | Not-determined |
| S3_extracted_k141_88551 | Not-determined |
| S3_extracted_k141_64621 | Not-determined |
| S3_extracted_k141_88557 | Not-determined |
| S3_extracted_k141_120312 | Not-determined |
| S3_extracted_k141_1067 | Not-determined |
| S3_extracted_k141_120313 | Not-determined |
| S3_extracted_k141_96961 | Low-quality |
| S3_extracted_k141_1075 | Low-quality |
| S3_extracted_k141_88565 | Not-determined |
| S3_extracted_k141_64623 | Low-quality |
| S3_extracted_k141_96967 | Not-determined |
| S3_extracted_k141_64627 | Not-determined |
| S3_extracted_k141_73309 | Not-determined |
| S3_extracted_k141_112444 | Not-determined |
| S3_extracted_k141_64629 | Not-determined |
| S3_extracted_k141_80886 | Low-quality |
| S3_extracted_k141_96971 | Not-determined |
| S3_extracted_k141_9611 | Low-quality |
| S3_extracted_k141_33243 | Not-determined |
| S3_extracted_k141_96973 | Not-determined |
| S3_extracted_k141_9613 | Not-determined |
| S3_extracted_k141_1084 | Not-determined |
| S3_extracted_k141_112452 | Not-determined |
| S3_extracted_k141_96975 | Not-determined |
| S3_extracted_k141_80887 | Low-quality |
| S3_extracted_k141_9621 | Not-determined |
| S3_extracted_k141_96980 | Low-quality |
| S3_extracted_k141_96982 | Low-quality |
| S3_extracted_k141_73317 | Not-determined |
| S3_extracted_k141_96984 | Not-determined |
| S3_extracted_k141_73319 | Not-determined |
| S3_extracted_k141_96985 | Not-determined |
| S3_extracted_k141_73323 | Not-determined |
| S3_extracted_k141_9629 | Not-determined |
| S3_extracted_k141_96989 | Not-determined |
| S3_extracted_k141_80906 | Not-determined |
| S3_extracted_k141_73331 | Not-determined |
| S3_extracted_k141_80907 | Not-determined |
| S3_extracted_k141_9638 | Not-determined |
| S3_extracted_k141_80909 | Not-determined |
| S3_extracted_k141_64634 | Not-determined |
| S3_extracted_k141_88590 | Not-determined |
| S3_extracted_k141_16672 | Not-determined |
| S3_extracted_k141_97013 | Not-determined |
| S3_extracted_k141_88599 | Not-determined |
| S3_extracted_k141_16676 | Not-determined |
| S3_extracted_k141_73351 | Not-determined |
| S3_extracted_k141_16682 | Not-determined |
| S3_extracted_k141_80917 | Not-determined |
| S3_extracted_k141_1124 | Not-determined |
| S3_extracted_k141_1127 | Not-determined |
| S3_extracted_k141_80920 | Not-determined |
| S3_extracted_k141_80921 | Not-determined |
| S3_extracted_k141_57074 | Not-determined |
| S3_extracted_k141_80932 | Not-determined |
| S3_extracted_k141_80934 | Not-determined |
| S3_extracted_k141_80938 | Not-determined |
| S3_extracted_k141_1131 | Low-quality |
| S3_extracted_k141_88625 | Not-determined |
| S3_extracted_k141_64649 | Not-determined |
| S3_extracted_k141_80946 | Low-quality |
| S3_extracted_k141_16698 | Not-determined |
| S3_extracted_k141_80957 | Not-determined |
| S3_extracted_k141_1142 | Not-determined |
| S3_extracted_k141_88636 | Low-quality |
| S3_extracted_k141_40958 | Not-determined |
| S3_extracted_k141_40961 | Not-determined |
| S3_extracted_k141_9659 | Low-quality |
| S3_extracted_k141_9660 | Not-determined |
| S3_extracted_k141_97015 | Not-determined |
| S3_extracted_k141_80970 | Not-determined |
| S3_extracted_k141_16712 | Low-quality |
| S3_extracted_k141_40970 | Not-determined |
| S3_extracted_k141_16715 | Not-determined |
| S3_extracted_k141_97021 | Low-quality |
| S3_extracted_k141_24935 | Not-determined |
| S3_extracted_k141_120327 | Not-determined |
| S3_extracted_k141_24939 | Not-determined |
| S3_extracted_k141_40978 | Not-determined |
| S3_extracted_k141_16726 | Low-quality |
| S3_extracted_k141_97026 | Not-determined |
| S3_extracted_k141_16733 | Low-quality |
| S3_extracted_k141_88643 | Not-determined |
| S3_extracted_k141_73363 | Not-determined |
| S3_extracted_k141_88652 | Not-determined |
| S3_extracted_k141_97033 | Not-determined |
| S3_extracted_k141_9662 | Not-determined |
| S3_extracted_k141_104669 | Not-determined |
| S3_extracted_k141_80995 | Low-quality |
| S3_extracted_k141_40989 | Not-determined |
| S3_extracted_k141_16741 | Not-determined |
| S3_extracted_k141_88658 | Not-determined |
| S3_extracted_k141_104675 | Not-determined |
| S3_extracted_k141_40991 | Not-determined |
| S3_extracted_k141_33264 | Not-determined |
| S3_extracted_k141_33265 | Not-determined |
| S3_extracted_k141_16746 | Not-determined |
| S3_extracted_k141_16752 | Not-determined |
| S3_extracted_k141_41000 | Not-determined |
| S3_extracted_k141_88673 | Not-determined |
| S3_extracted_k141_73391 | Low-quality |
| S3_extracted_k141_57107 | Not-determined |
| S3_extracted_k141_57110 | Not-determined |
| S3_extracted_k141_64674 | Not-determined |
| S3_extracted_k141_88683 | Not-determined |
| S3_extracted_k141_88684 | Not-determined |
| S3_extracted_k141_64680 | Not-determined |
| S3_extracted_k141_64682 | Not-determined |
| S3_extracted_k141_57117 | Not-determined |
| S3_extracted_k141_49333 | Low-quality |
| S3_extracted_k141_57120 | Not-determined |
| S3_extracted_k141_97047 | Not-determined |
| S3_extracted_k141_33282 | Not-determined |
| S3_extracted_k141_88699 | Low-quality |
| S3_extracted_k141_33293 | Not-determined |
| S3_extracted_k141_24978 | Low-quality |
| S3_extracted_k141_24982 | Not-determined |
| S3_extracted_k141_88706 | Not-determined |
| S3_extracted_k141_33303 | Not-determined |
| S3_extracted_k141_33308 | Low-quality |
| S3_extracted_k141_97053 | Not-determined |
| S3_extracted_k141_33310 | Not-determined |
| S3_extracted_k141_33312 | Low-quality |
| S3_extracted_k141_112476 | Not-determined |
| S3_extracted_k141_81004 | Low-quality |
| S3_extracted_k141_81006 | Not-determined |
| S3_extracted_k141_49343 | Not-determined |
| S3_extracted_k141_88711 | Low-quality |
| S3_extracted_k141_24993 | Not-determined |
| S3_extracted_k141_88713 | Not-determined |
| S3_extracted_k141_64686 | Not-determined |
| S3_extracted_k141_88714 | Low-quality |
| S3_extracted_k141_33326 | Not-determined |
| S3_extracted_k141_25005 | Not-determined |
| S3_extracted_k141_16781 | Not-determined |
| S3_extracted_k141_120349 | Not-determined |
| S3_extracted_k141_104699 | Not-determined |
| S3_extracted_k141_120351 | Not-determined |
| S3_extracted_k141_104701 | Not-determined |
| S3_extracted_k141_120358 | Low-quality |
| S3_extracted_k141_112481 | Not-determined |
| S3_extracted_k141_120360 | Not-determined |
| S3_extracted_k141_33327 | Not-determined |
| S3_extracted_k141_97070 | Not-determined |
| S3_extracted_k141_64699 | Low-quality |
| S3_extracted_k141_81031 | Not-determined |
| S3_extracted_k141_112492 | Not-determined |
| S3_extracted_k141_73425 | Not-determined |
| S3_extracted_k141_33342 | Low-quality |
| S3_extracted_k141_88726 | Not-determined |
| S3_extracted_k141_41045 | Not-determined |
| S3_extracted_k141_16799 | Not-determined |
| S3_extracted_k141_104707 | Not-determined |
| S3_extracted_k141_81050 | Low-quality |
| S3_extracted_k141_97081 | Not-determined |
| S3_extracted_k141_97082 | Not-determined |
| S3_extracted_k141_73442 | Not-determined |
| S3_extracted_k141_73444 | Not-determined |
| S3_extracted_k141_81069 | Not-determined |
| S3_extracted_k141_64711 | Not-determined |
| S3_extracted_k141_97091 | Not-determined |
| S3_extracted_k141_120380 | Not-determined |
| S3_extracted_k141_64716 | Not-determined |
| S3_extracted_k141_97096 | Not-determined |
| S3_extracted_k141_88753 | Not-determined |
| S3_extracted_k141_64721 | Not-determined |
| S3_extracted_k141_25030 | Not-determined |
| S3_extracted_k141_120393 | Low-quality |
| S3_extracted_k141_81091 | Low-quality |
| S3_extracted_k141_16813 | Low-quality |
| S3_extracted_k141_120401 | Not-determined |
| S3_extracted_k141_9685 | Low-quality |
| S3_extracted_k141_25037 | Not-determined |
| S3_extracted_k141_120402 | Low-quality |
| S3_extracted_k141_88765 | Not-determined |
| S3_extracted_k141_120403 | Not-determined |
| S3_extracted_k141_25043 | Low-quality |
| S3_extracted_k141_88768 | Low-quality |
| S3_extracted_k141_81097 | Low-quality |
| S3_extracted_k141_97107 | Not-determined |
| S3_extracted_k141_120405 | Not-determined |
| S3_extracted_k141_112518 | Not-determined |
| S3_extracted_k141_81103 | Not-determined |
| S3_extracted_k141_64725 | Low-quality |
| S3_extracted_k141_64726 | Not-determined |
| S3_extracted_k141_64730 | Not-determined |
| S3_extracted_k141_81106 | Not-determined |
| S3_extracted_k141_120409 | Not-determined |
| S3_extracted_k141_49360 | Not-determined |
| S3_extracted_k141_33385 | Not-determined |
| S3_extracted_k141_120421 | Not-determined |
| S3_extracted_k141_49361 | Low-quality |
| S3_extracted_k141_81110 | Low-quality |
| S3_extracted_k141_49366 | Not-determined |
| S3_extracted_k141_81117 | Low-quality |
| S3_extracted_k141_64743 | Not-determined |
| S3_extracted_k141_81118 | Not-determined |
| S3_extracted_k141_81120 | Not-determined |
| S3_extracted_k141_112527 | Not-determined |
| S3_extracted_k141_81121 | Not-determined |
| S3_extracted_k141_120433 | Low-quality |
| S3_extracted_k141_104715 | Not-determined |
| S3_extracted_k141_64751 | Not-determined |
| S3_extracted_k141_120444 | Low-quality |
| S3_extracted_k141_120449 | Not-determined |
| S3_extracted_k141_120450 | Not-determined |
| S3_extracted_k141_64773 | Not-determined |
| S3_extracted_k141_64775 | Not-determined |
| S3_extracted_k141_9695 | Not-determined |
| S16_vs_18440\|\|full | High-quality |
| S3_extracted_k141_9700 | Not-determined |
| S3_extracted_k141_1184 | Low-quality |
| S3_extracted_k141_81132 | Not-determined |
| S3_extracted_k141_64793 | Not-determined |
| S3_extracted_k141_81133 | Low-quality |
| S3_extracted_k141_104729 | Low-quality |
| S3_extracted_k141_112549 | Not-determined |
| S3_extracted_k141_1193 | Not-determined |
| S3_extracted_k141_64803 | Not-determined |
| S3_extracted_k141_1196 | Not-determined |
| S3_extracted_k141_120461 | Not-determined |
| S3_extracted_k141_112559 | Low-quality |
| S3_extracted_k141_1198 | Low-quality |
| S3_extracted_k141_16829 | Low-quality |
| S3_extracted_k141_81150 | Not-determined |
| S3_extracted_k141_41066 | Not-determined |
| S3_extracted_k141_16836 | Low-quality |
| S3_extracted_k141_33393 | Not-determined |
| S3_extracted_k141_120468 | Low-quality |
| S3_extracted_k141_16848 | Not-determined |
| S3_extracted_k141_112573 | Not-determined |
| S3_extracted_k141_120479 | Not-determined |
| S3_extracted_k141_1208 | Not-determined |
| S3_extracted_k141_9713 | Low-quality |
| S3_extracted_k141_9714 | Low-quality |
| S3_extracted_k141_64827 | Not-determined |
| S3_extracted_k141_104743 | Not-determined |
| S3_extracted_k141_9720 | Not-determined |
| S3_extracted_k141_64832 | Not-determined |
| S3_extracted_k141_9723 | Not-determined |
| S3_extracted_k141_9726 | Not-determined |
| S3_extracted_k141_112575 | Low-quality |
| S3_extracted_k141_112577 | Low-quality |
| S3_extracted_k141_104750 | Not-determined |
| S3_extracted_k141_88785 | Not-determined |
| S3_extracted_k141_88786 | Not-determined |
| S3_extracted_k141_88787 | Not-determined |
| S3_extracted_k141_64850 | Low-quality |
| S3_extracted_k141_16873 | Low-quality |
| S3_extracted_k141_88794 | Not-determined |
| S3_extracted_k141_64857 | Not-determined |
| S3_extracted_k141_112584 | Low-quality |
| S3_extracted_k141_64860 | Low-quality |
| S3_extracted_k141_112585 | Not-determined |
| S3_extracted_k141_41098 | Low-quality |
| S3_extracted_k141_16878 | Low-quality |
| S3_extracted_k141_81165 | Not-determined |
| S3_extracted_k141_112591 | Not-determined |
| S3_extracted_k141_88800 | Not-determined |
| S3_extracted_k141_9731 | Not-determined |
| S3_extracted_k141_112595 | Not-determined |
| S3_extracted_k141_33428 | Low-quality |
| S3_extracted_k141_81168 | Not-determined |
| S3_extracted_k141_88809 | Low-quality |
| S3_extracted_k141_64880 | Not-determined |
| S3_extracted_k141_81170 | Not-determined |
| S3_extracted_k141_33437 | Not-determined |
| S3_extracted_k141_112605 | Not-determined |
| S3_extracted_k141_64887 | Not-determined |
| S3_extracted_k141_49417 | Not-determined |
| S3_extracted_k141_104759 | Not-determined |
| S3_extracted_k141_33444 | Not-determined |
| S3_extracted_k141_64895 | Not-determined |
| S3_extracted_k141_73475 | Not-determined |
| S3_extracted_k141_64900 | Not-determined |
| S3_extracted_k141_73481 | Not-determined |
| S3_extracted_k141_88826 | Not-determined |
| S3_extracted_k141_1222 | Not-determined |
| S3_extracted_k141_1224 | Not-determined |
| S3_extracted_k141_49428 | Not-determined |
| S3_extracted_k141_1229 | Low-quality |
| S3_extracted_k141_49433 | Not-determined |
| S3_extracted_k141_49435 | Not-determined |
| S3_extracted_k141_1233 | Low-quality |
| S3_extracted_k141_73487 | Not-determined |
| S3_extracted_k141_97113 | Not-determined |
| S3_extracted_k141_97115 | Low-quality |
| S3_extracted_k141_73490 | Low-quality |
| S3_extracted_k141_49442 | Not-determined |
| S3_extracted_k141_16901 | Not-determined |
| S3_extracted_k141_49450 | Not-determined |
| S3_extracted_k141_120500 | Low-quality |

| | |
|---|---|
| S3_extracted_k141_120503 | Low-quality |
| S3_extracted_k141_33468 | Low-quality |
| S3_extracted_k141_97131 | Not-determined |
| S3_extracted_k141_81181 | Not-determined |
| S3_extracted_k141_33472 | Low-quality |
| S3_extracted_k141_41121 | Not-determined |
| S3_extracted_k141_41124 | Not-determined |
| S3_extracted_k141_41130 | Not-determined |
| S3_extracted_k141_81195 | Not-determined |
| S3_extracted_k141_104768 | Low-quality |
| S3_extracted_k141_112626 | Low-quality |
| S3_extracted_k141_41132 | Low-quality |
| S3_extracted_k141_41134 | Not-determined |
| S3_extracted_k141_88837 | Not-determined |
| S3_extracted_k141_41142 | Not-determined |
| S3_extracted_k141_33478 | Low-quality |
| S3_extracted_k141_120510 | Not-determined |
| S3_extracted_k141_120516 | Not-determined |
| S3_extracted_k141_33483 | Not-determined |
| S3_extracted_k141_97140 | Not-determined |
| S3_extracted_k141_16942 | Not-determined |
| S3_extracted_k141_33486 | Not-determined |
| S3_extracted_k141_120520 | Not-determined |
| S3_extracted_k141_97144 | Not-determined |
| S3_extracted_k141_73521 | Not-determined |
| S3_extracted_k141_73522 | Not-determined |
| S3_extracted_k141_1250 | Low-quality |
| S3_extracted_k141_1251 | Not-determined |
| S3_extracted_k141_16949 | Not-determined |
| S3_extracted_k141_33491 | Not-determined |
| S3_extracted_k141_97150 | Not-determined |
| S3_extracted_k141_16955 | Not-determined |
| S3_extracted_k141_16959 | Not-determined |
| S3_extracted_k141_57123 | Not-determined |
| S3_extracted_k141_97155 | Not-determined |
| S3_extracted_k141_73538 | Low-quality |
| S3_extracted_k141_73539 | Not-determined |
| S3_extracted_k141_81200 | Not-determined |
| S3_extracted_k141_1259 | Not-determined |
| S3_extracted_k141_57131 | Not-determined |
| S3_extracted_k141_57132 | Not-determined |
| S3_extracted_k141_73550 | Low-quality |
| S3_extracted_k141_33500 | Not-determined |
| S3_extracted_k141_57135 | Not-determined |
| S3_extracted_k141_81207 | Low-quality |
| S3_extracted_k141_16969 | Not-determined |
| S3_extracted_k141_120533 | Not-determined |
| S3_extracted_k141_41160 | Low-quality |
| S3_extracted_k141_64920 | Low-quality |
| S3_extracted_k141_41165 | Not-determined |
| S3_extracted_k141_1263 | Not-determined |
| S3_extracted_k141_104782 | Not-determined |
| S3_extracted_k141_1264 | Low-quality |
| S3_extracted_k141_41168 | Not-determined |
| S3_extracted_k141_104785 | Not-determined |
| S3_extracted_k141_16983 | Not-determined |
| S3_extracted_k141_104787 | Not-determined |
| S3_extracted_k141_81221 | Not-determined |
| S3_extracted_k141_81222 | Not-determined |
| S3_extracted_k141_9763 | Not-determined |
| S3_extracted_k141_64933 | Not-determined |
| S3_extracted_k141_1275 | Not-determined |
| S3_extracted_k141_1276 | Not-determined |
| S3_extracted_k141_1278 | Not-determined |
| S3_extracted_k141_33515 | Not-determined |
| S3_extracted_k141_9773 | Not-determined |
| S3_extracted_k141_33526 | Not-determined |
| S3_extracted_k141_1290 | Not-determined |
| S3_extracted_k141_49473 | Not-determined |
| S3_extracted_k141_49476 | Not-determined |
| S3_extracted_k141_104799 | Not-determined |
| S3_extracted_k141_88847 | Low-quality |
| S3_extracted_k141_81226 | Not-determined |
| S3_extracted_k141_49480 | Not-determined |
| S3_extracted_k141_112629 | Not-determined |
| S3_extracted_k141_73563 | Not-determined |
| S3_extracted_k141_88853 | Not-determined |
| S3_extracted_k141_88854 | Not-determined |
| S3_extracted_k141_81238 | Low-quality |
| S3_extracted_k141_49487 | Low-quality |
| S3_extracted_k141_1299 | Low-quality |
| S3_extracted_k141_1300 | Not-determined |
| S3_extracted_k141_1302 | Not-determined |
| S3_extracted_k141_49492 | Low-quality |
| S3_extracted_k141_88863 | Not-determined |
| S3_extracted_k141_104806 | Not-determined |
| S3_extracted_k141_1307 | Not-determined |
| S3_extracted_k141_49493 | Low-quality |
| S3_extracted_k141_112645 | Not-determined |
| S3_extracted_k141_49495 | Not-determined |
| S3_extracted_k141_112647 | Low-quality |
| S3_extracted_k141_88868 | Not-determined |
| S3_extracted_k141_57143 | Not-determined |
| S3_extracted_k141_120545 | Not-determined |
| S3_extracted_k141_57144 | Not-determined |
| S3_extracted_k141_112655 | Not-determined |
| S3_extracted_k141_73576 | Not-determined |
| S3_extracted_k141_81249 | Not-determined |
| S3_extracted_k141_57147 | Not-determined |
| S3_extracted_k141_1324 | Not-determined |
| S3_extracted_k141_57154 | Not-determined |
| S3_extracted_k141_112657 | Low-quality |
| S3_extracted_k141_120556 | Not-determined |
| S3_extracted_k141_49501 | Not-determined |
| S3_extracted_k141_49504 | Not-determined |
| S3_extracted_k141_104818 | Not-determined |
| S3_extracted_k141_41187 | Low-quality |
| S3_extracted_k141_104830 | Low-quality |
| S3_extracted_k141_64937 | Low-quality |
| S3_extracted_k141_64938 | Not-determined |
| S3_extracted_k141_104832 | Not-determined |
| S3_extracted_k141_1334 | Not-determined |
| S3_extracted_k141_49518 | Not-determined |
| S3_extracted_k141_104839 | Not-determined |
| S3_extracted_k141_104840 | Not-determined |
| S3_extracted_k141_97167 | Not-determined |
| S3_extracted_k141_33550 | Low-quality |
| S3_extracted_k141_49520 | Not-determined |
| S3_extracted_k141_1335 | Not-determined |
| S3_extracted_k141_1336 | Not-determined |
| S3_extracted_k141_1337 | Not-determined |
| S3_extracted_k141_49522 | Not-determined |
| S3_extracted_k141_104842 | Not-determined |
| S3_extracted_k141_1340 | Not-determined |
| S3_extracted_k141_64945 | Not-determined |
| S3_extracted_k141_33560 | Not-determined |
| S3_extracted_k141_64948 | Not-determined |
| S3_extracted_k141_104844 | Not-determined |
| S3_extracted_k141_1344 | Not-determined |
| S3_extracted_k141_104845 | Not-determined |
| S3_extracted_k141_41207 | Not-determined |
| S3_extracted_k141_120562 | Not-determined |
| S3_extracted_k141_64953 | Not-determined |
| S3_extracted_k141_41208 | Not-determined |
| S3_extracted_k141_73583 | Not-determined |
| S3_extracted_k141_64957 | Not-determined |
| S3_extracted_k141_9808 | Not-determined |
| S3_extracted_k141_41212 | Not-determined |
| S3_extracted_k141_57177 | Not-determined |
| S3_extracted_k141_73589 | Not-determined |
| S3_extracted_k141_64962 | Low-quality |
| S3_extracted_k141_73592 | Not-determined |
| S3_extracted_k141_97177 | Low-quality |
| S3_extracted_k141_1352 | Not-determined |
| S3_extracted_k141_33571 | Low-quality |
| S3_extracted_k141_49526 | Not-determined |
| S3_extracted_k141_33575 | Not-determined |
| S3_extracted_k141_57184 | Not-determined |
| S3_extracted_k141_73595 | Not-determined |
| S3_extracted_k141_41221 | Not-determined |
| S3_extracted_k141_41222 | Not-determined |
| S3_extracted_k141_120573 | Not-determined |
| S3_extracted_k141_41228 | Not-determined |
| S3_extracted_k141_33593 | Not-determined |
| S3_extracted_k141_120577 | Not-determined |
| S3_extracted_k141_112667 | Low-quality |
| S3_extracted_k141_33597 | Low-quality |
| S3_extracted_k141_57197 | Not-determined |
| S3_extracted_k141_41239 | Not-determined |
| S3_extracted_k141_57199 | Not-determined |
| S3_extracted_k141_9816 | Not-determined |
| S3_extracted_k141_49543 | Not-determined |
| S3_extracted_k141_104859 | Not-determined |
| S3_extracted_k141_104862 | Not-determined |
| S3_extracted_k141_33605 | Low-quality |
| S3_extracted_k141_49546 | Not-determined |
| S3_extracted_k141_57205 | Not-determined |
| S3_extracted_k141_104870 | Not-determined |
| S3_extracted_k141_9824 | Not-determined |
| S3_extracted_k141_9825 | Not-determined |
| S3_extracted_k141_9830 | Low-quality |
| S3_extracted_k141_17005 | Not-determined |
| S3_extracted_k141_81254 | Low-quality |
| S3_extracted_k141_57208 | Not-determined |
| S3_extracted_k141_81258 | Low-quality |
| S3_extracted_k141_41251 | Low-quality |
| S3_extracted_k141_9836 | Not-determined |
| S3_extracted_k141_33616 | Not-determined |
| S3_extracted_k141_9840 | Not-determined |
| S3_extracted_k141_81262 | Not-determined |
| S3_extracted_k141_17011 | Not-determined |
| S3_extracted_k141_9843 | Not-determined |
| S3_extracted_k141_81264 | Not-determined |
| S3_extracted_k141_64973 | Low-quality |
| S3_extracted_k141_17015 | Not-determined |
| S3_extracted_k141_64976 | Not-determined |
| S3_extracted_k141_81275 | Not-determined |
| S3_extracted_k141_104879 | Not-determined |
| S3_extracted_k141_120583 | Not-determined |
| S3_extracted_k141_49559 | Low-quality |
| S3_extracted_k141_64989 | Not-determined |
| S3_extracted_k141_49560 | Not-determined |
| S3_extracted_k141_97194 | Not-determined |
| S3_extracted_k141_120587 | Not-determined |
| S3_extracted_k141_104889 | Not-determined |
| S3_extracted_k141_97196 | Not-determined |
| S3_extracted_k141_120589 | Not-determined |
| S3_extracted_k141_25059 | Not-determined |
| S3_extracted_k141_81285 | Not-determined |
| S3_extracted_k141_57226 | Not-determined |
| S3_extracted_k141_81286 | Not-determined |
| S3_extracted_k141_81289 | Not-determined |
| S3_extracted_k141_57229 | Not-determined |
| S3_extracted_k141_112696 | Not-determined |
| S3_extracted_k141_112697 | Not-determined |
| S3_extracted_k141_81293 | Not-determined |
| S3_extracted_k141_81296 | Low-quality |
| S3_extracted_k141_81297 | Not-determined |
| S3_extracted_k141_25068 | Not-determined |
| S3_extracted_k141_25070 | Not-determined |
| S3_extracted_k141_81300 | Not-determined |
| S3_extracted_k141_81302 | Not-determined |
| S3_extracted_k141_25073 | Not-determined |
| S3_extracted_k141_57232 | Not-determined |
| S3_extracted_k141_57233 | Not-determined |
| S3_extracted_k141_57235 | Not-determined |
| S3_extracted_k141_57239 | Not-determined |
| S3_extracted_k141_33628 | Not-determined |
| S3_extracted_k141_81314 | Low-quality |
| S3_extracted_k141_1363 | Low-quality |
| S3_extracted_k141_25081 | Not-determined |
| S3_extracted_k141_97202 | Low-quality |
| S3_extracted_k141_65001 | Low-quality |
| S3_extracted_k141_81324 | Low-quality |
| S3_extracted_k141_81329 | Low-quality |
| S3_extracted_k141_81330 | Not-determined |
| S3_extracted_k141_1370 | Low-quality |
| S3_extracted_k141_49565 | Not-determined |
| S3_extracted_k141_17035 | Not-determined |
| S3_extracted_k141_65008 | Not-determined |
| S3_extracted_k141_49576 | Not-determined |
| S3_extracted_k141_33632 | Not-determined |
| S3_extracted_k141_25103 | Not-determined |
| S3_extracted_k141_25108 | Not-determined |
| S3_extracted_k141_33638 | Not-determined |
| S3_extracted_k141_49583 | Not-determined |
| S3_extracted_k141_33648 | Not-determined |
| S3_extracted_k141_25122 | Low-quality |
| S3_extracted_k141_25123 | Not-determined |
| S3_extracted_k141_81332 | Not-determined |
| S3_extracted_k141_49584 | Low-quality |
| S3_extracted_k141_120603 | Low-quality |
| S3_extracted_k141_1384 | Not-determined |
| S3_extracted_k141_1385 | Not-determined |
| S4_extracted_10506 | Complete |
| S3_extracted_k141_112725 | Not-determined |
| S3_extracted_k141_1390 | Not-determined |
| S3_extracted_k141_33664 | Not-determined |
| S3_extracted_k141_120611 | Not-determined |
| S3_extracted_k141_17053 | Not-determined |
| S3_extracted_k141_81343 | Not-determined |
| S3_extracted_k141_17054 | Not-determined |
| S3_extracted_k141_120614 | Not-determined |
| S3_extracted_k141_49602 | Not-determined |
| S3_extracted_k141_1401 | Not-determined |
| S3_extracted_k141_33672 | Not-determined |
| S3_extracted_k141_49607 | Not-determined |
| S3_extracted_k141_33673 | Not-determined |
| S3_extracted_k141_17063 | Not-determined |
| S3_extracted_k141_81348 | Not-determined |
| S3_extracted_k141_33676 | Not-determined |
| S3_extracted_k141_49611 | Not-determined |
| S3_extracted_k141_33678 | Not-determined |
| S3_extracted_k141_112734 | Not-determined |
| S3_extracted_k141_112737 | Not-determined |
| S3_extracted_k141_1414 | Not-determined |
| S3_extracted_k141_1417 | Not-determined |
| S3_extracted_k141_57274 | Not-determined |
| S3_extracted_k141_49619 | Low-quality |
| S3_extracted_k141_112741 | Not-determined |
| S3_extracted_k141_49620 | Not-determined |
| S3_extracted_k141_73611 | Not-determined |
| S3_extracted_k141_104918 | Not-determined |
| S3_extracted_k141_9854 | Not-determined |
| S3_extracted_k141_104920 | Not-determined |
| S3_extracted_k141_57280 | Not-determined |
| S3_extracted_k141_104925 | Not-determined |
| S3_extracted_k141_104927 | Not-determined |
| S3_extracted_k141_1423 | Not-determined |
| S3_extracted_k141_25130 | Not-determined |
| S3_extracted_k141_73617 | Not-determined |
| S3_extracted_k141_25131 | Not-determined |
| S3_extracted_k141_9866 | Not-determined |
| S3_extracted_k141_104935 | Low-quality |
| S3_extracted_k141_81368 | Not-determined |
| S3_extracted_k141_81370 | Not-determined |
| S3_extracted_k141_17092 | Not-determined |
| S3_extracted_k141_120620 | Not-determined |
| S3_extracted_k141_25136 | Not-determined |
| S3_extracted_k141_9871 | Not-determined |
| S3_extracted_k141_57295 | Not-determined |
| S3_extracted_k141_120626 | Not-determined |
| S3_extracted_k141_41255 | Not-determined |
| S3_extracted_k141_25145 | Low-quality |
| S3_extracted_k141_9884 | Not-determined |
| S3_extracted_k141_25151 | Not-determined |
| S3_extracted_k141_57299 | Not-determined |
| S3_extracted_k141_57302 | Not-determined |
| S3_extracted_k141_65051 | Not-determined |
| S3_extracted_k141_41264 | Not-determined |
| S3_extracted_k141_1442 | Not-determined |
| S3_extracted_k141_17101 | Not-determined |
| S3_extracted_k141_17103 | Not-determined |
| S3_extracted_k141_17107 | Not-determined |
| S3_extracted_k141_57312 | Low-quality |
| S3_extracted_k141_25161 | Not-determined |
| S3_extracted_k141_57318 | Not-determined |
| S3_extracted_k141_73652 | Not-determined |
| S3_extracted_k141_9898 | Low-quality |
| S3_extracted_k141_73653 | Not-determined |
| S3_extracted_k141_33696 | Not-determined |
| S3_extracted_k141_1450 | Not-determined |
| S3_extracted_k141_88911 | Not-determined |
| S3_extracted_k141_73657 | Not-determined |
| S3_extracted_k141_1456 | Not-determined |
| S3_extracted_k141_1461 | Low-quality |
| S3_extracted_k141_1462 | Low-quality |
| S3_extracted_k141_17119 | Not-determined |
| S3_extracted_k141_73666 | Not-determined |
| S3_extracted_k141_33708 | Not-determined |
| S3_extracted_k141_41273 | Not-determined |
| S3_extracted_k141_120640 | Not-determined |
| S3_extracted_k141_17124 | Not-determined |
| S3_extracted_k141_73680 | Low-quality |
| S3_extracted_k141_73681 | Low-quality |
| S3_extracted_k141_17128 | Low-quality |
| S3_extracted_k141_97242 | Not-determined |
| S3_extracted_k141_17130 | Not-determined |
| S3_extracted_k141_33728 | Not-determined |
| S3_extracted_k141_73687 | Low-quality |
| S3_extracted_k141_81384 | Not-determined |
| S3_extracted_k141_81385 | Not-determined |
| S3_extracted_k141_73691 | Not-determined |
| S3_extracted_k141_97244 | Low-quality |
| S3_extracted_k141_17142 | Not-determined |
| S3_extracted_k141_112756 | Not-determined |
| S3_extracted_k141_120650 | Not-determined |
| S3_extracted_k141_81401 | Not-determined |
| S3_extracted_k141_73700 | Low-quality |
| S3_extracted_k141_112768 | Low-quality |
| S3_extracted_k141_33739 | Not-determined |
| S3_extracted_k141_1486 | Not-determined |
| S3_extracted_k141_97250 | Not-determined |
| S3_extracted_k141_1489 | Not-determined |
| S3_extracted_k141_112777 | Not-determined |
| S3_extracted_k141_1492 | Not-determined |
| S3_extracted_k141_120661 | Not-determined |
| S3_extracted_k141_73718 | Not-determined |
| S3_extracted_k141_17150 | Not-determined |
| S3_extracted_k141_73720 | Not-determined |
| S3_extracted_k141_41292 | Not-determined |
| S3_extracted_k141_73724 | Not-determined |
| S3_extracted_k141_17157 | Not-determined |
| S3_extracted_k141_1496 | Not-determined |
| S3_extracted_k141_41300 | Not-determined |
| S3_extracted_k141_57336 | Low-quality |
| S3_extracted_k141_73732 | Not-determined |
| S3_extracted_k141_17163 | Not-determined |
| S3_extracted_k141_1499 | Not-determined |
| S3_extracted_k141_73734 | Not-determined |
| S3_extracted_k141_73735 | Not-determined |
| S3_extracted_k141_57340 | Not-determined |
| S3_extracted_k141_81409 | Low-quality |
| S3_extracted_k141_57346 | Not-determined |
| S3_extracted_k141_41310 | Not-determined |
| S3_extracted_k141_81416 | Not-determined |
| S3_extracted_k141_41317 | Low-quality |
| S3_extracted_k141_33751 | Low-quality |
| S3_extracted_k141_112785 | Not-determined |
| S3_extracted_k141_1510 | Not-determined |
| S3_extracted_k141_9927 | Not-determined |
| S3_extracted_k141_97271 | Low-quality |
| S3_extracted_k141_9931 | Not-determined |
| S3_extracted_k141_112794 | Not-determined |
| S3_extracted_k141_88934 | Not-determined |
| S3_extracted_k141_81426 | Not-determined |
| S3_extracted_k141_81427 | Not-determined |
| S3_extracted_k141_65085 | Not-determined |
| S3_extracted_k141_73739 | Not-determined |
| S3_extracted_k141_65090 | Not-determined |
| S3_extracted_k141_1527 | Not-determined |
| S3_extracted_k141_57356 | Not-determined |
| S3_extracted_k141_112799 | Not-determined |
| S3_extracted_k141_65092 | Not-determined |
| S3_extracted_k141_73742 | Not-determined |
| S3_extracted_k141_120672 | Not-determined |
| S3_extracted_k141_112802 | Not-determined |
| S3_extracted_k141_112804 | Low-quality |
| S3_extracted_k141_88945 | Not-determined |
| S3_extracted_k141_88947 | Not-determined |
| S3_extracted_k141_1539 | Not-determined |
| S3_extracted_k141_57359 | Not-determined |
| S3_extracted_k141_112809 | Not-determined |
| S3_extracted_k141_57360 | Not-determined |
| S3_extracted_k141_112811 | Not-determined |
| S3_extracted_k141_1543 | Not-determined |
| S3_extracted_k141_88952 | Not-determined |
| S3_extracted_k141_73746 | Not-determined |
| S3_extracted_k141_49638 | Not-determined |
| S3_extracted_k141_1552 | Low-quality |
| S3_extracted_k141_41334 | Not-determined |
| S3_extracted_k141_120685 | Not-determined |
| S3_extracted_k141_41336 | Low-quality |
| S3_extracted_k141_88966 | Low-quality |
| S3_extracted_k141_88971 | Low-quality |
| S3_extracted_k141_88980 | Low-quality |
| S3_extracted_k141_41347 | Not-determined |
| S3_extracted_k141_88985 | Not-determined |
| S3_extracted_k141_88986 | Not-determined |
| S3_extracted_k141_41348 | Not-determined |
| S3_extracted_k141_112834 | Not-determined |
| S3_extracted_k141_41359 | Low-quality |
| S3_extracted_k141_49667 | Not-determined |
| S3_extracted_k141_112838 | Not-determined |
| S3_extracted_k141_112840 | Not-determined |
| S3_extracted_k141_41365 | Low-quality |
| S3_extracted_k141_112842 | Not-determined |
| S3_extracted_k141_97294 | Not-determined |
| S3_extracted_k141_81440 | Not-determined |
| S3_extracted_k141_97296 | Low-quality |
| S3_extracted_k141_49679 | Not-determined |
| S3_extracted_k141_57386 | Not-determined |
| S3_extracted_k141_97297 | Low-quality |
| S3_extracted_k141_97298 | Not-determined |
| S3_extracted_k141_41373 | Not-determined |
| S3_extracted_k141_49680 | Not-determined |
| S3_extracted_k141_49681 | Not-determined |
| S3_extracted_k141_57390 | Not-determined |
| S3_extracted_k141_57392 | Not-determined |
| S3_extracted_k141_57396 | Not-determined |
| S3_extracted_k141_9963 | Low-quality |
| S3_extracted_k141_57398 | Not-determined |
| S3_extracted_k141_120690 | Not-determined |
| S3_extracted_k141_9968 | Not-determined |
| S4_vs_VIRSorter_10506-circular-cat | High-quality |
| S3_extracted_k141_57410 | Not-determined |
| S3_extracted_k141_65115 | Low-quality |
| S3_extracted_k141_120692 | Low-quality |
| S3_extracted_k141_120693 | Low-quality |
| S3_extracted_k141_9980 | Not-determined |
| S3_extracted_k141_9982 | Not-determined |
| S3_extracted_k141_65125 | Not-determined |
| S3_extracted_k141_9988 | Low-quality |
| S3_extracted_k141_25170 | Not-determined |
| S3_extracted_k141_9991 | Not-determined |
| S3_extracted_k141_25175 | Not-determined |
| S3_extracted_k141_9997 | Not-determined |
| S3_extracted_k141_73769 | Not-determined |
| S3_extracted_k141_104945 | Not-determined |
| S3_extracted_k141_104947 | Not-determined |
| S3_extracted_k141_57419 | Not-determined |
| S3_extracted_k141_57420 | Not-determined |
| S3_extracted_k141_97309 | Not-determined |
| S3_extracted_k141_9999 | Low-quality |
| S3_extracted_k141_97317 | Not-determined |
| S3_extracted_k141_10003 | Not-determined |
| S3_extracted_k141_81448 | Not-determined |
| S3_extracted_k141_25183 | Low-quality |
| S3_extracted_k141_10010 | Not-determined |
| S3_extracted_k141_104963 | Not-determined |
| S3_extracted_k141_104977 | Not-determined |
| S16_vs_6102\|\|full | High-quality |
| S3_extracted_k141_65127 | Not-determined |
| S3_extracted_k141_10026 | Not-determined |
| S3_extracted_k141_73779 | Not-determined |
| S3_extracted_k141_49694 | Not-determined |
| S3_extracted_k141_57436 | Not-determined |
| S3_extracted_k141_17169 | Not-determined |
| S3_extracted_k141_65137 | Not-determined |
| S3_extracted_k141_17171 | Not-determined |
| S3_extracted_k141_65143 | Low-quality |
| S3_extracted_k141_1578 | Not-determined |
| S3_extracted_k141_17177 | Not-determined |
| S3_extracted_k141_112867 | Not-determined |
| S3_extracted_k141_57453 | Not-determined |
| S3_extracted_k141_57455 | Not-determined |
| S3_extracted_k141_49708 | Not-determined |
| S3_extracted_k141_1585 | Low-quality |
| S3_extracted_k141_73784 | Not-determined |
| S3_extracted_k141_73785 | Not-determined |
| S3_extracted_k141_112877 | Not-determined |
| S3_extracted_k141_73792 | Not-determined |
| S3_extracted_k141_73794 | Not-determined |
| S3_extracted_k141_10052 | Not-determined |
| S3_extracted_k141_25203 | Low-quality |
| S3_extracted_k141_1593 | Not-determined |
| S3_extracted_k141_49714 | Not-determined |
| S3_extracted_k141_10062 | Low-quality |
| S3_extracted_k141_25206 | Not-determined |
| S3_extracted_k141_10065 | Not-determined |
| S3_extracted_k141_1596 | Low-quality |
| S3_extracted_k141_97346 | Not-determined |
| S3_extracted_k141_17189 | Low-quality |
| S3_extracted_k141_73808 | Low-quality |
| S3_extracted_k141_73809 | Not-determined |
| S3_extracted_k141_10074 | Not-determined |
| S3_extracted_k141_89048 | Low-quality |
| S3_extracted_k141_41423 | Not-determined |
| S3_extracted_k141_10076 | Not-determined |
| S3_extracted_k141_41426 | Not-determined |
| S3_extracted_k141_25223 | Not-determined |
| S3_extracted_k141_49731 | Low-quality |
| S3_extracted_k141_41427 | Not-determined |
| S3_extracted_k141_73814 | Not-determined |
| S3_extracted_k141_10081 | Low-quality |
| S3_extracted_k141_49735 | Not-determined |
| S3_extracted_k141_73828 | Low-quality |
| S3_extracted_k141_41431 | Not-determined |
| S3_extracted_k141_10085 | Low-quality |
| S3_extracted_k141_25233 | Not-determined |
| S3_extracted_k141_73833 | Not-determined |
| S3_extracted_k141_89052 | Not-determined |
| S3_extracted_k141_97354 | Not-determined |
| S3_extracted_k141_89057 | Not-determined |
| S3_extracted_k141_17198 | Low-quality |
| S3_extracted_k141_112905 | Not-determined |
| S3_extracted_k141_112906 | Not-determined |
| S3_extracted_k141_49744 | Not-determined |
| S3_extracted_k141_73847 | Not-determined |
| S3_extracted_k141_89068 | Not-determined |
| S3_extracted_k141_41435 | Low-quality |
| S3_extracted_k141_49757 | Not-determined |
| S3_extracted_k141_73855 | Not-determined |
| S3_extracted_k141_112923 | Not-determined |
| S3_extracted_k141_112924 | Not-determined |
| S3_extracted_k141_25241 | Not-determined |
| S3_extracted_k141_89074 | Not-determined |
| S3_extracted_k141_97368 | Low-quality |
| S3_extracted_k141_112932 | Not-determined |
| S3_extracted_k141_89083 | Not-determined |
| S3_extracted_k141_112934 | Low-quality |
| S3_extracted_k141_17212 | Low-quality |
| S3_extracted_k141_104992 | Low-quality |
| S3_extracted_k141_89090 | Not-determined |
| S3_extracted_k141_25253 | Not-determined |
| S3_extracted_k141_97376 | Not-determined |
| S3_extracted_k141_25254 | Not-determined |
| S3_extracted_k141_17216 | Not-determined |
| S3_extracted_k141_97377 | Not-determined |
| S3_extracted_k141_97379 | Not-determined |
| S3_extracted_k141_25257 | Not-determined |
| S3_extracted_k141_49772 | Not-determined |
| S3_extracted_k141_49774 | Not-determined |
| S3_extracted_k141_10111 | Not-determined |
| S3_extracted_k141_81464 | Low-quality |
| S3_extracted_k141_89097 | Not-determined |
| S3_extracted_k141_41440 | Low-quality |
| S3_extracted_k141_1625 | Not-determined |
| S3_extracted_k141_97385 | Low-quality |
| S3_extracted_k141_41447 | Not-determined |
| S3_extracted_k141_97388 | Not-determined |
| S3_extracted_k141_41449 | Not-determined |
| S3_extracted_k141_89105 | Not-determined |
| S3_extracted_k141_112948 | Low-quality |
| S3_extracted_k141_17230 | Not-determined |
| S3_extracted_k141_10130 | Not-determined |
| S3_extracted_k141_73893 | Not-determined |
| S3_extracted_k141_120747 | Not-determined |
| S3_extracted_k141_41453 | Not-determined |
| S3_extracted_k141_10137 | Not-determined |
| S3_extracted_k141_49782 | Low-quality |
| S3_extracted_k141_120751 | Not-determined |
| S3_extracted_k141_10139 | Not-determined |
| S3_extracted_k141_120755 | Not-determined |
| S3_extracted_k141_112956 | Low-quality |
| S3_extracted_k141_25276 | Not-determined |
| S3_extracted_k141_73899 | Not-determined |
| S3_extracted_k141_17246 | Not-determined |
| S3_extracted_k141_112960 | Not-determined |
| S3_extracted_k141_73903 | Not-determined |
| S3_extracted_k141_10146 | Low-quality |
| S3_extracted_k141_73906 | Low-quality |
| S3_extracted_k141_10148 | Not-determined |
| S3_extracted_k141_57473 | Not-determined |
| S3_extracted_k141_10153 | Low-quality |
| S3_extracted_k141_25287 | Not-determined |
| S3_extracted_k141_97406 | Not-determined |
| S3_extracted_k141_112984 | Not-determined |
| S3_extracted_k141_57481 | Not-determined |
| S3_extracted_k141_89126 | Low-quality |
| S3_extracted_k141_73914 | Not-determined |
| S3_extracted_k141_73916 | Not-determined |
| S3_extracted_k141_73918 | Low-quality |
| S3_extracted_k141_25292 | Low-quality |
| S3_extracted_k141_65166 | Not-determined |
| S3_extracted_k141_57490 | Not-determined |
| S3_extracted_k141_25294 | Not-determined |
| S3_extracted_k141_73922 | Not-determined |
| S3_extracted_k141_49805 | Not-determined |

| | |
|---|---|
| S3_extracted_k141_49808 | Not-determined |
| S3_extracted_k141_10179 | Not-determined |
| S3_extracted_k141_17255 | Not-determined |
| S3_extracted_k141_25304 | Not-determined |
| S3_extracted_k141_17259 | Not-determined |
| S3_extracted_k141_112987 | Low-quality |
| S3_extracted_k141_89146 | Not-determined |
| S3_extracted_k141_120794 | Not-determined |
| S3_extracted_k141_10183 | Not-determined |
| S3_extracted_k141_57505 | Low-quality |
| S3_extracted_k141_120799 | Not-determined |
| S3_extracted_k141_57506 | Low-quality |
| S3_extracted_k141_10192 | Not-determined |
| S3_extracted_k141_120806 | Not-determined |
| S3_extracted_k141_112999 | Not-determined |
| S3_extracted_k141_57512 | Not-determined |
| S3_extracted_k141_57514 | Not-determined |
| S3_extracted_k141_57515 | Not-determined |
| S3_extracted_k141_25321 | Not-determined |
| S3_extracted_k141_120817 | Not-determined |
| S3_extracted_k141_81485 | Not-determined |
| S3_extracted_k141_25324 | Not-determined |
| S3_extracted_k141_10205 | Not-determined |
| S3_extracted_k141_89163 | Not-determined |
| S3_extracted_k141_57530 | Not-determined |
| S3_extracted_k141_65171 | Not-determined |
| S3_extracted_k141_65176 | Not-determined |
| S3_extracted_k141_33772 | Not-determined |
| S3_extracted_k141_10213 | Not-determined |
| S3_extracted_k141_41466 | Not-determined |
| S3_extracted_k141_17266 | Not-determined |
| S3_extracted_k141_33783 | Not-determined |
| S3_extracted_k141_41469 | Not-determined |
| S3_extracted_k141_113020 | Not-determined |
| S3_extracted_k141_113035 | Not-determined |
| S3_extracted_k141_65182 | Not-determined |
| S3_extracted_k141_113037 | Not-determined |
| S3_extracted_k141_57540 | Not-determined |
| S3_extracted_k141_57541 | Not-determined |
| S3_extracted_k141_81494 | Not-determined |
| S3_extracted_k141_65186 | Not-determined |
| S3_extracted_k141_10220 | Not-determined |
| S3_extracted_k141_113043 | Not-determined |
| S3_extracted_k141_89169 | Not-determined |
| S3_extracted_k141_65190 | Not-determined |
| S3_extracted_k141_89178 | Not-determined |
| S3_extracted_k141_65196 | Low-quality |
| S3_extracted_k141_10239 | Not-determined |
| S3_extracted_k141_89183 | Not-determined |
| S3_extracted_k141_113046 | Low-quality |
| S3_extracted_k141_1668 | Not-determined |
| S3_extracted_k141_1670 | Not-determined |
| S3_extracted_k141_1671 | Low-quality |
| S3_extracted_k141_10254 | Not-determined |
| S3_extracted_k141_73933 | Not-determined |
| S3_extracted_k141_65207 | Not-determined |
| S3_extracted_k141_10258 | Not-determined |
| S3_extracted_k141_120822 | Not-determined |
| S3_extracted_k141_10262 | Not-determined |
| S3_extracted_k141_73946 | Not-determined |
| S3_extracted_k141_73948 | Not-determined |
| S3_extracted_k141_73949 | Not-determined |
| S3_extracted_k141_10271 | Not-determined |
| S3_extracted_k141_1680 | Not-determined |
| S3_extracted_k141_1681 | Not-determined |
| S3_extracted_k141_10273 | Not-determined |
| S3_extracted_k141_25331 | Not-determined |
| S3_extracted_k141_10281 | Not-determined |
| S3_extracted_k141_73966 | Not-determined |
| S3_extracted_k141_81510 | Not-determined |
| S3_extracted_k141_81514 | Not-determined |
| S3_extracted_k141_81515 | Not-determined |
| S3_extracted_k141_113053 | Not-determined |
| S3_extracted_k141_73973 | Not-determined |
| S3_extracted_k141_113056 | Not-determined |
| S3_extracted_k141_41481 | Not-determined |
| S3_extracted_k141_89196 | Not-determined |
| S3_extracted_k141_89198 | Low-quality |
| S3_extracted_k141_73975 | Not-determined |
| S3_extracted_k141_17280 | Not-determined |
| S3_extracted_k141_10301 | Not-determined |
| S3_extracted_k141_89208 | Not-determined |
| S3_extracted_k141_10309 | Not-determined |
| S3_extracted_k141_10312 | Not-determined |
| S3_extracted_k141_89209 | Not-determined |
| S3_extracted_k141_1706 | Not-determined |
| S3_extracted_k141_1708 | Not-determined |
| S3_extracted_k141_1709 | Not-determined |
| S3_extracted_k141_41491 | Low-quality |
| S3_extracted_k141_113065 | Low-quality |
| S3_extracted_k141_89210 | Low-quality |
| S3_extracted_k141_81529 | Not-determined |
| S3_extracted_k141_1718 | Not-determined |
| S3_extracted_k141_89217 | Not-determined |
| S3_extracted_k141_25345 | Low-quality |
| S3_extracted_k141_1720 | Not-determined |
| S3_extracted_k141_25351 | Low-quality |
| S3_extracted_k141_89224 | Not-determined |
| S3_extracted_k141_89225 | Not-determined |
| S3_extracted_k141_113074 | Low-quality |
| S3_extracted_k141_41508 | Not-determined |
| S3_extracted_k141_113075 | Not-determined |
| S3_extracted_k141_17284 | Not-determined |
| S3_extracted_k141_41510 | Not-determined |
| S3_extracted_k141_1733 | Low-quality |
| S3_extracted_k141_97436 | Not-determined |
| S3_extracted_k141_17290 | Low-quality |
| S3_extracted_k141_113079 | Not-determined |
| S3_extracted_k141_89241 | Not-determined |
| S3_extracted_k141_1743 | Not-determined |
| S3_extracted_k141_89246 | Not-determined |
| S3_extracted_k141_57570 | Low-quality |
| S3_extracted_k141_57572 | Low-quality |
| S3_extracted_k141_25360 | Low-quality |
| S3_extracted_k141_74010 | Not-determined |
| S3_extracted_k141_25362 | Not-determined |
| S3_extracted_k141_17301 | Not-determined |
| S3_extracted_k141_74016 | Not-determined |
| S3_extracted_k141_120851 | Not-determined |
| S3_extracted_k141_74017 | Low-quality |
| S3_extracted_k141_74018 | Low-quality |
| S3_extracted_k141_97449 | Low-quality |
| S3_extracted_k141_17316 | Not-determined |
| S3_extracted_k141_89263 | Not-determined |
| S3_extracted_k141_89267 | Not-determined |
| S3_extracted_k141_89268 | Not-determined |
| S3_extracted_k141_97453 | Not-determined |
| S3_extracted_k141_97454 | Not-determined |
| S3_extracted_k141_74033 | Not-determined |
| S3_extracted_k141_97457 | Low-quality |
| S3_extracted_k141_89275 | Not-determined |
| S3_extracted_k141_97459 | Not-determined |
| S3_extracted_k141_49837 | Low-quality |
| S3_extracted_k141_49839 | Not-determined |
| S3_extracted_k141_74037 | Not-determined |
| S3_extracted_k141_57599 | Low-quality |
| S3_extracted_k141_41523 | Low-quality |
| S3_extracted_k141_49843 | Low-quality |
| S3_extracted_k141_49844 | Low-quality |
| S3_extracted_k141_65217 | Not-determined |
| S3_extracted_k141_17331 | Not-determined |
| S3_extracted_k141_41530 | Not-determined |
| S3_extracted_k141_74051 | Low-quality |
| S3_extracted_k141_89281 | Not-determined |
| S3_extracted_k141_17340 | Not-determined |
| S3_extracted_k141_49855 | Not-determined |
| S3_extracted_k141_41542 | Not-determined |
| S3_extracted_k141_49856 | Not-determined |
| S3_extracted_k141_41544 | Not-determined |
| S3_extracted_k141_17345 | Not-determined |
| S3_extracted_k141_25380 | Not-determined |
| S3_extracted_k141_97477 | Low-quality |
| S3_extracted_k141_41550 | Not-determined |
| S3_extracted_k141_41553 | Not-determined |
| S3_extracted_k141_41554 | Not-determined |
| S3_extracted_k141_120872 | Low-quality |
| S3_extracted_k141_113101 | Not-determined |
| S3_extracted_k141_17365 | Not-determined |
| S3_extracted_k141_25384 | Low-quality |
| S3_extracted_k141_49866 | Not-determined |
| S3_extracted_k141_97488 | Low-quality |
| S3_extracted_k141_25389 | Low-quality |
| S3_extracted_k141_25390 | Low-quality |
| S3_extracted_k141_57623 | Not-determined |
| S3_extracted_k141_49873 | Not-determined |
| S3_extracted_k141_120880 | Not-determined |
| S3_extracted_k141_17369 | Not-determined |
| S3_extracted_k141_120886 | Not-determined |
| S3_extracted_k141_17374 | Low-quality |
| S3_extracted_k141_10354 | Not-determined |
| S3_extracted_k141_49894 | Low-quality |
| S3_extracted_k141_10364 | Not-determined |
| S3_extracted_k141_57632 | Not-determined |
| S3_extracted_k141_120898 | Low-quality |
| S3_extracted_k141_113139 | Low-quality |
| S3_extracted_k141_25410 | Not-determined |
| S3_extracted_k141_49904 | Not-determined |
| S3_extracted_k141_81538 | Not-determined |
| S3_extracted_k141_57643 | Not-determined |
| S3_extracted_k141_105014 | Not-determined |
| S3_extracted_k141_57647 | Not-determined |
| S3_extracted_k141_105023 | Not-determined |
| S3_extracted_k141_113153 | Not-determined |
| S3_extracted_k141_113154 | Not-determined |
| S3_extracted_k141_113155 | Not-determined |
| S3_extracted_k141_113156 | Low-quality |
| S3_extracted_k141_113160 | Low-quality |
| S3_extracted_k141_113162 | Not-determined |
| S3_extracted_k141_105036 | Not-determined |
| S3_extracted_k141_49926 | Not-determined |
| S3_extracted_k141_105045 | Not-determined |
| S3_extracted_k141_120903 | Not-determined |
| S3_extracted_k141_57656 | Not-determined |
| S3_extracted_k141_10372 | Not-determined |
| S3_extracted_k141_105051 | Not-determined |
| S3_extracted_k141_74064 | Low-quality |
| S3_extracted_k141_10377 | Not-determined |
| S3_extracted_k141_120908 | Not-determined |
| S3_extracted_k141_41573 | Not-determined |
| S3_extracted_k141_113178 | Not-determined |
| S3_extracted_k141_74066 | Not-determined |
| S3_extracted_k141_74070 | Not-determined |
| S3_extracted_k141_97509 | Not-determined |
| S3_extracted_k141_81568 | Not-determined |
| S3_extracted_k141_1751 | Not-determined |
| S3_extracted_k141_89299 | Not-determined |
| S3_extracted_k141_74079 | Low-quality |
| S3_extracted_k141_41590 | Not-determined |
| S3_extracted_k141_89307 | Not-determined |
| S3_extracted_k141_81574 | Not-determined |
| S3_extracted_k141_41592 | Not-determined |
| S3_extracted_k141_81579 | Not-determined |
| S3_extracted_k141_89316 | Not-determined |
| S8_vs_k141_185041\|\|full | High-quality |
| S3_extracted_k141_81584 | Not-determined |
| S3_extracted_k141_41599 | Not-determined |
| S3_extracted_k141_89318 | Not-determined |
| S3_extracted_k141_57677 | Not-determined |
| S3_extracted_k141_89321 | Not-determined |
| S3_extracted_k141_17384 | Low-quality |
| S3_extracted_k141_57681 | Low-quality |
| S3_extracted_k141_89323 | Not-determined |
| S3_extracted_k141_41604 | Low-quality |
| S3_extracted_k141_17394 | Not-determined |
| S3_extracted_k141_17399 | Not-determined |
| S3_extracted_k141_25440 | Not-determined |
| S3_extracted_k141_17400 | Low-quality |
| S3_extracted_k141_49949 | Low-quality |
| S3_extracted_k141_17401 | Not-determined |
| S3_extracted_k141_113200 | Not-determined |
| S3_extracted_k141_1778 | Not-determined |
| S3_extracted_k141_113201 | Not-determined |
| S3_extracted_k141_49951 | Not-determined |
| S3_extracted_k141_105091 | Not-determined |
| S3_extracted_k141_81592 | Not-determined |
| S3_extracted_k141_120941 | Not-determined |
| S3_extracted_k141_89338 | Low-quality |
| S3_extracted_k141_81598 | Not-determined |
| S3_extracted_k141_120949 | Not-determined |
| S3_extracted_k141_49965 | Not-determined |
| S3_extracted_k141_74088 | Low-quality |
| S3_extracted_k141_1794 | Not-determined |
| S3_extracted_k141_1795 | Not-determined |
| S3_extracted_k141_49969 | Low-quality |
| S3_extracted_k141_113223 | Low-quality |
| S3_extracted_k141_25450 | Not-determined |
| S3_extracted_k141_113227 | Not-determined |
| S3_extracted_k141_105102 | Not-determined |
| S3_extracted_k141_105105 | Not-determined |
| S3_extracted_k141_89358 | Not-determined |
| S3_extracted_k141_89360 | Not-determined |
| S3_extracted_k141_25458 | Low-quality |
| S3_extracted_k141_1808 | Not-determined |
| S3_extracted_k141_41625 | Not-determined |
| S3_extracted_k141_120958 | Not-determined |
| S3_extracted_k141_97528 | Not-determined |
| S3_extracted_k141_120964 | Low-quality |
| S3_extracted_k141_25462 | Low-quality |
| S3_extracted_k141_97532 | Not-determined |
| S3_extracted_k141_49984 | Not-determined |
| S3_extracted_k141_105109 | Low-quality |
| S3_extracted_k141_25471 | Not-determined |
| S3_extracted_k141_25473 | Not-determined |
| S3_extracted_k141_105111 | Not-determined |
| S3_extracted_k141_41639 | Not-determined |
| S3_extracted_k141_41640 | Not-determined |
| S3_extracted_k141_105117 | Not-determined |
| S3_extracted_k141_41646 | Not-determined |
| S3_extracted_k141_97533 | Low-quality |
| S3_extracted_k141_105122 | Not-determined |
| S3_extracted_k141_41654 | Not-determined |
| S3_extracted_k141_49996 | Not-determined |
| S3_extracted_k141_113245 | Low-quality |
| S3_extracted_k141_10428 | Low-quality |
| S3_extracted_k141_97538 | Not-determined |
| S3_extracted_k141_105130 | Not-determined |
| S3_extracted_k141_10430 | Not-determined |
| S3_extracted_k141_65223 | Not-determined |
| S3_extracted_k141_50002 | Not-determined |
| S3_extracted_k141_10432 | Not-determined |
| S3_extracted_k141_120969 | Not-determined |
| S3_extracted_k141_33869 | Not-determined |
| S3_extracted_k141_105131 | Not-determined |
| S3_extracted_k141_41665 | Not-determined |
| S3_extracted_k141_41670 | Not-determined |
| S3_extracted_k141_105139 | Not-determined |
| S3_extracted_k141_50005 | Not-determined |
| S3_extracted_k141_1832 | Not-determined |
| S3_extracted_k141_105143 | Not-determined |
| S3_extracted_k141_25494 | Not-determined |
| S3_extracted_k141_10437 | Low-quality |
| S3_extracted_k141_10438 | Not-determined |
| S3_extracted_k141_33879 | Not-determined |
| S3_extracted_k141_1835 | Not-determined |
| S3_extracted_k141_25497 | Not-determined |
| S3_extracted_k141_1837 | Not-determined |
| S3_extracted_k141_41682 | Low-quality |
| S3_extracted_k141_1843 | Not-determined |
| S3_extracted_k141_113254 | Not-determined |
| S3_extracted_k141_105157 | Not-determined |
| S3_extracted_k141_1846 | Not-determined |
| S3_extracted_k141_1850 | Not-determined |
| S3_extracted_k141_57720 | Not-determined |
| S3_extracted_k141_89376 | Low-quality |
| S3_extracted_k141_97548 | Low-quality |
| S3_extracted_k141_97549 | Not-determined |
| S3_extracted_k141_89381 | Not-determined |
| S3_extracted_k141_65251 | Not-determined |
| S3_extracted_k141_65253 | Low-quality |
| S3_extracted_k141_97553 | Not-determined |
| S3_extracted_k141_41697 | Not-determined |
| S3_extracted_k141_1865 | Not-determined |
| S3_extracted_k141_1866 | Not-determined |
| S3_extracted_k141_33884 | Not-determined |
| S3_extracted_k141_1868 | Not-determined |
| S3_extracted_k141_33888 | Not-determined |
| S3_extracted_k141_105168 | Not-determined |
| S3_extracted_k141_41703 | Not-determined |
| S3_extracted_k141_41704 | Not-determined |
| S3_extracted_k141_113258 | Not-determined |
| S3_extracted_k141_57736 | Not-determined |
| S3_extracted_k141_10466 | Low-quality |
| S3_extracted_k141_1871 | Not-determined |
| S3_extracted_k141_120985 | Not-determined |
| S3_extracted_k141_105175 | Not-determined |
| S3_extracted_k141_120987 | Not-determined |
| S3_extracted_k141_120988 | Not-determined |
| S3_extracted_k141_120992 | Low-quality |
| S3_extracted_k141_41712 | Low-quality |
| S3_extracted_k141_10490 | Not-determined |
| S3_extracted_k141_105181 | Not-determined |
| S3_extracted_k141_10495 | Low-quality |
| S3_extracted_k141_113275 | Not-determined |
| S3_extracted_k141_113276 | Not-determined |
| S3_extracted_k141_105191 | Not-determined |
| S3_extracted_k141_105192 | Not-determined |
| S3_extracted_k141_41721 | Not-determined |
| S3_extracted_k141_41722 | Not-determined |
| S3_extracted_k141_105194 | Not-determined |
| S3_extracted_k141_33893 | Not-determined |
| S3_extracted_k141_33894 | Not-determined |
| S3_extracted_k141_89386 | Not-determined |
| S3_extracted_k141_89387 | Not-determined |
| S3_extracted_k141_97561 | Not-determined |
| S3_extracted_k141_97562 | Not-determined |
| S3_extracted_k141_120998 | Not-determined |
| S3_extracted_k141_1888 | Low-quality |
| S3_extracted_k141_50053 | Not-determined |
| S3_extracted_k141_105203 | Not-determined |
| S3_extracted_k141_97567 | Not-determined |
| S3_extracted_k141_97568 | Not-determined |
| S3_extracted_k141_105211 | Not-determined |
| S3_extracted_k141_1895 | Low-quality |
| S3_extracted_k141_97572 | Low-quality |
| S3_extracted_k141_50057 | Not-determined |
| S3_extracted_k141_121012 | Not-determined |
| S3_extracted_k141_65282 | Not-determined |
| S3_extracted_k141_121015 | Not-determined |
| S3_extracted_k141_1905 | Not-determined |
| S3_extracted_k141_65286 | Not-determined |
| S3_extracted_k141_25513 | Not-determined |
| S3_extracted_k141_17434 | Not-determined |
| S3_extracted_k141_50069 | Not-determined |
| S3_extracted_k141_33901 | Low-quality |
| S3_extracted_k141_33903 | Not-determined |
| S3_extracted_k141_50074 | Not-determined |
| S3_extracted_k141_25521 | Not-determined |
| S3_extracted_k141_33909 | Low-quality |
| S3_extracted_k141_17444 | Low-quality |
| S3_extracted_k141_17446 | Not-determined |
| S3_extracted_k141_25526 | Low-quality |
| S3_extracted_k141_57772 | Not-determined |
| S3_extracted_k141_57774 | Not-determined |
| S3_extracted_k141_89407 | Low-quality |
| S3_extracted_k141_33918 | Low-quality |
| S3_extracted_k141_57780 | Not-determined |
| S3_extracted_k141_89412 | Not-determined |
| S3_extracted_k141_89416 | Low-quality |
| S3_extracted_k141_81620 | Not-determined |
| S3_extracted_k141_89418 | Not-determined |
| S3_extracted_k141_89426 | Not-determined |
| S3_extracted_k141_33923 | Not-determined |
| S3_extracted_k141_89429 | Not-determined |
| S3_extracted_k141_10539 | Not-determined |
| S3_extracted_k141_89435 | Not-determined |
| S3_extracted_k141_65300 | Not-determined |
| S3_extracted_k141_81634 | Not-determined |
| S3_extracted_k141_33935 | Not-determined |
| S3_extracted_k141_89450 | Not-determined |
| S3_extracted_k141_65310 | Low-quality |
| S3_extracted_k141_121037 | Low-quality |
| S3_extracted_k141_121040 | Low-quality |
| S3_extracted_k141_89461 | Low-quality |
| S3_extracted_k141_89463 | Not-determined |
| S3_extracted_k141_121052 | Low-quality |
| S3_extracted_k141_10552 | Not-determined |
| S3_extracted_k141_113302 | Low-quality |
| S3_extracted_k141_10557 | Not-determined |
| S3_extracted_k141_113316 | Not-determined |
| S3_extracted_k141_41725 | Not-determined |
| S3_extracted_k141_41726 | Not-determined |
| S3_extracted_k141_121059 | Low-quality |
| S3_extracted_k141_65325 | Low-quality |
| S3_extracted_k141_113330 | Not-determined |
| S3_extracted_k141_113331 | Not-determined |
| S3_extracted_k141_113332 | Not-determined |
| S3_extracted_k141_121064 | Low-quality |
| S3_extracted_k141_113335 | Not-determined |
| S3_extracted_k141_25557 | Not-determined |
| S3_extracted_k141_57795 | Not-determined |
| S3_extracted_k141_10583 | Not-determined |
| S3_extracted_k141_57797 | Not-determined |
| S3_extracted_k141_25560 | Not-determined |
| S3_extracted_k141_113343 | Not-determined |
| S3_extracted_k141_89471 | Low-quality |
| S3_extracted_k141_89473 | Not-determined |
| S3_extracted_k141_25564 | Not-determined |
| S3_extracted_k141_113346 | Low-quality |
| S3_extracted_k141_65336 | Not-determined |
| S3_extracted_k141_65337 | Not-determined |
| S3_extracted_k141_105220 | Low-quality |
| S3_extracted_k141_65341 | Not-determined |
| S3_extracted_k141_105223 | Not-determined |
| S3_extracted_k141_113348 | Low-quality |
| S3_extracted_k141_65348 | Not-determined |
| S3_extracted_k141_57811 | Not-determined |
| S3_extracted_k141_121080 | Not-determined |
| S3_extracted_k141_25579 | Not-determined |
| S3_extracted_k141_113350 | Low-quality |
| S3_extracted_k141_105235 | Not-determined |
| S3_extracted_k141_57817 | Not-determined |
| S3_extracted_k141_41730 | Not-determined |
| S3_extracted_k141_113352 | Not-determined |
| S3_extracted_k141_74099 | Not-determined |
| S3_extracted_k141_113361 | Not-determined |
| S3_extracted_k141_41743 | Not-determined |
| S3_extracted_k141_121091 | Not-determined |
| S3_extracted_k141_10621 | Not-determined |
| S3_extracted_k141_81648 | Not-determined |
| S3_extracted_k141_41755 | Not-determined |
| S3_extracted_k141_41757 | Not-determined |
| S3_extracted_k141_105256 | Low-quality |
| S3_extracted_k141_113382 | Not-determined |
| S3_extracted_k141_81664 | Not-determined |
| S3_extracted_k141_25585 | Not-determined |
| S3_extracted_k141_25588 | Not-determined |
| S3_extracted_k141_81669 | Not-determined |
| S3_extracted_k141_121111 | Not-determined |
| S3_extracted_k141_33953 | Not-determined |
| S3_extracted_k141_10639 | Not-determined |
| S3_extracted_k141_33958 | Not-determined |
| S3_extracted_k141_97588 | Not-determined |
| S3_extracted_k141_97592 | Not-determined |
| S3_extracted_k141_97594 | Not-determined |
| S3_extracted_k141_97599 | Low-quality |
| S3_extracted_k141_33975 | Not-determined |
| S3_extracted_k141_10655 | Not-determined |
| S3_extracted_k141_25593 | Not-determined |
| S3_extracted_k141_89494 | Not-determined |
| S3_extracted_k141_89495 | Not-determined |
| S3_extracted_k141_50083 | Not-determined |
| S3_extracted_k141_89499 | Not-determined |
| S3_extracted_k141_89500 | Not-determined |
| S3_extracted_k141_50085 | Low-quality |
| S3_extracted_k141_17469 | Not-determined |
| S3_extracted_k141_33994 | Not-determined |
| S3_extracted_k141_41759 | Low-quality |
| S3_extracted_k141_105288 | Not-determined |
| S3_extracted_k141_113402 | Not-determined |
| S3_extracted_k141_17471 | Not-determined |
| S3_extracted_k141_113404 | Not-determined |
| S3_extracted_k141_34000 | Not-determined |
| S3_extracted_k141_17473 | Not-determined |
| S3_extracted_k141_41765 | Not-determined |
| S3_extracted_k141_81693 | Not-determined |
| S3_extracted_k141_41767 | Not-determined |
| S3_extracted_k141_81697 | Not-determined |
| S3_extracted_k141_41772 | Not-determined |
| S3_extracted_k141_105298 | Low-quality |
| S3_extracted_k141_113411 | Not-determined |
| S3_extracted_k141_34005 | Not-determined |
| S3_extracted_k141_121121 | Not-determined |
| S3_extracted_k141_34008 | Not-determined |
| S3_extracted_k141_17480 | Not-determined |
| S3_extracted_k141_81702 | Low-quality |
| S3_extracted_k141_97605 | Not-determined |
| S3_extracted_k141_113420 | Not-determined |
| S3_extracted_k141_34009 | Not-determined |
| S3_extracted_k141_41786 | Low-quality |
| S3_extracted_k141_105305 | Not-determined |
| S3_extracted_k141_121133 | Not-determined |
| S3_extracted_k141_10672 | Not-determined |
| S3_extracted_k141_105316 | Low-quality |
| S3_extracted_k141_10678 | Low-quality |
| S3_extracted_k141_81711 | Not-determined |
| S3_extracted_k141_25616 | Not-determined |
| S3_extracted_k141_34017 | Not-determined |
| S3_extracted_k141_25618 | Not-determined |
| S3_extracted_k141_81714 | Not-determined |
| S3_extracted_k141_34019 | Low-quality |
| S3_extracted_k141_97610 | Not-determined |
| S3_extracted_k141_121136 | Low-quality |
| S3_extracted_k141_81720 | Not-determined |
| S3_extracted_k141_74146 | Not-determined |
| S3_extracted_k141_17487 | Not-determined |
| S3_extracted_k141_81726 | Not-determined |
| S3_extracted_k141_113441 | Not-determined |
| S3_extracted_k141_50097 | Not-determined |
| S3_extracted_k141_74153 | Not-determined |
| S3_extracted_k141_81731 | Low-quality |
| S3_extracted_k141_65359 | Not-determined |
| S3_extracted_k141_105334 | Not-determined |
| S3_extracted_k141_10696 | Not-determined |
| S3_extracted_k141_81748 | Not-determined |
| S3_extracted_k141_113463 | Low-quality |
| S3_extracted_k141_105342 | Not-determined |
| S3_extracted_k141_113469 | Not-determined |
| S3_extracted_k141_113471 | Not-determined |
| S3_extracted_k141_113473 | Not-determined |
| S3_extracted_k141_113477 | Not-determined |
| S3_extracted_k141_113479 | Low-quality |
| S3_extracted_k141_121167 | Not-determined |
| S3_extracted_k141_113481 | Low-quality |
| S3_extracted_k141_121169 | Not-determined |
| S3_extracted_k141_65363 | Not-determined |
| S3_extracted_k141_105358 | Not-determined |
| S3_extracted_k141_121172 | Low-quality |
| S3_extracted_k141_10717 | Not-determined |
| S3_extracted_k141_105366 | Not-determined |
| S3_extracted_k141_121185 | Not-determined |
| S3_extracted_k141_121188 | Not-determined |
| S3_extracted_k141_121192 | Not-determined |
| S3_extracted_k141_113498 | Not-determined |
| S3_extracted_k141_105373 | Not-determined |
| S3_extracted_k141_113499 | Not-determined |
| S3_extracted_k141_105375 | Low-quality |
| S3_extracted_k141_74183 | Low-quality |
| S3_extracted_k141_113505 | Not-determined |
| S3_extracted_k141_121204 | Not-determined |
| S3_extracted_k141_105389 | Not-determined |
| S3_extracted_k141_113508 | Not-determined |
| S3_extracted_k141_121209 | Not-determined |
| S3_extracted_k141_113511 | Not-determined |
| S3_extracted_k141_89562 | Not-determined |
| S3_extracted_k141_65379 | Not-determined |
| S3_extracted_k141_97642 | Not-determined |
| S3_extracted_k141_65384 | Not-determined |
| S3_extracted_k141_25639 | Not-determined |
| S3_extracted_k141_25641 | Not-determined |
| S3_extracted_k141_89572 | Not-determined |
| S3_extracted_k141_65388 | Not-determined |
| S3_extracted_k141_65393 | Low-quality |
| S3_extracted_k141_89574 | Not-determined |
| S3_extracted_k141_25651 | Not-determined |
| S3_extracted_k141_65400 | Not-determined |
| S3_extracted_k141_74198 | Not-determined |
| S3_extracted_k141_65413 | Not-determined |
| S3_extracted_k141_121233 | Not-determined |
| S3_extracted_k141_121235 | Low-quality |
| S3_extracted_k141_113535 | Not-determined |
| S3_extracted_k141_34069 | Low-quality |
| S3_extracted_k141_113544 | Not-determined |
| S3_extracted_k141_65421 | Low-quality |
| S3_extracted_k141_34075 | Not-determined |
| S3_extracted_k141_1915 | Not-determined |
| S3_extracted_k141_34082 | Not-determined |
| S3_extracted_k141_65431 | Not-determined |
| S3_extracted_k141_10746 | Not-determined |
| S3_extracted_k141_57829 | Low-quality |
| S3_extracted_k141_10748 | Not-determined |
| S3_extracted_k141_1923 | Not-determined |
| S3_extracted_k141_57835 | Not-determined |
| S3_extracted_k141_10753 | Not-determined |
| S3_extracted_k141_25660 | Not-determined |
| S3_extracted_k141_34096 | Not-determined |
| S3_extracted_k141_34099 | Not-determined |
| S3_extracted_k141_25664 | Not-determined |
| S3_extracted_k141_1940 | Not-determined |
| S3_extracted_k141_105427 | Not-determined |
| S3_extracted_k141_25667 | Not-determined |
| S3_extracted_k141_65443 | Not-determined |
| S3_extracted_k141_1942 | Not-determined |
| S3_extracted_k141_10763 | Not-determined |
| S3_extracted_k141_57853 | Not-determined |
| S3_extracted_k141_25673 | Not-determined |
| S3_extracted_k141_1947 | Not-determined |
| S3_extracted_k141_25677 | Not-determined |
| S3_extracted_k141_57858 | Low-quality |
| S3_extracted_k141_10771 | Not-determined |

| | |
|---|---|
| S3_extracted_k141_105445 | Not-determined |
| S3_extracted_k141_10772 | Low-quality |
| S3_extracted_k141_89577 | Low-quality |
| S3_extracted_k141_89578 | Not-determined |
| S3_extracted_k141_34111 | Low-quality |
| S3_extracted_k141_25686 | Not-determined |
| S3_extracted_k141_65461 | Not-determined |
| S3_extracted_k141_1956 | Not-determined |
| S3_extracted_k141_34118 | Low-quality |
| S3_extracted_k141_34123 | Not-determined |
| S3_extracted_k141_50124 | Low-quality |
| S3_extracted_k141_89591 | Not-determined |
| S3_extracted_k141_65476 | Not-determined |
| S3_extracted_k141_50127 | Not-determined |
| S3_extracted_k141_34131 | Not-determined |
| S3_extracted_k141_65480 | Not-determined |
| S3_extracted_k141_25694 | Not-determined |
| S3_extracted_k141_25695 | Not-determined |
| S3_extracted_k141_34137 | Not-determined |
| S3_extracted_k141_1973 | Not-determined |
| S3_extracted_k141_50137 | Not-determined |
| S3_extracted_k141_50138 | Not-determined |
| S3_extracted_k141_10785 | Low-quality |
| S3_extracted_k141_50146 | Low-quality |
| S3_extracted_k141_65491 | Not-determined |
| S3_extracted_k141_50149 | Not-determined |
| S3_extracted_k141_65492 | Low-quality |
| S3_extracted_k141_50154 | Not-determined |
| S3_extracted_k141_57871 | Low-quality |
| S3_extracted_k141_74223 | Not-determined |
| S3_extracted_k141_17534 | Not-determined |
| S3_extracted_k141_74226 | Not-determined |
| S3_extracted_k141_10793 | Low-quality |
| S3_extracted_k141_57879 | Not-determined |
| S3_extracted_k141_89597 | Not-determined |
| S3_extracted_k141_97659 | Not-determined |
| S3_extracted_k141_74234 | Low-quality |
| S3_extracted_k141_121262 | Low-quality |
| S3_extracted_k141_74238 | Not-determined |
| S3_extracted_k141_10807 | Not-determined |
| S3_extracted_k141_121268 | Not-determined |
| S10_vs_8854\|\|full | High-quality |
| S3_extracted_k141_34142 | Low-quality |
| S3_extracted_k141_34149 | Low-quality |
| S3_extracted_k141_121274 | Not-determined |
| S3_extracted_k141_25722 | Not-determined |
| S3_extracted_k141_2007 | Not-determined |
| S3_extracted_k141_2008 | Not-determined |
| S3_extracted_k141_34161 | Not-determined |
| S3_extracted_k141_10824 | Not-determined |
| S3_extracted_k141_34166 | Not-determined |
| S3_extracted_k141_89604 | Not-determined |
| S3_extracted_k141_89610 | Low-quality |
| S3_extracted_k141_65521 | Not-determined |
| S3_extracted_k141_97667 | Low-quality |
| S3_extracted_k141_10832 | Not-determined |
| S3_extracted_k141_2022 | Not-determined |
| S3_extracted_k141_97671 | Not-determined |
| S3_extracted_k141_97673 | Not-determined |
| S3_extracted_k141_89613 | Low-quality |
| S3_extracted_k141_57888 | Not-determined |
| S3_extracted_k141_41812 | Not-determined |
| S3_extracted_k141_2026 | Not-determined |
| S3_extracted_k141_121283 | Not-determined |
| S3_extracted_k141_34173 | Low-quality |
| S3_extracted_k141_57893 | Low-quality |
| S3_extracted_k141_25733 | Low-quality |
| S3_extracted_k141_57896 | Not-determined |
| S3_extracted_k141_10846 | Not-determined |
| S3_extracted_k141_25735 | Not-determined |
| S3_extracted_k141_34183 | Not-determined |
| S3_extracted_k141_121292 | Not-determined |
| S3_extracted_k141_2035 | Not-determined |
| S3_extracted_k141_41831 | Not-determined |
| S3_extracted_k141_57903 | Not-determined |
| S3_extracted_k141_10860 | Not-determined |
| S3_extracted_k141_10862 | Low-quality |
| S3_extracted_k141_2039 | Not-determined |
| S3_extracted_k141_57905 | Low-quality |
| S3_extracted_k141_17559 | Not-determined |
| S3_extracted_k141_57908 | Low-quality |
| S3_extracted_k141_41843 | Not-determined |
| S3_extracted_k141_113558 | Low-quality |
| S3_extracted_k141_113560 | Not-determined |
| S3_extracted_k141_57911 | Low-quality |
| S3_extracted_k141_41845 | Low-quality |
| S3_extracted_k141_57914 | Not-determined |
| S3_extracted_k141_17564 | Low-quality |
| S3_extracted_k141_113577 | Not-determined |
| S3_extracted_k141_34222 | Not-determined |
| S3_extracted_k141_34225 | Not-determined |
| S3_extracted_k141_10869 | Low-quality |
| S3_extracted_k141_10871 | Not-determined |
| S3_extracted_k141_97691 | Not-determined |
| S3_extracted_k141_2045 | Not-determined |
| S3_extracted_k141_10876 | Not-determined |
| S3_extracted_k141_17569 | Not-determined |
| S3_extracted_k141_17572 | Low-quality |
| S3_extracted_k141_10881 | Not-determined |
| S3_extracted_k141_25752 | Low-quality |
| S3_extracted_k141_34240 | Low-quality |
| S3_extracted_k141_25754 | Not-determined |
| S3_extracted_k141_41861 | Low-quality |
| S3_extracted_k141_34244 | Not-determined |
| S3_extracted_k141_10888 | Low-quality |
| S3_extracted_k141_34248 | Not-determined |
| S3_extracted_k141_10896 | Low-quality |
| S3_extracted_k141_25757 | Not-determined |
| S3_extracted_k141_34249 | Not-determined |
| S3_extracted_k141_121317 | Not-determined |
| S3_extracted_k141_34252 | Not-determined |
| S3_extracted_k141_10902 | Not-determined |
| S3_extracted_k141_34256 | Not-determined |
| S3_extracted_k141_121318 | Not-determined |
| S3_extracted_k141_41872 | Not-determined |
| S3_extracted_k141_10906 | Not-determined |
| S3_extracted_k141_2051 | Low-quality |
| S3_extracted_k141_121321 | Not-determined |
| S3_extracted_k141_121323 | Not-determined |
| S3_extracted_k141_121324 | Not-determined |
| S3_extracted_k141_121325 | Not-determined |
| S3_extracted_k141_57923 | Not-determined |
| S3_extracted_k141_34265 | Not-determined |
| S3_extracted_k141_121326 | Not-determined |
| S3_extracted_k141_17582 | Low-quality |
| S3_extracted_k141_2062 | Not-determined |
| S3_extracted_k141_34271 | Low-quality |
| S3_extracted_k141_17588 | Not-determined |
| S3_extracted_k141_34276 | Not-determined |
| S3_extracted_k141_74240 | Not-determined |
| S3_extracted_k141_121337 | Not-determined |
| S3_extracted_k141_2073 | Not-determined |
| S3_extracted_k141_57933 | Not-determined |
| S3_extracted_k141_2074 | Not-determined |
| S3_extracted_k141_2075 | Not-determined |
| S3_extracted_k141_57934 | Low-quality |
| S3_extracted_k141_17593 | Not-determined |
| S3_extracted_k141_34290 | Not-determined |
| S3_extracted_k141_34292 | Not-determined |
| S3_extracted_k141_10928 | Not-determined |
| S3_extracted_k141_17594 | Not-determined |
| S3_extracted_k141_10934 | Not-determined |
| S3_extracted_k141_17604 | Not-determined |
| S3_extracted_k141_74263 | Not-determined |
| S3_extracted_k141_89622 | Not-determined |
| S3_extracted_k141_57947 | Low-quality |
| S3_extracted_k141_17608 | Not-determined |
| S3_extracted_k141_105456 | Low-quality |
| S3_extracted_k141_89628 | Low-quality |
| S3_extracted_k141_57952 | Not-determined |
| S3_extracted_k141_17616 | Not-determined |
| S3_extracted_k141_41893 | Not-determined |
| S3_extracted_k141_17619 | Not-determined |
| S3_extracted_k141_89632 | Not-determined |
| S3_extracted_k141_17629 | Not-determined |
| S3_extracted_k141_10957 | Not-determined |
| S3_extracted_k141_57964 | Not-determined |
| S3_extracted_k141_57966 | Low-quality |
| S3_extracted_k141_10961 | Not-determined |
| S3_extracted_k141_10964 | Not-determined |
| S3_extracted_k141_17633 | Not-determined |
| S3_extracted_k141_121348 | Not-determined |
| S3_extracted_k141_25773 | Not-determined |
| S3_extracted_k141_25774 | Not-determined |
| S3_extracted_k141_10967 | Not-determined |
| S3_extracted_k141_65555 | Not-determined |
| S3_extracted_k141_121353 | Not-determined |
| S3_extracted_k141_10970 | Not-determined |
| S3_extracted_k141_121354 | Not-determined |
| S3_extracted_k141_25779 | Not-determined |
| S3_extracted_k141_65560 | Low-quality |
| S3_extracted_k141_41920 | Not-determined |
| S3_extracted_k141_34321 | Not-determined |
| S3_extracted_k141_34324 | Not-determined |
| S3_extracted_k141_10976 | Low-quality |
| S3_extracted_k141_65567 | Not-determined |
| S3_extracted_k141_25783 | Not-determined |
| S3_extracted_k141_65570 | Low-quality |
| S3_extracted_k141_17637 | Low-quality |
| S3_extracted_k141_74269 | Not-determined |
| S3_extracted_k141_89640 | Not-determined |
| S3_extracted_k141_57980 | Not-determined |
| S3_extracted_k141_105472 | Not-determined |
| S3_extracted_k141_74277 | Not-determined |
| S3_extracted_k141_41932 | Not-determined |
| S3_extracted_k141_57984 | Not-determined |
| S3_extracted_k141_105476 | Low-quality |
| S3_extracted_k141_89644 | Not-determined |
| S3_extracted_k141_105481 | Not-determined |
| S3_extracted_k141_105483 | Not-determined |
| S3_extracted_k141_121367 | Low-quality |
| S3_extracted_k141_97713 | Not-determined |
| S3_extracted_k141_121371 | Not-determined |
| S3_extracted_k141_25810 | Not-determined |
| S3_extracted_k141_89656 | Low-quality |
| S3_extracted_k141_89659 | Not-determined |
| S3_extracted_k141_97723 | Not-determined |
| S3_extracted_k141_57987 | Low-quality |
| S3_extracted_k141_41959 | Not-determined |
| S3_extracted_k141_97728 | Not-determined |
| S3_extracted_k141_34337 | Low-quality |
| S3_extracted_k141_121378 | Not-determined |
| S3_extracted_k141_57993 | Not-determined |
| S3_extracted_k141_34339 | Not-determined |
| S3_extracted_k141_34340 | Not-determined |
| S3_extracted_k141_97737 | Not-determined |
| S3_extracted_k141_58000 | Not-determined |
| S3_extracted_k141_113586 | Not-determined |
| S3_extracted_k141_105496 | Not-determined |
| S3_extracted_k141_34348 | Not-determined |
| S3_extracted_k141_25814 | Not-determined |
| S3_extracted_k141_25815 | Not-determined |
| S3_extracted_k141_58004 | Not-determined |
| S3_extracted_k141_81762 | Not-determined |
| S3_extracted_k141_41970 | Not-determined |
| S3_extracted_k141_113594 | Not-determined |
| S3_extracted_k141_121394 | Not-determined |
| S3_extracted_k141_113595 | Low-quality |
| S3_extracted_k141_81769 | Not-determined |
| S3_extracted_k141_74294 | Not-determined |
| S3_extracted_k141_74295 | Not-determined |
| S3_extracted_k141_113602 | Low-quality |
| S3_extracted_k141_25826 | Not-determined |
| S3_extracted_k141_41985 | Low-quality |
| S3_extracted_k141_113604 | Not-determined |
| S3_extracted_k141_2091 | Not-determined |
| S3_extracted_k141_113606 | Not-determined |
| S3_extracted_k141_81776 | Low-quality |
| S3_extracted_k141_81777 | Low-quality |
| S3_extracted_k141_113607 | Not-determined |
| S3_extracted_k141_25833 | Not-determined |
| S3_extracted_k141_65592 | Not-determined |
| S3_extracted_k141_25836 | Not-determined |
| S3_extracted_k141_89675 | Low-quality |
| S3_extracted_k141_65596 | Not-determined |
| S3_extracted_k141_17654 | Not-determined |
| S3_extracted_k141_42000 | Not-determined |
| S3_extracted_k141_58032 | Not-determined |
| S3_extracted_k141_34362 | Not-determined |
| S3_extracted_k141_50210 | Low-quality |
| S3_extracted_k141_113621 | Not-determined |
| S3_extracted_k141_34368 | Low-quality |
| S3_extracted_k141_65618 | Not-determined |
| S3_extracted_k141_42003 | Not-determined |
| S3_extracted_k141_105520 | Not-determined |
| S3_extracted_k141_113625 | Not-determined |
| S3_extracted_k141_105521 | Not-determined |
| S3_extracted_k141_25860 | Low-quality |
| S3_extracted_k141_34379 | Not-determined |
| S3_extracted_k141_25863 | Low-quality |
| S3_extracted_k141_89686 | Not-determined |
| S3_extracted_k141_121400 | Not-determined |
| S3_extracted_k141_113629 | Low-quality |
| S3_extracted_k141_105530 | Not-determined |
| S3_extracted_k141_121409 | Not-determined |
| S3_extracted_k141_121411 | Not-determined |
| S3_extracted_k141_89687 | Low-quality |
| S3_extracted_k141_89691 | Not-determined |
| S3_extracted_k141_65627 | Not-determined |
| S3_extracted_k141_105531 | Not-determined |
| S3_extracted_k141_105535 | Not-determined |
| S3_extracted_k141_42024 | Not-determined |
| S3_extracted_k141_105539 | Not-determined |
| S3_extracted_k141_97741 | Low-quality |
| S3_extracted_k141_42030 | Not-determined |
| S3_extracted_k141_89698 | Not-determined |
| S3_extracted_k141_97743 | Low-quality |
| S3_extracted_k141_97746 | Not-determined |
| S3_extracted_k141_89700 | Not-determined |
| S3_extracted_k141_89701 | Not-determined |
| S3_extracted_k141_58054 | Low-quality |
| S3_extracted_k141_58056 | Not-determined |
| S3_extracted_k141_113638 | Not-determined |
| S3_extracted_k141_89703 | Not-determined |
| S3_extracted_k141_58058 | Low-quality |
| S3_extracted_k141_113642 | Low-quality |
| S3_extracted_k141_105552 | Not-determined |
| S3_extracted_k141_97752 | Not-determined |
| S3_extracted_k141_34389 | Low-quality |
| S3_extracted_k141_113648 | Not-determined |
| S3_extracted_k141_11036 | Low-quality |
| S3_extracted_k141_17673 | Low-quality |
| S3_extracted_k141_11037 | Not-determined |
| S3_extracted_k141_113651 | Not-determined |
| S3_extracted_k141_11040 | Not-determined |
| S3_extracted_k141_65653 | Not-determined |
| S3_extracted_k141_34403 | Low-quality |
| S3_extracted_k141_50225 | Not-determined |
| S3_extracted_k141_89708 | Not-determined |
| S3_extracted_k141_34408 | Not-determined |
| S3_extracted_k141_113660 | Not-determined |
| S3_extracted_k141_65659 | Not-determined |
| S3_extracted_k141_58063 | Low-quality |
| S3_extracted_k141_50229 | Not-determined |
| S3_extracted_k141_58066 | Not-determined |
| S3_extracted_k141_58068 | Not-determined |
| S3_extracted_k141_11047 | Low-quality |
| S3_extracted_k141_58072 | Not-determined |
| S3_extracted_k141_11052 | Not-determined |
| S3_extracted_k141_58074 | Low-quality |
| S3_extracted_k141_113670 | Not-determined |
| S3_extracted_k141_11057 | Not-determined |
| S3_extracted_k141_58083 | Not-determined |
| S3_extracted_k141_65669 | Low-quality |
| S3_extracted_k141_113673 | Not-determined |
| S3_extracted_k141_105567 | Not-determined |
| S3_extracted_k141_105568 | Low-quality |
| S3_extracted_k141_113677 | Not-determined |
| S3_extracted_k141_105570 | Low-quality |
| S3_extracted_k141_89717 | Low-quality |
| S3_extracted_k141_50236 | Not-determined |
| S3_extracted_k141_113682 | Not-determined |
| S3_extracted_k141_89718 | Not-determined |
| S3_extracted_k141_11065 | Not-determined |
| S3_extracted_k141_58090 | Low-quality |
| S3_extracted_k141_58092 | Not-determined |
| S3_extracted_k141_65682 | Not-determined |
| S3_extracted_k141_34413 | Not-determined |
| S3_extracted_k141_11076 | Not-determined |
| S3_extracted_k141_34422 | Not-determined |
| S3_extracted_k141_17677 | Low-quality |
| S3_extracted_k141_113688 | Not-determined |
| S3_extracted_k141_113689 | Not-determined |
| S3_extracted_k141_17678 | Not-determined |
| S3_extracted_k141_113690 | Not-determined |
| S3_extracted_k141_65693 | Not-determined |
| S3_extracted_k141_65697 | Low-quality |
| S3_extracted_k141_105586 | Not-determined |
| S3_extracted_k141_89730 | Low-quality |
| S3_extracted_k141_17682 | Not-determined |
| S3_extracted_k141_113699 | Not-determined |
| S3_extracted_k141_58102 | Not-determined |
| S3_extracted_k141_65701 | Not-determined |
| S3_extracted_k141_113703 | Not-determined |
| S3_extracted_k141_2120 | Low-quality |
| S3_extracted_k141_74334 | Not-determined |
| S3_extracted_k141_105593 | Not-determined |
| S3_extracted_k141_58114 | Not-determined |
| S3_extracted_k141_58115 | Not-determined |
| S3_extracted_k141_81797 | Not-determined |
| S3_extracted_k141_113712 | Low-quality |
| S3_extracted_k141_105598 | Not-determined |
| S3_extracted_k141_113714 | Not-determined |
| S3_extracted_k141_42068 | Not-determined |
| S3_extracted_k141_81803 | Low-quality |
| S3_extracted_k141_11109 | Not-determined |
| S3_extracted_k141_121419 | Not-determined |
| S3_extracted_k141_113722 | Not-determined |
| S3_extracted_k141_121423 | Not-determined |
| S3_extracted_k141_121424 | Not-determined |
| S3_extracted_k141_34425 | Not-determined |
| S3_extracted_k141_11118 | Not-determined |
| S3_extracted_k141_105613 | Low-quality |
| S3_extracted_k141_11121 | Not-determined |
| S3_extracted_k141_105615 | Low-quality |
| S3_extracted_k141_105617 | Not-determined |
| S3_extracted_k141_65720 | Not-determined |
| S3_extracted_k141_81809 | Low-quality |
| S3_extracted_k141_113740 | Not-determined |
| S3_extracted_k141_89741 | Not-determined |
| S3_extracted_k141_113743 | Not-determined |
| S3_extracted_k141_65725 | Not-determined |
| S3_extracted_k141_89742 | Not-determined |
| S3_extracted_k141_58121 | Low-quality |
| S3_extracted_k141_50244 | Not-determined |
| S3_extracted_k141_113746 | Not-determined |
| S3_extracted_k141_17699 | Not-determined |
| S3_extracted_k141_113747 | Low-quality |
| S3_extracted_k141_58126 | Not-determined |
| S3_extracted_k141_17702 | Low-quality |
| S3_extracted_k141_2145 | Low-quality |
| S3_extracted_k141_81824 | Not-determined |
| S3_extracted_k141_105641 | Not-determined |
| S3_extracted_k141_17705 | Low-quality |
| S3_extracted_k141_74346 | Not-determined |
| S3_extracted_k141_74350 | Not-determined |
| S3_extracted_k141_105647 | Not-determined |
| S3_extracted_k141_11156 | Not-determined |
| S3_extracted_k141_11158 | Low-quality |
| S3_extracted_k141_58132 | Low-quality |
| S3_extracted_k141_25904 | Not-determined |
| S3_extracted_k141_121441 | Low-quality |
| S3_extracted_k141_11167 | Not-determined |
| S3_extracted_k141_34431 | Low-quality |
| S3_extracted_k141_74355 | Not-determined |
| S3_extracted_k141_50256 | Not-determined |
| S3_extracted_k141_58145 | Not-determined |
| S3_extracted_k141_65744 | Not-determined |
| S3_extracted_k141_105667 | Not-determined |
| S3_extracted_k141_50262 | Not-determined |
| S3_extracted_k141_74363 | Not-determined |
| S3_extracted_k141_25921 | Not-determined |
| S3_extracted_k141_11171 | Not-determined |
| S3_extracted_k141_17709 | Low-quality |
| S3_extracted_k141_105674 | Low-quality |
| S3_extracted_k141_65746 | Low-quality |
| S3_extracted_k141_65749 | Not-determined |
| S3_extracted_k141_17715 | Low-quality |
| S3_extracted_k141_74375 | Not-determined |
| S3_extracted_k141_81850 | Low-quality |
| S3_extracted_k141_81853 | Not-determined |
| S3_extracted_k141_11179 | Not-determined |
| S3_extracted_k141_25934 | Not-determined |
| S3_extracted_k141_34433 | Not-determined |
| S3_extracted_k141_42108 | Not-determined |
| S3_extracted_k141_50268 | Not-determined |
| S3_extracted_k141_58153 | Not-determined |
| S3_extracted_k141_34436 | Low-quality |
| S3_extracted_k141_17719 | Low-quality |
| S3_extracted_k141_17720 | Low-quality |
| S3_extracted_k141_65759 | Not-determined |
| S3_extracted_k141_81866 | Not-determined |
| S3_extracted_k141_50274 | Not-determined |
| S3_extracted_k141_11191 | Not-determined |
| S3_extracted_k141_97775 | Not-determined |
| S3_extracted_k141_25942 | Not-determined |
| S3_extracted_k141_58156 | Not-determined |
| S3_extracted_k141_97778 | Not-determined |
| S3_extracted_k141_58159 | Not-determined |
| S3_extracted_k141_42121 | Low-quality |
| S3_extracted_k141_65768 | Not-determined |
| S3_extracted_k141_58162 | Not-determined |
| S3_extracted_k141_105699 | Not-determined |
| S3_extracted_k141_50286 | Not-determined |
| S3_extracted_k141_65772 | Not-determined |
| S3_extracted_k141_11200 | Not-determined |
| S3_extracted_k141_25952 | Not-determined |
| S3_extracted_k141_65774 | Not-determined |
| S3_extracted_k141_50293 | Not-determined |
| S3_extracted_k141_11211 | Low-quality |
| S3_extracted_k141_58167 | Low-quality |
| S3_extracted_k141_34441 | Not-determined |
| S3_extracted_k141_97792 | Not-determined |
| S3_extracted_k141_58179 | Not-determined |
| S3_extracted_k141_65793 | Not-determined |
| S3_extracted_k141_58183 | Not-determined |
| S3_extracted_k141_105729 | Not-determined |
| S3_extracted_k141_89753 | Low-quality |
| S3_extracted_k141_25960 | Not-determined |
| S3_extracted_k141_89755 | Not-determined |
| S3_extracted_k141_25966 | Not-determined |
| S3_extracted_k141_50295 | Not-determined |
| S3_extracted_k141_50296 | Low-quality |
| S3_extracted_k141_74398 | Not-determined |
| S3_extracted_k141_97804 | Not-determined |
| S3_extracted_k141_34449 | Not-determined |
| S3_extracted_k141_81868 | Not-determined |
| S3_extracted_k141_58200 | Not-determined |
| S3_extracted_k141_58203 | Not-determined |
| S3_extracted_k141_50309 | Not-determined |
| S3_extracted_k141_25974 | Not-determined |
| S3_extracted_k141_89764 | Low-quality |
| S3_extracted_k141_65799 | Not-determined |
| S3_extracted_k141_65800 | Not-determined |
| S3_extracted_k141_50318 | Not-determined |
| S3_extracted_k141_65801 | Not-determined |
| S3_extracted_k141_121460 | Not-determined |
| S3_extracted_k141_97811 | Not-determined |
| S3_extracted_k141_2158 | Low-quality |
| S3_extracted_k141_25982 | Not-determined |
| S3_extracted_k141_89769 | Not-determined |
| S3_extracted_k141_58209 | Low-quality |
| S3_extracted_k141_17739 | Not-determined |
| S3_extracted_k141_121471 | Low-quality |
| S3_extracted_k141_74407 | Not-determined |
| S3_extracted_k141_50328 | Not-determined |
| S3_extracted_k141_97814 | Not-determined |
| S3_extracted_k141_65819 | Not-determined |
| S3_extracted_k141_65823 | Not-determined |
| S3_extracted_k141_74412 | Not-determined |
| S3_extracted_k141_89780 | Not-determined |
| S3_extracted_k141_58214 | Not-determined |
| S3_extracted_k141_50347 | Not-determined |
| S3_extracted_k141_113756 | Low-quality |
| S3_extracted_k141_34457 | Not-determined |
| S3_extracted_k141_58218 | Not-determined |
| S3_extracted_k141_34462 | Not-determined |
| S3_extracted_k141_121485 | Not-determined |
| S3_extracted_k141_58222 | Low-quality |
| S3_extracted_k141_105753 | Not-determined |
| S3_extracted_k141_34469 | Not-determined |
| S3_extracted_k141_58231 | Low-quality |
| S3_extracted_k141_74416 | Low-quality |
| S3_extracted_k141_74417 | Not-determined |
| S3_extracted_k141_74418 | Not-determined |
| S3_extracted_k141_105765 | Not-determined |
| S3_extracted_k141_74419 | Not-determined |
| S3_extracted_k141_113762 | Not-determined |
| S3_extracted_k141_34474 | Low-quality |
| S3_extracted_k141_50355 | Not-determined |
| S3_extracted_k141_50360 | Low-quality |
| S3_extracted_k141_65836 | Not-determined |
| S3_extracted_k141_11289 | Not-determined |
| S3_extracted_k141_89785 | Not-determined |
| S3_extracted_k141_65838 | Not-determined |
| S3_extracted_k141_121503 | Not-determined |
| S3_extracted_k141_121505 | Low-quality |
| S3_extracted_k141_65846 | Low-quality |
| S3_extracted_k141_89791 | Not-determined |
| S3_extracted_k141_81883 | Not-determined |
| S3_extracted_k141_50366 | Low-quality |
| S3_extracted_k141_89797 | Not-determined |
| S3_extracted_k141_11295 | Not-determined |
| S3_extracted_k141_81890 | Not-determined |
| S3_extracted_k141_81893 | Not-determined |
| S3_extracted_k141_65859 | Not-determined |
| S3_extracted_k141_65860 | Not-determined |
| S3_extracted_k141_34477 | Low-quality |
| S3_extracted_k141_11299 | Low-quality |
| S3_extracted_k141_34479 | Not-determined |
| S3_extracted_k141_58237 | Not-determined |
| S3_extracted_k141_2171 | Not-determined |
| S3_extracted_k141_65867 | Not-determined |
| S3_extracted_k141_89808 | Low-quality |
| S3_extracted_k141_58241 | Not-determined |
| S3_extracted_k141_2181 | Not-determined |
| S3_extracted_k141_105801 | Not-determined |
| S3_extracted_k141_105805 | Not-determined |
| S3_extracted_k141_42144 | Not-determined |
| S3_extracted_k141_65870 | Not-determined |
| S3_extracted_k141_89817 | Low-quality |
| S3_extracted_k141_105807 | Low-quality |
| S3_extracted_k141_65876 | Not-determined |
| S3_extracted_k141_113769 | Not-determined |
| S3_extracted_k141_105811 | Not-determined |
| S3_extracted_k141_121513 | Not-determined |
| S3_extracted_k141_11340 | Not-determined |
| S3_extracted_k141_65882 | Low-quality |
| S3_extracted_k141_74436 | Not-determined |
| S3_extracted_k141_50415 | Low-quality |
| S3_extracted_k141_50421 | Not-determined |
| S3_extracted_k141_105841 | Not-determined |
| S3_extracted_k141_65892 | Not-determined |
| S3_extracted_k141_65893 | Not-determined |
| S3_extracted_k141_121531 | Not-determined |
| S3_extracted_k141_81902 | Low-quality |
| S4_vs_VIRSorter_24603-circular-cat | High-quality |
| S3_extracted_k141_113777 | Not-determined |
| S3_extracted_k141_105848 | Not-determined |
| S3_extracted_k141_11391 | Low-quality |
| S3_extracted_k141_65904 | Not-determined |
| S3_extracted_k141_105850 | Low-quality |
| S3_extracted_k141_113780 | Not-determined |
| S3_extracted_k141_121541 | Not-determined |
| S3_extracted_k141_121545 | Not-determined |
| S3_extracted_k141_121547 | Not-determined |
| S3_extracted_k141_89858 | Not-determined |
| S3_extracted_k141_74441 | Low-quality |
| S3_extracted_k141_50444 | Low-quality |
| S16_extracted_47559 | Complete |
| S3_extracted_k141_50446 | Not-determined |
| S3_extracted_k141_50447 | Not-determined |
| S3_extracted_k141_74449 | Not-determined |
| S3_extracted_k141_113788 | Not-determined |
| S3_extracted_k141_89862 | Low-quality |
| S3_extracted_k141_17751 | Not-determined |
| S6_vs_k141_56496\|\|full | High-quality |
| S3_extracted_k141_89870 | Low-quality |
| S3_extracted_k141_113799 | Not-determined |
| S3_extracted_k141_74460 | Low-quality |
| S3_extracted_k141_2190 | Not-determined |
| S3_extracted_k141_74465 | Not-determined |
| S3_extracted_k141_89872 | Not-determined |
| S3_extracted_k141_50460 | Not-determined |
| S3_extracted_k141_113801 | Not-determined |
| S3_extracted_k141_113804 | Not-determined |
| S3_extracted_k141_74472 | Not-determined |
| S3_extracted_k141_89882 | Not-determined |
| S3_extracted_k141_74478 | Not-determined |
| S3_extracted_k141_50468 | Not-determined |
| S3_extracted_k141_121576 | Not-determined |
| S3_extracted_k141_121577 | Not-determined |
| S3_extracted_k141_97843 | Not-determined |
| S3_extracted_k141_121579 | Not-determined |
| S3_extracted_k141_74484 | Not-determined |
| S3_extracted_k141_113819 | Not-determined |
| S3_extracted_k141_113823 | Not-determined |
| S3_extracted_k141_17765 | Low-quality |
| S3_extracted_k141_65928 | Not-determined |
| S3_extracted_k141_65930 | Not-determined |
| S3_extracted_k141_89893 | Not-determined |
| S3_extracted_k141_121593 | Not-determined |
| S3_extracted_k141_25998 | Not-determined |
| S3_extracted_k141_26004 | Not-determined |
| S3_extracted_k141_121600 | Not-determined |
| S3_extracted_k141_65942 | Low-quality |
| S3_extracted_k141_121604 | Not-determined |
| S3_extracted_k141_58259 | Not-determined |
| S3_extracted_k141_121609 | Not-determined |

| | |
|---|---|
| S3_extracted_k141_121610 | Not-determined |
| S3_extracted_k141_105865 | Not-determined |
| S3_extracted_k141_121611 | Low-quality |
| S3_extracted_k141_74509 | Low-quality |
| S3_extracted_k141_105869 | Low-quality |
| S3_extracted_k141_17772 | Not-determined |
| S3_extracted_k141_74511 | Not-determined |
| S3_extracted_k141_105872 | Not-determined |
| S3_extracted_k141_105873 | Not-determined |
| S3_extracted_k141_42176 | Not-determined |
| S3_extracted_k141_42180 | Low-quality |
| S3_extracted_k141_26015 | Not-determined |
| S3_extracted_k141_58272 | Not-determined |
| S3_extracted_k141_50486 | Low-quality |
| S3_extracted_k141_26019 | Low-quality |
| S3_extracted_k141_58277 | Not-determined |
| S3_extracted_k141_113829 | Not-determined |
| S4_extracted_13353 | High-quality |
| S3_extracted_k141_42195 | Not-determined |
| S3_extracted_k141_26023 | Low-quality |
| S3_extracted_k141_121617 | Not-determined |
| S3_extracted_k141_97900 | Not-determined |
| S3_extracted_k141_113838 | Not-determined |
| S3_extracted_k141_26030 | Not-determined |
| S3_extracted_k141_42205 | Not-determined |
| S3_extracted_k141_42206 | Low-quality |
| S3_extracted_k141_81915 | Not-determined |
| S3_extracted_k141_81917 | Not-determined |
| S3_extracted_k141_81918 | Not-determined |
| S3_extracted_k141_58295 | Not-determined |
| S3_extracted_k141_121636 | Not-determined |
| S3_extracted_k141_58298 | Not-determined |
| S3_extracted_k141_26040 | Not-determined |
| S3_extracted_k141_121648 | Not-determined |
| S3_extracted_k141_42216 | Not-determined |
| S3_extracted_k141_58299 | Low-quality |
| S3_extracted_k141_105888 | Not-determined |
| S3_extracted_k141_81932 | Low-quality |
| S3_extracted_k141_26049 | Not-determined |
| S3_extracted_k141_81934 | Not-determined |
| S3_extracted_k141_11461 | Not-determined |
| S3_extracted_k141_11462 | Low-quality |
| S3_extracted_k141_50495 | Not-determined |
| S3_extracted_k141_81935 | Not-determined |
| S3_extracted_k141_17786 | Not-determined |
| S3_extracted_k141_26056 | Not-determined |
| S3_extracted_k141_17787 | Not-determined |
| S3_extracted_k141_89902 | Not-determined |
| S3_extracted_k141_11471 | Not-determined |
| S3_extracted_k141_89908 | Not-determined |
| S3_extracted_k141_42234 | Not-determined |
| S3_extracted_k141_11476 | Not-determined |
| S3_extracted_k141_113868 | Not-determined |
| S3_extracted_k141_17790 | Not-determined |
| S3_extracted_k141_17792 | Not-determined |
| S3_extracted_k141_26063 | Not-determined |
| S3_extracted_k141_11483 | Not-determined |
| S3_extracted_k141_42245 | Not-determined |
| S3_extracted_k141_17801 | Not-determined |
| S3_extracted_k141_74539 | Not-determined |
| S3_extracted_k141_42250 | Not-determined |
| S3_extracted_k141_97920 | Low-quality |
| S3_extracted_k141_26069 | Not-determined |
| S3_extracted_k141_11486 | Not-determined |
| S3_extracted_k141_26071 | Not-determined |
| S3_extracted_k141_97925 | Not-determined |
| S3_extracted_k141_74554 | Not-determined |
| S3_extracted_k141_74555 | Not-determined |
| S3_extracted_k141_42258 | Low-quality |
| S3_extracted_k141_74557 | Not-determined |
| S3_extracted_k141_11493 | Not-determined |
| S3_extracted_k141_42264 | Not-determined |
| S3_extracted_k141_97929 | Low-quality |
| S3_extracted_k141_97931 | Not-determined |
| S3_extracted_k141_58309 | Low-quality |
| S3_extracted_k141_11498 | Not-determined |
| S3_extracted_k141_42270 | Low-quality |
| S3_extracted_k141_11500 | Low-quality |
| S3_extracted_k141_105903 | Low-quality |
| S3_extracted_k141_58313 | Not-determined |
| S3_extracted_k141_50517 | Not-determined |
| S3_extracted_k141_97935 | Not-determined |
| S3_extracted_k141_50518 | Not-determined |
| S3_extracted_k141_11506 | Not-determined |
| S3_extracted_k141_74569 | Not-determined |
| S3_extracted_k141_81953 | Not-determined |
| S3_extracted_k141_81954 | Not-determined |
| S3_extracted_k141_74574 | Low-quality |
| S3_extracted_k141_81957 | Not-determined |
| S3_extracted_k141_26078 | Not-determined |
| S3_extracted_k141_74579 | Not-determined |
| S3_extracted_k141_74580 | Not-determined |
| S3_extracted_k141_113884 | Low-quality |
| S3_extracted_k141_113885 | Not-determined |
| S3_extracted_k141_74583 | Not-determined |
| S3_extracted_k141_11521 | Not-determined |
| S3_extracted_k141_74586 | Low-quality |
| S3_extracted_k141_74587 | Not-determined |
| S3_extracted_k141_42282 | Not-determined |
| S3_extracted_k141_74589 | Not-determined |
| S3_extracted_k141_89920 | Not-determined |
| S3_extracted_k141_89922 | Not-determined |
| S3_extracted_k141_42284 | Not-determined |
| S3_extracted_k141_2201 | Low-quality |
| S3_extracted_k141_81968 | Not-determined |
| S3_extracted_k141_81970 | Not-determined |
| S3_extracted_k141_42291 | Not-determined |
| S3_extracted_k141_26095 | Not-determined |
| S3_extracted_k141_65971 | Low-quality |
| S3_extracted_k141_74611 | Not-determined |
| S3_extracted_k141_74612 | Not-determined |
| S3_extracted_k141_81982 | Low-quality |
| S3_extracted_k141_50532 | Not-determined |
| S3_extracted_k141_105917 | Not-determined |
| S3_extracted_k141_50533 | Not-determined |
| S3_extracted_k141_81991 | Not-determined |
| S3_extracted_k141_50541 | Not-determined |
| S3_extracted_k141_11537 | Not-determined |
| S3_extracted_k141_81994 | Not-determined |
| S3_extracted_k141_89960 | Low-quality |
| S3_extracted_k141_105924 | Not-determined |
| S3_extracted_k141_42303 | Not-determined |
| S3_extracted_k141_89963 | Not-determined |
| S3_extracted_k141_89965 | Not-determined |
| S3_extracted_k141_89971 | Low-quality |
| S3_extracted_k141_26118 | Not-determined |
| S3_extracted_k141_42307 | Not-determined |
| S3_extracted_k141_113931 | Not-determined |
| S3_extracted_k141_105928 | Not-determined |
| S3_extracted_k141_42310 | Not-determined |
| S3_extracted_k141_113937 | Not-determined |
| S3_extracted_k141_113941 | Not-determined |
| S3_extracted_k141_74629 | Not-determined |
| S3_extracted_k141_26126 | Not-determined |
| S3_extracted_k141_105938 | Not-determined |
| S3_extracted_k141_105940 | Not-determined |
| S3_extracted_k141_11557 | Not-determined |
| S3_extracted_k141_74642 | Not-determined |
| S3_extracted_k141_26141 | Low-quality |
| S3_extracted_k141_34537 | Not-determined |
| S3_extracted_k141_89995 | Not-determined |
| S3_extracted_k141_26145 | Not-determined |
| S3_extracted_k141_34543 | Low-quality |
| S3_extracted_k141_82008 | Not-determined |
| S3_extracted_k141_82010 | Not-determined |
| S3_extracted_k141_105961 | Not-determined |
| S3_extracted_k141_58333 | Not-determined |
| S3_extracted_k141_50555 | Not-determined |
| S3_extracted_k141_105962 | Not-determined |
| S3_extracted_k141_105964 | Not-determined |
| S3_extracted_k141_82019 | Not-determined |
| S3_extracted_k141_82020 | Not-determined |
| S3_extracted_k141_2267 | Not-determined |
| S3_extracted_k141_34544 | Not-determined |
| S3_extracted_k141_82024 | Low-quality |
| S3_extracted_k141_42312 | Not-determined |
| S3_extracted_k141_34547 | Not-determined |
| S3_extracted_k141_90002 | Not-determined |
| S3_extracted_k141_42317 | Not-determined |
| S3_extracted_k141_82029 | Not-determined |
| S3_extracted_k141_42318 | Not-determined |
| S3_extracted_k141_90005 | Low-quality |
| S3_extracted_k141_42320 | Not-determined |
| S3_extracted_k141_11567 | Not-determined |
| S3_extracted_k141_90012 | Not-determined |
| S3_extracted_k141_121656 | Not-determined |
| S3_extracted_k141_121657 | Not-determined |
| S3_extracted_k141_121660 | Not-determined |
| S3_extracted_k141_2281 | Not-determined |
| S3_extracted_k141_42331 | Not-determined |
| S3_extracted_k141_82043 | Not-determined |
| S3_extracted_k141_97957 | Not-determined |
| S3_extracted_k141_42343 | Low-quality |
| S3_extracted_k141_42346 | Not-determined |
| S3_extracted_k141_97958 | Low-quality |
| S3_extracted_k141_26154 | Not-determined |
| S3_extracted_k141_97967 | Not-determined |
| S3_extracted_k141_11577 | Not-determined |
| S3_extracted_k141_82060 | Not-determined |
| S3_extracted_k141_2301 | Not-determined |
| S3_extracted_k141_97973 | Not-determined |
| S3_extracted_k141_105983 | Not-determined |
| S3_extracted_k141_113962 | Not-determined |
| S3_extracted_k141_26165 | Low-quality |
| S3_extracted_k141_74647 | Not-determined |
| S3_extracted_k141_105987 | Not-determined |
| S3_extracted_k141_105989 | Low-quality |
| S3_extracted_k141_26179 | Not-determined |
| S3_extracted_k141_105991 | Not-determined |
| S3_extracted_k141_74654 | Low-quality |
| S3_extracted_k141_121667 | Low-quality |
| S3_extracted_k141_2309 | Not-determined |
| S3_extracted_k141_11583 | Low-quality |
| S3_extracted_k141_11585 | Not-determined |
| S3_extracted_k141_11587 | Not-determined |
| S3_extracted_k141_11590 | Not-determined |
| S3_extracted_k141_121671 | Low-quality |
| S3_extracted_k141_11592 | Low-quality |
| S3_extracted_k141_113975 | Not-determined |
| S3_extracted_k141_11598 | Not-determined |
| S3_extracted_k141_113977 | Not-determined |
| S3_extracted_k141_11600 | Not-determined |
| S3_extracted_k141_90067 | Not-determined |
| S3_extracted_k141_58381 | Not-determined |
| S3_extracted_k141_50565 | Low-quality |
| S3_extracted_k141_26197 | Low-quality |
| S3_extracted_k141_90071 | Not-determined |
| S3_extracted_k141_26204 | Not-determined |
| S3_extracted_k141_2328 | Not-determined |
| S3_extracted_k141_97989 | Not-determined |
| S3_extracted_k141_11620 | Not-determined |
| S3_extracted_k141_2330 | Not-determined |
| S3_extracted_k141_106000 | Not-determined |
| S3_extracted_k141_58394 | Not-determined |
| S3_extracted_k141_97993 | Not-determined |
| S3_extracted_k141_106004 | Not-determined |
| S3_extracted_k141_90081 | Not-determined |
| S3_extracted_k141_11623 | Not-determined |
| S3_extracted_k141_74666 | Not-determined |
| S3_extracted_k141_2340 | Not-determined |
| S3_extracted_k141_2343 | Not-determined |
| S3_extracted_k141_74669 | Not-determined |
| S3_extracted_k141_2344 | Not-determined |
| S3_extracted_k141_2345 | Not-determined |
| S3_extracted_k141_11631 | Not-determined |
| S3_extracted_k141_2349 | Not-determined |
| S3_extracted_k141_42363 | Low-quality |
| S3_extracted_k141_11635 | Not-determined |
| S3_extracted_k141_106014 | Not-determined |
| S3_extracted_k141_2353 | Not-determined |
| S3_extracted_k141_11636 | Not-determined |
| S3_extracted_k141_42365 | Not-determined |
| S3_extracted_k141_42369 | Not-determined |
| S3_extracted_k141_42374 | Not-determined |
| S3_extracted_k141_42375 | Not-determined |
| S3_extracted_k141_82099 | Not-determined |
| S3_extracted_k141_11646 | Not-determined |
| S3_extracted_k141_98003 | Low-quality |
| S3_extracted_k141_42381 | Not-determined |
| S3_extracted_k141_11649 | Low-quality |
| S3_extracted_k141_98007 | Low-quality |
| S3_extracted_k141_11660 | Not-determined |
| S3_extracted_k141_11661 | Not-determined |
| S3_extracted_k141_2357 | Not-determined |
| S3_extracted_k141_50594 | Not-determined |
| S3_extracted_k141_50595 | Low-quality |
| S3_extracted_k141_58425 | Not-determined |
| S3_extracted_k141_82112 | Not-determined |
| S3_extracted_k141_2367 | Low-quality |
| S3_extracted_k141_90088 | Low-quality |
| S3_extracted_k141_42395 | Not-determined |
| S3_extracted_k141_82115 | Not-determined |
| S3_extracted_k141_2374 | Not-determined |
| S3_extracted_k141_113998 | Low-quality |
| S3_extracted_k141_26242 | Not-determined |
| S3_extracted_k141_26243 | Not-determined |
| S3_extracted_k141_42407 | Not-determined |
| S3_extracted_k141_17830 | Not-determined |
| S3_extracted_k141_42412 | Not-determined |
| S3_extracted_k141_2379 | Not-determined |
| S3_extracted_k141_26249 | Not-determined |
| S3_extracted_k141_42415 | Not-determined |
| S3_extracted_k141_26253 | Not-determined |
| S3_extracted_k141_42420 | Not-determined |
| S3_extracted_k141_90096 | Not-determined |
| S3_extracted_k141_114014 | Not-determined |
| S3_extracted_k141_98027 | Not-determined |
| S3_extracted_k141_50608 | Not-determined |
| S3_extracted_k141_106035 | Not-determined |
| S3_extracted_k141_90102 | Not-determined |
| S3_extracted_k141_90109 | Not-determined |
| S3_extracted_k141_90110 | Low-quality |
| S3_extracted_k141_11674 | Not-determined |
| S3_extracted_k141_82125 | Not-determined |
| S3_extracted_k141_42430 | Not-determined |
| S3_extracted_k141_114029 | Not-determined |
| S3_extracted_k141_17842 | Not-determined |
| S3_extracted_k141_17848 | Not-determined |
| S3_extracted_k141_114039 | Not-determined |
| S3_extracted_k141_42441 | Low-quality |
| S3_extracted_k141_11692 | Not-determined |
| S3_extracted_k141_121695 | Not-determined |
| S3_extracted_k141_74700 | Not-determined |
| S3_extracted_k141_50635 | Low-quality |
| S3_extracted_k141_50636 | Not-determined |
| S3_extracted_k141_74703 | Not-determined |
| S3_extracted_k141_74706 | Not-determined |
| S3_extracted_k141_82144 | Not-determined |
| S3_extracted_k141_42456 | Not-determined |
| S3_extracted_k141_42457 | Low-quality |
| S3_extracted_k141_50643 | Not-determined |
| S3_extracted_k141_74707 | Not-determined |
| S3_extracted_k141_74708 | Low-quality |
| S3_extracted_k141_17873 | Not-determined |
| S3_extracted_k141_74710 | Not-determined |
| S3_extracted_k141_74713 | Not-determined |
| S3_extracted_k141_26267 | Low-quality |
| S3_extracted_k141_82149 | Not-determined |
| S3_extracted_k141_17877 | Not-determined |
| S3_extracted_k141_26272 | Not-determined |
| S3_extracted_k141_17881 | Not-determined |
| S3_extracted_k141_17886 | Not-determined |
| S3_extracted_k141_98049 | Low-quality |
| S3_extracted_k141_11696 | Low-quality |
| S3_extracted_k141_26282 | Not-determined |
| S3_extracted_k141_98052 | Low-quality |
| S3_extracted_k141_34571 | Not-determined |
| S3_extracted_k141_17890 | Low-quality |
| S3_extracted_k141_50664 | Not-determined |
| S3_extracted_k141_11707 | Not-determined |
| S3_extracted_k141_82160 | Low-quality |
| S3_extracted_k141_98056 | Not-determined |
| S3_extracted_k141_34574 | Low-quality |
| S3_extracted_k141_90127 | Not-determined |
| S3_extracted_k141_82163 | Low-quality |
| S3_extracted_k141_34575 | Not-determined |
| S3_extracted_k141_34577 | Not-determined |
| S3_extracted_k141_50671 | Not-determined |
| S3_extracted_k141_34583 | Not-determined |
| S3_extracted_k141_98057 | Low-quality |
| S3_extracted_k141_106067 | Low-quality |
| S3_extracted_k141_98060 | Not-determined |
| S3_extracted_k141_17898 | Low-quality |
| S3_extracted_k141_106071 | Not-determined |
| S3_extracted_k141_82170 | Not-determined |
| S3_extracted_k141_17900 | Not-determined |
| S3_extracted_k141_114053 | Not-determined |
| S3_extracted_k141_74731 | Not-determined |
| S3_extracted_k141_17904 | Not-determined |
| S3_extracted_k141_58432 | Not-determined |
| S3_extracted_k141_2410 | Low-quality |
| S3_extracted_k141_2413 | Not-determined |
| S3_extracted_k141_34587 | Not-determined |
| S3_extracted_k141_34592 | Not-determined |
| S3_extracted_k141_17912 | Not-determined |
| S3_extracted_k141_26310 | Not-determined |
| S3_extracted_k141_82174 | Not-determined |
| S3_extracted_k141_82176 | Not-determined |
| S3_extracted_k141_82179 | Not-determined |
| S3_extracted_k141_82182 | Not-determined |
| S3_extracted_k141_50680 | Not-determined |
| S3_extracted_k141_58438 | Not-determined |
| S3_extracted_k141_82188 | Not-determined |
| S3_extracted_k141_17921 | Low-quality |
| S3_extracted_k141_90146 | Not-determined |
| S3_extracted_k141_98080 | Low-quality |
| S3_extracted_k141_98084 | Not-determined |
| S3_extracted_k141_121700 | Low-quality |
| S3_extracted_k141_121702 | Not-determined |
| S3_extracted_k141_17923 | Not-determined |
| S3_extracted_k141_74745 | Not-determined |
| S3_extracted_k141_2428 | Not-determined |
| S3_extracted_k141_82196 | Not-determined |
| S3_extracted_k141_74748 | Not-determined |
| S3_extracted_k141_74751 | Not-determined |
| S3_extracted_k141_58451 | Not-determined |
| S3_extracted_k141_121704 | Not-determined |
| S3_extracted_k141_82202 | Not-determined |
| S3_extracted_k141_26326 | Not-determined |
| S3_extracted_k141_42493 | Not-determined |
| S3_extracted_k141_42494 | Low-quality |
| S3_extracted_k141_26330 | Not-determined |
| S3_extracted_k141_98095 | Low-quality |
| S3_extracted_k141_42496 | Not-determined |
| S3_extracted_k141_2435 | Not-determined |
| S3_extracted_k141_106080 | Not-determined |
| S3_extracted_k141_98099 | Not-determined |
| S3_extracted_k141_114060 | Not-determined |
| S3_extracted_k141_26338 | Not-determined |
| S3_extracted_k141_90164 | Not-determined |
| S3_extracted_k141_42501 | Not-determined |
| S3_extracted_k141_106084 | Not-determined |
| S3_extracted_k141_106085 | Not-determined |
| S3_extracted_k141_17929 | Not-determined |
| S3_extracted_k141_34615 | Not-determined |
| S3_extracted_k141_106089 | Not-determined |
| S3_extracted_k141_26341 | Low-quality |
| S3_extracted_k141_90169 | Not-determined |
| S3_extracted_k141_114069 | Low-quality |
| S3_extracted_k141_90170 | Low-quality |
| S3_extracted_k141_58471 | Not-determined |
| S3_extracted_k141_34633 | Low-quality |
| S3_extracted_k141_34634 | Not-determined |
| S3_extracted_k141_106100 | Not-determined |
| S3_extracted_k141_2439 | Not-determined |
| S3_extracted_k141_2441 | Not-determined |
| S3_extracted_k141_58480 | Not-determined |
| S3_extracted_k141_90182 | Not-determined |
| S3_extracted_k141_114085 | Not-determined |
| S3_extracted_k141_58486 | Not-determined |
| S3_extracted_k141_114092 | Not-determined |
| S3_extracted_k141_106107 | Low-quality |
| S3_extracted_k141_58492 | Not-determined |
| S3_extracted_k141_42528 | Low-quality |
| S3_extracted_k141_98124 | Not-determined |
| S3_extracted_k141_98126 | Not-determined |
| S3_extracted_k141_58505 | Not-determined |
| S3_extracted_k141_121723 | Not-determined |
| S3_extracted_k141_106112 | Not-determined |
| S3_extracted_k141_82217 | Not-determined |
| S3_extracted_k141_90190 | Not-determined |
| S3_extracted_k141_82221 | Low-quality |
| S3_extracted_k141_98134 | Not-determined |
| S3_extracted_k141_98135 | Not-determined |
| S3_extracted_k141_26365 | Not-determined |
| S3_extracted_k141_90195 | Not-determined |
| S3_extracted_k141_34643 | Not-determined |
| S3_extracted_k141_82232 | Not-determined |
| S3_extracted_k141_98140 | Not-determined |
| S3_extracted_k141_90200 | Not-determined |
| S3_extracted_k141_34646 | Not-determined |
| S3_extracted_k141_90201 | Low-quality |
| S3_extracted_k141_90202 | Not-determined |
| S3_extracted_k141_90204 | Not-determined |
| S3_extracted_k141_26374 | Not-determined |
| S3_extracted_k141_34649 | Not-determined |
| S3_extracted_k141_34651 | Not-determined |
| S3_extracted_k141_58530 | Not-determined |
| S3_extracted_k141_58531 | Not-determined |
| S3_extracted_k141_114111 | Low-quality |
| S3_extracted_k141_58532 | Not-determined |
| S3_extracted_k141_90207 | Not-determined |
| S3_extracted_k141_34655 | Not-determined |
| S3_extracted_k141_90209 | Not-determined |
| S3_extracted_k141_74774 | Not-determined |
| S3_extracted_k141_82242 | Not-determined |
| S3_extracted_k141_34661 | Not-determined |
| S3_extracted_k141_50683 | Not-determined |
| S3_extracted_k141_106126 | Not-determined |
| S3_extracted_k141_50688 | Not-determined |
| S3_extracted_k141_121730 | Not-determined |
| S3_extracted_k141_58558 | Not-determined |
| S3_extracted_k141_34669 | Not-determined |
| S3_extracted_k141_26381 | Low-quality |
| S3_extracted_k141_58564 | Not-determined |
| S3_extracted_k141_90228 | Not-determined |
| S3_extracted_k141_90229 | Not-determined |
| S3_extracted_k141_58567 | Not-determined |
| S3_extracted_k141_58568 | Not-determined |
| S3_extracted_k141_74785 | Not-determined |
| S3_extracted_k141_50700 | Low-quality |
| S3_extracted_k141_17951 | Not-determined |
| S3_extracted_k141_90239 | Not-determined |
| S3_extracted_k141_82268 | Not-determined |
| S3_extracted_k141_74795 | Not-determined |
| S3_extracted_k141_121736 | Not-determined |
| S3_extracted_k141_114135 | Low-quality |
| S3_extracted_k141_2473 | Not-determined |
| S3_extracted_k141_114138 | Low-quality |
| S3_extracted_k141_34674 | Not-determined |
| S3_extracted_k141_42548 | Low-quality |
| S3_extracted_k141_2476 | Not-determined |
| S3_extracted_k141_74797 | Not-determined |
| S3_extracted_k141_58585 | Not-determined |
| S3_extracted_k141_2479 | Not-determined |
| S3_extracted_k141_42551 | Not-determined |
| S3_extracted_k141_98164 | Not-determined |
| S3_extracted_k141_106147 | Not-determined |
| S3_extracted_k141_58596 | Not-determined |
| S3_extracted_k141_42555 | Not-determined |
| S3_extracted_k141_42556 | Not-determined |
| S3_extracted_k141_42557 | Not-determined |
| S3_extracted_k141_90250 | Not-determined |
| S3_extracted_k141_42558 | Not-determined |
| S3_extracted_k141_34687 | Not-determined |
| S3_extracted_k141_58603 | Not-determined |
| S3_extracted_k141_42561 | Not-determined |
| S3_extracted_k141_2484 | Low-quality |
| S3_extracted_k141_50713 | Not-determined |
| S3_extracted_k141_26406 | Not-determined |
| S3_extracted_k141_90254 | Low-quality |
| S3_extracted_k141_17976 | Not-determined |
| S3_extracted_k141_82276 | Not-determined |
| S3_extracted_k141_42564 | Not-determined |
| S3_extracted_k141_82278 | Not-determined |
| S3_extracted_k141_50723 | Not-determined |
| S3_extracted_k141_121744 | Not-determined |
| S3_extracted_k141_26418 | Not-determined |
| S3_extracted_k141_58615 | Low-quality |
| S3_extracted_k141_82290 | Not-determined |
| S3_extracted_k141_98186 | Not-determined |
| S3_extracted_k141_50731 | Not-determined |
| S3_extracted_k141_98189 | Not-determined |
| S3_extracted_k141_90276 | Not-determined |
| S3_extracted_k141_90278 | Not-determined |
| S3_extracted_k141_90281 | Not-determined |
| S3_extracted_k141_98191 | Not-determined |
| S3_extracted_k141_82306 | Not-determined |
| S3_extracted_k141_90285 | Not-determined |
| S3_extracted_k141_90287 | Not-determined |
| S3_extracted_k141_50738 | Low-quality |
| S3_extracted_k141_2501 | Not-determined |
| S3_extracted_k141_26430 | Not-determined |
| S3_extracted_k141_50740 | Not-determined |
| S3_extracted_k141_106163 | Low-quality |
| S3_extracted_k141_106165 | Not-determined |
| S3_extracted_k141_26432 | Not-determined |
| S3_extracted_k141_106166 | Low-quality |
| S3_extracted_k141_90295 | Low-quality |
| S3_extracted_k141_26441 | Not-determined |
| S3_extracted_k141_58633 | Not-determined |
| S3_extracted_k141_2518 | Not-determined |
| S3_extracted_k141_58634 | Not-determined |
| S3_extracted_k141_82317 | Not-determined |
| S3_extracted_k141_106170 | Not-determined |
| S3_extracted_k141_106171 | Not-determined |
| S3_extracted_k141_2525 | Not-determined |
| S3_extracted_k141_11727 | Not-determined |
| S3_extracted_k141_98208 | Not-determined |
| S3_extracted_k141_50749 | Not-determined |
| S3_extracted_k141_82322 | Not-determined |
| S3_extracted_k141_121764 | Not-determined |
| S3_extracted_k141_58657 | Not-determined |
| S3_extracted_k141_98215 | Low-quality |
| S3_extracted_k141_2531 | Not-determined |
| S3_extracted_k141_98218 | Not-determined |
| S3_extracted_k141_121768 | Not-determined |
| S3_extracted_k141_98225 | Low-quality |
| S3_extracted_k141_82326 | Not-determined |
| S3_extracted_k141_121775 | Not-determined |
| S3_extracted_k141_2532 | Low-quality |
| S3_extracted_k141_74808 | Not-determined |
| S3_extracted_k141_2535 | Not-determined |
| S3_extracted_k141_121781 | Not-determined |
| S3_extracted_k141_114154 | Not-determined |
| S3_extracted_k141_114155 | Not-determined |
| S3_extracted_k141_98230 | Not-determined |
| S3_extracted_k141_121786 | Low-quality |
| S3_extracted_k141_106183 | Not-determined |
| S3_extracted_k141_74818 | Not-determined |
| S3_extracted_k141_74822 | Not-determined |
| S3_extracted_k141_114159 | Not-determined |
| S3_extracted_k141_90319 | Not-determined |
| S3_extracted_k141_58678 | Not-determined |
| S3_extracted_k141_98231 | Not-determined |
| S3_extracted_k141_90323 | Low-quality |
| S3_extracted_k141_121798 | Low-quality |
| S3_extracted_k141_98234 | Not-determined |
| S3_extracted_k141_58685 | Not-determined |
| S3_extracted_k141_106194 | Not-determined |
| S3_extracted_k141_121805 | Not-determined |
| S3_extracted_k141_106197 | Not-determined |
| S3_extracted_k141_74834 | Not-determined |
| S3_extracted_k141_106201 | Low-quality |
| S3_extracted_k141_121814 | Not-determined |
| S3_extracted_k141_2572 | Not-determined |
| S3_extracted_k141_74844 | Not-determined |
| S3_extracted_k141_98239 | Low-quality |
| S3_extracted_k141_74847 | Low-quality |
| S3_extracted_k141_82331 | Low-quality |
| S3_extracted_k141_2583 | Not-determined |
| S3_extracted_k141_2584 | Not-determined |
| S3_extracted_k141_26454 | Not-determined |
| S3_extracted_k141_17988 | Not-determined |
| S3_extracted_k141_2587 | Not-determined |
| S3_extracted_k141_11743 | Not-determined |
| S3_extracted_k141_26461 | Not-determined |
| S3_extracted_k141_65979 | Not-determined |
| S3_extracted_k141_26462 | Not-determined |
| S3_extracted_k141_42570 | Not-determined |
| S3_extracted_k141_42572 | Not-determined |
| S3_extracted_k141_11750 | Not-determined |
| S3_extracted_k141_2601 | Not-determined |
| S3_extracted_k141_50752 | Not-determined |
| S3_extracted_k141_98256 | Not-determined |
| S3_extracted_k141_65993 | Not-determined |
| S3_extracted_k141_42579 | Not-determined |
| S3_extracted_k141_66000 | Not-determined |
| S3_extracted_k141_82351 | Not-determined |
| S3_extracted_k141_11763 | Not-determined |
| S3_extracted_k141_42587 | Not-determined |
| S3_extracted_k141_42589 | Not-determined |
| S3_extracted_k141_26474 | Not-determined |
| S3_extracted_k141_11768 | Not-determined |
| S3_extracted_k141_26475 | Not-determined |
| S3_extracted_k141_34725 | Not-determined |
| S3_extracted_k141_11770 | Not-determined |
| S3_extracted_k141_98271 | Not-determined |
| S3_extracted_k141_58740 | Not-determined |
| S3_extracted_k141_26480 | Not-determined |
| S3_extracted_k141_98279 | Not-determined |
| S3_extracted_k141_18002 | Not-determined |
| S3_extracted_k141_74858 | Not-determined |
| S3_extracted_k141_26485 | Not-determined |
| S3_extracted_k141_11779 | Not-determined |
| S3_extracted_k141_18010 | Not-determined |
| S3_extracted_k141_74864 | Not-determined |
| S3_extracted_k141_18011 | Not-determined |
| S3_extracted_k141_34731 | Not-determined |

| | |
|---|---|
| S3_extracted_k141_34732 | Not-determined |
| S3_extracted_k141_2646 | Low-quality |
| S3_extracted_k141_66025 | Not-determined |
| S3_extracted_k141_34735 | Not-determined |
| S3_extracted_k141_18019 | Not-determined |
| S3_extracted_k141_2652 | Not-determined |
| S3_extracted_k141_98284 | Not-determined |
| S3_extracted_k141_34737 | Low-quality |
| S3_extracted_k141_50768 | Low-quality |
| S3_extracted_k141_11783 | Not-determined |
| S3_extracted_k141_34741 | Not-determined |
| S3_extracted_k141_26502 | Not-determined |
| S3_extracted_k141_2654 | Not-determined |
| S3_extracted_k141_42611 | Not-determined |
| S3_extracted_k141_66037 | Not-determined |
| S3_extracted_k141_11788 | Not-determined |
| S3_extracted_k141_58772 | Low-quality |
| S3_extracted_k141_58773 | Not-determined |
| S3_extracted_k141_11794 | Not-determined |
| S3_extracted_k141_34746 | Not-determined |
| S3_extracted_k141_34748 | Not-determined |
| S3_extracted_k141_50770 | Not-determined |
| S3_extracted_k141_42624 | Not-determined |
| S3_extracted_k141_66051 | Not-determined |
| S3_extracted_k141_42625 | Not-determined |
| S3_extracted_k141_34753 | Low-quality |
| S3_extracted_k141_42628 | Not-determined |
| S3_extracted_k141_58787 | Not-determined |
| S3_extracted_k141_2664 | Low-quality |
| S3_extracted_k141_2669 | Not-determined |
| S3_extracted_k141_42635 | Not-determined |
| S3_extracted_k141_34756 | Low-quality |
| S3_extracted_k141_66057 | Not-determined |
| S3_extracted_k141_11814 | Not-determined |
| S3_extracted_k141_50790 | Not-determined |
| S3_extracted_k141_50791 | Not-determined |
| S3_extracted_k141_42636 | Not-determined |
| S3_extracted_k141_74891 | Not-determined |
| S3_extracted_k141_2682 | Not-determined |
| S3_extracted_k141_2683 | Not-determined |
| S3_extracted_k141_50802 | Not-determined |
| S3_extracted_k141_66069 | Not-determined |
| S3_extracted_k141_2690 | Not-determined |
| S3_extracted_k141_66076 | Not-determined |
| S3_extracted_k141_106219 | Low-quality |
| S3_extracted_k141_50803 | Low-quality |
| S3_extracted_k141_50805 | Not-determined |
| S3_extracted_k141_106229 | Not-determined |
| S3_extracted_k141_34772 | Not-determined |
| S3_extracted_k141_114209 | Not-determined |
| S3_extracted_k141_74911 | Low-quality |
| S3_extracted_k141_106248 | Not-determined |
| S3_extracted_k141_74915 | Not-determined |
| S3_extracted_k141_121830 | Low-quality |
| S3_extracted_k141_74918 | Not-determined |
| S3_extracted_k141_121831 | Not-determined |
| S3_extracted_k141_18045 | Not-determined |
| S3_extracted_k141_74921 | Low-quality |
| S3_extracted_k141_74922 | Not-determined |
| S3_extracted_k141_26515 | Low-quality |
| S3_extracted_k141_11826 | Not-determined |
| S3_extracted_k141_34790 | Not-determined |
| S3_extracted_k141_18053 | Not-determined |
| S3_extracted_k141_26520 | Not-determined |
| S3_extracted_k141_58803 | Not-determined |
| S3_extracted_k141_74933 | Low-quality |
| S3_extracted_k141_74934 | Not-determined |
| S3_extracted_k141_2701 | Not-determined |
| S3_extracted_k141_42662 | Low-quality |
| S3_extracted_k141_42665 | Low-quality |
| S3_extracted_k141_74943 | Not-determined |
| S3_extracted_k141_11842 | Not-determined |
| S3_extracted_k141_82368 | Not-determined |
| S3_extracted_k141_82370 | Not-determined |
| S3_extracted_k141_66095 | Not-determined |
| S3_extracted_k141_50815 | Low-quality |
| S3_extracted_k141_106250 | Not-determined |
| S3_extracted_k141_121847 | Not-determined |
| S3_extracted_k141_74954 | Not-determined |
| S3_extracted_k141_106258 | Low-quality |
| S3_extracted_k141_50820 | Not-determined |
| S3_extracted_k141_106262 | Not-determined |
| S3_extracted_k141_42673 | Not-determined |
| S3_extracted_k141_106263 | Not-determined |
| S3_extracted_k141_106264 | Low-quality |
| S3_extracted_k141_26540 | Not-determined |
| S3_extracted_k141_26543 | Not-determined |
| S3_extracted_k141_106267 | Not-determined |
| S3_extracted_k141_34797 | Not-determined |
| S3_extracted_k141_26547 | Not-determined |
| S3_extracted_k141_74958 | Not-determined |
| S3_extracted_k141_42685 | Not-determined |
| S3_extracted_k141_11850 | Not-determined |
| S3_extracted_k141_74961 | Not-determined |
| S3_extracted_k141_34804 | Not-determined |
| S3_extracted_k141_66108 | Not-determined |
| S3_extracted_k141_2712 | Not-determined |
| S3_extracted_k141_98296 | Low-quality |
| S3_extracted_k141_42692 | Low-quality |
| S3_extracted_k141_50839 | Not-determined |
| S3_extracted_k141_50841 | Low-quality |
| S3_extracted_k141_50843 | Not-determined |
| S3_extracted_k141_74970 | Not-determined |
| S3_extracted_k141_82373 | Low-quality |
| S3_extracted_k141_26560 | Not-determined |
| S3_extracted_k141_50849 | Low-quality |
| S3_extracted_k141_26562 | Not-determined |
| S3_extracted_k141_74977 | Not-determined |
| S3_extracted_k141_58827 | Low-quality |
| S3_extracted_k141_82380 | Not-determined |
| S3_extracted_k141_82381 | Not-determined |
| S3_extracted_k141_58830 | Not-determined |
| S3_extracted_k141_58832 | Not-determined |
| S3_extracted_k141_114238 | Not-determined |
| S3_extracted_k141_114241 | Not-determined |
| S3_extracted_k141_42700 | Not-determined |
| S3_extracted_k141_66119 | Not-determined |
| S3_extracted_k141_42701 | Not-determined |
| S3_extracted_k141_34812 | Not-determined |
| S3_extracted_k141_66124 | Low-quality |
| S3_extracted_k141_50859 | Low-quality |
| S3_extracted_k141_34821 | Not-determined |
| S3_extracted_k141_50870 | Not-determined |
| S3_extracted_k141_2725 | Not-determined |
| S3_extracted_k141_42716 | Not-determined |
| S3_extracted_k141_50874 | Not-determined |
| S3_extracted_k141_26589 | Not-determined |
| S3_extracted_k141_106299 | Not-determined |
| S3_extracted_k141_114247 | Low-quality |
| S3_extracted_k141_98315 | Low-quality |
| S3_extracted_k141_121882 | Not-determined |
| S3_extracted_k141_106306 | Low-quality |
| S3_extracted_k141_98319 | Not-determined |
| S3_extracted_k141_42729 | Not-determined |
| S3_extracted_k141_114255 | Not-determined |
| S3_extracted_k141_58841 | Not-determined |
| S3_extracted_k141_98323 | Not-determined |
| S3_extracted_k141_42734 | Not-determined |
| S3_extracted_k141_58845 | Not-determined |
| S3_extracted_k141_74985 | Not-determined |
| S3_extracted_k141_42740 | Not-determined |
| S3_extracted_k141_74989 | Not-determined |
| S3_extracted_k141_74990 | Low-quality |
| S3_extracted_k141_42743 | Low-quality |
| S3_extracted_k141_42744 | Low-quality |
| S3_extracted_k141_18064 | Not-determined |
| S3_extracted_k141_121890 | Not-determined |
| S3_extracted_k141_58859 | Not-determined |
| S3_extracted_k141_42751 | Not-determined |
| S3_extracted_k141_66129 | Not-determined |
| S3_extracted_k141_98332 | Not-determined |
| S3_extracted_k141_66133 | Not-determined |
| S3_extracted_k141_114260 | Not-determined |
| S3_extracted_k141_98335 | Not-determined |
| S3_extracted_k141_11875 | Not-determined |
| S3_extracted_k141_2755 | Not-determined |
| S3_extracted_k141_2756 | Not-determined |
| S3_extracted_k141_18066 | Not-determined |
| S3_extracted_k141_2757 | Not-determined |
| S3_extracted_k141_11876 | Not-determined |
| S3_extracted_k141_114266 | Not-determined |
| S3_extracted_k141_66140 | Not-determined |
| S3_extracted_k141_18070 | Low-quality |
| S3_extracted_k141_58871 | Not-determined |
| S3_extracted_k141_82398 | Not-determined |
| S3_extracted_k141_50889 | Not-determined |
| S3_extracted_k141_26616 | Not-determined |
| S3_extracted_k141_18077 | Low-quality |
| S3_extracted_k141_42759 | Not-determined |
| S3_extracted_k141_106327 | Low-quality |
| S3_extracted_k141_42762 | Not-determined |
| S3_extracted_k141_82407 | Not-determined |
| S3_extracted_k141_42765 | Not-determined |
| S3_extracted_k141_50902 | Low-quality |
| S3_extracted_k141_82417 | Not-determined |
| S3_extracted_k141_82419 | Not-determined |
| S3_extracted_k141_82422 | Low-quality |
| S3_extracted_k141_50910 | Not-determined |
| S3_extracted_k141_82425 | Low-quality |
| S3_extracted_k141_82430 | Not-determined |
| S3_extracted_k141_11883 | Low-quality |
| S3_extracted_k141_75010 | Not-determined |
| S3_extracted_k141_75012 | Not-determined |
| S3_extracted_k141_58890 | Low-quality |
| S3_extracted_k141_58894 | Not-determined |
| S3_extracted_k141_114269 | Not-determined |
| S3_extracted_k141_42775 | Not-determined |
| S3_extracted_k141_106345 | Not-determined |
| S3_extracted_k141_42779 | Not-determined |
| S3_extracted_k141_58899 | Not-determined |
| S3_extracted_k141_42783 | Not-determined |
| S3_extracted_k141_82443 | Not-determined |
| S3_extracted_k141_106353 | Not-determined |
| S3_extracted_k141_42787 | Not-determined |
| S3_extracted_k141_121911 | Not-determined |
| S3_extracted_k141_106365 | Not-determined |
| S3_extracted_k141_82455 | Not-determined |
| S3_extracted_k141_82457 | Low-quality |
| S3_extracted_k141_75027 | Not-determined |
| S3_extracted_k141_75030 | Not-determined |
| S3_extracted_k141_121914 | Not-determined |
| S3_extracted_k141_2770 | Not-determined |
| S3_extracted_k141_121920 | Not-determined |
| S3_extracted_k141_82469 | Low-quality |
| S3_extracted_k141_106381 | Not-determined |
| S3_extracted_k141_106382 | Not-determined |
| S3_extracted_k141_98353 | Not-determined |
| S3_extracted_k141_11908 | Not-determined |
| S3_extracted_k141_90364 | Not-determined |
| S3_extracted_k141_42811 | Not-determined |
| S3_extracted_k141_121929 | Not-determined |
| S3_extracted_k141_82490 | Not-determined |
| S3_extracted_k141_82491 | Not-determined |
| S3_extracted_k141_98366 | Low-quality |
| S4_vs_VIRSorter_13353-cat_2 | High-quality |
| S3_extracted_k141_26640 | Not-determined |
| S3_extracted_k141_34832 | Not-determined |
| S3_extracted_k141_26643 | Not-determined |
| S3_extracted_k141_75046 | Not-determined |
| S3_extracted_k141_114274 | Not-determined |
| S3_extracted_k141_75051 | Not-determined |
| S3_extracted_k141_34846 | Not-determined |
| S3_extracted_k141_75057 | Not-determined |
| S3_extracted_k141_34847 | Not-determined |
| S3_extracted_k141_58913 | Not-determined |
| S3_extracted_k141_121939 | Not-determined |
| S3_extracted_k141_18082 | Not-determined |
| S3_extracted_k141_42825 | Not-determined |
| S3_extracted_k141_11919 | Not-determined |
| S3_extracted_k141_42827 | Not-determined |
| S3_extracted_k141_121940 | Not-determined |
| S3_extracted_k141_42832 | Not-determined |
| S3_extracted_k141_11929 | Not-determined |
| S3_extracted_k141_58923 | Not-determined |
| S3_extracted_k141_42836 | Not-determined |
| S3_extracted_k141_11934 | Not-determined |
| S3_extracted_k141_42844 | Not-determined |
| S3_extracted_k141_58929 | Not-determined |
| S3_extracted_k141_90378 | Low-quality |
| S3_extracted_k141_58930 | Low-quality |
| S3_extracted_k141_11949 | Not-determined |
| S3_extracted_k141_90380 | Not-determined |
| S3_extracted_k141_121947 | Not-determined |
| S3_extracted_k141_11952 | Low-quality |
| S3_extracted_k141_75066 | Not-determined |
| S3_extracted_k141_121948 | Not-determined |
| S3_extracted_k141_121953 | Not-determined |
| S3_extracted_k141_121954 | Not-determined |
| S3_extracted_k141_121956 | Not-determined |
| S3_extracted_k141_11956 | Low-quality |
| S3_extracted_k141_90390 | Low-quality |
| S3_extracted_k141_26656 | Not-determined |
| S3_extracted_k141_11959 | Low-quality |
| S3_extracted_k141_42868 | Not-determined |
| S3_extracted_k141_121965 | Not-determined |
| S3_extracted_k141_42872 | Not-determined |
| S3_extracted_k141_75069 | Not-determined |
| S3_extracted_k141_98391 | Not-determined |
| S16_vs_47559\|\|full | High-quality |
| S3_extracted_k141_121968 | Not-determined |
| S3_extracted_k141_34872 | Low-quality |
| S3_extracted_k141_11968 | Not-determined |
| S3_extracted_k141_90402 | Low-quality |
| S3_extracted_k141_121972 | Not-determined |
| S3_extracted_k141_121978 | Low-quality |
| S3_extracted_k141_98400 | Not-determined |
| S3_extracted_k141_106405 | Not-determined |
| S3_extracted_k141_34882 | Not-determined |
| S3_extracted_k141_50956 | Low-quality |
| S3_extracted_k141_75078 | Not-determined |
| S3_extracted_k141_114295 | Not-determined |
| S3_extracted_k141_42884 | Not-determined |
| S3_extracted_k141_106411 | Not-determined |
| S3_extracted_k141_121984 | Not-determined |
| S3_extracted_k141_42888 | Not-determined |
| S3_extracted_k141_75082 | Not-determined |
| S3_extracted_k141_11972 | Not-determined |
| S3_extracted_k141_106417 | Not-determined |
| S3_extracted_k141_42895 | Not-determined |
| S3_extracted_k141_75086 | Not-determined |
| S3_extracted_k141_42897 | Not-determined |
| S3_extracted_k141_50961 | Not-determined |
| S3_extracted_k141_75088 | Low-quality |
| S3_extracted_k141_58970 | Low-quality |
| S3_extracted_k141_121995 | Not-determined |
| S3_extracted_k141_82515 | Not-determined |
| S3_extracted_k141_121999 | Low-quality |
| S3_extracted_k141_82517 | Not-determined |
| S3_extracted_k141_26665 | Not-determined |
| S3_extracted_k141_98410 | Not-determined |
| S3_extracted_k141_82522 | Not-determined |
| S3_extracted_k141_26669 | Not-determined |
| S3_extracted_k141_34909 | Not-determined |
| S3_extracted_k141_82525 | Not-determined |
| S3_extracted_k141_122011 | Low-quality |
| S3_extracted_k141_50975 | Not-determined |
| S3_extracted_k141_34914 | Not-determined |
| S3_extracted_k141_114300 | Not-determined |
| S3_extracted_k141_90412 | Low-quality |
| S3_extracted_k141_34917 | Not-determined |
| S3_extracted_k141_82528 | Low-quality |
| S3_extracted_k141_114305 | Not-determined |
| S3_extracted_k141_82530 | Low-quality |
| S3_extracted_k141_90416 | Not-determined |
| S3_extracted_k141_90417 | Not-determined |
| S3_extracted_k141_75098 | Not-determined |
| S3_extracted_k141_90420 | Not-determined |
| S3_extracted_k141_75100 | Not-determined |
| S3_extracted_k141_75101 | Not-determined |
| S3_extracted_k141_90423 | Not-determined |
| S3_extracted_k141_90424 | Not-determined |
| S3_extracted_k141_114311 | Low-quality |
| S3_extracted_k141_75105 | Not-determined |
| S3_extracted_k141_122023 | Not-determined |
| S3_extracted_k141_75110 | Not-determined |
| S3_extracted_k141_11997 | Not-determined |
| S3_extracted_k141_11999 | Not-determined |
| S3_extracted_k141_122026 | Not-determined |
| S3_extracted_k141_106439 | Not-determined |
| S3_extracted_k141_59007 | Not-determined |
| S3_extracted_k141_12004 | Not-determined |
| S3_extracted_k141_75116 | Not-determined |
| S3_extracted_k141_122031 | Not-determined |
| S3_extracted_k141_75118 | Low-quality |
| S3_extracted_k141_90440 | Not-determined |
| S3_extracted_k141_90441 | Low-quality |
| S3_extracted_k141_75120 | Not-determined |
| S3_extracted_k141_59010 | Not-determined |
| S3_extracted_k141_90443 | Low-quality |
| S3_extracted_k141_50987 | Not-determined |
| S3_extracted_k141_12014 | Not-determined |
| S3_extracted_k141_90448 | Low-quality |
| S3_extracted_k141_98423 | Not-determined |
| S3_extracted_k141_106444 | Not-determined |
| S3_extracted_k141_90449 | Not-determined |
| S3_extracted_k141_12020 | Not-determined |
| S3_extracted_k141_50993 | Not-determined |
| S3_extracted_k141_106445 | Low-quality |
| S3_extracted_k141_82534 | Not-determined |
| S3_extracted_k141_82536 | Low-quality |
| S3_extracted_k141_26683 | Not-determined |
| S3_extracted_k141_82543 | Not-determined |
| S3_extracted_k141_90468 | Not-determined |
| S3_extracted_k141_98438 | Not-determined |
| S3_extracted_k141_98441 | Not-determined |
| S3_extracted_k141_82549 | Not-determined |
| S3_extracted_k141_51007 | Low-quality |
| S3_extracted_k141_82551 | Not-determined |
| S3_extracted_k141_98447 | Not-determined |
| S3_extracted_k141_82554 | Not-determined |
| S3_extracted_k141_2840 | Not-determined |
| S3_extracted_k141_75129 | Not-determined |
| S3_extracted_k141_51023 | Low-quality |
| S3_extracted_k141_26725 | Not-determined |
| S3_extracted_k141_75137 | Not-determined |
| S3_extracted_k141_66173 | Not-determined |
| S3_extracted_k141_59038 | Not-determined |
| S3_extracted_k141_75140 | Low-quality |
| S3_extracted_k141_75141 | Low-quality |
| S3_extracted_k141_98462 | Not-determined |
| S3_extracted_k141_106460 | Not-determined |
| S3_extracted_k141_59045 | Not-determined |
| S3_extracted_k141_106462 | Low-quality |
| S3_extracted_k141_106471 | Not-determined |
| S3_extracted_k141_82563 | Not-determined |
| S3_extracted_k141_114346 | Not-determined |
| S3_extracted_k141_26745 | Not-determined |
| S3_extracted_k141_106474 | Not-determined |
| S3_extracted_k141_26748 | Not-determined |
| S3_extracted_k141_114350 | Not-determined |
| S3_extracted_k141_34966 | Not-determined |
| S3_extracted_k141_106482 | Not-determined |
| S3_extracted_k141_75152 | Low-quality |
| S3_extracted_k141_34972 | Not-determined |
| S3_extracted_k141_106485 | Not-determined |
| S3_extracted_k141_114353 | Low-quality |
| S3_extracted_k141_59054 | Low-quality |
| S3_extracted_k141_59055 | Low-quality |
| S3_extracted_k141_59056 | Not-determined |
| S3_extracted_k141_114355 | Not-determined |
| S3_extracted_k141_114358 | Not-determined |
| S3_extracted_k141_59062 | Not-determined |
| S3_extracted_k141_59063 | Low-quality |
| S3_extracted_k141_114362 | Not-determined |
| S3_extracted_k141_114363 | Not-determined |
| S3_extracted_k141_2855 | Not-determined |
| S3_extracted_k141_114368 | Not-determined |
| S3_extracted_k141_90506 | Not-determined |
| S3_extracted_k141_90507 | Not-determined |
| S3_extracted_k141_90508 | Not-determined |
| S3_extracted_k141_18103 | Not-determined |
| S3_extracted_k141_106493 | Not-determined |
| S3_extracted_k141_18104 | Not-determined |
| S3_extracted_k141_34998 | Not-determined |
| S3_extracted_k141_34999 | Not-determined |
| S3_extracted_k141_35008 | Not-determined |
| S3_extracted_k141_114376 | Not-determined |
| S5_vs_k141_10428\|\|full | High-quality |
| S3_extracted_k141_59083 | Not-determined |
| S3_extracted_k141_26776 | Low-quality |
| S3_extracted_k141_51033 | Not-determined |
| S10_vs_87434\|\|full | High-quality |
| S3_extracted_k141_82572 | Not-determined |
| S3_extracted_k141_82573 | Not-determined |
| S3_extracted_k141_82575 | Not-determined |
| S3_extracted_k141_2865 | Not-determined |
| S3_extracted_k141_51038 | Not-determined |
| S3_extracted_k141_75168 | Not-determined |
| S3_extracted_k141_51040 | Low-quality |
| S3_extracted_k141_90524 | Low-quality |
| S3_extracted_k141_90525 | Not-determined |
| S3_extracted_k141_75171 | Not-determined |
| S3_extracted_k141_75175 | Not-determined |
| S3_extracted_k141_106509 | Not-determined |
| S3_extracted_k141_35026 | Not-determined |
| S3_extracted_k141_59110 | Not-determined |
| S3_extracted_k141_75182 | Not-determined |
| S3_extracted_k141_59111 | Not-determined |
| S3_extracted_k141_35035 | Not-determined |
| S3_extracted_k141_51044 | Not-determined |
| S3_extracted_k141_35039 | Not-determined |
| S3_extracted_k141_51045 | Low-quality |
| S3_extracted_k141_12025 | Not-determined |
| S3_extracted_k141_35045 | Not-determined |
| S3_extracted_k141_12029 | Not-determined |
| S3_extracted_k141_75196 | Not-determined |
| S3_extracted_k141_51054 | Low-quality |
| S3_extracted_k141_12033 | Not-determined |
| S3_extracted_k141_51055 | Low-quality |
| S3_extracted_k141_35059 | Not-determined |
| S3_extracted_k141_35060 | Not-determined |
| S3_extracted_k141_82595 | Not-determined |
| S3_extracted_k141_66190 | Not-determined |
| S3_extracted_k141_75201 | Not-determined |
| S3_extracted_k141_51060 | Not-determined |
| S3_extracted_k141_12037 | Low-quality |
| S3_extracted_k141_12039 | Not-determined |
| S3_extracted_k141_35065 | Not-determined |
| S3_extracted_k141_26782 | Not-determined |
| S3_extracted_k141_26783 | Not-determined |
| S3_extracted_k141_82609 | Not-determined |
| S3_extracted_k141_66206 | Low-quality |
| S3_extracted_k141_106523 | Low-quality |
| S3_extracted_k141_82614 | Not-determined |
| S3_extracted_k141_75214 | Not-determined |
| S3_extracted_k141_75215 | Not-determined |
| S3_extracted_k141_26796 | Not-determined |
| S3_extracted_k141_106526 | Not-determined |
| S3_extracted_k141_82615 | Not-determined |
| S3_extracted_k141_106528 | Not-determined |
| S3_extracted_k141_98485 | Not-determined |
| S3_extracted_k141_75223 | Low-quality |
| S3_extracted_k141_82623 | Not-determined |
| S3_extracted_k141_51090 | Not-determined |
| S3_extracted_k141_12049 | Not-determined |
| S3_extracted_k141_2873 | Low-quality |
| S3_extracted_k141_90535 | Not-determined |
| S3_extracted_k141_98491 | Not-determined |
| S3_extracted_k141_106535 | Not-determined |
| S3_extracted_k141_42904 | Low-quality |
| S3_extracted_k141_75231 | Not-determined |
| S3_extracted_k141_106540 | Not-determined |
| S3_extracted_k141_106545 | Not-determined |
| S3_extracted_k141_75238 | Not-determined |
| S3_extracted_k141_75239 | Not-determined |
| S3_extracted_k141_12060 | Not-determined |
| S3_extracted_k141_106551 | Not-determined |
| S3_extracted_k141_12061 | Not-determined |
| S3_extracted_k141_98499 | Low-quality |
| S3_extracted_k141_98502 | Not-determined |
| S3_extracted_k141_106558 | Not-determined |
| S3_extracted_k141_90550 | Low-quality |
| S3_extracted_k141_106563 | Not-determined |
| S3_extracted_k141_12068 | Not-determined |
| S3_extracted_k141_75247 | Not-determined |
| S3_extracted_k141_75248 | Not-determined |
| S3_extracted_k141_42916 | Not-determined |
| S3_extracted_k141_42917 | Not-determined |
| S3_extracted_k141_12071 | Not-determined |
| S3_extracted_k141_106566 | Not-determined |
| S3_extracted_k141_82636 | Not-determined |
| S3_extracted_k141_75252 | Not-determined |
| S3_extracted_k141_90564 | Not-determined |
| S3_extracted_k141_42922 | Not-determined |
| S3_extracted_k141_98509 | Not-determined |
| S3_extracted_k141_90566 | Not-determined |
| S3_extracted_k141_26817 | Not-determined |
| S3_extracted_k141_82647 | Not-determined |
| S3_extracted_k141_12080 | Not-determined |
| S3_extracted_k141_106571 | Not-determined |
| S3_extracted_k141_42932 | Not-determined |
| S3_extracted_k141_98516 | Not-determined |
| S3_extracted_k141_51112 | Not-determined |
| S3_extracted_k141_106579 | Not-determined |
| S3_extracted_k141_106580 | Not-determined |
| S3_extracted_k141_106581 | Not-determined |
| S3_extracted_k141_82659 | Not-determined |
| S3_extracted_k141_106583 | Not-determined |
| S3_extracted_k141_90585 | Not-determined |
| S3_extracted_k141_98532 | Not-determined |
| S3_extracted_k141_75280 | Not-determined |
| S3_extracted_k141_66236 | Not-determined |
| S3_extracted_k141_98535 | Not-determined |
| S3_extracted_k141_75283 | Not-determined |
| S3_extracted_k141_26830 | Low-quality |
| S3_extracted_k141_2891 | Not-determined |
| S3_extracted_k141_51124 | Low-quality |
| S10_vs_119298\|\|full | High-quality |
| S3_extracted_k141_66242 | Low-quality |
| S3_extracted_k141_66245 | Not-determined |
| S3_extracted_k141_51128 | Not-determined |
| S3_extracted_k141_90593 | Not-determined |
| S3_extracted_k141_35080 | Low-quality |
| S3_extracted_k141_82669 | Not-determined |
| S3_extracted_k141_2901 | Not-determined |
| S3_extracted_k141_35084 | Low-quality |
| S3_extracted_k141_35085 | Not-determined |
| S3_extracted_k141_2907 | Not-determined |
| S3_extracted_k141_51134 | Not-determined |
| S3_extracted_k141_18119 | Not-determined |
| S3_extracted_k141_42955 | Not-determined |
| S3_extracted_k141_18124 | Not-determined |
| S3_extracted_k141_90604 | Low-quality |
| S3_extracted_k141_18127 | Not-determined |
| S3_extracted_k141_75301 | Low-quality |
| S3_extracted_k141_35101 | Not-determined |
| S3_extracted_k141_75308 | Not-determined |
| S3_extracted_k141_75310 | Not-determined |
| S3_extracted_k141_2927 | Not-determined |
| S3_extracted_k141_82690 | Not-determined |
| S3_extracted_k141_51156 | Not-determined |
| S3_extracted_k141_35116 | Not-determined |
| S3_extracted_k141_51161 | Not-determined |
| S3_extracted_k141_75327 | Low-quality |
| S3_extracted_k141_35119 | Low-quality |
| S3_extracted_k141_12086 | Not-determined |
| S3_extracted_k141_18130 | Not-determined |
| S3_extracted_k141_35123 | Not-determined |
| S3_extracted_k141_35124 | Not-determined |
| S3_extracted_k141_18136 | Not-determined |
| S3_extracted_k141_12092 | Not-determined |
| S3_extracted_k141_75334 | Not-determined |
| S3_extracted_k141_18138 | Not-determined |
| S3_extracted_k141_75335 | Low-quality |
| S3_extracted_k141_12101 | Not-determined |
| S3_extracted_k141_12106 | Not-determined |
| S3_extracted_k141_51169 | Not-determined |
| S3_extracted_k141_12115 | Low-quality |
| S3_extracted_k141_26851 | Not-determined |
| S3_extracted_k141_90650 | Not-determined |
| S3_extracted_k141_51200 | Low-quality |
| S3_extracted_k141_66269 | Low-quality |
| S3_extracted_k141_51202 | Low-quality |
| S3_extracted_k141_122054 | Low-quality |
| S3_extracted_k141_51205 | Not-determined |
| S3_extracted_k141_82732 | Not-determined |
| S3_extracted_k141_26859 | Not-determined |
| S3_extracted_k141_122058 | Not-determined |
| S3_extracted_k141_82734 | Not-determined |
| S3_extracted_k141_12116 | Not-determined |
| S3_extracted_k141_90656 | Not-determined |
| S3_extracted_k141_12118 | Low-quality |
| S3_extracted_k141_106604 | Not-determined |
| S3_extracted_k141_82740 | Low-quality |
| S3_extracted_k141_82741 | Not-determined |
| S3_extracted_k141_43006 | Low-quality |
| S3_extracted_k141_106611 | Low-quality |
| S3_extracted_k141_106612 | Not-determined |
| S3_extracted_k141_66287 | Not-determined |
| S3_extracted_k141_51219 | Not-determined |
| S3_extracted_k141_51220 | Not-determined |
| S3_extracted_k141_82744 | Not-determined |
| S3_extracted_k141_43010 | Not-determined |
| S3_extracted_k141_106615 | Not-determined |
| S3_extracted_k141_98582 | Not-determined |
| S3_extracted_k141_75346 | Not-determined |
| S3_extracted_k141_66297 | Low-quality |
| S3_extracted_k141_12122 | Not-determined |
| S3_extracted_k141_66298 | Low-quality |
| S3_extracted_k141_66299 | Not-determined |
| S3_extracted_k141_114410 | Not-determined |
| S3_extracted_k141_98588 | Not-determined |
| S3_extracted_k141_75359 | Not-determined |
| S3_extracted_k141_114411 | Low-quality |
| S3_extracted_k141_51237 | Not-determined |
| S3_extracted_k141_51238 | Not-determined |
| S3_extracted_k141_12129 | Not-determined |
| S3_extracted_k141_12130 | Not-determined |
| S3_extracted_k141_98597 | Not-determined |

| | |
|---|---|
| S3_extracted_k141_114416 | Not-determined |
| S3_extracted_k141_114421 | Not-determined |
| S3_extracted_k141_12147 | Not-determined |
| S3_extracted_k141_12148 | Not-determined |
| S3_extracted_k141_106628 | Low-quality |
| S3_extracted_k141_122098 | Low-quality |
| S3_extracted_k141_51252 | Not-determined |
| S3_extracted_k141_26872 | Not-determined |
| S3_extracted_k141_82766 | Not-determined |
| S3_extracted_k141_18147 | Not-determined |
| S3_extracted_k141_51260 | Low-quality |
| S3_extracted_k141_82770 | Not-determined |
| S3_extracted_k141_35147 | Not-determined |
| S3_extracted_k141_43032 | Low-quality |
| S3_extracted_k141_51264 | Not-determined |
| S3_extracted_k141_59126 | Not-determined |
| S3_extracted_k141_82778 | Not-determined |
| S3_extracted_k141_114434 | Not-determined |
| S3_extracted_k141_59130 | Low-quality |
| S3_extracted_k141_12163 | Not-determined |
| S3_extracted_k141_12165 | Not-determined |
| S3_extracted_k141_2943 | Low-quality |
| S3_extracted_k141_114440 | Low-quality |
| S3_extracted_k141_106635 | Not-determined |
| S3_extracted_k141_2948 | Not-determined |
| S3_extracted_k141_106636 | Not-determined |
| S3_extracted_k141_26896 | Low-quality |
| S3_extracted_k141_59138 | Not-determined |
| S3_extracted_k141_106640 | Not-determined |
| S3_extracted_k141_82802 | Low-quality |
| S3_extracted_k141_59139 | Not-determined |
| S3_extracted_k141_59141 | Low-quality |
| S3_extracted_k141_12185 | Not-determined |
| S3_extracted_k141_12189 | Low-quality |
| S3_extracted_k141_2959 | Not-determined |
| S3_extracted_k141_18184 | Not-determined |
| S3_extracted_k141_43045 | Not-determined |
| S3_extracted_k141_106653 | Not-determined |
| S3_extracted_k141_2962 | Not-determined |
| S3_extracted_k141_18189 | Not-determined |
| S3_extracted_k141_82806 | Low-quality |
| S3_extracted_k141_82808 | Not-determined |
| S3_extracted_k141_122138 | Not-determined |
| S3_extracted_k141_114458 | Not-determined |
| S3_extracted_k141_2981 | Not-determined |
| S3_extracted_k141_2985 | Not-determined |
| S3_extracted_k141_26921 | Not-determined |
| S3_extracted_k141_26922 | Not-determined |
| S3_extracted_k141_2993 | Not-determined |
| S3_extracted_k141_2994 | Not-determined |
| S3_extracted_k141_122148 | Not-determined |
| S3_extracted_k141_2999 | Low-quality |
| S3_extracted_k141_43055 | Not-determined |
| S3_extracted_k141_12205 | Not-determined |
| S3_extracted_k141_106670 | Not-determined |
| S3_extracted_k141_12209 | Low-quality |
| S3_extracted_k141_3007 | Low-quality |
| S3_extracted_k141_82831 | Low-quality |
| S3_extracted_k141_98613 | Low-quality |
| S3_extracted_k141_82834 | Not-determined |
| S3_extracted_k141_106680 | Low-quality |
| S3_extracted_k141_3012 | Not-determined |
| S3_extracted_k141_114480 | Not-determined |
| S3_extracted_k141_3015 | Not-determined |
| S3_extracted_k141_114481 | Not-determined |
| S3_extracted_k141_114485 | Not-determined |
| S3_extracted_k141_98623 | Not-determined |
| S3_extracted_k141_106685 | Not-determined |
| S3_extracted_k141_3020 | Not-determined |
| S3_extracted_k141_35184 | Not-determined |
| S3_extracted_k141_75398 | Not-determined |
| S3_extracted_k141_75400 | Not-determined |
| S3_extracted_k141_98633 | Not-determined |
| S3_extracted_k141_75409 | Not-determined |
| S3_extracted_k141_75411 | Not-determined |
| S3_extracted_k141_43067 | Not-determined |
| S3_extracted_k141_114494 | Not-determined |
| S3_extracted_k141_114496 | Not-determined |
| S3_extracted_k141_98639 | Low-quality |
| S3_extracted_k141_51324 | Not-determined |
| S3_extracted_k141_43078 | Not-determined |
| S3_extracted_k141_106686 | Not-determined |
| S3_extracted_k141_66323 | Not-determined |
| S3_extracted_k141_66324 | Not-determined |
| S3_extracted_k141_51327 | Not-determined |
| S3_extracted_k141_114502 | Not-determined |
| S3_extracted_k141_3030 | Not-determined |
| S3_extracted_k141_114507 | Not-determined |
| S3_extracted_k141_98647 | Not-determined |
| S3_extracted_k141_35197 | Not-determined |
| S3_extracted_k141_122173 | Not-determined |
| S3_extracted_k141_98654 | Low-quality |
| S3_extracted_k141_75415 | Not-determined |
| S3_extracted_k141_51331 | Not-determined |
| S3_extracted_k141_75419 | Not-determined |
| S3_extracted_k141_51332 | Not-determined |
| S3_extracted_k141_12224 | Not-determined |
| S3_extracted_k141_75421 | Low-quality |
| S3_extracted_k141_35211 | Not-determined |
| S3_extracted_k141_114519 | Not-determined |
| S3_extracted_k141_35212 | Not-determined |
| S3_extracted_k141_43087 | Not-determined |
| S3_extracted_k141_43088 | Not-determined |
| S3_extracted_k141_106716 | Not-determined |
| S3_extracted_k141_90663 | Not-determined |
| S3_extracted_k141_90665 | Not-determined |
| S3_extracted_k141_90667 | Not-determined |
| S3_extracted_k141_98670 | Not-determined |
| S3_extracted_k141_90670 | Not-determined |
| S3_extracted_k141_12231 | Not-determined |
| S3_extracted_k141_106721 | Not-determined |
| S3_extracted_k141_66333 | Low-quality |
| S3_extracted_k141_59174 | Low-quality |
| S3_extracted_k141_3036 | Not-determined |
| S3_extracted_k141_66346 | Not-determined |
| S3_extracted_k141_66348 | Not-determined |
| S3_extracted_k141_35217 | Low-quality |
| S3_extracted_k141_3042 | Not-determined |
| S3_extracted_k141_26955 | Not-determined |
| S3_extracted_k141_106728 | Not-determined |
| S3_extracted_k141_90684 | Not-determined |
| S3_extracted_k141_3050 | Not-determined |
| S3_extracted_k141_26961 | Not-determined |
| S3_extracted_k141_43108 | Low-quality |
| S3_extracted_k141_43110 | Not-determined |
| S3_extracted_k141_35228 | Not-determined |
| S3_extracted_k141_43111 | Not-determined |
| S3_extracted_k141_26966 | Not-determined |
| S3_extracted_k141_43122 | Not-determined |
| S3_extracted_k141_59191 | Not-determined |
| S3_extracted_k141_66352 | Not-determined |
| S3_extracted_k141_75437 | Not-determined |
| S3_extracted_k141_75441 | Not-determined |
| S3_extracted_k141_75442 | Not-determined |
| S3_extracted_k141_35242 | Not-determined |
| S3_extracted_k141_106750 | Not-determined |
| S3_extracted_k141_26992 | Not-determined |
| S3_extracted_k141_75444 | Not-determined |
| S3_extracted_k141_106759 | Not-determined |
| S3_extracted_k141_27004 | Low-quality |
| S3_extracted_k141_90704 | Not-determined |
| S3_extracted_k141_35256 | Not-determined |
| S3_extracted_k141_106768 | Not-determined |
| S3_extracted_k141_27014 | Low-quality |
| S3_extracted_k141_90710 | Not-determined |
| S3_extracted_k141_59212 | Not-determined |
| S3_extracted_k141_59213 | Not-determined |
| S3_extracted_k141_35267 | Not-determined |
| S3_extracted_k141_82845 | Low-quality |
| S3_extracted_k141_82846 | Not-determined |
| S3_extracted_k141_27020 | Not-determined |
| S3_extracted_k141_75459 | Not-determined |
| S3_extracted_k141_18220 | Not-determined |
| S3_extracted_k141_27027 | Not-determined |
| S3_extracted_k141_75463 | Not-determined |
| S3_extracted_k141_106781 | Not-determined |
| S3_extracted_k141_106783 | Not-determined |
| S3_extracted_k141_18227 | Not-determined |
| S3_extracted_k141_82857 | Low-quality |
| S3_extracted_k141_90719 | Low-quality |
| S3_extracted_k141_18231 | Not-determined |
| S3_extracted_k141_18234 | Not-determined |
| S3_extracted_k141_35300 | Low-quality |
| S3_extracted_k141_35301 | Not-determined |
| S3_extracted_k141_27050 | Not-determined |
| S3_extracted_k141_27051 | Low-quality |
| S3_extracted_k141_35304 | Low-quality |
| S3_extracted_k141_59224 | Not-determined |
| S3_extracted_k141_114537 | Not-determined |
| S3_extracted_k141_106789 | Not-determined |
| S3_extracted_k141_90724 | Not-determined |
| S3_extracted_k141_90725 | Not-determined |
| S3_extracted_k141_114540 | Not-determined |
| S3_extracted_k141_18238 | Not-determined |
| S3_extracted_k141_114542 | Low-quality |
| S3_extracted_k141_66373 | Not-determined |
| S3_extracted_k141_27059 | Not-determined |
| S3_extracted_k141_18241 | Not-determined |
| S3_extracted_k141_106794 | Not-determined |
| S3_extracted_k141_90733 | Not-determined |
| S3_extracted_k141_27065 | Not-determined |
| S3_extracted_k141_43135 | Not-determined |
| S3_extracted_k141_90740 | Not-determined |
| S3_extracted_k141_114555 | Not-determined |
| S3_extracted_k141_75476 | Not-determined |
| S3_extracted_k141_12280 | Not-determined |
| S3_extracted_k141_106805 | Not-determined |
| S3_extracted_k141_82871 | Not-determined |
| S3_extracted_k141_59248 | Low-quality |
| S3_extracted_k141_75486 | Not-determined |
| S3_extracted_k141_82876 | Low-quality |
| S3_extracted_k141_35307 | Low-quality |
| S3_extracted_k141_3082 | Not-determined |
| S3_extracted_k141_35308 | Low-quality |
| S3_extracted_k141_106815 | Low-quality |
| S3_extracted_k141_98677 | Not-determined |
| S3_extracted_k141_3089 | Low-quality |
| S3_extracted_k141_98683 | Not-determined |
| S3_extracted_k141_114567 | Not-determined |
| S3_extracted_k141_12294 | Not-determined |
| S3_extracted_k141_3098 | Not-determined |
| S3_extracted_k141_75499 | Not-determined |
| S3_extracted_k141_51356 | Not-determined |
| S3_extracted_k141_75506 | Not-determined |
| S3_extracted_k141_51360 | Not-determined |
| S3_extracted_k141_51363 | Low-quality |
| S3_extracted_k141_35314 | Not-determined |
| S3_extracted_k141_35317 | Not-determined |
| S3_extracted_k141_114576 | Not-determined |
| S3_extracted_k141_66381 | Not-determined |
| S3_extracted_k141_114582 | Not-determined |
| S3_extracted_k141_82880 | Low-quality |
| S3_extracted_k141_35324 | Not-determined |
| S3_extracted_k141_106830 | Not-determined |
| S3_extracted_k141_59253 | Not-determined |
| S3_extracted_k141_59255 | Not-determined |
| S3_extracted_k141_90769 | Not-determined |
| S3_extracted_k141_114594 | Low-quality |
| S3_extracted_k141_75518 | Not-determined |
| S3_extracted_k141_35328 | Not-determined |
| S3_extracted_k141_27079 | Low-quality |
| S3_extracted_k141_98692 | Not-determined |
| S3_extracted_k141_75527 | Not-determined |
| S3_extracted_k141_27083 | Not-determined |
| S3_extracted_k141_114603 | Low-quality |
| S3_extracted_k141_35340 | Low-quality |
| S3_extracted_k141_98706 | Not-determined |
| S3_extracted_k141_12312 | Not-determined |
| S3_extracted_k141_82892 | Not-determined |
| S3_extracted_k141_43176 | Low-quality |
| S3_extracted_k141_75558 | Not-determined |
| S3_extracted_k141_59256 | Low-quality |
| S3_extracted_k141_59257 | Not-determined |
| S3_extracted_k141_90782 | Not-determined |
| S3_extracted_k141_51386 | Not-determined |
| S3_extracted_k141_75574 | Not-determined |
| S3_extracted_k141_90788 | Not-determined |
| S3_extracted_k141_51396 | Low-quality |
| S3_extracted_k141_51401 | Low-quality |
| S3_extracted_k141_106851 | Not-determined |
| S3_extracted_k141_106852 | Not-determined |
| S3_extracted_k141_106855 | Low-quality |
| S3_extracted_k141_43202 | Not-determined |
| S3_extracted_k141_59291 | Not-determined |
| S3_extracted_k141_59293 | Not-determined |
| S3_extracted_k141_43214 | Not-determined |
| S3_extracted_k141_59301 | Not-determined |
| S3_extracted_k141_66420 | Not-determined |
| S3_extracted_k141_59310 | Low-quality |
| S3_extracted_k141_27104 | Low-quality |
| S3_extracted_k141_59311 | Low-quality |
| S3_extracted_k141_27105 | Low-quality |
| S3_extracted_k141_114656 | Low-quality |
| S3_extracted_k141_35409 | Not-determined |
| S3_extracted_k141_51416 | Not-determined |
| S3_extracted_k141_43230 | Not-determined |
| S3_extracted_k141_98756 | Low-quality |
| S3_extracted_k141_75585 | Low-quality |
| S3_extracted_k141_98759 | Not-determined |
| S3_extracted_k141_51421 | Not-determined |
| S3_extracted_k141_43237 | Low-quality |
| S3_extracted_k141_66426 | Not-determined |
| S3_extracted_k141_122187 | Not-determined |
| S3_extracted_k141_122191 | Not-determined |
| S3_extracted_k141_51422 | Low-quality |
| S3_extracted_k141_122194 | Not-determined |
| S3_extracted_k141_3130 | Not-determined |
| S3_extracted_k141_106867 | Not-determined |
| S3_extracted_k141_3137 | Low-quality |
| S3_extracted_k141_3139 | Not-determined |
| S3_extracted_k141_59330 | Not-determined |
| S3_extracted_k141_59332 | Not-determined |
| S3_extracted_k141_75596 | Not-determined |
| S3_extracted_k141_35421 | Not-determined |
| S3_extracted_k141_75597 | Not-determined |
| S3_extracted_k141_75598 | Not-determined |
| S3_extracted_k141_3151 | Not-determined |
| S3_extracted_k141_35432 | Low-quality |
| S3_extracted_k141_75605 | Not-determined |
| S3_extracted_k141_106878 | Not-determined |
| S3_extracted_k141_98771 | Not-determined |
| S3_extracted_k141_75611 | Not-determined |
| S3_extracted_k141_122204 | Low-quality |
| S3_extracted_k141_122209 | Not-determined |
| S3_extracted_k141_75616 | Low-quality |
| S3_extracted_k141_75618 | Not-determined |
| S3_extracted_k141_59351 | Not-determined |
| S3_extracted_k141_35440 | Not-determined |
| S3_extracted_k141_75625 | Not-determined |
| S3_extracted_k141_106886 | Not-determined |
| S3_extracted_k141_106892 | Not-determined |
| S3_extracted_k141_75627 | Not-determined |
| S3_extracted_k141_59357 | Not-determined |
| S3_extracted_k141_75630 | Not-determined |
| S3_extracted_k141_75631 | Not-determined |
| S3_extracted_k141_106896 | Low-quality |
| S3_extracted_k141_114672 | Not-determined |
| S3_extracted_k141_75636 | Not-determined |
| S3_extracted_k141_114680 | Not-determined |
| S3_extracted_k141_75639 | Not-determined |
| S3_extracted_k141_59369 | Not-determined |
| S3_extracted_k141_75642 | Not-determined |
| S3_extracted_k141_114692 | Low-quality |
| S3_extracted_k141_75647 | Low-quality |
| S3_extracted_k141_114695 | Not-determined |
| S3_extracted_k141_75649 | Low-quality |
| S3_extracted_k141_18309 | Not-determined |
| S3_extracted_k141_35452 | Not-determined |
| S3_extracted_k141_35453 | Not-determined |
| S3_extracted_k141_18316 | Not-determined |
| S3_extracted_k141_75662 | Not-determined |
| S3_extracted_k141_75668 | Not-determined |
| S3_extracted_k141_35464 | Not-determined |
| S3_extracted_k141_18320 | Not-determined |
| S3_extracted_k141_90811 | Not-determined |
| S3_extracted_k141_12332 | Not-determined |
| S3_extracted_k141_12333 | Not-determined |
| S3_extracted_k141_51447 | Not-determined |
| S3_extracted_k141_98801 | Not-determined |
| S3_extracted_k141_98802 | Not-determined |
| S3_extracted_k141_122223 | Not-determined |
| S3_extracted_k141_122224 | Low-quality |
| S3_extracted_k141_51454 | Not-determined |
| S3_extracted_k141_106953 | Low-quality |
| S3_extracted_k141_90820 | Not-determined |
| S3_extracted_k141_122228 | Not-determined |
| S3_extracted_k141_122229 | Not-determined |
| S3_extracted_k141_18333 | Not-determined |
| S3_extracted_k141_90823 | Not-determined |
| S3_extracted_k141_12348 | Not-determined |
| S3_extracted_k141_90833 | Not-determined |
| S3_extracted_k141_35473 | Not-determined |
| S3_extracted_k141_12354 | Low-quality |
| S3_extracted_k141_12355 | Not-determined |
| S3_extracted_k141_35476 | Not-determined |
| S3_extracted_k141_51467 | Not-determined |
| S3_extracted_k141_90843 | Not-determined |
| S3_extracted_k141_106965 | Low-quality |
| S3_extracted_k141_12361 | Not-determined |
| S3_extracted_k141_51475 | Not-determined |
| S3_extracted_k141_51477 | Low-quality |
| S3_extracted_k141_12363 | Low-quality |
| S3_extracted_k141_12365 | Low-quality |
| S3_extracted_k141_35480 | Not-determined |
| S3_extracted_k141_90850 | Not-determined |
| S3_extracted_k141_51480 | Not-determined |
| S3_extracted_k141_114707 | Not-determined |
| S3_extracted_k141_75697 | Not-determined |
| S3_extracted_k141_35488 | Not-determined |
| S3_extracted_k141_75699 | Not-determined |
| S3_extracted_k141_51486 | Not-determined |
| S3_extracted_k141_122250 | Not-determined |
| S3_extracted_k141_12376 | Not-determined |
| S3_extracted_k141_51492 | Not-determined |
| S3_extracted_k141_35499 | Low-quality |
| S3_extracted_k141_35500 | Not-determined |
| S3_extracted_k141_106967 | Not-determined |
| S3_extracted_k141_122257 | Not-determined |
| S3_extracted_k141_90858 | Not-determined |
| S3_extracted_k141_12390 | Low-quality |
| S3_extracted_k141_106970 | Not-determined |
| S3_extracted_k141_90861 | Not-determined |
| S3_extracted_k141_12394 | Not-determined |
| S3_extracted_k141_12398 | Not-determined |
| S3_extracted_k141_12400 | Not-determined |
| S3_extracted_k141_12403 | Not-determined |
| S3_extracted_k141_12404 | Not-determined |
| S3_extracted_k141_51506 | Low-quality |
| S3_extracted_k141_12408 | Not-determined |
| S3_extracted_k141_43257 | Not-determined |
| S3_extracted_k141_98812 | Not-determined |
| S3_extracted_k141_75717 | Not-determined |
| S3_extracted_k141_90869 | Not-determined |
| S3_extracted_k141_106983 | Not-determined |
| S3_extracted_k141_35509 | Not-determined |
| S3_extracted_k141_35510 | Not-determined |
| S3_extracted_k141_98814 | Not-determined |
| S3_extracted_k141_114718 | Not-determined |
| S3_extracted_k141_98816 | Not-determined |
| S3_extracted_k141_12413 | Not-determined |
| S3_extracted_k141_75724 | Not-determined |
| S3_extracted_k141_12414 | Not-determined |
| S3_extracted_k141_114721 | Not-determined |
| S3_extracted_k141_35513 | Not-determined |
| S3_extracted_k141_35514 | Not-determined |
| S3_extracted_k141_106988 | Low-quality |
| S3_extracted_k141_114724 | Not-determined |
| S3_extracted_k141_66439 | Not-determined |
| S3_extracted_k141_90874 | Not-determined |
| S3_extracted_k141_43262 | Low-quality |
| S3_extracted_k141_75733 | Not-determined |
| S3_extracted_k141_35519 | Not-determined |
| S3_extracted_k141_98820 | Low-quality |
| S3_extracted_k141_35521 | Not-determined |
| S3_extracted_k141_18397 | Not-determined |
| S3_extracted_k141_114731 | Not-determined |
| S3_extracted_k141_66447 | Not-determined |
| S3_extracted_k141_98825 | Not-determined |
| S3_extracted_k141_114738 | Not-determined |
| S3_extracted_k141_18406 | Not-determined |
| S3_extracted_k141_98831 | Not-determined |
| S3_extracted_k141_98834 | Not-determined |
| S3_extracted_k141_66456 | Not-determined |
| S3_extracted_k141_106993 | Not-determined |
| S3_extracted_k141_90889 | Not-determined |
| S3_extracted_k141_90890 | Not-determined |
| S3_extracted_k141_114746 | Not-determined |
| S3_extracted_k141_12430 | Not-determined |
| S3_extracted_k141_12435 | Not-determined |
| S3_extracted_k141_12437 | Not-determined |
| S3_extracted_k141_18408 | Low-quality |
| S3_extracted_k141_75758 | Not-determined |
| S3_extracted_k141_75760 | Not-determined |
| S3_extracted_k141_107003 | Not-determined |
| S3_extracted_k141_59399 | Low-quality |
| S3_extracted_k141_59402 | Not-determined |
| S3_extracted_k141_59403 | Not-determined |
| S3_extracted_k141_18420 | Not-determined |
| S3_extracted_k141_59404 | Not-determined |
| S3_extracted_k141_18421 | Low-quality |
| S3_extracted_k141_75768 | Not-determined |
| S3_extracted_k141_59407 | Not-determined |
| S3_extracted_k141_59409 | Not-determined |
| S3_extracted_k141_12441 | Not-determined |
| S3_extracted_k141_75776 | Not-determined |
| S3_extracted_k141_75777 | Not-determined |
| S3_extracted_k141_51540 | Not-determined |
| S3_extracted_k141_98850 | Low-quality |
| S3_extracted_k141_51541 | Not-determined |
| S3_extracted_k141_107017 | Not-determined |
| S3_extracted_k141_114768 | Not-determined |
| S3_extracted_k141_90896 | Not-determined |
| S3_extracted_k141_90897 | Low-quality |
| S3_extracted_k141_18439 | Not-determined |
| S3_extracted_k141_90898 | Not-determined |
| S3_extracted_k141_90900 | Not-determined |
| S3_extracted_k141_51543 | Not-determined |
| S3_extracted_k141_98857 | Low-quality |
| S3_extracted_k141_107023 | Not-determined |
| S3_extracted_k141_12457 | Not-determined |
| S3_extracted_k141_90905 | Low-quality |
| S3_extracted_k141_90906 | Not-determined |
| S3_extracted_k141_107026 | Not-determined |
| S3_extracted_k141_107027 | Not-determined |
| S3_extracted_k141_98859 | Not-determined |
| S3_extracted_k141_98860 | Not-determined |
| S3_extracted_k141_107028 | Not-determined |
| S3_extracted_k141_66463 | Not-determined |
| S3_extracted_k141_18452 | Not-determined |
| S3_extracted_k141_75798 | Not-determined |
| S3_extracted_k141_75806 | Not-determined |
| S3_extracted_k141_66478 | Not-determined |
| S3_extracted_k141_35561 | Not-determined |
| S3_extracted_k141_35562 | Not-determined |
| S3_extracted_k141_51563 | Not-determined |
| S3_extracted_k141_75813 | Not-determined |
| S3_extracted_k141_18460 | Not-determined |
| S3_extracted_k141_107034 | Not-determined |
| S3_extracted_k141_90933 | Not-determined |
| S3_extracted_k141_90936 | Low-quality |
| S3_extracted_k141_75816 | Low-quality |
| S3_extracted_k141_107039 | Not-determined |
| S3_extracted_k141_90937 | Not-determined |
| S3_extracted_k141_12466 | Not-determined |
| S3_extracted_k141_90940 | Low-quality |
| S3_extracted_k141_66484 | Not-determined |
| S3_extracted_k141_3163 | Not-determined |
| S3_extracted_k141_107045 | Low-quality |
| S3_extracted_k141_59441 | Not-determined |
| S3_extracted_k141_59445 | Not-determined |
| S3_extracted_k141_98877 | Not-determined |
| S3_extracted_k141_98878 | Not-determined |
| S3_extracted_k141_114774 | Not-determined |
| S3_extracted_k141_35583 | Not-determined |
| S3_extracted_k141_3173 | Low-quality |
| S3_extracted_k141_35585 | Not-determined |
| S3_extracted_k141_27138 | Low-quality |
| S3_extracted_k141_114782 | Not-determined |
| S3_extracted_k141_59449 | Low-quality |
| S3_extracted_k141_27145 | Not-determined |
| S3_extracted_k141_66490 | Not-determined |
| S3_extracted_k141_27151 | Not-determined |
| S3_extracted_k141_107055 | Not-determined |
| S3_extracted_k141_114791 | Not-determined |
| S3_extracted_k141_3190 | Not-determined |
| S3_extracted_k141_114793 | Not-determined |
| S3_extracted_k141_51570 | Not-determined |
| S3_extracted_k141_18491 | Not-determined |
| S3_extracted_k141_35595 | Not-determined |
| S3_extracted_k141_51574 | Not-determined |
| S3_extracted_k141_98895 | Not-determined |
| S3_extracted_k141_98898 | Not-determined |
| S3_extracted_k141_27166 | Not-determined |
| S3_extracted_k141_18494 | Not-determined |
| S3_extracted_k141_122279 | Low-quality |
| S3_extracted_k141_82897 | Not-determined |
| S3_extracted_k141_18499 | Not-determined |
| S3_extracted_k141_122281 | Not-determined |
| S3_extracted_k141_82899 | Low-quality |
| S3_extracted_k141_98901 | Not-determined |
| S3_extracted_k141_59451 | Not-determined |
| S3_extracted_k141_122283 | Not-determined |
| S3_extracted_k141_3204 | Not-determined |
| S3_extracted_k141_98903 | Not-determined |
| S3_extracted_k141_107074 | Low-quality |
| S3_extracted_k141_27176 | Not-determined |
| S3_extracted_k141_59453 | Low-quality |
| S3_extracted_k141_18508 | Not-determined |
| S3_extracted_k141_3211 | Not-determined |
| S3_extracted_k141_3212 | Not-determined |
| S3_extracted_k141_3213 | Low-quality |
| S3_extracted_k141_98910 | Not-determined |
| S3_extracted_k141_82911 | Not-determined |
| S3_extracted_k141_98912 | Not-determined |
| S3_extracted_k141_66508 | Not-determined |
| S3_extracted_k141_82914 | Not-determined |
| S3_extracted_k141_82915 | Not-determined |
| S3_extracted_k141_43273 | Not-determined |
| S3_extracted_k141_114809 | Not-determined |
| S3_extracted_k141_75826 | Not-determined |
| S3_extracted_k141_27195 | Not-determined |
| S3_extracted_k141_27198 | Not-determined |
| S3_extracted_k141_3226 | Not-determined |
| S3_extracted_k141_27199 | Not-determined |
| S3_extracted_k141_122305 | Not-determined |
| S3_extracted_k141_122306 | Not-determined |
| S3_extracted_k141_122311 | Not-determined |
| S3_extracted_k141_12488 | Not-determined |
| S3_extracted_k141_51592 | Not-determined |
| S3_extracted_k141_35619 | Not-determined |
| S3_extracted_k141_43274 | Low-quality |
| S3_extracted_k141_114817 | Not-determined |
| S3_extracted_k141_35621 | Not-determined |
| S3_extracted_k141_43279 | Not-determined |
| S3_extracted_k141_18533 | Not-determined |
| S3_extracted_k141_27218 | Low-quality |
| S3_extracted_k141_27220 | Not-determined |
| S3_extracted_k141_35630 | Not-determined |
| S3_extracted_k141_82940 | Not-determined |
| S3_extracted_k141_35633 | Not-determined |
| S3_extracted_k141_35635 | Not-determined |
| S3_extracted_k141_98924 | Not-determined |
| S3_extracted_k141_43294 | Low-quality |
| S3_extracted_k141_122335 | Not-determined |
| S3_extracted_k141_90955 | Not-determined |
| S3_extracted_k141_114831 | Not-determined |
| S3_extracted_k141_51606 | Low-quality |
| S3_extracted_k141_59465 | Not-determined |
| S3_extracted_k141_43299 | Not-determined |
| S3_extracted_k141_114837 | Not-determined |
| S3_extracted_k141_51612 | Not-determined |
| S3_extracted_k141_18555 | Not-determined |
| S3_extracted_k141_43307 | Not-determined |
| S3_extracted_k141_35647 | Not-determined |
| S3_extracted_k141_35649 | Not-determined |
| S3_extracted_k141_12504 | Not-determined |
| S3_extracted_k141_90970 | Not-determined |
| S3_extracted_k141_18563 | Not-determined |
| S3_extracted_k141_27246 | Low-quality |
| S3_extracted_k141_12511 | Not-determined |
| S3_extracted_k141_59471 | Low-quality |
| S3_extracted_k141_66530 | Not-determined |
| S3_extracted_k141_51628 | Not-determined |
| S3_extracted_k141_27249 | Not-determined |
| S3_extracted_k141_122347 | Not-determined |
| S3_extracted_k141_66532 | Not-determined |
| S3_extracted_k141_66533 | Not-determined |
| S3_extracted_k141_75840 | Not-determined |
| S3_extracted_k141_59473 | Not-determined |
| S3_extracted_k141_122350 | Not-determined |
| S3_extracted_k141_27252 | Not-determined |
| S3_extracted_k141_107099 | Low-quality |
| S3_extracted_k141_27255 | Not-determined |
| S3_extracted_k141_27258 | Low-quality |
| S3_extracted_k141_51633 | Not-determined |
| S3_extracted_k141_27269 | Low-quality |
| S3_extracted_k141_27272 | Not-determined |
| S3_extracted_k141_59484 | Not-determined |
| S3_extracted_k141_43314 | Not-determined |
| S3_extracted_k141_66553 | Not-determined |
| S3_extracted_k141_27274 | Low-quality |
| S3_extracted_k141_27276 | Not-determined |
| S3_extracted_k141_114846 | Not-determined |
| S3_extracted_k141_90991 | Not-determined |
| S3_extracted_k141_59491 | Not-determined |
| S3_extracted_k141_98954 | Not-determined |
| S3_extracted_k141_43321 | Not-determined |
| S3_extracted_k141_114851 | Not-determined |
| S3_extracted_k141_43326 | Not-determined |

| | |
|---|---|
| S3_extracted_k141_27286 | Not-determined |
| S3_extracted_k141_27290 | Not-determined |
| S3_extracted_k141_43331 | Not-determined |
| S3_extracted_k141_91002 | Not-determined |
| S3_extracted_k141_91004 | Not-determined |
| S3_extracted_k141_91009 | Not-determined |
| S3_extracted_k141_35668 | Not-determined |
| S3_extracted_k141_12534 | Not-determined |
| S3_extracted_k141_114869 | Not-determined |
| S3_extracted_k141_12537 | Not-determined |
| S3_extracted_k141_91017 | Not-determined |
| S3_extracted_k141_114873 | Not-determined |
| S3_extracted_k141_43340 | Not-determined |
| S3_extracted_k141_122368 | Not-determined |
| S3_extracted_k141_122372 | Not-determined |
| S3_extracted_k141_122376 | Low-quality |
| S3_extracted_k141_43346 | Not-determined |
| S3_extracted_k141_51642 | Low-quality |
| S3_extracted_k141_122385 | Not-determined |
| S3_extracted_k141_107120 | Not-determined |
| S3_extracted_k141_91031 | Low-quality |
| S3_extracted_k141_122387 | Not-determined |
| S3_extracted_k141_114884 | Low-quality |
| S3_extracted_k141_91034 | Not-determined |
| S3_extracted_k141_114889 | Not-determined |
| S3_extracted_k141_27299 | Not-determined |
| S3_extracted_k141_51651 | Low-quality |
| S3_extracted_k141_59505 | Not-determined |
| S3_extracted_k141_27304 | Not-determined |
| S3_extracted_k141_98973 | Not-determined |
| S3_extracted_k141_91038 | Not-determined |
| S3_extracted_k141_107132 | Low-quality |
| S3_extracted_k141_114901 | Not-determined |
| S3_extracted_k141_122396 | Low-quality |
| S3_extracted_k141_27305 | Low-quality |
| S3_extracted_k141_82976 | Not-determined |
| S3_extracted_k141_122400 | Not-determined |
| S3_extracted_k141_82981 | Not-determined |
| S3_extracted_k141_82982 | Not-determined |
| S3_extracted_k141_82983 | Low-quality |
| S3_extracted_k141_98995 | Not-determined |
| S3_extracted_k141_27318 | Not-determined |
| S3_extracted_k141_114915 | Not-determined |
| S3_extracted_k141_114916 | Not-determined |
| S3_extracted_k141_107138 | Not-determined |
| S3_extracted_k141_91048 | Not-determined |
| S3_extracted_k141_59515 | Low-quality |
| S3_extracted_k141_91055 | Not-determined |
| S3_extracted_k141_99000 | Not-determined |
| S3_extracted_k141_59519 | Low-quality |
| S3_extracted_k141_107147 | Not-determined |
| S3_extracted_k141_18595 | Not-determined |
| S3_extracted_k141_107154 | Not-determined |
| S3_extracted_k141_18598 | Not-determined |
| S3_extracted_k141_59525 | Low-quality |
| S3_extracted_k141_59526 | Low-quality |
| S3_extracted_k141_91060 | Not-determined |
| S3_extracted_k141_114931 | Low-quality |
| S3_extracted_k141_114939 | Not-determined |
| S3_extracted_k141_114940 | Not-determined |
| S3_extracted_k141_99027 | Low-quality |
| S3_extracted_k141_18616 | Not-determined |
| S3_extracted_k141_83005 | Not-determined |
| S3_extracted_k141_99033 | Not-determined |
| S3_extracted_k141_83009 | Not-determined |
| S3_extracted_k141_114950 | Not-determined |
| S3_extracted_k141_18628 | Not-determined |
| S3_extracted_k141_43359 | Not-determined |
| S3_extracted_k141_43360 | Not-determined |
| S3_extracted_k141_35694 | Not-determined |
| S3_extracted_k141_91065 | Not-determined |
| S3_extracted_k141_83016 | Not-determined |
| S3_extracted_k141_35699 | Not-determined |
| S3_extracted_k141_99048 | Not-determined |
| S3_extracted_k141_122407 | Not-determined |
| S3_extracted_k141_75867 | Not-determined |
| S3_extracted_k141_27338 | Low-quality |
| S3_extracted_k141_66591 | Not-determined |
| S3_extracted_k141_99050 | Not-determined |
| S3_extracted_k141_91070 | Not-determined |
| S3_extracted_k141_91076 | Not-determined |
| S3_extracted_k141_114965 | Not-determined |
| S3_extracted_k141_114967 | Not-determined |
| S3_extracted_k141_12572 | Not-determined |
| S3_extracted_k141_83028 | Not-determined |
| S3_extracted_k141_18648 | Not-determined |
| S3_extracted_k141_12578 | Not-determined |
| S3_extracted_k141_12579 | Not-determined |
| S3_extracted_k141_83040 | Not-determined |
| S3_extracted_k141_122435 | Not-determined |
| S3_extracted_k141_107193 | Low-quality |
| S3_extracted_k141_18655 | Not-determined |
| S3_extracted_k141_114985 | Not-determined |
| S3_extracted_k141_91086 | Not-determined |
| S3_extracted_k141_83047 | Not-determined |
| S3_extracted_k141_122438 | Not-determined |
| S3_extracted_k141_91090 | Not-determined |
| S3_extracted_k141_75885 | Not-determined |
| S3_extracted_k141_91098 | Not-determined |
| S3_extracted_k141_35704 | Not-determined |
| S3_extracted_k141_122459 | Not-determined |
| S3_extracted_k141_3255 | Low-quality |
| S3_extracted_k141_59547 | Not-determined |
| S3_extracted_k141_3256 | Not-determined |
| S3_extracted_k141_3258 | Not-determined |
| S3_extracted_k141_12593 | Low-quality |
| S3_extracted_k141_114991 | Low-quality |
| S3_extracted_k141_114994 | Not-determined |
| S3_extracted_k141_107200 | Low-quality |
| S3_extracted_k141_122477 | Not-determined |
| S3_extracted_k141_83061 | Not-determined |
| S3_extracted_k141_83063 | Low-quality |
| S3_extracted_k141_27348 | Not-determined |
| S3_extracted_k141_122481 | Low-quality |
| S3_extracted_k141_114996 | Not-determined |
| S3_extracted_k141_12598 | Not-determined |
| S3_extracted_k141_83068 | Not-determined |
| S3_extracted_k141_12601 | Not-determined |
| S3_extracted_k141_75893 | Not-determined |
| S3_extracted_k141_75894 | Not-determined |
| S3_extracted_k141_83071 | Not-determined |
| S3_extracted_k141_115003 | Not-determined |
| S3_extracted_k141_12609 | Not-determined |
| S3_extracted_k141_27350 | Not-determined |
| S3_extracted_k141_66602 | Not-determined |
| S3_extracted_k141_75903 | Not-determined |
| S3_extracted_k141_75905 | Not-determined |
| S3_extracted_k141_51702 | Not-determined |
| S3_extracted_k141_91108 | Not-determined |
| S3_extracted_k141_122504 | Not-determined |
| S3_extracted_k141_12628 | Not-determined |
| S3_extracted_k141_107211 | Not-determined |
| S3_extracted_k141_122510 | Not-determined |
| S3_extracted_k141_12634 | Not-determined |
| S3_extracted_k141_3286 | Low-quality |
| S17_vs_k141_177203\|\|full | High-quality |
| S3_extracted_k141_12641 | Not-determined |
| S3_extracted_k141_3288 | Not-determined |
| S3_extracted_k141_12643 | Not-determined |
| S3_extracted_k141_83077 | Not-determined |
| S3_extracted_k141_122516 | Not-determined |
| S3_extracted_k141_43377 | Not-determined |
| S3_extracted_k141_122523 | Low-quality |
| S3_extracted_k141_107229 | Not-determined |
| S3_extracted_k141_27378 | Not-determined |
| S3_extracted_k141_99055 | Not-determined |
| S3_extracted_k141_107231 | Low-quality |
| S3_extracted_k141_27384 | Not-determined |
| S3_extracted_k141_43383 | Not-determined |
| S3_extracted_k141_83095 | Not-determined |
| S3_extracted_k141_27389 | Not-determined |
| S3_extracted_k141_122526 | Not-determined |
| S3_extracted_k141_43389 | Not-determined |
| S3_extracted_k141_99064 | Not-determined |
| S3_extracted_k141_43393 | Not-determined |
| S3_extracted_k141_35723 | Not-determined |
| S3_extracted_k141_122531 | Not-determined |
| S3_extracted_k141_3306 | Low-quality |
| S3_extracted_k141_99071 | Low-quality |
| S3_extracted_k141_51711 | Not-determined |
| S3_extracted_k141_83102 | Low-quality |
| S3_extracted_k141_99079 | Not-determined |
| S3_extracted_k141_107235 | Not-determined |
| S3_extracted_k141_83113 | Not-determined |
| S3_extracted_k141_66667 | Not-determined |
| S3_extracted_k141_18683 | Not-determined |
| S3_extracted_k141_99086 | Not-determined |
| S3_extracted_k141_107246 | Low-quality |
| S3_extracted_k141_99088 | Not-determined |
| S3_extracted_k141_3320 | Low-quality |
| S3_extracted_k141_18685 | Not-determined |
| S3_extracted_k141_107252 | Not-determined |
| S3_extracted_k141_43408 | Not-determined |
| S3_extracted_k141_27402 | Not-determined |
| S3_extracted_k141_107258 | Not-determined |
| S3_extracted_k141_18699 | Not-determined |
| S3_extracted_k141_43415 | Low-quality |
| S3_extracted_k141_35738 | Not-determined |
| S3_extracted_k141_99093 | Not-determined |
| S3_extracted_k141_35741 | Not-determined |
| S3_extracted_k141_51723 | Not-determined |
| S3_extracted_k141_35743 | Not-determined |
| S3_extracted_k141_18702 | Not-determined |
| S3_extracted_k141_43422 | Not-determined |
| S3_extracted_k141_27413 | Not-determined |
| S3_extracted_k141_115024 | Not-determined |
| S3_extracted_k141_83128 | Not-determined |
| S3_extracted_k141_99097 | Not-determined |
| S3_extracted_k141_27418 | Not-determined |
| S3_extracted_k141_27420 | Not-determined |
| S3_extracted_k141_27421 | Not-determined |
| S3_extracted_k141_83137 | Low-quality |
| S3_extracted_k141_12699 | Not-determined |
| S3_extracted_k141_91121 | Not-determined |
| S3_extracted_k141_83140 | Low-quality |
| S3_extracted_k141_107261 | Not-determined |
| S3_extracted_k141_12705 | Not-determined |
| S3_extracted_k141_51742 | Not-determined |
| S3_extracted_k141_107264 | Not-determined |
| S3_extracted_k141_115029 | Low-quality |
| S3_extracted_k141_122562 | Low-quality |
| S3_extracted_k141_66681 | Not-determined |
| S3_extracted_k141_27433 | Not-determined |
| S3_extracted_k141_107275 | Not-determined |
| S3_extracted_k141_43442 | Not-determined |
| S3_extracted_k141_43443 | Not-determined |
| S3_extracted_k141_12716 | Not-determined |
| S3_extracted_k141_59592 | Not-determined |
| S3_extracted_k141_12720 | Not-determined |
| S3_extracted_k141_122572 | Not-determined |
| S3_extracted_k141_115039 | Not-determined |
| S3_extracted_k141_122573 | Low-quality |
| S3_extracted_k141_66696 | Not-determined |
| S3_extracted_k141_27444 | Not-determined |
| S3_extracted_k141_59597 | Not-determined |
| S3_extracted_k141_66698 | Not-determined |
| S3_extracted_k141_66699 | Not-determined |
| S3_extracted_k141_43451 | Not-determined |
| S3_extracted_k141_51756 | Low-quality |
| S3_extracted_k141_99128 | Not-determined |
| S3_extracted_k141_99129 | Not-determined |
| S3_extracted_k141_59607 | Not-determined |
| S3_extracted_k141_27450 | Not-determined |
| S3_extracted_k141_51759 | Not-determined |
| S3_extracted_k141_35764 | Not-determined |
| S3_extracted_k141_59611 | Low-quality |
| S3_extracted_k141_35768 | Not-determined |
| S3_extracted_k141_99133 | Not-determined |
| S3_extracted_k141_51762 | Not-determined |
| S3_extracted_k141_83154 | Not-determined |
| S3_extracted_k141_51764 | Not-determined |
| S3_extracted_k141_51765 | Not-determined |
| S3_extracted_k141_51766 | Not-determined |
| S3_extracted_k141_115042 | Low-quality |
| S3_extracted_k141_115043 | Not-determined |
| S3_extracted_k141_83160 | Low-quality |
| S3_extracted_k141_35777 | Not-determined |
| S3_extracted_k141_115045 | Low-quality |
| S3_extracted_k141_115046 | Not-determined |
| S3_extracted_k141_43464 | Not-determined |
| S3_extracted_k141_59619 | Not-determined |
| S3_extracted_k141_115050 | Not-determined |
| S3_extracted_k141_115051 | Not-determined |
| S3_extracted_k141_107288 | Low-quality |
| S3_extracted_k141_27465 | Not-determined |
| S3_extracted_k141_115052 | Not-determined |
| S3_extracted_k141_51780 | Not-determined |
| S3_extracted_k141_51781 | Not-determined |
| S3_extracted_k141_66715 | Not-determined |
| S3_extracted_k141_91142 | Not-determined |
| S3_extracted_k141_27469 | Not-determined |
| S3_extracted_k141_51786 | Not-determined |
| S3_extracted_k141_91149 | Not-determined |
| S3_extracted_k141_122583 | Not-determined |
| S3_extracted_k141_18723 | Not-determined |
| S3_extracted_k141_75931 | Not-determined |
| S3_extracted_k141_75935 | Not-determined |
| S3_extracted_k141_91160 | Not-determined |
| S3_extracted_k141_91161 | Not-determined |
| S3_extracted_k141_35782 | Not-determined |
| S3_extracted_k141_107299 | Not-determined |
| S3_extracted_k141_91164 | Not-determined |
| S3_extracted_k141_91166 | Not-determined |
| S3_extracted_k141_3348 | Low-quality |
| S3_extracted_k141_18747 | Not-determined |
| S3_extracted_k141_83202 | Low-quality |
| S3_extracted_k141_107308 | Not-determined |
| S3_extracted_k141_3355 | Not-determined |
| S3_extracted_k141_27480 | Not-determined |
| S3_extracted_k141_115067 | Low-quality |
| S3_extracted_k141_51806 | Low-quality |
| S3_extracted_k141_59636 | Not-determined |
| S3_extracted_k141_107314 | Not-determined |
| S3_extracted_k141_59639 | Not-determined |
| S3_extracted_k141_27485 | Not-determined |
| S3_extracted_k141_51812 | Not-determined |
| S3_extracted_k141_59641 | Not-determined |
| S3_extracted_k141_115075 | Not-determined |
| S3_extracted_k141_91182 | Not-determined |
| S3_extracted_k141_115076 | Not-determined |
| S3_extracted_k141_91183 | Not-determined |
| S3_extracted_k141_43490 | Not-determined |
| S3_extracted_k141_3366 | Not-determined |
| S3_extracted_k141_75947 | Not-determined |
| S3_extracted_k141_107320 | Not-determined |
| S3_extracted_k141_91185 | Not-determined |
| S3_extracted_k141_3370 | Low-quality |
| S3_extracted_k141_91186 | Not-determined |
| S3_extracted_k141_51814 | Low-quality |
| S3_extracted_k141_83242 | Not-determined |
| S3_extracted_k141_91188 | Low-quality |
| S3_extracted_k141_51822 | Low-quality |
| S3_extracted_k141_83256 | Not-determined |
| S3_extracted_k141_91195 | Not-determined |
| S3_extracted_k141_59662 | Not-determined |
| S3_extracted_k141_83262 | Not-determined |
| S3_extracted_k141_107327 | Not-determined |
| S3_extracted_k141_27493 | Not-determined |
| S3_extracted_k141_59670 | Low-quality |
| S3_extracted_k141_91201 | Not-determined |
| S3_extracted_k141_107334 | Not-determined |
| S3_extracted_k141_59672 | Not-determined |
| S3_extracted_k141_43498 | Not-determined |
| S3_extracted_k141_59674 | Not-determined |
| S3_extracted_k141_122596 | Not-determined |
| S3_extracted_k141_91210 | Not-determined |
| S3_extracted_k141_12736 | Not-determined |
| S3_extracted_k141_91215 | Not-determined |
| S3_extracted_k141_43501 | Not-determined |
| S3_extracted_k141_43502 | Not-determined |
| S3_extracted_k141_122606 | Not-determined |
| S3_extracted_k141_83277 | Not-determined |
| S3_extracted_k141_12739 | Not-determined |
| S3_extracted_k141_59691 | Not-determined |
| S3_extracted_k141_107360 | Not-determined |
| S3_extracted_k141_107362 | Not-determined |
| S3_extracted_k141_99156 | Not-determined |
| S3_extracted_k141_51833 | Not-determined |
| S3_extracted_k141_59702 | Not-determined |
| S3_extracted_k141_12744 | Not-determined |
| S3_extracted_k141_43519 | Not-determined |
| S3_extracted_k141_75960 | Low-quality |
| S3_extracted_k141_122612 | Not-determined |
| S3_extracted_k141_122613 | Not-determined |
| S3_extracted_k141_43521 | Not-determined |
| S3_extracted_k141_83294 | Not-determined |
| S3_extracted_k141_66723 | Not-determined |
| S3_extracted_k141_99163 | Not-determined |
| S3_extracted_k141_115095 | Low-quality |
| S3_extracted_k141_27511 | Not-determined |
| S3_extracted_k141_99164 | Not-determined |
| S3_extracted_k141_83296 | Not-determined |
| S3_extracted_k141_43529 | Not-determined |
| S3_extracted_k141_66725 | Not-determined |
| S3_extracted_k141_43531 | Not-determined |
| S3_extracted_k141_75963 | Not-determined |
| S3_extracted_k141_91231 | Low-quality |
| S3_extracted_k141_122622 | Not-determined |
| S3_extracted_k141_66731 | Not-determined |
| S3_extracted_k141_43538 | Not-determined |
| S3_extracted_k141_27517 | Not-determined |
| S3_extracted_k141_51850 | Not-determined |
| S3_extracted_k141_99167 | Not-determined |
| S3_extracted_k141_91234 | Low-quality |
| S3_extracted_k141_99168 | Not-determined |
| S3_extracted_k141_83312 | Low-quality |
| S3_extracted_k141_59729 | Not-determined |
| S3_extracted_k141_59730 | Not-determined |
| S3_extracted_k141_3390 | Not-determined |
| S3_extracted_k141_91242 | Low-quality |
| S3_extracted_k141_91243 | Not-determined |
| S3_extracted_k141_115105 | Not-determined |
| S3_extracted_k141_66750 | Low-quality |
| S3_extracted_k141_12762 | Not-determined |
| S3_extracted_k141_43543 | Not-determined |
| S3_extracted_k141_12765 | Not-determined |
| S3_extracted_k141_115112 | Not-determined |
| S3_extracted_k141_18761 | Not-determined |
| S3_extracted_k141_43547 | Not-determined |
| S3_extracted_k141_122643 | Not-determined |
| S3_extracted_k141_12773 | Not-determined |
| S3_extracted_k141_43549 | Not-determined |
| S3_extracted_k141_59745 | Low-quality |
| S3_extracted_k141_12777 | Not-determined |
| S3_extracted_k141_122649 | Not-determined |
| S3_extracted_k141_122651 | Not-determined |
| S3_extracted_k141_43554 | Not-determined |
| S3_extracted_k141_115115 | Not-determined |
| S3_extracted_k141_27538 | Not-determined |
| S3_extracted_k141_51876 | Not-determined |
| S3_extracted_k141_12779 | Not-determined |
| S3_extracted_k141_91270 | Not-determined |
| S3_extracted_k141_99173 | Not-determined |
| S3_extracted_k141_91272 | Not-determined |
| S3_extracted_k141_91274 | Not-determined |
| S3_extracted_k141_59752 | Not-determined |
| S3_extracted_k141_3410 | Not-determined |
| S3_extracted_k141_3411 | Not-determined |
| S3_extracted_k141_122662 | Not-determined |
| S3_extracted_k141_115125 | Not-determined |
| S3_extracted_k141_115126 | Not-determined |
| S3_extracted_k141_35798 | Low-quality |
| S3_extracted_k141_3416 | Not-determined |
| S3_extracted_k141_27542 | Not-determined |
| S3_extracted_k141_35803 | Not-determined |
| S3_extracted_k141_59766 | Low-quality |
| S3_extracted_k141_59769 | Not-determined |
| S3_extracted_k141_66761 | Low-quality |
| S3_extracted_k141_91288 | Not-determined |
| S3_extracted_k141_43571 | Not-determined |
| S3_extracted_k141_91295 | Low-quality |
| S3_extracted_k141_91297 | Not-determined |
| S3_extracted_k141_12786 | Not-determined |
| S3_extracted_k141_35815 | Not-determined |
| S3_extracted_k141_66776 | Not-determined |
| S3_extracted_k141_51883 | Not-determined |
| S3_extracted_k141_122673 | Not-determined |
| S3_extracted_k141_83333 | Not-determined |
| S3_extracted_k141_83335 | Not-determined |
| S3_extracted_k141_59795 | Not-determined |
| S3_extracted_k141_59801 | Not-determined |
| S3_extracted_k141_3422 | Not-determined |
| S3_extracted_k141_83341 | Not-determined |
| S3_extracted_k141_59805 | Low-quality |
| S3_extracted_k141_83343 | Not-determined |
| S3_extracted_k141_99179 | Low-quality |
| S3_extracted_k141_66786 | Not-determined |
| S3_extracted_k141_83348 | Not-determined |
| S3_extracted_k141_99180 | Not-determined |
| S3_extracted_k141_83350 | Not-determined |
| S3_extracted_k141_122682 | Low-quality |
| S3_extracted_k141_83355 | Not-determined |
| S3_extracted_k141_91315 | Not-determined |
| S3_extracted_k141_122694 | Not-determined |
| S3_extracted_k141_122696 | Not-determined |
| S3_extracted_k141_91327 | Low-quality |
| S3_extracted_k141_12807 | Low-quality |
| S3_extracted_k141_122702 | Not-determined |
| S3_extracted_k141_91335 | Not-determined |
| S3_extracted_k141_27585 | Low-quality |
| S3_extracted_k141_59818 | Not-determined |
| S3_extracted_k141_35825 | Not-determined |
| S3_extracted_k141_115138 | Not-determined |
| S3_extracted_k141_12812 | Not-determined |
| S3_extracted_k141_51905 | Not-determined |
| S3_extracted_k141_51907 | Not-determined |
| S3_extracted_k141_115140 | Not-determined |
| S3_extracted_k141_76007 | Not-determined |
| S3_extracted_k141_91347 | Not-determined |
| S3_extracted_k141_51913 | Not-determined |
| S3_extracted_k141_3449 | Not-determined |
| S3_extracted_k141_115150 | Not-determined |
| S3_extracted_k141_115153 | Low-quality |
| S3_extracted_k141_99188 | Not-determined |
| S3_extracted_k141_59823 | Not-determined |
| S3_extracted_k141_3460 | Not-determined |
| S3_extracted_k141_115162 | Not-determined |
| S3_extracted_k141_59837 | Not-determined |
| S3_extracted_k141_3465 | Low-quality |
| S3_extracted_k141_83361 | Not-determined |
| S3_extracted_k141_115164 | Not-determined |
| S3_extracted_k141_91357 | Not-determined |
| S3_extracted_k141_35828 | Low-quality |
| S3_extracted_k141_91358 | Not-determined |
| S3_extracted_k141_35830 | Not-determined |
| S3_extracted_k141_91360 | Low-quality |
| S3_extracted_k141_122711 | Low-quality |
| S3_extracted_k141_51942 | Not-determined |
| S3_extracted_k141_51943 | Not-determined |
| S3_extracted_k141_59854 | Not-determined |
| S3_extracted_k141_66788 | Not-determined |
| S3_extracted_k141_3474 | Not-determined |
| S3_extracted_k141_51948 | Not-determined |
| S3_extracted_k141_3475 | Not-determined |
| S3_extracted_k141_59861 | Not-determined |
| S3_extracted_k141_3476 | Not-determined |
| S3_extracted_k141_3477 | Not-determined |
| S3_extracted_k141_115179 | Not-determined |
| S3_extracted_k141_83364 | Not-determined |
| S3_extracted_k141_76027 | Not-determined |
| S3_extracted_k141_115182 | Not-determined |
| S3_extracted_k141_115183 | Not-determined |
| S3_extracted_k141_3488 | Not-determined |
| S3_extracted_k141_76029 | Not-determined |
| S3_extracted_k141_115185 | Not-determined |
| S3_extracted_k141_3492 | Not-determined |
| S3_extracted_k141_51966 | Not-determined |
| S3_extracted_k141_51969 | Not-determined |
| S3_extracted_k141_76040 | Low-quality |
| S3_extracted_k141_51971 | Not-determined |
| S3_extracted_k141_76042 | Not-determined |
| S3_extracted_k141_76043 | Not-determined |
| S3_extracted_k141_3500 | Low-quality |
| S3_extracted_k141_99191 | Not-determined |
| S3_extracted_k141_76046 | Not-determined |
| S3_extracted_k141_35849 | Low-quality |
| S3_extracted_k141_35852 | Not-determined |
| S3_extracted_k141_122722 | Low-quality |
| S3_extracted_k141_3514 | Not-determined |
| S3_extracted_k141_122724 | Not-determined |
| S3_extracted_k141_91371 | Not-determined |
| S3_extracted_k141_66837 | Not-determined |
| S3_extracted_k141_115209 | Low-quality |
| S3_extracted_k141_115210 | Not-determined |
| S3_extracted_k141_66840 | Not-determined |
| S3_extracted_k141_12836 | Not-determined |
| S3_extracted_k141_66841 | Not-determined |
| S3_extracted_k141_91377 | Low-quality |
| S3_extracted_k141_66844 | Not-determined |
| S3_extracted_k141_12838 | Not-determined |
| S3_extracted_k141_66848 | Not-determined |
| S3_extracted_k141_83392 | Not-determined |
| S3_extracted_k141_99195 | Not-determined |
| S3_extracted_k141_115214 | Not-determined |
| S3_extracted_k141_66858 | Not-determined |
| S3_extracted_k141_76053 | Not-determined |
| S3_extracted_k141_91394 | Not-determined |
| S3_extracted_k141_91396 | Not-determined |
| S3_extracted_k141_76055 | Low-quality |
| S3_extracted_k141_76057 | Not-determined |
| S3_extracted_k141_115225 | Not-determined |
| S3_extracted_k141_18765 | Not-determined |
| S3_extracted_k141_122731 | Low-quality |
| S3_extracted_k141_76061 | Not-determined |
| S3_extracted_k141_115228 | Not-determined |
| S3_extracted_k141_122738 | Low-quality |
| S3_extracted_k141_18766 | Not-determined |
| S3_extracted_k141_122741 | Not-determined |
| S3_extracted_k141_52023 | Low-quality |
| S3_extracted_k141_76073 | Not-determined |
| S3_extracted_k141_12846 | Low-quality |
| S3_extracted_k141_12847 | Not-determined |
| S3_extracted_k141_99206 | Low-quality |
| S3_extracted_k141_76079 | Not-determined |
| S3_extracted_k141_3519 | Low-quality |
| S3_extracted_k141_18771 | Low-quality |
| S3_extracted_k141_52037 | Not-determined |
| S3_extracted_k141_3525 | Not-determined |
| S3_extracted_k141_52042 | Not-determined |
| S3_extracted_k141_115232 | Not-determined |
| S3_extracted_k141_18774 | Not-determined |
| S3_extracted_k141_52046 | Not-determined |
| S3_extracted_k141_122748 | Not-determined |
| S3_extracted_k141_52049 | Low-quality |
| S3_extracted_k141_18782 | Not-determined |
| S3_extracted_k141_83417 | Not-determined |
| S3_extracted_k141_18784 | Not-determined |
| S3_extracted_k141_3535 | Not-determined |
| S3_extracted_k141_43611 | Not-determined |
| S3_extracted_k141_66875 | Not-determined |
| S3_extracted_k141_122750 | Not-determined |
| S3_extracted_k141_27596 | Not-determined |
| S3_extracted_k141_52051 | Not-determined |
| S3_extracted_k141_27599 | Not-determined |
| S3_extracted_k141_66876 | Low-quality |
| S3_extracted_k141_3546 | Not-determined |
| S3_extracted_k141_35875 | Low-quality |
| S3_extracted_k141_3548 | Not-determined |
| S3_extracted_k141_83428 | Not-determined |
| S3_extracted_k141_35876 | Not-determined |
| S3_extracted_k141_122760 | Low-quality |
| S3_extracted_k141_27605 | Not-determined |
| S3_extracted_k141_3555 | Not-determined |
| S3_extracted_k141_18798 | Not-determined |
| S3_extracted_k141_35885 | Not-determined |
| S3_extracted_k141_43624 | Not-determined |
| S3_extracted_k141_91425 | Not-determined |
| S3_extracted_k141_18800 | Not-determined |
| S3_extracted_k141_52059 | Not-determined |
| S3_extracted_k141_91432 | Not-determined |
| S3_extracted_k141_12866 | Not-determined |
| S3_extracted_k141_27614 | Not-determined |
| S3_extracted_k141_43634 | Not-determined |
| S3_extracted_k141_52060 | Not-determined |
| S3_extracted_k141_91440 | Not-determined |
| S3_extracted_k141_18812 | Not-determined |
| S3_extracted_k141_18813 | Not-determined |
| S3_extracted_k141_83443 | Not-determined |
| S3_extracted_k141_91442 | Not-determined |
| S3_extracted_k141_52067 | Not-determined |
| S3_extracted_k141_83446 | Not-determined |
| S3_extracted_k141_115244 | Not-determined |
| S3_extracted_k141_83450 | Not-determined |
| S3_extracted_k141_52070 | Low-quality |
| S3_extracted_k141_52071 | Not-determined |
| S3_extracted_k141_52074 | Not-determined |
| S3_extracted_k141_115253 | Low-quality |
| S3_extracted_k141_3563 | Low-quality |
| S3_extracted_k141_35894 | Not-determined |
| S3_extracted_k141_52075 | Low-quality |
| S3_extracted_k141_35899 | Not-determined |
| S3_extracted_k141_122780 | Not-determined |
| S3_extracted_k141_3570 | Not-determined |
| S3_extracted_k141_115267 | Not-determined |
| S3_extracted_k141_43643 | Not-determined |
| S3_extracted_k141_43645 | Not-determined |
| S3_extracted_k141_122790 | Not-determined |
| S3_extracted_k141_115278 | Not-determined |
| S3_extracted_k141_59876 | Not-determined |
| S3_extracted_k141_83456 | Not-determined |
| S3_extracted_k141_83460 | Not-determined |
| S3_extracted_k141_52088 | Low-quality |
| S3_extracted_k141_83462 | Low-quality |
| S3_extracted_k141_52089 | Not-determined |
| S3_extracted_k141_12874 | Not-determined |
| S3_extracted_k141_76108 | Low-quality |
| S3_extracted_k141_76109 | Low-quality |
| S3_extracted_k141_52095 | Not-determined |
| S3_extracted_k141_59887 | Not-determined |

| | |
|---|---|
| S3_extracted_k141_59891 | Not-determined |
| S3_extracted_k141_59892 | Not-determined |
| S3_extracted_k141_83473 | Not-determined |
| S3_extracted_k141_43654 | Low-quality |
| S3_extracted_k141_59894 | Not-determined |
| S3_extracted_k141_43658 | Not-determined |
| S3_extracted_k141_66891 | Not-determined |
| S3_extracted_k141_52102 | Not-determined |
| S3_extracted_k141_43659 | Not-determined |
| S3_extracted_k141_76120 | Not-determined |
| S3_extracted_k141_83481 | Not-determined |
| S3_extracted_k141_76122 | Not-determined |
| S3_extracted_k141_66898 | Not-determined |
| S3_extracted_k141_43660 | Low-quality |
| S3_extracted_k141_3579 | Not-determined |
| S3_extracted_k141_83485 | Low-quality |
| S3_extracted_k141_59904 | Not-determined |
| S3_extracted_k141_35922 | Not-determined |
| S3_extracted_k141_43663 | Not-determined |
| S3_extracted_k141_83489 | Not-determined |
| S3_extracted_k141_52109 | Not-determined |
| S3_extracted_k141_43666 | Not-determined |
| S3_extracted_k141_12893 | Not-determined |
| S3_extracted_k141_27636 | Not-determined |
| S3_extracted_k141_3590 | Not-determined |
| S3_extracted_k141_91455 | Not-determined |
| S3_extracted_k141_83497 | Not-determined |
| S3_extracted_k141_59909 | Not-determined |
| S3_extracted_k141_35935 | Not-determined |
| S3_extracted_k141_35937 | Not-determined |
| S3_extracted_k141_3599 | Low-quality |
| S3_extracted_k141_59911 | Low-quality |
| S3_extracted_k141_52114 | Not-determined |
| S3_extracted_k141_12903 | Not-determined |
| S3_extracted_k141_27642 | Not-determined |
| S3_extracted_k141_91466 | Not-determined |
| S3_extracted_k141_122794 | Not-determined |
| S3_extracted_k141_35950 | Low-quality |
| S3_extracted_k141_122795 | Not-determined |
| S3_extracted_k141_35951 | Low-quality |
| S3_extracted_k141_12912 | Low-quality |
| S3_extracted_k141_107394 | Not-determined |
| S3_extracted_k141_3612 | Not-determined |
| S3_extracted_k141_59922 | Low-quality |
| S3_extracted_k141_83505 | Not-determined |
| S3_extracted_k141_35953 | Not-determined |
| S3_extracted_k141_12916 | Not-determined |
| S3_extracted_k141_91472 | Not-determined |
| S3_extracted_k141_91473 | Low-quality |
| S3_extracted_k141_91474 | Not-determined |
| S3_extracted_k141_43681 | Not-determined |
| S3_extracted_k141_52123 | Low-quality |
| S3_extracted_k141_76124 | Not-determined |
| S3_extracted_k141_115293 | Not-determined |
| S3_extracted_k141_83514 | Low-quality |
| S3_extracted_k141_43684 | Not-determined |
| S3_extracted_k141_107404 | Not-determined |
| S3_extracted_k141_12932 | Not-determined |
| S3_extracted_k141_91480 | Low-quality |
| S3_extracted_k141_52133 | Not-determined |
| S3_extracted_k141_12933 | Low-quality |
| S3_extracted_k141_66921 | Not-determined |
| S3_extracted_k141_76133 | Not-determined |
| S3_extracted_k141_43696 | Low-quality |
| S3_extracted_k141_66926 | Low-quality |
| S3_extracted_k141_66929 | Not-determined |
| S3_extracted_k141_91487 | Not-determined |
| S3_extracted_k141_91488 | Not-determined |
| S3_extracted_k141_76142 | Not-determined |
| S3_extracted_k141_35980 | Not-determined |
| S3_extracted_k141_52137 | Low-quality |
| S3_extracted_k141_52138 | Not-determined |
| S3_extracted_k141_52139 | Not-determined |
| S3_extracted_k141_35982 | Low-quality |
| S3_extracted_k141_83519 | Low-quality |
| S3_extracted_k141_35991 | Not-determined |
| S3_extracted_k141_76146 | Not-determined |
| S3_extracted_k141_36005 | Not-determined |
| S3_extracted_k141_36007 | Not-determined |
| S3_extracted_k141_76154 | Not-determined |
| S3_extracted_k141_76155 | Low-quality |
| S3_extracted_k141_36008 | Not-determined |
| S3_extracted_k141_36010 | Not-determined |
| S3_extracted_k141_12942 | Not-determined |
| S3_extracted_k141_36013 | Low-quality |
| S3_extracted_k141_12945 | Not-determined |
| S3_extracted_k141_43706 | Not-determined |
| S3_extracted_k141_12951 | Not-determined |
| S3_extracted_k141_36020 | Not-determined |
| S3_extracted_k141_91490 | Not-determined |
| S3_extracted_k141_52180 | Not-determined |
| S3_extracted_k141_12959 | Not-determined |
| S3_extracted_k141_43715 | Not-determined |
| S3_extracted_k141_91495 | Not-determined |
| S3_extracted_k141_43716 | Low-quality |
| S3_extracted_k141_52183 | Not-determined |
| S3_extracted_k141_12965 | Not-determined |
| S7_vs_24757\|\|full | High-quality |
| S3_extracted_k141_83538 | Not-determined |
| S3_extracted_k141_3616 | Low-quality |
| S3_extracted_k141_43719 | Low-quality |
| S16_vs_141218\|\|full | Medium-quality |
| S3_extracted_k141_36028 | Not-determined |
| S3_extracted_k141_36034 | Not-determined |
| S3_extracted_k141_76170 | Low-quality |
| S3_extracted_k141_83552 | Low-quality |
| S3_extracted_k141_52185 | Not-determined |
| S3_extracted_k141_43722 | Not-determined |
| S3_extracted_k141_43726 | Not-determined |
| S3_extracted_k141_36063 | Not-determined |
| S3_extracted_k141_27681 | Low-quality |
| S3_extracted_k141_27685 | Not-determined |
| S3_extracted_k141_76175 | Not-determined |
| S3_extracted_k141_91525 | Not-determined |
| S3_extracted_k141_52200 | Not-determined |
| S3_extracted_k141_91531 | Not-determined |
| S3_extracted_k141_83555 | Not-determined |
| S3_extracted_k141_59972 | Not-determined |
| S3_extracted_k141_52205 | Low-quality |
| S3_extracted_k141_52206 | Low-quality |
| S3_extracted_k141_76184 | Low-quality |
| S3_extracted_k141_36068 | Not-determined |
| S3_extracted_k141_36071 | Not-determined |
| S3_extracted_k141_91538 | Not-determined |
| S3_extracted_k141_91542 | Not-determined |
| S3_extracted_k141_59983 | Low-quality |
| S3_extracted_k141_91546 | Not-determined |
| S3_extracted_k141_36079 | Not-determined |
| S3_extracted_k141_36080 | Not-determined |
| S3_extracted_k141_27707 | Not-determined |
| S3_extracted_k141_91550 | Not-determined |
| S3_extracted_k141_3635 | Not-determined |
| S3_extracted_k141_27713 | Low-quality |
| S3_extracted_k141_3643 | Not-determined |
| S3_extracted_k141_36092 | Not-determined |
| S3_extracted_k141_107446 | Low-quality |
| S3_extracted_k141_83560 | Not-determined |
| S3_extracted_k141_91556 | Not-determined |
| S3_extracted_k141_83563 | Not-determined |
| S3_extracted_k141_43737 | Not-determined |
| S3_extracted_k141_107452 | Not-determined |
| S3_extracted_k141_52228 | Not-determined |
| S3_extracted_k141_59997 | Not-determined |
| S3_extracted_k141_107460 | Not-determined |
| S3_extracted_k141_60002 | Not-determined |
| S3_extracted_k141_107465 | Not-determined |
| S3_extracted_k141_91567 | Not-determined |
| S3_extracted_k141_3662 | Not-determined |
| S3_extracted_k141_83573 | Not-determined |
| S3_extracted_k141_43748 | Not-determined |
| S3_extracted_k141_83574 | Not-determined |
| S3_extracted_k141_66956 | Low-quality |
| S3_extracted_k141_27725 | Not-determined |
| S3_extracted_k141_66959 | Not-determined |
| S3_extracted_k141_91573 | Not-determined |
| S3_extracted_k141_3664 | Low-quality |
| S3_extracted_k141_52239 | Not-determined |
| S3_extracted_k141_91576 | Not-determined |
| S3_extracted_k141_91579 | Not-determined |
| S3_extracted_k141_83583 | Not-determined |
| S3_extracted_k141_91582 | Not-determined |
| S3_extracted_k141_66964 | Not-determined |
| S3_extracted_k141_83589 | Not-determined |
| S3_extracted_k141_18842 | Low-quality |
| S3_extracted_k141_27745 | Not-determined |
| S3_extracted_k141_27746 | Not-determined |
| S3_extracted_k141_8058 | High-quality |
| S3_extracted_k141_18849 | Low-quality |
| S3_extracted_k141_27750 | Not-determined |
| S3_extracted_k141_107475 | Not-determined |
| S3_extracted_k141_107477 | Not-determined |
| S3_extracted_k141_122836 | Not-determined |
| S3_extracted_k141_91592 | Not-determined |
| S3_extracted_k141_36093 | Not-determined |
| S3_extracted_k141_91594 | Not-determined |
| S3_extracted_k141_36097 | Not-determined |
| S3_extracted_k141_66976 | Not-determined |
| S3_extracted_k141_27756 | Not-determined |
| S3_extracted_k141_107482 | Not-determined |
| S3_extracted_k141_60036 | Not-determined |
| S3_extracted_k141_36101 | Not-determined |
| S3_extracted_k141_36104 | Not-determined |
| S3_extracted_k141_36105 | Not-determined |
| S3_extracted_k141_60045 | Not-determined |
| S3_extracted_k141_3681 | Not-determined |
| S3_extracted_k141_27765 | Not-determined |
| S3_extracted_k141_3682 | Not-determined |
| S3_extracted_k141_60053 | Not-determined |
| S3_extracted_k141_122856 | Not-determined |
| S3_extracted_k141_52260 | Not-determined |
| S3_extracted_k141_83603 | Not-determined |
| S3_extracted_k141_83604 | Not-determined |
| S3_extracted_k141_27772 | Low-quality |
| S3_extracted_k141_83607 | Not-determined |
| S3_extracted_k141_107504 | Not-determined |
| S3_extracted_k141_60063 | Not-determined |
| S3_extracted_k141_115329 | Not-determined |
| S3_extracted_k141_27776 | Not-determined |
| S3_extracted_k141_83611 | Not-determined |
| S3_extracted_k141_3692 | Not-determined |
| S3_extracted_k141_83613 | Low-quality |
| S3_extracted_k141_83615 | Not-determined |
| S3_extracted_k141_83616 | Not-determined |
| S3_extracted_k141_91604 | Not-determined |
| S3_extracted_k141_83622 | Not-determined |
| S3_extracted_k141_91609 | Low-quality |
| S3_extracted_k141_27785 | Not-determined |
| S3_extracted_k141_60071 | Not-determined |
| S3_extracted_k141_27790 | Not-determined |
| S3_extracted_k141_107512 | Not-determined |
| S3_extracted_k141_115343 | Not-determined |
| S3_extracted_k141_52279 | Not-determined |
| S3_extracted_k141_83625 | Not-determined |
| S3_extracted_k141_52283 | Not-determined |
| S3_extracted_k141_91613 | Low-quality |
| S3_extracted_k141_60075 | Not-determined |
| S3_extracted_k141_115346 | Low-quality |
| S3_extracted_k141_83634 | Not-determined |
| S3_extracted_k141_52286 | Not-determined |
| S3_extracted_k141_60083 | Not-determined |
| S3_extracted_k141_18860 | Not-determined |
| S3_extracted_k141_36120 | Not-determined |
| S3_extracted_k141_60085 | Not-determined |
| S3_extracted_k141_60086 | Not-determined |
| S3_extracted_k141_60087 | Not-determined |
| S3_extracted_k141_36125 | Not-determined |
| S3_extracted_k141_52298 | Not-determined |
| S3_extracted_k141_60089 | Low-quality |
| S3_extracted_k141_36128 | Not-determined |
| S3_extracted_k141_83646 | Not-determined |
| S3_extracted_k141_60094 | Not-determined |
| S3_extracted_k141_76199 | Not-determined |
| S3_extracted_k141_83648 | Low-quality |
| S3_extracted_k141_36130 | Low-quality |
| S3_extracted_k141_66984 | Low-quality |
| S3_extracted_k141_36132 | Not-determined |
| S3_extracted_k141_13008 | Not-determined |
| S3_extracted_k141_36137 | Not-determined |
| S3_extracted_k141_60095 | Not-determined |
| S3_extracted_k141_36142 | Not-determined |
| S3_extracted_k141_36157 | Not-determined |
| S3_extracted_k141_60105 | Not-determined |
| S3_extracted_k141_60112 | Not-determined |
| S3_extracted_k141_18880 | Not-determined |
| S3_extracted_k141_18881 | Low-quality |
| S3_extracted_k141_18882 | Not-determined |
| S3_extracted_k141_76204 | Low-quality |
| S3_extracted_k141_107523 | Not-determined |
| S3_extracted_k141_27799 | Not-determined |
| S3_extracted_k141_52337 | Not-determined |
| S3_extracted_k141_115363 | Not-determined |
| S3_extracted_k141_60119 | Not-determined |
| S3_extracted_k141_115365 | Not-determined |
| S3_extracted_k141_52344 | Not-determined |
| S3_extracted_k141_60121 | Not-determined |
| S3_extracted_k141_76218 | Not-determined |
| S3_extracted_k141_13024 | Not-determined |
| S3_extracted_k141_27810 | Not-determined |
| S3_extracted_k141_107535 | Low-quality |
| S3_extracted_k141_27813 | Not-determined |
| S3_extracted_k141_60132 | Not-determined |
| S3_extracted_k141_60133 | Not-determined |
| S3_extracted_k141_107537 | Not-determined |
| S3_extracted_k141_83699 | Not-determined |
| S3_extracted_k141_13027 | Not-determined |
| S3_extracted_k141_60136 | Low-quality |
| S3_extracted_k141_52356 | Not-determined |
| S3_extracted_k141_83703 | Low-quality |
| S3_extracted_k141_83705 | Low-quality |
| S3_extracted_k141_13035 | Not-determined |
| S3_extracted_k141_13036 | Not-determined |
| S3_extracted_k141_55571 | High-quality |
| S3_extracted_k141_99245 | Not-determined |
| S3_extracted_k141_76225 | Not-determined |
| S3_extracted_k141_52366 | Not-determined |
| S3_extracted_k141_107553 | Not-determined |
| S3_extracted_k141_107556 | Not-determined |
| S3_extracted_k141_83720 | Low-quality |
| S3_extracted_k141_13049 | Not-determined |
| S3_extracted_k141_13051 | Not-determined |
| S3_extracted_k141_107561 | Not-determined |
| S3_extracted_k141_36169 | Not-determined |
| S3_extracted_k141_3716 | Not-determined |
| S3_extracted_k141_76238 | Not-determined |
| S3_extracted_k141_107569 | Not-determined |
| S3_extracted_k141_3718 | Not-determined |
| S3_extracted_k141_52373 | Not-determined |
| S3_extracted_k141_3719 | Not-determined |
| S3_extracted_k141_3721 | Not-determined |
| S3_extracted_k141_36178 | Not-determined |
| S3_extracted_k141_36179 | Not-determined |
| S3_extracted_k141_76243 | Not-determined |
| S3_extracted_k141_99253 | Low-quality |
| S3_extracted_k141_76248 | Not-determined |
| S3_extracted_k141_76250 | Not-determined |
| S3_extracted_k141_36185 | Not-determined |
| S3_extracted_k141_107579 | Not-determined |
| S3_extracted_k141_91630 | Not-determined |
| S3_extracted_k141_107581 | Not-determined |
| S3_extracted_k141_83730 | Not-determined |
| S3_extracted_k141_91631 | Not-determined |
| S3_extracted_k141_115410 | Not-determined |
| S3_extracted_k141_27831 | Not-determined |
| S3_extracted_k141_52384 | Not-determined |
| S3_extracted_k141_107583 | Low-quality |
| S3_extracted_k141_91637 | Not-determined |
| S3_extracted_k141_13075 | Not-determined |
| S3_extracted_k141_107584 | Not-determined |
| S3_extracted_k141_13076 | Not-determined |
| S3_extracted_k141_52386 | Not-determined |
| S3_extracted_k141_18940 | Not-determined |
| S3_extracted_k141_18941 | Not-determined |
| S3_extracted_k141_52388 | Not-determined |
| S3_extracted_k141_91642 | Not-determined |
| S3_extracted_k141_52389 | Not-determined |
| S3_extracted_k141_18951 | Not-determined |
| S3_extracted_k141_52394 | Not-determined |
| S3_extracted_k141_76271 | Low-quality |
| S3_extracted_k141_99267 | Not-determined |
| S3_extracted_k141_60147 | Not-determined |
| S3_extracted_k141_60148 | Not-determined |
| S3_extracted_k141_52403 | Not-determined |
| S3_extracted_k141_66995 | Not-determined |
| S3_extracted_k141_13103 | Not-determined |
| S3_extracted_k141_3729 | Not-determined |
| S3_extracted_k141_60158 | Low-quality |
| S3_extracted_k141_3730 | Not-determined |
| S3_extracted_k141_52409 | Not-determined |
| S3_extracted_k141_13112 | Not-determined |
| S3_extracted_k141_115429 | Not-determined |
| S3_extracted_k141_60166 | Low-quality |
| S3_extracted_k141_43764 | Not-determined |
| S3_extracted_k141_76279 | Not-determined |
| S3_extracted_k141_36200 | Not-determined |
| S3_extracted_k141_52416 | Not-determined |
| S3_extracted_k141_18955 | Not-determined |
| S3_extracted_k141_52418 | Not-determined |
| S3_extracted_k141_122862 | Low-quality |
| S3_extracted_k141_43773 | Not-determined |
| S3_extracted_k141_91670 | Not-determined |
| S3_extracted_k141_99280 | Not-determined |
| S3_extracted_k141_122867 | Not-determined |
| S3_extracted_k141_91673 | Not-determined |
| S3_extracted_k141_3747 | Not-determined |
| S3_extracted_k141_27848 | Not-determined |
| S3_extracted_k141_3755 | Not-determined |
| S3_extracted_k141_99290 | Not-determined |
| S3_extracted_k141_99292 | Not-determined |
| S3_extracted_k141_91677 | Not-determined |
| S3_extracted_k141_27859 | Not-determined |
| S3_extracted_k141_27861 | Not-determined |
| S3_extracted_k141_122882 | Not-determined |
| S3_extracted_k141_91680 | Not-determined |
| S3_extracted_k141_91681 | Not-determined |
| S3_extracted_k141_91682 | Not-determined |
| S3_extracted_k141_99299 | Not-determined |
| S3_extracted_k141_18959 | Low-quality |
| S3_extracted_k141_52438 | Low-quality |
| S3_extracted_k141_43795 | Low-quality |
| S3_extracted_k141_43796 | Low-quality |
| S3_extracted_k141_99310 | Not-determined |
| S3_extracted_k141_99312 | Not-determined |
| S3_extracted_k141_107608 | Low-quality |
| S3_extracted_k141_67014 | Not-determined |
| S3_extracted_k141_3780 | Not-determined |
| S3_extracted_k141_13151 | Low-quality |
| S3_extracted_k141_122901 | Not-determined |
| S3_extracted_k141_18975 | Not-determined |
| S3_extracted_k141_43812 | Not-determined |
| S3_extracted_k141_107621 | Low-quality |
| S3_extracted_k141_43813 | Not-determined |
| S3_extracted_k141_99326 | Not-determined |
| S3_extracted_k141_76318 | Not-determined |
| S3_extracted_k141_107627 | Not-determined |
| S3_extracted_k141_115453 | Not-determined |
| S3_extracted_k141_107628 | Not-determined |
| S3_extracted_k141_91685 | Low-quality |
| S3_extracted_k141_99331 | Not-determined |
| S3_extracted_k141_27882 | Not-determined |
| S3_extracted_k141_76328 | Not-determined |
| S3_extracted_k141_13154 | Not-determined |
| S3_extracted_k141_76332 | Not-determined |
| S3_extracted_k141_76333 | Not-determined |
| S3_extracted_k141_3792 | Not-determined |
| S3_extracted_k141_76335 | Not-determined |
| S3_extracted_k141_52453 | Not-determined |
| S3_extracted_k141_3796 | Not-determined |
| S3_extracted_k141_43828 | Not-determined |
| S3_extracted_k141_115469 | Not-determined |
| S3_extracted_k141_115470 | Low-quality |
| S3_extracted_k141_60175 | Low-quality |
| S3_extracted_k141_76343 | Not-determined |
| S3_extracted_k141_60176 | Not-determined |
| S3_extracted_k141_27897 | Not-determined |
| S3_extracted_k141_115472 | Not-determined |
| S3_extracted_k141_18984 | Not-determined |
| S3_extracted_k141_60180 | Low-quality |
| S3_extracted_k141_43834 | Not-determined |
| S3_extracted_k141_52454 | Not-determined |
| S3_extracted_k141_60184 | Low-quality |
| S3_extracted_k141_18987 | Not-determined |
| S3_extracted_k141_27903 | Not-determined |
| S3_extracted_k141_76349 | Not-determined |
| S3_extracted_k141_18992 | Low-quality |
| S3_extracted_k141_52457 | Not-determined |
| S3_extracted_k141_3809 | Not-determined |
| S3_extracted_k141_67039 | Low-quality |
| S3_extracted_k141_67040 | Not-determined |
| S3_extracted_k141_3815 | Not-determined |
| S3_extracted_k141_52463 | Not-determined |
| S3_extracted_k141_19001 | Not-determined |
| S3_extracted_k141_76357 | Low-quality |
| S3_extracted_k141_60193 | Not-determined |
| S3_extracted_k141_76358 | Not-determined |
| S3_extracted_k141_107655 | Not-determined |
| S3_extracted_k141_76361 | Low-quality |
| S3_extracted_k141_67050 | Not-determined |
| S3_extracted_k141_43858 | Not-determined |
| S3_extracted_k141_19007 | Not-determined |
| S3_extracted_k141_115497 | Not-determined |
| S3_extracted_k141_3823 | Low-quality |
| S3_extracted_k141_60201 | Not-determined |
| S3_extracted_k141_3826 | Low-quality |
| S3_extracted_k141_107662 | Not-determined |
| S3_extracted_k141_3831 | Not-determined |
| S3_extracted_k141_76372 | Low-quality |
| S3_extracted_k141_27905 | Not-determined |
| S3_extracted_k141_27906 | Not-determined |
| S3_extracted_k141_91697 | Not-determined |
| S3_extracted_k141_36204 | Not-determined |
| S3_extracted_k141_91700 | Not-determined |
| S3_extracted_k141_43882 | Not-determined |
| S3_extracted_k141_91704 | Not-determined |
| S3_extracted_k141_91710 | Not-determined |
| S3_extracted_k141_60208 | Low-quality |
| S3_extracted_k141_3835 | Not-determined |
| S3_extracted_k141_3841 | Not-determined |
| S3_extracted_k141_3842 | Not-determined |
| S3_extracted_k141_60214 | Not-determined |
| S3_extracted_k141_3846 | Not-determined |
| S3_extracted_k141_27919 | Low-quality |
| S3_extracted_k141_3848 | Not-determined |
| S3_extracted_k141_91721 | Low-quality |
| S3_extracted_k141_76395 | Not-determined |
| S3_extracted_k141_60221 | Not-determined |
| S3_extracted_k141_99367 | Not-determined |
| S3_extracted_k141_27924 | Not-determined |
| S3_extracted_k141_60222 | Not-determined |
| S3_extracted_k141_107676 | Not-determined |
| S3_extracted_k141_107677 | Not-determined |
| S3_extracted_k141_99371 | Not-determined |
| S3_extracted_k141_99373 | Not-determined |
| S3_extracted_k141_67062 | Not-determined |
| S3_extracted_k141_76399 | Not-determined |
| S3_extracted_k141_76405 | Not-determined |
| S3_extracted_k141_27927 | Not-determined |
| S3_extracted_k141_76406 | Not-determined |
| S3_extracted_k141_107685 | Not-determined |
| S3_extracted_k141_3858 | Not-determined |
| S3_extracted_k141_115503 | Not-determined |
| S3_extracted_k141_91740 | Not-determined |
| S3_extracted_k141_3866 | Not-determined |
| S3_extracted_k141_91741 | Not-determined |
| S3_extracted_k141_3868 | Not-determined |
| S3_extracted_k141_122915 | Not-determined |
| S3_extracted_k141_122916 | Not-determined |
| S3_extracted_k141_52484 | Low-quality |
| S3_extracted_k141_36209 | Not-determined |
| S3_extracted_k141_13192 | Not-determined |
| S3_extracted_k141_19020 | Not-determined |
| S3_extracted_k141_36211 | Not-determined |
| S3_extracted_k141_13195 | Not-determined |
| S3_extracted_k141_52486 | Not-determined |
| S3_extracted_k141_36215 | Not-determined |
| S3_extracted_k141_52487 | Not-determined |
| S3_extracted_k141_122920 | Low-quality |
| S3_extracted_k141_19025 | Not-determined |
| S3_extracted_k141_52489 | Not-determined |
| S3_extracted_k141_36224 | Low-quality |
| S3_extracted_k141_13202 | Not-determined |
| S3_extracted_k141_13203 | Not-determined |
| S3_extracted_k141_76408 | Not-determined |
| S3_extracted_k141_13205 | Not-determined |
| S3_extracted_k141_67071 | Not-determined |
| S3_extracted_k141_67072 | Not-determined |
| S3_extracted_k141_3881 | Not-determined |
| S3_extracted_k141_91744 | Low-quality |
| S3_extracted_k141_60251 | Not-determined |
| S3_extracted_k141_36237 | Not-determined |
| S3_extracted_k141_60252 | Not-determined |
| S3_extracted_k141_122931 | Not-determined |
| S3_extracted_k141_60253 | Not-determined |
| S3_extracted_k141_3889 | Not-determined |
| S3_extracted_k141_67082 | Not-determined |
| S3_extracted_k141_36240 | Low-quality |
| S3_extracted_k141_76426 | Not-determined |
| S3_extracted_k141_52503 | Not-determined |
| S3_extracted_k141_76428 | Not-determined |
| S3_extracted_k141_76429 | Low-quality |
| S3_extracted_k141_60261 | Not-determined |
| S3_extracted_k141_52513 | Not-determined |
| S3_extracted_k141_76436 | Not-determined |
| S3_extracted_k141_91757 | Not-determined |
| S3_extracted_k141_60269 | Low-quality |
| S3_extracted_k141_43907 | Low-quality |
| S3_extracted_k141_67095 | Not-determined |
| S3_extracted_k141_43910 | Not-determined |
| S3_extracted_k141_43912 | Not-determined |
| S3_extracted_k141_19036 | Low-quality |
| S3_extracted_k141_3904 | Not-determined |
| S3_extracted_k141_99384 | Not-determined |
| S3_extracted_k141_67101 | Not-determined |
| S3_extracted_k141_99385 | Not-determined |
| S3_extracted_k141_3907 | Not-determined |
| S3_extracted_k141_122968 | Not-determined |
| S3_extracted_k141_91764 | Low-quality |
| S3_extracted_k141_43917 | Not-determined |
| S3_extracted_k141_43919 | Not-determined |
| S3_extracted_k141_99390 | Low-quality |
| S3_extracted_k141_91772 | Not-determined |
| S3_extracted_k141_43921 | Not-determined |
| S3_extracted_k141_43923 | Not-determined |
| S3_extracted_k141_13222 | Low-quality |
| S3_extracted_k141_43924 | Not-determined |
| S3_extracted_k141_3920 | Not-determined |
| S3_extracted_k141_91776 | Not-determined |
| S3_extracted_k141_122974 | Not-determined |
| S3_extracted_k141_60294 | Not-determined |
| S3_extracted_k141_3922 | Not-determined |
| S3_extracted_k141_19064 | Not-determined |
| S3_extracted_k141_19067 | Not-determined |
| S3_extracted_k141_19068 | Not-determined |
| S3_extracted_k141_67119 | Low-quality |
| S3_extracted_k141_67120 | Not-determined |
| S3_extracted_k141_76467 | Not-determined |
| S3_extracted_k141_122982 | Low-quality |
| S3_extracted_k141_91782 | Not-determined |
| S3_extracted_k141_13245 | Low-quality |
| S3_extracted_k141_67126 | Not-determined |
| S3_extracted_k141_67128 | Not-determined |
| S3_extracted_k141_19076 | Not-determined |
| S3_extracted_k141_76468 | Low-quality |
| S3_extracted_k141_60302 | Not-determined |
| S3_extracted_k141_13254 | Not-determined |
| S3_extracted_k141_3939 | Not-determined |
| S3_extracted_k141_67138 | Not-determined |
| S3_extracted_k141_76476 | Not-determined |
| S3_extracted_k141_107722 | Not-determined |
| S3_extracted_k141_67143 | Not-determined |
| S3_extracted_k141_115527 | Not-determined |
| S3_extracted_k141_122995 | Not-determined |
| S3_extracted_k141_107725 | Low-quality |
| S3_extracted_k141_3957 | Not-determined |
| S3_extracted_k141_67154 | Not-determined |
| S3_extracted_k141_91803 | Not-determined |
| S3_extracted_k141_27937 | Not-determined |
| S3_extracted_k141_3963 | Not-determined |
| S3_extracted_k141_91805 | Not-determined |
| S3_extracted_k141_19106 | Not-determined |
| S3_extracted_k141_76489 | Low-quality |
| S3_extracted_k141_27947 | Not-determined |
| S3_extracted_k141_36248 | Not-determined |
| S3_extracted_k141_115541 | Low-quality |
| S3_extracted_k141_27951 | Low-quality |
| S3_extracted_k141_19122 | Low-quality |
| S3_extracted_k141_27952 | Not-determined |
| S3_extracted_k141_115543 | Not-determined |
| S3_extracted_k141_115545 | Not-determined |
| S3_extracted_k141_19131 | Not-determined |
| S3_extracted_k141_3969 | Low-quality |
| S3_extracted_k141_123012 | Not-determined |
| S3_extracted_k141_67163 | Not-determined |
| S3_extracted_k141_107755 | Not-determined |
| S3_extracted_k141_13274 | Not-determined |
| S3_extracted_k141_19145 | Not-determined |
| S3_extracted_k141_60317 | Not-determined |
| S3_extracted_k141_43943 | Not-determined |
| S3_extracted_k141_107758 | Low-quality |
| S3_extracted_k141_3987 | Not-determined |
| S3_extracted_k141_13277 | Low-quality |
| S3_extracted_k141_43947 | Not-determined |
| S3_extracted_k141_13278 | Not-determined |
| S3_extracted_k141_76492 | Not-determined |
| S3_extracted_k141_76493 | Not-determined |
| S3_extracted_k141_123013 | Not-determined |
| S3_extracted_k141_123017 | Low-quality |
| S3_extracted_k141_115557 | Low-quality |
| S3_extracted_k141_107759 | Not-determined |
| S3_extracted_k141_60343 | Low-quality |

| | |
|---|---|
| S3_extracted_k141_76515 | Not-determined |
| S3_extracted_k141_91855 | Not-determined |
| S3_extracted_k141_43969 | Not-determined |
| S3_extracted_k141_91858 | Not-determined |
| S3_extracted_k141_43970 | Not-determined |
| S3_extracted_k141_67175 | Not-determined |
| S3_extracted_k141_43972 | Low-quality |
| S3_extracted_k141_27957 | Low-quality |
| S3_extracted_k141_27958 | Low-quality |
| S3_extracted_k141_43974 | Low-quality |
| S3_extracted_k141_76529 | Not-determined |
| S3_extracted_k141_67183 | Not-determined |
| S3_extracted_k141_76530 | Not-determined |
| S3_extracted_k141_27968 | Not-determined |
| S3_extracted_k141_76535 | Low-quality |
| S3_extracted_k141_99416 | Not-determined |
| S3_extracted_k141_67189 | Not-determined |
| S3_extracted_k141_123031 | Not-determined |
| S3_extracted_k141_60363 | Not-determined |
| S3_extracted_k141_123035 | Low-quality |
| S3_extracted_k141_13319 | Not-determined |
| S3_extracted_k141_13328 | Not-determined |
| S3_extracted_k141_60378 | Not-determined |
| S3_extracted_k141_91865 | Not-determined |
| S3_extracted_k141_107767 | Not-determined |
| S3_extracted_k141_13335 | Low-quality |
| S3_extracted_k141_107768 | Not-determined |
| S3_extracted_k141_13337 | Not-determined |
| S3_extracted_k141_19179 | Not-determined |
| S3_extracted_k141_19180 | Low-quality |
| S3_extracted_k141_19184 | Not-determined |
| S3_extracted_k141_36278 | Not-determined |
| S3_extracted_k141_60388 | Not-determined |
| S3_extracted_k141_115575 | Low-quality |
| S3_extracted_k141_115581 | Not-determined |
| S3_extracted_k141_107787 | Not-determined |
| S3_extracted_k141_67206 | Not-determined |
| S3_extracted_k141_60395 | Low-quality |
| S3_extracted_k141_13346 | Not-determined |
| S3_extracted_k141_67213 | Not-determined |
| S3_extracted_k141_13348 | Not-determined |
| S3_extracted_k141_36298 | Not-determined |
| S3_extracted_k141_36299 | Not-determined |
| S3_extracted_k141_60403 | Not-determined |
| S3_extracted_k141_52530 | Not-determined |
| S3_extracted_k141_107799 | Not-determined |
| S3_extracted_k141_13358 | Not-determined |
| S3_extracted_k141_52532 | Low-quality |
| S3_extracted_k141_60405 | Low-quality |
| S3_extracted_k141_4003 | Not-determined |
| S3_extracted_k141_52534 | Not-determined |
| S3_extracted_k141_13359 | Not-determined |
| S3_extracted_k141_115597 | Not-determined |
| S3_extracted_k141_67229 | Not-determined |
| S3_extracted_k141_60408 | Low-quality |
| S3_extracted_k141_19190 | Not-determined |
| S3_extracted_k141_99422 | Not-determined |
| S3_extracted_k141_13375 | Not-determined |
| S3_extracted_k141_19196 | Not-determined |
| S3_extracted_k141_4012 | Not-determined |
| S3_extracted_k141_91873 | Not-determined |
| S3_extracted_k141_115608 | Not-determined |
| S3_extracted_k141_4014 | Not-determined |
| S3_extracted_k141_99434 | Not-determined |
| S3_extracted_k141_99435 | Not-determined |
| S3_extracted_k141_4016 | Not-determined |
| S3_extracted_k141_91875 | Not-determined |
| S3_extracted_k141_4018 | Not-determined |
| S3_extracted_k141_91877 | Not-determined |
| S3_extracted_k141_115618 | Not-determined |
| S3_extracted_k141_115620 | Not-determined |
| S3_extracted_k141_4022 | Low-quality |
| S3_extracted_k141_91882 | Not-determined |
| S3_extracted_k141_115625 | Not-determined |
| S3_extracted_k141_67245 | Not-determined |
| S3_extracted_k141_27977 | Not-determined |
| S3_extracted_k141_13388 | Not-determined |
| S3_extracted_k141_115627 | Not-determined |
| S3_extracted_k141_107801 | Low-quality |
| S3_extracted_k141_123048 | Not-determined |
| S3_extracted_k141_27978 | Low-quality |
| S3_extracted_k141_67254 | Not-determined |
| S3_extracted_k141_13392 | Not-determined |
| S3_extracted_k141_27980 | Not-determined |
| S3_extracted_k141_36317 | Not-determined |
| S3_extracted_k141_107804 | Not-determined |
| S3_extracted_k141_107805 | Not-determined |
| S3_extracted_k141_4030 | Low-quality |
| S3_extracted_k141_4031 | Not-determined |
| S3_extracted_k141_52545 | Not-determined |
| S3_extracted_k141_52547 | Not-determined |
| S3_extracted_k141_115635 | Not-determined |
| S3_extracted_k141_99445 | Low-quality |
| S3_extracted_k141_36329 | Not-determined |
| S3_extracted_k141_99448 | Not-determined |
| S3_extracted_k141_115639 | Low-quality |
| S3_extracted_k141_115640 | Not-determined |
| S3_extracted_k141_60421 | Low-quality |
| S3_extracted_k141_123060 | Not-determined |
| S3_extracted_k141_27984 | Not-determined |
| S3_extracted_k141_123068 | Low-quality |
| S3_extracted_k141_60423 | Low-quality |
| S3_extracted_k141_115653 | Not-determined |
| S3_extracted_k141_19200 | Low-quality |
| S3_extracted_k141_4036 | Not-determined |
| S3_extracted_k141_99455 | Low-quality |
| S3_extracted_k141_4040 | Not-determined |
| S3_extracted_k141_123074 | Not-determined |
| S3_extracted_k141_99460 | Not-determined |
| S3_extracted_k141_91891 | Not-determined |
| S3_extracted_k141_99462 | Not-determined |
| S3_extracted_k141_13409 | Not-determined |
| S3_extracted_k141_99465 | Not-determined |
| S3_extracted_k141_13414 | Not-determined |
| S3_extracted_k141_123085 | Low-quality |
| S3_extracted_k141_67265 | Low-quality |
| S3_extracted_k141_60442 | Not-determined |
| S3_extracted_k141_52576 | Not-determined |
| S3_extracted_k141_76547 | Not-determined |
| S3_extracted_k141_76550 | Not-determined |
| S3_extracted_k141_99489 | Not-determined |
| S3_extracted_k141_76556 | Not-determined |
| S3_extracted_k141_67270 | Not-determined |
| S3_extracted_k141_36337 | Not-determined |
| S3_extracted_k141_67273 | Not-determined |
| S3_extracted_k141_13438 | Not-determined |
| S3_extracted_k141_4050 | Low-quality |
| S3_extracted_k141_115665 | Not-determined |
| S3_extracted_k141_60457 | Not-determined |
| S3_extracted_k141_115667 | Not-determined |
| S3_extracted_k141_13441 | Not-determined |
| S3_extracted_k141_76561 | Low-quality |
| S3_extracted_k141_13444 | Not-determined |
| S3_extracted_k141_36346 | Not-determined |
| S3_extracted_k141_52583 | Not-determined |
| S3_extracted_k141_13447 | Low-quality |
| S3_extracted_k141_36350 | Not-determined |
| S3_extracted_k141_13452 | Low-quality |
| S3_extracted_k141_76564 | Not-determined |
| S3_extracted_k141_36352 | Not-determined |
| S3_extracted_k141_99500 | Not-determined |
| S3_extracted_k141_60464 | Not-determined |
| S3_extracted_k141_123100 | Not-determined |
| S3_extracted_k141_44028 | Low-quality |
| S3_extracted_k141_123102 | Not-determined |
| S3_extracted_k141_60471 | Not-determined |
| S3_extracted_k141_123103 | Not-determined |
| S3_extracted_k141_19223 | Not-determined |
| S3_extracted_k141_123105 | Low-quality |
| S3_extracted_k141_123108 | Not-determined |
| S3_extracted_k141_19230 | Low-quality |
| S3_extracted_k141_13460 | Not-determined |
| S3_extracted_k141_115672 | Not-determined |
| S3_extracted_k141_13461 | Not-determined |
| S3_extracted_k141_4060 | Not-determined |
| S3_extracted_k141_52606 | Not-determined |
| S3_extracted_k141_67304 | Not-determined |
| S3_extracted_k141_52607 | Not-determined |
| S3_extracted_k141_123112 | Not-determined |
| S3_extracted_k141_67307 | Not-determined |
| S3_extracted_k141_19250 | Not-determined |
| S3_extracted_k141_76581 | Not-determined |
| S3_extracted_k141_76582 | Not-determined |
| S3_extracted_k141_36385 | Not-determined |
| S3_extracted_k141_19265 | Not-determined |
| S3_extracted_k141_4067 | Not-determined |
| S3_extracted_k141_67313 | Not-determined |
| S3_extracted_k141_107830 | Not-determined |
| S3_extracted_k141_4072 | Not-determined |
| S3_extracted_k141_107834 | Not-determined |
| S3_extracted_k141_115694 | Not-determined |
| S3_extracted_k141_123125 | Not-determined |
| S3_extracted_k141_123126 | Not-determined |
| S3_extracted_k141_107843 | Low-quality |
| S3_extracted_k141_36390 | Low-quality |
| S3_extracted_k141_123135 | Not-determined |
| S3_extracted_k141_36391 | Low-quality |
| S3_extracted_k141_76586 | Low-quality |
| S3_extracted_k141_67325 | Low-quality |
| S3_extracted_k141_99514 | Not-determined |
| S3_extracted_k141_107849 | Not-determined |
| S3_extracted_k141_123146 | Not-determined |
| S3_extracted_k141_115695 | Not-determined |
| S3_extracted_k141_13490 | Not-determined |
| S3_extracted_k141_76588 | Not-determined |
| S3_extracted_k141_107852 | Not-determined |
| S3_extracted_k141_83759 | Low-quality |
| S3_extracted_k141_4082 | Not-determined |
| S3_extracted_k141_123162 | Not-determined |
| S3_extracted_k141_115700 | Low-quality |
| S3_extracted_k141_36397 | Not-determined |
| S3_extracted_k141_99528 | Not-determined |
| S3_extracted_k141_4085 | Not-determined |
| S3_extracted_k141_19282 | Low-quality |
| S3_extracted_k141_4087 | Not-determined |
| S3_extracted_k141_83766 | Not-determined |
| S3_extracted_k141_19286 | Not-determined |
| S3_extracted_k141_44041 | Not-determined |
| S3_extracted_k141_36402 | Not-determined |
| S3_extracted_k141_52628 | Not-determined |
| S3_extracted_k141_52629 | Not-determined |
| S3_extracted_k141_52630 | Not-determined |
| S3_extracted_k141_13510 | Not-determined |
| S3_extracted_k141_44044 | Not-determined |
| S3_extracted_k141_19301 | Not-determined |
| S3_extracted_k141_123174 | Not-determined |
| S3_extracted_k141_52638 | Not-determined |
| S3_extracted_k141_60505 | Not-determined |
| S3_extracted_k141_60506 | Low-quality |
| S3_extracted_k141_123182 | Not-determined |
| S3_extracted_k141_115702 | Not-determined |
| S3_extracted_k141_13533 | Not-determined |
| S3_extracted_k141_13537 | Not-determined |
| S3_extracted_k141_115704 | Low-quality |
| S3_extracted_k141_13540 | Low-quality |
| S3_extracted_k141_13541 | Not-determined |
| S3_extracted_k141_99543 | Not-determined |
| S3_extracted_k141_52650 | Not-determined |
| S3_extracted_k141_107875 | Not-determined |
| S3_extracted_k141_52651 | Not-determined |
| S3_extracted_k141_99546 | Not-determined |
| S3_extracted_k141_99547 | Not-determined |
| S3_extracted_k141_60540 | Not-determined |
| S3_extracted_k141_60542 | Not-determined |
| S3_extracted_k141_99550 | Low-quality |
| S3_extracted_k141_76621 | Not-determined |
| S3_extracted_k141_60543 | Not-determined |
| S3_extracted_k141_44050 | Not-determined |
| S3_extracted_k141_4097 | Not-determined |
| S3_extracted_k141_107879 | Low-quality |
| S3_extracted_k141_107882 | Not-determined |
| S3_extracted_k141_123209 | Not-determined |
| S3_extracted_k141_107893 | Not-determined |
| S3_extracted_k141_123213 | Not-determined |
| S3_extracted_k141_76630 | Not-determined |
| S3_vs_k141_8058\|\|full | High-quality |
| S3_extracted_k141_115709 | Not-determined |
| S3_extracted_k141_91911 | Not-determined |
| S3_extracted_k141_76643 | Low-quality |
| S3_extracted_k141_123227 | Not-determined |
| S3_extracted_k141_60552 | Not-determined |
| S3_extracted_k141_52664 | Not-determined |
| S3_extracted_k141_91916 | Not-determined |
| S3_extracted_k141_83786 | Low-quality |
| S3_extracted_k141_123230 | Not-determined |
| S3_extracted_k141_52671 | Not-determined |
| S3_extracted_k141_60564 | Not-determined |
| S3_extracted_k141_52677 | Not-determined |
| S3_extracted_k141_13561 | Not-determined |
| S3_extracted_k141_52679 | Low-quality |
| S3_extracted_k141_13563 | Not-determined |
| S3_extracted_k141_91923 | Low-quality |
| S3_extracted_k141_13567 | Not-determined |
| S3_extracted_k141_107905 | Low-quality |
| S3_extracted_k141_13568 | Low-quality |
| S3_extracted_k141_123242 | Low-quality |
| S3_extracted_k141_44066 | Not-determined |
| S3_extracted_k141_44068 | Not-determined |
| S3_extracted_k141_123248 | Not-determined |
| S3_extracted_k141_27997 | Not-determined |
| S3_extracted_k141_52684 | Not-determined |
| S3_extracted_k141_13571 | Not-determined |
| S3_extracted_k141_52688 | Low-quality |
| S3_extracted_k141_60573 | Not-determined |
| S3_extracted_k141_60578 | Not-determined |
| S7_vs_134583\|\|full | High-quality |
| S3_extracted_k141_44073 | Not-determined |
| S3_extracted_k141_60581 | Not-determined |
| S3_extracted_k141_99567 | Not-determined |
| S3_extracted_k141_83802 | Low-quality |
| S3_extracted_k141_44082 | Not-determined |
| S3_extracted_k141_107920 | Not-determined |
| S3_extracted_k141_107921 | Not-determined |
| S3_extracted_k141_13587 | Not-determined |
| S3_extracted_k141_44089 | Low-quality |
| S3_extracted_k141_4111 | Not-determined |
| S3_extracted_k141_44092 | Low-quality |
| S3_extracted_k141_4116 | Not-determined |
| S3_extracted_k141_60602 | Low-quality |
| S3_extracted_k141_76681 | Not-determined |
| S3_extracted_k141_83804 | Low-quality |
| S3_extracted_k141_60607 | Not-determined |
| S3_extracted_k141_115733 | Not-determined |
| S3_extracted_k141_76686 | Not-determined |
| S3_extracted_k141_123255 | Not-determined |
| S3_extracted_k141_60610 | Not-determined |
| S3_extracted_k141_4132 | Not-determined |
| S3_extracted_k141_52709 | Not-determined |
| S3_extracted_k141_67374 | Not-determined |
| S3_extracted_k141_60619 | Not-determined |
| S3_extracted_k141_4138 | Not-determined |
| S3_extracted_k141_60620 | Not-determined |
| S3_extracted_k141_83808 | Not-determined |
| S3_extracted_k141_52715 | Not-determined |
| S3_extracted_k141_76698 | Not-determined |
| S3_extracted_k141_83811 | Not-determined |
| S3_extracted_k141_60626 | Low-quality |
| S3_extracted_k141_28017 | Not-determined |
| S3_extracted_k141_44102 | Not-determined |
| S3_extracted_k141_28018 | Not-determined |
| S3_extracted_k141_83814 | Not-determined |
| S3_extracted_k141_52722 | Low-quality |
| S3_extracted_k141_36418 | Low-quality |
| S3_extracted_k141_28024 | Not-determined |
| S3_extracted_k141_107930 | Low-quality |
| S3_extracted_k141_19314 | Not-determined |
| S3_extracted_k141_99572 | Not-determined |
| S3_extracted_k141_19315 | Not-determined |
| S3_extracted_k141_76707 | Not-determined |
| S3_extracted_k141_52728 | Not-determined |
| S3_extracted_k141_52729 | Not-determined |
| S3_extracted_k141_76713 | Not-determined |
| S3_extracted_k141_123276 | Not-determined |
| S3_extracted_k141_44104 | Not-determined |
| S3_extracted_k141_76714 | Not-determined |
| S3_extracted_k141_28037 | Low-quality |
| S3_extracted_k141_44107 | Low-quality |
| S3_extracted_k141_44108 | Not-determined |
| S3_extracted_k141_99580 | Low-quality |
| S3_extracted_k141_13601 | Low-quality |
| S3_extracted_k141_123284 | Not-determined |
| S3_extracted_k141_44113 | Low-quality |
| S3_extracted_k141_115757 | Not-determined |
| S3_extracted_k141_36425 | Not-determined |
| S3_extracted_k141_123286 | Not-determined |
| S3_extracted_k141_60645 | Not-determined |
| S3_extracted_k141_99583 | Low-quality |
| S3_extracted_k141_36427 | Not-determined |
| S3_extracted_k141_13604 | Not-determined |
| S3_extracted_k141_123291 | Not-determined |
| S3_extracted_k141_83825 | Not-determined |
| S3_extracted_k141_83826 | Not-determined |
| S3_extracted_k141_36437 | Not-determined |
| S3_extracted_k141_13611 | Not-determined |
| S3_extracted_k141_4149 | Low-quality |
| S3_extracted_k141_83828 | Not-determined |
| S3_extracted_k141_13613 | Not-determined |
| S3_extracted_k141_123294 | Not-determined |
| S3_extracted_k141_123295 | Not-determined |
| S3_extracted_k141_52751 | Not-determined |
| S3_extracted_k141_60657 | Not-determined |
| S3_extracted_k141_60659 | Not-determined |
| S3_extracted_k141_60660 | Not-determined |
| S3_extracted_k141_36439 | Low-quality |
| S3_extracted_k141_123303 | Not-determined |
| S3_extracted_k141_28058 | Not-determined |
| S3_extracted_k141_107936 | Not-determined |
| S3_extracted_k141_28059 | Not-determined |
| S3_extracted_k141_52758 | Not-determined |
| S3_extracted_k141_36442 | Low-quality |
| S3_extracted_k141_107940 | Not-determined |
| S3_extracted_k141_123306 | Not-determined |
| S3_extracted_k141_60663 | Not-determined |
| S3_extracted_k141_28070 | Not-determined |
| S3_extracted_k141_115772 | Not-determined |
| S3_extracted_k141_123309 | Low-quality |
| S3_extracted_k141_123310 | Not-determined |
| S3_extracted_k141_115778 | Not-determined |
| S3_extracted_k141_28079 | Not-determined |
| S3_extracted_k141_76726 | Low-quality |
| S3_extracted_k141_76727 | Not-determined |
| S3_extracted_k141_28082 | Not-determined |
| S3_extracted_k141_52766 | Not-determined |
| S3_extracted_k141_52767 | Not-determined |
| S3_extracted_k141_52770 | Not-determined |
| S3_extracted_k141_52771 | Not-determined |
| S3_extracted_k141_28089 | Low-quality |
| S3_extracted_k141_28093 | Not-determined |
| S3_extracted_k141_76729 | Not-determined |
| S3_extracted_k141_52772 | Not-determined |
| S3_extracted_k141_123321 | Not-determined |
| S3_extracted_k141_123323 | Low-quality |
| S3_extracted_k141_123326 | Not-determined |
| S3_extracted_k141_44119 | Not-determined |
| S3_extracted_k141_115792 | Not-determined |
| S3_extracted_k141_28100 | Not-determined |
| S3_extracted_k141_28102 | Not-determined |
| S3_extracted_k141_76742 | Not-determined |
| S3_extracted_k141_76743 | Not-determined |
| S3_extracted_k141_123329 | Not-determined |
| S3_extracted_k141_123330 | Not-determined |
| S3_extracted_k141_123333 | Not-determined |
| S3_extracted_k141_52780 | Low-quality |
| S3_extracted_k141_107952 | Not-determined |
| S3_extracted_k141_28112 | Not-determined |
| S3_extracted_k141_4161 | Low-quality |
| S3_extracted_k141_115804 | Not-determined |
| S3_extracted_k141_115806 | Low-quality |
| S3_extracted_k141_52786 | Not-determined |
| S3_extracted_k141_83839 | Not-determined |
| S3_extracted_k141_60678 | Not-determined |
| S3_extracted_k141_52790 | Not-determined |
| S3_extracted_k141_60682 | Not-determined |
| S3_extracted_k141_28122 | Not-determined |
| S3_extracted_k141_76755 | Low-quality |
| S3_extracted_k141_4170 | Not-determined |
| S3_extracted_k141_123342 | Low-quality |
| S3_extracted_k141_60687 | Not-determined |
| S3_extracted_k141_13630 | Not-determined |
| S3_extracted_k141_123348 | Not-determined |
| S3_extracted_k141_28129 | Not-determined |
| S3_extracted_k141_60692 | Not-determined |
| S3_extracted_k141_115815 | Not-determined |
| S3_extracted_k141_76762 | Not-determined |
| S3_extracted_k141_91942 | Not-determined |
| S3_extracted_k141_13633 | Not-determined |
| S3_extracted_k141_99608 | Not-determined |
| S3_extracted_k141_107959 | Not-determined |
| S3_extracted_k141_107960 | Not-determined |
| S3_extracted_k141_123357 | Not-determined |
| S3_extracted_k141_123358 | Not-determined |
| S3_extracted_k141_28143 | Low-quality |
| S3_extracted_k141_91949 | Not-determined |
| S3_extracted_k141_13640 | Low-quality |
| S3_extracted_k141_67387 | Low-quality |
| S3_extracted_k141_99615 | Not-determined |
| S3_extracted_k141_115824 | Low-quality |
| S3_extracted_k141_13644 | Not-determined |
| S3_extracted_k141_67391 | Not-determined |
| S3_extracted_k141_44142 | Not-determined |
| S3_extracted_k141_123364 | Not-determined |
| S3_extracted_k141_44143 | Not-determined |
| S3_extracted_k141_123365 | Not-determined |
| S3_extracted_k141_91959 | Not-determined |
| S3_extracted_k141_44145 | Not-determined |
| S3_extracted_k141_99622 | Not-determined |
| S3_extracted_k141_44148 | Low-quality |
| S3_extracted_k141_123374 | Not-determined |
| S3_extracted_k141_44158 | Not-determined |
| S3_extracted_k141_107977 | Low-quality |
| S3_extracted_k141_107978 | Not-determined |
| S3_extracted_k141_60694 | Low-quality |
| S3_extracted_k141_44163 | Not-determined |
| S3_extracted_k141_44165 | Not-determined |
| S3_extracted_k141_83847 | Low-quality |
| S3_extracted_k141_115844 | Not-determined |
| S3_extracted_k141_83858 | Not-determined |
| S3_vs_k141_55571\|\|full | High-quality |
| S3_extracted_k141_83861 | Not-determined |
| S3_extracted_k141_44173 | Not-determined |
| S3_extracted_k141_44177 | Not-determined |
| S3_extracted_k141_115847 | Low-quality |
| S3_extracted_k141_44184 | Low-quality |
| S3_extracted_k141_91971 | Not-determined |
| S3_extracted_k141_91972 | Not-determined |
| S3_extracted_k141_67400 | Not-determined |
| S3_extracted_k141_115854 | Not-determined |
| S3_extracted_k141_13654 | Not-determined |
| S3_extracted_k141_67403 | Not-determined |
| S3_extracted_k141_67404 | Not-determined |
| S3_extracted_k141_108001 | Not-determined |
| S3_extracted_k141_52847 | Not-determined |
| S3_extracted_k141_115861 | Not-determined |
| S3_extracted_k141_67416 | Not-determined |
| S3_extracted_k141_52848 | Low-quality |
| S3_extracted_k141_60701 | Not-determined |
| S3_extracted_k141_115866 | Low-quality |
| S3_extracted_k141_83874 | Not-determined |
| S3_extracted_k141_115867 | Not-determined |
| S3_extracted_k141_44208 | Not-determined |
| S3_extracted_k141_83878 | Not-determined |
| S3_extracted_k141_52852 | Low-quality |
| S3_extracted_k141_13664 | Not-determined |
| S3_extracted_k141_83880 | Not-determined |
| S3_extracted_k141_67424 | Not-determined |
| S2_extracted_k141_21630 | Complete |
| S3_extracted_k141_13668 | Not-determined |
| S3_extracted_k141_13670 | Not-determined |
| S3_extracted_k141_99628 | Not-determined |
| S3_extracted_k141_99630 | Not-determined |
| S3_extracted_k141_60711 | Not-determined |
| S3_extracted_k141_60713 | Not-determined |
| S3_extracted_k141_67428 | Not-determined |
| S3_extracted_k141_83899 | Not-determined |
| S3_extracted_k141_83900 | Not-determined |
| S3_extracted_k141_76777 | Low-quality |
| S3_extracted_k141_60720 | Not-determined |
| S3_extracted_k141_76780 | Not-determined |
| S3_extracted_k141_76785 | Not-determined |
| S3_extracted_k141_67436 | Not-determined |
| S3_extracted_k141_13673 | Not-determined |
| S3_extracted_k141_13676 | Not-determined |
| S3_extracted_k141_67437 | Low-quality |
| S3_extracted_k141_123409 | Not-determined |
| S3_extracted_k141_13677 | Low-quality |
| S3_extracted_k141_52882 | Not-determined |
| S3_extracted_k141_76800 | Low-quality |
| S3_extracted_k141_115869 | Not-determined |
| S3_extracted_k141_19358 | Not-determined |
| S3_extracted_k141_76801 | Not-determined |
| S3_extracted_k141_19359 | Not-determined |
| S3_extracted_k141_115872 | Not-determined |
| S3_extracted_k141_19361 | Not-determined |
| S3_extracted_k141_28159 | Not-determined |
| S3_extracted_k141_67441 | Not-determined |
| S3_extracted_k141_115878 | Not-determined |
| S3_extracted_k141_115879 | Not-determined |
| S3_extracted_k141_19368 | Not-determined |
| S3_extracted_k141_123425 | Not-determined |
| S3_extracted_k141_67451 | Not-determined |
| S3_extracted_k141_52899 | Low-quality |
| S3_extracted_k141_52900 | Low-quality |
| S3_extracted_k141_115885 | Not-determined |
| S3_extracted_k141_115886 | Not-determined |
| S3_extracted_k141_67455 | Not-determined |
| S3_extracted_k141_52909 | Not-determined |
| S3_extracted_k141_67461 | Not-determined |
| S3_extracted_k141_52911 | Not-determined |
| S3_extracted_k141_60732 | Low-quality |
| S3_extracted_k141_13698 | Low-quality |
| S3_extracted_k141_13700 | Not-determined |
| S3_extracted_k141_19372 | Not-determined |
| S3_extracted_k141_28171 | Not-determined |
| S3_extracted_k141_91998 | Not-determined |
| S3_extracted_k141_115901 | Not-determined |
| S3_extracted_k141_67472 | Low-quality |
| S3_extracted_k141_67473 | Low-quality |
| S3_extracted_k141_13703 | Low-quality |
| S3_extracted_k141_44228 | Not-determined |
| S3_extracted_k141_83902 | Low-quality |
| S3_extracted_k141_13715 | Not-determined |
| S3_extracted_k141_115908 | Not-determined |
| S3_extracted_k141_83909 | Low-quality |
| S3_extracted_k141_19392 | Not-determined |
| S3_extracted_k141_67485 | Low-quality |
| S3_extracted_k141_44238 | Not-determined |
| S3_extracted_k141_67488 | Not-determined |
| S3_extracted_k141_67491 | Low-quality |
| S3_extracted_k141_36485 | Not-determined |
| S3_extracted_k141_76818 | Not-determined |
| S3_extracted_k141_44244 | Not-determined |
| S3_extracted_k141_60733 | Not-determined |
| S3_extracted_k141_123462 | Not-determined |
| S3_extracted_k141_28188 | Not-determined |
| S3_extracted_k141_123467 | Not-determined |
| S3_extracted_k141_60740 | Not-determined |
| S3_extracted_k141_83910 | Not-determined |
| S3_extracted_k141_13733 | Not-determined |
| S3_extracted_k141_19402 | Not-determined |
| S3_extracted_k141_83913 | Not-determined |
| S3_extracted_k141_60748 | Not-determined |
| S3_extracted_k141_19406 | Not-determined |
| S3_extracted_k141_19408 | Not-determined |
| S3_extracted_k141_123481 | Low-quality |
| S3_extracted_k141_115934 | Not-determined |
| S3_extracted_k141_19414 | Not-determined |
| S3_extracted_k141_60749 | Low-quality |
| S3_extracted_k141_67507 | Not-determined |
| S3_extracted_k141_28196 | Not-determined |
| S3_extracted_k141_123485 | Not-determined |
| S3_extracted_k141_115939 | Low-quality |
| S3_extracted_k141_108037 | Not-determined |
| S3_extracted_k141_123488 | Not-determined |
| S3_extracted_k141_123490 | Low-quality |
| S3_extracted_k141_76832 | Low-quality |
| S3_extracted_k141_60755 | Low-quality |
| S3_extracted_k141_123500 | Not-determined |
| S3_extracted_k141_19430 | Not-determined |
| S3_extracted_k141_28197 | Low-quality |
| S3_extracted_k141_60758 | Not-determined |
| S3_extracted_k141_83919 | Not-determined |
| S3_extracted_k141_60759 | Not-determined |
| S3_extracted_k141_115941 | Low-quality |
| S3_extracted_k141_28200 | Not-determined |
| S3_extracted_k141_83921 | Low-quality |
| S3_extracted_k141_44265 | Not-determined |
| S3_extracted_k141_123503 | Not-determined |
| S3_extracted_k141_115950 | Low-quality |
| S3_extracted_k141_92019 | Not-determined |
| S3_extracted_k141_92020 | Low-quality |
| S3_extracted_k141_92026 | Not-determined |
| S3_extracted_k141_13758 | Not-determined |
| S3_extracted_k141_67532 | Not-determined |
| S3_extracted_k141_92033 | Not-determined |
| S3_extracted_k141_83931 | Not-determined |
| S3_extracted_k141_60765 | Low-quality |
| S3_extracted_k141_4213 | Not-determined |
| S3_extracted_k141_4214 | Not-determined |
| S3_extracted_k141_115961 | Not-determined |
| S3_extracted_k141_60767 | Low-quality |
| S3_extracted_k141_76845 | Not-determined |
| S3_extracted_k141_19450 | Not-determined |
| S3_extracted_k141_108049 | Not-determined |
| S3_extracted_k141_28213 | Low-quality |
| S3_extracted_k141_76853 | Not-determined |
| S3_extracted_k141_115965 | Low-quality |
| S3_extracted_k141_44276 | Not-determined |
| S3_extracted_k141_4225 | Not-determined |
| S3_extracted_k141_108060 | Not-determined |

| | |
|---|---|
| S3_extracted_k141_44279 | Not-determined |
| S3_extracted_k141_60779 | Not-determined |
| S3_extracted_k141_76863 | Not-determined |
| S3_extracted_k141_76865 | Not-determined |
| S3_extracted_k141_76866 | Low-quality |
| S3_extracted_k141_44289 | Not-determined |
| S3_extracted_k141_60787 | Not-determined |
| S3_extracted_k141_83937 | Not-determined |
| S3_extracted_k141_4241 | Not-determined |
| S3_extracted_k141_99655 | Not-determined |
| S3_extracted_k141_67556 | Not-determined |
| S3_extracted_k141_36509 | Not-determined |
| S3_extracted_k141_19469 | Not-determined |
| S3_extracted_k141_76870 | Not-determined |
| S3_extracted_k141_44299 | Not-determined |
| S3_extracted_k141_36514 | Not-determined |
| S3_extracted_k141_28217 | Not-determined |
| S3_extracted_k141_76876 | Not-determined |
| S3_extracted_k141_92042 | Not-determined |
| S3_extracted_k141_4248 | Not-determined |
| S3_extracted_k141_28219 | Low-quality |
| S3_extracted_k141_28222 | Not-determined |
| S3_extracted_k141_13776 | Not-determined |
| S3_extracted_k141_108071 | Not-determined |
| S3_extracted_k141_19486 | Not-determined |
| S3_extracted_k141_28224 | Low-quality |
| S3_extracted_k141_36530 | Not-determined |
| S3_extracted_k141_36532 | Not-determined |
| S3_extracted_k141_28226 | Not-determined |
| S3_extracted_k141_83974 | Not-determined |
| S3_extracted_k141_28229 | Not-determined |
| S3_extracted_k141_99679 | Not-determined |
| S3_extracted_k141_13791 | Not-determined |
| S3_extracted_k141_76883 | Not-determined |
| S3_extracted_k141_13795 | Not-determined |
| S3_extracted_k141_92045 | Not-determined |
| S3_extracted_k141_115990 | Low-quality |
| S3_extracted_k141_76886 | Not-determined |
| S3_extracted_k141_19488 | Not-determined |
| S3_extracted_k141_92047 | Not-determined |
| S3_extracted_k141_44316 | Not-determined |
| S3_extracted_k141_115994 | Not-determined |
| S3_extracted_k141_44322 | Not-determined |
| S3_extracted_k141_36542 | Not-determined |
| S3_extracted_k141_83984 | Not-determined |
| S3_extracted_k141_13804 | Not-determined |
| S3_extracted_k141_92053 | Not-determined |
| S3_extracted_k141_13807 | Not-determined |
| S3_extracted_k141_13810 | Not-determined |
| S3_extracted_k141_52947 | Not-determined |
| S3_extracted_k141_44333 | Not-determined |
| S3_extracted_k141_115998 | Low-quality |
| S3_extracted_k141_13813 | Not-determined |
| S3_extracted_k141_76893 | Not-determined |
| S3_extracted_k141_76894 | Not-determined |
| S3_extracted_k141_44339 | Not-determined |
| S3_extracted_k141_13818 | Not-determined |
| S3_extracted_k141_116002 | Not-determined |
| S3_extracted_k141_76896 | Not-determined |
| S3_extracted_k141_28244 | Not-determined |
| S3_extracted_k141_76898 | Low-quality |
| S3_extracted_k141_60814 | Not-determined |
| S3_extracted_k141_4254 | Not-determined |
| S3_extracted_k141_36558 | Not-determined |
| S3_extracted_k141_13825 | Not-determined |
| S3_extracted_k141_116005 | Not-determined |
| S3_extracted_k141_116006 | Not-determined |
| S3_extracted_k141_13827 | Not-determined |
| S3_extracted_k141_36563 | Not-determined |
| S3_extracted_k141_76910 | Low-quality |
| S3_extracted_k141_123523 | Low-quality |
| S3_extracted_k141_123527 | Low-quality |
| S3_extracted_k141_44350 | Not-determined |
| S3_extracted_k141_123532 | Not-determined |
| S3_extracted_k141_4270 | Low-quality |
| S3_extracted_k141_116017 | Low-quality |
| S3_extracted_k141_123537 | Low-quality |
| S3_extracted_k141_4271 | Not-determined |
| S3_extracted_k141_4272 | Not-determined |
| S3_extracted_k141_60824 | Not-determined |
| S3_extracted_k141_60826 | Not-determined |
| S3_extracted_k141_116023 | Not-determined |
| S3_extracted_k141_116024 | Not-determined |
| S3_extracted_k141_13857 | Not-determined |
| S3_extracted_k141_4280 | Not-determined |
| S3_extracted_k141_108083 | Not-determined |
| S3_extracted_k141_108085 | Low-quality |
| S3_extracted_k141_44354 | Not-determined |
| S3_extracted_k141_76931 | Low-quality |
| S3_extracted_k141_28277 | Not-determined |
| S3_extracted_k141_108087 | Not-determined |
| S3_extracted_k141_76939 | Not-determined |
| S3_extracted_k141_19519 | Low-quality |
| S3_extracted_k141_13870 | Not-determined |
| S3_extracted_k141_36577 | Not-determined |
| S3_extracted_k141_76944 | Not-determined |
| S3_extracted_k141_99696 | Low-quality |
| S3_extracted_k141_52972 | Not-determined |
| S3_extracted_k141_19526 | Not-determined |
| S3_extracted_k141_76954 | Not-determined |
| S3_extracted_k141_52981 | Not-determined |
| S3_extracted_k141_76957 | Not-determined |
| S3_extracted_k141_76965 | Not-determined |
| S3_extracted_k141_116050 | Not-determined |
| S3_extracted_k141_108098 | Not-determined |
| S3_extracted_k141_60833 | Not-determined |
| S3_extracted_k141_108103 | Not-determined |
| S3_extracted_k141_108106 | Not-determined |
| S3_extracted_k141_123544 | Not-determined |
| S3_extracted_k141_108114 | Not-determined |
| S3_extracted_k141_108115 | Not-determined |
| S3_extracted_k141_116056 | Not-determined |
| S3_extracted_k141_116058 | Not-determined |
| S3_extracted_k141_108117 | Not-determined |
| S3_extracted_k141_28292 | Low-quality |
| S3_extracted_k141_84024 | Not-determined |
| S3_extracted_k141_19536 | Not-determined |
| S3_extracted_k141_108120 | Not-determined |
| S3_extracted_k141_67578 | Low-quality |
| S3_extracted_k141_92082 | Low-quality |
| S3_extracted_k141_92083 | Not-determined |
| S3_extracted_k141_44383 | Not-determined |
| S3_extracted_k141_19539 | Not-determined |
| S3_extracted_k141_108122 | Not-determined |
| S3_extracted_k141_108123 | Low-quality |
| S3_extracted_k141_108124 | Not-determined |
| S3_extracted_k141_92087 | Not-determined |
| S3_extracted_k141_123552 | Low-quality |
| S3_extracted_k141_123553 | Low-quality |
| S3_extracted_k141_67581 | Low-quality |
| S3_extracted_k141_92088 | Low-quality |
| S3_extracted_k141_44389 | Not-determined |
| S3_extracted_k141_123555 | Not-determined |
| S3_extracted_k141_108127 | Not-determined |
| S3_extracted_k141_108128 | Not-determined |
| S3_extracted_k141_19544 | Not-determined |
| S3_extracted_k141_92095 | Not-determined |
| S3_extracted_k141_19547 | Low-quality |
| S3_extracted_k141_4287 | Not-determined |
| S3_extracted_k141_4288 | Not-determined |
| S3_extracted_k141_13915 | Not-determined |
| S3_extracted_k141_67590 | Not-determined |
| S3_extracted_k141_67592 | Not-determined |
| S3_extracted_k141_44397 | Not-determined |
| S3_extracted_k141_67593 | Not-determined |
| S3_extracted_k141_53005 | Not-determined |
| S3_extracted_k141_28306 | Not-determined |
| S3_extracted_k141_60842 | Low-quality |
| S3_extracted_k141_60845 | Not-determined |
| S3_extracted_k141_84037 | Not-determined |
| S3_extracted_k141_19556 | Low-quality |
| S3_extracted_k141_53019 | Low-quality |
| S3_extracted_k141_67596 | Low-quality |
| S3_extracted_k141_84042 | Low-quality |
| S3_extracted_k141_53021 | Not-determined |
| S3_extracted_k141_36593 | Low-quality |
| S3_extracted_k141_19562 | Low-quality |
| S3_extracted_k141_67601 | Not-determined |
| S3_extracted_k141_28309 | Not-determined |
| S3_extracted_k141_28311 | Not-determined |
| S3_extracted_k141_123565 | Not-determined |
| S3_extracted_k141_28313 | Not-determined |
| S3_extracted_k141_67603 | Low-quality |
| S3_extracted_k141_36598 | Not-determined |
| S3_extracted_k141_36602 | Not-determined |
| S3_extracted_k141_44400 | Not-determined |
| S3_extracted_k141_84049 | Low-quality |
| S3_extracted_k141_84052 | Low-quality |
| S3_extracted_k141_44403 | Not-determined |
| S3_extracted_k141_84054 | Not-determined |
| S3_extracted_k141_53038 | Not-determined |
| S3_extracted_k141_84056 | Not-determined |
| S3_extracted_k141_13936 | Not-determined |
| S3_extracted_k141_116086 | Not-determined |
| S3_extracted_k141_84059 | Not-determined |
| S3_extracted_k141_36610 | Not-determined |
| S3_extracted_k141_84060 | Not-determined |
| S3_extracted_k141_53044 | Not-determined |
| S3_extracted_k141_53048 | Not-determined |
| S3_extracted_k141_67611 | Not-determined |
| S3_extracted_k141_76980 | Low-quality |
| S3_extracted_k141_19577 | Not-determined |
| S3_extracted_k141_84066 | Not-determined |
| S3_extracted_k141_116094 | Not-determined |
| S3_extracted_k141_36618 | Not-determined |
| S3_extracted_k141_116096 | Not-determined |
| S3_extracted_k141_19583 | Not-determined |
| S3_extracted_k141_92111 | Not-determined |
| S3_extracted_k141_53053 | Not-determined |
| S3_extracted_k141_67621 | Low-quality |
| S3_extracted_k141_92112 | Low-quality |
| S3_extracted_k141_92113 | Not-determined |
| S3_extracted_k141_84076 | Not-determined |
| S3_extracted_k141_116103 | Not-determined |
| S3_extracted_k141_60861 | Not-determined |
| S3_extracted_k141_36623 | Not-determined |
| S3_extracted_k141_28320 | Not-determined |
| S3_extracted_k141_116105 | Not-determined |
| S3_extracted_k141_84081 | Low-quality |
| S3_extracted_k141_116106 | Low-quality |
| S3_extracted_k141_53062 | Not-determined |
| S3_extracted_k141_53063 | Low-quality |
| S3_extracted_k141_99705 | Not-determined |
| S3_extracted_k141_36627 | Low-quality |
| S3_extracted_k141_92122 | Not-determined |
| S3_extracted_k141_13961 | Not-determined |
| S3_extracted_k141_60872 | Not-determined |
| S3_extracted_k141_116120 | Not-determined |
| S3_extracted_k141_28326 | Not-determined |
| S3_extracted_k141_60881 | Not-determined |
| S3_extracted_k141_36637 | Not-determined |
| S3_extracted_k141_84092 | Low-quality |
| S3_extracted_k141_36641 | Not-determined |
| S3_extracted_k141_19600 | Not-determined |
| S3_extracted_k141_19601 | Not-determined |
| S3_extracted_k141_116135 | Not-determined |
| S3_extracted_k141_108156 | Not-determined |
| S3_extracted_k141_60890 | Not-determined |
| S3_extracted_k141_76996 | Not-determined |
| S3_extracted_k141_67627 | Not-determined |
| S3_extracted_k141_99709 | Not-determined |
| S3_extracted_k141_92135 | Not-determined |
| S3_extracted_k141_67631 | Not-determined |
| S3_extracted_k141_19611 | Not-determined |
| S3_extracted_k141_116142 | Not-determined |
| S3_extracted_k141_53084 | Not-determined |
| S3_extracted_k141_84102 | Not-determined |
| S3_extracted_k141_123575 | Not-determined |
| S3_extracted_k141_13985 | Not-determined |
| S3_extracted_k141_67637 | Not-determined |
| S3_extracted_k141_123580 | Not-determined |
| S3_extracted_k141_92142 | Not-determined |
| S3_extracted_k141_60891 | Not-determined |
| S3_extracted_k141_84109 | Not-determined |
| S3_extracted_k141_60892 | Not-determined |
| S3_extracted_k141_13989 | Not-determined |
| S3_extracted_k141_92144 | Not-determined |
| S3_extracted_k141_84111 | Not-determined |
| S3_extracted_k141_92147 | Not-determined |
| S3_extracted_k141_28342 | Not-determined |
| S3_extracted_k141_92149 | Not-determined |
| S3_extracted_k141_92150 | Not-determined |
| S3_extracted_k141_84115 | Not-determined |
| S3_extracted_k141_19621 | Low-quality |
| S3_extracted_k141_92155 | Not-determined |
| S3_extracted_k141_28345 | Not-determined |
| S11_vs_k141_104036\|\|full | High-quality |
| S3_extracted_k141_116155 | Not-determined |
| S3_extracted_k141_28348 | Not-determined |
| S3_extracted_k141_14002 | Not-determined |
| S3_extracted_k141_84122 | Not-determined |
| S3_extracted_k141_19627 | Low-quality |
| S3_extracted_k141_44410 | Not-determined |
| S3_extracted_k141_84126 | Not-determined |
| S3_extracted_k141_116160 | Low-quality |
| S3_extracted_k141_92163 | Not-determined |
| S3_extracted_k141_19634 | Not-determined |
| S3_extracted_k141_14010 | Not-determined |
| S3_extracted_k141_28353 | Not-determined |
| S3_extracted_k141_123584 | Not-determined |
| S3_extracted_k141_19636 | Not-determined |
| S3_extracted_k141_123587 | Not-determined |
| S3_extracted_k141_14015 | Not-determined |
| S3_extracted_k141_53103 | Not-determined |
| S3_extracted_k141_36646 | Not-determined |
| S3_extracted_k141_4337 | Not-determined |
| S3_extracted_k141_19637 | Not-determined |
| S3_extracted_k141_14018 | Low-quality |
| S3_extracted_k141_36652 | Not-determined |
| S3_extracted_k141_84137 | Low-quality |
| S3_extracted_k141_36653 | Not-determined |
| S3_extracted_k141_53108 | Not-determined |
| S3_extracted_k141_28358 | Not-determined |
| S3_extracted_k141_19643 | Not-determined |
| S3_extracted_k141_53109 | Not-determined |
| S3_extracted_k141_53110 | Not-determined |
| S3_extracted_k141_77021 | Not-determined |
| S3_extracted_k141_19646 | Low-quality |
| S3_extracted_k141_77023 | Not-determined |
| S3_extracted_k141_92174 | Not-determined |
| S3_extracted_k141_77026 | Not-determined |
| S3_extracted_k141_77030 | Not-determined |
| S3_extracted_k141_67657 | Low-quality |
| S3_extracted_k141_4355 | Not-determined |
| S3_extracted_k141_108165 | Not-determined |
| S3_extracted_k141_67664 | Not-determined |
| S3_extracted_k141_14026 | Not-determined |
| S3_extracted_k141_14029 | Low-quality |
| S3_extracted_k141_53120 | Not-determined |
| S3_extracted_k141_53121 | Not-determined |
| S3_extracted_k141_14031 | Not-determined |
| S3_extracted_k141_116187 | Low-quality |
| S3_extracted_k141_53124 | Not-determined |
| S3_extracted_k141_116188 | Not-determined |
| S3_extracted_k141_19668 | Not-determined |
| S3_extracted_k141_108170 | Not-determined |
| S3_extracted_k141_116190 | Not-determined |
| S3_extracted_k141_14037 | Not-determined |
| S3_extracted_k141_14042 | Low-quality |
| S3_extracted_k141_4359 | Not-determined |
| S3_extracted_k141_77045 | Not-determined |
| S3_extracted_k141_19678 | Not-determined |
| S3_extracted_k141_84166 | Low-quality |
| S3_extracted_k141_4363 | Not-determined |
| S3_extracted_k141_28381 | Low-quality |
| S3_extracted_k141_116201 | Low-quality |
| S3_extracted_k141_116204 | Not-determined |
| S3_extracted_k141_19683 | Low-quality |
| S3_extracted_k141_77061 | Not-determined |
| S3_extracted_k141_19686 | Not-determined |
| S3_extracted_k141_28389 | Not-determined |
| S3_extracted_k141_99732 | Not-determined |
| S3_extracted_k141_108174 | Not-determined |
| S3_extracted_k141_19688 | Low-quality |
| S3_extracted_k141_84179 | Not-determined |
| S3_extracted_k141_67675 | Not-determined |
| S3_extracted_k141_116209 | Low-quality |
| S3_extracted_k141_67676 | Not-determined |
| S3_extracted_k141_116210 | Not-determined |
| S3_extracted_k141_19693 | Not-determined |
| S3_extracted_k141_84189 | Not-determined |
| S3_extracted_k141_108189 | Not-determined |
| S3_extracted_k141_116219 | Not-determined |
| S3_extracted_k141_14060 | Not-determined |
| S3_extracted_k141_14061 | Not-determined |
| S3_extracted_k141_67689 | Not-determined |
| S3_extracted_k141_108191 | Low-quality |
| S3_extracted_k141_77081 | Not-determined |
| S3_extracted_k141_14067 | Not-determined |
| S3_extracted_k141_116226 | Not-determined |
| S3_extracted_k141_77083 | Not-determined |
| S3_extracted_k141_67692 | Not-determined |
| S3_extracted_k141_14072 | Not-determined |
| S3_extracted_k141_14074 | Low-quality |
| S3_extracted_k141_84197 | Not-determined |
| S3_extracted_k141_116235 | Not-determined |
| S3_extracted_k141_14078 | Not-determined |
| S3_extracted_k141_77086 | Low-quality |
| S3_extracted_k141_108205 | Not-determined |
| S3_extracted_k141_14083 | Not-determined |
| S3_extracted_k141_84200 | Not-determined |
| S3_extracted_k141_77093 | Low-quality |
| S3_extracted_k141_123612 | Not-determined |
| S3_extracted_k141_84203 | Not-determined |
| S3_extracted_k141_116241 | Not-determined |
| S3_extracted_k141_116242 | Not-determined |
| S3_extracted_k141_116244 | Not-determined |
| S3_extracted_k141_67702 | Not-determined |
| S3_extracted_k141_123616 | Not-determined |
| S3_extracted_k141_123617 | Not-determined |
| S3_extracted_k141_92218 | Not-determined |
| S3_extracted_k141_108222 | Not-determined |
| S3_extracted_k141_4391 | Not-determined |
| S3_extracted_k141_92225 | Not-determined |
| S3_extracted_k141_4393 | Not-determined |
| S3_extracted_k141_4394 | Not-determined |
| S3_extracted_k141_77117 | Not-determined |
| S3_extracted_k141_77123 | Not-determined |
| S3_extracted_k141_60926 | Not-determined |
| S3_extracted_k141_84214 | Not-determined |
| S3_extracted_k141_28400 | Not-determined |
| S3_extracted_k141_92234 | Not-determined |
| S3_extracted_k141_84224 | Low-quality |
| S3_extracted_k141_123631 | Not-determined |
| S3_extracted_k141_92238 | Not-determined |
| S3_extracted_k141_116279 | Not-determined |
| S3_extracted_k141_92241 | Not-determined |
| S3_extracted_k141_77134 | Not-determined |
| S12_vs_k141_66302\|\|full | High-quality |
| S3_extracted_k141_14130 | Not-determined |
| S3_extracted_k141_99735 | Low-quality |
| S3_extracted_k141_19728 | Not-determined |
| S3_extracted_k141_4403 | Low-quality |
| S3_extracted_k141_4404 | Not-determined |
| S3_extracted_k141_4406 | Not-determined |
| S3_extracted_k141_123648 | Not-determined |
| S3_extracted_k141_19734 | Not-determined |
| S3_extracted_k141_77154 | Not-determined |
| S3_extracted_k141_108236 | Not-determined |
| S3_extracted_k141_99750 | Not-determined |
| S3_extracted_k141_19745 | Not-determined |
| S3_extracted_k141_60947 | Not-determined |
| S3_extracted_k141_92263 | Not-determined |
| S3_extracted_k141_108241 | Not-determined |
| S3_extracted_k141_92273 | Not-determined |
| S3_extracted_k141_108247 | Not-determined |
| S3_extracted_k141_14147 | Not-determined |
| S3_extracted_k141_108249 | Low-quality |
| S3_extracted_k141_67731 | Not-determined |
| S3_extracted_k141_19762 | Not-determined |
| S3_extracted_k141_14153 | Not-determined |
| S3_extracted_k141_99774 | Not-determined |
| S3_extracted_k141_60966 | Not-determined |
| S3_extracted_k141_19767 | Not-determined |
| S3_extracted_k141_60967 | Low-quality |
| S3_extracted_k141_67738 | Not-determined |
| S3_extracted_k141_116281 | Not-determined |
| S3_extracted_k141_44424 | Not-determined |
| S3_extracted_k141_67742 | Not-determined |
| S3_extracted_k141_60972 | Not-determined |
| S3_extracted_k141_99785 | Not-determined |
| S3_extracted_k141_84235 | Not-determined |
| S3_extracted_k141_28404 | Not-determined |
| S3_extracted_k141_84238 | Not-determined |
| S3_extracted_k141_28407 | Not-determined |
| S3_extracted_k141_108251 | Low-quality |
| S3_extracted_k141_92306 | Not-determined |
| S3_extracted_k141_99795 | Not-determined |
| S3_extracted_k141_36683 | Not-determined |
| S3_extracted_k141_60989 | Not-determined |
| S3_extracted_k141_67749 | Low-quality |
| S3_extracted_k141_116292 | Not-determined |
| S3_extracted_k141_92312 | Not-determined |
| S3_extracted_k141_67751 | Not-determined |
| S3_extracted_k141_123657 | Not-determined |
| S3_extracted_k141_4408 | Not-determined |
| S3_extracted_k141_28419 | Not-determined |
| S3_extracted_k141_28420 | Not-determined |
| S3_extracted_k141_4411 | Not-determined |
| S3_extracted_k141_4412 | Not-determined |
| S3_extracted_k141_123658 | Not-determined |
| S3_extracted_k141_123659 | Low-quality |
| S3_extracted_k141_60995 | Not-determined |
| S3_extracted_k141_4415 | Not-determined |
| S3_extracted_k141_99805 | Not-determined |
| S3_extracted_k141_108261 | Low-quality |
| S3_extracted_k141_77170 | Not-determined |
| S3_extracted_k141_99809 | Not-determined |
| S3_extracted_k141_77176 | Not-determined |
| S3_extracted_k141_36690 | Not-determined |
| S3_extracted_k141_108270 | Not-determined |
| S3_extracted_k141_108272 | Not-determined |
| S3_extracted_k141_67771 | Not-determined |
| S3_extracted_k141_116300 | Not-determined |
| S3_extracted_k141_99818 | Not-determined |
| S3_extracted_k141_84267 | Not-determined |
| S3_extracted_k141_92331 | Low-quality |
| S3_extracted_k141_99820 | Not-determined |
| S3_extracted_k141_99821 | Not-determined |
| S3_extracted_k141_67772 | Not-determined |
| S3_extracted_k141_61009 | Not-determined |
| S3_extracted_k141_99824 | Not-determined |
| S3_extracted_k141_92336 | Not-determined |
| S3_extracted_k141_108290 | Not-determined |
| S3_extracted_k141_84281 | Low-quality |
| S3_extracted_k141_44431 | Not-determined |
| S3_extracted_k141_44432 | Not-determined |
| S3_extracted_k141_77191 | Not-determined |
| S3_extracted_k141_28439 | Not-determined |
| S3_extracted_k141_67783 | Not-determined |
| S3_extracted_k141_67784 | Not-determined |
| S3_extracted_k141_99834 | Not-determined |
| S3_extracted_k141_77197 | Not-determined |
| S3_extracted_k141_123679 | Not-determined |
| S3_extracted_k141_61020 | Not-determined |
| S3_extracted_k141_4428 | Not-determined |
| S3_extracted_k141_92340 | Not-determined |
| S3_extracted_k141_123682 | Not-determined |
| S3_extracted_k141_61026 | Not-determined |
| S3_extracted_k141_44443 | Not-determined |
| S3_extracted_k141_67786 | Not-determined |
| S3_extracted_k141_92348 | Not-determined |
| S3_extracted_k141_61030 | Low-quality |
| S3_extracted_k141_77204 | Not-determined |
| S3_extracted_k141_67792 | Not-determined |
| S3_extracted_k141_92350 | Not-determined |
| S3_extracted_k141_67796 | Not-determined |
| S3_extracted_k141_67797 | Not-determined |
| S3_extracted_k141_116313 | Not-determined |
| S3_extracted_k141_61040 | Not-determined |
| S3_extracted_k141_28455 | Low-quality |
| S3_extracted_k141_44445 | Not-determined |
| S3_extracted_k141_61045 | Not-determined |
| S3_extracted_k141_116316 | Not-determined |
| S3_extracted_k141_77211 | Not-determined |
| S3_extracted_k141_77212 | Not-determined |
| S3_extracted_k141_77215 | Not-determined |
| S3_extracted_k141_28457 | Not-determined |
| S3_extracted_k141_116323 | Not-determined |
| S3_extracted_k141_61051 | Not-determined |
| S3_extracted_k141_14175 | Low-quality |
| S3_extracted_k141_92361 | Not-determined |
| S3_extracted_k141_14181 | Not-determined |
| S3_extracted_k141_77224 | Not-determined |
| S3_extracted_k141_14186 | Not-determined |
| S3_extracted_k141_77227 | Not-determined |
| S3_extracted_k141_92370 | Low-quality |
| S3_extracted_k141_92371 | Not-determined |
| S3_extracted_k141_28466 | Not-determined |
| S3_extracted_k141_14188 | Not-determined |
| S3_extracted_k141_77230 | Not-determined |
| S3_extracted_k141_77234 | Not-determined |
| S3_extracted_k141_28485 | Not-determined |
| S3_extracted_k141_14196 | Low-quality |
| S3_extracted_k141_4440 | Not-determined |
| S3_extracted_k141_14202 | Not-determined |
| S3_extracted_k141_61062 | Not-determined |
| S3_extracted_k141_4444 | Not-determined |
| S3_extracted_k141_84313 | Not-determined |
| S3_extracted_k141_84316 | Not-determined |
| S3_extracted_k141_4449 | Not-determined |
| S3_extracted_k141_61074 | Not-determined |
| S3_extracted_k141_84319 | Not-determined |
| S3_extracted_k141_84320 | Not-determined |
| S3_extracted_k141_61076 | Not-determined |
| S3_extracted_k141_92378 | Not-determined |
| S3_extracted_k141_108311 | Not-determined |
| S3_extracted_k141_28500 | Low-quality |
| S3_extracted_k141_19774 | Not-determined |
| S3_extracted_k141_28505 | Not-determined |
| S3_extracted_k141_19778 | Not-determined |
| S3_extracted_k141_108325 | Low-quality |
| S3_extracted_k141_84325 | Low-quality |
| S3_extracted_k141_61099 | Low-quality |
| S3_extracted_k141_61100 | Not-determined |
| S3_extracted_k141_61101 | Not-determined |
| S3_extracted_k141_14213 | Not-determined |
| S3_extracted_k141_19782 | Low-quality |
| S3_extracted_k141_14216 | Not-determined |
| S3_extracted_k141_14219 | Low-quality |
| S3_extracted_k141_61104 | Not-determined |
| S3_extracted_k141_84333 | Not-determined |
| S3_extracted_k141_84334 | Not-determined |
| S3_extracted_k141_77242 | Not-determined |
| S3_extracted_k141_14221 | Low-quality |
| S3_extracted_k141_77245 | Not-determined |
| S3_extracted_k141_61108 | Not-determined |
| S3_extracted_k141_67821 | Not-determined |
| S3_extracted_k141_84339 | Not-determined |
| S3_extracted_k141_92391 | Not-determined |
| S3_extracted_k141_19792 | Not-determined |
| S3_extracted_k141_4458 | Not-determined |
| S3_extracted_k141_4463 | Not-determined |
| S3_extracted_k141_77267 | Not-determined |
| S3_extracted_k141_61114 | Low-quality |
| S3_extracted_k141_92405 | Not-determined |
| S3_extracted_k141_4465 | Not-determined |
| S3_extracted_k141_77281 | Not-determined |
| S3_extracted_k141_28514 | Low-quality |
| S3_extracted_k141_4473 | Not-determined |
| S3_extracted_k141_92412 | Not-determined |
| S4_extracted_15376 | Medium-quality |
| S3_extracted_k141_67830 | Not-determined |
| S3_extracted_k141_4476 | Not-determined |
| S3_extracted_k141_77284 | Low-quality |
| S3_extracted_k141_108344 | Not-determined |
| S3_extracted_k141_67832 | Not-determined |
| S3_extracted_k141_14238 | Not-determined |
| S3_extracted_k141_67833 | Low-quality |
| S3_extracted_k141_67834 | Not-determined |
| S3_extracted_k141_61134 | Low-quality |
| S3_extracted_k141_61135 | Not-determined |
| S3_extracted_k141_14241 | Not-determined |
| S3_extracted_k141_28525 | Not-determined |
| S3_extracted_k141_61140 | Not-determined |
| S3_extracted_k141_92419 | Not-determined |
| S3_extracted_k141_108357 | Not-determined |
| S3_extracted_k141_28536 | Low-quality |
| S3_extracted_k141_67842 | Low-quality |
| S3_extracted_k141_67843 | Not-determined |
| S3_extracted_k141_61153 | Not-determined |
| S3_extracted_k141_28541 | Not-determined |
| S3_extracted_k141_28542 | Low-quality |
| S3_extracted_k141_61158 | Not-determined |
| S3_extracted_k141_28549 | Not-determined |
| S3_extracted_k141_14267 | Not-determined |
| S3_extracted_k141_14273 | Not-determined |
| S3_extracted_k141_61167 | Not-determined |
| S3_extracted_k141_19820 | Not-determined |
| S3_extracted_k141_108362 | Low-quality |
| S3_extracted_k141_44466 | Not-determined |
| S3_extracted_k141_36693 | Not-determined |
| S3_extracted_k141_61183 | Not-determined |
| S3_extracted_k141_67863 | Low-quality |
| S3_extracted_k141_36696 | Not-determined |
| S3_extracted_k141_44470 | Not-determined |
| S3_extracted_k141_28580 | Low-quality |
| S3_extracted_k141_77289 | Not-determined |
| S3_extracted_k141_99857 | Not-determined |
| S3_extracted_k141_61191 | Not-determined |
| S3_extracted_k141_53143 | Not-determined |
| S3_extracted_k141_77294 | Low-quality |
| S3_extracted_k141_61199 | Not-determined |
| S3_extracted_k141_28591 | Not-determined |
| S3_extracted_k141_108367 | Not-determined |
| S3_extracted_k141_84385 | Not-determined |
| S3_extracted_k141_108368 | Not-determined |
| S3_extracted_k141_53151 | Not-determined |
| S3_extracted_k141_108369 | Not-determined |

| | |
|---|---|
| S3_extracted_k141_92448 | Not-determined |
| S3_extracted_k141_36713 | Low-quality |
| S3_extracted_k141_36715 | Not-determined |
| S3_extracted_k141_44486 | Not-determined |
| S3_extracted_k141_99866 | Not-determined |
| S3_extracted_k141_19843 | Not-determined |
| S3_extracted_k141_53157 | Not-determined |
| S3_extracted_k141_19849 | Not-determined |
| S3_extracted_k141_14292 | Not-determined |
| S3_extracted_k141_53168 | Not-determined |
| S3_extracted_k141_53169 | Not-determined |
| S3_extracted_k141_44497 | Low-quality |
| S3_extracted_k141_36729 | Not-determined |
| S3_extracted_k141_36734 | Not-determined |
| S3_extracted_k141_53178 | Low-quality |
| S3_extracted_k141_36738 | Not-determined |
| S3_extracted_k141_19858 | Not-determined |
| S3_extracted_k141_36739 | Not-determined |
| S3_extracted_k141_36741 | Not-determined |
| S3_extracted_k141_84389 | Not-determined |
| S3_extracted_k141_28616 | Not-determined |
| S3_extracted_k141_28617 | Not-determined |
| S3_extracted_k141_84390 | Low-quality |
| S3_extracted_k141_28619 | Not-determined |
| S3_extracted_k141_123726 | Low-quality |
| S3_extracted_k141_28623 | Not-determined |
| S3_extracted_k141_99881 | Low-quality |
| S3_extracted_k141_28625 | Not-determined |
| S3_extracted_k141_4497 | Not-determined |
| S3_extracted_k141_108377 | Not-determined |
| S3_extracted_k141_123731 | Not-determined |
| S3_extracted_k141_19874 | Not-determined |
| S3_extracted_k141_19877 | Not-determined |
| S3_extracted_k141_123735 | Not-determined |
| S3_extracted_k141_99895 | Not-determined |
| S3_extracted_k141_19881 | Not-determined |
| S3_extracted_k141_108383 | Not-determined |
| S3_extracted_k141_28636 | Not-determined |
| S3_extracted_k141_28637 | Not-determined |
| S3_extracted_k141_123742 | Not-determined |
| S3_extracted_k141_92464 | Low-quality |
| S3_extracted_k141_53194 | Not-determined |
| S3_extracted_k141_28642 | Not-determined |
| S3_extracted_k141_53197 | Not-determined |
| S3_extracted_k141_19888 | Not-determined |
| S3_extracted_k141_61239 | Not-determined |
| S3_extracted_k141_99908 | Not-determined |
| S3_extracted_k141_99915 | Low-quality |
| S3_extracted_k141_61247 | Not-determined |
| S3_extracted_k141_28654 | Low-quality |
| S3_extracted_k141_19899 | Not-determined |
| S3_extracted_k141_19903 | Not-determined |
| S3_extracted_k141_99919 | Not-determined |
| S3_extracted_k141_61255 | Not-determined |
| S3_extracted_k141_4509 | Not-determined |
| S3_extracted_k141_116346 | Low-quality |
| S3_extracted_k141_116347 | Not-determined |
| S3_extracted_k141_92481 | Low-quality |
| S3_extracted_k141_77305 | Not-determined |
| S3_extracted_k141_77306 | Not-determined |
| S3_extracted_k141_53208 | Low-quality |
| S3_extracted_k141_61273 | Not-determined |
| S3_extracted_k141_53210 | Not-determined |
| S3_extracted_k141_61277 | Not-determined |
| S3_extracted_k141_92484 | Not-determined |
| S3_extracted_k141_84456 | Not-determined |
| S3_extracted_k141_61278 | Not-determined |
| S3_extracted_k141_92487 | Not-determined |
| S3_extracted_k141_19918 | Not-determined |
| S3_extracted_k141_4527 | Low-quality |
| S3_extracted_k141_53225 | Low-quality |
| S3_extracted_k141_19925 | Not-determined |
| S3_extracted_k141_4537 | Not-determined |
| S3_extracted_k141_92501 | Not-determined |
| S3_extracted_k141_92502 | Not-determined |
| S3_extracted_k141_53235 | Low-quality |
| S3_extracted_k141_99941 | Not-determined |
| S3_extracted_k141_84480 | Low-quality |
| S3_extracted_k141_99942 | Not-determined |
| S3_extracted_k141_4544 | Not-determined |
| S3_extracted_k141_99944 | Not-determined |
| S3_extracted_k141_92509 | Not-determined |
| S3_extracted_k141_61294 | Not-determined |
| S3_extracted_k141_4545 | Not-determined |
| S3_extracted_k141_19938 | Not-determined |
| S3_extracted_k141_4547 | Not-determined |
| S3_extracted_k141_44521 | Not-determined |
| S3_extracted_k141_61299 | Not-determined |
| S3_extracted_k141_61302 | Low-quality |
| S3_extracted_k141_123768 | Low-quality |
| S3_extracted_k141_92512 | Not-determined |
| S3_extracted_k141_99961 | Not-determined |
| S3_extracted_k141_4560 | Low-quality |
| S3_extracted_k141_61317 | Not-determined |
| S3_extracted_k141_4564 | Low-quality |
| S3_extracted_k141_108403 | Not-determined |
| S3_extracted_k141_108410 | Not-determined |
| S3_extracted_k141_116361 | Low-quality |
| S3_extracted_k141_14336 | Not-determined |
| S3_extracted_k141_108412 | Low-quality |
| S3_extracted_k141_4574 | Not-determined |
| S3_extracted_k141_77322 | Not-determined |
| S3_extracted_k141_61332 | Not-determined |
| S3_extracted_k141_116362 | Not-determined |
| S3_extracted_k141_61333 | Not-determined |
| S3_extracted_k141_19949 | Low-quality |
| S3_extracted_k141_53264 | Not-determined |
| S3_extracted_k141_77331 | Not-determined |
| S3_extracted_k141_116377 | Not-determined |
| S3_extracted_k141_4585 | Not-determined |
| S3_extracted_k141_4586 | Not-determined |
| S3_extracted_k141_108416 | Not-determined |
| S3_extracted_k141_108419 | Not-determined |
| S3_extracted_k141_61345 | Low-quality |
| S3_extracted_k141_61349 | Not-determined |
| S3_extracted_k141_92517 | Not-determined |
| S3_extracted_k141_61351 | Not-determined |
| S3_extracted_k141_28663 | Low-quality |
| S3_extracted_k141_77346 | Not-determined |
| S3_extracted_k141_28667 | Not-determined |
| S3_extracted_k141_14361 | Not-determined |
| S3_extracted_k141_77351 | Not-determined |
| S3_extracted_k141_14362 | Not-determined |
| S3_extracted_k141_19962 | Low-quality |
| S3_extracted_k141_84502 | Not-determined |
| S3_extracted_k141_84503 | Not-determined |
| S3_extracted_k141_44543 | Not-determined |
| S3_extracted_k141_84508 | Not-determined |
| S3_extracted_k141_99988 | Not-determined |
| S3_extracted_k141_36751 | Not-determined |
| S3_extracted_k141_84510 | Not-determined |
| S3_extracted_k141_77364 | Not-determined |
| S3_extracted_k141_99990 | Not-determined |
| S3_extracted_k141_84516 | Not-determined |
| S3_extracted_k141_77365 | Not-determined |
| S3_extracted_k141_67894 | Not-determined |
| S3_extracted_k141_116386 | Not-determined |
| S3_extracted_k141_99992 | Not-determined |
| S3_extracted_k141_116388 | Not-determined |
| S3_extracted_k141_99994 | Not-determined |
| S3_extracted_k141_36761 | Not-determined |
| S3_extracted_k141_116392 | Not-determined |
| S3_extracted_k141_14378 | Not-determined |
| S3_extracted_k141_19974 | Low-quality |
| S3_extracted_k141_36768 | Not-determined |
| S3_extracted_k141_77375 | Not-determined |
| S3_extracted_k141_67903 | Not-determined |
| S3_extracted_k141_36776 | Not-determined |
| S3_extracted_k141_67904 | Low-quality |
| S3_extracted_k141_67906 | Not-determined |
| S3_extracted_k141_36782 | Not-determined |
| S3_extracted_k141_61380 | Not-determined |
| S3_extracted_k141_99999 | Not-determined |
| S3_extracted_k141_61382 | Not-determined |
| S3_extracted_k141_44555 | Not-determined |
| S3_extracted_k141_67909 | Not-determined |
| S3_extracted_k141_108434 | Low-quality |
| S3_extracted_k141_108435 | Not-determined |
| S3_extracted_k141_14385 | Low-quality |
| S3_extracted_k141_108444 | Not-determined |
| S3_extracted_k141_19987 | Not-determined |
| S3_extracted_k141_67920 | Not-determined |
| S3_extracted_k141_61397 | Not-determined |
| S3_extracted_k141_28687 | Not-determined |
| S3_extracted_k141_108452 | Low-quality |
| S3_extracted_k141_19990 | Not-determined |
| S3_extracted_k141_108456 | Not-determined |
| S3_extracted_k141_123797 | Not-determined |
| S3_extracted_k141_84520 | Not-determined |
| S3_extracted_k141_61407 | Not-determined |
| S3_extracted_k141_116394 | Not-determined |
| S3_extracted_k141_19993 | Not-determined |
| S3_extracted_k141_67926 | Not-determined |
| S3_extracted_k141_4592 | Not-determined |
| S3_extracted_k141_116396 | Not-determined |
| S3_extracted_k141_92531 | Low-quality |
| S3_extracted_k141_123799 | Not-determined |
| S3_extracted_k141_44565 | Low-quality |
| S3_extracted_k141_116400 | Not-determined |
| S3_extracted_k141_67933 | Not-determined |
| S3_extracted_k141_44572 | Not-determined |
| S3_extracted_k141_67940 | Not-determined |
| S3_extracted_k141_84522 | Low-quality |
| S3_extracted_k141_108459 | Not-determined |
| S3_extracted_k141_92538 | Low-quality |
| S3_extracted_k141_61424 | Not-determined |
| S3_extracted_k141_4598 | Not-determined |
| S3_extracted_k141_123809 | Not-determined |
| S3_extracted_k141_116418 | Not-determined |
| S3_extracted_k141_84541 | Not-determined |
| S3_extracted_k141_116419 | Not-determined |
| S3_extracted_k141_100008 | Not-determined |
| S3_extracted_k141_84544 | Not-determined |
| S3_extracted_k141_100009 | Not-determined |
| S3_extracted_k141_84545 | Low-quality |
| S3_extracted_k141_36800 | Not-determined |
| S3_extracted_k141_108470 | Not-determined |
| S3_extracted_k141_84548 | Not-determined |
| S3_extracted_k141_84549 | Not-determined |
| S3_extracted_k141_4618 | Not-determined |
| S3_extracted_k141_36808 | Not-determined |
| S3_extracted_k141_116428 | Not-determined |
| S3_extracted_k141_84559 | Not-determined |
| S3_extracted_k141_61455 | Low-quality |
| S3_extracted_k141_123827 | Not-determined |
| S3_extracted_k141_123828 | Low-quality |
| S3_extracted_k141_123832 | Not-determined |
| S3_extracted_k141_20024 | Low-quality |
| S3_extracted_k141_77410 | Not-determined |
| S3_extracted_k141_36821 | Not-determined |
| S3_extracted_k141_116439 | Not-determined |
| S3_extracted_k141_84564 | Not-determined |
| S3_extracted_k141_77417 | Not-determined |
| S3_extracted_k141_116442 | Not-determined |
| S3_extracted_k141_61458 | Low-quality |
| S3_extracted_k141_116444 | Not-determined |
| S3_extracted_k141_44597 | Not-determined |
| S3_extracted_k141_36831 | Not-determined |
| S3_extracted_k141_28699 | Not-determined |
| S3_extracted_k141_36835 | Not-determined |
| S3_extracted_k141_14391 | Low-quality |
| S3_extracted_k141_44603 | Not-determined |
| S3_extracted_k141_116448 | Not-determined |
| S3_extracted_k141_77426 | Not-determined |
| S3_extracted_k141_77428 | Low-quality |
| S3_extracted_k141_77429 | Not-determined |
| S3_extracted_k141_61466 | Not-determined |
| S3_extracted_k141_36845 | Not-determined |
| S3_extracted_k141_14399 | Not-determined |
| S3_extracted_k141_61471 | Not-determined |
| S3_extracted_k141_116458 | Not-determined |
| S3_extracted_k141_77438 | Not-determined |
| S3_extracted_k141_77439 | Not-determined |
| S3_extracted_k141_116461 | Not-determined |
| S3_extracted_k141_92554 | Not-determined |
| S3_extracted_k141_67955 | Low-quality |
| S3_extracted_k141_77448 | Not-determined |
| S3_extracted_k141_77449 | Not-determined |
| S3_extracted_k141_77450 | Not-determined |
| S3_extracted_k141_53273 | Not-determined |
| S3_extracted_k141_116468 | Not-determined |
| S3_extracted_k141_20053 | Not-determined |
| S3_extracted_k141_116471 | Not-determined |
| S3_extracted_k141_67958 | Not-determined |
| S3_extracted_k141_61489 | Not-determined |
| S3_extracted_k141_20060 | Not-determined |
| S3_extracted_k141_53283 | Not-determined |
| S3_extracted_k141_84585 | Not-determined |
| S3_extracted_k141_61492 | Not-determined |
| S3_extracted_k141_53285 | Not-determined |
| S3_extracted_k141_77465 | Not-determined |
| S3_extracted_k141_53295 | Not-determined |
| S3_extracted_k141_61496 | Low-quality |
| S3_extracted_k141_53296 | Not-determined |
| S3_extracted_k141_44616 | Not-determined |
| S3_extracted_k141_84594 | Not-determined |
| S3_extracted_k141_84597 | Not-determined |
| S3_extracted_k141_44617 | Low-quality |
| S3_extracted_k141_14429 | Not-determined |
| S3_extracted_k141_20071 | Not-determined |
| S3_extracted_k141_20072 | Not-determined |
| S3_extracted_k141_4622 | Low-quality |
| S3_extracted_k141_108487 | Not-determined |
| S3_extracted_k141_28704 | Not-determined |
| S3_extracted_k141_4628 | Not-determined |
| S3_extracted_k141_20079 | Not-determined |
| S3_extracted_k141_36886 | Not-determined |
| S3_extracted_k141_44637 | Not-determined |
| S3_extracted_k141_4636 | Low-quality |
| S3_extracted_k141_36899 | Not-determined |
| S3_extracted_k141_36901 | Not-determined |
| S3_extracted_k141_77483 | Not-determined |
| S3_extracted_k141_20086 | Not-determined |
| S3_extracted_k141_84602 | Not-determined |
| S3_extracted_k141_77486 | Not-determined |
| S3_extracted_k141_77489 | Not-determined |
| S3_extracted_k141_123846 | Low-quality |
| S3_extracted_k141_36907 | Not-determined |
| S3_extracted_k141_36910 | Not-determined |
| S3_extracted_k141_36911 | Not-determined |
| S3_extracted_k141_123850 | Low-quality |
| S3_extracted_k141_14444 | Not-determined |
| S3_extracted_k141_77495 | Not-determined |
| S3_extracted_k141_77496 | Low-quality |
| S3_extracted_k141_84613 | Low-quality |
| S3_extracted_k141_84614 | Not-determined |
| S3_extracted_k141_36917 | Low-quality |
| S3_extracted_k141_100026 | Not-determined |
| S3_extracted_k141_14453 | Not-determined |
| S3_extracted_k141_108490 | Low-quality |
| S3_extracted_k141_123861 | Not-determined |
| S3_extracted_k141_14456 | Low-quality |
| S3_extracted_k141_14458 | Not-determined |
| S3_extracted_k141_123864 | Low-quality |
| S3_extracted_k141_123865 | Not-determined |
| S3_extracted_k141_14465 | Not-determined |
| S3_extracted_k141_14467 | Not-determined |
| S3_extracted_k141_123870 | Low-quality |
| S3_extracted_k141_84621 | Low-quality |
| S3_extracted_k141_14471 | Not-determined |
| S3_extracted_k141_53310 | Not-determined |
| S3_extracted_k141_53311 | Low-quality |
| S3_extracted_k141_84632 | Not-determined |
| S3_extracted_k141_61517 | Not-determined |
| S3_extracted_k141_123878 | Not-determined |
| S3_extracted_k141_53322 | Not-determined |
| S3_extracted_k141_123880 | Not-determined |
| S3_extracted_k141_123883 | Not-determined |
| S3_extracted_k141_20107 | Not-determined |
| S3_extracted_k141_123885 | Not-determined |
| S3_extracted_k141_84638 | Not-determined |
| S3_extracted_k141_123886 | Not-determined |
| S3_extracted_k141_84648 | Not-determined |
| S3_extracted_k141_123888 | Not-determined |
| S3_extracted_k141_36932 | Not-determined |
| S3_extracted_k141_84653 | Not-determined |
| S3_extracted_k141_36936 | Not-determined |
| S3_extracted_k141_20116 | Not-determined |
| S3_extracted_k141_84656 | Not-determined |
| S3_extracted_k141_20117 | Low-quality |
| S3_extracted_k141_20118 | Not-determined |
| S3_extracted_k141_53325 | Low-quality |
| S3_extracted_k141_84659 | Not-determined |
| S3_extracted_k141_84660 | Not-determined |
| S3_extracted_k141_123890 | Not-determined |
| S3_extracted_k141_53332 | Not-determined |
| S3_extracted_k141_4646 | Not-determined |
| S3_extracted_k141_14473 | Low-quality |
| S3_extracted_k141_4649 | Low-quality |
| S3_extracted_k141_4653 | Not-determined |
| S3_extracted_k141_116478 | Not-determined |
| S3_extracted_k141_20131 | Not-determined |
| S3_extracted_k141_4659 | Low-quality |
| S3_extracted_k141_44669 | Not-determined |
| S3_extracted_k141_116484 | Low-quality |
| S3_extracted_k141_20140 | Not-determined |
| S3_extracted_k141_123902 | Low-quality |
| S3_extracted_k141_53354 | Not-determined |
| S3_extracted_k141_116488 | Not-determined |
| S3_extracted_k141_20145 | Low-quality |
| S3_extracted_k141_77527 | Not-determined |
| S3_extracted_k141_44676 | Not-determined |
| S3_extracted_k141_36950 | Low-quality |
| S3_extracted_k141_77528 | Low-quality |
| S3_extracted_k141_14489 | Not-determined |
| S3_extracted_k141_123913 | Low-quality |
| S3_extracted_k141_116490 | Not-determined |
| S3_extracted_k141_77536 | Not-determined |
| S3_extracted_k141_92584 | Not-determined |
| S3_extracted_k141_61540 | Not-determined |
| S3_extracted_k141_44691 | Not-determined |
| S3_extracted_k141_28718 | Not-determined |
| S3_extracted_k141_28719 | Not-determined |
| S3_extracted_k141_77539 | Not-determined |
| S3_extracted_k141_77540 | Not-determined |
| S3_extracted_k141_28722 | Not-determined |
| S3_extracted_k141_53357 | Not-determined |
| S3_extracted_k141_108519 | Not-determined |
| S3_extracted_k141_61548 | Low-quality |
| S3_extracted_k141_108521 | Low-quality |
| S3_extracted_k141_36965 | Not-determined |
| S3_extracted_k141_116512 | Not-determined |
| S3_extracted_k141_44702 | Not-determined |
| S3_extracted_k141_20161 | Not-determined |
| S3_extracted_k141_84672 | Not-determined |
| S3_extracted_k141_84673 | Not-determined |
| S3_extracted_k141_20170 | Not-determined |
| S3_extracted_k141_20172 | Not-determined |
| S3_extracted_k141_36973 | Not-determined |
| S3_extracted_k141_53373 | Low-quality |
| S3_extracted_k141_53375 | Not-determined |
| S3_extracted_k141_44715 | Not-determined |
| S3_extracted_k141_36976 | Not-determined |
| S3_extracted_k141_28731 | Not-determined |
| S3_extracted_k141_14497 | Not-determined |
| S3_extracted_k141_20176 | Not-determined |
| S3_extracted_k141_84680 | Not-determined |
| S3_extracted_k141_77562 | Not-determined |
| S3_extracted_k141_84684 | Not-determined |
| S3_extracted_k141_77564 | Not-determined |
| S3_extracted_k141_44720 | Not-determined |
| S3_extracted_k141_36980 | Not-determined |
| S3_extracted_k141_53386 | Not-determined |
| S3_extracted_k141_77567 | Not-determined |
| S3_extracted_k141_36982 | Not-determined |
| S3_extracted_k141_123929 | Not-determined |
| S3_extracted_k141_84690 | Not-determined |
| S3_extracted_k141_53390 | Not-determined |
| S3_extracted_k141_123930 | Not-determined |
| S3_extracted_k141_92599 | Low-quality |
| S3_extracted_k141_77578 | Not-determined |
| S3_extracted_k141_123937 | Not-determined |
| S3_extracted_k141_53399 | Not-determined |
| S3_extracted_k141_123942 | Not-determined |
| S3_extracted_k141_123943 | Not-determined |
| S3_extracted_k141_20198 | Not-determined |
| S3_extracted_k141_77585 | Not-determined |
| S3_extracted_k141_14510 | Not-determined |
| S3_extracted_k141_36998 | Not-determined |
| S3_extracted_k141_36999 | Not-determined |
| S3_extracted_k141_28744 | Not-determined |
| S3_extracted_k141_28746 | Not-determined |
| S3_extracted_k141_108536 | Not-determined |
| S3_extracted_k141_53402 | Not-determined |
| S3_extracted_k141_92615 | Not-determined |
| S3_extracted_k141_77597 | Low-quality |
| S3_extracted_k141_20202 | Low-quality |
| S3_extracted_k141_77598 | Not-determined |
| S3_extracted_k141_14529 | Not-determined |
| S3_extracted_k141_28753 | Not-determined |
| S3_extracted_k141_61564 | Not-determined |
| S3_extracted_k141_92621 | Low-quality |
| S3_extracted_k141_61566 | Not-determined |
| S3_extracted_k141_53410 | Not-determined |
| S3_extracted_k141_92632 | Not-determined |
| S3_extracted_k141_108546 | Not-determined |
| S3_extracted_k141_92634 | Not-determined |
| S3_extracted_k141_92636 | Not-determined |
| S3_extracted_k141_28754 | Not-determined |
| S3_extracted_k141_116532 | Low-quality |
| S3_extracted_k141_67988 | Low-quality |
| S3_extracted_k141_28758 | Low-quality |
| S3_extracted_k141_44767 | Not-determined |
| S3_extracted_k141_37010 | Low-quality |
| S3_extracted_k141_37011 | Not-determined |
| S3_extracted_k141_67992 | Not-determined |
| S3_extracted_k141_116537 | Low-quality |
| S3_extracted_k141_77615 | Not-determined |
| S3_extracted_k141_37014 | Not-determined |
| S3_extracted_k141_44771 | Not-determined |
| S3_extracted_k141_123964 | Low-quality |
| S3_extracted_k141_67998 | Low-quality |
| S3_extracted_k141_116544 | Not-determined |
| S3_extracted_k141_116545 | Not-determined |
| S3_extracted_k141_77623 | Not-determined |
| S3_extracted_k141_53433 | Low-quality |
| S3_extracted_k141_20214 | Not-determined |
| S3_extracted_k141_53435 | Low-quality |
| S3_extracted_k141_53436 | Not-determined |
| S3_extracted_k141_53439 | Not-determined |
| S3_extracted_k141_20220 | Not-determined |
| S3_extracted_k141_116549 | Not-determined |
| S3_extracted_k141_20224 | Not-determined |
| S3_extracted_k141_68009 | Not-determined |
| S3_extracted_k141_68010 | Not-determined |
| S3_extracted_k141_20227 | Not-determined |
| S3_extracted_k141_84701 | Not-determined |
| S3_extracted_k141_77634 | Not-determined |
| S3_extracted_k141_20231 | Not-determined |
| S3_extracted_k141_100048 | Not-determined |
| S3_extracted_k141_116558 | Low-quality |
| S3_extracted_k141_77638 | Not-determined |
| S3_extracted_k141_68015 | Not-determined |
| S3_extracted_k141_20233 | Not-determined |
| S3_extracted_k141_53446 | Not-determined |
| S3_extracted_k141_53447 | Not-determined |
| S3_extracted_k141_92664 | Not-determined |
| S3_extracted_k141_100058 | Not-determined |
| S3_extracted_k141_20242 | Low-quality |
| S3_extracted_k141_53453 | Not-determined |
| S3_extracted_k141_84722 | Not-determined |
| S3_extracted_k141_100063 | Not-determined |
| S3_extracted_k141_92673 | Not-determined |
| S3_extracted_k141_20248 | Not-determined |
| S3_extracted_k141_100068 | Not-determined |
| S3_extracted_k141_100071 | Not-determined |
| S3_extracted_k141_123978 | Not-determined |
| S3_extracted_k141_20264 | Not-determined |
| S3_extracted_k141_123980 | Not-determined |
| S3_extracted_k141_53462 | Not-determined |
| S3_extracted_k141_84737 | Not-determined |
| S3_extracted_k141_100076 | Not-determined |
| S3_extracted_k141_100077 | Not-determined |
| S3_extracted_k141_108555 | Not-determined |
| S3_extracted_k141_84742 | Low-quality |
| S3_extracted_k141_100080 | Not-determined |
| S3_extracted_k141_44803 | Not-determined |
| S3_extracted_k141_92684 | Low-quality |
| S3_extracted_k141_116579 | Not-determined |
| S3_extracted_k141_61586 | Not-determined |
| S3_extracted_k141_44805 | Not-determined |
| S3_extracted_k141_108562 | Not-determined |
| S3_extracted_k141_53473 | Low-quality |
| S3_extracted_k141_28767 | Not-determined |
| S3_extracted_k141_53475 | Not-determined |
| S3_extracted_k141_61590 | Not-determined |
| S3_extracted_k141_116587 | Not-determined |
| S3_extracted_k141_20285 | Not-determined |
| S3_extracted_k141_84756 | Low-quality |
| S3_extracted_k141_44806 | Low-quality |
| S3_extracted_k141_53485 | Not-determined |
| S3_extracted_k141_53487 | Not-determined |
| S3_extracted_k141_44813 | Not-determined |
| S3_extracted_k141_61597 | Not-determined |
| S3_extracted_k141_53492 | Not-determined |
| S3_extracted_k141_77675 | Not-determined |
| S3_extracted_k141_92696 | Not-determined |
| S3_extracted_k141_84766 | Low-quality |
| S3_extracted_k141_116601 | Not-determined |
| S3_extracted_k141_53496 | Not-determined |
| S3_extracted_k141_84772 | Not-determined |
| S3_extracted_k141_92700 | Low-quality |
| S3_extracted_k141_14546 | Low-quality |
| S3_extracted_k141_53501 | Not-determined |
| S3_extracted_k141_92703 | Low-quality |
| S3_extracted_k141_44830 | Not-determined |
| S3_extracted_k141_53503 | Not-determined |
| S3_extracted_k141_44832 | Not-determined |
| S3_extracted_k141_116610 | Not-determined |
| S3_extracted_k141_116611 | Not-determined |
| S3_extracted_k141_61613 | Not-determined |
| S3_extracted_k141_92713 | Not-determined |
| S3_extracted_k141_100111 | Low-quality |
| S3_extracted_k141_124004 | Not-determined |
| S3_extracted_k141_92716 | Not-determined |
| S3_extracted_k141_100116 | Not-determined |
| S3_extracted_k141_100118 | Not-determined |
| S3_extracted_k141_44847 | Not-determined |
| S3_extracted_k141_108579 | Not-determined |
| S3_extracted_k141_84793 | Not-determined |
| S3_extracted_k141_108587 | Not-determined |
| S3_extracted_k141_100129 | Low-quality |
| S3_extracted_k141_44860 | Not-determined |
| S3_extracted_k141_108594 | Not-determined |
| S3_extracted_k141_53512 | Not-determined |
| S3_extracted_k141_84800 | Low-quality |
| S3_extracted_k141_124012 | Not-determined |
| S3_extracted_k141_44865 | Not-determined |
| S3_extracted_k141_84804 | Not-determined |
| S3_extracted_k141_84805 | Not-determined |
| S3_extracted_k141_84807 | Low-quality |
| S3_extracted_k141_53521 | Not-determined |
| S3_extracted_k141_53526 | Not-determined |
| S3_extracted_k141_92736 | Not-determined |
| S3_extracted_k141_14571 | Not-determined |
| S3_extracted_k141_84813 | Not-determined |
| S3_extracted_k141_44892 | Not-determined |
| S3_extracted_k141_44896 | Not-determined |
| S3_extracted_k141_84820 | Not-determined |
| S3_extracted_k141_37071 | Not-determined |
| S3_extracted_k141_61625 | Not-determined |
| S3_extracted_k141_44904 | Not-determined |
| S3_extracted_k141_61633 | Not-determined |
| S3_extracted_k141_44913 | Not-determined |
| S3_extracted_k141_77684 | Not-determined |
| S3_extracted_k141_53541 | Not-determined |
| S3_extracted_k141_92757 | Not-determined |
| S3_extracted_k141_92760 | Not-determined |
| S3_extracted_k141_53546 | Not-determined |
| S3_extracted_k141_53549 | Low-quality |
| S3_extracted_k141_92767 | Not-determined |
| S3_extracted_k141_77696 | Not-determined |
| S3_extracted_k141_20323 | Low-quality |
| S3_extracted_k141_77708 | Not-determined |
| S3_extracted_k141_20325 | Not-determined |
| S3_extracted_k141_20328 | Not-determined |
| S4_vs_VIRSorter_15376-cat_3 | Medium-quality |
| S3_extracted_k141_68038 | Not-determined |
| S3_extracted_k141_124043 | Low-quality |
| S3_extracted_k141_92779 | Not-determined |
| S3_extracted_k141_124044 | Not-determined |
| S3_extracted_k141_20333 | Not-determined |
| S3_extracted_k141_20335 | Not-determined |
| S3_extracted_k141_124049 | Not-determined |
| S3_extracted_k141_14605 | Not-determined |
| S3_extracted_k141_84848 | Not-determined |
| S3_extracted_k141_53574 | Not-determined |
| S3_extracted_k141_14609 | Not-determined |
| S3_extracted_k141_53576 | Not-determined |
| S3_extracted_k141_84853 | Not-determined |
| S3_extracted_k141_53580 | Not-determined |
| S3_extracted_k141_124056 | Not-determined |
| S3_extracted_k141_124058 | Not-determined |
| S3_extracted_k141_14616 | Not-determined |
| S3_extracted_k141_124064 | Not-determined |
| S3_extracted_k141_28800 | Not-determined |
| S13_vs_26756\|\|full | High-quality |
| S3_extracted_k141_92788 | Not-determined |
| S3_extracted_k141_124070 | Not-determined |
| S3_extracted_k141_124071 | Not-determined |
| S3_extracted_k141_37079 | Not-determined |
| S3_extracted_k141_20372 | Low-quality |
| S3_extracted_k141_37087 | Not-determined |
| S3_extracted_k141_108597 | Not-determined |
| S3_extracted_k141_108598 | Not-determined |
| S3_extracted_k141_100165 | Low-quality |
| S3_extracted_k141_68055 | Not-determined |
| S3_extracted_k141_108599 | Not-determined |
| S3_extracted_k141_20375 | Not-determined |

| | |
|---|---|
| S3_extracted_k141_68057 | Not-determined |
| S3_extracted_k141_68059 | Not-determined |
| S3_extracted_k141_108605 | Low-quality |
| S3_extracted_k141_100173 | Low-quality |
| S3_extracted_k141_20385 | Low-quality |
| S3_extracted_k141_124077 | Not-determined |
| S3_extracted_k141_4690 | Not-determined |
| S3_extracted_k141_4697 | Not-determined |
| S3_extracted_k141_100192 | Not-determined |
| S3_extracted_k141_14646 | Not-determined |
| S3_extracted_k141_20391 | Low-quality |
| S3_extracted_k141_124092 | Low-quality |
| S3_extracted_k141_68086 | Not-determined |
| S3_extracted_k141_124098 | Not-determined |
| S3_extracted_k141_61658 | Not-determined |
| S3_extracted_k141_14655 | Not-determined |
| S3_extracted_k141_77743 | Low-quality |
| S3_extracted_k141_84866 | Not-determined |
| S3_extracted_k141_61665 | Low-quality |
| S3_extracted_k141_84873 | Not-determined |
| S3_extracted_k141_61670 | Not-determined |
| S3_extracted_k141_14665 | Low-quality |
| S3_extracted_k141_124115 | Not-determined |
| S3_extracted_k141_68110 | Not-determined |
| S3_extracted_k141_14669 | Not-determined |
| S3_extracted_k141_68111 | Not-determined |
| S3_extracted_k141_68114 | Not-determined |
| S3_extracted_k141_84892 | Not-determined |
| S3_extracted_k141_14672 | Not-determined |
| S3_extracted_k141_14678 | Low-quality |
| S8_extracted_k141_170430 | Complete |
| S3_extracted_k141_77746 | Not-determined |
| S3_extracted_k141_84895 | Low-quality |
| S3_extracted_k141_92792 | Not-determined |
| S3_extracted_k141_108630 | Not-determined |
| S3_extracted_k141_108633 | Not-determined |
| S3_extracted_k141_14682 | Not-determined |
| S3_extracted_k141_92794 | Low-quality |
| S3_extracted_k141_108636 | Not-determined |
| S3_extracted_k141_84904 | Not-determined |
| S3_extracted_k141_77749 | Low-quality |
| S3_extracted_k141_77750 | Not-determined |
| S3_extracted_k141_53599 | Low-quality |
| S3_extracted_k141_92798 | Not-determined |
| S3_extracted_k141_53607 | Not-determined |
| S3_extracted_k141_53609 | Not-determined |
| S3_extracted_k141_61699 | Not-determined |
| S3_extracted_k141_14698 | Not-determined |
| S3_extracted_k141_84923 | Not-determined |
| S3_extracted_k141_61703 | Not-determined |
| S3_extracted_k141_53617 | Not-determined |
| S3_extracted_k141_53620 | Not-determined |
| S3_extracted_k141_44933 | Not-determined |
| S3_extracted_k141_53621 | Low-quality |
| S3_extracted_k141_100206 | Not-determined |
| S3_extracted_k141_100211 | Not-determined |
| S3_extracted_k141_84935 | Not-determined |
| S3_extracted_k141_37106 | Not-determined |
| S3_extracted_k141_37107 | Not-determined |
| S3_extracted_k141_53627 | Not-determined |
| S3_extracted_k141_84937 | Not-determined |
| S3_extracted_k141_108641 | Not-determined |
| S3_extracted_k141_53635 | Not-determined |
| S3_extracted_k141_53637 | Not-determined |
| S3_extracted_k141_84939 | Not-determined |
| S3_extracted_k141_108642 | Low-quality |
| S3_extracted_k141_14720 | Not-determined |
| S3_extracted_k141_84943 | Not-determined |
| S3_extracted_k141_92829 | Not-determined |
| S3_extracted_k141_53642 | Not-determined |
| S3_extracted_k141_53643 | Not-determined |
| S3_extracted_k141_44950 | Not-determined |
| S3_extracted_k141_116615 | Not-determined |
| S3_extracted_k141_84946 | Not-determined |
| S3_extracted_k141_37123 | Not-determined |
| S3_extracted_k141_68124 | Low-quality |
| S3_extracted_k141_116619 | Not-determined |
| S3_extracted_k141_124145 | Not-determined |
| S3_extracted_k141_61712 | Not-determined |
| S3_extracted_k141_53649 | Not-determined |
| S3_extracted_k141_124147 | Not-determined |
| S3_extracted_k141_124148 | Not-determined |
| S3_extracted_k141_14732 | Low-quality |
| S3_extracted_k141_108646 | Not-determined |
| S3_extracted_k141_108649 | Not-determined |
| S3_extracted_k141_116631 | Not-determined |
| S3_extracted_k141_14740 | Not-determined |
| S3_extracted_k141_116648 | Not-determined |
| S3_extracted_k141_20403 | Not-determined |
| S3_extracted_k141_108662 | Not-determined |
| S3_extracted_k141_68139 | Low-quality |
| S3_extracted_k141_116651 | Not-determined |
| S3_extracted_k141_68141 | Not-determined |
| S3_extracted_k141_92836 | Not-determined |
| S3_extracted_k141_108673 | Not-determined |
| S3_extracted_k141_14754 | Not-determined |
| S3_extracted_k141_28817 | Not-determined |
| S3_extracted_k141_92842 | Not-determined |
| S3_extracted_k141_20413 | Not-determined |
| S3_extracted_k141_108677 | Not-determined |
| S3_extracted_k141_92844 | Not-determined |
| S3_extracted_k141_77758 | Low-quality |
| S3_extracted_k141_44954 | Low-quality |
| S3_extracted_k141_61731 | Not-determined |
| S3_extracted_k141_68157 | Not-determined |
| S3_extracted_k141_108690 | Not-determined |
| S3_extracted_k141_44972 | Not-determined |
| S3_extracted_k141_44973 | Not-determined |
| S3_extracted_k141_68177 | Not-determined |
| S3_extracted_k141_100271 | Low-quality |
| S3_extracted_k141_68184 | Low-quality |
| S3_extracted_k141_108693 | Not-determined |
| S3_extracted_k141_108698 | Low-quality |
| S3_extracted_k141_108701 | Not-determined |
| S3_extracted_k141_4712 | Not-determined |
| S3_extracted_k141_14778 | Low-quality |
| S3_extracted_k141_4715 | Low-quality |
| S3_extracted_k141_53665 | Low-quality |
| S3_extracted_k141_108710 | Not-determined |
| S3_extracted_k141_37165 | Not-determined |
| S3_extracted_k141_92855 | Not-determined |
| S3_extracted_k141_68193 | Not-determined |
| S3_extracted_k141_61742 | Not-determined |
| S3_extracted_k141_108719 | Low-quality |
| S3_extracted_k141_68200 | Not-determined |
| S3_extracted_k141_4723 | Not-determined |
| S3_extracted_k141_92863 | Not-determined |
| S3_extracted_k141_37180 | Not-determined |
| S3_extracted_k141_77773 | Low-quality |
| S3_extracted_k141_28827 | Not-determined |
| S3_extracted_k141_28828 | Not-determined |
| S3_extracted_k141_28831 | Not-determined |
| S3_extracted_k141_68210 | Not-determined |
| S3_extracted_k141_28838 | Low-quality |
| S3_extracted_k141_28840 | Not-determined |
| S3_extracted_k141_108724 | Low-quality |
| S3_extracted_k141_77783 | Low-quality |
| S3_extracted_k141_53679 | Low-quality |
| S3_extracted_k141_108727 | Not-determined |
| S3_extracted_k141_28844 | Not-determined |
| S3_extracted_k141_61756 | Not-determined |
| S3_extracted_k141_28846 | Not-determined |
| S3_extracted_k141_108732 | Low-quality |
| S3_extracted_k141_68219 | Not-determined |
| S3_extracted_k141_68220 | Not-determined |
| S3_extracted_k141_4741 | Not-determined |
| S3_extracted_k141_61761 | Not-determined |
| S3_extracted_k141_28854 | Low-quality |
| S3_extracted_k141_100293 | Low-quality |
| S3_extracted_k141_92883 | Not-determined |
| S3_extracted_k141_61767 | Not-determined |
| S3_extracted_k141_4744 | Low-quality |
| S3_extracted_k141_108745 | Not-determined |
| S3_extracted_k141_108750 | Not-determined |
| S3_extracted_k141_68233 | Not-determined |
| S3_extracted_k141_28862 | Not-determined |
| S3_extracted_k141_116667 | Not-determined |
| S3_extracted_k141_108757 | Not-determined |
| S3_extracted_k141_84970 | Not-determined |
| S3_extracted_k141_84973 | Not-determined |
| S3_extracted_k141_68240 | Not-determined |
| S3_extracted_k141_108761 | Not-determined |
| S3_extracted_k141_68242 | Not-determined |
| S3_extracted_k141_108763 | Not-determined |
| S3_extracted_k141_108768 | Not-determined |
| S3_extracted_k141_92899 | Not-determined |
| S3_extracted_k141_100324 | Not-determined |
| S3_extracted_k141_92901 | Low-quality |
| S3_extracted_k141_14796 | Not-determined |
| S3_extracted_k141_116683 | Not-determined |
| S3_extracted_k141_37200 | Low-quality |
| S3_extracted_k141_28868 | Not-determined |
| S3_extracted_k141_68248 | Not-determined |
| S3_extracted_k141_53716 | Not-determined |
| S3_extracted_k141_37208 | Not-determined |
| S3_extracted_k141_108772 | Not-determined |
| S3_extracted_k141_61777 | Not-determined |
| S3_extracted_k141_53723 | Not-determined |
| S3_extracted_k141_14809 | Not-determined |
| S3_extracted_k141_116692 | Not-determined |
| S3_extracted_k141_4759 | Not-determined |
| S3_extracted_k141_4761 | Not-determined |
| S3_extracted_k141_14814 | Not-determined |
| S3_extracted_k141_37210 | Low-quality |
| S3_extracted_k141_84988 | Not-determined |
| S3_extracted_k141_14815 | Not-determined |
| S3_extracted_k141_53734 | Not-determined |
| S3_extracted_k141_108781 | Not-determined |
| S3_extracted_k141_92910 | Low-quality |
| S3_extracted_k141_100329 | Low-quality |
| S3_extracted_k141_108783 | Not-determined |
| S3_extracted_k141_92911 | Not-determined |
| S3_extracted_k141_4775 | Not-determined |
| S3_extracted_k141_61795 | Not-determined |
| S3_extracted_k141_100337 | Not-determined |
| S3_extracted_k141_37217 | Low-quality |
| S3_extracted_k141_61805 | Low-quality |
| S3_extracted_k141_92916 | Low-quality |
| S3_extracted_k141_4783 | Not-determined |
| S3_extracted_k141_28876 | Not-determined |
| S3_extracted_k141_28877 | Low-quality |
| S3_extracted_k141_61809 | Not-determined |
| S3_extracted_k141_4785 | Not-determined |
| S3_vs_k141_43925\|\|full | High-quality |
| S3_extracted_k141_124180 | Not-determined |
| S3_extracted_k141_77793 | Not-determined |
| S3_extracted_k141_28881 | Low-quality |
| S3_extracted_k141_28882 | Not-determined |
| S3_extracted_k141_92927 | Not-determined |
| S3_extracted_k141_100342 | Not-determined |
| S3_extracted_k141_20443 | Low-quality |
| S3_extracted_k141_85003 | Not-determined |
| S3_extracted_k141_92931 | Low-quality |
| S3_extracted_k141_61821 | Not-determined |
| S3_extracted_k141_28889 | Low-quality |
| S3_extracted_k141_124184 | Not-determined |
| S3_extracted_k141_85013 | Not-determined |
| S3_extracted_k141_20448 | Not-determined |
| S3_extracted_k141_124185 | Low-quality |
| S3_extracted_k141_100353 | Not-determined |
| S3_extracted_k141_4797 | Not-determined |
| S3_extracted_k141_4798 | Not-determined |
| S3_extracted_k141_37218 | Not-determined |
| S3_extracted_k141_108807 | Not-determined |
| S3_extracted_k141_116704 | Not-determined |
| S3_extracted_k141_61834 | Not-determined |
| S3_extracted_k141_108810 | Not-determined |
| S3_extracted_k141_61836 | Not-determined |
| S3_extracted_k141_124197 | Not-determined |
| S3_extracted_k141_108813 | Low-quality |
| S3_extracted_k141_4800 | Low-quality |
| S3_extracted_k141_77811 | Not-determined |
| S3_extracted_k141_124200 | Not-determined |
| S3_extracted_k141_77812 | Not-determined |
| S3_extracted_k141_53740 | Not-determined |
| S3_extracted_k141_61838 | Low-quality |
| S3_extracted_k141_108823 | Not-determined |
| S3_extracted_k141_108824 | Not-determined |
| S3_extracted_k141_20466 | Not-determined |
| S3_extracted_k141_20467 | Not-determined |
| S3_extracted_k141_108827 | Not-determined |
| S3_extracted_k141_61856 | Not-determined |
| S3_extracted_k141_108830 | Low-quality |
| S3_extracted_k141_61858 | Not-determined |
| S3_extracted_k141_4813 | Not-determined |
| S3_extracted_k141_124206 | Not-determined |
| S3_extracted_k141_100376 | Not-determined |
| S3_extracted_k141_4816 | Not-determined |
| S3_extracted_k141_20482 | Not-determined |
| S3_extracted_k141_4832 | Low-quality |
| S3_extracted_k141_116714 | Not-determined |
| S3_extracted_k141_108841 | Not-determined |
| S3_extracted_k141_100385 | Not-determined |
| S3_extracted_k141_85039 | Not-determined |
| S3_extracted_k141_124220 | Not-determined |
| S3_extracted_k141_116724 | Low-quality |
| S3_extracted_k141_85043 | Not-determined |
| S3_extracted_k141_37242 | Not-determined |
| S3_extracted_k141_116727 | Low-quality |
| S3_extracted_k141_124227 | Not-determined |
| S3_extracted_k141_4849 | Not-determined |
| S3_extracted_k141_124230 | Not-determined |
| S3_extracted_k141_108845 | Not-determined |
| S3_extracted_k141_100400 | Low-quality |
| S3_extracted_k141_116736 | Not-determined |
| S3_extracted_k141_100407 | Not-determined |
| S3_extracted_k141_77826 | Not-determined |
| S3_extracted_k141_14849 | Not-determined |
| S3_extracted_k141_108856 | Not-determined |
| S3_extracted_k141_85052 | Not-determined |
| S3_extracted_k141_37252 | Not-determined |
| S3_extracted_k141_37253 | Not-determined |
| S3_extracted_k141_100414 | Not-determined |
| S3_extracted_k141_100415 | Not-determined |
| S3_extracted_k141_77831 | Not-determined |
| S3_extracted_k141_4864 | Not-determined |
| S3_extracted_k141_85060 | Not-determined |
| S3_extracted_k141_53764 | Not-determined |
| S3_extracted_k141_28910 | Not-determined |
| S3_extracted_k141_108865 | Not-determined |
| S3_extracted_k141_108868 | Not-determined |
| S3_extracted_k141_77840 | Not-determined |
| S3_extracted_k141_4884 | Not-determined |
| S3_extracted_k141_124246 | Not-determined |
| S3_extracted_k141_53779 | Low-quality |
| S3_extracted_k141_4886 | Low-quality |
| S3_extracted_k141_77844 | Not-determined |
| S3_extracted_k141_28914 | Not-determined |
| S3_extracted_k141_77845 | Not-determined |
| S3_extracted_k141_124250 | Low-quality |
| S3_extracted_k141_4891 | Not-determined |
| S3_extracted_k141_108891 | Not-determined |
| S3_extracted_k141_108896 | Not-determined |
| S3_extracted_k141_53788 | Not-determined |
| S3_extracted_k141_20502 | Not-determined |
| S3_extracted_k141_108897 | Not-determined |
| S3_extracted_k141_108904 | Not-determined |
| S3_extracted_k141_37272 | Not-determined |
| S3_extracted_k141_28936 | Not-determined |
| S3_extracted_k141_124257 | Not-determined |
| S3_extracted_k141_124261 | Not-determined |
| S3_extracted_k141_28944 | Not-determined |
| S3_extracted_k141_53799 | Not-determined |
| S3_extracted_k141_68292 | Not-determined |
| S3_extracted_k141_53806 | Not-determined |
| S3_extracted_k141_28948 | Not-determined |
| S3_extracted_k141_14858 | Not-determined |
| S3_extracted_k141_77851 | Not-determined |
| S3_extracted_k141_20511 | Not-determined |
| S3_extracted_k141_116769 | Not-determined |
| S3_extracted_k141_68297 | Low-quality |
| S3_extracted_k141_68298 | Not-determined |
| S3_extracted_k141_92974 | Low-quality |
| S3_extracted_k141_20518 | Not-determined |
| S3_extracted_k141_68301 | Low-quality |
| S3_extracted_k141_116775 | Not-determined |
| S7_vs_138173\|\|full | High-quality |
| S3_extracted_k141_108936 | Not-determined |
| S3_extracted_k141_61862 | Low-quality |
| S3_extracted_k141_68307 | Not-determined |
| S3_extracted_k141_61863 | Low-quality |
| S3_extracted_k141_108939 | Not-determined |
| S3_extracted_k141_116785 | Not-determined |
| S3_extracted_k141_68312 | Not-determined |
| S3_extracted_k141_68313 | Not-determined |
| S3_extracted_k141_68316 | Not-determined |
| S3_extracted_k141_92980 | Not-determined |
| S3_extracted_k141_68317 | Not-determined |
| S3_extracted_k141_85078 | Not-determined |
| S3_extracted_k141_85079 | Not-determined |
| S3_extracted_k141_61864 | Low-quality |
| S3_extracted_k141_116798 | Not-determined |
| S3_extracted_k141_4926 | Not-determined |
| S3_extracted_k141_14871 | Not-determined |
| S3_extracted_k141_14872 | Not-determined |
| S3_extracted_k141_108948 | Not-determined |
| S3_extracted_k141_68332 | Not-determined |
| S3_extracted_k141_4933 | Not-determined |
| S3_extracted_k141_28962 | Not-determined |
| S3_extracted_k141_4935 | Not-determined |
| S3_extracted_k141_61876 | Not-determined |
| S3_extracted_k141_53825 | Not-determined |
| S3_extracted_k141_93005 | Not-determined |
| S3_extracted_k141_53832 | Not-determined |
| S3_extracted_k141_20536 | Not-determined |
| S3_extracted_k141_4955 | Not-determined |
| S3_extracted_k141_4956 | Low-quality |
| S3_extracted_k141_20540 | Not-determined |
| S3_extracted_k141_20541 | Not-determined |
| S3_extracted_k141_53836 | Not-determined |
| S3_extracted_k141_68340 | Not-determined |
| S3_extracted_k141_77915 | Not-determined |
| S3_extracted_k141_93013 | Not-determined |
| S3_extracted_k141_77920 | Not-determined |
| S3_extracted_k141_93016 | Not-determined |
| S3_extracted_k141_53839 | Low-quality |
| S3_extracted_k141_93024 | Not-determined |
| S3_extracted_k141_116817 | Not-determined |
| S3_extracted_k141_28980 | Not-determined |
| S3_extracted_k141_28981 | Not-determined |
| S3_extracted_k141_116827 | Low-quality |
| S3_extracted_k141_116836 | Not-determined |
| S3_extracted_k141_116841 | Not-determined |
| S3_extracted_k141_28988 | Low-quality |
| S3_extracted_k141_85104 | Not-determined |
| S3_extracted_k141_37296 | Not-determined |
| S3_extracted_k141_28993 | Not-determined |
| S3_extracted_k141_28999 | Low-quality |
| S3_extracted_k141_85120 | Not-determined |
| S3_extracted_k141_29002 | Not-determined |
| S3_extracted_k141_29005 | Not-determined |
| S3_extracted_k141_85127 | Not-determined |
| S3_extracted_k141_4981 | Not-determined |
| S3_extracted_k141_4989 | Not-determined |
| S3_extracted_k141_29008 | Not-determined |
| S3_extracted_k141_4990 | Not-determined |
| S3_extracted_k141_4992 | Not-determined |
| S3_extracted_k141_68363 | Not-determined |
| S3_extracted_k141_4995 | Not-determined |
| S3_extracted_k141_29010 | Not-determined |
| S3_extracted_k141_68366 | Not-determined |
| S3_extracted_k141_93044 | Not-determined |
| S3_extracted_k141_53865 | Low-quality |
| S3_extracted_k141_53867 | Not-determined |
| S3_extracted_k141_68372 | Not-determined |
| S3_extracted_k141_61918 | Low-quality |
| S3_extracted_k141_124271 | Not-determined |
| S3_extracted_k141_124272 | Not-determined |
| S3_extracted_k141_5005 | Not-determined |
| S4_vs_VIRSorter_31993-circular-cat | High-quality |
| S3_extracted_k141_61920 | Not-determined |
| S3_extracted_k141_100443 | Not-determined |
| S3_extracted_k141_100446 | Not-determined |
| S3_extracted_k141_37312 | Not-determined |
| S3_extracted_k141_37314 | Not-determined |
| S3_extracted_k141_5018 | Not-determined |
| S3_extracted_k141_85144 | Not-determined |
| S3_extracted_k141_5021 | Not-determined |
| S3_extracted_k141_85149 | Not-determined |
| S3_extracted_k141_61938 | Not-determined |
| S3_extracted_k141_29034 | Not-determined |
| S4_vs_VIRSorter_31560-circular-cat | High-quality |
| S3_extracted_k141_124274 | Not-determined |
| S3_extracted_k141_5030 | Low-quality |
| S3_extracted_k141_61941 | Not-determined |
| S3_extracted_k141_37324 | Not-determined |
| S3_extracted_k141_61943 | Not-determined |
| S3_extracted_k141_68391 | Not-determined |
| S3_extracted_k141_53871 | Not-determined |
| S3_extracted_k141_14910 | Not-determined |
| S3_extracted_k141_108971 | Not-determined |
| S3_extracted_k141_14911 | Not-determined |
| S3_extracted_k141_14912 | Not-determined |
| S3_extracted_k141_68393 | Not-determined |
| S3_extracted_k141_61953 | Not-determined |
| S3_extracted_k141_61954 | Not-determined |
| S3_extracted_k141_85160 | Not-determined |
| S3_extracted_k141_108975 | Not-determined |
| S3_extracted_k141_68398 | Not-determined |
| S3_extracted_k141_5038 | Not-determined |
| S3_extracted_k141_29046 | Not-determined |
| S3_extracted_k141_77961 | Not-determined |
| S3_extracted_k141_29048 | Not-determined |
| S3_extracted_k141_61959 | Not-determined |
| S3_extracted_k141_68408 | Not-determined |
| S3_extracted_k141_61961 | Not-determined |
| S3_extracted_k141_37339 | Low-quality |
| S3_extracted_k141_37340 | Not-determined |
| S3_extracted_k141_68411 | Low-quality |
| S3_extracted_k141_37342 | Not-determined |
| S3_extracted_k141_5042 | Not-determined |
| S3_extracted_k141_116848 | Not-determined |
| S3_extracted_k141_85167 | Not-determined |
| S3_extracted_k141_77969 | Not-determined |
| S3_extracted_k141_5046 | Not-determined |
| S3_extracted_k141_77970 | Not-determined |
| S3_extracted_k141_85171 | Not-determined |
| S3_extracted_k141_100468 | Low-quality |
| S3_extracted_k141_100469 | Not-determined |
| S3_extracted_k141_68417 | Not-determined |
| S3_extracted_k141_53877 | Low-quality |
| S3_extracted_k141_77980 | Not-determined |
| S3_extracted_k141_77982 | Not-determined |
| S3_extracted_k141_116852 | Low-quality |
| S3_extracted_k141_124286 | Not-determined |
| S3_extracted_k141_85176 | Not-determined |
| S3_extracted_k141_45032 | Not-determined |
| S3_extracted_k141_116869 | Not-determined |
| S3_extracted_k141_53894 | Not-determined |
| S3_extracted_k141_68428 | Not-determined |
| S3_extracted_k141_124302 | Not-determined |
| S3_extracted_k141_45039 | Low-quality |
| S3_extracted_k141_68436 | Not-determined |
| S3_extracted_k141_29053 | Not-determined |
| S3_extracted_k141_124303 | Not-determined |
| S3_extracted_k141_68437 | Not-determined |
| S3_extracted_k141_29063 | Not-determined |
| S3_extracted_k141_29065 | Not-determined |
| S3_extracted_k141_108985 | Not-determined |
| S3_extracted_k141_53906 | Not-determined |
| S3_extracted_k141_5071 | Not-determined |
| S3_extracted_k141_29071 | Not-determined |
| S3_extracted_k141_5076 | Low-quality |
| S3_extracted_k141_124308 | Not-determined |
| S3_extracted_k141_124309 | Not-determined |
| S3_extracted_k141_37356 | Not-determined |
| S3_extracted_k141_78002 | Not-determined |
| S3_extracted_k141_68441 | Not-determined |
| S3_extracted_k141_20567 | Low-quality |
| S3_extracted_k141_68442 | Not-determined |
| S3_extracted_k141_124319 | Low-quality |
| S3_extracted_k141_124320 | Low-quality |
| S3_extracted_k141_78017 | Low-quality |
| S3_extracted_k141_85222 | Not-determined |
| S3_extracted_k141_85224 | Not-determined |
| S3_extracted_k141_85226 | Not-determined |
| S3_extracted_k141_116880 | Not-determined |
| S3_extracted_k141_78027 | Not-determined |
| S3_extracted_k141_109012 | Not-determined |
| S3_extracted_k141_62002 | Not-determined |
| S3_extracted_k141_20584 | Not-determined |
| S3_extracted_k141_62006 | Not-determined |
| S3_extracted_k141_5087 | Not-determined |
| S3_extracted_k141_85233 | Low-quality |
| S3_extracted_k141_5094 | Not-determined |
| S3_extracted_k141_5103 | Low-quality |
| S3_extracted_k141_116900 | Not-determined |
| S3_extracted_k141_109028 | Low-quality |
| S3_extracted_k141_5123 | Not-determined |
| S3_extracted_k141_109036 | Not-determined |
| S3_extracted_k141_62011 | Not-determined |
| S3_extracted_k141_37377 | Low-quality |
| S3_extracted_k141_20609 | Not-determined |
| S3_extracted_k141_5131 | Not-determined |
| S3_extracted_k141_109042 | Not-determined |
| S3_extracted_k141_109044 | Not-determined |
| S3_extracted_k141_116909 | Low-quality |
| S3_extracted_k141_20617 | Not-determined |
| S3_extracted_k141_5138 | Not-determined |
| S3_extracted_k141_85248 | Not-determined |
| S3_extracted_k141_5141 | Low-quality |
| S3_extracted_k141_100483 | Not-determined |
| S3_extracted_k141_116913 | Low-quality |
| S3_extracted_k141_109058 | Low-quality |
| S3_extracted_k141_109066 | Not-determined |
| S3_extracted_k141_68498 | Not-determined |
| S3_extracted_k141_68500 | Not-determined |
| S3_extracted_k141_109072 | Not-determined |
| S3_extracted_k141_100489 | Not-determined |
| S3_extracted_k141_100490 | Not-determined |
| S3_extracted_k141_100492 | Not-determined |
| S3_extracted_k141_85261 | Not-determined |
| S3_extracted_k141_53915 | Low-quality |
| S3_extracted_k141_78031 | Not-determined |
| S3_extracted_k141_78032 | Not-determined |
| S3_extracted_k141_85271 | Not-determined |
| S3_extracted_k141_85272 | Low-quality |
| S3_extracted_k141_53922 | Low-quality |
| S3_extracted_k141_37390 | Not-determined |
| S3_extracted_k141_53924 | Not-determined |
| S3_extracted_k141_45078 | Low-quality |
| S3_extracted_k141_37392 | Not-determined |
| S3_extracted_k141_37398 | Not-determined |
| S3_extracted_k141_68510 | Not-determined |
| S3_extracted_k141_5143 | Not-determined |
| S3_extracted_k141_45080 | Not-determined |
| S3_extracted_k141_68512 | Not-determined |
| S3_extracted_k141_124343 | Not-determined |
| S3_extracted_k141_5145 | Low-quality |
| S3_extracted_k141_5146 | Not-determined |
| S3_extracted_k141_124344 | Not-determined |
| S3_extracted_k141_53931 | Not-determined |
| S3_extracted_k141_53932 | Not-determined |
| S3_extracted_k141_62031 | Not-determined |
| S3_extracted_k141_53935 | Not-determined |
| S3_extracted_k141_45091 | Not-determined |
| S3_extracted_k141_45093 | Not-determined |
| S3_extracted_k141_78037 | Not-determined |
| S3_extracted_k141_85282 | Not-determined |
| S3_extracted_k141_45096 | Not-determined |
| S3_extracted_k141_78042 | Not-determined |
| S3_extracted_k141_109080 | Not-determined |
| S3_extracted_k141_62042 | Not-determined |
| S3_extracted_k141_53949 | Not-determined |
| S3_extracted_k141_29085 | Not-determined |
| S3_extracted_k141_29087 | Not-determined |
| S3_extracted_k141_62049 | Not-determined |
| S3_extracted_k141_29089 | Not-determined |
| S3_extracted_k141_68518 | Not-determined |
| S3_extracted_k141_62053 | Not-determined |
| S3_extracted_k141_20635 | Low-quality |
| S3_extracted_k141_14983 | Not-determined |
| S3_extracted_k141_37410 | Not-determined |
| S3_extracted_k141_109089 | Low-quality |
| S3_extracted_k141_109090 | Not-determined |
| S3_extracted_k141_109091 | Not-determined |
| S3_extracted_k141_85302 | Not-determined |
| S3_extracted_k141_62064 | Low-quality |
| S3_extracted_k141_85306 | Not-determined |
| S3_extracted_k141_14994 | Low-quality |
| S3_extracted_k141_20641 | Not-determined |
| S3_extracted_k141_29098 | Not-determined |
| S3_extracted_k141_37420 | Not-determined |
| S3_extracted_k141_68523 | Not-determined |
| S3_extracted_k141_29099 | Not-determined |
| S3_extracted_k141_29100 | Not-determined |
| S3_extracted_k141_116927 | Not-determined |
| S3_extracted_k141_29101 | Not-determined |
| S3_extracted_k141_78056 | Not-determined |
| S3_extracted_k141_109111 | Not-determined |
| S3_extracted_k141_109112 | Not-determined |
| S3_extracted_k141_29104 | Not-determined |
| S3_extracted_k141_53971 | Not-determined |
| S3_extracted_k141_15003 | Not-determined |
| S3_extracted_k141_29108 | Not-determined |
| S3_extracted_k141_85318 | Not-determined |
| S3_extracted_k141_109117 | Not-determined |
| S3_extracted_k141_68545 | Not-determined |
| S3_extracted_k141_85324 | Not-determined |
| S3_extracted_k141_85325 | Low-quality |
| S3_extracted_k141_62070 | Low-quality |
| S3_extracted_k141_5168 | Not-determined |
| S3_extracted_k141_62072 | Not-determined |
| S3_extracted_k141_68552 | Low-quality |
| S3_extracted_k141_85333 | Low-quality |
| S3_extracted_k141_68558 | Not-determined |
| S3_extracted_k141_15015 | Not-determined |
| S3_extracted_k141_124362 | Not-determined |
| S3_extracted_k141_109127 | Low-quality |

| | |
|---|---|
| S3_extracted_k141_109131 | Not-determined |
| S3_extracted_k141_29117 | Not-determined |
| S3_extracted_k141_62091 | Not-determined |
| S3_extracted_k141_109136 | Low-quality |
| S3_extracted_k141_15020 | Not-determined |
| S3_extracted_k141_109137 | Low-quality |
| S3_extracted_k141_124374 | Not-determined |
| S3_extracted_k141_109139 | Not-determined |
| S3_extracted_k141_116935 | Not-determined |
| S3_extracted_k141_109145 | Not-determined |
| S3_extracted_k141_124381 | Not-determined |
| S3_extracted_k141_109151 | Not-determined |
| S3_extracted_k141_124384 | Low-quality |
| S3_extracted_k141_116941 | Not-determined |
| S3_extracted_k141_109155 | Low-quality |
| S3_extracted_k141_54004 | Low-quality |
| S3_extracted_k141_54005 | Not-determined |
| S3_extracted_k141_116943 | Low-quality |
| S3_extracted_k141_15040 | Not-determined |
| S3_extracted_k141_5210 | Not-determined |
| S3_extracted_k141_5212 | Not-determined |
| S3_extracted_k141_15047 | Not-determined |
| S3_extracted_k141_15051 | Not-determined |
| S3_extracted_k141_116955 | Not-determined |
| S3_extracted_k141_5218 | Not-determined |
| S3_extracted_k141_15054 | Low-quality |
| S3_extracted_k141_15058 | Not-determined |
| S3_extracted_k141_116962 | Not-determined |
| S3_extracted_k141_116964 | Not-determined |
| S3_extracted_k141_5225 | Not-determined |
| S3_extracted_k141_85356 | Not-determined |
| S3_extracted_k141_124388 | Low-quality |
| S3_extracted_k141_109168 | Not-determined |
| S3_extracted_k141_15069 | Not-determined |
| S3_extracted_k141_124393 | Not-determined |
| S3_extracted_k141_62111 | Not-determined |
| S3_extracted_k141_15072 | Not-determined |
| S3_extracted_k141_124395 | Not-determined |
| S3_extracted_k141_109174 | Not-determined |
| S3_extracted_k141_68576 | Not-determined |
| S3_extracted_k141_68577 | Not-determined |
| S3_extracted_k141_109177 | Low-quality |
| S3_extracted_k141_15084 | Not-determined |
| S3_extracted_k141_85359 | Low-quality |
| S3_extracted_k141_15088 | Not-determined |
| S3_extracted_k141_15089 | Not-determined |
| S3_extracted_k141_116976 | Not-determined |
| S3_extracted_k141_62112 | Low-quality |
| S3_extracted_k141_29125 | Not-determined |
| S3_extracted_k141_62113 | Not-determined |
| S3_extracted_k141_62122 | Low-quality |
| S3_extracted_k141_116982 | Not-determined |
| S3_extracted_k141_15102 | Not-determined |
| S3_extracted_k141_62126 | Not-determined |
| S3_extracted_k141_116989 | Not-determined |
| S3_extracted_k141_15107 | Not-determined |
| S3_extracted_k141_29139 | Low-quality |
| S3_extracted_k141_29141 | Not-determined |
| S3_extracted_k141_93081 | Not-determined |
| S3_extracted_k141_29146 | Not-determined |
| S3_extracted_k141_62139 | Not-determined |
| S3_extracted_k141_29158 | Not-determined |
| S3_extracted_k141_93087 | Not-determined |
| S3_extracted_k141_85363 | Not-determined |
| S3_extracted_k141_15132 | Not-determined |
| S3_extracted_k141_62157 | Not-determined |
| S3_extracted_k141_124398 | Not-determined |
| S3_extracted_k141_68622 | Low-quality |
| S3_extracted_k141_5274 | Not-determined |
| S3_extracted_k141_68624 | Not-determined |
| S3_extracted_k141_29173 | Not-determined |
| S3_extracted_k141_37464 | Not-determined |
| S3_extracted_k141_29178 | Low-quality |
| S3_extracted_k141_37466 | Not-determined |
| S3_extracted_k141_45117 | Not-determined |
| S3_extracted_k141_45119 | Not-determined |
| S3_extracted_k141_37472 | Not-determined |
| S3_extracted_k141_93096 | Not-determined |
| S3_extracted_k141_62163 | Low-quality |
| S3_extracted_k141_93099 | Not-determined |
| S3_extracted_k141_15147 | Not-determined |
| S3_extracted_k141_29187 | Low-quality |
| S3_extracted_k141_93103 | Low-quality |
| S3_extracted_k141_15163 | Not-determined |
| S3_extracted_k141_15164 | Not-determined |
| S3_extracted_k141_78098 | Not-determined |
| S3_extracted_k141_15171 | Not-determined |
| S3_extracted_k141_78099 | Not-determined |
| S3_extracted_k141_15173 | Not-determined |
| S3_extracted_k141_85378 | Not-determined |
| S3_extracted_k141_68661 | Not-determined |
| S3_extracted_k141_37512 | Not-determined |
| S3_extracted_k141_109195 | Low-quality |
| S3_extracted_k141_109197 | Not-determined |
| S3_extracted_k141_62178 | Not-determined |
| S3_extracted_k141_62179 | Not-determined |
| S3_extracted_k141_109205 | Not-determined |
| S3_extracted_k141_124442 | Low-quality |
| S3_extracted_k141_68675 | Not-determined |
| S3_extracted_k141_68676 | Not-determined |
| S3_extracted_k141_37531 | Not-determined |
| S3_extracted_k141_62188 | Not-determined |
| S3_extracted_k141_37534 | Not-determined |
| S3_extracted_k141_37537 | Not-determined |
| S3_extracted_k141_109212 | Low-quality |
| S3_extracted_k141_68687 | Not-determined |
| S3_extracted_k141_78104 | Not-determined |
| S3_extracted_k141_62199 | Not-determined |
| S3_extracted_k141_109213 | Low-quality |
| S3_extracted_k141_5281 | Not-determined |
| S3_extracted_k141_5282 | Not-determined |
| S3_extracted_k141_68699 | Not-determined |
| S3_extracted_k141_62207 | Not-determined |
| S3_extracted_k141_62208 | Not-determined |
| S3_extracted_k141_109216 | Not-determined |
| S3_extracted_k141_5284 | Not-determined |
| S3_extracted_k141_62210 | Not-determined |
| S3_extracted_k141_37552 | Not-determined |
| S3_extracted_k141_37553 | Not-determined |
| S3_extracted_k141_37555 | Not-determined |
| S3_extracted_k141_5293 | Not-determined |
| S3_extracted_k141_37559 | Not-determined |
| S3_extracted_k141_29197 | Not-determined |
| S3_extracted_k141_37561 | Low-quality |
| S3_extracted_k141_62219 | Not-determined |
| S3_extracted_k141_68708 | Not-determined |
| S3_extracted_k141_62224 | Not-determined |
| S3_extracted_k141_116997 | Not-determined |
| S3_extracted_k141_29200 | Not-determined |
| S3_extracted_k141_116998 | Not-determined |
| S3_extracted_k141_68710 | Not-determined |
| S3_extracted_k141_62226 | Low-quality |
| S3_extracted_k141_85383 | Not-determined |
| S3_extracted_k141_37563 | Not-determined |
| S3_extracted_k141_15199 | Not-determined |
| S3_extracted_k141_109228 | Not-determined |
| S3_extracted_k141_29204 | Not-determined |
| S3_extracted_k141_68716 | Not-determined |
| S3_extracted_k141_15203 | Not-determined |
| S3_extracted_k141_68718 | Not-determined |
| S3_extracted_k141_78117 | Not-determined |
| S3_extracted_k141_85392 | Not-determined |
| S3_extracted_k141_85397 | Not-determined |
| S3_extracted_k141_62244 | Not-determined |
| S3_extracted_k141_20701 | Not-determined |
| S3_extracted_k141_37571 | Not-determined |
| S3_extracted_k141_29218 | Low-quality |
| S3_extracted_k141_29219 | Not-determined |
| S3_extracted_k141_124458 | Low-quality |
| S3_extracted_k141_124460 | Not-determined |
| S3_extracted_k141_124461 | Low-quality |
| S3_extracted_k141_29224 | Not-determined |
| S3_extracted_k141_85409 | Not-determined |
| S3_extracted_k141_109251 | Not-determined |
| S3_extracted_k141_68723 | Low-quality |
| S3_extracted_k141_20713 | Not-determined |
| S3_extracted_k141_124478 | Not-determined |
| S3_extracted_k141_15218 | Not-determined |
| S3_extracted_k141_109256 | Not-determined |
| S3_extracted_k141_124482 | Not-determined |
| S3_extracted_k141_5314 | Not-determined |
| S3_extracted_k141_93119 | Not-determined |
| S3_extracted_k141_109259 | Not-determined |
| S3_extracted_k141_15221 | Not-determined |
| S3_extracted_k141_109264 | Not-determined |
| S3_extracted_k141_93125 | Not-determined |
| S3_extracted_k141_124487 | Low-quality |
| S3_extracted_k141_109269 | Not-determined |
| S3_extracted_k141_109270 | Low-quality |
| S3_extracted_k141_5326 | Not-determined |
| S3_extracted_k141_5329 | Not-determined |
| S3_extracted_k141_15231 | Not-determined |
| S3_extracted_k141_93147 | Not-determined |
| S3_extracted_k141_93149 | Not-determined |
| S3_extracted_k141_124497 | Not-determined |
| S3_extracted_k141_85421 | Not-determined |
| S3_extracted_k141_127423 | Complete |
| S3_extracted_k141_124499 | Not-determined |
| S3_extracted_k141_15238 | Not-determined |
| S3_extracted_k141_20723 | Not-determined |
| S3_extracted_k141_93154 | Not-determined |
| S3_extracted_k141_5341 | Not-determined |
| S3_extracted_k141_5344 | Not-determined |
| S3_extracted_k141_20727 | Not-determined |
| S3_extracted_k141_5347 | Low-quality |
| S3_extracted_k141_68734 | Not-determined |
| S3_extracted_k141_124510 | Not-determined |
| S3_extracted_k141_5354 | Low-quality |
| S3_extracted_k141_124516 | Not-determined |
| S3_extracted_k141_117014 | Not-determined |
| S3_extracted_k141_78129 | Not-determined |
| S3_extracted_k141_85437 | Low-quality |
| S3_extracted_k141_37592 | Low-quality |
| S3_extracted_k141_100513 | Not-determined |
| S3_extracted_k141_5366 | Not-determined |
| S3_extracted_k141_5367 | Not-determined |
| S3_extracted_k141_100514 | Low-quality |
| S3_extracted_k141_85448 | Not-determined |
| S3_extracted_k141_5374 | Not-determined |
| S3_extracted_k141_100520 | Not-determined |
| S3_extracted_k141_93157 | Not-determined |
| S3_extracted_k141_93159 | Low-quality |
| S3_extracted_k141_93160 | Low-quality |
| S3_extracted_k141_5381 | Not-determined |
| S3_extracted_k141_5383 | Not-determined |
| S3_extracted_k141_109290 | Not-determined |
| S3_extracted_k141_100528 | Low-quality |
| S3_extracted_k141_68763 | Not-determined |
| S3_extracted_k141_68765 | Not-determined |
| S3_extracted_k141_68770 | Low-quality |
| S3_extracted_k141_109298 | Not-determined |
| S3_extracted_k141_78134 | Not-determined |
| S3_extracted_k141_68777 | Not-determined |
| S3_extracted_k141_37600 | Low-quality |
| S3_extracted_k141_85474 | Not-determined |
| S3_extracted_k141_117029 | Not-determined |
| S3_extracted_k141_62324 | Not-determined |
| S3_extracted_k141_62329 | Not-determined |
| S3_extracted_k141_5397 | Not-determined |
| S3_extracted_k141_117036 | Not-determined |
| S3_extracted_k141_117040 | Not-determined |
| S3_extracted_k141_62335 | Low-quality |
| S3_extracted_k141_85487 | Not-determined |
| S3_extracted_k141_29247 | Not-determined |
| S3_extracted_k141_54032 | Low-quality |
| S3_extracted_k141_117052 | Low-quality |
| S3_extracted_k141_29253 | Not-determined |
| S3_extracted_k141_5412 | Not-determined |
| S3_extracted_k141_100557 | Low-quality |
| S3_extracted_k141_54035 | Low-quality |
| S3_extracted_k141_100561 | Not-determined |
| S3_extracted_k141_45169 | Not-determined |
| S3_extracted_k141_93177 | Low-quality |
| S3_extracted_k141_29270 | Low-quality |
| S3_extracted_k141_62347 | Not-determined |
| S3_extracted_k141_93180 | Low-quality |
| S3_extracted_k141_93184 | Low-quality |
| S3_extracted_k141_45179 | Not-determined |
| S3_extracted_k141_85496 | Low-quality |
| S3_extracted_k141_45181 | Not-determined |
| S3_extracted_k141_37619 | Not-determined |
| S3_extracted_k141_37620 | Not-determined |
| S3_extracted_k141_20739 | Not-determined |
| S3_extracted_k141_93207 | Not-determined |
| S3_extracted_k141_15257 | Not-determined |
| S3_extracted_k141_45190 | Not-determined |
| S3_extracted_k141_45191 | Not-determined |
| S3_extracted_k141_78144 | Not-determined |
| S3_extracted_k141_117065 | Low-quality |
| S3_extracted_k141_68792 | Not-determined |
| S3_extracted_k141_45199 | Not-determined |
| S3_extracted_k141_117069 | Not-determined |
| S3_extracted_k141_45204 | Low-quality |
| S3_extracted_k141_93225 | Not-determined |
| S3_extracted_k141_68799 | Not-determined |
| S3_extracted_k141_93226 | Low-quality |
| S3_extracted_k141_85523 | Low-quality |
| S3_extracted_k141_85524 | Not-determined |
| S3_extracted_k141_93238 | Not-determined |
| S3_extracted_k141_15269 | Not-determined |
| S3_extracted_k141_109373 | Not-determined |
| S3_extracted_k141_20754 | Not-determined |
| S3_extracted_k141_78160 | Not-determined |
| S3_extracted_k141_20757 | Not-determined |
| S3_extracted_k141_109377 | Not-determined |
| S3_extracted_k141_37632 | Low-quality |
| S3_extracted_k141_37634 | Low-quality |
| S3_extracted_k141_109384 | Not-determined |
| S3_extracted_k141_109385 | Not-determined |
| S3_extracted_k141_37635 | Not-determined |
| S3_extracted_k141_45216 | Low-quality |
| S3_extracted_k141_15274 | Not-determined |
| S3_extracted_k141_85533 | Not-determined |
| S3_extracted_k141_20759 | Not-determined |
| S3_extracted_k141_93251 | Not-determined |
| S3_extracted_k141_20762 | Not-determined |
| S3_extracted_k141_93257 | Not-determined |
| S3_extracted_k141_85539 | Not-determined |
| S3_extracted_k141_109401 | Not-determined |
| S3_extracted_k141_68827 | Not-determined |
| S3_extracted_k141_37639 | Not-determined |
| S3_extracted_k141_109405 | Not-determined |
| S3_extracted_k141_68829 | Not-determined |
| S3_extracted_k141_93262 | Not-determined |
| S3_extracted_k141_78168 | Not-determined |
| S3_extracted_k141_109414 | Not-determined |
| S3_extracted_k141_15288 | Not-determined |
| S3_extracted_k141_109416 | Not-determined |
| S3_extracted_k141_100572 | Not-determined |
| S3_extracted_k141_109422 | Not-determined |
| S3_extracted_k141_15291 | Not-determined |
| S3_extracted_k141_124566 | Not-determined |
| S3_extracted_k141_93274 | Not-determined |
| S3_extracted_k141_37640 | Not-determined |
| S3_extracted_k141_37643 | Not-determined |
| S3_extracted_k141_93277 | Not-determined |
| S3_extracted_k141_93282 | Low-quality |
| S3_extracted_k141_93288 | Not-determined |
| S3_extracted_k141_109442 | Not-determined |
| S3_extracted_k141_29306 | Not-determined |
| S3_extracted_k141_54058 | Not-determined |
| S3_extracted_k141_29307 | Not-determined |
| S3_extracted_k141_117086 | Not-determined |
| S3_extracted_k141_29309 | Low-quality |
| S3_extracted_k141_100583 | Not-determined |
| S3_extracted_k141_117088 | Not-determined |
| S3_extracted_k141_100584 | Not-determined |
| S3_extracted_k141_29311 | Not-determined |
| S3_extracted_k141_85574 | Not-determined |
| S3_extracted_k141_109451 | Low-quality |
| S3_extracted_k141_109455 | Not-determined |
| S3_extracted_k141_29321 | Not-determined |
| S3_extracted_k141_37645 | Not-determined |
| S3_extracted_k141_15304 | Not-determined |
| S3_extracted_k141_15305 | Not-determined |
| S3_extracted_k141_45243 | Not-determined |
| S3_extracted_k141_15309 | Not-determined |
| S3_extracted_k141_109465 | Not-determined |
| S3_extracted_k141_100605 | Not-determined |
| S3_extracted_k141_100608 | Not-determined |
| S3_extracted_k141_124576 | Not-determined |
| S3_extracted_k141_29329 | Not-determined |
| S3_extracted_k141_109470 | Not-determined |
| S3_extracted_k141_78191 | Not-determined |
| S3_extracted_k141_117092 | Not-determined |
| S3_extracted_k141_29334 | Not-determined |
| S3_extracted_k141_117094 | Not-determined |
| S3_extracted_k141_100612 | Low-quality |
| S3_extracted_k141_20778 | Low-quality |
| S3_extracted_k141_20780 | Not-determined |
| S3_extracted_k141_109473 | Not-determined |
| S3_extracted_k141_100617 | Not-determined |
| S3_extracted_k141_109474 | Not-determined |
| S3_extracted_k141_100619 | Not-determined |
| S3_extracted_k141_37662 | Not-determined |
| S3_extracted_k141_37664 | Low-quality |
| S3_extracted_k141_29343 | Not-determined |
| S3_extracted_k141_109475 | Low-quality |
| S3_extracted_k141_37673 | Not-determined |
| S3_extracted_k141_45259 | Not-determined |
| S3_extracted_k141_100627 | Not-determined |
| S3_extracted_k141_5435 | Not-determined |
| S3_extracted_k141_78212 | Low-quality |
| S3_extracted_k141_5437 | Not-determined |
| S3_extracted_k141_100628 | Not-determined |
| S3_extracted_k141_20784 | Not-determined |
| S3_extracted_k141_62399 | Low-quality |
| S3_extracted_k141_54073 | Not-determined |
| S3_extracted_k141_100629 | Not-determined |
| S3_extracted_k141_37683 | Not-determined |
| S3_extracted_k141_37688 | Not-determined |
| S3_extracted_k141_68877 | Not-determined |
| S3_extracted_k141_15318 | Not-determined |
| S3_extracted_k141_15319 | Not-determined |
| S3_extracted_k141_45282 | Not-determined |
| S3_extracted_k141_29373 | Not-determined |
| S3_extracted_k141_29375 | Low-quality |
| S3_extracted_k141_62409 | Not-determined |
| S3_extracted_k141_100648 | Not-determined |
| S3_extracted_k141_93316 | Not-determined |
| S3_extracted_k141_5454 | Not-determined |
| S3_extracted_k141_85597 | Not-determined |
| S3_extracted_k141_100649 | Low-quality |
| S3_extracted_k141_85602 | Not-determined |
| S3_extracted_k141_5460 | Not-determined |
| S3_extracted_k141_85608 | Not-determined |
| S3_extracted_k141_29381 | Low-quality |
| S3_extracted_k141_45305 | Not-determined |
| S3_extracted_k141_68898 | Low-quality |
| S3_extracted_k141_45306 | Not-determined |
| S3_extracted_k141_54081 | Not-determined |
| S3_extracted_k141_45311 | Low-quality |
| S3_extracted_k141_54084 | Not-determined |
| S3_extracted_k141_117113 | Low-quality |
| S3_extracted_k141_100656 | Low-quality |
| S3_extracted_k141_100657 | Low-quality |
| S3_extracted_k141_29410 | Not-determined |
| S3_extracted_k141_93337 | Not-determined |
| S3_extracted_k141_93338 | Not-determined |
| S3_extracted_k141_20816 | Not-determined |
| S3_extracted_k141_85624 | Not-determined |
| S3_extracted_k141_20819 | Not-determined |
| S3_extracted_k141_29416 | Not-determined |
| S3_extracted_k141_78260 | Not-determined |
| S3_extracted_k141_85633 | Not-determined |
| S3_extracted_k141_78263 | Low-quality |
| S3_extracted_k141_78264 | Not-determined |
| S3_extracted_k141_93348 | Not-determined |
| S3_extracted_k141_93349 | Low-quality |
| S3_extracted_k141_109486 | Not-determined |
| S3_extracted_k141_109487 | Not-determined |
| S3_extracted_k141_62416 | Low-quality |
| S3_extracted_k141_20834 | Not-determined |
| S3_extracted_k141_29431 | Not-determined |
| S3_extracted_k141_45333 | Not-determined |
| S3_extracted_k141_29435 | Not-determined |
| S3_extracted_k141_45340 | Low-quality |
| S3_extracted_k141_15330 | Low-quality |
| S3_extracted_k141_117115 | Not-determined |
| S3_extracted_k141_109495 | Not-determined |
| S3_extracted_k141_100671 | Low-quality |
| S3_extracted_k141_109504 | Not-determined |
| S3_extracted_k141_54094 | Not-determined |
| S3_extracted_k141_78265 | Not-determined |
| S3_extracted_k141_54096 | Not-determined |
| S3_extracted_k141_54097 | Not-determined |
| S3_extracted_k141_85646 | Not-determined |
| S3_extracted_k141_100679 | Not-determined |
| S3_extracted_k141_62431 | Low-quality |
| S3_extracted_k141_124596 | Not-determined |
| S3_extracted_k141_124597 | Low-quality |
| S3_extracted_k141_124599 | Not-determined |
| S3_extracted_k141_62435 | Not-determined |
| S3_extracted_k141_124600 | Not-determined |
| S3_extracted_k141_15335 | Not-determined |
| S3_extracted_k141_29450 | Not-determined |
| S3_extracted_k141_78281 | Not-determined |
| S3_extracted_k141_62442 | Not-determined |
| S3_extracted_k141_78287 | Not-determined |
| S3_extracted_k141_124605 | Low-quality |
| S3_extracted_k141_100690 | Not-determined |
| S3_extracted_k141_124608 | Not-determined |
| S3_extracted_k141_78288 | Not-determined |
| S3_extracted_k141_62447 | Not-determined |
| S3_extracted_k141_78290 | Not-determined |
| S3_extracted_k141_29461 | Not-determined |
| S3_extracted_k141_62452 | Not-determined |
| S3_extracted_k141_78295 | Not-determined |
| S3_extracted_k141_85662 | Low-quality |
| S3_extracted_k141_78297 | Not-determined |
| S3_extracted_k141_15342 | Not-determined |
| S3_extracted_k141_29464 | Not-determined |
| S3_extracted_k141_93372 | Low-quality |
| S3_extracted_k141_124610 | Not-determined |
| S3_extracted_k141_20842 | Not-determined |
| S3_extracted_k141_20846 | Not-determined |
| S3_extracted_k141_20847 | Not-determined |
| S3_extracted_k141_124614 | Low-quality |
| S3_extracted_k141_117129 | Low-quality |
| S3_extracted_k141_124615 | Not-determined |
| S3_extracted_k141_124618 | Not-determined |
| S3_extracted_k141_15355 | Low-quality |
| S3_extracted_k141_62457 | Not-determined |
| S3_extracted_k141_100695 | Not-determined |
| S3_extracted_k141_78312 | Not-determined |
| S3_extracted_k141_15363 | Not-determined |
| S3_extracted_k141_117135 | Not-determined |
| S3_extracted_k141_109522 | Not-determined |
| S3_extracted_k141_29480 | Not-determined |
| S3_extracted_k141_15370 | Not-determined |
| S3_extracted_k141_45352 | Not-determined |
| S3_extracted_k141_5473 | Low-quality |
| S3_extracted_k141_5476 | Not-determined |
| S3_extracted_k141_93389 | Not-determined |
| S3_extracted_k141_54140 | Not-determined |
| S3_extracted_k141_100704 | Not-determined |
| S3_extracted_k141_85674 | Not-determined |
| S3_extracted_k141_117149 | Not-determined |
| S3_extracted_k141_109545 | Low-quality |
| S3_extracted_k141_109546 | Not-determined |
| S3_extracted_k141_93399 | Low-quality |
| S3_extracted_k141_109556 | Not-determined |
| S3_extracted_k141_37715 | Low-quality |
| S3_extracted_k141_15383 | Not-determined |
| S3_extracted_k141_15385 | Low-quality |
| S3_extracted_k141_109562 | Not-determined |
| S3_extracted_k141_93406 | Not-determined |
| S3_extracted_k141_100713 | Low-quality |
| S3_extracted_k141_109567 | Not-determined |
| S3_extracted_k141_100717 | Not-determined |
| S3_extracted_k141_93410 | Low-quality |
| S3_extracted_k141_109575 | Not-determined |
| S3_extracted_k141_85694 | Low-quality |
| S3_extracted_k141_45377 | Not-determined |
| S3_extracted_k141_100720 | Not-determined |
| S3_extracted_k141_100721 | Not-determined |
| S3_extracted_k141_100722 | Not-determined |
| S3_extracted_k141_93420 | Low-quality |
| S3_extracted_k141_109581 | Not-determined |
| S3_extracted_k141_100729 | Not-determined |
| S3_extracted_k141_29499 | Not-determined |
| S3_extracted_k141_15399 | Not-determined |
| S3_extracted_k141_45391 | Not-determined |
| S3_extracted_k141_100736 | Not-determined |
| S3_extracted_k141_109593 | Not-determined |
| S3_extracted_k141_54153 | Not-determined |
| S3_extracted_k141_45394 | Low-quality |
| S3_extracted_k141_124644 | Not-determined |
| S3_extracted_k141_100747 | Low-quality |
| S3_extracted_k141_109599 | Low-quality |
| S3_extracted_k141_5508 | Low-quality |
| S3_extracted_k141_109602 | Not-determined |
| S3_extracted_k141_109606 | Not-determined |
| S3_extracted_k141_78354 | Not-determined |
| S3_extracted_k141_29511 | Not-determined |
| S3_extracted_k141_100755 | Not-determined |
| S3_extracted_k141_109607 | Not-determined |
| S3_extracted_k141_93432 | Not-determined |
| S3_extracted_k141_109614 | Not-determined |
| S3_extracted_k141_93434 | Not-determined |
| S3_extracted_k141_78363 | Not-determined |
| S3_extracted_k141_78370 | Low-quality |
| S3_extracted_k141_5519 | Not-determined |
| S3_extracted_k141_78374 | Not-determined |
| S3_extracted_k141_78376 | Not-determined |
| S3_extracted_k141_29514 | Not-determined |
| S3_extracted_k141_93457 | Not-determined |
| S3_extracted_k141_93458 | Not-determined |
| S3_extracted_k141_93460 | Not-determined |
| S3_extracted_k141_109628 | Not-determined |
| S3_extracted_k141_109629 | Not-determined |
| S3_extracted_k141_20888 | Not-determined |
| S3_extracted_k141_100768 | Not-determined |
| S3_extracted_k141_93475 | Not-determined |
| S3_extracted_k141_20892 | Low-quality |
| S3_extracted_k141_93477 | Not-determined |
| S3_extracted_k141_45409 | Not-determined |
| S3_extracted_k141_124653 | Not-determined |
| S3_extracted_k141_100776 | Not-determined |
| S3_extracted_k141_29531 | Not-determined |
| S3_extracted_k141_93482 | Low-quality |
| S3_extracted_k141_29542 | Low-quality |
| S3_extracted_k141_78388 | Not-determined |
| S3_extracted_k141_109651 | Not-determined |
| S3_extracted_k141_20900 | Low-quality |
| S3_extracted_k141_20903 | Not-determined |
| S3_extracted_k141_85707 | Not-determined |
| S3_extracted_k141_124666 | Not-determined |
| S3_extracted_k141_93498 | Not-determined |
| S3_extracted_k141_109655 | Low-quality |
| S3_extracted_k141_78393 | Low-quality |
| S3_extracted_k141_109658 | Not-determined |
| S3_extracted_k141_124675 | Not-determined |
| S3_extracted_k141_45425 | Not-determined |
| S3_extracted_k141_54168 | Not-determined |
| S2_vs_k141_21630\|\|full | High-quality |
| S3_extracted_k141_5526 | Not-determined |
| S3_extracted_k141_124677 | Not-determined |
| S3_extracted_k141_45426 | Low-quality |
| S3_extracted_k141_85717 | Not-determined |
| S3_extracted_k141_124682 | Not-determined |
| S3_extracted_k141_54173 | Not-determined |
| S3_extracted_k141_54175 | Not-determined |
| S3_extracted_k141_124688 | Not-determined |
| S3_extracted_k141_78403 | Not-determined |
| S3_extracted_k141_124691 | Not-determined |
| S3_extracted_k141_124694 | Not-determined |
| S3_extracted_k141_78409 | Not-determined |
| S3_extracted_k141_5536 | Not-determined |
| S3_extracted_k141_93502 | Not-determined |
| S3_extracted_k141_5539 | Not-determined |
| S3_extracted_k141_45447 | Not-determined |
| S3_extracted_k141_15427 | Not-determined |
| S3_extracted_k141_78412 | Low-quality |
| S3_extracted_k141_29565 | Not-determined |
| S3_extracted_k141_5545 | Not-determined |
| S3_extracted_k141_109691 | Not-determined |
| S3_extracted_k141_85728 | Not-determined |
| S3_extracted_k141_54185 | Not-determined |
| S3_extracted_k141_29581 | Not-determined |
| S3_extracted_k141_5562 | Not-determined |
| S3_extracted_k141_93520 | Not-determined |
| S3_extracted_k141_45468 | Not-determined |
| S3_extracted_k141_45471 | Not-determined |
| S3_extracted_k141_117188 | Not-determined |
| S3_extracted_k141_85744 | Not-determined |
| S3_extracted_k141_54193 | Not-determined |
| S3_extracted_k141_117190 | Not-determined |
| S3_extracted_k141_85746 | Not-determined |
| S3_extracted_k141_29588 | Low-quality |
| S3_extracted_k141_5569 | Not-determined |
| S3_extracted_k141_117196 | Not-determined |
| S3_extracted_k141_117199 | Not-determined |
| S3_extracted_k141_124711 | Not-determined |
| S3_extracted_k141_5579 | Not-determined |
| S3_extracted_k141_78424 | Not-determined |
| S3_extracted_k141_78428 | Not-determined |
| S3_extracted_k141_45477 | Not-determined |
| S3_extracted_k141_45478 | Not-determined |
| S3_extracted_k141_45479 | Not-determined |
| S3_extracted_k141_45481 | Low-quality |
| S3_extracted_k141_5587 | Not-determined |
| S3_extracted_k141_100790 | Not-determined |
| S3_extracted_k141_20928 | Not-determined |
| S3_extracted_k141_93527 | Low-quality |
| S3_extracted_k141_45484 | Not-determined |
| S3_extracted_k141_54199 | Low-quality |
| S3_extracted_k141_78435 | Not-determined |
| S3_extracted_k141_54201 | Not-determined |

| | |
|---|---|
| S3_extracted_k141_93535 | Not-determined |
| S3_extracted_k141_5597 | Not-determined |
| S3_extracted_k141_5599 | Not-determined |
| S3_extracted_k141_54205 | Not-determined |
| S3_extracted_k141_78445 | Not-determined |
| S3_extracted_k141_93540 | Not-determined |
| S3_extracted_k141_45492 | Not-determined |
| S3_extracted_k141_100798 | Low-quality |
| S3_extracted_k141_5607 | Low-quality |
| S3_extracted_k141_45497 | Not-determined |
| S3_extracted_k141_117224 | Not-determined |
| S3_extracted_k141_45500 | Not-determined |
| S3_extracted_k141_117229 | Not-determined |
| S3_extracted_k141_117231 | Not-determined |
| S3_extracted_k141_29614 | Not-determined |
| S3_extracted_k141_45506 | Not-determined |
| S3_extracted_k141_5623 | Not-determined |
| S3_extracted_k141_85750 | Not-determined |
| S3_extracted_k141_45510 | Not-determined |
| S3_extracted_k141_109719 | Not-determined |
| S3_extracted_k141_124721 | Low-quality |
| S3_extracted_k141_109721 | Not-determined |
| S3_extracted_k141_78452 | Not-determined |
| S3_extracted_k141_109728 | Low-quality |
| S3_extracted_k141_78457 | Not-determined |
| S3_extracted_k141_109731 | Low-quality |
| S3_extracted_k141_29633 | Low-quality |
| S3_extracted_k141_93545 | Not-determined |
| S3_extracted_k141_100802 | Not-determined |
| S3_extracted_k141_109736 | Not-determined |
| S3_extracted_k141_93550 | Not-determined |
| S3_extracted_k141_29640 | Not-determined |
| S3_extracted_k141_109738 | Not-determined |
| S3_extracted_k141_78465 | Low-quality |
| S3_extracted_k141_45537 | Not-determined |
| S3_extracted_k141_93555 | Not-determined |
| S3_extracted_k141_117251 | Not-determined |
| S3_extracted_k141_78468 | Not-determined |
| S3_extracted_k141_45540 | Not-determined |
| S3_extracted_k141_78474 | Not-determined |
| S3_extracted_k141_78476 | Not-determined |
| S3_extracted_k141_29647 | Low-quality |
| S3_extracted_k141_45552 | Not-determined |
| S3_extracted_k141_117265 | Low-quality |
| S3_extracted_k141_45563 | Not-determined |
| S3_extracted_k141_100806 | Not-determined |
| S3_extracted_k141_117271 | Not-determined |
| S3_extracted_k141_45569 | Not-determined |
| S3_extracted_k141_117272 | Not-determined |
| S3_extracted_k141_100814 | Not-determined |
| S3_extracted_k141_124746 | Not-determined |
| S3_extracted_k141_45575 | Low-quality |
| S3_extracted_k141_85764 | Not-determined |
| S3_extracted_k141_37731 | Not-determined |
| S3_extracted_k141_37734 | Not-determined |
| S3_extracted_k141_93556 | Not-determined |
| S3_extracted_k141_37738 | Not-determined |
| S3_extracted_k141_15465 | Not-determined |
| S3_extracted_k141_93558 | Not-determined |
| S3_extracted_k141_37739 | Not-determined |
| S3_extracted_k141_93562 | Not-determined |
| S3_extracted_k141_45597 | Not-determined |
| S3_extracted_k141_15475 | Not-determined |
| S3_extracted_k141_29674 | Low-quality |
| S3_extracted_k141_15477 | Not-determined |
| S3_extracted_k141_20945 | Not-determined |
| S3_extracted_k141_15486 | Not-determined |
| S3_extracted_k141_93574 | Not-determined |
| S3_extracted_k141_20951 | Not-determined |
| S3_extracted_k141_20955 | Not-determined |
| S3_extracted_k141_29679 | Not-determined |
| S3_extracted_k141_62471 | Low-quality |
| S3_extracted_k141_15492 | Not-determined |
| S3_extracted_k141_5641 | Not-determined |
| S3_extracted_k141_5642 | Not-determined |
| S3_extracted_k141_15494 | Not-determined |
| S3_extracted_k141_37760 | Not-determined |
| S3_extracted_k141_15507 | Not-determined |
| S3_extracted_k141_37764 | Not-determined |
| S3_extracted_k141_15510 | Low-quality |
| S3_extracted_k141_20964 | Low-quality |
| S3_extracted_k141_20965 | Not-determined |
| S3_extracted_k141_20973 | Not-determined |
| S3_extracted_k141_15522 | Low-quality |
| S3_extracted_k141_62495 | Not-determined |
| S3_extracted_k141_20976 | Not-determined |
| S3_extracted_k141_62501 | Not-determined |
| S3_extracted_k141_124764 | Low-quality |
| S3_extracted_k141_62506 | Not-determined |
| S3_extracted_k141_62507 | Not-determined |
| S3_extracted_k141_93587 | Not-determined |
| S3_extracted_k141_20994 | Not-determined |
| S3_extracted_k141_93593 | Not-determined |
| S3_extracted_k141_93594 | Not-determined |
| S3_extracted_k141_54240 | Low-quality |
| S3_extracted_k141_117301 | Low-quality |
| S3_extracted_k141_124772 | Not-determined |
| S3_extracted_k141_93600 | Low-quality |
| S3_extracted_k141_37785 | Not-determined |
| S3_extracted_k141_93604 | Low-quality |
| S3_extracted_k141_20997 | Not-determined |
| S3_extracted_k141_93606 | Not-determined |
| S3_extracted_k141_100835 | Not-determined |
| S3_extracted_k141_68921 | Not-determined |
| S3_extracted_k141_68926 | Not-determined |
| S3_extracted_k141_45615 | Not-determined |
| S3_extracted_k141_37797 | Not-determined |
| S3_extracted_k141_45619 | Not-determined |
| S3_extracted_k141_85774 | Not-determined |
| S3_extracted_k141_85780 | Not-determined |
| S3_extracted_k141_100847 | Not-determined |
| S3_extracted_k141_5659 | Not-determined |
| S3_extracted_k141_124791 | Not-determined |
| S3_extracted_k141_62526 | Low-quality |
| S3_extracted_k141_62527 | Low-quality |
| S3_extracted_k141_37805 | Not-determined |
| S3_extracted_k141_68935 | Not-determined |
| S3_extracted_k141_29706 | Not-determined |
| S3_extracted_k141_85794 | Low-quality |
| S3_extracted_k141_124799 | Low-quality |
| S3_extracted_k141_54283 | Low-quality |
| S3_extracted_k141_54285 | Not-determined |
| S3_extracted_k141_15559 | Low-quality |
| S3_extracted_k141_21022 | Not-determined |
| S3_extracted_k141_37809 | Not-determined |
| S3_extracted_k141_93626 | Not-determined |
| S3_extracted_k141_124803 | Not-determined |
| S3_extracted_k141_62530 | Not-determined |
| S3_extracted_k141_93633 | Low-quality |
| S3_extracted_k141_62534 | Not-determined |
| S3_extracted_k141_93635 | Not-determined |
| S3_extracted_k141_68939 | Not-determined |
| S3_extracted_k141_62538 | Not-determined |
| S3_extracted_k141_85809 | Low-quality |
| S3_extracted_k141_45652 | Not-determined |
| S3_extracted_k141_78523 | Low-quality |
| S3_extracted_k141_85810 | Low-quality |
| S3_extracted_k141_124815 | Low-quality |
| S3_extracted_k141_45655 | Not-determined |
| S3_extracted_k141_109748 | Not-determined |
| S3_extracted_k141_78534 | Not-determined |
| S3_extracted_k141_93645 | Not-determined |
| S3_extracted_k141_78540 | Not-determined |
| S3_extracted_k141_68954 | Not-determined |
| S3_extracted_k141_68955 | Not-determined |
| S3_extracted_k141_68958 | Not-determined |
| S3_extracted_k141_109758 | Not-determined |
| S3_extracted_k141_15591 | Not-determined |
| S3_extracted_k141_85823 | Low-quality |
| S3_extracted_k141_29731 | Not-determined |
| S3_extracted_k141_85826 | Not-determined |
| S3_extracted_k141_45663 | Not-determined |
| S3_extracted_k141_85829 | Low-quality |
| S3_extracted_k141_68970 | Not-determined |
| S3_extracted_k141_85834 | Not-determined |
| S3_extracted_k141_78553 | Not-determined |
| S3_extracted_k141_15602 | Not-determined |
| S3_extracted_k141_15605 | Not-determined |
| S3_extracted_k141_45671 | Not-determined |
| S3_extracted_k141_15606 | Not-determined |
| S3_extracted_k141_37829 | Not-determined |
| S3_extracted_k141_85839 | Not-determined |
| S3_extracted_k141_68978 | Not-determined |
| S3_extracted_k141_85840 | Not-determined |
| S3_extracted_k141_85842 | Not-determined |
| S3_extracted_k141_85845 | Low-quality |
| S3_extracted_k141_15619 | Not-determined |
| S3_extracted_k141_78558 | Low-quality |
| S3_extracted_k141_85848 | Low-quality |
| S3_extracted_k141_68981 | Not-determined |
| S3_extracted_k141_68983 | Not-determined |
| S3_extracted_k141_15623 | Not-determined |
| S3_extracted_k141_124829 | Not-determined |
| S3_extracted_k141_15624 | Not-determined |
| S3_extracted_k141_68989 | Not-determined |
| S3_extracted_k141_21052 | Not-determined |
| S3_extracted_k141_124835 | Not-determined |
| S3_extracted_k141_78569 | Low-quality |
| S3_extracted_k141_21063 | Not-determined |
| S3_extracted_k141_68998 | Not-determined |
| S3_extracted_k141_124850 | Not-determined |
| S3_extracted_k141_124853 | Not-determined |
| S3_extracted_k141_124860 | Not-determined |
| S3_extracted_k141_69003 | Not-determined |
| S3_extracted_k141_85852 | Not-determined |
| S3_extracted_k141_85857 | Not-determined |
| S3_extracted_k141_69009 | Not-determined |
| S3_extracted_k141_124883 | Not-determined |
| S3_extracted_k141_124885 | Low-quality |
| S3_extracted_k141_45697 | Not-determined |
| S3_extracted_k141_93662 | Not-determined |
| S3_extracted_k141_29756 | Not-determined |
| S3_extracted_k141_78576 | Not-determined |
| S3_extracted_k141_93674 | Not-determined |
| S3_extracted_k141_93676 | Low-quality |
| S3_extracted_k141_85870 | Not-determined |
| S3_extracted_k141_85874 | Not-determined |
| S3_extracted_k141_15635 | Not-determined |
| S3_extracted_k141_85878 | Not-determined |
| S3_extracted_k141_45718 | Not-determined |
| S3_extracted_k141_37861 | Not-determined |
| S3_extracted_k141_124897 | Not-determined |
| S3_extracted_k141_93693 | Not-determined |
| S3_extracted_k141_85889 | Not-determined |
| S3_extracted_k141_124903 | Not-determined |
| S3_extracted_k141_78588 | Not-determined |
| S3_extracted_k141_54325 | Not-determined |
| S3_extracted_k141_54328 | Low-quality |
| S3_extracted_k141_37867 | Low-quality |
| S3_extracted_k141_37868 | Not-determined |
| S3_extracted_k141_54331 | Not-determined |
| S3_extracted_k141_124910 | Not-determined |
| S3_extracted_k141_124913 | Not-determined |
| S3_extracted_k141_29796 | Not-determined |
| S3_extracted_k141_54342 | Not-determined |
| S3_extracted_k141_21086 | Not-determined |
| S3_extracted_k141_124917 | Not-determined |
| S3_extracted_k141_45740 | Not-determined |
| S3_extracted_k141_78594 | Not-determined |
| S3_extracted_k141_29804 | Not-determined |
| S3_extracted_k141_45751 | Low-quality |
| S3_extracted_k141_117318 | Not-determined |
| S3_extracted_k141_45756 | Not-determined |
| S3_extracted_k141_45761 | Not-determined |
| S3_extracted_k141_21106 | Not-determined |
| S3_extracted_k141_5668 | Not-determined |
| S3_extracted_k141_5669 | Not-determined |
| S3_extracted_k141_124956 | Not-determined |
| S3_extracted_k141_93701 | Not-determined |
| S3_extracted_k141_93702 | Not-determined |
| S3_extracted_k141_21120 | Not-determined |
| S3_extracted_k141_117334 | Not-determined |
| S3_extracted_k141_124964 | Low-quality |
| S3_extracted_k141_37894 | Not-determined |
| S3_extracted_k141_37896 | Not-determined |
| S3_extracted_k141_21126 | Not-determined |
| S3_extracted_k141_85953 | Not-determined |
| S3_extracted_k141_5685 | Not-determined |
| S3_extracted_k141_21130 | Low-quality |
| S3_extracted_k141_62551 | Low-quality |
| S3_extracted_k141_62553 | Not-determined |
| S3_extracted_k141_93723 | Not-determined |
| S3_extracted_k141_85971 | Not-determined |
| S3_extracted_k141_62557 | Not-determined |
| S3_extracted_k141_69011 | Low-quality |
| S3_extracted_k141_93732 | Not-determined |
| S3_extracted_k141_69013 | Low-quality |
| S3_extracted_k141_21140 | Not-determined |
| S3_extracted_k141_69014 | Low-quality |
| S3_extracted_k141_15679 | Not-determined |
| S3_extracted_k141_85988 | Not-determined |
| S3_extracted_k141_124971 | Not-determined |
| S3_extracted_k141_5699 | Not-determined |
| S3_extracted_k141_5700 | Not-determined |
| S3_extracted_k141_78622 | Not-determined |
| S3_extracted_k141_78623 | Not-determined |
| S3_extracted_k141_78625 | Not-determined |
| S3_extracted_k141_100903 | Not-determined |
| S3_extracted_k141_100909 | Not-determined |
| S3_extracted_k141_100910 | Not-determined |
| S3_extracted_k141_62579 | Not-determined |
| S3_extracted_k141_78632 | Not-determined |
| S3_extracted_k141_37906 | Not-determined |
| S3_extracted_k141_100917 | Not-determined |
| S3_extracted_k141_15703 | Low-quality |
| S3_extracted_k141_62596 | Not-determined |
| S3_extracted_k141_109793 | Not-determined |
| S3_extracted_k141_62604 | Not-determined |
| S3_extracted_k141_37919 | Not-determined |
| S3_extracted_k141_62605 | Low-quality |
| S3_extracted_k141_62606 | Not-determined |
| S3_extracted_k141_37920 | Not-determined |
| S3_extracted_k141_5713 | Not-determined |
| S3_extracted_k141_109804 | Not-determined |
| S3_extracted_k141_21143 | Not-determined |
| S3_extracted_k141_109806 | Not-determined |
| S3_extracted_k141_37927 | Not-determined |
| S3_extracted_k141_21145 | Not-determined |
| S3_extracted_k141_93745 | Low-quality |
| S3_extracted_k141_124985 | Not-determined |
| S3_extracted_k141_93747 | Not-determined |
| S3_extracted_k141_124988 | Not-determined |
| S3_extracted_k141_37934 | Low-quality |
| S3_extracted_k141_78646 | Not-determined |
| S14_vs_k141_20193\|\|full | High-quality |
| S3_extracted_k141_15712 | Not-determined |
| S3_extracted_k141_21161 | Low-quality |
| S3_extracted_k141_21162 | Not-determined |
| S3_extracted_k141_29819 | Not-determined |
| S3_extracted_k141_5730 | Not-determined |
| S3_extracted_k141_45806 | Not-determined |
| S3_extracted_k141_78657 | Not-determined |
| S3_extracted_k141_54364 | Low-quality |
| S3_extracted_k141_62621 | Not-determined |
| S3_extracted_k141_45807 | Not-determined |
| S3_extracted_k141_117347 | Not-determined |
| S3_extracted_k141_29831 | Low-quality |
| S3_extracted_k141_15721 | Not-determined |
| S3_extracted_k141_62629 | Not-determined |
| S3_extracted_k141_45814 | Not-determined |
| S3_extracted_k141_62631 | Not-determined |
| S3_extracted_k141_5737 | Not-determined |
| S3_extracted_k141_15726 | Not-determined |
| S3_extracted_k141_5739 | Not-determined |
| S3_extracted_k141_29836 | Not-determined |
| S3_extracted_k141_45819 | Not-determined |
| S3_extracted_k141_15730 | Not-determined |
| S3_extracted_k141_62642 | Not-determined |
| S3_extracted_k141_93764 | Not-determined |
| S3_extracted_k141_100925 | Not-determined |
| S3_extracted_k141_100932 | Not-determined |
| S3_extracted_k141_86017 | Not-determined |
| S3_extracted_k141_124994 | Not-determined |
| S3_extracted_k141_117354 | Not-determined |
| S3_extracted_k141_117355 | Not-determined |
| S3_extracted_k141_15735 | Not-determined |
| S3_extracted_k141_5756 | Not-determined |
| S3_extracted_k141_15741 | Low-quality |
| S3_extracted_k141_124999 | Not-determined |
| S3_extracted_k141_5763 | Not-determined |
| S3_extracted_k141_29855 | Not-determined |
| S3_extracted_k141_125001 | Not-determined |
| S3_extracted_k141_62649 | Not-determined |
| S3_extracted_k141_29859 | Low-quality |
| S3_extracted_k141_62653 | Not-determined |
| S3_extracted_k141_29863 | Not-determined |
| S3_extracted_k141_100938 | Low-quality |
| S3_extracted_k141_100941 | Low-quality |
| S3_extracted_k141_109818 | Not-determined |
| S3_extracted_k141_69026 | Not-determined |
| S3_extracted_k141_29868 | Not-determined |
| S3_extracted_k141_15752 | Not-determined |
| S3_extracted_k141_86037 | Not-determined |
| S3_extracted_k141_62658 | Not-determined |
| S3_extracted_k141_86042 | Not-determined |
| S3_extracted_k141_62661 | Not-determined |
| S3_extracted_k141_29876 | Not-determined |
| S3_extracted_k141_69031 | Not-determined |
| S3_extracted_k141_62663 | Not-determined |
| S3_extracted_k141_54418 | Not-determined |
| S3_extracted_k141_109831 | Low-quality |
| S3_extracted_k141_29883 | Low-quality |
| S3_extracted_k141_93826 | Not-determined |
| S3_extracted_k141_93827 | Not-determined |
| S3_extracted_k141_117375 | Not-determined |
| S3_extracted_k141_54425 | Not-determined |
| S3_extracted_k141_62681 | Not-determined |
| S3_extracted_k141_109833 | Not-determined |
| S3_extracted_k141_86047 | Not-determined |
| S3_extracted_k141_93829 | Not-determined |
| S3_extracted_k141_62688 | Not-determined |
| S3_extracted_k141_86056 | Low-quality |
| S3_extracted_k141_69034 | Low-quality |
| S3_extracted_k141_62694 | Low-quality |
| S3_extracted_k141_62699 | Not-determined |
| S3_extracted_k141_45880 | Not-determined |
| S3_extracted_k141_69038 | Low-quality |
| S3_extracted_k141_54429 | Low-quality |
| S3_extracted_k141_86071 | Not-determined |
| S3_extracted_k141_69039 | Not-determined |
| S3_extracted_k141_15760 | Not-determined |
| S3_extracted_k141_37943 | Low-quality |
| S3_extracted_k141_37944 | Not-determined |
| S3_extracted_k141_21194 | Low-quality |
| S3_extracted_k141_93844 | Not-determined |
| S3_extracted_k141_45887 | Not-determined |
| S3_extracted_k141_54433 | Not-determined |
| S3_extracted_k141_54438 | Not-determined |
| S3_extracted_k141_69041 | Low-quality |
| S3_extracted_k141_117380 | Low-quality |
| S3_extracted_k141_15767 | Not-determined |
| S3_extracted_k141_54444 | Not-determined |
| S3_extracted_k141_37957 | Not-determined |
| S3_extracted_k141_54445 | Low-quality |
| S3_extracted_k141_109851 | Not-determined |
| S3_extracted_k141_69047 | Not-determined |
| S3_extracted_k141_37964 | Not-determined |
| S3_extracted_k141_21196 | Not-determined |
| S3_extracted_k141_5795 | Not-determined |
| S3_extracted_k141_54452 | Not-determined |
| S3_extracted_k141_21203 | Not-determined |
| S3_extracted_k141_21204 | Low-quality |
| S3_extracted_k141_109863 | Not-determined |
| S3_extracted_k141_5801 | Not-determined |
| S3_extracted_k141_54455 | Not-determined |
| S3_extracted_k141_125004 | Low-quality |
| S3_extracted_k141_78696 | Not-determined |
| S3_extracted_k141_21214 | Not-determined |
| S3_extracted_k141_21216 | Not-determined |
| S3_extracted_k141_37977 | Not-determined |
| S3_extracted_k141_21217 | Not-determined |
| S3_extracted_k141_15778 | Low-quality |
| S3_extracted_k141_37981 | Not-determined |
| S3_extracted_k141_21221 | Not-determined |
| S3_extracted_k141_45901 | Not-determined |
| S3_extracted_k141_45902 | Not-determined |
| S3_extracted_k141_117389 | Not-determined |
| S3_extracted_k141_117390 | Not-determined |
| S3_extracted_k141_29895 | Low-quality |
| S3_extracted_k141_86093 | Low-quality |
| S3_extracted_k141_29896 | Low-quality |
| S3_extracted_k141_45908 | Not-determined |
| S3_extracted_k141_117396 | Not-determined |
| S3_extracted_k141_45909 | Not-determined |
| S3_extracted_k141_15791 | Not-determined |
| S3_extracted_k141_117399 | Low-quality |
| S3_extracted_k141_29902 | Not-determined |
| S3_extracted_k141_93879 | Not-determined |
| S3_extracted_k141_37998 | Not-determined |
| S3_extracted_k141_45917 | Low-quality |
| S3_extracted_k141_93883 | Not-determined |
| S3_extracted_k141_93887 | Not-determined |
| S3_extracted_k141_93891 | Low-quality |
| S3_extracted_k141_69051 | Not-determined |
| S3_extracted_k141_69053 | Not-determined |
| S3_extracted_k141_117419 | Not-determined |
| S3_extracted_k141_93896 | Low-quality |
| S3_extracted_k141_54486 | Not-determined |
| S3_extracted_k141_109878 | Not-determined |
| S3_extracted_k141_69058 | Not-determined |
| S3_extracted_k141_54492 | Not-determined |
| S3_extracted_k141_109883 | Not-determined |
| S3_extracted_k141_109885 | Not-determined |
| S3_extracted_k141_38007 | Not-determined |
| S3_extracted_k141_117428 | Not-determined |
| S3_extracted_k141_15803 | Not-determined |
| S3_extracted_k141_69061 | Low-quality |
| S3_extracted_k141_62732 | Not-determined |
| S3_extracted_k141_69064 | Not-determined |
| S3_extracted_k141_38012 | Not-determined |
| S3_extracted_k141_38013 | Not-determined |
| S3_extracted_k141_86119 | Not-determined |
| S3_extracted_k141_93912 | Not-determined |
| S3_vs_k141_127423\|\|full | High-quality |
| S3_extracted_k141_117439 | Not-determined |
| S3_extracted_k141_125017 | Not-determined |
| S3_extracted_k141_117440 | Not-determined |
| S3_extracted_k141_117445 | Not-determined |
| S3_extracted_k141_117448 | Not-determined |
| S3_extracted_k141_54507 | Not-determined |
| S3_extracted_k141_69069 | Not-determined |
| S3_extracted_k141_125023 | Not-determined |
| S3_extracted_k141_45949 | Low-quality |
| S3_extracted_k141_45950 | Not-determined |
| S3_extracted_k141_45952 | Not-determined |
| S3_extracted_k141_45954 | Not-determined |
| S3_extracted_k141_38023 | Low-quality |
| S3_extracted_k141_45956 | Not-determined |
| S3_extracted_k141_125025 | Not-determined |
| S3_extracted_k141_45958 | Not-determined |
| S3_extracted_k141_109897 | Not-determined |
| S3_extracted_k141_15824 | Not-determined |
| S3_extracted_k141_109899 | Low-quality |
| S3_extracted_k141_125029 | Not-determined |
| S3_extracted_k141_38034 | Not-determined |
| S3_extracted_k141_54523 | Not-determined |
| S3_extracted_k141_54524 | Not-determined |
| S3_extracted_k141_54525 | Not-determined |
| S3_extracted_k141_29945 | Not-determined |
| S3_extracted_k141_15836 | Not-determined |
| S3_extracted_k141_69072 | Low-quality |
| S3_extracted_k141_29950 | Not-determined |
| S3_extracted_k141_109907 | Not-determined |
| S3_extracted_k141_69083 | Not-determined |
| S3_extracted_k141_54536 | Not-determined |
| S3_extracted_k141_86166 | Not-determined |
| S3_extracted_k141_86167 | Not-determined |
| S3_extracted_k141_54540 | Not-determined |
| S3_extracted_k141_125035 | Low-quality |
| S3_extracted_k141_38046 | Not-determined |
| S3_extracted_k141_38047 | Not-determined |
| S3_extracted_k141_54546 | Not-determined |
| S3_extracted_k141_5816 | Not-determined |
| S3_extracted_k141_109917 | Low-quality |
| S3_extracted_k141_38053 | Not-determined |
| S3_extracted_k141_117484 | Not-determined |
| S3_extracted_k141_21225 | Not-determined |
| S3_extracted_k141_15840 | Not-determined |
| S3_extracted_k141_54554 | Not-determined |
| S3_extracted_k141_54557 | Not-determined |
| S3_extracted_k141_117487 | Low-quality |
| S3_extracted_k141_15848 | Not-determined |
| S3_extracted_k141_54558 | Low-quality |
| S3_extracted_k141_38064 | Not-determined |
| S3_extracted_k141_117499 | Not-determined |
| S3_extracted_k141_117502 | Not-determined |
| S3_extracted_k141_21237 | Low-quality |
| S3_extracted_k141_69087 | Not-determined |
| S3_extracted_k141_62738 | Not-determined |
| S3_extracted_k141_86200 | Not-determined |
| S3_extracted_k141_38067 | Not-determined |
| S3_extracted_k141_38069 | Not-determined |
| S3_extracted_k141_117508 | Not-determined |
| S3_extracted_k141_38074 | Low-quality |
| S3_extracted_k141_62746 | Not-determined |
| S3_extracted_k141_46003 | Low-quality |
| S3_extracted_k141_62747 | Not-determined |
| S3_extracted_k141_86204 | Not-determined |
| S3_extracted_k141_78743 | Not-determined |
| S3_extracted_k141_46018 | Not-determined |
| S3_extracted_k141_38081 | Not-determined |
| S3_extracted_k141_30004 | Not-determined |
| S3_extracted_k141_21258 | Not-determined |
| S3_extracted_k141_78744 | Low-quality |
| S3_extracted_k141_38089 | Not-determined |
| S3_extracted_k141_21267 | Not-determined |
| S3_extracted_k141_62766 | Not-determined |
| S3_extracted_k141_30010 | Not-determined |
| S3_extracted_k141_78751 | Not-determined |
| S3_extracted_k141_30013 | Not-determined |
| S3_extracted_k141_21275 | Not-determined |
| S3_extracted_k141_21276 | Low-quality |
| S3_extracted_k141_30020 | Not-determined |
| S3_extracted_k141_46026 | Not-determined |
| S3_extracted_k141_21283 | Not-determined |
| S3_extracted_k141_78766 | Not-determined |
| S3_extracted_k141_78770 | Not-determined |
| S3_extracted_k141_109950 | Not-determined |
| S3_extracted_k141_109953 | Not-determined |
| S3_extracted_k141_21290 | Low-quality |
| S3_extracted_k141_15871 | Not-determined |
| S3_extracted_k141_21292 | Not-determined |
| S3_extracted_k141_62771 | Not-determined |
| S3_extracted_k141_62772 | Not-determined |
| S3_extracted_k141_46043 | Not-determined |
| S3_extracted_k141_78779 | Not-determined |
| S3_extracted_k141_117563 | Low-quality |
| S3_extracted_k141_86252 | Not-determined |
| S3_extracted_k141_125070 | Not-determined |
| S3_extracted_k141_38111 | Not-determined |
| S3_extracted_k141_117564 | Not-determined |
| S3_extracted_k141_78786 | Not-determined |
| S3_extracted_k141_21301 | Not-determined |
| S3_extracted_k141_86258 | Not-determined |
| S3_extracted_k141_38113 | Low-quality |
| S3_extracted_k141_15881 | Not-determined |
| S3_extracted_k141_21304 | Low-quality |
| S3_extracted_k141_86263 | Not-determined |
| S3_extracted_k141_93917 | Not-determined |
| S3_extracted_k141_93918 | Not-determined |
| S3_extracted_k141_78792 | Low-quality |
| S3_extracted_k141_125073 | Low-quality |
| S3_extracted_k141_86274 | Not-determined |
| S3_extracted_k141_117575 | Not-determined |
| S3_extracted_k141_125076 | Not-determined |
| S3_extracted_k141_125079 | Not-determined |
| S3_extracted_k141_86279 | Not-determined |
| S3_extracted_k141_86280 | Not-determined |
| S3_extracted_k141_117578 | Not-determined |
| S3_extracted_k141_69104 | Not-determined |
| S3_extracted_k141_117583 | Not-determined |
| S3_extracted_k141_30039 | Low-quality |
| S3_extracted_k141_78807 | Not-determined |
| S3_extracted_k141_78809 | Not-determined |
| S3_extracted_k141_117586 | Low-quality |
| S3_extracted_k141_125083 | Low-quality |
| S3_extracted_k141_46074 | Not-determined |
| S3_extracted_k141_30043 | Not-determined |
| S3_extracted_k141_93924 | Not-determined |
| S3_extracted_k141_93925 | Not-determined |
| S3_extracted_k141_125086 | Not-determined |
| S3_extracted_k141_38128 | Not-determined |
| S3_extracted_k141_38131 | Not-determined |
| S3_extracted_k141_38132 | Not-determined |
| S3_extracted_k141_62806 | Not-determined |
| S3_extracted_k141_86292 | Not-determined |
| S3_extracted_k141_15891 | Not-determined |
| S3_extracted_k141_30052 | Not-determined |
| S3_extracted_k141_125092 | Low-quality |
| S3_extracted_k141_30055 | Not-determined |
| S3_extracted_k141_78828 | Not-determined |
| S3_extracted_k141_125094 | Not-determined |
| S3_extracted_k141_86301 | Not-determined |
| S3_extracted_k141_125100 | Not-determined |
| S3_extracted_k141_93942 | Not-determined |
| S3_extracted_k141_78835 | Not-determined |
| S3_extracted_k141_62823 | Low-quality |
| S3_extracted_k141_86315 | Not-determined |
| S3_extracted_k141_125109 | Not-determined |
| S3_extracted_k141_109973 | Not-determined |
| S3_extracted_k141_5855 | Not-determined |
| S3_extracted_k141_109974 | Not-determined |
| S3_extracted_k141_86319 | Not-determined |
| S3_extracted_k141_5858 | Not-determined |
| S3_extracted_k141_62833 | Low-quality |
| S3_extracted_k141_125118 | Not-determined |
| S3_extracted_k141_117623 | Low-quality |
| S3_extracted_k141_109980 | Not-determined |
| S3_extracted_k141_125122 | Not-determined |

| | |
|---|---|
| S3_extracted_k141_93950 | Not-determined |
| S3_extracted_k141_5872 | Not-determined |
| S3_extracted_k141_86339 | Not-determined |
| S3_extracted_k141_93953 | Low-quality |
| S3_extracted_k141_93957 | Not-determined |
| S3_extracted_k141_109983 | Not-determined |
| S3_extracted_k141_30080 | Not-determined |
| S3_extracted_k141_109987 | Not-determined |
| S3_extracted_k141_117640 | Not-determined |
| S3_extracted_k141_117643 | Not-determined |
| S3_extracted_k141_30083 | Not-determined |
| S3_extracted_k141_109989 | Not-determined |
| S3_extracted_k141_109991 | Not-determined |
| S3_extracted_k141_125124 | Low-quality |
| S3_extracted_k141_46078 | Not-determined |
| S3_extracted_k141_109999 | Not-determined |
| S3_extracted_k141_62839 | Low-quality |
| S3_extracted_k141_110005 | Not-determined |
| S3_extracted_k141_93973 | Not-determined |
| S3_extracted_k141_93974 | Not-determined |
| S3_extracted_k141_110010 | Low-quality |
| S3_extracted_k141_110011 | Not-determined |
| S3_extracted_k141_78845 | Not-determined |
| S3_extracted_k141_78847 | Low-quality |
| S3_extracted_k141_110024 | Not-determined |
| S3_extracted_k141_78854 | Not-determined |
| S3_extracted_k141_117655 | Low-quality |
| S3_extracted_k141_117659 | Low-quality |
| S3_extracted_k141_5884 | Not-determined |
| S3_extracted_k141_5886 | Not-determined |
| S3_extracted_k141_5891 | Not-determined |
| S3_extracted_k141_5893 | Not-determined |
| S3_extracted_k141_15897 | Low-quality |
| S3_extracted_k141_15898 | Not-determined |
| S3_extracted_k141_125152 | Not-determined |
| S3_extracted_k141_62845 | Not-determined |
| S3_extracted_k141_15903 | Low-quality |
| S3_extracted_k141_15905 | Not-determined |
| S3_extracted_k141_46090 | Low-quality |
| S3_extracted_k141_125153 | Low-quality |
| S3_extracted_k141_110042 | Not-determined |
| S3_extracted_k141_46092 | Not-determined |
| S3_extracted_k141_46093 | Not-determined |
| S3_extracted_k141_93983 | Not-determined |
| S3_extracted_k141_78856 | Not-determined |
| S3_extracted_k141_110048 | Low-quality |
| S3_extracted_k141_117675 | Not-determined |
| S3_extracted_k141_30111 | Not-determined |
| S3_extracted_k141_110050 | Not-determined |
| S3_extracted_k141_46102 | Not-determined |
| S3_extracted_k141_86358 | Not-determined |
| S3_extracted_k141_54584 | Not-determined |
| S3_extracted_k141_15929 | Not-determined |
| S3_extracted_k141_78866 | Not-determined |
| S3_extracted_k141_54589 | Low-quality |
| S3_extracted_k141_110062 | Not-determined |
| S3_extracted_k141_30118 | Not-determined |
| S3_extracted_k141_117688 | Not-determined |
| S3_extracted_k141_54603 | Not-determined |
| S3_extracted_k141_117691 | Not-determined |
| S3_extracted_k141_46115 | Not-determined |
| S3_extracted_k141_110073 | Not-determined |
| S3_extracted_k141_62856 | Not-determined |
| S3_extracted_k141_110077 | Not-determined |
| S3_extracted_k141_110079 | Not-determined |
| S3_extracted_k141_62861 | Not-determined |
| S3_extracted_k141_78877 | Not-determined |
| S3_extracted_k141_94009 | Not-determined |
| S3_extracted_k141_110090 | Not-determined |
| S3_extracted_k141_54618 | Not-determined |
| S3_extracted_k141_30132 | Low-quality |
| S3_extracted_k141_94012 | Not-determined |
| S3_extracted_k141_117709 | Not-determined |
| S3_extracted_k141_94013 | Not-determined |
| S3_extracted_k141_125171 | Low-quality |
| S3_extracted_k141_30134 | Not-determined |
| S3_extracted_k141_94016 | Not-determined |
| S3_extracted_k141_125174 | Not-determined |
| S3_extracted_k141_94017 | Not-determined |
| S3_extracted_k141_62867 | Low-quality |
| S3_extracted_k141_110094 | Not-determined |
| S3_extracted_k141_94020 | Low-quality |
| S3_extracted_k141_54624 | Not-determined |
| S3_extracted_k141_38157 | Not-determined |
| S3_extracted_k141_54629 | Not-determined |
| S3_extracted_k141_62875 | Low-quality |
| S3_extracted_k141_38159 | Not-determined |
| S3_extracted_k141_54631 | Not-determined |
| S3_extracted_k141_110102 | Not-determined |
| S3_extracted_k141_54634 | Not-determined |
| S3_extracted_k141_62881 | Not-determined |
| S3_extracted_k141_86393 | Not-determined |
| S3_extracted_k141_110112 | Not-determined |
| S3_extracted_k141_62889 | Not-determined |
| S3_extracted_k141_46142 | Low-quality |
| S3_extracted_k141_69130 | Not-determined |
| S3_extracted_k141_110120 | Not-determined |
| S3_extracted_k141_86405 | Not-determined |
| S3_extracted_k141_94035 | Not-determined |
| S3_extracted_k141_86406 | Not-determined |
| S3_extracted_k141_62897 | Not-determined |
| S3_extracted_k141_62898 | Not-determined |
| S3_extracted_k141_94041 | Not-determined |
| S3_extracted_k141_110134 | Low-quality |
| S3_extracted_k141_94045 | Not-determined |
| S3_extracted_k141_117726 | Not-determined |
| S3_extracted_k141_62907 | Not-determined |
| S3_extracted_k141_62908 | Not-determined |
| S3_extracted_k141_54656 | Not-determined |
| S3_extracted_k141_46150 | Not-determined |
| S3_extracted_k141_54659 | Not-determined |
| S3_extracted_k141_117732 | Not-determined |
| S3_extracted_k141_30157 | Low-quality |
| S3_extracted_k141_100977 | Not-determined |
| S3_extracted_k141_125183 | Low-quality |
| S3_extracted_k141_94057 | Not-determined |
| S3_extracted_k141_46155 | Not-determined |
| S3_extracted_k141_117738 | Not-determined |
| S3_extracted_k141_30163 | Not-determined |
| S3_extracted_k141_21319 | Not-determined |
| S3_extracted_k141_21321 | Not-determined |
| S3_extracted_k141_78891 | Not-determined |
| S3_extracted_k141_125193 | Not-determined |
| S3_extracted_k141_21323 | Not-determined |
| S3_extracted_k141_21324 | Not-determined |
| S3_extracted_k141_54672 | Not-determined |
| S3_extracted_k141_21327 | Not-determined |
| S3_extracted_k141_78898 | Low-quality |
| S3_extracted_k141_46165 | Not-determined |
| S3_extracted_k141_86423 | Not-determined |
| S3_extracted_k141_100985 | Not-determined |
| S3_extracted_k141_117745 | Not-determined |
| S3_extracted_k141_21334 | Low-quality |
| S3_extracted_k141_54683 | Not-determined |
| S3_extracted_k141_110146 | Not-determined |
| S3_extracted_k141_38182 | Not-determined |
| S3_extracted_k141_94062 | Not-determined |
| S3_extracted_k141_5959 | Not-determined |
| S3_extracted_k141_69158 | Not-determined |
| S3_extracted_k141_62932 | Low-quality |
| S3_extracted_k141_62933 | Not-determined |
| S3_extracted_k141_30170 | Not-determined |
| S3_extracted_k141_86440 | Not-determined |
| S3_extracted_k141_94067 | Not-determined |
| S3_extracted_k141_78909 | Not-determined |
| S3_extracted_k141_46178 | Not-determined |
| S3_extracted_k141_94071 | Not-determined |
| S3_extracted_k141_21360 | Not-determined |
| S3_extracted_k141_21361 | Not-determined |
| S3_extracted_k141_86445 | Not-determined |
| S3_extracted_k141_78919 | Low-quality |
| S3_extracted_k141_78924 | Low-quality |
| S3_extracted_k141_94085 | Not-determined |
| S3_extracted_k141_86454 | Not-determined |
| S3_extracted_k141_69162 | Not-determined |
| S3_extracted_k141_69165 | Not-determined |
| S3_extracted_k141_38196 | Not-determined |
| S3_extracted_k141_117761 | Not-determined |
| S3_extracted_k141_125221 | Not-determined |
| S3_extracted_k141_30189 | Low-quality |
| S3_extracted_k141_117764 | Not-determined |
| S3_extracted_k141_30191 | Low-quality |
| S3_extracted_k141_94100 | Not-determined |
| S3_extracted_k141_54708 | Not-determined |
| S3_extracted_k141_100993 | Not-determined |
| S3_extracted_k141_100995 | Not-determined |
| S3_extracted_k141_46199 | Low-quality |
| S3_extracted_k141_117771 | Not-determined |
| S3_extracted_k141_46204 | Not-determined |
| S3_extracted_k141_62956 | Not-determined |
| S3_extracted_k141_86464 | Not-determined |
| S3_extracted_k141_30206 | Not-determined |
| S3_extracted_k141_117777 | Not-determined |
| S3_extracted_k141_86473 | Not-determined |
| S3_extracted_k141_125243 | Not-determined |
| S3_extracted_k141_86475 | Not-determined |
| S3_extracted_k141_94118 | Not-determined |
| S3_extracted_k141_86478 | Low-quality |
| S3_extracted_k141_38227 | Not-determined |
| S3_extracted_k141_125249 | Not-determined |
| S3_extracted_k141_30223 | Not-determined |
| S3_extracted_k141_5976 | Not-determined |
| S3_extracted_k141_110152 | Not-determined |
| S3_extracted_k141_46218 | Not-determined |
| S3_extracted_k141_62973 | Not-determined |
| S3_extracted_k141_54714 | Not-determined |
| S3_extracted_k141_38234 | Not-determined |
| S3_extracted_k141_110159 | Not-determined |
| S3_extracted_k141_117795 | Not-determined |
| S3_extracted_k141_78936 | Not-determined |
| S3_extracted_k141_30232 | Not-determined |
| S3_extracted_k141_78945 | Low-quality |
| S3_extracted_k141_38243 | Not-determined |
| S3_extracted_k141_46233 | Not-determined |
| S3_extracted_k141_62983 | Not-determined |
| S3_extracted_k141_38253 | Not-determined |
| S3_extracted_k141_100996 | Low-quality |
| S3_extracted_k141_62986 | Not-determined |
| S3_extracted_k141_117812 | Not-determined |
| S3_extracted_k141_38257 | Not-determined |
| S3_extracted_k141_30241 | Not-determined |
| S3_extracted_k141_38261 | Not-determined |
| S3_extracted_k141_38262 | Not-determined |
| S3_extracted_k141_62994 | Not-determined |
| S3_extracted_k141_78953 | Not-determined |
| S3_extracted_k141_62995 | Not-determined |
| S3_extracted_k141_117821 | Not-determined |
| S3_extracted_k141_62998 | Not-determined |
| S3_extracted_k141_117823 | Not-determined |
| S3_extracted_k141_38266 | Low-quality |
| S16_extracted_97279 | High-quality |
| S3_extracted_k141_54726 | Not-determined |
| S3_extracted_k141_125269 | Not-determined |
| S3_extracted_k141_78956 | Low-quality |
| S3_extracted_k141_78957 | Not-determined |
| S3_extracted_k141_69188 | Low-quality |
| S3_extracted_k141_125278 | Not-determined |
| S3_extracted_k141_69189 | Not-determined |
| S3_extracted_k141_63010 | Low-quality |
| S3_extracted_k141_125280 | Not-determined |
| S3_extracted_k141_30251 | Not-determined |
| S3_extracted_k141_6001 | Not-determined |
| S3_extracted_k141_6002 | Not-determined |
| S3_extracted_k141_69191 | Not-determined |
| S3_extracted_k141_69193 | Low-quality |
| S3_extracted_k141_46258 | Not-determined |
| S3_extracted_k141_54751 | Not-determined |
| S3_extracted_k141_30258 | Not-determined |
| S3_extracted_k141_117835 | Low-quality |
| S3_extracted_k141_78961 | Low-quality |
| S3_extracted_k141_69196 | Low-quality |
| S3_extracted_k141_110185 | Not-determined |
| S3_extracted_k141_46263 | Not-determined |
| S3_extracted_k141_117836 | Not-determined |
| S3_extracted_k141_54756 | Not-determined |
| S3_extracted_k141_86529 | Not-determined |
| S3_extracted_k141_38277 | Not-determined |
| S3_extracted_k141_63034 | Not-determined |
| S3_extracted_k141_117846 | Not-determined |
| S3_extracted_k141_117847 | Not-determined |
| S3_extracted_k141_46278 | Low-quality |
| S3_extracted_k141_110196 | Not-determined |
| S3_extracted_k141_54768 | Low-quality |
| S3_extracted_k141_101009 | Not-determined |
| S3_extracted_k141_94145 | Not-determined |
| S3_extracted_k141_110204 | Not-determined |
| S3_extracted_k141_117852 | Not-determined |
| S3_extracted_k141_63039 | Not-determined |
| S3_extracted_k141_117860 | Not-determined |
| S3_extracted_k141_54775 | Not-determined |
| S3_extracted_k141_54776 | Not-determined |
| S3_extracted_k141_125309 | Low-quality |
| S3_extracted_k141_117864 | Not-determined |
| S3_extracted_k141_21396 | Not-determined |
| S3_extracted_k141_86540 | Not-determined |
| S3_extracted_k141_110213 | Not-determined |
| S3_extracted_k141_117868 | Not-determined |
| S3_extracted_k141_38301 | Not-determined |
| S3_extracted_k141_78967 | Not-determined |
| S3_extracted_k141_63043 | Not-determined |
| S3_extracted_k141_110216 | Not-determined |
| S3_extracted_k141_110217 | Not-determined |
| S3_extracted_k141_110221 | Not-determined |
| S3_extracted_k141_86547 | Not-determined |
| S3_extracted_k141_110223 | Not-determined |
| S3_extracted_k141_86550 | Not-determined |
| S3_extracted_k141_94169 | Not-determined |
| S3_extracted_k141_63051 | Not-determined |
| S3_extracted_k141_78976 | Not-determined |
| S3_extracted_k141_117874 | Not-determined |
| S3_extracted_k141_117875 | Not-determined |
| S3_extracted_k141_125318 | Not-determined |
| S3_extracted_k141_117879 | Not-determined |
| S3_extracted_k141_38318 | Not-determined |
| S3_extracted_k141_78980 | Not-determined |
| S3_extracted_k141_21400 | Not-determined |
| S3_extracted_k141_46297 | Not-determined |
| S3_extracted_k141_30269 | Low-quality |
| S3_extracted_k141_125330 | Not-determined |
| S3_extracted_k141_125334 | Not-determined |
| S3_extracted_k141_94186 | Not-determined |
| S3_extracted_k141_101039 | Low-quality |
| S3_extracted_k141_86564 | Not-determined |
| S3_extracted_k141_30276 | Not-determined |
| S3_extracted_k141_110247 | Not-determined |
| S3_extracted_k141_86567 | Not-determined |
| S3_extracted_k141_63077 | Not-determined |
| S3_extracted_k141_63078 | Not-determined |
| S3_extracted_k141_6037 | Low-quality |
| S3_extracted_k141_30286 | Not-determined |
| S3_extracted_k141_54802 | Not-determined |
| S3_extracted_k141_38325 | Low-quality |
| S3_extracted_k141_30292 | Not-determined |
| S3_extracted_k141_6043 | Not-determined |
| S3_extracted_k141_63091 | Low-quality |
| S3_extracted_k141_86582 | Not-determined |
| S3_extracted_k141_69210 | Low-quality |
| S3_extracted_k141_54815 | Not-determined |
| S3_extracted_k141_54819 | Not-determined |
| S3_extracted_k141_69213 | Low-quality |
| S3_extracted_k141_69214 | Low-quality |
| S3_extracted_k141_54822 | Low-quality |
| S3_extracted_k141_69216 | Not-determined |
| S3_extracted_k141_21435 | Low-quality |
| S3_extracted_k141_63102 | Not-determined |
| S3_extracted_k141_94221 | Low-quality |
| S3_extracted_k141_63110 | Not-determined |
| S3_extracted_k141_38348 | Not-determined |
| S3_extracted_k141_38349 | Low-quality |
| S3_extracted_k141_110257 | Not-determined |
| S3_extracted_k141_21453 | Not-determined |
| S3_extracted_k141_110260 | Low-quality |
| S3_extracted_k141_46317 | Not-determined |
| S3_extracted_k141_101062 | Not-determined |
| S3_extracted_k141_63119 | Not-determined |
| S3_extracted_k141_101063 | Not-determined |
| S3_extracted_k141_63120 | Not-determined |
| S3_extracted_k141_101067 | Not-determined |
| S3_extracted_k141_94233 | Not-determined |
| S3_extracted_k141_63125 | Not-determined |
| S3_extracted_k141_110271 | Not-determined |
| S3_extracted_k141_101077 | Not-determined |
| S3_extracted_k141_54844 | Low-quality |
| S3_extracted_k141_63134 | Not-determined |
| S3_extracted_k141_63137 | Not-determined |
| S3_extracted_k141_101083 | Not-determined |
| S3_extracted_k141_69234 | Low-quality |
| S3_extracted_k141_69238 | Low-quality |
| S3_extracted_k141_54848 | Not-determined |
| S3_extracted_k141_21478 | Not-determined |
| S3_extracted_k141_94239 | Not-determined |
| S3_extracted_k141_125345 | Not-determined |
| S3_extracted_k141_38359 | Low-quality |
| S3_extracted_k141_101087 | Not-determined |
| S3_extracted_k141_125349 | Not-determined |
| S3_extracted_k141_125352 | Low-quality |
| S3_extracted_k141_6052 | Not-determined |
| S3_extracted_k141_54849 | Not-determined |
| S3_extracted_k141_125354 | Low-quality |
| S3_extracted_k141_94246 | Not-determined |
| S3_extracted_k141_101100 | Not-determined |
| S3_extracted_k141_125357 | Not-determined |
| S3_extracted_k141_6066 | Low-quality |
| S3_extracted_k141_86598 | Not-determined |
| S3_extracted_k141_6067 | Not-determined |
| S3_extracted_k141_125360 | Not-determined |
| S3_extracted_k141_6072 | Not-determined |
| S3_extracted_k141_110296 | Not-determined |
| S3_extracted_k141_46338 | Low-quality |
| S3_extracted_k141_86605 | Not-determined |
| S3_extracted_k141_54853 | Low-quality |
| S3_extracted_k141_21489 | Not-determined |
| S3_extracted_k141_21490 | Not-determined |
| S3_extracted_k141_86612 | Not-determined |
| S3_extracted_k141_38389 | Not-determined |
| S3_extracted_k141_86618 | Not-determined |
| S3_extracted_k141_101106 | Low-quality |
| S3_extracted_k141_6086 | Not-determined |
| S3_extracted_k141_30307 | Not-determined |
| S3_extracted_k141_30311 | Not-determined |
| S3_extracted_k141_30312 | Not-determined |
| S3_extracted_k141_69262 | Not-determined |
| S3_extracted_k141_110312 | Not-determined |
| S3_extracted_k141_54873 | Not-determined |
| S3_extracted_k141_86627 | Not-determined |
| S3_extracted_k141_69273 | Not-determined |
| S3_extracted_k141_38407 | Not-determined |
| S3_extracted_k141_6103 | Low-quality |
| S3_extracted_k141_6104 | Not-determined |
| S3_extracted_k141_21514 | Low-quality |
| S3_extracted_k141_21515 | Not-determined |
| S3_extracted_k141_6106 | Not-determined |
| S3_extracted_k141_21522 | Low-quality |
| S3_extracted_k141_110314 | Not-determined |
| S3_extracted_k141_110320 | Not-determined |
| S3_extracted_k141_110321 | Not-determined |
| S3_extracted_k141_125367 | Not-determined |
| S3_extracted_k141_125370 | Not-determined |
| S3_extracted_k141_38414 | Not-determined |
| S3_extracted_k141_6126 | Low-quality |
| S3_extracted_k141_101128 | Not-determined |
| S3_extracted_k141_6130 | Not-determined |
| S3_extracted_k141_46366 | Not-determined |
| S3_extracted_k141_46369 | Not-determined |
| S3_extracted_k141_86643 | Not-determined |
| S3_extracted_k141_86645 | Not-determined |
| S3_extracted_k141_21533 | Not-determined |
| S3_extracted_k141_38434 | Not-determined |
| S3_extracted_k141_38435 | Not-determined |
| S3_extracted_k141_6137 | Not-determined |
| S3_extracted_k141_21538 | Low-quality |
| S3_extracted_k141_6141 | Not-determined |
| S3_extracted_k141_69304 | Not-determined |
| S3_extracted_k141_6144 | Not-determined |
| S3_extracted_k141_101143 | Low-quality |
| S3_extracted_k141_69307 | Not-determined |
| S3_extracted_k141_46382 | Not-determined |
| S3_extracted_k141_38448 | Not-determined |
| S3_extracted_k141_54908 | Not-determined |
| S3_extracted_k141_69311 | Low-quality |
| S3_extracted_k141_46383 | Not-determined |
| S3_extracted_k141_21548 | Low-quality |
| S3_extracted_k141_38455 | Not-determined |
| S3_extracted_k141_21549 | Not-determined |
| S3_extracted_k141_86651 | Not-determined |
| S3_extracted_k141_86653 | Not-determined |
| S3_extracted_k141_38463 | Not-determined |
| S3_extracted_k141_30325 | Not-determined |
| S3_extracted_k141_86657 | Not-determined |
| S3_extracted_k141_30328 | Not-determined |
| S3_extracted_k141_21557 | Not-determined |
| S3_extracted_k141_38467 | Low-quality |
| S3_extracted_k141_21561 | Not-determined |
| S3_extracted_k141_6153 | Not-determined |
| S3_extracted_k141_94267 | Not-determined |
| S3_extracted_k141_94268 | Not-determined |
| S3_extracted_k141_86670 | Not-determined |
| S3_extracted_k141_101149 | Low-quality |
| S3_extracted_k141_54916 | Low-quality |
| S3_extracted_k141_38479 | Not-determined |
| S3_extracted_k141_38482 | Not-determined |
| S3_extracted_k141_6157 | Not-determined |
| S3_extracted_k141_79012 | Low-quality |
| S3_extracted_k141_6161 | Low-quality |
| S3_extracted_k141_69325 | Not-determined |
| S3_extracted_k141_63173 | Not-determined |
| S3_extracted_k141_54924 | Not-determined |
| S3_extracted_k141_38491 | Low-quality |
| S3_extracted_k141_54927 | Not-determined |
| S3_extracted_k141_38494 | Not-determined |
| S3_extracted_k141_6167 | Not-determined |
| S3_extracted_k141_86684 | Not-determined |
| S3_extracted_k141_46393 | Not-determined |
| S3_extracted_k141_110340 | Not-determined |
| S3_extracted_k141_101173 | Not-determined |
| S3_extracted_k141_110344 | Not-determined |
| S3_extracted_k141_21590 | Not-determined |
| S3_extracted_k141_38502 | Not-determined |
| S3_extracted_k141_54935 | Not-determined |
| S3_extracted_k141_38506 | Not-determined |
| S3_extracted_k141_21593 | Not-determined |
| S3_extracted_k141_110347 | Low-quality |
| S3_extracted_k141_21595 | Not-determined |
| S3_extracted_k141_6177 | Not-determined |
| S3_extracted_k141_101183 | Not-determined |
| S3_extracted_k141_21599 | Low-quality |
| S3_extracted_k141_21600 | Not-determined |
| S3_extracted_k141_46405 | Not-determined |
| S3_extracted_k141_21602 | Not-determined |
| S3_extracted_k141_6185 | Not-determined |
| S3_extracted_k141_6186 | Not-determined |
| S3_extracted_k141_21606 | Not-determined |
| S3_extracted_k141_101190 | Not-determined |
| S3_extracted_k141_101198 | Low-quality |
| S3_extracted_k141_21611 | Not-determined |
| S3_extracted_k141_101202 | Not-determined |
| S3_extracted_k141_79025 | Not-determined |
| S3_extracted_k141_63192 | Not-determined |
| S3_extracted_k141_38515 | Not-determined |
| S3_extracted_k141_69334 | Not-determined |
| S3_extracted_k141_21618 | Not-determined |
| S3_extracted_k141_6193 | Not-determined |
| S3_extracted_k141_110361 | Not-determined |
| S3_extracted_k141_63204 | Not-determined |
| S3_extracted_k141_21622 | Not-determined |
| S3_extracted_k141_110365 | Not-determined |
| S3_extracted_k141_21627 | Not-determined |
| S3_extracted_k141_63209 | Not-determined |
| S3_extracted_k141_117898 | Not-determined |
| S3_extracted_k141_101210 | Not-determined |
| S3_extracted_k141_101211 | Low-quality |
| S3_extracted_k141_110372 | Not-determined |
| S3_extracted_k141_86688 | Not-determined |
| S3_extracted_k141_38527 | Low-quality |
| S3_extracted_k141_125408 | Not-determined |
| S3_extracted_k141_54942 | Not-determined |
| S3_extracted_k141_86697 | Not-determined |
| S3_extracted_k141_101225 | Not-determined |
| S3_extracted_k141_54949 | Not-determined |
| S3_extracted_k141_54950 | Not-determined |
| S3_extracted_k141_94297 | Not-determined |
| S3_extracted_k141_54952 | Not-determined |
| S3_extracted_k141_117905 | Low-quality |
| S3_extracted_k141_117906 | Not-determined |
| S3_extracted_k141_38533 | Low-quality |
| S3_extracted_k141_38534 | Not-determined |
| S3_extracted_k141_54957 | Not-determined |
| S3_extracted_k141_54960 | Not-determined |
| S3_extracted_k141_117909 | Not-determined |
| S3_extracted_k141_54963 | Not-determined |
| S3_extracted_k141_69362 | Not-determined |
| S3_extracted_k141_21632 | Not-determined |
| S3_extracted_k141_21633 | Not-determined |
| S3_extracted_k141_117916 | Not-determined |
| S3_extracted_k141_125430 | Low-quality |
| S3_extracted_k141_38553 | Not-determined |
| S3_extracted_k141_125434 | Not-determined |
| S3_extracted_k141_125435 | Not-determined |
| S3_extracted_k141_69370 | Not-determined |
| S3_extracted_k141_63217 | Not-determined |
| S3_extracted_k141_117920 | Low-quality |
| S3_extracted_k141_46417 | Not-determined |
| S3_extracted_k141_101243 | Not-determined |
| S3_extracted_k141_46419 | Not-determined |
| S3_extracted_k141_46423 | Not-determined |
| S3_extracted_k141_69384 | Low-quality |
| S3_extracted_k141_69387 | Not-determined |
| S3_extracted_k141_86728 | Not-determined |
| S3_extracted_k141_86731 | Not-determined |
| S3_extracted_k141_63228 | Low-quality |
| S3_extracted_k141_69389 | Not-determined |
| S3_extracted_k141_79048 | Not-determined |
| S3_extracted_k141_30399 | Not-determined |
| S3_extracted_k141_21649 | Not-determined |
| S3_extracted_k141_63231 | Not-determined |
| S3_extracted_k141_21654 | Low-quality |
| S3_extracted_k141_86738 | Not-determined |
| S3_extracted_k141_86744 | Not-determined |
| S3_extracted_k141_125443 | Low-quality |
| S3_extracted_k141_101252 | Not-determined |
| S3_extracted_k141_101253 | Not-determined |
| S3_extracted_k141_86745 | Not-determined |
| S3_extracted_k141_63237 | Not-determined |
| S3_extracted_k141_117927 | Not-determined |
| S3_extracted_k141_30416 | Not-determined |
| S3_extracted_k141_117931 | Not-determined |
| S3_extracted_k141_101273 | Not-determined |
| S3_extracted_k141_30433 | Not-determined |
| S3_extracted_k141_110381 | Not-determined |
| S3_extracted_k141_110382 | Not-determined |
| S3_extracted_k141_30437 | Low-quality |
| S3_extracted_k141_110386 | Not-determined |
| S3_extracted_k141_110387 | Not-determined |
| S3_extracted_k141_30442 | Not-determined |
| S3_extracted_k141_110388 | Low-quality |
| S3_extracted_k141_69411 | Not-determined |
| S3_extracted_k141_101280 | Not-determined |
| S3_extracted_k141_117955 | Not-determined |
| S3_extracted_k141_86753 | Not-determined |
| S3_extracted_k141_110395 | Not-determined |
| S3_extracted_k141_101286 | Not-determined |
| S3_extracted_k141_54977 | Not-determined |
| S3_extracted_k141_54978 | Not-determined |
| S3_extracted_k141_86756 | Low-quality |
| S3_extracted_k141_63274 | Not-determined |
| S3_extracted_k141_86759 | Not-determined |
| S3_extracted_k141_94301 | Not-determined |
| S3_extracted_k141_54983 | Not-determined |
| S3_extracted_k141_69422 | Low-quality |
| S3_extracted_k141_63280 | Not-determined |
| S3_extracted_k141_69423 | Not-determined |
| S3_extracted_k141_54987 | Not-determined |
| S3_extracted_k141_94308 | Low-quality |
| S3_extracted_k141_94309 | Low-quality |
| S3_extracted_k141_86762 | Low-quality |
| S3_extracted_k141_6220 | Not-determined |
| S3_extracted_k141_86766 | Not-determined |
| S3_extracted_k141_101297 | Low-quality |
| S3_extracted_k141_86768 | Not-determined |
| S3_extracted_k141_101304 | Not-determined |
| S3_extracted_k141_6234 | Low-quality |
| S3_extracted_k141_30468 | Not-determined |
| S3_extracted_k141_54992 | Not-determined |
| S3_extracted_k141_21668 | Not-determined |
| S3_extracted_k141_110403 | Low-quality |
| S3_extracted_k141_117969 | Not-determined |
| S3_extracted_k141_117972 | Not-determined |
| S3_extracted_k141_54994 | Not-determined |
| S3_extracted_k141_117986 | Low-quality |
| S3_extracted_k141_55004 | Not-determined |
| S3_extracted_k141_55005 | Not-determined |
| S3_extracted_k141_101317 | Not-determined |
| S3_extracted_k141_125456 | Not-determined |
| S3_extracted_k141_117991 | Low-quality |
| S3_extracted_k141_6270 | Not-determined |
| S3_extracted_k141_21683 | Low-quality |
| S3_extracted_k141_21685 | Low-quality |
| S3_extracted_k141_118004 | Not-determined |
| S3_extracted_k141_125466 | Not-determined |
| S3_extracted_k141_86791 | Not-determined |
| S3_extracted_k141_86792 | Not-determined |
| S3_extracted_k141_118008 | Not-determined |
| S3_extracted_k141_125471 | Not-determined |
| S3_extracted_k141_125472 | Not-determined |
| S3_extracted_k141_125475 | Low-quality |
| S3_extracted_k141_118010 | Low-quality |
| S3_extracted_k141_118011 | Not-determined |
| S3_extracted_k141_118012 | Not-determined |
| S3_extracted_k141_118015 | Not-determined |

| | |
|---|---|
| S3_extracted_k141_101330 | Low-quality |
| S3_extracted_k141_118019 | Not-determined |
| S3_extracted_k141_101335 | Not-determined |
| S3_extracted_k141_6287 | Not-determined |
| S3_extracted_k141_110417 | Not-determined |
| S3_extracted_k141_55021 | Not-determined |
| S3_extracted_k141_63290 | Low-quality |
| S3_extracted_k141_118023 | Not-determined |
| S3_extracted_k141_110421 | Not-determined |
| S3_extracted_k141_118024 | Not-determined |
| S3_extracted_k141_21712 | Not-determined |
| S3_extracted_k141_118032 | Not-determined |
| S3_extracted_k141_110422 | Low-quality |
| S3_extracted_k141_110424 | Not-determined |
| S3_extracted_k141_125480 | Not-determined |
| S3_extracted_k141_125482 | Not-determined |
| S3_extracted_k141_125484 | Not-determined |
| S3_extracted_k141_6301 | Not-determined |
| S3_extracted_k141_101342 | Not-determined |
| S3_extracted_k141_6307 | Not-determined |
| S3_extracted_k141_6309 | Not-determined |
| S3_extracted_k141_118047 | Not-determined |
| S3_extracted_k141_118050 | Not-determined |
| S3_extracted_k141_125495 | Not-determined |
| S3_extracted_k141_125497 | Not-determined |
| S3_extracted_k141_6319 | Not-determined |
| S3_extracted_k141_101345 | Not-determined |
| S3_extracted_k141_118059 | Not-determined |
| S3_extracted_k141_125504 | Not-determined |
| S3_extracted_k141_55034 | Not-determined |
| S3_extracted_k141_55037 | Not-determined |
| S3_extracted_k141_101359 | Not-determined |
| S3_extracted_k141_6340 | Not-determined |
| S3_extracted_k141_6342 | Not-determined |
| S3_extracted_k141_55044 | Low-quality |
| S3_extracted_k141_125521 | Not-determined |
| S3_extracted_k141_110457 | Not-determined |
| S3_extracted_k141_94321 | Low-quality |
| S3_extracted_k141_94323 | Not-determined |
| S3_extracted_k141_94331 | Not-determined |
| S3_extracted_k141_101362 | Not-determined |
| S3_extracted_k141_110472 | Low-quality |
| S3_extracted_k141_6359 | Not-determined |
| S3_extracted_k141_110475 | Low-quality |
| S3_extracted_k141_101370 | Not-determined |
| S3_extracted_k141_101371 | Not-determined |
| S3_extracted_k141_101372 | Not-determined |
| S3_extracted_k141_21742 | Not-determined |
| S3_extracted_k141_101379 | Not-determined |
| S3_extracted_k141_94339 | Not-determined |
| S3_extracted_k141_94341 | Not-determined |
| S3_extracted_k141_125548 | Not-determined |
| S3_extracted_k141_125549 | Not-determined |
| S3_extracted_k141_110480 | Not-determined |
| S3_extracted_k141_125556 | Not-determined |
| S3_extracted_k141_63320 | Not-determined |
| S3_extracted_k141_118072 | Low-quality |
| S3_extracted_k141_94353 | Not-determined |
| S3_extracted_k141_101395 | Not-determined |
| S3_extracted_k141_46428 | Not-determined |
| S3_extracted_k141_118074 | Not-determined |
| S3_extracted_k141_46431 | Not-determined |
| S3_extracted_k141_63326 | Not-determined |
| S3_extracted_k141_94360 | Not-determined |
| S3_extracted_k141_69429 | Not-determined |
| S3_extracted_k141_55048 | Not-determined |
| S3_extracted_k141_55049 | Not-determined |
| S3_extracted_k141_101401 | Low-quality |
| S3_extracted_k141_6369 | Not-determined |
| S3_extracted_k141_110492 | Not-determined |
| S3_extracted_k141_6371 | Not-determined |
| S3_extracted_k141_110497 | Not-determined |
| S3_extracted_k141_21770 | Not-determined |
| S3_extracted_k141_21771 | Not-determined |
| S3_extracted_k141_101410 | Not-determined |
| S3_extracted_k141_55059 | Not-determined |
| S3_extracted_k141_55060 | Not-determined |
| S3_extracted_k141_55061 | Not-determined |
| S3_extracted_k141_21772 | Not-determined |
| S3_extracted_k141_55063 | Low-quality |
| S3_extracted_k141_69449 | Not-determined |
| S3_extracted_k141_110506 | Low-quality |
| S3_extracted_k141_6385 | Not-determined |
| S3_extracted_k141_55069 | Not-determined |
| S3_extracted_k141_125574 | Not-determined |
| S3_extracted_k141_55072 | Not-determined |
| S3_extracted_k141_110514 | Not-determined |
| S3_extracted_k141_55074 | Not-determined |
| S3_extracted_k141_6390 | Low-quality |
| S3_extracted_k141_86811 | Not-determined |
| S3_extracted_k141_21777 | Low-quality |
| S3_extracted_k141_6393 | Not-determined |
| S3_extracted_k141_118109 | Not-determined |
| S3_extracted_k141_21783 | Not-determined |
| S3_extracted_k141_46467 | Not-determined |
| S3_extracted_k141_21784 | Low-quality |
| S3_extracted_k141_125582 | Not-determined |
| S3_extracted_k141_30484 | Not-determined |
| S3_extracted_k141_86816 | Low-quality |
| S3_extracted_k141_55079 | Not-determined |
| S3_extracted_k141_6399 | Not-determined |
| S3_extracted_k141_46472 | Not-determined |
| S3_extracted_k141_101429 | Not-determined |
| S3_extracted_k141_55082 | Not-determined |
| S3_extracted_k141_101430 | Not-determined |
| S3_extracted_k141_94371 | Not-determined |
| S3_extracted_k141_69455 | Not-determined |
| S3_extracted_k141_55084 | Low-quality |
| S3_extracted_k141_118123 | Not-determined |
| S3_extracted_k141_94385 | Not-determined |
| S3_extracted_k141_55088 | Not-determined |
| S3_extracted_k141_101440 | Low-quality |
| S3_extracted_k141_21788 | Not-determined |
| S3_extracted_k141_55092 | Not-determined |
| S3_extracted_k141_94395 | Low-quality |
| S3_extracted_k141_6415 | Not-determined |
| S3_extracted_k141_69458 | Low-quality |
| S3_extracted_k141_21799 | Not-determined |
| S3_extracted_k141_46494 | Low-quality |
| S3_extracted_k141_94399 | Not-determined |
| S3_extracted_k141_69462 | Not-determined |
| S3_extracted_k141_46497 | Not-determined |
| S3_extracted_k141_94400 | Low-quality |
| S3_extracted_k141_69464 | Not-determined |
| S3_extracted_k141_125587 | Not-determined |
| S3_extracted_k141_69471 | Not-determined |
| S3_extracted_k141_55105 | Not-determined |
| S3_extracted_k141_86820 | Not-determined |
| S3_extracted_k141_125595 | Not-determined |
| S3_extracted_k141_118127 | Not-determined |
| S3_extracted_k141_125597 | Not-determined |
| S3_extracted_k141_6427 | Not-determined |
| S3_extracted_k141_6428 | Not-determined |
| S3_extracted_k141_86824 | Not-determined |
| S3_extracted_k141_6429 | Not-determined |
| S3_extracted_k141_110540 | Low-quality |
| S3_extracted_k141_110541 | Not-determined |
| S3_extracted_k141_110542 | Not-determined |
| S3_extracted_k141_55112 | Low-quality |
| S3_extracted_k141_110545 | Not-determined |
| S3_extracted_k141_110546 | Not-determined |
| S3_extracted_k141_55113 | Low-quality |
| S3_extracted_k141_63336 | Not-determined |
| S3_extracted_k141_94406 | Not-determined |
| S3_extracted_k141_69494 | Not-determined |
| S3_extracted_k141_21815 | Not-determined |
| S3_extracted_k141_38559 | Low-quality |
| S3_extracted_k141_30492 | Not-determined |
| S3_extracted_k141_55124 | Not-determined |
| S3_extracted_k141_21819 | Not-determined |
| S3_extracted_k141_63344 | Not-determined |
| S3_extracted_k141_86827 | Low-quality |
| S3_extracted_k141_86831 | Not-determined |
| S3_extracted_k141_110557 | Not-determined |
| S3_extracted_k141_38576 | Not-determined |
| S3_extracted_k141_38580 | Not-determined |
| S3_extracted_k141_86841 | Low-quality |
| S3_extracted_k141_125609 | Not-determined |
| S3_extracted_k141_63354 | Low-quality |
| S3_extracted_k141_118138 | Low-quality |
| S3_extracted_k141_86846 | Not-determined |
| S3_extracted_k141_21833 | Not-determined |
| S3_extracted_k141_30516 | Not-determined |
| S3_extracted_k141_110566 | Not-determined |
| S3_extracted_k141_118140 | Not-determined |
| S3_extracted_k141_94417 | Low-quality |
| S3_extracted_k141_125621 | Not-determined |
| S3_extracted_k141_21840 | Not-determined |
| S3_extracted_k141_94424 | Not-determined |
| S3_extracted_k141_86855 | Not-determined |
| S3_extracted_k141_30534 | Not-determined |
| S3_extracted_k141_110584 | Not-determined |
| S3_extracted_k141_94428 | Not-determined |
| S3_extracted_k141_110585 | Not-determined |
| S3_extracted_k141_21853 | Low-quality |
| S3_extracted_k141_125633 | Not-determined |
| S3_extracted_k141_21854 | Not-determined |
| S3_extracted_k141_118148 | Not-determined |
| S3_extracted_k141_86857 | Not-determined |
| S3_extracted_k141_79096 | Not-determined |
| S3_extracted_k141_118150 | Not-determined |
| S3_extracted_k141_94435 | Not-determined |
| S3_extracted_k141_118153 | Not-determined |
| S3_extracted_k141_79102 | Not-determined |
| S3_extracted_k141_79104 | Not-determined |
| S3_extracted_k141_94439 | Not-determined |
| S3_extracted_k141_69517 | Not-determined |
| S3_extracted_k141_30539 | Not-determined |
| S3_extracted_k141_101446 | Low-quality |
| S3_extracted_k141_21869 | Not-determined |
| S3_extracted_k141_30542 | Not-determined |
| S3_extracted_k141_79125 | Not-determined |
| S3_extracted_k141_79131 | Not-determined |
| S3_extracted_k141_94452 | Not-determined |
| S3_extracted_k141_46509 | Not-determined |
| S3_extracted_k141_30563 | Not-determined |
| S3_extracted_k141_46513 | Not-determined |
| S3_extracted_k141_86867 | Not-determined |
| S3_extracted_k141_110588 | Not-determined |
| S3_extracted_k141_69538 | Low-quality |
| S3_extracted_k141_46518 | Low-quality |
| S3_extracted_k141_38625 | Low-quality |
| S3_extracted_k141_86874 | Not-determined |
| S3_extracted_k141_21904 | Not-determined |
| S3_extracted_k141_30569 | Not-determined |
| S3_extracted_k141_21906 | Not-determined |
| S3_extracted_k141_21907 | Not-determined |
| S3_extracted_k141_38629 | Low-quality |
| S3_extracted_k141_21909 | Not-determined |
| S3_extracted_k141_46525 | Not-determined |
| S3_extracted_k141_46526 | Low-quality |
| S3_extracted_k141_79136 | Low-quality |
| S3_extracted_k141_86879 | Low-quality |
| S3_extracted_k141_21921 | Low-quality |
| S3_extracted_k141_6450 | Not-determined |
| S3_extracted_k141_6451 | Low-quality |
| S3_extracted_k141_101454 | Not-determined |
| S3_extracted_k141_6459 | Not-determined |
| S3_extracted_k141_86889 | Low-quality |
| S3_extracted_k141_101456 | Not-determined |
| S3_extracted_k141_21929 | Not-determined |
| S3_extracted_k141_79146 | Not-determined |
| S3_extracted_k141_46539 | Low-quality |
| S3_extracted_k141_79147 | Not-determined |
| S3_extracted_k141_21931 | Not-determined |
| S3_extracted_k141_63364 | Not-determined |
| S3_extracted_k141_69540 | Low-quality |
| S3_extracted_k141_101464 | Not-determined |
| S3_extracted_k141_101465 | Not-determined |
| S3_extracted_k141_38647 | Not-determined |
| S3_extracted_k141_21942 | Not-determined |
| S3_extracted_k141_63373 | Not-determined |
| S3_extracted_k141_21943 | Not-determined |
| S3_extracted_k141_30575 | Not-determined |
| S3_extracted_k141_69553 | Not-determined |
| S3_extracted_k141_101468 | Not-determined |
| S3_extracted_k141_6469 | Not-determined |
| S3_extracted_k141_63379 | Not-determined |
| S3_extracted_k141_101470 | Not-determined |
| S3_extracted_k141_63382 | Not-determined |
| S3_extracted_k141_6472 | Not-determined |
| S3_extracted_k141_86895 | Not-determined |
| S3_extracted_k141_86896 | Not-determined |
| S3_extracted_k141_94477 | Low-quality |
| S3_extracted_k141_101476 | Not-determined |
| S3_extracted_k141_69557 | Not-determined |
| S3_extracted_k141_101477 | Not-determined |
| S3_extracted_k141_46553 | Not-determined |
| S3_extracted_k141_69559 | Not-determined |
| S3_extracted_k141_86904 | Not-determined |
| S3_extracted_k141_63386 | Not-determined |
| S3_extracted_k141_86912 | Not-determined |
| S3_extracted_k141_38677 | Low-quality |
| S3_extracted_k141_86914 | Not-determined |
| S3_extracted_k141_86915 | Not-determined |
| S3_extracted_k141_21949 | Not-determined |
| S3_extracted_k141_21950 | Not-determined |
| S3_extracted_k141_86917 | Not-determined |
| S3_extracted_k141_86918 | Not-determined |
| S3_extracted_k141_63396 | Not-determined |
| S3_extracted_k141_79157 | Not-determined |
| S3_extracted_k141_69568 | Low-quality |
| S3_extracted_k141_79166 | Not-determined |
| S3_extracted_k141_38686 | Not-determined |
| S3_extracted_k141_46570 | Not-determined |
| S3_extracted_k141_38687 | Not-determined |
| S3_extracted_k141_38688 | Not-determined |
| S3_extracted_k141_46574 | Not-determined |
| S3_extracted_k141_79167 | Low-quality |
| S3_extracted_k141_86928 | Not-determined |
| S3_extracted_k141_63408 | Not-determined |
| S3_extracted_k141_69581 | Not-determined |
| S3_extracted_k141_6485 | Low-quality |
| S3_extracted_k141_94495 | Not-determined |
| S3_extracted_k141_21987 | Not-determined |
| S3_extracted_k141_79177 | Not-determined |
| S3_extracted_k141_69588 | Not-determined |
| S3_extracted_k141_94498 | Not-determined |
| S3_extracted_k141_69594 | Low-quality |
| S3_extracted_k141_118162 | Low-quality |
| S3_extracted_k141_118165 | Not-determined |
| S3_extracted_k141_118166 | Not-determined |
| S3_extracted_k141_86945 | Not-determined |
| S3_extracted_k141_110601 | Not-determined |
| S3_extracted_k141_30592 | Not-determined |
| S3_extracted_k141_30593 | Not-determined |
| S3_extracted_k141_22009 | Not-determined |
| S3_extracted_k141_69599 | Not-determined |
| S3_extracted_k141_46593 | Low-quality |
| S3_extracted_k141_69606 | Not-determined |
| S3_extracted_k141_69607 | Not-determined |
| S3_extracted_k141_69608 | Not-determined |
| S3_extracted_k141_79192 | Not-determined |
| S3_extracted_k141_79194 | Not-determined |
| S3_extracted_k141_86963 | Not-determined |
| S3_extracted_k141_125687 | Not-determined |
| S3_extracted_k141_86964 | Not-determined |
| S3_extracted_k141_125689 | Not-determined |
| S3_extracted_k141_22018 | Not-determined |
| S3_extracted_k141_79198 | Not-determined |
| S3_extracted_k141_101520 | Low-quality |
| S3_extracted_k141_118189 | Not-determined |
| S3_extracted_k141_30604 | Not-determined |
| S3_extracted_k141_101527 | Not-determined |
| S3_extracted_k141_55146 | Not-determined |
| S3_extracted_k141_30609 | Not-determined |
| S3_extracted_k141_6510 | Not-determined |
| S3_extracted_k141_38737 | Not-determined |
| S3_extracted_k141_55149 | Not-determined |
| S3_extracted_k141_63417 | Low-quality |
| S3_extracted_k141_63420 | Low-quality |
| S3_extracted_k141_55157 | Not-determined |
| S3_extracted_k141_22031 | Not-determined |
| S3_extracted_k141_118198 | Not-determined |
| S3_extracted_k141_110612 | Not-determined |
| S3_extracted_k141_94518 | Not-determined |
| S3_extracted_k141_38753 | Not-determined |
| S3_extracted_k141_55164 | Not-determined |
| S3_extracted_k141_79212 | Not-determined |
| S3_extracted_k141_101542 | Not-determined |
| S3_extracted_k141_94521 | Low-quality |
| S3_extracted_k141_101551 | Not-determined |
| S3_extracted_k141_87001 | Not-determined |
| S3_extracted_k141_110620 | Not-determined |
| S3_extracted_k141_38756 | Not-determined |
| S3_extracted_k141_125714 | Low-quality |
| S3_extracted_k141_110628 | Not-determined |
| S3_extracted_k141_110629 | Not-determined |
| S3_extracted_k141_79231 | Not-determined |
| S3_extracted_k141_79232 | Not-determined |
| S3_extracted_k141_110633 | Not-determined |
| S3_extracted_k141_79234 | Low-quality |
| S3_extracted_k141_55170 | Low-quality |
| S3_extracted_k141_110637 | Not-determined |
| S3_extracted_k141_94541 | Not-determined |
| S3_extracted_k141_6513 | Not-determined |
| S3_extracted_k141_55174 | Not-determined |
| S3_extracted_k141_79244 | Not-determined |
| S3_extracted_k141_55176 | Not-determined |
| S3_extracted_k141_79254 | Not-determined |
| S3_extracted_k141_118219 | Not-determined |
| S3_extracted_k141_69616 | Not-determined |
| S3_extracted_k141_69618 | Low-quality |
| S3_extracted_k141_118223 | Not-determined |
| S3_extracted_k141_94547 | Not-determined |
| S3_extracted_k141_6521 | Not-determined |
| S3_extracted_k141_6522 | Not-determined |
| S3_extracted_k141_79265 | Not-determined |
| S3_extracted_k141_118227 | Not-determined |
| S3_extracted_k141_6523 | Low-quality |
| S3_extracted_k141_118229 | Not-determined |
| S3_extracted_k141_69623 | Not-determined |
| S3_extracted_k141_55183 | Not-determined |
| S3_extracted_k141_38772 | Not-determined |
| S3_extracted_k141_38776 | Not-determined |
| S3_extracted_k141_38777 | Not-determined |
| S3_extracted_k141_79272 | Not-determined |
| S3_extracted_k141_79273 | Not-determined |
| S3_extracted_k141_46632 | Not-determined |
| S3_extracted_k141_46633 | Not-determined |
| S15_vs_76231\|\|full | High-quality |
| S3_extracted_k141_87014 | Not-determined |
| S3_extracted_k141_38782 | Low-quality |
| S3_extracted_k141_94552 | Not-determined |
| S3_extracted_k141_87017 | Not-determined |
| S3_extracted_k141_87019 | Not-determined |
| S3_extracted_k141_38784 | Not-determined |
| S3_extracted_k141_94555 | Not-determined |
| S3_extracted_k141_125723 | Not-determined |
| S3_extracted_k141_63459 | Not-determined |
| S3_extracted_k141_110651 | Low-quality |
| S3_extracted_k141_63461 | Not-determined |
| S3_extracted_k141_87027 | Not-determined |
| S3_extracted_k141_63463 | Not-determined |
| S3_extracted_k141_6536 | Not-determined |
| S3_extracted_k141_125734 | Not-determined |
| S3_extracted_k141_6539 | Not-determined |
| S3_extracted_k141_46636 | Low-quality |
| S3_extracted_k141_79279 | Not-determined |
| S3_extracted_k141_69641 | Low-quality |
| S3_extracted_k141_46640 | Not-determined |
| S3_extracted_k141_110660 | Not-determined |
| S3_extracted_k141_118247 | Not-determined |
| S3_extracted_k141_6550 | Not-determined |
| S3_extracted_k141_79286 | Low-quality |
| S3_extracted_k141_79287 | Not-determined |
| S3_extracted_k141_110664 | Not-determined |
| S3_extracted_k141_110666 | Not-determined |
| S3_extracted_k141_118253 | Not-determined |
| S3_extracted_k141_118258 | Not-determined |
| S3_extracted_k141_118259 | Not-determined |
| S3_extracted_k141_110671 | Not-determined |
| S3_extracted_k141_118260 | Not-determined |
| S3_extracted_k141_38789 | Not-determined |
| S3_extracted_k141_79296 | Not-determined |
| S3_extracted_k141_79297 | Not-determined |
| S3_extracted_k141_69674 | Not-determined |
| S3_extracted_k141_79304 | Not-determined |
| S3_extracted_k141_22071 | Not-determined |
| S3_extracted_k141_87047 | Low-quality |
| S3_extracted_k141_46652 | Not-determined |
| S3_extracted_k141_125738 | Not-determined |
| S3_extracted_k141_110679 | Not-determined |
| S3_extracted_k141_79306 | Not-determined |
| S3_extracted_k141_6557 | Not-determined |
| S3_extracted_k141_110683 | Not-determined |
| S3_extracted_k141_22079 | Low-quality |
| S3_extracted_k141_79312 | Low-quality |
| S3_extracted_k141_79313 | Not-determined |
| S3_extracted_k141_6559 | Not-determined |
| S3_extracted_k141_79315 | Not-determined |
| S3_extracted_k141_69681 | Not-determined |
| S3_extracted_k141_69682 | Not-determined |
| S3_extracted_k141_22087 | Not-determined |
| S3_extracted_k141_69683 | Low-quality |
| S3_extracted_k141_63511 | Not-determined |
| S3_extracted_k141_46655 | Not-determined |
| S3_extracted_k141_101597 | Not-determined |
| S3_extracted_k141_101598 | Not-determined |
| S3_extracted_k141_79327 | Not-determined |
| S3_extracted_k141_55194 | Not-determined |
| S3_extracted_k141_63519 | Not-determined |
| S3_extracted_k141_101603 | Low-quality |
| S3_extracted_k141_118296 | Not-determined |
| S3_extracted_k141_101607 | Not-determined |
| S3_extracted_k141_87061 | Not-determined |
| S3_extracted_k141_22093 | Not-determined |
| S3_extracted_k141_110713 | Not-determined |
| S3_extracted_k141_46670 | Not-determined |
| S3_extracted_k141_125752 | Not-determined |
| S3_extracted_k141_125753 | Not-determined |
| S3_extracted_k141_69697 | Low-quality |
| S3_extracted_k141_125754 | Not-determined |
| S3_extracted_k141_79342 | Low-quality |
| S3_extracted_k141_69700 | Not-determined |
| S3_extracted_k141_125755 | Low-quality |
| S3_extracted_k141_125756 | Low-quality |
| S3_extracted_k141_94585 | Not-determined |
| S3_extracted_k141_79354 | Not-determined |
| S3_extracted_k141_22098 | Not-determined |
| S3_extracted_k141_63540 | Not-determined |
| S3_extracted_k141_46685 | Not-determined |
| S3_extracted_k141_22101 | Not-determined |
| S3_extracted_k141_79360 | Not-determined |
| S3_extracted_k141_6572 | Not-determined |
| S3_extracted_k141_110721 | Not-determined |
| S3_extracted_k141_22106 | Not-determined |
| S3_extracted_k141_63546 | Not-determined |
| S3_extracted_k141_63547 | Not-determined |
| S3_extracted_k141_110724 | Not-determined |
| S3_extracted_k141_63549 | Not-determined |
| S3_extracted_k141_46694 | Not-determined |
| S3_extracted_k141_55205 | Low-quality |
| S3_extracted_k141_63552 | Not-determined |
| S3_extracted_k141_79371 | Not-determined |
| S3_extracted_k141_110730 | Low-quality |
| S3_extracted_k141_55208 | Not-determined |
| S3_extracted_k141_63557 | Not-determined |
| S3_extracted_k141_94600 | Low-quality |
| S3_extracted_k141_22120 | Not-determined |
| S3_extracted_k141_6594 | Low-quality |
| S3_extracted_k141_69728 | Low-quality |
| S3_extracted_k141_6595 | Not-determined |
| S3_extracted_k141_118320 | Not-determined |
| S3_extracted_k141_94604 | Not-determined |
| S3_extracted_k141_69731 | Not-determined |
| S3_extracted_k141_22125 | Not-determined |
| S3_extracted_k141_22127 | Low-quality |
| S3_extracted_k141_69732 | Low-quality |
| S3_extracted_k141_118322 | Not-determined |
| S3_extracted_k141_46703 | Low-quality |
| S3_extracted_k141_22133 | Not-determined |
| S3_extracted_k141_125778 | Not-determined |
| S3_extracted_k141_69737 | Not-determined |
| S3_extracted_k141_87081 | Low-quality |
| S3_extracted_k141_55214 | Not-determined |
| S3_extracted_k141_87084 | Low-quality |
| S3_extracted_k141_63577 | Not-determined |
| S3_extracted_k141_22145 | Not-determined |
| S3_extracted_k141_79387 | Low-quality |
| S3_extracted_k141_87091 | Not-determined |
| S3_extracted_k141_46723 | Not-determined |
| S3_extracted_k141_94616 | Low-quality |
| S3_extracted_k141_79391 | Low-quality |
| S3_extracted_k141_69755 | Not-determined |
| S3_extracted_k141_69756 | Not-determined |
| S3_extracted_k141_63585 | Low-quality |
| S3_extracted_k141_110734 | Low-quality |
| S3_extracted_k141_63587 | Not-determined |
| S3_extracted_k141_63590 | Not-determined |
| S3_extracted_k141_63595 | Not-determined |
| S3_extracted_k141_125788 | Not-determined |
| S3_extracted_k141_22158 | Not-determined |
| S15_extracted_6822 | Complete |
| S3_extracted_k141_79411 | Not-determined |
| S3_extracted_k141_63600 | Not-determined |
| S3_extracted_k141_63603 | Not-determined |
| S3_extracted_k141_22166 | Not-determined |
| S3_extracted_k141_69758 | Not-determined |
| S3_extracted_k141_69759 | Not-determined |
| S3_extracted_k141_87110 | Not-determined |
| S3_extracted_k141_118333 | Not-determined |
| S3_extracted_k141_79418 | Not-determined |
| S3_extracted_k141_94641 | Not-determined |
| S3_extracted_k141_79422 | Not-determined |
| S3_extracted_k141_79425 | Not-determined |
| S3_extracted_k141_118341 | Not-determined |
| S3_extracted_k141_87113 | Not-determined |
| S3_extracted_k141_22169 | Not-determined |
| S3_extracted_k141_94650 | Not-determined |
| S3_extracted_k141_125798 | Not-determined |
| S3_extracted_k141_46748 | Low-quality |
| S3_extracted_k141_46749 | Not-determined |
| S3_extracted_k141_46750 | Not-determined |
| S3_extracted_k141_94654 | Not-determined |
| S3_extracted_k141_110766 | Not-determined |
| S3_extracted_k141_46753 | Low-quality |
| S3_extracted_k141_87129 | Not-determined |
| S3_extracted_k141_79427 | Low-quality |
| S3_extracted_k141_79428 | Not-determined |
| S3_extracted_k141_125801 | Low-quality |
| S3_extracted_k141_125802 | Low-quality |
| S3_extracted_k141_125805 | Low-quality |
| S3_extracted_k141_79440 | Not-determined |
| S3_extracted_k141_94659 | Not-determined |
| S3_extracted_k141_94662 | Not-determined |
| S3_extracted_k141_55243 | Not-determined |
| S3_extracted_k141_118347 | Not-determined |
| S3_extracted_k141_55244 | Low-quality |
| S3_extracted_k141_55245 | Not-determined |
| S3_extracted_k141_87133 | Not-determined |
| S3_extracted_k141_22186 | Not-determined |
| S3_extracted_k141_55252 | Not-determined |
| S3_extracted_k141_55253 | Not-determined |
| S3_extracted_k141_94665 | Not-determined |
| S3_extracted_k141_22189 | Low-quality |
| S3_extracted_k141_94667 | Low-quality |
| S3_extracted_k141_125823 | Not-determined |
| S3_extracted_k141_22199 | Not-determined |
| S3_extracted_k141_46770 | Not-determined |
| S3_extracted_k141_46771 | Not-determined |
| S3_extracted_k141_125826 | Not-determined |
| S3_extracted_k141_125827 | Not-determined |
| S3_extracted_k141_94681 | Not-determined |
| S3_extracted_k141_118359 | Not-determined |
| S3_extracted_k141_22204 | Not-determined |
| S3_extracted_k141_94683 | Not-determined |
| S3_extracted_k141_94686 | Not-determined |
| S3_extracted_k141_94688 | Not-determined |
| S3_extracted_k141_125832 | Not-determined |
| S3_extracted_k141_94690 | Not-determined |
| S3_extracted_k141_94696 | Not-determined |
| S3_extracted_k141_101633 | Low-quality |
| S3_extracted_k141_63629 | Not-determined |
| S3_extracted_k141_101637 | Not-determined |
| S3_extracted_k141_101639 | Not-determined |
| S3_extracted_k141_101640 | Not-determined |
| S3_extracted_k141_63636 | Not-determined |
| S3_extracted_k141_46781 | Not-determined |
| S3_extracted_k141_101642 | Not-determined |
| S3_extracted_k141_46787 | Not-determined |
| S3_extracted_k141_46792 | Not-determined |
| S3_extracted_k141_118367 | Not-determined |
| S3_extracted_k141_101650 | Not-determined |
| S3_extracted_k141_30640 | Low-quality |
| S3_extracted_k141_94710 | Not-determined |
| S3_extracted_k141_46798 | Not-determined |
| S3_extracted_k141_55262 | Not-determined |
| S3_extracted_k141_110770 | Not-determined |
| S3_extracted_k141_63653 | Not-determined |
| S3_extracted_k141_63656 | Low-quality |
| S3_extracted_k141_55263 | Low-quality |
| S3_extracted_k141_101659 | Not-determined |
| S3_extracted_k141_94721 | Not-determined |
| S3_extracted_k141_101661 | Low-quality |
| S3_extracted_k141_101664 | Not-determined |
| S3_extracted_k141_69788 | Not-determined |
| S3_extracted_k141_69790 | Not-determined |
| S3_extracted_k141_101673 | Not-determined |
| S3_extracted_k141_94738 | Not-determined |
| S3_extracted_k141_94741 | Not-determined |
| S3_extracted_k141_38855 | Low-quality |
| S3_extracted_k141_101686 | Not-determined |
| S3_extracted_k141_6609 | Not-determined |
| S3_extracted_k141_38857 | Low-quality |
| S3_extracted_k141_38860 | Low-quality |
| S3_extracted_k141_118371 | Low-quality |
| S3_extracted_k141_118375 | Not-determined |
| S3_extracted_k141_87173 | Low-quality |

| | |
|---|---|
| S3_extracted_k141_110784 | Not-determined |
| S3_extracted_k141_6619 | Not-determined |
| S3_extracted_k141_22263 | Not-determined |
| S3_extracted_k141_6621 | Not-determined |
| S3_extracted_k141_69812 | Not-determined |
| S3_extracted_k141_69813 | Not-determined |
| S3_extracted_k141_69814 | Not-determined |
| S3_extracted_k141_22268 | Low-quality |
| S3_extracted_k141_46842 | Not-determined |
| S3_extracted_k141_87181 | Not-determined |
| S3_extracted_k141_101695 | Not-determined |
| S3_extracted_k141_118381 | Not-determined |
| S3_extracted_k141_69818 | Low-quality |
| S3_extracted_k141_69824 | Low-quality |
| S3_extracted_k141_69826 | Not-determined |
| S3_extracted_k141_69828 | Not-determined |
| S3_extracted_k141_101702 | Low-quality |
| S3_extracted_k141_110790 | Low-quality |
| S3_extracted_k141_22275 | Low-quality |
| S3_extracted_k141_30684 | Not-determined |
| S3_extracted_k141_55278 | Not-determined |
| S3_extracted_k141_38867 | Not-determined |
| S3_extracted_k141_6650 | Not-determined |
| S3_extracted_k141_101717 | Not-determined |
| S3_extracted_k141_55291 | Not-determined |
| S3_extracted_k141_22286 | Not-determined |
| S3_extracted_k141_6656 | Not-determined |
| S3_extracted_k141_6658 | Not-determined |
| S3_extracted_k141_55295 | Low-quality |
| S3_extracted_k141_6662 | Not-determined |
| S3_extracted_k141_38879 | Low-quality |
| S3_extracted_k141_6666 | Not-determined |
| S3_extracted_k141_38887 | Low-quality |
| S3_extracted_k141_101732 | Not-determined |
| S3_extracted_k141_87198 | Not-determined |
| S3_extracted_k141_87199 | Low-quality |
| S3_extracted_k141_101734 | Not-determined |
| S3_extracted_k141_55307 | Not-determined |
| S3_extracted_k141_22296 | Not-determined |
| S3_extracted_k141_22297 | Not-determined |
| S3_extracted_k141_87202 | Low-quality |
| S3_extracted_k141_30722 | Not-determined |
| S3_extracted_k141_46859 | Not-determined |
| S3_extracted_k141_46863 | Not-determined |
| S3_extracted_k141_38891 | Not-determined |
| S3_extracted_k141_94785 | Low-quality |
| S3_extracted_k141_110801 | Low-quality |
| S3_extracted_k141_87221 | Low-quality |
| S3_extracted_k141_110807 | Not-determined |
| S3_extracted_k141_94791 | Not-determined |
| S3_extracted_k141_30729 | Not-determined |
| S3_extracted_k141_69859 | Low-quality |
| S3_extracted_k141_69865 | Not-determined |
| S3_extracted_k141_30732 | Not-determined |
| S3_extracted_k141_38899 | Not-determined |
| S3_extracted_k141_38901 | Not-determined |
| S3_extracted_k141_94799 | Not-determined |
| S3_extracted_k141_94801 | Not-determined |
| S3_extracted_k141_94802 | Low-quality |
| S3_extracted_k141_125845 | Low-quality |
| S3_extracted_k141_46898 | Not-determined |
| S3_extracted_k141_125847 | Not-determined |
| S3_extracted_k141_6677 | Not-determined |
| S3_extracted_k141_38907 | Not-determined |
| S3_extracted_k141_101744 | Not-determined |
| S3_extracted_k141_22304 | Low-quality |
| S3_extracted_k141_101747 | Low-quality |
| S3_extracted_k141_87243 | Not-determined |
| S3_extracted_k141_69881 | Not-determined |
| S3_extracted_k141_69882 | Not-determined |
| S3_extracted_k141_30741 | Not-determined |
| S3_extracted_k141_125859 | Not-determined |
| S3_extracted_k141_101749 | Low-quality |
| S3_extracted_k141_125861 | Not-determined |
| S3_extracted_k141_87251 | Not-determined |
| S3_extracted_k141_110823 | Not-determined |
| S3_extracted_k141_94811 | Not-determined |
| S3_extracted_k141_125864 | Not-determined |
| S3_extracted_k141_46915 | Not-determined |
| S3_extracted_k141_46916 | Not-determined |
| S3_extracted_k141_87257 | Not-determined |
| S3_extracted_k141_101756 | Low-quality |
| S3_extracted_k141_46920 | Not-determined |
| S3_extracted_k141_101760 | Not-determined |
| S3_extracted_k141_118398 | Not-determined |
| S3_extracted_k141_125868 | Not-determined |
| S3_extracted_k141_101762 | Not-determined |
| S3_extracted_k141_101763 | Not-determined |
| S3_extracted_k141_6699 | Low-quality |
| S3_extracted_k141_125871 | Not-determined |
| S3_extracted_k141_30757 | Not-determined |
| S3_extracted_k141_22318 | Not-determined |
| S3_extracted_k141_125873 | Not-determined |
| S3_extracted_k141_30759 | Not-determined |
| S3_extracted_k141_30760 | Not-determined |
| S3_extracted_k141_69889 | Not-determined |
| S3_extracted_k141_69890 | Not-determined |
| S3_extracted_k141_101774 | Not-determined |
| S3_extracted_k141_101776 | Not-determined |
| S3_extracted_k141_118417 | Not-determined |
| S3_extracted_k141_55356 | Not-determined |
| S3_extracted_k141_94821 | Low-quality |
| S3_extracted_k141_69897 | Not-determined |
| S3_extracted_k141_110833 | Not-determined |
| S3_extracted_k141_101782 | Not-determined |
| S3_extracted_k141_87269 | Not-determined |
| S3_extracted_k141_38921 | Low-quality |
| S3_extracted_k141_101788 | Not-determined |
| S3_extracted_k141_22335 | Not-determined |
| S3_extracted_k141_118423 | Low-quality |
| S3_extracted_k141_30777 | Not-determined |
| S3_extracted_k141_94831 | Not-determined |
| S3_extracted_k141_30779 | Not-determined |
| S3_extracted_k141_118428 | Not-determined |
| S3_extracted_k141_125886 | Not-determined |
| S3_extracted_k141_22344 | Not-determined |
| S3_extracted_k141_22347 | Not-determined |
| S3_extracted_k141_69916 | Low-quality |
| S3_extracted_k141_94835 | Not-determined |
| S3_extracted_k141_94836 | Not-determined |
| S3_extracted_k141_101800 | Not-determined |
| S3_extracted_k141_94840 | Not-determined |
| S3_extracted_k141_125895 | Not-determined |
| S3_extracted_k141_69920 | Low-quality |
| S3_extracted_k141_125896 | Not-determined |
| S3_extracted_k141_79453 | Not-determined |
| S3_extracted_k141_79455 | Not-determined |
| S3_extracted_k141_94846 | Not-determined |
| S3_extracted_k141_22359 | Not-determined |
| S3_extracted_k141_69921 | Not-determined |
| S3_extracted_k141_118439 | Not-determined |
| S3_extracted_k141_79462 | Not-determined |
| S3_extracted_k141_118441 | Not-determined |
| S3_extracted_k141_94852 | Not-determined |
| S3_extracted_k141_79465 | Low-quality |
| S3_extracted_k141_46927 | Low-quality |
| S3_extracted_k141_125901 | Not-determined |
| S3_extracted_k141_101809 | Not-determined |
| S3_extracted_k141_118442 | Not-determined |
| S3_extracted_k141_22370 | Not-determined |
| S3_extracted_k141_69927 | Not-determined |
| S3_extracted_k141_79474 | Not-determined |
| S3_extracted_k141_22375 | Not-determined |
| S3_extracted_k141_30782 | Not-determined |
| S3_extracted_k141_101815 | Not-determined |
| S3_extracted_k141_110849 | Not-determined |
| S3_extracted_k141_94858 | Not-determined |
| S3_extracted_k141_79480 | Not-determined |
| S3_extracted_k141_6725 | Low-quality |
| S3_extracted_k141_94861 | Not-determined |
| S3_extracted_k141_125916 | Low-quality |
| S3_extracted_k141_101821 | Not-determined |
| S3_extracted_k141_125920 | Not-determined |
| S3_extracted_k141_94874 | Low-quality |
| S3_extracted_k141_6727 | Not-determined |
| S3_extracted_k141_125925 | Not-determined |
| S3_extracted_k141_69937 | Not-determined |
| S3_extracted_k141_38948 | Not-determined |
| S3_extracted_k141_69938 | Not-determined |
| S3_extracted_k141_55370 | Not-determined |
| S3_extracted_k141_69941 | Not-determined |
| S3_extracted_k141_110859 | Not-determined |
| S3_extracted_k141_94883 | Not-determined |
| S3_extracted_k141_46955 | Low-quality |
| S3_extracted_k141_30796 | Not-determined |
| S3_extracted_k141_87284 | Low-quality |
| S3_extracted_k141_118467 | Not-determined |
| S3_extracted_k141_118469 | Not-determined |
| S3_extracted_k141_79499 | Not-determined |
| S3_extracted_k141_79500 | Low-quality |
| S3_extracted_k141_87288 | Low-quality |
| S3_extracted_k141_79501 | Not-determined |
| S3_extracted_k141_125932 | Not-determined |
| S3_extracted_k141_30801 | Not-determined |
| S3_extracted_k141_30802 | Not-determined |
| S3_extracted_k141_79502 | Not-determined |
| S3_extracted_k141_30803 | Not-determined |
| S3_extracted_k141_55380 | Not-determined |
| S3_extracted_k141_55384 | Not-determined |
| S3_extracted_k141_101846 | Low-quality |
| S3_extracted_k141_101850 | Not-determined |
| S3_extracted_k141_6733 | Not-determined |
| S3_extracted_k141_63739 | Not-determined |
| S3_extracted_k141_22401 | Not-determined |
| S3_extracted_k141_94887 | Not-determined |
| S3_extracted_k141_6734 | Low-quality |
| S3_extracted_k141_63743 | Not-determined |
| S3_extracted_k141_22406 | Not-determined |
| S3_extracted_k141_30819 | Not-determined |
| S3_extracted_k141_110862 | Not-determined |
| S3_extracted_k141_30825 | Not-determined |
| S3_extracted_k141_55391 | Low-quality |
| S3_extracted_k141_38983 | Not-determined |
| S3_extracted_k141_46959 | Low-quality |
| S3_extracted_k141_55394 | Not-determined |
| S3_extracted_k141_38986 | Not-determined |
| S3_extracted_k141_63758 | Not-determined |
| S3_extracted_k141_101853 | Not-determined |
| S3_extracted_k141_118497 | Not-determined |
| S3_extracted_k141_55401 | Not-determined |
| S3_extracted_k141_46965 | Low-quality |
| S3_extracted_k141_110880 | Not-determined |
| S3_extracted_k141_118506 | Not-determined |
| S3_extracted_k141_55408 | Not-determined |
| S3_extracted_k141_38990 | Not-determined |
| S3_extracted_k141_46974 | Not-determined |
| S3_extracted_k141_125956 | Not-determined |
| S3_extracted_k141_46975 | Not-determined |
| S3_extracted_k141_118511 | Not-determined |
| S3_extracted_k141_55410 | Not-determined |
| S3_extracted_k141_101860 | Low-quality |
| S3_extracted_k141_69946 | Not-determined |
| S3_extracted_k141_69947 | Not-determined |
| S3_extracted_k141_69952 | Not-determined |
| S3_extracted_k141_69953 | Not-determined |
| S3_extracted_k141_6759 | Not-determined |
| S3_extracted_k141_79521 | Low-quality |
| S3_extracted_k141_79523 | Low-quality |
| S3_extracted_k141_125975 | Not-determined |
| S3_extracted_k141_69963 | Not-determined |
| S3_extracted_k141_6761 | Low-quality |
| S3_extracted_k141_125981 | Not-determined |
| S3_extracted_k141_30857 | Not-determined |
| S3_extracted_k141_101872 | Not-determined |
| S3_extracted_k141_101877 | Not-determined |
| S3_extracted_k141_101878 | Low-quality |
| S3_extracted_k141_6775 | Not-determined |
| S3_extracted_k141_38995 | Not-determined |
| S3_extracted_k141_6778 | Not-determined |
| S3_extracted_k141_39003 | Not-determined |
| S3_extracted_k141_118520 | Low-quality |
| S3_extracted_k141_30863 | Low-quality |
| S3_extracted_k141_30864 | Not-determined |
| S3_extracted_k141_39009 | Not-determined |
| S3_extracted_k141_6780 | Not-determined |
| S3_extracted_k141_30869 | Not-determined |
| S3_extracted_k141_94919 | Not-determined |
| S3_extracted_k141_118526 | Low-quality |
| S3_extracted_k141_30872 | Not-determined |
| S3_extracted_k141_118527 | Low-quality |
| S3_extracted_k141_69991 | Not-determined |
| S3_extracted_k141_118531 | Not-determined |
| S3_extracted_k141_46996 | Not-determined |
| S3_extracted_k141_69996 | Not-determined |
| S3_extracted_k141_94923 | Not-determined |
| S3_extracted_k141_22414 | Not-determined |
| S3_extracted_k141_47004 | Low-quality |
| S3_extracted_k141_47005 | Low-quality |
| S3_extracted_k141_55420 | Not-determined |
| S3_extracted_k141_94943 | Not-determined |
| S3_extracted_k141_55424 | Not-determined |
| S3_extracted_k141_55426 | Not-determined |
| S3_extracted_k141_70005 | Not-determined |
| S3_extracted_k141_125983 | Not-determined |
| S3_extracted_k141_55430 | Not-determined |
| S3_extracted_k141_47026 | Not-determined |
| S3_extracted_k141_55444 | Not-determined |
| S3_extracted_k141_126006 | Not-determined |
| S3_extracted_k141_126008 | Not-determined |
| S3_extracted_k141_70023 | Not-determined |
| S3_extracted_k141_6794 | Not-determined |
| S3_extracted_k141_94953 | Not-determined |
| S3_extracted_k141_22424 | Not-determined |
| S3_extracted_k141_94955 | Not-determined |
| S3_extracted_k141_22426 | Low-quality |
| S3_extracted_k141_39027 | Not-determined |
| S3_extracted_k141_55451 | Not-determined |
| S3_extracted_k141_126017 | Not-determined |
| S3_extracted_k141_22429 | Not-determined |
| S3_extracted_k141_94961 | Not-determined |
| S3_extracted_k141_47030 | Not-determined |
| S3_extracted_k141_110943 | Not-determined |
| S3_extracted_k141_94968 | Low-quality |
| S3_extracted_k141_6799 | Not-determined |
| S3_extracted_k141_6801 | Not-determined |
| S3_extracted_k141_55454 | Not-determined |
| S3_extracted_k141_70037 | Low-quality |
| S3_extracted_k141_94977 | Not-determined |
| S3_extracted_k141_47043 | Not-determined |
| S3_extracted_k141_6810 | Low-quality |
| S3_extracted_k141_70044 | Not-determined |
| S3_extracted_k141_70045 | Not-determined |
| S3_extracted_k141_110970 | Not-determined |
| S3_extracted_k141_94988 | Not-determined |
| S3_extracted_k141_47053 | Not-determined |
| S3_extracted_k141_94997 | Not-determined |
| S3_extracted_k141_118567 | Not-determined |
| S3_extracted_k141_79593 | Not-determined |
| S3_extracted_k141_118571 | Not-determined |
| S3_extracted_k141_126036 | Not-determined |
| S3_extracted_k141_55464 | Not-determined |
| S3_extracted_k141_118574 | Not-determined |
| S3_extracted_k141_70054 | Not-determined |
| S3_extracted_k141_110995 | Not-determined |
| S3_extracted_k141_101888 | Low-quality |
| S3_extracted_k141_70057 | Not-determined |
| S3_extracted_k141_126040 | Not-determined |
| S3_extracted_k141_70061 | Not-determined |
| S3_extracted_k141_39040 | Low-quality |
| S3_extracted_k141_55470 | Not-determined |
| S3_extracted_k141_70064 | Not-determined |
| S3_extracted_k141_101896 | Not-determined |
| S3_extracted_k141_95006 | Not-determined |
| S3_extracted_k141_101900 | Not-determined |
| S3_extracted_k141_126054 | Not-determined |
| S3_extracted_k141_101901 | Not-determined |
| S3_extracted_k141_47079 | Not-determined |
| S3_extracted_k141_6827 | Not-determined |
| S3_extracted_k141_111018 | Low-quality |
| S3_extracted_k141_47083 | Not-determined |
| S3_extracted_k141_55481 | Not-determined |
| S3_extracted_k141_55482 | Not-determined |
| S3_extracted_k141_101924 | Not-determined |
| S3_extracted_k141_55487 | Not-determined |
| S3_extracted_k141_6837 | Not-determined |
| S3_extracted_k141_111026 | Not-determined |
| S3_extracted_k141_55488 | Not-determined |
| S3_extracted_k141_55490 | Not-determined |
| S3_extracted_k141_39067 | Not-determined |
| S3_extracted_k141_118592 | Not-determined |
| S3_extracted_k141_111028 | Low-quality |
| S3_extracted_k141_95014 | Not-determined |
| S3_extracted_k141_79629 | Not-determined |
| S3_extracted_k141_30894 | Not-determined |
| S3_extracted_k141_79635 | Not-determined |
| S3_extracted_k141_39076 | Not-determined |
| S3_extracted_k141_118599 | Not-determined |
| S3_extracted_k141_39077 | Not-determined |
| S3_extracted_k141_22435 | Low-quality |
| S3_extracted_k141_39082 | Not-determined |
| S3_extracted_k141_22438 | Not-determined |
| S3_extracted_k141_118603 | Not-determined |
| S3_extracted_k141_79645 | Not-determined |
| S3_extracted_k141_30920 | Not-determined |
| S3_extracted_k141_126071 | Not-determined |
| S3_extracted_k141_111051 | Not-determined |
| S3_extracted_k141_126078 | Low-quality |
| S3_extracted_k141_70098 | Low-quality |
| S3_extracted_k141_126080 | Not-determined |
| S3_extracted_k141_126086 | Not-determined |
| S3_extracted_k141_126087 | Not-determined |
| S3_extracted_k141_126091 | Not-determined |
| S3_extracted_k141_111064 | Not-determined |
| S3_extracted_k141_126092 | Not-determined |
| S3_extracted_k141_95031 | Not-determined |
| S3_extracted_k141_95032 | Not-determined |
| S3_extracted_k141_39118 | Not-determined |
| S3_extracted_k141_95035 | Low-quality |
| S3_extracted_k141_70121 | Not-determined |
| S3_extracted_k141_118616 | Not-determined |
| S3_extracted_k141_95036 | Not-determined |
| S3_extracted_k141_55503 | Not-determined |
| S3_extracted_k141_39124 | Not-determined |
| S3_extracted_k141_95040 | Not-determined |
| S3_extracted_k141_118622 | Low-quality |
| S3_extracted_k141_39130 | Not-determined |
| S3_extracted_k141_55508 | Not-determined |
| S3_extracted_k141_6871 | Not-determined |
| S3_extracted_k141_55513 | Not-determined |
| S3_extracted_k141_47112 | Low-quality |
| S3_extracted_k141_126109 | Low-quality |
| S3_extracted_k141_118634 | Not-determined |
| S3_extracted_k141_39143 | Not-determined |
| S3_extracted_k141_55518 | Low-quality |
| S3_extracted_k141_111076 | Not-determined |
| S3_extracted_k141_39144 | Not-determined |
| S3_extracted_k141_47118 | Not-determined |
| S3_extracted_k141_101937 | Not-determined |
| S3_extracted_k141_126116 | Not-determined |
| S3_extracted_k141_126117 | Not-determined |
| S3_extracted_k141_39149 | Not-determined |
| S3_extracted_k141_39150 | Not-determined |
| S3_extracted_k141_126122 | Not-determined |
| S3_extracted_k141_79672 | Not-determined |
| S3_extracted_k141_47125 | Low-quality |
| S3_extracted_k141_30963 | Not-determined |
| S3_extracted_k141_79678 | Low-quality |
| S3_extracted_k141_126124 | Not-determined |
| S3_extracted_k141_22495 | Not-determined |
| S3_extracted_k141_118653 | Not-determined |
| S3_extracted_k141_95062 | Not-determined |
| S3_extracted_k141_111096 | Not-determined |
| S3_extracted_k141_70146 | Not-determined |
| S3_extracted_k141_30972 | Not-determined |
| S3_extracted_k141_22499 | Not-determined |
| S3_extracted_k141_111100 | Not-determined |
| S3_extracted_k141_118654 | Low-quality |
| S3_extracted_k141_111102 | Not-determined |
| S3_extracted_k141_118656 | Not-determined |
| S3_extracted_k141_126138 | Not-determined |
| S3_extracted_k141_126140 | Not-determined |
| S3_extracted_k141_126144 | Not-determined |
| S3_extracted_k141_111115 | Not-determined |
| S3_extracted_k141_126149 | Not-determined |
| S3_extracted_k141_22516 | Not-determined |
| S3_extracted_k141_39169 | Not-determined |
| S3_extracted_k141_39171 | Not-determined |
| S3_extracted_k141_126153 | Low-quality |
| S3_extracted_k141_39173 | Not-determined |
| S3_extracted_k141_79682 | Not-determined |
| S3_extracted_k141_126155 | Low-quality |
| S3_extracted_k141_55533 | Not-determined |
| S3_extracted_k141_47141 | Not-determined |
| S3_extracted_k141_126159 | Not-determined |
| S3_extracted_k141_87335 | Not-determined |
| S3_extracted_k141_39181 | Not-determined |
| S3_extracted_k141_47144 | Not-determined |
| S3_extracted_k141_87339 | Not-determined |
| S3_extracted_k141_22527 | Not-determined |
| S3_extracted_k141_6878 | Not-determined |
| S3_extracted_k141_87340 | Low-quality |
| S3_extracted_k141_22532 | Not-determined |
| S3_extracted_k141_47151 | Not-determined |
| S3_extracted_k141_118672 | Not-determined |
| S3_extracted_k141_55542 | Low-quality |
| S3_extracted_k141_39187 | Not-determined |
| S3_extracted_k141_87351 | Not-determined |
| S3_extracted_k141_111154 | Not-determined |
| S3_extracted_k141_101971 | Not-determined |
| S3_extracted_k141_87357 | Not-determined |
| S3_extracted_k141_22542 | Low-quality |
| S3_extracted_k141_22543 | Not-determined |
| S3_extracted_k141_79692 | Not-determined |
| S3_extracted_k141_70155 | Not-determined |
| S3_extracted_k141_39193 | Not-determined |
| S3_extracted_k141_79696 | Not-determined |
| S3_extracted_k141_22546 | Not-determined |
| S3_extracted_k141_70158 | Low-quality |
| S3_extracted_k141_118688 | Not-determined |
| S3_extracted_k141_126179 | Not-determined |
| S3_extracted_k141_22551 | Not-determined |
| S3_extracted_k141_39202 | Not-determined |
| S3_extracted_k141_118694 | Not-determined |
| S3_extracted_k141_70161 | Not-determined |
| S3_extracted_k141_118695 | Not-determined |
| S3_extracted_k141_6905 | Not-determined |
| S3_extracted_k141_118697 | Not-determined |
| S3_extracted_k141_39210 | Not-determined |
| S3_extracted_k141_22562 | Not-determined |
| S3_extracted_k141_111172 | Low-quality |
| S3_extracted_k141_111176 | Not-determined |
| S3_extracted_k141_87373 | Not-determined |
| S3_extracted_k141_111182 | Low-quality |
| S3_extracted_k141_22566 | Not-determined |
| S3_extracted_k141_118710 | Not-determined |
| S3_extracted_k141_6917 | Not-determined |
| S3_extracted_k141_118711 | Not-determined |
| S3_extracted_k141_95081 | Low-quality |
| S3_extracted_k141_30987 | Not-determined |
| S3_extracted_k141_126205 | Not-determined |
| S3_extracted_k141_118720 | Not-determined |
| S3_extracted_k141_111190 | Not-determined |
| S3_extracted_k141_70178 | Not-determined |
| S3_extracted_k141_22581 | Not-determined |
| S3_extracted_k141_6931 | Not-determined |
| S3_extracted_k141_87377 | Low-quality |
| S3_extracted_k141_30990 | Not-determined |
| S3_extracted_k141_111195 | Not-determined |
| S3_extracted_k141_95089 | Not-determined |
| S3_extracted_k141_39218 | Low-quality |
| S3_extracted_k141_30996 | Low-quality |
| S3_extracted_k141_95091 | Not-determined |
| S3_extracted_k141_70179 | Not-determined |
| S3_extracted_k141_126219 | Not-determined |
| S3_extracted_k141_30999 | Not-determined |
| S3_extracted_k141_118735 | Not-determined |
| S3_extracted_k141_31003 | Low-quality |
| S3_extracted_k141_39223 | Not-determined |
| S3_extracted_k141_47186 | Not-determined |
| S3_extracted_k141_87391 | Low-quality |
| S3_extracted_k141_39224 | Low-quality |
| S3_extracted_k141_118742 | Not-determined |
| S3_extracted_k141_126228 | Not-determined |
| S3_extracted_k141_126229 | Not-determined |
| S3_extracted_k141_22599 | Low-quality |
| S3_extracted_k141_118745 | Not-determined |
| S3_extracted_k141_111216 | Not-determined |
| S3_extracted_k141_111218 | Not-determined |
| S3_extracted_k141_39234 | Not-determined |
| S3_extracted_k141_95108 | Not-determined |
| S3_extracted_k141_101989 | Not-determined |
| S3_extracted_k141_111232 | Low-quality |
| S3_extracted_k141_22630 | Not-determined |
| S3_extracted_k141_126247 | Not-determined |
| S3_extracted_k141_39241 | Not-determined |
| S3_extracted_k141_111238 | Not-determined |
| S3_extracted_k141_126250 | Not-determined |
| S3_extracted_k141_126254 | Not-determined |
| S3_extracted_k141_70190 | Not-determined |
| S3_extracted_k141_111244 | Not-determined |
| S3_extracted_k141_39255 | Not-determined |
| S3_extracted_k141_126262 | Not-determined |
| S3_extracted_k141_39261 | Not-determined |
| S3_extracted_k141_126271 | Not-determined |
| S3_extracted_k141_39269 | Not-determined |
| S3_extracted_k141_39271 | Low-quality |
| S3_extracted_k141_47209 | Not-determined |
| S3_extracted_k141_87412 | Not-determined |
| S3_extracted_k141_118761 | Low-quality |
| S3_extracted_k141_87416 | Not-determined |
| S3_extracted_k141_111271 | Not-determined |
| S3_extracted_k141_111272 | Not-determined |
| S3_extracted_k141_87417 | Not-determined |
| S3_extracted_k141_111274 | Not-determined |
| S3_extracted_k141_70217 | Not-determined |
| S3_extracted_k141_87419 | Not-determined |
| S3_extracted_k141_70218 | Low-quality |
| S3_extracted_k141_47220 | Not-determined |
| S8_vs_k141_208127\|\|full | High-quality |
| S3_extracted_k141_55573 | Not-determined |
| S3_extracted_k141_87427 | Not-determined |
| S3_extracted_k141_111287 | Not-determined |
| S3_extracted_k141_102010 | Not-determined |
| S3_extracted_k141_47223 | Low-quality |
| S3_extracted_k141_31044 | Not-determined |
| S3_extracted_k141_55578 | Low-quality |
| S3_extracted_k141_70236 | Not-determined |
| S3_extracted_k141_87433 | Not-determined |
| S3_extracted_k141_102019 | Low-quality |
| S3_extracted_k141_95133 | Not-determined |
| S3_extracted_k141_102024 | Not-determined |
| S3_extracted_k141_118785 | Low-quality |
| S3_extracted_k141_55591 | Not-determined |
| S3_extracted_k141_47239 | Not-determined |
| S3_extracted_k141_95142 | Not-determined |
| S3_extracted_k141_31060 | Not-determined |
| S3_extracted_k141_87447 | Low-quality |
| S3_extracted_k141_111291 | Low-quality |
| S3_extracted_k141_31062 | Not-determined |
| S3_extracted_k141_31064 | Not-determined |
| S3_extracted_k141_102033 | Not-determined |
| S3_extracted_k141_47251 | Not-determined |
| S3_extracted_k141_87454 | Low-quality |
| S3_extracted_k141_55600 | Not-determined |
| S3_extracted_k141_55601 | Not-determined |
| S3_extracted_k141_102038 | Not-determined |
| S3_extracted_k141_55605 | Low-quality |
| S3_extracted_k141_95151 | Not-determined |
| S3_extracted_k141_47258 | Not-determined |
| S3_extracted_k141_31082 | Not-determined |
| S3_extracted_k141_87468 | Not-determined |
| S3_extracted_k141_31088 | Not-determined |
| S3_extracted_k141_111297 | Not-determined |
| S3_extracted_k141_87474 | Not-determined |
| S3_extracted_k141_55630 | Not-determined |
| S3_extracted_k141_87481 | Not-determined |
| S3_extracted_k141_31108 | Not-determined |
| S3_extracted_k141_111310 | Not-determined |
| S3_extracted_k141_70243 | Not-determined |
| S3_extracted_k141_87484 | Not-determined |
| S3_extracted_k141_95178 | Not-determined |
| S3_extracted_k141_47286 | Not-determined |
| S3_extracted_k141_87491 | Not-determined |
| S3_extracted_k141_47293 | Low-quality |
| S3_extracted_k141_70258 | Low-quality |
| S3_extracted_k141_111315 | Low-quality |
| S3_extracted_k141_31121 | Not-determined |
| S3_extracted_k141_111317 | Not-determined |
| S3_extracted_k141_87501 | Not-determined |
| S3_extracted_k141_111321 | Not-determined |
| S3_extracted_k141_95188 | Not-determined |
| S3_extracted_k141_55655 | Not-determined |
| S3_extracted_k141_111325 | Not-determined |
| S3_extracted_k141_31132 | Low-quality |
| S3_extracted_k141_111330 | Not-determined |
| S3_extracted_k141_47310 | Not-determined |
| S3_extracted_k141_111334 | Not-determined |
| S3_extracted_k141_95195 | Not-determined |
| S3_extracted_k141_6957 | Low-quality |
| S3_extracted_k141_47313 | Not-determined |
| S3_extracted_k141_6961 | Not-determined |
| S3_extracted_k141_126279 | Not-determined |
| S3_extracted_k141_87511 | Not-determined |
| S3_extracted_k141_87515 | Low-quality |
| S3_extracted_k141_126300 | Not-determined |
| S3_extracted_k141_95202 | Low-quality |
| S3_extracted_k141_126302 | Not-determined |
| S3_extracted_k141_126303 | Not-determined |
| S3_extracted_k141_126305 | Not-determined |
| S3_extracted_k141_126306 | Not-determined |
| S3_extracted_k141_126307 | Not-determined |
| S3_extracted_k141_126314 | Not-determined |
| S3_extracted_k141_31158 | Not-determined |
| S3_extracted_k141_47333 | Low-quality |
| S3_extracted_k141_70268 | Low-quality |
| S3_extracted_k141_47334 | Not-determined |
| S3_extracted_k141_111357 | Low-quality |
| S3_extracted_k141_47340 | Not-determined |
| S3_extracted_k141_47341 | Low-quality |

| | |
|---|---|
| S3_extracted_k141_126328 | Not-determined |
| S3_extracted_k141_111361 | Not-determined |
| S3_extracted_k141_118788 | Not-determined |
| S3_extracted_k141_87538 | Not-determined |
| S3_extracted_k141_111362 | Low-quality |
| S3_extracted_k141_126332 | Not-determined |
| S3_extracted_k141_55665 | Not-determined |
| S3_extracted_k141_39287 | Not-determined |
| S3_extracted_k141_87546 | Not-determined |
| S3_extracted_k141_47347 | Low-quality |
| S3_extracted_k141_95214 | Not-determined |
| S3_extracted_k141_111372 | Low-quality |
| S3_extracted_k141_87551 | Not-determined |
| S3_extracted_k141_39297 | Not-determined |
| S3_extracted_k141_95217 | Not-determined |
| S3_extracted_k141_126340 | Low-quality |
| S3_extracted_k141_111379 | Low-quality |
| S3_extracted_k141_87562 | Not-determined |
| S3_extracted_k141_55673 | Not-determined |
| S3_extracted_k141_39306 | Not-determined |
| S3_extracted_k141_87568 | Not-determined |
| S3_extracted_k141_111382 | Not-determined |
| S3_extracted_k141_118794 | Not-determined |
| S3_extracted_k141_118795 | Not-determined |
| S3_extracted_k141_87574 | Not-determined |
| S3_extracted_k141_39316 | Not-determined |
| S3_extracted_k141_31178 | Not-determined |
| S3_extracted_k141_102045 | Low-quality |
| S3_extracted_k141_6997 | Low-quality |
| S3_extracted_k141_102046 | Not-determined |
| S3_extracted_k141_102047 | Not-determined |
| S3_extracted_k141_118807 | Low-quality |
| S3_extracted_k141_7002 | Not-determined |
| S3_extracted_k141_102054 | Low-quality |
| S3_extracted_k141_111394 | Not-determined |
| S3_extracted_k141_31189 | Not-determined |
| S3_extracted_k141_111395 | Low-quality |
| S3_extracted_k141_87578 | Not-determined |
| S3_extracted_k141_31193 | Not-determined |
| S3_extracted_k141_87583 | Not-determined |
| S3_extracted_k141_87584 | Not-determined |
| S3_extracted_k141_39330 | Not-determined |
| S3_extracted_k141_39331 | Not-determined |
| S3_extracted_k141_55697 | Not-determined |
| S3_extracted_k141_31201 | Low-quality |
| S3_extracted_k141_118814 | Not-determined |
| S3_extracted_k141_55702 | Not-determined |
| S3_extracted_k141_118820 | Not-determined |
| S3_extracted_k141_70300 | Not-determined |
| S3_extracted_k141_118822 | Not-determined |
| S3_extracted_k141_111402 | Low-quality |
| S3_extracted_k141_31207 | Not-determined |
| S3_extracted_k141_111406 | Not-determined |
| S3_extracted_k141_126367 | Low-quality |
| S3_extracted_k141_95243 | Not-determined |
| S3_extracted_k141_55707 | Not-determined |
| S3_extracted_k141_111410 | Not-determined |
| S3_extracted_k141_70303 | Not-determined |
| S3_extracted_k141_70305 | Not-determined |
| S3_extracted_k141_111413 | Not-determined |
| S3_extracted_k141_126375 | Not-determined |
| S3_extracted_k141_31221 | Not-determined |
| S3_extracted_k141_22647 | Not-determined |
| S3_extracted_k141_47383 | Not-determined |
| S3_extracted_k141_70316 | Not-determined |
| S3_extracted_k141_70317 | Not-determined |
| S3_extracted_k141_118838 | Not-determined |
| S3_extracted_k141_55719 | Not-determined |
| S3_extracted_k141_126389 | Not-determined |
| S3_extracted_k141_118839 | Low-quality |
| S3_extracted_k141_39344 | Not-determined |
| S3_extracted_k141_102061 | Not-determined |
| S3_extracted_k141_70323 | Not-determined |
| S3_extracted_k141_55736 | Not-determined |
| S3_extracted_k141_102066 | Not-determined |
| S3_extracted_k141_22666 | Not-determined |
| S3_extracted_k141_31240 | Not-determined |
| S3_extracted_k141_70332 | Not-determined |
| S3_extracted_k141_22669 | Not-determined |
| S3_extracted_k141_87590 | Not-determined |
| S3_extracted_k141_102073 | Not-determined |
| S3_extracted_k141_126405 | Not-determined |
| S3_extracted_k141_126406 | Not-determined |
| S3_extracted_k141_126408 | Not-determined |
| S3_extracted_k141_126409 | Not-determined |
| S3_extracted_k141_102078 | Not-determined |
| S3_extracted_k141_102081 | Low-quality |
| S3_extracted_k141_31248 | Not-determined |
| S3_extracted_k141_102082 | Not-determined |
| S3_extracted_k141_118849 | Not-determined |
| S3_extracted_k141_102087 | Not-determined |
| S3_extracted_k141_22682 | Low-quality |
| S3_extracted_k141_39356 | Not-determined |
| S3_extracted_k141_31254 | Low-quality |
| S3_extracted_k141_47389 | Not-determined |
| S3_extracted_k141_102092 | Not-determined |
| S3_extracted_k141_55755 | Low-quality |
| S3_extracted_k141_47390 | Low-quality |
| S3_extracted_k141_55758 | Not-determined |
| S3_extracted_k141_22683 | Not-determined |
| S3_extracted_k141_31262 | Not-determined |
| S3_extracted_k141_95274 | Not-determined |
| S3_extracted_k141_126425 | Not-determined |
| S3_extracted_k141_126426 | Not-determined |
| S3_extracted_k141_55759 | Not-determined |
| S3_extracted_k141_55763 | Not-determined |
| S3_extracted_k141_87606 | Not-determined |
| S3_extracted_k141_126431 | Not-determined |
| S3_extracted_k141_126433 | Not-determined |
| S3_extracted_k141_39370 | Not-determined |
| S3_extracted_k141_126436 | Not-determined |
| S3_extracted_k141_87615 | Low-quality |
| S3_extracted_k141_87616 | Not-determined |
| S3_extracted_k141_111425 | Not-determined |
| S3_extracted_k141_87621 | Low-quality |
| S3_extracted_k141_31282 | Not-determined |
| S3_extracted_k141_111426 | Not-determined |
| S3_extracted_k141_47409 | Not-determined |
| S3_extracted_k141_55775 | Not-determined |
| S3_extracted_k141_70352 | Not-determined |
| S3_extracted_k141_70353 | Not-determined |
| S3_extracted_k141_39374 | Not-determined |
| S3_extracted_k141_87623 | Not-determined |
| S3_extracted_k141_87625 | Not-determined |
| S3_extracted_k141_95284 | Not-determined |
| S3_extracted_k141_102115 | Not-determined |
| S3_extracted_k141_87626 | Low-quality |
| S3_extracted_k141_111441 | Not-determined |
| S3_extracted_k141_111442 | Not-determined |
| S3_extracted_k141_87630 | Not-determined |
| S3_extracted_k141_102124 | Not-determined |
| S3_extracted_k141_31288 | Not-determined |
| S3_extracted_k141_70368 | Not-determined |
| S3_extracted_k141_39387 | Low-quality |
| S3_extracted_k141_7011 | Not-determined |
| S3_extracted_k141_102130 | Not-determined |
| S3_extracted_k141_39402 | Low-quality |
| S3_extracted_k141_95294 | Low-quality |
| S3_extracted_k141_95299 | Not-determined |
| S3_extracted_k141_126480 | Not-determined |
| S3_extracted_k141_47449 | Not-determined |
| S3_extracted_k141_47451 | Not-determined |
| S3_extracted_k141_39416 | Not-determined |
| S3_extracted_k141_95302 | Not-determined |
| S3_extracted_k141_95304 | Not-determined |
| S3_extracted_k141_95306 | Not-determined |
| S3_extracted_k141_102147 | Not-determined |
| S3_extracted_k141_7035 | Not-determined |
| S3_extracted_k141_111456 | Not-determined |
| S3_extracted_k141_47460 | Not-determined |
| S3_extracted_k141_87636 | Not-determined |
| S3_extracted_k141_111467 | Not-determined |
| S3_extracted_k141_87639 | Not-determined |
| S3_extracted_k141_47467 | Not-determined |
| S3_extracted_k141_47473 | Not-determined |
| S3_extracted_k141_31302 | Not-determined |
| S3_extracted_k141_87650 | Low-quality |
| S3_extracted_k141_102171 | Not-determined |
| S3_extracted_k141_39433 | Not-determined |
| S3_extracted_k141_70385 | Not-determined |
| S3_extracted_k141_31308 | Not-determined |
| S3_extracted_k141_22693 | Not-determined |
| S3_extracted_k141_102179 | Not-determined |
| S3_extracted_k141_70392 | Low-quality |
| S3_extracted_k141_47483 | Low-quality |
| S3_extracted_k141_39450 | Not-determined |
| S3_extracted_k141_39455 | Not-determined |
| S3_extracted_k141_87668 | Not-determined |
| S3_extracted_k141_39459 | Not-determined |
| S3_extracted_k141_95315 | Not-determined |
| S3_extracted_k141_47494 | Not-determined |
| S3_extracted_k141_47495 | Not-determined |
| S3_extracted_k141_70396 | Not-determined |
| S3_extracted_k141_31332 | Low-quality |
| S3_extracted_k141_31333 | Not-determined |
| S3_extracted_k141_111474 | Not-determined |
| S3_extracted_k141_39467 | Not-determined |
| S3_extracted_k141_111479 | Not-determined |
| S3_extracted_k141_111488 | Not-determined |
| S3_extracted_k141_31361 | Low-quality |
| S3_extracted_k141_22734 | Not-determined |
| S3_extracted_k141_47511 | Not-determined |
| S3_extracted_k141_47512 | Not-determined |
| S3_extracted_k141_118872 | Not-determined |
| S3_extracted_k141_47513 | Not-determined |
| S3_extracted_k141_118873 | Not-determined |
| S3_extracted_k141_39502 | Not-determined |
| S3_extracted_k141_39503 | Not-determined |
| S3_extracted_k141_31380 | Not-determined |
| S3_extracted_k141_118877 | Not-determined |
| S3_extracted_k141_39504 | Low-quality |
| S3_extracted_k141_31383 | Not-determined |
| S3_extracted_k141_39510 | Not-determined |
| S3_extracted_k141_31387 | Not-determined |
| S3_extracted_k141_47528 | Not-determined |
| S3_extracted_k141_31388 | Not-determined |
| S3_extracted_k141_95329 | Not-determined |
| S3_extracted_k141_31392 | Not-determined |
| S3_extracted_k141_31393 | Not-determined |
| S3_extracted_k141_31394 | Not-determined |
| S3_extracted_k141_70403 | Low-quality |
| S3_extracted_k141_7052 | Not-determined |
| S3_extracted_k141_118896 | Not-determined |
| S3_extracted_k141_7054 | Not-determined |
| S3_extracted_k141_7055 | Not-determined |
| S3_extracted_k141_7057 | Not-determined |
| S3_extracted_k141_95341 | Not-determined |
| S3_extracted_k141_70421 | Low-quality |
| S3_extracted_k141_47536 | Low-quality |
| S3_extracted_k141_70423 | Not-determined |
| S3_extracted_k141_126519 | Low-quality |
| S3_extracted_k141_111495 | Not-determined |
| S3_extracted_k141_126520 | Not-determined |
| S3_extracted_k141_111496 | Not-determined |
| S3_extracted_k141_31407 | Not-determined |
| S3_extracted_k141_70426 | Low-quality |
| S3_extracted_k141_126525 | Not-determined |
| S3_extracted_k141_7062 | Not-determined |
| S3_extracted_k141_95347 | Not-determined |
| S3_extracted_k141_7064 | Not-determined |
| S3_extracted_k141_70436 | Not-determined |
| S3_extracted_k141_70437 | Not-determined |
| S3_extracted_k141_31417 | Not-determined |
| S3_extracted_k141_70440 | Not-determined |
| S3_extracted_k141_31422 | Not-determined |
| S3_extracted_k141_111508 | Not-determined |
| S3_extracted_k141_31430 | Not-determined |
| S3_extracted_k141_111515 | Not-determined |
| S3_extracted_k141_111516 | Not-determined |
| S3_extracted_k141_39551 | Not-determined |
| S3_extracted_k141_47569 | Not-determined |
| S3_extracted_k141_39554 | Not-determined |
| S3_extracted_k141_95369 | Not-determined |
| S3_extracted_k141_102210 | Not-determined |
| S3_extracted_k141_47576 | Not-determined |
| S3_extracted_k141_126548 | Low-quality |
| S3_extracted_k141_39566 | Not-determined |
| S3_extracted_k141_39567 | Not-determined |
| S3_extracted_k141_95376 | Not-determined |
| S3_extracted_k141_22761 | Not-determined |
| S3_extracted_k141_39569 | Not-determined |
| S3_extracted_k141_47584 | Not-determined |
| S3_extracted_k141_39572 | Low-quality |
| S3_extracted_k141_22765 | Not-determined |
| S3_extracted_k141_22767 | Low-quality |
| S3_extracted_k141_111534 | Low-quality |
| S3_extracted_k141_126560 | Not-determined |
| S3_extracted_k141_31460 | Not-determined |
| S3_extracted_k141_126562 | Not-determined |
| S3_extracted_k141_70448 | Low-quality |
| S3_extracted_k141_70450 | Not-determined |
| S3_extracted_k141_95392 | Low-quality |
| S3_extracted_k141_39590 | Low-quality |
| S3_extracted_k141_111543 | Not-determined |
| S3_extracted_k141_39597 | Not-determined |
| S3_extracted_k141_47587 | Not-determined |
| S3_extracted_k141_111546 | Low-quality |
| S3_extracted_k141_22779 | Not-determined |
| S3_extracted_k141_31479 | Not-determined |
| S3_extracted_k141_31480 | Low-quality |
| S3_extracted_k141_22784 | Not-determined |
| S3_extracted_k141_111550 | Not-determined |
| S3_extracted_k141_70469 | Not-determined |
| S3_extracted_k141_22792 | Not-determined |
| S3_extracted_k141_70482 | Not-determined |
| S3_extracted_k141_111559 | Not-determined |
| S3_extracted_k141_111560 | Not-determined |
| S3_extracted_k141_111569 | Not-determined |
| S3_extracted_k141_7078 | Not-determined |
| S3_extracted_k141_111570 | Not-determined |
| S3_extracted_k141_102240 | Not-determined |
| S3_extracted_k141_70485 | Not-determined |
| S3_extracted_k141_111574 | Low-quality |
| S3_extracted_k141_95429 | Not-determined |
| S3_extracted_k141_102245 | Not-determined |
| S3_extracted_k141_47612 | Not-determined |
| S3_extracted_k141_111578 | Not-determined |
| S3_extracted_k141_47623 | Not-determined |
| S3_extracted_k141_111579 | Not-determined |
| S3_extracted_k141_39631 | Not-determined |
| S3_extracted_k141_39634 | Not-determined |
| S3_extracted_k141_39635 | Not-determined |
| S3_extracted_k141_47625 | Not-determined |
| S3_extracted_k141_39643 | Low-quality |
| S3_extracted_k141_7093 | Not-determined |
| S3_extracted_k141_39644 | Not-determined |
| S3_extracted_k141_102257 | Not-determined |
| S3_extracted_k141_7095 | Low-quality |
| S3_extracted_k141_7097 | Not-determined |
| S3_extracted_k141_22801 | Not-determined |
| S3_extracted_k141_95445 | Not-determined |
| S3_extracted_k141_47632 | Not-determined |
| S3_extracted_k141_95448 | Not-determined |
| S3_extracted_k141_22805 | Not-determined |
| S3_extracted_k141_126595 | Low-quality |
| S3_extracted_k141_102259 | Low-quality |
| S3_extracted_k141_47644 | Not-determined |
| S3_extracted_k141_126602 | Not-determined |
| S3_extracted_k141_95459 | Not-determined |
| S3_extracted_k141_95460 | Not-determined |
| S3_extracted_k141_39662 | Not-determined |
| S3_extracted_k141_102274 | Not-determined |
| S3_extracted_k141_22818 | Not-determined |
| S3_extracted_k141_126612 | Not-determined |
| S3_extracted_k141_126613 | Not-determined |
| S3_extracted_k141_7107 | Not-determined |
| S3_extracted_k141_22824 | Not-determined |
| S3_extracted_k141_126618 | Not-determined |
| S3_extracted_k141_95472 | Not-determined |
| S3_extracted_k141_22830 | Not-determined |
| S3_extracted_k141_39675 | Not-determined |
| S3_extracted_k141_22833 | Not-determined |
| S3_extracted_k141_22835 | Not-determined |
| S3_extracted_k141_126624 | Not-determined |
| S3_extracted_k141_126627 | Low-quality |
| S3_extracted_k141_102284 | Not-determined |
| S3_extracted_k141_39691 | Not-determined |
| S3_extracted_k141_39694 | Not-determined |
| S3_extracted_k141_7125 | Not-determined |
| S3_extracted_k141_95490 | Low-quality |
| S3_extracted_k141_95491 | Low-quality |
| S3_extracted_k141_39699 | Not-determined |
| S3_extracted_k141_126636 | Not-determined |
| S3_extracted_k141_126641 | Not-determined |
| S3_extracted_k141_95498 | Not-determined |
| S3_extracted_k141_95501 | Not-determined |
| S3_extracted_k141_95503 | Not-determined |
| S3_extracted_k141_126651 | Not-determined |
| S3_extracted_k141_126652 | Not-determined |
| S3_extracted_k141_126655 | Not-determined |
| S3_extracted_k141_102307 | Not-determined |
| S3_extracted_k141_102308 | Not-determined |
| S3_extracted_k141_39709 | Not-determined |
| S3_extracted_k141_47700 | Not-determined |
| S3_extracted_k141_102311 | Not-determined |
| S3_extracted_k141_47708 | Not-determined |
| S3_extracted_k141_22861 | Not-determined |
| S3_extracted_k141_39710 | Not-determined |
| S3_extracted_k141_47709 | Not-determined |
| S3_extracted_k141_39713 | Not-determined |
| S3_extracted_k141_7131 | Not-determined |
| S3_extracted_k141_47720 | Not-determined |
| S3_extracted_k141_95518 | Not-determined |
| S3_extracted_k141_22874 | Not-determined |
| S3_extracted_k141_70520 | Not-determined |
| S3_extracted_k141_102325 | Not-determined |
| S3_extracted_k141_22882 | Not-determined |
| S3_extracted_k141_102331 | Low-quality |
| S3_extracted_k141_47731 | Not-determined |
| S3_extracted_k141_31495 | Not-determined |
| S3_extracted_k141_31499 | Not-determined |
| S6_extracted_k141_56484 | Complete |
| S3_extracted_k141_7139 | Not-determined |
| S3_extracted_k141_7145 | Not-determined |
| S3_extracted_k141_7147 | Not-determined |
| S3_extracted_k141_22906 | Low-quality |
| S3_extracted_k141_70556 | Low-quality |
| S3_extracted_k141_126665 | Not-determined |
| S3_extracted_k141_126666 | Low-quality |
| S3_extracted_k141_126667 | Not-determined |
| S3_extracted_k141_126670 | Not-determined |
| S3_extracted_k141_102341 | Not-determined |
| S3_extracted_k141_126682 | Not-determined |
| S3_extracted_k141_39733 | Not-determined |
| S3_extracted_k141_7164 | Not-determined |
| S3_extracted_k141_39739 | Not-determined |
| S3_extracted_k141_7172 | Low-quality |
| S3_extracted_k141_102347 | Not-determined |
| S3_extracted_k141_7173 | Not-determined |
| S3_extracted_k141_7174 | Not-determined |
| S3_extracted_k141_39750 | Not-determined |
| S3_extracted_k141_31530 | Not-determined |
| S3_extracted_k141_22939 | Not-determined |
| S3_extracted_k141_39761 | Not-determined |
| S3_extracted_k141_47755 | Not-determined |
| S3_extracted_k141_22947 | Not-determined |
| S3_extracted_k141_31545 | Low-quality |
| S3_extracted_k141_7198 | Not-determined |
| S3_extracted_k141_22953 | Not-determined |
| S3_extracted_k141_47769 | Not-determined |
| S3_extracted_k141_31551 | Not-determined |
| S3_extracted_k141_102361 | Not-determined |
| S3_extracted_k141_70599 | Not-determined |
| S5_extracted_k141_110171 | High-quality |
| S3_extracted_k141_47796 | Not-determined |
| S3_extracted_k141_70607 | Not-determined |
| S3_extracted_k141_22975 | Not-determined |
| S3_extracted_k141_7220 | Not-determined |
| S3_extracted_k141_31563 | Not-determined |
| S3_extracted_k141_7225 | Not-determined |
| S3_extracted_k141_118901 | Not-determined |
| S3_extracted_k141_31570 | Not-determined |
| S3_extracted_k141_22984 | Not-determined |
| S3_extracted_k141_31573 | Not-determined |
| S3_extracted_k141_7236 | Not-determined |
| S3_extracted_k141_22988 | Low-quality |
| S3_extracted_k141_102390 | Not-determined |
| S3_extracted_k141_118908 | Not-determined |
| S3_extracted_k141_7251 | Low-quality |
| S3_extracted_k141_70633 | Not-determined |
| S3_extracted_k141_7256 | Not-determined |
| S3_extracted_k141_31582 | Not-determined |
| S3_extracted_k141_95533 | Low-quality |
| S3_extracted_k141_95536 | Not-determined |
| S3_extracted_k141_23008 | Not-determined |
| S3_extracted_k141_95542 | Not-determined |
| S3_extracted_k141_23010 | Not-determined |
| S3_extracted_k141_23013 | Not-determined |
| S3_extracted_k141_23014 | Not-determined |
| S3_extracted_k141_102406 | Low-quality |
| S3_extracted_k141_23019 | Not-determined |
| S3_extracted_k141_95555 | Not-determined |
| S3_extracted_k141_118917 | Not-determined |
| S3_extracted_k141_126729 | Not-determined |
| S3_extracted_k141_126730 | Not-determined |
| S3_extracted_k141_95557 | Low-quality |
| S3_extracted_k141_118920 | Not-determined |
| S3_extracted_k141_126731 | Low-quality |
| S3_extracted_k141_118922 | Not-determined |
| S3_extracted_k141_95560 | Not-determined |
| S3_extracted_k141_118924 | Low-quality |
| S3_extracted_k141_31597 | Low-quality |
| S3_extracted_k141_126733 | Not-determined |
| S3_extracted_k141_31600 | Not-determined |
| S3_extracted_k141_31603 | Not-determined |
| S3_extracted_k141_95563 | Not-determined |
| S3_extracted_k141_118935 | Not-determined |
| S3_extracted_k141_70650 | Not-determined |
| S3_extracted_k141_23036 | Not-determined |
| S3_extracted_k141_23037 | Not-determined |
| S3_extracted_k141_70655 | Not-determined |
| S3_extracted_k141_118945 | Not-determined |
| S3_extracted_k141_118946 | Low-quality |
| S3_extracted_k141_118948 | Not-determined |
| S3_extracted_k141_31616 | Not-determined |
| S3_extracted_k141_7277 | Not-determined |
| S3_extracted_k141_7282 | Not-determined |
| S3_extracted_k141_31622 | Not-determined |
| S3_extracted_k141_126736 | Not-determined |
| S3_extracted_k141_31628 | Low-quality |
| S3_extracted_k141_118959 | Not-determined |
| S3_extracted_k141_39796 | Low-quality |
| S3_extracted_k141_39798 | Not-determined |
| S3_extracted_k141_31634 | Not-determined |
| S3_extracted_k141_39803 | Not-determined |
| S3_extracted_k141_95576 | Not-determined |
| S3_extracted_k141_118965 | Not-determined |
| S14_vs_k141_101929\|\|full | Medium-quality |
| S3_extracted_k141_31647 | Not-determined |
| S3_extracted_k141_7295 | Low-quality |
| S3_extracted_k141_39806 | Not-determined |
| S3_extracted_k141_39807 | Not-determined |
| S3_extracted_k141_7298 | Not-determined |
| S3_extracted_k141_39808 | Not-determined |
| S3_extracted_k141_23043 | Low-quality |
| S3_extracted_k141_23046 | Not-determined |
| S3_extracted_k141_23050 | Low-quality |
| S3_extracted_k141_70681 | Not-determined |
| S3_extracted_k141_23058 | Not-determined |
| S3_extracted_k141_23059 | Not-determined |
| S3_extracted_k141_102433 | Low-quality |
| S3_extracted_k141_7311 | Not-determined |
| S3_extracted_k141_70688 | Not-determined |
| S3_extracted_k141_7315 | Not-determined |
| S3_extracted_k141_23065 | Low-quality |
| S3_extracted_k141_102443 | Low-quality |
| S3_extracted_k141_39818 | Not-determined |
| S3_extracted_k141_7321 | Low-quality |
| S3_extracted_k141_39823 | Not-determined |
| S3_extracted_k141_70701 | Not-determined |
| S3_extracted_k141_70702 | Not-determined |
| S3_extracted_k141_118971 | Not-determined |
| S3_extracted_k141_70708 | Not-determined |
| S3_extracted_k141_23078 | Low-quality |
| S3_extracted_k141_70715 | Not-determined |
| S3_extracted_k141_70717 | Not-determined |
| S3_extracted_k141_7333 | Not-determined |
| S3_extracted_k141_23080 | Not-determined |
| S3_extracted_k141_7336 | Low-quality |
| S3_extracted_k141_7337 | Not-determined |
| S3_extracted_k141_7340 | Not-determined |
| S3_extracted_k141_70726 | Low-quality |
| S3_extracted_k141_70728 | Not-determined |
| S3_extracted_k141_31684 | Not-determined |
| S3_extracted_k141_23089 | Not-determined |
| S3_extracted_k141_102462 | Not-determined |
| S3_extracted_k141_70731 | Not-determined |
| S3_extracted_k141_31688 | Not-determined |
| S3_extracted_k141_39843 | Not-determined |
| S3_extracted_k141_39845 | Low-quality |
| S3_extracted_k141_39846 | Not-determined |
| S3_extracted_k141_102466 | Not-determined |
| S3_extracted_k141_23095 | Not-determined |
| S3_extracted_k141_102468 | Not-determined |
| S3_extracted_k141_23097 | Not-determined |
| S3_extracted_k141_39849 | Low-quality |
| S3_extracted_k141_119007 | Not-determined |
| S3_extracted_k141_102472 | Not-determined |
| S3_extracted_k141_23100 | Not-determined |
| S3_extracted_k141_119010 | Not-determined |
| S3_extracted_k141_102477 | Not-determined |
| S3_extracted_k141_7355 | Not-determined |
| S3_extracted_k141_119024 | Not-determined |
| S3_extracted_k141_7357 | Not-determined |
| S3_extracted_k141_126760 | Not-determined |
| S3_extracted_k141_7364 | Low-quality |
| S3_extracted_k141_102496 | Low-quality |
| S3_extracted_k141_23110 | Not-determined |
| S3_extracted_k141_126765 | Not-determined |
| S3_extracted_k141_95598 | Not-determined |
| S3_extracted_k141_126768 | Not-determined |
| S3_extracted_k141_95601 | Not-determined |
| S3_extracted_k141_23124 | Not-determined |
| S3_extracted_k141_102509 | Low-quality |
| S3_extracted_k141_119032 | Not-determined |
| S3_extracted_k141_7377 | Not-determined |
| S3_extracted_k141_95608 | Low-quality |
| S3_extracted_k141_7380 | Low-quality |
| S3_extracted_k141_23130 | Not-determined |
| S3_extracted_k141_102514 | Not-determined |
| S3_extracted_k141_126790 | Not-determined |
| S3_extracted_k141_7394 | Not-determined |
| S3_extracted_k141_23141 | Not-determined |
| S3_extracted_k141_7405 | Not-determined |
| S3_extracted_k141_102546 | Not-determined |
| S3_extracted_k141_102555 | Not-determined |
| S3_extracted_k141_7424 | Not-determined |
| S5_vs_k141_110171\|\|full | High-quality |
| S3_extracted_k141_31730 | Not-determined |
| S3_extracted_k141_119042 | Not-determined |
| S3_extracted_k141_7446 | Not-determined |
| S3_extracted_k141_7450 | Not-determined |
| S3_extracted_k141_7458 | Not-determined |
| S3_extracted_k141_7462 | Not-determined |
| S3_extracted_k141_7466 | Not-determined |
| S3_extracted_k141_102589 | Not-determined |
| S3_extracted_k141_7467 | Not-determined |
| S3_extracted_k141_102597 | Not-determined |
| S3_extracted_k141_102603 | Not-determined |
| S3_extracted_k141_102605 | Not-determined |
| S3_extracted_k141_102606 | Not-determined |
| S3_extracted_k141_102609 | Not-determined |
| S3_extracted_k141_102613 | Not-determined |
| S3_extracted_k141_31760 | Low-quality |
| S3_extracted_k141_70762 | Not-determined |
| S3_extracted_k141_70764 | Not-determined |
| S3_extracted_k141_102626 | Not-determined |
| S3_extracted_k141_7489 | Low-quality |
| S3_extracted_k141_70770 | Not-determined |
| S3_extracted_k141_7497 | Not-determined |
| S3_extracted_k141_70776 | Not-determined |
| S3_extracted_k141_23197 | Low-quality |
| S3_extracted_k141_23200 | Not-determined |
| S3_extracted_k141_102640 | Low-quality |
| S3_extracted_k141_70786 | Not-determined |
| S3_extracted_k141_70788 | Low-quality |
| S3_extracted_k141_70791 | Not-determined |
| S3_extracted_k141_7510 | Not-determined |
| S3_extracted_k141_7511 | Not-determined |
| S3_extracted_k141_119063 | Not-determined |
| S3_extracted_k141_23204 | Low-quality |
| S3_extracted_k141_23205 | Not-determined |
| S3_extracted_k141_7519 | Not-determined |
| S3_extracted_k141_23209 | Not-determined |
| S3_extracted_k141_70810 | Not-determined |
| S3_extracted_k141_7529 | Not-determined |
| S3_extracted_k141_102655 | Not-determined |
| S3_extracted_k141_119073 | Low-quality |
| S3_extracted_k141_119077 | Not-determined |
| S3_extracted_k141_70828 | Not-determined |
| S3_extracted_k141_70830 | Not-determined |
| S3_extracted_k141_126804 | Low-quality |
| S3_extracted_k141_126805 | Not-determined |
| S3_extracted_k141_126810 | Not-determined |
| S3_extracted_k141_126812 | Not-determined |
| S3_extracted_k141_70844 | Not-determined |
| S3_extracted_k141_70849 | Not-determined |
| S3_extracted_k141_70855 | Low-quality |
| S3_extracted_k141_126815 | Not-determined |
| S3_extracted_k141_23235 | Not-determined |
| S3_extracted_k141_126819 | Not-determined |
| S3_extracted_k141_119086 | Not-determined |
| S3_extracted_k141_119089 | Not-determined |
| S3_extracted_k141_126823 | Not-determined |
| S3_extracted_k141_7537 | Not-determined |
| S3_extracted_k141_31775 | Not-determined |
| S3_extracted_k141_31781 | Not-determined |
| S3_extracted_k141_7541 | Not-determined |
| S3_extracted_k141_126833 | Not-determined |

| | |
|---|---|
| S3_extracted_k141_70875 | Not-determined |
| S3_extracted_k141_126840 | Not-determined |
| S3_extracted_k141_126841 | Not-determined |
| S3_extracted_k141_70883 | Not-determined |
| S3_extracted_k141_70884 | Not-determined |
| S3_extracted_k141_126847 | Low-quality |
| S3_extracted_k141_31804 | Not-determined |
| S3_extracted_k141_31806 | Not-determined |
| S3_extracted_k141_126853 | Not-determined |
| S3_extracted_k141_70896 | Not-determined |
| S3_extracted_k141_126861 | Not-determined |
| S3_extracted_k141_7556 | Not-determined |
| S3_extracted_k141_126864 | Not-determined |
| S3_extracted_k141_126868 | Not-determined |
| S3_extracted_k141_102703 | Not-determined |
| S3_extracted_k141_70912 | Not-determined |
| S3_extracted_k141_23253 | Not-determined |
| S3_extracted_k141_102713 | Not-determined |
| S3_extracted_k141_23269 | Not-determined |
| S3_extracted_k141_23270 | Not-determined |
| S3_extracted_k141_31831 | Not-determined |
| S3_extracted_k141_70923 | Not-determined |
| S3_extracted_k141_70926 | Not-determined |
| S3_extracted_k141_126871 | Not-determined |
| S3_extracted_k141_70935 | Not-determined |
| S3_extracted_k141_126877 | Not-determined |
| S3_extracted_k141_23278 | Low-quality |
| S3_extracted_k141_126881 | Not-determined |
| S3_extracted_k141_126882 | Not-determined |
| S3_extracted_k141_7573 | Low-quality |
| S3_extracted_k141_23283 | Not-determined |
| S3_extracted_k141_7575 | Low-quality |
| S3_extracted_k141_119111 | Not-determined |
| S3_extracted_k141_70943 | Not-determined |
| S3_extracted_k141_119114 | Low-quality |
| S3_extracted_k141_70946 | Not-determined |
| S3_extracted_k141_23288 | Not-determined |
| S3_extracted_k141_126900 | Not-determined |
| S3_extracted_k141_126901 | Not-determined |
| S3_extracted_k141_23292 | Not-determined |
| S3_extracted_k141_31862 | Not-determined |
| S3_extracted_k141_126904 | Not-determined |
| S3_extracted_k141_31863 | Low-quality |
| S3_extracted_k141_70961 | Not-determined |
| S3_extracted_k141_7591 | Not-determined |
| S3_extracted_k141_126914 | Not-determined |
| S3_extracted_k141_119131 | Not-determined |
| S3_extracted_k141_70969 | Not-determined |
| S3_extracted_k141_70972 | Not-determined |
| S3_extracted_k141_102747 | Low-quality |
| S3_extracted_k141_119148 | Not-determined |
| S3_extracted_k141_31868 | Low-quality |
| S3_extracted_k141_70980 | Not-determined |
| S3_extracted_k141_102756 | Not-determined |
| S3_extracted_k141_102758 | Not-determined |
| S3_extracted_k141_70983 | Not-determined |
| S3_extracted_k141_70998 | Not-determined |
| S3_extracted_k141_7597 | Not-determined |
| S3_extracted_k141_102768 | Not-determined |
| S3_extracted_k141_102770 | Not-determined |
| S3_extracted_k141_102774 | Not-determined |
| S3_extracted_k141_71002 | Not-determined |
| S3_extracted_k141_102780 | Low-quality |
| S3_extracted_k141_102782 | Not-determined |
| S3_extracted_k141_102783 | Not-determined |
| S3_extracted_k141_71007 | Not-determined |
| S3_extracted_k141_119154 | Low-quality |
| S3_extracted_k141_102785 | Not-determined |
| S3_extracted_k141_102787 | Not-determined |
| S3_extracted_k141_71014 | Not-determined |
| S3_extracted_k141_7637 | Not-determined |
| S3_extracted_k141_23351 | Not-determined |
| S3_extracted_k141_71026 | Not-determined |
| S3_extracted_k141_119167 | Not-determined |
| S3_extracted_k141_126944 | Not-determined |
| S3_extracted_k141_71040 | Not-determined |
| S3_extracted_k141_7663 | Not-determined |
| S3_extracted_k141_7675 | Not-determined |
| S3_extracted_k141_71061 | Not-determined |
| S3_extracted_k141_7682 | Not-determined |
| S3_extracted_k141_71063 | Not-determined |
| S3_extracted_k141_7683 | Not-determined |
| S3_extracted_k141_7688 | Not-determined |
| S3_extracted_k141_126956 | Not-determined |
| S3_extracted_k141_7692 | Not-determined |
| S3_extracted_k141_126958 | Low-quality |
| S3_extracted_k141_102807 | Not-determined |
| S3_extracted_k141_102823 | Not-determined |
| S3_extracted_k141_7721 | Not-determined |
| S3_extracted_k141_119212 | Not-determined |
| S3_extracted_k141_7728 | Not-determined |
| S3_extracted_k141_7731 | Low-quality |
| S3_extracted_k141_102835 | Not-determined |
| S3_extracted_k141_7732 | Low-quality |
| S3_extracted_k141_23380 | Not-determined |
| S3_extracted_k141_102839 | Not-determined |
| S3_extracted_k141_23383 | Not-determined |
| S3_extracted_k141_23385 | Not-determined |
| S3_extracted_k141_23387 | Not-determined |
| S3_extracted_k141_71112 | Low-quality |
| S3_extracted_k141_7766 | Not-determined |
| S3_extracted_k141_119229 | Not-determined |
| S3_extracted_k141_71122 | Not-determined |
| S3_extracted_k141_71123 | Not-determined |
| S3_extracted_k141_119237 | Not-determined |
| S3_extracted_k141_71125 | Low-quality |
| S3_extracted_k141_71126 | Not-determined |
| S3_extracted_k141_119239 | Low-quality |
| S3_extracted_k141_71129 | Not-determined |
| S3_extracted_k141_7773 | Not-determined |
| S3_extracted_k141_23434 | Low-quality |
| S3_extracted_k141_7777 | Not-determined |
| S3_extracted_k141_126998 | Not-determined |
| S3_extracted_k141_119246 | Not-determined |
| S3_extracted_k141_23442 | Not-determined |
| S3_extracted_k141_23444 | Not-determined |
| S3_extracted_k141_71132 | Not-determined |
| S3_extracted_k141_7796 | Not-determined |
| S3_extracted_k141_7797 | Not-determined |
| S3_extracted_k141_127019 | Not-determined |
| S3_extracted_k141_7804 | Not-determined |
| S3_extracted_k141_71139 | Not-determined |
| S3_extracted_k141_71140 | Low-quality |
| S3_extracted_k141_7817 | Not-determined |
| S3_extracted_k141_7818 | Not-determined |
| S3_extracted_k141_7822 | Not-determined |
| S3_extracted_k141_7825 | Not-determined |
| S3_extracted_k141_7828 | Low-quality |
| S3_extracted_k141_7830 | Low-quality |
| S3_extracted_k141_7833 | Not-determined |
| S3_extracted_k141_71153 | Low-quality |
| S3_extracted_k141_102859 | Not-determined |
| S3_extracted_k141_23467 | Not-determined |
| S3_extracted_k141_23469 | Low-quality |
| S3_extracted_k141_102866 | Not-determined |
| S3_extracted_k141_102867 | Not-determined |
| S3_extracted_k141_127039 | Not-determined |
| S3_extracted_k141_102873 | Not-determined |
| S3_extracted_k141_127051 | Not-determined |
| S3_extracted_k141_23489 | Not-determined |
| S3_extracted_k141_23490 | Not-determined |
| S3_extracted_k141_127058 | Not-determined |
| S3_extracted_k141_71165 | Not-determined |
| S3_extracted_k141_119276 | Not-determined |
| S3_extracted_k141_119278 | Not-determined |
| S3_extracted_k141_127064 | Low-quality |
| S3_extracted_k141_119283 | Not-determined |
| S3_extracted_k141_127074 | Not-determined |
| S3_extracted_k141_119288 | Not-determined |
| S3_extracted_k141_71183 | Not-determined |
| S3_extracted_k141_7870 | Not-determined |
| S3_extracted_k141_127085 | Not-determined |
| S3_extracted_k141_23510 | Not-determined |
| S3_extracted_k141_23513 | Not-determined |
| S3_extracted_k141_127091 | Not-determined |
| S3_extracted_k141_71213 | Not-determined |
| S3_extracted_k141_119293 | Low-quality |
| S3_extracted_k141_119296 | Not-determined |
| S3_extracted_k141_127100 | Not-determined |
| S3_extracted_k141_23531 | Low-quality |
| S3_extracted_k141_119303 | Not-determined |
| S3_extracted_k141_102919 | Not-determined |
| S3_extracted_k141_71219 | Not-determined |
| S3_extracted_k141_119307 | Not-determined |
| S3_extracted_k141_119308 | Not-determined |
| S3_extracted_k141_119315 | Not-determined |
| S3_extracted_k141_119318 | Not-determined |
| S3_extracted_k141_119324 | Not-determined |
| S3_extracted_k141_127107 | Not-determined |
| S3_extracted_k141_102933 | Low-quality |
| S3_extracted_k141_127111 | Not-determined |
| S3_extracted_k141_7887 | Not-determined |
| S3_extracted_k141_102936 | Low-quality |
| S3_extracted_k141_7895 | Not-determined |
| S3_extracted_k141_102941 | Not-determined |
| S3_extracted_k141_119347 | Not-determined |
| S3_extracted_k141_7912 | Low-quality |
| S3_extracted_k141_127131 | Not-determined |
| S3_extracted_k141_7913 | Low-quality |
| S3_extracted_k141_71254 | Not-determined |
| S3_extracted_k141_102945 | Low-quality |
| S3_extracted_k141_102947 | Not-determined |
| S3_extracted_k141_127132 | Not-determined |
| S3_extracted_k141_127135 | Not-determined |
| S3_extracted_k141_127140 | Low-quality |
| S3_extracted_k141_102953 | Low-quality |
| S3_extracted_k141_71260 | Not-determined |
| S3_extracted_k141_119360 | Not-determined |
| S3_extracted_k141_71263 | Not-determined |
| S3_extracted_k141_71264 | Low-quality |
| S3_extracted_k141_23538 | Not-determined |
| S3_extracted_k141_102962 | Not-determined |
| S3_extracted_k141_71273 | Not-determined |
| S3_extracted_k141_71278 | Not-determined |
| S3_extracted_k141_127157 | Not-determined |
| S3_extracted_k141_71292 | Not-determined |
| S3_extracted_k141_23559 | Not-determined |
| S3_extracted_k141_23560 | Not-determined |
| S3_extracted_k141_119377 | Low-quality |
| S3_extracted_k141_119379 | Low-quality |
| S3_extracted_k141_23571 | Not-determined |
| S3_extracted_k141_119383 | Not-determined |
| S3_extracted_k141_102986 | Low-quality |
| S3_extracted_k141_102991 | Not-determined |
| S3_extracted_k141_7919 | Not-determined |
| S3_extracted_k141_7922 | Low-quality |
| S3_extracted_k141_119402 | Low-quality |
| S3_extracted_k141_103007 | Low-quality |
| S15_vs_6822\|\|full | High-quality |
| S3_extracted_k141_127173 | Not-determined |
| S3_extracted_k141_127178 | Not-determined |
| S3_extracted_k141_103019 | Not-determined |
| S3_extracted_k141_23593 | Not-determined |
| S3_extracted_k141_103022 | Not-determined |
| S3_extracted_k141_127185 | Low-quality |
| S3_extracted_k141_119419 | Not-determined |
| S3_extracted_k141_103034 | Not-determined |
| S3_extracted_k141_127195 | Not-determined |
| S3_extracted_k141_119425 | Low-quality |
| S3_extracted_k141_71335 | Not-determined |
| S3_extracted_k141_103042 | Not-determined |
| S3_extracted_k141_71341 | Low-quality |
| S3_extracted_k141_23603 | Low-quality |
| S3_extracted_k141_23604 | Not-determined |
| S3_extracted_k141_71347 | Not-determined |
| S3_extracted_k141_71351 | Not-determined |
| S3_extracted_k141_23609 | Low-quality |
| S3_extracted_k141_23611 | Not-determined |
| S3_extracted_k141_23613 | Not-determined |
| S3_extracted_k141_119441 | Not-determined |
| S3_extracted_k141_103057 | Not-determined |
| S3_extracted_k141_71365 | Not-determined |
| S3_extracted_k141_103062 | Low-quality |
| S3_extracted_k141_103063 | Low-quality |
| S3_extracted_k141_103067 | Not-determined |
| S3_extracted_k141_71392 | Low-quality |
| S3_extracted_k141_23615 | Not-determined |
| S3_extracted_k141_71399 | Not-determined |
| S3_extracted_k141_103074 | Not-determined |
| S3_extracted_k141_103075 | Not-determined |
| S3_extracted_k141_23621 | Not-determined |
| S3_extracted_k141_71406 | Not-determined |
| S3_extracted_k141_71407 | Not-determined |
| S3_extracted_k141_71410 | Not-determined |
| S3_extracted_k141_23632 | Not-determined |
| S3_extracted_k141_23637 | Not-determined |
| S3_extracted_k141_103085 | Not-determined |
| S3_extracted_k141_103100 | Not-determined |
| S3_extracted_k141_23650 | Not-determined |
| S3_extracted_k141_103104 | Low-quality |
| S3_extracted_k141_23651 | Not-determined |
| S3_extracted_k141_127205 | Not-determined |
| S3_extracted_k141_127209 | Not-determined |
| S3_extracted_k141_127216 | Not-determined |
| S3_extracted_k141_127233 | Not-determined |
| S3_extracted_k141_23680 | Not-determined |
| S3_extracted_k141_127261 | Not-determined |
| S3_extracted_k141_71448 | Low-quality |
| S3_extracted_k141_23705 | Not-determined |
| S3_extracted_k141_103131 | Not-determined |
| S3_extracted_k141_23716 | Not-determined |
| S3_extracted_k141_71458 | Not-determined |
| S3_extracted_k141_23722 | Low-quality |
| S3_extracted_k141_103140 | Not-determined |
| S3_extracted_k141_127272 | Not-determined |
| S3_extracted_k141_23735 | Not-determined |
| S3_extracted_k141_71471 | Not-determined |
| S3_extracted_k141_71480 | Not-determined |
| S3_extracted_k141_127282 | Not-determined |
| S3_extracted_k141_71482 | Not-determined |
| S3_extracted_k141_103149 | Low-quality |
| S3_extracted_k141_127289 | Low-quality |
| S3_extracted_k141_127292 | Not-determined |
| S3_extracted_k141_119465 | Not-determined |
| S3_extracted_k141_103157 | Not-determined |
| S3_extracted_k141_119471 | Not-determined |
| S3_extracted_k141_103162 | Not-determined |
| S3_extracted_k141_103164 | Not-determined |
| S3_extracted_k141_103167 | Not-determined |
| S3_extracted_k141_71491 | Not-determined |
| S3_extracted_k141_127309 | Not-determined |
| S3_extracted_k141_103173 | Not-determined |
| S3_extracted_k141_127317 | Not-determined |
| S3_extracted_k141_103175 | Not-determined |
| S3_extracted_k141_119494 | Not-determined |
| S3_extracted_k141_71498 | Not-determined |
| S3_extracted_k141_127320 | Not-determined |
| S3_extracted_k141_127327 | Not-determined |
| S3_extracted_k141_127331 | Not-determined |
| S3_extracted_k141_71507 | Not-determined |
| S3_extracted_k141_23741 | Not-determined |
| S3_extracted_k141_23742 | Not-determined |
| S3_extracted_k141_23746 | Not-determined |
| S3_extracted_k141_71512 | Not-determined |
| S3_extracted_k141_103191 | Not-determined |
| S3_extracted_k141_127344 | Not-determined |
| S3_extracted_k141_103193 | Low-quality |
| S3_extracted_k141_127346 | Not-determined |
| S3_extracted_k141_23760 | Not-determined |
| S3_extracted_k141_127347 | Not-determined |
| S3_extracted_k141_71523 | Low-quality |
| S3_extracted_k141_71529 | Not-determined |
| S3_extracted_k141_71533 | Not-determined |
| S3_extracted_k141_103206 | Not-determined |
| S3_extracted_k141_127366 | Low-quality |
| S3_extracted_k141_127367 | Not-determined |
| S3_extracted_k141_119514 | Not-determined |
| S3_extracted_k141_71541 | Not-determined |
| S3_extracted_k141_71543 | Low-quality |
| S3_extracted_k141_23778 | Not-determined |
| S3_extracted_k141_103211 | Not-determined |
| S3_extracted_k141_103213 | Not-determined |
| S3_extracted_k141_127377 | Low-quality |
| S3_extracted_k141_23784 | Not-determined |
| S3_extracted_k141_23786 | Not-determined |
| S3_extracted_k141_71546 | Not-determined |
| S3_extracted_k141_119540 | Not-determined |
| S3_extracted_k141_71552 | Not-determined |
| S3_extracted_k141_23794 | Not-determined |
| S3_extracted_k141_71566 | Not-determined |
| S3_extracted_k141_71567 | Not-determined |
| S3_extracted_k141_23807 | Low-quality |
| S3_extracted_k141_71592 | Low-quality |
| S3_extracted_k141_103245 | Not-determined |
| S3_extracted_k141_23828 | Not-determined |
| S3_extracted_k141_23829 | Not-determined |
| S3_extracted_k141_23830 | Not-determined |
| S3_extracted_k141_23837 | Low-quality |
| S3_extracted_k141_23838 | Not-determined |
| S3_extracted_k141_103260 | Not-determined |
| S3_extracted_k141_103261 | Not-determined |
| S3_extracted_k141_103262 | Low-quality |
| S3_extracted_k141_103265 | Not-determined |
| S3_extracted_k141_103268 | Not-determined |
| S3_extracted_k141_103271 | Not-determined |
| S3_extracted_k141_23886 | Not-determined |
| S3_extracted_k141_23889 | Not-determined |
| S3_extracted_k141_23895 | Not-determined |
| S3_extracted_k141_23896 | Not-determined |
| S3_extracted_k141_23898 | Not-determined |
| S3_extracted_k141_103281 | Not-determined |
| S3_extracted_k141_103284 | Not-determined |
| S3_extracted_k141_23908 | Not-determined |
| S3_extracted_k141_71608 | Low-quality |
| S3_extracted_k141_103297 | Not-determined |
| S3_extracted_k141_103310 | Not-determined |
| S3_extracted_k141_71618 | Not-determined |
| S3_extracted_k141_103322 | Not-determined |
| S3_extracted_k141_103335 | Not-determined |
| S3_extracted_k141_71636 | Not-determined |
| S3_extracted_k141_103338 | Low-quality |
| S3_extracted_k141_71645 | Not-determined |
| S3_extracted_k141_71650 | Not-determined |
| S3_extracted_k141_71657 | Not-determined |
| S3_extracted_k141_103354 | Low-quality |
| S3_extracted_k141_71659 | Not-determined |
| S3_extracted_k141_71660 | Not-determined |
| S3_vs_k141_1143\|\|full | High-quality |
| S3_extracted_k141_71676 | Not-determined |
| S3_extracted_k141_71685 | Not-determined |
| S3_extracted_k141_71691 | Low-quality |
| S3_extracted_k141_103391 | Not-determined |
| S3_extracted_k141_71698 | Not-determined |
| S3_extracted_k141_71705 | Not-determined |
| S16_vs_97279\|\|full | High-quality |
| S3_extracted_k141_71711 | Not-determined |
| S3_extracted_k141_71721 | Not-determined |
| S3_extracted_k141_103413 | Not-determined |
| S3_extracted_k141_103431 | Not-determined |
| S3_extracted_k141_103432 | Not-determined |
| S3_extracted_k141_103435 | Not-determined |
| S3_extracted_k141_127398 | Low-quality |
| S3_extracted_k141_103449 | Not-determined |
| S3_extracted_k141_103451 | Not-determined |
| S3_extracted_k141_103455 | Not-determined |
| S3_extracted_k141_103460 | Not-determined |
| S8_extracted_k141_55476 | Complete |
| S6_vs_k141_56484\|\|full | High-quality |
| S3_extracted_k141_103466 | Low-quality |
| S3_extracted_k141_103491 | Not-determined |
| S3_extracted_k141_103493 | Not-determined |
| S3_extracted_k141_103494 | Not-determined |
| S3_extracted_k141_103495 | Low-quality |
| S3_extracted_k141_103509 | Not-determined |
| S3_extracted_k141_103516 | Not-determined |
| S7_vs_51332\|\|full | Medium-quality |
| S3_extracted_k141_103553 | Low-quality |
| S3_extracted_k141_103554 | Not-determined |
| S3_extracted_k141_103569 | Low-quality |
| S3_extracted_k141_103585 | Not-determined |
| S8_vs_k141_170430\|\|full | High-quality |
| S3_extracted_k141_103601 | Low-quality |
| S8_vs_k141_55476\|\|full | High-quality |
| S6_vs_k141_39065\|\|0_partial | Medium-quality |
| S11_vs_k141_92134\|\|full | Medium-quality |
| S3_extracted_k141_127414 | Low-quality |
| S3_extracted_k141_127415 | Not-determined |
| S3_extracted_k141_127417 | Not-determined |
| S3_extracted_k141_127420 | Low-quality |
| S12_extracted_k141_99317 | High-quality |
| S16_vs_86116\|\|full | High-quality |
| S3_extracted_k141_127427 | Not-determined |
| S3_vs_k141_89898\|\|full | High-quality |
| S3_extracted_k141_127437 | Not-determined |
| S3_extracted_k141_127438 | Not-determined |
| S3_extracted_k141_127446 | Not-determined |
| S12_vs_k141_99317\|\|full | High-quality |
| S12_vs_k141_1980\|\|full | High-quality |
| S3_extracted_k141_127462 | Not-determined |
| S3_extracted_k141_127464 | Not-determined |
| S3_extracted_k141_127471 | Not-determined |
| S3_extracted_k141_127474 | Not-determined |
| S3_extracted_k141_127481 | Not-determined |
| S3_extracted_k141_127482 | Not-determined |
| S3_extracted_k141_127483 | Not-determined |
| S3_extracted_k141_127488 | Not-determined |
| S3_extracted_k141_127489 | Low-quality |
| S3_extracted_k141_127493 | Low-quality |
| S3_extracted_k141_127497 | Not-determined |
| S3_extracted_k141_127506 | Not-determined |
| S3_extracted_k141_127509 | Not-determined |
| S3_extracted_k141_127515 | Not-determined |
| S3_extracted_k141_127517 | Not-determined |
| S3_vs_k141_79759\|\|full | Not-determined |
| S7_extracted_49007 | Medium-quality |
| S3_vs_k141_234\|\|full | Not-determined |
| S7_vs_49007\|\|full | Medium-quality |
| S2_vs_k141_7306\|\|full | High-quality |
| S3_extracted_k141_18080 | Medium-quality |
| S11_extracted_k141_89309 | Medium-quality |
| S8_extracted_k141_183045 | Complete |
| S14_extracted_k141_56130 | High-quality |
| S3_vs_k141_104754\|\|full | Low-quality |
| S3_vs_k141_9823\|\|full | Low-quality |
| S14_vs_k141_56130\|\|full | High-quality |
| S6_vs_k141_56491\|\|full | High-quality |
| S2_vs_k141_10289\|\|full | Medium-quality |
| S3_vs_k141_74111\|\|full | Low-quality |
| S8_vs_k141_183045\|\|full | High-quality |
| S16_vs_11649\|\|full | High-quality |
| S8_vs_k141_43212\|\|full | Medium-quality |
| S11_extracted_k141_95067 | Complete |
| S1_extracted_k141_107308 | Complete |
| S3_vs_k141_18080\|\|full | Medium-quality |
| S9_vs_k141_65087\|\|full | Medium-quality |
| S11_vs_k141_95067\|\|full | High-quality |
| S3_vs_k141_98315\|\|full | Low-quality |
| S3_extracted_k141_99734 | High-quality |
| S1_vs_k141_107308\|\|full | High-quality |
| S3_vs_k141_12120\|\|full | Not-determined |
| S3_vs_k141_43047\|\|full | Low-quality |
| S7_extracted_76463 | High-quality |
| S6_vs_k141_15813\|\|full | High-quality |
| S3_vs_k141_122810\|\|full | Not-determined |
| S9_vs_k141_88286\|\|full | High-quality |
| S12_extracted_k141_79666 | High-quality |
| S3_vs_k141_27821\|\|full | Low-quality |
| S11_vs_k141_36748\|\|full | High-quality |
| S7_vs_2599\|\|full | High-quality |
| S11_vs_k141_52322\|\|full | Medium-quality |
| S7_vs_76463\|\|full | High-quality |
| S3_vs_k141_84325\|\|full | Low-quality |
| S3_vs_k141_99734\|\|full | High-quality |
| S3_vs_k141_45010\|\|full | Low-quality |
| S5_extracted_k141_145588 | Complete |
| S6_extracted_k141_17627 | Complete |
| S12_vs_k141_79666\|\|full | High-quality |
| S9_vs_k141_90851\|\|full | High-quality |
| S5_vs_k141_145588\|\|full | High-quality |
| S6_vs_k141_17627\|\|full | High-quality |
| S7_vs_90915\|\|full | High-quality |
| S3_extracted_k141_17383 | Complete |
| S8_vs_k141_28311\|\|full | High-quality |
| S3_vs_k141_127398\|\|full | Low-quality |
| S8_vs_k141_132359\|\|full | High-quality |
| S6_vs_k141_56492\|\|full | High-quality |
| S8_vs_k141_206526\|\|full | High-quality |
| S3_vs_k141_17383\|\|full | High-quality |
| S3_vs_k141_127403\|\|full | Low-quality |
| S16_extracted_57310 | High-quality |
| S3_vs_k141_25896\|\|full | High-quality |
| S8_extracted_k141_201695 | Medium-quality |
| S8_vs_k141_201695\|\|full | Medium-quality |
| S11_vs_k141_82368\|\|full | High-quality |
| S5_extracted_k141_136076 | High-quality |
| S13_extracted_109235 | Complete |
| S3_extracted_k141_11428 | High-quality |
| S3_vs_k141_127412\|\|full | Not-determined |
| S3_vs_k141_11428\|\|full | High-quality |
| S8_vs_k141_99362\|\|full | High-quality |
| S3_vs_k141_127422\|\|full | Not-determined |
| S13_extracted_135369 | High-quality |
| S11_extracted_k141_84303 | Complete |
| S3_vs_k141_127428\|\|full | Not-determined |
| S3_vs_k141_127436\|\|full | Not-determined |
| S13_vs_135369\|\|full | High-quality |
| S5_vs_k141_136076\|\|full | High-quality |
| S13_vs_109235\|\|full | High-quality |
| S3_vs_k141_127448\|\|full | Not-determined |
| S3_vs_k141_127404\|\|full | High-quality |
| S3_vs_k141_63760\|\|full | Not-determined |
| S3_vs_k141_103613\|\|full | Low-quality |
| S3_vs_k141_15941\|\|full | Not-determined |
| S3_vs_k141_23918\|\|full | Low-quality |
| S3_vs_k141_103618\|\|full | Not-determined |
| S3_vs_k141_7979\|\|full | Low-quality |
| S3_vs_k141_0\|\|full | Not-determined |
| S3_vs_k141_95649\|\|full | Low-quality |
| S3_vs_k141_63768\|\|full | Not-determined |
| S3_vs_k141_119567\|\|full | Not-determined |
| S3_vs_k141_55817\|\|full | Low-quality |
| S3_vs_k141_15957\|\|full | Not-determined |
| S3_vs_k141_47838\|\|full | Low-quality |
| S3_vs_k141_63782\|\|full | Not-determined |
| S3_vs_k141_28\|\|full | Low-quality |
| S3_vs_k141_111595\|\|full | Not-determined |
| S3_vs_k141_71756\|\|full | Not-determined |
| S3_vs_k141_47858\|\|full | Not-determined |
| S3_vs_k141_63788\|\|full | Low-quality |
| S3_vs_k141_111607\|\|full | Low-quality |
| S3_vs_k141_47872\|\|full | Low-quality |
| S3_vs_k141_95685\|\|full | Low-quality |
| S3_vs_k141_47879\|\|full | Low-quality |
| S3_vs_k141_49\|\|full | Low-quality |
| S3_vs_k141_23996\|\|full | Not-determined |
| S3_vs_k141_57\|\|full | Low-quality |
| S3_vs_k141_47894\|\|full | Not-determined |
| S3_vs_k141_24012\|\|full | Not-determined |
| S3_vs_k141_103632\|\|full | Low-quality |
| S3_vs_k141_55847\|\|full | Low-quality |
| S3_vs_k141_47904\|\|full | Low-quality |
| S3_vs_k141_24018\|\|full | Low-quality |
| S3_vs_k141_111635\|\|full | Not-determined |
| S3_vs_k141_63814\|\|full | Not-determined |
| S3_vs_k141_71813\|\|full | Low-quality |
| S3_vs_k141_111644\|\|full | Low-quality |
| S3_vs_k141_55879\|\|full | Low-quality |
| S3_vs_k141_15993\|\|full | Low-quality |
| S3_vs_k141_55892\|\|full | Low-quality |
| S3_vs_k141_79766\|\|full | Not-determined |
| S3_vs_k141_92\|\|full | Not-determined |
| S3_vs_k141_111663\|\|full | Low-quality |
| S3_vs_k141_111667\|\|full | Low-quality |
| S3_vs_k141_24076\|\|full | Low-quality |
| S3_vs_k141_63844\|\|full | Not-determined |
| S3_vs_k141_119611\|\|full | Low-quality |
| S11_vs_k141_84303\|\|full | High-quality |
| S3_vs_k141_8059\|\|full | Low-quality |
| S3_vs_k141_103646\|\|full | Low-quality |
| S3_vs_k141_47994\|\|full | Low-quality |
| S3_vs_k141_55912\|\|full | Not-determined |
| S3_vs_k141_48005\|\|full | Low-quality |
| S3_vs_k141_71861\|\|full | Low-quality |
| S3_vs_k141_71869\|\|full | Low-quality |
| S3_vs_k141_48009\|\|full | Low-quality |
| S3_vs_k141_24105\|\|full | Not-determined |
| S3_vs_k141_55937\|\|full | Not-determined |
| S3_vs_k141_31953\|\|full | Not-determined |
| S3_vs_k141_48032\|\|full | Low-quality |
| S3_vs_k141_8094\|\|full | Low-quality |
| S3_vs_k141_24131\|\|full | Not-determined |
| S3_vs_k141_31956\|\|full | Low-quality |
| S3_vs_k141_24135\|\|full | Low-quality |
| S3_vs_k141_39930\|\|full | Not-determined |
| S3_vs_k141_24138\|\|full | Not-determined |
| S3_vs_k141_71895\|\|full | Low-quality |
| S3_vs_k141_71896\|\|full | Not-determined |
| S3_vs_k141_8123\|\|full | Low-quality |
| S3_vs_k141_39938\|\|full | Not-determined |
| S3_vs_k141_111717\|\|full | Low-quality |
| S3_vs_k141_87747\|\|full | Low-quality |
| S3_vs_k141_55954\|\|full | Low-quality |
| S3_vs_k141_142\|\|full | Low-quality |
| S3_vs_k141_71920\|\|full | Low-quality |
| S3_vs_k141_79808\|\|full | Low-quality |
| S3_vs_k141_71927\|\|full | Not-determined |
| S3_vs_k141_79810\|\|full | Low-quality |
| S4_extracted_2643 | Complete |
| S3_vs_k141_87771\|\|full | Not-determined |
| S3_vs_k141_63915\|\|full | Not-determined |
| S3_vs_k141_87777\|\|full | Low-quality |
| S3_vs_k141_56002\|\|full | Not-determined |
| S3_vs_k141_63919\|\|full | Not-determined |
| S3_vs_k141_63920\|\|full | Not-determined |
| S3_vs_k141_8157\|\|full | Not-determined |
| S3_vs_k141_63927\|\|full | Low-quality |
| S3_vs_k141_8161\|\|full | Not-determined |
| S6_extracted_k141_56482 | Complete |
| S3_vs_k141_103681\|\|full | Low-quality |

| | |
|---|---|
| S3_vs_k141_39945\|\|full | Not-determined |
| S3_vs_k141_79828\|\|full | Low-quality |
| S3_vs_k141_103686\|\|full | Not-determined |
| S3_vs_k141_87790\|\|full | Low-quality |
| S3_vs_k141_111727\|\|full | Low-quality |
| S3_vs_k141_63952\|\|full | Low-quality |
| S3_vs_k141_31993\|\|full | Low-quality |
| S3_vs_k141_63954\|\|full | Not-determined |
| S3_vs_k141_103693\|\|full | Not-determined |
| S3_vs_k141_24158\|\|full | Not-determined |
| S3_vs_k141_24167\|\|full | Low-quality |
| S3_vs_k141_63963\|\|full | Low-quality |
| S3_vs_k141_48131\|\|full | Low-quality |
| S3_vs_k141_32037\|\|full | Low-quality |
| S3_vs_k141_119694\|\|full | Not-determined |
| S3_vs_k141_79870\|\|full | Not-determined |
| S3_vs_k141_39971\|\|full | Not-determined |
| S16_vs_57310\|\|full | High-quality |
| S3_vs_k141_119695\|\|full | Not-determined |
| S3_vs_k141_8167\|\|full | Low-quality |
| S3_vs_k141_79878\|\|full | Low-quality |
| S3_vs_k141_111755\|\|full | Not-determined |
| S3_vs_k141_48147\|\|full | Not-determined |
| S3_vs_k141_39981\|\|full | Low-quality |
| S3_vs_k141_87806\|\|full | Low-quality |
| S3_vs_k141_8172\|\|full | Not-determined |
| S3_vs_k141_24177\|\|full | Low-quality |
| S3_vs_k141_32053\|\|full | Low-quality |
| S3_vs_k141_39992\|\|full | Not-determined |
| S3_vs_k141_24181\|\|full | Not-determined |
| S3_vs_k141_8176\|\|full | Not-determined |
| S3_vs_k141_95803\|\|full | Low-quality |
| S3_vs_k141_79907\|\|full | Low-quality |
| S3_vs_k141_87808\|\|full | Low-quality |
| S3_vs_k141_87816\|\|full | Low-quality |
| S3_vs_k141_79918\|\|full | Low-quality |
| S3_vs_k141_40007\|\|full | Low-quality |
| S3_vs_k141_111773\|\|full | Not-determined |
| S3_vs_k141_103772\|\|full | Low-quality |
| S3_vs_k141_8211\|\|full | Not-determined |
| S3_vs_k141_48198\|\|full | Not-determined |
| S3_vs_k141_56043\|\|full | Low-quality |
| S3_vs_k141_95863\|\|full | Not-determined |
| S3_vs_k141_32086\|\|full | Low-quality |
| S3_vs_k141_87825\|\|full | Low-quality |
| S3_vs_k141_71992\|\|full | Low-quality |
| S3_vs_k141_247\|\|full | Not-determined |
| S3_vs_k141_40013\|\|full | Low-quality |
| S3_vs_k141_40018\|\|full | Not-determined |
| S3_vs_k141_256\|\|full | Low-quality |
| S3_vs_k141_95884\|\|full | Low-quality |
| S3_vs_k141_40028\|\|full | Low-quality |
| S3_vs_k141_64034\|\|full | Low-quality |
| S3_vs_k141_32102\|\|full | Not-determined |
| S3_vs_k141_40040\|\|full | Low-quality |
| S3_vs_k141_32108\|\|full | Low-quality |
| S3_vs_k141_32110\|\|full | Low-quality |
| S3_vs_k141_95900\|\|full | Low-quality |
| S3_vs_k141_95901\|\|full | Not-determined |
| S3_vs_k141_40045\|\|full | Not-determined |
| S3_vs_k141_95903\|\|full | Low-quality |
| S3_vs_k141_64046\|\|full | Low-quality |
| S3_vs_k141_32121\|\|full | Low-quality |
| S3_vs_k141_56051\|\|full | Low-quality |
| S3_vs_k141_95918\|\|full | Low-quality |
| S3_vs_k141_8277\|\|full | Low-quality |
| S3_vs_k141_95945\|\|full | Not-determined |
| S3_vs_k141_16017\|\|full | Not-determined |
| S3_vs_k141_56082\|\|full | Low-quality |
| S3_vs_k141_8299\|\|full | Not-determined |
| S3_vs_k141_40071\|\|full | Low-quality |
| S3_vs_k141_40074\|\|full | Not-determined |
| S3_vs_k141_48250\|\|full | Not-determined |
| S3_vs_k141_48254\|\|full | Not-determined |
| S3_vs_k141_8308\|\|full | Low-quality |
| S3_vs_k141_48255\|\|full | Low-quality |
| S3_vs_k141_103805\|\|full | Not-determined |
| S3_vs_k141_8325\|\|full | Not-determined |
| S3_vs_k141_87855\|\|full | Not-determined |
| S3_vs_k141_103822\|\|full | Not-determined |
| S12_extracted_k141_128794 | Complete |
| S3_vs_k141_111777\|\|full | Not-determined |
| S3_vs_k141_24268\|\|full | Low-quality |
| S3_vs_k141_111778\|\|full | Low-quality |
| S3_vs_k141_48282\|\|full | Not-determined |
| S3_vs_k141_95983\|\|full | Not-determined |
| S3_vs_k141_87857\|\|full | Low-quality |
| S3_vs_k141_40121\|\|full | Low-quality |
| S3_vs_k141_119735\|\|full | Not-determined |
| S3_vs_k141_95986\|\|full | Low-quality |
| S3_vs_k141_64106\|\|full | Not-determined |
| S3_vs_k141_111792\|\|full | Low-quality |
| S3_vs_k141_111793\|\|full | Low-quality |
| S3_vs_k141_8357\|\|full | Low-quality |
| S3_vs_k141_8358\|\|full | Low-quality |
| S3_vs_k141_336\|\|full | Low-quality |
| S3_vs_k141_8361\|\|full | Low-quality |
| S3_vs_k141_95995\|\|full | Low-quality |
| S3_vs_k141_48341\|\|full | Low-quality |
| S3_vs_k141_96005\|\|full | Low-quality |
| S3_vs_k141_48351\|\|full | Low-quality |
| S3_vs_k141_96020\|\|full | Not-determined |
| S3_vs_k141_8400\|\|full | Low-quality |
| S3_vs_k141_372\|\|full | Not-determined |
| S3_vs_k141_56151\|\|full | Low-quality |
| S3_vs_k141_111819\|\|full | Low-quality |
| S3_vs_k141_96034\|\|full | Not-determined |
| S3_vs_k141_72060\|\|full | Low-quality |
| S3_vs_k141_96037\|\|full | Not-determined |
| S4_vs_VIRSorter_2643-circular-cat_ | High-quality |
| S3_vs_k141_96040\|\|full | Low-quality |
| S3_vs_k141_111827\|\|full | Not-determined |
| S3_vs_k141_8414\|\|full | Low-quality |
| S3_vs_k141_24306\|\|full | Low-quality |
| S3_vs_k141_111829\|\|full | Low-quality |
| S3_vs_k141_48365\|\|full | Not-determined |
| S3_vs_k141_8424\|\|full | Low-quality |
| S3_vs_k141_111837\|\|full | Low-quality |
| S3_vs_k141_40161\|\|full | Not-determined |
| S3_vs_k141_80022\|\|full | Low-quality |
| S6_vs_k141_56482\|\|full | High-quality |
| S3_vs_k141_8430\|\|full | Low-quality |
| S3_vs_k141_404\|\|full | Low-quality |
| S3_vs_k141_87897\|\|full | Not-determined |
| S3_vs_k141_96054\|\|full | Low-quality |
| S3_vs_k141_103908\|\|full | Low-quality |
| S3_vs_k141_103916\|\|full | Low-quality |
| S3_vs_k141_103927\|\|full | Not-determined |
| S3_vs_k141_40187\|\|full | Not-determined |
| S3_vs_k141_24359\|\|full | Low-quality |
| S3_vs_k141_412\|\|full | Low-quality |
| S3_vs_k141_40200\|\|full | Low-quality |
| S3_vs_k141_8474\|\|full | Low-quality |
| S3_vs_k141_111880\|\|full | Low-quality |
| S3_vs_k141_103951\|\|full | Not-determined |
| S3_vs_k141_119828\|\|full | Not-determined |
| S3_vs_k141_40239\|\|full | Low-quality |
| S3_vs_k141_429\|\|full | Not-determined |
| S3_vs_k141_103971\|\|full | Low-quality |
| S3_vs_k141_72110\|\|full | Low-quality |
| S3_vs_k141_40268\|\|full | Low-quality |
| S3_vs_k141_32295\|\|full | Low-quality |
| S3_vs_k141_48407\|\|full | Not-determined |
| S3_vs_k141_24401\|\|full | Not-determined |
| S3_vs_k141_72116\|\|full | Low-quality |
| S3_vs_k141_119870\|\|full | Low-quality |
| S3_vs_k141_103988\|\|full | Low-quality |
| S3_vs_k141_40278\|\|full | Not-determined |
| S3_vs_k141_119875\|\|full | Low-quality |
| S3_vs_k141_8532\|\|full | Low-quality |
| S3_vs_k141_24420\|\|full | Low-quality |
| S3_vs_k141_40280\|\|full | Low-quality |
| S3_vs_k141_24424\|\|full | Low-quality |
| S3_vs_k141_119885\|\|full | Low-quality |
| S3_vs_k141_24441\|\|full | Not-determined |
| S3_vs_k141_32358\|\|full | Low-quality |
| S3_vs_k141_87977\|\|full | Low-quality |
| S3_vs_k141_111919\|\|full | Low-quality |
| S3_vs_k141_8570\|\|full | Low-quality |
| S3_vs_k141_473\|\|full | Not-determined |
| S3_vs_k141_32375\|\|full | Low-quality |
| S3_vs_k141_87996\|\|full | Low-quality |
| S3_vs_k141_87998\|\|full | Low-quality |
| S3_vs_k141_119931\|\|full | Low-quality |
| S3_vs_k141_32391\|\|full | Low-quality |
| S3_vs_k141_8602\|\|full | Low-quality |
| S3_vs_k141_88004\|\|full | Low-quality |
| S3_vs_k141_111921\|\|full | Low-quality |
| S3_vs_k141_96148\|\|full | Not-determined |
| S3_vs_k141_32431\|\|full | Not-determined |
| S3_vs_k141_508\|\|full | Low-quality |
| S3_vs_k141_24482\|\|full | Low-quality |
| S3_vs_k141_104045\|\|full | Low-quality |
| S3_vs_k141_64203\|\|full | Low-quality |
| S3_vs_k141_32438\|\|full | Not-determined |
| S3_vs_k141_32441\|\|full | Not-determined |
| S3_vs_k141_104054\|\|full | Not-determined |
| S3_vs_k141_64209\|\|full | Low-quality |
| S3_vs_k141_119991\|\|full | Not-determined |
| S3_vs_k141_24501\|\|full | Low-quality |
| S3_vs_k141_48428\|\|full | Not-determined |
| S3_vs_k141_537\|\|full | Low-quality |
| S3_vs_k141_40381\|\|full | Low-quality |
| S3_extracted_k141_127409 | Complete |
| S3_vs_k141_72126\|\|full | Low-quality |
| S3_vs_k141_16171\|\|full | Not-determined |
| S3_vs_k141_24518\|\|full | Low-quality |
| S3_vs_k141_32474\|\|full | Not-determined |
| S3_vs_k141_88048\|\|full | Not-determined |
| S3_vs_k141_32486\|\|full | Low-quality |
| S3_vs_k141_16184\|\|full | Low-quality |
| S3_vs_k141_40392\|\|full | Low-quality |
| S3_vs_k141_88052\|\|full | Low-quality |
| S3_vs_k141_40395\|\|full | Not-determined |
| S3_vs_k141_88057\|\|full | Low-quality |
| S3_vs_k141_40405\|\|full | Low-quality |
| S3_vs_k141_72186\|\|full | Low-quality |
| S3_vs_k141_40417\|\|full | Low-quality |
| S3_vs_k141_88067\|\|full | Not-determined |
| S3_vs_k141_48474\|\|full | Low-quality |
| S3_vs_k141_32501\|\|full | Low-quality |
| S3_vs_k141_80179\|\|full | Not-determined |
| S3_vs_k141_104097\|\|full | Low-quality |
| S3_vs_k141_8720\|\|full | Low-quality |
| S3_vs_k141_72213\|\|full | Not-determined |
| S3_vs_k141_111974\|\|full | Low-quality |
| S3_vs_k141_602\|\|full | Not-determined |
| S3_vs_k141_111977\|\|full | Low-quality |
| S3_vs_k141_104112\|\|full | Low-quality |
| S3_vs_k141_8745\|\|full | Not-determined |
| S3_vs_k141_72249\|\|full | Low-quality |
| S3_vs_k141_96207\|\|full | Low-quality |
| S3_vs_k141_56284\|\|full | Low-quality |
| S3_vs_k141_56286\|\|full | Low-quality |
| S3_vs_k141_111987\|\|full | Not-determined |
| S3_vs_k141_80188\|\|full | Low-quality |
| S3_vs_k141_40451\|\|full | Low-quality |
| S3_vs_k141_48507\|\|full | Not-determined |
| S3_vs_k141_40461\|\|full | Low-quality |
| S3_vs_k141_112002\|\|full | Low-quality |
| S3_vs_k141_104138\|\|full | Not-determined |
| S3_vs_k141_689\|\|full | Not-determined |
| S3_vs_k141_8810\|\|full | Not-determined |
| S3_vs_k141_104152\|\|full | Low-quality |
| S3_vs_k141_8822\|\|full | Not-determined |
| S8_extracted_k141_97004 | High-quality |
| S8_vs_k141_97004\|\|full | High-quality |
| S3_vs_k141_119996\|\|full | Low-quality |
| S3_vs_k141_8859\|\|full | Low-quality |
| S3_vs_k141_56297\|\|full | Low-quality |
| S3_vs_k141_718\|\|full | Not-determined |
| S3_vs_k141_96281\|\|full | Not-determined |
| S3_vs_k141_16226\|\|full | Low-quality |
| S3_vs_k141_72360\|\|full | Low-quality |
| S3_vs_k141_32622\|\|full | Low-quality |
| S3_vs_k141_32624\|\|full | Low-quality |
| S3_vs_k141_104238\|\|full | Low-quality |
| S3_vs_k141_48591\|\|full | Not-determined |
| S3_vs_k141_96306\|\|full | Low-quality |
| S3_vs_k141_120013\|\|full | Low-quality |
| S3_vs_k141_88141\|\|full | Low-quality |
| S3_vs_k141_733\|\|full | Low-quality |
| S3_vs_k141_32665\|\|full | Low-quality |
| S3_vs_k141_40548\|\|full | Low-quality |
| S3_vs_k141_88149\|\|full | Not-determined |
| S3_vs_k141_104249\|\|full | Low-quality |
| S3_vs_k141_32677\|\|full | Not-determined |
| S3_vs_k141_96321\|\|full | Low-quality |
| S3_vs_k141_8892\|\|full | Low-quality |
| S3_vs_k141_32679\|\|full | Not-determined |
| S3_vs_k141_40567\|\|full | Low-quality |
| S3_vs_k141_80298\|\|full | Not-determined |
| S3_vs_k141_80301\|\|full | Not-determined |
| S12_vs_k141_128794\|\|full | High-quality |
| S3_vs_k141_72462\|\|full | Low-quality |
| S3_vs_k141_104263\|\|full | Low-quality |
| S3_vs_k141_8908\|\|full | Low-quality |
| S3_vs_k141_32689\|\|full | Low-quality |
| S3_vs_k141_112077\|\|full | Not-determined |
| S3_vs_k141_40587\|\|full | Low-quality |
| S3_vs_k141_48664\|\|full | Not-determined |
| S3_vs_k141_112078\|\|full | Not-determined |
| S3_vs_k141_72479\|\|full | Low-quality |
| S3_vs_k141_96353\|\|full | Low-quality |
| S3_vs_k141_64313\|\|full | Low-quality |
| S3_vs_k141_780\|\|full | Low-quality |
| S3_vs_k141_96357\|\|full | Low-quality |
| S3_vs_k141_64324\|\|full | Low-quality |
| S3_vs_k141_48675\|\|full | Not-determined |
| S3_vs_k141_112108\|\|full | Low-quality |
| S3_vs_k141_48682\|\|full | Low-quality |
| S3_vs_k141_80348\|\|full | Not-determined |
| S3_vs_k141_795\|\|full | Low-quality |
| S3_vs_k141_88202\|\|full | Not-determined |
| S3_vs_k141_796\|\|full | Not-determined |
| S3_vs_k141_16243\|\|full | Not-determined |
| S3_vs_k141_120070\|\|full | Not-determined |
| S3_vs_k141_32717\|\|full | Low-quality |
| S3_vs_k141_48702\|\|full | Not-determined |
| S3_vs_k141_24628\|\|full | Low-quality |
| S3_vs_k141_120071\|\|full | Low-quality |
| S3_vs_k141_16246\|\|full | Low-quality |
| S3_vs_k141_112160\|\|full | Not-determined |
| S3_vs_k141_48716\|\|full | Not-determined |
| S3_vs_k141_56459\|\|full | Low-quality |
| S3_vs_k141_104303\|\|full | Low-quality |
| S3_vs_k141_56466\|\|full | Low-quality |
| S3_vs_k141_120086\|\|full | Low-quality |
| S3_vs_k141_16256\|\|full | Low-quality |
| S3_vs_k141_96398\|\|full | Low-quality |
| S3_vs_k141_96404\|\|full | Not-determined |
| S3_vs_k141_72538\|\|full | Not-determined |
| S3_vs_k141_112186\|\|full | Low-quality |
| S3_vs_k141_104315\|\|full | Low-quality |
| S3_vs_k141_96425\|\|full | Not-determined |
| S3_vs_k141_104318\|\|full | Low-quality |
| S3_vs_k141_72557\|\|full | Low-quality |
| S3_vs_k141_72560\|\|full | Low-quality |
| S3_vs_k141_9030\|\|full | Not-determined |
| S3_vs_k141_16268\|\|full | Low-quality |
| S3_vs_k141_32772\|\|full | Not-determined |
| S3_vs_k141_72562\|\|full | Low-quality |
| S3_vs_k141_96451\|\|full | Low-quality |
| S3_vs_k141_72580\|\|full | Low-quality |
| S3_vs_k141_826\|\|full | Low-quality |
| S3_vs_k141_16286\|\|full | Low-quality |
| S3_vs_k141_72588\|\|full | Not-determined |
| S3_vs_k141_9052\|\|full | Low-quality |
| S3_vs_k141_80367\|\|full | Low-quality |
| S3_vs_k141_40669\|\|full | Low-quality |
| S3_vs_k141_9082\|\|full | Not-determined |
| S3_vs_k141_16314\|\|full | Not-determined |
| S3_vs_k141_120117\|\|full | Low-quality |
| S3_vs_k141_40680\|\|full | Low-quality |
| S3_vs_k141_48756\|\|full | Low-quality |
| S3_vs_k141_48763\|\|full | Not-determined |
| S3_vs_k141_72630\|\|full | Low-quality |
| S3_vs_k141_64335\|\|full | Not-determined |
| S3_vs_k141_96475\|\|full | Low-quality |
| S3_vs_k141_32839\|\|full | Not-determined |
| S3_vs_k141_72637\|\|full | Not-determined |
| S3_vs_k141_64342\|\|full | Low-quality |
| S3_vs_k141_96479\|\|full | Low-quality |
| S3_vs_k141_32846\|\|full | Low-quality |
| S3_vs_k141_80384\|\|full | Low-quality |
| S3_vs_k141_32853\|\|full | Not-determined |
| S3_vs_k141_72649\|\|full | Low-quality |
| S3_vs_k141_16337\|\|full | Low-quality |
| S3_vs_k141_120126\|\|full | Not-determined |
| S3_vs_k141_9122\|\|full | Not-determined |
| S3_vs_k141_72675\|\|full | Low-quality |
| S3_vs_k141_80406\|\|full | Low-quality |
| S3_vs_k141_16362\|\|full | Low-quality |
| S3_vs_k141_56580\|\|full | Not-determined |
| S3_vs_k141_56584\|\|full | Low-quality |
| S3_vs_k141_16365\|\|full | Low-quality |
| S3_vs_k141_40745\|\|full | Low-quality |
| S3_vs_k141_56590\|\|full | Low-quality |
| S3_vs_k141_96533\|\|full | Low-quality |
| S3_vs_k141_16372\|\|full | Low-quality |
| S3_vs_k141_9152\|\|full | Low-quality |
| S3_vs_k141_104355\|\|full | Low-quality |
| S3_vs_k141_40753\|\|full | Low-quality |
| S3_vs_k141_80424\|\|full | Not-determined |
| S3_vs_k141_40762\|\|full | Low-quality |
| S3_vs_k141_16381\|\|full | Low-quality |
| S3_vs_k141_104360\|\|full | Low-quality |
| S3_vs_k141_112274\|\|full | Low-quality |
| S3_vs_k141_64376\|\|full | Not-determined |
| S3_vs_k141_80439\|\|full | Not-determined |
| S3_vs_k141_80441\|\|full | Low-quality |
| S3_vs_k141_80446\|\|full | Not-determined |
| S3_vs_k141_64385\|\|full | Low-quality |
| S3_vs_k141_88306\|\|full | Not-determined |
| S3_vs_k141_16406\|\|full | Low-quality |
| S3_vs_k141_112283\|\|full | Not-determined |
| S3_vs_k141_48797\|\|full | Low-quality |
| S3_vs_k141_96541\|\|full | Low-quality |
| S3_vs_k141_104371\|\|full | Low-quality |
| S3_vs_k141_88313\|\|full | Low-quality |
| S3_vs_k141_104376\|\|full | Not-determined |
| S3_vs_k141_48813\|\|full | Not-determined |
| S3_vs_k141_40782\|\|full | Low-quality |
| S3_vs_k141_88322\|\|full | Not-determined |
| S3_vs_k141_72750\|\|full | Low-quality |
| S3_vs_k141_80470\|\|full | Low-quality |
| S3_vs_k141_96557\|\|full | Not-determined |
| S3_vs_k141_875\|\|full | Low-quality |
| S3_vs_k141_32914\|\|full | Not-determined |
| S3_vs_k141_96559\|\|full | Low-quality |
| S3_vs_k141_32915\|\|full | Not-determined |
| S3_vs_k141_96565\|\|full | Not-determined |
| S3_vs_k141_32937\|\|full | Not-determined |
| S3_vs_k141_88328\|\|full | Not-determined |
| S3_vs_k141_16432\|\|full | Low-quality |
| S3_vs_k141_889\|\|full | Not-determined |
| S3_vs_k141_104387\|\|full | Not-determined |
| S3_vs_k141_88338\|\|full | Low-quality |
| S3_vs_k141_48852\|\|full | Low-quality |
| S3_vs_k141_120142\|\|full | Low-quality |
| S8_vs_k141_43662\|\|full | High-quality |
| S3_vs_k141_32960\|\|full | Low-quality |
| S3_vs_k141_9164\|\|full | Low-quality |
| S3_vs_k141_48864\|\|full | Low-quality |
| S3_vs_k141_40822\|\|full | Not-determined |
| S3_vs_k141_32983\|\|full | Low-quality |
| S3_vs_k141_96592\|\|full | Not-determined |
| S3_vs_k141_48893\|\|full | Low-quality |
| S3_vs_k141_48905\|\|full | Low-quality |
| S3_vs_k141_48910\|\|full | Low-quality |
| S3_vs_k141_112316\|\|full | Low-quality |
| S3_vs_k141_88386\|\|full | Not-determined |
| S3_vs_k141_80559\|\|full | Low-quality |
| S3_vs_k141_40843\|\|full | Low-quality |
| S3_vs_k141_88388\|\|full | Low-quality |
| S3_vs_k141_88389\|\|full | Not-determined |
| S3_vs_k141_9194\|\|full | Not-determined |
| S3_vs_k141_104445\|\|full | Low-quality |
| S3_vs_k141_72863\|\|full | Not-determined |
| S3_vs_k141_112348\|\|full | Low-quality |
| S3_vs_k141_112352\|\|full | Low-quality |
| S10_extracted_18671 | Complete |
| S3_vs_k141_9207\|\|full | Low-quality |
| S3_vs_k141_88400\|\|full | Low-quality |
| S3_vs_k141_112353\|\|full | Low-quality |
| S3_vs_k141_33009\|\|full | Low-quality |
| S3_vs_k141_96663\|\|full | Low-quality |
| S3_vs_k141_88406\|\|full | Low-quality |
| S3_vs_k141_33014\|\|full | Not-determined |
| S3_vs_k141_33017\|\|full | Low-quality |
| S3_vs_k141_80565\|\|full | Low-quality |
| S3_vs_k141_72884\|\|full | Not-determined |
| S3_vs_k141_96674\|\|full | Low-quality |
| S3_vs_k141_33019\|\|full | Low-quality |
| S3_vs_k141_56735\|\|full | Low-quality |
| S3_vs_k141_88424\|\|full | Not-determined |
| S3_vs_k141_9260\|\|full | Not-determined |
| S3_vs_k141_24769\|\|full | Not-determined |
| S3_vs_k141_48980\|\|full | Not-determined |
| S3_vs_k141_80591\|\|full | Low-quality |
| S3_vs_k141_48981\|\|full | Low-quality |
| S3_vs_k141_56753\|\|full | Low-quality |
| S3_vs_k141_112370\|\|full | Not-determined |
| S3_vs_k141_88430\|\|full | Low-quality |
| S3_vs_k141_48992\|\|full | Low-quality |
| S3_vs_k141_80602\|\|full | Not-determined |
| S3_vs_k141_33050\|\|full | Not-determined |
| S3_vs_k141_40906\|\|full | Not-determined |
| S3_vs_k141_88451\|\|full | Low-quality |
| S3_vs_k141_72942\|\|full | Low-quality |
| S3_vs_k141_9284\|\|full | Not-determined |
| S3_vs_k141_24786\|\|full | Not-determined |
| S3_vs_k141_56778\|\|full | Low-quality |
| S3_vs_k141_88458\|\|full | Low-quality |
| S3_vs_k141_64477\|\|full | Not-determined |
| S3_vs_k141_33055\|\|full | Low-quality |
| S3_vs_k141_56789\|\|full | Not-determined |
| S3_vs_k141_88467\|\|full | Low-quality |
| S3_vs_k141_40930\|\|full | Low-quality |
| S3_vs_k141_64493\|\|full | Low-quality |
| S3_vs_k141_88471\|\|full | Low-quality |
| S3_vs_k141_947\|\|full | Not-determined |
| S3_vs_k141_33069\|\|full | Not-determined |
| S3_vs_k141_955\|\|full | Not-determined |
| S3_vs_k141_56794\|\|full | Low-quality |
| S3_vs_k141_9295\|\|full | Not-determined |
| S3_vs_k141_88493\|\|full | Low-quality |
| S3_vs_k141_24828\|\|full | Not-determined |
| S3_vs_k141_24835\|\|full | Not-determined |
| S3_vs_k141_33090\|\|full | Low-quality |
| S3_vs_k141_112391\|\|full | Low-quality |
| S3_vs_k141_9318\|\|full | Low-quality |
| S3_vs_k141_120200\|\|full | Low-quality |
| S3_vs_k141_969\|\|full | Low-quality |
| S3_vs_k141_64527\|\|full | Low-quality |
| S3_vs_k141_56811\|\|full | Low-quality |
| S3_vs_k141_80648\|\|full | Low-quality |
| S3_vs_k141_64533\|\|full | Not-determined |
| S3_vs_k141_80649\|\|full | Low-quality |
| S3_vs_k141_80651\|\|full | Low-quality |
| S3_vs_k141_80653\|\|full | Low-quality |
| S3_vs_k141_49053\|\|full | Not-determined |
| S3_vs_k141_991\|\|full | Low-quality |
| S3_vs_k141_88502\|\|full | Low-quality |
| S3_vs_k141_88515\|\|full | Not-determined |
| S3_vs_k141_56834\|\|full | Low-quality |
| S3_vs_k141_96742\|\|full | Low-quality |
| S3_vs_k141_104507\|\|full | Low-quality |
| S3_vs_k141_88526\|\|full | Low-quality |
| S3_vs_k141_49079\|\|full | Not-determined |
| S3_vs_k141_88530\|\|full | Not-determined |
| S3_vs_k141_9339\|\|full | Low-quality |
| S3_vs_k141_80675\|\|full | Not-determined |
| S3_vs_k141_73059\|\|full | Not-determined |
| S3_vs_k141_80678\|\|full | Low-quality |
| S3_vs_k141_9353\|\|full | Low-quality |
| S3_vs_k141_80679\|\|full | Not-determined |
| S3_vs_k141_49095\|\|full | Not-determined |
| S3_vs_k141_9373\|\|full | Not-determined |
| S3_vs_k141_49111\|\|full | Low-quality |
| S3_vs_k141_9375\|\|full | Low-quality |
| S3_vs_k141_24901\|\|full | Not-determined |
| S3_vs_k141_16524\|\|full | Low-quality |
| S3_vs_k141_16525\|\|full | Low-quality |
| S15_vs_43913\|\|full | Medium-quality |
| S3_vs_k141_24908\|\|full | Low-quality |
| S3_vs_k141_9402\|\|full | Low-quality |
| S3_vs_k141_56885\|\|full | Low-quality |
| S3_vs_k141_9414\|\|full | Low-quality |
| S3_vs_k141_16537\|\|full | Not-determined |
| S3_vs_k141_56895\|\|full | Not-determined |
| S3_vs_k141_73120\|\|full | Not-determined |
| S3_vs_k141_56902\|\|full | Low-quality |
| S3_vs_k141_73121\|\|full | Not-determined |
| S3_vs_k141_80725\|\|full | Low-quality |
| S3_vs_k141_16553\|\|full | Not-determined |
| S3_vs_k141_33118\|\|full | Low-quality |
| S3_vs_k141_56906\|\|full | Not-determined |
| S3_vs_k141_49173\|\|full | Not-determined |
| S3_vs_k141_104542\|\|full | Not-determined |
| S3_vs_k141_104545\|\|full | Low-quality |
| S3_vs_k141_16576\|\|full | Low-quality |
| S3_vs_k141_49192\|\|full | Not-determined |
| S3_vs_k141_80771\|\|full | Low-quality |
| S3_vs_k141_49200\|\|full | Not-determined |
| S3_vs_k141_49209\|\|full | Not-determined |
| S3_vs_k141_96818\|\|full | Low-quality |
| S3_vs_k141_9492\|\|full | Low-quality |
| S3_vs_k141_9498\|\|full | Not-determined |
| S3_vs_k141_16600\|\|full | Not-determined |
| S3_vs_k141_73188\|\|full | Low-quality |
| S3_vs_k141_9500\|\|full | Low-quality |
| S3_vs_k141_16609\|\|full | Low-quality |
| S3_vs_k141_49227\|\|full | Low-quality |
| S3_vs_k141_96849\|\|full | Not-determined |
| S3_vs_k141_96852\|\|full | Low-quality |
| S3_vs_k141_73208\|\|full | Not-determined |
| S3_vs_k141_73210\|\|full | Low-quality |
| S3_vs_k141_73225\|\|full | Not-determined |
| S3_vs_k141_73229\|\|full | Low-quality |
| S3_vs_k141_16642\|\|full | Low-quality |
| S3_vs_k141_9553\|\|full | Low-quality |
| S3_vs_k141_57012\|\|full | Not-determined |
| S3_vs_k141_64589\|\|full | Not-determined |
| S3_vs_k141_16670\|\|full | Low-quality |
| S3_vs_k141_73265\|\|full | Low-quality |
| S3_vs_k141_80823\|\|full | Low-quality |
| S3_vs_k141_73278\|\|full | Low-quality |
| S3_vs_k141_80832\|\|full | Not-determined |
| S3_vs_k141_57038\|\|full | Low-quality |
| S3_vs_k141_49283\|\|full | Low-quality |
| S3_vs_k141_33211\|\|full | Not-determined |
| S3_vs_k141_96937\|\|full | Not-determined |
| S3_vs_k141_49301\|\|full | Not-determined |
| S3_vs_k141_9582\|\|full | Not-determined |
| S3_vs_k141_49309\|\|full | Not-determined |
| S3_vs_k141_1051\|\|full | Not-determined |
| S3_vs_k141_33225\|\|full | Low-quality |
| S3_vs_k141_1055\|\|full | Not-determined |
| S3_vs_k141_120298\|\|full | Low-quality |
| S3_vs_k141_64617\|\|full | Not-determined |
| S3_vs_k141_120308\|\|full | Not-determined |
| S3_vs_k141_96960\|\|full | Low-quality |
| S3_vs_k141_96963\|\|full | Low-quality |
| S3_vs_k141_64623\|\|full | Low-quality |
| S3_vs_k141_73307\|\|full | Not-determined |
| S3_vs_k141_80887\|\|full | Low-quality |
| S3_vs_k141_1086\|\|full | Low-quality |
| S3_vs_k141_96977\|\|full | Low-quality |
| S3_vs_k141_73317\|\|full | Not-determined |
| S3_vs_k141_73336\|\|full | Low-quality |
| S3_vs_k141_97002\|\|full | Not-determined |
| S3_vs_k141_97005\|\|full | Low-quality |
| S3_vs_k141_57052\|\|full | Low-quality |
| S3_vs_k141_73348\|\|full | Not-determined |
| S3_vs_k141_80924\|\|full | Not-determined |
| S3_vs_k141_57067\|\|full | Low-quality |
| S3_vs_k141_57079\|\|full | Low-quality |
| S3_vs_k141_73357\|\|full | Low-quality |
| S3_vs_k141_88628\|\|full | Low-quality |
| S3_vs_k141_120316\|\|full | Low-quality |
| S3_vs_k141_57093\|\|full | Not-determined |
| S3_vs_k141_104660\|\|full | Low-quality |
| S3_vs_k141_80965\|\|full | Low-quality |
| S3_vs_k141_9659\|\|full | Low-quality |
| S3_vs_k141_104664\|\|full | Low-quality |
| S3_vs_k141_33249\|\|full | Not-determined |
| S3_vs_k141_120317\|\|full | Low-quality |
| S3_vs_k141_16712\|\|full | Low-quality |
| S3_vs_k141_80974\|\|full | Low-quality |
| S3_vs_k141_16729\|\|full | Low-quality |
| S3_vs_k141_40939\|\|0_partial | Medium-quality |
| S3_vs_k141_80985\|\|full | Low-quality |
| S3_vs_k141_97029\|\|full | Low-quality |
| S3_vs_k141_40987\|\|full | Low-quality |
| S3_vs_k141_9663\|\|full | Not-determined |
| S3_vs_k141_9664\|\|full | Low-quality |
| S3_vs_k141_73375\|\|full | Low-quality |
| S3_vs_k141_49329\|\|full | Low-quality |
| S3_vs_k141_88676\|\|full | Not-determined |
| S3_vs_k141_64674\|\|full | Not-determined |
| S3_vs_k141_64675\|\|full | Low-quality |
| S3_vs_k141_49333\|\|full | Low-quality |
| S10_extracted_24791 | High-quality |
| S3_vs_k141_88696\|\|full | Not-determined |
| S3_vs_k141_97050\|\|full | Low-quality |
| S3_vs_k141_1148\|\|full | Not-determined |
| S3_vs_k141_24967\|\|full | Low-quality |
| S3_vs_k141_9681\|\|full | Low-quality |

| | |
|---|---|
| S3_vs_k141_1158\|\|full | Low-quality |
| S3_vs_k141_33312\|\|full | Low-quality |
| S3_vs_k141_81004\|\|full | Low-quality |
| S3_vs_k141_81011\|\|full | Low-quality |
| S3_vs_k141_25003\|\|full | Not-determined |
| S3_vs_k141_112485\|\|full | Not-determined |
| S3_vs_k141_81027\|\|full | Not-determined |
| S3_vs_k141_33333\|\|full | Low-quality |
| S3_vs_k141_73430\|\|full | Low-quality |
| S3_vs_k141_104710\|\|full | Not-determined |
| S3_vs_k141_73440\|\|full | Low-quality |
| S3_vs_k141_73445\|\|full | Not-determined |
| S3_vs_k141_73451\|\|full | Low-quality |
| S3_vs_k141_64715\|\|full | Low-quality |
| S3_vs_k141_88744\|\|full | Low-quality |
| S3_vs_k141_16808\|\|full | Not-determined |
| S3_vs_k141_33367\|\|full | Low-quality |
| S3_vs_k141_120393\|\|full | Low-quality |
| S3_vs_k141_16813\|\|full | Low-quality |
| S3_vs_k141_9685\|\|full | Low-quality |
| S3_vs_k141_81097\|\|full | Low-quality |
| S3_vs_k141_88773\|\|full | Low-quality |
| S3_vs_k141_64725\|\|full | Low-quality |
| S3_vs_k141_25054\|\|full | Not-determined |
| S3_vs_k141_120417\|\|full | Not-determined |
| S3_vs_k141_49361\|\|full | Low-quality |
| S3_vs_k141_81110\|\|full | Low-quality |
| S3_vs_k141_81117\|\|full | Low-quality |
| S3_vs_k141_9691\|\|full | Not-determined |
| S3_vs_k141_112528\|\|full | Not-determined |
| S3_vs_k141_64757\|\|full | Not-determined |
| S3_vs_k141_81125\|\|full | Not-determined |
| S3_vs_k141_120452\|\|full | Low-quality |
| S3_vs_k141_49386\|\|full | Low-quality |
| S3_vs_k141_127409\|\|full | High-quality |
| S3_vs_k141_1184\|\|full | Low-quality |
| S3_vs_k141_41053\|\|full | Low-quality |
| S3_vs_k141_9701\|\|full | Low-quality |
| S3_vs_k141_1185\|\|full | Not-determined |
| S3_vs_k141_112539\|\|full | Low-quality |
| S3_vs_k141_104733\|\|full | Low-quality |
| S3_vs_k141_112559\|\|full | Low-quality |
| S3_vs_k141_1198\|\|full | Low-quality |
| S3_vs_k141_112563\|\|full | Not-determined |
| S3_vs_k141_33390\|\|full | Not-determined |
| S3_vs_k141_41070\|\|full | Not-determined |
| S3_vs_k141_49410\|\|full | Low-quality |
| S3_vs_k141_112575\|\|full | Low-quality |
| S3_vs_k141_41075\|\|full | Low-quality |
| S3_vs_k141_64850\|\|full | Low-quality |
| S3_vs_k141_16871\|\|full | Not-determined |
| S3_vs_k141_88796\|\|full | Low-quality |
| S3_vs_k141_64860\|\|full | Low-quality |
| S3_vs_k141_112585\|\|full | Not-determined |
| S3_vs_k141_16878\|\|full | Low-quality |
| S3_vs_k141_88798\|\|full | Low-quality |
| S3_vs_k141_33423\|\|full | Low-quality |
| S3_vs_k141_33429\|\|full | Low-quality |
| S3_vs_k141_112598\|\|full | Low-quality |
| S3_vs_k141_88809\|\|full | Low-quality |
| S3_vs_k141_41108\|\|full | Not-determined |
| S3_vs_k141_64882\|\|full | Low-quality |
| S3_vs_k141_104758\|\|full | Low-quality |
| S3_vs_k141_73473\|\|full | Not-determined |
| S3_vs_k141_73476\|\|full | Low-quality |
| S3_vs_k141_1223\|\|full | Low-quality |
| S3_vs_k141_73484\|\|full | Low-quality |
| S3_vs_k141_1230\|\|full | Low-quality |
| S3_vs_k141_49438\|\|full | Low-quality |
| S3_vs_k141_9757\|\|full | Not-determined |
| S3_vs_k141_73490\|\|full | Low-quality |
| S3_vs_k141_73492\|\|full | Not-determined |
| S3_vs_k141_120497\|\|full | Low-quality |
| S3_vs_k141_1238\|\|full | Low-quality |
| S3_vs_k141_73502\|\|full | Low-quality |
| S3_vs_k141_41121\|\|full | Not-determined |
| S3_vs_k141_104768\|\|full | Low-quality |
| S3_vs_k141_41132\|\|full | Low-quality |
| S3_vs_k141_33478\|\|full | Low-quality |
| S3_vs_k141_1247\|\|full | Low-quality |
| S3_vs_k141_73531\|\|full | Not-determined |
| S3_vs_k141_97153\|\|full | Low-quality |
| S3_vs_k141_57126\|\|full | Low-quality |
| S3_vs_k141_64917\|\|full | Low-quality |
| S3_vs_k141_64919\|\|full | Low-quality |
| S3_vs_k141_16973\|\|full | Not-determined |
| S3_vs_k141_64920\|\|full | Low-quality |
| S3_vs_k141_1274\|\|full | Low-quality |
| S3_vs_k141_16988\|\|full | Low-quality |
| S3_vs_k141_33513\|\|full | Low-quality |
| S3_vs_k141_33521\|\|full | Low-quality |
| S3_vs_k141_81229\|\|full | Low-quality |
| S3_vs_k141_88852\|\|full | Low-quality |
| S3_vs_k141_81238\|\|full | Low-quality |
| S3_vs_k141_49487\|\|full | Low-quality |
| S3_vs_k141_1299\|\|full | Low-quality |
| S3_vs_k141_49491\|\|full | Low-quality |
| S3_vs_k141_81241\|\|full | Not-determined |
| S3_vs_k141_1308\|\|full | Low-quality |
| S3_vs_k141_112647\|\|full | Low-quality |
| S3_vs_k141_49498\|\|full | Low-quality |
| S3_vs_k141_57148\|\|full | Not-determined |
| S3_vs_k141_1320\|\|full | Low-quality |
| S3_vs_k141_120548\|\|full | Low-quality |
| S3_vs_k141_49499\|\|full | Low-quality |
| S3_vs_k141_112657\|\|full | Low-quality |
| S3_vs_k141_1330\|\|full | Low-quality |
| S3_vs_k141_49509\|\|full | Low-quality |
| S3_vs_k141_64937\|\|full | Low-quality |
| S3_vs_k141_49515\|\|full | Low-quality |
| S3_vs_k141_49519\|\|full | Not-determined |
| S3_vs_k141_9802\|\|full | Low-quality |
| S3_vs_k141_33554\|\|full | Not-determined |
| S3_vs_k141_104845\|\|full | Not-determined |
| S3_vs_k141_41207\|\|full | Not-determined |
| S3_vs_k141_97175\|\|full | Low-quality |
| S3_vs_k141_64962\|\|full | Low-quality |
| S3_vs_k141_97177\|\|full | Low-quality |
| S3_vs_k141_33571\|\|full | Low-quality |
| S3_vs_k141_73594\|\|full | Not-determined |
| S3_vs_k141_33587\|\|full | Low-quality |
| S3_vs_k141_33597\|\|full | Low-quality |
| S3_vs_k141_104851\|\|full | Low-quality |
| S3_vs_k141_49541\|\|full | Low-quality |
| S3_vs_k141_33605\|\|full | Low-quality |
| S3_vs_k141_112672\|\|full | Low-quality |
| S3_vs_k141_9821\|\|full | Low-quality |
| S3_vs_k141_112676\|\|full | Low-quality |
| S3_vs_k141_104874\|\|full | Not-determined |
| S3_vs_k141_88898\|\|full | Not-determined |
| S3_vs_k141_17005\|\|full | Low-quality |
| S3_vs_k141_57208\|\|full | Not-determined |
| S3_vs_k141_81258\|\|full | Low-quality |
| S3_vs_k141_64968\|\|full | Low-quality |
| S3_vs_k141_81263\|\|full | Not-determined |
| S3_vs_k141_17015\|\|full | Low-quality |
| S3_vs_k141_64974\|\|full | Low-quality |
| S3_vs_k141_112688\|\|full | Not-determined |
| S3_vs_k141_49559\|\|full | Low-quality |
| S3_vs_k141_64989\|\|full | Not-determined |
| S3_vs_k141_1357\|\|full | Low-quality |
| S3_vs_k141_64993\|\|full | Low-quality |
| S3_vs_k141_57243\|\|full | Not-determined |
| S3_vs_k141_112710\|\|full | Low-quality |
| S3_vs_k141_25090\|\|full | Low-quality |
| S3_vs_k141_1370\|\|full | Low-quality |
| S3_vs_k141_112713\|\|full | Low-quality |
| S3_vs_k141_65010\|\|full | Low-quality |
| S3_vs_k141_25099\|\|full | Low-quality |
| S3_vs_k141_49578\|\|full | Not-determined |
| S3_vs_k141_104900\|\|full | Not-determined |
| S3_vs_k141_17044\|\|full | Low-quality |
| S3_vs_k141_112723\|\|full | Not-determined |
| S3_vs_k141_49584\|\|full | Low-quality |
| S3_vs_k141_25124\|\|full | Low-quality |
| S17_vs_k141_92866\|\|full | High-quality |
| S3_vs_k141_65031\|\|full | Low-quality |
| S3_vs_k141_17064\|\|full | Not-determined |
| S3_vs_k141_49619\|\|full | Low-quality |
| S3_vs_k141_1421\|\|full | Low-quality |
| S3_vs_k141_49634\|\|full | Not-determined |
| S3_vs_k141_73617\|\|full | Not-determined |
| S3_vs_k141_73622\|\|full | Low-quality |
| S3_vs_k141_17098\|\|full | Low-quality |
| S3_vs_k141_57297\|\|full | Low-quality |
| S3_vs_k141_57298\|\|full | Low-quality |
| S3_vs_k141_112743\|\|full | Low-quality |
| S3_vs_k141_57308\|\|full | Not-determined |
| S3_vs_k141_9898\|\|full | Low-quality |
| S3_vs_k141_120634\|\|full | Not-determined |
| S3_vs_k141_17117\|\|full | Not-determined |
| S3_vs_k141_1468\|\|full | Low-quality |
| S3_vs_k141_73670\|\|full | Not-determined |
| S3_vs_k141_97232\|\|full | Low-quality |
| S3_vs_k141_33713\|\|full | Low-quality |
| S3_vs_k141_1471\|\|full | Low-quality |
| S3_vs_k141_81383\|\|full | Not-determined |
| S3_vs_k141_73700\|\|full | Low-quality |
| S3_vs_k141_97255\|\|full | Low-quality |
| S3_vs_k141_57325\|\|full | Low-quality |
| S3_vs_k141_88927\|\|full | Low-quality |
| S3_vs_k141_1497\|\|full | Low-quality |
| S3_vs_k141_1502\|\|full | Low-quality |
| S3_vs_k141_81409\|\|full | Low-quality |
| S3_vs_k141_81417\|\|full | Low-quality |
| S3_vs_k141_57348\|\|full | Not-determined |
| S3_vs_k141_33751\|\|full | Low-quality |
| S3_vs_k141_112784\|\|full | Low-quality |
| S3_vs_k141_88929\|\|full | Low-quality |
| S3_vs_k141_57351\|\|full | Low-quality |
| S3_vs_k141_1518\|\|full | Low-quality |
| S3_vs_k141_1519\|\|full | Low-quality |
| S3_vs_k141_57358\|\|full | Not-determined |
| S3_vs_k141_1536\|\|full | Not-determined |
| S3_vs_k141_73743\|\|full | Low-quality |
| S3_vs_k141_120681\|\|full | Low-quality |
| S3_vs_k141_65097\|\|full | Low-quality |
| S3_vs_k141_112812\|\|full | Not-determined |
| S3_vs_k141_1551\|\|full | Low-quality |
| S3_vs_k141_88962\|\|full | Low-quality |
| S3_vs_k141_41336\|\|full | Low-quality |
| S3_vs_k141_41339\|\|full | Low-quality |
| S3_vs_k141_1563\|\|full | Not-determined |
| S3_vs_k141_112824\|\|full | Low-quality |
| S3_vs_k141_88999\|\|full | Not-determined |
| S3_vs_k141_41361\|\|full | Low-quality |
| S3_vs_k141_97296\|\|full | Low-quality |
| S3_vs_k141_97297\|\|full | Low-quality |
| S3_vs_k141_112853\|\|full | Low-quality |
| S3_vs_k141_9963\|\|full | Low-quality |
| S3_vs_k141_120692\|\|full | Low-quality |
| S3_vs_k141_57414\|\|full | Not-determined |
| S3_vs_k141_65124\|\|full | Low-quality |
| S3_vs_k141_120696\|\|full | Low-quality |
| S3_vs_k141_97307\|\|full | Not-determined |
| S3_vs_k141_25185\|\|full | Low-quality |
| S3_vs_k141_104964\|\|full | Low-quality |
| S10_vs_18671\|\|full | High-quality |
| S3_extracted_k141_80287 | Complete |
| S3_vs_k141_17173\|\|full | Low-quality |
| S3_vs_k141_57446\|\|full | Low-quality |
| S3_vs_k141_25193\|\|full | Low-quality |
| S3_vs_k141_25199\|\|full | Not-determined |
| S3_vs_k141_112879\|\|full | Not-determined |
| S3_vs_k141_41413\|\|full | Low-quality |
| S3_vs_k141_112884\|\|full | Low-quality |
| S3_vs_k141_10074\|\|full | Not-determined |
| S3_vs_k141_10081\|\|full | Low-quality |
| S3_vs_k141_10085\|\|full | Low-quality |
| S3_vs_k141_89051\|\|full | Low-quality |
| S3_vs_k141_17198\|\|full | Low-quality |
| S3_vs_k141_65147\|\|full | Low-quality |
| S3_vs_k141_81456\|\|full | Not-determined |
| S3_vs_k141_41435\|\|full | Low-quality |
| S3_vs_k141_49754\|\|full | Not-determined |
| S3_vs_k141_112921\|\|full | Not-determined |
| S3_vs_k141_112927\|\|full | Low-quality |
| S3_vs_k141_25242\|\|full | Not-determined |
| S3_vs_k141_112934\|\|full | Low-quality |
| S3_vs_k141_17207\|\|full | Low-quality |
| S3_vs_k141_49766\|\|full | Low-quality |
| S3_vs_k141_120725\|\|full | Low-quality |
| S3_vs_k141_104992\|\|full | Low-quality |
| S3_vs_k141_17221\|\|full | Not-determined |
| S3_vs_k141_81464\|\|full | Low-quality |
| S3_vs_k141_73889\|\|full | Not-determined |
| S3_vs_k141_97385\|\|full | Low-quality |
| S3_vs_k141_81471\|\|full | Not-determined |
| S3_vs_k141_41448\|\|full | Not-determined |
| S3_vs_k141_89104\|\|full | Not-determined |
| S3_vs_k141_89108\|\|full | Not-determined |
| S3_vs_k141_17237\|\|full | Not-determined |
| S3_vs_k141_112956\|\|full | Low-quality |
| S3_vs_k141_10146\|\|full | Low-quality |
| S3_vs_k141_97395\|\|full | Low-quality |
| S3_vs_k141_120763\|\|full | Not-determined |
| S3_vs_k141_112969\|\|full | Low-quality |
| S3_vs_k141_49787\|\|full | Not-determined |
| S3_vs_k141_49788\|\|full | Low-quality |
| S3_vs_k141_57481\|\|full | Not-determined |
| S3_vs_k141_25292\|\|full | Low-quality |
| S3_vs_k141_89131\|\|full | Low-quality |
| S3_vs_k141_57493\|\|full | Low-quality |
| S3_vs_k141_10172\|\|full | Low-quality |
| S3_vs_k141_65167\|\|full | Not-determined |
| S3_vs_k141_65168\|\|full | Not-determined |
| S3_vs_k141_49809\|\|full | Low-quality |
| S3_vs_k141_49812\|\|full | Not-determined |
| S3_vs_k141_120794\|\|full | Not-determined |
| S3_vs_k141_89148\|\|full | Not-determined |
| S3_vs_k141_113000\|\|full | Low-quality |
| S3_vs_k141_57522\|\|full | Not-determined |
| S3_vs_k141_25323\|\|full | Not-determined |
| S3_vs_k141_89158\|\|full | Not-determined |
| S3_vs_k141_113021\|\|full | Low-quality |
| S3_vs_k141_81502\|\|full | Low-quality |
| S3_vs_k141_1664\|\|full | Low-quality |
| S3_vs_k141_113046\|\|full | Low-quality |
| S3_vs_k141_1671\|\|full | Low-quality |
| S3_vs_k141_65206\|\|full | Low-quality |
| S3_vs_k141_73935\|\|full | Low-quality |
| S3_vs_k141_1676\|\|full | Low-quality |
| S3_vs_k141_73969\|\|full | Low-quality |
| S3_vs_k141_89196\|\|full | Not-determined |
| S3_vs_k141_17275\|\|full | Not-determined |
| S3_vs_k141_10294\|\|full | Not-determined |
| S3_vs_k141_41491\|\|full | Low-quality |
| S3_vs_k141_113065\|\|full | Low-quality |
| S3_vs_k141_89210\|\|full | Low-quality |
| S3_vs_k141_41505\|\|full | Not-determined |
| S3_vs_k141_41506\|\|full | Low-quality |
| S3_vs_k141_89222\|\|full | Low-quality |
| S3_vs_k141_113074\|\|full | Low-quality |
| S3_vs_k141_97435\|\|full | Not-determined |
| S3_vs_k141_74005\|\|full | Not-determined |
| S3_vs_k141_57578\|\|full | Low-quality |
| S3_vs_k141_74017\|\|full | Low-quality |
| S3_vs_k141_57582\|\|full | Low-quality |
| S3_vs_k141_97445\|\|full | Low-quality |
| S3_vs_k141_89256\|\|full | Not-determined |
| S3_vs_k141_113081\|\|full | Low-quality |
| S3_vs_k141_97449\|\|full | Low-quality |
| S3_vs_k141_57587\|\|full | Low-quality |
| S3_vs_k141_97452\|\|full | Low-quality |
| S3_vs_k141_89273\|\|full | Not-determined |
| S3_vs_k141_41522\|\|full | Low-quality |
| S3_vs_k141_49843\|\|full | Low-quality |
| S3_vs_k141_113095\|\|full | Not-determined |
| S3_vs_k141_120872\|\|full | Low-quality |
| S3_vs_k141_25387\|\|full | Not-determined |
| S3_vs_k141_49877\|\|full | Low-quality |
| S3_vs_k141_81536\|\|full | Low-quality |
| S3_vs_k141_113130\|\|full | Not-determined |
| S3_vs_k141_113139\|\|full | Low-quality |
| S3_vs_k141_105027\|\|full | Low-quality |
| S3_vs_k141_25421\|\|full | Not-determined |
| S3_vs_k141_113172\|\|full | Not-determined |
| S3_vs_k141_74064\|\|full | Low-quality |
| S3_vs_k141_81561\|\|full | Not-determined |
| S3_vs_k141_10381\|\|full | Low-quality |
| S3_vs_k141_81564\|\|full | Low-quality |
| S3_vs_k141_120910\|\|full | Low-quality |
| S3_vs_k141_113180\|\|full | Low-quality |
| S3_vs_k141_74066\|\|full | Not-determined |
| S3_vs_k141_74069\|\|full | Low-quality |
| S3_vs_k141_89298\|\|full | Low-quality |
| S3_vs_k141_17384\|\|full | Low-quality |
| S3_vs_k141_17400\|\|full | Low-quality |
| S3_vs_k141_49949\|\|full | Low-quality |
| S3_vs_k141_105089\|\|full | Low-quality |
| S3_vs_k141_81589\|\|full | Not-determined |
| S3_vs_k141_89336\|\|full | Low-quality |
| S3_vs_k141_10406\|\|full | Low-quality |
| S3_vs_k141_49962\|\|full | Low-quality |
| S3_vs_k141_25447\|\|full | Low-quality |
| S3_vs_k141_49968\|\|full | Low-quality |
| S3_vs_k141_41611\|\|full | Low-quality |
| S3_vs_k141_113230\|\|full | Low-quality |
| S3_vs_k141_25458\|\|full | Low-quality |
| S3_vs_k141_74091\|\|full | Low-quality |
| S3_vs_k141_97523\|\|full | Not-determined |
| S3_vs_k141_113237\|\|full | Low-quality |
| S3_vs_k141_49982\|\|full | Low-quality |
| S3_vs_k141_105108\|\|full | Low-quality |
| S3_vs_k141_10415\|\|full | Not-determined |
| S3_vs_k141_49990\|\|full | Not-determined |
| S3_vs_k141_105119\|\|full | Not-determined |
| S3_vs_k141_57692\|\|full | Low-quality |
| S3_vs_k141_33870\|\|full | Not-determined |
| S3_vs_k141_105131\|\|full | Not-determined |
| S3_vs_k141_1830\|\|full | Not-determined |
| S3_vs_k141_113247\|\|full | Low-quality |
| S3_vs_k141_57717\|\|full | Low-quality |
| S3_vs_k141_89373\|\|full | Low-quality |
| S3_vs_k141_105160\|\|full | Low-quality |
| S3_vs_k141_89376\|\|full | Low-quality |
| S3_vs_k141_41695\|\|full | Low-quality |
| S3_vs_k141_50023\|\|full | Not-determined |
| S3_vs_k141_1853\|\|full | Not-determined |
| S3_vs_k141_65253\|\|full | Low-quality |
| S3_vs_k141_97555\|\|full | Low-quality |
| S3_vs_k141_57734\|\|full | Low-quality |
| S3_vs_k141_10466\|\|full | Low-quality |
| S3_vs_k141_113263\|\|full | Low-quality |
| S3_vs_k141_10475\|\|full | Low-quality |
| S3_vs_k141_65263\|\|full | Low-quality |
| S3_vs_k141_17411\|\|full | Low-quality |
| S3_vs_k141_41711\|\|full | Low-quality |
| S3_vs_k141_10495\|\|full | Low-quality |
| S3_vs_k141_10496\|\|full | Low-quality |
| S3_vs_k141_50037\|\|full | Low-quality |
| S3_vs_k141_120994\|\|full | Low-quality |
| S3_vs_k141_121014\|\|full | Low-quality |
| S3_vs_k141_65285\|\|full | Low-quality |
| S3_vs_k141_10514\|\|full | Not-determined |
| S3_vs_k141_50070\|\|full | Not-determined |
| S3_vs_k141_57764\|\|full | Low-quality |
| S3_vs_k141_33909\|\|full | Low-quality |
| S3_vs_k141_57768\|\|full | Low-quality |
| S3_vs_k141_25526\|\|full | Low-quality |
| S3_vs_k141_89407\|\|full | Low-quality |
| S3_vs_k141_33918\|\|full | Low-quality |
| S3_vs_k141_25534\|\|full | Low-quality |
| S3_vs_k141_89411\|\|full | Not-determined |
| S3_vs_k141_97574\|\|full | Low-quality |
| S3_vs_k141_121025\|\|full | Not-determined |
| S3_vs_k141_57782\|\|full | Not-determined |
| S3_vs_k141_65306\|\|full | Low-quality |
| S3_vs_k141_10552\|\|full | Not-determined |
| S3_vs_k141_113302\|\|full | Low-quality |
| S3_vs_k141_113309\|\|full | Low-quality |
| S3_vs_k141_89464\|\|full | Low-quality |
| S3_vs_k141_65334\|\|full | Not-determined |
| S3_vs_k141_105220\|\|full | Low-quality |
| S3_vs_k141_113348\|\|full | Low-quality |
| S3_vs_k141_113350\|\|full | Low-quality |
| S3_vs_k141_121092\|\|full | Low-quality |
| S3_vs_k141_113368\|\|full | Not-determined |
| S3_vs_k141_33945\|\|full | Low-quality |
| S3_vs_k141_121096\|\|full | Low-quality |
| S3_vs_k141_33954\|\|full | Low-quality |
| S3_vs_k141_105274\|\|full | Not-determined |
| S3_vs_k141_113389\|\|full | Low-quality |
| S3_vs_k141_25604\|\|full | Low-quality |
| S3_vs_k141_41759\|\|full | Low-quality |
| S3_vs_k141_113406\|\|full | Low-quality |
| S3_vs_k141_41774\|\|full | Not-determined |
| S3_vs_k141_41786\|\|full | Low-quality |
| S3_vs_k141_105316\|\|full | Low-quality |
| S3_vs_k141_25612\|\|full | Not-determined |
| S3_vs_k141_81710\|\|full | Not-determined |
| S3_vs_k141_121135\|\|full | Low-quality |
| S3_vs_k141_121136\|\|full | Low-quality |
| S3_vs_k141_25626\|\|full | Not-determined |
| S3_vs_k141_81731\|\|full | Low-quality |
| S3_vs_k141_50099\|\|full | Not-determined |
| S3_vs_k141_105324\|\|full | Not-determined |
| S3_vs_k141_50100\|\|full | Low-quality |
| S3_vs_k141_121149\|\|full | Low-quality |
| S3_vs_k141_81745\|\|full | Not-determined |
| S3_vs_k141_105337\|\|full | Low-quality |
| S3_vs_k141_17492\|\|full | Low-quality |
| S3_vs_k141_105349\|\|full | Not-determined |
| S3_vs_k141_113479\|\|full | Low-quality |
| S3_vs_k141_105354\|\|full | Not-determined |
| S3_vs_k141_89530\|\|full | Not-determined |
| S3_vs_k141_89539\|\|full | Not-determined |
| S3_vs_k141_113504\|\|full | Low-quality |
| S3_vs_k141_25632\|\|full | Low-quality |
| S3_vs_k141_121206\|\|full | Not-determined |
| S3_vs_k141_89566\|\|full | Low-quality |
| S3_vs_k141_113523\|\|full | Low-quality |
| S3_vs_k141_113526\|\|full | Low-quality |
| S3_vs_k141_74203\|\|full | Low-quality |
| S3_vs_k141_105410\|\|full | Not-determined |
| S3_vs_k141_121237\|\|full | Low-quality |
| S3_vs_k141_113538\|\|full | Low-quality |
| S3_vs_k141_50120\|\|full | Not-determined |
| S3_vs_k141_10745\|\|full | Low-quality |
| S3_vs_k141_25654\|\|full | Low-quality |
| S3_vs_k141_105424\|\|full | Not-determined |
| S3_vs_k141_105425\|\|full | Low-quality |
| S3_vs_k141_113552\|\|full | Low-quality |
| S3_vs_k141_10761\|\|full | Not-determined |
| S3_vs_k141_34105\|\|full | Low-quality |
| S3_vs_k141_105443\|\|full | Not-determined |
| S3_vs_k141_121250\|\|full | Low-quality |
| S3_vs_k141_1950\|\|full | Low-quality |
| S3_vs_k141_34111\|\|full | Low-quality |
| S3_vs_k141_65457\|\|full | Not-determined |
| S3_vs_k141_34117\|\|full | Not-determined |
| S3_vs_k141_89581\|\|full | Not-determined |
| S3_vs_k141_50124\|\|full | Low-quality |
| S3_vs_k141_50135\|\|full | Not-determined |
| S3_vs_k141_10785\|\|full | Low-quality |
| S3_vs_k141_65492\|\|full | Low-quality |
| S3_vs_k141_57871\|\|full | Low-quality |
| S3_vs_k141_74223\|\|full | Not-determined |
| S3_vs_k141_74234\|\|full | Low-quality |
| S3_vs_k141_10800\|\|full | Not-determined |
| S3_vs_k141_121262\|\|full | Low-quality |
| S14_extracted_k141_20383 | High-quality |
| S3_vs_k141_10812\|\|full | Not-determined |
| S3_vs_k141_25725\|\|full | Not-determined |
| S3_vs_k141_89604\|\|full | Not-determined |
| S3_vs_k141_97667\|\|full | Low-quality |
| S3_vs_k141_89613\|\|full | Low-quality |
| S3_vs_k141_25733\|\|full | Low-quality |
| S3_vs_k141_34185\|\|full | Low-quality |
| S3_vs_k141_41836\|\|full | Low-quality |
| S3_vs_k141_10862\|\|full | Low-quality |
| S3_vs_k141_57905\|\|full | Low-quality |
| S3_vs_k141_41853\|\|full | Low-quality |
| S3_vs_k141_113570\|\|full | Low-quality |
| S3_vs_k141_17564\|\|full | Low-quality |
| S3_vs_k141_97692\|\|full | Low-quality |
| S3_vs_k141_97694\|\|full | Low-quality |
| S3_vs_k141_34242\|\|full | Not-determined |
| S3_vs_k141_34246\|\|full | Not-determined |
| S3_vs_k141_121312\|\|full | Low-quality |
| S3_vs_k141_10896\|\|full | Low-quality |
| S3_vs_k141_34249\|\|full | Not-determined |
| S3_vs_k141_121319\|\|full | Low-quality |
| S3_vs_k141_2051\|\|full | Low-quality |
| S3_vs_k141_57919\|\|full | Low-quality |
| S3_vs_k141_10908\|\|full | Low-quality |
| S3_vs_k141_25763\|\|full | Not-determined |
| S3_vs_k141_57931\|\|full | Low-quality |
| S3_vs_k141_57932\|\|full | Not-determined |
| S3_vs_k141_10914\|\|full | Not-determined |
| S3_vs_k141_57934\|\|full | Low-quality |
| S3_vs_k141_34289\|\|full | Low-quality |
| S3_vs_k141_97703\|\|full | Low-quality |
| S3_vs_k141_17600\|\|full | Not-determined |
| S3_vs_k141_10946\|\|full | Low-quality |
| S3_vs_k141_57951\|\|full | Low-quality |
| S3_vs_k141_89628\|\|full | Low-quality |
| S3_vs_k141_65551\|\|full | Not-determined |
| S3_vs_k141_17637\|\|full | Low-quality |
| S3_vs_k141_41941\|\|full | Low-quality |
| S3_vs_k141_97716\|\|full | Low-quality |
| S3_vs_k141_50191\|\|full | Not-determined |
| S3_vs_k141_41943\|\|full | Low-quality |
| S3_vs_k141_97723\|\|full | Not-determined |
| S3_vs_k141_57987\|\|full | Low-quality |
| S3_vs_k141_34337\|\|full | Low-quality |
| S3_vs_k141_121378\|\|full | Not-determined |
| S3_vs_k141_113595\|\|full | Low-quality |
| S3_vs_k141_89672\|\|full | Not-determined |
| S3_vs_k141_81776\|\|full | Low-quality |
| S3_vs_k141_81782\|\|full | Low-quality |
| S3_vs_k141_17658\|\|full | Low-quality |
| S3_vs_k141_105514\|\|full | Low-quality |
| S3_vs_k141_34361\|\|full | Not-determined |
| S3_vs_k141_58041\|\|full | Low-quality |
| S3_vs_k141_50210\|\|full | Low-quality |
| S3_vs_k141_34365\|\|full | Low-quality |
| S3_vs_k141_34369\|\|full | Low-quality |
| S3_vs_k141_105520\|\|full | Not-determined |
| S3_vs_k141_105526\|\|full | Low-quality |
| S3_vs_k141_74306\|\|full | Not-determined |
| S3_vs_k141_113629\|\|full | Low-quality |
| S3_vs_k141_25864\|\|full | Not-determined |
| S3_vs_k141_65628\|\|full | Not-determined |
| S3_vs_k141_97741\|\|full | Low-quality |
| S3_vs_k141_58054\|\|full | Low-quality |
| S3_vs_k141_89702\|\|full | Low-quality |
| S3_vs_k141_58058\|\|full | Low-quality |
| S3_vs_k141_97749\|\|full | Not-determined |
| S3_vs_k141_113642\|\|full | Low-quality |
| S3_vs_k141_34389\|\|full | Low-quality |
| S3_vs_k141_89706\|\|full | Not-determined |
| S3_vs_k141_17673\|\|full | Low-quality |
| S3_vs_k141_97756\|\|full | Low-quality |
| S3_vs_k141_25892\|\|full | Low-quality |
| S3_vs_k141_42038\|\|full | Low-quality |
| S3_vs_k141_65669\|\|full | Low-quality |
| S3_vs_k141_113680\|\|full | Low-quality |
| S3_vs_k141_42058\|\|full | Low-quality |
| S3_vs_k141_105580\|\|full | Not-determined |
| S3_vs_k141_17677\|\|full | Low-quality |
| S3_vs_k141_11079\|\|full | Not-determined |
| S3_vs_k141_17680\|\|full | Not-determined |
| S3_vs_k141_2120\|\|full | Low-quality |
| S3_vs_k141_11093\|\|full | Not-determined |
| S3_vs_k141_121413\|\|full | Low-quality |
| S3_vs_k141_81803\|\|full | Low-quality |
| S3_vs_k141_105613\|\|full | Low-quality |
| S3_vs_k141_81809\|\|full | Low-quality |
| S3_vs_k141_58121\|\|full | Low-quality |
| S3_vs_k141_81810\|\|full | Low-quality |
| S3_vs_k141_89745\|\|full | Not-determined |
| S3_vs_k141_113747\|\|full | Low-quality |
| S3_vs_k141_2139\|\|full | Not-determined |
| S3_vs_k141_17702\|\|full | Low-quality |
| S8_vs_k141_145173\|\|full | High-quality |
| S3_vs_k141_17705\|\|full | Low-quality |
| S3_vs_k141_42074\|\|full | Not-determined |
| S3_vs_k141_58133\|\|full | Low-quality |
| S3_vs_k141_81839\|\|full | Low-quality |
| S3_vs_k141_50252\|\|full | Low-quality |
| S3_vs_k141_121441\|\|full | Low-quality |
| S3_vs_k141_34431\|\|full | Low-quality |
| S3_vs_k141_58143\|\|full | Low-quality |
| S3_vs_k141_17708\|\|full | Not-determined |
| S3_vs_k141_97766\|\|full | Not-determined |
| S3_vs_k141_58148\|\|full | Low-quality |
| S3_vs_k141_11192\|\|full | Low-quality |
| S3_vs_k141_97773\|\|full | Low-quality |
| S3_vs_k141_58154\|\|full | Not-determined |
| S3_vs_k141_25941\|\|full | Low-quality |
| S3_vs_k141_105696\|\|full | Low-quality |
| S3_vs_k141_42121\|\|full | Low-quality |
| S3_vs_k141_65769\|\|full | Low-quality |
| S3_vs_k141_58167\|\|full | Low-quality |
| S3_vs_k141_121456\|\|full | Low-quality |
| S3_vs_k141_105722\|\|full | Not-determined |
| S3_vs_k141_25953\|\|full | Low-quality |
| S3_vs_k141_97802\|\|full | Low-quality |
| S4_vs_VIRSorter_7386-circular-cat_ | High-quality |
| S3_vs_k141_81867\|\|full | Low-quality |
| S3_vs_k141_11246\|\|full | Low-quality |
| S3_vs_k141_25975\|\|full | Low-quality |
| S3_vs_k141_2158\|\|full | Low-quality |
| S3_vs_k141_58209\|\|full | Low-quality |
| S3_vs_k141_2162\|\|full | Low-quality |
| S3_vs_k141_105738\|\|full | Low-quality |
| S3_vs_k141_89777\|\|full | Low-quality |
| S3_vs_k141_74410\|\|full | Low-quality |

S3_vs_k141_58212||full Low-quality
S3_vs_k141_105746||full Low-quality
S3_vs_k141_65828||full Not-determined
S3_vs_k141_113756||full Low-quality
S3_vs_k141_58222||full Low-quality
S3_vs_k141_121489||full Low-quality
S3_vs_k141_113762||full Not-determined
S3_vs_k141_105768||full Not-determined
S3_vs_k141_65834||full Low-quality
S3_vs_k141_74422||full Not-determined
S3_vs_k141_113764||full Not-determined
S3_vs_k141_74424||full Not-determined
S3_vs_k141_81881||full Not-determined
S3_vs_k141_11295||full Not-determined
S3_vs_k141_81891||full Low-quality
S3_vs_k141_50374||full Low-quality
S3_vs_k141_2168||full Not-determined
S3_vs_k141_11303||full Not-determined
S3_vs_k141_89808||full Low-quality
S3_vs_k141_42138||full Low-quality
S3_vs_k141_2177||full Not-determined
S3_vs_k141_42142||full Low-quality
S3_vs_k141_89817||full Low-quality
S3_vs_k141_74428||full Not-determined
S3_vs_k141_113771||full Not-determined
S3_vs_k141_121510||full Low-quality
S3_vs_k141_50415||full Low-quality
S3_vs_k141_65886||full Not-determined
S3_vs_k141_11364||full Low-quality
S3_vs_k141_121528||full Low-quality
S3_vs_k141_81902||full Low-quality
S3_vs_k141_74439||full Not-determined
S3_vs_k141_65915||full Low-quality
S3_vs_k141_80287||full High-quality
S3_vs_k141_74446||full Low-quality
S3_vs_k141_97829||full Low-quality
S3_vs_k141_89862||full Low-quality
S3_vs_k141_97831||full Not-determined
S3_vs_k141_89868||full Not-determined
S3_vs_k141_74460||full Low-quality
S3_vs_k141_50459||full Not-determined
S3_vs_k141_97834||full Low-quality
S3_vs_k141_17754||full Low-quality
S3_vs_k141_17758||full Low-quality
S3_vs_k141_105863||full Not-determined
S3_vs_k141_17765||full Low-quality
S3_vs_k141_121585||full Low-quality
S3_vs_k141_97863||full Low-quality
S3_vs_k141_74496||full Low-quality
S3_vs_k141_65941||full Not-determined
S3_vs_k141_74500||full Not-determined
S3_vs_k141_26007||full Low-quality
S3_vs_k141_42164||full Not-determined
S3_vs_k141_74507||full Not-determined
S3_vs_k141_74509||full Low-quality
S3_vs_k141_105877||full Low-quality
S3_vs_k141_50486||full Low-quality
S3_vs_k141_26019||full Low-quality
S11_vs_k141_40945||full High-quality
S3_vs_k141_26023||full Low-quality
S3_vs_k141_113837||full Not-determined
S3_vs_k141_42206||full Low-quality
S3_vs_k141_26033||full Low-quality
S3_vs_k141_81915||full Not-determined
S3_vs_k141_113839||full Low-quality
S3_vs_k141_42213||full Not-determined
S3_vs_k141_26044||full Low-quality
S3_vs_k141_81928||full Not-determined
S3_vs_k141_121650||full Not-determined
S3_vs_k141_58299||full Low-quality
S3_vs_k141_58300||full Low-quality
S3_vs_k141_74522||full Low-quality
S14_vs_k141_20383||full High-quality
S3_vs_k141_42231||full Low-quality
S3_vs_k141_105901||full Not-determined
S3_vs_k141_113871||full Low-quality
S3_vs_k141_97913||full Not-determined
S3_vs_k141_42258||full Low-quality
S3_vs_k141_97929||full Low-quality
S3_vs_k141_58309||full Low-quality
S3_vs_k141_34496||full Low-quality
S3_vs_k141_97935||full Not-determined
S3_vs_k141_74574||full Low-quality
S3_vs_k141_105910||full Low-quality
S3_vs_k141_74586||full Low-quality
S3_vs_k141_50522||full Not-determined
S3_vs_k141_42283||full Not-determined
S3_vs_k141_2201||full Low-quality
S3_vs_k141_34511||full Low-quality
S3_vs_k141_74611||full Not-determined
S3_vs_k141_89941||full Low-quality
S3_vs_k141_81982||full Low-quality
S3_vs_k141_113912||full Not-determined
S3_vs_k141_50542||full Low-quality
S3_vs_k141_113915||full Not-determined
S3_vs_k141_89960||full Low-quality
S3_vs_k141_113927||full Not-determined
S3_vs_k141_89977||full Not-determined
S3_vs_k141_34524||full Low-quality
S3_vs_k141_2231||full Low-quality
S3_vs_k141_11555||full Low-quality
S3_vs_k141_26141||full Not-determined
S3_vs_k141_89990||full Not-determined
S3_vs_k141_58326||full Low-quality
S3_vs_k141_90000||full Low-quality
S3_vs_k141_90026||full Not-determined
S3_vs_k141_42338||full Low-quality
S3_vs_k141_2290||full Low-quality
S3_vs_k141_105984||full Not-determined
S3_vs_k141_2303||full Low-quality
S3_vs_k141_26174||full Low-quality
S3_vs_k141_2306||full Not-determined
S3_vs_k141_2307||full Low-quality
S3_vs_k141_11583||full Low-quality
S1_vs_k141_23682||full High-quality
S3_vs_k141_42355||full Not-determined
S3_vs_k141_74657||full Not-determined
S3_vs_k141_50562||full Not-determined
S3_vs_k141_50563||full Not-determined
S3_vs_k141_50565||full Low-quality
S3_vs_k141_58388||full Low-quality
S3_vs_k141_97991||full Low-quality
S3_vs_k141_74661||full Low-quality
S3_vs_k141_74668||full Not-determined
S3_vs_k141_26216||full Low-quality
S3_vs_k141_26218||full Low-quality
S3_vs_k141_11640||full Not-determined
S3_vs_k141_98003||full Low-quality
S3_vs_k141_106019||full Not-determined
S3_vs_k141_26232||full Not-determined
S3_vs_k141_2367||full Low-quality
S3_vs_k141_90088||full Low-quality
S3_vs_k141_98024||full Low-quality
S3_vs_k141_114021||full Not-determined
S3_vs_k141_121685||full Low-quality
S3_vs_k141_98031||full Not-determined
S3_vs_k141_17850||full Low-quality
S3_vs_k141_2383||full Low-quality
S3_vs_k141_50630||full Not-determined
S3_vs_k141_74684||full Not-determined
S3_vs_k141_98043||full Low-quality
S3_vs_k141_42453||full Low-quality
S3_vs_k141_17876||full Not-determined
S3_vs_k141_50650||full Not-determined
S3_vs_k141_42463||full Low-quality
S3_vs_k141_11696||full Low-quality
S3_vs_k141_50660||full Not-determined
S3_vs_k141_34569||full Low-quality
S3_vs_k141_82160||full Low-quality
S3_vs_k141_82162||full Low-quality
S3_vs_k141_17892||full Low-quality
S3_vs_k141_11708||full Low-quality
S3_vs_k141_11711||full Low-quality
S3_vs_k141_74728||full Low-quality
S3_vs_k141_2410||full Low-quality
S3_vs_k141_26304||full Low-quality
S3_vs_k141_82173||full Not-determined
S3_vs_k141_17921||full Low-quality
S3_vs_k141_98080||full Low-quality
S3_vs_k141_121700||full Low-quality
S3_vs_k141_82190||full Not-determined
S3_vs_k141_74743||full Low-quality
S3_vs_k141_17922||full Low-quality
S3_vs_k141_98087||full Low-quality
S3_vs_k141_121703||full Not-determined
S3_vs_k141_98090||full Not-determined
S3_vs_k141_17926||full Low-quality
S3_vs_k141_34600||full Low-quality
S3_vs_k141_98102||full Not-determined
S3_vs_k141_90167||full Low-quality
S3_vs_k141_114069||full Low-quality
S3_vs_k141_90171||full Not-determined
S3_vs_k141_114081||full Not-determined
S3_vs_k141_106107||full Low-quality
S3_vs_k141_114095||full Low-quality
S3_vs_k141_114102||full Not-determined
S3_vs_k141_17932||full Low-quality
S3_vs_k141_26358||full Low-quality
S3_vs_k141_42535||full Not-determined
S3_vs_k141_58519||full Not-determined
S3_vs_k141_114108||full Low-quality
S3_vs_k141_2452||full Low-quality
S3_vs_k141_90226||full Not-determined
S3_vs_k141_26381||full Low-quality
S3_vs_k141_74782||full Low-quality
S3_vs_k141_98158||full Low-quality
S3_vs_k141_17945||full Low-quality
S3_vs_k141_58569||full Low-quality
S3_vs_k141_106130||full Low-quality
S3_vs_k141_82263||full Low-quality
S3_vs_k141_82265||full Low-quality
S3_vs_k141_50700||full Low-quality
S3_vs_k141_42548||full Low-quality
S3_vs_k141_58578||full Not-determined
S3_vs_k141_114139||full Low-quality
S3_vs_k141_26383||full Low-quality
S3_vs_k141_17965||full Low-quality
S3_vs_k141_74799||full Low-quality
S3_vs_k141_98178||full Low-quality
S3_vs_k141_17976||full Not-determined
S3_vs_k141_58610||full Not-determined
S3_vs_k141_17985||full Low-quality
S3_vs_k141_98185||full Not-determined
S3_vs_k141_98187||full Low-quality
S3_vs_k141_82294||full Not-determined
S3_vs_k141_2492||full Low-quality
S3_vs_k141_98190||full Low-quality
S3_vs_k141_58616||full Not-determined
S3_vs_k141_26427||full Not-determined
S3_vs_k141_90286||full Low-quality
S3_vs_k141_121753||full Not-determined
S3_vs_k141_58620||full Not-determined
S3_vs_k141_50738||full Low-quality
S3_vs_k141_90294||full Not-determined
S3_vs_k141_106166||full Low-quality
S3_vs_k141_90295||full Low-quality
S3_vs_k141_2522||full Low-quality
S3_vs_k141_58644||full Not-determined
S3_vs_k141_98202||full Not-determined
S3_vs_k141_106172||full Low-quality
S3_vs_k141_2528||full Low-quality
S3_vs_k141_98215||full Low-quality
S3_vs_k141_82324||full Not-determined
S3_vs_k141_2532||full Low-quality
S3_vs_k141_106180||full Not-determined
S3_vs_k141_121796||full Not-determined
S3_vs_k141_2557||full Low-quality
S3_vs_k141_106194||full Not-determined
S3_vs_k141_114167||full Low-quality
S3_vs_k141_34702||full Low-quality
S3_vs_k141_2562||full Low-quality
S3_vs_k141_114171||full Low-quality
S3_vs_k141_121808||full Low-quality
S3_vs_k141_2565||full Not-determined
S3_vs_k141_98239||full Low-quality
S3_vs_k141_74845||full Not-determined
S3_vs_k141_82331||full Low-quality
S3_vs_k141_121821||full Not-determined
S7_vs_48523||full High-quality
S3_vs_k141_11749||full Low-quality
S3_vs_k141_42576||full Not-determined
S3_vs_k141_58731||full Low-quality
S3_vs_k141_114193||full Not-determined
S3_vs_k141_26472||full Not-determined
S3_vs_k141_2623||full Low-quality
S3_vs_k141_2624||full Not-determined
S3_vs_k141_42597||full Low-quality
S3_vs_k141_26476||full Low-quality
S3_vs_k141_50756||full Low-quality
S3_vs_k141_50761||full Not-determined
S3_vs_k141_66023||full Low-quality
S3_vs_k141_58762||full Not-determined
S3_vs_k141_34737||full Low-quality
S3_vs_k141_11782||full Not-determined
S3_vs_k141_74869||full Not-determined
S3_vs_k141_98290||full Not-determined
S3_vs_k141_34745||full Low-quality
S3_vs_k141_50770||full Not-determined
S3_vs_k141_82363||full Not-determined
S3_vs_k141_34756||full Low-quality
S3_vs_k141_34761||full Low-quality
S3_vs_k141_74890||full Low-quality
S3_vs_k141_66062||full Not-determined
S3_vs_k141_50803||full Low-quality
S3_vs_k141_114204||full Not-determined
S3_vs_k141_66083||full Low-quality
S3_vs_k141_106243||full Low-quality
S3_vs_k141_74911||full Low-quality
S3_vs_k141_121830||full Low-quality
S3_vs_k141_58796||full Not-determined
S3_vs_k141_11829||full Low-quality
S3_vs_k141_114223||full Low-quality
S3_vs_k141_121834||full Low-quality
S3_vs_k141_26522||full Low-quality
S3_vs_k141_42668||full Not-determined
S3_vs_k141_50817||full Low-quality
S3_vs_k141_106258||full Low-quality
S3_vs_k141_42670||full Low-quality
S3_vs_k141_66103||full Low-quality
S3_vs_k141_90337||full Not-determined
S3_vs_k141_34796||full Low-quality
S3_vs_k141_74958||full Not-determined
S3_vs_k141_34799||full Not-determined
S3_vs_k141_11849||full Low-quality
S3_vs_k141_34806||full Not-determined
S3_vs_k141_82375||full Low-quality
S3_vs_k141_2717||full Not-determined
S3_vs_k141_58827||full Low-quality
S3_vs_k141_50859||full Low-quality
S3_vs_k141_114244||full Not-determined
S3_vs_k141_42714||full Low-quality
S3_vs_k141_2727||full Low-quality
S3_vs_k141_114247||full Low-quality
S3_vs_k141_82389||full Low-quality
S3_vs_k141_26593||full Not-determined
S3_vs_k141_98329||full Not-determined
S3_vs_k141_98338||full Not-determined
S3_vs_k141_50885||full Low-quality
S3_vs_k141_26611||full Low-quality
S3_vs_k141_106322||full Low-quality
S3_vs_k141_42766||full Low-quality
S3_vs_k141_106330||full Low-quality
S3_vs_k141_42767||full Not-determined
S3_vs_k141_42770||full Low-quality
S3_vs_k141_50918||full Low-quality
S3_vs_k141_106343||full Not-determined
S3_vs_k141_42782||full Low-quality
S3_vs_k141_82446||full Low-quality
S3_vs_k141_106369||full Not-determined
S3_vs_k141_11907||full Low-quality
S3_vs_k141_75042||full Low-quality
S3_vs_k141_11909||full Low-quality
S3_vs_k141_98361||full Low-quality
S3_vs_k141_98366||full Low-quality
S7_extracted_53960 Complete
S8_vs_k141_144183||full Medium-quality
S3_vs_k141_82495||full Not-determined
S3_vs_k141_90375||full Low-quality
S3_vs_k141_121932||full Low-quality
S3_vs_k141_34842||full Low-quality
S3_vs_k141_98369||full Low-quality
S3_vs_k141_58919||full Not-determined
S3_vs_k141_42834||full Low-quality
S3_vs_k141_2789||full Low-quality
S3_vs_k141_58924||full Low-quality
S3_vs_k141_50937||full Not-determined
S3_vs_k141_58930||full Low-quality
S3_vs_k141_58931||full Low-quality
S3_vs_k141_26651||full Low-quality
S3_vs_k141_11953||full Low-quality
S3_vs_k141_66154||full Low-quality
S3_vs_k141_11958||full Not-determined
S3_vs_k141_34870||full Low-quality
S11_vs_k141_104069||full High-quality
S3_vs_k141_34872||full Low-quality
S3_vs_k141_90402||full Low-quality
S3_vs_k141_98395||full Not-determined
S3_vs_k141_58941||full Low-quality
S3_vs_k141_90406||full Low-quality
S3_vs_k141_114293||full Low-quality
S3_vs_k141_50957||full Not-determined
S3_vs_k141_82501||full Low-quality
S3_vs_k141_75088||full Low-quality
S3_vs_k141_34889||full Low-quality
S3_vs_k141_121994||full Low-quality
S3_vs_k141_106427||full Low-quality
S3_vs_k141_82510||full Low-quality
S3_vs_k141_34899||full Not-determined
S3_vs_k141_11994||full Low-quality
S3_vs_k141_114301||full Low-quality
S3_vs_k141_50977||full Not-determined
S3_vs_k141_82528||full Low-quality
S3_vs_k141_11996||full Not-determined
S3_vs_k141_50979||full Low-quality
S3_vs_k141_2817||full Not-determined
S3_vs_k141_106440||full Low-quality
S3_vs_k141_59012||full Low-quality
S3_vs_k141_90447||full Low-quality
S3_vs_k141_98428||full Low-quality
S3_vs_k141_106445||full Low-quality
S3_vs_k141_82536||full Low-quality
S3_vs_k141_2833||full Low-quality
S3_vs_k141_90456||full Low-quality
S3_vs_k141_50999||full Not-determined
S3_vs_k141_98434||full Low-quality
S3_vs_k141_26689||full Low-quality
S3_vs_k141_34937||full Low-quality
S3_vs_k141_59024||full Low-quality
S3_vs_k141_90470||full Low-quality
S12_vs_k141_141337||full High-quality
S3_vs_k141_51008||full Low-quality
S3_vs_k141_98442||full Not-determined
S3_vs_k141_82550||full Not-determined
S3_vs_k141_75130||full Low-quality
S3_vs_k141_75141||full Low-quality
S3_vs_k141_2841||full Low-quality
S3_vs_k141_59051||full Low-quality
S3_vs_k141_59052||full Low-quality
S3_vs_k141_114351||full Not-determined
S3_vs_k141_59054||full Low-quality
S3_vs_k141_75155||full Not-determined
S3_vs_k141_26764||full Low-quality
S3_vs_k141_75162||full Low-quality
S3_vs_k141_114363||full Not-determined
S3_vs_k141_2857||full Low-quality
S7_vs_4956||full High-quality
S8_vs_k141_208118||full High-quality
S3_vs_k141_75170||full Not-determined
S3_vs_k141_75176||full Not-determined
S3_vs_k141_75183||full Low-quality
S3_vs_k141_59119||full Not-determined
S3_vs_k141_82585||full Not-determined
S3_vs_k141_51054||full Low-quality
S3_vs_k141_12037||full Low-quality
S3_vs_k141_82604||full Not-determined
S3_vs_k141_66207||full Low-quality
S3_vs_k141_26793||full Not-determined
S3_vs_k141_12046||full Not-determined
S3_vs_k141_82615||full Not-determined
S3_vs_k141_98487||full Low-quality
S3_vs_k141_82616||full Low-quality
S3_vs_k141_75223||full Low-quality
S3_vs_k141_12053||full Low-quality
S3_vs_k141_90546||full Low-quality
S3_vs_k141_51101||full Low-quality
S3_vs_k141_2878||full Low-quality
S3_vs_k141_82642||full Low-quality
S3_vs_k141_42929||full Not-determined
S3_vs_k141_51114||full Low-quality
S3_vs_k141_26826||full Not-determined
S3_vs_k141_66232||full Not-determined
S3_vs_k141_51124||full Low-quality
S14_vs_k141_112068||full High-quality
S3_vs_k141_66242||full Low-quality
S3_vs_k141_42933||full Low-quality
S3_vs_k141_75289||full Not-determined
S3_vs_k141_26836||full Not-determined
S3_vs_k141_35079||full Low-quality
S3_vs_k141_90593||full Not-determined
S3_vs_k141_98550||full Low-quality
S3_vs_k141_51163||full Not-determined
S3_vs_k141_106599||full Low-quality
S3_vs_k141_106603||full Low-quality
S3_vs_k141_51184||full Not-determined
S7_vs_53960||full High-quality
S3_vs_k141_114401||full Low-quality
S3_vs_k141_51199||full Low-quality
S3_vs_k141_66269||full Low-quality
S3_vs_k141_122055||full Low-quality
S3_vs_k141_106606||full Low-quality
S3_vs_k141_12119||full Low-quality
S3_vs_k141_66297||full Low-quality
S3_vs_k141_114411||full Low-quality
S3_vs_k141_51241||full Low-quality
S3_vs_k141_12137||full Not-determined
S3_vs_k141_51243||full Not-determined
S3_vs_k141_106628||full Low-quality
S3_vs_k141_122098||full Low-quality
S3_vs_k141_82762||full Not-determined
S3_vs_k141_35142||full Not-determined
S3_vs_k141_43032||full Low-quality
S3_vs_k141_18151||full Not-determined
S3_vs_k141_75376||full Low-quality
S3_vs_k141_51278||full Low-quality
S3_vs_k141_2943||full Low-quality
S3_vs_k141_114435||full Low-quality
S3_vs_k141_66308||full Low-quality
S3_vs_k141_51280||full Low-quality
S3_vs_k141_26896||full Low-quality
S3_vs_k141_66314||full Low-quality
S3_vs_k141_114443||full Not-determined
S3_vs_k141_122130||full Not-determined
S3_vs_k141_114453||full Low-quality
S3_vs_k141_75390||full Not-determined
S3_vs_k141_82817||full Low-quality
S3_vs_k141_2998||full Low-quality
S3_vs_k141_2999||full Low-quality
S3_vs_k141_106663||full Not-determined
S3_vs_k141_106665||full Low-quality
S3_vs_k141_26929||full Low-quality
S3_vs_k141_82821||full Low-quality
S3_vs_k141_98608||full Not-determined
S3_vs_k141_106672||full Low-quality
S3_vs_k141_12209||full Low-quality
S3_vs_k141_59142||full Low-quality
S3_vs_k141_82828||full Low-quality
S3_vs_k141_114477||full Not-determined
S3_vs_k141_114478||full Low-quality
S3_vs_k141_106683||full Low-quality
S3_vs_k141_98639||full Low-quality
S3_vs_k141_51321||full Low-quality
S3_vs_k141_114500||full Not-determined
S3_vs_k141_26932||full Low-quality
S3_vs_k141_3033||full Low-quality
S3_vs_k141_43080||full Not-determined
S3_vs_k141_26934||full Low-quality
S3_vs_k141_98647||full Not-determined
S3_vs_k141_3034||full Low-quality
S3_vs_k141_90657||full Low-quality
S3_vs_k141_35200||full Low-quality
S3_vs_k141_43084||full Not-determined
S3_vs_k141_12219||full Not-determined
S3_vs_k141_51330||full Low-quality
S3_vs_k141_98659||full Low-quality
S3_vs_k141_35213||full Not-determined
S3_vs_k141_26941||full Not-determined
S3_vs_k141_12229||full Not-determined
S3_vs_k141_98668||full Low-quality
S3_vs_k141_90666||full Not-determined
S3_vs_k141_106720||full Not-determined
S3_vs_k141_3035||full Low-quality
S3_vs_k141_12245||full Low-quality
S3_vs_k141_12252||full Low-quality
S3_vs_k141_59188||full Not-determined
S3_vs_k141_90695||full Low-quality
S3_vs_k141_12256||full Low-quality
S3_vs_k141_51342||full Low-quality
S3_vs_k141_82845||full Low-quality
S3_vs_k141_35285||full Low-quality
S3_vs_k141_82857||full Low-quality
S3_vs_k141_66362||full Not-determined
S3_vs_k141_90721||full Low-quality
S3_vs_k141_59221||full Not-determined
S3_vs_k141_66367||full Not-determined
S3_vs_k141_114538||full Low-quality
S3_vs_k141_18237||full Low-quality
S3_vs_k141_82863||full Low-quality
S3_vs_k141_90737||full Low-quality
S3_vs_k141_66375||full Not-determined
S3_vs_k141_12277||full Low-quality
S3_vs_k141_66376||full Low-quality
S3_vs_k141_82873||full Not-determined
S3_vs_k141_90742||full Not-determined
S3_vs_k141_18262||full Low-quality
S3_vs_k141_35307||full Low-quality
S3_vs_k141_106815||full Low-quality
S3_vs_k141_90745||full Not-determined
S3_vs_k141_3084||full Low-quality
S3_vs_k141_3097||full Not-determined
S3_vs_k141_18282||full Low-quality
S3_vs_k141_114580||full Low-quality
S3_vs_k141_90757||full Low-quality
S3_vs_k141_66384||full Low-quality
S3_vs_k141_90773||full Not-determined
S3_vs_k141_27079||full Low-quality
S3_vs_k141_35340||full Low-quality
S3_vs_k141_98704||full Low-quality
S3_vs_k141_51371||full Low-quality
S3_vs_k141_98713||full Low-quality
S3_vs_k141_114623||full Not-determined
S3_vs_k141_98720||full Low-quality
S3_vs_k141_90777||full Not-determined
S3_vs_k141_59256||full Low-quality
S3_vs_k141_114630||full Low-quality
S3_vs_k141_18292||full Not-determined
S3_vs_k141_43185||full Not-determined
S3_vs_k141_43186||full Low-quality
S3_vs_k141_75569||full Low-quality
S3_vs_k141_3108||full Low-quality
S3_vs_k141_3109||full Low-quality
S3_vs_k141_98730||full Low-quality
S3_vs_k141_90791||full Low-quality
S3_vs_k141_114647||full Not-determined
S3_vs_k141_114652||full Not-determined
S3_vs_k141_27104||full Low-quality
S3_vs_k141_59311||full Low-quality
S3_vs_k141_59312||full Not-determined
S3_vs_k141_122183||full Low-quality
S3_vs_k141_98756||full Low-quality
S3_vs_k141_98759||full Not-determined
S3_vs_k141_122188||full Low-quality
S3_vs_k141_51422||full Low-quality
S3_vs_k141_3131||full Low-quality
S3_vs_k141_122198||full Low-quality
S3_vs_k141_122204||full Low-quality
S3_vs_k141_98775||full Low-quality
S3_vs_k141_59362||full Not-determined
S3_vs_k141_98778||full Low-quality
S3_vs_k141_106922||full Low-quality
S3_vs_k141_75644||full Not-determined
S3_vs_k141_114692||full Low-quality
S3_vs_k141_75655||full Not-determined
S3_vs_k141_35460||full Not-determined
S3_vs_k141_18317||full Not-determined
S3_vs_k141_51439||full Not-determined
S3_vs_k141_106942||full Low-quality
S3_vs_k141_18326||full Low-quality
S3_vs_k141_90819||full Not-determined
S3_vs_k141_90820||full Not-determined
S3_vs_k141_90831||full Low-quality
S3_vs_k141_90833||full Not-determined
S3_vs_k141_18341||full Low-quality
S3_vs_k141_12358||full Not-determined
S3_vs_k141_122242||full Not-determined
S3_vs_k141_122245||full Low-quality
S3_vs_k141_122253||full Low-quality
S3_vs_k141_12383||full Low-quality
S3_vs_k141_106967||full Not-determined
S3_vs_k141_75706||full Low-quality
S3_vs_k141_75715||full Not-determined
S3_vs_k141_75716||full Not-determined
S3_vs_k141_90873||full Not-determined
S3_vs_k141_106988||full Low-quality
S3_vs_k141_18408||full Low-quality
S3_vs_k141_75772||full Not-determined
S3_vs_k141_51543||full Not-determined
S3_vs_k141_98857||full Low-quality
S3_vs_k141_75786||full Not-determined
S3_vs_k141_12460||full Low-quality

| | |
|---|---|
| S3_vs_k141_75799\|\|full | Low-quality |
| S3_vs_k141_98866\|\|full | Not-determined |
| S3_vs_k141_18461\|\|full | Low-quality |
| S3_vs_k141_75816\|\|full | Low-quality |
| S3_vs_k141_90937\|\|full | Not-determined |
| S3_vs_k141_90940\|\|full | Low-quality |
| S3_vs_k141_107043\|\|full | Low-quality |
| S3_vs_k141_35571\|\|full | Low-quality |
| S3_vs_k141_59440\|\|full | Not-determined |
| S3_vs_k141_98876\|\|full | Not-determined |
| S3_vs_k141_18465\|\|full | Low-quality |
| S3_vs_k141_27138\|\|full | Low-quality |
| S3_vs_k141_114781\|\|full | Not-determined |
| S3_vs_k141_114789\|\|full | Not-determined |
| S3_vs_k141_18484\|\|full | Not-determined |
| S3_vs_k141_107068\|\|full | Low-quality |
| S3_vs_k141_82897\|\|full | Not-determined |
| S3_vs_k141_3204\|\|full | Not-determined |
| S3_vs_k141_107074\|\|full | Low-quality |
| S3_vs_k141_107075\|\|full | Not-determined |
| S3_vs_k141_66500\|\|full | Low-quality |
| S3_vs_k141_35608\|\|full | Low-quality |
| S3_vs_k141_3213\|\|full | Low-quality |
| S3_vs_k141_12470\|\|full | Low-quality |
| S3_vs_k141_43272\|\|full | Low-quality |
| S3_vs_k141_114803\|\|full | Not-determined |
| S3_vs_k141_122296\|\|full | Not-determined |
| S3_vs_k141_82922\|\|full | Low-quality |
| S3_vs_k141_75829\|\|full | Low-quality |
| S3_vs_k141_35609\|\|full | Low-quality |
| S3_vs_k141_43274\|\|full | Low-quality |
| S3_vs_k141_114816\|\|full | Low-quality |
| S3_vs_k141_35620\|\|full | Not-determined |
| S3_vs_k141_12495\|\|full | Low-quality |
| S3_vs_k141_3232\|\|full | Low-quality |
| S3_vs_k141_18559\|\|full | Not-determined |
| S3_vs_k141_122343\|\|full | Not-determined |
| S3_vs_k141_43311\|\|full | Low-quality |
| S3_vs_k141_107099\|\|full | Low-quality |
| S3_vs_k141_90977\|\|full | Low-quality |
| S3_vs_k141_59483\|\|full | Not-determined |
| S3_vs_k141_98955\|\|full | Low-quality |
| S3_vs_k141_98963\|\|full | Low-quality |
| S3_vs_k141_35670\|\|full | Low-quality |
| S3_vs_k141_51642\|\|full | Low-quality |
| S3_vs_k141_114877\|\|full | Low-quality |
| S3_vs_k141_114884\|\|full | Low-quality |
| S3_vs_k141_114891\|\|full | Not-determined |
| S3_vs_k141_98974\|\|full | Low-quality |
| S3_vs_k141_98980\|\|full | Not-determined |
| S3_vs_k141_114899\|\|full | Not-determined |
| S3_vs_k141_18587\|\|full | Not-determined |
| S3_vs_k141_122396\|\|full | Low-quality |
| S3_vs_k141_122399\|\|full | Low-quality |
| S3_vs_k141_27305\|\|full | Low-quality |
| S3_vs_k141_35683\|\|full | Not-determined |
| S3_vs_k141_12560\|\|full | Low-quality |
| S3_vs_k141_51685\|\|full | Low-quality |
| S3_vs_k141_82994\|\|full | Low-quality |
| S3_vs_k141_107162\|\|full | Not-determined |
| S3_vs_k141_114930\|\|full | Low-quality |
| S3_vs_k141_114935\|\|full | Not-determined |
| S3_vs_k141_59536\|\|full | Low-quality |
| S3_vs_k141_18617\|\|full | Low-quality |
| S3_vs_k141_91068\|\|full | Low-quality |
| S3_vs_k141_75870\|\|full | Low-quality |
| S3_vs_k141_107181\|\|full | Low-quality |
| S3_vs_k141_83023\|\|full | Low-quality |
| S3_vs_k141_114978\|\|full | Low-quality |
| S3_vs_k141_75882\|\|full | Not-determined |
| S3_vs_k141_75883\|\|full | Not-determined |
| S3_vs_k141_12593\|\|full | Low-quality |
| S3_vs_k141_51696\|\|full | Not-determined |
| S3_vs_k141_114991\|\|full | Low-quality |
| S3_vs_k141_18669\|\|full | Not-determined |
| S3_vs_k141_83056\|\|full | Not-determined |
| S3_vs_k141_107200\|\|full | Low-quality |
| S3_vs_k141_83057\|\|full | Low-quality |
| S3_vs_k141_91099\|\|full | Low-quality |
| S3_vs_k141_122480\|\|full | Low-quality |
| S3_vs_k141_122484\|\|full | Low-quality |
| S3_vs_k141_91100\|\|full | Low-quality |
| S3_vs_k141_51707\|\|full | Not-determined |
| S3_vs_k141_18675\|\|full | Low-quality |
| S3_vs_k141_3286\|\|full | Low-quality |
| S6_extracted_k141_45618 | Complete |
| S3_vs_k141_3292\|\|full | Low-quality |
| S3_vs_k141_107229\|\|full | Not-determined |
| S3_vs_k141_12652\|\|full | Low-quality |
| S3_vs_k141_66629\|\|full | Low-quality |
| S3_vs_k141_3306\|\|full | Low-quality |
| S3_vs_k141_35725\|\|full | Low-quality |
| S3_vs_k141_12673\|\|full | Low-quality |
| S3_vs_k141_83102\|\|full | Low-quality |
| S3_vs_k141_12674\|\|full | Not-determined |
| S3_vs_k141_107235\|\|full | Not-determined |
| S3_vs_k141_66659\|\|full | Not-determined |
| S3_vs_k141_83117\|\|full | Low-quality |
| S3_vs_k141_107246\|\|full | Low-quality |
| S3_vs_k141_18684\|\|full | Not-determined |
| S3_vs_k141_12684\|\|full | Low-quality |
| S3_vs_k141_43403\|\|full | Low-quality |
| S3_vs_k141_27403\|\|full | Not-determined |
| S3_vs_k141_27410\|\|full | Low-quality |
| S3_vs_k141_122562\|\|full | Low-quality |
| S3_vs_k141_12717\|\|full | Not-determined |
| S3_vs_k141_59594\|\|full | Low-quality |
| S3_vs_k141_83149\|\|full | Not-determined |
| S3_vs_k141_122575\|\|full | Not-determined |
| S3_vs_k141_51758\|\|full | Low-quality |
| S3_vs_k141_83153\|\|full | Low-quality |
| S3_vs_k141_115042\|\|full | Low-quality |
| S3_vs_k141_83160\|\|full | Low-quality |
| S3_vs_k141_115045\|\|full | Low-quality |
| S3_vs_k141_51778\|\|full | Low-quality |
| S3_vs_k141_75916\|\|full | Low-quality |
| S3_vs_k141_107293\|\|full | Low-quality |
| S3_vs_k141_115056\|\|full | Not-determined |
| S3_vs_k141_83175\|\|full | Low-quality |
| S3_vs_k141_107294\|\|full | Not-determined |
| S3_vs_k141_91146\|\|full | Low-quality |
| S3_vs_k141_51783\|\|full | Not-determined |
| S3_vs_k141_66720\|\|full | Low-quality |
| S12_vs_k141_79785\|\|full | High-quality |
| S3_vs_k141_91152\|\|full | Low-quality |
| S3_vs_k141_91154\|\|full | Low-quality |
| S3_vs_k141_51793\|\|full | Low-quality |
| S3_vs_k141_35781\|\|full | Low-quality |
| S3_vs_k141_3348\|\|full | Low-quality |
| S3_vs_k141_83202\|\|full | Low-quality |
| S3_vs_k141_91179\|\|full | Low-quality |
| S3_vs_k141_107311\|\|full | Not-determined |
| S3_vs_k141_51808\|\|full | Not-determined |
| S3_vs_k141_27488\|\|full | Low-quality |
| S3_vs_k141_75949\|\|full | Low-quality |
| S3_vs_k141_51814\|\|full | Low-quality |
| S3_vs_k141_91188\|\|full | Low-quality |
| S3_vs_k141_51822\|\|full | Low-quality |
| S3_vs_k141_83254\|\|full | Not-determined |
| S3_vs_k141_59659\|\|full | Low-quality |
| S3_vs_k141_107330\|\|full | Not-determined |
| S16_vs_137222\|\|full | High-quality |
| S3_vs_k141_35785\|\|full | Low-quality |
| S3_vs_k141_83278\|\|full | Low-quality |
| S3_vs_k141_43509\|\|full | Low-quality |
| S3_vs_k141_107363\|\|full | Low-quality |
| S3_vs_k141_91224\|\|full | Not-determined |
| S3_vs_k141_83288\|\|full | Low-quality |
| S3_vs_k141_75959\|\|full | Low-quality |
| S3_vs_k141_12754\|\|full | Not-determined |
| S3_vs_k141_107372\|\|full | Not-determined |
| S3_vs_k141_75960\|\|full | Low-quality |
| S3_vs_k141_107380\|\|full | Not-determined |
| S3_vs_k141_43522\|\|full | Low-quality |
| S3_vs_k141_122618\|\|full | Not-determined |
| S3_vs_k141_99164\|\|full | Not-determined |
| S3_vs_k141_66732\|\|full | Not-determined |
| S3_vs_k141_83303\|\|full | Low-quality |
| S3_vs_k141_122631\|\|full | Low-quality |
| S3_vs_k141_59728\|\|full | Low-quality |
| S3_vs_k141_91242\|\|full | Low-quality |
| S3_vs_k141_122639\|\|full | Low-quality |
| S3_vs_k141_122641\|\|full | Low-quality |
| S3_vs_k141_51863\|\|full | Low-quality |
| S3_vs_k141_3404\|\|full | Low-quality |
| S3_vs_k141_43548\|\|full | Not-determined |
| S3_vs_k141_91263\|\|full | Low-quality |
| S3_vs_k141_91268\|\|full | Not-determined |
| S3_vs_k141_66752\|\|full | Not-determined |
| S3_vs_k141_75979\|\|full | Not-determined |
| S3_vs_k141_91277\|\|full | Low-quality |
| S3_vs_k141_3413\|\|full | Low-quality |
| S3_vs_k141_43562\|\|full | Not-determined |
| S3_vs_k141_59766\|\|full | Low-quality |
| S3_vs_k141_66761\|\|full | Low-quality |
| S3_vs_k141_59803\|\|full | Low-quality |
| S3_vs_k141_27564\|\|full | Low-quality |
| S3_vs_k141_75992\|\|full | Low-quality |
| S3_vs_k141_3428\|\|full | Not-determined |
| S3_vs_k141_59807\|\|full | Not-determined |
| S3_vs_k141_27570\|\|full | Low-quality |
| S3_vs_k141_51886\|\|full | Low-quality |
| S3_vs_k141_27572\|\|full | Low-quality |
| S3_vs_k141_12801\|\|full | Not-determined |
| S3_vs_k141_51889\|\|full | Low-quality |
| S3_vs_k141_27577\|\|full | Not-determined |
| S3_vs_k141_91324\|\|full | Low-quality |
| S3_vs_k141_122699\|\|full | Not-determined |
| S3_vs_k141_122701\|\|full | Low-quality |
| S3_vs_k141_27581\|\|full | Not-determined |
| S3_vs_k141_3433\|\|full | Low-quality |
| S3_vs_k141_27588\|\|full | Low-quality |
| S3_vs_k141_115139\|\|full | Not-determined |
| S3_vs_k141_115141\|\|full | Low-quality |
| S3_vs_k141_12817\|\|full | Not-determined |
| S3_vs_k141_3445\|\|full | Low-quality |
| S3_vs_k141_12819\|\|full | Low-quality |
| S3_vs_k141_3448\|\|full | Low-quality |
| S3_vs_k141_76013\|\|full | Low-quality |
| S3_vs_k141_115155\|\|full | Not-determined |
| S3_vs_k141_15450\|\|full | High-quality |
| S3_vs_k141_35828\|\|full | Low-quality |
| S8_vs_k141_208114\|\|full | High-quality |
| S3_vs_k141_91360\|\|full | Low-quality |
| S3_vs_k141_66789\|\|full | Low-quality |
| S3_vs_k141_83376\|\|full | Low-quality |
| S3_vs_k141_66810\|\|full | Low-quality |
| S3_vs_k141_3500\|\|full | Low-quality |
| S3_vs_k141_3509\|\|full | Not-determined |
| S3_vs_k141_115202\|\|full | Low-quality |
| S3_vs_k141_43584\|\|full | Low-quality |
| S3_vs_k141_91373\|\|full | Low-quality |
| S3_vs_k141_51989\|\|full | Not-determined |
| S3_vs_k141_51994\|\|full | Not-determined |
| S3_vs_k141_91390\|\|full | Low-quality |
| S3_vs_k141_115217\|\|full | Not-determined |
| S3_vs_k141_115220\|\|full | Low-quality |
| S3_vs_k141_122730\|\|full | Low-quality |
| S3_vs_k141_52010\|\|full | Low-quality |
| S3_vs_k141_115229\|\|full | Not-determined |
| S3_vs_k141_18770\|\|full | Low-quality |
| S3_vs_k141_3519\|\|full | Low-quality |
| S3_vs_k141_18771\|\|full | Low-quality |
| S3_vs_k141_18772\|\|full | Not-determined |
| S3_vs_k141_18773\|\|full | Not-determined |
| S3_vs_k141_66868\|\|full | Not-determined |
| S3_vs_k141_66876\|\|full | Low-quality |
| S3_vs_k141_27610\|\|full | Not-determined |
| S3_vs_k141_27612\|\|full | Not-determined |
| S3_vs_k141_83439\|\|full | Low-quality |
| S3_vs_k141_76101\|\|full | Not-determined |
| S3_vs_k141_18821\|\|full | Low-quality |
| S3_vs_k141_52070\|\|full | Low-quality |
| S3_vs_k141_27624\|\|full | Low-quality |
| S3_vs_k141_3563\|\|full | Low-quality |
| S3_vs_k141_52075\|\|full | Low-quality |
| S3_vs_k141_122779\|\|full | Low-quality |
| S3_vs_k141_83457\|\|full | Low-quality |
| S3_vs_k141_52088\|\|full | Low-quality |
| S3_vs_k141_35908\|\|full | Low-quality |
| S3_vs_k141_76108\|\|full | Low-quality |
| S3_vs_k141_66883\|\|full | Not-determined |
| S3_vs_k141_43654\|\|full | Low-quality |
| S3_vs_k141_35918\|\|full | Low-quality |
| S3_vs_k141_59900\|\|full | Low-quality |
| S3_vs_k141_27630\|\|full | Not-determined |
| S3_vs_k141_107387\|\|full | Low-quality |
| S3_vs_k141_43660\|\|full | Low-quality |
| S3_vs_k141_83485\|\|full | Low-quality |
| S3_vs_k141_91452\|\|full | Low-quality |
| S3_vs_k141_43671\|\|full | Not-determined |
| S3_vs_k141_43679\|\|full | Low-quality |
| S3_vs_k141_59911\|\|full | Low-quality |
| S3_vs_k141_52112\|\|full | Not-determined |
| S3_vs_k141_59916\|\|full | Not-determined |
| S3_vs_k141_52117\|\|full | Low-quality |
| S3_vs_k141_59922\|\|full | Low-quality |
| S3_vs_k141_99223\|\|full | Not-determined |
| S3_vs_k141_107409\|\|full | Low-quality |
| S3_vs_k141_76136\|\|full | Low-quality |
| S3_vs_k141_43696\|\|full | Low-quality |
| S3_vs_k141_35975\|\|full | Not-determined |
| S3_vs_k141_76140\|\|full | Low-quality |
| S3_vs_k141_99229\|\|full | Low-quality |
| S3_vs_k141_43699\|\|full | Not-determined |
| S3_vs_k141_52137\|\|full | Low-quality |
| S3_vs_k141_35981\|\|full | Not-determined |
| S3_vs_k141_99237\|\|full | Low-quality |
| S3_vs_k141_59926\|\|full | Low-quality |
| S3_vs_k141_52157\|\|full | Low-quality |
| S3_vs_k141_76149\|\|full | Low-quality |
| S3_vs_k141_52167\|\|full | Low-quality |
| S3_vs_k141_76155\|\|full | Low-quality |
| S3_vs_k141_12941\|\|full | Not-determined |
| S3_vs_k141_43707\|\|full | Not-determined |
| S3_vs_k141_83536\|\|full | Low-quality |
| S3_vs_k141_27660\|\|full | Low-quality |
| S3_vs_k141_43721\|\|full | Not-determined |
| S3_vs_k141_91500\|\|full | Not-determined |
| S3_vs_k141_27672\|\|full | Not-determined |
| S3_vs_k141_27676\|\|full | Low-quality |
| S3_vs_k141_18834\|\|full | Not-determined |
| S3_vs_k141_27678\|\|full | Not-determined |
| S5_vs_k141_850\|\|full | High-quality |
| S3_vs_k141_59969\|\|full | Low-quality |
| S3_vs_k141_52206\|\|full | Low-quality |
| S3_vs_k141_27693\|\|full | Low-quality |
| S3_vs_k141_59977\|\|full | Not-determined |
| S3_vs_k141_27704\|\|full | Not-determined |
| S3_vs_k141_107438\|\|full | Low-quality |
| S3_vs_k141_122823\|\|full | Not-determined |
| S3_vs_k141_122827\|\|full | Low-quality |
| S3_vs_k141_36091\|\|full | Low-quality |
| S3_vs_k141_91556\|\|full | Not-determined |
| S3_vs_k141_27721\|\|full | Low-quality |
| S3_vs_k141_83571\|\|full | Not-determined |
| S3_vs_k141_66956\|\|full | Low-quality |
| S3_vs_k141_43751\|\|full | Not-determined |
| S3_vs_k141_66962\|\|full | Not-determined |
| S3_vs_k141_18842\|\|full | Low-quality |
| S3_vs_k141_60011\|\|full | Low-quality |
| S3_vs_k141_27740\|\|full | Not-determined |
| S3_vs_k141_18849\|\|full | Low-quality |
| S3_vs_k141_107474\|\|full | Not-determined |
| S3_vs_k141_27753\|\|full | Low-quality |
| S3_vs_k141_36098\|\|full | Low-quality |
| S3_vs_k141_3670\|\|full | Low-quality |
| S3_vs_k141_107492\|\|full | Not-determined |
| S3_vs_k141_115324\|\|full | Not-determined |
| S3_vs_k141_107501\|\|full | Low-quality |
| S3_vs_k141_3688\|\|full | Not-determined |
| S3_vs_k141_60068\|\|full | Low-quality |
| S3_vs_k141_115341\|\|full | Low-quality |
| S3_vs_k141_91613\|\|full | Low-quality |
| S3_vs_k141_122860\|\|full | Not-determined |
| S3_vs_k141_115346\|\|full | Low-quality |
| S6_vs_k141_45618\|\|full | High-quality |
| S3_vs_k141_60077\|\|full | Not-determined |
| S3_vs_k141_83631\|\|full | Low-quality |
| S3_vs_k141_52285\|\|full | Not-determined |
| S5_vs_k141_145600\|\|full | High-quality |
| S3_vs_k141_66984\|\|full | Low-quality |
| S3_vs_k141_52327\|\|full | Low-quality |
| S3_vs_k141_115349\|\|full | Low-quality |
| S3_vs_k141_107529\|\|full | Low-quality |
| S3_vs_k141_115376\|\|full | Low-quality |
| S3_vs_k141_36160\|\|full | Not-determined |
| S4_vs_VIRSorter_31196-circular-cat | High-quality |
| S3_vs_k141_115391\|\|full | Low-quality |
| S3_vs_k141_107548\|\|full | Not-determined |
| S3_vs_k141_107552\|\|full | Not-determined |
| S3_vs_k141_115400\|\|full | Not-determined |
| S3_vs_k141_52371\|\|full | Low-quality |
| S3_vs_k141_52373\|\|full | Not-determined |
| S3_vs_k141_99253\|\|full | Low-quality |
| S3_vs_k141_107575\|\|full | Low-quality |
| S3_vs_k141_3726\|\|full | Low-quality |
| S3_vs_k141_60146\|\|full | Not-determined |
| S3_vs_k141_115414\|\|full | Not-determined |
| S3_vs_k141_76268\|\|full | Not-determined |
| S3_vs_k141_76269\|\|full | Low-quality |
| S3_vs_k141_3727\|\|full | Not-determined |
| S3_vs_k141_83743\|\|full | Low-quality |
| S3_vs_k141_60165\|\|full | Low-quality |
| S3_vs_k141_66997\|\|full | Not-determined |
| S3_vs_k141_115436\|\|full | Not-determined |
| S3_vs_k141_60166\|\|full | Low-quality |
| S3_vs_k141_76276\|\|full | Low-quality |
| S3_vs_k141_52415\|\|full | Not-determined |
| S3_vs_k141_3748\|\|full | Low-quality |
| S3_vs_k141_122869\|\|full | Not-determined |
| S3_vs_k141_52422\|\|full | Low-quality |
| S3_vs_k141_107595\|\|full | Not-determined |
| S3_vs_k141_27855\|\|full | Low-quality |
| S3_vs_k141_91683\|\|full | Not-determined |
| S3_vs_k141_18959\|\|full | Low-quality |
| S3_vs_k141_52438\|\|full | Low-quality |
| S3_vs_k141_43795\|\|full | Low-quality |
| S3_vs_k141_43797\|\|full | Not-determined |
| S3_vs_k141_13143\|\|full | Low-quality |
| S3_vs_k141_52447\|\|full | Low-quality |
| S3_vs_k141_27872\|\|full | Low-quality |
| S3_vs_k141_13147\|\|full | Not-determined |
| S3_vs_k141_76309\|\|full | Low-quality |
| S3_vs_k141_43810\|\|full | Low-quality |
| S3_vs_k141_67016\|\|full | Not-determined |
| S3_vs_k141_67017\|\|full | Low-quality |
| S3_vs_k141_115452\|\|full | Not-determined |
| S3_vs_k141_76322\|\|full | Not-determined |
| S3_vs_k141_91685\|\|full | Low-quality |
| S3_vs_k141_107631\|\|full | Not-determined |
| S3_vs_k141_91690\|\|full | Not-determined |
| S3_vs_k141_13157\|\|full | Not-determined |
| S3_vs_k141_3792\|\|full | Not-determined |
| S3_vs_k141_91692\|\|full | Low-quality |
| S3_vs_k141_122910\|\|full | Low-quality |
| S3_vs_k141_60177\|\|full | Low-quality |
| S3_vs_k141_60180\|\|full | Low-quality |
| S3_vs_k141_52454\|\|full | Not-determined |
| S3_vs_k141_67026\|\|full | Low-quality |
| S3_vs_k141_76347\|\|full | Not-determined |
| S3_vs_k141_52461\|\|full | Low-quality |
| S3_vs_k141_115485\|\|full | Low-quality |
| S3_vs_k141_107653\|\|full | Not-determined |
| S3_vs_k141_3823\|\|full | Low-quality |
| S3_vs_k141_43861\|\|full | Not-determined |
| S3_vs_k141_107661\|\|full | Low-quality |
| S3_vs_k141_3830\|\|full | Low-quality |
| S3_vs_k141_76372\|\|full | Low-quality |
| S3_vs_k141_91699\|\|full | Not-determined |
| S3_vs_k141_60206\|\|full | Low-quality |
| S3_vs_k141_60208\|\|full | Low-quality |
| S3_vs_k141_13172\|\|full | Low-quality |
| S3_vs_k141_3841\|\|full | Not-determined |
| S3_vs_k141_115500\|\|full | Not-determined |
| S3_vs_k141_43898\|\|full | Low-quality |
| S3_vs_k141_107678\|\|full | Not-determined |
| S3_vs_k141_76398\|\|full | Not-determined |
| S3_vs_k141_91731\|\|full | Not-determined |
| S3_vs_k141_115502\|\|full | Low-quality |
| S3_vs_k141_52484\|\|full | Low-quality |
| S3_vs_k141_19017\|\|full | Low-quality |
| S3_vs_k141_122920\|\|full | Low-quality |
| S3_vs_k141_67063\|\|full | Low-quality |
| S3_vs_k141_52490\|\|full | Low-quality |
| S3_vs_k141_36226\|\|full | Low-quality |
| S3_vs_k141_76411\|\|full | Not-determined |
| S3_vs_k141_122935\|\|full | Low-quality |
| S3_vs_k141_60258\|\|full | Low-quality |
| S3_vs_k141_76429\|\|full | Low-quality |
| S3_vs_k141_91746\|\|full | Low-quality |
| S3_vs_k141_67088\|\|full | Low-quality |
| S3_vs_k141_3899\|\|full | Low-quality |
| S3_vs_k141_43907\|\|full | Low-quality |
| S3_vs_k141_91759\|\|full | Low-quality |
| S3_vs_k141_91764\|\|full | Low-quality |
| S3_vs_k141_3913\|\|full | Low-quality |
| S3_vs_k141_99390\|\|full | Low-quality |
| S3_vs_k141_122973\|\|full | Low-quality |
| S3_vs_k141_76455\|\|full | Not-determined |
| S3_vs_k141_3921\|\|full | Not-determined |
| S3_vs_k141_122982\|\|full | Low-quality |
| S3_vs_k141_91782\|\|full | Not-determined |
| S3_vs_k141_19074\|\|full | Low-quality |
| S3_vs_k141_107719\|\|full | Low-quality |
| S3_vs_k141_91788\|\|full | Not-determined |
| S3_vs_k141_91791\|\|full | Low-quality |
| S3_vs_k141_67145\|\|full | Low-quality |
| S3_vs_k141_76489\|\|full | Low-quality |
| S3_vs_k141_3966\|\|full | Not-determined |
| S3_vs_k141_91808\|\|full | Low-quality |
| S3_vs_k141_115541\|\|full | Low-quality |
| S4_extracted_7318 | High-quality |
| S3_vs_k141_91811\|\|full | Not-determined |
| S3_vs_k141_91814\|\|full | Low-quality |
| S3_vs_k141_19133\|\|full | Not-determined |
| S3_vs_k141_3969\|\|full | Low-quality |
| S3_vs_k141_36251\|\|full | Not-determined |
| S3_vs_k141_107758\|\|full | Low-quality |
| S3_vs_k141_13277\|\|full | Low-quality |
| S3_vs_k141_123017\|\|full | Low-quality |
| S3_vs_k141_115556\|\|full | Low-quality |
| S3_vs_k141_115557\|\|full | Low-quality |
| S3_vs_k141_27954\|\|full | Not-determined |
| S3_vs_k141_107759\|\|full | Not-determined |
| S3_vs_k141_60343\|\|full | Low-quality |
| S3_vs_k141_76518\|\|full | Low-quality |
| S3_vs_k141_43971\|\|full | Low-quality |
| S3_vs_k141_27957\|\|full | Low-quality |
| S3_vs_k141_76527\|\|full | Low-quality |
| S3_vs_k141_91859\|\|full | Low-quality |
| S3_vs_k141_36258\|\|full | Low-quality |
| S3_vs_k141_13327\|\|full | Low-quality |
| S3_vs_k141_115571\|\|full | Not-determined |
| S3_vs_k141_19180\|\|full | Low-quality |
| S3_vs_k141_36272\|\|full | Not-determined |
| S3_vs_k141_91868\|\|full | Not-determined |
| S3_vs_k141_36282\|\|full | Low-quality |
| S3_vs_k141_3992\|\|full | Low-quality |
| S3_vs_k141_52519\|\|full | Low-quality |
| S3_vs_k141_115585\|\|full | Not-determined |
| S3_vs_k141_13350\|\|full | Low-quality |
| S3_vs_k141_36300\|\|full | Not-determined |
| S3_vs_k141_107799\|\|full | Not-determined |
| S3_vs_k141_115593\|\|full | Low-quality |
| S3_vs_k141_76544\|\|full | Not-determined |
| S3_vs_k141_91872\|\|full | Low-quality |
| S3_vs_k141_115626\|\|full | Low-quality |
| S3_vs_k141_4026\|\|full | Not-determined |
| S3_vs_k141_107801\|\|full | Low-quality |
| S3_vs_k141_27978\|\|full | Low-quality |
| S3_vs_k141_123051\|\|full | Low-quality |
| S3_vs_k141_99445\|\|full | Low-quality |
| S3_vs_k141_115637\|\|full | Low-quality |
| S3_vs_k141_123058\|\|full | Not-determined |
| S3_vs_k141_27984\|\|full | Not-determined |
| S3_vs_k141_44006\|\|full | Low-quality |
| S3_vs_k141_60423\|\|full | Low-quality |
| S3_vs_k141_36331\|\|full | Low-quality |
| S3_vs_k141_91885\|\|full | Not-determined |
| S3_vs_k141_123078\|\|full | Not-determined |
| S3_vs_k141_123085\|\|full | Low-quality |
| S3_vs_k141_52572\|\|full | Low-quality |
| S3_vs_k141_13419\|\|full | Not-determined |
| S3_vs_k141_44010\|\|full | Low-quality |
| S3_vs_k141_76549\|\|full | Not-determined |
| S3_vs_k141_13436\|\|full | Not-determined |
| S3_vs_k141_4050\|\|full | Low-quality |
| S3_vs_k141_91903\|\|full | Low-quality |
| S3_vs_k141_123094\|\|full | Low-quality |
| S3_vs_k141_76562\|\|full | Not-determined |
| S3_vs_k141_36351\|\|full | Low-quality |
| S3_vs_k141_67282\|\|full | Low-quality |
| S3_vs_k141_44031\|\|full | Not-determined |
| S3_vs_k141_67295\|\|full | Not-determined |
| S3_vs_k141_52602\|\|full | Not-determined |
| S3_vs_k141_52607\|\|full | Not-determined |
| S3_vs_k141_76574\|\|full | Low-quality |
| S3_vs_k141_36390\|\|full | Low-quality |
| S3_vs_k141_123138\|\|full | Low-quality |
| S3_vs_k141_36391\|\|full | Low-quality |
| S3_vs_k141_76586\|\|full | Low-quality |
| S3_vs_k141_13489\|\|full | Low-quality |
| S3_vs_k141_107857\|\|full | Not-determined |
| S3_vs_k141_52619\|\|full | Low-quality |
| S3_vs_k141_123161\|\|full | Not-determined |
| S3_vs_k141_115700\|\|full | Low-quality |
| S3_vs_k141_19282\|\|full | Low-quality |
| S3_vs_k141_52632\|\|full | Not-determined |
| S3_vs_k141_60496\|\|full | Low-quality |
| S3_vs_k141_13519\|\|full | Low-quality |
| S3_vs_k141_107865\|\|full | Low-quality |
| S3_vs_k141_13540\|\|full | Low-quality |
| S3_vs_k141_4095\|\|full | Low-quality |
| S3_vs_k141_76615\|\|full | Low-quality |
| S3_vs_k141_76625\|\|full | Not-determined |
| S3_vs_k141_107879\|\|full | Low-quality |
| S3_vs_k141_76629\|\|full | Low-quality |
| S4_vs_VIRSorter_7318-cat_2 | High-quality |
| S3_vs_k141_115708\|\|full | Not-determined |
| S3_vs_k141_83785\|\|full | Not-determined |
| S3_vs_k141_91912\|\|full | Not-determined |
| S3_vs_k141_83786\|\|full | Low-quality |
| S3_vs_k141_60563\|\|full | Low-quality |
| S3_vs_k141_52674\|\|full | Not-determined |
| S3_vs_k141_52678\|\|full | Low-quality |
| S3_vs_k141_52680\|\|full | Low-quality |
| S3_vs_k141_91927\|\|full | Low-quality |
| S3_vs_k141_83791\|\|full | Not-determined |
| S3_vs_k141_99563\|\|full | Not-determined |
| S3_vs_k141_91932\|\|full | Not-determined |
| S3_vs_k141_60579\|\|full | Not-determined |
| S3_vs_k141_123252\|\|full | Low-quality |
| S3_vs_k141_99568\|\|full | Not-determined |
| S3_vs_k141_67344\|\|full | Not-determined |
| S3_vs_k141_91933\|\|full | Not-determined |
| S3_vs_k141_76678\|\|full | Not-determined |
| S3_vs_k141_67367\|\|full | Not-determined |
| S3_vs_k141_60616\|\|full | Low-quality |
| S3_vs_k141_123263\|\|full | Not-determined |
| S3_vs_k141_60629\|\|full | Low-quality |
| S3_vs_k141_36418\|\|full | Low-quality |
| S3_vs_k141_123272\|\|full | Not-determined |
| S3_vs_k141_99577\|\|full | Low-quality |
| S3_vs_k141_99580\|\|full | Low-quality |
| S3_vs_k141_13601\|\|full | Low-quality |
| S3_vs_k141_13602\|\|full | Low-quality |
| S3_vs_k141_44113\|\|full | Low-quality |
| S3_vs_k141_36425\|\|full | Not-determined |
| S3_vs_k141_99583\|\|full | Low-quality |
| S3_vs_k141_13603\|\|full | Low-quality |
| S3_vs_k141_52743\|\|full | Low-quality |
| S3_vs_k141_99584\|\|full | Low-quality |
| S3_vs_k141_60652\|\|full | Not-determined |
| S3_vs_k141_36439\|\|full | Low-quality |
| S3_vs_k141_13616\|\|full | Low-quality |
| S3_vs_k141_123307\|\|full | Low-quality |
| S3_vs_k141_76719\|\|full | Low-quality |
| S3_vs_k141_107948\|\|full | Low-quality |
| S3_vs_k141_28089\|\|full | Low-quality |
| S3_vs_k141_123320\|\|full | Low-quality |
| S3_vs_k141_52772\|\|full | Not-determined |
| S3_vs_k141_28094\|\|full | Not-determined |
| S3_vs_k141_91936\|\|full | Not-determined |
| S3_vs_k141_19344\|\|full | Low-quality |
| S3_vs_k141_44122\|\|full | Low-quality |
| S3_vs_k141_83835\|\|full | Low-quality |
| S3_vs_k141_4161\|\|full | Low-quality |
| S3_vs_k141_44127\|\|full | Not-determined |
| S3_vs_k141_76750\|\|full | Low-quality |
| S3_vs_k141_83838\|\|full | Low-quality |
| S3_vs_k141_76755\|\|full | Low-quality |
| S3_vs_k141_115809\|\|full | Not-determined |
| S3_vs_k141_4178\|\|full | Low-quality |
| S3_vs_k141_28143\|\|full | Low-quality |
| S3_vs_k141_107961\|\|full | Low-quality |
| S3_vs_k141_13646\|\|full | Not-determined |
| S3_vs_k141_115828\|\|full | Low-quality |
| S3_vs_k141_4195\|\|full | Not-determined |
| S3_vs_k141_28149\|\|full | Low-quality |
| S3_vs_k141_4203\|\|full | Low-quality |
| S3_vs_k141_52809\|\|full | Not-determined |
| S9_vs_k141_90859\|\|full | High-quality |
| S3_vs_k141_123390\|\|full | Low-quality |
| S3_vs_k141_115847\|\|full | Low-quality |
| S3_vs_k141_91973\|\|full | Low-quality |
| S3_vs_k141_91974\|\|full | Low-quality |
| S3_vs_k141_115853\|\|full | Not-determined |
| S3_vs_k141_52848\|\|full | Low-quality |

| | |
|---|---|
| S3_vs_k141_83883\|\|full | Low-quality |
| S3_vs_k141_91983\|\|full | Not-determined |
| S3_vs_k141_13666\|\|full | Low-quality |
| S3_vs_k141_91986\|\|full | Not-determined |
| S3_vs_k141_76769\|\|full | Not-determined |
| S3_vs_k141_91987\|\|full | Not-determined |
| S3_vs_k141_52857\|\|full | Not-determined |
| S3_vs_k141_99632\|\|full | Low-quality |
| S3_vs_k141_76777\|\|full | Low-quality |
| S3_vs_k141_67437\|\|full | Low-quality |
| S3_vs_k141_13677\|\|full | Low-quality |
| S3_vs_k141_76793\|\|full | Low-quality |
| S3_vs_k141_52880\|\|full | Not-determined |
| S3_vs_k141_76800\|\|full | Low-quality |
| S3_vs_k141_28165\|\|full | Not-determined |
| S3_vs_k141_60732\|\|full | Low-quality |
| S3_vs_k141_44226\|\|full | Not-determined |
| S3_vs_k141_67472\|\|full | Low-quality |
| S3_vs_k141_92000\|\|full | Low-quality |
| S3_vs_k141_52923\|\|full | Not-determined |
| S3_vs_k141_92008\|\|full | Not-determined |
| S3_vs_k141_36487\|\|full | Not-determined |
| S3_vs_k141_115932\|\|full | Not-determined |
| S3_vs_k141_123481\|\|full | Low-quality |
| S3_vs_k141_60749\|\|full | Low-quality |
| S3_vs_k141_123490\|\|full | Low-quality |
| S3_vs_k141_99638\|\|full | Low-quality |
| S3_vs_k141_28197\|\|full | Low-quality |
| S3_vs_k141_115941\|\|full | Low-quality |
| S3_vs_k141_83921\|\|full | Low-quality |
| S3_vs_k141_67521\|\|full | Low-quality |
| S3_vs_k141_19433\|\|full | Low-quality |
| S5_vs_k141_38949\|\|full | High-quality |
| S3_vs_k141_92014\|\|full | Low-quality |
| S3_vs_k141_67522\|\|full | Not-determined |
| S3_vs_k141_52940\|\|full | Low-quality |
| S3_vs_k141_19435\|\|full | Low-quality |
| S3_vs_k141_76836\|\|full | Low-quality |
| S3_vs_k141_123509\|\|full | Not-determined |
| S3_vs_k141_60767\|\|full | Low-quality |
| S3_vs_k141_76849\|\|full | Low-quality |
| S3_vs_k141_115965\|\|full | Low-quality |
| S3_vs_k141_76863\|\|full | Not-determined |
| S3_vs_k141_19454\|\|full | Not-determined |
| S3_vs_k141_108062\|\|full | Not-determined |
| S3_vs_k141_4237\|\|full | Not-determined |
| S3_vs_k141_36513\|\|full | Low-quality |
| S3_vs_k141_76871\|\|full | Low-quality |
| S3_vs_k141_99667\|\|full | Low-quality |
| S3_vs_k141_28216\|\|full | Low-quality |
| S3_vs_k141_28219\|\|full | Low-quality |
| S3_vs_k141_36521\|\|full | Low-quality |
| S3_vs_k141_19485\|\|full | Low-quality |
| S3_vs_k141_28225\|\|full | Low-quality |
| S3_vs_k141_99678\|\|full | Low-quality |
| S3_vs_k141_36538\|\|full | Not-determined |
| S3_vs_k141_44326\|\|full | Low-quality |
| S3_vs_k141_123515\|\|full | Low-quality |
| S3_vs_k141_116000\|\|full | Low-quality |
| S3_vs_k141_123520\|\|full | Not-determined |
| S3_vs_k141_92070\|\|full | Low-quality |
| S3_vs_k141_84001\|\|full | Low-quality |
| S3_vs_k141_123521\|\|full | Low-quality |
| S3_vs_k141_116009\|\|full | Not-determined |
| S3_vs_k141_123523\|\|full | Low-quality |
| S3_vs_k141_123527\|\|full | Low-quality |
| S3_vs_k141_60818\|\|full | Low-quality |
| S3_vs_k141_4281\|\|full | Low-quality |
| S3_vs_k141_76931\|\|full | Low-quality |
| S3_vs_k141_108086\|\|full | Not-determined |
| S3_vs_k141_19519\|\|full | Low-quality |
| S3_vs_k141_52969\|\|full | Low-quality |
| S3_vs_k141_99698\|\|full | Low-quality |
| S3_vs_k141_76950\|\|full | Not-determined |
| S3_vs_k141_84009\|\|full | Low-quality |
| S3_vs_k141_52985\|\|full | Low-quality |
| S3_vs_k141_76971\|\|full | Low-quality |
| S3_vs_k141_44372\|\|full | Not-determined |
| S3_vs_k141_28287\|\|full | Low-quality |
| S3_vs_k141_84017\|\|full | Not-determined |
| S3_vs_k141_92081\|\|full | Low-quality |
| S3_vs_k141_92082\|\|full | Low-quality |
| S3_vs_k141_67582\|\|full | Not-determined |
| S3_vs_k141_44397\|\|full | Not-determined |
| S3_vs_k141_84030\|\|full | Low-quality |
| S3_vs_k141_84035\|\|full | Not-determined |
| S3_vs_k141_60842\|\|full | Low-quality |
| S3_vs_k141_36590\|\|full | Not-determined |
| S3_vs_k141_67596\|\|full | Low-quality |
| S3_vs_k141_36595\|\|full | Low-quality |
| S3_vs_k141_53025\|\|full | Low-quality |
| S3_vs_k141_36596\|\|full | Not-determined |
| S3_vs_k141_53027\|\|full | Low-quality |
| S3_vs_k141_19569\|\|full | Low-quality |
| S3_vs_k141_44402\|\|full | Not-determined |
| S3_vs_k141_19571\|\|full | Low-quality |
| S3_vs_k141_116082\|\|full | Low-quality |
| S3_vs_k141_76980\|\|full | Low-quality |
| S3_vs_k141_36613\|\|full | Low-quality |
| S3_vs_k141_19585\|\|full | Not-determined |
| S3_vs_k141_67621\|\|full | Low-quality |
| S3_vs_k141_92112\|\|full | Low-quality |
| S3_vs_k141_116103\|\|full | Not-determined |
| S3_vs_k141_67622\|\|full | Low-quality |
| S3_vs_k141_84079\|\|full | Low-quality |
| S3_vs_k141_19591\|\|full | Low-quality |
| S3_vs_k141_19596\|\|full | Low-quality |
| S3_vs_k141_116125\|\|full | Low-quality |
| S3_vs_k141_28333\|\|full | Not-determined |
| S3_vs_k141_19610\|\|full | Not-determined |
| S3_vs_k141_4313\|\|full | Not-determined |
| S3_vs_k141_116147\|\|full | Low-quality |
| S3_vs_k141_19621\|\|full | Low-quality |
| S16_vs_173137\|\|full | High-quality |
| S3_vs_k141_67643\|\|full | Low-quality |
| S3_vs_k141_19627\|\|full | Low-quality |
| S3_vs_k141_67645\|\|full | Low-quality |
| S3_vs_k141_99710\|\|full | Low-quality |
| S3_vs_k141_84137\|\|full | Low-quality |
| S3_vs_k141_92168\|\|full | Low-quality |
| S3_vs_k141_19642\|\|full | Low-quality |
| S3_vs_k141_53108\|\|full | Not-determined |
| S3_vs_k141_92172\|\|full | Low-quality |
| S3_vs_k141_4346\|\|full | Low-quality |
| S3_vs_k141_99719\|\|full | Low-quality |
| S3_vs_k141_67657\|\|full | Low-quality |
| S3_vs_k141_116184\|\|full | Low-quality |
| S3_vs_k141_36654\|\|full | Low-quality |
| S3_vs_k141_28381\|\|full | Low-quality |
| S3_vs_k141_4364\|\|full | Low-quality |
| S3_vs_k141_116201\|\|full | Low-quality |
| S3_vs_k141_77058\|\|full | Not-determined |
| S3_vs_k141_4366\|\|full | Low-quality |
| S3_vs_k141_19681\|\|full | Low-quality |
| S3_vs_k141_19688\|\|full | Low-quality |
| S3_vs_k141_84183\|\|full | Low-quality |
| S3_vs_k141_77076\|\|full | Low-quality |
| S3_vs_k141_108196\|\|full | Low-quality |
| S3_vs_k141_77086\|\|full | Low-quality |
| S3_vs_k141_92208\|\|full | Low-quality |
| S3_vs_k141_116238\|\|full | Not-determined |
| S3_vs_k141_84199\|\|full | Low-quality |
| S3_vs_k141_77091\|\|full | Low-quality |
| S3_vs_k141_84202\|\|full | Not-determined |
| S3_vs_k141_92217\|\|full | Low-quality |
| S3_vs_k141_92218\|\|full | Not-determined |
| S3_vs_k141_92226\|\|full | Low-quality |
| S3_vs_k141_92235\|\|full | Low-quality |
| S3_vs_k141_92236\|\|full | Not-determined |
| S5_vs_k141_81911\|\|full | High-quality |
| S3_vs_k141_67709\|\|full | Low-quality |
| S3_vs_k141_99735\|\|full | Low-quality |
| S3_vs_k141_4403\|\|full | Low-quality |
| S3_vs_k141_60942\|\|full | Low-quality |
| S3_vs_k141_108237\|\|full | Low-quality |
| S3_vs_k141_108238\|\|full | Not-determined |
| S3_vs_k141_67722\|\|full | Low-quality |
| S9_vs_k141_56354\|\|full | High-quality |
| S3_vs_k141_77163\|\|full | Not-determined |
| S3_vs_k141_67733\|\|full | Low-quality |
| S3_vs_k141_60967\|\|full | Low-quality |
| S3_vs_k141_60968\|\|full | Low-quality |
| S3_vs_k141_99796\|\|full | Low-quality |
| S3_vs_k141_14173\|\|full | Not-determined |
| S3_vs_k141_116288\|\|full | Not-determined |
| S3_vs_k141_99797\|\|full | Low-quality |
| S3_vs_k141_108261\|\|full | Low-quality |
| S3_vs_k141_92315\|\|full | Low-quality |
| S3_vs_k141_123664\|\|full | Low-quality |
| S3_vs_k141_92317\|\|full | Not-determined |
| S3_vs_k141_116303\|\|full | Low-quality |
| S3_vs_k141_77187\|\|full | Low-quality |
| S3_vs_k141_28434\|\|full | Low-quality |
| S3_vs_k141_61030\|\|full | Low-quality |
| S3_vs_k141_77222\|\|full | Low-quality |
| S3_vs_k141_92367\|\|full | Low-quality |
| S3_vs_k141_14191\|\|full | Low-quality |
| S3_vs_k141_123692\|\|full | Low-quality |
| S3_vs_k141_61060\|\|full | Not-determined |
| S3_vs_k141_123696\|\|full | Not-determined |
| S3_vs_k141_92377\|\|full | Low-quality |
| S3_vs_k141_92389\|\|full | Low-quality |
| S3_vs_k141_4460\|\|full | Low-quality |
| S3_vs_k141_77264\|\|full | Not-determined |
| S3_vs_k141_84349\|\|full | Not-determined |
| S3_vs_k141_28514\|\|full | Low-quality |
| S3_vs_k141_28515\|\|full | Low-quality |
| S3_vs_k141_77284\|\|full | Low-quality |
| S3_vs_k141_67833\|\|full | Low-quality |
| S3_vs_k141_19804\|\|full | Low-quality |
| S3_vs_k141_19810\|\|full | Not-determined |
| S3_vs_k141_14263\|\|full | Low-quality |
| S3_vs_k141_108362\|\|full | Low-quality |
| S3_vs_k141_61181\|\|full | Not-determined |
| S3_vs_k141_108364\|\|full | Not-determined |
| S3_vs_k141_28575\|\|full | Low-quality |
| S3_vs_k141_61189\|\|full | Not-determined |
| S3_vs_k141_36707\|\|full | Low-quality |
| S3_vs_k141_92447\|\|full | Not-determined |
| S3_vs_k141_36710\|\|full | Low-quality |
| S3_vs_k141_28598\|\|full | Low-quality |
| S3_vs_k141_67876\|\|full | Low-quality |
| S3_vs_k141_14290\|\|full | Low-quality |
| S3_vs_k141_99870\|\|full | Low-quality |
| S3_vs_k141_19857\|\|full | Low-quality |
| S3_vs_k141_28622\|\|full | Low-quality |
| S3_vs_k141_99881\|\|full | Low-quality |
| S3_vs_k141_99883\|\|full | Not-determined |
| S3_vs_k141_19873\|\|full | Low-quality |
| S3_vs_k141_28629\|\|full | Low-quality |
| S3_vs_k141_61227\|\|full | Not-determined |
| S3_vs_k141_92464\|\|full | Low-quality |
| S3_vs_k141_99905\|\|full | Not-determined |
| S3_vs_k141_84416\|\|full | Low-quality |
| S3_vs_k141_61238\|\|full | Low-quality |
| S3_vs_k141_19896\|\|full | Not-determined |
| S3_vs_k141_14319\|\|full | Low-quality |
| S3_vs_k141_61269\|\|full | Not-determined |
| S3_vs_k141_53208\|\|full | Low-quality |
| S3_vs_k141_77315\|\|full | Not-determined |
| S3_vs_k141_92488\|\|full | Not-determined |
| S3_vs_k141_77317\|\|full | Low-quality |
| S3_vs_k141_4527\|\|full | Low-quality |
| S3_vs_k141_53225\|\|full | Low-quality |
| S3_vs_k141_19932\|\|full | Low-quality |
| S3_vs_k141_99939\|\|full | Not-determined |
| S3_vs_k141_84481\|\|full | Not-determined |
| S3_vs_k141_61297\|\|full | Not-determined |
| S3_vs_k141_44523\|\|full | Not-determined |
| S3_vs_k141_4549\|\|full | Not-determined |
| S3_vs_k141_123768\|\|full | Low-quality |
| S3_vs_k141_53239\|\|full | Low-quality |
| S3_vs_k141_116361\|\|full | Low-quality |
| S3_vs_k141_99968\|\|full | Not-determined |
| S3_vs_k141_53255\|\|full | Low-quality |
| S3_vs_k141_123770\|\|full | Low-quality |
| S3_vs_k141_19949\|\|full | Low-quality |
| S3_vs_k141_14354\|\|full | Not-determined |
| S3_vs_k141_61340\|\|full | Not-determined |
| S3_vs_k141_28663\|\|full | Low-quality |
| S3_vs_k141_36748\|\|full | Not-determined |
| S3_vs_k141_61359\|\|full | Low-quality |
| S3_vs_k141_44544\|\|full | Not-determined |
| S3_vs_k141_67904\|\|full | Low-quality |
| S3_vs_k141_123783\|\|full | Not-determined |
| S3_vs_k141_44557\|\|full | Low-quality |
| S3_vs_k141_44565\|\|full | Low-quality |
| S3_vs_k141_84522\|\|full | Low-quality |
| S3_vs_k141_108459\|\|full | Not-determined |
| S3_vs_k141_92538\|\|full | Low-quality |
| S3_vs_k141_20007\|\|full | Not-determined |
| S3_vs_k141_44585\|\|full | Low-quality |
| S3_vs_k141_116409\|\|full | Not-determined |
| S3_vs_k141_84545\|\|full | Low-quality |
| S3_vs_k141_36815\|\|full | Not-determined |
| S3_vs_k141_20024\|\|full | Low-quality |
| S3_vs_k141_116432\|\|full | Not-determined |
| S3_vs_k141_116441\|\|full | Low-quality |
| S3_vs_k141_77419\|\|full | Low-quality |
| S3_vs_k141_36826\|\|full | Not-determined |
| S3_vs_k141_77420\|\|full | Low-quality |
| S3_vs_k141_116446\|\|full | Not-determined |
| S3_vs_k141_84571\|\|full | Not-determined |
| S3_vs_k141_20036\|\|full | Not-determined |
| S3_vs_k141_67955\|\|full | Low-quality |
| S3_vs_k141_67958\|\|full | Not-determined |
| S3_vs_k141_84582\|\|full | Not-determined |
| S3_vs_k141_84587\|\|full | Low-quality |
| S3_vs_k141_61493\|\|full | Not-determined |
| S3_vs_k141_77469\|\|full | Low-quality |
| S3_vs_k141_67962\|\|full | Not-determined |
| S3_vs_k141_108481\|\|full | Low-quality |
| S3_vs_k141_14434\|\|full | Low-quality |
| S4_extracted_12614 | Medium-quality |
| S3_vs_k141_20077\|\|full | Low-quality |
| S3_vs_k141_123843\|\|full | Not-determined |
| S3_vs_k141_123845\|\|full | Low-quality |
| S3_vs_k141_123846\|\|full | Low-quality |
| S3_vs_k141_20092\|\|full | Low-quality |
| S3_vs_k141_14456\|\|full | Low-quality |
| S3_vs_k141_14465\|\|full | Not-determined |
| S3_vs_k141_84621\|\|full | Low-quality |
| S3_vs_k141_53311\|\|full | Low-quality |
| S3_vs_k141_20099\|\|full | Not-determined |
| S3_vs_k141_61523\|\|full | Not-determined |
| S3_vs_k141_20108\|\|full | Low-quality |
| S3_vs_k141_61529\|\|full | Low-quality |
| S3_vs_k141_61534\|\|full | Not-determined |
| S3_vs_k141_123889\|\|full | Not-determined |
| S3_vs_k141_53325\|\|full | Low-quality |
| S3_vs_k141_4645\|\|full | Low-quality |
| S3_vs_k141_14473\|\|full | Low-quality |
| S3_vs_k141_4654\|\|full | Not-determined |
| S3_vs_k141_36942\|\|full | Not-determined |
| S3_vs_k141_44681\|\|full | Low-quality |
| S3_vs_k141_108516\|\|full | Low-quality |
| S3_vs_k141_61543\|\|full | Low-quality |
| S3_vs_k141_53359\|\|full | Low-quality |
| S3_vs_k141_53363\|\|full | Low-quality |
| S3_vs_k141_92587\|\|full | Not-determined |
| S3_vs_k141_36975\|\|full | Low-quality |
| S3_vs_k141_44716\|\|full | Low-quality |
| S3_vs_k141_53384\|\|full | Not-determined |
| S3_vs_k141_123933\|\|full | Low-quality |
| S3_vs_k141_14503\|\|full | Low-quality |
| S3_vs_k141_67976\|\|full | Not-determined |
| S3_vs_k141_67981\|\|full | Low-quality |
| S3_vs_k141_53407\|\|full | Low-quality |
| S3_vs_k141_20202\|\|full | Low-quality |
| S3_vs_k141_67988\|\|full | Low-quality |
| S3_vs_k141_67989\|\|full | Low-quality |
| S3_vs_k141_67991\|\|full | Not-determined |
| S3_vs_k141_116537\|\|full | Low-quality |
| S3_vs_k141_44768\|\|full | Not-determined |
| S3_vs_k141_92640\|\|full | Low-quality |
| S3_vs_k141_53425\|\|full | Not-determined |
| S3_vs_k141_123962\|\|full | Not-determined |
| S3_vs_k141_123965\|\|full | Not-determined |
| S3_vs_k141_77620\|\|full | Low-quality |
| S3_vs_k141_116546\|\|full | Low-quality |
| S3_vs_k141_100050\|\|full | Low-quality |
| S3_vs_k141_53444\|\|full | Not-determined |
| S3_vs_k141_68025\|\|full | Not-determined |
| S3_vs_k141_37027\|\|full | Low-quality |
| S3_vs_k141_20242\|\|full | Low-quality |
| S3_vs_k141_123973\|\|full | Not-determined |
| S3_vs_k141_20255\|\|full | Low-quality |
| S3_vs_k141_100072\|\|full | Low-quality |
| S3_vs_k141_84734\|\|full | Not-determined |
| S3_vs_k141_37033\|\|full | Not-determined |
| S3_vs_k141_100075\|\|full | Not-determined |
| S3_vs_k141_123981\|\|full | Not-determined |
| S3_vs_k141_37037\|\|full | Low-quality |
| S3_vs_k141_84742\|\|full | Low-quality |
| S3_vs_k141_53467\|\|full | Not-determined |
| S3_vs_k141_44801\|\|full | Low-quality |
| S3_vs_k141_100083\|\|full | Low-quality |
| S3_vs_k141_53473\|\|full | Low-quality |
| S3_vs_k141_116586\|\|full | Low-quality |
| S3_vs_k141_44806\|\|full | Low-quality |
| S3_vs_k141_20288\|\|full | Low-quality |
| S3_vs_k141_20292\|\|full | Low-quality |
| S3_vs_k141_28779\|\|full | Low-quality |
| S3_vs_k141_61599\|\|full | Low-quality |
| S3_vs_k141_84764\|\|full | Low-quality |
| S3_vs_k141_116603\|\|full | Low-quality |
| S3_vs_k141_92706\|\|full | Low-quality |
| S3_vs_k141_84792\|\|full | Low-quality |
| S3_vs_k141_14560\|\|full | Not-determined |
| S3_vs_k141_92724\|\|full | Low-quality |
| S3_vs_k141_92728\|\|full | Not-determined |
| S3_vs_k141_100132\|\|full | Not-determined |
| S3_vs_k141_84807\|\|full | Low-quality |
| S3_vs_k141_124019\|\|full | Not-determined |
| S3_vs_k141_100141\|\|full | Not-determined |
| S3_vs_k141_14583\|\|full | Low-quality |
| S3_vs_k141_77687\|\|full | Low-quality |
| S4_vs_VIRSorter_12614-cat_2 | Medium-quality |
| S3_vs_k141_124030\|\|full | Not-determined |
| S5_vs_k141_68254\|\|full | High-quality |
| S3_vs_k141_20323\|\|full | Low-quality |
| S11_vs_k141_104032\|\|full | High-quality |
| S3_vs_k141_100146\|\|full | Not-determined |
| S3_vs_k141_124049\|\|full | Not-determined |
| S3_vs_k141_124055\|\|full | Low-quality |
| S3_vs_k141_84859\|\|full | Low-quality |
| S3_vs_k141_28793\|\|full | Not-determined |
| S10_vs_24791\|\|full | High-quality |
| S3_vs_k141_37075\|\|full | Not-determined |
| S3_vs_k141_100157\|\|full | Not-determined |
| S3_vs_k141_108599\|\|full | Not-determined |
| S3_vs_k141_124074\|\|full | Low-quality |
| S3_vs_k141_108605\|\|full | Low-quality |
| S3_vs_k141_20385\|\|full | Low-quality |
| S3_vs_k141_100180\|\|full | Not-determined |
| S3_vs_k141_77715\|\|full | Low-quality |
| S3_vs_k141_100183\|\|full | Not-determined |
| S3_vs_k141_68077\|\|full | Low-quality |
| S3_vs_k141_84863\|\|full | Low-quality |
| S3_vs_k141_20391\|\|full | Low-quality |
| S3_vs_k141_124092\|\|full | Low-quality |
| S3_vs_k141_124096\|\|full | Low-quality |
| S3_vs_k141_77740\|\|full | Low-quality |
| S3_vs_k141_77742\|\|full | Low-quality |
| S3_vs_k141_61673\|\|full | Not-determined |
| S16_extracted_173106 | Complete |
| S3_vs_k141_92802\|\|full | Low-quality |
| S3_vs_k141_14704\|\|full | Low-quality |
| S3_vs_k141_84931\|\|full | Low-quality |
| S3_vs_k141_108642\|\|full | Low-quality |
| S3_vs_k141_84940\|\|full | Not-determined |
| S3_vs_k141_92826\|\|full | Not-determined |
| S3_vs_k141_53640\|\|full | Low-quality |
| S3_vs_k141_108645\|\|full | Low-quality |
| S3_vs_k141_116616\|\|full | Low-quality |
| S3_vs_k141_61711\|\|full | Low-quality |
| S3_vs_k141_100223\|\|full | Low-quality |
| S3_vs_k141_14732\|\|full | Low-quality |
| S3_vs_k141_108657\|\|full | Low-quality |
| S3_vs_k141_108658\|\|full | Not-determined |
| S3_vs_k141_108662\|\|full | Not-determined |
| S3_vs_k141_100260\|\|full | Low-quality |
| S3_vs_k141_61726\|\|full | Not-determined |
| S3_vs_k141_77758\|\|full | Low-quality |
| S3_vs_k141_44954\|\|full | Low-quality |
| S3_vs_k141_108685\|\|full | Not-determined |
| S3_vs_k141_37150\|\|full | Not-determined |
| S3_vs_k141_44962\|\|full | Low-quality |
| S3_vs_k141_14766\|\|full | Low-quality |
| S3_vs_k141_68173\|\|full | Low-quality |
| S3_vs_k141_61735\|\|full | Not-determined |
| S3_vs_k141_14778\|\|full | Low-quality |
| S3_vs_k141_100276\|\|full | Low-quality |
| S3_vs_k141_37159\|\|full | Not-determined |
| S3_vs_k141_37161\|\|full | Low-quality |
| S3_vs_k141_53666\|\|full | Not-determined |
| S3_vs_k141_4719\|\|full | Low-quality |
| S3_vs_k141_4720\|\|full | Low-quality |
| S3_vs_k141_68204\|\|full | Not-determined |
| S3_vs_k141_77773\|\|full | Low-quality |
| S3_vs_k141_53674\|\|full | Low-quality |
| S3_vs_k141_61751\|\|full | Low-quality |
| S3_vs_k141_108724\|\|full | Low-quality |
| S3_vs_k141_108737\|\|full | Low-quality |
| S3_vs_k141_28854\|\|full | Low-quality |
| S3_vs_k141_100297\|\|full | Low-quality |
| S3_vs_k141_20416\|\|full | Not-determined |
| S3_vs_k141_28857\|\|full | Not-determined |
| S3_vs_k141_84966\|\|full | Not-determined |
| S3_vs_k141_68237\|\|full | Low-quality |
| S3_vs_k141_4752\|\|full | Low-quality |
| S3_vs_k141_53711\|\|full | Low-quality |
| S3_vs_k141_20429\|\|full | Low-quality |
| S3_vs_k141_14795\|\|full | Low-quality |
| S3_vs_k141_45007\|\|full | Low-quality |
| S3_vs_k141_37197\|\|full | Low-quality |
| S3_vs_k141_53713\|\|full | Low-quality |
| S3_vs_k141_37204\|\|full | Not-determined |
| S3_vs_k141_14801\|\|full | Low-quality |
| S3_vs_k141_116686\|\|full | Not-determined |
| S3_vs_k141_20432\|\|full | Low-quality |
| S3_vs_k141_61775\|\|full | Low-quality |
| S3_vs_k141_68255\|\|full | Not-determined |
| S3_vs_k141_124151\|\|full | Not-determined |
| S3_vs_k141_124153\|\|full | Low-quality |
| S3_vs_k141_100339\|\|full | Low-quality |
| S3_vs_k141_92916\|\|full | Low-quality |
| S3_vs_k141_4783\|\|full | Not-determined |
| S8_vs_k141_51368\|\|full | High-quality |
| S3_vs_k141_77795\|\|full | Low-quality |
| S3_vs_k141_77797\|\|full | Not-determined |
| S3_vs_k141_124183\|\|full | Low-quality |
| S3_vs_k141_20447\|\|full | Not-determined |
| S3_vs_k141_20451\|\|full | Low-quality |
| S3_vs_k141_100357\|\|full | Not-determined |
| S3_vs_k141_108813\|\|full | Low-quality |
| S3_vs_k141_14842\|\|full | Not-determined |
| S3_vs_k141_61838\|\|full | Low-quality |
| S3_vs_k141_92940\|\|full | Low-quality |
| S3_vs_k141_92941\|\|full | Low-quality |
| S3_vs_k141_61850\|\|full | Low-quality |
| S3_vs_k141_124202\|\|full | Low-quality |
| S3_vs_k141_4810\|\|full | Low-quality |
| S3_vs_k141_108830\|\|full | Low-quality |
| S3_vs_k141_14848\|\|full | Not-determined |
| S3_vs_k141_85019\|\|full | Not-determined |
| S3_vs_k141_85028\|\|full | Not-determined |
| S3_vs_k141_116718\|\|full | Not-determined |
| S3_vs_k141_100390\|\|full | Not-determined |
| S3_vs_k141_116727\|\|full | Low-quality |
| S3_vs_k141_108852\|\|full | Low-quality |
| S3_vs_k141_77828\|\|full | Not-determined |
| S3_vs_k141_92945\|\|full | Low-quality |
| S3_vs_k141_28909\|\|full | Low-quality |
| S3_vs_k141_108863\|\|full | Not-determined |
| S3_vs_k141_53768\|\|full | Low-quality |
| S3_vs_k141_116740\|\|full | Low-quality |
| S3_vs_k141_20493\|\|full | Low-quality |
| S3_vs_k141_53787\|\|full | Not-determined |
| S3_vs_k141_4900\|\|full | Low-quality |
| S3_vs_k141_37261\|\|full | Low-quality |
| S3_vs_k141_37284\|\|full | Low-quality |
| S3_vs_k141_20509\|\|full | Low-quality |
| S3_vs_k141_108927\|\|full | Not-determined |
| S3_vs_k141_68297\|\|full | Low-quality |
| S3_vs_k141_100437\|\|full | Not-determined |
| S3_vs_k141_77864\|\|full | Low-quality |
| S3_vs_k141_53813\|\|full | Low-quality |
| S3_vs_k141_68319\|\|full | Not-determined |
| S3_vs_k141_61864\|\|full | Low-quality |
| S3_vs_k141_4930\|\|full | Not-determined |
| S3_vs_k141_28957\|\|full | Low-quality |
| S3_vs_k141_61868\|\|full | Not-determined |
| S3_vs_k141_20529\|\|full | Not-determined |
| S3_vs_k141_14890\|\|full | Low-quality |
| S3_vs_k141_77921\|\|full | Not-determined |
| S3_vs_k141_93022\|\|full | Low-quality |
| S3_vs_k141_20549\|\|full | Low-quality |
| S3_vs_k141_4969\|\|full | Not-determined |
| S3_vs_k141_4972\|\|full | Not-determined |
| S3_vs_k141_28985\|\|full | Low-quality |
| S3_vs_k141_68350\|\|full | Low-quality |
| S3_vs_k141_68351\|\|full | Not-determined |
| S3_vs_k141_29006\|\|full | Not-determined |
| S3_vs_k141_53853\|\|full | Not-determined |
| S3_vs_k141_4996\|\|full | Low-quality |
| S3_vs_k141_68364\|\|full | Not-determined |
| S3_vs_k141_68366\|\|full | Not-determined |
| S3_vs_k141_53865\|\|full | Low-quality |
| S3_vs_k141_61918\|\|full | Low-quality |
| S16_vs_173106\|\|full | High-quality |
| S3_vs_k141_93065\|\|full | Not-determined |
| S3_vs_k141_5021\|\|full | Not-determined |
| S3_vs_k141_29030\|\|full | Low-quality |
| S3_vs_k141_29033\|\|full | Low-quality |
| S3_vs_k141_5031\|\|full | Low-quality |
| S3_vs_k141_77947\|\|full | Low-quality |
| S3_vs_k141_5035\|\|full | Low-quality |
| S3_vs_k141_29045\|\|full | Low-quality |
| S3_vs_k141_37332\|\|full | Low-quality |
| S3_vs_k141_68412\|\|full | Low-quality |
| S3_vs_k141_53877\|\|full | Low-quality |
| S3_vs_k141_85175\|\|full | Low-quality |
| S3_vs_k141_116872\|\|full | Not-determined |
| S3_vs_k141_77998\|\|full | Not-determined |
| S3_vs_k141_124306\|\|full | Low-quality |
| S3_vs_k141_53905\|\|full | Low-quality |
| S3_vs_k141_14955\|\|full | Not-determined |
| S3_vs_k141_53907\|\|full | Low-quality |
| S3_vs_k141_20573\|\|full | Low-quality |
| S3_vs_k141_68445\|\|full | Low-quality |
| S3_vs_k141_20574\|\|full | Not-determined |
| S3_vs_k141_109009\|\|full | Not-determined |
| S3_vs_k141_14974\|\|full | Not-determined |
| S3_vs_k141_14978\|\|full | Low-quality |
| S3_vs_k141_109021\|\|full | Low-quality |
| S3_vs_k141_116887\|\|full | Not-determined |
| S3_vs_k141_45054\|\|full | Low-quality |
| S3_vs_k141_20591\|\|full | Not-determined |
| S3_vs_k141_68473\|\|full | Low-quality |
| S3_vs_k141_37376\|\|full | Not-determined |
| S3_vs_k141_116908\|\|full | Not-determined |
| S3_vs_k141_116909\|\|full | Low-quality |
| S3_vs_k141_109049\|\|full | Low-quality |
| S3_vs_k141_5141\|\|full | Low-quality |
| S3_vs_k141_85251\|\|full | Low-quality |
| S3_vs_k141_85252\|\|full | Not-determined |
| S3_vs_k141_116913\|\|full | Low-quality |
| S3_vs_k141_68497\|\|full | Not-determined |
| S3_vs_k141_85254\|\|full | Low-quality |
| S3_vs_k141_116919\|\|full | Low-quality |
| S3_vs_k141_124341\|\|full | Not-determined |
| S3_vs_k141_68507\|\|full | Not-determined |
| S3_vs_k141_53923\|\|full | Low-quality |
| S3_vs_k141_45078\|\|full | Low-quality |
| S3_vs_k141_5145\|\|full | Low-quality |
| S3_vs_k141_124345\|\|full | Not-determined |
| S3_vs_k141_85279\|\|full | Not-determined |
| S16_vs_173102\|\|full | High-quality |
| S3_vs_k141_85293\|\|full | Not-determined |
| S3_vs_k141_109089\|\|full | Low-quality |
| S3_vs_k141_14987\|\|full | Not-determined |
| S3_vs_k141_62064\|\|full | Low-quality |
| S3_vs_k141_85305\|\|full | Not-determined |
| S3_vs_k141_37415\|\|full | Low-quality |
| S3_vs_k141_68522\|\|full | Not-determined |
| S3_vs_k141_15002\|\|full | Low-quality |
| S3_vs_k141_68525\|\|full | Low-quality |
| S3_vs_k141_29102\|\|full | Low-quality |
| S3_vs_k141_68530\|\|full | Low-quality |
| S3_vs_k141_78067\|\|full | Not-determined |
| S3_vs_k141_78071\|\|full | Low-quality |
| S3_vs_k141_53985\|\|full | Not-determined |
| S3_vs_k141_68556\|\|full | Not-determined |
| S3_vs_k141_5172\|\|full | Not-determined |
| S3_vs_k141_5194\|\|full | Not-determined |
| S3_vs_k141_5211\|\|full | Not-determined |
| S3_vs_k141_5220\|\|full | Not-determined |
| S3_vs_k141_20683\|\|full | Low-quality |
| S3_vs_k141_116971\|\|full | Not-determined |
| S3_vs_k141_5241\|\|full | Low-quality |
| S3_vs_k141_109177\|\|full | Low-quality |
| S3_vs_k141_29119\|\|full | Not-determined |
| S3_vs_k141_68587\|\|full | Low-quality |
| S3_vs_k141_85359\|\|full | Low-quality |
| S3_vs_k141_62112\|\|full | Low-quality |
| S3_vs_k141_68590\|\|full | Low-quality |
| S3_vs_k141_68591\|\|full | Not-determined |
| S3_vs_k141_116979\|\|full | Not-determined |

| | |
|---|---|
| S3_vs_k141_62122\|\|full | Low-quality |
| S3_vs_k141_62129\|\|full | Not-determined |
| S3_vs_k141_29140\|\|full | Low-quality |
| S3_vs_k141_68607\|\|full | Low-quality |
| S3_vs_k141_15113\|\|full | Not-determined |
| S3_vs_k141_37447\|\|full | Not-determined |
| S3_vs_k141_93084\|\|full | Not-determined |
| S3_vs_k141_45111\|\|full | Low-quality |
| S3_vs_k141_93089\|\|full | Low-quality |
| S3_vs_k141_85361\|\|full | Low-quality |
| S3_vs_k141_37457\|\|full | Low-quality |
| S3_vs_k141_5276\|\|full | Not-determined |
| S3_vs_k141_124404\|\|full | Low-quality |
| S3_vs_k141_62158\|\|full | Not-determined |
| S3_vs_k141_29187\|\|full | Low-quality |
| S3_vs_k141_124412\|\|full | Not-determined |
| S3_vs_k141_93110\|\|full | Low-quality |
| S3_vs_k141_93112\|\|full | Not-determined |
| S3_vs_k141_15176\|\|full | Not-determined |
| S3_vs_k141_45143\|\|full | Low-quality |
| S3_vs_k141_109212\|\|full | Low-quality |
| S3_vs_k141_78103\|\|full | Low-quality |
| S3_vs_k141_68689\|\|full | Not-determined |
| S3_vs_k141_109215\|\|full | Low-quality |
| S3_vs_k141_37550\|\|full | Low-quality |
| S3_vs_k141_5287\|\|full | Not-determined |
| S3_vs_k141_62216\|\|full | Low-quality |
| S3_vs_k141_85380\|\|full | Low-quality |
| S3_vs_k141_68707\|\|full | Low-quality |
| S3_vs_k141_37562\|\|full | Low-quality |
| S3_vs_k141_68710\|\|full | Not-determined |
| S3_vs_k141_37567\|\|full | Not-determined |
| S3_vs_k141_62246\|\|full | Not-determined |
| S3_vs_k141_109247\|\|full | Low-quality |
| S3_vs_k141_62251\|\|full | Not-determined |
| S3_vs_k141_85410\|\|full | Not-determined |
| S3_vs_k141_15223\|\|full | Not-determined |
| S3_vs_k141_5323\|\|full | Low-quality |
| S3_vs_k141_85419\|\|full | Low-quality |
| S3_vs_k141_109277\|\|full | Low-quality |
| S3_vs_k141_5354\|\|full | Low-quality |
| S3_vs_k141_124515\|\|full | Low-quality |
| S3_vs_k141_85431\|\|full | Low-quality |
| S3_vs_k141_124521\|\|full | Not-determined |
| S3_vs_k141_85445\|\|full | Low-quality |
| S3_vs_k141_100514\|\|full | Low-quality |
| S3_vs_k141_5368\|\|full | Not-determined |
| S3_vs_k141_85456\|\|full | Not-determined |
| S3_vs_k141_85458\|\|full | Not-determined |
| S3_vs_k141_109291\|\|full | Not-determined |
| S3_vs_k141_100531\|\|full | Low-quality |
| S3_vs_k141_68767\|\|full | Not-determined |
| S3_vs_k141_93168\|\|full | Low-quality |
| S3_vs_k141_54303\|\|full | High-quality |
| S3_vs_k141_37600\|\|full | Low-quality |
| S3_vs_k141_85471\|\|full | Low-quality |
| S3_vs_k141_100535\|\|full | Low-quality |
| S3_vs_k141_117032\|\|full | Low-quality |
| S3_vs_k141_100544\|\|full | Not-determined |
| S3_vs_k141_100547\|\|full | Not-determined |
| S3_vs_k141_54035\|\|full | Low-quality |
| S3_vs_k141_100558\|\|full | Not-determined |
| S3_vs_k141_109320\|\|full | Not-determined |
| S3_vs_k141_54039\|\|full | Not-determined |
| S3_vs_k141_93180\|\|full | Low-quality |
| S3_vs_k141_109339\|\|full | Low-quality |
| S3_vs_k141_20738\|\|full | Not-determined |
| S3_vs_k141_100566\|\|full | Low-quality |
| S3_vs_k141_117065\|\|full | Low-quality |
| S3_vs_k141_117066\|\|full | Low-quality |
| S3_vs_k141_93223\|\|full | Not-determined |
| S3_vs_k141_117073\|\|full | Not-determined |
| S3_vs_k141_93226\|\|full | Low-quality |
| S3_vs_k141_85523\|\|full | Low-quality |
| S3_vs_k141_109358\|\|full | Low-quality |
| S3_vs_k141_37629\|\|full | Low-quality |
| S3_vs_k141_20759\|\|full | Not-determined |
| S3_vs_k141_117077\|\|full | Low-quality |
| S3_vs_k141_45230\|\|full | Low-quality |
| S3_vs_k141_124548\|\|full | Not-determined |
| S3_vs_k141_93258\|\|full | Low-quality |
| S3_vs_k141_29301\|\|full | Not-determined |
| S3_vs_k141_124568\|\|full | Not-determined |
| S3_vs_k141_68845\|\|full | Not-determined |
| S3_vs_k141_29309\|\|full | Low-quality |
| S3_vs_k141_100582\|\|full | Not-determined |
| S3_vs_k141_29310\|\|full | Not-determined |
| S3_vs_k141_20769\|\|full | Low-quality |
| S3_vs_k141_109451\|\|full | Low-quality |
| S3_vs_k141_29318\|\|full | Not-determined |
| S3_vs_k141_100598\|\|full | Not-determined |
| S3_vs_k141_29323\|\|full | Low-quality |
| S3_vs_k141_93293\|\|full | Low-quality |
| S3_vs_k141_29327\|\|full | Low-quality |
| S3_vs_k141_124574\|\|full | Not-determined |
| S3_vs_k141_93304\|\|full | Low-quality |
| S3_vs_k141_20773\|\|full | Low-quality |
| S3_vs_k141_85586\|\|full | Not-determined |
| S3_vs_k141_100612\|\|full | Low-quality |
| S3_vs_k141_109472\|\|full | Low-quality |
| S3_vs_k141_29341\|\|full | Low-quality |
| S3_vs_k141_124582\|\|full | Not-determined |
| S3_vs_k141_100624\|\|full | Low-quality |
| S3_vs_k141_78199\|\|full | Low-quality |
| S3_vs_k141_78212\|\|full | Low-quality |
| S3_vs_k141_62399\|\|full | Low-quality |
| S3_vs_k141_20785\|\|full | Low-quality |
| S3_vs_k141_29375\|\|full | Low-quality |
| S3_vs_k141_62409\|\|full | Not-determined |
| S3_vs_k141_45294\|\|full | Low-quality |
| S3_vs_k141_85604\|\|full | Not-determined |
| S3_vs_k141_93327\|\|full | Low-quality |
| S3_vs_k141_29387\|\|full | Not-determined |
| S3_vs_k141_54082\|\|full | Low-quality |
| S3_vs_k141_93333\|\|full | Low-quality |
| S3_vs_k141_117113\|\|full | Low-quality |
| S3_vs_k141_100656\|\|full | Low-quality |
| S12_extracted_k141_105542 | High-quality |
| S3_vs_k141_109485\|\|full | Low-quality |
| S3_vs_k141_117114\|\|full | Low-quality |
| S3_vs_k141_15330\|\|full | Low-quality |
| S3_vs_k141_109496\|\|full | Low-quality |
| S3_vs_k141_37703\|\|full | Low-quality |
| S3_vs_k141_124592\|\|full | Not-determined |
| S3_vs_k141_100671\|\|full | Low-quality |
| S3_vs_k141_124593\|\|full | Low-quality |
| S3_vs_k141_109507\|\|full | Low-quality |
| S3_vs_k141_100689\|\|full | Not-determined |
| S3_vs_k141_124605\|\|full | Low-quality |
| S3_vs_k141_62448\|\|full | Low-quality |
| S3_vs_k141_85661\|\|full | Low-quality |
| S3_vs_k141_15340\|\|full | Low-quality |
| S3_vs_k141_29462\|\|full | Not-determined |
| S3_vs_k141_15342\|\|full | Not-determined |
| S3_vs_k141_117122\|\|full | Not-determined |
| S3_vs_k141_54117\|\|full | Low-quality |
| S3_vs_k141_100691\|\|full | Low-quality |
| S3_vs_k141_124613\|\|full | Not-determined |
| S3_vs_k141_117130\|\|full | Low-quality |
| S3_vs_k141_78309\|\|full | Not-determined |
| S3_vs_k141_45342\|\|full | Low-quality |
| S3_vs_k141_54124\|\|full | Low-quality |
| S3_vs_k141_109514\|\|full | Not-determined |
| S3_vs_k141_45344\|\|full | Not-determined |
| S3_vs_k141_29477\|\|full | Low-quality |
| S3_vs_k141_124625\|\|full | Not-determined |
| S3_vs_k141_5473\|\|full | Low-quality |
| S3_vs_k141_117145\|\|full | Low-quality |
| S3_vs_k141_5478\|\|full | Not-determined |
| S3_vs_k141_45354\|\|full | Low-quality |
| S3_vs_k141_5480\|\|full | Low-quality |
| S3_vs_k141_93394\|\|full | Low-quality |
| S3_vs_k141_15382\|\|full | Not-determined |
| S3_vs_k141_54149\|\|full | Not-determined |
| S3_vs_k141_45367\|\|full | Not-determined |
| S3_vs_k141_93406\|\|full | Not-determined |
| S3_vs_k141_100713\|\|full | Low-quality |
| S3_vs_k141_5483\|\|full | Not-determined |
| S3_vs_k141_45371\|\|full | Not-determined |
| S3_vs_k141_93408\|\|full | Not-determined |
| S3_vs_k141_124632\|\|full | Low-quality |
| S3_vs_k141_85694\|\|full | Low-quality |
| S3_vs_k141_85699\|\|full | Low-quality |
| S3_vs_k141_20871\|\|full | Low-quality |
| S3_vs_k141_5495\|\|full | Not-determined |
| S3_vs_k141_45396\|\|full | Not-determined |
| S3_vs_k141_109599\|\|full | Low-quality |
| S3_vs_k141_5515\|\|full | Low-quality |
| S3_vs_k141_109623\|\|full | Not-determined |
| S3_vs_k141_93446\|\|full | Low-quality |
| S3_vs_k141_45406\|\|full | Low-quality |
| S3_vs_k141_93482\|\|full | Low-quality |
| S3_vs_k141_100779\|\|full | Not-determined |
| S3_vs_k141_45418\|\|full | Low-quality |
| S3_vs_k141_20900\|\|full | Low-quality |
| S3_vs_k141_93495\|\|full | Low-quality |
| S3_vs_k141_124665\|\|full | Not-determined |
| S3_vs_k141_117154\|\|full | Not-determined |
| S3_vs_k141_109655\|\|full | Low-quality |
| S3_vs_k141_78393\|\|full | Low-quality |
| S3_vs_k141_20908\|\|full | Low-quality |
| S12_vs_k141_105542\|\|full | High-quality |
| S3_vs_k141_45426\|\|full | Low-quality |
| S3_vs_k141_124683\|\|full | Low-quality |
| S3_vs_k141_85718\|\|full | Low-quality |
| S3_vs_k141_78400\|\|full | Not-determined |
| S3_vs_k141_109671\|\|full | Low-quality |
| S3_vs_k141_45435\|\|full | Low-quality |
| S3_vs_k141_78408\|\|full | Not-determined |
| S3_vs_k141_93501\|\|full | Low-quality |
| S3_vs_k141_78412\|\|full | Low-quality |
| S3_vs_k141_78415\|\|full | Not-determined |
| S3_vs_k141_78417\|\|full | Not-determined |
| S3_vs_k141_5540\|\|full | Low-quality |
| S3_vs_k141_5559\|\|full | Low-quality |
| S3_vs_k141_20913\|\|full | Not-determined |
| S3_vs_k141_20915\|\|full | Low-quality |
| S4_vs_VIRSorter_27132-circular-cat | High-quality |
| S3_vs_k141_109702\|\|full | Low-quality |
| S3_vs_k141_93527\|\|full | Low-quality |
| S3_vs_k141_5594\|\|full | Low-quality |
| S3_vs_k141_45497\|\|full | Not-determined |
| S3_vs_k141_124716\|\|full | Not-determined |
| S3_vs_k141_124721\|\|full | Low-quality |
| S3_vs_k141_85753\|\|full | Not-determined |
| S3_vs_k141_5632\|\|full | Not-determined |
| S3_vs_k141_29654\|\|full | Not-determined |
| S3_vs_k141_124735\|\|full | Low-quality |
| S3_vs_k141_100804\|\|full | Low-quality |
| S3_vs_k141_124743\|\|full | Low-quality |
| S4_vs_VIRSorter_8622-cat_2 | High-quality |
| S3_vs_k141_78481\|\|full | Low-quality |
| S3_vs_k141_29665\|\|full | Not-determined |
| S3_vs_k141_15457\|\|full | Low-quality |
| S3_vs_k141_117280\|\|full | Low-quality |
| S3_vs_k141_93558\|\|full | Not-determined |
| S3_vs_k141_37739\|\|full | Not-determined |
| S3_vs_k141_15467\|\|full | Low-quality |
| S3_vs_k141_29674\|\|full | Low-quality |
| S3_vs_k141_15487\|\|full | Not-determined |
| S3_vs_k141_62471\|\|full | Low-quality |
| S3_vs_k141_93575\|\|full | Low-quality |
| S3_vs_k141_20957\|\|full | Low-quality |
| S3_vs_k141_15509\|\|full | Low-quality |
| S3_vs_k141_20983\|\|full | Low-quality |
| S3_vs_k141_15537\|\|full | Not-determined |
| S3_vs_k141_117301\|\|full | Low-quality |
| S3_vs_k141_93603\|\|full | Low-quality |
| S3_vs_k141_78502\|\|full | Low-quality |
| S3_vs_k141_78504\|\|full | Not-determined |
| S3_vs_k141_29690\|\|full | Not-determined |
| S3_vs_k141_100844\|\|full | Low-quality |
| S3_vs_k141_124783\|\|full | Not-determined |
| S3_vs_k141_21012\|\|full | Low-quality |
| S3_vs_k141_124790\|\|full | Low-quality |
| S3_vs_k141_85794\|\|full | Low-quality |
| S3_vs_k141_15559\|\|full | Low-quality |
| S3_vs_k141_93634\|\|full | Not-determined |
| S3_vs_k141_85810\|\|full | Low-quality |
| S3_vs_k141_68946\|\|full | Low-quality |
| S3_vs_k141_68959\|\|full | Low-quality |
| S3_vs_k141_15591\|\|full | Not-determined |
| S3_vs_k141_29732\|\|full | Not-determined |
| S3_vs_k141_109761\|\|full | Low-quality |
| S3_vs_k141_85838\|\|full | Not-determined |
| S3_vs_k141_15615\|\|full | Low-quality |
| S3_vs_k141_78558\|\|full | Low-quality |
| S3_vs_k141_124828\|\|full | Low-quality |
| S17_vs_k141_1962\|\|full | High-quality |
| S3_vs_k141_124859\|\|full | Low-quality |
| S3_vs_k141_54307\|\|full | Not-determined |
| S3_vs_k141_69006\|\|full | Not-determined |
| S3_vs_k141_85854\|\|full | Low-quality |
| S3_vs_k141_93660\|\|full | Low-quality |
| S3_vs_k141_45708\|\|full | Low-quality |
| S3_vs_k141_85864\|\|full | Low-quality |
| S3_vs_k141_29780\|\|full | Low-quality |
| S3_vs_k141_85881\|\|full | Low-quality |
| S3_vs_k141_78578\|\|full | Low-quality |
| S3_vs_k141_85891\|\|full | Not-determined |
| S3_vs_k141_29790\|\|full | Low-quality |
| S3_vs_k141_15646\|\|full | Not-determined |
| S3_vs_k141_37864\|\|full | Low-quality |
| S3_vs_k141_85903\|\|full | Low-quality |
| S3_vs_k141_78597\|\|full | Low-quality |
| S3_vs_k141_93696\|\|full | Low-quality |
| S3_vs_k141_5667\|\|full | Not-determined |
| S3_vs_k141_124956\|\|full | Not-determined |
| S3_vs_k141_37889\|\|full | Not-determined |
| S3_vs_k141_37891\|\|full | Low-quality |
| S3_vs_k141_5680\|\|full | Low-quality |
| S3_vs_k141_124964\|\|full | Low-quality |
| S3_vs_k141_5684\|\|full | Low-quality |
| S3_vs_k141_21130\|\|full | Low-quality |
| S3_vs_k141_5687\|\|full | Not-determined |
| S3_vs_k141_21134\|\|full | Not-determined |
| S3_vs_k141_62551\|\|full | Low-quality |
| S3_vs_k141_21138\|\|full | Not-determined |
| S3_vs_k141_69011\|\|full | Low-quality |
| S3_vs_k141_69013\|\|full | Low-quality |
| S3_vs_k141_69014\|\|full | Low-quality |
| S3_vs_k141_93733\|\|full | Low-quality |
| S3_vs_k141_15678\|\|full | Not-determined |
| S3_vs_k141_93734\|\|full | Low-quality |
| S3_vs_k141_45799\|\|full | Low-quality |
| S3_vs_k141_15680\|\|full | Low-quality |
| S3_vs_k141_15695\|\|full | Low-quality |
| S3_vs_k141_15696\|\|full | Low-quality |
| S3_vs_k141_15702\|\|full | Low-quality |
| S3_vs_k141_5706\|\|full | Low-quality |
| S3_vs_k141_109798\|\|full | Low-quality |
| S3_vs_k141_62603\|\|full | Low-quality |
| S3_vs_k141_62605\|\|full | Low-quality |
| S3_vs_k141_124978\|\|full | Low-quality |
| S3_vs_k141_15708\|\|full | Not-determined |
| S3_vs_k141_78643\|\|full | Low-quality |
| S3_vs_k141_62613\|\|full | Not-determined |
| S3_vs_k141_37934\|\|full | Low-quality |
| S15_vs_1140\|\|full | High-quality |
| S3_vs_k141_15711\|\|full | Low-quality |
| S3_vs_k141_93751\|\|full | Low-quality |
| S3_vs_k141_29825\|\|full | Low-quality |
| S3_vs_k141_21174\|\|full | Low-quality |
| S3_vs_k141_29835\|\|full | Not-determined |
| S3_vs_k141_100928\|\|full | Low-quality |
| S3_vs_k141_21189\|\|full | Not-determined |
| S3_vs_k141_93773\|\|full | Low-quality |
| S3_vs_k141_124995\|\|full | Low-quality |
| S3_vs_k141_86023\|\|full | Not-determined |
| S3_vs_k141_15741\|\|full | Low-quality |
| S3_vs_k141_45832\|\|full | Low-quality |
| S3_vs_k141_117359\|\|full | Not-determined |
| S3_vs_k141_93793\|\|full | Not-determined |
| S3_vs_k141_100938\|\|full | Low-quality |
| S3_vs_k141_15749\|\|full | Low-quality |
| S3_vs_k141_54400\|\|full | Low-quality |
| S3_vs_k141_100941\|\|full | Low-quality |
| S3_vs_k141_69026\|\|full | Not-determined |
| S3_vs_k141_45852\|\|full | Low-quality |
| S3_vs_k141_54410\|\|full | Not-determined |
| S3_vs_k141_86056\|\|full | Low-quality |
| S3_vs_k141_69034\|\|full | Low-quality |
| S3_vs_k141_54429\|\|full | Low-quality |
| S3_vs_k141_45886\|\|full | Not-determined |
| S3_vs_k141_93846\|\|full | Low-quality |
| S3_vs_k141_69041\|\|full | Low-quality |
| S3_vs_k141_15778\|\|full | Low-quality |
| S3_vs_k141_5809\|\|full | Low-quality |
| S3_vs_k141_93867\|\|full | Not-determined |
| S3_vs_k141_54465\|\|full | Low-quality |
| S3_vs_k141_29898\|\|full | Not-determined |
| S3_vs_k141_100970\|\|full | Not-determined |
| S3_vs_k141_117408\|\|full | Low-quality |
| S3_vs_k141_45925\|\|full | Low-quality |
| S3_vs_k141_29907\|\|full | Not-determined |
| S3_vs_k141_54481\|\|full | Not-determined |
| S3_vs_k141_54483\|\|full | Not-determined |
| S3_vs_k141_93896\|\|full | Low-quality |
| S3_vs_k141_29909\|\|full | Not-determined |
| S3_vs_k141_38006\|\|full | Low-quality |
| S3_vs_k141_69059\|\|full | Not-determined |
| S8_vs_k141_54183\|\|full | High-quality |
| S3_vs_k141_86122\|\|full | Not-determined |
| S3_vs_k141_125017\|\|full | Not-determined |
| S3_vs_k141_29921\|\|full | Not-determined |
| S3_vs_k141_69069\|\|full | Not-determined |
| S3_vs_k141_45949\|\|full | Low-quality |
| S3_vs_k141_38023\|\|full | Low-quality |
| S3_vs_k141_125024\|\|full | Low-quality |
| S3_vs_k141_29941\|\|full | Not-determined |
| S3_vs_k141_15828\|\|full | Not-determined |
| S3_vs_k141_86150\|\|full | Not-determined |
| S3_vs_k141_45963\|\|full | Low-quality |
| S3_vs_k141_15830\|\|full | Low-quality |
| S3_vs_k141_69071\|\|full | Not-determined |
| S3_vs_k141_69074\|\|full | Not-determined |
| S3_vs_k141_117469\|\|full | Low-quality |
| S3_vs_k141_29966\|\|full | Low-quality |
| S3_vs_k141_109917\|\|full | Low-quality |
| S3_vs_k141_15843\|\|full | Not-determined |
| S3_vs_k141_117487\|\|full | Low-quality |
| S3_vs_k141_54558\|\|full | Low-quality |
| S3_vs_k141_86189\|\|full | Low-quality |
| S3_vs_k141_78732\|\|full | Low-quality |
| S3_vs_k141_86201\|\|full | Low-quality |
| S3_vs_k141_46003\|\|full | Low-quality |
| S3_vs_k141_78744\|\|full | Low-quality |
| S3_vs_k141_117532\|\|full | Not-determined |
| S3_vs_k141_62760\|\|full | Low-quality |
| S3_vs_k141_38094\|\|full | Low-quality |
| S3_vs_k141_86240\|\|full | Not-determined |
| S3_vs_k141_78756\|\|full | Low-quality |
| S3_vs_k141_38105\|\|full | Not-determined |
| S3_vs_k141_109944\|\|full | Not-determined |
| S3_vs_k141_109945\|\|full | Not-determined |
| S3_vs_k141_117551\|\|full | Low-quality |
| S3_vs_k141_21290\|\|full | Low-quality |
| S3_vs_k141_30029\|\|full | Low-quality |
| S3_vs_k141_86250\|\|full | Not-determined |
| S3_vs_k141_46045\|\|full | Low-quality |
| S3_vs_k141_21297\|\|full | Low-quality |
| S3_vs_k141_15876\|\|full | Low-quality |
| S3_vs_k141_38113\|\|full | Low-quality |
| S3_vs_k141_21302\|\|full | Low-quality |
| S3_vs_k141_69090\|\|full | Low-quality |
| S3_vs_k141_5841\|\|full | Low-quality |
| S3_vs_k141_125073\|\|full | Low-quality |
| S3_vs_k141_30037\|\|full | Not-determined |
| S3_vs_k141_86285\|\|full | Low-quality |
| S3_vs_k141_125083\|\|full | Low-quality |
| S3_vs_k141_30042\|\|full | Low-quality |
| S3_vs_k141_38129\|\|full | Not-determined |
| S3_vs_k141_62823\|\|full | Low-quality |
| S3_vs_k141_30057\|\|full | Low-quality |
| S3_vs_k141_86324\|\|full | Low-quality |
| S3_vs_k141_117623\|\|full | Low-quality |
| S3_vs_k141_86338\|\|full | Not-determined |
| S3_vs_k141_93953\|\|full | Low-quality |
| S3_vs_k141_117632\|\|full | Low-quality |
| S3_vs_k141_5876\|\|full | Not-determined |
| S3_vs_k141_125124\|\|full | Low-quality |
| S3_vs_k141_109998\|\|full | Low-quality |
| S3_vs_k141_125134\|\|full | Not-determined |
| S3_vs_k141_125138\|\|full | Not-determined |
| S3_vs_k141_117659\|\|full | Low-quality |
| S3_vs_k141_110041\|\|full | Low-quality |
| S3_vs_k141_15899\|\|full | Low-quality |
| S3_vs_k141_46090\|\|full | Low-quality |
| S3_vs_k141_117673\|\|full | Low-quality |
| S3_vs_k141_125153\|\|full | Low-quality |
| S3_vs_k141_117680\|\|full | Low-quality |
| S3_vs_k141_117681\|\|full | Low-quality |
| S3_vs_k141_93987\|\|full | Low-quality |
| S3_vs_k141_78868\|\|full | Not-determined |
| S3_vs_k141_117689\|\|full | Low-quality |
| S3_vs_k141_86375\|\|full | Not-determined |
| S3_vs_k141_78876\|\|full | Not-determined |
| S3_vs_k141_78879\|\|full | Low-quality |
| S3_vs_k141_30134\|\|full | Not-determined |
| S3_vs_k141_86379\|\|full | Low-quality |
| S3_vs_k141_62867\|\|full | Low-quality |
| S3_vs_k141_94021\|\|full | Low-quality |
| S3_vs_k141_110097\|\|full | Low-quality |
| S3_vs_k141_110104\|\|full | Low-quality |
| S3_vs_k141_62885\|\|full | Not-determined |
| S3_vs_k141_110122\|\|full | Low-quality |
| S3_vs_k141_30155\|\|full | Low-quality |
| S3_vs_k141_46149\|\|full | Low-quality |
| S3_vs_k141_69150\|\|full | Low-quality |
| S3_vs_k141_30157\|\|full | Low-quality |
| S3_vs_k141_117742\|\|full | Not-determined |
| S3_vs_k141_46179\|\|full | Low-quality |
| S3_vs_k141_69161\|\|full | Low-quality |
| S3_vs_k141_86443\|\|full | Not-determined |
| S3_vs_k141_86447\|\|full | Low-quality |
| S3_vs_k141_125209\|\|full | Not-determined |
| S3_vs_k141_100990\|\|full | Not-determined |
| S3_vs_k141_46198\|\|full | Low-quality |
| S3_vs_k141_94098\|\|full | Low-quality |
| S3_vs_k141_46199\|\|full | Low-quality |
| S3_vs_k141_125224\|\|full | Low-quality |
| S3_vs_k141_62956\|\|full | Not-determined |
| S3_vs_k141_46206\|\|full | Not-determined |
| S3_vs_k141_117776\|\|full | Low-quality |
| S3_vs_k141_78931\|\|full | Low-quality |
| S3_vs_k141_38219\|\|full | Low-quality |
| S3_vs_k141_94117\|\|full | Low-quality |
| S3_vs_k141_5975\|\|full | Not-determined |
| S3_vs_k141_54714\|\|full | Not-determined |
| S3_vs_k141_86492\|\|full | Not-determined |
| S3_vs_k141_100998\|\|full | Not-determined |
| S3_vs_k141_78953\|\|full | Not-determined |
| S3_vs_k141_38266\|\|full | Low-quality |
| S3_vs_k141_125266\|\|full | Low-quality |
| S3_vs_k141_69179\|\|full | Low-quality |
| S3_vs_k141_78956\|\|full | Low-quality |
| S3_vs_k141_63010\|\|full | Low-quality |
| S3_vs_k141_30250\|\|full | Low-quality |
| S3_vs_k141_117833\|\|full | Low-quality |
| S3_vs_k141_110177\|\|full | Low-quality |
| S3_vs_k141_69196\|\|full | Low-quality |
| S3_vs_k141_6017\|\|full | Not-determined |
| S3_vs_k141_54770\|\|full | Low-quality |
| S3_vs_k141_63036\|\|full | Low-quality |
| S3_vs_k141_125303\|\|full | Low-quality |
| S3_vs_k141_38295\|\|full | Not-determined |
| S3_vs_k141_117872\|\|full | Not-determined |
| S3_vs_k141_94171\|\|full | Not-determined |
| S3_vs_k141_94175\|\|full | Not-determined |
| S3_vs_k141_117884\|\|full | Low-quality |
| S3_vs_k141_101038\|\|full | Low-quality |
| S3_vs_k141_125329\|\|full | Low-quality |
| S3_vs_k141_63066\|\|full | Low-quality |
| S3_vs_k141_54800\|\|full | Low-quality |
| S3_vs_k141_6037\|\|full | Low-quality |
| S3_vs_k141_54801\|\|full | Not-determined |
| S3_vs_k141_63088\|\|full | Not-determined |
| S3_vs_k141_69210\|\|full | Low-quality |
| S3_vs_k141_110254\|\|full | Not-determined |
| S3_vs_k141_69211\|\|full | Not-determined |
| S3_vs_k141_54822\|\|full | Low-quality |
| S3_vs_k141_69216\|\|full | Not-determined |
| S3_vs_k141_63101\|\|full | Low-quality |
| S3_vs_k141_94221\|\|full | Low-quality |
| S3_vs_k141_38349\|\|full | Low-quality |
| S3_vs_k141_63113\|\|full | Low-quality |
| S3_vs_k141_69221\|\|full | Low-quality |
| S3_vs_k141_110260\|\|full | Low-quality |
| S3_vs_k141_54843\|\|full | Low-quality |
| S3_vs_k141_69234\|\|full | Low-quality |
| S3_vs_k141_38361\|\|full | Low-quality |
| S3_vs_k141_101091\|\|full | Low-quality |
| S14_extracted_k141_72335 | High-quality |
| S3_vs_k141_125356\|\|full | Low-quality |
| S3_vs_k141_38376\|\|full | Low-quality |
| S3_vs_k141_46338\|\|full | Low-quality |
| S3_vs_k141_54855\|\|full | Low-quality |
| S3_vs_k141_69256\|\|full | Low-quality |
| S3_vs_k141_54858\|\|full | Low-quality |
| S3_vs_k141_30302\|\|full | Not-determined |
| S3_vs_k141_110310\|\|full | Not-determined |
| S3_vs_k141_6086\|\|full | Not-determined |
| S3_vs_k141_54867\|\|full | Low-quality |
| S3_vs_k141_86620\|\|full | Low-quality |
| S3_vs_k141_21506\|\|full | Low-quality |
| S3_vs_k141_69275\|\|full | Low-quality |
| S3_vs_k141_21511\|\|full | Low-quality |
| S3_vs_k141_110325\|\|full | Not-determined |
| S3_vs_k141_6126\|\|full | Low-quality |
| S3_vs_k141_69292\|\|full | Not-determined |
| S3_vs_k141_54898\|\|full | Low-quality |
| S3_vs_k141_21529\|\|full | Low-quality |
| S3_vs_k141_101133\|\|full | Low-quality |
| S3_vs_k141_101134\|\|full | Low-quality |
| S3_vs_k141_69302\|\|full | Low-quality |
| S3_vs_k141_101143\|\|full | Low-quality |
| S3_vs_k141_69307\|\|full | Not-determined |
| S3_vs_k141_54904\|\|full | Not-determined |
| S3_vs_k141_69308\|\|full | Not-determined |
| S3_vs_k141_69311\|\|full | Low-quality |
| S3_vs_k141_21548\|\|full | Low-quality |
| S4_vs_VIRSorter_6022-circular-cat_ | High-quality |
| S3_vs_k141_63157\|\|full | Low-quality |
| S3_vs_k141_79004\|\|full | Low-quality |
| S3_vs_k141_21569\|\|full | Low-quality |
| S3_vs_k141_101149\|\|full | Low-quality |
| S3_vs_k141_110330\|\|full | Not-determined |
| S3_vs_k141_69320\|\|full | Low-quality |
| S3_vs_k141_6159\|\|full | Low-quality |
| S3_vs_k141_101159\|\|full | Not-determined |
| S3_vs_k141_38488\|\|full | Not-determined |
| S3_vs_k141_79015\|\|full | Low-quality |
| S3_vs_k141_63175\|\|full | Low-quality |
| S3_vs_k141_101171\|\|full | Low-quality |
| S3_vs_k141_6169\|\|full | Not-determined |
| S3_vs_k141_6174\|\|full | Not-determined |
| S3_vs_k141_110345\|\|full | Not-determined |
| S3_vs_k141_46397\|\|full | Low-quality |
| S3_vs_k141_6177\|\|full | Not-determined |
| S3_vs_k141_38507\|\|full | Not-determined |
| S3_vs_k141_101187\|\|full | Low-quality |
| S3_vs_k141_38510\|\|full | Low-quality |
| S3_vs_k141_101198\|\|full | Low-quality |
| S3_vs_k141_101199\|\|full | Low-quality |
| S3_vs_k141_69341\|\|full | Low-quality |
| S3_vs_k141_101211\|\|full | Low-quality |
| S3_vs_k141_38527\|\|full | Low-quality |
| S3_vs_k141_94290\|\|full | Low-quality |
| S3_vs_k141_69351\|\|full | Not-determined |
| S3_vs_k141_86703\|\|full | Low-quality |
| S3_vs_k141_117904\|\|full | Low-quality |
| S3_vs_k141_38528\|\|full | Low-quality |
| S3_vs_k141_54955\|\|full | Not-determined |
| S3_vs_k141_86712\|\|full | Low-quality |
| S3_vs_k141_117912\|\|full | Not-determined |
| S3_vs_k141_125430\|\|full | Low-quality |
| S3_vs_k141_125442\|\|full | Not-determined |
| S3_vs_k141_46414\|\|full | Not-determined |
| S3_vs_k141_101249\|\|full | Low-quality |
| S3_vs_k141_21642\|\|full | Low-quality |
| S3_vs_k141_86726\|\|full | Low-quality |
| S3_vs_k141_69384\|\|full | Low-quality |
| S3_vs_k141_63225\|\|full | Not-determined |
| S3_vs_k141_21646\|\|full | Low-quality |
| S3_vs_k141_21658\|\|full | Not-determined |
| S3_vs_k141_125443\|\|full | Low-quality |
| S3_vs_k141_63235\|\|full | Low-quality |
| S3_vs_k141_117944\|\|full | Low-quality |
| S3_vs_k141_69406\|\|full | Low-quality |
| S3_vs_k141_101277\|\|full | Not-determined |
| S3_vs_k141_69410\|\|full | Low-quality |
| S3_vs_k141_110390\|\|full | Low-quality |
| S3_vs_k141_86754\|\|full | Low-quality |
| S3_vs_k141_63281\|\|full | Not-determined |
| S3_vs_k141_94308\|\|full | Low-quality |
| S3_vs_k141_86762\|\|full | Low-quality |
| S3_vs_k141_101297\|\|full | Low-quality |
| S3_vs_k141_110403\|\|full | Low-quality |
| S3_vs_k141_30480\|\|full | Low-quality |
| S3_vs_k141_86775\|\|full | Low-quality |
| S3_vs_k141_117985\|\|full | Not-determined |
| S3_vs_k141_21685\|\|full | Low-quality |
| S3_vs_k141_118004\|\|full | Not-determined |
| S3_vs_k141_118010\|\|full | Low-quality |
| S3_vs_k141_101330\|\|full | Low-quality |
| S3_vs_k141_110408\|\|full | Not-determined |
| S3_vs_k141_55016\|\|full | Low-quality |
| S3_vs_k141_118022\|\|full | Not-determined |
| S3_vs_k141_55024\|\|full | Low-quality |

| | |
|---|---|
| S3_vs_k141_110422\|\|full | Low-quality |
| S3_vs_k141_125483\|\|full | Not-determined |
| S3_vs_k141_6334\|\|full | Low-quality |
| S3_vs_k141_101358\|\|full | Low-quality |
| S3_vs_k141_86799\|\|full | Low-quality |
| S3_vs_k141_55044\|\|full | Low-quality |
| S3_vs_k141_110455\|\|full | Low-quality |
| S3_vs_k141_94317\|\|full | Low-quality |
| S3_vs_k141_86804\|\|full | Not-determined |
| S3_vs_k141_101361\|\|full | Low-quality |
| S3_vs_k141_21745\|\|full | Low-quality |
| S3_vs_k141_110483\|\|full | Low-quality |
| S3_vs_k141_46425\|\|full | Not-determined |
| S3_vs_k141_101396\|\|full | Not-determined |
| S3_vs_k141_101397\|\|full | Low-quality |
| S3_vs_k141_6367\|\|full | Low-quality |
| S3_vs_k141_110495\|\|full | Low-quality |
| S3_vs_k141_55057\|\|full | Not-determined |
| S3_vs_k141_110506\|\|full | Low-quality |
| S3_vs_k141_125568\|\|full | Low-quality |
| S3_vs_k141_101414\|\|full | Low-quality |
| S3_vs_k141_55071\|\|full | Not-determined |
| S3_vs_k141_101440\|\|full | Low-quality |
| S3_vs_k141_101442\|\|full | Not-determined |
| S3_vs_k141_69458\|\|full | Low-quality |
| S3_vs_k141_86819\|\|full | Not-determined |
| S3_vs_k141_55108\|\|full | Not-determined |
| S3_vs_k141_110535\|\|full | Not-determined |
| S3_vs_k141_21810\|\|full | Not-determined |
| S3_vs_k141_94404\|\|full | Low-quality |
| S3_vs_k141_94407\|\|full | Not-determined |
| S3_vs_k141_69498\|\|full | Low-quality |
| S3_vs_k141_86827\|\|full | Low-quality |
| S3_vs_k141_69503\|\|full | Low-quality |
| S3_vs_k141_86832\|\|full | Low-quality |
| S3_vs_k141_38569\|\|full | Not-determined |
| S3_vs_k141_21825\|\|full | Low-quality |
| S3_vs_k141_86841\|\|full | Low-quality |
| S3_vs_k141_86842\|\|full | Not-determined |
| S3_vs_k141_21838\|\|full | Low-quality |
| S3_vs_k141_110569\|\|full | Not-determined |
| S3_vs_k141_86857\|\|full | Not-determined |
| S3_vs_k141_55137\|\|full | Low-quality |
| S3_vs_k141_38597\|\|full | Not-determined |
| S3_vs_k141_30551\|\|full | Low-quality |
| S3_vs_k141_46507\|\|full | Low-quality |
| S3_vs_k141_94462\|\|full | Not-determined |
| S3_vs_k141_94464\|\|full | Low-quality |
| S3_vs_k141_69534\|\|full | Not-determined |
| S3_vs_k141_125650\|\|full | Low-quality |
| S3_vs_k141_125654\|\|full | Low-quality |
| S3_vs_k141_86879\|\|full | Low-quality |
| S3_vs_k141_101451\|\|full | Low-quality |
| S3_vs_k141_46544\|\|full | Low-quality |
| S3_vs_k141_63384\|\|full | Low-quality |
| S3_vs_k141_46557\|\|full | Low-quality |
| S3_vs_k141_69562\|\|full | Low-quality |
| S3_vs_k141_101489\|\|full | Low-quality |
| S3_vs_k141_30581\|\|full | Low-quality |
| S3_vs_k141_101497\|\|full | Not-determined |
| S3_vs_k141_79167\|\|full | Low-quality |
| S3_vs_k141_118162\|\|full | Low-quality |
| S3_vs_k141_30588\|\|full | Low-quality |
| S3_vs_k141_110603\|\|full | Not-determined |
| S3_vs_k141_46591\|\|full | Low-quality |
| S3_vs_k141_118171\|\|full | Low-quality |
| S3_vs_k141_22012\|\|full | Not-determined |
| S3_vs_k141_46593\|\|full | Low-quality |
| S3_vs_k141_94505\|\|full | Low-quality |
| S3_vs_k141_86961\|\|full | Not-determined |
| S3_vs_k141_79194\|\|full | Not-determined |
| S3_vs_k141_94506\|\|full | Not-determined |
| S3_vs_k141_94507\|\|full | Not-determined |
| S3_vs_k141_79195\|\|full | Low-quality |
| S3_vs_k141_79202\|\|full | Not-determined |
| S3_vs_k141_125700\|\|full | Low-quality |
| S3_vs_k141_63417\|\|full | Low-quality |
| S3_vs_k141_101533\|\|full | Not-determined |
| S3_vs_k141_86992\|\|full | Low-quality |
| S3_vs_k141_94521\|\|full | Low-quality |
| S3_vs_k141_55166\|\|full | Not-determined |
| S3_vs_k141_110617\|\|full | Not-determined |
| S3_vs_k141_118210\|\|full | Not-determined |
| S3_vs_k141_125712\|\|full | Not-determined |
| S3_vs_k141_38757\|\|full | Not-determined |
| S3_vs_k141_63433\|\|full | Not-determined |
| S3_vs_k141_55170\|\|full | Low-quality |
| S3_vs_k141_110639\|\|full | Not-determined |
| S3_vs_k141_101558\|\|full | Low-quality |
| S3_vs_k141_94542\|\|full | Not-determined |
| S3_vs_k141_63445\|\|full | Not-determined |
| S3_vs_k141_101573\|\|full | Low-quality |
| S3_vs_k141_79263\|\|full | Not-determined |
| S3_vs_k141_55184\|\|full | Low-quality |
| S3_vs_k141_79268\|\|full | Low-quality |
| S3_vs_k141_6525\|\|full | Low-quality |
| S7_vs_83100\|\|full | High-quality |
| S3_vs_k141_87006\|\|full | Low-quality |
| S3_vs_k141_38781\|\|full | Not-determined |
| S3_vs_k141_69641\|\|full | Low-quality |
| S3_vs_k141_63474\|\|full | Low-quality |
| S3_vs_k141_22052\|\|full | Low-quality |
| S3_vs_k141_79286\|\|full | Low-quality |
| S3_vs_k141_22059\|\|full | Not-determined |
| S3_vs_k141_110668\|\|full | Low-quality |
| S3_vs_k141_94574\|\|full | Low-quality |
| S3_vs_k141_6553\|\|full | Low-quality |
| S3_vs_k141_79305\|\|full | Not-determined |
| S3_vs_k141_63508\|\|full | Low-quality |
| S3_vs_k141_69683\|\|full | Low-quality |
| S3_vs_k141_118287\|\|full | Not-determined |
| S3_vs_k141_87055\|\|full | Not-determined |
| S3_vs_k141_125749\|\|full | Low-quality |
| S8_extracted_k141_208126 | Complete |
| S3_vs_k141_101603\|\|full | Low-quality |
| S3_vs_k141_101610\|\|full | Low-quality |
| S3_vs_k141_110714\|\|full | Low-quality |
| S3_vs_k141_125756\|\|full | Low-quality |
| S3_vs_k141_94590\|\|full | Not-determined |
| S3_vs_k141_22104\|\|full | Low-quality |
| S3_vs_k141_79363\|\|full | Not-determined |
| S3_vs_k141_22108\|\|full | Low-quality |
| S3_vs_k141_55205\|\|full | Low-quality |
| S3_vs_k141_46702\|\|full | Low-quality |
| S3_vs_k141_94597\|\|full | Low-quality |
| S3_vs_k141_63560\|\|full | Low-quality |
| S3_vs_k141_46703\|\|full | Low-quality |
| S3_vs_k141_94612\|\|full | Not-determined |
| S3_vs_k141_87084\|\|full | Low-quality |
| S3_vs_k141_79379\|\|full | Low-quality |
| S3_vs_k141_94616\|\|full | Low-quality |
| S3_vs_k141_63581\|\|full | Low-quality |
| S3_vs_k141_87102\|\|full | Low-quality |
| S3_vs_k141_69757\|\|full | Low-quality |
| S3_vs_k141_46736\|\|full | Low-quality |
| S3_vs_k141_110749\|\|full | Not-determined |
| S3_vs_k141_46739\|\|full | Low-quality |
| S3_vs_k141_63609\|\|full | Not-determined |
| S3_vs_k141_87113\|\|full | Not-determined |
| S3_vs_k141_22171\|\|full | Low-quality |
| S3_vs_k141_125797\|\|full | Low-quality |
| S3_vs_k141_46748\|\|full | Low-quality |
| S3_vs_k141_79427\|\|full | Low-quality |
| S3_vs_k141_125801\|\|full | Low-quality |
| S3_vs_k141_118345\|\|full | Low-quality |
| S3_vs_k141_55244\|\|full | Low-quality |
| S3_vs_k141_118357\|\|full | Not-determined |
| S3_vs_k141_46764\|\|full | Low-quality |
| S3_vs_k141_94672\|\|full | Not-determined |
| S3_vs_k141_63614\|\|full | Low-quality |
| S3_vs_k141_63639\|\|full | Low-quality |
| S3_vs_k141_38840\|\|full | Not-determined |
| S3_vs_k141_46793\|\|full | Low-quality |
| S3_vs_k141_55263\|\|full | Low-quality |
| S3_vs_k141_63661\|\|full | Low-quality |
| S3_vs_k141_22238\|\|full | Low-quality |
| S3_vs_k141_22255\|\|full | Not-determined |
| S3_vs_k141_101681\|\|full | Low-quality |
| S3_vs_k141_30658\|\|full | Not-determined |
| S3_vs_k141_101687\|\|full | Low-quality |
| S13_vs_148294\|\|full | High-quality |
| S3_vs_k141_46826\|\|full | Low-quality |
| S3_vs_k141_6607\|\|full | Low-quality |
| S3_vs_k141_6609\|\|full | Not-determined |
| S3_vs_k141_46830\|\|full | Not-determined |
| S3_vs_k141_38857\|\|full | Low-quality |
| S3_vs_k141_118371\|\|full | Low-quality |
| S3_vs_k141_46840\|\|full | Not-determined |
| S3_vs_k141_30674\|\|full | Low-quality |
| S3_vs_k141_101702\|\|full | Low-quality |
| S3_vs_k141_94756\|\|full | Not-determined |
| S3_vs_k141_22275\|\|full | Low-quality |
| S3_vs_k141_30683\|\|full | Low-quality |
| S3_vs_k141_6644\|\|full | Not-determined |
| S3_vs_k141_69844\|\|full | Low-quality |
| S3_vs_k141_55301\|\|full | Not-determined |
| S3_vs_k141_87202\|\|full | Low-quality |
| S3_vs_k141_87209\|\|full | Not-determined |
| S3_vs_k141_94776\|\|full | Not-determined |
| S3_vs_k141_110801\|\|full | Low-quality |
| S3_vs_k141_69850\|\|full | Not-determined |
| S3_vs_k141_94793\|\|full | Not-determined |
| S3_vs_k141_125844\|\|full | Low-quality |
| S3_vs_k141_94795\|\|full | Not-determined |
| S3_vs_k141_38899\|\|full | Not-determined |
| S3_vs_k141_6671\|\|full | Not-determined |
| S3_vs_k141_125845\|\|full | Low-quality |
| S3_vs_k141_87231\|\|full | Not-determined |
| S3_vs_k141_101747\|\|full | Low-quality |
| S3_vs_k141_101748\|\|full | Not-determined |
| S3_vs_k141_94806\|\|full | Not-determined |
| S3_vs_k141_30745\|\|full | Low-quality |
| S3_vs_k141_30747\|\|full | Not-determined |
| S3_vs_k141_6699\|\|full | Low-quality |
| S3_vs_k141_22323\|\|full | Low-quality |
| S3_vs_k141_94818\|\|full | Not-determined |
| S3_vs_k141_63705\|\|full | Not-determined |
| S3_vs_k141_69893\|\|full | Low-quality |
| S3_vs_k141_94829\|\|full | Not-determined |
| S3_vs_k141_22338\|\|full | Not-determined |
| S3_vs_k141_94833\|\|full | Low-quality |
| S3_vs_k141_69920\|\|full | Low-quality |
| S3_vs_k141_69921\|\|full | Not-determined |
| S3_vs_k141_101804\|\|full | Not-determined |
| S3_vs_k141_79465\|\|full | Low-quality |
| S3_vs_k141_46927\|\|full | Low-quality |
| S3_vs_k141_79471\|\|full | Not-determined |
| S3_vs_k141_69930\|\|full | Not-determined |
| S3_vs_k141_6725\|\|full | Low-quality |
| S3_vs_k141_63713\|\|full | Not-determined |
| S3_vs_k141_30789\|\|full | Not-determined |
| S3_vs_k141_69944\|\|full | Low-quality |
| S3_vs_k141_118462\|\|full | Not-determined |
| S3_vs_k141_101834\|\|full | Low-quality |
| S3_vs_k141_87284\|\|full | Low-quality |
| S3_vs_k141_6729\|\|full | Low-quality |
| S3_vs_k141_30800\|\|full | Low-quality |
| S3_vs_k141_55378\|\|full | Not-determined |
| S3_vs_k141_22397\|\|full | Not-determined |
| S3_vs_k141_6734\|\|full | Low-quality |
| S3_vs_k141_30817\|\|full | Low-quality |
| S3_vs_k141_55391\|\|full | Low-quality |
| S3_vs_k141_125954\|\|full | Low-quality |
| S3_vs_k141_30828\|\|full | Low-quality |
| S3_vs_k141_46961\|\|full | Not-determined |
| S3_vs_k141_118501\|\|full | Not-determined |
| S3_vs_k141_6739\|\|full | Low-quality |
| S3_vs_k141_101860\|\|full | Low-quality |
| S3_vs_k141_46977\|\|full | Low-quality |
| S3_vs_k141_30841\|\|full | Not-determined |
| S3_vs_k141_110892\|\|full | Not-determined |
| S3_vs_k141_79512\|\|full | Not-determined |
| S3_vs_k141_125968\|\|full | Not-determined |
| S3_vs_k141_79523\|\|full | Low-quality |
| S3_vs_k141_6761\|\|full | Low-quality |
| S3_vs_k141_46990\|\|full | Not-determined |
| S3_vs_k141_118517\|\|full | Not-determined |
| S3_vs_k141_30863\|\|full | Low-quality |
| S3_vs_k141_94913\|\|full | Not-determined |
| S3_vs_k141_118526\|\|full | Low-quality |
| S3_vs_k141_118527\|\|full | Low-quality |
| S3_vs_k141_47003\|\|full | Not-determined |
| S3_vs_k141_94924\|\|full | Low-quality |
| S3_vs_k141_47005\|\|full | Low-quality |
| S3_vs_k141_70007\|\|full | Low-quality |
| S3_vs_k141_110910\|\|full | Not-determined |
| S3_vs_k141_94949\|\|full | Low-quality |
| S3_vs_k141_110912\|\|full | Not-determined |
| S3_vs_k141_55436\|\|full | Low-quality |
| S3_vs_k141_55442\|\|full | Low-quality |
| S3_vs_k141_79571\|\|full | Low-quality |
| S3_vs_k141_126004\|\|full | Not-determined |
| S3_vs_k141_22419\|\|full | Not-determined |
| S3_vs_k141_55453\|\|full | Not-determined |
| S3_vs_k141_79585\|\|full | Not-determined |
| S3_vs_k141_110959\|\|full | Not-determined |
| S3_vs_k141_110964\|\|full | Not-determined |
| S3_vs_k141_118553\|\|full | Low-quality |
| S3_vs_k141_110971\|\|full | Not-determined |
| S3_vs_k141_70046\|\|full | Not-determined |
| S3_vs_k141_47056\|\|full | Low-quality |
| S3_vs_k141_70049\|\|full | Not-determined |
| S3_vs_k141_70052\|\|full | Not-determined |
| S3_vs_k141_95002\|\|full | Not-determined |
| S3_vs_k141_39040\|\|full | Low-quality |
| S3_vs_k141_118581\|\|full | Not-determined |
| S3_vs_k141_118585\|\|full | Low-quality |
| S3_vs_k141_55475\|\|full | Not-determined |
| S3_vs_k141_6831\|\|full | Not-determined |
| S3_vs_k141_111023\|\|full | Low-quality |
| S3_vs_k141_6838\|\|full | Not-determined |
| S3_vs_k141_118591\|\|full | Low-quality |
| S3_vs_k141_111028\|\|full | Low-quality |
| S3_vs_k141_79636\|\|full | Not-determined |
| S3_vs_k141_39079\|\|full | Low-quality |
| S3_vs_k141_22435\|\|full | Low-quality |
| S3_vs_k141_118601\|\|full | Low-quality |
| S9_vs_k141_27813\|\|full | High-quality |
| S3_vs_k141_95030\|\|full | Not-determined |
| S3_vs_k141_111065\|\|full | Low-quality |
| S3_vs_k141_95035\|\|full | Low-quality |
| S3_vs_k141_111067\|\|full | Not-determined |
| S3_vs_k141_95036\|\|full | Not-determined |
| S3_vs_k141_70128\|\|full | Low-quality |
| S3_vs_k141_22468\|\|full | Low-quality |
| S3_vs_k141_47112\|\|full | Low-quality |
| S3_vs_k141_22473\|\|full | Low-quality |
| S3_vs_k141_95043\|\|full | Low-quality |
| S3_vs_k141_47117\|\|full | Not-determined |
| S3_vs_k141_22474\|\|full | Low-quality |
| S3_vs_k141_30955\|\|full | Low-quality |
| S3_vs_k141_39146\|\|full | Low-quality |
| S3_vs_k141_95045\|\|full | Low-quality |
| S3_vs_k141_47123\|\|full | Low-quality |
| S3_vs_k141_111086\|\|full | Not-determined |
| S3_vs_k141_95051\|\|full | Not-determined |
| S3_vs_k141_118644\|\|full | Not-determined |
| S3_vs_k141_118653\|\|full | Not-determined |
| S3_vs_k141_111094\|\|full | Not-determined |
| S3_vs_k141_118654\|\|full | Low-quality |
| S3_vs_k141_79679\|\|full | Low-quality |
| S3_vs_k141_101957\|\|full | Low-quality |
| S3_vs_k141_55536\|\|full | Not-determined |
| S3_vs_k141_87340\|\|full | Low-quality |
| S3_vs_k141_47153\|\|full | Not-determined |
| S3_vs_k141_55542\|\|full | Low-quality |
| S3_vs_k141_118675\|\|full | Not-determined |
| S3_vs_k141_118678\|\|full | Low-quality |
| S3_vs_k141_101970\|\|full | Low-quality |
| S3_vs_k141_70158\|\|full | Low-quality |
| S3_vs_k141_111170\|\|full | Low-quality |
| S3_vs_k141_87374\|\|full | Low-quality |
| S3_vs_k141_95081\|\|full | Low-quality |
| S3_vs_k141_111188\|\|full | Not-determined |
| S3_vs_k141_87376\|\|full | Low-quality |
| S3_vs_k141_87377\|\|full | Low-quality |
| S3_vs_k141_30989\|\|full | Low-quality |
| S3_vs_k141_6936\|\|full | Low-quality |
| S3_vs_k141_39220\|\|full | Not-determined |
| S3_vs_k141_118736\|\|full | Not-determined |
| S3_vs_k141_39224\|\|full | Low-quality |
| S3_vs_k141_111218\|\|full | Not-determined |
| S3_vs_k141_95102\|\|full | Low-quality |
| S3_vs_k141_95113\|\|full | Low-quality |
| S3_vs_k141_111241\|\|full | Not-determined |
| S3_vs_k141_111242\|\|full | Low-quality |
| S3_vs_k141_70191\|\|full | Not-determined |
| S3_vs_k141_31033\|\|full | Low-quality |
| S3_vs_k141_126263\|\|full | Low-quality |
| S3_vs_k141_39268\|\|full | Low-quality |
| S3_vs_k141_126275\|\|full | Low-quality |
| S3_vs_k141_111260\|\|full | Not-determined |
| S3_vs_k141_102000\|\|full | Low-quality |
| S3_vs_k141_70218\|\|full | Low-quality |
| S8_extracted_k141_28956 | Medium-quality |
| S3_vs_k141_95123\|\|full | Low-quality |
| S3_vs_k141_111288\|\|full | Not-determined |
| S3_vs_k141_31047\|\|full | Not-determined |
| S3_vs_k141_102026\|\|full | Low-quality |
| S3_vs_k141_118785\|\|full | Low-quality |
| S3_vs_k141_47238\|\|full | Low-quality |
| S3_vs_k141_95140\|\|full | Not-determined |
| S3_vs_k141_111291\|\|full | Low-quality |
| S3_vs_k141_95156\|\|full | Low-quality |
| S3_vs_k141_87469\|\|full | Low-quality |
| S3_vs_k141_95167\|\|full | Low-quality |
| S3_vs_k141_55626\|\|full | Low-quality |
| S3_vs_k141_47268\|\|full | Low-quality |
| S3_vs_k141_87484\|\|full | Not-determined |
| S3_vs_k141_47293\|\|full | Low-quality |
| S3_vs_k141_111315\|\|full | Low-quality |
| S3_vs_k141_95185\|\|full | Low-quality |
| S3_vs_k141_111330\|\|full | Not-determined |
| S3_vs_k141_47311\|\|full | Low-quality |
| S3_vs_k141_87515\|\|full | Low-quality |
| S3_vs_k141_111350\|\|full | Low-quality |
| S3_vs_k141_6981\|\|full | Not-determined |
| S3_vs_k141_31147\|\|full | Low-quality |
| S3_vs_k141_47333\|\|full | Low-quality |
| S3_vs_k141_70268\|\|full | Low-quality |
| S3_vs_k141_111357\|\|full | Low-quality |
| S3_vs_k141_55664\|\|full | Not-determined |
| S3_vs_k141_95212\|\|full | Not-determined |
| S3_vs_k141_111360\|\|full | Low-quality |
| S3_vs_k141_111362\|\|full | Low-quality |
| S3_vs_k141_55665\|\|full | Not-determined |
| S3_vs_k141_126334\|\|full | Low-quality |
| S3_vs_k141_111375\|\|full | Low-quality |
| S3_vs_k141_126340\|\|full | Low-quality |
| S3_vs_k141_111379\|\|full | Low-quality |
| S3_vs_k141_118802\|\|full | Not-determined |
| S3_vs_k141_6997\|\|full | Low-quality |
| S3_vs_k141_95223\|\|full | Low-quality |
| S3_vs_k141_87579\|\|full | Not-determined |
| S3_vs_k141_70294\|\|full | Low-quality |
| S3_vs_k141_70295\|\|full | Not-determined |
| S3_vs_k141_39328\|\|full | Not-determined |
| S3_vs_k141_39333\|\|full | Not-determined |
| S3_vs_k141_118813\|\|full | Low-quality |
| S3_vs_k141_118817\|\|full | Low-quality |
| S3_vs_k141_70299\|\|full | Not-determined |
| S3_vs_k141_111402\|\|full | Low-quality |
| S3_vs_k141_31206\|\|full | Not-determined |
| S3_vs_k141_126367\|\|full | Low-quality |
| S3_vs_k141_111411\|\|full | Not-determined |
| S3_vs_k141_70307\|\|full | Low-quality |
| S3_vs_k141_70311\|\|full | Not-determined |
| S3_vs_k141_47384\|\|full | Not-determined |
| S3_vs_k141_22653\|\|full | Not-determined |
| S3_vs_k141_118839\|\|full | Low-quality |
| S3_vs_k141_126412\|\|full | Not-determined |
| S3_vs_k141_47385\|\|full | Low-quality |
| S3_vs_k141_87595\|\|full | Low-quality |
| S3_vs_k141_22683\|\|full | Not-determined |
| S3_vs_k141_102105\|\|full | Low-quality |
| S3_vs_k141_55765\|\|full | Low-quality |
| S3_vs_k141_111419\|\|full | Low-quality |
| S3_vs_k141_47400\|\|full | Low-quality |
| S3_vs_k141_111423\|\|full | Low-quality |
| S3_vs_k141_55769\|\|full | Low-quality |
| S3_vs_k141_111428\|\|full | Not-determined |
| S3_vs_k141_31285\|\|full | Not-determined |
| S3_vs_k141_55777\|\|full | Not-determined |
| S3_vs_k141_55784\|\|full | Low-quality |
| S3_vs_k141_126464\|\|full | Low-quality |
| S3_vs_k141_39383\|\|full | Not-determined |
| S3_vs_k141_31295\|\|full | Low-quality |
| S3_vs_k141_7012\|\|full | Not-determined |
| S3_vs_k141_47443\|\|full | Low-quality |
| S3_vs_k141_47455\|\|full | Low-quality |
| S3_vs_k141_31299\|\|full | Low-quality |
| S3_vs_k141_87650\|\|full | Low-quality |
| S3_vs_k141_102182\|\|full | Low-quality |
| S3_vs_k141_87656\|\|full | Low-quality |
| S3_vs_k141_39460\|\|full | Not-determined |
| S3_vs_k141_22707\|\|full | Low-quality |
| S3_vs_k141_31324\|\|full | Low-quality |
| S3_vs_k141_47506\|\|full | Low-quality |
| S3_vs_k141_22738\|\|full | Low-quality |
| S3_vs_k141_118893\|\|full | Not-determined |
| S3_vs_k141_70403\|\|full | Low-quality |
| S3_vs_k141_7053\|\|full | Low-quality |
| S3_vs_k141_70426\|\|full | Low-quality |
| S3_vs_k141_31410\|\|full | Not-determined |
| S3_vs_k141_95345\|\|full | Low-quality |
| S3_vs_k141_39521\|\|full | Low-quality |
| S3_vs_k141_39525\|\|full | Not-determined |
| S3_vs_k141_111499\|\|full | Not-determined |
| S3_vs_k141_111500\|\|full | Low-quality |
| S3_vs_k141_111511\|\|full | Not-determined |
| S3_vs_k141_102202\|\|full | Not-determined |
| S3_vs_k141_47566\|\|full | Not-determined |
| S3_vs_k141_39555\|\|full | Low-quality |
| S3_vs_k141_102208\|\|full | Low-quality |
| S3_vs_k141_22753\|\|full | Not-determined |
| S3_vs_k141_39561\|\|full | Low-quality |
| S3_vs_k141_31449\|\|full | Not-determined |
| S3_vs_k141_126546\|\|full | Low-quality |
| S3_vs_k141_126548\|\|full | Low-quality |
| S3_vs_k141_126556\|\|full | Not-determined |
| S3_vs_k141_22764\|\|full | Low-quality |
| S3_vs_k141_95380\|\|full | Not-determined |
| S3_vs_k141_22767\|\|full | Low-quality |
| S3_vs_k141_31463\|\|full | Low-quality |
| S3_vs_k141_39590\|\|full | Low-quality |
| S3_vs_k141_70453\|\|full | Low-quality |
| S3_vs_k141_95399\|\|full | Low-quality |
| S3_vs_k141_22772\|\|full | Not-determined |
| S3_vs_k141_102219\|\|full | Not-determined |
| S3_vs_k141_111546\|\|full | Low-quality |
| S3_vs_k141_102229\|\|full | Not-determined |
| S3_vs_k141_70464\|\|full | Low-quality |
| S3_vs_k141_102232\|\|full | Not-determined |
| S3_vs_k141_102233\|\|full | Not-determined |
| S3_vs_k141_111558\|\|full | Not-determined |
| S3_vs_k141_95416\|\|full | Low-quality |
| S3_vs_k141_47602\|\|full | Low-quality |
| S3_vs_k141_102238\|\|full | Not-determined |
| S3_vs_k141_47604\|\|full | Low-quality |
| S3_vs_k141_111575\|\|full | Low-quality |
| S3_vs_k141_7085\|\|full | Low-quality |
| S3_vs_k141_95436\|\|full | Low-quality |
| S3_vs_k141_95440\|\|full | Low-quality |
| S3_vs_k141_47629\|\|full | Not-determined |
| S3_vs_k141_102259\|\|full | Low-quality |
| S3_vs_k141_22812\|\|full | Low-quality |
| S3_vs_k141_95453\|\|full | Low-quality |
| S3_vs_k141_126603\|\|full | Low-quality |
| S3_vs_k141_126608\|\|full | Not-determined |
| S3_vs_k141_39685\|\|full | Not-determined |
| S3_vs_k141_22838\|\|full | Low-quality |
| S3_vs_k141_126627\|\|full | Low-quality |
| S3_vs_k141_95490\|\|full | Low-quality |
| S3_vs_k141_22841\|\|full | Low-quality |
| S3_vs_k141_95494\|\|full | Not-determined |
| S3_vs_k141_102306\|\|full | Not-determined |
| S3_vs_k141_102310\|\|full | Low-quality |
| S3_vs_k141_22861\|\|full | Not-determined |
| S3_vs_k141_102314\|\|full | Not-determined |
| S12_vs_k141_58560\|\|full | Medium-quality |
| S3_vs_k141_102322\|\|full | Low-quality |
| S3_vs_k141_102327\|\|full | Not-determined |
| S3_vs_k141_31494\|\|full | Not-determined |
| S13_extracted_151755 | Complete |
| S3_vs_k141_7149\|\|full | Low-quality |
| S3_vs_k141_7155\|\|full | Low-quality |
| S3_vs_k141_22923\|\|full | Low-quality |
| S3_vs_k141_47748\|\|full | Not-determined |
| S3_vs_k141_22939\|\|full | Not-determined |
| S3_vs_k141_47749\|\|full | Not-determined |
| S3_vs_k141_102349\|\|full | Not-determined |
| S3_vs_k141_31539\|\|full | Low-quality |
| S3_vs_k141_102351\|\|full | Low-quality |
| S3_vs_k141_102367\|\|full | Low-quality |
| S3_vs_k141_22968\|\|full | Low-quality |
| S3_vs_k141_126696\|\|full | Not-determined |
| S3_vs_k141_95529\|\|full | Low-quality |
| S3_vs_k141_70605\|\|full | Low-quality |
| S3_vs_k141_22981\|\|full | Low-quality |
| S3_vs_k141_47814\|\|full | Not-determined |
| S3_vs_k141_22988\|\|full | Low-quality |
| S3_vs_k141_102391\|\|full | Not-determined |
| S3_vs_k141_95533\|\|full | Low-quality |
| S3_vs_k141_126726\|\|full | Not-determined |
| S3_vs_k141_95549\|\|full | Not-determined |
| S3_vs_k141_102406\|\|full | Low-quality |
| S3_vs_k141_102410\|\|full | Not-determined |
| S3_vs_k141_126731\|\|full | Low-quality |
| S3_vs_k141_118930\|\|full | Not-determined |
| S3_vs_k141_31615\|\|full | Low-quality |
| S8_extracted_k141_182875 | Complete |
| S11_vs_k141_23357\|\|full | High-quality |
| S3_vs_k141_31641\|\|full | Low-quality |
| S8_vs_k141_208126\|\|full | High-quality |
| S3_vs_k141_7295\|\|full | Low-quality |
| S3_vs_k141_23043\|\|full | Low-quality |
| S3_vs_k141_118967\|\|full | Not-determined |
| S3_vs_k141_70679\|\|full | Low-quality |
| S3_vs_k141_23050\|\|full | Low-quality |
| S3_vs_k141_70686\|\|full | Low-quality |
| S3_vs_k141_23066\|\|full | Not-determined |
| S3_vs_k141_39815\|\|full | Low-quality |
| S3_vs_k141_39819\|\|full | Low-quality |
| S3_vs_k141_7321\|\|full | Low-quality |
| S3_vs_k141_118970\|\|full | Not-determined |
| S3_vs_k141_102451\|\|full | Not-determined |
| S3_vs_k141_31671\|\|full | Low-quality |
| S3_vs_k141_7336\|\|full | Low-quality |
| S3_vs_k141_102456\|\|full | Not-determined |
| S3_vs_k141_39841\|\|full | Low-quality |
| S3_vs_k141_7345\|\|full | Low-quality |
| S3_vs_k141_102465\|\|full | Not-determined |
| S3_vs_k141_23101\|\|full | Low-quality |
| S3_vs_k141_31700\|\|full | Not-determined |
| S3_vs_k141_95592\|\|full | Low-quality |
| S3_vs_k141_7364\|\|full | Low-quality |
| S3_vs_k141_119029\|\|full | Low-quality |
| S3_vs_k141_95608\|\|full | Low-quality |
| S3_vs_k141_126782\|\|full | Not-determined |
| S3_vs_k141_7401\|\|full | Low-quality |
| S3_vs_k141_31720\|\|full | Not-determined |
| S1_extracted_k141_158961 | Medium-quality |
| S3_vs_k141_7437\|\|full | Not-determined |
| S3_vs_k141_70753\|\|full | Low-quality |
| S3_vs_k141_23167\|\|full | Low-quality |
| S3_vs_k141_102608\|\|full | Not-determined |
| S3_vs_k141_102625\|\|full | Low-quality |
| S3_vs_k141_7489\|\|full | Low-quality |
| S17_vs_k141_103009\|\|full | High-quality |
| S3_vs_k141_102640\|\|full | Low-quality |
| S3_vs_k141_7505\|\|full | Low-quality |
| S3_vs_k141_7508\|\|full | Not-determined |
| S3_vs_k141_119061\|\|full | Low-quality |
| S3_vs_k141_23204\|\|full | Low-quality |
| S3_vs_k141_119082\|\|full | Not-determined |
| S13_vs_60596\|\|full | High-quality |
| S3_vs_k141_31771\|\|full | Low-quality |
| S3_vs_k141_7543\|\|full | Low-quality |
| S3_vs_k141_31794\|\|full | Low-quality |
| S3_vs_k141_126842\|\|full | Not-determined |
| S3_vs_k141_126844\|\|full | Low-quality |
| S3_vs_k141_102684\|\|full | Not-determined |
| S3_vs_k141_119100\|\|full | Low-quality |
| S3_vs_k141_126856\|\|full | Not-determined |
| S3_vs_k141_70894\|\|full | Low-quality |
| S3_vs_k141_7553\|\|full | Not-determined |
| S3_vs_k141_31813\|\|full | Not-determined |
| S3_vs_k141_23258\|\|full | Low-quality |
| S3_vs_k141_102712\|\|full | Not-determined |
| S3_vs_k141_23265\|\|full | Low-quality |
| S3_vs_k141_126879\|\|full | Low-quality |
| S3_vs_k141_7573\|\|full | Low-quality |
| S3_vs_k141_31857\|\|full | Low-quality |
| S3_vs_k141_7584\|\|full | Low-quality |
| S3_vs_k141_31863\|\|full | Low-quality |
| S3_vs_k141_102734\|\|full | Low-quality |
| S3_vs_k141_31868\|\|full | Low-quality |
| S3_vs_k141_31869\|\|full | Low-quality |
| S3_vs_k141_102774\|\|full | Not-determined |
| S3_vs_k141_102779\|\|full | Low-quality |
| S3_vs_k141_7622\|\|full | Not-determined |
| S3_vs_k141_119154\|\|full | Low-quality |
| S3_vs_k141_119155\|\|full | Low-quality |
| S3_vs_k141_7632\|\|full | Not-determined |
| S3_vs_k141_7638\|\|full | Low-quality |
| S3_vs_k141_126935\|\|full | Low-quality |
| S3_vs_k141_119162\|\|full | Low-quality |
| S3_vs_k141_7652\|\|full | Low-quality |
| S3_vs_k141_71044\|\|full | Low-quality |

S3_vs_k141_71046||full Not-determined
S3_vs_k141_23364||full Not-determined
S3_vs_k141_126958||full Low-quality
S3_vs_k141_119193||full Low-quality
S3_vs_k141_23368||full Low-quality
S3_vs_k141_102812||full Low-quality
S3_vs_k141_119202||full Low-quality
S3_vs_k141_102831||full Low-quality
S3_vs_k141_7731||full Low-quality
S3_vs_k141_7736||full Low-quality
S3_vs_k141_102844||full Low-quality
S3_vs_k141_119216||full Not-determined
S3_vs_k141_126959||full Low-quality
S3_vs_k141_126961||full Not-determined
S3_vs_k141_71097||full Low-quality
S3_vs_k141_7754||full Low-quality
S3_vs_k141_7769||full Low-quality
S3_vs_k141_119228||full Low-quality
S3_vs_k141_126991||full Low-quality
S3_vs_k141_71128||full Low-quality
S3_vs_k141_127020||full Low-quality
S3_vs_k141_23454||full Low-quality
S3_vs_k141_119252||full Low-quality
S3_vs_k141_127030||full Low-quality
S3_vs_k141_119253||full Low-quality
S3_vs_k141_119266||full Low-quality
S3_vs_k141_23493||full Not-determined
S3_vs_k141_127068||full Low-quality
S3_vs_k141_102900||full Low-quality
S3_vs_k141_71186||full Low-quality
S3_vs_k141_127085||full Not-determined
S3_vs_k141_23508||full Not-determined
S3_vs_k141_127088||full Not-determined
S3_vs_k141_127093||full Not-determined
S3_vs_k141_7878||full Low-quality
S3_vs_k141_119306||full Not-determined
S3_vs_k141_119332||full Low-quality
S3_vs_k141_7913||full Low-quality
S3_vs_k141_102945||full Low-quality
S3_vs_k141_119354||full Low-quality
S3_vs_k141_71264||full Low-quality
S3_vs_k141_127145||full Not-determined
S3_vs_k141_23544||full Not-determined
S3_vs_k141_119377||full Low-quality
S3_vs_k141_119402||full Low-quality
S1_vs_k141_159958||full High-quality
S3_vs_k141_23586||full Low-quality
S3_vs_k141_7941||full Low-quality
S3_vs_k141_127192||full Not-determined
S3_vs_k141_71323||full Low-quality
S3_vs_k141_23594||full Low-quality
S3_vs_k141_7948||full Not-determined
S3_vs_k141_71331||full Not-determined
S3_vs_k141_23597||full Not-determined
S3_vs_k141_103047||full Low-quality
S3_vs_k141_71341||full Low-quality
S3_vs_k141_71342||full Low-quality
S3_vs_k141_23605||full Not-determined
S3_vs_k141_119441||full Not-determined
S3_vs_k141_119446||full Low-quality
S3_vs_k141_103061||full Not-determined
S3_vs_k141_103063||full Low-quality
S3_vs_k141_71389||full Low-quality
S3_vs_k141_103093||full Low-quality
S3_vs_k141_127211||full Low-quality
S3_vs_k141_23664||full Not-determined
S3_vs_k141_127233||full Not-determined
S3_vs_k141_71436||full Low-quality
S3_vs_k141_71439||full Not-determined
S3_vs_k141_71453||full Low-quality
S3_vs_k141_127264||full Low-quality
S3_vs_k141_71476||full Low-quality
S3_vs_k141_119479||full Not-determined
S3_vs_k141_71492||full Not-determined
S3_vs_k141_127319||full Not-determined
S3_vs_k141_71510||full Not-determined
S3_vs_k141_103193||full Low-quality
S3_vs_k141_127360||full Not-determined
S3_vs_k141_103204||full Low-quality
S3_vs_k141_127371||full Low-quality
S3_vs_k141_127377||full Low-quality
S3_vs_k141_23807||full Low-quality
S16_vs_173094||full High-quality
S3_vs_k141_127388||full Not-determined
S3_vs_k141_23831||full Not-determined
S3_vs_k141_103298||full Not-determined
S3_vs_k141_71620||full Not-determined
S3_vs_k141_103355||full Low-quality
S3_vs_k141_103361||full Low-quality
S3_vs_k141_103368||full Not-determined
S3_vs_k141_103383||full Low-quality
S3_vs_k141_71690||full Not-determined
S3_vs_k141_71693||full Low-quality
S3_vs_k141_103401||full Low-quality
S3_vs_k141_103411||full Not-determined
S3_vs_k141_103421||full Low-quality
S3_vs_k141_103426||full Low-quality
S3_vs_k141_103465||full Not-determined
S3_vs_k141_103466||full Low-quality
S3_vs_k141_103471||full Low-quality
S3_vs_k141_103498||full Low-quality
S3_vs_k141_103537||full Low-quality
S3_vs_k141_103557||full Low-quality
S3_vs_k141_15975||0_partial Low-quality
S17_vs_k141_157209||full High-quality
S3_vs_k141_104463||0_partial Not-determined
S3_vs_k141_16671||0_partial Not-determined
S11_vs_k141_10172||full High-quality
S3_vs_k141_40939||1_partial Not-determined
S3_vs_k141_65537||0_partial Low-quality
S3_vs_k141_81761||0_partial Not-determined
S3_vs_k141_82496||0_partial Low-quality
S3_vs_k141_122047||0_partial Not-determined
S3_vs_k141_82843||0_partial Not-determined
S3_vs_k141_122179||0_partial Low-quality
S3_vs_k141_82962||0_partial Not-determined
S3_vs_k141_3235||0_partial Low-quality
S3_vs_k141_66595||0_partial Low-quality
S3_vs_k141_3710||0_partial Not-determined
S3_vs_k141_43757||0_partial Not-determined
S3_vs_k141_19171||0_partial Not-determined
S3_vs_k141_99701||0_partial Low-quality
S3_vs_k141_36691||0_partial Not-determined
S3_vs_k141_14542||0_partial Low-quality
S3_vs_k141_68123||0_partial Not-determined
S3_vs_k141_85076||0_partial Not-determined
S3_vs_k141_124525||0_partial Not-determined
S5_vs_k141_27918||full High-quality
S3_vs_k141_6985||0_partial Low-quality
S3_vs_k141_71320||0_partial Not-determined
S3_vs_k141_103464||0_partial Not-determined
S3_vs_k141_47822||lt2gene Not-determined
S3_vs_k141_31885||lt2gene Low-quality
S3_vs_k141_13||lt2gene Low-quality
S3_vs_k141_103619||lt2gene Low-quality
S3_vs_k141_19||lt2gene Low-quality
S3_vs_k141_55814||lt2gene Not-determined
S3_vs_k141_119572||lt2gene Low-quality
S3_vs_k141_95671||lt2gene Low-quality
S3_vs_k141_55822||lt2gene Not-determined
S3_vs_k141_55828||lt2gene Not-determined
S3_vs_k141_37||lt2gene Low-quality
S3_vs_k141_39887||lt2gene Low-quality
S3_vs_k141_111613||lt2gene Low-quality
S3_vs_k141_95703||lt2gene Not-determined
S3_vs_k141_111615||lt2gene Not-determined
S3_vs_k141_55845||lt2gene Not-determined
S3_vs_k141_103633||lt2gene Not-determined
S3_vs_k141_60||lt2gene Not-determined
S3_vs_k141_47944||lt2gene Not-determined
S3_vs_k141_15983||lt2gene Low-quality
S3_vs_k141_71834||lt2gene Not-determined
S3_vs_k141_87703||lt2gene Low-quality
S3_vs_k141_87710||lt2gene Low-quality
S3_vs_k141_48002||lt2gene Low-quality
S3_vs_k141_48004||lt2gene Low-quality
S3_vs_k141_71858||lt2gene Not-determined
S3_vs_k141_48024||lt2gene Low-quality
S3_vs_k141_55947||lt2gene Low-quality
S3_vs_k141_111691||lt2gene Low-quality
S3_vs_k141_111702||lt2gene Low-quality
S3_vs_k141_87733||lt2gene Low-quality
S3_vs_k141_87746||lt2gene Low-quality
S3_vs_k141_111718||lt2gene Not-determined
S3_vs_k141_146||lt2gene Not-determined
S3_vs_k141_31998||lt2gene Not-determined
S3_vs_k141_39951||lt2gene Low-quality
S3_vs_k141_24166||lt2gene Low-quality
S3_vs_k141_56011||lt2gene Not-determined
S3_vs_k141_103741||lt2gene Low-quality
S3_vs_k141_63970||lt2gene Low-quality
S3_vs_k141_87810||lt2gene Low-quality
S3_vs_k141_95835||lt2gene Not-determined
S3_vs_k141_111767||lt2gene Low-quality
S3_vs_k141_71950||lt2gene Low-quality
S3_vs_k141_95842||lt2gene Not-determined
S3_vs_k141_79934||lt2gene Not-determined
S3_vs_k141_40019||lt2gene Low-quality
S3_vs_k141_64038||lt2gene Low-quality
S3_vs_k141_40047||lt2gene Low-quality
S3_vs_k141_40051||lt2gene Low-quality
S3_vs_k141_95923||lt2gene Low-quality
S3_vs_k141_95927||lt2gene Low-quality
S3_vs_k141_95928||lt2gene Low-quality
S3_vs_k141_72020||lt2gene Low-quality
S3_vs_k141_95936||lt2gene Low-quality
S3_vs_k141_8284||lt2gene Low-quality
S3_vs_k141_72040||lt2gene Low-quality
S3_vs_k141_16054||lt2gene Low-quality
S3_vs_k141_302||lt2gene Low-quality
S3_vs_k141_303||lt2gene Low-quality
S3_vs_k141_308||lt2gene Not-determined
S3_vs_k141_16071||lt2gene Not-determined
S3_vs_k141_119742||lt2gene Low-quality
S3_vs_k141_328||lt2gene Not-determined
S3_vs_k141_48332||lt2gene Low-quality
S3_vs_k141_80006||lt2gene Not-determined
S3_vs_k141_24296||lt2gene Low-quality
S3_vs_k141_119773||lt2gene Low-quality
S3_vs_k141_56164||lt2gene Low-quality
S3_vs_k141_64130||lt2gene Low-quality
S3_vs_k141_64132||lt2gene Low-quality
S3_vs_k141_24317||lt2gene Not-determined
S3_vs_k141_405||lt2gene Low-quality
S3_vs_k141_56186||lt2gene Low-quality
S3_vs_k141_8445||lt2gene Low-quality
S3_vs_k141_80037||lt2gene Low-quality
S3_vs_k141_80041||lt2gene Low-quality
S3_vs_k141_96070||lt2gene Low-quality
S3_vs_k141_111857||lt2gene Low-quality
S3_vs_k141_24361||lt2gene Low-quality
S3_vs_k141_32256||lt2gene Low-quality
S3_vs_k141_32260||lt2gene Low-quality
S3_vs_k141_32265||lt2gene Low-quality
S3_vs_k141_80087||lt2gene Low-quality
S3_vs_k141_40249||lt2gene Not-determined
S3_vs_k141_8499||lt2gene Low-quality
S3_vs_k141_48385||lt2gene Not-determined
S3_vs_k141_48390||lt2gene Low-quality
S3_vs_k141_24389||lt2gene Low-quality
S3_vs_k141_32331||lt2gene Low-quality
S3_vs_k141_64146||lt2gene Low-quality
S3_vs_k141_32344||lt2gene Not-determined
S3_vs_k141_64150||lt2gene Not-determined
S3_vs_k141_8556||lt2gene Low-quality
S3_vs_k141_111916||lt2gene Low-quality
S3_vs_k141_119916||lt2gene Low-quality
S3_vs_k141_119934||lt2gene Not-determined
S3_vs_k141_88010||lt2gene Low-quality
S3_vs_k141_32426||lt2gene Low-quality
S3_vs_k141_513||lt2gene Low-quality
S3_vs_k141_96171||lt2gene Low-quality
S3_vs_k141_96175||lt2gene Not-determined
S3_vs_k141_24506||lt2gene Not-determined
S3_vs_k141_111964||lt2gene Low-quality
S3_vs_k141_32478||lt2gene Not-determined
S3_vs_k141_32483||lt2gene Not-determined
S3_vs_k141_24529||lt2gene Low-quality
S3_vs_k141_16185||lt2gene Not-determined
S3_vs_k141_72164||lt2gene Low-quality
S3_vs_k141_88091||lt2gene Low-quality
S3_vs_k141_8735||lt2gene Low-quality
S3_vs_k141_48484||lt2gene Low-quality
S3_vs_k141_32519||lt2gene Low-quality
S3_vs_k141_663||lt2gene Low-quality
S3_vs_k141_72280||lt2gene Low-quality
S3_vs_k141_112018||lt2gene Low-quality
S3_vs_k141_8824||lt2gene Not-determined
S3_vs_k141_104174||lt2gene Not-determined
S3_vs_k141_112041||lt2gene Low-quality
S3_vs_k141_104190||lt2gene Low-quality
S3_vs_k141_8848||lt2gene Low-quality
S3_vs_k141_48532||lt2gene Low-quality
S3_vs_k141_40492||lt2gene Low-quality
S3_vs_k141_96257||lt2gene Not-determined
S3_vs_k141_56295||lt2gene Not-determined
S3_vs_k141_80233||lt2gene Low-quality
S3_vs_k141_72373||lt2gene Low-quality
S3_vs_k141_72384||lt2gene Low-quality
S3_vs_k141_56342||lt2gene Low-quality
S3_vs_k141_96304||lt2gene Not-determined
S3_vs_k141_32657||lt2gene Low-quality
S3_vs_k141_64281||lt2gene Low-quality
S3_vs_k141_88142||lt2gene Low-quality
S3_vs_k141_48636||lt2gene Low-quality
S3_vs_k141_24572||lt2gene Low-quality
S3_vs_k141_96322||lt2gene Low-quality
S3_vs_k141_8889||lt2gene Low-quality
S3_vs_k141_40553||lt2gene Not-determined
S3_vs_k141_8891||lt2gene Low-quality
S3_vs_k141_8900||lt2gene Not-determined
S3_vs_k141_8926||lt2gene Low-quality
S3_vs_k141_56385||lt2gene Low-quality
S3_vs_k141_48672||lt2gene Low-quality
S3_vs_k141_56403||lt2gene Low-quality
S3_vs_k141_80339||lt2gene Low-quality
S3_vs_k141_72501||lt2gene Low-quality
S3_vs_k141_104284||lt2gene Low-quality
S3_vs_k141_88208||lt2gene Low-quality
S3_vs_k141_88215||lt2gene Low-quality
S3_vs_k141_24623||lt2gene Low-quality
S3_vs_k141_112141||lt2gene Low-quality
S3_vs_k141_48707||lt2gene Low-quality
S3_vs_k141_112171||lt2gene Low-quality
S3_vs_k141_40630||lt2gene Low-quality
S3_vs_k141_120089||lt2gene Low-quality
S3_vs_k141_48729||lt2gene Low-quality
S3_vs_k141_56477||lt2gene Low-quality
S3_vs_k141_112202||lt2gene Low-quality
S3_vs_k141_96444||lt2gene Low-quality
S3_vs_k141_88247||lt2gene Low-quality
S3_vs_k141_56517||lt2gene Low-quality
S3_vs_k141_16283||lt2gene Low-quality
S3_vs_k141_56528||lt2gene Low-quality
S3_vs_k141_40652||lt2gene Low-quality
S3_vs_k141_56535||lt2gene Not-determined
S3_vs_k141_32814||lt2gene Not-determined
S3_vs_k141_40657||lt2gene Not-determined
S3_vs_k141_104341||lt2gene Low-quality
S3_vs_k141_842||lt2gene Low-quality
S3_vs_k141_40682||lt2gene Low-quality
S3_vs_k141_16325||lt2gene Low-quality
S3_vs_k141_16347||lt2gene Low-quality
S3_vs_k141_9132||lt2gene Low-quality
S3_vs_k141_32866||lt2gene Low-quality
S3_vs_k141_112266||lt2gene Not-determined
S3_vs_k141_40756||lt2gene Low-quality
S3_vs_k141_88295||lt2gene Low-quality
S3_vs_k141_48810||lt2gene Not-determined
S3_vs_k141_32919||lt2gene Low-quality
S3_vs_k141_48823||lt2gene Not-determined
S3_vs_k141_32934||lt2gene Low-quality
S3_vs_k141_32935||lt2gene Low-quality
S3_vs_k141_80478||lt2gene Low-quality
S3_vs_k141_88357||lt2gene Not-determined
S3_vs_k141_48862||lt2gene Low-quality
S3_vs_k141_912||lt2gene Low-quality
S3_vs_k141_72799||lt2gene Low-quality
S3_vs_k141_72808||lt2gene Not-determined
S3_vs_k141_80534||lt2gene Low-quality
S3_vs_k141_88364||lt2gene Low-quality
S3_vs_k141_56681||lt2gene Low-quality
S3_vs_k141_96593||lt2gene Low-quality
S3_vs_k141_96597||lt2gene Low-quality
S3_vs_k141_48902||lt2gene Not-determined
S3_vs_k141_48907||lt2gene Low-quality
S3_vs_k141_32989||lt2gene Low-quality
S3_vs_k141_96643||lt2gene Not-determined
S3_vs_k141_104440||lt2gene Low-quality
S3_vs_k141_40868||lt2gene Low-quality
S3_vs_k141_48935||lt2gene Low-quality
S3_vs_k141_96659||lt2gene Not-determined
S3_vs_k141_96678||lt2gene Low-quality
S3_vs_k141_96688||lt2gene Low-quality
S3_vs_k141_112364||lt2gene Not-determined
S3_vs_k141_112371||lt2gene Low-quality
S3_vs_k141_88428||lt2gene Low-quality
S3_vs_k141_33052||lt2gene Not-determined
S3_vs_k141_88454||lt2gene Low-quality
S3_vs_k141_56783||lt2gene Low-quality
S3_vs_k141_120178||lt2gene Low-quality
S3_vs_k141_40918||lt2gene Low-quality
S3_vs_k141_72956||lt2gene Low-quality
S3_vs_k141_64489||lt2gene Low-quality
S3_vs_k141_96709||lt2gene Not-determined
S3_vs_k141_56796||lt2gene Not-determined
S3_vs_k141_958||lt2gene Low-quality
S3_vs_k141_112381||lt2gene Low-quality
S3_vs_k141_24841||lt2gene Low-quality
S3_vs_k141_976||lt2gene Low-quality
S3_vs_k141_96727||lt2gene Low-quality
S3_vs_k141_72994||lt2gene Low-quality
S3_vs_k141_24865||lt2gene Low-quality
S3_vs_k141_88506||lt2gene Low-quality
S3_vs_k141_49073||lt2gene Not-determined
S3_vs_k141_9337||lt2gene Low-quality
S3_vs_k141_80670||lt2gene Not-determined
S3_vs_k141_1019||lt2gene Low-quality
S3_vs_k141_112409||lt2gene Not-determined
S3_vs_k141_80717||lt2gene Low-quality
S3_vs_k141_9399||lt2gene Not-determined
S3_vs_k141_33105||lt2gene Low-quality
S3_vs_k141_16564||lt2gene Low-quality
S3_vs_k141_33133||lt2gene Low-quality
S3_vs_k141_104569||lt2gene Low-quality
S3_vs_k141_56927||lt2gene Low-quality
S3_vs_k141_40935||lt2gene Low-quality
S3_vs_k141_56937||lt2gene Low-quality
S3_vs_k141_33140||lt2gene Low-quality
S3_vs_k141_9481||lt2gene Not-determined
S3_vs_k141_96812||lt2gene Low-quality
S3_vs_k141_49221||lt2gene Low-quality
S3_vs_k141_96842||lt2gene Low-quality
S3_vs_k141_9509||lt2gene Low-quality
S3_vs_k141_56978||lt2gene Not-determined
S3_vs_k141_73207||lt2gene Low-quality
S3_vs_k141_16627||lt2gene Not-determined
S3_vs_k141_49240||lt2gene Low-quality
S3_vs_k141_49246||lt2gene Low-quality
S3_vs_k141_16652||lt2gene Low-quality
S3_vs_k141_73281||lt2gene Low-quality
S3_vs_k141_73286||lt2gene Low-quality
S3_vs_k141_33221||lt2gene Low-quality
S3_vs_k141_73298||lt2gene Low-quality
S3_vs_k141_96961||lt2gene Low-quality
S3_vs_k141_9611||lt2gene Low-quality
S3_vs_k141_96980||lt2gene Low-quality
S3_vs_k141_57075||lt2gene Low-quality
S3_vs_k141_80935||lt2gene Not-determined
S3_vs_k141_57091||lt2gene Low-quality
S3_vs_k141_80946||lt2gene Low-quality
S3_vs_k141_88636||lt2gene Low-quality
S3_vs_k141_88671||lt2gene Low-quality
S3_vs_k141_73395||lt2gene Low-quality
S3_vs_k141_57117||lt2gene Not-determined
S3_vs_k141_88691||lt2gene Low-quality
S3_vs_k141_88699||lt2gene Low-quality
S3_vs_k141_49349||lt2gene Not-determined
S3_vs_k141_33340||lt2gene Low-quality
S3_vs_k141_33342||lt2gene Low-quality
S3_vs_k141_1181||lt2gene Not-determined
S3_vs_k141_81083||lt2gene Low-quality
S3_vs_k141_97093||lt2gene Low-quality
S3_vs_k141_88768||lt2gene Low-quality
S3_vs_k141_33386||lt2gene Low-quality
S3_vs_k141_64742||lt2gene Low-quality
S3_vs_k141_120433||lt2gene Low-quality
S3_vs_k141_64790||lt2gene Low-quality
S3_vs_k141_104729||lt2gene Low-quality
S3_vs_k141_16829||lt2gene Low-quality
S3_vs_k141_16833||lt2gene Low-quality
S3_vs_k141_16836||lt2gene Low-quality
S3_vs_k141_9725||lt2gene Low-quality
S3_vs_k141_41087||lt2gene Low-quality
S3_vs_k141_88792||lt2gene Not-determined
S3_vs_k141_16876||lt2gene Not-determined
S3_vs_k141_88808||lt2gene Low-quality
S3_vs_k141_73477||lt2gene Low-quality
S3_vs_k141_73480||lt2gene Low-quality
S3_vs_k141_1229||lt2gene Low-quality
S3_vs_k141_1233||lt2gene Low-quality
S3_vs_k141_49439||lt2gene Low-quality
S3_vs_k141_120500||lt2gene Low-quality
S3_vs_k141_120503||lt2gene Low-quality
S3_vs_k141_1240||lt2gene Low-quality
S3_vs_k141_81182||lt2gene Low-quality
S3_vs_k141_120529||lt2gene Low-quality
S3_vs_k141_81207||lt2gene Low-quality
S3_vs_k141_41165||lt2gene Not-determined
S3_vs_k141_1264||lt2gene Low-quality
S3_vs_k141_41168||lt2gene Not-determined
S3_vs_k141_41175||lt2gene Low-quality
S3_vs_k141_16991||lt2gene Low-quality
S3_vs_k141_73560||lt2gene Low-quality
S3_vs_k141_41177||lt2gene Not-determined
S3_vs_k141_49492||lt2gene Low-quality
S3_vs_k141_1303||lt2gene Low-quality
S3_vs_k141_104813||lt2gene Not-determined
S3_vs_k141_9787||lt2gene Low-quality
S3_vs_k141_41187||lt2gene Low-quality
S3_vs_k141_41192||lt2gene Low-quality
S3_vs_k141_104834||lt2gene Low-quality
S3_vs_k141_64947||lt2gene Low-quality
S3_vs_k141_57174||lt2gene Low-quality
S3_vs_k141_41213||lt2gene Low-quality
S3_vs_k141_104850||lt2gene Low-quality
S3_vs_k141_9832||lt2gene Low-quality
S3_vs_k141_64981||lt2gene Low-quality
S3_vs_k141_57237||lt2gene Not-determined
S3_vs_k141_81314||lt2gene Low-quality
S3_vs_k141_1363||lt2gene Low-quality
S3_vs_k141_112709||lt2gene Not-determined
S3_vs_k141_49563||lt2gene Low-quality
S3_vs_k141_33642||lt2gene Low-quality
S3_vs_k141_65023||lt2gene Low-quality
S3_vs_k141_49609||lt2gene Low-quality
S3_vs_k141_17075||lt2gene Low-quality
S3_vs_k141_104935||lt2gene Low-quality
S3_vs_k141_81377||lt2gene Low-quality
S3_vs_k141_41260||lt2gene Not-determined
S3_vs_k141_57306||lt2gene Not-determined
S3_vs_k141_33685||lt2gene Low-quality
S3_vs_k141_97233||lt2gene Not-determined
S3_vs_k141_17128||lt2gene Low-quality
S3_vs_k141_73687||lt2gene Low-quality
S3_vs_k141_17142||lt2gene Not-determined
S3_vs_k141_112770||lt2gene Low-quality
S3_vs_k141_41289||lt2gene Not-determined
S3_vs_k141_1539||lt2gene Not-determined
S3_vs_k141_88966||lt2gene Low-quality
S3_vs_k141_88971||lt2gene Low-quality
S3_vs_k141_41380||lt2gene Not-determined
S3_vs_k141_9985||lt2gene Low-quality
S3_vs_k141_1585||lt2gene Low-quality
S3_vs_k141_10062||lt2gene Low-quality
S3_vs_k141_17189||lt2gene Low-quality
S3_vs_k141_89048||lt2gene Low-quality
S3_vs_k141_73823||lt2gene Low-quality
S3_vs_k141_89059||lt2gene Low-quality
S3_vs_k141_120734||lt2gene Not-determined
S3_vs_k141_17226||lt2gene Low-quality
S3_vs_k141_10153||lt2gene Low-quality
S3_vs_k141_57475||lt2gene Low-quality
S3_vs_k141_112984||lt2gene Not-determined
S3_vs_k141_73918||lt2gene Low-quality
S3_vs_k141_57492||lt2gene Low-quality
S3_vs_k141_120800||lt2gene Low-quality
S3_vs_k141_57507||lt2gene Not-determined
S3_vs_k141_120806||lt2gene Not-determined
S3_vs_k141_81500||lt2gene Not-determined
S3_vs_k141_65198||lt2gene Not-determined
S3_vs_k141_81518||lt2gene Low-quality
S3_vs_k141_81521||lt2gene Low-quality
S3_vs_k141_89200||lt2gene Not-determined
S3_vs_k141_49830||lt2gene Low-quality
S3_vs_k141_73992||lt2gene Low-quality
S3_vs_k141_113067||lt2gene Low-quality
S3_vs_k141_89212||lt2gene Low-quality
S3_vs_k141_25351||lt2gene Low-quality
S3_vs_k141_89228||lt2gene Low-quality
S3_vs_k141_57573||lt2gene Not-determined
S3_vs_k141_33820||lt2gene Not-determined
S3_vs_k141_17336||lt2gene Low-quality
S3_vs_k141_49851||lt2gene Low-quality
S3_vs_k141_89285||lt2gene Low-quality
S3_vs_k141_89287||lt2gene Low-quality
S3_vs_k141_41544||lt2gene Not-determined
S3_vs_k141_97477||lt2gene Low-quality
S3_vs_k141_97482||lt2gene Not-determined
S3_vs_k141_97487||lt2gene Not-determined
S3_vs_k141_49868||lt2gene Low-quality
S3_vs_k141_25389||lt2gene Low-quality
S3_vs_k141_120898||lt2gene Low-quality
S3_vs_k141_81543||lt2gene Low-quality
S3_vs_k141_49908||lt2gene Low-quality
S3_vs_k141_105058||lt2gene Low-quality
S3_vs_k141_120909||lt2gene Low-quality
S3_vs_k141_74079||lt2gene Low-quality
S3_vs_k141_41593||lt2gene Low-quality
S3_vs_k141_57680||lt2gene Low-quality
S3_vs_k141_57681||lt2gene Low-quality
S3_vs_k141_41604||lt2gene Low-quality
S3_vs_k141_89326||lt2gene Not-determined
S3_vs_k141_25444||lt2gene Low-quality
S3_vs_k141_105095||lt2gene Not-determined
S3_vs_k141_120941||lt2gene Not-determined
S3_vs_k141_10408||lt2gene Low-quality
S3_vs_k141_74088||lt2gene Low-quality
S3_vs_k141_33847||lt2gene Low-quality
S3_vs_k141_105109||lt2gene Low-quality
S3_vs_k141_113244||lt2gene Low-quality
S3_vs_k141_97533||lt2gene Low-quality
S3_vs_k141_10428||lt2gene Low-quality
S3_vs_k141_10435||lt2gene Low-quality
S3_vs_k141_33876||lt2gene Low-quality
S3_vs_k141_50007||lt2gene Low-quality
S3_vs_k141_105144||lt2gene Low-quality
S3_vs_k141_41682||lt2gene Low-quality
S3_vs_k141_1862||lt2gene Low-quality
S3_vs_k141_120985||lt2gene Not-determined
S3_vs_k141_10468||lt2gene Low-quality
S3_vs_k141_17404||lt2gene Not-determined
S3_vs_k141_41718||lt2gene Low-quality
S3_vs_k141_10505||lt2gene Low-quality
S3_vs_k141_17429||lt2gene Not-determined
S3_vs_k141_97572||lt2gene Low-quality
S3_vs_k141_50061||lt2gene Low-quality
S3_vs_k141_121018||lt2gene Low-quality
S3_vs_k141_50069||lt2gene Not-determined
S3_vs_k141_57757||lt2gene Low-quality
S3_vs_k141_10528||lt2gene Low-quality
S3_vs_k141_17444||lt2gene Low-quality
S3_vs_k141_10538||lt2gene Not-determined
S3_vs_k141_33931||lt2gene Low-quality
S3_vs_k141_81639||lt2gene Low-quality
S3_vs_k141_121037||lt2gene Low-quality
S3_vs_k141_113328||lt2gene Low-quality
S3_vs_k141_113336||lt2gene Low-quality
S3_vs_k141_65330||lt2gene Not-determined
S3_vs_k141_25561||lt2gene Low-quality
S3_vs_k141_113346||lt2gene Low-quality
S3_vs_k141_10594||lt2gene Low-quality
S3_vs_k141_65342||lt2gene Low-quality
S3_vs_k141_113355||lt2gene Low-quality
S3_vs_k141_10614||lt2gene Low-quality
S3_vs_k141_74128||lt2gene Low-quality
S3_vs_k141_81676||lt2gene Not-determined
S3_vs_k141_97599||lt2gene Low-quality
S3_vs_k141_105298||lt2gene Low-quality
S3_vs_k141_41777||lt2gene Low-quality
S3_vs_k141_17485||lt2gene Not-determined
S3_vs_k141_41787||lt2gene Low-quality
S3_vs_k141_113438||lt2gene Not-determined
S3_vs_k141_34028||lt2gene Low-quality
S3_vs_k141_81719||lt2gene Low-quality
S3_vs_k141_74151||lt2gene Not-determined
S3_vs_k141_105325||lt2gene Low-quality
S3_vs_k141_105350||lt2gene Not-determined
S3_vs_k141_113481||lt2gene Low-quality
S3_vs_k141_121172||lt2gene Low-quality
S3_vs_k141_89544||lt2gene Low-quality
S3_vs_k141_113517||lt2gene Low-quality
S3_vs_k141_65407||lt2gene Low-quality
S3_vs_k141_74200||lt2gene Not-determined
S3_vs_k141_113527||lt2gene Low-quality
S3_vs_k141_121235||lt2gene Low-quality
S3_vs_k141_113544||lt2gene Not-determined
S3_vs_k141_113546||lt2gene Low-quality
S3_vs_k141_34090||lt2gene Low-quality

S3_vs_k141_34107||lt2gene Low-quality
S3_vs_k141_65462||lt2gene Not-determined
S3_vs_k141_1958||lt2gene Low-quality
S3_vs_k141_89585||lt2gene Low-quality
S3_vs_k141_50146||lt2gene Low-quality
S3_vs_k141_10793||lt2gene Low-quality
S3_vs_k141_89608||lt2gene Not-determined
S3_vs_k141_50162||lt2gene Not-determined
S3_vs_k141_17553||lt2gene Low-quality
S3_vs_k141_41845||lt2gene Low-quality
S3_vs_k141_97689||lt2gene Low-quality
S3_vs_k141_10881||lt2gene Not-determined
S3_vs_k141_34243||lt2gene Low-quality
S3_vs_k141_41866||lt2gene Low-quality
S3_vs_k141_121324||lt2gene Not-determined
S3_vs_k141_34268||lt2gene Low-quality
S3_vs_k141_34271||lt2gene Low-quality
S3_vs_k141_2075||lt2gene Not-determined
S3_vs_k141_105469||lt2gene Low-quality
S3_vs_k141_57955||lt2gene Low-quality
S3_vs_k141_34312||lt2gene Not-determined
S3_vs_k141_65560||lt2gene Low-quality
S3_vs_k141_65564||lt2gene Low-quality
S3_vs_k141_34325||lt2gene Not-determined
S3_vs_k141_105476||lt2gene Low-quality
S3_vs_k141_121367||lt2gene Low-quality
S3_vs_k141_50202||lt2gene Low-quality
S3_vs_k141_50203||lt2gene Low-quality
S3_vs_k141_34348||lt2gene Not-determined
S3_vs_k141_41978||lt2gene Not-determined
S3_vs_k141_81777||lt2gene Low-quality
S3_vs_k141_34366||lt2gene Low-quality
S3_vs_k141_11015||lt2gene Not-determined
S3_vs_k141_42004||lt2gene Low-quality
S3_vs_k141_105523||lt2gene Low-quality
S3_vs_k141_89693||lt2gene Low-quality
S3_vs_k141_97750||lt2gene Not-determined
S3_vs_k141_58063||lt2gene Low-quality
S3_vs_k141_11045||lt2gene Low-quality
S3_vs_k141_74309||lt2gene Low-quality
S3_vs_k141_58074||lt2gene Low-quality
S3_vs_k141_11059||lt2gene Not-determined
S3_vs_k141_105562||lt2gene Not-determined
S3_vs_k141_58090||lt2gene Low-quality
S3_vs_k141_42065||lt2gene Low-quality
S3_vs_k141_58110||lt2gene Low-quality
S3_vs_k141_42069||lt2gene Low-quality
S3_vs_k141_121423||lt2gene Not-determined
S3_vs_k141_74342||lt2gene Low-quality
S3_vs_k141_121431||lt2gene Low-quality
S3_vs_k141_58132||lt2gene Low-quality
S3_vs_k141_81850||lt2gene Low-quality
S3_vs_k141_58160||lt2gene Low-quality
S3_vs_k141_25950||lt2gene Not-determined
S3_vs_k141_42126||lt2gene Low-quality
S3_vs_k141_89753||lt2gene Low-quality
S3_vs_k141_11260||lt2gene Low-quality
S3_vs_k141_121483||lt2gene Not-determined
S3_vs_k141_58220||lt2gene Low-quality
S3_vs_k141_121498||lt2gene Low-quality
S3_vs_k141_65846||lt2gene Low-quality
S3_vs_k141_11297||lt2gene Low-quality
S3_vs_k141_65866||lt2gene Not-determined
S3_vs_k141_105792||lt2gene Low-quality
S3_vs_k141_89814||lt2gene Low-quality
S3_vs_k141_105801||lt2gene Not-determined
S3_vs_k141_50402||lt2gene Not-determined
S3_vs_k141_11362||lt2gene Not-determined
S3_vs_k141_113778||lt2gene Low-quality
S3_vs_k141_11391||lt2gene Low-quality
S3_vs_k141_121547||lt2gene Not-determined
S3_vs_k141_74441||lt2gene Low-quality
S3_vs_k141_97830||lt2gene Low-quality
S3_vs_k141_89870||lt2gene Low-quality
S3_vs_k141_121591||lt2gene Not-determined
S3_vs_k141_121611||lt2gene Low-quality
S3_vs_k141_11429||lt2gene Low-quality
S3_vs_k141_26025||lt2gene Low-quality
S3_vs_k141_11460||lt2gene Not-determined
S3_vs_k141_74524||lt2gene Not-determined
S3_vs_k141_11475||lt2gene Low-quality
S3_vs_k141_42241||lt2gene Low-quality
S3_vs_k141_97920||lt2gene Low-quality
S3_vs_k141_11487||lt2gene Not-determined
S3_vs_k141_74556||lt2gene Low-quality
S3_vs_k141_42259||lt2gene Low-quality
S3_vs_k141_34502||lt2gene Low-quality
S3_vs_k141_74569||lt2gene Not-determined
S3_vs_k141_74573||lt2gene Low-quality
S3_vs_k141_105914||lt2gene Low-quality
S3_vs_k141_89921||lt2gene Low-quality
S3_vs_k141_74598||lt2gene Low-quality
S3_vs_k141_89934||lt2gene Low-quality
S3_vs_k141_74608||lt2gene Low-quality
S3_vs_k141_74636||lt2gene Not-determined
S3_vs_k141_11561||lt2gene Low-quality
S3_vs_k141_105954||lt2gene Not-determined
S3_vs_k141_97964||lt2gene Low-quality
S3_vs_k141_42351||lt2gene Low-quality
S3_vs_k141_90033||lt2gene Low-quality
S3_vs_k141_82061||lt2gene Not-determined
S3_vs_k141_11594||lt2gene Low-quality
S3_vs_k141_58380||lt2gene Low-quality
S3_vs_k141_90074||lt2gene Low-quality
S3_vs_k141_90083||lt2gene Low-quality
S3_vs_k141_58424||lt2gene Low-quality
S3_vs_k141_50606||lt2gene Not-determined
S3_vs_k141_90110||lt2gene Low-quality
S3_vs_k141_11678||lt2gene Not-determined
S3_vs_k141_114031||lt2gene Low-quality
S3_vs_k141_11687||lt2gene Low-quality
S3_vs_k141_106051||lt2gene Low-quality
S3_vs_k141_106054||lt2gene Not-determined
S3_vs_k141_98052||lt2gene Low-quality
S3_vs_k141_106067||lt2gene Low-quality
S3_vs_k141_2414||lt2gene Low-quality
S3_vs_k141_26309||lt2gene Low-quality
S3_vs_k141_82174||lt2gene Not-determined
S3_vs_k141_58439||lt2gene Low-quality
S3_vs_k141_26325||lt2gene Low-quality
S3_vs_k141_90152||lt2gene Low-quality
S3_vs_k141_82202||lt2gene Not-determined
S3_vs_k141_42494||lt2gene Low-quality
S3_vs_k141_98104||lt2gene Low-quality
S3_vs_k141_42507||lt2gene Not-determined
S3_vs_k141_2440||lt2gene Low-quality
S3_vs_k141_26350||lt2gene Low-quality
S3_vs_k141_42534||lt2gene Low-quality
S3_vs_k141_26361||lt2gene Not-determined
S3_vs_k141_114111||lt2gene Low-quality
S3_vs_k141_34668||lt2gene Not-determined
S3_vs_k141_106134||lt2gene Low-quality
S3_vs_k141_58585||lt2gene Not-determined
S3_vs_k141_90254||lt2gene Low-quality
S3_vs_k141_26408||lt2gene Low-quality
S3_vs_k141_82281||lt2gene Low-quality
S3_vs_k141_90287||lt2gene Not-determined
S3_vs_k141_98209||lt2gene Low-quality
S3_vs_k141_98220||lt2gene Low-quality
S3_vs_k141_114154||lt2gene Not-determined
S3_vs_k141_106188||lt2gene Low-quality
S3_vs_k141_98233||lt2gene Low-quality
S3_vs_k141_98237||lt2gene Low-quality
S3_vs_k141_74847||lt2gene Low-quality
S3_vs_k141_42570||lt2gene Not-determined
S3_vs_k141_11757||lt2gene Low-quality
S3_vs_k141_2606||lt2gene Low-quality
S3_vs_k141_34723||lt2gene Low-quality
S3_vs_k141_66009||lt2gene Low-quality
S3_vs_k141_98272||lt2gene Not-determined
S3_vs_k141_26492||lt2gene Low-quality
S3_vs_k141_50768||lt2gene Low-quality
S3_vs_k141_58765||lt2gene Not-determined
S3_vs_k141_34767||lt2gene Low-quality
S3_vs_k141_106241||lt2gene Not-determined
S3_vs_k141_18040||lt2gene Low-quality
S3_vs_k141_74933||lt2gene Low-quality
S3_vs_k141_2703||lt2gene Low-quality
S3_vs_k141_42662||lt2gene Low-quality
S3_vs_k141_74963||lt2gene Not-determined
S3_vs_k141_121871||lt2gene Not-determined
S3_vs_k141_50866||lt2gene Not-determined
S3_vs_k141_106291||lt2gene Low-quality
S3_vs_k141_50879||lt2gene Low-quality
S3_vs_k141_121892||lt2gene Low-quality
S3_vs_k141_26610||lt2gene Low-quality
S3_vs_k141_11877||lt2gene Low-quality
S3_vs_k141_18070||lt2gene Low-quality
S3_vs_k141_82425||lt2gene Low-quality
S3_vs_k141_106340||lt2gene Low-quality
S3_vs_k141_106342||lt2gene Low-quality
S3_vs_k141_75007||lt2gene Low-quality
S3_vs_k141_58887||lt2gene Low-quality
S3_vs_k141_106352||lt2gene Low-quality
S3_vs_k141_82449||lt2gene Low-quality
S3_vs_k141_106378||lt2gene Low-quality
S3_vs_k141_106383||lt2gene Low-quality
S3_vs_k141_18085||lt2gene Low-quality
S3_vs_k141_50942||lt2gene Low-quality
S3_vs_k141_121955||lt2gene Low-quality
S3_vs_k141_50953||lt2gene Low-quality
S3_vs_k141_114295||lt2gene Not-determined
S3_vs_k141_58970||lt2gene Low-quality
S3_vs_k141_50966||lt2gene Low-quality
S3_vs_k141_58983||lt2gene Low-quality
S3_vs_k141_58989||lt2gene Low-quality
S3_vs_k141_90425||lt2gene Low-quality
S3_vs_k141_75118||lt2gene Low-quality
S3_vs_k141_42900||lt2gene Low-quality
S3_vs_k141_82538||lt2gene Not-determined
S3_vs_k141_51007||lt2gene Low-quality
S3_vs_k141_51023||lt2gene Low-quality
S3_vs_k141_59055||lt2gene Low-quality
S3_vs_k141_75156||lt2gene Not-determined
S3_vs_k141_35003||lt2gene Low-quality
S3_vs_k141_59088||lt2gene Low-quality
S3_vs_k141_51039||lt2gene Low-quality
S3_vs_k141_51040||lt2gene Low-quality
S3_vs_k141_59098||lt2gene Low-quality
S3_vs_k141_35061||lt2gene Low-quality
S3_vs_k141_106523||lt2gene Low-quality
S3_vs_k141_2873||lt2gene Low-quality
S3_vs_k141_42905||lt2gene Not-determined
S3_vs_k141_66217||lt2gene Low-quality
S3_vs_k141_42914||lt2gene Low-quality
S3_vs_k141_98509||lt2gene Not-determined
S3_vs_k141_42928||lt2gene Low-quality
S3_vs_k141_106570||lt2gene Not-determined
S3_vs_k141_51111||lt2gene Low-quality
S3_vs_k141_82655||lt2gene Not-determined
S3_vs_k141_75283||lt2gene Not-determined
S3_vs_k141_82669||lt2gene Not-determined
S3_vs_k141_98547||lt2gene Not-determined
S3_vs_k141_42943||lt2gene Low-quality
S3_vs_k141_90604||lt2gene Low-quality
S3_vs_k141_51144||lt2gene Low-quality
S3_vs_k141_90609||lt2gene Low-quality
S3_vs_k141_75327||lt2gene Low-quality
S3_vs_k141_82696||lt2gene Low-quality
S3_vs_k141_75335||lt2gene Low-quality
S3_vs_k141_51228||lt2gene Low-quality
S3_vs_k141_106623||lt2gene Low-quality
S3_vs_k141_43024||lt2gene Low-quality
S3_vs_k141_51251||lt2gene Low-quality
S3_vs_k141_51254||lt2gene Low-quality
S3_vs_k141_51281||lt2gene Not-determined
S3_vs_k141_114440||lt2gene Low-quality
S3_vs_k141_82801||lt2gene Not-determined
S3_vs_k141_106649||lt2gene Not-determined
S3_vs_k141_106653||lt2gene Not-determined
S3_vs_k141_18197||lt2gene Low-quality
S3_vs_k141_51302||lt2gene Not-determined
S3_vs_k141_75384||lt2gene Low-quality
S3_vs_k141_114464||lt2gene Low-quality
S3_vs_k141_82826||lt2gene Low-quality
S3_vs_k141_3007||lt2gene Low-quality
S3_vs_k141_82831||lt2gene Low-quality
S3_vs_k141_35177||lt2gene Low-quality
S3_vs_k141_43057||lt2gene Low-quality
S3_vs_k141_43068||lt2gene Low-quality
S3_vs_k141_12218||lt2gene Not-determined
S3_vs_k141_98654||lt2gene Low-quality
S3_vs_k141_75421||lt2gene Low-quality
S3_vs_k141_35215||lt2gene Low-quality
S3_vs_k141_66334||lt2gene Low-quality
S3_vs_k141_59184||lt2gene Not-determined
S3_vs_k141_26977||lt2gene Not-determined
S3_vs_k141_75452||lt2gene Not-determined
S3_vs_k141_18222||lt2gene Not-determined
S3_vs_k141_18223||lt2gene Low-quality
S3_vs_k141_82856||lt2gene Not-determined
S3_vs_k141_35300||lt2gene Low-quality
S3_vs_k141_35304||lt2gene Low-quality
S3_vs_k141_114542||lt2gene Low-quality
S3_vs_k141_18242||lt2gene Low-quality
S3_vs_k141_114550||lt2gene Low-quality
S3_vs_k141_35308||lt2gene Low-quality
S3_vs_k141_75499||lt2gene Not-determined
S3_vs_k141_51363||lt2gene Low-quality
S3_vs_k141_90753||lt2gene Not-determined
S3_vs_k141_82880||lt2gene Low-quality
S3_vs_k141_27093||lt2gene Low-quality
S3_vs_k141_27105||lt2gene Low-quality
S3_vs_k141_114656||lt2gene Low-quality
S3_vs_k141_98758||lt2gene Not-determined
S3_vs_k141_3137||lt2gene Low-quality
S3_vs_k141_106896||lt2gene Low-quality
S3_vs_k141_106914||lt2gene Low-quality
S3_vs_k141_90822||lt2gene Low-quality
S3_vs_k141_35478||lt2gene Low-quality
S3_vs_k141_106965||lt2gene Low-quality
S3_vs_k141_51477||lt2gene Low-quality
S3_vs_k141_51482||lt2gene Low-quality
S3_vs_k141_51506||lt2gene Low-quality
S3_vs_k141_18393||lt2gene Not-determined
S3_vs_k141_106989||lt2gene Low-quality
S3_vs_k141_98820||lt2gene Low-quality
S3_vs_k141_90880||lt2gene Low-quality
S3_vs_k141_66449||lt2gene Low-quality
S3_vs_k141_75745||lt2gene Not-determined
S3_vs_k141_75749||lt2gene Not-determined
S3_vs_k141_35528||lt2gene Low-quality
S3_vs_k141_12436||lt2gene Low-quality
S3_vs_k141_75774||lt2gene Not-determined
S3_vs_k141_18441||lt2gene Low-quality
S3_vs_k141_75792||lt2gene Low-quality
S3_vs_k141_3155||lt2gene Not-determined
S3_vs_k141_35587||lt2gene Low-quality
S3_vs_k141_122273||lt2gene Low-quality
S3_vs_k141_35594||lt2gene Low-quality
S3_vs_k141_122279||lt2gene Low-quality
S3_vs_k141_43269||lt2gene Low-quality
S3_vs_k141_82921||lt2gene Low-quality
S3_vs_k141_12477||lt2gene Not-determined
S3_vs_k141_3225||lt2gene Low-quality
S3_vs_k141_27210||lt2gene Low-quality
S3_vs_k141_35623||lt2gene Low-quality
S3_vs_k141_43289||lt2gene Not-determined
S3_vs_k141_82941||lt2gene Low-quality
S3_vs_k141_43294||lt2gene Low-quality
S3_vs_k141_82950||lt2gene Low-quality
S3_vs_k141_59471||lt2gene Low-quality
S3_vs_k141_66534||lt2gene Not-determined
S3_vs_k141_27293||lt2gene Low-quality
S3_vs_k141_98964||lt2gene Low-quality
S3_vs_k141_91031||lt2gene Low-quality
S3_vs_k141_51647||lt2gene Not-determined
S3_vs_k141_122387||lt2gene Not-determined
S3_vs_k141_51651||lt2gene Low-quality
S3_vs_k141_18582||lt2gene Not-determined
S3_vs_k141_27309||lt2gene Low-quality
S3_vs_k141_107141||lt2gene Low-quality
S3_vs_k141_75860||lt2gene Low-quality
S3_vs_k141_75864||lt2gene Low-quality
S3_vs_k141_83008||lt2gene Low-quality
S3_vs_k141_83030||lt2gene Not-determined
S3_vs_k141_107189||lt2gene Not-determined
S3_vs_k141_91091||lt2gene Low-quality
S3_vs_k141_122453||lt2gene Low-quality
S3_vs_k141_3272||lt2gene Low-quality
S3_vs_k141_122481||lt2gene Low-quality
S3_vs_k141_35714||lt2gene Low-quality
S3_vs_k141_27356||lt2gene Low-quality
S3_vs_k141_122505||lt2gene Not-determined
S3_vs_k141_12627||lt2gene Low-quality
S3_vs_k141_122517||lt2gene Low-quality
S3_vs_k141_35724||lt2gene Low-quality
S3_vs_k141_122543||lt2gene Low-quality
S3_vs_k141_99090||lt2gene Low-quality
S3_vs_k141_59574||lt2gene Low-quality
S3_vs_k141_66695||lt2gene Low-quality
S3_vs_k141_18714||lt2gene Low-quality
S3_vs_k141_107281||lt2gene Low-quality
S3_vs_k141_51773||lt2gene Not-determined
S3_vs_k141_107288||lt2gene Low-quality
S3_vs_k141_75919||lt2gene Low-quality
S3_vs_k141_51790||lt2gene Low-quality
S3_vs_k141_3351||lt2gene Low-quality
S3_vs_k141_3356||lt2gene Low-quality
S3_vs_k141_115067||lt2gene Low-quality
S3_vs_k141_83210||lt2gene Low-quality
S3_vs_k141_107314||lt2gene Not-determined
S3_vs_k141_43486||lt2gene Low-quality
S3_vs_k141_99147||lt2gene Not-determined
S3_vs_k141_59651||lt2gene Not-determined
S3_vs_k141_59670||lt2gene Low-quality
S3_vs_k141_91214||lt2gene Low-quality
S3_vs_k141_12743||lt2gene Low-quality
S3_vs_k141_12759||lt2gene Low-quality
S3_vs_k141_27531||lt2gene Not-determined
S3_vs_k141_59748||lt2gene Low-quality
S3_vs_k141_35798||lt2gene Low-quality
S3_vs_k141_59805||lt2gene Low-quality
S3_vs_k141_83346||lt2gene Not-determined
S3_vs_k141_83353||lt2gene Not-determined
S3_vs_k141_83355||lt2gene Not-determined
S3_vs_k141_115153||lt2gene Low-quality
S3_vs_k141_76021||lt2gene Not-determined
S3_vs_k141_59835||lt2gene Not-determined
S3_vs_k141_122710||lt2gene Low-quality
S3_vs_k141_51973||lt2gene Low-quality
S3_vs_k141_35855||lt2gene Low-quality
S3_vs_k141_3544||lt2gene Low-quality
S3_vs_k141_122760||lt2gene Low-quality
S3_vs_k141_76102||lt2gene Low-quality
S3_vs_k141_115253||lt2gene Low-quality
S3_vs_k141_35898||lt2gene Low-quality
S3_vs_k141_83462||lt2gene Low-quality
S3_vs_k141_76109||lt2gene Low-quality
S3_vs_k141_52097||lt2gene Low-quality
S3_vs_k141_66894||lt2gene Low-quality
S3_vs_k141_12892||lt2gene Not-determined
S3_vs_k141_27636||lt2gene Not-determined
S3_vs_k141_3597||lt2gene Low-quality
S3_vs_k141_3598||lt2gene Low-quality
S3_vs_k141_59915||lt2gene Low-quality
S3_vs_k141_35950||lt2gene Low-quality
S3_vs_k141_12912||lt2gene Low-quality
S3_vs_k141_91473||lt2gene Low-quality
S3_vs_k141_35958||lt2gene Low-quality
S3_vs_k141_115291||lt2gene Low-quality
S3_vs_k141_115295||lt2gene Not-determined
S3_vs_k141_35972||lt2gene Not-determined
S3_vs_k141_99236||lt2gene Low-quality
S3_vs_k141_52152||lt2gene Not-determined
S3_vs_k141_12944||lt2gene Low-quality
S3_vs_k141_36014||lt2gene Low-quality
S3_vs_k141_52172||lt2gene Low-quality
S3_vs_k141_43716||lt2gene Low-quality
S3_vs_k141_43719||lt2gene Low-quality
S3_vs_k141_76170||lt2gene Low-quality
S3_vs_k141_36050||lt2gene Not-determined
S3_vs_k141_36055||lt2gene Low-quality
S3_vs_k141_36061||lt2gene Not-determined
S3_vs_k141_76184||lt2gene Low-quality
S3_vs_k141_91544||lt2gene Low-quality
S3_vs_k141_91551||lt2gene Low-quality
S3_vs_k141_27713||lt2gene Low-quality
S3_vs_k141_3637||lt2gene Not-determined
S3_vs_k141_3646||lt2gene Low-quality
S3_vs_k141_107449||lt2gene Low-quality
S3_vs_k141_3653||lt2gene Low-quality
S3_vs_k141_91576||lt2gene Not-determined
S3_vs_k141_60016||lt2gene Low-quality
S3_vs_k141_60039||lt2gene Low-quality
S3_vs_k141_83613||lt2gene Low-quality
S3_vs_k141_52268||lt2gene Low-quality
S3_vs_k141_91609||lt2gene Low-quality
S3_vs_k141_60086||lt2gene Not-determined
S3_vs_k141_83648||lt2gene Low-quality
S3_vs_k141_83661||lt2gene Not-determined
S3_vs_k141_13019||lt2gene Low-quality
S3_vs_k141_27796||lt2gene Low-quality
S3_vs_k141_52335||lt2gene Low-quality
S3_vs_k141_27797||lt2gene Low-quality
S3_vs_k141_76212||lt2gene Not-determined
S3_vs_k141_52350||lt2gene Low-quality
S3_vs_k141_18909||lt2gene Low-quality
S3_vs_k141_99249||lt2gene Low-quality
S3_vs_k141_83718||lt2gene Low-quality
S3_vs_k141_91625||lt2gene Low-quality
S3_vs_k141_107558||lt2gene Low-quality
S3_vs_k141_83720||lt2gene Low-quality
S3_vs_k141_60143||lt2gene Not-determined
S3_vs_k141_27825||lt2gene Low-quality
S3_vs_k141_36178||lt2gene Not-determined
S3_vs_k141_52376||lt2gene Low-quality
S3_vs_k141_18921||lt2gene Low-quality
S3_vs_k141_76249||lt2gene Not-determined
S3_vs_k141_13077||lt2gene Low-quality
S3_vs_k141_76266||lt2gene Low-quality
S3_vs_k141_13087||lt2gene Low-quality
S3_vs_k141_52392||lt2gene Low-quality
S3_vs_k141_66996||lt2gene Low-quality
S3_vs_k141_60158||lt2gene Low-quality
S3_vs_k141_13129||lt2gene Not-determined
S3_vs_k141_13130||lt2gene Not-determined
S3_vs_k141_3754||lt2gene Low-quality
S3_vs_k141_60186||lt2gene Not-determined
S3_vs_k141_67032||lt2gene Low-quality
S3_vs_k141_76357||lt2gene Low-quality
S3_vs_k141_52469||lt2gene Low-quality
S3_vs_k141_67056||lt2gene Low-quality
S3_vs_k141_91726||lt2gene Low-quality
S3_vs_k141_60226||lt2gene Low-quality
S3_vs_k141_115503||lt2gene Not-determined
S3_vs_k141_13192||lt2gene Not-determined
S3_vs_k141_67078||lt2gene Low-quality
S3_vs_k141_122932||lt2gene Not-determined
S3_vs_k141_91754||lt2gene Low-quality
S3_vs_k141_43913||lt2gene Low-quality
S3_vs_k141_99383||lt2gene Low-quality
S3_vs_k141_3932||lt2gene Low-quality
S3_vs_k141_67141||lt2gene Not-determined
S3_vs_k141_115527||lt2gene Not-determined
S3_vs_k141_67151||lt2gene Low-quality
S3_vs_k141_19104||lt2gene Low-quality
S3_vs_k141_36247||lt2gene Low-quality
S3_vs_k141_19122||lt2gene Low-quality
S3_vs_k141_3975||lt2gene Low-quality
S3_vs_k141_27955||lt2gene Low-quality
S3_vs_k141_60339||lt2gene Low-quality
S3_vs_k141_43960||lt2gene Not-determined
S3_vs_k141_67173||lt2gene Low-quality
S3_vs_k141_43973||lt2gene Low-quality
S3_vs_k141_60353||lt2gene Low-quality
S3_vs_k141_76535||lt2gene Low-quality
S3_vs_k141_123035||lt2gene Low-quality
S3_vs_k141_43983||lt2gene Low-quality
S3_vs_k141_43986||lt2gene Not-determined
S3_vs_k141_13329||lt2gene Low-quality
S3_vs_k141_13338||lt2gene Not-determined
S3_vs_k141_115575||lt2gene Low-quality
S3_vs_k141_115581||lt2gene Not-determined
S3_vs_k141_67204||lt2gene Not-determined
S3_vs_k141_115591||lt2gene Low-quality
S3_vs_k141_60408||lt2gene Low-quality
S3_vs_k141_19194||lt2gene Low-quality
S3_vs_k141_99434||lt2gene Not-determined
S3_vs_k141_115625||lt2gene Not-determined
S3_vs_k141_107815||lt2gene Low-quality
S3_vs_k141_115639||lt2gene Low-quality
S3_vs_k141_60421||lt2gene Low-quality
S3_vs_k141_115653||lt2gene Not-determined
S3_vs_k141_115654||lt2gene Not-determined
S3_vs_k141_19200||lt2gene Low-quality
S3_vs_k141_91901||lt2gene Low-quality
S3_vs_k141_13452||lt2gene Low-quality
S3_vs_k141_123109||lt2gene Low-quality
S3_vs_k141_123134||lt2gene Low-quality
S3_vs_k141_60491||lt2gene Low-quality
S3_vs_k141_52630||lt2gene Not-determined
S3_vs_k141_60515||lt2gene Low-quality
S3_vs_k141_13530||lt2gene Low-quality
S3_vs_k141_123189||lt2gene Low-quality
S3_vs_k141_123196||lt2gene Low-quality
S3_vs_k141_123205||lt2gene Not-determined
S3_vs_k141_99560||lt2gene Low-quality
S3_vs_k141_123216||lt2gene Not-determined
S3_vs_k141_123228||lt2gene Not-determined
S3_vs_k141_13568||lt2gene Low-quality
S3_vs_k141_123242||lt2gene Low-quality
S3_vs_k141_44092||lt2gene Low-quality
S3_vs_k141_76674||lt2gene Low-quality
S3_vs_k141_76676||lt2gene Low-quality
S3_vs_k141_83804||lt2gene Low-quality
S3_vs_k141_60610||lt2gene Not-determined
S3_vs_k141_28014||lt2gene Not-determined
S3_vs_k141_76696||lt2gene Low-quality
S3_vs_k141_107930||lt2gene Low-quality
S3_vs_k141_99571||lt2gene Low-quality
S3_vs_k141_36424||lt2gene Low-quality
S3_vs_k141_83825||lt2gene Not-determined
S3_vs_k141_99588||lt2gene Low-quality
S3_vs_k141_123309||lt2gene Low-quality
S3_vs_k141_36462||lt2gene Low-quality
S3_vs_k141_123323||lt2gene Low-quality
S3_vs_k141_123347||lt2gene Not-determined
S3_vs_k141_123352||lt2gene Low-quality
S3_vs_k141_13640||lt2gene Low-quality
S3_vs_k141_115824||lt2gene Low-quality
S3_vs_k141_44139||lt2gene Low-quality
S3_vs_k141_91964||lt2gene Low-quality
S3_vs_k141_52814||lt2gene Not-determined
S3_vs_k141_83847||lt2gene Low-quality
S3_vs_k141_52818||lt2gene Not-determined
S3_vs_k141_107985||lt2gene Low-quality
S3_vs_k141_107998||lt2gene Low-quality
S3_vs_k141_91981||lt2gene Low-quality
S3_vs_k141_52867||lt2gene Low-quality
S3_vs_k141_28160||lt2gene Low-quality
S3_vs_k141_123426||lt2gene Low-quality
S3_vs_k141_13698||lt2gene Low-quality
S3_vs_k141_13703||lt2gene Low-quality
S3_vs_k141_67485||lt2gene Low-quality
S3_vs_k141_67502||lt2gene Low-quality
S3_vs_k141_92020||lt2gene Low-quality
S3_vs_k141_13768||lt2gene Not-determined
S3_vs_k141_92039||lt2gene Low-quality
S3_vs_k141_4232||lt2gene Not-determined
S3_vs_k141_60787||lt2gene Not-determined
S3_vs_k141_67561||lt2gene Low-quality
S3_vs_k141_36518||lt2gene Low-quality
S3_vs_k141_67565||lt2gene Low-quality
S3_vs_k141_28224||lt2gene Low-quality
S3_vs_k141_76898||lt2gene Low-quality
S3_vs_k141_4259||lt2gene Low-quality
S3_vs_k141_4270||lt2gene Low-quality
S3_vs_k141_123538||lt2gene Low-quality
S3_vs_k141_13839||lt2gene Not-determined
S3_vs_k141_116029||lt2gene Low-quality
S3_vs_k141_84012||lt2gene Low-quality
S3_vs_k141_28289||lt2gene Low-quality
S3_vs_k141_28292||lt2gene Low-quality
S3_vs_k141_44396||lt2gene Not-determined
S3_vs_k141_84042||lt2gene Low-quality
S3_vs_k141_36593||lt2gene Low-quality
S3_vs_k141_13935||lt2gene Low-quality
S3_vs_k141_13940||lt2gene Low-quality
S3_vs_k141_67619||lt2gene Low-quality
S3_vs_k141_60867||lt2gene Low-quality
S3_vs_k141_116117||lt2gene Low-quality
S3_vs_k141_116123||lt2gene Not-determined
S3_vs_k141_84088||lt2gene Low-quality
S3_vs_k141_84092||lt2gene Low-quality
S3_vs_k141_28334||lt2gene Not-determined
S3_vs_k141_76999||lt2gene Low-quality
S3_vs_k141_92146||lt2gene Low-quality
S3_vs_k141_4317||lt2gene Low-quality
S3_vs_k141_19628||lt2gene Low-quality
S3_vs_k141_84134||lt2gene Low-quality
S3_vs_k141_99717||lt2gene Low-quality
S3_vs_k141_14018||lt2gene Low-quality
S3_vs_k141_28359||lt2gene Low-quality
S3_vs_k141_92178||lt2gene Low-quality
S3_vs_k141_14040||lt2gene Low-quality
S3_vs_k141_116197||lt2gene Not-determined
S3_vs_k141_92192||lt2gene Low-quality
S3_vs_k141_19683||lt2gene Low-quality
S3_vs_k141_92197||lt2gene Low-quality
S3_vs_k141_108191||lt2gene Low-quality
S3_vs_k141_14072||lt2gene Not-determined
S3_vs_k141_19709||lt2gene Low-quality
S3_vs_k141_67702||lt2gene Not-determined
S3_vs_k141_116253||lt2gene Low-quality
S3_vs_k141_14103||lt2gene Not-determined
S3_vs_k141_123626||lt2gene Not-determined
S3_vs_k141_77133||lt2gene Low-quality
S3_vs_k141_92242||lt2gene Low-quality

S3_vs_k141_19754||lt2gene Low-quality
S3_vs_k141_4411||lt2gene Not-determined
S3_vs_k141_77178||lt2gene Not-determined
S3_vs_k141_84281||lt2gene Low-quality
S3_vs_k141_28437||lt2gene Low-quality
S3_vs_k141_92345||lt2gene Low-quality
S3_vs_k141_116327||lt2gene Low-quality
S3_vs_k141_84298||lt2gene Low-quality
S3_vs_k141_92370||lt2gene Low-quality
S3_vs_k141_4450||lt2gene Low-quality
S3_vs_k141_84326||lt2gene Low-quality
S3_vs_k141_14221||lt2gene Low-quality
S3_vs_k141_108350||lt2gene Low-quality
S3_vs_k141_61134||lt2gene Low-quality
S3_vs_k141_28524||lt2gene Low-quality
S3_vs_k141_14247||lt2gene Low-quality
S3_vs_k141_67854||lt2gene Low-quality
S3_vs_k141_67863||lt2gene Low-quality
S3_vs_k141_99852||lt2gene Not-determined
S3_vs_k141_77294||lt2gene Low-quality
S3_vs_k141_77295||lt2gene Not-determined
S3_vs_k141_28585||lt2gene Not-determined
S3_vs_k141_77298||lt2gene Low-quality
S3_vs_k141_108370||lt2gene Low-quality
S3_vs_k141_44483||lt2gene Low-quality
S3_vs_k141_36719||lt2gene Low-quality
S3_vs_k141_14306||lt2gene Low-quality
S3_vs_k141_108374||lt2gene Low-quality
S3_vs_k141_28621||lt2gene Low-quality
S3_vs_k141_53193||lt2gene Low-quality
S3_vs_k141_84427||lt2gene Low-quality
S3_vs_k141_61266||lt2gene Low-quality
S3_vs_k141_92481||lt2gene Low-quality
S3_vs_k141_77308||lt2gene Low-quality
S3_vs_k141_61285||lt2gene Low-quality
S3_vs_k141_14342||lt2gene Low-quality
S3_vs_k141_116366||lt2gene Low-quality
S3_vs_k141_84498||lt2gene Low-quality
S3_vs_k141_108421||lt2gene Low-quality
S3_vs_k141_116389||lt2gene Not-determined
S3_vs_k141_36766||lt2gene Low-quality
S3_vs_k141_36769||lt2gene Low-quality
S3_vs_k141_77391||lt2gene Low-quality
S3_vs_k141_116408||lt2gene Low-quality
S3_vs_k141_4611||lt2gene Not-determined
S3_vs_k141_123815||lt2gene Low-quality
S3_vs_k141_84552||lt2gene Low-quality
S3_vs_k141_28698||lt2gene Low-quality
S3_vs_k141_77428||lt2gene Low-quality
S3_vs_k141_77430||lt2gene Low-quality
S3_vs_k141_108476||lt2gene Low-quality
S3_vs_k141_44613||lt2gene Not-determined
S3_vs_k141_20059||lt2gene Low-quality
S3_vs_k141_61496||lt2gene Low-quality
S3_vs_k141_84601||lt2gene Low-quality
S3_vs_k141_36916||lt2gene Low-quality
S3_vs_k141_36917||lt2gene Low-quality
S3_vs_k141_77501||lt2gene Low-quality
S3_vs_k141_77521||lt2gene Low-quality
S3_vs_k141_36923||lt2gene Low-quality
S3_vs_k141_36950||lt2gene Low-quality
S3_vs_k141_36953||lt2gene Low-quality
S3_vs_k141_53356||lt2gene Not-determined
S3_vs_k141_20173||lt2gene Low-quality
S3_vs_k141_84675||lt2gene Low-quality
S3_vs_k141_14509||lt2gene Low-quality
S3_vs_k141_53402||lt2gene Not-determined
S3_vs_k141_14522||lt2gene Not-determined
S3_vs_k141_77597||lt2gene Low-quality
S3_vs_k141_108547||lt2gene Low-quality
S3_vs_k141_92651||lt2gene Low-quality
S3_vs_k141_53433||lt2gene Low-quality
S3_vs_k141_68012||lt2gene Low-quality
S3_vs_k141_68018||lt2gene Low-quality
S3_vs_k141_77652||lt2gene Not-determined
S3_vs_k141_92679||lt2gene Low-quality
S3_vs_k141_20259||lt2gene Low-quality
S3_vs_k141_37035||lt2gene Low-quality
S3_vs_k141_37038||lt2gene Not-determined
S3_vs_k141_84743||lt2gene Low-quality
S3_vs_k141_116584||lt2gene Low-quality
S3_vs_k141_37046||lt2gene Low-quality
S3_vs_k141_20289||lt2gene Not-determined
S3_vs_k141_53492||lt2gene Not-determined
S3_vs_k141_92700||lt2gene Low-quality
S3_vs_k141_14546||lt2gene Low-quality
S3_vs_k141_61612||lt2gene Low-quality
S3_vs_k141_44856||lt2gene Low-quality
S3_vs_k141_92735||lt2gene Not-determined
S3_vs_k141_84821||lt2gene Low-quality
S3_vs_k141_44903||lt2gene Low-quality
S3_vs_k141_53563||lt2gene Low-quality
S3_vs_k141_124043||lt2gene Low-quality
S3_vs_k141_20338||lt2gene Low-quality
S3_vs_k141_20362||lt2gene Low-quality
S3_vs_k141_124070||lt2gene Not-determined
S3_vs_k141_68061||lt2gene Low-quality
S3_vs_k141_124082||lt2gene Low-quality
S3_vs_k141_108621||lt2gene Low-quality
S3_vs_k141_100193||lt2gene Low-quality
S3_vs_k141_100196||lt2gene Not-determined
S3_vs_k141_92794||lt2gene Low-quality
S3_vs_k141_61692||lt2gene Low-quality
S3_vs_k141_53602||lt2gene Not-determined
S3_vs_k141_77754||lt2gene Not-determined
S3_vs_k141_92804||lt2gene Low-quality
S3_vs_k141_61706||lt2gene Low-quality
S3_vs_k141_53621||lt2gene Low-quality
S3_vs_k141_84932||lt2gene Not-determined
S3_vs_k141_14710||lt2gene Low-quality
S3_vs_k141_37112||lt2gene Low-quality
S3_vs_k141_108671||lt2gene Not-determined
S3_vs_k141_20410||lt2gene Not-determined
S3_vs_k141_37151||lt2gene Low-quality
S3_vs_k141_68162||lt2gene Low-quality
S3_vs_k141_108697||lt2gene Low-quality
S3_vs_k141_4715||lt2gene Low-quality
S3_vs_k141_53665||lt2gene Low-quality
S3_vs_k141_108719||lt2gene Low-quality
S3_vs_k141_100283||lt2gene Not-determined
S3_vs_k141_28837||lt2gene Low-quality
S3_vs_k141_28838||lt2gene Low-quality
S3_vs_k141_14784||lt2gene Low-quality
S3_vs_k141_53700||lt2gene Low-quality
S3_vs_k141_4744||lt2gene Low-quality
S3_vs_k141_28861||lt2gene Not-determined
S3_vs_k141_68252||lt2gene Low-quality
S3_vs_k141_68259||lt2gene Low-quality
S3_vs_k141_92910||lt2gene Low-quality
S3_vs_k141_14819||lt2gene Low-quality
S3_vs_k141_37217||lt2gene Low-quality
S3_vs_k141_20443||lt2gene Low-quality
S3_vs_k141_85003||lt2gene Not-determined
S3_vs_k141_92931||lt2gene Low-quality
S3_vs_k141_108807||lt2gene Not-determined
S3_vs_k141_53740||lt2gene Not-determined
S3_vs_k141_20480||lt2gene Low-quality
S3_vs_k141_124214||lt2gene Not-determined
S3_vs_k141_124222||lt2gene Low-quality
S3_vs_k141_85048||lt2gene Low-quality
S3_vs_k141_108845||lt2gene Not-determined
S3_vs_k141_116738||lt2gene Not-determined
S3_vs_k141_28910||lt2gene Not-determined
S3_vs_k141_4878||lt2gene Not-determined
S3_vs_k141_4879||lt2gene Low-quality
S3_vs_k141_28916||lt2gene Low-quality
S3_vs_k141_37278||lt2gene Low-quality
S3_vs_k141_124264||lt2gene Low-quality
S3_vs_k141_14854||lt2gene Not-determined
S3_vs_k141_53806||lt2gene Not-determined
S3_vs_k141_61862||lt2gene Low-quality
S3_vs_k141_108939||lt2gene Not-determined
S3_vs_k141_77866||lt2gene Low-quality
S3_vs_k141_4925||lt2gene Low-quality
S3_vs_k141_28992||lt2gene Low-quality
S3_vs_k141_61880||lt2gene Low-quality
S3_vs_k141_85133||lt2gene Low-quality
S3_vs_k141_14903||lt2gene Not-determined
S3_vs_k141_29029||lt2gene Low-quality
S3_vs_k141_85157||lt2gene Not-determined
S3_vs_k141_77962||lt2gene Low-quality
S3_vs_k141_68405||lt2gene Not-determined
S3_vs_k141_68407||lt2gene Low-quality
S3_vs_k141_37343||lt2gene Low-quality
S3_vs_k141_5064||lt2gene Not-determined
S3_vs_k141_108990||lt2gene Low-quality
S3_vs_k141_20567||lt2gene Low-quality
S3_vs_k141_20576||lt2gene Low-quality
S3_vs_k141_116884||lt2gene Low-quality
S3_vs_k141_14976||lt2gene Low-quality
S3_vs_k141_68457||lt2gene Low-quality
S3_vs_k141_109026||lt2gene Low-quality
S3_vs_k141_62014||lt2gene Low-quality
S3_vs_k141_37377||lt2gene Low-quality
S3_vs_k141_62016||lt2gene Low-quality
S3_vs_k141_100489||lt2gene Not-determined
S3_vs_k141_20631||lt2gene Low-quality
S3_vs_k141_85278||lt2gene Low-quality
S3_vs_k141_62037||lt2gene Not-determined
S3_vs_k141_78039||lt2gene Low-quality
S3_vs_k141_78049||lt2gene Low-quality
S3_vs_k141_85297||lt2gene Low-quality
S3_vs_k141_62058||lt2gene Low-quality
S3_vs_k141_20641||lt2gene Not-determined
S3_vs_k141_45105||lt2gene Low-quality
S3_vs_k141_62067||lt2gene Low-quality
S3_vs_k141_62070||lt2gene Low-quality
S3_vs_k141_5170||lt2gene Low-quality
S3_vs_k141_54004||lt2gene Low-quality
S3_vs_k141_116943||lt2gene Low-quality
S3_vs_k141_15051||lt2gene Not-determined
S3_vs_k141_15054||lt2gene Low-quality
S3_vs_k141_109179||lt2gene Low-quality
S3_vs_k141_29139||lt2gene Low-quality
S3_vs_k141_29178||lt2gene Low-quality
S3_vs_k141_109190||lt2gene Low-quality
S3_vs_k141_109195||lt2gene Low-quality
S3_vs_k141_109214||lt2gene Low-quality
S3_vs_k141_62226||lt2gene Low-quality
S3_vs_k141_29218||lt2gene Low-quality
S3_vs_k141_29219||lt2gene Not-determined
S3_vs_k141_124458||lt2gene Low-quality
S3_vs_k141_124462||lt2gene Low-quality
S3_vs_k141_5306||lt2gene Low-quality
S3_vs_k141_100502||lt2gene Low-quality
S3_vs_k141_124487||lt2gene Low-quality
S3_vs_k141_15226||lt2gene Low-quality
S3_vs_k141_109270||lt2gene Low-quality
S3_vs_k141_62272||lt2gene Low-quality
S3_vs_k141_5347||lt2gene Low-quality
S3_vs_k141_85427||lt2gene Low-quality
S3_vs_k141_93159||lt2gene Low-quality
S3_vs_k141_85461||lt2gene Low-quality
S3_vs_k141_100536||lt2gene Low-quality
S3_vs_k141_100542||lt2gene Low-quality
S3_vs_k141_62335||lt2gene Low-quality
S3_vs_k141_109306||lt2gene Not-determined
S3_vs_k141_54032||lt2gene Low-quality
S3_vs_k141_5409||lt2gene Not-determined
S3_vs_k141_62343||lt2gene Not-determined
S3_vs_k141_29259||lt2gene Not-determined
S3_vs_k141_62350||lt2gene Not-determined
S3_vs_k141_100565||lt2gene Low-quality
S3_vs_k141_93220||lt2gene Low-quality
S3_vs_k141_20753||lt2gene Low-quality
S3_vs_k141_37632||lt2gene Low-quality
S3_vs_k141_124544||lt2gene Low-quality
S3_vs_k141_93276||lt2gene Low-quality
S3_vs_k141_20790||lt2gene Low-quality
S3_vs_k141_45277||lt2gene Low-quality
S3_vs_k141_93320||lt2gene Low-quality
S3_vs_k141_93331||lt2gene Not-determined
S3_vs_k141_29428||lt2gene Low-quality
S3_vs_k141_45334||lt2gene Low-quality
S3_vs_k141_45340||lt2gene Low-quality
S3_vs_k141_29438||lt2gene Low-quality
S3_vs_k141_62424||lt2gene Low-quality
S3_vs_k141_100673||lt2gene Low-quality
S3_vs_k141_29444||lt2gene Low-quality
S3_vs_k141_54102||lt2gene Low-quality
S3_vs_k141_29447||lt2gene Low-quality
S3_vs_k141_78277||lt2gene Not-determined
S3_vs_k141_124619||lt2gene Low-quality
S3_vs_k141_62456||lt2gene Not-determined
S3_vs_k141_15355||lt2gene Low-quality
S3_vs_k141_62464||lt2gene Not-determined
S3_vs_k141_78312||lt2gene Not-determined
S3_vs_k141_15373||lt2gene Not-determined
S3_vs_k141_109545||lt2gene Low-quality
S3_vs_k141_45359||lt2gene Low-quality
S3_vs_k141_93410||lt2gene Low-quality
S3_vs_k141_15390||lt2gene Low-quality
S3_vs_k141_124634||lt2gene Low-quality
S3_vs_k141_45394||lt2gene Low-quality
S3_vs_k141_78350||lt2gene Not-determined
S3_vs_k141_100751||lt2gene Not-determined
S3_vs_k141_78361||lt2gene Low-quality
S3_vs_k141_109674||lt2gene Low-quality
S3_vs_k141_124703||lt2gene Low-quality
S3_vs_k141_54181||lt2gene Not-determined
S3_vs_k141_85729||lt2gene Low-quality
S3_vs_k141_117176||lt2gene Not-determined
S3_vs_k141_85731||lt2gene Low-quality
S3_vs_k141_29588||lt2gene Low-quality
S3_vs_k141_45481||lt2gene Low-quality
S3_vs_k141_29609||lt2gene Not-determined
S3_vs_k141_78444||lt2gene Low-quality
S3_vs_k141_54209||lt2gene Low-quality
S3_vs_k141_5620||lt2gene Low-quality
S3_vs_k141_29647||lt2gene Low-quality
S3_vs_k141_45564||lt2gene Low-quality
S3_vs_k141_100808||lt2gene Low-quality
S3_vs_k141_45571||lt2gene Low-quality
S3_vs_k141_93559||lt2gene Low-quality
S3_vs_k141_93568||lt2gene Low-quality
S3_vs_k141_37755||lt2gene Not-determined
S3_vs_k141_5639||lt2gene Low-quality
S3_vs_k141_78486||lt2gene Low-quality
S3_vs_k141_15498||lt2gene Low-quality
S3_vs_k141_15499||lt2gene Low-quality
S3_vs_k141_15522||lt2gene Low-quality
S3_vs_k141_29688||lt2gene Low-quality
S3_vs_k141_21009||lt2gene Not-determined
S3_vs_k141_62526||lt2gene Low-quality
S3_vs_k141_93633||lt2gene Low-quality
S3_vs_k141_54299||lt2gene Not-determined
S3_vs_k141_45652||lt2gene Not-determined
S3_vs_k141_15578||lt2gene Low-quality
S3_vs_k141_100888||lt2gene Not-determined
S3_vs_k141_45678||lt2gene Low-quality
S3_vs_k141_85848||lt2gene Low-quality
S3_vs_k141_54308||lt2gene Low-quality
S3_vs_k141_124877||lt2gene Not-determined
S3_vs_k141_29767||lt2gene Low-quality
S3_vs_k141_93676||lt2gene Low-quality
S3_vs_k141_124893||lt2gene Low-quality
S3_vs_k141_124900||lt2gene Low-quality
S3_vs_k141_78590||lt2gene Not-determined
S3_vs_k141_54335||lt2gene Low-quality
S3_vs_k141_15649||lt2gene Not-determined
S3_vs_k141_85912||lt2gene Low-quality
S3_vs_k141_117306||lt2gene Low-quality
S3_vs_k141_117319||lt2gene Not-determined
S3_vs_k141_117325||lt2gene Low-quality
S3_vs_k141_85958||lt2gene Not-determined
S3_vs_k141_15675||lt2gene Not-determined
S3_vs_k141_62564||lt2gene Low-quality
S3_vs_k141_15691||lt2gene Low-quality
S3_vs_k141_62594||lt2gene Low-quality
S3_vs_k141_21160||lt2gene Not-determined
S3_vs_k141_29818||lt2gene Low-quality
S3_vs_k141_54364||lt2gene Low-quality
S3_vs_k141_29831||lt2gene Low-quality
S3_vs_k141_86037||lt2gene Not-determined
S3_vs_k141_29871||lt2gene Low-quality
S3_vs_k141_29883||lt2gene Low-quality
S3_vs_k141_93821||lt2gene Low-quality
S3_vs_k141_62694||lt2gene Low-quality
S3_vs_k141_37943||lt2gene Low-quality
S3_vs_k141_15773||lt2gene Low-quality
S3_vs_k141_21204||lt2gene Low-quality
S3_vs_k141_78696||lt2gene Not-determined
S3_vs_k141_117386||lt2gene Low-quality
S3_vs_k141_37985||lt2gene Low-quality
S3_vs_k141_78702||lt2gene Low-quality
S3_vs_k141_86093||lt2gene Low-quality
S3_vs_k141_117397||lt2gene Low-quality
S3_vs_k141_54496||lt2gene Not-determined
S3_vs_k141_117449||lt2gene Low-quality
S3_vs_k141_86134||lt2gene Low-quality
S3_vs_k141_117457||lt2gene Low-quality
S3_vs_k141_86154||lt2gene Low-quality
S3_vs_k141_29947||lt2gene Low-quality
S3_vs_k141_69072||lt2gene Low-quality
S3_vs_k141_86158||lt2gene Low-quality
S3_vs_k141_29964||lt2gene Low-quality
S3_vs_k141_38051||lt2gene Low-quality
S3_vs_k141_117480||lt2gene Low-quality
S3_vs_k141_45999||lt2gene Low-quality
S3_vs_k141_109928||lt2gene Low-quality
S3_vs_k141_117540||lt2gene Not-determined
S3_vs_k141_21269||lt2gene Low-quality
S3_vs_k141_86229||lt2gene Low-quality
S3_vs_k141_38096||lt2gene Low-quality
S3_vs_k141_86243||lt2gene Low-quality
S3_vs_k141_46027||lt2gene Low-quality
S3_vs_k141_86276||lt2gene Low-quality
S3_vs_k141_117586||lt2gene Low-quality
S3_vs_k141_78814||lt2gene Not-determined
S3_vs_k141_93930||lt2gene Low-quality
S3_vs_k141_86291||lt2gene Low-quality
S3_vs_k141_125092||lt2gene Low-quality
S3_vs_k141_30084||lt2gene Low-quality
S3_vs_k141_109994||lt2gene Low-quality
S3_vs_k141_46084||lt2gene Low-quality
S3_vs_k141_30113||lt2gene Low-quality
S3_vs_k141_54589||lt2gene Low-quality
S3_vs_k141_46107||lt2gene Low-quality
S3_vs_k141_46118||lt2gene Low-quality
S3_vs_k141_125171||lt2gene Low-quality
S3_vs_k141_94020||lt2gene Low-quality
S3_vs_k141_69114||lt2gene Low-quality
S3_vs_k141_62876||lt2gene Low-quality
S3_vs_k141_5921||lt2gene Not-determined
S3_vs_k141_110125||lt2gene Low-quality
S3_vs_k141_110134||lt2gene Low-quality
S3_vs_k141_30158||lt2gene Low-quality
S3_vs_k141_54690||lt2gene Low-quality
S3_vs_k141_54693||lt2gene Not-determined
S3_vs_k141_94080||lt2gene Not-determined
S3_vs_k141_94083||lt2gene Not-determined
S3_vs_k141_86455||lt2gene Low-quality
S3_vs_k141_125212||lt2gene Low-quality
S3_vs_k141_54704||lt2gene Low-quality
S3_vs_k141_94106||lt2gene Low-quality
S3_vs_k141_62962||lt2gene Not-determined
S3_vs_k141_125247||lt2gene Low-quality
S3_vs_k141_117786||lt2gene Low-quality
S3_vs_k141_117800||lt2gene Not-determined
S3_vs_k141_46246||lt2gene Low-quality
S3_vs_k141_30246||lt2gene Low-quality
S3_vs_k141_46268||lt2gene Low-quality
S3_vs_k141_78973||lt2gene Low-quality
S3_vs_k141_63056||lt2gene Low-quality
S3_vs_k141_69214||lt2gene Low-quality
S3_vs_k141_94219||lt2gene Low-quality
S3_vs_k141_63110||lt2gene Not-determined
S3_vs_k141_46317||lt2gene Not-determined
S3_vs_k141_101072||lt2gene Low-quality
S3_vs_k141_69238||lt2gene Low-quality
S3_vs_k141_54848||lt2gene Not-determined
S3_vs_k141_6052||lt2gene Not-determined
S3_vs_k141_38388||lt2gene Not-determined
S3_vs_k141_6079||lt2gene Low-quality
S3_vs_k141_86626||lt2gene Low-quality
S3_vs_k141_69272||lt2gene Low-quality
S3_vs_k141_21523||lt2gene Low-quality
S3_vs_k141_6122||lt2gene Low-quality
S3_vs_k141_101122||lt2gene Low-quality
S3_vs_k141_46370||lt2gene Low-quality
S3_vs_k141_38477||lt2gene Not-determined
S3_vs_k141_79012||lt2gene Low-quality
S3_vs_k141_101163||lt2gene Low-quality
S3_vs_k141_21588||lt2gene Not-determined
S3_vs_k141_110339||lt2gene Low-quality
S3_vs_k141_30344||lt2gene Low-quality
S3_vs_k141_30373||lt2gene Not-determined
S3_vs_k141_117911||lt2gene Not-determined
S3_vs_k141_86716||lt2gene Low-quality
S3_vs_k141_101257||lt2gene Not-determined
S3_vs_k141_30437||lt2gene Low-quality
S3_vs_k141_63276||lt2gene Low-quality
S3_vs_k141_30449||lt2gene Low-quality
S3_vs_k141_30453||lt2gene Low-quality
S3_vs_k141_110401||lt2gene Low-quality
S3_vs_k141_54994||lt2gene Not-determined
S3_vs_k141_125459||lt2gene Low-quality
S3_vs_k141_86793||lt2gene Low-quality
S3_vs_k141_6302||lt2gene Low-quality
S3_vs_k141_21729||lt2gene Low-quality
S3_vs_k141_6333||lt2gene Low-quality
S3_vs_k141_6346||lt2gene Not-determined
S3_vs_k141_110475||lt2gene Low-quality
S3_vs_k141_125538||lt2gene Low-quality
S3_vs_k141_21753||lt2gene Low-quality
S3_vs_k141_94358||lt2gene Not-determined
S3_vs_k141_118081||lt2gene Low-quality
S3_vs_k141_101411||lt2gene Low-quality
S3_vs_k141_69444||lt2gene Not-determined
S3_vs_k141_118090||lt2gene Low-quality
S3_vs_k141_55066||lt2gene Not-determined
S3_vs_k141_46455||lt2gene Low-quality
S3_vs_k141_6390||lt2gene Low-quality
S3_vs_k141_21777||lt2gene Low-quality
S3_vs_k141_21784||lt2gene Low-quality
S3_vs_k141_21794||lt2gene Not-determined
S3_vs_k141_69473||lt2gene Low-quality
S3_vs_k141_110559||lt2gene Low-quality
S3_vs_k141_21845||lt2gene Low-quality
S3_vs_k141_21849||lt2gene Low-quality
S3_vs_k141_94428||lt2gene Not-determined
S3_vs_k141_94433||lt2gene Low-quality
S3_vs_k141_86859||lt2gene Low-quality
S3_vs_k141_118154||lt2gene Low-quality
S3_vs_k141_79129||lt2gene Low-quality
S3_vs_k141_46518||lt2gene Low-quality
S3_vs_k141_46526||lt2gene Low-quality
S3_vs_k141_21924||lt2gene Low-quality
S3_vs_k141_86889||lt2gene Low-quality
S3_vs_k141_6461||lt2gene Low-quality
S3_vs_k141_46539||lt2gene Low-quality
S3_vs_k141_69540||lt2gene Low-quality
S3_vs_k141_125664||lt2gene Low-quality
S3_vs_k141_101471||lt2gene Low-quality
S3_vs_k141_69558||lt2gene Low-quality
S3_vs_k141_6478||lt2gene Not-determined
S3_vs_k141_38677||lt2gene Low-quality
S3_vs_k141_86916||lt2gene Low-quality
S3_vs_k141_69564||lt2gene Low-quality
S3_vs_k141_46572||lt2gene Not-determined
S3_vs_k141_46577||lt2gene Low-quality
S3_vs_k141_6485||lt2gene Low-quality
S3_vs_k141_110593||lt2gene Not-determined
S3_vs_k141_55140||lt2gene Not-determined
S3_vs_k141_86985||lt2gene Low-quality
S3_vs_k141_94517||lt2gene Low-quality
S3_vs_k141_101543||lt2gene Not-determined
S3_vs_k141_46617||lt2gene Low-quality
S3_vs_k141_69624||lt2gene Low-quality
S3_vs_k141_55186||lt2gene Low-quality
S3_vs_k141_79275||lt2gene Low-quality
S3_vs_k141_63480||lt2gene Low-quality
S3_vs_k141_79301||lt2gene Low-quality
S3_vs_k141_38810||lt2gene Low-quality
S3_vs_k141_79308||lt2gene Low-quality
S3_vs_k141_22079||lt2gene Low-quality
S3_vs_k141_125740||lt2gene Not-determined
S3_vs_k141_46663||lt2gene Not-determined
S3_vs_k141_79334||lt2gene Low-quality
S3_vs_k141_87066||lt2gene Low-quality
S3_vs_k141_6566||lt2gene Low-quality
S3_vs_k141_79359||lt2gene Low-quality
S3_vs_k141_22123||lt2gene Low-quality
S3_vs_k141_125777||lt2gene Low-quality
S3_vs_k141_87081||lt2gene Low-quality
S3_vs_k141_6600||lt2gene Low-quality
S3_vs_k141_79391||lt2gene Low-quality
S3_vs_k141_79396||lt2gene Not-determined
S3_vs_k141_22152||lt2gene Low-quality
S3_vs_k141_110748||lt2gene Low-quality
S3_vs_k141_69775||lt2gene Low-quality
S3_vs_k141_55245||lt2gene Not-determined
S3_vs_k141_22185||lt2gene Low-quality
S3_vs_k141_22210||lt2gene Low-quality
S3_vs_k141_46774||lt2gene Low-quality
S3_vs_k141_22247||lt2gene Not-determined
S3_vs_k141_69796||lt2gene Low-quality
S3_vs_k141_101685||lt2gene Not-determined
S3_vs_k141_69818||lt2gene Low-quality
S3_vs_k141_6632||lt2gene Low-quality
S3_vs_k141_94766||lt2gene Low-quality
S3_vs_k141_55300||lt2gene Not-determined
S3_vs_k141_55313||lt2gene Low-quality
S3_vs_k141_30726||lt2gene Not-determined
S3_vs_k141_46883||lt2gene Low-quality
S3_vs_k141_125846||lt2gene Low-quality
S3_vs_k141_46906||lt2gene Low-quality
S3_vs_k141_118406||lt2gene Not-determined
S3_vs_k141_30768||lt2gene Not-determined
S3_vs_k141_110832||lt2gene Low-quality
S3_vs_k141_110835||lt2gene Low-quality
S3_vs_k141_118423||lt2gene Low-quality
S3_vs_k141_125887||lt2gene Low-quality
S3_vs_k141_69916||lt2gene Low-quality
S3_vs_k141_125892||lt2gene Not-determined
S3_vs_k141_101801||lt2gene Not-determined
S3_vs_k141_38927||lt2gene Low-quality
S3_vs_k141_101820||lt2gene Low-quality
S3_vs_k141_46950||lt2gene Low-quality
S3_vs_k141_125929||lt2gene Low-quality
S3_vs_k141_101835||lt2gene Low-quality
S3_vs_k141_55381||lt2gene Low-quality
S3_vs_k141_101846||lt2gene Low-quality
S3_vs_k141_38971||lt2gene Low-quality
S3_vs_k141_118491||lt2gene Not-determined
S3_vs_k141_118496||lt2gene Low-quality
S3_vs_k141_101855||lt2gene Not-determined
S3_vs_k141_46982||lt2gene Low-quality
S3_vs_k141_110893||lt2gene Low-quality
S3_vs_k141_30845||lt2gene Not-determined
S3_vs_k141_6767||lt2gene Low-quality
S3_vs_k141_79538||lt2gene Low-quality
S3_vs_k141_6779||lt2gene Not-determined
S3_vs_k141_69987||lt2gene Low-quality
S3_vs_k141_30873||lt2gene Not-determined
S3_vs_k141_79582||lt2gene Not-determined
S3_vs_k141_6806||lt2gene Not-determined
S3_vs_k141_55468||lt2gene Low-quality
S3_vs_k141_101909||lt2gene Not-determined
S3_vs_k141_111018||lt2gene Low-quality
S3_vs_k141_70086||lt2gene Low-quality
S3_vs_k141_22444||lt2gene Low-quality
S3_vs_k141_39092||lt2gene Low-quality
S3_vs_k141_111043||lt2gene Low-quality
S3_vs_k141_118612||lt2gene Low-quality
S3_vs_k141_111054||lt2gene Not-determined
S3_vs_k141_70098||lt2gene Low-quality
S3_vs_k141_70110||lt2gene Low-quality
S3_vs_k141_70113||lt2gene Low-quality
S3_vs_k141_39121||lt2gene Low-quality
S3_vs_k141_126109||lt2gene Low-quality
S3_vs_k141_95061||lt2gene Not-determined
S3_vs_k141_22507||lt2gene Low-quality
S3_vs_k141_70154||lt2gene Low-quality
S3_vs_k141_111164||lt2gene Low-quality
S3_vs_k141_111176||lt2gene Not-determined
S3_vs_k141_118721||lt2gene Low-quality
S3_vs_k141_39214||lt2gene Not-determined
S3_vs_k141_31003||lt2gene Low-quality
S3_vs_k141_111211||lt2gene Low-quality
S3_vs_k141_39229||lt2gene Not-determined
S3_vs_k141_111245||lt2gene Low-quality
S3_vs_k141_70192||lt2gene Low-quality
S3_vs_k141_39271||lt2gene Low-quality
S3_vs_k141_70207||lt2gene Not-determined
S3_vs_k141_47211||lt2gene Low-quality
S3_vs_k141_118760||lt2gene Low-quality
S3_vs_k141_111273||lt2gene Low-quality
S3_vs_k141_47226||lt2gene Not-determined
S3_vs_k141_55596||lt2gene Not-determined
S3_vs_k141_87447||lt2gene Low-quality
S3_vs_k141_87458||lt2gene Low-quality
S3_vs_k141_31072||lt2gene Low-quality
S3_vs_k141_95158||lt2gene Not-determined
S3_vs_k141_47287||lt2gene Low-quality
S3_vs_k141_70258||lt2gene Low-quality
S3_vs_k141_95186||lt2gene Low-quality
S3_vs_k141_6979||lt2gene Low-quality
S3_vs_k141_126325||lt2gene Low-quality
S3_vs_k141_111372||lt2gene Low-quality
S3_vs_k141_55670||lt2gene Low-quality
S3_vs_k141_87554||lt2gene Low-quality
S3_vs_k141_39297||lt2gene Not-determined
S3_vs_k141_126352||lt2gene Not-determined
S3_vs_k141_111391||lt2gene Not-determined
S3_vs_k141_102056||lt2gene Low-quality
S3_vs_k141_111401||lt2gene Low-quality
S3_vs_k141_22644||lt2gene Low-quality
S3_vs_k141_22667||lt2gene Not-determined

| | |
|---|---|
| S3_vs_k141_102081\|\|lt2gene | Low-quality |
| S3_vs_k141_55755\|\|lt2gene | Low-quality |
| S3_vs_k141_102099\|\|lt2gene | Low-quality |
| S3_vs_k141_39362\|\|lt2gene | Low-quality |
| S3_vs_k141_87609\|\|lt2gene | Not-determined |
| S3_vs_k141_7036\|\|lt2gene | Low-quality |
| S3_vs_k141_87651\|\|lt2gene | Low-quality |
| S3_vs_k141_47482\|\|lt2gene | Not-determined |
| S3_vs_k141_31361\|\|lt2gene | Low-quality |
| S3_vs_k141_31371\|\|lt2gene | Low-quality |
| S3_vs_k141_39505\|\|lt2gene | Low-quality |
| S3_vs_k141_39510\|\|lt2gene | Not-determined |
| S3_vs_k141_70419\|\|lt2gene | Low-quality |
| S3_vs_k141_47536\|\|lt2gene | Low-quality |
| S3_vs_k141_70429\|\|lt2gene | Low-quality |
| S3_vs_k141_7064\|\|lt2gene | Not-determined |
| S3_vs_k141_111501\|\|lt2gene | Low-quality |
| S3_vs_k141_126536\|\|lt2gene | Low-quality |
| S3_vs_k141_22763\|\|lt2gene | Not-determined |
| S3_vs_k141_111534\|\|lt2gene | Low-quality |
| S3_vs_k141_70466\|\|lt2gene | Low-quality |
| S3_vs_k141_39607\|\|lt2gene | Not-determined |
| S3_vs_k141_111569\|\|lt2gene | Not-determined |
| S3_vs_k141_102244\|\|lt2gene | Not-determined |
| S3_vs_k141_70494\|\|lt2gene | Low-quality |
| S3_vs_k141_39629\|\|lt2gene | Low-quality |
| S3_vs_k141_39643\|\|lt2gene | Low-quality |
| S3_vs_k141_102261\|\|lt2gene | Low-quality |
| S3_vs_k141_47648\|\|lt2gene | Low-quality |
| S3_vs_k141_95478\|\|lt2gene | Low-quality |
| S3_vs_k141_39693\|\|lt2gene | Low-quality |
| S3_vs_k141_95491\|\|lt2gene | Low-quality |
| S3_vs_k141_22885\|\|lt2gene | Not-determined |
| S3_vs_k141_102335\|\|lt2gene | Not-determined |
| S3_vs_k141_7141\|\|lt2gene | Low-quality |
| S3_vs_k141_70563\|\|lt2gene | Low-quality |
| S3_vs_k141_7172\|\|lt2gene | Low-quality |
| S3_vs_k141_39764\|\|lt2gene | Low-quality |
| S3_vs_k141_47769\|\|lt2gene | Not-determined |
| S3_vs_k141_22998\|\|lt2gene | Not-determined |
| S3_vs_k141_31597\|\|lt2gene | Low-quality |
| S3_vs_k141_95568\|\|lt2gene | Low-quality |
| S3_vs_k141_118935\|\|lt2gene | Not-determined |
| S3_vs_k141_70666\|\|lt2gene | Low-quality |
| S3_vs_k141_118953\|\|lt2gene | Low-quality |
| S3_vs_k141_7292\|\|lt2gene | Low-quality |
| S3_vs_k141_126742\|\|lt2gene | Low-quality |
| S3_vs_k141_39801\|\|lt2gene | Low-quality |
| S3_vs_k141_102413\|\|lt2gene | Low-quality |
| S3_vs_k141_95584\|\|lt2gene | Not-determined |
| S3_vs_k141_23065\|\|lt2gene | Low-quality |
| S3_vs_k141_70695\|\|lt2gene | Low-quality |
| S3_vs_k141_102443\|\|lt2gene | Low-quality |
| S3_vs_k141_31668\|\|lt2gene | Low-quality |
| S3_vs_k141_23078\|\|lt2gene | Low-quality |
| S3_vs_k141_70726\|\|lt2gene | Low-quality |
| S3_vs_k141_39845\|\|lt2gene | Low-quality |
| S3_vs_k141_102489\|\|lt2gene | Low-quality |
| S3_vs_k141_102496\|\|lt2gene | Low-quality |
| S3_vs_k141_95600\|\|lt2gene | Low-quality |
| S3_vs_k141_7380\|\|lt2gene | Low-quality |
| S3_vs_k141_7381\|\|lt2gene | Low-quality |
| S3_vs_k141_95622\|\|lt2gene | Low-quality |
| S3_vs_k141_31766\|\|lt2gene | Low-quality |
| S3_vs_k141_102636\|\|lt2gene | Low-quality |
| S3_vs_k141_70794\|\|lt2gene | Low-quality |
| S3_vs_k141_23218\|\|lt2gene | Low-quality |
| S3_vs_k141_70846\|\|lt2gene | Not-determined |
| S3_vs_k141_23238\|\|lt2gene | Low-quality |
| S3_vs_k141_31817\|\|lt2gene | Low-quality |
| S3_vs_k141_23278\|\|lt2gene | Low-quality |
| S3_vs_k141_126906\|\|lt2gene | Low-quality |
| S3_vs_k141_126910\|\|lt2gene | Low-quality |
| S3_vs_k141_70969\|\|lt2gene | Not-determined |
| S3_vs_k141_23314\|\|lt2gene | Low-quality |
| S3_vs_k141_70992\|\|lt2gene | Not-determined |
| S3_vs_k141_7609\|\|lt2gene | Low-quality |
| S3_vs_k141_102794\|\|lt2gene | Low-quality |
| S3_vs_k141_7640\|\|lt2gene | Low-quality |
| S3_vs_k141_119166\|\|lt2gene | Low-quality |
| S3_vs_k141_71063\|\|lt2gene | Not-determined |
| S3_vs_k141_7687\|\|lt2gene | Not-determined |
| S3_vs_k141_102842\|\|lt2gene | Low-quality |
| S3_vs_k141_23391\|\|lt2gene | Not-determined |
| S3_vs_k141_126963\|\|lt2gene | Low-quality |
| S3_vs_k141_71132\|\|lt2gene | Not-determined |
| S3_vs_k141_7828\|\|lt2gene | Low-quality |
| S3_vs_k141_23507\|\|lt2gene | Low-quality |
| S3_vs_k141_71196\|\|lt2gene | Low-quality |
| S3_vs_k141_71209\|\|lt2gene | Low-quality |
| S3_vs_k141_23526\|\|lt2gene | Low-quality |
| S3_vs_k141_23531\|\|lt2gene | Low-quality |
| S3_vs_k141_102936\|\|lt2gene | Low-quality |
| S3_vs_k141_23558\|\|lt2gene | Low-quality |
| S3_vs_k141_102986\|\|lt2gene | Low-quality |
| S3_vs_k141_71317\|\|lt2gene | Low-quality |
| S3_vs_k141_23590\|\|lt2gene | Low-quality |
| S3_vs_k141_119425\|\|lt2gene | Low-quality |
| S3_vs_k141_71340\|\|lt2gene | Low-quality |
| S3_vs_k141_103070\|\|lt2gene | Low-quality |
| S3_vs_k141_103079\|\|lt2gene | Not-determined |
| S3_vs_k141_103104\|\|lt2gene | Low-quality |
| S3_vs_k141_23660\|\|lt2gene | Not-determined |
| S3_vs_k141_23698\|\|lt2gene | Low-quality |
| S3_vs_k141_127289\|\|lt2gene | Low-quality |
| S3_vs_k141_71523\|\|lt2gene | Low-quality |
| S3_vs_k141_71543\|\|lt2gene | Low-quality |
| S3_vs_k141_127374\|\|lt2gene | Low-quality |
| S3_vs_k141_23810\|\|lt2gene | Not-determined |
| S3_vs_k141_71577\|\|lt2gene | Not-determined |
| S3_vs_k141_23825\|\|lt2gene | Low-quality |
| S3_vs_k141_71592\|\|lt2gene | Low-quality |
| S3_vs_k141_23837\|\|lt2gene | Low-quality |
| S3_vs_k141_103262\|\|lt2gene | Low-quality |
| S3_vs_k141_103279\|\|lt2gene | Low-quality |
| S3_vs_k141_103446\|\|lt2gene | Low-quality |
| S3_vs_k141_103569\|\|lt2gene | Low-quality |
| S3_vs_k141_103605\|\|lt2gene | Not-determined |
| S5_extracted_k141_3 | Not-determined |
| S5_extracted_k141_24289 | Not-determined |
| S5_extracted_k141_6076 | Not-determined |
| S5_extracted_k141_72860 | Not-determined |
| S5_extracted_k141_91080 | Low-quality |
| S5_extracted_k141_97148 | Low-quality |
| S5_extracted_k141_48578 | Not-determined |
| S5_extracted_k141_21 | Not-determined |
| S5_extracted_k141_24294 | Not-determined |
| S5_extracted_k141_127504 | Low-quality |
| S5_extracted_k141_48582 | Not-determined |
| S5_extracted_k141_91088 | Not-determined |
| S5_extracted_k141_133574 | Not-determined |
| S5_extracted_k141_121433 | Not-determined |
| S5_extracted_k141_36441 | Not-determined |
| S5_extracted_k141_29 | Not-determined |
| S5_extracted_k141_103226 | Not-determined |
| S5_extracted_k141_54650 | Not-determined |
| S5_extracted_k141_103228 | Not-determined |
| S5_extracted_k141_42524 | Not-determined |
| S5_extracted_k141_41 | Not-determined |
| S5_extracted_k141_42528 | Not-determined |
| S5_extracted_k141_54656 | Not-determined |
| S5_extracted_k141_36446 | Low-quality |
| S5_extracted_k141_42531 | Not-determined |
| S5_extracted_k141_36448 | Not-determined |
| S5_extracted_k141_91097 | Not-determined |
| S5_extracted_k141_121446 | Not-determined |
| S5_extracted_k141_97164 | Not-determined |
| S5_extracted_k141_97165 | Low-quality |
| S5_extracted_k141_66810 | Not-determined |
| S5_extracted_k141_133583 | Not-determined |
| S5_extracted_k141_54661 | Not-determined |
| S5_extracted_k141_133586 | Not-determined |
| S5_extracted_k141_121452 | Not-determined |
| S5_extracted_k141_49 | Not-determined |
| S5_extracted_k141_36451 | Low-quality |
| S5_extracted_k141_18240 | Low-quality |
| S5_extracted_k141_91113 | Not-determined |
| S5_extracted_k141_91114 | Not-determined |
| S5_extracted_k141_91118 | Not-determined |
| S5_extracted_k141_42547 | Not-determined |
| S5_extracted_k141_121467 | Not-determined |
| S5_extracted_k141_54667 | Not-determined |
| S5_extracted_k141_139675 | Not-determined |
| S5_extracted_k141_91124 | Not-determined |
| S5_extracted_k141_91125 | Low-quality |
| S5_extracted_k141_115375 | Not-determined |
| S5_extracted_k141_60773 | Not-determined |
| S5_extracted_k141_103242 | Not-determined |
| S5_extracted_k141_115377 | Low-quality |
| S5_extracted_k141_42553 | Not-determined |
| S5_extracted_k141_64 | Not-determined |
| S5_extracted_k141_139681 | Not-determined |
| S5_extracted_k141_133596 | Low-quality |
| S5_extracted_k141_91130 | Not-determined |
| S5_extracted_k141_121477 | Low-quality |
| S5_extracted_k141_85044 | Not-determined |
| S5_extracted_k141_121480 | Not-determined |
| S5_extracted_k141_85049 | Low-quality |
| S5_extracted_k141_36490 | Not-determined |
| S5_extracted_k141_103254 | Not-determined |
| S5_extracted_k141_36493 | Not-determined |
| S5_extracted_k141_103269 | Not-determined |
| S5_extracted_k141_42581 | Not-determined |
| S5_extracted_k141_42583 | Not-determined |
| S5_extracted_k141_127524 | Not-determined |
| S5_extracted_k141_89 | Not-determined |
| S5_extracted_k141_109325 | Low-quality |
| S5_extracted_k141_91153 | Not-determined |
| S5_extracted_k141_54682 | Low-quality |
| S5_extracted_k141_12176 | Not-determined |
| S5_extracted_k141_12177 | Not-determined |
| S5_extracted_k141_85063 | Not-determined |
| S5_extracted_k141_6116 | Not-determined |
| S5_extracted_k141_103279 | Not-determined |
| S5_extracted_k141_54688 | Not-determined |
| S5_extracted_k141_12183 | Low-quality |
| S5_extracted_k141_54692 | Not-determined |
| S5_extracted_k141_109335 | Low-quality |
| S5_extracted_k141_121509 | Not-determined |
| S5_extracted_k141_54697 | Not-determined |
| S5_extracted_k141_36503 | Not-determined |
| S5_extracted_k141_121513 | Not-determined |
| S5_extracted_k141_121515 | Not-determined |
| S5_extracted_k141_97172 | Low-quality |
| S5_extracted_k141_42597 | Not-determined |
| S5_extracted_k141_42599 | Not-determined |
| S5_extracted_k141_24324 | Not-determined |
| S5_extracted_k141_121529 | Not-determined |
| S5_extracted_k141_48650 | Not-determined |
| S5_extracted_k141_121530 | Not-determined |
| S5_extracted_k141_121531 | Not-determined |
| S5_extracted_k141_109354 | Low-quality |
| S5_extracted_k141_12197 | Not-determined |
| S5_extracted_k141_24330 | Not-determined |
| S5_extracted_k141_18302 | Not-determined |
| S5_extracted_k141_97192 | Not-determined |
| S5_extracted_k141_97193 | Not-determined |
| S5_extracted_k141_60817 | Not-determined |
| S5_extracted_k141_127556 | Low-quality |
| S5_extracted_k141_103301 | Not-determined |
| S5_extracted_k141_48664 | Low-quality |
| S5_extracted_k141_42610 | Low-quality |
| S5_extracted_k141_12211 | Not-determined |
| S5_extracted_k141_139717 | Low-quality |
| S5_extracted_k141_103318 | Not-determined |
| S5_extracted_k141_42618 | Low-quality |
| S5_extracted_k141_12219 | Low-quality |
| S5_extracted_k141_42624 | Not-determined |
| S5_extracted_k141_18326 | Low-quality |
| S5_extracted_k141_133615 | Not-determined |
| S5_extracted_k141_72906 | Not-determined |
| S5_extracted_k141_54719 | Not-determined |
| S5_extracted_k141_115395 | Not-determined |
| S5_extracted_k141_115396 | Not-determined |
| S5_extracted_k141_103342 | Not-determined |
| S5_extracted_k141_54724 | Low-quality |
| S5_extracted_k141_54727 | Not-determined |
| S5_extracted_k141_6186 | Low-quality |
| S5_extracted_k141_103351 | Not-determined |
| S5_extracted_k141_127592 | Low-quality |
| S5_extracted_k141_60854 | Not-determined |
| S5_extracted_k141_103355 | Low-quality |
| S5_extracted_k141_72917 | Not-determined |
| S5_extracted_k141_85071 | Not-determined |
| S5_extracted_k141_42663 | Not-determined |
| S5_extracted_k141_42665 | Not-determined |
| S5_extracted_k141_24348 | Not-determined |
| S5_extracted_k141_85074 | Low-quality |
| S5_extracted_k141_6195 | Not-determined |
| S5_extracted_k141_103378 | Not-determined |
| S5_extracted_k141_72942 | Not-determined |
| S5_extracted_k141_109382 | Not-determined |
| S5_extracted_k141_103386 | Not-determined |
| S5_extracted_k141_109383 | Low-quality |
| S5_extracted_k141_6212 | Not-determined |
| S5_extracted_k141_139755 | Not-determined |
| S5_extracted_k141_85089 | Low-quality |
| S5_extracted_k141_54749 | Not-determined |
| S5_extracted_k141_60888 | Not-determined |
| S5_extracted_k141_6220 | Not-determined |
| S5_extracted_k141_6221 | Not-determined |
| S5_extracted_k141_6222 | Not-determined |
| S5_extracted_k141_109398 | Not-determined |
| S5_extracted_k141_139765 | Not-determined |
| S5_extracted_k141_36533 | Not-determined |
| S5_extracted_k141_54756 | Not-determined |
| S5_extracted_k141_103406 | Low-quality |
| S5_extracted_k141_54757 | Low-quality |
| S5_extracted_k141_54759 | Low-quality |
| S5_extracted_k141_42691 | Not-determined |
| S5_extracted_k141_103413 | Low-quality |
| S5_extracted_k141_109410 | Not-determined |
| S5_extracted_k141_103414 | Not-determined |
| S5_extracted_k141_103415 | Not-determined |
| S5_extracted_k141_97239 | Not-determined |
| S5_extracted_k141_6233 | Not-determined |
| S5_extracted_k141_60906 | Not-determined |
| S5_extracted_k141_97240 | Low-quality |
| S5_extracted_k141_103422 | Not-determined |
| S5_extracted_k141_103423 | Low-quality |
| S5_extracted_k141_42702 | Not-determined |
| S5_extracted_k141_42704 | Not-determined |
| S5_extracted_k141_97249 | Not-determined |
| S5_extracted_k141_42707 | Not-determined |
| S5_extracted_k141_30421 | Not-determined |
| S5_extracted_k141_109423 | Low-quality |
| S5_extracted_k141_109424 | Not-determined |
| S5_extracted_k141_103436 | Not-determined |
| S5_extracted_k141_36542 | Not-determined |
| S5_extracted_k141_60923 | Not-determined |
| S5_extracted_k141_60924 | Not-determined |
| S5_extracted_k141_103438 | Not-determined |
| S5_extracted_k141_103439 | Not-determined |
| S5_extracted_k141_60928 | Not-determined |
| S5_extracted_k141_72962 | Low-quality |
| S5_extracted_k141_97251 | Not-determined |
| S5_extracted_k141_30430 | Low-quality |
| S5_extracted_k141_36548 | Not-determined |
| S5_extracted_k141_133630 | Not-determined |
| S5_extracted_k141_97264 | Not-determined |
| S5_extracted_k141_127617 | Low-quality |
| S5_extracted_k141_99 | Not-determined |
| S5_extracted_k141_100 | Not-determined |
| S5_extracted_k141_85140 | Not-determined |
| S5_extracted_k141_85143 | Low-quality |
| S5_extracted_k141_48678 | Not-determined |
| S5_extracted_k141_85146 | Not-determined |
| S5_extracted_k141_108 | Low-quality |
| S5_extracted_k141_30449 | Low-quality |
| S5_extracted_k141_103455 | Low-quality |
| S5_extracted_k141_127630 | Low-quality |
| S5_extracted_k141_103459 | Not-determined |
| S5_extracted_k141_121571 | Not-determined |
| S5_extracted_k141_78987 | Low-quality |
| S5_extracted_k141_127634 | Not-determined |
| S5_extracted_k141_48689 | Not-determined |
| S5_extracted_k141_48690 | Low-quality |
| S5_extracted_k141_6258 | Not-determined |
| S5_extracted_k141_48695 | Not-determined |
| S5_extracted_k141_121588 | Not-determined |
| S5_extracted_k141_42740 | Not-determined |
| S5_extracted_k141_127640 | Not-determined |
| S5_extracted_k141_127641 | Not-determined |
| S5_extracted_k141_36569 | Not-determined |
| S5_extracted_k141_66851 | Not-determined |
| S5_extracted_k141_66853 | Not-determined |
| S5_extracted_k141_36577 | Low-quality |
| S5_extracted_k141_18352 | Not-determined |
| S5_extracted_k141_66858 | Not-determined |
| S5_extracted_k141_66859 | Not-determined |
| S5_extracted_k141_97284 | Low-quality |
| S5_extracted_k141_6275 | Not-determined |
| S5_extracted_k141_18361 | Not-determined |
| S5_extracted_k141_118 | Not-determined |
| S5_extracted_k141_121 | Not-determined |
| S5_extracted_k141_97297 | Not-determined |
| S5_extracted_k141_42743 | Not-determined |
| S5_extracted_k141_109469 | Not-determined |
| S5_extracted_k141_121612 | Low-quality |
| S5_extracted_k141_121615 | Not-determined |
| S5_extracted_k141_60958 | Not-determined |
| S5_extracted_k141_133649 | Low-quality |
| S5_extracted_k141_139781 | Not-determined |
| S5_extracted_k141_133 | Not-determined |
| S5_extracted_k141_139787 | Not-determined |
| S5_extracted_k141_18377 | Not-determined |
| S5_extracted_k141_79002 | Not-determined |
| S5_extracted_k141_79004 | Not-determined |
| S5_extracted_k141_133653 | Not-determined |
| S5_extracted_k141_18379 | Not-determined |
| S5_extracted_k141_97303 | Not-determined |
| S5_extracted_k141_133656 | Not-determined |
| S5_extracted_k141_42778 | Not-determined |
| S5_extracted_k141_121630 | Low-quality |
| S5_extracted_k141_66898 | Low-quality |
| S5_extracted_k141_66904 | Not-determined |
| S5_extracted_k141_18392 | Not-determined |
| S5_extracted_k141_54806 | Not-determined |
| S5_extracted_k141_18394 | Not-determined |
| S5_extracted_k141_42792 | Not-determined |
| S5_extracted_k141_139804 | Not-determined |
| S5_extracted_k141_42796 | Not-determined |
| S5_extracted_k141_18410 | Not-determined |
| S5_extracted_k141_97312 | Not-determined |
| S5_extracted_k141_139813 | Low-quality |
| S5_extracted_k141_121656 | Not-determined |
| S5_extracted_k141_121657 | Not-determined |
| S5_extracted_k141_121658 | Not-determined |
| S5_extracted_k141_85182 | Not-determined |
| S5_extracted_k141_103482 | Low-quality |
| S5_extracted_k141_103484 | Not-determined |
| S5_extracted_k141_97315 | Low-quality |
| S5_extracted_k141_6305 | Not-determined |
| S5_extracted_k141_42805 | Not-determined |
| S5_extracted_k141_18424 | Not-determined |
| S5_extracted_k141_85191 | Not-determined |
| S5_extracted_k141_54812 | Low-quality |
| S5_extracted_k141_103490 | Not-determined |
| S5_extracted_k141_139829 | Low-quality |
| S5_extracted_k141_139830 | Not-determined |
| S5_extracted_k141_54818 | Low-quality |
| S5_extracted_k141_194 | Low-quality |
| S5_extracted_k141_48718 | Not-determined |
| S5_extracted_k141_42829 | Not-determined |
| S5_extracted_k141_202 | Not-determined |
| S5_extracted_k141_139850 | Low-quality |
| S5_extracted_k141_18449 | Low-quality |
| S5_extracted_k141_139853 | Low-quality |
| S5_extracted_k141_18452 | Not-determined |
| S5_extracted_k141_54837 | Not-determined |
| S5_extracted_k141_54838 | Not-determined |
| S5_extracted_k141_85207 | Not-determined |
| S5_extracted_k141_85209 | Not-determined |
| S5_extracted_k141_30492 | Not-determined |
| S5_extracted_k141_85211 | Not-determined |
| S5_extracted_k141_66939 | Not-determined |
| S5_extracted_k141_217 | Low-quality |
| S5_extracted_k141_85218 | Not-determined |
| S5_extracted_k141_60967 | Not-determined |
| S5_extracted_k141_85224 | Low-quality |
| S5_extracted_k141_60974 | Not-determined |
| S5_extracted_k141_60975 | Not-determined |
| S5_extracted_k141_139871 | Not-determined |
| S5_extracted_k141_6338 | Not-determined |
| S5_extracted_k141_115422 | Not-determined |
| S5_extracted_k141_121683 | Low-quality |
| S5_extracted_k141_85234 | Not-determined |
| S5_extracted_k141_24370 | Not-determined |
| S5_extracted_k141_240 | Not-determined |
| S5_extracted_k141_12268 | Not-determined |
| S5_extracted_k141_48735 | Not-determined |
| S5_extracted_k141_103524 | Not-determined |
| S5_extracted_k141_103525 | Not-determined |
| S5_extracted_k141_243 | Not-determined |
| S5_extracted_k141_139891 | Not-determined |
| S5_extracted_k141_139892 | Not-determined |
| S5_extracted_k141_85244 | Not-determined |
| S5_extracted_k141_48738 | Low-quality |
| S5_extracted_k141_85248 | Low-quality |
| S5_extracted_k141_60987 | Not-determined |
| S5_extracted_k141_252 | Not-determined |
| S5_extracted_k141_261 | Not-determined |
| S5_extracted_k141_72998 | Not-determined |
| S5_extracted_k141_6349 | Not-determined |
| S5_extracted_k141_121720 | Low-quality |
| S14_vs_k141_72335\|\|full | High-quality |
| S5_extracted_k141_85263 | Not-determined |
| S5_extracted_k141_48749 | Not-determined |
| S5_extracted_k141_139903 | Not-determined |
| S5_extracted_k141_127671 | Not-determined |
| S5_extracted_k141_24390 | Not-determined |
| S5_extracted_k141_42864 | Not-determined |
| S5_extracted_k141_30508 | Not-determined |
| S5_extracted_k141_127684 | Not-determined |
| S5_extracted_k141_282 | Not-determined |
| S5_extracted_k141_127686 | Not-determined |
| S5_extracted_k141_284 | Not-determined |
| S5_extracted_k141_139919 | Not-determined |
| S5_extracted_k141_127688 | Low-quality |
| S5_extracted_k141_6372 | Not-determined |
| S5_extracted_k141_121742 | Not-determined |
| S5_extracted_k141_85281 | Not-determined |
| S5_extracted_k141_139920 | Not-determined |
| S5_extracted_k141_287 | Low-quality |
| S5_extracted_k141_115452 | Not-determined |
| S5_extracted_k141_18473 | Not-determined |
| S5_extracted_k141_73010 | Low-quality |
| S5_extracted_k141_73011 | Not-determined |
| S5_extracted_k141_103542 | Low-quality |
| S5_extracted_k141_48753 | Not-determined |
| S5_extracted_k141_73014 | Not-determined |
| S5_extracted_k141_48758 | Not-determined |
| S5_extracted_k141_302 | Not-determined |
| S5_extracted_k141_48762 | Not-determined |
| S5_extracted_k141_85297 | Not-determined |
| S5_extracted_k141_6377 | Low-quality |
| S5_extracted_k141_115464 | Not-determined |
| S5_extracted_k141_18481 | Low-quality |
| S5_extracted_k141_103563 | Not-determined |
| S5_extracted_k141_30529 | Not-determined |
| S5_extracted_k141_103565 | Low-quality |
| S5_extracted_k141_73036 | Not-determined |
| S5_extracted_k141_60996 | Not-determined |
| S5_extracted_k141_115474 | Not-determined |
| S5_extracted_k141_61005 | Not-determined |
| S5_extracted_k141_73041 | Not-determined |
| S5_extracted_k141_61008 | Not-determined |
| S5_extracted_k141_61009 | Low-quality |
| S5_extracted_k141_18485 | Low-quality |
| S5_extracted_k141_115492 | Not-determined |
| S5_extracted_k141_18487 | Not-determined |
| S5_extracted_k141_24438 | Not-determined |
| S5_extracted_k141_6384 | Low-quality |
| S5_extracted_k141_42892 | Not-determined |
| S5_extracted_k141_73048 | Not-determined |
| S5_extracted_k141_61025 | Not-determined |
| S5_extracted_k141_115505 | Not-determined |
| S5_extracted_k141_139943 | Low-quality |
| S5_extracted_k141_91156 | Not-determined |
| S5_extracted_k141_42902 | Not-determined |
| S5_extracted_k141_91163 | Low-quality |
| S5_extracted_k141_121765 | Low-quality |
| S5_extracted_k141_121766 | Not-determined |
| S5_extracted_k141_121769 | Not-determined |
| S5_extracted_k141_121770 | Not-determined |
| S5_extracted_k141_103579 | Not-determined |
| S5_extracted_k141_42911 | Not-determined |
| S5_extracted_k141_42915 | Not-determined |
| S5_extracted_k141_18512 | Not-determined |
| S5_extracted_k141_61037 | Not-determined |
| S5_extracted_k141_103586 | Not-determined |
| S5_extracted_k141_133687 | Not-determined |
| S5_extracted_k141_133689 | Not-determined |
| S5_extracted_k141_36602 | Not-determined |
| S5_extracted_k141_103597 | Not-determined |
| S5_extracted_k141_115524 | Not-determined |
| S5_extracted_k141_121794 | Not-determined |
| S5_extracted_k141_18515 | Not-determined |
| S5_extracted_k141_109483 | Not-determined |
| S5_extracted_k141_61053 | Not-determined |
| S5_extracted_k141_127725 | Not-determined |
| S5_extracted_k141_115529 | Low-quality |
| S5_extracted_k141_97330 | Not-determined |
| S5_extracted_k141_61059 | Not-determined |
| S5_extracted_k141_79087 | Low-quality |
| S5_extracted_k141_97339 | Not-determined |
| S5_extracted_k141_121805 | Not-determined |
| S5_extracted_k141_6445 | Low-quality |
| S5_extracted_k141_42950 | Not-determined |
| S5_extracted_k141_121811 | Not-determined |
| S5_extracted_k141_18543 | Not-determined |
| S5_extracted_k141_109494 | Low-quality |
| S5_extracted_k141_97350 | Not-determined |
| S5_extracted_k141_121816 | Not-determined |
| S5_extracted_k141_109496 | Low-quality |
| S5_extracted_k141_18555 | Not-determined |
| S5_extracted_k141_121823 | Not-determined |
| S5_extracted_k141_6463 | Not-determined |
| S5_extracted_k141_115551 | Not-determined |
| S5_extracted_k141_6470 | Low-quality |
| S5_extracted_k141_73072 | Not-determined |
| S5_extracted_k141_73073 | Not-determined |
| S5_extracted_k141_344 | Not-determined |
| S5_extracted_k141_54862 | Not-determined |
| S5_extracted_k141_109506 | Not-determined |
| S5_extracted_k141_103640 | Not-determined |
| S5_extracted_k141_115564 | Not-determined |
| S5_extracted_k141_346 | Not-determined |
| S5_extracted_k141_103645 | Not-determined |
| S5_extracted_k141_127742 | Not-determined |
| S5_extracted_k141_103648 | Not-determined |
| S5_extracted_k141_36611 | Not-determined |
| S5_extracted_k141_91236 | Not-determined |
| S5_extracted_k141_368 | Not-determined |
| S5_extracted_k141_91257 | Low-quality |
| S5_extracted_k141_24451 | Not-determined |
| S5_extracted_k141_24453 | Not-determined |
| S5_extracted_k141_97360 | Not-determined |
| S5_extracted_k141_115584 | Not-determined |
| S5_extracted_k141_127761 | Low-quality |
| S5_extracted_k141_127762 | Low-quality |
| S5_extracted_k141_115589 | Not-determined |
| S5_extracted_k141_24460 | Not-determined |
| S5_extracted_k141_133708 | Not-determined |
| S5_extracted_k141_24462 | Not-determined |
| S5_extracted_k141_61074 | Not-determined |
| S5_extracted_k141_97370 | Not-determined |
| S5_extracted_k141_103656 | Not-determined |
| S5_extracted_k141_6480 | Low-quality |
| S5_extracted_k141_66976 | Not-determined |
| S5_extracted_k141_97376 | Not-determined |
| S5_extracted_k141_127776 | Not-determined |
| S5_extracted_k141_6487 | Not-determined |
| S5_extracted_k141_109518 | Not-determined |
| S5_extracted_k141_121865 | Not-determined |
| S5_extracted_k141_91287 | Not-determined |
| S5_extracted_k141_79117 | Not-determined |
| S5_extracted_k141_109523 | Low-quality |
| S5_extracted_k141_91291 | Not-determined |
| S5_extracted_k141_398 | Not-determined |
| S5_extracted_k141_121875 | Low-quality |
| S5_extracted_k141_30559 | Not-determined |
| S5_extracted_k141_66998 | Not-determined |
| S5_extracted_k141_66999 | Not-determined |
| S5_extracted_k141_133722 | Low-quality |
| S5_extracted_k141_48821 | Not-determined |
| S5_extracted_k141_67001 | Not-determined |
| S5_extracted_k141_48822 | Not-determined |
| S5_extracted_k141_91306 | Not-determined |
| S5_extracted_k141_103678 | Not-determined |
| S5_extracted_k141_6500 | Not-determined |
| S5_extracted_k141_411 | Not-determined |
| S5_extracted_k141_414 | Not-determined |
| S5_extracted_k141_91311 | Not-determined |
| S5_extracted_k141_127802 | Not-determined |
| S5_extracted_k141_121888 | Not-determined |
| S5_extracted_k141_419 | Not-determined |
| S5_extracted_k141_127810 | Not-determined |
| S5_extracted_k141_30589 | Not-determined |
| S5_extracted_k141_103693 | Not-determined |
| S5_extracted_k141_139959 | Not-determined |
| S5_extracted_k141_422 | Not-determined |
| S5_extracted_k141_42975 | Not-determined |
| S5_extracted_k141_424 | Not-determined |
| S5_extracted_k141_425 | Not-determined |
| S5_extracted_k141_91333 | Low-quality |
| S5_extracted_k141_91335 | Not-determined |
| S5_extracted_k141_42982 | Not-determined |
| S5_extracted_k141_434 | Low-quality |

| | |
|---|---|
| SS_extracted_k141_115607 | Not-determined |
| SS_extracted_k141_91341 | Not-determined |
| SS_extracted_k141_115610 | Not-determined |
| SS_extracted_k141_24471 | Not-determined |
| SS_extracted_k141_121896 | Low-quality |
| SS_extracted_k141_115611 | Low-quality |
| SS_extracted_k141_121900 | Not-determined |
| SS_extracted_k141_115614 | Not-determined |
| S13_vs_151755\|\|full | High-quality |
| SS_extracted_k141_115617 | Not-determined |
| SS_extracted_k141_103717 | Low-quality |
| SS_extracted_k141_115619 | Low-quality |
| SS_extracted_k141_36657 | Not-determined |
| SS_extracted_k141_447 | Not-determined |
| SS_extracted_k141_121909 | Not-determined |
| SS_extracted_k141_24489 | Not-determined |
| SS_extracted_k141_451 | Not-determined |
| SS_extracted_k141_30614 | Low-quality |
| SS_extracted_k141_139986 | Not-determined |
| SS_extracted_k141_30619 | Not-determined |
| SS_extracted_k141_6512 | Not-determined |
| SS_extracted_k141_139996 | Not-determined |
| SS_extracted_k141_73105 | Not-determined |
| SS_extracted_k141_115635 | Not-determined |
| SS_extracted_k141_103728 | Not-determined |
| SS_extracted_k141_115638 | Not-determined |
| SS_extracted_k141_121919 | Low-quality |
| SS_extracted_k141_43007 | Not-determined |
| SS_extracted_k141_73108 | Not-determined |
| SS_extracted_k141_115640 | Low-quality |
| SS_extracted_k141_30636 | Not-determined |
| SS_extracted_k141_115646 | Not-determined |
| SS_extracted_k141_467 | Not-determined |
| SS_extracted_k141_48846 | Not-determined |
| SS_extracted_k141_85340 | Not-determined |
| SS_extracted_k141_115656 | Low-quality |
| SS_extracted_k141_43022 | Not-determined |
| SS_extracted_k141_6528 | Low-quality |
| SS_extracted_k141_140008 | Low-quality |
| SS_extracted_k141_103737 | Not-determined |
| SS_extracted_k141_6531 | Not-determined |
| SS_extracted_k141_61123 | Low-quality |
| SS_extracted_k141_61124 | Low-quality |
| SS_extracted_k141_6534 | Not-determined |
| SS_extracted_k141_61127 | Not-determined |
| SS_extracted_k141_48857 | Not-determined |
| SS_extracted_k141_6536 | Not-determined |
| SS_extracted_k141_140011 | Not-determined |
| SS_extracted_k141_6537 | Not-determined |
| SS_extracted_k141_115658 | Low-quality |
| SS_extracted_k141_30654 | Not-determined |
| SS_extracted_k141_115663 | Not-determined |
| SS_extracted_k141_6551 | Low-quality |
| SS_extracted_k141_48875 | Not-determined |
| SS_extracted_k141_43036 | Low-quality |
| SS_extracted_k141_103751 | Not-determined |
| SS_extracted_k141_97396 | Not-determined |
| SS_extracted_k141_48879 | Not-determined |
| SS_extracted_k141_85342 | Low-quality |
| SS_extracted_k141_43039 | Not-determined |
| SS_extracted_k141_127819 | Not-determined |
| SS_extracted_k141_487 | Not-determined |
| SS_extracted_k141_121956 | Low-quality |
| SS_extracted_k141_48880 | Low-quality |
| SS_extracted_k141_97416 | Not-determined |
| SS_extracted_k141_127824 | Not-determined |
| SS_extracted_k141_30680 | Not-determined |
| SS_extracted_k141_6569 | Not-determined |
| SS_extracted_k141_48889 | Low-quality |
| SS_extracted_k141_85353 | Not-determined |
| SS_extracted_k141_127836 | Not-determined |
| SS_extracted_k141_85356 | Not-determined |
| SS_extracted_k141_115694 | Not-determined |
| SS_extracted_k141_48899 | Not-determined |
| SS_extracted_k141_85361 | Low-quality |
| SS_extracted_k141_54880 | Not-determined |
| SS_extracted_k141_43047 | Not-determined |
| SS_extracted_k141_133733 | Not-determined |
| SS_extracted_k141_30691 | Low-quality |
| SS_extracted_k141_43060 | Not-determined |
| SS_extracted_k141_115704 | Not-determined |
| SS_extracted_k141_133744 | Not-determined |
| SS_extracted_k141_133748 | Not-determined |
| SS_extracted_k141_30701 | Not-determined |
| SS_extracted_k141_48921 | Not-determined |
| SS_extracted_k141_133754 | Not-determined |
| SS_extracted_k141_48923 | Not-determined |
| SS_extracted_k141_115711 | Low-quality |
| SS_extracted_k141_97430 | Not-determined |
| SS_extracted_k141_528 | Not-determined |
| SS_extracted_k141_529 | Not-determined |
| SS_extracted_k141_103775 | Not-determined |
| SS_extracted_k141_12318 | Low-quality |
| SS_extracted_k141_97433 | Not-determined |
| SS_extracted_k141_97434 | Low-quality |
| SS_extracted_k141_54898 | Low-quality |
| SS_extracted_k141_48932 | Not-determined |
| SS_extracted_k141_537 | Low-quality |
| SS_extracted_k141_549 | Not-determined |
| SS_extracted_k141_48939 | Not-determined |
| SS_extracted_k141_552 | Not-determined |
| SS_extracted_k141_140019 | Not-determined |
| SS_extracted_k141_556 | Not-determined |
| SS_extracted_k141_43075 | Not-determined |
| SS_extracted_k141_43076 | Not-determined |
| SS_extracted_k141_91434 | Not-determined |
| SS_extracted_k141_48945 | Not-determined |
| SS_extracted_k141_91438 | Not-determined |
| SS_extracted_k141_121984 | Low-quality |
| SS_extracted_k141_12331 | Not-determined |
| SS_extracted_k141_36682 | Not-determined |
| SS_extracted_k141_115719 | Not-determined |
| SS_extracted_k141_140024 | Not-determined |
| SS_extracted_k141_43086 | Not-determined |
| SS_extracted_k141_54901 | Not-determined |
| SS_extracted_k141_43090 | Not-determined |
| SS_extracted_k141_140029 | Not-determined |
| SS_extracted_k141_115728 | Not-determined |
| SS_extracted_k141_30746 | Not-determined |
| SS_extracted_k141_12337 | Low-quality |
| SS_extracted_k141_127848 | Not-determined |
| SS_extracted_k141_79162 | Not-determined |
| SS_extracted_k141_127857 | Not-determined |
| SS_extracted_k141_97443 | Not-determined |
| SS_extracted_k141_36698 | Not-determined |
| SS_extracted_k141_582 | Low-quality |
| SS_extracted_k141_85381 | Not-determined |
| SS_extracted_k141_85382 | Low-quality |
| SS_extracted_k141_127865 | Low-quality |
| SS_extracted_k141_91458 | Not-determined |
| SS_extracted_k141_97456 | Low-quality |
| SS_extracted_k141_30761 | Not-determined |
| SS_extracted_k141_79167 | Not-determined |
| SS_extracted_k141_12362 | Low-quality |
| SS_extracted_k141_79172 | Not-determined |
| SS_extracted_k141_109545 | Not-determined |
| SS_extracted_k141_127881 | Low-quality |
| SS_extracted_k141_48958 | Low-quality |
| SS_extracted_k141_48959 | Not-determined |
| SS_extracted_k141_127885 | Not-determined |
| SS_extracted_k141_54924 | Not-determined |
| SS_extracted_k141_79174 | Not-determined |
| SS_extracted_k141_122026 | Not-determined |
| SS_extracted_k141_48965 | Not-determined |
| SS_extracted_k141_48967 | Not-determined |
| SS_extracted_k141_103778 | Not-determined |
| SS_extracted_k141_127890 | Low-quality |
| SS_extracted_k141_73128 | Not-determined |
| SS_extracted_k141_127891 | Not-determined |
| SS_extracted_k141_54928 | Low-quality |
| SS_extracted_k141_43124 | Not-determined |
| SS_extracted_k141_61144 | Not-determined |
| SS_extracted_k141_61145 | Not-determined |
| SS_extracted_k141_61148 | Not-determined |
| SS_extracted_k141_54933 | Not-determined |
| SS_extracted_k141_54934 | Low-quality |
| SS_extracted_k141_61150 | Not-determined |
| SS_extracted_k141_103793 | Not-determined |
| SS_extracted_k141_24517 | Not-determined |
| SS_extracted_k141_103794 | Not-determined |
| SS_extracted_k141_24519 | Not-determined |
| SS_extracted_k141_127913 | Not-determined |
| SS_extracted_k141_61159 | Not-determined |
| SS_extracted_k141_48979 | Not-determined |
| SS_extracted_k141_24524 | Not-determined |
| SS_extracted_k141_54940 | Not-determined |
| SS_extracted_k141_61161 | Not-determined |
| SS_extracted_k141_127916 | Low-quality |
| SS_extracted_k141_54942 | Not-determined |
| SS_extracted_k141_43131 | Not-determined |
| SS_extracted_k141_12377 | Not-determined |
| SS_extracted_k141_103801 | Not-determined |
| SS_extracted_k141_115752 | Low-quality |
| SS_extracted_k141_54945 | Low-quality |
| SS_extracted_k141_12382 | Low-quality |
| SS_extracted_k141_12383 | Not-determined |
| SS_extracted_k141_115757 | Not-determined |
| SS_extracted_k141_109589 | Not-determined |
| SS_extracted_k141_127922 | Not-determined |
| SS_extracted_k141_103811 | Low-quality |
| SS_extracted_k141_103813 | Low-quality |
| SS_extracted_k141_48994 | Not-determined |
| SS_extracted_k141_48998 | Not-determined |
| SS_extracted_k141_12389 | Low-quality |
| SS_extracted_k141_109591 | Low-quality |
| SS_extracted_k141_115767 | Not-determined |
| SS_extracted_k141_97484 | Low-quality |
| SS_extracted_k141_97485 | Low-quality |
| SS_extracted_k141_54950 | Not-determined |
| SS_extracted_k141_61185 | Not-determined |
| SS_extracted_k141_115775 | Not-determined |
| SS_extracted_k141_103824 | Not-determined |
| SS_extracted_k141_122077 | Low-quality |
| SS_extracted_k141_109601 | Not-determined |
| SS_extracted_k141_103829 | Not-determined |
| SS_extracted_k141_54957 | Low-quality |
| SS_extracted_k141_127947 | Not-determined |
| SS_extracted_k141_43142 | Not-determined |
| SS_extracted_k141_127948 | Not-determined |
| SS_extracted_k141_103836 | Not-determined |
| SS_extracted_k141_91492 | Not-determined |
| SS_extracted_k141_43151 | Not-determined |
| SS_extracted_k141_659 | Not-determined |
| SS_extracted_k141_97509 | Not-determined |
| SS_extracted_k141_97513 | Not-determined |
| SS_extracted_k141_91509 | Not-determined |
| SS_extracted_k141_97524 | Not-determined |
| SS_extracted_k141_54961 | Low-quality |
| SS_extracted_k141_43165 | Not-determined |
| SS_extracted_k141_43166 | Not-determined |
| SS_extracted_k141_677 | Not-determined |
| SS_extracted_k141_680 | Not-determined |
| SS_extracted_k141_91530 | Not-determined |
| SS_extracted_k141_91536 | Not-determined |
| SS_extracted_k141_30824 | Not-determined |
| SS_extracted_k141_122091 | Not-determined |
| SS_extracted_k141_700 | Low-quality |
| SS_extracted_k141_73147 | Low-quality |
| SS_extracted_k141_30830 | Not-determined |
| SS_extracted_k141_24559 | Low-quality |
| SS_extracted_k141_18597 | Not-determined |
| SS_extracted_k141_717 | Not-determined |
| SS_extracted_k141_49029 | Not-determined |
| SS_extracted_k141_97549 | Not-determined |
| SS_extracted_k141_49032 | Not-determined |
| SS_extracted_k141_18609 | Not-determined |
| SS_extracted_k141_733 | Not-determined |
| SS_extracted_k141_6598 | Low-quality |
| SS_extracted_k141_30841 | Low-quality |
| SS_extracted_k141_43186 | Not-determined |
| SS_extracted_k141_43188 | Not-determined |
| SS_extracted_k141_742 | Not-determined |
| SS_extracted_k141_122107 | Not-determined |
| SS_extracted_k141_109618 | Not-determined |
| SS_extracted_k141_18621 | Low-quality |
| SS_extracted_k141_18622 | Not-determined |
| SS_extracted_k141_61194 | Not-determined |
| SS_extracted_k141_109624 | Not-determined |
| SS_extracted_k141_749 | Not-determined |
| SS_extracted_k141_115779 | Low-quality |
| SS_extracted_k141_761 | Low-quality |
| SS_extracted_k141_30858 | Low-quality |
| SS_extracted_k141_30859 | Not-determined |
| SS_extracted_k141_115789 | Not-determined |
| SS_extracted_k141_85396 | Not-determined |
| SS_extracted_k141_115791 | Not-determined |
| SS_extracted_k141_115794 | Not-determined |
| SS_extracted_k141_43206 | Not-determined |
| SS_extracted_k141_30867 | Not-determined |
| SS_extracted_k141_85402 | Low-quality |
| SS_extracted_k141_18630 | Not-determined |
| SS_extracted_k141_768 | Low-quality |
| SS_extracted_k141_772 | Not-determined |
| SS_extracted_k141_109660 | Low-quality |
| SS_extracted_k141_30876 | Not-determined |
| SS_extracted_k141_30878 | Not-determined |
| SS_extracted_k141_109667 | Low-quality |
| SS_extracted_k141_12396 | Not-determined |
| SS_extracted_k141_122148 | Not-determined |
| SS_extracted_k141_109673 | Not-determined |
| SS_extracted_k141_54975 | Low-quality |
| SS_extracted_k141_18649 | Low-quality |
| SS_extracted_k141_12409 | Not-determined |
| SS_extracted_k141_109686 | Not-determined |
| SS_extracted_k141_122162 | Not-determined |
| SS_extracted_k141_85421 | Not-determined |
| SS_extracted_k141_18660 | Not-determined |
| SS_extracted_k141_85424 | Not-determined |
| SS_extracted_k141_115827 | Not-determined |
| SS_extracted_k141_6629 | Low-quality |
| SS_extracted_k141_6631 | Low-quality |
| SS_extracted_k141_24595 | Not-determined |
| SS_extracted_k141_122170 | Not-determined |
| SS_extracted_k141_109701 | Not-determined |
| SS_extracted_k141_122175 | Not-determined |
| SS_extracted_k141_779 | Not-determined |
| SS_extracted_k141_122177 | Not-determined |
| SS_extracted_k141_115837 | Not-determined |
| SS_extracted_k141_115842 | Not-determined |
| SS_extracted_k141_18665 | Not-determined |
| SS_extracted_k141_18667 | Not-determined |
| SS_extracted_k141_30898 | Not-determined |
| SS_extracted_k141_18673 | Not-determined |
| SS_extracted_k141_791 | Not-determined |
| SS_extracted_k141_36704 | Not-determined |
| SS_extracted_k141_36705 | Not-determined |
| SS_extracted_k141_18674 | Not-determined |
| SS_extracted_k141_43211 | Not-determined |
| SS_extracted_k141_109719 | Low-quality |
| SS_extracted_k141_61211 | Not-determined |
| SS_extracted_k141_49045 | Not-determined |
| SS_extracted_k141_127963 | Not-determined |
| SS_extracted_k141_30905 | Not-determined |
| SS_extracted_k141_115858 | Not-determined |
| SS_extracted_k141_43213 | Not-determined |
| SS_extracted_k141_61215 | Not-determined |
| SS_extracted_k141_109726 | Not-determined |
| SS_extracted_k141_49048 | Low-quality |
| SS_extracted_k141_30912 | Not-determined |
| SS_extracted_k141_800 | Low-quality |
| SS_extracted_k141_36719 | Not-determined |
| SS_extracted_k141_54990 | Not-determined |
| SS_extracted_k141_122195 | Not-determined |
| SS_extracted_k141_30915 | Low-quality |
| SS_extracted_k141_109731 | Not-determined |
| SS_extracted_k141_54996 | Not-determined |
| SS_extracted_k141_61232 | Not-determined |
| SS_extracted_k141_115869 | Not-determined |
| SS_extracted_k141_122201 | Not-determined |
| SS_extracted_k141_122202 | Not-determined |
| SS_extracted_k141_109738 | Low-quality |
| SS_extracted_k141_18679 | Not-determined |
| SS_extracted_k141_109740 | Not-determined |
| SS_extracted_k141_127972 | Not-determined |
| SS_extracted_k141_816 | Not-determined |
| SS_extracted_k141_109741 | Not-determined |
| SS_extracted_k141_109742 | Not-determined |
| SS_extracted_k141_820 | Not-determined |
| SS_extracted_k141_122212 | Not-determined |
| SS_extracted_k141_18683 | Not-determined |
| SS_extracted_k141_140041 | Not-determined |
| SS_extracted_k141_122216 | Not-determined |
| SS_extracted_k141_49059 | Low-quality |
| SS_extracted_k141_49061 | Not-determined |
| SS_extracted_k141_115881 | Not-determined |
| SS_extracted_k141_115887 | Not-determined |
| SS_extracted_k141_49063 | Not-determined |
| SS_extracted_k141_6659 | Not-determined |
| SS_extracted_k141_24623 | Not-determined |
| SS_extracted_k141_122225 | Not-determined |
| SS_extracted_k141_103844 | Not-determined |
| SS_extracted_k141_103846 | Not-determined |
| SS_extracted_k141_30920 | Not-determined |
| SS_extracted_k141_18694 | Not-determined |
| SS_extracted_k141_18695 | Not-determined |
| SS_extracted_k141_18696 | Not-determined |
| SS_extracted_k141_55016 | Not-determined |
| SS_extracted_k141_122240 | Not-determined |
| SS_extracted_k141_49072 | Not-determined |
| SS_extracted_k141_43266 | Not-determined |
| SS_extracted_k141_61259 | Low-quality |
| SS_extracted_k141_122248 | Not-determined |
| SS_extracted_k141_133815 | Low-quality |
| SS_extracted_k141_830 | Low-quality |
| SS_extracted_k141_36744 | Not-determined |
| SS_extracted_k141_30928 | Low-quality |
| SS_extracted_k141_49080 | Not-determined |
| SS_extracted_k141_36752 | Not-determined |
| SS_extracted_k141_133825 | Not-determined |
| SS_extracted_k141_6668 | Low-quality |
| SS_extracted_k141_43275 | Low-quality |
| SS_extracted_k141_133829 | Not-determined |
| SS_extracted_k141_6676 | Not-determined |
| SS_extracted_k141_73155 | Not-determined |
| SS_extracted_k141_49085 | Not-determined |
| SS_extracted_k141_30931 | Not-determined |
| SS_extracted_k141_43288 | Not-determined |
| SS_extracted_k141_109783 | Low-quality |
| SS_extracted_k141_122262 | Low-quality |
| SS_extracted_k141_49092 | Not-determined |
| SS_extracted_k141_85468 | Low-quality |
| SS_extracted_k141_122268 | Low-quality |
| SS_extracted_k141_73168 | Not-determined |
| SS_extracted_k141_73171 | Not-determined |
| SS_extracted_k141_122272 | Low-quality |
| SS_extracted_k141_30952 | Not-determined |
| SS_extracted_k141_30955 | Not-determined |
| SS_extracted_k141_36763 | Low-quality |
| SS_extracted_k141_36765 | Not-determined |
| SS_extracted_k141_12419 | Not-determined |
| SS_extracted_k141_109796 | Not-determined |
| SS_extracted_k141_127997 | Not-determined |
| SS_extracted_k141_24669 | Not-determined |
| SS_extracted_k141_109800 | Not-determined |
| SS_extracted_k141_85492 | Not-determined |
| SS_extracted_k141_73174 | Low-quality |
| SS_extracted_k141_36771 | Low-quality |
| SS_extracted_k141_122287 | Low-quality |
| SS_extracted_k141_109807 | Not-determined |
| SS_extracted_k141_140047 | Not-determined |
| SS_extracted_k141_30967 | Not-determined |
| SS_extracted_k141_49111 | Low-quality |
| SS_extracted_k141_24680 | Not-determined |
| SS_extracted_k141_24681 | Not-determined |
| SS_extracted_k141_30972 | Not-determined |
| SS_extracted_k141_122289 | Not-determined |
| SS_extracted_k141_109813 | Not-determined |
| SS_extracted_k141_109824 | Not-determined |
| SS_extracted_k141_49123 | Low-quality |
| SS_extracted_k141_122300 | Not-determined |
| SS_extracted_k141_24692 | Not-determined |
| SS_extracted_k141_122304 | Not-determined |
| SS_extracted_k141_122306 | Not-determined |
| SS_extracted_k141_73189 | Not-determined |
| SS_extracted_k141_122314 | Low-quality |
| SS_extracted_k141_122315 | Low-quality |
| SS_extracted_k141_36774 | Not-determined |
| SS_extracted_k141_73193 | Low-quality |
| SS_extracted_k141_103912 | Low-quality |
| SS_extracted_k141_67045 | Low-quality |
| SS_extracted_k141_73197 | Not-determined |
| SS_extracted_k141_67046 | Low-quality |
| SS_extracted_k141_24715 | Low-quality |
| SS_extracted_k141_85516 | Not-determined |
| SS_extracted_k141_18728 | Low-quality |
| SS_extracted_k141_67050 | Not-determined |
| SS_extracted_k141_122326 | Low-quality |
| SS_extracted_k141_115897 | Not-determined |
| SS_extracted_k141_43305 | Not-determined |
| SS_extracted_k141_43306 | Not-determined |
| SS_extracted_k141_24733 | Not-determined |
| SS_extracted_k141_855 | Not-determined |
| SS_extracted_k141_103930 | Not-determined |
| SS_extracted_k141_103931 | Not-determined |
| SS_extracted_k141_43310 | Not-determined |
| SS_extracted_k141_85520 | Not-determined |
| SS_extracted_k141_858 | Not-determined |
| SS_extracted_k141_85523 | Not-determined |
| SS_extracted_k141_91568 | Low-quality |
| SS_extracted_k141_85525 | Not-determined |
| SS_extracted_k141_133848 | Not-determined |
| SS_extracted_k141_128042 | Not-determined |
| SS_extracted_k141_31011 | Low-quality |
| SS_extracted_k141_133849 | Not-determined |
| SS_extracted_k141_85531 | Not-determined |
| SS_extracted_k141_36812 | Not-determined |
| SS_extracted_k141_859 | Low-quality |
| SS_extracted_k141_36821 | Low-quality |
| SS_extracted_k141_128050 | Low-quality |
| SS_extracted_k141_128051 | Not-determined |
| SS_extracted_k141_133859 | Not-determined |
| SS_extracted_k141_866 | Not-determined |
| SS_extracted_k141_128064 | Not-determined |
| SS_extracted_k141_128070 | Not-determined |
| SS_extracted_k141_18753 | Not-determined |
| SS_extracted_k141_36836 | Not-determined |
| SS_extracted_k141_49141 | Not-determined |
| SS_extracted_k141_49142 | Low-quality |
| SS_extracted_k141_36841 | Not-determined |
| SS_extracted_k141_49145 | Low-quality |
| SS_extracted_k141_18757 | Not-determined |
| SS_extracted_k141_18761 | Not-determined |
| SS_extracted_k141_85539 | Not-determined |
| SS_extracted_k141_115912 | Not-determined |
| SS_extracted_k141_36849 | Not-determined |
| SS_extracted_k141_6704 | Not-determined |
| SS_extracted_k141_85540 | Not-determined |
| SS_extracted_k141_115918 | Not-determined |
| SS_extracted_k141_67068 | Low-quality |
| SS_extracted_k141_67069 | Not-determined |
| SS_extracted_k141_49155 | Low-quality |
| SS_extracted_k141_128073 | Not-determined |
| SS_extracted_k141_12433 | Not-determined |
| SS_extracted_k141_128081 | Not-determined |
| SS_extracted_k141_49161 | Low-quality |
| SS_extracted_k141_49162 | Low-quality |
| SS_extracted_k141_36856 | Not-determined |
| SS_extracted_k141_905 | Not-determined |
| SS_extracted_k141_115930 | Low-quality |
| SS_extracted_k141_31040 | Not-determined |
| SS_extracted_k141_133874 | Not-determined |
| SS_extracted_k141_67082 | Not-determined |
| SS_extracted_k141_67084 | Not-determined |
| SS_extracted_k141_133877 | Not-determined |
| SS_extracted_k141_85548 | Not-determined |
| SS_extracted_k141_115936 | Not-determined |
| SS_extracted_k141_122352 | Low-quality |
| SS_extracted_k141_115945 | Not-determined |
| SS_extracted_k141_67094 | Not-determined |
| SS_extracted_k141_49186 | Low-quality |
| SS_extracted_k141_49187 | Low-quality |
| SS_extracted_k141_49188 | Not-determined |
| SS_extracted_k141_913 | Not-determined |
| SS_extracted_k141_916 | Not-determined |
| SS_extracted_k141_36876 | Not-determined |
| SS_extracted_k141_128100 | Not-determined |
| SS_extracted_k141_67103 | Not-determined |
| SS_extracted_k141_85567 | Not-determined |
| SS_extracted_k141_128106 | Not-determined |
| SS_extracted_k141_12462 | Not-determined |
| SS_extracted_k141_12464 | Not-determined |
| SS_extracted_k141_133891 | Not-determined |
| SS_extracted_k141_85568 | Low-quality |
| SS_extracted_k141_85571 | Not-determined |
| SS_extracted_k141_91577 | Not-determined |
| SS_extracted_k141_67118 | Not-determined |
| SS_extracted_k141_91580 | Not-determined |
| SS_extracted_k141_115964 | Low-quality |
| SS_extracted_k141_91589 | Not-determined |
| SS_extracted_k141_940 | Low-quality |
| SS_extracted_k141_67123 | Not-determined |
| SS_extracted_k141_122374 | Not-determined |
| SS_extracted_k141_67125 | Low-quality |
| SS_extracted_k141_133913 | Not-determined |
| SS_extracted_k141_941 | Low-quality |
| SS_extracted_k141_18801 | Not-determined |
| SS_extracted_k141_18803 | Not-determined |
| SS_extracted_k141_115972 | Not-determined |
| SS_extracted_k141_12499 | Not-determined |
| SS_extracted_k141_91601 | Not-determined |
| SS_extracted_k141_12507 | Not-determined |
| SS_extracted_k141_115980 | Low-quality |
| SS_extracted_k141_85573 | Not-determined |
| SS_extracted_k141_115985 | Low-quality |
| SS_extracted_k141_133923 | Low-quality |
| SS_extracted_k141_6718 | Low-quality |
| SS_extracted_k141_115989 | Low-quality |
| SS_extracted_k141_12515 | Not-determined |
| SS_extracted_k141_31062 | Not-determined |
| SS_extracted_k141_67145 | Not-determined |
| SS_extracted_k141_115996 | Not-determined |
| SS_extracted_k141_12525 | Not-determined |
| SS_extracted_k141_945 | Low-quality |
| SS_extracted_k141_67153 | Not-determined |
| SS_extracted_k141_91615 | Not-determined |
| SS_extracted_k141_49232 | Low-quality |
| SS_extracted_k141_12530 | Low-quality |
| SS_extracted_k141_91616 | Not-determined |
| SS_extracted_k141_91617 | Not-determined |
| SS_extracted_k141_67160 | Not-determined |
| SS_extracted_k141_116006 | Not-determined |
| SS_extracted_k141_49235 | Not-determined |
| SS_extracted_k141_31079 | Not-determined |
| SS_extracted_k141_122388 | Not-determined |
| SS_extracted_k141_43361 | Not-determined |
| SS_extracted_k141_49241 | Low-quality |
| SS_extracted_k141_43364 | Not-determined |
| SS_extracted_k141_128129 | Not-determined |
| SS_extracted_k141_122395 | Not-determined |
| SS_extracted_k141_43368 | Not-determined |
| SS_extracted_k141_12540 | Not-determined |
| SS_extracted_k141_128132 | Not-determined |
| SS_extracted_k141_128133 | Not-determined |
| SS_extracted_k141_955 | Not-determined |
| SS_extracted_k141_49248 | Low-quality |
| SS_extracted_k141_91631 | Not-determined |
| SS_extracted_k141_85605 | Not-determined |
| SS_extracted_k141_128140 | Not-determined |
| SS_extracted_k141_122409 | Not-determined |
| SS_extracted_k141_67182 | Not-determined |
| SS_extracted_k141_55066 | Not-determined |
| SS_extracted_k141_12550 | Not-determined |
| SS_extracted_k141_43385 | Low-quality |
| SS_extracted_k141_91651 | Not-determined |
| SS_extracted_k141_6730 | Not-determined |
| SS_extracted_k141_67186 | Not-determined |
| SS_extracted_k141_36902 | Not-determined |
| SS_extracted_k141_133929 | Not-determined |
| SS_extracted_k141_31114 | Low-quality |
| SS_extracted_k141_133936 | Not-determined |
| SS_extracted_k141_133943 | Low-quality |
| SS_extracted_k141_91659 | Not-determined |
| SS_extracted_k141_122418 | Not-determined |
| SS_extracted_k141_122420 | Low-quality |
| SS_extracted_k141_85622 | Not-determined |
| SS_extracted_k141_140059 | Low-quality |
| SS_extracted_k141_36920 | Not-determined |
| SS_extracted_k141_67206 | Not-determined |
| SS_extracted_k141_122425 | Not-determined |
| SS_extracted_k141_122426 | Not-determined |
| SS_extracted_k141_967 | Low-quality |
| SS_extracted_k141_18819 | Not-determined |
| SS_extracted_k141_31123 | Not-determined |
| SS_extracted_k141_122432 | Not-determined |
| SS_extracted_k141_18825 | Not-determined |
| SS_extracted_k141_973 | Low-quality |
| SS_extracted_k141_31127 | Not-determined |
| SS_extracted_k141_91679 | Not-determined |
| SS_extracted_k141_43411 | Not-determined |
| SS_extracted_k141_91681 | Not-determined |
| SS_extracted_k141_67215 | Not-determined |
| SS_extracted_k141_12557 | Not-determined |
| SS_extracted_k141_67217 | Not-determined |
| SS_extracted_k141_6752 | Low-quality |
| SS_extracted_k141_122438 | Not-determined |
| SS_extracted_k141_18830 | Not-determined |
| SS_extracted_k141_49252 | Not-determined |
| SS_extracted_k141_116048 | Not-determined |
| SS_extracted_k141_36937 | Not-determined |
| SS_extracted_k141_122441 | Not-determined |
| SS_extracted_k141_49255 | Not-determined |
| SS_extracted_k141_984 | Low-quality |
| SS_extracted_k141_67223 | Not-determined |
| SS_extracted_k141_49256 | Low-quality |
| SS_extracted_k141_36940 | Not-determined |
| SS_extracted_k141_67225 | Not-determined |
| SS_extracted_k141_31138 | Not-determined |
| SS_extracted_k141_31139 | Not-determined |
| SS_extracted_k141_128166 | Low-quality |

| | |
|---|---|
| SS_extracted_k141_991 | Not-determined |
| SS_extracted_k141_36956 | Not-determined |
| SS_extracted_k141_18837 | Low-quality |
| SS_extracted_k141_67236 | Not-determined |
| SS_extracted_k141_67238 | Low-quality |
| SS_extracted_k141_43422 | Not-determined |
| SS_extracted_k141_128172 | Not-determined |
| SS_extracted_k141_116060 | Not-determined |
| SS_extracted_k141_18841 | Not-determined |
| SS_extracted_k141_140082 | Low-quality |
| SS_extracted_k141_122458 | Not-determined |
| SS_extracted_k141_67242 | Not-determined |
| SS_extracted_k141_122460 | Not-determined |
| SS_extracted_k141_91704 | Not-determined |
| SS_extracted_k141_91705 | Not-determined |
| SS_extracted_k141_18846 | Not-determined |
| SS_extracted_k141_49275 | Not-determined |
| SS_extracted_k141_116064 | Not-determined |
| SS_extracted_k141_31154 | Not-determined |
| SS_extracted_k141_91710 | Not-determined |
| SS_extracted_k141_91711 | Not-determined |
| SS_extracted_k141_18854 | Low-quality |
| SS_extracted_k141_91717 | Not-determined |
| SS_extracted_k141_91719 | Low-quality |
| SS_extracted_k141_122476 | Not-determined |
| SS_extracted_k141_49286 | Not-determined |
| SS_extracted_k141_85650 | Not-determined |
| SS_extracted_k141_36969 | Not-determined |
| SS_extracted_k141_1012 | Low-quality |
| SS_extracted_k141_122485 | Not-determined |
| SS_extracted_k141_6769 | Not-determined |
| SS_extracted_k141_1015 | Not-determined |
| SS_extracted_k141_49307 | Not-determined |
| SS_extracted_k141_140107 | Not-determined |
| SS_extracted_k141_6777 | Low-quality |
| SS_extracted_k141_49310 | Not-determined |
| SS_extracted_k141_31178 | Low-quality |
| SS_extracted_k141_36979 | Low-quality |
| SS_extracted_k141_122492 | Not-determined |
| SS_extracted_k141_43437 | Not-determined |
| SS_extracted_k141_55101 | Not-determined |
| SS_extracted_k141_55103 | Not-determined |
| SS_extracted_k141_140115 | Not-determined |
| SS_extracted_k141_116095 | Not-determined |
| SS_extracted_k141_36989 | Not-determined |
| SS_extracted_k141_36990 | Not-determined |
| SS_extracted_k141_55118 | Not-determined |
| SS_extracted_k141_122499 | Not-determined |
| SS_extracted_k141_122501 | Not-determined |
| SS_extracted_k141_31181 | Low-quality |
| SS_extracted_k141_1035 | Not-determined |
| SS_extracted_k141_67253 | Not-determined |
| SS_extracted_k141_140119 | Low-quality |
| SS_extracted_k141_128203 | Not-determined |
| SS_extracted_k141_31187 | Not-determined |
| SS_extracted_k141_36998 | Not-determined |
| SS_extracted_k141_140122 | Not-determined |
| SS_extracted_k141_122513 | Not-determined |
| SS_extracted_k141_122514 | Not-determined |
| SS_extracted_k141_55133 | Not-determined |
| SS_extracted_k141_6794 | Not-determined |
| SS_extracted_k141_6795 | Not-determined |
| SS_extracted_k141_67265 | Not-determined |
| SS_extracted_k141_6798 | Low-quality |
| SS_extracted_k141_43460 | Low-quality |
| SS_extracted_k141_31202 | Not-determined |
| SS_extracted_k141_91727 | Low-quality |
| SS_extracted_k141_67270 | Not-determined |
| SS_extracted_k141_116115 | Not-determined |
| SS_extracted_k141_6803 | Not-determined |
| SS_extracted_k141_1059 | Low-quality |
| SS_extracted_k141_43471 | Not-determined |
| SS_extracted_k141_91741 | Low-quality |
| SS_extracted_k141_43481 | Not-determined |
| SS_extracted_k141_140130 | Low-quality |
| SS_extracted_k141_43491 | Not-determined |
| SS_extracted_k141_91752 | Not-determined |
| SS_extracted_k141_1084 | Not-determined |
| SS_extracted_k141_6815 | Not-determined |
| SS_extracted_k141_122536 | Not-determined |
| SS_extracted_k141_49322 | Not-determined |
| SS_extracted_k141_49323 | Not-determined |
| SS_extracted_k141_37020 | Low-quality |
| SS_extracted_k141_43497 | Low-quality |
| SS_extracted_k141_122541 | Not-determined |
| SS_extracted_k141_37023 | Not-determined |
| SS_extracted_k141_1089 | Not-determined |
| SS_extracted_k141_55147 | Not-determined |
| SS_extracted_k141_91775 | Not-determined |
| SS_extracted_k141_43505 | Not-determined |
| SS_extracted_k141_6836 | Not-determined |
| SS_extracted_k141_122558 | Low-quality |
| SS_extracted_k141_128217 | Low-quality |
| SS_extracted_k141_134005 | Not-determined |
| SS_extracted_k141_43519 | Not-determined |
| SS_extracted_k141_37029 | Not-determined |
| SS_extracted_k141_18871 | Not-determined |
| SS_extracted_k141_1109 | Not-determined |
| SS_extracted_k141_91796 | Not-determined |
| SS_extracted_k141_122566 | Not-determined |
| SS_extracted_k141_18874 | Low-quality |
| SS_extracted_k141_116123 | Low-quality |
| SS_extracted_k141_1116 | Not-determined |
| SS_extracted_k141_79194 | Low-quality |
| SS_extracted_k141_37039 | Not-determined |
| SS_extracted_k141_37040 | Not-determined |
| SS_extracted_k141_91809 | Not-determined |
| SS_extracted_k141_67281 | Not-determined |
| SS_extracted_k141_79203 | Not-determined |
| SS_extracted_k141_18884 | Not-determined |
| SS_extracted_k141_37051 | Not-determined |
| SS_extracted_k141_18885 | Not-determined |
| SS_extracted_k141_12571 | Not-determined |
| SS_extracted_k141_1137 | Not-determined |
| SS_extracted_k141_1139 | Not-determined |
| SS_extracted_k141_37058 | Not-determined |
| SS_extracted_k141_128233 | Not-determined |
| SS_extracted_k141_91816 | Low-quality |
| SS_extracted_k141_128240 | Not-determined |
| SS_extracted_k141_91817 | Not-determined |
| SS_extracted_k141_91819 | Not-determined |
| SS_extracted_k141_49360 | Not-determined |
| SS_extracted_k141_43540 | Not-determined |
| SS_extracted_k141_49361 | Not-determined |
| SS_extracted_k141_91825 | Low-quality |
| SS_extracted_k141_6874 | Not-determined |
| SS_extracted_k141_79216 | Not-determined |
| SS_extracted_k141_18901 | Not-determined |
| SS_extracted_k141_49367 | Not-determined |
| SS_extracted_k141_49370 | Not-determined |
| SS_extracted_k141_79220 | Low-quality |
| SS_extracted_k141_6878 | Not-determined |
| SS_extracted_k141_67304 | Not-determined |
| SS_extracted_k141_79223 | Low-quality |
| SS_extracted_k141_67306 | Not-determined |
| SS_extracted_k141_67309 | Low-quality |
| SS_extracted_k141_49375 | Not-determined |
| SS_extracted_k141_43547 | Not-determined |
| SS_extracted_k141_103936 | Not-determined |
| SS_extracted_k141_49380 | Low-quality |
| SS_extracted_k141_103939 | Not-determined |
| SS_extracted_k141_18904 | Not-determined |
| SS_extracted_k141_18906 | Not-determined |
| SS_extracted_k141_140153 | Low-quality |
| SS_extracted_k141_6883 | Not-determined |
| SS_extracted_k141_128246 | Low-quality |
| SS_extracted_k141_49387 | Not-determined |
| SS_extracted_k141_1166 | Not-determined |
| SS_extracted_k141_55167 | Low-quality |
| SS_extracted_k141_49391 | Low-quality |
| SS_extracted_k141_1173 | Not-determined |
| SS_extracted_k141_55175 | Not-determined |
| SS_extracted_k141_1175 | Not-determined |
| SS_extracted_k141_55178 | Not-determined |
| SS_extracted_k141_1178 | Not-determined |
| SS_extracted_k141_140166 | Low-quality |
| SS_extracted_k141_49397 | Low-quality |
| SS_extracted_k141_49399 | Low-quality |
| SS_extracted_k141_79234 | Not-determined |
| SS_extracted_k141_97565 | Not-determined |
| SS_extracted_k141_18933 | Not-determined |
| SS_extracted_k141_79237 | Not-determined |
| SS_extracted_k141_103956 | Not-determined |
| SB_vs_k141_182875\|\|full | High-quality |
| SS_extracted_k141_61277 | Not-determined |
| SS_extracted_k141_61280 | Not-determined |
| SS_extracted_k141_140185 | Not-determined |
| SS_extracted_k141_128270 | Not-determined |
| SS_extracted_k141_49415 | Low-quality |
| SS_extracted_k141_140186 | Not-determined |
| SS_extracted_k141_140187 | Not-determined |
| SS_extracted_k141_49418 | Not-determined |
| SS_extracted_k141_67357 | Low-quality |
| SS_extracted_k141_49421 | Not-determined |
| SS_extracted_k141_1201 | Not-determined |
| SS_extracted_k141_1202 | Not-determined |
| SS_extracted_k141_49427 | Not-determined |
| SS_extracted_k141_140196 | Low-quality |
| SS_extracted_k141_1208 | Not-determined |
| SS_extracted_k141_1212 | Not-determined |
| SS_extracted_k141_6911 | Not-determined |
| SS_extracted_k141_6913 | Low-quality |
| SS_extracted_k141_140205 | Not-determined |
| SS_extracted_k141_103980 | Not-determined |
| SS_extracted_k141_103981 | Not-determined |
| SS_extracted_k141_31248 | Not-determined |
| SS_extracted_k141_1216 | Not-determined |
| SS_extracted_k141_31261 | Not-determined |
| SS_extracted_k141_6932 | Not-determined |
| SS_extracted_k141_79251 | Not-determined |
| SS_extracted_k141_49438 | Not-determined |
| SS_extracted_k141_79255 | Not-determined |
| SS_extracted_k141_122596 | Not-determined |
| SS_extracted_k141_1235 | Not-determined |
| SS_extracted_k141_122597 | Not-determined |
| SS_extracted_k141_79259 | Not-determined |
| SS_extracted_k141_79263 | Low-quality |
| SS_extracted_k141_116196 | Not-determined |
| SS_extracted_k141_31286 | Not-determined |
| SS_extracted_k141_1248 | Not-determined |
| SS_extracted_k141_79273 | Not-determined |
| SS_extracted_k141_116208 | Not-determined |
| SS_extracted_k141_104004 | Low-quality |
| SS_extracted_k141_1263 | Not-determined |
| SS_extracted_k141_43610 | Low-quality |
| SS_extracted_k141_116217 | Not-determined |
| SS_extracted_k141_1266 | Not-determined |
| SS_extracted_k141_1274 | Not-determined |
| SS_extracted_k141_116226 | Low-quality |
| SS_extracted_k141_43630 | Not-determined |
| SB_vs_k141_28956\|\|full | Medium-quality |
| SS_extracted_k141_79277 | Not-determined |
| SS_extracted_k141_79278 | Not-determined |
| SS_extracted_k141_43637 | Not-determined |
| SS_extracted_k141_73208 | Not-determined |
| SS_extracted_k141_79285 | Not-determined |
| SS_extracted_k141_134051 | Not-determined |
| SS_extracted_k141_134052 | Low-quality |
| SS_extracted_k141_1287 | Not-determined |
| SS_extracted_k141_134057 | Low-quality |
| SS_extracted_k141_18944 | Not-determined |
| SS_extracted_k141_18946 | Not-determined |
| SS_extracted_k141_134059 | Low-quality |
| SS_extracted_k141_1294 | Low-quality |
| SS_extracted_k141_1296 | Not-determined |
| SS_extracted_k141_1302 | Not-determined |
| SS_extracted_k141_134068 | Not-determined |
| SS_extracted_k141_6954 | Not-determined |
| SS_extracted_k141_31317 | Not-determined |
| SS_extracted_k141_31324 | Not-determined |
| SS_extracted_k141_104025 | Not-determined |
| SS_extracted_k141_104029 | Low-quality |
| SS_extracted_k141_31333 | Low-quality |
| SS_extracted_k141_67360 | Not-determined |
| SS_extracted_k141_12593 | Not-determined |
| SS_extracted_k141_18960 | Not-determined |
| SS_extracted_k141_122622 | Low-quality |
| SS_extracted_k141_122624 | Not-determined |
| SS_extracted_k141_73234 | Not-determined |
| SS_extracted_k141_104039 | Not-determined |
| SS_extracted_k141_67368 | Low-quality |
| SS_extracted_k141_43661 | Not-determined |
| SS_extracted_k141_134076 | Not-determined |
| SS_extracted_k141_67375 | Low-quality |
| SS_extracted_k141_18979 | Not-determined |
| SS_extracted_k141_67378 | Not-determined |
| SS_extracted_k141_104053 | Not-determined |
| SS_extracted_k141_18980 | Not-determined |
| SS_extracted_k141_67379 | Not-determined |
| SS_extracted_k141_67392 | Low-quality |
| SS_extracted_k141_12608 | Not-determined |
| SS_extracted_k141_91837 | Not-determined |
| SS_extracted_k141_122626 | Not-determined |
| SS_extracted_k141_122628 | Not-determined |
| SS_extracted_k141_37079 | Not-determined |
| SS_extracted_k141_116236 | Not-determined |
| SS_extracted_k141_1327 | Low-quality |
| SS_extracted_k141_67406 | Not-determined |
| SS_extracted_k141_79331 | Not-determined |
| SS_extracted_k141_73260 | Not-determined |
| SS_extracted_k141_12619 | Not-determined |
| SS_extracted_k141_67421 | Not-determined |
| SS_extracted_k141_134080 | Not-determined |
| SS_extracted_k141_67424 | Not-determined |
| SS_extracted_k141_31364 | Not-determined |
| SS_extracted_k141_134087 | Not-determined |
| SS_extracted_k141_19002 | Not-determined |
| SS_extracted_k141_67435 | Not-determined |
| SS_extracted_k141_24790 | Not-determined |
| SS_extracted_k141_12641 | Not-determined |
| SS_extracted_k141_6992 | Low-quality |
| SS_extracted_k141_6995 | Not-determined |
| SS_extracted_k141_73265 | Not-determined |
| SS_extracted_k141_31377 | Not-determined |
| SS_extracted_k141_12646 | Low-quality |
| SS_extracted_k141_79345 | Not-determined |
| SS_extracted_k141_79347 | Low-quality |
| SS_extracted_k141_7005 | Not-determined |
| SS_extracted_k141_12653 | Low-quality |
| SS_extracted_k141_104075 | Not-determined |
| SS_extracted_k141_7012 | Not-determined |
| SS_extracted_k141_79354 | Not-determined |
| SS_extracted_k141_19016 | Not-determined |
| SS_extracted_k141_31393 | Low-quality |
| SS_extracted_k141_31395 | Not-determined |
| SS_extracted_k141_140226 | Not-determined |
| SS_extracted_k141_122669 | Not-determined |
| SS_extracted_k141_140233 | Not-determined |
| SS_extracted_k141_104090 | Low-quality |
| SS_extracted_k141_19031 | Low-quality |
| SS_extracted_k141_91852 | Not-determined |
| SS_extracted_k141_91856 | Not-determined |
| SS_extracted_k141_104100 | Low-quality |
| SS_extracted_k141_19036 | Not-determined |
| SS_extracted_k141_104103 | Not-determined |
| SS_extracted_k141_122691 | Not-determined |
| SS_extracted_k141_19040 | Not-determined |
| SS_extracted_k141_140243 | Not-determined |
| SS_extracted_k141_140247 | Not-determined |
| SS_extracted_k141_85694 | Not-determined |
| SS_extracted_k141_79365 | Not-determined |
| SS_extracted_k141_19053 | Low-quality |
| SS_extracted_k141_19056 | Not-determined |
| SS_extracted_k141_122705 | Low-quality |
| SS_extracted_k141_37087 | Not-determined |
| SS_extracted_k141_31415 | Not-determined |
| SS_extracted_k141_116253 | Not-determined |
| SS_extracted_k141_140259 | Not-determined |
| SS_extracted_k141_37092 | Low-quality |
| SS_extracted_k141_140261 | Not-determined |
| SS_extracted_k141_85702 | Not-determined |
| SS_extracted_k141_97578 | Not-determined |
| SS_extracted_k141_97579 | Not-determined |
| SS_extracted_k141_19063 | Not-determined |
| SS_extracted_k141_37098 | Not-determined |
| SS_extracted_k141_91874 | Not-determined |
| SS_extracted_k141_134129 | Not-determined |
| SS_extracted_k141_116262 | Not-determined |
| SS_extracted_k141_37101 | Not-determined |
| SS_extracted_k141_140268 | Low-quality |
| SS_extracted_k141_116263 | Low-quality |
| SS_extracted_k141_116266 | Not-determined |
| SS_extracted_k141_140271 | Not-determined |
| SS_extracted_k141_109832 | Not-determined |
| SS_extracted_k141_140272 | Not-determined |
| SS_extracted_k141_67469 | Low-quality |
| SS_extracted_k141_109834 | Low-quality |
| SS_extracted_k141_19080 | Not-determined |
| SS_extracted_k141_73278 | Not-determined |
| SS_extracted_k141_85719 | Low-quality |
| SS_extracted_k141_37117 | Not-determined |
| SS_extracted_k141_43694 | Not-determined |
| SS_extracted_k141_12690 | Not-determined |
| SS_extracted_k141_73281 | Low-quality |
| SS_extracted_k141_122748 | Not-determined |
| SS_extracted_k141_140274 | Not-determined |
| SS_extracted_k141_104117 | Not-determined |
| SS_extracted_k141_31433 | Not-determined |
| SS_extracted_k141_37131 | Low-quality |
| SS_extracted_k141_134130 | Not-determined |
| SS_extracted_k141_134131 | Low-quality |
| SS_extracted_k141_134133 | Not-determined |
| SS_extracted_k141_122762 | Not-determined |
| SS_extracted_k141_19107 | Not-determined |
| SS_extracted_k141_37133 | Not-determined |
| SS_extracted_k141_134138 | Not-determined |
| SS_extracted_k141_104124 | Not-determined |
| SS_extracted_k141_7051 | Not-determined |
| SS_extracted_k141_134142 | Not-determined |
| SS_extracted_k141_37139 | Not-determined |
| SS_extracted_k141_79371 | Not-determined |
| SS_extracted_k141_97604 | Not-determined |
| SS_extracted_k141_122771 | Not-determined |
| SS_extracted_k141_134148 | Not-determined |
| SS_extracted_k141_134150 | Not-determined |
| SS_extracted_k141_97612 | Not-determined |
| SS_extracted_k141_104128 | Not-determined |
| SS_extracted_k141_19122 | Not-determined |
| SS_extracted_k141_134155 | Not-determined |
| SS_extracted_k141_104132 | Not-determined |
| SS_extracted_k141_19125 | Not-determined |
| SS_extracted_k141_104135 | Not-determined |
| SS_extracted_k141_97616 | Not-determined |
| SS_extracted_k141_104138 | Not-determined |
| SS_extracted_k141_104141 | Low-quality |
| SS_extracted_k141_109849 | Low-quality |
| SS_extracted_k141_31460 | Low-quality |
| SS_extracted_k141_21869 | Complete |
| SS_extracted_k141_12720 | Not-determined |
| SS_extracted_k141_104144 | Low-quality |
| SS_extracted_k141_19139 | Not-determined |
| SS_extracted_k141_12729 | Not-determined |
| SS_extracted_k141_85739 | Not-determined |
| SS_extracted_k141_85745 | Not-determined |
| SS_extracted_k141_37163 | Low-quality |
| SS_extracted_k141_116314 | Not-determined |
| SS_extracted_k141_73296 | Not-determined |
| SS_extracted_k141_116320 | Not-determined |
| SS_extracted_k141_85751 | Low-quality |
| SS_extracted_k141_7084 | Not-determined |
| SS_extracted_k141_55209 | Not-determined |
| SS_extracted_k141_116322 | Not-determined |
| SS_extracted_k141_116323 | Not-determined |
| SS_extracted_k141_104151 | Not-determined |
| SS_extracted_k141_122802 | Low-quality |
| SS_extracted_k141_55215 | Not-determined |
| SS_extracted_k141_12747 | Not-determined |
| SS_extracted_k141_55229 | Not-determined |
| SS_extracted_k141_104167 | Not-determined |
| SS_extracted_k141_85761 | Not-determined |
| SS_extracted_k141_73299 | Not-determined |
| SS_extracted_k141_85767 | Not-determined |
| SS_extracted_k141_109882 | Not-determined |
| SS_extracted_k141_37180 | Not-determined |
| SS_extracted_k141_55246 | Low-quality |
| SS_extracted_k141_104173 | Not-determined |
| SS_extracted_k141_104174 | Not-determined |
| SS_extracted_k141_37184 | Not-determined |
| SS_extracted_k141_104177 | Not-determined |
| SS_extracted_k141_55249 | Not-determined |
| SS_extracted_k141_12786 | Not-determined |
| SS_extracted_k141_12788 | Low-quality |
| SS_extracted_k141_73307 | Low-quality |
| SS_extracted_k141_122831 | Low-quality |
| SS_extracted_k141_7103 | Not-determined |
| SS_extracted_k141_109901 | Not-determined |
| SS_extracted_k141_7112 | Not-determined |
| SS_extracted_k141_109909 | Not-determined |
| SS_extracted_k141_61288 | Not-determined |
| SS_extracted_k141_85802 | Not-determined |
| SS_extracted_k141_85808 | Low-quality |
| SS_extracted_k141_134195 | Not-determined |
| SS_extracted_k141_122856 | Not-determined |
| SS_extracted_k141_7117 | Not-determined |
| SS_extracted_k141_122858 | Not-determined |
| SS_extracted_k141_7120 | Low-quality |
| SS_extracted_k141_19161 | Low-quality |
| SS_extracted_k141_97626 | Not-determined |
| SS_extracted_k141_37202 | Low-quality |
| SS_extracted_k141_73318 | Not-determined |
| SS_extracted_k141_140290 | Not-determined |
| SS_extracted_k141_116406 | Not-determined |
| SS_extracted_k141_73322 | Not-determined |
| SS_extracted_k141_67510 | Not-determined |
| SS_extracted_k141_97652 | Low-quality |
| SS_extracted_k141_67516 | Not-determined |
| SS_extracted_k141_67519 | Not-determined |
| SS_extracted_k141_109918 | Low-quality |
| SS_extracted_k141_67521 | Not-determined |
| SS_extracted_k141_85832 | Not-determined |
| SS_extracted_k141_109922 | Not-determined |
| SS_extracted_k141_122878 | Low-quality |
| SS_extracted_k141_61311 | Not-determined |
| SS_extracted_k141_67534 | Not-determined |
| SS_extracted_k141_85848 | Low-quality |
| SS_extracted_k141_140306 | Low-quality |
| SS_extracted_k141_116425 | Not-determined |
| SS_extracted_k141_97662 | Not-determined |
| SS_extracted_k141_7140 | Not-determined |
| SS_extracted_k141_67544 | Not-determined |
| SS_extracted_k141_1397 | Not-determined |
| SS_extracted_k141_122882 | Not-determined |
| SS_extracted_k141_122884 | Not-determined |
| SS_extracted_k141_109950 | Not-determined |
| SS_extracted_k141_85872 | Not-determined |
| SS_extracted_k141_134209 | Not-determined |
| SS_extracted_k141_109952 | Not-determined |
| SS_extracted_k141_116427 | Low-quality |
| SS_extracted_k141_122887 | Not-determined |
| SS_extracted_k141_12803 | Low-quality |
| SS_extracted_k141_109960 | Not-determined |
| SS_extracted_k141_7161 | Not-determined |
| SS_extracted_k141_61333 | Not-determined |
| SS_extracted_k141_67569 | Not-determined |
| SS_extracted_k141_116457 | Low-quality |
| SS_extracted_k141_7166 | Not-determined |
| SS_extracted_k141_7171 | Not-determined |
| SS_extracted_k141_109970 | Not-determined |
| SS_extracted_k141_12828 | Not-determined |
| SS_extracted_k141_109973 | Not-determined |
| SS_extracted_k141_109974 | Not-determined |
| SS_extracted_k141_49465 | Low-quality |
| SS_extracted_k141_109978 | Not-determined |
| SS_extracted_k141_97681 | Low-quality |
| SS_extracted_k141_109982 | Not-determined |
| SS_extracted_k141_67576 | Low-quality |
| SS_extracted_k141_61340 | Low-quality |
| SS_extracted_k141_12839 | Not-determined |
| SS_extracted_k141_12843 | Not-determined |
| SS_extracted_k141_67581 | Not-determined |
| SS_extracted_k141_67582 | Not-determined |
| SS_extracted_k141_7185 | Not-determined |
| SS_extracted_k141_122911 | Low-quality |
| SS_extracted_k141_67595 | Not-determined |
| SS_extracted_k141_12854 | Not-determined |
| SS_extracted_k141_67604 | Not-determined |
| SS_extracted_k141_122917 | Low-quality |
| SS_extracted_k141_67608 | Not-determined |
| SS_extracted_k141_67609 | Not-determined |
| SS_extracted_k141_67614 | Low-quality |
| SS_extracted_k141_110013 | Not-determined |
| SS_extracted_k141_122921 | Not-determined |
| SS_extracted_k141_122922 | Not-determined |
| SS_extracted_k141_116475 | Not-determined |
| SS_extracted_k141_110021 | Not-determined |
| SS_extracted_k141_67619 | Not-determined |
| SS_extracted_k141_67620 | Not-determined |
| SS_extracted_k141_73344 | Low-quality |
| SS_extracted_k141_116482 | Not-determined |
| SS_extracted_k141_73349 | Not-determined |
| SS_extracted_k141_73352 | Not-determined |
| SS_extracted_k141_140354 | Not-determined |
| SS_extracted_k141_73355 | Not-determined |
| SS_extracted_k141_19189 | Not-determined |
| SS_extracted_k141_116487 | Low-quality |
| SS_extracted_k141_134232 | Not-determined |
| SS_extracted_k141_49484 | Not-determined |
| SS_extracted_k141_7198 | Low-quality |
| SS_extracted_k141_7199 | Low-quality |
| SS_extracted_k141_134241 | Not-determined |
| SS_extracted_k141_12859 | Not-determined |
| SS_extracted_k141_110051 | Not-determined |
| SS_extracted_k141_7207 | Not-determined |
| SS_extracted_k141_116502 | Not-determined |
| SS_extracted_k141_97727 | Not-determined |
| SS_extracted_k141_19220 | Not-determined |
| SS_extracted_k141_116505 | Not-determined |
| SS_extracted_k141_12869 | Low-quality |
| SS_extracted_k141_49495 | Not-determined |
| SS_extracted_k141_85894 | Not-determined |
| SS_extracted_k141_37228 | Not-determined |
| SS_extracted_k141_73358 | Not-determined |
| SS_extracted_k141_37229 | Not-determined |
| SS_extracted_k141_97732 | Not-determined |
| SS_extracted_k141_12872 | Not-determined |
| SS_extracted_k141_85899 | Not-determined |
| SS_extracted_k141_73375 | Low-quality |
| SS_extracted_k141_43749 | Not-determined |
| SS_extracted_k141_19225 | Low-quality |
| SS_extracted_k141_85914 | Low-quality |
| SS_extracted_k141_73385 | Not-determined |
| SS_extracted_k141_49513 | Not-determined |
| SS_extracted_k141_85918 | Not-determined |
| SS_extracted_k141_85922 | Not-determined |
| SS_extracted_k141_7255 | Not-determined |
| SS_extracted_k141_85923 | Low-quality |
| SS_extracted_k141_37247 | Not-determined |
| SS_extracted_k141_85925 | Not-determined |
| SS_extracted_k141_122928 | Low-quality |
| SS_extracted_k141_37256 | Not-determined |
| SS_extracted_k141_31559 | Not-determined |
| SS_extracted_k141_122932 | Not-determined |
| SS_extracted_k141_140372 | Not-determined |
| SS_extracted_k141_110073 | Not-determined |
| SS_extracted_k141_116522 | Not-determined |
| SS_extracted_k141_91900 | Not-determined |
| SS_extracted_k141_31563 | Low-quality |
| SS_extracted_k141_31568 | Not-determined |
| SS_extracted_k141_104233 | Not-determined |
| SS_extracted_k141_122939 | Low-quality |
| SS_extracted_k141_116530 | Not-determined |
| SS_extracted_k141_85936 | Not-determined |
| SS_extracted_k141_104239 | Not-determined |
| SS_extracted_k141_140377 | Low-quality |
| SS_extracted_k141_37273 | Not-determined |
| SS_extracted_k141_61372 | Low-quality |
| SS_extracted_k141_61380 | Not-determined |
| SS_extracted_k141_31586 | Not-determined |
| SS_extracted_k141_43780 | Not-determined |
| SS_extracted_k141_140391 | Not-determined |
| SS_extracted_k141_12902 | Low-quality |
| SS_extracted_k141_43785 | Not-determined |
| SS_extracted_k141_85955 | Not-determined |
| SS_extracted_k141_31591 | Not-determined |
| SS_extracted_k141_140406 | Not-determined |
| SS_extracted_k141_140409 | Not-determined |
| SS_extracted_k141_37304 | Not-determined |
| SS_extracted_k141_140411 | Not-determined |
| SS_extracted_k141_97763 | Not-determined |
| SS_extracted_k141_7302 | Not-determined |
| SS_extracted_k141_61411 | Not-determined |
| SS_extracted_k141_7303 | Not-determined |
| SS_extracted_k141_85976 | Not-determined |
| SS_extracted_k141_19288 | Not-determined |
| SS_extracted_k141_104255 | Not-determined |
| SS_extracted_k141_110118 | Low-quality |
| SS_extracted_k141_85985 | Low-quality |
| SS_extracted_k141_12934 | Not-determined |
| SS_extracted_k141_61431 | Not-determined |
| SS_extracted_k141_104263 | Not-determined |
| SS_extracted_k141_140436 | Not-determined |
| SS_extracted_k141_104271 | Not-determined |
| SS_extracted_k141_19303 | Not-determined |
| SS_extracted_k141_122975 | Not-determined |
| SS_extracted_k141_12939 | Not-determined |
| SS_extracted_k141_116567 | Not-determined |
| SS_extracted_k141_110141 | Not-determined |
| SS_extracted_k141_31616 | Not-determined |
| SS_extracted_k141_12942 | Not-determined |
| SS_extracted_k141_134272 | Not-determined |
| SS_extracted_k141_37339 | Low-quality |
| SS_extracted_k141_73414 | Not-determined |
| SS_extracted_k141_122990 | Not-determined |
| SS_extracted_k141_19313 | Low-quality |
| SS_extracted_k141_116577 | Low-quality |
| SS_extracted_k141_61455 | Low-quality |
| SS_extracted_k141_110155 | Not-determined |
| SS_extracted_k141_37349 | Not-determined |
| SS_extracted_k141_37351 | Not-determined |

| | |
|---|---|
| SS_extracted_k141_37353 | Not-determined |
| SS_extracted_k141_43806 | Not-determined |
| SS_extracted_k141_97783 | Not-determined |
| SS_extracted_k141_110159 | Low-quality |
| SS_extracted_k141_97787 | Not-determined |
| SS_extracted_k141_37363 | Not-determined |
| SS_extracted_k141_12960 | Low-quality |
| SS_extracted_k141_19320 | Not-determined |
| SS_extracted_k141_12965 | Not-determined |
| SS_extracted_k141_19322 | Not-determined |
| SS_extracted_k141_97799 | Not-determined |
| SS_extracted_k141_49537 | Not-determined |
| SS_extracted_k141_97800 | Not-determined |
| SS_extracted_k141_97801 | Not-determined |
| SS_extracted_k141_37371 | Not-determined |
| SS_extracted_k141_37374 | Not-determined |
| SS_extracted_k141_61479 | Not-determined |
| SS_extracted_k141_37376 | Not-determined |
| SS_extracted_k141_123004 | Low-quality |
| SS_extracted_k141_73420 | Low-quality |
| SS_extracted_k141_61482 | Not-determined |
| SS_extracted_k141_140457 | Low-quality |
| SS_extracted_k141_12972 | Low-quality |
| SS_extracted_k141_140460 | Not-determined |
| SS_extracted_k141_104308 | Low-quality |
| SS_extracted_k141_104309 | Not-determined |
| SS_extracted_k141_73435 | Not-determined |
| SS_extracted_k141_104312 | Not-determined |
| SS_extracted_k141_104313 | Not-determined |
| SS_extracted_k141_12986 | Not-determined |
| SS_extracted_k141_55266 | Not-determined |
| SS_extracted_k141_55272 | Not-determined |
| SS_extracted_k141_61495 | Not-determined |
| SS_extracted_k141_61498 | Low-quality |
| SS_extracted_k141_19335 | Not-determined |
| SS_extracted_k141_140478 | Not-determined |
| SS_extracted_k141_73451 | Not-determined |
| SS_extracted_k141_73452 | Not-determined |
| SS_extracted_k141_123016 | Not-determined |
| SS_extracted_k141_123019 | Not-determined |
| SS_extracted_k141_73459 | Not-determined |
| SS_extracted_k141_91915 | Not-determined |
| SS_extracted_k141_61512 | Low-quality |
| SS_extracted_k141_19350 | Low-quality |
| SS_extracted_k141_43835 | Not-determined |
| SS_extracted_k141_43841 | Not-determined |
| S6_extracted_k141_7268 | Complete |
| SS_extracted_k141_134307 | Not-determined |
| SS_extracted_k141_116608 | Not-determined |
| SS_extracted_k141_43845 | Not-determined |
| SS_extracted_k141_19357 | Not-determined |
| SS_extracted_k141_61527 | Not-determined |
| SS_extracted_k141_97806 | Low-quality |
| SS_extracted_k141_37394 | Not-determined |
| SS_extracted_k141_110170 | Not-determined |
| SS_extracted_k141_123034 | Not-determined |
| SS_extracted_k141_123035 | Not-determined |
| SS_extracted_k141_97814 | Low-quality |
| SS_extracted_k141_91928 | Not-determined |
| SS_extracted_k141_97820 | Not-determined |
| SS_extracted_k141_19369 | Not-determined |
| SS_vs_k141_145591\|\|full | High-quality |
| SS_extracted_k141_123050 | Not-determined |
| SS_extracted_k141_140504 | Not-determined |
| SS_extracted_k141_123053 | Not-determined |
| SS_extracted_k141_55298 | Not-determined |
| SS_extracted_k141_61560 | Not-determined |
| SS_extracted_k141_123055 | Low-quality |
| SS_extracted_k141_24796 | Not-determined |
| SS_extracted_k141_37407 | Not-determined |
| SS_extracted_k141_116618 | Not-determined |
| SS_extracted_k141_13016 | Not-determined |
| SS_extracted_k141_37421 | Low-quality |
| SS_extracted_k141_61566 | Not-determined |
| SS_extracted_k141_116634 | Not-determined |
| SS_extracted_k141_116635 | Low-quality |
| SS_extracted_k141_86034 | Not-determined |
| SS_extracted_k141_86037 | Not-determined |
| SS_extracted_k141_37438 | Low-quality |
| SS_extracted_k141_91960 | Not-determined |
| SS_extracted_k141_86042 | Low-quality |
| SS_extracted_k141_123080 | Not-determined |
| SS_extracted_k141_116644 | Not-determined |
| SS_extracted_k141_134335 | Not-determined |
| SS_extracted_k141_97839 | Not-determined |
| SS_extracted_k141_116649 | Not-determined |
| SS_extracted_k141_97840 | Low-quality |
| SS_extracted_k141_67665 | Not-determined |
| SS_extracted_k141_13041 | Low-quality |
| SS_extracted_k141_55308 | Not-determined |
| SS_extracted_k141_79385 | Low-quality |
| SS_extracted_k141_61586 | Not-determined |
| SS_extracted_k141_91981 | Not-determined |
| SS_extracted_k141_104337 | Not-determined |
| SS_extracted_k141_116657 | Low-quality |
| S4_extracted_4891 | Medium-quality |
| SS_extracted_k141_67670 | Low-quality |
| SS_extracted_k141_61606 | Not-determined |
| SS_extracted_k141_91998 | Not-determined |
| SS_extracted_k141_13049 | Not-determined |
| SS_extracted_k141_61610 | Not-determined |
| SS_extracted_k141_79389 | Low-quality |
| SS_extracted_k141_37460 | Not-determined |
| SS_extracted_k141_55320 | Not-determined |
| SS_extracted_k141_104351 | Not-determined |
| SS_extracted_k141_73502 | Not-determined |
| SS_extracted_k141_79394 | Not-determined |
| SS_extracted_k141_110184 | Low-quality |
| SS_extracted_k141_73511 | Not-determined |
| SS_extracted_k141_61616 | Not-determined |
| SS_extracted_k141_73512 | Not-determined |
| SS_extracted_k141_37472 | Not-determined |
| SS_extracted_k141_86062 | Not-determined |
| SS_extracted_k141_116667 | Not-determined |
| SS_extracted_k141_67685 | Not-determined |
| SS_extracted_k141_79405 | Not-determined |
| SS_extracted_k141_31670 | Low-quality |
| SS_extracted_k141_37478 | Not-determined |
| SS_extracted_k141_123088 | Not-determined |
| SS_extracted_k141_31674 | Not-determined |
| SS_extracted_k141_97860 | Not-determined |
| SS_extracted_k141_13071 | Not-determined |
| SS_extracted_k141_79413 | Not-determined |
| SS_extracted_k141_79414 | Not-determined |
| SS_extracted_k141_97865 | Not-determined |
| SS_extracted_k141_97872 | Not-determined |
| SS_extracted_k141_31681 | Not-determined |
| SS_extracted_k141_61631 | Not-determined |
| SS_extracted_k141_55323 | Low-quality |
| SS_extracted_k141_7313 | Not-determined |
| SS_extracted_k141_55332 | Not-determined |
| SS_extracted_k141_37488 | Not-determined |
| SS_extracted_k141_123104 | Not-determined |
| SS_extracted_k141_73541 | Not-determined |
| SS_extracted_k141_61648 | Not-determined |
| SS_extracted_k141_67708 | Not-determined |
| SS_extracted_k141_67710 | Not-determined |
| SS_extracted_k141_110202 | Not-determined |
| SS_extracted_k141_116677 | Low-quality |
| SS_extracted_k141_73550 | Not-determined |
| SS_extracted_k141_134351 | Not-determined |
| SS_extracted_k141_31697 | Not-determined |
| SS_extracted_k141_134352 | Not-determined |
| SS_extracted_k141_92009 | Not-determined |
| SS_extracted_k141_128294 | Not-determined |
| SS_extracted_k141_97883 | Not-determined |
| SS_extracted_k141_134359 | Not-determined |
| SS_extracted_k141_97884 | Low-quality |
| SS_extracted_k141_128299 | Low-quality |
| SS_extracted_k141_7325 | Not-determined |
| SS_extracted_k141_110218 | Not-determined |
| SS_extracted_k141_67719 | Not-determined |
| SS_extracted_k141_67720 | Not-determined |
| SS_extracted_k141_92015 | Low-quality |
| SS_extracted_k141_55343 | Not-determined |
| SS_extracted_k141_116692 | Low-quality |
| SS_extracted_k141_110225 | Not-determined |
| SS_extracted_k141_123126 | Not-determined |
| SS_extracted_k141_134363 | Low-quality |
| SS_extracted_k141_134365 | Not-determined |
| SS_extracted_k141_73559 | Not-determined |
| SS_extracted_k141_134366 | Not-determined |
| SS_extracted_k141_123129 | Not-determined |
| SS_extracted_k141_92027 | Low-quality |
| SS_extracted_k141_19435 | Not-determined |
| SS_extracted_k141_61670 | Not-determined |
| SS_extracted_k141_43876 | Not-determined |
| SS_extracted_k141_31721 | Low-quality |
| SS_extracted_k141_110236 | Not-determined |
| SS_extracted_k141_116703 | Not-determined |
| SS_extracted_k141_110239 | Not-determined |
| SS_extracted_k141_61682 | Not-determined |
| SS_extracted_k141_92035 | Low-quality |
| SS_extracted_k141_116707 | Not-determined |
| SS_extracted_k141_128307 | Not-determined |
| SS_extracted_k141_61684 | Not-determined |
| SS_extracted_k141_123141 | Low-quality |
| SS_extracted_k141_61694 | Low-quality |
| SS_extracted_k141_61696 | Not-determined |
| SS_extracted_k141_123149 | Not-determined |
| SS_extracted_k141_128315 | Not-determined |
| SS_extracted_k141_92042 | Not-determined |
| SS_extracted_k141_55354 | Not-determined |
| SS_extracted_k141_110252 | Not-determined |
| SS_extracted_k141_55359 | Not-determined |
| SS_extracted_k141_123157 | Not-determined |
| SS_extracted_k141_55363 | Not-determined |
| SS_extracted_k141_61703 | Low-quality |
| SS_extracted_k141_97893 | Not-determined |
| SS_extracted_k141_123160 | Not-determined |
| SS_extracted_k141_97895 | Not-determined |
| SS_extracted_k141_97896 | Not-determined |
| SS_extracted_k141_110262 | Not-determined |
| SS_extracted_k141_13103 | Not-determined |
| SS_extracted_k141_116726 | Not-determined |
| SS_extracted_k141_110266 | Low-quality |
| SS_extracted_k141_43893 | Not-determined |
| SS_extracted_k141_110268 | Not-determined |
| SS_extracted_k141_116728 | Low-quality |
| SS_extracted_k141_123162 | Low-quality |
| SS_extracted_k141_123165 | Not-determined |
| SS_extracted_k141_13111 | Low-quality |
| SS_extracted_k141_19467 | Not-determined |
| SS_extracted_k141_116740 | Low-quality |
| SS_extracted_k141_43913 | Not-determined |
| SS_extracted_k141_67734 | Not-determined |
| SS_extracted_k141_97902 | Not-determined |
| SS_extracted_k141_116744 | Not-determined |
| SS_extracted_k141_116746 | Not-determined |
| SS_extracted_k141_92052 | Not-determined |
| SS_extracted_k141_13122 | Not-determined |
| SS_extracted_k141_73580 | Low-quality |
| SS_extracted_k141_67745 | Not-determined |
| SS_extracted_k141_19491 | Not-determined |
| SS_extracted_k141_43928 | Not-determined |
| SS_extracted_k141_19506 | Not-determined |
| SS_extracted_k141_13144 | Not-determined |
| SS_extracted_k141_67757 | Low-quality |
| SS_extracted_k141_97916 | Low-quality |
| SS_extracted_k141_110321 | Low-quality |
| SS_extracted_k141_134380 | Not-determined |
| SS_extracted_k141_110328 | Not-determined |
| SS_extracted_k141_7347 | Not-determined |
| SS_extracted_k141_31763 | Not-determined |
| SS_extracted_k141_31764 | Not-determined |
| SS_extracted_k141_67767 | Not-determined |
| SS_extracted_k141_61710 | Not-determined |
| SS_extracted_k141_19510 | Not-determined |
| SS_extracted_k141_61712 | Low-quality |
| SS_extracted_k141_97921 | Not-determined |
| SS_extracted_k141_128321 | Low-quality |
| SS_extracted_k141_128322 | Not-determined |
| SS_extracted_k141_86084 | Not-determined |
| SS_extracted_k141_92087 | Not-determined |
| SS_extracted_k141_61722 | Low-quality |
| SS_extracted_k141_110344 | Not-determined |
| SS_extracted_k141_19518 | Not-determined |
| SS_extracted_k141_19519 | Not-determined |
| SS_extracted_k141_19520 | Not-determined |
| SS_extracted_k141_31772 | Low-quality |
| SS_extracted_k141_134400 | Not-determined |
| SS_extracted_k141_67786 | Not-determined |
| SS_extracted_k141_61727 | Not-determined |
| SS_extracted_k141_61728 | Not-determined |
| SS_extracted_k141_67787 | Not-determined |
| SS_extracted_k141_92102 | Not-determined |
| SS_extracted_k141_97925 | Not-determined |
| SS_extracted_k141_86095 | Not-determined |
| SS_extracted_k141_61735 | Not-determined |
| SS_extracted_k141_86100 | Not-determined |
| SS_extracted_k141_7365 | Not-determined |
| SS_extracted_k141_110354 | Not-determined |
| SS_extracted_k141_128345 | Not-determined |
| SS_extracted_k141_13158 | Not-determined |
| SS_extracted_k141_92108 | Not-determined |
| SS_extracted_k141_37513 | Not-determined |
| SS_extracted_k141_7370 | Not-determined |
| SS_extracted_k141_37521 | Not-determined |
| SS_extracted_k141_7372 | Not-determined |
| SS_extracted_k141_92113 | Not-determined |
| SS_extracted_k141_134415 | Low-quality |
| SS_extracted_k141_134419 | Low-quality |
| SS_extracted_k141_37528 | Not-determined |
| SS_extracted_k141_31791 | Not-determined |
| SS_extracted_k141_13162 | Not-determined |
| SS_extracted_k141_134425 | Not-determined |
| SS_extracted_k141_123207 | Low-quality |
| SS_extracted_k141_61737 | Not-determined |
| SS_extracted_k141_61738 | Not-determined |
| SS_extracted_k141_134439 | Not-determined |
| SS_extracted_k141_7387 | Not-determined |
| SS_extracted_k141_61747 | Low-quality |
| SS_extracted_k141_7392 | Low-quality |
| SS_extracted_k141_128371 | Not-determined |
| SS_extracted_k141_7393 | Not-determined |
| SS_extracted_k141_128372 | Not-determined |
| SS_extracted_k141_61753 | Not-determined |
| SS_extracted_k141_37545 | Not-determined |
| SS_extracted_k141_61756 | Not-determined |
| SS_extracted_k141_67800 | Low-quality |
| SS_extracted_k141_37555 | Low-quality |
| SS_extracted_k141_79466 | Not-determined |
| SS_extracted_k141_43953 | Low-quality |
| SS_extracted_k141_31812 | Not-determined |
| SS_extracted_k141_128381 | Not-determined |
| SS_extracted_k141_123225 | Not-determined |
| SS_extracted_k141_31816 | Low-quality |
| SS_extracted_k141_37565 | Not-determined |
| SS_extracted_k141_134454 | Not-determined |
| SS_extracted_k141_92153 | Not-determined |
| SS_extracted_k141_13199 | Not-determined |
| SS_extracted_k141_31821 | Not-determined |
| SS_extracted_k141_123237 | Low-quality |
| SS_extracted_k141_104368 | Not-determined |
| SS_extracted_k141_73617 | Not-determined |
| SS_extracted_k141_104372 | Not-determined |
| SS_extracted_k141_55387 | Not-determined |
| SS_extracted_k141_110378 | Not-determined |
| SS_extracted_k141_104377 | Not-determined |
| SS_extracted_k141_104380 | Not-determined |
| SS_extracted_k141_123252 | Not-determined |
| SS_extracted_k141_116827 | Not-determined |
| SS_extracted_k141_116838 | Not-determined |
| SS_extracted_k141_110389 | Not-determined |
| SS_extracted_k141_37582 | Not-determined |
| SS_extracted_k141_134463 | Not-determined |
| SS_extracted_k141_67828 | Not-determined |
| SS_extracted_k141_116844 | Not-determined |
| SS_extracted_k141_116845 | Not-determined |
| SS_extracted_k141_61797 | Low-quality |
| SS_extracted_k141_1474 | Not-determined |
| SS_extracted_k141_67840 | Not-determined |
| SS_extracted_k141_37595 | Low-quality |
| SS_extracted_k141_97968 | Not-determined |
| SS_extracted_k141_1477 | Not-determined |
| SS_extracted_k141_7408 | Not-determined |
| SS_extracted_k141_67846 | Not-determined |
| SS_extracted_k141_67847 | Not-determined |
| SS_extracted_k141_73637 | Not-determined |
| SS_extracted_k141_92169 | Not-determined |
| SS_extracted_k141_1483 | Low-quality |
| SS_extracted_k141_61818 | Not-determined |
| SS_extracted_k141_13214 | Low-quality |
| SS_extracted_k141_92175 | Not-determined |
| SS_extracted_k141_37604 | Not-determined |
| SS_extracted_k141_49568 | Not-determined |
| SS_extracted_k141_49569 | Not-determined |
| SS_extracted_k141_73651 | Not-determined |
| SS_extracted_k141_13225 | Not-determined |
| SS_extracted_k141_73656 | Not-determined |
| SS_extracted_k141_134484 | Not-determined |
| SS_extracted_k141_43993 | Not-determined |
| SS_extracted_k141_49578 | Low-quality |
| SS_extracted_k141_55402 | Not-determined |
| SS_extracted_k141_31832 | Low-quality |
| SS_extracted_k141_110423 | Low-quality |
| SS_extracted_k141_92193 | Not-determined |
| SS_extracted_k141_73670 | Not-determined |
| SS_extracted_k141_31841 | Not-determined |
| SS_extracted_k141_110429 | Not-determined |
| SS_extracted_k141_31847 | Not-determined |
| SS_extracted_k141_110434 | Low-quality |
| SS_extracted_k141_7437 | Not-determined |
| SS_extracted_k141_31856 | Not-determined |
| SS_extracted_k141_128412 | Low-quality |
| SS_extracted_k141_128413 | Not-determined |
| SS_extracted_k141_73689 | Not-determined |
| SS_extracted_k141_37615 | Low-quality |
| SS_extracted_k141_110447 | Not-determined |
| SS_extracted_k141_128422 | Not-determined |
| SS_extracted_k141_86116 | Not-determined |
| SS_extracted_k141_67868 | Not-determined |
| SS_extracted_k141_116914 | Not-determined |
| SS_extracted_k141_31880 | Not-determined |
| SS_extracted_k141_67871 | Not-determined |
| SS_extracted_k141_128435 | Not-determined |
| SS_extracted_k141_110457 | Not-determined |
| SS_extracted_k141_67875 | Not-determined |
| SS_extracted_k141_92219 | Not-determined |
| SS_extracted_k141_116929 | Not-determined |
| SS_extracted_k141_49616 | Not-determined |
| SS_extracted_k141_49619 | Not-determined |
| SS_extracted_k141_92226 | Not-determined |
| SS_extracted_k141_49622 | Not-determined |
| SS_extracted_k141_128451 | Not-determined |
| SS_extracted_k141_134513 | Low-quality |
| SS_extracted_k141_92241 | Not-determined |
| SS_extracted_k141_116938 | Not-determined |
| SS_extracted_k141_116939 | Not-determined |
| SS_extracted_k141_92243 | Low-quality |
| SS_extracted_k141_79511 | Not-determined |
| SS_extracted_k141_79514 | Not-determined |
| SS_extracted_k141_110479 | Low-quality |
| SS_extracted_k141_31895 | Not-determined |
| SS_extracted_k141_67884 | Not-determined |
| SS_extracted_k141_92259 | Low-quality |
| SS_extracted_k141_31901 | Not-determined |
| SS_extracted_k141_73729 | Not-determined |
| SS_extracted_k141_49655 | Not-determined |
| SS_extracted_k141_67892 | Low-quality |
| SS_extracted_k141_13255 | Not-determined |
| SS_extracted_k141_123261 | Not-determined |
| SS_extracted_k141_49660 | Low-quality |
| SS_extracted_k141_49663 | Not-determined |
| SS_extracted_k141_49666 | Low-quality |
| SS_extracted_k141_86160 | Not-determined |
| SS_extracted_k141_110496 | Not-determined |
| SS_extracted_k141_110498 | Not-determined |
| SS_extracted_k141_116965 | Not-determined |
| SS_extracted_k141_86165 | Not-determined |
| SS_extracted_k141_86169 | Not-determined |
| SS_extracted_k141_13268 | Not-determined |
| SS_extracted_k141_73733 | Not-determined |
| SS_extracted_k141_123274 | Not-determined |
| SS_extracted_k141_140553 | Not-determined |
| SS_extracted_k141_24864 | Not-determined |
| SS_extracted_k141_97981 | Not-determined |
| SS_extracted_k141_92289 | Not-determined |
| SS_extracted_k141_92290 | Not-determined |
| SS_extracted_k141_24868 | Not-determined |
| SS_extracted_k141_116975 | Not-determined |
| SS_extracted_k141_97987 | Not-determined |
| SS_extracted_k141_92294 | Not-determined |
| SS_extracted_k141_116977 | Low-quality |
| SS_extracted_k141_73740 | Low-quality |
| SS_extracted_k141_86177 | Not-determined |
| SS_extracted_k141_73744 | Not-determined |
| SS_extracted_k141_73748 | Not-determined |
| SS_extracted_k141_123283 | Not-determined |
| SS_extracted_k141_123286 | Not-determined |
| SS_extracted_k141_67905 | Not-determined |
| SS_extracted_k141_86183 | Low-quality |
| SS_extracted_k141_116996 | Not-determined |
| SS_extracted_k141_19538 | Low-quality |
| SS_extracted_k141_92326 | Not-determined |
| SS_extracted_k141_92330 | Low-quality |
| SS_extracted_k141_98001 | Low-quality |
| SS_extracted_k141_49672 | Not-determined |
| SS_extracted_k141_31907 | Not-determined |
| SS_extracted_k141_117008 | Not-determined |
| SS_extracted_k141_49675 | Low-quality |
| SS_extracted_k141_67919 | Not-determined |
| SS_extracted_k141_92341 | Not-determined |
| SS_extracted_k141_1531 | Not-determined |
| SS_extracted_k141_110500 | Low-quality |
| SS_extracted_k141_92343 | Low-quality |
| SS_extracted_k141_140561 | Not-determined |
| SS_extracted_k141_92344 | Not-determined |
| SS_extracted_k141_73770 | Not-determined |
| SS_extracted_k141_92347 | Not-determined |
| SS_extracted_k141_24888 | Low-quality |
| SS_extracted_k141_67923 | Not-determined |
| SS_extracted_k141_73775 | Low-quality |
| SS_extracted_k141_73776 | Not-determined |
| SS_extracted_k141_110510 | Low-quality |
| SS_extracted_k141_117028 | Low-quality |
| SS_extracted_k141_117030 | Low-quality |
| SS_extracted_k141_67930 | Not-determined |
| SS_extracted_k141_117032 | Not-determined |
| SS_extracted_k141_117033 | Not-determined |
| SS_extracted_k141_31928 | Not-determined |
| SS_extracted_k141_37652 | Low-quality |
| SS_extracted_k141_110521 | Not-determined |
| SS_extracted_k141_1558 | Low-quality |
| SS_extracted_k141_140573 | Not-determined |
| SS_extracted_k141_1560 | Not-determined |
| SS_extracted_k141_61832 | Low-quality |
| SS_extracted_k141_140577 | Not-determined |
| SS_extracted_k141_67937 | Low-quality |
| SS_extracted_k141_49690 | Low-quality |
| SS_extracted_k141_31948 | Low-quality |
| SS_extracted_k141_140582 | Not-determined |
| SS_extracted_k141_86196 | Not-determined |
| SS_extracted_k141_49692 | Not-determined |
| SS_extracted_k141_140585 | Not-determined |
| SS_extracted_k141_92376 | Low-quality |
| SS_extracted_k141_140588 | Not-determined |
| SS_extracted_k141_117045 | Not-determined |
| SS_extracted_k141_117046 | Not-determined |
| SS_extracted_k141_92384 | Not-determined |
| SS_extracted_k141_92385 | Not-determined |
| SS_extracted_k141_44021 | Not-determined |
| SS_extracted_k141_140599 | Not-determined |
| SS_extracted_k141_140600 | Not-determined |
| SS_extracted_k141_13295 | Not-determined |
| SS_extracted_k141_92400 | Not-determined |
| SS_extracted_k141_140611 | Not-determined |
| SS_extracted_k141_110545 | Not-determined |
| SS_extracted_k141_123327 | Not-determined |
| SS_extracted_k141_110547 | Not-determined |
| SS_extracted_k141_49716 | Not-determined |
| SS_extracted_k141_110548 | Low-quality |
| SS_extracted_k141_49719 | Not-determined |
| SS_extracted_k141_92411 | Not-determined |
| SS_extracted_k141_110556 | Not-determined |
| SS_extracted_k141_61873 | Not-determined |
| SS_extracted_k141_79526 | Not-determined |
| SS_extracted_k141_92420 | Low-quality |
| SS_extracted_k141_128473 | Not-determined |
| SS_extracted_k141_49728 | Not-determined |
| SS_extracted_k141_92428 | Not-determined |
| SS_extracted_k141_49729 | Not-determined |
| SS_extracted_k141_117059 | Not-determined |
| SS_extracted_k141_117060 | Not-determined |
| SS_extracted_k141_123335 | Low-quality |
| SS_extracted_k141_73804 | Not-determined |
| SS_extracted_k141_73806 | Not-determined |
| SS_extracted_k141_37663 | Not-determined |
| SS_extracted_k141_123337 | Low-quality |
| SS_extracted_k141_117065 | Not-determined |
| SS_extracted_k141_104437 | Not-determined |
| SS_extracted_k141_37665 | Not-determined |
| SS_extracted_k141_19576 | Low-quality |
| SS_extracted_k141_92445 | Not-determined |
| SS_extracted_k141_19581 | Not-determined |
| SS_extracted_k141_117072 | Not-determined |
| SS_extracted_k141_19583 | Low-quality |
| SS_extracted_k141_128479 | Not-determined |
| SS_extracted_k141_67961 | Not-determined |
| SS_extracted_k141_13319 | Low-quality |
| SS_extracted_k141_117080 | Not-determined |
| SS_extracted_k141_128483 | Not-determined |
| SS_extracted_k141_128484 | Not-determined |
| SS_extracted_k141_117083 | Not-determined |
| SS_extracted_k141_13323 | Not-determined |
| SS_extracted_k141_117084 | Not-determined |
| SS_extracted_k141_49759 | Not-determined |
| SS_extracted_k141_32001 | Not-determined |
| SS_extracted_k141_13327 | Not-determined |
| SS_extracted_k141_140641 | Not-determined |
| SS_extracted_k141_19587 | Low-quality |
| SS_extracted_k141_117088 | Not-determined |
| SS_extracted_k141_49765 | Not-determined |
| SS_extracted_k141_140643 | Not-determined |
| SS_extracted_k141_19590 | Not-determined |
| SS_extracted_k141_86238 | Not-determined |
| SS_extracted_k141_117092 | Not-determined |
| SS_extracted_k141_110573 | Not-determined |
| SS_extracted_k141_117094 | Low-quality |
| SS_extracted_k141_104457 | Not-determined |
| SS_extracted_k141_117095 | Not-determined |
| SS_extracted_k141_37691 | Not-determined |
| SS_extracted_k141_104458 | Not-determined |
| SS_extracted_k141_32007 | Not-determined |
| SS_extracted_k141_37693 | Low-quality |
| SS_extracted_k141_32011 | Not-determined |
| SS_extracted_k141_1578 | Not-determined |
| SS_extracted_k141_32018 | Not-determined |
| SS_extracted_k141_13333 | Not-determined |
| SS_extracted_k141_1583 | Low-quality |
| SS_extracted_k141_13336 | Not-determined |
| SS_extracted_k141_123368 | Not-determined |
| SS_extracted_k141_61892 | Not-determined |
| SS_extracted_k141_92472 | Not-determined |
| SS_extracted_k141_61897 | Not-determined |
| SS_extracted_k141_13338 | Low-quality |
| SS_extracted_k141_104469 | Not-determined |
| SS_extracted_k141_13341 | Not-determined |
| SS_extracted_k141_13342 | Low-quality |
| SS_extracted_k141_140663 | Not-determined |
| SS_extracted_k141_73847 | Not-determined |
| SS_extracted_k141_98043 | Low-quality |
| SS_extracted_k141_104481 | Low-quality |
| SS_extracted_k141_67978 | Not-determined |
| SS_extracted_k141_140672 | Low-quality |
| SS_extracted_k141_13357 | Not-determined |
| SS_extracted_k141_92489 | Not-determined |
| SS_extracted_k141_1597 | Not-determined |
| SS_extracted_k141_37711 | Not-determined |
| SS_extracted_k141_32037 | Not-determined |
| SS_extracted_k141_123375 | Not-determined |
| SS_extracted_k141_140681 | Low-quality |
| SS_extracted_k141_110608 | Not-determined |
| SS_extracted_k141_140688 | Not-determined |
| SS_extracted_k141_110611 | Low-quality |
| SS_extracted_k141_68002 | Not-determined |
| SS_extracted_k141_117136 | Not-determined |
| SS_extracted_k141_117139 | Not-determined |
| SS_extracted_k141_32046 | Not-determined |
| SS_extracted_k141_32051 | Not-determined |
| SS_extracted_k141_92498 | Not-determined |
| SS_extracted_k141_140705 | Not-determined |
| SS_extracted_k141_140706 | Not-determined |
| SS_extracted_k141_140707 | Low-quality |
| S4_vs_VIRSorter_4891-cat_3 | Medium-quality |
| SS_extracted_k141_140708 | Low-quality |
| SS_extracted_k141_117141 | Not-determined |
| SS_extracted_k141_92502 | Low-quality |
| SS_extracted_k141_19596 | Low-quality |
| SS_extracted_k141_44051 | Low-quality |
| SS_extracted_k141_32065 | Low-quality |
| SS_extracted_k141_19600 | Not-determined |
| SS_extracted_k141_55425 | Not-determined |
| SS_extracted_k141_92506 | Low-quality |
| SS_extracted_k141_104518 | Not-determined |
| SS_extracted_k141_104520 | Not-determined |
| SS_extracted_k141_117146 | Low-quality |
| SS_extracted_k141_104522 | Low-quality |
| SS_extracted_k141_44063 | Low-quality |
| SS_extracted_k141_44066 | Not-determined |
| SS_extracted_k141_92517 | Not-determined |
| SS_extracted_k141_44069 | Not-determined |
| SS_extracted_k141_104524 | Not-determined |
| SS_extracted_k141_7456 | Not-determined |
| SS_extracted_k141_92528 | Not-determined |
| SS_extracted_k141_44071 | Not-determined |
| SS_extracted_k141_104528 | Not-determined |
| SS_extracted_k141_19619 | Not-determined |
| SS_extracted_k141_117165 | Low-quality |

| | |
|---|---|
| SS_extracted_k141_117166 | Not-determined |
| SS_extracted_k141_134550 | Low-quality |
| SS_extracted_k141_32086 | Not-determined |
| SS_extracted_k141_123408 | Not-determined |
| SS_extracted_k141_37781 | Not-determined |
| SS_extracted_k141_110628 | Not-determined |
| SS_extracted_k141_68027 | Not-determined |
| SS_extracted_k141_55445 | Not-determined |
| SS_extracted_k141_44078 | Not-determined |
| SS_extracted_k141_68029 | Not-determined |
| SS_extracted_k141_79533 | Not-determined |
| SS_extracted_k141_140713 | Not-determined |
| SS_extracted_k141_1631 | Not-determined |
| SS_extracted_k141_140714 | Not-determined |
| SS_extracted_k141_7457 | Low-quality |
| SS_extracted_k141_1633 | Low-quality |
| SS_extracted_k141_1634 | Low-quality |
| SS_extracted_k141_68035 | Not-determined |
| SS_extracted_k141_92540 | Not-determined |
| SS_extracted_k141_32099 | Not-determined |
| SS_extracted_k141_92544 | Not-determined |
| SS_extracted_k141_32102 | Not-determined |
| SS_extracted_k141_128502 | Not-determined |
| SS_extracted_k141_24910 | Not-determined |
| SS_extracted_k141_128508 | Not-determined |
| SS_extracted_k141_7474 | Low-quality |
| SS_extracted_k141_1640 | Not-determined |
| SS_extracted_k141_128511 | Not-determined |
| SS_extracted_k141_104549 | Low-quality |
| SS_extracted_k141_140718 | Not-determined |
| SS_extracted_k141_1644 | Not-determined |
| SS_extracted_k141_128514 | Not-determined |
| SS_extracted_k141_1648 | Low-quality |
| SS_extracted_k141_1650 | Not-determined |
| SS_extracted_k141_98067 | Not-determined |
| SS_extracted_k141_128519 | Not-determined |
| SS_extracted_k141_13367 | Not-determined |
| SS_extracted_k141_55465 | Not-determined |
| SS_extracted_k141_104559 | Not-determined |
| SS_extracted_k141_110644 | Not-determined |
| SS_extracted_k141_110646 | Not-determined |
| SS_extracted_k141_55471 | Not-determined |
| SS_extracted_k141_49845 | Not-determined |
| SS_extracted_k141_1661 | Not-determined |
| SS_extracted_k141_13378 | Not-determined |
| SS_extracted_k141_128534 | Not-determined |
| SS_extracted_k141_61908 | Not-determined |
| SS_extracted_k141_19637 | Not-determined |
| SS_extracted_k141_61909 | Not-determined |
| SS_extracted_k141_110655 | Low-quality |
| SS_extracted_k141_1670 | Low-quality |
| SS_extracted_k141_128539 | Not-determined |
| SS_extracted_k141_98077 | Low-quality |
| SS_extracted_k141_61916 | Not-determined |
| SS_extracted_k141_104576 | Not-determined |
| SS_extracted_k141_128550 | Not-determined |
| SS_extracted_k141_140735 | Not-determined |
| SS_extracted_k141_19651 | Low-quality |
| SS_extracted_k141_13381 | Not-determined |
| SS_extracted_k141_61924 | Low-quality |
| SS_extracted_k141_110667 | Not-determined |
| SS_extracted_k141_140737 | Low-quality |
| SS_extracted_k141_32146 | Low-quality |
| SS_extracted_k141_32153 | Not-determined |
| SS_extracted_k141_1693 | Not-determined |
| SS_extracted_k141_55487 | Low-quality |
| SS_extracted_k141_104592 | Low-quality |
| SS_extracted_k141_32158 | Not-determined |
| SS_extracted_k141_128567 | Not-determined |
| SS_extracted_k141_49881 | Low-quality |
| SS_extracted_k141_44119 | Not-determined |
| SS_extracted_k141_128573 | Not-determined |
| SS_extracted_k141_49886 | Low-quality |
| SS_extracted_k141_128574 | Low-quality |
| SS_extracted_k141_1710 | Not-determined |
| SS_extracted_k141_1711 | Low-quality |
| SS_extracted_k141_7498 | Not-determined |
| SS_extracted_k141_7499 | Not-determined |
| SS_extracted_k141_134562 | Low-quality |
| SS_extracted_k141_110677 | Not-determined |
| SS_extracted_k141_44128 | Not-determined |
| SS_extracted_k141_61932 | Not-determined |
| SS_extracted_k141_134566 | Not-determined |
| SS_extracted_k141_7509 | Low-quality |
| SS_extracted_k141_44131 | Low-quality |
| SS_extracted_k141_128585 | Not-determined |
| SS_extracted_k141_92594 | Not-determined |
| SS_extracted_k141_7515 | Not-determined |
| SS_extracted_k141_86264 | Not-determined |
| SS_extracted_k141_55494 | Not-determined |
| SS_extracted_k141_7520 | Not-determined |
| SS_extracted_k141_55501 | Low-quality |
| SS_extracted_k141_55505 | Not-determined |
| SS_extracted_k141_55507 | Not-determined |
| SS_extracted_k141_128599 | Not-determined |
| SS_extracted_k141_92613 | Not-determined |
| SS_extracted_k141_55510 | Not-determined |
| SS_extracted_k141_128602 | Low-quality |
| SS_extracted_k141_92618 | Not-determined |
| SS_extracted_k141_128604 | Not-determined |
| SS_extracted_k141_73872 | Low-quality |
| SS_extracted_k141_86279 | Not-determined |
| SS_extracted_k141_61952 | Low-quality |
| SS_extracted_k141_92622 | Low-quality |
| SS_extracted_k141_7536 | Not-determined |
| SS_extracted_k141_92624 | Not-determined |
| SS_extracted_k141_55515 | Not-determined |
| SS_extracted_k141_32170 | Not-determined |
| SS_extracted_k141_68090 | Not-determined |
| SS_extracted_k141_68091 | Not-determined |
| SS_extracted_k141_110696 | Not-determined |
| SS_extracted_k141_24930 | Not-determined |
| SS_extracted_k141_128613 | Not-determined |
| SS_extracted_k141_24932 | Not-determined |
| SS_extracted_k141_68099 | Not-determined |
| SS_extracted_k141_117212 | Not-determined |
| SS_extracted_k141_24935 | Low-quality |
| SS_extracted_k141_92635 | Not-determined |
| SS_extracted_k141_110702 | Not-determined |
| SS_extracted_k141_49926 | Low-quality |
| SS_extracted_k141_117216 | Not-determined |
| SS_extracted_k141_68105 | Not-determined |
| SS_extracted_k141_61965 | Not-determined |
| SS_extracted_k141_55528 | Not-determined |
| SS_extracted_k141_104628 | Not-determined |
| SS_extracted_k141_7552 | Not-determined |
| SS_extracted_k141_117222 | Not-determined |
| SS_extracted_k141_49948 | Not-determined |
| SS_extracted_k141_7560 | Low-quality |
| SS_extracted_k141_117229 | Low-quality |
| SS_extracted_k141_104639 | Not-determined |
| SS_extracted_k141_110720 | Low-quality |
| SS_extracted_k141_44181 | Not-determined |
| SS_extracted_k141_92653 | Low-quality |
| SS_extracted_k141_79556 | Low-quality |
| SS_extracted_k141_73891 | Not-determined |
| SS_extracted_k141_49952 | Not-determined |
| SS_extracted_k141_92658 | Not-determined |
| SS_extracted_k141_104648 | Not-determined |
| SS_extracted_k141_79559 | Not-determined |
| SS_extracted_k141_44183 | Not-determined |
| SS_extracted_k141_123485 | Not-determined |
| SS_extracted_k141_98093 | Not-determined |
| SS_extracted_k141_110733 | Not-determined |
| SS_extracted_k141_44192 | Not-determined |
| SS_extracted_k141_86305 | Not-determined |
| SS_extracted_k141_104658 | Not-determined |
| SS_extracted_k141_98096 | Not-determined |
| SS_extracted_k141_110742 | Not-determined |
| SS_extracted_k141_98100 | Not-determined |
| SS_extracted_k141_134585 | Not-determined |
| SS_extracted_k141_98101 | Not-determined |
| SS_extracted_k141_110743 | Not-determined |
| SS_extracted_k141_98108 | Not-determined |
| SS_extracted_k141_117260 | Not-determined |
| SS_extracted_k141_98114 | Not-determined |
| SS_extracted_k141_123496 | Not-determined |
| SS_extracted_k141_110749 | Not-determined |
| SS_extracted_k141_13447 | Not-determined |
| SS_extracted_k141_13448 | Not-determined |
| SS_extracted_k141_86317 | Low-quality |
| SS_extracted_k141_13460 | Not-determined |
| SS_extracted_k141_13461 | Not-determined |
| SS_extracted_k141_104673 | Not-determined |
| SS_extracted_k141_104674 | Not-determined |
| SS_extracted_k141_128655 | Not-determined |
| SS_extracted_k141_49978 | Not-determined |
| SS_extracted_k141_92698 | Not-determined |
| SS_extracted_k141_110772 | Not-determined |
| SS_extracted_k141_19669 | Not-determined |
| SS_extracted_k141_110774 | Not-determined |
| SS_extracted_k141_61990 | Low-quality |
| SS_extracted_k141_1732 | Not-determined |
| SS_extracted_k141_128669 | Not-determined |
| SS_extracted_k141_128671 | Not-determined |
| SS_extracted_k141_128672 | Not-determined |
| SS_extracted_k141_37845 | Not-determined |
| SS_extracted_k141_55542 | Not-determined |
| SS_extracted_k141_1737 | Low-quality |
| SS_extracted_k141_1739 | Not-determined |
| SS_extracted_k141_92711 | Not-determined |
| SS_extracted_k141_128676 | Low-quality |
| SS_extracted_k141_49997 | Low-quality |
| SS_extracted_k141_68174 | Not-determined |
| SS_extracted_k141_92716 | Not-determined |
| SS_extracted_k141_117293 | Not-determined |
| SS_extracted_k141_55555 | Not-determined |
| SS_extracted_k141_50003 | Not-determined |
| SS_extracted_k141_13480 | Not-determined |
| SS_extracted_k141_117297 | Not-determined |
| SS_extracted_k141_117298 | Not-determined |
| SS_extracted_k141_117299 | Low-quality |
| SS_extracted_k141_92722 | Not-determined |
| SS_extracted_k141_68183 | Not-determined |
| SS_extracted_k141_117300 | Low-quality |
| SS_extracted_k141_110791 | Not-determined |
| SS_extracted_k141_55561 | Not-determined |
| SS_extracted_k141_50014 | Not-determined |
| SS_extracted_k141_140755 | Not-determined |
| SS_extracted_k141_140756 | Not-determined |
| SS_extracted_k141_68191 | Not-determined |
| SS_extracted_k141_128686 | Not-determined |
| SS_extracted_k141_110793 | Not-determined |
| SS_extracted_k141_68195 | Not-determined |
| SS_extracted_k141_73927 | Not-determined |
| SS_extracted_k141_50022 | Low-quality |
| SS_extracted_k141_128693 | Not-determined |
| SS_extracted_k141_50024 | Not-determined |
| SS_extracted_k141_68206 | Not-determined |
| SS_extracted_k141_140766 | Not-determined |
| SS_extracted_k141_128695 | Not-determined |
| SS_extracted_k141_140768 | Not-determined |
| SS_extracted_k141_50034 | Not-determined |
| SS_extracted_k141_104723 | Not-determined |
| SS_extracted_k141_68211 | Not-determined |
| SS_extracted_k141_134609 | Not-determined |
| SS_extracted_k141_1748 | Not-determined |
| SS_extracted_k141_134611 | Not-determined |
| SS_extracted_k141_128703 | Not-determined |
| SS_extracted_k141_1751 | Not-determined |
| SS_extracted_k141_1752 | Low-quality |
| SS_extracted_k141_110816 | Not-determined |
| SS_extracted_k141_128709 | Low-quality |
| SS_extracted_k141_140775 | Not-determined |
| SS_extracted_k141_55567 | Not-determined |
| SS_extracted_k141_55574 | Not-determined |
| SS_extracted_k141_55575 | Not-determined |
| SS_extracted_k141_140780 | Not-determined |
| SS_extracted_k141_24958 | Not-determined |
| SS_extracted_k141_134625 | Not-determined |
| SS_extracted_k141_140784 | Low-quality |
| SS_extracted_k141_79581 | Not-determined |
| SS_extracted_k141_55581 | Not-determined |
| SS_extracted_k141_50044 | Low-quality |
| SS_extracted_k141_79585 | Not-determined |
| SS_extracted_k141_44230 | Not-determined |
| SS_extracted_k141_79587 | Low-quality |
| SS_extracted_k141_44232 | Not-determined |
| SS_extracted_k141_79588 | Not-determined |
| SS_extracted_k141_1775 | Not-determined |
| SS_extracted_k141_140795 | Not-determined |
| SS_extracted_k141_55583 | Not-determined |
| SS_extracted_k141_134642 | Not-determined |
| SS_extracted_k141_79592 | Not-determined |
| SS_extracted_k141_86344 | Not-determined |
| SS_extracted_k141_110836 | Not-determined |
| SS_extracted_k141_86345 | Low-quality |
| SS_extracted_k141_37887 | Not-determined |
| SS_extracted_k141_86346 | Not-determined |
| SS_extracted_k141_128739 | Not-determined |
| SS_extracted_k141_32223 | Low-quality |
| SS_extracted_k141_44244 | Not-determined |
| SS_extracted_k141_104742 | Not-determined |
| SS_extracted_k141_140800 | Not-determined |
| SS_extracted_k141_79602 | Not-determined |
| SS_extracted_k141_140803 | Not-determined |
| SS_extracted_k141_86359 | Not-determined |
| SS_extracted_k141_104747 | Not-determined |
| SS_extracted_k141_110843 | Not-determined |
| SS_extracted_k141_1794 | Not-determined |
| SS_extracted_k141_123503 | Not-determined |
| SS_extracted_k141_24963 | Low-quality |
| SS_extracted_k141_123505 | Not-determined |
| SS_extracted_k141_24965 | Low-quality |
| SS_extracted_k141_13497 | Not-determined |
| SS_extracted_k141_62010 | Not-determined |
| SS_extracted_k141_50087 | Not-determined |
| SS_extracted_k141_86362 | Not-determined |
| SS_extracted_k141_73949 | Not-determined |
| SS_extracted_k141_13500 | Not-determined |
| SS_extracted_k141_32248 | Not-determined |
| SS_extracted_k141_13505 | Not-determined |
| SS_extracted_k141_62022 | Low-quality |
| SS_extracted_k141_140806 | Low-quality |
| SS_extracted_k141_50097 | Not-determined |
| SS_extracted_k141_13510 | Not-determined |
| SS_extracted_k141_37903 | Not-determined |
| SS_extracted_k141_37905 | Not-determined |
| SS_extracted_k141_24994 | Not-determined |
| SS_extracted_k141_44277 | Low-quality |
| SS_extracted_k141_44282 | Not-determined |
| SS_extracted_k141_44290 | Not-determined |
| SS_extracted_k141_92789 | Not-determined |
| SS_extracted_k141_50112 | Low-quality |
| SS_extracted_k141_86397 | Not-determined |
| SS_extracted_k141_86398 | Not-determined |
| SS_extracted_k141_92792 | Not-determined |
| SS_extracted_k141_140831 | Not-determined |
| SS_extracted_k141_13528 | Not-determined |
| SS_extracted_k141_1821 | Not-determined |
| SS_extracted_k141_128753 | Not-determined |
| SS_extracted_k141_92797 | Not-determined |
| SS_extracted_k141_123534 | Not-determined |
| SS_extracted_k141_25021 | Not-determined |
| SS_extracted_k141_128757 | Not-determined |
| SS_extracted_k141_92798 | Low-quality |
| SS_extracted_k141_1832 | Not-determined |
| SS_extracted_k141_1833 | Not-determined |
| SS_extracted_k141_37921 | Not-determined |
| SS_extracted_k141_37923 | Low-quality |
| SS_extracted_k141_92802 | Not-determined |
| SS_extracted_k141_86406 | Not-determined |
| SS_extracted_k141_92812 | Not-determined |
| SS_extracted_k141_32290 | Not-determined |
| SS_extracted_k141_37931 | Not-determined |
| SS_extracted_k141_50142 | Low-quality |
| SS_extracted_k141_128781 | Low-quality |
| SS_extracted_k141_86408 | Low-quality |
| SS_extracted_k141_128788 | Not-determined |
| SS_extracted_k141_62058 | Low-quality |
| SS_extracted_k141_128795 | Not-determined |
| SS_extracted_k141_62066 | Not-determined |
| SS_extracted_k141_128798 | Not-determined |
| SS_extracted_k141_92829 | Not-determined |
| SS_extracted_k141_134659 | Not-determined |
| SS_extracted_k141_92833 | Not-determined |
| SS_extracted_k141_13573 | Not-determined |
| SS_extracted_k141_62071 | Not-determined |
| SS_extracted_k141_7665 | Not-determined |
| SS_extracted_k141_44320 | Not-determined |
| SS_extracted_k141_37954 | Not-determined |
| SS_extracted_k141_44321 | Low-quality |
| SS_extracted_k141_37961 | Not-determined |
| SS_extracted_k141_13584 | Low-quality |
| SS_extracted_k141_37968 | Not-determined |
| SS_extracted_k141_128817 | Low-quality |
| SS_extracted_k141_128818 | Not-determined |
| SS_extracted_k141_25037 | Not-determined |
| SS_extracted_k141_98136 | Not-determined |
| SS_extracted_k141_32324 | Not-determined |
| SS_extracted_k141_32330 | Not-determined |
| SS_extracted_k141_128825 | Not-determined |
| SS_extracted_k141_128827 | Not-determined |
| SS_extracted_k141_37976 | Not-determined |
| SS_extracted_k141_86430 | Not-determined |
| SS_extracted_k141_37984 | Not-determined |
| SS_extracted_k141_7690 | Not-determined |
| SS_extracted_k141_7692 | Low-quality |
| SS_extracted_k141_86433 | Not-determined |
| SS_extracted_k141_98143 | Not-determined |
| SS_extracted_k141_1865 | Low-quality |
| SS_extracted_k141_50146 | Not-determined |
| SS_extracted_k141_98147 | Low-quality |
| SS_extracted_k141_1870 | Not-determined |
| SS_extracted_k141_1872 | Low-quality |
| SS_extracted_k141_1876 | Low-quality |
| SS_extracted_k141_104752 | Not-determined |
| SS_extracted_k141_50153 | Low-quality |
| SS_extracted_k141_68244 | Not-determined |
| SS_extracted_k141_117323 | Not-determined |
| SS_extracted_k141_32358 | Low-quality |
| SS_extracted_k141_1879 | Not-determined |
| SS_extracted_k141_117326 | Low-quality |
| SS_extracted_k141_25060 | Not-determined |
| SS_extracted_k141_86459 | Not-determined |
| SS_extracted_k141_37991 | Low-quality |
| SS_extracted_k141_68253 | Not-determined |
| SS_extracted_k141_32364 | Not-determined |
| SS_extracted_k141_37996 | Not-determined |
| SS_extracted_k141_13639 | Not-determined |
| SS_extracted_k141_50161 | Not-determined |
| SS_extracted_k141_79621 | Not-determined |
| SS_extracted_k141_25078 | Not-determined |
| SS_extracted_k141_62103 | Low-quality |
| SS_extracted_k141_44340 | Not-determined |
| SS_extracted_k141_19712 | Not-determined |
| SS_extracted_k141_86473 | Low-quality |
| SS_extracted_k141_62111 | Low-quality |
| SS_extracted_k141_79627 | Not-determined |
| SS_extracted_k141_86476 | Not-determined |
| SS_extracted_k141_123566 | Low-quality |
| SS_extracted_k141_117369 | Low-quality |
| SS_extracted_k141_1914 | Not-determined |
| SS_extracted_k141_44355 | Not-determined |
| SS_extracted_k141_25089 | Not-determined |
| SS_extracted_k141_44361 | Not-determined |
| S16_vs_59697\|\|full | High-quality |
| SS_extracted_k141_25093 | Not-determined |
| SS_extracted_k141_62122 | Not-determined |
| SS_extracted_k141_62123 | Not-determined |
| SS_extracted_k141_1926 | Low-quality |
| SS_extracted_k141_62125 | Low-quality |
| SS_extracted_k141_117383 | Not-determined |
| SS_extracted_k141_62129 | Not-determined |
| SS_extracted_k141_1936 | Low-quality |
| SS_extracted_k141_25102 | Low-quality |
| SS_extracted_k141_19721 | Not-determined |
| SS_extracted_k141_25103 | Not-determined |
| SS_extracted_k141_44373 | Low-quality |
| SS_extracted_k141_62138 | Not-determined |
| SS_extracted_k141_44375 | Low-quality |
| SS_extracted_k141_32384 | Low-quality |
| SS_extracted_k141_44380 | Not-determined |
| SS_extracted_k141_117402 | Low-quality |
| SS_extracted_k141_98175 | Not-determined |
| SS_extracted_k141_73998 | Not-determined |
| SS_extracted_k141_32392 | Not-determined |
| SS_extracted_k141_134689 | Not-determined |
| SS_extracted_k141_44388 | Not-determined |
| SS_extracted_k141_44389 | Not-determined |
| SS_extracted_k141_68259 | Not-determined |
| SS_extracted_k141_98182 | Not-determined |
| SS_extracted_k141_98183 | Not-determined |
| SS_extracted_k141_68262 | Not-determined |
| SS_extracted_k141_68264 | Low-quality |
| SS_extracted_k141_98188 | Not-determined |
| SS_extracted_k141_62160 | Not-determined |
| SS_extracted_k141_68266 | Not-determined |
| SS_extracted_k141_117415 | Not-determined |
| SS_extracted_k141_86488 | Not-determined |
| SS_extracted_k141_86489 | Not-determined |
| SS_extracted_k141_98195 | Low-quality |
| SS_extracted_k141_50170 | Not-determined |
| SS_extracted_k141_92846 | Not-determined |
| SS_extracted_k141_62168 | Not-determined |
| SS_extracted_k141_50175 | Not-determined |
| SS_extracted_k141_86498 | Low-quality |
| SS_extracted_k141_50176 | Not-determined |
| SS_extracted_k141_7729 | Not-determined |
| SS_extracted_k141_50178 | Not-determined |
| SS_extracted_k141_7731 | Not-determined |
| SS_extracted_k141_110857 | Not-determined |
| SS_extracted_k141_7734 | Not-determined |
| SS_extracted_k141_44406 | Not-determined |
| SS_extracted_k141_86505 | Not-determined |
| SS_extracted_k141_68278 | Not-determined |
| SS_extracted_k141_25120 | Not-determined |
| SS_extracted_k141_92855 | Not-determined |
| SS_extracted_k141_110875 | Not-determined |
| SS_extracted_k141_74006 | Not-determined |
| SS_extracted_k141_1964 | Not-determined |
| SS_extracted_k141_44422 | Low-quality |
| SS_extracted_k141_92857 | Low-quality |
| SS_extracted_k141_7747 | Not-determined |
| SS_extracted_k141_110878 | Low-quality |
| SS_extracted_k141_134703 | Not-determined |
| SS_extracted_k141_62196 | Low-quality |
| SS_extracted_k141_104813 | Low-quality |
| SS_extracted_k141_79654 | Not-determined |
| SS_extracted_k141_50207 | Not-determined |
| SS_extracted_k141_55622 | Not-determined |
| SS_extracted_k141_79656 | Not-determined |
| SS_extracted_k141_50209 | Low-quality |
| SS_extracted_k141_62201 | Not-determined |
| SS_extracted_k141_104817 | Low-quality |
| SS_extracted_k141_7761 | Not-determined |
| SS_extracted_k141_55632 | Not-determined |
| SS_extracted_k141_55634 | Not-determined |
| SS_extracted_k141_13686 | Not-determined |
| SS_extracted_k141_25137 | Not-determined |
| SS_extracted_k141_25138 | Low-quality |
| SS_extracted_k141_110891 | Low-quality |
| SS_extracted_k141_13689 | Low-quality |
| SS_extracted_k141_44437 | Not-determined |
| SS_extracted_k141_134715 | Low-quality |
| SS_extracted_k141_134717 | Low-quality |
| SS_extracted_k141_134725 | Low-quality |
| SS_extracted_k141_44450 | Not-determined |
| SS_extracted_k141_32430 | Low-quality |
| SS_extracted_k141_50232 | Not-determined |
| SS_extracted_k141_104842 | Low-quality |
| SS_extracted_k141_32436 | Not-determined |
| SS_extracted_k141_110896 | Low-quality |
| SS_extracted_k141_32443 | Low-quality |
| SS_extracted_k141_110899 | Not-determined |
| SS_extracted_k141_110901 | Not-determined |
| SS_extracted_k141_104859 | Not-determined |
| SS_extracted_k141_140868 | Low-quality |
| SS_extracted_k141_13716 | Not-determined |
| SS_extracted_k141_86521 | Low-quality |
| SS_extracted_k141_110905 | Low-quality |
| SS_extracted_k141_55648 | Not-determined |
| SS_extracted_k141_55649 | Not-determined |
| SS_extracted_k141_110909 | Not-determined |
| SS_extracted_k141_62218 | Low-quality |
| SS_extracted_k141_25147 | Not-determined |
| SS_extracted_k141_117432 | Not-determined |
| SS_extracted_k141_134742 | Not-determined |
| SS_extracted_k141_13731 | Not-determined |
| SS_extracted_k141_38067 | Not-determined |
| SS_extracted_k141_13743 | Not-determined |
| SS_extracted_k141_134762 | Not-determined |
| SS_extracted_k141_86541 | Not-determined |
| SS_extracted_k141_110935 | Not-determined |
| SS_extracted_k141_38076 | Not-determined |
| SS_extracted_k141_7788 | Not-determined |
| SS_extracted_k141_134774 | Not-determined |
| SS_extracted_k141_38086 | Not-determined |
| SS_extracted_k141_134778 | Not-determined |
| SS_extracted_k141_117450 | Not-determined |
| SS_extracted_k141_7807 | Low-quality |
| SS_extracted_k141_86551 | Not-determined |
| SS_extracted_k141_104888 | Not-determined |
| SS_extracted_k141_86553 | Not-determined |
| SS_extracted_k141_7812 | Not-determined |
| SS_extracted_k141_7813 | Not-determined |
| SS_extracted_k141_110960 | Not-determined |
| SS_extracted_k141_50269 | Not-determined |
| SS_extracted_k141_13767 | Not-determined |
| SS_extracted_k141_110963 | Low-quality |
| SS_extracted_k141_117457 | Low-quality |
| SS_extracted_k141_92861 | Not-determined |
| SS_extracted_k141_19737 | Not-determined |
| SS_extracted_k141_50273 | Low-quality |
| SS_extracted_k141_92866 | Not-determined |
| SS_extracted_k141_110970 | Low-quality |
| SS_extracted_k141_92867 | Not-determined |
| SS_extracted_k141_110971 | Not-determined |
| SS_extracted_k141_55676 | Not-determined |
| SS_extracted_k141_104896 | Not-determined |
| SS_extracted_k141_92870 | Not-determined |
| SS_extracted_k141_110972 | Low-quality |
| SS_extracted_k141_13772 | Not-determined |
| SS_extracted_k141_50279 | Not-determined |
| SS_extracted_k141_55685 | Not-determined |
| SS_extracted_k141_128877 | Not-determined |
| SS_extracted_k141_50282 | Low-quality |
| SS_extracted_k141_140924 | Not-determined |
| SS_extracted_k141_19751 | Not-determined |
| SS_extracted_k141_79671 | Not-determined |
| SS_extracted_k141_140931 | Low-quality |
| SS_extracted_k141_92879 | Low-quality |
| SS_extracted_k141_92881 | Low-quality |
| SS_extracted_k141_79679 | Not-determined |
| SS_extracted_k141_13792 | Not-determined |
| SS_extracted_k141_13795 | Low-quality |
| SS_extracted_k141_68296 | Low-quality |
| SS_extracted_k141_19766 | Not-determined |
| SS_extracted_k141_117467 | Not-determined |
| SS_extracted_k141_117468 | Low-quality |
| SS_extracted_k141_117469 | Not-determined |
| SS_extracted_k141_44477 | Low-quality |
| SS_extracted_k141_13802 | Low-quality |
| SS_extracted_k141_50298 | Not-determined |
| SS_extracted_k141_79691 | Low-quality |
| SS_extracted_k141_50303 | Not-determined |
| SS_extracted_k141_13808 | Low-quality |
| SS_extracted_k141_79699 | Not-determined |
| SS_extracted_k141_110992 | Not-determined |
| SS_extracted_k141_110993 | Low-quality |
| SS_extracted_k141_110996 | Not-determined |
| SS_extracted_k141_79702 | Not-determined |
| SS_extracted_k141_19778 | Not-determined |
| SS_extracted_k141_19779 | Not-determined |
| SS_extracted_k141_104918 | Not-determined |
| SS_extracted_k141_134800 | Not-determined |
| SS_extracted_k141_7846 | Not-determined |
| SS_extracted_k141_7847 | Not-determined |
| SS_extracted_k141_50314 | Low-quality |
| SS_extracted_k141_32475 | Not-determined |
| S9_vs_k141_55738\|\|full | Medium-quality |
| SS_extracted_k141_117489 | Not-determined |
| SS_extracted_k141_55691 | Low-quality |
| SS_extracted_k141_50316 | Not-determined |
| SS_extracted_k141_7860 | Low-quality |
| SS_extracted_k141_50317 | Not-determined |
| SS_extracted_k141_92887 | Not-determined |
| SS_extracted_k141_44480 | Not-determined |
| SS_extracted_k141_92890 | Not-determined |
| SS_extracted_k141_117495 | Not-determined |
| SS_extracted_k141_13826 | Low-quality |
| SS_extracted_k141_92894 | Not-determined |
| SS_extracted_k141_104932 | Low-quality |
| SS_extracted_k141_128897 | Not-determined |
| SS_extracted_k141_140939 | Not-determined |
| SS_extracted_k141_140940 | Not-determined |
| SS_extracted_k141_44488 | Not-determined |
| SS_extracted_k141_32492 | Not-determined |
| SS_extracted_k141_55705 | Not-determined |
| SS_extracted_k141_140942 | Low-quality |
| SS_extracted_k141_134816 | Not-determined |
| SS_extracted_k141_140944 | Not-determined |
| SS_extracted_k141_55707 | Not-determined |
| SS_extracted_k141_86574 | Not-determined |
| SS_extracted_k141_92906 | Low-quality |
| SS_extracted_k141_134818 | Not-determined |
| SS_extracted_k141_104939 | Not-determined |
| SS_extracted_k141_2025 | Not-determined |
| SS_extracted_k141_19792 | Not-determined |
| SS_extracted_k141_68319 | Not-determined |
| SS_extracted_k141_117506 | Low-quality |
| SS_extracted_k141_19797 | Not-determined |
| SS_extracted_k141_44496 | Not-determined |
| SS_extracted_k141_92914 | Not-determined |
| SS_extracted_k141_50345 | Not-determined |
| SS_extracted_k141_50346 | Not-determined |
| SS_extracted_k141_104957 | Not-determined |
| SS_extracted_k141_68336 | Not-determined |
| SS_extracted_k141_2035 | Not-determined |

| | |
|---|---|
| SS_extracted_k141_104962 | Not-determined |
| SS_extracted_k141_111014 | Not-determined |
| SS_extracted_k141_134836 | Not-determined |
| SS_extracted_k141_86588 | Not-determined |
| SS_extracted_k141_2041 | Not-determined |
| SS_extracted_k141_19808 | Low-quality |
| SS_extracted_k141_92929 | Low-quality |
| SS_extracted_k141_104967 | Not-determined |
| SS_extracted_k141_2048 | Low-quality |
| SS_extracted_k141_92937 | Not-determined |
| SS_extracted_k141_86590 | Not-determined |
| SS_extracted_k141_117523 | Not-determined |
| SS_extracted_k141_128935 | Not-determined |
| SS_extracted_k141_2054 | Not-determined |
| SS_extracted_k141_79721 | Low-quality |
| SS_extracted_k141_86594 | Not-determined |
| SS_extracted_k141_117530 | Not-determined |
| SS_extracted_k141_19809 | Low-quality |
| SS_extracted_k141_134847 | Not-determined |
| SS_extracted_k141_7900 | Not-determined |
| SS_extracted_k141_92948 | Not-determined |
| SS_extracted_k141_134850 | Not-determined |
| SS_extracted_k141_86605 | Not-determined |
| SS_extracted_k141_134853 | Low-quality |
| SS_extracted_k141_134854 | Not-determined |
| SS_extracted_k141_92951 | Not-determined |
| SS_extracted_k141_134859 | Not-determined |
| SS_extracted_k141_86610 | Not-determined |
| SS_extracted_k141_79728 | Not-determined |
| SS_extracted_k141_92954 | Low-quality |
| SS_extracted_k141_128950 | Not-determined |
| SS_extracted_k141_140950 | Not-determined |
| SS_extracted_k141_2074 | Low-quality |
| SS_extracted_k141_134867 | Not-determined |
| SS_extracted_k141_2077 | Not-determined |
| SS_extracted_k141_111028 | Not-determined |
| SS_extracted_k141_134869 | Not-determined |
| SS_extracted_k141_2083 | Low-quality |
| SS_extracted_k141_92958 | Not-determined |
| SS_extracted_k141_128956 | Not-determined |
| SS_extracted_k141_68347 | Not-determined |
| SS_extracted_k141_111034 | Not-determined |
| SS_extracted_k141_128965 | Low-quality |
| SS_extracted_k141_55719 | Not-determined |
| SS_extracted_k141_2093 | Not-determined |
| SS_extracted_k141_50365 | Not-determined |
| SS_extracted_k141_92968 | Not-determined |
| SS_extracted_k141_86640 | Not-determined |
| SS_extracted_k141_86641 | Not-determined |
| SS_extracted_k141_2109 | Not-determined |
| SS_extracted_k141_117550 | Low-quality |
| SS_extracted_k141_68358 | Low-quality |
| SS_extracted_k141_92977 | Not-determined |
| SS_extracted_k141_111048 | Not-determined |
| SS_extracted_k141_111049 | Not-determined |
| SS_extracted_k141_117556 | Low-quality |
| SS_extracted_k141_128981 | Not-determined |
| SS_extracted_k141_19814 | Not-determined |
| SS_extracted_k141_128984 | Not-determined |
| SS_extracted_k141_55736 | Not-determined |
| SS_extracted_k141_19820 | Low-quality |
| SS_extracted_k141_50379 | Not-determined |
| SS_extracted_k141_19824 | Not-determined |
| SS_extracted_k141_55738 | Not-determined |
| SS_extracted_k141_111062 | Low-quality |
| SS_extracted_k141_19825 | Not-determined |
| SS_extracted_k141_2134 | Not-determined |
| SS_extracted_k141_25202 | Not-determined |
| SS_extracted_k141_19827 | Not-determined |
| SS_extracted_k141_128988 | Low-quality |
| SS_extracted_k141_2138 | Low-quality |
| SS_extracted_k141_32500 | Not-determined |
| SS_extracted_k141_128997 | Not-determined |
| SS_extracted_k141_68384 | Not-determined |
| SS_extracted_k141_86657 | Low-quality |
| SS_extracted_k141_62221 | Not-determined |
| SS_extracted_k141_50395 | Not-determined |
| SS_extracted_k141_50398 | Not-determined |
| SS_extracted_k141_111068 | Low-quality |
| SS_extracted_k141_86659 | Not-determined |
| SS_extracted_k141_86660 | Not-determined |
| SS_extracted_k141_32506 | Not-determined |
| SS_extracted_k141_129001 | Not-determined |
| SS_extracted_k141_19838 | Low-quality |
| SS_extracted_k141_68390 | Not-determined |
| SS_extracted_k141_2159 | Low-quality |
| SS_extracted_k141_2160 | Not-determined |
| SS_extracted_k141_111073 | Low-quality |
| SS_extracted_k141_117566 | Low-quality |
| SS_extracted_k141_55751 | Not-determined |
| SS_extracted_k141_19842 | Not-determined |
| SS_extracted_k141_111075 | Not-determined |
| SS_extracted_k141_111078 | Not-determined |
| SS_extracted_k141_123594 | Not-determined |
| SS_extracted_k141_123597 | Not-determined |
| SS_extracted_k141_86663 | Not-determined |
| SS_extracted_k141_68404 | Not-determined |
| SS_extracted_k141_19858 | Low-quality |
| SS_extracted_k141_7985 | Low-quality |
| SS_extracted_k141_111095 | Not-determined |
| SS_extracted_k141_123613 | Not-determined |
| SS_extracted_k141_117582 | Low-quality |
| SS_extracted_k141_7987 | Not-determined |
| SS_extracted_k141_55759 | Not-determined |
| SS_extracted_k141_93003 | Low-quality |
| SS_extracted_k141_111107 | Low-quality |
| SS_extracted_k141_93007 | Not-determined |
| SS_extracted_k141_19874 | Not-determined |
| SS_extracted_k141_86685 | Not-determined |
| SS_extracted_k141_25244 | Low-quality |
| SS_extracted_k141_7995 | Low-quality |
| SS_extracted_k141_111117 | Low-quality |
| SS_extracted_k141_7997 | Not-determined |
| SS_extracted_k141_68420 | Not-determined |
| SS_extracted_k141_50414 | Not-determined |
| SS_extracted_k141_50415 | Low-quality |
| SS_extracted_k141_68422 | Not-determined |
| SS_extracted_k141_25253 | Not-determined |
| SS_extracted_k141_13843 | Not-determined |
| SS_extracted_k141_123632 | Not-determined |
| SS_extracted_k141_55775 | Not-determined |
| SS_extracted_k141_50424 | Not-determined |
| SS_extracted_k141_111130 | Not-determined |
| SS_extracted_k141_8021 | Not-determined |
| SS_extracted_k141_50427 | Low-quality |
| SS_extracted_k141_104979 | Low-quality |
| SS_extracted_k141_19889 | Not-determined |
| SS_extracted_k141_25264 | Not-determined |
| SS_extracted_k141_19898 | Not-determined |
| SS_extracted_k141_32529 | Not-determined |
| SS_extracted_k141_79759 | Low-quality |
| SS_extracted_k141_32532 | Not-determined |
| SS_extracted_k141_38107 | Not-determined |
| SS_extracted_k141_117613 | Not-determined |
| SS_extracted_k141_123651 | Low-quality |
| SS_extracted_k141_123652 | Not-determined |
| SS_extracted_k141_79761 | Low-quality |
| SS_extracted_k141_50433 | Not-determined |
| SS_extracted_k141_123655 | Not-determined |
| SS_extracted_k141_117622 | Not-determined |
| SS_extracted_k141_123661 | Not-determined |
| SS_extracted_k141_19914 | Low-quality |
| SS_extracted_k141_13854 | Not-determined |
| SS_extracted_k141_105002 | Low-quality |
| SS_extracted_k141_38124 | Not-determined |
| SS_extracted_k141_105005 | Not-determined |
| SS_extracted_k141_38130 | Not-determined |
| SS_extracted_k141_44515 | Not-determined |
| SS_extracted_k141_13869 | Not-determined |
| SS_extracted_k141_111141 | Not-determined |
| SS_extracted_k141_8035 | Not-determined |
| SS_extracted_k141_8036 | Not-determined |
| SS_extracted_k141_19924 | Not-determined |
| SS_extracted_k141_68444 | Low-quality |
| SS_extracted_k141_8040 | Not-determined |
| SS_extracted_k141_19930 | Not-determined |
| SS_extracted_k141_25284 | Not-determined |
| SS_extracted_k141_123670 | Not-determined |
| SS_extracted_k141_123675 | Not-determined |
| SS_extracted_k141_62242 | Low-quality |
| SS_extracted_k141_62244 | Low-quality |
| SS_extracted_k141_62245 | Not-determined |
| SS_extracted_k141_105019 | Not-determined |
| SS_extracted_k141_32540 | Not-determined |
| SS_extracted_k141_129042 | Low-quality |
| SS_extracted_k141_134913 | Not-determined |
| SS_extracted_k141_140975 | Low-quality |
| SS_extracted_k141_129047 | Not-determined |
| SS_extracted_k141_105023 | Not-determined |
| SS_extracted_k141_134916 | Not-determined |
| SS_extracted_k141_50438 | Not-determined |
| SS_extracted_k141_129057 | Low-quality |
| SS_extracted_k141_105031 | Not-determined |
| SS_extracted_k141_86726 | Not-determined |
| SS_extracted_k141_140978 | Not-determined |
| SS_extracted_k141_50444 | Not-determined |
| SS_extracted_k141_44534 | Not-determined |
| SS_extracted_k141_55797 | Not-determined |
| SS_extracted_k141_62257 | Not-determined |
| SS_extracted_k141_105043 | Not-determined |
| SS_extracted_k141_123683 | Not-determined |
| SS_extracted_k141_117632 | Not-determined |
| SS_extracted_k141_129061 | Low-quality |
| SS_extracted_k141_105048 | Low-quality |
| SS_extracted_k141_105050 | Not-determined |
| SS_extracted_k141_8044 | Low-quality |
| SS_extracted_k141_117637 | Not-determined |
| SS_extracted_k141_140991 | Not-determined |
| SS_extracted_k141_19944 | Not-determined |
| SS_extracted_k141_111147 | Not-determined |
| SS_extracted_k141_129066 | Not-determined |
| SS_extracted_k141_8050 | Low-quality |
| SS_extracted_k141_2243 | Not-determined |
| SS_extracted_k141_79780 | Not-determined |
| SS_extracted_k141_117647 | Not-determined |
| SS_extracted_k141_50467 | Not-determined |
| SS_extracted_k141_50470 | Not-determined |
| SS_extracted_k141_117652 | Not-determined |
| SS_extracted_k141_50472 | Not-determined |
| SS_extracted_k141_44550 | Not-determined |
| SS_extracted_k141_32551 | Low-quality |
| SS_extracted_k141_129088 | Low-quality |
| SS_extracted_k141_123697 | Low-quality |
| SS_extracted_k141_129090 | Low-quality |
| SS_extracted_k141_44556 | Not-determined |
| SS_extracted_k141_111162 | Low-quality |
| SS_extracted_k141_2264 | Not-determined |
| SS_extracted_k141_55822 | Not-determined |
| SS_extracted_k141_8061 | Not-determined |
| SS_extracted_k141_123703 | Not-determined |
| SS_extracted_k141_2268 | Not-determined |
| SS_extracted_k141_55825 | Not-determined |
| SS_extracted_k141_62284 | Not-determined |
| SS_extracted_k141_105058 | Not-determined |
| SS_extracted_k141_111173 | Not-determined |
| SS_extracted_k141_93015 | Not-determined |
| SS_extracted_k141_13915 | Not-determined |
| SS_extracted_k141_2275 | Not-determined |
| SS_extracted_k141_129113 | Not-determined |
| SS_extracted_k141_8064 | Low-quality |
| SS_extracted_k141_32568 | Not-determined |
| SS_extracted_k141_55843 | Low-quality |
| SS_extracted_k141_105071 | Not-determined |
| SS_extracted_k141_55847 | Not-determined |
| SS_extracted_k141_55848 | Low-quality |
| SS_extracted_k141_62298 | Low-quality |
| SS_extracted_k141_129121 | Not-determined |
| SS_extracted_k141_62301 | Not-determined |
| SS_extracted_k141_129123 | Low-quality |
| SS_extracted_k141_55851 | Not-determined |
| SS_extracted_k141_129125 | Not-determined |
| SS_extracted_k141_44572 | Not-determined |
| SS_extracted_k141_117689 | Not-determined |
| SS_extracted_k141_117690 | Not-determined |
| SS_extracted_k141_13932 | Not-determined |
| SS_extracted_k141_50484 | Not-determined |
| SS_extracted_k141_2303 | Not-determined |
| SS_extracted_k141_117696 | Not-determined |
| SS_extracted_k141_44579 | Not-determined |
| SS_extracted_k141_79802 | Not-determined |
| SS_extracted_k141_44582 | Low-quality |
| SS_extracted_k141_129132 | Not-determined |
| SS_extracted_k141_129133 | Not-determined |
| SS_extracted_k141_105085 | Not-determined |
| SS_extracted_k141_74053 | Not-determined |
| SS_extracted_k141_50496 | Not-determined |
| SS_extracted_k141_2311 | Not-determined |
| SS_extracted_k141_50502 | Not-determined |
| SS_extracted_k141_93021 | Not-determined |
| SS_extracted_k141_13945 | Not-determined |
| SS_extracted_k141_74057 | Not-determined |
| SS_extracted_k141_2319 | Not-determined |
| SS_extracted_k141_93025 | Not-determined |
| SS_extracted_k141_74064 | Not-determined |
| SS_extracted_k141_8100 | Not-determined |
| SS_extracted_k141_44601 | Not-determined |
| SS_extracted_k141_74070 | Low-quality |
| SS_extracted_k141_8104 | Not-determined |
| SS_extracted_k141_141048 | Not-determined |
| SS_extracted_k141_129158 | Not-determined |
| SS_extracted_k141_129165 | Not-determined |
| SS_extracted_k141_8110 | Low-quality |
| SS_extracted_k141_55865 | Not-determined |
| SS_extracted_k141_44617 | Not-determined |
| SS_extracted_k141_55868 | Not-determined |
| SS_extracted_k141_141056 | Not-determined |
| SS_extracted_k141_8113 | Not-determined |
| SS_extracted_k141_8114 | Not-determined |
| SS_extracted_k141_8115 | Not-determined |
| SS_extracted_k141_25308 | Not-determined |
| SS_extracted_k141_79826 | Not-determined |
| SS_extracted_k141_2344 | Not-determined |
| SS_extracted_k141_55878 | Not-determined |
| SS_extracted_k141_79828 | Low-quality |
| SS_extracted_k141_141076 | Not-determined |
| SS_extracted_k141_105113 | Low-quality |
| SS_extracted_k141_129188 | Not-determined |
| SS_extracted_k141_38144 | Not-determined |
| SS_extracted_k141_117733 | Not-determined |
| SS_extracted_k141_2349 | Not-determined |
| SS_extracted_k141_79835 | Not-determined |
| SS_extracted_k141_38148 | Not-determined |
| SS_extracted_k141_129195 | Not-determined |
| SS_extracted_k141_79839 | Not-determined |
| SS_extracted_k141_79841 | Not-determined |
| SS_extracted_k141_68453 | Not-determined |
| SS_extracted_k141_129202 | Not-determined |
| SS_extracted_k141_141092 | Not-determined |
| SS_extracted_k141_123725 | Not-determined |
| SS_extracted_k141_50508 | Not-determined |
| SS_extracted_k141_2364 | Not-determined |
| SS_extracted_k141_38162 | Not-determined |
| SS_extracted_k141_123730 | Low-quality |
| SS_extracted_k141_105137 | Not-determined |
| SS_extracted_k141_141098 | Low-quality |
| SS_extracted_k141_141099 | Not-determined |
| SS_extracted_k141_50519 | Not-determined |
| SS_extracted_k141_98247 | Low-quality |
| SS_extracted_k141_79842 | Not-determined |
| SS_extracted_k141_55892 | Not-determined |
| SS_extracted_k141_129208 | Not-determined |
| SS_extracted_k141_50522 | Not-determined |
| SS_extracted_k141_117755 | Not-determined |
| SS_extracted_k141_50523 | Low-quality |
| SS_extracted_k141_2378 | Not-determined |
| SS_extracted_k141_105159 | Not-determined |
| SS_extracted_k141_105162 | Not-determined |
| SS_extracted_k141_2382 | Low-quality |
| SS_extracted_k141_105165 | Not-determined |
| SS_extracted_k141_134921 | Not-determined |
| SS_extracted_k141_79862 | Not-determined |
| SS_extracted_k141_2387 | Not-determined |
| SS_extracted_k141_123758 | Not-determined |
| SS_extracted_k141_68478 | Not-determined |
| SS_extracted_k141_68480 | Low-quality |
| SS_extracted_k141_25322 | Low-quality |
| SS_extracted_k141_105176 | Not-determined |
| SS_extracted_k141_68484 | Low-quality |
| SS_extracted_k141_8135 | Not-determined |
| SS_extracted_k141_105180 | Not-determined |
| SS_extracted_k141_8142 | Not-determined |
| SS_extracted_k141_8147 | Not-determined |
| SS_extracted_k141_2401 | Not-determined |
| SS_extracted_k141_123764 | Not-determined |
| SS_extracted_k141_44640 | Not-determined |
| SS_extracted_k141_86756 | Low-quality |
| SS_extracted_k141_19977 | Not-determined |
| SS_extracted_k141_50553 | Not-determined |
| SS_extracted_k141_50554 | Not-determined |
| SS_extracted_k141_2406 | Not-determined |
| SS_extracted_k141_44642 | Low-quality |
| SS_extracted_k141_19982 | Not-determined |
| SS_extracted_k141_93039 | Not-determined |
| SS_extracted_k141_2410 | Not-determined |
| SS_extracted_k141_105202 | Not-determined |
| SS_extracted_k141_105203 | Not-determined |
| SS_extracted_k141_105206 | Not-determined |
| SS_extracted_k141_44659 | Not-determined |
| SS_extracted_k141_68499 | Not-determined |
| SS_extracted_k141_123778 | Not-determined |
| SS_extracted_k141_79881 | Not-determined |
| SS_extracted_k141_2417 | Low-quality |
| SS_extracted_k141_134942 | Not-determined |
| SS_extracted_k141_44661 | Not-determined |
| SS_extracted_k141_2418 | Not-determined |
| SS_extracted_k141_32580 | Low-quality |
| SS_extracted_k141_62317 | Not-determined |
| SS_extracted_k141_8168 | Not-determined |
| SS_extracted_k141_8170 | Not-determined |
| SS_extracted_k141_20002 | Low-quality |
| SS_extracted_k141_134947 | Not-determined |
| SS_extracted_k141_134950 | Low-quality |
| SS_extracted_k141_62324 | Not-determined |
| SS_extracted_k141_8183 | Not-determined |
| SS_extracted_k141_62331 | Low-quality |
| SS_extracted_k141_32598 | Not-determined |
| SS_extracted_k141_98275 | Not-determined |
| SS_extracted_k141_62335 | Not-determined |
| SS_extracted_k141_62342 | Not-determined |
| SS_extracted_k141_8200 | Not-determined |
| SS_extracted_k141_93089 | Not-determined |
| SS_extracted_k141_14024 | Low-quality |
| SS_extracted_k141_68528 | Low-quality |
| SS_extracted_k141_86788 | Not-determined |
| SS_extracted_k141_74082 | Not-determined |
| SS_extracted_k141_74083 | Not-determined |
| SS_extracted_k141_44689 | Not-determined |
| SS_extracted_k141_32606 | Low-quality |
| SS_extracted_k141_50582 | Not-determined |
| SS_extracted_k141_14035 | Low-quality |
| SS_extracted_k141_105265 | Low-quality |
| SS_extracted_k141_62351 | Not-determined |
| SS_extracted_k141_105274 | Not-determined |
| SS_extracted_k141_98291 | Low-quality |
| SS_extracted_k141_32623 | Not-determined |
| SS_extracted_k141_93101 | Low-quality |
| SS_extracted_k141_93102 | Not-determined |
| SS_extracted_k141_32627 | Low-quality |
| SS_extracted_k141_50600 | Not-determined |
| SS_extracted_k141_123821 | Low-quality |
| SS_extracted_k141_68532 | Low-quality |
| SS_extracted_k141_2447 | Not-determined |
| SS_extracted_k141_50603 | Low-quality |
| SS_extracted_k141_105296 | Low-quality |
| SS_extracted_k141_20014 | Not-determined |
| SS_extracted_k141_20016 | Not-determined |
| SS_extracted_k141_2453 | Not-determined |
| SS_extracted_k141_93113 | Not-determined |
| SS_extracted_k141_105300 | Not-determined |
| SS_extracted_k141_44725 | Not-determined |
| SS_extracted_k141_32645 | Not-determined |
| SS_extracted_k141_105303 | Low-quality |
| SS_extracted_k141_14073 | Not-determined |
| SS_extracted_k141_44730 | Not-determined |
| SS_extracted_k141_117806 | Low-quality |
| SS_extracted_k141_93118 | Low-quality |
| SS_extracted_k141_79910 | Low-quality |
| SS_extracted_k141_20025 | Not-determined |
| SS_extracted_k141_44733 | Not-determined |
| SS_extracted_k141_20027 | Not-determined |
| SS_extracted_k141_105309 | Low-quality |
| SS_extracted_k141_44740 | Not-determined |
| SS_extracted_k141_117815 | Not-determined |
| SS_extracted_k141_105315 | Not-determined |
| SS_extracted_k141_93125 | Not-determined |
| SS_extracted_k141_117817 | Not-determined |
| SS_extracted_k141_44745 | Low-quality |
| SS_extracted_k141_117820 | Not-determined |
| SS_extracted_k141_68549 | Low-quality |
| SS_extracted_k141_50628 | Not-determined |
| SS_extracted_k141_105323 | Low-quality |
| SS_extracted_k141_8215 | Not-determined |
| SS_extracted_k141_50632 | Not-determined |
| SS_extracted_k141_129233 | Not-determined |
| SS_extracted_k141_129236 | Not-determined |
| SS_extracted_k141_38207 | Not-determined |
| SS_extracted_k141_93141 | Not-determined |
| SS_extracted_k141_93142 | Not-determined |
| SS_extracted_k141_123845 | Not-determined |
| SS_extracted_k141_50650 | Low-quality |
| SS_extracted_k141_25340 | Not-determined |
| SS_extracted_k141_123846 | Not-determined |
| SS_extracted_k141_105341 | Not-determined |
| SS_extracted_k141_55904 | Not-determined |
| SS_extracted_k141_2485 | Low-quality |
| SS_extracted_k141_32678 | Not-determined |
| SS_extracted_k141_32680 | Not-determined |
| SS_extracted_k141_25348 | Low-quality |
| SS_extracted_k141_68566 | Low-quality |
| SS_extracted_k141_32685 | Not-determined |
| SS_extracted_k141_98310 | Low-quality |
| SS_extracted_k141_38216 | Not-determined |
| SS_extracted_k141_74118 | Low-quality |
| SS_extracted_k141_98311 | Low-quality |
| SS_extracted_k141_105351 | Not-determined |
| SS_extracted_k141_20057 | Not-determined |
| SS_extracted_k141_2490 | Not-determined |
| SS_extracted_k141_93158 | Not-determined |
| SS_extracted_k141_105360 | Not-determined |
| SS_extracted_k141_117867 | Not-determined |
| SS_extracted_k141_38222 | Not-determined |
| SS_extracted_k141_2498 | Low-quality |
| SS_extracted_k141_32700 | Not-determined |
| SS_extracted_k141_14106 | Not-determined |
| SS_extracted_k141_117877 | Not-determined |
| SS_extracted_k141_38231 | Low-quality |
| SS_extracted_k141_105375 | Low-quality |
| SS_extracted_k141_123875 | Not-determined |
| SS_extracted_k141_2507 | Low-quality |
| SS_extracted_k141_74123 | Not-determined |
| SS_extracted_k141_129270 | Low-quality |
| SS_extracted_k141_129271 | Not-determined |
| SS_extracted_k141_32714 | Not-determined |
| SS_extracted_k141_123878 | Not-determined |
| SS_extracted_k141_38240 | Low-quality |
| SS_extracted_k141_38241 | Not-determined |
| SS_extracted_k141_129274 | Not-determined |
| SS_extracted_k141_74132 | Not-determined |
| SS_extracted_k141_117894 | Not-determined |
| SS_extracted_k141_93184 | Low-quality |
| SS_extracted_k141_129279 | Not-determined |
| SS_vs_k141_21869\|\|full | High-quality |
| SS_extracted_k141_44747 | Not-determined |
| SS_extracted_k141_141129 | Not-determined |
| SS_extracted_k141_44752 | Not-determined |
| SS_extracted_k141_141134 | Not-determined |
| SS_extracted_k141_14130 | Not-determined |
| SS_extracted_k141_74147 | Not-determined |
| SS_extracted_k141_20076 | Low-quality |
| SS_extracted_k141_105404 | Not-determined |
| SS_extracted_k141_141147 | Not-determined |
| SS_extracted_k141_134981 | Not-determined |
| SS_extracted_k141_134982 | Low-quality |
| SS_extracted_k141_8222 | Not-determined |
| SS_extracted_k141_38266 | Not-determined |
| SS_extracted_k141_123902 | Not-determined |
| SS_extracted_k141_141155 | Not-determined |
| SS_extracted_k141_74151 | Low-quality |
| SS_extracted_k141_129284 | Not-determined |
| SS_extracted_k141_8244 | Low-quality |
| SS_extracted_k141_14153 | Not-determined |
| SS_extracted_k141_38270 | Not-determined |
| SS_extracted_k141_129294 | Low-quality |
| SS_extracted_k141_141158 | Not-determined |
| SS_extracted_k141_141159 | Not-determined |
| SS_extracted_k141_14158 | Not-determined |
| SS_extracted_k141_14160 | Not-determined |
| SS_extracted_k141_25390 | Not-determined |
| SS_extracted_k141_50696 | Low-quality |
| SS_extracted_k141_50697 | Not-determined |
| SS_extracted_k141_44773 | Not-determined |
| SS_extracted_k141_105417 | Not-determined |
| SS_extracted_k141_50699 | Low-quality |
| SS_extracted_k141_141171 | Not-determined |
| SS_extracted_k141_105419 | Not-determined |
| SS_extracted_k141_105420 | Not-determined |
| SS_extracted_k141_129303 | Not-determined |
| SS_extracted_k141_117902 | Not-determined |
| SS_extracted_k141_62379 | Not-determined |
| SS_extracted_k141_62380 | Low-quality |
| SS_extracted_k141_86834 | Not-determined |
| SS_extracted_k141_20102 | Not-determined |
| SS_extracted_k141_55950 | Not-determined |
| SS_extracted_k141_50716 | Low-quality |
| SS_extracted_k141_62388 | Not-determined |
| SS_extracted_k141_44785 | Not-determined |
| SS_extracted_k141_55957 | Not-determined |
| SS_extracted_k141_55958 | Low-quality |
| SS_extracted_k141_55960 | Not-determined |
| SS_extracted_k141_141199 | Not-determined |
| SS_extracted_k141_117921 | Low-quality |
| SS_extracted_k141_55962 | Not-determined |
| SS_extracted_k141_105424 | Not-determined |
| SS_extracted_k141_25400 | Low-quality |
| SS_extracted_k141_20121 | Low-quality |
| SS_extracted_k141_105427 | Not-determined |
| SS_extracted_k141_62399 | Not-determined |
| SS_extracted_k141_44795 | Not-determined |
| SS_extracted_k141_20124 | Low-quality |
| SS_extracted_k141_20127 | Low-quality |
| SS_extracted_k141_134999 | Not-determined |
| SS_extracted_k141_44801 | Low-quality |
| SS_extracted_k141_135008 | Low-quality |
| SS_extracted_k141_141211 | Not-determined |
| SS_extracted_k141_105445 | Not-determined |
| SS_extracted_k141_32750 | Low-quality |
| SS_extracted_k141_129334 | Not-determined |
| SS_extracted_k141_32755 | Not-determined |
| SS_extracted_k141_117932 | Not-determined |
| SS_extracted_k141_32762 | Low-quality |
| SS_extracted_k141_141224 | Low-quality |
| SS_extracted_k141_141225 | Low-quality |
| SS_extracted_k141_135012 | Not-determined |
| SS_extracted_k141_20144 | Low-quality |
| SS_extracted_k141_117941 | Not-determined |
| SS_extracted_k141_20148 | Not-determined |
| SS_extracted_k141_8302 | Not-determined |
| SS_extracted_k141_62420 | Not-determined |
| SS_extracted_k141_14195 | Not-determined |
| SS_extracted_k141_14201 | Not-determined |
| SS_extracted_k141_14202 | Not-determined |
| SS_extracted_k141_74194 | Not-determined |
| SS_extracted_k141_8308 | Low-quality |
| SS_extracted_k141_62431 | Not-determined |
| SS_extracted_k141_105462 | Not-determined |
| SS_extracted_k141_141232 | Low-quality |
| SS_extracted_k141_62434 | Not-determined |
| SS_extracted_k141_32774 | Low-quality |
| SS_extracted_k141_105469 | Not-determined |
| SS_extracted_k141_55974 | Not-determined |
| SS_extracted_k141_2537 | Not-determined |
| SS_extracted_k141_123952 | Not-determined |
| SS_extracted_k141_93189 | Not-determined |
| SS_extracted_k141_14214 | Low-quality |
| SS_extracted_k141_2542 | Not-determined |
| SS_extracted_k141_141244 | Not-determined |
| SS_extracted_k141_105478 | Not-determined |
| SS_extracted_k141_111203 | Low-quality |
| SS_extracted_k141_86854 | Low-quality |
| SS_extracted_k141_117959 | Not-determined |
| SS_extracted_k141_117960 | Not-determined |
| SS_extracted_k141_111205 | Not-determined |
| SS_extracted_k141_111209 | Not-determined |
| SS_extracted_k141_111213 | Not-determined |
| SS_extracted_k141_55983 | Not-determined |
| SS_extracted_k141_141252 | Not-determined |
| SS_extracted_k141_14228 | Not-determined |
| SS_extracted_k141_14232 | Not-determined |
| SS_extracted_k141_14233 | Not-determined |
| SS_extracted_k141_62451 | Not-determined |
| SS_extracted_k141_111217 | Low-quality |
| SS_extracted_k141_2558 | Low-quality |
| SS_extracted_k141_32795 | Not-determined |
| SS_extracted_k141_141266 | Not-determined |
| SS_extracted_k141_111219 | Not-determined |
| SS_extracted_k141_32799 | Not-determined |
| SS_extracted_k141_25419 | Not-determined |
| SS_extracted_k141_117979 | Not-determined |
| SS_extracted_k141_32801 | Not-determined |
| SS_extracted_k141_129363 | Not-determined |
| SS_extracted_k141_117983 | Not-determined |
| SS_extracted_k141_8341 | Not-determined |
| SS_extracted_k141_93206 | Not-determined |
| SS_extracted_k141_105503 | Not-determined |
| SS_extracted_k141_14239 | Not-determined |
| SS_extracted_k141_117987 | Not-determined |
| SS_extracted_k141_105507 | Low-quality |
| SS_extracted_k141_105509 | Low-quality |
| SS_extracted_k141_32808 | Not-determined |

| | |
|---|---|
| SS_extracted_k141_93213 | Not-determined |
| SS_extracted_k141_105516 | Not-determined |
| SS_extracted_k141_141281 | Low-quality |
| SS_extracted_k141_2571 | Not-determined |
| SS_extracted_k141_129385 | Not-determined |
| SS_extracted_k141_8344 | Low-quality |
| SS_extracted_k141_111235 | Not-determined |
| SS_extracted_k141_93227 | Not-determined |
| SS_extracted_k141_68614 | Not-determined |
| SS_extracted_k141_8354 | Not-determined |
| SS_extracted_k141_141296 | Not-determined |
| SS_extracted_k141_8356 | Not-determined |
| SS_extracted_k141_105523 | Not-determined |
| SS_extracted_k141_105524 | Not-determined |
| SS_extracted_k141_111242 | Not-determined |
| SS_extracted_k141_111243 | Not-determined |
| SS_extracted_k141_141300 | Not-determined |
| SS_extracted_k141_135036 | Not-determined |
| SS_extracted_k141_68621 | Not-determined |
| SS_extracted_k141_79931 | Low-quality |
| SS_extracted_k141_68622 | Not-determined |
| SS_extracted_k141_50744 | Not-determined |
| SS_extracted_k141_111245 | Not-determined |
| SS_extracted_k141_135043 | Not-determined |
| SS_extracted_k141_129398 | Low-quality |
| SS_extracted_k141_129399 | Low-quality |
| SS_extracted_k141_79935 | Low-quality |
| SS_extracted_k141_8372 | Not-determined |
| SS_extracted_k141_50747 | Not-determined |
| SS_extracted_k141_105545 | Not-determined |
| SS_extracted_k141_129409 | Low-quality |
| SS_extracted_k141_141315 | Not-determined |
| SS_extracted_k141_105548 | Low-quality |
| SS_extracted_k141_8379 | Not-determined |
| SS_extracted_k141_14283 | Not-determined |
| SS_extracted_k141_62454 | Not-determined |
| SS_extracted_k141_86896 | Not-determined |
| SS_extracted_k141_2594 | Not-determined |
| SS_extracted_k141_2599 | Not-determined |
| SS_extracted_k141_86898 | Not-determined |
| SS_extracted_k141_105565 | Not-determined |
| SS_extracted_k141_111255 | Not-determined |
| SS_extracted_k141_129418 | Not-determined |
| SS_extracted_k141_105566 | Low-quality |
| SS_extracted_k141_129425 | Not-determined |
| SS_extracted_k141_129426 | Not-determined |
| SS_extracted_k141_50759 | Not-determined |
| S6_vs_k141_7268\|\|full | High-quality |
| SS_extracted_k141_135065 | Low-quality |
| SS_extracted_k141_25424 | Low-quality |
| SS_extracted_k141_93261 | Not-determined |
| SS_extracted_k141_118054 | Not-determined |
| SS_extracted_k141_14311 | Not-determined |
| SS_extracted_k141_14313 | Not-determined |
| SS_extracted_k141_86911 | Not-determined |
| SS_extracted_k141_118058 | Not-determined |
| SS_extracted_k141_14318 | Not-determined |
| SS_extracted_k141_14321 | Not-determined |
| SS_extracted_k141_105589 | Low-quality |
| SS_extracted_k141_86917 | Not-determined |
| SS_vs_k141_130294\|\|full | High-quality |
| SS_extracted_k141_111265 | Not-determined |
| SS_extracted_k141_25437 | Not-determined |
| SS_extracted_k141_68631 | Not-determined |
| SS_extracted_k141_50772 | Not-determined |
| SS_extracted_k141_129437 | Low-quality |
| SS_extracted_k141_8396 | Low-quality |
| SS_extracted_k141_68636 | Low-quality |
| SS_extracted_k141_25447 | Not-determined |
| SS_extracted_k141_25452 | Not-determined |
| SS_extracted_k141_8403 | Low-quality |
| SS_extracted_k141_68637 | Not-determined |
| SS_extracted_k141_118077 | Not-determined |
| SS_extracted_k141_111284 | Not-determined |
| SS_extracted_k141_25459 | Not-determined |
| SS_extracted_k141_62477 | Not-determined |
| SS_extracted_k141_111289 | Not-determined |
| SS_extracted_k141_111290 | Not-determined |
| SS_extracted_k141_68651 | Not-determined |
| SS_extracted_k141_62483 | Not-determined |
| SS_extracted_k141_105628 | Not-determined |
| SS_extracted_k141_124011 | Not-determined |
| SS_extracted_k141_25483 | Not-determined |
| SS_extracted_k141_118087 | Not-determined |
| SS_extracted_k141_25488 | Not-determined |
| SS_extracted_k141_25489 | Low-quality |
| SS_extracted_k141_50798 | Not-determined |
| SS_extracted_k141_118097 | Not-determined |
| SS_extracted_k141_105643 | Not-determined |
| SS_extracted_k141_8422 | Not-determined |
| SS_extracted_k141_79972 | Not-determined |
| SS_extracted_k141_20193 | Not-determined |
| SS_extracted_k141_50813 | Not-determined |
| SS_extracted_k141_111316 | Low-quality |
| SS_extracted_k141_105652 | Not-determined |
| SS_extracted_k141_111318 | Low-quality |
| SS_extracted_k141_135073 | Not-determined |
| SS_extracted_k141_8432 | Not-determined |
| SS_extracted_k141_25505 | Not-determined |
| SS_extracted_k141_124034 | Not-determined |
| SS_extracted_k141_86929 | Not-determined |
| SS_extracted_k141_20206 | Low-quality |
| SS_extracted_k141_50814 | Low-quality |
| SS_extracted_k141_86932 | Not-determined |
| SS_extracted_k141_8439 | Not-determined |
| SS_extracted_k141_8444 | Not-determined |
| SS_extracted_k141_105663 | Not-determined |
| SS_extracted_k141_105666 | Not-determined |
| SS_extracted_k141_20214 | Not-determined |
| SS_extracted_k141_20215 | Not-determined |
| SS_extracted_k141_111335 | Low-quality |
| SS_extracted_k141_50824 | Not-determined |
| SS_extracted_k141_56008 | Not-determined |
| SS_extracted_k141_105667 | Low-quality |
| SS_extracted_k141_56009 | Not-determined |
| SS_extracted_k141_79985 | Not-determined |
| SS_extracted_k141_20222 | Low-quality |
| SS_extracted_k141_118127 | Not-determined |
| SS_extracted_k141_118128 | Low-quality |
| SS_extracted_k141_20225 | Not-determined |
| SS_extracted_k141_8463 | Not-determined |
| SS_extracted_k141_8467 | Not-determined |
| SS_extracted_k141_141339 | Not-determined |
| SS_extracted_k141_141341 | Not-determined |
| SS_extracted_k141_20236 | Not-determined |
| SS_extracted_k141_25523 | Not-determined |
| SS_extracted_k141_56012 | Low-quality |
| SS_extracted_k141_50854 | Not-determined |
| SS_extracted_k141_56014 | Not-determined |
| SS_extracted_k141_124053 | Not-determined |
| SS_extracted_k141_141359 | Not-determined |
| SS_extracted_k141_50858 | Not-determined |
| SS_extracted_k141_50863 | Not-determined |
| SS_extracted_k141_62493 | Not-determined |
| SS_extracted_k141_105703 | Not-determined |
| SS_extracted_k141_129448 | Not-determined |
| SS_extracted_k141_20253 | Not-determined |
| SS_extracted_k141_2633 | Not-determined |
| SS_extracted_k141_50867 | Not-determined |
| SS_extracted_k141_129451 | Low-quality |
| SS_extracted_k141_2647 | Low-quality |
| SS_extracted_k141_93285 | Not-determined |
| SS_extracted_k141_50878 | Not-determined |
| SS_extracted_k141_38304 | Not-determined |
| SS_extracted_k141_141374 | Not-determined |
| SS_extracted_k141_2652 | Low-quality |
| SS_extracted_k141_50883 | Low-quality |
| SS_extracted_k141_50888 | Not-determined |
| SS_extracted_k141_141382 | Not-determined |
| SS_extracted_k141_25532 | Not-determined |
| SS_extracted_k141_124075 | Not-determined |
| SS_extracted_k141_105719 | Not-determined |
| SS_extracted_k141_2668 | Not-determined |
| SS_extracted_k141_124084 | Not-determined |
| SS_extracted_k141_38318 | Not-determined |
| SS_extracted_k141_118137 | Not-determined |
| SS_extracted_k141_93304 | Low-quality |
| SS_extracted_k141_86983 | Not-determined |
| SS_extracted_k141_86984 | Not-determined |
| SS_extracted_k141_98332 | Not-determined |
| SS_extracted_k141_93315 | Not-determined |
| SS_extracted_k141_93318 | Not-determined |
| SS_extracted_k141_20271 | Not-determined |
| SS_extracted_k141_50910 | Not-determined |
| SS_extracted_k141_38323 | Not-determined |
| SS_extracted_k141_86999 | Not-determined |
| SS_extracted_k141_87000 | Not-determined |
| SS_extracted_k141_25560 | Not-determined |
| SS_extracted_k141_68668 | Not-determined |
| SS_extracted_k141_2683 | Not-determined |
| SS_extracted_k141_98350 | Not-determined |
| SS_extracted_k141_98351 | Not-determined |
| SS_extracted_k141_50926 | Not-determined |
| SS_extracted_k141_50930 | Not-determined |
| SS_extracted_k141_50932 | Not-determined |
| SS_extracted_k141_2696 | Low-quality |
| SS_extracted_k141_38332 | Not-determined |
| SS_extracted_k141_135080 | Not-determined |
| SS_extracted_k141_50938 | Not-determined |
| SS_extracted_k141_38335 | Not-determined |
| SS_extracted_k141_118160 | Not-determined |
| S11_vs_k141_6458\|\|full | Medium-quality |
| SS_extracted_k141_2708 | Not-determined |
| SS_extracted_k141_2710 | Low-quality |
| SS_extracted_k141_2711 | Low-quality |
| SS_extracted_k141_105737 | Not-determined |
| SS_extracted_k141_129467 | Not-determined |
| SS_extracted_k141_105742 | Not-determined |
| SS_extracted_k141_25591 | Not-determined |
| SS_extracted_k141_98362 | Not-determined |
| SS_extracted_k141_98370 | Not-determined |
| SS_extracted_k141_25601 | Low-quality |
| SS_extracted_k141_25602 | Not-determined |
| SS_extracted_k141_2733 | Not-determined |
| SS_extracted_k141_50955 | Not-determined |
| SS_extracted_k141_14356 | Not-determined |
| SS_extracted_k141_20293 | Not-determined |
| SS_extracted_k141_50962 | Low-quality |
| SS_extracted_k141_2737 | Not-determined |
| SS_extracted_k141_14361 | Not-determined |
| SS_extracted_k141_141398 | Not-determined |
| SS_extracted_k141_20297 | Low-quality |
| SS_extracted_k141_38349 | Low-quality |
| SS_extracted_k141_135105 | Not-determined |
| SS_extracted_k141_20300 | Low-quality |
| SS_extracted_k141_2747 | Not-determined |
| SS_extracted_k141_44829 | Not-determined |
| SS_extracted_k141_2748 | Not-determined |
| SS_extracted_k141_135115 | Not-determined |
| SS_extracted_k141_44830 | Low-quality |
| SS_extracted_k141_20306 | Not-determined |
| SS_extracted_k141_50981 | Not-determined |
| SS_extracted_k141_68680 | Not-determined |
| SS_extracted_k141_25636 | Not-determined |
| SS_extracted_k141_68688 | Not-determined |
| SS_extracted_k141_14377 | Not-determined |
| SS_extracted_k141_32834 | Low-quality |
| SS_extracted_k141_25638 | Low-quality |
| SS_extracted_k141_2760 | Not-determined |
| SS_extracted_k141_141410 | Not-determined |
| SS_extracted_k141_14382 | Not-determined |
| SS_extracted_k141_20308 | Not-determined |
| SS_extracted_k141_141413 | Not-determined |
| SS_extracted_k141_20310 | Not-determined |
| SS_extracted_k141_68693 | Low-quality |
| SS_extracted_k141_44844 | Low-quality |
| SS_extracted_k141_38378 | Low-quality |
| SS_extracted_k141_141420 | Not-determined |
| SS_extracted_k141_44847 | Low-quality |
| SS_extracted_k141_50996 | Low-quality |
| SS_extracted_k141_111357 | Not-determined |
| SS_extracted_k141_124133 | Not-determined |
| SS_extracted_k141_129483 | Low-quality |
| SS_extracted_k141_44857 | Not-determined |
| SS_extracted_k141_20321 | Low-quality |
| SS_extracted_k141_141436 | Not-determined |
| SS_extracted_k141_129487 | Not-determined |
| SS_extracted_k141_129493 | Not-determined |
| SS_extracted_k141_14394 | Not-determined |
| SS_extracted_k141_25662 | Not-determined |
| SS_extracted_k141_25665 | Not-determined |
| SS_extracted_k141_129498 | Not-determined |
| SS_extracted_k141_14401 | Not-determined |
| SS_extracted_k141_68719 | Not-determined |
| SS_extracted_k141_129504 | Not-determined |
| SS_extracted_k141_20330 | Not-determined |
| SS_extracted_k141_20331 | Not-determined |
| SS_extracted_k141_68726 | Not-determined |
| SS_extracted_k141_25672 | Not-determined |
| SS_extracted_k141_68734 | Not-determined |
| SS_extracted_k141_129523 | Not-determined |
| SS_extracted_k141_129525 | Not-determined |
| SS_extracted_k141_129529 | Not-determined |
| SS_extracted_k141_129533 | Not-determined |
| SS_extracted_k141_62534 | Not-determined |
| SS_extracted_k141_62535 | Not-determined |
| SS_extracted_k141_135142 | Not-determined |
| SS_extracted_k141_62538 | Not-determined |
| SS_extracted_k141_25688 | Not-determined |
| SS_extracted_k141_111381 | Not-determined |
| SS_extracted_k141_135151 | Low-quality |
| SS_extracted_k141_124178 | Not-determined |
| SS_extracted_k141_68750 | Not-determined |
| SS_extracted_k141_62543 | Not-determined |
| SS_extracted_k141_111388 | Not-determined |
| SS_extracted_k141_135158 | Not-determined |
| SS_extracted_k141_135160 | Not-determined |
| SS_extracted_k141_135161 | Not-determined |
| SS_extracted_k141_14416 | Not-determined |
| SS_extracted_k141_14420 | Not-determined |
| SS_extracted_k141_129561 | Not-determined |
| SS_extracted_k141_111400 | Not-determined |
| SS_extracted_k141_2792 | Not-determined |
| SS_extracted_k141_14421 | Not-determined |
| SS_extracted_k141_105787 | Low-quality |
| SS_extracted_k141_2798 | Not-determined |
| SS_extracted_k141_124187 | Not-determined |
| SS_extracted_k141_124191 | Not-determined |
| SS_extracted_k141_105795 | Not-determined |
| SS_extracted_k141_135168 | Not-determined |
| SS_extracted_k141_111415 | Not-determined |
| SS_extracted_k141_135171 | Not-determined |
| SS_extracted_k141_124201 | Not-determined |
| SS_extracted_k141_25725 | Low-quality |
| SS_extracted_k141_98406 | Not-determined |
| SS_extracted_k141_98409 | Not-determined |
| SS_extracted_k141_129567 | Low-quality |
| SS_extracted_k141_93330 | Not-determined |
| SS_extracted_k141_51017 | Low-quality |
| SS_extracted_k141_93334 | Not-determined |
| SS_extracted_k141_20344 | Low-quality |
| SS_extracted_k141_25734 | Low-quality |
| SS_extracted_k141_93338 | Low-quality |
| SS_extracted_k141_93340 | Not-determined |
| SS_extracted_k141_25746 | Not-determined |
| SS_extracted_k141_93346 | Not-determined |
| SS_extracted_k141_20353 | Low-quality |
| SS_extracted_k141_56031 | Not-determined |
| SS_extracted_k141_2828 | Not-determined |
| SS_extracted_k141_62589 | Not-determined |
| SS_extracted_k141_105822 | Low-quality |
| SS_extracted_k141_38404 | Not-determined |
| SS_extracted_k141_124221 | Low-quality |
| SS_extracted_k141_105826 | Not-determined |
| SS_extracted_k141_111437 | Not-determined |
| SS_extracted_k141_111439 | Low-quality |
| SS_extracted_k141_56037 | Not-determined |
| SS_extracted_k141_68772 | Low-quality |
| SS_extracted_k141_68773 | Not-determined |
| SS_extracted_k141_93365 | Low-quality |
| SS_extracted_k141_98426 | Low-quality |
| SS_extracted_k141_68776 | Low-quality |
| SS_extracted_k141_98430 | Not-determined |
| SS_extracted_k141_51046 | Low-quality |
| SS_extracted_k141_32877 | Not-determined |
| SS_extracted_k141_105845 | Not-determined |
| SS_extracted_k141_51052 | Low-quality |
| SS_extracted_k141_93379 | Not-determined |
| SS_extracted_k141_98437 | Low-quality |
| SS_extracted_k141_80073 | Not-determined |
| SS_extracted_k141_38437 | Not-determined |
| SS_extracted_k141_38439 | Not-determined |
| SS_extracted_k141_51063 | Not-determined |
| SS_extracted_k141_51066 | Low-quality |
| SS_extracted_k141_2857 | Not-determined |
| SS_extracted_k141_105861 | Low-quality |
| SS_extracted_k141_38453 | Not-determined |
| SS_extracted_k141_105862 | Not-determined |
| SS_extracted_k141_93389 | Not-determined |
| SS_extracted_k141_129606 | Not-determined |
| SS_extracted_k141_38457 | Not-determined |
| SS_extracted_k141_38460 | Not-determined |
| SS_extracted_k141_93396 | Not-determined |
| SS_extracted_k141_129612 | Not-determined |
| SS_extracted_k141_135188 | Low-quality |
| SS_extracted_k141_68789 | Not-determined |
| SS_extracted_k141_51075 | Not-determined |
| SS_extracted_k141_68791 | Low-quality |
| SS_extracted_k141_8515 | Low-quality |
| SS_extracted_k141_68792 | Not-determined |
| SS_extracted_k141_2871 | Not-determined |
| SS_extracted_k141_8517 | Low-quality |
| SS_extracted_k141_98455 | Low-quality |
| SS_extracted_k141_20393 | Not-determined |
| SS_extracted_k141_68796 | Not-determined |
| SS_extracted_k141_105878 | Not-determined |
| SS_extracted_k141_98458 | Not-determined |
| SS_extracted_k141_118179 | Low-quality |
| SS_extracted_k141_87011 | Not-determined |
| SS_extracted_k141_129630 | Not-determined |
| SS_extracted_k141_93405 | Not-determined |
| SS_extracted_k141_38482 | Not-determined |
| SS_extracted_k141_80081 | Not-determined |
| SS_extracted_k141_93410 | Not-determined |
| SS_extracted_k141_20411 | Not-determined |
| SS_extracted_k141_93413 | Not-determined |
| SS_extracted_k141_20412 | Low-quality |
| SS_extracted_k141_56039 | Low-quality |
| SS_extracted_k141_98464 | Not-determined |
| SS_extracted_k141_93424 | Not-determined |
| SS_extracted_k141_56046 | Not-determined |
| SS_extracted_k141_87039 | Low-quality |
| SS_extracted_k141_8532 | Low-quality |
| SS_extracted_k141_68807 | Not-determined |
| SS_extracted_k141_135200 | Low-quality |
| SS_extracted_k141_51092 | Not-determined |
| SS_extracted_k141_20429 | Not-determined |
| SS_extracted_k141_32891 | Low-quality |
| SS_extracted_k141_135205 | Not-determined |
| SS_extracted_k141_2881 | Not-determined |
| SS_extracted_k141_93432 | Not-determined |
| SS_extracted_k141_111458 | Not-determined |
| SS_extracted_k141_80091 | Not-determined |
| SS_extracted_k141_93437 | Low-quality |
| SS_extracted_k141_2886 | Not-determined |
| SS_extracted_k141_51114 | Not-determined |
| SS_extracted_k141_93443 | Not-determined |
| SS_extracted_k141_129669 | Not-determined |
| SS_extracted_k141_80100 | Not-determined |
| SS_extracted_k141_135209 | Not-determined |
| SS_extracted_k141_51119 | Low-quality |
| SS_extracted_k141_124254 | Not-determined |
| SS_extracted_k141_111466 | Low-quality |
| SS_extracted_k141_141452 | Not-determined |
| SS_extracted_k141_68846 | Not-determined |
| SS_extracted_k141_32901 | Not-determined |
| SS_extracted_k141_68848 | Not-determined |
| SS_extracted_k141_32903 | Not-determined |
| SS_extracted_k141_111470 | Not-determined |
| SS_extracted_k141_51127 | Low-quality |
| SS_extracted_k141_80104 | Not-determined |
| SS_extracted_k141_141459 | Not-determined |
| SS_extracted_k141_118207 | Low-quality |
| SS_extracted_k141_93478 | Not-determined |
| SS_extracted_k141_74211 | Not-determined |
| SS_extracted_k141_74212 | Not-determined |
| SS_extracted_k141_68858 | Not-determined |
| SS_extracted_k141_62607 | Low-quality |
| SS_extracted_k141_93494 | Not-determined |
| SS_extracted_k141_93495 | Not-determined |
| SS_extracted_k141_93496 | Not-determined |
| SS_extracted_k141_118219 | Not-determined |
| SS_extracted_k141_93504 | Not-determined |
| SS_extracted_k141_62613 | Not-determined |
| SS_extracted_k141_135221 | Not-determined |
| SS_extracted_k141_124285 | Low-quality |
| SS_extracted_k141_20450 | Not-determined |
| SS_extracted_k141_14453 | Not-determined |
| SS_extracted_k141_14455 | Not-determined |
| SS_extracted_k141_56074 | Not-determined |
| SS_extracted_k141_124294 | Not-determined |
| SS_extracted_k141_20454 | Not-determined |
| SS_extracted_k141_14460 | Not-determined |
| SS_extracted_k141_62615 | Low-quality |
| SS_extracted_k141_129673 | Not-determined |
| SS_extracted_k141_124299 | Not-determined |
| SS_extracted_k141_129675 | Not-determined |
| SS_extracted_k141_14465 | Not-determined |
| SS_extracted_k141_124302 | Not-determined |
| SS_extracted_k141_118231 | Not-determined |
| SS_extracted_k141_93520 | Not-determined |
| SS_extracted_k141_87056 | Low-quality |
| SS_extracted_k141_62622 | Low-quality |
| SS_extracted_k141_118237 | Not-determined |
| SS_extracted_k141_51129 | Not-determined |
| SS_extracted_k141_68888 | Low-quality |
| SS_extracted_k141_2935 | Not-determined |
| SS_extracted_k141_62628 | Not-determined |
| SS_extracted_k141_51130 | Low-quality |
| SS_extracted_k141_68891 | Not-determined |
| SS_extracted_k141_68893 | Not-determined |
| SS_extracted_k141_8603 | Not-determined |
| SS_extracted_k141_20466 | Not-determined |
| SS_extracted_k141_2947 | Not-determined |
| SS_extracted_k141_14473 | Not-determined |
| SS_extracted_k141_74250 | Not-determined |
| SS_extracted_k141_25780 | Not-determined |
| SS_extracted_k141_74261 | Not-determined |
| SS_extracted_k141_80125 | Not-determined |
| SS_extracted_k141_14487 | Not-determined |
| SS_extracted_k141_124333 | Not-determined |
| SS_extracted_k141_8630 | Not-determined |
| SS_extracted_k141_80135 | Not-determined |
| SS_extracted_k141_68902 | Not-determined |
| SS_extracted_k141_93566 | Not-determined |
| SS_extracted_k141_93567 | Not-determined |
| SS_extracted_k141_8641 | Not-determined |
| SS_extracted_k141_14500 | Not-determined |
| SS_extracted_k141_51144 | Low-quality |
| SS_extracted_k141_124346 | Not-determined |
| SS_extracted_k141_51145 | Low-quality |
| SS_extracted_k141_80141 | Low-quality |
| SS_extracted_k141_2976 | Not-determined |
| SS_extracted_k141_14506 | Low-quality |
| SS_extracted_k141_25790 | Low-quality |
| SS_extracted_k141_105886 | Low-quality |
| SS_extracted_k141_51151 | Not-determined |
| SS_extracted_k141_51157 | Not-determined |
| SS_extracted_k141_38489 | Low-quality |
| SS_extracted_k141_51160 | Not-determined |
| SS_extracted_k141_141484 | Not-determined |
| SS_extracted_k141_25792 | Not-determined |
| SS_extracted_k141_141486 | Low-quality |
| SS_extracted_k141_141487 | Not-determined |
| SS_extracted_k141_56086 | Low-quality |
| SS_extracted_k141_74285 | Not-determined |
| SS_extracted_k141_8666 | Not-determined |
| SS_extracted_k141_74290 | Not-determined |
| SS_extracted_k141_20499 | Not-determined |
| SS_extracted_k141_93584 | Not-determined |
| SS_extracted_k141_51169 | Not-determined |
| SS_extracted_k141_80146 | Not-determined |
| SS_extracted_k141_38505 | Not-determined |
| SS_extracted_k141_68929 | Not-determined |
| SS_extracted_k141_51174 | Not-determined |
| SS_extracted_k141_105922 | Not-determined |
| SS_extracted_k141_14527 | Not-determined |
| SS_extracted_k141_68935 | Low-quality |
| SS_extracted_k141_68936 | Low-quality |
| SS_extracted_k141_51180 | Low-quality |
| SS_extracted_k141_68941 | Low-quality |
| SS_extracted_k141_135253 | Not-determined |
| SS_extracted_k141_93599 | Not-determined |
| SS_extracted_k141_44864 | Not-determined |
| SS_extracted_k141_8675 | Not-determined |
| SS_extracted_k141_80154 | Not-determined |
| SS_extracted_k141_93609 | Not-determined |
| SS_extracted_k141_38525 | Low-quality |
| SS_extracted_k141_56091 | Not-determined |
| SS_extracted_k141_3013 | Not-determined |
| SS_extracted_k141_93615 | Not-determined |
| SS_extracted_k141_68946 | Not-determined |
| SS_extracted_k141_3026 | Not-determined |
| SS_extracted_k141_3028 | Low-quality |
| SS_extracted_k141_68950 | Not-determined |
| SS_extracted_k141_14546 | Not-determined |
| SS_extracted_k141_129726 | Low-quality |
| SS_extracted_k141_8694 | Not-determined |
| SS_extracted_k141_44869 | Not-determined |
| SS_extracted_k141_129733 | Not-determined |
| SS_extracted_k141_14555 | Low-quality |
| SS_extracted_k141_135262 | Not-determined |
| SS_extracted_k141_135263 | Not-determined |
| SS_extracted_k141_129737 | Not-determined |
| SS_extracted_k141_129738 | Low-quality |
| SS_extracted_k141_14560 | Low-quality |
| SS_extracted_k141_93625 | Low-quality |
| SS_extracted_k141_93627 | Not-determined |
| SS_extracted_k141_93630 | Not-determined |
| SS_extracted_k141_56115 | Not-determined |
| SS_extracted_k141_38547 | Not-determined |
| SS_extracted_k141_56121 | Not-determined |
| SS_extracted_k141_93636 | Low-quality |
| SS_extracted_k141_124387 | Not-determined |
| SS_extracted_k141_93637 | Not-determined |
| SS_extracted_k141_56123 | Not-determined |
| SS_extracted_k141_56124 | Not-determined |
| SS_extracted_k141_129757 | Not-determined |
| SS_extracted_k141_38552 | Not-determined |
| SS_extracted_k141_141507 | Not-determined |
| SS_extracted_k141_44891 | Not-determined |
| SS_extracted_k141_129760 | Low-quality |
| SS_extracted_k141_3046 | Not-determined |
| SS_extracted_k141_56132 | Low-quality |
| SS_extracted_k141_141512 | Not-determined |
| SS_extracted_k141_74316 | Low-quality |
| SS_extracted_k141_98471 | Not-determined |
| SS_extracted_k141_135265 | Not-determined |
| SS_extracted_k141_129765 | Not-determined |
| SS_extracted_k141_25819 | Not-determined |
| SS_extracted_k141_118325 | Not-determined |
| SS_extracted_k141_25822 | Low-quality |
| SS_extracted_k141_118329 | Not-determined |
| SS_extracted_k141_62688 | Low-quality |
| SS_extracted_k141_3070 | Low-quality |
| SS_extracted_k141_3071 | Not-determined |
| SS_extracted_k141_38556 | Not-determined |
| SS_extracted_k141_38557 | Not-determined |
| SS_extracted_k141_38558 | Not-determined |
| SS_extracted_k141_25831 | Not-determined |
| SS_extracted_k141_135275 | Not-determined |
| SS_extracted_k141_32950 | Low-quality |
| SS_extracted_k141_14566 | Low-quality |
| SS_extracted_k141_3077 | Low-quality |
| SS_extracted_k141_3078 | Not-determined |
| SS_extracted_k141_32953 | Not-determined |
| SS_extracted_k141_32956 | Not-determined |
| SS_extracted_k141_135282 | Not-determined |
| SS_extracted_k141_32958 | Not-determined |
| SS_extracted_k141_14580 | Not-determined |
| SS_extracted_k141_44928 | Not-determined |
| SS_extracted_k141_93660 | Not-determined |
| SS_extracted_k141_3089 | Not-determined |
| SS_extracted_k141_62710 | Low-quality |
| SS_extracted_k141_32965 | Not-determined |
| SS_extracted_k141_38563 | Low-quality |
| SS_extracted_k141_124408 | Not-determined |
| SS_extracted_k141_118352 | Low-quality |
| SS_extracted_k141_124411 | Not-determined |
| SS_extracted_k141_8721 | Not-determined |
| SS_extracted_k141_135289 | Low-quality |
| SS_extracted_k141_62718 | Not-determined |
| SS_extracted_k141_105987 | Not-determined |
| SS_extracted_k141_62719 | Low-quality |
| SS_extracted_k141_141533 | Low-quality |
| SS_extracted_k141_135302 | Not-determined |
| SS_extracted_k141_8731 | Not-determined |
| SS_extracted_k141_80202 | Not-determined |
| SS_extracted_k141_141536 | Not-determined |
| SS_extracted_k141_8735 | Not-determined |
| SS_extracted_k141_25837 | Not-determined |
| SS_extracted_k141_56147 | Not-determined |
| SS_extracted_k141_3108 | Not-determined |
| SS_extracted_k141_20512 | Not-determined |
| SS_extracted_k141_124427 | Low-quality |
| SS_extracted_k141_20522 | Not-determined |
| SS_extracted_k141_62735 | Not-determined |
| SS_extracted_k141_106020 | Not-determined |
| SS_extracted_k141_124439 | Not-determined |
| SS_extracted_k141_129785 | Low-quality |
| SS_extracted_k141_118360 | Not-determined |
| SS_extracted_k141_106027 | Not-determined |
| SS_extracted_k141_56158 | Not-determined |
| SS_extracted_k141_124447 | Not-determined |
| SS_extracted_k141_129792 | Not-determined |
| SS_extracted_k141_106033 | Not-determined |
| SS_extracted_k141_8754 | Not-determined |
| SS_extracted_k141_62749 | Not-determined |
| SS_extracted_k141_129800 | Not-determined |

| | |
|---|---|
| SS_extracted_k141_20544 | Low-quality |
| SS_extracted_k141_124456 | Not-determined |
| SS_extracted_k141_118373 | Not-determined |
| SS_extracted_k141_62752 | Low-quality |
| SS_extracted_k141_62753 | Low-quality |
| SS_extracted_k141_124458 | Low-quality |
| SS_extracted_k141_135325 | Not-determined |
| SS_extracted_k141_44959 | Not-determined |
| SS_extracted_k141_20558 | Not-determined |
| SS_extracted_k141_14612 | Not-determined |
| SS_extracted_k141_3147 | Not-determined |
| SS_extracted_k141_141547 | Not-determined |
| SS_extracted_k141_62763 | Not-determined |
| SS_extracted_k141_118379 | Not-determined |
| SS_extracted_k141_118380 | Low-quality |
| SS_extracted_k141_62771 | Not-determined |
| SS_extracted_k141_32994 | Not-determined |
| SS_extracted_k141_44969 | Not-determined |
| SS_extracted_k141_62776 | Not-determined |
| SS_extracted_k141_51191 | Not-determined |
| SS_extracted_k141_51193 | Not-determined |
| SS_extracted_k141_124470 | Not-determined |
| SS_extracted_k141_56179 | Not-determined |
| SS_extracted_k141_33006 | Low-quality |
| SS_extracted_k141_3160 | Not-determined |
| SS_extracted_k141_14629 | Not-determined |
| SS_extracted_k141_62782 | Not-determined |
| SS_extracted_k141_33014 | Not-determined |
| SS_extracted_k141_3163 | Low-quality |
| SS_extracted_k141_141557 | Low-quality |
| SS_extracted_k141_62783 | Not-determined |
| SS_extracted_k141_106047 | Not-determined |
| SS_extracted_k141_141560 | Not-determined |
| SS_extracted_k141_38592 | Not-determined |
| SS_extracted_k141_51202 | Not-determined |
| SS_extracted_k141_51203 | Low-quality |
| SS_extracted_k141_14640 | Not-determined |
| SS_extracted_k141_124473 | Low-quality |
| SS_extracted_k141_106056 | Not-determined |
| SS_extracted_k141_124478 | Not-determined |
| SS_extracted_k141_124481 | Not-determined |
| SS_extracted_k141_106060 | Not-determined |
| SS_extracted_k141_124483 | Not-determined |
| SS_extracted_k141_38606 | Not-determined |
| SS_extracted_k141_124486 | Not-determined |
| SS_extracted_k141_20564 | Not-determined |
| SS_extracted_k141_106062 | Not-determined |
| SS_extracted_k141_135331 | Not-determined |
| SS_extracted_k141_106063 | Low-quality |
| SS_extracted_k141_106065 | Not-determined |
| SS_extracted_k141_106068 | Low-quality |
| SS_extracted_k141_118406 | Not-determined |
| SS_extracted_k141_124490 | Not-determined |
| SS_extracted_k141_80242 | Not-determined |
| SS_extracted_k141_25861 | Not-determined |
| SS_extracted_k141_135348 | Low-quality |
| SS_extracted_k141_51219 | Low-quality |
| SS_extracted_k141_56192 | Low-quality |
| SS_extracted_k141_141577 | Not-determined |
| SS_extracted_k141_111510 | Not-determined |
| SS_extracted_k141_38629 | Low-quality |
| SS_extracted_k141_98507 | Not-determined |
| SS_extracted_k141_111514 | Low-quality |
| SS_extracted_k141_38634 | Low-quality |
| SS_extracted_k141_62811 | Low-quality |
| SS_extracted_k141_118429 | Not-determined |
| SS_extracted_k141_62813 | Not-determined |
| SS_extracted_k141_25880 | Not-determined |
| SS_extracted_k141_98520 | Not-determined |
| SS_extracted_k141_3198 | Not-determined |
| SS_extracted_k141_80245 | Not-determined |
| SS_extracted_k141_3201 | Not-determined |
| SS_extracted_k141_33045 | Not-determined |
| SS_extracted_k141_33047 | Not-determined |
| SS_extracted_k141_68967 | Not-determined |
| SS_extracted_k141_106093 | Not-determined |
| SS_extracted_k141_51239 | Not-determined |
| SS_extracted_k141_118440 | Not-determined |
| SS_extracted_k141_74361 | Low-quality |
| SS_extracted_k141_51244 | Not-determined |
| SS_extracted_k141_62842 | Not-determined |
| SS_extracted_k141_80258 | Not-determined |
| SS_extracted_k141_118448 | Not-determined |
| SS_extracted_k141_118450 | Not-determined |
| SS_extracted_k141_51264 | Not-determined |
| SS_extracted_k141_33075 | Not-determined |
| SS_extracted_k141_3219 | Not-determined |
| SS_extracted_k141_111535 | Low-quality |
| SS_extracted_k141_51271 | Not-determined |
| SS_extracted_k141_33082 | Low-quality |
| SS_extracted_k141_106118 | Not-determined |
| SS_extracted_k141_118464 | Not-determined |
| SS_extracted_k141_141608 | Low-quality |
| SS_extracted_k141_68990 | Not-determined |
| SS_extracted_k141_51276 | Low-quality |
| SS_extracted_k141_51277 | Low-quality |
| SS_extracted_k141_141617 | Not-determined |
| SS_extracted_k141_69000 | Not-determined |
| SS_extracted_k141_69002 | Not-determined |
| SS_extracted_k141_51293 | Low-quality |
| SS_extracted_k141_106126 | Not-determined |
| SS_extracted_k141_98533 | Low-quality |
| SS_extracted_k141_106129 | Not-determined |
| SS_extracted_k141_106133 | Not-determined |
| SS_extracted_k141_51304 | Not-determined |
| SS_extracted_k141_80280 | Not-determined |
| SS_extracted_k141_106136 | Not-determined |
| SS_extracted_k141_98541 | Low-quality |
| SS_extracted_k141_8775 | Low-quality |
| SS_extracted_k141_8778 | Not-determined |
| SS_extracted_k141_3260 | Not-determined |
| SS_extracted_k141_135351 | Low-quality |
| SS_extracted_k141_51318 | Not-determined |
| SS_extracted_k141_62884 | Not-determined |
| SS_extracted_k141_3270 | Not-determined |
| SS_extracted_k141_118482 | Low-quality |
| SS_extracted_k141_3274 | Not-determined |
| SS_extracted_k141_93723 | Low-quality |
| SS_extracted_k141_135358 | Low-quality |
| SS_extracted_k141_20587 | Not-determined |
| SS_extracted_k141_20588 | Not-determined |
| SS_extracted_k141_93725 | Low-quality |
| SS_extracted_k141_56194 | Not-determined |
| SS_extracted_k141_74410 | Not-determined |
| SS_extracted_k141_118488 | Not-determined |
| SS_extracted_k141_106153 | Low-quality |
| SS_extracted_k141_124528 | Not-determined |
| SS_extracted_k141_74412 | Not-determined |
| SS_extracted_k141_98551 | Not-determined |
| SS_extracted_k141_98552 | Not-determined |
| SS_extracted_k141_124535 | Not-determined |
| SS_extracted_k141_51328 | Low-quality |
| SS_extracted_k141_93738 | Not-determined |
| SS_extracted_k141_118501 | Not-determined |
| SS_extracted_k141_106163 | Not-determined |
| SS_extracted_k141_51331 | Not-determined |
| SS_extracted_k141_14705 | Not-determined |
| SS_extracted_k141_106165 | Low-quality |
| SS_extracted_k141_14709 | Not-determined |
| SS_extracted_k141_118509 | Not-determined |
| SS_extracted_k141_45040 | Not-determined |
| SS_extracted_k141_93748 | Low-quality |
| SS_extracted_k141_38655 | Not-determined |
| SS_extracted_k141_69020 | Not-determined |
| SS_extracted_k141_33102 | Low-quality |
| SS_extracted_k141_69023 | Not-determined |
| SS_extracted_k141_20605 | Not-determined |
| SS_extracted_k141_69024 | Not-determined |
| SS_extracted_k141_20608 | Not-determined |
| SS_extracted_k141_118516 | Not-determined |
| SS_extracted_k141_14722 | Not-determined |
| SS_extracted_k141_129862 | Low-quality |
| SS_extracted_k141_45044 | Not-determined |
| SS_extracted_k141_69037 | Not-determined |
| SS_extracted_k141_69038 | Not-determined |
| SS_extracted_k141_124552 | Low-quality |
| SS_extracted_k141_106175 | Not-determined |
| SS_extracted_k141_74426 | Not-determined |
| SS_extracted_k141_124557 | Not-determined |
| SS_extracted_k141_74427 | Not-determined |
| SS_extracted_k141_124560 | Not-determined |
| SS_extracted_k141_69054 | Not-determined |
| SS_extracted_k141_56236 | Not-determined |
| SS_extracted_k141_51364 | Not-determined |
| SS_extracted_k141_98563 | Not-determined |
| SS_extracted_k141_98564 | Not-determined |
| SS_extracted_k141_111547 | Low-quality |
| SS_extracted_k141_25887 | Low-quality |
| SS_extracted_k141_45066 | Not-determined |
| SS_extracted_k141_51367 | Not-determined |
| SS_extracted_k141_51368 | Low-quality |
| SS_extracted_k141_3317 | Not-determined |
| SS_extracted_k141_25897 | Not-determined |
| SS_extracted_k141_51375 | Not-determined |
| SS_extracted_k141_20632 | Not-determined |
| SS_extracted_k141_51376 | Not-determined |
| SS_extracted_k141_3322 | Not-determined |
| SS_extracted_k141_62900 | Low-quality |
| SS_extracted_k141_38672 | Not-determined |
| SS_extracted_k141_124592 | Not-determined |
| SS_extracted_k141_51385 | Not-determined |
| SS_extracted_k141_62902 | Not-determined |
| SS_extracted_k141_38676 | Not-determined |
| SS_extracted_k141_38677 | Not-determined |
| SS_extracted_k141_111559 | Not-determined |
| SS_extracted_k141_45091 | Not-determined |
| SS_extracted_k141_45098 | Low-quality |
| SS_extracted_k141_56264 | Not-determined |
| SS_extracted_k141_74437 | Not-determined |
| SS_extracted_k141_45102 | Not-determined |
| SS_extracted_k141_25914 | Low-quality |
| SS_extracted_k141_56268 | Not-determined |
| SS_extracted_k141_74442 | Not-determined |
| SS_extracted_k141_62908 | Not-determined |
| SS_extracted_k141_111577 | Not-determined |
| SS_extracted_k141_56282 | Not-determined |
| SS_extracted_k141_45116 | Not-determined |
| SS_extracted_k141_14742 | Not-determined |
| SS_extracted_k141_106221 | Not-determined |
| SS_extracted_k141_20642 | Not-determined |
| SS_extracted_k141_124627 | Low-quality |
| SS_extracted_k141_56298 | Not-determined |
| SS_extracted_k141_111593 | Not-determined |
| SS_extracted_k141_56300 | Not-determined |
| SS_extracted_k141_124633 | Not-determined |
| SS_extracted_k141_124635 | Not-determined |
| SS_extracted_k141_20648 | Low-quality |
| SS_extracted_k141_111599 | Not-determined |
| SS_extracted_k141_124637 | Not-determined |
| SS_extracted_k141_20654 | Not-determined |
| SS_extracted_k141_56306 | Not-determined |
| SS_extracted_k141_62924 | Low-quality |
| SS_extracted_k141_38707 | Low-quality |
| SS_extracted_k141_93770 | Low-quality |
| SS_extracted_k141_38710 | Not-determined |
| SS_extracted_k141_145604 | Complete |
| SS_extracted_k141_80316 | Not-determined |
| SS_extracted_k141_20669 | Low-quality |
| SS_extracted_k141_135384 | Not-determined |
| SS_extracted_k141_135386 | Not-determined |
| SS_extracted_k141_93783 | Low-quality |
| SS_extracted_k141_74460 | Not-determined |
| SS_extracted_k141_20673 | Not-determined |
| SS_extracted_k141_93790 | Not-determined |
| SS_extracted_k141_20677 | Not-determined |
| SS_extracted_k141_3365 | Not-determined |
| SS_extracted_k141_98604 | Not-determined |
| SS_extracted_k141_14766 | Not-determined |
| SS_extracted_k141_93798 | Not-determined |
| SS_extracted_k141_93801 | Not-determined |
| SS_extracted_k141_69087 | Not-determined |
| SS_extracted_k141_3383 | Not-determined |
| SS_extracted_k141_93808 | Not-determined |
| SS_extracted_k141_80330 | Not-determined |
| SS_extracted_k141_80334 | Not-determined |
| SS_extracted_k141_3392 | Not-determined |
| SS_extracted_k141_62963 | Not-determined |
| SS_extracted_k141_62968 | Not-determined |
| SS_extracted_k141_3396 | Not-determined |
| SS_extracted_k141_62970 | Not-determined |
| SS_extracted_k141_69111 | Not-determined |
| SS_extracted_k141_51411 | Not-determined |
| SS_extracted_k141_69115 | Not-determined |
| SS_extracted_k141_51413 | Not-determined |
| SS_extracted_k141_3410 | Not-determined |
| SS_extracted_k141_80342 | Not-determined |
| SS_extracted_k141_14806 | Not-determined |
| SS_extracted_k141_111634 | Not-determined |
| SS_extracted_k141_3420 | Not-determined |
| SS_extracted_k141_69129 | Not-determined |
| SS_extracted_k141_20679 | Not-determined |
| SS_extracted_k141_111643 | Not-determined |
| SS_extracted_k141_80349 | Not-determined |
| SS_extracted_k141_3426 | Not-determined |
| SS_extracted_k141_3427 | Not-determined |
| SS_extracted_k141_56337 | Not-determined |
| SS_extracted_k141_14830 | Not-determined |
| SS_extracted_k141_45119 | Not-determined |
| SS_extracted_k141_20685 | Low-quality |
| SS_extracted_k141_69139 | Low-quality |
| SS_extracted_k141_14840 | Not-determined |
| SS_extracted_k141_80361 | Not-determined |
| SS_extracted_k141_93816 | Low-quality |
| SS_extracted_k141_93817 | Not-determined |
| SS_extracted_k141_45132 | Not-determined |
| SS_extracted_k141_3447 | Not-determined |
| SS_extracted_k141_69147 | Low-quality |
| SS_extracted_k141_38759 | Not-determined |
| SS_extracted_k141_80368 | Low-quality |
| SS_extracted_k141_45146 | Not-determined |
| SS_extracted_k141_33122 | Not-determined |
| SS_extracted_k141_93832 | Not-determined |
| SS_extracted_k141_20689 | Not-determined |
| SS_extracted_k141_14859 | Low-quality |
| SS_extracted_k141_20693 | Not-determined |
| SS_extracted_k141_56356 | Not-determined |
| SS_extracted_k141_74486 | Not-determined |
| SS_extracted_k141_124651 | Low-quality |
| SS_extracted_k141_14865 | Low-quality |
| SS_extracted_k141_98623 | Low-quality |
| SS_extracted_k141_124655 | Not-determined |
| SS_extracted_k141_20698 | Not-determined |
| SS_extracted_k141_98628 | Not-determined |
| SS_extracted_k141_98631 | Not-determined |
| SS_extracted_k141_129932 | Low-quality |
| SS_extracted_k141_129933 | Low-quality |
| SS_extracted_k141_45159 | Low-quality |
| SS_extracted_k141_38790 | Not-determined |
| SS_extracted_k141_74504 | Low-quality |
| SS_extracted_k141_129938 | Low-quality |
| SS_extracted_k141_56366 | Not-determined |
| SS_extracted_k141_45170 | Not-determined |
| SS_extracted_k141_51426 | Low-quality |
| SS_extracted_k141_8864 | Not-determined |
| SS_extracted_k141_33152 | Not-determined |
| SS_extracted_k141_3494 | Low-quality |
| SS_extracted_k141_8865 | Low-quality |
| SS_extracted_k141_8867 | Low-quality |
| SS_extracted_k141_3499 | Not-determined |
| SS_extracted_k141_118576 | Not-determined |
| SS_extracted_k141_51438 | Not-determined |
| SS_extracted_k141_93842 | Low-quality |
| SS_extracted_k141_98650 | Not-determined |
| SS_extracted_k141_118594 | Not-determined |
| SS_extracted_k141_98651 | Low-quality |
| SS_extracted_k141_45181 | Not-determined |
| SS_extracted_k141_8875 | Not-determined |
| SS_extracted_k141_51452 | Not-determined |
| SS_extracted_k141_118602 | Not-determined |
| SS_extracted_k141_118603 | Not-determined |
| SS_extracted_k141_98669 | Not-determined |
| SS_extracted_k141_33171 | Not-determined |
| SS_extracted_k141_33176 | Low-quality |
| SS_extracted_k141_93850 | Not-determined |
| SS_extracted_k141_56390 | Not-determined |
| SS_extracted_k141_135439 | Low-quality |
| SS_extracted_k141_62995 | Not-determined |
| SS_extracted_k141_93855 | Not-determined |
| SS_extracted_k141_62997 | Not-determined |
| SS_extracted_k141_141684 | Not-determined |
| SS_extracted_k141_20736 | Not-determined |
| SS_extracted_k141_87063 | Low-quality |
| S4_vs_VIRSorter_31980-circular-cat | High-quality |
| SS_extracted_k141_87065 | Low-quality |
| SS_extracted_k141_14954 | Not-determined |
| SS_extracted_k141_20743 | Not-determined |
| SS_extracted_k141_129941 | Not-determined |
| SS_extracted_k141_69173 | Low-quality |
| SS_extracted_k141_69174 | Not-determined |
| SS_extracted_k141_69175 | Low-quality |
| SS_extracted_k141_93878 | Low-quality |
| SS_extracted_k141_87079 | Not-determined |
| SS_extracted_k141_45211 | Not-determined |
| SS_extracted_k141_135450 | Not-determined |
| SS_extracted_k141_141696 | Not-determined |
| SS_extracted_k141_20751 | Low-quality |
| SS_extracted_k141_45216 | Low-quality |
| SS_extracted_k141_14964 | Low-quality |
| SS_extracted_k141_141705 | Not-determined |
| SS_extracted_k141_129960 | Not-determined |
| SS_extracted_k141_3514 | Low-quality |
| SS_extracted_k141_129965 | Not-determined |
| SS_extracted_k141_51486 | Not-determined |
| SS_extracted_k141_20754 | Not-determined |
| SS_extracted_k141_93888 | Not-determined |
| SS_extracted_k141_14971 | Not-determined |
| SS_extracted_k141_141715 | Not-determined |
| SS_extracted_k141_118631 | Not-determined |
| SS_extracted_k141_3517 | Low-quality |
| SS_extracted_k141_141718 | Not-determined |
| SS_extracted_k141_118632 | Low-quality |
| SS_extracted_k141_69205 | Not-determined |
| SS_extracted_k141_69208 | Not-determined |
| SS_extracted_k141_93892 | Not-determined |
| SS_extracted_k141_56403 | Low-quality |
| SS_extracted_k141_51496 | Not-determined |
| SS_extracted_k141_69211 | Low-quality |
| SS_extracted_k141_87085 | Low-quality |
| SS_extracted_k141_93894 | Not-determined |
| SS_extracted_k141_63020 | Not-determined |
| SS_extracted_k141_69216 | Not-determined |
| SS_extracted_k141_69218 | Not-determined |
| SS_extracted_k141_8905 | Not-determined |
| SS_extracted_k141_141726 | Low-quality |
| SS_extracted_k141_111657 | Low-quality |
| SS_extracted_k141_45232 | Low-quality |
| SS_extracted_k141_56408 | Not-determined |
| SS_extracted_k141_135458 | Low-quality |
| SS_extracted_k141_33201 | Not-determined |
| SS_extracted_k141_87093 | Low-quality |
| SS_extracted_k141_51510 | Not-determined |
| SS_extracted_k141_8917 | Not-determined |
| SS_extracted_k141_51511 | Not-determined |
| SS_extracted_k141_20772 | Not-determined |
| SS_extracted_k141_141732 | Not-determined |
| SS_extracted_k141_51514 | Low-quality |
| SS_extracted_k141_141734 | Not-determined |
| SS_extracted_k141_98711 | Not-determined |
| SS_extracted_k141_141736 | Not-determined |
| SS_extracted_k141_98715 | Not-determined |
| SS_extracted_k141_63028 | Not-determined |
| SS_extracted_k141_51521 | Not-determined |
| SS_extracted_k141_56411 | Not-determined |
| SS_extracted_k141_141743 | Low-quality |
| SS_extracted_k141_141744 | Not-determined |
| SS_extracted_k141_69244 | Not-determined |
| SS_extracted_k141_20776 | Low-quality |
| SS_extracted_k141_63033 | Not-determined |
| SS_extracted_k141_87105 | Not-determined |
| SS_extracted_k141_20777 | Not-determined |
| SS_extracted_k141_51525 | Not-determined |
| SS_extracted_k141_20781 | Low-quality |
| SS_extracted_k141_51530 | Not-determined |
| SS_extracted_k141_87108 | Low-quality |
| SS_extracted_k141_135466 | Not-determined |
| SS_extracted_k141_15016 | Low-quality |
| SS_extracted_k141_69251 | Not-determined |
| SS_extracted_k141_45257 | Not-determined |
| SS_extracted_k141_56416 | Not-determined |
| SS_extracted_k141_15020 | Low-quality |
| SS_extracted_k141_20791 | Low-quality |
| SS_extracted_k141_93906 | Low-quality |
| SS_extracted_k141_51538 | Low-quality |
| SS_extracted_k141_15024 | Low-quality |
| SS_extracted_k141_3565 | Not-determined |
| SS_extracted_k141_141765 | Not-determined |
| SS_extracted_k141_15029 | Low-quality |
| SS_extracted_k141_111685 | Not-determined |
| SS_extracted_k141_20797 | Not-determined |
| SS_extracted_k141_8935 | Low-quality |
| SS_extracted_k141_111690 | Not-determined |
| SS_extracted_k141_51545 | Low-quality |
| SS_extracted_k141_51546 | Not-determined |
| SS_extracted_k141_51547 | Not-determined |
| SS_extracted_k141_111693 | Not-determined |
| SS_extracted_k141_87121 | Not-determined |
| SS_extracted_k141_87122 | Not-determined |
| SS_extracted_k141_8938 | Not-determined |
| SS_extracted_k141_87126 | Not-determined |
| SS_extracted_k141_3577 | Not-determined |
| SS_extracted_k141_69260 | Low-quality |
| SS_extracted_k141_51551 | Not-determined |
| SS_extracted_k141_111697 | Not-determined |
| SS_extracted_k141_129980 | Not-determined |
| SS_extracted_k141_69263 | Not-determined |
| SS_extracted_k141_129981 | Low-quality |
| SS_extracted_k141_3585 | Not-determined |
| SS_extracted_k141_87133 | Not-determined |
| SS_extracted_k141_63054 | Not-determined |
| SS_extracted_k141_141787 | Low-quality |
| SS_extracted_k141_141788 | Low-quality |
| SS_extracted_k141_141792 | Not-determined |
| SS_extracted_k141_69275 | Not-determined |
| SS_extracted_k141_141795 | Low-quality |
| SS_extracted_k141_51569 | Not-determined |
| SS_extracted_k141_20802 | Not-determined |
| SS_extracted_k141_141797 | Not-determined |
| SS_extracted_k141_8961 | Not-determined |
| SS_extracted_k141_51574 | Not-determined |
| SS_extracted_k141_51578 | Not-determined |
| SS_extracted_k141_111724 | Not-determined |
| SS_extracted_k141_63069 | Not-determined |
| SS_extracted_k141_111730 | Not-determined |
| SS_extracted_k141_111735 | Not-determined |
| SS_extracted_k141_51584 | Not-determined |
| SS_extracted_k141_63081 | Not-determined |
| SS_extracted_k141_20821 | Low-quality |
| SS_extracted_k141_106259 | Not-determined |
| SS_extracted_k141_106266 | Not-determined |
| SS_extracted_k141_141807 | Not-determined |
| SS_extracted_k141_106275 | Not-determined |
| SS_extracted_k141_106276 | Low-quality |
| SS_extracted_k141_20831 | Not-determined |
| SS_extracted_k141_20832 | Not-determined |
| SS_extracted_k141_15062 | Not-determined |
| SS_extracted_k141_118670 | Not-determined |
| SS_extracted_k141_118671 | Not-determined |
| SS_extracted_k141_118673 | Not-determined |
| SS_extracted_k141_118674 | Not-determined |
| SS_extracted_k141_45297 | Not-determined |
| SS_extracted_k141_20845 | Not-determined |
| SS_extracted_k141_20847 | Not-determined |
| SS_extracted_k141_118677 | Not-determined |
| SS_extracted_k141_87155 | Low-quality |
| SS_extracted_k141_15067 | Not-determined |
| SS_extracted_k141_87157 | Not-determined |
| SS_extracted_k141_87158 | Not-determined |
| SS_extracted_k141_106289 | Not-determined |
| SS_extracted_k141_74519 | Low-quality |
| SS_extracted_k141_45304 | Not-determined |
| SS_extracted_k141_141812 | Not-determined |
| SS_extracted_k141_20852 | Low-quality |
| S8_extracted_k141_72329 | Complete |
| SS_extracted_k141_20853 | Not-determined |
| SS_extracted_k141_106301 | Not-determined |
| SS_extracted_k141_51622 | Not-determined |
| SS_extracted_k141_45311 | Not-determined |
| SS_extracted_k141_111743 | Not-determined |
| SS_extracted_k141_111744 | Not-determined |
| SS_extracted_k141_69310 | Not-determined |
| SS_extracted_k141_80422 | Low-quality |
| SS_extracted_k141_51635 | Not-determined |
| S1_vs_k141_158961\|\|full | Medium-quality |
| SS_extracted_k141_130018 | Not-determined |
| SS_extracted_k141_98741 | Not-determined |
| SS_extracted_k141_38799 | Not-determined |
| SS_extracted_k141_69321 | Not-determined |
| SS_extracted_k141_118689 | Low-quality |
| SS_extracted_k141_118691 | Not-determined |
| SS_extracted_k141_118692 | Not-determined |
| SS_extracted_k141_51642 | Not-determined |
| SS_extracted_k141_8971 | Not-determined |
| SS_extracted_k141_20872 | Not-determined |
| SS_extracted_k141_111756 | Not-determined |
| SS_extracted_k141_111757 | Not-determined |
| SS_extracted_k141_63116 | Not-determined |
| SS_extracted_k141_51645 | Not-determined |
| SS_extracted_k141_106324 | Not-determined |
| SS_extracted_k141_45331 | Not-determined |
| SS_extracted_k141_118699 | Not-determined |
| SS_extracted_k141_98748 | Not-determined |
| SS_extracted_k141_3623 | Not-determined |
| SS_extracted_k141_111766 | Not-determined |
| SS_extracted_k141_111767 | Not-determined |
| SS_extracted_k141_111770 | Not-determined |
| SS_extracted_k141_69337 | Not-determined |
| SS_extracted_k141_8977 | Not-determined |
| SS_extracted_k141_118709 | Not-determined |
| SS_extracted_k141_69340 | Not-determined |
| SS_extracted_k141_98753 | Not-determined |
| SS_extracted_k141_98755 | Not-determined |
| SS_extracted_k141_69344 | Not-determined |
| SS_extracted_k141_141838 | Not-determined |
| SS_extracted_k141_111773 | Not-determined |
| SS_extracted_k141_98758 | Not-determined |
| SS_extracted_k141_63136 | Low-quality |
| SS_extracted_k141_98762 | Not-determined |
| SS_extracted_k141_141845 | Not-determined |
| SS_extracted_k141_141846 | Not-determined |
| SS_extracted_k141_33232 | Not-determined |
| SS_extracted_k141_33235 | Not-determined |
| SS_extracted_k141_20891 | Not-determined |
| SS_extracted_k141_98765 | Not-determined |
| SS_extracted_k141_20892 | Not-determined |
| SS_extracted_k141_33237 | Low-quality |
| SS_extracted_k141_74547 | Not-determined |
| SS_extracted_k141_38803 | Not-determined |
| SS_extracted_k141_20895 | Not-determined |
| SS_extracted_k141_80431 | Not-determined |
| SS_extracted_k141_80432 | Not-determined |
| SS_extracted_k141_15076 | Not-determined |
| SS_extracted_k141_63156 | Not-determined |
| SS_extracted_k141_69354 | Low-quality |
| SS_extracted_k141_69358 | Not-determined |
| SS_extracted_k141_51656 | Not-determined |
| SS_extracted_k141_63165 | Not-determined |
| SS_extracted_k141_106349 | Not-determined |
| SS_extracted_k141_15083 | Not-determined |
| SS_extracted_k141_33262 | Not-determined |
| SS_extracted_k141_63169 | Not-determined |
| SS_extracted_k141_106355 | Not-determined |
| SS_extracted_k141_56441 | Low-quality |
| SS_extracted_k141_38827 | Not-determined |
| SS_extracted_k141_87194 | Not-determined |
| SS_extracted_k141_38830 | Not-determined |
| SS_extracted_k141_106361 | Low-quality |
| SS_extracted_k141_69368 | Not-determined |
| SS_extracted_k141_3657 | Not-determined |
| SS_extracted_k141_3661 | Not-determined |
| SS_extracted_k141_38844 | Not-determined |
| SS_extracted_k141_38845 | Not-determined |
| SS_extracted_k141_98769 | Low-quality |
| SS_extracted_k141_141871 | Not-determined |
| SS_extracted_k141_20918 | Not-determined |
| SS_extracted_k141_38847 | Not-determined |
| SS_extracted_k141_33275 | Not-determined |
| SS_extracted_k141_20921 | Low-quality |
| SS_extracted_k141_69382 | Not-determined |
| SS_extracted_k141_87214 | Low-quality |
| SS_extracted_k141_106385 | Not-determined |
| SS_extracted_k141_38856 | Not-determined |
| SS_extracted_k141_38858 | Not-determined |
| SS_extracted_k141_141875 | Not-determined |
| SS_extracted_k141_38859 | Not-determined |
| SS_extracted_k141_38860 | Not-determined |
| SS_extracted_k141_130048 | Not-determined |
| SS_extracted_k141_106395 | Not-determined |
| SS_extracted_k141_141876 | Low-quality |
| SS_extracted_k141_106398 | Low-quality |
| SS_extracted_k141_141879 | Low-quality |
| SS_extracted_k141_38870 | Not-determined |
| SS_extracted_k141_20934 | Not-determined |
| SS_extracted_k141_111795 | Not-determined |
| SS_extracted_k141_111796 | Not-determined |
| SS_extracted_k141_38875 | Low-quality |
| SS_extracted_k141_130060 | Not-determined |
| SS_extracted_k141_38876 | Low-quality |
| SS_extracted_k141_63208 | Not-determined |
| SS_extracted_k141_20937 | Low-quality |
| SS_extracted_k141_106414 | Not-determined |
| SS_extracted_k141_130064 | Not-determined |
| SS_extracted_k141_130065 | Low-quality |
| SS_extracted_k141_111813 | Not-determined |
| SS_extracted_k141_63214 | Not-determined |
| SS_extracted_k141_141895 | Not-determined |
| SS_extracted_k141_56464 | Not-determined |
| SS_extracted_k141_111819 | Not-determined |
| SS_extracted_k141_33292 | Not-determined |
| SS_extracted_k141_20940 | Not-determined |

| | |
|---|---|
| SS_extracted_k141_15136 | Not-determined |
| SS_extracted_k141_20941 | Low-quality |
| SS_extracted_k141_33293 | Not-determined |
| SS_extracted_k141_141904 | Not-determined |
| SS_extracted_k141_33294 | Low-quality |
| SS_extracted_k141_141905 | Not-determined |
| SS_extracted_k141_15138 | Not-determined |
| SS_extracted_k141_56471 | Not-determined |
| SS_extracted_k141_124687 | Not-determined |
| SS_extracted_k141_106416 | Not-determined |
| SS_extracted_k141_3691 | Not-determined |
| SS_extracted_k141_111825 | Not-determined |
| SS_extracted_k141_141913 | Not-determined |
| SS_extracted_k141_93919 | Not-determined |
| SS_extracted_k141_63233 | Low-quality |
| SS_extracted_k141_98791 | Low-quality |
| SS_extracted_k141_98793 | Low-quality |
| SS_extracted_k141_124703 | Not-determined |
| SS_extracted_k141_15159 | Low-quality |
| SS_extracted_k141_141926 | Not-determined |
| SS_extracted_k141_98794 | Low-quality |
| SS_extracted_k141_3707 | Not-determined |
| SS_extracted_k141_87226 | Not-determined |
| SS_extracted_k141_124709 | Not-determined |
| SS_extracted_k141_33313 | Low-quality |
| SS_extracted_k141_111840 | Not-determined |
| SS_extracted_k141_38915 | Not-determined |
| SS_extracted_k141_63247 | Not-determined |
| SS_extracted_k141_93940 | Low-quality |
| SS_extracted_k141_124720 | Not-determined |
| SS_extracted_k141_3719 | Not-determined |
| SS_extracted_k141_20947 | Not-determined |
| SS_extracted_k141_111852 | Not-determined |
| SS_extracted_k141_51694 | Not-determined |
| SS_extracted_k141_56485 | Not-determined |
| SS_extracted_k141_51700 | Low-quality |
| SS_extracted_k141_98802 | Not-determined |
| SS_extracted_k141_130104 | Not-determined |
| SS_extracted_k141_56494 | Not-determined |
| SS_extracted_k141_124728 | Not-determined |
| SS_extracted_k141_33318 | Not-determined |
| SS_extracted_k141_111864 | Not-determined |
| SS_extracted_k141_69401 | Not-determined |
| SS_extracted_k141_63268 | Not-determined |
| SS_extracted_k141_118714 | Not-determined |
| SS_extracted_k141_127400 | Complete |
| SS_extracted_k141_106432 | Not-determined |
| SS_extracted_k141_111870 | Not-determined |
| SS_extracted_k141_69416 | Not-determined |
| SS_extracted_k141_20966 | Low-quality |
| SS_extracted_k141_106434 | Not-determined |
| SS_extracted_k141_118720 | Low-quality |
| SS_extracted_k141_118721 | Not-determined |
| SS_extracted_k141_51723 | Low-quality |
| SS_extracted_k141_69429 | Not-determined |
| SS_extracted_k141_33338 | Not-determined |
| SS_extracted_k141_80449 | Low-quality |
| SS_extracted_k141_51732 | Not-determined |
| SS_extracted_k141_63282 | Not-determined |
| SS_extracted_k141_69441 | Not-determined |
| SS_extracted_k141_118734 | Not-determined |
| SS_extracted_k141_69444 | Not-determined |
| SS_extracted_k141_141951 | Low-quality |
| SS_extracted_k141_38932 | Low-quality |
| SS_extracted_k141_130119 | Low-quality |
| SS_extracted_k141_141956 | Low-quality |
| SS_extracted_k141_38934 | Not-determined |
| SS_extracted_k141_63293 | Low-quality |
| SS_extracted_k141_130122 | Low-quality |
| SS_extracted_k141_33347 | Not-determined |
| SS_extracted_k141_130123 | Low-quality |
| SS_extracted_k141_93978 | Low-quality |
| SS_extracted_k141_130125 | Not-determined |
| SS_extracted_k141_118747 | Not-determined |
| SS_extracted_k141_3755 | Not-determined |
| SS_extracted_k141_33353 | Not-determined |
| SS_extracted_k141_69457 | Not-determined |
| SS_extracted_k141_141965 | Low-quality |
| SS_extracted_k141_93987 | Low-quality |
| SS_extracted_k141_130132 | Low-quality |
| SS_extracted_k141_124763 | Low-quality |
| SS_extracted_k141_3762 | Low-quality |
| SS_extracted_k141_124766 | Not-determined |
| SS_extracted_k141_69463 | Not-determined |
| SS_extracted_k141_3765 | Not-determined |
| SS_extracted_k141_80461 | Not-determined |
| SS_extracted_k141_141981 | Not-determined |
| SS_extracted_k141_3768 | Not-determined |
| SS_extracted_k141_51760 | Not-determined |
| SS_extracted_k141_3772 | Not-determined |
| SS_extracted_k141_111885 | Not-determined |
| SS_extracted_k141_20977 | Not-determined |
| SS_extracted_k141_51765 | Low-quality |
| SS_extracted_k141_20978 | Not-determined |
| SS_extracted_k141_3780 | Not-determined |
| SS_extracted_k141_106462 | Not-determined |
| SS_extracted_k141_130146 | Low-quality |
| SS_extracted_k141_15194 | Not-determined |
| SS_extracted_k141_106470 | Not-determined |
| SS_extracted_k141_111904 | Not-determined |
| SS_extracted_k141_106472 | Not-determined |
| SS_extracted_k141_20987 | Low-quality |
| SS_extracted_k141_15199 | Not-determined |
| SS_extracted_k141_15201 | Not-determined |
| SS_extracted_k141_3789 | Not-determined |
| SS_extracted_k141_135491 | Not-determined |
| SS_extracted_k141_15204 | Not-determined |
| SS_extracted_k141_124781 | Not-determined |
| SS_extracted_k141_111915 | Not-determined |
| SS_extracted_k141_124783 | Not-determined |
| SS_extracted_k141_33369 | Not-determined |
| SS_extracted_k141_56510 | Not-determined |
| SS_extracted_k141_33374 | Not-determined |
| SS_extracted_k141_15213 | Not-determined |
| SS_extracted_k141_118782 | Not-determined |
| SS_extracted_k141_15219 | Low-quality |
| SS_extracted_k141_25954 | Not-determined |
| SS_extracted_k141_51794 | Not-determined |
| SS_extracted_k141_69503 | Not-determined |
| SS_extracted_k141_63301 | Not-determined |
| SS_extracted_k141_124797 | Low-quality |
| SS_extracted_k141_21013 | Not-determined |
| SS_extracted_k141_142011 | Low-quality |
| SS_extracted_k141_15237 | Not-determined |
| SS_extracted_k141_69506 | Not-determined |
| SS_extracted_k141_3812 | Not-determined |
| SS_extracted_k141_51797 | Not-determined |
| SS_extracted_k141_51799 | Low-quality |
| SS_extracted_k141_33384 | Low-quality |
| SS_extracted_k141_118803 | Not-determined |
| SS_extracted_k141_33390 | Not-determined |
| SS_extracted_k141_21020 | Not-determined |
| SS_extracted_k141_142028 | Not-determined |
| SS_extracted_k141_142030 | Not-determined |
| SS_extracted_k141_56518 | Not-determined |
| SS_extracted_k141_38950 | Not-determined |
| SS_extracted_k141_142032 | Not-determined |
| SS_extracted_k141_56520 | Not-determined |
| SS_extracted_k141_56521 | Not-determined |
| SS_extracted_k141_21033 | Low-quality |
| SS_extracted_k141_130187 | Low-quality |
| SS_extracted_k141_80493 | Not-determined |
| SS_extracted_k141_21046 | Not-determined |
| SS_extracted_k141_111971 | Not-determined |
| SS_extracted_k141_142033 | Low-quality |
| SS_extracted_k141_63319 | Not-determined |
| SS_extracted_k141_25979 | Not-determined |
| SS_extracted_k141_142034 | Not-determined |
| SS_extracted_k141_106481 | Not-determined |
| SS_extracted_k141_63321 | Not-determined |
| SS_extracted_k141_130206 | Not-determined |
| SS_extracted_k141_15255 | Not-determined |
| SS_extracted_k141_135527 | Not-determined |
| SS_extracted_k141_118830 | Not-determined |
| SS_extracted_k141_111980 | Not-determined |
| SS_extracted_k141_118834 | Not-determined |
| SS_extracted_k141_33424 | Not-determined |
| SS_extracted_k141_33425 | Not-determined |
| SS_extracted_k141_9011 | Not-determined |
| SS_extracted_k141_9018 | Not-determined |
| SS_extracted_k141_15259 | Low-quality |
| SS_extracted_k141_33429 | Not-determined |
| SS_extracted_k141_56534 | Not-determined |
| SS_extracted_k141_33431 | Not-determined |
| SS_extracted_k141_130213 | Not-determined |
| SS_extracted_k141_118840 | Not-determined |
| SS_extracted_k141_56536 | Low-quality |
| SS_extracted_k141_33439 | Not-determined |
| SS_extracted_k141_130220 | Not-determined |
| SS_extracted_k141_51810 | Low-quality |
| SS_extracted_k141_98881 | Not-determined |
| SS_extracted_k141_124825 | Not-determined |
| SS_extracted_k141_9036 | Not-determined |
| SS_extracted_k141_112007 | Not-determined |
| SS_extracted_k141_130230 | Not-determined |
| SS_extracted_k141_26003 | Not-determined |
| SS_extracted_k141_124835 | Not-determined |
| SS_extracted_k141_118853 | Low-quality |
| SS_extracted_k141_112016 | Not-determined |
| SS_extracted_k141_94026 | Low-quality |
| SS_extracted_k141_106505 | Not-determined |
| SS_extracted_k141_112024 | Not-determined |
| SS_extracted_k141_3845 | Not-determined |
| SS_extracted_k141_135537 | Not-determined |
| SS_extracted_k141_63349 | Not-determined |
| SS_extracted_k141_98887 | Not-determined |
| SS_extracted_k141_124847 | Not-determined |
| SS_extracted_k141_3854 | Not-determined |
| SS_extracted_k141_106518 | Not-determined |
| SS_extracted_k141_63362 | Not-determined |
| SS_extracted_k141_33451 | Low-quality |
| SS_extracted_k141_106525 | Not-determined |
| SS_extracted_k141_94040 | Not-determined |
| SS_extracted_k141_3866 | Not-determined |
| SS_extracted_k141_94042 | Not-determined |
| SS_extracted_k141_26022 | Not-determined |
| SS_extracted_k141_118861 | Not-determined |
| SS_extracted_k141_106532 | Not-determined |
| SS_extracted_k141_51833 | Not-determined |
| SS_extracted_k141_15286 | Low-quality |
| SS_extracted_k141_112042 | Not-determined |
| SS_extracted_k141_118864 | Not-determined |
| SS_extracted_k141_94052 | Low-quality |
| SS_extracted_k141_51838 | Low-quality |
| SS_extracted_k141_9058 | Not-determined |
| SS_extracted_k141_130262 | Not-determined |
| SS_extracted_k141_112047 | Not-determined |
| SS_extracted_k141_87295 | Not-determined |
| SS_extracted_k141_87296 | Not-determined |
| SS_extracted_k141_118874 | Not-determined |
| SS_extracted_k141_9064 | Not-determined |
| SS_extracted_k141_87297 | Not-determined |
| SS_extracted_k141_56546 | Low-quality |
| SS_extracted_k141_56547 | Not-determined |
| SS_extracted_k141_106549 | Not-determined |
| SS_extracted_k141_112054 | Low-quality |
| SS_extracted_k141_33461 | Not-determined |
| SS_extracted_k141_130270 | Not-determined |
| SS_extracted_k141_51852 | Low-quality |
| SS_extracted_k141_130273 | Low-quality |
| SS_extracted_k141_130274 | Not-determined |
| SS_extracted_k141_21062 | Not-determined |
| SS_extracted_k141_130275 | Not-determined |
| SS_extracted_k141_63385 | Not-determined |
| SS_extracted_k141_51854 | Not-determined |
| SS_extracted_k141_112061 | Not-determined |
| SS_extracted_k141_106559 | Not-determined |
| SS_extracted_k141_3885 | Not-determined |
| SS_extracted_k141_106560 | Not-determined |
| SS_extracted_k141_106564 | Not-determined |
| SS_extracted_k141_94067 | Not-determined |
| SS_extracted_k141_21074 | Not-determined |
| SS_extracted_k141_106568 | Low-quality |
| SS_extracted_k141_21077 | Not-determined |
| SS_extracted_k141_142047 | Not-determined |
| SS_extracted_k141_87308 | Not-determined |
| SS_extracted_k141_142050 | Not-determined |
| SS_extracted_k141_21079 | Not-determined |
| SS_extracted_k141_45402 | Not-determined |
| SS_extracted_k141_9077 | Not-determined |
| SS_extracted_k141_21080 | Not-determined |
| SS_extracted_k141_51860 | Low-quality |
| SS_extracted_k141_135558 | Low-quality |
| SS_extracted_k141_21083 | Low-quality |
| SS_extracted_k141_51861 | Not-determined |
| SS_extracted_k141_45404 | Not-determined |
| SS_extracted_k141_63400 | Not-determined |
| SS_extracted_k141_63401 | Not-determined |
| SS_extracted_k141_51864 | Not-determined |
| SS_extracted_k141_33472 | Not-determined |
| SS_extracted_k141_87314 | Not-determined |
| SS_extracted_k141_69545 | Not-determined |
| SS_extracted_k141_98892 | Not-determined |
| SS_extracted_k141_15312 | Not-determined |
| SS_extracted_k141_63404 | Not-determined |
| SS_extracted_k141_3904 | Not-determined |
| SS_extracted_k141_98895 | Not-determined |
| SS_extracted_k141_74559 | Not-determined |
| SS_extracted_k141_94084 | Not-determined |
| SS_extracted_k141_112085 | Not-determined |
| SS_extracted_k141_112091 | Not-determined |
| SS_extracted_k141_118917 | Not-determined |
| SS_extracted_k141_51870 | Not-determined |
| SS_extracted_k141_112097 | Not-determined |
| SS_extracted_k141_98908 | Not-determined |
| SS_extracted_k141_63420 | Not-determined |
| SS_extracted_k141_94087 | Not-determined |
| SS_extracted_k141_87335 | Not-determined |
| SS_extracted_k141_9093 | Not-determined |
| SS_extracted_k141_51878 | Low-quality |
| SS_extracted_k141_94094 | Not-determined |
| SS_extracted_k141_9096 | Low-quality |
| SS_extracted_k141_45426 | Not-determined |
| SS_extracted_k141_9097 | Not-determined |
| SS_extracted_k141_45427 | Not-determined |
| SS_extracted_k141_51886 | Not-determined |
| SS_extracted_k141_118936 | Not-determined |
| SS_extracted_k141_51893 | Not-determined |
| SS_extracted_k141_94112 | Not-determined |
| SS_extracted_k141_3932 | Not-determined |
| SS_extracted_k141_87352 | Not-determined |
| SS_extracted_k141_51896 | Not-determined |
| SS_extracted_k141_106613 | Not-determined |
| SS_extracted_k141_74586 | Not-determined |
| SS_extracted_k141_26042 | Not-determined |
| SS_extracted_k141_26045 | Not-determined |
| SS_extracted_k141_106619 | Low-quality |
| SS_extracted_k141_69568 | Not-determined |
| SS_extracted_k141_69569 | Not-determined |
| SS_extracted_k141_94124 | Not-determined |
| SS_extracted_k141_74590 | Not-determined |
| SS_extracted_k141_51902 | Low-quality |
| SS_extracted_k141_98911 | Low-quality |
| SS_extracted_k141_106623 | Not-determined |
| SS_extracted_k141_69573 | Low-quality |
| SS_extracted_k141_118960 | Not-determined |
| SS_extracted_k141_142089 | Not-determined |
| SS_extracted_k141_15375 | Not-determined |
| SS_extracted_k141_15378 | Not-determined |
| SS_extracted_k141_56578 | Not-determined |
| SS_extracted_k141_112126 | Not-determined |
| SS_extracted_k141_63437 | Not-determined |
| SS_extracted_k141_51907 | Low-quality |
| SS_extracted_k141_3949 | Not-determined |
| SS_extracted_k141_112127 | Not-determined |
| SS_extracted_k141_142096 | Low-quality |
| SS_extracted_k141_15384 | Not-determined |
| SS_extracted_k141_106633 | Not-determined |
| SS_extracted_k141_106635 | Not-determined |
| SS_extracted_k141_106637 | Not-determined |
| SS_extracted_k141_45459 | Not-determined |
| SS_extracted_k141_142099 | Not-determined |
| SS_extracted_k141_142100 | Not-determined |
| SS_extracted_k141_69579 | Low-quality |
| SS_extracted_k141_106640 | Not-determined |
| SS_extracted_k141_3954 | Low-quality |
| SS_extracted_k141_9123 | Low-quality |
| SS_extracted_k141_94131 | Not-determined |
| SS_extracted_k141_142107 | Not-determined |
| SS_extracted_k141_124914 | Not-determined |
| SS_extracted_k141_118971 | Not-determined |
| SS_extracted_k141_51912 | Not-determined |
| SS_extracted_k141_87376 | Not-determined |
| SS_extracted_k141_94136 | Not-determined |
| SS_extracted_k141_3961 | Low-quality |
| SS_extracted_k141_106647 | Not-determined |
| SS_extracted_k141_118973 | Not-determined |
| SS_extracted_k141_63455 | Not-determined |
| SS_extracted_k141_94140 | Not-determined |
| SS_extracted_k141_56586 | Not-determined |
| SS_extracted_k141_94145 | Not-determined |
| SS_extracted_k141_124921 | Low-quality |
| SS_extracted_k141_51924 | Not-determined |
| SS_extracted_k141_118986 | Not-determined |
| SS_extracted_k141_98934 | Not-determined |
| SS_extracted_k141_130306 | Not-determined |
| SS_extracted_k141_130311 | Not-determined |
| SS_extracted_k141_38978 | Not-determined |
| SS_extracted_k141_130313 | Low-quality |
| SS_extracted_k141_45483 | Not-determined |
| SS_extracted_k141_51935 | Not-determined |
| SS_extracted_k141_142121 | Not-determined |
| SS_extracted_k141_15408 | Not-determined |
| SS_extracted_k141_118996 | Not-determined |
| SS_extracted_k141_45488 | Not-determined |
| SS_extracted_k141_112147 | Not-determined |
| SS_extracted_k141_112148 | Not-determined |
| SS_extracted_k141_130321 | Not-determined |
| SS_extracted_k141_63472 | Not-determined |
| SS_extracted_k141_63473 | Not-determined |
| SS_extracted_k141_15418 | Not-determined |
| SS_extracted_k141_130325 | Not-determined |
| SS_extracted_k141_112154 | Not-determined |
| SS_extracted_k141_15421 | Not-determined |
| SS_extracted_k141_63475 | Not-determined |
| SS_extracted_k141_51946 | Not-determined |
| SS_extracted_k141_94155 | Low-quality |
| SS_extracted_k141_9134 | Not-determined |
| SS_extracted_k141_45504 | Not-determined |
| SS_extracted_k141_124946 | Not-determined |
| SS_extracted_k141_3984 | Not-determined |
| SS_extracted_k141_26063 | Not-determined |
| SS_extracted_k141_130332 | Not-determined |
| SS_extracted_k141_87415 | Not-determined |
| SS_extracted_k141_87416 | Not-determined |
| SS_extracted_k141_87424 | Not-determined |
| SS_extracted_k141_142141 | Not-determined |
| SS_extracted_k141_45516 | Not-determined |
| SS_extracted_k141_39010 | Low-quality |
| SS_extracted_k141_45518 | Not-determined |
| SS_extracted_k141_63494 | Not-determined |
| SS_extracted_k141_56610 | Not-determined |
| SS_extracted_k141_33562 | Not-determined |
| SS_extracted_k141_9151 | Low-quality |
| SS_extracted_k141_106670 | Not-determined |
| SS_extracted_k141_39019 | Not-determined |
| SS_extracted_k141_135585 | Not-determined |
| SS_extracted_k141_106674 | Not-determined |
| SS_extracted_k141_106678 | Not-determined |
| SS_extracted_k141_124960 | Not-determined |
| SS_extracted_k141_87432 | Not-determined |
| SS_extracted_k141_56613 | Not-determined |
| SS_extracted_k141_124961 | Low-quality |
| SS_extracted_k141_130343 | Not-determined |
| SS_extracted_k141_87436 | Not-determined |
| SS_extracted_k141_106680 | Low-quality |
| SS_extracted_k141_9159 | Low-quality |
| SS_extracted_k141_106683 | Low-quality |
| SS_extracted_k141_124965 | Not-determined |
| SS_extracted_k141_106685 | Not-determined |
| SS_extracted_k141_33579 | Not-determined |
| SS_extracted_k141_45527 | Low-quality |
| SS_extracted_k141_135597 | Not-determined |
| SS_extracted_k141_33581 | Low-quality |
| SS_extracted_k141_63517 | Low-quality |
| SS_extracted_k141_51970 | Not-determined |
| SS_extracted_k141_39031 | Not-determined |
| SS_extracted_k141_39032 | Low-quality |
| SS_extracted_k141_39036 | Not-determined |
| SS_extracted_k141_26077 | Not-determined |
| SS_extracted_k141_112190 | Not-determined |
| SS_extracted_k141_124979 | Low-quality |
| SS_extracted_k141_9185 | Not-determined |
| SS_extracted_k141_124982 | Not-determined |
| SS_extracted_k141_3988 | Not-determined |
| SS_extracted_k141_9187 | Not-determined |
| SS_extracted_k141_39051 | Not-determined |
| SS_extracted_k141_106708 | Low-quality |
| SS_extracted_k141_98951 | Not-determined |
| SS_extracted_k141_39053 | Low-quality |
| SS_extracted_k141_9197 | Not-determined |
| SS_extracted_k141_106716 | Low-quality |
| SS_extracted_k141_106717 | Not-determined |
| SS_extracted_k141_15428 | Low-quality |
| SS_extracted_k141_51977 | Not-determined |
| SS_extracted_k141_15431 | Not-determined |
| SS_extracted_k141_9201 | Low-quality |
| SS_extracted_k141_4002 | Low-quality |
| SS_extracted_k141_135609 | Not-determined |
| SS_extracted_k141_9203 | Not-determined |
| SS_extracted_k141_94162 | Low-quality |
| SS_extracted_k141_130359 | Low-quality |
| SS_extracted_k141_9205 | Not-determined |
| SS_extracted_k141_9206 | Not-determined |
| SS_extracted_k141_4005 | Not-determined |
| SS_extracted_k141_87464 | Not-determined |
| SS_extracted_k141_87465 | Not-determined |
| SS_extracted_k141_106718 | Not-determined |
| SS_extracted_k141_15435 | Not-determined |
| SS_extracted_k141_130365 | Not-determined |
| SS_extracted_k141_130366 | Not-determined |
| SS_extracted_k141_106722 | Not-determined |
| SS_extracted_k141_112200 | Not-determined |
| SS_extracted_k141_21160 | Not-determined |
| SS_extracted_k141_51983 | Low-quality |
| SS_extracted_k141_94165 | Not-determined |
| SS_extracted_k141_106731 | Not-determined |
| SS_extracted_k141_124988 | Not-determined |
| SS_extracted_k141_63550 | Not-determined |
| SS_extracted_k141_4020 | Not-determined |
| SS_extracted_k141_63551 | Low-quality |
| SS_extracted_k141_51989 | Not-determined |
| SS_extracted_k141_9224 | Not-determined |
| SS_extracted_k141_4023 | Not-determined |
| SS_extracted_k141_124990 | Not-determined |
| SS_extracted_k141_15447 | Not-determined |
| SS_extracted_k141_130384 | Not-determined |
| SS_extracted_k141_15449 | Low-quality |
| SS_extracted_k141_106741 | Not-determined |
| SS_extracted_k141_142191 | Low-quality |
| SS_extracted_k141_9237 | Not-determined |
| SS_extracted_k141_63556 | Low-quality |
| SS_extracted_k141_63558 | Not-determined |
| SS_extracted_k141_106743 | Not-determined |
| SS_extracted_k141_94189 | Not-determined |
| SS_extracted_k141_94192 | Low-quality |
| SS_extracted_k141_15464 | Not-determined |
| SS_extracted_k141_9248 | Not-determined |
| SS_extracted_k141_106748 | Not-determined |
| SS_extracted_k141_63565 | Not-determined |
| SS_extracted_k141_15471 | Not-determined |
| SS_extracted_k141_125012 | Not-determined |
| SS_extracted_k141_63573 | Not-determined |
| SS_extracted_k141_106752 | Not-determined |
| SS_extracted_k141_52007 | Not-determined |
| SS_extracted_k141_94200 | Low-quality |
| SS_extracted_k141_33616 | Not-determined |
| SS_extracted_k141_4045 | Not-determined |
| SS_extracted_k141_33626 | Not-determined |
| SS_extracted_k141_87476 | Not-determined |
| SS_extracted_k141_9263 | Not-determined |
| SS_extracted_k141_94213 | Low-quality |
| SS_extracted_k141_33636 | Not-determined |
| SS_extracted_k141_4064 | Low-quality |
| SS_extracted_k141_135638 | Low-quality |
| SS_extracted_k141_94223 | Not-determined |
| SS_extracted_k141_125021 | Not-determined |
| SS_extracted_k141_21173 | Not-determined |
| SS_extracted_k141_112240 | Not-determined |
| SS_extracted_k141_9277 | Not-determined |
| SS_extracted_k141_21175 | Not-determined |
| SS_extracted_k141_112243 | Not-determined |
| SS_extracted_k141_106786 | Not-determined |
| SS_extracted_k141_52025 | Not-determined |
| SS_extracted_k141_130428 | Not-determined |
| SS_extracted_k141_135643 | Low-quality |
| SS_extracted_k141_39065 | Not-determined |
| SS_extracted_k141_9281 | Low-quality |
| SS_extracted_k141_94236 | Not-determined |
| SS_extracted_k141_112255 | Not-determined |
| SS_extracted_k141_135651 | Not-determined |
| SS_extracted_k141_9290 | Not-determined |
| SS_extracted_k141_52038 | Not-determined |
| SS_extracted_k141_112262 | Not-determined |
| SS_extracted_k141_52040 | Low-quality |
| SS_extracted_k141_21189 | Low-quality |
| SS_extracted_k141_9297 | Low-quality |
| SS_extracted_k141_63610 | Not-determined |
| SS_extracted_k141_94266 | Not-determined |
| SS_extracted_k141_112283 | Low-quality |
| SS_extracted_k141_112286 | Not-determined |
| SS_extracted_k141_52074 | Not-determined |
| SS_extracted_k141_33694 | Not-determined |
| SS_extracted_k141_135671 | Not-determined |
| SS_extracted_k141_63660 | Not-determined |
| SS_extracted_k141_130466 | Not-determined |
| SS_extracted_k141_125033 | Not-determined |
| SS_extracted_k141_9320 | Not-determined |
| SS_extracted_k141_94283 | Low-quality |
| SS_extracted_k141_94284 | Low-quality |
| SS_extracted_k141_33708 | Not-determined |
| SS_extracted_k141_39086 | Not-determined |
| SS_extracted_k141_98998 | Not-determined |
| SS_extracted_k141_112288 | Low-quality |
| SS_extracted_k141_9331 | Not-determined |
| SS_extracted_k141_135675 | Not-determined |
| SS_extracted_k141_125049 | Not-determined |
| SS_extracted_k141_106836 | Not-determined |
| SS_extracted_k141_39100 | Not-determined |
| SS_extracted_k141_125050 | Not-determined |
| SS_extracted_k141_106838 | Not-determined |
| SS_extracted_k141_106839 | Low-quality |
| SS_extracted_k141_9339 | Low-quality |
| SS_extracted_k141_9342 | Not-determined |
| SS_extracted_k141_33722 | Not-determined |
| SS_extracted_k141_106840 | Not-determined |
| SS_extracted_k141_45579 | Not-determined |
| SS_extracted_k141_135681 | Not-determined |
| SS_extracted_k141_9349 | Not-determined |
| SS_extracted_k141_9350 | Not-determined |
| SS_extracted_k141_52103 | Not-determined |
| SS_extracted_k141_119009 | Not-determined |
| SS_extracted_k141_26101 | Not-determined |
| SS_extracted_k141_125066 | Not-determined |
| SS_extracted_k141_106858 | Low-quality |
| SS_extracted_k141_106860 | Not-determined |
| SS_extracted_k141_39106 | Not-determined |
| SS_extracted_k141_9366 | Not-determined |
| SS_extracted_k141_125078 | Not-determined |
| SS_extracted_k141_135693 | Not-determined |
| SS_extracted_k141_142215 | Low-quality |
| SS_extracted_k141_106869 | Low-quality |
| SS_extracted_k141_9371 | Not-determined |
| SS_extracted_k141_45595 | Not-determined |
| SS_extracted_k141_9372 | Not-determined |
| SS_extracted_k141_125083 | Not-determined |
| SS_extracted_k141_142225 | Not-determined |
| SS_extracted_k141_9375 | Not-determined |
| SS_extracted_k141_106872 | Not-determined |
| SS_extracted_k141_106873 | Not-determined |
| SS_extracted_k141_106874 | Not-determined |
| SS_extracted_k141_52137 | Not-determined |
| SS_extracted_k141_125090 | Not-determined |
| SS_extracted_k141_52143 | Not-determined |
| SS_extracted_k141_45599 | Low-quality |
| SS_extracted_k141_26129 | Not-determined |
| SS_extracted_k141_45602 | Not-determined |
| SS_extracted_k141_52149 | Not-determined |
| SS_extracted_k141_39126 | Not-determined |
| SS_extracted_k141_52155 | Not-determined |
| SS_extracted_k141_39127 | Not-determined |
| SS_extracted_k141_142246 | Not-determined |
| SS_extracted_k141_112315 | Not-determined |
| SS_extracted_k141_112316 | Low-quality |
| SS_extracted_k141_142249 | Low-quality |
| SS_extracted_k141_33756 | Low-quality |
| SS_extracted_k141_80521 | Not-determined |
| SS_extracted_k141_106893 | Not-determined |
| SS_extracted_k141_94341 | Not-determined |
| SS_extracted_k141_99013 | Low-quality |
| SS_extracted_k141_125113 | Not-determined |
| SS_extracted_k141_125114 | Not-determined |
| SS_extracted_k141_52164 | Not-determined |
| SS_extracted_k141_130491 | Not-determined |
| SS_extracted_k141_125117 | Not-determined |
| SS_extracted_k141_4111 | Not-determined |
| SS_extracted_k141_45626 | Not-determined |
| SS_extracted_k141_87524 | Not-determined |
| SS_extracted_k141_142271 | Low-quality |
| SS_extracted_k141_106911 | Not-determined |
| SS_extracted_k141_130494 | Not-determined |
| SS_extracted_k141_26156 | Not-determined |
| SS_extracted_k141_130496 | Not-determined |
| SS_extracted_k141_39140 | Not-determined |
| SS_extracted_k141_130499 | Not-determined |
| SS_extracted_k141_39141 | Not-determined |
| SS_extracted_k141_142279 | Not-determined |
| SS_extracted_k141_39144 | Not-determined |
| SS_extracted_k141_87536 | Not-determined |

| | |
|---|---|
| SS_extracted_k141_87538 | Low-quality |
| SS_extracted_k141_4125 | Not-determined |
| SS_extracted_k141_87540 | Not-determined |
| SS_extracted_k141_52182 | Not-determined |
| SS_extracted_k141_4128 | Low-quality |
| SS_extracted_k141_52187 | Not-determined |
| SS_extracted_k141_9404 | Not-determined |
| SS_extracted_k141_4131 | Not-determined |
| SS_extracted_k141_112336 | Low-quality |
| SS_extracted_k141_39157 | Not-determined |
| SS_extracted_k141_87542 | Not-determined |
| SS_extracted_k141_106925 | Not-determined |
| SS_extracted_k141_125138 | Low-quality |
| SS_extracted_k141_56631 | Not-determined |
| SS_extracted_k141_135714 | Not-determined |
| SS_extracted_k141_74651 | Not-determined |
| SS_extracted_k141_142284 | Not-determined |
| SS_extracted_k141_106932 | Not-determined |
| SS_extracted_k141_142288 | Not-determined |
| SS_extracted_k141_52195 | Not-determined |
| SS_extracted_k141_80544 | Not-determined |
| SS_extracted_k141_26166 | Not-determined |
| SS_extracted_k141_26168 | Not-determined |
| SS_extracted_k141_106945 | Low-quality |
| SS_extracted_k141_39173 | Not-determined |
| SS_extracted_k141_125153 | Not-determined |
| SS_extracted_k141_106950 | Not-determined |
| SS_extracted_k141_125156 | Low-quality |
| SS_extracted_k141_106952 | Not-determined |
| SS_extracted_k141_80558 | Low-quality |
| SS_extracted_k141_39179 | Not-determined |
| SS_extracted_k141_125158 | Low-quality |
| SS_extracted_k141_74657 | Not-determined |
| SS_extracted_k141_106954 | Not-determined |
| SS_extracted_k141_87546 | Not-determined |
| SS_extracted_k141_87548 | Not-determined |
| SS_extracted_k141_87553 | Not-determined |
| SS_extracted_k141_4166 | Not-determined |
| SS_extracted_k141_21214 | Not-determined |
| SS_extracted_k141_106974 | Low-quality |
| SS_extracted_k141_142300 | Not-determined |
| SS_extracted_k141_142303 | Not-determined |
| SS_extracted_k141_106978 | Not-determined |
| SS_extracted_k141_26188 | Not-determined |
| SS_extracted_k141_112351 | Not-determined |
| SS_extracted_k141_26192 | Not-determined |
| SS_extracted_k141_112361 | Not-determined |
| SS_extracted_k141_142314 | Not-determined |
| SS_extracted_k141_135737 | Not-determined |
| SS_extracted_k141_106983 | Not-determined |
| SS_extracted_k141_106984 | Not-determined |
| SS_extracted_k141_4186 | Not-determined |
| SS_extracted_k141_119026 | Not-determined |
| SS_extracted_k141_87579 | Not-determined |
| SS_extracted_k141_80589 | Not-determined |
| SS_extracted_k141_52238 | Not-determined |
| SS_extracted_k141_142321 | Not-determined |
| SS_extracted_k141_99042 | Low-quality |
| SS_extracted_k141_4192 | Not-determined |
| SS_extracted_k141_4194 | Low-quality |
| SS_extracted_k141_52239 | Not-determined |
| SS_extracted_k141_87587 | Not-determined |
| SS_extracted_k141_39199 | Not-determined |
| SS_extracted_k141_119038 | Low-quality |
| SS_extracted_k141_119044 | Not-determined |
| SS_extracted_k141_142328 | Not-determined |
| SS_extracted_k141_4217 | Not-determined |
| SS_extracted_k141_45670 | Not-determined |
| SS_extracted_k141_33815 | Not-determined |
| SS_extracted_k141_33819 | Not-determined |
| SS_extracted_k141_125227 | Not-determined |
| SS_extracted_k141_142339 | Not-determined |
| SS_extracted_k141_125228 | Not-determined |
| SS_extracted_k141_33822 | Not-determined |
| SS_extracted_k141_87600 | Not-determined |
| SS_extracted_k141_39225 | Not-determined |
| SS_extracted_k141_4235 | Not-determined |
| SS_extracted_k141_130517 | Not-determined |
| SS_extracted_k141_26201 | Not-determined |
| SS_extracted_k141_26204 | Not-determined |
| SS_extracted_k141_87608 | Not-determined |
| SS_extracted_k141_142350 | Low-quality |
| SS_extracted_k141_142352 | Not-determined |
| SS_extracted_k141_87614 | Not-determined |
| SS_extracted_k141_4252 | Not-determined |
| SS_extracted_k141_99049 | Not-determined |
| SS_extracted_k141_142360 | Low-quality |
| SS_extracted_k141_119057 | Not-determined |
| SS_extracted_k141_39232 | Not-determined |
| SS_extracted_k141_142363 | Not-determined |
| SS_extracted_k141_52258 | Not-determined |
| SS_extracted_k141_130542 | Low-quality |
| SS_extracted_k141_112369 | Not-determined |
| SS_extracted_k141_119068 | Not-determined |
| SS_extracted_k141_119069 | Low-quality |
| SS_extracted_k141_142371 | Not-determined |
| SS_extracted_k141_4265 | Low-quality |
| SS_extracted_k141_39245 | Not-determined |
| SS_extracted_k141_142373 | Not-determined |
| SS_extracted_k141_39248 | Not-determined |
| SS_extracted_k141_39249 | Low-quality |
| SS_extracted_k141_130551 | Not-determined |
| SS_extracted_k141_130554 | Not-determined |
| SS_vs_k141_145604\|\|full | High-quality |
| SS_extracted_k141_106998 | Not-determined |
| SS_extracted_k141_106999 | Not-determined |
| SS_extracted_k141_107001 | Not-determined |
| SS_extracted_k141_99059 | Not-determined |
| SS_extracted_k141_130570 | Not-determined |
| SS_extracted_k141_99061 | Low-quality |
| SS_extracted_k141_4280 | Low-quality |
| SS_extracted_k141_80626 | Low-quality |
| SS_extracted_k141_33877 | Not-determined |
| SS_extracted_k141_4281 | Low-quality |
| SS_extracted_k141_130576 | Not-determined |
| SS_extracted_k141_107022 | Not-determined |
| SS_extracted_k141_119096 | Not-determined |
| SS_extracted_k141_142387 | Not-determined |
| SS_extracted_k141_107029 | Not-determined |
| SS_extracted_k141_45701 | Not-determined |
| SS_extracted_k141_125281 | Not-determined |
| SS_extracted_k141_39256 | Not-determined |
| SS_extracted_k141_80629 | Not-determined |
| SS_extracted_k141_4299 | Not-determined |
| SS_extracted_k141_107039 | Not-determined |
| SS_extracted_k141_4305 | Not-determined |
| SS_extracted_k141_4307 | Not-determined |
| SS_extracted_k141_26217 | Not-determined |
| SS_extracted_k141_87681 | Not-determined |
| SS_extracted_k141_125291 | Not-determined |
| SS_extracted_k141_33887 | Not-determined |
| SS_extracted_k141_80641 | Not-determined |
| SS_extracted_k141_107052 | Low-quality |
| SS_extracted_k141_26227 | Not-determined |
| SS_extracted_k141_80643 | Low-quality |
| SS_extracted_k141_125300 | Not-determined |
| SS_extracted_k141_130597 | Not-determined |
| SS_extracted_k141_80655 | Not-determined |
| SS_extracted_k141_52292 | Not-determined |
| SS_extracted_k141_26235 | Not-determined |
| SS_extracted_k141_80658 | Low-quality |
| SS_extracted_k141_33893 | Not-determined |
| SS_extracted_k141_45716 | Low-quality |
| SS_extracted_k141_99071 | Low-quality |
| SS_extracted_k141_33898 | Low-quality |
| SS_extracted_k141_74670 | Not-determined |
| SS_extracted_k141_142395 | Low-quality |
| SS_extracted_k141_33902 | Not-determined |
| SS_extracted_k141_63681 | Not-determined |
| SS_extracted_k141_135765 | Not-determined |
| SS_extracted_k141_125318 | Not-determined |
| SS_extracted_k141_107067 | Not-determined |
| SS_extracted_k141_45724 | Not-determined |
| SS_extracted_k141_69584 | Not-determined |
| SS_extracted_k141_125325 | Not-determined |
| SS_extracted_k141_125328 | Not-determined |
| SS_extracted_k141_33910 | Not-determined |
| SS_extracted_k141_33911 | Not-determined |
| SS_extracted_k141_9435 | Not-determined |
| SS_extracted_k141_26247 | Low-quality |
| SS_extracted_k141_125333 | Not-determined |
| SS_extracted_k141_69595 | Not-determined |
| SS_extracted_k141_142409 | Not-determined |
| SS_extracted_k141_33915 | Low-quality |
| SS_extracted_k141_69597 | Low-quality |
| SS_extracted_k141_142412 | Not-determined |
| SS_extracted_k141_45737 | Not-determined |
| SS_extracted_k141_9447 | Not-determined |
| SS_extracted_k141_87683 | Not-determined |
| SS_extracted_k141_63697 | Not-determined |
| SS_extracted_k141_107087 | Not-determined |
| SS_extracted_k141_107089 | Low-quality |
| SS_extracted_k141_74695 | Not-determined |
| SS_extracted_k141_45746 | Not-determined |
| SS_extracted_k141_130630 | Not-determined |
| SS_extracted_k141_87685 | Low-quality |
| SS_extracted_k141_130632 | Not-determined |
| SS_extracted_k141_112410 | Not-determined |
| SS_extracted_k141_112411 | Low-quality |
| SS_extracted_k141_4345 | Not-determined |
| SS_extracted_k141_26258 | Low-quality |
| SS_extracted_k141_130636 | Low-quality |
| SS_extracted_k141_142418 | Not-determined |
| SS_extracted_k141_130637 | Not-determined |
| SS_extracted_k141_130642 | Not-determined |
| SS_extracted_k141_107098 | Not-determined |
| SS_extracted_k141_87695 | Not-determined |
| SS_extracted_k141_142424 | Not-determined |
| SS_extracted_k141_142425 | Not-determined |
| SS_extracted_k141_26267 | Not-determined |
| SS_extracted_k141_135775 | Not-determined |
| SS_extracted_k141_130647 | Not-determined |
| SS_extracted_k141_112427 | Not-determined |
| SS_extracted_k141_130648 | Not-determined |
| SS_extracted_k141_130655 | Low-quality |
| SS_extracted_k141_33936 | Low-quality |
| SS_extracted_k141_45770 | Not-determined |
| SS_extracted_k141_45771 | Not-determined |
| SS_extracted_k141_130662 | Not-determined |
| SS_extracted_k141_80709 | Not-determined |
| SS_extracted_k141_45775 | Not-determined |
| SS_extracted_k141_87713 | Not-determined |
| SS_extracted_k141_135778 | Not-determined |
| SS_extracted_k141_142439 | Not-determined |
| SS_extracted_k141_135783 | Low-quality |
| SS_extracted_k141_4364 | Not-determined |
| SS_extracted_k141_45780 | Not-determined |
| SS_extracted_k141_80722 | Not-determined |
| SS_extracted_k141_130675 | Not-determined |
| SS_extracted_k141_87721 | Not-determined |
| SS_extracted_k141_63722 | Low-quality |
| SS_extracted_k141_9480 | Not-determined |
| SS_extracted_k141_9481 | Not-determined |
| SS_extracted_k141_125340 | Low-quality |
| SS_extracted_k141_33956 | Low-quality |
| SS_extracted_k141_52336 | Not-determined |
| SS_extracted_k141_4376 | Not-determined |
| SS_extracted_k141_74729 | Low-quality |
| SS_extracted_k141_33962 | Not-determined |
| SS_extracted_k141_4379 | Not-determined |
| SS_extracted_k141_112435 | Low-quality |
| SS_extracted_k141_4380 | Not-determined |
| SS_extracted_k141_130697 | Not-determined |
| SS_extracted_k141_52339 | Not-determined |
| SS_extracted_k141_130699 | Not-determined |
| SS_extracted_k141_4383 | Not-determined |
| SS_extracted_k141_33966 | Low-quality |
| SS_extracted_k141_9495 | Not-determined |
| SS_extracted_k141_63753 | Not-determined |
| SS_extracted_k141_130708 | Low-quality |
| SS_extracted_k141_52352 | Not-determined |
| SS_extracted_k141_119109 | Low-quality |
| SS_extracted_k141_74748 | Low-quality |
| SS_extracted_k141_87732 | Not-determined |
| SS_extracted_k141_107112 | Not-determined |
| SS_extracted_k141_9504 | Low-quality |
| SS_extracted_k141_63765 | Not-determined |
| SS_extracted_k141_87740 | Not-determined |
| SS_extracted_k141_130717 | Low-quality |
| SS_extracted_k141_69620 | Not-determined |
| SS_extracted_k141_119123 | Not-determined |
| SS_extracted_k141_130722 | Low-quality |
| SS_extracted_k141_63767 | Low-quality |
| SS_extracted_k141_69625 | Not-determined |
| SS_extracted_k141_63770 | Not-determined |
| SS_extracted_k141_69627 | Not-determined |
| SS_extracted_k141_4402 | Not-determined |
| SS_extracted_k141_26289 | Low-quality |
| SS_extracted_k141_99113 | Not-determined |
| SS_extracted_k141_34010 | Low-quality |
| SS_extracted_k141_69636 | Not-determined |
| SS_extracted_k141_99115 | Not-determined |
| SS_extracted_k141_130729 | Not-determined |
| SS_extracted_k141_52365 | Low-quality |
| SS_extracted_k141_9523 | Not-determined |
| SS_extracted_k141_130733 | Not-determined |
| SS_extracted_k141_9531 | Not-determined |
| SS_extracted_k141_119137 | Not-determined |
| SS_extracted_k141_4415 | Low-quality |
| SS_extracted_k141_4416 | Not-determined |
| SS_extracted_k141_119147 | Not-determined |
| SS_extracted_k141_45814 | Not-determined |
| SS_extracted_k141_69656 | Not-determined |
| SS_extracted_k141_107134 | Not-determined |
| SS_extracted_k141_45825 | Not-determined |
| SS_extracted_k141_52390 | Low-quality |
| SS_extracted_k141_34019 | Not-determined |
| SS_extracted_k141_87753 | Not-determined |
| SS_extracted_k141_45828 | Not-determined |
| SS_extracted_k141_69667 | Not-determined |
| SS_extracted_k141_4429 | Not-determined |
| SS_extracted_k141_69669 | Low-quality |
| SS_extracted_k141_107140 | Low-quality |
| SS_extracted_k141_26290 | Not-determined |
| SS_extracted_k141_130744 | Not-determined |
| SS_extracted_k141_69672 | Low-quality |
| SS_extracted_k141_26298 | Low-quality |
| SS_extracted_k141_125373 | Not-determined |
| SS_extracted_k141_52402 | Low-quality |
| SS_extracted_k141_34029 | Not-determined |
| SS_extracted_k141_135821 | Not-determined |
| SS_extracted_k141_130754 | Not-determined |
| SS_extracted_k141_130755 | Not-determined |
| SS_extracted_k141_130758 | Not-determined |
| SS_extracted_k141_135826 | Low-quality |
| SS_extracted_k141_26308 | Not-determined |
| SS_extracted_k141_125383 | Not-determined |
| SS_extracted_k141_130762 | Not-determined |
| SS_extracted_k141_45846 | Not-determined |
| SS_extracted_k141_4453 | Low-quality |
| SS_extracted_k141_125395 | Not-determined |
| SS_extracted_k141_45848 | Not-determined |
| SS_extracted_k141_4457 | Low-quality |
| SS_extracted_k141_15522 | Not-determined |
| SS_extracted_k141_107174 | Not-determined |
| SS_extracted_k141_45852 | Not-determined |
| SS_extracted_k141_107178 | Not-determined |
| SS_extracted_k141_99178 | Not-determined |
| SS_extracted_k141_107180 | Low-quality |
| SS_extracted_k141_107181 | Not-determined |
| SS_extracted_k141_15528 | Not-determined |
| SS_extracted_k141_107183 | Not-determined |
| SS_extracted_k141_87784 | Not-determined |
| SS_extracted_k141_119165 | Not-determined |
| SS_extracted_k141_119167 | Low-quality |
| SS_extracted_k141_135831 | Not-determined |
| SS_extracted_k141_125417 | Not-determined |
| SS_extracted_k141_4465 | Low-quality |
| SS_extracted_k141_4469 | Not-determined |
| SS_extracted_k141_4470 | Not-determined |
| SS_extracted_k141_87790 | Not-determined |
| SS_extracted_k141_34063 | Not-determined |
| SS_extracted_k141_15540 | Not-determined |
| SS_extracted_k141_4473 | Not-determined |
| SS_extracted_k141_34065 | Not-determined |
| SS_extracted_k141_4475 | Not-determined |
| SS_extracted_k141_45875 | Not-determined |
| SS_extracted_k141_107192 | Low-quality |
| SS_extracted_k141_135842 | Not-determined |
| SS_extracted_k141_45876 | Not-determined |
| SS_extracted_k141_4477 | Not-determined |
| SS_extracted_k141_34069 | Not-determined |
| SS_extracted_k141_125429 | Not-determined |
| SS_extracted_k141_130774 | Not-determined |
| SS_extracted_k141_130777 | Low-quality |
| SS_extracted_k141_99200 | Low-quality |
| SS_extracted_k141_45881 | Low-quality |
| SS_extracted_k141_119183 | Not-determined |
| SS_extracted_k141_45886 | Not-determined |
| SS_extracted_k141_130781 | Low-quality |
| SS_extracted_k141_45890 | Not-determined |
| SS_extracted_k141_87807 | Not-determined |
| SS_extracted_k141_142482 | Not-determined |
| SS_extracted_k141_119197 | Not-determined |
| SS_extracted_k141_34092 | Not-determined |
| SS_extracted_k141_119202 | Not-determined |
| SS_extracted_k141_142491 | Low-quality |
| SS_extracted_k141_142493 | Not-determined |
| SS_extracted_k141_52452 | Not-determined |
| SS_extracted_k141_125449 | Not-determined |
| SS_extracted_k141_142496 | Not-determined |
| SS_extracted_k141_34101 | Not-determined |
| S8_vs_k141_72329\|\|full | High-quality |
| SS_extracted_k141_142504 | Not-determined |
| SS_extracted_k141_39303 | Not-determined |
| SS_extracted_k141_9552 | Not-determined |
| SS_extracted_k141_52471 | Not-determined |
| SS_extracted_k141_39310 | Not-determined |
| SS_extracted_k141_94355 | Not-determined |
| SS_extracted_k141_125473 | Not-determined |
| SS_extracted_k141_119218 | Not-determined |
| SS_extracted_k141_119222 | Not-determined |
| SS_extracted_k141_112443 | Not-determined |
| SS_extracted_k141_45927 | Not-determined |
| SS_extracted_k141_39325 | Not-determined |
| SS_extracted_k141_125481 | Not-determined |
| SS_extracted_k141_99217 | Low-quality |
| SS_extracted_k141_34122 | Not-determined |
| SS_extracted_k141_107223 | Not-determined |
| SS_extracted_k141_119235 | Not-determined |
| SS_extracted_k141_125498 | Not-determined |
| SS_extracted_k141_125499 | Not-determined |
| SS_extracted_k141_107233 | Not-determined |
| SS_extracted_k141_119242 | Low-quality |
| SS_extracted_k141_107242 | Not-determined |
| SS_extracted_k141_99228 | Not-determined |
| SS_extracted_k141_45943 | Low-quality |
| SS_extracted_k141_45944 | Not-determined |
| SS_extracted_k141_52508 | Not-determined |
| SS_extracted_k141_99235 | Not-determined |
| SS_extracted_k141_87837 | Low-quality |
| SS_extracted_k141_125522 | Not-determined |
| SS_extracted_k141_107257 | Not-determined |
| SS_extracted_k141_130834 | Low-quality |
| SS_extracted_k141_39338 | Not-determined |
| SS_extracted_k141_125531 | Not-determined |
| SS_extracted_k141_107267 | Not-determined |
| SS_extracted_k141_45966 | Not-determined |
| SS_extracted_k141_125538 | Not-determined |
| SS_extracted_k141_112458 | Not-determined |
| SS_extracted_k141_80749 | Not-determined |
| SS_extracted_k141_69705 | Not-determined |
| SS_extracted_k141_69707 | Not-determined |
| SS_extracted_k141_99257 | Not-determined |
| SS_extracted_k141_52531 | Low-quality |
| SS_extracted_k141_125542 | Not-determined |
| SS_extracted_k141_125543 | Not-determined |
| SS_extracted_k141_39351 | Not-determined |
| SS_extracted_k141_80754 | Low-quality |
| SS_extracted_k141_39353 | Low-quality |
| SS_extracted_k141_69712 | Not-determined |
| SS_extracted_k141_69716 | Not-determined |
| SS_extracted_k141_99274 | Low-quality |
| SS_extracted_k141_34141 | Low-quality |
| SS_extracted_k141_135876 | Not-determined |
| SS_extracted_k141_52545 | Not-determined |
| SS_extracted_k141_125557 | Not-determined |
| SS_extracted_k141_135877 | Low-quality |
| SS_extracted_k141_80757 | Low-quality |
| SS_extracted_k141_34146 | Not-determined |
| SS_extracted_k141_9597 | Not-determined |
| SS_extracted_k141_45983 | Not-determined |
| SS_extracted_k141_125563 | Not-determined |
| SS_extracted_k141_34151 | Not-determined |
| SS_extracted_k141_9604 | Not-determined |
| SS_extracted_k141_21271 | Not-determined |
| SS_extracted_k141_15553 | Not-determined |
| SS_extracted_k141_46000 | Not-determined |
| SS_extracted_k141_125584 | Not-determined |
| SS_extracted_k141_21278 | Not-determined |
| SS_extracted_k141_99280 | Not-determined |
| SS_extracted_k141_125589 | Not-determined |
| SS_extracted_k141_99286 | Not-determined |
| SS_extracted_k141_119266 | Low-quality |
| SS_extracted_k141_52568 | Not-determined |
| SS_extracted_k141_125598 | Not-determined |
| SS_extracted_k141_34171 | Not-determined |
| SS_extracted_k141_63782 | Not-determined |
| SS_extracted_k141_34174 | Not-determined |
| SS_extracted_k141_125611 | Low-quality |
| SS_extracted_k141_39381 | Not-determined |
| SS_extracted_k141_46020 | Low-quality |
| SS_extracted_k141_63789 | Low-quality |
| SS_extracted_k141_63790 | Not-determined |
| SS_extracted_k141_99306 | Not-determined |
| SS_extracted_k141_69753 | Not-determined |
| SS_extracted_k141_69755 | Not-determined |
| SS_extracted_k141_46032 | Not-determined |
| SS_extracted_k141_107328 | Not-determined |
| SS_extracted_k141_69756 | Low-quality |
| SS_extracted_k141_135888 | Low-quality |
| SS_extracted_k141_63806 | Not-determined |
| SS_extracted_k141_69757 | Not-determined |
| SS_extracted_k141_46039 | Low-quality |
| SS_extracted_k141_107337 | Not-determined |
| SS_extracted_k141_107338 | Not-determined |
| SS_extracted_k141_112516 | Not-determined |
| SS_extracted_k141_99321 | Not-determined |
| SS_extracted_k141_63813 | Low-quality |
| SS_extracted_k141_99323 | Not-determined |
| SS_extracted_k141_34185 | Not-determined |
| SS_extracted_k141_119274 | Low-quality |
| SS_extracted_k141_125621 | Not-determined |
| SS_extracted_k141_119275 | Not-determined |
| SS_extracted_k141_119277 | Low-quality |
| SS_extracted_k141_135891 | Not-determined |
| SS_extracted_k141_135894 | Not-determined |
| SS_extracted_k141_99338 | Not-determined |
| SS_extracted_k141_34194 | Not-determined |
| SS_extracted_k141_39408 | Not-determined |
| SS_extracted_k141_69771 | Not-determined |
| SS_extracted_k141_112529 | Low-quality |
| SS_extracted_k141_107360 | Low-quality |
| SS_extracted_k141_107362 | Not-determined |
| SS_extracted_k141_69774 | Not-determined |
| SS_extracted_k141_39412 | Low-quality |
| SS_extracted_k141_87859 | Not-determined |
| SS_extracted_k141_39413 | Not-determined |
| SS_extracted_k141_34200 | Low-quality |
| SS_extracted_k141_112536 | Low-quality |
| SS_extracted_k141_39420 | Low-quality |
| SS_extracted_k141_125648 | Not-determined |
| SS_extracted_k141_125652 | Not-determined |
| SS_extracted_k141_69785 | Not-determined |
| SS_extracted_k141_125654 | Not-determined |
| SS_extracted_k141_74763 | Not-determined |
| S16_vs_97502\|\|full | Medium-quality |
| SS_extracted_k141_112546 | Not-determined |
| SS_extracted_k141_107372 | Not-determined |
| SS_extracted_k141_39433 | Not-determined |
| SS_extracted_k141_63835 | Low-quality |
| SS_extracted_k141_119303 | Low-quality |
| SS_extracted_k141_21306 | Not-determined |
| SS_extracted_k141_21308 | Not-determined |
| SS_extracted_k141_39440 | Not-determined |
| SS_extracted_k141_21310 | Not-determined |
| SS_extracted_k141_125659 | Not-determined |
| SS_extracted_k141_125662 | Not-determined |
| SS_extracted_k141_39447 | Not-determined |
| SS_extracted_k141_63848 | Not-determined |
| SS_extracted_k141_21316 | Not-determined |
| SS_extracted_k141_142549 | Not-determined |
| SS_extracted_k141_4517 | Not-determined |
| SS_extracted_k141_107389 | Not-determined |
| SS_extracted_k141_39455 | Not-determined |
| SS_extracted_k141_119312 | Not-determined |
| SS_extracted_k141_130843 | Not-determined |
| SS_extracted_k141_130844 | Not-determined |
| SS_extracted_k141_119315 | Not-determined |
| SS_extracted_k141_112555 | Not-determined |
| SS_extracted_k141_21320 | Not-determined |
| SS_extracted_k141_4527 | Not-determined |
| SS_extracted_k141_4531 | Not-determined |
| SS_extracted_k141_9627 | Low-quality |
| SS_extracted_k141_107397 | Low-quality |
| SS_extracted_k141_119328 | Not-determined |
| SS_extracted_k141_119330 | Not-determined |
| SS_extracted_k141_107401 | Not-determined |
| SS_extracted_k141_69804 | Not-determined |
| SS_extracted_k141_119334 | Not-determined |
| SS_extracted_k141_39467 | Low-quality |
| SS_extracted_k141_74785 | Not-determined |
| SS_extracted_k141_39470 | Not-determined |
| SS_extracted_k141_99359 | Not-determined |
| SS_extracted_k141_87911 | Not-determined |
| SS_extracted_k141_87917 | Not-determined |
| SS_extracted_k141_69816 | Not-determined |
| SS_extracted_k141_94389 | Not-determined |
| SS_extracted_k141_39479 | Not-determined |
| SS_extracted_k141_125677 | Not-determined |
| SS_extracted_k141_87928 | Not-determined |
| SS_extracted_k141_87929 | Not-determined |
| SS_extracted_k141_112572 | Not-determined |
| SS_extracted_k141_69820 | Not-determined |
| SS_extracted_k141_112574 | Not-determined |
| SS_extracted_k141_99370 | Low-quality |
| SS_extracted_k141_63871 | Not-determined |
| SS_extracted_k141_9653 | Not-determined |
| SS_extracted_k141_69824 | Not-determined |
| SS_extracted_k141_107428 | Not-determined |
| SS_extracted_k141_94392 | Low-quality |
| SS_extracted_k141_34211 | Not-determined |
| SS_extracted_k141_125701 | Not-determined |
| SS_extracted_k141_94405 | Not-determined |
| SS_extracted_k141_52598 | Not-determined |
| SS_extracted_k141_52603 | Not-determined |
| SS_extracted_k141_63880 | Not-determined |
| SS_extracted_k141_119366 | Low-quality |
| SS_extracted_k141_119369 | Not-determined |
| SS_extracted_k141_112603 | Not-determined |
| SS_extracted_k141_125709 | Not-determined |
| SS_extracted_k141_39493 | Not-determined |
| SS_extracted_k141_52611 | Not-determined |
| SS_extracted_k141_80771 | Not-determined |
| SS_extracted_k141_107443 | Low-quality |
| SS_extracted_k141_112621 | Not-determined |
| SS_extracted_k141_94422 | Not-determined |
| SS_extracted_k141_119396 | Not-determined |
| SS_extracted_k141_34239 | Not-determined |
| SS_extracted_k141_119398 | Not-determined |
| SS_extracted_k141_9669 | Not-determined |
| SS_extracted_k141_135948 | Low-quality |
| SS_extracted_k141_4555 | Not-determined |
| SS_extracted_k141_119405 | Low-quality |
| SS_extracted_k141_63890 | Not-determined |
| SS_extracted_k141_63891 | Not-determined |
| SS_extracted_k141_21350 | Not-determined |
| SS_extracted_k141_52620 | Not-determined |
| SS_extracted_k141_21353 | Not-determined |
| SS_extracted_k141_87970 | Not-determined |
| SS_extracted_k141_63899 | Not-determined |
| SS_extracted_k141_15593 | Not-determined |
| SS_extracted_k141_21354 | Low-quality |
| SS_extracted_k141_142587 | Not-determined |
| SS_extracted_k141_130903 | Not-determined |
| SS_extracted_k141_112648 | Not-determined |
| SS_extracted_k141_119419 | Not-determined |
| SS_extracted_k141_112651 | Not-determined |
| SS_extracted_k141_52633 | Not-determined |
| SS_extracted_k141_63904 | Not-determined |
| SS_extracted_k141_52635 | Not-determined |
| SS_extracted_k141_63905 | Not-determined |
| SS_extracted_k141_9688 | Not-determined |
| SS_extracted_k141_119425 | Low-quality |
| SS_extracted_k141_119426 | Not-determined |
| SS_extracted_k141_4571 | Low-quality |
| SS_extracted_k141_94445 | Low-quality |
| SS_extracted_k141_130910 | Not-determined |
| SS_extracted_k141_4572 | Not-determined |
| SS_extracted_k141_15599 | Low-quality |
| SS_extracted_k141_26328 | Low-quality |
| SS_extracted_k141_94448 | Not-determined |
| SS_extracted_k141_4577 | Not-determined |
| SS_extracted_k141_87985 | Not-determined |
| SS_extracted_k141_52650 | Low-quality |
| SS_extracted_k141_125734 | Not-determined |
| SS_extracted_k141_130915 | Not-determined |
| SS_extracted_k141_52654 | Not-determined |
| SS_extracted_k141_63919 | Low-quality |
| SS_extracted_k141_125743 | Not-determined |
| SS_extracted_k141_52663 | Not-determined |
| SS_extracted_k141_80781 | Low-quality |
| SS_extracted_k141_26343 | Not-determined |
| SS_extracted_k141_52666 | Not-determined |
| SS_extracted_k141_94466 | Low-quality |
| SS_extracted_k141_130926 | Not-determined |
| SS_extracted_k141_125749 | Not-determined |
| SS_extracted_k141_9714 | Low-quality |
| SS_extracted_k141_52678 | Not-determined |

| | |
|---|---|
| SS_extracted_k141_88004 | Not-determined |
| SS_extracted_k141_26351 | Not-determined |
| SS_extracted_k141_130933 | Not-determined |
| SS_extracted_k141_130938 | Not-determined |
| SS_extracted_k141_15616 | Not-determined |
| SS_extracted_k141_15617 | Not-determined |
| SS_extracted_k141_21392 | Not-determined |
| SS_extracted_k141_107476 | Not-determined |
| SS_extracted_k141_130943 | Not-determined |
| SS_extracted_k141_125761 | Not-determined |
| SS_extracted_k141_130949 | Not-determined |
| SS_extracted_k141_15630 | Not-determined |
| SS_extracted_k141_107485 | Not-determined |
| SS_extracted_k141_107489 | Not-determined |
| SS_extracted_k141_21416 | Not-determined |
| SS_extracted_k141_119449 | Low-quality |
| SS_extracted_k141_125770 | Not-determined |
| SS_extracted_k141_21425 | Low-quality |
| SS_extracted_k141_4604 | Low-quality |
| SS_extracted_k141_112681 | Low-quality |
| SS_extracted_k141_107517 | Not-determined |
| SS_extracted_k141_15645 | Not-determined |
| SS_extracted_k141_125777 | Not-determined |
| SS_extracted_k141_26366 | Not-determined |
| SS_extracted_k141_88025 | Low-quality |
| SS_extracted_k141_4619 | Low-quality |
| SS_extracted_k141_112688 | Low-quality |
| SS_extracted_k141_15659 | Not-determined |
| SS_extracted_k141_125791 | Not-determined |
| SS_extracted_k141_4624 | Not-determined |
| SS_extracted_k141_135988 | Not-determined |
| SS_extracted_k141_69851 | Low-quality |
| SS_extracted_k141_15673 | Not-determined |
| SS_extracted_k141_125803 | Low-quality |
| SS_extracted_k141_21435 | Low-quality |
| SS_extracted_k141_112704 | Not-determined |
| SS_extracted_k141_135991 | Not-determined |
| SS_extracted_k141_15681 | Not-determined |
| SS_extracted_k141_69864 | Not-determined |
| SS_extracted_k141_4642 | Not-determined |
| SS_extracted_k141_4643 | Not-determined |
| SS_extracted_k141_63927 | Not-determined |
| SS_extracted_k141_21453 | Not-determined |
| SS_extracted_k141_88055 | Not-determined |
| SS_extracted_k141_125824 | Not-determined |
| SS_extracted_k141_125826 | Not-determined |
| SS_extracted_k141_125827 | Low-quality |
| SS_extracted_k141_21458 | Low-quality |
| SS_extracted_k141_4651 | Not-determined |
| SS_extracted_k141_15695 | Not-determined |
| SS_extracted_k141_112711 | Low-quality |
| SS_extracted_k141_125837 | Not-determined |
| SS_extracted_k141_136015 | Not-determined |
| SS_extracted_k141_15705 | Not-determined |
| SS_extracted_k141_125840 | Not-determined |
| SS_extracted_k141_63956 | Not-determined |
| SS_extracted_k141_63957 | Not-determined |
| SS_extracted_k141_63958 | Not-determined |
| SS_extracted_k141_125842 | Not-determined |
| SS_extracted_k141_125843 | Low-quality |
| SS_extracted_k141_125844 | Low-quality |
| SS_extracted_k141_15711 | Low-quality |
| SS_extracted_k141_136019 | Not-determined |
| SS_extracted_k141_136020 | Not-determined |
| SS_extracted_k141_99424 | Low-quality |
| SS_extracted_k141_4663 | Not-determined |
| SS_extracted_k141_125850 | Not-determined |
| SS_extracted_k141_39496 | Low-quality |
| SS_extracted_k141_88077 | Not-determined |
| SS_extracted_k141_99431 | Not-determined |
| SS_extracted_k141_63964 | Not-determined |
| SS_extracted_k141_125854 | Not-determined |
| SS_extracted_k141_21475 | Not-determined |
| SS_extracted_k141_15721 | Not-determined |
| SS_extracted_k141_52727 | Not-determined |
| SS_extracted_k141_4672 | Low-quality |
| SS_extracted_k141_107539 | Not-determined |
| SS_extracted_k141_63975 | Not-determined |
| SS_extracted_k141_52733 | Not-determined |
| SS_extracted_k141_63977 | Not-determined |
| SS_extracted_k141_52735 | Low-quality |
| SS_extracted_k141_4680 | Not-determined |
| SS_extracted_k141_63981 | Not-determined |
| SS_extracted_k141_15726 | Not-determined |
| SS_extracted_k141_21485 | Not-determined |
| SS_extracted_k141_88090 | Not-determined |
| SS_extracted_k141_99447 | Not-determined |
| SS_extracted_k141_15738 | Low-quality |
| SS_extracted_k141_94537 | Low-quality |
| SS_extracted_k141_21493 | Not-determined |
| SS_extracted_k141_63991 | Not-determined |
| SS_extracted_k141_63992 | Not-determined |
| SS_extracted_k141_15741 | Not-determined |
| SS_extracted_k141_21497 | Not-determined |
| SS_extracted_k141_63994 | Not-determined |
| SS_extracted_k141_94540 | Low-quality |
| SS_extracted_k141_94541 | Not-determined |
| SS_extracted_k141_112730 | Low-quality |
| SS_extracted_k141_88110 | Not-determined |
| SS_extracted_k141_142644 | Low-quality |
| SS_extracted_k141_21507 | Not-determined |
| SS_extracted_k141_94546 | Not-determined |
| SS_extracted_k141_125875 | Not-determined |
| SS_extracted_k141_112735 | Not-determined |
| SS_extracted_k141_56663 | Not-determined |
| SS_extracted_k141_94553 | Not-determined |
| SS_extracted_k141_56666 | Not-determined |
| SS_extracted_k141_21511 | Low-quality |
| SS_extracted_k141_94562 | Not-determined |
| SS_extracted_k141_112745 | Not-determined |
| SS_extracted_k141_142665 | Not-determined |
| SS_extracted_k141_39556 | Not-determined |
| SS_extracted_k141_142668 | Not-determined |
| SS_extracted_k141_94572 | Not-determined |
| SS_extracted_k141_94573 | Not-determined |
| SS_extracted_k141_56682 | Not-determined |
| SS_extracted_k141_94576 | Not-determined |
| SS_extracted_k141_21528 | Not-determined |
| SS_extracted_k141_94577 | Not-determined |
| SS_extracted_k141_69891 | Not-determined |
| SS_extracted_k141_52770 | Low-quality |
| SS_extracted_k141_94579 | Low-quality |
| SS_extracted_k141_34329 | Not-determined |
| SS_extracted_k141_136043 | Not-determined |
| SS_extracted_k141_88123 | Not-determined |
| SS_extracted_k141_80794 | Low-quality |
| SS_extracted_k141_52774 | Low-quality |
| SS_extracted_k141_136047 | Not-determined |
| SS_extracted_k141_94583 | Not-determined |
| SS_extracted_k141_99485 | Not-determined |
| SS_extracted_k141_52785 | Not-determined |
| SS_extracted_k141_99489 | Not-determined |
| SS_extracted_k141_39565 | Not-determined |
| SS_extracted_k141_52789 | Not-determined |
| SS_extracted_k141_4712 | Low-quality |
| SS_extracted_k141_4723 | Not-determined |
| SS_extracted_k141_34353 | Not-determined |
| SS_extracted_k141_80817 | Low-quality |
| SS_extracted_k141_56706 | Not-determined |
| SS_extracted_k141_112814 | Not-determined |
| SS_extracted_k141_56712 | Not-determined |
| SS_extracted_k141_56713 | Not-determined |
| SS_extracted_k141_9767 | Not-determined |
| SS_extracted_k141_21558 | Not-determined |
| SS_extracted_k141_69925 | Not-determined |
| SS_extracted_k141_99516 | Not-determined |
| SS_extracted_k141_21562 | Not-determined |
| SS_extracted_k141_80825 | Not-determined |
| SS_extracted_k141_56731 | Not-determined |
| SS_extracted_k141_56732 | Not-determined |
| SS_extracted_k141_80829 | Not-determined |
| SS_extracted_k141_94613 | Not-determined |
| SS_extracted_k141_94616 | Low-quality |
| SS_extracted_k141_39607 | Low-quality |
| SS_extracted_k141_21571 | Low-quality |
| SS_extracted_k141_56740 | Not-determined |
| SS_extracted_k141_21572 | Not-determined |
| SS_extracted_k141_94620 | Not-determined |
| SS_extracted_k141_56741 | Not-determined |
| SS_extracted_k141_112816 | Low-quality |
| SS_extracted_k141_69933 | Low-quality |
| SS_extracted_k141_80833 | Low-quality |
| SS_extracted_k141_34369 | Not-determined |
| SS_extracted_k141_56743 | Low-quality |
| SS_extracted_k141_119460 | Not-determined |
| SS_extracted_k141_34371 | Not-determined |
| SS_extracted_k141_80835 | Low-quality |
| SS_extracted_k141_52795 | Not-determined |
| SS_extracted_k141_112829 | Not-determined |
| SS_extracted_k141_9792 | Not-determined |
| SS_extracted_k141_34376 | Not-determined |
| SS_extracted_k141_130965 | Not-determined |
| SS_extracted_k141_130968 | Not-determined |
| SS_extracted_k141_9799 | Not-determined |
| SS_extracted_k141_119475 | Not-determined |
| SS_extracted_k141_26376 | Low-quality |
| SS_extracted_k141_34383 | Not-determined |
| SS_extracted_k141_21589 | Not-determined |
| SS_extracted_k141_130972 | Low-quality |
| SS_extracted_k141_4742 | Not-determined |
| SS_extracted_k141_130974 | Not-determined |
| SS_extracted_k141_4743 | Not-determined |
| SS_extracted_k141_34387 | Not-determined |
| SS_extracted_k141_34391 | Not-determined |
| SS_extracted_k141_119480 | Not-determined |
| SS_extracted_k141_9805 | Not-determined |
| SS_extracted_k141_21597 | Low-quality |
| SS_extracted_k141_119481 | Not-determined |
| SS_extracted_k141_4751 | Not-determined |
| SS_extracted_k141_112850 | Not-determined |
| SS_extracted_k141_9809 | Not-determined |
| SS_extracted_k141_34398 | Not-determined |
| SS_extracted_k141_26387 | Not-determined |
| SS_extracted_k141_112862 | Not-determined |
| SS_extracted_k141_52804 | Low-quality |
| SS_extracted_k141_34407 | Not-determined |
| SS_extracted_k141_119492 | Not-determined |
| SS_extracted_k141_107584 | Not-determined |
| SS_extracted_k141_125977 | Not-determined |
| SS_extracted_k141_130996 | Not-determined |
| SS_extracted_k141_119499 | Not-determined |
| SS_extracted_k141_119500 | Not-determined |
| SS_extracted_k141_56759 | Not-determined |
| SS_extracted_k141_21619 | Low-quality |
| SS_extracted_k141_80858 | Not-determined |
| SS_extracted_k141_15754 | Not-determined |
| SS_extracted_k141_131015 | Not-determined |
| SS_extracted_k141_15756 | Not-determined |
| SS_extracted_k141_112887 | Not-determined |
| SS_extracted_k141_34419 | Low-quality |
| SS_extracted_k141_26404 | Not-determined |
| SS_extracted_k141_4794 | Low-quality |
| SS_extracted_k141_34424 | Not-determined |
| SS_extracted_k141_34425 | Not-determined |
| SS_extracted_k141_99544 | Not-determined |
| SS_extracted_k141_9835 | Low-quality |
| SS_extracted_k141_131023 | Low-quality |
| SS_extracted_k141_99546 | Not-determined |
| SS_extracted_k141_9839 | Not-determined |
| SS_extracted_k141_4802 | Not-determined |
| SS_extracted_k141_80884 | Not-determined |
| SS_extracted_k141_34430 | Not-determined |
| SS_extracted_k141_131033 | Low-quality |
| SS_extracted_k141_26422 | Not-determined |
| SS_extracted_k141_80893 | Not-determined |
| SS_extracted_k141_88231 | Not-determined |
| SS_extracted_k141_88233 | Not-determined |
| SS_extracted_k141_9853 | Low-quality |
| SS_extracted_k141_107606 | Low-quality |
| SS_extracted_k141_107608 | Low-quality |
| SS_extracted_k141_4830 | Low-quality |
| SS_extracted_k141_9858 | Not-determined |
| SS_extracted_k141_74814 | Not-determined |
| SS_extracted_k141_9863 | Not-determined |
| SS_extracted_k141_52824 | Not-determined |
| SS_extracted_k141_52825 | Not-determined |
| SS_extracted_k141_34438 | Not-determined |
| SS_extracted_k141_34440 | Not-determined |
| SS_extracted_k141_15770 | Not-determined |
| SS_extracted_k141_119558 | Low-quality |
| SS_extracted_k141_52832 | Low-quality |
| SS_extracted_k141_9872 | Not-determined |
| SS_extracted_k141_99558 | Not-determined |
| SS_extracted_k141_52835 | Not-determined |
| SS_extracted_k141_9873 | Not-determined |
| SS_extracted_k141_34448 | Not-determined |
| SS_extracted_k141_99561 | Low-quality |
| SS_extracted_k141_9876 | Low-quality |
| SS_extracted_k141_56792 | Not-determined |
| SS_extracted_k141_52843 | Not-determined |
| SS_extracted_k141_52845 | Not-determined |
| SS_extracted_k141_52846 | Not-determined |
| SS_extracted_k141_52848 | Not-determined |
| SS_extracted_k141_99566 | Low-quality |
| SS_extracted_k141_94639 | Not-determined |
| SS_extracted_k141_52849 | Not-determined |
| SS_extracted_k141_99567 | Not-determined |
| SS_extracted_k141_119568 | Low-quality |
| SS_extracted_k141_64025 | Not-determined |
| S17_vs_k141_94625\|\|full | High-quality |
| SS_extracted_k141_94647 | Low-quality |
| SS_extracted_k141_34461 | Not-determined |
| SS_extracted_k141_52856 | Not-determined |
| SS_extracted_k141_94648 | Low-quality |
| SS_extracted_k141_52858 | Low-quality |
| SS_extracted_k141_64028 | Low-quality |
| SS_extracted_k141_52865 | Not-determined |
| SS_extracted_k141_94656 | Not-determined |
| SS_extracted_k141_15780 | Not-determined |
| SS_extracted_k141_107635 | Low-quality |
| SS_extracted_k141_107636 | Not-determined |
| SS_extracted_k141_74834 | Not-determined |
| SS_extracted_k141_119578 | Not-determined |
| SS_extracted_k141_107639 | Not-determined |
| SS_extracted_k141_52875 | Not-determined |
| SS_extracted_k141_9880 | Not-determined |
| SS_extracted_k141_112909 | Not-determined |
| SS_extracted_k141_112910 | Not-determined |
| SS_extracted_k141_34477 | Not-determined |
| SS_extracted_k141_9886 | Not-determined |
| SS_extracted_k141_9889 | Not-determined |
| SS_extracted_k141_136085 | Not-determined |
| SS_extracted_k141_136088 | Not-determined |
| SS_extracted_k141_136090 | Low-quality |
| SS_extracted_k141_136093 | Not-determined |
| SS_extracted_k141_15801 | Low-quality |
| SS_extracted_k141_94671 | Not-determined |
| SS_extracted_k141_112928 | Not-determined |
| SS_extracted_k141_56821 | Not-determined |
| SS_extracted_k141_34503 | Not-determined |
| SS_extracted_k141_119586 | Low-quality |
| SS_extracted_k141_4872 | Not-determined |
| SS_extracted_k141_15809 | Not-determined |
| SS_extracted_k141_119588 | Not-determined |
| SS_extracted_k141_80913 | Not-determined |
| SS_extracted_k141_126016 | Low-quality |
| SS_extracted_k141_126020 | Not-determined |
| SS_extracted_k141_94688 | Not-determined |
| SS_extracted_k141_15816 | Not-determined |
| SS_extracted_k141_107650 | Not-determined |
| SS_extracted_k141_4880 | Not-determined |
| SS_extracted_k141_136108 | Not-determined |
| SS_extracted_k141_94691 | Not-determined |
| SS_extracted_k141_4885 | Not-determined |
| SS_extracted_k141_94697 | Not-determined |
| SS_extracted_k141_64048 | Not-determined |
| SS_extracted_k141_107656 | Low-quality |
| SS_extracted_k141_15824 | Low-quality |
| SS_extracted_k141_107659 | Low-quality |
| SS_extracted_k141_74858 | Low-quality |
| SS_extracted_k141_21643 | Not-determined |
| SS_extracted_k141_94703 | Low-quality |
| SS_extracted_k141_74863 | Not-determined |
| SS_extracted_k141_99594 | Not-determined |
| SS_extracted_k141_9908 | Low-quality |
| SS_extracted_k141_99595 | Not-determined |
| SS_extracted_k141_34529 | Not-determined |
| SS_extracted_k141_126027 | Not-determined |
| SS_extracted_k141_4897 | Not-determined |
| SS_extracted_k141_21646 | Low-quality |
| SS_extracted_k141_21647 | Not-determined |
| SS_extracted_k141_74867 | Low-quality |
| SS_extracted_k141_99600 | Low-quality |
| SS_extracted_k141_4899 | Low-quality |
| SS_extracted_k141_9913 | Not-determined |
| SS_extracted_k141_99604 | Not-determined |
| SS_extracted_k141_4904 | Not-determined |
| SS_extracted_k141_9919 | Not-determined |
| SS_extracted_k141_34535 | Not-determined |
| SS_extracted_k141_4906 | Not-determined |
| SS_extracted_k141_142706 | Not-determined |
| SS_extracted_k141_88245 | Not-determined |
| SS_extracted_k141_142714 | Not-determined |
| SS_extracted_k141_131100 | Not-determined |
| SS_extracted_k141_131103 | Low-quality |
| SS_extracted_k141_107676 | Not-determined |
| SS_extracted_k141_99618 | Not-determined |
| SS_extracted_k141_131107 | Not-determined |
| SS_extracted_k141_142719 | Not-determined |
| SS_extracted_k141_64068 | Low-quality |
| SS_extracted_k141_64071 | Low-quality |
| SS_extracted_k141_107678 | Not-determined |
| SS_extracted_k141_126055 | Low-quality |
| SS_extracted_k141_107680 | Low-quality |
| SS_extracted_k141_4930 | Not-determined |
| SS_extracted_k141_131116 | Not-determined |
| SS_extracted_k141_52936 | Not-determined |
| SS_extracted_k141_64077 | Not-determined |
| SS_extracted_k141_69963 | Not-determined |
| SS_extracted_k141_26429 | Low-quality |
| SS_extracted_k141_64079 | Not-determined |
| SS_extracted_k141_94728 | Not-determined |
| SS_extracted_k141_74882 | Low-quality |
| SS_extracted_k141_142733 | Not-determined |
| SS_extracted_k141_142734 | Not-determined |
| SS_extracted_k141_126059 | Not-determined |
| SS_extracted_k141_9933 | Low-quality |
| SS_extracted_k141_64086 | Low-quality |
| SS_extracted_k141_4948 | Low-quality |
| SS_extracted_k141_99641 | Low-quality |
| SS_extracted_k141_56835 | Not-determined |
| SS_extracted_k141_142744 | Not-determined |
| SS_extracted_k141_56837 | Not-determined |
| SS_extracted_k141_26439 | Low-quality |
| SS_extracted_k141_4955 | Not-determined |
| SS_extracted_k141_34575 | Not-determined |
| SS_extracted_k141_52947 | Not-determined |
| SS_extracted_k141_56842 | Not-determined |
| SS_extracted_k141_4961 | Not-determined |
| SS_extracted_k141_46048 | Not-determined |
| SS_extracted_k141_94742 | Not-determined |
| SS_extracted_k141_21700 | Not-determined |
| SS_extracted_k141_4972 | Low-quality |
| SS_extracted_k141_88288 | Not-determined |
| SS_extracted_k141_34586 | Not-determined |
| SS_extracted_k141_126065 | Low-quality |
| SS_extracted_k141_88293 | Not-determined |
| SS_extracted_k141_34590 | Not-determined |
| SS_extracted_k141_107705 | Not-determined |
| SS_extracted_k141_126068 | Not-determined |
| SS_extracted_k141_4979 | Not-determined |
| SS_extracted_k141_88300 | Not-determined |
| SS_extracted_k141_52972 | Low-quality |
| SS_extracted_k141_46052 | Not-determined |
| SS_extracted_k141_34601 | Not-determined |
| SS_extracted_k141_94753 | Not-determined |
| SS_extracted_k141_64112 | Not-determined |
| SS_extracted_k141_99653 | Low-quality |
| SS_extracted_k141_126078 | Not-determined |
| SS_extracted_k141_46055 | Not-determined |
| SS_extracted_k141_34607 | Low-quality |
| SS_extracted_k141_107718 | Not-determined |
| SS_extracted_k141_26451 | Not-determined |
| SS_extracted_k141_94760 | Not-determined |
| SS_extracted_k141_99659 | Not-determined |
| SS_extracted_k141_94764 | Not-determined |
| SS_extracted_k141_99664 | Not-determined |
| SS_extracted_k141_64119 | Not-determined |
| SS_extracted_k141_34614 | Not-determined |
| SS_extracted_k141_34615 | Low-quality |
| SS_extracted_k141_142784 | Not-determined |
| SS_extracted_k141_34617 | Low-quality |
| SS_extracted_k141_99667 | Low-quality |
| SS_extracted_k141_21713 | Not-determined |
| SS_extracted_k141_46061 | Not-determined |
| SS_extracted_k141_21716 | Not-determined |
| SS_extracted_k141_26462 | Not-determined |
| SS_extracted_k141_52991 | Low-quality |
| SS_extracted_k141_34623 | Low-quality |
| SS_extracted_k141_52993 | Not-determined |
| SS_extracted_k141_52994 | Not-determined |
| SS_extracted_k141_88334 | Not-determined |
| SS_extracted_k141_99671 | Low-quality |
| SS_extracted_k141_88335 | Not-determined |
| SS_extracted_k141_56869 | Not-determined |
| SS_extracted_k141_94781 | Not-determined |
| SS_extracted_k141_142798 | Not-determined |
| SS_extracted_k141_46074 | Not-determined |
| SS_extracted_k141_34636 | Not-determined |
| SS_extracted_k141_94786 | Not-determined |
| SS_extracted_k141_74908 | Not-determined |
| SS_extracted_k141_46085 | Low-quality |
| SS_extracted_k141_126095 | Not-determined |
| SS_extracted_k141_99691 | Not-determined |
| SS_extracted_k141_56877 | Not-determined |
| SS_extracted_k141_94788 | Not-determined |
| SS_extracted_k141_107722 | Not-determined |
| SS_extracted_k141_74918 | Low-quality |
| SS_extracted_k141_64136 | Low-quality |
| SS_extracted_k141_112985 | Not-determined |
| SS_extracted_k141_5044 | Not-determined |
| SS_extracted_k141_131124 | Not-determined |
| SS_extracted_k141_126104 | Not-determined |
| SS_extracted_k141_56883 | Low-quality |
| SS_extracted_k141_15876 | Not-determined |
| SS_extracted_k141_5049 | Not-determined |
| SS_extracted_k141_64139 | Not-determined |
| SS_extracted_k141_56886 | Not-determined |
| SS_extracted_k141_15878 | Not-determined |
| SS_extracted_k141_64141 | Not-determined |
| SS_extracted_k141_5053 | Low-quality |
| SS_extracted_k141_94803 | Not-determined |
| SS_extracted_k141_74927 | Not-determined |
| SS_extracted_k141_53040 | Not-determined |
| SS_extracted_k141_69986 | Low-quality |
| SS_extracted_k141_46105 | Not-determined |
| SS_extracted_k141_112989 | Low-quality |
| SS_extracted_k141_74932 | Not-determined |
| SS_extracted_k141_74933 | Not-determined |
| SS_extracted_k141_21737 | Not-determined |
| SS_extracted_k141_112994 | Low-quality |
| SS_extracted_k141_112995 | Not-determined |
| SS_extracted_k141_46110 | Not-determined |
| SS_extracted_k141_94815 | Low-quality |
| SS_extracted_k141_56902 | Not-determined |
| SS_extracted_k141_74938 | Not-determined |
| SS_extracted_k141_94818 | Not-determined |
| SS_extracted_k141_46113 | Low-quality |
| SS_extracted_k141_107744 | Not-determined |
| SS_extracted_k141_94821 | Low-quality |
| SS_extracted_k141_131149 | Not-determined |
| SS_extracted_k141_99698 | Not-determined |
| SS_extracted_k141_88363 | Not-determined |
| SS_extracted_k141_113000 | Not-determined |
| SS_extracted_k141_15900 | Not-determined |
| SS_extracted_k141_74942 | Low-quality |
| SS_extracted_k141_21747 | Not-determined |
| SS_extracted_k141_15903 | Not-determined |
| SS_extracted_k141_46118 | Not-determined |
| SS_extracted_k141_131154 | Not-determined |
| SS_extracted_k141_21748 | Not-determined |
| SS_extracted_k141_131163 | Not-determined |
| SS_extracted_k141_26491 | Low-quality |
| SS_extracted_k141_53067 | Low-quality |
| SS_extracted_k141_107757 | Not-determined |
| SS_extracted_k141_88373 | Not-determined |
| SS_extracted_k141_15917 | Not-determined |
| SS_extracted_k141_142821 | Not-determined |
| SS_extracted_k141_9965 | Not-determined |
| SS_extracted_k141_88374 | Not-determined |
| SS_extracted_k141_15921 | Not-determined |
| SS_extracted_k141_53075 | Not-determined |
| SS_extracted_k141_34672 | Not-determined |
| SS_extracted_k141_70008 | Not-determined |
| SS_extracted_k141_70009 | Not-determined |
| SS_extracted_k141_70011 | Not-determined |
| SS_extracted_k141_136130 | Not-determined |
| SS_extracted_k141_5068 | Not-determined |
| SS_extracted_k141_5070 | Not-determined |
| SS_extracted_k141_5071 | Not-determined |
| SS_extracted_k141_34679 | Low-quality |
| SS_extracted_k141_70022 | Low-quality |
| SS_extracted_k141_113026 | Not-determined |
| SS_extracted_k141_131174 | Not-determined |
| SS_extracted_k141_107776 | Not-determined |
| SS_extracted_k141_107777 | Not-determined |
| SS_extracted_k141_26502 | Not-determined |
| S9_vs_k141_90848\|\|full | Medium-quality |
| SS_extracted_k141_113030 | Not-determined |
| SS_extracted_k141_99720 | Not-determined |
| SS_extracted_k141_126140 | Not-determined |
| SS_extracted_k141_70031 | Not-determined |
| SS_extracted_k141_94832 | Not-determined |
| SS_extracted_k141_126147 | Not-determined |
| SS_extracted_k141_142829 | Low-quality |
| SS_extracted_k141_107788 | Low-quality |
| SS_extracted_k141_126149 | Not-determined |
| SS_extracted_k141_142837 | Low-quality |
| SS_extracted_k141_64147 | Not-determined |
| SS_extracted_k141_136149 | Not-determined |
| SS_extracted_k141_21767 | Not-determined |
| SS_extracted_k141_56933 | Not-determined |
| SS_extracted_k141_21770 | Not-determined |
| SS_extracted_k141_9973 | Not-determined |
| SS_extracted_k141_21773 | Not-determined |
| SS_extracted_k141_70041 | Not-determined |
| SS_extracted_k141_94846 | Not-determined |
| SS_extracted_k141_56942 | Low-quality |
| SS_extracted_k141_107795 | Not-determined |
| SS_extracted_k141_64161 | Not-determined |
| SS_extracted_k141_136158 | Not-determined |
| SS_extracted_k141_136159 | Not-determined |
| SS_extracted_k141_107802 | Not-determined |
| SS_extracted_k141_142839 | Low-quality |
| SS_extracted_k141_53095 | Not-determined |
| SS_extracted_k141_5086 | Low-quality |
| SS_extracted_k141_88410 | Low-quality |
| SS_extracted_k141_142844 | Low-quality |
| SS_extracted_k141_5094 | Not-determined |
| SS_extracted_k141_126167 | Not-determined |
| SS_extracted_k141_126168 | Not-determined |
| SS_extracted_k141_64175 | Not-determined |
| SS_extracted_k141_9995 | Low-quality |
| SS_extracted_k141_88418 | Low-quality |
| SS_extracted_k141_9999 | Not-determined |
| SS_extracted_k141_64181 | Not-determined |
| SS_extracted_k141_80989 | Not-determined |
| SS_extracted_k141_53100 | Not-determined |
| SS_extracted_k141_74973 | Not-determined |
| SS_extracted_k141_94855 | Not-determined |
| SS_extracted_k141_74974 | Low-quality |
| SS_extracted_k141_126171 | Not-determined |
| SS_extracted_k141_126172 | Not-determined |
| SS_extracted_k141_74975 | Not-determined |
| SS_extracted_k141_15954 | Low-quality |
| SS_extracted_k141_136175 | Not-determined |
| SS_extracted_k141_136176 | Not-determined |
| SS_extracted_k141_126178 | Not-determined |
| SS_extracted_k141_5110 | Not-determined |
| SS_extracted_k141_10003 | Not-determined |
| SS_extracted_k141_64193 | Not-determined |
| SS_extracted_k141_142868 | Not-determined |
| SS_extracted_k141_107820 | Not-determined |
| SS_extracted_k141_46130 | Not-determined |
| SS_extracted_k141_74982 | Not-determined |
| SS_extracted_k141_126191 | Not-determined |
| SS_extracted_k141_94874 | Not-determined |
| SS_extracted_k141_70062 | Not-determined |
| SS_extracted_k141_94876 | Not-determined |
| SS_extracted_k141_107831 | Not-determined |
| SS_extracted_k141_56963 | Not-determined |
| SS_extracted_k141_70065 | Low-quality |
| SS_extracted_k141_70066 | Not-determined |
| SS_extracted_k141_94886 | Not-determined |
| SS_extracted_k141_46144 | Low-quality |
| SS_extracted_k141_99736 | Low-quality |
| SS_extracted_k141_136198 | Not-determined |
| SS_extracted_k141_94893 | Not-determined |
| SS_extracted_k141_142888 | Not-determined |
| SS_extracted_k141_46150 | Not-determined |
| SS_extracted_k141_5123 | Not-determined |
| SS_extracted_k141_99743 | Low-quality |
| SS_extracted_k141_94900 | Low-quality |
| SS_extracted_k141_119612 | Not-determined |
| SS_extracted_k141_142895 | Not-determined |
| SS_extracted_k141_119622 | Not-determined |
| SS_extracted_k141_10040 | Not-determined |
| SS_extracted_k141_88457 | Not-determined |
| SS_extracted_k141_136200 | Not-determined |
| SS_extracted_k141_34766 | Not-determined |
| SS_extracted_k141_53133 | Not-determined |
| SS_extracted_k141_136204 | Low-quality |
| SS_extracted_k141_75002 | Not-determined |
| SS_extracted_k141_34771 | Not-determined |
| SS_extracted_k141_53140 | Not-determined |
| SS_extracted_k141_126214 | Not-determined |
| SS_extracted_k141_56978 | Not-determined |
| SS_extracted_k141_119634 | Not-determined |
| SS_extracted_k141_46165 | Low-quality |

| | |
|---|---|
| SS_extracted_k141_126218 | Not-determined |
| SS_extracted_k141_126221 | Low-quality |
| SS_extracted_k141_56983 | Not-determined |
| SS_extracted_k141_136215 | Low-quality |
| SS_extracted_k141_34779 | Not-determined |
| SS_extracted_k141_34781 | Not-determined |
| SS_extracted_k141_64219 | Not-determined |
| SS_extracted_k141_64221 | Not-determined |
| SS_extracted_k141_70100 | Low-quality |
| SS_extracted_k141_56990 | Not-determined |
| SS_extracted_k141_10059 | Not-determined |
| SS_extracted_k141_34784 | Low-quality |
| SS_extracted_k141_10063 | Not-determined |
| SS_extracted_k141_46174 | Not-determined |
| SS_extracted_k141_10065 | Not-determined |
| SS_extracted_k141_126234 | Not-determined |
| SS_extracted_k141_94933 | Not-determined |
| SS_extracted_k141_46178 | Not-determined |
| SS_extracted_k141_46181 | Not-determined |
| SS_extracted_k141_94937 | Not-determined |
| SS_extracted_k141_64223 | Not-determined |
| SS_extracted_k141_88476 | Low-quality |
| SS_extracted_k141_64225 | Not-determined |
| SS_extracted_k141_46186 | Low-quality |
| SS_extracted_k141_119654 | Not-determined |
| SS_extracted_k141_94948 | Not-determined |
| SS_extracted_k141_53154 | Low-quality |
| SS_extracted_k141_107877 | Not-determined |
| SS_extracted_k141_136225 | Not-determined |
| SS_extracted_k141_53157 | Not-determined |
| SS_extracted_k141_119661 | Not-determined |
| SS_extracted_k141_88490 | Not-determined |
| SS_extracted_k141_136230 | Not-determined |
| SS_extracted_k141_88494 | Low-quality |
| SS_extracted_k141_70128 | Not-determined |
| SS_extracted_k141_94964 | Low-quality |
| SS_extracted_k141_64239 | Low-quality |
| SS_extracted_k141_64240 | Low-quality |
| SS_extracted_k141_16013 | Low-quality |
| SS_extracted_k141_131212 | Not-determined |
| SS_extracted_k141_107897 | Not-determined |
| SS_extracted_k141_107902 | Not-determined |
| SS_extracted_k141_5175 | Low-quality |
| SS_extracted_k141_81008 | Not-determined |
| SS_extracted_k141_16022 | Not-determined |
| SS_extracted_k141_119679 | Not-determined |
| SS_extracted_k141_16028 | Not-determined |
| SS_extracted_k141_70141 | Not-determined |
| SS_extracted_k141_136240 | Not-determined |
| SS_extracted_k141_136242 | Not-determined |
| SS_extracted_k141_88508 | Low-quality |
| SS_extracted_k141_88510 | Not-determined |
| SS_extracted_k141_70146 | Low-quality |
| SS_extracted_k141_136246 | Low-quality |
| SS_extracted_k141_70155 | Not-determined |
| SS_extracted_k141_70157 | Not-determined |
| SS_extracted_k141_107910 | Not-determined |
| SS_extracted_k141_113059 | Not-determined |
| SS_extracted_k141_119692 | Not-determined |
| SS_extracted_k141_131224 | Not-determined |
| SS_extracted_k141_53178 | Not-determined |
| SS_extracted_k141_113062 | Not-determined |
| SS_extracted_k141_64251 | Not-determined |
| SS_extracted_k141_131229 | Not-determined |
| SS_extracted_k141_53187 | Not-determined |
| SS_extracted_k141_16036 | Not-determined |
| SS_extracted_k141_107917 | Low-quality |
| SS_extracted_k141_39656 | Not-determined |
| SS_extracted_k141_16042 | Not-determined |
| SS_extracted_k141_136263 | Not-determined |
| SS_extracted_k141_16048 | Not-determined |
| SS_extracted_k141_88534 | Low-quality |
| SS_extracted_k141_131248 | Not-determined |
| SS_extracted_k141_113077 | Not-determined |
| SS_extracted_k141_39671 | Not-determined |
| SS_extracted_k141_81034 | Not-determined |
| SS_extracted_k141_53207 | Not-determined |
| SS_extracted_k141_64269 | Not-determined |
| SS_extracted_k141_126289 | Not-determined |
| SS_extracted_k141_119716 | Not-determined |
| SS_extracted_k141_136266 | Not-determined |
| SS_extracted_k141_107924 | Not-determined |
| SS_extracted_k141_39680 | Not-determined |
| SS_extracted_k141_39682 | Not-determined |
| SS_extracted_k141_64272 | Not-determined |
| SS_extracted_k141_81049 | Low-quality |
| SS_extracted_k141_99771 | Not-determined |
| SS_extracted_k141_39685 | Low-quality |
| SS_extracted_k141_53225 | Not-determined |
| SS_extracted_k141_113089 | Not-determined |
| SS_extracted_k141_53233 | Not-determined |
| SS_extracted_k141_113091 | Not-determined |
| SS_extracted_k141_99780 | Not-determined |
| SS_extracted_k141_107932 | Not-determined |
| SS_extracted_k141_99783 | Not-determined |
| SS_extracted_k141_119739 | Not-determined |
| SS_extracted_k141_119745 | Not-determined |
| SS_extracted_k141_99787 | Not-determined |
| SS_extracted_k141_131279 | Not-determined |
| SS_extracted_k141_70192 | Not-determined |
| SS_extracted_k141_119747 | Not-determined |
| SS_extracted_k141_88551 | Not-determined |
| SS_extracted_k141_119756 | Not-determined |
| SS_extracted_k141_113114 | Not-determined |
| SS_extracted_k141_113115 | Not-determined |
| SS_extracted_k141_113116 | Not-determined |
| SS_extracted_k141_53248 | Not-determined |
| SS_extracted_k141_5252 | Low-quality |
| SS_extracted_k141_10092 | Not-determined |
| SS_extracted_k141_16078 | Low-quality |
| SS_extracted_k141_5257 | Not-determined |
| SS_extracted_k141_131294 | Not-determined |
| SS_extracted_k141_10095 | Not-determined |
| SS_extracted_k141_136311 | Not-determined |
| SS_extracted_k141_10102 | Not-determined |
| SS_extracted_k141_136313 | Low-quality |
| SS_extracted_k141_16087 | Not-determined |
| SS_extracted_k141_88568 | Not-determined |
| SS_extracted_k141_16092 | Not-determined |
| SS_extracted_k141_16094 | Not-determined |
| SS_extracted_k141_95032 | Low-quality |
| SS_extracted_k141_131317 | Not-determined |
| SS_extracted_k141_70219 | Low-quality |
| SS_extracted_k141_5269 | Not-determined |
| SS_extracted_k141_99809 | Not-determined |
| SS_extracted_k141_99810 | Not-determined |
| SS_extracted_k141_95034 | Low-quality |
| SS_extracted_k141_21782 | Not-determined |
| SS_extracted_k141_107956 | Low-quality |
| SS_extracted_k141_10120 | Not-determined |
| SS_extracted_k141_107960 | Not-determined |
| SS_extracted_k141_131322 | Low-quality |
| S14_vs_k141_9676\|\|full | High-quality |
| SS_extracted_k141_53273 | Not-determined |
| SS_extracted_k141_26590 | Low-quality |
| SS_extracted_k141_21784 | Not-determined |
| SS_extracted_k141_81073 | Not-determined |
| SS_extracted_k141_99817 | Not-determined |
| SS_extracted_k141_119783 | Not-determined |
| SS_extracted_k141_136326 | Not-determined |
| SS_extracted_k141_99820 | Not-determined |
| SS_extracted_k141_119784 | Low-quality |
| SS_extracted_k141_53282 | Not-determined |
| SS_extracted_k141_34849 | Not-determined |
| SS_extracted_k141_119786 | Not-determined |
| SS_extracted_k141_142923 | Not-determined |
| SS_extracted_k141_99822 | Low-quality |
| SS_extracted_k141_119789 | Low-quality |
| SS_extracted_k141_99823 | Not-determined |
| SS_extracted_k141_39725 | Not-determined |
| SS_extracted_k141_113154 | Not-determined |
| SS_extracted_k141_21801 | Not-determined |
| SS_extracted_k141_21804 | Not-determined |
| SS_extracted_k141_107973 | Low-quality |
| SS_extracted_k141_119795 | Low-quality |
| SS_extracted_k141_34854 | Not-determined |
| SS_extracted_k141_21809 | Not-determined |
| SS_extracted_k141_88596 | Not-determined |
| SS_extracted_k141_136334 | Not-determined |
| SS_extracted_k141_99834 | Not-determined |
| SS_extracted_k141_119800 | Not-determined |
| SS_extracted_k141_113164 | Low-quality |
| SS_extracted_k141_136337 | Not-determined |
| SS_extracted_k141_34858 | Not-determined |
| SS_extracted_k141_113169 | Not-determined |
| SS_extracted_k141_119806 | Not-determined |
| SS_extracted_k141_5294 | Not-determined |
| SS_extracted_k141_99841 | Low-quality |
| SS_extracted_k141_34866 | Not-determined |
| SS_extracted_k141_99842 | Low-quality |
| SS_extracted_k141_5297 | Not-determined |
| SS_extracted_k141_34867 | Not-determined |
| SS_extracted_k141_88619 | Low-quality |
| SS_extracted_k141_21828 | Not-determined |
| SS_extracted_k141_53310 | Not-determined |
| SS_extracted_k141_39748 | Not-determined |
| SS_extracted_k141_39750 | Low-quality |
| SS_extracted_k141_10141 | Not-determined |
| SS_extracted_k141_75015 | Not-determined |
| SS_extracted_k141_39762 | Not-determined |
| SS_extracted_k141_57007 | Not-determined |
| SS_extracted_k141_39763 | Not-determined |
| SS_extracted_k141_136349 | Low-quality |
| SS_extracted_k141_99865 | Not-determined |
| SS_extracted_k141_5312 | Low-quality |
| SS_extracted_k141_39766 | Not-determined |
| SS_extracted_k141_119820 | Low-quality |
| SS_extracted_k141_113194 | Not-determined |
| SS_extracted_k141_119824 | Not-determined |
| SS_extracted_k141_10156 | Not-determined |
| SS_extracted_k141_108010 | Not-determined |
| SS_extracted_k141_39773 | Not-determined |
| SS_extracted_k141_81126 | Not-determined |
| SS_extracted_k141_113199 | Not-determined |
| SS_extracted_k141_81128 | Not-determined |
| SS_extracted_k141_95074 | Not-determined |
| SS_extracted_k141_95075 | Not-determined |
| SS_extracted_k141_95076 | Not-determined |
| SS_extracted_k141_95079 | Not-determined |
| SS_extracted_k141_5322 | Low-quality |
| SS_extracted_k141_5323 | Low-quality |
| SS_extracted_k141_88641 | Low-quality |
| SS_extracted_k141_70256 | Not-determined |
| SS_extracted_k141_136366 | Not-determined |
| SS_extracted_k141_10159 | Low-quality |
| SS_extracted_k141_21852 | Not-determined |
| SS_extracted_k141_99874 | Not-determined |
| SS_extracted_k141_10161 | Not-determined |
| SS_extracted_k141_88649 | Not-determined |
| SS_extracted_k141_88650 | Not-determined |
| SS_extracted_k141_70260 | Not-determined |
| SS_extracted_k141_16114 | Not-determined |
| SS_extracted_k141_75032 | Not-determined |
| SS_extracted_k141_95089 | Not-determined |
| SS_extracted_k141_10169 | Low-quality |
| SS_extracted_k141_39794 | Not-determined |
| SS_extracted_k141_88659 | Low-quality |
| SS_extracted_k141_10171 | Not-determined |
| SS_extracted_k141_21861 | Low-quality |
| SS_extracted_k141_99886 | Low-quality |
| SS_extracted_k141_113224 | Not-determined |
| SS_extracted_k141_70272 | Low-quality |
| SS_extracted_k141_88662 | Not-determined |
| SS_extracted_k141_26611 | Not-determined |
| SS_extracted_k141_95104 | Not-determined |
| SS_extracted_k141_10181 | Not-determined |
| SS_extracted_k141_81162 | Not-determined |
| SS_extracted_k141_10182 | Not-determined |
| SS_extracted_k141_108024 | Not-determined |
| SS_extracted_k141_88667 | Low-quality |
| SS_extracted_k141_81165 | Not-determined |
| SS_extracted_k141_108027 | Not-determined |
| SS_extracted_k141_10185 | Not-determined |
| SS_extracted_k141_95112 | Not-determined |
| SS_extracted_k141_142932 | Not-determined |
| SS_extracted_k141_119858 | Not-determined |
| SS_extracted_k141_119859 | Not-determined |
| SS_extracted_k141_108034 | Not-determined |
| SS_extracted_k141_26612 | Low-quality |
| SS_extracted_k141_95119 | Not-determined |
| SS_extracted_k141_39817 | Not-determined |
| SS_extracted_k141_88676 | Not-determined |
| SS_extracted_k141_131351 | Not-determined |
| SS_extracted_k141_75050 | Not-determined |
| SS_extracted_k141_113250 | Not-determined |
| SS_extracted_k141_131354 | Low-quality |
| SS_extracted_k141_88683 | Not-determined |
| SS_extracted_k141_10190 | Low-quality |
| SS_extracted_k141_108039 | Low-quality |
| SS_extracted_k141_113253 | Not-determined |
| SS_extracted_k141_113258 | Not-determined |
| SS_extracted_k141_57024 | Not-determined |
| SS_extracted_k141_113262 | Not-determined |
| SS_extracted_k141_16155 | Not-determined |
| SS_extracted_k141_39829 | Not-determined |
| SS_extracted_k141_113267 | Not-determined |
| SS_extracted_k141_57036 | Not-determined |
| SS_extracted_k141_95134 | Not-determined |
| SS_extracted_k141_53348 | Not-determined |
| SS_extracted_k141_95138 | Not-determined |
| SS_extracted_k141_10201 | Not-determined |
| SS_extracted_k141_10202 | Not-determined |
| SS_extracted_k141_136375 | Low-quality |
| SS_extracted_k141_95142 | Not-determined |
| SS_extracted_k141_70316 | Not-determined |
| SS_extracted_k141_57045 | Not-determined |
| SS_extracted_k141_108046 | Not-determined |
| SS_extracted_k141_142946 | Not-determined |
| SS_extracted_k141_108048 | Low-quality |
| SS_extracted_k141_10204 | Not-determined |
| SS_extracted_k141_16167 | Low-quality |
| SS_extracted_k141_136384 | Not-determined |
| SS_extracted_k141_119887 | Low-quality |
| SS_extracted_k141_53356 | Not-determined |
| SS_extracted_k141_136389 | Not-determined |
| SS_extracted_k141_16172 | Low-quality |
| SS_extracted_k141_57052 | Not-determined |
| SS_extracted_k141_119897 | Not-determined |
| SS_extracted_k141_136402 | Not-determined |
| SS_extracted_k141_119900 | Not-determined |
| SS_extracted_k141_53363 | Low-quality |
| SS_extracted_k141_142957 | Low-quality |
| SS_extracted_k141_88705 | Not-determined |
| SS_extracted_k141_119906 | Low-quality |
| SS_extracted_k141_53367 | Not-determined |
| SS_extracted_k141_136415 | Not-determined |
| SS_extracted_k141_88715 | Not-determined |
| SS_extracted_k141_119911 | Not-determined |
| S3_vs_k141_127400\|\|full | High-quality |
| SS_extracted_k141_70339 | Not-determined |
| SS_extracted_k141_88721 | Not-determined |
| SS_extracted_k141_34911 | Not-determined |
| SS_extracted_k141_108081 | Not-determined |
| SS_extracted_k141_53381 | Not-determined |
| SS_extracted_k141_119922 | Not-determined |
| SS_extracted_k141_99919 | Not-determined |
| SS_extracted_k141_119923 | Not-determined |
| SS_extracted_k141_119925 | Low-quality |
| SS_extracted_k141_119926 | Not-determined |
| SS_extracted_k141_16194 | Low-quality |
| SS_extracted_k141_119928 | Not-determined |
| SS_extracted_k141_88729 | Not-determined |
| SS_extracted_k141_16198 | Low-quality |
| SS_extracted_k141_5395 | Not-determined |
| SS_extracted_k141_5396 | Not-determined |
| SS_extracted_k141_81181 | Not-determined |
| SS_extracted_k141_34929 | Not-determined |
| SS_extracted_k141_88737 | Not-determined |
| SS_extracted_k141_64295 | Not-determined |
| SS_extracted_k141_119938 | Low-quality |
| SS_extracted_k141_75075 | Not-determined |
| SS_extracted_k141_21884 | Low-quality |
| SS_extracted_k141_64297 | Not-determined |
| SS_extracted_k141_5409 | Not-determined |
| SS_extracted_k141_88743 | Not-determined |
| SS_extracted_k141_26637 | Not-determined |
| SS_extracted_k141_88745 | Not-determined |
| SS_extracted_k141_88748 | Low-quality |
| SS_extracted_k141_88750 | Low-quality |
| SS_extracted_k141_119950 | Not-determined |
| SS_extracted_k141_75086 | Not-determined |
| SS_extracted_k141_88755 | Not-determined |
| SS_extracted_k141_95178 | Low-quality |
| SS_extracted_k141_21892 | Low-quality |
| SS_extracted_k141_95180 | Not-determined |
| SS_extracted_k141_95183 | Not-determined |
| SS_extracted_k141_21896 | Low-quality |
| SS_extracted_k141_64308 | Not-determined |
| SS_extracted_k141_5425 | Not-determined |
| SS_extracted_k141_113288 | Low-quality |
| SS_extracted_k141_95186 | Not-determined |
| SS_extracted_k141_113290 | Not-determined |
| SS_extracted_k141_21903 | Not-determined |
| SS_extracted_k141_10259 | Not-determined |
| SS_extracted_k141_136435 | Not-determined |
| SS_extracted_k141_113292 | Not-determined |
| SS_extracted_k141_64316 | Low-quality |
| SS_extracted_k141_88769 | Not-determined |
| SS_extracted_k141_136443 | Not-determined |
| SS_extracted_k141_136444 | Not-determined |
| SS_extracted_k141_136446 | Not-determined |
| SS_extracted_k141_39875 | Low-quality |
| SS_extracted_k141_21913 | Not-determined |
| SS_extracted_k141_88775 | Not-determined |
| SS_extracted_k141_108100 | Not-determined |
| SS_extracted_k141_64334 | Not-determined |
| SS_extracted_k141_142983 | Not-determined |
| SS_extracted_k141_75107 | Not-determined |
| SS_extracted_k141_34970 | Not-determined |
| SS_extracted_k141_75109 | Not-determined |
| SS_extracted_k141_39883 | Low-quality |
| SS_extracted_k141_75110 | Not-determined |
| SS_extracted_k141_39884 | Not-determined |
| SS_extracted_k141_64340 | Not-determined |
| SS_extracted_k141_64347 | Not-determined |
| SS_extracted_k141_64348 | Not-determined |
| SS_extracted_k141_95216 | Not-determined |
| SS_extracted_k141_10296 | Not-determined |
| SS_extracted_k141_21935 | Low-quality |
| SS_extracted_k141_64368 | Not-determined |
| SS_extracted_k141_81235 | Not-determined |
| SS_extracted_k141_131409 | Not-determined |
| SS_extracted_k141_131410 | Low-quality |
| SS_extracted_k141_21938 | Low-quality |
| SS_extracted_k141_88786 | Low-quality |
| SS_extracted_k141_64374 | Not-determined |
| SS_extracted_k141_119974 | Low-quality |
| SS_extracted_k141_34996 | Not-determined |
| SS_extracted_k141_35000 | Not-determined |
| SS_extracted_k141_95237 | Low-quality |
| SS_extracted_k141_75124 | Low-quality |
| SS_extracted_k141_21949 | Not-determined |
| SS_extracted_k141_35008 | Not-determined |
| SS_extracted_k141_57101 | Not-determined |
| SS_extracted_k141_35013 | Not-determined |
| SS_extracted_k141_21954 | Not-determined |
| SS_extracted_k141_108126 | Not-determined |
| SS_extracted_k141_64384 | Not-determined |
| SS_extracted_k141_57105 | Not-determined |
| SS_extracted_k141_35020 | Not-determined |
| SS_extracted_k141_10327 | Not-determined |
| SS_extracted_k141_5465 | Not-determined |
| SS_extracted_k141_35024 | Not-determined |
| SS_extracted_k141_88808 | Low-quality |
| SS_extracted_k141_88809 | Low-quality |
| SS_extracted_k141_21966 | Not-determined |
| SS_extracted_k141_21967 | Not-determined |
| SS_extracted_k141_57108 | Not-determined |
| SS_extracted_k141_136476 | Not-determined |
| SS_extracted_k141_21977 | Not-determined |
| SS_extracted_k141_35043 | Not-determined |
| SS_extracted_k141_21978 | Low-quality |
| SS_extracted_k141_75150 | Not-determined |
| SS_extracted_k141_21981 | Not-determined |
| SS_extracted_k141_120014 | Not-determined |
| SS_extracted_k141_35049 | Not-determined |
| SS_extracted_k141_131436 | Low-quality |
| SS_extracted_k141_99938 | Not-determined |
| SS_extracted_k141_88840 | Low-quality |
| SS_extracted_k141_57115 | Not-determined |
| SS_extracted_k141_108134 | Low-quality |
| SS_extracted_k141_108135 | Low-quality |
| SS_extracted_k141_108136 | Low-quality |
| SS_extracted_k141_53441 | Not-determined |
| SS_extracted_k141_120045 | Not-determined |
| SS_extracted_k141_131447 | Low-quality |
| SS_extracted_k141_53449 | Not-determined |
| SS_extracted_k141_75161 | Low-quality |
| SS_extracted_k141_136485 | Not-determined |
| SS_extracted_k141_95277 | Not-determined |
| SS_extracted_k141_95278 | Not-determined |
| SS_extracted_k141_64419 | Low-quality |
| SS_extracted_k141_136489 | Not-determined |
| SS_extracted_k141_108143 | Low-quality |
| SS_extracted_k141_46209 | Not-determined |
| SS_extracted_k141_88853 | Not-determined |
| SS_extracted_k141_75172 | Not-determined |
| SS_extracted_k141_5517 | Low-quality |
| SS_extracted_k141_70357 | Not-determined |
| SS_extracted_k141_5520 | Low-quality |
| SS_extracted_k141_95285 | Not-determined |
| SS_extracted_k141_35083 | Not-determined |
| SS_extracted_k141_99962 | Not-determined |
| SS_extracted_k141_131456 | Not-determined |
| SS_extracted_k141_108151 | Not-determined |
| SS_extracted_k141_70362 | Not-determined |
| SS_extracted_k141_53462 | Not-determined |
| SS_extracted_k141_131460 | Not-determined |
| SS_extracted_k141_5531 | Not-determined |
| SS_extracted_k141_108156 | Not-determined |
| SS_extracted_k141_136503 | Low-quality |
| SS_extracted_k141_143010 | Low-quality |
| SS_extracted_k141_131466 | Low-quality |
| SS_extracted_k141_120060 | Not-determined |
| SS_extracted_k141_136504 | Not-determined |
| SS_extracted_k141_70373 | Not-determined |
| SS_extracted_k141_143014 | Not-determined |
| SS_extracted_k141_131471 | Low-quality |
| SS_extracted_k141_53465 | Not-determined |
| SS_extracted_k141_136512 | Not-determined |
| SS_extracted_k141_75192 | Not-determined |
| SS_extracted_k141_108172 | Not-determined |
| SS_extracted_k141_108173 | Not-determined |
| SS_extracted_k141_75197 | Not-determined |
| SS_extracted_k141_88870 | Not-determined |
| SS_extracted_k141_10360 | Not-determined |
| SS_extracted_k141_10361 | Not-determined |
| SS_extracted_k141_46229 | Not-determined |
| SS_extracted_k141_99975 | Not-determined |
| SS_extracted_k141_35103 | Not-determined |
| SS_extracted_k141_75201 | Not-determined |
| SS_extracted_k141_99978 | Not-determined |
| SS_extracted_k141_64445 | Not-determined |
| SS_extracted_k141_39949 | Not-determined |
| SS_extracted_k141_120063 | Low-quality |
| SS_extracted_k141_53479 | Not-determined |
| SS_extracted_k141_10370 | Not-determined |
| SS_extracted_k141_64451 | Not-determined |
| SS_extracted_k141_64452 | Not-determined |
| SS_extracted_k141_39952 | Not-determined |
| SS_extracted_k141_88875 | Not-determined |
| SS_extracted_k141_53484 | Low-quality |
| SS_extracted_k141_39958 | Low-quality |
| SS_extracted_k141_10377 | Not-determined |
| SS_extracted_k141_39959 | Not-determined |
| SS_extracted_k141_99995 | Not-determined |
| SS_extracted_k141_35118 | Not-determined |
| SS_extracted_k141_108184 | Not-determined |
| SS_extracted_k141_39964 | Low-quality |
| SS_extracted_k141_113388 | Not-determined |
| SS_extracted_k141_131491 | Low-quality |
| SS_extracted_k141_75212 | Not-determined |
| SS_extracted_k141_39969 | Not-determined |
| SS_extracted_k141_39970 | Not-determined |
| SS_extracted_k141_108187 | Not-determined |
| SS_extracted_k141_131503 | Low-quality |
| SS_extracted_k141_53494 | Not-determined |
| SS_extracted_k141_88886 | Not-determined |
| SS_extracted_k141_53499 | Low-quality |
| SS_extracted_k141_46242 | Low-quality |
| SS_extracted_k141_46243 | Not-determined |
| SS_extracted_k141_143033 | Not-determined |
| SS_extracted_k141_10390 | Low-quality |
| SS_extracted_k141_143035 | Not-determined |
| SS_extracted_k141_53506 | Not-determined |
| SS_extracted_k141_35127 | Not-determined |
| SS_extracted_k141_35128 | Not-determined |
| SS_extracted_k141_10393 | Not-determined |
| SS_extracted_k141_57160 | Not-determined |
| SS_extracted_k141_88903 | Low-quality |
| SS_extracted_k141_53513 | Not-determined |
| SS_extracted_k141_10396 | Not-determined |
| SS_extracted_k141_35140 | Not-determined |
| SS_extracted_k141_136518 | Not-determined |
| SS_extracted_k141_108214 | Not-determined |
| SS_extracted_k141_108215 | Not-determined |
| SS_extracted_k141_10397 | Not-determined |
| SS_extracted_k141_10398 | Not-determined |
| SS_extracted_k141_10400 | Not-determined |
| SS_extracted_k141_10402 | Not-determined |
| SS_extracted_k141_70433 | Not-determined |
| SS_extracted_k141_126327 | Not-determined |
| SS_extracted_k141_16233 | Low-quality |
| SS_extracted_k141_143052 | Low-quality |
| SS_extracted_k141_88915 | Not-determined |
| SS_extracted_k141_46247 | Not-determined |
| SS_extracted_k141_46248 | Low-quality |
| SS_extracted_k141_10410 | Low-quality |
| SS_extracted_k141_35156 | Low-quality |
| SS_extracted_k141_16242 | Not-determined |
| SS_extracted_k141_136534 | Not-determined |
| SS_extracted_k141_64485 | Low-quality |
| SS_extracted_k141_126330 | Not-determined |
| SS_extracted_k141_39977 | Low-quality |
| SS_extracted_k141_108223 | Not-determined |
| SS_extracted_k141_75221 | Not-determined |
| SS_extracted_k141_39980 | Low-quality |
| SS_extracted_k141_10418 | Not-determined |
| SS_extracted_k141_88923 | Not-determined |
| SS_extracted_k141_108227 | Not-determined |
| SS_extracted_k141_10420 | Not-determined |
| SS_extracted_k141_70462 | Not-determined |
| SS_extracted_k141_10422 | Not-determined |
| SS_extracted_k141_75231 | Not-determined |
| SS_extracted_k141_75233 | Not-determined |
| SS_extracted_k141_39985 | Not-determined |
| SS_extracted_k141_64493 | Low-quality |
| SS_extracted_k141_64494 | Not-determined |
| SS_extracted_k141_126341 | Low-quality |
| SS_extracted_k141_70466 | Not-determined |
| SS_extracted_k141_35166 | Not-determined |
| SS_extracted_k141_39988 | Not-determined |
| SS_extracted_k141_95320 | Not-determined |
| SS_extracted_k141_16264 | Not-determined |
| SS_extracted_k141_95321 | Not-determined |
| SS_extracted_k141_136542 | Not-determined |
| SS_extracted_k141_95323 | Not-determined |
| SS_extracted_k141_75239 | Not-determined |
| SS_extracted_k141_70473 | Low-quality |
| SS_extracted_k141_136549 | Not-determined |
| SS_extracted_k141_126354 | Not-determined |
| SS_extracted_k141_64501 | Not-determined |
| SS_extracted_k141_35172 | Low-quality |
| SS_extracted_k141_75248 | Low-quality |
| SS_extracted_k141_64510 | Not-determined |
| SS_extracted_k141_75256 | Not-determined |
| SS_extracted_k141_70488 | Not-determined |
| SS_extracted_k141_57173 | Not-determined |
| SS_extracted_k141_57174 | Not-determined |
| SS_extracted_k141_57177 | Not-determined |
| SS_extracted_k141_75263 | Not-determined |
| SS_extracted_k141_64515 | Not-determined |
| SS_extracted_k141_108251 | Not-determined |
| SS_extracted_k141_64519 | Low-quality |
| SS_extracted_k141_95336 | Not-determined |
| SS_extracted_k141_75273 | Not-determined |
| SS_extracted_k141_64521 | Not-determined |
| SS_extracted_k141_126374 | Not-determined |
| SS_extracted_k141_64524 | Low-quality |
| SS_extracted_k141_57185 | Low-quality |
| SS_extracted_k141_75281 | Not-determined |
| SS_extracted_k141_120110 | Not-determined |
| SS_extracted_k141_26720 | Not-determined |
| SS_extracted_k141_75285 | Not-determined |
| SS_extracted_k141_95355 | Not-determined |
| SS_extracted_k141_64541 | Not-determined |
| SS_extracted_k141_136558 | Low-quality |
| SS_extracted_k141_46273 | Not-determined |
| SS_extracted_k141_70511 | Not-determined |
| SS_extracted_k141_95361 | Not-determined |
| SS_extracted_k141_81248 | Low-quality |
| SS_extracted_k141_120120 | Not-determined |
| SS_extracted_k141_70517 | Low-quality |
| SS_extracted_k141_143065 | Not-determined |
| SS_extracted_k141_16298 | Not-determined |
| SS_extracted_k141_143067 | Not-determined |
| SS_extracted_k141_40019 | Not-determined |
| SS_extracted_k141_16299 | Not-determined |
| SS_extracted_k141_70521 | Not-determined |
| SS_extracted_k141_64547 | Not-determined |
| SS_extracted_k141_16301 | Not-determined |
| SS_extracted_k141_16302 | Not-determined |
| SS_extracted_k141_143076 | Not-determined |
| SS_extracted_k141_120132 | Low-quality |
| SS_extracted_k141_26740 | Not-determined |
| SS_extracted_k141_16304 | Not-determined |
| SS_extracted_k141_143079 | Low-quality |
| SS_extracted_k141_16310 | Low-quality |
| SS_extracted_k141_46287 | Not-determined |

| | |
|---|---|
| SS_extracted_k141_88941 | Not-determined |
| SS_extracted_k141_16313 | Not-determined |
| SS_extracted_k141_26758 | Not-determined |
| SS_extracted_k141_16316 | Not-determined |
| SS_extracted_k141_57202 | Not-determined |
| SS_extracted_k141_126403 | Low-quality |
| SS_extracted_k141_16319 | Not-determined |
| SS_extracted_k141_75314 | Not-determined |
| SS_extracted_k141_126408 | Not-determined |
| SS_extracted_k141_88946 | Not-determined |
| SS_extracted_k141_26761 | Low-quality |
| SS_extracted_k141_26764 | Not-determined |
| SS_extracted_k141_16325 | Low-quality |
| SS_extracted_k141_143081 | Not-determined |
| SS_extracted_k141_35219 | Not-determined |
| SS_extracted_k141_88950 | Low-quality |
| SS_extracted_k141_40046 | Not-determined |
| SS_extracted_k141_143083 | Not-determined |
| SS_extracted_k141_143084 | Not-determined |
| SS_extracted_k141_26767 | Not-determined |
| SS_extracted_k141_57208 | Not-determined |
| SS_extracted_k141_5545 | Not-determined |
| SS_extracted_k141_40050 | Low-quality |
| SS_extracted_k141_70522 | Low-quality |
| SS_extracted_k141_70523 | Not-determined |
| SS_extracted_k141_64561 | Not-determined |
| SS_extracted_k141_16334 | Not-determined |
| SS_extracted_k141_143094 | Low-quality |
| SS_extracted_k141_64573 | Not-determined |
| SS_extracted_k141_126428 | Not-determined |
| SS_extracted_k141_88961 | Low-quality |
| SS_extracted_k141_16345 | Not-determined |
| SS_extracted_k141_5566 | Not-determined |
| SS_extracted_k141_88967 | Not-determined |
| SS_extracted_k141_5573 | Not-determined |
| SS_extracted_k141_108290 | Not-determined |
| SS_extracted_k141_5575 | Not-determined |
| SS_extracted_k141_26782 | Not-determined |
| SS_extracted_k141_57226 | Not-determined |
| SS_extracted_k141_64583 | Not-determined |
| SS_extracted_k141_5580 | Not-determined |
| SS_extracted_k141_22030 | Not-determined |
| SS_extracted_k141_22031 | Not-determined |
| SS_extracted_k141_22034 | Not-determined |
| SS_extracted_k141_126439 | Not-determined |
| SS_extracted_k141_5591 | Not-determined |
| SS_extracted_k141_5595 | Not-determined |
| SS_extracted_k141_35249 | Not-determined |
| SS_extracted_k141_5596 | Not-determined |
| SS_extracted_k141_40091 | Not-determined |
| SS_extracted_k141_120148 | Not-determined |
| SS_extracted_k141_136583 | Not-determined |
| SS_extracted_k141_40093 | Low-quality |
| SS_extracted_k141_100003 | Not-determined |
| SS_extracted_k141_16351 | Low-quality |
| SS_extracted_k141_88990 | Not-determined |
| SS_extracted_k141_136589 | Low-quality |
| SS_extracted_k141_113402 | Not-determined |
| SS_extracted_k141_75333 | Not-determined |
| SS_extracted_k141_126450 | Not-determined |
| SS_extracted_k141_126454 | Not-determined |
| SS_extracted_k141_22045 | Low-quality |
| SS_extracted_k141_108314 | Not-determined |
| SS_extracted_k141_22047 | Not-determined |
| SS_extracted_k141_120159 | Low-quality |
| SS_extracted_k141_113413 | Low-quality |
| SS_extracted_k141_64608 | Not-determined |
| SS_extracted_k141_120162 | Not-determined |
| SS_extracted_k141_120163 | Not-determined |
| SS_extracted_k141_64611 | Not-determined |
| SS_extracted_k141_64614 | Low-quality |
| SS_extracted_k141_64616 | Not-determined |
| SS_extracted_k141_75347 | Not-determined |
| SS_extracted_k141_113417 | Not-determined |
| SS_extracted_k141_113419 | Not-determined |
| SS_extracted_k141_64623 | Not-determined |
| SS_extracted_k141_81266 | Not-determined |
| SS_extracted_k141_108332 | Not-determined |
| SS_extracted_k141_75352 | Not-determined |
| SS_extracted_k141_16371 | Not-determined |
| SS_extracted_k141_22070 | Not-determined |
| SS_extracted_k141_64626 | Low-quality |
| SS_extracted_k141_26813 | Not-determined |
| SS_extracted_k141_75354 | Not-determined |
| SS_extracted_k141_89011 | Not-determined |
| SS_extracted_k141_75356 | Not-determined |
| SS_extracted_k141_57242 | Not-determined |
| SS_extracted_k141_81270 | Not-determined |
| SS_extracted_k141_120173 | Not-determined |
| SS_extracted_k141_120174 | Not-determined |
| SS_extracted_k141_89016 | Not-determined |
| SS_extracted_k141_81271 | Low-quality |
| SS_extracted_k141_81272 | Not-determined |
| SS_extracted_k141_89020 | Not-determined |
| SS_extracted_k141_35282 | Not-determined |
| SS_extracted_k141_89021 | Not-determined |
| SS_extracted_k141_108344 | Not-determined |
| SS_extracted_k141_35285 | Low-quality |
| SS_extracted_k141_108355 | Not-determined |
| SS_extracted_k141_89029 | Low-quality |
| SS_extracted_k141_5603 | Not-determined |
| SS_extracted_k141_126479 | Not-determined |
| SS_extracted_k141_5608 | Not-determined |
| SS_extracted_k141_143136 | Not-determined |
| SS_extracted_k141_26819 | Not-determined |
| SS_extracted_k141_16396 | Not-determined |
| SS_extracted_k141_16398 | Low-quality |
| SS_extracted_k141_26825 | Not-determined |
| SS_extracted_k141_53536 | Not-determined |
| SS_extracted_k141_53538 | Low-quality |
| SS_extracted_k141_100018 | Not-determined |
| SS_extracted_k141_64638 | Not-determined |
| SS_extracted_k141_53540 | Not-determined |
| SS_extracted_k141_113433 | Low-quality |
| SS_extracted_k141_136605 | Not-determined |
| SS_extracted_k141_40126 | Low-quality |
| SS_extracted_k141_100021 | Not-determined |
| SS_extracted_k141_16406 | Not-determined |
| SS_extracted_k141_53545 | Not-determined |
| SS_extracted_k141_100023 | Not-determined |
| SS_extracted_k141_89041 | Not-determined |
| SS_extracted_k141_100030 | Not-determined |
| SS_extracted_k141_100031 | Not-determined |
| SS_extracted_k141_5628 | Not-determined |
| SS_extracted_k141_40132 | Not-determined |
| SS_extracted_k141_5630 | Low-quality |
| SS_extracted_k141_126506 | Not-determined |
| SS_extracted_k141_126507 | Not-determined |
| SS_extracted_k141_75380 | Not-determined |
| SS_extracted_k141_143156 | Not-determined |
| SS_extracted_k141_53555 | Low-quality |
| SS_extracted_k141_5636 | Low-quality |
| SS_extracted_k141_95401 | Not-determined |
| SS_extracted_k141_143162 | Low-quality |
| SS_extracted_k141_75391 | Not-determined |
| SS_extracted_k141_75392 | Not-determined |
| SS_extracted_k141_53567 | Not-determined |
| SS_extracted_k141_75394 | Not-determined |
| SS_extracted_k141_64654 | Low-quality |
| SS_extracted_k141_75396 | Not-determined |
| SS_extracted_k141_126515 | Not-determined |
| SS_extracted_k141_131520 | Low-quality |
| SS_extracted_k141_100044 | Not-determined |
| SS_extracted_k141_40158 | Not-determined |
| SS_extracted_k141_89056 | Not-determined |
| SS_extracted_k141_16427 | Not-determined |
| SS_extracted_k141_64666 | Low-quality |
| SS_extracted_k141_108373 | Not-determined |
| SS_extracted_k141_16431 | Not-determined |
| SS_extracted_k141_131528 | Low-quality |
| SS_extracted_k141_95416 | Low-quality |
| SS_extracted_k141_89062 | Not-determined |
| SS_extracted_k141_40164 | Not-determined |
| SS_extracted_k141_100057 | Low-quality |
| SS_extracted_k141_53579 | Not-determined |
| SS_extracted_k141_108381 | Not-determined |
| SS_extracted_k141_53584 | Not-determined |
| SS_extracted_k141_131538 | Low-quality |
| SS_extracted_k141_126530 | Not-determined |
| SS_extracted_k141_64677 | Low-quality |
| SS_extracted_k141_75413 | Not-determined |
| SS_extracted_k141_75415 | Not-determined |
| SS_extracted_k141_108389 | Not-determined |
| SS_extracted_k141_64679 | Low-quality |
| SS_extracted_k141_64681 | Not-determined |
| SS_extracted_k141_95426 | Not-determined |
| SS_extracted_k141_64682 | Low-quality |
| SS_extracted_k141_108393 | Not-determined |
| SS_extracted_k141_108396 | Not-determined |
| SS_extracted_k141_22088 | Not-determined |
| SS_extracted_k141_70553 | Not-determined |
| SS_extracted_k141_53590 | Not-determined |
| SS_extracted_k141_131553 | Low-quality |
| SS_extracted_k141_131555 | Not-determined |
| SS_extracted_k141_16460 | Not-determined |
| SS_extracted_k141_108404 | Not-determined |
| SS_extracted_k141_131560 | Not-determined |
| SS_extracted_k141_100075 | Low-quality |
| SS_extracted_k141_22101 | Not-determined |
| SS_extracted_k141_108411 | Not-determined |
| SS_extracted_k141_120176 | Not-determined |
| SS_extracted_k141_136633 | Not-determined |
| SS_extracted_k141_22106 | Not-determined |
| SS_extracted_k141_108417 | Not-determined |
| SS_extracted_k141_81336 | Not-determined |
| SS_extracted_k141_22112 | Not-determined |
| SS_extracted_k141_70565 | Not-determined |
| SS_extracted_k141_22116 | Not-determined |
| SS_extracted_k141_16481 | Not-determined |
| SS_extracted_k141_64709 | Not-determined |
| SS_extracted_k141_64710 | Not-determined |
| SS_extracted_k141_53607 | Not-determined |
| SS_extracted_k141_75436 | Not-determined |
| SS_extracted_k141_53609 | Not-determined |
| SS_extracted_k141_75443 | Not-determined |
| SS_extracted_k141_126566 | Not-determined |
| SS_extracted_k141_131576 | Not-determined |
| SS_extracted_k141_16494 | Not-determined |
| SS_extracted_k141_53618 | Not-determined |
| SS_extracted_k141_143203 | Not-determined |
| SS_extracted_k141_95454 | Not-determined |
| SS_extracted_k141_95455 | Not-determined |
| SS_extracted_k141_126567 | Low-quality |
| SS_extracted_k141_136648 | Not-determined |
| SS_extracted_k141_40189 | Not-determined |
| SS_extracted_k141_16504 | Not-determined |
| SS_extracted_k141_64722 | Not-determined |
| SS_extracted_k141_16505 | Not-determined |
| SS_extracted_k141_53628 | Not-determined |
| SS_extracted_k141_75458 | Not-determined |
| SS_extracted_k141_53629 | Not-determined |
| SS_extracted_k141_26901 | Not-determined |
| SS_extracted_k141_131589 | Not-determined |
| SS_extracted_k141_75462 | Not-determined |
| SS_extracted_k141_75464 | Not-determined |
| SS_extracted_k141_108440 | Not-determined |
| SS_extracted_k141_143209 | Not-determined |
| SS_extracted_k141_108441 | Not-determined |
| SS_extracted_k141_64725 | Not-determined |
| SS_extracted_k141_16513 | Not-determined |
| SS_extracted_k141_53635 | Not-determined |
| SS_extracted_k141_126576 | Not-determined |
| SS_extracted_k141_16516 | Not-determined |
| SS_extracted_k141_22145 | Not-determined |
| SS_extracted_k141_16517 | Not-determined |
| SS_extracted_k141_108448 | Low-quality |
| SS_extracted_k141_75474 | Low-quality |
| SS_extracted_k141_108452 | Not-determined |
| SS_extracted_k141_95472 | Not-determined |
| SS_extracted_k141_108456 | Not-determined |
| SS_extracted_k141_75479 | Not-determined |
| SS_extracted_k141_95475 | Not-determined |
| SS_extracted_k141_70605 | Not-determined |
| SS_extracted_k141_40210 | Not-determined |
| SS_extracted_k141_95477 | Not-determined |
| SS_extracted_k141_75484 | Not-determined |
| SS_extracted_k141_75485 | Not-determined |
| SS_extracted_k141_16535 | Not-determined |
| SS_extracted_k141_75488 | Low-quality |
| SS_extracted_k141_131607 | Not-determined |
| SS_extracted_k141_126595 | Not-determined |
| SS_extracted_k141_53657 | Not-determined |
| SS_extracted_k141_95483 | Not-determined |
| SS_extracted_k141_16538 | Not-determined |
| SS_extracted_k141_70612 | Not-determined |
| SS_extracted_k141_16539 | Not-determined |
| SS_extracted_k141_108476 | Not-determined |
| SS_extracted_k141_108482 | Low-quality |
| SS_extracted_k141_53668 | Not-determined |
| SS_extracted_k141_100113 | Low-quality |
| SS_extracted_k141_131615 | Low-quality |
| SS_extracted_k141_100118 | Low-quality |
| SS_extracted_k141_53674 | Low-quality |
| SS_extracted_k141_10431 | Low-quality |
| SS_extracted_k141_131621 | Not-determined |
| SS_extracted_k141_131622 | Not-determined |
| SS_extracted_k141_100121 | Not-determined |
| SS_extracted_k141_16542 | Not-determined |
| SS_extracted_k141_100123 | Not-determined |
| SS_extracted_k141_16543 | Not-determined |
| SS_extracted_k141_75519 | Not-determined |
| SS_extracted_k141_70615 | Not-determined |
| SS_extracted_k141_143217 | Low-quality |
| SS_extracted_k141_53682 | Not-determined |
| SS_extracted_k141_126615 | Not-determined |
| SS_extracted_k141_131630 | Not-determined |
| SS_extracted_k141_126623 | Not-determined |
| SS_extracted_k141_108501 | Not-determined |
| SS_extracted_k141_26903 | Not-determined |
| SS_extracted_k141_95496 | Low-quality |
| SS_extracted_k141_26907 | Not-determined |
| SS_extracted_k141_26908 | Not-determined |
| SS_extracted_k141_53697 | Not-determined |
| SS_extracted_k141_108505 | Low-quality |
| SS_extracted_k141_26917 | Not-determined |
| SS_extracted_k141_108510 | Low-quality |
| SS_extracted_k141_70639 | Not-determined |
| SS_extracted_k141_16555 | Not-determined |
| SS_extracted_k141_26919 | Low-quality |
| SS_extracted_k141_113481 | Not-determined |
| SS_extracted_k141_89109 | Not-determined |
| SS_extracted_k141_95499 | Low-quality |
| SS_extracted_k141_53708 | Not-determined |
| SS_extracted_k141_113488 | Not-determined |
| SS_extracted_k141_16560 | Not-determined |
| SS_extracted_k141_136669 | Not-determined |
| SS_extracted_k141_89111 | Not-determined |
| SS_extracted_k141_108522 | Not-determined |
| SS_extracted_k141_136671 | Not-determined |
| SS_extracted_k141_131657 | Not-determined |
| SS_extracted_k141_108524 | Not-determined |
| SS_extracted_k141_53713 | Not-determined |
| SS_extracted_k141_108528 | Not-determined |
| SS_extracted_k141_53715 | Low-quality |
| SS_extracted_k141_100164 | Low-quality |
| SS_extracted_k141_113498 | Low-quality |
| SS_extracted_k141_108531 | Not-determined |
| SS_extracted_k141_126638 | Not-determined |
| SS_extracted_k141_100167 | Not-determined |
| SS_extracted_k141_131664 | Low-quality |
| SS_extracted_k141_131668 | Not-determined |
| SS_extracted_k141_100169 | Not-determined |
| SS_extracted_k141_131671 | Low-quality |
| SS_extracted_k141_95514 | Low-quality |
| SS_extracted_k141_113507 | Low-quality |
| SS_extracted_k141_131676 | Low-quality |
| SS_extracted_k141_53731 | Low-quality |
| SS_extracted_k141_70664 | Not-determined |
| SS_extracted_k141_64732 | Not-determined |
| SS_extracted_k141_100184 | Not-determined |
| SS_extracted_k141_53737 | Not-determined |
| SS_extracted_k141_95527 | Not-determined |
| SS_extracted_k141_131689 | Not-determined |
| SS_extracted_k141_143249 | Not-determined |
| SS_extracted_k141_108562 | Low-quality |
| SS_extracted_k141_5685 | Not-determined |
| SS_extracted_k141_131696 | Not-determined |
| SS_extracted_k141_64743 | Not-determined |
| SS_extracted_k141_113527 | Not-determined |
| SS_extracted_k141_16604 | Not-determined |
| SS_extracted_k141_113530 | Not-determined |
| SS_extracted_k141_35327 | Not-determined |
| SS_extracted_k141_70677 | Low-quality |
| SS_extracted_k141_57251 | Not-determined |
| SS_extracted_k141_5701 | Not-determined |
| SS_extracted_k141_5702 | Not-determined |
| SS_extracted_k141_95549 | Low-quality |
| SS_extracted_k141_143266 | Not-determined |
| SS_extracted_k141_16616 | Not-determined |
| SS_extracted_k141_35341 | Not-determined |
| SS_extracted_k141_108582 | Low-quality |
| SS_extracted_k141_35342 | Not-determined |
| SS_extracted_k141_113543 | Not-determined |
| SS_extracted_k141_89155 | Not-determined |
| SS_extracted_k141_108588 | Not-determined |
| SS_extracted_k141_16627 | Low-quality |
| SS_extracted_k141_16628 | Not-determined |
| SS_extracted_k141_5719 | Not-determined |
| SS_extracted_k141_64770 | Not-determined |
| SS_extracted_k141_100209 | Not-determined |
| SS_extracted_k141_16635 | Not-determined |
| SS_extracted_k141_64772 | Low-quality |
| SS_extracted_k141_10458 | Not-determined |
| SS_extracted_k141_26943 | Not-determined |
| SS_extracted_k141_108602 | Low-quality |
| SS_extracted_k141_64777 | Not-determined |
| SS_extracted_k141_143279 | Low-quality |
| SS_extracted_k141_35356 | Not-determined |
| SS_extracted_k141_16644 | Not-determined |
| SS_extracted_k141_70699 | Not-determined |
| SS_extracted_k141_108611 | Not-determined |
| SS_extracted_k141_75538 | Not-determined |
| SS_extracted_k141_53766 | Not-determined |
| SS_extracted_k141_108617 | Not-determined |
| SS_extracted_k141_35367 | Low-quality |
| SS_extracted_k141_53770 | Not-determined |
| SS_extracted_k141_53771 | Low-quality |
| SS_extracted_k141_22163 | Not-determined |
| SS_extracted_k141_143297 | Not-determined |
| SS_extracted_k141_35371 | Not-determined |
| SS_extracted_k141_89178 | Low-quality |
| SS_extracted_k141_64803 | Not-determined |
| SS_extracted_k141_26962 | Low-quality |
| SS_extracted_k141_35376 | Not-determined |
| SS_extracted_k141_53779 | Not-determined |
| SS_extracted_k141_26968 | Not-determined |
| SS_extracted_k141_143303 | Not-determined |
| SS_extracted_k141_22168 | Not-determined |
| SS_extracted_k141_53781 | Low-quality |
| SS_extracted_k141_26973 | Low-quality |
| SS_extracted_k141_64813 | Not-determined |
| SS_extracted_k141_64819 | Not-determined |
| SS_extracted_k141_75573 | Low-quality |
| SS_extracted_k141_95581 | Not-determined |
| SS_extracted_k141_95582 | Low-quality |
| SS_extracted_k141_143320 | Not-determined |
| SS_extracted_k141_35390 | Not-determined |
| SS_extracted_k141_57256 | Low-quality |
| SS_extracted_k141_22173 | Not-determined |
| SS_extracted_k141_95594 | Low-quality |
| SS_extracted_k141_40241 | Not-determined |
| SS_extracted_k141_113594 | Not-determined |
| SS_extracted_k141_70733 | Not-determined |
| SS_extracted_k141_113596 | Low-quality |
| SS_extracted_k141_95600 | Not-determined |
| SS_extracted_k141_113598 | Not-determined |
| SS_extracted_k141_35396 | Not-determined |
| SS_extracted_k141_22188 | Low-quality |
| SS_extracted_k141_16710 | Not-determined |
| SS_extracted_k141_35406 | Not-determined |
| SS_extracted_k141_22198 | Low-quality |
| SS_extracted_k141_95612 | Not-determined |
| SS_extracted_k141_95615 | Not-determined |
| SS_extracted_k141_57273 | Not-determined |
| SS_extracted_k141_16718 | Not-determined |
| SS_extracted_k141_126675 | Not-determined |
| SS_extracted_k141_35416 | Not-determined |
| SS_extracted_k141_75587 | Not-determined |
| SS_extracted_k141_100236 | Not-determined |
| SS_extracted_k141_75598 | Low-quality |
| SS_extracted_k141_35430 | Not-determined |
| SS_extracted_k141_64830 | Not-determined |
| SS_extracted_k141_40254 | Low-quality |
| SS_extracted_k141_70767 | Not-determined |
| SS_extracted_k141_27008 | Not-determined |
| SS_extracted_k141_16725 | Not-determined |
| SS_extracted_k141_70769 | Low-quality |
| SS_extracted_k141_35433 | Not-determined |
| SS_extracted_k141_5737 | Not-determined |
| SS_extracted_k141_126687 | Low-quality |
| SS_extracted_k141_126688 | Not-determined |
| SS_extracted_k141_64838 | Not-determined |
| SS_extracted_k141_35435 | Not-determined |
| SS_extracted_k141_5738 | Low-quality |
| SS_extracted_k141_89182 | Low-quality |
| SS_extracted_k141_100263 | Not-determined |
| SS_extracted_k141_35440 | Not-determined |
| SS_extracted_k141_16736 | Not-determined |
| SS_extracted_k141_75611 | Low-quality |
| SS_extracted_k141_95634 | Not-determined |
| SS_extracted_k141_64850 | Not-determined |
| SS_extracted_k141_131722 | Not-determined |
| SS_extracted_k141_95639 | Not-determined |
| SS_extracted_k141_70773 | Not-determined |
| SS_extracted_k141_95640 | Not-determined |
| SS_extracted_k141_70777 | Low-quality |
| SS_extracted_k141_100279 | Low-quality |
| SS_extracted_k141_22212 | Low-quality |
| SS_extracted_k141_89201 | Not-determined |
| SS_extracted_k141_40289 | Not-determined |
| SS_extracted_k141_131728 | Low-quality |
| SS_extracted_k141_57292 | Low-quality |
| SS_extracted_k141_40293 | Not-determined |
| SS_extracted_k141_16743 | Not-determined |
| SS_extracted_k141_40298 | Low-quality |
| SS_extracted_k141_136715 | Low-quality |
| SS_extracted_k141_16744 | Not-determined |
| SS_extracted_k141_35459 | Not-determined |
| SS_extracted_k141_35462 | Not-determined |
| SS_extracted_k141_22217 | Not-determined |
| SS_extracted_k141_136720 | Not-determined |
| SS_extracted_k141_35464 | Not-determined |
| SS_extracted_k141_120190 | Not-determined |
| SS_extracted_k141_70790 | Not-determined |
| SS_extracted_k141_120192 | Not-determined |
| SS_extracted_k141_81350 | Low-quality |
| SS_extracted_k141_22219 | Not-determined |
| SS_extracted_k141_75634 | Not-determined |
| SS_extracted_k141_35477 | Not-determined |
| SS_extracted_k141_126707 | Not-determined |
| SS_extracted_k141_81361 | Not-determined |
| SS_extracted_k141_35481 | Not-determined |
| SS_extracted_k141_75638 | Not-determined |
| SS_extracted_k141_81364 | Not-determined |
| SS_extracted_k141_57308 | Not-determined |
| SS_extracted_k141_75642 | Not-determined |
| SS_extracted_k141_81374 | Not-determined |
| SS_extracted_k141_35489 | Not-determined |
| SS_extracted_k141_70809 | Not-determined |
| SS_extracted_k141_27024 | Low-quality |
| SS_extracted_k141_16786 | Not-determined |
| SS_extracted_k141_16788 | Not-determined |
| SS_extracted_k141_57311 | Not-determined |
| SS_extracted_k141_16797 | Not-determined |
| SS_extracted_k141_81385 | Not-determined |
| SS_extracted_k141_16799 | Not-determined |
| SS_extracted_k141_143347 | Low-quality |
| SS_extracted_k141_136729 | Not-determined |
| SS_extracted_k141_108633 | Low-quality |
| SS_extracted_k141_126737 | Not-determined |
| SS_extracted_k141_108635 | Not-determined |
| SS_extracted_k141_131762 | Not-determined |
| SS_extracted_k141_108637 | Not-determined |
| SS_extracted_k141_70833 | Not-determined |
| SS_extracted_k141_113667 | Low-quality |
| SS_extracted_k141_108640 | Low-quality |
| SS_extracted_k141_113674 | Not-determined |
| SS_extracted_k141_35515 | Not-determined |
| SS_extracted_k141_35516 | Not-determined |
| SS_extracted_k141_70851 | Not-determined |
| SS_extracted_k141_16812 | Not-determined |
| SS_extracted_k141_131778 | Not-determined |
| SS_extracted_k141_126744 | Not-determined |
| SS_extracted_k141_16818 | Not-determined |
| SS_extracted_k141_113681 | Not-determined |
| SS_extracted_k141_16819 | Not-determined |
| SS_extracted_k141_113683 | Not-determined |
| SS_extracted_k141_131784 | Not-determined |
| SS_extracted_k141_95662 | Not-determined |
| SS_extracted_k141_120217 | Not-determined |
| SS_extracted_k141_75675 | Not-determined |
| SS_extracted_k141_16827 | Not-determined |
| SS_extracted_k141_27040 | Not-determined |
| SS_extracted_k141_113692 | Not-determined |
| SS_extracted_k141_81419 | Not-determined |
| SS_extracted_k141_81422 | Not-determined |
| SS_extracted_k141_126748 | Low-quality |
| SS_extracted_k141_70859 | Not-determined |
| SS_extracted_k141_70860 | Not-determined |
| SS_extracted_k141_126753 | Low-quality |
| SS_extracted_k141_75688 | Not-determined |
| SS_extracted_k141_35531 | Low-quality |
| SS_extracted_k141_75690 | Not-determined |
| SS_extracted_k141_57336 | Low-quality |
| SS_extracted_k141_35537 | Low-quality |
| SS_extracted_k141_35538 | Not-determined |
| SS_extracted_k141_57341 | Not-determined |
| SS_extracted_k141_95666 | Not-determined |
| SS_extracted_k141_27063 | Not-determined |
| SS_extracted_k141_75695 | Low-quality |
| SS_extracted_k141_126767 | Low-quality |
| SS_extracted_k141_131804 | Not-determined |
| SS_extracted_k141_75705 | Not-determined |
| SS_extracted_k141_131807 | Not-determined |
| SS_extracted_k141_126777 | Low-quality |
| SS_extracted_k141_126780 | Not-determined |
| SS_extracted_k141_75706 | Low-quality |
| SS_extracted_k141_136746 | Not-determined |
| SS_extracted_k141_143367 | Not-determined |
| SS_extracted_k141_136747 | Not-determined |
| SS_extracted_k141_95676 | Not-determined |
| SS_extracted_k141_143370 | Not-determined |
| SS_extracted_k141_136751 | Not-determined |
| SS_extracted_k141_95679 | Not-determined |
| SS_extracted_k141_35550 | Not-determined |
| SS_extracted_k141_16848 | Not-determined |
| SS_extracted_k141_143383 | Not-determined |
| SS_extracted_k141_120259 | Low-quality |
| SS_extracted_k141_46313 | Low-quality |
| SS_extracted_k141_75722 | Not-determined |
| SS_extracted_k141_113718 | Not-determined |
| SS_extracted_k141_57363 | Not-determined |
| SS_extracted_k141_113719 | Low-quality |
| SS_extracted_k141_143386 | Not-determined |
| SS_extracted_k141_131829 | Not-determined |
| SS_extracted_k141_131830 | Not-determined |
| SS_extracted_k141_46322 | Low-quality |
| SS_extracted_k141_113728 | Not-determined |
| SS_extracted_k141_46328 | Low-quality |
| SS_extracted_k141_120269 | Not-determined |
| SS_extracted_k141_27073 | Low-quality |
| SS_extracted_k141_57372 | Low-quality |
| SS_extracted_k141_126807 | Not-determined |
| SS_extracted_k141_35574 | Low-quality |
| SS_extracted_k141_64888 | Not-determined |
| SS_extracted_k141_136767 | Not-determined |
| SS_extracted_k141_16867 | Not-determined |
| SS_extracted_k141_126810 | Low-quality |
| SS_extracted_k141_108664 | Low-quality |
| SS_extracted_k141_113749 | Low-quality |
| SS_extracted_k141_5749 | Low-quality |
| SS_extracted_k141_81462 | Not-determined |
| SS_extracted_k141_120293 | Not-determined |
| SS_extracted_k141_16885 | Not-determined |
| SS_extracted_k141_22235 | Not-determined |
| SS_extracted_k141_57387 | Low-quality |
| SS_extracted_k141_46349 | Not-determined |
| SS_extracted_k141_64918 | Low-quality |
| SS_extracted_k141_57390 | Low-quality |
| SS_extracted_k141_143401 | Not-determined |
| SS_extracted_k141_75743 | Not-determined |
| SS_extracted_k141_46354 | Not-determined |
| SS_extracted_k141_113768 | Not-determined |
| SS_extracted_k141_5751 | Not-determined |
| SS_extracted_k141_46364 | Not-determined |
| SS_extracted_k141_136773 | Not-determined |
| SS_extracted_k141_131847 | Not-determined |
| SS_extracted_k141_27095 | Not-determined |
| SS_extracted_k141_95724 | Not-determined |
| SS_extracted_k141_70874 | Not-determined |
| SS_extracted_k141_22241 | Low-quality |
| SS_extracted_k141_113776 | Not-determined |
| SS_extracted_k141_108693 | Low-quality |
| SS_extracted_k141_46377 | Low-quality |
| SS_extracted_k141_35598 | Low-quality |
| SS_extracted_k141_108696 | Not-determined |
| SS_extracted_k141_46386 | Not-determined |
| SS_extracted_k141_108697 | Not-determined |
| SS_extracted_k141_64933 | Low-quality |
| SS_extracted_k141_108700 | Not-determined |
| SS_extracted_k141_131852 | Low-quality |
| SS_extracted_k141_22254 | Not-determined |
| SS_extracted_k141_136783 | Not-determined |
| SS_extracted_k141_64941 | Not-determined |
| SS_extracted_k141_22260 | Low-quality |
| SS_extracted_k141_143407 | Not-determined |
| SS_extracted_k141_35609 | Not-determined |
| SS_extracted_k141_70881 | Low-quality |
| SS_extracted_k141_108707 | Not-determined |

| | |
|---|---|
| SS_extracted_k141_131866 | Low-quality |
| SS_extracted_k141_64947 | Not-determined |
| SS_extracted_k141_120345 | Low-quality |
| SS_extracted_k141_108714 | Not-determined |
| SS_extracted_k141_81465 | Low-quality |
| SS_extracted_k141_120353 | Not-determined |
| SS_extracted_k141_120354 | Not-determined |
| SS_extracted_k141_113783 | Not-determined |
| SS_extracted_k141_5763 | Low-quality |
| SS_extracted_k141_35615 | Not-determined |
| SS_extracted_k141_108720 | Not-determined |
| SS_extracted_k141_64956 | Low-quality |
| SS_extracted_k141_120372 | Not-determined |
| SS_extracted_k141_27113 | Not-determined |
| SS_extracted_k141_35624 | Not-determined |
| SS_extracted_k141_126822 | Not-determined |
| SS_extracted_k141_95765 | Low-quality |
| SS_extracted_k141_81484 | Low-quality |
| SS_extracted_k141_131889 | Not-determined |
| SS_extracted_k141_95770 | Low-quality |
| SS_extracted_k141_57416 | Not-determined |
| SS_extracted_k141_35633 | Low-quality |
| SS_extracted_k141_70912 | Not-determined |
| SS_extracted_k141_126831 | Not-determined |
| SS_extracted_k141_143432 | Not-determined |
| SS_extracted_k141_126839 | Not-determined |
| SS_extracted_k141_75778 | Not-determined |
| SS_extracted_k141_22281 | Not-determined |
| SS_extracted_k141_108741 | Low-quality |
| SS_extracted_k141_22287 | Low-quality |
| SS_extracted_k141_143444 | Low-quality |
| SS_extracted_k141_108751 | Not-determined |
| SS_extracted_k141_143448 | Not-determined |
| SS_extracted_k141_27123 | Low-quality |
| SS_extracted_k141_108755 | Not-determined |
| SS_extracted_k141_100297 | Not-determined |
| SS_extracted_k141_27127 | Not-determined |
| SS_extracted_k141_108757 | Not-determined |
| SS_extracted_k141_57426 | Not-determined |
| SS_extracted_k141_57430 | Not-determined |
| SS_extracted_k141_81505 | Low-quality |
| SS_extracted_k141_108763 | Low-quality |
| SS_extracted_k141_65004 | Not-determined |
| SS_extracted_k141_53799 | Not-determined |
| SS_extracted_k141_108765 | Not-determined |
| SS_extracted_k141_89212 | Low-quality |
| SS_extracted_k141_126872 | Not-determined |
| SS_extracted_k141_100311 | Not-determined |
| SS_extracted_k141_70927 | Not-determined |
| SS_extracted_k141_126876 | Low-quality |
| SS_extracted_k141_27147 | Low-quality |
| SS_extracted_k141_100316 | Not-determined |
| SS_extracted_k141_89218 | Not-determined |
| SS_extracted_k141_27149 | Not-determined |
| SS_extracted_k141_100319 | Not-determined |
| SS_extracted_k141_81523 | Not-determined |
| SS_extracted_k141_65014 | Not-determined |
| SS_extracted_k141_108770 | Not-determined |
| SS_extracted_k141_126887 | Not-determined |
| SS_extracted_k141_75799 | Not-determined |
| SS_extracted_k141_81525 | Low-quality |
| SS_extracted_k141_89224 | Not-determined |
| SS_extracted_k141_143472 | Not-determined |
| SS_extracted_k141_89225 | Not-determined |
| SS_extracted_k141_10499 | Not-determined |
| SS_extracted_k141_70935 | Not-determined |
| SS_extracted_k141_16904 | Not-determined |
| SS_extracted_k141_120405 | Not-determined |
| SS_extracted_k141_81532 | Not-determined |
| SS_extracted_k141_89232 | Not-determined |
| SS_extracted_k141_143476 | Not-determined |
| SS_extracted_k141_143478 | Not-determined |
| SS_extracted_k141_5800 | Not-determined |
| SS_extracted_k141_5802 | Not-determined |
| SS_extracted_k141_81539 | Not-determined |
| SS_extracted_k141_120416 | Not-determined |
| SS_extracted_k141_22316 | Not-determined |
| SS_extracted_k141_100328 | Not-determined |
| SS_extracted_k141_95795 | Not-determined |
| SS_extracted_k141_70945 | Not-determined |
| SS_extracted_k141_136813 | Not-determined |
| SS_extracted_k141_22322 | Not-determined |
| SS_extracted_k141_70950 | Low-quality |
| SS_extracted_k141_95799 | Low-quality |
| SS_extracted_k141_143488 | Low-quality |
| SS_extracted_k141_95801 | Not-determined |
| SS_extracted_k141_89243 | Low-quality |
| SS_extracted_k141_95802 | Not-determined |
| SS_extracted_k141_22327 | Not-determined |
| SS_extracted_k141_10511 | Not-determined |
| SS_extracted_k141_120424 | Not-determined |
| SS_extracted_k141_89244 | Not-determined |
| SS_extracted_k141_120425 | Not-determined |
| SS_extracted_k141_89245 | Low-quality |
| SS_extracted_k141_10516 | Not-determined |
| SS_extracted_k141_143491 | Low-quality |
| SS_extracted_k141_113833 | Not-determined |
| SS_extracted_k141_120432 | Not-determined |
| SS_extracted_k141_75820 | Low-quality |
| SS_extracted_k141_136821 | Not-determined |
| SS_extracted_k141_136822 | Not-determined |
| SS_extracted_k141_53817 | Not-determined |
| SS_extracted_k141_22339 | Low-quality |
| SS_extracted_k141_100331 | Low-quality |
| SS_extracted_k141_22343 | Not-determined |
| SS_extracted_k141_75825 | Not-determined |
| SS_extracted_k141_70966 | Low-quality |
| SS_extracted_k141_10522 | Not-determined |
| SS_extracted_k141_22350 | Not-determined |
| SS_extracted_k141_143501 | Low-quality |
| SS_extracted_k141_10527 | Not-determined |
| SS_extracted_k141_143505 | Not-determined |
| SS_extracted_k141_81567 | Not-determined |
| SS_extracted_k141_126910 | Not-determined |
| SS_extracted_k141_89251 | Not-determined |
| SS_extracted_k141_113846 | Not-determined |
| SS_extracted_k141_131905 | Not-determined |
| SS_extracted_k141_53830 | Not-determined |
| SS_extracted_k141_143514 | Not-determined |
| SS_extracted_k141_65043 | Not-determined |
| SS_extracted_k141_46414 | Not-determined |
| SS_extracted_k141_89263 | Not-determined |
| SS_extracted_k141_35651 | Not-determined |
| SS_extracted_k141_113863 | Not-determined |
| SS_extracted_k141_10546 | Not-determined |
| SS_extracted_k141_75848 | Not-determined |
| SS_extracted_k141_27185 | Low-quality |
| SS_extracted_k141_120460 | Low-quality |
| SS_extracted_k141_143525 | Low-quality |
| SS_extracted_k141_75853 | Not-determined |
| SS_extracted_k141_95847 | Not-determined |
| SS_extracted_k141_126931 | Not-determined |
| SS_extracted_k141_5836 | Low-quality |
| SS_extracted_k141_126932 | Not-determined |
| SS_extracted_k141_27196 | Not-determined |
| SS_extracted_k141_70994 | Not-determined |
| SS_extracted_k141_113883 | Not-determined |
| SS_extracted_k141_70995 | Not-determined |
| SS_extracted_k141_10562 | Not-determined |
| SS_extracted_k141_113885 | Not-determined |
| SS_extracted_k141_5849 | Not-determined |
| SS_extracted_k141_120480 | Not-determined |
| SS_extracted_k141_113888 | Not-determined |
| SS_extracted_k141_95860 | Not-determined |
| SS_extracted_k141_22364 | Not-determined |
| SS_extracted_k141_126941 | Not-determined |
| SS_extracted_k141_100354 | Not-determined |
| SS_extracted_k141_65079 | Not-determined |
| SS_extracted_k141_126943 | Not-determined |
| SS_extracted_k141_126948 | Not-determined |
| SS_extracted_k141_65085 | Not-determined |
| SS_extracted_k141_100361 | Not-determined |
| SS_extracted_k141_143535 | Not-determined |
| SS_extracted_k141_108779 | Not-determined |
| SS_extracted_k141_100363 | Low-quality |
| SS_extracted_k141_126952 | Not-determined |
| SS_extracted_k141_10573 | Not-determined |
| SS_extracted_k141_22376 | Low-quality |
| SS_extracted_k141_27215 | Not-determined |
| SS_extracted_k141_143543 | Not-determined |
| SS_extracted_k141_143545 | Not-determined |
| SS_extracted_k141_16912 | Not-determined |
| SS_extracted_k141_10586 | Not-determined |
| SS_extracted_k141_22382 | Not-determined |
| SS_extracted_k141_27219 | Low-quality |
| SS_extracted_k141_143551 | Not-determined |
| SS_extracted_k141_22383 | Not-determined |
| SS_extracted_k141_113912 | Not-determined |
| SS_extracted_k141_22384 | Not-determined |
| SS_extracted_k141_22385 | Not-determined |
| SS_extracted_k141_126965 | Low-quality |
| SS_extracted_k141_53857 | Low-quality |
| SS_extracted_k141_113918 | Low-quality |
| SS_extracted_k141_75887 | Low-quality |
| SS_extracted_k141_113922 | Not-determined |
| SS_extracted_k141_65105 | Not-determined |
| SS_extracted_k141_10605 | Not-determined |
| SS_extracted_k141_126975 | Low-quality |
| SS_extracted_k141_27227 | Not-determined |
| SS_extracted_k141_126976 | Not-determined |
| SS_extracted_k141_126978 | Low-quality |
| SS_extracted_k141_16934 | Not-determined |
| SS_extracted_k141_89306 | Not-determined |
| SS_extracted_k141_89307 | Not-determined |
| SS_extracted_k141_89308 | Not-determined |
| SS_extracted_k141_75897 | Not-determined |
| SS_extracted_k141_126982 | Not-determined |
| SS_extracted_k141_40346 | Not-determined |
| SS_extracted_k141_10607 | Low-quality |
| SS_extracted_k141_81589 | Not-determined |
| SS_extracted_k141_40351 | Not-determined |
| SS_extracted_k141_113932 | Not-determined |
| SS_extracted_k141_10614 | Not-determined |
| SS_extracted_k141_27243 | Not-determined |
| SS_extracted_k141_27244 | Not-determined |
| SS_extracted_k141_113934 | Not-determined |
| SS_extracted_k141_100397 | Not-determined |
| SS_extracted_k141_16947 | Not-determined |
| SS_extracted_k141_136869 | Not-determined |
| SS_extracted_k141_126998 | Low-quality |
| SS_extracted_k141_127000 | Low-quality |
| SS_extracted_k141_46444 | Not-determined |
| SS_extracted_k141_22405 | Not-determined |
| SS_extracted_k141_127003 | Not-determined |
| SS_extracted_k141_27254 | Not-determined |
| SS_extracted_k141_40364 | Not-determined |
| SS_extracted_k141_143582 | Not-determined |
| SS_extracted_k141_108802 | Not-determined |
| SS_extracted_k141_22408 | Not-determined |
| SS_extracted_k141_57441 | Not-determined |
| SS_extracted_k141_108806 | Not-determined |
| SS_extracted_k141_120505 | Low-quality |
| SS_extracted_k141_136884 | Not-determined |
| SS_extracted_k141_53867 | Low-quality |
| SS_extracted_k141_131928 | Not-determined |
| SS_extracted_k141_46446 | Not-determined |
| SS_extracted_k141_35714 | Not-determined |
| SS_extracted_k141_40384 | Not-determined |
| SS_extracted_k141_35716 | Not-determined |
| SS_extracted_k141_131937 | Not-determined |
| SS_extracted_k141_131938 | Not-determined |
| SS_extracted_k141_89333 | Low-quality |
| SS_extracted_k141_10642 | Not-determined |
| SS_extracted_k141_89336 | Not-determined |
| SS_extracted_k141_35723 | Not-determined |
| SS_extracted_k141_100404 | Not-determined |
| SS_extracted_k141_100407 | Not-determined |
| SS_extracted_k141_108819 | Not-determined |
| SS_extracted_k141_16986 | Not-determined |
| SS_extracted_k141_113955 | Low-quality |
| SS_extracted_k141_65156 | Low-quality |
| SS_extracted_k141_35731 | Not-determined |
| SS_extracted_k141_108824 | Not-determined |
| SS_extracted_k141_143604 | Not-determined |
| SS_extracted_k141_35742 | Not-determined |
| SS_extracted_k141_143609 | Not-determined |
| SS_extracted_k141_65164 | Low-quality |
| SS_extracted_k141_143611 | Not-determined |
| SS_extracted_k141_136888 | Not-determined |
| SS_extracted_k141_17001 | Not-determined |
| SS_extracted_k141_35753 | Not-determined |
| SS_extracted_k141_89352 | Not-determined |
| SS_extracted_k141_10668 | Not-determined |
| SS_extracted_k141_53888 | Not-determined |
| SS_extracted_k141_89354 | Not-determined |
| SS_extracted_k141_89355 | Not-determined |
| SS_extracted_k141_75936 | Low-quality |
| SS_extracted_k141_10670 | Not-determined |
| SS_extracted_k141_75938 | Low-quality |
| SS_extracted_k141_120536 | Not-determined |
| SS_extracted_k141_143621 | Not-determined |
| SS_extracted_k141_100421 | Low-quality |
| SS_extracted_k141_100422 | Low-quality |
| SS_extracted_k141_89362 | Not-determined |
| SS_extracted_k141_17012 | Not-determined |
| SS_extracted_k141_17013 | Not-determined |
| SS_extracted_k141_127025 | Not-determined |
| SS_extracted_k141_108845 | Low-quality |
| SS_extracted_k141_89367 | Not-determined |
| SS_extracted_k141_75945 | Not-determined |
| SS_extracted_k141_10680 | Not-determined |
| SS_extracted_k141_35766 | Not-determined |
| SS_extracted_k141_113980 | Low-quality |
| SS_extracted_k141_75948 | Not-determined |
| SS_extracted_k141_108853 | Not-determined |
| SS_extracted_k141_108854 | Not-determined |
| SS_extracted_k141_22446 | Not-determined |
| SS_extracted_k141_75951 | Low-quality |
| SS_extracted_k141_22448 | Not-determined |
| SS_extracted_k141_22449 | Not-determined |
| SS_extracted_k141_108858 | Low-quality |
| SS_extracted_k141_120547 | Not-determined |
| SS_extracted_k141_5858 | Low-quality |
| SS_extracted_k141_65177 | Not-determined |
| SS_extracted_k141_35773 | Not-determined |
| SS_extracted_k141_100442 | Not-determined |
| SS_extracted_k141_131977 | Low-quality |
| SS_extracted_k141_10694 | Not-determined |
| SS_extracted_k141_10695 | Not-determined |
| SS_extracted_k141_53916 | Low-quality |
| SS_extracted_k141_35782 | Not-determined |
| SS_extracted_k141_22457 | Low-quality |
| SS_extracted_k141_10704 | Low-quality |
| SS_extracted_k141_46496 | Not-determined |
| SS_extracted_k141_10706 | Not-determined |
| SS_extracted_k141_71029 | Not-determined |
| SS_extracted_k141_108869 | Not-determined |
| SS_extracted_k141_113990 | Not-determined |
| SS_extracted_k141_131986 | Not-determined |
| SS_extracted_k141_127038 | Not-determined |
| SS_extracted_k141_113993 | Not-determined |
| SS_extracted_k141_100460 | Not-determined |
| SS_extracted_k141_10714 | Not-determined |
| SS_extracted_k141_65199 | Low-quality |
| SS_extracted_k141_53931 | Not-determined |
| SS_extracted_k141_10717 | Not-determined |
| SS_extracted_k141_113996 | Not-determined |
| SS_extracted_k141_75973 | Not-determined |
| SS_extracted_k141_100465 | Not-determined |
| SS_extracted_k141_136894 | Not-determined |
| SS_extracted_k141_100467 | Not-determined |
| SS_extracted_k141_17026 | Low-quality |
| SS_extracted_k141_113999 | Not-determined |
| SS_extracted_k141_136896 | Not-determined |
| SS_extracted_k141_136897 | Low-quality |
| SS_extracted_k141_65203 | Not-determined |
| SS_extracted_k141_136899 | Not-determined |
| SS_extracted_k141_75976 | Not-determined |
| SS_extracted_k141_136900 | Not-determined |
| SS_extracted_k141_95911 | Low-quality |
| SS_extracted_k141_95912 | Not-determined |
| SS_extracted_k141_71047 | Not-determined |
| SS_extracted_k141_17030 | Low-quality |
| SS_extracted_k141_120579 | Not-determined |
| SS_extracted_k141_108890 | Not-determined |
| SS_extracted_k141_95913 | Low-quality |
| SS_extracted_k141_89372 | Not-determined |
| SS_extracted_k141_95922 | Not-determined |
| SS_extracted_k141_65209 | Not-determined |
| SS_extracted_k141_89383 | Not-determined |
| SS_extracted_k141_53956 | Not-determined |
| SS_extracted_k141_46527 | Not-determined |
| SS_extracted_k141_81646 | Not-determined |
| SS_extracted_k141_46532 | Not-determined |
| SS_extracted_k141_57448 | Low-quality |
| SS_extracted_k141_17060 | Not-determined |
| SS_extracted_k141_53971 | Not-determined |
| SS_extracted_k141_46537 | Not-determined |
| SS_extracted_k141_17063 | Not-determined |
| SS_extracted_k141_100504 | Not-determined |
| SS_extracted_k141_89394 | Not-determined |
| SS_extracted_k141_46541 | Not-determined |
| SS_extracted_k141_114024 | Low-quality |
| SS_extracted_k141_17070 | Not-determined |
| SS_extracted_k141_75999 | Not-determined |
| SS_extracted_k141_22485 | Not-determined |
| SS_extracted_k141_76004 | Not-determined |
| SS_extracted_k141_95925 | Not-determined |
| SS_extracted_k141_132014 | Not-determined |
| SS_extracted_k141_95930 | Not-determined |
| SS_extracted_k141_10742 | Not-determined |
| SS_extracted_k141_46556 | Not-determined |
| SS_extracted_k141_95931 | Low-quality |
| SS_extracted_k141_22492 | Not-determined |
| SS_extracted_k141_22494 | Not-determined |
| SS_extracted_k141_81657 | Low-quality |
| SS_extracted_k141_108897 | Low-quality |
| SS_extracted_k141_65238 | Not-determined |
| SS_extracted_k141_136933 | Not-determined |
| SS_extracted_k141_54004 | Not-determined |
| SS_extracted_k141_136936 | Low-quality |
| SS_extracted_k141_136937 | Not-determined |
| SS_extracted_k141_95944 | Not-determined |
| SS_extracted_k141_81662 | Not-determined |
| SS_extracted_k141_76015 | Not-determined |
| SS_extracted_k141_76017 | Not-determined |
| SS_extracted_k141_136940 | Not-determined |
| SS_extracted_k141_22504 | Not-determined |
| SS_extracted_k141_65246 | Not-determined |
| SS_extracted_k141_89402 | Low-quality |
| SS_extracted_k141_65253 | Low-quality |
| SS_extracted_k141_76030 | Not-determined |
| SS_extracted_k141_54015 | Not-determined |
| SS_extracted_k141_54016 | Low-quality |
| SS_extracted_k141_132047 | Not-determined |
| SS_extracted_k141_100511 | Not-determined |
| SS_extracted_k141_54025 | Not-determined |
| SS_extracted_k141_10766 | Not-determined |
| SS_extracted_k141_81678 | Not-determined |
| SS_extracted_k141_17105 | Low-quality |
| SS_extracted_k141_46588 | Not-determined |
| SS_extracted_k141_22521 | Not-determined |
| SS_extracted_k141_132060 | Low-quality |
| SS_extracted_k141_17107 | Not-determined |
| SS_extracted_k141_46589 | Not-determined |
| SS_extracted_k141_22523 | Low-quality |
| SS_extracted_k141_108904 | Not-determined |
| SS_extracted_k141_132065 | Not-determined |
| SS_extracted_k141_100526 | Low-quality |
| SS_extracted_k141_100528 | Not-determined |
| SS_extracted_k141_35835 | Not-determined |
| SS_extracted_k141_120593 | Not-determined |
| SS_extracted_k141_108911 | Not-determined |
| SS_extracted_k141_46596 | Low-quality |
| SS_extracted_k141_136946 | Not-determined |
| SS_extracted_k141_120596 | Not-determined |
| SS_extracted_k141_46604 | Not-determined |
| SS_extracted_k141_120600 | Not-determined |
| SS_extracted_k141_46606 | Not-determined |
| SS_extracted_k141_71059 | Low-quality |
| SS_extracted_k141_10784 | Low-quality |
| SS_extracted_k141_95974 | Not-determined |
| SS_extracted_k141_65275 | Not-determined |
| SS_extracted_k141_57485 | Not-determined |
| SS_extracted_k141_17137 | Not-determined |
| SS_extracted_k141_95985 | Not-determined |
| SS_extracted_k141_57490 | Low-quality |
| SS_extracted_k141_65281 | Low-quality |
| SS_extracted_k141_89453 | Not-determined |
| SS_extracted_k141_35862 | Not-determined |
| SS_extracted_k141_17139 | Low-quality |
| SS_extracted_k141_17140 | Not-determined |
| SS_extracted_k141_127068 | Not-determined |
| SS_extracted_k141_17142 | Not-determined |
| SS_extracted_k141_127070 | Not-determined |
| SS_extracted_k141_65288 | Not-determined |
| SS_extracted_k141_17144 | Not-determined |
| SS_extracted_k141_17146 | Not-determined |
| SS_extracted_k141_71073 | Not-determined |
| SS_extracted_k141_40416 | Not-determined |
| SS_extracted_k141_114066 | Not-determined |
| SS_extracted_k141_57504 | Not-determined |
| SS_extracted_k141_40422 | Not-determined |
| SS_extracted_k141_57510 | Not-determined |
| SS_extracted_k141_71082 | Not-determined |
| SS_extracted_k141_127077 | Not-determined |
| SS_extracted_k141_46627 | Not-determined |
| SS_extracted_k141_46628 | Not-determined |
| SS_extracted_k141_76052 | Not-determined |
| SS_extracted_k141_46630 | Not-determined |
| SS_extracted_k141_22601 | Not-determined |
| SS_extracted_k141_10796 | Not-determined |
| SS_extracted_k141_120636 | Not-determined |
| SS_extracted_k141_71087 | Not-determined |
| SS_extracted_k141_71088 | Not-determined |
| SS_extracted_k141_114077 | Not-determined |
| SS_extracted_k141_22607 | Not-determined |
| SS_extracted_k141_54062 | Not-determined |
| SS_extracted_k141_10806 | Not-determined |
| SS_extracted_k141_17167 | Not-determined |
| SS_extracted_k141_10807 | Not-determined |
| SS_extracted_k141_5953 | Not-determined |
| SS_extracted_k141_76071 | Not-determined |
| SS_extracted_k141_76073 | Not-determined |
| SS_extracted_k141_35866 | Low-quality |
| SS_extracted_k141_114083 | Not-determined |
| SS_extracted_k141_22615 | Not-determined |
| SS_extracted_k141_65311 | Not-determined |
| SS_extracted_k141_5965 | Not-determined |
| SS_extracted_k141_54081 | Not-determined |
| SS_extracted_k141_54082 | Not-determined |
| SS_extracted_k141_54086 | Low-quality |
| SS_extracted_k141_22626 | Not-determined |
| SS_extracted_k141_120644 | Low-quality |
| SS_extracted_k141_17182 | Not-determined |
| SS_extracted_k141_65326 | Not-determined |
| SS_extracted_k141_120646 | Not-determined |
| SS_extracted_k141_54094 | Not-determined |
| SS_extracted_k141_46650 | Not-determined |
| SS_extracted_k141_127100 | Low-quality |
| SS_extracted_k141_120648 | Not-determined |
| SS_extracted_k141_136980 | Not-determined |
| SS_extracted_k141_54102 | Not-determined |
| SS_extracted_k141_136984 | Not-determined |
| SS_extracted_k141_5984 | Not-determined |
| SS_extracted_k141_127106 | Low-quality |
| SS_extracted_k141_65337 | Not-determined |
| SS_extracted_k141_46659 | Not-determined |
| SS_extracted_k141_22642 | Not-determined |
| SS_extracted_k141_5986 | Not-determined |
| SS_extracted_k141_108948 | Not-determined |
| SS_extracted_k141_65341 | Not-determined |
| SS_extracted_k141_136996 | Not-determined |
| SS_extracted_k141_127117 | Low-quality |
| SS_extracted_k141_127119 | Not-determined |
| SS_extracted_k141_17202 | Not-determined |
| SS_extracted_k141_127120 | Not-determined |
| SS_extracted_k141_137000 | Not-determined |
| SS_extracted_k141_132111 | Not-determined |
| SS_extracted_k141_17207 | Not-determined |
| SS_extracted_k141_108958 | Not-determined |
| SS_extracted_k141_127124 | Not-determined |
| SS_extracted_k141_108960 | Not-determined |
| SS_extracted_k141_10824 | Low-quality |
| SS_extracted_k141_22650 | Not-determined |
| SS_extracted_k141_127128 | Not-determined |
| SS_extracted_k141_127130 | Not-determined |
| SS_extracted_k141_22655 | Not-determined |
| SS_extracted_k141_17214 | Not-determined |
| SS_extracted_k141_46670 | Not-determined |
| SS_extracted_k141_127132 | Low-quality |
| SS_extracted_k141_65354 | Not-determined |
| SS_extracted_k141_54124 | Low-quality |
| SS_extracted_k141_114133 | Not-determined |
| SS_extracted_k141_17228 | Not-determined |
| SS_extracted_k141_120664 | Not-determined |
| SS_extracted_k141_54130 | Not-determined |
| SS_extracted_k141_54131 | Not-determined |
| SS_extracted_k141_40476 | Not-determined |
| SS_extracted_k141_108980 | Not-determined |
| SS_extracted_k141_65370 | Not-determined |
| SS_extracted_k141_108981 | Not-determined |
| SS_extracted_k141_40479 | Not-determined |
| SS_extracted_k141_120669 | Not-determined |
| SS_extracted_k141_127154 | Not-determined |
| SS_extracted_k141_96025 | Not-determined |
| SS_extracted_k141_132131 | Low-quality |
| SS_extracted_k141_108989 | Not-determined |
| SS_extracted_k141_89496 | Not-determined |
| SS_extracted_k141_108995 | Low-quality |
| SS_extracted_k141_54147 | Not-determined |
| SS_extracted_k141_71109 | Not-determined |
| SS_extracted_k141_46684 | Not-determined |
| SS_extracted_k141_6029 | Not-determined |
| SS_extracted_k141_109008 | Not-determined |
| SS_extracted_k141_89506 | Low-quality |
| SS_extracted_k141_100560 | Low-quality |
| SS_extracted_k141_57519 | Not-determined |
| SS_extracted_k141_22699 | Not-determined |
| SS_extracted_k141_27262 | Low-quality |
| SS_extracted_k141_137035 | Not-determined |
| SS_extracted_k141_54163 | Not-determined |
| SS_extracted_k141_17253 | Not-determined |
| SS_extracted_k141_57526 | Low-quality |
| SS_extracted_k141_57527 | Not-determined |
| SS_extracted_k141_40489 | Not-determined |
| SS_extracted_k141_46711 | Not-determined |
| SS_extracted_k141_22721 | Not-determined |
| SS_extracted_k141_40500 | Low-quality |
| SS_extracted_k141_35899 | Not-determined |
| SS_extracted_k141_27284 | Not-determined |
| SS_extracted_k141_109038 | Not-determined |
| SS_extracted_k141_40505 | Not-determined |
| SS_extracted_k141_76093 | Low-quality |
| SS_extracted_k141_109042 | Not-determined |
| SS_extracted_k141_109044 | Not-determined |
| SS_extracted_k141_10831 | Not-determined |
| SS_extracted_k141_132153 | Not-determined |
| SS_extracted_k141_96038 | Not-determined |
| SS_extracted_k141_96039 | Not-determined |
| SS_extracted_k141_132158 | Not-determined |
| SS_extracted_k141_132161 | Not-determined |
| SS_extracted_k141_40512 | Not-determined |
| SS_extracted_k141_57532 | Low-quality |
| SS_extracted_k141_17282 | Not-determined |
| SS_extracted_k141_17283 | Not-determined |
| SS_extracted_k141_109063 | Not-determined |
| SS_extracted_k141_114163 | Not-determined |
| SS_extracted_k141_40515 | Low-quality |
| SS_extracted_k141_35920 | Low-quality |
| SB_extracted_k141_106946 | Complete |
| SS_extracted_k141_6033 | Not-determined |
| SS_extracted_k141_81712 | Not-determined |
| SS_extracted_k141_143637 | Low-quality |
| SS_extracted_k141_143639 | Not-determined |
| SS_extracted_k141_143640 | Not-determined |
| SS_extracted_k141_54201 | Not-determined |
| SS_extracted_k141_54203 | Not-determined |
| SS_extracted_k141_46724 | Not-determined |
| SS_extracted_k141_96066 | Not-determined |
| SS_extracted_k141_57539 | Not-determined |
| SS_extracted_k141_96069 | Low-quality |
| SS_extracted_k141_120707 | Not-determined |
| SS_extracted_k141_54212 | Not-determined |
| SS_extracted_k141_46733 | Not-determined |
| SS_extracted_k141_54214 | Low-quality |
| SS_extracted_k141_120718 | Not-determined |
| SS_extracted_k141_46738 | Not-determined |
| SS_extracted_k141_10837 | Low-quality |
| SS_extracted_k141_40541 | Not-determined |
| SS_extracted_k141_40543 | Low-quality |
| SS_extracted_k141_81732 | Not-determined |
| SS_extracted_k141_17301 | Not-determined |
| SS_extracted_k141_132170 | Not-determined |
| SS_extracted_k141_54228 | Not-determined |
| SS_extracted_k141_120733 | Low-quality |
| SS_extracted_k141_35938 | Not-determined |
| SS_extracted_k141_40553 | Not-determined |
| SS_extracted_k141_27302 | Not-determined |
| SS_extracted_k141_46772 | Not-determined |
| SS_extracted_k141_57548 | Not-determined |
| SS_extracted_k141_71130 | Not-determined |
| SS_extracted_k141_17309 | Low-quality |
| SS_extracted_k141_40562 | Not-determined |
| SS_extracted_k141_17313 | Low-quality |
| SS_extracted_k141_120749 | Not-determined |
| SS_extracted_k141_114217 | Not-determined |
| SS_extracted_k141_120751 | Not-determined |
| SS_extracted_k141_114219 | Not-determined |
| SS_extracted_k141_10848 | Low-quality |
| SS_extracted_k141_120759 | Not-determined |
| SS_extracted_k141_40570 | Not-determined |
| SS_extracted_k141_10851 | Low-quality |
| SS_extracted_k141_114225 | Low-quality |
| SS_extracted_k141_40577 | Not-determined |
| SS_extracted_k141_40579 | Low-quality |
| SS_extracted_k141_27315 | Not-determined |
| SS_extracted_k141_27317 | Not-determined |
| SS_extracted_k141_81746 | Low-quality |

| | |
|---|---|
| SS_extracted_k141_120766 | Not-determined |
| SS_extracted_k141_132184 | Not-determined |
| SS_extracted_k141_120767 | Low-quality |
| SS_extracted_k141_71150 | Not-determined |
| SS_extracted_k141_96100 | Not-determined |
| SS_extracted_k141_114243 | Not-determined |
| SS_extracted_k141_17327 | Not-determined |
| SS_extracted_k141_54246 | Not-determined |
| SS_extracted_k141_96111 | Not-determined |
| SS_extracted_k141_40607 | Not-determined |
| SS_extracted_k141_40609 | Not-determined |
| SS_extracted_k141_96113 | Low-quality |
| SS_extracted_k141_40612 | Not-determined |
| SS_extracted_k141_40613 | Not-determined |
| SS_extracted_k141_132196 | Not-determined |
| SS_extracted_k141_40615 | Not-determined |
| SS_extracted_k141_10853 | Not-determined |
| SS_extracted_k141_109088 | Not-determined |
| SS_extracted_k141_96119 | Not-determined |
| SS_extracted_k141_10854 | Not-determined |
| SS_extracted_k141_89545 | Not-determined |
| SS_extracted_k141_40619 | Not-determined |
| SS_extracted_k141_71163 | Not-determined |
| SS_extracted_k141_76116 | Not-determined |
| SS_extracted_k141_40622 | Not-determined |
| SS_extracted_k141_96126 | Not-determined |
| SS_extracted_k141_143645 | Low-quality |
| SS_extracted_k141_71177 | Not-determined |
| SS_extracted_k141_96134 | Not-determined |
| SS_extracted_k141_109113 | Not-determined |
| SS_extracted_k141_71185 | Not-determined |
| SS_extracted_k141_89555 | Not-determined |
| SS_extracted_k141_89557 | Not-determined |
| SS_extracted_k141_120797 | Low-quality |
| SS_extracted_k141_35994 | Not-determined |
| SS_extracted_k141_35997 | Not-determined |
| SS_extracted_k141_40645 | Not-determined |
| SS_extracted_k141_40646 | Not-determined |
| SS_extracted_k141_10873 | Not-determined |
| SS_extracted_k141_89566 | Not-determined |
| SS_extracted_k141_120809 | Not-determined |
| SS_extracted_k141_36005 | Not-determined |
| SS_extracted_k141_71212 | Low-quality |
| SS_extracted_k141_27353 | Not-determined |
| SS_extracted_k141_40657 | Low-quality |
| SS_extracted_k141_36019 | Not-determined |
| SS_extracted_k141_40658 | Low-quality |
| SS_extracted_k141_17374 | Not-determined |
| SS_extracted_k141_109122 | Not-determined |
| SS_extracted_k141_89579 | Low-quality |
| SS_extracted_k141_89580 | Not-determined |
| SS_extracted_k141_89584 | Low-quality |
| SS_extracted_k141_65410 | Not-determined |
| SS_extracted_k141_137052 | Low-quality |
| SS_extracted_k141_132220 | Low-quality |
| SS_extracted_k141_71226 | Low-quality |
| S12_vs_k141_167155\|\|full | High-quality |
| SS_extracted_k141_36040 | Not-determined |
| SS_extracted_k141_54302 | Low-quality |
| SS_extracted_k141_132226 | Not-determined |
| SS_extracted_k141_46822 | Not-determined |
| SS_extracted_k141_46823 | Not-determined |
| SS_extracted_k141_54306 | Low-quality |
| SS_extracted_k141_89598 | Not-determined |
| SS_extracted_k141_17391 | Not-determined |
| SS_extracted_k141_114255 | Not-determined |
| SS_extracted_k141_40681 | Not-determined |
| SS_extracted_k141_27375 | Not-determined |
| SS_extracted_k141_114259 | Not-determined |
| SS_extracted_k141_76180 | Not-determined |
| SS_extracted_k141_120862 | Low-quality |
| SS_extracted_k141_40687 | Not-determined |
| SS_extracted_k141_89611 | Low-quality |
| SS_extracted_k141_109149 | Not-determined |
| SS_extracted_k141_76184 | Not-determined |
| SS_extracted_k141_54313 | Not-determined |
| SS_extracted_k141_89612 | Not-determined |
| SS_extracted_k141_17409 | Not-determined |
| SS_extracted_k141_17411 | Not-determined |
| SS_extracted_k141_46846 | Not-determined |
| SS_extracted_k141_54320 | Low-quality |
| SS_extracted_k141_109156 | Not-determined |
| SS_extracted_k141_40696 | Low-quality |
| SS_extracted_k141_46851 | Not-determined |
| SS_extracted_k141_36055 | Not-determined |
| SS_extracted_k141_114271 | Not-determined |
| SS_extracted_k141_137073 | Not-determined |
| SS_extracted_k141_36057 | Not-determined |
| SS_extracted_k141_36058 | Not-determined |
| SS_extracted_k141_89631 | Not-determined |
| SS_extracted_k141_89632 | Not-determined |
| SS_extracted_k141_143664 | Not-determined |
| SS_extracted_k141_54330 | Not-determined |
| SS_extracted_k141_143666 | Not-determined |
| SS_extracted_k141_54331 | Not-determined |
| SS_extracted_k141_22751 | Not-determined |
| SS_extracted_k141_17427 | Not-determined |
| SS_extracted_k141_81812 | Not-determined |
| SS_extracted_k141_114279 | Not-determined |
| SS_extracted_k141_22753 | Not-determined |
| SS_extracted_k141_143670 | Not-determined |
| SS_extracted_k141_57570 | Not-determined |
| SS_extracted_k141_76199 | Not-determined |
| SS_extracted_k141_81813 | Low-quality |
| SS_extracted_k141_81814 | Not-determined |
| SS_extracted_k141_46876 | Low-quality |
| SS_extracted_k141_114288 | Not-determined |
| SS_extracted_k141_57573 | Not-determined |
| SS_extracted_k141_57574 | Not-determined |
| SS_extracted_k141_120887 | Not-determined |
| SS_extracted_k141_114294 | Not-determined |
| SS_extracted_k141_120891 | Not-determined |
| SS_extracted_k141_22758 | Low-quality |
| SS_extracted_k141_81818 | Low-quality |
| SS_extracted_k141_46888 | Not-determined |
| SS_extracted_k141_109175 | Not-determined |
| SS_extracted_k141_120895 | Not-determined |
| SS_extracted_k141_22762 | Low-quality |
| SS_extracted_k141_54356 | Low-quality |
| SS_extracted_k141_22768 | Not-determined |
| SS_extracted_k141_54357 | Low-quality |
| SS_extracted_k141_54358 | Not-determined |
| SS_extracted_k141_132243 | Low-quality |
| SS_extracted_k141_132246 | Not-determined |
| SS_extracted_k141_114310 | Not-determined |
| SS_extracted_k141_17462 | Not-determined |
| SS_extracted_k141_46911 | Not-determined |
| SS_extracted_k141_81831 | Not-determined |
| SS_extracted_k141_22771 | Low-quality |
| SS_extracted_k141_114317 | Low-quality |
| SS_extracted_k141_137077 | Not-determined |
| SS_extracted_k141_40745 | Not-determined |
| SS_extracted_k141_46922 | Not-determined |
| SS_extracted_k141_57583 | Not-determined |
| SS_extracted_k141_137083 | Not-determined |
| SS_extracted_k141_120915 | Not-determined |
| SS_extracted_k141_46925 | Not-determined |
| SS_extracted_k141_109195 | Not-determined |
| SS_extracted_k141_120917 | Not-determined |
| SS_extracted_k141_57592 | Not-determined |
| SS_extracted_k141_137090 | Low-quality |
| SS_extracted_k141_57594 | Low-quality |
| SS_extracted_k141_137094 | Not-determined |
| SS_extracted_k141_10911 | Not-determined |
| SS_extracted_k141_54372 | Not-determined |
| SS_extracted_k141_76251 | Not-determined |
| SS_extracted_k141_109205 | Low-quality |
| SS_extracted_k141_114336 | Not-determined |
| SS_extracted_k141_17476 | Not-determined |
| SS_extracted_k141_10917 | Low-quality |
| SS_extracted_k141_10920 | Not-determined |
| SS_extracted_k141_120924 | Low-quality |
| SS_extracted_k141_114342 | Not-determined |
| SS_extracted_k141_137102 | Low-quality |
| SS_extracted_k141_65432 | Low-quality |
| SS_extracted_k141_120929 | Not-determined |
| SS_extracted_k141_22796 | Not-determined |
| SS_extracted_k141_46941 | Low-quality |
| SS_extracted_k141_40750 | Not-determined |
| SS_extracted_k141_40754 | Low-quality |
| SS_extracted_k141_114351 | Not-determined |
| SS_extracted_k141_76270 | Not-determined |
| SS_extracted_k141_65448 | Not-determined |
| SS_extracted_k141_54386 | Not-determined |
| SS_extracted_k141_76276 | Not-determined |
| SS_extracted_k141_65451 | Not-determined |
| SS_extracted_k141_89667 | Not-determined |
| SS_extracted_k141_114360 | Not-determined |
| SS_extracted_k141_54400 | Not-determined |
| SS_extracted_k141_17489 | Not-determined |
| SS_extracted_k141_46970 | Not-determined |
| SS_extracted_k141_46971 | Not-determined |
| SS_extracted_k141_46975 | Not-determined |
| SS_extracted_k141_40773 | Not-determined |
| SS_extracted_k141_57597 | Not-determined |
| SS_extracted_k141_114372 | Not-determined |
| SS_extracted_k141_65465 | Low-quality |
| SS_extracted_k141_96139 | Not-determined |
| SS_extracted_k141_81853 | Not-determined |
| SS_extracted_k141_65469 | Not-determined |
| SS_extracted_k141_46983 | Low-quality |
| SS_extracted_k141_17505 | Low-quality |
| SS_extracted_k141_40784 | Not-determined |
| SS_extracted_k141_109215 | Not-determined |
| SS_extracted_k141_76298 | Not-determined |
| SS_extracted_k141_114389 | Not-determined |
| SS_extracted_k141_109224 | Low-quality |
| SS_extracted_k141_22846 | Not-determined |
| SS_extracted_k141_81856 | Not-determined |
| SS_extracted_k141_81857 | Not-determined |
| SS_extracted_k141_109227 | Not-determined |
| SS_extracted_k141_89702 | Not-determined |
| SS_extracted_k141_109229 | Not-determined |
| SS_extracted_k141_89704 | Not-determined |
| SS_extracted_k141_100576 | Not-determined |
| SS_extracted_k141_100577 | Not-determined |
| SS_extracted_k141_81862 | Not-determined |
| SS_extracted_k141_22854 | Not-determined |
| SS_extracted_k141_27403 | Not-determined |
| SS_extracted_k141_109232 | Low-quality |
| SS_extracted_k141_109233 | Not-determined |
| SS_extracted_k141_22860 | Not-determined |
| SS_extracted_k141_114409 | Not-determined |
| SS_extracted_k141_100586 | Not-determined |
| SS_extracted_k141_114412 | Not-determined |
| SS_extracted_k141_114414 | Low-quality |
| SS_extracted_k141_47019 | Low-quality |
| SS_extracted_k141_22864 | Low-quality |
| SS_extracted_k141_47022 | Not-determined |
| SS_extracted_k141_114417 | Low-quality |
| SS_extracted_k141_109244 | Not-determined |
| SS_extracted_k141_89722 | Low-quality |
| SS_extracted_k141_114418 | Not-determined |
| SS_extracted_k141_114424 | Not-determined |
| SS_extracted_k141_114429 | Not-determined |
| SS_extracted_k141_27428 | Low-quality |
| SS_extracted_k141_71229 | Not-determined |
| SS_extracted_k141_114437 | Not-determined |
| SS_extracted_k141_36072 | Not-determined |
| SS_extracted_k141_36074 | Low-quality |
| SS_extracted_k141_36075 | Low-quality |
| SS_extracted_k141_22887 | Not-determined |
| SS_extracted_k141_96152 | Low-quality |
| SS_extracted_k141_114446 | Low-quality |
| SS_extracted_k141_27438 | Not-determined |
| SS_extracted_k141_36083 | Not-determined |
| SS_extracted_k141_71241 | Not-determined |
| SS_extracted_k141_109261 | Not-determined |
| SS_extracted_k141_114453 | Low-quality |
| SS_extracted_k141_76310 | Not-determined |
| SS_extracted_k141_17536 | Low-quality |
| SS_extracted_k141_120953 | Low-quality |
| SS_extracted_k141_17540 | Low-quality |
| SS_extracted_k141_76319 | Low-quality |
| SS_extracted_k141_109275 | Not-determined |
| SS_extracted_k141_71254 | Not-determined |
| SS_extracted_k141_36097 | Not-determined |
| SS_extracted_k141_109277 | Not-determined |
| SS_extracted_k141_22913 | Not-determined |
| SS_extracted_k141_120960 | Not-determined |
| SS_extracted_k141_132261 | Low-quality |
| SS_extracted_k141_120964 | Not-determined |
| SS_extracted_k141_120965 | Low-quality |
| SS_extracted_k141_89728 | Not-determined |
| SS_extracted_k141_76327 | Not-determined |
| SS_extracted_k141_76328 | Not-determined |
| SS_extracted_k141_114469 | Low-quality |
| SS_extracted_k141_47046 | Not-determined |
| SS_extracted_k141_114470 | Not-determined |
| SS_extracted_k141_71261 | Not-determined |
| SS_extracted_k141_132270 | Not-determined |
| SS_extracted_k141_81880 | Low-quality |
| SS_extracted_k141_132276 | Not-determined |
| SS_extracted_k141_143698 | Not-determined |
| SS_extracted_k141_71267 | Not-determined |
| SS_extracted_k141_143702 | Low-quality |
| SS_extracted_k141_132286 | Not-determined |
| SS_extracted_k141_132289 | Not-determined |
| SS_extracted_k141_47064 | Not-determined |
| SS_extracted_k141_36118 | Not-determined |
| SS_extracted_k141_81890 | Not-determined |
| SS_extracted_k141_47068 | Not-determined |
| SS_extracted_k141_22925 | Not-determined |
| SS_extracted_k141_47069 | Not-determined |
| SS_extracted_k141_47070 | Not-determined |
| SS_extracted_k141_143711 | Not-determined |
| SS_extracted_k141_96165 | Not-determined |
| SS_extracted_k141_17572 | Not-determined |
| SS_extracted_k141_76362 | Not-determined |
| SS_extracted_k141_76370 | Low-quality |
| SS_extracted_k141_143719 | Low-quality |
| SS_extracted_k141_120999 | Not-determined |
| SS_extracted_k141_22935 | Not-determined |
| SS_extracted_k141_76372 | Not-determined |
| SS_extracted_k141_76374 | Low-quality |
| SS_extracted_k141_143722 | Low-quality |
| SS_extracted_k141_22939 | Not-determined |
| SS_extracted_k141_76378 | Not-determined |
| SS_extracted_k141_54478 | Low-quality |
| SS_extracted_k141_10957 | Not-determined |
| SS_extracted_k141_143726 | Not-determined |
| SS_extracted_k141_132310 | Not-determined |
| SS_extracted_k141_71278 | Not-determined |
| SS_extracted_k141_89762 | Not-determined |
| SS_extracted_k141_143732 | Not-determined |
| SS_extracted_k141_47093 | Low-quality |
| SS_extracted_k141_54485 | Not-determined |
| SS_extracted_k141_143737 | Not-determined |
| SS_extracted_k141_143739 | Not-determined |
| SS_extracted_k141_143741 | Not-determined |
| SS_extracted_k141_81895 | Not-determined |
| SS_extracted_k141_17602 | Not-determined |
| SS_extracted_k141_114504 | Not-determined |
| SS_extracted_k141_96200 | Low-quality |
| SS_extracted_k141_143745 | Not-determined |
| SS_extracted_k141_132318 | Not-determined |
| SS_extracted_k141_132321 | Low-quality |
| SS_extracted_k141_96205 | Low-quality |
| SS_extracted_k141_81903 | Low-quality |
| SS_extracted_k141_96208 | Not-determined |
| SS_extracted_k141_54497 | Low-quality |
| SS_extracted_k141_54498 | Not-determined |
| SS_extracted_k141_121019 | Not-determined |
| SS_extracted_k141_132326 | Not-determined |
| SS_extracted_k141_47113 | Not-determined |
| SS_extracted_k141_132330 | Low-quality |
| SS_extracted_k141_96216 | Low-quality |
| SS_extracted_k141_17614 | Not-determined |
| SS_extracted_k141_17616 | Low-quality |
| SS_extracted_k141_27467 | Low-quality |
| SS_extracted_k141_40789 | Not-determined |
| SS_extracted_k141_40790 | Low-quality |
| SS_extracted_k141_22975 | Low-quality |
| SS_extracted_k141_76418 | Low-quality |
| SS_extracted_k141_27473 | Not-determined |
| SS_extracted_k141_89800 | Low-quality |
| SS_extracted_k141_127163 | Not-determined |
| SS_extracted_k141_89804 | Not-determined |
| SS_extracted_k141_76426 | Not-determined |
| SS_extracted_k141_121030 | Low-quality |
| SS_extracted_k141_127169 | Not-determined |
| SS_extracted_k141_47149 | Not-determined |
| SS_extracted_k141_76433 | Not-determined |
| SS_extracted_k141_76434 | Not-determined |
| SS_extracted_k141_127177 | Not-determined |
| SS_extracted_k141_76441 | Low-quality |
| SS_extracted_k141_89812 | Not-determined |
| SS_extracted_k141_47155 | Not-determined |
| SS_extracted_k141_137161 | Low-quality |
| SS_extracted_k141_132345 | Not-determined |
| SS_extracted_k141_137167 | Not-determined |
| SS_extracted_k141_76445 | Not-determined |
| SS_extracted_k141_36171 | Not-determined |
| SS_extracted_k141_47166 | Not-determined |
| SS_extracted_k141_36174 | Low-quality |
| SS_extracted_k141_76450 | Low-quality |
| SS_extracted_k141_27479 | Not-determined |
| SS_extracted_k141_89830 | Low-quality |
| SS_extracted_k141_17651 | Not-determined |
| SS_extracted_k141_89837 | Not-determined |
| SS_extracted_k141_17658 | Not-determined |
| SS_extracted_k141_89839 | Low-quality |
| SS_extracted_k141_81914 | Low-quality |
| SS_extracted_k141_89845 | Not-determined |
| SS_extracted_k141_23007 | Low-quality |
| SS_extracted_k141_54528 | Not-determined |
| SS_extracted_k141_17661 | Not-determined |
| SS_extracted_k141_89851 | Low-quality |
| SS_extracted_k141_57628 | Not-determined |
| SS_extracted_k141_27496 | Not-determined |
| SS_extracted_k141_27497 | Not-determined |
| SS_extracted_k141_100620 | Not-determined |
| SS_extracted_k141_76475 | Not-determined |
| SS_extracted_k141_100622 | Low-quality |
| SS_extracted_k141_76482 | Not-determined |
| SS_extracted_k141_47191 | Not-determined |
| SS_extracted_k141_76488 | Not-determined |
| SS_extracted_k141_81938 | Low-quality |
| SS_extracted_k141_54556 | Not-determined |
| SS_extracted_k141_10976 | Not-determined |
| SS_extracted_k141_114535 | Not-determined |
| SS_extracted_k141_10982 | Not-determined |
| SS_extracted_k141_121069 | Low-quality |
| SS_extracted_k141_47208 | Not-determined |
| SS_extracted_k141_36222 | Not-determined |
| SS_extracted_k141_36224 | Low-quality |
| SS_extracted_k141_81955 | Not-determined |
| SS_extracted_k141_47216 | Low-quality |
| SS_extracted_k141_127224 | Low-quality |
| SS_extracted_k141_54579 | Not-determined |
| SS_extracted_k141_127230 | Not-determined |
| SS_extracted_k141_114548 | Not-determined |
| SS_extracted_k141_89878 | Not-determined |
| SS_extracted_k141_47221 | Not-determined |
| SS_extracted_k141_57652 | Not-determined |
| SS_extracted_k141_132368 | Not-determined |
| SS_extracted_k141_36235 | Not-determined |
| SS_extracted_k141_36236 | Not-determined |
| SS_extracted_k141_114555 | Not-determined |
| SS_extracted_k141_89886 | Not-determined |
| SS_extracted_k141_11006 | Not-determined |
| SS_extracted_k141_127236 | Low-quality |
| SS_extracted_k141_143796 | Low-quality |
| SS_extracted_k141_100663 | Not-determined |
| SS_extracted_k141_143797 | Not-determined |
| SS_extracted_k141_54595 | Not-determined |
| SS_extracted_k141_89895 | Not-determined |
| SS_extracted_k141_23041 | Not-determined |
| SS_extracted_k141_96226 | Not-determined |
| SS_extracted_k141_47231 | Not-determined |
| SS_extracted_k141_36244 | Not-determined |
| SS_extracted_k141_137203 | Not-determined |
| SS_extracted_k141_36249 | Not-determined |
| SS_extracted_k141_36250 | Not-determined |
| SS_extracted_k141_121079 | Not-determined |
| SS_extracted_k141_137208 | Not-determined |
| SS_extracted_k141_96231 | Low-quality |
| SS_extracted_k141_11011 | Not-determined |
| SS_extracted_k141_54598 | Low-quality |
| SS_extracted_k141_11013 | Not-determined |
| SS_extracted_k141_54599 | Not-determined |
| SS_extracted_k141_96246 | Not-determined |
| SS_extracted_k141_100675 | Not-determined |
| SS_extracted_k141_23049 | Not-determined |
| SS_extracted_k141_143811 | Not-determined |
| SS_extracted_k141_23054 | Not-determined |
| SS_extracted_k141_81976 | Not-determined |
| SS_extracted_k141_27528 | Not-determined |
| SS_extracted_k141_81977 | Not-determined |
| SS_extracted_k141_47248 | Low-quality |
| SS_extracted_k141_96254 | Not-determined |
| SS_extracted_k141_40839 | Low-quality |
| SS_extracted_k141_96258 | Not-determined |
| SS_extracted_k141_47258 | Not-determined |
| SS_extracted_k141_40840 | Low-quality |
| SS_extracted_k141_17764 | Not-determined |
| SS_extracted_k141_23062 | Not-determined |
| SS_extracted_k141_47263 | Not-determined |
| SS_extracted_k141_40841 | Low-quality |
| SS_extracted_k141_96261 | Low-quality |
| SS_extracted_k141_36276 | Not-determined |
| SS_extracted_k141_143825 | Not-determined |
| SS_extracted_k141_143827 | Not-determined |
| SS_extracted_k141_23071 | Not-determined |
| SS_extracted_k141_100689 | Not-determined |
| SS_extracted_k141_114581 | Not-determined |
| SS_extracted_k141_127271 | Not-determined |
| SS_extracted_k141_127272 | Not-determined |
| SS_extracted_k141_96266 | Not-determined |
| SS_extracted_k141_143833 | Low-quality |
| SS_extracted_k141_100695 | Not-determined |
| SS_extracted_k141_89946 | Not-determined |
| SS_extracted_k141_36285 | Not-determined |
| SS_extracted_k141_127273 | Low-quality |
| SS_extracted_k141_100700 | Not-determined |
| SS_extracted_k141_17783 | Low-quality |
| SS_extracted_k141_143840 | Not-determined |
| SS_extracted_k141_23078 | Not-determined |
| SS_extracted_k141_89950 | Not-determined |
| SS_extracted_k141_127281 | Not-determined |
| SS_extracted_k141_137232 | Low-quality |
| SS_extracted_k141_127284 | Not-determined |
| SS_extracted_k141_65511 | Not-determined |
| SS_extracted_k141_23080 | Low-quality |
| SS_extracted_k141_143849 | Not-determined |
| SS_extracted_k141_96271 | Not-determined |
| SS_extracted_k141_137236 | Not-determined |
| SS_extracted_k141_121108 | Not-determined |
| SS_extracted_k141_96277 | Low-quality |
| SS_extracted_k141_143862 | Not-determined |
| SS_extracted_k141_36317 | Not-determined |
| SS_extracted_k141_132391 | Not-determined |
| SS_extracted_k141_36318 | Low-quality |
| SS_extracted_k141_96285 | Low-quality |
| SS_extracted_k141_36322 | Not-determined |
| SS_extracted_k141_65535 | Low-quality |
| SS_extracted_k141_11034 | Not-determined |
| SS_extracted_k141_127297 | Not-determined |
| SS_extracted_k141_132395 | Not-determined |
| SS_extracted_k141_23094 | Not-determined |
| SS_extracted_k141_23095 | Not-determined |
| SS_extracted_k141_121121 | Low-quality |
| SS_extracted_k141_65543 | Low-quality |
| SS_extracted_k141_71325 | Not-determined |
| SS_extracted_k141_65544 | Not-determined |
| SS_extracted_k141_132403 | Not-determined |
| SS_extracted_k141_114613 | Not-determined |
| SS_extracted_k141_114615 | Not-determined |
| SS_extracted_k141_100732 | Not-determined |
| SS_extracted_k141_82015 | Low-quality |
| SS_extracted_k141_57676 | Not-determined |
| SS_extracted_k141_71329 | Low-quality |
| SS_extracted_k141_89980 | Not-determined |
| SS_extracted_k141_121134 | Low-quality |
| SS_extracted_k141_132410 | Not-determined |
| SS_extracted_k141_127306 | Not-determined |
| SS_extracted_k141_57679 | Not-determined |
| SS_extracted_k141_137240 | Not-determined |
| SS_extracted_k141_96299 | Not-determined |
| SS_extracted_k141_127311 | Not-determined |
| SS_extracted_k141_132421 | Not-determined |
| SS_extracted_k141_36353 | Not-determined |
| SS_extracted_k141_65557 | Not-determined |
| SS_extracted_k141_23108 | Not-determined |
| SS_extracted_k141_27550 | Not-determined |
| SS_extracted_k141_23113 | Not-determined |
| SS_extracted_k141_57697 | Not-determined |
| SS_extracted_k141_17817 | Not-determined |
| SS_extracted_k141_127324 | Not-determined |
| SS_extracted_k141_127327 | Not-determined |
| SS_extracted_k141_132438 | Not-determined |
| SS_extracted_k141_100759 | Low-quality |
| SS_extracted_k141_17827 | Low-quality |
| SS_extracted_k141_96318 | Not-determined |
| SS_extracted_k141_127336 | Not-determined |
| SS_extracted_k141_114629 | Not-determined |
| SS_extracted_k141_132452 | Not-determined |
| SS_extracted_k141_114631 | Not-determined |
| SS_extracted_k141_27560 | Not-determined |
| SS_extracted_k141_47303 | Not-determined |
| SS_extracted_k141_96330 | Low-quality |
| SS_extracted_k141_100765 | Low-quality |
| SS_extracted_k141_114641 | Not-determined |
| SS_extracted_k141_23134 | Low-quality |
| SS_extracted_k141_114645 | Not-determined |
| SS_extracted_k141_100772 | Low-quality |
| SS_extracted_k141_82034 | Not-determined |
| SS_extracted_k141_47315 | Not-determined |
| SS_extracted_k141_127346 | Low-quality |
| SS_extracted_k141_65579 | Not-determined |
| SS_extracted_k141_47331 | Not-determined |
| SS_extracted_k141_114651 | Low-quality |
| SS_extracted_k141_11041 | Not-determined |
| SS_extracted_k141_57722 | Not-determined |
| SS_extracted_k141_47335 | Not-determined |
| SS_extracted_k141_27565 | Not-determined |
| SS_extracted_k141_47337 | Not-determined |
| SS_extracted_k141_47341 | Not-determined |
| SS_extracted_k141_47342 | Not-determined |
| SS_extracted_k141_36371 | Low-quality |
| SS_extracted_k141_121200 | Not-determined |
| SS_extracted_k141_17864 | Not-determined |
| SS_extracted_k141_143884 | Low-quality |
| SS_extracted_k141_96349 | Not-determined |
| SS_extracted_k141_121210 | Not-determined |
| SS_extracted_k141_23144 | Not-determined |
| SS_extracted_k141_65594 | Not-determined |
| SS_extracted_k141_96352 | Not-determined |
| SS_extracted_k141_57745 | Not-determined |
| SS_extracted_k141_143889 | Not-determined |
| SS_extracted_k141_143890 | Not-determined |
| SS_extracted_k141_143892 | Low-quality |
| SS_extracted_k141_27574 | Not-determined |
| SS_extracted_k141_82053 | Not-determined |
| SS_extracted_k141_36386 | Not-determined |
| SS_extracted_k141_23158 | Not-determined |
| SS_extracted_k141_71355 | Not-determined |
| SS_extracted_k141_143902 | Not-determined |
| S11_vs_k141_6878\|\|full | High-quality |
| SS_extracted_k141_132460 | Not-determined |
| SS_extracted_k141_132461 | Not-determined |
| SS_extracted_k141_71362 | Not-determined |
| SS_extracted_k141_137263 | Not-determined |
| SS_extracted_k141_71364 | Not-determined |
| SS_extracted_k141_57760 | Not-determined |
| SS_extracted_k141_114677 | Not-determined |
| SS_extracted_k141_143904 | Not-determined |
| SS_extracted_k141_132472 | Low-quality |
| SS_extracted_k141_143911 | Not-determined |
| SS_extracted_k141_143912 | Not-determined |
| SS_extracted_k141_114685 | Not-determined |
| SS_extracted_k141_100792 | Not-determined |
| SS_extracted_k141_71379 | Not-determined |
| SS_extracted_k141_137271 | Not-determined |
| SS_extracted_k141_71380 | Not-determined |
| SS_extracted_k141_100794 | Not-determined |
| SS_extracted_k141_36408 | Not-determined |
| SS_extracted_k141_71381 | Not-determined |
| SS_extracted_k141_23175 | Not-determined |
| SS_extracted_k141_11068 | Not-determined |
| SS_extracted_k141_137281 | Not-determined |
| SS_extracted_k141_27594 | Not-determined |
| SS_extracted_k141_127408 | Low-quality |
| SS_extracted_k141_23180 | Not-determined |
| SS_extracted_k141_71391 | Not-determined |
| SS_extracted_k141_121271 | Not-determined |
| SS_extracted_k141_121273 | Not-determined |
| SS_extracted_k141_121274 | Not-determined |
| SS_extracted_k141_127413 | Not-determined |
| SS_extracted_k141_137289 | Not-determined |
| SS_extracted_k141_71400 | Not-determined |
| SS_extracted_k141_23185 | Not-determined |
| SS_extracted_k141_114695 | Not-determined |
| SS_extracted_k141_100800 | Low-quality |
| SS_extracted_k141_47359 | Not-determined |
| SS_extracted_k141_100802 | Not-determined |
| SS_extracted_k141_36426 | Not-determined |
| SS_extracted_k141_132488 | Not-determined |
| SS_extracted_k141_71406 | Not-determined |
| SS_extracted_k141_100813 | Not-determined |
| SS_extracted_k141_121282 | Low-quality |
| SS_extracted_k141_137291 | Not-determined |
| SS_extracted_k141_121284 | Not-determined |
| SS_extracted_k141_121285 | Not-determined |
| SS_extracted_k141_11076 | Not-determined |
| SS_extracted_k141_17889 | Not-determined |
| SS_extracted_k141_121294 | Not-determined |
| SS_extracted_k141_89999 | Not-determined |
| SS_extracted_k141_137298 | Not-determined |

| | |
|---|---|
| SS_extracted_k141_121302 | Not-determined |
| SS_extracted_k141_17897 | Not-determined |
| SS_extracted_k141_127443 | Not-determined |
| SS_extracted_k141_114728 | Not-determined |
| SS_extracted_k141_71435 | Not-determined |
| SS_extracted_k141_17901 | Not-determined |
| SS_extracted_k141_137300 | Low-quality |
| SS_extracted_k141_47396 | Not-determined |
| SS_extracted_k141_90006 | Not-determined |
| SS_extracted_k141_143931 | Not-determined |
| SS_extracted_k141_132513 | Not-determined |
| SS_extracted_k141_121322 | Not-determined |
| SS_extracted_k141_47398 | Low-quality |
| SS_extracted_k141_47399 | Not-determined |
| SS_extracted_k141_121326 | Low-quality |
| SS_extracted_k141_71442 | Low-quality |
| SS_extracted_k141_96365 | Not-determined |
| SS_extracted_k141_47404 | Not-determined |
| SS_extracted_k141_132524 | Not-determined |
| SS_extracted_k141_82093 | Not-determined |
| SS_extracted_k141_143941 | Not-determined |
| SS_extracted_k141_100838 | Not-determined |
| SS_extracted_k141_23198 | Not-determined |
| SS_extracted_k141_127467 | Low-quality |
| SS_extracted_k141_132534 | Not-determined |
| SS_extracted_k141_96378 | Not-determined |
| SS_extracted_k141_17917 | Low-quality |
| SS_extracted_k141_143946 | Not-determined |
| SS_extracted_k141_71455 | Not-determined |
| SS_extracted_k141_23208 | Not-determined |
| SS_extracted_k141_96382 | Low-quality |
| SS_extracted_k141_71458 | Not-determined |
| SS_extracted_k141_23212 | Not-determined |
| SS_extracted_k141_11109 | Low-quality |
| SS_extracted_k141_143953 | Not-determined |
| SS_extracted_k141_143954 | Not-determined |
| SS_extracted_k141_114754 | Not-determined |
| SS_extracted_k141_11111 | Low-quality |
| SS_extracted_k141_90018 | Low-quality |
| SS_extracted_k141_96391 | Low-quality |
| SS_extracted_k141_71464 | Low-quality |
| SS_extracted_k141_96395 | Not-determined |
| SS_extracted_k141_96397 | Low-quality |
| SS_extracted_k141_71465 | Not-determined |
| SS_extracted_k141_47445 | Not-determined |
| SS_extracted_k141_71470 | Not-determined |
| SS_extracted_k141_47448 | Low-quality |
| SS_extracted_k141_127497 | Not-determined |
| SS_extracted_k141_47455 | Not-determined |
| SS_extracted_k141_71480 | Not-determined |
| SS_extracted_k141_90048 | Not-determined |
| SS_extracted_k141_47456 | Low-quality |
| SS_extracted_k141_132569 | Not-determined |
| SS_extracted_k141_47465 | Low-quality |
| SS_extracted_k141_47474 | Not-determined |
| SS_extracted_k141_96425 | Not-determined |
| SS_extracted_k141_47475 | Not-determined |
| SS_extracted_k141_71482 | Low-quality |
| SS_extracted_k141_96429 | Not-determined |
| SS_extracted_k141_96430 | Not-determined |
| SS_extracted_k141_96431 | Not-determined |
| SS_extracted_k141_47482 | Not-determined |
| SS_extracted_k141_96437 | Low-quality |
| SS_extracted_k141_132588 | Not-determined |
| SS_extracted_k141_90056 | Not-determined |
| SS_extracted_k141_132592 | Not-determined |
| SS_extracted_k141_27629 | Low-quality |
| SS_extracted_k141_47500 | Low-quality |
| SS_extracted_k141_27632 | Not-determined |
| SS_extracted_k141_23228 | Not-determined |
| SS_extracted_k141_90067 | Not-determined |
| SS_extracted_k141_96452 | Low-quality |
| SS_extracted_k141_47505 | Not-determined |
| SS_extracted_k141_90070 | Not-determined |
| SS_extracted_k141_71501 | Not-determined |
| SS_extracted_k141_96454 | Not-determined |
| SS_extracted_k141_47510 | Not-determined |
| SS_extracted_k141_90072 | Not-determined |
| SS_extracted_k141_90074 | Not-determined |
| SS_extracted_k141_90076 | Not-determined |
| SS_extracted_k141_90078 | Not-determined |
| SS_extracted_k141_47516 | Not-determined |
| SS_extracted_k141_90079 | Not-determined |
| SS_extracted_k141_90080 | Not-determined |
| SS_extracted_k141_71511 | Low-quality |
| SS_extracted_k141_96458 | Low-quality |
| SS_extracted_k141_90082 | Low-quality |
| SS_extracted_k141_90086 | Not-determined |
| SS_extracted_k141_47531 | Not-determined |
| SS_extracted_k141_96464 | Not-determined |
| SS_extracted_k141_71520 | Not-determined |
| SS_extracted_k141_47539 | Not-determined |
| SS_extracted_k141_96468 | Not-determined |
| SS_extracted_k141_96469 | Low-quality |
| SS_extracted_k141_96470 | Low-quality |
| SS_extracted_k141_57797 | Low-quality |
| SS_extracted_k141_47541 | Not-determined |
| SS_extracted_k141_47543 | Not-determined |
| SS_extracted_k141_27652 | Low-quality |
| SS_extracted_k141_17929 | Not-determined |
| SS_extracted_k141_143969 | Not-determined |
| SS_extracted_k141_27660 | Not-determined |
| SS_extracted_k141_47550 | Not-determined |
| SS_extracted_k141_71537 | Not-determined |
| SS_extracted_k141_100860 | Not-determined |
| SS_extracted_k141_40862 | Not-determined |
| SS_extracted_k141_96481 | Low-quality |
| SS_extracted_k141_47558 | Not-determined |
| SS_extracted_k141_96486 | Not-determined |
| SS_extracted_k141_132620 | Not-determined |
| SS_extracted_k141_121352 | Not-determined |
| SS_extracted_k141_96497 | Low-quality |
| SS_extracted_k141_143979 | Not-determined |
| SS_extracted_k141_71551 | Not-determined |
| SS_extracted_k141_121358 | Not-determined |
| SS_extracted_k141_132632 | Not-determined |
| SS_extracted_k141_96504 | Not-determined |
| SS_extracted_k141_71556 | Not-determined |
| SS_extracted_k141_132636 | Not-determined |
| SS_extracted_k141_100873 | Not-determined |
| SS_extracted_k141_132638 | Not-determined |
| SS_extracted_k141_143989 | Not-determined |
| SS_extracted_k141_100874 | Not-determined |
| SS_extracted_k141_71562 | Not-determined |
| SS_extracted_k141_121369 | Not-determined |
| SS_extracted_k141_57810 | Not-determined |
| SS_extracted_k141_100881 | Not-determined |
| SS_extracted_k141_71574 | Not-determined |
| SS_extracted_k141_144000 | Not-determined |
| SS_extracted_k141_71578 | Not-determined |
| SS_extracted_k141_71580 | Not-determined |
| SS_extracted_k141_17967 | Low-quality |
| SS_extracted_k141_71587 | Not-determined |
| SS_extracted_k141_100893 | Not-determined |
| SS_extracted_k141_17976 | Not-determined |
| SS_extracted_k141_100899 | Not-determined |
| SS_extracted_k141_137325 | Low-quality |
| SS_extracted_k141_27685 | Low-quality |
| SS_extracted_k141_27686 | Not-determined |
| SS_extracted_k141_96530 | Not-determined |
| SS_extracted_k141_96532 | Not-determined |
| SS_extracted_k141_121382 | Not-determined |
| SS_extracted_k141_121388 | Not-determined |
| SS_extracted_k141_17985 | Not-determined |
| SS_extracted_k141_132658 | Low-quality |
| SS_extracted_k141_47604 | Not-determined |
| SS_extracted_k141_71605 | Not-determined |
| SS_extracted_k141_17987 | Not-determined |
| SS_extracted_k141_71611 | Not-determined |
| SS_extracted_k141_47609 | Not-determined |
| SS_extracted_k141_71614 | Not-determined |
| SS_extracted_k141_47612 | Not-determined |
| SS_extracted_k141_137335 | Not-determined |
| SS_extracted_k141_47618 | Not-determined |
| SS_extracted_k141_23247 | Low-quality |
| SS_extracted_k141_47626 | Not-determined |
| SS_extracted_k141_137340 | Low-quality |
| SS_extracted_k141_27711 | Not-determined |
| SS_extracted_k141_96561 | Not-determined |
| SS_extracted_k141_96562 | Not-determined |
| SS_extracted_k141_23255 | Not-determined |
| SS_extracted_k141_121402 | Not-determined |
| SS_extracted_k141_18011 | Not-determined |
| SS_extracted_k141_11122 | Not-determined |
| SS_extracted_k141_121408 | Not-determined |
| SS_extracted_k141_47630 | Not-determined |
| SS_extracted_k141_121409 | Not-determined |
| SS_extracted_k141_23264 | Low-quality |
| SS_extracted_k141_18015 | Not-determined |
| SS_extracted_k141_23278 | Low-quality |
| SS_extracted_k141_71637 | Low-quality |
| SS_extracted_k141_23282 | Not-determined |
| SS_extracted_k141_47650 | Low-quality |
| SS_extracted_k141_47655 | Not-determined |
| SS_extracted_k141_47660 | Not-determined |
| SS_extracted_k141_47661 | Not-determined |
| SS_extracted_k141_23295 | Not-determined |
| SS_extracted_k141_23296 | Low-quality |
| SS_extracted_k141_90098 | Low-quality |
| SS_extracted_k141_90100 | Not-determined |
| SS_extracted_k141_137352 | Low-quality |
| SS_extracted_k141_27744 | Not-determined |
| SS_extracted_k141_90104 | Not-determined |
| SS_extracted_k141_47678 | Not-determined |
| SS_extracted_k141_47680 | Not-determined |
| SS_extracted_k141_23312 | Not-determined |
| SS_extracted_k141_82134 | Low-quality |
| SS_extracted_k141_90110 | Not-determined |
| SS_extracted_k141_114758 | Not-determined |
| SS_extracted_k141_90117 | Not-determined |
| SS_extracted_k141_100914 | Not-determined |
| SS_extracted_k141_132731 | Not-determined |
| SS_extracted_k141_137369 | Low-quality |
| SS_extracted_k141_100915 | Not-determined |
| SS_extracted_k141_100916 | Not-determined |
| SS_extracted_k141_71656 | Not-determined |
| SS_extracted_k141_144048 | Low-quality |
| SS_extracted_k141_71658 | Low-quality |
| SS_extracted_k141_23321 | Not-determined |
| SS_extracted_k141_71662 | Not-determined |
| SS_extracted_k141_90131 | Not-determined |
| SS_extracted_k141_71664 | Low-quality |
| SS_extracted_k141_27752 | Not-determined |
| SS_extracted_k141_144057 | Not-determined |
| SS_extracted_k141_71669 | Not-determined |
| SS_extracted_k141_23329 | Low-quality |
| SS_extracted_k141_71672 | Not-determined |
| SS_extracted_k141_11144 | Low-quality |
| SS_extracted_k141_132751 | Not-determined |
| SS_extracted_k141_100926 | Not-determined |
| SS_extracted_k141_11147 | Low-quality |
| SS_extracted_k141_23336 | Low-quality |
| SS_extracted_k141_82146 | Not-determined |
| SS_extracted_k141_23339 | Low-quality |
| SS_extracted_k141_132759 | Not-determined |
| SS_extracted_k141_27773 | Not-determined |
| SS_extracted_k141_27775 | Not-determined |
| SS_extracted_k141_71690 | Not-determined |
| SS_extracted_k141_132766 | Not-determined |
| SS_extracted_k141_27786 | Not-determined |
| SS_extracted_k141_47710 | Not-determined |
| SS_extracted_k141_47711 | Not-determined |
| SS_extracted_k141_40899 | Not-determined |
| SS_extracted_k141_11163 | Low-quality |
| SS_extracted_k141_40901 | Not-determined |
| SS_extracted_k141_47713 | Not-determined |
| SS_extracted_k141_132774 | Not-determined |
| SS_extracted_k141_76514 | Low-quality |
| SS_extracted_k141_57838 | Not-determined |
| SS_extracted_k141_144073 | Not-determined |
| SS_extracted_k141_96588 | Low-quality |
| SS_extracted_k141_132780 | Not-determined |
| SS_extracted_k141_76518 | Low-quality |
| SS_extracted_k141_96589 | Low-quality |
| SS_extracted_k141_76524 | Not-determined |
| SS_extracted_k141_76526 | Not-determined |
| SS_extracted_k141_18087 | Not-determined |
| SS_extracted_k141_18089 | Low-quality |
| SS_extracted_k141_18091 | Not-determined |
| SS_extracted_k141_96595 | Not-determined |
| SS_extracted_k141_76530 | Not-determined |
| SS_extracted_k141_76531 | Not-determined |
| SS_extracted_k141_132789 | Low-quality |
| SS_extracted_k141_132790 | Low-quality |
| SS_extracted_k141_27804 | Not-determined |
| SS_extracted_k141_90145 | Not-determined |
| SS_extracted_k141_144087 | Not-determined |
| SS_extracted_k141_71699 | Low-quality |
| SS_extracted_k141_71701 | Low-quality |
| SS_extracted_k141_18102 | Not-determined |
| SS_extracted_k141_47739 | Not-determined |
| SS_extracted_k141_18103 | Not-determined |
| SS_extracted_k141_132794 | Low-quality |
| SS_extracted_k141_132795 | Not-determined |
| SS_extracted_k141_132796 | Not-determined |
| SS_extracted_k141_71703 | Not-determined |
| SS_extracted_k141_96609 | Not-determined |
| SS_extracted_k141_144097 | Not-determined |
| SS_extracted_k141_137385 | Low-quality |
| SS_extracted_k141_71716 | Not-determined |
| SS_extracted_k141_47763 | Low-quality |
| SS_extracted_k141_90152 | Not-determined |
| SS_extracted_k141_71720 | Not-determined |
| SS_extracted_k141_132812 | Low-quality |
| SS_extracted_k141_40939 | Not-determined |
| SS_extracted_k141_144107 | Not-determined |
| SS_extracted_k141_71729 | Not-determined |
| SS_extracted_k141_144111 | Low-quality |
| SS_extracted_k141_40942 | Low-quality |
| SS_extracted_k141_132823 | Low-quality |
| SS_extracted_k141_76539 | Not-determined |
| SS_extracted_k141_132825 | Not-determined |
| SS_extracted_k141_90160 | Not-determined |
| SS_extracted_k141_76542 | Low-quality |
| SS_extracted_k141_114815 | Low-quality |
| SS_extracted_k141_144119 | Not-determined |
| SS_extracted_k141_144121 | Not-determined |
| SS_extracted_k141_144124 | Not-determined |
| SS_extracted_k141_100956 | Low-quality |
| SS_extracted_k141_144126 | Low-quality |
| SS_extracted_k141_100958 | Not-determined |
| SS_extracted_k141_76556 | Not-determined |
| SS_extracted_k141_76557 | Not-determined |
| SS_extracted_k141_144135 | Not-determined |
| SS_extracted_k141_90168 | Not-determined |
| SS_extracted_k141_132840 | Not-determined |
| SS_extracted_k141_71756 | Not-determined |
| SS_extracted_k141_71759 | Not-determined |
| SS_extracted_k141_71761 | Not-determined |
| SS_extracted_k141_71763 | Not-determined |
| SS_extracted_k141_47796 | Low-quality |
| SS_extracted_k141_76579 | Not-determined |
| SS_extracted_k141_47797 | Low-quality |
| SS_extracted_k141_47801 | Not-determined |
| SS_extracted_k141_71772 | Not-determined |
| SS_extracted_k141_90181 | Not-determined |
| SS_extracted_k141_47807 | Not-determined |
| SS_extracted_k141_82187 | Not-determined |
| SS_extracted_k141_90196 | Low-quality |
| SS_extracted_k141_27844 | Not-determined |
| SS_extracted_k141_82201 | Not-determined |
| SS_extracted_k141_90203 | Low-quality |
| SS_extracted_k141_114825 | Low-quality |
| SS_extracted_k141_100979 | Not-determined |
| SS_extracted_k141_90212 | Not-determined |
| SS_extracted_k141_65646 | Low-quality |
| SS_extracted_k141_90215 | Not-determined |
| SS_extracted_k141_27866 | Low-quality |
| SS_extracted_k141_100993 | Low-quality |
| SS_extracted_k141_18141 | Low-quality |
| SS_extracted_k141_76625 | Low-quality |
| SS_extracted_k141_132878 | Not-determined |
| SS_extracted_k141_27872 | Low-quality |
| SS_extracted_k141_27875 | Not-determined |
| SS_extracted_k141_132889 | Low-quality |
| SS_extracted_k141_114866 | Not-determined |
| SS_extracted_k141_90234 | Not-determined |
| SS_extracted_k141_18158 | Not-determined |
| SS_extracted_k141_132898 | Low-quality |
| SS_extracted_k141_47833 | Not-determined |
| SS_extracted_k141_114877 | Not-determined |
| SS_extracted_k141_114879 | Not-determined |
| SS_extracted_k141_76634 | Not-determined |
| SS_extracted_k141_96635 | Not-determined |
| SS_extracted_k141_96636 | Not-determined |
| SS_extracted_k141_96641 | Not-determined |
| SS_extracted_k141_90242 | Not-determined |
| SS_extracted_k141_23392 | Low-quality |
| SS_extracted_k141_114898 | Not-determined |
| SS_extracted_k141_71789 | Low-quality |
| SS_extracted_k141_90252 | Not-determined |
| SS_extracted_k141_65681 | Not-determined |
| SS_extracted_k141_76649 | Not-determined |
| SS_extracted_k141_47855 | Not-determined |
| SS_extracted_k141_71795 | Not-determined |
| SS_extracted_k141_76653 | Not-determined |
| SS_extracted_k141_90258 | Not-determined |
| SS_extracted_k141_47860 | Not-determined |
| SS_extracted_k141_65688 | Not-determined |
| SS_extracted_k141_65690 | Low-quality |
| SS_extracted_k141_23405 | Low-quality |
| SS_extracted_k141_114909 | Not-determined |
| SS_extracted_k141_47865 | Not-determined |
| SS_extracted_k141_47866 | Not-determined |
| SS_extracted_k141_23409 | Not-determined |
| SS_extracted_k141_47873 | Low-quality |
| SS_extracted_k141_23411 | Not-determined |
| SS_extracted_k141_132930 | Not-determined |
| SS_extracted_k141_27906 | Not-determined |
| SS_extracted_k141_90276 | Not-determined |
| SS_extracted_k141_96668 | Not-determined |
| SS_extracted_k141_90278 | Low-quality |
| SS_extracted_k141_96669 | Low-quality |
| SS_extracted_k141_132939 | Not-determined |
| SS_extracted_k141_40976 | Low-quality |
| SS_extracted_k141_114926 | Low-quality |
| SS_extracted_k141_65706 | Not-determined |
| SS_extracted_k141_40982 | Not-determined |
| SS_extracted_k141_65712 | Not-determined |
| SS_extracted_k141_90290 | Low-quality |
| SS_extracted_k141_23417 | Not-determined |
| SS_extracted_k141_114935 | Not-determined |
| SS_extracted_k141_40989 | Not-determined |
| SS_extracted_k141_137441 | Not-determined |
| SS_extracted_k141_96687 | Not-determined |
| SS_extracted_k141_11180 | Not-determined |
| SS_extracted_k141_90298 | Not-determined |
| SS_extracted_k141_96693 | Not-determined |
| SS_extracted_k141_137447 | Not-determined |
| SS_extracted_k141_96694 | Not-determined |
| SS_extracted_k141_11184 | Not-determined |
| SS_extracted_k141_132957 | Not-determined |
| SS_extracted_k141_137450 | Not-determined |
| SS_extracted_k141_96699 | Not-determined |
| SS_extracted_k141_132961 | Not-determined |
| SS_extracted_k141_132964 | Not-determined |
| SS_extracted_k141_23426 | Not-determined |
| SS_extracted_k141_65728 | Low-quality |
| SS_extracted_k141_132967 | Not-determined |
| SS_extracted_k141_76675 | Not-determined |
| SS_extracted_k141_76678 | Not-determined |
| SS_extracted_k141_47885 | Low-quality |
| SS_extracted_k141_132969 | Not-determined |
| SS_extracted_k141_23432 | Not-determined |
| SS_extracted_k141_23436 | Not-determined |
| SS_extracted_k141_90310 | Low-quality |
| SS_extracted_k141_137462 | Not-determined |
| SS_extracted_k141_96708 | Low-quality |
| SS_extracted_k141_23440 | Not-determined |
| SS_extracted_k141_96709 | Not-determined |
| SS_extracted_k141_96710 | Low-quality |
| SS_extracted_k141_23442 | Not-determined |
| SS_extracted_k141_65743 | Not-determined |
| SS_extracted_k141_137469 | Low-quality |
| SS_extracted_k141_96718 | Not-determined |
| SS_extracted_k141_132979 | Not-determined |
| SS_extracted_k141_65748 | Not-determined |
| SS_extracted_k141_137473 | Not-determined |
| SS_extracted_k141_96728 | Not-determined |
| SS_extracted_k141_137477 | Not-determined |
| SS_extracted_k141_41005 | Not-determined |
| SS_extracted_k141_137485 | Not-determined |
| SS_extracted_k141_96739 | Not-determined |
| SS_extracted_k141_96740 | Not-determined |
| SS_extracted_k141_11221 | Low-quality |
| SS_extracted_k141_76706 | Not-determined |
| SS_extracted_k141_41013 | Not-determined |
| SS_extracted_k141_65770 | Not-determined |
| SS_extracted_k141_133015 | Not-determined |
| SS_extracted_k141_90341 | Low-quality |
| SS_extracted_k141_65773 | Not-determined |
| SS_extracted_k141_96744 | Not-determined |
| SS_extracted_k141_76715 | Not-determined |
| SS_extracted_k141_90344 | Low-quality |
| SS_extracted_k141_96751 | Not-determined |
| SS_extracted_k141_23482 | Not-determined |
| SS_extracted_k141_96754 | Not-determined |
| SS_extracted_k141_90346 | Not-determined |
| SS_extracted_k141_96756 | Not-determined |
| SS_extracted_k141_71875 | Not-determined |
| SS_extracted_k141_71876 | Not-determined |
| SS_extracted_k141_57855 | Not-determined |
| SS_extracted_k141_133017 | Low-quality |
| SS_extracted_k141_96763 | Not-determined |
| SS_extracted_k141_144156 | Not-determined |
| SS_extracted_k141_133026 | Not-determined |
| SS_extracted_k141_23492 | Not-determined |
| SS_extracted_k141_96769 | Not-determined |
| SS_extracted_k141_114958 | Not-determined |
| SS_extracted_k141_144159 | Not-determined |
| SS_extracted_k141_133037 | Low-quality |
| SS_extracted_k141_133038 | Not-determined |
| SS_extracted_k141_57876 | Not-determined |
| SS_extracted_k141_133040 | Not-determined |
| SS_extracted_k141_90375 | Low-quality |
| SS_extracted_k141_90378 | Not-determined |
| SS_extracted_k141_90385 | Not-determined |
| SS_extracted_k141_23503 | Low-quality |
| SS_extracted_k141_23507 | Not-determined |
| SS_extracted_k141_90391 | Low-quality |
| SS_extracted_k141_41059 | Low-quality |
| SS_extracted_k141_41065 | Not-determined |
| SS_extracted_k141_76719 | Not-determined |
| SS_extracted_k141_82263 | Not-determined |
| SS_extracted_k141_18196 | Low-quality |
| SS_extracted_k141_23514 | Not-determined |
| SS_extracted_k141_133051 | Not-determined |
| SS_extracted_k141_18198 | Not-determined |
| SS_extracted_k141_23533 | Low-quality |
| SS_extracted_k141_96778 | Low-quality |
| SS_extracted_k141_18202 | Not-determined |
| SS_extracted_k141_82276 | Not-determined |
| SS_extracted_k141_82279 | Not-determined |
| SS_extracted_k141_82283 | Not-determined |
| SS_extracted_k141_23544 | Not-determined |
| SS_extracted_k141_57894 | Not-determined |
| SS_extracted_k141_82290 | Not-determined |
| SS_extracted_k141_23552 | Not-determined |
| SS_extracted_k141_96797 | Low-quality |
| SS_extracted_k141_23556 | Not-determined |
| SS_extracted_k141_18211 | Not-determined |
| SS_extracted_k141_23566 | Not-determined |
| SS_extracted_k141_18212 | Not-determined |
| SS_extracted_k141_96806 | Not-determined |
| SS_extracted_k141_90403 | Not-determined |
| SS_extracted_k141_23581 | Not-determined |
| SS_extracted_k141_23584 | Not-determined |
| SS_extracted_k141_144188 | Low-quality |
| SS_extracted_k141_65815 | Not-determined |
| SS_extracted_k141_133075 | Low-quality |
| SS_extracted_k141_90410 | Not-determined |
| SS_extracted_k141_82305 | Not-determined |
| SS_extracted_k141_90414 | Not-determined |
| SS_extracted_k141_144201 | Not-determined |
| SS_extracted_k141_144205 | Not-determined |
| SS_extracted_k141_144210 | Not-determined |
| SS_extracted_k141_41138 | Not-determined |
| SS_extracted_k141_90423 | Not-determined |
| SS_extracted_k141_144217 | Not-determined |
| SS_extracted_k141_41142 | Not-determined |
| SS_extracted_k141_23613 | Low-quality |
| SS_extracted_k141_82317 | Not-determined |
| SS_extracted_k141_144227 | Not-determined |
| SS_extracted_k141_144228 | Not-determined |
| SS_extracted_k141_65848 | Not-determined |
| SS_extracted_k141_65854 | Not-determined |
| SS_extracted_k141_47908 | Not-determined |
| SS_extracted_k141_82324 | Low-quality |
| SS_extracted_k141_47915 | Not-determined |
| SS_extracted_k141_11268 | Not-determined |
| SS_extracted_k141_11269 | Not-determined |
| SS_extracted_k141_47919 | Not-determined |
| SS_extracted_k141_71892 | Not-determined |
| SS_extracted_k141_82329 | Not-determined |
| SS_extracted_k141_47927 | Not-determined |
| SS_extracted_k141_71902 | Not-determined |
| SS_extracted_k141_115008 | Not-determined |
| SS_extracted_k141_82337 | Not-determined |
| SS_extracted_k141_82340 | Not-determined |
| SS_extracted_k141_115012 | Not-determined |
| SS_extracted_k141_90440 | Low-quality |
| SS_extracted_k141_27926 | Not-determined |
| SS_extracted_k141_144234 | Not-determined |
| SS_extracted_k141_115036 | Not-determined |
| SS_extracted_k141_27932 | Not-determined |
| SS_extracted_k141_90452 | Not-determined |
| SS_extracted_k141_144238 | Low-quality |
| SS_extracted_k141_133135 | Not-determined |
| SS_extracted_k141_71934 | Not-determined |
| SS_extracted_k141_71936 | Not-determined |
| SS_extracted_k141_115054 | Low-quality |
| SS_extracted_k141_115055 | Not-determined |
| SS_extracted_k141_101002 | Not-determined |
| SS_extracted_k141_27953 | Not-determined |
| SS_extracted_k141_71946 | Not-determined |
| SS_extracted_k141_47941 | Not-determined |
| SS_extracted_k141_47944 | Not-determined |
| SS_extracted_k141_47945 | Not-determined |
| SS_extracted_k141_27961 | Not-determined |
| SS_extracted_k141_47946 | Not-determined |
| SS_extracted_k141_90471 | Not-determined |
| SS_extracted_k141_133163 | Low-quality |
| SS_extracted_k141_133170 | Not-determined |
| SS_extracted_k141_90483 | Not-determined |
| SS_extracted_k141_47952 | Low-quality |
| SS_extracted_k141_41153 | Not-determined |
| SS_extracted_k141_115098 | Not-determined |
| SS_extracted_k141_47959 | Low-quality |
| SS_extracted_k141_101009 | Not-determined |
| SS_extracted_k141_96859 | Not-determined |
| SS_extracted_k141_115104 | Low-quality |
| SS_extracted_k141_96865 | Low-quality |
| SS_extracted_k141_96866 | Not-determined |
| SS_extracted_k141_115114 | Not-determined |
| SS_extracted_k141_96869 | Not-determined |
| SS_extracted_k141_47973 | Low-quality |
| SS_extracted_k141_133192 | Not-determined |
| SS_extracted_k141_90500 | Not-determined |
| SS_extracted_k141_133194 | Low-quality |
| SS_extracted_k141_41167 | Not-determined |
| SS_extracted_k141_90504 | Not-determined |
| SS_extracted_k141_133199 | Not-determined |
| SS_extracted_k141_76751 | Not-determined |
| SS_extracted_k141_90506 | Not-determined |
| SS_extracted_k141_41171 | Not-determined |
| SS_extracted_k141_96880 | Not-determined |
| SS_extracted_k141_41172 | Not-determined |
| SS_extracted_k141_96882 | Not-determined |
| SS_extracted_k141_96883 | Not-determined |
| SS_extracted_k141_76755 | Not-determined |
| SS_extracted_k141_133207 | Not-determined |
| SS_extracted_k141_76760 | Not-determined |
| SS_extracted_k141_82362 | Not-determined |
| SS_extracted_k141_96897 | Low-quality |
| SS_extracted_k141_76769 | Not-determined |
| SS_extracted_k141_41189 | Not-determined |
| SS_extracted_k141_41196 | Not-determined |
| SS_extracted_k141_47984 | Low-quality |
| SS_extracted_k141_47985 | Low-quality |
| SS_extracted_k141_47986 | Low-quality |
| SS_extracted_k141_133225 | Not-determined |
| SS_extracted_k141_96902 | Low-quality |
| SS_extracted_k141_90529 | Low-quality |
| SS_extracted_k141_47990 | Not-determined |
| SS_extracted_k141_76779 | Low-quality |
| SS_extracted_k141_115147 | Not-determined |
| SS_extracted_k141_115148 | Not-determined |
| SS_extracted_k141_76782 | Not-determined |
| SS_extracted_k141_82383 | Not-determined |
| SS_extracted_k141_76786 | Not-determined |
| SS_extracted_k141_133232 | Not-determined |
| SS_extracted_k141_96907 | Low-quality |
| SS_extracted_k141_71974 | Not-determined |
| SS_extracted_k141_96914 | Low-quality |
| SS_extracted_k141_71976 | Not-determined |
| SS_extracted_k141_76792 | Low-quality |
| SS_extracted_k141_96925 | Low-quality |
| SS_extracted_k141_137536 | Not-determined |
| SS_extracted_k141_96928 | Not-determined |
| SS_extracted_k141_41215 | Not-determined |
| SS_extracted_k141_96938 | Not-determined |
| SS_extracted_k141_71988 | Not-determined |
| SS_extracted_k141_48012 | Low-quality |
| SS_extracted_k141_82403 | Not-determined |
| SS_extracted_k141_23628 | Low-quality |
| SS_extracted_k141_48019 | Not-determined |
| SS_extracted_k141_96947 | Not-determined |
| SS_extracted_k141_72001 | Low-quality |
| SS_extracted_k141_27993 | Low-quality |

| | |
|---|---|
| SS_extracted_k141_41227 | Low-quality |
| SS_extracted_k141_137553 | Low-quality |
| SS_extracted_k141_27995 | Not-determined |
| SS_extracted_k141_23639 | Low-quality |
| SS_extracted_k141_27996 | Not-determined |
| SS_extracted_k141_96956 | Not-determined |
| SS_extracted_k141_82419 | Not-determined |
| SS_extracted_k141_23643 | Low-quality |
| SS_extracted_k141_90544 | Not-determined |
| SS_extracted_k141_137559 | Not-determined |
| SS_extracted_k141_23648 | Not-determined |
| SS_extracted_k141_90550 | Not-determined |
| SS_extracted_k141_72017 | Not-determined |
| SS_extracted_k141_23659 | Not-determined |
| SS_extracted_k141_41240 | Not-determined |
| SS_extracted_k141_90568 | Not-determined |
| SS_extracted_k141_23665 | Not-determined |
| SS_extracted_k141_11315 | Not-determined |
| SS_extracted_k141_72029 | Not-determined |
| SS_extracted_k141_11333 | Low-quality |
| SS_extracted_k141_11334 | Not-determined |
| SS_extracted_k141_11336 | Not-determined |
| SS_extracted_k141_72045 | Not-determined |
| SS_extracted_k141_27998 | Not-determined |
| SS_extracted_k141_11345 | Not-determined |
| SS_extracted_k141_72049 | Low-quality |
| SS_extracted_k141_76814 | Not-determined |
| SS_extracted_k141_72059 | Not-determined |
| SS_extracted_k141_90601 | Low-quality |
| SS_extracted_k141_115186 | Not-determined |
| SS_extracted_k141_115189 | Not-determined |
| SS_extracted_k141_11361 | Not-determined |
| SS_extracted_k141_115194 | Low-quality |
| SS_extracted_k141_82451 | Not-determined |
| SS_extracted_k141_90612 | Not-determined |
| SS_extracted_k141_97018 | Low-quality |
| SS_extracted_k141_133259 | Not-determined |
| SS_extracted_k141_11367 | Not-determined |
| SS_extracted_k141_97031 | Not-determined |
| SS_extracted_k141_133264 | Not-determined |
| SS_extracted_k141_23686 | Not-determined |
| SS_extracted_k141_76832 | Not-determined |
| SS_extracted_k141_133273 | Not-determined |
| SS_extracted_k141_76835 | Not-determined |
| SS_extracted_k141_72082 | Not-determined |
| SS_extracted_k141_72085 | Not-determined |
| SS_extracted_k141_115207 | Not-determined |
| SS_extracted_k141_90645 | Not-determined |
| SS_extracted_k141_115209 | Not-determined |
| SS_extracted_k141_57941 | Not-determined |
| SS_extracted_k141_90650 | Not-determined |
| SS_extracted_k141_57942 | Not-determined |
| SS_extracted_k141_115210 | Low-quality |
| SS_extracted_k141_11381 | Not-determined |
| SS_extracted_k141_115211 | Not-determined |
| SS_extracted_k141_72098 | Not-determined |
| SS_extracted_k141_11387 | Not-determined |
| SS_extracted_k141_57951 | Not-determined |
| SS_extracted_k141_76852 | Not-determined |
| SS_extracted_k141_133285 | Not-determined |
| SS_extracted_k141_144303 | Not-determined |
| SS_extracted_k141_144307 | Not-determined |
| SS_extracted_k141_144308 | Not-determined |
| SS_extracted_k141_144309 | Not-determined |
| SS_extracted_k141_144311 | Not-determined |
| SS_extracted_k141_72113 | Not-determined |
| SS_extracted_k141_115230 | Low-quality |
| SS_extracted_k141_133306 | Not-determined |
| SS_extracted_k141_133308 | Not-determined |
| SS_extracted_k141_133311 | Not-determined |
| SS_extracted_k141_133312 | Not-determined |
| SS_extracted_k141_115237 | Not-determined |
| SS_extracted_k141_72117 | Not-determined |
| SS_extracted_k141_133315 | Not-determined |
| SS_extracted_k141_133316 | Not-determined |
| SS_extracted_k141_97056 | Not-determined |
| SS_extracted_k141_115243 | Not-determined |
| SS_extracted_k141_144324 | Not-determined |
| SS_extracted_k141_72122 | Not-determined |
| SS_extracted_k141_11414 | Not-determined |
| SS_extracted_k141_133319 | Low-quality |
| SS_extracted_k141_65875 | Not-determined |
| SS_extracted_k141_97075 | Not-determined |
| SS_extracted_k141_115265 | Not-determined |
| SS_extracted_k141_11428 | Not-determined |
| SS_extracted_k141_133331 | Not-determined |
| SS_extracted_k141_72135 | Low-quality |
| SS_extracted_k141_115268 | Not-determined |
| SS_extracted_k141_115269 | Not-determined |
| SS_extracted_k141_65889 | Not-determined |
| SS_extracted_k141_97092 | Low-quality |
| SS_extracted_k141_72140 | Not-determined |
| SS_extracted_k141_133341 | Not-determined |
| SS_extracted_k141_76864 | Not-determined |
| SS_extracted_k141_133348 | Not-determined |
| SS_extracted_k141_133355 | Not-determined |
| SS_extracted_k141_72159 | Not-determined |
| SS_extracted_k141_133356 | Not-determined |
| SS_extracted_k141_72160 | Not-determined |
| SS_extracted_k141_41282 | Not-determined |
| SS_extracted_k141_41284 | Not-determined |
| SS_extracted_k141_41285 | Not-determined |
| SS_extracted_k141_65900 | Low-quality |
| SS_extracted_k141_97115 | Low-quality |
| SS_extracted_k141_90691 | Not-determined |
| SS_extracted_k141_57987 | Low-quality |
| SS_extracted_k141_115286 | Not-determined |
| SS_extracted_k141_76891 | Not-determined |
| SS_extracted_k141_101052 | Low-quality |
| SS_extracted_k141_57993 | Not-determined |
| SS_extracted_k141_115290 | Low-quality |
| SS_extracted_k141_133377 | Low-quality |
| SS_extracted_k141_133378 | Low-quality |
| SS_extracted_k141_133383 | Not-determined |
| SS_extracted_k141_90699 | Low-quality |
| SS_extracted_k141_90702 | Low-quality |
| SS_extracted_k141_90704 | Low-quality |
| SS_extracted_k141_115303 | Not-determined |
| SS_extracted_k141_76900 | Not-determined |
| SS_extracted_k141_133395 | Not-determined |
| SS_extracted_k141_115305 | Not-determined |
| SS_extracted_k141_137608 | Not-determined |
| SS_extracted_k141_115308 | Not-determined |
| SS_extracted_k141_133404 | Not-determined |
| SS_extracted_k141_28008 | Not-determined |
| SS_extracted_k141_115314 | Not-determined |
| SS_extracted_k141_65914 | Low-quality |
| SS_extracted_k141_28011 | Not-determined |
| SS_extracted_k141_72193 | Not-determined |
| SS_extracted_k141_11446 | Not-determined |
| SS_extracted_k141_28022 | Not-determined |
| SS_extracted_k141_115334 | Not-determined |
| SS_extracted_k141_41297 | Not-determined |
| SS_extracted_k141_48028 | Not-determined |
| SS_extracted_k141_101060 | Not-determined |
| SS_extracted_k141_48029 | Not-determined |
| SS_extracted_k141_115346 | Not-determined |
| SS_extracted_k141_101065 | Not-determined |
| SS_extracted_k141_72211 | Low-quality |
| SS_extracted_k141_115350 | Low-quality |
| SS_extracted_k141_90729 | Not-determined |
| SS_extracted_k141_133427 | Not-determined |
| SS_extracted_k141_72216 | Not-determined |
| SS_extracted_k141_41306 | Not-determined |
| SS_extracted_k141_41310 | Not-determined |
| SS_extracted_k141_72222 | Not-determined |
| SS_extracted_k141_76933 | Not-determined |
| SS_extracted_k141_65928 | Not-determined |
| SS_extracted_k141_133437 | Low-quality |
| SS_extracted_k141_48043 | Low-quality |
| SS_extracted_k141_76939 | Low-quality |
| SS_extracted_k141_133440 | Not-determined |
| SS_extracted_k141_90742 | Not-determined |
| SS_extracted_k141_65935 | Not-determined |
| SS_extracted_k141_90744 | Not-determined |
| S17_vs_k141_43307\|\|full | Medium-quality |
| SS_extracted_k141_133446 | Low-quality |
| SS_extracted_k141_23720 | Not-determined |
| SS_extracted_k141_23721 | Low-quality |
| SS_extracted_k141_23724 | Not-determined |
| SS_extracted_k141_144330 | Low-quality |
| SS_extracted_k141_144332 | Low-quality |
| SS_extracted_k141_72240 | Low-quality |
| S8_vs_k141_106946\|\|full | High-quality |
| SS_extracted_k141_76959 | Not-determined |
| SS_extracted_k141_144340 | Not-determined |
| SS_extracted_k141_72248 | Not-determined |
| SS_extracted_k141_48073 | Not-determined |
| SS_extracted_k141_144344 | Not-determined |
| SS_extracted_k141_144345 | Not-determined |
| SS_extracted_k141_76969 | Not-determined |
| SS_extracted_k141_137640 | Not-determined |
| SS_extracted_k141_11456 | Not-determined |
| SS_extracted_k141_48077 | Not-determined |
| SS_extracted_k141_76972 | Low-quality |
| SS_extracted_k141_11459 | Not-determined |
| SS_extracted_k141_76975 | Not-determined |
| SS_extracted_k141_11460 | Not-determined |
| SS_extracted_k141_133452 | Not-determined |
| SS_extracted_k141_41328 | Not-determined |
| SS_extracted_k141_133457 | Low-quality |
| SS_extracted_k141_133461 | Not-determined |
| SS_extracted_k141_23740 | Not-determined |
| SS_extracted_k141_133466 | Not-determined |
| SS_extracted_k141_137645 | Not-determined |
| SS_extracted_k141_133470 | Low-quality |
| SS_extracted_k141_137647 | Not-determined |
| SS_extracted_k141_23751 | Low-quality |
| SS_extracted_k141_23757 | Not-determined |
| SS_extracted_k141_133481 | Not-determined |
| SS_extracted_k141_58015 | Not-determined |
| SS_extracted_k141_41356 | Low-quality |
| SS_extracted_k141_72270 | Low-quality |
| SS_extracted_k141_101092 | Not-determined |
| SS_extracted_k141_41359 | Low-quality |
| SS_extracted_k141_23765 | Low-quality |
| SS_extracted_k141_65990 | Not-determined |
| SS_extracted_k141_11499 | Not-determined |
| SS_extracted_k141_137653 | Not-determined |
| SS_extracted_k141_48113 | Low-quality |
| SS_extracted_k141_77012 | Not-determined |
| SS_extracted_k141_48116 | Not-determined |
| SS_extracted_k141_66004 | Not-determined |
| SS_extracted_k141_144380 | Not-determined |
| SS_extracted_k141_23783 | Not-determined |
| SS_extracted_k141_144384 | Low-quality |
| SS_extracted_k141_72286 | Not-determined |
| SS_extracted_k141_23785 | Not-determined |
| SS_extracted_k141_66009 | Not-determined |
| SS_extracted_k141_137668 | Not-determined |
| SS_extracted_k141_144410 | Not-determined |
| SS_extracted_k141_41377 | Low-quality |
| SS_extracted_k141_133490 | Not-determined |
| SS_extracted_k141_133493 | Not-determined |
| SS_extracted_k141_23820 | Not-determined |
| SS_extracted_k141_101104 | Not-determined |
| SS_extracted_k141_133496 | Low-quality |
| SS_extracted_k141_23840 | Not-determined |
| SS_extracted_k141_101106 | Low-quality |
| SS_extracted_k141_23846 | Not-determined |
| SS_extracted_k141_23850 | Not-determined |
| SS_extracted_k141_101111 | Not-determined |
| SS_extracted_k141_133516 | Low-quality |
| SS_extracted_k141_101114 | Low-quality |
| SS_extracted_k141_144416 | Low-quality |
| SS_extracted_k141_72320 | Low-quality |
| SS_extracted_k141_101117 | Not-determined |
| SS_extracted_k141_133520 | Not-determined |
| SS_extracted_k141_133521 | Not-determined |
| SS_extracted_k141_48163 | Not-determined |
| SS_extracted_k141_82505 | Not-determined |
| SS_extracted_k141_48166 | Low-quality |
| SS_extracted_k141_48167 | Not-determined |
| SS_extracted_k141_48168 | Not-determined |
| SS_extracted_k141_133527 | Not-determined |
| SS_extracted_k141_11521 | Not-determined |
| SS_extracted_k141_82517 | Low-quality |
| SS_vs_k141_145599\|\|full | High-quality |
| SS_extracted_k141_11534 | Low-quality |
| SS_extracted_k141_144430 | Not-determined |
| SS_extracted_k141_48179 | Not-determined |
| SS_extracted_k141_48183 | Not-determined |
| SS_extracted_k141_23870 | Not-determined |
| SS_extracted_k141_144436 | Not-determined |
| SS_extracted_k141_48190 | Not-determined |
| SS_extracted_k141_23874 | Low-quality |
| SS_extracted_k141_66044 | Not-determined |
| SS_extracted_k141_133551 | Low-quality |
| SS_extracted_k141_133559 | Not-determined |
| SS_extracted_k141_48206 | Not-determined |
| SS_extracted_k141_28087 | Low-quality |
| SS_extracted_k141_28089 | Low-quality |
| SS_extracted_k141_137708 | Not-determined |
| SS_extracted_k141_11541 | Low-quality |
| SS_extracted_k141_23902 | Not-determined |
| SS_extracted_k141_133565 | Not-determined |
| SS_extracted_k141_48230 | Low-quality |
| SS_extracted_k141_11556 | Not-determined |
| SS_extracted_k141_77060 | Not-determined |
| SS_extracted_k141_144443 | Not-determined |
| SS_extracted_k141_66068 | Not-determined |
| SS_extracted_k141_77064 | Not-determined |
| SS_extracted_k141_77066 | Not-determined |
| SS_extracted_k141_144452 | Low-quality |
| SS_extracted_k141_77067 | Not-determined |
| SS_extracted_k141_144454 | Not-determined |
| SS_extracted_k141_23919 | Low-quality |
| SS_extracted_k141_48237 | Not-determined |
| SS_extracted_k141_48244 | Not-determined |
| SS_extracted_k141_77077 | Not-determined |
| SS_extracted_k141_137713 | Not-determined |
| SS_extracted_k141_144463 | Not-determined |
| SS_extracted_k141_41416 | Not-determined |
| SS_extracted_k141_11565 | Not-determined |
| SS_extracted_k141_137717 | Not-determined |
| SS_extracted_k141_101151 | Not-determined |
| SS_extracted_k141_41428 | Not-determined |
| SS_extracted_k141_82553 | Low-quality |
| SS_extracted_k141_41433 | Not-determined |
| SS_extracted_k141_11575 | Not-determined |
| SS_extracted_k141_144472 | Not-determined |
| SS_extracted_k141_66122 | Not-determined |
| SS_extracted_k141_77092 | Not-determined |
| SS_extracted_k141_48290 | Not-determined |
| SS_extracted_k141_28114 | Not-determined |
| SS_extracted_k141_48295 | Not-determined |
| SS_extracted_k141_101177 | Not-determined |
| SS_extracted_k141_41455 | Low-quality |
| SS_extracted_k141_41457 | Not-determined |
| SS_extracted_k141_137732 | Low-quality |
| SS_extracted_k141_11580 | Not-determined |
| SS_extracted_k141_137736 | Not-determined |
| SS_extracted_k141_77106 | Not-determined |
| SS_extracted_k141_48313 | Low-quality |
| SS_extracted_k141_11595 | Not-determined |
| SS_extracted_k141_137738 | Not-determined |
| SS_extracted_k141_77119 | Not-determined |
| SS_extracted_k141_77126 | Not-determined |
| SS_extracted_k141_48338 | Not-determined |
| SS_extracted_k141_82576 | Not-determined |
| SS_extracted_k141_82582 | Not-determined |
| SS_extracted_k141_48349 | Not-determined |
| SS_extracted_k141_144475 | Not-determined |
| SS_extracted_k141_144481 | Not-determined |
| SS_extracted_k141_72371 | Not-determined |
| SS_extracted_k141_48354 | Not-determined |
| SS_extracted_k141_144484 | Not-determined |
| SS_extracted_k141_41472 | Not-determined |
| SS_extracted_k141_48357 | Not-determined |
| SS_extracted_k141_72378 | Not-determined |
| SS_extracted_k141_28146 | Low-quality |
| SS_extracted_k141_144491 | Low-quality |
| SS_extracted_k141_72383 | Not-determined |
| SS_extracted_k141_137754 | Not-determined |
| SS_extracted_k141_137757 | Low-quality |
| SS_extracted_k141_11620 | Not-determined |
| SS_extracted_k141_41483 | Not-determined |
| SS_extracted_k141_48375 | Not-determined |
| SS_extracted_k141_41489 | Not-determined |
| SS_extracted_k141_48377 | Not-determined |
| SS_extracted_k141_23990 | Not-determined |
| SS_extracted_k141_41493 | Not-determined |
| SS_extracted_k141_41495 | Not-determined |
| SS_extracted_k141_66176 | Not-determined |
| SS_extracted_k141_23993 | Not-determined |
| SS_extracted_k141_137770 | Low-quality |
| SS_extracted_k141_77155 | Not-determined |
| SS_extracted_k141_41501 | Not-determined |
| SS_extracted_k141_11629 | Not-determined |
| SS_extracted_k141_82590 | Low-quality |
| SS_extracted_k141_101249 | Not-determined |
| SS_extracted_k141_144516 | Not-determined |
| SS_extracted_k141_82592 | Low-quality |
| SS_extracted_k141_144523 | Low-quality |
| SS_extracted_k141_137781 | Not-determined |
| SS_extracted_k141_66185 | Not-determined |
| SS_extracted_k141_77167 | Not-determined |
| SS_extracted_k141_24022 | Not-determined |
| SS_extracted_k141_28163 | Low-quality |
| SS_extracted_k141_24025 | Not-determined |
| SS_extracted_k141_77172 | Low-quality |
| SS_extracted_k141_11654 | Not-determined |
| SS_extracted_k141_144546 | Not-determined |
| SS_extracted_k141_11656 | Not-determined |
| SS_extracted_k141_24032 | Not-determined |
| SS_extracted_k141_144554 | Not-determined |
| SS_extracted_k141_72406 | Not-determined |
| SS_extracted_k141_90769 | Not-determined |
| SS_extracted_k141_24041 | Not-determined |
| SS_extracted_k141_41537 | Not-determined |
| SS_extracted_k141_82608 | Not-determined |
| SS_extracted_k141_72409 | Not-determined |
| SS_extracted_k141_41545 | Not-determined |
| SS_extracted_k141_72414 | Low-quality |
| SS_extracted_k141_82614 | Low-quality |
| SS_extracted_k141_72417 | Not-determined |
| SS_extracted_k141_90778 | Not-determined |
| SS_extracted_k141_24055 | Not-determined |
| SS_extracted_k141_82619 | Not-determined |
| SS_extracted_k141_24061 | Not-determined |
| SS_extracted_k141_28182 | Not-determined |
| SS_extracted_k141_66189 | Not-determined |
| SS_extracted_k141_144569 | Not-determined |
| SS_extracted_k141_72437 | Low-quality |
| SS_extracted_k141_72441 | Low-quality |
| SS_extracted_k141_77208 | Not-determined |
| SS_extracted_k141_77209 | Not-determined |
| SS_extracted_k141_28192 | Not-determined |
| SS_extracted_k141_90782 | Not-determined |
| SS_extracted_k141_41557 | Not-determined |
| SS_extracted_k141_58041 | Not-determined |
| SS_extracted_k141_28197 | Not-determined |
| SS_extracted_k141_28198 | Not-determined |
| SS_extracted_k141_72447 | Not-determined |
| SS_extracted_k141_24069 | Not-determined |
| SS_extracted_k141_28202 | Not-determined |
| SS_extracted_k141_77224 | Not-determined |
| SS_extracted_k141_28205 | Not-determined |
| SS_extracted_k141_28206 | Not-determined |
| SS_extracted_k141_77231 | Not-determined |
| SS_extracted_k141_24075 | Not-determined |
| SS_extracted_k141_41573 | Not-determined |
| SS_extracted_k141_24078 | Low-quality |
| SS_extracted_k141_77238 | Not-determined |
| SS_extracted_k141_24081 | Not-determined |
| SS_extracted_k141_90802 | Not-determined |
| SS_extracted_k141_66203 | Not-determined |
| SS_extracted_k141_82632 | Not-determined |
| SS_extracted_k141_101275 | Not-determined |
| SS_extracted_k141_137802 | Not-determined |
| SS_extracted_k141_137803 | Low-quality |
| SS_extracted_k141_58071 | Not-determined |
| SS_extracted_k141_137805 | Not-determined |
| SS_extracted_k141_28244 | Not-determined |
| SS_extracted_k141_144626 | Not-determined |
| SS_extracted_k141_137814 | Not-determined |
| SS_extracted_k141_137817 | Not-determined |
| SS_extracted_k141_28250 | Not-determined |
| SS_extracted_k141_144638 | Not-determined |
| SS_extracted_k141_48436 | Not-determined |
| SS_extracted_k141_137822 | Low-quality |
| SS_extracted_k141_58088 | Not-determined |
| SS_extracted_k141_82660 | Not-determined |
| SS_extracted_k141_41607 | Not-determined |
| SS_extracted_k141_41611 | Not-determined |
| SS_extracted_k141_58099 | Low-quality |
| SS_extracted_k141_101300 | Not-determined |
| SS_extracted_k141_48455 | Not-determined |
| SS_extracted_k141_72482 | Not-determined |
| SS_extracted_k141_58103 | Low-quality |
| SS_extracted_k141_137835 | Not-determined |
| SS_extracted_k141_137838 | Not-determined |
| SS_extracted_k141_41620 | Low-quality |
| SS_extracted_k141_48459 | Low-quality |
| SS_extracted_k141_72483 | Not-determined |
| SS_extracted_k141_77264 | Low-quality |
| SS_extracted_k141_82672 | Not-determined |
| SS_extracted_k141_41623 | Not-determined |
| SS_extracted_k141_41629 | Not-determined |
| SS_extracted_k141_82675 | Not-determined |
| SS_extracted_k141_82676 | Low-quality |
| SS_extracted_k141_41637 | Not-determined |
| SS_extracted_k141_82678 | Not-determined |
| SS_extracted_k141_90815 | Not-determined |
| SS_extracted_k141_41652 | Low-quality |
| SS_extracted_k141_144645 | Not-determined |
| SS_extracted_k141_41656 | Low-quality |
| SS_extracted_k141_72521 | Not-determined |
| SS_extracted_k141_137846 | Not-determined |
| SS_extracted_k141_24100 | Not-determined |
| SS_extracted_k141_41660 | Not-determined |
| SS_extracted_k141_66239 | Not-determined |
| SS_extracted_k141_66240 | Not-determined |
| SS_extracted_k141_90832 | Not-determined |
| SS_extracted_k141_66241 | Not-determined |
| SS_extracted_k141_72527 | Not-determined |
| SS_extracted_k141_144653 | Not-determined |
| SS_extracted_k141_48466 | Not-determined |
| SS_extracted_k141_144662 | Not-determined |
| SS_extracted_k141_77269 | Not-determined |
| SS_extracted_k141_82692 | Low-quality |
| SS_extracted_k141_28272 | Not-determined |
| SS_extracted_k141_144669 | Not-determined |
| SS_extracted_k141_77277 | Not-determined |
| SS_extracted_k141_82697 | Low-quality |
| SS_extracted_k141_144673 | Not-determined |
| SS_extracted_k141_24117 | Not-determined |
| SS_extracted_k141_66265 | Not-determined |
| SS_extracted_k141_24119 | Not-determined |
| SS_extracted_k141_66269 | Not-determined |
| SS_extracted_k141_137858 | Not-determined |
| SS_extracted_k141_66278 | Not-determined |
| SS_extracted_k141_137866 | Not-determined |
| SS_extracted_k141_101323 | Not-determined |
| SS_extracted_k141_24126 | Low-quality |
| SS_extracted_k141_66286 | Not-determined |
| SS_extracted_k141_90859 | Not-determined |
| SS_extracted_k141_66293 | Not-determined |
| SS_extracted_k141_90865 | Low-quality |
| SS_extracted_k141_90869 | Not-determined |
| SS_extracted_k141_24154 | Not-determined |
| SS_extracted_k141_66308 | Not-determined |
| SS_extracted_k141_48490 | Low-quality |
| SS_extracted_k141_90880 | Not-determined |
| SS_extracted_k141_66309 | Low-quality |
| SS_extracted_k141_137878 | Not-determined |
| SS_extracted_k141_58137 | Not-determined |
| SS_extracted_k141_24161 | Not-determined |
| SS_extracted_k141_58142 | Not-determined |
| SS_extracted_k141_66314 | Not-determined |
| SS_extracted_k141_66316 | Not-determined |
| SS_extracted_k141_48497 | Low-quality |
| SS_extracted_k141_144698 | Not-determined |
| SS_extracted_k141_137884 | Not-determined |
| SS_extracted_k141_48501 | Not-determined |
| SS_extracted_k141_101326 | Low-quality |
| SS_extracted_k141_90893 | Not-determined |
| SS_extracted_k141_82725 | Low-quality |
| SS_extracted_k141_58151 | Low-quality |
| SS_extracted_k141_90906 | Low-quality |
| SS_extracted_k141_144715 | Not-determined |
| SS_extracted_k141_48508 | Low-quality |
| SS_extracted_k141_101335 | Not-determined |
| SS_extracted_k141_144717 | Low-quality |
| SS_extracted_k141_28315 | Not-determined |
| SS_extracted_k141_144720 | Not-determined |
| SS_extracted_k141_28319 | Not-determined |
| SS_extracted_k141_58162 | Low-quality |
| SS_extracted_k141_101347 | Not-determined |
| SS_extracted_k141_144722 | Not-determined |
| SS_extracted_k141_144723 | Not-determined |
| SS_extracted_k141_82733 | Not-determined |
| SS_extracted_k141_144726 | Not-determined |
| SS_extracted_k141_48518 | Not-determined |
| SS_extracted_k141_58165 | Low-quality |
| SS_extracted_k141_90929 | Low-quality |
| SS_extracted_k141_144731 | Not-determined |
| SS_extracted_k141_48531 | Not-determined |
| SS_extracted_k141_24214 | Low-quality |
| SS_extracted_k141_66336 | Low-quality |
| SS_extracted_k141_144741 | Not-determined |
| SS_extracted_k141_41676 | Not-determined |
| SS_extracted_k141_144742 | Low-quality |
| SS_extracted_k141_66337 | Low-quality |
| SS_extracted_k141_41682 | Not-determined |
| SS_extracted_k141_101351 | Not-determined |
| SS_extracted_k141_24229 | Not-determined |
| SS_extracted_k141_48541 | Not-determined |
| SS_extracted_k141_24232 | Not-determined |
| SS_extracted_k141_48545 | Not-determined |
| SS_extracted_k141_82751 | Not-determined |
| SS_extracted_k141_58178 | Low-quality |
| SS_extracted_k141_90942 | Not-determined |
| SS_extracted_k141_101354 | Not-determined |
| SS_extracted_k141_58182 | Not-determined |
| SS_extracted_k141_90945 | Not-determined |
| SS_extracted_k141_72560 | Not-determined |
| SS_extracted_k141_41688 | Low-quality |
| SS_extracted_k141_90949 | Not-determined |
| SS_extracted_k141_90952 | Low-quality |
| SS_extracted_k141_82771 | Low-quality |
| SS_extracted_k141_144744 | Low-quality |
| SS_extracted_k141_41700 | Low-quality |
| SS_extracted_k141_82777 | Not-determined |
| SS_extracted_k141_90968 | Not-determined |
| SS_extracted_k141_58201 | Not-determined |
| SS_extracted_k141_72579 | Not-determined |
| SS_extracted_k141_66358 | Not-determined |
| SS_extracted_k141_90976 | Low-quality |
| SS_extracted_k141_58210 | Not-determined |
| SS_extracted_k141_28334 | Not-determined |
| SS_extracted_k141_90982 | Not-determined |
| SS_extracted_k141_66361 | Not-determined |
| SS_extracted_k141_24240 | Low-quality |
| SS_extracted_k141_90986 | Low-quality |
| SS_extracted_k141_28339 | Not-determined |
| SS_extracted_k141_41716 | Not-determined |
| SS_extracted_k141_24244 | Not-determined |
| SS_extracted_k141_24247 | Low-quality |
| SS_extracted_k141_82794 | Not-determined |
| SS_extracted_k141_90993 | Not-determined |
| SS_extracted_k141_72599 | Not-determined |
| SS_extracted_k141_24267 | Not-determined |
| SS_extracted_k141_41732 | Not-determined |
| SS_extracted_k141_101362 | Low-quality |
| SS_extracted_k141_28347 | Not-determined |
| SS_extracted_k141_58232 | Not-determined |
| SS_extracted_k141_58233 | Not-determined |
| SS_extracted_k141_144755 | Low-quality |
| SS_extracted_k141_28354 | Low-quality |
| SS_extracted_k141_82815 | Not-determined |
| SS_extracted_k141_101373 | Not-determined |
| SS_extracted_k141_82817 | Not-determined |
| SS_extracted_k141_58241 | Low-quality |
| SS_extracted_k141_82819 | Not-determined |
| SS_extracted_k141_101380 | Not-determined |
| SS_extracted_k141_101381 | Low-quality |
| SS_extracted_k141_66370 | Not-determined |
| SS_extracted_k141_28368 | Low-quality |
| SS_extracted_k141_91023 | Not-determined |
| SS_extracted_k141_28372 | Not-determined |
| SS_extracted_k141_28378 | Not-determined |
| SS_extracted_k141_41756 | Not-determined |
| SS_extracted_k141_82831 | Low-quality |
| SS_extracted_k141_144768 | Low-quality |
| SS_extracted_k141_82832 | Not-determined |
| SS_extracted_k141_91027 | Not-determined |
| SS_extracted_k141_41765 | Not-determined |
| SS_extracted_k141_58265 | Not-determined |
| SS_extracted_k141_28384 | Not-determined |
| SS_extracted_k141_28389 | Not-determined |
| SS_extracted_k141_72639 | Low-quality |
| SS_extracted_k141_82838 | Not-determined |
| SS_extracted_k141_58272 | Not-determined |
| SS_extracted_k141_144788 | Not-determined |
| SS_extracted_k141_28392 | Not-determined |
| SS_extracted_k141_72643 | Not-determined |
| SS_extracted_k141_72647 | Low-quality |
| SS_extracted_k141_82846 | Low-quality |
| SS_extracted_k141_28397 | Low-quality |
| SS_extracted_k141_41778 | Not-determined |
| SS_extracted_k141_41785 | Not-determined |
| SS_extracted_k141_66387 | Not-determined |
| SS_extracted_k141_144800 | Not-determined |
| SS_extracted_k141_28415 | Not-determined |
| SS_extracted_k141_58279 | Not-determined |
| SS_extracted_k141_58282 | Not-determined |
| SS_extracted_k141_41795 | Not-determined |
| SS_extracted_k141_41799 | Not-determined |

| | |
|---|---|
| S5_extracted_k141_41808 | Not-determined |
| S5_extracted_k141_28429 | Low-quality |
| S5_extracted_k141_28436 | Not-determined |
| S5_extracted_k141_137925 | Not-determined |
| S5_extracted_k141_72671 | Not-determined |
| S5_extracted_k141_72672 | Not-determined |
| S5_extracted_k141_72674 | Not-determined |
| S5_extracted_k141_72679 | Not-determined |
| S5_extracted_k141_41840 | Not-determined |
| S5_extracted_k141_82904 | Low-quality |
| S5_extracted_k141_58286 | Not-determined |
| S5_extracted_k141_28461 | Not-determined |
| S5_extracted_k141_41853 | Not-determined |
| S5_extracted_k141_72687 | Not-determined |
| S5_extracted_k141_101421 | Not-determined |
| S5_extracted_k141_28471 | Not-determined |
| S5_extracted_k141_82910 | Not-determined |
| S5_extracted_k141_72694 | Not-determined |
| S5_extracted_k141_82913 | Low-quality |
| S5_extracted_k141_66397 | Not-determined |
| S5_extracted_k141_41872 | Not-determined |
| S5_extracted_k141_28488 | Not-determined |
| S5_extracted_k141_77288 | Not-determined |
| S5_extracted_k141_82921 | Not-determined |
| S5_extracted_k141_41882 | Not-determined |
| S5_extracted_k141_137935 | Not-determined |
| S5_extracted_k141_41888 | Low-quality |
| S5_extracted_k141_28510 | Not-determined |
| S5_extracted_k141_66406 | Not-determined |
| S5_extracted_k141_66409 | Low-quality |
| S5_extracted_k141_72722 | Not-determined |
| S5_extracted_k141_144833 | Not-determined |
| S5_extracted_k141_58305 | Not-determined |
| S5_extracted_k141_137950 | Not-determined |
| S5_extracted_k141_82951 | Low-quality |
| S5_extracted_k141_77297 | Not-determined |
| S5_extracted_k141_72743 | Not-determined |
| S5_extracted_k141_58324 | Not-determined |
| S5_extracted_k141_72750 | Not-determined |
| S5_extracted_k141_144861 | Low-quality |
| S5_extracted_k141_144862 | Not-determined |
| S5_extracted_k141_66452 | Not-determined |
| S5_extracted_k141_72760 | Not-determined |
| S5_extracted_k141_28565 | Not-determined |
| S5_extracted_k141_77321 | Not-determined |
| S5_extracted_k141_72773 | Not-determined |
| S5_extracted_k141_82973 | Not-determined |
| S5_extracted_k141_41920 | Low-quality |
| S5_extracted_k141_41922 | Not-determined |
| S5_extracted_k141_66472 | Not-determined |
| S5_extracted_k141_77325 | Not-determined |
| S5_extracted_k141_41927 | Not-determined |
| S5_extracted_k141_41941 | Not-determined |
| S5_extracted_k141_77339 | Not-determined |
| S5_extracted_k141_66490 | Not-determined |
| S5_extracted_k141_77344 | Not-determined |
| S5_extracted_k141_66496 | Low-quality |
| S5_extracted_k141_137984 | Not-determined |
| S5_extracted_k141_144910 | Not-determined |
| S5_extracted_k141_137990 | Not-determined |
| S5_extracted_k141_41945 | Not-determined |
| S5_extracted_k141_144915 | Not-determined |
| S5_extracted_k141_66511 | Not-determined |
| S5_extracted_k141_66513 | Not-determined |
| S5_extracted_k141_144922 | Not-determined |
| S5_extracted_k141_144924 | Not-determined |
| S5_extracted_k141_144925 | Low-quality |
| S5_extracted_k141_144926 | Not-determined |
| S5_extracted_k141_144927 | Not-determined |
| S5_extracted_k141_28590 | Not-determined |
| S5_extracted_k141_144932 | Not-determined |
| S5_extracted_k141_28595 | Not-determined |
| S5_extracted_k141_144938 | Low-quality |
| S5_extracted_k141_144945 | Not-determined |
| S5_extracted_k141_83018 | Not-determined |
| S5_extracted_k141_58358 | Not-determined |
| S5_extracted_k141_41968 | Not-determined |
| S5_extracted_k141_83023 | Not-determined |
| S5_extracted_k141_83026 | Not-determined |
| S5_extracted_k141_77412 | Not-determined |
| S5_extracted_k141_144954 | Not-determined |
| S5_extracted_k141_77418 | Not-determined |
| S5_extracted_k141_58385 | Not-determined |
| S5_extracted_k141_144964 | Not-determined |
| S5_extracted_k141_58388 | Low-quality |
| S5_extracted_k141_58389 | Not-determined |
| S5_extracted_k141_58390 | Not-determined |
| S5_extracted_k141_77428 | Not-determined |
| S5_extracted_k141_28609 | Not-determined |
| S5_extracted_k141_41985 | Low-quality |
| S5_extracted_k141_41988 | Not-determined |
| S5_extracted_k141_83073 | Low-quality |
| S5_extracted_k141_41997 | Low-quality |
| S5_extracted_k141_42007 | Not-determined |
| S5_extracted_k141_42012 | Not-determined |
| S5_extracted_k141_42013 | Not-determined |
| S5_extracted_k141_83084 | Not-determined |
| S5_extracted_k141_83091 | Not-determined |
| S5_extracted_k141_72784 | Not-determined |
| S5_extracted_k141_72786 | Not-determined |
| S5_extracted_k141_77440 | Not-determined |
| S5_extracted_k141_42043 | Not-determined |
| S5_extracted_k141_58424 | Not-determined |
| S5_extracted_k141_72790 | Not-determined |
| S5_extracted_k141_83113 | Low-quality |
| S5_extracted_k141_28629 | Not-determined |
| S5_extracted_k141_72803 | Low-quality |
| S5_extracted_k141_77459 | Not-determined |
| S5_extracted_k141_72811 | Not-determined |
| S5_extracted_k141_28640 | Not-determined |
| S5_extracted_k141_42075 | Not-determined |
| S5_extracted_k141_77471 | Low-quality |
| S5_extracted_k141_77472 | Not-determined |
| S5_extracted_k141_28647 | Not-determined |
| S5_extracted_k141_42076 | Not-determined |
| S5_extracted_k141_72821 | Not-determined |
| S5_extracted_k141_77476 | Not-determined |
| S5_extracted_k141_42083 | Low-quality |
| S5_extracted_k141_42086 | Not-determined |
| S5_extracted_k141_42088 | Not-determined |
| S5_extracted_k141_58428 | Low-quality |
| S5_extracted_k141_83161 | Not-determined |
| S5_extracted_k141_42103 | Not-determined |
| S5_extracted_k141_28653 | Not-determined |
| S5_extracted_k141_83172 | Not-determined |
| S5_extracted_k141_83174 | Low-quality |
| S5_extracted_k141_83177 | Not-determined |
| S5_extracted_k141_42121 | Not-determined |
| S5_extracted_k141_28664 | Not-determined |
| S5_extracted_k141_83182 | Not-determined |
| S5_extracted_k141_58446 | Not-determined |
| S5_extracted_k141_72855 | Not-determined |
| S5_extracted_k141_83188 | Low-quality |
| S5_extracted_k141_58453 | Not-determined |
| S5_extracted_k141_58454 | Not-determined |
| S5_extracted_k141_83202 | Low-quality |
| S5_extracted_k141_83204 | Low-quality |
| S5_extracted_k141_83215 | Low-quality |
| S5_extracted_k141_58477 | Low-quality |
| S5_extracted_k141_77512 | Not-determined |
| S5_extracted_k141_83220 | Not-determined |
| S5_extracted_k141_77521 | Not-determined |
| S5_extracted_k141_145005 | Not-determined |
| S5_extracted_k141_83224 | Low-quality |
| S5_extracted_k141_145013 | Low-quality |
| S5_extracted_k141_58510 | Not-determined |
| S5_extracted_k141_77548 | Not-determined |
| S5_extracted_k141_77562 | Not-determined |
| S5_extracted_k141_58531 | Not-determined |
| S5_extracted_k141_77572 | Not-determined |
| S5_extracted_k141_77585 | Not-determined |
| S5_extracted_k141_77586 | Not-determined |
| S5_extracted_k141_77587 | Not-determined |
| S5_extracted_k141_77589 | Not-determined |
| S5_extracted_k141_42135 | Not-determined |
| S5_extracted_k141_42151 | Not-determined |
| S5_extracted_k141_42156 | Not-determined |
| S5_extracted_k141_28695 | Not-determined |
| S5_extracted_k141_66527 | Low-quality |
| S5_extracted_k141_42163 | Not-determined |
| S5_extracted_k141_58572 | Not-determined |
| S5_extracted_k141_137998 | Not-determined |
| S5_extracted_k141_145033 | Not-determined |
| S5_extracted_k141_58586 | Not-determined |
| S5_extracted_k141_83260 | Not-determined |
| S5_extracted_k141_145041 | Low-quality |
| S5_extracted_k141_145044 | Not-determined |
| S5_extracted_k141_138025 | Not-determined |
| S5_extracted_k141_138027 | Not-determined |
| S5_extracted_k141_83271 | Not-determined |
| S5_extracted_k141_58596 | Not-determined |
| S5_extracted_k141_42176 | Not-determined |
| S5_extracted_k141_42177 | Not-determined |
| S5_extracted_k141_42181 | Not-determined |
| S5_extracted_k141_42188 | Not-determined |
| S5_extracted_k141_28719 | Not-determined |
| S5_extracted_k141_28722 | Not-determined |
| S5_extracted_k141_83287 | Not-determined |
| S5_extracted_k141_83295 | Not-determined |
| S5_extracted_k141_42203 | Not-determined |
| S5_extracted_k141_138034 | Not-determined |
| S5_extracted_k141_42212 | Low-quality |
| S5_extracted_k141_42214 | Low-quality |
| S5_extracted_k141_83302 | Not-determined |
| S5_extracted_k141_83304 | Not-determined |
| S5_extracted_k141_145055 | Not-determined |
| S5_extracted_k141_42229 | Not-determined |
| S5_extracted_k141_145068 | Not-determined |
| S5_extracted_k141_145069 | Low-quality |
| S5_extracted_k141_42239 | Not-determined |
| S5_extracted_k141_83323 | Not-determined |
| S5_extracted_k141_138039 | Low-quality |
| S5_extracted_k141_138041 | Not-determined |
| S5_extracted_k141_42243 | Not-determined |
| S5_extracted_k141_42247 | Not-determined |
| S5_extracted_k141_28736 | Not-determined |
| S5_extracted_k141_28737 | Not-determined |
| S5_extracted_k141_138063 | Low-quality |
| S5_extracted_k141_145083 | Not-determined |
| S5_extracted_k141_145084 | Not-determined |
| S5_extracted_k141_101438 | Not-determined |
| S5_extracted_k141_42258 | Not-determined |
| S5_extracted_k141_138077 | Not-determined |
| S5_extracted_k141_145100 | Not-determined |
| S5_extracted_k141_58662 | Not-determined |
| S5_extracted_k141_101457 | Not-determined |
| S5_extracted_k141_58673 | Not-determined |
| S5_extracted_k141_58674 | Not-determined |
| S5_extracted_k141_42271 | Not-determined |
| S5_extracted_k141_66553 | Not-determined |
| S5_extracted_k141_42274 | Low-quality |
| S5_extracted_k141_145129 | Not-determined |
| S5_extracted_k141_42277 | Not-determined |
| S5_extracted_k141_138093 | Not-determined |
| S5_extracted_k141_138094 | Not-determined |
| S5_extracted_k141_66557 | Not-determined |
| S5_extracted_k141_42284 | Low-quality |
| S5_extracted_k141_101470 | Not-determined |
| S5_extracted_k141_77594 | Not-determined |
| S5_extracted_k141_101472 | Not-determined |
| S5_extracted_k141_42288 | Low-quality |
| S5_extracted_k141_42291 | Not-determined |
| S5_extracted_k141_145151 | Not-determined |
| S5_extracted_k141_42300 | Not-determined |
| S5_extracted_k141_101484 | Low-quality |
| S5_extracted_k141_83390 | Low-quality |
| S5_extracted_k141_138119 | Low-quality |
| S5_extracted_k141_42311 | Not-determined |
| S5_extracted_k141_66594 | Not-determined |
| S5_extracted_k141_77611 | Not-determined |
| S5_extracted_k141_77617 | Low-quality |
| S5_extracted_k141_66600 | Low-quality |
| S5_extracted_k141_58699 | Not-determined |
| S5_extracted_k141_28779 | Not-determined |
| S5_extracted_k141_138131 | Not-determined |
| S5_extracted_k141_101494 | Not-determined |
| S5_extracted_k141_101497 | Not-determined |
| S5_extracted_k141_101498 | Low-quality |
| S5_extracted_k141_66617 | Not-determined |
| S5_extracted_k141_58731 | Not-determined |
| S5_extracted_k141_83409 | Not-determined |
| S5_extracted_k141_42352 | Low-quality |
| S5_extracted_k141_42359 | Low-quality |
| S5_extracted_k141_28799 | Not-determined |
| S5_extracted_k141_42369 | Not-determined |
| S5_extracted_k141_66663 | Not-determined |
| S5_extracted_k141_42376 | Not-determined |
| S5_extracted_k141_83428 | Not-determined |
| S5_extracted_k141_42377 | Not-determined |
| S5_extracted_k141_83431 | Not-determined |
| S5_extracted_k141_138159 | Not-determined |
| S5_extracted_k141_11665 | Not-determined |
| S5_extracted_k141_11667 | Not-determined |
| S5_extracted_k141_138175 | Not-determined |
| S5_extracted_k141_138177 | Low-quality |
| S5_extracted_k141_42406 | Not-determined |
| S5_extracted_k141_42410 | Not-determined |
| S5_extracted_k141_66692 | Not-determined |
| S5_extracted_k141_83453 | Not-determined |
| S5_extracted_k141_77679 | Not-determined |
| S5_extracted_k141_66698 | Not-determined |
| S5_extracted_k141_77680 | Not-determined |
| S5_extracted_k141_28818 | Not-determined |
| S5_extracted_k141_42420 | Not-determined |
| S5_extracted_k141_42425 | Not-determined |
| S5_extracted_k141_66706 | Not-determined |
| S5_extracted_k141_42430 | Not-determined |
| S5_extracted_k141_28827 | Not-determined |
| S5_extracted_k141_138207 | Not-determined |
| S5_extracted_k141_138208 | Not-determined |
| S5_extracted_k141_138221 | Not-determined |
| S5_extracted_k141_66728 | Not-determined |
| S5_extracted_k141_145230 | Low-quality |
| S5_extracted_k141_66732 | Low-quality |
| S5_extracted_k141_101507 | Not-determined |
| S5_extracted_k141_145237 | Low-quality |
| S5_extracted_k141_145238 | Not-determined |
| S5_extracted_k141_145245 | Not-determined |
| S5_extracted_k141_83473 | Not-determined |
| S5_extracted_k141_42456 | Not-determined |
| S5_extracted_k141_42457 | Not-determined |
| S8_vs_k141_88013\|\|full | High-quality |
| S5_extracted_k141_66763 | Not-determined |
| S5_extracted_k141_83491 | Not-determined |
| S5_extracted_k141_83522 | Not-determined |
| S5_extracted_k141_11688 | Not-determined |
| S5_extracted_k141_66783 | Not-determined |
| S5_extracted_k141_66784 | Not-determined |
| S5_extracted_k141_145259 | Not-determined |
| S5_extracted_k141_42464 | Not-determined |
| S5_extracted_k141_145262 | Not-determined |
| S5_extracted_k141_145265 | Not-determined |
| S5_extracted_k141_145271 | Not-determined |
| S5_extracted_k141_145273 | Not-determined |
| S5_extracted_k141_145275 | Not-determined |
| S5_extracted_k141_28874 | Not-determined |
| S5_extracted_k141_101531 | Not-determined |
| S5_extracted_k141_83562 | Not-determined |
| S5_extracted_k141_58807 | Not-determined |
| S5_extracted_k141_138272 | Not-determined |
| S5_extracted_k141_145290 | Not-determined |
| S5_extracted_k141_42483 | Not-determined |
| S5_extracted_k141_42484 | Not-determined |
| S5_extracted_k141_58813 | Low-quality |
| S5_extracted_k141_28885 | Not-determined |
| S5_extracted_k141_145296 | Not-determined |
| S5_extracted_k141_11697 | Not-determined |
| S5_extracted_k141_145298 | Low-quality |
| S5_extracted_k141_138281 | Not-determined |
| S5_extracted_k141_28894 | Low-quality |
| S5_extracted_k141_138284 | Low-quality |
| S5_extracted_k141_138286 | Low-quality |
| S5_extracted_k141_11704 | Not-determined |
| S5_extracted_k141_42494 | Low-quality |
| S5_extracted_k141_42495 | Not-determined |
| S5_extracted_k141_83583 | Not-determined |
| S5_extracted_k141_58829 | Not-determined |
| S5_extracted_k141_58831 | Not-determined |
| S5_extracted_k141_83601 | Not-determined |
| S5_extracted_k141_145329 | Not-determined |
| S5_extracted_k141_138304 | Not-determined |
| S5_extracted_k141_58840 | Low-quality |
| S5_extracted_k141_83610 | Not-determined |
| S5_extracted_k141_58841 | Low-quality |
| S5_extracted_k141_138313 | Not-determined |
| S5_extracted_k141_28909 | Low-quality |
| S5_extracted_k141_138317 | Not-determined |
| S5_extracted_k141_138324 | Not-determined |
| S5_extracted_k141_138325 | Low-quality |
| S5_extracted_k141_145357 | Not-determined |
| S5_extracted_k141_28911 | Low-quality |
| S5_extracted_k141_145361 | Not-determined |
| S5_extracted_k141_77738 | Low-quality |
| S5_extracted_k141_145363 | Not-determined |
| S5_extracted_k141_145365 | Not-determined |
| S5_extracted_k141_28923 | Not-determined |
| S5_extracted_k141_83633 | Not-determined |
| S5_extracted_k141_145373 | Not-determined |
| S5_extracted_k141_83641 | Not-determined |
| S5_extracted_k141_28930 | Not-determined |
| S5_extracted_k141_28932 | Not-determined |
| S5_extracted_k141_11711 | Not-determined |
| S5_extracted_k141_11712 | Low-quality |
| S5_extracted_k141_58856 | Low-quality |
| S5_extracted_k141_58857 | Not-determined |
| S5_extracted_k141_83661 | Not-determined |
| S5_extracted_k141_83676 | Not-determined |
| S5_extracted_k141_101541 | Not-determined |
| S5_extracted_k141_28954 | Not-determined |
| S5_extracted_k141_77748 | Not-determined |
| S5_extracted_k141_138350 | Not-determined |
| S5_extracted_k141_77755 | Not-determined |
| S5_extracted_k141_28968 | Not-determined |
| S5_extracted_k141_58871 | Not-determined |
| S5_extracted_k141_138355 | Low-quality |
| S5_extracted_k141_77761 | Not-determined |
| S5_extracted_k141_138357 | Low-quality |
| S5_extracted_k141_28971 | Low-quality |
| S5_extracted_k141_11723 | Low-quality |
| S5_extracted_k141_145401 | Not-determined |
| S5_extracted_k141_145403 | Low-quality |
| S5_extracted_k141_77775 | Not-determined |
| S5_extracted_k141_58878 | Low-quality |
| S5_extracted_k141_58880 | Not-determined |
| S5_extracted_k141_138374 | Low-quality |
| S5_extracted_k141_83686 | Not-determined |
| S5_extracted_k141_11736 | Not-determined |
| S5_extracted_k141_145417 | Not-determined |
| S5_extracted_k141_138377 | Low-quality |
| S5_extracted_k141_145427 | Not-determined |
| S5_extracted_k141_101547 | Not-determined |
| S5_extracted_k141_145433 | Not-determined |
| S5_extracted_k141_58889 | Not-determined |
| S5_extracted_k141_83694 | Not-determined |
| S5_extracted_k141_58891 | Not-determined |
| S5_extracted_k141_58895 | Low-quality |
| S5_extracted_k141_11745 | Low-quality |
| S5_extracted_k141_11749 | Not-determined |
| S5_extracted_k141_145445 | Not-determined |
| S5_extracted_k141_138388 | Not-determined |
| S5_extracted_k141_145451 | Not-determined |
| S5_extracted_k141_138398 | Not-determined |
| S5_extracted_k141_138399 | Not-determined |
| S5_extracted_k141_58913 | Not-determined |
| S5_extracted_k141_28994 | Not-determined |
| S5_extracted_k141_101556 | Low-quality |
| S5_extracted_k141_11768 | Not-determined |
| S5_extracted_k141_145481 | Not-determined |
| S5_extracted_k141_101580 | Not-determined |
| S5_extracted_k141_11791 | Not-determined |
| S5_extracted_k141_145487 | Not-determined |
| S5_extracted_k141_145489 | Not-determined |
| S5_extracted_k141_138422 | Low-quality |
| S5_extracted_k141_58931 | Low-quality |
| S5_extracted_k141_138430 | Not-determined |
| S5_extracted_k141_138431 | Not-determined |
| S5_extracted_k141_145507 | Low-quality |
| S5_extracted_k141_29011 | Not-determined |
| S5_extracted_k141_101594 | Not-determined |
| S5_extracted_k141_145510 | Not-determined |
| S5_extracted_k141_145513 | Low-quality |
| S5_extracted_k141_29019 | Low-quality |
| S5_extracted_k141_101602 | Not-determined |
| S5_extracted_k141_58941 | Not-determined |
| S5_extracted_k141_29025 | Low-quality |
| S5_extracted_k141_138447 | Not-determined |
| S5_extracted_k141_101608 | Low-quality |
| S5_extracted_k141_101609 | Not-determined |
| S5_extracted_k141_138464 | Not-determined |
| S5_extracted_k141_138475 | Not-determined |
| S5_extracted_k141_138477 | Low-quality |
| S5_extracted_k141_29041 | Not-determined |
| S5_extracted_k141_101623 | Low-quality |
| S5_extracted_k141_145522 | Not-determined |
| S5_extracted_k141_101625 | Low-quality |
| S5_extracted_k141_145524 | Low-quality |
| S5_extracted_k141_101630 | Not-determined |
| S5_extracted_k141_101631 | Not-determined |
| S5_extracted_k141_138485 | Not-determined |
| S5_extracted_k141_101640 | Not-determined |
| S5_extracted_k141_138490 | Low-quality |
| S5_extracted_k141_83741 | Not-determined |
| S5_extracted_k141_145544 | Not-determined |
| S5_extracted_k141_145545 | Not-determined |
| S5_extracted_k141_101649 | Not-determined |
| S5_extracted_k141_58975 | Not-determined |
| S5_extracted_k141_145557 | Not-determined |
| S5_extracted_k141_29057 | Not-determined |
| S5_extracted_k141_29059 | Not-determined |
| S5_extracted_k141_58982 | Not-determined |
| S5_extracted_k141_83772 | Not-determined |
| S5_extracted_k141_83773 | Not-determined |
| S5_extracted_k141_138502 | Not-determined |
| S5_extracted_k141_11871 | Low-quality |
| S5_extracted_k141_58991 | Not-determined |
| S5_extracted_k141_83779 | Not-determined |
| S5_extracted_k141_83797 | Not-determined |
| S5_extracted_k141_29068 | Low-quality |
| S5_extracted_k141_29072 | Not-determined |
| S5_extracted_k141_83843 | Not-determined |
| S5_extracted_k141_83844 | Low-quality |
| S5_extracted_k141_83851 | Not-determined |
| S5_extracted_k141_29094 | Low-quality |
| S5_extracted_k141_145571 | Not-determined |
| S5_extracted_k141_145575 | Not-determined |
| S5_extracted_k141_101653 | Low-quality |
| S5_extracted_k141_101656 | Not-determined |
| S5_extracted_k141_101658 | Not-determined |
| S5_extracted_k141_101664 | Not-determined |
| S5_extracted_k141_59043 | Not-determined |
| S5_extracted_k141_101666 | Low-quality |
| S5_extracted_k141_138527 | Not-determined |
| S5_extracted_k141_101674 | Not-determined |
| S5_extracted_k141_59055 | Not-determined |
| S5_extracted_k141_101676 | Not-determined |
| S5_extracted_k141_59059 | Not-determined |
| S5_extracted_k141_138563 | Low-quality |
| S5_extracted_k141_59064 | Low-quality |
| S5_extracted_k141_59065 | Not-determined |
| S5_extracted_k141_29126 | Not-determined |
| S5_extracted_k141_138571 | Not-determined |
| S5_extracted_k141_101683 | Not-determined |
| S5_extracted_k141_138576 | Not-determined |
| S5_extracted_k141_83876 | Not-determined |
| S5_extracted_k141_59076 | Not-determined |
| S5_extracted_k141_59077 | Not-determined |
| S5_extracted_k141_101695 | Low-quality |
| S5_extracted_k141_101697 | Not-determined |
| S5_extracted_k141_59111 | Not-determined |
| S5_extracted_k141_101699 | Not-determined |
| S5_extracted_k141_59127 | Not-determined |
| S5_extracted_k141_101702 | Low-quality |
| S5_extracted_k141_101705 | Low-quality |
| S5_extracted_k141_59140 | Low-quality |
| S5_extracted_k141_59144 | Low-quality |
| S5_extracted_k141_29144 | Low-quality |
| S5_extracted_k141_29151 | Not-determined |
| S5_extracted_k141_59154 | Not-determined |
| S5_extracted_k141_59167 | Low-quality |
| S5_extracted_k141_83912 | Low-quality |
| S5_extracted_k141_59185 | Low-quality |
| S5_extracted_k141_59188 | Not-determined |
| S5_extracted_k141_59190 | Not-determined |
| S5_extracted_k141_59198 | Low-quality |
| S5_extracted_k141_101745 | Not-determined |
| S5_extracted_k141_101769 | Not-determined |
| S5_extracted_k141_101783 | Low-quality |
| S5_extracted_k141_59215 | Not-determined |
| S5_extracted_k141_59216 | Low-quality |
| S5_extracted_k141_59221 | Not-determined |
| S5_extracted_k141_59224 | Low-quality |
| S14_extracted_k141_115736 | Complete |
| S5_extracted_k141_83963 | Low-quality |
| S5_extracted_k141_101799 | Low-quality |
| S5_extracted_k141_83968 | Not-determined |
| S5_extracted_k141_83972 | Not-determined |
| S5_extracted_k141_83973 | Not-determined |
| S5_extracted_k141_83983 | Not-determined |
| S5_extracted_k141_59249 | Not-determined |
| S5_extracted_k141_101811 | Not-determined |
| S5_extracted_k141_59264 | Not-determined |
| S5_extracted_k141_29211 | Not-determined |
| S5_extracted_k141_59267 | Not-determined |
| S5_extracted_k141_101815 | Low-quality |
| S5_extracted_k141_84003 | Not-determined |
| S5_extracted_k141_77851 | Not-determined |
| S5_extracted_k141_101826 | Not-determined |
| S5_extracted_k141_29222 | Not-determined |
| S5_extracted_k141_29223 | Not-determined |
| S5_extracted_k141_59287 | Not-determined |
| S5_extracted_k141_77862 | Low-quality |
| S5_extracted_k141_77866 | Low-quality |
| S5_extracted_k141_29232 | Not-determined |
| S5_extracted_k141_59290 | Low-quality |
| S5_extracted_k141_84035 | Not-determined |
| S5_extracted_k141_84042 | Not-determined |
| S5_extracted_k141_101838 | Not-determined |
| S5_extracted_k141_101840 | Low-quality |
| S5_extracted_k141_101842 | Not-determined |
| S5_extracted_k141_101843 | Not-determined |
| S7_vs_138109\|\|full | High-quality |
| S5_extracted_k141_101850 | Not-determined |
| S5_extracted_k141_84071 | Not-determined |
| S5_extracted_k141_29259 | Not-determined |
| S5_extracted_k141_59308 | Not-determined |
| S5_extracted_k141_84086 | Low-quality |
| S5_extracted_k141_77916 | Not-determined |
| S5_extracted_k141_59321 | Not-determined |
| S4_extracted_31979 | Complete |
| S5_extracted_k141_77919 | Low-quality |
| S5_extracted_k141_101876 | Not-determined |
| S5_extracted_k141_77922 | Not-determined |
| S5_extracted_k141_77925 | Low-quality |
| S5_extracted_k141_77930 | Not-determined |
| S5_extracted_k141_77936 | Not-determined |
| S5_extracted_k141_77942 | Not-determined |
| S5_extracted_k141_77945 | Not-determined |
| S5_extracted_k141_84119 | Not-determined |
| S5_extracted_k141_59353 | Not-determined |
| S5_extracted_k141_11880 | Not-determined |
| S5_extracted_k141_77953 | Not-determined |
| S5_extracted_k141_11881 | Not-determined |
| S5_extracted_k141_77969 | Low-quality |
| S5_extracted_k141_29288 | Not-determined |
| S5_extracted_k141_77970 | Not-determined |
| S5_extracted_k141_29300 | Low-quality |
| S5_extracted_k141_84170 | Not-determined |
| S5_extracted_k141_101897 | Not-determined |
| S5_extracted_k141_101902 | Not-determined |
| S5_extracted_k141_59385 | Not-determined |
| S5_extracted_k141_29317 | Low-quality |
| S5_extracted_k141_77984 | Not-determined |
| S5_extracted_k141_59387 | Not-determined |
| S5_extracted_k141_84193 | Not-determined |
| S5_extracted_k141_59391 | Not-determined |
| S5_extracted_k141_59393 | Not-determined |
| S5_extracted_k141_77999 | Not-determined |
| S5_extracted_k141_101911 | Not-determined |
| S5_extracted_k141_78006 | Not-determined |
| S5_extracted_k141_78009 | Not-determined |
| S5_extracted_k141_78013 | Not-determined |
| S5_extracted_k141_78021 | Not-determined |
| S5_extracted_k141_11937 | Not-determined |
| S5_extracted_k141_59429 | Low-quality |
| S5_extracted_k141_11938 | Low-quality |
| S5_extracted_k141_11944 | Not-determined |
| S5_extracted_k141_78054 | Low-quality |
| S5_extracted_k141_78056 | Not-determined |
| S5_extracted_k141_11951 | Not-determined |
| S5_extracted_k141_11958 | Not-determined |
| S5_extracted_k141_101916 | Not-determined |
| S5_extracted_k141_78077 | Not-determined |
| S8_extracted_k141_117796 | High-quality |
| S8_vs_k141_117796\|\|full | High-quality |
| S5_extracted_k141_59472 | Low-quality |
| S5_extracted_k141_78104 | Not-determined |
| S5_extracted_k141_11968 | Not-determined |
| S5_extracted_k141_78109 | Not-determined |
| S5_extracted_k141_29336 | Not-determined |
| S5_extracted_k141_101926 | Not-determined |
| S5_extracted_k141_78124 | Not-determined |
| S5_extracted_k141_29352 | Not-determined |
| S5_extracted_k141_101936 | Low-quality |
| S5_extracted_k141_78136 | Not-determined |
| S5_extracted_k141_138629 | Not-determined |
| S5_extracted_k141_78142 | Not-determined |
| S5_extracted_k141_11979 | Low-quality |
| S5_extracted_k141_59508 | Not-determined |
| S5_extracted_k141_59512 | Not-determined |

| | |
|---|---|
| S5_extracted_k141_11990 | Not-determined |
| S5_extracted_k141_59514 | Not-determined |
| S5_extracted_k141_101941 | Not-determined |
| S5_extracted_k141_84226 | Not-determined |
| S5_extracted_k141_138657 | Low-quality |
| S5_extracted_k141_84229 | Not-determined |
| S5_extracted_k141_59529 | Low-quality |
| S5_extracted_k141_101955 | Low-quality |
| S5_extracted_k141_101965 | Not-determined |
| S5_extracted_k141_101970 | Not-determined |
| S5_extracted_k141_78189 | Low-quality |
| S5_extracted_k141_84255 | Not-determined |
| S5_extracted_k141_84257 | Low-quality |
| S5_extracted_k141_59557 | Low-quality |
| S5_extracted_k141_78201 | Low-quality |
| S5_extracted_k141_78207 | Not-determined |
| S5_extracted_k141_11996 | Not-determined |
| S5_extracted_k141_78213 | Low-quality |
| S10_vs_89054\|\|full | High-quality |
| S5_extracted_k141_101972 | Not-determined |
| S5_extracted_k141_78219 | Not-determined |
| S5_extracted_k141_84279 | Not-determined |
| S5_extracted_k141_78227 | Not-determined |
| S5_extracted_k141_84287 | Low-quality |
| S5_extracted_k141_138714 | Not-determined |
| S5_extracted_k141_138717 | Not-determined |
| S5_extracted_k141_78230 | Low-quality |
| S5_extracted_k141_12000 | Not-determined |
| S5_extracted_k141_12005 | Not-determined |
| S5_extracted_k141_84302 | Not-determined |
| S5_extracted_k141_12013 | Low-quality |
| S5_extracted_k141_138743 | Not-determined |
| S5_extracted_k141_12020 | Low-quality |
| S5_extracted_k141_138760 | Not-determined |
| S8_extracted_k141_72773 | Complete |
| S5_extracted_k141_138764 | Not-determined |
| S5_extracted_k141_78249 | Not-determined |
| S5_extracted_k141_59618 | Low-quality |
| S5_extracted_k141_102017 | Not-determined |
| S5_extracted_k141_102020 | Not-determined |
| S5_extracted_k141_78268 | Not-determined |
| S5_extracted_k141_138777 | Not-determined |
| S5_extracted_k141_138779 | Not-determined |
| S5_extracted_k141_12039 | Not-determined |
| S5_extracted_k141_138784 | Not-determined |
| S5_extracted_k141_78277 | Low-quality |
| S5_extracted_k141_138789 | Not-determined |
| S5_extracted_k141_102040 | Not-determined |
| S5_extracted_k141_12043 | Not-determined |
| S5_extracted_k141_78286 | Not-determined |
| S5_extracted_k141_78297 | Low-quality |
| S5_extracted_k141_102041 | Not-determined |
| S5_extracted_k141_138808 | Not-determined |
| S5_extracted_k141_138816 | Not-determined |
| S5_extracted_k141_59623 | Not-determined |
| S5_extracted_k141_102047 | Not-determined |
| S5_extracted_k141_59631 | Not-determined |
| S5_extracted_k141_102048 | Not-determined |
| S5_extracted_k141_59632 | Low-quality |
| S14_vs_k141_115736\|\|full | High-quality |
| S5_extracted_k141_78315 | Low-quality |
| S5_extracted_k141_138835 | Not-determined |
| S5_extracted_k141_138838 | Low-quality |
| S5_extracted_k141_78323 | Not-determined |
| S5_extracted_k141_78339 | Not-determined |
| S5_extracted_k141_138846 | Not-determined |
| S4_vs_VIRSorter_31979-circular-cat | High-quality |
| S5_extracted_k141_145589 | Not-determined |
| S5_extracted_k141_138857 | Not-determined |
| S5_extracted_k141_138869 | Not-determined |
| S5_extracted_k141_138873 | Not-determined |
| S5_extracted_k141_12075 | Not-determined |
| S5_extracted_k141_78360 | Low-quality |
| S5_extracted_k141_84318 | Not-determined |
| S5_extracted_k141_78376 | Not-determined |
| S5_extracted_k141_78378 | Low-quality |
| S5_extracted_k141_102074 | Not-determined |
| S5_extracted_k141_78386 | Low-quality |
| S5_extracted_k141_102075 | Low-quality |
| S5_extracted_k141_84326 | Low-quality |
| S5_extracted_k141_78398 | Low-quality |
| S5_extracted_k141_102098 | Low-quality |
| S5_extracted_k141_102107 | Not-determined |
| S5_extracted_k141_12102 | Low-quality |
| S5_extracted_k141_78425 | Low-quality |
| S5_extracted_k141_78430 | Not-determined |
| S5_extracted_k141_78438 | Not-determined |
| S5_extracted_k141_102123 | Not-determined |
| S5_extracted_k141_102125 | Not-determined |
| S5_extracted_k141_138882 | Not-determined |
| S5_extracted_k141_102148 | Low-quality |
| S5_extracted_k141_138907 | Not-determined |
| S5_extracted_k141_84375 | Not-determined |
| S5_extracted_k141_84376 | Not-determined |
| S5_extracted_k141_102166 | Not-determined |
| S5_extracted_k141_102169 | Not-determined |
| S5_extracted_k141_102171 | Not-determined |
| S5_extracted_k141_84396 | Not-determined |
| S5_extracted_k141_102176 | Not-determined |
| S5_extracted_k141_84403 | Not-determined |
| S5_extracted_k141_102181 | Not-determined |
| S5_extracted_k141_138931 | Not-determined |
| S5_extracted_k141_138942 | Not-determined |
| S5_extracted_k141_138943 | Not-determined |
| S5_extracted_k141_78451 | Not-determined |
| S5_extracted_k141_84439 | Not-determined |
| S5_extracted_k141_78461 | Not-determined |
| S5_extracted_k141_138957 | Not-determined |
| S5_extracted_k141_78470 | Not-determined |
| S5_extracted_k141_78476 | Not-determined |
| S5_extracted_k141_78481 | Low-quality |
| S5_extracted_k141_78488 | Not-determined |
| S5_extracted_k141_138964 | Not-determined |
| S5_extracted_k141_59646 | Low-quality |
| S5_extracted_k141_138969 | Low-quality |
| S5_extracted_k141_78503 | Not-determined |
| S5_extracted_k141_59674 | Not-determined |
| S5_extracted_k141_78518 | Not-determined |
| S5_extracted_k141_59675 | Low-quality |
| S5_extracted_k141_78533 | Not-determined |
| S5_extracted_k141_59683 | Not-determined |
| S5_extracted_k141_78568 | Not-determined |
| S5_extracted_k141_59699 | Not-determined |
| S5_extracted_k141_29424 | Not-determined |
| S5_extracted_k141_138986 | Not-determined |
| S5_extracted_k141_29428 | Not-determined |
| S5_extracted_k141_78617 | Not-determined |
| S5_extracted_k141_78619 | Not-determined |
| S5_extracted_k141_29439 | Not-determined |
| S5_extracted_k141_78625 | Not-determined |
| S5_extracted_k141_29442 | Not-determined |
| S5_extracted_k141_78630 | Not-determined |
| S5_extracted_k141_78641 | Not-determined |
| S5_extracted_k141_78645 | Low-quality |
| S5_extracted_k141_29456 | Not-determined |
| S5_extracted_k141_139011 | Not-determined |
| S5_extracted_k141_29463 | Not-determined |
| S5_extracted_k141_29469 | Not-determined |
| S5_extracted_k141_139022 | Not-determined |
| S5_extracted_k141_78666 | Not-determined |
| S5_extracted_k141_78669 | Not-determined |
| S5_extracted_k141_29473 | Not-determined |
| S5_extracted_k141_29474 | Not-determined |
| S5_extracted_k141_59746 | Not-determined |
| S5_extracted_k141_29486 | Not-determined |
| S5_extracted_k141_59751 | Not-determined |
| S5_extracted_k141_29488 | Not-determined |
| S5_extracted_k141_139044 | Not-determined |
| S5_extracted_k141_102194 | Not-determined |
| S5_extracted_k141_139057 | Low-quality |
| S5_extracted_k141_59764 | Not-determined |
| S5_extracted_k141_29501 | Not-determined |
| S5_extracted_k141_59766 | Not-determined |
| S5_extracted_k141_139065 | Low-quality |
| S5_extracted_k141_139072 | Not-determined |
| S5_extracted_k141_29509 | Not-determined |
| S5_extracted_k141_59775 | Low-quality |
| S5_extracted_k141_78681 | Not-determined |
| S5_extracted_k141_78683 | Not-determined |
| S5_extracted_k141_139078 | Not-determined |
| S5_extracted_k141_78692 | Not-determined |
| S5_extracted_k141_29535 | Not-determined |
| S5_extracted_k141_78706 | Not-determined |
| S5_extracted_k141_102243 | Low-quality |
| S5_extracted_k141_102245 | Low-quality |
| S5_extracted_k141_78717 | Not-determined |
| S5_extracted_k141_78726 | Not-determined |
| S5_extracted_k141_102253 | Not-determined |
| S5_extracted_k141_78743 | Not-determined |
| S5_extracted_k141_102262 | Not-determined |
| S5_extracted_k141_102269 | Not-determined |
| S5_extracted_k141_59802 | Not-determined |
| S5_extracted_k141_78790 | Not-determined |
| S5_extracted_k141_78801 | Not-determined |
| S5_extracted_k141_59808 | Low-quality |
| S5_extracted_k141_102293 | Not-determined |
| S5_extracted_k141_29575 | Not-determined |
| S5_extracted_k141_29577 | Not-determined |
| S5_extracted_k141_102300 | Not-determined |
| S5_extracted_k141_102302 | Not-determined |
| S5_extracted_k141_59813 | Not-determined |
| S5_extracted_k141_59814 | Not-determined |
| S5_extracted_k141_78825 | Not-determined |
| S5_extracted_k141_102318 | Low-quality |
| S5_extracted_k141_78835 | Not-determined |
| S5_extracted_k141_102335 | Low-quality |
| S5_extracted_k141_29602 | Not-determined |
| S5_extracted_k141_102341 | Not-determined |
| S5_extracted_k141_29617 | Not-determined |
| S5_extracted_k141_102346 | Not-determined |
| S5_extracted_k141_102361 | Low-quality |
| S5_extracted_k141_84553 | Not-determined |
| S5_extracted_k141_59856 | Not-determined |
| S5_extracted_k141_102372 | Not-determined |
| S5_extracted_k141_84565 | Not-determined |
| S5_extracted_k141_84568 | Not-determined |
| S5_extracted_k141_78845 | Not-determined |
| S5_extracted_k141_59864 | Not-determined |
| S5_extracted_k141_102383 | Not-determined |
| S5_extracted_k141_102384 | Not-determined |
| S5_extracted_k141_102386 | Not-determined |
| S5_extracted_k141_78863 | Not-determined |
| S5_extracted_k141_139097 | Not-determined |
| S5_extracted_k141_139098 | Not-determined |
| S5_extracted_k141_139099 | Low-quality |
| S5_extracted_k141_59873 | Not-determined |
| S5_extracted_k141_78866 | Low-quality |
| S5_extracted_k141_78878 | Not-determined |
| S5_extracted_k141_78880 | Not-determined |
| S12_extracted_k141_188826 | Complete |
| S5_extracted_k141_78883 | Not-determined |
| S5_extracted_k141_139108 | Not-determined |
| S5_extracted_k141_139111 | Not-determined |
| S5_extracted_k141_102400 | Low-quality |
| S5_extracted_k141_139114 | Low-quality |
| S5_extracted_k141_139117 | Not-determined |
| S5_extracted_k141_139118 | Low-quality |
| S5_extracted_k141_78893 | Not-determined |
| S5_extracted_k141_84579 | Not-determined |
| S5_extracted_k141_84585 | Not-determined |
| S5_extracted_k141_139126 | Not-determined |
| S5_extracted_k141_84589 | Not-determined |
| S5_extracted_k141_78901 | Low-quality |
| S5_extracted_k141_29688 | Not-determined |
| S5_extracted_k141_84608 | Not-determined |
| S5_extracted_k141_78911 | Not-determined |
| S5_extracted_k141_102413 | Not-determined |
| S5_extracted_k141_29720 | Not-determined |
| S5_extracted_k141_102420 | Not-determined |
| S5_extracted_k141_84630 | Not-determined |
| S5_extracted_k141_84635 | Low-quality |
| S5_extracted_k141_29730 | Not-determined |
| S5_extracted_k141_29734 | Not-determined |
| S5_extracted_k141_59920 | Not-determined |
| S5_extracted_k141_139136 | Not-determined |
| S5_extracted_k141_59935 | Low-quality |
| S5_extracted_k141_84650 | Not-determined |
| S5_extracted_k141_84654 | Not-determined |
| S5_extracted_k141_59947 | Low-quality |
| S5_extracted_k141_59948 | Not-determined |
| S5_extracted_k141_145613 | Not-determined |
| S5_extracted_k141_145619 | Low-quality |
| S5_extracted_k141_102442 | Not-determined |
| S5_extracted_k141_102443 | Not-determined |
| S5_extracted_k141_84663 | Not-determined |
| S5_extracted_k141_59961 | Not-determined |
| S5_extracted_k141_84666 | Not-determined |
| S5_extracted_k141_145623 | Low-quality |
| S5_extracted_k141_84681 | Not-determined |
| S5_extracted_k141_139185 | Not-determined |
| S5_extracted_k141_84684 | Not-determined |
| S5_extracted_k141_145628 | Not-determined |
| S5_extracted_k141_59976 | Not-determined |
| S5_extracted_k141_102465 | Low-quality |
| S5_extracted_k141_59998 | Not-determined |
| S5_extracted_k141_102468 | Not-determined |
| S5_extracted_k141_145638 | Not-determined |
| S5_extracted_k141_60005 | Not-determined |
| S5_extracted_k141_102483 | Not-determined |
| S5_extracted_k141_102484 | Not-determined |
| S5_extracted_k141_145645 | Not-determined |
| S5_extracted_k141_84705 | Low-quality |
| S5_extracted_k141_60015 | Not-determined |
| S5_extracted_k141_145657 | Not-determined |
| S5_extracted_k141_84714 | Not-determined |
| S5_extracted_k141_145664 | Not-determined |
| S5_extracted_k141_102491 | Not-determined |
| S5_extracted_k141_145675 | Not-determined |
| S5_extracted_k141_102494 | Low-quality |
| S5_extracted_k141_60025 | Not-determined |
| S5_extracted_k141_60026 | Not-determined |
| S5_extracted_k141_145686 | Not-determined |
| S5_extracted_k141_145688 | Not-determined |
| S5_extracted_k141_145693 | Not-determined |
| S5_extracted_k141_145701 | Not-determined |
| S5_extracted_k141_60035 | Not-determined |
| S5_extracted_k141_60036 | Low-quality |
| S5_extracted_k141_29843 | Not-determined |
| S5_extracted_k141_84744 | Low-quality |
| S5_extracted_k141_84750 | Not-determined |
| S5_extracted_k141_29847 | Not-determined |
| S5_extracted_k141_84785 | Not-determined |
| S5_extracted_k141_139255 | Not-determined |
| S5_extracted_k141_84791 | Not-determined |
| S5_extracted_k141_84793 | Not-determined |
| S5_extracted_k141_84801 | Not-determined |
| S5_extracted_k141_29860 | Low-quality |
| S5_extracted_k141_102557 | Not-determined |
| S5_extracted_k141_84811 | Not-determined |
| S5_extracted_k141_29868 | Not-determined |
| S5_extracted_k141_60068 | Not-determined |
| S5_extracted_k141_60078 | Not-determined |
| S5_extracted_k141_29885 | Not-determined |
| S5_extracted_k141_84820 | Not-determined |
| S5_extracted_k141_102570 | Not-determined |
| S5_extracted_k141_29899 | Not-determined |
| S5_extracted_k141_29900 | Not-determined |
| S5_extracted_k141_29904 | Not-determined |
| S5_extracted_k141_29905 | Not-determined |
| S5_extracted_k141_84832 | Not-determined |
| S5_extracted_k141_60093 | Low-quality |
| S5_extracted_k141_29910 | Low-quality |
| S5_extracted_k141_84837 | Not-determined |
| S5_extracted_k141_102579 | Not-determined |
| S5_extracted_k141_102581 | Not-determined |
| S5_extracted_k141_102586 | Not-determined |
| S5_extracted_k141_102587 | Not-determined |
| S5_extracted_k141_29922 | Not-determined |
| S5_extracted_k141_102592 | Low-quality |
| S5_extracted_k141_29937 | Not-determined |
| S5_extracted_k141_84841 | Not-determined |
| S5_extracted_k141_102618 | Not-determined |
| S5_extracted_k141_102623 | Not-determined |
| S5_extracted_k141_84855 | Not-determined |
| S5_extracted_k141_84862 | Not-determined |
| S5_extracted_k141_29959 | Low-quality |
| S5_extracted_k141_29964 | Low-quality |
| S5_extracted_k141_29967 | Not-determined |
| S5_extracted_k141_29971 | Low-quality |
| S5_extracted_k141_29980 | Not-determined |
| S5_extracted_k141_30009 | Not-determined |
| S5_extracted_k141_102662 | Not-determined |
| S5_extracted_k141_102683 | Not-determined |
| S5_extracted_k141_84895 | Not-determined |
| S5_extracted_k141_84896 | Not-determined |
| S5_extracted_k141_84902 | Low-quality |
| S5_extracted_k141_60139 | Not-determined |
| S5_extracted_k141_84916 | Low-quality |
| S5_extracted_k141_84918 | Not-determined |
| S5_extracted_k141_84920 | Not-determined |
| S5_extracted_k141_30042 | Not-determined |
| S5_extracted_k141_102698 | Not-determined |
| S5_extracted_k141_84937 | Low-quality |
| S5_extracted_k141_84938 | Not-determined |
| S5_extracted_k141_60147 | Not-determined |
| S5_extracted_k141_102715 | Not-determined |
| S5_extracted_k141_102717 | Not-determined |
| S5_extracted_k141_60155 | Low-quality |
| S5_extracted_k141_102734 | Not-determined |
| S5_extracted_k141_30073 | Not-determined |
| S5_extracted_k141_139293 | Low-quality |
| S5_extracted_k141_30080 | Low-quality |
| S5_extracted_k141_30083 | Not-determined |
| S5_extracted_k141_30092 | Not-determined |
| S5_extracted_k141_60168 | Not-determined |
| S5_extracted_k141_139312 | Not-determined |
| S5_extracted_k141_60177 | Not-determined |
| S5_extracted_k141_102751 | Low-quality |
| S5_extracted_k141_102755 | Not-determined |
| S5_extracted_k141_84954 | Low-quality |
| S5_extracted_k141_60182 | Not-determined |
| S5_extracted_k141_30108 | Not-determined |
| S5_extracted_k141_84960 | Not-determined |
| S5_extracted_k141_84962 | Not-determined |
| S5_extracted_k141_84972 | Not-determined |
| S5_extracted_k141_60215 | Low-quality |
| S5_extracted_k141_102778 | Not-determined |
| S5_extracted_k141_102782 | Low-quality |
| S5_extracted_k141_102794 | Not-determined |
| S5_extracted_k141_30162 | Not-determined |
| S5_extracted_k141_60234 | Not-determined |
| S5_extracted_k141_30168 | Not-determined |
| S5_extracted_k141_30182 | Low-quality |
| S5_extracted_k141_102818 | Low-quality |
| S5_extracted_k141_30189 | Not-determined |
| S5_extracted_k141_102824 | Not-determined |
| S5_extracted_k141_30191 | Not-determined |
| S5_extracted_k141_102827 | Low-quality |
| S5_extracted_k141_30195 | Low-quality |
| S5_extracted_k141_85000 | Not-determined |
| S5_extracted_k141_30205 | Not-determined |
| S5_extracted_k141_30209 | Not-determined |
| S5_extracted_k141_30217 | Not-determined |
| S5_extracted_k141_102852 | Not-determined |
| S5_extracted_k141_102856 | Not-determined |
| S5_extracted_k141_102858 | Not-determined |
| S5_extracted_k141_139344 | Not-determined |
| S5_extracted_k141_30236 | Not-determined |
| S5_extracted_k141_30242 | Low-quality |
| S5_extracted_k141_60243 | Not-determined |
| S5_extracted_k141_102900 | Not-determined |
| S5_extracted_k141_102904 | Not-determined |
| S5_extracted_k141_102905 | Not-determined |
| S5_extracted_k141_102906 | Not-determined |
| S5_extracted_k141_102910 | Not-determined |
| S5_extracted_k141_102914 | Not-determined |
| S5_extracted_k141_60250 | Not-determined |
| S5_extracted_k141_30261 | Not-determined |
| S5_extracted_k141_139356 | Not-determined |
| S5_extracted_k141_60262 | Low-quality |
| S5_extracted_k141_60266 | Not-determined |
| S5_extracted_k141_60273 | Not-determined |
| S5_extracted_k141_60277 | Low-quality |
| S5_extracted_k141_60279 | Not-determined |
| S5_extracted_k141_30285 | Not-determined |
| S5_extracted_k141_30286 | Not-determined |
| S5_extracted_k141_139391 | Not-determined |
| S5_extracted_k141_139394 | Not-determined |
| S5_extracted_k141_60292 | Not-determined |
| S5_extracted_k141_30301 | Low-quality |
| S5_extracted_k141_102972 | Low-quality |
| S5_extracted_k141_30303 | Low-quality |
| S5_extracted_k141_102974 | Low-quality |
| S5_extracted_k141_30307 | Not-determined |
| S5_extracted_k141_102979 | Not-determined |
| S5_extracted_k141_102992 | Not-determined |
| S5_extracted_k141_102998 | Not-determined |
| S5_extracted_k141_60320 | Not-determined |
| S5_extracted_k141_60325 | Not-determined |
| S5_extracted_k141_30328 | Low-quality |
| S5_extracted_k141_30329 | Not-determined |
| S5_extracted_k141_30331 | Low-quality |
| S5_extracted_k141_30332 | Not-determined |
| S5_extracted_k141_60330 | Not-determined |
| S5_extracted_k141_30340 | Not-determined |
| S5_extracted_k141_30343 | Not-determined |
| S5_extracted_k141_60344 | Low-quality |
| S5_extracted_k141_103023 | Not-determined |
| S5_extracted_k141_103029 | Not-determined |
| S5_extracted_k141_60363 | Low-quality |
| S5_extracted_k141_139448 | Not-determined |
| S5_extracted_k141_60371 | Not-determined |
| S5_extracted_k141_60373 | Not-determined |
| S5_extracted_k141_60378 | Low-quality |
| S5_extracted_k141_103057 | Not-determined |
| S5_extracted_k141_103061 | Not-determined |
| S5_extracted_k141_139461 | Low-quality |
| S5_extracted_k141_139464 | Not-determined |
| S5_extracted_k141_139472 | Not-determined |
| S5_extracted_k141_60405 | Low-quality |
| S5_extracted_k141_103096 | Not-determined |
| S5_extracted_k141_60409 | Low-quality |
| S5_extracted_k141_139497 | Not-determined |
| S5_extracted_k141_103105 | Not-determined |
| S5_extracted_k141_139502 | Low-quality |
| S5_extracted_k141_103119 | Not-determined |
| S5_extracted_k141_139506 | Low-quality |
| S5_extracted_k141_139508 | Not-determined |
| S5_extracted_k141_103125 | Not-determined |
| S5_extracted_k141_60415 | Not-determined |
| S5_extracted_k141_139519 | Not-determined |
| S5_extracted_k141_60419 | Not-determined |
| S5_extracted_k141_60422 | Not-determined |
| S5_extracted_k141_103128 | Low-quality |
| S5_extracted_k141_103130 | Low-quality |
| S5_extracted_k141_139536 | Not-determined |
| S5_extracted_k141_139540 | Not-determined |
| S5_extracted_k141_139545 | Not-determined |
| S5_extracted_k141_103152 | Not-determined |
| S5_extracted_k141_103156 | Low-quality |
| S5_extracted_k141_103157 | Not-determined |
| S5_extracted_k141_103164 | Low-quality |
| S5_extracted_k141_139558 | Not-determined |
| S5_extracted_k141_139563 | Not-determined |
| S5_extracted_k141_103173 | Not-determined |
| S5_extracted_k141_103177 | Not-determined |
| S5_extracted_k141_103184 | Low-quality |
| S5_extracted_k141_103186 | Low-quality |
| S5_extracted_k141_103194 | Low-quality |
| S5_extracted_k141_139586 | Low-quality |
| S5_extracted_k141_103201 | Not-determined |
| S5_extracted_k141_139587 | Not-determined |
| S5_extracted_k141_139591 | Not-determined |
| S9_vs_k141_32838\|\|full | High-quality |
| S5_extracted_k141_60474 | Not-determined |
| S5_extracted_k141_139639 | Not-determined |
| S5_extracted_k141_60476 | Low-quality |
| S5_extracted_k141_60486 | Not-determined |
| S5_extracted_k141_60488 | Low-quality |
| S5_extracted_k141_60495 | Not-determined |
| S5_extracted_k141_60496 | Not-determined |
| S5_extracted_k141_60506 | Not-determined |
| S5_extracted_k141_60508 | Not-determined |
| S5_extracted_k141_60510 | Not-determined |
| S5_extracted_k141_60517 | Not-determined |
| S5_extracted_k141_60520 | Not-determined |
| S5_extracted_k141_60521 | Not-determined |
| S5_extracted_k141_60539 | Not-determined |
| S5_extracted_k141_60553 | Not-determined |
| S5_extracted_k141_60581 | Not-determined |
| S5_extracted_k141_60582 | Not-determined |
| S5_extracted_k141_60593 | Not-determined |
| S5_vs_k141_145602\|\|full | High-quality |
| S5_extracted_k141_60599 | Not-determined |
| S5_extracted_k141_60629 | Not-determined |
| S5_extracted_k141_60640 | Not-determined |
| S5_extracted_k141_60652 | Not-determined |
| S5_extracted_k141_60672 | Not-determined |
| S5_extracted_k141_60700 | Low-quality |
| S5_extracted_k141_60712 | Not-determined |
| S5_vs_k141_121506\|\|full | Not-determined |
| S5_vs_k141_60959\|\|full | Not-determined |
| S3_extracted_k141_37073 | High-quality |
| S5_vs_k141_666\|\|full | Not-determined |
| S3_vs_k141_37073\|\|full | High-quality |
| S12_extracted_k141_55175 | Complete |
| S12_extracted_k141_48278 | Complete |
| S3_extracted_k141_127394 | Complete |
| S5_vs_k141_19188\|\|full | Low-quality |
| S8_vs_k141_72773\|\|full | High-quality |
| S4_vs_VIRSorter_15837-cat_2 | High-quality |
| S10_vs_18901\|\|full | High-quality |
| S1_extracted_k141_22149 | High-quality |
| S5_vs_k141_123766\|\|full | Not-determined |
| S12_vs_k141_188826\|\|full | High-quality |
| S5_vs_k141_129343\|\|full | Not-determined |
| S3_vs_k141_50557\|\|full | High-quality |
| S5_vs_k141_118434\|\|full | Not-determined |
| S6_vs_k141_15390\|\|full | Medium-quality |
| S5_vs_k141_130256\|\|full | Low-quality |
| S5_vs_k141_87335\|\|full | Not-determined |
| S5_vs_k141_21164\|\|full | Not-determined |
| S1_vs_k141_22149\|\|full | High-quality |
| S12_vs_k141_55175\|\|full | High-quality |
| S12_vs_k141_48278\|\|full | High-quality |
| S5_vs_k141_39984\|\|full | Not-determined |
| S5_vs_k141_40202\|\|full | Not-determined |
| S3_vs_k141_127394\|\|full | High-quality |
| S5_vs_k141_145601\|\|full | High-quality |
| S5_vs_k141_113616\|\|full | Not-determined |
| S5_vs_k141_35522\|\|full | Not-determined |
| S7_extracted_60968 | Complete |
| S8_extracted_k141_38785 | High-quality |
| S8_vs_k141_38785\|\|full | High-quality |
| S7_vs_36656\|\|full | Medium-quality |
| S7_vs_95276\|\|full | High-quality |
| S7_vs_60968\|\|full | High-quality |
| S3_vs_k141_127421\|\|full | High-quality |
| S7_extracted_134023 | Complete |
| S5_vs_k141_82785\|\|full | Not-determined |
| S8_vs_k141_208113\|\|full | High-quality |
| S11_vs_k141_84848\|\|full | High-quality |
| S12_vs_k141_86928\|\|full | Medium-quality |
| S4_vs_VIRSorter_31916-circular-cat | High-quality |
| S11_vs_k141_68348\|\|full | High-quality |
| S7_vs_134023\|\|full | High-quality |
| S13_extracted_44871 | Complete |
| S1_extracted_k141_22963 | Complete |
| S7_vs_135679\|\|full | High-quality |
| S5_vs_k141_145026\|\|full | High-quality |
| S12_vs_k141_75098\|\|full | Medium-quality |
| S1_extracted_k141_6955 | High-quality |
| S7_vs_27530\|\|full | High-quality |
| S17_vs_k141_167589\|\|full | Medium-quality |
| S5_vs_k141_6478\|\|full | High-quality |
| S5_vs_k141_145594\|\|full | Not-determined |
| S5_vs_k141_145595\|\|full | Low-quality |
| S14_vs_k141_39797\|\|full | High-quality |
| S16_vs_20060\|\|full | High-quality |
| S13_vs_44871\|\|full | High-quality |
| S1_vs_k141_22963\|\|full | High-quality |
| S9_extracted_k141_19284 | Complete |
| S8_vs_k141_3224\|\|full | High-quality |
| S9_vs_k141_37437\|\|full | High-quality |
| S16_vs_75108\|\|full | High-quality |
| S9_vs_k141_19284\|\|full | High-quality |
| S15_vs_35445\|\|full | Medium-quality |
| S8_vs_k141_35308\|\|full | High-quality |
| S5_vs_k141_145612\|\|full | Low-quality |
| S5_vs_k141_145621\|\|full | Not-determined |
| S5_vs_k141_145627\|\|full | Low-quality |
| S7_vs_40259\|\|full | Medium-quality |
| S6_extracted_k141_25710 | High-quality |
| S5_vs_k141_133569\|\|full | Not-determined |
| S5_vs_k141_36438\|\|full | Not-determined |
| S5_vs_k141_121428\|\|full | Low-quality |
| S5_vs_k141_36439\|\|full | Low-quality |
| S5_vs_k141_121435\|\|full | Low-quality |
| S5_vs_k141_6079\|\|full | Low-quality |
| S5_vs_k141_42521\|\|full | Low-quality |
| S5_vs_k141_40\|\|full | Low-quality |
| S5_vs_k141_121438\|\|full | Low-quality |
| S5_vs_k141_36446\|\|full | Low-quality |
| S5_vs_k141_18231\|\|full | Low-quality |
| S5_vs_k141_97162\|\|full | Not-determined |
| S5_vs_k141_54660\|\|full | Low-quality |
| S5_vs_k141_42534\|\|full | Low-quality |
| S5_vs_k141_60751\|\|full | Not-determined |
| S5_vs_k141_42537\|\|full | Low-quality |
| S5_vs_k141_49\|\|full | Not-determined |
| S5_vs_k141_6084\|\|full | Not-determined |
| S5_vs_k141_115367\|\|full | Low-quality |
| S5_vs_k141_18246\|\|full | Low-quality |
| S5_vs_k141_24312\|\|full | Not-determined |
| S5_vs_k141_53\|\|full | Low-quality |
| S5_vs_k141_72867\|\|full | Low-quality |
| S5_vs_k141_48605\|\|full | Not-determined |
| S5_vs_k141_18251\|\|full | Not-determined |
| S5_vs_k141_85031\|\|full | Low-quality |

| | |
|---|---|
| SS_vs_k141_139680\|\|full | Low-quality |
| SS_vs_k141_139684\|\|full | Low-quality |
| SS_vs_k141_69\|\|full | Not-determined |
| SS_vs_k141_91134\|\|full | Not-determined |
| SS_vs_k141_48611\|\|full | Not-determined |
| SS_vs_k141_6091\|\|full | Not-determined |
| SS_vs_k141_139690\|\|full | Low-quality |
| SS_vs_k141_85060\|\|full | Low-quality |
| SS_vs_k141_127521\|\|full | Low-quality |
| SS_vs_k141_72878\|\|full | Low-quality |
| SS_vs_k141_91149\|\|full | Low-quality |
| SS_vs_k141_109324\|\|full | Not-determined |
| SS_vs_k141_60798\|\|full | Not-determined |
| SS_vs_k141_127536\|\|full | Low-quality |
| SS_vs_k141_127539\|\|full | Not-determined |
| SS_vs_k141_12185\|\|full | Not-determined |
| SS_vs_k141_36510\|\|full | Low-quality |
| SS_vs_k141_24324\|\|full | Not-determined |
| SS_vs_k141_18299\|\|full | Not-determined |
| SS_vs_k141_97186\|\|full | Not-determined |
| SS_vs_k141_127546\|\|full | Not-determined |
| SS_vs_k141_12197\|\|full | Low-quality |
| SS_vs_k141_127549\|\|full | Low-quality |
| SS_vs_k141_97188\|\|full | Low-quality |
| SS_vs_k141_6138\|\|full | Low-quality |
| SS_vs_k141_121532\|\|full | Not-determined |
| SS_vs_k141_97194\|\|full | Low-quality |
| SS_vs_k141_6140\|\|full | Low-quality |
| SS_vs_k141_139705\|\|full | Low-quality |
| SS_vs_k141_48655\|\|full | Low-quality |
| SS_vs_k141_48657\|\|full | Low-quality |
| SS_vs_k141_127556\|\|full | Low-quality |
| SS_vs_k141_127558\|\|full | Not-determined |
| SS_vs_k141_42610\|\|full | Low-quality |
| SS_vs_k141_60827\|\|full | Low-quality |
| SS_vs_k141_6145\|\|full | Low-quality |
| SS_vs_k141_6155\|\|full | Not-determined |
| SS_vs_k141_18322\|\|full | Not-determined |
| SS_vs_k141_6172\|\|full | Low-quality |
| SS_vs_k141_18327\|\|full | Low-quality |
| SS_vs_k141_18328\|\|full | Not-determined |
| SS_vs_k141_18331\|\|full | Not-determined |
| SS_vs_k141_103332\|\|full | Not-determined |
| SS_vs_k141_54724\|\|full | Low-quality |
| SS_vs_k141_127576\|\|full | Low-quality |
| SS_vs_k141_133618\|\|full | Not-determined |
| SS_vs_k141_139743\|\|full | Not-determined |
| SS_vs_k141_54728\|\|full | Not-determined |
| SS_vs_k141_60841\|\|full | Low-quality |
| SS_vs_k141_42647\|\|full | Not-determined |
| S6_vs_k141_25710\|\|full | High-quality |
| SS_vs_k141_60846\|\|full | Not-determined |
| SS_vs_k141_36514\|\|full | Not-determined |
| SS_vs_k141_60849\|\|full | Low-quality |
| SS_vs_k141_54734\|\|full | Not-determined |
| SS_vs_k141_24337\|\|full | Low-quality |
| SS_vs_k141_85067\|\|full | Not-determined |
| SS_vs_k141_60859\|\|full | Low-quality |
| SS_vs_k141_60873\|\|full | Low-quality |
| SS_vs_k141_97230\|\|full | Low-quality |
| SS_vs_k141_97232\|\|full | Low-quality |
| SS_vs_k141_72943\|\|full | Low-quality |
| SS_vs_k141_109383\|\|full | Low-quality |
| SS_vs_k141_60882\|\|full | Not-determined |
| SS_vs_k141_6214\|\|full | Low-quality |
| SS_vs_k141_85089\|\|full | Low-quality |
| SS_vs_k141_109392\|\|full | Not-determined |
| SS_vs_k141_103400\|\|full | Low-quality |
| SS_vs_k141_42683\|\|full | Not-determined |
| SS_vs_k141_103404\|\|full | Not-determined |
| SS_vs_k141_54757\|\|full | Low-quality |
| SS_vs_k141_6229\|\|full | Not-determined |
| SS_vs_k141_60905\|\|full | Not-determined |
| SS_vs_k141_97240\|\|full | Low-quality |
| SS_vs_k141_42695\|\|full | Low-quality |
| SS_vs_k141_42698\|\|full | Low-quality |
| SS_vs_k141_78961\|\|full | Not-determined |
| SS_vs_k141_60921\|\|full | Not-determined |
| SS_vs_k141_72962\|\|full | Low-quality |
| SS_vs_k141_97253\|\|full | Not-determined |
| SS_vs_k141_30439\|\|full | Low-quality |
| SS_vs_k141_78972\|\|full | Low-quality |
| SS_vs_k141_127618\|\|full | Not-determined |
| SS_vs_k141_30445\|\|full | Low-quality |
| SS_vs_k141_30449\|\|full | Low-quality |
| SS_vs_k141_103455\|\|full | Low-quality |
| SS_vs_k141_36563\|\|full | Not-determined |
| SS_vs_k141_30453\|\|full | Not-determined |
| SS_vs_k141_48686\|\|full | Low-quality |
| SS_vs_k141_139777\|\|full | Not-determined |
| SS_vs_k141_30457\|\|full | Not-determined |
| SS_vs_k141_66839\|\|full | Not-determined |
| SS_vs_k141_30472\|\|full | Low-quality |
| SS_vs_k141_48696\|\|full | Not-determined |
| SS_vs_k141_109453\|\|full | Not-determined |
| SS_vs_k141_121591\|\|full | Low-quality |
| SS_vs_k141_121592\|\|full | Not-determined |
| SS_vs_k141_30476\|\|full | Low-quality |
| SS_vs_k141_6264\|\|full | Low-quality |
| SS_vs_k141_30480\|\|full | Low-quality |
| SS_vs_k141_97280\|\|full | Low-quality |
| SS_vs_k141_72979\|\|full | Not-determined |
| SS_vs_k141_18353\|\|full | Not-determined |
| SS_vs_k141_54789\|\|full | Not-determined |
| SS_vs_k141_42741\|\|full | Low-quality |
| SS_vs_k141_30484\|\|full | Low-quality |
| SS_vs_k141_60952\|\|full | Not-determined |
| SS_vs_k141_121617\|\|full | Low-quality |
| SS_vs_k141_133649\|\|full | Low-quality |
| SS_vs_k141_18376\|\|full | Not-determined |
| SS_vs_k141_97302\|\|full | Low-quality |
| SS_vs_k141_138\|\|full | Low-quality |
| SS_vs_k141_18378\|\|full | Low-quality |
| SS_vs_k141_79007\|\|full | Not-determined |
| SS_vs_k141_139790\|\|full | Low-quality |
| SS_vs_k141_42781\|\|full | Low-quality |
| SS_vs_k141_66898\|\|full | Low-quality |
| SS_vs_k141_42782\|\|full | Not-determined |
| SS_vs_k141_156\|\|full | Low-quality |
| SS_vs_k141_121642\|\|full | Not-determined |
| SS_vs_k141_85174\|\|full | Not-determined |
| SS_vs_k141_42797\|\|full | Low-quality |
| SS_vs_k141_121655\|\|full | Low-quality |
| SS_vs_k141_97313\|\|full | Low-quality |
| SS_vs_k141_97317\|\|full | Not-determined |
| SS_vs_k141_121664\|\|full | Low-quality |
| SS_vs_k141_139827\|\|full | Low-quality |
| SS_vs_k141_6313\|\|full | Low-quality |
| SS_vs_k141_42811\|\|full | Low-quality |
| SS_vs_k141_54818\|\|full | Low-quality |
| SS_vs_k141_48708\|\|full | Low-quality |
| SS_vs_k141_184\|\|full | Low-quality |
| SS_vs_k141_18443\|\|full | Low-quality |
| SS_vs_k141_54823\|\|full | Low-quality |
| SS_vs_k141_48713\|\|full | Low-quality |
| SS_vs_k141_85197\|\|full | Low-quality |
| SS_vs_k141_79015\|\|full | Low-quality |
| SS_vs_k141_6335\|\|full | Low-quality |
| SS_vs_k141_133665\|\|full | Low-quality |
| SS_vs_k141_103493\|\|full | Low-quality |
| S1_vs_k141_6955\|\|full | High-quality |
| SS_vs_k141_66933\|\|full | Low-quality |
| SS_vs_k141_133678\|\|full | Low-quality |
| SS_vs_k141_211\|\|full | Low-quality |
| SS_vs_k141_213\|\|full | Low-quality |
| SS_vs_k141_12260\|\|full | Low-quality |
| SS_vs_k141_60964\|\|full | Low-quality |
| SS_vs_k141_139866\|\|full | Low-quality |
| SS_vs_k141_103509\|\|full | Not-determined |
| SS_vs_k141_121683\|\|full | Low-quality |
| SS_vs_k141_115423\|\|full | Not-determined |
| SS_vs_k141_60977\|\|full | Low-quality |
| SS_vs_k141_42837\|\|full | Low-quality |
| SS_vs_k141_103522\|\|full | Low-quality |
| SS_vs_k141_139885\|\|full | Not-determined |
| SS_vs_k141_79025\|\|full | Low-quality |
| SS_vs_k141_48738\|\|full | Low-quality |
| SS_vs_k141_103531\|\|full | Low-quality |
| SS_vs_k141_121714\|\|full | Low-quality |
| SS_vs_k141_139897\|\|full | Low-quality |
| S12_vs_k141_188825\|\|full | Medium-quality |
| SS_vs_k141_85260\|\|full | Not-determined |
| SS_vs_k141_139902\|\|full | Low-quality |
| SS_vs_k141_139912\|\|full | Low-quality |
| SS_vs_k141_115448\|\|full | Low-quality |
| SS_vs_k141_79052\|\|full | Low-quality |
| SS_vs_k141_30520\|\|full | Low-quality |
| SS_vs_k141_292\|\|full | Low-quality |
| SS_vs_k141_103542\|\|full | Low-quality |
| SS_vs_k141_73015\|\|full | Low-quality |
| SS_vs_k141_6377\|\|full | Low-quality |
| SS_vs_k141_121753\|\|full | Low-quality |
| SS_vs_k141_115464\|\|full | Not-determined |
| SS_vs_k141_42881\|\|full | Low-quality |
| SS_vs_k141_66966\|\|full | Low-quality |
| SS_vs_k141_322\|\|full | Low-quality |
| SS_vs_k141_18489\|\|full | Low-quality |
| SS_vs_k141_61018\|\|full | Not-determined |
| SS_vs_k141_42889\|\|full | Not-determined |
| SS_vs_k141_139943\|\|full | Low-quality |
| SS_vs_k141_139948\|\|full | Not-determined |
| SS_vs_k141_18510\|\|full | Not-determined |
| SS_vs_k141_42921\|\|full | Low-quality |
| SS_vs_k141_91175\|\|full | Low-quality |
| SS_vs_k141_103594\|\|full | Not-determined |
| SS_vs_k141_42930\|\|full | Low-quality |
| SS_vs_k141_115528\|\|full | Low-quality |
| SS_vs_k141_109484\|\|full | Not-determined |
| SS_vs_k141_115529\|\|full | Low-quality |
| SS_vs_k141_115531\|\|full | Not-determined |
| SS_vs_k141_127726\|\|full | Low-quality |
| SS_vs_k141_79087\|\|full | Low-quality |
| SS_vs_k141_6438\|\|full | Low-quality |
| SS_vs_k141_79094\|\|full | Low-quality |
| SS_vs_k141_109494\|\|full | Low-quality |
| SS_vs_k141_127737\|\|full | Low-quality |
| SS_vs_k141_91196\|\|full | Low-quality |
| SS_vs_k141_18561\|\|full | Low-quality |
| SS_vs_k141_121830\|\|full | Low-quality |
| SS_vs_k141_54852\|\|full | Low-quality |
| SS_vs_k141_342\|\|full | Low-quality |
| SS_vs_k141_115563\|\|full | Low-quality |
| SS_vs_k141_115566\|\|full | Low-quality |
| SS_vs_k141_103646\|\|full | Low-quality |
| SS_vs_k141_79107\|\|full | Low-quality |
| SS_vs_k141_91233\|\|full | Low-quality |
| SS_vs_k141_351\|\|full | Low-quality |
| SS_vs_k141_133698\|\|full | Low-quality |
| SS_vs_k141_115581\|\|full | Low-quality |
| SS_vs_k141_36618\|\|full | Low-quality |
| SS_vs_k141_115590\|\|full | Not-determined |
| SS_vs_k141_79110\|\|full | Low-quality |
| SS_vs_k141_61075\|\|full | Low-quality |
| SS_vs_k141_127770\|\|full | Not-determined |
| SS_vs_k141_121856\|\|full | Not-determined |
| SS_vs_k141_79112\|\|full | Low-quality |
| SS_vs_k141_48798\|\|full | Low-quality |
| S3_extracted_k141_101611 | Medium-quality |
| SS_vs_k141_61077\|\|full | Not-determined |
| SS_vs_k141_91279\|\|full | Low-quality |
| SS_vs_k141_109513\|\|full | Not-determined |
| SS_vs_k141_36630\|\|full | Not-determined |
| SS_vs_k141_121860\|\|full | Low-quality |
| SS_vs_k141_6486\|\|full | Low-quality |
| SS_vs_k141_121869\|\|full | Low-quality |
| SS_vs_k141_393\|\|full | Low-quality |
| SS_vs_k141_48809\|\|full | Low-quality |
| SS_vs_k141_133716\|\|full | Low-quality |
| SS_vs_k141_48811\|\|full | Low-quality |
| SS_vs_k141_79126\|\|full | Low-quality |
| SS_vs_k141_66996\|\|full | Not-determined |
| SS_vs_k141_399\|\|full | Not-determined |
| SS_vs_k141_79129\|\|full | Low-quality |
| SS_vs_k141_103681\|\|full | Low-quality |
| SS_vs_k141_48827\|\|full | Low-quality |
| SS_vs_k141_103686\|\|full | Low-quality |
| SS_vs_k141_416\|\|full | Low-quality |
| SS_vs_k141_420\|\|full | Low-quality |
| SS_vs_k141_79142\|\|full | Low-quality |
| SS_vs_k141_73100\|\|full | Low-quality |
| SS_vs_k141_36646\|\|full | Low-quality |
| SS_vs_k141_127813\|\|full | Low-quality |
| SS_vs_k141_115611\|\|full | Low-quality |
| SS_vs_k141_6506\|\|full | Low-quality |
| SS_vs_k141_30610\|\|full | Not-determined |
| SS_vs_k141_115625\|\|full | Low-quality |
| SS_vs_k141_454\|\|full | Not-determined |
| SS_vs_k141_115631\|\|full | Low-quality |
| SS_vs_k141_6523\|\|full | Not-determined |
| SS_vs_k141_115640\|\|full | Low-quality |
| SS_vs_k141_73110\|\|full | Not-determined |
| SS_vs_k141_121923\|\|full | Not-determined |
| SS_vs_k141_121927\|\|full | Not-determined |
| SS_vs_k141_115658\|\|full | Low-quality |
| SS_vs_k141_48861\|\|full | Low-quality |
| SS_vs_k141_115659\|\|full | Not-determined |
| SS_vs_k141_91372\|\|full | Low-quality |
| SS_vs_k141_48876\|\|full | Not-determined |
| SS_vs_k141_6555\|\|full | Low-quality |
| SS_vs_k141_6560\|\|full | Not-determined |
| SS_vs_k141_85342\|\|full | Low-quality |
| SS_vs_k141_73119\|\|full | Not-determined |
| S11_vs_k141_79831\|\|full | Medium-quality |
| SS_vs_k141_30666\|\|full | Not-determined |
| SS_vs_k141_30670\|\|full | Low-quality |
| SS_vs_k141_121956\|\|full | Low-quality |
| SS_vs_k141_48880\|\|full | Low-quality |
| SS_vs_k141_54877\|\|full | Low-quality |
| SS_vs_k141_48898\|\|full | Low-quality |
| SS_vs_k141_504\|\|full | Low-quality |
| SS_vs_k141_85360\|\|full | Not-determined |
| SS_vs_k141_103764\|\|full | Not-determined |
| SS_vs_k141_43053\|\|full | Low-quality |
| SS_vs_k141_115701\|\|full | Low-quality |
| SS_vs_k141_30691\|\|full | Low-quality |
| SS_vs_k141_36671\|\|full | Not-determined |
| SS_vs_k141_133744\|\|full | Not-determined |
| SS_vs_k141_127840\|\|full | Low-quality |
| SS_vs_k141_30697\|\|full | Low-quality |
| SS_vs_k141_48919\|\|full | Not-determined |
| SS_vs_k141_36680\|\|full | Low-quality |
| SS_vs_k141_43074\|\|full | Low-quality |
| SS_vs_k141_48943\|\|full | Not-determined |
| SS_vs_k141_48947\|\|full | Low-quality |
| SS_vs_k141_133756\|\|full | Low-quality |
| SS_vs_k141_12337\|\|full | Low-quality |
| SS_vs_k141_79159\|\|full | Not-determined |
| SS_vs_k141_122002\|\|full | Low-quality |
| SS_vs_k141_582\|\|full | Low-quality |
| SS_vs_k141_97447\|\|full | Low-quality |
| SS_vs_k141_122011\|\|full | Low-quality |
| S3_vs_k141_18858\|\|full | Medium-quality |
| SS_vs_k141_30749\|\|full | Low-quality |
| SS_vs_k141_127865\|\|full | Low-quality |
| SS_vs_k141_97456\|\|full | Low-quality |
| SS_vs_k141_127868\|\|full | Not-determined |
| SS_vs_k141_122014\|\|full | Low-quality |
| SS_vs_k141_109537\|\|full | Low-quality |
| SS_vs_k141_79164\|\|full | Low-quality |
| SS_vs_k141_127881\|\|full | Low-quality |
| SS_vs_k141_24509\|\|full | Low-quality |
| SS_vs_k141_61131\|\|full | Low-quality |
| SS_vs_k141_109555\|\|full | Low-quality |
| SS_vs_k141_43122\|\|full | Low-quality |
| SS_vs_k141_91473\|\|full | Low-quality |
| SS_vs_k141_626\|\|full | Not-determined |
| SS_vs_k141_127899\|\|full | Low-quality |
| SS_vs_k141_122042\|\|full | Low-quality |
| SS_vs_k141_109565\|\|full | Low-quality |
| SS_vs_k141_48970\|\|full | Low-quality |
| SS_vs_k141_115747\|\|full | Low-quality |
| SS_vs_k141_103797\|\|full | Low-quality |
| SS_vs_k141_127916\|\|full | Low-quality |
| SS_vs_k141_127918\|\|full | Low-quality |
| SS_vs_k141_115752\|\|full | Low-quality |
| SS_vs_k141_43137\|\|full | Low-quality |
| SS_vs_k141_61174\|\|full | Not-determined |
| SS_vs_k141_30787\|\|full | Not-determined |
| SS_vs_k141_12389\|\|full | Low-quality |
| SS_vs_k141_109591\|\|full | Low-quality |
| SS_vs_k141_97483\|\|full | Not-determined |
| SS_vs_k141_43140\|\|full | Low-quality |
| SS_vs_k141_103820\|\|full | Low-quality |
| SS_vs_k141_122077\|\|full | Low-quality |
| SS_vs_k141_73137\|\|full | Not-determined |
| SS_vs_k141_79184\|\|full | Low-quality |
| SS_vs_k141_97502\|\|full | Low-quality |
| SS_vs_k141_661\|\|full | Not-determined |
| SS_vs_k141_6582\|\|full | Not-determined |
| SS_vs_k141_79191\|\|full | Low-quality |
| SS_vs_k141_97522\|\|full | Not-determined |
| SS_vs_k141_54961\|\|full | Low-quality |
| S1_extracted_k141_6272 | Complete |
| SS_vs_k141_91547\|\|full | Not-determined |
| SS_vs_k141_43175\|\|full | Not-determined |
| SS_vs_k141_43180\|\|full | Not-determined |
| SS_vs_k141_18599\|\|full | Not-determined |
| SS_vs_k141_733\|\|full | Not-determined |
| SS_vs_k141_18610\|\|full | Low-quality |
| SS_vs_k141_122106\|\|full | Low-quality |
| SS_vs_k141_30842\|\|full | Not-determined |
| SS_vs_k141_109623\|\|full | Low-quality |
| SS_vs_k141_115779\|\|full | Low-quality |
| SS_vs_k141_109627\|\|full | Not-determined |
| SS_vs_k141_30851\|\|full | Low-quality |
| SS_vs_k141_115782\|\|full | Low-quality |
| SS_vs_k141_761\|\|full | Low-quality |
| SS_vs_k141_30858\|\|full | Low-quality |
| SS_vs_k141_43200\|\|full | Low-quality |
| SS_vs_k141_18626\|\|full | Not-determined |
| SS_vs_k141_18628\|\|full | Low-quality |
| SS_vs_k141_109669\|\|full | Low-quality |
| SS_vs_k141_54972\|\|full | Low-quality |
| SS_vs_k141_18654\|\|full | Low-quality |
| SS_vs_k141_18659\|\|full | Low-quality |
| SS_vs_k141_6629\|\|full | Low-quality |
| SS_vs_k141_122165\|\|full | Low-quality |
| SS_vs_k141_776\|\|full | Not-determined |
| SS_vs_k141_778\|\|full | Not-determined |
| SS_vs_k141_30886\|\|full | Not-determined |
| SS_vs_k141_115835\|\|full | Low-quality |
| SS_vs_k141_30887\|\|full | Low-quality |
| SS_vs_k141_122179\|\|full | Low-quality |
| SS_vs_k141_789\|\|full | Not-determined |
| S7_vs_78507\|\|full | Medium-quality |
| SS_vs_k141_18674\|\|full | Not-determined |
| SS_vs_k141_85432\|\|full | Not-determined |
| SS_vs_k141_49044\|\|full | Low-quality |
| SS_vs_k141_115861\|\|full | Not-determined |
| SS_vs_k141_800\|\|full | Low-quality |
| SS_vs_k141_36720\|\|full | Not-determined |
| SS_vs_k141_803\|\|full | Low-quality |
| SS_vs_k141_30916\|\|full | Not-determined |
| SS_vs_k141_85442\|\|full | Not-determined |
| SS_vs_k141_30918\|\|full | Not-determined |
| SS_vs_k141_24613\|\|full | Low-quality |
| SS_vs_k141_18681\|\|full | Low-quality |
| SS_vs_k141_821\|\|full | Low-quality |
| SS_vs_k141_140040\|\|full | Not-determined |
| SS_vs_k141_43235\|\|full | Low-quality |
| SS_vs_k141_85450\|\|full | Low-quality |
| SS_vs_k141_49059\|\|full | Low-quality |
| SS_vs_k141_133803\|\|full | Low-quality |
| SS_vs_k141_6652\|\|full | Low-quality |
| SS_vs_k141_43243\|\|full | Low-quality |
| SS_vs_k141_127979\|\|full | Not-determined |
| SS_vs_k141_55002\|\|full | Not-determined |
| SS_vs_k141_122229\|\|full | Low-quality |
| SS_vs_k141_67019\|\|full | Low-quality |
| SS_vs_k141_6666\|\|full | Not-determined |
| SS_vs_k141_122234\|\|full | Low-quality |
| SS_vs_k141_36742\|\|full | Low-quality |
| SS_vs_k141_133813\|\|full | Low-quality |
| SS_vs_k141_826\|\|full | Low-quality |
| SS_vs_k141_55023\|\|full | Not-determined |
| SS_vs_k141_830\|\|full | Low-quality |
| SS_vs_k141_49077\|\|full | Not-determined |
| SS_vs_k141_833\|\|full | Not-determined |
| SS_vs_k141_103869\|\|full | Low-quality |
| SS_vs_k141_49083\|\|full | Low-quality |
| SS_vs_k141_109780\|\|full | Low-quality |
| SS_vs_k141_109782\|\|full | Not-determined |
| SS_vs_k141_85468\|\|full | Low-quality |
| SS_vs_k141_24639\|\|full | Low-quality |
| SS_vs_k141_122268\|\|full | Low-quality |
| SS_vs_k141_30944\|\|full | Low-quality |
| SS_vs_k141_122272\|\|full | Low-quality |
| SS_vs_k141_36759\|\|full | Low-quality |
| SS_vs_k141_12417\|\|full | Not-determined |
| SS_vs_k141_848\|\|full | Not-determined |
| SS_vs_k141_36770\|\|full | Low-quality |
| SS_vs_k141_73174\|\|full | Low-quality |
| SS_vs_k141_24671\|\|full | Not-determined |
| SS_vs_k141_49100\|\|full | Low-quality |
| SS_vs_k141_18703\|\|full | Not-determined |
| SS_vs_k141_85504\|\|full | Low-quality |
| SS_vs_k141_30974\|\|full | Low-quality |
| SS_vs_k141_18713\|\|full | Low-quality |
| SS_vs_k141_128008\|\|full | Low-quality |
| SS_vs_k141_73179\|\|full | Not-determined |
| SS_vs_k141_122314\|\|full | Low-quality |
| SS_vs_k141_73193\|\|full | Low-quality |
| SS_vs_k141_122317\|\|full | Not-determined |
| SS_vs_k141_103912\|\|full | Low-quality |
| SS_vs_k141_67046\|\|full | Low-quality |
| SS_vs_k141_30982\|\|full | Not-determined |
| SS_vs_k141_24718\|\|full | Low-quality |
| SS_vs_k141_73198\|\|full | Low-quality |
| SS_vs_k141_24728\|\|full | Low-quality |
| SS_vs_k141_134874\|\|full | High-quality |
| SS_vs_k141_91568\|\|full | Low-quality |
| SS_vs_k141_36802\|\|full | Low-quality |
| SS_vs_k141_31009\|\|full | Low-quality |
| SS_vs_k141_36803\|\|full | Not-determined |
| SS_vs_k141_85527\|\|full | Not-determined |
| SS_vs_k141_115906\|\|full | Low-quality |
| SS_vs_k141_85533\|\|full | Low-quality |
| SS_vs_k141_6694\|\|full | Not-determined |
| SS_vs_k141_6696\|\|full | Low-quality |
| SS_vs_k141_6703\|\|full | Not-determined |
| SS_vs_k141_36849\|\|full | Not-determined |
| SS_vs_k141_67072\|\|full | Not-determined |
| SS_vs_k141_36852\|\|full | Not-determined |
| SS_vs_k141_115922\|\|full | Not-determined |
| SS_vs_k141_49161\|\|full | Low-quality |
| SS_vs_k141_49166\|\|full | Low-quality |
| SS_vs_k141_31038\|\|full | Not-determined |
| SS_vs_k141_6711\|\|full | Low-quality |
| SS_vs_k141_12443\|\|full | Low-quality |
| SS_vs_k141_115937\|\|full | Low-quality |
| SS_vs_k141_36870\|\|full | Not-determined |
| SS_vs_k141_49186\|\|full | Low-quality |
| SS_vs_k141_12455\|\|full | Low-quality |
| SS_vs_k141_49195\|\|full | Low-quality |
| SS_vs_k141_128107\|\|full | Low-quality |
| SS_vs_k141_133890\|\|full | Not-determined |
| SS_vs_k141_85568\|\|full | Low-quality |
| SS_vs_k141_67115\|\|full | Not-determined |
| SS_vs_k141_122363\|\|full | Not-determined |
| SS_vs_k141_12482\|\|full | Low-quality |
| SS_vs_k141_18792\|\|full | Low-quality |
| SS_vs_k141_122368\|\|full | Low-quality |
| SS_vs_k141_940\|\|full | Low-quality |
| SS_vs_k141_941\|\|full | Low-quality |
| SS_vs_k141_12497\|\|full | Not-determined |
| SS_vs_k141_43334\|\|full | Low-quality |
| SS_vs_k141_6720\|\|full | Low-quality |
| SS_vs_k141_946\|\|full | Not-determined |
| SS_vs_k141_122381\|\|full | Low-quality |
| SS_vs_k141_91620\|\|full | Low-quality |
| SS_vs_k141_122390\|\|full | Low-quality |
| SS_vs_k141_49241\|\|full | Low-quality |
| SS_vs_k141_12539\|\|full | Not-determined |
| SS_vs_k141_122400\|\|full | Low-quality |
| SS_vs_k141_961\|\|full | Not-determined |
| SS_vs_k141_116016\|\|full | Low-quality |
| SS_vs_k141_122413\|\|full | Low-quality |
| SS_vs_k141_67187\|\|full | Low-quality |
| SS_vs_k141_43389\|\|full | Low-quality |
| SS_vs_k141_91652\|\|full | Low-quality |
| SS_vs_k141_122418\|\|full | Not-determined |
| SS_vs_k141_140059\|\|full | Low-quality |
| SS_vs_k141_67203\|\|full | Not-determined |
| SS_vs_k141_967\|\|full | Low-quality |
| SS_vs_k141_133950\|\|full | Not-determined |
| SS_vs_k141_140062\|\|full | Not-determined |
| SS_vs_k141_43410\|\|full | Not-determined |
| SS_vs_k141_36926\|\|full | Not-determined |
| SS_vs_k141_85626\|\|full | Not-determined |
| SS_vs_k141_43413\|\|full | Low-quality |
| SS_vs_k141_67218\|\|full | Low-quality |
| SS_vs_k141_18831\|\|full | Low-quality |
| SS_vs_k141_36939\|\|full | Low-quality |
| SS_vs_k141_43416\|\|full | Low-quality |
| SS_vs_k141_91689\|\|full | Not-determined |
| SS_vs_k141_128166\|\|full | Low-quality |
| SS_vs_k141_18837\|\|full | Low-quality |
| SS_vs_k141_49268\|\|full | Low-quality |
| SS_vs_k141_116062\|\|full | Low-quality |
| SS_vs_k141_116064\|\|full | Not-determined |
| SS_vs_k141_91708\|\|full | Low-quality |
| SS_vs_k141_31160\|\|full | Low-quality |
| SS_vs_k141_31164\|\|full | Not-determined |
| SS_vs_k141_49297\|\|full | Low-quality |
| SS_vs_k141_1012\|\|full | Low-quality |
| SS_vs_k141_85659\|\|full | Not-determined |
| SS_vs_k141_1018\|\|full | Low-quality |
| SS_vs_k141_55093\|\|full | Low-quality |
| SS_vs_k141_122493\|\|full | Low-quality |
| S14_vs_k141_115455\|\|full | Medium-quality |
| SS_vs_k141_1027\|\|full | Not-determined |
| SS_vs_k141_36987\|\|full | Not-determined |
| SS_vs_k141_133971\|\|full | Not-determined |
| SS_vs_k141_43448\|\|full | Not-determined |
| SS_vs_k141_140119\|\|full | Low-quality |
| SS_vs_k141_1043\|\|full | Low-quality |
| SS_vs_k141_43452\|\|full | Not-determined |
| SS_vs_k141_31187\|\|full | Low-quality |
| SS_vs_k141_6794\|\|full | Not-determined |
| SS_vs_k141_55134\|\|full | Low-quality |
| SS_vs_k141_6796\|\|full | Low-quality |
| SS_vs_k141_43460\|\|full | Low-quality |
| SS_vs_k141_128211\|\|full | Low-quality |
| SS_vs_k141_1059\|\|full | Low-quality |
| SS_vs_k141_116119\|\|full | Not-determined |
| SS_vs_k141_1064\|\|full | Low-quality |
| SS_vs_k141_31206\|\|full | Low-quality |
| SS_vs_k141_1065\|\|full | Not-determined |
| SS_vs_k141_122530\|\|full | Low-quality |
| SS_vs_k141_140130\|\|full | Low-quality |
| SS_vs_k141_6814\|\|full | Low-quality |
| SS_vs_k141_43496\|\|full | Low-quality |
| SS_vs_k141_55149\|\|full | Not-determined |
| SS_vs_k141_122558\|\|full | Low-quality |
| SS_vs_k141_128221\|\|full | Low-quality |
| SS_vs_k141_116123\|\|full | Low-quality |
| S1_vs_k141_6272\|\|full | High-quality |
| SS_vs_k141_79194\|\|full | Low-quality |
| SS_vs_k141_1118\|\|full | Not-determined |
| SS_vs_k141_49347\|\|full | Not-determined |
| SS_vs_k141_18882\|\|full | Low-quality |
| SS_vs_k141_1139\|\|full | Low-quality |
| SS_vs_k141_128231\|\|full | Low-quality |
| SS_vs_k141_49357\|\|full | Low-quality |
| SS_vs_k141_116132\|\|full | Not-determined |
| SS_vs_k141_43538\|\|full | Low-quality |
| SS_vs_k141_18893\|\|full | Not-determined |
| SS_vs_k141_43543\|\|full | Low-quality |
| SS_vs_k141_1146\|\|full | Low-quality |
| SS_vs_k141_43546\|\|full | Low-quality |
| SS_vs_k141_140148\|\|full | Not-determined |
| SS_vs_k141_43551\|\|full | Not-determined |
| SS_vs_k141_103938\|\|full | Low-quality |
| SS_vs_k141_140154\|\|full | Low-quality |
| SS_vs_k141_67322\|\|full | Not-determined |
| SS_vs_k141_55167\|\|full | Low-quality |
| SS_vs_k141_140167\|\|full | Not-determined |
| SS_vs_k141_67333\|\|full | Low-quality |
| SS_vs_k141_43573\|\|full | Low-quality |
| SS_vs_k141_1182\|\|full | Not-determined |
| SS_vs_k141_61275\|\|full | Low-quality |
| SS_vs_k141_1192\|\|full | Not-determined |
| SS_vs_k141_122580\|\|full | Low-quality |
| SS_vs_k141_1210\|\|full | Low-quality |
| SS_vs_k141_1215\|\|full | Low-quality |
| SS_vs_k141_116178\|\|full | Low-quality |
| SS_vs_k141_1221\|\|full | Not-determined |
| SS_vs_k141_31262\|\|full | Low-quality |
| SS_vs_k141_116181\|\|full | Low-quality |
| SS_vs_k141_79263\|\|full | Low-quality |
| SS_vs_k141_122607\|\|full | Not-determined |
| SS_vs_k141_122613\|\|full | Low-quality |
| SS_vs_k141_31292\|\|full | Low-quality |
| SS_vs_k141_43603\|\|full | Low-quality |
| SS_vs_k141_43608\|\|full | Not-determined |
| SS_vs_k141_117770\|\|full | Medium-quality |
| SS_vs_k141_134055\|\|full | Low-quality |
| SS_vs_k141_134057\|\|full | Low-quality |
| SS_vs_k141_79291\|\|full | Not-determined |
| SS_vs_k141_134059\|\|full | Low-quality |
| SS_vs_k141_43645\|\|full | Low-quality |
| SS_vs_k141_1297\|\|full | Low-quality |
| SS_vs_k141_43650\|\|full | Low-quality |
| SS_vs_k141_12579\|\|full | Not-determined |
| SS_vs_k141_31315\|\|full | Low-quality |
| SS_vs_k141_6957\|\|full | Low-quality |
| SS_vs_k141_67363\|\|full | Low-quality |
| SS_vs_k141_6958\|\|full | Low-quality |
| SS_vs_k141_79300\|\|full | Not-determined |

| | |
|---|---|
| S5_vs_k141_134071\|\|full | Low-quality |
| S5_vs_k141_104045\|\|full | Not-determined |
| S5_vs_k141_12596\|\|full | Low-quality |
| S5_vs_k141_104048\|\|full | Not-determined |
| S5_vs_k141_67375\|\|full | Low-quality |
| S5_vs_k141_122627\|\|full | Low-quality |
| S5_vs_k141_67397\|\|full | Low-quality |
| S14_vs_k141_117158\|\|full | High-quality |
| S5_vs_k141_18995\|\|full | Low-quality |
| S5_vs_k141_1335\|\|full | Low-quality |
| S5_vs_k141_79332\|\|full | Low-quality |
| S5_vs_k141_79337\|\|full | Low-quality |
| S3_vs_k141_101611\|\|full | Medium-quality |
| S5_vs_k141_19004\|\|full | Low-quality |
| S5_vs_k141_104062\|\|full | Not-determined |
| S5_vs_k141_79339\|\|full | Low-quality |
| S5_vs_k141_31370\|\|full | Not-determined |
| S5_vs_k141_43665\|\|full | Low-quality |
| S5_vs_k141_104069\|\|full | Low-quality |
| S5_vs_k141_6999\|\|full | Not-determined |
| S5_vs_k141_43667\|\|full | Low-quality |
| S5_vs_k141_19014\|\|full | Not-determined |
| S5_vs_k141_134108\|\|full | Not-determined |
| S5_vs_k141_79356\|\|full | Low-quality |
| S5_vs_k141_122674\|\|full | Not-determined |
| S5_vs_k141_104090\|\|full | Low-quality |
| S5_vs_k141_122678\|\|full | Low-quality |
| S5_vs_k141_91854\|\|full | Not-determined |
| S5_vs_k141_73268\|\|full | Low-quality |
| S5_vs_k141_145603\|\|full | Medium-quality |
| S5_vs_k141_104104\|\|full | Low-quality |
| S5_vs_k141_79359\|\|full | Low-quality |
| S5_vs_k141_85690\|\|full | Low-quality |
| S5_vs_k141_140262\|\|full | Not-determined |
| S5_vs_k141_122712\|\|full | Low-quality |
| S5_vs_k141_19069\|\|full | Not-determined |
| S5_vs_k141_140268\|\|full | Low-quality |
| S5_vs_k141_97589\|\|full | Low-quality |
| S5_vs_k141_109830\|\|full | Low-quality |
| S5_vs_k141_140270\|\|full | Not-determined |
| S5_vs_k141_128281\|\|full | Not-determined |
| S5_vs_k141_73271\|\|full | Not-determined |
| S5_vs_k141_116282\|\|full | Low-quality |
| S5_vs_k141_109835\|\|full | Low-quality |
| S5_vs_k141_85719\|\|full | Low-quality |
| S5_vs_k141_1372\|\|full | Low-quality |
| S5_vs_k141_73281\|\|full | Low-quality |
| S5_vs_k141_31438\|\|full | Low-quality |
| S5_vs_k141_140279\|\|full | Low-quality |
| S5_vs_k141_97601\|\|full | Not-determined |
| S5_vs_k141_79370\|\|full | Low-quality |
| S5_vs_k141_122770\|\|full | Low-quality |
| S5_vs_k141_85726\|\|full | Low-quality |
| S5_vs_k141_104141\|\|full | Low-quality |
| S5_vs_k141_122787\|\|full | Not-determined |
| S5_vs_k141_109849\|\|full | Low-quality |
| S5_vs_k141_116300\|\|full | Low-quality |
| S5_vs_k141_104144\|\|full | Low-quality |
| S5_vs_k141_122793\|\|full | Not-determined |
| S5_vs_k141_37158\|\|full | Low-quality |
| S5_vs_k141_19142\|\|full | Low-quality |
| S5_vs_k141_19145\|\|full | Low-quality |
| S5_vs_k141_85751\|\|full | Low-quality |
| S5_vs_k141_104149\|\|full | Low-quality |
| S5_vs_k141_7091\|\|full | Not-determined |
| S5_vs_k141_134175\|\|full | Not-determined |
| S5_vs_k141_104159\|\|full | Not-determined |
| S5_vs_k141_116331\|\|full | Not-determined |
| S5_vs_k141_134182\|\|full | Low-quality |
| S5_vs_k141_116341\|\|full | Low-quality |
| S5_vs_k141_104172\|\|full | Not-determined |
| S5_vs_k141_12782\|\|full | Not-determined |
| S5_vs_k141_116352\|\|full | Low-quality |
| S5_vs_k141_7098\|\|full | Low-quality |
| S5_vs_k141_12789\|\|full | Not-determined |
| S5_vs_k141_73307\|\|full | Low-quality |
| S5_vs_k141_109895\|\|full | Low-quality |
| S5_vs_k141_7104\|\|full | Low-quality |
| S5_vs_k141_116373\|\|full | Low-quality |
| S5_vs_k141_67487\|\|full | Not-determined |
| S5_vs_k141_122847\|\|full | Low-quality |
| S5_vs_k141_67490\|\|full | Not-determined |
| S5_vs_k141_122849\|\|full | Not-determined |
| S5_vs_k141_116379\|\|full | Low-quality |
| S5_vs_k141_55252\|\|full | Low-quality |
| S5_vs_k141_55253\|\|full | Not-determined |
| S5_vs_k141_122858\|\|full | Not-determined |
| S5_vs_k141_7120\|\|full | Low-quality |
| S5_vs_k141_67503\|\|full | Low-quality |
| S5_vs_k141_134197\|\|full | Not-determined |
| S5_vs_k141_85817\|\|full | Not-determined |
| S5_vs_k141_19161\|\|full | Low-quality |
| S5_vs_k141_97632\|\|full | Not-determined |
| S5_vs_k141_140289\|\|full | Low-quality |
| S5_vs_k141_97645\|\|full | Low-quality |
| S5_vs_k141_97654\|\|full | Low-quality |
| S5_vs_k141_31498\|\|full | Not-determined |
| S5_vs_k141_61305\|\|full | Not-determined |
| S5_vs_k141_109918\|\|full | Low-quality |
| S5_vs_k141_31505\|\|full | Low-quality |
| S5_vs_k141_116411\|\|full | Not-determined |
| S5_vs_k141_19175\|\|full | Low-quality |
| S5_vs_k141_67527\|\|full | Low-quality |
| S5_vs_k141_134204\|\|full | Low-quality |
| S5_vs_k141_109932\|\|full | Low-quality |
| S5_vs_k141_61320\|\|full | Low-quality |
| S5_vs_k141_31520\|\|full | Low-quality |
| S5_vs_k141_109944\|\|full | Not-determined |
| S5_vs_k141_85867\|\|full | Low-quality |
| S5_vs_k141_67554\|\|full | Not-determined |
| S5_vs_k141_12803\|\|full | Low-quality |
| S5_vs_k141_67568\|\|full | Not-determined |
| S5_vs_k141_122898\|\|full | Not-determined |
| S5_vs_k141_12829\|\|full | Low-quality |
| S5_vs_k141_49470\|\|full | Not-determined |
| S5_vs_k141_67574\|\|full | Not-determined |
| S5_vs_k141_67576\|\|full | Low-quality |
| S5_vs_k141_61338\|\|full | Low-quality |
| S5_vs_k141_109997\|\|full | Not-determined |
| S5_vs_k141_122913\|\|full | Low-quality |
| S5_vs_k141_61346\|\|full | Low-quality |
| S5_vs_k141_122916\|\|full | Low-quality |
| S5_vs_k141_97710\|\|full | Low-quality |
| S5_vs_k141_134217\|\|full | Not-determined |
| S5_vs_k141_140330\|\|full | Not-determined |
| S5_vs_k141_140333\|\|full | Not-determined |
| S5_vs_k141_116471\|\|full | Low-quality |
| S5_vs_k141_67617\|\|full | Low-quality |
| S5_vs_k141_140343\|\|full | Low-quality |
| S5_vs_k141_73344\|\|full | Low-quality |
| S5_vs_k141_140346\|\|full | Low-quality |
| S5_vs_k141_104183\|\|full | Low-quality |
| S5_vs_k141_97718\|\|full | Low-quality |
| S5_vs_k141_134223\|\|full | Low-quality |
| S11_extracted_k141_104034 | Complete |
| S5_vs_k141_37210\|\|full | Not-determined |
| S5_vs_k141_104195\|\|full | Not-determined |
| S5_vs_k141_19211\|\|full | Low-quality |
| S5_vs_k141_116495\|\|full | Low-quality |
| S5_vs_k141_85892\|\|full | Not-determined |
| S5_vs_k141_85893\|\|full | Not-determined |
| S5_vs_k141_85895\|\|full | Low-quality |
| S5_vs_k141_7219\|\|full | Not-determined |
| S5_vs_k141_31538\|\|full | Low-quality |
| S5_vs_k141_73375\|\|full | Low-quality |
| S5_vs_k141_49510\|\|full | Low-quality |
| S5_vs_k141_49514\|\|full | Low-quality |
| S5_vs_k141_19242\|\|full | Low-quality |
| S11_extracted_k141_104052 | Complete |
| S5_vs_k141_140370\|\|full | Not-determined |
| S5_vs_k141_85934\|\|full | Not-determined |
| S5_vs_k141_140377\|\|full | Low-quality |
| S5_vs_k141_104240\|\|full | Low-quality |
| S5_vs_k141_110083\|\|full | Low-quality |
| S5_vs_k141_19245\|\|full | Low-quality |
| S5_vs_k141_110092\|\|full | Low-quality |
| S5_vs_k141_31587\|\|full | Low-quality |
| S5_vs_k141_31595\|\|full | Not-determined |
| S5_vs_k141_140405\|\|full | Low-quality |
| S5_vs_k141_31598\|\|full | Not-determined |
| S5_vs_k141_12914\|\|full | Not-determined |
| S5_vs_k141_7298\|\|full | Low-quality |
| S5_vs_k141_37309\|\|full | Not-determined |
| S5_vs_k141_7302\|\|full | Not-determined |
| S5_vs_k141_104256\|\|full | Not-determined |
| S5_vs_k141_12936\|\|full | Low-quality |
| S5_vs_k141_104259\|\|full | Not-determined |
| S5_vs_k141_85988\|\|full | Not-determined |
| S5_vs_k141_19309\|\|full | Not-determined |
| S5_vs_k141_61445\|\|full | Low-quality |
| S5_vs_k141_116577\|\|full | Low-quality |
| S5_vs_k141_61455\|\|full | Low-quality |
| S5_vs_k141_43805\|\|full | Low-quality |
| S5_vs_k141_110159\|\|full | Low-quality |
| S5_vs_k141_12960\|\|full | Low-quality |
| S5_vs_k141_140457\|\|full | Low-quality |
| S5_vs_k141_73427\|\|full | Low-quality |
| S5_vs_k141_134286\|\|full | Low-quality |
| S5_vs_k141_12990\|\|full | Low-quality |
| S5_vs_k141_86000\|\|full | Low-quality |
| S5_vs_k141_140471\|\|full | Low-quality |
| S5_vs_k141_91909\|\|full | Low-quality |
| S5_vs_k141_61500\|\|full | Low-quality |
| S5_vs_k141_19336\|\|full | Low-quality |
| S5_vs_k141_55284\|\|full | Low-quality |
| S5_vs_k141_19341\|\|full | Not-determined |
| S5_vs_k141_86014\|\|full | Not-determined |
| S5_vs_k141_37385\|\|full | Low-quality |
| S7_extracted_117697 | High-quality |
| S5_vs_k141_91925\|\|full | Not-determined |
| S5_vs_k141_97814\|\|full | Low-quality |
| S5_vs_k141_37397\|\|full | Not-determined |
| S5_vs_k141_31634\|\|full | Not-determined |
| S16_vs_140709\|\|full | High-quality |
| S5_vs_k141_73470\|\|full | Low-quality |
| S5_vs_k141_13007\|\|full | Not-determined |
| S5_vs_k141_31659\|\|full | Low-quality |
| S5_vs_k141_13013\|\|full | Low-quality |
| S5_vs_k141_24795\|\|full | Not-determined |
| S5_vs_k141_116615\|\|full | Low-quality |
| S5_vs_k141_61564\|\|full | Not-determined |
| S5_vs_k141_123070\|\|full | Not-determined |
| S5_vs_k141_73484\|\|full | Low-quality |
| S5_vs_k141_140519\|\|full | Not-determined |
| S5_vs_k141_86027\|\|full | Low-quality |
| S5_vs_k141_61575\|\|full | Low-quality |
| S5_vs_k141_91956\|\|full | Low-quality |
| S5_vs_k141_37441\|\|full | Low-quality |
| S5_vs_k141_97840\|\|full | Low-quality |
| S5_vs_k141_7309\|\|full | Low-quality |
| S5_vs_k141_13041\|\|full | Low-quality |
| S5_vs_k141_19411\|\|full | Not-determined |
| S5_vs_k141_73497\|\|full | Low-quality |
| S5_vs_k141_73498\|\|full | Not-determined |
| S5_vs_k141_67670\|\|full | Low-quality |
| S5_vs_k141_19417\|\|full | Low-quality |
| S5_vs_k141_37459\|\|full | Low-quality |
| S5_vs_k141_79389\|\|full | Low-quality |
| S5_vs_k141_43855\|\|full | Low-quality |
| S5_vs_k141_61612\|\|full | Low-quality |
| S5_vs_k141_97851\|\|full | Not-determined |
| S5_vs_k141_134340\|\|full | Low-quality |
| S5_vs_k141_79398\|\|full | Low-quality |
| S5_vs_k141_116663\|\|full | Low-quality |
| S5_vs_k141_79404\|\|full | Low-quality |
| S5_vs_k141_31666\|\|full | Low-quality |
| S5_vs_k141_13065\|\|full | Low-quality |
| S5_vs_k141_123095\|\|full | Not-determined |
| S5_vs_k141_37485\|\|full | Not-determined |
| S5_vs_k141_67693\|\|full | Low-quality |
| S5_vs_k141_134347\|\|full | Low-quality |
| S5_vs_k141_55323\|\|full | Low-quality |
| S5_vs_k141_73531\|\|full | Low-quality |
| S5_vs_k141_31686\|\|full | Not-determined |
| S5_vs_k141_61638\|\|full | Not-determined |
| S5_vs_k141_31691\|\|full | Not-determined |
| S5_vs_k141_37490\|\|full | Not-determined |
| S5_vs_k141_55335\|\|full | Low-quality |
| S5_vs_k141_116677\|\|full | Low-quality |
| S5_vs_k141_37498\|\|full | Not-determined |
| S5_vs_k141_92007\|\|full | Low-quality |
| S5_vs_k141_92015\|\|full | Low-quality |
| S5_vs_k141_116692\|\|full | Low-quality |
| S5_vs_k141_97890\|\|full | Not-determined |
| S5_vs_k141_1446\|\|full | Not-determined |
| S5_vs_k141_43877\|\|full | Not-determined |
| S5_vs_k141_92035\|\|full | Low-quality |
| S5_vs_k141_123137\|\|full | Not-determined |
| S5_vs_k141_128307\|\|full | Not-determined |
| S7_vs_65719\|\|full | Medium-quality |
| S5_vs_k141_92039\|\|full | Low-quality |
| S5_vs_k141_55372\|\|full | Low-quality |
| S5_vs_k141_123162\|\|full | Low-quality |
| S5_vs_k141_13111\|\|full | Low-quality |
| S5_vs_k141_67730\|\|full | Not-determined |
| S5_vs_k141_13116\|\|full | Low-quality |
| S5_vs_k141_110294\|\|full | Low-quality |
| S5_vs_k141_110297\|\|full | Low-quality |
| S5_vs_k141_67742\|\|full | Low-quality |
| S5_vs_k141_110303\|\|full | Not-determined |
| S5_vs_k141_7341\|\|full | Low-quality |
| S5_vs_k141_97916\|\|full | Low-quality |
| S5_vs_k141_134374\|\|full | Not-determined |
| S5_vs_k141_134375\|\|full | Not-determined |
| S5_vs_k141_92068\|\|full | Low-quality |
| S5_vs_k141_24841\|\|full | Not-determined |
| S5_vs_k141_24842\|\|full | Low-quality |
| S5_vs_k141_13150\|\|full | Not-determined |
| S5_vs_k141_7352\|\|full | Low-quality |
| S5_vs_k141_67781\|\|full | Low-quality |
| S5_vs_k141_92096\|\|full | Not-determined |
| S5_vs_k141_19522\|\|full | Not-determined |
| S5_vs_k141_24852\|\|full | Low-quality |
| S5_vs_k141_1454\|\|full | Not-determined |
| S5_vs_k141_73603\|\|full | Not-determined |
| S5_vs_k141_92106\|\|full | Low-quality |
| S5_vs_k141_116807\|\|full | Not-determined |
| S5_vs_k141_128346\|\|full | Low-quality |
| S5_vs_k141_92112\|\|full | Low-quality |
| S5_vs_k141_128347\|\|full | Not-determined |
| S5_vs_k141_134421\|\|full | Not-determined |
| S5_vs_k141_134424\|\|full | Low-quality |
| S5_vs_k141_123206\|\|full | Low-quality |
| S5_vs_k141_61736\|\|full | Not-determined |
| S5_vs_k141_123207\|\|full | Low-quality |
| S5_vs_k141_13173\|\|full | Low-quality |
| S5_vs_k141_7382\|\|full | Low-quality |
| S5_vs_k141_61748\|\|full | Low-quality |
| S5_vs_k141_79462\|\|full | Low-quality |
| S5_vs_k141_134446\|\|full | Low-quality |
| S5_vs_k141_67805\|\|full | Low-quality |
| S5_vs_k141_43953\|\|full | Low-quality |
| S5_vs_k141_134447\|\|full | Low-quality |
| S5_vs_k141_123223\|\|full | Low-quality |
| S5_vs_k141_37565\|\|full | Not-determined |
| S5_vs_k141_123238\|\|full | Low-quality |
| S5_vs_k141_92160\|\|full | Low-quality |
| S5_vs_k141_61788\|\|full | Low-quality |
| S5_vs_k141_61794\|\|full | Low-quality |
| S5_vs_k141_37580\|\|full | Low-quality |
| S5_vs_k141_92163\|\|full | Not-determined |
| S5_vs_k141_67830\|\|full | Low-quality |
| S5_vs_k141_37589\|\|full | Low-quality |
| S5_vs_k141_92164\|\|full | Low-quality |
| S5_vs_k141_134472\|\|full | Low-quality |
| S5_vs_k141_37592\|\|full | Low-quality |
| S5_vs_k141_73635\|\|full | Not-determined |
| S5_vs_k141_110404\|\|full | Low-quality |
| S5_vs_k141_61812\|\|full | Low-quality |
| S5_vs_k141_1480\|\|full | Not-determined |
| S5_vs_k141_116867\|\|full | Low-quality |
| S5_vs_k141_110408\|\|full | Low-quality |
| S16_vs_172827\|\|full | Medium-quality |
| S5_vs_k141_13210\|\|full | Low-quality |
| S5_vs_k141_110413\|\|full | Not-determined |
| S5_vs_k141_128403\|\|full | Not-determined |
| S5_vs_k141_73645\|\|full | Not-determined |
| S5_vs_k141_49566\|\|full | Low-quality |
| S5_vs_k141_13225\|\|full | Not-determined |
| S5_vs_k141_31832\|\|full | Low-quality |
| S5_vs_k141_116882\|\|full | Low-quality |
| S5_vs_k141_110423\|\|full | Low-quality |
| S5_vs_k141_31838\|\|full | Not-determined |
| S5_vs_k141_73676\|\|full | Not-determined |
| S5_vs_k141_73680\|\|full | Low-quality |
| S5_vs_k141_134497\|\|full | Not-determined |
| S5_vs_k141_73690\|\|full | Low-quality |
| S5_vs_k141_37623\|\|full | Low-quality |
| S5_vs_k141_86118\|\|full | Low-quality |
| S5_vs_k141_116918\|\|full | Not-determined |
| S5_vs_k141_110456\|\|full | Low-quality |
| S5_vs_k141_134500\|\|full | Low-quality |
| S5_vs_k141_49639\|\|full | Not-determined |
| S5_vs_k141_110476\|\|full | Not-determined |
| S5_vs_k141_110479\|\|full | Low-quality |
| S5_vs_k141_73724\|\|full | Low-quality |
| S11_vs_k141_104034\|\|full | High-quality |
| S11_vs_k141_104052\|\|full | High-quality |
| S5_vs_k141_13262\|\|full | Low-quality |
| S5_vs_k141_73732\|\|full | Low-quality |
| S5_vs_k141_73736\|\|full | Low-quality |
| S5_vs_k141_86174\|\|full | Low-quality |
| S5_vs_k141_116981\|\|full | Not-determined |
| S5_vs_k141_97999\|\|full | Not-determined |
| S5_vs_k141_92312\|\|full | Low-quality |
| S5_vs_k141_67908\|\|full | Low-quality |
| S5_vs_k141_134534\|\|full | Not-determined |
| S5_vs_k141_92314\|\|full | Low-quality |
| S5_vs_k141_1514\|\|full | Low-quality |
| S5_vs_k141_116999\|\|full | Low-quality |
| S5_vs_k141_98001\|\|full | Low-quality |
| S5_vs_k141_49672\|\|full | Not-determined |
| S5_vs_k141_31905\|\|full | Not-determined |
| S5_vs_k141_1522\|\|full | Not-determined |
| S5_vs_k141_31907\|\|full | Not-determined |
| S5_vs_k141_49675\|\|full | Low-quality |
| S5_vs_k141_98006\|\|full | Not-determined |
| S5_vs_k141_73766\|\|full | Not-determined |
| S5_vs_k141_1530\|\|full | Not-determined |
| S5_vs_k141_110501\|\|full | Low-quality |
| S5_vs_k141_24888\|\|full | Low-quality |
| S5_vs_k141_117024\|\|full | Not-determined |
| S5_vs_k141_110514\|\|full | Low-quality |
| S5_vs_k141_37647\|\|full | Not-determined |
| S5_vs_k141_134540\|\|full | Not-determined |
| S5_vs_k141_140564\|\|full | Low-quality |
| S5_vs_k141_110519\|\|full | Low-quality |
| S5_vs_k141_1552\|\|full | Not-determined |
| S5_vs_k141_67935\|\|full | Low-quality |
| S5_vs_k141_61832\|\|full | Low-quality |
| S5_vs_k141_61835\|\|full | Low-quality |
| S5_vs_k141_49689\|\|full | Not-determined |
| S5_vs_k141_61841\|\|full | Low-quality |
| S5_vs_k141_104397\|\|full | Low-quality |
| S5_vs_k141_86198\|\|full | Low-quality |
| S5_vs_k141_140588\|\|full | Not-determined |
| S5_vs_k141_104399\|\|full | Not-determined |
| S5_vs_k141_61857\|\|full | Not-determined |
| S5_vs_k141_86201\|\|full | Low-quality |
| S5_vs_k141_140597\|\|full | Not-determined |
| S5_vs_k141_49695\|\|full | Not-determined |
| S5_vs_k141_104409\|\|full | Low-quality |
| S5_vs_k141_49702\|\|full | Low-quality |
| S5_vs_k141_44022\|\|full | Low-quality |
| S5_vs_k141_61861\|\|full | Low-quality |
| S5_vs_k141_110537\|\|full | Not-determined |
| S5_vs_k141_13303\|\|full | High-quality |
| S5_vs_k141_24897\|\|full | Not-determined |
| S5_vs_k141_110543\|\|full | Not-determined |
| S5_vs_k141_110548\|\|full | Low-quality |
| S5_vs_k141_110552\|\|full | Low-quality |
| S5_vs_k141_92420\|\|full | Low-quality |
| S5_vs_k141_92424\|\|full | Not-determined |
| S5_vs_k141_49727\|\|full | Low-quality |
| S5_vs_k141_73802\|\|full | Not-determined |
| S5_vs_k141_110559\|\|full | Not-determined |
| S5_vs_k141_117076\|\|full | Low-quality |
| S5_vs_k141_86223\|\|full | Low-quality |
| S5_vs_k141_122618\|\|full | High-quality |
| S5_vs_k141_13319\|\|full | Low-quality |
| S5_vs_k141_37682\|\|full | Low-quality |
| S5_vs_k141_67965\|\|full | Low-quality |
| S5_vs_k141_110560\|\|full | Low-quality |
| S5_vs_k141_19587\|\|full | Low-quality |
| S5_vs_k141_86231\|\|full | Low-quality |
| S5_vs_k141_49764\|\|full | Low-quality |
| S5_vs_k141_37690\|\|full | Low-quality |
| S5_vs_k141_37693\|\|full | Low-quality |
| S5_vs_k141_104461\|\|full | Not-determined |
| S5_vs_k141_92466\|\|full | Low-quality |
| S5_vs_k141_61891\|\|full | Low-quality |
| S5_vs_k141_13342\|\|full | Low-quality |
| S5_vs_k141_67971\|\|full | Not-determined |
| S5_vs_k141_37704\|\|full | Low-quality |
| S5_vs_k141_49783\|\|full | Low-quality |
| S5_vs_k141_49784\|\|full | Low-quality |
| S5_vs_k141_32032\|\|full | Not-determined |
| S5_vs_k141_49799\|\|full | Low-quality |
| S5_vs_k141_140681\|\|full | Low-quality |
| S5_vs_k141_123377\|\|full | Low-quality |
| S5_vs_k141_110611\|\|full | Low-quality |
| S5_vs_k141_117133\|\|full | Low-quality |
| S5_vs_k141_110613\|\|full | Not-determined |
| S5_vs_k141_32046\|\|full | Not-determined |
| S5_vs_k141_49808\|\|full | Not-determined |
| S5_vs_k141_44034\|\|full | Low-quality |
| S5_vs_k141_13363\|\|full | Low-quality |
| S5_vs_k141_68008\|\|full | Low-quality |
| S9_vs_k141_90849\|\|full | High-quality |
| S5_vs_k141_140708\|\|full | Low-quality |
| S5_vs_k141_117140\|\|full | Low-quality |
| S5_vs_k141_19596\|\|full | Low-quality |
| S5_vs_k141_110620\|\|full | Low-quality |
| S5_vs_k141_49827\|\|full | Not-determined |
| S5_vs_k141_55427\|\|full | Low-quality |
| S5_vs_k141_98059\|\|full | Low-quality |
| S5_vs_k141_104525\|\|full | Not-determined |
| S5_vs_k141_32081\|\|full | Low-quality |
| S5_vs_k141_134549\|\|full | Low-quality |
| S5_vs_k141_92533\|\|full | Not-determined |
| S5_vs_k141_55440\|\|full | Not-determined |
| S5_vs_k141_44072\|\|full | Low-quality |
| S5_vs_k141_117173\|\|full | Not-determined |
| S5_vs_k141_92535\|\|full | Low-quality |
| S5_vs_k141_37784\|\|full | Low-quality |
| S5_vs_k141_55446\|\|full | Low-quality |
| S7_extracted_9334 | Complete |
| S5_vs_k141_79537\|\|full | Low-quality |
| S5_vs_k141_128499\|\|full | Not-determined |
| S5_vs_k141_24906\|\|full | Low-quality |
| S5_vs_k141_98061\|\|full | Not-determined |
| S5_vs_k141_128512\|\|full | Low-quality |
| S5_vs_k141_104549\|\|full | Low-quality |
| S5_vs_k141_140718\|\|full | Not-determined |
| S5_vs_k141_140719\|\|full | Low-quality |
| S5_vs_k141_13373\|\|full | Not-determined |
| S5_vs_k141_110642\|\|full | Low-quality |
| S5_vs_k141_123411\|\|full | Not-determined |
| S5_vs_k141_55470\|\|full | Low-quality |
| S5_vs_k141_68051\|\|full | Low-quality |
| S5_vs_k141_61904\|\|full | Not-determined |
| S5_vs_k141_49850\|\|full | Not-determined |
| S5_vs_k141_140723\|\|full | Low-quality |
| S5_vs_k141_98073\|\|full | Low-quality |
| S5_vs_k141_110656\|\|full | Not-determined |
| S5_vs_k141_61918\|\|full | Low-quality |
| S5_vs_k141_19651\|\|full | Low-quality |
| S5_vs_k141_55482\|\|full | Not-determined |
| S5_vs_k141_128558\|\|full | Not-determined |
| S5_vs_k141_104592\|\|full | Low-quality |
| S5_vs_k141_68068\|\|full | Low-quality |
| S5_vs_k141_32157\|\|full | Not-determined |
| S5_vs_k141_92572\|\|full | Not-determined |
| S5_vs_k141_117184\|\|full | Low-quality |
| S5_vs_k141_134553\|\|full | Low-quality |
| S5_vs_k141_128574\|\|full | Low-quality |
| S5_vs_k141_134558\|\|full | Low-quality |
| S5_vs_k141_117191\|\|full | Not-determined |
| S5_vs_k141_49890\|\|full | Low-quality |
| S5_vs_k141_92588\|\|full | Not-determined |
| S5_vs_k141_79552\|\|full | Low-quality |
| S5_vs_k141_44131\|\|full | Low-quality |
| S5_vs_k141_49905\|\|full | Not-determined |
| S5_vs_k141_61948\|\|full | Low-quality |
| S5_vs_k141_24919\|\|full | Not-determined |
| S5_vs_k141_55512\|\|full | Not-determined |
| S5_vs_k141_55513\|\|full | Low-quality |
| S5_vs_k141_86282\|\|full | Low-quality |
| S5_vs_k141_128608\|\|full | Not-determined |
| S5_vs_k141_44158\|\|full | Low-quality |
| S5_vs_k141_32176\|\|full | Low-quality |
| S5_vs_k141_68105\|\|full | Not-determined |
| S5_vs_k141_44163\|\|full | Not-determined |
| S5_vs_k141_49935\|\|full | Low-quality |
| S5_vs_k141_86288\|\|full | Low-quality |
| S5_vs_k141_73890\|\|full | Low-quality |
| S5_vs_k141_110714\|\|full | Low-quality |
| S5_vs_k141_7560\|\|full | Low-quality |
| S5_vs_k141_123473\|\|full | Low-quality |
| S5_vs_k141_123476\|\|full | Not-determined |
| S5_vs_k141_61972\|\|full | Low-quality |
| S5_vs_k141_49951\|\|full | Not-determined |
| S5_vs_k141_92664\|\|full | Low-quality |
| S5_vs_k141_92667\|\|full | Low-quality |
| S5_vs_k141_134579\|\|full | Not-determined |
| S5_vs_k141_13434\|\|full | Not-determined |
| S5_vs_k141_92684\|\|full | Low-quality |
| S5_vs_k141_123497\|\|full | Low-quality |
| S5_vs_k141_134600\|\|full | Low-quality |
| S5_vs_k141_117263\|\|full | Not-determined |
| S5_vs_k141_86317\|\|full | Low-quality |
| S5_vs_k141_117266\|\|full | Low-quality |
| S5_vs_k141_68159\|\|full | Low-quality |
| S5_vs_k141_19665\|\|full | Low-quality |
| S5_vs_k141_61990\|\|full | Low-quality |
| S5_vs_k141_7566\|\|full | Low-quality |
| S5_vs_k141_1731\|\|full | Not-determined |
| S5_vs_k141_55544\|\|full | Low-quality |
| S5_vs_k141_68177\|\|full | Low-quality |
| S5_vs_k141_110789\|\|full | Low-quality |
| S5_vs_k141_117300\|\|full | Low-quality |
| S5_vs_k141_140754\|\|full | Low-quality |
| S5_vs_k141_104707\|\|full | Low-quality |
| S5_vs_k141_68194\|\|full | Low-quality |
| S5_vs_k141_13482\|\|full | Not-determined |
| S5_vs_k141_50020\|\|full | Not-determined |
| S5_vs_k141_68201\|\|full | Low-quality |
| S5_vs_k141_104716\|\|full | Low-quality |
| S14_vs_k141_4887\|\|full | High-quality |
| S5_vs_k141_7598\|\|full | Not-determined |
| S5_vs_k141_68210\|\|full | Low-quality |
| S5_vs_k141_55564\|\|full | Low-quality |
| S5_vs_k141_140774\|\|full | Not-determined |
| S5_vs_k141_128710\|\|full | Not-determined |
| S5_vs_k141_140777\|\|full | Not-determined |
| S5_vs_k141_134618\|\|full | Not-determined |
| S5_vs_k141_140779\|\|full | Not-determined |
| S5_vs_k141_86335\|\|full | Low-quality |
| S5_vs_k141_1761\|\|full | Low-quality |
| S5_vs_k141_86337\|\|full | Low-quality |
| S5_vs_k141_50041\|\|full | Low-quality |
| S5_vs_k141_79581\|\|full | Not-determined |
| S5_vs_k141_55578\|\|full | Low-quality |
| S5_vs_k141_32210\|\|full | Low-quality |
| S5_vs_k141_37884\|\|full | Not-determined |
| S5_vs_k141_86345\|\|full | Low-quality |
| S5_vs_k141_86350\|\|full | Low-quality |
| S5_vs_k141_134648\|\|full | Low-quality |
| S5_vs_k141_37893\|\|full | Low-quality |
| S5_vs_k141_1792\|\|full | Low-quality |
| S5_vs_k141_13490\|\|full | Not-determined |
| S5_vs_k141_123508\|\|full | Low-quality |
| S5_vs_k141_50077\|\|full | Not-determined |
| S5_vs_k141_62005\|\|full | Low-quality |
| S5_vs_k141_24973\|\|full | Not-determined |
| S5_vs_k141_24974\|\|full | Low-quality |
| S5_vs_k141_86364\|\|full | Not-determined |
| S5_vs_k141_140805\|\|full | Low-quality |
| S5_vs_k141_79604\|\|full | Low-quality |
| S5_vs_k141_123514\|\|full | Not-determined |
| S5_vs_k141_32255\|\|full | Not-determined |
| S5_vs_k141_44283\|\|full | Low-quality |
| S5_vs_k141_7628\|\|full | Not-determined |
| S5_vs_k141_50112\|\|full | Low-quality |
| S5_vs_k141_13526\|\|full | Not-determined |
| S5_vs_k141_123532\|\|full | Not-determined |
| S5_vs_k141_7632\|\|full | Not-determined |
| S5_vs_k141_92798\|\|full | Low-quality |
| S5_vs_k141_13534\|\|full | Not-determined |
| S5_vs_k141_37925\|\|full | Not-determined |
| S5_vs_k141_62040\|\|full | Not-determined |
| S5_vs_k141_50137\|\|full | Low-quality |
| S5_vs_k141_37928\|\|full | Low-quality |
| S5_vs_k141_44304\|\|full | Not-determined |
| S5_vs_k141_50142\|\|full | Low-quality |
| S5_vs_k141_128781\|\|full | Low-quality |
| S5_vs_k141_50144\|\|full | Low-quality |
| S5_vs_k141_62058\|\|full | Low-quality |
| S5_vs_k141_32301\|\|full | Low-quality |
| S5_vs_k141_7657\|\|full | Not-determined |
| S5_vs_k141_13573\|\|full | Not-determined |
| S5_vs_k141_32305\|\|full | Low-quality |
| S5_vs_k141_13581\|\|full | Not-determined |
| S5_vs_k141_134660\|\|full | Not-determined |
| S5_vs_k141_7675\|\|full | Not-determined |
| S5_vs_k141_32333\|\|full | Not-determined |
| S5_vs_k141_62086\|\|full | Low-quality |
| S5_vs_k141_7692\|\|full | Low-quality |
| S5_vs_k141_32344\|\|full | Low-quality |

| | |
|---|---|
| S5_vs_k141_1865\|\|full | Low-quality |
| S5_vs_k141_32349\|\|full | Low-quality |
| S5_vs_k141_128832\|\|full | Low-quality |
| S17_vs_k141_127360\|\|full | High-quality |
| S5_vs_k141_134664\|\|full | Not-determined |
| S5_vs_k141_98155\|\|full | Not-determined |
| S5_vs_k141_25057\|\|full | Low-quality |
| S5_vs_k141_13624\|\|full | Not-determined |
| S5_vs_k141_32360\|\|full | Low-quality |
| S5_vs_k141_37991\|\|full | Low-quality |
| S5_vs_k141_104755\|\|full | Low-quality |
| S5_vs_k141_117328\|\|full | Not-determined |
| S5_vs_k141_62096\|\|full | Not-determined |
| S5_vs_k141_117354\|\|full | Not-determined |
| S5_vs_k141_13642\|\|full | Not-determined |
| S5_vs_k141_86466\|\|full | Low-quality |
| S5_vs_k141_86468\|\|full | Not-determined |
| S5_vs_k141_1897\|\|full | Not-determined |
| S5_vs_k141_19715\|\|full | Not-determined |
| S5_vs_k141_86473\|\|full | Low-quality |
| S5_vs_k141_104756\|\|full | Low-quality |
| S5_vs_k141_13649\|\|full | Low-quality |
| S5_vs_k141_123566\|\|full | Low-quality |
| S5_vs_k141_117369\|\|full | Low-quality |
| S5_vs_k141_19719\|\|full | Low-quality |
| S5_vs_k141_117372\|\|full | Low-quality |
| S7_vs_117697\|\|full | High-quality |
| S5_vs_k141_44362\|\|full | Not-determined |
| S5_vs_k141_117378\|\|full | Low-quality |
| S5_vs_k141_32370\|\|full | Low-quality |
| S5_vs_k141_44377\|\|full | Low-quality |
| S5_vs_k141_62141\|\|full | Low-quality |
| S5_vs_k141_7713\|\|full | Low-quality |
| S5_vs_k141_117395\|\|full | Low-quality |
| S5_vs_k141_7718\|\|full | Low-quality |
| S5_vs_k141_104786\|\|full | Low-quality |
| S5_vs_k141_7722\|\|full | Not-determined |
| S5_vs_k141_62152\|\|full | Not-determined |
| S5_vs_k141_68264\|\|full | Low-quality |
| S5_vs_k141_25113\|\|full | Low-quality |
| S5_vs_k141_55598\|\|full | Not-determined |
| S5_vs_k141_86498\|\|full | Low-quality |
| S5_vs_k141_104793\|\|full | Low-quality |
| S5_vs_k141_7729\|\|full | Not-determined |
| S5_vs_k141_38047\|\|full | Low-quality |
| S5_vs_k141_68272\|\|full | Not-determined |
| S5_vs_k141_13669\|\|full | Low-quality |
| S5_vs_k141_92856\|\|full | Low-quality |
| S4_vs_VIRSorter_16493-cat_2 | High-quality |
| S5_vs_k141_1973\|\|full | Not-determined |
| S5_vs_k141_7756\|\|full | Low-quality |
| S5_vs_k141_104817\|\|full | Low-quality |
| S5_vs_k141_79661\|\|full | Low-quality |
| S5_vs_k141_32419\|\|full | Not-determined |
| S5_vs_k141_140856\|\|full | Not-determined |
| S5_vs_k141_50216\|\|full | Not-determined |
| S5_vs_k141_79668\|\|full | Low-quality |
| S5_vs_k141_1990\|\|full | Low-quality |
| S5_vs_k141_62216\|\|full | Not-determined |
| S5_vs_k141_62217\|\|full | Low-quality |
| S5_vs_k141_110899\|\|full | Not-determined |
| S5_vs_k141_117424\|\|full | Low-quality |
| S5_vs_k141_140866\|\|full | Low-quality |
| S5_vs_k141_32444\|\|full | Low-quality |
| S5_vs_k141_32448\|\|full | Low-quality |
| S5_vs_k141_13736\|\|full | Not-determined |
| S5_vs_k141_38074\|\|full | Not-determined |
| S5_vs_k141_7785\|\|full | Not-determined |
| S5_vs_k141_74027\|\|full | Low-quality |
| S5_vs_k141_7792\|\|full | Low-quality |
| S5_vs_k141_134776\|\|full | Low-quality |
| S5_vs_k141_128854\|\|full | Not-determined |
| S5_vs_k141_13759\|\|full | Low-quality |
| S5_vs_k141_128855\|\|full | Not-determined |
| S5_vs_k141_104884\|\|full | Low-quality |
| S5_vs_k141_55657\|\|full | Not-determined |
| S5_vs_k141_140910\|\|full | Low-quality |
| S5_vs_k141_104893\|\|full | Low-quality |
| S5_vs_k141_25165\|\|full | Low-quality |
| S5_vs_k141_55666\|\|full | Not-determined |
| S5_vs_k141_140919\|\|full | Low-quality |
| S5_vs_k141_117457\|\|full | Low-quality |
| S5_vs_k141_19741\|\|full | Low-quality |
| S5_vs_k141_2004\|\|full | Low-quality |
| S5_vs_k141_74039\|\|full | Low-quality |
| S5_vs_k141_13778\|\|full | Not-determined |
| S5_vs_k141_79671\|\|full | Not-determined |
| S5_vs_k141_92879\|\|full | Low-quality |
| S5_vs_k141_134795\|\|full | Not-determined |
| S5_vs_k141_79685\|\|full | Not-determined |
| S5_vs_k141_117468\|\|full | Low-quality |
| S5_vs_k141_44477\|\|full | Low-quality |
| S5_vs_k141_79691\|\|full | Low-quality |
| S5_vs_k141_19769\|\|full | Low-quality |
| S5_vs_k141_68305\|\|full | Not-determined |
| S5_vs_k141_50305\|\|full | Not-determined |
| S5_vs_k141_68314\|\|full | Low-quality |
| S5_vs_k141_104917\|\|full | Low-quality |
| S5_vs_k141_68316\|\|full | Not-determined |
| S5_vs_k141_25183\|\|full | Not-determined |
| S5_vs_k141_13816\|\|full | Low-quality |
| S13_extracted_24991 | Medium-quality |
| S5_vs_k141_134804\|\|full | Low-quality |
| S7_vs_9334\|\|full | High-quality |
| S5_vs_k141_92882\|\|full | Not-determined |
| S5_vs_k141_55688\|\|full | Low-quality |
| S11_extracted_k141_68693 | Medium-quality |
| S5_vs_k141_13823\|\|full | Low-quality |
| S5_vs_k141_55692\|\|full | Low-quality |
| S5_vs_k141_117501\|\|full | Low-quality |
| S5_vs_k141_2017\|\|full | Low-quality |
| S5_vs_k141_111002\|\|full | Low-quality |
| S5_vs_k141_86575\|\|full | Low-quality |
| S5_vs_k141_104937\|\|full | Low-quality |
| S5_vs_k141_92906\|\|full | Low-quality |
| S5_vs_k141_104940\|\|full | Low-quality |
| S5_vs_k141_68319\|\|full | Not-determined |
| S5_vs_k141_104949\|\|full | Not-determined |
| S5_vs_k141_117515\|\|full | Low-quality |
| S5_vs_k141_19802\|\|full | Low-quality |
| S5_vs_k141_111011\|\|full | Not-determined |
| S5_vs_k141_134834\|\|full | Low-quality |
| S5_vs_k141_2037\|\|full | Not-determined |
| S5_vs_k141_86586\|\|full | Low-quality |
| S5_vs_k141_19808\|\|full | Low-quality |
| S5_vs_k141_134837\|\|full | Low-quality |
| S5_vs_k141_2048\|\|full | Low-quality |
| S5_vs_k141_79720\|\|full | Not-determined |
| S5_vs_k141_2055\|\|full | Low-quality |
| S5_vs_k141_7898\|\|full | Not-determined |
| S5_vs_k141_92942\|\|full | Low-quality |
| S5_extracted_k141_74661 | High-quality |
| S5_vs_k141_2075\|\|full | Low-quality |
| S5_vs_k141_111026\|\|full | Low-quality |
| S5_vs_k141_128952\|\|full | Not-determined |
| S5_vs_k141_2083\|\|full | Low-quality |
| S5_vs_k141_92962\|\|full | Low-quality |
| S5_vs_k141_86624\|\|full | Low-quality |
| S5_vs_k141_55715\|\|full | Low-quality |
| S5_vs_k141_50361\|\|full | Low-quality |
| S5_vs_k141_111038\|\|full | Not-determined |
| S5_vs_k141_68353\|\|full | Not-determined |
| S5_vs_k141_7931\|\|full | Low-quality |
| S5_vs_k141_68361\|\|full | Low-quality |
| S5_vs_k141_117553\|\|full | Not-determined |
| S5_vs_k141_123578\|\|full | Not-determined |
| S5_vs_k141_92979\|\|full | Not-determined |
| S5_vs_k141_50368\|\|full | Not-determined |
| S5_vs_k141_68364\|\|full | Low-quality |
| S5_vs_k141_68374\|\|full | Low-quality |
| S5_vs_k141_2132\|\|full | Low-quality |
| S5_vs_k141_92996\|\|full | Low-quality |
| S5_vs_k141_2133\|\|full | Low-quality |
| S5_vs_k141_128994\|\|full | Not-determined |
| S5_vs_k141_7958\|\|full | Low-quality |
| S5_vs_k141_86657\|\|full | Low-quality |
| S5_vs_k141_25212\|\|full | Not-determined |
| S5_vs_k141_62222\|\|full | Low-quality |
| S5_vs_k141_25214\|\|full | Not-determined |
| S5_vs_k141_2157\|\|full | Low-quality |
| S5_vs_k141_123589\|\|full | Low-quality |
| S5_vs_k141_111085\|\|full | Not-determined |
| S5_vs_k141_2165\|\|full | Low-quality |
| S5_vs_k141_86663\|\|full | Not-determined |
| S5_vs_k141_62229\|\|full | Low-quality |
| S5_vs_k141_68411\|\|full | Low-quality |
| S5_vs_k141_86672\|\|full | Low-quality |
| S8_extracted_k141_7243 | Medium-quality |
| S5_vs_k141_93003\|\|full | Low-quality |
| S5_vs_k141_19875\|\|full | Low-quality |
| S5_vs_k141_19878\|\|full | Low-quality |
| S5_vs_k141_111120\|\|full | Low-quality |
| S5_vs_k141_8004\|\|full | Low-quality |
| S5_vs_k141_38093\|\|full | Not-determined |
| S5_vs_k141_50422\|\|full | Not-determined |
| S5_vs_k141_117601\|\|full | Low-quality |
| S5_vs_k141_129028\|\|full | Low-quality |
| S5_vs_k141_123636\|\|full | Low-quality |
| S5_vs_k141_38098\|\|full | Low-quality |
| S5_vs_k141_8024\|\|full | Not-determined |
| S5_vs_k141_104979\|\|full | Low-quality |
| S5_vs_k141_117606\|\|full | Low-quality |
| S5_vs_k141_111134\|\|full | Not-determined |
| S5_vs_k141_104987\|\|full | Low-quality |
| S5_vs_k141_123651\|\|full | Low-quality |
| S5_vs_k141_117617\|\|full | Low-quality |
| S5_vs_k141_134883\|\|full | Not-determined |
| S5_vs_k141_25279\|\|full | Not-determined |
| S5_vs_k141_13862\|\|full | Low-quality |
| S5_vs_k141_105013\|\|full | Low-quality |
| S5_vs_k141_134892\|\|full | Low-quality |
| S5_vs_k141_86705\|\|full | Low-quality |
| S5_vs_k141_86714\|\|full | Not-determined |
| S5_vs_k141_140975\|\|full | Low-quality |
| S5_vs_k141_2216\|\|full | Not-determined |
| S5_vs_k141_129053\|\|full | Not-determined |
| S5_vs_k141_44528\|\|full | Low-quality |
| S5_vs_k141_62250\|\|full | Low-quality |
| S5_vs_k141_129061\|\|full | Low-quality |
| S16_extracted_52513 | High-quality |
| S5_vs_k141_140990\|\|full | Low-quality |
| S5_vs_k141_105052\|\|full | Not-determined |
| S5_vs_k141_123691\|\|full | Not-determined |
| S5_vs_k141_129065\|\|full | Not-determined |
| S5_vs_k141_117643\|\|full | Low-quality |
| S5_vs_k141_129075\|\|full | Not-determined |
| S5_vs_k141_13893\|\|full | Low-quality |
| S5_vs_k141_86735\|\|full | Low-quality |
| S5_vs_k141_123696\|\|full | Not-determined |
| S5_vs_k141_129090\|\|full | Low-quality |
| S5_vs_k141_32555\|\|full | Not-determined |
| S5_vs_k141_86738\|\|full | Not-determined |
| S5_vs_k141_62278\|\|full | Not-determined |
| S5_vs_k141_129105\|\|full | Low-quality |
| S5_vs_k141_62298\|\|full | Low-quality |
| S5_vs_k141_8067\|\|full | Low-quality |
| S5_vs_k141_13925\|\|full | Not-determined |
| S5_vs_k141_13926\|\|full | Low-quality |
| S5_vs_k141_44573\|\|full | Not-determined |
| S5_vs_k141_8088\|\|full | Low-quality |
| S5_vs_k141_74059\|\|full | Low-quality |
| S5_vs_k141_44599\|\|full | Low-quality |
| S5_vs_k141_79810\|\|full | Not-determined |
| S5_vs_k141_79812\|\|full | Low-quality |
| S5_vs_k141_13948\|\|full | Not-determined |
| S5_vs_k141_2326\|\|full | Low-quality |
| S5_vs_k141_2329\|\|full | Low-quality |
| S5_vs_k141_2338\|\|full | Low-quality |
| S5_vs_k141_117726\|\|full | Not-determined |
| S5_vs_k141_13960\|\|full | Low-quality |
| S5_vs_k141_123723\|\|full | Not-determined |
| S5_vs_k141_50507\|\|full | Low-quality |
| S5_vs_k141_2358\|\|full | Low-quality |
| S5_vs_k141_123730\|\|full | Low-quality |
| S5_vs_k141_141098\|\|full | Low-quality |
| S5_vs_k141_98246\|\|full | Not-determined |
| S5_vs_k141_79844\|\|full | Low-quality |
| S5_vs_k141_68464\|\|full | Not-determined |
| S5_vs_k141_38167\|\|full | Low-quality |
| S5_vs_k141_13980\|\|full | Not-determined |
| S5_vs_k141_13986\|\|full | Not-determined |
| S16_vs_52513\|\|full | High-quality |
| S5_vs_k141_50535\|\|full | Not-determined |
| S5_vs_k141_50538\|\|full | Low-quality |
| S5_vs_k141_79872\|\|full | Not-determined |
| S5_vs_k141_2397\|\|full | Low-quality |
| S5_vs_k141_86758\|\|full | Not-determined |
| S5_vs_k141_44642\|\|full | Low-quality |
| S5_vs_k141_93039\|\|full | Not-determined |
| S5_vs_k141_19987\|\|full | Low-quality |
| S5_vs_k141_19990\|\|full | Low-quality |
| S5_vs_k141_105209\|\|full | Low-quality |
| S5_vs_k141_2412\|\|full | Low-quality |
| S5_vs_k141_79879\|\|full | Low-quality |
| S5_vs_k141_14008\|\|full | Not-determined |
| S5_vs_k141_2416\|\|full | Low-quality |
| S5_vs_k141_62312\|\|full | Not-determined |
| S5_vs_k141_86770\|\|full | Low-quality |
| S5_vs_k141_19999\|\|full | Not-determined |
| S5_vs_k141_134946\|\|full | Not-determined |
| S5_vs_k141_86782\|\|full | Low-quality |
| S16_vs_136935\|\|full | Medium-quality |
| S5_vs_k141_2427\|\|full | Low-quality |
| S5_vs_k141_93073\|\|full | Not-determined |
| S5_vs_k141_117774\|\|full | Low-quality |
| S5_vs_k141_117777\|\|full | Not-determined |
| S5_vs_k141_68521\|\|full | Low-quality |
| S5_vs_k141_14024\|\|full | Low-quality |
| S5_vs_k141_93095\|\|full | Not-determined |
| S5_vs_k141_117789\|\|full | Low-quality |
| S5_vs_k141_62346\|\|full | Low-quality |
| S5_vs_k141_2436\|\|full | Not-determined |
| S5_vs_k141_14043\|\|full | Low-quality |
| S5_vs_k141_93101\|\|full | Low-quality |
| S5_vs_k141_123815\|\|full | Low-quality |
| S5_vs_k141_62359\|\|full | Not-determined |
| S5_vs_k141_62360\|\|full | Not-determined |
| S5_vs_k141_123825\|\|full | Low-quality |
| S5_vs_k141_68538\|\|full | Not-determined |
| S5_vs_k141_93108\|\|full | Low-quality |
| S5_vs_k141_68544\|\|full | Not-determined |
| S5_vs_k141_32643\|\|full | Not-determined |
| S5_vs_k141_50610\|\|full | Low-quality |
| S5_vs_k141_117798\|\|full | Low-quality |
| S5_vs_k141_105303\|\|full | Low-quality |
| S5_vs_k141_79910\|\|full | Low-quality |
| S5_vs_k141_117809\|\|full | Low-quality |
| S5_vs_k141_38184\|\|full | Not-determined |
| S5_vs_k141_134972\|\|full | Low-quality |
| S5_vs_k141_38186\|\|full | Low-quality |
| S4_extracted_8925 | Medium-quality |
| S5_vs_k141_32662\|\|full | Low-quality |
| S5_vs_k141_117816\|\|full | Low-quality |
| S5_vs_k141_8209\|\|full | Low-quality |
| S5_vs_k141_134974\|\|full | Low-quality |
| S5_vs_k141_38193\|\|full | Not-determined |
| S5_vs_k141_2480\|\|full | Low-quality |
| S5_vs_k141_32666\|\|full | Low-quality |
| S5_vs_k141_105324\|\|full | Not-determined |
| S5_vs_k141_123834\|\|full | Not-determined |
| S5_vs_k141_38199\|\|full | Low-quality |
| S5_vs_k141_129240\|\|full | Low-quality |
| S5_vs_k141_68558\|\|full | Low-quality |
| S5_vs_k141_105338\|\|full | Not-determined |
| S5_vs_k141_2485\|\|full | Low-quality |
| S5_vs_k141_93147\|\|full | Not-determined |
| S5_vs_k141_68566\|\|full | Low-quality |
| S5_vs_k141_20051\|\|full | Not-determined |
| S5_vs_k141_123865\|\|full | Low-quality |
| S5_vs_k141_68569\|\|full | Low-quality |
| S5_vs_k141_129266\|\|full | Low-quality |
| S5_vs_k141_38220\|\|full | Not-determined |
| S5_vs_k141_38231\|\|full | Low-quality |
| S5_vs_k141_74121\|\|full | Not-determined |
| S5_vs_k141_129270\|\|full | Low-quality |
| S5_vs_k141_2510\|\|full | Not-determined |
| S5_vs_k141_93184\|\|full | Low-quality |
| S4_vs_VIRSorter_8925-cat_2 | Medium-quality |
| S5_vs_k141_117897\|\|full | Low-quality |
| S5_vs_k141_74137\|\|full | Low-quality |
| S5_vs_k141_44751\|\|full | Not-determined |
| S5_vs_k141_38249\|\|full | Low-quality |
| S5_vs_k141_20076\|\|full | Low-quality |
| S5_vs_k141_134980\|\|full | Not-determined |
| S5_vs_k141_38263\|\|full | Low-quality |
| S5_vs_k141_68585\|\|full | Low-quality |
| S5_vs_k141_14135\|\|full | Low-quality |
| S5_vs_k141_38264\|\|full | Not-determined |
| S5_vs_k141_38265\|\|full | Not-determined |
| S5_vs_k141_8242\|\|full | Low-quality |
| S5_vs_k141_141156\|\|full | Low-quality |
| S5_vs_k141_129297\|\|full | Low-quality |
| S5_vs_k141_25389\|\|full | Low-quality |
| S5_vs_k141_50696\|\|full | Low-quality |
| S5_vs_k141_129300\|\|full | Not-determined |
| S5_vs_k141_129301\|\|full | Not-determined |
| S5_vs_k141_68600\|\|full | Low-quality |
| S5_vs_k141_123923\|\|full | Low-quality |
| S5_vs_k141_38281\|\|full | Not-determined |
| S5_vs_k141_20106\|\|full | Low-quality |
| S5_vs_k141_129312\|\|full | Low-quality |
| S5_vs_k141_62392\|\|full | Low-quality |
| S5_vs_k141_20121\|\|full | Low-quality |
| S5_vs_k141_134996\|\|full | Low-quality |
| S5_vs_k141_20127\|\|full | Low-quality |
| S5_vs_k141_134998\|\|full | Low-quality |
| S5_vs_k141_38295\|\|full | Low-quality |
| S5_vs_k141_44801\|\|full | Low-quality |
| S5_vs_k141_135008\|\|full | Low-quality |
| S5_vs_k141_105444\|\|full | Low-quality |
| S5_vs_k141_32750\|\|full | Low-quality |
| S5_vs_k141_141219\|\|full | Not-determined |
| S5_vs_k141_20136\|\|full | Low-quality |
| S5_vs_k141_117930\|\|full | Low-quality |
| S5_vs_k141_20139\|\|full | Low-quality |
| S5_vs_k141_20140\|\|full | Low-quality |
| S5_vs_k141_141225\|\|full | Low-quality |
| S5_vs_k141_74197\|\|full | Not-determined |
| S5_vs_k141_141232\|\|full | Low-quality |
| S5_vs_k141_55972\|\|full | Low-quality |
| S5_vs_k141_32774\|\|full | Low-quality |
| S5_vs_k141_14214\|\|full | Low-quality |
| S5_vs_k141_141244\|\|full | Not-determined |
| S5_vs_k141_62440\|\|full | Not-determined |
| S5_vs_k141_86854\|\|full | Low-quality |
| S5_vs_k141_105481\|\|full | Not-determined |
| S5_vs_k141_2547\|\|full | Not-determined |
| S5_vs_k141_117964\|\|full | Not-determined |
| S5_vs_k141_32790\|\|full | Not-determined |
| S5_vs_k141_111216\|\|full | Low-quality |
| S5_vs_k141_111217\|\|full | Low-quality |
| S5_vs_k141_141263\|\|full | Low-quality |
| S5_vs_k141_2558\|\|full | Low-quality |
| S5_vs_k141_93204\|\|full | Not-determined |
| S5_vs_k141_117981\|\|full | Not-determined |
| S5_vs_k141_93206\|\|full | Not-determined |
| S5_vs_k141_2564\|\|full | Low-quality |
| S5_vs_k141_32809\|\|full | Not-determined |
| S5_vs_k141_117992\|\|full | Low-quality |
| S5_vs_k141_141280\|\|full | Low-quality |
| S5_vs_k141_93217\|\|full | Not-determined |
| S5_vs_k141_141282\|\|full | Not-determined |
| S5_vs_k141_2572\|\|full | Low-quality |
| S5_vs_k141_111228\|\|full | Low-quality |
| S5_vs_k141_111230\|\|full | Not-determined |
| S5_vs_k141_32818\|\|full | Not-determined |
| S5_vs_k141_8343\|\|full | Not-determined |
| S5_vs_k141_135027\|\|full | Low-quality |
| S5_vs_k141_68613\|\|full | Low-quality |
| S5_vs_k141_93228\|\|full | Not-determined |
| S5_vs_k141_8357\|\|full | Low-quality |
| S5_vs_k141_79931\|\|full | Low-quality |
| S5_vs_k141_105527\|\|full | Not-determined |
| S5_vs_k141_68623\|\|full | Low-quality |
| S5_vs_k141_123981\|\|full | Not-determined |
| S5_vs_k141_79946\|\|full | Low-quality |
| S5_vs_k141_50748\|\|full | Low-quality |
| S5_vs_k141_129408\|\|full | Low-quality |
| S5_vs_k141_62453\|\|full | Low-quality |
| S5_vs_k141_141325\|\|full | Not-determined |
| S5_vs_k141_118033\|\|full | Not-determined |
| S5_vs_k141_111251\|\|full | Not-determined |
| S5_vs_k141_118041\|\|full | Low-quality |
| S5_vs_k141_8388\|\|full | Not-determined |
| S15_vs_7103\|\|full | High-quality |
| S5_vs_k141_14309\|\|full | Not-determined |
| S5_vs_k141_8391\|\|full | Not-determined |
| S5_vs_k141_25425\|\|full | Low-quality |
| S5_vs_k141_25432\|\|full | Low-quality |
| S8_vs_k141_9114\|\|full | High-quality |
| S5_vs_k141_93267\|\|full | Low-quality |
| S5_vs_k141_8402\|\|full | Not-determined |
| S5_vs_k141_105596\|\|full | Low-quality |
| S5_vs_k141_62465\|\|full | Low-quality |
| S5_vs_k141_105607\|\|full | Not-determined |
| S5_vs_k141_105614\|\|full | Not-determined |
| S5_vs_k141_105632\|\|full | Low-quality |
| S5_vs_k141_124019\|\|full | Not-determined |
| S5_vs_k141_8425\|\|full | Low-quality |
| S5_vs_k141_124024\|\|full | Low-quality |
| S5_vs_k141_135068\|\|full | Low-quality |
| S5_vs_k141_111316\|\|full | Low-quality |
| S5_vs_k141_124032\|\|full | Not-determined |
| S5_vs_k141_20205\|\|full | Low-quality |
| S5_vs_k141_118116\|\|full | Low-quality |
| S5_vs_k141_118117\|\|full | Low-quality |
| S5_vs_k141_20213\|\|full | Low-quality |
| S5_vs_k141_50828\|\|full | Low-quality |
| S14_vs_k141_117165\|\|full | High-quality |
| S5_vs_k141_50835\|\|full | Not-determined |
| S5_vs_k141_141328\|\|full | Not-determined |
| S5_vs_k141_141331\|\|full | Low-quality |
| S5_vs_k141_50838\|\|full | Low-quality |
| S5_vs_k141_25516\|\|full | Low-quality |
| S5_vs_k141_93278\|\|full | Low-quality |
| S5_vs_k141_20234\|\|full | Low-quality |
| S5_vs_k141_20238\|\|full | Low-quality |
| S5_vs_k141_25525\|\|full | Not-determined |
| S5_vs_k141_20244\|\|full | Not-determined |
| S5_vs_k141_124061\|\|full | Not-determined |
| S5_vs_k141_141366\|\|full | Low-quality |
| S5_vs_k141_141367\|\|full | Low-quality |
| S5_vs_k141_129451\|\|full | Low-quality |
| S5_vs_k141_141370\|\|full | Low-quality |
| S5_vs_k141_93292\|\|full | Not-determined |
| S5_vs_k141_38307\|\|full | Not-determined |
| S5_vs_k141_2656\|\|full | Not-determined |
| S5_vs_k141_62502\|\|full | Low-quality |
| S5_vs_k141_93304\|\|full | Low-quality |
| S5_vs_k141_25547\|\|full | Not-determined |
| S12_extracted_k141_174374 | High-quality |
| S5_vs_k141_124105\|\|full | Low-quality |
| S5_vs_k141_32823\|\|full | Low-quality |
| S5_vs_k141_50928\|\|full | Low-quality |
| S5_vs_k141_2696\|\|full | Low-quality |
| S5_vs_k141_32833\|\|full | Low-quality |
| S5_vs_k141_38334\|\|full | Not-determined |
| S13_vs_24991\|\|full | Medium-quality |
| S5_vs_k141_20280\|\|full | Low-quality |
| S5_vs_k141_25601\|\|full | Low-quality |
| S5_vs_k141_2723\|\|full | Low-quality |
| S5_vs_k141_20287\|\|full | Low-quality |
| S5_vs_k141_38347\|\|full | Not-determined |
| S5_vs_k141_38349\|\|full | Low-quality |
| S5_vs_k141_2743\|\|full | Not-determined |
| S3_extracted_k141_19349 | High-quality |
| S5_vs_k141_135116\|\|full | Low-quality |
| S5_vs_k141_44830\|\|full | Low-quality |
| S5_vs_k141_14371\|\|full | Low-quality |
| S5_vs_k141_25628\|\|full | Low-quality |
| S5_vs_k141_135117\|\|full | Not-determined |
| S5_vs_k141_32834\|\|full | Low-quality |
| S5_vs_k141_50984\|\|full | Not-determined |
| S5_vs_k141_44839\|\|full | Not-determined |
| S5_vs_k141_68693\|\|full | Low-quality |
| S5_vs_k141_44843\|\|full | Low-quality |
| S5_vs_k141_124130\|\|full | Low-quality |
| S5_vs_k141_98380\|\|full | Low-quality |
| S5_vs_k141_20321\|\|full | Low-quality |
| S5_vs_k141_98383\|\|full | Low-quality |
| S5_vs_k141_141439\|\|full | Not-determined |
| S5_vs_k141_14399\|\|full | Not-determined |
| S5_vs_k141_129499\|\|full | Low-quality |
| S12_vs_k141_174374\|\|full | High-quality |
| S5_vs_k141_124154\|\|full | Low-quality |
| S5_vs_k141_111369\|\|full | Low-quality |
| S5_vs_k141_98396\|\|full | Low-quality |
| S5_vs_k141_20332\|\|full | Not-determined |
| S5_vs_k141_129524\|\|full | Not-determined |
| S5_vs_k141_124165\|\|full | Low-quality |
| S5_vs_k141_14415\|\|full | Not-determined |
| S5_vs_k141_62536\|\|full | Not-determined |
| S5_vs_k141_25687\|\|full | Low-quality |
| S5_vs_k141_25701\|\|full | Low-quality |
| S5_vs_k141_135157\|\|full | Low-quality |
| S5_vs_k141_80021\|\|full | Low-quality |
| S5_vs_k141_25704\|\|full | Not-determined |
| S5_vs_k141_25713\|\|full | Low-quality |
| S5_vs_k141_105787\|\|full | Low-quality |
| S5_vs_k141_80023\|\|full | Not-determined |
| S5_vs_k141_25716\|\|full | Low-quality |
| S5_vs_k141_105791\|\|full | Not-determined |
| S5_vs_k141_124197\|\|full | Low-quality |
| S5_vs_k141_51016\|\|full | Low-quality |
| S5_vs_k141_62565\|\|full | Low-quality |
| S5_vs_k141_135172\|\|full | Low-quality |
| S5_vs_k141_25730\|\|full | Low-quality |
| S5_vs_k141_105800\|\|full | Low-quality |
| S5_vs_k141_20344\|\|full | Low-quality |
| S5_vs_k141_25734\|\|full | Low-quality |
| S5_vs_k141_135182\|\|full | Low-quality |
| S5_vs_k141_2822\|\|full | Low-quality |
| S5_vs_k141_105807\|\|full | Low-quality |
| S5_vs_k141_56029\|\|full | Not-determined |
| S5_vs_k141_80043\|\|full | Low-quality |
| S5_vs_k141_105818\|\|full | Low-quality |
| S5_vs_k141_80047\|\|full | Not-determined |
| S5_vs_k141_93348\|\|full | Not-determined |
| S5_vs_k141_105821\|\|full | Low-quality |
| S5_vs_k141_2831\|\|full | Not-determined |
| S5_vs_k141_111430\|\|full | Low-quality |
| S5_vs_k141_80051\|\|full | Not-determined |
| S5_vs_k141_62592\|\|full | Low-quality |
| S5_vs_k141_111435\|\|full | Low-quality |
| S5_vs_k141_20364\|\|full | Not-determined |
| S5_vs_k141_68772\|\|full | Low-quality |
| S5_vs_k141_98426\|\|full | Low-quality |
| S5_vs_k141_2844\|\|full | Not-determined |
| S5_vs_k141_68776\|\|full | Low-quality |
| S5_vs_k141_51046\|\|full | Low-quality |
| S5_vs_k141_105849\|\|full | Not-determined |
| S5_vs_k141_98439\|\|full | Not-determined |
| S5_vs_k141_105859\|\|full | Not-determined |
| S5_vs_k141_51072\|\|full | Not-determined |
| S5_vs_k141_59798\|\|full | Medium-quality |
| S5_vs_k141_98450\|\|full | Low-quality |
| S5_vs_k141_2869\|\|full | Low-quality |
| S5_vs_k141_135188\|\|full | Low-quality |
| S5_vs_k141_20390\|\|full | Not-determined |
| S5_vs_k141_98455\|\|full | Low-quality |
| S5_vs_k141_20412\|\|full | Low-quality |
| S5_vs_k141_56039\|\|full | Low-quality |
| S5_vs_k141_124252\|\|full | Not-determined |
| S5_vs_k141_56044\|\|full | Not-determined |
| S5_vs_k141_135196\|\|full | Low-quality |
| S5_vs_k141_8532\|\|full | Low-quality |
| S8_vs_k141_71212\|\|full | High-quality |
| S5_vs_k141_74661\|\|full | High-quality |
| S5_vs_k141_68817\|\|full | Low-quality |
| S5_vs_k141_2878\|\|full | Low-quality |
| S5_vs_k141_111454\|\|full | Low-quality |
| S5_vs_k141_68818\|\|full | Not-determined |
| S5_vs_k141_2883\|\|full | Low-quality |
| S5_vs_k141_2888\|\|full | Low-quality |
| S5_vs_k141_68821\|\|full | Not-determined |
| S5_vs_k141_93457\|\|full | Not-determined |
| S5_vs_k141_2895\|\|full | Low-quality |
| S5_vs_k141_20434\|\|full | Low-quality |
| S5_vs_k141_51127\|\|full | Low-quality |
| S5_vs_k141_62605\|\|full | Low-quality |
| S5_vs_k141_74213\|\|full | Low-quality |
| S5_vs_k141_14440\|\|full | Low-quality |
| S5_vs_k141_62608\|\|full | Low-quality |
| S5_vs_k141_118220\|\|full | Not-determined |
| S5_vs_k141_62614\|\|full | Not-determined |
| S5_vs_k141_124285\|\|full | Low-quality |
| S5_vs_k141_8569\|\|full | Not-determined |
| S5_vs_k141_14448\|\|full | Low-quality |
| S5_vs_k141_68869\|\|full | Low-quality |
| S5_vs_k141_135225\|\|full | Not-determined |
| S5_vs_k141_124288\|\|full | Low-quality |
| S5_vs_k141_129671\|\|full | Low-quality |
| S5_vs_k141_93512\|\|full | Low-quality |
| S5_vs_k141_2920\|\|full | Low-quality |
| S5_vs_k141_62615\|\|full | Low-quality |
| S5_vs_k141_68882\|\|full | Low-quality |
| S5_vs_k141_129676\|\|full | Low-quality |
| S5_vs_k141_93521\|\|full | Not-determined |
| S5_vs_k141_62622\|\|full | Low-quality |
| S5_vs_k141_68888\|\|full | Low-quality |
| S5_vs_k141_124308\|\|full | Low-quality |
| S5_vs_k141_51130\|\|full | Low-quality |
| S5_vs_k141_129684\|\|full | Low-quality |
| S5_vs_k141_124313\|\|full | Low-quality |
| S5_vs_k141_20464\|\|full | Not-determined |
| S5_vs_k141_2941\|\|full | Not-determined |
| S5_vs_k141_20469\|\|full | Not-determined |
| S5_vs_k141_2948\|\|full | Not-determined |
| S5_vs_k141_129687\|\|full | Low-quality |

| | |
|---|---|
| S5_vs_k141_2952\|\|full | Not-determined |
| S5_vs_k141_62654\|\|full | Low-quality |
| S5_vs_k141_135237\|\|full | Low-quality |
| S5_vs_k141_93553\|\|full | Low-quality |
| S5_vs_k141_14497\|\|full | Low-quality |
| S5_vs_k141_68903\|\|full | Low-quality |
| S5_vs_k141_2970\|\|full | Low-quality |
| S5_vs_k141_25790\|\|full | Low-quality |
| S5_vs_k141_2981\|\|full | Not-determined |
| S5_vs_k141_14513\|\|full | Low-quality |
| S5_vs_k141_145598\|\|full | Medium-quality |
| S5_vs_k141_124358\|\|full | Low-quality |
| S5_vs_k141_105904\|\|full | Not-determined |
| S5_vs_k141_93581\|\|full | Low-quality |
| S5_vs_k141_14519\|\|full | Low-quality |
| S5_vs_k141_14520\|\|full | Low-quality |
| S5_vs_k141_51173\|\|full | Low-quality |
| S5_vs_k141_129707\|\|full | Low-quality |
| S5_vs_k141_8670\|\|full | Low-quality |
| S5_vs_k141_38520\|\|full | Low-quality |
| S5_vs_k141_129713\|\|full | Not-determined |
| S5_vs_k141_105929\|\|full | Low-quality |
| S5_vs_k141_8684\|\|full | Low-quality |
| S5_vs_k141_124369\|\|full | Not-determined |
| S5_vs_k141_56100\|\|full | Low-quality |
| S5_vs_k141_44872\|\|full | Low-quality |
| S5_vs_k141_38536\|\|full | Low-quality |
| S5_vs_k141_14558\|\|full | Not-determined |
| S5_vs_k141_80165\|\|full | Low-quality |
| S5_vs_k141_38540\|\|full | Not-determined |
| S5_vs_k141_8702\|\|full | Low-quality |
| S5_vs_k141_44877\|\|full | Not-determined |
| S5_vs_k141_129748\|\|full | Not-determined |
| S5_vs_k141_8703\|\|full | Not-determined |
| S5_vs_k141_44883\|\|full | Low-quality |
| S5_vs_k141_80178\|\|full | Low-quality |
| S5_vs_k141_38550\|\|full | Low-quality |
| S5_vs_k141_93642\|\|full | Not-determined |
| S5_vs_k141_74316\|\|full | Low-quality |
| S5_vs_k141_105945\|\|full | Low-quality |
| S5_vs_k141_68958\|\|full | Low-quality |
| S5_vs_k141_25827\|\|full | Low-quality |
| S5_vs_k141_62702\|\|full | Not-determined |
| S5_vs_k141_105968\|\|full | Low-quality |
| S5_vs_k141_62710\|\|full | Low-quality |
| S5_vs_k141_38563\|\|full | Low-quality |
| S5_vs_k141_44935\|\|full | Low-quality |
| S5_vs_k141_118352\|\|full | Low-quality |
| S5_vs_k141_3093\|\|full | Not-determined |
| S5_vs_k141_105980\|\|full | Low-quality |
| S14_vs_k141_79505\|\|full | Medium-quality |
| S5_vs_k141_106005\|\|full | Not-determined |
| S5_vs_k141_38569\|\|full | Low-quality |
| S5_vs_k141_25844\|\|full | Not-determined |
| S5_vs_k141_80213\|\|full | Not-determined |
| S5_vs_k141_8750\|\|full | Low-quality |
| S5_vs_k141_141538\|\|full | Low-quality |
| S5_vs_k141_56155\|\|full | Not-determined |
| S5_vs_k141_56156\|\|full | Low-quality |
| S5_vs_k141_3118\|\|full | Low-quality |
| S5_vs_k141_44949\|\|full | Low-quality |
| S5_vs_k141_129802\|\|full | Low-quality |
| S5_vs_k141_62752\|\|full | Low-quality |
| S5_vs_k141_3135\|\|full | Low-quality |
| S5_vs_k141_14607\|\|full | Not-determined |
| S5_vs_k141_141543\|\|full | Not-determined |
| S5_vs_k141_32980\|\|full | Low-quality |
| S5_vs_k141_141546\|\|full | Low-quality |
| S5_vs_k141_3151\|\|full | Low-quality |
| S5_vs_k141_44964\|\|full | Low-quality |
| S5_vs_k141_3163\|\|full | Low-quality |
| S5_vs_k141_141557\|\|full | Low-quality |
| S5_vs_k141_33016\|\|full | Low-quality |
| S5_vs_k141_38599\|\|full | Low-quality |
| S5_vs_k141_124473\|\|full | Low-quality |
| S5_vs_k141_124475\|\|full | Low-quality |
| S5_vs_k141_141567\|\|full | Not-determined |
| S5_vs_k141_124485\|\|full | Low-quality |
| S5_vs_k141_106061\|\|full | Low-quality |
| S5_vs_k141_135332\|\|full | Low-quality |
| S5_vs_k141_135334\|\|full | Not-determined |
| S5_vs_k141_106069\|\|full | Not-determined |
| S5_vs_k141_118408\|\|full | Not-determined |
| S5_vs_k141_25861\|\|full | Low-quality |
| S5_vs_k141_135346\|\|full | Low-quality |
| S5_vs_k141_25864\|\|full | Not-determined |
| S5_vs_k141_51217\|\|full | Not-determined |
| S5_vs_k141_98501\|\|full | Not-determined |
| S5_vs_k141_56192\|\|full | Low-quality |
| S5_vs_k141_51222\|\|full | Low-quality |
| S5_vs_k141_38629\|\|full | Low-quality |
| S3_vs_k141_19349\|\|full | High-quality |
| S5_vs_k141_25872\|\|full | Low-quality |
| S5_vs_k141_111514\|\|full | Low-quality |
| S5_vs_k141_124502\|\|full | Not-determined |
| S5_vs_k141_38634\|\|full | Low-quality |
| S5_vs_k141_51231\|\|full | Not-determined |
| S5_vs_k141_51239\|\|full | Not-determined |
| S5_vs_k141_98521\|\|full | Low-quality |
| S5_vs_k141_74368\|\|full | Not-determined |
| S5_vs_k141_93698\|\|full | Low-quality |
| S5_vs_k141_62849\|\|full | Low-quality |
| S5_vs_k141_111530\|\|full | Low-quality |
| S5_vs_k141_3215\|\|full | Low-quality |
| S5_vs_k141_33074\|\|full | Low-quality |
| S5_vs_k141_106113\|\|full | Not-determined |
| S5_vs_k141_68987\|\|full | Low-quality |
| S5_vs_k141_141608\|\|full | Low-quality |
| S5_vs_k141_124521\|\|full | Not-determined |
| S5_vs_k141_80272\|\|full | Not-determined |
| S5_vs_k141_51293\|\|full | Low-quality |
| S5_vs_k141_51295\|\|full | Low-quality |
| S5_vs_k141_80273\|\|full | Not-determined |
| S5_vs_k141_98533\|\|full | Low-quality |
| S5_vs_k141_74381\|\|full | Low-quality |
| S5_vs_k141_45002\|\|full | Not-determined |
| S5_vs_k141_98541\|\|full | Low-quality |
| S5_vs_k141_98543\|\|full | Low-quality |
| S5_vs_k141_106139\|\|full | Low-quality |
| S5_vs_k141_80289\|\|full | Low-quality |
| S5_vs_k141_69006\|\|full | Not-determined |
| S7_extracted_138225 | Complete |
| S5_vs_k141_118482\|\|full | Low-quality |
| S5_vs_k141_106149\|\|full | Low-quality |
| S5_vs_k141_62890\|\|full | Not-determined |
| S5_vs_k141_8783\|\|full | Low-quality |
| S5_vs_k141_93723\|\|full | Low-quality |
| S5_vs_k141_141628\|\|full | Low-quality |
| S5_vs_k141_106151\|\|full | Not-determined |
| S5_vs_k141_98549\|\|full | Not-determined |
| S5_vs_k141_93730\|\|full | Low-quality |
| S5_vs_k141_51328\|\|full | Low-quality |
| S5_vs_k141_69010\|\|full | Low-quality |
| S5_vs_k141_118499\|\|full | Low-quality |
| S5_vs_k141_129851\|\|full | Not-determined |
| S5_vs_k141_56203\|\|full | Not-determined |
| S5_vs_k141_93748\|\|full | Low-quality |
| S5_vs_k141_20600\|\|full | Not-determined |
| S5_vs_k141_98558\|\|full | Low-quality |
| S5_vs_k141_33102\|\|full | Low-quality |
| S5_vs_k141_129858\|\|full | Low-quality |
| S5_vs_k141_3294\|\|full | Not-determined |
| S5_vs_k141_124552\|\|full | Low-quality |
| S5_vs_k141_45047\|\|full | Low-quality |
| S5_vs_k141_56224\|\|full | Not-determined |
| S5_vs_k141_124557\|\|full | Not-determined |
| S5_vs_k141_3306\|\|full | Low-quality |
| S5_vs_k141_3308\|\|full | Not-determined |
| S5_vs_k141_106183\|\|full | Low-quality |
| S5_vs_k141_51368\|\|full | Low-quality |
| S5_vs_k141_69055\|\|full | Not-determined |
| S5_vs_k141_129875\|\|full | Low-quality |
| S5_vs_k141_38667\|\|full | Low-quality |
| S5_vs_k141_51378\|\|full | Not-determined |
| S5_vs_k141_45082\|\|full | Low-quality |
| S5_vs_k141_62900\|\|full | Low-quality |
| S5_vs_k141_80304\|\|full | Low-quality |
| S5_vs_k141_3327\|\|full | Low-quality |
| S5_vs_k141_45098\|\|full | Low-quality |
| S5_vs_k141_74436\|\|full | Low-quality |
| S5_vs_k141_25914\|\|full | Low-quality |
| S5_vs_k141_111569\|\|full | Not-determined |
| S5_vs_k141_129899\|\|full | Not-determined |
| S5_vs_k141_74440\|\|full | Not-determined |
| S5_vs_k141_45109\|\|full | Not-determined |
| S5_vs_k141_69075\|\|full | Not-determined |
| S5_vs_k141_98585\|\|full | Low-quality |
| S5_vs_k141_38688\|\|full | Not-determined |
| S5_vs_k141_111588\|\|full | Low-quality |
| S5_vs_k141_56289\|\|full | Low-quality |
| S5_vs_k141_20642\|\|full | Not-determined |
| S5_vs_k141_74448\|\|full | Not-determined |
| S5_vs_k141_56294\|\|full | Low-quality |
| S5_vs_k141_124627\|\|full | Low-quality |
| S5_vs_k141_3349\|\|full | Not-determined |
| S5_vs_k141_93756\|\|full | Not-determined |
| S5_vs_k141_111597\|\|full | Low-quality |
| S5_vs_k141_56302\|\|full | Not-determined |
| S5_vs_k141_98589\|\|full | Not-determined |
| S5_vs_k141_56307\|\|full | Low-quality |
| S5_vs_k141_124638\|\|full | Low-quality |
| S5_vs_k141_135380\|\|full | Not-determined |
| S5_vs_k141_62924\|\|full | Low-quality |
| S5_vs_k141_111606\|\|full | Not-determined |
| S14_vs_k141_61490\|\|full | Medium-quality |
| S5_vs_k141_38715\|\|full | Low-quality |
| S5_vs_k141_3354\|\|full | Not-determined |
| S5_vs_k141_93783\|\|full | Low-quality |
| S5_vs_k141_3357\|\|full | Not-determined |
| S5_vs_k141_3358\|\|full | Low-quality |
| S5_vs_k141_3359\|\|full | Low-quality |
| S5_vs_k141_3366\|\|full | Low-quality |
| S3_extracted_k141_5525 | High-quality |
| S5_vs_k141_111610\|\|full | Not-determined |
| S5_vs_k141_129920\|\|full | Not-determined |
| S5_vs_k141_69085\|\|full | Not-determined |
| S5_vs_k141_111616\|\|full | Not-determined |
| S5_vs_k141_14775\|\|full | Low-quality |
| S5_vs_k141_74464\|\|full | Low-quality |
| S5_vs_k141_14803\|\|full | Low-quality |
| S5_vs_k141_56326\|\|full | Not-determined |
| S5_vs_k141_80344\|\|full | Low-quality |
| S5_vs_k141_33105\|\|full | Low-quality |
| S5_vs_k141_8830\|\|full | Not-determined |
| S5_vs_k141_80360\|\|full | Low-quality |
| S5_vs_k141_45126\|\|full | Not-determined |
| S5_vs_k141_38756\|\|full | Low-quality |
| S5_vs_k141_56350\|\|full | Low-quality |
| S5_vs_k141_80364\|\|full | Low-quality |
| S5_vs_k141_69147\|\|full | Low-quality |
| S5_vs_k141_74476\|\|full | Not-determined |
| S5_vs_k141_38759\|\|full | Not-determined |
| S5_vs_k141_3454\|\|full | Not-determined |
| S5_vs_k141_74478\|\|full | Not-determined |
| S5_vs_k141_69150\|\|full | Not-determined |
| S5_vs_k141_74482\|\|full | Low-quality |
| S5_vs_k141_80368\|\|full | Low-quality |
| S3_extracted_k141_69175 | Complete |
| S5_vs_k141_38778\|\|full | Not-determined |
| S5_vs_k141_20699\|\|full | Not-determined |
| S5_vs_k141_124657\|\|full | Low-quality |
| S5_vs_k141_38785\|\|full | Not-determined |
| S5_vs_k141_69158\|\|full | Not-determined |
| S6_vs_k141_56485\|\|full | High-quality |
| S5_vs_k141_129934\|\|full | Low-quality |
| S5_vs_k141_98641\|\|full | Low-quality |
| S17_extracted_k141_177199 | Complete |
| S5_vs_k141_56365\|\|full | Not-determined |
| S5_vs_k141_8854\|\|full | Low-quality |
| S5_vs_k141_8865\|\|full | Low-quality |
| S5_vs_k141_51430\|\|full | Low-quality |
| S5_vs_k141_74507\|\|full | Low-quality |
| S5_vs_k141_51431\|\|full | Low-quality |
| S5_vs_k141_145611\|\|full | High-quality |
| S5_vs_k141_20709\|\|full | Low-quality |
| S5_vs_k141_93840\|\|full | Not-determined |
| S5_vs_k141_20710\|\|full | Not-determined |
| S5_vs_k141_14917\|\|full | Low-quality |
| S5_vs_k141_62983\|\|full | Low-quality |
| S5_vs_k141_51455\|\|full | Low-quality |
| S5_vs_k141_98664\|\|full | Low-quality |
| S5_vs_k141_118613\|\|full | Low-quality |
| S5_vs_k141_141668\|\|full | Low-quality |
| S5_vs_k141_141672\|\|full | Low-quality |
| S5_vs_k141_33176\|\|full | Low-quality |
| S5_vs_k141_20728\|\|full | Not-determined |
| S5_vs_k141_135439\|\|full | Low-quality |
| S5_vs_k141_14945\|\|full | Low-quality |
| S5_vs_k141_141688\|\|full | Low-quality |
| S5_vs_k141_80389\|\|full | Low-quality |
| S5_vs_k141_8125\|\|full | High-quality |
| S12_vs_k141_93357\|\|full | Medium-quality |
| S5_vs_k141_93878\|\|full | Low-quality |
| S5_vs_k141_63007\|\|full | Not-determined |
| S5_vs_k141_141693\|\|full | Not-determined |
| S5_vs_k141_129951\|\|full | Not-determined |
| S5_vs_k141_69187\|\|full | Low-quality |
| S5_vs_k141_93888\|\|full | Not-determined |
| S5_vs_k141_20755\|\|full | Low-quality |
| S5_vs_k141_33187\|\|full | Not-determined |
| S5_vs_k141_3516\|\|full | Low-quality |
| S5_vs_k141_3517\|\|full | Low-quality |
| S5_vs_k141_93891\|\|full | Low-quality |
| S5_vs_k141_56403\|\|full | Low-quality |
| S5_vs_k141_87085\|\|full | Low-quality |
| S5_vs_k141_80397\|\|full | Low-quality |
| S5_vs_k141_118645\|\|full | Low-quality |
| S5_vs_k141_135457\|\|full | Low-quality |
| S5_vs_k141_141726\|\|full | Low-quality |
| S5_vs_k141_111657\|\|full | Low-quality |
| S5_vs_k141_56408\|\|full | Not-determined |
| S5_vs_k141_141729\|\|full | Low-quality |
| S5_vs_k141_63025\|\|full | Not-determined |
| S5_vs_k141_141730\|\|full | Low-quality |
| S5_vs_k141_3541\|\|full | Low-quality |
| S5_vs_k141_8922\|\|full | Not-determined |
| S5_vs_k141_69243\|\|full | Low-quality |
| S5_vs_k141_141743\|\|full | Low-quality |
| S5_vs_k141_111679\|\|full | Low-quality |
| S5_vs_k141_87108\|\|full | Low-quality |
| S5_vs_k141_45251\|\|full | Low-quality |
| S5_vs_k141_15027\|\|full | Low-quality |
| S5_vs_k141_20793\|\|full | Not-determined |
| S5_vs_k141_20795\|\|full | Not-determined |
| S5_vs_k141_69259\|\|full | Low-quality |
| S3_vs_k141_5525\|\|full | High-quality |
| S5_vs_k141_51545\|\|full | Low-quality |
| S5_vs_k141_15035\|\|full | Low-quality |
| S5_vs_k141_87119\|\|full | Low-quality |
| S5_vs_k141_69260\|\|full | Low-quality |
| S5_vs_k141_111697\|\|full | Not-determined |
| S5_vs_k141_15042\|\|full | Low-quality |
| S5_vs_k141_3580\|\|full | Low-quality |
| S5_vs_k141_69264\|\|full | Low-quality |
| S5_vs_k141_129981\|\|full | Low-quality |
| S5_vs_k141_69265\|\|full | Low-quality |
| S5_vs_k141_111704\|\|full | Low-quality |
| S5_vs_k141_63055\|\|full | Not-determined |
| S5_vs_k141_87135\|\|full | Low-quality |
| S5_vs_k141_141787\|\|full | Low-quality |
| S5_vs_k141_69273\|\|full | Not-determined |
| S5_vs_k141_145578\|\|full | High-quality |
| S5_vs_k141_63060\|\|full | Not-determined |
| S5_vs_k141_141795\|\|full | Low-quality |
| S5_vs_k141_63076\|\|full | Low-quality |
| S5_vs_k141_118648\|\|full | Low-quality |
| S5_vs_k141_69292\|\|full | Not-determined |
| S5_vs_k141_87139\|\|full | Low-quality |
| S5_vs_k141_106276\|\|full | Low-quality |
| S5_vs_k141_118659\|\|full | Not-determined |
| S5_vs_k141_20831\|\|full | Not-determined |
| S5_vs_k141_87150\|\|full | Not-determined |
| S5_vs_k141_87153\|\|full | Low-quality |
| S5_vs_k141_63097\|\|full | Low-quality |
| S5_vs_k141_106289\|\|full | Not-determined |
| S5_vs_k141_129998\|\|full | Not-determined |
| S5_vs_k141_20850\|\|full | Low-quality |
| S5_vs_k141_20852\|\|full | Low-quality |
| S7_vs_138225\|\|full | High-quality |
| S5_vs_k141_74525\|\|full | Low-quality |
| S5_vs_k141_111745\|\|full | Not-determined |
| S5_vs_k141_3604\|\|full | Not-determined |
| S3_vs_k141_69175\|\|full | High-quality |
| S5_vs_k141_98742\|\|full | Low-quality |
| S5_vs_k141_111760\|\|full | Not-determined |
| S5_vs_k141_141830\|\|full | Low-quality |
| S5_vs_k141_63136\|\|full | Low-quality |
| S5_vs_k141_45348\|\|full | Low-quality |
| S5_vs_k141_3629\|\|full | Low-quality |
| S5_vs_k141_63140\|\|full | Low-quality |
| S5_vs_k141_20889\|\|full | Low-quality |
| S5_vs_k141_69346\|\|full | Low-quality |
| S5_vs_k141_106339\|\|full | Low-quality |
| S17_vs_k141_177199\|\|full | High-quality |
| S5_vs_k141_45370\|\|full | Not-determined |
| S5_vs_k141_20897\|\|full | Low-quality |
| S5_vs_k141_33253\|\|full | Low-quality |
| S5_vs_k141_106346\|\|full | Low-quality |
| S5_vs_k141_69361\|\|full | Not-determined |
| S5_vs_k141_63172\|\|full | Low-quality |
| S5_vs_k141_106361\|\|full | Low-quality |
| S5_vs_k141_3658\|\|full | Low-quality |
| S5_vs_k141_3659\|\|full | Not-determined |
| S5_vs_k141_106368\|\|full | Low-quality |
| S5_vs_k141_69372\|\|full | Low-quality |
| S5_vs_k141_106369\|\|full | Low-quality |
| S12_vs_k141_87088\|\|full | High-quality |
| S5_vs_k141_3665\|\|full | Not-determined |
| S5_vs_k141_87211\|\|full | Low-quality |
| S5_vs_k141_69385\|\|full | Low-quality |
| S5_vs_k141_15117\|\|full | Low-quality |
| S5_vs_k141_106389\|\|full | Low-quality |
| S5_vs_k141_15121\|\|full | Low-quality |
| S5_vs_k141_141876\|\|full | Low-quality |
| S5_vs_k141_106398\|\|full | Low-quality |
| S5_vs_k141_111788\|\|full | Not-determined |
| S5_vs_k141_141881\|\|full | Not-determined |
| S5_vs_k141_38876\|\|full | Low-quality |
| S5_vs_k141_106405\|\|full | Low-quality |
| S5_vs_k141_124680\|\|full | Not-determined |
| S5_vs_k141_63211\|\|full | Not-determined |
| S5_vs_k141_15128\|\|full | Low-quality |
| S5_vs_k141_63213\|\|full | Low-quality |
| S5_vs_k141_141894\|\|full | Not-determined |
| S5_vs_k141_111816\|\|full | Low-quality |
| S5_vs_k141_141900\|\|full | Not-determined |
| S5_vs_k141_63218\|\|full | Low-quality |
| S5_vs_k141_141907\|\|full | Low-quality |
| S5_vs_k141_141908\|\|full | Not-determined |
| S5_vs_k141_33298\|\|full | Not-determined |
| S5_vs_k141_33302\|\|full | Low-quality |
| S5_vs_k141_141916\|\|full | Low-quality |
| S5_vs_k141_63233\|\|full | Low-quality |
| S5_vs_k141_98791\|\|full | Low-quality |
| S5_vs_k141_141921\|\|full | Low-quality |
| S5_vs_k141_3701\|\|full | Low-quality |
| S5_vs_k141_33311\|\|full | Not-determined |
| S5_vs_k141_33313\|\|full | Low-quality |
| S5_vs_k141_63239\|\|full | Low-quality |
| S5_vs_k141_51688\|\|full | Not-determined |
| S5_vs_k141_15176\|\|full | Not-determined |
| S5_vs_k141_33314\|\|full | Low-quality |
| S5_vs_k141_56492\|\|full | Low-quality |
| S5_vs_k141_124725\|\|full | Low-quality |
| S5_vs_k141_3729\|\|full | Not-determined |
| S5_vs_k141_20959\|\|full | Low-quality |
| S5_vs_k141_51708\|\|full | Not-determined |
| S8_vs_k141_7243\|\|full | Medium-quality |
| S5_vs_k141_20963\|\|full | Not-determined |
| S5_vs_k141_3735\|\|full | Low-quality |
| S5_vs_k141_63271\|\|full | Low-quality |
| S5_vs_k141_33328\|\|full | Low-quality |
| S5_vs_k141_20966\|\|full | Low-quality |
| S5_vs_k141_33330\|\|full | Not-determined |
| S5_vs_k141_141941\|\|full | Low-quality |
| S5_vs_k141_118716\|\|full | Low-quality |
| S5_vs_k141_80449\|\|full | Low-quality |
| S5_vs_k141_130114\|\|full | Low-quality |
| S5_vs_k141_141951\|\|full | Low-quality |
| S5_vs_k141_106446\|\|full | Low-quality |
| S5_vs_k141_130119\|\|full | Low-quality |
| S5_vs_k141_33345\|\|full | Low-quality |
| S5_vs_k141_98819\|\|full | Low-quality |
| S5_vs_k141_51741\|\|full | Not-determined |
| S5_vs_k141_69452\|\|full | Low-quality |
| S5_vs_k141_141965\|\|full | Low-quality |
| S5_vs_k141_3762\|\|full | Low-quality |
| S5_vs_k141_38943\|\|full | Not-determined |
| S5_vs_k141_3764\|\|full | Low-quality |
| S5_vs_k141_118755\|\|full | Not-determined |
| S5_vs_k141_106453\|\|full | Not-determined |
| S5_vs_k141_80466\|\|full | Not-determined |
| S5_vs_k141_98833\|\|full | Not-determined |
| S5_vs_k141_141985\|\|full | Low-quality |
| S5_vs_k141_15189\|\|full | Low-quality |
| S5_vs_k141_20980\|\|full | Low-quality |
| S5_vs_k141_15191\|\|full | Not-determined |
| S5_vs_k141_106468\|\|full | Low-quality |
| S5_vs_k141_111897\|\|full | Low-quality |
| S5_vs_k141_33359\|\|full | Low-quality |
| S5_vs_k141_111902\|\|full | Low-quality |
| S5_vs_k141_80471\|\|full | Low-quality |
| S7_vs_94738\|\|full | High-quality |
| S5_vs_k141_80474\|\|full | Not-determined |
| S5_vs_k141_80479\|\|full | Low-quality |
| S5_vs_k141_135494\|\|full | Low-quality |
| S5_vs_k141_130158\|\|full | Low-quality |
| S5_vs_k141_51786\|\|full | Low-quality |
| S5_vs_k141_124784\|\|full | Low-quality |
| S5_vs_k141_3802\|\|full | Low-quality |
| S5_vs_k141_51790\|\|full | Low-quality |
| S5_vs_k141_3805\|\|full | Low-quality |
| S5_vs_k141_98858\|\|full | Low-quality |
| S5_vs_k141_142011\|\|full | Low-quality |
| S5_vs_k141_142018\|\|full | Low-quality |
| S5_vs_k141_3818\|\|full | Low-quality |
| S5_vs_k141_25973\|\|full | Not-determined |
| S5_vs_k141_130187\|\|full | Low-quality |
| S5_vs_k141_80491\|\|full | Low-quality |
| S5_vs_k141_3825\|\|full | Not-determined |
| S5_vs_k141_21042\|\|full | Not-determined |
| S5_vs_k141_111971\|\|full | Not-determined |
| S5_vs_k141_142033\|\|full | Low-quality |
| S5_vs_k141_130204\|\|full | Low-quality |
| S5_vs_k141_118824\|\|full | Low-quality |
| S5_vs_k141_111975\|\|full | Low-quality |
| S5_vs_k141_87255\|\|full | Not-determined |
| S5_vs_k141_3835\|\|full | Low-quality |
| S5_vs_k141_142039\|\|full | Not-determined |
| S5_vs_k141_9010\|\|full | Low-quality |
| S5_vs_k141_15259\|\|full | Low-quality |
| S5_vs_k141_87266\|\|full | Low-quality |
| S5_vs_k141_106495\|\|full | Not-determined |
| S5_vs_k141_112000\|\|full | Low-quality |
| S5_vs_k141_51810\|\|full | Low-quality |
| S5_vs_k141_130221\|\|full | Not-determined |
| S5_vs_k141_15262\|\|full | Low-quality |
| S5_vs_k141_9038\|\|full | Low-quality |
| S5_vs_k141_124831\|\|full | Low-quality |
| S5_vs_k141_63343\|\|full | Low-quality |
| S5_vs_k141_15270\|\|full | Not-determined |
| S5_vs_k141_106501\|\|full | Low-quality |
| S5_vs_k141_94026\|\|full | Low-quality |
| S5_vs_k141_26008\|\|full | Not-determined |
| S5_vs_k141_118854\|\|full | Low-quality |
| S5_vs_k141_9046\|\|full | Not-determined |
| S5_vs_k141_51812\|\|full | Not-determined |
| S5_vs_k141_26013\|\|full | Low-quality |
| S5_vs_k141_112031\|\|full | Not-determined |
| S5_vs_k141_26018\|\|full | Not-determined |
| S5_vs_k141_15281\|\|full | Low-quality |
| S5_vs_k141_63372\|\|full | Not-determined |
| S5_vs_k141_130258\|\|full | Low-quality |
| S5_vs_k141_94052\|\|full | Low-quality |
| S5_vs_k141_51838\|\|full | Low-quality |
| S5_vs_k141_56546\|\|full | Low-quality |
| S5_vs_k141_112052\|\|full | Not-determined |
| S5_vs_k141_45376\|\|full | Low-quality |
| S5_vs_k141_112054\|\|full | Low-quality |
| S5_vs_k141_118882\|\|full | Low-quality |
| S5_vs_k141_130269\|\|full | Low-quality |
| S5_vs_k141_33466\|\|full | Low-quality |
| S5_vs_k141_9065\|\|full | Low-quality |
| S5_vs_k141_3883\|\|full | Low-quality |
| S5_vs_k141_135554\|\|full | Not-determined |
| S5_vs_k141_106570\|\|full | Low-quality |
| S5_vs_k141_130285\|\|full | Low-quality |
| S5_vs_k141_33468\|\|full | Not-determined |
| S5_vs_k141_15305\|\|full | Low-quality |
| S5_vs_k141_51859\|\|full | Not-determined |
| S5_vs_k141_45403\|\|full | Not-determined |
| S5_vs_k141_51860\|\|full | Low-quality |
| S5_vs_k141_135558\|\|full | Low-quality |
| S5_vs_k141_124865\|\|full | Not-determined |
| S5_vs_k141_142054\|\|full | Low-quality |
| S5_vs_k141_51866\|\|full | Low-quality |
| S5_vs_k141_45407\|\|full | Low-quality |
| S5_vs_k141_118913\|\|full | Not-determined |
| S5_vs_k141_63407\|\|full | Low-quality |
| S5_vs_k141_94085\|\|full | Not-determined |
| S5_vs_k141_51869\|\|full | Low-quality |
| S5_vs_k141_33489\|\|full | Low-quality |
| S5_vs_k141_124872\|\|full | Not-determined |
| S5_vs_k141_74565\|\|full | Not-determined |
| S5_vs_k141_74569\|\|full | Not-determined |
| S5_vs_k141_112100\|\|full | Low-quality |
| S5_vs_k141_94087\|\|full | Not-determined |
| S5_vs_k141_3910\|\|full | Not-determined |
| S5_vs_k141_94088\|\|full | Not-determined |
| S5_vs_k141_51878\|\|full | Low-quality |
| S5_vs_k141_106595\|\|full | Low-quality |
| S5_vs_k141_112104\|\|full | Not-determined |
| S5_vs_k141_63421\|\|full | Low-quality |
| S5_vs_k141_33506\|\|full | Not-determined |
| S5_vs_k141_142072\|\|full | Not-determined |
| S5_vs_k141_124889\|\|full | Not-determined |
| S5_vs_k141_106601\|\|full | Low-quality |
| S5_vs_k141_142076\|\|full | Low-quality |
| S5_vs_k141_124891\|\|full | Low-quality |
| S5_vs_k141_45440\|\|full | Not-determined |
| S5_vs_k141_94117\|\|full | Not-determined |
| S5_vs_k141_112110\|\|full | Low-quality |
| S5_vs_k141_124898\|\|full | Not-determined |
| S5_vs_k141_94120\|\|full | Low-quality |
| S5_vs_k141_3941\|\|full | Not-determined |
| S5_vs_k141_69572\|\|full | Not-determined |
| S5_vs_k141_69574\|\|full | Not-determined |
| S5_vs_k141_106630\|\|full | Low-quality |
| S5_vs_k141_142096\|\|full | Low-quality |
| S5_vs_k141_98915\|\|full | Low-quality |
| S5_vs_k141_69579\|\|full | Low-quality |
| S5_vs_k141_15386\|\|full | Low-quality |
| S5_vs_k141_9121\|\|full | Low-quality |
| S5_vs_k141_87372\|\|full | Low-quality |
| S5_vs_k141_124913\|\|full | Not-determined |
| S5_vs_k141_118969\|\|full | Low-quality |
| S5_vs_k141_51908\|\|full | Low-quality |
| S3_vs_k141_119660\|\|full | Medium-quality |
| S5_vs_k141_98931\|\|full | Not-determined |
| S5_vs_k141_56587\|\|full | Not-determined |
| S5_vs_k141_98938\|\|full | Not-determined |
| S5_vs_k141_51934\|\|full | Low-quality |
| S5_vs_k141_33540\|\|full | Not-determined |
| S5_vs_k141_106667\|\|full | Low-quality |
| S5_vs_k141_56588\|\|full | Low-quality |
| S5_vs_k141_119001\|\|full | Low-quality |
| S5_vs_k141_56590\|\|full | Not-determined |
| S5_vs_k141_142130\|\|full | Not-determined |
| S5_vs_k141_112165\|\|full | Low-quality |
| S5_vs_k141_63483\|\|full | Not-determined |
| S5_vs_k141_33549\|\|full | Low-quality |
| S5_vs_k141_130336\|\|full | Low-quality |
| S5_vs_k141_9143\|\|full | Low-quality |
| S5_vs_k141_9147\|\|full | Low-quality |
| S5_vs_k141_124957\|\|full | Not-determined |
| S5_vs_k141_9150\|\|full | Low-quality |
| S5_vs_k141_142149\|\|full | Low-quality |
| S5_vs_k141_9155\|\|full | Low-quality |
| S5_vs_k141_33570\|\|full | Not-determined |
| S5_vs_k141_106680\|\|full | Low-quality |
| S5_vs_k141_106683\|\|full | Low-quality |
| S5_vs_k141_130344\|\|full | Not-determined |
| S5_vs_k141_51967\|\|full | Low-quality |
| S5_vs_k141_87445\|\|full | Low-quality |
| S5_vs_k141_39027\|\|full | Not-determined |
| S5_vs_k141_21134\|\|full | Low-quality |
| S5_vs_k141_106708\|\|full | Low-quality |
| S5_vs_k141_106711\|\|full | Low-quality |
| S5_vs_k141_21145\|\|full | Not-determined |
| S5_vs_k141_15428\|\|full | Low-quality |
| S5_vs_k141_3998\|\|full | Low-quality |
| S5_vs_k141_94159\|\|full | Not-determined |
| S5_vs_k141_21159\|\|full | Low-quality |
| S5_vs_k141_112206\|\|full | Not-determined |
| S5_vs_k141_51987\|\|full | Not-determined |
| S5_vs_k141_112209\|\|full | Low-quality |
| S5_vs_k141_98952\|\|full | Not-determined |
| S5_vs_k141_98957\|\|full | Low-quality |
| S5_vs_k141_51996\|\|full | Low-quality |
| S5_vs_k141_63555\|\|full | Low-quality |
| S5_vs_k141_63556\|\|full | Low-quality |
| S5_vs_k141_124991\|\|full | Not-determined |
| S5_vs_k141_15477\|\|full | Low-quality |
| S5_vs_k141_4044\|\|full | Low-quality |
| S5_vs_k141_112215\|\|full | Low-quality |
| S5_vs_k141_9252\|\|full | Not-determined |
| S5_vs_k141_33625\|\|full | Not-determined |
| S5_vs_k141_63585\|\|full | Low-quality |
| S5_vs_k141_125013\|\|full | Low-quality |

| | |
|---|---|
| S5_vs_k141_135638\|\|full | Low-quality |
| S5_vs_k141_33641\|\|full | Not-determined |
| S5_vs_k141_67274\|\|full | High-quality |
| S5_vs_k141_9281\|\|full | Low-quality |
| S5_vs_k141_87496\|\|full | Not-determined |
| S5_vs_k141_52034\|\|full | Low-quality |
| S5_vs_k141_63595\|\|full | Low-quality |
| S5_vs_k141_106807\|\|full | Not-determined |
| S5_vs_k141_94259\|\|full | Low-quality |
| S5_vs_k141_9308\|\|full | Not-determined |
| S5_vs_k141_45551\|\|full | Low-quality |
| S5_vs_k141_45552\|\|full | Not-determined |
| S5_vs_k141_9317\|\|full | Low-quality |
| S5_vs_k141_52075\|\|full | Not-determined |
| S5_vs_k141_63671\|\|full | Not-determined |
| S5_vs_k141_94283\|\|full | Low-quality |
| S5_vs_k141_106815\|\|full | Low-quality |
| S5_vs_k141_39086\|\|full | Not-determined |
| S5_vs_k141_135673\|\|full | Low-quality |
| S5_vs_k141_112288\|\|full | Low-quality |
| S5_vs_k141_135675\|\|full | Not-determined |
| S5_vs_k141_94301\|\|full | Not-determined |
| S5_vs_k141_130471\|\|full | Not-determined |
| S3_extracted_k141_106401 | Medium-quality |
| S5_vs_k141_125051\|\|full | Low-quality |
| S5_vs_k141_135677\|\|full | Low-quality |
| S3_vs_k141_106401\|\|full | Medium-quality |
| S5_vs_k141_125060\|\|full | Not-determined |
| S5_vs_k141_142200\|\|full | Low-quality |
| S5_vs_k141_94315\|\|full | Low-quality |
| S5_vs_k141_74629\|\|full | Low-quality |
| S5_vs_k141_142213\|\|full | Not-determined |
| S5_vs_k141_106869\|\|full | Low-quality |
| S5_vs_k141_106871\|\|full | Low-quality |
| S5_vs_k141_39118\|\|full | Not-determined |
| S11_vs_k141_68693\|\|full | Medium-quality |
| S5_vs_k141_45599\|\|full | Low-quality |
| S5_vs_k141_9380\|\|full | Low-quality |
| S5_vs_k141_9383\|\|full | Low-quality |
| S5_vs_k141_94328\|\|full | Low-quality |
| S5_vs_k141_80514\|\|full | Low-quality |
| S5_vs_k141_106881\|\|full | Low-quality |
| S5_vs_k141_125099\|\|full | Not-determined |
| S5_vs_k141_130481\|\|full | Low-quality |
| S5_vs_k141_4101\|\|full | Not-determined |
| S5_vs_k141_99013\|\|full | Low-quality |
| S5_vs_k141_45618\|\|full | Not-determined |
| S5_vs_k141_142271\|\|full | Low-quality |
| S5_vs_k141_112329\|\|full | Not-determined |
| S5_vs_k141_52180\|\|full | Not-determined |
| S5_vs_k141_87538\|\|full | Low-quality |
| S5_vs_k141_80530\|\|full | Low-quality |
| S5_vs_k141_56627\|\|full | Not-determined |
| S5_vs_k141_106923\|\|full | Not-determined |
| S5_vs_k141_125138\|\|full | Low-quality |
| S5_vs_k141_9412\|\|full | Not-determined |
| S5_vs_k141_106933\|\|full | Not-determined |
| S5_vs_k141_99024\|\|full | Low-quality |
| S5_vs_k141_125151\|\|full | Low-quality |
| S5_vs_k141_106943\|\|full | Not-determined |
| S5_vs_k141_39181\|\|full | Not-determined |
| S5_vs_k141_125167\|\|full | Low-quality |
| S5_vs_k141_4153\|\|full | Low-quality |
| S5_vs_k141_99026\|\|full | Not-determined |
| S5_vs_k141_106971\|\|full | Low-quality |
| S5_vs_k141_4161\|\|full | Not-determined |
| S5_vs_k141_45654\|\|full | Not-determined |
| S5_vs_k141_21211\|\|full | Low-quality |
| S5_vs_k141_142295\|\|full | Not-determined |
| S5_vs_k141_37659\|\|full | Medium-quality |
| S5_vs_k141_99027\|\|full | Low-quality |
| S5_vs_k141_4170\|\|full | Low-quality |
| S5_vs_k141_125184\|\|full | Low-quality |
| S5_vs_k141_4171\|\|full | Not-determined |
| S5_vs_k141_106979\|\|full | Low-quality |
| S5_vs_k141_106982\|\|full | Not-determined |
| S5_vs_k141_45657\|\|full | Not-determined |
| S5_vs_k141_4181\|\|full | Not-determined |
| S5_vs_k141_26196\|\|full | Not-determined |
| S5_vs_k141_26197\|\|full | Not-determined |
| S5_vs_k141_45660\|\|full | Low-quality |
| S5_vs_k141_119028\|\|full | Low-quality |
| S5_vs_k141_45666\|\|full | Low-quality |
| S5_vs_k141_142326\|\|full | Not-determined |
| S5_vs_k141_4212\|\|full | Not-determined |
| S5_vs_k141_45670\|\|full | Not-determined |
| S5_vs_k141_87590\|\|full | Not-determined |
| S5_vs_k141_119054\|\|full | Low-quality |
| S5_vs_k141_125226\|\|full | Low-quality |
| S5_vs_k141_33827\|\|full | Not-determined |
| S5_vs_k141_130524\|\|full | Low-quality |
| S5_vs_k141_142361\|\|full | Not-determined |
| S5_vs_k141_33846\|\|full | Not-determined |
| S5_vs_k141_130541\|\|full | Not-determined |
| S5_vs_k141_130542\|\|full | Low-quality |
| S5_vs_k141_125237\|\|full | Low-quality |
| S5_vs_k141_52262\|\|full | Not-determined |
| S5_vs_k141_4262\|\|full | Low-quality |
| S5_vs_k141_112374\|\|full | Low-quality |
| S16_vs_78429\|\|full | High-quality |
| S5_vs_k141_80608\|\|full | Low-quality |
| S11_vs_k141_84627\|\|full | High-quality |
| S5_vs_k141_107005\|\|full | Not-determined |
| S5_vs_k141_87644\|\|full | Not-determined |
| S5_vs_k141_125267\|\|full | Not-determined |
| S5_vs_k141_33873\|\|full | Low-quality |
| S5_vs_k141_87647\|\|full | Low-quality |
| S5_vs_k141_125269\|\|full | Low-quality |
| S5_vs_k141_99062\|\|full | Low-quality |
| S5_vs_k141_80626\|\|full | Low-quality |
| S5_vs_k141_45693\|\|full | Low-quality |
| S5_vs_k141_45695\|\|full | Low-quality |
| S5_vs_k141_4289\|\|full | Low-quality |
| S5_vs_k141_112395\|\|full | Not-determined |
| S5_vs_k141_52285\|\|full | Not-determined |
| S5_vs_k141_107028\|\|full | Low-quality |
| S5_vs_k141_4298\|\|full | Not-determined |
| S5_vs_k141_142395\|\|full | Low-quality |
| S5_vs_k141_4320\|\|full | Low-quality |
| S5_vs_k141_107063\|\|full | Low-quality |
| S5_vs_k141_26241\|\|full | Low-quality |
| S5_vs_k141_130609\|\|full | Low-quality |
| S5_vs_k141_63688\|\|full | Low-quality |
| S5_vs_k141_9434\|\|full | Low-quality |
| S5_vs_k141_45732\|\|full | Low-quality |
| S5_vs_k141_69597\|\|full | Low-quality |
| S5_vs_k141_130619\|\|full | Low-quality |
| S5_vs_k141_9450\|\|full | Low-quality |
| S5_vs_k141_112411\|\|full | Low-quality |
| S5_vs_k141_26258\|\|full | Low-quality |
| S5_vs_k141_74706\|\|full | Low-quality |
| S5_vs_k141_142429\|\|full | Not-determined |
| S5_vs_k141_9455\|\|full | Low-quality |
| S5_vs_k141_33937\|\|full | Low-quality |
| S5_vs_k141_142430\|\|full | Not-determined |
| S5_vs_k141_33938\|\|full | Low-quality |
| S5_vs_k141_63712\|\|full | Not-determined |
| S5_vs_k141_33952\|\|full | Low-quality |
| S5_vs_k141_4368\|\|full | Not-determined |
| S5_vs_k141_39274\|\|full | Not-determined |
| S5_vs_k141_63722\|\|full | Low-quality |
| S5_vs_k141_125340\|\|full | Low-quality |
| S5_vs_k141_33956\|\|full | Low-quality |
| S5_vs_k141_74726\|\|full | Not-determined |
| S5_vs_k141_33957\|\|full | Low-quality |
| S5_vs_k141_74729\|\|full | Low-quality |
| S5_vs_k141_112435\|\|full | Low-quality |
| S5_vs_k141_33966\|\|full | Low-quality |
| S11_vs_k141_55144\|\|full | High-quality |
| S3_extracted_k141_99626 | Complete |
| S5_vs_k141_74738\|\|full | Not-determined |
| S5_vs_k141_33971\|\|full | Low-quality |
| S5_vs_k141_135795\|\|full | Not-determined |
| S5_vs_k141_74741\|\|full | Low-quality |
| S5_vs_k141_87728\|\|full | Low-quality |
| S5_vs_k141_74748\|\|full | Low-quality |
| S5_vs_k141_52355\|\|full | Low-quality |
| S5_vs_k141_4387\|\|full | Low-quality |
| S5_vs_k141_130722\|\|full | Low-quality |
| S5_vs_k141_130724\|\|full | Not-determined |
| S5_vs_k141_130727\|\|full | Low-quality |
| S5_vs_k141_34010\|\|full | Low-quality |
| S5_vs_k141_45799\|\|full | Low-quality |
| S5_vs_k141_99116\|\|full | Not-determined |
| S5_vs_k141_99122\|\|full | Low-quality |
| S5_vs_k141_107129\|\|full | Not-determined |
| S5_vs_k141_4415\|\|full | Low-quality |
| S5_vs_k141_45813\|\|full | Not-determined |
| S5_vs_k141_135811\|\|full | Not-determined |
| S5_vs_k141_135812\|\|full | Not-determined |
| S5_vs_k141_135815\|\|full | Low-quality |
| S5_vs_k141_69671\|\|full | Low-quality |
| S5_vs_k141_69672\|\|full | Low-quality |
| S5_vs_k141_107144\|\|full | Not-determined |
| S5_vs_k141_52402\|\|full | Low-quality |
| S5_vs_k141_69675\|\|full | Low-quality |
| S5_vs_k141_130752\|\|full | Low-quality |
| S5_vs_k141_15505\|\|full | Not-determined |
| S5_vs_k141_15507\|\|full | Low-quality |
| S5_vs_k141_26314\|\|full | Low-quality |
| S5_vs_k141_15508\|\|full | Not-determined |
| S5_vs_k141_99157\|\|full | Low-quality |
| S5_vs_k141_99160\|\|full | Low-quality |
| S5_vs_k141_87766\|\|full | Not-determined |
| S5_vs_k141_125400\|\|full | Low-quality |
| S5_vs_k141_4457\|\|full | Low-quality |
| S5_vs_k141_4462\|\|full | Low-quality |
| S5_vs_k141_52429\|\|full | Not-determined |
| S5_vs_k141_99185\|\|full | Not-determined |
| S5_vs_k141_119167\|\|full | Low-quality |
| S5_vs_k141_4465\|\|full | Low-quality |
| S5_vs_k141_107190\|\|full | Low-quality |
| S5_vs_k141_130765\|\|full | Not-determined |
| S5_vs_k141_45881\|\|full | Low-quality |
| S5_vs_k141_130781\|\|full | Low-quality |
| S5_vs_k141_34082\|\|full | Low-quality |
| S5_vs_k141_52453\|\|full | Not-determined |
| S5_vs_k141_142497\|\|full | Low-quality |
| S16_extracted_134705 | High-quality |
| S5_vs_k141_9549\|\|full | Low-quality |
| S5_vs_k141_34111\|\|full | Low-quality |
| S5_vs_k141_130798\|\|full | Low-quality |
| S5_vs_k141_45920\|\|full | Low-quality |
| S5_vs_k141_130805\|\|full | Low-quality |
| S5_vs_k141_52488\|\|full | Not-determined |
| S5_vs_k141_34121\|\|full | Low-quality |
| S5_vs_k141_80731\|\|full | Not-determined |
| S5_vs_k141_119227\|\|full | Not-determined |
| S5_vs_k141_112450\|\|full | Not-determined |
| S5_vs_k141_99222\|\|full | Low-quality |
| S5_vs_k141_119230\|\|full | Low-quality |
| S5_vs_k141_130813\|\|full | Low-quality |
| S5_vs_k141_99224\|\|full | Not-determined |
| S5_vs_k141_135844\|\|full | Low-quality |
| S5_vs_k141_94362\|\|full | Low-quality |
| S5_vs_k141_87830\|\|full | Not-determined |
| S5_vs_k141_130823\|\|full | Low-quality |
| S5_vs_k141_45943\|\|full | Low-quality |
| S5_vs_k141_125508\|\|full | Low-quality |
| S5_vs_k141_9564\|\|full | Low-quality |
| S5_vs_k141_94372\|\|full | Low-quality |
| S5_vs_k141_87837\|\|full | Low-quality |
| S5_vs_k141_45950\|\|full | Low-quality |
| S5_vs_k141_52518\|\|full | Low-quality |
| S5_vs_k141_99240\|\|full | Low-quality |
| S5_vs_k141_9570\|\|full | Low-quality |
| S5_vs_k141_135864\|\|full | Low-quality |
| S5_vs_k141_39339\|\|full | Not-determined |
| S1_vs_k141_143929\|\|full | High-quality |
| S5_vs_k141_80744\|\|full | Not-determined |
| S5_vs_k141_69705\|\|full | Not-determined |
| S5_vs_k141_52531\|\|full | Low-quality |
| S5_vs_k141_9591\|\|full | Low-quality |
| S5_vs_k141_119249\|\|full | Low-quality |
| S5_vs_k141_39353\|\|full | Low-quality |
| S5_vs_k141_45972\|\|full | Low-quality |
| S5_vs_k141_125546\|\|full | Low-quality |
| S5_vs_k141_135866\|\|full | Low-quality |
| S5_vs_k141_112470\|\|full | Not-determined |
| S5_vs_k141_80757\|\|full | Low-quality |
| S5_vs_k141_69736\|\|full | Low-quality |
| S5_vs_k141_45991\|\|full | Low-quality |
| S5_vs_k141_125594\|\|full | Not-determined |
| S5_vs_k141_112496\|\|full | Not-determined |
| S5_vs_k141_34170\|\|full | Low-quality |
| S5_vs_k141_69751\|\|full | Not-determined |
| S5_vs_k141_74756\|\|full | Low-quality |
| S5_vs_k141_107329\|\|full | Not-determined |
| S5_vs_k141_135888\|\|full | Low-quality |
| S5_vs_k141_112515\|\|full | Low-quality |
| S5_vs_k141_87841\|\|full | Low-quality |
| S5_vs_k141_119274\|\|full | Low-quality |
| S5_vs_k141_63818\|\|full | Low-quality |
| S5_vs_k141_135893\|\|full | Low-quality |
| S5_vs_k141_21288\|\|full | Low-quality |
| S5_vs_k141_125630\|\|full | Low-quality |
| S5_vs_k141_135898\|\|full | Low-quality |
| S5_vs_k141_39410\|\|full | Low-quality |
| S5_vs_k141_87860\|\|full | Not-determined |
| S7_vs_138140\|\|full | High-quality |
| S5_vs_k141_99342\|\|full | Not-determined |
| S5_vs_k141_63835\|\|full | Low-quality |
| S5_vs_k141_4509\|\|full | Low-quality |
| S5_vs_k141_119310\|\|full | Low-quality |
| S5_vs_k141_87878\|\|full | Not-determined |
| S5_vs_k141_9615\|\|full | Low-quality |
| S5_vs_k141_21318\|\|full | Low-quality |
| S5_vs_k141_9621\|\|full | Low-quality |
| S5_vs_k141_112556\|\|full | Not-determined |
| S5_vs_k141_135902\|\|full | Low-quality |
| S5_vs_k141_9627\|\|full | Low-quality |
| S5_vs_k141_142559\|\|full | Low-quality |
| S5_vs_k141_9629\|\|full | Low-quality |
| S5_vs_k141_130860\|\|full | Low-quality |
| S5_vs_k141_21331\|\|full | Low-quality |
| S5_vs_k141_107406\|\|full | Low-quality |
| S5_vs_k141_107409\|\|full | Not-determined |
| S5_vs_k141_99362\|\|full | Not-determined |
| S5_vs_k141_9649\|\|full | Not-determined |
| S5_vs_k141_125682\|\|full | Low-quality |
| S5_vs_k141_99370\|\|full | Low-quality |
| S5_vs_k141_63875\|\|full | Low-quality |
| S5_vs_k141_94392\|\|full | Low-quality |
| S5_vs_k141_112589\|\|full | Not-determined |
| S5_vs_k141_99380\|\|full | Low-quality |
| S5_vs_k141_69830\|\|full | Not-determined |
| S5_vs_k141_135919\|\|full | Not-determined |
| S5_vs_k141_94404\|\|full | Low-quality |
| S5_vs_k141_69833\|\|full | Low-quality |
| S5_vs_k141_87930\|\|full | Not-determined |
| S5_vs_k141_135929\|\|full | Low-quality |
| S5_vs_k141_112610\|\|full | Not-determined |
| S5_vs_k141_87945\|\|full | Low-quality |
| S5_vs_k141_9660\|\|full | Not-determined |
| S5_vs_k141_119387\|\|full | Not-determined |
| S5_vs_k141_107441\|\|full | Low-quality |
| S5_vs_k141_135938\|\|full | Not-determined |
| S5_vs_k141_135950\|\|full | Low-quality |
| S5_vs_k141_107451\|\|full | Not-determined |
| S5_vs_k141_63886\|\|full | Not-determined |
| S5_vs_k141_87961\|\|full | Low-quality |
| S5_vs_k141_52617\|\|full | Not-determined |
| S5_vs_k141_9679\|\|full | Not-determined |
| S5_vs_k141_9682\|\|full | Low-quality |
| S5_vs_k141_4558\|\|full | Low-quality |
| S5_vs_k141_15596\|\|full | Not-determined |
| S5_vs_k141_94445\|\|full | Low-quality |
| S5_vs_k141_142596\|\|full | Low-quality |
| S5_vs_k141_125736\|\|full | Low-quality |
| S5_vs_k141_52657\|\|full | Not-determined |
| S5_vs_k141_80781\|\|full | Low-quality |
| S5_vs_k141_130924\|\|full | Low-quality |
| S5_vs_k141_130925\|\|full | Low-quality |
| S5_vs_k141_107465\|\|full | Low-quality |
| S5_vs_k141_15615\|\|full | Low-quality |
| S5_vs_k141_125762\|\|full | Not-determined |
| S5_vs_k141_135970\|\|full | Low-quality |
| S5_vs_k141_88018\|\|full | Low-quality |
| S5_vs_k141_88024\|\|full | Not-determined |
| S5_vs_k141_94497\|\|full | Not-determined |
| S5_vs_k141_52696\|\|full | Not-determined |
| S5_vs_k141_112688\|\|full | Low-quality |
| S5_vs_k141_94505\|\|full | Not-determined |
| S5_vs_k141_135986\|\|full | Not-determined |
| S5_vs_k141_69851\|\|full | Low-quality |
| S5_vs_k141_34258\|\|full | Not-determined |
| S5_vs_k141_4635\|\|full | Low-quality |
| S5_vs_k141_21440\|\|full | Low-quality |
| S5_vs_k141_34260\|\|full | Not-determined |
| S5_vs_k141_69865\|\|full | Low-quality |
| S17_vs_k141_177202\|\|full | High-quality |
| S5_vs_k141_9744\|\|full | Low-quality |
| S5_vs_k141_4644\|\|full | Low-quality |
| S5_vs_k141_34263\|\|full | Not-determined |
| S5_vs_k141_9745\|\|full | Not-determined |
| S5_vs_k141_88054\|\|full | Low-quality |
| S5_vs_k141_4647\|\|full | Low-quality |
| S5_vs_k141_125828\|\|full | Not-determined |
| S5_vs_k141_4649\|\|full | Low-quality |
| S5_vs_k141_69869\|\|full | Not-determined |
| S5_vs_k141_125837\|\|full | Not-determined |
| S5_vs_k141_4655\|\|full | Low-quality |
| S5_vs_k141_130955\|\|full | Low-quality |
| S5_vs_k141_112715\|\|full | Low-quality |
| S5_vs_k141_125843\|\|full | Low-quality |
| S5_vs_k141_63960\|\|full | Low-quality |
| S5_vs_k141_107532\|\|full | Not-determined |
| S5_vs_k141_4665\|\|full | Low-quality |
| S5_vs_k141_107534\|\|full | Not-determined |
| S5_vs_k141_52727\|\|full | Not-determined |
| S5_vs_k141_136025\|\|full | Not-determined |
| S5_vs_k141_99438\|\|full | Low-quality |
| S5_vs_k141_15722\|\|full | Low-quality |
| S5_vs_k141_21480\|\|full | Not-determined |
| S5_vs_k141_63981\|\|full | Not-determined |
| S5_vs_k141_15733\|\|full | Not-determined |
| S5_vs_k141_52745\|\|full | Low-quality |
| S5_vs_k141_94536\|\|full | Not-determined |
| S5_vs_k141_21493\|\|full | Not-determined |
| S5_vs_k141_52746\|\|full | Low-quality |
| S5_vs_k141_112721\|\|full | Low-quality |
| S5_vs_k141_99449\|\|full | Not-determined |
| S5_vs_k141_94544\|\|full | Not-determined |
| S5_vs_k141_142645\|\|full | Low-quality |
| S5_vs_k141_39551\|\|full | Not-determined |
| S5_vs_k141_21511\|\|full | Low-quality |
| S5_vs_k141_56668\|\|full | Low-quality |
| S5_vs_k141_107553\|\|full | Low-quality |
| S3_vs_k141_99626\|\|full | High-quality |
| S5_vs_k141_99473\|\|full | Low-quality |
| S5_vs_k141_56677\|\|full | Not-determined |
| S5_vs_k141_34322\|\|full | Not-determined |
| S5_vs_k141_130959\|\|full | Low-quality |
| S5_vs_k141_112755\|\|full | Low-quality |
| S5_vs_k141_94579\|\|full | Low-quality |
| S5_vs_k141_4686\|\|full | Not-determined |
| S5_vs_k141_21533\|\|full | Low-quality |
| S5_vs_k141_52774\|\|full | Low-quality |
| S5_vs_k141_94581\|\|full | Low-quality |
| S5_vs_k141_125894\|\|full | Low-quality |
| S16_vs_134705\|\|full | High-quality |
| S5_vs_k141_39566\|\|full | Not-determined |
| S5_vs_k141_39570\|\|full | Not-determined |
| S5_vs_k141_99502\|\|full | Low-quality |
| S5_vs_k141_69896\|\|full | Low-quality |
| S5_vs_k141_56699\|\|full | Low-quality |
| S5_vs_k141_125899\|\|full | Low-quality |
| S5_vs_k141_99506\|\|full | Low-quality |
| S5_vs_k141_125903\|\|full | Low-quality |
| S5_vs_k141_99510\|\|full | Not-determined |
| S5_vs_k141_136056\|\|full | Low-quality |
| S5_vs_k141_94616\|\|full | Low-quality |
| S5_vs_k141_112816\|\|full | Low-quality |
| S5_vs_k141_94621\|\|full | Low-quality |
| S5_vs_k141_56743\|\|full | Low-quality |
| S5_vs_k141_52793\|\|full | Not-determined |
| S5_vs_k141_34370\|\|full | Low-quality |
| S5_vs_k141_99525\|\|full | Low-quality |
| S5_vs_k141_80835\|\|full | Low-quality |
| S5_vs_k141_34375\|\|full | Not-determined |
| S5_vs_k141_9797\|\|full | Low-quality |
| S5_vs_k141_26375\|\|full | Low-quality |
| S5_vs_k141_39630\|\|full | Not-determined |
| S5_vs_k141_80841\|\|full | Not-determined |
| S3_vs_k141_127401\|\|full | High-quality |
| S5_vs_k141_21588\|\|full | Low-quality |
| S5_vs_k141_130971\|\|full | Not-determined |
| S5_vs_k141_130972\|\|full | Low-quality |
| S5_vs_k141_119480\|\|full | Not-determined |
| S5_vs_k141_112847\|\|full | Low-quality |
| S5_vs_k141_88187\|\|full | Low-quality |
| S5_vs_k141_52796\|\|full | Not-determined |
| S5_vs_k141_34393\|\|full | Not-determined |
| S5_vs_k141_26382\|\|full | Low-quality |
| S5_vs_k141_4754\|\|full | Low-quality |
| S5_vs_k141_4758\|\|full | Not-determined |
| S5_vs_k141_125973\|\|full | Low-quality |
| S5_vs_k141_80850\|\|full | Low-quality |
| S5_vs_k141_125976\|\|full | Not-determined |
| S5_vs_k141_125986\|\|full | Low-quality |
| S5_vs_k141_9822\|\|full | Not-determined |
| S5_vs_k141_21625\|\|full | Low-quality |
| S5_vs_k141_34414\|\|full | Low-quality |
| S5_vs_k141_88207\|\|full | Not-determined |
| S5_vs_k141_107597\|\|full | Low-quality |
| S5_vs_k141_26404\|\|full | Not-determined |
| S5_vs_k141_4792\|\|full | Low-quality |
| S5_vs_k141_74804\|\|full | Low-quality |
| S5_vs_k141_4795\|\|full | Low-quality |
| S5_vs_k141_39654\|\|full | Low-quality |
| S5_vs_k141_131023\|\|full | Low-quality |
| S5_vs_k141_26409\|\|full | Low-quality |
| S5_vs_k141_9843\|\|full | Low-quality |
| S5_vs_k141_34429\|\|full | Not-determined |
| S5_vs_k141_131033\|\|full | Low-quality |
| S5_vs_k141_88221\|\|full | Not-determined |
| S5_vs_k141_69949\|\|full | Low-quality |
| S5_vs_k141_112902\|\|full | Low-quality |
| S5_vs_k141_131042\|\|full | Low-quality |
| S5_vs_k141_26423\|\|full | Low-quality |
| S5_vs_k141_119537\|\|full | Low-quality |
| S5_vs_k141_107606\|\|full | Low-quality |
| S5_vs_k141_131055\|\|full | Low-quality |
| S5_vs_k141_64011\|\|full | Low-quality |
| S5_vs_k141_74824\|\|full | Not-determined |
| S5_vs_k141_52832\|\|full | Low-quality |
| S5_vs_k141_9871\|\|full | Not-determined |
| S5_vs_k141_99559\|\|full | Low-quality |
| S5_vs_k141_94631\|\|full | Not-determined |
| S5_vs_k141_9873\|\|full | Not-determined |
| S5_vs_k141_99561\|\|full | Low-quality |
| S5_vs_k141_4852\|\|full | Low-quality |
| S5_vs_k141_34454\|\|full | Low-quality |
| S5_vs_k141_64019\|\|full | Low-quality |
| S5_vs_k141_99567\|\|full | Not-determined |
| S5_vs_k141_56804\|\|full | Low-quality |
| S5_vs_k141_107625\|\|full | Low-quality |
| S4_vs_VIRSorter_21643-cat_2 | High-quality |
| S5_vs_k141_94651\|\|full | Not-determined |
| S5_vs_k141_64026\|\|full | Not-determined |
| S5_vs_k141_119572\|\|full | Not-determined |
| S5_vs_k141_52863\|\|full | Not-determined |
| S5_vs_k141_34472\|\|full | Not-determined |
| S5_vs_k141_64033\|\|full | Low-quality |
| S5_vs_k141_64034\|\|full | Not-determined |
| S5_vs_k141_34475\|\|full | Low-quality |
| S5_vs_k141_107642\|\|full | Not-determined |
| S5_vs_k141_34476\|\|full | Low-quality |
| S5_vs_k141_9887\|\|full | Low-quality |
| S5_vs_k141_21631\|\|full | Not-determined |
| S5_vs_k141_64041\|\|full | Not-determined |
| S5_vs_k141_131082\|\|full | Low-quality |
| S5_vs_k141_112920\|\|full | Low-quality |
| S10_vs_120541\|\|full | High-quality |
| S5_vs_k141_119586\|\|full | Low-quality |
| S5_vs_k141_52890\|\|full | Low-quality |
| S5_vs_k141_126016\|\|full | Low-quality |
| S5_vs_k141_112929\|\|full | Not-determined |
| S5_vs_k141_80916\|\|full | Not-determined |
| S5_vs_k141_74854\|\|full | Low-quality |
| S5_vs_k141_4888\|\|full | Low-quality |
| S5_vs_k141_142684\|\|full | Not-determined |
| S5_vs_k141_112938\|\|full | Low-quality |
| S5_vs_k141_107658\|\|full | Not-determined |
| S5_vs_k141_94703\|\|full | Low-quality |
| S5_vs_k141_74867\|\|full | Low-quality |
| S5_vs_k141_94709\|\|full | Not-determined |
| S5_vs_k141_142700\|\|full | Low-quality |
| S5_vs_k141_56829\|\|full | Low-quality |
| S5_vs_k141_142708\|\|full | Low-quality |
| S5_vs_k141_126040\|\|full | Low-quality |
| S5_vs_k141_99616\|\|full | Not-determined |
| S5_vs_k141_21670\|\|full | Low-quality |
| S5_vs_k141_142724\|\|full | Low-quality |
| S5_vs_k141_64074\|\|full | Not-determined |
| S5_vs_k141_64080\|\|full | Low-quality |
| S5_vs_k141_34563\|\|full | Not-determined |
| S5_vs_k141_88262\|\|full | Low-quality |
| S5_vs_k141_99637\|\|full | Low-quality |
| S5_vs_k141_64086\|\|full | Low-quality |
| S5_vs_k141_21681\|\|full | Not-determined |
| S5_vs_k141_56840\|\|full | Low-quality |
| S5_vs_k141_21692\|\|full | Low-quality |
| S5_vs_k141_34577\|\|full | Low-quality |
| S5_vs_k141_107694\|\|full | Low-quality |
| S5_vs_k141_88283\|\|full | Not-determined |
| S5_vs_k141_88284\|\|full | Low-quality |
| S5_vs_k141_88291\|\|full | Low-quality |
| S5_vs_k141_107708\|\|full | Not-determined |
| S5_vs_k141_74889\|\|full | Low-quality |
| S5_vs_k141_56849\|\|full | Not-determined |
| S5_vs_k141_52984\|\|full | Low-quality |
| S5_vs_k141_142778\|\|full | Low-quality |
| S5_vs_k141_34617\|\|full | Low-quality |
| S5_vs_k141_46060\|\|full | Not-determined |
| S5_vs_k141_88318\|\|full | Low-quality |
| S5_vs_k141_52987\|\|full | Low-quality |
| S5_vs_k141_21725\|\|full | Low-quality |
| S5_vs_k141_15850\|\|full | Low-quality |
| S5_vs_k141_56864\|\|full | Low-quality |
| S5_vs_k141_99671\|\|full | Low-quality |
| S5_vs_k141_74898\|\|full | Not-determined |
| S5_vs_k141_15854\|\|full | Low-quality |
| S5_vs_k141_64132\|\|full | Low-quality |
| S5_vs_k141_112984\|\|full | Not-determined |
| S5_vs_k141_56883\|\|full | Low-quality |
| S8_extracted_k141_197061 | Complete |
| S5_vs_k141_69981\|\|full | Not-determined |
| S5_vs_k141_94805\|\|full | Low-quality |
| S5_vs_k141_112989\|\|full | Low-quality |
| S5_vs_k141_46106\|\|full | Low-quality |
| S5_vs_k141_53048\|\|full | Not-determined |
| S5_vs_k141_112994\|\|full | Low-quality |
| S5_vs_k141_15892\|\|full | Low-quality |
| S5_vs_k141_126110\|\|full | Low-quality |
| S5_vs_k141_131144\|\|full | Low-quality |
| S5_vs_k141_74937\|\|full | Low-quality |
| S5_vs_k141_88360\|\|full | Low-quality |
| S5_vs_k141_26477\|\|full | Low-quality |
| S5_vs_k141_131148\|\|full | Low-quality |
| S5_vs_k141_21745\|\|full | Low-quality |
| S5_vs_k141_126117\|\|full | Low-quality |
| S5_vs_k141_74947\|\|full | Low-quality |
| S5_vs_k141_107751\|\|full | Low-quality |
| S5_vs_k141_126121\|\|full | Not-determined |
| S5_vs_k141_34659\|\|full | Not-determined |
| S5_vs_k141_56911\|\|full | Not-determined |
| S5_vs_k141_70005\|\|full | Low-quality |
| S5_vs_k141_88375\|\|full | Low-quality |
| S5_vs_k141_9970\|\|full | Not-determined |
| S5_vs_k141_126129\|\|full | Not-determined |
| S5_vs_k141_126131\|\|full | Not-determined |
| S5_vs_k141_113028\|\|full | Not-determined |
| S4_vs_VIRSorter_5698-cat_2 | High-quality |
| S5_vs_k141_70025\|\|full | Low-quality |
| S5_vs_k141_34689\|\|full | Low-quality |
| S5_vs_k141_107781\|\|full | Low-quality |
| S5_vs_k141_113041\|\|full | Low-quality |
| S5_vs_k141_99725\|\|full | Not-determined |
| S5_vs_k141_131184\|\|full | Not-determined |
| S5_vs_k141_56922\|\|full | Low-quality |
| S5_vs_k141_94840\|\|full | Low-quality |
| S5_vs_k141_142837\|\|full | Low-quality |
| S5_vs_k141_5077\|\|full | Not-determined |
| S5_vs_k141_9971\|\|full | Low-quality |
| S5_vs_k141_142839\|\|full | Low-quality |
| S5_vs_k141_64164\|\|full | Low-quality |
| S5_vs_k141_88410\|\|full | Low-quality |
| S5_vs_k141_64168\|\|full | Not-determined |
| S5_vs_k141_34717\|\|full | Low-quality |
| S5_vs_k141_126164\|\|full | Low-quality |
| S5_vs_k141_126165\|\|full | Not-determined |
| S5_vs_k141_15936\|\|full | Low-quality |
| S5_vs_k141_26511\|\|full | Not-determined |
| S5_vs_k141_88418\|\|full | Low-quality |
| S5_vs_k141_142848\|\|full | Low-quality |
| S5_vs_k141_5104\|\|full | Low-quality |
| S5_vs_k141_34722\|\|full | Not-determined |
| S5_vs_k141_107814\|\|full | Low-quality |
| S5_vs_k141_46125\|\|full | Low-quality |
| S5_vs_k141_70058\|\|full | Low-quality |
| S5_vs_k141_94871\|\|full | Low-quality |
| S5_vs_k141_64195\|\|full | Not-determined |
| S5_vs_k141_136185\|\|full | Not-determined |
| S5_vs_k141_46133\|\|full | Low-quality |
| S5_vs_k141_107831\|\|full | Not-determined |
| S5_vs_k141_15976\|\|full | Low-quality |
| S5_vs_k141_70065\|\|full | Low-quality |
| S5_vs_k141_53113\|\|full | Not-determined |

| | |
|---|---|
| S5_vs_k141_46142\|\|full | Not-determined |
| S5_vs_k141_88442\|\|full | Low-quality |
| S5_vs_k141_15989\|\|full | Low-quality |
| S5_vs_k141_99743\|\|full | Low-quality |
| S5_vs_k141_94900\|\|full | Low-quality |
| S5_vs_k141_126207\|\|full | Low-quality |
| S5_vs_k141_119610\|\|full | Low-quality |
| S5_vs_k141_70073\|\|full | Low-quality |
| S5_vs_k141_46155\|\|full | Not-determined |
| S5_vs_k141_74995\|\|full | Low-quality |
| S5_vs_k141_10040\|\|full | Not-determined |
| S5_vs_k141_56971\|\|full | Not-determined |
| S5_vs_k141_88457\|\|full | Not-determined |
| S5_vs_k141_136200\|\|full | Not-determined |
| S5_vs_k141_136201\|\|full | Low-quality |
| S5_vs_k141_56973\|\|full | Not-determined |
| S5_vs_k141_34770\|\|full | Low-quality |
| S5_vs_k141_64211\|\|full | Low-quality |
| S5_vs_k141_5144\|\|full | Low-quality |
| S5_vs_k141_46165\|\|full | Low-quality |
| S5_vs_k141_70096\|\|full | Not-determined |
| S5_vs_k141_5153\|\|full | Not-determined |
| S5_vs_k141_136215\|\|full | Low-quality |
| S5_vs_k141_70100\|\|full | Low-quality |
| S5_vs_k141_136216\|\|full | Low-quality |
| S5_vs_k141_94934\|\|full | Not-determined |
| S5_vs_k141_46183\|\|full | Not-determined |
| S5_vs_k141_53144\|\|full | Low-quality |
| S5_vs_k141_5159\|\|full | Low-quality |
| S5_vs_k141_99752\|\|full | Low-quality |
| S5_vs_k141_5161\|\|full | Low-quality |
| S5_vs_k141_88477\|\|full | Low-quality |
| S5_vs_k141_131209\|\|full | Not-determined |
| S5_vs_k141_136229\|\|full | Low-quality |
| S5_vs_k141_10073\|\|full | Not-determined |
| S5_vs_k141_88494\|\|full | Low-quality |
| S5_vs_k141_64239\|\|full | Low-quality |
| S5_vs_k141_119666\|\|full | Low-quality |
| S5_vs_k141_70133\|\|full | Low-quality |
| S5_vs_k141_26551\|\|full | Not-determined |
| S5_vs_k141_26555\|\|full | Low-quality |
| S5_vs_k141_16023\|\|full | Low-quality |
| S5_vs_k141_88508\|\|full | Low-quality |
| S5_vs_k141_88516\|\|full | Low-quality |
| S5_vs_k141_94974\|\|full | Low-quality |
| S5_vs_k141_131227\|\|full | Low-quality |
| S5_vs_k141_107913\|\|full | Low-quality |
| S5_vs_k141_5207\|\|full | Low-quality |
| S5_vs_k141_113074\|\|full | Low-quality |
| S5_vs_k141_126284\|\|full | Low-quality |
| S5_vs_k141_70162\|\|full | Not-determined |
| S5_vs_k141_142899\|\|full | Not-determined |
| S5_vs_k141_94986\|\|full | Not-determined |
| S5_vs_k141_70165\|\|full | Not-determined |
| S5_vs_k141_107919\|\|full | Not-determined |
| S5_vs_k141_39675\|\|full | Low-quality |
| S5_vs_k141_113084\|\|full | Not-determined |
| S5_vs_k141_107928\|\|full | Low-quality |
| S5_vs_k141_53224\|\|full | Not-determined |
| S5_vs_k141_34841\|\|full | Low-quality |
| S5_vs_k141_88541\|\|full | Not-determined |
| S5_vs_k141_107933\|\|full | Low-quality |
| S5_vs_k141_136291\|\|full | Low-quality |
| S5_vs_k141_5232\|\|full | Low-quality |
| S5_vs_k141_16073\|\|full | Low-quality |
| S5_vs_k141_88543\|\|full | Not-determined |
| S7_vs_20200\|\|full | High-quality |
| S5_vs_k141_99794\|\|full | Not-determined |
| S5_vs_k141_16078\|\|full | Low-quality |
| S13_extracted_85097 | High-quality |
| S5_vs_k141_119763\|\|full | Not-determined |
| S5_vs_k141_53253\|\|full | Low-quality |
| S5_vs_k141_136316\|\|full | Not-determined |
| S6_vs_k141_20970\|\|full | Medium-quality |
| S5_vs_k141_10109\|\|full | Not-determined |
| S5_vs_k141_10113\|\|full | Low-quality |
| S5_vs_k141_88573\|\|full | Low-quality |
| S5_vs_k141_21781\|\|full | Not-determined |
| S5_vs_k141_113126\|\|full | Low-quality |
| S5_vs_k141_131322\|\|full | Low-quality |
| S11_vs_k141_38860\|\|full | High-quality |
| S5_vs_k141_39721\|\|full | Not-determined |
| S5_vs_k141_107968\|\|full | Not-determined |
| S5_vs_k141_5276\|\|full | Low-quality |
| S5_vs_k141_99822\|\|full | Low-quality |
| S5_vs_k141_119790\|\|full | Low-quality |
| S5_vs_k141_136333\|\|full | Not-determined |
| S5_vs_k141_119802\|\|full | Low-quality |
| S5_vs_k141_119804\|\|full | Low-quality |
| S5_vs_k141_107982\|\|full | Low-quality |
| S5_vs_k141_39741\|\|full | Not-determined |
| S5_vs_k141_88619\|\|full | Low-quality |
| S5_vs_k141_53306\|\|full | Low-quality |
| S5_vs_k141_88621\|\|full | Low-quality |
| S5_vs_k141_107997\|\|full | Low-quality |
| S5_vs_k141_136342\|\|full | Not-determined |
| S5_vs_k141_5305\|\|full | Low-quality |
| S5_vs_k141_113187\|\|full | Not-determined |
| S5_vs_k141_99862\|\|full | Not-determined |
| S5_vs_k141_57006\|\|full | Low-quality |
| S5_vs_k141_88628\|\|full | Low-quality |
| S5_vs_k141_21844\|\|full | Low-quality |
| S5_vs_k141_119820\|\|full | Low-quality |
| S5_vs_k141_136351\|\|full | Low-quality |
| S5_vs_k141_119821\|\|full | Low-quality |
| S5_vs_k141_5314\|\|full | Low-quality |
| S5_vs_k141_136355\|\|full | Low-quality |
| S5_vs_k141_75023\|\|full | Low-quality |
| S5_vs_k141_10159\|\|full | Low-quality |
| S5_vs_k141_108016\|\|full | Low-quality |
| S5_vs_k141_39790\|\|full | Not-determined |
| S5_vs_k141_99881\|\|full | Low-quality |
| S5_vs_k141_21855\|\|full | Not-determined |
| S5_vs_k141_113216\|\|full | Not-determined |
| S5_vs_k141_21861\|\|full | Low-quality |
| S5_vs_k141_75038\|\|full | Low-quality |
| S5_vs_k141_99886\|\|full | Low-quality |
| S5_vs_k141_99888\|\|full | Low-quality |
| S5_vs_k141_70272\|\|full | Low-quality |
| S5_vs_k141_99889\|\|full | Not-determined |
| S5_vs_k141_113228\|\|full | Low-quality |
| S5_vs_k141_108021\|\|full | Low-quality |
| S5_vs_k141_95108\|\|full | Not-determined |
| S5_vs_k141_10183\|\|full | Low-quality |
| S5_vs_k141_119866\|\|full | Not-determined |
| S5_vs_k141_16142\|\|full | Not-determined |
| S5_vs_k141_131354\|\|full | Low-quality |
| S5_vs_k141_70288\|\|full | Low-quality |
| S5_vs_k141_108040\|\|full | Not-determined |
| S5_vs_k141_75054\|\|full | Low-quality |
| S5_vs_k141_34890\|\|full | Low-quality |
| S5_vs_k141_88687\|\|full | Low-quality |
| S5_vs_k141_99905\|\|full | Low-quality |
| S5_vs_k141_136375\|\|full | Low-quality |
| S5_vs_k141_57043\|\|full | Low-quality |
| S5_vs_k141_75067\|\|full | Not-determined |
| S5_vs_k141_119887\|\|full | Low-quality |
| S5_vs_k141_136387\|\|full | Not-determined |
| S5_vs_k141_142954\|\|full | Not-determined |
| S5_vs_k141_95155\|\|full | Not-determined |
| S5_vs_k141_53361\|\|full | Low-quality |
| S5_vs_k141_88697\|\|full | Low-quality |
| S5_vs_k141_34900\|\|full | Low-quality |
| S5_vs_k141_53363\|\|full | Low-quality |
| S5_vs_k141_5381\|\|full | Low-quality |
| S5_vs_k141_131374\|\|full | Low-quality |
| S5_vs_k141_88716\|\|full | Low-quality |
| S5_vs_k141_34914\|\|full | Not-determined |
| S5_vs_k141_5389\|\|full | Low-quality |
| S5_vs_k141_26627\|\|full | Low-quality |
| S5_vs_k141_88726\|\|full | Not-determined |
| S8_vs_k141_136922\|\|full | Medium-quality |
| S5_vs_k141_26633\|\|full | Low-quality |
| S5_vs_k141_88736\|\|full | Low-quality |
| S5_vs_k141_21884\|\|full | Low-quality |
| S5_vs_k141_21887\|\|full | Low-quality |
| S5_vs_k141_10244\|\|full | Not-determined |
| S5_vs_k141_39855\|\|full | Low-quality |
| S5_vs_k141_88750\|\|full | Low-quality |
| S5_vs_k141_131399\|\|full | Low-quality |
| S5_vs_k141_21892\|\|full | Low-quality |
| S5_vs_k141_81198\|\|full | Low-quality |
| S5_vs_k141_39861\|\|full | Low-quality |
| S5_vs_k141_39864\|\|full | Low-quality |
| S5_vs_k141_81205\|\|full | Low-quality |
| S5_extracted_k141_114249 | Complete |
| S5_vs_k141_113291\|\|full | Low-quality |
| S5_vs_k141_26651\|\|full | Not-determined |
| S5_vs_k141_10266\|\|full | Low-quality |
| S5_vs_k141_39870\|\|full | Low-quality |
| S5_vs_k141_81208\|\|full | Not-determined |
| S5_vs_k141_75095\|\|full | Low-quality |
| S5_vs_k141_39875\|\|full | Low-quality |
| S5_vs_k141_142972\|\|full | Low-quality |
| S5_vs_k141_88774\|\|full | Low-quality |
| S5_vs_k141_88777\|\|full | Low-quality |
| S5_vs_k141_81215\|\|full | Not-determined |
| S5_vs_k141_5450\|\|full | Not-determined |
| S5_vs_k141_75111\|\|full | Not-determined |
| S5_vs_k141_113313\|\|full | Low-quality |
| S5_vs_k141_34980\|\|full | Low-quality |
| S5_vs_k141_113319\|\|full | Low-quality |
| S5_vs_k141_81231\|\|full | Not-determined |
| S5_vs_k141_39889\|\|full | Not-determined |
| S5_vs_k141_131404\|\|full | Low-quality |
| S5_vs_k141_95224\|\|full | Low-quality |
| S5_vs_k141_10297\|\|full | Low-quality |
| S5_vs_k141_88786\|\|full | Low-quality |
| S5_vs_k141_57088\|\|full | Not-determined |
| S5_vs_k141_131412\|\|full | Not-determined |
| S5_vs_k141_88794\|\|full | Not-determined |
| S5_vs_k141_95233\|\|full | Not-determined |
| S5_vs_k141_57094\|\|full | Not-determined |
| S5_vs_k141_75123\|\|full | Low-quality |
| S5_vs_k141_75124\|\|full | Low-quality |
| S5_vs_k141_64380\|\|full | Not-determined |
| S5_vs_k141_10314\|\|full | Not-determined |
| S5_vs_k141_88805\|\|full | Not-determined |
| S5_vs_k141_10323\|\|full | Low-quality |
| S5_vs_k141_35022\|\|full | Low-quality |
| S5_vs_k141_95239\|\|full | Low-quality |
| S5_vs_k141_88808\|\|full | Low-quality |
| S5_vs_k141_57107\|\|full | Low-quality |
| S5_vs_k141_21965\|\|full | Not-determined |
| S5_vs_k141_21969\|\|full | Low-quality |
| S5_vs_k141_21978\|\|full | Low-quality |
| S5_vs_k141_131429\|\|full | Not-determined |
| S5_vs_k141_64400\|\|full | Low-quality |
| S5_vs_k141_21989\|\|full | Not-determined |
| S5_vs_k141_95264\|\|full | Not-determined |
| S5_vs_k141_5500\|\|full | Not-determined |
| S5_vs_k141_108138\|\|full | Low-quality |
| S5_vs_k141_35072\|\|full | Not-determined |
| S5_vs_k141_75161\|\|full | Low-quality |
| S5_vs_k141_64415\|\|full | Not-determined |
| S5_vs_k141_75162\|\|full | Not-determined |
| S5_vs_k141_113379\|\|full | Low-quality |
| S5_vs_k141_108143\|\|full | Low-quality |
| S5_vs_k141_120052\|\|full | Low-quality |
| S5_vs_k141_5521\|\|full | Not-determined |
| S5_vs_k141_108150\|\|full | Not-determined |
| S5_vs_k141_113385\|\|full | Low-quality |
| S5_vs_k141_136503\|\|full | Low-quality |
| S5_vs_k141_143010\|\|full | Low-quality |
| S5_vs_k141_46221\|\|full | Low-quality |
| S5_vs_k141_99970\|\|full | Not-determined |
| S5_vs_k141_120062\|\|full | Not-determined |
| S5_vs_k141_39945\|\|full | Not-determined |
| S5_vs_k141_35102\|\|full | Low-quality |
| S5_vs_k141_46230\|\|full | Low-quality |
| S5_vs_k141_120063\|\|full | Low-quality |
| S5_vs_k141_108179\|\|full | Not-determined |
| S5_vs_k141_70390\|\|full | Low-quality |
| S5_vs_k141_53480\|\|full | Not-determined |
| S5_vs_k141_53482\|\|full | Low-quality |
| S5_vs_k141_57128\|\|full | Not-determined |
| S5_vs_k141_88878\|\|full | Not-determined |
| S5_vs_k141_32885\|\|full | Medium-quality |
| S5_vs_k141_39964\|\|full | Low-quality |
| S5_vs_k141_53486\|\|full | Low-quality |
| S5_vs_k141_113389\|\|full | Not-determined |
| S5_vs_k141_113395\|\|full | Low-quality |
| S5_vs_k141_126304\|\|full | Low-quality |
| S5_vs_k141_10389\|\|full | Not-determined |
| S5_vs_k141_64474\|\|full | Low-quality |
| S5_vs_k141_10395\|\|full | Low-quality |
| S5_vs_k141_108212\|\|full | Low-quality |
| S7_vs_69630\|\|full | High-quality |
| S5_vs_k141_108213\|\|full | Low-quality |
| S5_vs_k141_95310\|\|full | Low-quality |
| S5_vs_k141_16233\|\|full | Low-quality |
| S5_vs_k141_88910\|\|full | Not-determined |
| S5_vs_k141_143050\|\|full | Not-determined |
| S5_vs_k141_64479\|\|full | Low-quality |
| S5_vs_k141_46248\|\|full | Low-quality |
| S5_vs_k141_10410\|\|full | Low-quality |
| S5_vs_k141_35156\|\|full | Low-quality |
| S5_vs_k141_108220\|\|full | Low-quality |
| S5_vs_k141_75219\|\|full | Not-determined |
| S5_vs_k141_88920\|\|full | Low-quality |
| S5_vs_k141_136536\|\|full | Not-determined |
| S5_vs_k141_64493\|\|full | Low-quality |
| S5_vs_k141_35164\|\|full | Low-quality |
| S5_vs_k141_16258\|\|full | Not-determined |
| S5_vs_k141_35166\|\|full | Low-quality |
| S5_vs_k141_126344\|\|full | Low-quality |
| S5_vs_k141_64497\|\|full | Low-quality |
| S5_vs_k141_35172\|\|full | Low-quality |
| S5_vs_k141_70482\|\|full | Low-quality |
| S5_vs_k141_64511\|\|full | Low-quality |
| S5_vs_k141_136552\|\|full | Not-determined |
| S5_vs_k141_95332\|\|full | Low-quality |
| S5_vs_k141_39996\|\|full | Not-determined |
| S5_vs_k141_64513\|\|full | Not-determined |
| S5_vs_k141_120101\|\|full | Low-quality |
| S5_vs_k141_95340\|\|full | Low-quality |
| S5_vs_k141_120108\|\|full | Not-determined |
| S5_vs_k141_75284\|\|full | Not-determined |
| S5_vs_k141_95356\|\|full | Not-determined |
| S5_vs_k141_40009\|\|full | Low-quality |
| S5_vs_k141_26723\|\|full | Not-determined |
| S5_vs_k141_46274\|\|full | Low-quality |
| S5_vs_k141_35195\|\|full | Low-quality |
| S5_vs_k141_108264\|\|full | Low-quality |
| S5_vs_k141_70516\|\|full | Low-quality |
| S5_vs_k141_143066\|\|full | Not-determined |
| S5_vs_k141_136564\|\|full | Low-quality |
| S5_vs_k141_120133\|\|full | Low-quality |
| S5_vs_k141_88931\|\|full | Not-determined |
| S5_vs_k141_46285\|\|full | Low-quality |
| S5_vs_k141_75306\|\|full | Low-quality |
| S5_vs_k141_35206\|\|full | Low-quality |
| S5_vs_k141_120136\|\|full | Not-determined |
| S5_vs_k141_143081\|\|full | Not-determined |
| S5_vs_k141_57203\|\|full | Not-determined |
| S5_vs_k141_75315\|\|full | Low-quality |
| S5_vs_k141_40048\|\|full | Low-quality |
| S5_vs_k141_57207\|\|full | Not-determined |
| S5_vs_k141_40050\|\|full | Low-quality |
| S5_vs_k141_16333\|\|full | Low-quality |
| S5_vs_k141_5553\|\|full | Low-quality |
| S5_vs_k141_88954\|\|full | Low-quality |
| S5_vs_k141_5563\|\|full | Low-quality |
| S5_vs_k141_26777\|\|full | Not-determined |
| S5_vs_k141_16348\|\|full | Low-quality |
| S5_vs_k141_22019\|\|full | Not-determined |
| S5_vs_k141_40070\|\|full | Not-determined |
| S5_vs_k141_88980\|\|full | Low-quality |
| S5_vs_k141_126430\|\|full | Not-determined |
| S5_vs_k141_108291\|\|full | Low-quality |
| S5_vs_k141_40078\|\|full | Low-quality |
| S5_vs_k141_120145\|\|full | Not-determined |
| S5_vs_k141_126449\|\|full | Not-determined |
| S5_vs_k141_75320\|\|full | Low-quality |
| S5_vs_k141_22040\|\|full | Not-determined |
| S5_vs_k141_40093\|\|full | Low-quality |
| S5_vs_k141_120152\|\|full | Low-quality |
| S15_vs_71968\|\|full | Medium-quality |
| S5_vs_k141_16351\|\|full | Low-quality |
| S5_vs_k141_136589\|\|full | Low-quality |
| S5_vs_k141_26787\|\|full | Not-determined |
| S5_vs_k141_64605\|\|full | Low-quality |
| S5_vs_k141_120159\|\|full | Low-quality |
| S5_vs_k141_16360\|\|full | Low-quality |
| S5_vs_k141_75344\|\|full | Not-determined |
| S5_vs_k141_120166\|\|full | Low-quality |
| S5_vs_k141_108342\|\|full | Low-quality |
| S5_vs_k141_81275\|\|full | Low-quality |
| S5_vs_k141_53533\|\|full | Low-quality |
| S5_vs_k141_89026\|\|full | Low-quality |
| S5_vs_k141_75370\|\|full | Low-quality |
| S5_vs_k141_40112\|\|full | Low-quality |
| S5_vs_k141_40114\|\|full | Not-determined |
| S5_vs_k141_126486\|\|full | Not-determined |
| S5_vs_k141_5609\|\|full | Low-quality |
| S5_vs_k141_143143\|\|full | Not-determined |
| S5_vs_k141_143144\|\|full | Low-quality |
| S5_vs_k141_40126\|\|full | Low-quality |
| S5_vs_k141_126492\|\|full | Not-determined |
| S5_vs_k141_143148\|\|full | Not-determined |
| S5_vs_k141_26835\|\|full | Low-quality |
| S5_vs_k141_53552\|\|full | Low-quality |
| S5_vs_k141_35298\|\|full | Low-quality |
| S5_vs_k141_89054\|\|full | Low-quality |
| S5_vs_k141_5634\|\|full | Not-determined |
| S5_extracted_k141_145586 | Complete |
| S5_vs_k141_53560\|\|full | Low-quality |
| S5_vs_k141_143162\|\|full | Low-quality |
| S5_vs_k141_126513\|\|full | Low-quality |
| S5_vs_k141_75395\|\|full | Not-determined |
| S5_vs_k141_35301\|\|full | Not-determined |
| S5_vs_k141_126517\|\|full | Low-quality |
| S5_vs_k141_81299\|\|full | Low-quality |
| S5_vs_k141_64666\|\|full | Low-quality |
| S5_vs_k141_131528\|\|full | Low-quality |
| S5_vs_k141_53577\|\|full | Low-quality |
| S5_vs_k141_126525\|\|full | Low-quality |
| S5_vs_k141_53579\|\|full | Not-determined |
| S5_vs_k141_16437\|\|full | Low-quality |
| S5_vs_k141_126528\|\|full | Not-determined |
| S5_vs_k141_81307\|\|full | Low-quality |
| S5_vs_k141_53587\|\|full | Not-determined |
| S5_vs_k141_81309\|\|full | Low-quality |
| S5_vs_k141_64679\|\|full | Low-quality |
| S5_vs_k141_75417\|\|full | Not-determined |
| S5_vs_k141_126541\|\|full | Low-quality |
| S5_vs_k141_143179\|\|full | Not-determined |
| S5_vs_k141_53591\|\|full | Low-quality |
| S5_vs_k141_100075\|\|full | Low-quality |
| S5_vs_k141_126543\|\|full | Not-determined |
| S4_vs_VIRSorter_31977-circular-cat | High-quality |
| S5_vs_k141_53599\|\|full | Not-determined |
| S5_vs_k141_53600\|\|full | Not-determined |
| S5_vs_k141_136639\|\|full | Low-quality |
| S5_vs_k141_95449\|\|full | Not-determined |
| S5_vs_k141_53610\|\|full | Low-quality |
| S5_vs_k141_126562\|\|full | Low-quality |
| S5_vs_k141_26889\|\|full | Low-quality |
| S5_vs_k141_126567\|\|full | Low-quality |
| S5_vs_k141_64722\|\|full | Not-determined |
| S5_vs_k141_22142\|\|full | Low-quality |
| S5_vs_k141_75473\|\|full | Low-quality |
| S5_vs_k141_108448\|\|full | Low-quality |
| S5_vs_k141_75474\|\|full | Low-quality |
| S5_vs_k141_53649\|\|full | Not-determined |
| S5_vs_k141_75482\|\|full | Low-quality |
| S5_vs_k141_53651\|\|full | Low-quality |
| S5_vs_k141_75492\|\|full | Low-quality |
| S5_vs_k141_40215\|\|full | Low-quality |
| S5_vs_k141_95486\|\|full | Not-determined |
| S5_vs_k141_126600\|\|full | Not-determined |
| S5_vs_k141_95492\|\|full | Not-determined |
| S5_vs_k141_113450\|\|full | Low-quality |
| S5_vs_k141_108490\|\|full | Not-determined |
| S5_vs_k141_53682\|\|full | Not-determined |
| S5_vs_k141_126614\|\|full | Low-quality |
| S5_vs_k141_126616\|\|full | Not-determined |
| S5_vs_k141_10444\|\|full | Low-quality |
| S5_vs_k141_95495\|\|full | Low-quality |
| S5_vs_k141_113475\|\|full | Low-quality |
| S5_vs_k141_113476\|\|full | Low-quality |
| S5_vs_k141_131642\|\|full | Not-determined |
| S5_vs_k141_26919\|\|full | Low-quality |
| S5_vs_k141_16557\|\|full | Not-determined |
| S5_vs_k141_53705\|\|full | Low-quality |
| S5_vs_k141_95499\|\|full | Low-quality |
| S5_vs_k141_126634\|\|full | Low-quality |
| S5_vs_k141_16558\|\|full | Not-determined |
| S5_vs_k141_126635\|\|full | Not-determined |
| S5_vs_k141_108533\|\|full | Low-quality |
| S5_vs_k141_16565\|\|full | Low-quality |
| S5_vs_k141_131671\|\|full | Low-quality |
| S5_vs_k141_89116\|\|full | Low-quality |
| S5_vs_k141_113504\|\|full | Not-determined |
| S5_vs_k141_113507\|\|full | Low-quality |
| S5_vs_k141_5662\|\|full | Not-determined |
| S5_vs_k141_131680\|\|full | Not-determined |
| S5_vs_k141_16577\|\|full | Not-determined |
| S5_vs_k141_53731\|\|full | Low-quality |
| S5_vs_k141_89129\|\|full | Low-quality |
| S5_vs_k141_113514\|\|full | Low-quality |
| S5_vs_k141_95526\|\|full | Low-quality |
| S5_vs_k141_108554\|\|full | Low-quality |
| S5_vs_k141_131698\|\|full | Low-quality |
| S5_vs_k141_100205\|\|full | Low-quality |
| S5_vs_k141_53756\|\|full | Low-quality |
| S5_vs_k141_64755\|\|full | Not-determined |
| S5_vs_k141_53758\|\|full | Low-quality |
| S5_vs_k141_113538\|\|full | Low-quality |
| S5_vs_k141_143268\|\|full | Not-determined |
| S5_vs_k141_108582\|\|full | Low-quality |
| S5_vs_k141_70691\|\|full | Low-quality |
| S5_vs_k141_131707\|\|full | Low-quality |
| S5_vs_k141_108586\|\|full | Low-quality |
| S5_vs_k141_16627\|\|full | Low-quality |
| S5_vs_k141_100206\|\|full | Low-quality |
| S5_vs_k141_26939\|\|full | Low-quality |
| S5_vs_k141_143273\|\|full | Not-determined |
| S3_extracted_k141_44925 | Medium-quality |
| S5_vs_k141_89162\|\|full | Not-determined |
| S5_vs_k141_108607\|\|full | Not-determined |
| S5_vs_k141_100218\|\|full | Not-determined |
| S5_vs_k141_35362\|\|full | Low-quality |
| S5_vs_k141_53769\|\|full | Low-quality |
| S5_vs_k141_16662\|\|full | Low-quality |
| S5_vs_k141_143298\|\|full | Low-quality |
| S5_vs_k141_95556\|\|full | Low-quality |
| S5_vs_k141_113575\|\|full | Not-determined |
| S5_vs_k141_53777\|\|full | Low-quality |
| S5_vs_k141_10485\|\|full | Low-quality |
| S5_vs_k141_70707\|\|full | Not-determined |
| S5_vs_k141_53781\|\|full | Low-quality |
| S5_vs_k141_75564\|\|full | Low-quality |
| S5_vs_k141_70714\|\|full | Low-quality |
| S5_vs_k141_95582\|\|full | Low-quality |
| S5_vs_k141_26983\|\|full | Low-quality |
| S5_vs_k141_143324\|\|full | Not-determined |
| S5_vs_k141_95588\|\|full | Not-determined |
| S5_vs_k141_70722\|\|full | Low-quality |
| S5_vs_k141_143327\|\|full | Low-quality |
| S5_vs_k141_35395\|\|full | Not-determined |
| S5_vs_k141_22198\|\|full | Low-quality |
| S5_vs_k141_113611\|\|full | Not-determined |
| S5_vs_k141_57272\|\|full | Not-determined |
| S5_vs_k141_57273\|\|full | Not-determined |
| S5_vs_k141_22202\|\|full | Low-quality |
| S5_vs_k141_22203\|\|full | Low-quality |
| S5_vs_k141_26997\|\|full | Not-determined |
| S5_vs_k141_126677\|\|full | Low-quality |
| S5_vs_k141_35431\|\|full | Not-determined |
| S5_vs_k141_5739\|\|full | Low-quality |
| S5_vs_k141_22209\|\|full | Not-determined |
| S5_vs_k141_75606\|\|full | Low-quality |
| S5_vs_k141_136704\|\|full | Not-determined |
| S5_vs_k141_27012\|\|full | Low-quality |
| S5_vs_k141_75611\|\|full | Low-quality |
| S5_vs_k141_40269\|\|full | Not-determined |
| S5_vs_k141_70770\|\|full | Low-quality |
| S5_vs_k141_95637\|\|full | Not-determined |
| S5_vs_k141_40281\|\|full | Not-determined |
| S5_vs_k141_75624\|\|full | Low-quality |
| S5_vs_k141_22210\|\|full | Not-determined |
| S5_vs_k141_40285\|\|full | Not-determined |
| S5_vs_k141_57284\|\|full | Not-determined |
| S5_vs_k141_100279\|\|full | Low-quality |
| S5_vs_k141_22212\|\|full | Low-quality |
| S5_vs_k141_70778\|\|full | Low-quality |
| S5_vs_k141_131726\|\|full | Low-quality |
| S5_vs_k141_70787\|\|full | Not-determined |
| S5_vs_k141_75630\|\|full | Low-quality |
| S5_vs_k141_81352\|\|full | Low-quality |
| S5_vs_k141_81357\|\|full | Low-quality |
| S5_vs_k141_126707\|\|full | Not-determined |
| S5_vs_k141_70803\|\|full | Low-quality |
| S5_vs_k141_16773\|\|full | Low-quality |
| S5_vs_k141_35482\|\|full | Low-quality |
| S5_vs_k141_70804\|\|full | Not-determined |
| S5_vs_k141_113653\|\|full | Low-quality |
| S5_vs_k141_113655\|\|full | Not-determined |
| S5_vs_k141_126726\|\|full | Not-determined |
| S5_vs_k141_75644\|\|full | Not-determined |
| S5_vs_k141_131759\|\|full | Not-determined |
| S5_vs_k141_113667\|\|full | Low-quality |
| S5_vs_k141_136736\|\|full | Low-quality |
| S5_vs_k141_35519\|\|full | Not-determined |
| S5_vs_k141_113676\|\|full | Low-quality |
| S5_vs_k141_113678\|\|full | Not-determined |
| S5_vs_k141_113679\|\|full | Not-determined |
| S5_vs_k141_120215\|\|full | Low-quality |
| S5_vs_k141_40308\|\|full | Low-quality |
| S5_vs_k141_131792\|\|full | Not-determined |
| S5_vs_k141_126748\|\|full | Low-quality |
| S5_vs_k141_120228\|\|full | Not-determined |
| S5_vs_k141_57333\|\|full | Low-quality |
| S5_vs_k141_120239\|\|full | Low-quality |
| S5_vs_k141_75695\|\|full | Low-quality |
| S5_vs_k141_27065\|\|full | Low-quality |
| S5_vs_k141_113706\|\|full | Not-determined |
| S5_vs_k141_70863\|\|full | Low-quality |
| S5_vs_k141_16839\|\|full | Low-quality |
| S5_vs_k141_75706\|\|full | Low-quality |
| S5_vs_k141_35546\|\|full | Not-determined |
| S5_vs_k141_57351\|\|full | Not-determined |
| S5_vs_k141_46306\|\|full | Not-determined |
| S5_vs_k141_113716\|\|full | Low-quality |
| S5_vs_k141_46313\|\|full | Low-quality |
| S5_vs_k141_113719\|\|full | Low-quality |
| S5_vs_k141_143388\|\|full | Low-quality |
| S5_vs_k141_35559\|\|full | Low-quality |
| S5_vs_k141_46328\|\|full | Low-quality |
| S5_vs_k141_131840\|\|full | Low-quality |
| S5_vs_k141_35570\|\|full | Low-quality |
| S5_vs_k141_95697\|\|full | Not-determined |
| S5_vs_k141_57378\|\|full | Not-determined |
| S5_vs_k141_120287\|\|full | Low-quality |
| S5_vs_k141_64903\|\|full | Low-quality |
| S5_vs_k141_126813\|\|full | Not-determined |
| S5_vs_k141_120297\|\|full | Not-determined |
| S5_vs_k141_64918\|\|full | Low-quality |
| S5_vs_k141_46350\|\|full | Not-determined |
| S5_vs_k141_57392\|\|full | Not-determined |
| S5_vs_k141_95717\|\|full | Low-quality |
| S5_vs_k141_136774\|\|full | Low-quality |
| S5_vs_k141_75754\|\|full | Low-quality |
| S5_vs_k141_22241\|\|full | Low-quality |
| S5_vs_k141_35598\|\|full | Low-quality |
| S5_vs_k141_113780\|\|full | Not-determined |
| S5_vs_k141_143405\|\|full | Not-determined |
| S5_vs_k141_22260\|\|full | Low-quality |
| S5_vs_k141_131866\|\|full | Low-quality |
| S5_vs_k141_113784\|\|full | Low-quality |
| S5_vs_k141_95759\|\|full | Low-quality |
| S5_vs_k141_131887\|\|full | Not-determined |
| S5_vs_k141_35626\|\|full | Not-determined |
| S5_vs_k141_5772\|\|full | Not-determined |
| S5_vs_k141_22266\|\|full | Low-quality |
| S5_vs_k141_70906\|\|full | Low-quality |
| S5_vs_k141_35633\|\|full | Low-quality |
| S5_vs_k141_131894\|\|full | Low-quality |
| S5_vs_k141_75776\|\|full | Low-quality |
| S5_vs_k141_136792\|\|full | Low-quality |
| S5_vs_k141_27123\|\|full | Low-quality |
| S5_vs_k141_81505\|\|full | Low-quality |
| S5_vs_k141_70920\|\|full | Low-quality |
| S5_vs_k141_5781\|\|full | Low-quality |
| S5_vs_k141_100308\|\|full | Low-quality |
| S5_vs_k141_89212\|\|full | Low-quality |
| S5_vs_k141_53800\|\|full | Low-quality |
| S5_vs_k141_100310\|\|full | Not-determined |
| S5_vs_k141_70928\|\|full | Low-quality |
| S5_vs_k141_27147\|\|full | Low-quality |
| S5_vs_k141_81525\|\|full | Low-quality |
| S5_vs_k141_16899\|\|full | Low-quality |
| S5_vs_k141_143471\|\|full | Not-determined |
| S5_vs_k141_10499\|\|full | Not-determined |
| S5_vs_k141_70933\|\|full | Low-quality |
| S5_vs_k141_5803\|\|full | Low-quality |
| S5_vs_k141_114249\|\|full | High-quality |
| S5_vs_k141_113821\|\|full | Low-quality |
| S5_vs_k141_136814\|\|full | Low-quality |
| S5_vs_k141_143488\|\|full | Low-quality |
| S5_vs_k141_53809\|\|full | Low-quality |
| S5_vs_k141_136818\|\|full | Not-determined |
| S5_vs_k141_46404\|\|full | Low-quality |
| S5_vs_k141_75820\|\|full | Low-quality |
| S5_vs_k141_131901\|\|full | Not-determined |
| S5_vs_k141_46405\|\|full | Low-quality |
| S5_vs_k141_53827\|\|full | Not-determined |
| S5_vs_k141_143512\|\|full | Not-determined |

| | |
|---|---|
| SS_vs_k141_5813\|\|full | Low-quality |
| SS_vs_k141_35648\|\|full | Low-quality |
| SS_vs_k141_5823\|\|full | Not-determined |
| SS_vs_k141_75843\|\|full | Not-determined |
| SS_vs_k141_143521\|\|full | Not-determined |
| SS_vs_k141_27182\|\|full | Not-determined |
| SS_vs_k141_65055\|\|full | Low-quality |
| SS_vs_k141_35653\|\|full | Low-quality |
| SS_vs_k141_120462\|\|full | Low-quality |
| SS_vs_k141_126927\|\|full | Not-determined |
| SS_vs_k141_89270\|\|full | Low-quality |
| SS_vs_k141_5841\|\|full | Not-determined |
| SS_vs_k141_22353\|\|full | Low-quality |
| SS_vs_k141_70994\|\|full | Not-determined |
| SS_vs_k141_5846\|\|full | Not-determined |
| SS_vs_k141_27197\|\|full | Not-determined |
| SS_vs_k141_22359\|\|full | Low-quality |
| SS_vs_k141_89278\|\|full | Low-quality |
| SS_vs_k141_89280\|\|full | Low-quality |
| SS_vs_k141_113895\|\|full | Low-quality |
| SS_vs_k141_75866\|\|full | Not-determined |
| SS_vs_k141_22372\|\|full | Low-quality |
| SS_vs_k141_65089\|\|full | Low-quality |
| SS_vs_k141_35672\|\|full | Low-quality |
| SS_vs_k141_22380\|\|full | Not-determined |
| SS_vs_k141_100369\|\|full | Not-determined |
| SS_vs_k141_22384\|\|full | Not-determined |
| SS_vs_k141_53864\|\|full | Low-quality |
| SS_vs_k141_46436\|\|full | Not-determined |
| SS_vs_k141_10607\|\|full | Low-quality |
| SS_vs_k141_100392\|\|full | Not-determined |
| SS_vs_k141_22401\|\|full | Not-determined |
| SS_vs_k141_143572\|\|full | Not-determined |
| SS_vs_k141_89317\|\|full | Not-determined |
| SS_vs_k141_126998\|\|full | Low-quality |
| SS_vs_k141_27251\|\|full | Not-determined |
| SS_vs_k141_65125\|\|full | Not-determined |
| SS_vs_k141_40362\|\|full | Not-determined |
| SS_vs_k141_108799\|\|full | Low-quality |
| SS_vs_k141_81598\|\|full | Not-determined |
| SS_vs_k141_120504\|\|full | Low-quality |
| SS_vs_k141_22410\|\|full | Low-quality |
| SS_vs_k141_40377\|\|full | Not-determined |
| SS_vs_k141_75912\|\|full | Low-quality |
| SS_vs_k141_89333\|\|full | Low-quality |
| SS_vs_k141_65156\|\|full | Low-quality |
| SS_vs_k141_46462\|\|full | Low-quality |
| SS_vs_k141_16990\|\|full | Low-quality |
| SS_vs_k141_71009\|\|full | Low-quality |
| SS_vs_k141_22431\|\|full | Low-quality |
| SS_vs_k141_71012\|\|full | Low-quality |
| SS_vs_k141_136889\|\|full | Low-quality |
| SS_vs_k141_108843\|\|full | Low-quality |
| SS_vs_k141_53898\|\|full | Low-quality |
| SS_vs_k141_5854\|\|full | Low-quality |
| SS_vs_k141_75951\|\|full | Low-quality |
| SS_vs_k141_120547\|\|full | Not-determined |
| SS_vs_k141_131977\|\|full | Low-quality |
| SS_vs_k141_100445\|\|full | Low-quality |
| SS_vs_k141_75963\|\|full | Low-quality |
| SS_vs_k141_46497\|\|full | Low-quality |
| SS_vs_k141_5872\|\|full | Low-quality |
| SS_vs_k141_71030\|\|full | Low-quality |
| SS_vs_k141_35789\|\|full | Not-determined |
| SS_vs_k141_100459\|\|full | Not-determined |
| SS_vs_k141_120566\|\|full | Not-determined |
| SS_vs_k141_136897\|\|full | Low-quality |
| SS_vs_k141_95911\|\|full | Low-quality |
| SS_vs_k141_100476\|\|full | Low-quality |
| SS_vs_k141_5887\|\|full | Low-quality |
| SS_vs_k141_114002\|\|full | Not-determined |
| SS_vs_k141_75980\|\|full | Not-determined |
| SS_vs_k141_95913\|\|full | Low-quality |
| SS_vs_k141_95918\|\|full | Not-determined |
| SS_vs_k141_35796\|\|full | Not-determined |
| SS_vs_k141_136926\|\|full | Not-determined |
| SS_vs_k141_114015\|\|full | Low-quality |
| SS_vs_k141_57448\|\|full | Low-quality |
| SS_vs_k141_46538\|\|full | Not-determined |
| SS_vs_k141_46551\|\|full | Not-determined |
| SS_vs_k141_17078\|\|full | Not-determined |
| SS_vs_k141_17081\|\|full | Low-quality |
| SS_vs_k141_65233\|\|full | Low-quality |
| SS_vs_k141_10744\|\|full | Low-quality |
| SS_vs_k141_136936\|\|full | Low-quality |
| SS_vs_k141_65242\|\|full | Low-quality |
| SS_vs_k141_89402\|\|full | Low-quality |
| SS_vs_k141_17092\|\|full | Low-quality |
| SS_vs_k141_10760\|\|full | Not-determined |
| SS_vs_k141_76031\|\|full | Low-quality |
| SS_vs_k141_35819\|\|full | Not-determined |
| SS_vs_k141_132060\|\|full | Low-quality |
| SS_vs_k141_17108\|\|full | Low-quality |
| SS_vs_k141_35827\|\|full | Low-quality |
| SS_vs_k141_100526\|\|full | Low-quality |
| SS_vs_k141_57470\|\|full | Low-quality |
| SS_vs_k141_10776\|\|full | Not-determined |
| SS_vs_k141_5904\|\|full | Low-quality |
| SS_vs_k141_22537\|\|full | Not-determined |
| SS_vs_k141_46599\|\|full | Not-determined |
| SS_vs_k141_10779\|\|full | Low-quality |
| SS_vs_k141_71059\|\|full | Low-quality |
| SS_vs_k141_76043\|\|full | Not-determined |
| SS_vs_k141_46620\|\|full | Not-determined |
| SS_vs_k141_95973\|\|full | Not-determined |
| SS_vs_k141_76048\|\|full | Low-quality |
| SS_vs_k141_35856\|\|full | Not-determined |
| SS_vs_k141_120612\|\|full | Not-determined |
| SS_vs_k141_95985\|\|full | Not-determined |
| SS_vs_k141_35859\|\|full | Not-determined |
| SS_vs_k141_65281\|\|full | Low-quality |
| SS_vs_k141_17139\|\|full | Low-quality |
| SS_vs_k141_132089\|\|full | Not-determined |
| SS_vs_k141_22587\|\|full | Low-quality |
| SS_vs_k141_10793\|\|full | Low-quality |
| SS_vs_k141_40431\|\|full | Not-determined |
| SS_vs_k141_5941\|\|full | Not-determined |
| SS_vs_k141_22606\|\|full | Not-determined |
| SS_vs_k141_76063\|\|full | Not-determined |
| SS_vs_k141_54061\|\|full | Low-quality |
| SS_vs_k141_114082\|\|full | Not-determined |
| SS_vs_k141_46638\|\|full | Low-quality |
| SS_vs_k141_136969\|\|full | Low-quality |
| SS_vs_k141_10808\|\|full | Low-quality |
| SS_vs_k141_54086\|\|full | Low-quality |
| SS_vs_k141_114093\|\|full | Low-quality |
| SS_vs_k141_136973\|\|full | Not-determined |
| SS_vs_k141_17180\|\|full | Not-determined |
| SS_vs_k141_114094\|\|full | Low-quality |
| SS_vs_k141_127097\|\|full | Not-determined |
| SS_vs_k141_17190\|\|full | Low-quality |
| SS_vs_k141_114105\|\|full | Low-quality |
| SS_vs_k141_108947\|\|full | Low-quality |
| SS_vs_k141_127117\|\|full | Low-quality |
| SS_vs_k141_40455\|\|full | Low-quality |
| SS_vs_k141_132109\|\|full | Low-quality |
| SS_vs_k141_22647\|\|full | Low-quality |
| SS_vs_k141_22652\|\|full | Not-determined |
| SS_vs_k141_46669\|\|full | Not-determined |
| SS_vs_k141_89482\|\|full | Not-determined |
| SS_vs_k141_108965\|\|full | Low-quality |
| SS_vs_k141_127132\|\|full | Low-quality |
| SS_vs_k141_100540\|\|full | Low-quality |
| SS_vs_k141_54124\|\|full | Low-quality |
| SS_vs_k141_22662\|\|full | Low-quality |
| SS_vs_k141_127141\|\|full | Low-quality |
| SS_vs_k141_40473\|\|full | Low-quality |
| SS_vs_k141_96021\|\|full | Not-determined |
| SS_vs_k141_100549\|\|full | Not-determined |
| S3_vs_k141_108076\|\|full | High-quality |
| SS_vs_k141_81687\|\|full | Not-determined |
| SS_vs_k141_6021\|\|full | Not-determined |
| SS_vs_k141_22685\|\|full | Not-determined |
| SS_vs_k141_89502\|\|full | Not-determined |
| SS_vs_k141_6030\|\|full | Low-quality |
| SS_vs_k141_100560\|\|full | Low-quality |
| SS_vs_k141_114150\|\|full | Low-quality |
| SS_vs_k141_54170\|\|full | Not-determined |
| SS_vs_k141_76091\|\|full | Not-determined |
| SS_vs_k141_132148\|\|full | Low-quality |
| SS_vs_k141_35910\|\|full | Low-quality |
| SS_vs_k141_114161\|\|full | Not-determined |
| SS_vs_k141_57532\|\|full | Low-quality |
| SS_vs_k141_40515\|\|full | Low-quality |
| SS_vs_k141_36605\|\|full | High-quality |
| SS_vs_k141_143637\|\|full | Low-quality |
| SS_vs_k141_114168\|\|full | Not-determined |
| SS_vs_k141_6038\|\|full | Low-quality |
| SS_vs_k141_46727\|\|full | Low-quality |
| SS_vs_k141_46734\|\|full | Low-quality |
| SS_vs_k141_109078\|\|full | Not-determined |
| SS_vs_k141_10837\|\|full | Low-quality |
| SS_vs_k141_120724\|\|full | Not-determined |
| SS_vs_k141_40547\|\|full | Low-quality |
| SS_vs_k141_27300\|\|full | Not-determined |
| SS_vs_k141_40556\|\|full | Not-determined |
| SS_vs_k141_71125\|\|full | Low-quality |
| SS_vs_k141_114205\|\|full | Not-determined |
| SS_vs_k141_71128\|\|full | Low-quality |
| SS_vs_k141_17308\|\|full | Low-quality |
| SS_vs_k141_17309\|\|full | Low-quality |
| SS_vs_k141_114217\|\|full | Low-quality |
| SS_vs_k141_17315\|\|full | Low-quality |
| SS_vs_k141_81744\|\|full | Low-quality |
| SS_vs_k141_120765\|\|full | Low-quality |
| SS_vs_k141_17317\|\|full | Low-quality |
| SS_vs_k141_114241\|\|full | Not-determined |
| SS_vs_k141_71154\|\|full | Low-quality |
| SS_vs_k141_71156\|\|full | Low-quality |
| S3_vs_k141_44925\|\|full | Medium-quality |
| SS_vs_k141_17337\|\|full | Low-quality |
| SS_vs_k141_40617\|\|full | Not-determined |
| SS_vs_k141_89547\|\|full | Not-determined |
| SS_vs_k141_17348\|\|full | Low-quality |
| SS_vs_k141_109104\|\|full | Low-quality |
| SS_vs_k141_96132\|\|full | Low-quality |
| SS_vs_k141_81764\|\|full | Low-quality |
| SS_vs_k141_89551\|\|full | Not-determined |
| SS_vs_k141_40635\|\|full | Not-determined |
| SS_vs_k141_89554\|\|full | Not-determined |
| SS_vs_k141_65378\|\|full | Not-determined |
| SS_vs_k141_10868\|\|full | Low-quality |
| SS_vs_k141_120797\|\|full | Low-quality |
| SS_vs_k141_89563\|\|full | Low-quality |
| SS_vs_k141_46797\|\|full | Low-quality |
| SS_vs_k141_132206\|\|full | Low-quality |
| SS_vs_k141_36002\|\|full | Not-determined |
| SS_vs_k141_71211\|\|full | Not-determined |
| SS_vs_k141_40657\|\|full | Low-quality |
| SS_vs_k141_89579\|\|full | Low-quality |
| SS_vs_k141_36022\|\|full | Low-quality |
| SS_vs_k141_40663\|\|full | Not-determined |
| SS_vs_k141_76162\|\|full | Low-quality |
| SS_vs_k141_89584\|\|full | Low-quality |
| SS_vs_k141_137052\|\|full | Low-quality |
| SS_vs_k141_76165\|\|full | Not-determined |
| SS_vs_k141_132227\|\|full | Not-determined |
| SS_vs_k141_137058\|\|full | Low-quality |
| SS_vs_k141_137059\|\|full | Not-determined |
| S3_extracted_k141_127405 | Complete |
| SS_vs_k141_17401\|\|full | Low-quality |
| SS_vs_k141_120862\|\|full | Low-quality |
| SS_vs_k141_137066\|\|full | Low-quality |
| SS_vs_k141_89611\|\|full | Low-quality |
| SS_vs_k141_109148\|\|full | Not-determined |
| SS_vs_k141_46839\|\|full | Low-quality |
| SS_vs_k141_17408\|\|full | Low-quality |
| SS_vs_k141_143658\|\|full | Not-determined |
| SS_vs_k141_36054\|\|full | Low-quality |
| SS_vs_k141_76194\|\|full | Not-determined |
| SS_vs_k141_76197\|\|full | Not-determined |
| SS_vs_k141_114283\|\|full | Not-determined |
| SS_vs_k141_46876\|\|full | Low-quality |
| SS_vs_k141_46877\|\|full | Not-determined |
| SS_vs_k141_54348\|\|full | Low-quality |
| SS_vs_k141_81818\|\|full | Low-quality |
| SS_vs_k141_54356\|\|full | Low-quality |
| SS_vs_k141_54357\|\|full | Low-quality |
| SS_vs_k141_46904\|\|full | Not-determined |
| SS_vs_k141_40738\|\|full | Low-quality |
| SS_vs_k141_22771\|\|full | Low-quality |
| SS_vs_k141_137080\|\|full | Low-quality |
| SS_vs_k141_54367\|\|full | Low-quality |
| SS_vs_k141_137090\|\|full | Low-quality |
| SS_vs_k141_76251\|\|full | Not-determined |
| SS_vs_k141_65427\|\|full | Low-quality |
| SS_vs_k141_114338\|\|full | Not-determined |
| SS_vs_k141_10917\|\|full | Low-quality |
| SS_vs_k141_120924\|\|full | Low-quality |
| SS_vs_k141_40751\|\|full | Not-determined |
| SS_vs_k141_114345\|\|full | Low-quality |
| SS_vs_k141_22801\|\|full | Not-determined |
| SS_vs_k141_22806\|\|full | Low-quality |
| SS_vs_k141_81841\|\|full | Not-determined |
| SS_vs_k141_22814\|\|full | Low-quality |
| SS_vs_k141_65464\|\|full | Not-determined |
| SS_vs_k141_65465\|\|full | Low-quality |
| SS_vs_k141_76285\|\|full | Not-determined |
| SS_vs_k141_17501\|\|full | Low-quality |
| SS_vs_k141_137118\|\|full | Low-quality |
| SS_vs_k141_89690\|\|full | Not-determined |
| SS_vs_k141_76299\|\|full | Low-quality |
| SS_vs_k141_76300\|\|full | Not-determined |
| SS_vs_k141_89697\|\|full | Low-quality |
| SS_vs_k141_10944\|\|full | Low-quality |
| SS_vs_k141_109232\|\|full | Low-quality |
| SS_vs_k141_89709\|\|full | Low-quality |
| SS_vs_k141_54433\|\|full | Not-determined |
| SS_vs_k141_47014\|\|full | Low-quality |
| SS_vs_k141_114414\|\|full | Low-quality |
| SS_vs_k141_114417\|\|full | Low-quality |
| SS_vs_k141_89722\|\|full | Low-quality |
| SS_vs_k141_27427\|\|full | Not-determined |
| SS_vs_k141_71228\|\|full | Low-quality |
| SS_vs_k141_22886\|\|full | Low-quality |
| SS_vs_k141_96152\|\|full | Low-quality |
| SS_vs_k141_114446\|\|full | Low-quality |
| SS_vs_k141_145586\|\|full | High-quality |
| SS_vs_k141_36086\|\|full | Low-quality |
| SS_vs_k141_109265\|\|full | Low-quality |
| SS_vs_k141_17530\|\|full | Low-quality |
| SS_vs_k141_17539\|\|full | Low-quality |
| SS_vs_k141_120953\|\|full | Low-quality |
| SS_vs_k141_22907\|\|full | Not-determined |
| SS_vs_k141_114465\|\|full | Not-determined |
| SS_vs_k141_114466\|\|full | Low-quality |
| SS_vs_k141_109284\|\|full | Not-determined |
| SS_vs_k141_22914\|\|full | Not-determined |
| SS_vs_k141_36112\|\|full | Not-determined |
| SS_vs_k141_143702\|\|full | Low-quality |
| SS_vs_k141_81888\|\|full | Low-quality |
| SS_vs_k141_132291\|\|full | Not-determined |
| SS_vs_k141_96164\|\|full | Low-quality |
| SS_vs_k141_132299\|\|full | Not-determined |
| SS_vs_k141_89744\|\|full | Low-quality |
| SS_vs_k141_143719\|\|full | Low-quality |
| SS_vs_k141_36135\|\|full | Low-quality |
| SS_vs_k141_17582\|\|full | Not-determined |
| SS_vs_k141_54478\|\|full | Low-quality |
| SS_vs_k141_143723\|\|full | Low-quality |
| SS_vs_k141_132308\|\|full | Low-quality |
| SS_vs_k141_10960\|\|full | Not-determined |
| SS_vs_k141_71275\|\|full | Not-determined |
| SS_vs_k141_10961\|\|full | Low-quality |
| SS_vs_k141_54481\|\|full | Not-determined |
| SS_vs_k141_47093\|\|full | Low-quality |
| SS_vs_k141_121005\|\|full | Low-quality |
| SS_vs_k141_54486\|\|full | Low-quality |
| SS_vs_k141_114502\|\|full | Not-determined |
| SS_vs_k141_81895\|\|full | Not-determined |
| SS_vs_k141_54490\|\|full | Not-determined |
| SS_vs_k141_47106\|\|full | Low-quality |
| SS_vs_k141_36150\|\|full | Not-determined |
| SS_vs_k141_76407\|\|full | Low-quality |
| SS_vs_k141_71294\|\|full | Low-quality |
| SS_vs_k141_132330\|\|full | Low-quality |
| SS_vs_k141_71295\|\|full | Low-quality |
| SS_vs_k141_96216\|\|full | Low-quality |
| SS_vs_k141_22951\|\|full | Not-determined |
| SS_vs_k141_17613\|\|full | Low-quality |
| SS_vs_k141_47135\|\|full | Low-quality |
| SS_vs_k141_76418\|\|full | Low-quality |
| SS_vs_k141_40793\|\|full | Low-quality |
| S3_vs_k141_3501\|\|full | Medium-quality |
| SS_vs_k141_121030\|\|full | Low-quality |
| SS_vs_k141_71311\|\|full | Not-determined |
| SS_vs_k141_127185\|\|full | Low-quality |
| SS_vs_k141_137166\|\|full | Not-determined |
| SS_vs_k141_36168\|\|full | Low-quality |
| SS_vs_k141_47165\|\|full | Low-quality |
| SS_vs_k141_36172\|\|full | Not-determined |
| SS_vs_k141_27475\|\|full | Low-quality |
| SS_vs_k141_76450\|\|full | Low-quality |
| SS_vs_k141_22996\|\|full | Low-quality |
| SS_vs_k141_17654\|\|full | Low-quality |
| S3_vs_k141_50924\|\|full | High-quality |
| SS_vs_k141_89851\|\|full | Low-quality |
| SS_vs_k141_121052\|\|full | Not-determined |
| SS_vs_k141_76474\|\|full | Not-determined |
| SS_vs_k141_89866\|\|full | Low-quality |
| SS_vs_k141_27506\|\|full | Low-quality |
| SS_vs_k141_17679\|\|full | Not-determined |
| SS_vs_k141_54554\|\|full | Not-determined |
| SS_vs_k141_23021\|\|full | Not-determined |
| SS_vs_k141_121069\|\|full | Low-quality |
| SS_vs_k141_47216\|\|full | Low-quality |
| SS_vs_k141_89877\|\|full | Low-quality |
| SS_vs_k141_127231\|\|full | Not-determined |
| SS_vs_k141_127232\|\|full | Low-quality |
| SS_vs_k141_47226\|\|full | Low-quality |
| SS_vs_k141_54589\|\|full | Low-quality |
| SS_vs_k141_127236\|\|full | Low-quality |
| SS_vs_k141_143796\|\|full | Low-quality |
| SS_vs_k141_127241\|\|full | Not-determined |
| SS_vs_k141_17722\|\|full | Not-determined |
| SS_vs_k141_121074\|\|full | Not-determined |
| SS_vs_k141_27519\|\|full | Not-determined |
| SS_vs_k141_47233\|\|full | Low-quality |
| SS_vs_k141_89901\|\|full | Low-quality |
| SS_vs_k141_114558\|\|full | Not-determined |
| SS_vs_k141_54598\|\|full | Low-quality |
| SS_vs_k141_17746\|\|full | Not-determined |
| SS_vs_k141_40839\|\|full | Low-quality |
| SS_vs_k141_40840\|\|full | Low-quality |
| SS_vs_k141_47264\|\|full | Low-quality |
| SS_vs_k141_114577\|\|full | Low-quality |
| SS_vs_k141_17769\|\|full | Low-quality |
| SS_vs_k141_47267\|\|full | Not-determined |
| SS_vs_k141_27534\|\|full | Low-quality |
| SS_vs_k141_17774\|\|full | Not-determined |
| SS_vs_k141_27541\|\|full | Not-determined |
| SS_vs_k141_114590\|\|full | Not-determined |
| SS_vs_k141_127273\|\|full | Low-quality |
| SS_vs_k141_127274\|\|full | Low-quality |
| SS_vs_k141_23080\|\|full | Low-quality |
| SS_vs_k141_89955\|\|full | Not-determined |
| SS_vs_k141_71313\|\|full | Not-determined |
| SS_vs_k141_96277\|\|full | Low-quality |
| SS_vs_k141_65532\|\|full | Low-quality |
| SS_vs_k141_132393\|\|full | Low-quality |
| SS_vs_k141_17796\|\|full | Not-determined |
| SS_vs_k141_132395\|\|full | Not-determined |
| SS_vs_k141_82010\|\|full | Not-determined |
| SS_vs_k141_132403\|\|full | Not-determined |
| SS_vs_k141_100728\|\|full | Not-determined |
| SS_vs_k141_127298\|\|full | Low-quality |
| SS_vs_k141_82015\|\|full | Low-quality |
| SS_vs_k141_114618\|\|full | Low-quality |
| SS_vs_k141_82017\|\|full | Low-quality |
| SS_vs_k141_36342\|\|full | Low-quality |
| SS_vs_k141_82024\|\|full | Low-quality |
| SS_vs_k141_71334\|\|full | Not-determined |
| SS_vs_k141_132436\|\|full | Low-quality |
| SS_vs_k141_17827\|\|full | Low-quality |
| SS_vs_k141_114626\|\|full | Not-determined |
| SS_vs_k141_17834\|\|full | Not-determined |
| SS_vs_k141_132447\|\|full | Not-determined |
| SS_vs_k141_36355\|\|full | Low-quality |
| SS_vs_k141_96327\|\|full | Not-determined |
| SS_vs_k141_82031\|\|full | Low-quality |
| SS_vs_k141_100771\|\|full | Low-quality |
| SS_vs_k141_17844\|\|full | Not-determined |
| SS_vs_k141_17854\|\|full | Low-quality |
| SS_vs_k141_11043\|\|full | Low-quality |
| SS_vs_k141_65589\|\|full | Low-quality |
| SS_vs_k141_121196\|\|full | Low-quality |
| SS_vs_k141_23139\|\|full | Low-quality |
| SS_vs_k141_143884\|\|full | Low-quality |
| SS_vs_k141_47348\|\|full | Low-quality |
| SS_vs_k141_96350\|\|full | Not-determined |
| SS_vs_k141_114653\|\|full | Low-quality |
| SS_vs_k141_100774\|\|full | Low-quality |
| SS_vs_k141_127373\|\|full | Low-quality |
| SS_vs_k141_100777\|\|full | Not-determined |
| SS_vs_k141_143893\|\|full | Low-quality |
| SS_vs_k141_127382\|\|full | Not-determined |
| S3_extracted_k141_12212 | Medium-quality |
| SS_vs_k141_27582\|\|full | Not-determined |
| SS_vs_k141_137261\|\|full | Low-quality |
| SS_vs_k141_23161\|\|full | Low-quality |
| SS_vs_k141_71365\|\|full | Low-quality |
| SS_vs_k141_127394\|\|full | Low-quality |
| SS_vs_k141_132472\|\|full | Low-quality |
| SS_vs_k141_36399\|\|full | Low-quality |
| SS_vs_k141_114682\|\|full | Low-quality |
| SS_vs_k141_137273\|\|full | Low-quality |
| SS_vs_k141_71386\|\|full | Low-quality |
| SS_vs_k141_71389\|\|full | Low-quality |
| SS_vs_k141_100800\|\|full | Low-quality |
| SS_vs_k141_127419\|\|full | Not-determined |
| SS_vs_k141_143919\|\|full | Low-quality |
| SS_vs_k141_71409\|\|full | Not-determined |
| SS_vs_k141_27616\|\|full | Not-determined |
| SS_vs_k141_82080\|\|full | Low-quality |
| SS_vs_k141_89987\|\|full | Not-determined |
| SS_vs_k141_114715\|\|full | Low-quality |
| SS_vs_k141_121281\|\|full | Low-quality |
| SS_vs_k141_96358\|\|full | Low-quality |
| SS_vs_k141_47377\|\|full | Low-quality |
| SS_vs_k141_100814\|\|full | Low-quality |
| SS_vs_k141_121291\|\|full | Not-determined |
| SS_vs_k141_17888\|\|full | Low-quality |
| SS_vs_k141_127436\|\|full | Low-quality |
| SS_vs_k141_121295\|\|full | Low-quality |
| SS_vs_k141_90002\|\|full | Low-quality |
| SS_vs_k141_137300\|\|full | Low-quality |
| SS_vs_k141_90004\|\|full | Not-determined |
| SS_vs_k141_143923\|\|full | Not-determined |
| SS_vs_k141_11080\|\|full | Not-determined |
| SS_vs_k141_127455\|\|full | Not-determined |
| SS_vs_k141_121324\|\|full | Low-quality |
| SS_vs_k141_47398\|\|full | Low-quality |
| SS_vs_k141_96363\|\|full | Low-quality |
| SS_vs_k141_127460\|\|full | Not-determined |
| SS_vs_k141_23192\|\|full | Low-quality |
| SS_vs_k141_23197\|\|full | Not-determined |
| SS_vs_k141_127467\|\|full | Low-quality |
| SS_vs_k141_82103\|\|full | Low-quality |
| SS_vs_k141_127468\|\|full | Low-quality |
| SS_vs_k141_132554\|\|full | Not-determined |
| SS_vs_k141_132555\|\|full | Not-determined |
| SS_vs_k141_71464\|\|full | Low-quality |
| SS_vs_k141_82123\|\|full | Low-quality |
| SS_vs_k141_71465\|\|full | Not-determined |
| SS_vs_k141_47448\|\|full | Low-quality |
| SS_vs_k141_90048\|\|full | Not-determined |
| SS_vs_k141_132565\|\|full | Low-quality |
| SS_vs_k141_47480\|\|full | Not-determined |
| SS_vs_k141_96437\|\|full | Low-quality |
| SS_vs_k141_90054\|\|full | Low-quality |
| SS_vs_k141_11114\|\|full | Low-quality |
| SS_vs_k141_90071\|\|full | Low-quality |
| SS_vs_k141_71511\|\|full | Low-quality |
| SS_vs_k141_96458\|\|full | Low-quality |
| SS_vs_k141_47536\|\|full | Low-quality |
| SS_vs_k141_57796\|\|full | Not-determined |
| SS_vs_k141_27652\|\|full | Low-quality |
| SS_vs_k141_47546\|\|full | Low-quality |
| S3_vs_k141_12212\|\|full | Medium-quality |
| SS_vs_k141_71528\|\|full | Low-quality |
| SS_vs_k141_71530\|\|full | Low-quality |
| SS_vs_k141_17933\|\|full | Low-quality |
| SS_vs_k141_96478\|\|full | Low-quality |
| SS_vs_k141_47555\|\|full | Low-quality |
| SS_vs_k141_47559\|\|full | Not-determined |
| SS_vs_k141_143980\|\|full | Low-quality |
| SS_vs_k141_47567\|\|full | Low-quality |
| SS_vs_k141_96502\|\|full | Low-quality |
| SS_vs_k141_27683\|\|full | Low-quality |
| SS_vs_k141_96511\|\|full | Not-determined |
| SS_vs_k141_57818\|\|full | Low-quality |
| SS_vs_k141_17967\|\|full | Low-quality |
| SS_vs_k141_137325\|\|full | Low-quality |
| SS_vs_k141_27685\|\|full | Low-quality |
| SS_vs_k141_17979\|\|full | Not-determined |
| SS_vs_k141_27700\|\|full | Not-determined |
| SS_vs_k141_23247\|\|full | Low-quality |
| SS_vs_k141_144003\|\|full | Low-quality |
| SS_vs_k141_27705\|\|full | Low-quality |
| SS_vs_k141_137339\|\|full | Not-determined |
| SS_vs_k141_132682\|\|full | Not-determined |
| SS_vs_k141_18002\|\|full | Low-quality |
| SS_vs_k141_121399\|\|full | Low-quality |
| SS_vs_k141_132689\|\|full | Low-quality |
| SS_vs_k141_132690\|\|full | Not-determined |
| SS_vs_k141_71637\|\|full | Low-quality |
| SS_vs_k141_47647\|\|full | Not-determined |
| SS_vs_k141_23283\|\|full | Low-quality |
| SS_vs_k141_132710\|\|full | Low-quality |
| SS_vs_k141_47663\|\|full | Not-determined |
| SS_vs_k141_132720\|\|full | Low-quality |
| SS_vs_k141_71649\|\|full | Low-quality |
| SS_vs_k141_132731\|\|full | Low-quality |
| SS_vs_k141_47687\|\|full | Low-quality |
| SS_vs_k141_18040\|\|full | Low-quality |
| SS_vs_k141_90124\|\|full | Not-determined |
| SS_vs_k141_144053\|\|full | Not-determined |
| SS_vs_k141_132748\|\|full | Low-quality |
| SS_vs_k141_27762\|\|full | Not-determined |
| SS_vs_k141_144059\|\|full | Low-quality |
| SS_vs_k141_11145\|\|full | Low-quality |
| SS_vs_k141_71680\|\|full | Not-determined |
| SS_vs_k141_18062\|\|full | Low-quality |
| SS_vs_k141_18066\|\|full | Low-quality |
| SS_vs_k141_47708\|\|full | Not-determined |
| SS_vs_k141_57833\|\|full | Not-determined |
| SS_vs_k141_11163\|\|full | Low-quality |
| SS_vs_k141_18080\|\|full | Low-quality |
| SS_vs_k141_18084\|\|full | Low-quality |
| SS_vs_k141_18085\|\|full | Low-quality |
| SS_vs_k141_96589\|\|full | Low-quality |
| SS_vs_k141_27796\|\|full | Not-determined |
| SS_vs_k141_47722\|\|full | Not-determined |
| SS_vs_k141_144077\|\|full | Low-quality |
| SS_vs_k141_144078\|\|full | Low-quality |
| SS_vs_k141_27806\|\|full | Low-quality |
| SS_vs_k141_18106\|\|full | Not-determined |
| SS_vs_k141_82157\|\|full | Low-quality |
| SS_vs_k141_40937\|\|full | Low-quality |
| SS_vs_k141_90149\|\|full | Low-quality |
| SS_vs_k141_132812\|\|full | Low-quality |
| SS_vs_k141_144103\|\|full | Not-determined |
| SS_vs_k141_114802\|\|full | Low-quality |
| SS_vs_k141_132815\|\|full | Low-quality |
| SS_vs_k141_76537\|\|full | Low-quality |
| SS_vs_k141_132823\|\|full | Low-quality |
| SS_vs_k141_144126\|\|full | Low-quality |
| SS_vs_k141_40953\|\|full | Not-determined |
| SS_vs_k141_71750\|\|full | Not-determined |
| SS_vs_k141_71755\|\|full | Low-quality |
| SS_vs_k141_90169\|\|full | Not-determined |
| SS_vs_k141_47796\|\|full | Low-quality |
| SS_vs_k141_76580\|\|full | Low-quality |
| SS_vs_k141_47799\|\|full | Low-quality |
| SS_vs_k141_47804\|\|full | Low-quality |
| SS_vs_k141_82184\|\|full | Low-quality |
| SS_vs_k141_76591\|\|full | Not-determined |
| SS_vs_k141_82192\|\|full | Low-quality |
| SS_vs_k141_27838\|\|full | Low-quality |
| SS_vs_k141_90192\|\|full | Low-quality |
| SS_vs_k141_114825\|\|full | Low-quality |
| SS_vs_k141_100985\|\|full | Low-quality |
| SS_vs_k141_47828\|\|full | Low-quality |
| SS_vs_k141_100990\|\|full | Low-quality |
| SS_vs_k141_96618\|\|full | Low-quality |
| SS_vs_k141_18141\|\|full | Low-quality |
| SS_vs_k141_27872\|\|full | Low-quality |
| SS_vs_k141_76627\|\|full | Not-determined |
| SS_vs_k141_132889\|\|full | Low-quality |
| SS_vs_k141_137416\|\|full | Not-determined |
| SS_vs_k141_23382\|\|full | Low-quality |
| SS_vs_k141_96637\|\|full | Low-quality |
| SS_vs_k141_27891\|\|full | Not-determined |
| SS_vs_k141_132910\|\|full | Low-quality |
| SS_vs_k141_137418\|\|full | Not-determined |
| SS_vs_k141_65679\|\|full | Low-quality |
| SS_vs_k141_71789\|\|full | Low-quality |
| SS_vs_k141_23397\|\|full | Low-quality |
| SS_vs_k141_65681\|\|full | Not-determined |
| SS_vs_k141_40956\|\|full | Not-determined |
| SS_vs_k141_65682\|\|full | Low-quality |
| SS_vs_k141_27899\|\|full | Low-quality |
| SS_vs_k141_23408\|\|full | Not-determined |
| SS_vs_k141_65703\|\|full | Low-quality |
| SS_vs_k141_114926\|\|full | Low-quality |
| SS_vs_k141_23414\|\|full | Not-determined |
| SS_vs_k141_132945\|\|full | Low-quality |
| SS_vs_k141_90295\|\|full | Not-determined |
| SS_vs_k141_65718\|\|full | Low-quality |

| | |
|---|---|
| S5_vs_k141_71820\|\|full | Low-quality |
| S5_vs_k141_65726\|\|full | Not-determined |
| S5_vs_k141_11189\|\|full | Low-quality |
| S5_vs_k141_47883\|\|full | Not-determined |
| S5_vs_k141_76680\|\|full | Low-quality |
| S5_vs_k141_65739\|\|full | Low-quality |
| S5_vs_k141_96711\|\|full | Low-quality |
| S5_vs_k141_137467\|\|full | Low-quality |
| S5_vs_k141_82238\|\|full | Low-quality |
| S5_vs_k141_76693\|\|full | Low-quality |
| S5_vs_k141_96731\|\|full | Low-quality |
| S5_vs_k141_71846\|\|full | Low-quality |
| S5_vs_k141_41005\|\|full | Not-determined |
| S5_vs_k141_96733\|\|full | Not-determined |
| S5_vs_k141_133004\|\|full | Low-quality |
| S5_vs_k141_76704\|\|full | Low-quality |
| S5_vs_k141_71858\|\|full | Not-determined |
| S5_vs_k141_11225\|\|full | Not-determined |
| S5_vs_k141_114947\|\|full | Low-quality |
| S5_vs_k141_90344\|\|full | Low-quality |
| S5_vs_k141_57851\|\|full | Low-quality |
| S5_vs_k141_114951\|\|full | Not-determined |
| S5_vs_k141_144145\|\|full | Low-quality |
| S5_vs_k141_23483\|\|full | Low-quality |
| S5_vs_k141_133017\|\|full | Low-quality |
| S5_vs_k141_23486\|\|full | Low-quality |
| S5_vs_k141_96767\|\|full | Low-quality |
| S5_vs_k141_65778\|\|full | Not-determined |
| S5_vs_k141_144159\|\|full | Not-determined |
| S5_vs_k141_114965\|\|full | Low-quality |
| S5_vs_k141_57874\|\|full | Low-quality |
| S5_vs_k141_137496\|\|full | Low-quality |
| S5_vs_k141_90391\|\|full | Low-quality |
| S5_vs_k141_11227\|\|full | Not-determined |
| S5_vs_k141_41067\|\|full | Not-determined |
| S5_vs_k141_23526\|\|full | Not-determined |
| S5_vs_k141_133050\|\|full | Low-quality |
| S5_vs_k141_133055\|\|full | Not-determined |
| S5_vs_k141_18205\|\|full | Not-determined |
| S5_vs_k141_57880\|\|full | Low-quality |
| S5_vs_k141_41089\|\|full | Not-determined |
| S5_vs_k141_18208\|\|full | Low-quality |
| S5_vs_k141_18210\|\|full | Low-quality |
| S5_vs_k141_144183\|\|full | Not-determined |
| S5_vs_k141_65815\|\|full | Not-determined |
| S5_vs_k141_90404\|\|full | Low-quality |
| S5_vs_k141_133075\|\|full | Low-quality |
| S5_vs_k141_144193\|\|full | Not-determined |
| S5_vs_k141_41129\|\|full | Low-quality |
| S5_vs_k141_82307\|\|full | Not-determined |
| S5_vs_k141_65842\|\|full | Not-determined |
| S5_vs_k141_82321\|\|full | Not-determined |
| S5_vs_k141_115032\|\|full | Low-quality |
| S5_vs_k141_27930\|\|full | Low-quality |
| S5_vs_k141_144240\|\|full | Low-quality |
| S5_vs_k141_133136\|\|full | Low-quality |
| S5_vs_k141_11282\|\|full | Low-quality |
| S5_vs_k141_115056\|\|full | Not-determined |
| S5_vs_k141_76725\|\|full | Not-determined |
| S5_vs_k141_76730\|\|full | Low-quality |
| S5_vs_k141_133163\|\|full | Low-quality |
| S5_vs_k141_27969\|\|full | Low-quality |
| S5_vs_k141_96849\|\|full | Low-quality |
| S5_vs_k141_76731\|\|full | Low-quality |
| S5_vs_k141_96853\|\|full | Not-determined |
| S5_vs_k141_115100\|\|full | Low-quality |
| S5_vs_k141_96857\|\|full | Low-quality |
| S5_vs_k141_47969\|\|full | Low-quality |
| S5_vs_k141_96865\|\|full | Low-quality |
| S5_vs_k141_41161\|\|full | Not-determined |
| S5_vs_k141_47973\|\|full | Low-quality |
| S5_vs_k141_90501\|\|full | Low-quality |
| S5_vs_k141_11288\|\|full | Not-determined |
| S5_vs_k141_76758\|\|full | Low-quality |
| S5_vs_k141_115138\|\|full | Not-determined |
| S5_vs_k141_47984\|\|full | Low-quality |
| S5_vs_k141_71970\|\|full | Low-quality |
| S5_vs_k141_115150\|\|full | Not-determined |
| S5_vs_k141_96921\|\|full | Low-quality |
| S5_vs_k141_48002\|\|full | Low-quality |
| S5_vs_k141_41210\|\|full | Low-quality |
| S5_vs_k141_41211\|\|full | Not-determined |
| S5_vs_k141_96935\|\|full | Low-quality |
| S5_vs_k141_76805\|\|full | Not-determined |
| S5_vs_k141_11307\|\|full | Low-quality |
| S5_vs_k141_76806\|\|full | Low-quality |
| S5_vs_k141_48012\|\|full | Low-quality |
| S5_vs_k141_27989\|\|full | Low-quality |
| S5_vs_k141_72001\|\|full | Low-quality |
| S5_vs_k141_137551\|\|full | Not-determined |
| S5_vs_k141_23635\|\|full | Low-quality |
| S5_vs_k141_23642\|\|full | Not-determined |
| S5_vs_k141_72020\|\|full | Not-determined |
| S5_vs_k141_137565\|\|full | Low-quality |
| S5_vs_k141_137580\|\|full | Not-determined |
| S5_vs_k141_96988\|\|full | Not-determined |
| S5_vs_k141_101032\|\|full | Low-quality |
| S5_vs_k141_72036\|\|full | Not-determined |
| S5_vs_k141_72044\|\|full | Low-quality |
| S5_vs_k141_27998\|\|full | Not-determined |
| S5_vs_k141_11353\|\|full | Low-quality |
| S5_vs_k141_82456\|\|full | Not-determined |
| S5_vs_k141_97018\|\|full | Low-quality |
| S5_vs_k141_97026\|\|full | Low-quality |
| S5_vs_k141_11368\|\|full | Not-determined |
| S5_vs_k141_97034\|\|full | Low-quality |
| S5_vs_k141_76831\|\|full | Low-quality |
| S5_vs_k141_72077\|\|full | Low-quality |
| S5_vs_k141_11374\|\|full | Low-quality |
| S5_vs_k141_115209\|\|full | Not-determined |
| S5_vs_k141_82478\|\|full | Low-quality |
| S5_vs_k141_90651\|\|full | Low-quality |
| S5_vs_k141_115210\|\|full | Low-quality |
| S5_vs_k141_144301\|\|full | Low-quality |
| S5_vs_k141_133291\|\|full | Low-quality |
| S5_vs_k141_90659\|\|full | Not-determined |
| S5_vs_k141_65868\|\|full | Low-quality |
| S5_vs_k141_90665\|\|full | Not-determined |
| S5_vs_k141_90667\|\|full | Low-quality |
| S5_vs_k141_115260\|\|full | Low-quality |
| S5_vs_k141_90670\|\|full | Not-determined |
| S5_vs_k141_133328\|\|full | Not-determined |
| S5_vs_k141_97100\|\|full | Not-determined |
| S5_vs_k141_137599\|\|full | Low-quality |
| S5_vs_k141_76881\|\|full | Not-determined |
| S5_vs_k141_57989\|\|full | Low-quality |
| S5_vs_k141_101052\|\|full | Low-quality |
| S5_vs_k141_133377\|\|full | Low-quality |
| S5_vs_k141_115304\|\|full | Low-quality |
| S5_vs_k141_28009\|\|full | Low-quality |
| S5_vs_k141_133407\|\|full | Low-quality |
| S5_vs_k141_65914\|\|full | Low-quality |
| S5_vs_k141_72187\|\|full | Low-quality |
| S5_vs_k141_76909\|\|full | Low-quality |
| S5_vs_k141_28020\|\|full | Not-determined |
| S5_vs_k141_28022\|\|full | Not-determined |
| S5_vs_k141_90722\|\|full | Low-quality |
| S5_vs_k141_133418\|\|full | Not-determined |
| S5_vs_k141_41305\|\|full | Not-determined |
| S5_vs_k141_76935\|\|full | Low-quality |
| S5_vs_k141_72228\|\|full | Low-quality |
| S7_vs_13799\|\|full | High-quality |
| S5_vs_k141_133446\|\|full | Low-quality |
| S5_vs_k141_48051\|\|full | Low-quality |
| S5_vs_k141_48057\|\|full | Low-quality |
| S5_vs_k141_144329\|\|full | Low-quality |
| S5_vs_k141_101072\|\|full | Low-quality |
| S5_vs_k141_48064\|\|full | Low-quality |
| S5_vs_k141_23741\|\|full | Low-quality |
| S5_vs_k141_133467\|\|full | Low-quality |
| S5_vs_k141_48090\|\|full | Low-quality |
| S5_vs_k141_23751\|\|full | Low-quality |
| S5_vs_k141_72270\|\|full | Low-quality |
| S5_vs_k141_28040\|\|full | Low-quality |
| S5_vs_k141_65989\|\|full | Low-quality |
| S5_vs_k141_48114\|\|full | Low-quality |
| S5_vs_k141_65999\|\|full | Not-determined |
| S5_vs_k141_11503\|\|full | Low-quality |
| S5_vs_k141_66015\|\|full | Not-determined |
| S5_vs_k141_77020\|\|full | Low-quality |
| S5_vs_k141_48126\|\|full | Low-quality |
| S5_vs_k141_23806\|\|full | Not-determined |
| S5_vs_k141_77027\|\|full | Low-quality |
| S5_vs_k141_72298\|\|full | Not-determined |
| S5_vs_k141_48137\|\|full | Not-determined |
| S5_vs_k141_41377\|\|full | Low-quality |
| S5_vs_k141_101103\|\|full | Not-determined |
| S5_vs_k141_23817\|\|full | Not-determined |
| S5_vs_k141_23833\|\|full | Low-quality |
| S5_vs_k141_101106\|\|full | Low-quality |
| S5_vs_k141_23856\|\|full | Low-quality |
| S5_vs_k141_133530\|\|full | Not-determined |
| S5_vs_k141_144419\|\|full | Not-determined |
| S5_vs_k141_144420\|\|full | Low-quality |
| S5_vs_k141_82515\|\|full | Low-quality |
| S5_vs_k141_144424\|\|full | Not-determined |
| S5_vs_k141_11538\|\|full | Low-quality |
| S5_vs_k141_133550\|\|full | Low-quality |
| S5_vs_k141_48230\|\|full | Low-quality |
| S5_vs_k141_82545\|\|full | Not-determined |
| S5_vs_k141_77059\|\|full | Low-quality |
| S5_vs_k141_48232\|\|full | Not-determined |
| S5_vs_k141_23910\|\|full | Not-determined |
| S5_vs_k141_144446\|\|full | Not-determined |
| S5_vs_k141_66070\|\|full | Low-quality |
| S5_vs_k141_144452\|\|full | Low-quality |
| S5_vs_k141_41401\|\|full | Low-quality |
| S5_vs_k141_28099\|\|full | Low-quality |
| S5_vs_k141_66088\|\|full | Not-determined |
| S5_vs_k141_77080\|\|full | Low-quality |
| S5_vs_k141_82553\|\|full | Low-quality |
| S5_vs_k141_66107\|\|full | Not-determined |
| S5_vs_k141_137719\|\|full | Not-determined |
| S5_vs_k141_23949\|\|full | Not-determined |
| S5_vs_k141_66113\|\|full | Low-quality |
| S5_vs_k141_137730\|\|full | Not-determined |
| S5_vs_k141_66132\|\|full | Not-determined |
| S5_vs_k141_48306\|\|full | Not-determined |
| S5_vs_k141_137732\|\|full | Low-quality |
| S5_vs_k141_48313\|\|full | Low-quality |
| S5_vs_k141_11598\|\|full | Low-quality |
| S5_vs_k141_23954\|\|full | Not-determined |
| S5_vs_k141_137738\|\|full | Not-determined |
| S5_vs_k141_66159\|\|full | Low-quality |
| S5_vs_k141_48344\|\|full | Low-quality |
| S5_vs_k141_137747\|\|full | Low-quality |
| S5_vs_k141_48346\|\|full | Low-quality |
| S5_vs_k141_41463\|\|full | Low-quality |
| S5_vs_k141_28147\|\|full | Low-quality |
| S5_vs_k141_23987\|\|full | Low-quality |
| S5_vs_k141_90754\|\|full | Low-quality |
| S5_vs_k141_66177\|\|full | Low-quality |
| S5_vs_k141_11630\|\|full | Low-quality |
| S5_vs_k141_82590\|\|full | Low-quality |
| S5_vs_k141_23997\|\|full | Low-quality |
| S5_vs_k141_82592\|\|full | Low-quality |
| S5_vs_k141_11634\|\|full | Not-determined |
| S5_vs_k141_66183\|\|full | Not-determined |
| S5_vs_k141_77165\|\|full | Not-determined |
| S5_vs_k141_41513\|\|full | Low-quality |
| S5_vs_k141_137782\|\|full | Low-quality |
| S5_vs_k141_137788\|\|full | Not-determined |
| S5_vs_k141_77176\|\|full | Low-quality |
| S5_vs_k141_82602\|\|full | Low-quality |
| S5_vs_k141_24033\|\|full | Low-quality |
| S5_vs_k141_77180\|\|full | Low-quality |
| S5_vs_k141_90769\|\|full | Not-determined |
| S5_vs_k141_24045\|\|full | Low-quality |
| S5_vs_k141_77192\|\|full | Not-determined |
| S5_vs_k141_82614\|\|full | Low-quality |
| S5_vs_k141_24064\|\|full | Not-determined |
| S5_vs_k141_101260\|\|full | Low-quality |
| S5_vs_k141_41554\|\|full | Not-determined |
| S5_vs_k141_41556\|\|full | Low-quality |
| S5_vs_k141_28196\|\|full | Low-quality |
| S5_vs_k141_77222\|\|full | Not-determined |
| S5_vs_k141_41562\|\|full | Not-determined |
| S5_vs_k141_72451\|\|full | Not-determined |
| S5_vs_k141_90792\|\|full | Low-quality |
| S5_vs_k141_58057\|\|full | Low-quality |
| S5_vs_k141_77242\|\|full | Not-determined |
| S5_vs_k141_82629\|\|full | Not-determined |
| S5_vs_k141_82636\|\|full | Low-quality |
| S5_vs_k141_28221\|\|full | Low-quality |
| S5_vs_k141_58070\|\|full | Low-quality |
| S5_vs_k141_144626\|\|full | Not-determined |
| S5_vs_k141_41599\|\|full | Low-quality |
| S5_vs_k141_41600\|\|full | Low-quality |
| S5_vs_k141_101289\|\|full | Not-determined |
| S5_vs_k141_48458\|\|full | Not-determined |
| S5_vs_k141_77264\|\|full | Low-quality |
| S5_vs_k141_82671\|\|full | Low-quality |
| S5_vs_k141_82676\|\|full | Low-quality |
| S5_vs_k141_66234\|\|full | Not-determined |
| S5_vs_k141_82682\|\|full | Low-quality |
| S5_vs_k141_66237\|\|full | Not-determined |
| S5_vs_k141_144649\|\|full | Low-quality |
| S14_extracted_k141_117150 | Complete |
| S5_vs_k141_66244\|\|full | Not-determined |
| S5_vs_k141_24111\|\|full | Not-determined |
| S5_vs_k141_144667\|\|full | Low-quality |
| S5_vs_k141_82696\|\|full | Low-quality |
| S5_vs_k141_66264\|\|full | Low-quality |
| S5_vs_k141_28288\|\|full | Not-determined |
| S5_vs_k141_144683\|\|full | Low-quality |
| S5_vs_k141_66309\|\|full | Low-quality |
| S5_vs_k141_82717\|\|full | Not-determined |
| S5_vs_k141_82721\|\|full | Not-determined |
| S5_vs_k141_90902\|\|full | Low-quality |
| S5_vs_k141_101329\|\|full | Low-quality |
| S5_vs_k141_90907\|\|full | Low-quality |
| S5_vs_k141_58153\|\|full | Not-determined |
| S5_vs_k141_28313\|\|full | Low-quality |
| S5_vs_k141_144717\|\|full | Low-quality |
| S5_vs_k141_24190\|\|full | Low-quality |
| S5_vs_k141_90913\|\|full | Not-determined |
| S5_vs_k141_82733\|\|full | Not-determined |
| S5_vs_k141_24200\|\|full | Low-quality |
| S6_vs_k141_12945\|\|full | Medium-quality |
| S5_vs_k141_58165\|\|full | Low-quality |
| S5_vs_k141_144729\|\|full | Low-quality |
| S3_vs_k141_127405\|\|full | High-quality |
| S5_vs_k141_90932\|\|full | Not-determined |
| S5_vs_k141_66336\|\|full | Low-quality |
| S5_vs_k141_66337\|\|full | Low-quality |
| S5_vs_k141_58172\|\|full | Low-quality |
| S5_vs_k141_82749\|\|full | Low-quality |
| S5_vs_k141_72554\|\|full | Low-quality |
| S5_vs_k141_144744\|\|full | Low-quality |
| S5_vs_k141_90956\|\|full | Low-quality |
| S5_vs_k141_90976\|\|full | Low-quality |
| S5_vs_k141_58209\|\|full | Low-quality |
| S5_vs_k141_66359\|\|full | Low-quality |
| S5_vs_k141_66360\|\|full | Not-determined |
| S5_vs_k141_82787\|\|full | Low-quality |
| S5_vs_k141_90992\|\|full | Low-quality |
| S5_vs_k141_28341\|\|full | Low-quality |
| S5_vs_k141_72607\|\|full | Low-quality |
| S5_vs_k141_24269\|\|full | Not-determined |
| S5_vs_k141_144748\|\|full | Low-quality |
| S5_vs_k141_144749\|\|full | Not-determined |
| S5_vs_k141_41734\|\|full | Low-quality |
| S5_vs_k141_82818\|\|full | Not-determined |
| S5_vs_k141_58244\|\|full | Low-quality |
| S5_vs_k141_144763\|\|full | Low-quality |
| S5_vs_k141_91025\|\|full | Low-quality |
| S5_vs_k141_24287\|\|full | Low-quality |
| S5_vs_k141_72631\|\|full | Not-determined |
| S5_vs_k141_28380\|\|full | Not-determined |
| S5_vs_k141_41761\|\|full | Low-quality |
| S5_vs_k141_28390\|\|full | Low-quality |
| S5_vs_k141_41776\|\|full | Not-determined |
| S5_vs_k141_82858\|\|full | Low-quality |
| S5_vs_k141_101388\|\|full | Low-quality |
| S5_vs_k141_91034\|\|full | Low-quality |
| S5_vs_k141_144802\|\|full | Low-quality |
| S5_vs_k141_101398\|\|full | Low-quality |
| S5_vs_k141_91059\|\|full | Not-determined |
| S5_vs_k141_82909\|\|full | Not-determined |
| S5_vs_k141_28491\|\|full | Not-determined |
| S5_vs_k141_28515\|\|full | Not-determined |
| S5_vs_k141_145590\|\|full | High-quality |
| S5_vs_k141_41898\|\|full | Not-determined |
| S5_vs_k141_137941\|\|full | Low-quality |
| S5_vs_k141_137943\|\|full | Low-quality |
| S5_vs_k141_82932\|\|full | Low-quality |
| S5_vs_k141_72732\|\|full | Low-quality |
| S5_vs_k141_144846\|\|full | Not-determined |
| S5_vs_k141_58314\|\|full | Low-quality |
| S5_vs_k141_144855\|\|full | Not-determined |
| S5_vs_k141_58317\|\|full | Not-determined |
| S5_vs_k141_72750\|\|full | Not-determined |
| S5_vs_k141_82956\|\|full | Low-quality |
| S5_vs_k141_137962\|\|full | Low-quality |
| S5_vs_k141_82969\|\|full | Not-determined |
| S5_vs_k141_66468\|\|full | Not-determined |
| S5_vs_k141_82979\|\|full | Low-quality |
| S5_vs_k141_137979\|\|full | Not-determined |
| S5_vs_k141_66496\|\|full | Low-quality |
| S5_vs_k141_82997\|\|full | Low-quality |
| S5_vs_k141_144916\|\|full | Low-quality |
| S5_vs_k141_58345\|\|full | Low-quality |
| S5_vs_k141_41951\|\|full | Low-quality |
| S5_vs_k141_66517\|\|full | Not-determined |
| S5_vs_k141_72774\|\|full | Not-determined |
| S5_vs_k141_144942\|\|full | Low-quality |
| S5_vs_k141_83036\|\|full | Low-quality |
| S5_vs_k141_144953\|\|full | Low-quality |
| S5_vs_k141_83048\|\|full | Not-determined |
| S5_vs_k141_58385\|\|full | Not-determined |
| S5_vs_k141_28602\|\|full | Not-determined |
| S5_vs_k141_28604\|\|full | Low-quality |
| S5_vs_k141_144970\|\|full | Not-determined |
| S5_vs_k141_58398\|\|full | Low-quality |
| S5_vs_k141_83071\|\|full | Not-determined |
| S5_vs_k141_83075\|\|full | Not-determined |
| S5_vs_k141_28626\|\|full | Not-determined |
| S5_vs_k141_58407\|\|full | Not-determined |
| S5_vs_k141_83106\|\|full | Low-quality |
| S5_vs_k141_83124\|\|full | Low-quality |
| S5_vs_k141_77460\|\|full | Low-quality |
| S5_vs_k141_77467\|\|full | Low-quality |
| S5_vs_k141_83158\|\|full | Low-quality |
| S5_vs_k141_58428\|\|full | Low-quality |
| S5_vs_k141_72838\|\|full | Low-quality |
| S5_vs_k141_83167\|\|full | Not-determined |
| S5_vs_k141_77497\|\|full | Not-determined |
| S5_vs_k141_42109\|\|full | Not-determined |
| S5_vs_k141_28654\|\|full | Low-quality |
| S5_vs_k141_83175\|\|full | Not-determined |
| S5_vs_k141_77499\|\|full | Low-quality |
| S5_vs_k141_77503\|\|full | Not-determined |
| S5_vs_k141_83224\|\|full | Low-quality |
| S5_vs_k141_77531\|\|full | Low-quality |
| S5_vs_k141_58512\|\|full | Not-determined |
| S5_vs_k141_77564\|\|full | Not-determined |
| S5_vs_k141_145025\|\|full | Low-quality |
| S5_vs_k141_28690\|\|full | Not-determined |
| S5_vs_k141_42162\|\|full | Low-quality |
| S5_vs_k141_83240\|\|full | Low-quality |
| S7_vs_47050\|\|full | High-quality |
| S5_vs_k141_138004\|\|full | Not-determined |
| S5_vs_k141_101425\|\|full | Low-quality |
| S5_vs_k141_145036\|\|full | Not-determined |
| S5_vs_k141_145038\|\|full | Not-determined |
| S5_vs_k141_28717\|\|full | Not-determined |
| S5_vs_k141_138036\|\|full | Low-quality |
| S5_vs_k141_145069\|\|full | Low-quality |
| S5_vs_k141_83320\|\|full | Low-quality |
| S5_vs_k141_138039\|\|full | Low-quality |
| S5_vs_k141_28743\|\|full | Low-quality |
| S5_vs_k141_101465\|\|full | Low-quality |
| S5_vs_k141_66559\|\|full | Low-quality |
| S7_vs_73280\|\|full | High-quality |
| S5_vs_k141_42296\|\|full | Not-determined |
| S5_vs_k141_66581\|\|full | Not-determined |
| S5_vs_k141_42301\|\|full | Not-determined |
| S5_vs_k141_145158\|\|full | Low-quality |
| S5_vs_k141_66587\|\|full | Low-quality |
| S5_vs_k141_58688\|\|full | Low-quality |
| S5_vs_k141_138123\|\|full | Not-determined |
| S5_vs_k141_83402\|\|full | Low-quality |
| S5_vs_k141_28771\|\|full | Low-quality |
| S5_vs_k141_77626\|\|full | Not-determined |
| S5_vs_k141_138130\|\|full | Not-determined |
| S5_vs_k141_138134\|\|full | Low-quality |
| S5_vs_k141_66618\|\|full | Low-quality |
| S5_vs_k141_58726\|\|full | Not-determined |
| S5_vs_k141_77646\|\|full | Not-determined |
| S5_vs_k141_42343\|\|full | Low-quality |
| S5_vs_k141_138145\|\|full | Low-quality |
| S5_vs_k141_66646\|\|full | Low-quality |
| S5_vs_k141_42370\|\|full | Low-quality |
| S5_vs_k141_83419\|\|full | Low-quality |
| S5_vs_k141_28804\|\|full | Low-quality |
| S5_vs_k141_66660\|\|full | Low-quality |
| S5_vs_k141_83434\|\|full | Not-determined |
| S5_vs_k141_83435\|\|full | Low-quality |
| S5_vs_k141_83439\|\|full | Low-quality |
| S5_vs_k141_42398\|\|full | Low-quality |
| S5_vs_k141_58744\|\|full | Not-determined |
| S5_vs_k141_83453\|\|full | Not-determined |
| S5_vs_k141_42418\|\|full | Not-determined |
| S5_vs_k141_77686\|\|full | Not-determined |
| S5_vs_k141_28825\|\|full | Low-quality |
| S5_vs_k141_77696\|\|full | Low-quality |
| S5_vs_k141_66720\|\|full | Low-quality |
| S5_vs_k141_138210\|\|full | Low-quality |
| S5_vs_k141_138218\|\|full | Low-quality |
| S5_vs_k141_145230\|\|full | Low-quality |
| S5_vs_k141_138231\|\|full | Not-determined |
| S5_vs_k141_42455\|\|full | Not-determined |
| S5_vs_k141_83482\|\|full | Low-quality |
| S5_vs_k141_66757\|\|full | Low-quality |
| S12_vs_k141_94042\|\|full | Medium-quality |
| S5_vs_k141_77705\|\|full | Not-determined |
| S5_vs_k141_83499\|\|full | Not-determined |
| S14_vs_k141_117150\|\|full | High-quality |
| S5_vs_k141_58773\|\|full | Not-determined |
| S5_vs_k141_58775\|\|full | Not-determined |
| S5_vs_k141_145267\|\|full | Not-determined |
| S5_vs_k141_28861\|\|full | Not-determined |
| S5_vs_k141_138270\|\|full | Low-quality |
| S5_vs_k141_83567\|\|full | Low-quality |
| S5_vs_k141_58813\|\|full | Low-quality |
| S5_vs_k141_138276\|\|full | Not-determined |
| S5_vs_k141_145298\|\|full | Low-quality |
| S5_vs_k141_42494\|\|full | Low-quality |
| S5_vs_k141_58827\|\|full | Low-quality |
| S5_vs_k141_83591\|\|full | Not-determined |
| S5_vs_k141_28901\|\|full | Not-determined |
| S5_vs_k141_42501\|\|full | Not-determined |
| S5_vs_k141_145332\|\|full | Low-quality |
| S5_vs_k141_58841\|\|full | Low-quality |
| S5_vs_k141_145346\|\|full | Low-quality |
| S5_vs_k141_145348\|\|full | Low-quality |
| S5_vs_k141_83622\|\|full | Low-quality |
| S5_vs_k141_77737\|\|full | Low-quality |
| S5_vs_k141_83626\|\|full | Low-quality |
| S5_vs_k141_58856\|\|full | Low-quality |
| S5_vs_k141_28941\|\|full | Low-quality |
| S5_vs_k141_101536\|\|full | Low-quality |
| S5_vs_k141_11718\|\|full | Low-quality |
| S14_extracted_k141_117155 | Medium-quality |
| S9_extracted_k141_73311 | Medium-quality |
| S5_vs_k141_77757\|\|full | Low-quality |
| S5_vs_k141_28972\|\|full | Not-determined |
| S5_vs_k141_145398\|\|full | Not-determined |
| S5_vs_k141_11723\|\|full | Low-quality |
| S5_vs_k141_138377\|\|full | Low-quality |
| S5_vs_k141_58895\|\|full | Low-quality |
| S5_vs_k141_77820\|\|full | Not-determined |
| S5_vs_k141_11753\|\|full | Low-quality |
| S5_vs_k141_77828\|\|full | Not-determined |
| S5_vs_k141_11764\|\|full | Low-quality |
| S5_vs_k141_28993\|\|full | Not-determined |
| S5_vs_k141_77834\|\|full | Low-quality |
| S5_vs_k141_101556\|\|full | Low-quality |
| S5_vs_k141_138400\|\|full | Low-quality |
| S5_vs_k141_145471\|\|full | Not-determined |
| S5_vs_k141_11792\|\|full | Low-quality |
| S5_vs_k141_101592\|\|full | Low-quality |
| S5_vs_k141_145507\|\|full | Low-quality |
| S5_vs_k141_101598\|\|full | Low-quality |
| S5_vs_k141_29027\|\|full | Not-determined |
| S5_vs_k141_138450\|\|full | Not-determined |
| S5_vs_k141_58948\|\|full | Low-quality |
| S5_vs_k141_11816\|\|full | Low-quality |
| S5_vs_k141_138466\|\|full | Low-quality |
| S5_vs_k141_83740\|\|full | Not-determined |
| S5_vs_k141_138490\|\|full | Low-quality |
| S5_vs_k141_11859\|\|full | Low-quality |
| S5_vs_k141_145551\|\|full | Low-quality |
| S5_vs_k141_11863\|\|full | Low-quality |
| S5_vs_k141_58989\|\|full | Not-determined |
| S5_vs_k141_83797\|\|full | Not-determined |
| S5_vs_k141_29094\|\|full | Low-quality |
| S5_vs_k141_145569\|\|full | Not-determined |
| S5_vs_k141_138511\|\|full | Not-determined |
| S5_vs_k141_101663\|\|full | Not-determined |
| S5_vs_k141_59042\|\|full | Low-quality |
| S5_vs_k141_101680\|\|full | Low-quality |
| S5_vs_k141_138537\|\|full | Low-quality |
| S5_vs_k141_59061\|\|full | Low-quality |
| S5_vs_k141_29113\|\|full | Not-determined |
| S5_vs_k141_59064\|\|full | Low-quality |
| S5_vs_k141_101688\|\|full | Low-quality |
| S5_vs_k141_101695\|\|full | Low-quality |
| S5_vs_k141_59097\|\|full | Low-quality |
| S5_vs_k141_145581\|\|full | High-quality |
| S5_vs_k141_101702\|\|full | Low-quality |
| S5_vs_k141_59144\|\|full | Low-quality |
| S5_vs_k141_83912\|\|full | Low-quality |
| S5_vs_k141_101736\|\|full | Not-determined |
| S5_vs_k141_101741\|\|full | Low-quality |
| S5_vs_k141_101766\|\|full | Low-quality |
| S5_vs_k141_59217\|\|full | Low-quality |
| S5_vs_k141_77843\|\|full | Not-determined |
| S5_vs_k141_59254\|\|full | Low-quality |
| S5_vs_k141_83993\|\|full | Low-quality |
| S5_vs_k141_83998\|\|full | Not-determined |
| S5_vs_k141_84004\|\|full | Low-quality |
| S5_vs_k141_101829\|\|full | Not-determined |
| S5_vs_k141_59281\|\|full | Not-determined |
| S5_vs_k141_59290\|\|full | Low-quality |
| S5_vs_k141_77874\|\|full | Low-quality |
| S5_vs_k141_29243\|\|full | Low-quality |
| S5_vs_k141_77875\|\|full | Not-determined |
| S5_vs_k141_101843\|\|full | Not-determined |
| S5_vs_k141_101845\|\|full | Low-quality |
| S5_vs_k141_77887\|\|full | Not-determined |
| S13_vs_85097\|\|full | High-quality |
| S5_vs_k141_29262\|\|full | Low-quality |
| S5_vs_k141_77920\|\|full | Not-determined |
| S5_vs_k141_77926\|\|full | Low-quality |
| S5_vs_k141_101887\|\|full | Low-quality |
| S5_vs_k141_11890\|\|full | Not-determined |
| S5_vs_k141_29300\|\|full | Low-quality |
| S5_vs_k141_11909\|\|full | Low-quality |
| S5_vs_k141_11915\|\|full | Not-determined |
| S5_vs_k141_84187\|\|full | Not-determined |
| S5_vs_k141_101905\|\|full | Low-quality |
| S5_vs_k141_59388\|\|full | Low-quality |
| S5_vs_k141_78027\|\|full | Not-determined |
| S5_vs_k141_59417\|\|full | Not-determined |
| S5_vs_k141_11938\|\|full | Low-quality |
| S5_vs_k141_78053\|\|full | Low-quality |
| S5_vs_k141_59440\|\|full | Low-quality |
| S5_vs_k141_78070\|\|full | Low-quality |
| S5_vs_k141_78076\|\|full | Low-quality |
| S5_vs_k141_78086\|\|full | Low-quality |
| S5_extracted_k141_145583 | Complete |
| S5_vs_k141_101924\|\|full | Low-quality |
| S5_vs_k141_78117\|\|full | Not-determined |
| S5_vs_k141_29351\|\|full | Low-quality |
| S5_vs_k141_138645\|\|full | Low-quality |
| S5_vs_k141_78161\|\|full | Low-quality |
| S5_vs_k141_78164\|\|full | Low-quality |
| S5_vs_k141_101947\|\|full | Low-quality |
| S5_vs_k141_138657\|\|full | Low-quality |
| S5_vs_k141_59529\|\|full | Low-quality |
| S5_vs_k141_78173\|\|full | Not-determined |
| S5_vs_k141_84245\|\|full | Low-quality |
| S5_vs_k141_138675\|\|full | Low-quality |
| S5_vs_k141_59557\|\|full | Low-quality |
| S5_vs_k141_59560\|\|full | Low-quality |
| S5_vs_k141_29368\|\|full | Not-determined |
| S5_vs_k141_78220\|\|full | Low-quality |
| S5_vs_k141_138709\|\|full | Low-quality |
| S5_vs_k141_78230\|\|full | Low-quality |
| S5_vs_k141_78240\|\|full | Not-determined |
| S5_vs_k141_29394\|\|full | Not-determined |
| S5_vs_k141_138745\|\|full | Not-determined |
| S5_vs_k141_59598\|\|full | Not-determined |
| S5_vs_k141_59613\|\|full | Low-quality |
| S5_vs_k141_59618\|\|full | Low-quality |
| S5_vs_k141_102030\|\|full | Low-quality |
| S5_vs_k141_12046\|\|full | Low-quality |
| S5_vs_k141_59625\|\|full | Low-quality |
| S5_vs_k141_138819\|\|full | Not-determined |
| S5_vs_k141_138822\|\|full | Low-quality |
| S5_vs_k141_78315\|\|full | Low-quality |
| S5_vs_k141_138876\|\|full | Low-quality |
| S5_vs_k141_84316\|\|full | Low-quality |
| S5_vs_k141_102075\|\|full | Low-quality |
| S5_vs_k141_84328\|\|full | Low-quality |
| S5_vs_k141_84333\|\|full | Not-determined |
| S5_vs_k141_78425\|\|full | Low-quality |
| S5_vs_k141_102115\|\|full | Low-quality |

| | |
|---|---|
| S5_vs_k141_84356\|\|full | Low-quality |
| S5_vs_k141_78434\|\|full | Low-quality |
| S5_vs_k141_78445\|\|full | Not-determined |
| S5_vs_k141_102151\|\|full | Low-quality |
| S5_vs_k141_138904\|\|full | Low-quality |
| S5_vs_k141_138919\|\|full | Not-determined |
| S5_vs_k141_138930\|\|full | Not-determined |
| S5_vs_k141_78481\|\|full | Low-quality |
| S5_vs_k141_59647\|\|full | Low-quality |
| S5_vs_k141_78501\|\|full | Low-quality |
| S5_vs_k141_59668\|\|full | Low-quality |
| S5_vs_k141_59675\|\|full | Low-quality |
| S5_vs_k141_84491\|\|full | Low-quality |
| S5_vs_k141_29405\|\|full | Not-determined |
| S5_vs_k141_29406\|\|full | Low-quality |
| S14_vs_k141_117155\|\|full | Medium-quality |
| S5_vs_k141_29438\|\|full | Low-quality |
| S5_vs_k141_138997\|\|full | Low-quality |
| S5_vs_k141_84543\|\|full | Low-quality |
| S5_vs_k141_29444\|\|full | Not-determined |
| S5_vs_k141_29454\|\|full | Low-quality |
| S5_vs_k141_78650\|\|full | Low-quality |
| S5_vs_k141_78660\|\|full | Not-determined |
| S5_vs_k141_59719\|\|full | Low-quality |
| S5_vs_k141_29470\|\|full | Low-quality |
| S5_vs_k141_59728\|\|full | Low-quality |
| S5_vs_k141_139037\|\|full | Low-quality |
| S5_vs_k141_139041\|\|full | Low-quality |
| S5_vs_k141_102199\|\|full | Low-quality |
| S5_vs_k141_139061\|\|full | Not-determined |
| S5_vs_k141_29506\|\|full | Not-determined |
| S5_vs_k141_139074\|\|full | Low-quality |
| S5_vs_k141_139083\|\|full | Low-quality |
| S5_vs_k141_78699\|\|full | Low-quality |
| S5_vs_k141_59789\|\|full | Low-quality |
| S5_vs_k141_102232\|\|full | Low-quality |
| S5_vs_k141_29555\|\|full | Low-quality |
| S9_vs_k141_73311\|\|full | Medium-quality |
| S5_vs_k141_29562\|\|full | Low-quality |
| S5_vs_k141_29567\|\|full | Not-determined |
| S5_vs_k141_59808\|\|full | Low-quality |
| S5_vs_k141_78804\|\|full | Low-quality |
| S5_vs_k141_102318\|\|full | Low-quality |
| S5_vs_k141_59826\|\|full | Not-determined |
| S5_vs_k141_29590\|\|full | Low-quality |
| S5_vs_k141_59834\|\|full | Low-quality |
| S5_vs_k141_78841\|\|full | Low-quality |
| S5_vs_k141_102337\|\|full | Not-determined |
| S5_vs_k141_29622\|\|full | Not-determined |
| S5_vs_k141_102354\|\|full | Low-quality |
| S5_vs_k141_29624\|\|full | Low-quality |
| S5_vs_k141_29626\|\|full | Low-quality |
| S5_vs_k141_102365\|\|full | Low-quality |
| S5_vs_k141_59854\|\|full | Not-determined |
| S5_vs_k141_102382\|\|full | Not-determined |
| S5_vs_k141_102384\|\|full | Not-determined |
| S5_vs_k141_59871\|\|full | Low-quality |
| S5_vs_k141_59875\|\|full | Low-quality |
| S5_vs_k141_78886\|\|full | Low-quality |
| S5_vs_k141_59890\|\|full | Not-determined |
| S5_vs_k141_78891\|\|full | Low-quality |
| S5_vs_k141_29669\|\|full | Low-quality |
| S7_vs_50194\|\|full | High-quality |
| S5_vs_k141_78892\|\|full | Not-determined |
| S5_vs_k141_84580\|\|full | Not-determined |
| S5_vs_k141_59898\|\|full | Not-determined |
| S5_vs_k141_84600\|\|full | Low-quality |
| S5_vs_k141_102408\|\|full | Not-determined |
| S5_vs_k141_84609\|\|full | Low-quality |
| S5_vs_k141_78916\|\|full | Low-quality |
| S5_vs_k141_84612\|\|full | Low-quality |
| S5_vs_k141_139130\|\|full | Not-determined |
| S5_vs_k141_29740\|\|full | Low-quality |
| S5_vs_k141_139135\|\|full | Low-quality |
| S5_vs_k141_84649\|\|full | Low-quality |
| S5_vs_k141_59946\|\|full | Low-quality |
| S5_vs_k141_139145\|\|full | Low-quality |
| S5_vs_k141_29754\|\|full | Low-quality |
| S5_vs_k141_29760\|\|full | Not-determined |
| S5_vs_k141_139170\|\|full | Low-quality |
| S5_vs_k141_102460\|\|full | Low-quality |
| S5_vs_k141_102465\|\|full | Low-quality |
| S5_vs_k141_102488\|\|full | Not-determined |
| S5_vs_k141_84708\|\|full | Low-quality |
| S5_vs_k141_60016\|\|full | Not-determined |
| S5_vs_k141_60028\|\|full | Not-determined |
| S5_vs_k141_84736\|\|full | Low-quality |
| S5_vs_k141_60045\|\|full | Low-quality |
| S5_vs_k141_29860\|\|full | Low-quality |
| S5_vs_k141_102558\|\|full | Not-determined |
| S5_vs_k141_102560\|\|full | Low-quality |
| S5_vs_k141_84819\|\|full | Not-determined |
| S5_vs_k141_29911\|\|full | Not-determined |
| S5_vs_k141_84840\|\|full | Low-quality |
| S5_vs_k141_84843\|\|full | Low-quality |
| S5_vs_k141_102627\|\|full | Not-determined |
| S5_vs_k141_29954\|\|full | Low-quality |
| S5_vs_k141_29959\|\|full | Low-quality |
| S5_vs_k141_29971\|\|full | Low-quality |
| S5_vs_k141_102670\|\|full | Not-determined |
| S5_vs_k141_102676\|\|full | Low-quality |
| S5_vs_k141_84884\|\|full | Not-determined |
| S5_vs_k141_84888\|\|full | Low-quality |
| S5_vs_k141_84901\|\|full | Low-quality |
| S5_vs_k141_84903\|\|full | Low-quality |
| S5_vs_k141_60134\|\|full | Low-quality |
| S5_vs_k141_60141\|\|full | Low-quality |
| S5_vs_k141_30063\|\|full | Low-quality |
| S5_vs_k141_60160\|\|full | Low-quality |
| S5_vs_k141_30074\|\|full | Not-determined |
| S5_vs_k141_102735\|\|full | Low-quality |
| S5_vs_k141_60162\|\|full | Low-quality |
| S5_vs_k141_60163\|\|full | Not-determined |
| S5_vs_k141_30088\|\|full | Low-quality |
| S5_vs_k141_139308\|\|full | Low-quality |
| S9_vs_k141_90850\|\|full | High-quality |
| S5_vs_k141_30094\|\|full | Not-determined |
| S5_vs_k141_102751\|\|full | Low-quality |
| S5_vs_k141_84954\|\|full | Low-quality |
| S5_vs_k141_30110\|\|full | Low-quality |
| S5_vs_k141_30117\|\|full | Low-quality |
| S5_vs_k141_84982\|\|full | Not-determined |
| S5_vs_k141_102781\|\|full | Not-determined |
| S5_vs_k141_60230\|\|full | Not-determined |
| S5_vs_k141_30182\|\|full | Low-quality |
| S5_vs_k141_102827\|\|full | Low-quality |
| S5_vs_k141_84996\|\|full | Not-determined |
| S5_vs_k141_139329\|\|full | Low-quality |
| S5_vs_k141_139332\|\|full | Low-quality |
| S5_vs_k141_30226\|\|full | Low-quality |
| S5_vs_k141_102881\|\|full | Low-quality |
| S5_vs_k141_102885\|\|full | Low-quality |
| S5_vs_k141_30249\|\|full | Low-quality |
| S5_vs_k141_102902\|\|full | Low-quality |
| S5_vs_k141_102935\|\|full | Low-quality |
| S5_vs_k141_102937\|\|full | Not-determined |
| S5_vs_k141_102939\|\|full | Low-quality |
| S5_vs_k141_60280\|\|full | Not-determined |
| S5_vs_k141_30292\|\|full | Low-quality |
| S5_vs_k141_30301\|\|full | Low-quality |
| S5_vs_k141_30303\|\|full | Low-quality |
| S5_vs_k141_102974\|\|full | Low-quality |
| S15_extracted_41956 | Medium-quality |
| S5_vs_k141_139423\|\|full | Low-quality |
| S5_vs_k141_60343\|\|full | Not-determined |
| S5_vs_k141_103035\|\|full | Low-quality |
| S5_vs_k141_103055\|\|full | Not-determined |
| S5_vs_k141_139454\|\|full | Not-determined |
| S5_vs_k141_103073\|\|full | Low-quality |
| S5_vs_k141_60405\|\|full | Low-quality |
| S5_vs_k141_60409\|\|full | Low-quality |
| S5_vs_k141_139502\|\|full | Low-quality |
| S5_vs_k141_139506\|\|full | Low-quality |
| S5_vs_k141_139521\|\|full | Low-quality |
| S5_vs_k141_103128\|\|full | Low-quality |
| S5_vs_k141_139530\|\|full | Not-determined |
| S5_vs_k141_139533\|\|full | Low-quality |
| S5_vs_k141_103140\|\|full | Low-quality |
| S5_vs_k141_103156\|\|full | Low-quality |
| S5_vs_k141_139554\|\|full | Not-determined |
| S5_vs_k141_139555\|\|full | Not-determined |
| S5_vs_k141_103170\|\|full | Low-quality |
| S11_vs_k141_14882\|\|full | Medium-quality |
| S5_vs_k141_60458\|\|full | Low-quality |
| S5_vs_k141_60461\|\|full | Low-quality |
| S5_vs_k141_139624\|\|full | Low-quality |
| S5_vs_k141_60475\|\|full | Low-quality |
| S5_vs_k141_139636\|\|full | Not-determined |
| S5_vs_k141_60476\|\|full | Low-quality |
| S5_vs_k141_60502\|\|full | Low-quality |
| S14_vs_k141_117166\|\|full | High-quality |
| S5_vs_k141_60532\|\|full | Low-quality |
| S5_vs_k141_60647\|\|full | Not-determined |
| S5_vs_k141_60667\|\|full | Not-determined |
| S5_vs_k141_60672\|\|full | Not-determined |
| S5_vs_k141_60691\|\|full | Not-determined |
| S5_vs_k141_60700\|\|full | Low-quality |
| S5_vs_k141_24363\|\|0_partial | Not-determined |
| S5_vs_k141_91154\|\|0_partial | Low-quality |
| S5_vs_k141_48777\|\|0_partial | Low-quality |
| S5_vs_k141_133796\|\|0_partial | Low-quality |
| S5_vs_k141_36890\|\|0_partial | Low-quality |
| S5_vs_k141_55140\|\|0_partial | Not-determined |
| S5_vs_k141_61284\|\|0_partial | Not-determined |
| S5_vs_k141_122923\|\|0_partial | Low-quality |
| S5_vs_k141_37506\|\|0_partial | Low-quality |
| S5_vs_k141_44025\|\|0_partial | Low-quality |
| S5_vs_k141_64146\|\|full | High-quality |
| S5_vs_k141_24905\|\|0_partial | Not-determined |
| S5_vs_k141_86243\|\|0_partial | Low-quality |
| S5_vs_k141_61991\|\|0_partial | Not-determined |
| S5_vs_k141_25023\|\|0_partial | Not-determined |
| S5_vs_k141_117564\|\|0_partial | Not-determined |
| S5_vs_k141_117625\|\|0_partial | Not-determined |
| S5_vs_k141_38139\|\|0_partial | Low-quality |
| S5_vs_k141_86751\|\|0_partial | Not-determined |
| S5_vs_k141_141116\|\|0_partial | Low-quality |
| S5_vs_k141_111186\|\|0_partial | Low-quality |
| S5_vs_k141_32820\|\|0_partial | Not-determined |
| S5_vs_k141_38380\|\|0_partial | Not-determined |
| S6_vs_k141_56497\|\|full | High-quality |
| S5_vs_k141_8767\|\|0_partial | Not-determined |
| S5_vs_k141_8767\|\|1_partial | Low-quality |
| S5_vs_k141_93704\|\|0_partial | Not-determined |
| S5_vs_k141_51394\|\|0_partial | Not-determined |
| S5_vs_k141_33103\|\|0_partial | Low-quality |
| S5_vs_k141_141661\|\|0_partial | Not-determined |
| S5_vs_k141_8988\|\|0_partial | Low-quality |
| S5_vs_k141_21058\|\|0_partial | Not-determined |
| S5_vs_k141_15478\|\|0_partial | Low-quality |
| S5_vs_k141_4539\|\|0_partial | Low-quality |
| S5_vs_k141_46047\|\|0_partial | Low-quality |
| S5_vs_k141_99996\|\|0_partial | Low-quality |
| S5_vs_k141_46296\|\|0_partial | Low-quality |
| S5_vs_k141_108624\|\|0_partial | Not-determined |
| S5_vs_k141_27258\|\|0_partial | Not-determined |
| S5_vs_k141_145583\|\|full | High-quality |
| S5_vs_k141_127162\|\|1_partial | Low-quality |
| S5_vs_k141_132364\|\|0_partial | Low-quality |
| S5_vs_k141_130032\|\|full | Medium-quality |
| S5_vs_k141_96355\|\|0_partial | Low-quality |
| S5_vs_k141_23242\|\|0_partial | Low-quality |
| S3_vs_k141_97827\|\|full | Medium-quality |
| S5_vs_k141_66521\|\|0_partial | Low-quality |
| S5_vs_k141_72859\|\|lt2gene | Low-quality |
| S5_vs_k141_48579\|\|lt2gene | Not-determined |
| S5_vs_k141_103223\|\|lt2gene | Low-quality |
| S5_vs_k141_48592\|\|lt2gene | Low-quality |
| S5_vs_k141_42528\|\|lt2gene | Not-determined |
| S5_vs_k141_18230\|\|lt2gene | Low-quality |
| S5_vs_k141_44\|\|lt2gene | Low-quality |
| S5_vs_k141_121453\|\|lt2gene | Low-quality |
| S5_vs_k141_18240\|\|lt2gene | Low-quality |
| S5_vs_k141_115382\|\|lt2gene | Low-quality |
| S5_vs_k141_60783\|\|lt2gene | Low-quality |
| S5_vs_k141_91141\|\|lt2gene | Low-quality |
| S5_vs_k141_127520\|\|lt2gene | Low-quality |
| S5_vs_k141_127522\|\|lt2gene | Not-determined |
| S5_vs_k141_42582\|\|lt2gene | Low-quality |
| S5_vs_k141_85064\|\|lt2gene | Low-quality |
| S5_vs_k141_109332\|\|lt2gene | Low-quality |
| S5_vs_k141_109334\|\|lt2gene | Low-quality |
| S5_vs_k141_109335\|\|lt2gene | Low-quality |
| S5_vs_k141_72883\|\|lt2gene | Low-quality |
| S5_vs_k141_24321\|\|lt2gene | Low-quality |
| S5_vs_k141_121519\|\|lt2gene | Not-determined |
| S5_vs_k141_18305\|\|lt2gene | Not-determined |
| S5_vs_k141_18306\|\|lt2gene | Not-determined |
| S5_vs_k141_60819\|\|lt2gene | Not-determined |
| S5_vs_k141_127557\|\|lt2gene | Not-determined |
| S5_vs_k141_115387\|\|lt2gene | Not-determined |
| S5_vs_k141_6176\|\|lt2gene | Low-quality |
| S5_vs_k141_72912\|\|lt2gene | Low-quality |
| S5_vs_k141_42645\|\|lt2gene | Not-determined |
| S5_vs_k141_127581\|\|lt2gene | Not-determined |
| S5_vs_k141_103348\|\|lt2gene | Low-quality |
| S5_vs_k141_103365\|\|lt2gene | Not-determined |
| S5_vs_k141_85074\|\|lt2gene | Low-quality |
| S5_vs_k141_109381\|\|lt2gene | Low-quality |
| S5_vs_k141_60885\|\|lt2gene | Low-quality |
| S5_vs_k141_103406\|\|lt2gene | Low-quality |
| S5_vs_k141_72946\|\|lt2gene | Low-quality |
| S5_vs_k141_54762\|\|lt2gene | Not-determined |
| S5_vs_k141_103417\|\|lt2gene | Low-quality |
| S5_vs_k141_30407\|\|lt2gene | Not-determined |
| S5_vs_k141_109423\|\|lt2gene | Low-quality |
| S5_vs_k141_133621\|\|lt2gene | Low-quality |
| S5_vs_k141_133627\|\|lt2gene | Low-quality |
| S5_vs_k141_85132\|\|lt2gene | Low-quality |
| S5_vs_k141_109426\|\|lt2gene | Low-quality |
| S5_vs_k141_127621\|\|lt2gene | Low-quality |
| S5_vs_k141_42714\|\|lt2gene | Low-quality |
| S5_vs_k141_36564\|\|lt2gene | Low-quality |
| S5_vs_k141_121569\|\|lt2gene | Not-determined |
| S5_vs_k141_60939\|\|lt2gene | Low-quality |
| S5_vs_k141_48690\|\|lt2gene | Low-quality |
| S5_vs_k141_121585\|\|lt2gene | Low-quality |
| S5_vs_k141_60949\|\|lt2gene | Low-quality |
| S5_vs_k141_72981\|\|lt2gene | Low-quality |
| S5_vs_k141_121607\|\|lt2gene | Low-quality |
| S5_vs_k141_97287\|\|lt2gene | Low-quality |
| S5_vs_k141_109461\|\|lt2gene | Not-determined |
| S5_vs_k141_127662\|\|lt2gene | Low-quality |
| S5_vs_k141_139782\|\|lt2gene | Low-quality |
| S5_vs_k141_97304\|\|lt2gene | Low-quality |
| S5_vs_k141_66911\|\|lt2gene | Low-quality |
| S5_vs_k141_173\|\|lt2gene | Not-determined |
| S5_vs_k141_18419\|\|lt2gene | Not-determined |
| S5_vs_k141_18428\|\|lt2gene | Low-quality |
| S5_vs_k141_54813\|\|lt2gene | Low-quality |
| S5_vs_k141_18429\|\|lt2gene | Low-quality |
| S5_vs_k141_66919\|\|lt2gene | Low-quality |
| S5_vs_k141_48714\|\|lt2gene | Low-quality |
| S5_vs_k141_139849\|\|lt2gene | Low-quality |
| S5_vs_k141_60970\|\|lt2gene | Low-quality |
| S5_vs_k141_85234\|\|lt2gene | Not-determined |
| S5_vs_k141_121698\|\|lt2gene | Low-quality |
| S5_vs_k141_121713\|\|lt2gene | Low-quality |
| S5_vs_k141_121720\|\|lt2gene | Low-quality |
| S5_vs_k141_79045\|\|lt2gene | Low-quality |
| S5_vs_k141_6354\|\|lt2gene | Low-quality |
| S5_vs_k141_24398\|\|lt2gene | Low-quality |
| S5_vs_k141_287\|\|lt2gene | Low-quality |
| S5_vs_k141_115467\|\|lt2gene | Low-quality |
| S5_vs_k141_18481\|\|lt2gene | Low-quality |
| S5_vs_k141_61025\|\|lt2gene | Not-determined |
| S5_vs_k141_18497\|\|lt2gene | Low-quality |
| S5_vs_k141_42900\|\|lt2gene | Low-quality |
| S5_vs_k141_61028\|\|lt2gene | Low-quality |
| S5_vs_k141_91163\|\|lt2gene | Low-quality |
| S5_vs_k141_121767\|\|lt2gene | Low-quality |
| S5_vs_k141_42920\|\|lt2gene | Low-quality |
| S5_vs_k141_42932\|\|lt2gene | Low-quality |
| S5_vs_k141_6445\|\|lt2gene | Low-quality |
| S5_vs_k141_115547\|\|lt2gene | Low-quality |
| S5_vs_k141_109496\|\|lt2gene | Low-quality |
| S5_vs_k141_6465\|\|lt2gene | Low-quality |
| S5_vs_k141_6470\|\|lt2gene | Low-quality |
| S5_vs_k141_73075\|\|lt2gene | Low-quality |
| S5_vs_k141_12276\|\|lt2gene | Low-quality |
| S5_vs_k141_127740\|\|lt2gene | Not-determined |
| S5_vs_k141_348\|\|lt2gene | Not-determined |
| S5_vs_k141_48784\|\|lt2gene | Low-quality |
| S5_vs_k141_127762\|\|lt2gene | Low-quality |
| S5_vs_k141_377\|\|lt2gene | Low-quality |
| S5_vs_k141_30545\|\|lt2gene | Low-quality |
| S5_vs_k141_127776\|\|lt2gene | Not-determined |
| S5_vs_k141_61079\|\|lt2gene | Low-quality |
| S5_vs_k141_30553\|\|lt2gene | Not-determined |
| S5_vs_k141_109523\|\|lt2gene | Low-quality |
| S5_vs_k141_121875\|\|lt2gene | Low-quality |
| S5_vs_k141_133720\|\|lt2gene | Low-quality |
| S5_vs_k141_404\|\|lt2gene | Low-quality |
| S5_vs_k141_36637\|\|lt2gene | Low-quality |
| S5_vs_k141_73092\|\|lt2gene | Low-quality |
| S5_vs_k141_91339\|\|lt2gene | Low-quality |
| S5_vs_k141_434\|\|lt2gene | Low-quality |
| S5_vs_k141_91344\|\|lt2gene | Low-quality |
| S5_vs_k141_30614\|\|lt2gene | Low-quality |
| S5_vs_k141_30617\|\|lt2gene | Low-quality |
| S5_vs_k141_61111\|\|lt2gene | Low-quality |
| S5_vs_k141_458\|\|lt2gene | Low-quality |
| S5_vs_k141_43010\|\|lt2gene | Low-quality |
| S5_vs_k141_115656\|\|lt2gene | Low-quality |
| S5_vs_k141_6528\|\|lt2gene | Low-quality |
| S5_vs_k141_61123\|\|lt2gene | Low-quality |
| S5_vs_k141_61124\|\|lt2gene | Low-quality |
| S5_vs_k141_30645\|\|lt2gene | Low-quality |
| S5_vs_k141_481\|\|lt2gene | Low-quality |
| S5_vs_k141_6551\|\|lt2gene | Low-quality |
| S5_vs_k141_121951\|\|lt2gene | Low-quality |
| S5_vs_k141_103749\|\|lt2gene | Low-quality |
| S5_vs_k141_43036\|\|lt2gene | Low-quality |
| S5_vs_k141_6567\|\|lt2gene | Low-quality |
| S5_vs_k141_6570\|\|lt2gene | Low-quality |
| S5_vs_k141_48889\|\|lt2gene | Low-quality |
| S5_vs_k141_48891\|\|lt2gene | Low-quality |
| S5_vs_k141_85361\|\|lt2gene | Low-quality |
| S5_vs_k141_12300\|\|lt2gene | Low-quality |
| S5_vs_k141_115706\|\|lt2gene | Low-quality |
| S5_vs_k141_12308\|\|lt2gene | Low-quality |
| S5_vs_k141_12318\|\|lt2gene | Low-quality |
| S5_vs_k141_97434\|\|lt2gene | Low-quality |
| S5_vs_k141_91427\|\|lt2gene | Low-quality |
| S5_vs_k141_30730\|\|lt2gene | Low-quality |
| S5_vs_k141_91443\|\|lt2gene | Low-quality |
| S5_vs_k141_91446\|\|lt2gene | Low-quality |
| S5_vs_k141_36697\|\|lt2gene | Low-quality |
| S5_vs_k141_593\|\|lt2gene | Not-determined |
| S5_vs_k141_97465\|\|lt2gene | Low-quality |
| S5_vs_k141_48957\|\|lt2gene | Low-quality |
| S5_vs_k141_54926\|\|lt2gene | Not-determined |
| S5_vs_k141_122047\|\|lt2gene | Low-quality |
| S5_vs_k141_54941\|\|lt2gene | Low-quality |
| S5_vs_k141_115754\|\|lt2gene | Low-quality |
| S5_vs_k141_97484\|\|lt2gene | Low-quality |
| S5_vs_k141_97487\|\|lt2gene | Low-quality |
| S5_vs_k141_97497\|\|lt2gene | Not-determined |
| S5_vs_k141_54957\|\|lt2gene | Low-quality |
| S5_vs_k141_91496\|\|lt2gene | Not-determined |
| S5_vs_k141_109607\|\|lt2gene | Not-determined |
| S5_vs_k141_24546\|\|lt2gene | Low-quality |
| S5_vs_k141_73147\|\|lt2gene | Low-quality |
| S5_vs_k141_737\|\|lt2gene | Not-determined |
| S5_vs_k141_61194\|\|lt2gene | Not-determined |
| S5_vs_k141_115781\|\|lt2gene | Low-quality |
| S5_vs_k141_85402\|\|lt2gene | Low-quality |
| S5_vs_k141_109654\|\|lt2gene | Low-quality |
| S5_vs_k141_18636\|\|lt2gene | Not-determined |
| S5_vs_k141_85413\|\|lt2gene | Low-quality |
| S5_vs_k141_85416\|\|lt2gene | Low-quality |
| S5_vs_k141_115822\|\|lt2gene | Low-quality |
| S5_vs_k141_85422\|\|lt2gene | Not-determined |
| S5_vs_k141_85424\|\|lt2gene | Not-determined |
| S5_vs_k141_109695\|\|lt2gene | Not-determined |
| S5_vs_k141_122171\|\|lt2gene | Not-determined |
| S5_vs_k141_782\|\|lt2gene | Low-quality |
| S5_vs_k141_24604\|\|lt2gene | Low-quality |
| S5_vs_k141_109719\|\|lt2gene | Low-quality |
| S5_vs_k141_61214\|\|lt2gene | Low-quality |
| S5_vs_k141_49048\|\|lt2gene | Low-quality |
| S5_vs_k141_85447\|\|lt2gene | Low-quality |
| S5_vs_k141_24621\|\|lt2gene | Low-quality |
| S5_vs_k141_67017\|\|lt2gene | Not-determined |
| S5_vs_k141_67018\|\|lt2gene | Low-quality |
| S5_vs_k141_24630\|\|lt2gene | Low-quality |
| S5_vs_k141_43255\|\|lt2gene | Low-quality |
| S5_vs_k141_24636\|\|lt2gene | Low-quality |
| S5_vs_k141_61255\|\|lt2gene | Low-quality |
| S5_vs_k141_49076\|\|lt2gene | Low-quality |
| S5_vs_k141_133815\|\|lt2gene | Low-quality |
| S5_vs_k141_61261\|\|lt2gene | Low-quality |
| S5_vs_k141_133820\|\|lt2gene | Low-quality |
| S5_vs_k141_43275\|\|lt2gene | Low-quality |
| S5_vs_k141_109783\|\|lt2gene | Low-quality |
| S5_vs_k141_24650\|\|lt2gene | Low-quality |
| S5_vs_k141_36763\|\|lt2gene | Low-quality |
| S5_vs_k141_85486\|\|lt2gene | Low-quality |
| S5_vs_k141_109797\|\|lt2gene | Low-quality |
| S5_vs_k141_36771\|\|lt2gene | Low-quality |
| S5_vs_k141_140045\|\|lt2gene | Low-quality |
| S5_vs_k141_30965\|\|lt2gene | Low-quality |
| S5_vs_k141_122290\|\|lt2gene | Low-quality |
| S5_vs_k141_109818\|\|lt2gene | Low-quality |
| S5_vs_k141_49123\|\|lt2gene | Low-quality |
| S5_vs_k141_122306\|\|lt2gene | Not-determined |
| S5_vs_k141_67045\|\|lt2gene | Low-quality |
| S5_vs_k141_103920\|\|lt2gene | Low-quality |
| S5_vs_k141_122320\|\|lt2gene | Low-quality |
| S5_vs_k141_36783\|\|lt2gene | Low-quality |
| S5_vs_k141_122332\|\|lt2gene | Low-quality |
| S5_vs_k141_36813\|\|lt2gene | Low-quality |
| S5_vs_k141_36823\|\|lt2gene | Low-quality |
| S5_vs_k141_128064\|\|lt2gene | Not-determined |
| S5_vs_k141_49142\|\|lt2gene | Low-quality |
| S5_vs_k141_49144\|\|lt2gene | Not-determined |
| S5_vs_k141_18763\|\|lt2gene | Low-quality |
| S5_vs_k141_67068\|\|lt2gene | Low-quality |
| S5_vs_k141_115933\|\|lt2gene | Low-quality |
| S5_vs_k141_49184\|\|lt2gene | Not-determined |
| S5_vs_k141_115947\|\|lt2gene | Not-determined |
| S5_vs_k141_85564\|\|lt2gene | Low-quality |
| S5_vs_k141_67105\|\|lt2gene | Low-quality |
| S5_vs_k141_36886\|\|lt2gene | Not-determined |
| S5_vs_k141_67118\|\|lt2gene | Not-determined |
| S5_vs_k141_12479\|\|lt2gene | Low-quality |
| S5_vs_k141_115964\|\|lt2gene | Low-quality |
| S5_vs_k141_122369\|\|lt2gene | Low-quality |
| S5_vs_k141_115989\|\|lt2gene | Low-quality |
| S5_vs_k141_24759\|\|lt2gene | Not-determined |
| S5_vs_k141_43339\|\|lt2gene | Low-quality |
| S5_vs_k141_43343\|\|lt2gene | Low-quality |
| S5_vs_k141_49232\|\|lt2gene | Low-quality |
| S5_vs_k141_12530\|\|lt2gene | Low-quality |
| S5_vs_k141_12533\|\|lt2gene | Not-determined |
| S5_vs_k141_67160\|\|lt2gene | Not-determined |
| S5_vs_k141_85599\|\|lt2gene | Low-quality |
| S5_vs_k141_49248\|\|lt2gene | Low-quality |
| S5_vs_k141_67178\|\|lt2gene | Not-determined |
| S5_vs_k141_67181\|\|lt2gene | Low-quality |
| S5_vs_k141_140055\|\|lt2gene | Low-quality |
| S5_vs_k141_133943\|\|lt2gene | Low-quality |
| S5_vs_k141_133949\|\|lt2gene | Not-determined |
| S5_vs_k141_122435\|\|lt2gene | Not-determined |
| S5_vs_k141_116043\|\|lt2gene | Low-quality |
| S5_vs_k141_6752\|\|lt2gene | Low-quality |
| S5_vs_k141_31129\|\|lt2gene | Not-determined |
| S5_vs_k141_140074\|\|lt2gene | Low-quality |
| S5_vs_k141_67238\|\|lt2gene | Low-quality |
| S5_vs_k141_18839\|\|lt2gene | Low-quality |
| S5_vs_k141_140083\|\|lt2gene | Low-quality |
| S5_vs_k141_116068\|\|lt2gene | Low-quality |
| S5_vs_k141_91719\|\|lt2gene | Low-quality |
| S5_vs_k141_36973\|\|lt2gene | Low-quality |
| S5_vs_k141_140106\|\|lt2gene | Low-quality |
| S5_vs_k141_140107\|\|lt2gene | Not-determined |
| S5_vs_k141_49308\|\|lt2gene | Low-quality |
| S5_vs_k141_6777\|\|lt2gene | Low-quality |
| S5_vs_k141_6778\|\|lt2gene | Low-quality |
| S5_vs_k141_55117\|\|lt2gene | Low-quality |
| S5_vs_k141_128205\|\|lt2gene | Low-quality |
| S5_vs_k141_6797\|\|lt2gene | Low-quality |
| S5_vs_k141_6798\|\|lt2gene | Low-quality |
| S5_vs_k141_91727\|\|lt2gene | Low-quality |
| S5_vs_k141_43497\|\|lt2gene | Low-quality |
| S5_vs_k141_122552\|\|lt2gene | Low-quality |
| S5_vs_k141_91796\|\|lt2gene | Not-determined |
| S5_vs_k141_122566\|\|lt2gene | Not-determined |
| S5_vs_k141_18874\|\|lt2gene | Low-quality |
| S5_vs_k141_37033\|\|lt2gene | Low-quality |
| S5_vs_k141_1124\|\|lt2gene | Low-quality |
| S5_vs_k141_37044\|\|lt2gene | Low-quality |
| S5_vs_k141_91813\|\|lt2gene | Low-quality |
| S5_vs_k141_79223\|\|lt2gene | Low-quality |
| S5_vs_k141_67309\|\|lt2gene | Low-quality |
| S5_vs_k141_49380\|\|lt2gene | Low-quality |
| S5_vs_k141_18906\|\|lt2gene | Not-determined |
| S5_vs_k141_140153\|\|lt2gene | Low-quality |
| S5_vs_k141_43562\|\|lt2gene | Low-quality |
| S5_vs_k141_140159\|\|lt2gene | Not-determined |
| S5_vs_k141_49397\|\|lt2gene | Low-quality |
| S5_vs_k141_49412\|\|lt2gene | Low-quality |
| S5_vs_k141_49415\|\|lt2gene | Low-quality |
| S5_vs_k141_49418\|\|lt2gene | Not-determined |
| S5_vs_k141_43587\|\|lt2gene | Low-quality |
| S5_vs_k141_140208\|\|lt2gene | Low-quality |
| S5_vs_k141_116171\|\|lt2gene | Low-quality |
| S5_vs_k141_31259\|\|lt2gene | Low-quality |
| S5_vs_k141_31261\|\|lt2gene | Not-determined |
| S5_vs_k141_103998\|\|lt2gene | Low-quality |
| S5_vs_k141_1249\|\|lt2gene | Not-determined |
| S5_vs_k141_116220\|\|lt2gene | Low-quality |
| S5_vs_k141_1281\|\|lt2gene | Low-quality |
| S5_vs_k141_43628\|\|lt2gene | Low-quality |
| S5_vs_k141_43630\|\|lt2gene | Not-determined |
| S5_vs_k141_79285\|\|lt2gene | Not-determined |
| S5_vs_k141_134052\|\|lt2gene | Low-quality |
| S5_vs_k141_79293\|\|lt2gene | Low-quality |
| S5_vs_k141_79305\|\|lt2gene | Low-quality |
| S5_vs_k141_31344\|\|lt2gene | Low-quality |
| S5_vs_k141_12601\|\|lt2gene | Not-determined |
| S5_vs_k141_1324\|\|lt2gene | Low-quality |
| S5_vs_k141_67408\|\|lt2gene | Low-quality |
| S5_vs_k141_73252\|\|lt2gene | Low-quality |
| S5_vs_k141_31364\|\|lt2gene | Not-determined |
| S5_vs_k141_12636\|\|lt2gene | Not-determined |
| S5_vs_k141_31375\|\|lt2gene | Not-determined |
| S5_vs_k141_79347\|\|lt2gene | Low-quality |
| S5_vs_k141_122662\|\|lt2gene | Low-quality |
| S5_vs_k141_104079\|\|lt2gene | Low-quality |
| S5_vs_k141_19020\|\|lt2gene | Low-quality |
| S5_vs_k141_43673\|\|lt2gene | Not-determined |
| S5_vs_k141_140239\|\|lt2gene | Low-quality |
| S5_vs_k141_140243\|\|lt2gene | Not-determined |
| S5_vs_k141_122706\|\|lt2gene | Low-quality |
| S5_vs_k141_31418\|\|lt2gene | Low-quality |
| S5_vs_k141_37092\|\|lt2gene | Low-quality |
| S5_vs_k141_37093\|\|lt2gene | Low-quality |
| S5_vs_k141_1358\|\|lt2gene | Low-quality |
| S5_vs_k141_7025\|\|lt2gene | Low-quality |
| S5_vs_k141_109834\|\|lt2gene | Low-quality |
| S5_vs_k141_37107\|\|lt2gene | Not-determined |
| S5_vs_k141_67478\|\|lt2gene | Low-quality |
| S5_vs_k141_19092\|\|lt2gene | Low-quality |
| S5_vs_k141_122789\|\|lt2gene | Low-quality |
| S5_vs_k141_19134\|\|lt2gene | Low-quality |
| S5_vs_k141_31463\|\|lt2gene | Low-quality |
| S5_vs_k141_73291\|\|lt2gene | Low-quality |
| S5_vs_k141_31477\|\|lt2gene | Low-quality |
| S5_vs_k141_122802\|\|lt2gene | Low-quality |
| S5_vs_k141_55218\|\|lt2gene | Low-quality |
| S5_vs_k141_85760\|\|lt2gene | Low-quality |
| S5_vs_k141_12768\|\|lt2gene | Low-quality |
| S5_vs_k141_55246\|\|lt2gene | Low-quality |
| S5_vs_k141_73304\|\|lt2gene | Low-quality |
| S5_vs_k141_116361\|\|lt2gene | Low-quality |
| S5_vs_k141_122831\|\|lt2gene | Low-quality |
| S5_vs_k141_85801\|\|lt2gene | Low-quality |
| S5_vs_k141_85808\|\|lt2gene | Low-quality |
| S5_vs_k141_19163\|\|lt2gene | Not-determined |
| S5_vs_k141_122864\|\|lt2gene | Low-quality |
| S5_vs_k141_31504\|\|lt2gene | Low-quality |
| S5_vs_k141_31511\|\|lt2gene | Low-quality |
| S5_vs_k141_85842\|\|lt2gene | Low-quality |
| S5_vs_k141_67528\|\|lt2gene | Low-quality |
| S5_vs_k141_7147\|\|lt2gene | Low-quality |
| S5_vs_k141_140313\|\|lt2gene | Not-determined |
| S5_vs_k141_140316\|\|lt2gene | Not-determined |
| S5_vs_k141_67555\|\|lt2gene | Not-determined |
| S5_vs_k141_1404\|\|lt2gene | Low-quality |
| S5_vs_k141_116427\|\|lt2gene | Low-quality |
| S5_vs_k141_12805\|\|lt2gene | Low-quality |
| S5_vs_k141_12812\|\|lt2gene | Low-quality |
| S5_vs_k141_12823\|\|lt2gene | Low-quality |
| S5_vs_k141_109990\|\|lt2gene | Low-quality |
| S5_vs_k141_109992\|\|lt2gene | Not-determined |
| S5_vs_k141_122917\|\|lt2gene | Low-quality |
| S5_vs_k141_140340\|\|lt2gene | Low-quality |
| S5_vs_k141_116476\|\|lt2gene | Low-quality |
| S5_vs_k141_19190\|\|lt2gene | Low-quality |
| S5_vs_k141_134231\|\|lt2gene | Not-determined |
| S5_vs_k141_116487\|\|lt2gene | Low-quality |
| S5_vs_k141_116490\|\|lt2gene | Low-quality |
| S5_vs_k141_7198\|\|lt2gene | Low-quality |
| S5_vs_k141_37215\|\|lt2gene | Low-quality |
| S5_vs_k141_134243\|\|lt2gene | Low-quality |
| S5_vs_k141_104198\|\|lt2gene | Low-quality |
| S5_vs_k141_43727\|\|lt2gene | Not-determined |
| S5_vs_k141_12869\|\|lt2gene | Low-quality |

SS_vs_k141_43737||lt2gene Low-quality
SS_vs_k141_7229||lt2gene Low-quality
SS_vs_k141_49506||lt2gene Low-quality
SS_vs_k141_104225||lt2gene Low-quality
SS_vs_k141_12877||lt2gene Low-quality
SS_vs_k141_31585||lt2gene Not-determined
SS_vs_k141_140396||lt2gene Low-quality
SS_vs_k141_43786||lt2gene Low-quality
SS_vs_k141_12913||lt2gene Low-quality
SS_vs_k141_37301||lt2gene Low-quality
SS_vs_k141_110118||lt2gene Low-quality
SS_vs_k141_85985||lt2gene Low-quality
SS_vs_k141_61426||lt2gene Low-quality
SS_vs_k141_140431||lt2gene Low-quality
SS_vs_k141_104272||lt2gene Low-quality
SS_vs_k141_37339||lt2gene Low-quality
SS_vs_k141_12952||lt2gene Low-quality
SS_vs_k141_61454||lt2gene Not-determined
SS_vs_k141_140451||lt2gene Not-determined
SS_vs_k141_37371||lt2gene Not-determined
SS_vs_k141_134276||lt2gene Not-determined
SS_vs_k141_37375||lt2gene Not-determined
SS_vs_k141_73422||lt2gene Not-determined
SS_vs_k141_73428||lt2gene Low-quality
SS_vs_k141_104305||lt2gene Not-determined
SS_vs_k141_140465||lt2gene Low-quality
SS_vs_k141_134291||lt2gene Low-quality
SS_vs_k141_55292||lt2gene Low-quality
SS_vs_k141_43847||lt2gene Low-quality
SS_vs_k141_134311||lt2gene Not-determined
SS_vs_k141_97806||lt2gene Low-quality
SS_vs_k141_134312||lt2gene Low-quality
SS_vs_k141_61546||lt2gene Not-determined
SS_vs_k141_19379||lt2gene Low-quality
SS_vs_k141_61552||lt2gene Low-quality
SS_vs_k141_123055||lt2gene Low-quality
SS_vs_k141_37409||lt2gene Not-determined
SS_vs_k141_13029||lt2gene Not-determined
SS_vs_k141_61578||lt2gene Not-determined
SS_vs_k141_79385||lt2gene Low-quality
SS_vs_k141_104350||lt2gene Low-quality
SS_vs_k141_24818||lt2gene Low-quality
SS_vs_k141_37464||lt2gene Low-quality
SS_vs_k141_67679||lt2gene Low-quality
SS_vs_k141_73511||lt2gene Not-determined
SS_vs_k141_73522||lt2gene Low-quality
SS_vs_k141_134345||lt2gene Not-determined
SS_vs_k141_97872||lt2gene Not-determined
SS_vs_k141_123113||lt2gene Low-quality
SS_vs_k141_128295||lt2gene Not-determined
SS_vs_k141_92027||lt2gene Low-quality
SS_vs_k141_31721||lt2gene Low-quality
SS_vs_k141_116738||lt2gene Low-quality
SS_vs_k141_19467||lt2gene Not-determined
SS_vs_k141_116760||lt2gene Low-quality
SS_vs_k141_19501||lt2gene Low-quality
SS_vs_k141_24835||lt2gene Low-quality
SS_vs_k141_31758||lt2gene Low-quality
SS_vs_k141_110329||lt2gene Low-quality
SS_vs_k141_128321||lt2gene Low-quality
SS_vs_k141_92087||lt2gene Not-determined
SS_vs_k141_37508||lt2gene Low-quality
SS_vs_k141_92091||lt2gene Not-determined
SS_vs_k141_67784||lt2gene Low-quality
SS_vs_k141_61728||lt2gene Not-determined
SS_vs_k141_31778||lt2gene Low-quality
SS_vs_k141_134403||lt2gene Not-determined
SS_vs_k141_116796||lt2gene Low-quality
SS_vs_k141_7366||lt2gene Low-quality
SS_vs_k141_116804||lt2gene Low-quality
SS_vs_k141_79442||lt2gene Not-determined
SS_vs_k141_134413||lt2gene Low-quality
SS_vs_k141_134415||lt2gene Low-quality
SS_vs_k141_134419||lt2gene Low-quality
SS_vs_k141_7377||lt2gene Low-quality
SS_vs_k141_7383||lt2gene Low-quality
SS_vs_k141_61747||lt2gene Low-quality
SS_vs_k141_61756||lt2gene Not-determined
SS_vs_k141_31808||lt2gene Low-quality
SS_vs_k141_128394||lt2gene Not-determined
SS_vs_k141_61783||lt2gene Low-quality
SS_vs_k141_110403||lt2gene Low-quality
SS_vs_k141_61805||lt2gene Low-quality
SS_vs_k141_13214||lt2gene Low-quality
SS_vs_k141_128405||lt2gene Not-determined
SS_vs_k141_73662||lt2gene Low-quality
SS_vs_k141_49577||lt2gene Not-determined
SS_vs_k141_116880||lt2gene Low-quality
SS_vs_k141_49578||lt2gene Low-quality
SS_vs_k141_73666||lt2gene Not-determined
SS_vs_k141_110434||lt2gene Low-quality
SS_vs_k141_128415||lt2gene Low-quality
SS_vs_k141_110443||lt2gene Low-quality
SS_vs_k141_31864||lt2gene Low-quality
SS_vs_k141_73698||lt2gene Low-quality
SS_vs_k141_73699||lt2gene Not-determined
SS_vs_k141_67872||lt2gene Low-quality
SS_vs_k141_134513||lt2gene Low-quality
SS_vs_k141_92238||lt2gene Low-quality
SS_vs_k141_86131||lt2gene Low-quality
SS_vs_k141_116951||lt2gene Low-quality
SS_vs_k141_86143||lt2gene Not-determined
SS_vs_k141_92259||lt2gene Low-quality
SS_vs_k141_67891||lt2gene Low-quality
SS_vs_k141_86149||lt2gene Low-quality
SS_vs_k141_92290||lt2gene Not-determined
SS_vs_k141_24869||lt2gene Low-quality
SS_vs_k141_92295||lt2gene Not-determined
SS_vs_k141_97991||lt2gene Low-quality
SS_vs_k141_86185||lt2gene Low-quality
SS_vs_k141_92330||lt2gene Low-quality
SS_vs_k141_31906||lt2gene Low-quality
SS_vs_k141_117021||lt2gene Low-quality
SS_vs_k141_73775||lt2gene Low-quality
SS_vs_k141_37652||lt2gene Low-quality
SS_vs_k141_140568||lt2gene Low-quality
SS_vs_k141_1555||lt2gene Low-quality
SS_vs_k141_61830||lt2gene Low-quality
SS_vs_k141_1558||lt2gene Low-quality
SS_vs_k141_67937||lt2gene Low-quality
SS_vs_k141_92376||lt2gene Low-quality
SS_vs_k141_92384||lt2gene Not-determined
SS_vs_k141_31975||lt2gene Low-quality
SS_vs_k141_61879||lt2gene Low-quality
SS_vs_k141_128477||lt2gene Low-quality
SS_vs_k141_123335||lt2gene Low-quality
SS_vs_k141_123337||lt2gene Low-quality
SS_vs_k141_19574||lt2gene Low-quality
SS_vs_k141_19576||lt2gene Low-quality
SS_vs_k141_117071||lt2gene Low-quality
SS_vs_k141_86222||lt2gene Low-quality
SS_vs_k141_140629||lt2gene Low-quality
SS_vs_k141_140631||lt2gene Low-quality
SS_vs_k141_98019||lt2gene Low-quality
SS_vs_k141_32004||lt2gene Not-determined
SS_vs_k141_98025||lt2gene Not-determined
SS_vs_k141_117094||lt2gene Low-quality
SS_vs_k141_92460||lt2gene Low-quality
SS_vs_k141_73838||lt2gene Low-quality
SS_vs_k141_61896||lt2gene Not-determined
SS_vs_k141_104471||lt2gene Low-quality
SS_vs_k141_32026||lt2gene Low-quality
SS_vs_k141_140665||lt2gene Not-determined
SS_vs_k141_92480||lt2gene Low-quality
SS_vs_k141_104487||lt2gene Low-quality
SS_vs_k141_49801||lt2gene Low-quality
SS_vs_k141_67996||lt2gene Low-quality
SS_vs_k141_140689||lt2gene Low-quality
SS_vs_k141_44039||lt2gene Low-quality
SS_vs_k141_32058||lt2gene Low-quality
SS_vs_k141_110619||lt2gene Low-quality
SS_vs_k141_92502||lt2gene Low-quality
SS_vs_k141_37739||lt2gene Not-determined
SS_vs_k141_32065||lt2gene Low-quality
SS_vs_k141_49824||lt2gene Not-determined
SS_vs_k141_92506||lt2gene Low-quality
SS_vs_k141_44063||lt2gene Low-quality
SS_vs_k141_110624||lt2gene Low-quality
SS_vs_k141_117157||lt2gene Low-quality
SS_vs_k141_49837||lt2gene Low-quality
SS_vs_k141_55438||lt2gene Low-quality
SS_vs_k141_104527||lt2gene Low-quality
SS_vs_k141_104534||lt2gene Not-determined
SS_vs_k141_37775||lt2gene Not-determined
SS_vs_k141_128500||lt2gene Low-quality
SS_vs_k141_32104||lt2gene Not-determined
SS_vs_k141_123417||lt2gene Low-quality
SS_vs_k141_123418||lt2gene Low-quality
SS_vs_k141_61906||lt2gene Low-quality
SS_vs_k141_1664||lt2gene Low-quality
SS_vs_k141_19639||lt2gene Low-quality
SS_vs_k141_44105||lt2gene Low-quality
SS_vs_k141_61924||lt2gene Low-quality
SS_vs_k141_55487||lt2gene Low-quality
SS_vs_k141_86250||lt2gene Low-quality
SS_vs_k141_49875||lt2gene Low-quality
SS_vs_k141_44119||lt2gene Not-determined
SS_vs_k141_49886||lt2gene Low-quality
SS_vs_k141_44128||lt2gene Not-determined
SS_vs_k141_123467||lt2gene Low-quality
SS_vs_k141_44137||lt2gene Low-quality
SS_vs_k141_44138||lt2gene Low-quality
SS_vs_k141_92610||lt2gene Low-quality
SS_vs_k141_73872||lt2gene Low-quality
SS_vs_k141_7537||lt2gene Low-quality
SS_vs_k141_73877||lt2gene Not-determined
SS_vs_k141_49926||lt2gene Low-quality
SS_vs_k141_49934||lt2gene Low-quality
SS_vs_k141_128624||lt2gene Low-quality
SS_vs_k141_128625||lt2gene Low-quality
SS_vs_k141_44181||lt2gene Not-determined
SS_vs_k141_92654||lt2gene Low-quality
SS_vs_k141_117238||lt2gene Low-quality
SS_vs_k141_92678||lt2gene Low-quality
SS_vs_k141_68148||lt2gene Low-quality
SS_vs_k141_79560||lt2gene Low-quality
SS_vs_k141_110760||lt2gene Low-quality
SS_vs_k141_110765||lt2gene Not-determined
SS_vs_k141_110775||lt2gene Low-quality
SS_vs_k141_1737||lt2gene Low-quality
SS_vs_k141_68173||lt2gene Low-quality
SS_vs_k141_49997||lt2gene Low-quality
SS_vs_k141_19686||lt2gene Low-quality
SS_vs_k141_37856||lt2gene Low-quality
SS_vs_k141_7602||lt2gene Low-quality
SS_vs_k141_7603||lt2gene Low-quality
SS_vs_k141_19710||lt2gene Low-quality
SS_vs_k141_55574||lt2gene Not-determined
SS_vs_k141_140782||lt2gene Not-determined
SS_vs_k141_140784||lt2gene Low-quality
SS_vs_k141_32205||lt2gene Low-quality
SS_vs_k141_134633||lt2gene Low-quality
SS_vs_k141_37881||lt2gene Low-quality
SS_vs_k141_50056||lt2gene Low-quality
SS_vs_k141_44243||lt2gene Low-quality
SS_vs_k141_55591||lt2gene Low-quality
SS_vs_k141_24963||lt2gene Low-quality
SS_vs_k141_73938||lt2gene Low-quality
SS_vs_k141_32237||lt2gene Low-quality
SS_vs_k141_24965||lt2gene Low-quality
SS_vs_k141_62026||lt2gene Low-quality
SS_vs_k141_13516||lt2gene Low-quality
SS_vs_k141_32279||lt2gene Low-quality
SS_vs_k141_68223||lt2gene Low-quality
SS_vs_k141_86408||lt2gene Low-quality
SS_vs_k141_7647||lt2gene Low-quality
SS_vs_k141_128799||lt2gene Low-quality
SS_vs_k141_44321||lt2gene Low-quality
SS_vs_k141_32312||lt2gene Low-quality
SS_vs_k141_25031||lt2gene Low-quality
SS_vs_k141_128817||lt2gene Low-quality
SS_vs_k141_86427||lt2gene Low-quality
SS_vs_k141_13610||lt2gene Low-quality
SS_vs_k141_1876||lt2gene Low-quality
SS_vs_k141_25066||lt2gene Low-quality
SS_vs_k141_25067||lt2gene Low-quality
SS_vs_k141_50165||lt2gene Not-determined
SS_vs_k141_73982||lt2gene Not-determined
SS_vs_k141_86471||lt2gene Not-determined
SS_vs_k141_1908||lt2gene Low-quality
SS_vs_k141_86479||lt2gene Not-determined
SS_vs_k141_1926||lt2gene Low-quality
SS_vs_k141_117380||lt2gene Low-quality
SS_vs_k141_62127||lt2gene Low-quality
SS_vs_k141_1936||lt2gene Low-quality
SS_vs_k141_25102||lt2gene Low-quality
SS_vs_k141_44373||lt2gene Low-quality
SS_vs_k141_1942||lt2gene Low-quality
SS_vs_k141_32384||lt2gene Low-quality
SS_vs_k141_44381||lt2gene Not-determined
SS_vs_k141_117402||lt2gene Low-quality
SS_vs_k141_7726||lt2gene Low-quality
SS_vs_k141_98195||lt2gene Low-quality
SS_vs_k141_38048||lt2gene Low-quality
SS_vs_k141_86503||lt2gene Low-quality
SS_vs_k141_110875||lt2gene Not-determined
SS_vs_k141_44422||lt2gene Low-quality
SS_vs_k141_32409||lt2gene Low-quality
SS_vs_k141_7748||lt2gene Low-quality
SS_vs_k141_32412||lt2gene Low-quality
SS_vs_k141_44433||lt2gene Low-quality
SS_vs_k141_79663||lt2gene Low-quality
SS_vs_k141_50224||lt2gene Low-quality
SS_vs_k141_104838||lt2gene Low-quality
SS_vs_k141_134751||lt2gene Not-determined
SS_vs_k141_110926||lt2gene Low-quality
SS_vs_k141_7807||lt2gene Low-quality
SS_vs_k141_110961||lt2gene Low-quality
SS_vs_k141_55667||lt2gene Not-determined
SS_vs_k141_110963||lt2gene Low-quality
SS_vs_k141_50274||lt2gene Low-quality
SS_vs_k141_110970||lt2gene Low-quality
SS_vs_k141_92880||lt2gene Low-quality
SS_vs_k141_79694||lt2gene Low-quality
SS_vs_k141_25182||lt2gene Low-quality
SS_vs_k141_7859||lt2gene Low-quality
SS_vs_k141_55691||lt2gene Low-quality
SS_vs_k141_50345||lt2gene Not-determined
SS_vs_k141_92929||lt2gene Low-quality
SS_vs_k141_117524||lt2gene Low-quality
SS_vs_k141_128937||lt2gene Low-quality
SS_vs_k141_117530||lt2gene Not-determined
SS_vs_k141_134850||lt2gene Not-determined
SS_vs_k141_134853||lt2gene Low-quality
SS_vs_k141_140947||lt2gene Low-quality
SS_vs_k141_111024||lt2gene Low-quality
SS_vs_k141_92954||lt2gene Low-quality
SS_vs_k141_140952||lt2gene Low-quality
SS_vs_k141_86619||lt2gene Low-quality
SS_vs_k141_128965||lt2gene Low-quality
SS_vs_k141_111046||lt2gene Low-quality
SS_vs_k141_92980||lt2gene Low-quality
SS_vs_k141_128988||lt2gene Low-quality
SS_vs_k141_79740||lt2gene Low-quality
SS_vs_k141_128996||lt2gene Low-quality
SS_vs_k141_32505||lt2gene Low-quality
SS_vs_k141_2159||lt2gene Low-quality
SS_vs_k141_117566||lt2gene Low-quality
SS_vs_k141_19852||lt2gene Not-determined
SS_vs_k141_68405||lt2gene Not-determined
SS_vs_k141_68407||lt2gene Not-determined
SS_vs_k141_19858||lt2gene Low-quality
SS_vs_k141_7984||lt2gene Low-quality
SS_vs_k141_13833||lt2gene Not-determined
SS_vs_k141_111123||lt2gene Not-determined
SS_vs_k141_25266||lt2gene Low-quality
SS_vs_k141_79759||lt2gene Low-quality
SS_vs_k141_2207||lt2gene Low-quality
SS_vs_k141_105001||lt2gene Low-quality
SS_vs_k141_105002||lt2gene Low-quality
SS_vs_k141_13865||lt2gene Low-quality
SS_vs_k141_8034||lt2gene Low-quality
SS_vs_k141_68444||lt2gene Low-quality
SS_vs_k141_2215||lt2gene Not-determined
SS_vs_k141_98231||lt2gene Low-quality
SS_vs_k141_32538||lt2gene Low-quality
SS_vs_k141_25296||lt2gene Low-quality
SS_vs_k141_129049||lt2gene Low-quality
SS_vs_k141_62254||lt2gene Low-quality
SS_vs_k141_8043||lt2gene Not-determined
SS_vs_k141_140993||lt2gene Low-quality
SS_vs_k141_50476||lt2gene Low-quality
SS_vs_k141_123697||lt2gene Low-quality
SS_vs_k141_105062||lt2gene Low-quality
SS_vs_k141_8064||lt2gene Low-quality
SS_vs_k141_55845||lt2gene Low-quality
SS_vs_k141_55848||lt2gene Low-quality
SS_vs_k141_8075||lt2gene Low-quality
SS_vs_k141_44577||lt2gene Low-quality
SS_vs_k141_44582||lt2gene Low-quality
SS_vs_k141_93019||lt2gene Not-determined
SS_vs_k141_93021||lt2gene Not-determined
SS_vs_k141_93029||lt2gene Low-quality
SS_vs_k141_141069||lt2gene Low-quality
SS_vs_k141_79828||lt2gene Low-quality
SS_vs_k141_105113||lt2gene Low-quality
SS_vs_k141_129199||lt2gene Low-quality
SS_vs_k141_68452||lt2gene Low-quality
SS_vs_k141_68459||lt2gene Low-quality
SS_vs_k141_13973||lt2gene Low-quality
SS_vs_k141_13981||lt2gene Low-quality
SS_vs_k141_134918||lt2gene Not-determined
SS_vs_k141_105158||lt2gene Low-quality
SS_vs_k141_117765||lt2gene Low-quality
SS_vs_k141_68480||lt2gene Low-quality
SS_vs_k141_25322||lt2gene Low-quality
SS_vs_k141_105176||lt2gene Not-determined
SS_vs_k141_68484||lt2gene Low-quality
SS_vs_k141_68486||lt2gene Low-quality
SS_vs_k141_86756||lt2gene Low-quality
SS_vs_k141_86762||lt2gene Not-determined
SS_vs_k141_105208||lt2gene Low-quality
SS_vs_k141_44656||lt2gene Low-quality
SS_vs_k141_105210||lt2gene Low-quality
SS_vs_k141_44659||lt2gene Not-determined
SS_vs_k141_32575||lt2gene Low-quality
SS_vs_k141_2417||lt2gene Low-quality
SS_vs_k141_134945||lt2gene Low-quality
SS_vs_k141_20002||lt2gene Low-quality
SS_vs_k141_50573||lt2gene Not-determined
SS_vs_k141_62331||lt2gene Low-quality
SS_vs_k141_62332||lt2gene Low-quality
SS_vs_k141_32608||lt2gene Not-determined
SS_vs_k141_74091||lt2gene Low-quality
SS_vs_k141_50582||lt2gene Not-determined
SS_vs_k141_32617||lt2gene Not-determined
SS_vs_k141_14042||lt2gene Low-quality
SS_vs_k141_32623||lt2gene Not-determined
SS_vs_k141_50594||lt2gene Low-quality
SS_vs_k141_32625||lt2gene Low-quality
SS_vs_k141_14058||lt2gene Not-determined
SS_vs_k141_2446||lt2gene Low-quality
SS_vs_k141_44717||lt2gene Not-determined
SS_vs_k141_105296||lt2gene Low-quality
SS_vs_k141_129224||lt2gene Not-determined
SS_vs_k141_44732||lt2gene Low-quality
SS_vs_k141_105309||lt2gene Low-quality
SS_vs_k141_117812||lt2gene Low-quality
SS_vs_k141_98311||lt2gene Low-quality
SS_vs_k141_117857||lt2gene Low-quality
SS_vs_k141_14094||lt2gene Low-quality
SS_vs_k141_2498||lt2gene Low-quality
SS_vs_k141_68571||lt2gene Low-quality
SS_vs_k141_105369||lt2gene Low-quality
SS_vs_k141_2500||lt2gene Low-quality
SS_vs_k141_50679||lt2gene Low-quality
SS_vs_k141_32713||lt2gene Low-quality
SS_vs_k141_38240||lt2gene Low-quality
SS_vs_k141_68580||lt2gene Low-quality
SS_vs_k141_44758||lt2gene Low-quality
SS_vs_k141_123888||lt2gene Low-quality
SS_vs_k141_14134||lt2gene Low-quality
SS_vs_k141_98326||lt2gene Low-quality
SS_vs_k141_8221||lt2gene Low-quality
SS_vs_k141_8225||lt2gene Low-quality
SS_vs_k141_55913||lt2gene Low-quality
SS_vs_k141_8252||lt2gene Not-determined
SS_vs_k141_14157||lt2gene Low-quality
SS_vs_k141_8259||lt2gene Low-quality
SS_vs_k141_50699||lt2gene Low-quality
SS_vs_k141_117905||lt2gene Low-quality
SS_vs_k141_117907||lt2gene Not-determined
SS_vs_k141_123926||lt2gene Low-quality
SS_vs_k141_50716||lt2gene Low-quality
SS_vs_k141_123931||lt2gene Low-quality
SS_vs_k141_55958||lt2gene Low-quality
SS_vs_k141_20124||lt2gene Low-quality
SS_vs_k141_105438||lt2gene Low-quality
SS_vs_k141_141211||lt2gene Not-determined
SS_vs_k141_20144||lt2gene Low-quality
SS_vs_k141_62416||lt2gene Low-quality
SS_vs_k141_2537||lt2gene Not-determined
SS_vs_k141_62439||lt2gene Not-determined
SS_vs_k141_141249||lt2gene Low-quality
SS_vs_k141_111213||lt2gene Not-determined
SS_vs_k141_14225||lt2gene Not-determined
SS_vs_k141_2554||lt2gene Low-quality
SS_vs_k141_86881||lt2gene Low-quality
SS_vs_k141_135043||lt2gene Not-determined
SS_vs_k141_79935||lt2gene Low-quality
SS_vs_k141_14275||lt2gene Low-quality
SS_vs_k141_79945||lt2gene Not-determined
SS_vs_k141_105540||lt2gene Low-quality
SS_vs_k141_105548||lt2gene Low-quality
SS_vs_k141_50774||lt2gene Low-quality
SS_vs_k141_129437||lt2gene Low-quality
SS_vs_k141_8396||lt2gene Low-quality
SS_vs_k141_55993||lt2gene Low-quality
SS_vs_k141_105603||lt2gene Low-quality
SS_vs_k141_68652||lt2gene Not-determined
SS_vs_k141_68657||lt2gene Low-quality
SS_vs_k141_25474||lt2gene Low-quality
SS_vs_k141_25489||lt2gene Low-quality
SS_vs_k141_111318||lt2gene Low-quality
SS_vs_k141_105667||lt2gene Low-quality
SS_vs_k141_20222||lt2gene Low-quality
SS_vs_k141_50837||lt2gene Not-determined
SS_vs_k141_20230||lt2gene Low-quality
SS_vs_k141_20233||lt2gene Low-quality
SS_vs_k141_2622||lt2gene Low-quality
SS_vs_k141_8484||lt2gene Low-quality
SS_vs_k141_141362||lt2gene Not-determined
SS_vs_k141_129453||lt2gene Not-determined
SS_vs_k141_141383||lt2gene Low-quality
SS_vs_k141_20267||lt2gene Low-quality
SS_vs_k141_124086||lt2gene Low-quality
SS_vs_k141_25542||lt2gene Low-quality
SS_vs_k141_124103||lt2gene Low-quality
SS_vs_k141_86999||lt2gene Not-determined
SS_vs_k141_118146||lt2gene Not-determined
SS_vs_k141_50926||lt2gene Not-determined
SS_vs_k141_14343||lt2gene Low-quality
SS_vs_k141_98355||lt2gene Low-quality
SS_vs_k141_135090||lt2gene Low-quality
SS_vs_k141_105739||lt2gene Low-quality
SS_vs_k141_50962||lt2gene Low-quality
SS_vs_k141_111350||lt2gene Low-quality
SS_vs_k141_32842||lt2gene Not-determined
SS_vs_k141_50996||lt2gene Low-quality
SS_vs_k141_68699||lt2gene Low-quality
SS_vs_k141_141428||lt2gene Not-determined
SS_vs_k141_38381||lt2gene Low-quality
SS_vs_k141_25681||lt2gene Low-quality
SS_vs_k141_135144||lt2gene Low-quality
SS_vs_k141_68743||lt2gene Low-quality
SS_vs_k141_68745||lt2gene Low-quality
SS_vs_k141_135151||lt2gene Low-quality
SS_vs_k141_2803||lt2gene Low-quality
SS_vs_k141_135168||lt2gene Not-determined
SS_vs_k141_105796||lt2gene Low-quality
SS_vs_k141_98409||lt2gene Not-determined
SS_vs_k141_93338||lt2gene Low-quality
SS_vs_k141_93343||lt2gene Low-quality
SS_vs_k141_135183||lt2gene Low-quality
SS_vs_k141_93346||lt2gene Not-determined
SS_vs_k141_2825||lt2gene Low-quality
SS_vs_k141_20353||lt2gene Low-quality
SS_vs_k141_62589||lt2gene Not-determined
SS_vs_k141_105822||lt2gene Low-quality
SS_vs_k141_111441||lt2gene Low-quality
SS_vs_k141_98424||lt2gene Low-quality
SS_vs_k141_93365||lt2gene Low-quality
SS_vs_k141_105842||lt2gene Low-quality
SS_vs_k141_51052||lt2gene Low-quality
SS_vs_k141_93374||lt2gene Low-quality
SS_vs_k141_98437||lt2gene Low-quality
SS_vs_k141_93380||lt2gene Low-quality
SS_vs_k141_93397||lt2gene Low-quality
SS_vs_k141_51082||lt2gene Low-quality
SS_vs_k141_56056||lt2gene Not-determined
SS_vs_k141_93437||lt2gene Low-quality
SS_vs_k141_80095||lt2gene Low-quality
SS_vs_k141_56058||lt2gene Low-quality
SS_vs_k141_68839||lt2gene Not-determined
SS_vs_k141_93456||lt2gene Low-quality
SS_vs_k141_124261||lt2gene Not-determined
SS_vs_k141_2899||lt2gene Low-quality
SS_vs_k141_124273||lt2gene Low-quality
SS_vs_k141_74212||lt2gene Not-determined
SS_vs_k141_62607||lt2gene Low-quality
SS_vs_k141_124280||lt2gene Low-quality
SS_vs_k141_56072||lt2gene Not-determined
SS_vs_k141_20449||lt2gene Low-quality
SS_vs_k141_124292||lt2gene Low-quality
SS_vs_k141_124293||lt2gene Low-quality
SS_vs_k141_56077||lt2gene Low-quality
SS_vs_k141_20457||lt2gene Low-quality
SS_vs_k141_68897||lt2gene Low-quality
SS_vs_k141_135228||lt2gene Low-quality
SS_vs_k141_62656||lt2gene Low-quality
SS_vs_k141_62657||lt2gene Not-determined
SS_vs_k141_2965||lt2gene Low-quality
SS_vs_k141_141479||lt2gene Low-quality
SS_vs_k141_14500||lt2gene Not-determined
SS_vs_k141_124342||lt2gene Not-determined
SS_vs_k141_80141||lt2gene Low-quality
SS_vs_k141_25788||lt2gene Low-quality
SS_vs_k141_14508||lt2gene Low-quality
SS_vs_k141_141486||lt2gene Low-quality
SS_vs_k141_56086||lt2gene Low-quality
SS_vs_k141_38495||lt2gene Low-quality
SS_vs_k141_124357||lt2gene Low-quality
SS_vs_k141_68935||lt2gene Low-quality
SS_vs_k141_68936||lt2gene Low-quality
SS_vs_k141_38525||lt2gene Low-quality
SS_vs_k141_38528||lt2gene Low-quality
SS_vs_k141_93612||lt2gene Low-quality
SS_vs_k141_56092||lt2gene Low-quality
SS_vs_k141_105940||lt2gene Low-quality
SS_vs_k141_93619||lt2gene Low-quality
SS_vs_k141_129730||lt2gene Low-quality
SS_vs_k141_93641||lt2gene Low-quality
SS_vs_k141_56131||lt2gene Low-quality
SS_vs_k141_56132||lt2gene Low-quality
SS_vs_k141_118313||lt2gene Low-quality
SS_vs_k141_135269||lt2gene Low-quality
SS_vs_k141_118338||lt2gene Not-determined
SS_vs_k141_105977||lt2gene Not-determined
SS_vs_k141_3094||lt2gene Low-quality
SS_vs_k141_118353||lt2gene Not-determined
SS_vs_k141_8724||lt2gene Not-determined
SS_vs_k141_135293||lt2gene Low-quality
SS_vs_k141_93675||lt2gene Low-quality
SS_vs_k141_62719||lt2gene Low-quality
SS_vs_k141_141533||lt2gene Low-quality
SS_vs_k141_105999||lt2gene Not-determined
SS_vs_k141_80210||lt2gene Low-quality
SS_vs_k141_124426||lt2gene Low-quality
SS_vs_k141_8752||lt2gene Low-quality
SS_vs_k141_129784||lt2gene Low-quality
SS_vs_k141_62742||lt2gene Not-determined
SS_vs_k141_62747||lt2gene Low-quality
SS_vs_k141_20544||lt2gene Low-quality
SS_vs_k141_124455||lt2gene Not-determined
SS_vs_k141_124458||lt2gene Low-quality
SS_vs_k141_141564||lt2gene Not-determined
SS_vs_k141_74328||lt2gene Low-quality
SS_vs_k141_129819||lt2gene Low-quality
SS_vs_k141_106041||lt2gene Low-quality
SS_vs_k141_33006||lt2gene Low-quality
SS_vs_k141_98494||lt2gene Low-quality
SS_vs_k141_38593||lt2gene Not-determined
SS_vs_k141_98495||lt2gene Low-quality
SS_vs_k141_106073||lt2gene Low-quality
SS_vs_k141_51219||lt2gene Low-quality
SS_vs_k141_111509||lt2gene Low-quality
SS_vs_k141_118424||lt2gene Low-quality
SS_vs_k141_3184||lt2gene Low-quality
SS_vs_k141_51236||lt2gene Low-quality
SS_vs_k141_51254||lt2gene Low-quality
SS_vs_k141_111535||lt2gene Low-quality
SS_vs_k141_106117||lt2gene Not-determined
SS_vs_k141_106119||lt2gene Low-quality
SS_vs_k141_51276||lt2gene Low-quality
SS_vs_k141_44992||lt2gene Low-quality
SS_vs_k141_3243||lt2gene Not-determined
SS_vs_k141_3252||lt2gene Low-quality
SS_vs_k141_51305||lt2gene Not-determined
SS_vs_k141_8778||lt2gene Not-determined
SS_vs_k141_69005||lt2gene Low-quality
SS_vs_k141_14698||lt2gene Low-quality
SS_vs_k141_93725||lt2gene Low-quality
SS_vs_k141_118498||lt2gene Low-quality
SS_vs_k141_14707||lt2gene Low-quality
SS_vs_k141_69022||lt2gene Low-quality
SS_vs_k141_20607||lt2gene Not-determined
SS_vs_k141_129862||lt2gene Low-quality
SS_vs_k141_69041||lt2gene Not-determined
SS_vs_k141_111547||lt2gene Low-quality

| | |
|---|---|
| SS_vs_k141_38677\|\|lt2gene | Not-determined |
| SS_vs_k141_124608\|\|lt2gene | Not-determined |
| SS_vs_k141_20643\|\|lt2gene | Not-determined |
| SS_vs_k141_56300\|\|lt2gene | Not-determined |
| SS_vs_k141_124633\|\|lt2gene | Not-determined |
| SS_vs_k141_14753\|\|lt2gene | Low-quality |
| SS_vs_k141_56308\|\|lt2gene | Low-quality |
| SS_vs_k141_56314\|\|lt2gene | Low-quality |
| SS_vs_k141_111611\|\|lt2gene | Low-quality |
| SS_vs_k141_135400\|\|lt2gene | Low-quality |
| SS_vs_k141_135401\|\|lt2gene | Low-quality |
| SS_vs_k141_80335\|\|lt2gene | Low-quality |
| SS_vs_k141_14796\|\|lt2gene | Low-quality |
| SS_vs_k141_14806\|\|lt2gene | Not-determined |
| SS_vs_k141_80347\|\|lt2gene | Not-determined |
| SS_vs_k141_111640\|\|lt2gene | Low-quality |
| SS_vs_k141_3422\|\|lt2gene | Low-quality |
| SS_vs_k141_45125\|\|lt2gene | Not-determined |
| SS_vs_k141_45135\|\|lt2gene | Low-quality |
| SS_vs_k141_45148\|\|lt2gene | Low-quality |
| SS_vs_k141_98623\|\|lt2gene | Low-quality |
| SS_vs_k141_129931\|\|lt2gene | Low-quality |
| SS_vs_k141_98646\|\|lt2gene | Low-quality |
| SS_vs_k141_74513\|\|lt2gene | Low-quality |
| SS_vs_k141_93842\|\|lt2gene | Low-quality |
| SS_vs_k141_118595\|\|lt2gene | Low-quality |
| SS_vs_k141_98665\|\|lt2gene | Low-quality |
| SS_vs_k141_87063\|\|lt2gene | Low-quality |
| SS_vs_k141_8883\|\|lt2gene | Low-quality |
| SS_vs_k141_45198\|\|lt2gene | Low-quality |
| SS_vs_k141_87065\|\|lt2gene | Low-quality |
| SS_vs_k141_80391\|\|lt2gene | Low-quality |
| SS_vs_k141_69175\|\|lt2gene | Low-quality |
| SS_vs_k141_56394\|\|lt2gene | Low-quality |
| SS_vs_k141_118631\|\|lt2gene | Not-determined |
| SS_vs_k141_14977\|\|lt2gene | Low-quality |
| SS_vs_k141_45219\|\|lt2gene | Low-quality |
| SS_vs_k141_69206\|\|lt2gene | Low-quality |
| SS_vs_k141_69211\|\|lt2gene | Low-quality |
| SS_vs_k141_8907\|\|lt2gene | Low-quality |
| SS_vs_k141_98705\|\|lt2gene | Not-determined |
| SS_vs_k141_135458\|\|lt2gene | Low-quality |
| SS_vs_k141_111665\|\|lt2gene | Low-quality |
| SS_vs_k141_135463\|\|lt2gene | Not-determined |
| SS_vs_k141_15002\|\|lt2gene | Low-quality |
| SS_vs_k141_141747\|\|lt2gene | Low-quality |
| SS_vs_k141_93908\|\|lt2gene | Low-quality |
| SS_vs_k141_106235\|\|lt2gene | Low-quality |
| SS_vs_k141_45270\|\|lt2gene | Not-determined |
| SS_vs_k141_8938\|\|lt2gene | Not-determined |
| SS_vs_k141_69262\|\|lt2gene | Low-quality |
| SS_vs_k141_45282\|\|lt2gene | Low-quality |
| SS_vs_k141_3589\|\|lt2gene | Low-quality |
| SS_vs_k141_69284\|\|lt2gene | Low-quality |
| SS_vs_k141_20813\|\|lt2gene | Low-quality |
| SS_vs_k141_63086\|\|lt2gene | Low-quality |
| SS_vs_k141_87155\|\|lt2gene | Low-quality |
| SS_vs_k141_74519\|\|lt2gene | Low-quality |
| SS_vs_k141_74521\|\|lt2gene | Low-quality |
| SS_vs_k141_130015\|\|lt2gene | Low-quality |
| SS_vs_k141_69320\|\|lt2gene | Low-quality |
| SS_vs_k141_45327\|\|lt2gene | Low-quality |
| SS_vs_k141_118700\|\|lt2gene | Not-determined |
| SS_vs_k141_20876\|\|lt2gene | Low-quality |
| SS_vs_k141_69334\|\|lt2gene | Low-quality |
| SS_vs_k141_3637\|\|lt2gene | Low-quality |
| SS_vs_k141_45369\|\|lt2gene | Low-quality |
| SS_vs_k141_69354\|\|lt2gene | Low-quality |
| SS_vs_k141_33252\|\|lt2gene | Not-determined |
| SS_vs_k141_38810\|\|lt2gene | Low-quality |
| SS_vs_k141_38816\|\|lt2gene | Not-determined |
| SS_vs_k141_20905\|\|lt2gene | Not-determined |
| SS_vs_k141_15097\|\|lt2gene | Not-determined |
| SS_vs_k141_3654\|\|lt2gene | Not-determined |
| SS_vs_k141_38847\|\|lt2gene | Not-determined |
| SS_vs_k141_20930\|\|lt2gene | Low-quality |
| SS_vs_k141_111802\|\|lt2gene | Low-quality |
| SS_vs_k141_20941\|\|lt2gene | Low-quality |
| SS_vs_k141_130071\|\|lt2gene | Low-quality |
| SS_vs_k141_98794\|\|lt2gene | Low-quality |
| SS_vs_k141_141932\|\|lt2gene | Low-quality |
| SS_vs_k141_93940\|\|lt2gene | Low-quality |
| SS_vs_k141_20948\|\|lt2gene | Low-quality |
| SS_vs_k141_51697\|\|lt2gene | Low-quality |
| SS_vs_k141_3732\|\|lt2gene | Low-quality |
| SS_vs_k141_33324\|\|lt2gene | Low-quality |
| SS_vs_k141_20965\|\|lt2gene | Low-quality |
| SS_vs_k141_20967\|\|lt2gene | Low-quality |
| SS_vs_k141_20970\|\|lt2gene | Low-quality |
| SS_vs_k141_63281\|\|lt2gene | Not-determined |
| SS_vs_k141_63291\|\|lt2gene | Low-quality |
| SS_vs_k141_130123\|\|lt2gene | Low-quality |
| SS_vs_k141_141960\|\|lt2gene | Low-quality |
| SS_vs_k141_93987\|\|lt2gene | Low-quality |
| SS_vs_k141_130132\|\|lt2gene | Low-quality |
| SS_vs_k141_111882\|\|lt2gene | Low-quality |
| SS_vs_k141_3776\|\|lt2gene | Low-quality |
| SS_vs_k141_118771\|\|lt2gene | Low-quality |
| SS_vs_k141_15194\|\|lt2gene | Not-determined |
| SS_vs_k141_80470\|\|lt2gene | Low-quality |
| SS_vs_k141_15219\|\|lt2gene | Low-quality |
| SS_vs_k141_135509\|\|lt2gene | Low-quality |
| SS_vs_k141_124797\|\|lt2gene | Low-quality |
| SS_vs_k141_51799\|\|lt2gene | Low-quality |
| SS_vs_k141_33384\|\|lt2gene | Low-quality |
| SS_vs_k141_21016\|\|lt2gene | Low-quality |
| SS_vs_k141_33397\|\|lt2gene | Not-determined |
| SS_vs_k141_87262\|\|lt2gene | Low-quality |
| SS_vs_k141_87263\|\|lt2gene | Low-quality |
| SS_vs_k141_56528\|\|lt2gene | Low-quality |
| SS_vs_k141_63329\|\|lt2gene | Low-quality |
| SS_vs_k141_56536\|\|lt2gene | Low-quality |
| SS_vs_k141_15272\|\|lt2gene | Low-quality |
| SS_vs_k141_106507\|\|lt2gene | Low-quality |
| SS_vs_k141_51816\|\|lt2gene | Not-determined |
| SS_vs_k141_106525\|\|lt2gene | Not-determined |
| SS_vs_k141_15278\|\|lt2gene | Low-quality |
| SS_vs_k141_15286\|\|lt2gene | Low-quality |
| SS_vs_k141_112041\|\|lt2gene | Low-quality |
| SS_vs_k141_94053\|\|lt2gene | Not-determined |
| SS_vs_k141_15288\|\|lt2gene | Low-quality |
| SS_vs_k141_94071\|\|lt2gene | Low-quality |
| SS_vs_k141_9083\|\|lt2gene | Low-quality |
| SS_vs_k141_15309\|\|lt2gene | Low-quality |
| SS_vs_k141_63406\|\|lt2gene | Low-quality |
| SS_vs_k141_112090\|\|lt2gene | Low-quality |
| SS_vs_k141_98905\|\|lt2gene | Not-determined |
| SS_vs_k141_98907\|\|lt2gene | Low-quality |
| SS_vs_k141_106592\|\|lt2gene | Low-quality |
| SS_vs_k141_142068\|\|lt2gene | Low-quality |
| SS_vs_k141_9097\|\|lt2gene | Not-determined |
| SS_vs_k141_106600\|\|lt2gene | Not-determined |
| SS_vs_k141_3924\|\|lt2gene | Low-quality |
| SS_vs_k141_118957\|\|lt2gene | Not-determined |
| SS_vs_k141_33522\|\|lt2gene | Low-quality |
| SS_vs_k141_106625\|\|lt2gene | Low-quality |
| SS_vs_k141_9123\|\|lt2gene | Low-quality |
| SS_vs_k141_124914\|\|lt2gene | Not-determined |
| SS_vs_k141_94139\|\|lt2gene | Low-quality |
| SS_vs_k141_51921\|\|lt2gene | Low-quality |
| SS_vs_k141_118985\|\|lt2gene | Low-quality |
| SS_vs_k141_118986\|\|lt2gene | Not-determined |
| SS_vs_k141_45480\|\|lt2gene | Low-quality |
| SS_vs_k141_38985\|\|lt2gene | Low-quality |
| SS_vs_k141_21107\|\|lt2gene | Low-quality |
| SS_vs_k141_130330\|\|lt2gene | Low-quality |
| SS_vs_k141_3985\|\|lt2gene | Low-quality |
| SS_vs_k141_9141\|\|lt2gene | Low-quality |
| SS_vs_k141_39006\|\|lt2gene | Low-quality |
| SS_vs_k141_124961\|\|lt2gene | Low-quality |
| SS_vs_k141_45527\|\|lt2gene | Low-quality |
| SS_vs_k141_112176\|\|lt2gene | Low-quality |
| SS_vs_k141_33581\|\|lt2gene | Low-quality |
| SS_vs_k141_21135\|\|lt2gene | Low-quality |
| SS_vs_k141_130353\|\|lt2gene | Low-quality |
| SS_vs_k141_9201\|\|lt2gene | Low-quality |
| SS_vs_k141_4002\|\|lt2gene | Low-quality |
| SS_vs_k141_130359\|\|lt2gene | Low-quality |
| SS_vs_k141_4008\|\|lt2gene | Low-quality |
| SS_vs_k141_51983\|\|lt2gene | Low-quality |
| SS_vs_k141_98956\|\|lt2gene | Not-determined |
| SS_vs_k141_142188\|\|lt2gene | Low-quality |
| SS_vs_k141_51997\|\|lt2gene | Low-quality |
| SS_vs_k141_63564\|\|lt2gene | Low-quality |
| SS_vs_k141_98968\|\|lt2gene | Low-quality |
| SS_vs_k141_63589\|\|lt2gene | Low-quality |
| SS_vs_k141_94213\|\|lt2gene | Low-quality |
| SS_vs_k141_106777\|\|lt2gene | Low-quality |
| SS_vs_k141_39063\|\|lt2gene | Not-determined |
| SS_vs_k141_21189\|\|lt2gene | Low-quality |
| SS_vs_k141_94272\|\|lt2gene | Low-quality |
| SS_vs_k141_52073\|\|lt2gene | Low-quality |
| SS_vs_k141_94284\|\|lt2gene | Low-quality |
| SS_vs_k141_135674\|\|lt2gene | Low-quality |
| SS_vs_k141_99002\|\|lt2gene | Not-determined |
| SS_vs_k141_94298\|\|lt2gene | Low-quality |
| SS_vs_k141_9336\|\|lt2gene | Low-quality |
| SS_vs_k141_52121\|\|lt2gene | Low-quality |
| SS_vs_k141_45587\|\|lt2gene | Low-quality |
| SS_vs_k141_125077\|\|lt2gene | Low-quality |
| SS_vs_k141_52127\|\|lt2gene | Low-quality |
| SS_vs_k141_94334\|\|lt2gene | Low-quality |
| SS_vs_k141_4096\|\|lt2gene | Not-determined |
| SS_vs_k141_142249\|\|lt2gene | Low-quality |
| SS_vs_k141_94343\|\|lt2gene | Low-quality |
| SS_vs_k141_130488\|\|lt2gene | Low-quality |
| SS_vs_k141_94346\|\|lt2gene | Low-quality |
| SS_vs_k141_142265\|\|lt2gene | Not-determined |
| SS_vs_k141_87524\|\|lt2gene | Not-determined |
| SS_vs_k141_80525\|\|lt2gene | Low-quality |
| SS_vs_k141_142273\|\|lt2gene | Not-determined |
| SS_vs_k141_33770\|\|lt2gene | Not-determined |
| SS_vs_k141_87532\|\|lt2gene | Low-quality |
| SS_vs_k141_106913\|\|lt2gene | Low-quality |
| SS_vs_k141_142279\|\|lt2gene | Not-determined |
| SS_vs_k141_106920\|\|lt2gene | Low-quality |
| SS_vs_k141_52195\|\|lt2gene | Not-determined |
| SS_vs_k141_106941\|\|lt2gene | Low-quality |
| SS_vs_k141_106945\|\|lt2gene | Low-quality |
| SS_vs_k141_26179\|\|lt2gene | Low-quality |
| SS_vs_k141_135738\|\|lt2gene | Low-quality |
| SS_vs_k141_87578\|\|lt2gene | Not-determined |
| SS_vs_k141_99042\|\|lt2gene | Low-quality |
| SS_vs_k141_4209\|\|lt2gene | Low-quality |
| SS_vs_k141_39204\|\|lt2gene | Low-quality |
| SS_vs_k141_125222\|\|lt2gene | Low-quality |
| SS_vs_k141_142350\|\|lt2gene | Low-quality |
| SS_vs_k141_142360\|\|lt2gene | Low-quality |
| SS_vs_k141_130532\|\|lt2gene | Not-determined |
| SS_vs_k141_87619\|\|lt2gene | Low-quality |
| SS_vs_k141_119069\|\|lt2gene | Low-quality |
| SS_vs_k141_4265\|\|lt2gene | Low-quality |
| SS_vs_k141_112375\|\|lt2gene | Not-determined |
| SS_vs_k141_99054\|\|lt2gene | Not-determined |
| SS_vs_k141_99057\|\|lt2gene | Low-quality |
| SS_vs_k141_119094\|\|lt2gene | Low-quality |
| SS_vs_k141_4293\|\|lt2gene | Low-quality |
| SS_vs_k141_112403\|\|lt2gene | Low-quality |
| SS_vs_k141_4304\|\|lt2gene | Low-quality |
| SS_vs_k141_125288\|\|lt2gene | Low-quality |
| SS_vs_k141_80643\|\|lt2gene | Low-quality |
| SS_vs_k141_26235\|\|lt2gene | Not-determined |
| SS_vs_k141_45716\|\|lt2gene | Low-quality |
| SS_vs_k141_99071\|\|lt2gene | Low-quality |
| SS_vs_k141_74671\|\|lt2gene | Low-quality |
| SS_vs_k141_135768\|\|lt2gene | Low-quality |
| SS_vs_k141_4329\|\|lt2gene | Low-quality |
| SS_vs_k141_26247\|\|lt2gene | Low-quality |
| SS_vs_k141_69594\|\|lt2gene | Low-quality |
| SS_vs_k141_4334\|\|lt2gene | Low-quality |
| SS_vs_k141_9443\|\|lt2gene | Low-quality |
| SS_vs_k141_107095\|\|lt2gene | Low-quality |
| SS_vs_k141_33927\|\|lt2gene | Low-quality |
| SS_vs_k141_107101\|\|lt2gene | Low-quality |
| SS_vs_k141_130655\|\|lt2gene | Low-quality |
| SS_vs_k141_9459\|\|lt2gene | Low-quality |
| SS_vs_k141_33936\|\|lt2gene | Low-quality |
| SS_vs_k141_99092\|\|lt2gene | Low-quality |
| SS_vs_k141_142442\|\|lt2gene | Low-quality |
| SS_vs_k141_80723\|\|lt2gene | Low-quality |
| SS_vs_k141_52334\|\|lt2gene | Low-quality |
| SS_vs_k141_52337\|\|lt2gene | Not-determined |
| SS_vs_k141_142451\|\|lt2gene | Low-quality |
| SS_vs_k141_130708\|\|lt2gene | Low-quality |
| SS_vs_k141_119109\|\|lt2gene | Low-quality |
| SS_vs_k141_87736\|\|lt2gene | Low-quality |
| SS_vs_k141_69624\|\|lt2gene | Low-quality |
| SS_vs_k141_99106\|\|lt2gene | Low-quality |
| SS_vs_k141_99107\|\|lt2gene | Low-quality |
| SS_vs_k141_26289\|\|lt2gene | Low-quality |
| SS_vs_k141_69647\|\|lt2gene | Low-quality |
| SS_vs_k141_135810\|\|lt2gene | Low-quality |
| SS_vs_k141_52389\|\|lt2gene | Low-quality |
| SS_vs_k141_52390\|\|lt2gene | Low-quality |
| SS_vs_k141_87752\|\|lt2gene | Low-quality |
| SS_vs_k141_69669\|\|lt2gene | Low-quality |
| SS_vs_k141_52396\|\|lt2gene | Low-quality |
| SS_vs_k141_34025\|\|lt2gene | Low-quality |
| SS_vs_k141_45838\|\|lt2gene | Low-quality |
| SS_vs_k141_69674\|\|lt2gene | Low-quality |
| SS_vs_k141_99147\|\|lt2gene | Low-quality |
| SS_vs_k141_130759\|\|lt2gene | Low-quality |
| SS_vs_k141_4444\|\|lt2gene | Low-quality |
| SS_vs_k141_99158\|\|lt2gene | Not-determined |
| SS_vs_k141_4453\|\|lt2gene | Low-quality |
| SS_vs_k141_52415\|\|lt2gene | Low-quality |
| SS_vs_k141_125403\|\|lt2gene | Not-determined |
| SS_vs_k141_15529\|\|lt2gene | Low-quality |
| SS_vs_k141_135830\|\|lt2gene | Low-quality |
| SS_vs_k141_119175\|\|lt2gene | Low-quality |
| SS_vs_k141_34071\|\|lt2gene | Low-quality |
| SS_vs_k141_130777\|\|lt2gene | Low-quality |
| SS_vs_k141_125433\|\|lt2gene | Low-quality |
| SS_vs_k141_99202\|\|lt2gene | Low-quality |
| SS_vs_k141_130785\|\|lt2gene | Low-quality |
| SS_vs_k141_142486\|\|lt2gene | Low-quality |
| SS_vs_k141_4501\|\|lt2gene | Low-quality |
| SS_vs_k141_34102\|\|lt2gene | Not-determined |
| SS_vs_k141_94354\|\|lt2gene | Low-quality |
| SS_vs_k141_45928\|\|lt2gene | Low-quality |
| SS_vs_k141_99217\|\|lt2gene | Low-quality |
| SS_vs_k141_125484\|\|lt2gene | Not-determined |
| SS_vs_k141_34126\|\|lt2gene | Not-determined |
| SS_vs_k141_107227\|\|lt2gene | Low-quality |
| SS_vs_k141_69687\|\|lt2gene | Not-determined |
| SS_vs_k141_52514\|\|lt2gene | Not-determined |
| SS_vs_k141_125523\|\|lt2gene | Low-quality |
| SS_vs_k141_107266\|\|lt2gene | Low-quality |
| SS_vs_k141_99249\|\|lt2gene | Low-quality |
| SS_vs_k141_45968\|\|lt2gene | Low-quality |
| SS_vs_k141_9582\|\|lt2gene | Low-quality |
| SS_vs_k141_69708\|\|lt2gene | Not-determined |
| SS_vs_k141_52534\|\|lt2gene | Low-quality |
| SS_vs_k141_125544\|\|lt2gene | Low-quality |
| SS_vs_k141_107287\|\|lt2gene | Not-determined |
| SS_vs_k141_34141\|\|lt2gene | Low-quality |
| SS_vs_k141_52545\|\|lt2gene | Not-determined |
| SS_vs_k141_125559\|\|lt2gene | Low-quality |
| SS_vs_k141_21262\|\|lt2gene | Low-quality |
| SS_vs_k141_21263\|\|lt2gene | Not-determined |
| SS_vs_k141_119265\|\|lt2gene | Low-quality |
| SS_vs_k141_119266\|\|lt2gene | Low-quality |
| SS_vs_k141_125600\|\|lt2gene | Not-determined |
| SS_vs_k141_63781\|\|lt2gene | Not-determined |
| SS_vs_k141_39381\|\|lt2gene | Not-determined |
| SS_vs_k141_46020\|\|lt2gene | Low-quality |
| SS_vs_k141_99304\|\|lt2gene | Not-determined |
| SS_vs_k141_46029\|\|lt2gene | Low-quality |
| SS_vs_k141_107330\|\|lt2gene | Low-quality |
| SS_vs_k141_63811\|\|lt2gene | Low-quality |
| SS_vs_k141_63813\|\|lt2gene | Low-quality |
| SS_vs_k141_112518\|\|lt2gene | Low-quality |
| SS_vs_k141_39399\|\|lt2gene | Not-determined |
| SS_vs_k141_119277\|\|lt2gene | Low-quality |
| SS_vs_k141_63820\|\|lt2gene | Low-quality |
| SS_vs_k141_107358\|\|lt2gene | Low-quality |
| SS_vs_k141_107364\|\|lt2gene | Low-quality |
| SS_vs_k141_112550\|\|lt2gene | Low-quality |
| SS_vs_k141_107385\|\|lt2gene | Low-quality |
| SS_vs_k141_87884\|\|lt2gene | Low-quality |
| SS_vs_k141_135903\|\|lt2gene | Low-quality |
| SS_vs_k141_87901\|\|lt2gene | Low-quality |
| SS_vs_k141_9630\|\|lt2gene | Low-quality |
| SS_vs_k141_99366\|\|lt2gene | Low-quality |
| SS_vs_k141_9653\|\|lt2gene | Not-determined |
| SS_vs_k141_94396\|\|lt2gene | Low-quality |
| SS_vs_k141_125699\|\|lt2gene | Low-quality |
| SS_vs_k141_135932\|\|lt2gene | Low-quality |
| SS_vs_k141_135933\|\|lt2gene | Low-quality |
| SS_vs_k141_34232\|\|lt2gene | Low-quality |
| SS_vs_k141_112624\|\|lt2gene | Low-quality |
| SS_vs_k141_87956\|\|lt2gene | Low-quality |
| SS_vs_k141_107455\|\|lt2gene | Low-quality |
| SS_vs_k141_34246\|\|lt2gene | Low-quality |
| SS_vs_k141_119405\|\|lt2gene | Low-quality |
| SS_vs_k141_135955\|\|lt2gene | Low-quality |
| SS_vs_k141_63896\|\|lt2gene | Not-determined |
| SS_vs_k141_52624\|\|lt2gene | Not-determined |
| SS_vs_k141_125721\|\|lt2gene | Low-quality |
| SS_vs_k141_119420\|\|lt2gene | Low-quality |
| SS_vs_k141_4571\|\|lt2gene | Low-quality |
| SS_vs_k141_9700\|\|lt2gene | Low-quality |
| SS_vs_k141_21369\|\|lt2gene | Low-quality |
| SS_vs_k141_9704\|\|lt2gene | Low-quality |
| SS_vs_k141_63919\|\|lt2gene | Low-quality |
| SS_vs_k141_99409\|\|lt2gene | Not-determined |
| SS_vs_k141_4590\|\|lt2gene | Low-quality |
| SS_vs_k141_21420\|\|lt2gene | Low-quality |
| SS_vs_k141_112681\|\|lt2gene | Low-quality |
| SS_vs_k141_142606\|\|lt2gene | Low-quality |
| SS_vs_k141_94508\|\|lt2gene | Low-quality |
| SS_vs_k141_34255\|\|lt2gene | Low-quality |
| SS_vs_k141_125803\|\|lt2gene | Low-quality |
| SS_vs_k141_4643\|\|lt2gene | Not-determined |
| SS_vs_k141_125827\|\|lt2gene | Low-quality |
| SS_vs_k141_21458\|\|lt2gene | Low-quality |
| SS_vs_k141_125832\|\|lt2gene | Not-determined |
| SS_vs_k141_130957\|\|lt2gene | Low-quality |
| SS_vs_k141_63956\|\|lt2gene | Not-determined |
| SS_vs_k141_15711\|\|lt2gene | Low-quality |
| SS_vs_k141_136020\|\|lt2gene | Not-determined |
| SS_vs_k141_99424\|\|lt2gene | Low-quality |
| SS_vs_k141_63970\|\|lt2gene | Low-quality |
| SS_vs_k141_4672\|\|lt2gene | Low-quality |
| SS_vs_k141_107543\|\|lt2gene | Low-quality |
| SS_vs_k141_15738\|\|lt2gene | Low-quality |
| SS_vs_k141_88102\|\|lt2gene | Not-determined |
| SS_vs_k141_142644\|\|lt2gene | Low-quality |
| SS_vs_k141_136026\|\|lt2gene | Not-determined |
| SS_vs_k141_112748\|\|lt2gene | Low-quality |
| SS_vs_k141_80790\|\|lt2gene | Not-determined |
| SS_vs_k141_112762\|\|lt2gene | Low-quality |
| SS_vs_k141_52777\|\|lt2gene | Low-quality |
| SS_vs_k141_99493\|\|lt2gene | Low-quality |
| SS_vs_k141_4712\|\|lt2gene | Low-quality |
| SS_vs_k141_94591\|\|lt2gene | Low-quality |
| SS_vs_k141_39575\|\|lt2gene | Low-quality |
| SS_vs_k141_112788\|\|lt2gene | Not-determined |
| SS_vs_k141_69905\|\|lt2gene | Low-quality |
| SS_vs_k141_21549\|\|lt2gene | Low-quality |
| SS_vs_k141_94620\|\|lt2gene | Not-determined |
| SS_vs_k141_9791\|\|lt2gene | Not-determined |
| SS_vs_k141_130967\|\|lt2gene | Low-quality |
| SS_vs_k141_26376\|\|lt2gene | Low-quality |
| SS_vs_k141_21597\|\|lt2gene | Low-quality |
| SS_vs_k141_21600\|\|lt2gene | Low-quality |
| SS_vs_k141_119486\|\|lt2gene | Low-quality |
| SS_vs_k141_112856\|\|lt2gene | Low-quality |
| SS_vs_k141_112862\|\|lt2gene | Not-determined |
| SS_vs_k141_21611\|\|lt2gene | Low-quality |
| SS_vs_k141_4767\|\|lt2gene | Low-quality |
| SS_vs_k141_4776\|\|lt2gene | Not-determined |
| SS_vs_k141_26400\|\|lt2gene | Low-quality |
| SS_vs_k141_88217\|\|lt2gene | Not-determined |
| SS_vs_k141_80893\|\|lt2gene | Not-determined |
| SS_vs_k141_107608\|\|lt2gene | Low-quality |
| SS_vs_k141_119558\|\|lt2gene | Low-quality |
| SS_vs_k141_9876\|\|lt2gene | Low-quality |
| SS_vs_k141_94647\|\|lt2gene | Low-quality |
| SS_vs_k141_94648\|\|lt2gene | Low-quality |
| SS_vs_k141_52857\|\|lt2gene | Low-quality |
| SS_vs_k141_52864\|\|lt2gene | Low-quality |
| SS_vs_k141_9882\|\|lt2gene | Low-quality |
| SS_vs_k141_125998\|\|lt2gene | Low-quality |
| SS_vs_k141_94667\|\|lt2gene | Not-determined |
| SS_vs_k141_34488\|\|lt2gene | Low-quality |
| SS_vs_k141_112925\|\|lt2gene | Low-quality |
| SS_vs_k141_4883\|\|lt2gene | Low-quality |
| SS_vs_k141_94694\|\|lt2gene | Low-quality |
| SS_vs_k141_4885\|\|lt2gene | Not-determined |
| SS_vs_k141_112936\|\|lt2gene | Low-quality |
| SS_vs_k141_34517\|\|lt2gene | Low-quality |
| SS_vs_k141_80923\|\|lt2gene | Not-determined |
| SS_vs_k141_74861\|\|lt2gene | Low-quality |
| SS_vs_k141_34529\|\|lt2gene | Not-determined |
| SS_vs_k141_21646\|\|lt2gene | Low-quality |
| SS_vs_k141_99605\|\|lt2gene | Low-quality |
| SS_vs_k141_34542\|\|lt2gene | Low-quality |
| SS_vs_k141_131097\|\|lt2gene | Low-quality |
| SS_vs_k141_69955\|\|lt2gene | Low-quality |
| SS_vs_k141_99613\|\|lt2gene | Low-quality |
| SS_vs_k141_131103\|\|lt2gene | Low-quality |
| SS_vs_k141_88254\|\|lt2gene | Not-determined |
| SS_vs_k141_64068\|\|lt2gene | Low-quality |
| SS_vs_k141_142723\|\|lt2gene | Low-quality |
| SS_vs_k141_131114\|\|lt2gene | Low-quality |
| SS_vs_k141_26429\|\|lt2gene | Low-quality |
| SS_vs_k141_9933\|\|lt2gene | Low-quality |
| SS_vs_k141_88263\|\|lt2gene | Not-determined |
| SS_vs_k141_142744\|\|lt2gene | Not-determined |
| SS_vs_k141_88287\|\|lt2gene | Low-quality |
| SS_vs_k141_64107\|\|lt2gene | Low-quality |
| SS_vs_k141_34607\|\|lt2gene | Low-quality |
| SS_vs_k141_69979\|\|lt2gene | Low-quality |
| SS_vs_k141_99667\|\|lt2gene | Low-quality |
| SS_vs_k141_34622\|\|lt2gene | Low-quality |
| SS_vs_k141_21727\|\|lt2gene | Low-quality |
| SS_vs_k141_74897\|\|lt2gene | Not-determined |
| SS_vs_k141_56868\|\|lt2gene | Low-quality |
| SS_vs_k141_142797\|\|lt2gene | Low-quality |
| SS_vs_k141_99682\|\|lt2gene | Low-quality |
| SS_vs_k141_46085\|\|lt2gene | Low-quality |
| SS_vs_k141_112985\|\|lt2gene | Not-determined |
| SS_vs_k141_15878\|\|lt2gene | Not-determined |
| SS_vs_k141_5053\|\|lt2gene | Low-quality |
| SS_vs_k141_46113\|\|lt2gene | Low-quality |
| SS_vs_k141_74942\|\|lt2gene | Low-quality |
| SS_vs_k141_34663\|\|lt2gene | Low-quality |
| SS_vs_k141_15925\|\|lt2gene | Not-determined |
| SS_vs_k141_88384\|\|lt2gene | Low-quality |
| SS_vs_k141_34691\|\|lt2gene | Low-quality |
| SS_vs_k141_70033\|\|lt2gene | Low-quality |
| SS_vs_k141_142829\|\|lt2gene | Low-quality |
| SS_vs_k141_136154\|\|lt2gene | Low-quality |
| SS_vs_k141_56940\|\|lt2gene | Low-quality |
| SS_vs_k141_56942\|\|lt2gene | Low-quality |
| SS_vs_k141_53085\|\|lt2gene | Low-quality |
| SS_vs_k141_5086\|\|lt2gene | Low-quality |
| SS_vs_k141_142844\|\|lt2gene | Low-quality |
| SS_vs_k141_80983\|\|lt2gene | Low-quality |
| SS_vs_k141_15950\|\|lt2gene | Low-quality |
| SS_vs_k141_88437\|\|lt2gene | Low-quality |
| SS_vs_k141_34731\|\|lt2gene | Low-quality |
| SS_vs_k141_94876\|\|lt2gene | Not-determined |
| SS_vs_k141_46144\|\|lt2gene | Low-quality |
| SS_vs_k141_99739\|\|lt2gene | Low-quality |
| SS_vs_k141_15994\|\|lt2gene | Low-quality |
| SS_vs_k141_15997\|\|lt2gene | Low-quality |
| SS_vs_k141_94899\|\|lt2gene | Low-quality |
| SS_vs_k141_70075\|\|lt2gene | Low-quality |
| SS_vs_k141_53132\|\|lt2gene | Low-quality |
| SS_vs_k141_64208\|\|lt2gene | Low-quality |
| SS_vs_k141_34767\|\|lt2gene | Not-determined |
| SS_vs_k141_107853\|\|lt2gene | Not-determined |
| SS_vs_k141_5148\|\|lt2gene | Not-determined |
| SS_vs_k141_64217\|\|lt2gene | Low-quality |
| SS_vs_k141_126221\|\|lt2gene | Low-quality |
| SS_vs_k141_10064\|\|lt2gene | Low-quality |
| SS_vs_k141_94933\|\|lt2gene | Not-determined |
| SS_vs_k141_46180\|\|lt2gene | Low-quality |
| SS_vs_k141_5165\|\|lt2gene | Low-quality |
| SS_vs_k141_119658\|\|lt2gene | Low-quality |
| SS_vs_k141_126263\|\|lt2gene | Not-determined |
| SS_vs_k141_64240\|\|lt2gene | Low-quality |
| SS_vs_k141_70137\|\|lt2gene | Not-determined |
| SS_vs_k141_26553\|\|lt2gene | Low-quality |
| SS_vs_k141_34793\|\|lt2gene | Low-quality |
| SS_vs_k141_88512\|\|lt2gene | Low-quality |
| SS_vs_k141_136246\|\|lt2gene | Low-quality |
| SS_vs_k141_53184\|\|lt2gene | Low-quality |
| SS_vs_k141_136260\|\|lt2gene | Low-quality |
| SS_vs_k141_88534\|\|lt2gene | Low-quality |
| SS_vs_k141_26576\|\|lt2gene | Low-quality |
| SS_vs_k141_119724\|\|lt2gene | Low-quality |
| SS_vs_k141_113086\|\|lt2gene | Not-determined |
| SS_vs_k141_34839\|\|lt2gene | Low-quality |
| SS_vs_k141_70176\|\|lt2gene | Low-quality |
| SS_vs_k141_53234\|\|lt2gene | Low-quality |
| SS_vs_k141_131283\|\|lt2gene | Low-quality |
| SS_vs_k141_131286\|\|lt2gene | Low-quality |
| SS_vs_k141_10091\|\|lt2gene | Not-determined |
| SS_vs_k141_70210\|\|lt2gene | Not-determined |
| SS_vs_k141_70214\|\|lt2gene | Low-quality |
| SS_vs_k141_95032\|\|lt2gene | Low-quality |
| SS_vs_k141_107952\|\|lt2gene | Not-determined |
| SS_vs_k141_70219\|\|lt2gene | Low-quality |
| SS_vs_k141_10117\|\|lt2gene | Low-quality |
| SS_vs_k141_39715\|\|lt2gene | Low-quality |
| SS_vs_k141_26590\|\|lt2gene | Low-quality |
| SS_vs_k141_113140\|\|lt2gene | Low-quality |
| SS_vs_k141_136328\|\|lt2gene | Low-quality |
| SS_vs_k141_10123\|\|lt2gene | Not-determined |
| SS_vs_k141_119789\|\|lt2gene | Low-quality |
| SS_vs_k141_53291\|\|lt2gene | Not-determined |
| SS_vs_k141_81088\|\|lt2gene | Low-quality |
| SS_vs_k141_99839\|\|lt2gene | Not-determined |
| SS_vs_k141_99842\|\|lt2gene | Low-quality |
| SS_vs_k141_75017\|\|lt2gene | Low-quality |
| SS_vs_k141_81126\|\|lt2gene | Not-determined |
| SS_vs_k141_95071\|\|lt2gene | Low-quality |
| SS_vs_k141_16120\|\|lt2gene | Low-quality |
| SS_vs_k141_39800\|\|lt2gene | Low-quality |
| SS_vs_k141_39804\|\|lt2gene | Low-quality |
| SS_vs_k141_39809\|\|lt2gene | Low-quality |
| SS_vs_k141_142935\|\|lt2gene | Low-quality |
| SS_vs_k141_88677\|\|lt2gene | Low-quality |
| SS_vs_k141_108039\|\|lt2gene | Low-quality |
| SS_vs_k141_34888\|\|lt2gene | Low-quality |
| SS_vs_k141_57034\|\|lt2gene | Low-quality |
| SS_vs_k141_57038\|\|lt2gene | Low-quality |
| SS_vs_k141_5351\|\|lt2gene | Not-determined |
| SS_vs_k141_88692\|\|lt2gene | Low-quality |
| SS_vs_k141_70315\|\|lt2gene | Low-quality |
| SS_vs_k141_108048\|\|lt2gene | Low-quality |
| SS_vs_k141_95148\|\|lt2gene | Low-quality |
| SS_vs_k141_136385\|\|lt2gene | Low-quality |
| SS_vs_k141_136386\|\|lt2gene | Low-quality |
| SS_vs_k141_5377\|\|lt2gene | Not-determined |
| SS_vs_k141_5387\|\|lt2gene | Low-quality |
| SS_vs_k141_53377\|\|lt2gene | Low-quality |
| SS_vs_k141_119925\|\|lt2gene | Low-quality |
| SS_vs_k141_10234\|\|lt2gene | Not-determined |
| SS_vs_k141_113275\|\|lt2gene | Low-quality |
| SS_vs_k141_88748\|\|lt2gene | Low-quality |
| SS_vs_k141_136427\|\|lt2gene | Low-quality |
| SS_vs_k141_21896\|\|lt2gene | Low-quality |
| SS_vs_k141_21900\|\|lt2gene | Low-quality |
| SS_vs_k141_57075\|\|lt2gene | Low-quality |
| SS_vs_k141_113288\|\|lt2gene | Low-quality |
| SS_vs_k141_64316\|\|lt2gene | Low-quality |
| SS_vs_k141_95192\|\|lt2gene | Low-quality |
| SS_vs_k141_119965\|\|lt2gene | Low-quality |
| SS_vs_k141_88768\|\|lt2gene | Not-determined |
| SS_vs_k141_136443\|\|lt2gene | Not-determined |
| SS_vs_k141_34974\|\|lt2gene | Low-quality |
| SS_vs_k141_136455\|\|lt2gene | Low-quality |
| SS_vs_k141_108117\|\|lt2gene | Low-quality |
| SS_vs_k141_64378\|\|lt2gene | Low-quality |
| SS_vs_k141_35018\|\|lt2gene | Low-quality |
| SS_vs_k141_136471\|\|lt2gene | Low-quality |
| SS_vs_k141_136473\|\|lt2gene | Not-determined |
| SS_vs_k141_35038\|\|lt2gene | Low-quality |
| SS_vs_k141_57116\|\|lt2gene | Low-quality |
| SS_vs_k141_108136\|\|lt2gene | Low-quality |
| SS_vs_k141_5517\|\|lt2gene | Low-quality |
| SS_vs_k141_5520\|\|lt2gene | Low-quality |
| SS_vs_k141_99962\|\|lt2gene | Not-determined |
| SS_vs_k141_70362\|\|lt2gene | Not-determined |
| SS_vs_k141_108155\|\|lt2gene | Low-quality |
| SS_vs_k141_143009\|\|lt2gene | Low-quality |
| SS_vs_k141_131466\|\|lt2gene | Low-quality |
| SS_vs_k141_131471\|\|lt2gene | Low-quality |
| SS_vs_k141_39958\|\|lt2gene | Low-quality |
| SS_vs_k141_53487\|\|lt2gene | Low-quality |
| SS_vs_k141_131503\|\|lt2gene | Low-quality |
| SS_vs_k141_131504\|\|lt2gene | Not-determined |
| SS_vs_k141_53499\|\|lt2gene | Low-quality |
| SS_vs_k141_10390\|\|lt2gene | Low-quality |
| SS_vs_k141_16209\|\|lt2gene | Low-quality |
| SS_vs_k141_35127\|\|lt2gene | Not-determined |
| SS_vs_k141_126318\|\|lt2gene | Low-quality |
| SS_vs_k141_35150\|\|lt2gene | Low-quality |
| SS_vs_k141_143057\|\|lt2gene | Low-quality |
| SS_vs_k141_64485\|\|lt2gene | Low-quality |
| SS_vs_k141_108224\|\|lt2gene | Not-determined |
| SS_vs_k141_64491\|\|lt2gene | Low-quality |
| SS_vs_k141_75242\|\|lt2gene | Low-quality |
| SS_vs_k141_39999\|\|lt2gene | Low-quality |
| SS_vs_k141_35187\|\|lt2gene | Low-quality |

SS_vs_k141_64519||lt2gene Low-quality
SS_vs_k141_64524||lt2gene Low-quality
SS_vs_k141_57185||lt2gene Low-quality
SS_vs_k141_40016||lt2gene Low-quality
SS_vs_k141_81248||lt2gene Low-quality
SS_vs_k141_120120||lt2gene Not-determined
SS_vs_k141_143064||lt2gene Not-determined
SS_vs_k141_70517||lt2gene Low-quality
SS_vs_k141_40030||lt2gene Low-quality
SS_vs_k141_81258||lt2gene Low-quality
SS_vs_k141_126403||lt2gene Low-quality
SS_vs_k141_40041||lt2gene Low-quality
SS_vs_k141_88945||lt2gene Low-quality
SS_vs_k141_16325||lt2gene Low-quality
SS_vs_k141_88950||lt2gene Low-quality
SS_vs_k141_5551||lt2gene Low-quality
SS_vs_k141_143094||lt2gene Low-quality
SS_vs_k141_16347||lt2gene Not-determined
SS_vs_k141_35237||lt2gene Low-quality
SS_vs_k141_143102||lt2gene Low-quality
SS_vs_k141_99997||lt2gene Not-determined
SS_vs_k141_26804||lt2gene Low-quality
SS_vs_k141_57237||lt2gene Low-quality
SS_vs_k141_64626||lt2gene Low-quality
SS_vs_k141_35278||lt2gene Not-determined
SS_vs_k141_120174||lt2gene Not-determined
SS_vs_k141_81271||lt2gene Low-quality
SS_vs_k141_81272||lt2gene Not-determined
SS_vs_k141_136605||lt2gene Not-determined
SS_vs_k141_53555||lt2gene Low-quality
SS_vs_k141_5636||lt2gene Low-quality
SS_vs_k141_40136||lt2gene Not-determined
SS_vs_k141_64645||lt2gene Low-quality
SS_vs_k141_64654||lt2gene Low-quality
SS_vs_k141_40161||lt2gene Low-quality
SS_vs_k141_40175||lt2gene Low-quality
SS_vs_k141_64682||lt2gene Low-quality
SS_vs_k141_131566||lt2gene Not-determined
SS_vs_k141_131570||lt2gene Low-quality
SS_vs_k141_81338||lt2gene Low-quality
SS_vs_k141_100084||lt2gene Low-quality
SS_vs_k141_16509||lt2gene Low-quality
SS_vs_k141_126572||lt2gene Not-determined
SS_vs_k141_64727||lt2gene Low-quality
SS_vs_k141_131605||lt2gene Not-determined
SS_vs_k141_16535||lt2gene Not-determined
SS_vs_k141_75488||lt2gene Low-quality
SS_vs_k141_108482||lt2gene Low-quality
SS_vs_k141_100113||lt2gene Low-quality
SS_vs_k141_10431||lt2gene Low-quality
SS_vs_k141_131621||lt2gene Not-determined
SS_vs_k141_16556||lt2gene Low-quality
SS_vs_k141_113486||lt2gene Low-quality
SS_vs_k141_100164||lt2gene Low-quality
SS_vs_k141_131662||lt2gene Not-determined
SS_vs_k141_70658||lt2gene Low-quality
SS_vs_k141_53722||lt2gene Low-quality
SS_vs_k141_95514||lt2gene Low-quality
SS_vs_k141_5669||lt2gene Low-quality
SS_vs_k141_16589||lt2gene Low-quality
SS_vs_k141_16593||lt2gene Low-quality
SS_vs_k141_113523||lt2gene Low-quality
SS_vs_k141_70677||lt2gene Low-quality
SS_vs_k141_95549||lt2gene Low-quality
SS_vs_k141_89149||lt2gene Low-quality
SS_vs_k141_16638||lt2gene Low-quality
SS_vs_k141_26947||lt2gene Low-quality
SS_vs_k141_143276||lt2gene Not-determined
SS_vs_k141_16672||lt2gene Low-quality
SS_vs_k141_113577||lt2gene Low-quality
SS_vs_k141_143312||lt2gene Low-quality
SS_vs_k141_16690||lt2gene Low-quality
SS_vs_k141_113580||lt2gene Low-quality
SS_vs_k141_22178||lt2gene Low-quality
SS_vs_k141_143329||lt2gene Low-quality
SS_vs_k141_35392||lt2gene Low-quality
SS_vs_k141_40243||lt2gene Not-determined
SS_vs_k141_113598||lt2gene Not-determined
SS_vs_k141_75579||lt2gene Low-quality
SS_vs_k141_100246||lt2gene Low-quality
SS_vs_k141_40254||lt2gene Low-quality
SS_vs_k141_113626||lt2gene Not-determined
SS_vs_k141_113637||lt2gene Not-determined
SS_vs_k141_40275||lt2gene Low-quality
SS_vs_k141_70777||lt2gene Low-quality
SS_vs_k141_131728||lt2gene Low-quality
SS_vs_k141_40293||lt2gene Not-determined
SS_vs_k141_16753||lt2gene Not-determined
SS_vs_k141_16762||lt2gene Not-determined
SS_vs_k141_143332||lt2gene Low-quality
SS_vs_k141_143347||lt2gene Low-quality
SS_vs_k141_108640||lt2gene Low-quality
SS_vs_k141_113673||lt2gene Low-quality
SS_vs_k141_108649||lt2gene Not-determined
SS_vs_k141_113687||lt2gene Low-quality
SS_vs_k141_27045||lt2gene Low-quality
SS_vs_k141_126753||lt2gene Low-quality
SS_vs_k141_35531||lt2gene Low-quality
SS_vs_k141_35537||lt2gene Low-quality
SS_vs_k141_126767||lt2gene Low-quality
SS_vs_k141_131812||lt2gene Not-determined
SS_vs_k141_143387||lt2gene Low-quality
SS_vs_k141_35561||lt2gene Low-quality
SS_vs_k141_131839||lt2gene Low-quality
SS_vs_k141_35568||lt2gene Low-quality
SS_vs_k141_27073||lt2gene Low-quality
SS_vs_k141_35574||lt2gene Low-quality
SS_vs_k141_136766||lt2gene Low-quality
SS_vs_k141_126810||lt2gene Low-quality
SS_vs_k141_120291||lt2gene Low-quality
SS_vs_k141_113759||lt2gene Not-determined
SS_vs_k141_136777||lt2gene Low-quality
SS_vs_k141_113774||lt2gene Low-quality
SS_vs_k141_120328||lt2gene Low-quality
SS_vs_k141_108703||lt2gene Low-quality
SS_vs_k141_81465||lt2gene Low-quality
SS_vs_k141_5777||lt2gene Low-quality
SS_vs_k141_81489||lt2gene Low-quality
SS_vs_k141_126842||lt2gene Low-quality
SS_vs_k141_108744||lt2gene Low-quality
SS_vs_k141_108763||lt2gene Low-quality
SS_vs_k141_126876||lt2gene Low-quality
SS_vs_k141_89220||lt2gene Low-quality
SS_vs_k141_27152||lt2gene Not-determined
SS_vs_k141_81536||lt2gene Not-determined
SS_vs_k141_65027||lt2gene Low-quality
SS_vs_k141_81539||lt2gene Not-determined
SS_vs_k141_89243||lt2gene Low-quality
SS_vs_k141_95805||lt2gene Low-quality
SS_vs_k141_113837||lt2gene Low-quality
SS_vs_k141_120446||lt2gene Low-quality
SS_vs_k141_131910||lt2gene Not-determined
SS_vs_k141_70986||lt2gene Low-quality
SS_vs_k141_143525||lt2gene Low-quality
SS_vs_k141_89271||lt2gene Low-quality
SS_vs_k141_100349||lt2gene Low-quality
SS_vs_k141_113891||lt2gene Low-quality
SS_vs_k141_108778||lt2gene Low-quality
SS_vs_k141_108784||lt2gene Low-quality
SS_vs_k141_22382||lt2gene Not-determined
SS_vs_k141_27219||lt2gene Low-quality
SS_vs_k141_113911||lt2gene Low-quality
SS_vs_k141_75884||lt2gene Low-quality
SS_vs_k141_22387||lt2gene Low-quality
SS_vs_k141_53857||lt2gene Low-quality
SS_vs_k141_113918||lt2gene Low-quality
SS_vs_k141_65098||lt2gene Low-quality
SS_vs_k141_75890||lt2gene Not-determined
SS_vs_k141_126992||lt2gene Not-determined
SS_vs_k141_27243||lt2gene Not-determined
SS_vs_k141_10616||lt2gene Low-quality
SS_vs_k141_136865||lt2gene Low-quality
SS_vs_k141_136873||lt2gene Low-quality
SS_vs_k141_127000||lt2gene Low-quality
SS_vs_k141_120505||lt2gene Low-quality
SS_vs_k141_16961||lt2gene Low-quality
SS_vs_k141_65145||lt2gene Low-quality
SS_vs_k141_143598||lt2gene Low-quality
SS_vs_k141_113956||lt2gene Not-determined
SS_vs_k141_120529||lt2gene Low-quality
SS_vs_k141_22436||lt2gene Low-quality
SS_vs_k141_89359||lt2gene Not-determined
SS_vs_k141_108846||lt2gene Not-determined
SS_vs_k141_113980||lt2gene Low-quality
SS_vs_k141_65172||lt2gene Low-quality
SS_vs_k141_5852||lt2gene Low-quality
SS_vs_k141_127029||lt2gene Low-quality
SS_vs_k141_10704||lt2gene Low-quality
SS_vs_k141_113995||lt2gene Low-quality
SS_vs_k141_113998||lt2gene Low-quality
SS_vs_k141_17026||lt2gene Low-quality
SS_vs_k141_127048||lt2gene Low-quality
SS_vs_k141_100473||lt2gene Not-determined
SS_vs_k141_100475||lt2gene Low-quality
SS_vs_k141_100477||lt2gene Low-quality
SS_vs_k141_22462||lt2gene Low-quality
SS_vs_k141_75987||lt2gene Low-quality
SS_vs_k141_100487||lt2gene Low-quality
SS_vs_k141_100493||lt2gene Not-determined
SS_vs_k141_53959||lt2gene Not-determined
SS_vs_k141_114016||lt2gene Low-quality
SS_vs_k141_114024||lt2gene Low-quality
SS_vs_k141_75996||lt2gene Not-determined
SS_vs_k141_53984||lt2gene Low-quality
SS_vs_k141_53986||lt2gene Low-quality
SS_vs_k141_22492||lt2gene Not-determined
SS_vs_k141_5892||lt2gene Low-quality
SS_vs_k141_108897||lt2gene Low-quality
SS_vs_k141_22499||lt2gene Not-determined
SS_vs_k141_35811||lt2gene Low-quality
SS_vs_k141_95955||lt2gene Low-quality
SS_vs_k141_100509||lt2gene Low-quality
SS_vs_k141_95958||lt2gene Low-quality
SS_vs_k141_89412||lt2gene Low-quality
SS_vs_k141_10767||lt2gene Low-quality
SS_vs_k141_40402||lt2gene Low-quality
SS_vs_k141_108907||lt2gene Low-quality
SS_vs_k141_35838||lt2gene Low-quality
SS_vs_k141_22542||lt2gene Low-quality
SS_vs_k141_10784||lt2gene Low-quality
SS_vs_k141_76047||lt2gene Low-quality
SS_vs_k141_46627||lt2gene Not-determined
SS_vs_k141_46628||lt2gene Not-determined
SS_vs_k141_89460||lt2gene Low-quality
SS_vs_k141_35866||lt2gene Low-quality
SS_vs_k141_54075||lt2gene Low-quality
SS_vs_k141_54087||lt2gene Low-quality
SS_vs_k141_120644||lt2gene Low-quality
SS_vs_k141_5983||lt2gene Low-quality
SS_vs_k141_22643||lt2gene Low-quality
SS_vs_k141_22648||lt2gene Low-quality
SS_vs_k141_114114||lt2gene Low-quality
SS_vs_k141_132113||lt2gene Low-quality
SS_vs_k141_10824||lt2gene Low-quality
SS_vs_k141_127130||lt2gene Not-determined
SS_vs_k141_40463||lt2gene Not-determined
SS_vs_k141_46671||lt2gene Low-quality
SS_vs_k141_127138||lt2gene Low-quality
SS_vs_k141_114132||lt2gene Low-quality
SS_vs_k141_108976||lt2gene Not-determined
SS_vs_k141_127153||lt2gene Low-quality
SS_vs_k141_132131||lt2gene Low-quality
SS_vs_k141_108990||lt2gene Low-quality
SS_vs_k141_71101||lt2gene Not-determined
SS_vs_k141_108993||lt2gene Low-quality
SS_vs_k141_71102||lt2gene Low-quality
SS_vs_k141_54149||lt2gene Low-quality
SS_vs_k141_109001||lt2gene Low-quality
SS_vs_k141_35872||lt2gene Not-determined
SS_vs_k141_27260||lt2gene Low-quality
SS_vs_k141_89506||lt2gene Low-quality
SS_vs_k141_109013||lt2gene Low-quality
SS_vs_k141_137034||lt2gene Low-quality
SS_vs_k141_89516||lt2gene Low-quality
SS_vs_k141_109019||lt2gene Low-quality
SS_vs_k141_96028||lt2gene Low-quality
SS_vs_k141_27279||lt2gene Low-quality
SS_vs_k141_54185||lt2gene Low-quality
SS_vs_k141_17271||lt2gene Low-quality
SS_vs_k141_96044||lt2gene Not-determined
SS_vs_k141_81709||lt2gene Not-determined
SS_vs_k141_96069||lt2gene Low-quality
SS_vs_k141_54214||lt2gene Low-quality
SS_vs_k141_46739||lt2gene Low-quality
SS_vs_k141_46743||lt2gene Not-determined
SS_vs_k141_132169||lt2gene Not-determined
SS_vs_k141_35953||lt2gene Not-determined
SS_vs_k141_46788||lt2gene Low-quality
SS_vs_k141_114228||lt2gene Low-quality
SS_vs_k141_132184||lt2gene Not-determined
SS_vs_k141_54250||lt2gene Low-quality
SS_vs_k141_132195||lt2gene Low-quality
SS_vs_k141_22741||lt2gene Low-quality
SS_vs_k141_109098||lt2gene Not-determined
SS_vs_k141_40622||lt2gene Not-determined
SS_vs_k141_96131||lt2gene Low-quality
SS_vs_k141_35983||lt2gene Low-quality
SS_vs_k141_81763||lt2gene Not-determined
SS_vs_k141_54272||lt2gene Low-quality
SS_vs_k141_120807||lt2gene Low-quality
SS_vs_k141_71216||lt2gene Low-quality
SS_vs_k141_57561||lt2gene Low-quality
SS_vs_k141_40658||lt2gene Low-quality
SS_vs_k141_132214||lt2gene Low-quality
SS_vs_k141_54289||lt2gene Not-determined
SS_vs_k141_132220||lt2gene Low-quality
SS_vs_k141_132222||lt2gene Low-quality
SS_vs_k141_54302||lt2gene Low-quality
SS_vs_k141_27372||lt2gene Not-determined
SS_vs_k141_54306||lt2gene Low-quality
SS_vs_k141_114253||lt2gene Low-quality
SS_vs_k141_109153||lt2gene Low-quality
SS_vs_k141_54320||lt2gene Low-quality
SS_vs_k141_109157||lt2gene Low-quality
SS_vs_k141_17427||lt2gene Not-determined
SS_vs_k141_143669||lt2gene Not-determined
SS_vs_k141_22754||lt2gene Low-quality
SS_vs_k141_10902||lt2gene Not-determined
SS_vs_k141_22758||lt2gene Low-quality
SS_vs_k141_81820||lt2gene Low-quality
SS_vs_k141_22762||lt2gene Low-quality
SS_vs_k141_120903||lt2gene Not-determined
SS_vs_k141_114317||lt2gene Low-quality
SS_vs_k141_22782||lt2gene Not-determined
SS_vs_k141_89646||lt2gene Low-quality
SS_vs_k141_109205||lt2gene Low-quality
SS_vs_k141_137102||lt2gene Low-quality
SS_vs_k141_46941||lt2gene Low-quality
SS_vs_k141_137104||lt2gene Low-quality
SS_vs_k141_89668||lt2gene Low-quality
SS_vs_k141_114358||lt2gene Low-quality
SS_vs_k141_65456||lt2gene Low-quality
SS_vs_k141_114372||lt2gene Not-determined
SS_vs_k141_46984||lt2gene Low-quality
SS_vs_k141_17519||lt2gene Low-quality
SS_vs_k141_109224||lt2gene Low-quality
SS_vs_k141_109238||lt2gene Low-quality
SS_vs_k141_27428||lt2gene Low-quality
SS_vs_k141_36074||lt2gene Low-quality
SS_vs_k141_114450||lt2gene Low-quality
SS_vs_k141_114453||lt2gene Low-quality
SS_vs_k141_17536||lt2gene Low-quality
SS_vs_k141_109270||lt2gene Low-quality
SS_vs_k141_17540||lt2gene Low-quality
SS_vs_k141_114467||lt2gene Low-quality
SS_vs_k141_120965||lt2gene Low-quality
SS_vs_k141_114469||lt2gene Low-quality
SS_vs_k141_120997||lt2gene Low-quality
SS_vs_k141_76370||lt2gene Low-quality
SS_vs_k141_114490||lt2gene Not-determined
SS_vs_k141_76374||lt2gene Low-quality
SS_vs_k141_22946||lt2gene Low-quality
SS_vs_k141_96188||lt2gene Not-determined
SS_vs_k141_76386||lt2gene Low-quality
SS_vs_k141_96200||lt2gene Low-quality
SS_vs_k141_96205||lt2gene Low-quality
SS_vs_k141_81903||lt2gene Low-quality
SS_vs_k141_36151||lt2gene Low-quality
SS_vs_k141_121019||lt2gene Not-determined
SS_vs_k141_22973||lt2gene Low-quality
SS_vs_k141_76421||lt2gene Low-quality
SS_vs_k141_137156||lt2gene Low-quality
SS_vs_k141_89804||lt2gene Not-determined
SS_vs_k141_22980||lt2gene Low-quality
SS_vs_k141_127178||lt2gene Low-quality
SS_vs_k141_137167||lt2gene Not-determined
SS_vs_k141_76444||lt2gene Low-quality
SS_vs_k141_121032||lt2gene Low-quality
SS_vs_k141_89839||lt2gene Low-quality
SS_vs_k141_47177||lt2gene Low-quality
SS_vs_k141_23007||lt2gene Low-quality
SS_vs_k141_47181||lt2gene Low-quality
SS_vs_k141_27503||lt2gene Low-quality
SS_vs_k141_23019||lt2gene Low-quality
SS_vs_k141_100629||lt2gene Not-determined
SS_vs_k141_47197||lt2gene Low-quality
SS_vs_k141_40810||lt2gene Not-determined
SS_vs_k141_17680||lt2gene Low-quality
SS_vs_k141_54560||lt2gene Low-quality
SS_vs_k141_89868||lt2gene Low-quality
SS_vs_k141_27512||lt2gene Low-quality
SS_vs_k141_23034||lt2gene Low-quality
SS_vs_k141_143795||lt2gene Low-quality
SS_vs_k141_17726||lt2gene Low-quality
SS_vs_k141_47247||lt2gene Low-quality
SS_vs_k141_96254||lt2gene Not-determined
SS_vs_k141_17761||lt2gene Low-quality
SS_vs_k141_114581||lt2gene Not-determined
SS_vs_k141_137229||lt2gene Low-quality
SS_vs_k141_127285||lt2gene Low-quality
SS_vs_k141_36304||lt2gene Low-quality
SS_vs_k141_23085||lt2gene Low-quality
SS_vs_k141_121106||lt2gene Low-quality
SS_vs_k141_143865||lt2gene Low-quality
SS_vs_k141_96285||lt2gene Low-quality
SS_vs_k141_11035||lt2gene Not-determined
SS_vs_k141_89981||lt2gene Low-quality
SS_vs_k141_121134||lt2gene Low-quality
SS_vs_k141_57680||lt2gene Low-quality
SS_vs_k141_71340||lt2gene Not-determined
SS_vs_k141_23106||lt2gene Low-quality
SS_vs_k141_23108||lt2gene Not-determined
SS_vs_k141_54633||lt2gene Low-quality
SS_vs_k141_27561||lt2gene Low-quality
SS_vs_k141_127346||lt2gene Low-quality
SS_vs_k141_143876||lt2gene Low-quality
SS_vs_k141_96351||lt2gene Not-determined
SS_vs_k141_143892||lt2gene Low-quality
SS_vs_k141_143896||lt2gene Low-quality
SS_vs_k141_71359||lt2gene Not-determined
SS_vs_k141_11052||lt2gene Low-quality
SS_vs_k141_36423||lt2gene Low-quality
SS_vs_k141_47369||lt2gene Low-quality
SS_vs_k141_127442||lt2gene Low-quality
SS_vs_k141_114728||lt2gene Not-determined
SS_vs_k141_47397||lt2gene Not-determined
SS_vs_k141_127452||lt2gene Low-quality
SS_vs_k141_143933||lt2gene Low-quality
SS_vs_k141_143945||lt2gene Low-quality
SS_vs_k141_82100||lt2gene Low-quality
SS_vs_k141_132534||lt2gene Not-determined
SS_vs_k141_96382||lt2gene Low-quality
SS_vs_k141_11111||lt2gene Low-quality
SS_vs_k141_23221||lt2gene Low-quality
SS_vs_k141_90024||lt2gene Low-quality
SS_vs_k141_96391||lt2gene Low-quality
SS_vs_k141_47440||lt2gene Low-quality
SS_vs_k141_137314||lt2gene Low-quality
SS_vs_k141_47465||lt2gene Low-quality
SS_vs_k141_57786||lt2gene Not-determined
SS_vs_k141_27621||lt2gene Low-quality
SS_vs_k141_27629||lt2gene Low-quality
SS_vs_k141_27634||lt2gene Not-determined
SS_vs_k141_90078||lt2gene Not-determined
SS_vs_k141_96459||lt2gene Low-quality
SS_vs_k141_90082||lt2gene Low-quality
SS_vs_k141_71520||lt2gene Not-determined
SS_vs_k141_100862||lt2gene Low-quality
SS_vs_k141_96481||lt2gene Low-quality
SS_vs_k141_17938||lt2gene Low-quality
SS_vs_k141_96497||lt2gene Low-quality
SS_vs_k141_57804||lt2gene Low-quality
SS_vs_k141_40877||lt2gene Low-quality
SS_vs_k141_47602||lt2gene Low-quality
SS_vs_k141_96545||lt2gene Not-determined
SS_vs_k141_137340||lt2gene Low-quality
SS_vs_k141_47629||lt2gene Low-quality
SS_vs_k141_11125||lt2gene Low-quality
SS_vs_k141_11129||lt2gene Low-quality
SS_vs_k141_47653||lt2gene Not-determined
SS_vs_k141_90095||lt2gene Low-quality
SS_vs_k141_27745||lt2gene Not-determined
SS_vs_k141_23307||lt2gene Low-quality
SS_vs_k141_82134||lt2gene Low-quality
SS_vs_k141_144048||lt2gene Low-quality
SS_vs_k141_11142||lt2gene Not-determined
SS_vs_k141_27758||lt2gene Low-quality
SS_vs_k141_23339||lt2gene Low-quality
SS_vs_k141_121416||lt2gene Low-quality
SS_vs_k141_40891||lt2gene Low-quality
SS_vs_k141_96577||lt2gene Not-determined
SS_vs_k141_100942||lt2gene Not-determined
SS_vs_k141_76518||lt2gene Low-quality
SS_vs_k141_40926||lt2gene Not-determined
SS_vs_k141_71705||lt2gene Low-quality
SS_vs_k141_137389||lt2gene Low-quality
SS_vs_k141_27831||lt2gene Low-quality
SS_vs_k141_40942||lt2gene Low-quality
SS_vs_k141_100956||lt2gene Low-quality
SS_vs_k141_71751||lt2gene Low-quality
SS_vs_k141_47797||lt2gene Low-quality
SS_vs_k141_71766||lt2gene Not-determined
SS_vs_k141_132858||lt2gene Low-quality
SS_vs_k141_82186||lt2gene Low-quality
SS_vs_k141_82201||lt2gene Not-determined
SS_vs_k141_27850||lt2gene Not-determined
SS_vs_k141_100980||lt2gene Not-determined
SS_vs_k141_100993||lt2gene Low-quality
SS_vs_k141_137409||lt2gene Low-quality
SS_vs_k141_18156||lt2gene Low-quality
SS_vs_k141_65668||lt2gene Low-quality
SS_vs_k141_114886||lt2gene Not-determined
SS_vs_k141_82217||lt2gene Low-quality
SS_vs_k141_47862||lt2gene Low-quality
SS_vs_k141_23405||lt2gene Low-quality
SS_vs_k141_114915||lt2gene Low-quality
SS_vs_k141_23413||lt2gene Low-quality
SS_vs_k141_96669||lt2gene Low-quality
SS_vs_k141_40976||lt2gene Low-quality
SS_vs_k141_132941||lt2gene Low-quality
SS_vs_k141_40981||lt2gene Low-quality
SS_vs_k141_132949||lt2gene Low-quality
SS_vs_k141_90297||lt2gene Not-determined
SS_vs_k141_71824||lt2gene Low-quality
SS_vs_k141_96715||lt2gene Not-determined
SS_vs_k141_137469||lt2gene Low-quality
SS_vs_k141_71837||lt2gene Low-quality
SS_vs_k141_23461||lt2gene Low-quality
SS_vs_k141_11221||lt2gene Low-quality
SS_vs_k141_137494||lt2gene Low-quality
SS_vs_k141_90353||lt2gene Not-determined
SS_vs_k141_23491||lt2gene Not-determined
SS_vs_k141_65786||lt2gene Low-quality
SS_vs_k141_114976||lt2gene Low-quality
SS_vs_k141_41042||lt2gene Not-determined
SS_vs_k141_90375||lt2gene Low-quality
SS_vs_k141_23503||lt2gene Low-quality
SS_vs_k141_18203||lt2gene Low-quality
SS_vs_k141_57881||lt2gene Low-quality
SS_vs_k141_144184||lt2gene Low-quality
SS_vs_k141_65811||lt2gene Low-quality
SS_vs_k141_23597||lt2gene Low-quality
SS_vs_k141_65828||lt2gene Low-quality
SS_vs_k141_65832||lt2gene Low-quality
SS_vs_k141_133091||lt2gene Not-determined
SS_vs_k141_133093||lt2gene Not-determined
SS_vs_k141_23611||lt2gene Low-quality
SS_vs_k141_23613||lt2gene Low-quality
SS_vs_k141_144224||lt2gene Low-quality
SS_vs_k141_71891||lt2gene Low-quality
SS_vs_k141_47920||lt2gene Low-quality
SS_vs_k141_115035||lt2gene Low-quality
SS_vs_k141_115040||lt2gene Low-quality
SS_vs_k141_144238||lt2gene Low-quality
SS_vs_k141_90457||lt2gene Low-quality
SS_vs_k141_144246||lt2gene Not-determined
SS_vs_k141_133166||lt2gene Low-quality
SS_vs_k141_133170||lt2gene Not-determined
SS_vs_k141_47960||lt2gene Low-quality
SS_vs_k141_96866||lt2gene Not-determined
SS_vs_k141_101018||lt2gene Low-quality
SS_vs_k141_144282||lt2gene Not-determined
SS_vs_k141_71963||lt2gene Low-quality
SS_vs_k141_76765||lt2gene Low-quality
SS_vs_k141_47985||lt2gene Low-quality
SS_vs_k141_90528||lt2gene Low-quality
SS_vs_k141_90529||lt2gene Low-quality
SS_vs_k141_47996||lt2gene Low-quality
SS_vs_k141_96907||lt2gene Low-quality
SS_vs_k141_96913||lt2gene Low-quality
SS_vs_k141_96914||lt2gene Low-quality
SS_vs_k141_96924||lt2gene Not-determined
SS_vs_k141_82403||lt2gene Not-determined
SS_vs_k141_82408||lt2gene Low-quality
SS_vs_k141_82414||lt2gene Not-determined
SS_vs_k141_137553||lt2gene Low-quality
SS_vs_k141_23639||lt2gene Low-quality
SS_vs_k141_137556||lt2gene Low-quality
SS_vs_k141_96957||lt2gene Not-determined
SS_vs_k141_82421||lt2gene Low-quality
SS_vs_k141_23643||lt2gene Low-quality
SS_vs_k141_41232||lt2gene Low-quality
SS_vs_k141_96978||lt2gene Low-quality
SS_vs_k141_41247||lt2gene Low-quality
SS_vs_k141_23673||lt2gene Low-quality
SS_vs_k141_72049||lt2gene Low-quality
SS_vs_k141_101036||lt2gene Low-quality
SS_vs_k141_11356||lt2gene Low-quality
SS_vs_k141_82451||lt2gene Not-determined
SS_vs_k141_72086||lt2gene Low-quality
SS_vs_k141_23703||lt2gene Low-quality
SS_vs_k141_97062||lt2gene Low-quality
SS_vs_k141_133319||lt2gene Low-quality
SS_vs_k141_97065||lt2gene Low-quality
SS_vs_k141_97066||lt2gene Low-quality
SS_vs_k141_65889||lt2gene Not-determined
SS_vs_k141_97092||lt2gene Low-quality
SS_vs_k141_72142||lt2gene Low-quality
SS_vs_k141_137601||lt2gene Not-determined
SS_vs_k141_65900||lt2gene Low-quality
SS_vs_k141_115283||lt2gene Low-quality
SS_vs_k141_76893||lt2gene Low-quality
SS_vs_k141_133378||lt2gene Low-quality
SS_vs_k141_133385||lt2gene Low-quality
SS_vs_k141_115349||lt2gene Low-quality
SS_vs_k141_72211||lt2gene Low-quality
SS_vs_k141_137619||lt2gene Not-determined
SS_vs_k141_48043||lt2gene Low-quality
SS_vs_k141_76939||lt2gene Low-quality
SS_vs_k141_90740||lt2gene Not-determined
SS_vs_k141_41316||lt2gene Low-quality
SS_vs_k141_90746||lt2gene Low-quality
SS_vs_k141_144332||lt2gene Low-quality
SS_vs_k141_48060||lt2gene Low-quality
SS_vs_k141_72240||lt2gene Low-quality
SS_vs_k141_65952||lt2gene Low-quality
SS_vs_k141_48080||lt2gene Low-quality
SS_vs_k141_48081||lt2gene Low-quality
SS_vs_k141_41331||lt2gene Low-quality
SS_vs_k141_133470||lt2gene Low-quality
SS_vs_k141_11498||lt2gene Not-determined
SS_vs_k141_144384||lt2gene Low-quality
SS_vs_k141_28057||lt2gene Not-determined
SS_vs_k141_72296||lt2gene Low-quality
SS_vs_k141_23840||lt2gene Not-determined
SS_vs_k141_101118||lt2gene Not-determined
SS_vs_k141_82517||lt2gene Low-quality
SS_vs_k141_28064||lt2gene Low-quality
SS_vs_k141_48182||lt2gene Not-determined
SS_vs_k141_23869||lt2gene Low-quality
SS_vs_k141_48192||lt2gene Low-quality
SS_vs_k141_48205||lt2gene Not-determined
SS_vs_k141_66071||lt2gene Low-quality
SS_vs_k141_23919||lt2gene Low-quality
SS_vs_k141_48240||lt2gene Not-determined
SS_vs_k141_66108||lt2gene Low-quality
SS_vs_k141_144468||lt2gene Low-quality
SS_vs_k141_41455||lt2gene Low-quality
SS_vs_k141_137733||lt2gene Low-quality
SS_vs_k141_11580||lt2gene Not-determined
SS_vs_k141_77104||lt2gene Low-quality
SS_vs_k141_72357||lt2gene Not-determined
SS_vs_k141_72359||lt2gene Not-determined
SS_vs_k141_144483||lt2gene Low-quality
SS_vs_k141_48357||lt2gene Not-determined
SS_vs_k141_137750||lt2gene Low-quality
SS_vs_k141_41478||lt2gene Low-quality
SS_vs_k141_41497||lt2gene Low-quality
SS_vs_k141_77153||lt2gene Low-quality
SS_vs_k141_28163||lt2gene Low-quality
SS_vs_k141_24027||lt2gene Low-quality
SS_vs_k141_77191||lt2gene Low-quality
SS_vs_k141_72414||lt2gene Low-quality
SS_vs_k141_24059||lt2gene Low-quality
SS_vs_k141_66193||lt2gene Low-quality
SS_vs_k141_66195||lt2gene Low-quality
SS_vs_k141_90790||lt2gene Low-quality
SS_vs_k141_41574||lt2gene Not-determined
SS_vs_k141_137803||lt2gene Low-quality

SS_vs_k141_137822||lt2gene Low-quality
SS_vs_k141_58099||lt2gene Low-quality
SS_vs_k141_48456||lt2gene Low-quality
SS_vs_k141_41620||lt2gene Low-quality
SS_vs_k141_48460||lt2gene Not-determined
SS_vs_k141_48461||lt2gene Not-determined
SS_vs_k141_77269||lt2gene Not-determined
SS_vs_k141_66256||lt2gene Low-quality
SS_vs_k141_82697||lt2gene Low-quality
SS_vs_k141_66268||lt2gene Low-quality
SS_vs_k141_66272||lt2gene Low-quality
SS_vs_k141_66292||lt2gene Low-quality
SS_vs_k141_90865||lt2gene Low-quality
SS_vs_k141_58126||lt2gene Low-quality
SS_vs_k141_24156||lt2gene Low-quality
SS_vs_k141_90891||lt2gene Not-determined
SS_vs_k141_144706||lt2gene Low-quality
SS_vs_k141_82725||lt2gene Low-quality
SS_vs_k141_58151||lt2gene Low-quality
SS_vs_k141_24214||lt2gene Low-quality
SS_vs_k141_82771||lt2gene Low-quality
SS_vs_k141_66344||lt2gene Low-quality
SS_vs_k141_41702||lt2gene Low-quality
SS_vs_k141_41708||lt2gene Not-determined
SS_vs_k141_24240||lt2gene Low-quality
SS_vs_k141_24247||lt2gene Low-quality
SS_vs_k141_24252||lt2gene Low-quality
SS_vs_k141_24255||lt2gene Low-quality
SS_vs_k141_41738||lt2gene Low-quality
SS_vs_k141_72624||lt2gene Low-quality
SS_vs_k141_28367||lt2gene Low-quality
SS_vs_k141_41747||lt2gene Low-quality
SS_vs_k141_82831||lt2gene Low-quality
SS_vs_k141_144768||lt2gene Low-quality
SS_vs_k141_58259||lt2gene Low-quality
SS_vs_k141_72639||lt2gene Low-quality
SS_vs_k141_82841||lt2gene Low-quality
SS_vs_k141_58273||lt2gene Low-quality
SS_vs_k141_72647||lt2gene Low-quality
SS_vs_k141_82846||lt2gene Low-quality
SS_vs_k141_28397||lt2gene Low-quality
SS_vs_k141_72650||lt2gene Low-quality
SS_vs_k141_41793||lt2gene Low-quality
SS_vs_k141_82885||lt2gene Not-determined
SS_vs_k141_41833||lt2gene Low-quality
SS_vs_k141_82904||lt2gene Low-quality
SS_vs_k141_82913||lt2gene Low-quality
SS_vs_k141_28521||lt2gene Low-quality
SS_vs_k141_77296||lt2gene Low-quality
SS_vs_k141_66437||lt2gene Not-determined
SS_vs_k141_82965||lt2gene Low-quality
SS_vs_k141_41943||lt2gene Low-quality
SS_vs_k141_82994||lt2gene Not-determined
SS_vs_k141_137990||lt2gene Not-determined
SS_vs_k141_144919||lt2gene Low-quality
SS_vs_k141_58351||lt2gene Low-quality
SS_vs_k141_83011||lt2gene Low-quality
SS_vs_k141_144938||lt2gene Low-quality
SS_vs_k141_77392||lt2gene Not-determined
SS_vs_k141_83046||lt2gene Not-determined
SS_vs_k141_58388||lt2gene Low-quality
SS_vs_k141_41981||lt2gene Low-quality
SS_vs_k141_41985||lt2gene Low-quality
SS_vs_k141_41994||lt2gene Low-quality
SS_vs_k141_41997||lt2gene Low-quality
SS_vs_k141_72803||lt2gene Low-quality
SS_vs_k141_42059||lt2gene Not-determined
SS_vs_k141_77476||lt2gene Not-determined
SS_vs_k141_77478||lt2gene Low-quality
SS_vs_k141_58438||lt2gene Low-quality
SS_vs_k141_83204||lt2gene Low-quality
SS_vs_k141_83211||lt2gene Low-quality
SS_vs_k141_83213||lt2gene Low-quality
SS_vs_k141_83215||lt2gene Low-quality
SS_vs_k141_58478||lt2gene Low-quality
SS_vs_k141_77521||lt2gene Not-determined
SS_vs_k141_58497||lt2gene Low-quality
SS_vs_k141_66523||lt2gene Low-quality
SS_vs_k141_28697||lt2gene Low-quality
SS_vs_k141_138020||lt2gene Not-determined
SS_vs_k141_42200||lt2gene Low-quality
SS_vs_k141_58617||lt2gene Low-quality
SS_vs_k141_42212||lt2gene Low-quality
SS_vs_k141_42214||lt2gene Low-quality
SS_vs_k141_83306||lt2gene Low-quality
SS_vs_k141_42244||lt2gene Low-quality
SS_vs_k141_42255||lt2gene Not-determined
SS_vs_k141_101449||lt2gene Low-quality
SS_vs_k141_83363||lt2gene Low-quality
SS_vs_k141_145136||lt2gene Low-quality
SS_vs_k141_42288||lt2gene Low-quality
SS_vs_k141_42295||lt2gene Low-quality
SS_vs_k141_101479||lt2gene Not-determined
SS_vs_k141_101484||lt2gene Low-quality
SS_vs_k141_66600||lt2gene Low-quality
SS_vs_k141_28782||lt2gene Low-quality
SS_vs_k141_83416||lt2gene Low-quality
SS_vs_k141_83431||lt2gene Not-determined
SS_vs_k141_83449||lt2gene Not-determined
SS_vs_k141_42404||lt2gene Not-determined
SS_vs_k141_138182||lt2gene Not-determined
SS_vs_k141_138234||lt2gene Low-quality
SS_vs_k141_66765||lt2gene Low-quality
SS_vs_k141_58788||lt2gene Low-quality
SS_vs_k141_28869||lt2gene Low-quality
SS_vs_k141_145287||lt2gene Low-quality
SS_vs_k141_83568||lt2gene Not-determined
SS_vs_k141_138284||lt2gene Low-quality
SS_vs_k141_138286||lt2gene Low-quality
SS_vs_k141_83604||lt2gene Not-determined
SS_vs_k141_77722||lt2gene Low-quality
SS_vs_k141_28911||lt2gene Low-quality
SS_vs_k141_83674||lt2gene Low-quality
SS_vs_k141_101540||lt2gene Not-determined
SS_vs_k141_28971||lt2gene Low-quality
SS_vs_k141_77766||lt2gene Low-quality
SS_vs_k141_58875||lt2gene Low-quality
SS_vs_k141_145403||lt2gene Low-quality
SS_vs_k141_58880||lt2gene Not-determined
SS_vs_k141_138374||lt2gene Low-quality
SS_vs_k141_11745||lt2gene Low-quality
SS_vs_k141_11757||lt2gene Not-determined
SS_vs_k141_11772||lt2gene Not-determined
SS_vs_k141_145486||lt2gene Low-quality
SS_vs_k141_138422||lt2gene Low-quality
SS_vs_k141_101587||lt2gene Low-quality
SS_vs_k141_58931||lt2gene Low-quality
SS_vs_k141_101596||lt2gene Low-quality
SS_vs_k141_145512||lt2gene Low-quality
SS_vs_k141_29019||lt2gene Low-quality
SS_vs_k141_29025||lt2gene Low-quality
SS_vs_k141_58959||lt2gene Low-quality
SS_vs_k141_145524||lt2gene Low-quality
SS_vs_k141_58973||lt2gene Low-quality
SS_vs_k141_58975||lt2gene Not-determined
SS_vs_k141_58990||lt2gene Low-quality
SS_vs_k141_83868||lt2gene Low-quality
SS_vs_k141_101653||lt2gene Low-quality
SS_vs_k141_59030||lt2gene Low-quality
SS_vs_k141_59066||lt2gene Low-quality
SS_vs_k141_59095||lt2gene Low-quality
SS_vs_k141_83889||lt2gene Low-quality
SS_vs_k141_59140||lt2gene Low-quality
SS_vs_k141_29144||lt2gene Low-quality
SS_vs_k141_29150||lt2gene Low-quality
SS_vs_k141_29160||lt2gene Low-quality
SS_vs_k141_29175||lt2gene Low-quality
SS_vs_k141_59198||lt2gene Low-quality
SS_vs_k141_101769||lt2gene Not-determined
SS_vs_k141_101770||lt2gene Not-determined
SS_vs_k141_59209||lt2gene Not-determined
SS_vs_k141_101783||lt2gene Low-quality
SS_vs_k141_29198||lt2gene Low-quality
SS_vs_k141_83959||lt2gene Low-quality
SS_vs_k141_29201||lt2gene Low-quality
SS_vs_k141_29212||lt2gene Low-quality
SS_vs_k141_59291||lt2gene Not-determined
SS_vs_k141_101840||lt2gene Low-quality
SS_vs_k141_59306||lt2gene Not-determined
SS_vs_k141_84056||lt2gene Not-determined
SS_vs_k141_84068||lt2gene Low-quality
SS_vs_k141_84091||lt2gene Low-quality
SS_vs_k141_77919||lt2gene Low-quality
SS_vs_k141_77921||lt2gene Not-determined
SS_vs_k141_77933||lt2gene Not-determined
SS_vs_k141_59350||lt2gene Low-quality
SS_vs_k141_11919||lt2gene Not-determined
SS_vs_k141_84184||lt2gene Low-quality
SS_vs_k141_59386||lt2gene Low-quality
SS_vs_k141_59429||lt2gene Low-quality
SS_vs_k141_59449||lt2gene Not-determined
SS_vs_k141_59466||lt2gene Low-quality
SS_vs_k141_29333||lt2gene Not-determined
SS_vs_k141_29356||lt2gene Low-quality
SS_vs_k141_101955||lt2gene Low-quality
SS_vs_k141_78189||lt2gene Low-quality
SS_vs_k141_138674||lt2gene Low-quality
SS_vs_k141_78213||lt2gene Low-quality
SS_vs_k141_12013||lt2gene Low-quality
SS_vs_k141_12020||lt2gene Low-quality
SS_vs_k141_138790||lt2gene Not-determined
SS_vs_k141_78297||lt2gene Low-quality
SS_vs_k141_59626||lt2gene Not-determined
SS_vs_k141_138841||lt2gene Not-determined
SS_vs_k141_78339||lt2gene Not-determined
SS_vs_k141_138849||lt2gene Not-determined
SS_vs_k141_138870||lt2gene Not-determined
SS_vs_k141_78360||lt2gene Low-quality
SS_vs_k141_102065||lt2gene Not-determined
SS_vs_k141_84326||lt2gene Low-quality
SS_vs_k141_12129||lt2gene Low-quality
SS_vs_k141_84430||lt2gene Low-quality
SS_vs_k141_84439||lt2gene Not-determined
SS_vs_k141_84459||lt2gene Low-quality
SS_vs_k141_78492||lt2gene Not-determined
SS_vs_k141_78545||lt2gene Low-quality
SS_vs_k141_78558||lt2gene Low-quality
SS_vs_k141_138992||lt2gene Low-quality
SS_vs_k141_138995||lt2gene Low-quality
SS_vs_k141_139065||lt2gene Low-quality
SS_vs_k141_59775||lt2gene Low-quality
SS_vs_k141_78736||lt2gene Low-quality
SS_vs_k141_78748||lt2gene Low-quality
SS_vs_k141_102271||lt2gene Low-quality
SS_vs_k141_59801||lt2gene Low-quality
SS_vs_k141_78794||lt2gene Low-quality
SS_vs_k141_59833||lt2gene Low-quality
SS_vs_k141_102329||lt2gene Low-quality
SS_vs_k141_102335||lt2gene Low-quality
SS_vs_k141_102383||lt2gene Not-determined
SS_vs_k141_78866||lt2gene Low-quality
SS_vs_k141_29662||lt2gene Low-quality
SS_vs_k141_59888||lt2gene Not-determined
SS_vs_k141_102403||lt2gene Low-quality
SS_vs_k141_139114||lt2gene Low-quality
SS_vs_k141_139125||lt2gene Not-determined
SS_vs_k141_29705||lt2gene Not-determined
SS_vs_k141_29753||lt2gene Low-quality
SS_vs_k141_29759||lt2gene Low-quality
SS_vs_k141_29772||lt2gene Not-determined
SS_vs_k141_59999||lt2gene Low-quality
SS_vs_k141_102469||lt2gene Low-quality
SS_vs_k141_60004||lt2gene Low-quality
SS_vs_k141_29825||lt2gene Not-determined
SS_vs_k141_60036||lt2gene Low-quality
SS_vs_k141_84756||lt2gene Low-quality
SS_vs_k141_102528||lt2gene Low-quality
SS_vs_k141_139252||lt2gene Low-quality
SS_vs_k141_60055||lt2gene Low-quality
SS_vs_k141_29876||lt2gene Not-determined
SS_vs_k141_60082||lt2gene Low-quality
SS_vs_k141_60089||lt2gene Low-quality
SS_vs_k141_60093||lt2gene Low-quality
SS_vs_k141_29914||lt2gene Low-quality
SS_vs_k141_102592||lt2gene Low-quality
SS_vs_k141_84864||lt2gene Not-determined
SS_vs_k141_84868||lt2gene Low-quality
SS_vs_k141_30006||lt2gene Low-quality
SS_vs_k141_60130||lt2gene Low-quality
SS_vs_k141_60132||lt2gene Low-quality
SS_vs_k141_84916||lt2gene Low-quality
SS_vs_k141_30036||lt2gene Low-quality
SS_vs_k141_139279||lt2gene Low-quality
SS_vs_k141_102710||lt2gene Not-determined
SS_vs_k141_60212||lt2gene Not-determined
SS_vs_k141_30134||lt2gene Low-quality
SS_vs_k141_30139||lt2gene Not-determined
SS_vs_k141_30184||lt2gene Low-quality
SS_vs_k141_102818||lt2gene Low-quality
SS_vs_k141_102823||lt2gene Low-quality
SS_vs_k141_102847||lt2gene Low-quality
SS_vs_k141_30222||lt2gene Not-determined
SS_vs_k141_102865||lt2gene Low-quality
SS_vs_k141_30242||lt2gene Low-quality
SS_vs_k141_102916||lt2gene Low-quality
SS_vs_k141_139361||lt2gene Not-determined
SS_vs_k141_60266||lt2gene Not-determined
SS_vs_k141_60273||lt2gene Not-determined
SS_vs_k141_139380||lt2gene Low-quality
SS_vs_k141_102990||lt2gene Low-quality
SS_vs_k141_30328||lt2gene Low-quality
SS_vs_k141_30331||lt2gene Low-quality
SS_vs_k141_30334||lt2gene Low-quality
SS_vs_k141_60339||lt2gene Low-quality
SS_vs_k141_103032||lt2gene Low-quality
SS_vs_k141_60363||lt2gene Low-quality
SS_vs_k141_139452||lt2gene Not-determined
SS_vs_k141_103103||lt2gene Low-quality
SS_vs_k141_139526||lt2gene Not-determined
SS_vs_k141_139564||lt2gene Low-quality
SS_vs_k141_139567||lt2gene Low-quality
SS_vs_k141_103197||lt2gene Low-quality
SS_vs_k141_139583||lt2gene Low-quality
SS_vs_k141_139586||lt2gene Low-quality
SS_vs_k141_60477||lt2gene Not-determined
SS_vs_k141_60503||lt2gene Low-quality
SS_vs_k141_60616||lt2gene Low-quality
SS_vs_k141_60655||lt2gene Low-quality
S7_extracted_9 Not-determined
S7_extracted_11 Not-determined
S7_extracted_26 Not-determined
S7_extracted_27 Not-determined
S7_extracted_30 Not-determined
S7_extracted_34 Low-quality
S7_extracted_39 Not-determined
S7_extracted_77 Not-determined
S7_extracted_81 Not-determined
S7_extracted_103 Not-determined
S7_extracted_105 Not-determined
S7_extracted_117 Not-determined
S7_extracted_123 Not-determined
S7_extracted_125 Not-determined
S7_extracted_132 Not-determined
S7_extracted_138 Not-determined
S7_extracted_159 Low-quality
S7_extracted_162 Not-determined
S7_extracted_172 Not-determined
S7_extracted_178 Not-determined
S7_extracted_191 Low-quality
S7_extracted_196 Not-determined
S7_extracted_210 Not-determined
S7_extracted_219 Low-quality
S7_extracted_220 Not-determined
S7_extracted_249 Low-quality
S7_extracted_271 Not-determined
S7_extracted_295 Not-determined
S7_extracted_296 Not-determined
S7_extracted_304 Not-determined
S7_extracted_336 Low-quality
S7_extracted_364 Not-determined
S7_extracted_370 Not-determined
S7_extracted_380 Low-quality
S7_extracted_388 Not-determined
S7_extracted_430 Not-determined
S7_extracted_446 Not-determined
S7_extracted_453 Not-determined
S7_extracted_454 Low-quality
S7_extracted_462 Not-determined
S7_extracted_483 Not-determined
S7_extracted_485 Not-determined
S7_extracted_488 Not-determined
S7_extracted_504 Not-determined
S7_extracted_510 Not-determined
S7_extracted_514 Low-quality
S7_extracted_548 Not-determined
S7_extracted_574 Low-quality
S7_extracted_575 Not-determined
S7_extracted_578 Not-determined
S7_extracted_587 Not-determined
S7_extracted_590 Not-determined
S7_extracted_595 Low-quality
S7_extracted_632 Not-determined
S7_extracted_658 Not-determined
S7_extracted_674 Low-quality
S7_extracted_684 Not-determined
S7_extracted_687 Low-quality
S7_extracted_688 Not-determined
S7_extracted_689 Low-quality
S7_extracted_700 Not-determined
S7_extracted_705 Not-determined
S7_extracted_726 Not-determined
S7_extracted_741 Not-determined
S7_extracted_742 Low-quality
S7_extracted_748 Not-determined
S7_extracted_751 Low-quality
S7_extracted_771 Not-determined
S7_extracted_785 Low-quality
S7_extracted_809 Not-determined
S7_extracted_829 Low-quality
S7_extracted_842 Low-quality
S7_extracted_844 Not-determined
S7_extracted_877 Low-quality
S7_extracted_898 Not-determined
S7_extracted_902 Not-determined
S7_extracted_916 Not-determined
S7_extracted_933 Not-determined
S7_extracted_942 Not-determined
S7_extracted_954 Not-determined
S7_extracted_980 Not-determined
S7_extracted_1003 Low-quality
S7_extracted_1013 Not-determined
S7_extracted_1039 Low-quality
S7_extracted_1054 Not-determined
S7_extracted_1060 Not-determined
S7_extracted_1065 Not-determined
S7_extracted_1066 Not-determined
S7_extracted_1082 Low-quality
S7_extracted_1108 Not-determined
S7_extracted_1116 Not-determined
S7_extracted_1124 Not-determined
S7_extracted_1132 Low-quality
S7_extracted_1146 Low-quality
S7_extracted_1168 Not-determined
S7_extracted_1181 Not-determined
S7_extracted_1194 Not-determined
S7_extracted_1198 Not-determined
S7_extracted_1207 Not-determined
S7_extracted_1218 Not-determined
S7_extracted_1236 Not-determined
S7_extracted_1239 Not-determined
S7_extracted_1248 Not-determined
S7_extracted_1266 Low-quality
S7_extracted_1314 Not-determined
S7_extracted_1317 Not-determined
S7_extracted_1319 Not-determined
S7_extracted_1324 Not-determined
S7_extracted_1336 Not-determined
S7_extracted_1337 Not-determined
S7_extracted_1348 Not-determined
S7_extracted_1356 Not-determined
S7_extracted_1415 Not-determined
S7_extracted_1422 Not-determined
S7_extracted_1426 Not-determined
S7_extracted_1432 Not-determined
S7_extracted_1470 Not-determined
S7_extracted_1483 Not-determined
S7_extracted_1493 Not-determined
S7_extracted_1495 Low-quality
S7_extracted_1507 Not-determined
S7_extracted_1511 Not-determined
S7_extracted_1516 Not-determined
S7_extracted_1540 Low-quality
S7_extracted_1541 Not-determined
S7_extracted_1548 Low-quality
S7_extracted_1551 Low-quality
S7_extracted_1571 Not-determined
S7_extracted_1572 Not-determined
S7_extracted_1596 Not-determined
S7_extracted_1611 Not-determined
S7_extracted_1629 Not-determined
S7_extracted_1642 Not-determined
S7_extracted_1646 Not-determined
S7_extracted_1649 Not-determined
S7_extracted_1652 Not-determined
S7_extracted_1656 Not-determined
S7_extracted_1660 Not-determined
S7_extracted_1670 Not-determined
S7_extracted_1680 Not-determined
S7_extracted_1693 Not-determined
S7_extracted_1710 Not-determined
S7_extracted_1719 Not-determined
S7_extracted_1729 Not-determined
S7_extracted_1737 Low-quality
S7_extracted_1744 Low-quality
S7_extracted_1774 Low-quality
S7_extracted_1780 Not-determined
S7_extracted_1784 Not-determined
S7_extracted_1788 Not-determined
S7_extracted_1792 Not-determined
S7_extracted_1811 Not-determined
S7_extracted_1814 Not-determined
S7_extracted_1829 Not-determined
S7_extracted_1833 Not-determined
S7_extracted_1887 Not-determined
S7_extracted_1893 Not-determined
S7_extracted_1899 Not-determined
S7_extracted_1902 Not-determined
S7_extracted_1945 Low-quality
S7_extracted_1960 Not-determined
S7_extracted_1994 Not-determined
S7_extracted_2009 Not-determined
S7_extracted_2017 Not-determined
S7_extracted_2035 Low-quality
S7_extracted_2043 Not-determined
S7_extracted_2050 Low-quality
S7_extracted_2069 Not-determined
S7_extracted_2081 Not-determined
S7_extracted_2082 Not-determined
S7_extracted_2083 Not-determined
S7_extracted_2084 Not-determined
S7_extracted_2096 Not-determined
S7_extracted_2105 Not-determined
S7_extracted_2107 Not-determined
S7_extracted_2127 Not-determined
S7_extracted_2137 Not-determined
S7_extracted_2140 Not-determined
S7_extracted_2146 Not-determined
S7_extracted_2192 Not-determined
S7_extracted_2212 Low-quality
S7_extracted_2213 Not-determined
S7_extracted_2215 Low-quality
S7_extracted_2222 Not-determined
S7_extracted_2234 Not-determined
S7_extracted_2237 Low-quality
S7_extracted_2246 Not-determined
S7_extracted_2254 Low-quality
S7_extracted_2257 Low-quality
S7_extracted_2272 Not-determined
S7_extracted_2289 Low-quality
S7_extracted_2315 Not-determined
S7_extracted_2334 Not-determined
S7_extracted_2337 Not-determined
S7_extracted_2345 Not-determined
S7_extracted_2346 Not-determined
S7_extracted_2347 Not-determined
S7_extracted_2357 Not-determined
S7_extracted_2371 Not-determined
S7_extracted_2374 Not-determined
S7_extracted_2381 Not-determined
S7_extracted_2385 Not-determined
S7_extracted_2390 Not-determined
S7_extracted_2394 Not-determined
S7_extracted_2400 Not-determined
S7_extracted_2405 Not-determined
S7_extracted_2415 Not-determined
S7_extracted_2425 Not-determined
S7_extracted_2439 Not-determined
S7_extracted_2446 Not-determined
S7_extracted_2452 Not-determined
S7_extracted_2453 Not-determined
S7_extracted_2459 Not-determined
S7_extracted_2469 Not-determined
S7_extracted_2472 Not-determined
S7_extracted_2481 Not-determined
S7_extracted_2502 Low-quality
S7_extracted_2526 Low-quality
S7_extracted_2528 Not-determined
S7_extracted_2539 Not-determined
S7_extracted_2545 Not-determined
S7_extracted_2554 Low-quality
S7_extracted_2560 Not-determined
S7_extracted_2561 Low-quality
S7_extracted_2564 Not-determined
S7_extracted_2568 Not-determined
S7_extracted_2584 Not-determined
S7_extracted_2600 Not-determined
S7_extracted_2604 Not-determined
S7_extracted_2620 Not-determined
S7_extracted_2633 Not-determined
S7_extracted_2647 Not-determined
S7_extracted_2652 Not-determined
S7_extracted_2658 Not-determined
S7_extracted_2689 Not-determined
S7_extracted_2691 Not-determined
S7_extracted_2703 Not-determined
S7_extracted_2704 Not-determined
S7_extracted_2713 Not-determined
S7_extracted_2721 Low-quality
S7_extracted_2744 Not-determined
S7_extracted_2761 Not-determined
S7_extracted_2763 Not-determined
S7_extracted_2777 Not-determined
S7_extracted_2778 Not-determined
S7_extracted_2780 Not-determined
S7_extracted_2794 Not-determined
S7_extracted_2795 Not-determined
S7_extracted_2806 Low-quality
S7_extracted_2816 Not-determined
S7_extracted_2820 Not-determined
S7_extracted_2837 Low-quality
S7_extracted_2839 Low-quality
S7_extracted_2840 Not-determined
S7_extracted_2846 Not-determined
S7_extracted_2873 Not-determined
S7_extracted_2876 Not-determined
S7_extracted_2890 Low-quality
S7_extracted_2903 Not-determined
S7_extracted_2913 Not-determined
S7_extracted_2930 Low-quality
S7_extracted_2936 Not-determined
S7_extracted_2937 Not-determined
S7_extracted_2943 Not-determined
S7_extracted_2963 Not-determined
S7_extracted_2967 Not-determined
S7_extracted_2983 Not-determined
S7_extracted_2997 Not-determined
S7_extracted_3006 Low-quality
S7_extracted_3015 Not-determined
S7_extracted_3016 Not-determined
S7_extracted_3027 Not-determined
S7_extracted_3040 Not-determined
S7_extracted_3041 Not-determined
S7_extracted_3052 Low-quality
S7_extracted_3068 Low-quality
S7_extracted_3069 Low-quality
S7_extracted_3074 Not-determined
S7_extracted_3085 Not-determined
S7_extracted_3086 Not-determined
S7_extracted_3095 Not-determined
S7_extracted_3103 Not-determined
S7_extracted_3105 Not-determined
S7_extracted_3106 Not-determined
S7_extracted_3110 Low-quality
S7_extracted_3118 Not-determined
S7_extracted_3120 Not-determined
S7_extracted_3121 Low-quality
S7_extracted_3127 Low-quality
S7_extracted_3129 Not-determined
S7_extracted_3132 Not-determined
S7_extracted_3144 Low-quality
S7_extracted_3149 Low-quality
S7_extracted_3152 Not-determined
S7_extracted_3160 Not-determined
S7_extracted_3167 Not-determined
S7_extracted_3176 Not-determined
S7_extracted_3181 Not-determined
S7_extracted_3191 Not-determined
S7_extracted_3200 Not-determined
S7_extracted_3206 Not-determined
S7_extracted_3214 Not-determined
S7_extracted_3221 Not-determined
S7_extracted_3255 Not-determined
S7_extracted_3272 Not-determined
S7_extracted_3292 Not-determined
S7_extracted_3298 Not-determined
S7_extracted_3344 Not-determined
S7_extracted_3359 Not-determined
S7_extracted_3375 Not-determined
S7_extracted_3397 Not-determined

| | |
|---|---|
| S7_extracted_3404 | Not-determined |
| S7_extracted_3426 | Not-determined |
| S7_extracted_3429 | Not-determined |
| S7_extracted_3454 | Not-determined |
| S7_extracted_3455 | Not-determined |
| S7_extracted_3472 | Low-quality |
| S7_extracted_3476 | Not-determined |
| S7_extracted_3479 | Not-determined |
| S7_extracted_3482 | Not-determined |
| S7_extracted_3487 | Not-determined |
| S7_extracted_3490 | Low-quality |
| S7_extracted_3506 | Low-quality |
| S7_extracted_3519 | Not-determined |
| S7_extracted_3524 | Not-determined |
| S7_extracted_3534 | Low-quality |
| S7_extracted_3538 | Not-determined |
| S7_extracted_3542 | Not-determined |
| S7_extracted_3549 | Low-quality |
| S7_extracted_3551 | Low-quality |
| S7_extracted_3557 | Not-determined |
| S7_extracted_3584 | Not-determined |
| S7_extracted_3587 | Low-quality |
| S7_extracted_3593 | Not-determined |
| S7_extracted_3617 | Not-determined |
| S7_extracted_3629 | Not-determined |
| S7_extracted_3639 | Low-quality |
| S7_extracted_3640 | Not-determined |
| S7_extracted_3650 | Low-quality |
| S7_extracted_3653 | Not-determined |
| S7_extracted_3663 | Not-determined |
| S7_extracted_3687 | Not-determined |
| S7_extracted_3714 | Not-determined |
| S7_extracted_3717 | Not-determined |
| S7_extracted_3719 | Not-determined |
| S7_extracted_3752 | Low-quality |
| S7_extracted_3771 | Not-determined |
| S7_extracted_3785 | Not-determined |
| S7_extracted_3786 | Not-determined |
| S7_extracted_3790 | Not-determined |
| S7_extracted_3812 | Not-determined |
| S7_extracted_3815 | Not-determined |
| S7_extracted_3853 | Not-determined |
| S7_extracted_3857 | Not-determined |
| S7_extracted_3871 | Not-determined |
| S7_extracted_3880 | Not-determined |
| S7_extracted_3893 | Low-quality |
| S7_extracted_3895 | Not-determined |
| S7_extracted_3914 | Not-determined |
| S7_extracted_3928 | Low-quality |
| S7_extracted_3929 | Not-determined |
| S7_extracted_3955 | Not-determined |
| S7_extracted_3956 | Not-determined |
| S7_extracted_3960 | Not-determined |
| S7_extracted_3961 | Not-determined |
| S7_extracted_3987 | Not-determined |
| S7_extracted_4035 | Not-determined |
| S7_extracted_4072 | Not-determined |
| S7_extracted_4084 | Low-quality |
| S7_extracted_4085 | Not-determined |
| S7_extracted_4087 | Not-determined |
| S7_extracted_4093 | Not-determined |
| S7_extracted_4100 | Not-determined |
| S7_extracted_4120 | Not-determined |
| S7_extracted_4127 | Not-determined |
| S7_extracted_4135 | Not-determined |
| S7_extracted_4137 | Not-determined |
| S7_extracted_4146 | Not-determined |
| S7_extracted_4168 | Not-determined |
| S7_extracted_4175 | Not-determined |
| S7_extracted_4179 | Not-determined |
| S7_extracted_4190 | Low-quality |
| S7_extracted_4202 | Not-determined |
| S7_extracted_4213 | Not-determined |
| S7_extracted_4228 | Low-quality |
| S7_extracted_4230 | Not-determined |
| S7_extracted_4243 | Not-determined |
| S7_extracted_4247 | Not-determined |
| S7_extracted_4265 | Not-determined |
| S7_extracted_4289 | Not-determined |
| S7_extracted_4294 | Not-determined |
| S7_extracted_4303 | Not-determined |
| S7_extracted_4315 | Not-determined |
| S7_extracted_4317 | Not-determined |
| S7_extracted_4336 | Not-determined |
| S7_extracted_4343 | Not-determined |
| S7_extracted_4361 | Not-determined |
| S7_extracted_4363 | Not-determined |
| S7_extracted_4364 | Not-determined |
| S7_extracted_4398 | Not-determined |
| S7_extracted_4400 | Not-determined |
| S7_extracted_4407 | Not-determined |
| S7_extracted_4409 | Low-quality |
| S7_extracted_4417 | Not-determined |
| S7_extracted_4418 | Not-determined |
| S7_extracted_4419 | Not-determined |
| S7_extracted_4421 | Not-determined |
| S7_extracted_4433 | Not-determined |
| S7_extracted_4446 | Not-determined |
| S7_extracted_4459 | Not-determined |
| S7_extracted_4480 | Not-determined |
| S7_extracted_4499 | Not-determined |
| S7_extracted_4500 | Not-determined |
| S7_extracted_4540 | Not-determined |
| S7_extracted_4549 | Not-determined |
| S7_extracted_4560 | Low-quality |
| S7_extracted_4583 | Not-determined |
| S7_extracted_4634 | Not-determined |
| S7_extracted_4666 | Not-determined |
| S7_extracted_4668 | Not-determined |
| S7_extracted_4674 | Low-quality |
| S7_extracted_4679 | Not-determined |
| S7_extracted_4688 | Not-determined |
| S7_extracted_4707 | Not-determined |
| S7_extracted_4717 | Not-determined |
| S7_extracted_4720 | Not-determined |
| S7_extracted_4724 | Not-determined |
| S7_extracted_4737 | Not-determined |
| S7_extracted_4738 | Not-determined |
| S7_extracted_4740 | Low-quality |
| S7_extracted_4748 | Low-quality |
| S7_extracted_4750 | Not-determined |
| S7_extracted_4757 | Not-determined |
| S7_extracted_4767 | Not-determined |
| S7_extracted_4786 | Not-determined |
| S7_extracted_4803 | Not-determined |
| S7_extracted_4805 | Low-quality |
| S7_extracted_4808 | Not-determined |
| S7_extracted_4815 | Not-determined |
| S7_extracted_4819 | Not-determined |
| S7_extracted_4829 | Low-quality |
| S7_extracted_4834 | Not-determined |
| S7_extracted_4845 | Not-determined |
| S7_extracted_4853 | Not-determined |
| S7_extracted_4856 | Not-determined |
| S7_extracted_4861 | Not-determined |
| S7_extracted_4864 | Not-determined |
| S7_extracted_4869 | Not-determined |
| S7_extracted_4872 | Low-quality |
| S7_extracted_4881 | Not-determined |
| S7_extracted_4884 | Not-determined |
| S7_extracted_4887 | Not-determined |
| S7_extracted_4891 | Not-determined |
| S7_extracted_4930 | Not-determined |
| S7_extracted_4949 | Not-determined |
| S7_extracted_4966 | Not-determined |
| S7_extracted_4978 | Low-quality |
| S7_extracted_4997 | Not-determined |
| S7_extracted_4998 | Not-determined |
| S7_extracted_4999 | Not-determined |
| S7_extracted_5004 | Not-determined |
| S7_extracted_5008 | Not-determined |
| S7_extracted_5034 | Not-determined |
| S7_extracted_5040 | Not-determined |
| S7_extracted_5049 | Not-determined |
| S7_extracted_5066 | Low-quality |
| S7_extracted_5068 | Not-determined |
| S7_extracted_5091 | Not-determined |
| S7_extracted_5108 | Low-quality |
| S7_extracted_5111 | Not-determined |
| S7_extracted_5123 | Not-determined |
| S7_extracted_5164 | Low-quality |
| S7_extracted_5165 | Not-determined |
| S7_extracted_5177 | Low-quality |
| S7_extracted_5188 | Not-determined |
| S7_extracted_5192 | Not-determined |
| S7_extracted_5194 | Not-determined |
| S7_extracted_5201 | Low-quality |
| S7_extracted_5216 | Not-determined |
| S7_extracted_5225 | Not-determined |
| S7_extracted_5233 | Not-determined |
| S7_extracted_5253 | Not-determined |
| S7_extracted_5259 | Not-determined |
| S7_extracted_5273 | Low-quality |
| S7_extracted_5281 | Not-determined |
| S7_extracted_5288 | Low-quality |
| S7_extracted_5307 | Not-determined |
| S7_extracted_5314 | Not-determined |
| S7_extracted_5329 | Low-quality |
| S7_extracted_5342 | Not-determined |
| S7_extracted_5345 | Not-determined |
| S7_extracted_5361 | Not-determined |
| S7_extracted_5375 | Not-determined |
| S7_extracted_5383 | Not-determined |
| S7_extracted_5389 | Not-determined |
| S7_extracted_5391 | Low-quality |
| S7_extracted_5398 | Not-determined |
| S7_extracted_5404 | Low-quality |
| S7_extracted_5416 | Not-determined |
| S7_extracted_5418 | Not-determined |
| S7_extracted_5436 | Not-determined |
| S3_vs_k141_49835\|\|full | High-quality |
| S7_extracted_5440 | Not-determined |
| S7_extracted_5462 | Not-determined |
| S7_extracted_5464 | Not-determined |
| S7_extracted_5470 | Not-determined |
| S7_extracted_5478 | Not-determined |
| S7_extracted_5479 | Low-quality |
| S7_extracted_5489 | Not-determined |
| S7_extracted_5496 | Not-determined |
| S7_extracted_5502 | Not-determined |
| S7_extracted_5507 | Not-determined |
| S7_extracted_5508 | Not-determined |
| S7_extracted_5512 | Not-determined |
| S7_extracted_5519 | Low-quality |
| S7_extracted_5533 | Not-determined |
| S7_extracted_5540 | Not-determined |
| S7_extracted_5560 | Not-determined |
| S7_extracted_5564 | Not-determined |
| S7_extracted_5582 | Not-determined |
| S7_extracted_5584 | Not-determined |
| S7_extracted_5613 | Not-determined |
| S7_extracted_5616 | Not-determined |
| S7_extracted_5638 | Not-determined |
| S7_extracted_5645 | Not-determined |
| S7_extracted_5646 | Not-determined |
| S7_extracted_5652 | Not-determined |
| S7_extracted_5665 | Not-determined |
| S7_extracted_5671 | Low-quality |
| S7_extracted_5686 | Not-determined |
| S7_extracted_5731 | Not-determined |
| S7_extracted_5733 | Not-determined |
| S7_extracted_5738 | Not-determined |
| S7_extracted_5750 | Not-determined |
| S7_extracted_5757 | Low-quality |
| S7_extracted_5758 | Not-determined |
| S7_extracted_5762 | Not-determined |
| S7_extracted_5770 | Low-quality |
| S5_vs_k141_121411\|\|full | High-quality |
| S7_extracted_5818 | Low-quality |
| S7_extracted_5827 | Not-determined |
| S7_extracted_5840 | Not-determined |
| S5_vs_k141_8203\|\|full | Medium-quality |
| S7_extracted_5849 | Not-determined |
| S7_extracted_5877 | Not-determined |
| S7_extracted_5895 | Not-determined |
| S7_extracted_5919 | Not-determined |
| S7_extracted_5921 | Not-determined |
| S7_extracted_5933 | Not-determined |
| S7_extracted_5940 | Not-determined |
| S7_extracted_5944 | Not-determined |
| S7_extracted_5956 | Not-determined |
| S7_extracted_5978 | Not-determined |
| S7_extracted_5988 | Low-quality |
| S7_extracted_6034 | Not-determined |
| S7_extracted_6037 | Low-quality |
| S7_extracted_6047 | Low-quality |
| S7_extracted_6058 | Not-determined |
| S7_extracted_6092 | Low-quality |
| S7_extracted_6093 | Low-quality |
| S7_extracted_6103 | Not-determined |
| S7_extracted_6111 | Low-quality |
| S7_extracted_6116 | Not-determined |
| S7_extracted_6122 | Low-quality |
| S7_extracted_6134 | Not-determined |
| S7_extracted_6138 | Not-determined |
| S7_extracted_6165 | Not-determined |
| S7_extracted_6168 | Low-quality |
| S7_extracted_6197 | Not-determined |
| S7_extracted_6216 | Low-quality |
| S7_extracted_6226 | Low-quality |
| S7_extracted_6250 | Not-determined |
| S7_extracted_6264 | Not-determined |
| S7_extracted_6270 | Not-determined |
| S7_extracted_6277 | Not-determined |
| S7_extracted_6287 | Not-determined |
| S7_extracted_6291 | Not-determined |
| S7_extracted_6317 | Not-determined |
| S7_extracted_6324 | Not-determined |
| S7_extracted_6338 | Low-quality |
| S7_extracted_6362 | Not-determined |
| S7_extracted_6372 | Not-determined |
| S7_extracted_6386 | Not-determined |
| S7_extracted_6398 | Not-determined |
| S7_extracted_6407 | Not-determined |
| S7_extracted_6410 | Not-determined |
| S7_extracted_6422 | Low-quality |
| S7_extracted_6423 | Not-determined |
| S7_extracted_6456 | Low-quality |
| S7_extracted_6461 | Not-determined |
| S7_extracted_6464 | Not-determined |
| S7_extracted_6479 | Not-determined |
| S7_extracted_6494 | Not-determined |
| S7_extracted_6507 | Not-determined |
| S7_extracted_6508 | Not-determined |
| S7_extracted_6520 | Not-determined |
| S7_extracted_6523 | Not-determined |
| S7_extracted_6524 | Not-determined |
| S7_extracted_6536 | Not-determined |
| S7_extracted_6559 | Not-determined |
| S7_extracted_6561 | Not-determined |
| S7_extracted_6566 | Not-determined |
| S7_extracted_6591 | Low-quality |
| S7_extracted_6611 | Low-quality |
| S7_extracted_6623 | Not-determined |
| S7_extracted_6632 | Not-determined |
| S7_extracted_6650 | Not-determined |
| S7_extracted_6658 | Not-determined |
| S15_vs_41956\|\|full | Medium-quality |
| S7_extracted_6670 | Low-quality |
| S7_extracted_6673 | Not-determined |
| S7_extracted_6679 | Not-determined |
| S7_extracted_6691 | Not-determined |
| S7_extracted_6699 | Not-determined |
| S7_extracted_6712 | Not-determined |
| S7_extracted_6728 | Not-determined |
| S7_extracted_6732 | Low-quality |
| S7_extracted_6734 | Not-determined |
| S7_extracted_6737 | Not-determined |
| S7_extracted_6743 | Not-determined |
| S7_extracted_6758 | Not-determined |
| S7_extracted_6774 | Not-determined |
| S7_extracted_6779 | Not-determined |
| S7_extracted_6786 | Not-determined |
| S7_extracted_6808 | Not-determined |
| S7_extracted_6814 | Not-determined |
| S7_extracted_6816 | Not-determined |
| S7_extracted_6826 | Not-determined |
| S7_extracted_6835 | Not-determined |
| S7_extracted_6840 | Not-determined |
| S7_extracted_6856 | Not-determined |
| S7_extracted_6878 | Not-determined |
| S7_extracted_6903 | Not-determined |
| S7_extracted_6906 | Not-determined |
| S7_extracted_6920 | Not-determined |
| S7_extracted_6922 | Not-determined |
| S7_extracted_6943 | Not-determined |
| S7_extracted_6957 | Not-determined |
| S7_extracted_6961 | Not-determined |
| S7_extracted_7002 | Not-determined |
| S7_extracted_7003 | Not-determined |
| S7_extracted_7024 | Low-quality |
| S7_extracted_7031 | Not-determined |
| S7_extracted_7037 | Not-determined |
| S7_extracted_7051 | Low-quality |
| S7_extracted_7068 | Not-determined |
| S7_extracted_7072 | Not-determined |
| S7_extracted_7087 | Low-quality |
| S7_extracted_7116 | Low-quality |
| S7_extracted_7148 | Not-determined |
| S7_extracted_7170 | Low-quality |
| S7_extracted_7174 | Low-quality |
| S7_extracted_7175 | Not-determined |
| S7_extracted_7185 | Not-determined |
| S7_extracted_7212 | Low-quality |
| S7_extracted_7217 | Not-determined |
| S7_extracted_7227 | Not-determined |
| S7_extracted_7233 | Low-quality |
| S7_extracted_7240 | Low-quality |
| S7_extracted_7247 | Not-determined |
| S7_extracted_7274 | Not-determined |
| S7_extracted_7288 | Low-quality |
| S7_extracted_7323 | Not-determined |
| S7_extracted_7325 | Not-determined |
| S7_extracted_7327 | Not-determined |
| S7_extracted_7346 | Not-determined |
| S7_extracted_7349 | Not-determined |
| S7_extracted_7356 | Not-determined |
| S7_extracted_7369 | Not-determined |
| S7_extracted_7405 | Low-quality |
| S7_extracted_7414 | Not-determined |
| S7_extracted_7433 | Not-determined |
| S7_extracted_7441 | Low-quality |
| S7_extracted_7442 | Not-determined |
| S7_extracted_7447 | Not-determined |
| S7_extracted_7448 | Low-quality |
| S7_extracted_7496 | Not-determined |
| S7_extracted_7526 | Not-determined |
| S7_extracted_7539 | Not-determined |
| S7_extracted_7541 | Low-quality |
| S7_extracted_7544 | Low-quality |
| S7_extracted_7574 | Not-determined |
| S7_extracted_7583 | Not-determined |
| S7_extracted_7590 | Low-quality |
| S7_extracted_7599 | Not-determined |
| S7_extracted_7604 | Not-determined |
| S7_extracted_7605 | Not-determined |
| S7_extracted_7642 | Not-determined |
| S7_extracted_7644 | Not-determined |
| S7_extracted_7647 | Not-determined |
| S7_extracted_7660 | Not-determined |
| S7_extracted_7663 | Low-quality |
| S7_extracted_7665 | Low-quality |
| S7_extracted_7666 | Not-determined |
| S7_extracted_7669 | Not-determined |
| S7_extracted_7688 | Low-quality |
| S7_extracted_7705 | Low-quality |
| S7_extracted_7708 | Not-determined |
| S7_extracted_7711 | Not-determined |
| S7_extracted_7716 | Not-determined |
| S7_extracted_7718 | Low-quality |
| S7_extracted_7727 | Not-determined |
| S7_extracted_7732 | Not-determined |
| S7_extracted_7741 | Low-quality |
| S7_extracted_7742 | Not-determined |
| S7_extracted_7760 | Not-determined |
| S7_extracted_7769 | Not-determined |
| S7_extracted_7775 | Not-determined |
| S7_extracted_7795 | Not-determined |
| S7_extracted_7808 | Not-determined |
| S7_extracted_7814 | Not-determined |
| S7_extracted_7839 | Not-determined |
| S7_extracted_7851 | Not-determined |
| S7_extracted_7863 | Not-determined |
| S7_extracted_7891 | Not-determined |
| S7_extracted_7908 | Not-determined |
| S7_extracted_7913 | Not-determined |
| S7_extracted_7946 | Low-quality |
| S7_extracted_7961 | Not-determined |
| S7_extracted_7966 | Not-determined |
| S7_extracted_7969 | Not-determined |
| S7_extracted_7980 | Not-determined |
| S7_extracted_7981 | Not-determined |
| S7_extracted_7988 | Not-determined |
| S7_extracted_7990 | Not-determined |
| S7_extracted_8005 | Low-quality |
| S7_extracted_8020 | Not-determined |
| S7_extracted_8023 | Not-determined |
| S7_extracted_8038 | Not-determined |
| S7_extracted_8065 | Not-determined |
| S7_extracted_8069 | Not-determined |
| S7_extracted_8072 | Not-determined |
| S7_extracted_8098 | Not-determined |
| S7_extracted_8104 | Not-determined |
| S7_extracted_8118 | Low-quality |
| S7_extracted_8137 | Low-quality |
| S7_extracted_8144 | Low-quality |
| S7_extracted_8155 | Not-determined |
| S7_extracted_8188 | Not-determined |
| S7_extracted_8191 | Not-determined |
| S7_extracted_8208 | Not-determined |
| S7_extracted_8211 | Not-determined |
| S7_extracted_8212 | Not-determined |
| S7_extracted_8219 | Not-determined |
| S7_extracted_8224 | Not-determined |
| S7_extracted_8234 | Not-determined |
| S7_extracted_8245 | Low-quality |
| S7_extracted_8256 | Low-quality |
| S7_extracted_8287 | Not-determined |
| S7_extracted_8298 | Not-determined |
| S7_extracted_8309 | Not-determined |
| S7_extracted_8331 | Not-determined |
| S6_vs_k141_32886\|\|full | High-quality |
| S7_extracted_8371 | Not-determined |
| S7_extracted_8377 | Not-determined |
| S7_extracted_8384 | Not-determined |
| S7_extracted_8398 | Not-determined |
| S7_extracted_8416 | Not-determined |
| S7_extracted_8446 | Not-determined |
| S7_extracted_8449 | Not-determined |
| S7_extracted_8486 | Not-determined |
| S7_extracted_8499 | Not-determined |
| S7_extracted_8518 | Not-determined |
| S7_extracted_8519 | Not-determined |
| S7_extracted_8521 | Not-determined |
| S7_extracted_8522 | Not-determined |
| S7_extracted_8523 | Not-determined |
| S7_extracted_8527 | Not-determined |
| S7_extracted_8529 | Not-determined |
| S7_extracted_8538 | Not-determined |
| S7_extracted_8541 | Not-determined |
| S7_extracted_8557 | Not-determined |
| S7_extracted_8563 | Not-determined |
| S7_extracted_8570 | Not-determined |
| S7_extracted_8575 | Not-determined |
| S7_extracted_8578 | Not-determined |
| S7_extracted_8582 | Not-determined |
| S7_extracted_8602 | Low-quality |
| S7_extracted_8604 | Not-determined |
| S7_extracted_8606 | Low-quality |
| S7_extracted_8607 | Not-determined |
| S7_extracted_8613 | Not-determined |
| S7_extracted_8616 | Not-determined |
| S7_extracted_8646 | Low-quality |
| S7_extracted_8648 | Not-determined |
| S7_extracted_8649 | Low-quality |
| S7_extracted_8677 | Not-determined |
| S7_extracted_8679 | Not-determined |
| S7_extracted_8680 | Not-determined |
| S7_extracted_8709 | Not-determined |
| S7_extracted_8720 | Not-determined |
| S7_extracted_8735 | Not-determined |
| S7_extracted_8746 | Not-determined |
| S7_extracted_8749 | Not-determined |
| S7_extracted_8782 | Not-determined |
| S7_extracted_8786 | Not-determined |
| S7_extracted_8798 | Not-determined |
| S7_extracted_8819 | Not-determined |
| S7_extracted_8847 | Low-quality |
| S7_extracted_8849 | Not-determined |
| S7_extracted_8851 | Not-determined |
| S7_extracted_8861 | Not-determined |
| S7_extracted_8866 | Not-determined |
| S7_extracted_8881 | Low-quality |
| S7_extracted_8903 | Not-determined |
| S7_extracted_8905 | Not-determined |
| S7_extracted_8906 | Not-determined |
| S7_extracted_8909 | Not-determined |
| S7_extracted_8922 | Not-determined |
| S7_extracted_8930 | Not-determined |
| S7_extracted_8941 | Not-determined |
| S7_extracted_8956 | Not-determined |
| S7_extracted_8957 | Not-determined |
| S7_extracted_8958 | Not-determined |
| S7_extracted_8959 | Not-determined |
| S7_extracted_8986 | Low-quality |
| S7_extracted_8997 | Not-determined |
| S7_extracted_8998 | Not-determined |
| S7_extracted_9003 | Not-determined |
| S7_extracted_9011 | Not-determined |
| S7_extracted_9019 | Not-determined |
| S7_extracted_9023 | Not-determined |
| S7_extracted_9030 | Low-quality |
| S7_extracted_9047 | Not-determined |
| S7_extracted_9057 | Not-determined |
| S7_extracted_9058 | Low-quality |
| S7_extracted_9060 | Not-determined |
| S7_extracted_9061 | Not-determined |
| S7_extracted_9079 | Not-determined |
| S7_extracted_9130 | Not-determined |
| S7_extracted_9148 | Low-quality |
| S7_extracted_9212 | Not-determined |
| S7_extracted_9218 | Not-determined |
| S7_extracted_9226 | Not-determined |
| S7_extracted_9227 | Not-determined |
| S7_extracted_9231 | Not-determined |
| S7_extracted_9232 | Low-quality |
| S7_extracted_9246 | Not-determined |
| S7_extracted_9250 | Not-determined |
| S7_extracted_9252 | Not-determined |
| S7_extracted_9261 | Low-quality |
| S7_extracted_9292 | Not-determined |
| S7_extracted_9298 | Not-determined |
| S7_extracted_9312 | Low-quality |
| S7_extracted_9316 | Low-quality |
| S7_extracted_9324 | Not-determined |
| S7_extracted_9332 | Low-quality |
| S5_extracted_k141_77701 | Medium-quality |
| S7_extracted_9350 | Not-determined |
| S7_extracted_9379 | Not-determined |
| S7_extracted_9383 | Not-determined |
| S7_extracted_9392 | Low-quality |
| S7_extracted_9393 | Not-determined |
| S7_extracted_9397 | Not-determined |
| S7_extracted_9442 | Not-determined |
| S7_extracted_9445 | Not-determined |
| S7_extracted_9447 | Not-determined |
| S7_extracted_9469 | Not-determined |
| S7_extracted_9489 | Not-determined |
| S7_extracted_9492 | Not-determined |
| S7_extracted_9494 | Not-determined |
| S7_extracted_9502 | Not-determined |
| S7_extracted_9509 | Low-quality |
| S7_extracted_9528 | Not-determined |
| S7_extracted_9538 | Not-determined |
| S7_extracted_9544 | Not-determined |
| S7_extracted_9566 | Not-determined |
| S7_extracted_9577 | Not-determined |
| S7_extracted_9589 | Not-determined |
| S7_extracted_9597 | Not-determined |
| S7_extracted_9601 | Not-determined |
| S7_extracted_9608 | Not-determined |
| S7_extracted_9624 | Not-determined |
| S7_extracted_9633 | Not-determined |
| S7_extracted_9636 | Not-determined |
| S7_extracted_9647 | Low-quality |
| S7_extracted_9666 | Not-determined |
| S7_extracted_9671 | Low-quality |
| S7_extracted_9695 | Not-determined |
| S7_extracted_9703 | Not-determined |
| S7_extracted_9710 | Not-determined |
| S7_extracted_9711 | Not-determined |
| S7_extracted_9717 | Not-determined |
| S7_extracted_9727 | Not-determined |
| S7_extracted_9733 | Not-determined |
| S7_extracted_9736 | Low-quality |
| S7_extracted_9738 | Not-determined |
| S7_extracted_9761 | Not-determined |
| S7_extracted_9766 | Low-quality |
| S7_extracted_9773 | Not-determined |
| S7_extracted_9809 | Not-determined |
| S7_extracted_9819 | Not-determined |
| S7_extracted_9820 | Not-determined |
| S7_extracted_9821 | Not-determined |
| S7_extracted_9833 | Not-determined |
| S7_extracted_9836 | Not-determined |
| S7_extracted_9873 | Not-determined |
| S7_extracted_9877 | Not-determined |
| S7_extracted_9904 | Not-determined |
| S7_extracted_9929 | Not-determined |

| | |
|---|---|
| S7_extracted_9937 | Not-determined |
| S7_extracted_9946 | Low-quality |
| S7_extracted_9958 | Not-determined |
| S7_extracted_9963 | Not-determined |
| S7_extracted_9976 | Not-determined |
| S7_extracted_9977 | Not-determined |
| S7_extracted_9996 | Not-determined |
| S7_extracted_10007 | Not-determined |
| S7_extracted_10021 | Not-determined |
| S7_extracted_10040 | Not-determined |
| S7_extracted_10054 | Not-determined |
| S7_extracted_10061 | Not-determined |
| S7_extracted_10079 | Not-determined |
| S7_extracted_10088 | Not-determined |
| S7_extracted_10095 | Not-determined |
| S7_extracted_10101 | Not-determined |
| S7_extracted_10119 | Not-determined |
| S7_extracted_10127 | Not-determined |
| S7_extracted_10162 | Not-determined |
| S7_extracted_10174 | Not-determined |
| S7_extracted_10182 | Not-determined |
| S7_extracted_10230 | Not-determined |
| S7_extracted_10240 | Not-determined |
| S7_extracted_10252 | Not-determined |
| S7_extracted_10258 | Not-determined |
| S7_extracted_10285 | Not-determined |
| S7_extracted_10286 | Not-determined |
| S7_extracted_10298 | Not-determined |
| S7_extracted_10304 | Not-determined |
| S7_extracted_10315 | Not-determined |
| S7_extracted_10317 | Not-determined |
| S7_extracted_10318 | Not-determined |
| S7_extracted_10325 | Low-quality |
| S7_extracted_10334 | Not-determined |
| S7_extracted_10347 | Not-determined |
| S7_extracted_10348 | Not-determined |
| S7_extracted_10371 | Low-quality |
| S7_extracted_10376 | Not-determined |
| S7_extracted_10381 | Not-determined |
| S7_extracted_10398 | Not-determined |
| S7_extracted_10407 | Not-determined |
| S7_extracted_10434 | Not-determined |
| S7_extracted_10457 | Low-quality |
| S7_extracted_10465 | Not-determined |
| S7_extracted_10468 | Not-determined |
| S7_extracted_10471 | Not-determined |
| S7_extracted_10492 | Low-quality |
| S7_extracted_10501 | Not-determined |
| S7_extracted_10502 | Not-determined |
| S7_extracted_10503 | Not-determined |
| S7_extracted_10505 | Low-quality |
| S7_extracted_10522 | Not-determined |
| S7_extracted_10531 | Not-determined |
| S7_extracted_10536 | Not-determined |
| S7_extracted_10540 | Low-quality |
| S7_extracted_10544 | Low-quality |
| S7_extracted_10549 | Not-determined |
| S7_extracted_10552 | Not-determined |
| S7_extracted_10559 | Not-determined |
| S7_extracted_10562 | Low-quality |
| S7_extracted_10582 | Not-determined |
| S7_extracted_10600 | Not-determined |
| S7_extracted_10602 | Not-determined |
| S7_extracted_10603 | Not-determined |
| S7_extracted_10632 | Not-determined |
| S7_extracted_10636 | Not-determined |
| S7_extracted_10678 | Low-quality |
| S7_extracted_10686 | Not-determined |
| S7_extracted_10717 | Not-determined |
| S7_extracted_10739 | Low-quality |
| S7_extracted_10783 | Low-quality |
| S7_extracted_10799 | Not-determined |
| S7_extracted_10823 | Not-determined |
| S7_extracted_10827 | Not-determined |
| S7_extracted_10828 | Not-determined |
| S7_extracted_10854 | Not-determined |
| S7_extracted_10876 | Low-quality |
| S7_extracted_10877 | Low-quality |
| S7_extracted_10901 | Not-determined |
| S7_extracted_10930 | Not-determined |
| S7_extracted_10932 | Not-determined |
| S7_extracted_10956 | Not-determined |
| S7_extracted_10967 | Not-determined |
| S7_extracted_10999 | Not-determined |
| S7_extracted_11011 | Low-quality |
| S7_extracted_11020 | Not-determined |
| S7_extracted_11021 | Not-determined |
| S7_extracted_11023 | Low-quality |
| S7_extracted_11062 | Low-quality |
| S7_extracted_11084 | Low-quality |
| S7_extracted_11087 | Not-determined |
| S7_extracted_11093 | Low-quality |
| S7_extracted_11118 | Not-determined |
| S7_extracted_11128 | Not-determined |
| S7_extracted_11133 | Not-determined |
| S7_extracted_11134 | Not-determined |
| S7_extracted_11162 | Not-determined |
| S7_extracted_11170 | Not-determined |
| S7_extracted_11174 | Not-determined |
| S7_extracted_11181 | Not-determined |
| S7_extracted_11185 | Low-quality |
| S7_extracted_11203 | Not-determined |
| S7_extracted_11207 | Not-determined |
| S7_extracted_11215 | Low-quality |
| S7_extracted_11236 | Not-determined |
| S7_extracted_11238 | Not-determined |
| S7_extracted_11261 | Not-determined |
| S7_extracted_11266 | Not-determined |
| S7_extracted_11275 | Not-determined |
| S7_extracted_11278 | Low-quality |
| S7_extracted_11280 | Not-determined |
| S7_extracted_11291 | Not-determined |
| S7_extracted_11293 | Not-determined |
| S7_extracted_11308 | Low-quality |
| S7_extracted_11328 | Not-determined |
| S7_extracted_11329 | Not-determined |
| S7_extracted_11332 | Not-determined |
| S7_extracted_11338 | Not-determined |
| S7_extracted_11349 | Low-quality |
| S7_extracted_11385 | Not-determined |
| S7_extracted_11390 | Low-quality |
| S7_extracted_11404 | Not-determined |
| S7_extracted_11414 | Not-determined |
| S7_extracted_11415 | Not-determined |
| S7_extracted_11417 | Not-determined |
| S7_extracted_11421 | Not-determined |
| S7_extracted_11429 | Low-quality |
| S7_extracted_11444 | Not-determined |
| S7_extracted_11453 | Not-determined |
| S7_extracted_11458 | Not-determined |
| S7_extracted_11460 | Not-determined |
| S7_extracted_11481 | Not-determined |
| S7_extracted_11492 | Low-quality |
| S7_extracted_11499 | Not-determined |
| S7_extracted_11515 | Not-determined |
| S7_extracted_11517 | Low-quality |
| S7_extracted_11538 | Low-quality |
| S7_extracted_11550 | Not-determined |
| S7_extracted_11562 | Not-determined |
| S7_extracted_11575 | Not-determined |
| S7_extracted_11606 | Not-determined |
| S7_extracted_11620 | Not-determined |
| S7_extracted_11627 | Not-determined |
| S7_extracted_11633 | Low-quality |
| S7_extracted_11640 | Not-determined |
| S7_extracted_11658 | Not-determined |
| S7_extracted_11659 | Low-quality |
| S7_extracted_11697 | Not-determined |
| S7_extracted_11702 | Not-determined |
| S7_extracted_11707 | Not-determined |
| S7_extracted_11714 | Not-determined |
| S7_extracted_11726 | Not-determined |
| S7_extracted_11734 | Not-determined |
| S7_extracted_11741 | Not-determined |
| S17_vs_k141_177196\|\|full | High-quality |
| S7_extracted_11757 | Not-determined |
| S7_extracted_11763 | Not-determined |
| S7_extracted_11771 | Not-determined |
| S7_extracted_11785 | Not-determined |
| S7_extracted_11795 | Not-determined |
| S7_extracted_11809 | Not-determined |
| S7_extracted_11813 | Not-determined |
| S7_extracted_11821 | Not-determined |
| S7_extracted_11823 | Low-quality |
| S7_extracted_11824 | Low-quality |
| S7_extracted_11833 | Not-determined |
| S7_extracted_11849 | Low-quality |
| S7_extracted_11856 | Not-determined |
| S7_extracted_11870 | Not-determined |
| S7_extracted_11898 | Not-determined |
| S7_extracted_11902 | Not-determined |
| S7_extracted_11906 | Not-determined |
| S7_extracted_11936 | Not-determined |
| S7_extracted_11971 | Low-quality |
| S7_extracted_11996 | Not-determined |
| S7_extracted_11998 | Not-determined |
| S7_extracted_12001 | Not-determined |
| S7_extracted_12002 | Not-determined |
| S7_extracted_12010 | Not-determined |
| S7_extracted_12016 | Not-determined |
| S7_extracted_12018 | Not-determined |
| S7_extracted_12058 | Not-determined |
| S7_extracted_12075 | Not-determined |
| S7_extracted_12099 | Not-determined |
| S7_extracted_12102 | Low-quality |
| S7_extracted_12123 | Low-quality |
| S7_extracted_12130 | Not-determined |
| S7_extracted_12133 | Low-quality |
| S7_extracted_12148 | Not-determined |
| S7_extracted_12159 | Not-determined |
| S7_extracted_12164 | Not-determined |
| S7_extracted_12165 | Not-determined |
| S7_extracted_12169 | Not-determined |
| S7_extracted_12171 | Low-quality |
| S7_extracted_12173 | Not-determined |
| S7_extracted_12185 | Not-determined |
| S7_extracted_12193 | Not-determined |
| S7_extracted_12206 | Not-determined |
| S7_extracted_12212 | Not-determined |
| S7_extracted_12225 | Not-determined |
| S7_extracted_12229 | Low-quality |
| S7_extracted_12254 | Not-determined |
| S7_extracted_12259 | Not-determined |
| S7_extracted_12273 | Not-determined |
| S7_extracted_12280 | Not-determined |
| S7_extracted_12324 | Not-determined |
| S7_extracted_12356 | Not-determined |
| S7_extracted_12378 | Not-determined |
| S7_extracted_12382 | Low-quality |
| S7_extracted_12386 | Low-quality |
| S7_extracted_12394 | Not-determined |
| S7_extracted_12403 | Not-determined |
| S7_extracted_12408 | Not-determined |
| S7_extracted_12421 | Not-determined |
| S7_extracted_12452 | Not-determined |
| S7_extracted_12453 | Not-determined |
| S7_extracted_12462 | Not-determined |
| S7_extracted_12485 | Not-determined |
| S7_extracted_12506 | Not-determined |
| S7_extracted_12512 | Not-determined |
| S7_extracted_12530 | Not-determined |
| S7_extracted_12531 | Low-quality |
| S7_extracted_12575 | Low-quality |
| S7_extracted_12576 | Not-determined |
| S7_extracted_12584 | Not-determined |
| S7_extracted_12595 | Not-determined |
| S7_extracted_12621 | Not-determined |
| S7_extracted_12624 | Not-determined |
| S7_extracted_12626 | Not-determined |
| S7_extracted_12637 | Low-quality |
| S7_extracted_12643 | Not-determined |
| S7_extracted_12682 | Low-quality |
| S7_extracted_12709 | Low-quality |
| S7_extracted_12722 | Not-determined |
| S7_extracted_12738 | Not-determined |
| S7_extracted_12741 | Not-determined |
| S7_extracted_12742 | Not-determined |
| S7_extracted_12744 | Not-determined |
| S7_extracted_12789 | Not-determined |
| S7_extracted_12801 | Not-determined |
| S7_extracted_12809 | Low-quality |
| S7_extracted_12813 | Not-determined |
| S7_extracted_12818 | Not-determined |
| S7_extracted_12827 | Low-quality |
| S7_extracted_12841 | Not-determined |
| S7_extracted_12843 | Low-quality |
| S7_extracted_12844 | Not-determined |
| S7_extracted_12855 | Not-determined |
| S7_extracted_12857 | Not-determined |
| S7_extracted_12881 | Low-quality |
| S7_extracted_12883 | Low-quality |
| S7_extracted_12887 | Not-determined |
| S7_extracted_12892 | Low-quality |
| S7_extracted_12902 | Not-determined |
| S7_extracted_12907 | Not-determined |
| S7_extracted_12919 | Not-determined |
| S7_extracted_12924 | Not-determined |
| S7_extracted_12927 | Low-quality |
| S7_extracted_12932 | Not-determined |
| S7_extracted_12940 | Not-determined |
| S7_extracted_12959 | Not-determined |
| S7_extracted_12961 | Not-determined |
| S7_extracted_12968 | Not-determined |
| S7_extracted_12998 | Not-determined |
| S7_extracted_13005 | Not-determined |
| S7_extracted_13009 | Low-quality |
| S7_extracted_13017 | Low-quality |
| S7_extracted_13026 | Not-determined |
| S7_extracted_13029 | Not-determined |
| S7_extracted_13044 | Not-determined |
| S7_extracted_13074 | Not-determined |
| S7_extracted_13117 | Low-quality |
| S7_extracted_13120 | Not-determined |
| S7_extracted_13123 | Not-determined |
| S7_extracted_13133 | Low-quality |
| S7_extracted_13177 | Not-determined |
| S7_extracted_13192 | Not-determined |
| S7_extracted_13226 | Not-determined |
| S7_extracted_13269 | Not-determined |
| S7_extracted_13273 | Not-determined |
| S7_extracted_13308 | Not-determined |
| S7_extracted_13368 | Not-determined |
| S7_extracted_13371 | Not-determined |
| S7_extracted_13376 | Not-determined |
| S7_extracted_13394 | Low-quality |
| S7_extracted_13396 | Not-determined |
| S7_extracted_13406 | Not-determined |
| S7_extracted_13410 | Not-determined |
| S7_extracted_13425 | Not-determined |
| S7_extracted_13427 | Not-determined |
| S7_extracted_13449 | Not-determined |
| S7_extracted_13455 | Not-determined |
| S7_extracted_13463 | Not-determined |
| S7_extracted_13475 | Not-determined |
| S7_extracted_13483 | Not-determined |
| S7_extracted_13520 | Not-determined |
| S7_extracted_13529 | Low-quality |
| S7_extracted_13536 | Low-quality |
| S7_extracted_13537 | Not-determined |
| S7_extracted_13545 | Not-determined |
| S7_extracted_13559 | Not-determined |
| S7_extracted_13571 | Not-determined |
| S7_extracted_13576 | Not-determined |
| S7_extracted_13579 | Not-determined |
| S7_extracted_13581 | Not-determined |
| S7_extracted_13595 | Not-determined |
| S16_vs_173100\|\|full | High-quality |
| S7_extracted_13624 | Not-determined |
| S7_extracted_13641 | Not-determined |
| S7_extracted_13672 | Not-determined |
| S7_extracted_13674 | Not-determined |
| S7_extracted_13676 | Not-determined |
| S7_extracted_13687 | Not-determined |
| S7_extracted_13694 | Not-determined |
| S7_extracted_13715 | Not-determined |
| S7_extracted_13736 | Not-determined |
| S7_extracted_13738 | Not-determined |
| S7_extracted_13740 | Not-determined |
| S7_extracted_13745 | Not-determined |
| S7_extracted_13761 | Low-quality |
| S7_extracted_13765 | Low-quality |
| S7_extracted_13778 | Not-determined |
| S7_extracted_13828 | Low-quality |
| S7_extracted_13834 | Not-determined |
| S7_extracted_13844 | Not-determined |
| S7_extracted_13845 | Not-determined |
| S7_extracted_13858 | Not-determined |
| S7_extracted_13872 | Not-determined |
| S7_extracted_13875 | Low-quality |
| S7_extracted_13911 | Not-determined |
| S7_extracted_13914 | Not-determined |
| S7_extracted_13917 | Low-quality |
| S7_extracted_13925 | Not-determined |
| S6_vs_k141_43762\|\|full | Medium-quality |
| S7_extracted_13963 | Not-determined |
| S7_extracted_13965 | Not-determined |
| S7_extracted_13983 | Not-determined |
| S7_extracted_13992 | Not-determined |
| S7_extracted_14013 | Not-determined |
| S7_extracted_14027 | Not-determined |
| S7_extracted_14034 | Low-quality |
| S7_extracted_14070 | Low-quality |
| S7_extracted_14073 | Not-determined |
| S7_extracted_14090 | Not-determined |
| S7_extracted_14092 | Not-determined |
| S7_extracted_14097 | Not-determined |
| S7_extracted_14103 | Not-determined |
| S7_extracted_14107 | Low-quality |
| S7_extracted_14111 | Low-quality |
| S7_extracted_14114 | Low-quality |
| S7_extracted_14141 | Low-quality |
| S7_extracted_14151 | Not-determined |
| S7_extracted_14156 | Not-determined |
| S7_extracted_14158 | Low-quality |
| S7_extracted_14159 | Not-determined |
| S7_extracted_14162 | Not-determined |
| S7_extracted_14171 | Not-determined |
| S7_extracted_14181 | Not-determined |
| S7_extracted_14195 | Not-determined |
| S7_extracted_14214 | Not-determined |
| S7_extracted_14215 | Not-determined |
| S7_extracted_14233 | Not-determined |
| S7_extracted_14239 | Low-quality |
| S7_extracted_14249 | Not-determined |
| S7_extracted_14260 | Not-determined |
| S7_extracted_14293 | Not-determined |
| S7_extracted_14294 | Not-determined |
| S7_extracted_14297 | Not-determined |
| S7_extracted_14301 | Not-determined |
| S7_extracted_14315 | Not-determined |
| S7_extracted_14329 | Not-determined |
| S7_extracted_14335 | Low-quality |
| S7_extracted_14347 | Not-determined |
| S7_extracted_14355 | Low-quality |
| S7_extracted_14357 | Not-determined |
| S7_extracted_14361 | Not-determined |
| S7_extracted_14368 | Not-determined |
| S7_extracted_14372 | Low-quality |
| S7_extracted_14403 | Not-determined |
| S7_extracted_14411 | Not-determined |
| S7_extracted_14412 | Not-determined |
| S7_extracted_14420 | Not-determined |
| S7_extracted_14422 | Low-quality |
| S7_extracted_14432 | Not-determined |
| S7_extracted_14454 | Low-quality |
| S7_extracted_14460 | Not-determined |
| S7_extracted_14468 | Not-determined |
| S7_extracted_14486 | Not-determined |
| S7_extracted_14488 | Not-determined |
| S7_extracted_14495 | Not-determined |
| S7_extracted_14498 | Low-quality |
| S7_extracted_14499 | Not-determined |
| S7_extracted_14505 | Low-quality |
| S7_extracted_14517 | Not-determined |
| S7_extracted_14520 | Not-determined |
| S7_extracted_14533 | Not-determined |
| S7_extracted_14555 | Low-quality |
| S7_extracted_14558 | Not-determined |
| S7_extracted_14593 | Not-determined |
| S7_extracted_14619 | Not-determined |
| S7_extracted_14624 | Low-quality |
| S7_extracted_14641 | Not-determined |
| S7_extracted_14650 | Not-determined |
| S7_extracted_14660 | Not-determined |
| S7_extracted_14667 | Not-determined |
| S7_extracted_14670 | Low-quality |
| S7_extracted_14672 | Not-determined |
| S7_extracted_14677 | Low-quality |
| S7_extracted_14694 | Not-determined |
| S7_extracted_14695 | Not-determined |
| S7_extracted_14707 | Not-determined |
| S7_extracted_14716 | Not-determined |
| S7_extracted_14727 | Not-determined |
| S7_extracted_14735 | Low-quality |
| S7_extracted_14737 | Not-determined |
| S7_extracted_14743 | Not-determined |
| S7_extracted_14752 | Low-quality |
| S7_extracted_14760 | Not-determined |
| S7_extracted_14765 | Low-quality |
| S7_extracted_14767 | Not-determined |
| S7_extracted_14771 | Not-determined |
| S7_extracted_14774 | Not-determined |
| S7_extracted_14778 | Not-determined |
| S7_extracted_14795 | Not-determined |
| S7_extracted_14818 | Low-quality |
| S7_extracted_14823 | Not-determined |
| S7_extracted_14854 | Not-determined |
| S7_extracted_14877 | Not-determined |
| S7_extracted_14885 | Not-determined |
| S7_extracted_14887 | Not-determined |
| S7_extracted_14901 | Not-determined |
| S7_extracted_14905 | Not-determined |
| S7_extracted_14909 | Not-determined |
| S7_extracted_14913 | Not-determined |
| S7_extracted_14922 | Not-determined |
| S7_extracted_14936 | Not-determined |
| S7_extracted_14950 | Not-determined |
| S7_extracted_14951 | Not-determined |
| S7_extracted_14973 | Not-determined |
| S7_extracted_14992 | Not-determined |
| S7_extracted_15000 | Not-determined |
| S7_extracted_15002 | Low-quality |
| S7_extracted_15015 | Not-determined |
| S7_extracted_15019 | Low-quality |
| S7_extracted_15028 | Not-determined |
| S7_extracted_15035 | Not-determined |
| S7_extracted_15038 | Not-determined |
| S7_extracted_15051 | Not-determined |
| S7_extracted_15052 | Not-determined |
| S7_extracted_15055 | Not-determined |
| S7_extracted_15056 | Not-determined |
| S7_extracted_15070 | Not-determined |
| S7_extracted_15074 | Low-quality |
| S7_extracted_15075 | Not-determined |
| S7_extracted_15091 | Not-determined |
| S7_extracted_15108 | Not-determined |
| S7_extracted_15114 | Not-determined |
| S7_extracted_15117 | Not-determined |
| S7_extracted_15138 | Not-determined |
| S7_extracted_15167 | Not-determined |
| S7_extracted_15183 | Not-determined |
| S7_extracted_15186 | Not-determined |
| S7_extracted_15242 | Low-quality |
| S7_extracted_15244 | Not-determined |
| S7_extracted_15251 | Not-determined |
| S7_extracted_15252 | Not-determined |
| S7_extracted_15283 | Not-determined |
| S7_extracted_15286 | Not-determined |
| S7_extracted_15290 | Not-determined |
| S7_extracted_15316 | Not-determined |
| S7_extracted_15320 | Not-determined |
| S7_extracted_15335 | Not-determined |
| S7_extracted_15364 | Not-determined |
| S7_extracted_15367 | Not-determined |
| S7_extracted_15373 | Not-determined |
| S7_extracted_15379 | Not-determined |
| S7_extracted_15391 | Not-determined |
| S7_extracted_15397 | Low-quality |
| S7_extracted_15447 | Not-determined |
| S7_extracted_15449 | Not-determined |
| S7_extracted_15454 | Not-determined |
| S7_extracted_15476 | Low-quality |
| S7_extracted_15481 | Not-determined |
| S7_extracted_15499 | Not-determined |
| S7_extracted_15512 | Low-quality |
| S7_extracted_15528 | Not-determined |
| S7_extracted_15547 | Low-quality |
| S7_extracted_15557 | Low-quality |
| S7_extracted_15567 | Low-quality |
| S7_extracted_15576 | Not-determined |
| S7_extracted_15597 | Not-determined |
| S7_extracted_15599 | Not-determined |
| S7_extracted_15617 | Not-determined |
| S7_extracted_15647 | Not-determined |
| S7_extracted_15649 | Low-quality |
| S7_extracted_15651 | Not-determined |
| S7_extracted_15669 | Not-determined |
| S7_extracted_15673 | Not-determined |
| S7_extracted_15686 | Low-quality |
| S7_extracted_15703 | Not-determined |
| S7_extracted_15710 | Not-determined |
| S7_extracted_15726 | Not-determined |
| S7_extracted_15731 | Not-determined |
| S7_extracted_15755 | Not-determined |
| S7_extracted_15759 | Low-quality |
| S7_extracted_15780 | Not-determined |
| S7_extracted_15800 | Not-determined |
| S7_extracted_15802 | Not-determined |
| S7_extracted_15807 | Not-determined |
| S7_extracted_15809 | Not-determined |
| S7_extracted_15829 | Not-determined |
| S7_extracted_15838 | Low-quality |
| S7_extracted_15859 | Low-quality |
| S7_extracted_15866 | Not-determined |
| S7_extracted_15884 | Not-determined |
| S7_extracted_15886 | Low-quality |
| S7_extracted_15889 | Not-determined |
| S7_extracted_15893 | Low-quality |
| S7_extracted_15896 | Not-determined |
| S7_extracted_15898 | Not-determined |
| S7_extracted_15900 | Not-determined |
| S7_extracted_15907 | Low-quality |
| S7_extracted_15909 | Not-determined |
| S7_extracted_15911 | Not-determined |
| S7_extracted_15916 | Not-determined |
| S7_extracted_15917 | Not-determined |
| S7_extracted_15922 | Not-determined |
| S7_extracted_15923 | Low-quality |
| S7_extracted_15931 | Not-determined |
| S7_extracted_15980 | Not-determined |
| S7_extracted_16018 | Not-determined |
| S7_extracted_16019 | Not-determined |
| S7_extracted_16034 | Not-determined |
| S7_extracted_16045 | Low-quality |
| S7_extracted_16061 | Not-determined |
| S7_extracted_16073 | Low-quality |
| S7_extracted_16076 | Not-determined |
| S7_extracted_16078 | Not-determined |
| S7_extracted_16100 | Not-determined |
| S7_extracted_16131 | Not-determined |
| S7_extracted_16135 | Not-determined |
| S7_extracted_16137 | Low-quality |
| S7_extracted_16138 | Not-determined |
| S7_extracted_16141 | Not-determined |
| S7_extracted_16146 | Not-determined |
| S7_extracted_16161 | Low-quality |
| S7_extracted_16165 | Low-quality |
| S7_extracted_16167 | Low-quality |
| S7_extracted_16178 | Not-determined |
| S7_extracted_16199 | Not-determined |
| S7_extracted_16204 | Not-determined |
| S7_extracted_16212 | Not-determined |
| S7_extracted_16218 | Not-determined |
| S7_extracted_16224 | Low-quality |
| S7_extracted_16241 | Not-determined |
| S7_extracted_16246 | Not-determined |
| S7_extracted_16254 | Not-determined |
| S7_extracted_16273 | Not-determined |
| S7_extracted_16274 | Low-quality |
| S7_extracted_16293 | Not-determined |
| S7_extracted_16295 | Not-determined |
| S7_extracted_16305 | Not-determined |
| S7_extracted_16361 | Low-quality |
| S7_extracted_16364 | Not-determined |
| S7_extracted_16375 | Not-determined |
| S7_extracted_16376 | Not-determined |
| S7_extracted_16409 | Not-determined |
| S7_extracted_16419 | Not-determined |
| S7_extracted_16439 | Not-determined |
| S7_extracted_16450 | Not-determined |
| S7_extracted_16454 | Low-quality |
| S7_extracted_16456 | Not-determined |
| S7_extracted_16470 | Low-quality |
| S7_extracted_16473 | Not-determined |
| S7_extracted_16474 | Not-determined |
| S7_extracted_16484 | Not-determined |
| S7_extracted_16487 | Not-determined |
| S7_extracted_16495 | Not-determined |
| S7_extracted_16526 | Not-determined |
| S7_extracted_16529 | Low-quality |
| S7_extracted_16533 | Not-determined |
| S7_extracted_16537 | Not-determined |
| S7_extracted_16547 | Not-determined |
| S7_extracted_16584 | Not-determined |
| S7_extracted_16596 | Not-determined |
| S7_extracted_16600 | Not-determined |
| S7_extracted_16632 | Not-determined |
| S7_extracted_16638 | Not-determined |
| S7_extracted_16645 | Low-quality |

| | |
|---|---|
| S7_extracted_16651 | Not-determined |
| S7_extracted_16670 | Not-determined |
| S7_extracted_16685 | Not-determined |
| S7_extracted_16700 | Not-determined |
| S7_extracted_16704 | Not-determined |
| S7_extracted_16707 | Low-quality |
| S7_extracted_16723 | Not-determined |
| S7_extracted_16739 | Low-quality |
| S7_extracted_16755 | Not-determined |
| S7_extracted_16758 | Low-quality |
| S7_extracted_16759 | Not-determined |
| S7_extracted_16766 | Not-determined |
| S7_extracted_16767 | Low-quality |
| S7_extracted_16785 | Not-determined |
| S7_extracted_16786 | Not-determined |
| S7_extracted_16788 | Not-determined |
| S7_extracted_16801 | Not-determined |
| S7_extracted_16803 | Not-determined |
| S7_extracted_16831 | Not-determined |
| S7_extracted_16834 | Not-determined |
| S7_extracted_16835 | Low-quality |
| S7_extracted_16842 | Low-quality |
| S7_extracted_16849 | Low-quality |
| S7_extracted_16852 | Not-determined |
| S7_extracted_16865 | Not-determined |
| S7_extracted_16869 | Low-quality |
| S7_extracted_16885 | Not-determined |
| S7_extracted_16888 | Not-determined |
| S7_extracted_16894 | Not-determined |
| S7_extracted_16900 | Low-quality |
| S7_extracted_16904 | Not-determined |
| S7_extracted_16906 | Not-determined |
| S7_extracted_16922 | Low-quality |
| S7_extracted_16927 | Not-determined |
| S7_extracted_16954 | Low-quality |
| S7_extracted_16960 | Not-determined |
| S7_extracted_16978 | Not-determined |
| S7_extracted_16983 | Not-determined |
| S7_extracted_17023 | Not-determined |
| S7_extracted_17025 | Not-determined |
| S7_extracted_17029 | Not-determined |
| S7_extracted_17031 | Not-determined |
| S7_extracted_17051 | Not-determined |
| S7_extracted_17067 | Not-determined |
| S7_extracted_17079 | Not-determined |
| S7_extracted_17080 | Low-quality |
| S7_extracted_17098 | Not-determined |
| S7_extracted_17127 | Not-determined |
| S7_extracted_17161 | Low-quality |
| S7_extracted_17172 | Not-determined |
| S7_extracted_17175 | Not-determined |
| S7_extracted_17193 | Not-determined |
| S7_extracted_17200 | Low-quality |
| S7_extracted_17202 | Not-determined |
| S7_extracted_17205 | Not-determined |
| S7_extracted_17209 | Low-quality |
| S7_extracted_17214 | Not-determined |
| S7_extracted_17219 | Low-quality |
| S7_extracted_17224 | Not-determined |
| S7_extracted_17238 | Not-determined |
| S7_extracted_17241 | Not-determined |
| S7_extracted_17264 | Not-determined |
| S7_extracted_17270 | Not-determined |
| S7_extracted_17277 | Not-determined |
| S7_extracted_17279 | Not-determined |
| S7_extracted_17287 | Not-determined |
| S7_extracted_17297 | Low-quality |
| S7_extracted_17304 | Not-determined |
| S7_extracted_17334 | Not-determined |
| S7_extracted_17346 | Not-determined |
| S7_extracted_17347 | Not-determined |
| S7_extracted_17353 | Not-determined |
| S7_extracted_17364 | Low-quality |
| S7_extracted_17367 | Not-determined |
| S7_extracted_17397 | Not-determined |
| S7_extracted_17417 | Low-quality |
| S7_extracted_17419 | Not-determined |
| S7_extracted_17430 | Not-determined |
| S7_extracted_17449 | Low-quality |
| S7_extracted_17455 | Not-determined |
| S7_extracted_17462 | Not-determined |
| S7_extracted_17470 | Not-determined |
| S7_extracted_17474 | Not-determined |
| S7_extracted_17516 | Not-determined |
| S7_extracted_17523 | Not-determined |
| S7_extracted_17526 | Not-determined |
| S7_extracted_17532 | Not-determined |
| S7_extracted_17541 | Not-determined |
| S7_extracted_17549 | Not-determined |
| S7_extracted_17554 | Low-quality |
| S7_extracted_17557 | Low-quality |
| S7_extracted_17558 | Not-determined |
| S7_extracted_17570 | Not-determined |
| S7_extracted_17575 | Not-determined |
| S7_extracted_17581 | Low-quality |
| S7_extracted_17612 | Not-determined |
| S7_extracted_17615 | Low-quality |
| S7_extracted_17619 | Not-determined |
| S7_extracted_17622 | Low-quality |
| S7_extracted_17628 | Not-determined |
| S7_extracted_17652 | Not-determined |
| S7_extracted_17677 | Not-determined |
| S7_extracted_17692 | Low-quality |
| S7_extracted_17706 | Low-quality |
| S7_extracted_17712 | Not-determined |
| S7_extracted_17721 | Not-determined |
| S7_extracted_17724 | Low-quality |
| S7_extracted_17731 | Not-determined |
| S7_extracted_17736 | Not-determined |
| S7_extracted_17749 | Not-determined |
| S7_extracted_17764 | Not-determined |
| S7_extracted_17773 | Not-determined |
| S7_extracted_17808 | Not-determined |
| S7_extracted_17812 | Not-determined |
| S7_extracted_17834 | Low-quality |
| S7_extracted_17836 | Not-determined |
| S7_extracted_17838 | Not-determined |
| S7_extracted_17851 | Not-determined |
| S7_extracted_17856 | Low-quality |
| S7_extracted_17897 | Not-determined |
| S7_extracted_17901 | Not-determined |
| S7_extracted_17926 | Not-determined |
| S7_extracted_17929 | Not-determined |
| S7_extracted_17935 | Not-determined |
| S7_extracted_17938 | Not-determined |
| S7_extracted_17949 | Not-determined |
| S7_extracted_17967 | Not-determined |
| S7_extracted_18002 | Not-determined |
| S7_extracted_18009 | Not-determined |
| S7_extracted_18018 | Not-determined |
| S7_extracted_18021 | Not-determined |
| S7_extracted_18033 | Not-determined |
| S7_extracted_18034 | Not-determined |
| S7_extracted_18039 | Low-quality |
| S7_extracted_18041 | Low-quality |
| S7_extracted_18062 | Low-quality |
| S7_extracted_18063 | Not-determined |
| S7_extracted_18066 | Not-determined |
| S7_extracted_18069 | Low-quality |
| S7_extracted_18070 | Not-determined |
| S7_extracted_18071 | Not-determined |
| S7_extracted_18095 | Not-determined |
| S7_extracted_18100 | Not-determined |
| S7_extracted_18112 | Low-quality |
| S7_extracted_18130 | Low-quality |
| S7_extracted_18141 | Not-determined |
| S7_extracted_18164 | Low-quality |
| S7_extracted_18183 | Not-determined |
| S7_extracted_18199 | Not-determined |
| S7_extracted_18214 | Not-determined |
| S7_extracted_18215 | Not-determined |
| S7_extracted_18216 | Not-determined |
| S7_extracted_18230 | Low-quality |
| S7_extracted_18243 | Low-quality |
| S7_extracted_18244 | Not-determined |
| S7_extracted_18260 | Not-determined |
| S7_extracted_18261 | Not-determined |
| S7_extracted_18269 | Low-quality |
| S7_extracted_18294 | Not-determined |
| S7_extracted_18299 | Not-determined |
| S7_extracted_18330 | Not-determined |
| S7_extracted_18350 | Not-determined |
| S7_extracted_18353 | Not-determined |
| S7_extracted_18366 | Not-determined |
| S7_extracted_18376 | Not-determined |
| S7_extracted_18398 | Not-determined |
| S7_extracted_18402 | Low-quality |
| S7_extracted_18409 | Low-quality |
| S7_extracted_18425 | Not-determined |
| S7_extracted_18449 | Not-determined |
| S7_extracted_18454 | Not-determined |
| S7_extracted_18461 | Not-determined |
| S7_extracted_18462 | Low-quality |
| S7_extracted_18494 | Not-determined |
| S7_extracted_18513 | Not-determined |
| S7_extracted_18525 | Low-quality |
| S7_extracted_18549 | Not-determined |
| S7_extracted_18551 | Not-determined |
| S7_extracted_18559 | Low-quality |
| S7_extracted_18562 | Low-quality |
| S7_extracted_18580 | Not-determined |
| S7_extracted_18596 | Not-determined |
| S7_extracted_18605 | Not-determined |
| S7_extracted_18614 | Not-determined |
| S7_extracted_18619 | Not-determined |
| S7_extracted_18620 | Not-determined |
| S7_extracted_18625 | Not-determined |
| S7_extracted_18635 | Low-quality |
| S7_extracted_18644 | Low-quality |
| S7_extracted_18647 | Not-determined |
| S7_extracted_18662 | Low-quality |
| S7_extracted_18668 | Not-determined |
| S7_extracted_18680 | Not-determined |
| S7_extracted_18681 | Not-determined |
| S7_extracted_18697 | Not-determined |
| S7_extracted_18708 | Low-quality |
| S7_extracted_18769 | Not-determined |
| S7_extracted_18772 | Not-determined |
| S7_extracted_18786 | Not-determined |
| S7_extracted_18810 | Not-determined |
| S7_extracted_18822 | Not-determined |
| S7_extracted_18827 | Not-determined |
| S7_extracted_18842 | Not-determined |
| S7_extracted_18847 | Low-quality |
| S7_extracted_18849 | Not-determined |
| S7_extracted_18851 | Not-determined |
| S7_extracted_18858 | Low-quality |
| S7_extracted_18866 | Not-determined |
| S7_extracted_18871 | Not-determined |
| S7_extracted_18872 | Not-determined |
| S7_extracted_18880 | Not-determined |
| S7_extracted_18900 | Low-quality |
| S7_extracted_18918 | Not-determined |
| S7_extracted_18920 | Not-determined |
| S7_extracted_18924 | Low-quality |
| S7_extracted_18935 | Not-determined |
| S7_extracted_18960 | Not-determined |
| S7_extracted_18970 | Not-determined |
| S7_extracted_18980 | Not-determined |
| S7_extracted_18987 | Low-quality |
| S7_extracted_18988 | Not-determined |
| S7_extracted_18989 | Not-determined |
| S7_extracted_19010 | Not-determined |
| S7_extracted_19040 | Not-determined |
| S7_extracted_19083 | Not-determined |
| S7_extracted_19086 | Not-determined |
| S7_extracted_19109 | Not-determined |
| S7_extracted_19110 | Low-quality |
| S7_extracted_19118 | Not-determined |
| S7_extracted_19122 | Not-determined |
| S7_extracted_19129 | Not-determined |
| S7_extracted_19140 | Not-determined |
| S7_extracted_19205 | Not-determined |
| S7_extracted_19214 | Not-determined |
| S7_extracted_19217 | Not-determined |
| S7_extracted_19219 | Not-determined |
| S7_extracted_19224 | Not-determined |
| S7_extracted_19254 | Not-determined |
| S7_extracted_19301 | Not-determined |
| S7_extracted_19307 | Not-determined |
| S7_extracted_19313 | Low-quality |
| S7_extracted_19364 | Not-determined |
| S7_extracted_19376 | Not-determined |
| S7_extracted_19378 | Not-determined |
| S7_extracted_19381 | Not-determined |
| S7_extracted_19384 | Low-quality |
| S7_extracted_19386 | Low-quality |
| S7_extracted_19390 | Not-determined |
| S7_extracted_19415 | Not-determined |
| S7_extracted_19431 | Low-quality |
| S7_extracted_19438 | Low-quality |
| S7_extracted_19485 | Not-determined |
| S7_extracted_19486 | Not-determined |
| S7_extracted_19508 | Not-determined |
| S7_extracted_19510 | Not-determined |
| S7_extracted_19515 | Not-determined |
| S7_extracted_19524 | Not-determined |
| S7_extracted_19527 | Not-determined |
| S7_extracted_19528 | Not-determined |
| S7_extracted_19534 | Not-determined |
| S7_extracted_19536 | Not-determined |
| S7_extracted_19547 | Not-determined |
| S7_extracted_19553 | Not-determined |
| S7_extracted_19565 | Not-determined |
| S7_extracted_19583 | Not-determined |
| S7_extracted_19585 | Not-determined |
| S7_extracted_19589 | Not-determined |
| S7_extracted_19592 | Not-determined |
| S7_extracted_19633 | Not-determined |
| S7_extracted_19634 | Not-determined |
| S7_extracted_19645 | Not-determined |
| S7_extracted_19664 | Not-determined |
| S7_extracted_19672 | Not-determined |
| S7_extracted_19687 | Not-determined |
| S7_extracted_19692 | Not-determined |
| S7_extracted_19693 | Low-quality |
| S7_extracted_19727 | Not-determined |
| S7_extracted_19735 | Not-determined |
| S7_extracted_19737 | Not-determined |
| S7_extracted_19743 | Low-quality |
| S7_extracted_19750 | Not-determined |
| S7_extracted_19758 | Not-determined |
| S7_extracted_19762 | Not-determined |
| S7_extracted_19765 | Low-quality |
| S7_extracted_19788 | Not-determined |
| S7_extracted_19795 | Low-quality |
| S7_extracted_19797 | Not-determined |
| S7_extracted_19799 | Low-quality |
| S7_extracted_19814 | Low-quality |
| S7_extracted_19820 | Not-determined |
| S7_extracted_19823 | Not-determined |
| S7_extracted_19824 | Not-determined |
| S7_extracted_19834 | Not-determined |
| S7_extracted_19850 | Not-determined |
| S7_extracted_19851 | Not-determined |
| S11_vs_k141_104049\|\|full | High-quality |
| S7_extracted_19911 | Not-determined |
| S7_extracted_19921 | Not-determined |
| S7_extracted_19923 | Not-determined |
| S7_extracted_19931 | Not-determined |
| S7_extracted_19943 | Low-quality |
| S7_extracted_19971 | Not-determined |
| S7_extracted_20001 | Not-determined |
| S7_extracted_20005 | Not-determined |
| S7_extracted_20009 | Low-quality |
| S7_extracted_20013 | Not-determined |
| S7_extracted_20017 | Not-determined |
| S7_extracted_20027 | Not-determined |
| S7_extracted_20043 | Not-determined |
| S7_extracted_20044 | Not-determined |
| S7_extracted_20050 | Not-determined |
| S7_extracted_20070 | Not-determined |
| S7_extracted_20087 | Not-determined |
| S7_extracted_20090 | Not-determined |
| S7_extracted_20113 | Not-determined |
| S7_extracted_20123 | Not-determined |
| S6_vs_k141_16135\|\|full | High-quality |
| S7_extracted_20156 | Not-determined |
| S7_extracted_20180 | Low-quality |
| S7_extracted_20182 | Not-determined |
| S7_extracted_20207 | Not-determined |
| S7_extracted_20214 | Not-determined |
| S7_extracted_20242 | Not-determined |
| S7_extracted_20245 | Not-determined |
| S7_extracted_20246 | Low-quality |
| S7_extracted_20266 | Not-determined |
| S7_extracted_20293 | Not-determined |
| S7_extracted_20297 | Low-quality |
| S7_extracted_20304 | Not-determined |
| S7_extracted_20328 | Not-determined |
| S7_extracted_20351 | Not-determined |
| S7_extracted_20368 | Not-determined |
| S7_extracted_20369 | Low-quality |
| S7_extracted_20376 | Not-determined |
| S7_extracted_20391 | Not-determined |
| S7_extracted_20401 | Low-quality |
| S7_extracted_20429 | Not-determined |
| S7_extracted_20436 | Not-determined |
| S7_extracted_20471 | Not-determined |
| S7_extracted_20498 | Not-determined |
| S7_extracted_20501 | Not-determined |
| S7_extracted_20513 | Not-determined |
| S7_extracted_20514 | Not-determined |
| S7_extracted_20523 | Not-determined |
| S7_extracted_20549 | Not-determined |
| S7_extracted_20559 | Not-determined |
| S7_extracted_20567 | Not-determined |
| S7_extracted_20569 | Not-determined |
| S7_extracted_20621 | Not-determined |
| S7_extracted_20622 | Low-quality |
| S7_extracted_20624 | Not-determined |
| S7_extracted_20634 | Not-determined |
| S7_extracted_20638 | Not-determined |
| S7_extracted_20645 | Not-determined |
| S7_extracted_20650 | Not-determined |
| S7_extracted_20655 | Not-determined |
| S7_extracted_20667 | Not-determined |
| S7_extracted_20679 | Not-determined |
| S7_extracted_20680 | Low-quality |
| S7_extracted_20685 | Not-determined |
| S7_extracted_20686 | Not-determined |
| S7_extracted_20689 | Not-determined |
| S7_extracted_20697 | Not-determined |
| S7_extracted_20701 | Not-determined |
| S7_extracted_20702 | Not-determined |
| S7_extracted_20724 | Low-quality |
| S7_extracted_20728 | Not-determined |
| S7_extracted_20734 | Not-determined |
| S7_extracted_20747 | Not-determined |
| S7_extracted_20748 | Not-determined |
| S7_extracted_20776 | Not-determined |
| S7_extracted_20779 | Not-determined |
| S7_extracted_20784 | Not-determined |
| S7_extracted_20806 | Low-quality |
| S7_extracted_20808 | Not-determined |
| S7_extracted_20834 | Not-determined |
| S7_extracted_20835 | Low-quality |
| S7_extracted_20840 | Not-determined |
| S7_extracted_20850 | Not-determined |
| S7_extracted_20861 | Not-determined |
| S7_extracted_20869 | Not-determined |
| S7_extracted_20894 | Not-determined |
| S7_extracted_20899 | Low-quality |
| S7_extracted_20903 | Low-quality |
| S7_extracted_20928 | Not-determined |
| S7_extracted_20932 | Not-determined |
| S7_extracted_20938 | Not-determined |
| S7_extracted_20953 | Low-quality |
| S7_extracted_20957 | Not-determined |
| S7_extracted_20973 | Not-determined |
| S7_extracted_20998 | Not-determined |
| S7_extracted_21007 | Low-quality |
| S7_extracted_21011 | Not-determined |
| S7_extracted_21019 | Low-quality |
| S7_extracted_21025 | Not-determined |
| S7_extracted_21028 | Not-determined |
| S7_extracted_21057 | Not-determined |
| S7_extracted_21060 | Low-quality |
| S7_extracted_21061 | Not-determined |
| S7_extracted_21066 | Not-determined |
| S7_extracted_21076 | Not-determined |
| S7_extracted_21097 | Low-quality |
| S7_extracted_21155 | Low-quality |
| S7_extracted_21164 | Not-determined |
| S7_extracted_21189 | Not-determined |
| S7_extracted_21196 | Not-determined |
| S7_extracted_21201 | Not-determined |
| S7_extracted_21205 | Not-determined |
| S7_extracted_21206 | Low-quality |
| S7_extracted_21217 | Not-determined |
| S7_extracted_21239 | Low-quality |
| S7_extracted_21270 | Not-determined |
| S7_extracted_21297 | Low-quality |
| S7_extracted_21319 | Not-determined |
| S7_extracted_21321 | Low-quality |
| S7_extracted_21339 | Not-determined |
| S7_extracted_21353 | Low-quality |
| S7_extracted_21360 | Low-quality |
| S7_extracted_21383 | Not-determined |
| S7_extracted_21386 | Low-quality |
| S7_extracted_21387 | Not-determined |
| S7_extracted_21410 | Not-determined |
| S7_extracted_21427 | Low-quality |
| S7_extracted_21446 | Not-determined |
| S7_extracted_21451 | Not-determined |
| S7_extracted_21463 | Not-determined |
| S7_extracted_21498 | Low-quality |
| S7_extracted_21514 | Not-determined |
| S7_extracted_21529 | Not-determined |
| S7_extracted_21530 | Not-determined |
| S7_extracted_21539 | Low-quality |
| S7_extracted_21543 | Not-determined |
| S7_extracted_21548 | Not-determined |
| S7_extracted_21557 | Not-determined |
| S7_extracted_21564 | Not-determined |
| S7_extracted_21574 | Not-determined |
| S7_extracted_21588 | Low-quality |
| S7_extracted_21593 | Not-determined |
| S7_extracted_21603 | Not-determined |
| S7_extracted_21611 | Not-determined |
| S7_extracted_21624 | Not-determined |
| S7_extracted_21625 | Not-determined |
| S7_extracted_21640 | Low-quality |
| S7_extracted_21652 | Not-determined |
| S7_extracted_21656 | Not-determined |
| S7_extracted_21661 | Not-determined |
| S7_extracted_21662 | Low-quality |
| S7_extracted_21668 | Not-determined |
| S7_extracted_21671 | Not-determined |
| S7_extracted_21702 | Not-determined |
| S7_extracted_21703 | Not-determined |
| S7_extracted_21710 | Not-determined |
| S7_extracted_21715 | Not-determined |
| S7_extracted_21733 | Not-determined |
| S7_extracted_21758 | Not-determined |
| S7_extracted_21765 | Not-determined |
| S7_extracted_21767 | Not-determined |
| S7_extracted_21793 | Not-determined |
| S7_extracted_21803 | Not-determined |
| S7_extracted_21812 | Low-quality |
| S7_extracted_21815 | Low-quality |
| S7_extracted_21831 | Low-quality |
| S7_extracted_21833 | Not-determined |
| S7_extracted_21843 | Not-determined |
| S7_extracted_21846 | Not-determined |
| S7_extracted_21849 | Not-determined |
| S7_extracted_21856 | Not-determined |
| S7_extracted_21872 | Low-quality |
| S7_extracted_21876 | Not-determined |
| S7_extracted_21892 | Not-determined |
| S7_extracted_21924 | Not-determined |
| S7_extracted_21932 | Low-quality |
| S7_extracted_21935 | Not-determined |
| S7_extracted_21954 | Not-determined |
| S7_extracted_21956 | Not-determined |
| S7_extracted_21966 | Not-determined |
| S7_extracted_21974 | Not-determined |
| S14_vs_k141_5414\|\|full | Medium-quality |
| S7_extracted_21987 | Not-determined |
| S7_extracted_21996 | Not-determined |
| S7_extracted_22000 | Low-quality |
| S7_extracted_22017 | Not-determined |
| S7_extracted_22030 | Not-determined |
| S7_extracted_22031 | Not-determined |
| S7_extracted_22062 | Low-quality |
| S7_extracted_22065 | Not-determined |
| S7_extracted_22086 | Low-quality |
| S7_extracted_22096 | Not-determined |
| S7_extracted_22106 | Not-determined |
| S7_extracted_22111 | Not-determined |
| S7_extracted_22121 | Low-quality |
| S7_extracted_22135 | Not-determined |
| S7_extracted_22138 | Not-determined |
| S7_extracted_22151 | Not-determined |
| S7_extracted_22167 | Low-quality |
| S7_extracted_22176 | Low-quality |
| S7_extracted_22201 | Not-determined |
| S7_extracted_22207 | Not-determined |
| S7_extracted_22213 | Not-determined |
| S7_extracted_22220 | Not-determined |
| S7_extracted_22223 | Not-determined |
| S7_extracted_22240 | Not-determined |
| S7_extracted_22241 | Not-determined |
| S7_extracted_22264 | Not-determined |
| S7_extracted_22302 | Not-determined |
| S7_extracted_22312 | Not-determined |
| S7_extracted_22314 | Low-quality |
| S7_extracted_22370 | Low-quality |
| S7_extracted_22378 | Low-quality |
| S7_extracted_22388 | Not-determined |
| S7_extracted_22400 | Not-determined |
| S7_extracted_22423 | Low-quality |
| S7_extracted_22447 | Low-quality |
| S7_extracted_22456 | Not-determined |
| S7_extracted_22466 | Not-determined |
| S7_extracted_22469 | Low-quality |
| S7_extracted_22493 | Low-quality |
| S7_extracted_22494 | Not-determined |
| S7_extracted_22496 | Not-determined |
| S7_extracted_22508 | Not-determined |
| S7_extracted_22509 | Not-determined |
| S7_extracted_22512 | Not-determined |
| S7_extracted_22532 | Low-quality |
| S7_extracted_22538 | Not-determined |
| S7_extracted_22541 | Not-determined |
| S7_extracted_22559 | Not-determined |
| S7_extracted_22572 | Not-determined |
| S7_extracted_22583 | Not-determined |
| S7_extracted_22584 | Not-determined |
| S7_extracted_22598 | Not-determined |
| S7_extracted_22617 | Not-determined |
| S7_extracted_22626 | Not-determined |
| S7_extracted_22639 | Not-determined |
| S7_extracted_22656 | Low-quality |
| S7_extracted_22663 | Not-determined |
| S7_extracted_22682 | Not-determined |
| S7_extracted_22691 | Low-quality |
| S7_extracted_22720 | Not-determined |
| S7_extracted_22724 | Not-determined |
| S7_extracted_22731 | Not-determined |
| S7_extracted_22739 | Not-determined |
| S7_extracted_22754 | Low-quality |
| S7_extracted_22757 | Low-quality |
| S7_extracted_22758 | Not-determined |
| S7_extracted_22766 | Not-determined |
| S7_extracted_22781 | Not-determined |
| S7_extracted_22785 | Not-determined |
| S7_extracted_22792 | Low-quality |
| S7_extracted_22794 | Not-determined |
| S7_extracted_22814 | Not-determined |
| S7_extracted_22819 | Low-quality |
| S7_extracted_22832 | Not-determined |
| S7_extracted_22835 | Not-determined |
| S7_extracted_22841 | Not-determined |
| S7_extracted_22842 | Not-determined |
| S7_extracted_22852 | Not-determined |
| S7_extracted_22870 | Not-determined |
| S7_extracted_22880 | Not-determined |
| S7_extracted_22884 | Not-determined |
| S7_extracted_22887 | Low-quality |
| S7_extracted_22889 | Not-determined |
| S7_extracted_22897 | Not-determined |
| S7_extracted_22898 | Low-quality |
| S7_extracted_22904 | Low-quality |
| S7_extracted_22914 | Not-determined |
| S7_extracted_22951 | Not-determined |
| S7_extracted_22953 | Low-quality |
| S7_extracted_22962 | Not-determined |
| S7_extracted_22963 | Not-determined |
| S7_extracted_22966 | Not-determined |
| S7_extracted_22981 | Low-quality |
| S7_extracted_22984 | Not-determined |
| S7_extracted_22991 | Not-determined |
| S7_extracted_23009 | Not-determined |
| S7_extracted_23013 | Not-determined |
| S7_extracted_23052 | Not-determined |
| S7_extracted_23059 | Not-determined |
| S7_extracted_23061 | Not-determined |
| S7_extracted_23090 | Not-determined |
| S7_extracted_23104 | Not-determined |
| S7_extracted_23121 | Not-determined |
| S7_extracted_23138 | Low-quality |
| S7_extracted_23154 | Not-determined |
| S7_extracted_23208 | Not-determined |
| S7_extracted_23215 | Low-quality |
| S7_extracted_23216 | Not-determined |
| S7_extracted_23237 | Not-determined |
| S7_extracted_23238 | Not-determined |
| S7_extracted_23241 | Not-determined |

| | |
|---|---|
| S7_extracted_23261 | Not-determined |
| S7_extracted_23280 | Not-determined |
| S7_extracted_23284 | Not-determined |
| S7_extracted_23288 | Not-determined |
| S7_extracted_23305 | Low-quality |
| S7_extracted_23306 | Not-determined |
| S7_extracted_23311 | Low-quality |
| S7_extracted_23333 | Not-determined |
| S7_extracted_23339 | Not-determined |
| S7_extracted_23372 | Not-determined |
| S7_extracted_23388 | Low-quality |
| S7_extracted_23389 | Not-determined |
| S7_extracted_23413 | Low-quality |
| S7_extracted_23415 | Not-determined |
| S7_extracted_23431 | Not-determined |
| S7_extracted_23447 | Not-determined |
| S7_extracted_23451 | Not-determined |
| S7_extracted_23453 | Not-determined |
| S7_extracted_23459 | Not-determined |
| S7_extracted_23467 | Not-determined |
| S7_extracted_23471 | Not-determined |
| S7_extracted_23481 | Not-determined |
| S7_extracted_23497 | Not-determined |
| S7_extracted_23504 | Not-determined |
| S7_extracted_23505 | Low-quality |
| S7_extracted_23507 | Not-determined |
| S7_extracted_23565 | Low-quality |
| S7_extracted_23581 | Not-determined |
| S7_extracted_23593 | Low-quality |
| S7_extracted_23595 | Not-determined |
| S7_extracted_23601 | Not-determined |
| S7_extracted_23603 | Not-determined |
| S7_extracted_23613 | Not-determined |
| S7_extracted_23621 | Not-determined |
| S7_extracted_23656 | Not-determined |
| S7_extracted_23657 | Not-determined |
| S7_extracted_23665 | Not-determined |
| S7_extracted_23687 | Not-determined |
| S7_extracted_23689 | Not-determined |
| S7_extracted_23695 | Not-determined |
| S7_extracted_23698 | Not-determined |
| S7_extracted_23717 | Not-determined |
| S7_extracted_23719 | Not-determined |
| S7_extracted_23748 | Not-determined |
| S7_extracted_23750 | Low-quality |
| S7_extracted_23766 | Not-determined |
| S7_extracted_23779 | Not-determined |
| S7_extracted_23780 | Not-determined |
| S7_extracted_23792 | Low-quality |
| S7_extracted_23801 | Not-determined |
| S7_extracted_23814 | Not-determined |
| S7_extracted_23820 | Not-determined |
| S7_extracted_23823 | Not-determined |
| S7_extracted_23826 | Not-determined |
| S7_extracted_23839 | Not-determined |
| S7_extracted_23873 | Low-quality |
| S7_extracted_23881 | Not-determined |
| S7_extracted_23903 | Low-quality |
| S7_extracted_23911 | Not-determined |
| S7_extracted_23913 | Low-quality |
| S7_extracted_23914 | Not-determined |
| S7_extracted_23918 | Low-quality |
| S7_extracted_23932 | Not-determined |
| S7_extracted_23947 | Not-determined |
| S7_extracted_23949 | Not-determined |
| S7_extracted_23950 | Not-determined |
| S7_extracted_23953 | Low-quality |
| S7_extracted_23955 | Not-determined |
| S7_extracted_23967 | Low-quality |
| S7_extracted_23987 | Not-determined |
| S7_extracted_24009 | Not-determined |
| S7_extracted_24010 | Not-determined |
| S7_extracted_24029 | Not-determined |
| S7_extracted_24042 | Low-quality |
| S7_extracted_24047 | Not-determined |
| S7_extracted_24059 | Not-determined |
| S7_extracted_24086 | Low-quality |
| S7_extracted_24088 | Low-quality |
| S7_extracted_24100 | Not-determined |
| S7_extracted_24105 | Not-determined |
| S7_extracted_24112 | Low-quality |
| S7_extracted_24124 | Not-determined |
| S7_extracted_24136 | Low-quality |
| S7_extracted_24179 | Not-determined |
| S7_extracted_24209 | Low-quality |
| S7_extracted_24276 | Not-determined |
| S7_extracted_24278 | Not-determined |
| S7_extracted_24296 | Low-quality |
| S7_extracted_24308 | Not-determined |
| S7_extracted_24317 | Not-determined |
| S7_extracted_24324 | Not-determined |
| S7_extracted_24329 | Low-quality |
| S7_extracted_24333 | Not-determined |
| S7_extracted_24334 | Not-determined |
| S7_extracted_24345 | Not-determined |
| S7_extracted_24346 | Not-determined |
| S7_extracted_24350 | Not-determined |
| S7_extracted_24356 | Not-determined |
| S7_extracted_24372 | Not-determined |
| S7_extracted_24379 | Low-quality |
| S7_extracted_24385 | Not-determined |
| S7_extracted_24395 | Not-determined |
| S7_extracted_24404 | Not-determined |
| S7_extracted_24411 | Not-determined |
| S7_extracted_24438 | Low-quality |
| S7_extracted_24442 | Low-quality |
| S7_extracted_24462 | Not-determined |
| S7_extracted_24467 | Not-determined |
| S7_extracted_24485 | Low-quality |
| S7_extracted_24499 | Not-determined |
| S7_extracted_24505 | Low-quality |
| S7_extracted_24509 | Not-determined |
| S7_extracted_24516 | Not-determined |
| S7_extracted_24528 | Not-determined |
| S7_extracted_24582 | Not-determined |
| S7_extracted_24611 | Not-determined |
| S7_extracted_24618 | Not-determined |
| S7_extracted_24662 | Not-determined |
| S7_extracted_24672 | Not-determined |
| S7_extracted_24680 | Not-determined |
| S7_extracted_24699 | Not-determined |
| S7_extracted_24702 | Not-determined |
| S7_extracted_24724 | Not-determined |
| S7_extracted_24729 | Not-determined |
| S7_extracted_24754 | Not-determined |
| S7_extracted_24759 | Low-quality |
| S7_extracted_24791 | Not-determined |
| S7_extracted_24801 | Not-determined |
| S7_extracted_24820 | Not-determined |
| S7_extracted_24821 | Not-determined |
| S7_extracted_24847 | Low-quality |
| S7_extracted_24856 | Not-determined |
| S7_extracted_24861 | Not-determined |
| S7_extracted_24867 | Low-quality |
| S7_extracted_24868 | Not-determined |
| S7_extracted_24891 | Not-determined |
| S7_extracted_24915 | Low-quality |
| S7_extracted_24919 | Low-quality |
| S7_extracted_24934 | Not-determined |
| S7_extracted_24938 | Not-determined |
| S7_extracted_24939 | Not-determined |
| S7_extracted_24942 | Not-determined |
| S7_extracted_24970 | Low-quality |
| S7_extracted_24999 | Not-determined |
| S7_extracted_25063 | Not-determined |
| S7_extracted_25077 | Low-quality |
| S7_extracted_25084 | Not-determined |
| S7_extracted_25090 | Not-determined |
| S7_extracted_25111 | Not-determined |
| S7_extracted_25131 | Not-determined |
| S7_extracted_25182 | Low-quality |
| S7_extracted_25227 | Not-determined |
| S7_extracted_25239 | Low-quality |
| S7_extracted_25245 | Not-determined |
| S7_extracted_25246 | Not-determined |
| S7_extracted_25249 | Low-quality |
| S7_extracted_25250 | Not-determined |
| S7_extracted_25258 | Not-determined |
| S7_extracted_25288 | Not-determined |
| S7_extracted_25302 | Low-quality |
| S7_extracted_25307 | Not-determined |
| S7_extracted_25310 | Not-determined |
| S7_extracted_25321 | Not-determined |
| S7_extracted_25323 | Not-determined |
| S7_extracted_25363 | Not-determined |
| S7_extracted_25376 | Low-quality |
| S7_extracted_25383 | Not-determined |
| S7_extracted_25395 | Not-determined |
| S7_extracted_25408 | Low-quality |
| S7_extracted_25409 | Not-determined |
| S7_extracted_25447 | Not-determined |
| S7_extracted_25451 | Low-quality |
| S7_extracted_25456 | Low-quality |
| S7_extracted_25484 | Not-determined |
| S7_extracted_25487 | Low-quality |
| S7_extracted_25491 | Not-determined |
| S7_extracted_25499 | Not-determined |
| S7_extracted_25504 | Not-determined |
| S7_extracted_25513 | Not-determined |
| S7_extracted_25522 | Not-determined |
| S7_extracted_25527 | Not-determined |
| S7_extracted_25533 | Not-determined |
| S7_extracted_25534 | Not-determined |
| S7_extracted_25544 | Not-determined |
| S7_extracted_25552 | Low-quality |
| S7_extracted_25563 | Not-determined |
| S7_extracted_25570 | Not-determined |
| S7_extracted_25584 | Not-determined |
| S7_extracted_25593 | Low-quality |
| S7_extracted_25617 | Not-determined |
| S7_extracted_25631 | Not-determined |
| S7_extracted_25651 | Not-determined |
| S7_extracted_25684 | Not-determined |
| S7_extracted_25688 | Not-determined |
| S7_extracted_25706 | Not-determined |
| S7_extracted_25708 | Not-determined |
| S7_extracted_25717 | Not-determined |
| S7_extracted_25721 | Not-determined |
| S7_extracted_25723 | Not-determined |
| S7_extracted_25752 | Not-determined |
| S7_extracted_25761 | Low-quality |
| S7_extracted_25773 | Low-quality |
| S7_extracted_25794 | Not-determined |
| S7_extracted_25801 | Not-determined |
| S7_extracted_25810 | Not-determined |
| S7_extracted_25818 | Low-quality |
| S7_extracted_25820 | Not-determined |
| S7_extracted_25836 | Not-determined |
| S7_extracted_25841 | Not-determined |
| S7_extracted_25843 | Not-determined |
| S7_extracted_25844 | Not-determined |
| S7_extracted_25868 | Low-quality |
| S7_extracted_25897 | Low-quality |
| S7_extracted_25905 | Low-quality |
| S7_extracted_25911 | Not-determined |
| S7_extracted_25916 | Low-quality |
| S7_extracted_25922 | Not-determined |
| S7_extracted_25947 | Not-determined |
| S7_extracted_25970 | Not-determined |
| S7_extracted_25977 | Not-determined |
| S7_extracted_25980 | Not-determined |
| S7_extracted_25997 | Not-determined |
| S7_extracted_26001 | Not-determined |
| S7_extracted_26022 | Not-determined |
| S7_extracted_26050 | Not-determined |
| S7_extracted_26066 | Not-determined |
| S7_extracted_26092 | Not-determined |
| S7_extracted_26111 | Not-determined |
| S7_extracted_26113 | Low-quality |
| S7_extracted_26131 | Not-determined |
| S7_extracted_26144 | Low-quality |
| S7_extracted_26159 | Not-determined |
| S7_extracted_26198 | Not-determined |
| S7_extracted_26199 | Not-determined |
| S7_extracted_26219 | Not-determined |
| S7_extracted_26222 | Low-quality |
| S7_extracted_26225 | Not-determined |
| S7_extracted_26227 | Not-determined |
| S7_extracted_26238 | Not-determined |
| S7_extracted_26240 | Not-determined |
| S7_extracted_26243 | Not-determined |
| S7_extracted_26248 | Not-determined |
| S7_extracted_26287 | Not-determined |
| S7_extracted_26306 | Low-quality |
| S7_extracted_26312 | Not-determined |
| S7_extracted_26314 | Not-determined |
| S7_extracted_26339 | Not-determined |
| S7_extracted_26351 | Not-determined |
| S7_extracted_26368 | Not-determined |
| S7_extracted_26410 | Not-determined |
| S7_extracted_26421 | Not-determined |
| S7_extracted_26430 | Not-determined |
| S7_extracted_26442 | Not-determined |
| S7_extracted_26445 | Not-determined |
| S7_extracted_26453 | Not-determined |
| S7_extracted_26455 | Not-determined |
| S7_extracted_26460 | Low-quality |
| S7_extracted_26473 | Not-determined |
| S7_extracted_26478 | Not-determined |
| S7_extracted_26496 | Not-determined |
| S7_extracted_26508 | Low-quality |
| S7_extracted_26518 | Not-determined |
| S7_extracted_26560 | Not-determined |
| S7_extracted_26568 | Not-determined |
| S7_extracted_26584 | Low-quality |
| S7_extracted_26587 | Not-determined |
| S7_extracted_26589 | Not-determined |
| S7_extracted_26595 | Not-determined |
| S7_extracted_26600 | Low-quality |
| S7_extracted_26621 | Not-determined |
| S7_extracted_26639 | Not-determined |
| S7_extracted_26645 | Not-determined |
| S7_extracted_26654 | Low-quality |
| S7_extracted_26670 | Not-determined |
| S7_extracted_26672 | Low-quality |
| S7_extracted_26683 | Not-determined |
| S7_extracted_26697 | Low-quality |
| S7_extracted_26699 | Low-quality |
| S7_extracted_26709 | Not-determined |
| S7_extracted_26719 | Not-determined |
| S7_extracted_26721 | Low-quality |
| S7_extracted_26725 | Not-determined |
| S7_extracted_26738 | Not-determined |
| S7_extracted_26761 | Not-determined |
| S7_extracted_26773 | Not-determined |
| S7_extracted_26787 | Not-determined |
| S7_extracted_26796 | Not-determined |
| S7_extracted_26821 | Not-determined |
| S7_extracted_26860 | Not-determined |
| S7_extracted_26877 | Low-quality |
| S7_extracted_26883 | Not-determined |
| S7_extracted_26893 | Not-determined |
| S7_extracted_26905 | Not-determined |
| S7_extracted_26919 | Not-determined |
| S7_extracted_26933 | Not-determined |
| S7_extracted_26937 | Low-quality |
| S7_extracted_26941 | Not-determined |
| S7_extracted_26959 | Not-determined |
| S7_extracted_26973 | Not-determined |
| S7_extracted_26978 | Not-determined |
| S7_extracted_26983 | Not-determined |
| S7_extracted_27004 | Not-determined |
| S7_extracted_27005 | Not-determined |
| S7_extracted_27016 | Not-determined |
| S7_extracted_27037 | Not-determined |
| S7_extracted_27058 | Not-determined |
| S7_extracted_27083 | Not-determined |
| S7_extracted_27085 | Not-determined |
| S7_extracted_27111 | Not-determined |
| S7_extracted_27132 | Not-determined |
| S7_extracted_27158 | Not-determined |
| S7_extracted_27165 | Not-determined |
| S7_extracted_27177 | Not-determined |
| S7_extracted_27194 | Not-determined |
| S7_extracted_27203 | Not-determined |
| S7_extracted_27207 | Low-quality |
| S7_extracted_27261 | Not-determined |
| S7_extracted_27262 | Low-quality |
| S7_extracted_27271 | Not-determined |
| S7_extracted_27272 | Not-determined |
| S7_extracted_27278 | Low-quality |
| S7_extracted_49485 | Medium-quality |
| S7_extracted_27321 | Not-determined |
| S7_extracted_27326 | Not-determined |
| S7_extracted_27327 | Not-determined |
| S7_extracted_27336 | Not-determined |
| S7_extracted_27337 | Not-determined |
| S7_extracted_27352 | Low-quality |
| S7_extracted_27360 | Not-determined |
| S7_extracted_27370 | Not-determined |
| S7_extracted_27378 | Not-determined |
| S7_extracted_27381 | Not-determined |
| S7_extracted_27388 | Not-determined |
| S7_extracted_27401 | Not-determined |
| S7_extracted_27405 | Not-determined |
| S7_extracted_27410 | Low-quality |
| S7_extracted_27430 | Low-quality |
| S8_vs_k141_197061\|\|full | High-quality |
| S7_extracted_27440 | Not-determined |
| S7_extracted_27456 | Low-quality |
| S7_extracted_27469 | Not-determined |
| S7_extracted_27474 | Not-determined |
| S7_extracted_27482 | Not-determined |
| S7_extracted_27493 | Not-determined |
| S7_extracted_27502 | Not-determined |
| S7_extracted_27517 | Not-determined |
| S7_extracted_27527 | Not-determined |
| S7_extracted_27549 | Not-determined |
| S7_extracted_27558 | Not-determined |
| S7_extracted_27561 | Not-determined |
| S7_extracted_27565 | Not-determined |
| S7_extracted_27577 | Not-determined |
| S7_extracted_27588 | Not-determined |
| S7_extracted_27601 | Low-quality |
| S7_extracted_27603 | Not-determined |
| S7_extracted_27607 | Not-determined |
| S7_extracted_27611 | Not-determined |
| S7_extracted_27614 | Not-determined |
| S7_extracted_27617 | Low-quality |
| S7_extracted_27639 | Not-determined |
| S7_extracted_27657 | Not-determined |
| S7_extracted_27662 | Not-determined |
| S7_extracted_27695 | Not-determined |
| S7_extracted_27700 | Low-quality |
| S7_extracted_27721 | Not-determined |
| S7_extracted_27723 | Not-determined |
| S7_extracted_27750 | Not-determined |
| S7_extracted_27757 | Not-determined |
| S7_extracted_27767 | Not-determined |
| S7_extracted_27775 | Not-determined |
| S7_extracted_27780 | Not-determined |
| S7_extracted_27797 | Low-quality |
| S7_extracted_27818 | Not-determined |
| S7_extracted_27823 | Not-determined |
| S7_extracted_27826 | Not-determined |
| S7_extracted_27827 | Low-quality |
| S7_extracted_27841 | Not-determined |
| S7_extracted_27859 | Not-determined |
| S7_extracted_27873 | Not-determined |
| S7_extracted_27905 | Not-determined |
| S7_extracted_27937 | Not-determined |
| S7_extracted_27953 | Not-determined |
| S7_extracted_27958 | Not-determined |
| S7_extracted_27993 | Not-determined |
| S7_extracted_28005 | Not-determined |
| S7_extracted_28007 | Low-quality |
| S7_extracted_28029 | Not-determined |
| S7_extracted_28031 | Not-determined |
| S7_extracted_28037 | Not-determined |
| S7_extracted_28038 | Not-determined |
| S7_extracted_28041 | Low-quality |
| S7_extracted_28044 | Not-determined |
| S7_extracted_28046 | Not-determined |
| S7_extracted_28064 | Not-determined |
| S7_extracted_28071 | Not-determined |
| S7_extracted_28077 | Not-determined |
| S7_extracted_28100 | Not-determined |
| S7_extracted_28107 | Low-quality |
| S7_extracted_28111 | Not-determined |
| S7_extracted_28119 | Not-determined |
| S7_extracted_28120 | Not-determined |
| S7_extracted_28127 | Not-determined |
| S7_extracted_28132 | Not-determined |
| S7_extracted_28145 | Not-determined |
| S7_extracted_28168 | Low-quality |
| S7_extracted_28189 | Low-quality |
| S7_extracted_28194 | Not-determined |
| S7_extracted_28205 | Low-quality |
| S7_extracted_28218 | Not-determined |
| S7_extracted_28220 | Low-quality |
| S7_extracted_28225 | Not-determined |
| S7_extracted_28231 | Not-determined |
| S7_extracted_28245 | Not-determined |
| S7_extracted_28255 | Not-determined |
| S7_extracted_28259 | Not-determined |
| S7_extracted_28263 | Not-determined |
| S7_extracted_28272 | Not-determined |
| S7_extracted_28282 | Not-determined |
| S7_extracted_28290 | Not-determined |
| S7_extracted_28313 | Not-determined |
| S7_extracted_28322 | Low-quality |
| S7_extracted_28332 | Not-determined |
| S7_extracted_28345 | Low-quality |
| S7_extracted_28352 | Low-quality |
| S7_extracted_28354 | Not-determined |
| S7_extracted_28373 | Not-determined |
| S7_extracted_28377 | Low-quality |
| S7_extracted_28380 | Not-determined |
| S7_extracted_28404 | Not-determined |
| S7_extracted_28423 | Not-determined |
| S7_extracted_28428 | Not-determined |
| S13_extracted_87873 | Medium-quality |
| S7_extracted_28431 | Not-determined |
| S7_extracted_28439 | Not-determined |
| S7_extracted_28442 | Not-determined |
| S7_extracted_28443 | Not-determined |
| S7_extracted_28470 | Not-determined |
| S7_extracted_28473 | Low-quality |
| S7_extracted_28475 | Not-determined |
| S7_extracted_28494 | Not-determined |
| S7_extracted_28500 | Not-determined |
| S7_extracted_28501 | Not-determined |
| S7_extracted_28504 | Not-determined |
| S7_extracted_28506 | Not-determined |
| S7_extracted_28526 | Not-determined |
| S7_extracted_28529 | Not-determined |
| S7_extracted_28539 | Not-determined |
| S7_extracted_28559 | Not-determined |
| S7_extracted_28570 | Not-determined |
| S7_extracted_28574 | Low-quality |
| S7_extracted_28577 | Low-quality |
| S7_extracted_28617 | Not-determined |
| S7_extracted_28631 | Not-determined |
| S7_extracted_28635 | Not-determined |
| S7_extracted_28642 | Not-determined |
| S7_extracted_28653 | Not-determined |
| S7_extracted_28658 | Not-determined |
| S7_extracted_28666 | Low-quality |
| S7_extracted_28677 | Not-determined |
| S7_extracted_28683 | Low-quality |
| S7_extracted_28691 | Not-determined |
| S7_extracted_28693 | Not-determined |
| S7_extracted_28766 | Not-determined |
| S7_extracted_28771 | Not-determined |
| S7_extracted_28794 | Not-determined |
| S7_extracted_28797 | Not-determined |
| S7_extracted_28799 | Not-determined |
| S7_extracted_28804 | Low-quality |
| S7_extracted_28810 | Low-quality |
| S7_extracted_28827 | Not-determined |
| S7_extracted_28835 | Low-quality |
| S7_extracted_28841 | Not-determined |
| S7_extracted_28843 | Not-determined |
| S7_extracted_28874 | Not-determined |
| S7_extracted_28880 | Low-quality |
| S7_extracted_28895 | Low-quality |
| S7_extracted_28906 | Not-determined |
| S7_extracted_28907 | Low-quality |
| S7_extracted_28922 | Not-determined |
| S7_extracted_28931 | Low-quality |
| S7_extracted_28970 | Not-determined |
| S7_extracted_28978 | Low-quality |
| S7_extracted_28985 | Not-determined |
| S7_extracted_28994 | Not-determined |
| S7_extracted_28998 | Not-determined |
| S7_extracted_29018 | Not-determined |
| S7_extracted_29019 | Not-determined |
| S7_extracted_29023 | Not-determined |
| S7_extracted_29024 | Not-determined |
| S7_extracted_29026 | Not-determined |
| S7_extracted_29028 | Not-determined |
| S7_extracted_29054 | Not-determined |
| S7_extracted_29064 | Not-determined |
| S7_extracted_29065 | Not-determined |
| S7_extracted_29082 | Low-quality |
| S7_extracted_29085 | Low-quality |
| S7_extracted_29091 | Not-determined |
| S7_extracted_29104 | Not-determined |
| S7_extracted_29122 | Not-determined |
| S7_extracted_29125 | Not-determined |
| S7_extracted_29134 | Not-determined |
| S7_extracted_29182 | Not-determined |
| S7_extracted_29185 | Not-determined |
| S7_extracted_29194 | Not-determined |
| S13_vs_87873\|\|full | Medium-quality |
| S7_extracted_29222 | Not-determined |
| S7_extracted_29247 | Not-determined |
| S7_extracted_29250 | Not-determined |
| S7_extracted_29252 | Low-quality |
| S7_extracted_29260 | Not-determined |
| S7_extracted_29274 | Low-quality |
| S7_extracted_29282 | Not-determined |
| S7_extracted_29283 | Not-determined |
| S7_extracted_29300 | Not-determined |
| S7_extracted_29305 | Not-determined |
| S7_extracted_29307 | Not-determined |
| S7_extracted_29309 | Low-quality |
| S7_extracted_29312 | Not-determined |
| S7_extracted_29323 | Not-determined |
| S7_extracted_29325 | Not-determined |
| S7_extracted_29334 | Low-quality |
| S7_extracted_29336 | Low-quality |
| S7_extracted_29344 | Low-quality |
| S7_extracted_29361 | Not-determined |
| S7_extracted_29364 | Low-quality |
| S7_extracted_29378 | Not-determined |
| S7_extracted_29406 | Not-determined |
| S7_extracted_29407 | Not-determined |
| S7_extracted_29408 | Not-determined |
| S7_extracted_29409 | Not-determined |
| S7_extracted_29426 | Not-determined |
| S7_extracted_29437 | Not-determined |
| S7_extracted_29438 | Not-determined |
| S7_extracted_29439 | Not-determined |
| S7_extracted_29441 | Not-determined |
| S7_extracted_29442 | Not-determined |
| S7_extracted_29444 | Not-determined |
| S7_extracted_29455 | Not-determined |
| S7_extracted_29465 | Not-determined |
| S7_extracted_29466 | Low-quality |
| S7_extracted_29467 | Not-determined |
| S7_extracted_29482 | Not-determined |
| S7_extracted_29500 | Not-determined |
| S7_extracted_29505 | Not-determined |
| S7_extracted_29510 | Not-determined |
| S7_extracted_29515 | Low-quality |
| S7_extracted_29543 | Low-quality |
| S7_extracted_29548 | Not-determined |
| S7_extracted_29549 | Not-determined |
| S7_extracted_29575 | Not-determined |
| S7_extracted_29576 | Not-determined |
| S7_extracted_29606 | Not-determined |
| S7_extracted_29607 | Not-determined |
| S7_extracted_29610 | Not-determined |
| S7_extracted_29611 | Not-determined |
| S7_extracted_29622 | Not-determined |
| S7_extracted_29638 | Not-determined |
| S7_extracted_29642 | Low-quality |
| S7_extracted_29645 | Not-determined |
| S7_extracted_29646 | Low-quality |
| S7_extracted_29650 | Low-quality |
| S7_extracted_29683 | Not-determined |
| S7_extracted_29687 | Not-determined |
| S7_extracted_29688 | Not-determined |
| S7_extracted_29694 | Not-determined |
| S7_extracted_29742 | Low-quality |
| S7_extracted_29747 | Not-determined |
| S7_extracted_29751 | Low-quality |
| S7_extracted_29758 | Not-determined |
| S7_extracted_29773 | Not-determined |
| S7_extracted_29777 | Not-determined |
| S7_extracted_29783 | Low-quality |
| S7_extracted_29785 | Not-determined |
| S7_extracted_29808 | Not-determined |
| S7_extracted_29815 | Not-determined |
| S7_extracted_29832 | Not-determined |
| S7_extracted_29835 | Not-determined |
| S7_extracted_29848 | Low-quality |
| S7_extracted_29852 | Not-determined |
| S7_extracted_29853 | Not-determined |
| S7_extracted_29856 | Low-quality |
| S7_extracted_29858 | Low-quality |
| S7_extracted_29879 | Not-determined |
| S7_extracted_29891 | Low-quality |
| S7_extracted_29892 | Low-quality |
| S7_extracted_29894 | Low-quality |
| S7_extracted_29900 | Not-determined |
| S7_extracted_29919 | Not-determined |
| S7_extracted_29935 | Not-determined |

| | |
|---|---|
| S7_extracted_29938 | Not-determined |
| S4_extracted_28601 | High-quality |
| S7_extracted_29953 | Not-determined |
| S7_extracted_29975 | Not-determined |
| S7_extracted_29983 | Not-determined |
| S7_extracted_29997 | Not-determined |
| S7_extracted_29999 | Not-determined |
| S7_extracted_30020 | Low-quality |
| S7_extracted_30025 | Not-determined |
| S7_extracted_30034 | Low-quality |
| S7_extracted_30040 | Not-determined |
| S7_extracted_30044 | Not-determined |
| S7_extracted_30045 | Not-determined |
| S7_extracted_30052 | Low-quality |
| S7_extracted_30054 | Low-quality |
| S7_extracted_30057 | Not-determined |
| S7_extracted_30067 | Not-determined |
| S7_extracted_30077 | Not-determined |
| S7_extracted_30083 | Not-determined |
| S7_extracted_30089 | Not-determined |
| S7_extracted_30095 | Not-determined |
| S7_extracted_30111 | Not-determined |
| S7_extracted_30131 | Not-determined |
| S7_extracted_30139 | Not-determined |
| S7_extracted_30173 | Not-determined |
| S7_extracted_30189 | Not-determined |
| S7_extracted_30210 | Not-determined |
| S7_extracted_30211 | Low-quality |
| S7_extracted_30222 | Not-determined |
| S7_extracted_30230 | Not-determined |
| S7_extracted_30235 | Not-determined |
| S7_extracted_30244 | Not-determined |
| S7_extracted_30260 | Not-determined |
| S7_extracted_30291 | Not-determined |
| S7_extracted_30301 | Not-determined |
| S7_extracted_30350 | Not-determined |
| S7_extracted_30359 | Low-quality |
| S7_extracted_30371 | Low-quality |
| S7_extracted_30384 | Not-determined |
| S7_extracted_30389 | Not-determined |
| S7_extracted_30405 | Not-determined |
| S7_extracted_30409 | Not-determined |
| S7_extracted_30417 | Not-determined |
| S7_extracted_30449 | Low-quality |
| S7_extracted_30460 | Not-determined |
| S7_extracted_30466 | Not-determined |
| S7_extracted_30467 | Not-determined |
| S7_extracted_30484 | Not-determined |
| S7_extracted_30485 | Not-determined |
| S7_extracted_30491 | Not-determined |
| S7_extracted_30494 | Not-determined |
| S7_extracted_30496 | Not-determined |
| S7_extracted_30503 | Not-determined |
| S7_extracted_30518 | Not-determined |
| S7_extracted_30529 | Not-determined |
| S7_extracted_30532 | Low-quality |
| S7_extracted_30546 | Not-determined |
| S7_extracted_30556 | Not-determined |
| S7_extracted_30565 | Not-determined |
| S7_extracted_30575 | Not-determined |
| S7_extracted_30580 | Not-determined |
| S7_extracted_30584 | Not-determined |
| S7_extracted_30592 | Not-determined |
| S7_extracted_30593 | Low-quality |
| S7_extracted_30619 | Not-determined |
| S7_extracted_30630 | Not-determined |
| S7_extracted_30646 | Not-determined |
| S7_extracted_30647 | Not-determined |
| S7_extracted_30650 | Not-determined |
| S7_extracted_30671 | Not-determined |
| S7_extracted_30672 | Not-determined |
| S7_extracted_30675 | Not-determined |
| S7_extracted_30676 | Not-determined |
| S7_extracted_30681 | Low-quality |
| S7_extracted_30691 | Low-quality |
| S7_extracted_30699 | Not-determined |
| S7_extracted_30709 | Not-determined |
| S7_extracted_30716 | Not-determined |
| S7_extracted_30719 | Not-determined |
| S7_extracted_30725 | Low-quality |
| S7_extracted_30729 | Not-determined |
| S7_extracted_30734 | Not-determined |
| S7_extracted_30737 | Not-determined |
| S7_extracted_30749 | Not-determined |
| S7_extracted_30752 | Not-determined |
| S7_extracted_30762 | Not-determined |
| S7_extracted_30773 | Not-determined |
| S7_extracted_30780 | Not-determined |
| S7_extracted_30787 | Low-quality |
| S7_extracted_30789 | Low-quality |
| S7_extracted_30794 | Low-quality |
| S7_extracted_30796 | Not-determined |
| S7_extracted_30812 | Not-determined |
| S7_extracted_30870 | Not-determined |
| S7_extracted_30881 | Not-determined |
| S7_extracted_30903 | Not-determined |
| S7_extracted_30910 | Not-determined |
| S7_extracted_30920 | Not-determined |
| S7_extracted_30925 | Not-determined |
| S7_extracted_30926 | Not-determined |
| S7_extracted_30963 | Not-determined |
| S7_extracted_30966 | Not-determined |
| S7_extracted_31025 | Not-determined |
| S7_extracted_31035 | Low-quality |
| S7_extracted_31046 | Low-quality |
| S7_extracted_31051 | Low-quality |
| S7_extracted_31066 | Not-determined |
| S7_extracted_31095 | Not-determined |
| S7_extracted_31110 | Not-determined |
| S7_extracted_31126 | Not-determined |
| S7_extracted_31132 | Low-quality |
| S7_extracted_31140 | Not-determined |
| S7_extracted_31155 | Low-quality |
| S7_extracted_31252 | Low-quality |
| S7_extracted_31255 | Not-determined |
| S7_extracted_31264 | Not-determined |
| S7_extracted_31276 | Not-determined |
| S7_extracted_31299 | Low-quality |
| S7_extracted_31301 | Not-determined |
| S7_extracted_31333 | Not-determined |
| S7_extracted_31340 | Low-quality |
| S7_extracted_31341 | Low-quality |
| S7_extracted_31344 | Not-determined |
| S7_extracted_31358 | Not-determined |
| S7_extracted_31374 | Not-determined |
| S7_extracted_31380 | Low-quality |
| S7_extracted_31393 | Not-determined |
| S7_extracted_31418 | Not-determined |
| S7_extracted_31433 | Not-determined |
| S7_extracted_31434 | Not-determined |
| S7_extracted_31435 | Not-determined |
| S7_extracted_31449 | Not-determined |
| S7_extracted_31455 | Not-determined |
| S7_extracted_31462 | Not-determined |
| S7_extracted_31470 | Not-determined |
| S7_extracted_31474 | Not-determined |
| S7_extracted_31480 | Low-quality |
| S7_extracted_31499 | Not-determined |
| S7_extracted_31500 | Not-determined |
| S7_extracted_31506 | Not-determined |
| S7_extracted_31546 | Not-determined |
| S7_extracted_31547 | Not-determined |
| S7_extracted_31549 | Low-quality |
| S7_extracted_31550 | Low-quality |
| S7_extracted_31555 | Not-determined |
| S7_extracted_31583 | Not-determined |
| S7_extracted_31628 | Not-determined |
| S7_extracted_31633 | Not-determined |
| S7_extracted_31654 | Not-determined |
| S7_extracted_31659 | Not-determined |
| S7_extracted_31663 | Not-determined |
| S7_extracted_31673 | Not-determined |
| S7_extracted_31686 | Not-determined |
| S7_extracted_31689 | Low-quality |
| S7_extracted_31692 | Not-determined |
| S7_extracted_31713 | Not-determined |
| S7_extracted_31715 | Not-determined |
| S7_extracted_31729 | Not-determined |
| S7_extracted_31741 | Not-determined |
| S7_extracted_31745 | Not-determined |
| S7_extracted_31753 | Not-determined |
| S7_extracted_31762 | Low-quality |
| S7_extracted_31763 | Not-determined |
| S7_extracted_31801 | Not-determined |
| S7_extracted_31806 | Not-determined |
| S7_extracted_31816 | Not-determined |
| S7_extracted_31833 | Not-determined |
| S7_extracted_31851 | Not-determined |
| S7_extracted_31856 | Not-determined |
| S7_extracted_31857 | Not-determined |
| S7_extracted_31868 | Low-quality |
| S7_extracted_31879 | Not-determined |
| S7_extracted_31881 | Not-determined |
| S7_extracted_31897 | Low-quality |
| S7_extracted_31899 | Low-quality |
| S7_extracted_31901 | Not-determined |
| S7_extracted_31903 | Not-determined |
| S7_extracted_31952 | Not-determined |
| S7_extracted_31958 | Not-determined |
| S7_extracted_31964 | Not-determined |
| S7_extracted_31980 | Low-quality |
| S7_extracted_31995 | Low-quality |
| S7_extracted_31998 | Not-determined |
| S7_extracted_32001 | Not-determined |
| S7_extracted_32014 | Not-determined |
| S7_extracted_32016 | Not-determined |
| S7_extracted_32040 | Not-determined |
| S7_extracted_32050 | Low-quality |
| S7_extracted_32054 | Not-determined |
| S7_extracted_32114 | Not-determined |
| S7_extracted_32115 | Not-determined |
| S7_extracted_32120 | Low-quality |
| S7_extracted_32127 | Low-quality |
| S7_extracted_32136 | Not-determined |
| S7_extracted_32140 | Not-determined |
| S7_extracted_32150 | Not-determined |
| S7_extracted_32182 | Not-determined |
| S7_extracted_32192 | Not-determined |
| S7_extracted_32196 | Not-determined |
| S7_extracted_32203 | Not-determined |
| S7_extracted_32215 | Not-determined |
| S7_extracted_32233 | Not-determined |
| S7_extracted_32237 | Not-determined |
| S7_extracted_32290 | Not-determined |
| S7_extracted_32294 | Low-quality |
| S7_extracted_32301 | Low-quality |
| S7_extracted_32309 | Not-determined |
| S7_extracted_32315 | Not-determined |
| S7_extracted_32325 | Not-determined |
| S7_extracted_32340 | Not-determined |
| S7_extracted_32350 | Not-determined |
| S7_extracted_32361 | Not-determined |
| S7_extracted_32372 | Not-determined |
| S7_extracted_32373 | Not-determined |
| S4_vs_VIRSorter_28601-cat_2 | High-quality |
| S7_extracted_32405 | Not-determined |
| S7_extracted_32412 | Not-determined |
| S7_extracted_32427 | Not-determined |
| S7_extracted_32431 | Not-determined |
| S7_extracted_32440 | Not-determined |
| S7_extracted_32451 | Not-determined |
| S7_extracted_32470 | Not-determined |
| S7_extracted_32475 | Not-determined |
| S7_extracted_32499 | Not-determined |
| S7_extracted_32510 | Not-determined |
| S7_extracted_32519 | Not-determined |
| S7_extracted_32534 | Not-determined |
| S7_extracted_32554 | Not-determined |
| S3_vs_k141_127395\|\|full | High-quality |
| S7_extracted_32594 | Not-determined |
| S7_extracted_32602 | Not-determined |
| S7_extracted_32631 | Not-determined |
| S7_extracted_32656 | Not-determined |
| S7_extracted_32676 | Not-determined |
| S7_extracted_32693 | Not-determined |
| S7_extracted_32697 | Not-determined |
| S7_extracted_32725 | Not-determined |
| S7_extracted_32738 | Not-determined |
| S7_extracted_32749 | Low-quality |
| S7_extracted_32752 | Not-determined |
| S7_extracted_32755 | Low-quality |
| S7_extracted_32764 | Not-determined |
| S7_extracted_32773 | Not-determined |
| S7_extracted_32774 | Not-determined |
| S7_extracted_32790 | Not-determined |
| S7_extracted_32805 | Not-determined |
| S7_extracted_32806 | Not-determined |
| S7_extracted_32817 | Low-quality |
| S7_extracted_32865 | Low-quality |
| S7_extracted_32872 | Not-determined |
| S7_extracted_32874 | Not-determined |
| S7_extracted_32878 | Not-determined |
| S7_extracted_32881 | Not-determined |
| S7_extracted_32895 | Low-quality |
| S7_extracted_32902 | Not-determined |
| S7_extracted_32906 | Not-determined |
| S7_extracted_32929 | Not-determined |
| S7_extracted_32930 | Not-determined |
| S7_extracted_32938 | Not-determined |
| S7_extracted_33002 | Low-quality |
| S7_extracted_33027 | Not-determined |
| S7_extracted_33037 | Not-determined |
| S7_extracted_33038 | Low-quality |
| S7_extracted_33046 | Not-determined |
| S7_extracted_33111 | Not-determined |
| S7_extracted_33122 | Low-quality |
| S7_extracted_33126 | Low-quality |
| S7_extracted_33138 | Not-determined |
| S7_extracted_33154 | Low-quality |
| S7_extracted_33166 | Not-determined |
| S7_extracted_33188 | Not-determined |
| S7_extracted_33194 | Not-determined |
| S7_extracted_33196 | Not-determined |
| S7_extracted_33208 | Not-determined |
| S7_extracted_33221 | Not-determined |
| S7_extracted_33228 | Not-determined |
| S7_extracted_33242 | Not-determined |
| S7_extracted_33251 | Not-determined |
| S7_extracted_33253 | Not-determined |
| S7_extracted_33264 | Not-determined |
| S7_extracted_33274 | Not-determined |
| S7_extracted_33276 | Not-determined |
| S7_extracted_33287 | Not-determined |
| S7_extracted_33298 | Not-determined |
| S7_extracted_33302 | Not-determined |
| S7_extracted_33309 | Not-determined |
| S7_extracted_33332 | Not-determined |
| S7_extracted_33346 | Not-determined |
| S7_extracted_33359 | Not-determined |
| S7_extracted_33399 | Not-determined |
| S7_extracted_33427 | Not-determined |
| S7_extracted_33433 | Not-determined |
| S7_extracted_33438 | Not-determined |
| S7_extracted_33447 | Not-determined |
| S7_extracted_33449 | Low-quality |
| S7_extracted_33459 | Not-determined |
| S7_extracted_33483 | Not-determined |
| S7_extracted_33486 | Not-determined |
| S7_extracted_33492 | Not-determined |
| S7_extracted_33501 | Not-determined |
| S7_extracted_33506 | Not-determined |
| S7_extracted_33522 | Low-quality |
| S7_extracted_33541 | Not-determined |
| S7_extracted_33546 | Not-determined |
| S7_extracted_33565 | Not-determined |
| S7_extracted_33593 | Not-determined |
| S7_extracted_33594 | Not-determined |
| S7_extracted_33598 | Not-determined |
| S7_extracted_33604 | Not-determined |
| S7_extracted_33609 | Not-determined |
| S7_extracted_33627 | Not-determined |
| S7_extracted_33650 | Not-determined |
| S7_extracted_33656 | Not-determined |
| S7_extracted_33669 | Not-determined |
| S7_extracted_33671 | Low-quality |
| S7_extracted_33672 | Not-determined |
| S7_extracted_33673 | Not-determined |
| S7_extracted_33674 | Not-determined |
| S7_extracted_33677 | Low-quality |
| S7_extracted_33700 | Low-quality |
| S7_extracted_33702 | Not-determined |
| S7_extracted_33704 | Not-determined |
| S7_extracted_33706 | Low-quality |
| S7_extracted_33716 | Not-determined |
| S7_extracted_33756 | Not-determined |
| S7_extracted_33766 | Not-determined |
| S7_extracted_33780 | Low-quality |
| S7_extracted_33808 | Not-determined |
| S7_extracted_33813 | Not-determined |
| S7_extracted_33829 | Not-determined |
| S7_extracted_33838 | Not-determined |
| S7_extracted_33853 | Not-determined |
| S7_extracted_33864 | Low-quality |
| S7_extracted_33902 | Not-determined |
| S7_extracted_33903 | Not-determined |
| S7_extracted_33908 | Not-determined |
| S7_extracted_33914 | Not-determined |
| S7_extracted_33924 | Not-determined |
| S7_extracted_33931 | Low-quality |
| S7_extracted_33937 | Not-determined |
| S7_extracted_33944 | Not-determined |
| S7_extracted_33946 | Not-determined |
| S7_extracted_33969 | Not-determined |
| S7_extracted_33980 | Not-determined |
| S7_extracted_33994 | Not-determined |
| S7_extracted_34004 | Not-determined |
| S7_extracted_34005 | Not-determined |
| S7_extracted_34008 | Not-determined |
| S7_extracted_34013 | Not-determined |
| S7_extracted_34024 | Not-determined |
| S7_extracted_34032 | Low-quality |
| S7_extracted_34050 | Not-determined |
| S7_extracted_34059 | Not-determined |
| S7_extracted_34060 | Not-determined |
| S7_extracted_34064 | Not-determined |
| S7_extracted_34084 | Not-determined |
| S7_extracted_34087 | Low-quality |
| S7_extracted_34092 | Not-determined |
| S7_extracted_34101 | Low-quality |
| S7_extracted_34117 | Not-determined |
| S7_extracted_34122 | Low-quality |
| S7_extracted_34126 | Not-determined |
| S7_extracted_34133 | Not-determined |
| S7_extracted_34136 | Not-determined |
| S7_extracted_34145 | Low-quality |
| S7_extracted_34152 | Not-determined |
| S7_extracted_34154 | Not-determined |
| S7_extracted_34158 | Low-quality |
| S7_extracted_34191 | Not-determined |
| S7_extracted_34197 | Low-quality |
| S7_extracted_34209 | Not-determined |
| S7_extracted_34236 | Not-determined |
| S7_extracted_34242 | Not-determined |
| S7_extracted_34253 | Not-determined |
| S7_extracted_34262 | Not-determined |
| S7_extracted_34271 | Not-determined |
| S7_extracted_34302 | Not-determined |
| S7_extracted_34306 | Not-determined |
| S7_extracted_34315 | Not-determined |
| S7_extracted_34318 | Not-determined |
| S7_extracted_34334 | Not-determined |
| S7_extracted_34350 | Low-quality |
| S7_extracted_34362 | Not-determined |
| S7_extracted_34373 | Not-determined |
| S7_extracted_34375 | Not-determined |
| S7_extracted_34384 | Not-determined |
| S7_extracted_34398 | Low-quality |
| S7_extracted_34445 | Not-determined |
| S7_extracted_34455 | Not-determined |
| S7_extracted_34481 | Low-quality |
| S7_extracted_34486 | Not-determined |
| S7_extracted_34490 | Low-quality |
| S7_extracted_34493 | Not-determined |
| S5_vs_k141_77701\|\|full | Medium-quality |
| S7_extracted_34530 | Low-quality |
| S7_extracted_34536 | Not-determined |
| S7_extracted_34561 | Low-quality |
| S7_extracted_34605 | Not-determined |
| S7_extracted_34613 | Not-determined |
| S7_extracted_34620 | Not-determined |
| S7_extracted_34623 | Not-determined |
| S7_extracted_34649 | Low-quality |
| S7_extracted_34679 | Not-determined |
| S7_extracted_34686 | Low-quality |
| S7_extracted_34690 | Not-determined |
| S7_extracted_34706 | Low-quality |
| S7_extracted_34736 | Not-determined |
| S7_extracted_34738 | Not-determined |
| S7_extracted_34752 | Not-determined |
| S7_extracted_34758 | Low-quality |
| S7_extracted_34770 | Not-determined |
| S7_extracted_34772 | Low-quality |
| S7_extracted_34812 | Low-quality |
| S7_extracted_34821 | Not-determined |
| S7_extracted_34830 | Not-determined |
| S7_extracted_34831 | Not-determined |
| S7_extracted_34833 | Not-determined |
| S7_extracted_34843 | Not-determined |
| S7_extracted_34863 | Not-determined |
| S7_extracted_34871 | Not-determined |
| S7_extracted_34877 | Not-determined |
| S7_extracted_34905 | Not-determined |
| S7_extracted_34911 | Low-quality |
| S7_extracted_34912 | Not-determined |
| S7_extracted_34920 | Low-quality |
| S7_extracted_34922 | Not-determined |
| S7_extracted_34942 | Not-determined |
| S7_extracted_34951 | Not-determined |
| S7_extracted_34967 | Not-determined |
| S7_extracted_35019 | Not-determined |
| S7_extracted_35033 | Not-determined |
| S7_extracted_35045 | Not-determined |
| S7_extracted_35064 | Not-determined |
| S7_extracted_35069 | Not-determined |
| S7_extracted_35100 | Low-quality |
| S7_extracted_35102 | Not-determined |
| S7_extracted_35103 | Not-determined |
| S7_extracted_35119 | Low-quality |
| S7_extracted_35123 | Low-quality |
| S7_extracted_35133 | Not-determined |
| S7_extracted_35142 | Not-determined |
| S7_extracted_35153 | Low-quality |
| S7_extracted_35169 | Low-quality |
| S7_extracted_35198 | Not-determined |
| S7_extracted_35199 | Not-determined |
| S7_extracted_35215 | Not-determined |
| S7_extracted_35217 | Not-determined |
| S7_extracted_35219 | Not-determined |
| S7_extracted_35220 | Not-determined |
| S7_extracted_35235 | Low-quality |
| S7_extracted_35257 | Not-determined |
| S7_extracted_35263 | Not-determined |
| S7_extracted_35292 | Not-determined |
| S7_extracted_35294 | Not-determined |
| S7_extracted_35303 | Not-determined |
| S7_extracted_35324 | Not-determined |
| S7_extracted_35350 | Low-quality |
| S7_extracted_35356 | Not-determined |
| S7_extracted_35363 | Not-determined |
| S7_extracted_35387 | Not-determined |
| S7_extracted_35392 | Not-determined |
| S7_extracted_35406 | Not-determined |
| S7_extracted_35413 | Not-determined |
| S7_extracted_35423 | Low-quality |
| S7_extracted_35428 | Not-determined |
| S7_extracted_35453 | Not-determined |
| S7_extracted_35464 | Not-determined |
| S7_extracted_35477 | Not-determined |
| S7_extracted_35496 | Not-determined |
| S7_extracted_35502 | Low-quality |
| S7_extracted_35528 | Not-determined |
| S7_extracted_35533 | Not-determined |
| S7_extracted_35536 | Not-determined |
| S7_extracted_35541 | Not-determined |
| S7_extracted_35546 | Not-determined |
| S7_extracted_35552 | Not-determined |
| S7_extracted_35568 | Not-determined |
| S7_extracted_35569 | Not-determined |
| S7_extracted_35570 | Not-determined |
| S7_extracted_35584 | Not-determined |
| S7_extracted_35594 | Low-quality |
| S7_extracted_35599 | Not-determined |
| S7_extracted_35605 | Not-determined |
| S7_extracted_35607 | Low-quality |
| S7_extracted_35615 | Low-quality |
| S7_extracted_35618 | Not-determined |
| S7_extracted_35622 | Not-determined |
| S7_extracted_35626 | Not-determined |
| S7_extracted_35638 | Low-quality |
| S7_extracted_35650 | Not-determined |
| S7_extracted_35665 | Low-quality |
| S7_extracted_35672 | Low-quality |
| S7_extracted_35674 | Not-determined |
| S7_extracted_35682 | Low-quality |
| S7_extracted_35688 | Not-determined |
| S7_extracted_35690 | Not-determined |
| S7_extracted_35701 | Not-determined |
| S7_extracted_35705 | Low-quality |
| S7_extracted_35706 | Low-quality |
| S7_extracted_35726 | Not-determined |
| S7_extracted_35730 | Low-quality |
| S7_extracted_35737 | Not-determined |
| S7_extracted_35775 | Not-determined |
| S7_extracted_35794 | Not-determined |
| S7_extracted_35795 | Low-quality |
| S7_extracted_35808 | Not-determined |
| S7_extracted_35812 | Not-determined |
| S7_extracted_35815 | Low-quality |
| S7_extracted_35821 | Not-determined |
| S7_extracted_35828 | Not-determined |
| S7_extracted_35843 | Not-determined |
| S7_extracted_35860 | Not-determined |
| S7_extracted_35862 | Not-determined |
| S7_extracted_35866 | Not-determined |
| S7_extracted_35890 | Not-determined |
| S7_extracted_35933 | Low-quality |
| S7_extracted_35943 | Not-determined |
| S7_extracted_35945 | Not-determined |
| S7_extracted_35984 | Not-determined |
| S7_extracted_36017 | Not-determined |
| S7_extracted_36031 | Not-determined |
| S7_extracted_36044 | Not-determined |
| S7_extracted_36057 | Not-determined |
| S7_extracted_36081 | Low-quality |
| S7_extracted_36085 | Low-quality |
| S7_extracted_36090 | Not-determined |
| S7_extracted_36098 | Not-determined |
| S7_extracted_36115 | Not-determined |
| S7_extracted_36131 | Not-determined |
| S7_extracted_36154 | Not-determined |
| S7_extracted_36166 | Not-determined |
| S7_extracted_36204 | Low-quality |
| S7_extracted_36226 | Low-quality |
| S7_extracted_36249 | Not-determined |
| S7_extracted_36260 | Not-determined |
| S7_extracted_36264 | Not-determined |
| S7_extracted_36277 | Not-determined |
| S7_extracted_36281 | Not-determined |
| S7_extracted_36290 | Not-determined |
| S7_extracted_36301 | Not-determined |
| S7_extracted_36307 | Low-quality |
| S7_extracted_36308 | Not-determined |
| S7_extracted_36312 | Low-quality |
| S7_extracted_36333 | Not-determined |
| S7_extracted_36335 | Not-determined |
| S7_extracted_36337 | Not-determined |
| S7_extracted_36390 | Not-determined |
| S7_extracted_36392 | Not-determined |
| S7_extracted_36418 | Not-determined |
| S7_extracted_36431 | Not-determined |
| S7_extracted_36440 | Not-determined |
| S7_extracted_36449 | Not-determined |
| S7_extracted_36455 | Low-quality |
| S7_extracted_36457 | Not-determined |
| S7_extracted_36459 | Not-determined |
| S7_extracted_36469 | Not-determined |
| S7_extracted_36482 | Not-determined |
| S7_extracted_36503 | Low-quality |
| S7_extracted_36519 | Not-determined |
| S7_extracted_36527 | Low-quality |
| S7_extracted_36538 | Not-determined |
| S7_extracted_36542 | Not-determined |
| S7_extracted_36564 | Not-determined |
| S7_extracted_36601 | Not-determined |
| S7_extracted_36602 | Not-determined |
| S7_extracted_36618 | Not-determined |
| S7_extracted_36619 | Not-determined |
| S5_extracted_k141_28061 | Complete |
| S7_extracted_36630 | Low-quality |
| S7_extracted_36645 | Not-determined |
| S7_extracted_36673 | Low-quality |
| S7_extracted_36682 | Not-determined |
| S7_extracted_36686 | Not-determined |
| S7_extracted_36690 | Low-quality |
| S7_extracted_36704 | Not-determined |
| S7_extracted_36714 | Not-determined |
| S7_extracted_36716 | Not-determined |
| S7_extracted_36737 | Low-quality |
| S7_extracted_36766 | Low-quality |
| S7_extracted_36773 | Low-quality |
| S7_extracted_36775 | Not-determined |
| S7_extracted_36776 | Low-quality |
| S7_extracted_36779 | Low-quality |
| S7_extracted_36780 | Not-determined |
| S7_extracted_36783 | Low-quality |
| S7_extracted_36787 | Not-determined |
| S7_extracted_36808 | Low-quality |
| S7_extracted_36829 | Not-determined |
| S7_extracted_36843 | Not-determined |

| | |
|---|---|
| S7_extracted_36844 | Not-determined |
| S7_extracted_36886 | Not-determined |
| S7_extracted_36899 | Not-determined |
| S7_extracted_36933 | Not-determined |
| S7_extracted_36949 | Not-determined |
| S7_extracted_36953 | Low-quality |
| S7_extracted_36954 | Not-determined |
| S7_extracted_36960 | Not-determined |
| S7_extracted_36972 | Not-determined |
| S7_extracted_36988 | Low-quality |
| S7_extracted_37020 | Not-determined |
| S7_extracted_37024 | Not-determined |
| S7_extracted_37030 | Not-determined |
| S7_extracted_37037 | Not-determined |
| S7_extracted_37040 | Not-determined |
| S7_extracted_37042 | Not-determined |
| S7_extracted_37043 | Not-determined |
| S7_extracted_37045 | Not-determined |
| S7_extracted_37049 | Not-determined |
| S7_extracted_37062 | Not-determined |
| S7_extracted_37066 | Not-determined |
| S7_extracted_37074 | Low-quality |
| S7_extracted_37082 | Low-quality |
| S7_extracted_37083 | Low-quality |
| S7_extracted_37084 | Not-determined |
| S7_extracted_37090 | Not-determined |
| S7_extracted_37091 | Not-determined |
| S7_extracted_37099 | Not-determined |
| S7_extracted_37101 | Not-determined |
| S7_extracted_37118 | Not-determined |
| S7_extracted_37132 | Not-determined |
| S7_extracted_37142 | Not-determined |
| S7_extracted_37174 | Not-determined |
| S7_extracted_37207 | Not-determined |
| S7_extracted_37211 | Not-determined |
| S7_extracted_37225 | Not-determined |
| S7_extracted_37253 | Not-determined |
| S7_extracted_37258 | Low-quality |
| S7_extracted_37260 | Not-determined |
| S7_extracted_37270 | Low-quality |
| S6_vs_k141_26816\|\|full | High-quality |
| S5_vs_k141_28061\|\|full | Medium-quality |
| S7_extracted_37330 | Not-determined |
| S7_extracted_37365 | Not-determined |
| S7_extracted_37397 | Not-determined |
| S7_extracted_37426 | Not-determined |
| S7_extracted_37431 | Not-determined |
| S7_extracted_37439 | Not-determined |
| S7_extracted_37454 | Not-determined |
| S7_extracted_37459 | Not-determined |
| S7_extracted_37466 | Not-determined |
| S7_extracted_37469 | Not-determined |
| S7_extracted_37478 | Not-determined |
| S7_extracted_37505 | Low-quality |
| S7_extracted_37520 | Low-quality |
| S7_extracted_37526 | Low-quality |
| S7_extracted_37532 | Not-determined |
| S7_extracted_37538 | Not-determined |
| S7_extracted_37547 | Not-determined |
| S7_extracted_37554 | Not-determined |
| S7_extracted_37583 | Not-determined |
| S7_extracted_37594 | Not-determined |
| S7_extracted_37602 | Not-determined |
| S7_extracted_37621 | Low-quality |
| S7_extracted_37630 | Not-determined |
| S7_extracted_37642 | Not-determined |
| S7_extracted_37670 | Not-determined |
| S7_extracted_37691 | Not-determined |
| S7_extracted_37720 | Not-determined |
| S7_extracted_37727 | Not-determined |
| S7_extracted_37729 | Low-quality |
| S7_extracted_37741 | Not-determined |
| S7_extracted_37742 | Not-determined |
| S7_extracted_37757 | Not-determined |
| S7_extracted_37768 | Not-determined |
| S7_extracted_37774 | Not-determined |
| S7_extracted_37789 | Low-quality |
| S7_extracted_37798 | Not-determined |
| S7_extracted_37810 | Not-determined |
| S7_extracted_37826 | Not-determined |
| S7_extracted_37834 | Not-determined |
| S7_extracted_37846 | Not-determined |
| S7_extracted_37863 | Low-quality |
| S7_extracted_37868 | Not-determined |
| S7_extracted_37872 | Not-determined |
| S7_extracted_37875 | Not-determined |
| S7_extracted_37888 | Low-quality |
| S7_extracted_37903 | Not-determined |
| S7_extracted_37905 | Low-quality |
| S7_extracted_37909 | Not-determined |
| S7_extracted_37920 | Not-determined |
| S7_extracted_37936 | Not-determined |
| S7_extracted_37962 | Not-determined |
| S7_extracted_37964 | Low-quality |
| S7_extracted_37969 | Not-determined |
| S7_extracted_37984 | Not-determined |
| S7_extracted_38012 | Not-determined |
| S7_extracted_38037 | Not-determined |
| S7_extracted_38043 | Not-determined |
| S7_extracted_38051 | Not-determined |
| S7_extracted_38060 | Not-determined |
| S7_extracted_38082 | Low-quality |
| S7_extracted_38085 | Not-determined |
| S7_extracted_38095 | Not-determined |
| S7_extracted_38101 | Not-determined |
| S7_extracted_38107 | Not-determined |
| S7_extracted_38115 | Low-quality |
| S7_extracted_38126 | Not-determined |
| S7_extracted_38144 | Not-determined |
| S7_extracted_38147 | Not-determined |
| S7_extracted_38163 | Not-determined |
| S7_extracted_38173 | Low-quality |
| S7_extracted_38226 | Not-determined |
| S7_extracted_38253 | Not-determined |
| S7_extracted_38262 | Low-quality |
| S7_extracted_38265 | Not-determined |
| S7_extracted_38296 | Not-determined |
| S7_extracted_38300 | Low-quality |
| S7_extracted_38306 | Low-quality |
| S7_extracted_38339 | Low-quality |
| S7_extracted_38348 | Not-determined |
| S7_extracted_38377 | Not-determined |
| S7_extracted_38415 | Not-determined |
| S7_extracted_38428 | Not-determined |
| S7_extracted_38462 | Not-determined |
| S7_extracted_38469 | Not-determined |
| S7_extracted_38476 | Low-quality |
| S7_extracted_38480 | Low-quality |
| S7_extracted_38484 | Low-quality |
| S7_extracted_38492 | Not-determined |
| S7_extracted_38494 | Not-determined |
| S7_extracted_38496 | Low-quality |
| S7_extracted_38503 | Not-determined |
| S7_extracted_38509 | Not-determined |
| S7_extracted_38517 | Not-determined |
| S7_extracted_38539 | Not-determined |
| S7_extracted_38545 | Not-determined |
| S7_extracted_38546 | Not-determined |
| S7_extracted_38554 | Not-determined |
| S7_extracted_38558 | Low-quality |
| S7_extracted_38574 | Not-determined |
| S7_extracted_38589 | Not-determined |
| S7_extracted_38602 | Low-quality |
| S7_extracted_38620 | Not-determined |
| S7_extracted_38631 | Not-determined |
| S7_extracted_38635 | Low-quality |
| S7_extracted_38679 | Not-determined |
| S7_extracted_38688 | Not-determined |
| S9_vs_k141_90846\|\|full | High-quality |
| S7_extracted_38699 | Not-determined |
| S7_extracted_38700 | Not-determined |
| S7_extracted_38707 | Not-determined |
| S7_extracted_38722 | Not-determined |
| S7_extracted_38748 | Not-determined |
| S7_extracted_38788 | Not-determined |
| S7_extracted_38793 | Not-determined |
| S7_extracted_38819 | Not-determined |
| S7_extracted_38824 | Not-determined |
| S7_extracted_38835 | Low-quality |
| S7_extracted_38837 | Not-determined |
| S7_extracted_38843 | Low-quality |
| S7_extracted_38846 | Low-quality |
| S7_extracted_38867 | Not-determined |
| S7_extracted_38904 | Not-determined |
| S7_extracted_38924 | Not-determined |
| S7_extracted_38941 | Not-determined |
| S7_extracted_38947 | Not-determined |
| S7_extracted_38960 | Low-quality |
| S7_extracted_38971 | Not-determined |
| S7_extracted_38978 | Not-determined |
| S7_extracted_38981 | Not-determined |
| S7_extracted_38984 | Not-determined |
| S7_extracted_38987 | Not-determined |
| S7_extracted_39005 | Not-determined |
| S7_extracted_39014 | Low-quality |
| S7_extracted_39025 | Low-quality |
| S7_extracted_39026 | Low-quality |
| S7_extracted_39038 | Not-determined |
| S7_extracted_39057 | Not-determined |
| S7_extracted_39065 | Not-determined |
| S7_extracted_39079 | Not-determined |
| S7_extracted_39085 | Not-determined |
| S7_extracted_39092 | Not-determined |
| S7_extracted_39124 | Not-determined |
| S7_extracted_39146 | Low-quality |
| S7_extracted_39148 | Low-quality |
| S7_extracted_39165 | Not-determined |
| S7_extracted_39167 | Not-determined |
| S7_extracted_39168 | Not-determined |
| S7_extracted_39172 | Not-determined |
| S7_extracted_39190 | Not-determined |
| S7_extracted_39222 | Not-determined |
| S7_extracted_39232 | Not-determined |
| S7_extracted_39240 | Not-determined |
| S7_extracted_39255 | Not-determined |
| S7_extracted_39272 | Not-determined |
| S7_extracted_39287 | Not-determined |
| S7_extracted_39299 | Not-determined |
| S7_extracted_39302 | Not-determined |
| S7_extracted_39305 | Not-determined |
| S7_extracted_39311 | Not-determined |
| S7_extracted_39334 | Not-determined |
| S7_extracted_39338 | Not-determined |
| S7_extracted_39354 | Low-quality |
| S7_extracted_39355 | Low-quality |
| S7_extracted_39372 | Not-determined |
| S7_extracted_39379 | Not-determined |
| S7_extracted_39384 | Low-quality |
| S7_extracted_39391 | Not-determined |
| S7_extracted_39410 | Not-determined |
| S7_extracted_39417 | Low-quality |
| S7_extracted_39442 | Low-quality |
| S7_extracted_39443 | Not-determined |
| S7_extracted_39450 | Low-quality |
| S7_extracted_39486 | Low-quality |
| S7_extracted_39488 | Not-determined |
| S7_extracted_39492 | Not-determined |
| S7_extracted_39507 | Not-determined |
| S7_extracted_39516 | Not-determined |
| S7_extracted_39518 | Not-determined |
| S7_extracted_39522 | Not-determined |
| S7_extracted_39528 | Not-determined |
| S7_extracted_39530 | Low-quality |
| S7_extracted_39534 | Not-determined |
| S7_extracted_39546 | Not-determined |
| S7_extracted_39548 | Low-quality |
| S7_extracted_39562 | Not-determined |
| S7_extracted_39563 | Not-determined |
| S7_extracted_39568 | Not-determined |
| S7_extracted_39603 | Not-determined |
| S7_extracted_39612 | Not-determined |
| S7_extracted_39639 | Not-determined |
| S7_extracted_39652 | Not-determined |
| S7_extracted_39658 | Not-determined |
| S7_extracted_39664 | Not-determined |
| S7_extracted_39680 | Not-determined |
| S7_extracted_39699 | Not-determined |
| S14_vs_k141_112345\|\|full | Medium-quality |
| S7_extracted_39727 | Not-determined |
| S7_extracted_39731 | Not-determined |
| S7_extracted_39745 | Not-determined |
| S7_extracted_39755 | Not-determined |
| S7_extracted_39785 | Not-determined |
| S7_extracted_39797 | Not-determined |
| S7_extracted_39849 | Not-determined |
| S7_extracted_39854 | Not-determined |
| S7_extracted_39855 | Not-determined |
| S7_extracted_39859 | Not-determined |
| S7_extracted_39873 | Not-determined |
| S7_extracted_39874 | Low-quality |
| S7_extracted_39875 | Low-quality |
| S7_extracted_39876 | Not-determined |
| S7_extracted_39879 | Not-determined |
| S7_extracted_39900 | Low-quality |
| S7_extracted_39904 | Not-determined |
| S7_extracted_39908 | Not-determined |
| S7_extracted_39918 | Not-determined |
| S7_extracted_39926 | Not-determined |
| S7_extracted_39935 | Not-determined |
| S7_extracted_39939 | Not-determined |
| S7_extracted_39941 | Not-determined |
| S7_extracted_39948 | Not-determined |
| S7_extracted_39962 | Not-determined |
| S7_extracted_39967 | Not-determined |
| S7_extracted_39972 | Not-determined |
| S7_extracted_40004 | Low-quality |
| S7_extracted_40011 | Low-quality |
| S7_extracted_40018 | Not-determined |
| S7_extracted_40030 | Not-determined |
| S7_extracted_40036 | Not-determined |
| S7_extracted_40046 | Not-determined |
| S7_extracted_40055 | Low-quality |
| S7_extracted_40062 | Low-quality |
| S7_extracted_40073 | Not-determined |
| S7_extracted_40089 | Not-determined |
| S7_extracted_40112 | Not-determined |
| S7_extracted_40117 | Low-quality |
| S7_extracted_40123 | Not-determined |
| S7_extracted_40126 | Low-quality |
| S7_extracted_40131 | Not-determined |
| S7_extracted_40136 | Low-quality |
| S7_extracted_40139 | Not-determined |
| S7_extracted_40143 | Not-determined |
| S7_extracted_40148 | Not-determined |
| S7_extracted_40158 | Low-quality |
| S7_extracted_40170 | Not-determined |
| S7_extracted_40204 | Not-determined |
| S7_extracted_40207 | Not-determined |
| S7_extracted_40222 | Not-determined |
| S7_extracted_40227 | Not-determined |
| S7_extracted_40229 | Not-determined |
| S7_extracted_40241 | Not-determined |
| S7_extracted_40265 | Not-determined |
| S7_extracted_40267 | Not-determined |
| S7_extracted_40273 | Not-determined |
| S7_extracted_40279 | Not-determined |
| S7_extracted_40280 | Not-determined |
| S7_extracted_40288 | Not-determined |
| S7_extracted_40297 | Not-determined |
| S7_extracted_40304 | Low-quality |
| S7_extracted_40315 | Not-determined |
| S7_extracted_40319 | Not-determined |
| S7_extracted_40325 | Not-determined |
| S7_extracted_40334 | Not-determined |
| S7_extracted_40346 | Not-determined |
| S7_extracted_40364 | Not-determined |
| S7_extracted_40376 | Not-determined |
| S7_extracted_40382 | Not-determined |
| S7_extracted_40389 | Not-determined |
| S7_extracted_40402 | Not-determined |
| S7_extracted_40407 | Not-determined |
| S7_extracted_40411 | Not-determined |
| S7_extracted_40416 | Not-determined |
| S7_extracted_40420 | Low-quality |
| S7_extracted_40427 | Low-quality |
| S7_extracted_40432 | Not-determined |
| S7_extracted_40433 | Not-determined |
| S7_extracted_40441 | Not-determined |
| S7_extracted_40443 | Not-determined |
| S7_extracted_40480 | Not-determined |
| S7_extracted_40502 | Low-quality |
| S7_extracted_40506 | Not-determined |
| S7_extracted_40514 | Not-determined |
| S7_extracted_40525 | Not-determined |
| S7_extracted_40527 | Not-determined |
| S7_extracted_40532 | Not-determined |
| S7_extracted_40533 | Not-determined |
| S7_extracted_40537 | Not-determined |
| S7_extracted_40548 | Not-determined |
| S7_extracted_40558 | Not-determined |
| S7_extracted_40559 | Not-determined |
| S7_extracted_40561 | Low-quality |
| S7_extracted_40563 | Not-determined |
| S7_extracted_40571 | Not-determined |
| S7_extracted_40598 | Not-determined |
| S7_extracted_40603 | Not-determined |
| S7_extracted_40604 | Not-determined |
| S7_extracted_40613 | Not-determined |
| S7_extracted_40620 | Not-determined |
| S7_extracted_40628 | Low-quality |
| S7_extracted_40632 | Not-determined |
| S7_extracted_40635 | Not-determined |
| S7_extracted_40648 | Not-determined |
| S7_extracted_40651 | Not-determined |
| S7_extracted_40663 | Not-determined |
| S7_extracted_40672 | Not-determined |
| S7_extracted_40674 | Not-determined |
| S7_extracted_40683 | Not-determined |
| S7_extracted_40712 | Low-quality |
| S7_extracted_40748 | Low-quality |
| S7_extracted_40752 | Not-determined |
| S7_extracted_40759 | Not-determined |
| S7_extracted_40770 | Not-determined |
| S7_extracted_40798 | Not-determined |
| S7_extracted_40808 | Not-determined |
| S7_extracted_40816 | Not-determined |
| S7_extracted_40832 | Not-determined |
| S7_extracted_40853 | Not-determined |
| S7_extracted_40865 | Not-determined |
| S7_extracted_40879 | Low-quality |
| S7_extracted_40908 | Low-quality |
| S7_extracted_40912 | Not-determined |
| S7_extracted_40915 | Not-determined |
| S7_extracted_40959 | Not-determined |
| S7_extracted_40980 | Low-quality |
| S7_extracted_40991 | Not-determined |
| S7_extracted_40999 | Low-quality |
| S7_extracted_41028 | Low-quality |
| S7_extracted_41030 | Not-determined |
| S7_extracted_41031 | Not-determined |
| S7_extracted_41041 | Not-determined |
| S7_extracted_41060 | Low-quality |
| S7_extracted_41091 | Not-determined |
| S7_extracted_41099 | Not-determined |
| S7_extracted_41101 | Not-determined |
| S7_extracted_41107 | Low-quality |
| S7_extracted_41114 | Not-determined |
| S7_extracted_41129 | Not-determined |
| S7_extracted_41145 | Not-determined |
| S7_extracted_41169 | Not-determined |
| S7_extracted_41171 | Not-determined |
| S7_extracted_41215 | Not-determined |
| S7_extracted_41227 | Not-determined |
| S7_extracted_41247 | Not-determined |
| S7_extracted_41252 | Not-determined |
| S7_extracted_41259 | Low-quality |
| S7_extracted_41267 | Not-determined |
| S7_extracted_41272 | Not-determined |
| S7_extracted_41288 | Low-quality |
| S7_extracted_41293 | Not-determined |
| S7_extracted_41307 | Not-determined |
| S7_extracted_41333 | Not-determined |
| S7_extracted_41351 | Not-determined |
| S7_extracted_41386 | Not-determined |
| S7_extracted_41392 | Not-determined |
| S7_extracted_41402 | Not-determined |
| S7_extracted_41413 | Not-determined |
| S7_extracted_41448 | Not-determined |
| S7_extracted_41449 | Not-determined |
| S7_extracted_41480 | Not-determined |
| S7_extracted_41494 | Not-determined |
| S7_extracted_41496 | Not-determined |
| S7_extracted_41510 | Not-determined |
| S7_extracted_41513 | Low-quality |
| S7_extracted_41521 | Not-determined |
| S7_extracted_41524 | Not-determined |
| S7_extracted_41535 | Not-determined |
| S7_extracted_41540 | Not-determined |
| S7_extracted_41542 | Low-quality |
| S7_extracted_41545 | Low-quality |
| S7_extracted_41578 | Not-determined |
| S7_extracted_41579 | Not-determined |
| S7_extracted_41591 | Not-determined |
| S7_extracted_41610 | Not-determined |
| S7_extracted_41655 | Not-determined |
| S7_extracted_41672 | Low-quality |
| S7_extracted_41682 | Not-determined |
| S7_extracted_41691 | Not-determined |
| S7_extracted_41726 | Not-determined |
| S7_extracted_41742 | Not-determined |
| S7_extracted_41758 | Not-determined |
| S7_extracted_41771 | Not-determined |
| S7_extracted_41780 | Not-determined |
| S7_extracted_41781 | Not-determined |
| S7_extracted_41798 | Not-determined |
| S7_extracted_41813 | Not-determined |
| S7_extracted_41825 | Low-quality |
| S7_extracted_41829 | Not-determined |
| S7_extracted_41852 | Not-determined |
| S7_extracted_41865 | Low-quality |
| S7_extracted_41886 | Not-determined |
| S7_extracted_41895 | Low-quality |
| S7_extracted_41910 | Not-determined |
| S7_extracted_41918 | Not-determined |
| S7_extracted_41930 | Not-determined |
| S7_extracted_41988 | Not-determined |
| S7_extracted_42010 | Low-quality |
| S7_extracted_42020 | Low-quality |
| S7_extracted_42040 | Not-determined |
| S7_extracted_42047 | Not-determined |
| S7_extracted_42060 | Not-determined |
| S7_extracted_42083 | Not-determined |
| S7_extracted_42100 | Not-determined |
| S7_extracted_42106 | Low-quality |
| S7_extracted_42115 | Not-determined |
| S7_extracted_42121 | Not-determined |
| S7_extracted_42133 | Low-quality |
| S7_extracted_42135 | Low-quality |
| S7_extracted_42162 | Not-determined |
| S7_extracted_42174 | Low-quality |
| S7_extracted_42177 | Low-quality |
| S7_extracted_42184 | Not-determined |
| S7_extracted_42189 | Not-determined |
| S7_extracted_42208 | Not-determined |
| S7_extracted_42209 | Low-quality |
| S7_extracted_42215 | Not-determined |
| S7_extracted_42222 | Not-determined |
| S7_extracted_42229 | Not-determined |
| S7_extracted_42234 | Not-determined |
| S7_extracted_42251 | Not-determined |
| S7_extracted_42265 | Not-determined |
| S7_extracted_42269 | Not-determined |
| S7_extracted_42271 | Not-determined |
| S7_extracted_42294 | Not-determined |
| S7_extracted_42304 | Not-determined |
| S7_extracted_42307 | Not-determined |
| S7_extracted_42318 | Not-determined |
| S7_extracted_42330 | Not-determined |
| S7_extracted_42335 | Not-determined |
| S7_extracted_42336 | Not-determined |
| S7_extracted_42363 | Not-determined |
| S7_extracted_42397 | Low-quality |
| S7_extracted_42398 | Not-determined |
| S7_extracted_42418 | Not-determined |
| S7_extracted_42432 | Low-quality |
| S7_extracted_42434 | Not-determined |
| S7_extracted_42452 | Not-determined |
| S7_extracted_42468 | Not-determined |
| S7_extracted_42475 | Not-determined |
| S7_extracted_42477 | Not-determined |
| S7_extracted_42493 | Not-determined |
| S7_extracted_42521 | Not-determined |
| S7_extracted_42534 | Not-determined |
| S7_extracted_42545 | Low-quality |
| S7_extracted_42574 | Not-determined |
| S7_extracted_42597 | Not-determined |
| S7_extracted_42600 | Not-determined |
| S7_extracted_42607 | Low-quality |
| S7_extracted_42608 | Not-determined |
| S7_extracted_42621 | Low-quality |
| S7_extracted_42629 | Not-determined |
| S7_extracted_42645 | Low-quality |
| S7_extracted_42656 | Not-determined |
| S7_extracted_42680 | Not-determined |
| S7_extracted_42691 | Not-determined |
| S7_extracted_42702 | Not-determined |
| S7_extracted_42704 | Not-determined |
| S7_extracted_42710 | Not-determined |
| S7_extracted_42732 | Not-determined |
| S7_extracted_42738 | Not-determined |
| S7_extracted_42739 | Not-determined |
| S7_extracted_42742 | Not-determined |
| S7_extracted_42766 | Not-determined |
| S7_extracted_42773 | Low-quality |
| S7_extracted_42779 | Not-determined |
| S7_extracted_42783 | Low-quality |
| S7_extracted_42785 | Not-determined |
| S7_extracted_42789 | Not-determined |
| S7_extracted_42793 | Not-determined |
| S7_extracted_42800 | Not-determined |
| S7_extracted_42803 | Not-determined |
| S7_extracted_42813 | Not-determined |
| S7_extracted_42821 | Low-quality |
| S7_extracted_42833 | Not-determined |
| S7_extracted_42839 | Not-determined |
| S7_extracted_42847 | Not-determined |
| S7_extracted_42854 | Not-determined |
| S7_extracted_42855 | Not-determined |
| S7_extracted_42871 | Not-determined |
| S7_extracted_42873 | Not-determined |
| S7_extracted_42880 | Not-determined |
| S7_extracted_42885 | Not-determined |
| S7_extracted_42899 | Not-determined |
| S7_extracted_42908 | Not-determined |
| S7_extracted_42913 | Not-determined |
| S7_extracted_42920 | Not-determined |
| S7_extracted_42931 | Low-quality |
| S7_extracted_42943 | Not-determined |
| S7_extracted_42946 | Not-determined |
| S7_extracted_42956 | Not-determined |
| S7_extracted_42966 | Not-determined |
| S7_extracted_42970 | Not-determined |
| S7_extracted_42977 | Low-quality |
| S7_extracted_42978 | Not-determined |
| S7_extracted_42989 | Not-determined |
| S7_extracted_42997 | Not-determined |
| S7_extracted_43001 | Not-determined |
| S7_extracted_43008 | Low-quality |
| S7_extracted_43012 | Not-determined |
| S7_extracted_43022 | Not-determined |
| S7_extracted_43059 | Not-determined |
| S7_extracted_43065 | Not-determined |
| S7_extracted_43068 | Not-determined |
| S7_extracted_43071 | Not-determined |
| S7_extracted_43073 | Low-quality |
| S7_extracted_43074 | Not-determined |
| S7_extracted_43075 | Not-determined |
| S7_extracted_43079 | Not-determined |
| S7_extracted_43093 | Not-determined |
| S7_extracted_43094 | Not-determined |
| S7_extracted_43117 | Low-quality |
| S7_extracted_43132 | Not-determined |
| S7_extracted_43168 | Low-quality |
| S7_extracted_43170 | Not-determined |
| S7_extracted_43192 | Not-determined |
| S7_extracted_43206 | Low-quality |
| S7_extracted_43229 | Not-determined |
| S7_extracted_43232 | Not-determined |
| S7_extracted_43233 | Not-determined |
| S7_extracted_43234 | Not-determined |
| S7_extracted_43237 | Not-determined |
| S7_extracted_43240 | Not-determined |
| S7_extracted_43251 | Low-quality |
| S7_extracted_43255 | Not-determined |
| S7_extracted_43256 | Not-determined |
| S7_extracted_43296 | Not-determined |
| S7_extracted_43328 | Low-quality |
| S7_extracted_43359 | Not-determined |
| S7_extracted_43375 | Not-determined |
| S7_extracted_43393 | Not-determined |
| S7_extracted_43394 | Not-determined |
| S7_extracted_43406 | Low-quality |
| S7_extracted_43414 | Not-determined |
| S7_extracted_43415 | Not-determined |
| S7_extracted_43420 | Low-quality |
| S7_extracted_43425 | Not-determined |
| S7_extracted_43441 | Not-determined |
| S7_extracted_43471 | Not-determined |
| S7_extracted_43489 | Not-determined |
| S7_extracted_43512 | Not-determined |
| S7_extracted_43524 | Not-determined |
| S7_extracted_43549 | Not-determined |
| S7_extracted_43560 | Not-determined |
| S7_extracted_43581 | Not-determined |
| S7_extracted_43593 | Low-quality |
| S7_extracted_43604 | Not-determined |
| S7_extracted_43610 | Not-determined |
| S7_extracted_43615 | Not-determined |
| S7_extracted_43648 | Not-determined |

| | |
|---|---|
| S7_extracted_43662 | Not-determined |
| S7_extracted_43663 | Not-determined |
| S7_extracted_43707 | Not-determined |
| S7_extracted_43716 | Not-determined |
| S7_extracted_43725 | Not-determined |
| S7_extracted_43741 | Not-determined |
| S7_extracted_43764 | Not-determined |
| S7_extracted_43794 | Low-quality |
| S7_extracted_43798 | Not-determined |
| S7_extracted_43807 | Low-quality |
| S7_extracted_43809 | Low-quality |
| S7_extracted_43818 | Not-determined |
| S7_extracted_43823 | Not-determined |
| S7_extracted_43831 | Not-determined |
| S7_extracted_43835 | Not-determined |
| S7_extracted_43837 | Not-determined |
| S7_extracted_43847 | Not-determined |
| S7_extracted_43862 | Low-quality |
| S7_extracted_43865 | Low-quality |
| S7_extracted_43889 | Not-determined |
| S7_extracted_43893 | Not-determined |
| S7_extracted_43896 | Not-determined |
| S7_extracted_43909 | Low-quality |
| S7_extracted_43920 | Not-determined |
| S7_extracted_43922 | Not-determined |
| S7_extracted_43925 | Not-determined |
| S7_extracted_43931 | Not-determined |
| S7_extracted_43936 | Not-determined |
| S7_extracted_43942 | Not-determined |
| S7_extracted_43955 | Low-quality |
| S7_extracted_43961 | Low-quality |
| S7_extracted_43983 | Not-determined |
| S7_extracted_43984 | Not-determined |
| S7_extracted_43994 | Not-determined |
| S7_extracted_44074 | Not-determined |
| S7_extracted_44087 | Low-quality |
| S7_extracted_44094 | Low-quality |
| S7_extracted_44100 | Low-quality |
| S7_extracted_44101 | Not-determined |
| S7_extracted_44131 | Not-determined |
| S7_extracted_44150 | Not-determined |
| S7_extracted_44153 | Not-determined |
| S7_extracted_44170 | Not-determined |
| S7_extracted_44181 | Not-determined |
| S7_extracted_44182 | Not-determined |
| S7_extracted_44195 | Not-determined |
| S7_extracted_44202 | Not-determined |
| S7_extracted_44203 | Not-determined |
| S7_extracted_44212 | Not-determined |
| S7_extracted_44230 | Not-determined |
| S7_extracted_44232 | Not-determined |
| S7_extracted_44243 | Not-determined |
| S7_extracted_44249 | Low-quality |
| S7_extracted_44269 | Not-determined |
| S7_extracted_44276 | Not-determined |
| S7_extracted_44286 | Not-determined |
| S7_extracted_44293 | Not-determined |
| S7_extracted_44309 | Not-determined |
| S7_extracted_44312 | Not-determined |
| S7_extracted_44325 | Not-determined |
| S7_extracted_44333 | Low-quality |
| S7_extracted_44337 | Low-quality |
| S7_extracted_44339 | Not-determined |
| S7_extracted_44344 | Not-determined |
| S7_extracted_44346 | Not-determined |
| S7_extracted_44348 | Not-determined |
| S7_extracted_44352 | Not-determined |
| S7_extracted_44372 | Not-determined |
| S7_extracted_44387 | Not-determined |
| S7_extracted_44391 | Not-determined |
| S7_extracted_44398 | Not-determined |
| S7_extracted_44408 | Not-determined |
| S7_extracted_44417 | Not-determined |
| S7_extracted_44429 | Low-quality |
| S7_extracted_44447 | Low-quality |
| S7_extracted_44454 | Not-determined |
| S7_extracted_44456 | Not-determined |
| S7_extracted_44531 | Low-quality |
| S7_extracted_44553 | Not-determined |
| S7_extracted_44580 | Not-determined |
| S7_extracted_44590 | Not-determined |
| S7_extracted_44594 | Not-determined |
| S7_extracted_44597 | Not-determined |
| S7_extracted_44604 | Not-determined |
| S7_extracted_44606 | Not-determined |
| S7_extracted_44611 | Low-quality |
| S7_extracted_44613 | Low-quality |
| S7_extracted_44625 | Not-determined |
| S7_extracted_44634 | Not-determined |
| S7_extracted_44636 | Not-determined |
| S7_extracted_44638 | Not-determined |
| S7_extracted_44674 | Not-determined |
| S7_extracted_44675 | Low-quality |
| S7_extracted_44676 | Low-quality |
| S7_extracted_44714 | Not-determined |
| S7_extracted_44720 | Not-determined |
| S7_extracted_44721 | Not-determined |
| S7_extracted_44723 | Not-determined |
| S7_extracted_44734 | Low-quality |
| S7_extracted_44737 | Not-determined |
| S7_extracted_44748 | Low-quality |
| S7_extracted_44773 | Not-determined |
| S7_extracted_44774 | Not-determined |
| S7_extracted_44775 | Not-determined |
| S7_extracted_44778 | Not-determined |
| S7_extracted_44812 | Low-quality |
| S7_extracted_44834 | Low-quality |
| S7_extracted_44837 | Not-determined |
| S7_extracted_44843 | Not-determined |
| S7_extracted_44872 | Not-determined |
| S7_extracted_44883 | Not-determined |
| S7_extracted_44885 | Not-determined |
| S7_extracted_44916 | Not-determined |
| S7_extracted_44940 | Not-determined |
| S7_extracted_44960 | Not-determined |
| S7_extracted_44963 | Not-determined |
| S7_extracted_44968 | Not-determined |
| S7_extracted_44970 | Not-determined |
| S7_extracted_44984 | Not-determined |
| S7_extracted_45007 | Not-determined |
| S7_extracted_45011 | Low-quality |
| S7_extracted_45044 | Not-determined |
| S7_extracted_45048 | Not-determined |
| S7_extracted_45059 | Not-determined |
| S7_extracted_45061 | Not-determined |
| S7_extracted_45120 | Not-determined |
| S7_extracted_45139 | Low-quality |
| S7_extracted_45191 | Not-determined |
| S7_extracted_45198 | Not-determined |
| S7_extracted_45205 | Not-determined |
| S7_extracted_45239 | Not-determined |
| S7_extracted_45241 | Not-determined |
| S7_extracted_45297 | Not-determined |
| S7_extracted_45314 | Not-determined |
| S7_extracted_45344 | Not-determined |
| S7_extracted_45387 | Not-determined |
| S7_extracted_45416 | Not-determined |
| S7_extracted_45418 | Not-determined |
| S7_extracted_45422 | Not-determined |
| S7_extracted_45423 | Not-determined |
| S7_extracted_45428 | Low-quality |
| S7_extracted_45465 | Not-determined |
| S7_extracted_45467 | Not-determined |
| S7_extracted_45475 | Low-quality |
| S7_extracted_45486 | Not-determined |
| S7_extracted_45514 | Low-quality |
| S7_extracted_45517 | Low-quality |
| S7_extracted_45521 | Not-determined |
| S7_extracted_45532 | Not-determined |
| S7_extracted_45538 | Not-determined |
| S7_extracted_45564 | Not-determined |
| S7_extracted_45565 | Not-determined |
| S7_extracted_45570 | Not-determined |
| S7_extracted_45572 | Not-determined |
| S7_extracted_45581 | Low-quality |
| S7_extracted_45615 | Not-determined |
| S7_extracted_45617 | Not-determined |
| S7_extracted_45630 | Not-determined |
| S7_extracted_45631 | Not-determined |
| S7_extracted_45639 | Low-quality |
| S7_extracted_45656 | Not-determined |
| S7_extracted_45661 | Not-determined |
| S7_extracted_45662 | Not-determined |
| S7_extracted_45666 | Low-quality |
| S7_extracted_45677 | Not-determined |
| S7_extracted_45684 | Low-quality |
| S7_extracted_45700 | Not-determined |
| S7_extracted_45709 | Not-determined |
| S7_extracted_45710 | Low-quality |
| S7_extracted_45738 | Not-determined |
| S7_extracted_45744 | Not-determined |
| S7_extracted_45749 | Not-determined |
| S7_extracted_45752 | Not-determined |
| S7_extracted_45759 | Not-determined |
| S7_extracted_45772 | Not-determined |
| S7_extracted_45777 | Not-determined |
| S7_extracted_45781 | Not-determined |
| S7_extracted_45802 | Not-determined |
| S7_extracted_45814 | Not-determined |
| S7_extracted_45832 | Not-determined |
| S7_extracted_45841 | Not-determined |
| S7_extracted_45855 | Low-quality |
| S7_extracted_45859 | Not-determined |
| S7_extracted_45867 | Not-determined |
| S7_extracted_45884 | Not-determined |
| S7_extracted_45896 | Not-determined |
| S7_extracted_45897 | Not-determined |
| S7_extracted_45907 | Not-determined |
| S7_extracted_45920 | Not-determined |
| S7_extracted_45925 | Not-determined |
| S7_extracted_45938 | Not-determined |
| S7_extracted_45940 | Not-determined |
| S7_extracted_45947 | Not-determined |
| S7_extracted_45952 | Low-quality |
| S7_extracted_45975 | Not-determined |
| S7_extracted_45980 | Not-determined |
| S7_extracted_45993 | Not-determined |
| S7_extracted_45999 | Not-determined |
| S7_extracted_46004 | Not-determined |
| S7_extracted_46008 | Not-determined |
| S7_extracted_46015 | Not-determined |
| S7_extracted_46043 | Low-quality |
| S7_extracted_46056 | Not-determined |
| S7_extracted_46073 | Not-determined |
| S7_extracted_46080 | Not-determined |
| S7_extracted_46085 | Low-quality |
| S7_extracted_46093 | Not-determined |
| S7_extracted_46102 | Not-determined |
| S7_extracted_46110 | Not-determined |
| S7_extracted_46119 | Low-quality |
| S7_extracted_46165 | Not-determined |
| S7_extracted_46180 | Not-determined |
| S7_extracted_46197 | Not-determined |
| S7_extracted_46203 | Not-determined |
| S7_extracted_46219 | Not-determined |
| S7_extracted_46221 | Not-determined |
| S7_extracted_46255 | Not-determined |
| S7_extracted_46277 | Not-determined |
| S7_extracted_46284 | Not-determined |
| S7_extracted_46288 | Low-quality |
| S7_extracted_46312 | Low-quality |
| S7_extracted_46314 | Not-determined |
| S7_extracted_46328 | Not-determined |
| S7_extracted_46342 | Not-determined |
| S7_extracted_46346 | Not-determined |
| S7_extracted_46347 | Low-quality |
| S7_extracted_46364 | Not-determined |
| S7_extracted_46383 | Low-quality |
| S7_extracted_46393 | Not-determined |
| S7_extracted_46398 | Not-determined |
| S7_extracted_46407 | Not-determined |
| S7_extracted_46443 | Low-quality |
| S7_extracted_46451 | Not-determined |
| S7_extracted_46461 | Not-determined |
| S7_extracted_46471 | Not-determined |
| S7_extracted_46475 | Not-determined |
| S7_extracted_46483 | Not-determined |
| S7_extracted_46489 | Low-quality |
| S7_extracted_46491 | Low-quality |
| S7_extracted_46509 | Not-determined |
| S7_extracted_46514 | Not-determined |
| S7_extracted_46515 | Not-determined |
| S7_extracted_46523 | Not-determined |
| S7_extracted_46536 | Not-determined |
| S7_extracted_46542 | Not-determined |
| S7_extracted_46559 | Not-determined |
| S7_extracted_46585 | Not-determined |
| S7_extracted_46588 | Not-determined |
| S7_extracted_46594 | Low-quality |
| S7_extracted_46599 | Low-quality |
| S7_extracted_46608 | Not-determined |
| S7_extracted_46616 | Not-determined |
| S7_extracted_46626 | Not-determined |
| S7_extracted_46630 | Low-quality |
| S7_extracted_46658 | Low-quality |
| S7_extracted_46675 | Low-quality |
| S7_extracted_46683 | Not-determined |
| S7_extracted_46692 | Not-determined |
| S7_extracted_46698 | Not-determined |
| S7_extracted_46705 | Not-determined |
| S7_extracted_46713 | Not-determined |
| S7_extracted_46714 | Not-determined |
| S7_extracted_46715 | Not-determined |
| S7_extracted_46736 | Not-determined |
| S7_extracted_46747 | Not-determined |
| S7_extracted_46753 | Not-determined |
| S7_extracted_46765 | Not-determined |
| S7_extracted_46786 | Not-determined |
| S7_extracted_46788 | Not-determined |
| S7_extracted_46806 | Not-determined |
| S7_extracted_46818 | Not-determined |
| S7_extracted_46843 | Low-quality |
| S7_extracted_46845 | Low-quality |
| S7_extracted_46860 | Not-determined |
| S7_extracted_46868 | Not-determined |
| S7_extracted_46879 | Not-determined |
| S7_extracted_46889 | Not-determined |
| S7_extracted_46895 | Not-determined |
| S7_extracted_46923 | Low-quality |
| S7_extracted_46924 | Not-determined |
| S7_extracted_46935 | Not-determined |
| S7_extracted_46952 | Not-determined |
| S7_extracted_46959 | Not-determined |
| S7_extracted_46969 | Not-determined |
| S7_extracted_46978 | Not-determined |
| S7_extracted_46986 | Not-determined |
| S7_extracted_47024 | Not-determined |
| S7_extracted_47025 | Not-determined |
| S7_extracted_47026 | Not-determined |
| S7_extracted_47035 | Not-determined |
| S7_extracted_47073 | Not-determined |
| S7_extracted_47099 | Not-determined |
| S7_extracted_47108 | Not-determined |
| S7_extracted_47111 | Not-determined |
| S7_extracted_47151 | Low-quality |
| S7_extracted_47171 | Not-determined |
| S7_extracted_47172 | Not-determined |
| S7_extracted_47179 | Not-determined |
| S7_extracted_47181 | Not-determined |
| S7_extracted_47186 | Not-determined |
| S7_extracted_47188 | Low-quality |
| S7_extracted_47191 | Low-quality |
| S7_extracted_47203 | Not-determined |
| S7_extracted_47235 | Low-quality |
| S7_extracted_47251 | Low-quality |
| S7_extracted_47252 | Not-determined |
| S7_extracted_47257 | Low-quality |
| S7_extracted_47264 | Not-determined |
| S7_extracted_47266 | Not-determined |
| S7_extracted_47269 | Not-determined |
| S7_extracted_47306 | Not-determined |
| S7_extracted_47309 | Not-determined |
| S7_extracted_47317 | Not-determined |
| S7_extracted_47328 | Low-quality |
| S7_extracted_47341 | Not-determined |
| S14_vs_k141_32999\|\|full | High-quality |
| S7_extracted_47355 | Not-determined |
| S7_extracted_47356 | Not-determined |
| S7_extracted_47360 | Not-determined |
| S7_extracted_47368 | Not-determined |
| S7_extracted_47374 | Low-quality |
| S7_extracted_47382 | Not-determined |
| S7_extracted_47391 | Not-determined |
| S7_extracted_47394 | Not-determined |
| S7_extracted_47398 | Low-quality |
| S7_extracted_47403 | Low-quality |
| S7_extracted_47413 | Not-determined |
| S7_extracted_47439 | Not-determined |
| S7_extracted_47442 | Not-determined |
| S7_extracted_47453 | Not-determined |
| S7_extracted_47476 | Not-determined |
| S7_extracted_47502 | Not-determined |
| S7_extracted_47511 | Not-determined |
| S7_extracted_47520 | Not-determined |
| S7_extracted_47523 | Not-determined |
| S7_extracted_47526 | Not-determined |
| S7_extracted_47533 | Not-determined |
| S7_extracted_47557 | Not-determined |
| S7_extracted_47561 | Low-quality |
| S7_extracted_47571 | Low-quality |
| S7_extracted_47600 | Not-determined |
| S7_extracted_47604 | Not-determined |
| S7_extracted_47605 | Not-determined |
| S7_extracted_47609 | Not-determined |
| S7_extracted_47616 | Not-determined |
| S7_extracted_47631 | Not-determined |
| S7_extracted_47641 | Not-determined |
| S7_extracted_47644 | Not-determined |
| S7_extracted_47654 | Not-determined |
| S7_extracted_47670 | Not-determined |
| S7_extracted_47682 | Not-determined |
| S7_extracted_47684 | Not-determined |
| S7_extracted_47691 | Not-determined |
| S7_extracted_47696 | Not-determined |
| S7_extracted_47713 | Not-determined |
| S7_extracted_47720 | Not-determined |
| S7_extracted_47742 | Not-determined |
| S7_extracted_47753 | Not-determined |
| S7_extracted_47769 | Not-determined |
| S7_extracted_47772 | Not-determined |
| S7_extracted_47777 | Not-determined |
| S7_extracted_47825 | Not-determined |
| S7_extracted_47836 | Not-determined |
| S7_extracted_47849 | Not-determined |
| S7_extracted_47853 | Low-quality |
| S7_extracted_47866 | Not-determined |
| S7_extracted_47873 | Not-determined |
| S7_extracted_47883 | Not-determined |
| S7_extracted_47895 | Not-determined |
| S7_extracted_47898 | Not-determined |
| S7_extracted_47901 | Not-determined |
| S7_extracted_47907 | Not-determined |
| S7_extracted_47908 | Not-determined |
| S7_extracted_47954 | Low-quality |
| S7_extracted_47959 | Low-quality |
| S7_extracted_47966 | Not-determined |
| S7_extracted_47997 | Low-quality |
| S7_extracted_48005 | Not-determined |
| S7_extracted_48019 | Low-quality |
| S7_extracted_48033 | Not-determined |
| S7_extracted_48042 | Not-determined |
| S7_extracted_48046 | Not-determined |
| S7_extracted_48061 | Not-determined |
| S7_extracted_48079 | Not-determined |
| S7_extracted_48083 | Not-determined |
| S7_extracted_48089 | Not-determined |
| S7_extracted_48099 | Not-determined |
| S7_extracted_48117 | Not-determined |
| S7_extracted_48118 | Low-quality |
| S7_extracted_48137 | Not-determined |
| S7_extracted_48141 | Not-determined |
| S7_extracted_48145 | Not-determined |
| S7_extracted_48151 | Not-determined |
| S7_extracted_48168 | Not-determined |
| S7_extracted_48183 | Not-determined |
| S7_extracted_48199 | Not-determined |
| S7_extracted_48200 | Not-determined |
| S7_extracted_48218 | Not-determined |
| S7_extracted_48260 | Low-quality |
| S7_extracted_48271 | Not-determined |
| S7_extracted_48275 | Not-determined |
| S7_extracted_48288 | Not-determined |
| S7_extracted_48290 | Not-determined |
| S7_extracted_48304 | Not-determined |
| S7_extracted_48309 | Not-determined |
| S7_extracted_48316 | Not-determined |
| S7_extracted_48331 | Not-determined |
| S7_extracted_48348 | Not-determined |
| S7_extracted_48350 | Low-quality |
| S7_extracted_48352 | Low-quality |
| S7_extracted_48400 | Low-quality |
| S7_extracted_48413 | Not-determined |
| S7_extracted_48455 | Not-determined |
| S7_extracted_48463 | Not-determined |
| S7_extracted_48464 | Low-quality |
| S7_extracted_48472 | Not-determined |
| S7_extracted_48476 | Not-determined |
| S7_extracted_48486 | Low-quality |
| S7_extracted_48488 | Not-determined |
| S7_extracted_48504 | Not-determined |
| S7_extracted_48532 | Not-determined |
| S7_extracted_48536 | Not-determined |
| S7_extracted_48547 | Not-determined |
| S7_extracted_48557 | Not-determined |
| S7_extracted_48559 | Not-determined |
| S7_extracted_48574 | Not-determined |
| S7_extracted_48577 | Low-quality |
| S7_extracted_48585 | Not-determined |
| S7_extracted_48586 | Not-determined |
| S7_extracted_48593 | Not-determined |
| S7_extracted_48594 | Low-quality |
| S7_extracted_48611 | Low-quality |
| S7_extracted_48632 | Low-quality |
| S7_extracted_48635 | Low-quality |
| S7_extracted_48644 | Low-quality |
| S7_extracted_48663 | Not-determined |
| S7_extracted_48679 | Not-determined |
| S7_extracted_48701 | Not-determined |
| S7_extracted_48702 | Low-quality |
| S7_extracted_48727 | Not-determined |
| S7_extracted_48731 | Not-determined |
| S7_extracted_48735 | Low-quality |
| S7_extracted_48752 | Not-determined |
| S7_extracted_48754 | Low-quality |
| S7_extracted_48777 | Not-determined |
| S7_extracted_48786 | Not-determined |
| S7_extracted_48834 | Not-determined |
| S7_extracted_48837 | Not-determined |
| S7_extracted_48842 | Not-determined |
| S7_extracted_48861 | Not-determined |
| S7_extracted_48867 | Not-determined |
| S7_extracted_48875 | Not-determined |
| S7_extracted_48877 | Not-determined |
| S7_extracted_48879 | Low-quality |
| S7_extracted_48883 | Not-determined |
| S7_extracted_48887 | Not-determined |
| S7_extracted_48908 | Not-determined |
| S7_extracted_48918 | Not-determined |
| S7_extracted_48933 | Not-determined |
| S7_extracted_48945 | Low-quality |
| S7_extracted_48951 | Not-determined |
| S7_extracted_48953 | Not-determined |
| S7_extracted_48970 | Not-determined |
| S7_extracted_48978 | Not-determined |
| S7_extracted_48983 | Not-determined |
| S7_extracted_48985 | Low-quality |
| S5_vs_k141_143651\|\|full | High-quality |
| S7_extracted_49011 | Not-determined |
| S7_extracted_49017 | Not-determined |
| S7_extracted_49043 | Not-determined |
| S7_extracted_49057 | Low-quality |
| S7_extracted_49061 | Not-determined |
| S7_extracted_49065 | Not-determined |
| S7_extracted_49070 | Not-determined |
| S7_extracted_49073 | Not-determined |
| S7_extracted_49098 | Not-determined |
| S7_extracted_49106 | Low-quality |
| S7_extracted_49107 | Not-determined |
| S7_extracted_49132 | Not-determined |
| S7_extracted_49140 | Low-quality |
| S7_extracted_49164 | Not-determined |
| S7_extracted_49170 | Not-determined |
| S7_extracted_49179 | Not-determined |
| S7_extracted_49207 | Not-determined |
| S7_extracted_49239 | Not-determined |
| S7_extracted_49242 | Not-determined |
| S7_extracted_49250 | Not-determined |
| S7_extracted_49257 | Low-quality |
| S7_extracted_49267 | Not-determined |
| S7_extracted_49276 | Not-determined |
| S7_extracted_49287 | Not-determined |
| S7_extracted_49324 | Not-determined |
| S7_extracted_49327 | Not-determined |
| S7_extracted_49332 | Low-quality |
| S7_extracted_49349 | Not-determined |
| S7_extracted_49353 | Low-quality |
| S7_extracted_49354 | Low-quality |
| S7_extracted_49357 | Low-quality |
| S7_extracted_49365 | Not-determined |
| S7_extracted_49394 | Low-quality |
| S7_extracted_49406 | Not-determined |
| S7_extracted_49410 | Low-quality |
| S7_extracted_49420 | Not-determined |
| S7_extracted_49431 | Not-determined |
| S7_extracted_49434 | Not-determined |
| S7_extracted_49435 | Not-determined |
| S7_extracted_49437 | Not-determined |
| S7_extracted_49445 | Not-determined |
| S7_extracted_49454 | Not-determined |
| S7_extracted_49473 | Not-determined |
| S7_extracted_49478 | Not-determined |
| S7_extracted_49481 | Not-determined |
| S5_vs_k141_120079\|\|full | Medium-quality |
| S7_extracted_49529 | Not-determined |
| S7_extracted_49541 | Not-determined |
| S7_extracted_49547 | Not-determined |
| S7_extracted_49565 | Not-determined |
| S7_extracted_49579 | Not-determined |
| S7_extracted_49627 | Not-determined |
| S7_extracted_49634 | Not-determined |
| S7_extracted_49660 | Not-determined |
| S7_extracted_49664 | Not-determined |
| S7_extracted_49666 | Not-determined |
| S7_extracted_49668 | Not-determined |
| S7_extracted_49691 | Low-quality |
| S7_extracted_49718 | Not-determined |
| S7_extracted_49726 | Not-determined |
| S7_extracted_49733 | Not-determined |
| S7_extracted_49738 | Not-determined |
| S7_extracted_49742 | Not-determined |
| S7_extracted_49753 | Not-determined |
| S7_extracted_49761 | Not-determined |
| S7_extracted_49764 | Low-quality |
| S7_extracted_49776 | Not-determined |
| S7_extracted_49791 | Not-determined |
| S7_extracted_49800 | Low-quality |
| S7_extracted_49816 | Not-determined |
| S7_extracted_49823 | Not-determined |
| S7_extracted_49835 | Not-determined |
| S7_extracted_49850 | Not-determined |
| S7_extracted_49911 | Not-determined |
| S7_extracted_49916 | Not-determined |
| S7_extracted_49919 | Not-determined |
| S7_extracted_49920 | Not-determined |
| S7_extracted_49932 | Not-determined |
| S7_extracted_49941 | Not-determined |
| S7_extracted_49943 | Not-determined |
| S7_extracted_49954 | Not-determined |
| S7_extracted_49960 | Low-quality |
| S7_extracted_49964 | Not-determined |
| S7_extracted_49972 | Not-determined |
| S7_extracted_49977 | Not-determined |
| S7_extracted_49987 | Not-determined |
| S7_extracted_49989 | Not-determined |
| S7_extracted_50001 | Not-determined |
| S7_extracted_50015 | Not-determined |
| S7_extracted_50017 | Not-determined |
| S7_extracted_50023 | Not-determined |
| S7_extracted_50025 | Not-determined |
| S7_extracted_50064 | Not-determined |
| S7_extracted_50077 | Not-determined |
| S7_extracted_50091 | Not-determined |
| S7_extracted_50094 | Not-determined |
| S7_extracted_50115 | Not-determined |
| S7_extracted_50123 | Low-quality |
| S7_extracted_50126 | Not-determined |
| S7_extracted_50136 | Low-quality |
| S7_extracted_50144 | Not-determined |
| S7_extracted_50175 | Not-determined |
| S7_extracted_50177 | Low-quality |
| S7_extracted_50185 | Low-quality |
| S7_extracted_50200 | Low-quality |
| S7_extracted_50201 | Not-determined |
| S7_extracted_50222 | Not-determined |
| S7_extracted_50233 | Low-quality |
| S7_extracted_50234 | Not-determined |
| S7_extracted_50250 | Not-determined |
| S7_extracted_50254 | Not-determined |
| S7_extracted_50261 | Not-determined |
| S7_extracted_50278 | Not-determined |
| S7_extracted_50283 | Not-determined |
| S7_extracted_50284 | Not-determined |
| S7_extracted_50285 | Low-quality |
| S7_extracted_50289 | Low-quality |
| S7_extracted_50290 | Low-quality |
| S7_extracted_50297 | Not-determined |
| S7_extracted_50360 | Not-determined |
| S7_extracted_50364 | Low-quality |
| S7_extracted_50380 | Not-determined |

| | |
|---|---|
| S7_extracted_S0393 | Low-quality |
| S7_extracted_S0398 | Not-determined |
| S7_extracted_S0410 | Not-determined |
| S7_extracted_S0416 | Not-determined |
| S7_extracted_S0425 | Low-quality |
| S7_extracted_S0436 | Low-quality |
| S7_extracted_S0468 | Not-determined |
| S7_extracted_S0491 | Not-determined |
| S7_extracted_S0494 | Not-determined |
| S7_extracted_S0505 | Not-determined |
| S7_extracted_S0516 | Not-determined |
| S7_extracted_S0530 | Low-quality |
| S7_extracted_S0536 | Not-determined |
| S7_extracted_S0537 | Not-determined |
| S7_extracted_S0538 | Not-determined |
| S7_extracted_S0555 | Not-determined |
| S7_extracted_S0572 | Not-determined |
| S7_extracted_S0591 | Not-determined |
| S7_extracted_S0594 | Not-determined |
| S7_extracted_S0632 | Not-determined |
| S7_extracted_S0641 | Not-determined |
| S7_extracted_S0642 | Not-determined |
| S7_extracted_S0655 | Not-determined |
| S7_extracted_S0670 | Low-quality |
| S7_extracted_S0678 | Not-determined |
| S7_extracted_S0686 | Not-determined |
| S7_extracted_S0690 | Not-determined |
| S7_extracted_S0695 | Not-determined |
| S7_extracted_S0701 | Not-determined |
| S7_extracted_S0705 | Not-determined |
| S7_extracted_S0707 | Not-determined |
| S7_extracted_S0716 | Not-determined |
| S7_extracted_S0723 | Not-determined |
| S7_extracted_S0730 | Low-quality |
| S7_extracted_S0731 | Not-determined |
| S7_extracted_S0747 | Not-determined |
| S7_extracted_S0752 | Not-determined |
| S7_extracted_S0754 | Not-determined |
| S7_extracted_S0764 | Not-determined |
| S7_extracted_S0768 | Low-quality |
| S7_extracted_S0770 | Not-determined |
| S7_extracted_S0797 | Not-determined |
| S7_extracted_S0804 | Low-quality |
| S7_extracted_S0812 | Not-determined |
| S7_extracted_S0852 | Not-determined |
| S7_extracted_S0867 | Not-determined |
| S7_extracted_S0869 | Low-quality |
| S7_extracted_S0899 | Not-determined |
| S7_extracted_S0907 | Not-determined |
| S7_extracted_S0927 | Not-determined |
| S7_extracted_S0936 | Not-determined |
| S7_extracted_S0943 | Not-determined |
| S7_extracted_S0948 | Not-determined |
| S7_extracted_S0958 | Low-quality |
| S7_extracted_S0967 | Not-determined |
| S7_extracted_S0978 | Not-determined |
| S7_extracted_S0982 | Not-determined |
| S7_extracted_S0990 | Not-determined |
| S7_extracted_S0996 | Not-determined |
| S7_extracted_S1005 | Not-determined |
| S7_extracted_S1006 | Not-determined |
| S7_extracted_S1016 | Not-determined |
| S7_extracted_S1035 | Not-determined |
| S7_extracted_S1048 | Not-determined |
| S7_extracted_S1053 | Not-determined |
| S7_extracted_S1073 | Not-determined |
| S7_extracted_S1093 | Not-determined |
| S7_extracted_S1094 | Not-determined |
| S7_extracted_S1124 | Not-determined |
| S7_extracted_S1126 | Not-determined |
| S7_extracted_S1139 | Not-determined |
| S7_extracted_S1146 | Not-determined |
| S7_extracted_S1149 | Low-quality |
| S7_extracted_S1167 | Not-determined |
| S7_extracted_S1179 | Not-determined |
| S7_extracted_S1187 | Not-determined |
| S7_extracted_S1203 | Not-determined |
| S7_extracted_S1205 | Low-quality |
| S7_extracted_S1207 | Low-quality |
| S7_extracted_S1222 | Not-determined |
| S7_extracted_S1236 | Not-determined |
| S7_extracted_S1237 | Not-determined |
| S7_extracted_S1250 | Not-determined |
| S7_extracted_S1268 | Not-determined |
| S7_extracted_S1290 | Low-quality |
| S7_extracted_S1305 | Not-determined |
| S7_extracted_S1306 | Not-determined |
| S7_extracted_S1315 | Not-determined |
| S7_extracted_S1323 | Not-determined |
| S7_extracted_S1327 | Not-determined |
| S7_extracted_S1356 | Not-determined |
| S7_extracted_S1359 | Low-quality |
| S7_extracted_S1363 | Not-determined |
| S7_extracted_S1375 | Not-determined |
| S7_extracted_S1385 | Not-determined |
| S7_extracted_S1397 | Not-determined |
| S7_extracted_S1408 | Not-determined |
| S7_extracted_S1417 | Not-determined |
| S7_extracted_S1422 | Not-determined |
| S7_extracted_S1439 | Not-determined |
| S7_extracted_S1440 | Low-quality |
| S7_extracted_S1451 | Not-determined |
| S7_extracted_S1464 | Not-determined |
| S7_extracted_S1470 | Not-determined |
| S7_extracted_S1495 | Not-determined |
| S7_extracted_S1542 | Not-determined |
| S7_extracted_S1562 | Not-determined |
| S7_extracted_S1573 | Not-determined |
| S7_extracted_S1575 | Low-quality |
| S7_extracted_S1581 | Not-determined |
| S7_extracted_S1582 | Low-quality |
| S7_extracted_S1596 | Not-determined |
| S7_extracted_S1598 | Not-determined |
| S7_extracted_S1599 | Not-determined |
| S7_extracted_S1606 | Not-determined |
| S7_extracted_S1632 | Not-determined |
| S7_extracted_S1653 | Not-determined |
| S7_extracted_S1658 | Low-quality |
| S7_extracted_S1688 | Not-determined |
| S7_extracted_S1698 | Not-determined |
| S7_extracted_S1699 | Not-determined |
| S7_extracted_S1700 | Not-determined |
| S7_extracted_S1707 | Low-quality |
| S7_extracted_S1721 | Not-determined |
| S7_extracted_S1727 | Not-determined |
| S7_extracted_S1730 | Not-determined |
| S7_extracted_S1736 | Not-determined |
| S7_extracted_S1737 | Not-determined |
| S7_extracted_S1750 | Low-quality |
| S7_extracted_S1776 | Not-determined |
| S7_extracted_S1780 | Not-determined |
| S7_extracted_S1785 | Not-determined |
| S7_extracted_S1815 | Not-determined |
| S7_extracted_S1839 | Not-determined |
| S7_extracted_S1852 | Low-quality |
| S7_extracted_S1873 | Low-quality |
| S7_extracted_S1886 | Not-determined |
| S7_extracted_S1918 | Not-determined |
| S7_extracted_S1928 | Not-determined |
| S7_extracted_S1941 | Not-determined |
| S7_extracted_S1947 | Not-determined |
| S7_extracted_S1951 | Not-determined |
| S7_extracted_S1976 | Not-determined |
| S7_extracted_S1987 | Not-determined |
| S3_extracted_k141_44405 | Medium-quality |
| S7_extracted_S2021 | Not-determined |
| S7_extracted_S2023 | Not-determined |
| S7_extracted_S2032 | Not-determined |
| S7_extracted_S2041 | Low-quality |
| S7_extracted_S2062 | Not-determined |
| S7_extracted_S2069 | Not-determined |
| S7_extracted_S2070 | Low-quality |
| S7_extracted_S2073 | Not-determined |
| S7_extracted_S2078 | Not-determined |
| S7_extracted_S2083 | Not-determined |
| S7_extracted_S2096 | Not-determined |
| S7_extracted_S2112 | Not-determined |
| S7_extracted_S2114 | Not-determined |
| S7_extracted_S2117 | Not-determined |
| S7_extracted_S2144 | Not-determined |
| S7_extracted_S2152 | Not-determined |
| S7_extracted_S2153 | Not-determined |
| S7_extracted_S2163 | Not-determined |
| S7_extracted_S2192 | Low-quality |
| S7_extracted_S2197 | Low-quality |
| S7_extracted_S2198 | Not-determined |
| S7_extracted_S2222 | Not-determined |
| S7_extracted_S2225 | Not-determined |
| S7_extracted_S2231 | Not-determined |
| S7_extracted_S2232 | Not-determined |
| S7_extracted_S2252 | Not-determined |
| S7_extracted_S2272 | Not-determined |
| S7_extracted_S2291 | Low-quality |
| S7_extracted_S2293 | Not-determined |
| S7_extracted_S2308 | Not-determined |
| S7_extracted_S2325 | Not-determined |
| S7_extracted_S2339 | Not-determined |
| S7_extracted_S2369 | Not-determined |
| S7_extracted_S2380 | Not-determined |
| S7_extracted_S2417 | Not-determined |
| S7_extracted_S2426 | Not-determined |
| S7_extracted_S2450 | Not-determined |
| S7_extracted_S2451 | Not-determined |
| S7_extracted_S2452 | Not-determined |
| S7_extracted_S2482 | Not-determined |
| S7_extracted_S2485 | Not-determined |
| S7_extracted_S2490 | Not-determined |
| S7_extracted_S2497 | Low-quality |
| S7_extracted_S2509 | Not-determined |
| S7_extracted_S2523 | Not-determined |
| S7_extracted_S2536 | Not-determined |
| S7_extracted_S2560 | Low-quality |
| S7_extracted_S2569 | Not-determined |
| S7_extracted_S2578 | Not-determined |
| S7_extracted_S2595 | Not-determined |
| S7_extracted_S2596 | Not-determined |
| S7_extracted_S2597 | Low-quality |
| S7_extracted_S2603 | Not-determined |
| S7_extracted_S2607 | Not-determined |
| S7_extracted_S2611 | Not-determined |
| S7_extracted_S2628 | Not-determined |
| S7_extracted_S2631 | Not-determined |
| S7_extracted_S2637 | Not-determined |
| S7_extracted_S2639 | Low-quality |
| S7_extracted_S2640 | Not-determined |
| S7_extracted_S2646 | Not-determined |
| S7_extracted_S2658 | Not-determined |
| S7_extracted_S2676 | Not-determined |
| S7_extracted_S2686 | Not-determined |
| S7_extracted_S2688 | Low-quality |
| S7_extracted_S2696 | Not-determined |
| S7_extracted_S2710 | Not-determined |
| S7_extracted_S2720 | Not-determined |
| S7_extracted_S2722 | Not-determined |
| S7_extracted_S2745 | Not-determined |
| S7_extracted_S2758 | Not-determined |
| S7_extracted_S2759 | Low-quality |
| S7_extracted_S2785 | Not-determined |
| S7_extracted_S2792 | Low-quality |
| S7_extracted_S2794 | Low-quality |
| S7_extracted_S2802 | Low-quality |
| S7_extracted_S2829 | Low-quality |
| S7_extracted_S2842 | Not-determined |
| S7_extracted_S2846 | Not-determined |
| S7_extracted_S2856 | Not-determined |
| S7_extracted_S2858 | Not-determined |
| S7_extracted_S2868 | Not-determined |
| S7_extracted_S2893 | Not-determined |
| S7_extracted_S2895 | Not-determined |
| S7_extracted_S2906 | Not-determined |
| S7_extracted_S2911 | Not-determined |
| S7_extracted_S2912 | Not-determined |
| S7_extracted_S2916 | Not-determined |
| S7_extracted_S2942 | Not-determined |
| S7_extracted_S2945 | Not-determined |
| S7_extracted_S2977 | Not-determined |
| S7_extracted_S2981 | Not-determined |
| S7_extracted_S3008 | Not-determined |
| S7_extracted_S3025 | Not-determined |
| S7_extracted_S3035 | Low-quality |
| S7_extracted_S3036 | Not-determined |
| S7_extracted_S3039 | Low-quality |
| S7_extracted_S3045 | Not-determined |
| S7_extracted_S3050 | Not-determined |
| S7_extracted_S3051 | Low-quality |
| S7_extracted_S3064 | Low-quality |
| S7_extracted_S3095 | Not-determined |
| S7_extracted_S3100 | Not-determined |
| S7_extracted_S3105 | Not-determined |
| S7_extracted_S3112 | Not-determined |
| S7_extracted_S3128 | Not-determined |
| S7_extracted_S3136 | Low-quality |
| S7_extracted_S3199 | Not-determined |
| S7_extracted_S3200 | Low-quality |
| S7_extracted_S3213 | Not-determined |
| S7_extracted_S3218 | Low-quality |
| S7_extracted_S3244 | Low-quality |
| S7_extracted_S3284 | Not-determined |
| S7_extracted_S3297 | Not-determined |
| S7_extracted_S3301 | Low-quality |
| S7_extracted_S3310 | Not-determined |
| S7_extracted_S3317 | Not-determined |
| S7_extracted_S3342 | Not-determined |
| S7_extracted_S3351 | Not-determined |
| S7_extracted_S3356 | Low-quality |
| S7_extracted_S3358 | Not-determined |
| S7_extracted_S3365 | Not-determined |
| S7_extracted_S3373 | Not-determined |
| S7_extracted_S3390 | Not-determined |
| S7_extracted_S3391 | Not-determined |
| S7_extracted_S3406 | Low-quality |
| S7_extracted_S3414 | Low-quality |
| S7_extracted_S3422 | Not-determined |
| S7_extracted_S3433 | Not-determined |
| S7_extracted_S3441 | Not-determined |
| S7_extracted_S3448 | Not-determined |
| S7_extracted_S3467 | Low-quality |
| S7_extracted_S3473 | Not-determined |
| S7_extracted_S3474 | Low-quality |
| S7_extracted_S3476 | Not-determined |
| S7_extracted_S3491 | Not-determined |
| S7_extracted_S3496 | Not-determined |
| S7_extracted_S3500 | Not-determined |
| S7_extracted_S3507 | Low-quality |
| S7_extracted_S3516 | Not-determined |
| S7_extracted_S3546 | Not-determined |
| S7_extracted_S3551 | Not-determined |
| S7_extracted_S3552 | Low-quality |
| S7_extracted_S3554 | Not-determined |
| S7_extracted_S3558 | Not-determined |
| S7_extracted_S3564 | Not-determined |
| S7_extracted_S3566 | Not-determined |
| S7_extracted_S3576 | Not-determined |
| S7_extracted_S3581 | Not-determined |
| S7_extracted_S3608 | Not-determined |
| S7_extracted_S3612 | Not-determined |
| S7_extracted_S3618 | Not-determined |
| S7_extracted_S3622 | Not-determined |
| S7_extracted_S3679 | Not-determined |
| S7_extracted_S3694 | Not-determined |
| S7_extracted_S3718 | Low-quality |
| S7_extracted_S3725 | Not-determined |
| S7_extracted_S3726 | Not-determined |
| S7_extracted_S3736 | Not-determined |
| S7_extracted_S3737 | Not-determined |
| S7_extracted_S3750 | Not-determined |
| S7_extracted_S3755 | Low-quality |
| S7_extracted_S3779 | Not-determined |
| S7_extracted_S3781 | Not-determined |
| S7_extracted_S3790 | Not-determined |
| S7_extracted_S3791 | Low-quality |
| S7_extracted_S3797 | Not-determined |
| S7_extracted_S3806 | Not-determined |
| S7_extracted_S3816 | Not-determined |
| S7_extracted_S3832 | Not-determined |
| S7_extracted_S3835 | Not-determined |
| S7_extracted_S3841 | Not-determined |
| S7_extracted_S3849 | Not-determined |
| S7_extracted_S3860 | Not-determined |
| S7_extracted_S3867 | Low-quality |
| S7_extracted_S3868 | Not-determined |
| S7_extracted_S3871 | Low-quality |
| S7_extracted_S3879 | Low-quality |
| S7_extracted_S3905 | Low-quality |
| S7_extracted_S3916 | Not-determined |
| S7_extracted_S3935 | Not-determined |
| S7_extracted_S3941 | Not-determined |
| S7_extracted_S3947 | Not-determined |
| S5_vs_k141_51416\|\|full | High-quality |
| S7_extracted_S3975 | Not-determined |
| S7_extracted_S3987 | Not-determined |
| S7_extracted_S3990 | Not-determined |
| S7_extracted_S4002 | Not-determined |
| S7_extracted_S4008 | Low-quality |
| S7_extracted_S4011 | Not-determined |
| S7_extracted_S4021 | Not-determined |
| S7_extracted_S4030 | Not-determined |
| S7_extracted_S4035 | Not-determined |
| S7_extracted_S4038 | Not-determined |
| S7_extracted_S4041 | Not-determined |
| S7_extracted_S4046 | Not-determined |
| S7_extracted_S4049 | Not-determined |
| S7_extracted_S4053 | Not-determined |
| S7_extracted_S4056 | Not-determined |
| S7_extracted_S4058 | Low-quality |
| S7_extracted_S4074 | Not-determined |
| S7_extracted_S4085 | Low-quality |
| S7_extracted_S4090 | Low-quality |
| S7_extracted_S4101 | Not-determined |
| S7_extracted_S4111 | Not-determined |
| S7_extracted_S4123 | Not-determined |
| S7_extracted_S4148 | Not-determined |
| S7_extracted_S4153 | Low-quality |
| S7_extracted_S4162 | Not-determined |
| S7_extracted_S4171 | Not-determined |
| S7_extracted_S4187 | Low-quality |
| S7_extracted_S4196 | Not-determined |
| S7_extracted_S4217 | Not-determined |
| S7_extracted_S4235 | Not-determined |
| S7_extracted_S4240 | Low-quality |
| S7_extracted_S4245 | Not-determined |
| S7_extracted_S4258 | Not-determined |
| S7_extracted_S4260 | Not-determined |
| S7_extracted_S4278 | Low-quality |
| S7_extracted_S4281 | Not-determined |
| S7_extracted_S4303 | Not-determined |
| S7_extracted_S4305 | Not-determined |
| S7_extracted_S4313 | Not-determined |
| S7_extracted_S4318 | Not-determined |
| S7_extracted_S4321 | Not-determined |
| S7_extracted_S4335 | Low-quality |
| S7_extracted_S4364 | Low-quality |
| S7_extracted_S4387 | Low-quality |
| S7_extracted_S4388 | Not-determined |
| S7_extracted_S4404 | Not-determined |
| S7_extracted_S4405 | Not-determined |
| S7_extracted_S4409 | Not-determined |
| S7_extracted_S4412 | Low-quality |
| S7_extracted_S4423 | Not-determined |
| S7_extracted_S4434 | Not-determined |
| S7_extracted_S4442 | Low-quality |
| S7_extracted_S4449 | Not-determined |
| S7_extracted_S4462 | Not-determined |
| S7_extracted_S4471 | Not-determined |
| S7_extracted_S4499 | Low-quality |
| S7_extracted_S4525 | Low-quality |
| S7_extracted_S4572 | Not-determined |
| S7_extracted_S4584 | Not-determined |
| S7_extracted_S4591 | Not-determined |
| S7_extracted_S4592 | Not-determined |
| S7_extracted_S4593 | Low-quality |
| S7_extracted_S4603 | Not-determined |
| S7_extracted_S4616 | Not-determined |
| S7_extracted_S4627 | Not-determined |
| S7_extracted_S4636 | Not-determined |
| S7_extracted_S4653 | Not-determined |
| S7_extracted_S4677 | Not-determined |
| S7_extracted_S4687 | Not-determined |
| S7_extracted_S4718 | Not-determined |
| S7_extracted_S4728 | Not-determined |
| S7_extracted_S4730 | Not-determined |
| S7_extracted_S4739 | Not-determined |
| S7_extracted_S4754 | Low-quality |
| S7_extracted_S4773 | Not-determined |
| S7_extracted_S4783 | Low-quality |
| S7_extracted_S4823 | Not-determined |
| S7_extracted_S4830 | Not-determined |
| S7_extracted_S4833 | Not-determined |
| S7_extracted_S4837 | Not-determined |
| S7_extracted_S4859 | Low-quality |
| S7_extracted_S4867 | Not-determined |
| S7_extracted_S4871 | Not-determined |
| S7_extracted_S4875 | Not-determined |
| S7_extracted_S4890 | Not-determined |
| S7_extracted_S4911 | Not-determined |
| S7_extracted_S4939 | Not-determined |
| S7_extracted_S4950 | Not-determined |
| S7_extracted_S4965 | Not-determined |
| S7_extracted_S4977 | Not-determined |
| S7_extracted_S4978 | Not-determined |
| S7_extracted_S4983 | Not-determined |
| S7_extracted_S4989 | Not-determined |
| S7_extracted_S5008 | Low-quality |
| S7_extracted_S5027 | Not-determined |
| S7_extracted_S5030 | Low-quality |
| S7_extracted_S5034 | Not-determined |
| S7_extracted_S5041 | Not-determined |
| S7_extracted_S5056 | Low-quality |
| S7_extracted_S5073 | Low-quality |
| S7_extracted_S5106 | Not-determined |
| S7_extracted_S5112 | Not-determined |
| S7_extracted_S5120 | Not-determined |
| S7_extracted_S5127 | Not-determined |
| S7_extracted_S5131 | Not-determined |
| S7_extracted_S5147 | Low-quality |
| S7_extracted_S5149 | Not-determined |
| S7_extracted_S5159 | Low-quality |
| S7_extracted_S5171 | Low-quality |
| S7_extracted_S5175 | Not-determined |
| S7_extracted_S5205 | Low-quality |
| S7_extracted_S5206 | Low-quality |
| S7_extracted_S5219 | Not-determined |
| S7_extracted_S5221 | Not-determined |
| S7_extracted_S5242 | Not-determined |
| S7_extracted_S5259 | Not-determined |
| S7_extracted_S5272 | Not-determined |
| S7_extracted_S5282 | Not-determined |
| S7_extracted_S5321 | Not-determined |
| S7_extracted_S5336 | Not-determined |
| S7_extracted_S5343 | Not-determined |
| S7_extracted_S5352 | Not-determined |
| S7_extracted_S5362 | Not-determined |
| S7_extracted_S5371 | Not-determined |
| S7_extracted_S5375 | Not-determined |
| S7_extracted_S5386 | Low-quality |
| S7_extracted_S5401 | Not-determined |
| S7_extracted_S5431 | Not-determined |
| S7_extracted_S5434 | Not-determined |
| S7_extracted_S5445 | Not-determined |
| S7_extracted_S5469 | Not-determined |
| S7_extracted_S5481 | Not-determined |
| S7_extracted_S5502 | Not-determined |
| S7_extracted_S5519 | Not-determined |
| S7_extracted_S5557 | Not-determined |
| S7_extracted_S5559 | Not-determined |
| S7_extracted_S5561 | Not-determined |
| S7_extracted_S5562 | Not-determined |
| S7_extracted_S5584 | Not-determined |
| S7_extracted_S5597 | Not-determined |
| S7_extracted_S5605 | Not-determined |
| S7_extracted_S5606 | Low-quality |
| S7_extracted_S5613 | Not-determined |
| S7_extracted_S5616 | Low-quality |
| S7_extracted_S5633 | Low-quality |
| S7_extracted_S5638 | Not-determined |
| S7_extracted_S5643 | Not-determined |
| S7_extracted_S5663 | Not-determined |
| S7_extracted_S5666 | Not-determined |
| S7_extracted_S5672 | Not-determined |
| S7_extracted_S5714 | Not-determined |
| S7_extracted_S5718 | Not-determined |
| S7_extracted_S5729 | Not-determined |
| S7_extracted_S5745 | Not-determined |
| S7_extracted_S5794 | Not-determined |
| S7_extracted_S5804 | Not-determined |
| S7_extracted_S5823 | Low-quality |
| S7_extracted_S5829 | Low-quality |
| S7_extracted_S5836 | Low-quality |
| S7_extracted_S5838 | Not-determined |
| S7_extracted_S5844 | Not-determined |
| S7_extracted_S5854 | Low-quality |
| S7_extracted_S5856 | Low-quality |
| S7_extracted_S5857 | Not-determined |
| S7_extracted_S5879 | Not-determined |
| S7_extracted_S5919 | Low-quality |
| S7_extracted_S5954 | Not-determined |
| S7_extracted_S5961 | Not-determined |
| S7_extracted_S5963 | Not-determined |
| S7_extracted_S5976 | Not-determined |
| S7_extracted_S5980 | Low-quality |
| S7_extracted_S5991 | Low-quality |
| S7_extracted_S5993 | Not-determined |
| S7_extracted_S6002 | Not-determined |
| S7_extracted_S6024 | Not-determined |
| S7_extracted_S6030 | Not-determined |
| S7_extracted_S6044 | Not-determined |
| S7_extracted_S6070 | Not-determined |
| S7_extracted_S6076 | Not-determined |
| S7_extracted_S6099 | Not-determined |
| S7_extracted_S6107 | Not-determined |
| S7_extracted_S6111 | Low-quality |
| S7_extracted_S6112 | Not-determined |
| S7_extracted_S6113 | Not-determined |
| S7_extracted_S6116 | Not-determined |
| S7_extracted_S6175 | Not-determined |
| S7_extracted_S6190 | Low-quality |
| S7_extracted_S6213 | Not-determined |
| S7_extracted_S6226 | Not-determined |
| S7_extracted_S6240 | Not-determined |
| S7_extracted_S6268 | Not-determined |
| S7_extracted_S6271 | Not-determined |
| S7_extracted_S6323 | Not-determined |
| S7_extracted_S6328 | Low-quality |
| S7_extracted_S6348 | Not-determined |
| S7_extracted_S6377 | Not-determined |
| S7_extracted_S6381 | Low-quality |
| S7_extracted_S6382 | Not-determined |
| S7_extracted_S6395 | Not-determined |
| S7_extracted_S6411 | Low-quality |
| S7_extracted_S6423 | Low-quality |
| S7_extracted_S6436 | Not-determined |
| S7_extracted_S6439 | Not-determined |
| S7_extracted_S6445 | Not-determined |
| S7_extracted_S6456 | Not-determined |
| S7_extracted_S6484 | Low-quality |
| S7_extracted_S6489 | Not-determined |
| S7_extracted_S6513 | Not-determined |
| S7_extracted_S6528 | Not-determined |
| S7_extracted_S6538 | Not-determined |
| S7_extracted_S6544 | Not-determined |
| S7_extracted_S6553 | Low-quality |
| S7_extracted_S6557 | Not-determined |
| S7_extracted_S6572 | Not-determined |
| S7_extracted_S6584 | Not-determined |
| S7_extracted_S6595 | Not-determined |
| S7_extracted_S6600 | Not-determined |
| S7_extracted_S6602 | Not-determined |
| S7_extracted_S6609 | Not-determined |
| S7_extracted_S6612 | Not-determined |
| S7_extracted_S6618 | Not-determined |
| S7_extracted_S6623 | Not-determined |
| S7_extracted_S6649 | Not-determined |
| S7_extracted_S6651 | Not-determined |
| S7_extracted_S6654 | Not-determined |
| S7_extracted_S6663 | Not-determined |
| S7_extracted_S6664 | Not-determined |
| S7_extracted_S6689 | Not-determined |
| S7_extracted_S6694 | Low-quality |
| S7_extracted_S6710 | Not-determined |
| S7_extracted_S6712 | Low-quality |
| S7_extracted_S6721 | Low-quality |
| S7_extracted_S6734 | Not-determined |
| S7_extracted_S6757 | Not-determined |
| S7_extracted_S6770 | Not-determined |
| S7_extracted_S6782 | Low-quality |
| S7_extracted_S6814 | Not-determined |
| S7_extracted_S6855 | Low-quality |
| S7_extracted_S6884 | Not-determined |
| S7_extracted_S6891 | Not-determined |
| S7_extracted_S6922 | Low-quality |
| S7_extracted_S6929 | Not-determined |
| S7_extracted_S6951 | Not-determined |
| S7_extracted_S6955 | Not-determined |
| S7_extracted_S6956 | Low-quality |
| S7_extracted_S6962 | Low-quality |
| S7_extracted_S6968 | Not-determined |
| S7_extracted_S7009 | Not-determined |
| S7_extracted_S7040 | Not-determined |
| S7_extracted_S7082 | Not-determined |
| S7_extracted_S7095 | Not-determined |
| S7_extracted_S7096 | Not-determined |
| S7_extracted_S7097 | Not-determined |
| S7_extracted_S7112 | Not-determined |
| S7_extracted_S7115 | Not-determined |
| S7_extracted_S7116 | Not-determined |
| S7_extracted_S7120 | Not-determined |
| S7_extracted_S7136 | Not-determined |
| S7_extracted_S7140 | Low-quality |
| S7_extracted_S7147 | Not-determined |

| | |
|---|---|
| S7_extracted_57149 | Not-determined |
| S7_extracted_57159 | Low-quality |
| S7_extracted_57203 | Not-determined |
| S7_extracted_57205 | Not-determined |
| S7_extracted_57206 | Not-determined |
| S7_extracted_57207 | Low-quality |
| S7_extracted_57209 | Not-determined |
| S7_extracted_57216 | Low-quality |
| S7_extracted_57234 | Low-quality |
| S7_extracted_57237 | Not-determined |
| S7_extracted_57239 | Not-determined |
| S7_extracted_57249 | Low-quality |
| S7_extracted_57261 | Not-determined |
| S7_extracted_57274 | Low-quality |
| S7_extracted_57276 | Not-determined |
| S7_extracted_57283 | Not-determined |
| S7_extracted_57284 | Not-determined |
| S7_extracted_57296 | Not-determined |
| S7_extracted_57307 | Not-determined |
| S7_extracted_57321 | Low-quality |
| S7_extracted_57337 | Low-quality |
| S7_extracted_57350 | Not-determined |
| S7_extracted_57389 | Not-determined |
| S7_extracted_57406 | Not-determined |
| S7_extracted_57412 | Not-determined |
| S7_extracted_57425 | Not-determined |
| S7_extracted_57447 | Not-determined |
| S7_extracted_57454 | Not-determined |
| S7_extracted_57455 | Not-determined |
| S7_extracted_57458 | Not-determined |
| S7_extracted_57462 | Not-determined |
| S7_extracted_57464 | Not-determined |
| S7_extracted_57472 | Not-determined |
| S7_extracted_57484 | Not-determined |
| S7_extracted_57496 | Not-determined |
| S7_extracted_57497 | Not-determined |
| S7_extracted_57505 | Not-determined |
| S7_extracted_57507 | Not-determined |
| S7_extracted_57521 | Not-determined |
| S7_extracted_57525 | Not-determined |
| S7_extracted_57526 | Not-determined |
| S7_extracted_57550 | Not-determined |
| S7_extracted_57575 | Not-determined |
| S7_extracted_57579 | Not-determined |
| S7_extracted_57583 | Not-determined |
| S7_extracted_57597 | Not-determined |
| S7_extracted_57599 | Not-determined |
| S7_extracted_57619 | Not-determined |
| S7_extracted_57623 | Low-quality |
| S7_extracted_57630 | Not-determined |
| S7_extracted_57644 | Not-determined |
| S7_extracted_57645 | Not-determined |
| S7_extracted_57660 | Low-quality |
| S7_extracted_57669 | Not-determined |
| S7_extracted_57687 | Low-quality |
| S7_extracted_57699 | Low-quality |
| S7_extracted_57727 | Not-determined |
| S7_extracted_57729 | Not-determined |
| S7_extracted_57737 | Not-determined |
| S7_extracted_57748 | Not-determined |
| S7_extracted_57752 | Not-determined |
| S7_extracted_57764 | Not-determined |
| S7_extracted_57768 | Not-determined |
| S7_extracted_57787 | Not-determined |
| S7_extracted_57802 | Not-determined |
| S7_extracted_57807 | Not-determined |
| S7_extracted_57824 | Low-quality |
| S7_extracted_57831 | Not-determined |
| S7_extracted_57843 | Low-quality |
| S7_extracted_57844 | Not-determined |
| S7_extracted_57845 | Not-determined |
| S7_extracted_57852 | Not-determined |
| S7_extracted_57854 | Not-determined |
| S7_extracted_57863 | Not-determined |
| S7_extracted_57869 | Not-determined |
| S7_extracted_57880 | Not-determined |
| S7_extracted_57893 | Low-quality |
| S7_extracted_57924 | Not-determined |
| S7_extracted_57929 | Not-determined |
| S7_extracted_57944 | Low-quality |
| S7_extracted_57948 | Not-determined |
| S7_extracted_57949 | Not-determined |
| S7_extracted_57952 | Not-determined |
| S7_extracted_57954 | Not-determined |
| S7_extracted_57958 | Not-determined |
| S7_extracted_57961 | Not-determined |
| S7_extracted_57965 | Not-determined |
| S7_extracted_57995 | Not-determined |
| S7_extracted_58012 | Not-determined |
| S7_extracted_58035 | Low-quality |
| S7_extracted_58053 | Not-determined |
| S7_extracted_58074 | Low-quality |
| S7_extracted_58081 | Not-determined |
| S7_extracted_58097 | Not-determined |
| S7_extracted_58136 | Low-quality |
| S7_extracted_58141 | Not-determined |
| S7_extracted_58161 | Not-determined |
| S7_extracted_58170 | Low-quality |
| S7_extracted_58191 | Not-determined |
| S7_extracted_58202 | Not-determined |
| S7_extracted_58221 | Not-determined |
| S7_extracted_58247 | Not-determined |
| S7_extracted_58249 | Not-determined |
| S7_extracted_58258 | Not-determined |
| S7_extracted_58265 | Not-determined |
| S7_extracted_58271 | Low-quality |
| S7_extracted_58291 | Not-determined |
| S7_extracted_58313 | Not-determined |
| S7_extracted_58321 | Not-determined |
| S7_extracted_58330 | Not-determined |
| S7_extracted_58401 | Not-determined |
| S7_extracted_58423 | Not-determined |
| S7_extracted_58428 | Low-quality |
| S7_extracted_58486 | Not-determined |
| S7_extracted_58494 | Not-determined |
| S7_extracted_58517 | Low-quality |
| S7_extracted_58539 | Not-determined |
| S7_extracted_58549 | Low-quality |
| S7_extracted_58591 | Low-quality |
| S7_extracted_58592 | Not-determined |
| S7_extracted_58624 | Not-determined |
| S7_extracted_58634 | Not-determined |
| S7_extracted_58644 | Not-determined |
| S7_extracted_58660 | Not-determined |
| S7_extracted_58665 | Low-quality |
| S7_extracted_58674 | Not-determined |
| S7_extracted_58683 | Not-determined |
| S7_extracted_58684 | Low-quality |
| S7_extracted_58700 | Not-determined |
| S7_extracted_58706 | Not-determined |
| S7_extracted_58710 | Not-determined |
| S7_extracted_58744 | Not-determined |
| S7_extracted_58745 | Not-determined |
| S3_vs_k141_100785\|\|full | High-quality |
| S7_extracted_58757 | Not-determined |
| S7_extracted_58759 | Not-determined |
| S7_extracted_58772 | Not-determined |
| S7_extracted_58807 | Low-quality |
| S7_extracted_58818 | Not-determined |
| S7_extracted_58826 | Not-determined |
| S7_extracted_58827 | Not-determined |
| S7_extracted_58835 | Not-determined |
| S7_extracted_58843 | Not-determined |
| S7_extracted_58857 | Not-determined |
| S7_extracted_58875 | Low-quality |
| S7_extracted_58900 | Not-determined |
| S7_extracted_58912 | Low-quality |
| S7_extracted_58930 | Not-determined |
| S7_extracted_58933 | Not-determined |
| S7_extracted_58958 | Low-quality |
| S7_extracted_58963 | Low-quality |
| S7_extracted_58969 | Not-determined |
| S7_extracted_58970 | Not-determined |
| S7_extracted_58987 | Not-determined |
| S7_extracted_58988 | Not-determined |
| S7_extracted_58997 | Not-determined |
| S7_extracted_59000 | Not-determined |
| S7_extracted_59017 | Not-determined |
| S7_extracted_59019 | Not-determined |
| S7_extracted_59031 | Low-quality |
| S7_extracted_59040 | Not-determined |
| S7_extracted_59050 | Not-determined |
| S7_extracted_59051 | Not-determined |
| S7_extracted_59074 | Not-determined |
| S7_extracted_59081 | Not-determined |
| S7_extracted_59087 | Low-quality |
| S7_extracted_59097 | Not-determined |
| S7_extracted_59099 | Low-quality |
| S7_extracted_59100 | Not-determined |
| S7_extracted_59109 | Not-determined |
| S7_extracted_59112 | Not-determined |
| S7_extracted_59116 | Not-determined |
| S7_extracted_59127 | Not-determined |
| S7_extracted_59131 | Not-determined |
| S7_extracted_59141 | Not-determined |
| S7_extracted_59150 | Not-determined |
| S7_extracted_59159 | Not-determined |
| S7_extracted_59163 | Not-determined |
| S7_extracted_59182 | Not-determined |
| S7_extracted_59189 | Not-determined |
| S7_extracted_59193 | Low-quality |
| S7_extracted_59199 | Not-determined |
| S7_extracted_59200 | Not-determined |
| S7_extracted_59208 | Not-determined |
| S7_extracted_59213 | Low-quality |
| S7_extracted_59215 | Not-determined |
| S7_extracted_59257 | Low-quality |
| S7_extracted_59265 | Not-determined |
| S7_extracted_59278 | Not-determined |
| S7_extracted_59314 | Low-quality |
| S7_extracted_59317 | Not-determined |
| S7_extracted_59340 | Not-determined |
| S7_extracted_59357 | Low-quality |
| S7_extracted_59374 | Not-determined |
| S7_extracted_59390 | Not-determined |
| S7_extracted_59415 | Not-determined |
| S7_extracted_59421 | Low-quality |
| S7_extracted_59435 | Not-determined |
| S7_extracted_59444 | Low-quality |
| S7_extracted_59448 | Not-determined |
| S7_extracted_59454 | Not-determined |
| S7_extracted_59478 | Not-determined |
| S7_extracted_59516 | Not-determined |
| S7_extracted_59549 | Not-determined |
| S7_extracted_59560 | Not-determined |
| S7_extracted_59597 | Low-quality |
| S7_extracted_59610 | Low-quality |
| S7_extracted_59614 | Not-determined |
| S7_extracted_59620 | Not-determined |
| S7_extracted_59633 | Not-determined |
| S7_extracted_59640 | Not-determined |
| S7_extracted_59658 | Low-quality |
| S7_extracted_59674 | Not-determined |
| S7_extracted_59704 | Low-quality |
| S7_extracted_59717 | Not-determined |
| S7_extracted_59721 | Not-determined |
| S7_extracted_59742 | Not-determined |
| S7_extracted_59746 | Low-quality |
| S7_extracted_59756 | Not-determined |
| S7_extracted_59759 | Not-determined |
| S7_extracted_59764 | Not-determined |
| S7_extracted_59766 | Low-quality |
| S7_extracted_59772 | Not-determined |
| S7_extracted_59791 | Not-determined |
| S7_extracted_59806 | Low-quality |
| S7_extracted_59821 | Low-quality |
| S7_extracted_59833 | Not-determined |
| S7_extracted_59834 | Not-determined |
| S7_extracted_59840 | Low-quality |
| S7_extracted_59851 | Not-determined |
| S7_extracted_59854 | Not-determined |
| S7_extracted_59855 | Not-determined |
| S7_extracted_59859 | Not-determined |
| S7_extracted_59885 | Not-determined |
| S7_extracted_59892 | Not-determined |
| S7_extracted_59898 | Not-determined |
| S7_extracted_59907 | Low-quality |
| S7_extracted_59929 | Not-determined |
| S7_extracted_59946 | Not-determined |
| S7_extracted_59947 | Not-determined |
| S7_extracted_59952 | Not-determined |
| S7_extracted_59972 | Not-determined |
| S7_extracted_59976 | Not-determined |
| S7_extracted_59989 | Not-determined |
| S7_extracted_59992 | Not-determined |
| S7_extracted_60011 | Not-determined |
| S7_extracted_60014 | Low-quality |
| S7_extracted_60016 | Not-determined |
| S7_extracted_60020 | Not-determined |
| S7_extracted_60031 | Low-quality |
| S7_extracted_60038 | Not-determined |
| S7_extracted_60047 | Not-determined |
| S7_extracted_60061 | Not-determined |
| S7_extracted_60067 | Not-determined |
| S7_extracted_60076 | Not-determined |
| S7_extracted_60090 | Not-determined |
| S7_extracted_60097 | Not-determined |
| S7_extracted_60100 | Not-determined |
| S7_extracted_60118 | Low-quality |
| S7_extracted_60130 | Not-determined |
| S7_extracted_60133 | Not-determined |
| S7_extracted_60134 | Low-quality |
| S7_extracted_60158 | Not-determined |
| S7_extracted_60164 | Not-determined |
| S7_extracted_60166 | Not-determined |
| S7_extracted_60171 | Low-quality |
| S7_extracted_60175 | Not-determined |
| S7_extracted_60179 | Not-determined |
| S7_extracted_60188 | Not-determined |
| S7_extracted_60206 | Low-quality |
| S7_extracted_60248 | Low-quality |
| S7_extracted_60250 | Not-determined |
| S7_extracted_60270 | Not-determined |
| S7_extracted_60273 | Not-determined |
| S7_extracted_60310 | Not-determined |
| S7_extracted_60337 | Low-quality |
| S7_extracted_60340 | Not-determined |
| S7_extracted_60347 | Not-determined |
| S7_extracted_60378 | Low-quality |
| S7_extracted_60400 | Low-quality |
| S7_extracted_60412 | Not-determined |
| S7_extracted_60427 | Not-determined |
| S7_extracted_60441 | Not-determined |
| S7_extracted_60454 | Not-determined |
| S7_extracted_60468 | Not-determined |
| S7_extracted_60489 | Not-determined |
| S7_extracted_60492 | Not-determined |
| S7_extracted_60498 | Low-quality |
| S7_extracted_60514 | Not-determined |
| S7_extracted_60523 | Not-determined |
| S7_extracted_60524 | Not-determined |
| S7_extracted_60540 | Not-determined |
| S7_extracted_60567 | Not-determined |
| S7_extracted_60573 | Not-determined |
| S7_extracted_60595 | Not-determined |
| S7_extracted_60604 | Low-quality |
| S7_extracted_60641 | Not-determined |
| S7_extracted_60644 | Not-determined |
| S7_extracted_60657 | Not-determined |
| S7_extracted_60675 | Not-determined |
| S7_extracted_60684 | Not-determined |
| S7_extracted_60685 | Not-determined |
| S7_extracted_60692 | Low-quality |
| S7_extracted_60701 | Not-determined |
| S7_extracted_60713 | Not-determined |
| S7_extracted_60756 | Not-determined |
| S7_extracted_60767 | Low-quality |
| S7_extracted_60792 | Not-determined |
| S7_extracted_60794 | Low-quality |
| S7_extracted_60813 | Not-determined |
| S7_extracted_60817 | Not-determined |
| S7_extracted_60818 | Not-determined |
| S7_extracted_60823 | Not-determined |
| S7_extracted_60826 | Not-determined |
| S7_extracted_60828 | Not-determined |
| S7_extracted_60853 | Not-determined |
| S7_extracted_60866 | Not-determined |
| S7_extracted_60966 | Not-determined |
| S9_extracted_k141_90856 | Complete |
| S7_extracted_60972 | Not-determined |
| S7_extracted_60975 | Not-determined |
| S7_extracted_60976 | Not-determined |
| S7_extracted_60983 | Not-determined |
| S7_extracted_60993 | Not-determined |
| S7_extracted_60997 | Not-determined |
| S7_extracted_60998 | Not-determined |
| S7_extracted_61011 | Low-quality |
| S7_extracted_61022 | Not-determined |
| S7_extracted_61024 | Not-determined |
| S7_extracted_61029 | Not-determined |
| S7_extracted_61047 | Not-determined |
| S7_extracted_61057 | Not-determined |
| S7_extracted_61084 | Not-determined |
| S7_extracted_61085 | Not-determined |
| S7_extracted_61087 | Not-determined |
| S7_extracted_61095 | Not-determined |
| S7_extracted_61139 | Not-determined |
| S7_extracted_61140 | Low-quality |
| S7_extracted_61155 | Not-determined |
| S7_extracted_61161 | Not-determined |
| S7_extracted_61166 | Not-determined |
| S7_extracted_61175 | Low-quality |
| S7_extracted_61188 | Not-determined |
| S7_extracted_61199 | Not-determined |
| S7_extracted_61236 | Not-determined |
| S7_extracted_61256 | Not-determined |
| S7_extracted_61271 | Not-determined |
| S7_extracted_61279 | Not-determined |
| S7_extracted_61283 | Not-determined |
| S7_extracted_61294 | Not-determined |
| S7_extracted_61300 | Not-determined |
| S7_extracted_61311 | Not-determined |
| S7_extracted_61313 | Not-determined |
| S7_extracted_61333 | Not-determined |
| S7_extracted_61343 | Not-determined |
| S7_extracted_61345 | Not-determined |
| S7_extracted_61350 | Not-determined |
| S7_extracted_61356 | Not-determined |
| S7_extracted_61376 | Not-determined |
| S7_extracted_61384 | Not-determined |
| S7_extracted_61403 | Low-quality |
| S7_extracted_61416 | Not-determined |
| S7_extracted_61418 | Low-quality |
| S7_extracted_61436 | Not-determined |
| S7_extracted_61437 | Not-determined |
| S7_extracted_61445 | Not-determined |
| S7_extracted_61460 | Not-determined |
| S7_extracted_61465 | Low-quality |
| S7_extracted_61473 | Not-determined |
| S7_extracted_61474 | Low-quality |
| S7_extracted_61487 | Not-determined |
| S7_extracted_61515 | Not-determined |
| S7_extracted_61520 | Not-determined |
| S7_extracted_61523 | Not-determined |
| S7_extracted_61533 | Not-determined |
| S7_extracted_61544 | Low-quality |
| S7_extracted_61547 | Not-determined |
| S7_extracted_61553 | Not-determined |
| S7_extracted_61559 | Not-determined |
| S7_extracted_61595 | Low-quality |
| S7_extracted_61596 | Not-determined |
| S7_extracted_61606 | Not-determined |
| S7_extracted_61611 | Not-determined |
| S7_extracted_61623 | Not-determined |
| S7_extracted_61626 | Not-determined |
| S7_extracted_61636 | Not-determined |
| S7_extracted_61646 | Not-determined |
| S7_extracted_61657 | Not-determined |
| S7_extracted_61663 | Low-quality |
| S7_extracted_61671 | Not-determined |
| S7_extracted_61675 | Not-determined |
| S7_extracted_61685 | Not-determined |
| S7_extracted_61694 | Not-determined |
| S7_extracted_61720 | Not-determined |
| S7_extracted_61722 | Not-determined |
| S7_extracted_61730 | Not-determined |
| S7_extracted_61737 | Not-determined |
| S7_extracted_61744 | Not-determined |
| S7_extracted_61785 | Not-determined |
| S7_extracted_61803 | Not-determined |
| S7_extracted_61817 | Not-determined |
| S7_extracted_61818 | Low-quality |
| S7_extracted_61820 | Not-determined |
| S7_extracted_61822 | Not-determined |
| S7_extracted_61830 | Not-determined |
| S7_extracted_61832 | Not-determined |
| S7_extracted_61834 | Not-determined |
| S7_extracted_61848 | Not-determined |
| S7_extracted_61857 | Not-determined |
| S7_extracted_61870 | Not-determined |
| S7_extracted_61890 | Not-determined |
| S7_extracted_61900 | Low-quality |
| S7_extracted_61902 | Not-determined |
| S7_extracted_61907 | Not-determined |
| S7_extracted_61912 | Not-determined |
| S7_extracted_61920 | Not-determined |
| S7_extracted_61922 | Not-determined |
| S7_extracted_61953 | Not-determined |
| S7_extracted_61962 | Not-determined |
| S7_extracted_61977 | Not-determined |
| S7_extracted_61980 | Low-quality |
| S7_extracted_61998 | Low-quality |
| S7_extracted_62008 | Not-determined |
| S7_extracted_62031 | Not-determined |
| S7_extracted_62042 | Low-quality |
| S7_extracted_62043 | Not-determined |
| S7_extracted_62053 | Not-determined |
| S7_extracted_62075 | Low-quality |
| S7_extracted_62080 | Not-determined |
| S7_extracted_62090 | Not-determined |
| S7_extracted_62117 | Not-determined |
| S7_extracted_62169 | Not-determined |
| S7_extracted_62190 | Not-determined |
| S7_extracted_62209 | Not-determined |
| S7_extracted_62211 | Not-determined |
| S7_extracted_62226 | Not-determined |
| S7_extracted_62233 | Not-determined |
| S7_extracted_62235 | Not-determined |
| S7_extracted_62238 | Low-quality |
| S7_extracted_62241 | Low-quality |
| S7_extracted_62250 | Low-quality |
| S7_extracted_62251 | Not-determined |
| S7_extracted_62255 | Low-quality |
| S7_extracted_62263 | Low-quality |
| S7_extracted_62265 | Not-determined |
| S7_extracted_62277 | Not-determined |
| S7_extracted_62287 | Low-quality |
| S7_extracted_62288 | Not-determined |
| S7_extracted_62294 | Low-quality |
| S7_extracted_62302 | Low-quality |
| S7_extracted_62303 | Not-determined |
| S7_extracted_62317 | Not-determined |
| S7_extracted_62324 | Not-determined |
| S7_extracted_62361 | Not-determined |
| S7_extracted_62373 | Not-determined |
| S7_extracted_62387 | Not-determined |
| S7_extracted_62392 | Not-determined |
| S7_extracted_62393 | Not-determined |
| S7_extracted_62399 | Low-quality |
| S7_extracted_62409 | Low-quality |
| S7_extracted_62422 | Not-determined |
| S7_extracted_62467 | Not-determined |
| S7_extracted_62469 | Not-determined |
| S7_extracted_62479 | Not-determined |
| S7_extracted_62480 | Not-determined |
| S7_extracted_62485 | Low-quality |
| S7_extracted_62487 | Low-quality |
| S7_extracted_62495 | Not-determined |
| S7_extracted_62560 | Not-determined |
| S7_extracted_62574 | Not-determined |
| S7_extracted_62581 | Not-determined |
| S7_extracted_62590 | Low-quality |
| S7_extracted_62611 | Not-determined |
| S7_extracted_62618 | Not-determined |
| S7_extracted_62619 | Low-quality |
| S7_extracted_62634 | Not-determined |
| S7_extracted_62663 | Not-determined |
| S7_extracted_62680 | Low-quality |
| S7_extracted_62698 | Low-quality |
| S7_extracted_62713 | Not-determined |
| S7_extracted_62716 | Not-determined |
| S7_extracted_62717 | Not-determined |
| S7_extracted_62724 | Low-quality |
| S7_extracted_62761 | Not-determined |
| S7_extracted_62762 | Not-determined |
| S7_extracted_62769 | Not-determined |
| S7_extracted_62794 | Not-determined |
| S7_extracted_62800 | Not-determined |
| S7_extracted_62816 | Not-determined |
| S7_extracted_62817 | Low-quality |
| S7_extracted_62830 | Not-determined |
| S7_extracted_62847 | Not-determined |
| S7_extracted_62863 | Not-determined |
| S7_extracted_62866 | Not-determined |
| S7_extracted_62869 | Not-determined |
| S7_extracted_62892 | Not-determined |
| S7_extracted_62901 | Low-quality |
| S7_extracted_62913 | Not-determined |
| S7_extracted_62916 | Not-determined |
| S7_extracted_62917 | Not-determined |
| S7_extracted_62927 | Low-quality |
| S7_extracted_62959 | Not-determined |
| S7_extracted_62972 | Not-determined |
| S7_extracted_62976 | Not-determined |
| S7_extracted_62981 | Not-determined |
| S7_extracted_62990 | Not-determined |
| S7_extracted_63009 | Not-determined |
| S7_extracted_63038 | Not-determined |
| S7_extracted_63051 | Not-determined |
| S7_extracted_63056 | Not-determined |
| S7_extracted_63059 | Not-determined |
| S7_extracted_63063 | Low-quality |
| S7_extracted_63068 | Low-quality |
| S7_extracted_63073 | Not-determined |
| S7_extracted_63079 | Not-determined |
| S7_extracted_63091 | Low-quality |
| S7_extracted_63093 | Not-determined |
| S7_extracted_63109 | Not-determined |
| S7_extracted_63116 | Low-quality |
| S7_extracted_63120 | Not-determined |
| S7_extracted_63121 | Not-determined |
| S7_extracted_63123 | Not-determined |
| S7_extracted_63154 | Low-quality |
| S7_extracted_63165 | Low-quality |
| S7_extracted_63181 | Low-quality |
| S7_extracted_63227 | Not-determined |
| S7_extracted_63244 | Low-quality |
| S7_extracted_63270 | Not-determined |
| S7_extracted_63271 | Not-determined |
| S7_extracted_63291 | Not-determined |
| S7_extracted_63298 | Not-determined |
| S7_extracted_63300 | Not-determined |
| S7_extracted_63308 | Not-determined |
| S7_extracted_63312 | Not-determined |
| S7_extracted_63322 | Not-determined |
| S7_extracted_63366 | Not-determined |
| S7_extracted_63375 | Low-quality |
| S7_extracted_63398 | Not-determined |
| S7_extracted_63402 | Not-determined |
| S7_extracted_63405 | Not-determined |
| S7_extracted_63415 | Low-quality |
| S7_extracted_63418 | Not-determined |
| S7_extracted_63460 | Not-determined |
| S7_extracted_63486 | Not-determined |
| S7_extracted_63498 | Not-determined |
| S7_extracted_63513 | Not-determined |
| S7_extracted_63562 | Not-determined |
| S7_extracted_63564 | Not-determined |
| S7_extracted_63585 | Not-determined |
| S7_extracted_63606 | Not-determined |
| S7_extracted_63625 | Not-determined |
| S7_extracted_63628 | Low-quality |
| S7_extracted_63631 | Not-determined |
| S7_extracted_63639 | Not-determined |
| S7_extracted_63648 | Not-determined |
| S7_extracted_63650 | Not-determined |
| S7_extracted_63652 | Not-determined |
| S7_extracted_63659 | Not-determined |
| S7_extracted_63666 | Not-determined |
| S7_extracted_63683 | Not-determined |
| S7_extracted_63690 | Not-determined |
| S7_extracted_63694 | Not-determined |
| S7_extracted_63699 | Not-determined |
| S7_extracted_63723 | Not-determined |
| S7_extracted_63731 | Not-determined |
| S7_extracted_63738 | Not-determined |
| S7_extracted_63749 | Low-quality |
| S7_extracted_63776 | Not-determined |
| S7_extracted_63799 | Not-determined |
| S7_extracted_63804 | Low-quality |
| S7_extracted_63805 | Not-determined |
| S7_extracted_63815 | Low-quality |
| S7_extracted_63820 | Not-determined |
| S7_extracted_63831 | Not-determined |
| S7_extracted_63859 | Not-determined |
| S7_extracted_63861 | Not-determined |
| S7_extracted_63869 | Not-determined |
| S7_extracted_63883 | Not-determined |
| S7_extracted_63887 | Not-determined |
| S7_extracted_63888 | Low-quality |
| S14_vs_k141_117154\|\|full | Medium-quality |
| S7_extracted_63903 | Not-determined |
| S7_extracted_63904 | Not-determined |
| S7_extracted_63919 | Not-determined |
| S7_extracted_63941 | Not-determined |
| S7_extracted_63953 | Not-determined |
| S7_extracted_63955 | Not-determined |
| S7_extracted_63965 | Not-determined |
| S7_extracted_64021 | Not-determined |

| | |
|---|---|
| S7_extracted_64030 | Not-determined |
| S7_extracted_64128 | Not-determined |
| S7_extracted_64130 | Not-determined |
| S7_extracted_64140 | Not-determined |
| S7_extracted_64146 | Low-quality |
| S7_extracted_64151 | Low-quality |
| S7_extracted_64154 | Low-quality |
| S7_extracted_64189 | Not-determined |
| S7_extracted_64194 | Not-determined |
| S7_extracted_64201 | Not-determined |
| S7_extracted_64202 | Not-determined |
| S7_extracted_64256 | Not-determined |
| S7_extracted_64257 | Low-quality |
| S7_extracted_64259 | Low-quality |
| S7_extracted_64262 | Not-determined |
| S7_extracted_64277 | Not-determined |
| S7_extracted_64320 | Not-determined |
| S7_extracted_64330 | Not-determined |
| S7_extracted_64338 | Not-determined |
| S7_extracted_64356 | Not-determined |
| S7_extracted_64375 | Not-determined |
| S7_extracted_64407 | Not-determined |
| S7_extracted_64413 | Not-determined |
| S7_extracted_64422 | Not-determined |
| S7_extracted_64429 | Not-determined |
| S7_extracted_64436 | Not-determined |
| S7_extracted_64461 | Not-determined |
| S7_extracted_64478 | Not-determined |
| S7_extracted_64485 | Not-determined |
| S7_extracted_64486 | Not-determined |
| S7_extracted_64498 | Not-determined |
| S7_extracted_64511 | Low-quality |
| S7_extracted_64530 | Not-determined |
| S7_extracted_64551 | Not-determined |
| S7_extracted_64567 | Not-determined |
| S7_extracted_64570 | Not-determined |
| S7_extracted_64571 | Not-determined |
| S7_extracted_64577 | Not-determined |
| S7_extracted_64578 | Not-determined |
| S7_extracted_64586 | Not-determined |
| S7_extracted_64595 | Not-determined |
| S7_extracted_64596 | Not-determined |
| S7_extracted_64599 | Not-determined |
| S7_extracted_64609 | Not-determined |
| S7_extracted_64620 | Not-determined |
| S7_extracted_64661 | Not-determined |
| S7_extracted_64671 | Not-determined |
| S7_extracted_64687 | Not-determined |
| S7_extracted_64705 | Not-determined |
| S7_extracted_64739 | Low-quality |
| S7_extracted_64741 | Not-determined |
| S7_extracted_64742 | Not-determined |
| S7_extracted_64746 | Not-determined |
| S7_extracted_64752 | Not-determined |
| S7_extracted_64784 | Not-determined |
| S7_extracted_64788 | Not-determined |
| S7_extracted_64806 | Not-determined |
| S7_extracted_64823 | Not-determined |
| S7_extracted_64825 | Not-determined |
| S7_extracted_64843 | Not-determined |
| S7_extracted_64857 | Not-determined |
| S7_extracted_64887 | Not-determined |
| S7_extracted_64888 | Low-quality |
| S7_extracted_64897 | Not-determined |
| S7_extracted_64904 | Low-quality |
| S7_extracted_64912 | Not-determined |
| S7_extracted_64914 | Not-determined |
| S7_extracted_64918 | Not-determined |
| S7_extracted_64929 | Not-determined |
| S7_extracted_64954 | Not-determined |
| S7_extracted_64960 | Low-quality |
| S7_extracted_64965 | Not-determined |
| S7_extracted_64967 | Low-quality |
| S7_extracted_64995 | Not-determined |
| S7_extracted_64996 | Not-determined |
| S7_extracted_65000 | Not-determined |
| S7_extracted_65017 | Low-quality |
| S7_extracted_65019 | Not-determined |
| S7_extracted_65023 | Not-determined |
| S7_extracted_65027 | Not-determined |
| S7_extracted_65058 | Not-determined |
| S7_extracted_65061 | Not-determined |
| S7_extracted_65070 | Not-determined |
| S7_extracted_65124 | Not-determined |
| S7_extracted_65129 | Low-quality |
| S7_extracted_65131 | Not-determined |
| S7_extracted_65144 | Not-determined |
| S7_extracted_65162 | Not-determined |
| S7_extracted_65175 | Not-determined |
| S7_extracted_65198 | Not-determined |
| S7_extracted_65221 | Not-determined |
| S7_extracted_65226 | Not-determined |
| S7_extracted_65233 | Not-determined |
| S7_extracted_65254 | Not-determined |
| S7_extracted_65261 | Low-quality |
| S7_extracted_65266 | Not-determined |
| S7_extracted_65294 | Not-determined |
| S7_extracted_65296 | Low-quality |
| S7_extracted_65300 | Not-determined |
| S7_extracted_65317 | Not-determined |
| S7_extracted_65325 | Not-determined |
| S7_extracted_65336 | Not-determined |
| S7_extracted_65346 | Not-determined |
| S7_extracted_65352 | Low-quality |
| S7_extracted_65354 | Not-determined |
| S7_extracted_65357 | Not-determined |
| S7_extracted_65369 | Low-quality |
| S7_extracted_65380 | Not-determined |
| S7_extracted_65391 | Not-determined |
| S7_extracted_65402 | Low-quality |
| S7_extracted_65414 | Not-determined |
| S7_extracted_65435 | Not-determined |
| S7_extracted_65456 | Low-quality |
| S7_extracted_65463 | Not-determined |
| S7_extracted_65468 | Not-determined |
| S7_extracted_65487 | Low-quality |
| S7_extracted_65498 | Not-determined |
| S7_extracted_65499 | Not-determined |
| S7_extracted_65516 | Not-determined |
| S7_extracted_65542 | Not-determined |
| S7_extracted_65549 | Not-determined |
| S7_extracted_65563 | Not-determined |
| S7_extracted_65569 | Not-determined |
| S7_extracted_65575 | Not-determined |
| S7_extracted_65585 | Not-determined |
| S7_extracted_65600 | Not-determined |
| S7_extracted_65608 | Not-determined |
| S7_extracted_65609 | Not-determined |
| S7_extracted_65615 | Not-determined |
| S7_extracted_65617 | Not-determined |
| S7_extracted_65630 | Not-determined |
| S7_extracted_65644 | Not-determined |
| S7_extracted_65660 | Not-determined |
| S7_extracted_65662 | Not-determined |
| S7_extracted_65676 | Low-quality |
| S7_extracted_65680 | Not-determined |
| S7_extracted_65687 | Low-quality |
| S7_extracted_65688 | Not-determined |
| S7_extracted_65689 | Not-determined |
| S7_extracted_65690 | Not-determined |
| S7_extracted_65702 | Not-determined |
| S7_extracted_65716 | Low-quality |
| S7_extracted_65727 | Low-quality |
| S7_extracted_65732 | Not-determined |
| S7_extracted_65733 | Not-determined |
| S7_extracted_65738 | Not-determined |
| S7_extracted_65747 | Low-quality |
| S7_extracted_65755 | Low-quality |
| S7_extracted_65759 | Not-determined |
| S7_extracted_65761 | Not-determined |
| S7_extracted_65780 | Low-quality |
| S7_extracted_65794 | Not-determined |
| S7_extracted_65811 | Not-determined |
| S7_extracted_65814 | Not-determined |
| S7_extracted_65816 | Not-determined |
| S7_extracted_65823 | Low-quality |
| S7_extracted_65826 | Not-determined |
| S7_extracted_65832 | Not-determined |
| S7_extracted_65853 | Not-determined |
| S7_extracted_65855 | Not-determined |
| S7_extracted_65857 | Not-determined |
| S7_extracted_65867 | Low-quality |
| S7_extracted_65873 | Not-determined |
| S7_extracted_65891 | Not-determined |
| S7_extracted_65897 | Not-determined |
| S7_extracted_65908 | Low-quality |
| S7_extracted_65935 | Not-determined |
| S7_extracted_65936 | Not-determined |
| S7_extracted_65937 | Low-quality |
| S7_extracted_65948 | Not-determined |
| S7_extracted_65949 | Low-quality |
| S7_extracted_65951 | Not-determined |
| S7_extracted_65986 | Not-determined |
| S7_extracted_66032 | Not-determined |
| S7_extracted_66044 | Not-determined |
| S7_extracted_66052 | Not-determined |
| S7_extracted_66057 | Not-determined |
| S7_extracted_66069 | Not-determined |
| S7_extracted_66072 | Low-quality |
| S7_extracted_66094 | Not-determined |
| S7_extracted_66133 | Not-determined |
| S7_extracted_66134 | Not-determined |
| S7_extracted_66140 | Not-determined |
| S7_extracted_66160 | Not-determined |
| S7_extracted_66166 | Not-determined |
| S7_extracted_66175 | Not-determined |
| S7_vs_49485\|\|full | Medium-quality |
| S7_extracted_66182 | Not-determined |
| S7_extracted_66212 | Not-determined |
| S7_extracted_66215 | Low-quality |
| S7_extracted_66223 | Not-determined |
| S7_extracted_66230 | Not-determined |
| S7_extracted_66237 | Not-determined |
| S7_extracted_66240 | Not-determined |
| S7_extracted_66246 | Not-determined |
| S7_extracted_66252 | Not-determined |
| S7_extracted_66262 | Not-determined |
| S7_extracted_66274 | Not-determined |
| S7_extracted_66330 | Not-determined |
| S7_extracted_66361 | Not-determined |
| S7_extracted_66363 | Low-quality |
| S7_extracted_66371 | Low-quality |
| S7_extracted_66376 | Not-determined |
| S7_extracted_66380 | Low-quality |
| S7_extracted_66397 | Not-determined |
| S7_extracted_66401 | Not-determined |
| S7_extracted_66409 | Not-determined |
| S7_extracted_66411 | Low-quality |
| S7_extracted_66420 | Not-determined |
| S7_extracted_66438 | Not-determined |
| S7_extracted_66452 | Not-determined |
| S7_extracted_66463 | Not-determined |
| S7_extracted_66465 | Not-determined |
| S7_extracted_66469 | Not-determined |
| S7_extracted_66480 | Not-determined |
| S7_extracted_66489 | Not-determined |
| S7_extracted_66492 | Not-determined |
| S7_extracted_66501 | Not-determined |
| S7_extracted_66506 | Not-determined |
| S7_extracted_66522 | Not-determined |
| S7_extracted_66525 | Not-determined |
| S7_extracted_66541 | Not-determined |
| S7_extracted_66568 | Not-determined |
| S7_extracted_66575 | Not-determined |
| S7_extracted_66578 | Not-determined |
| S7_extracted_66601 | Low-quality |
| S7_extracted_66604 | Not-determined |
| S7_extracted_66607 | Not-determined |
| S7_extracted_66616 | Low-quality |
| S7_extracted_66627 | Not-determined |
| S7_extracted_66629 | Not-determined |
| S7_extracted_66634 | Not-determined |
| S7_extracted_66643 | Not-determined |
| S7_extracted_66646 | Not-determined |
| S7_extracted_66657 | Low-quality |
| S7_extracted_66661 | Not-determined |
| S7_extracted_66672 | Not-determined |
| S7_extracted_66678 | Not-determined |
| S7_extracted_66682 | Low-quality |
| S7_extracted_66693 | Not-determined |
| S7_extracted_66698 | Not-determined |
| S7_extracted_66712 | Not-determined |
| S7_extracted_66735 | Not-determined |
| S7_extracted_66746 | Not-determined |
| S7_extracted_66755 | Not-determined |
| S7_extracted_66761 | Not-determined |
| S7_extracted_66764 | Not-determined |
| S7_extracted_66769 | Not-determined |
| S7_extracted_66774 | Not-determined |
| S7_extracted_66776 | Not-determined |
| S7_extracted_66777 | Not-determined |
| S7_extracted_66784 | Not-determined |
| S7_extracted_66786 | Not-determined |
| S7_extracted_66790 | Not-determined |
| S7_extracted_66804 | Not-determined |
| S7_extracted_66806 | Not-determined |
| S7_extracted_66820 | Not-determined |
| S7_extracted_66833 | Not-determined |
| S7_extracted_66854 | Low-quality |
| S7_extracted_66858 | Not-determined |
| S7_extracted_66870 | Not-determined |
| S7_extracted_66879 | Not-determined |
| S7_extracted_66889 | Not-determined |
| S7_extracted_66906 | Not-determined |
| S7_extracted_66914 | Low-quality |
| S7_extracted_66920 | Not-determined |
| S7_extracted_66921 | Low-quality |
| S7_extracted_66931 | Not-determined |
| S7_extracted_66943 | Not-determined |
| S7_extracted_66955 | Not-determined |
| S7_extracted_66956 | Not-determined |
| S7_extracted_66958 | Not-determined |
| S7_extracted_66961 | Not-determined |
| S7_extracted_66998 | Not-determined |
| S7_extracted_67013 | Not-determined |
| S7_extracted_67020 | Not-determined |
| S7_extracted_67022 | Not-determined |
| S7_extracted_67033 | Not-determined |
| S7_extracted_67073 | Not-determined |
| S7_extracted_67091 | Low-quality |
| S7_extracted_67092 | Not-determined |
| S7_extracted_67109 | Low-quality |
| S7_extracted_67116 | Low-quality |
| S7_extracted_67158 | Low-quality |
| S7_extracted_67172 | Not-determined |
| S7_extracted_67202 | Not-determined |
| S7_extracted_67225 | Not-determined |
| S7_extracted_67231 | Not-determined |
| S7_extracted_67234 | Not-determined |
| S7_extracted_67240 | Low-quality |
| S7_extracted_67244 | Not-determined |
| S7_extracted_67246 | Not-determined |
| S7_extracted_67256 | Not-determined |
| S7_extracted_67278 | Not-determined |
| S7_extracted_67284 | Low-quality |
| S7_extracted_67285 | Not-determined |
| S7_extracted_67289 | Not-determined |
| S7_extracted_67290 | Low-quality |
| S7_extracted_67302 | Not-determined |
| S7_extracted_67315 | Not-determined |
| S7_extracted_67317 | Not-determined |
| S7_extracted_67318 | Low-quality |
| S7_extracted_67323 | Not-determined |
| S7_extracted_67332 | Not-determined |
| S7_extracted_67348 | Not-determined |
| S7_extracted_67361 | Not-determined |
| S7_extracted_67363 | Not-determined |
| S7_extracted_67393 | Not-determined |
| S7_extracted_67406 | Not-determined |
| S7_extracted_67421 | Not-determined |
| S7_extracted_67425 | Not-determined |
| S7_extracted_67431 | Low-quality |
| S7_extracted_67436 | Not-determined |
| S7_extracted_67454 | Low-quality |
| S7_extracted_67465 | Low-quality |
| S7_extracted_67470 | Low-quality |
| S7_extracted_67488 | Low-quality |
| S7_extracted_67489 | Low-quality |
| S7_extracted_67492 | Not-determined |
| S7_extracted_67493 | Not-determined |
| S7_extracted_67494 | Low-quality |
| S7_extracted_67512 | Not-determined |
| S7_extracted_67519 | Low-quality |
| S7_extracted_67525 | Not-determined |
| S7_extracted_67538 | Not-determined |
| S7_extracted_67550 | Not-determined |
| S7_extracted_67552 | Not-determined |
| S7_extracted_67554 | Not-determined |
| S7_extracted_67566 | Low-quality |
| S7_extracted_67576 | Not-determined |
| S7_extracted_67586 | Low-quality |
| S7_extracted_67590 | Low-quality |
| S7_extracted_67592 | Not-determined |
| S7_extracted_67602 | Not-determined |
| S7_extracted_67603 | Not-determined |
| S7_extracted_67615 | Not-determined |
| S7_extracted_67617 | Not-determined |
| S7_extracted_67625 | Not-determined |
| S7_extracted_67630 | Low-quality |
| S7_extracted_67643 | Not-determined |
| S7_extracted_67650 | Not-determined |
| S7_extracted_67651 | Not-determined |
| S7_extracted_67676 | Low-quality |
| S7_extracted_67677 | Not-determined |
| S7_extracted_67682 | Not-determined |
| S7_extracted_67686 | Not-determined |
| S7_extracted_67693 | Not-determined |
| S7_extracted_67695 | Low-quality |
| S7_extracted_67702 | Not-determined |
| S7_extracted_67707 | Not-determined |
| S7_extracted_67710 | Not-determined |
| S7_extracted_67724 | Not-determined |
| S7_extracted_67730 | Not-determined |
| S7_extracted_67731 | Not-determined |
| S7_extracted_67735 | Not-determined |
| S7_extracted_67748 | Low-quality |
| S7_extracted_67753 | Not-determined |
| S7_extracted_67759 | Not-determined |
| S7_extracted_67766 | Not-determined |
| S7_extracted_67777 | Not-determined |
| S7_extracted_67789 | Not-determined |
| S7_extracted_67801 | Not-determined |
| S7_extracted_67802 | Low-quality |
| S7_extracted_67821 | Not-determined |
| S7_extracted_67833 | Not-determined |
| S7_extracted_67840 | Not-determined |
| S7_extracted_67862 | Low-quality |
| S7_extracted_67863 | Not-determined |
| S7_extracted_67872 | Not-determined |
| S14_vs_k141_78164\|\|full | Medium-quality |
| S7_extracted_67930 | Low-quality |
| S7_extracted_67964 | Not-determined |
| S7_extracted_67968 | Not-determined |
| S7_extracted_67981 | Low-quality |
| S7_extracted_68014 | Not-determined |
| S7_extracted_68018 | Not-determined |
| S7_extracted_68029 | Low-quality |
| S7_extracted_68036 | Not-determined |
| S7_extracted_68046 | Not-determined |
| S7_extracted_68050 | Not-determined |
| S7_extracted_68070 | Not-determined |
| S7_extracted_68080 | Not-determined |
| S7_extracted_68083 | Not-determined |
| S7_extracted_68094 | Low-quality |
| S7_extracted_68095 | Not-determined |
| S7_extracted_68109 | Not-determined |
| S7_extracted_68124 | Not-determined |
| S7_extracted_68134 | Not-determined |
| S7_extracted_68137 | Not-determined |
| S7_extracted_68141 | Not-determined |
| S7_extracted_68142 | Not-determined |
| S7_extracted_68153 | Not-determined |
| S7_extracted_68171 | Not-determined |
| S7_extracted_68181 | Not-determined |
| S7_extracted_68220 | Not-determined |
| S7_extracted_68231 | Not-determined |
| S7_extracted_68238 | Not-determined |
| S5_vs_k141_145592\|\|full | Medium-quality |
| S7_extracted_68243 | Low-quality |
| S7_extracted_68249 | Not-determined |
| S7_extracted_68250 | Not-determined |
| S7_extracted_68256 | Not-determined |
| S7_extracted_68262 | Not-determined |
| S7_extracted_68269 | Not-determined |
| S7_extracted_68270 | Not-determined |
| S7_extracted_68272 | Not-determined |
| S7_extracted_68274 | Not-determined |
| S7_extracted_68279 | Low-quality |
| S7_extracted_68282 | Low-quality |
| S7_extracted_68293 | Not-determined |
| S7_extracted_68299 | Not-determined |
| S7_extracted_68304 | Not-determined |
| S7_extracted_68318 | Not-determined |
| S7_extracted_68325 | Not-determined |
| S7_extracted_68327 | Not-determined |
| S7_extracted_68329 | Not-determined |
| S7_extracted_68342 | Not-determined |
| S7_extracted_68352 | Low-quality |
| S7_extracted_68403 | Not-determined |
| S7_extracted_68404 | Low-quality |
| S7_extracted_68411 | Not-determined |
| S7_extracted_68438 | Not-determined |
| S7_extracted_68448 | Not-determined |
| S7_extracted_68449 | Not-determined |
| S7_extracted_68507 | Not-determined |
| S7_extracted_68511 | Not-determined |
| S7_extracted_68526 | Not-determined |
| S7_extracted_68533 | Not-determined |
| S7_extracted_68535 | Low-quality |
| S6_vs_k141_56489\|\|full | High-quality |
| S7_extracted_68556 | Not-determined |
| S7_extracted_68571 | Not-determined |
| S7_extracted_68579 | Not-determined |
| S7_extracted_68593 | Not-determined |
| S7_extracted_68611 | Not-determined |
| S7_extracted_68618 | Not-determined |
| S7_extracted_68621 | Not-determined |
| S7_extracted_68631 | Not-determined |
| S7_extracted_68647 | Not-determined |
| S7_extracted_68649 | Not-determined |
| S7_extracted_68656 | Not-determined |
| S7_extracted_68667 | Not-determined |
| S7_extracted_68670 | Not-determined |
| S7_extracted_68679 | Not-determined |
| S7_extracted_68680 | Low-quality |
| S7_extracted_68684 | Not-determined |
| S7_extracted_68696 | Not-determined |
| S7_extracted_68737 | Low-quality |
| S7_extracted_68750 | Not-determined |
| S7_extracted_68781 | Not-determined |
| S7_extracted_68788 | Not-determined |
| S7_extracted_68802 | Not-determined |
| S7_extracted_68806 | Not-determined |
| S7_extracted_68814 | Not-determined |
| S7_extracted_68815 | Not-determined |
| S7_extracted_68819 | Not-determined |
| S7_extracted_68850 | Not-determined |
| S7_extracted_68852 | Low-quality |
| S7_extracted_68867 | Not-determined |
| S7_extracted_68880 | Not-determined |
| S7_extracted_68883 | Not-determined |
| S7_extracted_68894 | Not-determined |
| S7_extracted_68897 | Not-determined |
| S7_extracted_68905 | Not-determined |
| S7_extracted_68937 | Not-determined |
| S7_extracted_68951 | Low-quality |
| S7_extracted_68958 | Low-quality |
| S7_extracted_68964 | Low-quality |
| S7_extracted_68967 | Low-quality |
| S7_extracted_68970 | Not-determined |
| S7_extracted_68980 | Not-determined |
| S7_extracted_68988 | Not-determined |
| S7_extracted_68993 | Not-determined |
| S7_extracted_68994 | Not-determined |
| S7_extracted_69001 | Low-quality |
| S7_extracted_69016 | Not-determined |
| S7_extracted_69027 | Not-determined |
| S7_extracted_69068 | Not-determined |
| S7_extracted_69071 | Low-quality |
| S7_extracted_69074 | Not-determined |
| S7_extracted_69094 | Not-determined |
| S7_extracted_69126 | Not-determined |
| S7_extracted_69129 | Not-determined |
| S7_extracted_69132 | Low-quality |
| S7_extracted_69152 | Not-determined |
| S7_extracted_69153 | Not-determined |
| S7_extracted_69156 | Not-determined |
| S7_extracted_69180 | Low-quality |
| S7_extracted_69193 | Not-determined |
| S7_extracted_69220 | Not-determined |
| S7_extracted_69227 | Low-quality |
| S7_extracted_69273 | Not-determined |
| S7_extracted_69289 | Low-quality |
| S7_extracted_69293 | Not-determined |
| S7_extracted_69299 | Not-determined |
| S7_extracted_69305 | Not-determined |
| S7_extracted_69308 | Low-quality |
| S7_extracted_69322 | Not-determined |
| S7_extracted_69325 | Not-determined |
| S7_extracted_69330 | Low-quality |
| S7_extracted_69343 | Not-determined |
| S5_vs_k141_67958\|\|full | Medium-quality |
| S7_extracted_69358 | Not-determined |
| S7_extracted_69385 | Not-determined |
| S7_extracted_69386 | Not-determined |
| S7_extracted_69404 | Not-determined |
| S7_extracted_69405 | Not-determined |
| S7_extracted_69466 | Low-quality |
| S7_extracted_69480 | Not-determined |
| S7_extracted_69496 | Not-determined |
| S7_extracted_69502 | Not-determined |
| S7_extracted_69506 | Not-determined |
| S7_extracted_69514 | Not-determined |
| S7_extracted_69515 | Not-determined |
| S7_extracted_69522 | Not-determined |
| S7_extracted_69527 | Low-quality |
| S7_extracted_69551 | Not-determined |
| S7_extracted_69561 | Not-determined |
| S7_extracted_69574 | Low-quality |
| S7_extracted_69605 | Low-quality |
| S7_extracted_69629 | Not-determined |
| S7_extracted_69651 | Not-determined |
| S7_extracted_69694 | Not-determined |
| S7_extracted_69702 | Not-determined |
| S7_extracted_69712 | Not-determined |
| S7_extracted_69730 | Low-quality |
| S7_extracted_69738 | Not-determined |
| S7_extracted_69744 | Not-determined |
| S7_extracted_69753 | Not-determined |
| S7_extracted_69761 | Not-determined |
| S7_extracted_69797 | Not-determined |
| S7_extracted_69801 | Not-determined |
| S7_extracted_69815 | Low-quality |
| S7_extracted_69830 | Not-determined |
| S7_extracted_69836 | Not-determined |
| S7_extracted_69843 | Not-determined |
| S7_extracted_69851 | Not-determined |
| S7_extracted_69855 | Not-determined |
| S7_extracted_69870 | Not-determined |
| S7_extracted_69891 | Low-quality |
| S7_extracted_69898 | Not-determined |
| S7_extracted_69899 | Not-determined |
| S7_extracted_69908 | Not-determined |
| S7_extracted_69918 | Not-determined |
| S7_extracted_69931 | Not-determined |
| S7_extracted_69932 | Not-determined |
| S7_extracted_69934 | Low-quality |
| S7_extracted_69935 | Not-determined |
| S7_extracted_69965 | Not-determined |
| S7_extracted_70005 | Not-determined |
| S7_extracted_70019 | Not-determined |
| S7_extracted_70056 | Not-determined |
| S7_extracted_70073 | Not-determined |
| S7_extracted_70076 | Not-determined |
| S7_extracted_70097 | Not-determined |
| S7_extracted_70123 | Not-determined |
| S7_extracted_70142 | Not-determined |
| S7_extracted_70143 | Not-determined |
| S7_extracted_70158 | Not-determined |
| S7_extracted_70178 | Not-determined |
| S7_extracted_70193 | Low-quality |
| S7_extracted_70207 | Not-determined |
| S7_extracted_70213 | Not-determined |
| S7_extracted_70252 | Not-determined |
| S7_extracted_70257 | Low-quality |
| S7_extracted_70266 | Not-determined |
| S7_extracted_70267 | Not-determined |
| S7_extracted_70294 | Not-determined |
| S7_extracted_70299 | Not-determined |
| S7_extracted_70321 | Not-determined |
| S7_extracted_70347 | Not-determined |
| S7_extracted_70352 | Not-determined |
| S7_extracted_70369 | Not-determined |
| S7_extracted_70387 | Not-determined |
| S7_extracted_70391 | Not-determined |
| S7_extracted_70401 | Not-determined |
| S7_extracted_70407 | Not-determined |
| S7_extracted_70415 | Not-determined |
| S7_extracted_70426 | Not-determined |
| S7_extracted_70428 | Not-determined |
| S7_extracted_70448 | Not-determined |
| S7_extracted_70482 | Not-determined |
| S7_extracted_70486 | Low-quality |
| S7_extracted_70487 | Not-determined |
| S7_extracted_70491 | Not-determined |
| S7_extracted_70506 | Not-determined |
| S7_extracted_70508 | Not-determined |
| S7_extracted_70511 | Not-determined |
| S7_extracted_70514 | Not-determined |

| | |
|---|---|
| S7_extracted_70552 | Not-determined |
| S7_extracted_70581 | Not-determined |
| S7_extracted_70583 | Not-determined |
| S7_extracted_70587 | Not-determined |
| S7_extracted_70596 | Not-determined |
| S7_extracted_70613 | Not-determined |
| S7_extracted_70617 | Not-determined |
| S7_extracted_70626 | Not-determined |
| S7_extracted_70629 | Not-determined |
| S7_extracted_70644 | Not-determined |
| S7_extracted_70667 | Not-determined |
| S7_extracted_70676 | Not-determined |
| S7_extracted_70690 | Not-determined |
| S7_extracted_70704 | Low-quality |
| S7_extracted_70707 | Low-quality |
| S7_extracted_70735 | Not-determined |
| S7_extracted_70736 | Not-determined |
| S7_extracted_70748 | Not-determined |
| S7_extracted_70750 | Not-determined |
| S7_extracted_70770 | Not-determined |
| S7_extracted_70831 | Not-determined |
| S7_extracted_70837 | Not-determined |
| S7_extracted_70857 | Not-determined |
| S7_extracted_70869 | Not-determined |
| S7_extracted_70873 | Not-determined |
| S7_extracted_70877 | Low-quality |
| S7_extracted_70893 | Not-determined |
| S7_extracted_70902 | Not-determined |
| S7_extracted_70929 | Not-determined |
| S7_extracted_70942 | Not-determined |
| S7_extracted_70949 | Not-determined |
| S7_extracted_70959 | Not-determined |
| S7_extracted_70966 | Low-quality |
| S7_extracted_70975 | Not-determined |
| S7_extracted_70987 | Not-determined |
| S7_extracted_70990 | Not-determined |
| S7_extracted_71002 | Low-quality |
| S7_extracted_71009 | Not-determined |
| S7_extracted_71022 | Not-determined |
| S7_extracted_71025 | Not-determined |
| S7_extracted_71028 | Not-determined |
| S7_extracted_71071 | Not-determined |
| S7_extracted_71087 | Not-determined |
| S7_extracted_71089 | Low-quality |
| S7_extracted_71097 | Not-determined |
| S7_extracted_71099 | Not-determined |
| S7_extracted_71103 | Not-determined |
| S7_extracted_71107 | Not-determined |
| S7_extracted_71109 | Not-determined |
| S7_extracted_71141 | Not-determined |
| S7_extracted_71166 | Not-determined |
| S7_extracted_71184 | Not-determined |
| S7_extracted_71188 | Not-determined |
| S7_extracted_71192 | Not-determined |
| S7_extracted_71195 | Not-determined |
| S7_extracted_71203 | Not-determined |
| S7_extracted_71217 | Not-determined |
| S7_extracted_71249 | Not-determined |
| S7_extracted_71264 | Not-determined |
| S7_extracted_71268 | Not-determined |
| S7_extracted_71274 | Not-determined |
| S7_extracted_71299 | Not-determined |
| S7_extracted_71308 | Not-determined |
| S7_extracted_71321 | Not-determined |
| S7_extracted_71333 | Not-determined |
| S7_extracted_71336 | Not-determined |
| S7_extracted_71337 | Not-determined |
| S7_extracted_71341 | Not-determined |
| S7_extracted_71344 | Not-determined |
| S7_extracted_71346 | Not-determined |
| S7_extracted_71373 | Low-quality |
| S7_extracted_71378 | Not-determined |
| S7_extracted_71385 | Not-determined |
| S7_extracted_71395 | Not-determined |
| S7_extracted_71405 | Not-determined |
| S7_extracted_71410 | Not-determined |
| S7_extracted_71420 | Not-determined |
| S7_extracted_71429 | Low-quality |
| S7_extracted_71439 | Not-determined |
| S7_extracted_71450 | Not-determined |
| S7_extracted_71452 | Not-determined |
| S7_extracted_71458 | Not-determined |
| S7_extracted_71490 | Low-quality |
| S7_extracted_71516 | Not-determined |
| S14_extracted_k141_72953 | High-quality |
| S7_extracted_71537 | Low-quality |
| S7_extracted_71545 | Not-determined |
| S7_extracted_71551 | Low-quality |
| S7_extracted_71558 | Not-determined |
| S7_extracted_71589 | Low-quality |
| S7_extracted_71623 | Not-determined |
| S7_extracted_71624 | Not-determined |
| S7_extracted_71637 | Low-quality |
| S7_extracted_71641 | Not-determined |
| S7_extracted_71680 | Not-determined |
| S7_extracted_71705 | Not-determined |
| S7_extracted_71717 | Not-determined |
| S7_extracted_71766 | Not-determined |
| S7_extracted_71777 | Low-quality |
| S7_extracted_71778 | Low-quality |
| S7_extracted_71781 | Not-determined |
| S7_extracted_71788 | Not-determined |
| S7_extracted_71805 | Not-determined |
| S7_extracted_71811 | Not-determined |
| S7_extracted_71817 | Low-quality |
| S7_extracted_71833 | Low-quality |
| S7_extracted_71857 | Low-quality |
| S7_extracted_71860 | Not-determined |
| S7_extracted_71869 | Low-quality |
| S7_extracted_71882 | Low-quality |
| S7_extracted_71885 | Not-determined |
| S7_extracted_71886 | Not-determined |
| S7_extracted_71898 | Not-determined |
| S7_extracted_71912 | Not-determined |
| S7_extracted_71913 | Not-determined |
| S7_extracted_71925 | Low-quality |
| S7_extracted_71931 | Not-determined |
| S7_extracted_71938 | Not-determined |
| S7_extracted_71963 | Low-quality |
| S7_extracted_71967 | Not-determined |
| S7_extracted_71972 | Not-determined |
| S7_extracted_71981 | Not-determined |
| S7_extracted_71982 | Not-determined |
| S7_extracted_72023 | Not-determined |
| S7_extracted_72048 | Not-determined |
| S7_extracted_72065 | Not-determined |
| S7_extracted_72070 | Not-determined |
| S7_extracted_72074 | Low-quality |
| S7_extracted_72125 | Low-quality |
| S7_extracted_72157 | Low-quality |
| S7_extracted_72161 | Not-determined |
| S7_extracted_72166 | Not-determined |
| S7_extracted_72204 | Not-determined |
| S7_extracted_72216 | Not-determined |
| S7_extracted_72231 | Not-determined |
| S7_extracted_72247 | Not-determined |
| S7_extracted_72251 | Not-determined |
| S7_extracted_72257 | Low-quality |
| S7_extracted_72277 | Low-quality |
| S7_extracted_72280 | Low-quality |
| S7_extracted_72285 | Not-determined |
| S7_extracted_72293 | Not-determined |
| S7_extracted_72320 | Not-determined |
| S7_extracted_72325 | Not-determined |
| S7_extracted_72330 | Not-determined |
| S7_extracted_72353 | Low-quality |
| S7_extracted_72360 | Not-determined |
| S7_extracted_72365 | Not-determined |
| S7_extracted_72366 | Not-determined |
| S7_extracted_72378 | Not-determined |
| S7_extracted_72385 | Not-determined |
| S7_extracted_72396 | Not-determined |
| S7_extracted_72400 | Not-determined |
| S7_extracted_72407 | Low-quality |
| S7_extracted_72418 | Not-determined |
| S7_extracted_72439 | Not-determined |
| S7_extracted_72446 | Not-determined |
| S7_extracted_72450 | Not-determined |
| S7_extracted_72471 | Not-determined |
| S7_extracted_72487 | Low-quality |
| S10_vs_67471\|\|full | Medium-quality |
| S7_extracted_72511 | Not-determined |
| S7_extracted_72517 | Not-determined |
| S7_extracted_72527 | Low-quality |
| S7_extracted_72544 | Not-determined |
| S7_extracted_72559 | Not-determined |
| S7_extracted_72570 | Not-determined |
| S7_extracted_72575 | Not-determined |
| S7_extracted_72608 | Not-determined |
| S7_extracted_72623 | Not-determined |
| S7_extracted_72682 | Not-determined |
| S7_extracted_72683 | Not-determined |
| S7_extracted_72687 | Not-determined |
| S7_extracted_72691 | Not-determined |
| S7_extracted_72703 | Low-quality |
| S7_extracted_72704 | Low-quality |
| S7_extracted_72705 | Not-determined |
| S7_extracted_72720 | Low-quality |
| S7_extracted_72726 | Not-determined |
| S7_extracted_72733 | Not-determined |
| S7_extracted_72734 | Low-quality |
| S7_extracted_72736 | Not-determined |
| S7_extracted_72750 | Not-determined |
| S7_extracted_72753 | Not-determined |
| S7_extracted_72776 | Not-determined |
| S7_extracted_72777 | Not-determined |
| S7_extracted_72780 | Low-quality |
| S7_extracted_72788 | Low-quality |
| S7_extracted_72806 | Not-determined |
| S7_extracted_72817 | Not-determined |
| S7_extracted_72824 | Not-determined |
| S7_extracted_72849 | Not-determined |
| S7_extracted_72865 | Not-determined |
| S7_extracted_72876 | Not-determined |
| S7_extracted_72887 | Not-determined |
| S7_extracted_72912 | Not-determined |
| S7_extracted_72915 | Not-determined |
| S7_extracted_72916 | Not-determined |
| S7_extracted_72917 | Not-determined |
| S7_extracted_72918 | Not-determined |
| S7_extracted_72922 | Not-determined |
| S7_extracted_72955 | Low-quality |
| S7_extracted_72965 | Low-quality |
| S7_extracted_72969 | Not-determined |
| S7_extracted_72988 | Low-quality |
| S7_extracted_72998 | Low-quality |
| S7_extracted_73001 | Not-determined |
| S7_extracted_73014 | Low-quality |
| S7_extracted_73022 | Not-determined |
| S7_extracted_73031 | Not-determined |
| S7_extracted_73033 | Not-determined |
| S7_extracted_73045 | Not-determined |
| S7_extracted_73070 | Not-determined |
| S7_extracted_73108 | Not-determined |
| S7_extracted_73131 | Not-determined |
| S7_extracted_73141 | Not-determined |
| S7_extracted_73151 | Not-determined |
| S7_extracted_73156 | Not-determined |
| S7_extracted_73164 | Not-determined |
| S7_extracted_73168 | Not-determined |
| S7_extracted_73172 | Not-determined |
| S7_extracted_73180 | Low-quality |
| S7_extracted_73192 | Low-quality |
| S7_extracted_73195 | Not-determined |
| S7_extracted_73203 | Not-determined |
| S7_extracted_73205 | Not-determined |
| S7_extracted_73212 | Not-determined |
| S7_extracted_73218 | Low-quality |
| S7_extracted_73235 | Not-determined |
| S7_extracted_73239 | Not-determined |
| S7_extracted_73240 | Not-determined |
| S7_extracted_73242 | Low-quality |
| S7_extracted_73257 | Not-determined |
| S7_extracted_73261 | Not-determined |
| S7_extracted_73264 | Not-determined |
| S7_extracted_73266 | Not-determined |
| S7_extracted_73279 | Not-determined |
| S7_extracted_73286 | Not-determined |
| S7_extracted_73299 | Not-determined |
| S7_extracted_73311 | Not-determined |
| S7_extracted_73315 | Not-determined |
| S7_extracted_73317 | Low-quality |
| S7_extracted_73323 | Not-determined |
| S7_extracted_73327 | Not-determined |
| S7_extracted_73332 | Not-determined |
| S7_extracted_73335 | Not-determined |
| S7_extracted_73337 | Low-quality |
| S7_extracted_73381 | Not-determined |
| S7_extracted_73382 | Not-determined |
| S7_extracted_73383 | Not-determined |
| S7_extracted_73417 | Low-quality |
| S7_extracted_73456 | Not-determined |
| S7_extracted_73460 | Low-quality |
| S7_extracted_73497 | Not-determined |
| S7_extracted_73506 | Not-determined |
| S7_extracted_73507 | Not-determined |
| S7_extracted_73544 | Low-quality |
| S7_extracted_73556 | Not-determined |
| S7_extracted_73559 | Not-determined |
| S7_extracted_73560 | Not-determined |
| S7_extracted_73576 | Not-determined |
| S7_extracted_73591 | Not-determined |
| S7_extracted_73597 | Not-determined |
| S7_extracted_73599 | Low-quality |
| S7_extracted_73614 | Not-determined |
| S7_extracted_73617 | Not-determined |
| S7_extracted_73640 | Not-determined |
| S7_extracted_73681 | Not-determined |
| S7_extracted_73719 | Not-determined |
| S7_extracted_73730 | Low-quality |
| S7_extracted_73733 | Low-quality |
| S7_extracted_73777 | Low-quality |
| S7_extracted_73782 | Not-determined |
| S7_extracted_73808 | Not-determined |
| S7_extracted_73816 | Not-determined |
| S7_extracted_73839 | Low-quality |
| S7_extracted_73883 | Not-determined |
| S7_extracted_73893 | Not-determined |
| S7_extracted_73911 | Low-quality |
| S7_extracted_73955 | Not-determined |
| S7_extracted_73983 | Low-quality |
| S7_extracted_73994 | Not-determined |
| S7_extracted_73998 | Not-determined |
| S7_extracted_74000 | Not-determined |
| S7_extracted_74015 | Not-determined |
| S7_extracted_74019 | Low-quality |
| S7_extracted_74066 | Not-determined |
| S7_extracted_74108 | Not-determined |
| S7_extracted_74126 | Not-determined |
| S7_extracted_74148 | Not-determined |
| S7_extracted_74194 | Not-determined |
| S7_extracted_74202 | Not-determined |
| S7_extracted_74211 | Not-determined |
| S7_extracted_74230 | Low-quality |
| S7_extracted_74232 | Not-determined |
| S7_extracted_74247 | Not-determined |
| S7_extracted_74257 | Not-determined |
| S7_extracted_74267 | Not-determined |
| S7_extracted_74272 | Not-determined |
| S7_extracted_74282 | Not-determined |
| S7_extracted_74290 | Not-determined |
| S7_extracted_74296 | Low-quality |
| S7_extracted_74320 | Low-quality |
| S7_extracted_74328 | Low-quality |
| S7_extracted_74340 | Not-determined |
| S7_extracted_74350 | Not-determined |
| S7_extracted_74361 | Not-determined |
| S7_extracted_74386 | Not-determined |
| S7_extracted_74414 | Low-quality |
| S7_extracted_74420 | Not-determined |
| S7_extracted_74430 | Not-determined |
| S7_extracted_74440 | Not-determined |
| S7_extracted_74450 | Not-determined |
| S7_extracted_74457 | Low-quality |
| S7_extracted_74458 | Not-determined |
| S7_extracted_74464 | Not-determined |
| S7_extracted_74466 | Low-quality |
| S7_extracted_74475 | Not-determined |
| S7_extracted_74481 | Not-determined |
| S7_extracted_74494 | Not-determined |
| S7_extracted_74507 | Not-determined |
| S7_extracted_74510 | Not-determined |
| S7_extracted_74574 | Not-determined |
| S7_extracted_74594 | Not-determined |
| S7_extracted_74599 | Not-determined |
| S7_extracted_74605 | Not-determined |
| S7_extracted_74607 | Not-determined |
| S7_extracted_74613 | Not-determined |
| S7_extracted_74615 | Not-determined |
| S7_extracted_74640 | Low-quality |
| S7_extracted_74653 | Not-determined |
| S7_extracted_74664 | Not-determined |
| S7_extracted_74665 | Not-determined |
| S7_extracted_74666 | Not-determined |
| S7_extracted_74669 | Not-determined |
| S7_extracted_74673 | Not-determined |
| S7_extracted_74678 | Low-quality |
| S7_extracted_74725 | Not-determined |
| S7_extracted_74755 | Not-determined |
| S7_extracted_74770 | Not-determined |
| S7_extracted_74778 | Not-determined |
| S7_extracted_74795 | Low-quality |
| S7_extracted_74799 | Not-determined |
| S7_extracted_74800 | Not-determined |
| S7_extracted_74805 | Not-determined |
| S7_extracted_74806 | Not-determined |
| S7_extracted_74808 | Not-determined |
| S7_extracted_74829 | Low-quality |
| S7_extracted_74832 | Not-determined |
| S7_extracted_74842 | Low-quality |
| S7_extracted_74850 | Not-determined |
| S7_extracted_74869 | Not-determined |
| S7_extracted_74899 | Low-quality |
| S7_extracted_74903 | Not-determined |
| S7_extracted_74913 | Not-determined |
| S7_extracted_74919 | Not-determined |
| S7_extracted_74943 | Not-determined |
| S7_extracted_74953 | Low-quality |
| S7_extracted_74962 | Not-determined |
| S7_extracted_74982 | Not-determined |
| S7_extracted_74995 | Not-determined |
| S7_extracted_75001 | Not-determined |
| S7_extracted_75023 | Low-quality |
| S7_extracted_75030 | Not-determined |
| S7_extracted_75039 | Low-quality |
| S7_extracted_75040 | Not-determined |
| S7_extracted_75066 | Low-quality |
| S7_extracted_75090 | Low-quality |
| S7_extracted_75111 | Not-determined |
| S7_extracted_75137 | Not-determined |
| S7_extracted_75160 | Not-determined |
| S7_extracted_75170 | Low-quality |
| S7_extracted_75179 | Not-determined |
| S7_extracted_75233 | Not-determined |
| S7_extracted_75243 | Not-determined |
| S7_extracted_75267 | Not-determined |
| S7_extracted_75289 | Low-quality |
| S7_extracted_75298 | Not-determined |
| S7_extracted_75304 | Not-determined |
| S7_extracted_75310 | Not-determined |
| S7_extracted_75312 | Not-determined |
| S7_extracted_75315 | Not-determined |
| S7_extracted_75322 | Not-determined |
| S7_extracted_75339 | Not-determined |
| S7_extracted_75349 | Not-determined |
| S7_extracted_75356 | Not-determined |
| S7_extracted_75357 | Not-determined |
| S7_extracted_75392 | Low-quality |
| S7_extracted_75395 | Not-determined |
| S7_extracted_75396 | Not-determined |
| S7_extracted_75425 | Low-quality |
| S7_extracted_75434 | Low-quality |
| S7_extracted_75438 | Not-determined |
| S7_extracted_75446 | Not-determined |
| S7_extracted_75450 | Not-determined |
| S7_extracted_75453 | Low-quality |
| S7_extracted_75471 | Not-determined |
| S7_extracted_75487 | Not-determined |
| S7_extracted_75489 | Low-quality |
| S7_extracted_75500 | Not-determined |
| S7_extracted_75509 | Not-determined |
| S7_extracted_75523 | Low-quality |
| S7_extracted_75536 | Not-determined |
| S7_extracted_75538 | Not-determined |
| S7_extracted_75548 | Not-determined |
| S7_extracted_75574 | Low-quality |
| S7_extracted_75585 | Low-quality |
| S7_extracted_75594 | Not-determined |
| S7_extracted_75595 | Not-determined |
| S7_extracted_75598 | Not-determined |
| S7_extracted_75610 | Low-quality |
| S7_extracted_75616 | Low-quality |
| S7_extracted_75622 | Not-determined |
| S7_extracted_75660 | Not-determined |
| S7_extracted_75661 | Not-determined |
| S7_extracted_75673 | Not-determined |
| S7_extracted_75679 | Not-determined |
| S7_extracted_75690 | Not-determined |
| S7_extracted_75701 | Not-determined |
| S7_extracted_75705 | Not-determined |
| S7_extracted_75717 | Not-determined |
| S7_extracted_75718 | Not-determined |
| S7_extracted_75740 | Not-determined |
| S7_extracted_75744 | Not-determined |
| S7_extracted_75750 | Low-quality |
| S7_extracted_75759 | Low-quality |
| S7_extracted_75779 | Low-quality |
| S7_extracted_75780 | Not-determined |
| S7_extracted_75793 | Not-determined |
| S7_extracted_75819 | Not-determined |
| S7_extracted_75821 | Not-determined |
| S7_extracted_75829 | Not-determined |
| S7_extracted_75836 | Not-determined |
| S7_extracted_75838 | Not-determined |
| S7_extracted_75843 | Low-quality |
| S7_extracted_75875 | Low-quality |
| S7_extracted_75880 | Low-quality |
| S7_extracted_75887 | Not-determined |
| S7_extracted_75899 | Not-determined |
| S7_extracted_75904 | Not-determined |
| S7_extracted_75909 | Not-determined |
| S7_extracted_75911 | Not-determined |
| S7_extracted_75912 | Not-determined |
| S7_extracted_75915 | Low-quality |
| S7_extracted_75932 | Not-determined |
| S7_extracted_75961 | Not-determined |
| S7_extracted_75967 | Low-quality |
| S7_extracted_75973 | Not-determined |
| S7_extracted_75974 | Not-determined |
| S7_extracted_76026 | Not-determined |
| S7_extracted_76029 | Low-quality |
| S7_extracted_76059 | Low-quality |
| S7_extracted_76066 | Not-determined |
| S7_extracted_76080 | Not-determined |
| S7_extracted_76098 | Not-determined |
| S7_extracted_76101 | Not-determined |
| S7_extracted_76115 | Low-quality |
| S7_extracted_76124 | Not-determined |
| S7_extracted_76158 | Low-quality |
| S7_extracted_76168 | Low-quality |
| S7_extracted_76197 | Not-determined |
| S7_extracted_76209 | Not-determined |
| S7_extracted_76225 | Low-quality |
| S7_extracted_76234 | Low-quality |
| S7_extracted_76236 | Not-determined |
| S7_extracted_76238 | Not-determined |
| S7_extracted_76239 | Low-quality |
| S7_extracted_76243 | Low-quality |
| S7_extracted_76250 | Not-determined |
| S7_extracted_76257 | Low-quality |
| S7_extracted_76279 | Not-determined |
| S7_extracted_76294 | Not-determined |
| S7_extracted_76306 | Not-determined |
| S7_extracted_76307 | Not-determined |
| S7_extracted_76317 | Not-determined |
| S7_extracted_76327 | Not-determined |
| S7_extracted_76328 | Not-determined |
| S7_extracted_76330 | Low-quality |
| S7_extracted_76339 | Low-quality |
| S7_extracted_76344 | Not-determined |
| S7_extracted_76346 | Not-determined |
| S7_extracted_76365 | Not-determined |
| S7_extracted_76369 | Not-determined |
| S7_extracted_76377 | Not-determined |
| S8_vs_k141_4153\|\|full | Medium-quality |
| S7_extracted_76430 | Not-determined |
| S7_extracted_76433 | Not-determined |
| S7_extracted_76435 | Low-quality |
| S7_extracted_76447 | Low-quality |
| S7_extracted_76449 | Not-determined |
| S3_vs_k141_44405\|\|full | Medium-quality |
| S7_extracted_76483 | Not-determined |
| S7_extracted_76493 | Not-determined |
| S7_extracted_76500 | Low-quality |
| S7_extracted_76503 | Not-determined |
| S7_extracted_76504 | Low-quality |
| S7_extracted_76513 | Not-determined |
| S7_extracted_76517 | Not-determined |
| S7_extracted_76522 | Low-quality |
| S7_extracted_76524 | Not-determined |
| S7_extracted_76529 | Not-determined |
| S7_extracted_76533 | Not-determined |
| S7_extracted_76547 | Not-determined |
| S7_extracted_76572 | Low-quality |
| S7_extracted_76577 | Low-quality |
| S7_extracted_76586 | Not-determined |
| S7_extracted_76588 | Low-quality |
| S7_extracted_76591 | Not-determined |
| S7_extracted_76597 | Not-determined |
| S7_extracted_76604 | Not-determined |
| S7_extracted_76607 | Not-determined |
| S7_extracted_76619 | Not-determined |
| S7_extracted_76625 | Not-determined |
| S7_extracted_76632 | Not-determined |
| S7_extracted_76660 | Not-determined |
| S7_extracted_76673 | Not-determined |
| S7_extracted_76675 | Not-determined |
| S7_extracted_76681 | Not-determined |
| S7_extracted_76682 | Low-quality |
| S7_extracted_76711 | Not-determined |
| S7_extracted_76719 | Not-determined |
| S7_extracted_76735 | Low-quality |
| S7_extracted_76738 | Low-quality |
| S7_extracted_76741 | Not-determined |
| S7_extracted_76766 | Not-determined |
| S7_extracted_76767 | Not-determined |
| S7_extracted_76789 | Not-determined |
| S7_extracted_76791 | Low-quality |
| S7_extracted_76808 | Not-determined |
| S7_extracted_76819 | Low-quality |
| S7_extracted_76825 | Not-determined |
| S7_extracted_76848 | Not-determined |
| S7_extracted_76850 | Not-determined |
| S7_extracted_76881 | Not-determined |
| S7_extracted_76882 | Not-determined |
| S7_extracted_76883 | Low-quality |
| S7_extracted_76885 | Low-quality |
| S7_extracted_76889 | Not-determined |
| S7_extracted_76891 | Low-quality |
| S7_extracted_76905 | Low-quality |
| S7_extracted_76921 | Not-determined |
| S7_extracted_76925 | Not-determined |
| S7_extracted_76932 | Not-determined |
| S7_extracted_76934 | Not-determined |
| S7_extracted_76942 | Not-determined |
| S7_extracted_76947 | Not-determined |
| S7_extracted_76953 | Not-determined |
| S7_extracted_76982 | Not-determined |
| S7_extracted_77012 | Not-determined |
| S7_extracted_77014 | Not-determined |
| S7_extracted_77015 | Not-determined |
| S7_extracted_77028 | Low-quality |
| S7_extracted_77061 | Not-determined |
| S7_extracted_77066 | Not-determined |
| S7_extracted_77075 | Not-determined |
| S7_extracted_77076 | Not-determined |
| S7_extracted_77085 | Not-determined |
| S7_extracted_77104 | Not-determined |
| S7_extracted_77135 | Not-determined |
| S7_extracted_77147 | Not-determined |
| S7_extracted_77155 | Not-determined |
| S7_extracted_77163 | Not-determined |
| S7_extracted_77165 | Not-determined |
| S7_extracted_77175 | Not-determined |
| S7_extracted_77188 | Not-determined |
| S7_extracted_77190 | Not-determined |
| S7_extracted_77212 | Low-quality |
| S7_extracted_77220 | Not-determined |
| S7_extracted_77243 | Not-determined |
| S7_extracted_77246 | Not-determined |
| S7_extracted_77256 | Not-determined |
| S7_extracted_77285 | Not-determined |
| S7_extracted_77286 | Low-quality |
| S7_extracted_77300 | Not-determined |
| S7_extracted_77308 | Not-determined |
| S7_extracted_77329 | Not-determined |
| S7_extracted_77332 | Not-determined |
| S7_extracted_77348 | Not-determined |
| S7_extracted_77363 | Not-determined |
| S7_extracted_77366 | Not-determined |
| S7_extracted_77394 | Not-determined |
| S7_extracted_77408 | Not-determined |
| S7_extracted_77425 | Low-quality |
| S7_extracted_77436 | Not-determined |
| S7_extracted_77439 | Low-quality |
| S7_extracted_77452 | Not-determined |
| S7_extracted_77464 | Not-determined |
| S7_extracted_77469 | Not-determined |
| S7_extracted_77479 | Not-determined |

| | |
|---|---|
| S7_extracted_77492 | Not-determined |
| S7_extracted_77507 | Not-determined |
| S7_extracted_77518 | Not-determined |
| S7_extracted_77520 | Not-determined |
| S7_extracted_77530 | Not-determined |
| S7_extracted_77536 | Not-determined |
| S7_extracted_77568 | Not-determined |
| S7_extracted_77613 | Not-determined |
| S7_extracted_77614 | Not-determined |
| S7_extracted_77619 | Not-determined |
| S7_extracted_77628 | Not-determined |
| S7_extracted_77660 | Not-determined |
| S7_extracted_77673 | Not-determined |
| S7_extracted_77681 | Not-determined |
| S7_extracted_77698 | Not-determined |
| S7_extracted_77699 | Not-determined |
| S7_extracted_77704 | Not-determined |
| S7_extracted_77719 | Not-determined |
| S7_extracted_77725 | Not-determined |
| S7_extracted_77726 | Not-determined |
| S7_extracted_77738 | Not-determined |
| S7_extracted_77756 | Not-determined |
| S7_extracted_77762 | Low-quality |
| S7_extracted_77765 | Low-quality |
| S7_extracted_77777 | Not-determined |
| S7_extracted_77797 | Low-quality |
| S7_extracted_77819 | Low-quality |
| S7_extracted_77824 | Low-quality |
| S7_extracted_77829 | Not-determined |
| S7_extracted_77848 | Not-determined |
| S7_extracted_77864 | Not-determined |
| S7_extracted_77871 | Not-determined |
| S7_extracted_77875 | Not-determined |
| S7_extracted_77885 | Not-determined |
| S7_extracted_77888 | Not-determined |
| S7_extracted_77922 | Not-determined |
| S7_extracted_77932 | Not-determined |
| S7_extracted_77943 | Not-determined |
| S7_extracted_77948 | Not-determined |
| S7_extracted_77978 | Not-determined |
| S7_extracted_77998 | Low-quality |
| S7_extracted_78008 | Not-determined |
| S7_extracted_78040 | Low-quality |
| S7_extracted_78089 | Not-determined |
| S7_extracted_78093 | Not-determined |
| S7_extracted_78110 | Not-determined |
| S7_extracted_78121 | Not-determined |
| S7_extracted_78122 | Not-determined |
| S7_extracted_78148 | Not-determined |
| S7_extracted_78149 | Not-determined |
| S7_extracted_78155 | Not-determined |
| S7_extracted_78163 | Not-determined |
| S7_extracted_78177 | Not-determined |
| S7_extracted_78224 | Not-determined |
| S7_extracted_78238 | Not-determined |
| S7_extracted_78240 | Not-determined |
| S7_extracted_78248 | Not-determined |
| S7_extracted_78251 | Low-quality |
| S7_extracted_78277 | Not-determined |
| S7_extracted_78291 | Low-quality |
| S7_extracted_78306 | Low-quality |
| S7_extracted_78327 | Not-determined |
| S7_extracted_78336 | Not-determined |
| S7_extracted_78350 | Low-quality |
| S7_extracted_78354 | Not-determined |
| S7_extracted_78396 | Not-determined |
| S7_extracted_78398 | Not-determined |
| S7_extracted_78403 | Not-determined |
| S7_extracted_78409 | Not-determined |
| S7_extracted_78413 | Not-determined |
| S7_extracted_78441 | Not-determined |
| S7_extracted_78469 | Not-determined |
| S7_extracted_78479 | Not-determined |
| S7_extracted_78501 | Not-determined |
| S7_extracted_78504 | Not-determined |
| S7_extracted_78526 | Low-quality |
| S7_extracted_78545 | Low-quality |
| S7_extracted_78555 | Low-quality |
| S7_extracted_78559 | Not-determined |
| S7_extracted_78566 | Not-determined |
| S7_extracted_78587 | Not-determined |
| S7_extracted_78590 | Not-determined |
| S7_extracted_78598 | Low-quality |
| S7_extracted_78617 | Not-determined |
| S7_extracted_78625 | Low-quality |
| S7_extracted_78640 | Low-quality |
| S7_extracted_78644 | Not-determined |
| S7_extracted_78648 | Not-determined |
| S7_extracted_78649 | Not-determined |
| S7_extracted_78651 | Not-determined |
| S7_extracted_78656 | Not-determined |
| S7_extracted_78667 | Not-determined |
| S7_extracted_78683 | Not-determined |
| S7_extracted_78699 | Not-determined |
| S7_extracted_78726 | Not-determined |
| S7_extracted_78730 | Not-determined |
| S7_extracted_78739 | Not-determined |
| S7_extracted_78740 | Not-determined |
| S7_extracted_78762 | Not-determined |
| S7_extracted_78787 | Not-determined |
| S7_extracted_78805 | Not-determined |
| S7_extracted_78815 | Not-determined |
| S7_extracted_78817 | Not-determined |
| S7_extracted_78818 | Not-determined |
| S7_extracted_78829 | Not-determined |
| S7_extracted_78854 | Low-quality |
| S7_extracted_78861 | Not-determined |
| S7_extracted_78868 | Not-determined |
| S7_extracted_78874 | Not-determined |
| S7_extracted_78881 | Not-determined |
| S7_extracted_78893 | Low-quality |
| S7_extracted_78938 | Low-quality |
| S7_extracted_78947 | Not-determined |
| S7_extracted_78958 | Not-determined |
| S7_extracted_78963 | Not-determined |
| S7_extracted_78968 | Not-determined |
| S7_extracted_78976 | Low-quality |
| S7_extracted_78997 | Not-determined |
| S7_extracted_79018 | Not-determined |
| S7_extracted_79032 | Low-quality |
| S7_extracted_79034 | Not-determined |
| S7_extracted_79038 | Not-determined |
| S7_extracted_79056 | Low-quality |
| S7_extracted_79058 | Not-determined |
| S7_extracted_79072 | Not-determined |
| S7_extracted_79089 | Not-determined |
| S7_extracted_79100 | Not-determined |
| S7_extracted_79113 | Not-determined |
| S7_extracted_79117 | Not-determined |
| S7_extracted_79127 | Not-determined |
| S7_extracted_79138 | Not-determined |
| S7_extracted_79154 | Low-quality |
| S7_extracted_79165 | Not-determined |
| S7_extracted_79173 | Not-determined |
| S7_extracted_79178 | Low-quality |
| S7_extracted_79190 | Low-quality |
| S7_extracted_79199 | Not-determined |
| S7_extracted_79201 | Not-determined |
| S7_extracted_79207 | Not-determined |
| S7_extracted_79231 | Not-determined |
| S7_extracted_79234 | Low-quality |
| S7_extracted_79255 | Not-determined |
| S7_extracted_79285 | Not-determined |
| S7_extracted_79310 | Not-determined |
| S7_extracted_79311 | Not-determined |
| S7_extracted_79313 | Not-determined |
| S7_extracted_79325 | Low-quality |
| S7_extracted_79349 | Not-determined |
| S7_extracted_79351 | Not-determined |
| S7_extracted_79356 | Low-quality |
| S7_extracted_79358 | Low-quality |
| S7_extracted_79365 | Not-determined |
| S7_extracted_79367 | Low-quality |
| S7_extracted_79371 | Not-determined |
| S7_extracted_79375 | Not-determined |
| S7_extracted_79376 | Not-determined |
| S7_extracted_79379 | Not-determined |
| S7_extracted_79381 | Not-determined |
| S7_extracted_79414 | Not-determined |
| S7_extracted_79428 | Not-determined |
| S7_extracted_79437 | Low-quality |
| S7_extracted_79443 | Not-determined |
| S7_extracted_79449 | Not-determined |
| S7_extracted_79471 | Not-determined |
| S7_extracted_79487 | Not-determined |
| S7_extracted_79538 | Low-quality |
| S7_extracted_79560 | Not-determined |
| S7_extracted_79561 | Low-quality |
| S7_extracted_79638 | Not-determined |
| S7_extracted_79674 | Not-determined |
| S7_extracted_79680 | Not-determined |
| S7_extracted_79712 | Not-determined |
| S7_extracted_79726 | Low-quality |
| S7_extracted_79732 | Not-determined |
| S7_extracted_79739 | Not-determined |
| S7_extracted_79744 | Not-determined |
| S7_extracted_79753 | Not-determined |
| S7_extracted_79769 | Low-quality |
| S7_extracted_79789 | Not-determined |
| S7_extracted_79813 | Not-determined |
| S7_extracted_79829 | Not-determined |
| S7_extracted_79830 | Not-determined |
| S7_extracted_79863 | Not-determined |
| S7_extracted_79870 | Not-determined |
| S7_extracted_79888 | Not-determined |
| S7_extracted_79890 | Not-determined |
| S7_extracted_79911 | Not-determined |
| S7_extracted_79920 | Not-determined |
| S7_extracted_79929 | Not-determined |
| S7_extracted_79930 | Not-determined |
| S7_extracted_79934 | Not-determined |
| S7_extracted_79938 | Not-determined |
| S7_extracted_79944 | Not-determined |
| S7_extracted_79967 | Low-quality |
| S7_extracted_79974 | Not-determined |
| S7_extracted_79994 | Not-determined |
| S7_extracted_80008 | Not-determined |
| S7_extracted_80011 | Not-determined |
| S7_extracted_80020 | Low-quality |
| S7_extracted_80030 | Not-determined |
| S7_extracted_80039 | Not-determined |
| S7_extracted_80054 | Not-determined |
| S7_extracted_80067 | Not-determined |
| S7_extracted_80077 | Not-determined |
| S7_extracted_80083 | Not-determined |
| S7_extracted_80089 | Not-determined |
| S7_extracted_80090 | Low-quality |
| S7_extracted_80106 | Not-determined |
| S7_extracted_80109 | Not-determined |
| S7_extracted_80117 | Not-determined |
| S7_extracted_80129 | Not-determined |
| S7_extracted_80149 | Not-determined |
| S7_extracted_80163 | Not-determined |
| S7_extracted_80186 | Not-determined |
| S7_extracted_80201 | Low-quality |
| S7_extracted_80206 | Not-determined |
| S7_extracted_80207 | Low-quality |
| S7_extracted_80219 | Not-determined |
| S7_extracted_80258 | Not-determined |
| S7_extracted_80281 | Low-quality |
| S7_extracted_80286 | Low-quality |
| S7_extracted_80310 | Not-determined |
| S7_extracted_80318 | Not-determined |
| S7_extracted_80323 | Not-determined |
| S7_extracted_80326 | Not-determined |
| S7_extracted_80337 | Not-determined |
| S7_extracted_80343 | Not-determined |
| S7_extracted_80365 | Not-determined |
| S7_extracted_80366 | Not-determined |
| S7_extracted_80372 | Not-determined |
| S7_extracted_80391 | Not-determined |
| S7_extracted_80394 | Not-determined |
| S7_extracted_80405 | Not-determined |
| S7_extracted_80422 | Not-determined |
| S7_extracted_80435 | Not-determined |
| S7_extracted_80440 | Not-determined |
| S7_extracted_80444 | Not-determined |
| S7_extracted_80458 | Not-determined |
| S7_extracted_80469 | Not-determined |
| S7_extracted_80472 | Not-determined |
| S7_extracted_80490 | Not-determined |
| S7_extracted_80495 | Not-determined |
| S7_extracted_80496 | Not-determined |
| S7_extracted_80519 | Low-quality |
| S7_extracted_80520 | Not-determined |
| S7_extracted_80551 | Not-determined |
| S7_extracted_80555 | Not-determined |
| S7_extracted_80593 | Not-determined |
| S7_extracted_80598 | Not-determined |
| S7_extracted_80631 | Not-determined |
| S7_extracted_80634 | Not-determined |
| S7_extracted_80695 | Low-quality |
| S7_extracted_80696 | Not-determined |
| S7_extracted_80712 | Not-determined |
| S7_extracted_80718 | Not-determined |
| S7_extracted_80725 | Low-quality |
| S7_extracted_80754 | Not-determined |
| S7_extracted_80768 | Not-determined |
| S7_extracted_80788 | Low-quality |
| S7_extracted_80792 | Not-determined |
| S7_extracted_80811 | Not-determined |
| S7_extracted_80842 | Not-determined |
| S7_extracted_80845 | Low-quality |
| S7_extracted_80859 | Low-quality |
| S7_extracted_80876 | Not-determined |
| S7_extracted_80885 | Not-determined |
| S7_extracted_80891 | Not-determined |
| S7_extracted_80931 | Not-determined |
| S7_extracted_80940 | Low-quality |
| S7_extracted_80953 | Low-quality |
| S7_extracted_80967 | Not-determined |
| S7_extracted_80979 | Not-determined |
| S7_extracted_80992 | Not-determined |
| S7_extracted_81017 | Low-quality |
| S7_extracted_81043 | Not-determined |
| S7_extracted_81046 | Not-determined |
| S7_extracted_81053 | Not-determined |
| S7_extracted_81092 | Not-determined |
| S7_extracted_81116 | Not-determined |
| S7_extracted_81137 | Not-determined |
| S7_extracted_81170 | Not-determined |
| S7_extracted_81200 | Not-determined |
| S7_extracted_81209 | Not-determined |
| S7_extracted_81213 | Not-determined |
| S7_extracted_81219 | Not-determined |
| S7_extracted_81241 | Low-quality |
| S7_extracted_81254 | Not-determined |
| S7_extracted_81265 | Not-determined |
| S7_extracted_81276 | Low-quality |
| S7_extracted_81282 | Not-determined |
| S7_extracted_81298 | Not-determined |
| S7_extracted_81300 | Not-determined |
| S7_extracted_81306 | Not-determined |
| S7_extracted_81310 | Not-determined |
| S7_extracted_81327 | Low-quality |
| S7_extracted_81335 | Not-determined |
| S7_extracted_81349 | Low-quality |
| S7_extracted_81351 | Not-determined |
| S7_extracted_81357 | Not-determined |
| S7_extracted_81359 | Not-determined |
| S7_extracted_81368 | Not-determined |
| S7_extracted_81374 | Not-determined |
| S7_extracted_81388 | Not-determined |
| S7_extracted_81392 | Not-determined |
| S7_extracted_81398 | Not-determined |
| S7_extracted_81412 | Low-quality |
| S7_extracted_81425 | Not-determined |
| S7_extracted_81431 | Low-quality |
| S7_extracted_81435 | Not-determined |
| S7_extracted_81443 | Not-determined |
| S7_extracted_81457 | Not-determined |
| S7_extracted_81458 | Low-quality |
| S7_extracted_81463 | Not-determined |
| S7_extracted_81484 | Not-determined |
| S7_extracted_81524 | Not-determined |
| S7_extracted_81532 | Low-quality |
| S7_extracted_81534 | Not-determined |
| S7_extracted_81535 | Low-quality |
| S7_extracted_81579 | Not-determined |
| S7_extracted_81588 | Low-quality |
| S7_extracted_81595 | Not-determined |
| S7_extracted_81602 | Not-determined |
| S7_extracted_81623 | Not-determined |
| S7_extracted_81635 | Low-quality |
| S7_extracted_81666 | Not-determined |
| S7_extracted_81684 | Not-determined |
| S7_extracted_81696 | Not-determined |
| S7_extracted_81699 | Low-quality |
| S7_extracted_81700 | Not-determined |
| S7_extracted_81710 | Not-determined |
| S7_extracted_81715 | Low-quality |
| S7_extracted_81733 | Not-determined |
| S7_extracted_81734 | Not-determined |
| S7_extracted_81738 | Not-determined |
| S7_extracted_81790 | Low-quality |
| S7_extracted_81799 | Not-determined |
| S7_extracted_81801 | Low-quality |
| S7_extracted_81824 | Not-determined |
| S7_extracted_81836 | Not-determined |
| S7_extracted_81838 | Not-determined |
| S7_extracted_81840 | Low-quality |
| S7_extracted_81843 | Low-quality |
| S7_extracted_81853 | Not-determined |
| S7_extracted_81864 | Low-quality |
| S7_extracted_81868 | Low-quality |
| S7_extracted_81886 | Not-determined |
| S7_extracted_81947 | Not-determined |
| S7_extracted_81949 | Not-determined |
| S7_extracted_81959 | Not-determined |
| S7_extracted_81969 | Not-determined |
| S7_extracted_81970 | Low-quality |
| S7_extracted_81971 | Not-determined |
| S7_extracted_81972 | Not-determined |
| S7_extracted_81976 | Low-quality |
| S7_extracted_82016 | Not-determined |
| S7_extracted_82027 | Not-determined |
| S7_extracted_82036 | Not-determined |
| S7_extracted_82040 | Not-determined |
| S7_extracted_82045 | Not-determined |
| S7_extracted_82046 | Low-quality |
| S7_extracted_82059 | Not-determined |
| S7_extracted_82065 | Not-determined |
| S7_extracted_82072 | Not-determined |
| S7_extracted_82073 | Low-quality |
| S7_extracted_82079 | Not-determined |
| S7_extracted_82088 | Not-determined |
| S7_extracted_82096 | Not-determined |
| S7_extracted_82100 | Not-determined |
| S7_extracted_82107 | Low-quality |
| S7_extracted_82111 | Not-determined |
| S7_extracted_82116 | Not-determined |
| S7_extracted_82128 | Not-determined |
| S7_extracted_82163 | Not-determined |
| S7_extracted_82179 | Not-determined |
| S7_extracted_82188 | Not-determined |
| S7_extracted_82216 | Not-determined |
| S7_extracted_82218 | Not-determined |
| S7_extracted_82223 | Not-determined |
| S7_extracted_82231 | Not-determined |
| S7_extracted_82232 | Not-determined |
| S7_extracted_82233 | Not-determined |
| S7_extracted_82234 | Not-determined |
| S7_extracted_82235 | Not-determined |
| S7_extracted_82236 | Not-determined |
| S7_extracted_82242 | Not-determined |
| S7_extracted_82247 | Not-determined |
| S7_extracted_82259 | Not-determined |
| S7_extracted_82270 | Not-determined |
| S7_extracted_82277 | Not-determined |
| S7_extracted_82281 | Not-determined |
| S7_extracted_82285 | Low-quality |
| S7_extracted_82289 | Not-determined |
| S7_extracted_82297 | Not-determined |
| S7_extracted_82307 | Not-determined |
| S7_extracted_82330 | Not-determined |
| S7_extracted_82335 | Low-quality |
| S7_extracted_82354 | Not-determined |
| S7_extracted_82356 | Not-determined |
| S7_extracted_82359 | Not-determined |
| S7_extracted_82367 | Not-determined |
| S7_extracted_82375 | Not-determined |
| S7_extracted_82377 | Not-determined |
| S7_extracted_82381 | Not-determined |
| S7_extracted_82386 | Low-quality |
| S7_extracted_82388 | Not-determined |
| S7_extracted_82391 | Not-determined |
| S7_extracted_82418 | Low-quality |
| S7_extracted_82437 | Not-determined |
| S7_extracted_82451 | Not-determined |
| S7_extracted_82453 | Not-determined |
| S7_extracted_82458 | Not-determined |
| S7_extracted_82459 | Not-determined |
| S7_extracted_82460 | Low-quality |
| S7_extracted_82494 | Not-determined |
| S7_extracted_82555 | Not-determined |
| S7_extracted_82562 | Low-quality |
| S7_extracted_82577 | Low-quality |
| S7_extracted_82578 | Low-quality |
| S7_extracted_82581 | Not-determined |
| S7_extracted_82584 | Not-determined |
| S7_extracted_82589 | Low-quality |
| S7_extracted_82590 | Not-determined |
| S7_extracted_82594 | Not-determined |
| S7_extracted_82607 | Not-determined |
| S7_extracted_82635 | Not-determined |
| S7_extracted_82679 | Not-determined |
| S7_extracted_82717 | Low-quality |
| S7_extracted_82724 | Not-determined |
| S7_extracted_82762 | Not-determined |
| S7_extracted_82771 | Not-determined |
| S7_extracted_82774 | Not-determined |
| S7_extracted_82784 | Not-determined |
| S7_extracted_82790 | Not-determined |
| S7_extracted_82797 | Not-determined |
| S1_extracted_k141_17085 | Medium-quality |
| S7_extracted_82810 | Not-determined |
| S7_extracted_82834 | Not-determined |
| S7_extracted_82835 | Not-determined |
| S7_extracted_82843 | Not-determined |
| S7_extracted_82862 | Not-determined |
| S7_extracted_82882 | Not-determined |
| S7_extracted_82899 | Not-determined |
| S7_extracted_82906 | Not-determined |
| S7_extracted_82914 | Not-determined |
| S7_extracted_82920 | Low-quality |
| S7_extracted_82930 | Low-quality |
| S7_extracted_82948 | Not-determined |
| S7_extracted_82950 | Not-determined |
| S7_extracted_82968 | Not-determined |
| S7_extracted_82971 | Low-quality |
| S7_extracted_82972 | Not-determined |
| S7_extracted_82979 | Not-determined |
| S7_extracted_82990 | Not-determined |
| S7_extracted_82991 | Not-determined |
| S7_extracted_83002 | Not-determined |
| S7_extracted_83012 | Not-determined |
| S7_extracted_83014 | Not-determined |
| S7_extracted_83015 | Not-determined |
| S7_extracted_83025 | Not-determined |
| S7_extracted_83033 | Low-quality |
| S7_extracted_83038 | Not-determined |
| S7_extracted_83046 | Not-determined |
| S7_extracted_83086 | Not-determined |
| S7_extracted_83087 | Not-determined |
| S7_extracted_83095 | Not-determined |
| S7_extracted_83104 | Not-determined |
| S7_extracted_83119 | Not-determined |
| S7_extracted_83124 | Not-determined |
| S7_extracted_83167 | Low-quality |
| S7_extracted_83169 | Not-determined |
| S7_extracted_83183 | Not-determined |
| S7_extracted_83186 | Not-determined |
| S7_extracted_83194 | Low-quality |
| S7_extracted_83221 | Not-determined |
| S7_extracted_83224 | Low-quality |
| S7_extracted_83231 | Low-quality |
| S7_extracted_83237 | Low-quality |
| S7_extracted_83241 | Not-determined |
| S7_extracted_83254 | Not-determined |
| S7_extracted_83308 | Not-determined |
| S7_extracted_83315 | Not-determined |
| S7_extracted_83319 | Not-determined |
| S7_extracted_83329 | Not-determined |
| S7_extracted_83331 | Not-determined |
| S7_extracted_83343 | Not-determined |
| S7_extracted_83380 | Not-determined |
| S7_extracted_83383 | Low-quality |
| S7_extracted_83396 | Not-determined |
| S7_extracted_83399 | Not-determined |
| S7_extracted_83406 | Not-determined |
| S7_extracted_83417 | Not-determined |
| S7_extracted_83449 | Not-determined |
| S7_extracted_83454 | Not-determined |
| S7_extracted_83458 | Not-determined |
| S7_extracted_83463 | Not-determined |
| S7_extracted_83466 | Not-determined |
| S7_extracted_83478 | Not-determined |
| S7_extracted_83480 | Not-determined |
| S7_extracted_83490 | Not-determined |
| S7_extracted_83492 | Low-quality |
| S7_extracted_83495 | Not-determined |
| S7_extracted_83522 | Not-determined |
| S7_extracted_83531 | Not-determined |
| S7_extracted_83539 | Not-determined |
| S7_extracted_83543 | Not-determined |
| S7_extracted_83546 | Not-determined |
| S7_extracted_83562 | Not-determined |
| S7_extracted_83600 | Not-determined |
| S7_extracted_83606 | Low-quality |
| S13_extracted_66508 | High-quality |
| S7_extracted_83632 | Not-determined |
| S7_extracted_83642 | Not-determined |
| S7_extracted_83650 | Not-determined |
| S7_extracted_83653 | Low-quality |
| S7_extracted_83654 | Not-determined |
| S7_extracted_83656 | Not-determined |
| S7_extracted_83664 | Not-determined |
| S7_extracted_83704 | Not-determined |
| S7_extracted_83719 | Not-determined |
| S7_extracted_83729 | Not-determined |
| S7_extracted_83746 | Not-determined |
| S7_extracted_83750 | Not-determined |
| S7_extracted_83780 | Not-determined |
| S7_extracted_83782 | Not-determined |
| S7_extracted_83808 | Not-determined |
| S7_extracted_83814 | Not-determined |
| S7_extracted_83834 | Low-quality |
| S7_extracted_83836 | Low-quality |
| S7_extracted_83847 | Not-determined |
| S7_extracted_83852 | Not-determined |
| S7_extracted_83858 | Not-determined |
| S7_extracted_83875 | Not-determined |
| S7_extracted_83876 | Not-determined |
| S7_extracted_83881 | Not-determined |
| S7_extracted_83883 | Not-determined |
| S7_extracted_83895 | Not-determined |
| S7_extracted_83905 | Not-determined |
| S7_extracted_83912 | Not-determined |
| S7_extracted_83917 | Not-determined |
| S7_extracted_83922 | Not-determined |
| S7_extracted_83926 | Low-quality |
| S7_extracted_83927 | Not-determined |
| S7_extracted_83990 | Not-determined |
| S7_extracted_84015 | Not-determined |
| S7_extracted_84038 | Not-determined |
| S7_extracted_84039 | Not-determined |
| S7_extracted_84048 | Not-determined |
| S7_extracted_84055 | Low-quality |
| S7_extracted_84065 | Not-determined |
| S7_extracted_84085 | Not-determined |
| S7_extracted_84108 | Not-determined |
| S7_extracted_84111 | Not-determined |
| S7_extracted_84118 | Low-quality |
| S7_extracted_84126 | Not-determined |
| S7_extracted_84128 | Not-determined |
| S7_extracted_84134 | Not-determined |
| S7_extracted_84158 | Low-quality |
| S7_extracted_84161 | Not-determined |
| S7_extracted_84167 | Not-determined |
| S7_extracted_84168 | Not-determined |
| S7_extracted_84169 | Not-determined |
| S7_extracted_84186 | Low-quality |
| S7_extracted_84190 | Low-quality |
| S7_extracted_84235 | Low-quality |
| S7_extracted_84240 | Not-determined |
| S7_extracted_84273 | Not-determined |
| S7_extracted_84298 | Low-quality |
| S7_extracted_84302 | Not-determined |
| S7_extracted_84314 | Not-determined |
| S7_extracted_84321 | Not-determined |
| S7_extracted_84323 | Low-quality |
| S7_extracted_84328 | Not-determined |
| S7_extracted_84334 | Low-quality |
| S7_extracted_84360 | Not-determined |
| S7_extracted_84368 | Not-determined |
| S7_extracted_84384 | Not-determined |
| S7_extracted_84391 | Not-determined |
| S7_extracted_84400 | Not-determined |
| S7_extracted_84431 | Not-determined |
| S7_extracted_84432 | Not-determined |
| S7_extracted_84437 | Not-determined |
| S7_extracted_84471 | Low-quality |
| S7_extracted_84472 | Not-determined |
| S7_extracted_84502 | Low-quality |
| S7_extracted_84514 | Not-determined |
| S7_extracted_84533 | Not-determined |
| S7_extracted_84553 | Not-determined |
| S7_extracted_84554 | Not-determined |
| S7_extracted_84557 | Not-determined |

| | |
|---|---|
| S7_extracted_84563 | Not-determined |
| S7_extracted_84568 | Not-determined |
| S7_extracted_84570 | Low-quality |
| S7_extracted_84575 | Not-determined |
| S7_extracted_84578 | Not-determined |
| S7_extracted_84584 | Low-quality |
| S7_extracted_84597 | Low-quality |
| S7_extracted_84605 | Not-determined |
| S7_extracted_84610 | Not-determined |
| S7_extracted_84623 | Low-quality |
| S7_extracted_84633 | Not-determined |
| S7_extracted_84641 | Not-determined |
| S7_extracted_84642 | Not-determined |
| S7_extracted_84652 | Not-determined |
| S7_extracted_84655 | Not-determined |
| S7_extracted_84656 | Not-determined |
| S7_extracted_84662 | Not-determined |
| S7_extracted_84686 | Not-determined |
| S7_extracted_84689 | Not-determined |
| S7_extracted_84714 | Not-determined |
| S7_extracted_84757 | Not-determined |
| S7_extracted_84774 | Not-determined |
| S7_extracted_84780 | Not-determined |
| S7_extracted_84782 | Not-determined |
| S7_extracted_84786 | Not-determined |
| S7_extracted_84792 | Not-determined |
| S7_extracted_84801 | Not-determined |
| S7_extracted_84805 | Not-determined |
| S7_extracted_84816 | Not-determined |
| S7_extracted_84849 | Not-determined |
| S7_extracted_84854 | Not-determined |
| S7_extracted_84865 | Not-determined |
| S7_extracted_84872 | Not-determined |
| S7_extracted_84878 | Not-determined |
| S7_extracted_84894 | Not-determined |
| S7_extracted_84897 | Not-determined |
| S7_extracted_84906 | Not-determined |
| S7_extracted_84924 | Not-determined |
| S7_extracted_84930 | Not-determined |
| S7_extracted_84933 | Not-determined |
| S7_extracted_84940 | Not-determined |
| S7_extracted_84949 | Low-quality |
| S7_extracted_84953 | Not-determined |
| S7_extracted_84954 | Low-quality |
| S7_extracted_84968 | Low-quality |
| S9_vs_k141_90856\|\|full | High-quality |
| S7_extracted_85002 | Not-determined |
| S7_extracted_85027 | Low-quality |
| S7_extracted_85028 | Not-determined |
| S7_extracted_85031 | Not-determined |
| S7_extracted_85033 | Not-determined |
| S7_extracted_85043 | Not-determined |
| S7_extracted_85060 | Not-determined |
| S7_extracted_85062 | Not-determined |
| S7_extracted_85081 | Low-quality |
| S7_extracted_85090 | Not-determined |
| S7_extracted_85112 | Not-determined |
| S7_extracted_85118 | Not-determined |
| S7_extracted_85141 | Not-determined |
| S7_extracted_85160 | Not-determined |
| S7_extracted_85178 | Not-determined |
| S7_extracted_85182 | Not-determined |
| S7_extracted_85195 | Low-quality |
| S7_extracted_85199 | Not-determined |
| S7_extracted_85227 | Low-quality |
| S7_extracted_85229 | Not-determined |
| S7_extracted_85245 | Not-determined |
| S7_extracted_85249 | Not-determined |
| S7_extracted_85252 | Not-determined |
| S7_extracted_85272 | Not-determined |
| S7_extracted_85324 | Not-determined |
| S7_extracted_85340 | Low-quality |
| S7_extracted_85341 | Low-quality |
| S7_extracted_85367 | Not-determined |
| S7_extracted_85383 | Not-determined |
| S7_extracted_85390 | Not-determined |
| S7_extracted_85411 | Not-determined |
| S7_extracted_85421 | Not-determined |
| S7_extracted_85441 | Not-determined |
| S7_extracted_85453 | Not-determined |
| S7_extracted_85459 | Not-determined |
| S7_extracted_85466 | Low-quality |
| S7_extracted_85468 | Not-determined |
| S7_extracted_85472 | Not-determined |
| S7_extracted_85492 | Not-determined |
| S7_extracted_85497 | Low-quality |
| S7_extracted_85501 | Low-quality |
| S7_extracted_85520 | Not-determined |
| S7_extracted_85525 | Not-determined |
| S7_extracted_85526 | Not-determined |
| S7_extracted_85545 | Not-determined |
| S7_extracted_85551 | Not-determined |
| S7_extracted_85562 | Not-determined |
| S7_extracted_85571 | Not-determined |
| S7_extracted_85572 | Not-determined |
| S7_extracted_85583 | Not-determined |
| S7_extracted_85603 | Not-determined |
| S7_extracted_85623 | Not-determined |
| S7_extracted_85625 | Not-determined |
| S7_extracted_85663 | Not-determined |
| S7_extracted_85675 | Not-determined |
| S7_extracted_85678 | Not-determined |
| S7_extracted_85686 | Not-determined |
| S7_extracted_85687 | Not-determined |
| S7_extracted_85699 | Not-determined |
| S7_extracted_85700 | Not-determined |
| S7_extracted_85728 | Not-determined |
| S7_extracted_85778 | Not-determined |
| S7_extracted_85788 | Low-quality |
| S7_extracted_85789 | Not-determined |
| S7_extracted_85801 | Not-determined |
| S7_extracted_85802 | Low-quality |
| S7_extracted_85824 | Not-determined |
| S7_extracted_85844 | Low-quality |
| S7_extracted_85860 | Not-determined |
| S7_extracted_85861 | Not-determined |
| S7_extracted_85866 | Low-quality |
| S7_extracted_85889 | Not-determined |
| S7_extracted_85910 | Not-determined |
| S7_extracted_85920 | Not-determined |
| S7_extracted_85921 | Not-determined |
| S7_extracted_85927 | Not-determined |
| S7_extracted_85948 | Not-determined |
| S7_extracted_85960 | Not-determined |
| S7_extracted_85961 | Not-determined |
| S7_extracted_85993 | Not-determined |
| S7_extracted_85994 | Not-determined |
| S7_extracted_85997 | Not-determined |
| S7_extracted_86010 | Not-determined |
| S7_extracted_86015 | Not-determined |
| S7_extracted_86034 | Not-determined |
| S7_extracted_86050 | Not-determined |
| S7_extracted_86136 | Not-determined |
| S7_extracted_86146 | Low-quality |
| S7_extracted_86165 | Not-determined |
| S7_extracted_86182 | Not-determined |
| S7_extracted_86185 | Low-quality |
| S7_extracted_86189 | Not-determined |
| S7_extracted_86218 | Not-determined |
| S7_extracted_86226 | Not-determined |
| S7_extracted_86227 | Not-determined |
| S7_extracted_86229 | Not-determined |
| S7_extracted_86234 | Not-determined |
| S7_extracted_86241 | Not-determined |
| S7_extracted_86242 | Low-quality |
| S7_extracted_86252 | Low-quality |
| S7_extracted_86260 | Not-determined |
| S7_extracted_86283 | Not-determined |
| S7_extracted_86287 | Not-determined |
| S7_extracted_86306 | Not-determined |
| S7_extracted_86321 | Not-determined |
| S7_extracted_86337 | Not-determined |
| S7_extracted_86338 | Not-determined |
| S7_extracted_86339 | Not-determined |
| S7_extracted_86345 | Not-determined |
| S7_extracted_86350 | Not-determined |
| S7_extracted_86355 | Not-determined |
| S7_extracted_86378 | Not-determined |
| S7_extracted_86386 | Not-determined |
| S7_extracted_86406 | Low-quality |
| S7_extracted_86410 | Low-quality |
| S7_extracted_86415 | Not-determined |
| S7_extracted_86424 | Not-determined |
| S7_extracted_86433 | Not-determined |
| S7_extracted_86437 | Low-quality |
| S7_extracted_86444 | Not-determined |
| S7_extracted_86448 | Not-determined |
| S7_extracted_86450 | Low-quality |
| S7_extracted_86462 | Not-determined |
| S7_extracted_86464 | Low-quality |
| S7_extracted_86492 | Not-determined |
| S7_extracted_86517 | Not-determined |
| S7_extracted_86545 | Not-determined |
| S7_extracted_86547 | Low-quality |
| S7_extracted_86578 | Not-determined |
| S7_extracted_86589 | Low-quality |
| S7_extracted_86597 | Not-determined |
| S7_extracted_86598 | Not-determined |
| S7_extracted_86604 | Low-quality |
| S7_extracted_86605 | Not-determined |
| S7_extracted_86615 | Not-determined |
| S7_extracted_86630 | Not-determined |
| S7_extracted_86660 | Not-determined |
| S7_extracted_86666 | Not-determined |
| S7_extracted_86674 | Not-determined |
| S7_extracted_86679 | Low-quality |
| S7_extracted_86702 | Not-determined |
| S7_extracted_86711 | Not-determined |
| S7_extracted_86715 | Not-determined |
| S7_extracted_86721 | Not-determined |
| S7_extracted_86729 | Low-quality |
| S7_extracted_86743 | Not-determined |
| S7_extracted_86750 | Not-determined |
| S7_extracted_86762 | Not-determined |
| S7_extracted_86775 | Not-determined |
| S7_extracted_86779 | Low-quality |
| S7_extracted_86783 | Not-determined |
| S7_extracted_86803 | Not-determined |
| S7_extracted_86817 | Not-determined |
| S7_extracted_86844 | Low-quality |
| S7_extracted_86864 | Not-determined |
| S7_extracted_86872 | Not-determined |
| S7_extracted_86879 | Not-determined |
| S7_extracted_86891 | Low-quality |
| S7_extracted_86909 | Low-quality |
| S7_extracted_86925 | Not-determined |
| S7_extracted_86926 | Low-quality |
| S7_extracted_86927 | Not-determined |
| S7_extracted_86940 | Low-quality |
| S7_extracted_86977 | Not-determined |
| S7_extracted_86993 | Not-determined |
| S7_extracted_86997 | Not-determined |
| S7_extracted_87016 | Not-determined |
| S7_extracted_87020 | Not-determined |
| S7_extracted_87028 | Not-determined |
| S7_extracted_87035 | Low-quality |
| S7_extracted_87051 | Not-determined |
| S7_extracted_87078 | Not-determined |
| S7_extracted_87085 | Not-determined |
| S7_extracted_87092 | Not-determined |
| S7_extracted_87121 | Not-determined |
| S7_extracted_87130 | Not-determined |
| S7_extracted_87137 | Not-determined |
| S7_extracted_87143 | Not-determined |
| S7_extracted_87148 | Not-determined |
| S7_extracted_87156 | Not-determined |
| S7_extracted_87182 | Low-quality |
| S7_extracted_87187 | Not-determined |
| S7_extracted_87191 | Not-determined |
| S7_extracted_87202 | Low-quality |
| S7_extracted_87209 | Low-quality |
| S7_extracted_87229 | Low-quality |
| S7_extracted_87250 | Not-determined |
| S7_extracted_87258 | Low-quality |
| S7_extracted_87276 | Not-determined |
| S7_extracted_87307 | Not-determined |
| S7_extracted_87308 | Not-determined |
| S7_extracted_87310 | Not-determined |
| S7_extracted_87338 | Not-determined |
| S7_extracted_87342 | Not-determined |
| S7_extracted_87354 | Not-determined |
| S7_extracted_87356 | Not-determined |
| S7_extracted_87366 | Not-determined |
| S7_extracted_87375 | Not-determined |
| S7_extracted_87386 | Low-quality |
| S7_extracted_87421 | Not-determined |
| S7_extracted_87435 | Not-determined |
| S7_extracted_87458 | Not-determined |
| S7_extracted_87464 | Low-quality |
| S7_extracted_87481 | Low-quality |
| S7_extracted_87491 | Not-determined |
| S7_extracted_87493 | Not-determined |
| S7_extracted_87529 | Not-determined |
| S7_extracted_87534 | Not-determined |
| S7_extracted_87535 | Not-determined |
| S7_extracted_87536 | Not-determined |
| S7_extracted_87538 | Not-determined |
| S7_extracted_87541 | Not-determined |
| S7_extracted_87546 | Low-quality |
| S7_extracted_87547 | Not-determined |
| S7_extracted_87551 | Not-determined |
| S7_extracted_87556 | Not-determined |
| S7_extracted_87563 | Low-quality |
| S7_extracted_87566 | Not-determined |
| S7_extracted_87569 | Not-determined |
| S7_extracted_87611 | Low-quality |
| S7_extracted_87626 | Not-determined |
| S7_extracted_87630 | Not-determined |
| S7_extracted_87633 | Not-determined |
| S7_extracted_87634 | Not-determined |
| S7_extracted_87636 | Not-determined |
| S7_extracted_87637 | Not-determined |
| S7_extracted_87654 | Low-quality |
| S7_extracted_87656 | Not-determined |
| S7_extracted_87671 | Not-determined |
| S7_extracted_87672 | Low-quality |
| S7_extracted_87680 | Not-determined |
| S7_extracted_87686 | Not-determined |
| S7_extracted_87691 | Not-determined |
| S7_extracted_87692 | Not-determined |
| S7_extracted_87694 | Not-determined |
| S7_extracted_87707 | Not-determined |
| S7_extracted_87718 | Not-determined |
| S7_extracted_87720 | Not-determined |
| S7_extracted_87744 | Not-determined |
| S7_extracted_87811 | Not-determined |
| S7_extracted_87816 | Not-determined |
| S7_extracted_87821 | Not-determined |
| S7_extracted_87823 | Not-determined |
| S7_extracted_87826 | Not-determined |
| S7_extracted_87833 | Not-determined |
| S7_extracted_87846 | Not-determined |
| S7_extracted_87853 | Not-determined |
| S7_extracted_87878 | Not-determined |
| S7_extracted_87884 | Not-determined |
| S7_extracted_87888 | Not-determined |
| S7_extracted_87889 | Not-determined |
| S7_extracted_87897 | Not-determined |
| S7_extracted_87927 | Low-quality |
| S7_extracted_87942 | Not-determined |
| S7_extracted_87945 | Not-determined |
| S7_extracted_87950 | Not-determined |
| S7_extracted_87957 | Not-determined |
| S7_extracted_87978 | Not-determined |
| S7_extracted_87988 | Not-determined |
| S7_extracted_88017 | Not-determined |
| S7_extracted_88020 | Not-determined |
| S7_extracted_88027 | Not-determined |
| S7_extracted_88041 | Not-determined |
| S7_extracted_88047 | Not-determined |
| S7_extracted_88083 | Not-determined |
| S7_extracted_88092 | Not-determined |
| S7_extracted_88095 | Not-determined |
| S7_extracted_88099 | Not-determined |
| S7_extracted_88105 | Not-determined |
| S7_extracted_88112 | Low-quality |
| S7_extracted_88115 | Not-determined |
| S7_extracted_88119 | Low-quality |
| S7_extracted_88127 | Not-determined |
| S7_extracted_88176 | Not-determined |
| S7_extracted_88184 | Not-determined |
| S7_extracted_88189 | Low-quality |
| S7_extracted_88211 | Not-determined |
| S7_extracted_88212 | Not-determined |
| S7_extracted_88222 | Not-determined |
| S7_extracted_88227 | Low-quality |
| S7_extracted_88229 | Not-determined |
| S7_extracted_88248 | Low-quality |
| S7_extracted_88249 | Not-determined |
| S7_extracted_88251 | Low-quality |
| S7_extracted_88256 | Not-determined |
| S7_extracted_88260 | Low-quality |
| S7_extracted_88269 | Low-quality |
| S7_extracted_88271 | Low-quality |
| S7_extracted_88279 | Not-determined |
| S7_extracted_88291 | Not-determined |
| S7_extracted_88355 | Not-determined |
| S7_extracted_88372 | Not-determined |
| S7_extracted_88385 | Not-determined |
| S7_extracted_88387 | Not-determined |
| S7_extracted_88390 | Not-determined |
| S7_extracted_88419 | Not-determined |
| S7_extracted_88429 | Not-determined |
| S7_extracted_88438 | Low-quality |
| S7_extracted_88442 | Not-determined |
| S7_extracted_88444 | Not-determined |
| S7_extracted_88455 | Low-quality |
| S7_extracted_88464 | Not-determined |
| S7_extracted_88475 | Not-determined |
| S7_extracted_88478 | Not-determined |
| S7_extracted_88495 | Not-determined |
| S7_extracted_88498 | Low-quality |
| S7_extracted_88500 | Not-determined |
| S7_extracted_88501 | Not-determined |
| S7_extracted_88512 | Not-determined |
| S7_extracted_88517 | Not-determined |
| S7_extracted_88529 | Not-determined |
| S7_extracted_88530 | Not-determined |
| S7_extracted_88559 | Not-determined |
| S7_extracted_88564 | Not-determined |
| S7_extracted_88573 | Not-determined |
| S7_extracted_88576 | Not-determined |
| S7_extracted_88586 | Not-determined |
| S7_extracted_88600 | Not-determined |
| S7_extracted_88604 | Not-determined |
| S7_extracted_88611 | Not-determined |
| S7_extracted_88617 | Not-determined |
| S7_extracted_88630 | Not-determined |
| S7_extracted_88664 | Not-determined |
| S7_extracted_88691 | Not-determined |
| S7_extracted_88701 | Not-determined |
| S7_extracted_88729 | Not-determined |
| S7_extracted_88749 | Not-determined |
| S7_extracted_88765 | Low-quality |
| S7_extracted_88795 | Not-determined |
| S7_extracted_88829 | Not-determined |
| S7_extracted_88838 | Not-determined |
| S7_extracted_88846 | Not-determined |
| S7_extracted_88851 | Not-determined |
| S7_extracted_88858 | Not-determined |
| S7_extracted_88877 | Not-determined |
| S7_extracted_88880 | Not-determined |
| S7_extracted_88902 | Not-determined |
| S7_extracted_88919 | Not-determined |
| S7_extracted_88926 | Not-determined |
| S7_extracted_88939 | Low-quality |
| S7_extracted_88943 | Low-quality |
| S7_extracted_88952 | Not-determined |
| S7_extracted_88989 | Not-determined |
| S7_extracted_89018 | Not-determined |
| S7_extracted_89025 | Not-determined |
| S7_extracted_89042 | Not-determined |
| S7_extracted_89043 | Not-determined |
| S7_extracted_89049 | Not-determined |
| S7_extracted_89057 | Not-determined |
| S7_extracted_89061 | Not-determined |
| S7_extracted_89069 | Low-quality |
| S7_extracted_89075 | Not-determined |
| S7_extracted_89082 | Not-determined |
| S7_extracted_89094 | Not-determined |
| S7_extracted_89099 | Low-quality |
| S7_extracted_89110 | Low-quality |
| S7_extracted_89116 | Low-quality |
| S7_extracted_89118 | Not-determined |
| S7_extracted_89132 | Low-quality |
| S7_extracted_89134 | Not-determined |
| S7_extracted_89143 | Not-determined |
| S7_extracted_89144 | Not-determined |
| S7_extracted_89157 | Not-determined |
| S7_extracted_89184 | Not-determined |
| S14_vs_k141_72953\|\|full | High-quality |
| S7_extracted_89233 | Not-determined |
| S7_extracted_89236 | Not-determined |
| S7_extracted_89242 | Not-determined |
| S7_extracted_89258 | Low-quality |
| S7_extracted_89264 | Not-determined |
| S7_extracted_89266 | Low-quality |
| S7_extracted_89275 | Not-determined |
| S7_extracted_89311 | Not-determined |
| S7_extracted_89344 | Not-determined |
| S7_extracted_89387 | Not-determined |
| S7_extracted_89391 | Not-determined |
| S7_extracted_89398 | Not-determined |
| S7_extracted_89403 | Not-determined |
| S7_extracted_89413 | Not-determined |
| S7_extracted_89420 | Not-determined |
| S7_extracted_89431 | Not-determined |
| S7_extracted_89435 | Low-quality |
| S7_extracted_89474 | Not-determined |
| S7_extracted_89493 | Not-determined |
| S7_extracted_89507 | Not-determined |
| S7_extracted_89534 | Low-quality |
| S7_extracted_89546 | Not-determined |
| S7_extracted_89551 | Not-determined |
| S7_extracted_89564 | Not-determined |
| S7_extracted_89565 | Not-determined |
| S7_extracted_89577 | Not-determined |
| S7_extracted_89579 | Not-determined |
| S7_extracted_89597 | Not-determined |
| S7_extracted_89603 | Not-determined |
| S7_extracted_89609 | Low-quality |
| S7_extracted_89611 | Not-determined |
| S7_extracted_89613 | Low-quality |
| S7_extracted_89633 | Not-determined |
| S7_extracted_89640 | Not-determined |
| S7_extracted_89655 | Not-determined |
| S7_extracted_89656 | Not-determined |
| S7_extracted_89666 | Low-quality |
| S5_vs_k141_43207\|\|full | High-quality |
| S7_extracted_89677 | Not-determined |
| S7_extracted_89680 | Not-determined |
| S7_extracted_89708 | Not-determined |
| S7_extracted_89730 | Not-determined |
| S7_extracted_89733 | Not-determined |
| S7_extracted_89742 | Not-determined |
| S7_extracted_89749 | Not-determined |
| S7_extracted_89750 | Not-determined |
| S7_extracted_89776 | Not-determined |
| S7_extracted_89792 | Not-determined |
| S7_extracted_89796 | Not-determined |
| S7_extracted_89799 | Not-determined |
| S7_extracted_89804 | Not-determined |
| S7_extracted_89813 | Not-determined |
| S7_extracted_89835 | Not-determined |
| S7_extracted_89842 | Not-determined |
| S7_extracted_89880 | Not-determined |
| S7_extracted_89881 | Low-quality |
| S7_extracted_89904 | Not-determined |
| S7_extracted_89921 | Not-determined |
| S7_extracted_89977 | Not-determined |
| S7_extracted_90005 | Not-determined |
| S7_extracted_90032 | Not-determined |
| S7_extracted_90042 | Not-determined |
| S7_extracted_90053 | Not-determined |
| S7_extracted_90054 | Low-quality |
| S7_extracted_90060 | Low-quality |
| S7_extracted_90064 | Not-determined |
| S7_extracted_90106 | Not-determined |
| S7_extracted_90111 | Not-determined |
| S7_extracted_90114 | Not-determined |
| S7_extracted_90141 | Low-quality |
| S7_extracted_90147 | Not-determined |
| S7_extracted_90157 | Low-quality |
| S7_extracted_90193 | Not-determined |
| S7_extracted_90203 | Low-quality |
| S7_extracted_90206 | Not-determined |
| S7_extracted_90209 | Not-determined |
| S7_extracted_90216 | Not-determined |
| S7_extracted_90232 | Not-determined |
| S7_extracted_90239 | Not-determined |
| S7_extracted_90249 | Not-determined |
| S7_extracted_90252 | Not-determined |
| S7_extracted_90255 | Low-quality |
| S7_extracted_90272 | Not-determined |
| S7_extracted_90273 | Not-determined |
| S7_extracted_90275 | Low-quality |
| S7_extracted_90285 | Low-quality |
| S7_extracted_90294 | Not-determined |
| S7_extracted_90295 | Low-quality |
| S7_extracted_90298 | Not-determined |
| S7_extracted_90308 | Not-determined |
| S7_extracted_90329 | Not-determined |
| S7_extracted_90336 | Not-determined |
| S7_extracted_90341 | Not-determined |
| S7_extracted_90349 | Not-determined |
| S7_extracted_90356 | Not-determined |
| S7_extracted_90365 | Not-determined |
| S7_extracted_90395 | Not-determined |
| S7_extracted_90400 | Low-quality |
| S7_extracted_90420 | Not-determined |
| S7_extracted_90428 | Low-quality |
| S7_extracted_90434 | Not-determined |
| S7_extracted_90479 | Not-determined |
| S7_extracted_90500 | Not-determined |
| S7_extracted_90510 | Not-determined |
| S7_extracted_90539 | Not-determined |
| S7_extracted_90554 | Not-determined |
| S7_extracted_90555 | Not-determined |
| S7_extracted_90561 | Not-determined |
| S7_extracted_90569 | Not-determined |
| S7_extracted_90579 | Not-determined |
| S7_extracted_90612 | Not-determined |
| S7_extracted_90633 | Not-determined |
| S7_extracted_90636 | Not-determined |
| S7_extracted_90641 | Not-determined |
| S7_extracted_90677 | Not-determined |
| S7_extracted_90689 | Low-quality |
| S7_extracted_90698 | Not-determined |
| S7_extracted_90718 | Not-determined |
| S7_extracted_90722 | Low-quality |
| S7_extracted_90725 | Not-determined |
| S7_extracted_90744 | Not-determined |
| S7_extracted_90747 | Not-determined |
| S7_extracted_90748 | Not-determined |
| S7_extracted_90759 | Not-determined |
| S7_extracted_90804 | Not-determined |
| S7_extracted_90840 | Not-determined |
| S7_extracted_90850 | Low-quality |
| S7_extracted_90861 | Not-determined |
| S7_extracted_90886 | Not-determined |
| S7_extracted_90924 | Not-determined |
| S7_extracted_90968 | Low-quality |
| S7_extracted_90970 | Not-determined |
| S7_extracted_90989 | Not-determined |
| S7_extracted_90991 | Not-determined |
| S7_extracted_91005 | Not-determined |
| S7_extracted_91007 | Not-determined |
| S7_extracted_91008 | Not-determined |
| S7_extracted_91015 | Not-determined |
| S7_extracted_91017 | Not-determined |
| S7_extracted_91025 | Low-quality |
| S7_extracted_91033 | Not-determined |
| S7_extracted_91050 | Not-determined |
| S7_extracted_91077 | Low-quality |
| S7_extracted_91080 | Not-determined |
| S7_extracted_91089 | Not-determined |
| S7_extracted_91099 | Not-determined |
| S7_extracted_91154 | Low-quality |
| S7_extracted_91164 | Not-determined |
| S7_extracted_91167 | Not-determined |
| S7_extracted_91171 | Not-determined |
| S7_extracted_91189 | Not-determined |
| S7_extracted_91199 | Not-determined |
| S7_extracted_91208 | Not-determined |
| S7_extracted_91219 | Not-determined |
| S7_extracted_91227 | Not-determined |
| S7_extracted_91230 | Low-quality |
| S7_extracted_91233 | Not-determined |
| S7_extracted_91235 | Not-determined |
| S7_extracted_91237 | Low-quality |
| S7_extracted_91242 | Not-determined |
| S7_extracted_91249 | Not-determined |
| S7_extracted_91252 | Not-determined |
| S7_extracted_91254 | Not-determined |
| S7_extracted_91256 | Not-determined |
| S7_extracted_91282 | Not-determined |
| S7_extracted_91291 | Not-determined |
| S7_extracted_91298 | Low-quality |
| S7_extracted_91317 | Not-determined |
| S7_extracted_91321 | Not-determined |
| S7_extracted_91326 | Not-determined |
| S7_extracted_91327 | Not-determined |
| S7_extracted_91343 | Not-determined |
| S7_extracted_91349 | Not-determined |
| S7_extracted_91358 | Not-determined |
| S7_extracted_91370 | Low-quality |
| S7_extracted_91379 | Not-determined |
| S7_extracted_91386 | Low-quality |
| S7_extracted_91393 | Not-determined |

| | |
|---|---|
| S7_extracted_91401 | Not-determined |
| S7_extracted_91402 | Not-determined |
| S7_extracted_91408 | Not-determined |
| S7_extracted_91411 | Low-quality |
| S7_extracted_91417 | Not-determined |
| S7_extracted_91419 | Not-determined |
| S7_extracted_91420 | Not-determined |
| S7_extracted_91421 | Low-quality |
| S7_extracted_91438 | Not-determined |
| S7_extracted_91490 | Not-determined |
| S7_extracted_91505 | Not-determined |
| S7_extracted_91512 | Not-determined |
| S7_extracted_91519 | Not-determined |
| S7_extracted_91521 | Not-determined |
| S7_extracted_91523 | Low-quality |
| S7_extracted_91528 | Not-determined |
| S7_extracted_91546 | Not-determined |
| S7_extracted_91558 | Not-determined |
| S7_extracted_91561 | Not-determined |
| S7_extracted_91577 | Not-determined |
| S7_extracted_91589 | Low-quality |
| S7_extracted_91591 | Not-determined |
| S7_extracted_91600 | Not-determined |
| S7_extracted_91607 | Not-determined |
| S7_extracted_91646 | Not-determined |
| S7_extracted_91654 | Not-determined |
| S7_extracted_91663 | Not-determined |
| S7_extracted_91670 | Not-determined |
| S7_extracted_91696 | Not-determined |
| S7_extracted_91715 | Low-quality |
| S7_extracted_91720 | Low-quality |
| S7_extracted_91734 | Not-determined |
| S7_extracted_91762 | Not-determined |
| S7_extracted_91771 | Not-determined |
| S7_extracted_91774 | Not-determined |
| S7_extracted_91777 | Not-determined |
| S7_extracted_91815 | Not-determined |
| S7_extracted_91819 | Not-determined |
| S7_extracted_91831 | Not-determined |
| S7_extracted_91833 | Not-determined |
| S7_extracted_91844 | Not-determined |
| S7_extracted_91903 | Low-quality |
| S7_extracted_91952 | Low-quality |
| S7_extracted_91959 | Not-determined |
| S7_extracted_91968 | Low-quality |
| S7_extracted_91970 | Low-quality |
| S7_extracted_91976 | Not-determined |
| S7_extracted_91993 | Not-determined |
| S7_extracted_92011 | Low-quality |
| S7_extracted_92049 | Not-determined |
| S7_extracted_92060 | Low-quality |
| S7_extracted_92073 | Not-determined |
| S7_extracted_92090 | Low-quality |
| S7_extracted_92098 | Not-determined |
| S7_extracted_92122 | Not-determined |
| S7_extracted_92129 | Not-determined |
| S7_extracted_92144 | Low-quality |
| S7_extracted_92147 | Low-quality |
| S7_extracted_92156 | Not-determined |
| S7_extracted_92161 | Not-determined |
| S7_extracted_92176 | Not-determined |
| S7_extracted_92191 | Not-determined |
| S7_extracted_92193 | Not-determined |
| S7_extracted_92213 | Low-quality |
| S7_extracted_92220 | Not-determined |
| S7_extracted_92224 | Not-determined |
| S7_extracted_92233 | Low-quality |
| S7_extracted_92237 | Not-determined |
| S7_extracted_92259 | Not-determined |
| S7_extracted_92262 | Not-determined |
| S7_extracted_92265 | Low-quality |
| S7_extracted_92275 | Not-determined |
| S7_extracted_92290 | Not-determined |
| S7_extracted_92310 | Not-determined |
| S7_extracted_92313 | Not-determined |
| S7_extracted_92327 | Not-determined |
| S7_extracted_92348 | Not-determined |
| S7_extracted_92361 | Low-quality |
| S7_extracted_92379 | Not-determined |
| S7_extracted_92388 | Not-determined |
| S7_extracted_92415 | Low-quality |
| S7_extracted_92416 | Low-quality |
| S7_extracted_92420 | Not-determined |
| S7_extracted_92423 | Not-determined |
| S7_extracted_92432 | Not-determined |
| S7_extracted_92448 | Not-determined |
| S7_extracted_92450 | Low-quality |
| S7_extracted_92455 | Not-determined |
| S7_extracted_92456 | Not-determined |
| S7_extracted_92461 | Not-determined |
| S7_extracted_92471 | Not-determined |
| S7_extracted_92475 | Not-determined |
| S7_extracted_92513 | Low-quality |
| S7_extracted_92518 | Not-determined |
| S7_extracted_92527 | Not-determined |
| S7_extracted_92534 | Not-determined |
| S7_extracted_92538 | Not-determined |
| S7_extracted_92564 | Not-determined |
| S7_extracted_92584 | Low-quality |
| S7_extracted_92598 | Not-determined |
| S7_extracted_92602 | Not-determined |
| S7_extracted_92608 | Not-determined |
| S7_extracted_92611 | Not-determined |
| S7_extracted_92621 | Not-determined |
| S7_extracted_92632 | Not-determined |
| S7_extracted_92642 | Not-determined |
| S7_extracted_92651 | Not-determined |
| S7_extracted_92656 | Not-determined |
| S7_extracted_92684 | Not-determined |
| S7_extracted_92695 | Low-quality |
| S7_extracted_92746 | Not-determined |
| S7_extracted_92754 | Not-determined |
| S7_extracted_92759 | Not-determined |
| S7_extracted_92768 | Not-determined |
| S7_extracted_92771 | Not-determined |
| S7_extracted_92792 | Not-determined |
| S7_extracted_92813 | Not-determined |
| S7_extracted_92819 | Not-determined |
| S7_extracted_92829 | Not-determined |
| S7_extracted_92843 | Not-determined |
| S7_extracted_92845 | Not-determined |
| S7_extracted_92851 | Not-determined |
| S7_extracted_92875 | Not-determined |
| S7_extracted_92886 | Not-determined |
| S7_extracted_92887 | Not-determined |
| S7_extracted_92889 | Not-determined |
| S7_extracted_92903 | Not-determined |
| S7_extracted_92909 | Not-determined |
| S7_extracted_92914 | Not-determined |
| S7_extracted_92921 | Not-determined |
| S7_extracted_92922 | Not-determined |
| S7_extracted_92934 | Not-determined |
| S7_extracted_92941 | Not-determined |
| S7_extracted_92973 | Not-determined |
| S7_extracted_92985 | Not-determined |
| S7_extracted_93005 | Low-quality |
| S7_extracted_93013 | Not-determined |
| S7_extracted_93019 | Low-quality |
| S7_extracted_93052 | Low-quality |
| S7_extracted_93068 | Not-determined |
| S7_extracted_93069 | Not-determined |
| S7_extracted_93091 | Not-determined |
| S7_extracted_93093 | Low-quality |
| S1_vs_k141_17085\|\|full | Medium-quality |
| S7_extracted_93101 | Not-determined |
| S7_extracted_93111 | Not-determined |
| S7_extracted_93113 | Low-quality |
| S7_extracted_93159 | Low-quality |
| S7_extracted_93165 | Not-determined |
| S7_extracted_93174 | Not-determined |
| S7_extracted_93180 | Not-determined |
| S7_extracted_93205 | Not-determined |
| S7_extracted_93207 | Low-quality |
| S7_extracted_93208 | Not-determined |
| S7_extracted_93222 | Not-determined |
| S7_extracted_93225 | Not-determined |
| S7_extracted_93228 | Not-determined |
| S7_extracted_93261 | Not-determined |
| S7_extracted_93268 | Not-determined |
| S7_extracted_93286 | Not-determined |
| S7_extracted_93305 | Low-quality |
| S7_extracted_93309 | Not-determined |
| S7_extracted_93318 | Low-quality |
| S7_extracted_93335 | Not-determined |
| S7_extracted_93336 | Not-determined |
| S7_extracted_93337 | Not-determined |
| S7_extracted_93356 | Low-quality |
| S7_extracted_93361 | Not-determined |
| S7_extracted_93366 | Not-determined |
| S7_extracted_93423 | Not-determined |
| S7_extracted_93441 | Not-determined |
| S7_extracted_93444 | Not-determined |
| S7_extracted_93466 | Low-quality |
| S7_extracted_93480 | Not-determined |
| S7_extracted_93486 | Not-determined |
| S7_extracted_93492 | Not-determined |
| S7_extracted_93499 | Not-determined |
| S7_extracted_93509 | Not-determined |
| S7_extracted_93512 | Low-quality |
| S7_extracted_93526 | Not-determined |
| S7_extracted_93527 | Not-determined |
| S7_extracted_93532 | Not-determined |
| S7_extracted_93551 | Low-quality |
| S7_extracted_93552 | Not-determined |
| S7_extracted_93553 | Not-determined |
| S7_extracted_93560 | Not-determined |
| S7_extracted_93566 | Not-determined |
| S7_extracted_93595 | Not-determined |
| S7_extracted_93611 | Not-determined |
| S7_extracted_93632 | Not-determined |
| S7_extracted_93638 | Not-determined |
| S7_extracted_93644 | Not-determined |
| S7_extracted_93652 | Not-determined |
| S7_extracted_93655 | Low-quality |
| S7_extracted_93673 | Not-determined |
| S7_extracted_93681 | Not-determined |
| S7_extracted_93718 | Not-determined |
| S7_extracted_93724 | Low-quality |
| S7_extracted_93726 | Not-determined |
| S7_extracted_93746 | Low-quality |
| S7_extracted_93764 | Low-quality |
| S7_extracted_93767 | Not-determined |
| S7_extracted_93781 | Not-determined |
| S7_extracted_93788 | Low-quality |
| S7_extracted_93789 | Not-determined |
| S7_extracted_93791 | Not-determined |
| S7_extracted_93805 | Not-determined |
| S7_extracted_93807 | Not-determined |
| S7_extracted_93827 | Not-determined |
| S7_extracted_93830 | Not-determined |
| S7_extracted_93834 | Not-determined |
| S7_extracted_93842 | Not-determined |
| S7_extracted_93849 | Low-quality |
| S7_extracted_93871 | Not-determined |
| S7_extracted_93876 | Not-determined |
| S7_extracted_93901 | Not-determined |
| S7_extracted_93913 | Low-quality |
| S7_extracted_93927 | Not-determined |
| S7_extracted_93933 | Not-determined |
| S7_extracted_93943 | Not-determined |
| S7_extracted_93994 | Not-determined |
| S7_extracted_93997 | Not-determined |
| S7_extracted_93999 | Not-determined |
| S7_extracted_94013 | Not-determined |
| S7_extracted_94029 | Not-determined |
| S7_extracted_94110 | Not-determined |
| S7_extracted_94119 | Not-determined |
| S7_extracted_94120 | Not-determined |
| S7_extracted_94149 | Not-determined |
| S7_extracted_94150 | Not-determined |
| S7_extracted_94162 | Not-determined |
| S7_extracted_94167 | Not-determined |
| S7_extracted_94176 | Not-determined |
| S7_extracted_94202 | Not-determined |
| S7_extracted_94204 | Not-determined |
| S7_extracted_94209 | Not-determined |
| S7_extracted_94227 | Not-determined |
| S7_extracted_94228 | Not-determined |
| S7_extracted_94236 | Not-determined |
| S7_extracted_94240 | Not-determined |
| S7_extracted_94241 | Low-quality |
| S7_extracted_94243 | Not-determined |
| S7_extracted_94244 | Not-determined |
| S7_extracted_94266 | Not-determined |
| S7_extracted_94277 | Not-determined |
| S7_extracted_94281 | Not-determined |
| S7_extracted_94290 | Not-determined |
| S7_extracted_94337 | Not-determined |
| S7_extracted_94349 | Not-determined |
| S7_extracted_94356 | Not-determined |
| S7_extracted_94390 | Not-determined |
| S7_extracted_94421 | Not-determined |
| S7_extracted_94432 | Not-determined |
| S7_extracted_94436 | Not-determined |
| S7_extracted_94437 | Not-determined |
| S7_extracted_94441 | Low-quality |
| S7_extracted_94443 | Low-quality |
| S7_extracted_94455 | Not-determined |
| S7_extracted_94467 | Not-determined |
| S7_extracted_94474 | Not-determined |
| S7_extracted_94476 | Low-quality |
| S7_extracted_94477 | Not-determined |
| S7_extracted_94487 | Not-determined |
| S7_extracted_94492 | Not-determined |
| S7_extracted_94498 | Not-determined |
| S7_extracted_94514 | Low-quality |
| S7_extracted_94518 | Not-determined |
| S7_extracted_94519 | Not-determined |
| S7_extracted_94535 | Not-determined |
| S7_extracted_94536 | Not-determined |
| S7_extracted_94547 | Not-determined |
| S7_extracted_94555 | Low-quality |
| S7_extracted_94561 | Not-determined |
| S7_extracted_94570 | Not-determined |
| S7_extracted_94628 | Not-determined |
| S7_extracted_94662 | Not-determined |
| S7_extracted_94676 | Not-determined |
| S7_extracted_94681 | Not-determined |
| S7_extracted_94689 | Not-determined |
| S7_extracted_94690 | Not-determined |
| S7_extracted_94691 | Not-determined |
| S7_extracted_94696 | Low-quality |
| S7_extracted_94704 | Not-determined |
| S7_extracted_94742 | Not-determined |
| S7_extracted_94747 | Low-quality |
| S7_extracted_94753 | Not-determined |
| S7_extracted_94775 | Not-determined |
| S7_extracted_94796 | Not-determined |
| S7_extracted_94839 | Not-determined |
| S7_extracted_94849 | Not-determined |
| S7_extracted_94864 | Not-determined |
| S7_extracted_94873 | Low-quality |
| S7_extracted_94879 | Not-determined |
| S7_extracted_94892 | Not-determined |
| S7_extracted_94895 | Not-determined |
| S7_extracted_94907 | Low-quality |
| S7_extracted_94935 | Not-determined |
| S7_extracted_94941 | Low-quality |
| S7_extracted_94949 | Low-quality |
| S7_extracted_94954 | Not-determined |
| S7_extracted_94957 | Not-determined |
| S7_extracted_94958 | Not-determined |
| S7_extracted_94960 | Not-determined |
| S7_extracted_94964 | Not-determined |
| S7_extracted_94980 | Not-determined |
| S7_extracted_94996 | Not-determined |
| S7_extracted_94997 | Not-determined |
| S7_extracted_95010 | Low-quality |
| S7_extracted_95013 | Not-determined |
| S7_extracted_95025 | Not-determined |
| S7_extracted_95029 | Not-determined |
| S7_extracted_95037 | Not-determined |
| S7_extracted_95044 | Not-determined |
| S7_extracted_95065 | Low-quality |
| S7_extracted_95070 | Low-quality |
| S7_extracted_95079 | Low-quality |
| S7_extracted_95089 | Not-determined |
| S7_extracted_95103 | Not-determined |
| S7_extracted_95106 | Not-determined |
| S7_extracted_95118 | Not-determined |
| S7_extracted_95128 | Not-determined |
| S7_extracted_95132 | Low-quality |
| S7_extracted_95140 | Low-quality |
| S7_extracted_95158 | Low-quality |
| S7_extracted_95161 | Not-determined |
| S7_extracted_95162 | Not-determined |
| S7_extracted_95179 | Not-determined |
| S7_extracted_95194 | Low-quality |
| S7_extracted_95213 | Low-quality |
| S7_extracted_95220 | Not-determined |
| S7_extracted_95233 | Not-determined |
| S7_extracted_95234 | Not-determined |
| S7_extracted_95259 | Not-determined |
| S7_extracted_95273 | Not-determined |
| S7_extracted_95277 | Low-quality |
| S7_extracted_95289 | Not-determined |
| S7_extracted_95292 | Low-quality |
| S7_extracted_95306 | Not-determined |
| S7_extracted_95309 | Not-determined |
| S7_extracted_95315 | Not-determined |
| S7_extracted_95380 | Not-determined |
| S7_extracted_95386 | Not-determined |
| S7_extracted_95407 | Low-quality |
| S7_extracted_95408 | Not-determined |
| S7_extracted_95417 | Not-determined |
| S7_extracted_95440 | Not-determined |
| S7_extracted_95443 | Not-determined |
| S7_extracted_95456 | Not-determined |
| S7_extracted_95463 | Not-determined |
| S7_extracted_95472 | Low-quality |
| S7_extracted_95520 | Not-determined |
| S7_extracted_95524 | Not-determined |
| S7_extracted_95531 | Not-determined |
| S7_extracted_95535 | Not-determined |
| S7_extracted_95544 | Low-quality |
| S7_extracted_95545 | Not-determined |
| S7_extracted_95550 | Not-determined |
| S7_extracted_95551 | Not-determined |
| S7_extracted_95580 | Not-determined |
| S7_extracted_95599 | Not-determined |
| S7_extracted_95604 | Not-determined |
| S7_extracted_95606 | Low-quality |
| S7_extracted_95612 | Not-determined |
| S7_extracted_95619 | Low-quality |
| S7_extracted_95620 | Not-determined |
| S7_extracted_95636 | Not-determined |
| S7_extracted_95638 | Not-determined |
| S7_extracted_95645 | Not-determined |
| S7_extracted_95652 | Low-quality |
| S7_extracted_95673 | Not-determined |
| S7_extracted_95682 | Low-quality |
| S7_extracted_95690 | Not-determined |
| S7_extracted_95691 | Not-determined |
| S7_extracted_95692 | Not-determined |
| S7_extracted_95709 | Not-determined |
| S7_extracted_95714 | Not-determined |
| S7_extracted_95722 | Not-determined |
| S7_extracted_95765 | Not-determined |
| S7_extracted_95777 | Not-determined |
| S7_extracted_95780 | Low-quality |
| S7_extracted_95801 | Not-determined |
| S7_extracted_95810 | Not-determined |
| S7_extracted_95811 | Not-determined |
| S7_extracted_95812 | Not-determined |
| S7_extracted_95819 | Low-quality |
| S7_extracted_95838 | Not-determined |
| S7_extracted_95843 | Not-determined |
| S7_extracted_95856 | Not-determined |
| S7_extracted_95861 | Not-determined |
| S7_extracted_95873 | Low-quality |
| S7_extracted_95891 | Not-determined |
| S7_extracted_95899 | Not-determined |
| S7_extracted_95921 | Not-determined |
| S7_extracted_95957 | Not-determined |
| S7_extracted_95958 | Low-quality |
| S7_extracted_95959 | Not-determined |
| S7_extracted_95980 | Not-determined |
| S7_extracted_96043 | Not-determined |
| S7_extracted_96052 | Not-determined |
| S7_extracted_96067 | Not-determined |
| S7_extracted_96083 | Not-determined |
| S7_extracted_96084 | Not-determined |
| S7_extracted_96086 | Low-quality |
| S7_extracted_96088 | Not-determined |
| S7_extracted_96089 | Not-determined |
| S7_extracted_96118 | Low-quality |
| S7_extracted_96122 | Not-determined |
| S7_extracted_96133 | Not-determined |
| S7_extracted_96135 | Low-quality |
| S7_extracted_96137 | Not-determined |
| S7_extracted_96139 | Not-determined |
| S7_extracted_96149 | Not-determined |
| S7_extracted_96171 | Not-determined |
| S7_extracted_96187 | Low-quality |
| S7_extracted_96194 | Not-determined |
| S7_extracted_96207 | Not-determined |
| S7_extracted_96223 | Not-determined |
| S7_extracted_96226 | Low-quality |
| S7_extracted_96237 | Not-determined |
| S7_extracted_96249 | Not-determined |
| S7_extracted_96252 | Not-determined |
| S7_extracted_96253 | Not-determined |
| S7_extracted_96257 | Not-determined |
| S7_extracted_96297 | Not-determined |
| S7_extracted_96323 | Not-determined |
| S7_extracted_96334 | Low-quality |
| S7_extracted_96361 | Low-quality |
| S7_extracted_96368 | Not-determined |
| S7_extracted_96375 | Not-determined |
| S7_extracted_96385 | Not-determined |
| S7_extracted_96386 | Not-determined |
| S7_extracted_96387 | Not-determined |
| S7_extracted_96388 | Not-determined |
| S7_extracted_96395 | Not-determined |
| S7_extracted_96407 | Not-determined |
| S7_extracted_96409 | Not-determined |
| S7_extracted_96411 | Not-determined |
| S7_extracted_96428 | Not-determined |
| S7_extracted_96433 | Low-quality |
| S7_extracted_96439 | Low-quality |
| S7_extracted_96440 | Not-determined |
| S7_extracted_96445 | Not-determined |
| S7_extracted_96448 | Not-determined |
| S7_extracted_96450 | Low-quality |
| S7_extracted_96451 | Not-determined |
| S7_extracted_96462 | Not-determined |
| S7_extracted_96472 | Not-determined |
| S7_extracted_96476 | Not-determined |
| S7_extracted_96483 | Not-determined |
| S7_extracted_96491 | Not-determined |
| S7_extracted_96492 | Not-determined |
| S7_extracted_96506 | Low-quality |
| S7_extracted_96510 | Low-quality |
| S7_extracted_96520 | Not-determined |
| S7_extracted_96523 | Not-determined |
| S7_extracted_96527 | Not-determined |
| S7_extracted_96579 | Not-determined |
| S7_extracted_96616 | Not-determined |
| S7_extracted_96622 | Low-quality |
| S7_extracted_96637 | Not-determined |
| S7_extracted_96649 | Low-quality |
| S7_extracted_96657 | Not-determined |
| S7_extracted_96659 | Not-determined |
| S7_extracted_96667 | Not-determined |
| S7_extracted_96670 | Not-determined |
| S7_extracted_96686 | Not-determined |
| S7_extracted_96693 | Not-determined |
| S7_extracted_96701 | Not-determined |
| S7_extracted_96727 | Not-determined |
| S7_extracted_96735 | Not-determined |
| S7_extracted_96748 | Not-determined |
| S7_extracted_96749 | Not-determined |
| S7_extracted_96770 | Not-determined |
| S7_extracted_96799 | Not-determined |
| S7_extracted_96803 | Not-determined |
| S7_extracted_96823 | Low-quality |
| S7_extracted_96827 | Low-quality |
| S7_extracted_96828 | Not-determined |
| S7_extracted_96838 | Not-determined |
| S7_extracted_96883 | Not-determined |
| S7_extracted_96894 | Not-determined |
| S7_extracted_96948 | Not-determined |
| S7_extracted_96952 | Low-quality |
| S7_extracted_96981 | Not-determined |
| S7_extracted_97000 | Low-quality |
| S7_extracted_97003 | Not-determined |
| S7_extracted_97012 | Low-quality |
| S7_extracted_97037 | Not-determined |
| S7_extracted_97047 | Not-determined |
| S7_extracted_97055 | Not-determined |
| S7_extracted_97057 | Not-determined |
| S7_extracted_97059 | Not-determined |
| S7_extracted_97067 | Not-determined |
| S7_extracted_97086 | Not-determined |
| S7_extracted_97087 | Not-determined |
| S7_extracted_97096 | Not-determined |
| S7_extracted_97103 | Low-quality |
| S7_extracted_97108 | Not-determined |
| S7_extracted_97113 | Not-determined |
| S7_extracted_97115 | Not-determined |
| S7_extracted_97117 | Not-determined |
| S7_extracted_97123 | Not-determined |
| S7_extracted_97125 | Not-determined |
| S7_extracted_97133 | Not-determined |
| S7_extracted_97138 | Not-determined |
| S7_extracted_97165 | Not-determined |
| S7_extracted_97166 | Not-determined |
| S7_extracted_97169 | Not-determined |
| S7_extracted_97171 | Not-determined |
| S7_extracted_97172 | Low-quality |
| S7_extracted_97183 | Not-determined |
| S7_extracted_97186 | Not-determined |
| S7_extracted_97198 | Not-determined |
| S7_extracted_97200 | Not-determined |
| S7_extracted_97202 | Not-determined |
| S7_extracted_97225 | Not-determined |
| S7_extracted_97236 | Not-determined |
| S7_extracted_97240 | Not-determined |
| S7_extracted_97244 | Not-determined |
| S7_extracted_97255 | Not-determined |
| S7_extracted_97262 | Not-determined |
| S7_extracted_97268 | Not-determined |
| S7_extracted_97308 | Not-determined |
| S7_extracted_97315 | Not-determined |
| S7_extracted_97318 | Not-determined |
| S7_extracted_97346 | Not-determined |
| S7_extracted_97349 | Not-determined |
| S7_extracted_97364 | Low-quality |
| S7_extracted_97388 | Not-determined |
| S7_extracted_97397 | Not-determined |
| S7_extracted_97401 | Not-determined |
| S7_extracted_97417 | Not-determined |
| S7_extracted_97437 | Not-determined |
| S7_extracted_97451 | Not-determined |
| S7_extracted_97468 | Not-determined |
| S7_extracted_97489 | Not-determined |
| S7_extracted_97508 | Not-determined |
| S7_extracted_97523 | Low-quality |
| S7_extracted_97528 | Not-determined |
| S7_extracted_97552 | Not-determined |
| S7_extracted_97553 | Not-determined |
| S7_extracted_97567 | Not-determined |
| S7_extracted_97570 | Not-determined |
| S7_extracted_97577 | Not-determined |
| S7_extracted_97585 | Not-determined |
| S7_extracted_97596 | Not-determined |
| S7_extracted_97616 | Not-determined |
| S7_extracted_97617 | Not-determined |
| S7_extracted_97619 | Low-quality |
| S7_extracted_97622 | Not-determined |
| S7_extracted_97637 | Not-determined |
| S7_extracted_97640 | Not-determined |
| S7_extracted_97642 | Not-determined |
| S7_extracted_97666 | Low-quality |
| S7_extracted_97673 | Not-determined |
| S7_extracted_97714 | Not-determined |
| S7_extracted_97736 | Not-determined |
| S7_extracted_97739 | Not-determined |
| S7_extracted_97741 | Low-quality |
| S7_extracted_97766 | Low-quality |
| S7_extracted_97791 | Not-determined |
| S7_extracted_97797 | Not-determined |
| S7_extracted_97835 | Not-determined |
| S7_extracted_97843 | Not-determined |
| S7_extracted_97846 | Not-determined |
| S7_extracted_97885 | Low-quality |
| S7_extracted_97887 | Not-determined |
| S7_extracted_97902 | Not-determined |
| S7_extracted_97911 | Not-determined |
| S7_extracted_97923 | Not-determined |
| S7_extracted_97961 | Not-determined |
| S7_extracted_97963 | Not-determined |
| S7_extracted_97966 | Not-determined |
| S7_extracted_97988 | Low-quality |
| S7_extracted_97999 | Not-determined |
| S7_extracted_98001 | Not-determined |
| S7_extracted_98014 | Not-determined |
| S7_extracted_98027 | Not-determined |
| S7_extracted_98030 | Not-determined |
| S7_extracted_98032 | Low-quality |
| S7_extracted_98035 | Not-determined |
| S7_extracted_98043 | Not-determined |
| S7_extracted_98052 | Not-determined |
| S7_extracted_98100 | Low-quality |
| S7_extracted_98160 | Not-determined |
| S7_extracted_98181 | Not-determined |
| S7_extracted_98184 | Not-determined |
| S7_extracted_98187 | Low-quality |
| S7_extracted_98193 | Not-determined |

| | |
|---|---|
| S7_extracted_98221 | Not-determined |
| S7_extracted_98246 | Low-quality |
| S7_extracted_98249 | Not-determined |
| S7_extracted_98260 | Not-determined |
| S7_extracted_98292 | Not-determined |
| S7_extracted_98296 | Not-determined |
| S7_extracted_98313 | Low-quality |
| S7_extracted_98321 | Not-determined |
| S7_extracted_98373 | Low-quality |
| S7_extracted_98377 | Not-determined |
| S7_extracted_98380 | Not-determined |
| S7_extracted_98383 | Not-determined |
| S7_extracted_98386 | Not-determined |
| S7_extracted_98389 | Not-determined |
| S7_extracted_98391 | Not-determined |
| S7_extracted_98405 | Not-determined |
| S7_extracted_98418 | Not-determined |
| S7_extracted_98427 | Not-determined |
| S7_extracted_98434 | Low-quality |
| S7_extracted_98448 | Low-quality |
| S7_extracted_98462 | Not-determined |
| S7_extracted_98467 | Low-quality |
| S7_extracted_98473 | Low-quality |
| S7_extracted_98478 | Low-quality |
| S7_extracted_98482 | Not-determined |
| S7_extracted_98484 | Not-determined |
| S7_extracted_98486 | Not-determined |
| S7_extracted_98493 | Not-determined |
| S7_extracted_98503 | Not-determined |
| S7_extracted_98514 | Not-determined |
| S7_extracted_98528 | Not-determined |
| S7_extracted_98529 | Low-quality |
| S7_extracted_98530 | Not-determined |
| S7_extracted_98542 | Not-determined |
| S7_extracted_98551 | Not-determined |
| S7_extracted_98568 | Not-determined |
| S7_extracted_98579 | Low-quality |
| S7_extracted_98580 | Not-determined |
| S7_extracted_98582 | Not-determined |
| S7_extracted_98613 | Not-determined |
| S7_extracted_98616 | Not-determined |
| S7_extracted_98634 | Not-determined |
| S7_extracted_98649 | Not-determined |
| S7_extracted_98651 | Not-determined |
| S7_extracted_98666 | Not-determined |
| S7_extracted_98677 | Low-quality |
| S7_extracted_98681 | Low-quality |
| S7_extracted_98682 | Not-determined |
| S7_extracted_98711 | Not-determined |
| S7_extracted_98716 | Not-determined |
| S7_extracted_98718 | Not-determined |
| S7_extracted_98719 | Low-quality |
| S7_extracted_98726 | Low-quality |
| S7_extracted_98740 | Not-determined |
| S7_extracted_98743 | Low-quality |
| S7_extracted_98759 | Not-determined |
| S7_extracted_98770 | Not-determined |
| S7_extracted_98793 | Not-determined |
| S7_extracted_98802 | Not-determined |
| S7_extracted_98805 | Not-determined |
| S7_extracted_98807 | Not-determined |
| S7_extracted_98813 | Not-determined |
| S7_extracted_98819 | Not-determined |
| S7_extracted_98828 | Not-determined |
| S7_extracted_98838 | Not-determined |
| S7_extracted_98881 | Not-determined |
| S7_extracted_98885 | Not-determined |
| S7_extracted_98906 | Not-determined |
| S7_extracted_98912 | Not-determined |
| S7_extracted_98920 | Low-quality |
| S7_extracted_98922 | Not-determined |
| S7_extracted_98934 | Not-determined |
| S7_extracted_98948 | Not-determined |
| S7_extracted_98963 | Not-determined |
| S7_extracted_98970 | Low-quality |
| S7_extracted_98975 | Not-determined |
| S7_extracted_98976 | Not-determined |
| S7_extracted_98978 | Not-determined |
| S7_extracted_98987 | Not-determined |
| S7_extracted_99005 | Not-determined |
| S7_extracted_99019 | Not-determined |
| S7_extracted_99021 | Not-determined |
| S7_extracted_99026 | Low-quality |
| S7_extracted_99035 | Low-quality |
| S7_extracted_99036 | Not-determined |
| S7_extracted_99038 | Not-determined |
| S7_extracted_99052 | Not-determined |
| S7_extracted_99061 | Low-quality |
| S7_extracted_99062 | Not-determined |
| S7_extracted_99067 | Low-quality |
| S7_extracted_99069 | Not-determined |
| S7_extracted_99070 | Not-determined |
| S7_extracted_99074 | Low-quality |
| S7_extracted_99107 | Not-determined |
| S7_extracted_99116 | Not-determined |
| S7_extracted_99125 | Low-quality |
| S7_extracted_99132 | Low-quality |
| S7_extracted_99155 | Not-determined |
| S7_extracted_99156 | Not-determined |
| S7_extracted_99159 | Not-determined |
| S7_extracted_99175 | Not-determined |
| S7_extracted_99185 | Not-determined |
| S7_extracted_99194 | Not-determined |
| S7_extracted_99197 | Low-quality |
| S7_extracted_99204 | Not-determined |
| S7_extracted_99222 | Not-determined |
| S7_extracted_99223 | Not-determined |
| S7_extracted_99245 | Not-determined |
| S7_extracted_99258 | Not-determined |
| S7_extracted_99259 | Not-determined |
| S7_extracted_99284 | Not-determined |
| S7_extracted_99315 | Not-determined |
| S7_extracted_99324 | Not-determined |
| S7_extracted_99326 | Not-determined |
| S7_extracted_99330 | Not-determined |
| S7_extracted_99331 | Not-determined |
| S7_extracted_99347 | Low-quality |
| S7_extracted_99360 | Low-quality |
| S7_extracted_99362 | Low-quality |
| S7_extracted_99371 | Not-determined |
| S7_extracted_99389 | Not-determined |
| S7_extracted_99390 | Not-determined |
| S7_extracted_99400 | Not-determined |
| S7_extracted_99404 | Low-quality |
| S7_extracted_99411 | Not-determined |
| S7_extracted_99413 | Not-determined |
| S7_extracted_99420 | Not-determined |
| S7_extracted_99423 | Not-determined |
| S7_extracted_99431 | Not-determined |
| S7_extracted_99440 | Low-quality |
| S7_extracted_99446 | Not-determined |
| S7_extracted_99464 | Low-quality |
| S7_extracted_99499 | Not-determined |
| S7_extracted_99500 | Not-determined |
| S7_extracted_99501 | Not-determined |
| S7_extracted_99535 | Not-determined |
| S7_extracted_99540 | Not-determined |
| S7_extracted_99542 | Not-determined |
| S7_extracted_99546 | Not-determined |
| S7_extracted_99601 | Not-determined |
| S7_extracted_99603 | Not-determined |
| S7_extracted_99612 | Not-determined |
| S7_extracted_99613 | Not-determined |
| S7_extracted_99619 | Low-quality |
| S7_extracted_99627 | Not-determined |
| S7_extracted_99633 | Not-determined |
| S7_extracted_99635 | Not-determined |
| S7_extracted_99647 | Not-determined |
| S7_extracted_99679 | Not-determined |
| S7_extracted_99683 | Not-determined |
| S7_extracted_99685 | Not-determined |
| S7_extracted_99710 | Not-determined |
| S7_extracted_99725 | Not-determined |
| S7_extracted_99729 | Not-determined |
| S7_extracted_99733 | Not-determined |
| S7_extracted_99755 | Not-determined |
| S7_extracted_99758 | Not-determined |
| S7_extracted_99771 | Low-quality |
| S7_extracted_99806 | Not-determined |
| S7_extracted_99811 | Low-quality |
| S7_extracted_99814 | Not-determined |
| S7_extracted_99816 | Low-quality |
| S7_extracted_99823 | Not-determined |
| S7_extracted_99825 | Not-determined |
| S7_extracted_99839 | Not-determined |
| S7_extracted_99859 | Not-determined |
| S7_extracted_99861 | Not-determined |
| S7_extracted_99887 | Not-determined |
| S7_extracted_99890 | Not-determined |
| S7_extracted_99896 | Not-determined |
| S7_extracted_99913 | Not-determined |
| S7_extracted_99919 | Not-determined |
| S7_extracted_99923 | Not-determined |
| S7_extracted_99931 | Not-determined |
| S7_extracted_99934 | Not-determined |
| S7_extracted_99950 | Not-determined |
| S7_extracted_99960 | Not-determined |
| S7_extracted_99969 | Not-determined |
| S7_extracted_99975 | Not-determined |
| S7_extracted_99976 | Not-determined |
| S7_extracted_99977 | Not-determined |
| S7_extracted_99978 | Not-determined |
| S7_extracted_99991 | Not-determined |
| S7_extracted_99999 | Not-determined |
| S7_extracted_100003 | Not-determined |
| S7_extracted_100008 | Not-determined |
| S7_extracted_100045 | Low-quality |
| S7_extracted_100072 | Not-determined |
| S7_extracted_100090 | Not-determined |
| S7_extracted_100101 | Not-determined |
| S7_extracted_100103 | Not-determined |
| S7_extracted_100112 | Low-quality |
| S7_extracted_100117 | Not-determined |
| S7_extracted_100122 | Not-determined |
| S7_extracted_100137 | Not-determined |
| S7_extracted_100139 | Not-determined |
| S7_extracted_100143 | Not-determined |
| S7_extracted_100162 | Not-determined |
| S7_extracted_100197 | Low-quality |
| S7_extracted_100211 | Low-quality |
| S7_extracted_100229 | Not-determined |
| S7_extracted_100236 | Not-determined |
| S7_extracted_100240 | Not-determined |
| S7_extracted_100280 | Not-determined |
| S7_extracted_100289 | Not-determined |
| S7_extracted_100293 | Not-determined |
| S7_extracted_100313 | Not-determined |
| S7_extracted_100321 | Not-determined |
| S7_extracted_100323 | Not-determined |
| S7_extracted_100346 | Low-quality |
| S7_extracted_100372 | Not-determined |
| S7_extracted_100380 | Not-determined |
| S7_extracted_100390 | Not-determined |
| S7_extracted_100408 | Low-quality |
| S7_extracted_100412 | Not-determined |
| S7_extracted_100414 | Not-determined |
| S7_extracted_100422 | Not-determined |
| S7_extracted_100430 | Not-determined |
| S7_extracted_100442 | Not-determined |
| S7_extracted_100447 | Not-determined |
| S7_extracted_100449 | Low-quality |
| S7_extracted_100470 | Not-determined |
| S7_extracted_100475 | Not-determined |
| S7_extracted_100481 | Not-determined |
| S7_extracted_100494 | Low-quality |
| S7_extracted_100499 | Not-determined |
| S7_extracted_100514 | Not-determined |
| S7_extracted_100528 | Not-determined |
| S7_extracted_100559 | Not-determined |
| S7_extracted_100562 | Not-determined |
| S7_extracted_100577 | Low-quality |
| S7_extracted_100596 | Not-determined |
| S7_extracted_100598 | Not-determined |
| S7_extracted_100604 | Not-determined |
| S7_extracted_100610 | Not-determined |
| S7_extracted_100614 | Not-determined |
| S7_extracted_100631 | Not-determined |
| S7_extracted_100649 | Low-quality |
| S7_extracted_100664 | Low-quality |
| S7_extracted_100683 | Not-determined |
| S7_extracted_100685 | Not-determined |
| S7_extracted_100690 | Not-determined |
| S7_extracted_100711 | Not-determined |
| S7_extracted_100739 | Not-determined |
| S7_extracted_100740 | Not-determined |
| S7_extracted_100742 | Not-determined |
| S7_extracted_100747 | Not-determined |
| S7_extracted_100750 | Not-determined |
| S7_extracted_100775 | Not-determined |
| S7_extracted_100778 | Not-determined |
| S7_extracted_100809 | Not-determined |
| S7_extracted_100816 | Not-determined |
| S7_extracted_100818 | Not-determined |
| S7_extracted_100834 | Not-determined |
| S7_extracted_100840 | Low-quality |
| S7_extracted_100847 | Not-determined |
| S7_extracted_100850 | Not-determined |
| S7_extracted_100857 | Not-determined |
| S7_extracted_100859 | Not-determined |
| S7_extracted_100863 | Not-determined |
| S7_extracted_100870 | Not-determined |
| S7_extracted_100883 | Low-quality |
| S7_extracted_100888 | Not-determined |
| S7_extracted_100889 | Low-quality |
| S7_extracted_100901 | Not-determined |
| S7_extracted_100932 | Not-determined |
| S7_extracted_100935 | Low-quality |
| S7_extracted_100943 | Not-determined |
| S7_extracted_100947 | Not-determined |
| S7_extracted_100955 | Not-determined |
| S7_extracted_100956 | Not-determined |
| S7_extracted_100965 | Not-determined |
| S7_extracted_101000 | Not-determined |
| S7_extracted_101013 | Not-determined |
| S7_extracted_101039 | Not-determined |
| S7_extracted_101042 | Not-determined |
| S7_extracted_101048 | Not-determined |
| S7_extracted_101058 | Not-determined |
| S7_extracted_101059 | Not-determined |
| S7_extracted_101079 | Low-quality |
| S7_extracted_101081 | Low-quality |
| S7_extracted_101108 | Not-determined |
| S7_extracted_101114 | Not-determined |
| S7_extracted_101115 | Not-determined |
| S7_extracted_101116 | Not-determined |
| S7_extracted_101119 | Not-determined |
| S7_extracted_101134 | Not-determined |
| S7_extracted_101151 | Not-determined |
| S7_extracted_101156 | Not-determined |
| S7_extracted_101170 | Not-determined |
| S7_extracted_101179 | Not-determined |
| S7_extracted_101185 | Not-determined |
| S7_extracted_101189 | Not-determined |
| S7_extracted_101212 | Not-determined |
| S7_extracted_101226 | Not-determined |
| S7_extracted_101236 | Not-determined |
| S7_extracted_101240 | Not-determined |
| S7_extracted_101241 | Not-determined |
| S7_extracted_101245 | Not-determined |
| S7_extracted_101250 | Not-determined |
| S7_extracted_101258 | Not-determined |
| S7_extracted_101262 | Low-quality |
| S7_extracted_101268 | Low-quality |
| S7_extracted_101285 | Low-quality |
| S7_extracted_101290 | Not-determined |
| S7_extracted_101293 | Low-quality |
| S7_extracted_101297 | Not-determined |
| S7_extracted_101305 | Not-determined |
| S7_extracted_101307 | Low-quality |
| S7_extracted_101312 | Low-quality |
| S7_extracted_101320 | Not-determined |
| S7_extracted_101325 | Not-determined |
| S7_extracted_101349 | Low-quality |
| S7_extracted_101360 | Not-determined |
| S7_extracted_101364 | Not-determined |
| S7_extracted_101366 | Not-determined |
| S7_extracted_101370 | Not-determined |
| S7_extracted_101371 | Not-determined |
| S7_extracted_101380 | Not-determined |
| S7_extracted_101389 | Not-determined |
| S7_extracted_101391 | Not-determined |
| S7_extracted_101400 | Not-determined |
| S7_extracted_101432 | Not-determined |
| S7_extracted_101439 | Not-determined |
| S7_extracted_101448 | Not-determined |
| S7_extracted_101455 | Low-quality |
| S7_extracted_101457 | Not-determined |
| S7_extracted_101462 | Low-quality |
| S7_extracted_101485 | Not-determined |
| S7_extracted_101496 | Not-determined |
| S7_extracted_101497 | Not-determined |
| S7_extracted_101498 | Not-determined |
| S7_extracted_101530 | Not-determined |
| S7_extracted_101538 | Not-determined |
| S7_extracted_101575 | Low-quality |
| S7_extracted_101577 | Not-determined |
| S7_extracted_101591 | Not-determined |
| S7_extracted_101619 | Not-determined |
| S7_extracted_101629 | Not-determined |
| S7_extracted_101639 | Not-determined |
| S7_extracted_101640 | Not-determined |
| S7_extracted_101644 | Not-determined |
| S7_extracted_101645 | Not-determined |
| S7_extracted_101648 | Not-determined |
| S7_extracted_101658 | Not-determined |
| S7_extracted_101660 | Not-determined |
| S7_extracted_101669 | Not-determined |
| S7_extracted_101676 | Not-determined |
| S7_extracted_101680 | Not-determined |
| S7_extracted_101685 | Low-quality |
| S7_extracted_101686 | Not-determined |
| S7_extracted_101691 | Not-determined |
| S7_extracted_101718 | Low-quality |
| S7_extracted_101720 | Not-determined |
| S7_extracted_101724 | Not-determined |
| S7_extracted_101749 | Not-determined |
| S7_extracted_101773 | Not-determined |
| S7_extracted_101778 | Not-determined |
| S7_extracted_101786 | Not-determined |
| S7_extracted_101798 | Not-determined |
| S7_extracted_101813 | Not-determined |
| S7_extracted_101827 | Not-determined |
| S7_extracted_101839 | Not-determined |
| S7_extracted_101845 | Not-determined |
| S7_extracted_101878 | Not-determined |
| S7_extracted_101900 | Not-determined |
| S7_extracted_101906 | Not-determined |
| S7_extracted_101910 | Not-determined |
| S7_extracted_101918 | Low-quality |
| S7_extracted_101926 | Not-determined |
| S7_extracted_101935 | Not-determined |
| S7_extracted_101942 | Low-quality |
| S7_extracted_101971 | Not-determined |
| S7_extracted_101975 | Not-determined |
| S7_extracted_101984 | Not-determined |
| S7_extracted_101985 | Not-determined |
| S7_extracted_101988 | Not-determined |
| S7_extracted_101998 | Not-determined |
| S7_extracted_102032 | Not-determined |
| S7_extracted_102033 | Not-determined |
| S7_extracted_102040 | Low-quality |
| S7_extracted_102070 | Not-determined |
| S7_extracted_102141 | Not-determined |
| S7_extracted_102148 | Not-determined |
| S7_extracted_102151 | Not-determined |
| S7_extracted_102185 | Not-determined |
| S7_extracted_102192 | Not-determined |
| S7_extracted_102193 | Low-quality |
| S7_extracted_102196 | Not-determined |
| S7_extracted_102198 | Not-determined |
| S7_extracted_102205 | Not-determined |
| S7_extracted_102218 | Not-determined |
| S7_extracted_102255 | Low-quality |
| S7_extracted_102256 | Low-quality |
| S7_extracted_102261 | Not-determined |
| S7_extracted_102269 | Not-determined |
| S7_extracted_102340 | Not-determined |
| S7_extracted_102344 | Not-determined |
| S7_extracted_102356 | Not-determined |
| S7_extracted_102359 | Not-determined |
| S7_extracted_102369 | Not-determined |
| S7_extracted_102390 | Not-determined |
| S7_extracted_102394 | Not-determined |
| S7_extracted_102414 | Not-determined |
| S7_extracted_102420 | Not-determined |
| S7_extracted_102457 | Not-determined |
| S7_extracted_102478 | Not-determined |
| S7_extracted_102508 | Not-determined |
| S7_extracted_102509 | Not-determined |
| S7_extracted_102524 | Low-quality |
| S7_extracted_102532 | Not-determined |
| S7_extracted_102577 | Not-determined |
| S7_extracted_102594 | Not-determined |
| S7_extracted_102636 | Not-determined |
| S7_extracted_102643 | Not-determined |
| S7_extracted_102644 | Not-determined |
| S7_extracted_102646 | Not-determined |
| S7_extracted_102650 | Not-determined |
| S7_extracted_102656 | Not-determined |
| S7_extracted_102658 | Low-quality |
| S7_extracted_102667 | Not-determined |
| S7_extracted_102679 | Not-determined |
| S7_extracted_102705 | Not-determined |
| S7_extracted_102726 | Not-determined |
| S7_extracted_102727 | Low-quality |
| S7_extracted_102729 | Not-determined |
| S7_extracted_102791 | Low-quality |
| S7_extracted_102796 | Not-determined |
| S7_extracted_102809 | Not-determined |
| S7_extracted_102821 | Not-determined |
| S7_extracted_102827 | Not-determined |
| S7_extracted_102842 | Low-quality |
| S7_extracted_102847 | Not-determined |
| S7_extracted_102848 | Not-determined |
| S7_extracted_102893 | Not-determined |
| S7_extracted_102910 | Not-determined |
| S7_extracted_102921 | Not-determined |
| S7_extracted_102950 | Not-determined |
| S7_extracted_102977 | Not-determined |
| S7_extracted_103026 | Not-determined |
| S7_extracted_103027 | Not-determined |
| S7_extracted_103047 | Low-quality |
| S7_extracted_103054 | Not-determined |
| S7_extracted_103060 | Not-determined |
| S7_extracted_103064 | Not-determined |
| S7_extracted_103074 | Not-determined |
| S7_extracted_103084 | Not-determined |
| S7_extracted_103112 | Not-determined |
| S7_extracted_103115 | Not-determined |
| S7_extracted_103122 | Not-determined |
| S7_extracted_103124 | Not-determined |
| S7_extracted_103145 | Low-quality |
| S7_extracted_103160 | Not-determined |
| S7_extracted_103169 | Not-determined |
| S7_extracted_103201 | Not-determined |
| S7_extracted_103202 | Not-determined |
| S7_extracted_103214 | Not-determined |
| S7_extracted_103216 | Not-determined |
| S7_extracted_103219 | Not-determined |
| S7_extracted_103234 | Not-determined |
| S7_extracted_103235 | Not-determined |
| S7_extracted_103242 | Not-determined |
| S7_extracted_103248 | Not-determined |
| S7_extracted_103249 | Not-determined |
| S7_extracted_103251 | Not-determined |
| S7_extracted_103267 | Not-determined |
| S7_extracted_103269 | Not-determined |
| S7_extracted_103278 | Not-determined |
| S7_extracted_103291 | Not-determined |
| S7_extracted_103326 | Not-determined |
| S7_extracted_103340 | Not-determined |
| S7_extracted_103344 | Not-determined |
| S7_extracted_103349 | Not-determined |
| S7_extracted_103359 | Low-quality |
| S7_extracted_103363 | Low-quality |
| S7_extracted_103373 | Low-quality |
| S7_extracted_103392 | Not-determined |
| S7_extracted_103403 | Not-determined |
| S7_extracted_103409 | Not-determined |
| S7_extracted_103414 | Low-quality |
| S7_extracted_103425 | Low-quality |
| S7_extracted_103430 | Not-determined |
| S7_extracted_103437 | Not-determined |
| S7_extracted_103438 | Not-determined |
| S7_extracted_103459 | Not-determined |
| S7_extracted_103498 | Not-determined |
| S7_extracted_103511 | Not-determined |
| S7_extracted_103516 | Low-quality |
| S7_extracted_103529 | Not-determined |
| S7_extracted_103555 | Low-quality |
| S7_extracted_103580 | Not-determined |
| S7_extracted_103582 | Not-determined |
| S7_extracted_103626 | Not-determined |
| S7_extracted_103632 | Not-determined |
| S7_extracted_103636 | Not-determined |
| S7_extracted_103650 | Not-determined |
| S7_extracted_103663 | Not-determined |
| S7_extracted_103672 | Not-determined |
| S7_extracted_103684 | Not-determined |
| S7_extracted_103685 | Not-determined |
| S7_extracted_103690 | Low-quality |
| S7_extracted_103694 | Not-determined |
| S7_extracted_103696 | Not-determined |
| S7_extracted_103727 | Not-determined |
| S7_extracted_103756 | Not-determined |
| S7_extracted_103764 | Not-determined |
| S7_extracted_103766 | Not-determined |
| S7_extracted_103767 | Not-determined |
| S7_extracted_103783 | Low-quality |
| S7_extracted_103801 | Not-determined |
| S7_extracted_103802 | Not-determined |
| S7_extracted_103807 | Not-determined |
| S7_extracted_103843 | Low-quality |
| S7_extracted_103853 | Not-determined |
| S7_extracted_103904 | Not-determined |
| S7_extracted_103906 | Not-determined |
| S7_extracted_103911 | Not-determined |
| S7_extracted_103920 | Not-determined |
| S7_extracted_103924 | Low-quality |
| S7_extracted_103936 | Not-determined |
| S7_extracted_103945 | Low-quality |
| S7_extracted_103954 | Low-quality |
| S7_extracted_103960 | Low-quality |
| S7_extracted_103964 | Not-determined |
| S7_extracted_104021 | Not-determined |
| S7_extracted_104038 | Not-determined |
| S7_extracted_104048 | Not-determined |
| S7_extracted_104053 | Not-determined |
| S7_extracted_104064 | Not-determined |
| S7_extracted_104067 | Not-determined |
| S7_extracted_104068 | Not-determined |
| S7_extracted_104099 | Not-determined |
| S7_extracted_104100 | Not-determined |
| S7_extracted_104107 | Not-determined |
| S7_extracted_104136 | Low-quality |
| S7_extracted_104164 | Not-determined |
| S7_extracted_104181 | Not-determined |
| S7_extracted_104196 | Not-determined |
| S7_extracted_104198 | Not-determined |
| S7_extracted_104203 | Not-determined |
| S7_extracted_104213 | Not-determined |
| S7_extracted_104268 | Not-determined |
| S7_extracted_104283 | Not-determined |
| S7_extracted_104300 | Not-determined |
| S7_extracted_104315 | Not-determined |
| S7_extracted_104317 | Not-determined |
| S7_extracted_104320 | Not-determined |
| S7_extracted_104328 | Not-determined |
| S7_extracted_104329 | Not-determined |
| S7_extracted_104330 | Low-quality |
| S7_extracted_104337 | Not-determined |
| S7_extracted_104338 | Not-determined |
| S7_extracted_104388 | Not-determined |
| S7_extracted_104391 | Not-determined |
| S7_extracted_104422 | Not-determined |
| S7_extracted_104424 | Low-quality |
| S7_extracted_104426 | Not-determined |
| S7_extracted_104431 | Not-determined |
| S7_extracted_104446 | Not-determined |
| S7_extracted_104467 | Not-determined |
| S7_extracted_104468 | Not-determined |
| S7_extracted_104475 | Not-determined |
| S7_extracted_104477 | Not-determined |
| S7_extracted_104514 | Not-determined |
| S7_extracted_104565 | Not-determined |
| S7_extracted_104574 | Not-determined |
| S7_extracted_104581 | Low-quality |
| S7_extracted_104607 | Not-determined |
| S7_extracted_104608 | Low-quality |
| S7_extracted_104617 | Not-determined |
| S7_extracted_104620 | Not-determined |
| S7_extracted_104653 | Not-determined |
| S7_extracted_104654 | Not-determined |
| S7_extracted_104661 | Low-quality |
| S7_extracted_104691 | Not-determined |
| S7_extracted_104694 | Not-determined |
| S7_extracted_104698 | Not-determined |
| S7_extracted_104714 | Low-quality |
| S7_extracted_104723 | Not-determined |
| S7_extracted_104729 | Not-determined |
| S7_extracted_104746 | Not-determined |
| S7_extracted_104773 | Not-determined |
| S7_extracted_104774 | Not-determined |
| S7_extracted_104784 | Not-determined |
| S7_extracted_104786 | Not-determined |
| S7_extracted_104808 | Not-determined |
| S7_extracted_104809 | Not-determined |
| S7_extracted_104816 | Not-determined |
| S7_extracted_104817 | Not-determined |

| | |
|---|---|
| S7_extracted_104833 | Not-determined |
| S7_extracted_104835 | Not-determined |
| S7_extracted_104847 | Not-determined |
| S7_extracted_104880 | Not-determined |
| S7_extracted_104890 | Not-determined |
| S7_extracted_104899 | Not-determined |
| S7_extracted_104902 | Not-determined |
| S7_extracted_104904 | Not-determined |
| S7_extracted_104918 | Not-determined |
| S7_extracted_104920 | Not-determined |
| S7_extracted_104929 | Low-quality |
| S7_extracted_104933 | Not-determined |
| S7_extracted_104939 | Not-determined |
| S7_extracted_104941 | Not-determined |
| S7_extracted_104942 | Not-determined |
| S7_extracted_104954 | Not-determined |
| S7_extracted_104969 | Low-quality |
| S7_extracted_104978 | Not-determined |
| S7_extracted_104995 | Not-determined |
| S7_extracted_104996 | Not-determined |
| S7_extracted_105015 | Not-determined |
| S7_extracted_105026 | Not-determined |
| S7_extracted_105027 | Not-determined |
| S7_extracted_105053 | Low-quality |
| S7_extracted_105061 | Not-determined |
| S7_extracted_105069 | Not-determined |
| S7_extracted_105071 | Not-determined |
| S7_extracted_105079 | Not-determined |
| S7_extracted_105083 | Not-determined |
| S7_extracted_105102 | Not-determined |
| S7_extracted_105115 | Not-determined |
| S7_extracted_105116 | Not-determined |
| S7_extracted_105120 | Not-determined |
| S7_extracted_105121 | Not-determined |
| S7_extracted_105142 | Not-determined |
| S7_extracted_105157 | Not-determined |
| S7_extracted_105163 | Not-determined |
| S7_extracted_105172 | Not-determined |
| S7_extracted_105244 | Not-determined |
| S7_extracted_105254 | Not-determined |
| S7_extracted_105263 | Not-determined |
| S7_extracted_105278 | Low-quality |
| S7_extracted_105280 | Not-determined |
| S7_extracted_105284 | Not-determined |
| S7_extracted_105286 | Not-determined |
| S7_extracted_105287 | Not-determined |
| S7_extracted_105291 | Not-determined |
| S7_extracted_105292 | Not-determined |
| S7_extracted_105299 | Not-determined |
| S7_extracted_105311 | Not-determined |
| S7_extracted_105312 | Not-determined |
| S7_extracted_105340 | Not-determined |
| S7_extracted_105362 | Not-determined |
| S7_extracted_105366 | Not-determined |
| S7_extracted_105380 | Not-determined |
| S7_extracted_105386 | Not-determined |
| S7_extracted_105391 | Not-determined |
| S7_extracted_105408 | Not-determined |
| S7_extracted_105411 | Not-determined |
| S7_extracted_105412 | Low-quality |
| S7_extracted_105421 | Not-determined |
| S7_extracted_105441 | Not-determined |
| S7_extracted_105443 | Not-determined |
| S7_extracted_105445 | Not-determined |
| S7_extracted_105450 | Not-determined |
| S7_extracted_105468 | Low-quality |
| S7_extracted_105472 | Low-quality |
| S7_extracted_105474 | Not-determined |
| S7_extracted_105483 | Not-determined |
| S7_extracted_105491 | Low-quality |
| S7_extracted_105499 | Not-determined |
| S7_extracted_105504 | Not-determined |
| S7_extracted_105517 | Not-determined |
| S7_extracted_105553 | Low-quality |
| S7_extracted_105559 | Not-determined |
| S7_extracted_105577 | Not-determined |
| S7_extracted_105579 | Not-determined |
| S7_extracted_105597 | Low-quality |
| S7_extracted_105631 | Not-determined |
| S7_extracted_105664 | Not-determined |
| S7_extracted_105681 | Low-quality |
| S7_extracted_105689 | Not-determined |
| S7_extracted_105717 | Not-determined |
| S7_extracted_105727 | Not-determined |
| S7_extracted_105780 | Not-determined |
| S7_extracted_105792 | Not-determined |
| S7_extracted_105816 | Low-quality |
| S7_extracted_105819 | Not-determined |
| S7_extracted_105831 | Not-determined |
| S7_extracted_105840 | Not-determined |
| S7_extracted_105865 | Not-determined |
| S7_extracted_105874 | Not-determined |
| S7_extracted_105877 | Not-determined |
| S7_extracted_105878 | Not-determined |
| S7_extracted_105890 | Low-quality |
| S7_extracted_105903 | Not-determined |
| S7_extracted_105963 | Not-determined |
| S7_extracted_105981 | Low-quality |
| S7_extracted_105984 | Not-determined |
| S7_extracted_105986 | Not-determined |
| S7_extracted_105997 | Not-determined |
| S7_extracted_106054 | Not-determined |
| S7_extracted_106066 | Not-determined |
| S7_extracted_106068 | Not-determined |
| S7_extracted_106070 | Not-determined |
| S7_extracted_106093 | Not-determined |
| S7_extracted_106102 | Not-determined |
| S7_extracted_106104 | Not-determined |
| S7_extracted_106107 | Not-determined |
| S7_extracted_106116 | Low-quality |
| S7_extracted_106153 | Low-quality |
| S7_extracted_106157 | Not-determined |
| S7_extracted_106190 | Low-quality |
| S7_extracted_106192 | Not-determined |
| S7_extracted_106198 | Not-determined |
| S7_extracted_106215 | Not-determined |
| S7_extracted_106226 | Not-determined |
| S7_extracted_106258 | Not-determined |
| S7_extracted_106260 | Low-quality |
| S7_extracted_106271 | Not-determined |
| S7_extracted_106276 | Not-determined |
| S7_extracted_106279 | Low-quality |
| S7_extracted_106288 | Not-determined |
| S7_extracted_106291 | Not-determined |
| S7_extracted_106318 | Not-determined |
| S7_extracted_106347 | Not-determined |
| S7_extracted_106354 | Not-determined |
| S7_extracted_106373 | Low-quality |
| S7_extracted_106380 | Not-determined |
| S7_extracted_106381 | Not-determined |
| S7_extracted_106395 | Not-determined |
| S7_extracted_106417 | Not-determined |
| S7_extracted_106423 | Not-determined |
| S7_extracted_106438 | Not-determined |
| S7_extracted_106446 | Not-determined |
| S7_extracted_106449 | Not-determined |
| S7_extracted_106450 | Not-determined |
| S7_extracted_106456 | Not-determined |
| S7_extracted_106471 | Not-determined |
| S7_extracted_106477 | Not-determined |
| S7_extracted_106497 | Not-determined |
| S7_extracted_106499 | Not-determined |
| S7_extracted_106520 | Not-determined |
| S7_extracted_106528 | Not-determined |
| S7_extracted_106533 | Not-determined |
| S7_extracted_106535 | Not-determined |
| S7_extracted_106537 | Low-quality |
| S7_extracted_106539 | Not-determined |
| S7_extracted_106551 | Low-quality |
| S7_extracted_106557 | Not-determined |
| S7_extracted_106567 | Not-determined |
| S7_extracted_106574 | Not-determined |
| S7_extracted_106575 | Not-determined |
| S7_extracted_106576 | Not-determined |
| S7_extracted_106587 | Not-determined |
| S7_extracted_106600 | Not-determined |
| S7_extracted_106642 | Low-quality |
| S7_extracted_106647 | Not-determined |
| S7_extracted_106649 | Not-determined |
| S7_extracted_106656 | Not-determined |
| S7_extracted_106698 | Not-determined |
| S7_extracted_106703 | Not-determined |
| S7_extracted_106712 | Not-determined |
| S7_extracted_106718 | Not-determined |
| S7_extracted_106728 | Low-quality |
| S7_extracted_106737 | Not-determined |
| S7_extracted_106739 | Not-determined |
| S7_extracted_106745 | Not-determined |
| S7_extracted_106749 | Not-determined |
| S7_extracted_106753 | Low-quality |
| S7_extracted_106777 | Not-determined |
| S7_extracted_106786 | Not-determined |
| S7_extracted_106797 | Not-determined |
| S7_extracted_106798 | Not-determined |
| S7_extracted_106799 | Low-quality |
| S7_extracted_106815 | Not-determined |
| S7_extracted_106825 | Not-determined |
| S7_extracted_106830 | Not-determined |
| S7_extracted_106859 | Not-determined |
| S7_extracted_106860 | Not-determined |
| S7_extracted_106863 | Not-determined |
| S7_extracted_106868 | Not-determined |
| S7_extracted_106869 | Not-determined |
| S7_extracted_106873 | Not-determined |
| S7_extracted_106876 | Not-determined |
| S7_extracted_106901 | Low-quality |
| S7_extracted_106912 | Not-determined |
| S7_extracted_106918 | Low-quality |
| S7_extracted_106921 | Not-determined |
| S7_extracted_106934 | Not-determined |
| S7_extracted_106938 | Low-quality |
| S7_extracted_106939 | Low-quality |
| S7_extracted_106949 | Low-quality |
| S7_extracted_106950 | Not-determined |
| S7_extracted_106952 | Not-determined |
| S7_extracted_106956 | Not-determined |
| S7_extracted_106958 | Not-determined |
| S7_extracted_106974 | Not-determined |
| S7_extracted_106978 | Low-quality |
| S7_extracted_106981 | Not-determined |
| S7_extracted_106990 | Not-determined |
| S7_extracted_106999 | Not-determined |
| S7_extracted_107001 | Not-determined |
| S7_extracted_107025 | Not-determined |
| S7_extracted_107029 | Low-quality |
| S7_extracted_107046 | Low-quality |
| S7_extracted_107068 | Not-determined |
| S7_extracted_107076 | Not-determined |
| S7_extracted_107111 | Low-quality |
| S7_extracted_107116 | Not-determined |
| S7_extracted_107124 | Not-determined |
| S7_extracted_107125 | Not-determined |
| S7_extracted_107128 | Not-determined |
| S7_extracted_107143 | Low-quality |
| S7_extracted_107144 | Not-determined |
| S7_extracted_107158 | Not-determined |
| S7_extracted_107170 | Not-determined |
| S7_extracted_107178 | Low-quality |
| S7_extracted_107195 | Not-determined |
| S7_extracted_107205 | Not-determined |
| S7_extracted_107208 | Low-quality |
| S7_extracted_107217 | Not-determined |
| S7_extracted_107235 | Not-determined |
| S7_extracted_107239 | Not-determined |
| S7_extracted_107255 | Low-quality |
| S7_extracted_107291 | Not-determined |
| S7_extracted_107303 | Not-determined |
| S7_extracted_107310 | Not-determined |
| S7_extracted_107314 | Not-determined |
| S7_extracted_107328 | Not-determined |
| S7_extracted_107337 | Low-quality |
| S7_extracted_107338 | Not-determined |
| S7_extracted_107340 | Not-determined |
| S7_extracted_107366 | Not-determined |
| S7_extracted_107375 | Not-determined |
| S7_extracted_107394 | Not-determined |
| S7_extracted_107403 | Not-determined |
| S7_extracted_107405 | Not-determined |
| S7_extracted_107410 | Not-determined |
| S7_extracted_107424 | Not-determined |
| S7_extracted_107432 | Not-determined |
| S7_extracted_107438 | Not-determined |
| S7_extracted_107449 | Not-determined |
| S7_extracted_107463 | Not-determined |
| S7_extracted_107467 | Low-quality |
| S7_extracted_107469 | Not-determined |
| S7_extracted_107481 | Not-determined |
| S7_extracted_107494 | Not-determined |
| S7_extracted_107509 | Not-determined |
| S7_extracted_107512 | Not-determined |
| S7_extracted_107520 | Not-determined |
| S7_extracted_107523 | Not-determined |
| S7_extracted_107533 | Not-determined |
| S7_extracted_107538 | Not-determined |
| S7_extracted_107549 | Not-determined |
| S7_extracted_107553 | Not-determined |
| S7_extracted_107572 | Not-determined |
| S7_extracted_107582 | Not-determined |
| S7_extracted_107588 | Not-determined |
| S7_extracted_107590 | Not-determined |
| S7_extracted_107609 | Not-determined |
| S7_extracted_107625 | Low-quality |
| S7_extracted_107634 | Low-quality |
| S7_extracted_107641 | Not-determined |
| S7_extracted_107643 | Not-determined |
| S7_extracted_107650 | Not-determined |
| S7_extracted_107654 | Not-determined |
| S7_extracted_107656 | Not-determined |
| S7_extracted_107657 | Not-determined |
| S7_extracted_107672 | Not-determined |
| S7_extracted_107685 | Not-determined |
| S7_extracted_107690 | Not-determined |
| S7_extracted_107698 | Not-determined |
| S7_extracted_107703 | Not-determined |
| S7_extracted_107713 | Low-quality |
| S7_extracted_107720 | Not-determined |
| S7_extracted_107733 | Not-determined |
| S7_extracted_107739 | Not-determined |
| S7_extracted_107754 | Not-determined |
| S7_extracted_107770 | Not-determined |
| S7_extracted_107771 | Not-determined |
| S7_extracted_107781 | Not-determined |
| S7_extracted_107782 | Low-quality |
| S7_extracted_107784 | Low-quality |
| S7_extracted_107786 | Not-determined |
| S7_extracted_107792 | Not-determined |
| S7_extracted_107801 | Not-determined |
| S7_extracted_107832 | Not-determined |
| S7_extracted_107837 | Not-determined |
| S7_extracted_107871 | Not-determined |
| S7_extracted_107882 | Not-determined |
| S7_extracted_107887 | Not-determined |
| S7_extracted_107889 | Low-quality |
| S7_extracted_107892 | Not-determined |
| S7_extracted_107893 | Not-determined |
| S7_extracted_107916 | Low-quality |
| S7_extracted_107923 | Not-determined |
| S7_extracted_107942 | Not-determined |
| S7_extracted_107949 | Not-determined |
| S7_extracted_107956 | Not-determined |
| S7_extracted_107983 | Not-determined |
| S7_extracted_107993 | Not-determined |
| S7_extracted_108009 | Not-determined |
| S7_extracted_108027 | Not-determined |
| S7_extracted_108066 | Not-determined |
| S7_extracted_108102 | Low-quality |
| S7_extracted_108110 | Not-determined |
| S7_extracted_108119 | Not-determined |
| S7_extracted_108123 | Not-determined |
| S7_extracted_108134 | Not-determined |
| S7_extracted_108137 | Not-determined |
| S7_extracted_108152 | Not-determined |
| S7_extracted_108155 | Not-determined |
| S7_extracted_108160 | Not-determined |
| S7_extracted_108174 | Not-determined |
| S7_extracted_108178 | Not-determined |
| S7_extracted_108199 | Low-quality |
| S7_extracted_108211 | Low-quality |
| S7_extracted_108215 | Not-determined |
| S7_extracted_108220 | Not-determined |
| S7_extracted_108244 | Not-determined |
| S7_extracted_108265 | Not-determined |
| S7_extracted_108272 | Not-determined |
| S7_extracted_108276 | Not-determined |
| S7_extracted_108308 | Not-determined |
| S7_extracted_108312 | Not-determined |
| S7_extracted_108318 | Not-determined |
| S7_extracted_108324 | Not-determined |
| S7_extracted_108334 | Not-determined |
| S7_extracted_108337 | Not-determined |
| S7_extracted_108364 | Low-quality |
| S7_extracted_108382 | Not-determined |
| S7_extracted_108393 | Not-determined |
| S7_extracted_108394 | Not-determined |
| S7_extracted_108397 | Low-quality |
| S7_extracted_108403 | Low-quality |
| S7_extracted_108422 | Not-determined |
| S7_extracted_108424 | Low-quality |
| S7_extracted_108442 | Low-quality |
| S7_extracted_108481 | Not-determined |
| S7_extracted_108482 | Not-determined |
| S7_extracted_108499 | Not-determined |
| S7_extracted_108503 | Not-determined |
| S7_extracted_108504 | Not-determined |
| S7_extracted_108511 | Low-quality |
| S7_extracted_108526 | Low-quality |
| S7_extracted_108546 | Not-determined |
| S7_extracted_108563 | Not-determined |
| S7_extracted_108566 | Not-determined |
| S7_extracted_108569 | Not-determined |
| S7_extracted_108571 | Not-determined |
| S7_extracted_108576 | Low-quality |
| S7_extracted_108583 | Not-determined |
| S7_extracted_108603 | Not-determined |
| S7_extracted_108605 | Not-determined |
| S7_extracted_108613 | Not-determined |
| S7_extracted_108624 | Not-determined |
| S7_extracted_108632 | Low-quality |
| S7_extracted_108648 | Low-quality |
| S7_extracted_108653 | Low-quality |
| S7_extracted_108663 | Not-determined |
| S7_extracted_108692 | Not-determined |
| S7_extracted_108695 | Not-determined |
| S7_extracted_108699 | Not-determined |
| S7_extracted_108709 | Not-determined |
| S7_extracted_108716 | Low-quality |
| S7_extracted_108745 | Not-determined |
| S7_extracted_108754 | Low-quality |
| S7_extracted_108755 | Low-quality |
| S7_extracted_108757 | Not-determined |
| S7_extracted_108763 | Not-determined |
| S7_extracted_108765 | Not-determined |
| S7_extracted_108772 | Not-determined |
| S7_extracted_108778 | Not-determined |
| S7_extracted_108786 | Not-determined |
| S7_extracted_108805 | Not-determined |
| S7_extracted_108806 | Not-determined |
| S7_extracted_108842 | Not-determined |
| S7_extracted_108846 | Low-quality |
| S7_extracted_108851 | Not-determined |
| S7_extracted_108856 | Not-determined |
| S7_extracted_108871 | Not-determined |
| S7_extracted_108883 | Not-determined |
| S7_extracted_108908 | Not-determined |
| S7_extracted_108939 | Not-determined |
| S7_extracted_108940 | Not-determined |
| S7_extracted_108962 | Not-determined |
| S7_extracted_108970 | Low-quality |
| S7_extracted_108976 | Not-determined |
| S7_extracted_108989 | Not-determined |
| S7_extracted_108993 | Not-determined |
| S7_extracted_109010 | Not-determined |
| S7_extracted_109011 | Not-determined |
| S7_extracted_109013 | Not-determined |
| S7_extracted_109022 | Not-determined |
| S7_extracted_109035 | Not-determined |
| S7_extracted_109037 | Not-determined |
| S7_extracted_109054 | Not-determined |
| S7_extracted_109065 | Not-determined |
| S7_extracted_109073 | Not-determined |
| S7_extracted_109093 | Not-determined |
| S7_extracted_109101 | Low-quality |
| S7_extracted_109109 | Not-determined |
| S7_extracted_109126 | Low-quality |
| S7_extracted_109128 | Low-quality |
| S7_extracted_109130 | Not-determined |
| S7_extracted_109134 | Not-determined |
| S7_extracted_109156 | Not-determined |
| S7_extracted_109194 | Not-determined |
| S7_extracted_109200 | Not-determined |
| S7_extracted_109214 | Not-determined |
| S7_extracted_109219 | Not-determined |
| S7_extracted_109220 | Not-determined |
| S7_extracted_109231 | Not-determined |
| S7_extracted_109239 | Not-determined |
| S7_extracted_109246 | Not-determined |
| S7_extracted_109255 | Low-quality |
| S7_extracted_109266 | Not-determined |
| S7_extracted_109278 | Not-determined |
| S7_extracted_109286 | Not-determined |
| S7_extracted_109291 | Not-determined |
| S7_extracted_109309 | Not-determined |
| S7_extracted_109320 | Not-determined |
| S7_extracted_109321 | Low-quality |
| S7_extracted_109324 | Not-determined |
| S7_extracted_109346 | Not-determined |
| S7_extracted_109347 | Not-determined |
| S7_extracted_109359 | Not-determined |
| S7_extracted_109364 | Not-determined |
| S7_extracted_109381 | Not-determined |
| S7_extracted_109408 | Not-determined |
| S7_extracted_109409 | Not-determined |
| S7_extracted_109410 | Not-determined |
| S7_extracted_109417 | Not-determined |
| S7_extracted_109436 | Not-determined |
| S7_extracted_109438 | Not-determined |
| S7_extracted_109444 | Not-determined |
| S7_extracted_109447 | Not-determined |
| S7_extracted_109458 | Not-determined |
| S7_extracted_109468 | Not-determined |
| S7_extracted_109488 | Not-determined |
| S7_extracted_109499 | Not-determined |
| S7_extracted_109508 | Not-determined |
| S7_extracted_109509 | Not-determined |
| S7_extracted_109510 | Not-determined |
| S7_extracted_109515 | Low-quality |
| S7_extracted_109550 | Not-determined |
| S7_extracted_109553 | Not-determined |
| S7_extracted_109561 | Not-determined |
| S7_extracted_109603 | Low-quality |
| S7_extracted_109631 | Low-quality |
| S7_extracted_109650 | Not-determined |
| S7_extracted_109663 | Not-determined |
| S7_extracted_109675 | Low-quality |
| S7_extracted_109681 | Low-quality |
| S7_extracted_109688 | Not-determined |
| S7_extracted_109692 | Not-determined |
| S7_extracted_109717 | Not-determined |
| S7_extracted_109749 | Low-quality |
| S7_extracted_109755 | Low-quality |
| S7_extracted_109765 | Not-determined |
| S7_extracted_109770 | Not-determined |
| S7_extracted_109779 | Not-determined |
| S7_extracted_109802 | Not-determined |
| S7_extracted_109803 | Not-determined |
| S7_extracted_109814 | Not-determined |
| S7_extracted_109815 | Not-determined |
| S7_extracted_109821 | Not-determined |
| S7_extracted_109833 | Not-determined |
| S7_extracted_109851 | Not-determined |
| S7_extracted_109869 | Not-determined |
| S7_extracted_109880 | Low-quality |
| S7_extracted_109893 | Not-determined |
| S7_extracted_109905 | Not-determined |
| S7_extracted_109906 | Not-determined |
| S7_extracted_109926 | Not-determined |
| S7_extracted_109938 | Not-determined |
| S7_extracted_109949 | Not-determined |
| S7_extracted_109955 | Not-determined |
| S7_extracted_109964 | Not-determined |
| S7_extracted_109975 | Not-determined |
| S7_extracted_110002 | Not-determined |
| S7_extracted_110011 | Not-determined |
| S7_extracted_110013 | Not-determined |
| S7_extracted_110016 | Low-quality |
| S7_extracted_110018 | Not-determined |
| S7_extracted_110025 | Not-determined |
| S7_extracted_110044 | Low-quality |
| S7_extracted_110050 | Not-determined |
| S7_extracted_110063 | Not-determined |
| S7_extracted_110075 | Not-determined |
| S7_extracted_110077 | Low-quality |
| S7_extracted_110078 | Not-determined |
| S7_extracted_110115 | Not-determined |
| S7_extracted_110125 | Not-determined |
| S7_extracted_110126 | Not-determined |
| S7_extracted_110148 | Not-determined |
| S7_extracted_110206 | Low-quality |
| S7_extracted_110231 | Not-determined |
| S7_extracted_110254 | Not-determined |
| S7_extracted_110271 | Not-determined |
| S7_extracted_110276 | Low-quality |
| S7_extracted_110329 | Low-quality |
| S7_extracted_110342 | Not-determined |
| S7_extracted_110345 | Not-determined |
| S7_extracted_110363 | Not-determined |
| S7_extracted_110366 | Not-determined |
| S7_extracted_110382 | Not-determined |
| S7_extracted_110384 | Not-determined |
| S7_extracted_110390 | Not-determined |
| S7_extracted_110410 | Not-determined |
| S7_extracted_110415 | Not-determined |
| S7_extracted_110442 | Not-determined |
| S7_extracted_110460 | Low-quality |
| S7_extracted_110480 | Not-determined |
| S7_extracted_110484 | Not-determined |
| S7_extracted_110503 | Low-quality |
| S7_extracted_110520 | Not-determined |
| S7_extracted_110534 | Not-determined |
| S7_extracted_110555 | Low-quality |
| S7_extracted_110581 | Not-determined |
| S7_extracted_110590 | Low-quality |
| S7_extracted_110596 | Not-determined |
| S7_extracted_110603 | Not-determined |
| S7_extracted_110616 | Low-quality |
| S7_extracted_110621 | Low-quality |
| S7_extracted_110627 | Not-determined |
| S7_extracted_110634 | Not-determined |
| S7_extracted_110643 | Low-quality |
| S7_extracted_110652 | Low-quality |
| S7_extracted_110663 | Not-determined |
| S7_extracted_110665 | Not-determined |
| S7_extracted_110668 | Low-quality |
| S7_extracted_110674 | Low-quality |
| S7_extracted_110680 | Not-determined |
| S7_extracted_110681 | Not-determined |
| S7_extracted_110715 | Low-quality |
| S7_extracted_110717 | Not-determined |
| S7_extracted_110719 | Low-quality |
| S7_extracted_110730 | Not-determined |
| S7_extracted_110771 | Not-determined |
| S7_extracted_110815 | Not-determined |
| S7_extracted_110828 | Not-determined |
| S7_extracted_110829 | Not-determined |
| S7_extracted_110841 | Not-determined |
| S7_extracted_110846 | Not-determined |
| S7_extracted_110862 | Not-determined |
| S7_extracted_110867 | Not-determined |
| S7_extracted_110881 | Not-determined |
| S7_extracted_110883 | Not-determined |
| S7_extracted_110888 | Low-quality |
| S7_extracted_110892 | Not-determined |
| S7_extracted_110919 | Not-determined |
| S7_extracted_110965 | Not-determined |
| S7_extracted_110969 | Not-determined |
| S7_extracted_110972 | Low-quality |
| S7_extracted_111002 | Low-quality |
| S7_extracted_111005 | Not-determined |
| S7_extracted_111028 | Not-determined |
| S7_extracted_111040 | Not-determined |
| S7_extracted_111058 | Not-determined |
| S7_extracted_111074 | Not-determined |
| S7_extracted_111084 | Not-determined |
| S7_extracted_111129 | Not-determined |
| S7_extracted_111132 | Not-determined |
| S7_extracted_111141 | Not-determined |
| S7_extracted_111152 | Not-determined |
| S7_extracted_111173 | Not-determined |
| S7_extracted_111190 | Not-determined |
| S7_extracted_111191 | Not-determined |
| S7_extracted_111197 | Low-quality |
| S7_extracted_111208 | Low-quality |
| S7_extracted_111217 | Not-determined |
| S7_extracted_111220 | Not-determined |
| S7_extracted_111221 | Not-determined |
| S7_extracted_111222 | Not-determined |
| S7_extracted_111226 | Not-determined |
| S7_extracted_111242 | Low-quality |
| S7_extracted_111257 | Low-quality |
| S7_extracted_111265 | Not-determined |
| S7_extracted_111269 | Not-determined |
| S7_extracted_111280 | Low-quality |
| S7_extracted_111290 | Not-determined |
| S7_extracted_111328 | Low-quality |
| S7_extracted_111329 | Low-quality |
| S7_extracted_111348 | Not-determined |
| S7_extracted_111350 | Not-determined |
| S7_extracted_111367 | Not-determined |
| S7_extracted_111375 | Not-determined |

| | |
|---|---|
| S7_extracted_111396 | Not-determined |
| S7_extracted_111414 | Low-quality |
| S7_extracted_111417 | Not-determined |
| S7_extracted_111429 | Not-determined |
| S7_extracted_111436 | Not-determined |
| S7_extracted_111440 | Not-determined |
| S7_extracted_111452 | Not-determined |
| S7_extracted_111459 | Low-quality |
| S7_extracted_111487 | Not-determined |
| S7_extracted_111506 | Not-determined |
| S7_extracted_111507 | Not-determined |
| S7_extracted_111509 | Not-determined |
| S7_extracted_111510 | Not-determined |
| S7_extracted_111522 | Not-determined |
| S7_extracted_111525 | Not-determined |
| S7_extracted_111529 | Not-determined |
| S7_extracted_111532 | Low-quality |
| S7_extracted_111536 | Not-determined |
| S7_extracted_111549 | Not-determined |
| S7_extracted_111564 | Low-quality |
| S7_extracted_111570 | Low-quality |
| S7_extracted_111578 | Not-determined |
| S7_extracted_111607 | Not-determined |
| S7_extracted_111609 | Not-determined |
| S7_extracted_111639 | Not-determined |
| S7_extracted_111642 | Not-determined |
| S7_extracted_111654 | Not-determined |
| S7_extracted_111655 | Not-determined |
| S7_extracted_111663 | Not-determined |
| S7_extracted_111668 | Not-determined |
| S7_extracted_111678 | Not-determined |
| S7_extracted_111681 | Low-quality |
| S7_extracted_111717 | Low-quality |
| S7_extracted_111772 | Not-determined |
| S7_extracted_111789 | Not-determined |
| S7_extracted_111815 | Not-determined |
| S7_extracted_111832 | Not-determined |
| S7_extracted_111870 | Not-determined |
| S7_extracted_111877 | Low-quality |
| S7_extracted_111878 | Not-determined |
| S7_extracted_111900 | Not-determined |
| S7_extracted_111932 | Not-determined |
| S7_extracted_111938 | Low-quality |
| S7_extracted_111993 | Not-determined |
| S7_extracted_111998 | Not-determined |
| S7_extracted_112001 | Not-determined |
| S7_extracted_112004 | Not-determined |
| S7_extracted_112009 | Not-determined |
| S7_extracted_112012 | Not-determined |
| S7_extracted_112022 | Not-determined |
| S7_extracted_112029 | Not-determined |
| S7_extracted_112031 | Low-quality |
| S7_extracted_112037 | Not-determined |
| S7_extracted_112043 | Not-determined |
| S7_extracted_112050 | Low-quality |
| S7_extracted_112053 | Not-determined |
| S7_extracted_112057 | Not-determined |
| S7_extracted_112059 | Not-determined |
| S7_extracted_112068 | Not-determined |
| S7_extracted_112100 | Low-quality |
| S7_extracted_112115 | Not-determined |
| S7_extracted_112134 | Not-determined |
| S7_extracted_112150 | Not-determined |
| S7_extracted_112181 | Low-quality |
| S7_extracted_112192 | Low-quality |
| S7_extracted_112210 | Not-determined |
| S7_extracted_112225 | Not-determined |
| S7_extracted_112226 | Not-determined |
| S7_extracted_112229 | Not-determined |
| S7_extracted_112237 | Not-determined |
| S7_extracted_112241 | Not-determined |
| S7_extracted_112242 | Not-determined |
| S7_extracted_112246 | Not-determined |
| S7_extracted_112260 | Not-determined |
| S7_extracted_112263 | Not-determined |
| S7_extracted_112265 | Not-determined |
| S7_extracted_112291 | Not-determined |
| S7_extracted_112293 | Not-determined |
| S7_extracted_112299 | Not-determined |
| S7_extracted_112319 | Not-determined |
| S7_extracted_112321 | Low-quality |
| S7_extracted_112334 | Not-determined |
| S7_extracted_112339 | Not-determined |
| S7_extracted_112341 | Not-determined |
| S7_extracted_112356 | Not-determined |
| S7_extracted_112360 | Not-determined |
| S7_extracted_112425 | Low-quality |
| S7_extracted_112426 | Not-determined |
| S7_extracted_112430 | Not-determined |
| S7_extracted_112431 | Low-quality |
| S7_extracted_112438 | Not-determined |
| S7_extracted_112441 | Low-quality |
| S7_extracted_112465 | Not-determined |
| S7_extracted_112466 | Low-quality |
| S7_extracted_112518 | Not-determined |
| S7_extracted_112535 | Not-determined |
| S7_extracted_112566 | Not-determined |
| S7_extracted_112570 | Not-determined |
| S7_extracted_112582 | Low-quality |
| S7_extracted_112614 | Low-quality |
| S7_extracted_112633 | Not-determined |
| S7_extracted_112640 | Not-determined |
| S7_extracted_112655 | Not-determined |
| S7_extracted_112656 | Not-determined |
| S7_extracted_112662 | Not-determined |
| S7_extracted_112681 | Low-quality |
| S7_extracted_112683 | Not-determined |
| S7_extracted_112722 | Not-determined |
| S7_extracted_112728 | Not-determined |
| S7_extracted_112757 | Not-determined |
| S7_extracted_112774 | Not-determined |
| S7_extracted_112787 | Not-determined |
| S7_extracted_112803 | Not-determined |
| S7_extracted_112810 | Not-determined |
| S7_extracted_112811 | Not-determined |
| S7_extracted_112817 | Not-determined |
| S7_extracted_112819 | Not-determined |
| S7_extracted_112822 | Not-determined |
| S7_extracted_112836 | Not-determined |
| S13_vs_66508\|\|full | High-quality |
| S7_extracted_112926 | Not-determined |
| S7_extracted_112928 | Not-determined |
| S7_extracted_112939 | Not-determined |
| S7_extracted_112943 | Low-quality |
| S7_extracted_112946 | Not-determined |
| S7_extracted_112967 | Not-determined |
| S7_extracted_112988 | Not-determined |
| S7_extracted_113008 | Not-determined |
| S7_extracted_113015 | Not-determined |
| S7_extracted_113019 | Low-quality |
| S7_extracted_113070 | Not-determined |
| S7_extracted_113075 | Not-determined |
| S7_extracted_113084 | Low-quality |
| S7_extracted_113097 | Not-determined |
| S7_extracted_113123 | Low-quality |
| S7_extracted_113126 | Not-determined |
| S7_extracted_113140 | Low-quality |
| S7_extracted_113149 | Not-determined |
| S7_extracted_113155 | Not-determined |
| S7_extracted_113158 | Not-determined |
| S7_extracted_113199 | Not-determined |
| S7_extracted_113218 | Not-determined |
| S7_extracted_113219 | Not-determined |
| S7_extracted_113231 | Not-determined |
| S7_extracted_113236 | Not-determined |
| S7_extracted_113271 | Not-determined |
| S7_extracted_113292 | Not-determined |
| S7_extracted_113296 | Not-determined |
| S7_extracted_113303 | Low-quality |
| S7_extracted_113314 | Low-quality |
| S7_extracted_113333 | Not-determined |
| S7_extracted_113360 | Not-determined |
| S7_extracted_113362 | Low-quality |
| S7_extracted_113374 | Low-quality |
| S7_extracted_113377 | Low-quality |
| S7_extracted_113378 | Not-determined |
| S7_extracted_113380 | Not-determined |
| S7_extracted_113386 | Not-determined |
| S7_extracted_113393 | Low-quality |
| S7_extracted_113418 | Low-quality |
| S7_extracted_113445 | Not-determined |
| S7_extracted_113450 | Not-determined |
| S7_extracted_113453 | Not-determined |
| S7_extracted_113456 | Low-quality |
| S7_extracted_113457 | Not-determined |
| S7_extracted_113474 | Not-determined |
| S7_extracted_113478 | Not-determined |
| S7_extracted_113501 | Not-determined |
| S7_extracted_113504 | Not-determined |
| S7_extracted_113509 | Not-determined |
| S7_extracted_113516 | Not-determined |
| S7_extracted_113523 | Not-determined |
| S7_extracted_113530 | Low-quality |
| S7_extracted_113535 | Not-determined |
| S7_extracted_113538 | Not-determined |
| S7_extracted_113545 | Not-determined |
| S7_extracted_113563 | Low-quality |
| S7_extracted_113564 | Not-determined |
| S7_extracted_113566 | Not-determined |
| S7_extracted_113572 | Low-quality |
| S7_extracted_113583 | Not-determined |
| S7_extracted_113588 | Not-determined |
| S7_extracted_113589 | Not-determined |
| S7_extracted_113611 | Not-determined |
| S7_extracted_113636 | Not-determined |
| S7_extracted_113638 | Low-quality |
| S7_extracted_113642 | Not-determined |
| S7_extracted_113643 | Not-determined |
| S7_extracted_113644 | Not-determined |
| S7_extracted_113657 | Not-determined |
| S7_extracted_113658 | Low-quality |
| S7_extracted_113669 | Not-determined |
| S7_extracted_113683 | Not-determined |
| S7_extracted_113702 | Not-determined |
| S7_extracted_113714 | Not-determined |
| S7_extracted_113723 | Not-determined |
| S7_extracted_113732 | Not-determined |
| S7_extracted_113741 | Not-determined |
| S7_extracted_113744 | Not-determined |
| S7_extracted_113756 | Not-determined |
| S7_extracted_113764 | Not-determined |
| S7_extracted_113765 | Low-quality |
| S7_extracted_113766 | Low-quality |
| S7_extracted_113773 | Low-quality |
| S7_extracted_113775 | Low-quality |
| S7_extracted_113790 | Not-determined |
| S7_extracted_113791 | Not-determined |
| S7_extracted_113801 | Not-determined |
| S7_extracted_113804 | Not-determined |
| S7_extracted_113810 | Not-determined |
| S7_extracted_113820 | Not-determined |
| S7_extracted_113821 | Low-quality |
| S7_extracted_113830 | Low-quality |
| S7_extracted_113831 | Not-determined |
| S7_extracted_113847 | Not-determined |
| S7_extracted_113858 | Low-quality |
| S7_extracted_113864 | Not-determined |
| S7_extracted_113866 | Not-determined |
| S7_extracted_113881 | Not-determined |
| S7_extracted_113891 | Not-determined |
| S7_extracted_113913 | Not-determined |
| S7_extracted_113949 | Not-determined |
| S7_extracted_113962 | Not-determined |
| S7_extracted_113977 | Not-determined |
| S7_extracted_113982 | Low-quality |
| S7_extracted_113986 | Not-determined |
| S7_extracted_113993 | Not-determined |
| S7_extracted_114008 | Low-quality |
| S7_extracted_114019 | Not-determined |
| S7_extracted_114020 | Not-determined |
| S7_extracted_114061 | Not-determined |
| S7_extracted_114068 | Not-determined |
| S7_extracted_114074 | Not-determined |
| S7_extracted_114075 | Not-determined |
| S7_extracted_114081 | Not-determined |
| S7_extracted_114082 | Not-determined |
| S7_extracted_114094 | Not-determined |
| S7_extracted_114097 | Not-determined |
| S7_extracted_114100 | Not-determined |
| S7_extracted_114120 | Low-quality |
| S7_extracted_114128 | Not-determined |
| S7_extracted_114142 | Not-determined |
| S7_extracted_114160 | Not-determined |
| S7_extracted_114164 | Not-determined |
| S7_extracted_114200 | Not-determined |
| S7_extracted_114206 | Not-determined |
| S7_extracted_114208 | Low-quality |
| S7_extracted_114213 | Not-determined |
| S7_extracted_114220 | Not-determined |
| S7_extracted_114233 | Not-determined |
| S7_extracted_114246 | Low-quality |
| S7_extracted_114249 | Not-determined |
| S7_extracted_114251 | Not-determined |
| S7_extracted_114263 | Not-determined |
| S7_extracted_114277 | Low-quality |
| S7_extracted_114280 | Not-determined |
| S7_extracted_114281 | Not-determined |
| S7_extracted_114297 | Not-determined |
| S7_extracted_114345 | Low-quality |
| S7_extracted_114361 | Not-determined |
| S7_extracted_114370 | Not-determined |
| S7_extracted_114378 | Not-determined |
| S7_extracted_114389 | Not-determined |
| S7_extracted_114393 | Not-determined |
| S7_extracted_114395 | Not-determined |
| S7_extracted_114417 | Not-determined |
| S7_extracted_114440 | Low-quality |
| S7_extracted_114442 | Not-determined |
| S7_extracted_114445 | Not-determined |
| S7_extracted_114449 | Not-determined |
| S7_extracted_114455 | Not-determined |
| S7_extracted_114456 | Not-determined |
| S7_extracted_114461 | Not-determined |
| S7_vs_48832\|\|full | High-quality |
| S7_extracted_114478 | Low-quality |
| S7_extracted_114479 | Not-determined |
| S7_extracted_114480 | Not-determined |
| S7_extracted_114520 | Not-determined |
| S7_extracted_114545 | Not-determined |
| S7_extracted_114547 | Low-quality |
| S7_extracted_114549 | Not-determined |
| S7_extracted_114550 | Not-determined |
| S7_extracted_114555 | Not-determined |
| S7_extracted_114576 | Not-determined |
| S7_extracted_114587 | Not-determined |
| S7_extracted_114600 | Not-determined |
| S7_extracted_114614 | Not-determined |
| S7_extracted_114615 | Not-determined |
| S7_extracted_114629 | Not-determined |
| S7_extracted_114659 | Not-determined |
| S7_extracted_114665 | Not-determined |
| S7_extracted_114672 | Not-determined |
| S7_extracted_114686 | Not-determined |
| S7_extracted_114728 | Not-determined |
| S7_extracted_114737 | Not-determined |
| S7_extracted_114750 | Not-determined |
| S7_extracted_114771 | Not-determined |
| S7_extracted_114781 | Not-determined |
| S7_extracted_114807 | Not-determined |
| S7_extracted_114808 | Low-quality |
| S7_extracted_114828 | Not-determined |
| S7_extracted_114829 | Not-determined |
| S7_extracted_114833 | Low-quality |
| S7_extracted_114835 | Not-determined |
| S7_extracted_114839 | Not-determined |
| S7_extracted_114842 | Not-determined |
| S7_extracted_114845 | Low-quality |
| S7_extracted_114853 | Not-determined |
| S7_extracted_114877 | Not-determined |
| S7_extracted_114900 | Low-quality |
| S7_extracted_114921 | Not-determined |
| S7_extracted_114934 | Not-determined |
| S7_extracted_114947 | Not-determined |
| S7_extracted_114961 | Not-determined |
| S7_extracted_114965 | Low-quality |
| S7_extracted_114976 | Not-determined |
| S7_extracted_114978 | Not-determined |
| S7_extracted_115022 | Not-determined |
| S7_extracted_115050 | Not-determined |
| S7_extracted_115054 | Not-determined |
| S7_extracted_115066 | Not-determined |
| S7_extracted_115073 | Not-determined |
| S7_extracted_115076 | Not-determined |
| S7_extracted_115096 | Not-determined |
| S7_extracted_115115 | Low-quality |
| S7_extracted_115126 | Low-quality |
| S7_extracted_115133 | Not-determined |
| S7_extracted_115143 | Not-determined |
| S7_extracted_115173 | Low-quality |
| S7_extracted_115176 | Not-determined |
| S7_extracted_115195 | Not-determined |
| S7_extracted_115216 | Not-determined |
| S7_extracted_115231 | Not-determined |
| S7_extracted_115232 | Not-determined |
| S7_extracted_115235 | Low-quality |
| S7_extracted_115250 | Not-determined |
| S7_extracted_115252 | Not-determined |
| S7_extracted_115258 | Not-determined |
| S7_extracted_115263 | Not-determined |
| S7_extracted_115266 | Low-quality |
| S7_extracted_115275 | Not-determined |
| S7_extracted_115288 | Not-determined |
| S7_extracted_115291 | Not-determined |
| S7_extracted_115300 | Low-quality |
| S7_extracted_115314 | Not-determined |
| S7_extracted_115352 | Not-determined |
| S7_extracted_115359 | Not-determined |
| S7_extracted_115386 | Low-quality |
| S7_extracted_115389 | Not-determined |
| S7_extracted_115394 | Not-determined |
| S7_extracted_115396 | Not-determined |
| S7_extracted_115397 | Not-determined |
| S7_extracted_115408 | Not-determined |
| S7_extracted_115409 | Not-determined |
| S7_extracted_115430 | Low-quality |
| S7_extracted_115433 | Not-determined |
| S7_extracted_115436 | Not-determined |
| S7_extracted_115455 | Low-quality |
| S7_extracted_115485 | Not-determined |
| S7_extracted_115512 | Not-determined |
| S7_extracted_115521 | Not-determined |
| S7_extracted_115536 | Not-determined |
| S7_extracted_115562 | Not-determined |
| S7_extracted_115565 | Not-determined |
| S7_extracted_115576 | Low-quality |
| S7_extracted_115578 | Low-quality |
| S7_extracted_115582 | Low-quality |
| S7_extracted_115588 | Low-quality |
| S7_extracted_115607 | Not-determined |
| S7_extracted_115609 | Low-quality |
| S7_extracted_115616 | Not-determined |
| S7_extracted_115631 | Not-determined |
| S7_extracted_115654 | Not-determined |
| S7_extracted_115657 | Not-determined |
| S7_extracted_115664 | Not-determined |
| S7_extracted_115684 | Not-determined |
| S7_extracted_115688 | Not-determined |
| S7_extracted_115694 | Not-determined |
| S7_extracted_115722 | Low-quality |
| S7_extracted_115724 | Not-determined |
| S7_extracted_115744 | Not-determined |
| S7_extracted_115745 | Not-determined |
| S7_extracted_115750 | Not-determined |
| S7_extracted_115758 | Low-quality |
| S7_extracted_115769 | Low-quality |
| S7_extracted_115791 | Not-determined |
| S7_extracted_115796 | Low-quality |
| S7_extracted_115802 | Not-determined |
| S7_extracted_115822 | Not-determined |
| S7_extracted_115826 | Not-determined |
| S7_extracted_115840 | Not-determined |
| S7_extracted_115860 | Not-determined |
| S7_extracted_115867 | Not-determined |
| S7_extracted_115890 | Not-determined |
| S7_extracted_115901 | Not-determined |
| S7_extracted_115904 | Not-determined |
| S7_extracted_115911 | Not-determined |
| S7_extracted_115913 | Not-determined |
| S7_extracted_115914 | Low-quality |
| S7_extracted_115920 | Not-determined |
| S7_extracted_115923 | Not-determined |
| S7_extracted_115933 | Not-determined |
| S7_extracted_115953 | Not-determined |
| S7_extracted_115969 | Not-determined |
| S7_extracted_115978 | Low-quality |
| S7_extracted_115981 | Low-quality |
| S7_extracted_115989 | Not-determined |
| S7_extracted_116003 | Low-quality |
| S7_extracted_116012 | Not-determined |
| S7_extracted_116020 | Not-determined |
| S7_extracted_116022 | Not-determined |
| S7_extracted_116036 | Not-determined |
| S7_extracted_116039 | Not-determined |
| S7_extracted_116041 | Not-determined |
| S7_extracted_116055 | Not-determined |
| S7_extracted_116060 | Not-determined |
| S7_extracted_116066 | Not-determined |
| S7_extracted_116081 | Not-determined |
| S7_extracted_116110 | Not-determined |
| S7_extracted_116117 | Not-determined |
| S7_extracted_116118 | Not-determined |
| S7_extracted_116122 | Not-determined |
| S7_extracted_116126 | Not-determined |
| S7_extracted_116133 | Not-determined |
| S7_extracted_116136 | Not-determined |
| S7_extracted_116143 | Not-determined |
| S7_extracted_116148 | Not-determined |
| S7_extracted_116149 | Not-determined |
| S7_extracted_116160 | Not-determined |
| S7_extracted_116166 | Not-determined |
| S7_extracted_116170 | Not-determined |
| S7_extracted_116182 | Not-determined |
| S7_extracted_116199 | Not-determined |
| S7_extracted_116227 | Not-determined |
| S7_extracted_116229 | Not-determined |
| S7_extracted_116231 | Low-quality |
| S7_extracted_116266 | Not-determined |
| S7_extracted_116284 | Not-determined |
| S7_extracted_116291 | Not-determined |
| S7_extracted_116295 | Not-determined |
| S7_extracted_116298 | Low-quality |
| S7_extracted_116302 | Not-determined |
| S7_extracted_116308 | Not-determined |
| S7_extracted_116318 | Not-determined |
| S7_extracted_116336 | Not-determined |
| S7_extracted_116372 | Low-quality |
| S7_extracted_116382 | Not-determined |
| S7_extracted_116384 | Low-quality |
| S7_extracted_116387 | Not-determined |
| S7_extracted_116432 | Not-determined |
| S7_extracted_116434 | Not-determined |
| S7_extracted_116435 | Low-quality |
| S7_extracted_116440 | Not-determined |
| S7_extracted_116441 | Not-determined |
| S7_extracted_116454 | Not-determined |
| S7_extracted_116475 | Not-determined |
| S7_extracted_116487 | Not-determined |
| S7_extracted_116493 | Not-determined |
| S7_extracted_116497 | Not-determined |
| S7_extracted_116501 | Not-determined |
| S7_extracted_116536 | Not-determined |
| S7_extracted_116566 | Not-determined |
| S7_extracted_116572 | Not-determined |
| S7_extracted_116578 | Low-quality |
| S7_extracted_116585 | Not-determined |
| S7_extracted_116616 | Not-determined |
| S7_extracted_116617 | Low-quality |
| S7_extracted_116620 | Low-quality |
| S7_extracted_116626 | Low-quality |
| S7_extracted_116627 | Not-determined |
| S7_extracted_116646 | Not-determined |
| S7_extracted_116650 | Not-determined |
| S7_extracted_116657 | Not-determined |
| S7_extracted_116668 | Not-determined |
| S7_extracted_116671 | Not-determined |
| S7_extracted_116686 | Not-determined |
| S7_extracted_116690 | Not-determined |
| S7_extracted_116703 | Not-determined |
| S7_extracted_116709 | Low-quality |
| S7_extracted_116742 | Not-determined |
| S7_extracted_116750 | Low-quality |
| S7_extracted_116759 | Low-quality |
| S7_extracted_116779 | Not-determined |
| S7_extracted_116785 | Low-quality |
| S7_extracted_116786 | Not-determined |
| S7_extracted_116788 | Not-determined |
| S7_extracted_116793 | Not-determined |
| S7_extracted_116810 | Low-quality |
| S7_extracted_116825 | Not-determined |
| S7_extracted_116830 | Not-determined |
| S7_extracted_116838 | Not-determined |
| S7_extracted_116840 | Not-determined |
| S7_extracted_116842 | Not-determined |
| S7_extracted_116873 | Low-quality |
| S7_extracted_116883 | Not-determined |
| S7_extracted_116889 | Not-determined |
| S7_extracted_116890 | Not-determined |
| S7_extracted_116899 | Not-determined |
| S7_extracted_116901 | Not-determined |
| S7_extracted_116905 | Not-determined |
| S7_extracted_116913 | Not-determined |
| S7_extracted_116929 | Not-determined |
| S7_extracted_116940 | Low-quality |
| S7_extracted_116941 | Not-determined |
| S7_extracted_116962 | Not-determined |
| S7_extracted_116994 | Not-determined |
| S7_extracted_116995 | Not-determined |
| S7_extracted_117009 | Not-determined |
| S7_extracted_117012 | Not-determined |
| S7_extracted_117042 | Not-determined |
| S7_extracted_117047 | Not-determined |
| S7_extracted_117081 | Low-quality |
| S7_extracted_117084 | Low-quality |
| S7_extracted_117101 | Not-determined |
| S7_extracted_117113 | Not-determined |
| S7_extracted_117126 | Not-determined |
| S7_extracted_117127 | Not-determined |
| S7_extracted_117132 | Not-determined |
| S7_extracted_117149 | Low-quality |
| S7_extracted_117181 | Low-quality |
| S7_extracted_117200 | Not-determined |
| S7_extracted_117201 | Not-determined |
| S7_extracted_117207 | Low-quality |
| S7_extracted_117209 | Not-determined |
| S7_extracted_117215 | Not-determined |
| S7_extracted_117216 | Not-determined |
| S7_extracted_117249 | Not-determined |
| S7_extracted_117252 | Not-determined |
| S7_extracted_117253 | Not-determined |
| S7_extracted_117315 | Not-determined |
| S7_extracted_117325 | Not-determined |
| S7_extracted_117328 | Low-quality |
| S7_extracted_117347 | Not-determined |
| S7_extracted_117364 | Low-quality |
| S7_extracted_117386 | Not-determined |
| S7_extracted_117392 | Not-determined |
| S7_extracted_117395 | Low-quality |
| S7_extracted_117413 | Not-determined |
| S7_extracted_117428 | Not-determined |
| S7_extracted_117429 | Low-quality |
| S7_extracted_117432 | Not-determined |
| S7_extracted_117436 | Low-quality |
| S7_extracted_117438 | Not-determined |
| S7_extracted_117453 | Not-determined |
| S7_extracted_117478 | Not-determined |
| S7_extracted_117480 | Not-determined |
| S7_extracted_117483 | Not-determined |
| S7_extracted_117490 | Not-determined |
| S7_extracted_117495 | Low-quality |
| S7_extracted_117498 | Not-determined |
| S7_extracted_117507 | Low-quality |
| S7_extracted_117511 | Not-determined |
| S7_extracted_117552 | Not-determined |
| S7_extracted_117562 | Not-determined |
| S7_extracted_117580 | Not-determined |
| S7_extracted_117607 | Not-determined |
| S7_extracted_117609 | Not-determined |
| S7_extracted_117624 | Low-quality |
| S7_extracted_117631 | Not-determined |
| S7_extracted_117647 | Low-quality |
| S7_extracted_117649 | Not-determined |
| S7_extracted_117657 | Not-determined |
| S7_extracted_117658 | Not-determined |
| S7_extracted_117693 | Low-quality |
| S7_vs_59465\|\|full | Medium-quality |
| S7_extracted_117718 | Low-quality |
| S7_extracted_117732 | Not-determined |
| S7_extracted_117734 | Not-determined |
| S7_extracted_117738 | Not-determined |
| S7_extracted_117755 | Not-determined |
| S7_extracted_117784 | Low-quality |
| S7_extracted_117791 | Not-determined |
| S7_extracted_117802 | Low-quality |
| S7_extracted_117812 | Low-quality |
| S7_extracted_117823 | Not-determined |
| S7_extracted_117831 | Not-determined |
| S7_extracted_117832 | Not-determined |
| S7_extracted_117834 | Not-determined |
| S7_extracted_117854 | Not-determined |
| S7_extracted_117864 | Not-determined |
| S7_extracted_117890 | Not-determined |
| S7_extracted_117892 | Not-determined |
| S7_extracted_117898 | Not-determined |
| S7_extracted_117899 | Not-determined |
| S7_extracted_117956 | Not-determined |
| S7_extracted_117960 | Not-determined |
| S7_extracted_117984 | Low-quality |
| S7_extracted_118002 | Low-quality |
| S7_extracted_118017 | Low-quality |

| | |
|---|---|
| S7_extracted_118048 | Not-determined |
| S7_extracted_118051 | Not-determined |
| S7_extracted_118065 | Not-determined |
| S7_extracted_118069 | Not-determined |
| S7_extracted_118074 | Low-quality |
| S7_extracted_118079 | Not-determined |
| S7_extracted_118082 | Not-determined |
| S7_extracted_118105 | Not-determined |
| S7_extracted_118106 | Not-determined |
| S7_extracted_118110 | Not-determined |
| S7_extracted_118114 | Not-determined |
| S7_extracted_118122 | Not-determined |
| S7_extracted_118127 | Not-determined |
| S7_extracted_118132 | Not-determined |
| S7_extracted_118150 | Not-determined |
| S7_extracted_118152 | Low-quality |
| S7_extracted_118158 | Not-determined |
| S7_extracted_118171 | Not-determined |
| S7_extracted_118172 | Not-determined |
| S7_extracted_118192 | Not-determined |
| S7_extracted_118252 | Not-determined |
| S7_extracted_118255 | Not-determined |
| S7_extracted_118261 | Not-determined |
| S7_extracted_118298 | Not-determined |
| S7_extracted_118306 | Not-determined |
| S7_extracted_118312 | Not-determined |
| S7_extracted_118325 | Not-determined |
| S7_extracted_118346 | Not-determined |
| S7_extracted_118353 | Not-determined |
| S7_extracted_118356 | Not-determined |
| S7_extracted_118357 | Low-quality |
| S7_extracted_118361 | Not-determined |
| S7_extracted_118379 | Not-determined |
| S7_extracted_118384 | Low-quality |
| S7_extracted_118386 | Not-determined |
| S7_extracted_118388 | Not-determined |
| S7_extracted_118389 | Low-quality |
| S7_extracted_118392 | Not-determined |
| S7_extracted_118394 | Not-determined |
| S7_extracted_118401 | Not-determined |
| S7_extracted_118410 | Low-quality |
| S7_extracted_118411 | Not-determined |
| S7_extracted_118418 | Not-determined |
| S7_extracted_118441 | Not-determined |
| S7_extracted_118444 | Not-determined |
| S7_extracted_118482 | Not-determined |
| S7_extracted_118483 | Not-determined |
| S7_extracted_118505 | Not-determined |
| S7_extracted_118508 | Not-determined |
| S7_extracted_118516 | Not-determined |
| S7_extracted_118553 | Low-quality |
| S7_extracted_118561 | Not-determined |
| S7_extracted_118585 | Not-determined |
| S7_extracted_118603 | Not-determined |
| S7_extracted_118614 | Not-determined |
| S7_extracted_118615 | Low-quality |
| S7_extracted_118621 | Not-determined |
| S7_extracted_118627 | Not-determined |
| S7_extracted_118632 | Not-determined |
| S7_extracted_118656 | Not-determined |
| S7_extracted_118667 | Not-determined |
| S7_extracted_118668 | Not-determined |
| S7_extracted_118682 | Not-determined |
| S7_extracted_118692 | Not-determined |
| S7_extracted_118695 | Not-determined |
| S7_extracted_118697 | Not-determined |
| S7_extracted_118701 | Not-determined |
| S7_extracted_118704 | Low-quality |
| S7_extracted_118727 | Not-determined |
| S7_extracted_118739 | Not-determined |
| S7_extracted_118747 | Low-quality |
| S7_extracted_118780 | Low-quality |
| S7_extracted_118815 | Not-determined |
| S7_extracted_118853 | Not-determined |
| S7_extracted_118855 | Not-determined |
| S7_extracted_118863 | Not-determined |
| S7_extracted_118869 | Not-determined |
| S7_extracted_118870 | Not-determined |
| S7_extracted_118882 | Not-determined |
| S7_extracted_118913 | Low-quality |
| S7_extracted_118936 | Not-determined |
| S7_extracted_118961 | Not-determined |
| S7_extracted_118966 | Not-determined |
| S7_extracted_118972 | Low-quality |
| S7_extracted_118976 | Low-quality |
| S7_extracted_119020 | Not-determined |
| S7_extracted_119031 | Not-determined |
| S7_extracted_119052 | Low-quality |
| S7_extracted_119070 | Low-quality |
| S7_extracted_119093 | Low-quality |
| S7_extracted_119094 | Not-determined |
| S7_extracted_119106 | Not-determined |
| S7_extracted_119107 | Not-determined |
| S7_extracted_119109 | Low-quality |
| S7_extracted_119111 | Not-determined |
| S7_extracted_119115 | Low-quality |
| S7_extracted_119138 | Not-determined |
| S7_extracted_119144 | Low-quality |
| S7_extracted_119167 | Low-quality |
| S7_extracted_119169 | Low-quality |
| S7_extracted_119179 | Not-determined |
| S7_extracted_119211 | Not-determined |
| S7_extracted_119226 | Low-quality |
| S7_extracted_119249 | Not-determined |
| S7_extracted_119255 | Not-determined |
| S7_extracted_119257 | Not-determined |
| S7_extracted_119273 | Not-determined |
| S7_extracted_119276 | Low-quality |
| S7_extracted_119295 | Low-quality |
| S7_extracted_119319 | Not-determined |
| S7_extracted_119334 | Not-determined |
| S7_extracted_119336 | Low-quality |
| S7_extracted_119354 | Not-determined |
| S7_extracted_119359 | Not-determined |
| S7_extracted_119367 | Not-determined |
| S7_extracted_119392 | Low-quality |
| S7_extracted_119403 | Not-determined |
| S7_extracted_119404 | Not-determined |
| S7_extracted_119410 | Not-determined |
| S7_extracted_119425 | Low-quality |
| S7_extracted_119428 | Low-quality |
| S7_extracted_119464 | Not-determined |
| S7_extracted_119486 | Low-quality |
| S7_extracted_119501 | Not-determined |
| S7_extracted_119536 | Not-determined |
| S7_extracted_119538 | Low-quality |
| S7_extracted_119554 | Not-determined |
| S7_extracted_119564 | Not-determined |
| S7_extracted_119573 | Not-determined |
| S7_extracted_119590 | Low-quality |
| S7_extracted_119622 | Not-determined |
| S7_extracted_119643 | Not-determined |
| S7_extracted_119646 | Not-determined |
| S7_extracted_119647 | Low-quality |
| S7_extracted_119651 | Not-determined |
| S7_extracted_119676 | Not-determined |
| S7_extracted_119686 | Not-determined |
| S7_extracted_119691 | Not-determined |
| S7_extracted_119694 | Not-determined |
| S7_extracted_119719 | Not-determined |
| S7_extracted_119725 | Low-quality |
| S7_extracted_119727 | Not-determined |
| S7_extracted_119746 | Not-determined |
| S7_extracted_119748 | Not-determined |
| S7_extracted_119749 | Not-determined |
| S7_extracted_119752 | Not-determined |
| S7_extracted_119757 | Low-quality |
| S7_extracted_119760 | Not-determined |
| S7_extracted_119786 | Not-determined |
| S7_extracted_119793 | Not-determined |
| S7_extracted_119802 | Not-determined |
| S7_extracted_119805 | Not-determined |
| S7_extracted_119817 | Not-determined |
| S7_extracted_119823 | Not-determined |
| S7_extracted_119828 | Low-quality |
| S7_extracted_119830 | Not-determined |
| S7_extracted_119839 | Not-determined |
| S7_extracted_119847 | Not-determined |
| S7_extracted_119885 | Not-determined |
| S7_extracted_119893 | Not-determined |
| S7_extracted_119897 | Not-determined |
| S7_extracted_119900 | Not-determined |
| S7_extracted_119915 | Not-determined |
| S7_extracted_119924 | Low-quality |
| S7_extracted_119943 | Not-determined |
| S7_extracted_119961 | Not-determined |
| S7_extracted_119965 | Not-determined |
| S7_extracted_119973 | Not-determined |
| S7_extracted_119974 | Not-determined |
| S7_extracted_119981 | Not-determined |
| S7_extracted_119999 | Not-determined |
| S7_extracted_120009 | Not-determined |
| S7_extracted_120015 | Low-quality |
| S7_extracted_120032 | Not-determined |
| S7_extracted_120076 | Not-determined |
| S7_extracted_120085 | Not-determined |
| S7_extracted_120110 | Low-quality |
| S7_extracted_120123 | Not-determined |
| S7_extracted_120135 | Not-determined |
| S7_extracted_120137 | Not-determined |
| S7_extracted_120141 | Not-determined |
| S7_extracted_120144 | Not-determined |
| S7_extracted_120156 | Low-quality |
| S7_extracted_120157 | Not-determined |
| S7_extracted_120172 | Not-determined |
| S7_extracted_120177 | Not-determined |
| S7_extracted_120178 | Not-determined |
| S7_extracted_120188 | Not-determined |
| S7_extracted_120196 | Not-determined |
| S7_extracted_120215 | Low-quality |
| S7_extracted_120229 | Low-quality |
| S7_extracted_120242 | Low-quality |
| S7_extracted_120274 | Not-determined |
| S7_extracted_120283 | Not-determined |
| S7_extracted_120286 | Not-determined |
| S7_extracted_120297 | Not-determined |
| S7_extracted_120302 | Low-quality |
| S7_extracted_120323 | Not-determined |
| S7_extracted_120336 | Not-determined |
| S7_extracted_120354 | Low-quality |
| S7_extracted_120368 | Low-quality |
| S7_extracted_120372 | Not-determined |
| S7_extracted_120380 | Not-determined |
| S7_extracted_120385 | Not-determined |
| S7_extracted_120388 | Not-determined |
| S7_extracted_120422 | Not-determined |
| S7_extracted_120434 | Not-determined |
| S7_extracted_120440 | Not-determined |
| S7_extracted_120451 | Not-determined |
| S7_extracted_120452 | Not-determined |
| S7_extracted_120459 | Not-determined |
| S7_extracted_120478 | Not-determined |
| S7_extracted_120483 | Not-determined |
| S7_extracted_120505 | Low-quality |
| S7_extracted_120511 | Low-quality |
| S7_extracted_120514 | Low-quality |
| S7_extracted_120523 | Not-determined |
| S7_extracted_120527 | Not-determined |
| S7_extracted_120534 | Not-determined |
| S7_extracted_120556 | Not-determined |
| S7_extracted_120576 | Not-determined |
| S7_extracted_120579 | Not-determined |
| S7_extracted_120596 | Not-determined |
| S7_extracted_120617 | Not-determined |
| S7_extracted_120632 | Not-determined |
| S7_extracted_120647 | Not-determined |
| S7_extracted_120650 | Not-determined |
| S7_extracted_120658 | Not-determined |
| S7_extracted_120661 | Not-determined |
| S7_extracted_120671 | Low-quality |
| S7_extracted_120679 | Not-determined |
| S7_extracted_120688 | Not-determined |
| S7_extracted_120699 | Not-determined |
| S7_extracted_120701 | Not-determined |
| S7_extracted_120715 | Not-determined |
| S7_extracted_120738 | Not-determined |
| S7_extracted_120743 | Low-quality |
| S7_extracted_120757 | Not-determined |
| S7_extracted_120759 | Not-determined |
| S7_extracted_120763 | Not-determined |
| S7_extracted_120764 | Not-determined |
| S7_extracted_120791 | Not-determined |
| S7_extracted_120807 | Not-determined |
| S7_extracted_120831 | Not-determined |
| S7_extracted_120838 | Not-determined |
| S7_extracted_120842 | Low-quality |
| S7_extracted_120846 | Not-determined |
| S7_extracted_120848 | Not-determined |
| S7_extracted_120866 | Not-determined |
| S7_extracted_120896 | Not-determined |
| S7_extracted_120899 | Not-determined |
| S7_extracted_120921 | Not-determined |
| S7_extracted_120936 | Not-determined |
| S7_extracted_120953 | Not-determined |
| S7_extracted_120958 | Not-determined |
| S7_extracted_120969 | Not-determined |
| S7_extracted_120971 | Not-determined |
| S7_extracted_120977 | Not-determined |
| S7_extracted_120999 | Not-determined |
| S7_extracted_121018 | Not-determined |
| S7_extracted_121025 | Not-determined |
| S7_extracted_121026 | Not-determined |
| S7_extracted_121029 | Not-determined |
| S7_extracted_121045 | Not-determined |
| S7_extracted_121046 | Not-determined |
| S7_extracted_121058 | Not-determined |
| S7_extracted_121086 | Not-determined |
| S7_extracted_121091 | Not-determined |
| S7_extracted_121109 | Low-quality |
| S7_extracted_121124 | Low-quality |
| S7_extracted_121127 | Not-determined |
| S7_extracted_121132 | Not-determined |
| S7_extracted_121142 | Not-determined |
| S7_extracted_121156 | Not-determined |
| S7_extracted_121157 | Low-quality |
| S7_extracted_121158 | Not-determined |
| S7_extracted_121167 | Low-quality |
| S7_extracted_121168 | Not-determined |
| S7_extracted_121170 | Not-determined |
| S7_extracted_121190 | Not-determined |
| S7_extracted_121202 | Not-determined |
| S7_extracted_121207 | Not-determined |
| S7_extracted_121224 | Low-quality |
| S7_extracted_121273 | Low-quality |
| S7_extracted_121274 | Not-determined |
| S7_extracted_121283 | Low-quality |
| S7_extracted_121284 | Not-determined |
| S7_extracted_121287 | Not-determined |
| S7_extracted_121289 | Not-determined |
| S7_extracted_121292 | Not-determined |
| S7_extracted_121296 | Not-determined |
| S7_extracted_121301 | Not-determined |
| S7_extracted_121334 | Not-determined |
| S7_extracted_121350 | Not-determined |
| S7_extracted_121353 | Not-determined |
| S7_extracted_121355 | Not-determined |
| S7_extracted_121363 | Not-determined |
| S7_extracted_121373 | Low-quality |
| S7_extracted_121392 | Not-determined |
| S7_extracted_121420 | Not-determined |
| S7_extracted_121438 | Low-quality |
| S7_extracted_121441 | Not-determined |
| S7_extracted_121442 | Not-determined |
| S7_extracted_121495 | Not-determined |
| S7_extracted_121511 | Not-determined |
| S7_extracted_121522 | Not-determined |
| S7_extracted_121530 | Not-determined |
| S7_extracted_121537 | Low-quality |
| S7_extracted_121540 | Low-quality |
| S7_extracted_121579 | Not-determined |
| S7_extracted_121600 | Low-quality |
| S7_extracted_121612 | Not-determined |
| S7_extracted_121617 | Not-determined |
| S7_extracted_121626 | Not-determined |
| S7_extracted_121640 | Low-quality |
| S7_extracted_121645 | Low-quality |
| S7_extracted_121648 | Not-determined |
| S7_extracted_121659 | Not-determined |
| S7_extracted_121665 | Not-determined |
| S7_extracted_121676 | Not-determined |
| S7_extracted_121702 | Not-determined |
| S7_extracted_121720 | Low-quality |
| S7_extracted_121773 | Not-determined |
| S7_extracted_121785 | Not-determined |
| S7_extracted_121787 | Not-determined |
| S7_extracted_121790 | Not-determined |
| S7_extracted_121803 | Not-determined |
| S7_extracted_121808 | Not-determined |
| S7_extracted_121815 | Not-determined |
| S7_extracted_121826 | Not-determined |
| S7_extracted_121832 | Not-determined |
| S7_extracted_121848 | Not-determined |
| S7_extracted_121861 | Low-quality |
| S7_extracted_121866 | Not-determined |
| S7_extracted_121869 | Not-determined |
| S7_extracted_121871 | Not-determined |
| S7_extracted_121874 | Not-determined |
| S7_extracted_121909 | Not-determined |
| S7_extracted_121928 | Not-determined |
| S7_extracted_121932 | Not-determined |
| S7_extracted_121952 | Not-determined |
| S7_extracted_121958 | Not-determined |
| S7_extracted_121960 | Not-determined |
| S7_extracted_121970 | Low-quality |
| S7_extracted_121981 | Not-determined |
| S7_extracted_121992 | Not-determined |
| S7_extracted_122000 | Not-determined |
| S7_extracted_122001 | Not-determined |
| S7_extracted_122010 | Not-determined |
| S7_extracted_122015 | Not-determined |
| S7_extracted_122016 | Not-determined |
| S7_extracted_122017 | Not-determined |
| S7_extracted_122058 | Not-determined |
| S7_extracted_122063 | Not-determined |
| S7_extracted_122078 | Not-determined |
| S7_extracted_122088 | Not-determined |
| S7_extracted_122094 | Not-determined |
| S7_extracted_122096 | Not-determined |
| S7_extracted_122102 | Low-quality |
| S7_extracted_122127 | Not-determined |
| S7_extracted_122131 | Not-determined |
| S7_extracted_122132 | Not-determined |
| S7_extracted_122133 | Low-quality |
| S7_extracted_122141 | Not-determined |
| S7_extracted_122146 | Not-determined |
| S7_extracted_122150 | Not-determined |
| S7_extracted_122157 | Not-determined |
| S7_extracted_122167 | Not-determined |
| S7_extracted_122173 | Not-determined |
| S7_extracted_122180 | Not-determined |
| S7_extracted_122208 | Not-determined |
| S7_extracted_122231 | Not-determined |
| S7_extracted_122249 | Low-quality |
| S7_extracted_122250 | Not-determined |
| S7_extracted_122277 | Not-determined |
| S7_extracted_122293 | Not-determined |
| S7_extracted_122296 | Low-quality |
| S7_extracted_122304 | Low-quality |
| S7_extracted_122325 | Not-determined |
| S7_extracted_122330 | Not-determined |
| S7_extracted_122361 | Not-determined |
| S7_extracted_122365 | Not-determined |
| S7_extracted_122370 | Not-determined |
| S7_extracted_122371 | Not-determined |
| S7_extracted_122372 | Not-determined |
| S7_extracted_122376 | Not-determined |
| S7_extracted_122380 | Not-determined |
| S7_extracted_122387 | Not-determined |
| S7_extracted_122391 | Not-determined |
| S7_extracted_122403 | Not-determined |
| S7_extracted_122406 | Low-quality |
| S7_extracted_122436 | Low-quality |
| S7_extracted_122444 | Not-determined |
| S7_extracted_122467 | Not-determined |
| S7_extracted_122484 | Not-determined |
| S7_extracted_122486 | Not-determined |
| S7_extracted_122492 | Not-determined |
| S7_extracted_122505 | Not-determined |
| S7_extracted_122512 | Not-determined |
| S7_extracted_122518 | Not-determined |
| S7_extracted_122545 | Not-determined |
| S7_extracted_122547 | Low-quality |
| S7_extracted_122574 | Not-determined |
| S7_extracted_122576 | Not-determined |
| S7_extracted_122585 | Not-determined |
| S7_extracted_122614 | Not-determined |
| S7_extracted_122619 | Not-determined |
| S7_extracted_122625 | Not-determined |
| S7_extracted_122637 | Not-determined |
| S7_extracted_122651 | Low-quality |
| S7_extracted_122665 | Not-determined |
| S7_extracted_122673 | Low-quality |
| S7_extracted_122685 | Not-determined |
| S7_extracted_122690 | Low-quality |
| S7_extracted_122698 | Not-determined |
| S7_extracted_122706 | Low-quality |
| S7_extracted_122713 | Not-determined |
| S7_extracted_122718 | Not-determined |
| S7_extracted_122747 | Not-determined |
| S7_extracted_122751 | Not-determined |
| S7_extracted_122753 | Not-determined |
| S7_extracted_122760 | Not-determined |
| S7_extracted_122777 | Not-determined |
| S7_extracted_122790 | Not-determined |
| S7_extracted_122811 | Not-determined |
| S7_extracted_122812 | Low-quality |
| S7_extracted_122820 | Not-determined |
| S7_extracted_122830 | Not-determined |
| S7_extracted_122839 | Not-determined |
| S7_extracted_122867 | Not-determined |
| S7_extracted_122881 | Not-determined |
| S7_extracted_122889 | Not-determined |
| S7_extracted_122892 | Not-determined |
| S7_extracted_122895 | Not-determined |
| S7_extracted_122904 | Low-quality |
| S7_extracted_122906 | Low-quality |
| S7_extracted_122917 | Not-determined |
| S7_extracted_122949 | Not-determined |
| S7_extracted_122964 | Low-quality |
| S7_extracted_122966 | Not-determined |
| S7_extracted_122973 | Low-quality |
| S7_extracted_122978 | Not-determined |
| S7_extracted_122989 | Not-determined |
| S7_extracted_122997 | Not-determined |
| S7_extracted_123001 | Not-determined |
| S7_extracted_123064 | Low-quality |
| S7_extracted_123072 | Low-quality |
| S7_extracted_123080 | Not-determined |
| S7_extracted_123088 | Not-determined |
| S7_extracted_123090 | Not-determined |
| S7_extracted_123117 | Low-quality |
| S7_extracted_123121 | Not-determined |
| S7_extracted_123140 | Low-quality |
| S7_extracted_123152 | Low-quality |
| S7_extracted_123173 | Low-quality |
| S7_extracted_123190 | Low-quality |
| S7_extracted_123192 | Not-determined |
| S7_extracted_123193 | Not-determined |
| S7_extracted_123210 | Not-determined |
| S7_extracted_123212 | Not-determined |
| S7_extracted_123234 | Not-determined |
| S7_extracted_123267 | Low-quality |
| S7_extracted_123268 | Not-determined |
| S7_extracted_123282 | Not-determined |
| S7_extracted_123286 | Not-determined |
| S7_extracted_123299 | Not-determined |
| S7_extracted_123316 | Not-determined |
| S7_extracted_123324 | Low-quality |
| S7_extracted_123354 | Not-determined |
| S7_extracted_123355 | Not-determined |
| S7_extracted_123360 | Not-determined |
| S7_extracted_123374 | Low-quality |
| S7_extracted_123395 | Low-quality |
| S7_extracted_123400 | Not-determined |
| S7_extracted_123401 | Not-determined |
| S7_extracted_123407 | Not-determined |
| S7_extracted_123412 | Not-determined |
| S7_extracted_123420 | Not-determined |
| S7_extracted_123436 | Low-quality |
| S7_extracted_123462 | Not-determined |
| S7_extracted_123469 | Not-determined |
| S7_extracted_123473 | Low-quality |
| S7_extracted_123495 | Not-determined |
| S7_extracted_123498 | Not-determined |
| S7_extracted_123499 | Low-quality |
| S7_extracted_123515 | Low-quality |
| S7_extracted_123521 | Not-determined |
| S7_extracted_123526 | Not-determined |
| S7_extracted_123529 | Not-determined |
| S7_extracted_123541 | Not-determined |
| S7_extracted_123542 | Not-determined |
| S7_extracted_123561 | Low-quality |
| S7_extracted_123588 | Not-determined |
| S7_extracted_123594 | Not-determined |
| S7_extracted_123606 | Not-determined |
| S7_extracted_123607 | Not-determined |
| S7_extracted_123646 | Low-quality |
| S7_extracted_123672 | Not-determined |
| S7_extracted_123676 | Not-determined |
| S7_extracted_123700 | Not-determined |
| S7_extracted_123707 | Not-determined |
| S7_extracted_123732 | Not-determined |
| S7_extracted_123739 | Low-quality |
| S7_extracted_123743 | Not-determined |
| S7_extracted_123751 | Not-determined |
| S7_extracted_123755 | Not-determined |
| S7_extracted_123757 | Not-determined |
| S7_extracted_123776 | Low-quality |
| S7_extracted_123784 | Not-determined |
| S7_extracted_123795 | Not-determined |
| S7_extracted_123815 | Not-determined |
| S7_extracted_123822 | Not-determined |
| S7_extracted_123846 | Not-determined |
| S7_extracted_123847 | Not-determined |
| S7_extracted_123913 | Not-determined |
| S7_extracted_123919 | Not-determined |
| S7_extracted_123935 | Not-determined |
| S7_extracted_123937 | Low-quality |
| S7_extracted_123946 | Not-determined |
| S7_extracted_123961 | Not-determined |
| S7_extracted_123969 | Not-determined |
| S7_extracted_123971 | Not-determined |
| S7_extracted_123981 | Not-determined |
| S7_extracted_123986 | Not-determined |
| S7_extracted_123996 | Low-quality |
| S7_extracted_124004 | Not-determined |
| S7_extracted_124018 | Not-determined |
| S7_extracted_124020 | Not-determined |
| S7_extracted_124022 | Low-quality |
| S7_extracted_124024 | Not-determined |
| S7_extracted_124030 | Not-determined |
| S7_extracted_124040 | Low-quality |
| S7_extracted_124044 | Not-determined |
| S7_extracted_124048 | Not-determined |
| S7_extracted_124057 | Not-determined |
| S7_extracted_124091 | Not-determined |
| S7_extracted_124097 | Not-determined |
| S7_extracted_124134 | Not-determined |
| S7_extracted_124142 | Low-quality |
| S7_extracted_124157 | Not-determined |
| S7_extracted_124178 | Not-determined |
| S7_extracted_124190 | Not-determined |
| S7_extracted_124204 | Low-quality |
| S7_extracted_124211 | Not-determined |
| S7_extracted_124220 | Low-quality |
| S7_extracted_124240 | Not-determined |
| S7_extracted_124254 | Not-determined |
| S7_extracted_124275 | Not-determined |
| S7_extracted_124276 | Not-determined |
| S7_extracted_124288 | Not-determined |
| S7_extracted_124307 | Not-determined |
| S7_extracted_124312 | Low-quality |
| S7_extracted_124318 | Not-determined |
| S7_extracted_124319 | Not-determined |
| S7_extracted_124326 | Low-quality |
| S7_extracted_124335 | Not-determined |
| S7_extracted_124338 | Not-determined |
| S7_extracted_124342 | Low-quality |
| S7_extracted_124360 | Low-quality |
| S7_extracted_124367 | Not-determined |
| S7_extracted_124376 | Not-determined |
| S7_extracted_124382 | Low-quality |
| S7_extracted_124407 | Low-quality |
| S7_extracted_124412 | Not-determined |
| S7_extracted_124434 | Not-determined |
| S7_extracted_124438 | Not-determined |
| S7_extracted_124439 | Not-determined |
| S7_extracted_124441 | Low-quality |
| S7_extracted_124450 | Not-determined |
| S7_extracted_124463 | Not-determined |
| S7_extracted_124490 | Not-determined |
| S7_extracted_124491 | Low-quality |
| S7_extracted_124494 | Not-determined |
| S5_vs_k141_121419\|\|full | High-quality |
| S7_extracted_124521 | Not-determined |
| S7_extracted_124537 | Not-determined |
| S7_extracted_124547 | Not-determined |
| S7_extracted_124551 | Not-determined |
| S7_extracted_124553 | Not-determined |
| S7_extracted_124564 | Low-quality |
| S7_extracted_124575 | Low-quality |
| S7_extracted_124576 | Not-determined |
| S7_extracted_124586 | Not-determined |
| S7_extracted_124587 | Not-determined |
| S7_extracted_124593 | Not-determined |
| S7_extracted_124602 | Not-determined |
| S7_extracted_124604 | Not-determined |
| S7_extracted_124634 | Not-determined |
| S7_extracted_124655 | Not-determined |

| | |
|---|---|
| S7_extracted_124667 | Not-determined |
| S7_extracted_124698 | Not-determined |
| S7_extracted_124775 | Not-determined |
| S7_extracted_124777 | Not-determined |
| S7_extracted_124778 | Not-determined |
| S7_extracted_124801 | Not-determined |
| S7_extracted_124807 | Not-determined |
| S7_extracted_124864 | Low-quality |
| S7_extracted_124866 | Not-determined |
| S7_extracted_124876 | Low-quality |
| S7_extracted_124888 | Not-determined |
| S7_extracted_124900 | Not-determined |
| S7_extracted_124903 | Not-determined |
| S7_extracted_124908 | Not-determined |
| S7_extracted_124914 | Not-determined |
| S7_extracted_124917 | Not-determined |
| S7_extracted_124929 | Not-determined |
| S7_extracted_124941 | Not-determined |
| S7_extracted_124942 | Not-determined |
| S7_extracted_124945 | Not-determined |
| S7_extracted_124957 | Not-determined |
| S7_extracted_124980 | Not-determined |
| S7_extracted_124982 | Not-determined |
| S7_extracted_124986 | Low-quality |
| S7_extracted_124990 | Not-determined |
| S7_extracted_124991 | Not-determined |
| S7_extracted_124997 | Low-quality |
| S7_extracted_125002 | Not-determined |
| S7_extracted_125035 | Not-determined |
| S7_extracted_125037 | Low-quality |
| S7_extracted_125045 | Low-quality |
| S7_extracted_125102 | Low-quality |
| S7_extracted_125117 | Not-determined |
| S7_extracted_125145 | Not-determined |
| S7_extracted_125150 | Low-quality |
| S7_extracted_125152 | Not-determined |
| S7_extracted_125160 | Not-determined |
| S7_extracted_125166 | Not-determined |
| S7_extracted_125183 | Not-determined |
| S7_extracted_125189 | Not-determined |
| S7_extracted_125208 | Low-quality |
| S7_extracted_125218 | Not-determined |
| S7_extracted_125223 | Not-determined |
| S7_extracted_125248 | Not-determined |
| S7_extracted_125259 | Not-determined |
| S7_extracted_125299 | Not-determined |
| S7_extracted_125330 | Not-determined |
| S7_extracted_125341 | Not-determined |
| S7_extracted_125352 | Not-determined |
| S7_extracted_125354 | Low-quality |
| S7_extracted_125357 | Not-determined |
| S7_extracted_125361 | Not-determined |
| S7_extracted_125362 | Low-quality |
| S7_extracted_125371 | Low-quality |
| S7_extracted_125377 | Not-determined |
| S7_extracted_125417 | Not-determined |
| S7_extracted_125431 | Not-determined |
| S7_extracted_125437 | Not-determined |
| S7_extracted_125442 | Not-determined |
| S7_extracted_125443 | Low-quality |
| S7_extracted_125445 | Not-determined |
| S7_extracted_125476 | Not-determined |
| S7_extracted_125481 | Low-quality |
| S7_extracted_125482 | Not-determined |
| S7_extracted_125486 | Not-determined |
| S7_extracted_125492 | Not-determined |
| S7_extracted_125502 | Not-determined |
| S7_extracted_125510 | Not-determined |
| S7_extracted_125525 | Not-determined |
| S7_extracted_125535 | Not-determined |
| S7_extracted_125537 | Not-determined |
| S7_extracted_125547 | Low-quality |
| S7_extracted_125574 | Not-determined |
| S7_extracted_125601 | Not-determined |
| S7_extracted_125602 | Not-determined |
| S7_extracted_125610 | Not-determined |
| S7_extracted_125613 | Not-determined |
| S7_extracted_125660 | Not-determined |
| S7_extracted_125674 | Not-determined |
| S7_extracted_125677 | Not-determined |
| S7_extracted_125708 | Low-quality |
| S7_extracted_125734 | Not-determined |
| S7_extracted_125750 | Not-determined |
| S7_extracted_125752 | Not-determined |
| S7_extracted_125769 | Not-determined |
| S7_extracted_125784 | Not-determined |
| S7_extracted_125793 | Not-determined |
| S7_extracted_125812 | Not-determined |
| S7_extracted_125816 | Not-determined |
| S7_extracted_125823 | Not-determined |
| S7_extracted_125825 | Low-quality |
| S7_extracted_125837 | Low-quality |
| S7_extracted_125845 | Low-quality |
| S7_extracted_125847 | Not-determined |
| S7_extracted_125851 | Low-quality |
| S7_extracted_125856 | Not-determined |
| S7_extracted_125864 | Low-quality |
| S7_extracted_125883 | Not-determined |
| S7_extracted_125896 | Not-determined |
| S7_extracted_125903 | Not-determined |
| S7_extracted_125913 | Not-determined |
| S7_extracted_125923 | Not-determined |
| S7_extracted_125930 | Not-determined |
| S7_extracted_125941 | Not-determined |
| S7_extracted_125945 | Not-determined |
| S7_extracted_125947 | Not-determined |
| S7_extracted_125948 | Not-determined |
| S7_extracted_125953 | Not-determined |
| S7_extracted_125960 | Not-determined |
| S7_extracted_125979 | Not-determined |
| S7_extracted_125985 | Low-quality |
| S7_extracted_126006 | Low-quality |
| S7_extracted_126019 | Not-determined |
| S7_extracted_126035 | Not-determined |
| S7_extracted_126051 | Not-determined |
| S7_extracted_126078 | Not-determined |
| S7_extracted_126090 | Not-determined |
| S7_extracted_126092 | Not-determined |
| S7_extracted_126097 | Not-determined |
| S7_extracted_126114 | Not-determined |
| S7_extracted_126121 | Not-determined |
| S7_extracted_126122 | Not-determined |
| S7_extracted_126144 | Not-determined |
| S7_extracted_126150 | Not-determined |
| S7_extracted_126152 | Not-determined |
| S7_extracted_126153 | Not-determined |
| S7_extracted_126165 | Not-determined |
| S7_extracted_126177 | Not-determined |
| S7_extracted_126205 | Not-determined |
| S7_extracted_126209 | Not-determined |
| S7_extracted_126214 | Not-determined |
| S7_extracted_126232 | Not-determined |
| S7_extracted_126242 | Not-determined |
| S7_extracted_126243 | Low-quality |
| S7_extracted_126254 | Not-determined |
| S7_extracted_126276 | Low-quality |
| S7_extracted_126279 | Not-determined |
| S7_extracted_126288 | Low-quality |
| S7_extracted_126289 | Not-determined |
| S7_extracted_126307 | Not-determined |
| S7_extracted_126317 | Not-determined |
| S7_extracted_126326 | Not-determined |
| S7_extracted_126327 | Not-determined |
| S7_extracted_126348 | Not-determined |
| S7_extracted_126351 | Not-determined |
| S7_extracted_126353 | Not-determined |
| S7_extracted_126355 | Not-determined |
| S7_extracted_126356 | Not-determined |
| S7_extracted_126369 | Not-determined |
| S7_extracted_126374 | Not-determined |
| S7_extracted_126399 | Not-determined |
| S7_extracted_126403 | Low-quality |
| S7_extracted_126408 | Not-determined |
| S7_extracted_126410 | Not-determined |
| S7_extracted_126417 | Not-determined |
| S7_extracted_126420 | Not-determined |
| S7_extracted_126434 | Not-determined |
| S7_extracted_126443 | Not-determined |
| S7_extracted_126447 | Not-determined |
| S7_extracted_126466 | Not-determined |
| S7_extracted_126473 | Low-quality |
| S7_extracted_126476 | Not-determined |
| S7_extracted_126478 | Not-determined |
| S7_extracted_126481 | Low-quality |
| S7_extracted_126497 | Low-quality |
| S7_extracted_126522 | Not-determined |
| S7_extracted_126535 | Low-quality |
| S7_extracted_126542 | Not-determined |
| S7_extracted_126545 | Not-determined |
| S7_extracted_126554 | Not-determined |
| S7_extracted_126567 | Not-determined |
| S7_extracted_126568 | Not-determined |
| S7_extracted_126571 | Not-determined |
| S7_extracted_126574 | Not-determined |
| S7_extracted_126575 | Not-determined |
| S7_extracted_126606 | Not-determined |
| S7_extracted_126617 | Not-determined |
| S7_extracted_126630 | Not-determined |
| S7_extracted_126635 | Low-quality |
| S7_extracted_126645 | Not-determined |
| S7_extracted_126646 | Low-quality |
| S7_extracted_126657 | Not-determined |
| S7_extracted_126663 | Not-determined |
| S7_extracted_126665 | Not-determined |
| S7_extracted_126677 | Low-quality |
| S7_extracted_126700 | Low-quality |
| S7_extracted_126709 | Not-determined |
| S7_extracted_126719 | Not-determined |
| S7_extracted_126725 | Not-determined |
| S7_extracted_126732 | Not-determined |
| S7_extracted_126754 | Not-determined |
| S7_extracted_126757 | Not-determined |
| S7_extracted_126763 | Not-determined |
| S7_extracted_126774 | Not-determined |
| S7_extracted_126787 | Not-determined |
| S7_extracted_126790 | Not-determined |
| S7_extracted_126846 | Not-determined |
| S7_extracted_126863 | Not-determined |
| S7_extracted_126871 | Not-determined |
| S7_extracted_126873 | Not-determined |
| S7_extracted_126884 | Not-determined |
| S7_extracted_126887 | Low-quality |
| S7_extracted_126895 | Low-quality |
| S7_extracted_126921 | Not-determined |
| S7_extracted_126960 | Not-determined |
| S7_extracted_126990 | Not-determined |
| S7_extracted_127020 | Not-determined |
| S7_extracted_127030 | Low-quality |
| S7_extracted_127051 | Low-quality |
| S7_extracted_127056 | Not-determined |
| S7_extracted_127081 | Low-quality |
| S7_extracted_127082 | Not-determined |
| S7_extracted_127087 | Not-determined |
| S7_extracted_127092 | Not-determined |
| S7_extracted_127099 | Not-determined |
| S7_extracted_127100 | Not-determined |
| S7_extracted_127103 | Not-determined |
| S7_extracted_127130 | Low-quality |
| S7_extracted_127141 | Low-quality |
| S7_extracted_127146 | Not-determined |
| S7_extracted_127148 | Not-determined |
| S7_extracted_127150 | Not-determined |
| S7_extracted_127164 | Not-determined |
| S7_extracted_127175 | Not-determined |
| S7_extracted_127182 | Not-determined |
| S7_extracted_127191 | Not-determined |
| S7_extracted_127194 | Not-determined |
| S7_extracted_127208 | Not-determined |
| S7_extracted_127246 | Low-quality |
| S7_extracted_127248 | Not-determined |
| S7_extracted_127253 | Low-quality |
| S7_extracted_127255 | Not-determined |
| S7_extracted_127266 | Not-determined |
| S7_extracted_127276 | Not-determined |
| S7_extracted_127282 | Not-determined |
| S7_extracted_127286 | Not-determined |
| S7_extracted_127290 | Not-determined |
| S7_extracted_127303 | Not-determined |
| S7_extracted_127306 | Not-determined |
| S7_extracted_127308 | Not-determined |
| S7_extracted_127323 | Not-determined |
| S7_extracted_127325 | Not-determined |
| S7_extracted_127326 | Not-determined |
| S7_extracted_127327 | Not-determined |
| S7_extracted_127340 | Low-quality |
| S7_extracted_127351 | Not-determined |
| S7_extracted_127369 | Not-determined |
| S7_extracted_127375 | Not-determined |
| S7_extracted_127392 | Not-determined |
| S7_extracted_127398 | Not-determined |
| S7_extracted_127410 | Not-determined |
| S7_extracted_127420 | Not-determined |
| S7_extracted_127438 | Not-determined |
| S7_extracted_127456 | Not-determined |
| S7_extracted_127457 | Not-determined |
| S7_extracted_127483 | Not-determined |
| S7_extracted_127486 | Not-determined |
| S7_extracted_127529 | Not-determined |
| S7_extracted_127540 | Not-determined |
| S7_extracted_127554 | Not-determined |
| S7_extracted_127556 | Not-determined |
| S7_extracted_127572 | Not-determined |
| S7_extracted_127580 | Not-determined |
| S7_extracted_127589 | Not-determined |
| S7_extracted_127599 | Not-determined |
| S7_extracted_127602 | Not-determined |
| S7_extracted_127608 | Not-determined |
| S7_extracted_127623 | Not-determined |
| S7_extracted_127658 | Low-quality |
| S7_extracted_127673 | Not-determined |
| S7_extracted_127689 | Not-determined |
| S7_extracted_127690 | Not-determined |
| S7_extracted_127715 | Not-determined |
| S7_extracted_127717 | Not-determined |
| S7_extracted_127718 | Not-determined |
| S7_extracted_127719 | Not-determined |
| S7_extracted_127721 | Not-determined |
| S7_extracted_127724 | Low-quality |
| S7_extracted_127725 | Not-determined |
| S7_extracted_127727 | Not-determined |
| S7_extracted_127747 | Low-quality |
| S7_extracted_127766 | Not-determined |
| S7_extracted_127785 | Not-determined |
| S7_extracted_127801 | Not-determined |
| S7_extracted_127828 | Not-determined |
| S7_extracted_127829 | Not-determined |
| S7_extracted_127844 | Low-quality |
| S7_extracted_127851 | Low-quality |
| S7_extracted_127859 | Low-quality |
| S7_extracted_127860 | Not-determined |
| S7_extracted_127862 | Not-determined |
| S7_extracted_127871 | Not-determined |
| S7_extracted_127872 | Low-quality |
| S7_extracted_127889 | Not-determined |
| S7_extracted_127908 | Not-determined |
| S7_extracted_127925 | Not-determined |
| S7_extracted_127937 | Not-determined |
| S7_extracted_127939 | Not-determined |
| S7_extracted_127957 | Not-determined |
| S7_extracted_127970 | Not-determined |
| S7_extracted_127976 | Not-determined |
| S7_extracted_127985 | Not-determined |
| S7_extracted_128002 | Low-quality |
| S7_extracted_128006 | Low-quality |
| S7_extracted_128034 | Not-determined |
| S7_extracted_128040 | Low-quality |
| S7_extracted_128052 | Not-determined |
| S7_extracted_128053 | Not-determined |
| S7_extracted_128056 | Not-determined |
| S7_extracted_128061 | Not-determined |
| S7_extracted_128062 | Not-determined |
| S7_extracted_128081 | Not-determined |
| S7_extracted_128097 | Not-determined |
| S7_extracted_128105 | Not-determined |
| S7_extracted_128108 | Not-determined |
| S7_extracted_128114 | Not-determined |
| S7_extracted_128117 | Not-determined |
| S7_extracted_128120 | Not-determined |
| S7_extracted_128130 | Not-determined |
| S7_extracted_128133 | Low-quality |
| S7_extracted_128141 | Not-determined |
| S7_extracted_128142 | Not-determined |
| S7_extracted_128150 | Not-determined |
| S7_extracted_128165 | Not-determined |
| S7_extracted_128187 | Low-quality |
| S7_extracted_128194 | Not-determined |
| S7_extracted_128212 | Low-quality |
| S7_extracted_128232 | Low-quality |
| S7_extracted_128238 | Not-determined |
| S7_extracted_128259 | Not-determined |
| S7_extracted_128265 | Not-determined |
| S7_extracted_128309 | Not-determined |
| S7_extracted_128314 | Low-quality |
| S7_extracted_128317 | Not-determined |
| S7_extracted_128320 | Not-determined |
| S7_extracted_128344 | Not-determined |
| S7_extracted_128350 | Not-determined |
| S7_extracted_128372 | Not-determined |
| S7_extracted_128376 | Low-quality |
| S7_extracted_128382 | Not-determined |
| S7_extracted_128393 | Not-determined |
| S7_extracted_128428 | Not-determined |
| S7_extracted_128435 | Not-determined |
| S7_extracted_128450 | Low-quality |
| S7_extracted_128456 | Not-determined |
| S7_extracted_128477 | Low-quality |
| S7_extracted_128478 | Not-determined |
| S7_extracted_128499 | Not-determined |
| S7_extracted_128511 | Not-determined |
| S7_extracted_128512 | Not-determined |
| S7_extracted_128533 | Not-determined |
| S7_extracted_128537 | Not-determined |
| S7_extracted_128541 | Not-determined |
| S7_extracted_128566 | Low-quality |
| S7_extracted_128569 | Not-determined |
| S7_extracted_128572 | Not-determined |
| S7_extracted_128602 | Not-determined |
| S7_extracted_128627 | Not-determined |
| S7_extracted_128632 | Low-quality |
| S7_extracted_128684 | Not-determined |
| S7_extracted_128687 | Low-quality |
| S7_extracted_128688 | Low-quality |
| S7_extracted_128697 | Low-quality |
| S7_extracted_128710 | Not-determined |
| S7_extracted_128739 | Not-determined |
| S7_extracted_128742 | Not-determined |
| S7_extracted_128779 | Not-determined |
| S7_extracted_128780 | Not-determined |
| S7_extracted_128789 | Not-determined |
| S7_extracted_128795 | Not-determined |
| S7_extracted_128805 | Not-determined |
| S7_extracted_128809 | Not-determined |
| S7_extracted_128819 | Not-determined |
| S7_extracted_128827 | Not-determined |
| S7_extracted_128834 | Not-determined |
| S7_extracted_128841 | Not-determined |
| S7_extracted_128860 | Not-determined |
| S7_extracted_128861 | Not-determined |
| S7_extracted_128884 | Not-determined |
| S7_extracted_128891 | Not-determined |
| S7_extracted_128896 | Not-determined |
| S7_extracted_128918 | Not-determined |
| S7_extracted_128940 | Not-determined |
| S7_extracted_128959 | Not-determined |
| S7_extracted_128963 | Not-determined |
| S7_extracted_128969 | Not-determined |
| S7_extracted_128977 | Not-determined |
| S7_extracted_129008 | Not-determined |
| S7_extracted_129016 | Not-determined |
| S7_extracted_129020 | Low-quality |
| S7_extracted_129027 | Not-determined |
| S7_extracted_129033 | Not-determined |
| S7_extracted_129038 | Not-determined |
| S7_extracted_129039 | Not-determined |
| S7_extracted_129045 | Not-determined |
| S7_extracted_129051 | Low-quality |
| S7_extracted_129071 | Not-determined |
| S7_extracted_129094 | Not-determined |
| S7_extracted_129108 | Not-determined |
| S7_extracted_129111 | Not-determined |
| S7_extracted_129119 | Not-determined |
| S7_extracted_129123 | Not-determined |
| S7_extracted_129129 | Not-determined |
| S7_extracted_129135 | Not-determined |
| S7_extracted_129152 | Not-determined |
| S7_extracted_129193 | Not-determined |
| S7_extracted_129216 | Not-determined |
| S7_extracted_129219 | Not-determined |
| S7_extracted_129232 | Not-determined |
| S7_extracted_129236 | Low-quality |
| S7_extracted_129238 | Not-determined |
| S7_extracted_129245 | Not-determined |
| S7_extracted_129249 | Not-determined |
| S7_extracted_129252 | Not-determined |
| S7_extracted_129258 | Not-determined |
| S7_extracted_129274 | Not-determined |
| S7_extracted_129279 | Low-quality |
| S7_extracted_129294 | Not-determined |
| S7_extracted_129296 | Low-quality |
| S7_extracted_129300 | Not-determined |
| S7_extracted_129303 | Not-determined |
| S7_extracted_129305 | Not-determined |
| S7_extracted_129308 | Not-determined |
| S7_extracted_129325 | Not-determined |
| S7_extracted_129340 | Not-determined |
| S7_extracted_129342 | Low-quality |
| S7_extracted_129390 | Not-determined |
| S7_extracted_129399 | Low-quality |
| S7_extracted_129409 | Low-quality |
| S7_extracted_129442 | Not-determined |
| S7_extracted_129448 | Not-determined |
| S7_extracted_129451 | Not-determined |
| S7_extracted_129457 | Not-determined |
| S7_extracted_129474 | Low-quality |
| S7_extracted_129493 | Not-determined |
| S7_extracted_129499 | Not-determined |
| S7_extracted_129533 | Not-determined |
| S7_extracted_129535 | Not-determined |
| S7_extracted_129536 | Not-determined |
| S7_extracted_129541 | Low-quality |
| S7_extracted_129544 | Low-quality |
| S7_extracted_129547 | Not-determined |
| S7_extracted_129552 | Not-determined |
| S7_extracted_129570 | Not-determined |
| S7_extracted_129575 | Low-quality |
| S7_extracted_129598 | Not-determined |
| S7_extracted_129604 | Not-determined |
| S7_extracted_129608 | Not-determined |
| S7_extracted_129617 | Not-determined |
| S7_extracted_129624 | Not-determined |
| S7_extracted_129627 | Not-determined |
| S7_extracted_129632 | Low-quality |
| S7_extracted_129647 | Not-determined |
| S7_extracted_129651 | Not-determined |
| S7_extracted_129661 | Not-determined |
| S7_extracted_129664 | Not-determined |
| S7_extracted_129669 | Low-quality |
| S7_extracted_129687 | Not-determined |
| S7_extracted_129689 | Not-determined |
| S7_extracted_129693 | Low-quality |
| S7_extracted_129694 | Low-quality |
| S7_extracted_129708 | Low-quality |
| S7_extracted_129735 | Low-quality |
| S7_extracted_129737 | Not-determined |
| S7_extracted_129738 | Not-determined |
| S7_extracted_129747 | Not-determined |
| S7_extracted_129769 | Not-determined |
| S7_extracted_129772 | Low-quality |
| S7_extracted_129776 | Not-determined |
| S7_extracted_129781 | Not-determined |
| S7_extracted_129788 | Not-determined |
| S7_extracted_129799 | Not-determined |
| S7_extracted_129804 | Not-determined |
| S7_extracted_129806 | Not-determined |
| S7_extracted_129816 | Low-quality |
| S7_extracted_129822 | Not-determined |
| S7_extracted_129824 | Not-determined |
| S7_extracted_129831 | Not-determined |
| S7_extracted_129856 | Not-determined |
| S7_extracted_129887 | Not-determined |
| S7_extracted_129914 | Not-determined |
| S7_extracted_129922 | Low-quality |
| S7_extracted_129923 | Not-determined |
| S7_extracted_129927 | Not-determined |
| S7_extracted_129929 | Not-determined |
| S7_extracted_129963 | Not-determined |
| S7_extracted_129964 | Not-determined |
| S7_extracted_129973 | Not-determined |
| S7_extracted_129977 | Not-determined |
| S7_extracted_129981 | Not-determined |
| S7_extracted_129994 | Not-determined |
| S7_extracted_130018 | Not-determined |
| S7_extracted_130019 | Not-determined |
| S7_extracted_130020 | Not-determined |
| S7_extracted_130031 | Not-determined |
| S7_extracted_130044 | Not-determined |
| S7_extracted_130058 | Low-quality |
| S7_extracted_130066 | Not-determined |
| S7_extracted_130070 | Not-determined |
| S7_extracted_130073 | Not-determined |
| S7_extracted_130077 | Not-determined |
| S7_extracted_130079 | Not-determined |
| S7_extracted_130096 | Not-determined |
| S7_extracted_130100 | Not-determined |
| S7_extracted_130174 | Not-determined |
| S7_extracted_130178 | Not-determined |
| S7_extracted_130180 | Low-quality |
| S7_extracted_130181 | Not-determined |
| S7_extracted_130187 | Not-determined |
| S7_extracted_130190 | Not-determined |
| S7_extracted_130204 | Low-quality |
| S7_extracted_130212 | Low-quality |
| S7_extracted_130237 | Not-determined |
| S7_extracted_130257 | Not-determined |
| S7_extracted_130267 | Low-quality |
| S7_extracted_130281 | Not-determined |
| S7_extracted_130298 | Not-determined |
| S7_extracted_130302 | Not-determined |
| S7_extracted_130344 | Not-determined |
| S7_extracted_130351 | Low-quality |
| S7_extracted_130362 | Not-determined |
| S7_extracted_130437 | Not-determined |
| S7_extracted_130459 | Not-determined |
| S7_extracted_130462 | Not-determined |
| S7_extracted_130465 | Not-determined |
| S7_extracted_130477 | Low-quality |
| S7_extracted_130481 | Not-determined |
| S7_extracted_130485 | Not-determined |
| S7_extracted_130491 | Not-determined |
| S7_extracted_130505 | Not-determined |
| S7_extracted_130515 | Not-determined |
| S7_extracted_130522 | Low-quality |
| S7_extracted_130524 | Low-quality |
| S7_extracted_130526 | Not-determined |
| S7_extracted_130545 | Not-determined |
| S7_extracted_130557 | Not-determined |
| S7_extracted_130564 | Not-determined |
| S7_extracted_130566 | Not-determined |
| S7_extracted_130581 | Low-quality |
| S7_extracted_130587 | Not-determined |
| S7_extracted_130588 | Not-determined |
| S7_extracted_130599 | Not-determined |
| S7_extracted_130604 | Low-quality |
| S7_extracted_130605 | Not-determined |
| S7_extracted_130607 | Not-determined |
| S7_extracted_130616 | Not-determined |
| S7_extracted_130619 | Not-determined |
| S7_extracted_130620 | Not-determined |
| S7_extracted_130628 | Not-determined |
| S7_extracted_130643 | Not-determined |
| S7_extracted_130650 | Not-determined |
| S7_extracted_130660 | Low-quality |
| S7_extracted_130664 | Not-determined |
| S7_extracted_130680 | Low-quality |
| S7_extracted_130681 | Not-determined |
| S7_extracted_130689 | Not-determined |
| S7_extracted_130699 | Not-determined |
| S7_extracted_130700 | Not-determined |
| S7_extracted_130703 | Low-quality |
| S7_extracted_130720 | Low-quality |
| S7_extracted_130723 | Not-determined |
| S7_extracted_130729 | Low-quality |
| S7_extracted_130732 | Not-determined |
| S7_extracted_130734 | Not-determined |
| S7_extracted_130763 | Not-determined |
| S7_extracted_130766 | Not-determined |
| S7_extracted_130768 | Not-determined |
| S7_extracted_130780 | Low-quality |
| S7_extracted_130798 | Not-determined |
| S7_extracted_130801 | Not-determined |
| S7_extracted_130812 | Low-quality |
| S7_extracted_130818 | Not-determined |
| S7_extracted_130824 | Not-determined |
| S7_extracted_130829 | Not-determined |
| S7_extracted_130862 | Not-determined |
| S7_extracted_130883 | Not-determined |
| S7_extracted_130919 | Not-determined |
| S7_extracted_130933 | Low-quality |
| S7_extracted_130935 | Not-determined |
| S7_extracted_130941 | Not-determined |
| S7_extracted_130945 | Not-determined |
| S7_extracted_130947 | Not-determined |
| S7_extracted_130954 | Not-determined |
| S7_extracted_130985 | Not-determined |
| S7_extracted_130995 | Low-quality |
| S7_extracted_131003 | Low-quality |
| S7_extracted_131004 | Not-determined |
| S7_extracted_131007 | Not-determined |
| S7_extracted_131061 | Not-determined |

| | |
|---|---|
| S7_extracted_131062 | Low-quality |
| S7_extracted_131067 | Not-determined |
| S7_extracted_131074 | Not-determined |
| S7_extracted_131107 | Not-determined |
| S7_extracted_131122 | Low-quality |
| S7_extracted_131128 | Not-determined |
| S7_extracted_131130 | Not-determined |
| S7_extracted_131135 | Not-determined |
| S7_extracted_131150 | Not-determined |
| S7_extracted_131157 | Not-determined |
| S7_extracted_131176 | Not-determined |
| S7_extracted_131181 | Not-determined |
| S7_extracted_131184 | Not-determined |
| S7_extracted_131189 | Low-quality |
| S7_extracted_131193 | Not-determined |
| S7_extracted_131197 | Not-determined |
| S7_extracted_131213 | Not-determined |
| S7_extracted_131247 | Not-determined |
| S7_extracted_131272 | Not-determined |
| S7_extracted_131276 | Low-quality |
| S7_extracted_131278 | Not-determined |
| S7_extracted_131324 | Not-determined |
| S7_extracted_131327 | Not-determined |
| S7_extracted_131353 | Not-determined |
| S7_extracted_131359 | Not-determined |
| S7_extracted_131370 | Not-determined |
| S7_extracted_131376 | Not-determined |
| S7_extracted_131398 | Not-determined |
| S7_extracted_131425 | Not-determined |
| S7_extracted_131442 | Not-determined |
| S7_extracted_131446 | Not-determined |
| S7_extracted_131464 | Not-determined |
| S7_extracted_131480 | Low-quality |
| S7_extracted_131491 | Not-determined |
| S7_extracted_131502 | Not-determined |
| S7_extracted_131522 | Not-determined |
| S7_extracted_131536 | Not-determined |
| S7_extracted_131537 | Low-quality |
| S7_extracted_131545 | Not-determined |
| S7_extracted_131582 | Not-determined |
| S7_extracted_131583 | Not-determined |
| S7_extracted_131590 | Not-determined |
| S7_extracted_131597 | Not-determined |
| S7_extracted_131599 | Not-determined |
| S7_extracted_131619 | Not-determined |
| S7_extracted_131637 | Not-determined |
| S7_extracted_131664 | Not-determined |
| S7_extracted_131665 | Not-determined |
| S7_extracted_131685 | Not-determined |
| S7_extracted_131712 | Not-determined |
| S7_extracted_131736 | Low-quality |
| S7_extracted_131739 | Not-determined |
| S7_extracted_131742 | Not-determined |
| S7_extracted_131747 | Not-determined |
| S7_extracted_131768 | Not-determined |
| S7_extracted_131774 | Not-determined |
| S7_extracted_131775 | Not-determined |
| S7_extracted_131778 | Not-determined |
| S7_extracted_131804 | Not-determined |
| S7_extracted_131811 | Not-determined |
| S7_extracted_131816 | Not-determined |
| S7_extracted_131824 | Not-determined |
| S7_extracted_131825 | Not-determined |
| S7_extracted_131826 | Not-determined |
| S7_extracted_131835 | Not-determined |
| S7_extracted_131842 | Not-determined |
| S7_extracted_131845 | Not-determined |
| S7_extracted_131861 | Low-quality |
| S7_extracted_131864 | Not-determined |
| S7_extracted_131869 | Low-quality |
| S7_extracted_131897 | Low-quality |
| S7_extracted_131903 | Not-determined |
| S7_extracted_131905 | Not-determined |
| S7_extracted_131909 | Not-determined |
| S7_extracted_131914 | Low-quality |
| S7_extracted_131916 | Low-quality |
| S7_extracted_131952 | Not-determined |
| S7_extracted_131960 | Not-determined |
| S7_extracted_131962 | Not-determined |
| S7_extracted_131968 | Not-determined |
| S7_extracted_131977 | Not-determined |
| S7_extracted_131998 | Not-determined |
| S7_extracted_132023 | Not-determined |
| S7_extracted_132037 | Not-determined |
| S7_extracted_132043 | Not-determined |
| S7_extracted_132055 | Not-determined |
| S7_extracted_132064 | Not-determined |
| S7_extracted_132067 | Not-determined |
| S7_extracted_132069 | Not-determined |
| S7_extracted_132077 | Low-quality |
| S7_extracted_132079 | Not-determined |
| S7_extracted_132090 | Not-determined |
| S7_extracted_132099 | Not-determined |
| S7_extracted_132112 | Not-determined |
| S7_extracted_132129 | Not-determined |
| S7_extracted_132135 | Not-determined |
| S7_extracted_132137 | Not-determined |
| S7_extracted_132138 | Not-determined |
| S7_extracted_132160 | Not-determined |
| S7_extracted_132189 | Not-determined |
| S7_extracted_132208 | Not-determined |
| S7_extracted_132220 | Not-determined |
| S7_extracted_132228 | Not-determined |
| S7_extracted_132231 | Not-determined |
| S7_extracted_132234 | Not-determined |
| S7_extracted_132248 | Not-determined |
| S7_extracted_132259 | Not-determined |
| S7_extracted_132260 | Low-quality |
| S7_extracted_132269 | Not-determined |
| S7_extracted_132271 | Low-quality |
| S7_extracted_132272 | Not-determined |
| S7_extracted_132280 | Not-determined |
| S7_extracted_132283 | Not-determined |
| S7_extracted_132299 | Not-determined |
| S7_extracted_132317 | Not-determined |
| S7_extracted_132321 | Not-determined |
| S7_extracted_132342 | Not-determined |
| S7_extracted_132344 | Not-determined |
| S7_extracted_132348 | Low-quality |
| S7_extracted_132376 | Low-quality |
| S7_extracted_132377 | Low-quality |
| S7_extracted_132390 | Not-determined |
| S7_extracted_132392 | Low-quality |
| S7_extracted_132416 | Not-determined |
| S7_extracted_132432 | Not-determined |
| S7_extracted_132437 | Not-determined |
| S7_extracted_132457 | Not-determined |
| S7_extracted_132459 | Not-determined |
| S7_extracted_132467 | Not-determined |
| S7_extracted_132475 | Not-determined |
| S7_extracted_132482 | Not-determined |
| S7_extracted_132505 | Low-quality |
| S7_extracted_132547 | Not-determined |
| S7_extracted_132548 | Not-determined |
| S7_extracted_132562 | Not-determined |
| S7_extracted_132583 | Not-determined |
| S7_extracted_132585 | Not-determined |
| S7_extracted_132601 | Not-determined |
| S7_extracted_132610 | Not-determined |
| S7_extracted_132611 | Not-determined |
| S7_extracted_132615 | Not-determined |
| S7_extracted_132631 | Not-determined |
| S7_extracted_132636 | Low-quality |
| S7_extracted_132644 | Not-determined |
| S7_extracted_132649 | Not-determined |
| S7_extracted_132655 | Not-determined |
| S7_extracted_132656 | Not-determined |
| S7_extracted_132678 | Not-determined |
| S7_extracted_132682 | Not-determined |
| S7_extracted_132705 | Not-determined |
| S7_extracted_132710 | Not-determined |
| S7_extracted_132713 | Not-determined |
| S7_extracted_132715 | Not-determined |
| S7_extracted_132718 | Low-quality |
| S7_extracted_132721 | Low-quality |
| S7_extracted_132727 | Not-determined |
| S7_extracted_132730 | Not-determined |
| S7_extracted_132732 | Low-quality |
| S7_extracted_132733 | Not-determined |
| S7_extracted_132743 | Low-quality |
| S7_extracted_132748 | Not-determined |
| S7_extracted_132773 | Not-determined |
| S7_extracted_132774 | Not-determined |
| S7_extracted_132779 | Not-determined |
| S7_extracted_132781 | Not-determined |
| S7_extracted_132807 | Not-determined |
| S7_extracted_132825 | Not-determined |
| S7_extracted_132829 | Low-quality |
| S7_extracted_132835 | Not-determined |
| S7_extracted_132856 | Not-determined |
| S7_extracted_132857 | Low-quality |
| S7_extracted_132860 | Not-determined |
| S7_extracted_132863 | Not-determined |
| S7_extracted_132866 | Low-quality |
| S7_extracted_132882 | Low-quality |
| S7_extracted_132883 | Low-quality |
| S7_extracted_132886 | Low-quality |
| S7_extracted_132891 | Low-quality |
| S7_extracted_132892 | Not-determined |
| S7_extracted_132913 | Not-determined |
| S7_extracted_132919 | Not-determined |
| S7_extracted_132941 | Not-determined |
| S7_extracted_132950 | Not-determined |
| S7_extracted_132961 | Low-quality |
| S7_extracted_132968 | Not-determined |
| S7_extracted_132975 | Not-determined |
| S7_extracted_132976 | Not-determined |
| S7_extracted_132985 | Not-determined |
| S7_extracted_132987 | Not-determined |
| S7_extracted_132988 | Not-determined |
| S7_extracted_132990 | Not-determined |
| S7_extracted_133007 | Not-determined |
| S7_extracted_133020 | Not-determined |
| S7_extracted_133025 | Low-quality |
| S7_extracted_133026 | Not-determined |
| S7_extracted_133035 | Not-determined |
| S7_extracted_133040 | Low-quality |
| S7_extracted_133053 | Not-determined |
| S7_extracted_133056 | Not-determined |
| S7_extracted_133064 | Not-determined |
| S7_extracted_133067 | Not-determined |
| S7_extracted_133070 | Not-determined |
| S7_extracted_133082 | Not-determined |
| S7_extracted_133083 | Not-determined |
| S7_extracted_133089 | Not-determined |
| S7_extracted_133096 | Low-quality |
| S7_extracted_133098 | Not-determined |
| S7_extracted_133101 | Low-quality |
| S7_extracted_133105 | Not-determined |
| S7_extracted_133129 | Not-determined |
| S7_extracted_133131 | Low-quality |
| S7_extracted_133139 | Low-quality |
| S7_extracted_133145 | Not-determined |
| S7_extracted_133156 | Not-determined |
| S7_extracted_133163 | Not-determined |
| S7_extracted_133166 | Not-determined |
| S7_extracted_133173 | Not-determined |
| S7_extracted_133176 | Not-determined |
| S7_extracted_133180 | Not-determined |
| S7_extracted_133196 | Not-determined |
| S7_extracted_133205 | Not-determined |
| S7_extracted_133208 | Not-determined |
| S7_extracted_133234 | Low-quality |
| S7_extracted_133258 | Low-quality |
| S7_extracted_133276 | Not-determined |
| S7_extracted_133287 | Not-determined |
| S7_extracted_133296 | Not-determined |
| S7_extracted_133314 | Not-determined |
| S7_extracted_133324 | Not-determined |
| S7_extracted_133328 | Not-determined |
| S7_extracted_133337 | Low-quality |
| S7_extracted_133347 | Not-determined |
| S7_extracted_133350 | Not-determined |
| S7_extracted_133362 | Not-determined |
| S7_extracted_133364 | Not-determined |
| S7_extracted_133371 | Not-determined |
| S7_extracted_133378 | Not-determined |
| S7_extracted_133384 | Not-determined |
| S7_extracted_133385 | Not-determined |
| S7_extracted_133394 | Not-determined |
| S7_extracted_133410 | Not-determined |
| S7_extracted_133442 | Not-determined |
| S7_extracted_133460 | Not-determined |
| S7_extracted_133463 | Not-determined |
| S7_extracted_133487 | Not-determined |
| S7_extracted_133489 | Not-determined |
| S7_extracted_133493 | Not-determined |
| S7_extracted_133508 | Not-determined |
| S7_extracted_133516 | Not-determined |
| S7_extracted_133520 | Not-determined |
| S7_extracted_133523 | Not-determined |
| S7_extracted_133527 | Not-determined |
| S7_extracted_133528 | Not-determined |
| S7_extracted_133530 | Not-determined |
| S7_extracted_133540 | Not-determined |
| S7_extracted_133554 | Not-determined |
| S7_extracted_133556 | Not-determined |
| S7_extracted_133557 | Low-quality |
| S7_extracted_133558 | Low-quality |
| S7_extracted_133577 | Not-determined |
| S7_extracted_133578 | Not-determined |
| S7_extracted_133585 | Not-determined |
| S7_extracted_133594 | Not-determined |
| S7_extracted_133603 | Not-determined |
| S7_extracted_133605 | Not-determined |
| S7_extracted_133607 | Not-determined |
| S7_extracted_133613 | Not-determined |
| S7_extracted_133615 | Not-determined |
| S7_extracted_133624 | Not-determined |
| S7_extracted_133628 | Not-determined |
| S7_extracted_133636 | Not-determined |
| S7_extracted_133679 | Not-determined |
| S7_extracted_133696 | Not-determined |
| S7_extracted_133706 | Low-quality |
| S7_extracted_133722 | Not-determined |
| S7_extracted_133724 | Not-determined |
| S7_extracted_133730 | Low-quality |
| S7_extracted_133746 | Not-determined |
| S7_extracted_133761 | Not-determined |
| S7_extracted_133767 | Low-quality |
| S7_extracted_133769 | Low-quality |
| S7_extracted_133778 | Not-determined |
| S7_extracted_133784 | Not-determined |
| S7_extracted_133788 | Low-quality |
| S7_extracted_133794 | Not-determined |
| S7_extracted_133844 | Low-quality |
| S7_extracted_133849 | Not-determined |
| S7_extracted_133854 | Not-determined |
| S7_extracted_133861 | Not-determined |
| S7_extracted_133863 | Low-quality |
| S7_extracted_133878 | Not-determined |
| S7_extracted_133882 | Not-determined |
| S7_extracted_133884 | Low-quality |
| S7_extracted_133894 | Low-quality |
| S7_extracted_133909 | Not-determined |
| S7_extracted_133946 | Not-determined |
| S7_extracted_133967 | Not-determined |
| S7_extracted_133982 | Not-determined |
| S7_extracted_133991 | Not-determined |
| S7_extracted_134004 | Not-determined |
| S7_extracted_134005 | Not-determined |
| S7_extracted_134014 | Not-determined |
| S5_vs_k141_145388\|\|full | High-quality |
| S7_extracted_134033 | Not-determined |
| S7_extracted_134038 | Low-quality |
| S7_extracted_134058 | Not-determined |
| S7_extracted_134061 | Not-determined |
| S7_extracted_134077 | Not-determined |
| S7_extracted_134089 | Not-determined |
| S7_extracted_134123 | Not-determined |
| S7_extracted_134142 | Low-quality |
| S7_extracted_134185 | Not-determined |
| S7_extracted_134186 | Not-determined |
| S7_extracted_134187 | Low-quality |
| S7_extracted_134196 | Not-determined |
| S7_extracted_134201 | Not-determined |
| S7_extracted_134203 | Low-quality |
| S7_extracted_134215 | Low-quality |
| S7_extracted_134231 | Not-determined |
| S7_extracted_134237 | Not-determined |
| S7_extracted_134238 | Low-quality |
| S7_extracted_134247 | Low-quality |
| S7_extracted_134259 | Not-determined |
| S7_extracted_134270 | Not-determined |
| S7_extracted_134288 | Not-determined |
| S7_extracted_134303 | Low-quality |
| S7_extracted_134310 | Not-determined |
| S7_extracted_134314 | Not-determined |
| S7_extracted_134317 | Not-determined |
| S7_extracted_134325 | Not-determined |
| S7_extracted_134348 | Not-determined |
| S7_extracted_134378 | Not-determined |
| S7_extracted_134381 | Not-determined |
| S7_extracted_134396 | Not-determined |
| S7_extracted_134404 | Not-determined |
| S7_extracted_134409 | Low-quality |
| S7_extracted_134412 | Not-determined |
| S7_extracted_134428 | Not-determined |
| S7_extracted_134434 | Not-determined |
| S7_extracted_134435 | Not-determined |
| S7_extracted_134436 | Low-quality |
| S7_extracted_134438 | Not-determined |
| S7_extracted_134444 | Low-quality |
| S7_extracted_134486 | Not-determined |
| S7_extracted_134488 | Not-determined |
| S7_extracted_134521 | Not-determined |
| S7_extracted_134528 | Not-determined |
| S7_extracted_134540 | Not-determined |
| S7_extracted_134542 | Low-quality |
| S7_extracted_134556 | Low-quality |
| S7_extracted_134568 | Low-quality |
| S7_extracted_134569 | Low-quality |
| S7_extracted_134595 | Not-determined |
| S7_extracted_134597 | Not-determined |
| S7_extracted_134599 | Not-determined |
| S7_extracted_134600 | Not-determined |
| S7_extracted_134603 | Not-determined |
| S7_extracted_134618 | Not-determined |
| S7_extracted_134634 | Low-quality |
| S7_extracted_134653 | Not-determined |
| S7_extracted_134654 | Not-determined |
| S7_extracted_134662 | Low-quality |
| S7_extracted_134670 | Low-quality |
| S7_extracted_134673 | Not-determined |
| S7_extracted_134695 | Low-quality |
| S7_extracted_134706 | Not-determined |
| S7_extracted_134709 | Not-determined |
| S7_extracted_134718 | Not-determined |
| S7_extracted_134755 | Not-determined |
| S7_extracted_134756 | Low-quality |
| S7_extracted_134793 | Not-determined |
| S7_extracted_134808 | Not-determined |
| S7_extracted_134810 | Not-determined |
| S7_extracted_134841 | Low-quality |
| S7_extracted_134847 | Low-quality |
| S7_extracted_134848 | Not-determined |
| S7_extracted_134850 | Not-determined |
| S7_extracted_134872 | Low-quality |
| S7_extracted_134876 | Not-determined |
| S7_extracted_134887 | Not-determined |
| S7_extracted_134928 | Not-determined |
| S7_extracted_134929 | Low-quality |
| S7_extracted_134949 | Not-determined |
| S7_extracted_134957 | Not-determined |
| S7_extracted_135036 | Not-determined |
| S7_extracted_135039 | Not-determined |
| S7_extracted_135056 | Not-determined |
| S7_extracted_135063 | Low-quality |
| S7_extracted_135081 | Not-determined |
| S7_extracted_135084 | Not-determined |
| S7_extracted_135092 | Low-quality |
| S7_extracted_135099 | Not-determined |
| S7_extracted_135116 | Not-determined |
| S7_extracted_135136 | Low-quality |
| S7_extracted_135140 | Not-determined |
| S7_extracted_135142 | Not-determined |
| S7_extracted_135144 | Not-determined |
| S7_extracted_135170 | Not-determined |
| S7_extracted_135182 | Not-determined |
| S7_extracted_135183 | Not-determined |
| S7_extracted_135190 | Not-determined |
| S7_extracted_135197 | Not-determined |
| S7_extracted_135218 | Not-determined |
| S7_extracted_135245 | Not-determined |
| S7_extracted_135264 | Not-determined |
| S7_extracted_135267 | Not-determined |
| S7_extracted_135276 | Not-determined |
| S7_extracted_135277 | Not-determined |
| S7_extracted_135280 | Not-determined |
| S7_extracted_135281 | Not-determined |
| S7_extracted_135306 | Not-determined |
| S7_extracted_135309 | Not-determined |
| S7_extracted_135314 | Not-determined |
| S7_extracted_135317 | Not-determined |
| S7_extracted_135339 | Not-determined |
| S7_extracted_135357 | Not-determined |
| S7_extracted_135358 | Low-quality |
| S7_extracted_135368 | Not-determined |
| S7_extracted_135378 | Not-determined |
| S7_extracted_135385 | Not-determined |
| S7_extracted_135386 | Not-determined |
| S7_extracted_135392 | Not-determined |
| S7_extracted_135427 | Not-determined |
| S7_extracted_135444 | Not-determined |
| S7_extracted_135445 | Not-determined |
| S7_extracted_135451 | Not-determined |
| S7_extracted_135454 | Not-determined |
| S7_extracted_135455 | Not-determined |
| S7_extracted_135462 | Not-determined |
| S7_extracted_135467 | Not-determined |
| S7_extracted_135491 | Not-determined |
| S7_extracted_135497 | Not-determined |
| S7_extracted_135515 | Low-quality |
| S7_extracted_135518 | Low-quality |
| S7_extracted_135520 | Not-determined |
| S7_extracted_135529 | Not-determined |
| S7_extracted_135531 | Not-determined |
| S7_extracted_135540 | Not-determined |
| S7_extracted_135555 | Not-determined |
| S7_extracted_135562 | Not-determined |
| S7_extracted_135569 | Not-determined |
| S7_extracted_135571 | Not-determined |
| S7_extracted_135572 | Not-determined |
| S7_extracted_135575 | Not-determined |
| S7_extracted_135580 | Not-determined |
| S7_extracted_135586 | Not-determined |
| S7_extracted_135590 | Not-determined |
| S7_extracted_135605 | Not-determined |
| S7_extracted_135612 | Low-quality |
| S7_extracted_135631 | Not-determined |
| S7_extracted_135634 | Not-determined |
| S7_extracted_135641 | Low-quality |
| S7_extracted_135650 | Not-determined |
| S7_extracted_135656 | Low-quality |
| S7_extracted_135666 | Not-determined |
| S7_extracted_135678 | Not-determined |
| S7_extracted_135688 | Not-determined |
| S7_extracted_135689 | Not-determined |
| S7_extracted_135705 | Not-determined |
| S7_extracted_135730 | Not-determined |
| S7_extracted_135733 | Not-determined |
| S7_extracted_135737 | Not-determined |
| S7_extracted_135767 | Not-determined |
| S7_extracted_135777 | Low-quality |
| S9_vs_k141_63485\|\|full | High-quality |
| S7_extracted_135804 | Not-determined |
| S7_extracted_135810 | Not-determined |
| S7_extracted_135820 | Not-determined |
| S7_extracted_135831 | Not-determined |
| S7_extracted_135837 | Not-determined |
| S7_extracted_135841 | Not-determined |
| S7_extracted_135843 | Not-determined |
| S7_extracted_135847 | Not-determined |
| S7_extracted_135860 | Not-determined |
| S7_extracted_135862 | Low-quality |
| S7_extracted_135876 | Not-determined |
| S7_extracted_135877 | Not-determined |
| S7_extracted_135897 | Not-determined |
| S7_extracted_135907 | Low-quality |
| S7_extracted_135909 | Not-determined |
| S7_extracted_135910 | Not-determined |
| S7_extracted_135911 | Not-determined |
| S7_extracted_135932 | Not-determined |
| S7_extracted_135947 | Not-determined |
| S7_extracted_135948 | Not-determined |
| S7_extracted_135949 | Not-determined |
| S7_extracted_135980 | Not-determined |
| S7_extracted_135988 | Not-determined |
| S7_extracted_135995 | Not-determined |
| S7_extracted_135996 | Not-determined |
| S7_extracted_136018 | Not-determined |
| S7_extracted_136021 | Not-determined |
| S7_extracted_136023 | Not-determined |
| S7_extracted_136041 | Not-determined |
| S7_extracted_136043 | Not-determined |
| S7_extracted_136074 | Not-determined |
| S7_extracted_136082 | Low-quality |
| S7_extracted_136094 | Not-determined |
| S7_extracted_136125 | Not-determined |
| S7_extracted_136128 | Low-quality |
| S7_extracted_136152 | Not-determined |
| S7_extracted_136165 | Not-determined |
| S7_extracted_136168 | Not-determined |
| S7_extracted_136199 | Not-determined |
| S7_extracted_136200 | Not-determined |
| S7_extracted_136222 | Not-determined |
| S7_extracted_136224 | Not-determined |
| S7_extracted_136229 | Not-determined |
| S7_extracted_136231 | Not-determined |
| S7_extracted_136242 | Not-determined |
| S7_extracted_136243 | Not-determined |
| S7_extracted_136248 | Not-determined |
| S7_extracted_136266 | Not-determined |
| S7_extracted_136282 | Not-determined |
| S7_extracted_136296 | Not-determined |
| S7_extracted_136299 | Not-determined |
| S7_extracted_136323 | Low-quality |
| S7_extracted_136327 | Low-quality |
| S7_extracted_136341 | Not-determined |
| S7_extracted_136348 | Not-determined |
| S7_extracted_136351 | Not-determined |
| S7_extracted_136362 | Not-determined |
| S7_extracted_136369 | Not-determined |
| S7_extracted_136373 | Not-determined |
| S7_extracted_136384 | Not-determined |
| S7_extracted_136386 | Low-quality |
| S7_extracted_136395 | Low-quality |
| S7_extracted_136414 | Not-determined |
| S7_extracted_136418 | Not-determined |
| S7_extracted_136419 | Low-quality |
| S7_extracted_136427 | Not-determined |
| S7_extracted_136434 | Not-determined |
| S7_extracted_136442 | Not-determined |
| S7_extracted_136469 | Not-determined |
| S7_extracted_136474 | Not-determined |
| S7_extracted_136476 | Low-quality |
| S7_extracted_136481 | Not-determined |
| S7_extracted_136489 | Not-determined |
| S7_extracted_136506 | Not-determined |
| S7_extracted_136512 | Not-determined |
| S7_extracted_136514 | Not-determined |
| S7_extracted_136544 | Not-determined |
| S7_extracted_136552 | Not-determined |
| S7_extracted_136556 | Not-determined |
| S7_extracted_136561 | Not-determined |
| S7_extracted_136564 | Not-determined |
| S7_extracted_136581 | Not-determined |
| S7_extracted_136588 | Not-determined |
| S7_extracted_136592 | Not-determined |
| S7_extracted_136608 | Low-quality |
| S7_extracted_136613 | Not-determined |
| S7_extracted_136623 | Not-determined |
| S7_extracted_136629 | Not-determined |
| S7_extracted_136632 | Not-determined |
| S7_extracted_136641 | Not-determined |
| S7_extracted_136645 | Not-determined |
| S7_extracted_136649 | Not-determined |
| S7_extracted_136660 | Not-determined |
| S7_extracted_136664 | Not-determined |
| S7_extracted_136678 | Not-determined |
| S7_extracted_136709 | Not-determined |
| S7_extracted_136734 | Not-determined |
| S7_extracted_136737 | Not-determined |
| S7_extracted_136776 | Low-quality |
| S7_extracted_136777 | Not-determined |
| S7_extracted_136795 | Not-determined |
| S5_vs_k141_86562\|\|full | High-quality |
| S7_extracted_136813 | Not-determined |
| S7_extracted_136815 | Not-determined |
| S7_extracted_136837 | Not-determined |
| S7_extracted_136846 | Not-determined |
| S7_extracted_136854 | Not-determined |
| S7_extracted_136858 | Not-determined |
| S7_extracted_136860 | Not-determined |
| S7_extracted_136865 | Not-determined |
| S7_extracted_136886 | Not-determined |
| S7_extracted_136896 | Not-determined |
| S7_extracted_136903 | Not-determined |
| S7_extracted_136905 | Low-quality |
| S7_extracted_136920 | Not-determined |
| S7_extracted_136927 | Not-determined |
| S7_extracted_136936 | Low-quality |
| S7_extracted_136949 | Not-determined |
| S7_extracted_136972 | Low-quality |
| S7_extracted_136974 | Not-determined |
| S7_extracted_136983 | Not-determined |
| S7_extracted_136984 | Low-quality |
| S7_extracted_136985 | Not-determined |
| S7_extracted_136987 | Not-determined |

| | |
|---|---|
| S7_extracted_137001 | Low-quality |
| S7_extracted_137006 | Not-determined |
| S7_extracted_137008 | Not-determined |
| S7_extracted_137019 | Not-determined |
| S7_extracted_137023 | Not-determined |
| S7_extracted_137028 | Low-quality |
| S7_extracted_137042 | Not-determined |
| S7_extracted_137051 | Low-quality |
| S7_extracted_137071 | Low-quality |
| S7_extracted_137095 | Not-determined |
| S7_extracted_137097 | Not-determined |
| S7_extracted_137114 | Not-determined |
| S7_extracted_137130 | Low-quality |
| S7_extracted_137139 | Not-determined |
| S7_extracted_137146 | Low-quality |
| S7_extracted_137201 | Low-quality |
| S7_extracted_137206 | Not-determined |
| S7_extracted_137210 | Low-quality |
| S7_extracted_137217 | Not-determined |
| S7_extracted_137238 | Not-determined |
| S7_extracted_137246 | Not-determined |
| S7_extracted_137247 | Not-determined |
| S7_extracted_137263 | Not-determined |
| S7_extracted_137284 | Not-determined |
| S7_extracted_137293 | Not-determined |
| S7_extracted_137294 | Not-determined |
| S7_extracted_137300 | Not-determined |
| S7_extracted_137303 | Not-determined |
| S7_extracted_137321 | Not-determined |
| S7_extracted_137332 | Low-quality |
| S7_extracted_137342 | Not-determined |
| S7_extracted_137345 | Not-determined |
| S7_extracted_137347 | Low-quality |
| S7_extracted_137356 | Not-determined |
| S7_extracted_137376 | Not-determined |
| S7_extracted_137377 | Not-determined |
| S7_extracted_137385 | Not-determined |
| S7_extracted_137391 | Low-quality |
| S7_extracted_137394 | Low-quality |
| S7_extracted_137399 | Not-determined |
| S7_extracted_137402 | Not-determined |
| S7_extracted_137435 | Not-determined |
| S7_extracted_137454 | Not-determined |
| S7_extracted_137464 | Low-quality |
| S7_extracted_137474 | Not-determined |
| S7_extracted_137476 | Not-determined |
| S7_extracted_137491 | Not-determined |
| S7_extracted_137500 | Not-determined |
| S7_extracted_137518 | Not-determined |
| S7_extracted_137523 | Not-determined |
| S7_extracted_137534 | Not-determined |
| S7_extracted_137540 | Not-determined |
| S7_extracted_137559 | Not-determined |
| S7_extracted_137572 | Not-determined |
| S7_extracted_137573 | Not-determined |
| S7_extracted_137575 | Not-determined |
| S7_extracted_137580 | Not-determined |
| S7_extracted_137581 | Not-determined |
| S7_extracted_137586 | Not-determined |
| S7_extracted_137599 | Not-determined |
| S7_extracted_137607 | Not-determined |
| S7_extracted_137616 | Low-quality |
| S7_extracted_137622 | Not-determined |
| S7_extracted_137632 | Not-determined |
| S7_extracted_137647 | Not-determined |
| S7_extracted_137648 | Low-quality |
| S7_extracted_137652 | Not-determined |
| S7_extracted_137658 | Not-determined |
| S7_extracted_137660 | Not-determined |
| S7_extracted_137670 | Not-determined |
| S7_extracted_137671 | Not-determined |
| S7_extracted_137682 | Not-determined |
| S7_extracted_137689 | Low-quality |
| S7_extracted_137695 | Not-determined |
| S7_extracted_137699 | Low-quality |
| S7_extracted_137726 | Not-determined |
| S7_extracted_137742 | Not-determined |
| S7_extracted_137743 | Low-quality |
| S7_extracted_137744 | Not-determined |
| S7_extracted_137768 | Not-determined |
| S7_extracted_137773 | Low-quality |
| S7_extracted_137788 | Not-determined |
| S7_extracted_137792 | Not-determined |
| S7_extracted_137797 | Not-determined |
| S7_extracted_137815 | Not-determined |
| S7_extracted_137817 | Not-determined |
| S7_extracted_137833 | Not-determined |
| S7_extracted_137839 | Not-determined |
| S7_extracted_137841 | Not-determined |
| S7_extracted_137864 | Low-quality |
| S7_extracted_137880 | Not-determined |
| S7_extracted_137884 | Not-determined |
| S7_extracted_137896 | Low-quality |
| S7_extracted_137903 | Not-determined |
| S7_extracted_137911 | Not-determined |
| S7_extracted_137918 | Not-determined |
| S7_extracted_137933 | Low-quality |
| S7_extracted_137936 | Low-quality |
| S7_extracted_137939 | Not-determined |
| S7_extracted_137950 | Low-quality |
| S7_extracted_137953 | Not-determined |
| S7_extracted_137962 | Not-determined |
| S7_extracted_137974 | Not-determined |
| S7_extracted_137977 | Not-determined |
| S7_extracted_137989 | Not-determined |
| S7_extracted_138003 | Not-determined |
| S7_extracted_138008 | Not-determined |
| S7_extracted_138017 | Not-determined |
| S5_vs_k141_136679\|\|full | High-quality |
| S7_extracted_138036 | Not-determined |
| S7_extracted_138038 | Not-determined |
| S7_extracted_138039 | Not-determined |
| S7_extracted_138045 | Not-determined |
| S16_extracted_12551 | High-quality |
| S7_extracted_138078 | Low-quality |
| S7_extracted_138080 | Not-determined |
| S7_extracted_138088 | Not-determined |
| S5_extracted_k141_34842 | High-quality |
| S7_extracted_138104 | Low-quality |
| S7_extracted_138106 | Not-determined |
| S7_extracted_138114 | Not-determined |
| S7_extracted_138119 | Not-determined |
| S7_extracted_138127 | Low-quality |
| S7_extracted_138143 | Not-determined |
| S7_extracted_138149 | Not-determined |
| S7_extracted_138190 | Not-determined |
| S7_extracted_138193 | Not-determined |
| S7_extracted_138198 | Not-determined |
| S7_extracted_138199 | Not-determined |
| S7_extracted_138201 | Not-determined |
| S7_extracted_138210 | Low-quality |
| S7_extracted_138217 | Low-quality |
| S5_vs_k141_15069\|\|full | Medium-quality |
| S7_extracted_138237 | Not-determined |
| S7_extracted_138245 | Not-determined |
| S7_extracted_138246 | Not-determined |
| S7_extracted_138250 | Not-determined |
| S7_extracted_138258 | Not-determined |
| S7_extracted_138275 | Not-determined |
| S7_extracted_138278 | Not-determined |
| S7_extracted_138282 | Not-determined |
| S7_extracted_138290 | Not-determined |
| S7_extracted_138293 | Not-determined |
| S7_extracted_138295 | Not-determined |
| S7_extracted_138301 | Not-determined |
| S7_extracted_138302 | Not-determined |
| S7_extracted_138304 | Not-determined |
| S7_extracted_138305 | Not-determined |
| S4_vs_VIRSorter_31972-circular-cat | High-quality |
| S7_vs_42563\|\|full | High-quality |
| S7_vs_2692\|\|full | Not-determined |
| S16_vs_118029\|\|full | High-quality |
| S5_vs_k141_4737\|\|full | Medium-quality |
| S5_vs_k141_34842\|\|full | High-quality |
| S3_vs_k141_30318\|\|full | High-quality |
| S16_vs_78361\|\|full | High-quality |
| S8_vs_k141_148389\|\|full | Medium-quality |
| S16_vs_172749\|\|full | High-quality |
| S7_vs_14275\|\|full | High-quality |
| S9_vs_k141_4343\|\|full | High-quality |
| S7_vs_17785\|\|full | Low-quality |
| S1_vs_k141_133577\|\|full | High-quality |
| S7_vs_25868\|\|full | Low-quality |
| S4_vs_VIRSorter_12831-circular-cat | High-quality |
| S5_vs_k141_116235\|\|full | High-quality |
| S11_extracted_k141_10907 | Complete |
| S9_vs_k141_90857\|\|full | High-quality |
| S5_vs_k141_1572\|\|full | Medium-quality |
| S3_vs_k141_23194\|\|full | Medium-quality |
| S5_extracted_k141_29332 | High-quality |
| S4_extracted_31682 | Complete |
| S16_vs_12551\|\|full | High-quality |
| S7_vs_51110\|\|full | Not-determined |
| S12_vs_k141_150812\|\|full | Medium-quality |
| S11_vs_k141_10907\|\|full | Medium-quality |
| S11_extracted_k141_104048 | Complete |
| S6_extracted_k141_52601 | Complete |
| S5_vs_k141_29332\|\|full | High-quality |
| S7_vs_74969\|\|full | Not-determined |
| S7_vs_75613\|\|full | Not-determined |
| S7_vs_90087\|\|full | Low-quality |
| S7_vs_91589\|\|full | Low-quality |
| S16_vs_51901\|\|full | High-quality |
| S14_vs_k141_90934\|\|full | High-quality |
| S12_vs_k141_81729\|\|full | High-quality |
| S7_vs_106604\|\|full | Not-determined |
| S4_vs_VIRSorter_31682-circular-cat | High-quality |
| S14_vs_k141_632\|\|full | Medium-quality |
| S8_extracted_k141_16172 | Medium-quality |
| S8_vs_k141_16172\|\|full | Medium-quality |
| S14_vs_k141_39266\|\|full | Medium-quality |
| S13_extracted_54201 | Medium-quality |
| S7_extracted_39724 | High-quality |
| S13_vs_54201\|\|full | Medium-quality |
| S4_vs_VIRSorter_19324-cat_2 | High-quality |
| S11_vs_k141_41107\|\|full | Medium-quality |
| S6_vs_k141_52601\|\|full | High-quality |
| S9_extracted_k141_62658 | High-quality |
| S8_vs_k141_134805\|\|full | High-quality |
| S9_vs_k141_62658\|\|full | High-quality |
| S17_extracted_k141_1614 | High-quality |
| S6_vs_k141_20670\|\|full | Medium-quality |
| S17_vs_k141_1614\|\|full | High-quality |
| S5_vs_k141_127162\|\|0_partial | Medium-quality |
| S11_vs_k141_104042\|\|full | Medium-quality |
| S17_vs_k141_41845\|\|full | High-quality |
| S11_vs_k141_3277\|\|full | Medium-quality |
| S14_vs_k141_117153\|\|full | High-quality |
| S7_vs_138193\|\|full | Not-determined |
| S7_vs_138201\|\|full | Not-determined |
| S5_vs_k141_145596\|\|full | High-quality |
| S7_vs_138237\|\|full | Not-determined |
| S7_vs_138246\|\|full | Not-determined |
| S7_vs_138260\|\|full | Not-determined |
| S7_vs_37\|\|full | Low-quality |
| S7_vs_85\|\|full | Not-determined |
| S7_vs_119\|\|full | Not-determined |
| S7_vs_129\|\|full | Not-determined |
| S7_vs_149\|\|full | Not-determined |
| S7_vs_151\|\|full | Not-determined |
| S7_vs_152\|\|full | Not-determined |
| S7_vs_191\|\|full | Low-quality |
| S7_vs_231\|\|full | Not-determined |
| S7_vs_249\|\|full | Low-quality |
| S7_vs_277\|\|full | Not-determined |
| S7_vs_282\|\|full | Not-determined |
| S7_vs_299\|\|full | Low-quality |
| S7_vs_318\|\|full | Low-quality |
| S7_vs_340\|\|full | Not-determined |
| S7_vs_347\|\|full | Low-quality |
| S7_vs_352\|\|full | Low-quality |
| S7_vs_366\|\|full | Not-determined |
| S7_vs_380\|\|full | Low-quality |
| S7_vs_407\|\|full | Low-quality |
| S7_vs_445\|\|full | Low-quality |
| S7_vs_469\|\|full | Not-determined |
| S7_vs_489\|\|full | Low-quality |
| S7_vs_497\|\|full | Low-quality |
| S7_vs_521\|\|full | Low-quality |
| S7_vs_522\|\|full | Low-quality |
| S7_vs_532\|\|full | Not-determined |
| S7_vs_544\|\|full | Low-quality |
| S7_vs_561\|\|full | Low-quality |
| S7_vs_571\|\|full | Low-quality |
| S7_vs_596\|\|full | Low-quality |
| S7_vs_619\|\|full | Not-determined |
| S7_vs_631\|\|full | Low-quality |
| S7_vs_654\|\|full | Low-quality |
| S7_vs_660\|\|full | Not-determined |
| S7_vs_687\|\|full | Low-quality |
| S7_vs_785\|\|full | Low-quality |
| S7_vs_828\|\|full | Not-determined |
| S7_vs_829\|\|full | Low-quality |
| S7_vs_854\|\|full | Low-quality |
| S7_vs_859\|\|full | Not-determined |
| S7_vs_875\|\|full | Low-quality |
| S7_vs_895\|\|full | Not-determined |
| S7_vs_922\|\|full | Low-quality |
| S7_vs_934\|\|full | Not-determined |
| S8_vs_k141_173961\|\|full | Medium-quality |
| S7_vs_959\|\|full | Low-quality |
| S7_vs_965\|\|full | Low-quality |
| S7_vs_1020\|\|full | Low-quality |
| S7_vs_1069\|\|full | Not-determined |
| S7_vs_1082\|\|full | Low-quality |
| S7_vs_1102\|\|full | Low-quality |
| S7_vs_1108\|\|full | Not-determined |
| S7_vs_1117\|\|full | Not-determined |
| S7_vs_1137\|\|full | Low-quality |
| S7_vs_1146\|\|full | Low-quality |
| S7_vs_1266\|\|full | Low-quality |
| S7_vs_1268\|\|full | Low-quality |
| S7_vs_1280\|\|full | Low-quality |
| S7_vs_1345\|\|full | Low-quality |
| S7_vs_1421\|\|full | Not-determined |
| S7_vs_1439\|\|full | Not-determined |
| S7_vs_1468\|\|full | Low-quality |
| S7_vs_1518\|\|full | Low-quality |
| S7_vs_1547\|\|full | Low-quality |
| S7_vs_1609\|\|full | Not-determined |
| S7_vs_1626\|\|full | Low-quality |
| S7_vs_1637\|\|full | Low-quality |
| S7_vs_1662\|\|full | Not-determined |
| S7_vs_1667\|\|full | Low-quality |
| S7_vs_1677\|\|full | Low-quality |
| S7_vs_1679\|\|full | Low-quality |
| S7_vs_1727\|\|full | Not-determined |
| S7_vs_1744\|\|full | Low-quality |
| S7_vs_1792\|\|full | Not-determined |
| S7_vs_1854\|\|full | Not-determined |
| S7_vs_1896\|\|full | Low-quality |
| S7_vs_1906\|\|full | Low-quality |
| S7_vs_1907\|\|full | Not-determined |
| S7_vs_1930\|\|full | Not-determined |
| S7_vs_1945\|\|full | Low-quality |
| S7_vs_2012\|\|full | Low-quality |
| S7_vs_2050\|\|full | Low-quality |
| S7_vs_2075\|\|full | Low-quality |
| S7_vs_2112\|\|full | Not-determined |
| S7_vs_2133\|\|full | Not-determined |
| S7_vs_2153\|\|full | Low-quality |
| S7_vs_2159\|\|full | Low-quality |
| S7_vs_2212\|\|full | Low-quality |
| S7_vs_2215\|\|full | Low-quality |
| S7_vs_2257\|\|full | Low-quality |
| S7_vs_2295\|\|full | Low-quality |
| S7_vs_2297\|\|full | Not-determined |
| S7_vs_2354\|\|full | Low-quality |
| S7_vs_2406\|\|full | Not-determined |
| S7_vs_2420\|\|full | Not-determined |
| S7_vs_2456\|\|full | Not-determined |
| S7_vs_2490\|\|full | Not-determined |
| S7_vs_2492\|\|full | Low-quality |
| S7_vs_2526\|\|full | Low-quality |
| S7_vs_2559\|\|full | Not-determined |
| S7_vs_2561\|\|full | Low-quality |
| S7_vs_2583\|\|full | Low-quality |
| S7_vs_2589\|\|full | Not-determined |
| S7_vs_2604\|\|full | Not-determined |
| S7_vs_2611\|\|full | Low-quality |
| S7_vs_2632\|\|full | Not-determined |
| S7_vs_2656\|\|full | Low-quality |
| S7_vs_2666\|\|full | Low-quality |
| S7_vs_2686\|\|full | Low-quality |
| S7_vs_2687\|\|full | Low-quality |
| S7_vs_2688\|\|full | Low-quality |
| S7_vs_2696\|\|full | Low-quality |
| S7_vs_2697\|\|full | Low-quality |
| S7_vs_2699\|\|full | Low-quality |
| S7_vs_2747\|\|full | Low-quality |
| S7_vs_2830\|\|full | Not-determined |
| S7_vs_2837\|\|full | Low-quality |
| S7_vs_2839\|\|full | Low-quality |
| S7_vs_2909\|\|full | Not-determined |
| S7_vs_2928\|\|full | Not-determined |
| S7_vs_2939\|\|full | Low-quality |
| S7_vs_2952\|\|full | Low-quality |
| S7_vs_3014\|\|full | Low-quality |
| S7_vs_3040\|\|full | Not-determined |
| S7_vs_3044\|\|full | Low-quality |
| S7_vs_3052\|\|full | Low-quality |
| S7_vs_3094\|\|full | Low-quality |
| S7_vs_3110\|\|full | Low-quality |
| S7_vs_3121\|\|full | Low-quality |
| S7_vs_3127\|\|full | Low-quality |
| S7_vs_3144\|\|full | Low-quality |
| S7_vs_3149\|\|full | Low-quality |
| S7_vs_3197\|\|full | Low-quality |
| S7_vs_3209\|\|full | Not-determined |
| S7_vs_3246\|\|full | Not-determined |
| S7_vs_3284\|\|full | Low-quality |
| S7_vs_3287\|\|full | Not-determined |
| S7_vs_3391\|\|full | Not-determined |
| S7_vs_3425\|\|full | Not-determined |
| S7_vs_3494\|\|full | Low-quality |
| S7_vs_3574\|\|full | Low-quality |
| S7_vs_3604\|\|full | Low-quality |
| S7_vs_3624\|\|full | Low-quality |
| S7_vs_3626\|\|full | Low-quality |
| S7_vs_3685\|\|full | Not-determined |
| S7_vs_3701\|\|full | Low-quality |
| S7_vs_3729\|\|full | Not-determined |
| S7_vs_3739\|\|full | Not-determined |
| S7_vs_3752\|\|full | Low-quality |
| S7_vs_3756\|\|full | Low-quality |
| S7_vs_3761\|\|full | Low-quality |
| S7_vs_3797\|\|full | Not-determined |
| S7_vs_3829\|\|full | Not-determined |
| S7_vs_3893\|\|full | Low-quality |
| S7_vs_3928\|\|full | Low-quality |
| S7_vs_3942\|\|full | Low-quality |
| S7_vs_3952\|\|full | Not-determined |
| S7_vs_3957\|\|full | Not-determined |
| S7_vs_3981\|\|full | Low-quality |
| S7_vs_4017\|\|full | Low-quality |
| S7_vs_4091\|\|full | Low-quality |
| S9_vs_k141_21850\|\|full | Medium-quality |
| S7_vs_4129\|\|full | Not-determined |
| S7_vs_4131\|\|full | Low-quality |
| S7_vs_4151\|\|full | Not-determined |
| S7_vs_4152\|\|full | Not-determined |
| S7_vs_4158\|\|full | Not-determined |
| S7_vs_4164\|\|full | Not-determined |
| S7_vs_4165\|\|full | Not-determined |
| S7_vs_4189\|\|full | Not-determined |
| S7_vs_4200\|\|full | Not-determined |
| S7_vs_4208\|\|full | Low-quality |
| S7_vs_4269\|\|full | Low-quality |
| S7_vs_4277\|\|full | Low-quality |
| S7_vs_4335\|\|full | Low-quality |
| S7_vs_4352\|\|full | Low-quality |
| S7_vs_4410\|\|full | Not-determined |
| S7_vs_4412\|\|full | Not-determined |
| S7_vs_4451\|\|full | Low-quality |
| S7_vs_4495\|\|full | Low-quality |
| S7_vs_4529\|\|full | Low-quality |
| S7_vs_4556\|\|full | Not-determined |
| S7_vs_4560\|\|full | Low-quality |
| S7_vs_4564\|\|full | Not-determined |
| S7_vs_4695\|\|full | Low-quality |
| S7_vs_4708\|\|full | Low-quality |
| S7_vs_4709\|\|full | Not-determined |
| S7_vs_4729\|\|full | Low-quality |
| S7_vs_4740\|\|full | Low-quality |
| S7_vs_4745\|\|full | Not-determined |
| S7_vs_4768\|\|full | Low-quality |
| S7_vs_4804\|\|full | Low-quality |
| S7_vs_4807\|\|full | Low-quality |
| S7_vs_4825\|\|full | Low-quality |
| S7_vs_4935\|\|full | Low-quality |
| S7_vs_4941\|\|full | Not-determined |
| S7_vs_4944\|\|full | Low-quality |
| S7_vs_4970\|\|full | Low-quality |
| S7_vs_4982\|\|full | Not-determined |
| S7_vs_5041\|\|full | Low-quality |
| S7_vs_5048\|\|full | Low-quality |
| S7_vs_5077\|\|full | Low-quality |
| S7_vs_5105\|\|full | Low-quality |
| S7_vs_5108\|\|full | Low-quality |
| S7_vs_5148\|\|full | Low-quality |
| S7_vs_5257\|\|full | Low-quality |
| S7_vs_5267\|\|full | Low-quality |
| S7_vs_5273\|\|full | Low-quality |
| S7_vs_5283\|\|full | Not-determined |
| S7_vs_5287\|\|full | Low-quality |
| S7_vs_5291\|\|full | Low-quality |
| S7_vs_5299\|\|full | Low-quality |
| S7_vs_5428\|\|full | Low-quality |
| S7_vs_5443\|\|full | Low-quality |
| S7_vs_5446\|\|full | Low-quality |
| S7_vs_5479\|\|full | Low-quality |
| S7_vs_5485\|\|full | Low-quality |
| S7_vs_5493\|\|full | Not-determined |
| S7_vs_5496\|\|full | Not-determined |
| S7_vs_5511\|\|full | Low-quality |
| S7_vs_5550\|\|full | Not-determined |
| S7_vs_5578\|\|full | Low-quality |
| S7_vs_5579\|\|full | Low-quality |
| S7_vs_5627\|\|full | Low-quality |
| S7_vs_5645\|\|full | Not-determined |
| S7_vs_5668\|\|full | Not-determined |
| S7_vs_5671\|\|full | Low-quality |
| S7_vs_5673\|\|full | Low-quality |
| S7_vs_5708\|\|full | Not-determined |
| S7_vs_5717\|\|full | Low-quality |
| S7_vs_5723\|\|full | Low-quality |
| S7_vs_5757\|\|full | Low-quality |
| S7_vs_5767\|\|full | Low-quality |
| S7_vs_5770\|\|full | Low-quality |
| S7_vs_5826\|\|full | Low-quality |
| S12_vs_k141_5857\|\|full | Medium-quality |
| S7_vs_5850\|\|full | Not-determined |
| S7_vs_5855\|\|full | Low-quality |
| S7_vs_5930\|\|full | Not-determined |
| S7_vs_5966\|\|full | Low-quality |
| S7_vs_5974\|\|full | Low-quality |
| S7_vs_6037\|\|full | Low-quality |
| S7_vs_6044\|\|full | Low-quality |
| S7_vs_6047\|\|full | Low-quality |
| S7_vs_6056\|\|full | Low-quality |
| S7_vs_6077\|\|full | Low-quality |
| S7_vs_6138\|\|full | Not-determined |
| S7_vs_6146\|\|full | Low-quality |
| S7_vs_6168\|\|full | Low-quality |
| S7_vs_6196\|\|full | Low-quality |
| S7_vs_6218\|\|full | Low-quality |
| S7_vs_6298\|\|full | Not-determined |
| S7_vs_6308\|\|full | Low-quality |
| S7_vs_6330\|\|full | Low-quality |
| S7_vs_6332\|\|full | Low-quality |
| S7_vs_6334\|\|full | Not-determined |
| S7_vs_6377\|\|full | Not-determined |
| S7_vs_6428\|\|full | Not-determined |
| S7_vs_6501\|\|full | Low-quality |
| S7_vs_6504\|\|full | Low-quality |
| S7_vs_6555\|\|full | Not-determined |
| S7_vs_6581\|\|full | Low-quality |
| S7_vs_6591\|\|full | Low-quality |
| S7_vs_6611\|\|full | Low-quality |
| S3_vs_k141_123834\|\|full | High-quality |
| S7_vs_6638\|\|full | Not-determined |
| S7_vs_6652\|\|full | Low-quality |
| S7_vs_6669\|\|full | Not-determined |
| S7_vs_6670\|\|full | Low-quality |
| S7_vs_6672\|\|full | Low-quality |
| S7_vs_6713\|\|full | Low-quality |
| S7_vs_6753\|\|full | Not-determined |
| S7_vs_6762\|\|full | Low-quality |
| S7_vs_6785\|\|full | Not-determined |
| S7_vs_6800\|\|full | Not-determined |
| S7_vs_6806\|\|full | Low-quality |
| S7_vs_6850\|\|full | Low-quality |
| S7_vs_6851\|\|full | Low-quality |
| S7_vs_6860\|\|full | Low-quality |
| S7_vs_6925\|\|full | Low-quality |
| S7_vs_6963\|\|full | Low-quality |
| S7_vs_7038\|\|full | Not-determined |
| S7_vs_7045\|\|full | Low-quality |
| S7_vs_7065\|\|full | Low-quality |
| S7_vs_7071\|\|full | Not-determined |
| S7_vs_7109\|\|full | Low-quality |
| S7_vs_7135\|\|full | Not-determined |
| S7_vs_7170\|\|full | Low-quality |
| S7_vs_7174\|\|full | Low-quality |
| S7_vs_7197\|\|full | Low-quality |
| S7_vs_7202\|\|full | Not-determined |
| S7_vs_7213\|\|full | Low-quality |
| S7_vs_7233\|\|full | Low-quality |
| S7_vs_7240\|\|full | Low-quality |
| S7_vs_7267\|\|full | Low-quality |
| S7_vs_7281\|\|full | Low-quality |
| S7_vs_7293\|\|full | Low-quality |
| S7_vs_7331\|\|full | Not-determined |
| S7_vs_7378\|\|full | Not-determined |
| S7_vs_7441\|\|full | Low-quality |
| S7_vs_7590\|\|full | Low-quality |
| S7_vs_7610\|\|full | Not-determined |
| S7_vs_7665\|\|full | Low-quality |
| S7_vs_7741\|\|full | Low-quality |
| S7_vs_7761\|\|full | Not-determined |
| S7_vs_7850\|\|full | Low-quality |
| S7_vs_7944\|\|full | Not-determined |
| S7_vs_7953\|\|full | Low-quality |
| S7_vs_7969\|\|full | Not-determined |
| S7_vs_7997\|\|full | Not-determined |
| S7_vs_8005\|\|full | Low-quality |
| S7_vs_8054\|\|full | Low-quality |
| S7_vs_8056\|\|full | Low-quality |
| S7_vs_8076\|\|full | Low-quality |
| S7_vs_8083\|\|full | Not-determined |
| S7_vs_8084\|\|full | Not-determined |
| S7_vs_8105\|\|full | Low-quality |
| S7_vs_8118\|\|full | Low-quality |
| S7_vs_8121\|\|full | Not-determined |
| S7_vs_8130\|\|full | Not-determined |
| S7_vs_8137\|\|full | Low-quality |
| S7_vs_8140\|\|full | Low-quality |
| S7_vs_8146\|\|full | Low-quality |
| S7_vs_8162\|\|full | Low-quality |
| S7_vs_8163\|\|full | Not-determined |
| S7_vs_8202\|\|full | Low-quality |
| S7_vs_8224\|\|full | Not-determined |
| S7_vs_8229\|\|full | Low-quality |
| S7_vs_8255\|\|full | Low-quality |
| S7_vs_8256\|\|full | Low-quality |
| S7_vs_8281\|\|full | Low-quality |
| S7_vs_8292\|\|full | Not-determined |
| S7_vs_557\|\|full | High-quality |
| S7_vs_8351\|\|full | Low-quality |
| S7_vs_8367\|\|full | Low-quality |
| S7_vs_8413\|\|full | Not-determined |
| S7_vs_8453\|\|full | Not-determined |
| S7_vs_8467\|\|full | Not-determined |
| S7_vs_8492\|\|full | Low-quality |
| S7_vs_8507\|\|full | Not-determined |
| S7_vs_8510\|\|full | Not-determined |
| S7_vs_8555\|\|full | Low-quality |
| S7_vs_8557\|\|full | Not-determined |
| S7_vs_8596\|\|full | Low-quality |
| S7_vs_8597\|\|full | Low-quality |
| S7_vs_8625\|\|full | Low-quality |
| S7_vs_8632\|\|full | Not-determined |
| S7_vs_8646\|\|full | Low-quality |
| S7_vs_8667\|\|full | Not-determined |
| S7_vs_8672\|\|full | Not-determined |
| S7_vs_8681\|\|full | Low-quality |
| S7_vs_8697\|\|full | Not-determined |
| S7_vs_8853\|\|full | Low-quality |
| S7_vs_8892\|\|full | Not-determined |
| S7_vs_8893\|\|full | Low-quality |
| S7_vs_8913\|\|full | Low-quality |
| S7_vs_8921\|\|full | Low-quality |
| S7_vs_8925\|\|full | Low-quality |
| S7_vs_8977\|\|full | Not-determined |
| S7_vs_8986\|\|full | Low-quality |
| S7_vs_8996\|\|full | Low-quality |
| S7_vs_9020\|\|full | Not-determined |
| S7_vs_9127\|\|full | Low-quality |
| S7_vs_9136\|\|full | Not-determined |
| S7_vs_9156\|\|full | Low-quality |
| S7_vs_9171\|\|full | Low-quality |
| S7_vs_9203\|\|full | Not-determined |
| S7_vs_9232\|\|full | Low-quality |
| S7_vs_9256\|\|full | Low-quality |
| S7_vs_9261\|\|full | Low-quality |
| S7_vs_9310\|\|full | Low-quality |
| S7_vs_9312\|\|full | Low-quality |
| S7_vs_9316\|\|full | Low-quality |
| S7_vs_9317\|\|full | Low-quality |

S7_vs_9344||full Not-determined
S7_vs_9361||full Low-quality
S7_vs_9364||full Not-determined
S7_vs_9365||full Not-determined
S7_vs_9413||full Low-quality
S7_vs_9418||full Low-quality
S7_vs_9562||full Low-quality
S7_vs_9587||full Low-quality
S7_vs_9647||full Low-quality
S7_vs_9658||full Low-quality
S7_vs_9659||full Not-determined
S7_vs_9668||full Not-determined
S7_vs_9687||full Not-determined
S7_vs_9759||full Low-quality
S7_vs_9934||full Low-quality
S7_vs_9946||full Low-quality
S7_vs_10000||full Not-determined
S7_vs_10102||full Low-quality
S7_vs_10145||full Not-determined
S7_vs_10149||full Low-quality
S7_vs_10199||full Not-determined
S7_vs_10258||full Not-determined
S7_vs_10294||full Not-determined
S7_vs_10396||full Not-determined
S7_vs_10477||full Not-determined
S7_vs_10480||full Not-determined
S7_vs_10491||full Low-quality
S7_vs_10499||full Not-determined
S7_vs_10544||full Low-quality
S7_vs_10547||full Not-determined
S7_vs_10561||full Low-quality
S7_vs_10586||full Low-quality
S7_vs_10657||full Not-determined
S7_vs_10663||full Low-quality
S7_vs_10676||full Not-determined
S7_vs_10722||full Not-determined
S7_vs_10739||full Low-quality
S7_vs_10771||full Not-determined
S7_vs_10778||full Low-quality
S7_vs_10798||full Low-quality
S7_vs_10805||full Low-quality
S7_vs_10832||full Not-determined
S7_vs_10885||full Not-determined
S7_vs_10888||full Low-quality
S7_vs_10926||full Low-quality
S7_vs_10943||full Low-quality
S7_vs_10950||full Not-determined
S7_vs_10986||full Not-determined
S7_vs_11007||full Low-quality
S7_vs_11011||full Low-quality
S7_vs_11014||full Not-determined
S7_vs_11070||full Low-quality
S8_vs_k141_10200||full High-quality
S7_vs_11095||full Low-quality
S7_vs_11138||full Not-determined
S7_vs_11153||full Low-quality
S7_vs_11173||full Low-quality
S7_vs_11278||full Low-quality
S7_vs_11284||full Not-determined
S7_vs_11308||full Low-quality
S7_vs_11314||full Not-determined
S7_vs_11324||full Low-quality
S7_vs_11352||full Not-determined
S7_vs_11355||full Low-quality
S7_vs_11390||full Low-quality
S7_vs_11418||full Low-quality
S7_vs_11433||full Low-quality
S7_vs_11483||full Low-quality
S7_vs_11517||full Low-quality
S7_vs_11519||full Low-quality
S7_vs_11593||full Not-determined
S7_vs_11633||full Low-quality
S7_vs_11647||full Low-quality
S7_vs_11657||full Low-quality
S7_vs_11659||full Low-quality
S7_vs_11661||full Low-quality
S7_vs_11668||full Not-determined
S7_vs_11701||full Low-quality
S11_vs_k141_104048||full High-quality
S7_vs_11752||full Low-quality
S7_vs_11769||full Low-quality
S7_vs_11794||full Not-determined
S7_vs_11834||full Not-determined
S7_vs_11849||full Low-quality
S7_vs_11888||full Low-quality
S7_vs_11901||full Not-determined
S7_vs_11918||full Not-determined
S7_vs_11971||full Low-quality
S7_vs_12035||full Low-quality
S7_vs_12051||full Low-quality
S7_vs_12063||full Low-quality
S7_vs_12102||full Low-quality
S7_vs_12132||full Not-determined
S7_vs_12133||full Low-quality
S7_vs_12171||full Low-quality
S7_vs_12212||full Not-determined
S7_vs_12213||full Low-quality
S7_vs_12224||full Not-determined
S7_vs_12229||full Low-quality
S7_vs_12231||full Low-quality
S7_vs_12232||full Low-quality
S7_vs_12235||full Low-quality
S7_vs_12246||full Low-quality
S7_vs_12292||full Low-quality
S7_vs_12298||full Not-determined
S7_vs_12384||full Not-determined
S7_vs_12385||full Low-quality
S7_vs_12405||full Not-determined
S7_vs_12428||full Not-determined
S7_vs_12482||full Not-determined
S7_vs_12500||full Not-determined
S7_vs_12531||full Low-quality
S7_vs_12553||full Low-quality
S7_vs_12575||full Low-quality
S7_vs_12597||full Not-determined
S7_vs_12599||full Low-quality
S7_vs_12606||full Low-quality
S7_vs_12626||full Not-determined
S7_vs_12720||full Low-quality
S7_vs_12747||full Not-determined
S7_vs_12754||full Low-quality
S7_vs_12884||full Low-quality
S7_vs_12921||full Low-quality
S7_vs_12934||full Low-quality
S7_vs_13001||full Low-quality
S17_vs_k141_160502||full Medium-quality
S7_vs_13059||full Low-quality
S7_vs_13081||full Low-quality
S7_vs_13110||full Low-quality
S7_vs_13119||full Low-quality
S7_vs_13154||full Not-determined
S7_vs_13217||full Not-determined
S7_vs_13297||full Not-determined
S7_vs_134391||full Medium-quality
S7_vs_13386||full Low-quality
S7_vs_13388||full Not-determined
S7_vs_13390||full Not-determined
S7_vs_13397||full Not-determined
S7_vs_13406||full Not-determined
S7_vs_13425||full Not-determined
S7_vs_13436||full Low-quality
S7_vs_13454||full Not-determined
S7_vs_13480||full Low-quality
S7_vs_13501||full Not-determined
S7_vs_13505||full Low-quality
S7_vs_13543||full Low-quality
S7_vs_13549||full Not-determined
S7_vs_13578||full Not-determined
S7_vs_13601||full Low-quality
S7_vs_13603||full Not-determined
S7_vs_13636||full Not-determined
S7_vs_13640||full Not-determined
S7_vs_13653||full Not-determined
S7_vs_13698||full Low-quality
S7_vs_13716||full Low-quality
S7_vs_13726||full Low-quality
S7_vs_13761||full Low-quality
S7_vs_13765||full Low-quality
S7_vs_13773||full Not-determined
S9_extracted_k141_90871 Complete
S7_vs_13804||full Low-quality
S7_vs_13842||full Low-quality
S7_vs_13867||full Low-quality
S7_vs_13875||full Low-quality
S14_extracted_k141_117162 Complete
S7_vs_13906||full Low-quality
S7_vs_13949||full Low-quality
S7_vs_13955||full Not-determined
S7_vs_13966||full Low-quality
S7_vs_13986||full Not-determined
S7_vs_14020||full Low-quality
S7_vs_14034||full Low-quality
S7_vs_14037||full Not-determined
S7_vs_14040||full Not-determined
S7_vs_14056||full Low-quality
S7_vs_14061||full Not-determined
S7_vs_14063||full Low-quality
S7_vs_14107||full Low-quality
S7_vs_14108||full Not-determined
S7_vs_14140||full Not-determined
S7_vs_14141||full Low-quality
S7_vs_14143||full Low-quality
S7_vs_14145||full Low-quality
S7_vs_14199||full Not-determined
S7_vs_14232||full Not-determined
S7_vs_14239||full Low-quality
S7_vs_14242||full Not-determined
S5_vs_k141_80911||full High-quality
S7_vs_14424||full Low-quality
S7_vs_14455||full Low-quality
S7_vs_14466||full Not-determined
S7_vs_14469||full Not-determined
S7_vs_14505||full Low-quality
S7_vs_14581||full Low-quality
S7_vs_14606||full Low-quality
S7_vs_14618||full Not-determined
S7_vs_14645||full Not-determined
S7_vs_14669||full Not-determined
S7_vs_14671||full Low-quality
S7_vs_14679||full Low-quality
S7_vs_14689||full Not-determined
S7_vs_14698||full Not-determined
S7_vs_14804||full Not-determined
S7_vs_14857||full Low-quality
S7_vs_14867||full Low-quality
S7_vs_14917||full Low-quality
S7_vs_14935||full Low-quality
S7_vs_14945||full Low-quality
S7_vs_14952||full Low-quality
S7_vs_14959||full Low-quality
S7_vs_14964||full Low-quality
S7_vs_14969||full Not-determined
S7_vs_15017||full Low-quality
S7_vs_15045||full Not-determined
S7_vs_15061||full Low-quality
S7_vs_15071||full Low-quality
S7_vs_15074||full Low-quality
S7_vs_15077||full Low-quality
S7_vs_15101||full Not-determined
S7_vs_15212||full Low-quality
S7_vs_15215||full Low-quality
S7_vs_15236||full Not-determined
S7_vs_15273||full Low-quality
S7_vs_15285||full Not-determined
S7_vs_15288||full Not-determined
S7_vs_15332||full Not-determined
S7_vs_15343||full Low-quality
S7_vs_15411||full Not-determined
S7_vs_15473||full Low-quality
S7_vs_15478||full Low-quality
S7_vs_15512||full Low-quality
S7_vs_15522||full Low-quality
S7_vs_15526||full Low-quality
S7_vs_15547||full Low-quality
S7_vs_15549||full Low-quality
S7_vs_15563||full Low-quality
S7_vs_15620||full Low-quality
S7_vs_15675||full Not-determined
S7_vs_15694||full Not-determined
S7_vs_15819||full Low-quality
S7_vs_15838||full Low-quality
S7_vs_15859||full Low-quality
S7_vs_15860||full Low-quality
S7_vs_15878||full Not-determined
S7_vs_15920||full Not-determined
S7_vs_15991||full Low-quality
S7_vs_16038||full Not-determined
S7_vs_16045||full Low-quality
S7_vs_16073||full Low-quality
S7_vs_16081||full Low-quality
S7_vs_16087||full Low-quality
S7_vs_16113||full Low-quality
S7_vs_16130||full Not-determined
S7_vs_16161||full Low-quality
S7_vs_16226||full Low-quality
S7_vs_16246||full Not-determined
S7_vs_16274||full Low-quality
S7_vs_16289||full Not-determined
S7_vs_16295||full Not-determined
S7_vs_16328||full Not-determined
S7_vs_16334||full Not-determined
S7_vs_16337||full Low-quality
S16_extracted_5809 Medium-quality
S7_vs_16363||full Low-quality
S7_vs_16368||full Low-quality
S7_vs_16381||full Not-determined
S7_vs_16409||full Not-determined
S7_vs_16432||full Low-quality
S7_vs_16497||full Low-quality
S7_vs_16529||full Low-quality
S7_vs_16531||full Not-determined
S7_vs_16610||full Low-quality
S7_vs_16635||full Not-determined
S7_vs_16739||full Low-quality
S7_vs_16747||full Not-determined
S7_vs_16758||full Low-quality
S7_vs_16771||full Not-determined
S7_vs_16789||full Low-quality
S7_vs_16814||full Low-quality
S7_vs_16823||full Not-determined
S7_vs_16839||full Low-quality
S7_vs_16855||full Not-determined
S7_vs_16863||full Low-quality
S7_vs_16869||full Low-quality
S7_vs_16884||full Low-quality
S7_vs_16900||full Low-quality
S7_vs_16937||full Low-quality
S7_vs_16971||full Low-quality
S7_vs_17076||full Not-determined
S7_vs_17080||full Low-quality
S7_vs_17092||full Low-quality
S7_vs_17118||full Not-determined
S7_vs_17170||full Low-quality
S7_vs_17181||full Not-determined
S7_vs_17200||full Low-quality
S7_vs_17209||full Low-quality
S7_vs_17231||full Low-quality
S7_vs_17278||full Low-quality
S7_vs_17306||full Low-quality
S7_vs_17363||full Low-quality
S7_vs_17364||full Low-quality
S7_vs_17370||full Low-quality
S7_vs_17371||full Not-determined
S7_vs_17405||full Not-determined
S7_vs_17410||full Low-quality
S7_vs_17422||full Not-determined
S7_vs_17508||full Low-quality
S7_vs_17557||full Low-quality
S7_vs_17570||full Not-determined
S7_vs_17574||full Low-quality
S7_vs_17581||full Low-quality
S7_vs_17595||full Not-determined
S7_vs_17639||full Not-determined
S7_vs_17655||full Not-determined
S7_vs_17675||full Low-quality
S7_vs_17695||full Not-determined
S7_vs_17696||full Not-determined
S7_vs_17706||full Low-quality
S7_vs_17719||full Not-determined
S7_vs_17743||full Not-determined
S7_vs_17755||full Low-quality
S7_vs_17784||full Low-quality
S7_vs_17856||full Low-quality
S7_vs_17874||full Not-determined
S7_vs_17952||full Not-determined
S7_vs_17974||full Not-determined
S7_vs_17978||full Not-determined
S7_vs_18014||full Not-determined
S7_vs_18040||full Low-quality
S7_vs_18043||full Not-determined
S7_vs_18067||full Low-quality
S7_vs_18130||full Low-quality
S7_vs_18137||full Not-determined
S7_vs_18159||full Low-quality
S7_vs_18164||full Low-quality
S7_vs_18177||full Low-quality
S7_vs_18193||full Not-determined
S7_vs_18220||full Low-quality
S7_vs_18225||full Low-quality
S7_vs_18230||full Low-quality
S7_vs_18234||full Not-determined
S7_vs_18240||full Low-quality
S7_vs_18333||full Low-quality
S7_vs_18367||full Not-determined
S7_vs_18402||full Low-quality
S7_vs_18417||full Low-quality
S7_vs_18520||full Low-quality
S7_vs_18525||full Low-quality
S7_vs_18537||full Low-quality
S7_vs_18545||full Not-determined
S7_vs_18562||full Low-quality
S7_vs_18588||full Low-quality
S7_vs_18620||full Not-determined
S7_vs_18644||full Low-quality
S7_vs_18688||full Low-quality
S7_vs_18708||full Low-quality
S7_vs_18709||full Not-determined
S7_vs_18802||full Not-determined
S7_vs_18812||full Low-quality
S7_vs_18815||full Low-quality
S7_vs_18837||full Not-determined
S7_vs_18838||full Low-quality
S7_vs_18847||full Low-quality
S7_vs_18889||full Not-determined
S7_vs_18908||full Not-determined
S7_vs_18924||full Low-quality
S7_vs_18925||full Not-determined
S7_vs_19001||full Not-determined
S7_vs_19007||full Low-quality
S7_vs_19016||full Not-determined
S7_vs_19022||full Low-quality
S7_vs_19058||full Low-quality
S7_vs_19073||full Low-quality
S7_vs_19074||full Low-quality
S7_vs_19075||full Low-quality
S7_vs_19079||full Low-quality
S7_vs_19088||full Not-determined
S7_vs_19100||full Low-quality
S7_vs_19113||full Low-quality
S7_vs_19118||full Not-determined
S7_vs_19130||full Low-quality
S7_vs_19168||full Low-quality
S7_vs_19221||full Low-quality
S7_vs_19229||full Low-quality
S7_vs_19302||full Not-determined
S7_vs_19303||full Not-determined
S7_vs_19316||full Low-quality
S7_vs_19337||full Low-quality
S7_vs_19371||full Not-determined
S7_vs_19376||full Not-determined
S7_vs_19384||full Low-quality
S7_vs_19412||full Low-quality
S7_vs_19427||full Low-quality
S7_vs_19431||full Low-quality
S7_vs_19438||full Low-quality
S7_vs_19466||full Not-determined
S7_vs_19481||full Not-determined
S7_vs_19501||full Low-quality
S7_vs_19511||full Not-determined
S7_vs_19542||full Low-quality
S7_vs_19557||full Not-determined
S7_vs_19569||full Not-determined
S7_vs_19680||full Not-determined
S7_vs_19693||full Low-quality
S7_vs_19742||full Not-determined
S7_vs_19748||full Low-quality
S7_vs_19752||full Not-determined
S7_vs_19765||full Low-quality
S7_vs_19766||full Low-quality
S7_vs_19771||full Low-quality
S7_vs_19777||full Not-determined
S7_vs_19787||full Low-quality
S7_vs_19803||full Low-quality
S7_vs_19805||full Not-determined
S7_vs_19814||full Low-quality
S7_vs_19838||full Not-determined
S16_vs_5809||full Medium-quality
S7_vs_19896||full Low-quality
S7_vs_20038||full Low-quality
S7_vs_20046||full Not-determined
S7_vs_20067||full Not-determined
S13_vs_98591||full Medium-quality
S7_vs_20148||full Low-quality
S9_vs_k141_90871||full High-quality
S7_vs_20217||full Not-determined
S7_vs_20240||full Not-determined
S7_vs_20253||full Not-determined
S7_vs_20286||full Not-determined
S7_vs_20311||full Low-quality
S7_vs_20401||full Low-quality
S7_vs_20411||full Low-quality
S7_vs_20413||full Low-quality
S7_vs_20420||full Low-quality
S7_vs_20427||full Low-quality
S7_vs_20432||full Low-quality
S7_vs_20455||full Low-quality
S7_vs_20511||full Not-determined
S7_vs_20516||full Low-quality
S7_vs_20532||full Not-determined
S7_vs_20680||full Low-quality
S7_vs_20717||full Low-quality
S7_vs_20724||full Low-quality
S7_vs_20737||full Not-determined
S7_vs_20744||full Low-quality
S7_vs_20781||full Not-determined
S7_vs_20794||full Low-quality
S7_vs_20847||full Not-determined
S7_vs_20850||full Not-determined
S7_vs_20858||full Not-determined
S7_vs_20867||full Low-quality
S7_vs_20871||full Low-quality
S7_vs_20898||full Low-quality
S7_vs_20931||full Low-quality
S7_vs_21019||full Low-quality
S7_vs_21056||full Not-determined
S7_vs_21116||full Low-quality
S7_vs_21118||full Low-quality
S7_vs_21155||full Low-quality
S7_vs_21163||full Low-quality
S7_vs_21167||full Low-quality
S7_vs_21206||full Low-quality
S7_vs_21208||full Low-quality
S7_vs_21264||full Low-quality
S7_vs_21301||full Low-quality
S7_vs_21321||full Low-quality
S7_vs_21353||full Low-quality
S7_vs_21358||full Low-quality
S7_vs_21364||full Low-quality
S7_vs_21425||full Not-determined
S7_vs_21450||full Not-determined
S7_vs_21479||full Not-determined
S7_vs_21498||full Low-quality
S7_vs_21516||full Not-determined
S14_vs_k141_117162||full High-quality
S7_vs_21555||full Not-determined
S7_vs_21596||full Low-quality
S7_vs_21613||full Not-determined
S7_vs_21618||full Not-determined
S7_vs_21638||full Low-quality
S7_vs_21666||full Not-determined
S7_vs_21686||full Low-quality
S7_vs_21723||full Low-quality
S7_vs_21781||full Low-quality
S7_vs_21784||full Low-quality
S7_vs_21815||full Low-quality
S7_vs_21831||full Low-quality
S7_vs_21835||full Low-quality
S7_vs_21872||full Low-quality
S7_vs_21901||full Not-determined
S7_vs_21902||full Not-determined
S7_vs_21912||full Low-quality
S7_vs_21915||full Low-quality
S7_vs_21920||full Not-determined
S7_vs_21932||full Low-quality
S7_vs_21938||full Low-quality
S14_vs_k141_88666||full High-quality
S7_vs_21985||full Not-determined
S7_vs_21990||full Low-quality
S7_vs_21998||full Low-quality
S7_vs_22000||full Low-quality
S7_vs_22026||full Low-quality
S7_vs_22037||full Not-determined
S7_vs_22121||full Low-quality
S7_vs_22151||full Not-determined
S7_vs_22168||full Low-quality
S7_vs_22172||full Not-determined
S7_vs_22176||full Low-quality
S7_vs_22199||full Low-quality
S7_vs_22243||full Not-determined
S7_vs_22271||full Low-quality
S7_vs_22277||full Not-determined
S7_vs_22315||full Not-determined
S7_vs_22325||full Not-determined
S7_vs_22335||full Low-quality
S7_vs_22337||full Not-determined
S7_vs_22348||full Not-determined
S7_vs_22383||full Not-determined
S7_vs_22404||full Low-quality
S7_vs_22441||full Low-quality
S7_vs_22443||full Not-determined
S7_vs_22454||full Low-quality
S7_vs_22469||full Low-quality
S7_vs_22503||full Not-determined
S7_vs_22506||full Low-quality
S7_vs_22558||full Low-quality
S7_vs_22591||full Low-quality
S7_vs_22631||full Low-quality
S7_vs_22656||full Low-quality
S7_vs_22679||full Low-quality
S7_vs_22691||full Low-quality
S7_vs_22759||full Not-determined
S7_vs_22768||full Not-determined
S7_vs_22791||full Not-determined
S7_vs_22854||full Not-determined
S7_vs_22864||full Not-determined
S7_vs_22954||full Not-determined
S7_vs_23011||full Not-determined
S7_vs_23021||full Low-quality
S7_vs_23029||full Low-quality
S7_vs_23033||full Not-determined
S7_vs_23066||full Not-determined
S7_vs_23123||full Low-quality
S7_vs_23198||full Low-quality
S7_vs_23210||full Low-quality
S7_vs_23211||full Low-quality
S7_vs_23238||full Not-determined
S7_vs_23282||full Low-quality
S7_vs_23291||full Not-determined
S7_vs_23316||full Low-quality
S7_vs_23336||full Low-quality
S7_vs_23346||full Not-determined
S7_vs_23348||full Low-quality
S7_vs_23364||full Low-quality
S7_vs_23390||full Low-quality
S7_vs_23402||full Low-quality
S7_vs_23436||full Low-quality
S7_vs_23473||full Low-quality
S7_vs_23501||full Low-quality
S7_vs_23503||full Not-determined
S7_vs_23505||full Low-quality
S7_vs_23557||full Low-quality
S7_vs_23564||full Low-quality
S7_vs_23565||full Low-quality
S7_vs_23603||full Not-determined
S7_vs_23653||full Low-quality
S7_vs_23747||full Not-determined
S7_vs_23750||full Low-quality
S7_vs_23759||full Low-quality
S5_vs_k141_122344||full High-quality
S7_vs_23781||full Not-determined
S7_vs_23790||full Low-quality
S7_vs_23799||full Not-determined
S7_vs_23873||full Low-quality
S7_vs_23900||full Low-quality
S7_vs_23903||full Low-quality
S7_vs_23922||full Not-determined
S7_vs_23942||full Low-quality
S7_vs_23953||full Low-quality
S7_vs_23971||full Low-quality
S7_vs_23981||full Not-determined
S7_vs_24026||full Low-quality
S7_vs_24030||full Low-quality
S7_vs_24070||full Not-determined
S7_vs_24087||full Low-quality
S7_vs_24186||full Low-quality
S7_vs_24188||full Low-quality
S7_vs_24190||full Low-quality
S7_vs_24199||full Not-determined
S7_vs_24284||full Low-quality

| | |
|---|---|
| S7_vs_24295\|\|full | Low-quality |
| S7_vs_24320\|\|full | Not-determined |
| S7_vs_24344\|\|full | Low-quality |
| S7_vs_24352\|\|full | Not-determined |
| S7_vs_24438\|\|full | Low-quality |
| S7_vs_24465\|\|full | Not-determined |
| S7_vs_24481\|\|full | Low-quality |
| S7_vs_24505\|\|full | Low-quality |
| S7_vs_24529\|\|full | Low-quality |
| S7_vs_24532\|\|full | Low-quality |
| S7_vs_24542\|\|full | Low-quality |
| S7_vs_24546\|\|full | Not-determined |
| S7_vs_24550\|\|full | Not-determined |
| S7_vs_24566\|\|full | Low-quality |
| S7_vs_24607\|\|full | Low-quality |
| S7_vs_24632\|\|full | Low-quality |
| S7_vs_24685\|\|full | Low-quality |
| S7_vs_24695\|\|full | Low-quality |
| S7_vs_24714\|\|full | Not-determined |
| S7_vs_24724\|\|full | Not-determined |
| S7_vs_24759\|\|full | Low-quality |
| S7_vs_24805\|\|full | Not-determined |
| S7_vs_24806\|\|full | Low-quality |
| S7_vs_24816\|\|full | Low-quality |
| S7_vs_24843\|\|full | Not-determined |
| S7_vs_24851\|\|full | Low-quality |
| S7_vs_24867\|\|full | Low-quality |
| S7_vs_24868\|\|full | Not-determined |
| S7_vs_24874\|\|full | Low-quality |
| S7_vs_24919\|\|full | Low-quality |
| S7_vs_24923\|\|full | Low-quality |
| S7_vs_24942\|\|full | Not-determined |
| S7_vs_25008\|\|full | Low-quality |
| S7_vs_25035\|\|full | Not-determined |
| S7_vs_25055\|\|full | Not-determined |
| S7_vs_25056\|\|full | Low-quality |
| S7_vs_25078\|\|full | Not-determined |
| S7_vs_25166\|\|full | Not-determined |
| S7_vs_25169\|\|full | Low-quality |
| S7_vs_25200\|\|full | Low-quality |
| S7_vs_25215\|\|full | Not-determined |
| S7_vs_25223\|\|full | Low-quality |
| S7_vs_25228\|\|full | Not-determined |
| S7_vs_25269\|\|full | Not-determined |
| S7_vs_25281\|\|full | Not-determined |
| S7_vs_25293\|\|full | Low-quality |
| S7_vs_25296\|\|full | Low-quality |
| S7_vs_25330\|\|full | Low-quality |
| S7_vs_25343\|\|full | Low-quality |
| S7_vs_25362\|\|full | Low-quality |
| S7_vs_25387\|\|full | Low-quality |
| S7_vs_25405\|\|full | Not-determined |
| S7_vs_25426\|\|full | Not-determined |
| S7_vs_25428\|\|full | Not-determined |
| S7_vs_25466\|\|full | Not-determined |
| S7_vs_25468\|\|full | Not-determined |
| S7_vs_25469\|\|full | Low-quality |
| S7_vs_25471\|\|full | Not-determined |
| S7_vs_25552\|\|full | Low-quality |
| S7_vs_25565\|\|full | Not-determined |
| S7_vs_25616\|\|full | Low-quality |
| S7_vs_25618\|\|full | Not-determined |
| S7_vs_25620\|\|full | Low-quality |
| S7_vs_25656\|\|full | Low-quality |
| S7_vs_25695\|\|full | Low-quality |
| S7_vs_25746\|\|full | Not-determined |
| S7_vs_25757\|\|full | Not-determined |
| S7_vs_25759\|\|full | Not-determined |
| S7_vs_25763\|\|full | Low-quality |
| S7_vs_25773\|\|full | Low-quality |
| S7_vs_25811\|\|full | Low-quality |
| S7_vs_25892\|\|full | Not-determined |
| S7_vs_25897\|\|full | Low-quality |
| S7_vs_25933\|\|full | Not-determined |
| S7_vs_25941\|\|full | Low-quality |
| S7_vs_25952\|\|full | Low-quality |
| S7_vs_25991\|\|full | Not-determined |
| S7_vs_26007\|\|full | Not-determined |
| S7_vs_26041\|\|full | Low-quality |
| S7_vs_26047\|\|full | Low-quality |
| S7_vs_26048\|\|full | Not-determined |
| S7_vs_26075\|\|full | Low-quality |
| S7_vs_26113\|\|full | Low-quality |
| S7_vs_26116\|\|full | Low-quality |
| S7_vs_26144\|\|full | Low-quality |
| S7_vs_26222\|\|full | Low-quality |
| S7_vs_55031\|\|full | Medium-quality |
| S7_vs_26253\|\|full | Low-quality |
| S7_vs_26320\|\|full | Low-quality |
| S7_vs_26330\|\|full | Not-determined |
| S7_vs_26400\|\|full | Low-quality |
| S7_vs_26487\|\|full | Not-determined |
| S7_vs_26517\|\|full | Low-quality |
| S7_vs_26569\|\|full | Low-quality |
| S7_vs_26571\|\|full | Not-determined |
| S7_vs_26608\|\|full | Not-determined |
| S7_vs_26647\|\|full | Low-quality |
| S7_vs_26672\|\|full | Low-quality |
| S7_vs_26697\|\|full | Low-quality |
| S7_vs_26714\|\|full | Not-determined |
| S7_vs_26724\|\|full | Not-determined |
| S7_vs_26730\|\|full | Not-determined |
| S7_vs_26748\|\|full | Low-quality |
| S5_vs_k141_109085\|\|full | High-quality |
| S7_vs_26799\|\|full | Low-quality |
| S7_vs_26813\|\|full | Low-quality |
| S7_vs_26817\|\|full | Low-quality |
| S7_vs_26842\|\|full | Not-determined |
| S7_vs_26863\|\|full | Low-quality |
| S7_vs_26865\|\|full | Low-quality |
| S7_vs_26877\|\|full | Low-quality |
| S7_vs_27047\|\|full | Low-quality |
| S7_vs_27132\|\|full | Not-determined |
| S7_vs_27142\|\|full | Low-quality |
| S7_vs_27154\|\|full | Low-quality |
| S7_vs_27222\|\|full | Low-quality |
| S7_vs_27262\|\|full | Low-quality |
| S7_vs_27278\|\|full | Low-quality |
| S7_vs_27285\|\|full | Low-quality |
| S7_vs_27292\|\|full | Not-determined |
| S7_vs_27343\|\|full | Low-quality |
| S7_vs_27355\|\|full | Not-determined |
| S7_vs_27361\|\|full | Not-determined |
| S14_extracted_k141_58919 | High-quality |
| S7_vs_27458\|\|full | Not-determined |
| S7_vs_27471\|\|full | Not-determined |
| S7_vs_27495\|\|full | Low-quality |
| S7_vs_27501\|\|full | Low-quality |
| S7_vs_27544\|\|full | Low-quality |
| S7_vs_27580\|\|full | Low-quality |
| S7_vs_27590\|\|full | Not-determined |
| S7_vs_27598\|\|full | Not-determined |
| S7_vs_27661\|\|full | Low-quality |
| S7_vs_27675\|\|full | Not-determined |
| S7_vs_27677\|\|full | Not-determined |
| S7_vs_27741\|\|full | Low-quality |
| S7_vs_27795\|\|full | Low-quality |
| S7_vs_27801\|\|full | Low-quality |
| S7_vs_27821\|\|full | Low-quality |
| S7_vs_27882\|\|full | Not-determined |
| S7_vs_27892\|\|full | Not-determined |
| S7_vs_27895\|\|full | Low-quality |
| S7_vs_27917\|\|full | Low-quality |
| S7_vs_27927\|\|full | Not-determined |
| S7_vs_27932\|\|full | Not-determined |
| S7_vs_27933\|\|full | Low-quality |
| S7_vs_27965\|\|full | Not-determined |
| S7_vs_28000\|\|full | Low-quality |
| S7_vs_28007\|\|full | Low-quality |
| S7_vs_28009\|\|full | Not-determined |
| S7_vs_28072\|\|full | Not-determined |
| S7_vs_28099\|\|full | Low-quality |
| S7_vs_28107\|\|full | Low-quality |
| S7_vs_28137\|\|full | Not-determined |
| S14_vs_k141_59347\|\|full | High-quality |
| S7_vs_28158\|\|full | Low-quality |
| S7_vs_28189\|\|full | Low-quality |
| S7_vs_28277\|\|full | Not-determined |
| S7_vs_28285\|\|full | Not-determined |
| S7_vs_28305\|\|full | Low-quality |
| S7_vs_28338\|\|full | Low-quality |
| S7_vs_28372\|\|full | Not-determined |
| S7_vs_28377\|\|full | Low-quality |
| S7_vs_28458\|\|full | Not-determined |
| S7_vs_28463\|\|full | Not-determined |
| S7_vs_28478\|\|full | Low-quality |
| S7_vs_28536\|\|full | Not-determined |
| S7_vs_28541\|\|full | Low-quality |
| S7_vs_28543\|\|full | Low-quality |
| S7_vs_28563\|\|full | Low-quality |
| S7_vs_28647\|\|full | Low-quality |
| S7_vs_28648\|\|full | Not-determined |
| S7_vs_28654\|\|full | Low-quality |
| S7_vs_28657\|\|full | Not-determined |
| S7_vs_28666\|\|full | Low-quality |
| S8_extracted_k141_45776 | High-quality |
| S7_vs_28818\|\|full | Low-quality |
| S7_vs_28907\|\|full | Low-quality |
| S7_vs_28911\|\|full | Low-quality |
| S7_vs_28955\|\|full | Low-quality |
| S7_vs_28972\|\|full | Low-quality |
| S7_vs_28973\|\|full | Not-determined |
| S7_vs_28986\|\|full | Low-quality |
| S7_vs_28996\|\|full | Not-determined |
| S7_vs_29001\|\|full | Not-determined |
| S7_vs_29067\|\|full | Low-quality |
| S7_vs_29074\|\|full | Low-quality |
| S7_vs_29189\|\|full | Low-quality |
| S8_vs_k141_45776\|\|full | High-quality |
| S7_vs_29197\|\|full | Not-determined |
| S7_vs_29311\|\|full | Low-quality |
| S7_vs_29336\|\|full | Low-quality |
| S7_vs_29349\|\|full | Low-quality |
| S7_vs_29378\|\|full | Not-determined |
| S7_vs_29413\|\|full | Not-determined |
| S7_vs_29425\|\|full | Low-quality |
| S7_vs_29430\|\|full | Low-quality |
| S7_vs_29476\|\|full | Low-quality |
| S7_vs_29511\|\|full | Low-quality |
| S7_vs_29541\|\|full | Low-quality |
| S7_vs_29543\|\|full | Low-quality |
| S7_vs_29590\|\|full | Low-quality |
| S4_vs_VIRSorter_1135-cat_2 | Medium-quality |
| S7_vs_29598\|\|full | Low-quality |
| S7_vs_29602\|\|full | Low-quality |
| S7_vs_29610\|\|full | Not-determined |
| S7_vs_29623\|\|full | Low-quality |
| S7_vs_29632\|\|full | Low-quality |
| S7_vs_29646\|\|full | Low-quality |
| S7_vs_29662\|\|full | Low-quality |
| S7_vs_29673\|\|full | Low-quality |
| S7_vs_29684\|\|full | Low-quality |
| S7_vs_29707\|\|full | Not-determined |
| S7_vs_29718\|\|full | Not-determined |
| S7_vs_29759\|\|full | Low-quality |
| S7_vs_29783\|\|full | Low-quality |
| S7_vs_29844\|\|full | Not-determined |
| S7_vs_29871\|\|full | Low-quality |
| S7_vs_29891\|\|full | Low-quality |
| S7_vs_29892\|\|full | Low-quality |
| S7_vs_29912\|\|full | Not-determined |
| S7_vs_29928\|\|full | Low-quality |
| S8_vs_k141_193843\|\|full | High-quality |
| S7_vs_29972\|\|full | Low-quality |
| S7_vs_30021\|\|full | Not-determined |
| S7_vs_30054\|\|full | Low-quality |
| S7_vs_30074\|\|full | Not-determined |
| S7_vs_30140\|\|full | Low-quality |
| S7_vs_30149\|\|full | Low-quality |
| S7_vs_30156\|\|full | Not-determined |
| S7_vs_30183\|\|full | Low-quality |
| S7_vs_30328\|\|full | Low-quality |
| S7_vs_30348\|\|full | Low-quality |
| S7_vs_30360\|\|full | Low-quality |
| S7_vs_30371\|\|full | Low-quality |
| S7_vs_30426\|\|full | Low-quality |
| S7_vs_30429\|\|full | Low-quality |
| S7_vs_30435\|\|full | Low-quality |
| S7_vs_30448\|\|full | Low-quality |
| S7_vs_30474\|\|full | Not-determined |
| S7_vs_30500\|\|full | Low-quality |
| S7_vs_30505\|\|full | Not-determined |
| S7_vs_30532\|\|full | Low-quality |
| S7_vs_30548\|\|full | Not-determined |
| S7_vs_30559\|\|full | Low-quality |
| S7_vs_30591\|\|full | Not-determined |
| S7_vs_30605\|\|full | Low-quality |
| S17_extracted_k141_177200 | Complete |
| S7_vs_30663\|\|full | Low-quality |
| S16_extracted_172952 | Complete |
| S7_vs_30678\|\|full | Low-quality |
| S7_vs_30726\|\|full | Low-quality |
| S7_vs_30771\|\|full | Not-determined |
| S7_vs_30773\|\|full | Not-determined |
| S7_vs_30789\|\|full | Low-quality |
| S7_vs_30827\|\|full | Low-quality |
| S7_vs_30961\|\|full | Low-quality |
| S7_vs_31035\|\|full | Low-quality |
| S7_vs_31046\|\|full | Low-quality |
| S7_vs_31102\|\|full | Low-quality |
| S7_vs_31113\|\|full | Low-quality |
| S7_vs_31121\|\|full | Low-quality |
| S7_vs_31151\|\|full | Not-determined |
| S7_vs_31163\|\|full | Not-determined |
| S7_vs_31203\|\|full | Not-determined |
| S7_vs_31206\|\|full | Low-quality |
| S7_vs_31262\|\|full | Low-quality |
| S7_vs_31271\|\|full | Not-determined |
| S7_vs_31280\|\|full | Low-quality |
| S7_vs_31288\|\|full | Low-quality |
| S7_vs_31304\|\|full | Low-quality |
| S7_vs_31309\|\|full | Low-quality |
| S7_vs_31341\|\|full | Not-determined |
| S7_vs_31350\|\|full | Low-quality |
| S7_vs_31431\|\|full | Not-determined |
| S7_vs_31466\|\|full | Low-quality |
| S7_vs_31473\|\|full | Not-determined |
| S7_vs_31480\|\|full | Low-quality |
| S7_vs_31488\|\|full | Not-determined |
| S7_vs_31535\|\|full | Not-determined |
| S7_vs_31549\|\|full | Low-quality |
| S7_vs_31552\|\|full | Low-quality |
| S7_vs_31553\|\|full | Low-quality |
| S7_vs_31554\|\|full | Low-quality |
| S7_vs_31557\|\|full | Low-quality |
| S7_vs_31558\|\|full | Low-quality |
| S7_vs_31594\|\|full | Low-quality |
| S7_vs_31613\|\|full | Not-determined |
| S7_vs_39724\|\|full | High-quality |
| S7_vs_31669\|\|full | Low-quality |
| S7_vs_31689\|\|full | Low-quality |
| S7_vs_31711\|\|full | Low-quality |
| S7_vs_31714\|\|full | Not-determined |
| S7_vs_31721\|\|full | Not-determined |
| S7_vs_31730\|\|full | Low-quality |
| S7_vs_31762\|\|full | Low-quality |
| S7_vs_31763\|\|full | Not-determined |
| S7_vs_31781\|\|full | Not-determined |
| S7_vs_31786\|\|full | Low-quality |
| S7_vs_31790\|\|full | Low-quality |
| S7_vs_31807\|\|full | Not-determined |
| S7_vs_31832\|\|full | Low-quality |
| S7_vs_31843\|\|full | Not-determined |
| S7_vs_31848\|\|full | Not-determined |
| S7_vs_31857\|\|full | Not-determined |
| S7_vs_31862\|\|full | Low-quality |
| S7_vs_31868\|\|full | Low-quality |
| S7_vs_31876\|\|full | Not-determined |
| S7_vs_31890\|\|full | Not-determined |
| S7_vs_31991\|\|full | Low-quality |
| S7_vs_32018\|\|full | Low-quality |
| S7_vs_32030\|\|full | Low-quality |
| S7_vs_32043\|\|full | Not-determined |
| S7_vs_32050\|\|full | Low-quality |
| S7_vs_32068\|\|full | Low-quality |
| S7_vs_32096\|\|full | Not-determined |
| S7_vs_32124\|\|full | Low-quality |
| S7_vs_32127\|\|full | Low-quality |
| S7_vs_32138\|\|full | Low-quality |
| S7_vs_32161\|\|full | Low-quality |
| S7_vs_32184\|\|full | Not-determined |
| S7_vs_32186\|\|full | Low-quality |
| S7_vs_32199\|\|full | Not-determined |
| S7_vs_32211\|\|full | Low-quality |
| S7_vs_32256\|\|full | Not-determined |
| S7_vs_32294\|\|full | Low-quality |
| S7_vs_32308\|\|full | Not-determined |
| S7_vs_32335\|\|full | Low-quality |
| S7_vs_60775\|\|full | High-quality |
| S7_vs_32397\|\|full | Not-determined |
| S7_vs_32413\|\|full | Low-quality |
| S7_vs_32452\|\|full | Low-quality |
| S7_vs_32454\|\|full | Low-quality |
| S4_vs_VIRSorter_6812-circular-cat_ | High-quality |
| S7_vs_32610\|\|full | Low-quality |
| S7_vs_32626\|\|full | Low-quality |
| S7_vs_32655\|\|full | Low-quality |
| S7_vs_32718\|\|full | Low-quality |
| S7_vs_32732\|\|full | Low-quality |
| S7_vs_32817\|\|full | Low-quality |
| S7_vs_32822\|\|full | Not-determined |
| S7_vs_32840\|\|full | Not-determined |
| S7_vs_32964\|\|full | Not-determined |
| S7_vs_32973\|\|full | Not-determined |
| S7_vs_32990\|\|full | Low-quality |
| S7_vs_33038\|\|full | Low-quality |
| S7_vs_33040\|\|full | Low-quality |
| S7_vs_33055\|\|full | Low-quality |
| S7_vs_33067\|\|full | Not-determined |
| S7_vs_33070\|\|full | Not-determined |
| S7_vs_33073\|\|full | Low-quality |
| S7_vs_33076\|\|full | Not-determined |
| S7_vs_33085\|\|full | Not-determined |
| S7_vs_33091\|\|full | Not-determined |
| S7_vs_33137\|\|full | Low-quality |
| S7_vs_33150\|\|full | Low-quality |
| S7_vs_33169\|\|full | Not-determined |
| S7_vs_33180\|\|full | Not-determined |
| S7_vs_33244\|\|full | Low-quality |
| S7_vs_33250\|\|full | Low-quality |
| S7_vs_33312\|\|full | Low-quality |
| S7_vs_33339\|\|full | Low-quality |
| S7_vs_33361\|\|full | Low-quality |
| S7_vs_33363\|\|full | Low-quality |
| S7_vs_33414\|\|full | Not-determined |
| S7_vs_33431\|\|full | Low-quality |
| S7_vs_33462\|\|full | Low-quality |
| S7_vs_33478\|\|full | Low-quality |
| S7_vs_33514\|\|full | Low-quality |
| S7_vs_33516\|\|full | Not-determined |
| S7_vs_33522\|\|full | Low-quality |
| S7_vs_33559\|\|full | Low-quality |
| S7_vs_33566\|\|full | Low-quality |
| S7_vs_33600\|\|full | Low-quality |
| S7_vs_33603\|\|full | Low-quality |
| S7_vs_33730\|\|full | Not-determined |
| S7_vs_33738\|\|full | Not-determined |
| S7_vs_33819\|\|full | Low-quality |
| S7_vs_33840\|\|full | Low-quality |
| S7_vs_33904\|\|full | Not-determined |
| S7_vs_33916\|\|full | Low-quality |
| S7_vs_33931\|\|full | Low-quality |
| S7_vs_33966\|\|full | Low-quality |
| S7_vs_33971\|\|full | Low-quality |
| S7_vs_34032\|\|full | Low-quality |
| S7_vs_34056\|\|full | Low-quality |
| S7_vs_34069\|\|full | Low-quality |
| S7_vs_34099\|\|full | Not-determined |
| S7_vs_34124\|\|full | Not-determined |
| S7_vs_34130\|\|full | Not-determined |
| S7_vs_34149\|\|full | Low-quality |
| S7_vs_34159\|\|full | Low-quality |
| S7_vs_34176\|\|full | Low-quality |
| S7_vs_34197\|\|full | Low-quality |
| S7_vs_34226\|\|full | Not-determined |
| S7_vs_34304\|\|full | Low-quality |
| S7_vs_34305\|\|full | Low-quality |
| S7_vs_34339\|\|full | Low-quality |
| S7_vs_34351\|\|full | Low-quality |
| S7_vs_34357\|\|full | Low-quality |
| S7_vs_34405\|\|full | Low-quality |
| S7_vs_34413\|\|full | Not-determined |
| S7_vs_34465\|\|full | Low-quality |
| S7_vs_34481\|\|full | Low-quality |
| S7_vs_34485\|\|full | Low-quality |
| S7_vs_34490\|\|full | Low-quality |
| S7_vs_34495\|\|full | Not-determined |
| S7_vs_34529\|\|full | Low-quality |
| S7_vs_34542\|\|full | Not-determined |
| S7_vs_34561\|\|full | Low-quality |
| S7_vs_34596\|\|full | Low-quality |
| S7_vs_34597\|\|full | Low-quality |
| S7_vs_34665\|\|full | Low-quality |
| S7_vs_34729\|\|full | Low-quality |
| S7_vs_34735\|\|full | Low-quality |
| S7_vs_34787\|\|full | Not-determined |
| S7_vs_34835\|\|full | Low-quality |
| S7_vs_34848\|\|full | Low-quality |
| S7_vs_34895\|\|full | Low-quality |
| S7_vs_34904\|\|full | Low-quality |
| S7_vs_34907\|\|full | Low-quality |
| S7_vs_34911\|\|full | Low-quality |
| S7_vs_34920\|\|full | Low-quality |
| S7_vs_34948\|\|full | Low-quality |
| S7_vs_34954\|\|full | Low-quality |
| S7_vs_34966\|\|full | Not-determined |
| S7_vs_34983\|\|full | Not-determined |
| S7_vs_34997\|\|full | Not-determined |
| S7_vs_35002\|\|full | Low-quality |
| S7_vs_35020\|\|full | Not-determined |
| S7_vs_35051\|\|full | Low-quality |
| S7_vs_35053\|\|full | Low-quality |
| S7_vs_35091\|\|full | Not-determined |
| S7_vs_35094\|\|full | Low-quality |
| S7_vs_35121\|\|full | Not-determined |
| S7_vs_35137\|\|full | Low-quality |
| S7_vs_35156\|\|full | Low-quality |
| S7_vs_35183\|\|full | Low-quality |
| S7_vs_35189\|\|full | Low-quality |
| S7_vs_35244\|\|full | Low-quality |
| S7_vs_35263\|\|full | Not-determined |
| S7_vs_35283\|\|full | Low-quality |
| S7_vs_35295\|\|full | Not-determined |
| S7_vs_35300\|\|full | Low-quality |
| S7_vs_35351\|\|full | Not-determined |
| S7_vs_35393\|\|full | Not-determined |
| S7_vs_35420\|\|full | Not-determined |
| S7_vs_35425\|\|full | Low-quality |
| S7_vs_35453\|\|full | Not-determined |
| S7_vs_35513\|\|full | Not-determined |
| S7_vs_35521\|\|full | Not-determined |
| S7_vs_35569\|\|full | Not-determined |
| S7_vs_35582\|\|full | Not-determined |
| S7_vs_35607\|\|full | Low-quality |
| S7_vs_35639\|\|full | Low-quality |
| S7_vs_35676\|\|full | Low-quality |
| S7_vs_35682\|\|full | Low-quality |
| S7_vs_35700\|\|full | Low-quality |
| S7_vs_35714\|\|full | Low-quality |
| S7_vs_35718\|\|full | Not-determined |
| S7_vs_35740\|\|full | Not-determined |
| S7_vs_35777\|\|full | Not-determined |
| S7_vs_35788\|\|full | Low-quality |
| S7_vs_35798\|\|full | Low-quality |
| S7_vs_35883\|\|full | Low-quality |
| S7_vs_35888\|\|full | Not-determined |
| S7_vs_35941\|\|full | Low-quality |
| S7_vs_35988\|\|full | Low-quality |
| S7_vs_36014\|\|full | Not-determined |
| S7_vs_36032\|\|full | Low-quality |
| S7_vs_36054\|\|full | Low-quality |
| S7_vs_36084\|\|full | Low-quality |
| S7_vs_36109\|\|full | Low-quality |
| S7_vs_36164\|\|full | Not-determined |
| S7_vs_36185\|\|full | Low-quality |
| S7_vs_36199\|\|full | Low-quality |
| S7_vs_36226\|\|full | Low-quality |
| S7_vs_36235\|\|full | Not-determined |
| S7_vs_36237\|\|full | Low-quality |
| S7_vs_36257\|\|full | Low-quality |
| S7_vs_36296\|\|full | Low-quality |
| S7_vs_36358\|\|full | Not-determined |
| S7_vs_36415\|\|full | Low-quality |
| S7_vs_36434\|\|full | Low-quality |
| S7_vs_36435\|\|full | Not-determined |
| S7_vs_36451\|\|full | Not-determined |
| S7_vs_36455\|\|full | Low-quality |
| S7_vs_36503\|\|full | Low-quality |
| S7_vs_36555\|\|full | Low-quality |
| S7_vs_36556\|\|full | Low-quality |
| S7_vs_36582\|\|full | Low-quality |
| S7_vs_36592\|\|full | Low-quality |
| S7_vs_36596\|\|full | Low-quality |
| S7_vs_36610\|\|full | Not-determined |
| S16_vs_33281\|\|full | Medium-quality |
| S7_vs_36630\|\|full | Low-quality |
| S7_vs_36665\|\|full | Low-quality |
| S7_vs_36705\|\|full | Low-quality |
| S7_vs_36722\|\|full | Low-quality |
| S7_vs_36751\|\|full | Not-determined |
| S7_vs_36773\|\|full | Low-quality |
| S7_vs_36776\|\|full | Low-quality |
| S7_vs_36783\|\|full | Low-quality |
| S7_vs_36802\|\|full | Low-quality |
| S7_vs_36820\|\|full | Not-determined |
| S7_vs_36821\|\|full | Low-quality |
| S7_vs_36876\|\|full | Low-quality |
| S7_vs_36901\|\|full | Low-quality |
| S7_vs_36922\|\|full | Not-determined |
| S7_vs_36964\|\|full | Low-quality |
| S7_vs_37009\|\|full | Low-quality |
| S7_vs_37111\|\|full | Low-quality |
| S7_vs_37137\|\|full | Low-quality |
| S7_vs_37152\|\|full | Low-quality |
| S7_vs_37168\|\|full | Low-quality |
| S7_vs_37190\|\|full | Low-quality |
| S7_vs_37201\|\|full | Low-quality |
| S7_vs_37202\|\|full | Not-determined |
| S7_vs_37218\|\|full | Low-quality |
| S7_vs_37233\|\|full | Not-determined |
| S4_vs_VIRSorter_4959-cat_2 | Medium-quality |
| S7_vs_37317\|\|full | Not-determined |
| S7_vs_37373\|\|full | Low-quality |
| S7_vs_37434\|\|full | Low-quality |
| S7_vs_37438\|\|full | Low-quality |
| S7_vs_37453\|\|full | Low-quality |
| S7_vs_37471\|\|full | Low-quality |
| S7_vs_37489\|\|full | Not-determined |
| S7_vs_37492\|\|full | Not-determined |
| S7_vs_37556\|\|full | Low-quality |
| S7_vs_37570\|\|full | Not-determined |
| S7_vs_37616\|\|full | Not-determined |
| S7_vs_37697\|\|full | Low-quality |
| S7_vs_37728\|\|full | Not-determined |
| S7_vs_37769\|\|full | Low-quality |
| S7_vs_37776\|\|full | Not-determined |
| S7_vs_37801\|\|full | Not-determined |
| S7_vs_37811\|\|full | Low-quality |
| S7_vs_37827\|\|full | Low-quality |
| S7_vs_37851\|\|full | Low-quality |
| S7_vs_37855\|\|full | Low-quality |
| S7_vs_37877\|\|full | Low-quality |
| S7_vs_37889\|\|full | Low-quality |
| S7_vs_37962\|\|full | Not-determined |
| S7_vs_37964\|\|full | Low-quality |
| S7_vs_38028\|\|full | Low-quality |
| S7_vs_38047\|\|full | Low-quality |
| S7_vs_38106\|\|full | Low-quality |
| S11_extracted_k141_104071 | Complete |
| S7_vs_38123\|\|full | Not-determined |
| S7_vs_38164\|\|full | Low-quality |
| S7_vs_38224\|\|full | Not-determined |
| S7_vs_38254\|\|full | Not-determined |
| S7_vs_38295\|\|full | Low-quality |
| S7_vs_38300\|\|full | Low-quality |
| S7_vs_38308\|\|full | Not-determined |
| S7_vs_38322\|\|full | Not-determined |
| S7_vs_38339\|\|full | Low-quality |
| S7_vs_38386\|\|full | Not-determined |
| S7_vs_38408\|\|full | Not-determined |
| S7_vs_38423\|\|full | Low-quality |
| S7_vs_38437\|\|full | Not-determined |
| S7_vs_38446\|\|full | Low-quality |
| S7_vs_38451\|\|full | Low-quality |
| S7_vs_38476\|\|full | Low-quality |
| S7_vs_38497\|\|full | Not-determined |
| S7_vs_38500\|\|full | Not-determined |
| S7_vs_38565\|\|full | Low-quality |
| S7_vs_38579\|\|full | Low-quality |
| S7_vs_38598\|\|full | Not-determined |
| S7_vs_38603\|\|full | Not-determined |
| S7_vs_38605\|\|full | Not-determined |
| S7_vs_38653\|\|full | Low-quality |
| S7_vs_38675\|\|full | Not-determined |
| S7_vs_38676\|\|full | Not-determined |
| S7_vs_38680\|\|full | Low-quality |
| S7_vs_38683\|\|full | Low-quality |
| S7_vs_38720\|\|full | Not-determined |
| S7_vs_38725\|\|full | Low-quality |
| S7_vs_38728\|\|full | Low-quality |
| S7_vs_38810\|\|full | Not-determined |
| S7_vs_38833\|\|full | Low-quality |
| S9_vs_k141_40761\|\|full | High-quality |
| S7_vs_38838\|\|full | Not-determined |
| S7_vs_38843\|\|full | Low-quality |
| S7_vs_38880\|\|full | Not-determined |
| S7_vs_38915\|\|full | Not-determined |
| S7_vs_38926\|\|full | Low-quality |
| S7_vs_38956\|\|full | Low-quality |

| | |
|---|---|
| S7_vs_38985\|\|full | Not-determined |
| S7_vs_39012\|\|full | Low-quality |
| S7_vs_39023\|\|full | Low-quality |
| S7_vs_39034\|\|full | Low-quality |
| S7_vs_39072\|\|full | Low-quality |
| S7_vs_39076\|\|full | Low-quality |
| S7_vs_39120\|\|full | Not-determined |
| S7_vs_39124\|\|full | Not-determined |
| S7_vs_39127\|\|full | Low-quality |
| S7_vs_39143\|\|full | Low-quality |
| S7_vs_39222\|\|full | Not-determined |
| S7_vs_39246\|\|full | Not-determined |
| S7_vs_39255\|\|full | Not-determined |
| S7_vs_39262\|\|full | Low-quality |
| S7_vs_39314\|\|full | Low-quality |
| S7_vs_39344\|\|full | Low-quality |
| S7_vs_39356\|\|full | Not-determined |
| S7_vs_39373\|\|full | Not-determined |
| S7_vs_39382\|\|full | Not-determined |
| S7_vs_39393\|\|full | Not-determined |
| S7_vs_39400\|\|full | Not-determined |
| S7_vs_39417\|\|full | Low-quality |
| S7_vs_39422\|\|full | Low-quality |
| S7_vs_39430\|\|full | Low-quality |
| S7_vs_39456\|\|full | Not-determined |
| S7_vs_39503\|\|full | Low-quality |
| S7_vs_39525\|\|full | Low-quality |
| S7_vs_39548\|\|full | Low-quality |
| S7_vs_39555\|\|full | Not-determined |
| S7_vs_39631\|\|full | Not-determined |
| S7_vs_39632\|\|full | Low-quality |
| S7_vs_39647\|\|full | Low-quality |
| S7_vs_39673\|\|full | Not-determined |
| S7_vs_39678\|\|full | Low-quality |
| S17_vs_k141_177200\|\|full | High-quality |
| S7_vs_39740\|\|full | Not-determined |
| S7_vs_39792\|\|full | Low-quality |
| S7_vs_39802\|\|full | Not-determined |
| S7_vs_39826\|\|full | Low-quality |
| S7_vs_39856\|\|full | Low-quality |
| S7_vs_39862\|\|full | Low-quality |
| S7_vs_39882\|\|full | Low-quality |
| S7_vs_39966\|\|full | Not-determined |
| S7_vs_39984\|\|full | Low-quality |
| S7_vs_40004\|\|full | Low-quality |
| S7_vs_40050\|\|full | Not-determined |
| S7_vs_40055\|\|full | Low-quality |
| S7_vs_40063\|\|full | Low-quality |
| S7_vs_40104\|\|full | Low-quality |
| S7_vs_40185\|\|full | Low-quality |
| S7_vs_40189\|\|full | Low-quality |
| S7_vs_40197\|\|full | Not-determined |
| S7_vs_40202\|\|full | Low-quality |
| S16_vs_172952\|\|full | High-quality |
| S7_vs_40296\|\|full | Low-quality |
| S7_vs_40299\|\|full | Low-quality |
| S7_vs_40304\|\|full | Low-quality |
| S7_vs_40307\|\|full | Low-quality |
| S7_vs_40328\|\|full | Not-determined |
| S7_vs_40464\|\|full | Not-determined |
| S7_vs_40504\|\|full | Low-quality |
| S7_vs_40520\|\|full | Low-quality |
| S7_vs_40521\|\|full | Not-determined |
| S7_vs_40552\|\|full | Not-determined |
| S7_vs_40557\|\|full | Not-determined |
| S7_vs_40561\|\|full | Low-quality |
| S7_vs_40574\|\|full | Not-determined |
| S7_vs_40576\|\|full | Low-quality |
| S7_vs_40590\|\|full | Low-quality |
| S7_vs_40591\|\|full | Not-determined |
| S7_vs_40601\|\|full | Low-quality |
| S7_vs_40626\|\|full | Not-determined |
| S7_vs_40648\|\|full | Not-determined |
| S7_vs_40712\|\|full | Low-quality |
| S7_vs_40736\|\|full | Low-quality |
| S7_vs_40748\|\|full | Low-quality |
| S7_vs_40769\|\|full | Not-determined |
| S7_vs_40855\|\|full | Low-quality |
| S7_vs_40892\|\|full | Not-determined |
| S7_vs_40908\|\|full | Low-quality |
| S7_vs_40930\|\|full | Not-determined |
| S7_vs_40961\|\|full | Low-quality |
| S7_vs_40973\|\|full | Not-determined |
| S7_vs_40985\|\|full | Low-quality |
| S7_vs_41060\|\|full | Low-quality |
| S7_vs_41071\|\|full | Low-quality |
| S16_extracted_172897 | Complete |
| S7_vs_41198\|\|full | Not-determined |
| S7_vs_41259\|\|full | Low-quality |
| S7_vs_41272\|\|full | Not-determined |
| S7_vs_41273\|\|full | Not-determined |
| S7_vs_41275\|\|full | Not-determined |
| S7_vs_41397\|\|full | Not-determined |
| S7_vs_41417\|\|full | Low-quality |
| S7_vs_41423\|\|full | Low-quality |
| S7_vs_41487\|\|full | Low-quality |
| S7_vs_41520\|\|full | Low-quality |
| S11_vs_k141_104071\|\|full | Medium-quality |
| S7_vs_41537\|\|full | Not-determined |
| S7_vs_41545\|\|full | Low-quality |
| S7_vs_41622\|\|full | Not-determined |
| S7_vs_41672\|\|full | Low-quality |
| S7_vs_41724\|\|full | Low-quality |
| S7_vs_41805\|\|full | Low-quality |
| S7_vs_41816\|\|full | Not-determined |
| S7_vs_41841\|\|full | Low-quality |
| S7_vs_41849\|\|full | Low-quality |
| S7_vs_41865\|\|full | Low-quality |
| S7_vs_41900\|\|full | Low-quality |
| S7_vs_41934\|\|full | Low-quality |
| S7_vs_41959\|\|full | Low-quality |
| S7_vs_41989\|\|full | Not-determined |
| S7_vs_42010\|\|full | Low-quality |
| S7_vs_42026\|\|full | Low-quality |
| S7_vs_42030\|\|full | Not-determined |
| S7_vs_42048\|\|full | Not-determined |
| S7_vs_42051\|\|full | Not-determined |
| S7_vs_42090\|\|full | Not-determined |
| S7_vs_42134\|\|full | Low-quality |
| S7_vs_42135\|\|full | Low-quality |
| S7_vs_42177\|\|full | Low-quality |
| S7_vs_42180\|\|full | Low-quality |
| S7_vs_42188\|\|full | Low-quality |
| S7_vs_42270\|\|full | Low-quality |
| S7_vs_42284\|\|full | Low-quality |
| S7_vs_42287\|\|full | Low-quality |
| S7_vs_42299\|\|full | Not-determined |
| S7_vs_42327\|\|full | Low-quality |
| S7_vs_42341\|\|full | Low-quality |
| S7_vs_42372\|\|full | Not-determined |
| S7_vs_42386\|\|full | Not-determined |
| S7_vs_42419\|\|full | Not-determined |
| S7_vs_42453\|\|full | Not-determined |
| S7_vs_42457\|\|full | Low-quality |
| S7_vs_42464\|\|full | Low-quality |
| S7_vs_42483\|\|full | Low-quality |
| S7_vs_42567\|\|full | Low-quality |
| S7_vs_42645\|\|full | Low-quality |
| S7_vs_42650\|\|full | Low-quality |
| S7_vs_42668\|\|full | Low-quality |
| S7_vs_42713\|\|full | Not-determined |
| S7_vs_42724\|\|full | Low-quality |
| S7_vs_42762\|\|full | Low-quality |
| S7_vs_42799\|\|full | Low-quality |
| S7_vs_42807\|\|full | Low-quality |
| S7_vs_42824\|\|full | Not-determined |
| S7_vs_42840\|\|full | Not-determined |
| S7_vs_42907\|\|full | Low-quality |
| S7_vs_42931\|\|full | Low-quality |
| S7_vs_42976\|\|full | Low-quality |
| S7_vs_43023\|\|full | Low-quality |
| S7_vs_43027\|\|full | Low-quality |
| S7_vs_43073\|\|full | Low-quality |
| S7_vs_43153\|\|full | Low-quality |
| S7_vs_43173\|\|full | Low-quality |
| S7_vs_43229\|\|full | Not-determined |
| S7_vs_43233\|\|full | Not-determined |
| S7_vs_43251\|\|full | Low-quality |
| S7_vs_43272\|\|full | Not-determined |
| S7_vs_43330\|\|full | Not-determined |
| S7_vs_43362\|\|full | Low-quality |
| S7_vs_43385\|\|full | Not-determined |
| S7_vs_43391\|\|full | Low-quality |
| S7_vs_43406\|\|full | Low-quality |
| S7_vs_43515\|\|full | Not-determined |
| S7_vs_43517\|\|full | Low-quality |
| S11_vs_k141_20225\|\|full | High-quality |
| S7_vs_43538\|\|full | Not-determined |
| S7_vs_43545\|\|full | Not-determined |
| S7_vs_43551\|\|full | Not-determined |
| S7_vs_43575\|\|full | Low-quality |
| S7_vs_43601\|\|full | Not-determined |
| S7_vs_43640\|\|full | Low-quality |
| S7_vs_43807\|\|full | Low-quality |
| S7_vs_43809\|\|full | Low-quality |
| S7_vs_43832\|\|full | Not-determined |
| S7_vs_43857\|\|full | Low-quality |
| S7_vs_43869\|\|full | Not-determined |
| S7_vs_43876\|\|full | Not-determined |
| S7_vs_43884\|\|full | Low-quality |
| S7_vs_43909\|\|full | Low-quality |
| S7_vs_43928\|\|full | Low-quality |
| S7_vs_43961\|\|full | Low-quality |
| S7_vs_44012\|\|full | Not-determined |
| S7_vs_44030\|\|full | Low-quality |
| S7_vs_44079\|\|full | Not-determined |
| S7_vs_44106\|\|full | Not-determined |
| S7_vs_44107\|\|full | Not-determined |
| S7_vs_44191\|\|full | Not-determined |
| S7_vs_44201\|\|full | Low-quality |
| S7_vs_44220\|\|full | Low-quality |
| S7_vs_44250\|\|full | Low-quality |
| S7_vs_44265\|\|full | Not-determined |
| S7_vs_44301\|\|full | Not-determined |
| S7_vs_44365\|\|full | Low-quality |
| S7_vs_44390\|\|full | Low-quality |
| S7_vs_44393\|\|full | Not-determined |
| S7_vs_44412\|\|full | Low-quality |
| S7_vs_44423\|\|full | Not-determined |
| S7_vs_44447\|\|full | Low-quality |
| S7_vs_44448\|\|full | Not-determined |
| S7_vs_44546\|\|full | Not-determined |
| S7_vs_44556\|\|full | Low-quality |
| S7_vs_44611\|\|full | Low-quality |
| S7_vs_44612\|\|full | Not-determined |
| S7_vs_44613\|\|full | Low-quality |
| S7_vs_44629\|\|full | Not-determined |
| S7_vs_44675\|\|full | Low-quality |
| S7_vs_44731\|\|full | Not-determined |
| S7_vs_44748\|\|full | Low-quality |
| S7_vs_44806\|\|full | Low-quality |
| S7_vs_44909\|\|full | Not-determined |
| S7_vs_44914\|\|full | Low-quality |
| S7_vs_44921\|\|full | Not-determined |
| S7_vs_44937\|\|full | Low-quality |
| S7_vs_44943\|\|full | Not-determined |
| S7_vs_45011\|\|full | Low-quality |
| S7_vs_45028\|\|full | Low-quality |
| S7_vs_45033\|\|full | Not-determined |
| S7_vs_45085\|\|full | Low-quality |
| S7_vs_45096\|\|full | Low-quality |
| S7_vs_45128\|\|full | Low-quality |
| S7_vs_45166\|\|full | Low-quality |
| S7_vs_45206\|\|full | Low-quality |
| S7_vs_45318\|\|full | Not-determined |
| S7_vs_45447\|\|full | Low-quality |
| S7_vs_45482\|\|full | Low-quality |
| S7_vs_45494\|\|full | Low-quality |
| S7_vs_45505\|\|full | Not-determined |
| S7_vs_45525\|\|full | Low-quality |
| S7_vs_45545\|\|full | Low-quality |
| S7_vs_45576\|\|full | Low-quality |
| S7_vs_45577\|\|full | Low-quality |
| S7_vs_45582\|\|full | Low-quality |
| S7_vs_45605\|\|full | Not-determined |
| S7_vs_45635\|\|full | Not-determined |
| S7_vs_45690\|\|full | Not-determined |
| S7_vs_45741\|\|full | Not-determined |
| S7_vs_45773\|\|full | Low-quality |
| S7_vs_45790\|\|full | Low-quality |
| S7_vs_45866\|\|full | Not-determined |
| S7_vs_45904\|\|full | Not-determined |
| S7_vs_45908\|\|full | Not-determined |
| S7_vs_45965\|\|full | Not-determined |
| S7_vs_45987\|\|full | Low-quality |
| S7_vs_45990\|\|full | Low-quality |
| S7_vs_46039\|\|full | Low-quality |
| S7_vs_46051\|\|full | Not-determined |
| S7_vs_46064\|\|full | Not-determined |
| S7_vs_46123\|\|full | Not-determined |
| S7_vs_46134\|\|full | Low-quality |
| S7_vs_46141\|\|full | Low-quality |
| S7_vs_46142\|\|full | Not-determined |
| S7_vs_46145\|\|full | Not-determined |
| S7_vs_46182\|\|full | Not-determined |
| S7_vs_46212\|\|full | Low-quality |
| S7_vs_46246\|\|full | Low-quality |
| S7_vs_46247\|\|full | Low-quality |
| S7_vs_46270\|\|full | Not-determined |
| S7_vs_46284\|\|full | Not-determined |
| S7_vs_46288\|\|full | Low-quality |
| S7_vs_46312\|\|full | Low-quality |
| S7_vs_46328\|\|full | Not-determined |
| S7_vs_46330\|\|full | Low-quality |
| S7_vs_46351\|\|full | Not-determined |
| S7_vs_46369\|\|full | Low-quality |
| S7_vs_46372\|\|full | Low-quality |
| S7_vs_46373\|\|full | Low-quality |
| S7_vs_46382\|\|full | Low-quality |
| S7_vs_46384\|\|full | Not-determined |
| S7_vs_46406\|\|full | Not-determined |
| S7_vs_46436\|\|full | Not-determined |
| S7_vs_46443\|\|full | Low-quality |
| S7_vs_46466\|\|full | Low-quality |
| S7_vs_46489\|\|full | Low-quality |
| S7_vs_46510\|\|full | Low-quality |
| S7_vs_46520\|\|full | Low-quality |
| S7_vs_46599\|\|full | Low-quality |
| S7_vs_46630\|\|full | Low-quality |
| S7_vs_46639\|\|full | Low-quality |
| S7_vs_46643\|\|full | Low-quality |
| S7_vs_46650\|\|full | Low-quality |
| S7_vs_46658\|\|full | Low-quality |
| S7_vs_46680\|\|full | Not-determined |
| S7_vs_46733\|\|full | Not-determined |
| S7_vs_46759\|\|full | Not-determined |
| S7_vs_46769\|\|full | Not-determined |
| S7_vs_46787\|\|full | Not-determined |
| S7_vs_46807\|\|full | Low-quality |
| S7_vs_46828\|\|full | Low-quality |
| S7_vs_46838\|\|full | Not-determined |
| S7_vs_46839\|\|full | Low-quality |
| S7_vs_46860\|\|full | Not-determined |
| S7_vs_46950\|\|full | Not-determined |
| S7_vs_46987\|\|full | Not-determined |
| S7_vs_47010\|\|full | Low-quality |
| S7_vs_47042\|\|full | Not-determined |
| S3_extracted_k141_8164 | High-quality |
| S7_vs_47055\|\|full | Not-determined |
| S7_vs_47104\|\|full | Low-quality |
| S7_vs_47118\|\|full | Low-quality |
| S7_vs_47149\|\|full | Low-quality |
| S7_vs_47183\|\|full | Not-determined |
| S7_vs_47191\|\|full | Low-quality |
| S7_vs_47228\|\|full | Low-quality |
| S7_vs_47241\|\|full | Low-quality |
| S7_vs_47251\|\|full | Low-quality |
| S7_vs_47288\|\|full | Not-determined |
| S7_vs_47305\|\|full | Low-quality |
| S7_vs_47328\|\|full | Low-quality |
| S3_vs_k141_8164\|\|full | High-quality |
| S7_vs_47348\|\|full | Low-quality |
| S7_vs_47353\|\|full | Low-quality |
| S7_vs_47411\|\|full | Low-quality |
| S7_vs_47415\|\|full | Low-quality |
| S7_vs_47504\|\|full | Not-determined |
| S7_vs_47519\|\|full | Low-quality |
| S7_vs_47554\|\|full | Low-quality |
| S7_vs_47561\|\|full | Low-quality |
| S7_vs_47569\|\|full | Low-quality |
| S7_vs_47615\|\|full | Low-quality |
| S7_vs_47656\|\|full | Not-determined |
| S7_vs_47673\|\|full | Not-determined |
| S7_vs_47685\|\|full | Low-quality |
| S7_vs_47775\|\|full | Low-quality |
| S7_vs_47846\|\|full | Low-quality |
| S7_vs_47853\|\|full | Low-quality |
| S7_vs_47861\|\|full | Low-quality |
| S7_vs_47899\|\|full | Low-quality |
| S7_vs_47909\|\|full | Not-determined |
| S7_vs_47922\|\|full | Not-determined |
| S7_vs_47954\|\|full | Low-quality |
| S7_vs_47961\|\|full | Low-quality |
| S7_vs_47980\|\|full | Low-quality |
| S7_vs_48009\|\|full | Low-quality |
| S7_vs_48010\|\|full | Not-determined |
| S7_vs_48013\|\|full | Low-quality |
| S7_vs_48014\|\|full | Low-quality |
| S7_vs_48019\|\|full | Low-quality |
| S7_vs_48054\|\|full | Low-quality |
| S7_vs_48070\|\|full | Low-quality |
| S7_vs_48080\|\|full | Low-quality |
| S7_vs_48085\|\|full | Low-quality |
| S7_vs_48129\|\|full | Low-quality |
| S7_vs_48133\|\|full | Low-quality |
| S7_vs_48150\|\|full | Not-determined |
| S7_vs_48174\|\|full | Not-determined |
| S7_vs_48247\|\|full | Low-quality |
| S7_vs_48278\|\|full | Low-quality |
| S7_vs_48287\|\|full | Low-quality |
| S7_vs_48328\|\|full | Not-determined |
| S7_vs_48334\|\|full | Not-determined |
| S7_vs_48373\|\|full | Low-quality |
| S7_vs_48379\|\|full | Low-quality |
| S7_vs_48455\|\|full | Not-determined |
| S7_vs_48456\|\|full | Not-determined |
| S7_vs_48493\|\|full | Not-determined |
| S5_vs_k141_130958\|\|full | Medium-quality |
| S7_vs_48558\|\|full | Low-quality |
| S7_vs_48592\|\|full | Low-quality |
| S7_vs_48597\|\|full | Low-quality |
| S7_vs_48644\|\|full | Low-quality |
| S7_vs_48660\|\|full | Low-quality |
| S7_vs_48670\|\|full | Not-determined |
| S7_vs_48671\|\|full | Not-determined |
| S7_vs_48678\|\|full | Not-determined |
| S7_vs_48680\|\|full | Not-determined |
| S7_vs_48687\|\|full | Low-quality |
| S7_vs_48702\|\|full | Low-quality |
| S8_extracted_k141_102508 | High-quality |
| S7_vs_48711\|\|full | Low-quality |
| S7_vs_48735\|\|full | Low-quality |
| S7_vs_48803\|\|full | Low-quality |
| S7_vs_48830\|\|full | Not-determined |
| S9_vs_k141_45812\|\|full | Medium-quality |
| S7_vs_48843\|\|full | Low-quality |
| S7_vs_48876\|\|full | Low-quality |
| S7_vs_48927\|\|full | Low-quality |
| S7_vs_48963\|\|full | Not-determined |
| S13_vs_14794\|\|full | Medium-quality |
| S7_vs_49009\|\|full | Low-quality |
| S7_vs_49033\|\|full | Not-determined |
| S7_vs_49085\|\|full | Not-determined |
| S7_vs_49088\|\|full | Not-determined |
| S7_vs_49187\|\|full | Low-quality |
| S7_vs_49195\|\|full | Not-determined |
| S7_vs_49257\|\|full | Low-quality |
| S7_vs_49278\|\|full | Not-determined |
| S7_vs_49354\|\|full | Low-quality |
| S7_vs_49357\|\|full | Low-quality |
| S7_vs_49425\|\|full | Not-determined |
| S10_vs_59228\|\|full | Medium-quality |
| S7_vs_49440\|\|full | Not-determined |
| S7_vs_49465\|\|full | Low-quality |
| S7_vs_49467\|\|full | Low-quality |
| S17_vs_k141_39515\|\|full | Medium-quality |
| S7_vs_49512\|\|full | Low-quality |
| S7_vs_49529\|\|full | Not-determined |
| S7_vs_49537\|\|full | Low-quality |
| S7_vs_49556\|\|full | Not-determined |
| S7_vs_49602\|\|full | Not-determined |
| S7_vs_49637\|\|full | Low-quality |
| S7_vs_49651\|\|full | Low-quality |
| S7_vs_49663\|\|full | Low-quality |
| S7_vs_49691\|\|full | Low-quality |
| S7_vs_49693\|\|full | Low-quality |
| S7_vs_49694\|\|full | Not-determined |
| S7_vs_49798\|\|full | Low-quality |
| S7_vs_49801\|\|full | Low-quality |
| S7_vs_49848\|\|full | Low-quality |
| S7_vs_49926\|\|full | Low-quality |
| S7_vs_49978\|\|full | Not-determined |
| S7_vs_50031\|\|full | Low-quality |
| S7_vs_50036\|\|full | Not-determined |
| S7_vs_50073\|\|full | Low-quality |
| S7_vs_50090\|\|full | Low-quality |
| S7_vs_50123\|\|full | Low-quality |
| S7_vs_50175\|\|full | Not-determined |
| S7_vs_50185\|\|full | Low-quality |
| S14_extracted_k141_48775 | Medium-quality |
| S7_vs_50240\|\|full | Low-quality |
| S7_vs_50272\|\|full | Low-quality |
| S7_vs_50279\|\|full | Low-quality |
| S7_vs_50285\|\|full | Low-quality |
| S7_vs_50329\|\|full | Low-quality |
| S7_vs_50406\|\|full | Low-quality |
| S7_vs_50425\|\|full | Low-quality |
| S7_vs_50435\|\|full | Low-quality |
| S7_vs_50438\|\|full | Not-determined |
| S7_vs_50445\|\|full | Low-quality |
| S7_vs_50448\|\|full | Not-determined |
| S7_vs_50465\|\|full | Low-quality |
| S7_vs_50489\|\|full | Not-determined |
| S7_vs_50588\|\|full | Not-determined |
| S7_vs_50730\|\|full | Low-quality |
| S7_vs_50776\|\|full | Low-quality |
| S7_vs_50781\|\|full | Low-quality |
| S7_vs_50834\|\|full | Low-quality |
| S7_vs_50836\|\|full | Not-determined |
| S7_vs_50853\|\|full | Low-quality |
| S7_vs_50865\|\|full | Not-determined |
| S7_vs_50871\|\|full | Low-quality |
| S7_vs_50875\|\|full | Not-determined |
| S7_vs_50886\|\|full | Low-quality |
| S7_vs_50917\|\|full | Not-determined |
| S7_vs_50932\|\|full | Low-quality |
| S7_vs_50940\|\|full | Not-determined |
| S7_vs_50950\|\|full | Low-quality |
| S7_vs_50974\|\|full | Low-quality |
| S7_vs_50981\|\|full | Low-quality |
| S7_vs_50999\|\|full | Low-quality |
| S7_vs_51000\|\|full | Not-determined |
| S7_vs_51015\|\|full | Low-quality |
| S7_vs_51165\|\|full | Low-quality |
| S7_vs_51190\|\|full | Not-determined |
| S7_vs_51246\|\|full | Low-quality |
| S7_vs_51273\|\|full | Low-quality |
| S7_vs_51295\|\|full | Low-quality |
| S7_vs_51312\|\|full | Low-quality |
| S7_vs_51329\|\|full | Not-determined |
| S16_vs_172897\|\|full | High-quality |
| S7_vs_51348\|\|full | Low-quality |
| S7_vs_51365\|\|full | Not-determined |
| S7_vs_51393\|\|full | Not-determined |
| S7_vs_51416\|\|full | Low-quality |
| S7_vs_51504\|\|full | Low-quality |
| S7_vs_51535\|\|full | Low-quality |
| S7_vs_51575\|\|full | Low-quality |
| S7_vs_51582\|\|full | Low-quality |
| S7_vs_51603\|\|full | Not-determined |
| S7_vs_51658\|\|full | Low-quality |
| S7_vs_51669\|\|full | Not-determined |
| S7_vs_51679\|\|full | Low-quality |
| S7_vs_51753\|\|full | Low-quality |
| S7_vs_51761\|\|full | Not-determined |
| S7_vs_51772\|\|full | Low-quality |
| S7_vs_51783\|\|full | Low-quality |
| S7_vs_51855\|\|full | Not-determined |
| S7_vs_51873\|\|full | Low-quality |
| S7_vs_51905\|\|full | Low-quality |
| S7_vs_51939\|\|full | Low-quality |
| S7_vs_51980\|\|full | Not-determined |
| S7_vs_51997\|\|full | Low-quality |
| S9_vs_k141_66877\|\|full | High-quality |
| S7_vs_52041\|\|full | Low-quality |
| S7_vs_52053\|\|full | Not-determined |
| S7_vs_52069\|\|full | Not-determined |
| S7_vs_52070\|\|full | Low-quality |
| S7_vs_52146\|\|full | Low-quality |
| S7_vs_52160\|\|full | Low-quality |
| S7_vs_52197\|\|full | Low-quality |
| S7_vs_52264\|\|full | Low-quality |
| S7_vs_52283\|\|full | Low-quality |
| S7_vs_52291\|\|full | Low-quality |
| S7_vs_52295\|\|full | Low-quality |
| S7_vs_52309\|\|full | Low-quality |
| S7_vs_52331\|\|full | Low-quality |
| S7_vs_52414\|\|full | Low-quality |
| S7_vs_52502\|\|full | Low-quality |
| S7_vs_52515\|\|full | Low-quality |
| S7_vs_52584\|\|full | Low-quality |
| S7_vs_52634\|\|full | Not-determined |
| S7_vs_52699\|\|full | Not-determined |
| S7_vs_52722\|\|full | Not-determined |
| S7_vs_52737\|\|full | Low-quality |
| S7_vs_52759\|\|full | Low-quality |
| S7_vs_52795\|\|full | Low-quality |
| S7_vs_52829\|\|full | Low-quality |
| S7_vs_52837\|\|full | Low-quality |
| S7_vs_52913\|\|full | Low-quality |
| S7_vs_52918\|\|full | Not-determined |
| S7_vs_52987\|\|full | Low-quality |
| S7_vs_52994\|\|full | Low-quality |
| S7_vs_52995\|\|full | Not-determined |
| S7_vs_53044\|\|full | Low-quality |
| S7_vs_53064\|\|full | Low-quality |
| S7_vs_53065\|\|full | Not-determined |
| S7_vs_53092\|\|full | Not-determined |
| S7_vs_53106\|\|full | Not-determined |
| S7_vs_53134\|\|full | Not-determined |
| S7_vs_53135\|\|full | Low-quality |
| S7_vs_53141\|\|full | Low-quality |
| S7_vs_53200\|\|full | Low-quality |
| S7_vs_53226\|\|full | Low-quality |
| S7_vs_53229\|\|full | Not-determined |
| S8_vs_k141_102508\|\|full | High-quality |
| S7_vs_53284\|\|full | Not-determined |
| S7_vs_53357\|\|full | Low-quality |
| S7_vs_53406\|\|full | Low-quality |
| S7_vs_53456\|\|full | Not-determined |
| S7_vs_53591\|\|full | Not-determined |
| S7_vs_53600\|\|full | Low-quality |
| S7_vs_53604\|\|full | Not-determined |
| S7_vs_53609\|\|full | Not-determined |
| S7_vs_53826\|\|full | Low-quality |
| S7_vs_53852\|\|full | Not-determined |
| S7_vs_53871\|\|full | Low-quality |
| S7_vs_53875\|\|full | Not-determined |
| S7_vs_53897\|\|full | Not-determined |
| S7_vs_53921\|\|full | Not-determined |
| S7_vs_53926\|\|full | Not-determined |
| S7_vs_53966\|\|full | Low-quality |
| S7_vs_54033\|\|full | Not-determined |
| S7_vs_54071\|\|full | Low-quality |
| S7_vs_54072\|\|full | Low-quality |
| S7_vs_54085\|\|full | Low-quality |
| S7_vs_54094\|\|full | Low-quality |
| S7_vs_54138\|\|full | Not-determined |
| S7_vs_54146\|\|full | Not-determined |
| S7_vs_54151\|\|full | Not-determined |
| S7_vs_54187\|\|full | Low-quality |
| S7_vs_54249\|\|full | Not-determined |
| S7_vs_54271\|\|full | Low-quality |
| S7_vs_54283\|\|full | Low-quality |
| S7_vs_54364\|\|full | Low-quality |
| S7_vs_54413\|\|full | Low-quality |
| S7_vs_54433\|\|full | Low-quality |
| S7_vs_54453\|\|full | Not-determined |
| S7_vs_54457\|\|full | Not-determined |
| S7_vs_54467\|\|full | Low-quality |
| S7_vs_54499\|\|full | Low-quality |
| S7_vs_54507\|\|full | Not-determined |
| S7_vs_54544\|\|full | Low-quality |
| S7_vs_54625\|\|full | Not-determined |
| S7_vs_54658\|\|full | Not-determined |
| S7_vs_54706\|\|full | Not-determined |
| S7_vs_54712\|\|full | Low-quality |
| S7_vs_54716\|\|full | Low-quality |
| S7_vs_54738\|\|full | Not-determined |
| S7_vs_54761\|\|full | Low-quality |
| S7_vs_54859\|\|full | Low-quality |
| S7_vs_54897\|\|full | Low-quality |
| S7_vs_54901\|\|full | Low-quality |
| S7_vs_54950\|\|full | Not-determined |
| S7_vs_54954\|\|full | Low-quality |
| S7_vs_54974\|\|full | Not-determined |
| S7_vs_55008\|\|full | Low-quality |
| S7_vs_55014\|\|full | Not-determined |
| S7_vs_55030\|\|full | Low-quality |
| S5_vs_k141_84574\|\|full | High-quality |
| S7_vs_55073\|\|full | Low-quality |
| S7_vs_55097\|\|full | Not-determined |
| S7_vs_55124\|\|full | Low-quality |
| S7_vs_55176\|\|full | Low-quality |
| S7_vs_55205\|\|full | Low-quality |
| S7_vs_55206\|\|full | Low-quality |
| S7_vs_55314\|\|full | Low-quality |
| S7_vs_55379\|\|full | Low-quality |
| S7_vs_55393\|\|full | Not-determined |

| | |
|---|---|
| S11_vs_k141_8456\|\|full | High-quality |
| S7_vs_55433\|\|full | Low-quality |
| S7_vs_55506\|\|full | Low-quality |
| S7_vs_55513\|\|full | Not-determined |
| S7_vs_55523\|\|full | Not-determined |
| S11_vs_k141_81775\|\|full | High-quality |
| S7_vs_55530\|\|full | Not-determined |
| S7_vs_55535\|\|full | Not-determined |
| S7_vs_55565\|\|full | Not-determined |
| S7_vs_55587\|\|full | Low-quality |
| S7_vs_55624\|\|full | Not-determined |
| S7_vs_55647\|\|full | Not-determined |
| S7_vs_55667\|\|full | Not-determined |
| S7_vs_55673\|\|full | Low-quality |
| S7_vs_55685\|\|full | Not-determined |
| S7_vs_55694\|\|full | Low-quality |
| S7_vs_55701\|\|full | Low-quality |
| S7_vs_55723\|\|full | Low-quality |
| S7_vs_55740\|\|full | Low-quality |
| S7_vs_55747\|\|full | Not-determined |
| S7_vs_55769\|\|full | Not-determined |
| S7_vs_55789\|\|full | Not-determined |
| S7_vs_55813\|\|full | Not-determined |
| S7_vs_55814\|\|full | Low-quality |
| S7_vs_55818\|\|full | Low-quality |
| S7_vs_55829\|\|full | Low-quality |
| S7_vs_55853\|\|full | Low-quality |
| S7_vs_55866\|\|full | Low-quality |
| S7_vs_55898\|\|full | Low-quality |
| S7_vs_55917\|\|full | Low-quality |
| S7_vs_55926\|\|full | Not-determined |
| S7_vs_55984\|\|full | Low-quality |
| S7_vs_56004\|\|full | Not-determined |
| S7_vs_56080\|\|full | Low-quality |
| S7_vs_56104\|\|full | Low-quality |
| S7_vs_56111\|\|full | Low-quality |
| S7_vs_56116\|\|full | Not-determined |
| S7_vs_56123\|\|full | Low-quality |
| S7_vs_56176\|\|full | Not-determined |
| S7_vs_56190\|\|full | Low-quality |
| S7_vs_56260\|\|full | Not-determined |
| S7_vs_56267\|\|full | Low-quality |
| S7_vs_56270\|\|full | Low-quality |
| S7_vs_56286\|\|full | Not-determined |
| S7_vs_56310\|\|full | Low-quality |
| S7_vs_56322\|\|full | Not-determined |
| S7_vs_56335\|\|full | Not-determined |
| S7_vs_56402\|\|full | Low-quality |
| S7_vs_56423\|\|full | Low-quality |
| S7_vs_56484\|\|full | Low-quality |
| S7_vs_56500\|\|full | Low-quality |
| S7_vs_56527\|\|full | Low-quality |
| S14_extracted_k141_31122 | Medium-quality |
| S7_vs_56531\|\|full | Low-quality |
| S7_vs_56535\|\|full | Low-quality |
| S7_vs_56584\|\|full | Not-determined |
| S7_vs_56602\|\|full | Not-determined |
| S7_vs_56616\|\|full | Low-quality |
| S7_vs_56668\|\|full | Not-determined |
| S7_vs_56677\|\|full | Low-quality |
| S7_vs_56769\|\|full | Low-quality |
| S7_vs_56781\|\|full | Low-quality |
| S7_vs_56795\|\|full | Low-quality |
| S7_vs_56798\|\|full | Low-quality |
| S7_vs_56899\|\|full | Low-quality |
| S7_vs_56922\|\|full | Low-quality |
| S7_vs_56984\|\|full | Not-determined |
| S7_vs_57064\|\|full | Low-quality |
| S7_vs_57074\|\|full | Low-quality |
| S7_vs_57139\|\|full | Low-quality |
| S7_vs_57149\|\|full | Not-determined |
| S7_vs_57152\|\|full | Not-determined |
| S7_vs_57153\|\|full | Low-quality |
| S7_vs_57182\|\|full | Low-quality |
| S7_vs_57194\|\|full | Low-quality |
| S7_vs_57199\|\|full | Low-quality |
| S7_vs_57207\|\|full | Low-quality |
| S7_vs_57210\|\|full | Low-quality |
| S7_vs_57225\|\|full | Low-quality |
| S7_vs_57301\|\|full | Low-quality |
| S7_vs_57321\|\|full | Low-quality |
| S7_vs_57349\|\|full | Not-determined |
| S7_vs_57359\|\|full | Low-quality |
| S7_vs_57428\|\|full | Not-determined |
| S7_vs_57495\|\|full | Low-quality |
| S7_vs_57554\|\|full | Not-determined |
| S7_vs_57566\|\|full | Low-quality |
| S7_vs_57592\|\|full | Low-quality |
| S7_vs_57623\|\|full | Low-quality |
| S7_vs_57625\|\|full | Low-quality |
| S7_vs_57702\|\|full | Not-determined |
| S7_vs_57715\|\|full | Not-determined |
| S7_vs_57716\|\|full | Not-determined |
| S7_vs_57731\|\|full | Not-determined |
| S7_vs_57816\|\|full | Not-determined |
| S7_vs_57825\|\|full | Low-quality |
| S7_vs_57833\|\|full | Low-quality |
| S7_vs_57843\|\|full | Low-quality |
| S7_vs_57866\|\|full | Not-determined |
| S7_vs_57890\|\|full | Not-determined |
| S7_vs_57893\|\|full | Low-quality |
| S7_vs_57920\|\|full | Low-quality |
| S7_vs_57944\|\|full | Low-quality |
| S7_vs_57981\|\|full | Low-quality |
| S7_vs_58035\|\|full | Low-quality |
| S7_vs_58065\|\|full | Not-determined |
| S7_vs_58149\|\|full | Low-quality |
| S7_vs_58170\|\|full | Low-quality |
| S7_vs_58186\|\|full | Low-quality |
| S7_vs_58200\|\|full | Not-determined |
| S7_vs_58318\|\|full | Low-quality |
| S7_vs_58325\|\|full | Low-quality |
| S7_vs_58335\|\|full | Not-determined |
| S7_vs_58338\|\|full | Low-quality |
| S7_vs_58368\|\|full | Not-determined |
| S7_vs_58393\|\|full | Low-quality |
| S7_vs_58453\|\|full | Low-quality |
| S7_vs_58458\|\|full | Low-quality |
| S7_vs_58462\|\|full | Not-determined |
| S7_vs_58487\|\|full | Not-determined |
| S7_vs_58517\|\|full | Low-quality |
| S7_vs_58557\|\|full | Low-quality |
| S7_vs_58561\|\|full | Low-quality |
| S7_vs_58569\|\|full | Not-determined |
| S7_vs_58588\|\|full | Low-quality |
| S7_vs_58591\|\|full | Low-quality |
| S7_vs_58623\|\|full | Low-quality |
| S7_vs_58626\|\|full | Not-determined |
| S7_vs_58631\|\|full | Low-quality |
| S7_vs_58632\|\|full | Low-quality |
| S7_vs_58652\|\|full | Low-quality |
| S7_vs_58664\|\|full | Low-quality |
| S7_vs_58665\|\|full | Low-quality |
| S7_vs_58691\|\|full | Low-quality |
| S7_vs_58733\|\|full | Not-determined |
| S3_vs_k141_20315\|\|full | Medium-quality |
| S7_vs_58771\|\|full | Not-determined |
| S7_vs_58813\|\|full | Low-quality |
| S7_vs_58838\|\|full | Low-quality |
| S7_vs_58850\|\|full | Low-quality |
| S7_vs_58881\|\|full | Not-determined |
| S7_vs_58905\|\|full | Low-quality |
| S7_vs_58906\|\|full | Low-quality |
| S7_vs_58912\|\|full | Low-quality |
| S7_vs_58933\|\|full | Not-determined |
| S7_vs_58956\|\|full | Not-determined |
| S7_vs_58958\|\|full | Low-quality |
| S7_vs_58960\|\|full | Low-quality |
| S7_vs_58990\|\|full | Low-quality |
| S7_vs_58997\|\|full | Low-quality |
| S7_vs_59006\|\|full | Low-quality |
| S7_vs_59015\|\|full | Not-determined |
| S7_vs_59031\|\|full | Low-quality |
| S7_vs_59065\|\|full | Low-quality |
| S7_vs_59081\|\|full | Not-determined |
| S7_vs_59101\|\|full | Low-quality |
| S7_vs_59112\|\|full | Not-determined |
| S7_vs_59120\|\|full | Low-quality |
| S7_vs_59212\|\|full | Low-quality |
| S7_vs_59221\|\|full | Not-determined |
| S7_vs_59283\|\|full | Low-quality |
| S7_vs_59290\|\|full | Low-quality |
| S7_vs_59314\|\|full | Low-quality |
| S7_vs_59353\|\|full | Not-determined |
| S7_vs_59372\|\|full | Not-determined |
| S7_vs_59416\|\|full | Low-quality |
| S7_vs_59447\|\|full | Low-quality |
| S7_vs_59453\|\|full | Low-quality |
| S5_extracted_k141_134040 | Medium-quality |
| S7_vs_59482\|\|full | Low-quality |
| S7_vs_59532\|\|full | Low-quality |
| S7_vs_59573\|\|full | Low-quality |
| S7_vs_59657\|\|full | Low-quality |
| S7_vs_59670\|\|full | Low-quality |
| S7_vs_59704\|\|full | Low-quality |
| S7_vs_59708\|\|full | Not-determined |
| S7_vs_59792\|\|full | Low-quality |
| S7_vs_59821\|\|full | Low-quality |
| S7_vs_59881\|\|full | Low-quality |
| S7_vs_59914\|\|full | Low-quality |
| S7_vs_59924\|\|full | Low-quality |
| S7_vs_59954\|\|full | Not-determined |
| S7_vs_59960\|\|full | Not-determined |
| S7_vs_60013\|\|full | Not-determined |
| S7_vs_60014\|\|full | Low-quality |
| S7_vs_60023\|\|full | Not-determined |
| S7_vs_60032\|\|full | Not-determined |
| S7_vs_60036\|\|full | Low-quality |
| S7_vs_60072\|\|full | Not-determined |
| S7_vs_60143\|\|full | Low-quality |
| S7_vs_60171\|\|full | Low-quality |
| S7_vs_60187\|\|full | Low-quality |
| S7_vs_60196\|\|full | Not-determined |
| S7_vs_60201\|\|full | Not-determined |
| S7_vs_60207\|\|full | Not-determined |
| S7_vs_60234\|\|full | Low-quality |
| S7_vs_60248\|\|full | Low-quality |
| S7_vs_60258\|\|full | Not-determined |
| S7_vs_60300\|\|full | Not-determined |
| S7_vs_60320\|\|full | Not-determined |
| S7_vs_60348\|\|full | Low-quality |
| S7_vs_60358\|\|full | Not-determined |
| S7_vs_60378\|\|full | Low-quality |
| S7_vs_60379\|\|full | Not-determined |
| S7_vs_60386\|\|full | Not-determined |
| S7_vs_60387\|\|full | Low-quality |
| S7_vs_60391\|\|full | Low-quality |
| S7_vs_60415\|\|full | Low-quality |
| S7_vs_60416\|\|full | Low-quality |
| S7_vs_60463\|\|full | Low-quality |
| S7_vs_60498\|\|full | Low-quality |
| S7_vs_60602\|\|full | Low-quality |
| S7_vs_60604\|\|full | Low-quality |
| S7_vs_60692\|\|full | Low-quality |
| S7_vs_60764\|\|full | Not-determined |
| S7_vs_60779\|\|full | Not-determined |
| S7_vs_60786\|\|full | Low-quality |
| S7_vs_60802\|\|full | Not-determined |
| S7_vs_60814\|\|full | Low-quality |
| S7_vs_60829\|\|full | Low-quality |
| S7_vs_60843\|\|full | Low-quality |
| S7_vs_60872\|\|full | Low-quality |
| S7_vs_60881\|\|full | Low-quality |
| S7_vs_60904\|\|full | Not-determined |
| S7_vs_60947\|\|full | Not-determined |
| S7_vs_60969\|\|full | Low-quality |
| S7_vs_61037\|\|full | Low-quality |
| S7_vs_61041\|\|full | Low-quality |
| S7_vs_61050\|\|full | Low-quality |
| S7_vs_61096\|\|full | Low-quality |
| S7_vs_61140\|\|full | Low-quality |
| S7_vs_61147\|\|full | Not-determined |
| S7_vs_61162\|\|full | Not-determined |
| S7_vs_61175\|\|full | Low-quality |
| S7_vs_61176\|\|full | Low-quality |
| S7_vs_61195\|\|full | Low-quality |
| S7_vs_61241\|\|full | Low-quality |
| S7_vs_61262\|\|full | Not-determined |
| S7_vs_61265\|\|full | Low-quality |
| S7_vs_61286\|\|full | Low-quality |
| S7_vs_61296\|\|full | Not-determined |
| S7_vs_61319\|\|full | Low-quality |
| S7_vs_61353\|\|full | Low-quality |
| S7_vs_61389\|\|full | Low-quality |
| S7_vs_61418\|\|full | Low-quality |
| S7_vs_61448\|\|full | Low-quality |
| S7_vs_61469\|\|full | Not-determined |
| S7_vs_61501\|\|full | Low-quality |
| S7_vs_61522\|\|full | Not-determined |
| S7_vs_61546\|\|full | Low-quality |
| S7_vs_61548\|\|full | Not-determined |
| S7_vs_61550\|\|full | Not-determined |
| S7_vs_61601\|\|full | Not-determined |
| S7_vs_61610\|\|full | Not-determined |
| S7_vs_61611\|\|full | Not-determined |
| S7_vs_61612\|\|full | Low-quality |
| S7_vs_61614\|\|full | Not-determined |
| S7_vs_61635\|\|full | Not-determined |
| S7_vs_61649\|\|full | Low-quality |
| S7_vs_61686\|\|full | Low-quality |
| S7_vs_61853\|\|full | Not-determined |
| S7_vs_61866\|\|full | Not-determined |
| S7_vs_61939\|\|full | Not-determined |
| S7_vs_61987\|\|full | Low-quality |
| S7_vs_61998\|\|full | Low-quality |
| S7_vs_62015\|\|full | Not-determined |
| S7_vs_62025\|\|full | Not-determined |
| S7_vs_62056\|\|full | Not-determined |
| S7_vs_62060\|\|full | Not-determined |
| S7_vs_62061\|\|full | Low-quality |
| S7_vs_62062\|\|full | Not-determined |
| S7_vs_62064\|\|full | Low-quality |
| S7_vs_62068\|\|full | Not-determined |
| S7_vs_62085\|\|full | Low-quality |
| S7_vs_62125\|\|full | Not-determined |
| S7_vs_62137\|\|full | Not-determined |
| S7_vs_62158\|\|full | Not-determined |
| S7_vs_62175\|\|full | Low-quality |
| S7_vs_62182\|\|full | Not-determined |
| S7_vs_62238\|\|full | Low-quality |
| S7_vs_62284\|\|full | Low-quality |
| S7_vs_62287\|\|full | Low-quality |
| S7_vs_62294\|\|full | Low-quality |
| S7_vs_62304\|\|full | Not-determined |
| S7_vs_62320\|\|full | Low-quality |
| S7_vs_62327\|\|full | Low-quality |
| S7_vs_62337\|\|full | Low-quality |
| S7_vs_62339\|\|full | Not-determined |
| S11_vs_k141_104041\|\|full | High-quality |
| S7_vs_62458\|\|full | Not-determined |
| S7_vs_62470\|\|full | Not-determined |
| S7_vs_62483\|\|full | Low-quality |
| S7_vs_62590\|\|full | Low-quality |
| S7_vs_62603\|\|full | Low-quality |
| S7_vs_62619\|\|full | Low-quality |
| S7_vs_62640\|\|full | Low-quality |
| S7_vs_62641\|\|full | Low-quality |
| S7_vs_62647\|\|full | Low-quality |
| S7_vs_62677\|\|full | Not-determined |
| S7_vs_62692\|\|full | Not-determined |
| S7_vs_62733\|\|full | Not-determined |
| S7_vs_62750\|\|full | Low-quality |
| S7_vs_62775\|\|full | Not-determined |
| S7_vs_62785\|\|full | Low-quality |
| S7_vs_62787\|\|full | Low-quality |
| S7_vs_62791\|\|full | Low-quality |
| S7_vs_62824\|\|full | Low-quality |
| S7_vs_62840\|\|full | Low-quality |
| S7_vs_62906\|\|full | Low-quality |
| S7_vs_62919\|\|full | Low-quality |
| S7_vs_62927\|\|full | Low-quality |
| S7_vs_62955\|\|full | Low-quality |
| S7_vs_62996\|\|full | Not-determined |
| S7_vs_63001\|\|full | Low-quality |
| S7_vs_63031\|\|full | Low-quality |
| S7_vs_63077\|\|full | Not-determined |
| S7_vs_63090\|\|full | Low-quality |
| S7_vs_63144\|\|full | Not-determined |
| S7_vs_63194\|\|full | Low-quality |
| S7_vs_63203\|\|full | Not-determined |
| S7_vs_63218\|\|full | Low-quality |
| S7_vs_63238\|\|full | Not-determined |
| S7_vs_63263\|\|full | Not-determined |
| S7_vs_63284\|\|full | Not-determined |
| S7_vs_63302\|\|full | Low-quality |
| S7_vs_63314\|\|full | Low-quality |
| S7_vs_63339\|\|full | Not-determined |
| S7_vs_63341\|\|full | Not-determined |
| S7_vs_63373\|\|full | Not-determined |
| S7_vs_63393\|\|full | Not-determined |
| S7_vs_63408\|\|full | Not-determined |
| S7_vs_63415\|\|full | Low-quality |
| S7_vs_63486\|\|full | Not-determined |
| S7_vs_63532\|\|full | Not-determined |
| S7_vs_63565\|\|full | Not-determined |
| S7_vs_63566\|\|full | Low-quality |
| S7_vs_63635\|\|full | Low-quality |
| S7_vs_63664\|\|full | Not-determined |
| S7_vs_63685\|\|full | Not-determined |
| S7_vs_63696\|\|full | Low-quality |
| S7_vs_63719\|\|full | Low-quality |
| S7_vs_63725\|\|full | Low-quality |
| S7_vs_63731\|\|full | Not-determined |
| S7_vs_63743\|\|full | Not-determined |
| S7_vs_63781\|\|full | Low-quality |
| S7_vs_63786\|\|full | Low-quality |
| S7_vs_63793\|\|full | Low-quality |
| S7_vs_63804\|\|full | Low-quality |
| S7_vs_63811\|\|full | Low-quality |
| S7_vs_63841\|\|full | Not-determined |
| S7_vs_63849\|\|full | Low-quality |
| S7_vs_63882\|\|full | Not-determined |
| S11_extracted_k141_104070 | Complete |
| S7_vs_63953\|\|full | Not-determined |
| S7_vs_63961\|\|full | Not-determined |
| S7_vs_63990\|\|full | Low-quality |
| S7_vs_64017\|\|full | Not-determined |
| S7_vs_64081\|\|full | Low-quality |
| S7_vs_64098\|\|full | Low-quality |
| S7_vs_64112\|\|full | Low-quality |
| S7_vs_64118\|\|full | Low-quality |
| S7_vs_64165\|\|full | Not-determined |
| S7_vs_64203\|\|full | Low-quality |
| S7_vs_64252\|\|full | Low-quality |
| S7_vs_64280\|\|full | Not-determined |
| S7_vs_64391\|\|full | Low-quality |
| S7_vs_64399\|\|full | Low-quality |
| S7_vs_64417\|\|full | Low-quality |
| S7_vs_64421\|\|full | Low-quality |
| S7_vs_64529\|\|full | Low-quality |
| S7_vs_64585\|\|full | Low-quality |
| S7_vs_64598\|\|full | Low-quality |
| S7_vs_64603\|\|full | Low-quality |
| S7_vs_64616\|\|full | Not-determined |
| S7_vs_64648\|\|full | Not-determined |
| S7_vs_64669\|\|full | Low-quality |
| S7_vs_64683\|\|full | Low-quality |
| S7_vs_64688\|\|full | Not-determined |
| S7_vs_64796\|\|full | Not-determined |
| S7_vs_64801\|\|full | Low-quality |
| S7_vs_64849\|\|full | Low-quality |
| S7_vs_64858\|\|full | Low-quality |
| S7_vs_64872\|\|full | Low-quality |
| S7_vs_64876\|\|full | Low-quality |
| S7_vs_64888\|\|full | Low-quality |
| S7_vs_64985\|\|full | Not-determined |
| S7_vs_65061\|\|full | Not-determined |
| S7_vs_65077\|\|full | Not-determined |
| S7_vs_65098\|\|full | Low-quality |
| S7_vs_65128\|\|full | Low-quality |
| S7_vs_65174\|\|full | Not-determined |
| S7_vs_65205\|\|full | Low-quality |
| S7_vs_65211\|\|full | Not-determined |
| S7_vs_65238\|\|full | Low-quality |
| S7_vs_65258\|\|full | Low-quality |
| S7_vs_65310\|\|full | Not-determined |
| S7_vs_65331\|\|full | Low-quality |
| S7_vs_65351\|\|full | Low-quality |
| S7_vs_65352\|\|full | Low-quality |
| S7_vs_65362\|\|full | Not-determined |
| S7_vs_65369\|\|full | Low-quality |
| S7_vs_65438\|\|full | Not-determined |
| S7_vs_65465\|\|full | Not-determined |
| S7_vs_65471\|\|full | Not-determined |
| S7_vs_65518\|\|full | Low-quality |
| S7_vs_65557\|\|full | Not-determined |
| S7_vs_65590\|\|full | Low-quality |
| S7_vs_65591\|\|full | Low-quality |
| S7_vs_65620\|\|full | Low-quality |
| S7_vs_65641\|\|full | Low-quality |
| S7_vs_65649\|\|full | Low-quality |
| S7_vs_65650\|\|full | Low-quality |
| S7_vs_65681\|\|full | Low-quality |
| S7_vs_65712\|\|full | Low-quality |
| S7_vs_65747\|\|full | Low-quality |
| S7_vs_65753\|\|full | Not-determined |
| S7_vs_65755\|\|full | Low-quality |
| S7_vs_65780\|\|full | Low-quality |
| S7_vs_65867\|\|full | Low-quality |
| S7_vs_65908\|\|full | Low-quality |
| S7_vs_65970\|\|full | Low-quality |
| S7_vs_65992\|\|full | Low-quality |
| S7_vs_66017\|\|full | Low-quality |
| S7_vs_66085\|\|full | Not-determined |
| S7_vs_66086\|\|full | Not-determined |
| S7_vs_66174\|\|full | Low-quality |
| S3_vs_k141_89485\|\|full | High-quality |
| S7_vs_66211\|\|full | Not-determined |
| S7_vs_66215\|\|full | Low-quality |
| S7_vs_66229\|\|full | Not-determined |
| S7_vs_66305\|\|full | Low-quality |
| S7_vs_66319\|\|full | Not-determined |
| S7_vs_66345\|\|full | Not-determined |
| S7_vs_66371\|\|full | Low-quality |
| S7_vs_66380\|\|full | Low-quality |
| S7_vs_66404\|\|full | Not-determined |
| S7_vs_66486\|\|full | Low-quality |
| S7_vs_66491\|\|full | Low-quality |
| S7_vs_66545\|\|full | Low-quality |
| S7_vs_66573\|\|full | Not-determined |
| S7_vs_66574\|\|full | Not-determined |
| S7_vs_66593\|\|full | Not-determined |
| S7_vs_66598\|\|full | Not-determined |
| S7_vs_66599\|\|full | Low-quality |
| S7_vs_66608\|\|full | Low-quality |
| S7_vs_66618\|\|full | Not-determined |
| S7_vs_66620\|\|full | Low-quality |
| S7_vs_66631\|\|full | Not-determined |
| S7_vs_66639\|\|full | Low-quality |
| S7_vs_66651\|\|full | Not-determined |
| S7_vs_66655\|\|full | Not-determined |
| S7_vs_66659\|\|full | Low-quality |
| S7_vs_66682\|\|full | Low-quality |
| S7_vs_66685\|\|full | Low-quality |
| S7_vs_66713\|\|full | Not-determined |
| S7_vs_66750\|\|full | Not-determined |
| S7_vs_66760\|\|full | Low-quality |
| S7_vs_66763\|\|full | Low-quality |
| S7_vs_66813\|\|full | Low-quality |
| S7_vs_66815\|\|full | Low-quality |
| S7_vs_66838\|\|full | Not-determined |
| S7_vs_66845\|\|full | Low-quality |
| S7_vs_66854\|\|full | Low-quality |
| S7_vs_66865\|\|full | Not-determined |
| S7_vs_66921\|\|full | Low-quality |
| S7_vs_66979\|\|full | Not-determined |
| S7_vs_67042\|\|full | Low-quality |
| S7_vs_67065\|\|full | Not-determined |
| S7_vs_67069\|\|full | Low-quality |
| S7_vs_67090\|\|full | Low-quality |
| S7_vs_67116\|\|full | Low-quality |
| S7_vs_67128\|\|full | Not-determined |
| S7_vs_67152\|\|full | Not-determined |
| S7_vs_67209\|\|full | Not-determined |
| S7_vs_67231\|\|full | Not-determined |
| S7_vs_67286\|\|full | Low-quality |
| S7_vs_67318\|\|full | Low-quality |
| S7_vs_67326\|\|full | Low-quality |
| S7_vs_67334\|\|full | Low-quality |
| S7_vs_67378\|\|full | Low-quality |
| S7_vs_67435\|\|full | Low-quality |
| S7_vs_67462\|\|full | Low-quality |
| S7_vs_67487\|\|full | Low-quality |
| S7_vs_67489\|\|full | Low-quality |
| S7_vs_67491\|\|full | Low-quality |
| S7_vs_67494\|\|full | Low-quality |
| S7_vs_67503\|\|full | Low-quality |
| S7_vs_67534\|\|full | Not-determined |
| S7_vs_67540\|\|full | Low-quality |
| S7_vs_67565\|\|full | Low-quality |
| S7_vs_67566\|\|full | Low-quality |
| S7_vs_67596\|\|full | Low-quality |
| S7_vs_67619\|\|full | Not-determined |
| S7_vs_67642\|\|full | Not-determined |
| S7_vs_67644\|\|full | Low-quality |
| S7_vs_67652\|\|full | Not-determined |
| S7_vs_67661\|\|full | Not-determined |
| S7_vs_67705\|\|full | Low-quality |
| S7_vs_67713\|\|full | Low-quality |
| S7_vs_67748\|\|full | Low-quality |
| S7_vs_67765\|\|full | Low-quality |
| S7_vs_67782\|\|full | Low-quality |
| S7_vs_67804\|\|full | Low-quality |
| S7_vs_67846\|\|full | Not-determined |
| S7_vs_67862\|\|full | Low-quality |
| S7_vs_67887\|\|full | Not-determined |
| S8_vs_k141_176604\|\|full | High-quality |
| S7_vs_67911\|\|full | Low-quality |
| S7_vs_67945\|\|full | Not-determined |
| S7_vs_67965\|\|full | Low-quality |
| S7_vs_67978\|\|full | Low-quality |
| S7_vs_67981\|\|full | Low-quality |
| S7_vs_68063\|\|full | Low-quality |
| S7_vs_68074\|\|full | Low-quality |
| S7_vs_68094\|\|full | Low-quality |
| S7_vs_68174\|\|full | Not-determined |
| S7_vs_68175\|\|full | Not-determined |
| S7_vs_68218\|\|full | Low-quality |
| S13_extracted_36198 | Medium-quality |
| S7_vs_68243\|\|full | Low-quality |
| S7_vs_68252\|\|full | Low-quality |
| S7_vs_68278\|\|full | Not-determined |
| S7_vs_68279\|\|full | Low-quality |
| S7_vs_68294\|\|full | Not-determined |
| S7_vs_68307\|\|full | Low-quality |
| S7_vs_68326\|\|full | Low-quality |
| S7_vs_68390\|\|full | Low-quality |
| S7_vs_68404\|\|full | Low-quality |
| S7_vs_68431\|\|full | Low-quality |
| S7_vs_68451\|\|full | Low-quality |
| S7_vs_68453\|\|full | Not-determined |
| S7_vs_68454\|\|full | Not-determined |
| S7_vs_68477\|\|full | Low-quality |
| S7_vs_68507\|\|full | Not-determined |
| S7_vs_68516\|\|full | Low-quality |
| S8_vs_k141_175235\|\|full | High-quality |
| S7_vs_68566\|\|full | Low-quality |
| S7_vs_68572\|\|full | Not-determined |
| S7_vs_68630\|\|full | Low-quality |
| S7_vs_68643\|\|full | Not-determined |
| S7_vs_68666\|\|full | Low-quality |
| S7_vs_68680\|\|full | Low-quality |
| S7_vs_68688\|\|full | Low-quality |
| S7_vs_68702\|\|full | Low-quality |
| S7_vs_68731\|\|full | Low-quality |
| S7_vs_68737\|\|full | Low-quality |
| S7_vs_68754\|\|full | Low-quality |
| S7_vs_68791\|\|full | Not-determined |
| S7_vs_68805\|\|full | Low-quality |
| S7_vs_68808\|\|full | Not-determined |
| S7_vs_68852\|\|full | Low-quality |
| S7_vs_68889\|\|full | Low-quality |
| S7_vs_68969\|\|full | Low-quality |
| S7_vs_69024\|\|full | Low-quality |
| S7_vs_69046\|\|full | Low-quality |
| S7_vs_69072\|\|full | Not-determined |
| S7_vs_69095\|\|full | Not-determined |
| S7_vs_69113\|\|full | Low-quality |
| S7_vs_69149\|\|full | Low-quality |
| S7_vs_69157\|\|full | Not-determined |
| S7_vs_69167\|\|full | Low-quality |
| S7_vs_69180\|\|full | Low-quality |
| S7_vs_69227\|\|full | Low-quality |
| S7_vs_69238\|\|full | Not-determined |
| S7_vs_69326\|\|full | Not-determined |
| S7_vs_69330\|\|full | Not-determined |
| S7_vs_69333\|\|full | Low-quality |
| S6_vs_k141_33711\|\|full | High-quality |
| S7_vs_69389\|\|full | Not-determined |
| S7_vs_69414\|\|full | Low-quality |
| S7_vs_69415\|\|full | Not-determined |
| S7_vs_69435\|\|full | Not-determined |
| S7_vs_69438\|\|full | Low-quality |
| S7_vs_69477\|\|full | Low-quality |
| S7_vs_69527\|\|full | Low-quality |
| S7_vs_69534\|\|full | Low-quality |
| S7_vs_69544\|\|full | Low-quality |
| S7_vs_69555\|\|full | Low-quality |
| S7_vs_69559\|\|full | Not-determined |
| S7_vs_69628\|\|full | Low-quality |
| S16_extracted_96481 | Medium-quality |
| S7_vs_69644\|\|full | Not-determined |
| S7_vs_69664\|\|full | Not-determined |
| S7_vs_69678\|\|full | Low-quality |
| S7_vs_69717\|\|full | Not-determined |
| S7_vs_69728\|\|full | Low-quality |
| S7_vs_69752\|\|full | Not-determined |
| S7_vs_69815\|\|full | Low-quality |
| S7_vs_69868\|\|full | Low-quality |

| | |
|---|---|
| S7_vs_69877\|\|full | Low-quality |
| S7_vs_69901\|\|full | Not-determined |
| S7_vs_69934\|\|full | Low-quality |
| S7_vs_69947\|\|full | Not-determined |
| S7_vs_69951\|\|full | Not-determined |
| S7_vs_69970\|\|full | Not-determined |
| S7_vs_70016\|\|full | Low-quality |
| S7_vs_70034\|\|full | Low-quality |
| S7_vs_70047\|\|full | Low-quality |
| S7_vs_70065\|\|full | Low-quality |
| S7_vs_70068\|\|full | Not-determined |
| S7_vs_70070\|\|full | Low-quality |
| S7_vs_70085\|\|full | Low-quality |
| S7_vs_70112\|\|full | Low-quality |
| S7_vs_70139\|\|full | Low-quality |
| S7_vs_70146\|\|full | Low-quality |
| S7_vs_70171\|\|full | Low-quality |
| S7_vs_70181\|\|full | Low-quality |
| S7_vs_70225\|\|full | Low-quality |
| S7_vs_70240\|\|full | Low-quality |
| S13_vs_36198\|\|full | Medium-quality |
| S7_vs_70269\|\|full | Low-quality |
| S7_vs_70290\|\|full | Low-quality |
| S7_vs_70306\|\|full | Low-quality |
| S7_vs_70311\|\|full | Low-quality |
| S7_vs_70319\|\|full | Not-determined |
| S7_vs_70342\|\|full | Low-quality |
| S7_vs_70343\|\|full | Low-quality |
| S7_vs_70374\|\|full | Not-determined |
| S7_vs_70385\|\|full | Low-quality |
| S7_vs_70457\|\|full | Low-quality |
| S7_vs_70467\|\|full | Low-quality |
| S7_vs_70498\|\|full | Low-quality |
| S7_vs_70533\|\|full | Low-quality |
| S7_vs_70537\|\|full | Not-determined |
| S7_vs_70630\|\|full | Low-quality |
| S7_vs_70638\|\|full | Low-quality |
| S7_vs_70641\|\|full | Low-quality |
| S7_vs_70659\|\|full | Not-determined |
| S7_vs_70665\|\|full | Not-determined |
| S7_vs_70704\|\|full | Low-quality |
| S7_vs_70716\|\|full | Not-determined |
| S7_vs_70725\|\|full | Not-determined |
| S7_vs_70803\|\|full | Not-determined |
| S7_vs_70894\|\|full | Low-quality |
| S7_vs_70898\|\|full | Low-quality |
| S7_vs_71002\|\|full | Low-quality |
| S7_vs_71084\|\|full | Low-quality |
| S7_vs_71101\|\|full | Low-quality |
| S7_vs_71126\|\|full | Low-quality |
| S7_vs_71170\|\|full | Not-determined |
| S7_vs_71215\|\|full | Low-quality |
| S7_vs_71253\|\|full | Low-quality |
| S7_vs_71310\|\|full | Low-quality |
| S7_vs_71326\|\|full | Low-quality |
| S7_vs_71327\|\|full | Low-quality |
| S7_vs_71356\|\|full | Low-quality |
| S7_vs_71363\|\|full | Low-quality |
| S7_vs_71375\|\|full | Low-quality |
| S7_vs_71392\|\|full | Low-quality |
| S7_vs_71400\|\|full | Not-determined |
| S7_vs_71433\|\|full | Low-quality |
| S7_vs_71434\|\|full | Low-quality |
| S7_vs_71435\|\|full | Low-quality |
| S7_vs_71490\|\|full | Low-quality |
| S7_vs_71521\|\|full | Not-determined |
| S7_vs_71523\|\|full | Low-quality |
| S7_vs_71526\|\|full | Not-determined |
| S7_vs_4114\|\|full | Medium-quality |
| S7_vs_71577\|\|full | Low-quality |
| S7_vs_71601\|\|full | Low-quality |
| S7_vs_71614\|\|full | Not-determined |
| S7_vs_71711\|\|full | Low-quality |
| S7_vs_71762\|\|full | Low-quality |
| S7_vs_71836\|\|full | Not-determined |
| S7_vs_71854\|\|full | Not-determined |
| S7_vs_71868\|\|full | Low-quality |
| S7_vs_71911\|\|full | Low-quality |
| S7_vs_71925\|\|full | Low-quality |
| S7_vs_71971\|\|full | Low-quality |
| S7_vs_71979\|\|full | Low-quality |
| S7_vs_72007\|\|full | Not-determined |
| S7_vs_72012\|\|full | Not-determined |
| S7_vs_72036\|\|full | Low-quality |
| S7_vs_72170\|\|full | Low-quality |
| S7_vs_72189\|\|full | Low-quality |
| S7_vs_72203\|\|full | Not-determined |
| S7_vs_72213\|\|full | Not-determined |
| S7_vs_72242\|\|full | Not-determined |
| S7_vs_72280\|\|full | Low-quality |
| S7_vs_72336\|\|full | Not-determined |
| S7_vs_72415\|\|full | Not-determined |
| S7_vs_72421\|\|full | Not-determined |
| S7_vs_72455\|\|full | Low-quality |
| S7_vs_72487\|\|full | Low-quality |
| S8_extracted_k141_13709 | Complete |
| S7_vs_72504\|\|full | Not-determined |
| S7_vs_72508\|\|full | Low-quality |
| S7_vs_72510\|\|full | Low-quality |
| S7_vs_72550\|\|full | Not-determined |
| S7_vs_72557\|\|full | Low-quality |
| S7_vs_72563\|\|full | Not-determined |
| S7_vs_72610\|\|full | Low-quality |
| S7_vs_72613\|\|full | Low-quality |
| S7_vs_72628\|\|full | Not-determined |
| S7_vs_72703\|\|full | Low-quality |
| S7_vs_72720\|\|full | Low-quality |
| S7_vs_72724\|\|full | Low-quality |
| S7_vs_72743\|\|full | Low-quality |
| S7_vs_72855\|\|full | Low-quality |
| S7_vs_72858\|\|full | Not-determined |
| S7_vs_72863\|\|full | Not-determined |
| S7_vs_72921\|\|full | Low-quality |
| S7_vs_72927\|\|full | Not-determined |
| S7_vs_72930\|\|full | Not-determined |
| S7_vs_72952\|\|full | Low-quality |
| S7_vs_72965\|\|full | Low-quality |
| S7_vs_73002\|\|full | Low-quality |
| S7_vs_73028\|\|full | Not-determined |
| S7_vs_73106\|\|full | Low-quality |
| S7_vs_73172\|\|full | Not-determined |
| S7_vs_73173\|\|full | Low-quality |
| S7_vs_73218\|\|full | Low-quality |
| S7_vs_73256\|\|full | Low-quality |
| S7_vs_73263\|\|full | Low-quality |
| S13_extracted_22175 | High-quality |
| S7_vs_73286\|\|full | Not-determined |
| S7_vs_73287\|\|full | Not-determined |
| S7_vs_73305\|\|full | Not-determined |
| S7_vs_73331\|\|full | Low-quality |
| S7_vs_73401\|\|full | Low-quality |
| S7_vs_73437\|\|full | Not-determined |
| S7_vs_73472\|\|full | Low-quality |
| S7_vs_73475\|\|full | Not-determined |
| S7_vs_73531\|\|full | Low-quality |
| S7_vs_73543\|\|full | Low-quality |
| S7_vs_73628\|\|full | Not-determined |
| S7_vs_73647\|\|full | Not-determined |
| S7_vs_73653\|\|full | Low-quality |
| S7_vs_73656\|\|full | Low-quality |
| S7_vs_73733\|\|full | Low-quality |
| S7_vs_73746\|\|full | Not-determined |
| S7_vs_73758\|\|full | Not-determined |
| S7_vs_73769\|\|full | Not-determined |
| S7_vs_73774\|\|full | Not-determined |
| S7_vs_73804\|\|full | Not-determined |
| S7_vs_73827\|\|full | Not-determined |
| S7_vs_73839\|\|full | Low-quality |
| S7_vs_73867\|\|full | Not-determined |
| S7_vs_73892\|\|full | Not-determined |
| S7_vs_73898\|\|full | Not-determined |
| S7_vs_73974\|\|full | Not-determined |
| S7_vs_73984\|\|full | Low-quality |
| S7_vs_74001\|\|full | Low-quality |
| S7_vs_74004\|\|full | Low-quality |
| S7_vs_74011\|\|full | Not-determined |
| S7_vs_74095\|\|full | Low-quality |
| S7_vs_74120\|\|full | Not-determined |
| S7_vs_74157\|\|full | Not-determined |
| S7_vs_74158\|\|full | Not-determined |
| S7_vs_74217\|\|full | Not-determined |
| S7_vs_74226\|\|full | Low-quality |
| S7_vs_74229\|\|full | Not-determined |
| S7_vs_74230\|\|full | Low-quality |
| S7_vs_74244\|\|full | Low-quality |
| S7_vs_74293\|\|full | Not-determined |
| S7_vs_74294\|\|full | Low-quality |
| S7_vs_74327\|\|full | Low-quality |
| S7_vs_74328\|\|full | Low-quality |
| S7_vs_74342\|\|full | Low-quality |
| S7_vs_74349\|\|full | Not-determined |
| S7_vs_74388\|\|full | Not-determined |
| S7_vs_74395\|\|full | Low-quality |
| S7_vs_74431\|\|full | Not-determined |
| S7_vs_74457\|\|full | Low-quality |
| S7_vs_74511\|\|full | Not-determined |
| S7_vs_74545\|\|full | Not-determined |
| S7_vs_74583\|\|full | Not-determined |
| S7_vs_74670\|\|full | Low-quality |
| S7_vs_74678\|\|full | Low-quality |
| S7_vs_74783\|\|full | Low-quality |
| S7_vs_74874\|\|full | Not-determined |
| S7_vs_74888\|\|full | Not-determined |
| S7_vs_74899\|\|full | Low-quality |
| S7_vs_74956\|\|full | Low-quality |
| S7_vs_74985\|\|full | Not-determined |
| S7_vs_74998\|\|full | Not-determined |
| S7_vs_75023\|\|full | Low-quality |
| S7_vs_75057\|\|full | Not-determined |
| S7_vs_75058\|\|full | Low-quality |
| S7_vs_75071\|\|full | Low-quality |
| S7_vs_75084\|\|full | Low-quality |
| S7_vs_75087\|\|full | Low-quality |
| S7_vs_75106\|\|full | Not-determined |
| S7_vs_75108\|\|full | Low-quality |
| S7_vs_75111\|\|full | Not-determined |
| S7_vs_75148\|\|full | Low-quality |
| S7_vs_75153\|\|full | Low-quality |
| S7_vs_75192\|\|full | Low-quality |
| S7_vs_75220\|\|full | Low-quality |
| S7_vs_75321\|\|full | Low-quality |
| S7_vs_75368\|\|full | Low-quality |
| S7_vs_75392\|\|full | Low-quality |
| S7_vs_75496\|\|full | Low-quality |
| S7_vs_75537\|\|full | Low-quality |
| S7_vs_75557\|\|full | Low-quality |
| S7_vs_75579\|\|full | Low-quality |
| S7_vs_75657\|\|full | Low-quality |
| S7_vs_75680\|\|full | Not-determined |
| S7_vs_75727\|\|full | Low-quality |
| S7_vs_75759\|\|full | Low-quality |
| S7_vs_75771\|\|full | Low-quality |
| S7_vs_75790\|\|full | Not-determined |
| S7_vs_75813\|\|full | Not-determined |
| S7_vs_75875\|\|full | Low-quality |
| S7_vs_75915\|\|full | Low-quality |
| S7_vs_75967\|\|full | Low-quality |
| S7_vs_75974\|\|full | Not-determined |
| S7_vs_76024\|\|full | Not-determined |
| S7_vs_76029\|\|full | Low-quality |
| S7_vs_76052\|\|full | Not-determined |
| S7_vs_76059\|\|full | Low-quality |
| S7_vs_76122\|\|full | Low-quality |
| S7_vs_76150\|\|full | Not-determined |
| S7_vs_76151\|\|full | Low-quality |
| S7_vs_76196\|\|full | Low-quality |
| S7_vs_76257\|\|full | Low-quality |
| S7_vs_76275\|\|full | Low-quality |
| S7_vs_76284\|\|full | Low-quality |
| S7_vs_76301\|\|full | Low-quality |
| S7_vs_76339\|\|full | Low-quality |
| S7_vs_76359\|\|full | Not-determined |
| S14_vs_k141_58919\|\|full | High-quality |
| S7_vs_76401\|\|full | Low-quality |
| S7_vs_76402\|\|full | Low-quality |
| S7_vs_76409\|\|full | Low-quality |
| S7_vs_76435\|\|full | Low-quality |
| S7_vs_76441\|\|full | Low-quality |
| S5_extracted_k141_105728 | Medium-quality |
| S7_vs_76464\|\|full | Not-determined |
| S7_vs_76470\|\|full | Low-quality |
| S7_vs_76486\|\|full | Not-determined |
| S7_vs_76500\|\|full | Low-quality |
| S7_vs_76504\|\|full | Low-quality |
| S7_vs_76569\|\|full | Low-quality |
| S7_vs_76588\|\|full | Low-quality |
| S7_vs_76590\|\|full | Low-quality |
| S7_vs_76641\|\|full | Not-determined |
| S7_vs_76693\|\|full | Low-quality |
| S7_vs_76699\|\|full | Low-quality |
| S7_vs_76704\|\|full | Low-quality |
| S7_vs_76712\|\|full | Not-determined |
| S7_vs_76757\|\|full | Not-determined |
| S7_vs_76778\|\|full | Low-quality |
| S7_vs_76811\|\|full | Not-determined |
| S7_vs_76819\|\|full | Low-quality |
| S7_vs_76859\|\|full | Low-quality |
| S7_vs_76880\|\|full | Low-quality |
| S7_vs_76886\|\|full | Not-determined |
| S7_vs_76914\|\|full | Low-quality |
| S7_vs_76917\|\|full | Low-quality |
| S7_vs_76932\|\|full | Not-determined |
| S7_vs_77003\|\|full | Low-quality |
| S7_vs_77051\|\|full | Low-quality |
| S7_vs_77082\|\|full | Low-quality |
| S7_vs_77100\|\|full | Low-quality |
| S7_vs_77132\|\|full | Not-determined |
| S7_vs_77137\|\|full | Not-determined |
| S7_vs_77149\|\|full | Not-determined |
| S7_vs_77365\|\|full | Low-quality |
| S7_vs_77425\|\|full | Low-quality |
| S7_vs_77451\|\|full | Low-quality |
| S7_vs_77494\|\|full | Not-determined |
| S7_vs_77594\|\|full | Low-quality |
| S7_vs_77645\|\|full | Low-quality |
| S7_vs_77746\|\|full | Not-determined |
| S7_vs_77756\|\|full | Not-determined |
| S7_vs_77781\|\|full | Low-quality |
| S7_vs_77797\|\|full | Low-quality |
| S7_vs_77819\|\|full | Low-quality |
| S7_vs_77834\|\|full | Low-quality |
| S7_vs_77842\|\|full | Not-determined |
| S7_vs_77869\|\|full | Low-quality |
| S7_vs_77878\|\|full | Low-quality |
| S7_vs_77904\|\|full | Low-quality |
| S7_vs_77941\|\|full | Not-determined |
| S7_vs_77963\|\|full | Low-quality |
| S7_vs_77982\|\|full | Not-determined |
| S7_vs_78034\|\|full | Not-determined |
| S7_vs_78050\|\|full | Low-quality |
| S7_vs_78102\|\|full | Low-quality |
| S7_vs_78114\|\|full | Low-quality |
| S7_vs_78201\|\|full | Low-quality |
| S7_vs_78210\|\|full | Low-quality |
| S7_vs_78231\|\|full | Low-quality |
| S7_vs_78243\|\|full | Low-quality |
| S7_vs_78249\|\|full | Low-quality |
| S7_vs_78253\|\|full | Low-quality |
| S7_vs_78284\|\|full | Not-determined |
| S7_vs_78392\|\|full | Low-quality |
| S7_vs_78453\|\|full | Not-determined |
| S7_vs_78496\|\|full | Low-quality |
| S5_vs_k141_105728\|\|full | Medium-quality |
| S7_vs_78536\|\|full | Not-determined |
| S7_vs_78585\|\|full | Low-quality |
| S7_vs_78626\|\|full | Low-quality |
| S7_vs_78650\|\|full | Low-quality |
| S7_vs_78677\|\|full | Low-quality |
| S7_vs_78680\|\|full | Low-quality |
| S7_vs_78685\|\|full | Low-quality |
| S7_vs_78693\|\|full | Not-determined |
| S7_vs_78704\|\|full | Not-determined |
| S7_vs_78749\|\|full | Low-quality |
| S7_vs_78752\|\|full | Low-quality |
| S7_vs_78763\|\|full | Low-quality |
| S7_vs_78774\|\|full | Low-quality |
| S7_vs_78782\|\|full | Low-quality |
| S7_vs_78825\|\|full | Low-quality |
| S7_vs_78828\|\|full | Low-quality |
| S7_vs_78830\|\|full | Low-quality |
| S7_vs_78888\|\|full | Not-determined |
| S7_vs_78893\|\|full | Low-quality |
| S7_vs_78907\|\|full | Low-quality |
| S7_vs_78950\|\|full | Not-determined |
| S7_vs_79004\|\|full | Low-quality |
| S7_vs_79032\|\|full | Low-quality |
| S7_vs_79045\|\|full | Low-quality |
| S7_vs_79056\|\|full | Low-quality |
| S7_vs_79136\|\|full | Low-quality |
| S7_vs_79139\|\|full | Low-quality |
| S7_vs_79154\|\|full | Low-quality |
| S7_vs_79178\|\|full | Low-quality |
| S7_vs_79179\|\|full | Low-quality |
| S7_vs_79181\|\|full | Low-quality |
| S7_vs_79190\|\|full | Low-quality |
| S7_vs_79226\|\|full | Low-quality |
| S7_vs_79228\|\|full | Low-quality |
| S7_vs_79279\|\|full | Low-quality |
| S11_vs_k141_104070\|\|full | High-quality |
| S7_vs_79327\|\|full | Low-quality |
| S7_vs_79358\|\|full | Low-quality |
| S7_vs_79463\|\|full | Low-quality |
| S7_vs_79482\|\|full | Low-quality |
| S7_vs_79511\|\|full | Low-quality |
| S7_vs_79526\|\|full | Not-determined |
| S7_vs_79578\|\|full | Low-quality |
| S7_vs_79587\|\|full | Low-quality |
| S7_vs_79633\|\|full | Low-quality |
| S7_vs_79636\|\|full | Not-determined |
| S7_vs_79795\|\|full | Not-determined |
| S7_vs_79815\|\|full | Not-determined |
| S7_vs_79906\|\|full | Not-determined |
| S7_vs_79919\|\|full | Low-quality |
| S7_vs_79953\|\|full | Low-quality |
| S7_vs_80006\|\|full | Not-determined |
| S7_vs_80009\|\|full | Not-determined |
| S7_vs_80010\|\|full | Not-determined |
| S7_vs_80021\|\|full | Low-quality |
| S7_vs_80048\|\|full | Low-quality |
| S7_vs_80078\|\|full | Low-quality |
| S7_vs_80090\|\|full | Low-quality |
| S7_vs_80095\|\|full | Not-determined |
| S7_vs_80101\|\|full | Low-quality |
| S7_vs_80183\|\|full | Low-quality |
| S7_vs_80196\|\|full | Low-quality |
| S7_vs_80208\|\|full | Low-quality |
| S7_vs_80214\|\|full | Not-determined |
| S7_vs_80216\|\|full | Low-quality |
| S7_vs_80226\|\|full | Low-quality |
| S7_vs_80234\|\|full | Low-quality |
| S7_vs_80281\|\|full | Low-quality |
| S7_vs_80286\|\|full | Low-quality |
| S7_vs_80315\|\|full | Not-determined |
| S7_vs_80386\|\|full | Low-quality |
| S7_vs_80432\|\|full | Not-determined |
| S7_vs_80464\|\|full | Low-quality |
| S7_vs_80517\|\|full | Low-quality |
| S7_vs_80530\|\|full | Low-quality |
| S7_vs_80583\|\|full | Low-quality |
| S7_vs_80609\|\|full | Low-quality |
| S7_vs_80652\|\|full | Not-determined |
| S7_vs_80674\|\|full | Low-quality |
| S7_vs_80695\|\|full | Low-quality |
| S7_vs_80725\|\|full | Low-quality |
| S7_vs_80749\|\|full | Low-quality |
| S7_vs_80779\|\|full | Low-quality |
| S7_vs_80826\|\|full | Not-determined |
| S7_vs_80835\|\|full | Not-determined |
| S7_vs_80843\|\|full | Low-quality |
| S7_vs_80853\|\|full | Low-quality |
| S7_vs_80878\|\|full | Low-quality |
| S7_vs_80887\|\|full | Low-quality |
| S7_vs_80892\|\|full | Not-determined |
| S7_vs_80919\|\|full | Not-determined |
| S7_vs_80986\|\|full | Low-quality |
| S7_vs_81027\|\|full | Low-quality |
| S7_vs_81106\|\|full | Low-quality |
| S7_vs_81123\|\|full | Not-determined |
| S7_vs_81135\|\|full | Low-quality |
| S7_vs_81141\|\|full | Not-determined |
| S7_vs_81187\|\|full | Low-quality |
| S7_vs_81202\|\|full | Low-quality |
| S7_vs_81215\|\|full | Low-quality |
| S7_vs_81234\|\|full | Low-quality |
| S7_vs_81268\|\|full | Not-determined |
| S7_vs_81293\|\|full | Low-quality |
| S7_vs_81322\|\|full | Low-quality |
| S7_vs_81326\|\|full | Low-quality |
| S7_vs_81343\|\|full | Low-quality |
| S7_vs_81391\|\|full | Not-determined |
| S7_vs_81458\|\|full | Low-quality |
| S7_vs_81498\|\|full | Not-determined |
| S7_vs_81551\|\|full | Not-determined |
| S7_vs_81552\|\|full | Low-quality |
| S7_vs_81553\|\|full | Not-determined |
| S7_vs_81607\|\|full | Low-quality |
| S7_vs_81626\|\|full | Low-quality |
| S7_vs_81677\|\|full | Not-determined |
| S7_vs_81705\|\|full | Low-quality |
| S7_vs_81712\|\|full | Not-determined |
| S7_vs_81715\|\|full | Low-quality |
| S7_vs_81746\|\|full | Low-quality |
| S7_vs_81784\|\|full | Not-determined |
| S7_vs_81823\|\|full | Not-determined |
| S7_vs_81828\|\|full | Low-quality |
| S7_vs_81843\|\|full | Low-quality |
| S7_vs_81853\|\|full | Not-determined |
| S7_vs_81871\|\|full | Not-determined |
| S7_vs_81880\|\|full | Low-quality |
| S7_vs_81890\|\|full | Not-determined |
| S7_vs_81893\|\|full | Low-quality |
| S7_vs_81902\|\|full | Low-quality |
| S7_vs_81913\|\|full | Not-determined |
| S7_vs_81935\|\|full | Low-quality |
| S7_vs_81950\|\|full | Low-quality |
| S7_vs_81952\|\|full | Not-determined |
| S7_vs_81970\|\|full | Low-quality |
| S7_vs_81990\|\|full | Not-determined |
| S7_vs_82018\|\|full | Not-determined |
| S7_vs_82078\|\|full | Low-quality |
| S7_vs_82092\|\|full | Not-determined |
| S7_vs_82100\|\|full | Not-determined |
| S7_vs_82101\|\|full | Not-determined |
| S7_vs_82117\|\|full | Not-determined |
| S7_vs_82119\|\|full | Low-quality |
| S7_vs_82125\|\|full | Low-quality |
| S7_vs_82138\|\|full | Low-quality |
| S13_vs_22175\|\|full | High-quality |
| S7_vs_82191\|\|full | Low-quality |
| S7_vs_82308\|\|full | Not-determined |
| S7_vs_82383\|\|full | Not-determined |
| S7_vs_82404\|\|full | Low-quality |
| S7_vs_82411\|\|full | Low-quality |
| S7_vs_82430\|\|full | Low-quality |
| S7_vs_82439\|\|full | Low-quality |
| S7_vs_82460\|\|full | Low-quality |
| S7_vs_82471\|\|full | Not-determined |
| S7_vs_82490\|\|full | Low-quality |
| S7_vs_82534\|\|full | Not-determined |
| S7_vs_82562\|\|full | Low-quality |
| S7_vs_82563\|\|full | Low-quality |
| S7_vs_82576\|\|full | Low-quality |
| S7_vs_82595\|\|full | Low-quality |
| S7_vs_82610\|\|full | Not-determined |
| S7_vs_82655\|\|full | Low-quality |
| S7_vs_82658\|\|full | Not-determined |
| S7_vs_82663\|\|full | Not-determined |
| S7_vs_82687\|\|full | Low-quality |
| S7_vs_82705\|\|full | Low-quality |
| S7_vs_82712\|\|full | Low-quality |
| S7_vs_82720\|\|full | Not-determined |
| S7_vs_82742\|\|full | Low-quality |
| S7_vs_82747\|\|full | Not-determined |
| S7_vs_82753\|\|full | Not-determined |
| S7_vs_82758\|\|full | Not-determined |
| S7_vs_82769\|\|full | Low-quality |
| S14_vs_k141_31122\|\|full | Medium-quality |
| S7_vs_82854\|\|full | Low-quality |
| S7_vs_82884\|\|full | Not-determined |
| S7_vs_82971\|\|full | Low-quality |
| S7_vs_82977\|\|full | Low-quality |
| S7_vs_83042\|\|full | Low-quality |
| S7_vs_83089\|\|full | Low-quality |
| S11_vs_k141_34137\|\|full | High-quality |
| S7_vs_83166\|\|full | Not-determined |
| S7_vs_83194\|\|full | Low-quality |
| S7_vs_83227\|\|full | Not-determined |
| S7_vs_83289\|\|full | Low-quality |
| S7_vs_83292\|\|full | Not-determined |
| S7_vs_83305\|\|full | Low-quality |
| S7_vs_83322\|\|full | Not-determined |
| S7_vs_83337\|\|full | Low-quality |
| S7_vs_83339\|\|full | Not-determined |
| S7_vs_83346\|\|full | Low-quality |
| S7_vs_83347\|\|full | Low-quality |
| S7_vs_83370\|\|full | Not-determined |
| S7_vs_83390\|\|full | Low-quality |
| S7_vs_83433\|\|full | Not-determined |
| S7_vs_112384\|\|full | Medium-quality |
| S7_vs_83498\|\|full | Low-quality |
| S7_vs_83540\|\|full | Low-quality |
| S7_vs_83598\|\|full | Not-determined |
| S7_vs_83618\|\|full | Not-determined |
| S3_vs_k141_127413\|\|full | Medium-quality |
| S7_vs_83636\|\|full | Low-quality |
| S7_vs_83653\|\|full | Low-quality |
| S7_vs_83658\|\|full | Low-quality |
| S7_vs_83664\|\|full | Not-determined |
| S7_vs_83683\|\|full | Low-quality |
| S7_vs_83727\|\|full | Low-quality |
| S7_vs_83730\|\|full | Not-determined |
| S7_vs_83778\|\|full | Not-determined |
| S7_vs_83797\|\|full | Low-quality |
| S7_vs_83817\|\|full | Not-determined |
| S7_vs_83834\|\|full | Low-quality |
| S7_vs_83836\|\|full | Low-quality |
| S7_vs_83840\|\|full | Low-quality |
| S7_vs_83867\|\|full | Low-quality |
| S7_vs_83910\|\|full | Not-determined |
| S7_vs_83957\|\|full | Not-determined |
| S7_vs_83983\|\|full | Not-determined |
| S7_vs_83990\|\|full | Not-determined |
| S7_vs_84142\|\|full | Low-quality |
| S7_vs_84168\|\|full | Not-determined |
| S7_vs_84173\|\|full | Low-quality |
| S7_vs_84186\|\|full | Low-quality |
| S7_vs_84235\|\|full | Low-quality |
| S7_vs_84247\|\|full | Not-determined |
| S7_vs_84289\|\|full | Not-determined |
| S7_vs_84290\|\|full | Not-determined |
| S7_vs_84298\|\|full | Low-quality |
| S7_vs_84346\|\|full | Not-determined |
| S7_vs_84414\|\|full | Low-quality |
| S7_vs_84456\|\|full | Not-determined |
| S7_vs_84471\|\|full | Low-quality |
| S7_vs_84483\|\|full | Not-determined |
| S7_vs_84513\|\|full | Low-quality |
| S7_vs_84519\|\|full | Low-quality |
| S7_vs_84526\|\|full | Low-quality |
| S7_vs_84532\|\|full | Low-quality |
| S7_vs_84540\|\|full | Low-quality |
| S7_vs_84570\|\|full | Low-quality |
| S7_vs_84583\|\|full | Not-determined |
| S7_vs_84584\|\|full | Low-quality |
| S7_vs_84606\|\|full | Low-quality |
| S7_vs_84611\|\|full | Low-quality |
| S7_vs_84612\|\|full | Not-determined |
| S7_vs_84618\|\|full | Not-determined |
| S7_vs_84623\|\|full | Low-quality |
| S7_vs_84636\|\|full | Low-quality |
| S7_vs_84652\|\|full | Not-determined |
| S7_vs_84669\|\|full | Low-quality |
| S7_vs_84670\|\|full | Low-quality |
| S7_vs_84676\|\|full | Low-quality |
| S7_vs_84701\|\|full | Low-quality |
| S7_vs_84752\|\|full | Not-determined |
| S7_vs_84773\|\|full | Low-quality |
| S7_vs_84779\|\|full | Not-determined |
| S7_vs_84859\|\|full | Low-quality |
| S7_vs_84870\|\|full | Low-quality |
| S7_vs_84898\|\|full | Low-quality |
| S7_vs_84954\|\|full | Low-quality |
| S11_extracted_k141_34224 | Complete |
| S7_vs_85001\|\|full | Low-quality |
| S7_vs_85011\|\|full | Not-determined |
| S7_vs_85068\|\|full | Not-determined |
| S7_vs_85077\|\|full | Not-determined |
| S7_vs_85078\|\|full | Not-determined |
| S7_vs_85081\|\|full | Low-quality |
| S7_vs_85154\|\|full | Not-determined |
| S7_vs_85175\|\|full | Low-quality |
| S7_vs_85184\|\|full | Low-quality |
| S7_vs_85186\|\|full | Low-quality |
| S7_vs_85227\|\|full | Low-quality |
| S7_vs_85236\|\|full | Low-quality |
| S7_vs_85254\|\|full | Not-determined |
| S7_vs_85287\|\|full | Low-quality |
| S7_vs_85288\|\|full | Low-quality |
| S7_vs_85321\|\|full | Not-determined |
| S7_vs_85382\|\|full | Low-quality |
| S7_vs_85407\|\|full | Low-quality |
| S7_vs_85440\|\|full | Not-determined |
| S7_vs_85461\|\|full | Low-quality |
| S7_vs_85470\|\|full | Low-quality |
| S7_vs_85488\|\|full | Not-determined |
| S7_vs_85505\|\|full | Low-quality |
| S7_vs_85596\|\|full | Not-determined |
| S7_vs_85628\|\|full | Low-quality |
| S7_vs_85675\|\|full | Not-determined |

| | |
|---|---|
| S7_vs_85688\|\|full | Not-determined |
| S7_vs_85761\|\|full | Low-quality |
| S7_vs_85762\|\|full | Low-quality |
| S7_vs_85764\|\|full | Not-determined |
| S7_vs_85802\|\|full | Low-quality |
| S7_vs_85816\|\|full | Low-quality |
| S7_vs_85844\|\|full | Low-quality |
| S7_vs_85862\|\|full | Not-determined |
| S7_vs_85866\|\|full | Low-quality |
| S7_vs_85869\|\|full | Low-quality |
| S7_vs_85878\|\|full | Low-quality |
| S7_vs_85922\|\|full | Not-determined |
| S7_vs_85923\|\|full | Low-quality |
| S7_vs_86099\|\|full | Low-quality |
| S7_vs_86106\|\|full | Low-quality |
| S7_vs_86219\|\|full | Not-determined |
| S7_vs_86235\|\|full | Low-quality |
| S7_vs_86323\|\|full | Low-quality |
| S7_vs_86339\|\|full | Not-determined |
| S7_vs_86367\|\|full | Not-determined |
| S7_extracted_138097 | Complete |
| S7_vs_86420\|\|full | Low-quality |
| S7_vs_86435\|\|full | Low-quality |
| S7_vs_86437\|\|full | Low-quality |
| S7_vs_86440\|\|full | Not-determined |
| S7_vs_86450\|\|full | Low-quality |
| S7_vs_86461\|\|full | Low-quality |
| S7_vs_86464\|\|full | Low-quality |
| S7_vs_86469\|\|full | Low-quality |
| S7_vs_86496\|\|full | Low-quality |
| S7_vs_86497\|\|full | Low-quality |
| S7_vs_86528\|\|full | Low-quality |
| S7_vs_86551\|\|full | Low-quality |
| S7_vs_86635\|\|full | Low-quality |
| S7_vs_86675\|\|full | Low-quality |
| S7_vs_86688\|\|full | Not-determined |
| S7_vs_86704\|\|full | Low-quality |
| S7_vs_86818\|\|full | Not-determined |
| S7_vs_86840\|\|full | Low-quality |
| S7_vs_86858\|\|full | Not-determined |
| S7_vs_86891\|\|full | Low-quality |
| S7_vs_86942\|\|full | Low-quality |
| S7_vs_86976\|\|full | Not-determined |
| S7_vs_87035\|\|full | Low-quality |
| S7_vs_87071\|\|full | Low-quality |
| S7_vs_87108\|\|full | Low-quality |
| S7_vs_87116\|\|full | Not-determined |
| S7_vs_87122\|\|full | Low-quality |
| S7_vs_87153\|\|full | Low-quality |
| S7_vs_87223\|\|full | Low-quality |
| S7_vs_87229\|\|full | Low-quality |
| S7_vs_87258\|\|full | Low-quality |
| S7_vs_87269\|\|full | Not-determined |
| S8_vs_k141_102608\|\|full | Medium-quality |
| S7_vs_87280\|\|full | Not-determined |
| S7_vs_87313\|\|full | Low-quality |
| S7_vs_87329\|\|full | Low-quality |
| S7_vs_87335\|\|full | Low-quality |
| S7_vs_87341\|\|full | Not-determined |
| S7_vs_87386\|\|full | Low-quality |
| S7_vs_87422\|\|full | Low-quality |
| S7_vs_87426\|\|full | Low-quality |
| S8_vs_k141_13709\|\|full | High-quality |
| S7_vs_87443\|\|full | Low-quality |
| S7_vs_87447\|\|full | Not-determined |
| S7_vs_87481\|\|full | Low-quality |
| S7_vs_87497\|\|full | Low-quality |
| S7_vs_87512\|\|full | Not-determined |
| S7_vs_87526\|\|full | Low-quality |
| S7_vs_87563\|\|full | Low-quality |
| S7_vs_87587\|\|full | Low-quality |
| S7_vs_87594\|\|full | Low-quality |
| S7_vs_87599\|\|full | Low-quality |
| S7_vs_87619\|\|full | Not-determined |
| S7_vs_87621\|\|full | Low-quality |
| S7_vs_87651\|\|full | Low-quality |
| S7_vs_87660\|\|full | Not-determined |
| S7_vs_87668\|\|full | Low-quality |
| S7_vs_87824\|\|full | Not-determined |
| S7_vs_87865\|\|full | Low-quality |
| S7_vs_87866\|\|full | Not-determined |
| S7_vs_87881\|\|full | Low-quality |
| S7_vs_87898\|\|full | Low-quality |
| S7_vs_87919\|\|full | Low-quality |
| S7_vs_87933\|\|full | Not-determined |
| S7_vs_87946\|\|full | Low-quality |
| S7_vs_87982\|\|full | Not-determined |
| S7_vs_88046\|\|full | Low-quality |
| S7_vs_88160\|\|full | Low-quality |
| S7_vs_88168\|\|full | Low-quality |
| S7_vs_88169\|\|full | Low-quality |
| S7_vs_88195\|\|full | Low-quality |
| S7_vs_88206\|\|full | Not-determined |
| S7_vs_88223\|\|full | Low-quality |
| S7_vs_88236\|\|full | Low-quality |
| S7_vs_88266\|\|full | Not-determined |
| S7_vs_88271\|\|full | Low-quality |
| S7_vs_88274\|\|full | Low-quality |
| S7_vs_88303\|\|full | Low-quality |
| S7_vs_88306\|\|full | Low-quality |
| S7_vs_88328\|\|full | Low-quality |
| S7_vs_88344\|\|full | Not-determined |
| S7_vs_88361\|\|full | Low-quality |
| S7_vs_88373\|\|full | Not-determined |
| S7_vs_88438\|\|full | Low-quality |
| S7_vs_88498\|\|full | Low-quality |
| S7_vs_88506\|\|full | Not-determined |
| S7_vs_88514\|\|full | Low-quality |
| S7_vs_88516\|\|full | Not-determined |
| S7_vs_88569\|\|full | Low-quality |
| S7_vs_88598\|\|full | Not-determined |
| S7_vs_88635\|\|full | Not-determined |
| S7_vs_88653\|\|full | Not-determined |
| S7_vs_88700\|\|full | Low-quality |
| S7_vs_88709\|\|full | Not-determined |
| S7_vs_88727\|\|full | Low-quality |
| S7_vs_88738\|\|full | Low-quality |
| S7_vs_88765\|\|full | Low-quality |
| S7_vs_88779\|\|full | Not-determined |
| S7_vs_88782\|\|full | Not-determined |
| S7_vs_88789\|\|full | Not-determined |
| S7_vs_88873\|\|full | Not-determined |
| S7_vs_88901\|\|full | Low-quality |
| S7_vs_88912\|\|full | Not-determined |
| S7_vs_88938\|\|full | Low-quality |
| S7_vs_88939\|\|full | Low-quality |
| S7_vs_88943\|\|full | Low-quality |
| S7_vs_88984\|\|full | Not-determined |
| S7_vs_89006\|\|full | Low-quality |
| S7_vs_89067\|\|full | Not-determined |
| S7_vs_89080\|\|full | Not-determined |
| S7_vs_89090\|\|full | Not-determined |
| S7_vs_89110\|\|full | Low-quality |
| S7_vs_89143\|\|full | Not-determined |
| S7_vs_89176\|\|full | Not-determined |
| S5_vs_k141_134040\|\|full | Medium-quality |
| S7_vs_89258\|\|full | Low-quality |
| S7_vs_89296\|\|full | Not-determined |
| S7_vs_89361\|\|full | Low-quality |
| S7_vs_89383\|\|full | Low-quality |
| S7_vs_89419\|\|full | Low-quality |
| S7_vs_89428\|\|full | Low-quality |
| S7_vs_89431\|\|full | Not-determined |
| S7_vs_89438\|\|full | Not-determined |
| S7_vs_89439\|\|full | Low-quality |
| S7_vs_89441\|\|full | Low-quality |
| S7_vs_89454\|\|full | Not-determined |
| S7_vs_89473\|\|full | Low-quality |
| S7_vs_89479\|\|full | Low-quality |
| S7_vs_89489\|\|full | Low-quality |
| S7_vs_89534\|\|full | Low-quality |
| S7_vs_89593\|\|full | Low-quality |
| S7_vs_89601\|\|full | Not-determined |
| S7_vs_89606\|\|full | Not-determined |
| S7_vs_89618\|\|full | Not-determined |
| S7_vs_89621\|\|full | Low-quality |
| S7_extracted_68239 | High-quality |
| S7_vs_89731\|\|full | Low-quality |
| S7_vs_89746\|\|full | Not-determined |
| S7_vs_89758\|\|full | Low-quality |
| S7_vs_89818\|\|full | Low-quality |
| S7_vs_89853\|\|full | Low-quality |
| S7_vs_89896\|\|full | Low-quality |
| S7_vs_89945\|\|full | Not-determined |
| S7_vs_89980\|\|full | Low-quality |
| S7_vs_90008\|\|full | Not-determined |
| S7_vs_90009\|\|full | Low-quality |
| S7_vs_90017\|\|full | Low-quality |
| S7_vs_90038\|\|full | Low-quality |
| S7_vs_90049\|\|full | Low-quality |
| S7_vs_90083\|\|full | Low-quality |
| S7_vs_90128\|\|full | Not-determined |
| S7_vs_90134\|\|full | Low-quality |
| S7_vs_90137\|\|full | Low-quality |
| S7_vs_90143\|\|full | Low-quality |
| S7_vs_90146\|\|full | Low-quality |
| S7_vs_90197\|\|full | Low-quality |
| S7_vs_90208\|\|full | Low-quality |
| S7_vs_90214\|\|full | Low-quality |
| S7_vs_90253\|\|full | Low-quality |
| S7_vs_90274\|\|full | Not-determined |
| S7_vs_90328\|\|full | Low-quality |
| S7_vs_90341\|\|full | Not-determined |
| S7_vs_90373\|\|full | Low-quality |
| S7_vs_90391\|\|full | Not-determined |
| S7_vs_90468\|\|full | Not-determined |
| S7_vs_90501\|\|full | Not-determined |
| S7_vs_90565\|\|full | Not-determined |
| S7_vs_90586\|\|full | Low-quality |
| S7_vs_90627\|\|full | Not-determined |
| S7_vs_90681\|\|full | Low-quality |
| S7_vs_90763\|\|full | Not-determined |
| S7_vs_90774\|\|full | Low-quality |
| S7_vs_90794\|\|full | Low-quality |
| S7_vs_90810\|\|full | Low-quality |
| S7_vs_90842\|\|full | Low-quality |
| S7_vs_90891\|\|full | Low-quality |
| S7_vs_90911\|\|full | Not-determined |
| S15_vs_18274\|\|full | High-quality |
| S7_vs_90935\|\|full | Low-quality |
| S7_vs_90957\|\|full | Low-quality |
| S7_vs_90963\|\|full | Low-quality |
| S7_vs_90968\|\|full | Low-quality |
| S7_vs_90970\|\|full | Not-determined |
| S7_vs_90974\|\|full | Low-quality |
| S7_vs_91036\|\|full | Not-determined |
| S7_vs_91134\|\|full | Not-determined |
| S7_vs_91153\|\|full | Low-quality |
| S7_vs_91185\|\|full | Low-quality |
| S7_vs_91191\|\|full | Not-determined |
| S7_vs_91214\|\|full | Low-quality |
| S7_vs_91262\|\|full | Low-quality |
| S7_vs_91284\|\|full | Not-determined |
| S7_vs_91298\|\|full | Low-quality |
| S7_vs_91304\|\|full | Low-quality |
| S7_vs_91332\|\|full | Not-determined |
| S7_vs_91359\|\|full | Low-quality |
| S7_vs_91386\|\|full | Low-quality |
| S7_vs_91445\|\|full | Not-determined |
| S7_vs_91455\|\|full | Low-quality |
| S7_vs_91464\|\|full | Not-determined |
| S7_vs_91496\|\|full | Not-determined |
| S7_vs_91523\|\|full | Low-quality |
| S7_vs_91583\|\|full | Not-determined |
| S7_vs_91588\|\|full | Low-quality |
| S7_vs_91726\|\|full | Low-quality |
| S7_vs_91754\|\|full | Low-quality |
| S7_vs_91822\|\|full | Low-quality |
| S7_vs_91894\|\|full | Not-determined |
| S7_vs_91903\|\|full | Low-quality |
| S7_vs_91920\|\|full | Low-quality |
| S7_vs_91928\|\|full | Not-determined |
| S7_vs_91970\|\|full | Low-quality |
| S7_vs_91984\|\|full | Low-quality |
| S7_vs_91997\|\|full | Low-quality |
| S7_vs_92011\|\|full | Low-quality |
| S7_vs_92014\|\|full | Not-determined |
| S7_vs_92020\|\|full | Low-quality |
| S7_vs_92041\|\|full | Low-quality |
| S7_vs_92108\|\|full | Low-quality |
| S7_vs_92109\|\|full | Not-determined |
| S7_vs_92128\|\|full | Low-quality |
| S7_vs_92137\|\|full | Not-determined |
| S7_vs_92144\|\|full | Low-quality |
| S7_vs_92219\|\|full | Low-quality |
| S7_vs_92233\|\|full | Low-quality |
| S7_vs_92253\|\|full | Low-quality |
| S7_vs_92284\|\|full | Not-determined |
| S7_vs_92287\|\|full | Low-quality |
| S7_vs_92303\|\|full | Not-determined |
| S7_vs_92321\|\|full | Low-quality |
| S7_vs_92374\|\|full | Low-quality |
| S7_vs_92415\|\|full | Low-quality |
| S7_vs_92446\|\|full | Low-quality |
| S7_vs_92498\|\|full | Low-quality |
| S7_vs_92500\|\|full | Not-determined |
| S7_vs_92547\|\|full | Low-quality |
| S7_vs_92551\|\|full | Not-determined |
| S7_vs_92560\|\|full | Not-determined |
| S7_vs_92581\|\|full | Low-quality |
| S7_vs_92596\|\|full | Low-quality |
| S7_vs_92641\|\|full | Not-determined |
| S7_vs_92659\|\|full | Low-quality |
| S7_vs_92674\|\|full | Low-quality |
| S7_vs_92695\|\|full | Low-quality |
| S7_vs_92699\|\|full | Low-quality |
| S7_vs_92736\|\|full | Low-quality |
| S7_vs_92753\|\|full | Low-quality |
| S7_vs_92811\|\|full | Low-quality |
| S7_vs_92812\|\|full | Low-quality |
| S7_vs_92826\|\|full | Not-determined |
| S7_vs_92923\|\|full | Not-determined |
| S7_vs_92930\|\|full | Not-determined |
| S7_vs_92999\|\|full | Low-quality |
| S7_vs_93022\|\|full | Low-quality |
| S7_vs_93037\|\|full | Not-determined |
| S7_vs_93051\|\|full | Low-quality |
| S7_vs_93072\|\|full | Low-quality |
| S7_vs_93093\|\|full | Low-quality |
| S7_vs_93096\|\|full | Low-quality |
| S11_vs_k141_34224\|\|full | High-quality |
| S7_vs_93098\|\|full | Not-determined |
| S7_vs_93103\|\|full | Not-determined |
| S7_vs_93105\|\|full | Not-determined |
| S7_vs_93109\|\|full | Not-determined |
| S7_vs_93139\|\|full | Not-determined |
| S7_vs_93159\|\|full | Low-quality |
| S7_vs_93203\|\|full | Not-determined |
| S7_vs_93270\|\|full | Not-determined |
| S7_vs_93305\|\|full | Low-quality |
| S7_vs_93318\|\|full | Low-quality |
| S7_vs_93513\|\|full | Not-determined |
| S7_vs_93545\|\|full | Low-quality |
| S7_vs_93577\|\|full | Low-quality |
| S7_vs_93581\|\|full | Low-quality |
| S7_vs_93604\|\|full | Not-determined |
| S7_vs_93634\|\|full | Low-quality |
| S7_vs_93647\|\|full | Low-quality |
| S7_vs_93650\|\|full | Not-determined |
| S7_vs_93687\|\|full | Low-quality |
| S7_vs_93715\|\|full | Low-quality |
| S7_vs_93719\|\|full | Low-quality |
| S7_vs_93724\|\|full | Low-quality |
| S7_vs_93734\|\|full | Low-quality |
| S7_vs_93746\|\|full | Low-quality |
| S7_vs_93782\|\|full | Low-quality |
| S7_vs_93788\|\|full | Low-quality |
| S7_vs_93948\|\|full | Not-determined |
| S7_vs_93975\|\|full | Not-determined |
| S7_vs_94006\|\|full | Low-quality |
| S7_vs_94023\|\|full | Low-quality |
| S7_vs_94036\|\|full | Low-quality |
| S7_vs_94106\|\|full | Low-quality |
| S7_vs_94107\|\|full | Not-determined |
| S7_vs_94108\|\|full | Low-quality |
| S7_vs_94118\|\|full | Low-quality |
| S7_vs_94144\|\|full | Low-quality |
| S7_vs_94149\|\|full | Not-determined |
| S7_vs_94157\|\|full | Low-quality |
| S7_vs_94174\|\|full | Low-quality |
| S7_vs_94199\|\|full | Low-quality |
| S7_vs_94221\|\|full | Low-quality |
| S7_vs_94222\|\|full | Not-determined |
| S7_vs_94236\|\|full | Not-determined |
| S7_vs_94296\|\|full | Low-quality |
| S7_vs_94334\|\|full | Low-quality |
| S7_vs_94382\|\|full | Low-quality |
| S7_vs_94444\|\|full | Low-quality |
| S7_vs_94461\|\|full | Low-quality |
| S7_vs_94481\|\|full | Low-quality |
| S7_vs_94509\|\|full | Low-quality |
| S7_vs_94514\|\|full | Low-quality |
| S7_vs_94528\|\|full | Low-quality |
| S7_vs_94543\|\|full | Not-determined |
| S7_vs_94550\|\|full | Low-quality |
| S7_vs_94649\|\|full | Not-determined |
| S7_vs_94654\|\|full | Low-quality |
| S7_vs_94722\|\|full | Low-quality |
| S7_vs_94736\|\|full | Not-determined |
| S7_vs_94744\|\|full | Low-quality |
| S7_vs_94746\|\|full | Low-quality |
| S7_vs_94761\|\|full | Not-determined |
| S7_vs_94856\|\|full | Low-quality |
| S7_vs_94907\|\|full | Low-quality |
| S7_vs_94929\|\|full | Low-quality |
| S7_vs_94938\|\|full | Not-determined |
| S7_vs_94949\|\|full | Low-quality |
| S7_vs_94961\|\|full | Low-quality |
| S7_vs_94969\|\|full | Not-determined |
| S7_vs_95006\|\|full | Low-quality |
| S7_vs_95072\|\|full | Not-determined |
| S7_vs_95091\|\|full | Low-quality |
| S7_vs_95122\|\|full | Low-quality |
| S7_vs_95132\|\|full | Low-quality |
| S7_vs_95140\|\|full | Low-quality |
| S7_vs_95184\|\|full | Not-determined |
| S7_vs_95186\|\|full | Not-determined |
| S7_vs_95194\|\|full | Low-quality |
| S7_vs_95213\|\|full | Low-quality |
| S7_vs_95220\|\|full | Not-determined |
| S7_vs_95256\|\|full | Low-quality |
| S7_vs_95274\|\|full | Not-determined |
| S7_vs_95330\|\|full | Low-quality |
| S7_vs_95413\|\|full | Not-determined |
| S7_vs_95438\|\|full | Not-determined |
| S7_vs_95619\|\|full | Low-quality |
| S7_vs_95623\|\|full | Low-quality |
| S7_vs_95624\|\|full | Low-quality |
| S7_vs_95652\|\|full | Low-quality |
| S7_vs_95713\|\|full | Low-quality |
| S7_vs_95740\|\|full | Low-quality |
| S7_vs_95749\|\|full | Low-quality |
| S7_vs_95761\|\|full | Not-determined |
| S7_vs_95780\|\|full | Low-quality |
| S7_vs_95794\|\|full | Not-determined |
| S7_vs_95873\|\|full | Low-quality |
| S7_vs_95907\|\|full | Low-quality |
| S7_vs_95938\|\|full | Low-quality |
| S7_vs_95962\|\|full | Not-determined |
| S7_vs_95982\|\|full | Low-quality |
| S7_vs_96036\|\|full | Not-determined |
| S7_vs_96038\|\|full | Not-determined |
| S7_vs_96042\|\|full | Low-quality |
| S7_vs_96135\|\|full | Low-quality |
| S7_vs_96187\|\|full | Low-quality |
| S7_vs_96214\|\|full | Low-quality |
| S7_vs_96307\|\|full | Low-quality |
| S7_vs_96332\|\|full | Not-determined |
| S7_vs_96346\|\|full | Low-quality |
| S7_vs_96361\|\|full | Low-quality |
| S7_vs_96422\|\|full | Not-determined |
| S7_vs_96433\|\|full | Low-quality |
| S7_vs_96439\|\|full | Low-quality |
| S7_vs_96490\|\|full | Low-quality |
| S7_vs_96508\|\|full | Low-quality |
| S7_vs_96521\|\|full | Low-quality |
| S7_vs_96531\|\|full | Low-quality |
| S7_vs_96549\|\|full | Not-determined |
| S7_vs_96582\|\|full | Not-determined |
| S7_vs_96736\|\|full | Not-determined |
| S7_vs_96740\|\|full | Low-quality |
| S7_vs_96792\|\|full | Low-quality |
| S7_vs_96808\|\|full | Low-quality |
| S7_vs_96823\|\|full | Low-quality |
| S7_vs_96846\|\|full | Not-determined |
| S7_vs_96906\|\|full | Low-quality |
| S7_vs_97000\|\|full | Low-quality |
| S7_vs_97012\|\|full | Low-quality |
| S7_vs_97017\|\|full | Not-determined |
| S7_vs_97060\|\|full | Low-quality |
| S7_vs_97061\|\|full | Low-quality |
| S7_vs_97091\|\|full | Low-quality |
| S7_vs_97112\|\|full | Low-quality |
| S7_vs_97204\|\|full | Not-determined |
| S7_vs_97221\|\|full | Not-determined |
| S7_vs_97244\|\|full | Not-determined |
| S7_vs_97312\|\|full | Not-determined |
| S7_vs_97356\|\|full | Low-quality |
| S7_vs_97361\|\|full | Low-quality |
| S7_vs_97443\|\|full | Not-determined |
| S7_vs_97447\|\|full | Not-determined |
| S7_vs_97459\|\|full | Not-determined |
| S7_vs_97481\|\|full | Low-quality |
| S7_vs_97485\|\|full | Not-determined |
| S7_vs_97507\|\|full | Not-determined |
| S7_vs_97542\|\|full | Low-quality |
| S7_vs_97560\|\|full | Low-quality |
| S7_vs_97635\|\|full | Low-quality |
| S7_vs_97652\|\|full | Low-quality |
| S7_vs_97666\|\|full | Low-quality |
| S7_vs_97695\|\|full | Low-quality |
| S7_vs_97705\|\|full | Not-determined |
| S11_vs_k141_23188\|\|full | Medium-quality |
| S7_vs_97772\|\|full | Low-quality |
| S7_vs_97779\|\|full | Low-quality |
| S7_vs_97785\|\|full | Low-quality |
| S7_vs_97799\|\|full | Not-determined |
| S7_vs_97822\|\|full | Low-quality |
| S7_vs_97874\|\|full | Not-determined |
| S7_vs_97884\|\|full | Not-determined |
| S7_vs_97885\|\|full | Low-quality |
| S7_vs_97893\|\|full | Not-determined |
| S7_vs_97913\|\|full | Low-quality |
| S7_vs_97938\|\|full | Not-determined |
| S7_vs_97954\|\|full | Low-quality |
| S7_vs_97982\|\|full | Low-quality |
| S7_vs_97990\|\|full | Not-determined |
| S7_vs_98023\|\|full | Low-quality |
| S7_vs_98038\|\|full | Low-quality |
| S7_vs_98054\|\|full | Low-quality |
| S7_vs_98129\|\|full | Low-quality |
| S7_vs_98142\|\|full | Not-determined |
| S7_vs_98161\|\|full | Low-quality |
| S7_vs_98179\|\|full | Not-determined |
| S7_vs_98220\|\|full | Low-quality |
| S7_vs_98237\|\|full | Not-determined |
| S7_vs_98313\|\|full | Low-quality |
| S7_vs_98434\|\|full | Low-quality |
| S7_vs_98447\|\|full | Low-quality |
| S7_vs_98448\|\|full | Low-quality |
| S7_vs_98467\|\|full | Low-quality |
| S7_vs_98473\|\|full | Low-quality |
| S7_vs_98567\|\|full | Low-quality |
| S7_vs_98579\|\|full | Low-quality |
| S7_vs_98637\|\|full | Low-quality |
| S7_vs_98638\|\|full | Not-determined |
| S7_vs_98642\|\|full | Not-determined |
| S7_vs_98674\|\|full | Low-quality |
| S7_vs_98719\|\|full | Low-quality |
| S7_vs_98721\|\|full | Low-quality |
| S7_vs_98857\|\|full | Low-quality |
| S7_vs_98863\|\|full | Low-quality |
| S7_vs_98887\|\|full | Not-determined |
| S7_vs_98916\|\|full | Low-quality |
| S7_vs_98920\|\|full | Not-determined |
| S7_vs_98924\|\|full | Low-quality |
| S7_vs_98994\|\|full | Low-quality |
| S7_vs_99003\|\|full | Low-quality |
| S7_vs_99014\|\|full | Not-determined |
| S7_vs_99015\|\|full | Low-quality |
| S7_vs_99026\|\|full | Low-quality |
| S7_vs_99054\|\|full | Not-determined |
| S7_vs_99067\|\|full | Low-quality |
| S7_vs_99125\|\|full | Low-quality |
| S7_vs_99160\|\|full | Low-quality |
| S7_vs_99189\|\|full | Not-determined |
| S7_vs_99275\|\|full | Not-determined |
| S7_vs_99310\|\|full | Not-determined |
| S7_vs_99340\|\|full | Low-quality |
| S7_vs_99361\|\|full | Not-determined |
| S7_vs_99413\|\|full | Not-determined |
| S7_vs_99448\|\|full | Low-quality |
| S7_vs_99488\|\|full | Low-quality |
| S7_vs_99556\|\|full | Low-quality |
| S7_vs_99591\|\|full | Low-quality |
| S7_vs_99598\|\|full | Not-determined |
| S7_vs_99634\|\|full | Low-quality |
| S7_vs_99674\|\|full | Not-determined |
| S7_vs_99676\|\|full | Low-quality |
| S7_vs_99743\|\|full | Low-quality |
| S7_vs_99784\|\|full | Low-quality |
| S7_vs_99841\|\|full | Low-quality |
| S7_vs_99889\|\|full | Not-determined |
| S7_vs_99896\|\|full | Not-determined |
| S7_vs_99907\|\|full | Low-quality |
| S7_vs_99910\|\|full | Not-determined |
| S7_vs_99918\|\|full | Low-quality |
| S7_vs_99993\|\|full | Low-quality |
| S7_vs_100021\|\|full | Low-quality |
| S7_vs_100045\|\|full | Low-quality |
| S7_vs_100049\|\|full | Low-quality |
| S7_vs_100120\|\|full | Low-quality |
| S7_vs_100194\|\|full | Not-determined |
| S7_vs_100242\|\|full | Low-quality |
| S7_vs_100268\|\|full | Low-quality |
| S7_vs_100278\|\|full | Low-quality |
| S7_vs_100285\|\|full | Not-determined |
| S7_vs_100394\|\|full | Low-quality |
| S7_vs_100413\|\|full | Not-determined |
| S7_vs_100449\|\|full | Low-quality |
| S7_vs_100520\|\|full | Not-determined |
| S7_vs_100535\|\|full | Low-quality |
| S7_vs_100555\|\|full | Low-quality |
| S7_vs_100564\|\|full | Low-quality |
| S7_vs_100577\|\|full | Low-quality |
| S7_vs_100609\|\|full | Not-determined |
| S7_vs_138097\|\|full | High-quality |
| S7_vs_100647\|\|full | Low-quality |
| S7_vs_100649\|\|full | Low-quality |
| S7_vs_100659\|\|full | Low-quality |
| S7_vs_100664\|\|full | Low-quality |
| S7_vs_100677\|\|full | Not-determined |
| S7_vs_100691\|\|full | Low-quality |
| S7_vs_100695\|\|full | Low-quality |
| S7_vs_100741\|\|full | Low-quality |
| S7_vs_100745\|\|full | Not-determined |
| S7_vs_100760\|\|full | Not-determined |
| S7_vs_100805\|\|full | Low-quality |
| S7_vs_100814\|\|full | Low-quality |
| S7_vs_100830\|\|full | Not-determined |
| S7_vs_100889\|\|full | Low-quality |
| S7_vs_100891\|\|full | Low-quality |
| S7_vs_100895\|\|full | Low-quality |
| S7_vs_100935\|\|full | Low-quality |
| S7_vs_100949\|\|full | Not-determined |
| S7_vs_100966\|\|full | Low-quality |
| S7_vs_100977\|\|full | Low-quality |
| S7_vs_100992\|\|full | Low-quality |
| S7_vs_101006\|\|full | Not-determined |
| S7_vs_101069\|\|full | Low-quality |
| S7_vs_101078\|\|full | Not-determined |
| S7_vs_101079\|\|full | Low-quality |
| S7_vs_101084\|\|full | Low-quality |
| S7_vs_101104\|\|full | Not-determined |
| S7_vs_101128\|\|full | Not-determined |
| S7_vs_101141\|\|full | Low-quality |
| S7_vs_101221\|\|full | Not-determined |
| S7_vs_101225\|\|full | Low-quality |
| S7_vs_101231\|\|full | Not-determined |
| S7_vs_101248\|\|full | Low-quality |
| S7_vs_101259\|\|full | Low-quality |
| S7_vs_101263\|\|full | Not-determined |
| S7_vs_101267\|\|full | Not-determined |
| S7_vs_101285\|\|full | Low-quality |
| S7_vs_101303\|\|full | Low-quality |
| S7_vs_101317\|\|full | Low-quality |
| S7_vs_101433\|\|full | Low-quality |
| S7_vs_101455\|\|full | Low-quality |
| S7_vs_101456\|\|full | Not-determined |
| S7_vs_101469\|\|full | Low-quality |
| S7_vs_101474\|\|full | Not-determined |
| S7_vs_101544\|\|full | Low-quality |
| S7_vs_101554\|\|full | Low-quality |
| S7_vs_101564\|\|full | Not-determined |
| S7_vs_101586\|\|full | Low-quality |
| S7_vs_101589\|\|full | Low-quality |
| S7_vs_101606\|\|full | Low-quality |
| S7_vs_101607\|\|full | Low-quality |
| S7_vs_101616\|\|full | Not-determined |
| S7_vs_101631\|\|full | Low-quality |
| S7_vs_101654\|\|full | Low-quality |
| S7_vs_101685\|\|full | Low-quality |
| S7_vs_101689\|\|full | Low-quality |
| S7_vs_101714\|\|full | Not-determined |
| S7_vs_101718\|\|full | Low-quality |
| S7_vs_101728\|\|full | Not-determined |
| S5_vs_k141_84946\|\|full | High-quality |
| S7_vs_101812\|\|full | Low-quality |
| S7_vs_101815\|\|full | Not-determined |

| | |
|---|---|
| S7_vs_101903\|\|full | Low-quality |
| S7_vs_101919\|\|full | Not-determined |
| S7_vs_101942\|\|full | Low-quality |
| S7_vs_101979\|\|full | Low-quality |
| S7_vs_101993\|\|full | Low-quality |
| S7_vs_102067\|\|full | Low-quality |
| S7_vs_102072\|\|full | Not-determined |
| S7_vs_102087\|\|full | Low-quality |
| S7_vs_102125\|\|full | Not-determined |
| S7_vs_102151\|\|full | Not-determined |
| S7_vs_102181\|\|full | Low-quality |
| S7_vs_102256\|\|full | Low-quality |
| S7_vs_102283\|\|full | Not-determined |
| S7_vs_102327\|\|full | Low-quality |
| S7_vs_102374\|\|full | Low-quality |
| S7_vs_102386\|\|full | Low-quality |
| S7_vs_102388\|\|full | Not-determined |
| S7_vs_102394\|\|full | Not-determined |
| S7_vs_102401\|\|full | Low-quality |
| S7_vs_102486\|\|full | Not-determined |
| S7_vs_102499\|\|full | Not-determined |
| S7_vs_102524\|\|full | Low-quality |
| S7_vs_102598\|\|full | Not-determined |
| S7_vs_102671\|\|full | Low-quality |
| S7_vs_102700\|\|full | Low-quality |
| S7_vs_102738\|\|full | Low-quality |
| S7_vs_102751\|\|full | Not-determined |
| S7_vs_102763\|\|full | Low-quality |
| S7_vs_102772\|\|full | Low-quality |
| S7_vs_102789\|\|full | Low-quality |
| S7_vs_102802\|\|full | Not-determined |
| S7_vs_102807\|\|full | Not-determined |
| S7_vs_102842\|\|full | Low-quality |
| S7_vs_102844\|\|full | Not-determined |
| S7_vs_102855\|\|full | Low-quality |
| S7_vs_102895\|\|full | Low-quality |
| S7_vs_102924\|\|full | Low-quality |
| S7_vs_103011\|\|full | Not-determined |
| S7_vs_103045\|\|full | Low-quality |
| S7_vs_103054\|\|full | Not-determined |
| S7_vs_103071\|\|full | Not-determined |
| S7_vs_103093\|\|full | Not-determined |
| S7_vs_103172\|\|full | Not-determined |
| S7_vs_103205\|\|full | Not-determined |
| S7_vs_103208\|\|full | Not-determined |
| S7_vs_103336\|\|full | Low-quality |
| S7_vs_103373\|\|full | Low-quality |
| S7_vs_103404\|\|full | Low-quality |
| S7_vs_103425\|\|full | Low-quality |
| S7_vs_103428\|\|full | Not-determined |
| S7_vs_103450\|\|full | Low-quality |
| S7_vs_103555\|\|full | Low-quality |
| S7_vs_103593\|\|full | Low-quality |
| S7_vs_103612\|\|full | Low-quality |
| S7_vs_103637\|\|full | Low-quality |
| S7_vs_103674\|\|full | Low-quality |
| S7_vs_103690\|\|full | Low-quality |
| S7_vs_103726\|\|full | Low-quality |
| S7_vs_103783\|\|full | Low-quality |
| S7_vs_103812\|\|full | Not-determined |
| S7_vs_103833\|\|full | Not-determined |
| S7_vs_103838\|\|full | Low-quality |
| S7_vs_103880\|\|full | Not-determined |
| S7_vs_103896\|\|full | Low-quality |
| S7_vs_103942\|\|full | Not-determined |
| S7_vs_103974\|\|full | Low-quality |
| S7_vs_104013\|\|full | Low-quality |
| S7_vs_104033\|\|full | Low-quality |
| S7_vs_104036\|\|full | Not-determined |
| S7_vs_104038\|\|full | Not-determined |
| S7_vs_104101\|\|full | Not-determined |
| S7_vs_104131\|\|full | Low-quality |
| S7_vs_104161\|\|full | Low-quality |
| S7_vs_104193\|\|full | Not-determined |
| S7_vs_104195\|\|full | Not-determined |
| S7_vs_104204\|\|full | Not-determined |
| S7_vs_104312\|\|full | Not-determined |
| S7_vs_104344\|\|full | Not-determined |
| S7_vs_104346\|\|full | Low-quality |
| S7_vs_104348\|\|full | Low-quality |
| S7_vs_104370\|\|full | Low-quality |
| S7_vs_104392\|\|full | Not-determined |
| S7_vs_104493\|\|full | Not-determined |
| S7_vs_104517\|\|full | Low-quality |
| S7_vs_104533\|\|full | Not-determined |
| S7_vs_104555\|\|full | Not-determined |
| S7_vs_104557\|\|full | Not-determined |
| S7_vs_104572\|\|full | Low-quality |
| S7_vs_104576\|\|full | Low-quality |
| S7_vs_104590\|\|full | Low-quality |
| S7_vs_104650\|\|full | Low-quality |
| S7_vs_104661\|\|full | Low-quality |
| S7_vs_104695\|\|full | Low-quality |
| S7_vs_104765\|\|full | Low-quality |
| S7_vs_104779\|\|full | Not-determined |
| S7_vs_104825\|\|full | Low-quality |
| S7_vs_104864\|\|full | Not-determined |
| S7_vs_104969\|\|full | Low-quality |
| S7_vs_104986\|\|full | Not-determined |
| S7_vs_105009\|\|full | Not-determined |
| S7_vs_105053\|\|full | Low-quality |
| S7_vs_105065\|\|full | Low-quality |
| S7_vs_105147\|\|full | Not-determined |
| S7_vs_105150\|\|full | Low-quality |
| S7_vs_105201\|\|full | Low-quality |
| S7_vs_105210\|\|full | Not-determined |
| S7_vs_105216\|\|full | Low-quality |
| S7_vs_105245\|\|full | Not-determined |
| S7_vs_105278\|\|full | Low-quality |
| S7_vs_105288\|\|full | Low-quality |
| S7_vs_105290\|\|full | Not-determined |
| S7_vs_105297\|\|full | Low-quality |
| S7_vs_105393\|\|full | Not-determined |
| S7_vs_105402\|\|full | Not-determined |
| S7_vs_105470\|\|full | Low-quality |
| S7_vs_105472\|\|full | Low-quality |
| S7_vs_105498\|\|full | Low-quality |
| S7_vs_105515\|\|full | Low-quality |
| S7_vs_105546\|\|full | Not-determined |
| S7_vs_105671\|\|full | Low-quality |
| S7_vs_105752\|\|full | Not-determined |
| S7_vs_105754\|\|full | Low-quality |
| S7_vs_105784\|\|full | Low-quality |
| S7_vs_105809\|\|full | Low-quality |
| S7_vs_105816\|\|full | Low-quality |
| S7_vs_105855\|\|full | Not-determined |
| S7_vs_105870\|\|full | Low-quality |
| S7_vs_105916\|\|full | Low-quality |
| S7_vs_105921\|\|full | Not-determined |
| S7_vs_105936\|\|full | Low-quality |
| S7_vs_105946\|\|full | Low-quality |
| S7_vs_105962\|\|full | Low-quality |
| S7_vs_105965\|\|full | Not-determined |
| S7_vs_105989\|\|full | Low-quality |
| S7_vs_106001\|\|full | Low-quality |
| S7_vs_106077\|\|full | Low-quality |
| S7_vs_106088\|\|full | Low-quality |
| S7_vs_106108\|\|full | Not-determined |
| S7_vs_106116\|\|full | Low-quality |
| S7_vs_106117\|\|full | Low-quality |
| S7_vs_106166\|\|full | Low-quality |
| S7_vs_106248\|\|full | Not-determined |
| S7_vs_106286\|\|full | Not-determined |
| S7_vs_106314\|\|full | Low-quality |
| S7_vs_106315\|\|full | Low-quality |
| S7_vs_106350\|\|full | Low-quality |
| S7_vs_106377\|\|full | Not-determined |
| S7_vs_106454\|\|full | Not-determined |
| S7_vs_106464\|\|full | Not-determined |
| S7_vs_106487\|\|full | Low-quality |
| S7_vs_106493\|\|full | Low-quality |
| S7_vs_106506\|\|full | Low-quality |
| S7_vs_106518\|\|full | Low-quality |
| S7_vs_106529\|\|full | Low-quality |
| S7_vs_106551\|\|full | Low-quality |
| S7_vs_106568\|\|full | Not-determined |
| S7_vs_106578\|\|full | Low-quality |
| S7_vs_106597\|\|full | Not-determined |
| S7_vs_106605\|\|full | Not-determined |
| S7_vs_106651\|\|full | Not-determined |
| S7_vs_106680\|\|full | Low-quality |
| S7_vs_106704\|\|full | Not-determined |
| S7_vs_106740\|\|full | Not-determined |
| S7_vs_106752\|\|full | Low-quality |
| S7_vs_106822\|\|full | Not-determined |
| S7_vs_106871\|\|full | Low-quality |
| S7_vs_106875\|\|full | Low-quality |
| S7_vs_106897\|\|full | Low-quality |
| S7_vs_106912\|\|full | Not-determined |
| S7_vs_106918\|\|full | Low-quality |
| S7_vs_106924\|\|full | Not-determined |
| S7_vs_106960\|\|full | Low-quality |
| S7_vs_106963\|\|full | Not-determined |
| S7_vs_106978\|\|full | Low-quality |
| S7_vs_107015\|\|full | Not-determined |
| S7_vs_107043\|\|full | Not-determined |
| S7_vs_107073\|\|full | Low-quality |
| S7_vs_107104\|\|full | Not-determined |
| S7_vs_107107\|\|full | Not-determined |
| S7_vs_107152\|\|full | Not-determined |
| S7_vs_107164\|\|full | Not-determined |
| S7_vs_107206\|\|full | Not-determined |
| S7_vs_107211\|\|full | Low-quality |
| S7_vs_107250\|\|full | Not-determined |
| S7_vs_107280\|\|full | Low-quality |
| S7_vs_107314\|\|full | Not-determined |
| S7_vs_107343\|\|full | Low-quality |
| S7_vs_107348\|\|full | Not-determined |
| S7_vs_107369\|\|full | Not-determined |
| S7_vs_107391\|\|full | Low-quality |
| S7_vs_107467\|\|full | Low-quality |
| S7_vs_107485\|\|full | Low-quality |
| S7_vs_107497\|\|full | Not-determined |
| S7_vs_107526\|\|full | Not-determined |
| S7_vs_107572\|\|full | Not-determined |
| S7_vs_107595\|\|full | Low-quality |
| S7_vs_107659\|\|full | Not-determined |
| S7_vs_107691\|\|full | Low-quality |
| S7_vs_107715\|\|full | Not-determined |
| S7_vs_107731\|\|full | Low-quality |
| S7_vs_107735\|\|full | Not-determined |
| S7_vs_107778\|\|full | Low-quality |
| S7_vs_107785\|\|full | Low-quality |
| S7_vs_107792\|\|full | Not-determined |
| S7_vs_107811\|\|full | Not-determined |
| S7_vs_107820\|\|full | Not-determined |
| S7_vs_107862\|\|full | Low-quality |
| S7_vs_107866\|\|full | Low-quality |
| S14_vs_k141_14737\|\|full | High-quality |
| S7_vs_107881\|\|full | Low-quality |
| S7_vs_107889\|\|full | Low-quality |
| S7_vs_107906\|\|full | Low-quality |
| S7_vs_107933\|\|full | Low-quality |
| S7_vs_107940\|\|full | Low-quality |
| S7_vs_107990\|\|full | Low-quality |
| S7_vs_107999\|\|full | Low-quality |
| S7_vs_108000\|\|full | Not-determined |
| S7_vs_108010\|\|full | Low-quality |
| S7_vs_108075\|\|full | Not-determined |
| S7_vs_108113\|\|full | Not-determined |
| S7_vs_108155\|\|full | Not-determined |
| S7_vs_108179\|\|full | Low-quality |
| S7_vs_108191\|\|full | Low-quality |
| S7_vs_108199\|\|full | Low-quality |
| S7_vs_108211\|\|full | Low-quality |
| S7_vs_108239\|\|full | Low-quality |
| S7_vs_108304\|\|full | Low-quality |
| S7_vs_108345\|\|full | Not-determined |
| S7_vs_108368\|\|full | Low-quality |
| S7_vs_108378\|\|full | Not-determined |
| S7_vs_108395\|\|full | Low-quality |
| S7_vs_108397\|\|full | Low-quality |
| S7_vs_108398\|\|full | Low-quality |
| S7_vs_108433\|\|full | Low-quality |
| S7_vs_108436\|\|full | Low-quality |
| S7_vs_108442\|\|full | Low-quality |
| S7_vs_108450\|\|full | Low-quality |
| S7_vs_108478\|\|full | Not-determined |
| S7_vs_108511\|\|full | Low-quality |
| S7_vs_108514\|\|full | Low-quality |
| S7_vs_108526\|\|full | Not-determined |
| S7_vs_108544\|\|full | Low-quality |
| S7_vs_108580\|\|full | Low-quality |
| S7_vs_108610\|\|full | Low-quality |
| S7_vs_108612\|\|full | Not-determined |
| S7_vs_108664\|\|full | Low-quality |
| S9_vs_k141_62803\|\|full | High-quality |
| S7_vs_108686\|\|full | Not-determined |
| S7_vs_108708\|\|full | Low-quality |
| S7_vs_108795\|\|full | Low-quality |
| S7_vs_108833\|\|full | Not-determined |
| S7_vs_108851\|\|full | Not-determined |
| S7_vs_108865\|\|full | Not-determined |
| S7_vs_108887\|\|full | Not-determined |
| S7_vs_108892\|\|full | Low-quality |
| S7_vs_108919\|\|full | Low-quality |
| S7_vs_108931\|\|full | Low-quality |
| S7_vs_108949\|\|full | Low-quality |
| S7_vs_108958\|\|full | Not-determined |
| S7_vs_109019\|\|full | Not-determined |
| S7_vs_109090\|\|full | Low-quality |
| S7_vs_109114\|\|full | Low-quality |
| S7_vs_109126\|\|full | Low-quality |
| S7_vs_109180\|\|full | Low-quality |
| S7_vs_109186\|\|full | Low-quality |
| S7_vs_109191\|\|full | Low-quality |
| S7_vs_109197\|\|full | Low-quality |
| S7_vs_109229\|\|full | Low-quality |
| S7_vs_109354\|\|full | Not-determined |
| S7_vs_109474\|\|full | Low-quality |
| S7_vs_109486\|\|full | Not-determined |
| S7_vs_109489\|\|full | Not-determined |
| S5_vs_k141_63924\|\|full | High-quality |
| S7_vs_109558\|\|full | Low-quality |
| S7_vs_109582\|\|full | Not-determined |
| S7_vs_109603\|\|full | Low-quality |
| S7_vs_109649\|\|full | Low-quality |
| S7_vs_109651\|\|full | Low-quality |
| S7_vs_109656\|\|full | Not-determined |
| S7_vs_109694\|\|full | Low-quality |
| S7_vs_109696\|\|full | Low-quality |
| S7_vs_109755\|\|full | Low-quality |
| S7_vs_109773\|\|full | Not-determined |
| S7_vs_109817\|\|full | Not-determined |
| S7_vs_109837\|\|full | Low-quality |
| S7_vs_109847\|\|full | Low-quality |
| S7_vs_109860\|\|full | Low-quality |
| S7_vs_109880\|\|full | Low-quality |
| S7_vs_109887\|\|full | Not-determined |
| S7_vs_109895\|\|full | Low-quality |
| S7_vs_109901\|\|full | Low-quality |
| S7_vs_109915\|\|full | Low-quality |
| S7_vs_109919\|\|full | Not-determined |
| S7_vs_109990\|\|full | Low-quality |
| S7_vs_110016\|\|full | Low-quality |
| S7_vs_110024\|\|full | Not-determined |
| S7_vs_110039\|\|full | Low-quality |
| S7_vs_110046\|\|full | Low-quality |
| S7_vs_110074\|\|full | Not-determined |
| S7_vs_110104\|\|full | Low-quality |
| S7_vs_110112\|\|full | Low-quality |
| S7_vs_110141\|\|full | Not-determined |
| S7_vs_110152\|\|full | Low-quality |
| S7_vs_110164\|\|full | Not-determined |
| S7_vs_110184\|\|full | Low-quality |
| S7_vs_110206\|\|full | Low-quality |
| S7_vs_110215\|\|full | Low-quality |
| S7_vs_110258\|\|full | Not-determined |
| S7_vs_110291\|\|full | Not-determined |
| S7_vs_110318\|\|full | Low-quality |
| S7_vs_110321\|\|full | Not-determined |
| S7_vs_110362\|\|full | Low-quality |
| S7_vs_110371\|\|full | Low-quality |
| S7_vs_110383\|\|full | Low-quality |
| S7_vs_110421\|\|full | Low-quality |
| S7_vs_110430\|\|full | Low-quality |
| S7_vs_110505\|\|full | Low-quality |
| S7_vs_110511\|\|full | Low-quality |
| S7_vs_110667\|\|full | Low-quality |
| S7_vs_110668\|\|full | Low-quality |
| S7_vs_110715\|\|full | Low-quality |
| S7_vs_110722\|\|full | Low-quality |
| S7_vs_110748\|\|full | Low-quality |
| S7_vs_110761\|\|full | Low-quality |
| S7_vs_110778\|\|full | Not-determined |
| S7_vs_110826\|\|full | Not-determined |
| S7_vs_110914\|\|full | Low-quality |
| S7_vs_110926\|\|full | Low-quality |
| S7_vs_110932\|\|full | Not-determined |
| S7_vs_110952\|\|full | Low-quality |
| S7_vs_110998\|\|full | Not-determined |
| S7_vs_111011\|\|full | Low-quality |
| S7_vs_111035\|\|full | Low-quality |
| S7_vs_111049\|\|full | Low-quality |
| S7_vs_111073\|\|full | Low-quality |
| S7_vs_111077\|\|full | Not-determined |
| S7_vs_111085\|\|full | Not-determined |
| S7_vs_111176\|\|full | Low-quality |
| S7_vs_111184\|\|full | Low-quality |
| S7_vs_111205\|\|full | Not-determined |
| S7_vs_111248\|\|full | Low-quality |
| S7_vs_111300\|\|full | Low-quality |
| S7_vs_111317\|\|full | Low-quality |
| S7_vs_111328\|\|full | Low-quality |
| S7_vs_111329\|\|full | Low-quality |
| S7_vs_111346\|\|full | Low-quality |
| S7_vs_111532\|\|full | Low-quality |
| S7_vs_111545\|\|full | Not-determined |
| S7_vs_111553\|\|full | Low-quality |
| S7_vs_111555\|\|full | Low-quality |
| S7_vs_111614\|\|full | Low-quality |
| S7_vs_111617\|\|full | Not-determined |
| S7_vs_111619\|\|full | Not-determined |
| S7_vs_111655\|\|full | Not-determined |
| S7_vs_111681\|\|full | Low-quality |
| S7_vs_111735\|\|full | Low-quality |
| S16_vs_96481\|\|full | Medium-quality |
| S7_vs_111863\|\|full | Not-determined |
| S7_vs_111879\|\|full | Not-determined |
| S7_vs_111893\|\|full | Low-quality |
| S7_vs_111896\|\|full | Low-quality |
| S7_vs_112015\|\|full | Not-determined |
| S7_vs_112026\|\|full | Low-quality |
| S7_vs_112051\|\|full | Low-quality |
| S7_vs_112117\|\|full | Low-quality |
| S7_vs_112192\|\|full | Low-quality |
| S7_vs_112203\|\|full | Low-quality |
| S7_vs_112213\|\|full | Low-quality |
| S7_vs_112228\|\|full | Low-quality |
| S7_vs_112239\|\|full | Low-quality |
| S7_vs_112321\|\|full | Low-quality |
| S7_vs_112328\|\|full | Not-determined |
| S7_vs_112332\|\|full | Low-quality |
| S10_vs_77004\|\|full | High-quality |
| S7_vs_112407\|\|full | Not-determined |
| S7_vs_112419\|\|full | Not-determined |
| S7_vs_112431\|\|full | Low-quality |
| S7_vs_112436\|\|full | Low-quality |
| S7_vs_112437\|\|full | Not-determined |
| S7_vs_112452\|\|full | Low-quality |
| S7_vs_112460\|\|full | Low-quality |
| S7_vs_112466\|\|full | Low-quality |
| S7_vs_112478\|\|full | Low-quality |
| S7_vs_112490\|\|full | Low-quality |
| S7_vs_112498\|\|full | Not-determined |
| S7_vs_112526\|\|full | Low-quality |
| S7_vs_112571\|\|full | Low-quality |
| S7_vs_112574\|\|full | Low-quality |
| S7_vs_112582\|\|full | Low-quality |
| S7_vs_112607\|\|full | Low-quality |
| S7_vs_112689\|\|full | Not-determined |
| S7_vs_112762\|\|full | Not-determined |
| S7_vs_112765\|\|full | Low-quality |
| S7_vs_112808\|\|full | Low-quality |
| S7_vs_112830\|\|full | Not-determined |
| S7_vs_112831\|\|full | Low-quality |
| S7_vs_112865\|\|full | Low-quality |
| S7_vs_112877\|\|full | Not-determined |
| S7_vs_112910\|\|full | Low-quality |
| S11_vs_k141_1937\|\|full | Medium-quality |
| S7_vs_112917\|\|full | Low-quality |
| S7_vs_112931\|\|full | Not-determined |
| S7_vs_112958\|\|full | Not-determined |
| S7_vs_112960\|\|full | Not-determined |
| S7_vs_112968\|\|full | Low-quality |
| S7_vs_112992\|\|full | Low-quality |
| S7_vs_113013\|\|full | Low-quality |
| S7_vs_113034\|\|full | Not-determined |
| S7_vs_113070\|\|full | Not-determined |
| S7_vs_113077\|\|full | Low-quality |
| S7_vs_113096\|\|full | Low-quality |
| S7_vs_113128\|\|full | Low-quality |
| S7_vs_113142\|\|full | Not-determined |
| S7_vs_113157\|\|full | Not-determined |
| S7_vs_113193\|\|full | Low-quality |
| S7_vs_113221\|\|full | Not-determined |
| S7_vs_113240\|\|full | Low-quality |
| S7_vs_113314\|\|full | Low-quality |
| S7_vs_113318\|\|full | Low-quality |
| S7_vs_113329\|\|full | Low-quality |
| S7_vs_113345\|\|full | Not-determined |
| S7_vs_113356\|\|full | Low-quality |
| S7_vs_113362\|\|full | Low-quality |
| S7_vs_113393\|\|full | Low-quality |
| S7_vs_113434\|\|full | Low-quality |
| S7_vs_113447\|\|full | Not-determined |
| S7_vs_113456\|\|full | Low-quality |
| S7_vs_113482\|\|full | Not-determined |
| S7_vs_113484\|\|full | Low-quality |
| S7_vs_113497\|\|full | Low-quality |
| S7_vs_113547\|\|full | Not-determined |
| S7_vs_113563\|\|full | Low-quality |
| S7_vs_113579\|\|full | Low-quality |
| S7_vs_113643\|\|full | Not-determined |
| S7_vs_113708\|\|full | Low-quality |
| S7_vs_113728\|\|full | Not-determined |
| S7_vs_113750\|\|full | Not-determined |
| S7_vs_113775\|\|full | Low-quality |
| S7_vs_113812\|\|full | Low-quality |
| S7_vs_113813\|\|full | Low-quality |
| S7_vs_113889\|\|full | Not-determined |
| S7_vs_113906\|\|full | Not-determined |
| S7_vs_113927\|\|full | Not-determined |
| S7_vs_113959\|\|full | Low-quality |
| S7_vs_113961\|\|full | Not-determined |
| S7_vs_113971\|\|full | Low-quality |
| S7_vs_114008\|\|full | Low-quality |
| S7_vs_114024\|\|full | Low-quality |
| S7_vs_114083\|\|full | Low-quality |
| S7_vs_114238\|\|full | Not-determined |
| S7_vs_114246\|\|full | Low-quality |
| S7_vs_114260\|\|full | Low-quality |
| S7_vs_114293\|\|full | Low-quality |
| S7_vs_114300\|\|full | Low-quality |
| S7_vs_114308\|\|full | Low-quality |
| S7_vs_114310\|\|full | Low-quality |
| S7_vs_114338\|\|full | Low-quality |
| S7_vs_114378\|\|full | Not-determined |
| S7_vs_114424\|\|full | Low-quality |
| S7_vs_129271\|\|full | Medium-quality |
| S7_vs_114492\|\|full | Not-determined |
| S7_vs_114502\|\|full | Low-quality |
| S7_vs_114506\|\|full | Not-determined |
| S7_vs_114549\|\|full | Not-determined |
| S7_vs_114555\|\|full | Not-determined |
| S7_vs_114588\|\|full | Not-determined |
| S7_vs_114590\|\|full | Not-determined |
| S7_vs_114633\|\|full | Not-determined |
| S7_vs_114700\|\|full | Not-determined |
| S7_vs_114722\|\|full | Low-quality |
| S7_vs_114746\|\|full | Not-determined |
| S7_vs_114763\|\|full | Not-determined |
| S7_vs_114775\|\|full | Low-quality |
| S7_vs_114786\|\|full | Low-quality |
| S7_vs_114787\|\|full | Low-quality |
| S7_vs_114805\|\|full | Low-quality |
| S7_vs_114808\|\|full | Low-quality |
| S7_vs_114809\|\|full | Not-determined |
| S7_vs_114815\|\|full | Low-quality |
| S7_vs_114845\|\|full | Low-quality |
| S7_vs_114884\|\|full | Not-determined |
| S7_vs_114917\|\|full | Low-quality |
| S7_vs_114936\|\|full | Low-quality |
| S7_vs_114943\|\|full | Not-determined |
| S7_vs_114945\|\|full | Low-quality |
| S7_vs_114947\|\|full | Not-determined |
| S7_vs_114949\|\|full | Low-quality |
| S7_vs_114994\|\|full | Low-quality |
| S7_vs_114995\|\|full | Not-determined |
| S7_vs_115095\|\|full | Not-determined |
| S7_vs_115098\|\|full | Not-determined |
| S7_vs_115102\|\|full | Not-determined |
| S7_vs_115117\|\|full | Low-quality |
| S7_vs_115139\|\|full | Not-determined |
| S7_vs_115172\|\|full | Low-quality |
| S7_vs_115186\|\|full | Not-determined |
| S7_vs_115235\|\|full | Low-quality |
| S7_vs_115251\|\|full | Low-quality |
| S7_vs_115285\|\|full | Low-quality |
| S7_vs_115295\|\|full | Low-quality |
| S7_vs_115320\|\|full | Low-quality |
| S7_vs_115377\|\|full | Low-quality |
| S7_vs_115386\|\|full | Low-quality |
| S7_vs_115418\|\|full | Low-quality |
| S7_vs_115421\|\|full | Not-determined |
| S7_vs_115422\|\|full | Low-quality |
| S7_vs_115430\|\|full | Low-quality |
| S7_vs_115455\|\|full | Low-quality |
| S7_vs_115457\|\|full | Low-quality |
| S7_vs_115525\|\|full | Low-quality |
| S7_vs_115582\|\|full | Low-quality |
| S7_vs_115595\|\|full | Not-determined |
| S7_vs_115641\|\|full | Not-determined |
| S7_vs_115643\|\|full | Low-quality |
| S7_vs_115653\|\|full | Not-determined |
| S7_vs_115681\|\|full | Low-quality |
| S7_vs_115692\|\|full | Low-quality |
| S7_vs_115722\|\|full | Low-quality |
| S7_vs_115743\|\|full | Low-quality |
| S7_vs_115772\|\|full | Low-quality |
| S7_vs_115787\|\|full | Low-quality |
| S7_vs_115900\|\|full | Not-determined |
| S7_vs_115989\|\|full | Not-determined |
| S7_vs_115992\|\|full | Low-quality |
| S7_vs_116010\|\|full | Low-quality |
| S7_vs_116018\|\|full | Not-determined |
| S7_vs_116044\|\|full | Low-quality |
| S7_vs_116053\|\|full | Low-quality |
| S7_vs_116070\|\|full | Low-quality |
| S7_vs_116146\|\|full | Not-determined |
| S7_vs_116187\|\|full | Low-quality |
| S7_vs_116192\|\|full | Low-quality |
| S7_vs_116238\|\|full | Low-quality |
| S7_vs_116353\|\|full | Not-determined |
| S7_vs_116378\|\|full | Not-determined |
| S7_vs_116446\|\|full | Not-determined |
| S7_vs_116447\|\|full | Not-determined |
| S7_vs_116462\|\|full | Not-determined |
| S7_vs_116465\|\|full | Low-quality |
| S7_vs_116468\|\|full | Low-quality |
| S7_vs_116485\|\|full | Low-quality |
| S7_vs_116510\|\|full | Low-quality |
| S7_vs_116551\|\|full | Low-quality |
| S7_vs_116553\|\|full | Not-determined |
| S7_vs_116560\|\|full | Low-quality |
| S7_vs_116567\|\|full | Low-quality |
| S7_vs_116588\|\|full | Low-quality |
| S7_vs_116644\|\|full | Low-quality |
| S7_vs_116704\|\|full | Not-determined |
| S7_vs_116765\|\|full | Low-quality |
| S7_vs_116771\|\|full | Low-quality |
| S7_vs_116778\|\|full | Not-determined |
| S7_vs_116780\|\|full | Low-quality |
| S7_vs_116793\|\|full | Not-determined |
| S7_vs_116801\|\|full | Not-determined |
| S7_vs_116810\|\|full | Low-quality |
| S7_vs_116816\|\|full | Not-determined |
| S7_vs_116879\|\|full | Low-quality |
| S7_vs_116884\|\|full | Low-quality |
| S7_vs_116965\|\|full | Not-determined |
| S7_vs_116967\|\|full | Low-quality |
| S7_vs_116987\|\|full | Low-quality |
| S7_vs_117006\|\|full | Low-quality |
| S7_vs_117038\|\|full | Not-determined |
| S7_vs_117084\|\|full | Low-quality |
| S7_vs_117104\|\|full | Not-determined |
| S7_vs_117140\|\|full | Not-determined |
| S7_vs_117189\|\|full | Low-quality |
| S7_vs_117224\|\|full | Not-determined |
| S7_vs_117230\|\|full | Low-quality |
| S7_vs_117236\|\|full | Not-determined |
| S7_vs_117246\|\|full | Low-quality |
| S7_vs_117304\|\|full | Not-determined |
| S7_vs_117328\|\|full | Low-quality |
| S7_vs_117336\|\|full | Low-quality |
| S7_vs_117341\|\|full | Low-quality |
| S7_vs_117352\|\|full | Low-quality |
| S7_vs_117378\|\|full | Not-determined |
| S7_vs_117395\|\|full | Low-quality |
| S7_vs_117406\|\|full | Low-quality |
| S7_vs_117429\|\|full | Low-quality |
| S7_vs_117442\|\|full | Low-quality |
| S7_vs_117452\|\|full | Low-quality |
| S7_vs_117474\|\|full | Not-determined |
| S7_vs_117520\|\|full | Not-determined |
| S7_vs_117570\|\|full | Not-determined |
| S7_vs_117574\|\|full | Not-determined |
| S7_vs_117612\|\|full | Low-quality |
| S7_vs_117621\|\|full | Low-quality |

| | |
|---|---|
| S7_vs_117659\|\|full | Low-quality |
| S7_vs_117667\|\|full | Low-quality |
| S7_vs_117686\|\|full | Not-determined |
| S5_vs_k141_65490\|\|0_partial | High-quality |
| S7_vs_117717\|\|full | Low-quality |
| S7_vs_117784\|\|full | Low-quality |
| S7_vs_117795\|\|full | Low-quality |
| S7_vs_117804\|\|full | Not-determined |
| S7_vs_117875\|\|full | Not-determined |
| S7_vs_117897\|\|full | Low-quality |
| S7_vs_117998\|\|full | Low-quality |
| S7_vs_118006\|\|full | Low-quality |
| S7_vs_118026\|\|full | Low-quality |
| S7_vs_118033\|\|full | Low-quality |
| S7_vs_118062\|\|full | Low-quality |
| S7_vs_118073\|\|full | Not-determined |
| S7_vs_118075\|\|full | Low-quality |
| S7_vs_118089\|\|full | Low-quality |
| S7_vs_118106\|\|full | Not-determined |
| S7_vs_118133\|\|full | Low-quality |
| S7_vs_118167\|\|full | Low-quality |
| S7_vs_118181\|\|full | Low-quality |
| S7_vs_118226\|\|full | Low-quality |
| S7_vs_118229\|\|full | Not-determined |
| S7_vs_118277\|\|full | Low-quality |
| S11_vs_k141_79275\|\|full | High-quality |
| S7_vs_118304\|\|full | Low-quality |
| S7_vs_118330\|\|full | Not-determined |
| S7_vs_118384\|\|full | Low-quality |
| S7_vs_118389\|\|full | Low-quality |
| S7_vs_118402\|\|full | Low-quality |
| S7_vs_118432\|\|full | Low-quality |
| S7_vs_118478\|\|full | Low-quality |
| S7_vs_118507\|\|full | Low-quality |
| S7_vs_118538\|\|full | Not-determined |
| S7_vs_118595\|\|full | Low-quality |
| S7_vs_118648\|\|full | Not-determined |
| S7_vs_118650\|\|full | Low-quality |
| S7_vs_118676\|\|full | Not-determined |
| S7_vs_118689\|\|full | Low-quality |
| S7_vs_118705\|\|full | Not-determined |
| S7_vs_118718\|\|full | Low-quality |
| S7_vs_118782\|\|full | Low-quality |
| S7_vs_118826\|\|full | Low-quality |
| S7_vs_118831\|\|full | Low-quality |
| S7_vs_118871\|\|full | Low-quality |
| S7_vs_118875\|\|full | Not-determined |
| S7_vs_118876\|\|full | Low-quality |
| S7_vs_118879\|\|full | Low-quality |
| S7_vs_118898\|\|full | Low-quality |
| S7_vs_118935\|\|full | Low-quality |
| S7_vs_118959\|\|full | Not-determined |
| S7_vs_118976\|\|full | Low-quality |
| S7_vs_119070\|\|full | Low-quality |
| S7_vs_119093\|\|full | Low-quality |
| S7_vs_119097\|\|full | Not-determined |
| S7_vs_119103\|\|full | Not-determined |
| S7_vs_119115\|\|full | Low-quality |
| S7_vs_119130\|\|full | Low-quality |
| S7_vs_119144\|\|full | Low-quality |
| S7_vs_119162\|\|full | Low-quality |
| S7_vs_119169\|\|full | Low-quality |
| S7_vs_119191\|\|full | Not-determined |
| S7_vs_119199\|\|full | Low-quality |
| S7_vs_119217\|\|full | Low-quality |
| S7_vs_119250\|\|full | Low-quality |
| S7_vs_119289\|\|full | Not-determined |
| S7_vs_119295\|\|full | Low-quality |
| S7_vs_119345\|\|full | Low-quality |
| S7_vs_119428\|\|full | Low-quality |
| S7_vs_119469\|\|full | Not-determined |
| S7_vs_119473\|\|full | Low-quality |
| S7_vs_119486\|\|full | Low-quality |
| S7_vs_68239\|\|full | High-quality |
| S7_vs_119494\|\|full | Low-quality |
| S7_vs_119495\|\|full | Low-quality |
| S7_vs_119507\|\|full | Not-determined |
| S7_vs_119647\|\|full | Low-quality |
| S10_vs_58014\|\|full | Medium-quality |
| S7_vs_119685\|\|full | Low-quality |
| S7_vs_119725\|\|full | Low-quality |
| S7_vs_119759\|\|full | Not-determined |
| S7_vs_119779\|\|full | Not-determined |
| S7_vs_119787\|\|full | Low-quality |
| S7_vs_119808\|\|full | Low-quality |
| S7_vs_119825\|\|full | Low-quality |
| S7_vs_119853\|\|full | Low-quality |
| S7_vs_119867\|\|full | Low-quality |
| S7_vs_119908\|\|full | Not-determined |
| S7_vs_119916\|\|full | Not-determined |
| S7_vs_119930\|\|full | Not-determined |
| S7_vs_119932\|\|full | Low-quality |
| S7_vs_119980\|\|full | Low-quality |
| S7_vs_120015\|\|full | Low-quality |
| S7_vs_120037\|\|full | Low-quality |
| S7_vs_120047\|\|full | Low-quality |
| S7_vs_120123\|\|full | Not-determined |
| S7_vs_120164\|\|full | Low-quality |
| S7_vs_120186\|\|full | Not-determined |
| S7_vs_120190\|\|full | Low-quality |
| S7_vs_120207\|\|full | Not-determined |
| S7_vs_120210\|\|full | Not-determined |
| S7_vs_120215\|\|full | Low-quality |
| S7_vs_120229\|\|full | Low-quality |
| S7_vs_120264\|\|full | Low-quality |
| S7_vs_120302\|\|full | Low-quality |
| S7_vs_120371\|\|full | Not-determined |
| S7_vs_120383\|\|full | Low-quality |
| S7_vs_120402\|\|full | Not-determined |
| S7_vs_120441\|\|full | Low-quality |
| S7_vs_120462\|\|full | Low-quality |
| S7_vs_120520\|\|full | Not-determined |
| S7_vs_120600\|\|full | Not-determined |
| S7_vs_120605\|\|full | Not-determined |
| S7_vs_120641\|\|full | Low-quality |
| S7_vs_120675\|\|full | Not-determined |
| S7_vs_120725\|\|full | Low-quality |
| S7_vs_120818\|\|full | Low-quality |
| S7_vs_120896\|\|full | Not-determined |
| S7_vs_120951\|\|full | Not-determined |
| S7_vs_120953\|\|full | Not-determined |
| S7_vs_120954\|\|full | Low-quality |
| S7_vs_120988\|\|full | Not-determined |
| S7_vs_121109\|\|full | Low-quality |
| S7_vs_121153\|\|full | Not-determined |
| S7_vs_121167\|\|full | Low-quality |
| S7_vs_121185\|\|full | Low-quality |
| S7_vs_121217\|\|full | Not-determined |
| S7_vs_121224\|\|full | Low-quality |
| S7_vs_121238\|\|full | Low-quality |
| S7_vs_121348\|\|full | Low-quality |
| S7_vs_121350\|\|full | Not-determined |
| S7_vs_121373\|\|full | Low-quality |
| S7_vs_121375\|\|full | Low-quality |
| S7_vs_121417\|\|full | Low-quality |
| S7_vs_121438\|\|full | Low-quality |
| S7_vs_121444\|\|full | Not-determined |
| S7_vs_121493\|\|full | Low-quality |
| S7_vs_121520\|\|full | Low-quality |
| S7_vs_121530\|\|full | Not-determined |
| S7_vs_121568\|\|full | Not-determined |
| S7_vs_121569\|\|full | Not-determined |
| S7_vs_121577\|\|full | Low-quality |
| S7_vs_121609\|\|full | Not-determined |
| S7_vs_121611\|\|full | Not-determined |
| S7_vs_121638\|\|full | Low-quality |
| S7_vs_121645\|\|full | Low-quality |
| S7_vs_121663\|\|full | Low-quality |
| S7_vs_121706\|\|full | Not-determined |
| S7_vs_121718\|\|full | Not-determined |
| S7_vs_121720\|\|full | Low-quality |
| S7_vs_121764\|\|full | Not-determined |
| S7_vs_121861\|\|full | Low-quality |
| S7_vs_121865\|\|full | Not-determined |
| S7_vs_121942\|\|full | Low-quality |
| S7_vs_121964\|\|full | Not-determined |
| S7_vs_122002\|\|full | Low-quality |
| S7_vs_122067\|\|full | Low-quality |
| S7_vs_122121\|\|full | Low-quality |
| S7_vs_122133\|\|full | Low-quality |
| S6_vs_k141_56490\|\|full | High-quality |
| S7_vs_122153\|\|full | Not-determined |
| S7_vs_122160\|\|full | Low-quality |
| S7_vs_122165\|\|full | Not-determined |
| S7_vs_122169\|\|full | Not-determined |
| S7_vs_122171\|\|full | Low-quality |
| S7_vs_122172\|\|full | Low-quality |
| S7_vs_122197\|\|full | Low-quality |
| S7_vs_122225\|\|full | Low-quality |
| S7_vs_122236\|\|full | Low-quality |
| S7_vs_122296\|\|full | Low-quality |
| S7_vs_122334\|\|full | Not-determined |
| S7_vs_122351\|\|full | Low-quality |
| S7_vs_122367\|\|full | Low-quality |
| S7_vs_122369\|\|full | Low-quality |
| S7_vs_122405\|\|full | Low-quality |
| S7_vs_122424\|\|full | Not-determined |
| S7_vs_122483\|\|full | Low-quality |
| S7_vs_122511\|\|full | Low-quality |
| S7_vs_122522\|\|full | Low-quality |
| S7_vs_122559\|\|full | Low-quality |
| S7_vs_122594\|\|full | Low-quality |
| S7_vs_122599\|\|full | Not-determined |
| S7_vs_122603\|\|full | Not-determined |
| S7_vs_122627\|\|full | Not-determined |
| S7_vs_122653\|\|full | Not-determined |
| S7_vs_122660\|\|full | Not-determined |
| S7_vs_122673\|\|full | Low-quality |
| S7_vs_122761\|\|full | Not-determined |
| S7_vs_122812\|\|full | Low-quality |
| S7_vs_122816\|\|full | Not-determined |
| S7_vs_122820\|\|full | Not-determined |
| S7_vs_122837\|\|full | Low-quality |
| S7_vs_122897\|\|full | Low-quality |
| S7_vs_122961\|\|full | Low-quality |
| S7_vs_122964\|\|full | Low-quality |
| S7_vs_122990\|\|full | Low-quality |
| S7_vs_123018\|\|full | Low-quality |
| S7_vs_123055\|\|full | Low-quality |
| S7_vs_123070\|\|full | Not-determined |
| S7_vs_123075\|\|full | Low-quality |
| S7_vs_123077\|\|full | Not-determined |
| S7_vs_123096\|\|full | Low-quality |
| S7_vs_123110\|\|full | Not-determined |
| S7_vs_123117\|\|full | Low-quality |
| S7_vs_123129\|\|full | Not-determined |
| S7_vs_123152\|\|full | Low-quality |
| S7_vs_123161\|\|full | Low-quality |
| S7_vs_123202\|\|full | Not-determined |
| S7_vs_123216\|\|full | Not-determined |
| S7_vs_123245\|\|full | Not-determined |
| S7_vs_123265\|\|full | Not-determined |
| S7_vs_123266\|\|full | Not-determined |
| S7_vs_123292\|\|full | Low-quality |
| S7_vs_123308\|\|full | Low-quality |
| S7_vs_123324\|\|full | Low-quality |
| S7_vs_123348\|\|full | Not-determined |
| S7_vs_123368\|\|full | Low-quality |
| S7_vs_123395\|\|full | Low-quality |
| S7_vs_123435\|\|full | Not-determined |
| S7_vs_123439\|\|full | Low-quality |
| S7_vs_123473\|\|full | Low-quality |
| S7_vs_123515\|\|full | Low-quality |
| S7_vs_123541\|\|full | Not-determined |
| S7_vs_123556\|\|full | Low-quality |
| S7_vs_123563\|\|full | Low-quality |
| S7_vs_123614\|\|full | Low-quality |
| S7_vs_123646\|\|full | Low-quality |
| S7_vs_123665\|\|full | Low-quality |
| S7_vs_123671\|\|full | Low-quality |
| S7_vs_123761\|\|full | Not-determined |
| S7_vs_123762\|\|full | Low-quality |
| S7_vs_123802\|\|full | Not-determined |
| S7_vs_123807\|\|full | Not-determined |
| S7_vs_123810\|\|full | Low-quality |
| S7_vs_123848\|\|full | Not-determined |
| S7_vs_123940\|\|full | Not-determined |
| S7_vs_123960\|\|full | Not-determined |
| S7_vs_124003\|\|full | Low-quality |
| S7_vs_124016\|\|full | Not-determined |
| S7_vs_124094\|\|full | Not-determined |
| S7_vs_124172\|\|full | Not-determined |
| S7_vs_124176\|\|full | Low-quality |
| S7_vs_124220\|\|full | Low-quality |
| S7_vs_124230\|\|full | Not-determined |
| S7_vs_124242\|\|full | Low-quality |
| S7_vs_124320\|\|full | Low-quality |
| S7_vs_124376\|\|full | Not-determined |
| S7_vs_124383\|\|full | Not-determined |
| S7_vs_124387\|\|full | Low-quality |
| S7_vs_124407\|\|full | Low-quality |
| S7_vs_124415\|\|full | Low-quality |
| S7_vs_124460\|\|full | Not-determined |
| S7_vs_124461\|\|full | Not-determined |
| S7_vs_124493\|\|full | Low-quality |
| S7_vs_124501\|\|full | Not-determined |
| S5_vs_k141_145584\|\|full | High-quality |
| S7_vs_124575\|\|full | Low-quality |
| S7_vs_124583\|\|full | Not-determined |
| S7_vs_124601\|\|full | Low-quality |
| S7_vs_124608\|\|full | Not-determined |
| S7_vs_124631\|\|full | Not-determined |
| S7_vs_124650\|\|full | Not-determined |
| S7_vs_124687\|\|full | Not-determined |
| S7_vs_124737\|\|full | Not-determined |
| S7_vs_124793\|\|full | Not-determined |
| S7_vs_124796\|\|full | Low-quality |
| S7_vs_124799\|\|full | Low-quality |
| S7_vs_124818\|\|full | Low-quality |
| S7_vs_124822\|\|full | Not-determined |
| S7_vs_124823\|\|full | Not-determined |
| S7_vs_124844\|\|full | Low-quality |
| S7_vs_124864\|\|full | Low-quality |
| S7_vs_124930\|\|full | Not-determined |
| S7_vs_124936\|\|full | Not-determined |
| S7_vs_124950\|\|full | Low-quality |
| S7_vs_124967\|\|full | Not-determined |
| S7_vs_124975\|\|full | Low-quality |
| S7_vs_124986\|\|full | Low-quality |
| S7_vs_124997\|\|full | Low-quality |
| S7_vs_125016\|\|full | Low-quality |
| S7_vs_125017\|\|full | Not-determined |
| S7_vs_125045\|\|full | Low-quality |
| S7_vs_125063\|\|full | Low-quality |
| S7_vs_125064\|\|full | Low-quality |
| S7_vs_125094\|\|full | Low-quality |
| S7_vs_125101\|\|full | Not-determined |
| S7_vs_125102\|\|full | Low-quality |
| S7_vs_125166\|\|full | Not-determined |
| S7_vs_125201\|\|full | Low-quality |
| S7_vs_125208\|\|full | Low-quality |
| S7_vs_125219\|\|full | Low-quality |
| S7_vs_125290\|\|full | Low-quality |
| S7_vs_125296\|\|full | Not-determined |
| S7_vs_125355\|\|full | Not-determined |
| S7_vs_125443\|\|full | Low-quality |
| S7_vs_125474\|\|full | Not-determined |
| S7_vs_125478\|\|full | Not-determined |
| S7_vs_125527\|\|full | Low-quality |
| S7_vs_125533\|\|full | Low-quality |
| S7_vs_125546\|\|full | Not-determined |
| S7_vs_125548\|\|full | Low-quality |
| S7_vs_125564\|\|full | Low-quality |
| S7_vs_125653\|\|full | Not-determined |
| S7_vs_125655\|\|full | Low-quality |
| S7_vs_125711\|\|full | Low-quality |
| S7_vs_125802\|\|full | Low-quality |
| S7_vs_125808\|\|full | Not-determined |
| S7_vs_125837\|\|full | Low-quality |
| S7_vs_125845\|\|full | Low-quality |
| S7_vs_125889\|\|full | Not-determined |
| S7_vs_125900\|\|full | Not-determined |
| S7_vs_125907\|\|full | Low-quality |
| S7_vs_125910\|\|full | Low-quality |
| S7_vs_125924\|\|full | Low-quality |
| S7_vs_125942\|\|full | Low-quality |
| S7_vs_125946\|\|full | Low-quality |
| S7_vs_125993\|\|full | Low-quality |
| S7_vs_126001\|\|full | Low-quality |
| S7_vs_126006\|\|full | Low-quality |
| S7_vs_126075\|\|full | Not-determined |
| S7_vs_126138\|\|full | Not-determined |
| S7_vs_126141\|\|full | Low-quality |
| S7_vs_126173\|\|full | Low-quality |
| S7_vs_126180\|\|full | Low-quality |
| S7_vs_126203\|\|full | Low-quality |
| S7_vs_126237\|\|full | Low-quality |
| S7_vs_126240\|\|full | Not-determined |
| S7_vs_126248\|\|full | Low-quality |
| S7_vs_126260\|\|full | Low-quality |
| S7_vs_126288\|\|full | Low-quality |
| S7_vs_126299\|\|full | Not-determined |
| S7_vs_126362\|\|full | Low-quality |
| S7_vs_126373\|\|full | Low-quality |
| S7_vs_126377\|\|full | Low-quality |
| S7_vs_126379\|\|full | Low-quality |
| S7_vs_126568\|\|full | Not-determined |
| S7_vs_126604\|\|full | Not-determined |
| S7_vs_126646\|\|full | Low-quality |
| S7_vs_126702\|\|full | Low-quality |
| S7_vs_126743\|\|full | Low-quality |
| S7_vs_126786\|\|full | Not-determined |
| S7_vs_126808\|\|full | Not-determined |
| S7_vs_126879\|\|full | Not-determined |
| S7_vs_126883\|\|full | Not-determined |
| S7_vs_126885\|\|full | Low-quality |
| S7_vs_126891\|\|full | Low-quality |
| S7_vs_126993\|\|full | Low-quality |
| S7_vs_127051\|\|full | Low-quality |
| S7_vs_127089\|\|full | Not-determined |
| S7_vs_127130\|\|full | Low-quality |
| S7_vs_127141\|\|full | Low-quality |
| S7_vs_127192\|\|full | Not-determined |
| S7_vs_127246\|\|full | Low-quality |
| S7_vs_127257\|\|full | Not-determined |
| S7_vs_127316\|\|full | Low-quality |
| S7_vs_127340\|\|full | Low-quality |
| S7_vs_127366\|\|full | Not-determined |
| S7_vs_127461\|\|full | Not-determined |
| S7_vs_127477\|\|full | Low-quality |
| S7_vs_127506\|\|full | Low-quality |
| S7_vs_127586\|\|full | Low-quality |
| S7_vs_127603\|\|full | Low-quality |
| S7_vs_127658\|\|full | Low-quality |
| S7_vs_127667\|\|full | Low-quality |
| S7_vs_127737\|\|full | Not-determined |
| S7_vs_127769\|\|full | Not-determined |
| S7_vs_127778\|\|full | Low-quality |
| S7_vs_127796\|\|full | Low-quality |
| S7_vs_127887\|\|full | Not-determined |
| S7_vs_127944\|\|full | Not-determined |
| S7_vs_128006\|\|full | Low-quality |
| S7_vs_128029\|\|full | Not-determined |
| S7_vs_128047\|\|full | Not-determined |
| S7_vs_128067\|\|full | Low-quality |
| S7_vs_128069\|\|full | Low-quality |
| S7_vs_128108\|\|full | Not-determined |
| S7_vs_128118\|\|full | Low-quality |
| S7_vs_128162\|\|full | Not-determined |
| S7_vs_128173\|\|full | Low-quality |
| S7_vs_128181\|\|full | Low-quality |
| S7_vs_128227\|\|full | Low-quality |
| S7_vs_128232\|\|full | Low-quality |
| S7_vs_128266\|\|full | Low-quality |
| S7_vs_128280\|\|full | Low-quality |
| S7_vs_128301\|\|full | Low-quality |
| S7_vs_128307\|\|full | Not-determined |
| S7_vs_128359\|\|full | Not-determined |
| S7_vs_128375\|\|full | Not-determined |
| S7_vs_128387\|\|full | Not-determined |
| S7_vs_128447\|\|full | Not-determined |
| S7_vs_128459\|\|full | Not-determined |
| S7_vs_128477\|\|full | Low-quality |
| S7_vs_128504\|\|full | Not-determined |
| S7_vs_128509\|\|full | Not-determined |
| S7_vs_128527\|\|full | Not-determined |
| S7_vs_128590\|\|full | Low-quality |
| S7_vs_128623\|\|full | Not-determined |
| S7_vs_128631\|\|full | Low-quality |
| S7_vs_128643\|\|full | Not-determined |
| S7_vs_128687\|\|full | Low-quality |
| S7_vs_128694\|\|full | Low-quality |
| S7_vs_128697\|\|full | Low-quality |
| S7_vs_128700\|\|full | Not-determined |
| S7_vs_128752\|\|full | Not-determined |
| S7_vs_128815\|\|full | Low-quality |
| S7_vs_128887\|\|full | Low-quality |
| S7_vs_128926\|\|full | Low-quality |
| S7_vs_128966\|\|full | Low-quality |
| S7_vs_129084\|\|full | Not-determined |
| S7_vs_129183\|\|full | Low-quality |
| S7_vs_129275\|\|full | Low-quality |
| S7_vs_129296\|\|full | Low-quality |
| S7_vs_129297\|\|full | Not-determined |
| S7_vs_129330\|\|full | Low-quality |
| S7_vs_129344\|\|full | Not-determined |
| S7_vs_129364\|\|full | Low-quality |
| S7_vs_129399\|\|full | Low-quality |
| S7_vs_129544\|\|full | Low-quality |
| S7_vs_129575\|\|full | Low-quality |
| S7_vs_129606\|\|full | Low-quality |
| S7_vs_129609\|\|full | Low-quality |
| S7_vs_129641\|\|full | Low-quality |
| S7_vs_129730\|\|full | Low-quality |
| S7_vs_129735\|\|full | Low-quality |
| S7_vs_129747\|\|full | Not-determined |
| S7_vs_129750\|\|full | Not-determined |
| S7_vs_129772\|\|full | Low-quality |
| S7_vs_129816\|\|full | Low-quality |
| S7_vs_129821\|\|full | Not-determined |
| S7_vs_129848\|\|full | Not-determined |
| S14_extracted_k141_117147 | Complete |
| S7_vs_129901\|\|full | Low-quality |
| S7_vs_129960\|\|full | Not-determined |
| S7_vs_130060\|\|full | Low-quality |
| S7_vs_130081\|\|full | Low-quality |
| S7_vs_130086\|\|full | Low-quality |
| S7_vs_130179\|\|full | Low-quality |
| S7_vs_130180\|\|full | Low-quality |
| S7_vs_130221\|\|full | Not-determined |
| S7_vs_130232\|\|full | Low-quality |
| S7_vs_130315\|\|full | Low-quality |
| S7_vs_130335\|\|full | Not-determined |
| S7_vs_130463\|\|full | Not-determined |
| S7_vs_130493\|\|full | Not-determined |
| S7_vs_130522\|\|full | Low-quality |
| S7_vs_130549\|\|full | Low-quality |
| S7_vs_130567\|\|full | Not-determined |
| S7_vs_130586\|\|full | Not-determined |
| S7_vs_130592\|\|full | Low-quality |
| S7_vs_130678\|\|full | Not-determined |
| S7_vs_130716\|\|full | Not-determined |
| S7_vs_130773\|\|full | Not-determined |
| S7_vs_130784\|\|full | Not-determined |
| S7_vs_130802\|\|full | Not-determined |
| S7_vs_130810\|\|full | Not-determined |
| S7_vs_130811\|\|full | Low-quality |
| S7_vs_130812\|\|full | Low-quality |
| S7_vs_130845\|\|full | Low-quality |
| S7_vs_130901\|\|full | Not-determined |
| S7_vs_130918\|\|full | Not-determined |
| S7_vs_130925\|\|full | Not-determined |
| S7_vs_130930\|\|full | Low-quality |
| S7_vs_130938\|\|full | Low-quality |
| S7_vs_130956\|\|full | Low-quality |
| S7_vs_131040\|\|full | Not-determined |
| S7_vs_131053\|\|full | Not-determined |
| S7_vs_131074\|\|full | Not-determined |
| S7_vs_131100\|\|full | Low-quality |
| S7_vs_131189\|\|full | Low-quality |
| S7_vs_131217\|\|full | Not-determined |
| S7_vs_131218\|\|full | Low-quality |
| S7_vs_131252\|\|full | Low-quality |
| S14_vs_k141_34449\|\|full | High-quality |
| S7_vs_131330\|\|full | Not-determined |
| S7_vs_131367\|\|full | Low-quality |
| S7_vs_131416\|\|full | Not-determined |
| S7_vs_131443\|\|full | Not-determined |
| S7_vs_131447\|\|full | Low-quality |
| S7_vs_131466\|\|full | Low-quality |
| S7_vs_131489\|\|full | Not-determined |
| S7_vs_131553\|\|full | Not-determined |
| S7_vs_131561\|\|full | Not-determined |
| S7_vs_131566\|\|full | Low-quality |
| S7_vs_131572\|\|full | Low-quality |
| S7_vs_131573\|\|full | Not-determined |
| S7_vs_131608\|\|full | Not-determined |
| S7_vs_131655\|\|full | Low-quality |
| S7_vs_131698\|\|full | Not-determined |
| S7_vs_131753\|\|full | Low-quality |
| S7_vs_131803\|\|full | Low-quality |
| S7_vs_131817\|\|full | Low-quality |
| S7_vs_131831\|\|full | Low-quality |
| S7_vs_131854\|\|full | Low-quality |
| S7_vs_131859\|\|full | Not-determined |
| S7_vs_131869\|\|full | Low-quality |
| S7_vs_131910\|\|full | Not-determined |
| S7_vs_131914\|\|full | Low-quality |
| S7_vs_131933\|\|full | Low-quality |
| S7_vs_131946\|\|full | Low-quality |
| S7_vs_131959\|\|full | Not-determined |
| S7_vs_132006\|\|full | Low-quality |
| S7_vs_132018\|\|full | Not-determined |
| S7_vs_132026\|\|full | Not-determined |
| S7_vs_132070\|\|full | Low-quality |
| S7_vs_132086\|\|full | Low-quality |
| S7_vs_132098\|\|full | Not-determined |
| S7_vs_132139\|\|full | Low-quality |
| S7_vs_132157\|\|full | Low-quality |
| S7_vs_132201\|\|full | Low-quality |
| S7_vs_132245\|\|full | Low-quality |
| S7_vs_132305\|\|full | Low-quality |
| S7_vs_132310\|\|full | Not-determined |
| S7_vs_132330\|\|full | Low-quality |
| S7_vs_132347\|\|full | Not-determined |
| S7_vs_132357\|\|full | Not-determined |
| S7_vs_132399\|\|full | Not-determined |
| S7_vs_132412\|\|full | Not-determined |
| S7_vs_132454\|\|full | Not-determined |
| S7_vs_132463\|\|full | Not-determined |
| S7_vs_132489\|\|full | Low-quality |
| S7_vs_132505\|\|full | Low-quality |
| S7_vs_132555\|\|full | Not-determined |
| S7_vs_132562\|\|full | Not-determined |
| S7_vs_132643\|\|full | Low-quality |
| S7_vs_132650\|\|full | Low-quality |
| S7_vs_132679\|\|full | Low-quality |
| S7_vs_132680\|\|full | Not-determined |
| S7_vs_132687\|\|full | Low-quality |
| S7_vs_132710\|\|full | Not-determined |
| S7_vs_132718\|\|full | Low-quality |
| S7_vs_132721\|\|full | Low-quality |
| S7_vs_132828\|\|full | Low-quality |
| S7_vs_132857\|\|full | Low-quality |
| S7_vs_132865\|\|full | Not-determined |
| S7_vs_132882\|\|full | Low-quality |
| S7_vs_132893\|\|full | Low-quality |
| S7_vs_132921\|\|full | Low-quality |
| S7_vs_132941\|\|full | Not-determined |
| S7_vs_132959\|\|full | Low-quality |
| S7_vs_132961\|\|full | Low-quality |
| S7_vs_132972\|\|full | Low-quality |
| S7_vs_133040\|\|full | Low-quality |
| S7_vs_133051\|\|full | Low-quality |
| S7_vs_133086\|\|full | Low-quality |
| S7_vs_133101\|\|full | Low-quality |
| S7_vs_133103\|\|full | Low-quality |
| S7_vs_133131\|\|full | Low-quality |
| S7_vs_133144\|\|full | Low-quality |
| S7_vs_133216\|\|full | Not-determined |
| S7_vs_133258\|\|full | Low-quality |
| S7_vs_133263\|\|full | Low-quality |
| S7_vs_133275\|\|full | Not-determined |
| S7_vs_133288\|\|full | Low-quality |
| S7_vs_133315\|\|full | Low-quality |
| S7_vs_133378\|\|full | Not-determined |
| S7_vs_133427\|\|full | Low-quality |
| S7_vs_133472\|\|full | Not-determined |
| S7_vs_133474\|\|full | Low-quality |
| S7_vs_133477\|\|full | Low-quality |
| S7_vs_133501\|\|full | Low-quality |
| S7_vs_133511\|\|full | Low-quality |
| S7_vs_133557\|\|full | Low-quality |
| S7_vs_133756\|\|full | Low-quality |
| S7_vs_133787\|\|full | Low-quality |
| S7_vs_133793\|\|full | Low-quality |
| S7_vs_133838\|\|full | Not-determined |
| S7_vs_133861\|\|full | Not-determined |
| S7_vs_133884\|\|full | Low-quality |
| S7_vs_133935\|\|full | Not-determined |
| S7_vs_133943\|\|full | Not-determined |
| S7_vs_133950\|\|full | Low-quality |
| S7_vs_134024\|\|full | Low-quality |
| S7_vs_134037\|\|full | Low-quality |
| S7_vs_134038\|\|full | Low-quality |
| S7_vs_134050\|\|full | Low-quality |
| S7_vs_134053\|\|full | Low-quality |
| S7_vs_134069\|\|full | Low-quality |
| S7_vs_134070\|\|full | Not-determined |
| S7_vs_134144\|\|full | Not-determined |
| S7_vs_134157\|\|full | Low-quality |
| S7_vs_134173\|\|full | Low-quality |
| S7_vs_134248\|\|full | Low-quality |
| S7_vs_134250\|\|full | Low-quality |
| S7_vs_134289\|\|full | Low-quality |
| S7_vs_134299\|\|full | Not-determined |
| S7_vs_134337\|\|full | Not-determined |
| S7_vs_134343\|\|full | Not-determined |
| S7_vs_134365\|\|full | Low-quality |

| | |
|---|---|
| S7_vs_134374\|\|full | Low-quality |
| S7_vs_134429\|\|full | Low-quality |
| S7_vs_134439\|\|full | Low-quality |
| S7_vs_134450\|\|full | Not-determined |
| S7_vs_134464\|\|full | Not-determined |
| S7_vs_134493\|\|full | Not-determined |
| S7_vs_134509\|\|full | Not-determined |
| S7_vs_134529\|\|full | Low-quality |
| S7_vs_134542\|\|full | Low-quality |
| S7_vs_134556\|\|full | Low-quality |
| S7_vs_134562\|\|full | Low-quality |
| S7_vs_134568\|\|full | Low-quality |
| S7_vs_134569\|\|full | Low-quality |
| S12_vs_k141_99092\|\|full | High-quality |
| S7_vs_134636\|\|full | Not-determined |
| S7_vs_134638\|\|full | Not-determined |
| S7_vs_134647\|\|full | Low-quality |
| S7_vs_134701\|\|full | Low-quality |
| S7_vs_134704\|\|full | Low-quality |
| S7_vs_134722\|\|full | Not-determined |
| S7_vs_134771\|\|full | Not-determined |
| S7_vs_134819\|\|full | Low-quality |
| S7_vs_134886\|\|full | Low-quality |
| S7_vs_134898\|\|full | Low-quality |
| S7_vs_134906\|\|full | Low-quality |
| S7_vs_134950\|\|full | Low-quality |
| S7_vs_134990\|\|full | Low-quality |
| S7_vs_135002\|\|full | Low-quality |
| S7_vs_135009\|\|full | Not-determined |
| S7_vs_135111\|\|full | Low-quality |
| S7_vs_135117\|\|full | Not-determined |
| S7_vs_135134\|\|full | Low-quality |
| S7_vs_135142\|\|full | Not-determined |
| S7_vs_135183\|\|full | Not-determined |
| S7_vs_135200\|\|full | Low-quality |
| S7_vs_135239\|\|full | Low-quality |
| S7_vs_135257\|\|full | Not-determined |
| S7_vs_135284\|\|full | Low-quality |
| S7_vs_135313\|\|full | Low-quality |
| S7_vs_135405\|\|full | Low-quality |
| S7_vs_135416\|\|full | Not-determined |
| S7_vs_135483\|\|full | Low-quality |
| S7_vs_135525\|\|full | Not-determined |
| S7_vs_135586\|\|full | Not-determined |
| S7_vs_135656\|\|full | Low-quality |
| S7_vs_135657\|\|full | Low-quality |
| S7_vs_135777\|\|full | Low-quality |
| S7_vs_135821\|\|full | Low-quality |
| S7_vs_135867\|\|full | Low-quality |
| S7_vs_135907\|\|full | Low-quality |
| S7_vs_135920\|\|full | Low-quality |
| S7_vs_135928\|\|full | Not-determined |
| S7_vs_135962\|\|full | Low-quality |
| S7_vs_135991\|\|full | Not-determined |
| S7_vs_136008\|\|full | Low-quality |
| S7_vs_136088\|\|full | Low-quality |
| S7_vs_136090\|\|full | Not-determined |
| S7_vs_136096\|\|full | Not-determined |
| S7_vs_136128\|\|full | Low-quality |
| S7_vs_136139\|\|full | Not-determined |
| S7_vs_136142\|\|full | Low-quality |
| S7_vs_136181\|\|full | Not-determined |
| S7_vs_136182\|\|full | Low-quality |
| S7_vs_136207\|\|full | Not-determined |
| S7_vs_136216\|\|full | Low-quality |
| S7_vs_136258\|\|full | Not-determined |
| S7_vs_136273\|\|full | Low-quality |
| S7_vs_136295\|\|full | Not-determined |
| S7_vs_136308\|\|full | Not-determined |
| S7_vs_136311\|\|full | Not-determined |
| S7_vs_136316\|\|full | Not-determined |
| S7_vs_136327\|\|full | Low-quality |
| S7_vs_136358\|\|full | Not-determined |
| S7_vs_136381\|\|full | Low-quality |
| S7_vs_136399\|\|full | Low-quality |
| S7_vs_136408\|\|full | Low-quality |
| S7_vs_136428\|\|full | Low-quality |
| S7_vs_136443\|\|full | Low-quality |
| S7_vs_136447\|\|full | Not-determined |
| S7_vs_136460\|\|full | Low-quality |
| S7_vs_136538\|\|full | Low-quality |
| S7_vs_136540\|\|full | Not-determined |
| S7_vs_136635\|\|full | Low-quality |
| S7_vs_136668\|\|full | Low-quality |
| S7_vs_136670\|\|full | Low-quality |
| S7_vs_136749\|\|full | Low-quality |
| S7_vs_136759\|\|full | Low-quality |
| S7_vs_136776\|\|full | Low-quality |
| S7_vs_31644\|\|full | Medium-quality |
| S7_vs_136816\|\|full | Not-determined |
| S7_vs_136851\|\|full | Low-quality |
| S7_vs_136873\|\|full | Not-determined |
| S7_vs_136929\|\|full | Low-quality |
| S7_vs_136945\|\|full | Not-determined |
| S7_vs_136965\|\|full | Not-determined |
| S7_vs_136979\|\|full | Not-determined |
| S7_vs_137028\|\|full | Low-quality |
| S7_vs_137036\|\|full | Low-quality |
| S7_vs_137062\|\|full | Not-determined |
| S7_vs_137108\|\|full | Low-quality |
| S7_vs_137130\|\|full | Low-quality |
| S7_vs_137149\|\|full | Low-quality |
| S7_vs_137152\|\|full | Not-determined |
| S7_vs_137190\|\|full | Not-determined |
| S7_vs_137197\|\|full | Not-determined |
| S7_vs_137201\|\|full | Low-quality |
| S7_vs_137203\|\|full | Not-determined |
| S7_vs_137210\|\|full | Low-quality |
| S7_vs_137221\|\|full | Not-determined |
| S7_vs_137229\|\|full | Not-determined |
| S7_vs_137240\|\|full | Not-determined |
| S7_vs_137256\|\|full | Not-determined |
| S7_vs_137273\|\|full | Low-quality |
| S7_vs_137282\|\|full | Not-determined |
| S7_vs_137283\|\|full | Not-determined |
| S7_vs_137338\|\|full | Not-determined |
| S7_vs_137420\|\|full | Low-quality |
| S7_vs_137427\|\|full | Low-quality |
| S7_vs_137429\|\|full | Low-quality |
| S7_vs_137439\|\|full | Not-determined |
| S7_vs_137466\|\|full | Not-determined |
| S7_vs_137484\|\|full | Not-determined |
| S7_vs_137486\|\|full | Not-determined |
| S7_vs_137498\|\|full | Low-quality |
| S7_vs_137553\|\|full | Not-determined |
| S7_vs_137561\|\|full | Low-quality |
| S7_vs_137574\|\|full | Not-determined |
| S7_vs_137612\|\|full | Not-determined |
| S7_vs_137613\|\|full | Not-determined |
| S7_vs_137616\|\|full | Low-quality |
| S7_vs_137639\|\|full | Not-determined |
| S7_vs_137699\|\|full | Low-quality |
| S7_vs_137702\|\|full | Not-determined |
| S7_vs_137739\|\|full | Not-determined |
| S7_vs_137773\|\|full | Low-quality |
| S7_vs_137793\|\|full | Not-determined |
| S7_vs_137836\|\|full | Not-determined |
| S7_vs_137863\|\|full | Not-determined |
| S7_vs_137864\|\|full | Low-quality |
| S7_vs_137866\|\|full | Not-determined |
| S7_vs_137867\|\|full | Not-determined |
| S7_vs_137872\|\|full | Low-quality |
| S7_vs_137896\|\|full | Low-quality |
| S7_vs_137913\|\|full | Not-determined |
| S7_vs_137935\|\|full | Low-quality |
| S7_vs_137936\|\|full | Low-quality |
| S7_vs_137967\|\|full | Low-quality |
| S7_vs_137993\|\|full | Not-determined |
| S7_vs_138000\|\|full | Low-quality |
| S7_vs_138081\|\|full | Low-quality |
| S7_vs_138210\|\|full | Low-quality |
| S7_vs_138217\|\|full | Low-quality |
| S7_vs_4472\|\|0_partial | Low-quality |
| S7_vs_12150\|\|0_partial | Not-determined |
| S7_vs_15170\|\|0_partial | Not-determined |
| S7_vs_23373\|\|0_partial | Low-quality |
| S7_vs_45454\|\|0_partial | Not-determined |
| S7_vs_47488\|\|0_partial | Not-determined |
| S7_vs_47978\|\|0_partial | Not-determined |
| S7_vs_52351\|\|0_partial | Low-quality |
| S14_vs_k141_117147\|\|full | High-quality |
| S7_vs_72746\|\|0_partial | Low-quality |
| S7_vs_85914\|\|0_partial | Low-quality |
| S7_vs_104214\|\|0_partial | Not-determined |
| S7_vs_118374\|\|0_partial | Not-determined |
| S7_vs_120623\|\|0_partial | Low-quality |
| S7_vs_34\|\|lt2gene | Low-quality |
| S7_vs_219\|\|lt2gene | Low-quality |
| S7_vs_421\|\|lt2gene | Low-quality |
| S7_vs_454\|\|lt2gene | Low-quality |
| S7_vs_488\|\|lt2gene | Not-determined |
| S7_vs_492\|\|lt2gene | Low-quality |
| S7_vs_494\|\|lt2gene | Low-quality |
| S7_vs_514\|\|lt2gene | Low-quality |
| S7_vs_613\|\|lt2gene | Low-quality |
| S7_vs_629\|\|lt2gene | Low-quality |
| S7_vs_692\|\|lt2gene | Low-quality |
| S7_vs_731\|\|lt2gene | Low-quality |
| S7_vs_855\|\|lt2gene | Low-quality |
| S7_vs_883\|\|lt2gene | Low-quality |
| S7_vs_988\|\|lt2gene | Low-quality |
| S7_vs_999\|\|lt2gene | Not-determined |
| S7_vs_1003\|\|lt2gene | Low-quality |
| S7_vs_1112\|\|lt2gene | Low-quality |
| S7_vs_1283\|\|lt2gene | Low-quality |
| S7_vs_1301\|\|lt2gene | Low-quality |
| S7_vs_1339\|\|lt2gene | Low-quality |
| S7_vs_1369\|\|lt2gene | Not-determined |
| S7_vs_1395\|\|lt2gene | Low-quality |
| S7_vs_1551\|\|lt2gene | Low-quality |
| S7_vs_1651\|\|lt2gene | Low-quality |
| S7_vs_1672\|\|lt2gene | Low-quality |
| S7_vs_1750\|\|lt2gene | Low-quality |
| S7_vs_1764\|\|lt2gene | Low-quality |
| S7_vs_1842\|\|lt2gene | Low-quality |
| S7_vs_1910\|\|lt2gene | Not-determined |
| S7_vs_1916\|\|lt2gene | Low-quality |
| S7_vs_2023\|\|lt2gene | Not-determined |
| S7_vs_2152\|\|lt2gene | Low-quality |
| S7_vs_2189\|\|lt2gene | Low-quality |
| S7_vs_2205\|\|lt2gene | Low-quality |
| S7_vs_2216\|\|lt2gene | Low-quality |
| S7_vs_2357\|\|lt2gene | Not-determined |
| S7_vs_2440\|\|lt2gene | Low-quality |
| S7_vs_2603\|\|lt2gene | Low-quality |
| S7_vs_2694\|\|lt2gene | Low-quality |
| S7_vs_2887\|\|lt2gene | Low-quality |
| S7_vs_2890\|\|lt2gene | Low-quality |
| S7_vs_2913\|\|lt2gene | Not-determined |
| S7_vs_2924\|\|lt2gene | Not-determined |
| S7_vs_2925\|\|lt2gene | Low-quality |
| S7_vs_2930\|\|lt2gene | Low-quality |
| S7_vs_2945\|\|lt2gene | Not-determined |
| S7_vs_2950\|\|lt2gene | Low-quality |
| S7_vs_2967\|\|lt2gene | Not-determined |
| S7_vs_3003\|\|lt2gene | Low-quality |
| S7_vs_3059\|\|lt2gene | Low-quality |
| S7_vs_3069\|\|lt2gene | Low-quality |
| S7_vs_3295\|\|lt2gene | Low-quality |
| S7_vs_3344\|\|lt2gene | Not-determined |
| S7_vs_3373\|\|lt2gene | Low-quality |
| S7_vs_3551\|\|lt2gene | Low-quality |
| S7_vs_3617\|\|lt2gene | Not-determined |
| S7_vs_3654\|\|lt2gene | Not-determined |
| S7_vs_3755\|\|lt2gene | Low-quality |
| S7_vs_3802\|\|lt2gene | Low-quality |
| S7_vs_3857\|\|lt2gene | Not-determined |
| S7_vs_4144\|\|lt2gene | Low-quality |
| S7_vs_4145\|\|lt2gene | Low-quality |
| S7_vs_4156\|\|lt2gene | Low-quality |
| S7_vs_4214\|\|lt2gene | Not-determined |
| S7_vs_4238\|\|lt2gene | Low-quality |
| S7_vs_4299\|\|lt2gene | Low-quality |
| S7_vs_4320\|\|lt2gene | Low-quality |
| S7_vs_4349\|\|lt2gene | Not-determined |
| S7_vs_4590\|\|lt2gene | Low-quality |
| S7_vs_4595\|\|lt2gene | Low-quality |
| S7_vs_4658\|\|lt2gene | Low-quality |
| S7_vs_4674\|\|lt2gene | Low-quality |
| S7_vs_4857\|\|lt2gene | Low-quality |
| S7_vs_4877\|\|lt2gene | Low-quality |
| S7_vs_4901\|\|lt2gene | Low-quality |
| S7_vs_5002\|\|lt2gene | Not-determined |
| S7_vs_5020\|\|lt2gene | Low-quality |
| S7_vs_5026\|\|lt2gene | Low-quality |
| S7_vs_5066\|\|lt2gene | Low-quality |
| S7_vs_5113\|\|lt2gene | Low-quality |
| S7_vs_5138\|\|lt2gene | Low-quality |
| S7_vs_5168\|\|lt2gene | Low-quality |
| S7_vs_5219\|\|lt2gene | Low-quality |
| S7_vs_5227\|\|lt2gene | Low-quality |
| S7_vs_5254\|\|lt2gene | Not-determined |
| S7_vs_5339\|\|lt2gene | Not-determined |
| S7_vs_5355\|\|lt2gene | Low-quality |
| S7_vs_5396\|\|lt2gene | Low-quality |
| S7_vs_5545\|\|lt2gene | Not-determined |
| S7_vs_5641\|\|lt2gene | Low-quality |
| S7_vs_5677\|\|lt2gene | Low-quality |
| S7_vs_5744\|\|lt2gene | Low-quality |
| S7_vs_5751\|\|lt2gene | Low-quality |
| S7_vs_5818\|\|lt2gene | Low-quality |
| S7_vs_5982\|\|lt2gene | Not-determined |
| S7_vs_6093\|\|lt2gene | Low-quality |
| S7_vs_6139\|\|lt2gene | Not-determined |
| S7_vs_6173\|\|lt2gene | Low-quality |
| S7_vs_6222\|\|lt2gene | Low-quality |
| S7_vs_6310\|\|lt2gene | Not-determined |
| S7_vs_6319\|\|lt2gene | Low-quality |
| S7_vs_6363\|\|lt2gene | Low-quality |
| S7_vs_6382\|\|lt2gene | Low-quality |
| S7_vs_6417\|\|lt2gene | Low-quality |
| S7_vs_6510\|\|lt2gene | Low-quality |
| S7_vs_6651\|\|lt2gene | Low-quality |
| S7_vs_6661\|\|lt2gene | Low-quality |
| S7_vs_6847\|\|lt2gene | Low-quality |
| S7_vs_6865\|\|lt2gene | Low-quality |
| S7_vs_6959\|\|lt2gene | Low-quality |
| S7_vs_7024\|\|lt2gene | Low-quality |
| S7_vs_7047\|\|lt2gene | Low-quality |
| S7_vs_7054\|\|lt2gene | Not-determined |
| S7_vs_7288\|\|lt2gene | Low-quality |
| S7_vs_7297\|\|lt2gene | Low-quality |
| S7_vs_7300\|\|lt2gene | Low-quality |
| S7_vs_7327\|\|lt2gene | Not-determined |
| S7_vs_7341\|\|lt2gene | Low-quality |
| S7_vs_7347\|\|lt2gene | Not-determined |
| S7_vs_7426\|\|lt2gene | Low-quality |
| S7_vs_7437\|\|lt2gene | Low-quality |
| S7_vs_7476\|\|lt2gene | Low-quality |
| S7_vs_7542\|\|lt2gene | Low-quality |
| S7_vs_7544\|\|lt2gene | Low-quality |
| S7_vs_7598\|\|lt2gene | Low-quality |
| S7_vs_7679\|\|lt2gene | Not-determined |
| S7_vs_7883\|\|lt2gene | Low-quality |
| S7_vs_7907\|\|lt2gene | Low-quality |
| S7_vs_7958\|\|lt2gene | Low-quality |
| S7_vs_7975\|\|lt2gene | Low-quality |
| S7_vs_8159\|\|lt2gene | Low-quality |
| S7_vs_8200\|\|lt2gene | Low-quality |
| S7_vs_8205\|\|lt2gene | Not-determined |
| S7_vs_8288\|\|lt2gene | Low-quality |
| S7_vs_8434\|\|lt2gene | Low-quality |
| S7_vs_8560\|\|lt2gene | Low-quality |
| S7_vs_8601\|\|lt2gene | Low-quality |
| S7_vs_8602\|\|lt2gene | Low-quality |
| S7_vs_8612\|\|lt2gene | Low-quality |
| S7_vs_8642\|\|lt2gene | Low-quality |
| S7_vs_8723\|\|lt2gene | Low-quality |
| S7_vs_8737\|\|lt2gene | Low-quality |
| S7_vs_8847\|\|lt2gene | Low-quality |
| S7_vs_8883\|\|lt2gene | Not-determined |
| S7_vs_8964\|\|lt2gene | Low-quality |
| S7_vs_9030\|\|lt2gene | Low-quality |
| S7_vs_9086\|\|lt2gene | Not-determined |
| S7_vs_9229\|\|lt2gene | Low-quality |
| S7_vs_9286\|\|lt2gene | Not-determined |
| S7_vs_9327\|\|lt2gene | Low-quality |
| S7_vs_9484\|\|lt2gene | Low-quality |
| S7_vs_9548\|\|lt2gene | Not-determined |
| S7_vs_9599\|\|lt2gene | Not-determined |
| S7_vs_9652\|\|lt2gene | Low-quality |
| S7_vs_9766\|\|lt2gene | Low-quality |
| S7_vs_9805\|\|lt2gene | Low-quality |
| S7_vs_9848\|\|lt2gene | Not-determined |
| S7_vs_9927\|\|lt2gene | Low-quality |
| S7_vs_10099\|\|lt2gene | Low-quality |
| S7_vs_10303\|\|lt2gene | Low-quality |
| S7_vs_10306\|\|lt2gene | Not-determined |
| S7_vs_10371\|\|lt2gene | Low-quality |
| S7_vs_10378\|\|lt2gene | Low-quality |
| S7_vs_10379\|\|lt2gene | Low-quality |
| S7_vs_10479\|\|lt2gene | Low-quality |
| S7_vs_10508\|\|lt2gene | Low-quality |
| S7_vs_10514\|\|lt2gene | Not-determined |
| S7_vs_10565\|\|lt2gene | Low-quality |
| S7_vs_10570\|\|lt2gene | Not-determined |
| S7_vs_10662\|\|lt2gene | Low-quality |
| S7_vs_10678\|\|lt2gene | Low-quality |
| S7_vs_10877\|\|lt2gene | Low-quality |
| S7_vs_10878\|\|lt2gene | Low-quality |
| S7_vs_11084\|\|lt2gene | Low-quality |
| S7_vs_11141\|\|lt2gene | Not-determined |
| S7_vs_11213\|\|lt2gene | Low-quality |
| S7_vs_11215\|\|lt2gene | Low-quality |
| S7_vs_11372\|\|lt2gene | Not-determined |
| S7_vs_11492\|\|lt2gene | Low-quality |
| S7_vs_11596\|\|lt2gene | Not-determined |
| S7_vs_11640\|\|lt2gene | Not-determined |
| S7_vs_11671\|\|lt2gene | Low-quality |
| S7_vs_11699\|\|lt2gene | Not-determined |
| S7_vs_11745\|\|lt2gene | Low-quality |
| S7_vs_11749\|\|lt2gene | Not-determined |
| S7_vs_11783\|\|lt2gene | Low-quality |
| S7_vs_11800\|\|lt2gene | Low-quality |
| S7_vs_12018\|\|lt2gene | Not-determined |
| S7_vs_12116\|\|lt2gene | Low-quality |
| S7_vs_12122\|\|lt2gene | Low-quality |
| S7_vs_12167\|\|lt2gene | Not-determined |
| S7_vs_12242\|\|lt2gene | Low-quality |
| S7_vs_12301\|\|lt2gene | Low-quality |
| S7_vs_12333\|\|lt2gene | Low-quality |
| S7_vs_12487\|\|lt2gene | Not-determined |
| S7_vs_12569\|\|lt2gene | Low-quality |
| S7_vs_12624\|\|lt2gene | Not-determined |
| S7_vs_12634\|\|lt2gene | Not-determined |
| S7_vs_12709\|\|lt2gene | Low-quality |
| S7_vs_12721\|\|lt2gene | Low-quality |
| S7_vs_12809\|\|lt2gene | Low-quality |
| S7_vs_13018\|\|lt2gene | Low-quality |
| S7_vs_13039\|\|lt2gene | Not-determined |
| S7_vs_13040\|\|lt2gene | Not-determined |
| S7_vs_13049\|\|lt2gene | Low-quality |
| S7_vs_13076\|\|lt2gene | Low-quality |
| S7_vs_13140\|\|lt2gene | Low-quality |
| S7_vs_13185\|\|lt2gene | Low-quality |
| S7_vs_13374\|\|lt2gene | Low-quality |
| S7_vs_13420\|\|lt2gene | Not-determined |
| S7_vs_13529\|\|lt2gene | Low-quality |
| S7_vs_13536\|\|lt2gene | Low-quality |
| S7_vs_13618\|\|lt2gene | Not-determined |
| S7_vs_13625\|\|lt2gene | Low-quality |
| S7_vs_13642\|\|lt2gene | Not-determined |
| S7_vs_13668\|\|lt2gene | Low-quality |
| S7_vs_13689\|\|lt2gene | Low-quality |
| S7_vs_13707\|\|lt2gene | Not-determined |
| S7_vs_13713\|\|lt2gene | Not-determined |
| S7_vs_13848\|\|lt2gene | Not-determined |
| S7_vs_13859\|\|lt2gene | Low-quality |
| S7_vs_13865\|\|lt2gene | Not-determined |
| S7_vs_14134\|\|lt2gene | Low-quality |
| S7_vs_14298\|\|lt2gene | Low-quality |
| S7_vs_14307\|\|lt2gene | Low-quality |
| S7_vs_14335\|\|lt2gene | Low-quality |
| S7_vs_14346\|\|lt2gene | Low-quality |
| S7_vs_14355\|\|lt2gene | Low-quality |
| S7_vs_14555\|\|lt2gene | Low-quality |
| S7_vs_14562\|\|lt2gene | Low-quality |
| S7_vs_14585\|\|lt2gene | Not-determined |
| S7_vs_14670\|\|lt2gene | Low-quality |
| S7_vs_14818\|\|lt2gene | Low-quality |
| S7_vs_14834\|\|lt2gene | Not-determined |
| S7_vs_14906\|\|lt2gene | Low-quality |
| S7_vs_15205\|\|lt2gene | Low-quality |
| S7_vs_15211\|\|lt2gene | Low-quality |
| S7_vs_15233\|\|lt2gene | Low-quality |
| S7_vs_15429\|\|lt2gene | Not-determined |
| S7_vs_15541\|\|lt2gene | Low-quality |
| S7_vs_15557\|\|lt2gene | Low-quality |
| S7_vs_15577\|\|lt2gene | Low-quality |
| S7_vs_15591\|\|lt2gene | Low-quality |
| S7_vs_15603\|\|lt2gene | Not-determined |
| S7_vs_15649\|\|lt2gene | Low-quality |
| S7_vs_15661\|\|lt2gene | Low-quality |
| S7_vs_15718\|\|lt2gene | Low-quality |
| S7_vs_15907\|\|lt2gene | Low-quality |
| S7_vs_16196\|\|lt2gene | Low-quality |
| S7_vs_16198\|\|lt2gene | Low-quality |
| S7_vs_16221\|\|lt2gene | Not-determined |
| S7_vs_16231\|\|lt2gene | Low-quality |
| S7_vs_16238\|\|lt2gene | Low-quality |
| S7_vs_16252\|\|lt2gene | Low-quality |
| S7_vs_16288\|\|lt2gene | Not-determined |
| S7_vs_16429\|\|lt2gene | Low-quality |
| S7_vs_16448\|\|lt2gene | Low-quality |
| S7_vs_16470\|\|lt2gene | Low-quality |
| S7_vs_16645\|\|lt2gene | Low-quality |
| S7_vs_16714\|\|lt2gene | Low-quality |
| S7_vs_16849\|\|lt2gene | Low-quality |
| S7_vs_16954\|\|lt2gene | Low-quality |
| S7_vs_17058\|\|lt2gene | Low-quality |
| S7_vs_17097\|\|lt2gene | Low-quality |
| S7_vs_17267\|\|lt2gene | Low-quality |
| S7_vs_17279\|\|lt2gene | Not-determined |
| S7_vs_17449\|\|lt2gene | Low-quality |
| S7_vs_17475\|\|lt2gene | Low-quality |
| S7_vs_17556\|\|lt2gene | Not-determined |
| S7_vs_17615\|\|lt2gene | Low-quality |
| S7_vs_17622\|\|lt2gene | Low-quality |
| S7_vs_17810\|\|lt2gene | Not-determined |
| S7_vs_17941\|\|lt2gene | Low-quality |
| S7_vs_17981\|\|lt2gene | Not-determined |
| S7_vs_17996\|\|lt2gene | Not-determined |
| S7_vs_18000\|\|lt2gene | Low-quality |
| S7_vs_18212\|\|lt2gene | Low-quality |
| S7_vs_18238\|\|lt2gene | Low-quality |
| S7_vs_18400\|\|lt2gene | Low-quality |
| S7_vs_18403\|\|lt2gene | Not-determined |
| S7_vs_18434\|\|lt2gene | Not-determined |
| S7_vs_18478\|\|lt2gene | Low-quality |
| S7_vs_18544\|\|lt2gene | Low-quality |
| S7_vs_18641\|\|lt2gene | Low-quality |
| S7_vs_18890\|\|lt2gene | Low-quality |
| S7_vs_19047\|\|lt2gene | Low-quality |
| S7_vs_19289\|\|lt2gene | Low-quality |
| S7_vs_19325\|\|lt2gene | Low-quality |
| S7_vs_19354\|\|lt2gene | Low-quality |
| S7_vs_19381\|\|lt2gene | Not-determined |
| S7_vs_19471\|\|lt2gene | Not-determined |
| S7_vs_19581\|\|lt2gene | Low-quality |
| S7_vs_19641\|\|lt2gene | Not-determined |
| S7_vs_19718\|\|lt2gene | Low-quality |
| S7_vs_19792\|\|lt2gene | Low-quality |
| S7_vs_19808\|\|lt2gene | Low-quality |
| S7_vs_20009\|\|lt2gene | Low-quality |
| S7_vs_20106\|\|lt2gene | Low-quality |
| S7_vs_20117\|\|lt2gene | Low-quality |
| S7_vs_20192\|\|lt2gene | Low-quality |
| S7_vs_20194\|\|lt2gene | Low-quality |
| S7_vs_20250\|\|lt2gene | Low-quality |
| S7_vs_20260\|\|lt2gene | Low-quality |
| S7_vs_20346\|\|lt2gene | Not-determined |
| S7_vs_20361\|\|lt2gene | Low-quality |
| S7_vs_20373\|\|lt2gene | Not-determined |
| S7_vs_20477\|\|lt2gene | Low-quality |
| S7_vs_20572\|\|lt2gene | Low-quality |
| S7_vs_20807\|\|lt2gene | Not-determined |
| S7_vs_20857\|\|lt2gene | Low-quality |
| S7_vs_20900\|\|lt2gene | Low-quality |
| S7_vs_20953\|\|lt2gene | Low-quality |
| S7_vs_20968\|\|lt2gene | Not-determined |
| S7_vs_20989\|\|lt2gene | Low-quality |
| S7_vs_21098\|\|lt2gene | Low-quality |
| S7_vs_21156\|\|lt2gene | Low-quality |
| S7_vs_21268\|\|lt2gene | Low-quality |
| S7_vs_21274\|\|lt2gene | Not-determined |
| S7_vs_21539\|\|lt2gene | Low-quality |
| S7_vs_21699\|\|lt2gene | Low-quality |
| S7_vs_21713\|\|lt2gene | Low-quality |
| S7_vs_21904\|\|lt2gene | Low-quality |
| S7_vs_21962\|\|lt2gene | Not-determined |
| S7_vs_21984\|\|lt2gene | Low-quality |
| S7_vs_22062\|\|lt2gene | Low-quality |
| S7_vs_22080\|\|lt2gene | Low-quality |
| S7_vs_22086\|\|lt2gene | Low-quality |
| S7_vs_22183\|\|lt2gene | Low-quality |
| S7_vs_22207\|\|lt2gene | Not-determined |
| S7_vs_22225\|\|lt2gene | Low-quality |
| S7_vs_22231\|\|lt2gene | Not-determined |
| S7_vs_22548\|\|lt2gene | Not-determined |
| S7_vs_22671\|\|lt2gene | Low-quality |
| S7_vs_22713\|\|lt2gene | Low-quality |
| S7_vs_22819\|\|lt2gene | Low-quality |
| S7_vs_22887\|\|lt2gene | Low-quality |
| S7_vs_22920\|\|lt2gene | Low-quality |
| S7_vs_22970\|\|lt2gene | Low-quality |
| S7_vs_22973\|\|lt2gene | Low-quality |
| S7_vs_22981\|\|lt2gene | Low-quality |
| S7_vs_23058\|\|lt2gene | Low-quality |
| S7_vs_23107\|\|lt2gene | Low-quality |
| S7_vs_23149\|\|lt2gene | Low-quality |
| S7_vs_23220\|\|lt2gene | Low-quality |
| S7_vs_23311\|\|lt2gene | Low-quality |
| S7_vs_23323\|\|lt2gene | Low-quality |
| S7_vs_23411\|\|lt2gene | Low-quality |
| S7_vs_23440\|\|lt2gene | Low-quality |
| S7_vs_23485\|\|lt2gene | Low-quality |
| S7_vs_23663\|\|lt2gene | Low-quality |
| S7_vs_23736\|\|lt2gene | Low-quality |
| S7_vs_23820\|\|lt2gene | Not-determined |
| S7_vs_23914\|\|lt2gene | Not-determined |
| S7_vs_23967\|\|lt2gene | Low-quality |
| S7_vs_24025\|\|lt2gene | Low-quality |
| S7_vs_24086\|\|lt2gene | Low-quality |
| S7_vs_24096\|\|lt2gene | Low-quality |
| S7_vs_24102\|\|lt2gene | Low-quality |
| S7_vs_24204\|\|lt2gene | Low-quality |
| S7_vs_24209\|\|lt2gene | Low-quality |
| S7_vs_24257\|\|lt2gene | Not-determined |
| S7_vs_24524\|\|lt2gene | Low-quality |
| S7_vs_24631\|\|lt2gene | Low-quality |
| S7_vs_24704\|\|lt2gene | Low-quality |
| S7_vs_24794\|\|lt2gene | Not-determined |
| S7_vs_24931\|\|lt2gene | Low-quality |
| S7_vs_24949\|\|lt2gene | Low-quality |
| S7_vs_24970\|\|lt2gene | Low-quality |
| S7_vs_25105\|\|lt2gene | Low-quality |
| S7_vs_25147\|\|lt2gene | Low-quality |
| S7_vs_25164\|\|lt2gene | Low-quality |
| S7_vs_25291\|\|lt2gene | Low-quality |
| S7_vs_25445\|\|lt2gene | Not-determined |
| S7_vs_25802\|\|lt2gene | Not-determined |
| S7_vs_25877\|\|lt2gene | Low-quality |
| S7_vs_25916\|\|lt2gene | Low-quality |
| S7_vs_25975\|\|lt2gene | Not-determined |
| S7_vs_26128\|\|lt2gene | Low-quality |
| S7_vs_26171\|\|lt2gene | Low-quality |
| S7_vs_26232\|\|lt2gene | Not-determined |
| S7_vs_26381\|\|lt2gene | Low-quality |
| S7_vs_26472\|\|lt2gene | Low-quality |
| S7_vs_26508\|\|lt2gene | Low-quality |
| S7_vs_26520\|\|lt2gene | Low-quality |
| S7_vs_26525\|\|lt2gene | Low-quality |
| S7_vs_26631\|\|lt2gene | Low-quality |
| S7_vs_26646\|\|lt2gene | Not-determined |
| S7_vs_26802\|\|lt2gene | Low-quality |
| S7_vs_26915\|\|lt2gene | Low-quality |
| S7_vs_26937\|\|lt2gene | Low-quality |
| S7_vs_26982\|\|lt2gene | Not-determined |
| S7_vs_27121\|\|lt2gene | Low-quality |
| S7_vs_27141\|\|lt2gene | Low-quality |
| S7_vs_27456\|\|lt2gene | Low-quality |
| S7_vs_27477\|\|lt2gene | Low-quality |
| S7_vs_27583\|\|lt2gene | Not-determined |
| S7_vs_27663\|\|lt2gene | Low-quality |
| S7_vs_27975\|\|lt2gene | Low-quality |
| S7_vs_28011\|\|lt2gene | Low-quality |
| S7_vs_28041\|\|lt2gene | Low-quality |
| S7_vs_28188\|\|lt2gene | Not-determined |
| S7_vs_28220\|\|lt2gene | Low-quality |
| S7_vs_28414\|\|lt2gene | Not-determined |
| S7_vs_28473\|\|lt2gene | Low-quality |
| S7_vs_28578\|\|lt2gene | Low-quality |
| S7_vs_28608\|\|lt2gene | Low-quality |
| S7_vs_28672\|\|lt2gene | Low-quality |
| S7_vs_28839\|\|lt2gene | Low-quality |
| S7_vs_28842\|\|lt2gene | Not-determined |
| S7_vs_28876\|\|lt2gene | Low-quality |
| S7_vs_28916\|\|lt2gene | Low-quality |
| S7_vs_28930\|\|lt2gene | Low-quality |
| S7_vs_28953\|\|lt2gene | Not-determined |
| S7_vs_29024\|\|lt2gene | Not-determined |
| S7_vs_29093\|\|lt2gene | Low-quality |
| S7_vs_29097\|\|lt2gene | Low-quality |
| S7_vs_29117\|\|lt2gene | Low-quality |
| S7_vs_29184\|\|lt2gene | Low-quality |
| S7_vs_29251\|\|lt2gene | Not-determined |
| S7_vs_29252\|\|lt2gene | Low-quality |
| S7_vs_29283\|\|lt2gene | Not-determined |
| S7_vs_29338\|\|lt2gene | Not-determined |

S7_vs_29342||lt2gene Not-determined
S7_vs_29344||lt2gene Low-quality
S7_vs_29473||lt2gene Not-determined
S7_vs_29515||lt2gene Low-quality
S7_vs_29561||lt2gene Low-quality
S7_vs_29637||lt2gene Low-quality
S7_vs_29649||lt2gene Not-determined
S7_vs_29680||lt2gene Low-quality
S7_vs_29745||lt2gene Low-quality
S7_vs_29766||lt2gene Low-quality
S7_vs_29794||lt2gene Low-quality
S7_vs_29797||lt2gene Not-determined
S7_vs_29858||lt2gene Low-quality
S7_vs_29999||lt2gene Not-determined
S7_vs_30120||lt2gene Low-quality
S7_vs_30191||lt2gene Low-quality
S7_vs_30212||lt2gene Low-quality
S7_vs_30307||lt2gene Low-quality
S7_vs_30331||lt2gene Low-quality
S7_vs_30359||lt2gene Low-quality
S7_vs_30422||lt2gene Low-quality
S7_vs_30593||lt2gene Low-quality
S7_vs_30657||lt2gene Not-determined
S7_vs_30742||lt2gene Low-quality
S7_vs_30745||lt2gene Not-determined
S7_vs_30753||lt2gene Low-quality
S7_vs_30774||lt2gene Low-quality
S7_vs_30837||lt2gene Not-determined
S7_vs_30841||lt2gene Low-quality
S7_vs_30875||lt2gene Low-quality
S7_vs_30888||lt2gene Low-quality
S7_vs_30996||lt2gene Low-quality
S7_vs_31051||lt2gene Low-quality
S7_vs_31062||lt2gene Low-quality
S7_vs_31066||lt2gene Not-determined
S7_vs_31152||lt2gene Not-determined
S7_vs_31155||lt2gene Low-quality
S7_vs_31178||lt2gene Low-quality
S7_vs_31252||lt2gene Low-quality
S7_vs_31299||lt2gene Low-quality
S7_vs_31340||lt2gene Low-quality
S7_vs_31418||lt2gene Not-determined
S7_vs_31578||lt2gene Low-quality
S7_vs_31715||lt2gene Not-determined
S7_vs_31827||lt2gene Low-quality
S7_vs_31897||lt2gene Low-quality
S7_vs_31990||lt2gene Low-quality
S7_vs_31995||lt2gene Low-quality
S7_vs_32131||lt2gene Low-quality
S7_vs_32207||lt2gene Not-determined
S7_vs_32299||lt2gene Low-quality
S7_vs_32412||lt2gene Not-determined
S7_vs_32464||lt2gene Low-quality
S7_vs_32476||lt2gene Not-determined
S7_vs_32478||lt2gene Low-quality
S7_vs_32497||lt2gene Low-quality
S7_vs_32597||lt2gene Low-quality
S7_vs_32627||lt2gene Not-determined
S7_vs_32782||lt2gene Low-quality
S7_vs_32893||lt2gene Low-quality
S7_vs_32986||lt2gene Not-determined
S7_vs_32991||lt2gene Not-determined
S7_vs_33520||lt2gene Not-determined
S7_vs_33526||lt2gene Low-quality
S7_vs_33545||lt2gene Low-quality
S7_vs_33671||lt2gene Low-quality
S7_vs_33677||lt2gene Low-quality
S7_vs_33864||lt2gene Low-quality
S7_vs_33889||lt2gene Low-quality
S7_vs_33958||lt2gene Low-quality
S7_vs_34094||lt2gene Low-quality
S7_vs_34101||lt2gene Low-quality
S7_vs_34210||lt2gene Not-determined
S7_vs_34229||lt2gene Low-quality
S7_vs_34345||lt2gene Not-determined
S7_vs_34392||lt2gene Low-quality
S7_vs_34734||lt2gene Low-quality
S7_vs_35104||lt2gene Low-quality
S7_vs_35163||lt2gene Low-quality
S7_vs_35165||lt2gene Low-quality
S7_vs_35350||lt2gene Low-quality
S7_vs_35372||lt2gene Low-quality
S7_vs_35379||lt2gene Low-quality
S7_vs_35481||lt2gene Low-quality
S7_vs_35626||lt2gene Not-determined
S7_vs_35654||lt2gene Not-determined
S7_vs_35657||lt2gene Low-quality
S7_vs_35668||lt2gene Low-quality
S7_vs_35730||lt2gene Low-quality
S7_vs_35761||lt2gene Low-quality
S7_vs_35792||lt2gene Low-quality
S7_vs_35795||lt2gene Low-quality
S7_vs_35836||lt2gene Low-quality
S7_vs_35882||lt2gene Low-quality
S7_vs_35930||lt2gene Low-quality
S7_vs_36012||lt2gene Low-quality
S7_vs_36119||lt2gene Low-quality
S7_vs_36177||lt2gene Low-quality
S7_vs_36179||lt2gene Low-quality
S7_vs_36262||lt2gene Low-quality
S7_vs_36312||lt2gene Low-quality
S7_vs_36441||lt2gene Not-determined
S7_vs_36487||lt2gene Low-quality
S7_vs_36488||lt2gene Low-quality
S7_vs_36515||lt2gene Low-quality
S7_vs_36542||lt2gene Not-determined
S7_vs_36545||lt2gene Low-quality
S7_vs_36568||lt2gene Low-quality
S7_vs_36571||lt2gene Low-quality
S7_vs_36667||lt2gene Low-quality
S7_vs_36720||lt2gene Low-quality
S7_vs_36759||lt2gene Low-quality
S7_vs_36793||lt2gene Low-quality
S7_vs_36939||lt2gene Low-quality
S7_vs_36996||lt2gene Low-quality
S7_vs_37024||lt2gene Not-determined
S7_vs_37052||lt2gene Not-determined
S7_vs_37077||lt2gene Low-quality
S7_vs_37082||lt2gene Low-quality
S7_vs_37134||lt2gene Not-determined
S7_vs_37235||lt2gene Low-quality
S7_vs_37417||lt2gene Low-quality
S7_vs_37451||lt2gene Low-quality
S7_vs_37513||lt2gene Low-quality
S7_vs_37618||lt2gene Low-quality
S7_vs_37676||lt2gene Low-quality
S7_vs_37695||lt2gene Not-determined
S7_vs_37702||lt2gene Low-quality
S7_vs_37729||lt2gene Low-quality
S7_vs_37767||lt2gene Not-determined
S7_vs_37791||lt2gene Not-determined
S7_vs_37916||lt2gene Low-quality
S7_vs_37943||lt2gene Not-determined
S7_vs_37987||lt2gene Low-quality
S7_vs_38135||lt2gene Low-quality
S7_vs_38173||lt2gene Low-quality
S7_vs_38205||lt2gene Low-quality
S7_vs_38233||lt2gene Low-quality
S7_vs_38262||lt2gene Low-quality
S7_vs_38362||lt2gene Not-determined
S7_vs_38375||lt2gene Low-quality
S7_vs_38435||lt2gene Low-quality
S7_vs_38483||lt2gene Low-quality
S7_vs_38496||lt2gene Low-quality
S7_vs_38580||lt2gene Low-quality
S7_vs_38710||lt2gene Low-quality
S7_vs_38722||lt2gene Not-determined
S7_vs_38736||lt2gene Low-quality
S7_vs_38884||lt2gene Low-quality
S7_vs_38900||lt2gene Low-quality
S7_vs_39067||lt2gene Not-determined
S7_vs_39110||lt2gene Low-quality
S7_vs_39121||lt2gene Not-determined
S7_vs_39146||lt2gene Low-quality
S7_vs_39148||lt2gene Low-quality
S7_vs_39153||lt2gene Low-quality
S7_vs_39191||lt2gene Not-determined
S7_vs_39193||lt2gene Not-determined
S7_vs_39267||lt2gene Low-quality
S7_vs_39268||lt2gene Low-quality
S7_vs_39302||lt2gene Not-determined
S7_vs_39319||lt2gene Low-quality
S7_vs_39354||lt2gene Low-quality
S7_vs_39384||lt2gene Low-quality
S7_vs_39428||lt2gene Low-quality
S7_vs_39541||lt2gene Low-quality
S7_vs_39686||lt2gene Low-quality
S7_vs_39816||lt2gene Not-determined
S7_vs_39864||lt2gene Low-quality
S7_vs_39867||lt2gene Low-quality
S7_vs_39924||lt2gene Low-quality
S7_vs_39930||lt2gene Not-determined
S7_vs_39945||lt2gene Not-determined
S7_vs_40036||lt2gene Not-determined
S7_vs_40090||lt2gene Low-quality
S7_vs_40136||lt2gene Low-quality
S7_vs_40293||lt2gene Low-quality
S7_vs_40337||lt2gene Low-quality
S7_vs_40420||lt2gene Low-quality
S7_vs_40474||lt2gene Low-quality
S7_vs_40550||lt2gene Low-quality
S7_vs_40551||lt2gene Low-quality
S7_vs_40578||lt2gene Low-quality
S7_vs_40647||lt2gene Low-quality
S7_vs_40693||lt2gene Low-quality
S7_vs_40732||lt2gene Low-quality
S7_vs_40830||lt2gene Low-quality
S7_vs_40917||lt2gene Not-determined
S7_vs_41193||lt2gene Not-determined
S7_vs_41365||lt2gene Low-quality
S7_vs_41389||lt2gene Low-quality
S7_vs_41403||lt2gene Low-quality
S7_vs_41412||lt2gene Low-quality
S7_vs_41503||lt2gene Not-determined
S7_vs_41542||lt2gene Low-quality
S7_vs_41597||lt2gene Low-quality
S7_vs_41626||lt2gene Low-quality
S7_vs_41671||lt2gene Low-quality
S7_vs_41681||lt2gene Low-quality
S7_vs_41721||lt2gene Low-quality
S7_vs_41722||lt2gene Low-quality
S7_vs_41733||lt2gene Not-determined
S7_vs_41837||lt2gene Not-determined
S7_vs_41964||lt2gene Low-quality
S7_vs_41974||lt2gene Low-quality
S7_vs_42023||lt2gene Low-quality
S7_vs_42112||lt2gene Low-quality
S7_vs_42120||lt2gene Not-determined
S7_vs_42126||lt2gene Low-quality
S7_vs_42170||lt2gene Low-quality
S7_vs_42338||lt2gene Low-quality
S7_vs_42405||lt2gene Not-determined
S7_vs_42409||lt2gene Low-quality
S7_vs_42544||lt2gene Low-quality
S7_vs_42621||lt2gene Low-quality
S7_vs_42699||lt2gene Low-quality
S7_vs_42732||lt2gene Not-determined
S7_vs_42733||lt2gene Low-quality
S7_vs_42773||lt2gene Low-quality
S7_vs_43025||lt2gene Low-quality
S7_vs_43101||lt2gene Low-quality
S7_vs_43155||lt2gene Low-quality
S7_vs_43224||lt2gene Low-quality
S7_vs_43236||lt2gene Not-determined
S7_vs_43314||lt2gene Low-quality
S7_vs_43325||lt2gene Not-determined
S7_vs_43336||lt2gene Low-quality
S7_vs_43428||lt2gene Not-determined
S7_vs_43639||lt2gene Low-quality
S7_vs_43642||lt2gene Not-determined
S7_vs_43717||lt2gene Low-quality
S7_vs_43738||lt2gene Low-quality
S7_vs_43764||lt2gene Not-determined
S7_vs_43786||lt2gene Low-quality
S7_vs_43903||lt2gene Low-quality
S7_vs_43955||lt2gene Low-quality
S7_vs_44035||lt2gene Low-quality
S7_vs_44046||lt2gene Low-quality
S7_vs_44062||lt2gene Low-quality
S7_vs_44094||lt2gene Low-quality
S7_vs_44132||lt2gene Low-quality
S7_vs_44207||lt2gene Not-determined
S7_vs_44253||lt2gene Low-quality
S7_vs_44407||lt2gene Low-quality
S7_vs_44435||lt2gene Not-determined
S7_vs_44450||lt2gene Not-determined
S7_vs_44473||lt2gene Not-determined
S7_vs_44507||lt2gene Not-determined
S7_vs_44570||lt2gene Low-quality
S7_vs_44635||lt2gene Low-quality
S7_vs_44673||lt2gene Low-quality
S7_vs_44779||lt2gene Low-quality
S7_vs_44877||lt2gene Low-quality
S7_vs_44912||lt2gene Low-quality
S7_vs_45270||lt2gene Not-determined
S7_vs_45295||lt2gene Low-quality
S7_vs_45327||lt2gene Low-quality
S7_vs_45410||lt2gene Low-quality
S7_vs_45460||lt2gene Not-determined
S7_vs_45475||lt2gene Low-quality
S7_vs_45514||lt2gene Low-quality
S7_vs_45548||lt2gene Low-quality
S7_vs_45550||lt2gene Low-quality
S7_vs_45559||lt2gene Low-quality
S7_vs_45562||lt2gene Low-quality
S7_vs_45721||lt2gene Not-determined
S7_vs_45800||lt2gene Low-quality
S7_vs_45804||lt2gene Not-determined
S7_vs_45888||lt2gene Low-quality
S7_vs_46001||lt2gene Low-quality
S7_vs_46043||lt2gene Low-quality
S7_vs_46119||lt2gene Low-quality
S7_vs_46179||lt2gene Low-quality
S7_vs_46396||lt2gene Low-quality
S7_vs_46482||lt2gene Low-quality
S7_vs_46486||lt2gene Low-quality
S7_vs_46511||lt2gene Low-quality
S7_vs_46544||lt2gene Low-quality
S7_vs_46551||lt2gene Low-quality
S7_vs_46572||lt2gene Not-determined
S7_vs_46614||lt2gene Not-determined
S7_vs_46646||lt2gene Low-quality
S7_vs_46671||lt2gene Low-quality
S7_vs_46716||lt2gene Low-quality
S7_vs_46742||lt2gene Not-determined
S7_vs_46746||lt2gene Not-determined
S7_vs_46758||lt2gene Low-quality
S7_vs_46813||lt2gene Low-quality
S7_vs_46870||lt2gene Low-quality
S7_vs_46880||lt2gene Low-quality
S7_vs_46936||lt2gene Not-determined
S7_vs_46962||lt2gene Not-determined
S7_vs_46964||lt2gene Low-quality
S7_vs_46978||lt2gene Not-determined
S7_vs_46990||lt2gene Low-quality
S7_vs_47028||lt2gene Low-quality
S7_vs_47193||lt2gene Low-quality
S7_vs_47235||lt2gene Low-quality
S7_vs_47257||lt2gene Low-quality
S7_vs_47270||lt2gene Low-quality
S7_vs_47379||lt2gene Low-quality
S7_vs_47413||lt2gene Not-determined
S7_vs_47417||lt2gene Low-quality
S7_vs_47642||lt2gene Low-quality
S7_vs_47655||lt2gene Not-determined
S7_vs_47901||lt2gene Not-determined
S7_vs_47997||lt2gene Low-quality
S7_vs_48096||lt2gene Not-determined
S7_vs_48123||lt2gene Low-quality
S7_vs_48243||lt2gene Low-quality
S7_vs_48363||lt2gene Low-quality
S7_vs_48400||lt2gene Low-quality
S7_vs_48401||lt2gene Low-quality
S7_vs_48464||lt2gene Low-quality
S7_vs_48662||lt2gene Not-determined
S7_vs_48682||lt2gene Not-determined
S7_vs_48879||lt2gene Low-quality
S7_vs_48975||lt2gene Not-determined
S7_vs_48982||lt2gene Not-determined
S7_vs_49067||lt2gene Not-determined
S7_vs_49131||lt2gene Low-quality
S7_vs_49157||lt2gene Low-quality
S7_vs_49212||lt2gene Not-determined
S7_vs_49282||lt2gene Low-quality
S7_vs_49349||lt2gene Not-determined
S7_vs_49353||lt2gene Low-quality
S7_vs_49410||lt2gene Low-quality
S7_vs_49585||lt2gene Low-quality
S7_vs_49592||lt2gene Low-quality
S7_vs_49635||lt2gene Low-quality
S7_vs_49686||lt2gene Low-quality
S7_vs_49690||lt2gene Low-quality
S7_vs_49781||lt2gene Low-quality
S7_vs_49885||lt2gene Low-quality
S7_vs_49950||lt2gene Low-quality
S7_vs_49959||lt2gene Low-quality
S7_vs_49990||lt2gene Low-quality
S7_vs_49999||lt2gene Low-quality
S7_vs_50121||lt2gene Low-quality
S7_vs_50436||lt2gene Low-quality
S7_vs_50633||lt2gene Low-quality
S7_vs_50664||lt2gene Low-quality
S7_vs_50735||lt2gene Not-determined
S7_vs_50768||lt2gene Low-quality
S7_vs_50958||lt2gene Low-quality
S7_vs_51099||lt2gene Not-determined
S7_vs_51115||lt2gene Not-determined
S7_vs_51149||lt2gene Low-quality
S7_vs_51170||lt2gene Low-quality
S7_vs_51223||lt2gene Low-quality
S7_vs_51300||lt2gene Low-quality
S7_vs_51335||lt2gene Not-determined
S7_vs_51413||lt2gene Low-quality
S7_vs_51470||lt2gene Not-determined
S7_vs_51500||lt2gene Not-determined
S7_vs_51518||lt2gene Low-quality
S7_vs_51542||lt2gene Not-determined
S7_vs_51547||lt2gene Low-quality
S7_vs_51650||lt2gene Not-determined
S7_vs_51811||lt2gene Low-quality
S7_vs_51846||lt2gene Not-determined
S7_vs_51889||lt2gene Low-quality
S7_vs_51914||lt2gene Low-quality
S7_vs_52079||lt2gene Low-quality
S7_vs_52166||lt2gene Not-determined
S7_vs_52215||lt2gene Not-determined
S7_vs_52221||lt2gene Low-quality
S7_vs_52296||lt2gene Low-quality
S7_vs_52348||lt2gene Low-quality
S7_vs_52405||lt2gene Low-quality
S7_vs_52442||lt2gene Not-determined
S7_vs_52573||lt2gene Low-quality
S7_vs_52601||lt2gene Not-determined
S7_vs_52636||lt2gene Not-determined
S7_vs_52793||lt2gene Not-determined
S7_vs_52794||lt2gene Low-quality
S7_vs_53029||lt2gene Low-quality
S7_vs_53084||lt2gene Low-quality
S7_vs_53138||lt2gene Low-quality
S7_vs_53163||lt2gene Low-quality
S7_vs_53188||lt2gene Low-quality
S7_vs_53301||lt2gene Low-quality
S7_vs_53374||lt2gene Low-quality
S7_vs_53450||lt2gene Low-quality
S7_vs_53462||lt2gene Low-quality
S7_vs_53502||lt2gene Not-determined
S7_vs_53618||lt2gene Not-determined
S7_vs_53666||lt2gene Low-quality
S7_vs_53726||lt2gene Not-determined
S7_vs_53776||lt2gene Low-quality
S7_vs_53812||lt2gene Not-determined
S7_vs_53837||lt2gene Low-quality
S7_vs_53872||lt2gene Low-quality
S7_vs_53928||lt2gene Low-quality
S7_vs_53975||lt2gene Not-determined
S7_vs_53985||lt2gene Low-quality
S7_vs_53988||lt2gene Low-quality
S7_vs_54008||lt2gene Low-quality
S7_vs_54048||lt2gene Low-quality
S7_vs_54058||lt2gene Low-quality
S7_vs_54090||lt2gene Low-quality
S7_vs_54194||lt2gene Low-quality
S7_vs_54273||lt2gene Low-quality
S7_vs_54278||lt2gene Low-quality
S7_vs_54325||lt2gene Not-determined
S7_vs_54406||lt2gene Not-determined
S7_vs_54420||lt2gene Low-quality
S7_vs_54525||lt2gene Low-quality
S7_vs_54529||lt2gene Low-quality
S7_vs_54543||lt2gene Low-quality
S7_vs_54605||lt2gene Not-determined
S7_vs_54662||lt2gene Not-determined
S7_vs_54783||lt2gene Low-quality
S7_vs_54812||lt2gene Low-quality
S7_vs_54853||lt2gene Low-quality
S7_vs_54929||lt2gene Not-determined
S7_vs_55074||lt2gene Low-quality
S7_vs_55086||lt2gene Not-determined
S7_vs_55100||lt2gene Low-quality
S7_vs_55103||lt2gene Low-quality
S7_vs_55117||lt2gene Low-quality
S7_vs_55147||lt2gene Low-quality
S7_vs_55204||lt2gene Not-determined
S7_vs_55254||lt2gene Low-quality
S7_vs_55278||lt2gene Low-quality
S7_vs_55319||lt2gene Low-quality
S7_vs_55320||lt2gene Low-quality
S7_vs_55480||lt2gene Low-quality
S7_vs_55547||lt2gene Low-quality
S7_vs_55650||lt2gene Low-quality
S7_vs_55713||lt2gene Low-quality
S7_vs_55720||lt2gene Not-determined
S7_vs_55920||lt2gene Low-quality
S7_vs_55924||lt2gene Not-determined
S7_vs_56002||lt2gene Not-determined
S7_vs_56192||lt2gene Low-quality
S7_vs_56255||lt2gene Low-quality
S7_vs_56293||lt2gene Not-determined
S7_vs_56299||lt2gene Low-quality
S7_vs_56301||lt2gene Low-quality
S7_vs_56348||lt2gene Not-determined
S7_vs_56381||lt2gene Low-quality
S7_vs_56459||lt2gene Low-quality
S7_vs_56525||lt2gene Not-determined
S7_vs_56595||lt2gene Not-determined
S7_vs_56613||lt2gene Not-determined
S7_vs_56652||lt2gene Low-quality
S7_vs_56670||lt2gene Low-quality
S7_vs_56694||lt2gene Low-quality
S7_vs_56868||lt2gene Low-quality
S7_vs_56890||lt2gene Low-quality
S7_vs_56918||lt2gene Low-quality
S7_vs_56962||lt2gene Low-quality
S7_vs_56994||lt2gene Low-quality
S7_vs_57056||lt2gene Low-quality
S7_vs_57075||lt2gene Low-quality
S7_vs_57177||lt2gene Low-quality
S7_vs_57252||lt2gene Low-quality
S7_vs_57449||lt2gene Low-quality
S7_vs_57450||lt2gene Low-quality
S7_vs_57624||lt2gene Low-quality
S7_vs_57660||lt2gene Low-quality
S7_vs_57918||lt2gene Not-determined
S7_vs_57969||lt2gene Low-quality
S7_vs_58009||lt2gene Low-quality
S7_vs_58271||lt2gene Low-quality
S7_vs_58276||lt2gene Low-quality
S7_vs_58317||lt2gene Low-quality
S7_vs_58336||lt2gene Not-determined
S7_vs_58357||lt2gene Low-quality
S7_vs_58436||lt2gene Low-quality
S7_vs_58459||lt2gene Low-quality
S7_vs_58586||lt2gene Low-quality
S7_vs_58799||lt2gene Low-quality
S7_vs_58830||lt2gene Low-quality
S7_vs_58954||lt2gene Low-quality
S7_vs_59013||lt2gene Low-quality
S7_vs_59045||lt2gene Low-quality
S7_vs_59093||lt2gene Not-determined
S7_vs_59107||lt2gene Low-quality
S7_vs_59201||lt2gene Not-determined
S7_vs_59204||lt2gene Not-determined
S7_vs_59223||lt2gene Low-quality
S7_vs_59332||lt2gene Low-quality
S7_vs_59357||lt2gene Low-quality
S7_vs_59431||lt2gene Low-quality
S7_vs_59440||lt2gene Not-determined
S7_vs_59444||lt2gene Low-quality
S7_vs_59464||lt2gene Low-quality
S7_vs_59551||lt2gene Low-quality
S7_vs_59561||lt2gene Low-quality
S7_vs_59575||lt2gene Low-quality
S7_vs_59610||lt2gene Low-quality
S7_vs_59625||lt2gene Low-quality
S7_vs_59766||lt2gene Low-quality
S7_vs_59782||lt2gene Low-quality
S7_vs_59977||lt2gene Low-quality
S7_vs_59987||lt2gene Low-quality
S7_vs_60069||lt2gene Low-quality
S7_vs_60108||lt2gene Not-determined
S7_vs_60189||lt2gene Low-quality
S7_vs_60284||lt2gene Low-quality
S7_vs_60337||lt2gene Low-quality
S7_vs_60478||lt2gene Low-quality
S7_vs_60521||lt2gene Low-quality
S7_vs_60588||lt2gene Not-determined
S7_vs_60633||lt2gene Not-determined
S7_vs_60639||lt2gene Low-quality
S7_vs_60894||lt2gene Low-quality
S7_vs_60896||lt2gene Low-quality
S7_vs_61011||lt2gene Low-quality
S7_vs_61046||lt2gene Low-quality
S7_vs_61107||lt2gene Not-determined
S7_vs_61250||lt2gene Low-quality
S7_vs_61344||lt2gene Not-determined
S7_vs_61382||lt2gene Low-quality
S7_vs_61398||lt2gene Low-quality
S7_vs_61402||lt2gene Low-quality
S7_vs_61403||lt2gene Low-quality
S7_vs_61428||lt2gene Low-quality
S7_vs_61553||lt2gene Not-determined
S7_vs_61594||lt2gene Low-quality
S7_vs_61595||lt2gene Low-quality
S7_vs_61735||lt2gene Low-quality
S7_vs_61800||lt2gene Low-quality
S7_vs_61844||lt2gene Low-quality
S7_vs_61886||lt2gene Not-determined
S7_vs_61980||lt2gene Low-quality
S7_vs_61990||lt2gene Not-determined
S7_vs_62013||lt2gene Not-determined
S7_vs_62014||lt2gene Low-quality
S7_vs_62042||lt2gene Low-quality
S7_vs_62048||lt2gene Low-quality
S7_vs_62075||lt2gene Low-quality
S7_vs_62241||lt2gene Low-quality
S7_vs_62262||lt2gene Low-quality
S7_vs_62333||lt2gene Low-quality
S7_vs_62335||lt2gene Low-quality
S7_vs_62369||lt2gene Low-quality
S7_vs_62397||lt2gene Not-determined
S7_vs_62409||lt2gene Low-quality
S7_vs_62538||lt2gene Low-quality
S7_vs_62680||lt2gene Low-quality
S7_vs_62698||lt2gene Low-quality
S7_vs_62724||lt2gene Low-quality
S7_vs_62833||lt2gene Low-quality
S7_vs_62901||lt2gene Low-quality
S7_vs_62988||lt2gene Not-determined
S7_vs_63022||lt2gene Low-quality
S7_vs_63065||lt2gene Not-determined
S7_vs_63222||lt2gene Low-quality
S7_vs_63225||lt2gene Low-quality
S7_vs_63275||lt2gene Not-determined
S7_vs_63375||lt2gene Low-quality
S7_vs_63409||lt2gene Low-quality
S7_vs_63427||lt2gene Low-quality
S7_vs_63440||lt2gene Low-quality
S7_vs_63443||lt2gene Low-quality
S7_vs_63477||lt2gene Low-quality
S7_vs_63546||lt2gene Low-quality
S7_vs_63560||lt2gene Not-determined
S7_vs_63598||lt2gene Low-quality
S7_vs_63614||lt2gene Low-quality
S7_vs_63628||lt2gene Low-quality
S7_vs_63724||lt2gene Low-quality
S7_vs_63749||lt2gene Low-quality
S7_vs_63836||lt2gene Low-quality
S7_vs_63848||lt2gene Low-quality
S7_vs_63885||lt2gene Low-quality
S7_vs_64154||lt2gene Low-quality
S7_vs_64307||lt2gene Low-quality
S7_vs_64371||lt2gene Low-quality
S7_vs_64443||lt2gene Low-quality
S7_vs_64445||lt2gene Low-quality
S7_vs_64511||lt2gene Low-quality
S7_vs_64593||lt2gene Low-quality
S7_vs_64606||lt2gene Low-quality
S7_vs_64618||lt2gene Low-quality
S7_vs_64661||lt2gene Not-determined
S7_vs_64678||lt2gene Low-quality
S7_vs_64835||lt2gene Low-quality
S7_vs_64844||lt2gene Low-quality
S7_vs_64904||lt2gene Low-quality
S7_vs_64967||lt2gene Low-quality
S7_vs_65076||lt2gene Low-quality
S7_vs_65160||lt2gene Low-quality
S7_vs_65286||lt2gene Not-determined
S7_vs_65304||lt2gene Low-quality
S7_vs_65372||lt2gene Not-determined

| | |
|---|---|
| S7_vs_65377\|\|lt2gene | Low-quality |
| S7_vs_65491\|\|lt2gene | Low-quality |
| S7_vs_65515\|\|lt2gene | Not-determined |
| S7_vs_65538\|\|lt2gene | Not-determined |
| S7_vs_65571\|\|lt2gene | Low-quality |
| S7_vs_65615\|\|lt2gene | Not-determined |
| S7_vs_65624\|\|lt2gene | Low-quality |
| S7_vs_65676\|\|lt2gene | Low-quality |
| S7_vs_65687\|\|lt2gene | Low-quality |
| S7_vs_65942\|\|lt2gene | Not-determined |
| S7_vs_65995\|\|lt2gene | Not-determined |
| S7_vs_66009\|\|lt2gene | Not-determined |
| S7_vs_66116\|\|lt2gene | Low-quality |
| S7_vs_66129\|\|lt2gene | Not-determined |
| S7_vs_66143\|\|lt2gene | Low-quality |
| S7_vs_66191\|\|lt2gene | Low-quality |
| S7_vs_66337\|\|lt2gene | Low-quality |
| S7_vs_66427\|\|lt2gene | Not-determined |
| S7_vs_66430\|\|lt2gene | Not-determined |
| S7_vs_66492\|\|lt2gene | Not-determined |
| S7_vs_66601\|\|lt2gene | Low-quality |
| S7_vs_66626\|\|lt2gene | Low-quality |
| S7_vs_66732\|\|lt2gene | Low-quality |
| S7_vs_66874\|\|lt2gene | Low-quality |
| S7_vs_66971\|\|lt2gene | Not-determined |
| S7_vs_67035\|\|lt2gene | Low-quality |
| S7_vs_67053\|\|lt2gene | Low-quality |
| S7_vs_67109\|\|lt2gene | Low-quality |
| S7_vs_67397\|\|lt2gene | Not-determined |
| S7_vs_67454\|\|lt2gene | Low-quality |
| S7_vs_67470\|\|lt2gene | Low-quality |
| S7_vs_67502\|\|lt2gene | Low-quality |
| S7_vs_67630\|\|lt2gene | Low-quality |
| S7_vs_67759\|\|lt2gene | Not-determined |
| S7_vs_67776\|\|lt2gene | Low-quality |
| S7_vs_67838\|\|lt2gene | Low-quality |
| S7_vs_68132\|\|lt2gene | Low-quality |
| S7_vs_68267\|\|lt2gene | Low-quality |
| S7_vs_68288\|\|lt2gene | Low-quality |
| S7_vs_68298\|\|lt2gene | Not-determined |
| S7_vs_68330\|\|lt2gene | Not-determined |
| S7_vs_68448\|\|lt2gene | Not-determined |
| S7_vs_68465\|\|lt2gene | Low-quality |
| S7_vs_68515\|\|lt2gene | Not-determined |
| S7_vs_68519\|\|lt2gene | Not-determined |
| S7_vs_68525\|\|lt2gene | Low-quality |
| S7_vs_68589\|\|lt2gene | Low-quality |
| S7_vs_68803\|\|lt2gene | Not-determined |
| S7_vs_68816\|\|lt2gene | Low-quality |
| S7_vs_68818\|\|lt2gene | Low-quality |
| S7_vs_68890\|\|lt2gene | Low-quality |
| S7_vs_68933\|\|lt2gene | Low-quality |
| S7_vs_68942\|\|lt2gene | Low-quality |
| S7_vs_68951\|\|lt2gene | Low-quality |
| S7_vs_69010\|\|lt2gene | Low-quality |
| S7_vs_69240\|\|lt2gene | Low-quality |
| S7_vs_69308\|\|lt2gene | Low-quality |
| S7_vs_69324\|\|lt2gene | Low-quality |
| S7_vs_69378\|\|lt2gene | Low-quality |
| S7_vs_69382\|\|lt2gene | Low-quality |
| S7_vs_69466\|\|lt2gene | Low-quality |
| S7_vs_69467\|\|lt2gene | Low-quality |
| S7_vs_69502\|\|lt2gene | Not-determined |
| S7_vs_69869\|\|lt2gene | Low-quality |
| S7_vs_69948\|\|lt2gene | Low-quality |
| S7_vs_69954\|\|lt2gene | Low-quality |
| S7_vs_70066\|\|lt2gene | Low-quality |
| S7_vs_70073\|\|lt2gene | Not-determined |
| S7_vs_70132\|\|lt2gene | Not-determined |
| S7_vs_70164\|\|lt2gene | Low-quality |
| S7_vs_70168\|\|lt2gene | Low-quality |
| S7_vs_70193\|\|lt2gene | Low-quality |
| S7_vs_70236\|\|lt2gene | Low-quality |
| S7_vs_70298\|\|lt2gene | Low-quality |
| S7_vs_70320\|\|lt2gene | Low-quality |
| S7_vs_70486\|\|lt2gene | Low-quality |
| S7_vs_70543\|\|lt2gene | Not-determined |
| S7_vs_70564\|\|lt2gene | Not-determined |
| S7_vs_70673\|\|lt2gene | Low-quality |
| S7_vs_70691\|\|lt2gene | Not-determined |
| S7_vs_70866\|\|lt2gene | Low-quality |
| S7_vs_70966\|\|lt2gene | Low-quality |
| S7_vs_71038\|\|lt2gene | Low-quality |
| S7_vs_71105\|\|lt2gene | Low-quality |
| S7_vs_71111\|\|lt2gene | Low-quality |
| S7_vs_71113\|\|lt2gene | Low-quality |
| S7_vs_71159\|\|lt2gene | Low-quality |
| S7_vs_71164\|\|lt2gene | Low-quality |
| S7_vs_71248\|\|lt2gene | Low-quality |
| S7_vs_71269\|\|lt2gene | Low-quality |
| S7_vs_71273\|\|lt2gene | Not-determined |
| S7_vs_71311\|\|lt2gene | Low-quality |
| S7_vs_71367\|\|lt2gene | Low-quality |
| S7_vs_71466\|\|lt2gene | Not-determined |
| S7_vs_71477\|\|lt2gene | Low-quality |
| S7_vs_71537\|\|lt2gene | Low-quality |
| S7_vs_71643\|\|lt2gene | Low-quality |
| S7_vs_71649\|\|lt2gene | Low-quality |
| S7_vs_71670\|\|lt2gene | Not-determined |
| S7_vs_71700\|\|lt2gene | Low-quality |
| S7_vs_71778\|\|lt2gene | Low-quality |
| S7_vs_71786\|\|lt2gene | Not-determined |
| S7_vs_71815\|\|lt2gene | Low-quality |
| S7_vs_71842\|\|lt2gene | Low-quality |
| S7_vs_71988\|\|lt2gene | Low-quality |
| S7_vs_72006\|\|lt2gene | Low-quality |
| S7_vs_72064\|\|lt2gene | Not-determined |
| S7_vs_72072\|\|lt2gene | Not-determined |
| S7_vs_72113\|\|lt2gene | Low-quality |
| S7_vs_72147\|\|lt2gene | Low-quality |
| S7_vs_72277\|\|lt2gene | Low-quality |
| S7_vs_72347\|\|lt2gene | Low-quality |
| S7_vs_72353\|\|lt2gene | Low-quality |
| S7_vs_72393\|\|lt2gene | Low-quality |
| S7_vs_72407\|\|lt2gene | Low-quality |
| S7_vs_72456\|\|lt2gene | Low-quality |
| S7_vs_72467\|\|lt2gene | Low-quality |
| S7_vs_72545\|\|lt2gene | Low-quality |
| S7_vs_72651\|\|lt2gene | Not-determined |
| S7_vs_72657\|\|lt2gene | Low-quality |
| S7_vs_72669\|\|lt2gene | Not-determined |
| S7_vs_72742\|\|lt2gene | Not-determined |
| S7_vs_72758\|\|lt2gene | Low-quality |
| S7_vs_72780\|\|lt2gene | Low-quality |
| S7_vs_72883\|\|lt2gene | Low-quality |
| S7_vs_72973\|\|lt2gene | Low-quality |
| S7_vs_73007\|\|lt2gene | Low-quality |
| S7_vs_73413\|\|lt2gene | Low-quality |
| S7_vs_73417\|\|lt2gene | Low-quality |
| S7_vs_73516\|\|lt2gene | Low-quality |
| S7_vs_73544\|\|lt2gene | Low-quality |
| S7_vs_73553\|\|lt2gene | Low-quality |
| S7_vs_73674\|\|lt2gene | Low-quality |
| S7_vs_73777\|\|lt2gene | Low-quality |
| S7_vs_73965\|\|lt2gene | Low-quality |
| S7_vs_73983\|\|lt2gene | Low-quality |
| S7_vs_73997\|\|lt2gene | Low-quality |
| S7_vs_74037\|\|lt2gene | Low-quality |
| S7_vs_74048\|\|lt2gene | Low-quality |
| S7_vs_74079\|\|lt2gene | Low-quality |
| S7_vs_74098\|\|lt2gene | Low-quality |
| S7_vs_74253\|\|lt2gene | Low-quality |
| S7_vs_74296\|\|lt2gene | Low-quality |
| S7_vs_74316\|\|lt2gene | Low-quality |
| S7_vs_74338\|\|lt2gene | Low-quality |
| S7_vs_74361\|\|lt2gene | Not-determined |
| S7_vs_74414\|\|lt2gene | Low-quality |
| S7_vs_74509\|\|lt2gene | Not-determined |
| S7_vs_74514\|\|lt2gene | Low-quality |
| S7_vs_74550\|\|lt2gene | Not-determined |
| S7_vs_74592\|\|lt2gene | Low-quality |
| S7_vs_74640\|\|lt2gene | Low-quality |
| S7_vs_74749\|\|lt2gene | Low-quality |
| S7_vs_75066\|\|lt2gene | Low-quality |
| S7_vs_75122\|\|lt2gene | Low-quality |
| S7_vs_75151\|\|lt2gene | Low-quality |
| S7_vs_75228\|\|lt2gene | Low-quality |
| S7_vs_75434\|\|lt2gene | Low-quality |
| S7_vs_75515\|\|lt2gene | Low-quality |
| S7_vs_75523\|\|lt2gene | Low-quality |
| S7_vs_75539\|\|lt2gene | Low-quality |
| S7_vs_75610\|\|lt2gene | Low-quality |
| S7_vs_75665\|\|lt2gene | Not-determined |
| S7_vs_75696\|\|lt2gene | Low-quality |
| S7_vs_75734\|\|lt2gene | Low-quality |
| S7_vs_75862\|\|lt2gene | Low-quality |
| S7_vs_75987\|\|lt2gene | Low-quality |
| S7_vs_76053\|\|lt2gene | Low-quality |
| S7_vs_76103\|\|lt2gene | Low-quality |
| S7_vs_76109\|\|lt2gene | Low-quality |
| S7_vs_76158\|\|lt2gene | Low-quality |
| S7_vs_76168\|\|lt2gene | Low-quality |
| S7_vs_76239\|\|lt2gene | Low-quality |
| S7_vs_76262\|\|lt2gene | Low-quality |
| S7_vs_76447\|\|lt2gene | Low-quality |
| S7_vs_76498\|\|lt2gene | Low-quality |
| S7_vs_76563\|\|lt2gene | Low-quality |
| S7_vs_76619\|\|lt2gene | Not-determined |
| S7_vs_76685\|\|lt2gene | Low-quality |
| S7_vs_76721\|\|lt2gene | Not-determined |
| S7_vs_76791\|\|lt2gene | Low-quality |
| S7_vs_76792\|\|lt2gene | Low-quality |
| S7_vs_76891\|\|lt2gene | Low-quality |
| S7_vs_76901\|\|lt2gene | Low-quality |
| S7_vs_76905\|\|lt2gene | Low-quality |
| S7_vs_76918\|\|lt2gene | Low-quality |
| S7_vs_77071\|\|lt2gene | Low-quality |
| S7_vs_77122\|\|lt2gene | Low-quality |
| S7_vs_77162\|\|lt2gene | Not-determined |
| S7_vs_77168\|\|lt2gene | Low-quality |
| S7_vs_77310\|\|lt2gene | Low-quality |
| S7_vs_77401\|\|lt2gene | Not-determined |
| S7_vs_77413\|\|lt2gene | Low-quality |
| S7_vs_77448\|\|lt2gene | Low-quality |
| S7_vs_77460\|\|lt2gene | Low-quality |
| S7_vs_77506\|\|lt2gene | Low-quality |
| S7_vs_77602\|\|lt2gene | Low-quality |
| S7_vs_77610\|\|lt2gene | Low-quality |
| S7_vs_77652\|\|lt2gene | Not-determined |
| S7_vs_77724\|\|lt2gene | Low-quality |
| S7_vs_77764\|\|lt2gene | Low-quality |
| S7_vs_77921\|\|lt2gene | Not-determined |
| S7_vs_78315\|\|lt2gene | Not-determined |
| S7_vs_78494\|\|lt2gene | Low-quality |
| S7_vs_78498\|\|lt2gene | Low-quality |
| S7_vs_78526\|\|lt2gene | Low-quality |
| S7_vs_78545\|\|lt2gene | Low-quality |
| S7_vs_78631\|\|lt2gene | Low-quality |
| S7_vs_78644\|\|lt2gene | Not-determined |
| S7_vs_78690\|\|lt2gene | Low-quality |
| S7_vs_78777\|\|lt2gene | Low-quality |
| S7_vs_78837\|\|lt2gene | Low-quality |
| S7_vs_78858\|\|lt2gene | Not-determined |
| S7_vs_78875\|\|lt2gene | Not-determined |
| S7_vs_78983\|\|lt2gene | Low-quality |
| S7_vs_79039\|\|lt2gene | Not-determined |
| S7_vs_79244\|\|lt2gene | Not-determined |
| S7_vs_79286\|\|lt2gene | Not-determined |
| S7_vs_79291\|\|lt2gene | Low-quality |
| S7_vs_79401\|\|lt2gene | Not-determined |
| S7_vs_79438\|\|lt2gene | Not-determined |
| S7_vs_79481\|\|lt2gene | Not-determined |
| S7_vs_79500\|\|lt2gene | Low-quality |
| S7_vs_79557\|\|lt2gene | Low-quality |
| S7_vs_79561\|\|lt2gene | Low-quality |
| S7_vs_79714\|\|lt2gene | Not-determined |
| S7_vs_79853\|\|lt2gene | Low-quality |
| S7_vs_79859\|\|lt2gene | Not-determined |
| S7_vs_79886\|\|lt2gene | Not-determined |
| S7_vs_79925\|\|lt2gene | Low-quality |
| S7_vs_79967\|\|lt2gene | Low-quality |
| S7_vs_80002\|\|lt2gene | Low-quality |
| S7_vs_80179\|\|lt2gene | Low-quality |
| S7_vs_80207\|\|lt2gene | Low-quality |
| S7_vs_80298\|\|lt2gene | Low-quality |
| S7_vs_80303\|\|lt2gene | Low-quality |
| S7_vs_80308\|\|lt2gene | Low-quality |
| S7_vs_80378\|\|lt2gene | Not-determined |
| S7_vs_80416\|\|lt2gene | Low-quality |
| S7_vs_80519\|\|lt2gene | Low-quality |
| S7_vs_80534\|\|lt2gene | Not-determined |
| S7_vs_80579\|\|lt2gene | Low-quality |
| S7_vs_80585\|\|lt2gene | Not-determined |
| S7_vs_80607\|\|lt2gene | Low-quality |
| S7_vs_80745\|\|lt2gene | Low-quality |
| S7_vs_80859\|\|lt2gene | Low-quality |
| S7_vs_80894\|\|lt2gene | Not-determined |
| S7_vs_80912\|\|lt2gene | Not-determined |
| S7_vs_80950\|\|lt2gene | Not-determined |
| S7_vs_80962\|\|lt2gene | Low-quality |
| S7_vs_80996\|\|lt2gene | Not-determined |
| S7_vs_81041\|\|lt2gene | Not-determined |
| S7_vs_81157\|\|lt2gene | Low-quality |
| S7_vs_81246\|\|lt2gene | Low-quality |
| S7_vs_81276\|\|lt2gene | Low-quality |
| S7_vs_81315\|\|lt2gene | Low-quality |
| S7_vs_81327\|\|lt2gene | Low-quality |
| S7_vs_81348\|\|lt2gene | Low-quality |
| S7_vs_81376\|\|lt2gene | Low-quality |
| S7_vs_81460\|\|lt2gene | Low-quality |
| S7_vs_81503\|\|lt2gene | Low-quality |
| S7_vs_81532\|\|lt2gene | Low-quality |
| S7_vs_81539\|\|lt2gene | Low-quality |
| S7_vs_81578\|\|lt2gene | Low-quality |
| S7_vs_81829\|\|lt2gene | Low-quality |
| S7_vs_81864\|\|lt2gene | Low-quality |
| S7_vs_81892\|\|lt2gene | Low-quality |
| S7_vs_81998\|\|lt2gene | Low-quality |
| S7_vs_82024\|\|lt2gene | Low-quality |
| S7_vs_82050\|\|lt2gene | Low-quality |
| S7_vs_82051\|\|lt2gene | Not-determined |
| S7_vs_82179\|\|lt2gene | Not-determined |
| S7_vs_82190\|\|lt2gene | Not-determined |
| S7_vs_82212\|\|lt2gene | Not-determined |
| S7_vs_82271\|\|lt2gene | Low-quality |
| S7_vs_82285\|\|lt2gene | Low-quality |
| S7_vs_82338\|\|lt2gene | Low-quality |
| S7_vs_82386\|\|lt2gene | Low-quality |
| S7_vs_82395\|\|lt2gene | Low-quality |
| S7_vs_82403\|\|lt2gene | Low-quality |
| S7_vs_82448\|\|lt2gene | Low-quality |
| S7_vs_82504\|\|lt2gene | Low-quality |
| S7_vs_82530\|\|lt2gene | Low-quality |
| S7_vs_82572\|\|lt2gene | Low-quality |
| S7_vs_82634\|\|lt2gene | Not-determined |
| S7_vs_82639\|\|lt2gene | Low-quality |
| S7_vs_82684\|\|lt2gene | Low-quality |
| S7_vs_82745\|\|lt2gene | Not-determined |
| S7_vs_82784\|\|lt2gene | Not-determined |
| S7_vs_82820\|\|lt2gene | Low-quality |
| S7_vs_82904\|\|lt2gene | Low-quality |
| S7_vs_82920\|\|lt2gene | Low-quality |
| S7_vs_82930\|\|lt2gene | Low-quality |
| S7_vs_82969\|\|lt2gene | Low-quality |
| S7_vs_82974\|\|lt2gene | Not-determined |
| S7_vs_83054\|\|lt2gene | Low-quality |
| S7_vs_83055\|\|lt2gene | Low-quality |
| S7_vs_83087\|\|lt2gene | Not-determined |
| S7_vs_83206\|\|lt2gene | Low-quality |
| S7_vs_83212\|\|lt2gene | Low-quality |
| S7_vs_83224\|\|lt2gene | Low-quality |
| S7_vs_83237\|\|lt2gene | Low-quality |
| S7_vs_83310\|\|lt2gene | Not-determined |
| S7_vs_83481\|\|lt2gene | Low-quality |
| S7_vs_83604\|\|lt2gene | Low-quality |
| S7_vs_83606\|\|lt2gene | Low-quality |
| S7_vs_83667\|\|lt2gene | Not-determined |
| S7_vs_83863\|\|lt2gene | Low-quality |
| S7_vs_83881\|\|lt2gene | Not-determined |
| S7_vs_83897\|\|lt2gene | Low-quality |
| S7_vs_84012\|\|lt2gene | Low-quality |
| S7_vs_84049\|\|lt2gene | Low-quality |
| S7_vs_84169\|\|lt2gene | Not-determined |
| S7_vs_84228\|\|lt2gene | Low-quality |
| S7_vs_84264\|\|lt2gene | Not-determined |
| S7_vs_84317\|\|lt2gene | Low-quality |
| S7_vs_84323\|\|lt2gene | Low-quality |
| S7_vs_84330\|\|lt2gene | Low-quality |
| S7_vs_84382\|\|lt2gene | Not-determined |
| S7_vs_84425\|\|lt2gene | Low-quality |
| S7_vs_84436\|\|lt2gene | Low-quality |
| S7_vs_84593\|\|lt2gene | Low-quality |
| S7_vs_84707\|\|lt2gene | Low-quality |
| S7_vs_84737\|\|lt2gene | Low-quality |
| S7_vs_84809\|\|lt2gene | Low-quality |
| S7_vs_84941\|\|lt2gene | Low-quality |
| S7_vs_84942\|\|lt2gene | Low-quality |
| S7_vs_84968\|\|lt2gene | Low-quality |
| S7_vs_84973\|\|lt2gene | Low-quality |
| S7_vs_85004\|\|lt2gene | Low-quality |
| S7_vs_85089\|\|lt2gene | Low-quality |
| S7_vs_85111\|\|lt2gene | Not-determined |
| S7_vs_85119\|\|lt2gene | Low-quality |
| S7_vs_85125\|\|lt2gene | Low-quality |
| S7_vs_85149\|\|lt2gene | Low-quality |
| S7_vs_85224\|\|lt2gene | Low-quality |
| S7_vs_85333\|\|lt2gene | Low-quality |
| S7_vs_85497\|\|lt2gene | Low-quality |
| S7_vs_85640\|\|lt2gene | Low-quality |
| S7_vs_85741\|\|lt2gene | Low-quality |
| S7_vs_85857\|\|lt2gene | Low-quality |
| S7_vs_85931\|\|lt2gene | Low-quality |
| S7_vs_85985\|\|lt2gene | Not-determined |
| S7_vs_85999\|\|lt2gene | Low-quality |
| S7_vs_86065\|\|lt2gene | Low-quality |
| S7_vs_86076\|\|lt2gene | Not-determined |
| S7_vs_86121\|\|lt2gene | Not-determined |
| S7_vs_86139\|\|lt2gene | Not-determined |
| S7_vs_86146\|\|lt2gene | Low-quality |
| S7_vs_86197\|\|lt2gene | Not-determined |
| S7_vs_86267\|\|lt2gene | Low-quality |
| S7_vs_86311\|\|lt2gene | Low-quality |
| S7_vs_86320\|\|lt2gene | Not-determined |
| S7_vs_86396\|\|lt2gene | Low-quality |
| S7_vs_86406\|\|lt2gene | Low-quality |
| S7_vs_86410\|\|lt2gene | Low-quality |
| S7_vs_86479\|\|lt2gene | Low-quality |
| S7_vs_86526\|\|lt2gene | Not-determined |
| S7_vs_86679\|\|lt2gene | Low-quality |
| S7_vs_86709\|\|lt2gene | Low-quality |
| S7_vs_86719\|\|lt2gene | Low-quality |
| S7_vs_86779\|\|lt2gene | Low-quality |
| S7_vs_86813\|\|lt2gene | Low-quality |
| S7_vs_86815\|\|lt2gene | Low-quality |
| S7_vs_86925\|\|lt2gene | Not-determined |
| S7_vs_86940\|\|lt2gene | Low-quality |
| S7_vs_86975\|\|lt2gene | Low-quality |
| S7_vs_87009\|\|lt2gene | Not-determined |
| S7_vs_87036\|\|lt2gene | Low-quality |
| S7_vs_87048\|\|lt2gene | Low-quality |
| S7_vs_87134\|\|lt2gene | Low-quality |
| S7_vs_87182\|\|lt2gene | Low-quality |
| S7_vs_87303\|\|lt2gene | Low-quality |
| S7_vs_87365\|\|lt2gene | Low-quality |
| S7_vs_87369\|\|lt2gene | Low-quality |
| S7_vs_87464\|\|lt2gene | Low-quality |
| S7_vs_87546\|\|lt2gene | Low-quality |
| S7_vs_87625\|\|lt2gene | Not-determined |
| S7_vs_87648\|\|lt2gene | Low-quality |
| S7_vs_87672\|\|lt2gene | Low-quality |
| S7_vs_87700\|\|lt2gene | Low-quality |
| S7_vs_87766\|\|lt2gene | Low-quality |
| S7_vs_87772\|\|lt2gene | Not-determined |
| S7_vs_87872\|\|lt2gene | Low-quality |
| S7_vs_87927\|\|lt2gene | Low-quality |
| S7_vs_88036\|\|lt2gene | Low-quality |
| S7_vs_88133\|\|lt2gene | Low-quality |
| S7_vs_88192\|\|lt2gene | Low-quality |
| S7_vs_88227\|\|lt2gene | Low-quality |
| S7_vs_88280\|\|lt2gene | Low-quality |
| S7_vs_88333\|\|lt2gene | Low-quality |
| S7_vs_88334\|\|lt2gene | Low-quality |
| S7_vs_88335\|\|lt2gene | Low-quality |
| S7_vs_88541\|\|lt2gene | Low-quality |
| S7_vs_88588\|\|lt2gene | Low-quality |
| S7_vs_88677\|\|lt2gene | Low-quality |
| S7_vs_88683\|\|lt2gene | Low-quality |
| S7_vs_88748\|\|lt2gene | Low-quality |
| S7_vs_88783\|\|lt2gene | Low-quality |
| S7_vs_88845\|\|lt2gene | Low-quality |
| S7_vs_88876\|\|lt2gene | Not-determined |
| S7_vs_89046\|\|lt2gene | Not-determined |
| S7_vs_89047\|\|lt2gene | Not-determined |
| S7_vs_89116\|\|lt2gene | Low-quality |
| S7_vs_89127\|\|lt2gene | Low-quality |
| S7_vs_89150\|\|lt2gene | Low-quality |
| S7_vs_89211\|\|lt2gene | Not-determined |
| S7_vs_89294\|\|lt2gene | Low-quality |
| S7_vs_89429\|\|lt2gene | Low-quality |
| S7_vs_89442\|\|lt2gene | Low-quality |
| S7_vs_89464\|\|lt2gene | Low-quality |
| S7_vs_89609\|\|lt2gene | Low-quality |
| S7_vs_89666\|\|lt2gene | Low-quality |
| S7_vs_89673\|\|lt2gene | Low-quality |
| S7_vs_89821\|\|lt2gene | Low-quality |
| S7_vs_89847\|\|lt2gene | Low-quality |
| S7_vs_89916\|\|lt2gene | Low-quality |
| S7_vs_89927\|\|lt2gene | Low-quality |
| S7_vs_90000\|\|lt2gene | Not-determined |
| S7_vs_90003\|\|lt2gene | Low-quality |
| S7_vs_90371\|\|lt2gene | Low-quality |
| S7_vs_90443\|\|lt2gene | Low-quality |
| S7_vs_90513\|\|lt2gene | Low-quality |
| S7_vs_90558\|\|lt2gene | Low-quality |
| S7_vs_90573\|\|lt2gene | Low-quality |
| S7_vs_90689\|\|lt2gene | Low-quality |
| S7_vs_90711\|\|lt2gene | Not-determined |
| S7_vs_90804\|\|lt2gene | Not-determined |
| S7_vs_90820\|\|lt2gene | Low-quality |
| S7_vs_90840\|\|lt2gene | Not-determined |
| S7_vs_90846\|\|lt2gene | Low-quality |
| S7_vs_90939\|\|lt2gene | Low-quality |
| S7_vs_91025\|\|lt2gene | Low-quality |
| S7_vs_91112\|\|lt2gene | Not-determined |
| S7_vs_91155\|\|lt2gene | Low-quality |
| S7_vs_91162\|\|lt2gene | Not-determined |
| S7_vs_91168\|\|lt2gene | Low-quality |
| S7_vs_91261\|\|lt2gene | Low-quality |
| S7_vs_91292\|\|lt2gene | Not-determined |
| S7_vs_91349\|\|lt2gene | Not-determined |
| S7_vs_91357\|\|lt2gene | Not-determined |
| S7_vs_91367\|\|lt2gene | Low-quality |
| S7_vs_91390\|\|lt2gene | Not-determined |
| S7_vs_91411\|\|lt2gene | Low-quality |
| S7_vs_91448\|\|lt2gene | Low-quality |
| S7_vs_91751\|\|lt2gene | Low-quality |
| S7_vs_91841\|\|lt2gene | Low-quality |
| S7_vs_91847\|\|lt2gene | Low-quality |
| S7_vs_91925\|\|lt2gene | Not-determined |
| S7_vs_91935\|\|lt2gene | Low-quality |
| S7_vs_91949\|\|lt2gene | Not-determined |
| S7_vs_91966\|\|lt2gene | Low-quality |
| S7_vs_91968\|\|lt2gene | Low-quality |
| S7_vs_92006\|\|lt2gene | Low-quality |
| S7_vs_92060\|\|lt2gene | Low-quality |
| S7_vs_92145\|\|lt2gene | Low-quality |
| S7_vs_92207\|\|lt2gene | Low-quality |
| S7_vs_92245\|\|lt2gene | Low-quality |
| S7_vs_92265\|\|lt2gene | Low-quality |
| S7_vs_92270\|\|lt2gene | Low-quality |
| S7_vs_92330\|\|lt2gene | Low-quality |
| S7_vs_92341\|\|lt2gene | Not-determined |
| S7_vs_92396\|\|lt2gene | Low-quality |
| S7_vs_92430\|\|lt2gene | Not-determined |
| S7_vs_92488\|\|lt2gene | Low-quality |
| S7_vs_92501\|\|lt2gene | Low-quality |
| S7_vs_92534\|\|lt2gene | Not-determined |
| S7_vs_92557\|\|lt2gene | Low-quality |
| S7_vs_92561\|\|lt2gene | Low-quality |
| S7_vs_92593\|\|lt2gene | Low-quality |
| S7_vs_92612\|\|lt2gene | Low-quality |
| S7_vs_92634\|\|lt2gene | Low-quality |
| S7_vs_92637\|\|lt2gene | Not-determined |
| S7_vs_92878\|\|lt2gene | Low-quality |
| S7_vs_93113\|\|lt2gene | Low-quality |
| S7_vs_93207\|\|lt2gene | Low-quality |
| S7_vs_93356\|\|lt2gene | Low-quality |
| S7_vs_93409\|\|lt2gene | Not-determined |
| S7_vs_93501\|\|lt2gene | Not-determined |
| S7_vs_93626\|\|lt2gene | Low-quality |
| S7_vs_93792\|\|lt2gene | Not-determined |
| S7_vs_93849\|\|lt2gene | Low-quality |
| S7_vs_93904\|\|lt2gene | Low-quality |
| S7_vs_93913\|\|lt2gene | Low-quality |
| S7_vs_93952\|\|lt2gene | Low-quality |
| S7_vs_93955\|\|lt2gene | Low-quality |
| S7_vs_93974\|\|lt2gene | Not-determined |
| S7_vs_94443\|\|lt2gene | Low-quality |
| S7_vs_94530\|\|lt2gene | Not-determined |
| S7_vs_94590\|\|lt2gene | Not-determined |
| S7_vs_94705\|\|lt2gene | Low-quality |
| S7_vs_94756\|\|lt2gene | Not-determined |
| S7_vs_94798\|\|lt2gene | Low-quality |
| S7_vs_94845\|\|lt2gene | Low-quality |
| S7_vs_94852\|\|lt2gene | Not-determined |
| S7_vs_94941\|\|lt2gene | Low-quality |
| S7_vs_94943\|\|lt2gene | Low-quality |
| S7_vs_94945\|\|lt2gene | Low-quality |
| S7_vs_94952\|\|lt2gene | Low-quality |
| S7_vs_95010\|\|lt2gene | Low-quality |
| S7_vs_95038\|\|lt2gene | Low-quality |
| S7_vs_95039\|\|lt2gene | Low-quality |
| S7_vs_95079\|\|lt2gene | Low-quality |
| S7_vs_95362\|\|lt2gene | Low-quality |
| S7_vs_95388\|\|lt2gene | Low-quality |
| S7_vs_95407\|\|lt2gene | Low-quality |
| S7_vs_95606\|\|lt2gene | Low-quality |
| S7_vs_95654\|\|lt2gene | Low-quality |
| S7_vs_95868\|\|lt2gene | Low-quality |
| S7_vs_96004\|\|lt2gene | Low-quality |
| S7_vs_96083\|\|lt2gene | Not-determined |
| S7_vs_96117\|\|lt2gene | Low-quality |
| S7_vs_96147\|\|lt2gene | Low-quality |
| S7_vs_96184\|\|lt2gene | Low-quality |
| S7_vs_96196\|\|lt2gene | Not-determined |
| S7_vs_96510\|\|lt2gene | Low-quality |
| S7_vs_96574\|\|lt2gene | Low-quality |
| S7_vs_96595\|\|lt2gene | Low-quality |
| S7_vs_96621\|\|lt2gene | Not-determined |
| S7_vs_96671\|\|lt2gene | Not-determined |
| S7_vs_96822\|\|lt2gene | Low-quality |
| S7_vs_96832\|\|lt2gene | Low-quality |
| S7_vs_96882\|\|lt2gene | Low-quality |
| S7_vs_96974\|\|lt2gene | Low-quality |
| S7_vs_97004\|\|lt2gene | Not-determined |
| S7_vs_97106\|\|lt2gene | Not-determined |
| S7_vs_97151\|\|lt2gene | Low-quality |
| S7_vs_97172\|\|lt2gene | Low-quality |
| S7_vs_97196\|\|lt2gene | Low-quality |
| S7_vs_97206\|\|lt2gene | Low-quality |
| S7_vs_97207\|\|lt2gene | Low-quality |
| S7_vs_97307\|\|lt2gene | Low-quality |
| S7_vs_97364\|\|lt2gene | Low-quality |
| S7_vs_97366\|\|lt2gene | Not-determined |
| S7_vs_97472\|\|lt2gene | Low-quality |
| S7_vs_97520\|\|lt2gene | Low-quality |
| S7_vs_97553\|\|lt2gene | Not-determined |
| S7_vs_97569\|\|lt2gene | Not-determined |
| S7_vs_97710\|\|lt2gene | Not-determined |
| S7_vs_97766\|\|lt2gene | Low-quality |
| S7_vs_97828\|\|lt2gene | Not-determined |
| S7_vs_97843\|\|lt2gene | Not-determined |
| S7_vs_98015\|\|lt2gene | Low-quality |
| S7_vs_98059\|\|lt2gene | Not-determined |
| S7_vs_98100\|\|lt2gene | Low-quality |
| S7_vs_98241\|\|lt2gene | Low-quality |
| S7_vs_98263\|\|lt2gene | Low-quality |
| S7_vs_98280\|\|lt2gene | Low-quality |
| S7_vs_98320\|\|lt2gene | Low-quality |
| S7_vs_98328\|\|lt2gene | Low-quality |
| S7_vs_98350\|\|lt2gene | Not-determined |
| S7_vs_98541\|\|lt2gene | Not-determined |
| S7_vs_98677\|\|lt2gene | Low-quality |
| S7_vs_98734\|\|lt2gene | Not-determined |
| S7_vs_98763\|\|lt2gene | Low-quality |
| S7_vs_98874\|\|lt2gene | Not-determined |
| S7_vs_98970\|\|lt2gene | Low-quality |
| S7_vs_99035\|\|lt2gene | Low-quality |
| S7_vs_99177\|\|lt2gene | Low-quality |
| S7_vs_99197\|\|lt2gene | Low-quality |
| S7_vs_99224\|\|lt2gene | Not-determined |
| S7_vs_99270\|\|lt2gene | Not-determined |
| S7_vs_99283\|\|lt2gene | Low-quality |
| S7_vs_99289\|\|lt2gene | Not-determined |
| S7_vs_99679\|\|lt2gene | Not-determined |
| S7_vs_99777\|\|lt2gene | Low-quality |
| S7_vs_99799\|\|lt2gene | Low-quality |
| S7_vs_99816\|\|lt2gene | Low-quality |
| S7_vs_99862\|\|lt2gene | Low-quality |
| S7_vs_99865\|\|lt2gene | Low-quality |
| S7_vs_99901\|\|lt2gene | Low-quality |
| S7_vs_100091\|\|lt2gene | Not-determined |
| S7_vs_100157\|\|lt2gene | Low-quality |
| S7_vs_100233\|\|lt2gene | Low-quality |
| S7_vs_100245\|\|lt2gene | Low-quality |
| S7_vs_100374\|\|lt2gene | Low-quality |
| S7_vs_100464\|\|lt2gene | Low-quality |
| S7_vs_100473\|\|lt2gene | Not-determined |
| S7_vs_100488\|\|lt2gene | Not-determined |
| S7_vs_100667\|\|lt2gene | Low-quality |
| S7_vs_100704\|\|lt2gene | Low-quality |
| S7_vs_100792\|\|lt2gene | Not-determined |
| S7_vs_100978\|\|lt2gene | Low-quality |
| S7_vs_101031\|\|lt2gene | Low-quality |
| S7_vs_101081\|\|lt2gene | Low-quality |
| S7_vs_101087\|\|lt2gene | Not-determined |
| S7_vs_101293\|\|lt2gene | Low-quality |
| S7_vs_101416\|\|lt2gene | Low-quality |
| S7_vs_101529\|\|lt2gene | Low-quality |

| | |
|---|---|
| S7_vs_101738\|\|lt2gene | Low-quality |
| S7_vs_101753\|\|lt2gene | Not-determined |
| S7_vs_101884\|\|lt2gene | Low-quality |
| S7_vs_101948\|\|lt2gene | Low-quality |
| S7_vs_102036\|\|lt2gene | Low-quality |
| S7_vs_102040\|\|lt2gene | Low-quality |
| S7_vs_102245\|\|lt2gene | Low-quality |
| S7_vs_102392\|\|lt2gene | Low-quality |
| S7_vs_102416\|\|lt2gene | Not-determined |
| S7_vs_102501\|\|lt2gene | Not-determined |
| S7_vs_102601\|\|lt2gene | Low-quality |
| S7_vs_102727\|\|lt2gene | Low-quality |
| S7_vs_102755\|\|lt2gene | Low-quality |
| S7_vs_103047\|\|lt2gene | Low-quality |
| S7_vs_103056\|\|lt2gene | Not-determined |
| S7_vs_103146\|\|lt2gene | Low-quality |
| S7_vs_103194\|\|lt2gene | Low-quality |
| S7_vs_103213\|\|lt2gene | Low-quality |
| S7_vs_103313\|\|lt2gene | Low-quality |
| S7_vs_103328\|\|lt2gene | Low-quality |
| S7_vs_103382\|\|lt2gene | Not-determined |
| S7_vs_103398\|\|lt2gene | Low-quality |
| S7_vs_103500\|\|lt2gene | Low-quality |
| S7_vs_103516\|\|lt2gene | Low-quality |
| S7_vs_103751\|\|lt2gene | Low-quality |
| S7_vs_103772\|\|lt2gene | Low-quality |
| S7_vs_103924\|\|lt2gene | Low-quality |
| S7_vs_104014\|\|lt2gene | Low-quality |
| S7_vs_104117\|\|lt2gene | Low-quality |
| S7_vs_104263\|\|lt2gene | Low-quality |
| S7_vs_104380\|\|lt2gene | Low-quality |
| S7_vs_104456\|\|lt2gene | Low-quality |
| S7_vs_104562\|\|lt2gene | Low-quality |
| S7_vs_104581\|\|lt2gene | Low-quality |
| S7_vs_104589\|\|lt2gene | Low-quality |
| S7_vs_104646\|\|lt2gene | Not-determined |
| S7_vs_104712\|\|lt2gene | Low-quality |
| S7_vs_104801\|\|lt2gene | Low-quality |
| S7_vs_104804\|\|lt2gene | Not-determined |
| S7_vs_104837\|\|lt2gene | Low-quality |
| S7_vs_104854\|\|lt2gene | Low-quality |
| S7_vs_104904\|\|lt2gene | Not-determined |
| S7_vs_104925\|\|lt2gene | Not-determined |
| S7_vs_104983\|\|lt2gene | Low-quality |
| S7_vs_105031\|\|lt2gene | Not-determined |
| S7_vs_105115\|\|lt2gene | Not-determined |
| S7_vs_105158\|\|lt2gene | Not-determined |
| S7_vs_105161\|\|lt2gene | Not-determined |
| S7_vs_105162\|\|lt2gene | Low-quality |
| S7_vs_105262\|\|lt2gene | Low-quality |
| S7_vs_105339\|\|lt2gene | Low-quality |
| S7_vs_105373\|\|lt2gene | Not-determined |
| S7_vs_105388\|\|lt2gene | Low-quality |
| S7_vs_105389\|\|lt2gene | Low-quality |
| S7_vs_105438\|\|lt2gene | Low-quality |
| S7_vs_105451\|\|lt2gene | Not-determined |
| S7_vs_105492\|\|lt2gene | Not-determined |
| S7_vs_105602\|\|lt2gene | Low-quality |
| S7_vs_105655\|\|lt2gene | Not-determined |
| S7_vs_105750\|\|lt2gene | Low-quality |
| S7_vs_105850\|\|lt2gene | Not-determined |
| S7_vs_105872\|\|lt2gene | Low-quality |
| S7_vs_105890\|\|lt2gene | Low-quality |
| S7_vs_105941\|\|lt2gene | Low-quality |
| S7_vs_105969\|\|lt2gene | Low-quality |
| S7_vs_106000\|\|lt2gene | Low-quality |
| S7_vs_106059\|\|lt2gene | Low-quality |
| S7_vs_106137\|\|lt2gene | Low-quality |
| S7_vs_106139\|\|lt2gene | Not-determined |
| S7_vs_106147\|\|lt2gene | Low-quality |
| S7_vs_106153\|\|lt2gene | Low-quality |
| S7_vs_106249\|\|lt2gene | Low-quality |
| S7_vs_106272\|\|lt2gene | Low-quality |
| S7_vs_106275\|\|lt2gene | Low-quality |
| S7_vs_106279\|\|lt2gene | Low-quality |
| S7_vs_106285\|\|lt2gene | Not-determined |
| S7_vs_106328\|\|lt2gene | Low-quality |
| S7_vs_106394\|\|lt2gene | Low-quality |
| S7_vs_106547\|\|lt2gene | Low-quality |
| S7_vs_106567\|\|lt2gene | Not-determined |
| S7_vs_106576\|\|lt2gene | Not-determined |
| S7_vs_106630\|\|lt2gene | Not-determined |
| S7_vs_106687\|\|lt2gene | Low-quality |
| S7_vs_106697\|\|lt2gene | Not-determined |
| S7_vs_106713\|\|lt2gene | Low-quality |
| S7_vs_106753\|\|lt2gene | Low-quality |
| S7_vs_106859\|\|lt2gene | Not-determined |
| S7_vs_106906\|\|lt2gene | Low-quality |
| S7_vs_106962\|\|lt2gene | Low-quality |
| S7_vs_106967\|\|lt2gene | Low-quality |
| S7_vs_107013\|\|lt2gene | Low-quality |
| S7_vs_107111\|\|lt2gene | Low-quality |
| S7_vs_107143\|\|lt2gene | Low-quality |
| S7_vs_107237\|\|lt2gene | Low-quality |
| S7_vs_107245\|\|lt2gene | Low-quality |
| S7_vs_107255\|\|lt2gene | Low-quality |
| S7_vs_107283\|\|lt2gene | Low-quality |
| S7_vs_107352\|\|lt2gene | Low-quality |
| S7_vs_107420\|\|lt2gene | Low-quality |
| S7_vs_107522\|\|lt2gene | Not-determined |
| S7_vs_107570\|\|lt2gene | Low-quality |
| S7_vs_107573\|\|lt2gene | Low-quality |
| S7_vs_107638\|\|lt2gene | Low-quality |
| S7_vs_107640\|\|lt2gene | Low-quality |
| S7_vs_107648\|\|lt2gene | Not-determined |
| S7_vs_107661\|\|lt2gene | Low-quality |
| S7_vs_107796\|\|lt2gene | Low-quality |
| S7_vs_107802\|\|lt2gene | Low-quality |
| S7_vs_107847\|\|lt2gene | Low-quality |
| S7_vs_107941\|\|lt2gene | Low-quality |
| S7_vs_107950\|\|lt2gene | Not-determined |
| S7_vs_107954\|\|lt2gene | Low-quality |
| S7_vs_107979\|\|lt2gene | Low-quality |
| S7_vs_107985\|\|lt2gene | Low-quality |
| S7_vs_108086\|\|lt2gene | Low-quality |
| S7_vs_108102\|\|lt2gene | Low-quality |
| S7_vs_108184\|\|lt2gene | Low-quality |
| S7_vs_108226\|\|lt2gene | Low-quality |
| S7_vs_108323\|\|lt2gene | Low-quality |
| S7_vs_108371\|\|lt2gene | Low-quality |
| S7_vs_108504\|\|lt2gene | Not-determined |
| S7_vs_108570\|\|lt2gene | Low-quality |
| S7_vs_108576\|\|lt2gene | Low-quality |
| S7_vs_108722\|\|lt2gene | Low-quality |
| S7_vs_108746\|\|lt2gene | Low-quality |
| S7_vs_108754\|\|lt2gene | Low-quality |
| S7_vs_108755\|\|lt2gene | Low-quality |
| S7_vs_108774\|\|lt2gene | Low-quality |
| S7_vs_108828\|\|lt2gene | Not-determined |
| S7_vs_108971\|\|lt2gene | Low-quality |
| S7_vs_109034\|\|lt2gene | Low-quality |
| S7_vs_109115\|\|lt2gene | Low-quality |
| S7_vs_109128\|\|lt2gene | Low-quality |
| S7_vs_109141\|\|lt2gene | Low-quality |
| S7_vs_109168\|\|lt2gene | Low-quality |
| S7_vs_109309\|\|lt2gene | Not-determined |
| S7_vs_109453\|\|lt2gene | Low-quality |
| S7_vs_109465\|\|lt2gene | Low-quality |
| S7_vs_109483\|\|lt2gene | Not-determined |
| S7_vs_109571\|\|lt2gene | Low-quality |
| S7_vs_109675\|\|lt2gene | Low-quality |
| S7_vs_109863\|\|lt2gene | Low-quality |
| S7_vs_110064\|\|lt2gene | Low-quality |
| S7_vs_110072\|\|lt2gene | Not-determined |
| S7_vs_110276\|\|lt2gene | Low-quality |
| S7_vs_110404\|\|lt2gene | Low-quality |
| S7_vs_110503\|\|lt2gene | Low-quality |
| S7_vs_110557\|\|lt2gene | Low-quality |
| S7_vs_110570\|\|lt2gene | Low-quality |
| S7_vs_110590\|\|lt2gene | Low-quality |
| S7_vs_110616\|\|lt2gene | Low-quality |
| S7_vs_110677\|\|lt2gene | Low-quality |
| S7_vs_110702\|\|lt2gene | Low-quality |
| S7_vs_110705\|\|lt2gene | Not-determined |
| S7_vs_110735\|\|lt2gene | Low-quality |
| S7_vs_110850\|\|lt2gene | Not-determined |
| S7_vs_110881\|\|lt2gene | Not-determined |
| S7_vs_111095\|\|lt2gene | Low-quality |
| S7_vs_111114\|\|lt2gene | Low-quality |
| S7_vs_111174\|\|lt2gene | Not-determined |
| S7_vs_111242\|\|lt2gene | Low-quality |
| S7_vs_111257\|\|lt2gene | Low-quality |
| S7_vs_111263\|\|lt2gene | Not-determined |
| S7_vs_111405\|\|lt2gene | Low-quality |
| S7_vs_111426\|\|lt2gene | Low-quality |
| S7_vs_111477\|\|lt2gene | Not-determined |
| S7_vs_111527\|\|lt2gene | Not-determined |
| S7_vs_111546\|\|lt2gene | Low-quality |
| S7_vs_111664\|\|lt2gene | Low-quality |
| S7_vs_111846\|\|lt2gene | Not-determined |
| S7_vs_111921\|\|lt2gene | Low-quality |
| S7_vs_111938\|\|lt2gene | Low-quality |
| S7_vs_112035\|\|lt2gene | Low-quality |
| S7_vs_112067\|\|lt2gene | Not-determined |
| S7_vs_112095\|\|lt2gene | Not-determined |
| S7_vs_112231\|\|lt2gene | Low-quality |
| S7_vs_112255\|\|lt2gene | Low-quality |
| S7_vs_112295\|\|lt2gene | Low-quality |
| S7_vs_112370\|\|lt2gene | Low-quality |
| S7_vs_112445\|\|lt2gene | Low-quality |
| S7_vs_112614\|\|lt2gene | Low-quality |
| S7_vs_112674\|\|lt2gene | Low-quality |
| S7_vs_112738\|\|lt2gene | Low-quality |
| S7_vs_112793\|\|lt2gene | Low-quality |
| S7_vs_112828\|\|lt2gene | Not-determined |
| S7_vs_112976\|\|lt2gene | Not-determined |
| S7_vs_113031\|\|lt2gene | Low-quality |
| S7_vs_113039\|\|lt2gene | Not-determined |
| S7_vs_113084\|\|lt2gene | Low-quality |
| S7_vs_113130\|\|lt2gene | Low-quality |
| S7_vs_113152\|\|lt2gene | Low-quality |
| S7_vs_113186\|\|lt2gene | Not-determined |
| S7_vs_113232\|\|lt2gene | Low-quality |
| S7_vs_113237\|\|lt2gene | Low-quality |
| S7_vs_113245\|\|lt2gene | Not-determined |
| S7_vs_113336\|\|lt2gene | Low-quality |
| S7_vs_113374\|\|lt2gene | Low-quality |
| S7_vs_113529\|\|lt2gene | Low-quality |
| S7_vs_113562\|\|lt2gene | Low-quality |
| S7_vs_113597\|\|lt2gene | Low-quality |
| S7_vs_113623\|\|lt2gene | Low-quality |
| S7_vs_113624\|\|lt2gene | Not-determined |
| S7_vs_113685\|\|lt2gene | Not-determined |
| S7_vs_114007\|\|lt2gene | Low-quality |
| S7_vs_114044\|\|lt2gene | Low-quality |
| S7_vs_114106\|\|lt2gene | Low-quality |
| S7_vs_114169\|\|lt2gene | Low-quality |
| S7_vs_114277\|\|lt2gene | Low-quality |
| S7_vs_114319\|\|lt2gene | Low-quality |
| S7_vs_114323\|\|lt2gene | Low-quality |
| S7_vs_114440\|\|lt2gene | Low-quality |
| S7_vs_114638\|\|lt2gene | Low-quality |
| S7_vs_114663\|\|lt2gene | Low-quality |
| S7_vs_114729\|\|lt2gene | Low-quality |
| S7_vs_114731\|\|lt2gene | Low-quality |
| S7_vs_114801\|\|lt2gene | Low-quality |
| S7_vs_114863\|\|lt2gene | Not-determined |
| S7_vs_114869\|\|lt2gene | Low-quality |
| S7_vs_114965\|\|lt2gene | Low-quality |
| S7_vs_114971\|\|lt2gene | Not-determined |
| S7_vs_115063\|\|lt2gene | Low-quality |
| S7_vs_115115\|\|lt2gene | Low-quality |
| S7_vs_115151\|\|lt2gene | Low-quality |
| S7_vs_115173\|\|lt2gene | Low-quality |
| S7_vs_115177\|\|lt2gene | Not-determined |
| S7_vs_115233\|\|lt2gene | Low-quality |
| S7_vs_115272\|\|lt2gene | Not-determined |
| S7_vs_115354\|\|lt2gene | Low-quality |
| S7_vs_115461\|\|lt2gene | Low-quality |
| S7_vs_115578\|\|lt2gene | Low-quality |
| S7_vs_115609\|\|lt2gene | Low-quality |
| S7_vs_115770\|\|lt2gene | Not-determined |
| S7_vs_115857\|\|lt2gene | Low-quality |
| S7_vs_115861\|\|lt2gene | Low-quality |
| S7_vs_115935\|\|lt2gene | Low-quality |
| S7_vs_115947\|\|lt2gene | Not-determined |
| S7_vs_115950\|\|lt2gene | Low-quality |
| S7_vs_116076\|\|lt2gene | Low-quality |
| S7_vs_116121\|\|lt2gene | Low-quality |
| S7_vs_116142\|\|lt2gene | Low-quality |
| S7_vs_116155\|\|lt2gene | Low-quality |
| S7_vs_116189\|\|lt2gene | Low-quality |
| S7_vs_116275\|\|lt2gene | Low-quality |
| S7_vs_116336\|\|lt2gene | Not-determined |
| S7_vs_116372\|\|lt2gene | Low-quality |
| S7_vs_116496\|\|lt2gene | Not-determined |
| S7_vs_116622\|\|lt2gene | Low-quality |
| S7_vs_116651\|\|lt2gene | Low-quality |
| S7_vs_116741\|\|lt2gene | Low-quality |
| S7_vs_116785\|\|lt2gene | Low-quality |
| S7_vs_116868\|\|lt2gene | Low-quality |
| S7_vs_116918\|\|lt2gene | Low-quality |
| S7_vs_116940\|\|lt2gene | Low-quality |
| S7_vs_116951\|\|lt2gene | Not-determined |
| S7_vs_117024\|\|lt2gene | Low-quality |
| S7_vs_117044\|\|lt2gene | Low-quality |
| S7_vs_117182\|\|lt2gene | Not-determined |
| S7_vs_117303\|\|lt2gene | Low-quality |
| S7_vs_117354\|\|lt2gene | Not-determined |
| S7_vs_117360\|\|lt2gene | Low-quality |
| S7_vs_117414\|\|lt2gene | Low-quality |
| S7_vs_117525\|\|lt2gene | Not-determined |
| S7_vs_117526\|\|lt2gene | Low-quality |
| S7_vs_117788\|\|lt2gene | Not-determined |
| S7_vs_117798\|\|lt2gene | Low-quality |
| S7_vs_117830\|\|lt2gene | Low-quality |
| S7_vs_117888\|\|lt2gene | Low-quality |
| S7_vs_117919\|\|lt2gene | Low-quality |
| S7_vs_117983\|\|lt2gene | Low-quality |
| S7_vs_118011\|\|lt2gene | Low-quality |
| S7_vs_118152\|\|lt2gene | Low-quality |
| S7_vs_118160\|\|lt2gene | Low-quality |
| S7_vs_118250\|\|lt2gene | Low-quality |
| S7_vs_118262\|\|lt2gene | Low-quality |
| S7_vs_118464\|\|lt2gene | Low-quality |
| S7_vs_118599\|\|lt2gene | Low-quality |
| S7_vs_118710\|\|lt2gene | Not-determined |
| S7_vs_118972\|\|lt2gene | Low-quality |
| S7_vs_119069\|\|lt2gene | Low-quality |
| S7_vs_119134\|\|lt2gene | Low-quality |
| S7_vs_119266\|\|lt2gene | Not-determined |
| S7_vs_119402\|\|lt2gene | Low-quality |
| S7_vs_119697\|\|lt2gene | Low-quality |
| S7_vs_119713\|\|lt2gene | Not-determined |
| S7_vs_119826\|\|lt2gene | Low-quality |
| S7_vs_119837\|\|lt2gene | Low-quality |
| S7_vs_119941\|\|lt2gene | Low-quality |
| S7_vs_119952\|\|lt2gene | Low-quality |
| S7_vs_120110\|\|lt2gene | Low-quality |
| S7_vs_120244\|\|lt2gene | Low-quality |
| S7_vs_120443\|\|lt2gene | Low-quality |
| S7_vs_120446\|\|lt2gene | Low-quality |
| S7_vs_120450\|\|lt2gene | Not-determined |
| S7_vs_120573\|\|lt2gene | Low-quality |
| S7_vs_120634\|\|lt2gene | Low-quality |
| S7_vs_120698\|\|lt2gene | Not-determined |
| S7_vs_120743\|\|lt2gene | Low-quality |
| S7_vs_120876\|\|lt2gene | Low-quality |
| S7_vs_120883\|\|lt2gene | Not-determined |
| S7_vs_120938\|\|lt2gene | Not-determined |
| S7_vs_120986\|\|lt2gene | Low-quality |
| S7_vs_121040\|\|lt2gene | Low-quality |
| S7_vs_121086\|\|lt2gene | Not-determined |
| S7_vs_121124\|\|lt2gene | Low-quality |
| S7_vs_121144\|\|lt2gene | Low-quality |
| S7_vs_121160\|\|lt2gene | Low-quality |
| S7_vs_121273\|\|lt2gene | Low-quality |
| S7_vs_121309\|\|lt2gene | Low-quality |
| S7_vs_121402\|\|lt2gene | Low-quality |
| S7_vs_121409\|\|lt2gene | Low-quality |
| S7_vs_121540\|\|lt2gene | Low-quality |
| S7_vs_121559\|\|lt2gene | Not-determined |
| S7_vs_121600\|\|lt2gene | Low-quality |
| S7_vs_121647\|\|lt2gene | Low-quality |
| S7_vs_121683\|\|lt2gene | Low-quality |
| S7_vs_121688\|\|lt2gene | Low-quality |
| S7_vs_121805\|\|lt2gene | Low-quality |
| S7_vs_121810\|\|lt2gene | Low-quality |
| S7_vs_121963\|\|lt2gene | Low-quality |
| S7_vs_121967\|\|lt2gene | Not-determined |
| S7_vs_122102\|\|lt2gene | Low-quality |
| S7_vs_122122\|\|lt2gene | Low-quality |
| S7_vs_122202\|\|lt2gene | Low-quality |
| S7_vs_122249\|\|lt2gene | Low-quality |
| S7_vs_122254\|\|lt2gene | Low-quality |
| S7_vs_122266\|\|lt2gene | Low-quality |
| S7_vs_122406\|\|lt2gene | Low-quality |
| S7_vs_122477\|\|lt2gene | Low-quality |
| S7_vs_122491\|\|lt2gene | Not-determined |
| S7_vs_122613\|\|lt2gene | Low-quality |
| S7_vs_122616\|\|lt2gene | Low-quality |
| S7_vs_122618\|\|lt2gene | Low-quality |
| S7_vs_122642\|\|lt2gene | Low-quality |
| S7_vs_122690\|\|lt2gene | Low-quality |
| S7_vs_122706\|\|lt2gene | Low-quality |
| S7_vs_122836\|\|lt2gene | Low-quality |
| S7_vs_122904\|\|lt2gene | Low-quality |
| S7_vs_123140\|\|lt2gene | Low-quality |
| S7_vs_123192\|\|lt2gene | Not-determined |
| S7_vs_123310\|\|lt2gene | Not-determined |
| S7_vs_123314\|\|lt2gene | Low-quality |
| S7_vs_123575\|\|lt2gene | Not-determined |
| S7_vs_123584\|\|lt2gene | Low-quality |
| S7_vs_123674\|\|lt2gene | Low-quality |
| S7_vs_123719\|\|lt2gene | Low-quality |
| S7_vs_123739\|\|lt2gene | Low-quality |
| S7_vs_123776\|\|lt2gene | Low-quality |
| S7_vs_123863\|\|lt2gene | Low-quality |
| S7_vs_123884\|\|lt2gene | Low-quality |
| S7_vs_124142\|\|lt2gene | Low-quality |
| S7_vs_124180\|\|lt2gene | Low-quality |
| S7_vs_124281\|\|lt2gene | Low-quality |
| S7_vs_124299\|\|lt2gene | Low-quality |
| S7_vs_124312\|\|lt2gene | Low-quality |
| S7_vs_124507\|\|lt2gene | Not-determined |
| S7_vs_124564\|\|lt2gene | Low-quality |
| S7_vs_124665\|\|lt2gene | Low-quality |
| S7_vs_124699\|\|lt2gene | Low-quality |
| S7_vs_124811\|\|lt2gene | Not-determined |
| S7_vs_124878\|\|lt2gene | Not-determined |
| S7_vs_124955\|\|lt2gene | Not-determined |
| S7_vs_125065\|\|lt2gene | Not-determined |
| S7_vs_125080\|\|lt2gene | Not-determined |
| S7_vs_125111\|\|lt2gene | Low-quality |
| S7_vs_125146\|\|lt2gene | Not-determined |
| S7_vs_125150\|\|lt2gene | Low-quality |
| S7_vs_125169\|\|lt2gene | Low-quality |
| S7_vs_125217\|\|lt2gene | Not-determined |
| S7_vs_125244\|\|lt2gene | Low-quality |
| S7_vs_125301\|\|lt2gene | Low-quality |
| S7_vs_125340\|\|lt2gene | Low-quality |
| S7_vs_125354\|\|lt2gene | Low-quality |
| S7_vs_125362\|\|lt2gene | Low-quality |
| S7_vs_125365\|\|lt2gene | Low-quality |
| S7_vs_125392\|\|lt2gene | Low-quality |
| S7_vs_125464\|\|lt2gene | Low-quality |
| S7_vs_125483\|\|lt2gene | Not-determined |
| S7_vs_125562\|\|lt2gene | Low-quality |
| S7_vs_125610\|\|lt2gene | Not-determined |
| S7_vs_125905\|\|lt2gene | Low-quality |
| S7_vs_125932\|\|lt2gene | Low-quality |
| S7_vs_126029\|\|lt2gene | Low-quality |
| S7_vs_126085\|\|lt2gene | Not-determined |
| S7_vs_126113\|\|lt2gene | Not-determined |
| S7_vs_126114\|\|lt2gene | Not-determined |
| S7_vs_126137\|\|lt2gene | Not-determined |
| S7_vs_126378\|\|lt2gene | Low-quality |
| S7_vs_126399\|\|lt2gene | Not-determined |
| S7_vs_126420\|\|lt2gene | Not-determined |
| S7_vs_126424\|\|lt2gene | Low-quality |
| S7_vs_126481\|\|lt2gene | Low-quality |
| S7_vs_126635\|\|lt2gene | Low-quality |
| S7_vs_126804\|\|lt2gene | Not-determined |
| S7_vs_126864\|\|lt2gene | Low-quality |
| S7_vs_126915\|\|lt2gene | Low-quality |
| S7_vs_126972\|\|lt2gene | Low-quality |
| S7_vs_127033\|\|lt2gene | Not-determined |
| S7_vs_127278\|\|lt2gene | Low-quality |
| S7_vs_127283\|\|lt2gene | Low-quality |
| S7_vs_127286\|\|lt2gene | Not-determined |
| S7_vs_127318\|\|lt2gene | Not-determined |
| S7_vs_127433\|\|lt2gene | Low-quality |
| S7_vs_127465\|\|lt2gene | Low-quality |
| S7_vs_127549\|\|lt2gene | Low-quality |
| S7_vs_127564\|\|lt2gene | Low-quality |
| S7_vs_127565\|\|lt2gene | Low-quality |
| S7_vs_127614\|\|lt2gene | Low-quality |
| S7_vs_127657\|\|lt2gene | Not-determined |
| S7_vs_127673\|\|lt2gene | Not-determined |
| S7_vs_127681\|\|lt2gene | Not-determined |
| S7_vs_127781\|\|lt2gene | Not-determined |
| S7_vs_127811\|\|lt2gene | Low-quality |
| S7_vs_127822\|\|lt2gene | Low-quality |
| S7_vs_127836\|\|lt2gene | Low-quality |
| S7_vs_127844\|\|lt2gene | Low-quality |
| S7_vs_128042\|\|lt2gene | Low-quality |
| S7_vs_128076\|\|lt2gene | Low-quality |
| S7_vs_128154\|\|lt2gene | Low-quality |
| S7_vs_128157\|\|lt2gene | Low-quality |
| S7_vs_128244\|\|lt2gene | Low-quality |
| S7_vs_128310\|\|lt2gene | Low-quality |
| S7_vs_128376\|\|lt2gene | Low-quality |
| S7_vs_128430\|\|lt2gene | Low-quality |
| S7_vs_128457\|\|lt2gene | Not-determined |
| S7_vs_128469\|\|lt2gene | Low-quality |
| S7_vs_128674\|\|lt2gene | Not-determined |
| S7_vs_128681\|\|lt2gene | Low-quality |
| S7_vs_128688\|\|lt2gene | Low-quality |
| S7_vs_128736\|\|lt2gene | Low-quality |
| S7_vs_128847\|\|lt2gene | Not-determined |
| S7_vs_128870\|\|lt2gene | Low-quality |
| S7_vs_128958\|\|lt2gene | Low-quality |
| S7_vs_128970\|\|lt2gene | Not-determined |
| S7_vs_129005\|\|lt2gene | Not-determined |
| S7_vs_129180\|\|lt2gene | Low-quality |
| S7_vs_129217\|\|lt2gene | Low-quality |
| S7_vs_129272\|\|lt2gene | Not-determined |
| S7_vs_129282\|\|lt2gene | Low-quality |
| S7_vs_129376\|\|lt2gene | Low-quality |
| S7_vs_129467\|\|lt2gene | Low-quality |
| S7_vs_129632\|\|lt2gene | Low-quality |
| S7_vs_129784\|\|lt2gene | Low-quality |
| S7_vs_129870\|\|lt2gene | Low-quality |
| S7_vs_129922\|\|lt2gene | Low-quality |
| S7_vs_130019\|\|lt2gene | Not-determined |
| S7_vs_130189\|\|lt2gene | Low-quality |
| S7_vs_130273\|\|lt2gene | Low-quality |
| S7_vs_130280\|\|lt2gene | Low-quality |
| S7_vs_130318\|\|lt2gene | Low-quality |
| S7_vs_130378\|\|lt2gene | Low-quality |
| S7_vs_130440\|\|lt2gene | Low-quality |
| S7_vs_130477\|\|lt2gene | Low-quality |
| S7_vs_130505\|\|lt2gene | Not-determined |
| S7_vs_130524\|\|lt2gene | Low-quality |
| S7_vs_130680\|\|lt2gene | Low-quality |
| S7_vs_130683\|\|lt2gene | Low-quality |
| S7_vs_130695\|\|lt2gene | Low-quality |
| S7_vs_130837\|\|lt2gene | Low-quality |
| S7_vs_130851\|\|lt2gene | Low-quality |
| S7_vs_130994\|\|lt2gene | Not-determined |
| S7_vs_130995\|\|lt2gene | Low-quality |
| S7_vs_131003\|\|lt2gene | Low-quality |
| S7_vs_131201\|\|lt2gene | Low-quality |
| S7_vs_131214\|\|lt2gene | Low-quality |
| S7_vs_131225\|\|lt2gene | Not-determined |
| S7_vs_131227\|\|lt2gene | Low-quality |
| S7_vs_131256\|\|lt2gene | Low-quality |
| S7_vs_131329\|\|lt2gene | Low-quality |
| S7_vs_131357\|\|lt2gene | Low-quality |
| S7_vs_131480\|\|lt2gene | Low-quality |
| S7_vs_131683\|\|lt2gene | Not-determined |
| S7_vs_131781\|\|lt2gene | Low-quality |
| S7_vs_131825\|\|lt2gene | Not-determined |
| S7_vs_131892\|\|lt2gene | Not-determined |
| S7_vs_131919\|\|lt2gene | Low-quality |
| S7_vs_131971\|\|lt2gene | Low-quality |
| S7_vs_131975\|\|lt2gene | Low-quality |
| S7_vs_132115\|\|lt2gene | Not-determined |
| S7_vs_132264\|\|lt2gene | Not-determined |
| S7_vs_132348\|\|lt2gene | Low-quality |
| S7_vs_132377\|\|lt2gene | Low-quality |
| S7_vs_132392\|\|lt2gene | Low-quality |
| S7_vs_132537\|\|lt2gene | Not-determined |
| S7_vs_132564\|\|lt2gene | Not-determined |
| S7_vs_132590\|\|lt2gene | Low-quality |
| S7_vs_132688\|\|lt2gene | Low-quality |
| S7_vs_132734\|\|lt2gene | Low-quality |
| S7_vs_132756\|\|lt2gene | Low-quality |
| S7_vs_132833\|\|lt2gene | Low-quality |
| S7_vs_132861\|\|lt2gene | Not-determined |
| S7_vs_132866\|\|lt2gene | Low-quality |
| S7_vs_132891\|\|lt2gene | Low-quality |
| S7_vs_132901\|\|lt2gene | Low-quality |
| S7_vs_132919\|\|lt2gene | Not-determined |
| S7_vs_132930\|\|lt2gene | Low-quality |
| S7_vs_132967\|\|lt2gene | Low-quality |
| S7_vs_132998\|\|lt2gene | Not-determined |
| S7_vs_133030\|\|lt2gene | Low-quality |
| S7_vs_133054\|\|lt2gene | Low-quality |
| S7_vs_133068\|\|lt2gene | Low-quality |
| S7_vs_133150\|\|lt2gene | Low-quality |
| S7_vs_133334\|\|lt2gene | Not-determined |
| S7_vs_133355\|\|lt2gene | Low-quality |
| S7_vs_133383\|\|lt2gene | Low-quality |
| S7_vs_133432\|\|lt2gene | Low-quality |
| S7_vs_133452\|\|lt2gene | Low-quality |
| S7_vs_133533\|\|lt2gene | Not-determined |
| S7_vs_133627\|\|lt2gene | Low-quality |
| S7_vs_133667\|\|lt2gene | Low-quality |
| S7_vs_133828\|\|lt2gene | Not-determined |
| S7_vs_133937\|\|lt2gene | Low-quality |
| S7_vs_133986\|\|lt2gene | Low-quality |
| S7_vs_134043\|\|lt2gene | Low-quality |
| S7_vs_134056\|\|lt2gene | Low-quality |
| S7_vs_134063\|\|lt2gene | Low-quality |
| S7_vs_134187\|\|lt2gene | Low-quality |
| S7_vs_134215\|\|lt2gene | Low-quality |
| S7_vs_134237\|\|lt2gene | Not-determined |
| S7_vs_134278\|\|lt2gene | Low-quality |
| S7_vs_134336\|\|lt2gene | Not-determined |
| S7_vs_134436\|\|lt2gene | Low-quality |
| S7_vs_134473\|\|lt2gene | Low-quality |
| S7_vs_134508\|\|lt2gene | Not-determined |
| S7_vs_134649\|\|lt2gene | Low-quality |
| S7_vs_134653\|\|lt2gene | Not-determined |
| S7_vs_134683\|\|lt2gene | Low-quality |
| S7_vs_134695\|\|lt2gene | Low-quality |
| S7_vs_134775\|\|lt2gene | Low-quality |
| S7_vs_134847\|\|lt2gene | Low-quality |
| S7_vs_135063\|\|lt2gene | Low-quality |
| S7_vs_135068\|\|lt2gene | Not-determined |
| S7_vs_135123\|\|lt2gene | Low-quality |
| S7_vs_135136\|\|lt2gene | Low-quality |
| S7_vs_135289\|\|lt2gene | Low-quality |
| S7_vs_135584\|\|lt2gene | Low-quality |
| S7_vs_135811\|\|lt2gene | Low-quality |
| S7_vs_135843\|\|lt2gene | Not-determined |
| S7_vs_135855\|\|lt2gene | Not-determined |
| S7_vs_135989\|\|lt2gene | Not-determined |
| S7_vs_136078\|\|lt2gene | Low-quality |
| S7_vs_136107\|\|lt2gene | Low-quality |
| S7_vs_136204\|\|lt2gene | Low-quality |
| S7_vs_136275\|\|lt2gene | Low-quality |
| S7_vs_136309\|\|lt2gene | Low-quality |
| S7_vs_136319\|\|lt2gene | Low-quality |
| S7_vs_136385\|\|lt2gene | Low-quality |
| S7_vs_136394\|\|lt2gene | Low-quality |
| S7_vs_136476\|\|lt2gene | Low-quality |
| S7_vs_136491\|\|lt2gene | Low-quality |
| S7_vs_136493\|\|lt2gene | Low-quality |
| S7_vs_136674\|\|lt2gene | Low-quality |
| S7_vs_136689\|\|lt2gene | Low-quality |
| S7_vs_136750\|\|lt2gene | Low-quality |
| S7_vs_136799\|\|lt2gene | Low-quality |
| S7_vs_136918\|\|lt2gene | Not-determined |
| S7_vs_136935\|\|lt2gene | Low-quality |
| S7_vs_136936\|\|lt2gene | Low-quality |
| S7_vs_136958\|\|lt2gene | Not-determined |
| S7_vs_137024\|\|lt2gene | Low-quality |
| S7_vs_137050\|\|lt2gene | Low-quality |
| S7_vs_137100\|\|lt2gene | Low-quality |
| S7_vs_137133\|\|lt2gene | Low-quality |
| S7_vs_137146\|\|lt2gene | Low-quality |
| S7_vs_137233\|\|lt2gene | Low-quality |
| S7_vs_137269\|\|lt2gene | Not-determined |
| S7_vs_137421\|\|lt2gene | Not-determined |
| S7_vs_137434\|\|lt2gene | Not-determined |
| S7_vs_137571\|\|lt2gene | Low-quality |
| S7_vs_137680\|\|lt2gene | Low-quality |
| S7_vs_137734\|\|lt2gene | Low-quality |
| S7_vs_137743\|\|lt2gene | Low-quality |
| S7_vs_137929\|\|lt2gene | Low-quality |
| S7_vs_137933\|\|lt2gene | Low-quality |
| S7_vs_137950\|\|lt2gene | Low-quality |
| S7_vs_137969\|\|lt2gene | Low-quality |
| S7_vs_138037\|\|lt2gene | Low-quality |
| S7_vs_138067\|\|lt2gene | Not-determined |
| S7_vs_138127\|\|lt2gene | Low-quality |
| NC1_vf_k141_1575 | Not-determined |
| NC1_vf_k141_3374 | Not-determined |
| NC1_vf_k141_2700 | Not-determined |
| NC1_vf_k141_3150 | Not-determined |
| NC1_vf_k141_900 | Not-determined |
| NC1_vf_k141_225 | Not-determined |
| NC1_vf_k141_2925 | Not-determined |
| NC1_vf_k141_1800 | Not-determined |
| NC1_vf_k141_2025 | Not-determined |
| NC1_vf_k141_2475 | Not-determined |
| NC1_vf_k141_1350 | Not-determined |

| | |
|---|---|
| NC1_vf_k141_0 | Not-determined |
| NC1_vf_k141_450 | Not-determined |
| NC1_vf_k141_3375 | Not-determined |
| NC1_vf_k141_2250 | Not-determined |
| NC1_vf_k141_1125 | Not-determined |
| NC1_vf_k141_901 | Not-determined |
| NC1_vf_k141_675 | Not-determined |
| NC1_vf_k141_1576 | Not-determined |
| NC1_vf_k141_2926 | Not-determined |
| NC1_vf_k141_226 | Not-determined |
| NC1_vf_k141_1 | Not-determined |
| NC1_vf_k141_2026 | Not-determined |
| NC1_vf_k141_2701 | Not-determined |
| NC1_vf_k141_3151 | Not-determined |
| NC1_vf_k141_1801 | Not-determined |
| NC1_vf_k141_2476 | Not-determined |
| NC1_vf_k141_451 | Not-determined |
| NC1_vf_k141_1126 | Not-determined |
| NC1_vf_k141_1351 | Not-determined |
| NC1_vf_k141_3376 | Not-determined |
| NC1_vf_k141_1577 | Not-determined |
| NC1_vf_k141_2 | Not-determined |
| NC1_vf_k141_227 | Not-determined |
| NC1_vf_k141_2477 | Not-determined |
| NC1_vf_k141_2027 | Not-determined |
| NC1_vf_k141_676 | Not-determined |
| NC1_vf_k141_3152 | Not-determined |
| NC1_vf_k141_1802 | Not-determined |
| NC1_vf_k141_1127 | Not-determined |
| NC1_vf_k141_2251 | Not-determined |
| NC1_vf_k141_3 | Not-determined |
| NC1_vf_k141_1352 | Not-determined |
| NC1_vf_k141_2927 | Not-determined |
| NC1_vf_k141_452 | Not-determined |
| NC1_vf_k141_902 | Not-determined |
| NC1_vf_k141_228 | Not-determined |
| NC1_vf_k141_677 | Not-determined |
| NC1_vf_k141_1578 | Not-determined |
| NC1_vf_k141_2028 | Not-determined |
| NC1_vf_k141_4 | Not-determined |
| NC1_vf_k141_3153 | Not-determined |
| NC1_vf_k141_1803 | Not-determined |
| NC1_vf_k141_3377 | Low-quality |
| NC1_vf_k141_2702 | Not-determined |
| NC1_vf_k141_229 | Not-determined |
| NC1_vf_k141_5 | Not-determined |
| NC1_vf_k141_678 | Not-determined |
| NC1_vf_k141_453 | Not-determined |
| NC1_vf_k141_2252 | Not-determined |
| NC1_vf_k141_903 | Not-determined |
| NC1_vf_k141_1353 | Not-determined |
| NC1_vf_k141_1128 | Not-determined |
| NC1_vf_k141_2029 | Not-determined |
| NC1_vf_k141_2478 | Not-determined |
| NC1_vf_k141_2928 | Not-determined |
| NC1_vf_k141_1579 | Not-determined |
| NC1_vf_k141_6 | Not-determined |
| NC1_vf_k141_679 | Not-determined |
| NC1_vf_k141_454 | Low-quality |
| NC1_vf_k141_1804 | Not-determined |
| NC1_vf_k141_1354 | Not-determined |
| NC1_vf_k141_3154 | Not-determined |
| NC1_vf_k141_2253 | Not-determined |
| NC1_vf_k141_230 | Not-determined |
| NC1_vf_k141_1580 | Not-determined |
| NC1_vf_k141_2703 | Not-determined |
| NC1_vf_k141_3378 | Not-determined |
| NC1_vf_k141_904 | Not-determined |
| NC1_vf_k141_2030 | Not-determined |
| NC1_vf_k141_2479 | Not-determined |
| NC1_vf_k141_2929 | Not-determined |
| NC1_vf_k141_680 | Not-determined |
| NC1_vf_k141_1355 | Not-determined |
| NC1_vf_k141_455 | Not-determined |
| NC1_vf_k141_7 | Not-determined |
| NC1_vf_k141_905 | Not-determined |
| NC1_vf_k141_3379 | Low-quality |
| NC1_vf_k141_1805 | Not-determined |
| NC1_vf_k141_2480 | Not-determined |
| NC1_vf_k141_231 | Not-determined |
| NC1_vf_k141_8 | Not-determined |
| NC1_vf_k141_2254 | Not-determined |
| NC1_vf_k141_2704 | Not-determined |
| NC1_vf_k141_456 | Not-determined |
| NC1_vf_k141_2031 | Not-determined |
| NC1_vf_k141_3155 | Not-determined |
| NC1_vf_k141_681 | Not-determined |
| NC1_vf_k141_2930 | Not-determined |
| NC1_vf_k141_1356 | Not-determined |
| NC1_vf_k141_1581 | Not-determined |
| NC1_vf_k141_3380 | Not-determined |
| NC1_vf_k141_2481 | Not-determined |
| NC1_vf_k141_9 | Not-determined |
| NC1_vf_k141_2705 | Not-determined |
| NC1_vf_k141_1129 | Low-quality |
| NC1_vf_k141_2255 | Not-determined |
| NC1_vf_k141_3156 | Not-determined |
| NC1_vf_k141_457 | Not-determined |
| NC1_vf_k141_2931 | Low-quality |
| NC1_vf_k141_3381 | Not-determined |
| NC1_vf_k141_2482 | Not-determined |
| NC1_vf_k141_1806 | Not-determined |
| NC1_vf_k141_10 | Not-determined |
| NC1_vf_k141_1582 | Not-determined |
| NC1_vf_k141_232 | Not-determined |
| NC1_vf_k141_1130 | Not-determined |
| NC1_vf_k141_2256 | Not-determined |
| NC1_vf_k141_906 | Not-determined |
| NC1_vf_k141_2032 | Low-quality |
| NC1_vf_k141_682 | Not-determined |
| NC1_vf_k141_2483 | Not-determined |
| NC1_vf_k141_1583 | Not-determined |
| NC1_vf_k141_1807 | Not-determined |
| NC1_vf_k141_233 | Not-determined |
| NC1_vf_k141_2706 | Not-determined |
| NC1_vf_k141_2932 | Not-determined |
| NC1_vf_k141_458 | Not-determined |
| NC1_vf_k141_2257 | Not-determined |
| NC1_vf_k141_1584 | Not-determined |
| NC1_vf_k141_907 | Not-determined |
| NC1_vf_k141_11 | Not-determined |
| NC1_vf_k141_1131 | Not-determined |
| NC1_vf_k141_2484 | Not-determined |
| NC1_vf_k141_1808 | Not-determined |
| NC1_vf_k141_3382 | Low-quality |
| NC1_vf_k141_683 | Not-determined |
| NC1_vf_k141_234 | Not-determined |
| NC1_vf_k141_1357 | Low-quality |
| NC1_vf_k141_12 | Not-determined |
| NC1_vf_k141_2707 | Not-determined |
| NC1_vf_k141_908 | Not-determined |
| NC1_vf_k141_3383 | Not-determined |
| NC1_vf_k141_235 | Not-determined |
| NC1_vf_k141_684 | Not-determined |
| NC1_vf_k141_2485 | Not-determined |
| NC1_vf_k141_1585 | Not-determined |
| NC1_vf_k141_459 | Not-determined |
| NC1_vf_k141_2033 | Not-determined |
| NC1_vf_k141_1809 | Not-determined |
| NC1_vf_k141_909 | Not-determined |
| NC1_vf_k141_13 | Not-determined |
| NC1_vf_k141_1358 | Not-determined |
| NC1_vf_k141_2933 | Not-determined |
| NC1_vf_k141_1132 | Not-determined |
| NC1_vf_k141_685 | Not-determined |
| NC1_vf_k141_3157 | Not-determined |
| NC1_vf_k141_2034 | Not-determined |
| NC1_vf_k141_3384 | Not-determined |
| NC1_vf_k141_2708 | Not-determined |
| NC1_vf_k141_2258 | Low-quality |
| NC1_vf_k141_1810 | Not-determined |
| NC1_vf_k141_1586 | Not-determined |
| NC1_vf_k141_14 | Not-determined |
| NC1_vf_k141_2934 | Not-determined |
| NC1_vf_k141_460 | Not-determined |
| NC1_vf_k141_1359 | Not-determined |
| NC1_vf_k141_236 | Not-determined |
| NC1_vf_k141_686 | Not-determined |
| NC1_vf_k141_1133 | Not-determined |
| NC1_vf_k141_2259 | Not-determined |
| NC1_vf_k141_3158 | Not-determined |
| NC1_vf_k141_1811 | Not-determined |
| NC1_vf_k141_1587 | Not-determined |
| NC1_vf_k141_2486 | Not-determined |
| NC1_vf_k141_15 | Not-determined |
| NC1_vf_k141_2035 | Not-determined |
| NC1_vf_k141_2709 | Not-determined |
| NC1_vf_k141_3385 | Not-determined |
| NC1_vf_k141_687 | Not-determined |
| NC1_vf_k141_1134 | Not-determined |
| NC1_vf_k141_461 | Not-determined |
| NC1_vf_k141_1588 | Not-determined |
| NC1_vf_k141_910 | Not-determined |
| NC1_vf_k141_1360 | Not-determined |
| NC1_vf_k141_2935 | Not-determined |
| NC1_vf_k141_2487 | Not-determined |
| NC1_vf_k141_16 | Not-determined |
| NC1_vf_k141_3159 | Not-determined |
| NC1_vf_k141_237 | Not-determined |
| NC1_vf_k141_2260 | Not-determined |
| NC1_vf_k141_1589 | Not-determined |
| NC1_vf_k141_2036 | Not-determined |
| NC1_vf_k141_3386 | Not-determined |
| NC1_vf_k141_17 | Not-determined |
| NC1_vf_k141_462 | Not-determined |
| NC1_vf_k141_2710 | Not-determined |
| NC1_vf_k141_1135 | Not-determined |
| NC1_vf_k141_688 | Not-determined |
| NC1_vf_k141_1812 | Not-determined |
| NC1_vf_k141_1361 | Not-determined |
| NC1_vf_k141_2936 | Not-determined |
| NC1_vf_k141_2488 | Not-determined |
| NC1_vf_k141_2261 | Not-determined |
| NC1_vf_k141_1590 | Not-determined |
| NC1_vf_k141_3160 | Not-determined |
| NC1_vf_k141_463 | Not-determined |
| NC1_vf_k141_1813 | Not-determined |
| NC1_vf_k141_18 | Not-determined |
| NC1_vf_k141_2037 | Not-determined |
| NC1_vf_k141_911 | Not-determined |
| NC1_vf_k141_1362 | Low-quality |
| NC1_vf_k141_3387 | Low-quality |
| NC1_vf_k141_238 | Not-determined |
| NC1_vf_k141_464 | Not-determined |
| NC1_vf_k141_1591 | Not-determined |
| NC1_vf_k141_3161 | Not-determined |
| NC1_vf_k141_2262 | Not-determined |
| NC1_vf_k141_2038 | Not-determined |
| NC1_vf_k141_1136 | Not-determined |
| NC1_vf_k141_2937 | Not-determined |
| NC1_vf_k141_1814 | Not-determined |
| NC1_vf_k141_465 | Not-determined |
| NC1_vf_k141_912 | Not-determined |
| NC1_vf_k141_1592 | Not-determined |
| NC1_vf_k141_689 | Not-determined |
| NC1_vf_k141_19 | Not-determined |
| NC1_vf_k141_2263 | Not-determined |
| NC1_vf_k141_1363 | Not-determined |
| NC1_vf_k141_3388 | Not-determined |
| NC1_vf_k141_2489 | Not-determined |
| NC1_vf_k141_3162 | Not-determined |
| NC1_vf_k141_1815 | Not-determined |
| NC1_vf_k141_2938 | Not-determined |
| NC1_vf_k141_1137 | Not-determined |
| NC1_vf_k141_20 | Not-determined |
| NC1_vf_k141_913 | Not-determined |
| NC1_vf_k141_2711 | Not-determined |
| NC1_vf_k141_2264 | Not-determined |
| NC1_vf_k141_239 | Not-determined |
| NC1_vf_k141_690 | Not-determined |
| NC1_vf_k141_466 | Not-determined |
| NC1_vf_k141_3163 | Not-determined |
| NC1_vf_k141_21 | Not-determined |
| NC1_vf_k141_3389 | Not-determined |
| NC1_vf_k141_914 | Not-determined |
| NC1_vf_k141_1593 | Not-determined |
| NC1_vf_k141_240 | Not-determined |
| NC1_vf_k141_1816 | Not-determined |
| NC1_vf_k141_2490 | Not-determined |
| NC1_vf_k141_2712 | Not-determined |
| NC1_vf_k141_2939 | Not-determined |
| NC1_vf_k141_2265 | Not-determined |
| NC1_vf_k141_1364 | Not-determined |
| NC1_vf_k141_22 | Not-determined |
| NC1_vf_k141_1138 | Not-determined |
| NC1_vf_k141_467 | Not-determined |
| NC1_vf_k141_1817 | Not-determined |
| NC1_vf_k141_241 | Not-determined |
| NC1_vf_k141_3164 | Not-determined |
| NC1_vf_k141_915 | Not-determined |
| NC1_vf_k141_2266 | Not-determined |
| NC1_vf_k141_23 | Not-determined |
| NC1_vf_k141_691 | Not-determined |
| NC1_vf_k141_2713 | Not-determined |
| NC1_vf_k141_2940 | Low-quality |
| NC1_vf_k141_1365 | Not-determined |
| NC1_vf_k141_242 | Not-determined |
| NC1_vf_k141_1139 | Not-determined |
| NC1_vf_k141_2267 | Not-determined |
| NC1_vf_k141_3390 | Not-determined |
| NC1_vf_k141_916 | Not-determined |
| NC1_vf_k141_3165 | Not-determined |
| NC1_vf_k141_2491 | Not-determined |
| NC1_vf_k141_468 | Not-determined |
| NC1_vf_k141_1366 | Low-quality |
| NC1_vf_k141_692 | Not-determined |
| NC1_vf_k141_2039 | Low-quality |
| NC1_vf_k141_2492 | Not-determined |
| NC1_vf_k141_2268 | Not-determined |
| NC1_vf_k141_24 | Not-determined |
| NC1_vf_k141_3391 | Not-determined |
| NC1_vf_k141_917 | Not-determined |
| NC1_vf_k141_1818 | Not-determined |
| NC1_vf_k141_3166 | Not-determined |
| NC1_vf_k141_1367 | Not-determined |
| NC1_vf_k141_243 | Low-quality |
| NC1_vf_k141_1140 | Not-determined |
| NC1_vf_k141_693 | Not-determined |
| NC1_vf_k141_25 | Not-determined |
| NC1_vf_k141_2493 | Not-determined |
| NC1_vf_k141_2040 | Not-determined |
| NC1_vf_k141_3392 | Not-determined |
| NC1_vf_k141_2269 | Not-determined |
| NC1_vf_k141_3167 | Not-determined |
| NC1_vf_k141_918 | Not-determined |
| NC1_vf_k141_2941 | Not-determined |
| NC1_vf_k141_1819 | Not-determined |
| NC1_vf_k141_244 | Not-determined |
| NC1_vf_k141_2041 | Not-determined |
| NC1_vf_k141_26 | Not-determined |
| NC1_vf_k141_2494 | Not-determined |
| NC1_vf_k141_1368 | Not-determined |
| NC1_vf_k141_3168 | Not-determined |
| NC1_vf_k141_1141 | Not-determined |
| NC1_vf_k141_1594 | Not-determined |
| NC1_vf_k141_3393 | Not-determined |
| NC1_vf_k141_694 | Not-determined |
| NC1_vf_k141_2270 | Not-determined |
| NC1_vf_k141_2042 | Not-determined |
| NC1_vf_k141_919 | Not-determined |
| NC1_vf_k141_2495 | Low-quality |
| NC1_vf_k141_27 | Not-determined |
| NC1_vf_k141_1369 | Not-determined |
| NC1_vf_k141_3169 | Not-determined |
| NC1_vf_k141_2942 | Not-determined |
| NC1_vf_k141_245 | Not-determined |
| NC1_vf_k141_1595 | Not-determined |
| NC1_vf_k141_1142 | Not-determined |
| NC1_vf_k141_3394 | Not-determined |
| NC1_vf_k141_2043 | Not-determined |
| NC1_vf_k141_28 | Not-determined |
| NC1_vf_k141_920 | Not-determined |
| NC1_vf_k141_2496 | Not-determined |
| NC1_vf_k141_2271 | Not-determined |
| NC1_vf_k141_1596 | Not-determined |
| NC1_vf_k141_695 | Not-determined |
| NC1_vf_k141_1820 | Not-determined |
| NC1_vf_k141_3395 | Not-determined |
| NC1_vf_k141_2943 | Not-determined |
| NC1_vf_k141_3170 | Not-determined |
| NC1_vf_k141_1143 | Not-determined |
| NC1_vf_k141_1597 | Not-determined |
| NC1_vf_k141_469 | Not-determined |
| NC1_vf_k141_2497 | Not-determined |
| NC1_vf_k141_2272 | Not-determined |
| NC1_vf_k141_696 | Not-determined |
| NC1_vf_k141_29 | Not-determined |
| NC1_vf_k141_1370 | Not-determined |
| NC1_vf_k141_2044 | Not-determined |
| NC1_vf_k141_921 | Not-determined |
| NC1_vf_k141_246 | Not-determined |
| NC1_vf_k141_1144 | Not-determined |
| NC1_vf_k141_3171 | Not-determined |
| NC1_vf_k141_30 | Not-determined |
| NC1_vf_k141_922 | Low-quality |
| NC1_vf_k141_247 | Not-determined |
| NC1_vf_k141_2944 | Not-determined |
| NC1_vf_k141_1598 | Not-determined |
| NC1_vf_k141_1145 | Not-determined |
| NC1_vf_k141_697 | Not-determined |
| NC1_vf_k141_2045 | Not-determined |
| NC1_vf_k141_1371 | Not-determined |
| NC1_vf_k141_3396 | Not-determined |
| NC1_vf_k141_1821 | Not-determined |
| NC1_vf_k141_923 | Not-determined |
| NC1_vf_k141_31 | Not-determined |
| NC1_vf_k141_248 | Not-determined |
| NC1_vf_k141_2498 | Low-quality |
| NC1_vf_k141_2945 | Not-determined |
| NC1_vf_k141_1146 | Not-determined |
| NC1_vf_k141_1599 | Not-determined |
| NC1_vf_k141_2046 | Not-determined |
| NC1_vf_k141_2273 | Not-determined |
| NC1_vf_k141_3172 | Not-determined |
| NC1_vf_k141_3397 | Not-determined |
| NC1_vf_k141_924 | Not-determined |
| NC1_vf_k141_470 | Not-determined |
| NC1_vf_k141_698 | Not-determined |
| NC1_vf_k141_32 | Low-quality |
| NC1_vf_k141_1822 | Not-determined |
| NC1_vf_k141_2499 | Not-determined |
| NC1_vf_k141_2946 | Not-determined |
| NC1_vf_k141_1600 | Not-determined |
| NC1_vf_k141_1147 | Not-determined |
| NC1_vf_k141_3173 | Not-determined |
| NC1_vf_k141_925 | Not-determined |
| NC1_vf_k141_699 | Not-determined |
| NC1_vf_k141_33 | Not-determined |
| NC1_vf_k141_1823 | Not-determined |
| NC1_vf_k141_3398 | Not-determined |
| NC1_vf_k141_249 | Low-quality |
| NC1_vf_k141_2047 | Not-determined |
| NC1_vf_k141_471 | Not-determined |
| NC1_vf_k141_1148 | Not-determined |
| NC1_vf_k141_2500 | Not-determined |
| NC1_vf_k141_2947 | Not-determined |
| NC1_vf_k141_3174 | Not-determined |
| NC1_vf_k141_34 | Not-determined |
| NC1_vf_k141_1601 | Not-determined |
| NC1_vf_k141_926 | Not-determined |
| NC1_vf_k141_1824 | Not-determined |
| NC1_vf_k141_1372 | Not-determined |
| NC1_vf_k141_700 | Not-determined |
| NC1_vf_k141_472 | Not-determined |
| NC1_vf_k141_2048 | Not-determined |
| NC1_vf_k141_2501 | Not-determined |
| NC1_vf_k141_1149 | Not-determined |
| NC1_vf_k141_250 | Not-determined |
| NC1_vf_k141_35 | Not-determined |
| NC1_vf_k141_2714 | Low-quality |
| NC1_vf_k141_1602 | Not-determined |
| NC1_vf_k141_3175 | Not-determined |
| NC1_vf_k141_2948 | Not-determined |
| NC1_vf_k141_1373 | Not-determined |
| NC1_vf_k141_3399 | Not-determined |
| NC1_vf_k141_1825 | Not-determined |
| NC1_vf_k141_701 | Not-determined |
| NC1_vf_k141_36 | Not-determined |
| NC1_vf_k141_1603 | Not-determined |
| NC1_vf_k141_2949 | Not-determined |
| NC1_vf_k141_251 | Not-determined |
| NC1_vf_k141_473 | Not-determined |
| NC1_vf_k141_2715 | Not-determined |
| NC1_vf_k141_3400 | Not-determined |
| NC1_vf_k141_927 | Not-determined |
| NC1_vf_k141_2502 | Not-determined |
| NC1_vf_k141_3176 | Not-determined |
| NC1_vf_k141_2049 | Low-quality |
| NC1_vf_k141_1374 | Not-determined |
| NC1_vf_k141_1826 | Not-determined |
| NC1_vf_k141_1604 | Not-determined |
| NC1_vf_k141_1150 | Not-determined |
| NC1_vf_k141_2503 | Not-determined |
| NC1_vf_k141_928 | Not-determined |
| NC1_vf_k141_37 | Not-determined |
| NC1_vf_k141_3401 | Not-determined |
| NC1_vf_k141_474 | Not-determined |
| NC1_vf_k141_1605 | Not-determined |
| NC1_vf_k141_2716 | Not-determined |
| NC1_vf_k141_3177 | Not-determined |
| NC1_vf_k141_2950 | Not-determined |
| NC1_vf_k141_1375 | Not-determined |
| NC1_vf_k141_702 | Not-determined |
| NC1_vf_k141_2050 | Not-determined |
| NC1_vf_k141_929 | Not-determined |
| NC1_vf_k141_3402 | Not-determined |
| NC1_vf_k141_1151 | Not-determined |
| NC1_vf_k141_1606 | Not-determined |
| NC1_vf_k141_475 | Not-determined |
| NC1_vf_k141_38 | Not-determined |
| NC1_vf_k141_1827 | Not-determined |
| NC1_vf_k141_2504 | Not-determined |
| NC1_vf_k141_703 | Not-determined |
| NC1_vf_k141_2717 | Not-determined |
| NC1_vf_k141_1376 | Not-determined |
| NC1_vf_k141_2951 | Not-determined |
| NC1_vf_k141_930 | Not-determined |
| NC1_vf_k141_252 | Not-determined |
| NC1_vf_k141_1607 | Not-determined |
| NC1_vf_k141_1152 | Not-determined |
| NC1_vf_k141_3403 | Not-determined |
| NC1_vf_k141_2718 | Not-determined |
| NC1_vf_k141_1828 | Not-determined |
| NC1_vf_k141_2505 | Not-determined |
| NC1_vf_k141_1608 | Not-determined |
| NC1_vf_k141_1377 | Not-determined |
| NC1_vf_k141_704 | Not-determined |
| NC1_vf_k141_2952 | Not-determined |
| NC1_vf_k141_253 | Low-quality |
| NC1_vf_k141_931 | Not-determined |
| NC1_vf_k141_39 | Not-determined |
| NC1_vf_k141_2051 | Not-determined |
| NC1_vf_k141_476 | Not-determined |
| NC1_vf_k141_705 | Not-determined |
| NC1_vf_k141_1609 | Low-quality |
| NC1_vf_k141_2506 | Not-determined |
| NC1_vf_k141_932 | Not-determined |
| NC1_vf_k141_1153 | Not-determined |
| NC1_vf_k141_2953 | Not-determined |
| NC1_vf_k141_1610 | Not-determined |
| NC1_vf_k141_933 | Not-determined |
| NC1_vf_k141_706 | Not-determined |
| NC1_vf_k141_1378 | Not-determined |
| NC1_vf_k141_254 | Not-determined |
| NC1_vf_k141_2052 | Not-determined |
| NC1_vf_k141_477 | Not-determined |
| NC1_vf_k141_707 | Not-determined |
| NC1_vf_k141_1154 | Not-determined |
| NC1_vf_k141_934 | Not-determined |
| NC1_vf_k141_2274 | Low-quality |
| NC1_vf_k141_1829 | Low-quality |
| NC1_vf_k141_2507 | Not-determined |
| NC1_vf_k141_40 | Not-determined |
| NC1_vf_k141_1611 | Not-determined |
| NC1_vf_k141_1379 | Not-determined |
| NC1_vf_k141_3178 | Not-determined |
| NC1_vf_k141_708 | Not-determined |
| NC1_vf_k141_478 | Not-determined |
| NC1_vf_k141_935 | Low-quality |
| NC1_vf_k141_255 | Not-determined |
| NC1_vf_k141_1155 | Not-determined |
| NC1_vf_k141_41 | Not-determined |
| NC1_vf_k141_1612 | Not-determined |
| NC1_vf_k141_2508 | Not-determined |
| NC1_vf_k141_2275 | Not-determined |
| NC1_vf_k141_2719 | Not-determined |
| NC1_vf_k141_2053 | Not-determined |
| NC1_vf_k141_3179 | Not-determined |
| NC1_vf_k141_936 | Not-determined |
| NC1_vf_k141_2509 | Not-determined |
| NC1_vf_k141_479 | Not-determined |
| NC1_vf_k141_3404 | Not-determined |
| NC1_vf_k141_1830 | Not-determined |
| NC1_vf_k141_256 | Not-determined |
| NC1_vf_k141_709 | Not-determined |
| NC1_vf_k141_42 | Not-determined |
| NC1_vf_k141_1380 | Not-determined |
| NC1_vf_k141_1156 | Not-determined |
| NC1_vf_k141_1613 | Not-determined |
| NC1_vf_k141_3180 | Not-determined |
| NC1_vf_k141_2510 | Not-determined |
| NC1_vf_k141_937 | Not-determined |
| NC1_vf_k141_480 | Not-determined |
| NC1_vf_k141_2054 | Not-determined |
| NC1_vf_k141_710 | Not-determined |
| NC1_vf_k141_43 | Not-determined |
| NC1_vf_k141_1831 | Not-determined |
| NC1_vf_k141_2720 | Not-determined |
| NC1_vf_k141_3405 | Not-determined |
| NC1_vf_k141_1614 | Not-determined |
| NC1_vf_k141_257 | Not-determined |
| NC1_vf_k141_3181 | Not-determined |
| NC1_vf_k141_2511 | Not-determined |
| NC1_vf_k141_1157 | Not-determined |
| NC1_vf_k141_2055 | Not-determined |
| NC1_vf_k141_44 | Not-determined |
| NC1_vf_k141_1615 | Not-determined |
| NC1_vf_k141_3406 | Not-determined |
| NC1_vf_k141_258 | Not-determined |
| NC1_vf_k141_2512 | Not-determined |
| NC1_vf_k141_481 | Not-determined |
| NC1_vf_k141_1158 | Not-determined |
| NC1_vf_k141_938 | Not-determined |
| NC1_vf_k141_3182 | Not-determined |
| NC1_vf_k141_2056 | Not-determined |
| NC1_vf_k141_1616 | Not-determined |
| NC1_vf_k141_2276 | Not-determined |
| NC1_vf_k141_711 | Not-determined |
| NC1_vf_k141_2721 | Not-determined |
| NC1_vf_k141_1381 | Not-determined |
| NC1_vf_k141_45 | Not-determined |
| NC1_vf_k141_259 | Not-determined |
| NC1_vf_k141_482 | Not-determined |
| NC1_vf_k141_1617 | Not-determined |
| NC1_vf_k141_939 | Not-determined |
| NC1_vf_k141_1832 | Not-determined |
| NC1_vf_k141_3407 | Not-determined |
| NC1_vf_k141_3183 | Not-determined |
| NC1_vf_k141_2513 | Not-determined |
| NC1_vf_k141_2277 | Not-determined |
| NC1_vf_k141_1382 | Not-determined |
| NC1_vf_k141_1833 | Not-determined |
| NC1_vf_k141_2954 | Low-quality |
| NC1_vf_k141_3184 | Not-determined |
| NC1_vf_k141_1618 | Not-determined |
| NC1_vf_k141_2722 | Not-determined |
| NC1_vf_k141_712 | Not-determined |
| NC1_vf_k141_46 | Not-determined |
| NC1_vf_k141_260 | Not-determined |
| NC1_vf_k141_940 | Low-quality |
| NC1_vf_k141_3408 | Not-determined |
| NC1_vf_k141_483 | Not-determined |
| NC1_vf_k141_2514 | Not-determined |
| NC1_vf_k141_1834 | Not-determined |
| NC1_vf_k141_1619 | Not-determined |
| NC1_vf_k141_713 | Not-determined |
| NC1_vf_k141_2278 | Not-determined |
| NC1_vf_k141_1383 | Not-determined |
| NC1_vf_k141_2057 | Not-determined |
| NC1_vf_k141_2955 | Not-determined |
| NC1_vf_k141_3409 | Not-determined |
| NC1_vf_k141_2723 | Not-determined |
| NC1_vf_k141_261 | Low-quality |
| NC1_vf_k141_484 | Not-determined |
| NC1_vf_k141_47 | Not-determined |
| NC1_vf_k141_1620 | Not-determined |
| NC1_vf_k141_3185 | Not-determined |
| NC1_vf_k141_941 | Not-determined |
| NC1_vf_k141_2279 | Not-determined |
| NC1_vf_k141_1835 | Not-determined |
| NC1_vf_k141_1621 | Not-determined |
| NC1_vf_k141_2515 | Not-determined |
| NC1_vf_k141_2956 | Not-determined |
| NC1_vf_k141_1384 | Not-determined |
| NC1_vf_k141_48 | Not-determined |
| NC1_vf_k141_3186 | Not-determined |
| NC1_vf_k141_942 | Not-determined |
| NC1_vf_k141_714 | Not-determined |
| NC1_vf_k141_2724 | Not-determined |
| NC1_vf_k141_2280 | Not-determined |
| NC1_vf_k141_1622 | Not-determined |
| NC1_vf_k141_2058 | Not-determined |
| NC1_vf_k141_262 | Not-determined |
| NC1_vf_k141_1836 | Not-determined |
| NC1_vf_k141_3410 | Not-determined |
| NC1_vf_k141_2516 | Not-determined |
| NC1_vf_k141_49 | Not-determined |
| NC1_vf_k141_1385 | Not-determined |
| NC1_vf_k141_2281 | Not-determined |

| | |
|---|---|
| NC1_vf_k141_2725 | Not-determined |
| NC1_vf_k141_715 | Not-determined |
| NC1_vf_k141_2059 | Not-determined |
| NC1_vf_k141_2957 | Low-quality |
| NC1_vf_k141_485 | Not-determined |
| NC1_vf_k141_2517 | Not-determined |
| NC1_vf_k141_50 | Not-determined |
| NC1_vf_k141_1623 | Not-determined |
| NC1_vf_k141_1837 | Not-determined |
| NC1_vf_k141_3187 | Not-determined |
| NC1_vf_k141_3411 | Not-determined |
| NC1_vf_k141_2282 | Not-determined |
| NC1_vf_k141_51 | Not-determined |
| NC1_vf_k141_716 | Not-determined |
| NC1_vf_k141_2726 | Not-determined |
| NC1_vf_k141_2518 | Not-determined |
| NC1_vf_k141_1624 | Not-determined |
| NC1_vf_k141_1386 | Not-determined |
| NC1_vf_k141_2060 | Not-determined |
| NC1_vf_k141_3188 | Not-determined |
| NC1_vf_k141_2283 | Not-determined |
| NC1_vf_k141_3412 | Not-determined |
| NC1_vf_k141_943 | Not-determined |
| NC1_vf_k141_717 | Not-determined |
| NC1_vf_k141_1625 | Not-determined |
| NC1_vf_k141_1387 | Not-determined |
| NC1_vf_k141_2519 | Not-determined |
| NC1_vf_k141_2958 | Not-determined |
| NC1_vf_k141_1838 | Not-determined |
| NC1_vf_k141_3413 | Not-determined |
| NC1_vf_k141_2061 | Not-determined |
| NC1_vf_k141_486 | Not-determined |
| NC1_vf_k141_2284 | Not-determined |
| NC1_vf_k141_1388 | Not-determined |
| NC1_vf_k141_944 | Not-determined |
| NC1_vf_k141_3414 | Not-determined |
| NC1_vf_k141_2062 | Not-determined |
| NC1_vf_k141_52 | Not-determined |
| NC1_vf_k141_2727 | Not-determined |
| NC1_vf_k141_1626 | Not-determined |
| NC1_vf_k141_718 | Not-determined |
| NC1_vf_k141_1839 | Not-determined |
| NC1_vf_k141_2520 | Not-determined |
| NC1_vf_k141_1389 | Not-determined |
| NC1_vf_k141_3415 | Not-determined |
| NC1_vf_k141_2285 | Not-determined |
| NC1_vf_k141_945 | Not-determined |
| NC1_vf_k141_2959 | Low-quality |
| NC1_vf_k141_53 | Not-determined |
| NC1_vf_k141_3189 | Not-determined |
| NC1_vf_k141_2728 | Not-determined |
| NC1_vf_k141_719 | Not-determined |
| NC1_vf_k141_1840 | Not-determined |
| NC1_vf_k141_2063 | Not-determined |
| NC1_vf_k141_3416 | Not-determined |
| NC1_vf_k141_1390 | Not-determined |
| NC1_vf_k141_2286 | Not-determined |
| NC1_vf_k141_3190 | Not-determined |
| NC1_vf_k141_1627 | Not-determined |
| NC1_vf_k141_54 | Not-determined |
| NC1_vf_k141_2729 | Not-determined |
| NC1_vf_k141_720 | Not-determined |
| NC1_vf_k141_946 | Not-determined |
| NC1_vf_k141_2960 | Not-determined |
| NC1_vf_k141_1841 | Not-determined |
| NC1_vf_k141_2064 | Low-quality |
| NC1_vf_k141_2521 | Not-determined |
| NC1_vf_k141_55 | Not-determined |
| NC1_vf_k141_2287 | Not-determined |
| NC1_vf_k141_2730 | Not-determined |
| NC1_vf_k141_3191 | Not-determined |
| NC1_vf_k141_263 | Not-determined |
| NC1_vf_k141_1628 | Not-determined |
| NC1_vf_k141_947 | Not-determined |
| NC1_vf_k141_721 | Not-determined |
| NC1_vf_k141_1391 | Not-determined |
| NC1_vf_k141_2961 | Not-determined |
| NC1_vf_k141_3417 | Not-determined |
| NC1_vf_k141_2065 | Not-determined |
| NC1_vf_k141_56 | Not-determined |
| NC1_vf_k141_1842 | Not-determined |
| NC1_vf_k141_2522 | Low-quality |
| NC1_vf_k141_2731 | Not-determined |
| NC1_vf_k141_948 | Not-determined |
| NC1_vf_k141_1629 | Not-determined |
| NC1_vf_k141_57 | Not-determined |
| NC1_vf_k141_1392 | Not-determined |
| NC1_vf_k141_3192 | Not-determined |
| NC1_vf_k141_1843 | Not-determined |
| NC1_vf_k141_722 | Not-determined |
| NC1_vf_k141_2066 | Not-determined |
| NC1_vf_k141_2523 | Not-determined |
| NC1_vf_k141_487 | Not-determined |
| NC1_vf_k141_2732 | Not-determined |
| NC1_vf_k141_949 | Not-determined |
| NC1_vf_k141_2962 | Not-determined |
| NC1_vf_k141_723 | Not-determined |
| NC1_vf_k141_3193 | Not-determined |
| NC1_vf_k141_2288 | Not-determined |
| NC1_vf_k141_3418 | Not-determined |
| NC1_vf_k141_1630 | Not-determined |
| NC1_vf_k141_1393 | Not-determined |
| NC1_vf_k141_264 | Not-determined |
| NC1_vf_k141_58 | Not-determined |
| NC1_vf_k141_2067 | Not-determined |
| NC1_vf_k141_724 | Not-determined |
| NC1_vf_k141_488 | Not-determined |
| NC1_vf_k141_2733 | Not-determined |
| NC1_vf_k141_2289 | Not-determined |
| NC1_vf_k141_2963 | Not-determined |
| NC1_vf_k141_2524 | Not-determined |
| NC1_vf_k141_1394 | Not-determined |
| NC1_vf_k141_950 | Not-determined |
| NC1_vf_k141_59 | Not-determined |
| NC1_vf_k141_2068 | Not-determined |
| NC1_vf_k141_489 | Not-determined |
| NC1_vf_k141_1631 | Not-determined |
| NC1_vf_k141_2290 | Not-determined |
| NC1_vf_k141_725 | Not-determined |
| NC1_vf_k141_265 | Low-quality |
| NC1_vf_k141_3194 | Not-determined |
| NC1_vf_k141_3419 | Low-quality |
| NC1_vf_k141_60 | Not-determined |
| NC1_vf_k141_2734 | Not-determined |
| NC1_vf_k141_951 | Not-determined |
| NC1_vf_k141_2964 | Not-determined |
| NC1_vf_k141_2525 | Not-determined |
| NC1_vf_k141_1632 | Not-determined |
| NC1_vf_k141_726 | Not-determined |
| NC1_vf_k141_3420 | Low-quality |
| NC1_vf_k141_266 | Not-determined |
| NC1_vf_k141_2735 | Not-determined |
| NC1_vf_k141_3195 | Not-determined |
| NC1_vf_k141_2965 | Not-determined |
| NC1_vf_k141_1159 | Low-quality |
| NC1_vf_k141_1395 | Not-determined |
| NC1_vf_k141_1633 | Not-determined |
| NC1_vf_k141_61 | Not-determined |
| NC1_vf_k141_3421 | Not-determined |
| NC1_vf_k141_952 | Not-determined |
| NC1_vf_k141_267 | Not-determined |
| NC1_vf_k141_3196 | Not-determined |
| NC1_vf_k141_490 | Not-determined |
| NC1_vf_k141_727 | Not-determined |
| NC1_vf_k141_2526 | Not-determined |
| NC1_vf_k141_62 | Low-quality |
| NC1_vf_k141_2736 | Not-determined |
| NC1_vf_k141_1396 | Not-determined |
| NC1_vf_k141_953 | Not-determined |
| NC1_vf_k141_2291 | Not-determined |
| NC1_vf_k141_268 | Not-determined |
| NC1_vf_k141_2069 | Not-determined |
| NC1_vf_k141_2737 | Not-determined |
| NC1_vf_k141_728 | Not-determined |
| NC1_vf_k141_3197 | Not-determined |
| NC1_vf_k141_491 | Not-determined |
| NC1_vf_k141_1634 | Not-determined |
| NC1_vf_k141_1397 | Not-determined |
| NC1_vf_k141_2527 | Not-determined |
| NC1_vf_k141_2292 | Low-quality |
| NC1_vf_k141_1160 | Not-determined |
| NC1_vf_k141_1844 | Not-determined |
| NC1_vf_k141_729 | Not-determined |
| NC1_vf_k141_492 | Not-determined |
| NC1_vf_k141_269 | Not-determined |
| NC1_vf_k141_3422 | Not-determined |
| NC1_vf_k141_1635 | Not-determined |
| NC1_vf_k141_2528 | Not-determined |
| NC1_vf_k141_63 | Not-determined |
| NC1_vf_k141_954 | Not-determined |
| NC1_vf_k141_3198 | Low-quality |
| NC1_vf_k141_1845 | Not-determined |
| NC1_vf_k141_493 | Not-determined |
| NC1_vf_k141_3423 | Not-determined |
| NC1_vf_k141_270 | Not-determined |
| NC1_vf_k141_2738 | Not-determined |
| NC1_vf_k141_1161 | Not-determined |
| NC1_vf_k141_2070 | Not-determined |
| NC1_vf_k141_64 | Not-determined |
| NC1_vf_k141_3199 | Not-determined |
| NC1_vf_k141_2293 | Not-determined |
| NC1_vf_k141_730 | Not-determined |
| NC1_vf_k141_1636 | Not-determined |
| NC1_vf_k141_494 | Not-determined |
| NC1_vf_k141_2529 | Not-determined |
| NC1_vf_k141_1398 | Not-determined |
| NC1_vf_k141_65 | Not-determined |
| NC1_vf_k141_271 | Not-determined |
| NC1_vf_k141_1637 | Not-determined |
| NC1_vf_k141_955 | Not-determined |
| NC1_vf_k141_1846 | Not-determined |
| NC1_vf_k141_2071 | Not-determined |
| NC1_vf_k141_3424 | Not-determined |
| NC1_vf_k141_1399 | Not-determined |
| NC1_vf_k141_1162 | Not-determined |
| NC1_vf_k141_2739 | Not-determined |
| NC1_vf_k141_2294 | Not-determined |
| NC1_vf_k141_2966 | Not-determined |
| NC1_vf_k141_495 | Not-determined |
| NC1_vf_k141_731 | Not-determined |
| NC1_vf_k141_2530 | Not-determined |
| NC1_vf_k141_272 | Not-determined |
| NC1_vf_k141_2072 | Not-determined |
| NC1_vf_k141_956 | Low-quality |
| NC1_vf_k141_2967 | Not-determined |
| NC1_vf_k141_1638 | Not-determined |
| NC1_vf_k141_3425 | Not-determined |
| NC1_vf_k141_2740 | Not-determined |
| NC1_vf_k141_1163 | Not-determined |
| NC1_vf_k141_2295 | Not-determined |
| NC1_vf_k141_1847 | Not-determined |
| NC1_vf_k141_66 | Low-quality |
| NC1_vf_k141_1400 | Not-determined |
| NC1_vf_k141_2073 | Not-determined |
| NC1_vf_k141_1639 | Not-determined |
| NC1_vf_k141_273 | Not-determined |
| NC1_vf_k141_732 | Not-determined |
| NC1_vf_k141_957 | Not-determined |
| NC1_vf_k141_67 | Not-determined |
| NC1_vf_k141_2296 | Not-determined |
| NC1_vf_k141_3426 | Not-determined |
| NC1_vf_k141_2531 | Not-determined |
| NC1_vf_k141_2741 | Low-quality |
| NC1_vf_k141_1164 | Not-determined |
| NC1_vf_k141_2968 | Not-determined |
| NC1_vf_k141_2074 | Not-determined |
| NC1_vf_k141_1640 | Not-determined |
| NC1_vf_k141_274 | Low-quality |
| NC1_vf_k141_1848 | Not-determined |
| NC1_vf_k141_1401 | Not-determined |
| NC1_vf_k141_496 | Low-quality |
| NC1_vf_k141_958 | Not-determined |
| NC1_vf_k141_68 | Not-determined |
| NC1_vf_k141_733 | Not-determined |
| NC1_vf_k141_2532 | Not-determined |
| NC1_vf_k141_1641 | Not-determined |
| NC1_vf_k141_2297 | Not-determined |
| NC1_vf_k141_3427 | Not-determined |
| NC1_vf_k141_2075 | Not-determined |
| NC1_vf_k141_275 | Not-determined |
| NC1_vf_k141_1849 | Not-determined |
| NC1_vf_k141_69 | Not-determined |
| NC1_vf_k141_2742 | Low-quality |
| NC1_vf_k141_959 | Not-determined |
| NC1_vf_k141_497 | Not-determined |
| NC1_vf_k141_1642 | Not-determined |
| NC1_vf_k141_276 | Not-determined |
| NC1_vf_k141_2969 | Not-determined |
| NC1_vf_k141_2533 | Not-determined |
| NC1_vf_k141_734 | Not-determined |
| NC1_vf_k141_2076 | Not-determined |
| NC1_vf_k141_3200 | Not-determined |
| NC1_vf_k141_3428 | Not-determined |
| NC1_vf_k141_1850 | Not-determined |
| NC1_vf_k141_498 | Not-determined |
| NC1_vf_k141_1165 | Not-determined |
| NC1_vf_k141_1643 | Not-determined |
| NC1_vf_k141_2743 | Not-determined |
| NC1_vf_k141_2298 | Not-determined |
| NC1_vf_k141_2534 | Not-determined |
| NC1_vf_k141_735 | Not-determined |
| NC1_vf_k141_2077 | Not-determined |
| NC1_vf_k141_3429 | Not-determined |
| NC1_vf_k141_3201 | Not-determined |
| NC1_vf_k141_499 | Not-determined |
| NC1_vf_k141_70 | Not-determined |
| NC1_vf_k141_1644 | Not-determined |
| NC1_vf_k141_277 | Low-quality |
| NC1_vf_k141_960 | Not-determined |
| NC1_vf_k141_2299 | Not-determined |
| NC1_vf_k141_736 | Not-determined |
| NC1_vf_k141_1851 | Not-determined |
| NC1_vf_k141_2078 | Not-determined |
| NC1_vf_k141_1166 | Not-determined |
| NC1_vf_k141_1645 | Not-determined |
| NC1_vf_k141_3202 | Not-determined |
| NC1_vf_k141_500 | Not-determined |
| NC1_vf_k141_3430 | Not-determined |
| NC1_vf_k141_278 | Low-quality |
| NC1_vf_k141_2535 | Not-determined |
| NC1_vf_k141_2744 | Not-determined |
| NC1_vf_k141_961 | Not-determined |
| NC1_vf_k141_1646 | Not-determined |
| NC1_vf_k141_71 | Not-determined |
| NC1_vf_k141_737 | Not-determined |
| NC1_vf_k141_1167 | Not-determined |
| NC1_vf_k141_501 | Not-determined |
| NC1_vf_k141_2970 | Not-determined |
| NC1_vf_k141_279 | Not-determined |
| NC1_vf_k141_2079 | Not-determined |
| NC1_vf_k141_2536 | Not-determined |
| NC1_vf_k141_1647 | Not-determined |
| NC1_vf_k141_962 | Not-determined |
| NC1_vf_k141_1852 | Not-determined |
| NC1_vf_k141_2745 | Not-determined |
| NC1_vf_k141_1402 | Low-quality |
| NC1_vf_k141_2300 | Not-determined |
| NC1_vf_k141_738 | Not-determined |
| NC1_vf_k141_1168 | Not-determined |
| NC1_vf_k141_72 | Low-quality |
| NC1_vf_k141_502 | Not-determined |
| NC1_vf_k141_2080 | Not-determined |
| NC1_vf_k141_1648 | Not-determined |
| NC1_vf_k141_2971 | Not-determined |
| NC1_vf_k141_280 | Not-determined |
| NC1_vf_k141_3203 | Not-determined |
| NC1_vf_k141_1169 | Not-determined |
| NC1_vf_k141_2537 | Not-determined |
| NC1_vf_k141_2746 | Not-determined |
| NC1_vf_k141_1649 | Not-determined |
| NC1_vf_k141_2081 | Not-determined |
| NC1_vf_k141_739 | Not-determined |
| NC1_vf_k141_1403 | Not-determined |
| NC1_vf_k141_963 | Not-determined |
| NC1_vf_k141_73 | Not-determined |
| NC1_vf_k141_2972 | Not-determined |
| NC1_vf_k141_1853 | Low-quality |
| NC1_vf_k141_3431 | Not-determined |
| NC1_vf_k141_1170 | Not-determined |
| NC1_vf_k141_2301 | Not-determined |
| NC1_vf_k141_2538 | Not-determined |
| NC1_vf_k141_2082 | Not-determined |
| NC1_vf_k141_503 | Not-determined |
| NC1_vf_k141_1404 | Not-determined |
| NC1_vf_k141_3204 | Not-determined |
| NC1_vf_k141_281 | Not-determined |
| NC1_vf_k141_1650 | Not-determined |
| NC1_vf_k141_1854 | Not-determined |
| NC1_vf_k141_2539 | Not-determined |
| NC1_vf_k141_74 | Not-determined |
| NC1_vf_k141_2083 | Not-determined |
| NC1_vf_k141_1171 | Not-determined |
| NC1_vf_k141_3432 | Not-determined |
| NC1_vf_k141_964 | Not-determined |
| NC1_vf_k141_3205 | Not-determined |
| NC1_vf_k141_1651 | Not-determined |
| NC1_vf_k141_2302 | Not-determined |
| NC1_vf_k141_75 | Not-determined |
| NC1_vf_k141_1405 | Not-determined |
| NC1_vf_k141_1855 | Not-determined |
| NC1_vf_k141_282 | Not-determined |
| NC1_vf_k141_3433 | Not-determined |
| NC1_vf_k141_2084 | Not-determined |
| NC1_vf_k141_504 | Not-determined |
| NC1_vf_k141_3206 | Not-determined |
| NC1_vf_k141_2540 | Not-determined |
| NC1_vf_k141_2973 | Not-determined |
| NC1_vf_k141_1856 | Not-determined |
| NC1_vf_k141_76 | Not-determined |
| NC1_vf_k141_1652 | Not-determined |
| NC1_vf_k141_1172 | Not-determined |
| NC1_vf_k141_965 | Not-determined |
| NC1_vf_k141_2303 | Not-determined |
| NC1_vf_k141_283 | Not-determined |
| NC1_vf_k141_505 | Not-determined |
| NC1_vf_k141_2974 | Not-determined |
| NC1_vf_k141_3207 | Not-determined |
| NC1_vf_k141_77 | Not-determined |
| NC1_vf_k141_1857 | Low-quality |
| NC1_vf_k141_2085 | Not-determined |
| NC1_vf_k141_1406 | Not-determined |
| NC1_vf_k141_1653 | Not-determined |
| NC1_vf_k141_966 | Not-determined |
| NC1_vf_k141_2304 | Not-determined |
| NC1_vf_k141_740 | Not-determined |
| NC1_vf_k141_2541 | Not-determined |
| NC1_vf_k141_284 | Not-determined |
| NC1_vf_k141_506 | Not-determined |
| NC1_vf_k141_3434 | Not-determined |
| NC1_vf_k141_2975 | Not-determined |
| NC1_vf_k141_1654 | Not-determined |
| NC1_vf_k141_78 | Not-determined |
| NC1_vf_k141_1173 | Not-determined |
| NC1_vf_k141_2305 | Not-determined |
| NC1_vf_k141_1407 | Not-determined |
| NC1_vf_k141_285 | Not-determined |
| NC1_vf_k141_2086 | Low-quality |
| NC1_vf_k141_741 | Not-determined |
| NC1_vf_k141_507 | Not-determined |
| NC1_vf_k141_79 | Not-determined |
| NC1_vf_k141_1655 | Not-determined |
| NC1_vf_k141_3435 | Not-determined |
| NC1_vf_k141_2976 | Not-determined |
| NC1_vf_k141_2542 | Not-determined |
| NC1_vf_k141_80 | Not-determined |
| NC1_vf_k141_3208 | Not-determined |
| NC1_vf_k141_1656 | Not-determined |
| NC1_vf_k141_2087 | Not-determined |
| NC1_vf_k141_1408 | Not-determined |
| NC1_vf_k141_286 | Not-determined |
| NC1_vf_k141_967 | Not-determined |
| NC1_vf_k141_81 | Not-determined |
| NC1_vf_k141_1657 | Not-determined |
| NC1_vf_k141_2977 | Not-determined |
| NC1_vf_k141_2306 | Not-determined |
| NC1_vf_k141_3436 | Not-determined |
| NC1_vf_k141_1174 | Not-determined |
| NC1_vf_k141_742 | Not-determined |
| NC1_vf_k141_508 | Not-determined |
| NC1_vf_k141_82 | Not-determined |
| NC1_vf_k141_1658 | Not-determined |
| NC1_vf_k141_2543 | Not-determined |
| NC1_vf_k141_287 | Not-determined |
| NC1_vf_k141_2978 | Not-determined |
| NC1_vf_k141_1858 | Not-determined |
| NC1_vf_k141_2088 | Not-determined |
| NC1_vf_k141_83 | Not-determined |
| NC1_vf_k141_2307 | Not-determined |
| NC1_vf_k141_1175 | Not-determined |
| NC1_vf_k141_2544 | Not-determined |
| NC1_vf_k141_968 | Not-determined |
| NC1_vf_k141_1659 | Low-quality |
| NC1_vf_k141_1409 | Not-determined |
| NC1_vf_k141_288 | Not-determined |
| NC1_vf_k141_84 | Not-determined |
| NC1_vf_k141_2979 | Not-determined |
| NC1_vf_k141_3209 | Not-determined |
| NC1_vf_k141_3437 | Not-determined |
| NC1_vf_k141_509 | Not-determined |
| NC1_vf_k141_1660 | Not-determined |
| NC1_vf_k141_85 | Not-determined |
| NC1_vf_k141_969 | Not-determined |
| NC1_vf_k141_1859 | Not-determined |
| NC1_vf_k141_3438 | Not-determined |
| NC1_vf_k141_2308 | Not-determined |
| NC1_vf_k141_2980 | Not-determined |
| NC1_vf_k141_289 | Not-determined |
| NC1_vf_k141_1176 | Not-determined |
| NC1_vf_k141_510 | Not-determined |
| NC1_vf_k141_2089 | Not-determined |
| NC1_vf_k141_970 | Not-determined |
| NC1_vf_k141_86 | Not-determined |
| NC1_vf_k141_1661 | Not-determined |
| NC1_vf_k141_3210 | Not-determined |
| NC1_vf_k141_1410 | Not-determined |
| NC1_vf_k141_2309 | Not-determined |
| NC1_vf_k141_2545 | Not-determined |
| NC1_vf_k141_1177 | Not-determined |
| NC1_vf_k141_290 | Not-determined |
| NC1_vf_k141_3439 | Not-determined |
| NC1_vf_k141_511 | Not-determined |
| NC1_vf_k141_1662 | Not-determined |
| NC1_vf_k141_3211 | Not-determined |
| NC1_vf_k141_2090 | Not-determined |
| NC1_vf_k141_1860 | Not-determined |
| NC1_vf_k141_971 | Not-determined |
| NC1_vf_k141_1411 | Not-determined |
| NC1_vf_k141_2546 | Not-determined |
| NC1_vf_k141_87 | Not-determined |
| NC1_vf_k141_291 | Not-determined |
| NC1_vf_k141_2981 | Not-determined |
| NC1_vf_k141_1178 | Not-determined |
| NC1_vf_k141_1412 | Not-determined |
| NC1_vf_k141_1663 | Not-determined |
| NC1_vf_k141_2091 | Not-determined |
| NC1_vf_k141_2310 | Not-determined |
| NC1_vf_k141_1861 | Not-determined |
| NC1_vf_k141_2547 | Not-determined |
| NC1_vf_k141_1179 | Not-determined |
| NC1_vf_k141_292 | Not-determined |
| NC1_vf_k141_88 | Not-determined |
| NC1_vf_k141_512 | Not-determined |
| NC1_vf_k141_972 | Not-determined |
| NC1_vf_k141_1664 | Not-determined |
| NC1_vf_k141_3440 | Not-determined |
| NC1_vf_k141_2092 | Not-determined |
| NC1_vf_k141_1413 | Not-determined |
| NC1_vf_k141_3212 | Not-determined |
| NC1_vf_k141_2982 | Not-determined |
| NC1_vf_k141_1665 | Not-determined |
| NC1_vf_k141_2548 | Not-determined |
| NC1_vf_k141_89 | Not-determined |
| NC1_vf_k141_293 | Not-determined |
| NC1_vf_k141_1862 | Not-determined |
| NC1_vf_k141_3441 | Not-determined |
| NC1_vf_k141_2311 | Not-determined |
| NC1_vf_k141_973 | Not-determined |
| NC1_vf_k141_90 | Not-determined |
| NC1_vf_k141_1180 | Not-determined |
| NC1_vf_k141_2983 | Not-determined |
| NC1_vf_k141_2549 | Not-determined |
| NC1_vf_k141_1666 | Not-determined |
| NC1_vf_k141_513 | Not-determined |
| NC1_vf_k141_2093 | Not-determined |
| NC1_vf_k141_1863 | Not-determined |
| NC1_vf_k141_294 | Not-determined |
| NC1_vf_k141_3213 | Not-determined |
| NC1_vf_k141_1414 | Not-determined |
| NC1_vf_k141_2312 | Not-determined |
| NC1_vf_k141_974 | Not-determined |
| NC1_vf_k141_1181 | Not-determined |
| NC1_vf_k141_1667 | Not-determined |
| NC1_vf_k141_2984 | Low-quality |
| NC1_vf_k141_2094 | Low-quality |
| NC1_vf_k141_91 | Not-determined |
| NC1_vf_k141_3442 | Not-determined |
| NC1_vf_k141_1864 | Not-determined |
| NC1_vf_k141_2313 | Not-determined |
| NC1_vf_k141_1415 | Not-determined |
| NC1_vf_k141_3214 | Not-determined |
| NC1_vf_k141_975 | Not-determined |
| NC1_vf_k141_2550 | Not-determined |
| NC1_vf_k141_514 | Not-determined |
| NC1_vf_k141_92 | Not-determined |
| NC1_vf_k141_1182 | Not-determined |
| NC1_vf_k141_295 | Not-determined |
| NC1_vf_k141_3443 | Not-determined |
| NC1_vf_k141_2985 | Not-determined |
| NC1_vf_k141_1865 | Not-determined |
| NC1_vf_k141_976 | Not-determined |
| NC1_vf_k141_3215 | Not-determined |
| NC1_vf_k141_2314 | Not-determined |
| NC1_vf_k141_2551 | Not-determined |
| NC1_vf_k141_296 | Not-determined |
| NC1_vf_k141_93 | Not-determined |
| NC1_vf_k141_1183 | Low-quality |
| NC1_vf_k141_1668 | Not-determined |
| NC1_vf_k141_2986 | Not-determined |
| NC1_vf_k141_2095 | Not-determined |
| NC1_vf_k141_1866 | Not-determined |
| NC1_vf_k141_3216 | Not-determined |
| NC1_vf_k141_94 | Not-determined |
| NC1_vf_k141_2315 | Not-determined |
| NC1_vf_k141_743 | Not-determined |
| NC1_vf_k141_1669 | Not-determined |
| NC1_vf_k141_297 | Not-determined |
| NC1_vf_k141_1416 | Not-determined |
| NC1_vf_k141_2747 | Low-quality |
| NC1_vf_k141_1184 | Not-determined |
| NC1_vf_k141_2987 | Not-determined |
| NC1_vf_k141_2096 | Not-determined |
| NC1_vf_k141_744 | Not-determined |
| NC1_vf_k141_3217 | Low-quality |
| NC1_vf_k141_515 | Not-determined |
| NC1_vf_k141_298 | Not-determined |
| NC1_vf_k141_2316 | Not-determined |
| NC1_vf_k141_977 | Not-determined |
| NC1_vf_k141_1867 | Not-determined |
| NC1_vf_k141_1417 | Not-determined |
| NC1_vf_k141_2988 | Low-quality |
| NC1_vf_k141_745 | Low-quality |
| NC1_vf_k141_2097 | Not-determined |
| NC1_vf_k141_95 | Not-determined |
| NC1_vf_k141_299 | Not-determined |
| NC1_vf_k141_1185 | Not-determined |
| NC1_vf_k141_2748 | Not-determined |
| NC1_vf_k141_516 | Not-determined |
| NC1_vf_k141_2989 | Not-determined |
| NC1_vf_k141_978 | Not-determined |
| NC1_vf_k141_1418 | Not-determined |
| NC1_vf_k141_2317 | Not-determined |
| NC1_vf_k141_2098 | Not-determined |
| NC1_vf_k141_1186 | Not-determined |
| NC1_vf_k141_2749 | Not-determined |
| NC1_vf_k141_96 | Not-determined |
| NC1_vf_k141_979 | Not-determined |
| NC1_vf_k141_300 | Not-determined |
| NC1_vf_k141_1670 | Not-determined |
| NC1_vf_k141_1868 | Not-determined |
| NC1_vf_k141_2099 | Not-determined |
| NC1_vf_k141_2990 | Not-determined |
| NC1_vf_k141_517 | Not-determined |
| NC1_vf_k141_2318 | Not-determined |
| NC1_vf_k141_97 | Not-determined |
| NC1_vf_k141_746 | Not-determined |
| NC1_vf_k141_2750 | Not-determined |
| NC1_vf_k141_1671 | Not-determined |
| NC1_vf_k141_3219 | Not-determined |
| NC1_vf_k141_980 | Not-determined |
| NC1_vf_k141_2991 | Not-determined |
| NC1_vf_k141_3444 | Not-determined |
| NC1_vf_k141_2100 | Not-determined |
| NC1_vf_k141_2319 | Not-determined |
| NC1_vf_k141_301 | Not-determined |
| NC1_vf_k141_518 | Not-determined |
| NC1_vf_k141_747 | Not-determined |
| NC1_vf_k141_1672 | Not-determined |
| NC1_vf_k141_2992 | Not-determined |
| NC1_vf_k141_3220 | Not-determined |
| NC1_vf_k141_2320 | Not-determined |
| NC1_vf_k141_98 | Not-determined |
| NC1_vf_k141_1419 | Not-determined |
| NC1_vf_k141_981 | Not-determined |
| NC1_vf_k141_2101 | Not-determined |
| NC1_vf_k141_1673 | Not-determined |
| NC1_vf_k141_1869 | Low-quality |
| NC1_vf_k141_3445 | Not-determined |
| NC1_vf_k141_748 | Not-determined |
| NC1_vf_k141_302 | Not-determined |

| | |
|---|---|
| NC1_vf_k141_1187 | Not-determined |
| NC1_vf_k141_2321 | Not-determined |
| NC1_vf_k141_2751 | Not-determined |
| NC1_vf_k141_3221 | Not-determined |
| NC1_vf_k141_519 | Not-determined |
| NC1_vf_k141_1674 | Not-determined |
| NC1_vf_k141_982 | Not-determined |
| NC1_vf_k141_1420 | Not-determined |
| NC1_vf_k141_2102 | Not-determined |
| NC1_vf_k141_749 | Not-determined |
| NC1_vf_k141_1188 | Not-determined |
| NC1_vf_k141_303 | Not-determined |
| NC1_vf_k141_2752 | Not-determined |
| NC1_vf_k141_3446 | Not-determined |
| NC1_vf_k141_1870 | Not-determined |
| NC1_vf_k141_1675 | Not-determined |
| NC1_vf_k141_983 | Not-determined |
| NC1_vf_k141_2103 | Not-determined |
| NC1_vf_k141_750 | Not-determined |
| NC1_vf_k141_1189 | Not-determined |
| NC1_vf_k141_1421 | Not-determined |
| NC1_vf_k141_520 | Not-determined |
| NC1_vf_k141_3447 | Not-determined |
| NC1_vf_k141_304 | Not-determined |
| NC1_vf_k141_2322 | Not-determined |
| NC1_vf_k141_1676 | Not-determined |
| NC1_vf_k141_751 | Not-determined |
| NC1_vf_k141_984 | Not-determined |
| NC1_vf_k141_2104 | Not-determined |
| NC1_vf_k141_3448 | Not-determined |
| NC1_vf_k141_3222 | Not-determined |
| NC1_vf_k141_2753 | Not-determined |
| NC1_vf_k141_305 | Not-determined |
| NC1_vf_k141_1677 | Not-determined |
| NC1_vf_k141_2552 | Not-determined |
| NC1_vf_k141_752 | Not-determined |
| NC1_vf_k141_2993 | Not-determined |
| NC1_vf_k141_1871 | Not-determined |
| NC1_vf_k141_3223 | Not-determined |
| NC1_vf_k141_521 | Not-determined |
| NC1_vf_k141_1190 | Not-determined |
| NC1_vf_k141_2105 | Not-determined |
| NC1_vf_k141_3449 | Not-determined |
| NC1_vf_k141_1422 | Not-determined |
| NC1_vf_k141_2754 | Not-determined |
| NC1_vf_k141_985 | Not-determined |
| NC1_vf_k141_1678 | Not-determined |
| NC1_vf_k141_306 | Not-determined |
| NC1_vf_k141_3224 | Not-determined |
| NC1_vf_k141_3450 | Not-determined |
| NC1_vf_k141_2106 | Not-determined |
| NC1_vf_k141_1679 | Not-determined |
| NC1_vf_k141_1872 | Not-determined |
| NC1_vf_k141_753 | Not-determined |
| NC1_vf_k141_307 | Low-quality |
| NC1_vf_k141_2755 | Not-determined |
| NC1_vf_k141_2553 | Low-quality |
| NC1_vf_k141_522 | Not-determined |
| NC1_vf_k141_1423 | Not-determined |
| NC1_vf_k141_1191 | Low-quality |
| NC1_vf_k141_2323 | Low-quality |
| NC1_vf_k141_2994 | Not-determined |
| NC1_vf_k141_1680 | Not-determined |
| NC1_vf_k141_986 | Not-determined |
| NC1_vf_k141_3225 | Not-determined |
| NC1_vf_k141_754 | Not-determined |
| NC1_vf_k141_1873 | Not-determined |
| NC1_vf_k141_2107 | Not-determined |
| NC1_vf_k141_3451 | Not-determined |
| NC1_vf_k141_1424 | Not-determined |
| NC1_vf_k141_1192 | Not-determined |
| NC1_vf_k141_3226 | Not-determined |
| NC1_vf_k141_1681 | Not-determined |
| NC1_vf_k141_755 | Not-determined |
| NC1_vf_k141_2554 | Not-determined |
| NC1_vf_k141_2324 | Not-determined |
| NC1_vf_k141_1874 | Not-determined |
| NC1_vf_k141_523 | Not-determined |
| NC1_vf_k141_2995 | Not-determined |
| NC1_vf_k141_987 | Not-determined |
| NC1_vf_k141_1425 | Not-determined |
| NC1_vf_k141_1193 | Not-determined |
| NC1_vf_k141_2756 | Low-quality |
| NC1_vf_k141_2108 | Not-determined |
| NC1_vf_k141_3227 | Not-determined |
| NC1_vf_k141_2996 | Not-determined |
| NC1_vf_k141_756 | Not-determined |
| NC1_vf_k141_308 | Not-determined |
| NC1_vf_k141_1682 | Not-determined |
| NC1_vf_k141_2555 | Not-determined |
| NC1_vf_k141_1875 | Not-determined |
| NC1_vf_k141_1194 | Not-determined |
| NC1_vf_k141_988 | Not-determined |
| NC1_vf_k141_1426 | Not-determined |
| NC1_vf_k141_524 | Not-determined |
| NC1_vf_k141_3452 | Not-determined |
| NC1_vf_k141_2757 | Not-determined |
| NC1_vf_k141_2109 | Not-determined |
| NC1_vf_k141_309 | Not-determined |
| NC1_vf_k141_757 | Not-determined |
| NC1_vf_k141_2556 | Not-determined |
| NC1_vf_k141_525 | Not-determined |
| NC1_vf_k141_2997 | Not-determined |
| NC1_vf_k141_1195 | Not-determined |
| NC1_vf_k141_1683 | Not-determined |
| NC1_vf_k141_3228 | Not-determined |
| NC1_vf_k141_758 | Not-determined |
| NC1_vf_k141_2758 | Not-determined |
| NC1_vf_k141_2110 | Not-determined |
| NC1_vf_k141_2325 | Not-determined |
| NC1_vf_k141_1427 | Not-determined |
| NC1_vf_k141_310 | Not-determined |
| NC1_vf_k141_2998 | Not-determined |
| NC1_vf_k141_3453 | Not-determined |
| NC1_vf_k141_2557 | Not-determined |
| NC1_vf_k141_1684 | Not-determined |
| NC1_vf_k141_526 | Not-determined |
| NC1_vf_k141_989 | Not-determined |
| NC1_vf_k141_1876 | Not-determined |
| NC1_vf_k141_3229 | Not-determined |
| NC1_vf_k141_759 | Not-determined |
| NC1_vf_k141_3454 | Not-determined |
| NC1_vf_k141_2759 | Not-determined |
| NC1_vf_k141_2111 | Not-determined |
| NC1_vf_k141_2558 | Not-determined |
| NC1_vf_k141_1685 | Not-determined |
| NC1_vf_k141_527 | Not-determined |
| NC1_vf_k141_99 | Low-quality |
| NC1_vf_k141_2999 | Not-determined |
| NC1_vf_k141_990 | Not-determined |
| NC1_vf_k141_1428 | Not-determined |
| NC1_vf_k141_760 | Low-quality |
| NC1_vf_k141_3230 | Not-determined |
| NC1_vf_k141_1877 | Not-determined |
| NC1_vf_k141_2760 | Not-determined |
| NC1_vf_k141_1686 | Not-determined |
| NC1_vf_k141_2326 | Not-determined |
| NC1_vf_k141_100 | Not-determined |
| NC1_vf_k141_991 | Not-determined |
| NC1_vf_k141_2112 | Not-determined |
| NC1_vf_k141_528 | Not-determined |
| NC1_vf_k141_3455 | Not-determined |
| NC1_vf_k141_2559 | Not-determined |
| NC1_vf_k141_3000 | Not-determined |
| NC1_vf_k141_311 | Not-determined |
| NC1_vf_k141_761 | Not-determined |
| NC1_vf_k141_1878 | Not-determined |
| NC1_vf_k141_2327 | Not-determined |
| NC1_vf_k141_2761 | Not-determined |
| NC1_vf_k141_992 | Not-determined |
| NC1_vf_k141_3231 | Not-determined |
| NC1_vf_k141_1429 | Not-determined |
| NC1_vf_k141_529 | Not-determined |
| NC1_vf_k141_2113 | Not-determined |
| NC1_vf_k141_762 | Not-determined |
| NC1_vf_k141_1687 | Not-determined |
| NC1_vf_k141_101 | Not-determined |
| NC1_vf_k141_1879 | Not-determined |
| NC1_vf_k141_2560 | Not-determined |
| NC1_vf_k141_2762 | Not-determined |
| NC1_vf_k141_312 | Not-determined |
| NC1_vf_k141_1688 | Not-determined |
| NC1_vf_k141_3232 | Not-determined |
| NC1_vf_k141_1430 | Not-determined |
| NC1_vf_k141_2114 | Not-determined |
| NC1_vf_k141_1880 | Not-determined |
| NC1_vf_k141_1196 | Low-quality |
| NC1_vf_k141_993 | Not-determined |
| NC1_vf_k141_2561 | Not-determined |
| NC1_vf_k141_1689 | Not-determined |
| NC1_vf_k141_3001 | Not-determined |
| NC1_vf_k141_102 | Not-determined |
| NC1_vf_k141_2763 | Not-determined |
| NC1_vf_k141_1431 | Not-determined |
| NC1_vf_k141_1881 | Not-determined |
| NC1_vf_k141_313 | Not-determined |
| NC1_vf_k141_530 | Low-quality |
| NC1_vf_k141_763 | Not-determined |
| NC1_vf_k141_3233 | Not-determined |
| NC1_vf_k141_2328 | Not-determined |
| NC1_vf_k141_3456 | Not-determined |
| NC1_vf_k141_1690 | Not-determined |
| NC1_vf_k141_1197 | Not-determined |
| NC1_vf_k141_994 | Not-determined |
| NC1_vf_k141_3002 | Not-determined |
| NC1_vf_k141_1432 | Not-determined |
| NC1_vf_k141_531 | Not-determined |
| NC1_vf_k141_764 | Not-determined |
| NC1_vf_k141_2115 | Low-quality |
| NC1_vf_k141_1691 | Not-determined |
| NC1_vf_k141_1882 | Not-determined |
| NC1_vf_k141_2764 | Not-determined |
| NC1_vf_k141_3003 | Not-determined |
| NC1_vf_k141_3234 | Not-determined |
| NC1_vf_k141_2329 | Not-determined |
| NC1_vf_k141_3457 | Not-determined |
| NC1_vf_k141_1692 | Not-determined |
| NC1_vf_k141_1433 | Not-determined |
| NC1_vf_k141_2765 | Not-determined |
| NC1_vf_k141_1198 | Not-determined |
| NC1_vf_k141_2562 | Not-determined |
| NC1_vf_k141_1883 | Not-determined |
| NC1_vf_k141_3458 | Not-determined |
| NC1_vf_k141_532 | Not-determined |
| NC1_vf_k141_765 | Not-determined |
| NC1_vf_k141_1693 | Not-determined |
| NC1_vf_k141_3235 | Low-quality |
| NC1_vf_k141_2330 | Not-determined |
| NC1_vf_k141_314 | Low-quality |
| NC1_vf_k141_995 | Low-quality |
| NC1_vf_k141_1694 | Not-determined |
| NC1_vf_k141_1199 | Not-determined |
| NC1_vf_k141_1434 | Not-determined |
| NC1_vf_k141_3004 | Low-quality |
| NC1_vf_k141_2766 | Not-determined |
| NC1_vf_k141_103 | Not-determined |
| NC1_vf_k141_2116 | Not-determined |
| NC1_vf_k141_3459 | Not-determined |
| NC1_vf_k141_3236 | Not-determined |
| NC1_vf_k141_1884 | Not-determined |
| NC1_vf_k141_996 | Not-determined |
| NC1_vf_k141_2331 | Not-determined |
| NC1_vf_k141_2563 | Not-determined |
| NC1_vf_k141_766 | Not-determined |
| NC1_vf_k141_1695 | Not-determined |
| NC1_vf_k141_1435 | Not-determined |
| NC1_vf_k141_533 | Not-determined |
| NC1_vf_k141_1200 | Not-determined |
| NC1_vf_k141_3005 | Not-determined |
| NC1_vf_k141_104 | Not-determined |
| NC1_vf_k141_3460 | Not-determined |
| NC1_vf_k141_2117 | Not-determined |
| NC1_vf_k141_315 | Not-determined |
| NC1_vf_k141_997 | Not-determined |
| NC1_vf_k141_1696 | Not-determined |
| NC1_vf_k141_534 | Not-determined |
| NC1_vf_k141_1885 | Low-quality |
| NC1_vf_k141_1436 | Not-determined |
| NC1_vf_k141_2564 | Not-determined |
| NC1_vf_k141_2332 | Not-determined |
| NC1_vf_k141_2767 | Not-determined |
| NC1_vf_k141_3006 | Not-determined |
| NC1_vf_k141_3237 | Not-determined |
| NC1_vf_k141_535 | Not-determined |
| NC1_vf_k141_998 | Not-determined |
| NC1_vf_k141_1201 | Not-determined |
| NC1_vf_k141_3461 | Not-determined |
| NC1_vf_k141_316 | Not-determined |
| NC1_vf_k141_2118 | Not-determined |
| NC1_vf_k141_2333 | Not-determined |
| NC1_vf_k141_767 | Not-determined |
| NC1_vf_k141_1886 | Not-determined |
| NC1_vf_k141_2768 | Not-determined |
| NC1_vf_k141_1697 | Not-determined |
| NC1_vf_k141_536 | Not-determined |
| NC1_vf_k141_1437 | Not-determined |
| NC1_vf_k141_1202 | Not-determined |
| NC1_vf_k141_999 | Not-determined |
| NC1_vf_k141_3238 | Not-determined |
| NC1_vf_k141_3462 | Not-determined |
| NC1_vf_k141_2565 | Not-determined |
| NC1_vf_k141_768 | Not-determined |
| NC1_vf_k141_2119 | Not-determined |
| NC1_vf_k141_317 | Not-determined |
| NC1_vf_k141_1887 | Not-determined |
| NC1_vf_k141_2334 | Not-determined |
| NC1_vf_k141_3007 | Not-determined |
| NC1_vf_k141_1438 | Not-determined |
| NC1_vf_k141_1698 | Not-determined |
| NC1_vf_k141_105 | Not-determined |
| NC1_vf_k141_3463 | Low-quality |
| NC1_vf_k141_2769 | Not-determined |
| NC1_vf_k141_537 | Not-determined |
| NC1_vf_k141_3239 | Not-determined |
| NC1_vf_k141_1000 | Not-determined |
| NC1_vf_k141_769 | Not-determined |
| NC1_vf_k141_318 | Not-determined |
| NC1_vf_k141_2335 | Not-determined |
| NC1_vf_k141_1439 | Not-determined |
| NC1_vf_k141_1699 | Not-determined |
| NC1_vf_k141_2770 | Not-determined |
| NC1_vf_k141_3008 | Not-determined |
| NC1_vf_k141_1888 | Not-determined |
| NC1_vf_k141_3240 | Not-determined |
| NC1_vf_k141_2566 | Not-determined |
| NC1_vf_k141_106 | Not-determined |
| NC1_vf_k141_2120 | Not-determined |
| NC1_vf_k141_1700 | Not-determined |
| NC1_vf_k141_319 | Not-determined |
| NC1_vf_k141_770 | Not-determined |
| NC1_vf_k141_1440 | Not-determined |
| NC1_vf_k141_3009 | Not-determined |
| NC1_vf_k141_2336 | Not-determined |
| NC1_vf_k141_1889 | Not-determined |
| NC1_vf_k141_1701 | Not-determined |
| NC1_vf_k141_2121 | Not-determined |
| NC1_vf_k141_3241 | Not-determined |
| NC1_vf_k141_2771 | Not-determined |
| NC1_vf_k141_320 | Low-quality |
| NC1_vf_k141_2567 | Not-determined |
| NC1_vf_k141_1203 | Not-determined |
| NC1_vf_k141_107 | Not-determined |
| NC1_vf_k141_2337 | Not-determined |
| NC1_vf_k141_771 | Not-determined |
| NC1_vf_k141_1441 | Not-determined |
| NC1_vf_k141_1702 | Not-determined |
| NC1_vf_k141_3010 | Not-determined |
| NC1_vf_k141_3464 | Not-determined |
| NC1_vf_k141_2772 | Not-determined |
| NC1_vf_k141_108 | Not-determined |
| NC1_vf_k141_538 | Not-determined |
| NC1_vf_k141_2568 | Not-determined |
| NC1_vf_k141_1890 | Not-determined |
| NC1_vf_k141_321 | Not-determined |
| NC1_vf_k141_1204 | Not-determined |
| NC1_vf_k141_2122 | Not-determined |
| NC1_vf_k141_1001 | Low-quality |
| NC1_vf_k141_2338 | Not-determined |
| NC1_vf_k141_109 | Not-determined |
| NC1_vf_k141_3465 | Not-determined |
| NC1_vf_k141_772 | Not-determined |
| NC1_vf_k141_3242 | Not-determined |
| NC1_vf_k141_2773 | Not-determined |
| NC1_vf_k141_1442 | Not-determined |
| NC1_vf_k141_3011 | Not-determined |
| NC1_vf_k141_1703 | Not-determined |
| NC1_vf_k141_539 | Not-determined |
| NC1_vf_k141_2123 | Not-determined |
| NC1_vf_k141_1205 | Not-determined |
| NC1_vf_k141_322 | Not-determined |
| NC1_vf_k141_3466 | Not-determined |
| NC1_vf_k141_2569 | Not-determined |
| NC1_vf_k141_1704 | Not-determined |
| NC1_vf_k141_110 | Not-determined |
| NC1_vf_k141_3012 | Not-determined |
| NC1_vf_k141_2339 | Not-determined |
| NC1_vf_k141_1002 | Not-determined |
| NC1_vf_k141_2774 | Not-determined |
| NC1_vf_k141_773 | Not-determined |
| NC1_vf_k141_2124 | Not-determined |
| NC1_vf_k141_3243 | Not-determined |
| NC1_vf_k141_1705 | Not-determined |
| NC1_vf_k141_2570 | Not-determined |
| NC1_vf_k141_540 | Not-determined |
| NC1_vf_k141_111 | Not-determined |
| NC1_vf_k141_1443 | Not-determined |
| NC1_vf_k141_323 | Not-determined |
| NC1_vf_k141_3467 | Low-quality |
| NC1_vf_k141_1206 | Not-determined |
| NC1_vf_k141_3013 | Not-determined |
| NC1_vf_k141_1003 | Not-determined |
| NC1_vf_k141_541 | Not-determined |
| NC1_vf_k141_1706 | Not-determined |
| NC1_vf_k141_3244 | Not-determined |
| NC1_vf_k141_2340 | Not-determined |
| NC1_vf_k141_774 | Low-quality |
| NC1_vf_k141_2125 | Not-determined |
| NC1_vf_k141_1444 | Not-determined |
| NC1_vf_k141_3468 | Not-determined |
| NC1_vf_k141_2571 | Not-determined |
| NC1_vf_k141_3014 | Not-determined |
| NC1_vf_k141_1891 | Not-determined |
| NC1_vf_k141_1207 | Not-determined |
| NC1_vf_k141_324 | Not-determined |
| NC1_vf_k141_2775 | Not-determined |
| NC1_vf_k141_1707 | Not-determined |
| NC1_vf_k141_1445 | Not-determined |
| NC1_vf_k141_3469 | Not-determined |
| NC1_vf_k141_2341 | Not-determined |
| NC1_vf_k141_3015 | Not-determined |
| NC1_vf_k141_1208 | Not-determined |
| NC1_vf_k141_325 | Not-determined |
| NC1_vf_k141_1708 | Not-determined |
| NC1_vf_k141_1004 | Low-quality |
| NC1_vf_k141_2572 | Not-determined |
| NC1_vf_k141_3470 | Not-determined |
| NC1_vf_k141_1446 | Not-determined |
| NC1_vf_k141_775 | Not-determined |
| NC1_vf_k141_542 | Not-determined |
| NC1_vf_k141_3016 | Not-determined |
| NC1_vf_k141_1892 | Not-determined |
| NC1_vf_k141_3245 | Low-quality |
| NC1_vf_k141_1209 | Not-determined |
| NC1_vf_k141_1709 | Low-quality |
| NC1_vf_k141_2342 | Not-determined |
| NC1_vf_k141_326 | Not-determined |
| NC1_vf_k141_1005 | Not-determined |
| NC1_vf_k141_1447 | Not-determined |
| NC1_vf_k141_2776 | Not-determined |
| NC1_vf_k141_3017 | Not-determined |
| NC1_vf_k141_1210 | Not-determined |
| NC1_vf_k141_1893 | Not-determined |
| NC1_vf_k141_543 | Not-determined |
| NC1_vf_k141_1710 | Not-determined |
| NC1_vf_k141_2126 | Not-determined |
| NC1_vf_k141_2343 | Not-determined |
| NC1_vf_k141_1006 | Not-determined |
| NC1_vf_k141_2777 | Not-determined |
| NC1_vf_k141_3471 | Not-determined |
| NC1_vf_k141_327 | Not-determined |
| NC1_vf_k141_544 | Not-determined |
| NC1_vf_k141_776 | Not-determined |
| NC1_vf_k141_1211 | Not-determined |
| NC1_vf_k141_3018 | Not-determined |
| NC1_vf_k141_1711 | Not-determined |
| NC1_vf_k141_1007 | Not-determined |
| NC1_vf_k141_2778 | Not-determined |
| NC1_vf_k141_2127 | Not-determined |
| NC1_vf_k141_2344 | Not-determined |
| NC1_vf_k141_545 | Not-determined |
| NC1_vf_k141_777 | Not-determined |
| NC1_vf_k141_1212 | Not-determined |
| NC1_vf_k141_3472 | Not-determined |
| NC1_vf_k141_112 | Not-determined |
| NC1_vf_k141_1712 | Not-determined |
| NC1_vf_k141_328 | Not-determined |
| NC1_vf_k141_2779 | Not-determined |
| NC1_vf_k141_2128 | Not-determined |
| NC1_vf_k141_3019 | Not-determined |
| NC1_vf_k141_2345 | Not-determined |
| NC1_vf_k141_1713 | Not-determined |
| NC1_vf_k141_1448 | Not-determined |
| NC1_vf_k141_778 | Not-determined |
| NC1_vf_k141_1894 | Not-determined |
| NC1_vf_k141_1008 | Not-determined |
| NC1_vf_k141_113 | Not-determined |
| NC1_vf_k141_1213 | Not-determined |
| NC1_vf_k141_546 | Not-determined |
| NC1_vf_k141_2573 | Low-quality |
| NC1_vf_k141_1714 | Not-determined |
| NC1_vf_k141_2129 | Not-determined |
| NC1_vf_k141_329 | Not-determined |
| NC1_vf_k141_114 | Not-determined |
| NC1_vf_k141_1895 | Not-determined |
| NC1_vf_k141_3246 | Not-determined |
| NC1_vf_k141_1214 | Not-determined |
| NC1_vf_k141_547 | Not-determined |
| NC1_vf_k141_2780 | Not-determined |
| NC1_vf_k141_2346 | Not-determined |
| NC1_vf_k141_115 | Not-determined |
| NC1_vf_k141_3020 | Not-determined |
| NC1_vf_k141_330 | Not-determined |
| NC1_vf_k141_2574 | Low-quality |
| NC1_vf_k141_1896 | Not-determined |
| NC1_vf_k141_1449 | Not-determined |
| NC1_vf_k141_3247 | Not-determined |
| NC1_vf_k141_548 | Not-determined |
| NC1_vf_k141_2130 | Not-determined |
| NC1_vf_k141_1715 | Not-determined |
| NC1_vf_k141_1215 | Not-determined |
| NC1_vf_k141_779 | Not-determined |
| NC1_vf_k141_3473 | Not-determined |
| NC1_vf_k141_116 | Not-determined |
| NC1_vf_k141_1716 | Not-determined |
| NC1_vf_k141_2347 | Not-determined |
| NC1_vf_k141_3021 | Not-determined |
| NC1_vf_k141_2575 | Not-determined |
| NC1_vf_k141_2781 | Not-determined |
| NC1_vf_k141_1897 | Not-determined |
| NC1_vf_k141_2131 | Not-determined |
| NC1_vf_k141_1009 | Not-determined |
| NC1_vf_k141_3248 | Not-determined |
| NC1_vf_k141_549 | Not-determined |
| NC1_vf_k141_3474 | Not-determined |
| NC1_vf_k141_1717 | Not-determined |
| NC1_vf_k141_2576 | Not-determined |
| NC1_vf_k141_3022 | Not-determined |
| NC1_vf_k141_2132 | Not-determined |
| NC1_vf_k141_780 | Not-determined |
| NC1_vf_k141_2782 | Not-determined |
| NC1_vf_k141_1216 | Not-determined |
| NC1_vf_k141_331 | Not-determined |
| NC1_vf_k141_3249 | Not-determined |
| NC1_vf_k141_117 | Not-determined |
| NC1_vf_k141_2348 | Not-determined |
| NC1_vf_k141_550 | Not-determined |
| NC1_vf_k141_2577 | Not-determined |
| NC1_vf_k141_1898 | Not-determined |
| NC1_vf_k141_3023 | Not-determined |
| NC1_vf_k141_1450 | Not-determined |
| NC1_vf_k141_1217 | Not-determined |
| NC1_vf_k141_1718 | Not-determined |
| NC1_vf_k141_3475 | Not-determined |
| NC1_vf_k141_332 | Low-quality |
| NC1_vf_k141_2783 | Not-determined |
| NC1_vf_k141_2349 | Not-determined |
| NC1_vf_k141_3024 | Not-determined |
| NC1_vf_k141_2133 | Not-determined |
| NC1_vf_k141_781 | Not-determined |
| NC1_vf_k141_551 | Not-determined |
| NC1_vf_k141_3250 | Not-determined |
| NC1_vf_k141_1218 | Not-determined |
| NC1_vf_k141_1719 | Not-determined |
| NC1_vf_k141_1010 | Not-determined |
| NC1_vf_k141_1451 | Not-determined |
| NC1_vf_k141_118 | Not-determined |
| NC1_vf_k141_2784 | Not-determined |
| NC1_vf_k141_333 | Not-determined |
| NC1_vf_k141_1899 | Not-determined |
| NC1_vf_k141_3476 | Not-determined |
| NC1_vf_k141_2350 | Not-determined |
| NC1_vf_k141_1219 | Not-determined |
| NC1_vf_k141_2578 | Not-determined |
| NC1_vf_k141_3025 | Not-determined |
| NC1_vf_k141_2785 | Not-determined |
| NC1_vf_k141_119 | Not-determined |
| NC1_vf_k141_1011 | Not-determined |
| NC1_vf_k141_3251 | Not-determined |
| NC1_vf_k141_1900 | Not-determined |
| NC1_vf_k141_1452 | Not-determined |
| NC1_vf_k141_552 | Not-determined |
| NC1_vf_k141_782 | Not-determined |
| NC1_vf_k141_1720 | Not-determined |
| NC1_vf_k141_2351 | Not-determined |
| NC1_vf_k141_1220 | Not-determined |
| NC1_vf_k141_2134 | Not-determined |
| NC1_vf_k141_2786 | Not-determined |
| NC1_vf_k141_1721 | Not-determined |
| NC1_vf_k141_2352 | Not-determined |
| NC1_vf_k141_120 | Low-quality |
| NC1_vf_k141_3477 | Not-determined |
| NC1_vf_k141_783 | Low-quality |
| NC1_vf_k141_1453 | Not-determined |
| NC1_vf_k141_553 | Not-determined |
| NC1_vf_k141_1901 | Not-determined |
| NC1_vf_k141_1722 | Not-determined |
| NC1_vf_k141_121 | Not-determined |
| NC1_vf_k141_2135 | Not-determined |
| NC1_vf_k141_3478 | Not-determined |
| NC1_vf_k141_1221 | Not-determined |
| NC1_vf_k141_1012 | Not-determined |
| NC1_vf_k141_784 | Not-determined |
| NC1_vf_k141_1723 | Not-determined |
| NC1_vf_k141_1454 | Not-determined |
| NC1_vf_k141_554 | Not-determined |
| NC1_vf_k141_122 | Not-determined |
| NC1_vf_k141_1902 | Low-quality |
| NC1_vf_k141_3479 | Not-determined |
| NC1_vf_k141_2136 | Not-determined |
| NC1_vf_k141_3026 | Low-quality |
| NC1_vf_k141_2353 | Not-determined |
| NC1_vf_k141_2579 | Not-determined |
| NC1_vf_k141_2787 | Not-determined |
| NC1_vf_k141_1724 | Not-determined |
| NC1_vf_k141_3252 | Not-determined |
| NC1_vf_k141_785 | Not-determined |
| NC1_vf_k141_123 | Not-determined |
| NC1_vf_k141_1222 | Not-determined |
| NC1_vf_k141_1013 | Not-determined |
| NC1_vf_k141_2137 | Not-determined |
| NC1_vf_k141_1903 | Not-determined |
| NC1_vf_k141_2354 | Not-determined |
| NC1_vf_k141_555 | Not-determined |
| NC1_vf_k141_1725 | Not-determined |
| NC1_vf_k141_124 | Not-determined |
| NC1_vf_k141_3253 | Not-determined |
| NC1_vf_k141_3480 | Not-determined |
| NC1_vf_k141_2580 | Not-determined |
| NC1_vf_k141_334 | Low-quality |
| NC1_vf_k141_2138 | Not-determined |
| NC1_vf_k141_2788 | Not-determined |
| NC1_vf_k141_786 | Not-determined |
| NC1_vf_k141_1223 | Low-quality |
| NC1_vf_k141_1455 | Not-determined |
| NC1_vf_k141_125 | Not-determined |
| NC1_vf_k141_1014 | Not-determined |
| NC1_vf_k141_3254 | Not-determined |
| NC1_vf_k141_787 | Not-determined |
| NC1_vf_k141_2139 | Not-determined |
| NC1_vf_k141_2355 | Not-determined |
| NC1_vf_k141_2581 | Not-determined |
| NC1_vf_k141_556 | Not-determined |
| NC1_vf_k141_2789 | Not-determined |
| NC1_vf_k141_1904 | Not-determined |
| NC1_vf_k141_1015 | Not-determined |
| NC1_vf_k141_3481 | Not-determined |
| NC1_vf_k141_1224 | Not-determined |
| NC1_vf_k141_2140 | Not-determined |
| NC1_vf_k141_1456 | Not-determined |
| NC1_vf_k141_1726 | Not-determined |
| NC1_vf_k141_788 | Not-determined |
| NC1_vf_k141_3255 | Not-determined |
| NC1_vf_k141_2356 | Not-determined |
| NC1_vf_k141_557 | Not-determined |
| NC1_vf_k141_1016 | Not-determined |
| NC1_vf_k141_3482 | Not-determined |
| NC1_vf_k141_335 | Not-determined |
| NC1_vf_k141_1905 | Not-determined |
| NC1_vf_k141_1225 | Not-determined |

| | |
|---|---|
| NC1_vf_k141_1727 | Not-determined |
| NC1_vf_k141_2790 | Not-determined |
| NC1_vf_k141_3256 | Not-determined |
| NC1_vf_k141_126 | Not-determined |
| NC1_vf_k141_2357 | Not-determined |
| NC1_vf_k141_2141 | Not-determined |
| NC1_vf_k141_3483 | Not-determined |
| NC1_vf_k141_1457 | Not-determined |
| NC1_vf_k141_1017 | Not-determined |
| NC1_vf_k141_558 | Not-determined |
| NC1_vf_k141_2582 | Not-determined |
| NC1_vf_k141_336 | Not-determined |
| NC1_vf_k141_789 | Not-determined |
| NC1_vf_k141_1226 | Not-determined |
| NC1_vf_k141_1728 | Not-determined |
| NC1_vf_k141_2358 | Not-determined |
| NC1_vf_k141_3484 | Not-determined |
| NC1_vf_k141_127 | Not-determined |
| NC1_vf_k141_1458 | Not-determined |
| NC1_vf_k141_3027 | Low-quality |
| NC1_vf_k141_1906 | Not-determined |
| NC1_vf_k141_337 | Not-determined |
| NC1_vf_k141_2142 | Not-determined |
| NC1_vf_k141_1729 | Not-determined |
| NC1_vf_k141_3257 | Not-determined |
| NC1_vf_k141_128 | Not-determined |
| NC1_vf_k141_790 | Not-determined |
| NC1_vf_k141_2359 | Not-determined |
| NC1_vf_k141_3485 | Not-determined |
| NC1_vf_k141_2583 | Not-determined |
| NC1_vf_k141_1227 | Not-determined |
| NC1_vf_k141_2143 | Not-determined |
| NC1_vf_k141_3028 | Not-determined |
| NC1_vf_k141_1459 | Not-determined |
| NC1_vf_k141_3258 | Not-determined |
| NC1_vf_k141_1018 | Not-determined |
| NC1_vf_k141_1907 | Not-determined |
| NC1_vf_k141_129 | Not-determined |
| NC1_vf_k141_1730 | Not-determined |
| NC1_vf_k141_559 | Not-determined |
| NC1_vf_k141_791 | Not-determined |
| NC1_vf_k141_2584 | Not-determined |
| NC1_vf_k141_1228 | Not-determined |
| NC1_vf_k141_338 | Not-determined |
| NC1_vf_k141_2144 | Not-determined |
| NC1_vf_k141_1460 | Not-determined |
| NC1_vf_k141_1019 | Not-determined |
| NC1_vf_k141_3029 | Low-quality |
| NC1_vf_k141_3259 | Not-determined |
| NC1_vf_k141_2791 | Not-determined |
| NC1_vf_k141_1908 | Not-determined |
| NC1_vf_k141_2360 | Not-determined |
| NC1_vf_k141_339 | Not-determined |
| NC1_vf_k141_130 | Not-determined |
| NC1_vf_k141_1020 | Not-determined |
| NC1_vf_k141_560 | Low-quality |
| NC1_vf_k141_1731 | Not-determined |
| NC1_vf_k141_3260 | Not-determined |
| NC1_vf_k141_1461 | Not-determined |
| NC1_vf_k141_1909 | Not-determined |
| NC1_vf_k141_792 | Not-determined |
| NC1_vf_k141_3030 | Not-determined |
| NC1_vf_k141_3486 | Not-determined |
| NC1_vf_k141_2792 | Not-determined |
| NC1_vf_k141_561 | Not-determined |
| NC1_vf_k141_1732 | Not-determined |
| NC1_vf_k141_131 | Not-determined |
| NC1_vf_k141_340 | Not-determined |
| NC1_vf_k141_2145 | Not-determined |
| NC1_vf_k141_2361 | Not-determined |
| NC1_vf_k141_132 | Not-determined |
| NC1_vf_k141_793 | Not-determined |
| NC1_vf_k141_562 | Not-determined |
| NC1_vf_k141_1733 | Not-determined |
| NC1_vf_k141_2793 | Not-determined |
| NC1_vf_k141_2585 | Not-determined |
| NC1_vf_k141_341 | Not-determined |
| NC1_vf_k141_133 | Not-determined |
| NC1_vf_k141_1910 | Not-determined |
| NC1_vf_k141_1462 | Not-determined |
| NC1_vf_k141_3031 | Not-determined |
| NC1_vf_k141_1021 | Not-determined |
| NC1_vf_k141_2146 | Not-determined |
| NC1_vf_k141_134 | Not-determined |
| NC1_vf_k141_2362 | Not-determined |
| NC1_vf_k141_794 | Not-determined |
| NC1_vf_k141_2586 | Not-determined |
| NC1_vf_k141_1734 | Not-determined |
| NC1_vf_k141_342 | Not-determined |
| NC1_vf_k141_563 | Not-determined |
| NC1_vf_k141_1911 | Not-determined |
| NC1_vf_k141_135 | Not-determined |
| NC1_vf_k141_3487 | Not-determined |
| NC1_vf_k141_2794 | Not-determined |
| NC1_vf_k141_2147 | Not-determined |
| NC1_vf_k141_3032 | Not-determined |
| NC1_vf_k141_1022 | Not-determined |
| NC1_vf_k141_343 | Not-determined |
| NC1_vf_k141_795 | Not-determined |
| NC1_vf_k141_1735 | Low-quality |
| NC1_vf_k141_136 | Not-determined |
| NC1_vf_k141_564 | Not-determined |
| NC1_vf_k141_2587 | Not-determined |
| NC1_vf_k141_3261 | Low-quality |
| NC1_vf_k141_1912 | Not-determined |
| NC1_vf_k141_3488 | Not-determined |
| NC1_vf_k141_1463 | Not-determined |
| NC1_vf_k141_1736 | Not-determined |
| NC1_vf_k141_2148 | Not-determined |
| NC1_vf_k141_137 | Not-determined |
| NC1_vf_k141_2795 | Not-determined |
| NC1_vf_k141_3033 | Not-determined |
| NC1_vf_k141_796 | Not-determined |
| NC1_vf_k141_1023 | Not-determined |
| NC1_vf_k141_2363 | Not-determined |
| NC1_vf_k141_565 | Not-determined |
| NC1_vf_k141_1737 | Not-determined |
| NC1_vf_k141_344 | Not-determined |
| NC1_vf_k141_3262 | Not-determined |
| NC1_vf_k141_2588 | Not-determined |
| NC1_vf_k141_3034 | Not-determined |
| NC1_vf_k141_2149 | Low-quality |
| NC1_vf_k141_1913 | Not-determined |
| NC1_vf_k141_1738 | Not-determined |
| NC1_vf_k141_138 | Not-determined |
| NC1_vf_k141_797 | Not-determined |
| NC1_vf_k141_1024 | Low-quality |
| NC1_vf_k141_2796 | Not-determined |
| NC1_vf_k141_1464 | Not-determined |
| NC1_vf_k141_2364 | Not-determined |
| NC1_vf_k141_1914 | Not-determined |
| NC1_vf_k141_1739 | Not-determined |
| NC1_vf_k141_1025 | Not-determined |
| NC1_vf_k141_2150 | Not-determined |
| NC1_vf_k141_3035 | Not-determined |
| NC1_vf_k141_2589 | Not-determined |
| NC1_vf_k141_3263 | Not-determined |
| NC1_vf_k141_798 | Not-determined |
| NC1_vf_k141_345 | Not-determined |
| NC1_vf_k141_2797 | Not-determined |
| NC1_vf_k141_2365 | Not-determined |
| NC1_vf_k141_139 | Not-determined |
| NC1_vf_k141_3489 | Not-determined |
| NC1_vf_k141_1915 | Not-determined |
| NC1_vf_k141_1026 | Not-determined |
| NC1_vf_k141_1465 | Not-determined |
| NC1_vf_k141_2151 | Not-determined |
| NC1_vf_k141_3264 | Not-determined |
| NC1_vf_k141_2366 | Not-determined |
| NC1_vf_k141_1740 | Not-determined |
| NC1_vf_k141_2590 | Not-determined |
| NC1_vf_k141_3036 | Low-quality |
| NC1_vf_k141_2798 | Not-determined |
| NC1_vf_k141_346 | Not-determined |
| NC1_vf_k141_3490 | Not-determined |
| NC1_vf_k141_140 | Not-determined |
| NC1_vf_k141_1916 | Not-determined |
| NC1_vf_k141_1027 | Not-determined |
| NC1_vf_k141_1741 | Not-determined |
| NC1_vf_k141_2591 | Not-determined |
| NC1_vf_k141_3037 | Not-determined |
| NC1_vf_k141_3265 | Not-determined |
| NC1_vf_k141_141 | Not-determined |
| NC1_vf_k141_1466 | Not-determined |
| NC1_vf_k141_347 | Not-determined |
| NC1_vf_k141_799 | Not-determined |
| NC1_vf_k141_1742 | Not-determined |
| NC1_vf_k141_1917 | Not-determined |
| NC1_vf_k141_1028 | Not-determined |
| NC1_vf_k141_142 | Not-determined |
| NC1_vf_k141_2799 | Not-determined |
| NC1_vf_k141_2152 | Not-determined |
| NC1_vf_k141_2592 | Not-determined |
| NC1_vf_k141_1229 | Not-determined |
| NC1_vf_k141_3266 | Not-determined |
| NC1_vf_k141_1467 | Not-determined |
| NC1_vf_k141_3491 | Not-determined |
| NC1_vf_k141_3038 | Not-determined |
| NC1_vf_k141_1918 | Not-determined |
| NC1_vf_k141_800 | Not-determined |
| NC1_vf_k141_2367 | Not-determined |
| NC1_vf_k141_143 | Not-determined |
| NC1_vf_k141_2800 | Not-determined |
| NC1_vf_k141_566 | Low-quality |
| NC1_vf_k141_3267 | Not-determined |
| NC1_vf_k141_2153 | Not-determined |
| NC1_vf_k141_1230 | Not-determined |
| NC1_vf_k141_348 | Not-determined |
| NC1_vf_k141_1029 | Not-determined |
| NC1_vf_k141_1743 | Not-determined |
| NC1_vf_k141_1468 | Not-determined |
| NC1_vf_k141_3492 | Not-determined |
| NC1_vf_k141_2593 | Not-determined |
| NC1_vf_k141_2368 | Not-determined |
| NC1_vf_k141_1919 | Not-determined |
| NC1_vf_k141_144 | Not-determined |
| NC1_vf_k141_3039 | Not-determined |
| NC1_vf_k141_2154 | Not-determined |
| NC1_vf_k141_2801 | Not-determined |
| NC1_vf_k141_801 | Not-determined |
| NC1_vf_k141_1231 | Not-determined |
| NC1_vf_k141_3268 | Not-determined |
| NC1_vf_k141_1030 | Not-determined |
| NC1_vf_k141_349 | Not-determined |
| NC1_vf_k141_567 | Not-determined |
| NC1_vf_k141_2369 | Not-determined |
| NC1_vf_k141_1920 | Not-determined |
| NC1_vf_k141_145 | Low-quality |
| NC1_vf_k141_3040 | Not-determined |
| NC1_vf_k141_2802 | Not-determined |
| NC1_vf_k141_1232 | Not-determined |
| NC1_vf_k141_3269 | Not-determined |
| NC1_vf_k141_802 | Not-determined |
| NC1_vf_k141_1031 | Not-determined |
| NC1_vf_k141_2155 | Not-determined |
| NC1_vf_k141_3493 | Not-determined |
| NC1_vf_k141_2803 | Not-determined |
| NC1_vf_k141_1469 | Low-quality |
| NC1_vf_k141_146 | Not-determined |
| NC1_vf_k141_1744 | Not-determined |
| NC1_vf_k141_350 | Not-determined |
| NC1_vf_k141_568 | Not-determined |
| NC1_vf_k141_2594 | Not-determined |
| NC1_vf_k141_3041 | Not-determined |
| NC1_vf_k141_1032 | Not-determined |
| NC1_vf_k141_1233 | Not-determined |
| NC1_vf_k141_1921 | Not-determined |
| NC1_vf_k141_3270 | Not-determined |
| NC1_vf_k141_2156 | Low-quality |
| NC1_vf_k141_1745 | Not-determined |
| NC1_vf_k141_3494 | Not-determined |
| NC1_vf_k141_147 | Not-determined |
| NC1_vf_k141_1470 | Not-determined |
| NC1_vf_k141_569 | Not-determined |
| NC1_vf_k141_803 | Not-determined |
| NC1_vf_k141_2595 | Not-determined |
| NC1_vf_k141_2804 | Not-determined |
| NC1_vf_k141_1033 | Not-determined |
| NC1_vf_k141_3271 | Not-determined |
| NC1_vf_k141_1234 | Not-determined |
| NC1_vf_k141_351 | Not-determined |
| NC1_vf_k141_148 | Not-determined |
| NC1_vf_k141_1746 | Not-determined |
| NC1_vf_k141_1471 | Not-determined |
| NC1_vf_k141_570 | Not-determined |
| NC1_vf_k141_3042 | Not-determined |
| NC1_vf_k141_3495 | Not-determined |
| NC1_vf_k141_2157 | Not-determined |
| NC1_vf_k141_1034 | Not-determined |
| NC1_vf_k141_149 | Not-determined |
| NC1_vf_k141_1747 | Not-determined |
| NC1_vf_k141_2805 | Not-determined |
| NC1_vf_k141_1235 | Not-determined |
| NC1_vf_k141_804 | Not-determined |
| NC1_vf_k141_2596 | Not-determined |
| NC1_vf_k141_150 | Not-determined |
| NC1_vf_k141_352 | Not-determined |
| NC1_vf_k141_2370 | Not-determined |
| NC1_vf_k141_3496 | Not-determined |
| NC1_vf_k141_2158 | Not-determined |
| NC1_vf_k141_3272 | Not-determined |
| NC1_vf_k141_1472 | Not-determined |
| NC1_vf_k141_3043 | Low-quality |
| NC1_vf_k141_571 | Not-determined |
| NC1_vf_k141_1035 | Not-determined |
| NC1_vf_k141_1236 | Not-determined |
| NC1_vf_k141_2597 | Not-determined |
| NC1_vf_k141_2806 | Not-determined |
| NC1_vf_k141_805 | Not-determined |
| NC1_vf_k141_151 | Not-determined |
| NC1_vf_k141_353 | Not-determined |
| NC1_vf_k141_2371 | Not-determined |
| NC1_vf_k141_572 | Low-quality |
| NC1_vf_k141_3273 | Not-determined |
| NC1_vf_k141_2598 | Not-determined |
| NC1_vf_k141_2159 | Not-determined |
| NC1_vf_k141_1473 | Not-determined |
| NC1_vf_k141_1237 | Not-determined |
| NC1_vf_k141_2807 | Low-quality |
| NC1_vf_k141_3044 | Not-determined |
| NC1_vf_k141_152 | Not-determined |
| NC1_vf_k141_1036 | Not-determined |
| NC1_vf_k141_354 | Not-determined |
| NC1_vf_k141_2372 | Not-determined |
| NC1_vf_k141_573 | Not-determined |
| NC1_vf_k141_2160 | Not-determined |
| NC1_vf_k141_2808 | Not-determined |
| NC1_vf_k141_153 | Not-determined |
| NC1_vf_k141_3274 | Not-determined |
| NC1_vf_k141_3497 | Not-determined |
| NC1_vf_k141_806 | Not-determined |
| NC1_vf_k141_3045 | Not-determined |
| NC1_vf_k141_1238 | Not-determined |
| NC1_vf_k141_2599 | Not-determined |
| NC1_vf_k141_1474 | Not-determined |
| NC1_vf_k141_355 | Not-determined |
| NC1_vf_k141_1037 | Not-determined |
| NC1_vf_k141_2809 | Not-determined |
| NC1_vf_k141_154 | Not-determined |
| NC1_vf_k141_807 | Not-determined |
| NC1_vf_k141_2161 | Not-determined |
| NC1_vf_k141_3046 | Not-determined |
| NC1_vf_k141_3498 | Not-determined |
| NC1_vf_k141_356 | Not-determined |
| NC1_vf_k141_1038 | Not-determined |
| NC1_vf_k141_574 | Low-quality |
| NC1_vf_k141_3275 | Not-determined |
| NC1_vf_k141_155 | Not-determined |
| NC1_vf_k141_1922 | Low-quality |
| NC1_vf_k141_2810 | Not-determined |
| NC1_vf_k141_2600 | Not-determined |
| NC1_vf_k141_1239 | Not-determined |
| NC1_vf_k141_3047 | Not-determined |
| NC1_vf_k141_2162 | Not-determined |
| NC1_vf_k141_1475 | Not-determined |
| NC1_vf_k141_357 | Not-determined |
| NC1_vf_k141_3499 | Not-determined |
| NC1_vf_k141_575 | Not-determined |
| NC1_vf_k141_3048 | Not-determined |
| NC1_vf_k141_3276 | Low-quality |
| NC1_vf_k141_1923 | Not-determined |
| NC1_vf_k141_156 | Not-determined |
| NC1_vf_k141_2601 | Low-quality |
| NC1_vf_k141_808 | Not-determined |
| NC1_vf_k141_2163 | Not-determined |
| NC1_vf_k141_1476 | Not-determined |
| NC1_vf_k141_358 | Not-determined |
| NC1_vf_k141_1039 | Not-determined |
| NC1_vf_k141_1924 | Not-determined |
| NC1_vf_k141_2164 | Not-determined |
| NC1_vf_k141_3049 | Not-determined |
| NC1_vf_k141_2602 | Not-determined |
| NC1_vf_k141_359 | Not-determined |
| NC1_vf_k141_3277 | Not-determined |
| NC1_vf_k141_2811 | Not-determined |
| NC1_vf_k141_576 | Not-determined |
| NC1_vf_k141_3500 | Not-determined |
| NC1_vf_k141_157 | Low-quality |
| NC1_vf_k141_1040 | Not-determined |
| NC1_vf_k141_1925 | Not-determined |
| NC1_vf_k141_1240 | Not-determined |
| NC1_vf_k141_2165 | Not-determined |
| NC1_vf_k141_360 | Not-determined |
| NC1_vf_k141_2603 | Not-determined |
| NC1_vf_k141_158 | Low-quality |
| NC1_vf_k141_577 | Not-determined |
| NC1_vf_k141_1477 | Not-determined |
| NC1_vf_k141_2812 | Not-determined |
| NC1_vf_k141_809 | Low-quality |
| NC1_vf_k141_1041 | Not-determined |
| NC1_vf_k141_2373 | Not-determined |
| NC1_vf_k141_1241 | Not-determined |
| NC1_vf_k141_3501 | Not-determined |
| NC1_vf_k141_3278 | Not-determined |
| NC1_vf_k141_2166 | Not-determined |
| NC1_vf_k141_361 | Not-determined |
| NC1_vf_k141_159 | Not-determined |
| NC1_vf_k141_1042 | Not-determined |
| NC1_vf_k141_1478 | Not-determined |
| NC1_vf_k141_2374 | Not-determined |
| NC1_vf_k141_3502 | Not-determined |
| NC1_vf_k141_810 | Not-determined |
| NC1_vf_k141_2604 | Not-determined |
| NC1_vf_k141_2167 | Not-determined |
| NC1_vf_k141_578 | Not-determined |
| NC1_vf_k141_3279 | Not-determined |
| NC1_vf_k141_160 | Not-determined |
| NC1_vf_k141_1242 | Not-determined |
| NC1_vf_k141_362 | Not-determined |
| NC1_vf_k141_1043 | Not-determined |
| NC1_vf_k141_2375 | Not-determined |
| NC1_vf_k141_2605 | Not-determined |
| NC1_vf_k141_3503 | Not-determined |
| NC1_vf_k141_811 | Not-determined |
| NC1_vf_k141_1479 | Not-determined |
| NC1_vf_k141_2813 | Not-determined |
| NC1_vf_k141_161 | Not-determined |
| NC1_vf_k141_3280 | Not-determined |
| NC1_vf_k141_2168 | Not-determined |
| NC1_vf_k141_1044 | Not-determined |
| NC1_vf_k141_3050 | Not-determined |
| NC1_vf_k141_162 | Not-determined |
| NC1_vf_k141_1480 | Not-determined |
| NC1_vf_k141_1243 | Not-determined |
| NC1_vf_k141_579 | Not-determined |
| NC1_vf_k141_2814 | Not-determined |
| NC1_vf_k141_1045 | Not-determined |
| NC1_vf_k141_2169 | Not-determined |
| NC1_vf_k141_3281 | Not-determined |
| NC1_vf_k141_3504 | Not-determined |
| NC1_vf_k141_812 | Not-determined |
| NC1_vf_k141_163 | Not-determined |
| NC1_vf_k141_1926 | Not-determined |
| NC1_vf_k141_580 | Not-determined |
| NC1_vf_k141_2376 | Not-determined |
| NC1_vf_k141_1481 | Not-determined |
| NC1_vf_k141_1244 | Not-determined |
| NC1_vf_k141_363 | Not-determined |
| NC1_vf_k141_3505 | Not-determined |
| NC1_vf_k141_3051 | Not-determined |
| NC1_vf_k141_164 | Not-determined |
| NC1_vf_k141_2170 | Not-determined |
| NC1_vf_k141_2815 | Not-determined |
| NC1_vf_k141_3282 | Not-determined |
| NC1_vf_k141_2377 | Not-determined |
| NC1_vf_k141_1046 | Not-determined |
| NC1_vf_k141_1482 | Not-determined |
| NC1_vf_k141_1245 | Not-determined |
| NC1_vf_k141_1927 | Not-determined |
| NC1_vf_k141_3506 | Not-determined |
| NC1_vf_k141_3052 | Not-determined |
| NC1_vf_k141_165 | Not-determined |
| NC1_vf_k141_364 | Not-determined |
| NC1_vf_k141_1047 | Not-determined |
| NC1_vf_k141_1928 | Not-determined |
| NC1_vf_k141_3283 | Not-determined |
| NC1_vf_k141_3053 | Not-determined |
| NC1_vf_k141_813 | Low-quality |
| NC1_vf_k141_2378 | Not-determined |
| NC1_vf_k141_2171 | Not-determined |
| NC1_vf_k141_2606 | Not-determined |
| NC1_vf_k141_1246 | Not-determined |
| NC1_vf_k141_365 | Not-determined |
| NC1_vf_k141_166 | Not-determined |
| NC1_vf_k141_2816 | Not-determined |
| NC1_vf_k141_1048 | Not-determined |
| NC1_vf_k141_581 | Not-determined |
| NC1_vf_k141_1929 | Not-determined |
| NC1_vf_k141_3054 | Not-determined |
| NC1_vf_k141_2172 | Not-determined |
| NC1_vf_k141_814 | Not-determined |
| NC1_vf_k141_1483 | Not-determined |
| NC1_vf_k141_3507 | Not-determined |
| NC1_vf_k141_167 | Not-determined |
| NC1_vf_k141_366 | Not-determined |
| NC1_vf_k141_1049 | Not-determined |
| NC1_vf_k141_3284 | Not-determined |
| NC1_vf_k141_1484 | Not-determined |
| NC1_vf_k141_1930 | Not-determined |
| NC1_vf_k141_3055 | Not-determined |
| NC1_vf_k141_2817 | Not-determined |
| NC1_vf_k141_2607 | Low-quality |
| NC1_vf_k141_2379 | Not-determined |
| NC1_vf_k141_168 | Not-determined |
| NC1_vf_k141_3508 | Not-determined |
| NC1_vf_k141_815 | Not-determined |
| NC1_vf_k141_2173 | Not-determined |
| NC1_vf_k141_367 | Not-determined |
| NC1_vf_k141_1050 | Not-determined |
| NC1_vf_k141_1485 | Not-determined |
| NC1_vf_k141_1931 | Not-determined |
| NC1_vf_k141_169 | Not-determined |
| NC1_vf_k141_2818 | Not-determined |
| NC1_vf_k141_582 | Not-determined |
| NC1_vf_k141_1247 | Not-determined |
| NC1_vf_k141_3285 | Not-determined |
| NC1_vf_k141_3056 | Not-determined |
| NC1_vf_k141_368 | Not-determined |
| NC1_vf_k141_3509 | Not-determined |
| NC1_vf_k141_1486 | Not-determined |
| NC1_vf_k141_2608 | Not-determined |
| NC1_vf_k141_583 | Not-determined |
| NC1_vf_k141_2380 | Not-determined |
| NC1_vf_k141_816 | Not-determined |
| NC1_vf_k141_2174 | Not-determined |
| NC1_vf_k141_170 | Not-determined |
| NC1_vf_k141_2819 | Not-determined |
| NC1_vf_k141_3057 | Not-determined |
| NC1_vf_k141_1487 | Not-determined |
| NC1_vf_k141_3286 | Not-determined |
| NC1_vf_k141_1932 | Not-determined |
| NC1_vf_k141_584 | Not-determined |
| NC1_vf_k141_2381 | Not-determined |
| NC1_vf_k141_1248 | Not-determined |
| NC1_vf_k141_3510 | Not-determined |
| NC1_vf_k141_2609 | Not-determined |
| NC1_vf_k141_817 | Not-determined |
| NC1_vf_k141_2820 | Not-determined |
| NC1_vf_k141_369 | Not-determined |
| NC1_vf_k141_1051 | Not-determined |
| NC1_vf_k141_3058 | Not-determined |
| NC1_vf_k141_2175 | Not-determined |
| NC1_vf_k141_171 | Low-quality |
| NC1_vf_k141_3287 | Not-determined |
| NC1_vf_k141_2382 | Not-determined |
| NC1_vf_k141_585 | Not-determined |
| NC1_vf_k141_3511 | Low-quality |
| NC1_vf_k141_2610 | Not-determined |
| NC1_vf_k141_1933 | Not-determined |
| NC1_vf_k141_172 | Not-determined |
| NC1_vf_k141_3059 | Not-determined |
| NC1_vf_k141_370 | Not-determined |
| NC1_vf_k141_2821 | Not-determined |
| NC1_vf_k141_2383 | Not-determined |
| NC1_vf_k141_3288 | Not-determined |
| NC1_vf_k141_2611 | Not-determined |
| NC1_vf_k141_1488 | Not-determined |
| NC1_vf_k141_3512 | Not-determined |
| NC1_vf_k141_173 | Not-determined |
| NC1_vf_k141_2176 | Not-determined |
| NC1_vf_k141_818 | Not-determined |
| NC1_vf_k141_1249 | Not-determined |
| NC1_vf_k141_371 | Not-determined |
| NC1_vf_k141_2822 | Not-determined |
| NC1_vf_k141_1934 | Not-determined |
| NC1_vf_k141_3060 | Not-determined |
| NC1_vf_k141_1489 | Not-determined |
| NC1_vf_k141_174 | Not-determined |
| NC1_vf_k141_819 | Not-determined |
| NC1_vf_k141_1250 | Not-determined |
| NC1_vf_k141_3289 | Not-determined |
| NC1_vf_k141_1052 | Not-determined |
| NC1_vf_k141_2823 | Not-determined |
| NC1_vf_k141_175 | Not-determined |
| NC1_vf_k141_1935 | Not-determined |
| NC1_vf_k141_3061 | Not-determined |
| NC1_vf_k141_2612 | Not-determined |
| NC1_vf_k141_2384 | Not-determined |
| NC1_vf_k141_1490 | Not-determined |
| NC1_vf_k141_3513 | Not-determined |
| NC1_vf_k141_820 | Not-determined |
| NC1_vf_k141_176 | Not-determined |
| NC1_vf_k141_1251 | Not-determined |
| NC1_vf_k141_372 | Not-determined |
| NC1_vf_k141_1053 | Not-determined |
| NC1_vf_k141_1936 | Not-determined |
| NC1_vf_k141_2824 | Not-determined |
| NC1_vf_k141_2385 | Not-determined |
| NC1_vf_k141_1748 | Low-quality |
| NC1_vf_k141_2613 | Not-determined |
| NC1_vf_k141_373 | Not-determined |
| NC1_vf_k141_177 | Not-determined |
| NC1_vf_k141_821 | Not-determined |
| NC1_vf_k141_2177 | Not-determined |
| NC1_vf_k141_3290 | Low-quality |
| NC1_vf_k141_1491 | Not-determined |
| NC1_vf_k141_3062 | Not-determined |
| NC1_vf_k141_1054 | Not-determined |
| NC1_vf_k141_1252 | Not-determined |
| NC1_vf_k141_2825 | Not-determined |
| NC1_vf_k141_3514 | Not-determined |
| NC1_vf_k141_1937 | Not-determined |
| NC1_vf_k141_2178 | Not-determined |
| NC1_vf_k141_2614 | Not-determined |
| NC1_vf_k141_1492 | Not-determined |
| NC1_vf_k141_3291 | Not-determined |
| NC1_vf_k141_2826 | Not-determined |
| NC1_vf_k141_3063 | Not-determined |
| NC1_vf_k141_374 | Not-determined |
| NC1_vf_k141_178 | Not-determined |
| NC1_vf_k141_1055 | Not-determined |
| NC1_vf_k141_1253 | Not-determined |
| NC1_vf_k141_1749 | Not-determined |
| NC1_vf_k141_2179 | Low-quality |
| NC1_vf_k141_3515 | Not-determined |
| NC1_vf_k141_1938 | Not-determined |
| NC1_vf_k141_3292 | Not-determined |
| NC1_vf_k141_2615 | Not-determined |
| NC1_vf_k141_1493 | Not-determined |
| NC1_vf_k141_3064 | Not-determined |
| NC1_vf_k141_822 | Not-determined |
| NC1_vf_k141_1254 | Not-determined |
| NC1_vf_k141_375 | Not-determined |
| NC1_vf_k141_1750 | Not-determined |
| NC1_vf_k141_1056 | Not-determined |
| NC1_vf_k141_3293 | Not-determined |
| NC1_vf_k141_3065 | Not-determined |
| NC1_vf_k141_1494 | Not-determined |
| NC1_vf_k141_2180 | Not-determined |
| NC1_vf_k141_2616 | Not-determined |
| NC1_vf_k141_1939 | Not-determined |
| NC1_vf_k141_2386 | Not-determined |
| NC1_vf_k141_3516 | Not-determined |
| NC1_vf_k141_1255 | Not-determined |
| NC1_vf_k141_823 | Not-determined |
| NC1_vf_k141_2827 | Not-determined |
| NC1_vf_k141_1751 | Not-determined |
| NC1_vf_k141_3294 | Not-determined |
| NC1_vf_k141_3066 | Not-determined |
| NC1_vf_k141_1057 | Not-determined |
| NC1_vf_k141_1940 | Not-determined |
| NC1_vf_k141_2617 | Not-determined |
| NC1_vf_k141_1495 | Not-determined |
| NC1_vf_k141_376 | Not-determined |
| NC1_vf_k141_1256 | Not-determined |
| NC1_vf_k141_2387 | Low-quality |
| NC1_vf_k141_1752 | Not-determined |
| NC1_vf_k141_2181 | Not-determined |
| NC1_vf_k141_824 | Not-determined |

| | |
|---|---|
| NC1_vf_k141_1058 | Not-determined |
| NC1_vf_k141_2828 | Low-quality |
| NC1_vf_k141_2618 | Not-determined |
| NC1_vf_k141_1941 | Not-determined |
| NC1_vf_k141_1257 | Not-determined |
| NC1_vf_k141_377 | Not-determined |
| NC1_vf_k141_1496 | Not-determined |
| NC1_vf_k141_3517 | Not-determined |
| NC1_vf_k141_1753 | Not-determined |
| NC1_vf_k141_2388 | Not-determined |
| NC1_vf_k141_825 | Not-determined |
| NC1_vf_k141_2619 | Not-determined |
| NC1_vf_k141_1942 | Not-determined |
| NC1_vf_k141_3295 | Not-determined |
| NC1_vf_k141_2182 | Not-determined |
| NC1_vf_k141_1258 | Not-determined |
| NC1_vf_k141_1754 | Not-determined |
| NC1_vf_k141_1497 | Not-determined |
| NC1_vf_k141_826 | Not-determined |
| NC1_vf_k141_3518 | Not-determined |
| NC1_vf_k141_3296 | Not-determined |
| NC1_vf_k141_179 | Low-quality |
| NC1_vf_k141_2389 | Not-determined |
| NC1_vf_k141_1943 | Not-determined |
| NC1_vf_k141_378 | Not-determined |
| NC1_vf_k141_1498 | Not-determined |
| NC1_vf_k141_2829 | Not-determined |
| NC1_vf_k141_1259 | Not-determined |
| NC1_vf_k141_2620 | Not-determined |
| NC1_vf_k141_180 | Not-determined |
| NC1_vf_k141_1755 | Not-determined |
| NC1_vf_k141_3519 | Not-determined |
| NC1_vf_k141_3297 | Not-determined |
| NC1_vf_k141_1499 | Not-determined |
| NC1_vf_k141_2183 | Not-determined |
| NC1_vf_k141_2830 | Not-determined |
| NC1_vf_k141_1059 | Not-determined |
| NC1_vf_k141_1756 | Not-determined |
| NC1_vf_k141_1944 | Not-determined |
| NC1_vf_k141_1260 | Not-determined |
| NC1_vf_k141_379 | Not-determined |
| NC1_vf_k141_3298 | Not-determined |
| NC1_vf_k141_2390 | Not-determined |
| NC1_vf_k141_2831 | Not-determined |
| NC1_vf_k141_1757 | Not-determined |
| NC1_vf_k141_3520 | Not-determined |
| NC1_vf_k141_1500 | Low-quality |
| NC1_vf_k141_1945 | Not-determined |
| NC1_vf_k141_2621 | Not-determined |
| NC1_vf_k141_2184 | Not-determined |
| NC1_vf_k141_1261 | Not-determined |
| NC1_vf_k141_1060 | Not-determined |
| NC1_vf_k141_2832 | Not-determined |
| NC1_vf_k141_380 | Not-determined |
| NC1_vf_k141_1946 | Not-determined |
| NC1_vf_k141_3521 | Not-determined |
| NC1_vf_k141_3299 | Not-determined |
| NC1_vf_k141_1501 | Not-determined |
| NC1_vf_k141_1262 | Not-determined |
| NC1_vf_k141_2391 | Not-determined |
| NC1_vf_k141_1758 | Not-determined |
| NC1_vf_k141_381 | Not-determined |
| NC1_vf_k141_2622 | Not-determined |
| NC1_vf_k141_3522 | Not-determined |
| NC1_vf_k141_1947 | Not-determined |
| NC1_vf_k141_3300 | Not-determined |
| NC1_vf_k141_1061 | Not-determined |
| NC1_vf_k141_2185 | Not-determined |
| NC1_vf_k141_1759 | Not-determined |
| NC1_vf_k141_181 | Not-determined |
| NC1_vf_k141_1502 | Low-quality |
| NC1_vf_k141_2623 | Not-determined |
| NC1_vf_k141_382 | Not-determined |
| NC1_vf_k141_2833 | Low-quality |
| NC1_vf_k141_1263 | Not-determined |
| NC1_vf_k141_1062 | Not-determined |
| NC1_vf_k141_182 | Low-quality |
| NC1_vf_k141_3523 | Not-determined |
| NC1_vf_k141_1760 | Not-determined |
| NC1_vf_k141_3301 | Not-determined |
| NC1_vf_k141_2186 | Not-determined |
| NC1_vf_k141_2392 | Not-determined |
| NC1_vf_k141_3067 | Not-determined |
| NC1_vf_k141_383 | Not-determined |
| NC1_vf_k141_183 | Not-determined |
| NC1_vf_k141_1761 | Not-determined |
| NC1_vf_k141_1948 | Not-determined |
| NC1_vf_k141_1264 | Not-determined |
| NC1_vf_k141_2834 | Not-determined |
| NC1_vf_k141_2624 | Not-determined |
| NC1_vf_k141_1503 | Not-determined |
| NC1_vf_k141_3068 | Not-determined |
| NC1_vf_k141_1762 | Not-determined |
| NC1_vf_k141_827 | Not-determined |
| NC1_vf_k141_1949 | Low-quality |
| NC1_vf_k141_2835 | Not-determined |
| NC1_vf_k141_1063 | Not-determined |
| NC1_vf_k141_1265 | Not-determined |
| NC1_vf_k141_1504 | Not-determined |
| NC1_vf_k141_2393 | Not-determined |
| NC1_vf_k141_3069 | Not-determined |
| NC1_vf_k141_2187 | Not-determined |
| NC1_vf_k141_1763 | Not-determined |
| NC1_vf_k141_184 | Not-determined |
| NC1_vf_k141_828 | Not-determined |
| NC1_vf_k141_3302 | Not-determined |
| NC1_vf_k141_384 | Not-determined |
| NC1_vf_k141_1064 | Not-determined |
| NC1_vf_k141_2625 | Not-determined |
| NC1_vf_k141_1505 | Not-determined |
| NC1_vf_k141_1266 | Not-determined |
| NC1_vf_k141_2394 | Not-determined |
| NC1_vf_k141_2188 | Not-determined |
| NC1_vf_k141_2836 | Not-determined |
| NC1_vf_k141_185 | Not-determined |
| NC1_vf_k141_1764 | Not-determined |
| NC1_vf_k141_1065 | Not-determined |
| NC1_vf_k141_385 | Not-determined |
| NC1_vf_k141_3303 | Not-determined |
| NC1_vf_k141_1506 | Not-determined |
| NC1_vf_k141_2626 | Not-determined |
| NC1_vf_k141_3070 | Low-quality |
| NC1_vf_k141_1950 | Not-determined |
| NC1_vf_k141_2189 | Not-determined |
| NC1_vf_k141_186 | Not-determined |
| NC1_vf_k141_2837 | Not-determined |
| NC1_vf_k141_1765 | Not-determined |
| NC1_vf_k141_829 | Not-determined |
| NC1_vf_k141_2627 | Not-determined |
| NC1_vf_k141_1507 | Not-determined |
| NC1_vf_k141_386 | Not-determined |
| NC1_vf_k141_1066 | Not-determined |
| NC1_vf_k141_187 | Not-determined |
| NC1_vf_k141_3071 | Not-determined |
| NC1_vf_k141_2838 | Not-determined |
| NC1_vf_k141_1267 | Not-determined |
| NC1_vf_k141_2190 | Not-determined |
| NC1_vf_k141_1951 | Not-determined |
| NC1_vf_k141_3304 | Not-determined |
| NC1_vf_k141_1508 | Not-determined |
| NC1_vf_k141_830 | Not-determined |
| NC1_vf_k141_387 | Not-determined |
| NC1_vf_k141_1067 | Not-determined |
| NC1_vf_k141_2628 | Not-determined |
| NC1_vf_k141_1268 | Not-determined |
| NC1_vf_k141_2191 | Low-quality |
| NC1_vf_k141_188 | Not-determined |
| NC1_vf_k141_2839 | Not-determined |
| NC1_vf_k141_1509 | Low-quality |
| NC1_vf_k141_1269 | Not-determined |
| NC1_vf_k141_2629 | Not-determined |
| NC1_vf_k141_3524 | Not-determined |
| NC1_vf_k141_3305 | Not-determined |
| NC1_vf_k141_831 | Not-determined |
| NC1_vf_k141_388 | Not-determined |
| NC1_vf_k141_2395 | Not-determined |
| NC1_vf_k141_189 | Not-determined |
| NC1_vf_k141_3072 | Not-determined |
| NC1_vf_k141_1068 | Not-determined |
| NC1_vf_k141_1270 | Not-determined |
| NC1_vf_k141_1510 | Not-determined |
| NC1_vf_k141_2192 | Not-determined |
| NC1_vf_k141_1952 | Low-quality |
| NC1_vf_k141_3525 | Not-determined |
| NC1_vf_k141_2840 | Not-determined |
| NC1_vf_k141_190 | Not-determined |
| NC1_vf_k141_2630 | Not-determined |
| NC1_vf_k141_389 | Not-determined |
| NC1_vf_k141_1271 | Low-quality |
| NC1_vf_k141_2396 | Not-determined |
| NC1_vf_k141_832 | Not-determined |
| NC1_vf_k141_1766 | Not-determined |
| NC1_vf_k141_1511 | Not-determined |
| NC1_vf_k141_1069 | Not-determined |
| NC1_vf_k141_3306 | Not-determined |
| NC1_vf_k141_2193 | Not-determined |
| NC1_vf_k141_390 | Not-determined |
| NC1_vf_k141_1953 | Not-determined |
| NC1_vf_k141_1767 | Low-quality |
| NC1_vf_k141_2631 | Low-quality |
| NC1_vf_k141_2841 | Not-determined |
| NC1_vf_k141_191 | Not-determined |
| NC1_vf_k141_1512 | Not-determined |
| NC1_vf_k141_3307 | Not-determined |
| NC1_vf_k141_3073 | Not-determined |
| NC1_vf_k141_1272 | Not-determined |
| NC1_vf_k141_2194 | Not-determined |
| NC1_vf_k141_1768 | Not-determined |
| NC1_vf_k141_3526 | Not-determined |
| NC1_vf_k141_2397 | Not-determined |
| NC1_vf_k141_2842 | Not-determined |
| NC1_vf_k141_2632 | Not-determined |
| NC1_vf_k141_192 | Not-determined |
| NC1_vf_k141_1513 | Not-determined |
| NC1_vf_k141_3074 | Low-quality |
| NC1_vf_k141_1769 | Not-determined |
| NC1_vf_k141_1273 | Not-determined |
| NC1_vf_k141_3527 | Low-quality |
| NC1_vf_k141_2633 | Not-determined |
| NC1_vf_k141_1954 | Not-determined |
| NC1_vf_k141_833 | Not-determined |
| NC1_vf_k141_1274 | Not-determined |
| NC1_vf_k141_2398 | Not-determined |
| NC1_vf_k141_1070 | Not-determined |
| NC1_vf_k141_3075 | Not-determined |
| NC1_vf_k141_2195 | Not-determined |
| NC1_vf_k141_2843 | Not-determined |
| NC1_vf_k141_1770 | Not-determined |
| NC1_vf_k141_193 | Not-determined |
| NC1_vf_k141_391 | Not-determined |
| NC1_vf_k141_3528 | Not-determined |
| NC1_vf_k141_1955 | Not-determined |
| NC1_vf_k141_834 | Not-determined |
| NC1_vf_k141_1771 | Not-determined |
| NC1_vf_k141_1275 | Not-determined |
| NC1_vf_k141_2634 | Not-determined |
| NC1_vf_k141_2196 | Not-determined |
| NC1_vf_k141_3076 | Not-determined |
| NC1_vf_k141_3308 | Not-determined |
| NC1_vf_k141_2844 | Not-determined |
| NC1_vf_k141_392 | Not-determined |
| NC1_vf_k141_1071 | Not-determined |
| NC1_vf_k141_194 | Not-determined |
| NC1_vf_k141_1772 | Not-determined |
| NC1_vf_k141_3529 | Not-determined |
| NC1_vf_k141_835 | Low-quality |
| NC1_vf_k141_1514 | Not-determined |
| NC1_vf_k141_3077 | Not-determined |
| NC1_vf_k141_2635 | Not-determined |
| NC1_vf_k141_2197 | Not-determined |
| NC1_vf_k141_3309 | Not-determined |
| NC1_vf_k141_393 | Not-determined |
| NC1_vf_k141_836 | Not-determined |
| NC1_vf_k141_1773 | Not-determined |
| NC1_vf_k141_1276 | Not-determined |
| NC1_vf_k141_2399 | Not-determined |
| NC1_vf_k141_195 | Not-determined |
| NC1_vf_k141_1515 | Not-determined |
| NC1_vf_k141_1072 | Not-determined |
| NC1_vf_k141_2636 | Not-determined |
| NC1_vf_k141_1956 | Not-determined |
| NC1_vf_k141_3530 | Not-determined |
| NC1_vf_k141_3310 | Not-determined |
| NC1_vf_k141_1774 | Not-determined |
| NC1_vf_k141_837 | Not-determined |
| NC1_vf_k141_196 | Not-determined |
| NC1_vf_k141_394 | Not-determined |
| NC1_vf_k141_1073 | Not-determined |
| NC1_vf_k141_3531 | Not-determined |
| NC1_vf_k141_2400 | Not-determined |
| NC1_vf_k141_1516 | Low-quality |
| NC1_vf_k141_3311 | Not-determined |
| NC1_vf_k141_197 | Not-determined |
| NC1_vf_k141_1277 | Not-determined |
| NC1_vf_k141_838 | Not-determined |
| NC1_vf_k141_2198 | Not-determined |
| NC1_vf_k141_395 | Not-determined |
| NC1_vf_k141_2845 | Not-determined |
| NC1_vf_k141_2637 | Not-determined |
| NC1_vf_k141_3532 | Not-determined |
| NC1_vf_k141_1775 | Not-determined |
| NC1_vf_k141_198 | Not-determined |
| NC1_vf_k141_2401 | Not-determined |
| NC1_vf_k141_3312 | Not-determined |
| NC1_vf_k141_839 | Not-determined |
| NC1_vf_k141_1278 | Not-determined |
| NC1_vf_k141_1517 | Not-determined |
| NC1_vf_k141_396 | Not-determined |
| NC1_vf_k141_3533 | Not-determined |
| NC1_vf_k141_1074 | Not-determined |
| NC1_vf_k141_199 | Not-determined |
| NC1_vf_k141_1957 | Low-quality |
| NC1_vf_k141_2638 | Not-determined |
| NC1_vf_k141_2402 | Not-determined |
| NC1_vf_k141_3313 | Not-determined |
| NC1_vf_k141_2846 | Not-determined |
| NC1_vf_k141_1518 | Not-determined |
| NC1_vf_k141_200 | Not-determined |
| NC1_vf_k141_1776 | Not-determined |
| NC1_vf_k141_840 | Not-determined |
| NC1_vf_k141_2199 | Not-determined |
| NC1_vf_k141_3314 | Not-determined |
| NC1_vf_k141_1075 | Not-determined |
| NC1_vf_k141_2639 | Not-determined |
| NC1_vf_k141_397 | Not-determined |
| NC1_vf_k141_1279 | Not-determined |
| NC1_vf_k141_1777 | Not-determined |
| NC1_vf_k141_1519 | Not-determined |
| NC1_vf_k141_2403 | Not-determined |
| NC1_vf_k141_3078 | Not-determined |
| NC1_vf_k141_201 | Not-determined |
| NC1_vf_k141_2200 | Not-determined |
| NC1_vf_k141_1958 | Not-determined |
| NC1_vf_k141_1076 | Not-determined |
| NC1_vf_k141_2847 | Not-determined |
| NC1_vf_k141_3315 | Not-determined |
| NC1_vf_k141_1778 | Not-determined |
| NC1_vf_k141_1520 | Not-determined |
| NC1_vf_k141_1280 | Not-determined |
| NC1_vf_k141_2404 | Not-determined |
| NC1_vf_k141_398 | Not-determined |
| NC1_vf_k141_2848 | Not-determined |
| NC1_vf_k141_2640 | Not-determined |
| NC1_vf_k141_1779 | Not-determined |
| NC1_vf_k141_3079 | Not-determined |
| NC1_vf_k141_841 | Low-quality |
| NC1_vf_k141_1959 | Not-determined |
| NC1_vf_k141_1077 | Not-determined |
| NC1_vf_k141_1521 | Not-determined |
| NC1_vf_k141_1281 | Not-determined |
| NC1_vf_k141_3316 | Not-determined |
| NC1_vf_k141_2405 | Not-determined |
| NC1_vf_k141_202 | Not-determined |
| NC1_vf_k141_3080 | Not-determined |
| NC1_vf_k141_2641 | Not-determined |
| NC1_vf_k141_3534 | Not-determined |
| NC1_vf_k141_2849 | Not-determined |
| NC1_vf_k141_1078 | Not-determined |
| NC1_vf_k141_1780 | Low-quality |
| NC1_vf_k141_1960 | Not-determined |
| NC1_vf_k141_203 | Not-determined |
| NC1_vf_k141_1282 | Not-determined |
| NC1_vf_k141_2201 | Not-determined |
| NC1_vf_k141_1522 | Low-quality |
| NC1_vf_k141_399 | Not-determined |
| NC1_vf_k141_3317 | Not-determined |
| NC1_vf_k141_2406 | Not-determined |
| NC1_vf_k141_3535 | Not-determined |
| NC1_vf_k141_842 | Not-determined |
| NC1_vf_k141_1781 | Not-determined |
| NC1_vf_k141_3081 | Not-determined |
| NC1_vf_k141_1079 | Not-determined |
| NC1_vf_k141_1283 | Not-determined |
| NC1_vf_k141_400 | Not-determined |
| NC1_vf_k141_2642 | Not-determined |
| NC1_vf_k141_204 | Not-determined |
| NC1_vf_k141_2202 | Not-determined |
| NC1_vf_k141_1782 | Not-determined |
| NC1_vf_k141_843 | Not-determined |
| NC1_vf_k141_3082 | Not-determined |
| NC1_vf_k141_3318 | Not-determined |
| NC1_vf_k141_2407 | Not-determined |
| NC1_vf_k141_1284 | Not-determined |
| NC1_vf_k141_2850 | Low-quality |
| NC1_vf_k141_1961 | Not-determined |
| NC1_vf_k141_1080 | Not-determined |
| NC1_vf_k141_401 | Not-determined |
| NC1_vf_k141_205 | Not-determined |
| NC1_vf_k141_3083 | Low-quality |
| NC1_vf_k141_2643 | Not-determined |
| NC1_vf_k141_844 | Not-determined |
| NC1_vf_k141_1783 | Not-determined |
| NC1_vf_k141_1081 | Not-determined |
| NC1_vf_k141_206 | Not-determined |
| NC1_vf_k141_402 | Not-determined |
| NC1_vf_k141_1285 | Not-determined |
| NC1_vf_k141_2203 | Not-determined |
| NC1_vf_k141_2851 | Not-determined |
| NC1_vf_k141_3536 | Not-determined |
| NC1_vf_k141_1784 | Not-determined |
| NC1_vf_k141_1962 | Not-determined |
| NC1_vf_k141_845 | Not-determined |
| NC1_vf_k141_2644 | Not-determined |
| NC1_vf_k141_3084 | Not-determined |
| NC1_vf_k141_1286 | Not-determined |
| NC1_vf_k141_2204 | Not-determined |
| NC1_vf_k141_1785 | Not-determined |
| NC1_vf_k141_403 | Not-determined |
| NC1_vf_k141_3537 | Not-determined |
| NC1_vf_k141_1082 | Not-determined |
| NC1_vf_k141_1963 | Not-determined |
| NC1_vf_k141_3319 | Low-quality |
| NC1_vf_k141_2408 | Not-determined |
| NC1_vf_k141_2645 | Not-determined |
| NC1_vf_k141_2852 | Not-determined |
| NC1_vf_k141_1786 | Not-determined |
| NC1_vf_k141_3085 | Not-determined |
| NC1_vf_k141_2205 | Not-determined |
| NC1_vf_k141_1083 | Not-determined |
| NC1_vf_k141_404 | Not-determined |
| NC1_vf_k141_2646 | Not-determined |
| NC1_vf_k141_1787 | Not-determined |
| NC1_vf_k141_1287 | Not-determined |
| NC1_vf_k141_2853 | Not-determined |
| NC1_vf_k141_846 | Not-determined |
| NC1_vf_k141_3320 | Not-determined |
| NC1_vf_k141_3538 | Not-determined |
| NC1_vf_k141_1084 | Not-determined |
| NC1_vf_k141_1523 | Low-quality |
| NC1_vf_k141_2409 | Not-determined |
| NC1_vf_k141_1788 | Not-determined |
| NC1_vf_k141_1288 | Not-determined |
| NC1_vf_k141_2647 | Not-determined |
| NC1_vf_k141_3539 | Not-determined |
| NC1_vf_k141_847 | Not-determined |
| NC1_vf_k141_2206 | Not-determined |
| NC1_vf_k141_405 | Not-determined |
| NC1_vf_k141_3086 | Not-determined |
| NC1_vf_k141_3321 | Not-determined |
| NC1_vf_k141_207 | Not-determined |
| NC1_vf_k141_1789 | Not-determined |
| NC1_vf_k141_1289 | Not-determined |
| NC1_vf_k141_2410 | Not-determined |
| NC1_vf_k141_208 | Not-determined |
| NC1_vf_k141_848 | Not-determined |
| NC1_vf_k141_2648 | Not-determined |
| NC1_vf_k141_406 | Not-determined |
| NC1_vf_k141_1790 | Not-determined |
| NC1_vf_k141_3322 | Not-determined |
| NC1_vf_k141_2207 | Not-determined |
| NC1_vf_k141_1085 | Not-determined |
| NC1_vf_k141_2411 | Not-determined |
| NC1_vf_k141_1290 | Not-determined |
| NC1_vf_k141_209 | Not-determined |
| NC1_vf_k141_1791 | Not-determined |
| NC1_vf_k141_407 | Not-determined |
| NC1_vf_k141_849 | Not-determined |
| NC1_vf_k141_2412 | Not-determined |
| NC1_vf_k141_1291 | Not-determined |
| NC1_vf_k141_1792 | Not-determined |
| NC1_vf_k141_3540 | Not-determined |
| NC1_vf_k141_2208 | Not-determined |
| NC1_vf_k141_3087 | Not-determined |
| NC1_vf_k141_2854 | Not-determined |
| NC1_vf_k141_1086 | Not-determined |
| NC1_vf_k141_408 | Not-determined |
| NC1_vf_k141_1793 | Not-determined |
| NC1_vf_k141_3323 | Not-determined |
| NC1_vf_k141_2649 | Not-determined |
| NC1_vf_k141_1292 | Not-determined |
| NC1_vf_k141_2209 | Not-determined |
| NC1_vf_k141_1524 | Not-determined |
| NC1_vf_k141_3541 | Not-determined |
| NC1_vf_k141_210 | Not-determined |
| NC1_vf_k141_2413 | Not-determined |
| NC1_vf_k141_3088 | Not-determined |
| NC1_vf_k141_1794 | Not-determined |
| NC1_vf_k141_409 | Not-determined |
| NC1_vf_k141_1087 | Not-determined |
| NC1_vf_k141_2210 | Not-determined |
| NC1_vf_k141_1964 | Low-quality |
| NC1_vf_k141_1525 | Not-determined |
| NC1_vf_k141_2650 | Not-determined |
| NC1_vf_k141_211 | Low-quality |
| NC1_vf_k141_1293 | Not-determined |
| NC1_vf_k141_1795 | Not-determined |
| NC1_vf_k141_3089 | Not-determined |
| NC1_vf_k141_1088 | Not-determined |
| NC1_vf_k141_3324 | Not-determined |
| NC1_vf_k141_2414 | Not-determined |
| NC1_vf_k141_410 | Not-determined |
| NC1_vf_k141_1796 | Not-determined |
| NC1_vf_k141_2211 | Not-determined |
| NC1_vf_k141_1294 | Not-determined |
| NC1_vf_k141_1526 | Not-determined |
| NC1_vf_k141_2415 | Not-determined |
| NC1_vf_k141_2651 | Low-quality |
| NC1_vf_k141_1797 | Not-determined |
| NC1_vf_k141_1965 | Not-determined |
| NC1_vf_k141_411 | Not-determined |
| NC1_vf_k141_1089 | Not-determined |
| NC1_vf_k141_1527 | Not-determined |
| NC1_vf_k141_1295 | Not-determined |
| NC1_vf_k141_2212 | Not-determined |
| NC1_vf_k141_1798 | Not-determined |
| NC1_vf_k141_2416 | Not-determined |
| NC1_vf_k141_1966 | Not-determined |
| NC1_vf_k141_3090 | Low-quality |
| NC1_vf_k141_1528 | Not-determined |
| NC1_vf_k141_586 | Low-quality |
| NC1_vf_k141_1799 | Not-determined |
| NC1_vf_k141_1296 | Not-determined |
| NC1_vf_k141_3542 | Not-determined |
| NC1_vf_k141_1529 | Not-determined |
| NC1_vf_k141_1090 | Not-determined |
| NC1_vf_k141_412 | Not-determined |
| NC1_vf_k141_1967 | Not-determined |
| NC1_vf_k141_587 | Not-determined |
| NC1_vf_k141_3091 | Not-determined |
| NC1_vf_k141_2417 | Not-determined |
| NC1_vf_k141_2213 | Not-determined |
| NC1_vf_k141_2652 | Not-determined |
| NC1_vf_k141_3543 | Not-determined |
| NC1_vf_k141_413 | Not-determined |
| NC1_vf_k141_1968 | Not-determined |
| NC1_vf_k141_588 | Not-determined |
| NC1_vf_k141_1297 | Not-determined |
| NC1_vf_k141_212 | Not-determined |
| NC1_vf_k141_1530 | Not-determined |
| NC1_vf_k141_1091 | Not-determined |
| NC1_vf_k141_2214 | Not-determined |
| NC1_vf_k141_414 | Not-determined |
| NC1_vf_k141_589 | Not-determined |
| NC1_vf_k141_1969 | Not-determined |
| NC1_vf_k141_2418 | Not-determined |
| NC1_vf_k141_3544 | Low-quality |
| NC1_vf_k141_1298 | Not-determined |
| NC1_vf_k141_2215 | Not-determined |
| NC1_vf_k141_1531 | Not-determined |
| NC1_vf_k141_1092 | Not-determined |
| NC1_vf_k141_213 | Not-determined |
| NC1_vf_k141_1970 | Not-determined |
| NC1_vf_k141_2653 | Not-determined |
| NC1_vf_k141_2419 | Not-determined |
| NC1_vf_k141_1299 | Not-determined |
| NC1_vf_k141_590 | Not-determined |
| NC1_vf_k141_1093 | Not-determined |
| NC1_vf_k141_214 | Not-determined |
| NC1_vf_k141_1532 | Not-determined |
| NC1_vf_k141_3325 | Low-quality |
| NC1_vf_k141_1300 | Not-determined |
| NC1_vf_k141_2420 | Low-quality |
| NC1_vf_k141_215 | Not-determined |
| NC1_vf_k141_3545 | Low-quality |
| NC1_vf_k141_2216 | Not-determined |
| NC1_vf_k141_1971 | Not-determined |
| NC1_vf_k141_2654 | Not-determined |
| NC1_vf_k141_850 | Not-determined |
| NC1_vf_k141_415 | Not-determined |
| NC1_vf_k141_3326 | Not-determined |
| NC1_vf_k141_591 | Not-determined |
| NC1_vf_k141_216 | Not-determined |
| NC1_vf_k141_3546 | Not-determined |
| NC1_vf_k141_2655 | Not-determined |
| NC1_vf_k141_851 | Not-determined |
| NC1_vf_k141_3327 | Not-determined |
| NC1_vf_k141_217 | Not-determined |
| NC1_vf_k141_592 | Not-determined |
| NC1_vf_k141_1094 | Not-determined |
| NC1_vf_k141_2217 | Not-determined |
| NC1_vf_k141_416 | Low-quality |
| NC1_vf_k141_1301 | Not-determined |
| NC1_vf_k141_218 | Not-determined |
| NC1_vf_k141_1533 | Not-determined |
| NC1_vf_k141_2421 | Not-determined |
| NC1_vf_k141_3328 | Not-determined |
| NC1_vf_k141_593 | Not-determined |
| NC1_vf_k141_2855 | Not-determined |
| NC1_vf_k141_2656 | Not-determined |
| NC1_vf_k141_852 | Not-determined |
| NC1_vf_k141_1095 | Not-determined |
| NC1_vf_k141_417 | Not-determined |
| NC1_vf_k141_2218 | Not-determined |
| NC1_vf_k141_219 | Not-determined |
| NC1_vf_k141_3547 | Not-determined |
| NC1_vf_k141_2422 | Not-determined |
| NC1_vf_k141_1302 | Not-determined |
| NC1_vf_k141_1534 | Not-determined |
| NC1_vf_k141_594 | Not-determined |
| NC1_vf_k141_3329 | Not-determined |
| NC1_vf_k141_1972 | Not-determined |
| NC1_vf_k141_2856 | Not-determined |
| NC1_vf_k141_220 | Low-quality |
| NC1_vf_k141_418 | Not-determined |
| NC1_vf_k141_2219 | Not-determined |
| NC1_vf_k141_1096 | Not-determined |
| NC1_vf_k141_3548 | Not-determined |
| NC1_vf_k141_3330 | Not-determined |
| NC1_vf_k141_2657 | Not-determined |
| NC1_vf_k141_221 | Not-determined |
| NC1_vf_k141_1303 | Not-determined |
| NC1_vf_k141_2423 | Not-determined |
| NC1_vf_k141_3092 | Low-quality |
| NC1_vf_k141_853 | Not-determined |
| NC1_vf_k141_2220 | Not-determined |
| NC1_vf_k141_1973 | Not-determined |
| NC1_vf_k141_595 | Not-determined |
| NC1_vf_k141_1535 | Not-determined |
| NC1_vf_k141_3331 | Low-quality |
| NC1_vf_k141_222 | Not-determined |
| NC1_vf_k141_2857 | Not-determined |
| NC1_vf_k141_2658 | Not-determined |
| NC1_vf_k141_3093 | Not-determined |
| NC1_vf_k141_854 | Not-determined |
| NC1_vf_k141_3549 | Not-determined |
| NC1_vf_k141_1304 | Not-determined |
| NC1_vf_k141_1097 | Not-determined |
| NC1_vf_k141_223 | Not-determined |
| NC1_vf_k141_1536 | Not-determined |
| NC1_vf_k141_1974 | Not-determined |
| NC1_vf_k141_3332 | Low-quality |
| NC1_vf_k141_2659 | Not-determined |
| NC1_vf_k141_419 | Not-determined |
| NC1_vf_k141_855 | Not-determined |
| NC1_vf_k141_2858 | Not-determined |
| NC1_vf_k141_596 | Not-determined |
| NC1_vf_k141_1537 | Not-determined |
| NC1_vf_k141_2424 | Not-determined |
| NC1_vf_k141_2660 | Not-determined |

| | |
|---|---|
| NC1_vf_k141_2221 | Not-determined |
| NC1_vf_k141_224 | Low-quality |
| NC1_vf_k141_1098 | Not-determined |
| NC1_vf_k141_1305 | Not-determined |
| NC1_vf_k141_3333 | Not-determined |
| NC1_vf_k141_1975 | Not-determined |
| NC1_vf_k141_1538 | Not-determined |
| NC1_vf_k141_2859 | Not-determined |
| NC1_vf_k141_3550 | Not-determined |
| NC1_vf_k141_3094 | Not-determined |
| NC1_vf_k141_2222 | Not-determined |
| NC1_vf_k141_597 | Not-determined |
| NC1_vf_k141_1306 | Not-determined |
| NC1_vf_k141_2661 | Not-determined |
| NC1_vf_k141_1099 | Not-determined |
| NC1_vf_k141_856 | Not-determined |
| NC1_vf_k141_1976 | Not-determined |
| NC1_vf_k141_3334 | Not-determined |
| NC1_vf_k141_3551 | Not-determined |
| NC1_vf_k141_2860 | Not-determined |
| NC1_vf_k141_2223 | Not-determined |
| NC1_vf_k141_1539 | Not-determined |
| NC1_vf_k141_3095 | Not-determined |
| NC1_vf_k141_1100 | Not-determined |
| NC1_vf_k141_598 | Not-determined |
| NC1_vf_k141_2662 | Low-quality |
| NC1_vf_k141_1977 | Not-determined |
| NC1_vf_k141_2425 | Not-determined |
| NC1_vf_k141_2861 | Not-determined |
| NC1_vf_k141_2224 | Not-determined |
| NC1_vf_k141_3335 | Not-determined |
| NC1_vf_k141_857 | Not-determined |
| NC1_vf_k141_3552 | Not-determined |
| NC1_vf_k141_1540 | Not-determined |
| NC1_vf_k141_1307 | Not-determined |
| NC1_vf_k141_3096 | Not-determined |
| NC1_vf_k141_1978 | Not-determined |
| NC1_vf_k141_2426 | Not-determined |
| NC1_vf_k141_1101 | Not-determined |
| NC1_vf_k141_858 | Not-determined |
| NC1_vf_k141_2862 | Not-determined |
| NC1_vf_k141_2663 | Not-determined |
| NC1_vf_k141_2225 | Low-quality |
| NC1_vf_k141_3336 | Not-determined |
| NC1_vf_k141_3553 | Not-determined |
| NC1_vf_k141_1979 | Not-determined |
| NC1_vf_k141_2427 | Not-determined |
| NC1_vf_k141_420 | Low-quality |
| NC1_vf_k141_3097 | Not-determined |
| NC1_vf_k141_2863 | Not-determined |
| NC1_vf_k141_1541 | Not-determined |
| NC1_vf_k141_3337 | Not-determined |
| NC1_vf_k141_1308 | Not-determined |
| NC1_vf_k141_599 | Not-determined |
| NC1_vf_k141_2664 | Not-determined |
| NC1_vf_k141_421 | Not-determined |
| NC1_vf_k141_3554 | Not-determined |
| NC1_vf_k141_2428 | Not-determined |
| NC1_vf_k141_1980 | Not-determined |
| NC1_vf_k141_2226 | Not-determined |
| NC1_vf_k141_3338 | Not-determined |
| NC1_vf_k141_859 | Not-determined |
| NC1_vf_k141_2864 | Not-determined |
| NC1_vf_k141_3098 | Not-determined |
| NC1_vf_k141_600 | Not-determined |
| NC1_vf_k141_1542 | Not-determined |
| NC1_vf_k141_2429 | Not-determined |
| NC1_vf_k141_2665 | Not-determined |
| NC1_vf_k141_2227 | Not-determined |
| NC1_vf_k141_2865 | Not-determined |
| NC1_vf_k141_601 | Not-determined |
| NC1_vf_k141_1543 | Not-determined |
| NC1_vf_k141_860 | Not-determined |
| NC1_vf_k141_2666 | Not-determined |
| NC1_vf_k141_3339 | Not-determined |
| NC1_vf_k141_2430 | Not-determined |
| NC1_vf_k141_2228 | Not-determined |
| NC1_vf_k141_3099 | Low-quality |
| NC1_vf_k141_2866 | Low-quality |
| NC1_vf_k141_1981 | Not-determined |
| NC1_vf_k141_1309 | Low-quality |
| NC1_vf_k141_1544 | Not-determined |
| NC1_vf_k141_2229 | Low-quality |
| NC1_vf_k141_861 | Not-determined |
| NC1_vf_k141_2867 | Not-determined |
| NC1_vf_k141_2667 | Not-determined |
| NC1_vf_k141_3340 | Not-determined |
| NC1_vf_k141_3100 | Not-determined |
| NC1_vf_k141_2431 | Not-determined |
| NC1_vf_k141_1310 | Not-determined |
| NC1_vf_k141_1982 | Low-quality |
| NC1_vf_k141_2868 | Not-determined |
| NC1_vf_k141_2668 | Not-determined |
| NC1_vf_k141_422 | Not-determined |
| NC1_vf_k141_3101 | Not-determined |
| NC1_vf_k141_3341 | Not-determined |
| NC1_vf_k141_2230 | Not-determined |
| NC1_vf_k141_1311 | Not-determined |
| NC1_vf_k141_2432 | Not-determined |
| NC1_vf_k141_602 | Not-determined |
| NC1_vf_k141_862 | Not-determined |
| NC1_vf_k141_1545 | Not-determined |
| NC1_vf_k141_1312 | Not-determined |
| NC1_vf_k141_2669 | Not-determined |
| NC1_vf_k141_423 | Not-determined |
| NC1_vf_k141_3102 | Not-determined |
| NC1_vf_k141_2231 | Not-determined |
| NC1_vf_k141_2869 | Low-quality |
| NC1_vf_k141_3342 | Not-determined |
| NC1_vf_k141_2433 | Not-determined |
| NC1_vf_k141_603 | Not-determined |
| NC1_vf_k141_1546 | Not-determined |
| NC1_vf_k141_1313 | Not-determined |
| NC1_vf_k141_3103 | Low-quality |
| NC1_vf_k141_2232 | Not-determined |
| NC1_vf_k141_2870 | Not-determined |
| NC1_vf_k141_3343 | Not-determined |
| NC1_vf_k141_424 | Not-determined |
| NC1_vf_k141_1983 | Low-quality |
| NC1_vf_k141_863 | Not-determined |
| NC1_vf_k141_1314 | Not-determined |
| NC1_vf_k141_1547 | Low-quality |
| NC1_vf_k141_3104 | Not-determined |
| NC1_vf_k141_2233 | Not-determined |
| NC1_vf_k141_3344 | Low-quality |
| NC1_vf_k141_604 | Not-determined |
| NC1_vf_k141_2871 | Low-quality |
| NC1_vf_k141_864 | Not-determined |
| NC1_vf_k141_1548 | Not-determined |
| NC1_vf_k141_425 | Not-determined |
| NC1_vf_k141_2434 | Not-determined |
| NC1_vf_k141_1984 | Not-determined |
| NC1_vf_k141_1315 | Not-determined |
| NC1_vf_k141_2234 | Not-determined |
| NC1_vf_k141_2872 | Not-determined |
| NC1_vf_k141_3345 | Not-determined |
| NC1_vf_k141_865 | Not-determined |
| NC1_vf_k141_3105 | Not-determined |
| NC1_vf_k141_2235 | Not-determined |
| NC1_vf_k141_1102 | Low-quality |
| NC1_vf_k141_1985 | Not-determined |
| NC1_vf_k141_3555 | Low-quality |
| NC1_vf_k141_1316 | Not-determined |
| NC1_vf_k141_426 | Not-determined |
| NC1_vf_k141_2873 | Not-determined |
| NC1_vf_k141_1549 | Not-determined |
| NC1_vf_k141_605 | Not-determined |
| NC1_vf_k141_2435 | Not-determined |
| NC1_vf_k141_1103 | Low-quality |
| NC1_vf_k141_866 | Not-determined |
| NC1_vf_k141_3346 | Not-determined |
| NC1_vf_k141_3106 | Not-determined |
| NC1_vf_k141_2236 | Not-determined |
| NC1_vf_k141_1986 | Not-determined |
| NC1_vf_k141_427 | Not-determined |
| NC1_vf_k141_3556 | Low-quality |
| NC1_vf_k141_1317 | Not-determined |
| NC1_vf_k141_1550 | Not-determined |
| NC1_vf_k141_606 | Not-determined |
| NC1_vf_k141_3107 | Not-determined |
| NC1_vf_k141_2670 | Low-quality |
| NC1_vf_k141_2237 | Low-quality |
| NC1_vf_k141_1104 | Not-determined |
| NC1_vf_k141_3557 | Not-determined |
| NC1_vf_k141_1318 | Not-determined |
| NC1_vf_k141_1987 | Not-determined |
| NC1_vf_k141_428 | Not-determined |
| NC1_vf_k141_2436 | Not-determined |
| NC1_vf_k141_607 | Not-determined |
| NC1_vf_k141_867 | Not-determined |
| NC1_vf_k141_1551 | Not-determined |
| NC1_vf_k141_3347 | Not-determined |
| NC1_vf_k141_2671 | Not-determined |
| NC1_vf_k141_3558 | Not-determined |
| NC1_vf_k141_3108 | Not-determined |
| NC1_vf_k141_429 | Low-quality |
| NC1_vf_k141_608 | Not-determined |
| NC1_vf_k141_868 | Not-determined |
| NC1_vf_k141_3348 | Not-determined |
| NC1_vf_k141_1105 | Not-determined |
| NC1_vf_k141_1319 | Not-determined |
| NC1_vf_k141_2238 | Not-determined |
| NC1_vf_k141_1552 | Not-determined |
| NC1_vf_k141_3559 | Low-quality |
| NC1_vf_k141_3109 | Not-determined |
| NC1_vf_k141_2437 | Not-determined |
| NC1_vf_k141_2672 | Not-determined |
| NC1_vf_k141_3349 | Not-determined |
| NC1_vf_k141_430 | Not-determined |
| NC1_vf_k141_1106 | Low-quality |
| NC1_vf_k141_609 | Not-determined |
| NC1_vf_k141_1320 | Not-determined |
| NC1_vf_k141_1988 | Not-determined |
| NC1_vf_k141_2874 | Low-quality |
| NC1_vf_k141_2239 | Not-determined |
| NC1_vf_k141_869 | Not-determined |
| NC1_vf_k141_431 | Not-determined |
| NC1_vf_k141_3110 | Not-determined |
| NC1_vf_k141_3560 | Low-quality |
| NC1_vf_k141_2673 | Not-determined |
| NC1_vf_k141_1107 | Not-determined |
| NC1_vf_k141_1553 | Not-determined |
| NC1_vf_k141_2438 | Not-determined |
| NC1_vf_k141_610 | Not-determined |
| NC1_vf_k141_1989 | Not-determined |
| NC1_vf_k141_2875 | Not-determined |
| NC1_vf_k141_2240 | Not-determined |
| NC1_vf_k141_3350 | Low-quality |
| NC1_vf_k141_432 | Not-determined |
| NC1_vf_k141_3561 | Low-quality |
| NC1_vf_k141_1108 | Not-determined |
| NC1_vf_k141_2674 | Not-determined |
| NC1_vf_k141_3111 | Not-determined |
| NC1_vf_k141_870 | Not-determined |
| NC1_vf_k141_611 | Not-determined |
| NC1_vf_k141_2241 | Not-determined |
| NC1_vf_k141_1554 | Not-determined |
| NC1_vf_k141_1321 | Not-determined |
| NC1_vf_k141_3562 | Not-determined |
| NC1_vf_k141_2439 | Not-determined |
| NC1_vf_k141_1990 | Not-determined |
| NC1_vf_k141_2876 | Not-determined |
| NC1_vf_k141_3351 | Not-determined |
| NC1_vf_k141_433 | Not-determined |
| NC1_vf_k141_1109 | Not-determined |
| NC1_vf_k141_2675 | Not-determined |
| NC1_vf_k141_2242 | Not-determined |
| NC1_vf_k141_871 | Not-determined |
| NC1_vf_k141_3563 | Not-determined |
| NC1_vf_k141_1322 | Not-determined |
| NC1_vf_k141_434 | Not-determined |
| NC1_vf_k141_1555 | Not-determined |
| NC1_vf_k141_2440 | Not-determined |
| NC1_vf_k141_612 | Not-determined |
| NC1_vf_k141_1991 | Not-determined |
| NC1_vf_k141_3352 | Not-determined |
| NC1_vf_k141_2243 | Not-determined |
| NC1_vf_k141_3564 | Not-determined |
| NC1_vf_k141_2877 | Not-determined |
| NC1_vf_k141_1110 | Not-determined |
| NC1_vf_k141_613 | Not-determined |
| NC1_vf_k141_2676 | Not-determined |
| NC1_vf_k141_1556 | Not-determined |
| NC1_vf_k141_435 | Not-determined |
| NC1_vf_k141_2244 | Not-determined |
| NC1_vf_k141_3565 | Not-determined |
| NC1_vf_k141_2441 | Not-determined |
| NC1_vf_k141_2878 | Not-determined |
| NC1_vf_k141_3353 | Not-determined |
| NC1_vf_k141_1111 | Not-determined |
| NC1_vf_k141_1323 | Not-determined |
| NC1_vf_k141_2677 | Not-determined |
| NC1_vf_k141_1992 | Not-determined |
| NC1_vf_k141_436 | Not-determined |
| NC1_vf_k141_614 | Not-determined |
| NC1_vf_k141_1557 | Not-determined |
| NC1_vf_k141_3566 | Not-determined |
| NC1_vf_k141_2245 | Not-determined |
| NC1_vf_k141_2442 | Not-determined |
| NC1_vf_k141_872 | Not-determined |
| NC1_vf_k141_2879 | Not-determined |
| NC1_vf_k141_1558 | Not-determined |
| NC1_vf_k141_2678 | Not-determined |
| NC1_vf_k141_3354 | Not-determined |
| NC1_vf_k141_437 | Not-determined |
| NC1_vf_k141_615 | Not-determined |
| NC1_vf_k141_1993 | Not-determined |
| NC1_vf_k141_3567 | Not-determined |
| NC1_vf_k141_2246 | Not-determined |
| NC1_vf_k141_1324 | Not-determined |
| NC1_vf_k141_873 | Not-determined |
| NC1_vf_k141_1112 | Not-determined |
| NC1_vf_k141_2443 | Not-determined |
| NC1_vf_k141_3355 | Not-determined |
| NC1_vf_k141_3568 | Not-determined |
| NC1_vf_k141_1559 | Not-determined |
| NC1_vf_k141_2880 | Not-determined |
| NC1_vf_k141_1113 | Not-determined |
| NC1_vf_k141_1994 | Not-determined |
| NC1_vf_k141_2679 | Not-determined |
| NC1_vf_k141_3569 | Not-determined |
| NC1_vf_k141_438 | Not-determined |
| NC1_vf_k141_3356 | Not-determined |
| NC1_vf_k141_874 | Not-determined |
| NC1_vf_k141_1995 | Not-determined |
| NC1_vf_k141_1114 | Not-determined |
| NC1_vf_k141_3570 | Not-determined |
| NC1_vf_k141_2680 | Not-determined |
| NC1_vf_k141_2881 | Not-determined |
| NC1_vf_k141_875 | Not-determined |
| NC1_vf_k141_439 | Not-determined |
| NC1_vf_k141_1996 | Not-determined |
| NC1_vf_k141_1115 | Not-determined |
| NC1_vf_k141_3571 | Not-determined |
| NC1_vf_k141_3357 | Not-determined |
| NC1_vf_k141_3112 | Low-quality |
| NC1_vf_k141_2444 | Not-determined |
| NC1_vf_k141_2882 | Not-determined |
| NC1_vf_k141_616 | Not-determined |
| NC1_vf_k141_876 | Not-determined |
| NC1_vf_k141_3572 | Not-determined |
| NC1_vf_k141_3358 | Not-determined |
| NC1_vf_k141_1997 | Not-determined |
| NC1_vf_k141_1116 | Not-determined |
| NC1_vf_k141_3113 | Not-determined |
| NC1_vf_k141_1325 | Not-determined |
| NC1_vf_k141_2883 | Not-determined |
| NC1_vf_k141_440 | Not-determined |
| NC1_vf_k141_2445 | Not-determined |
| NC1_vf_k141_1560 | Not-determined |
| NC1_vf_k141_3359 | Not-determined |
| NC1_vf_k141_617 | Not-determined |
| NC1_vf_k141_1117 | Not-determined |
| NC1_vf_k141_3573 | Not-determined |
| NC1_vf_k141_3114 | Not-determined |
| NC1_vf_k141_877 | Not-determined |
| NC1_vf_k141_1326 | Not-determined |
| NC1_vf_k141_2446 | Not-determined |
| NC1_vf_k141_1998 | Not-determined |
| NC1_vf_k141_441 | Not-determined |
| NC1_vf_k141_1118 | Not-determined |
| NC1_vf_k141_3360 | Not-determined |
| NC1_vf_k141_3115 | Not-determined |
| NC1_vf_k141_1561 | Not-determined |
| NC1_vf_k141_2447 | Not-determined |
| NC1_vf_k141_2884 | Not-determined |
| NC1_vf_k141_1327 | Not-determined |
| NC1_vf_k141_878 | Not-determined |
| NC1_vf_k141_1119 | Not-determined |
| NC1_vf_k141_2681 | Not-determined |
| NC1_vf_k141_3116 | Not-determined |
| NC1_vf_k141_618 | Not-determined |
| NC1_vf_k141_1999 | Not-determined |
| NC1_vf_k141_2885 | Not-determined |
| NC1_vf_k141_442 | Not-determined |
| NC1_vf_k141_1562 | Not-determined |
| NC1_vf_k141_3361 | Not-determined |
| NC1_vf_k141_879 | Not-determined |
| NC1_vf_k141_1120 | Not-determined |
| NC1_vf_k141_2682 | Not-determined |
| NC1_vf_k141_2448 | Not-determined |
| NC1_vf_k141_3574 | Not-determined |
| NC1_vf_k141_2000 | Not-determined |
| NC1_vf_k141_1328 | Not-determined |
| NC1_vf_k141_880 | Not-determined |
| NC1_vf_k141_2683 | Low-quality |
| NC1_vf_k141_1121 | Not-determined |
| NC1_vf_k141_619 | Not-determined |
| NC1_vf_k141_2449 | Not-determined |
| NC1_vf_k141_2001 | Not-determined |
| NC1_vf_k141_1563 | Not-determined |
| NC1_vf_k141_1329 | Not-determined |
| NC1_vf_k141_443 | Low-quality |
| NC1_vf_k141_3117 | Not-determined |
| NC1_vf_k141_1122 | Not-determined |
| NC1_vf_k141_2886 | Not-determined |
| NC1_vf_k141_2684 | Not-determined |
| NC1_vf_k141_881 | Not-determined |
| NC1_vf_k141_3575 | Low-quality |
| NC1_vf_k141_2450 | Not-determined |
| NC1_vf_k141_3362 | Not-determined |
| NC1_vf_k141_1564 | Not-determined |
| NC1_vf_k141_3118 | Not-determined |
| NC1_vf_k141_444 | Not-determined |
| NC1_vf_k141_2685 | Not-determined |
| NC1_vf_k141_3576 | Not-determined |
| NC1_vf_k141_2002 | Not-determined |
| NC1_vf_k141_2887 | Not-determined |
| NC1_vf_k141_1565 | Not-determined |
| NC1_vf_k141_2451 | Not-determined |
| NC1_vf_k141_620 | Not-determined |
| NC1_vf_k141_1123 | Not-determined |
| NC1_vf_k141_882 | Not-determined |
| NC1_vf_k141_3119 | Not-determined |
| NC1_vf_k141_1330 | Not-determined |
| NC1_vf_k141_2003 | Not-determined |
| NC1_vf_k141_1566 | Not-determined |
| NC1_vf_k141_2888 | Not-determined |
| NC1_vf_k141_2686 | Not-determined |
| NC1_vf_k141_3577 | Not-determined |
| NC1_vf_k141_445 | Not-determined |
| NC1_vf_k141_1124 | Not-determined |
| NC1_vf_k141_3120 | Not-determined |
| NC1_vf_k141_1331 | Not-determined |
| NC1_vf_k141_2452 | Not-determined |
| NC1_vf_k141_621 | Not-determined |
| NC1_vf_k141_2004 | Not-determined |
| NC1_vf_k141_883 | Not-determined |
| NC1_vf_k141_2687 | Not-determined |
| NC1_vf_k141_3578 | Not-determined |
| NC1_vf_k141_3363 | Not-determined |
| NC1_vf_k141_3121 | Not-determined |
| NC1_vf_k141_446 | Not-determined |
| NC1_vf_k141_2453 | Not-determined |
| NC1_vf_k141_1332 | Not-determined |
| NC1_vf_k141_2889 | Not-determined |
| NC1_vf_k141_2005 | Not-determined |
| NC1_vf_k141_2247 | Not-determined |
| NC1_vf_k141_622 | Not-determined |
| NC1_vf_k141_1567 | Low-quality |
| NC1_vf_k141_3579 | Not-determined |
| NC1_vf_k141_2688 | Low-quality |
| NC1_vf_k141_3122 | Not-determined |
| NC1_vf_k141_1333 | Not-determined |
| NC1_vf_k141_884 | Not-determined |
| NC1_vf_k141_2890 | Not-determined |
| NC1_vf_k141_447 | Not-determined |
| NC1_vf_k141_2006 | Low-quality |
| NC1_vf_k141_3580 | Low-quality |
| NC1_vf_k141_1568 | Low-quality |
| NC1_vf_k141_3123 | Not-determined |
| NC1_vf_k141_2689 | Not-determined |
| NC1_vf_k141_885 | Not-determined |
| NC1_vf_k141_623 | Not-determined |
| NC1_vf_k141_1334 | Not-determined |
| NC1_vf_k141_2454 | Not-determined |
| NC1_vf_k141_2248 | Not-determined |
| NC1_vf_k141_448 | Not-determined |
| NC1_vf_k141_1569 | Not-determined |
| NC1_vf_k141_886 | Not-determined |
| NC1_vf_k141_3581 | Not-determined |
| NC1_vf_k141_2690 | Not-determined |
| NC1_vf_k141_2249 | Not-determined |
| NC1_vf_k141_2891 | Not-determined |
| NC1_vf_k141_1335 | Not-determined |
| NC1_vf_k141_449 | Not-determined |
| NC1_vf_k141_624 | Not-determined |
| NC1_vf_k141_3124 | Not-determined |
| NC1_vf_k141_887 | Not-determined |
| NC1_vf_k141_2691 | Not-determined |
| NC1_vf_k141_3582 | Not-determined |
| NC1_vf_k141_1336 | Not-determined |
| NC1_vf_k141_2455 | Not-determined |
| NC1_vf_k141_2007 | Not-determined |
| NC1_vf_k141_3125 | Not-determined |
| NC1_vf_k141_625 | Not-determined |
| NC1_vf_k141_2892 | Not-determined |
| NC1_vf_k141_1570 | Not-determined |
| NC1_vf_k141_2692 | Not-determined |
| NC1_vf_k141_3583 | Not-determined |
| NC1_vf_k141_1337 | Not-determined |
| NC1_vf_k141_888 | Not-determined |
| NC1_vf_k141_2693 | Not-determined |
| NC1_vf_k141_2008 | Not-determined |
| NC1_vf_k141_1571 | Not-determined |
| NC1_vf_k141_3126 | Not-determined |
| NC1_vf_k141_3584 | Not-determined |
| NC1_vf_k141_626 | Not-determined |
| NC1_vf_k141_2893 | Not-determined |
| NC1_vf_k141_2456 | Not-determined |
| NC1_vf_k141_3364 | Not-determined |
| NC1_vf_k141_1338 | Not-determined |
| NC1_vf_k141_889 | Not-determined |
| NC1_vf_k141_2009 | Not-determined |
| NC1_vf_k141_2694 | Not-determined |
| NC1_vf_k141_3127 | Not-determined |
| NC1_vf_k141_2457 | Not-determined |
| NC1_vf_k141_3585 | Not-determined |
| NC1_vf_k141_2894 | Not-determined |
| NC1_vf_k141_627 | Not-determined |
| NC1_vf_k141_3365 | Not-determined |
| NC1_vf_k141_890 | Not-determined |
| NC1_vf_k141_1339 | Not-determined |
| NC1_vf_k141_2695 | Not-determined |
| NC1_vf_k141_2458 | Not-determined |
| NC1_vf_k141_628 | Not-determined |
| NC1_vf_k141_3586 | Not-determined |
| NC1_vf_k141_3128 | Not-determined |
| NC1_vf_k141_3366 | Not-determined |
| NC1_vf_k141_1572 | Not-determined |
| NC1_vf_k141_2696 | Not-determined |
| NC1_vf_k141_2895 | Not-determined |
| NC1_vf_k141_2010 | Low-quality |
| NC1_vf_k141_891 | Not-determined |
| NC1_vf_k141_3129 | Not-determined |
| NC1_vf_k141_2697 | Not-determined |
| NC1_vf_k141_1573 | Not-determined |
| NC1_vf_k141_1340 | Not-determined |
| NC1_vf_k141_3587 | Not-determined |
| NC1_vf_k141_3367 | Not-determined |
| NC1_vf_k141_3130 | Not-determined |
| NC1_vf_k141_2011 | Not-determined |
| NC1_vf_k141_2698 | Not-determined |
| NC1_vf_k141_629 | Not-determined |
| NC1_vf_k141_2896 | Not-determined |
| NC1_vf_k141_892 | Not-determined |
| NC1_vf_k141_1341 | Not-determined |
| NC1_vf_k141_3588 | Not-determined |
| NC1_vf_k141_3368 | Not-determined |
| NC1_vf_k141_2699 | Not-determined |
| NC1_vf_k141_3131 | Low-quality |
| NC1_vf_k141_2897 | Not-determined |
| NC1_vf_k141_1342 | Not-determined |
| NC1_vf_k141_3589 | Not-determined |
| NC1_vf_k141_2012 | Not-determined |
| NC1_vf_k141_630 | Not-determined |
| NC1_vf_k141_1574 | Not-determined |
| NC1_vf_k141_3590 | Not-determined |
| NC1_vf_k141_3132 | Not-determined |
| NC1_vf_k141_893 | Not-determined |
| NC1_vf_k141_3369 | Not-determined |
| NC1_vf_k141_2013 | Not-determined |
| NC1_vf_k141_631 | Not-determined |
| NC1_vf_k141_3370 | Not-determined |
| NC1_vf_k141_894 | Not-determined |
| NC1_vf_k141_2014 | Not-determined |
| NC1_vf_k141_3591 | Not-determined |
| NC1_vf_k141_2898 | Not-determined |
| NC1_vf_k141_1343 | Not-determined |
| NC1_vf_k141_3371 | Not-determined |
| NC1_vf_k141_895 | Not-determined |
| NC1_vf_k141_2015 | Not-determined |
| NC1_vf_k141_3592 | Not-determined |
| NC1_vf_k141_632 | Not-determined |
| NC1_vf_k141_3133 | Not-determined |
| NC1_vf_k141_2899 | Not-determined |
| NC1_vf_k141_3593 | Not-determined |
| NC1_vf_k141_1344 | Not-determined |
| NC1_vf_k141_2016 | Not-determined |
| NC1_vf_k141_633 | Not-determined |
| NC1_vf_k141_3134 | Not-determined |
| NC1_vf_k141_3372 | Not-determined |
| NC1_vf_k141_3594 | Not-determined |
| NC1_vf_k141_634 | Not-determined |
| NC1_vf_k141_2017 | Not-determined |
| NC1_vf_k141_896 | Low-quality |
| NC1_vf_k141_3373 | Not-determined |
| NC1_vf_k141_3135 | Not-determined |
| NC1_vf_k141_1345 | Not-determined |
| NC1_vf_k141_3595 | Not-determined |
| NC1_vf_k141_635 | Not-determined |
| NC1_vf_k141_2900 | Not-determined |
| NC1_vf_k141_3136 | Not-determined |
| NC1_vf_k141_2018 | Not-determined |
| NC1_vf_k141_897 | Not-determined |
| NC1_vf_k141_1346 | Not-determined |
| NC1_vf_k141_3137 | Not-determined |
| NC1_vf_k141_636 | Not-determined |
| NC1_vf_k141_2019 | Not-determined |
| NC1_vf_k141_2901 | Not-determined |
| NC1_vf_k141_1347 | Not-determined |
| NC1_vf_k141_898 | Not-determined |
| NC1_vf_k141_3596 | Not-determined |
| NC1_vf_k141_3138 | Not-determined |
| NC1_vf_k141_2020 | Not-determined |
| NC1_vf_k141_637 | Not-determined |
| NC1_vf_k141_2902 | Not-determined |
| NC1_vf_k141_899 | Not-determined |
| NC1_vf_k141_1348 | Not-determined |
| NC1_vf_k141_3597 | Low-quality |
| NC1_vf_k141_638 | Not-determined |
| NC1_vf_k141_2903 | Not-determined |
| NC1_vf_k141_1349 | Low-quality |
| NC1_vf_k141_3139 | Not-determined |
| NC1_vf_k141_2021 | Not-determined |
| NC1_vf_k141_3140 | Low-quality |
| NC1_vf_k141_639 | Not-determined |
| NC1_vf_k141_2904 | Not-determined |
| NC1_vf_k141_3141 | Not-determined |
| NC1_vf_k141_640 | Not-determined |
| NC1_vf_k141_2022 | Not-determined |
| NC1_vf_k141_2905 | Not-determined |
| NC1_vf_k141_2906 | Low-quality |
| NC1_vf_k141_2023 | Not-determined |
| NC1_vf_k141_3142 | Low-quality |
| NC1_vf_k141_2907 | Not-determined |
| NC1_vf_k141_641 | Not-determined |
| NC1_vf_k141_3143 | Not-determined |
| NC1_vf_k141_2908 | Not-determined |
| NC1_vf_k141_2024 | Not-determined |
| NC1_vf_k141_3144 | Not-determined |
| NC1_vf_k141_642 | Not-determined |
| NC1_vf_k141_2909 | Not-determined |
| NC1_vf_k141_3145 | Not-determined |
| NC1_vf_k141_2910 | Not-determined |
| NC1_vf_k141_643 | Not-determined |
| NC1_vf_k141_644 | Not-determined |
| NC1_vf_k141_3146 | Not-determined |
| NC1_vf_k141_2911 | Not-determined |
| NC1_vf_k141_645 | Not-determined |
| NC1_vf_k141_2912 | Not-determined |
| NC1_vf_k141_3147 | Not-determined |
| NC1_vf_k141_646 | Not-determined |
| NC1_vf_k141_2913 | Not-determined |
| NC1_vf_k141_3148 | Not-determined |
| NC1_vf_k141_2914 | Not-determined |
| NC1_vf_k141_647 | Not-determined |
| NC1_vf_k141_2915 | Not-determined |
| NC1_vf_k141_3149 | Not-determined |

| | |
|---|---|
| NC1_vf_k141_648 | Not-determined |
| NC1_vf_k141_2916 | Not-determined |
| NC1_vf_k141_649 | Not-determined |
| NC1_vf_k141_650 | Not-determined |
| NC1_vf_k141_2917 | Not-determined |
| NC1_vf_k141_651 | Not-determined |
| NC1_vf_k141_2918 | Not-determined |
| NC1_vf_k141_2919 | Not-determined |
| NC1_vf_k141_2920 | Low-quality |
| NC1_vf_k141_652 | Not-determined |
| NC1_vf_k141_2921 | Not-determined |
| NC1_vf_k141_653 | Not-determined |
| NC1_vf_k141_654 | Not-determined |
| NC1_vf_k141_2922 | Not-determined |
| NC1_vf_k141_2923 | Low-quality |
| NC1_vf_k141_655 | Low-quality |
| NC1_vf_k141_2924 | Not-determined |
| NC1_vf_k141_656 | Not-determined |
| NC1_vf_k141_657 | Not-determined |
| NC1_vf_k141_658 | Not-determined |
| NC1_vf_k141_659 | Low-quality |
| NC1_vf_k141_2459 | Low-quality |
| NC1_vf_k141_660 | Not-determined |
| NC1_vf_k141_2460 | Low-quality |
| NC1_vf_k141_661 | Not-determined |
| NC1_vf_k141_2461 | Not-determined |
| NC1_vf_k141_662 | Not-determined |
| NC1_vf_k141_663 | Not-determined |
| NC1_vf_k141_664 | Not-determined |
| NC1_vf_k141_665 | Not-determined |
| NC1_vf_k141_666 | Not-determined |
| NC1_vf_k141_667 | Not-determined |
| NC1_vf_k141_668 | Not-determined |
| NC1_vf_k141_669 | Not-determined |
| NC1_vf_k141_670 | Not-determined |
| NC1_vf_k141_671 | Not-determined |
| NC1_vf_k141_2462 | Not-determined |
| NC1_vf_k141_672 | Low-quality |
| NC1_vf_k141_2463 | Not-determined |
| NC1_vf_k141_673 | Not-determined |
| NC1_vf_k141_2464 | Not-determined |
| NC1_vf_k141_674 | Not-determined |
| NC1_vf_k141_2465 | Not-determined |
| NC1_vf_k141_2466 | Not-determined |
| NC1_vf_k141_2467 | Not-determined |
| NC1_vf_k141_2468 | Not-determined |
| NC1_vf_k141_2469 | Not-determined |
| NC1_vf_k141_2470 | Not-determined |
| NC1_vf_k141_2471 | Not-determined |
| NC1_vf_k141_2472 | Not-determined |
| NC1_vf_k141_2473 | Not-determined |
| NC1_vf_k141_2474 | Not-determined |
| NC1_vs_k141_902\|\|full | Not-determined |
| NC1_vs_k141_3157\|\|full | Not-determined |
| NC1_vs_k141_2258\|\|full | Low-quality |
| NC1_vs_k141_243\|\|full | Low-quality |
| NC1_vs_k141_1820\|\|full | Not-determined |
| NC1_vs_k141_2714\|\|full | Low-quality |
| NC1_vs_k141_2274\|\|full | Low-quality |
| NC1_vs_k141_2954\|\|full | Low-quality |
| NC1_vs_k141_1159\|\|full | Low-quality |
| NC1_vs_k141_956\|\|full | Low-quality |
| NC1_vs_k141_2742\|\|full | Low-quality |
| NC1_vs_k141_1402\|\|full | Low-quality |
| NC1_vs_k141_2747\|\|full | Low-quality |
| NC1_vs_k141_2552\|\|full | Not-determined |
| NC1_vs_k141_99\|\|full | Low-quality |
| NC1_vs_k141_2115\|\|full | Low-quality |
| NC1_vs_k141_2573\|\|full | Low-quality |
| NC1_vs_k141_120\|\|full | Low-quality |
| NC1_vs_k141_334\|\|full | Low-quality |
| NC1_vs_k141_126\|\|full | Not-determined |
| NC1_vs_k141_348\|\|full | Not-determined |
| NC1_vs_k141_354\|\|full | Not-determined |
| NC1_vs_k141_809\|\|full | Low-quality |
| NC1_vs_k141_3050\|\|full | Not-determined |
| NC1_vs_k141_818\|\|full | Not-determined |
| NC1_vs_k141_1748\|\|full | Low-quality |
| NC1_vs_k141_3290\|\|full | Low-quality |
| NC1_vs_k141_3065\|\|full | Not-determined |
| NC1_vs_k141_179\|\|full | Low-quality |
| NC1_vs_k141_1952\|\|full | Low-quality |
| NC1_vs_k141_1523\|\|full | Low-quality |
| NC1_vs_k141_1964\|\|full | Low-quality |
| NC1_vs_k141_586\|\|full | Low-quality |
| NC1_vs_k141_3325\|\|full | Low-quality |
| NC1_vs_k141_3545\|\|full | Low-quality |
| NC1_vs_k141_2855\|\|full | Not-determined |
| NC1_vs_k141_420\|\|full | Low-quality |
| NC1_vs_k141_1983\|\|full | Low-quality |
| NC1_vs_k141_1102\|\|full | Low-quality |
| NC1_vs_k141_3555\|\|full | Low-quality |
| NC1_vs_k141_3112\|\|full | Low-quality |
| NC1_vs_k141_3575\|\|full | Low-quality |
| NC1_vs_k141_3364\|\|full | Not-determined |
| NC1_vs_k141_2459\|\|full | Low-quality |
| NC1_vs_k141_3377\|\|lt2gene | Low-quality |
| NC1_vs_k141_454\|\|lt2gene | Low-quality |
| NC1_vs_k141_3379\|\|lt2gene | Low-quality |
| NC1_vs_k141_2931\|\|lt2gene | Low-quality |
| NC1_vs_k141_1357\|\|lt2gene | Low-quality |
| NC1_vs_k141_2940\|\|lt2gene | Low-quality |
| NC1_vs_k141_253\|\|lt2gene | Low-quality |
| NC1_vs_k141_1829\|\|lt2gene | Low-quality |
| NC1_vs_k141_2064\|\|lt2gene | Low-quality |
| NC1_vs_k141_3419\|\|lt2gene | Low-quality |
| NC1_vs_k141_2741\|\|lt2gene | Low-quality |
| NC1_vs_k141_277\|\|lt2gene | Low-quality |
| NC1_vs_k141_3217\|\|lt2gene | Low-quality |
| NC1_vs_k141_2754\|\|lt2gene | Not-determined |
| NC1_vs_k141_760\|\|lt2gene | Low-quality |
| NC1_vs_k141_530\|\|lt2gene | Low-quality |
| NC1_vs_k141_995\|\|lt2gene | Low-quality |
| NC1_vs_k141_3004\|\|lt2gene | Low-quality |
| NC1_vs_k141_320\|\|lt2gene | Low-quality |
| NC1_vs_k141_3467\|\|lt2gene | Low-quality |
| NC1_vs_k141_774\|\|lt2gene | Low-quality |
| NC1_vs_k141_3245\|\|lt2gene | Low-quality |
| NC1_vs_k141_2779\|\|lt2gene | Not-determined |
| NC1_vs_k141_2574\|\|lt2gene | Low-quality |
| NC1_vs_k141_1735\|\|lt2gene | Low-quality |
| NC1_vs_k141_2598\|\|lt2gene | Not-determined |
| NC1_vs_k141_158\|\|lt2gene | Low-quality |
| NC1_vs_k141_3501\|\|lt2gene | Not-determined |
| NC1_vs_k141_813\|\|lt2gene | Low-quality |
| NC1_vs_k141_3511\|\|lt2gene | Low-quality |
| NC1_vs_k141_1491\|\|lt2gene | Not-determined |
| NC1_vs_k141_384\|\|lt2gene | Not-determined |
| NC1_vs_k141_1516\|\|lt2gene | Low-quality |
| NC1_vs_k141_1522\|\|lt2gene | Low-quality |
| NC1_vs_k141_206\|\|lt2gene | Not-determined |
| NC1_vs_k141_1785\|\|lt2gene | Not-determined |
| NC1_vs_k141_2651\|\|lt2gene | Low-quality |
| NC1_vs_k141_224\|\|lt2gene | Low-quality |
| NC1_vs_k141_3103\|\|lt2gene | Low-quality |
| NC1_vs_k141_2871\|\|lt2gene | Low-quality |
| NC1_vs_k141_2237\|\|lt2gene | Low-quality |
| NC1_vs_k141_2688\|\|lt2gene | Low-quality |
| NC1_vs_k141_3582\|\|lt2gene | Not-determined |
| NC1_vs_k141_2460\|\|lt2gene | Low-quality |
| NC2_vf_NC2_1 | Not-determined |
| NC2_vf_NC2_2 | Not-determined |
| NC2_vf_NC2_3 | Not-determined |
| NC2_vf_NC2_4 | Not-determined |
| NC2_vf_NC2_5 | Not-determined |
| NC2_vf_NC2_6 | Not-determined |
| NC2_vf_NC2_7 | Low-quality |
| NC2_vf_NC2_8 | Low-quality |
| NC2_vf_NC2_9 | Not-determined |
| NC2_vf_NC2_10 | Low-quality |
| NC2_vf_NC2_11 | Not-determined |
| NC2_vf_NC2_12 | Low-quality |
| NC2_vf_NC2_13 | Not-determined |
| NC2_vf_NC2_14 | Not-determined |
| NC2_vf_NC2_15 | Low-quality |
| NC2_vf_NC2_16 | Not-determined |
| NC2_vf_NC2_17 | Not-determined |
| NC2_vf_NC2_18 | Low-quality |
| NC2_vf_NC2_19 | Not-determined |
| NC3_vf_k141_0 | Not-determined |
| NC3_vf_k141_532 | Not-determined |
| NC3_vf_k141_1596 | Not-determined |
| NC3_vf_k141_1862 | Not-determined |
| NC3_vf_k141_665 | Not-determined |
| NC3_vf_k141_133 | Not-determined |
| NC3_vf_k141_1463 | Not-determined |
| NC3_vf_k141_1 | Not-determined |
| NC3_vf_k141_1729 | Not-determined |
| NC3_vf_k141_266 | Not-determined |
| NC3_vf_k141_1995 | Not-determined |
| NC3_vf_k141_931 | Not-determined |
| NC3_vf_k141_1863 | Not-determined |
| NC3_vf_k141_399 | Not-determined |
| NC3_vf_k141_666 | Not-determined |
| NC3_vf_k141_2 | Low-quality |
| NC3_vf_k141_134 | Not-determined |
| NC3_vf_k141_533 | Not-determined |
| NC3_vf_k141_1064 | Not-determined |
| NC3_vf_k141_267 | Not-determined |
| NC3_vf_k141_1730 | Not-determined |
| NC3_vf_k141_1330 | Not-determined |
| NC3_vf_k141_1597 | Not-determined |
| NC3_vf_k141_1197 | Not-determined |
| NC3_vf_k141_1996 | Not-determined |
| NC3_vf_k141_1864 | Not-determined |
| NC3_vf_k141_400 | Not-determined |
| NC3_vf_k141_3 | Not-determined |
| NC3_vf_k141_932 | Not-determined |
| NC3_vf_k141_135 | Not-determined |
| NC3_vf_k141_534 | Low-quality |
| NC3_vf_k141_667 | Not-determined |
| NC3_vf_k141_268 | Not-determined |
| NC3_vf_k141_1464 | Not-determined |
| NC3_vf_k141_1598 | Not-determined |
| NC3_vf_k141_1997 | Not-determined |
| NC3_vf_k141_4 | Not-determined |
| NC3_vf_k141_1731 | Low-quality |
| NC3_vf_k141_1331 | Not-determined |
| NC3_vf_k141_1198 | Not-determined |
| NC3_vf_k141_269 | Not-determined |
| NC3_vf_k141_1465 | Not-determined |
| NC3_vf_k141_535 | Not-determined |
| NC3_vf_k141_1599 | Not-determined |
| NC3_vf_k141_668 | Not-determined |
| NC3_vf_k141_401 | Not-determined |
| NC3_vf_k141_5 | Not-determined |
| NC3_vf_k141_1865 | Not-determined |
| NC3_vf_k141_933 | Not-determined |
| NC3_vf_k141_1065 | Low-quality |
| NC3_vf_k141_798 | Not-determined |
| NC3_vf_k141_1466 | Not-determined |
| NC3_vf_k141_1199 | Not-determined |
| NC3_vf_k141_1732 | Not-determined |
| NC3_vf_k141_270 | Not-determined |
| NC3_vf_k141_6 | Not-determined |
| NC3_vf_k141_1998 | Low-quality |
| NC3_vf_k141_536 | Not-determined |
| NC3_vf_k141_402 | Not-determined |
| NC3_vf_k141_934 | Not-determined |
| NC3_vf_k141_669 | Not-determined |
| NC3_vf_k141_1733 | Not-determined |
| NC3_vf_k141_1467 | Not-determined |
| NC3_vf_k141_271 | Low-quality |
| NC3_vf_k141_799 | Not-determined |
| NC3_vf_k141_537 | Not-determined |
| NC3_vf_k141_7 | Not-determined |
| NC3_vf_k141_1468 | Not-determined |
| NC3_vf_k141_272 | Not-determined |
| NC3_vf_k141_1866 | Not-determined |
| NC3_vf_k141_1600 | Not-determined |
| NC3_vf_k141_800 | Low-quality |
| NC3_vf_k141_1734 | Not-determined |
| NC3_vf_k141_8 | Not-determined |
| NC3_vf_k141_538 | Not-determined |
| NC3_vf_k141_935 | Not-determined |
| NC3_vf_k141_403 | Not-determined |
| NC3_vf_k141_670 | Not-determined |
| NC3_vf_k141_273 | Not-determined |
| NC3_vf_k141_1867 | Not-determined |
| NC3_vf_k141_1200 | Low-quality |
| NC3_vf_k141_136 | Low-quality |
| NC3_vf_k141_9 | Not-determined |
| NC3_vf_k141_1735 | Not-determined |
| NC3_vf_k141_1066 | Low-quality |
| NC3_vf_k141_1999 | Low-quality |
| NC3_vf_k141_1868 | Not-determined |
| NC3_vf_k141_801 | Not-determined |
| NC3_vf_k141_1469 | Not-determined |
| NC3_vf_k141_137 | Not-determined |
| NC3_vf_k141_936 | Not-determined |
| NC3_vf_k141_10 | Not-determined |
| NC3_vf_k141_539 | Not-determined |
| NC3_vf_k141_274 | Not-determined |
| NC3_vf_k141_1201 | Not-determined |
| NC3_vf_k141_1601 | Not-determined |
| NC3_vf_k141_671 | Not-determined |
| NC3_vf_k141_1736 | Not-determined |
| NC3_vf_k141_404 | Not-determined |
| NC3_vf_k141_2000 | Not-determined |
| NC3_vf_k141_1737 | Not-determined |
| NC3_vf_k141_11 | Not-determined |
| NC3_vf_k141_540 | Not-determined |
| NC3_vf_k141_138 | Not-determined |
| NC3_vf_k141_672 | Not-determined |
| NC3_vf_k141_1332 | Low-quality |
| NC3_vf_k141_1869 | Not-determined |
| NC3_vf_k141_1202 | Not-determined |
| NC3_vf_k141_405 | Not-determined |
| NC3_vf_k141_1738 | Not-determined |
| NC3_vf_k141_1470 | Not-determined |
| NC3_vf_k141_139 | Not-determined |
| NC3_vf_k141_1602 | Not-determined |
| NC3_vf_k141_1870 | Not-determined |
| NC3_vf_k141_1739 | Not-determined |
| NC3_vf_k141_673 | Not-determined |
| NC3_vf_k141_541 | Not-determined |
| NC3_vf_k141_937 | Not-determined |
| NC3_vf_k141_1067 | Not-determined |
| NC3_vf_k141_1871 | Not-determined |
| NC3_vf_k141_1333 | Not-determined |
| NC3_vf_k141_12 | Not-determined |
| NC3_vf_k141_1471 | Not-determined |
| NC3_vf_k141_1203 | Not-determined |
| NC3_vf_k141_1603 | Not-determined |
| NC3_vf_k141_542 | Not-determined |
| NC3_vf_k141_674 | Not-determined |
| NC3_vf_k141_406 | Not-determined |
| NC3_vf_k141_938 | Not-determined |
| NC3_vf_k141_1068 | Not-determined |
| NC3_vf_k141_140 | Not-determined |
| NC3_vf_k141_13 | Not-determined |
| NC3_vf_k141_1872 | Low-quality |
| NC3_vf_k141_1740 | Not-determined |
| NC3_vf_k141_1472 | Not-determined |
| NC3_vf_k141_1204 | Not-determined |
| NC3_vf_k141_141 | Not-determined |
| NC3_vf_k141_1069 | Not-determined |
| NC3_vf_k141_1873 | Not-determined |
| NC3_vf_k141_407 | Not-determined |
| NC3_vf_k141_1334 | Not-determined |
| NC3_vf_k141_675 | Not-determined |
| NC3_vf_k141_1473 | Not-determined |
| NC3_vf_k141_1741 | Not-determined |
| NC3_vf_k141_543 | Not-determined |
| NC3_vf_k141_142 | Not-determined |
| NC3_vf_k141_1070 | Not-determined |
| NC3_vf_k141_802 | Low-quality |
| NC3_vf_k141_1874 | Not-determined |
| NC3_vf_k141_1205 | Not-determined |
| NC3_vf_k141_14 | Not-determined |
| NC3_vf_k141_1474 | Not-determined |
| NC3_vf_k141_1742 | Not-determined |
| NC3_vf_k141_1604 | Low-quality |
| NC3_vf_k141_544 | Not-determined |
| NC3_vf_k141_676 | Not-determined |
| NC3_vf_k141_15 | Not-determined |
| NC3_vf_k141_1875 | Not-determined |
| NC3_vf_k141_1071 | Low-quality |
| NC3_vf_k141_1743 | Not-determined |
| NC3_vf_k141_1475 | Not-determined |
| NC3_vf_k141_1335 | Not-determined |
| NC3_vf_k141_1605 | Not-determined |
| NC3_vf_k141_16 | Not-determined |
| NC3_vf_k141_408 | Not-determined |
| NC3_vf_k141_1876 | Not-determined |
| NC3_vf_k141_275 | Low-quality |
| NC3_vf_k141_1476 | Not-determined |
| NC3_vf_k141_545 | Not-determined |
| NC3_vf_k141_803 | Not-determined |
| NC3_vf_k141_1744 | Not-determined |
| NC3_vf_k141_17 | Not-determined |
| NC3_vf_k141_1606 | Not-determined |
| NC3_vf_k141_1477 | Low-quality |
| NC3_vf_k141_1072 | Not-determined |
| NC3_vf_k141_276 | Not-determined |
| NC3_vf_k141_1336 | Not-determined |
| NC3_vf_k141_1877 | Not-determined |
| NC3_vf_k141_677 | Not-determined |
| NC3_vf_k141_1607 | Not-determined |
| NC3_vf_k141_1478 | Not-determined |
| NC3_vf_k141_18 | Not-determined |
| NC3_vf_k141_1745 | Not-determined |
| NC3_vf_k141_409 | Not-determined |
| NC3_vf_k141_1337 | Not-determined |
| NC3_vf_k141_277 | Not-determined |
| NC3_vf_k141_1608 | Not-determined |
| NC3_vf_k141_1073 | Not-determined |
| NC3_vf_k141_1878 | Not-determined |
| NC3_vf_k141_143 | Not-determined |
| NC3_vf_k141_19 | Not-determined |
| NC3_vf_k141_1479 | Not-determined |
| NC3_vf_k141_278 | Not-determined |
| NC3_vf_k141_410 | Not-determined |
| NC3_vf_k141_1879 | Not-determined |
| NC3_vf_k141_1074 | Not-determined |
| NC3_vf_k141_1746 | Not-determined |
| NC3_vf_k141_678 | Not-determined |
| NC3_vf_k141_804 | Low-quality |
| NC3_vf_k141_546 | Not-determined |
| NC3_vf_k141_279 | Not-determined |
| NC3_vf_k141_1880 | Not-determined |
| NC3_vf_k141_144 | Not-determined |
| NC3_vf_k141_1609 | Not-determined |
| NC3_vf_k141_1747 | Not-determined |
| NC3_vf_k141_411 | Not-determined |
| NC3_vf_k141_1480 | Not-determined |
| NC3_vf_k141_1338 | Low-quality |
| NC3_vf_k141_679 | Not-determined |
| NC3_vf_k141_20 | Not-determined |
| NC3_vf_k141_805 | Not-determined |
| NC3_vf_k141_1881 | Not-determined |
| NC3_vf_k141_145 | Not-determined |
| NC3_vf_k141_1075 | Not-determined |
| NC3_vf_k141_1748 | Not-determined |
| NC3_vf_k141_1610 | Not-determined |
| NC3_vf_k141_547 | Low-quality |
| NC3_vf_k141_412 | Not-determined |
| NC3_vf_k141_1481 | Low-quality |
| NC3_vf_k141_146 | Not-determined |
| NC3_vf_k141_1882 | Not-determined |
| NC3_vf_k141_939 | Low-quality |
| NC3_vf_k141_280 | Low-quality |
| NC3_vf_k141_2001 | Not-determined |
| NC3_vf_k141_1339 | Not-determined |
| NC3_vf_k141_21 | Not-determined |
| NC3_vf_k141_1611 | Not-determined |
| NC3_vf_k141_1076 | Not-determined |
| NC3_vf_k141_680 | Not-determined |
| NC3_vf_k141_806 | Not-determined |
| NC3_vf_k141_940 | Not-determined |
| NC3_vf_k141_1749 | Not-determined |
| NC3_vf_k141_1482 | Not-determined |
| NC3_vf_k141_1883 | Not-determined |
| NC3_vf_k141_413 | Not-determined |
| NC3_vf_k141_548 | Not-determined |
| NC3_vf_k141_2002 | Low-quality |
| NC3_vf_k141_1612 | Not-determined |
| NC3_vf_k141_147 | Low-quality |
| NC3_vf_k141_22 | Not-determined |
| NC3_vf_k141_1884 | Not-determined |
| NC3_vf_k141_1483 | Not-determined |
| NC3_vf_k141_549 | Not-determined |
| NC3_vf_k141_2003 | Not-determined |
| NC3_vf_k141_1077 | Not-determined |
| NC3_vf_k141_1750 | Not-determined |
| NC3_vf_k141_148 | Not-determined |
| NC3_vf_k141_1613 | Not-determined |
| NC3_vf_k141_941 | Not-determined |
| NC3_vf_k141_807 | Not-determined |
| NC3_vf_k141_1340 | Low-quality |
| NC3_vf_k141_681 | Not-determined |
| NC3_vf_k141_23 | Not-determined |
| NC3_vf_k141_281 | Low-quality |
| NC3_vf_k141_1484 | Not-determined |
| NC3_vf_k141_550 | Not-determined |
| NC3_vf_k141_414 | Not-determined |
| NC3_vf_k141_1078 | Not-determined |
| NC3_vf_k141_2004 | Not-determined |
| NC3_vf_k141_942 | Not-determined |
| NC3_vf_k141_1751 | Not-determined |
| NC3_vf_k141_1885 | Not-determined |
| NC3_vf_k141_1485 | Not-determined |
| NC3_vf_k141_1341 | Not-determined |
| NC3_vf_k141_808 | Not-determined |
| NC3_vf_k141_282 | Not-determined |
| NC3_vf_k141_1752 | Not-determined |
| NC3_vf_k141_2005 | Not-determined |
| NC3_vf_k141_1614 | Not-determined |
| NC3_vf_k141_1486 | Not-determined |
| NC3_vf_k141_943 | Not-determined |
| NC3_vf_k141_283 | Not-determined |
| NC3_vf_k141_24 | Low-quality |
| NC3_vf_k141_415 | Not-determined |
| NC3_vf_k141_1753 | Not-determined |
| NC3_vf_k141_2006 | Not-determined |
| NC3_vf_k141_551 | Not-determined |
| NC3_vf_k141_1342 | Not-determined |
| NC3_vf_k141_682 | Not-determined |
| NC3_vf_k141_1886 | Low-quality |
| NC3_vf_k141_1079 | Low-quality |
| NC3_vf_k141_944 | Not-determined |
| NC3_vf_k141_1754 | Not-determined |
| NC3_vf_k141_809 | Low-quality |
| NC3_vf_k141_1487 | Not-determined |
| NC3_vf_k141_1615 | Not-determined |
| NC3_vf_k141_552 | Not-determined |
| NC3_vf_k141_25 | Low-quality |
| NC3_vf_k141_1080 | Not-determined |
| NC3_vf_k141_1488 | Not-determined |
| NC3_vf_k141_2007 | Not-determined |
| NC3_vf_k141_810 | Not-determined |
| NC3_vf_k141_945 | Not-determined |
| NC3_vf_k141_149 | Not-determined |
| NC3_vf_k141_553 | Not-determined |
| NC3_vf_k141_416 | Not-determined |
| NC3_vf_k141_1887 | Not-determined |
| NC3_vf_k141_2008 | Not-determined |
| NC3_vf_k141_1343 | Not-determined |
| NC3_vf_k141_683 | Not-determined |
| NC3_vf_k141_1081 | Not-determined |
| NC3_vf_k141_26 | Not-determined |
| NC3_vf_k141_1489 | Not-determined |
| NC3_vf_k141_1616 | Not-determined |
| NC3_vf_k141_1755 | Not-determined |
| NC3_vf_k141_811 | Low-quality |
| NC3_vf_k141_150 | Not-determined |
| NC3_vf_k141_1888 | Not-determined |
| NC3_vf_k141_1082 | Not-determined |
| NC3_vf_k141_554 | Not-determined |
| NC3_vf_k141_2009 | Not-determined |
| NC3_vf_k141_1490 | Not-determined |
| NC3_vf_k141_684 | Not-determined |
| NC3_vf_k141_284 | Low-quality |
| NC3_vf_k141_1756 | Not-determined |
| NC3_vf_k141_27 | Not-determined |
| NC3_vf_k141_1617 | Not-determined |
| NC3_vf_k141_1344 | Not-determined |
| NC3_vf_k141_946 | Low-quality |
| NC3_vf_k141_812 | Not-determined |
| NC3_vf_k141_1083 | Not-determined |
| NC3_vf_k141_555 | Not-determined |
| NC3_vf_k141_1491 | Not-determined |
| NC3_vf_k141_1757 | Not-determined |
| NC3_vf_k141_151 | Not-determined |
| NC3_vf_k141_28 | Not-determined |
| NC3_vf_k141_1345 | Not-determined |
| NC3_vf_k141_1618 | Not-determined |
| NC3_vf_k141_2010 | Not-determined |
| NC3_vf_k141_1889 | Not-determined |
| NC3_vf_k141_1084 | Not-determined |
| NC3_vf_k141_685 | Not-determined |
| NC3_vf_k141_285 | Not-determined |
| NC3_vf_k141_556 | Not-determined |
| NC3_vf_k141_152 | Not-determined |
| NC3_vf_k141_1758 | Not-determined |
| NC3_vf_k141_813 | Not-determined |
| NC3_vf_k141_286 | Not-determined |
| NC3_vf_k141_1890 | Low-quality |
| NC3_vf_k141_29 | Not-determined |
| NC3_vf_k141_1346 | Low-quality |
| NC3_vf_k141_1492 | Not-determined |
| NC3_vf_k141_686 | Not-determined |
| NC3_vf_k141_153 | Not-determined |
| NC3_vf_k141_1619 | Not-determined |
| NC3_vf_k141_2011 | Not-determined |
| NC3_vf_k141_1891 | Not-determined |
| NC3_vf_k141_287 | Not-determined |
| NC3_vf_k141_30 | Not-determined |
| NC3_vf_k141_417 | Not-determined |
| NC3_vf_k141_1085 | Not-determined |
| NC3_vf_k141_1347 | Not-determined |
| NC3_vf_k141_947 | Not-determined |
| NC3_vf_k141_1493 | Not-determined |
| NC3_vf_k141_154 | Not-determined |
| NC3_vf_k141_814 | Not-determined |
| NC3_vf_k141_1620 | Not-determined |
| NC3_vf_k141_1892 | Not-determined |
| NC3_vf_k141_687 | Not-determined |
| NC3_vf_k141_1494 | Not-determined |
| NC3_vf_k141_1348 | Not-determined |
| NC3_vf_k141_557 | Not-determined |
| NC3_vf_k141_1759 | Not-determined |
| NC3_vf_k141_1086 | Not-determined |
| NC3_vf_k141_288 | Low-quality |
| NC3_vf_k141_1893 | Not-determined |
| NC3_vf_k141_31 | Not-determined |
| NC3_vf_k141_155 | Not-determined |
| NC3_vf_k141_1621 | Low-quality |
| NC3_vf_k141_558 | Not-determined |
| NC3_vf_k141_1760 | Not-determined |
| NC3_vf_k141_688 | Not-determined |
| NC3_vf_k141_1349 | Not-determined |
| NC3_vf_k141_1894 | Not-determined |
| NC3_vf_k141_289 | Not-determined |
| NC3_vf_k141_156 | Not-determined |
| NC3_vf_k141_1622 | Not-determined |
| NC3_vf_k141_1761 | Not-determined |
| NC3_vf_k141_559 | Not-determined |
| NC3_vf_k141_948 | Low-quality |
| NC3_vf_k141_1495 | Not-determined |
| NC3_vf_k141_2012 | Not-determined |
| NC3_vf_k141_32 | Not-determined |
| NC3_vf_k141_1350 | Not-determined |
| NC3_vf_k141_1762 | Not-determined |
| NC3_vf_k141_1895 | Not-determined |
| NC3_vf_k141_560 | Not-determined |
| NC3_vf_k141_157 | Not-determined |
| NC3_vf_k141_815 | Low-quality |
| NC3_vf_k141_1087 | Not-determined |
| NC3_vf_k141_33 | Not-determined |
| NC3_vf_k141_2013 | Not-determined |
| NC3_vf_k141_418 | Not-determined |
| NC3_vf_k141_290 | Not-determined |
| NC3_vf_k141_689 | Low-quality |
| NC3_vf_k141_158 | Not-determined |
| NC3_vf_k141_1623 | Low-quality |
| NC3_vf_k141_1351 | Not-determined |
| NC3_vf_k141_561 | Not-determined |
| NC3_vf_k141_1088 | Not-determined |
| NC3_vf_k141_2014 | Not-determined |
| NC3_vf_k141_949 | Not-determined |
| NC3_vf_k141_1896 | Not-determined |
| NC3_vf_k141_34 | Not-determined |
| NC3_vf_k141_159 | Not-determined |
| NC3_vf_k141_816 | Not-determined |
| NC3_vf_k141_1624 | Not-determined |
| NC3_vf_k141_1763 | Not-determined |
| NC3_vf_k141_1352 | Not-determined |
| NC3_vf_k141_419 | Not-determined |
| NC3_vf_k141_2015 | Not-determined |
| NC3_vf_k141_1089 | Not-determined |
| NC3_vf_k141_1897 | Not-determined |
| NC3_vf_k141_950 | Not-determined |
| NC3_vf_k141_562 | Not-determined |
| NC3_vf_k141_817 | Not-determined |
| NC3_vf_k141_1090 | Not-determined |
| NC3_vf_k141_1764 | Not-determined |
| NC3_vf_k141_35 | Low-quality |
| NC3_vf_k141_2016 | Not-determined |
| NC3_vf_k141_291 | Low-quality |
| NC3_vf_k141_1898 | Not-determined |
| NC3_vf_k141_160 | Not-determined |
| NC3_vf_k141_1353 | Not-determined |
| NC3_vf_k141_690 | Not-determined |
| NC3_vf_k141_1091 | Not-determined |
| NC3_vf_k141_420 | Not-determined |
| NC3_vf_k141_36 | Not-determined |
| NC3_vf_k141_2017 | Not-determined |
| NC3_vf_k141_951 | Not-determined |

| | |
|---|---|
| NC3_vf_k141_818 | Not-determined |
| NC3_vf_k141_161 | Not-determined |
| NC3_vf_k141_691 | Not-determined |
| NC3_vf_k141_1206 | Low-quality |
| NC3_vf_k141_37 | Not-determined |
| NC3_vf_k141_1625 | Not-determined |
| NC3_vf_k141_563 | Low-quality |
| NC3_vf_k141_1354 | Not-determined |
| NC3_vf_k141_1496 | Low-quality |
| NC3_vf_k141_819 | Not-determined |
| NC3_vf_k141_2018 | Not-determined |
| NC3_vf_k141_162 | Not-determined |
| NC3_vf_k141_952 | Not-determined |
| NC3_vf_k141_1092 | Not-determined |
| NC3_vf_k141_292 | Not-determined |
| NC3_vf_k141_1207 | Not-determined |
| NC3_vf_k141_692 | Not-determined |
| NC3_vf_k141_1765 | Not-determined |
| NC3_vf_k141_1626 | Low-quality |
| NC3_vf_k141_564 | Not-determined |
| NC3_vf_k141_163 | Low-quality |
| NC3_vf_k141_2019 | Not-determined |
| NC3_vf_k141_820 | Not-determined |
| NC3_vf_k141_693 | Not-determined |
| NC3_vf_k141_565 | Not-determined |
| NC3_vf_k141_38 | Not-determined |
| NC3_vf_k141_1093 | Not-determined |
| NC3_vf_k141_2020 | Not-determined |
| NC3_vf_k141_1497 | Not-determined |
| NC3_vf_k141_1355 | Not-determined |
| NC3_vf_k141_164 | Not-determined |
| NC3_vf_k141_1208 | Not-determined |
| NC3_vf_k141_1766 | Not-determined |
| NC3_vf_k141_1899 | Low-quality |
| NC3_vf_k141_1627 | Not-determined |
| NC3_vf_k141_293 | Not-determined |
| NC3_vf_k141_2021 | Not-determined |
| NC3_vf_k141_1498 | Not-determined |
| NC3_vf_k141_39 | Not-determined |
| NC3_vf_k141_953 | Not-determined |
| NC3_vf_k141_165 | Not-determined |
| NC3_vf_k141_694 | Not-determined |
| NC3_vf_k141_1356 | Not-determined |
| NC3_vf_k141_294 | Not-determined |
| NC3_vf_k141_1767 | Not-determined |
| NC3_vf_k141_1628 | Not-determined |
| NC3_vf_k141_821 | Not-determined |
| NC3_vf_k141_2022 | Not-determined |
| NC3_vf_k141_1094 | Not-determined |
| NC3_vf_k141_1209 | Not-determined |
| NC3_vf_k141_1499 | Not-determined |
| NC3_vf_k141_166 | Not-determined |
| NC3_vf_k141_40 | Not-determined |
| NC3_vf_k141_1900 | Not-determined |
| NC3_vf_k141_954 | Not-determined |
| NC3_vf_k141_695 | Not-determined |
| NC3_vf_k141_1629 | Not-determined |
| NC3_vf_k141_1095 | Not-determined |
| NC3_vf_k141_1500 | Not-determined |
| NC3_vf_k141_167 | Not-determined |
| NC3_vf_k141_1768 | Low-quality |
| NC3_vf_k141_566 | Not-determined |
| NC3_vf_k141_822 | Low-quality |
| NC3_vf_k141_1630 | Not-determined |
| NC3_vf_k141_2023 | Not-determined |
| NC3_vf_k141_955 | Low-quality |
| NC3_vf_k141_1901 | Not-determined |
| NC3_vf_k141_295 | Not-determined |
| NC3_vf_k141_1096 | Not-determined |
| NC3_vf_k141_1357 | Not-determined |
| NC3_vf_k141_696 | Not-determined |
| NC3_vf_k141_567 | Not-determined |
| NC3_vf_k141_823 | Not-determined |
| NC3_vf_k141_1097 | Not-determined |
| NC3_vf_k141_1210 | Low-quality |
| NC3_vf_k141_2024 | Not-determined |
| NC3_vf_k141_1631 | Not-determined |
| NC3_vf_k141_568 | Not-determined |
| NC3_vf_k141_1358 | Not-determined |
| NC3_vf_k141_1769 | Low-quality |
| NC3_vf_k141_697 | Low-quality |
| NC3_vf_k141_1902 | Not-determined |
| NC3_vf_k141_1098 | Not-determined |
| NC3_vf_k141_1632 | Not-determined |
| NC3_vf_k141_1501 | Not-determined |
| NC3_vf_k141_296 | Not-determined |
| NC3_vf_k141_824 | Not-determined |
| NC3_vf_k141_1211 | Not-determined |
| NC3_vf_k141_41 | Not-determined |
| NC3_vf_k141_1903 | Low-quality |
| NC3_vf_k141_1099 | Not-determined |
| NC3_vf_k141_1502 | Not-determined |
| NC3_vf_k141_168 | Not-determined |
| NC3_vf_k141_297 | Not-determined |
| NC3_vf_k141_421 | Low-quality |
| NC3_vf_k141_569 | Not-determined |
| NC3_vf_k141_698 | Not-determined |
| NC3_vf_k141_825 | Not-determined |
| NC3_vf_k141_2025 | Low-quality |
| NC3_vf_k141_1359 | Not-determined |
| NC3_vf_k141_1503 | Not-determined |
| NC3_vf_k141_169 | Not-determined |
| NC3_vf_k141_422 | Not-determined |
| NC3_vf_k141_570 | Not-determined |
| NC3_vf_k141_1100 | Not-determined |
| NC3_vf_k141_956 | Low-quality |
| NC3_vf_k141_2026 | Not-determined |
| NC3_vf_k141_699 | Not-determined |
| NC3_vf_k141_1770 | Not-determined |
| NC3_vf_k141_170 | Not-determined |
| NC3_vf_k141_1360 | Not-determined |
| NC3_vf_k141_826 | Not-determined |
| NC3_vf_k141_1633 | Low-quality |
| NC3_vf_k141_2027 | Not-determined |
| NC3_vf_k141_423 | Not-determined |
| NC3_vf_k141_298 | Low-quality |
| NC3_vf_k141_571 | Not-determined |
| NC3_vf_k141_1101 | Not-determined |
| NC3_vf_k141_171 | Not-determined |
| NC3_vf_k141_1771 | Low-quality |
| NC3_vf_k141_1361 | Not-determined |
| NC3_vf_k141_42 | Not-determined |
| NC3_vf_k141_700 | Not-determined |
| NC3_vf_k141_1504 | Not-determined |
| NC3_vf_k141_2028 | Not-determined |
| NC3_vf_k141_1102 | Not-determined |
| NC3_vf_k141_572 | Not-determined |
| NC3_vf_k141_1634 | Not-determined |
| NC3_vf_k141_172 | Low-quality |
| NC3_vf_k141_43 | Not-determined |
| NC3_vf_k141_1772 | Not-determined |
| NC3_vf_k141_1505 | Not-determined |
| NC3_vf_k141_424 | Not-determined |
| NC3_vf_k141_827 | Not-determined |
| NC3_vf_k141_573 | Not-determined |
| NC3_vf_k141_1362 | Not-determined |
| NC3_vf_k141_1103 | Not-determined |
| NC3_vf_k141_173 | Not-determined |
| NC3_vf_k141_701 | Not-determined |
| NC3_vf_k141_44 | Not-determined |
| NC3_vf_k141_1635 | Not-determined |
| NC3_vf_k141_1506 | Not-determined |
| NC3_vf_k141_2029 | Not-determined |
| NC3_vf_k141_425 | Low-quality |
| NC3_vf_k141_174 | Not-determined |
| NC3_vf_k141_957 | Low-quality |
| NC3_vf_k141_1104 | Not-determined |
| NC3_vf_k141_1212 | Not-determined |
| NC3_vf_k141_1363 | Not-determined |
| NC3_vf_k141_1773 | Not-determined |
| NC3_vf_k141_2030 | Low-quality |
| NC3_vf_k141_702 | Not-determined |
| NC3_vf_k141_1636 | Not-determined |
| NC3_vf_k141_574 | Not-determined |
| NC3_vf_k141_299 | Not-determined |
| NC3_vf_k141_1507 | Low-quality |
| NC3_vf_k141_426 | Not-determined |
| NC3_vf_k141_828 | Low-quality |
| NC3_vf_k141_1105 | Not-determined |
| NC3_vf_k141_958 | Not-determined |
| NC3_vf_k141_175 | Not-determined |
| NC3_vf_k141_1774 | Low-quality |
| NC3_vf_k141_1637 | Not-determined |
| NC3_vf_k141_300 | Not-determined |
| NC3_vf_k141_575 | Low-quality |
| NC3_vf_k141_1508 | Not-determined |
| NC3_vf_k141_1213 | Low-quality |
| NC3_vf_k141_2031 | Low-quality |
| NC3_vf_k141_1364 | Not-determined |
| NC3_vf_k141_301 | Low-quality |
| NC3_vf_k141_1638 | Not-determined |
| NC3_vf_k141_45 | Not-determined |
| NC3_vf_k141_959 | Not-determined |
| NC3_vf_k141_176 | Not-determined |
| NC3_vf_k141_703 | Low-quality |
| NC3_vf_k141_1214 | Not-determined |
| NC3_vf_k141_576 | Not-determined |
| NC3_vf_k141_1639 | Not-determined |
| NC3_vf_k141_302 | Not-determined |
| NC3_vf_k141_427 | Low-quality |
| NC3_vf_k141_1365 | Not-determined |
| NC3_vf_k141_177 | Not-determined |
| NC3_vf_k141_1775 | Not-determined |
| NC3_vf_k141_1509 | Low-quality |
| NC3_vf_k141_829 | Low-quality |
| NC3_vf_k141_1106 | Low-quality |
| NC3_vf_k141_2032 | Not-determined |
| NC3_vf_k141_1215 | Not-determined |
| NC3_vf_k141_1510 | Not-determined |
| NC3_vf_k141_46 | Not-determined |
| NC3_vf_k141_428 | Not-determined |
| NC3_vf_k141_704 | Low-quality |
| NC3_vf_k141_960 | Not-determined |
| NC3_vf_k141_1776 | Low-quality |
| NC3_vf_k141_1366 | Not-determined |
| NC3_vf_k141_577 | Not-determined |
| NC3_vf_k141_830 | Not-determined |
| NC3_vf_k141_178 | Not-determined |
| NC3_vf_k141_47 | Low-quality |
| NC3_vf_k141_1511 | Not-determined |
| NC3_vf_k141_1777 | Not-determined |
| NC3_vf_k141_1640 | Low-quality |
| NC3_vf_k141_1904 | Not-determined |
| NC3_vf_k141_1107 | Low-quality |
| NC3_vf_k141_429 | Not-determined |
| NC3_vf_k141_578 | Not-determined |
| NC3_vf_k141_1367 | Not-determined |
| NC3_vf_k141_1778 | Not-determined |
| NC3_vf_k141_1512 | Not-determined |
| NC3_vf_k141_831 | Not-determined |
| NC3_vf_k141_179 | Not-determined |
| NC3_vf_k141_1216 | Not-determined |
| NC3_vf_k141_961 | Not-determined |
| NC3_vf_k141_1905 | Low-quality |
| NC3_vf_k141_48 | Not-determined |
| NC3_vf_k141_303 | Low-quality |
| NC3_vf_k141_430 | Not-determined |
| NC3_vf_k141_705 | Not-determined |
| NC3_vf_k141_579 | Not-determined |
| NC3_vf_k141_180 | Not-determined |
| NC3_vf_k141_2033 | Not-determined |
| NC3_vf_k141_832 | Not-determined |
| NC3_vf_k141_1108 | Not-determined |
| NC3_vf_k141_49 | Low-quality |
| NC3_vf_k141_1368 | Not-determined |
| NC3_vf_k141_304 | Not-determined |
| NC3_vf_k141_962 | Not-determined |
| NC3_vf_k141_1906 | Not-determined |
| NC3_vf_k141_431 | Not-determined |
| NC3_vf_k141_1217 | Low-quality |
| NC3_vf_k141_181 | Not-determined |
| NC3_vf_k141_1641 | Not-determined |
| NC3_vf_k141_2034 | Not-determined |
| NC3_vf_k141_1907 | Not-determined |
| NC3_vf_k141_50 | Not-determined |
| NC3_vf_k141_833 | Not-determined |
| NC3_vf_k141_305 | Not-determined |
| NC3_vf_k141_706 | Not-determined |
| NC3_vf_k141_963 | Not-determined |
| NC3_vf_k141_432 | Not-determined |
| NC3_vf_k141_1218 | Not-determined |
| NC3_vf_k141_2035 | Not-determined |
| NC3_vf_k141_306 | Not-determined |
| NC3_vf_k141_1908 | Not-determined |
| NC3_vf_k141_1779 | Not-determined |
| NC3_vf_k141_580 | Not-determined |
| NC3_vf_k141_182 | Not-determined |
| NC3_vf_k141_51 | Not-determined |
| NC3_vf_k141_1109 | Not-determined |
| NC3_vf_k141_834 | Not-determined |
| NC3_vf_k141_1909 | Not-determined |
| NC3_vf_k141_1642 | Not-determined |
| NC3_vf_k141_307 | Not-determined |
| NC3_vf_k141_433 | Not-determined |
| NC3_vf_k141_1369 | Not-determined |
| NC3_vf_k141_183 | Not-determined |
| NC3_vf_k141_1780 | Not-determined |
| NC3_vf_k141_1110 | Not-determined |
| NC3_vf_k141_1643 | Not-determined |
| NC3_vf_k141_2036 | Low-quality |
| NC3_vf_k141_184 | Not-determined |
| NC3_vf_k141_1219 | Not-determined |
| NC3_vf_k141_1781 | Not-determined |
| NC3_vf_k141_52 | Not-determined |
| NC3_vf_k141_581 | Not-determined |
| NC3_vf_k141_434 | Not-determined |
| NC3_vf_k141_835 | Not-determined |
| NC3_vf_k141_1910 | Not-determined |
| NC3_vf_k141_185 | Not-determined |
| NC3_vf_k141_1782 | Not-determined |
| NC3_vf_k141_1644 | Not-determined |
| NC3_vf_k141_435 | Not-determined |
| NC3_vf_k141_1220 | Not-determined |
| NC3_vf_k141_53 | Not-determined |
| NC3_vf_k141_1370 | Low-quality |
| NC3_vf_k141_2037 | Not-determined |
| NC3_vf_k141_186 | Not-determined |
| NC3_vf_k141_1111 | Not-determined |
| NC3_vf_k141_1911 | Not-determined |
| NC3_vf_k141_836 | Not-determined |
| NC3_vf_k141_2038 | Low-quality |
| NC3_vf_k141_54 | Not-determined |
| NC3_vf_k141_1221 | Not-determined |
| NC3_vf_k141_1783 | Not-determined |
| NC3_vf_k141_1645 | Not-determined |
| NC3_vf_k141_582 | Low-quality |
| NC3_vf_k141_436 | Not-determined |
| NC3_vf_k141_1112 | Not-determined |
| NC3_vf_k141_1371 | Not-determined |
| NC3_vf_k141_837 | Not-determined |
| NC3_vf_k141_55 | Not-determined |
| NC3_vf_k141_1646 | Low-quality |
| NC3_vf_k141_1912 | Not-determined |
| NC3_vf_k141_1784 | Not-determined |
| NC3_vf_k141_707 | Low-quality |
| NC3_vf_k141_437 | Not-determined |
| NC3_vf_k141_56 | Not-determined |
| NC3_vf_k141_583 | Not-determined |
| NC3_vf_k141_1372 | Not-determined |
| NC3_vf_k141_1222 | Not-determined |
| NC3_vf_k141_838 | Not-determined |
| NC3_vf_k141_1913 | Not-determined |
| NC3_vf_k141_57 | Not-determined |
| NC3_vf_k141_584 | Not-determined |
| NC3_vf_k141_438 | Not-determined |
| NC3_vf_k141_308 | Not-determined |
| NC3_vf_k141_1373 | Not-determined |
| NC3_vf_k141_839 | Not-determined |
| NC3_vf_k141_1914 | Not-determined |
| NC3_vf_k141_1223 | Not-determined |
| NC3_vf_k141_1113 | Low-quality |
| NC3_vf_k141_58 | Not-determined |
| NC3_vf_k141_1785 | Not-determined |
| NC3_vf_k141_964 | Low-quality |
| NC3_vf_k141_1374 | Not-determined |
| NC3_vf_k141_1915 | Low-quality |
| NC3_vf_k141_59 | Not-determined |
| NC3_vf_k141_1224 | Not-determined |
| NC3_vf_k141_1114 | Not-determined |
| NC3_vf_k141_585 | Not-determined |
| NC3_vf_k141_840 | Not-determined |
| NC3_vf_k141_1786 | Low-quality |
| NC3_vf_k141_439 | Not-determined |
| NC3_vf_k141_60 | Not-determined |
| NC3_vf_k141_708 | Low-quality |
| NC3_vf_k141_1115 | Not-determined |
| NC3_vf_k141_1787 | Not-determined |
| NC3_vf_k141_61 | Not-determined |
| NC3_vf_k141_440 | Not-determined |
| NC3_vf_k141_1116 | Not-determined |
| NC3_vf_k141_1225 | Low-quality |
| NC3_vf_k141_841 | Not-determined |
| NC3_vf_k141_965 | Not-determined |
| NC3_vf_k141_709 | Not-determined |
| NC3_vf_k141_309 | Not-determined |
| NC3_vf_k141_62 | Not-determined |
| NC3_vf_k141_586 | Not-determined |
| NC3_vf_k141_1117 | Not-determined |
| NC3_vf_k141_710 | Not-determined |
| NC3_vf_k141_2039 | Not-determined |
| NC3_vf_k141_310 | Not-determined |
| NC3_vf_k141_1788 | Not-determined |
| NC3_vf_k141_441 | Not-determined |
| NC3_vf_k141_63 | Not-determined |
| NC3_vf_k141_187 | Not-determined |
| NC3_vf_k141_966 | Low-quality |
| NC3_vf_k141_2040 | Low-quality |
| NC3_vf_k141_311 | Not-determined |
| NC3_vf_k141_1118 | Not-determined |
| NC3_vf_k141_711 | Not-determined |
| NC3_vf_k141_1789 | Not-determined |
| NC3_vf_k141_442 | Not-determined |
| NC3_vf_k141_188 | Not-determined |
| NC3_vf_k141_2041 | Low-quality |
| NC3_vf_k141_1647 | Not-determined |
| NC3_vf_k141_967 | Not-determined |
| NC3_vf_k141_312 | Not-determined |
| NC3_vf_k141_1119 | Not-determined |
| NC3_vf_k141_1226 | Not-determined |
| NC3_vf_k141_842 | Not-determined |
| NC3_vf_k141_1790 | Not-determined |
| NC3_vf_k141_189 | Not-determined |
| NC3_vf_k141_64 | Not-determined |
| NC3_vf_k141_1513 | Not-determined |
| NC3_vf_k141_1375 | Not-determined |
| NC3_vf_k141_313 | Not-determined |
| NC3_vf_k141_712 | Not-determined |
| NC3_vf_k141_1648 | Not-determined |
| NC3_vf_k141_1120 | Not-determined |
| NC3_vf_k141_968 | Not-determined |
| NC3_vf_k141_190 | Not-determined |
| NC3_vf_k141_65 | Not-determined |
| NC3_vf_k141_1791 | Low-quality |
| NC3_vf_k141_1514 | Low-quality |
| NC3_vf_k141_1916 | Not-determined |
| NC3_vf_k141_1649 | Low-quality |
| NC3_vf_k141_314 | Low-quality |
| NC3_vf_k141_66 | Not-determined |
| NC3_vf_k141_1376 | Not-determined |
| NC3_vf_k141_1227 | Not-determined |
| NC3_vf_k141_713 | Not-determined |
| NC3_vf_k141_1515 | Not-determined |
| NC3_vf_k141_843 | Low-quality |
| NC3_vf_k141_2042 | Not-determined |
| NC3_vf_k141_315 | Not-determined |
| NC3_vf_k141_191 | Not-determined |
| NC3_vf_k141_1650 | Not-determined |
| NC3_vf_k141_1121 | Low-quality |
| NC3_vf_k141_1917 | Not-determined |
| NC3_vf_k141_714 | Not-determined |
| NC3_vf_k141_969 | Not-determined |
| NC3_vf_k141_67 | Not-determined |
| NC3_vf_k141_1792 | Not-determined |
| NC3_vf_k141_1377 | Not-determined |
| NC3_vf_k141_1516 | Not-determined |
| NC3_vf_k141_316 | Not-determined |
| NC3_vf_k141_2043 | Not-determined |
| NC3_vf_k141_1918 | Not-determined |
| NC3_vf_k141_844 | Not-determined |
| NC3_vf_k141_1651 | Not-determined |
| NC3_vf_k141_68 | Not-determined |
| NC3_vf_k141_192 | Not-determined |
| NC3_vf_k141_1793 | Not-determined |
| NC3_vf_k141_1378 | Not-determined |
| NC3_vf_k141_1122 | Not-determined |
| NC3_vf_k141_715 | Not-determined |
| NC3_vf_k141_1517 | Not-determined |
| NC3_vf_k141_587 | Not-determined |
| NC3_vf_k141_1228 | Not-determined |
| NC3_vf_k141_1919 | Not-determined |
| NC3_vf_k141_317 | Not-determined |
| NC3_vf_k141_970 | Not-determined |
| NC3_vf_k141_69 | Not-determined |
| NC3_vf_k141_845 | Not-determined |
| NC3_vf_k141_1123 | Not-determined |
| NC3_vf_k141_588 | Not-determined |
| NC3_vf_k141_716 | Not-determined |
| NC3_vf_k141_193 | Low-quality |
| NC3_vf_k141_1518 | Not-determined |
| NC3_vf_k141_443 | Low-quality |
| NC3_vf_k141_1920 | Not-determined |
| NC3_vf_k141_318 | Not-determined |
| NC3_vf_k141_1652 | Low-quality |
| NC3_vf_k141_1379 | Not-determined |
| NC3_vf_k141_1124 | Not-determined |
| NC3_vf_k141_70 | Not-determined |
| NC3_vf_k141_589 | Not-determined |
| NC3_vf_k141_846 | Not-determined |
| NC3_vf_k141_1794 | Low-quality |
| NC3_vf_k141_971 | Not-determined |
| NC3_vf_k141_194 | Not-determined |
| NC3_vf_k141_444 | Not-determined |
| NC3_vf_k141_319 | Not-determined |
| NC3_vf_k141_1125 | Not-determined |
| NC3_vf_k141_1795 | Low-quality |
| NC3_vf_k141_1519 | Not-determined |
| NC3_vf_k141_1921 | Not-determined |
| NC3_vf_k141_717 | Not-determined |
| NC3_vf_k141_1229 | Not-determined |
| NC3_vf_k141_1653 | Not-determined |
| NC3_vf_k141_590 | Not-determined |
| NC3_vf_k141_195 | Low-quality |
| NC3_vf_k141_320 | Not-determined |
| NC3_vf_k141_1796 | Not-determined |
| NC3_vf_k141_1126 | Not-determined |
| NC3_vf_k141_591 | Not-determined |
| NC3_vf_k141_1922 | Not-determined |
| NC3_vf_k141_718 | Not-determined |
| NC3_vf_k141_1520 | Not-determined |
| NC3_vf_k141_445 | Low-quality |
| NC3_vf_k141_2044 | Low-quality |
| NC3_vf_k141_847 | Not-determined |
| NC3_vf_k141_321 | Low-quality |
| NC3_vf_k141_196 | Not-determined |
| NC3_vf_k141_1923 | Not-determined |
| NC3_vf_k141_1380 | Low-quality |
| NC3_vf_k141_1230 | Not-determined |
| NC3_vf_k141_1654 | Not-determined |
| NC3_vf_k141_972 | Low-quality |
| NC3_vf_k141_1521 | Not-determined |
| NC3_vf_k141_322 | Not-determined |
| NC3_vf_k141_446 | Low-quality |
| NC3_vf_k141_719 | Low-quality |
| NC3_vf_k141_2045 | Not-determined |
| NC3_vf_k141_1127 | Low-quality |
| NC3_vf_k141_848 | Low-quality |
| NC3_vf_k141_71 | Low-quality |
| NC3_vf_k141_1522 | Not-determined |
| NC3_vf_k141_1924 | Low-quality |
| NC3_vf_k141_592 | Not-determined |
| NC3_vf_k141_1231 | Not-determined |
| NC3_vf_k141_197 | Not-determined |
| NC3_vf_k141_1381 | Not-determined |
| NC3_vf_k141_973 | Not-determined |
| NC3_vf_k141_323 | Not-determined |
| NC3_vf_k141_1128 | Not-determined |
| NC3_vf_k141_1797 | Not-determined |
| NC3_vf_k141_1523 | Not-determined |
| NC3_vf_k141_72 | Not-determined |
| NC3_vf_k141_1655 | Not-determined |
| NC3_vf_k141_1232 | Not-determined |
| NC3_vf_k141_593 | Not-determined |
| NC3_vf_k141_1925 | Not-determined |
| NC3_vf_k141_1524 | Not-determined |
| NC3_vf_k141_849 | Not-determined |
| NC3_vf_k141_1129 | Not-determined |
| NC3_vf_k141_720 | Not-determined |
| NC3_vf_k141_198 | Not-determined |
| NC3_vf_k141_1656 | Low-quality |
| NC3_vf_k141_1798 | Not-determined |
| NC3_vf_k141_1926 | Not-determined |
| NC3_vf_k141_1233 | Not-determined |
| NC3_vf_k141_594 | Not-determined |
| NC3_vf_k141_324 | Low-quality |
| NC3_vf_k141_447 | Not-determined |
| NC3_vf_k141_199 | Not-determined |
| NC3_vf_k141_974 | Not-determined |
| NC3_vf_k141_850 | Not-determined |
| NC3_vf_k141_1130 | Not-determined |
| NC3_vf_k141_1799 | Low-quality |
| NC3_vf_k141_1657 | Not-determined |
| NC3_vf_k141_1927 | Not-determined |
| NC3_vf_k141_1525 | Not-determined |
| NC3_vf_k141_721 | Not-determined |
| NC3_vf_k141_595 | Not-determined |
| NC3_vf_k141_1234 | Not-determined |
| NC3_vf_k141_73 | Not-determined |
| NC3_vf_k141_1131 | Not-determined |
| NC3_vf_k141_1526 | Not-determined |
| NC3_vf_k141_448 | Not-determined |
| NC3_vf_k141_1928 | Not-determined |
| NC3_vf_k141_200 | Not-determined |
| NC3_vf_k141_325 | Low-quality |
| NC3_vf_k141_1800 | Not-determined |
| NC3_vf_k141_1382 | Not-determined |
| NC3_vf_k141_1132 | Not-determined |
| NC3_vf_k141_74 | Not-determined |
| NC3_vf_k141_1658 | Low-quality |
| NC3_vf_k141_1235 | Low-quality |
| NC3_vf_k141_596 | Not-determined |
| NC3_vf_k141_1929 | Not-determined |
| NC3_vf_k141_326 | Not-determined |
| NC3_vf_k141_1527 | Not-determined |
| NC3_vf_k141_1133 | Not-determined |
| NC3_vf_k141_75 | Not-determined |
| NC3_vf_k141_201 | Not-determined |
| NC3_vf_k141_722 | Not-determined |
| NC3_vf_k141_975 | Low-quality |
| NC3_vf_k141_1383 | Not-determined |
| NC3_vf_k141_1801 | Not-determined |
| NC3_vf_k141_1930 | Not-determined |
| NC3_vf_k141_1134 | Not-determined |
| NC3_vf_k141_1528 | Not-determined |
| NC3_vf_k141_851 | Low-quality |
| NC3_vf_k141_76 | Not-determined |
| NC3_vf_k141_597 | Not-determined |
| NC3_vf_k141_449 | Low-quality |
| NC3_vf_k141_1659 | Not-determined |
| NC3_vf_k141_723 | Not-determined |
| NC3_vf_k141_976 | Not-determined |
| NC3_vf_k141_1931 | Not-determined |
| NC3_vf_k141_77 | Not-determined |
| NC3_vf_k141_1802 | Not-determined |
| NC3_vf_k141_1236 | Low-quality |
| NC3_vf_k141_1529 | Not-determined |
| NC3_vf_k141_2046 | Not-determined |
| NC3_vf_k141_1384 | Not-determined |
| NC3_vf_k141_1135 | Not-determined |
| NC3_vf_k141_724 | Low-quality |
| NC3_vf_k141_1932 | Not-determined |
| NC3_vf_k141_1660 | Not-determined |
| NC3_vf_k141_977 | Not-determined |
| NC3_vf_k141_1803 | Not-determined |
| NC3_vf_k141_202 | Low-quality |
| NC3_vf_k141_852 | Not-determined |
| NC3_vf_k141_450 | Not-determined |
| NC3_vf_k141_78 | Not-determined |
| NC3_vf_k141_1933 | Not-determined |
| NC3_vf_k141_1237 | Not-determined |
| NC3_vf_k141_1661 | Not-determined |
| NC3_vf_k141_1385 | Not-determined |
| NC3_vf_k141_1530 | Low-quality |
| NC3_vf_k141_978 | Not-determined |
| NC3_vf_k141_1136 | Not-determined |
| NC3_vf_k141_203 | Not-determined |
| NC3_vf_k141_1804 | Not-determined |
| NC3_vf_k141_79 | Not-determined |
| NC3_vf_k141_327 | Not-determined |
| NC3_vf_k141_1662 | Not-determined |
| NC3_vf_k141_1137 | Low-quality |
| NC3_vf_k141_1238 | Not-determined |
| NC3_vf_k141_979 | Not-determined |
| NC3_vf_k141_1386 | Not-determined |
| NC3_vf_k141_80 | Not-determined |
| NC3_vf_k141_1805 | Not-determined |
| NC3_vf_k141_1663 | Not-determined |
| NC3_vf_k141_451 | Not-determined |
| NC3_vf_k141_1531 | Not-determined |
| NC3_vf_k141_725 | Not-determined |
| NC3_vf_k141_1138 | Low-quality |
| NC3_vf_k141_204 | Not-determined |
| NC3_vf_k141_1806 | Not-determined |
| NC3_vf_k141_1387 | Not-determined |
| NC3_vf_k141_598 | Low-quality |
| NC3_vf_k141_81 | Not-determined |
| NC3_vf_k141_205 | Not-determined |
| NC3_vf_k141_1934 | Low-quality |
| NC3_vf_k141_2047 | Not-determined |
| NC3_vf_k141_1139 | Not-determined |

| | |
|---|---|
| NC3_vf_k141_328 | Low-quality |
| NC3_vf_k141_452 | Low-quality |
| NC3_vf_k141_726 | Not-determined |
| NC3_vf_k141_599 | Not-determined |
| NC3_vf_k141_980 | Not-determined |
| NC3_vf_k141_1664 | Not-determined |
| NC3_vf_k141_1388 | Not-determined |
| NC3_vf_k141_1807 | Not-determined |
| NC3_vf_k141_1239 | Not-determined |
| NC3_vf_k141_82 | Not-determined |
| NC3_vf_k141_2048 | Not-determined |
| NC3_vf_k141_1935 | Low-quality |
| NC3_vf_k141_853 | Not-determined |
| NC3_vf_k141_329 | Not-determined |
| NC3_vf_k141_1808 | Low-quality |
| NC3_vf_k141_83 | Not-determined |
| NC3_vf_k141_727 | Not-determined |
| NC3_vf_k141_1140 | Low-quality |
| NC3_vf_k141_206 | Not-determined |
| NC3_vf_k141_1936 | Not-determined |
| NC3_vf_k141_1532 | Not-determined |
| NC3_vf_k141_453 | Low-quality |
| NC3_vf_k141_1389 | Not-determined |
| NC3_vf_k141_1665 | Not-determined |
| NC3_vf_k141_2049 | Not-determined |
| NC3_vf_k141_1809 | Not-determined |
| NC3_vf_k141_84 | Not-determined |
| NC3_vf_k141_1240 | Not-determined |
| NC3_vf_k141_1937 | Not-determined |
| NC3_vf_k141_1533 | Not-determined |
| NC3_vf_k141_330 | Not-determined |
| NC3_vf_k141_854 | Not-determined |
| NC3_vf_k141_1141 | Not-determined |
| NC3_vf_k141_2050 | Not-determined |
| NC3_vf_k141_981 | Low-quality |
| NC3_vf_k141_728 | Not-determined |
| NC3_vf_k141_207 | Not-determined |
| NC3_vf_k141_1810 | Not-determined |
| NC3_vf_k141_85 | Not-determined |
| NC3_vf_k141_454 | Not-determined |
| NC3_vf_k141_1534 | Not-determined |
| NC3_vf_k141_1241 | Low-quality |
| NC3_vf_k141_331 | Not-determined |
| NC3_vf_k141_1666 | Not-determined |
| NC3_vf_k141_1142 | Not-determined |
| NC3_vf_k141_600 | Not-determined |
| NC3_vf_k141_2051 | Not-determined |
| NC3_vf_k141_208 | Not-determined |
| NC3_vf_k141_855 | Not-determined |
| NC3_vf_k141_332 | Not-determined |
| NC3_vf_k141_1938 | Not-determined |
| NC3_vf_k141_1535 | Not-determined |
| NC3_vf_k141_1143 | Not-determined |
| NC3_vf_k141_1811 | Low-quality |
| NC3_vf_k141_86 | Not-determined |
| NC3_vf_k141_455 | Not-determined |
| NC3_vf_k141_1242 | Not-determined |
| NC3_vf_k141_601 | Not-determined |
| NC3_vf_k141_1667 | Not-determined |
| NC3_vf_k141_729 | Not-determined |
| NC3_vf_k141_1939 | Not-determined |
| NC3_vf_k141_2052 | Not-determined |
| NC3_vf_k141_1812 | Not-determined |
| NC3_vf_k141_333 | Not-determined |
| NC3_vf_k141_1144 | Not-determined |
| NC3_vf_k141_856 | Low-quality |
| NC3_vf_k141_602 | Not-determined |
| NC3_vf_k141_1668 | Not-determined |
| NC3_vf_k141_87 | Not-determined |
| NC3_vf_k141_1940 | Not-determined |
| NC3_vf_k141_209 | Not-determined |
| NC3_vf_k141_730 | Not-determined |
| NC3_vf_k141_2053 | Not-determined |
| NC3_vf_k141_603 | Not-determined |
| NC3_vf_k141_1813 | Not-determined |
| NC3_vf_k141_1536 | Not-determined |
| NC3_vf_k141_857 | Not-determined |
| NC3_vf_k141_88 | Not-determined |
| NC3_vf_k141_1145 | Low-quality |
| NC3_vf_k141_456 | Not-determined |
| NC3_vf_k141_2054 | Not-determined |
| NC3_vf_k141_1537 | Not-determined |
| NC3_vf_k141_731 | Not-determined |
| NC3_vf_k141_1814 | Not-determined |
| NC3_vf_k141_604 | Not-determined |
| NC3_vf_k141_1146 | Not-determined |
| NC3_vf_k141_1669 | Not-determined |
| NC3_vf_k141_1941 | Not-determined |
| NC3_vf_k141_1538 | Not-determined |
| [illegible] | [illegible] |
| NC3_vf_k141_224 | Not-determined |
| NC3_vf_k141_100 | Not-determined |
| NC3_vf_k141_866 | Not-determined |
| NC3_vf_k141_1550 | Not-determined |
| NC3_vf_k141_1824 | Not-determined |
| NC3_vf_k141_1154 | Low-quality |
| NC3_vf_k141_1403 | Not-determined |
| NC3_vf_k141_1953 | Not-determined |
| NC3_vf_k141_346 | Not-determined |
| NC3_vf_k141_462 | Not-determined |
| NC3_vf_k141_986 | Low-quality |
| NC3_vf_k141_1551 | Not-determined |
| NC3_vf_k141_225 | Not-determined |
| NC3_vf_k141_613 | Not-determined |
| NC3_vf_k141_1155 | Not-determined |
| NC3_vf_k141_1825 | Not-determined |
| NC3_vf_k141_1675 | Low-quality |
| NC3_vf_k141_101 | Not-determined |
| NC3_vf_k141_2067 | Not-determined |
| NC3_vf_k141_614 | Not-determined |
| NC3_vf_k141_1156 | Not-determined |
| NC3_vf_k141_1552 | Not-determined |
| NC3_vf_k141_987 | Not-determined |
| NC3_vf_k141_734 | Not-determined |
| NC3_vf_k141_867 | Not-determined |
| NC3_vf_k141_1954 | Low-quality |
| NC3_vf_k141_463 | Not-determined |
| NC3_vf_k141_102 | Not-determined |
| NC3_vf_k141_226 | Not-determined |
| NC3_vf_k141_1676 | Not-determined |
| NC3_vf_k141_2068 | Not-determined |
| NC3_vf_k141_1826 | Not-determined |
| NC3_vf_k141_615 | Not-determined |
| NC3_vf_k141_1404 | Low-quality |
| NC3_vf_k141_1157 | Not-determined |
| NC3_vf_k141_1553 | Low-quality |
| NC3_vf_k141_103 | Not-determined |
| NC3_vf_k141_1955 | Not-determined |
| NC3_vf_k141_347 | Low-quality |
| NC3_vf_k141_227 | Not-determined |
| NC3_vf_k141_1827 | Not-determined |
| NC3_vf_k141_616 | Not-determined |
| NC3_vf_k141_868 | Not-determined |
| NC3_vf_k141_1248 | Low-quality |
| NC3_vf_k141_464 | Not-determined |
| NC3_vf_k141_348 | Not-determined |
| NC3_vf_k141_1158 | Not-determined |
| NC3_vf_k141_104 | Not-determined |
| NC3_vf_k141_1554 | Not-determined |
| NC3_vf_k141_988 | Not-determined |
| NC3_vf_k141_1677 | Not-determined |
| NC3_vf_k141_1956 | Not-determined |
| NC3_vf_k141_617 | Not-determined |
| NC3_vf_k141_1405 | Not-determined |
| NC3_vf_k141_465 | Not-determined |
| NC3_vf_k141_105 | Low-quality |
| NC3_vf_k141_1555 | Not-determined |
| NC3_vf_k141_1159 | Not-determined |
| NC3_vf_k141_618 | Not-determined |
| NC3_vf_k141_1678 | Not-determined |
| NC3_vf_k141_349 | Not-determined |
| NC3_vf_k141_466 | Not-determined |
| NC3_vf_k141_106 | Not-determined |
| NC3_vf_k141_1828 | Low-quality |
| NC3_vf_k141_1160 | Not-determined |
| NC3_vf_k141_1556 | Not-determined |
| NC3_vf_k141_350 | Not-determined |
| NC3_vf_k141_1249 | Not-determined |
| NC3_vf_k141_2069 | Low-quality |
| NC3_vf_k141_869 | Not-determined |
| NC3_vf_k141_1161 | Not-determined |
| NC3_vf_k141_1829 | Not-determined |
| NC3_vf_k141_1679 | Not-determined |
| NC3_vf_k141_107 | Not-determined |
| NC3_vf_k141_1250 | Not-determined |
| NC3_vf_k141_351 | Low-quality |
| NC3_vf_k141_1830 | Not-determined |
| NC3_vf_k141_1162 | Not-determined |
| NC3_vf_k141_228 | Not-determined |
| NC3_vf_k141_1680 | Not-determined |
| NC3_vf_k141_467 | Not-determined |
| NC3_vf_k141_1957 | Low-quality |
| NC3_vf_k141_870 | Not-determined |
| NC3_vf_k141_1831 | Not-determined |
| NC3_vf_k141_1251 | Not-determined |
| NC3_vf_k141_735 | Not-determined |
| NC3_vf_k141_229 | Low-quality |
| NC3_vf_k141_352 | Not-determined |
| NC3_vf_k141_1406 | Not-determined |
| NC3_vf_k141_1681 | Not-determined |
| NC3_vf_k141_1958 | Not-determined |
| NC3_vf_k141_1557 | Not-determined |
| [illegible] | [illegible] |

| | |
|---|---|
| NC3_vf_k141_744 | Not-determined |
| NC3_vf_k141_1182 | Not-determined |
| NC3_vf_k141_884 | Low-quality |
| NC3_vf_k141_639 | Not-determined |
| NC3_vf_k141_375 | Not-determined |
| NC3_vf_k141_1855 | Not-determined |
| NC3_vf_k141_252 | Low-quality |
| NC3_vf_k141_1576 | Not-determined |
| NC3_vf_k141_1420 | Not-determined |
| NC3_vf_k141_1266 | Not-determined |
| NC3_vf_k141_1705 | Not-determined |
| NC3_vf_k141_640 | Not-determined |
| NC3_vf_k141_1183 | Not-determined |
| NC3_vf_k141_483 | Not-determined |
| NC3_vf_k141_1856 | Not-determined |
| NC3_vf_k141_745 | Not-determined |
| NC3_vf_k141_885 | Not-determined |
| NC3_vf_k141_376 | Not-determined |
| NC3_vf_k141_2091 | Not-determined |
| NC3_vf_k141_1975 | Not-determined |
| NC3_vf_k141_1184 | Not-determined |
| NC3_vf_k141_1577 | Not-determined |
| NC3_vf_k141_1706 | Not-determined |
| NC3_vf_k141_253 | Not-determined |
| NC3_vf_k141_1421 | Not-determined |
| NC3_vf_k141_1267 | Not-determined |
| NC3_vf_k141_377 | Low-quality |
| NC3_vf_k141_2092 | Not-determined |
| NC3_vf_k141_641 | Not-determined |
| NC3_vf_k141_998 | Low-quality |
| NC3_vf_k141_1857 | Not-determined |
| NC3_vf_k141_1185 | Not-determined |
| NC3_vf_k141_1976 | Not-determined |
| NC3_vf_k141_1707 | Not-determined |
| NC3_vf_k141_746 | Not-determined |
| NC3_vf_k141_123 | Low-quality |
| NC3_vf_k141_484 | Not-determined |
| NC3_vf_k141_378 | Not-determined |
| NC3_vf_k141_2093 | Not-determined |
| NC3_vf_k141_886 | Not-determined |
| NC3_vf_k141_1186 | Not-determined |
| NC3_vf_k141_1977 | Not-determined |
| NC3_vf_k141_1578 | Low-quality |
| NC3_vf_k141_1422 | Low-quality |
| NC3_vf_k141_124 | Not-determined |
| NC3_vf_k141_747 | Not-determined |
| NC3_vf_k141_1708 | Not-determined |
| NC3_vf_k141_999 | Low-quality |
| NC3_vf_k141_1579 | Not-determined |
| NC3_vf_k141_1187 | Not-determined |
| NC3_vf_k141_379 | Not-determined |
| NC3_vf_k141_2094 | Not-determined |
| NC3_vf_k141_485 | Not-determined |
| NC3_vf_k141_1709 | Not-determined |
| NC3_vf_k141_1423 | Not-determined |
| NC3_vf_k141_1978 | Not-determined |
| NC3_vf_k141_125 | Not-determined |
| NC3_vf_k141_1268 | Not-determined |
| NC3_vf_k141_2095 | Not-determined |
| NC3_vf_k141_642 | Low-quality |
| NC3_vf_k141_1188 | Not-determined |
| NC3_vf_k141_1000 | Not-determined |
| NC3_vf_k141_1710 | Not-determined |
| NC3_vf_k141_126 | Not-determined |
| NC3_vf_k141_1580 | Not-determined |
| NC3_vf_k141_1424 | Not-determined |
| NC3_vf_k141_1858 | Low-quality |
| NC3_vf_k141_643 | Low-quality |
| NC3_vf_k141_1269 | Not-determined |
| NC3_vf_k141_254 | Low-quality |
| NC3_vf_k141_2096 | Low-quality |
| NC3_vf_k141_1189 | Not-determined |
| NC3_vf_k141_1979 | Not-determined |
| NC3_vf_k141_887 | Not-determined |
| NC3_vf_k141_255 | Not-determined |
| NC3_vf_k141_2097 | Not-determined |
| NC3_vf_k141_486 | Not-determined |
| NC3_vf_k141_127 | Low-quality |
| NC3_vf_k141_748 | Low-quality |
| NC3_vf_k141_1711 | Not-determined |
| NC3_vf_k141_1859 | Not-determined |
| NC3_vf_k141_1425 | Low-quality |
| NC3_vf_k141_1001 | Not-determined |
| NC3_vf_k141_1980 | Not-determined |
| NC3_vf_k141_256 | Not-determined |
| NC3_vf_k141_2098 | Not-determined |
| NC3_vf_k141_1712 | Not-determined |
| NC3_vf_k141_644 | Not-determined |
| NC3_vf_k141_380 | Low-quality |
| NC3_vf_k141_487 | Not-determined |
| NC3_vf_k141_1981 | Not-determined |
| NC3_vf_k141_128 | Not-determined |
| NC3_vf_k141_1860 | Not-determined |
| NC3_vf_k141_257 | Not-determined |
| NC3_vf_k141_2099 | Not-determined |
| NC3_vf_k141_1002 | Not-determined |
| NC3_vf_k141_1581 | Low-quality |
| NC3_vf_k141_1270 | Low-quality |
| NC3_vf_k141_645 | Not-determined |
| NC3_vf_k141_129 | Not-determined |
| NC3_vf_k141_749 | Not-determined |
| NC3_vf_k141_1982 | Not-determined |
| NC3_vf_k141_1861 | Not-determined |
| NC3_vf_k141_258 | Low-quality |
| NC3_vf_k141_1713 | Low-quality |
| NC3_vf_k141_1582 | Not-determined |
| NC3_vf_k141_888 | Not-determined |
| NC3_vf_k141_381 | Not-determined |
| NC3_vf_k141_2100 | Not-determined |
| NC3_vf_k141_646 | Not-determined |
| NC3_vf_k141_1983 | Not-determined |
| NC3_vf_k141_1426 | Not-determined |
| NC3_vf_k141_488 | Not-determined |
| NC3_vf_k141_130 | Not-determined |
| NC3_vf_k141_259 | Not-determined |
| NC3_vf_k141_1583 | Not-determined |
| NC3_vf_k141_1714 | Not-determined |
| NC3_vf_k141_1003 | Low-quality |
| NC3_vf_k141_382 | Not-determined |
| NC3_vf_k141_1984 | Not-determined |
| NC3_vf_k141_1715 | Not-determined |
| NC3_vf_k141_889 | Not-determined |
| NC3_vf_k141_647 | Not-determined |
| NC3_vf_k141_383 | Not-determined |
| NC3_vf_k141_1584 | Not-determined |
| NC3_vf_k141_1985 | Not-determined |
| NC3_vf_k141_2101 | Not-determined |
| NC3_vf_k141_750 | Not-determined |
| NC3_vf_k141_1271 | Not-determined |
| NC3_vf_k141_260 | Not-determined |
| NC3_vf_k141_1716 | Not-determined |
| NC3_vf_k141_489 | Not-determined |
| NC3_vf_k141_648 | Not-determined |
| NC3_vf_k141_1585 | Not-determined |
| NC3_vf_k141_1004 | Not-determined |
| NC3_vf_k141_1427 | Not-determined |
| NC3_vf_k141_2102 | Not-determined |
| NC3_vf_k141_384 | Not-determined |
| NC3_vf_k141_890 | Not-determined |
| NC3_vf_k141_1586 | Not-determined |
| NC3_vf_k141_261 | Not-determined |
| NC3_vf_k141_1717 | Not-determined |
| NC3_vf_k141_649 | Not-determined |
| NC3_vf_k141_751 | Not-determined |
| NC3_vf_k141_131 | Low-quality |
| NC3_vf_k141_2103 | Not-determined |
| NC3_vf_k141_490 | Not-determined |
| NC3_vf_k141_1428 | Not-determined |
| NC3_vf_k141_650 | Low-quality |
| NC3_vf_k141_385 | Not-determined |
| NC3_vf_k141_1986 | Not-determined |
| NC3_vf_k141_1718 | Not-determined |
| NC3_vf_k141_1272 | Not-determined |
| NC3_vf_k141_2104 | Not-determined |
| NC3_vf_k141_752 | Not-determined |
| NC3_vf_k141_491 | Not-determined |
| NC3_vf_k141_386 | Not-determined |
| NC3_vf_k141_651 | Not-determined |
| NC3_vf_k141_1587 | Not-determined |
| NC3_vf_k141_1429 | Not-determined |
| NC3_vf_k141_132 | Not-determined |
| NC3_vf_k141_1719 | Not-determined |
| NC3_vf_k141_1987 | Not-determined |
| NC3_vf_k141_652 | Not-determined |
| NC3_vf_k141_1430 | Not-determined |
| NC3_vf_k141_1588 | Not-determined |
| NC3_vf_k141_1988 | Not-determined |
| NC3_vf_k141_262 | Not-determined |
| NC3_vf_k141_387 | Not-determined |
| NC3_vf_k141_653 | Not-determined |
| NC3_vf_k141_1720 | Not-determined |
| NC3_vf_k141_1273 | Not-determined |
| NC3_vf_k141_1005 | Not-determined |
| NC3_vf_k141_1589 | Not-determined |
| NC3_vf_k141_263 | Not-determined |
| NC3_vf_k141_1989 | Not-determined |
| NC3_vf_k141_388 | Not-determined |
| NC3_vf_k141_1431 | Not-determined |
| NC3_vf_k141_891 | Not-determined |
| NC3_vf_k141_753 | Low-quality |
| NC3_vf_k141_2105 | Not-determined |
| NC3_vf_k141_1590 | Not-determined |
| NC3_vf_k141_1721 | Not-determined |
| NC3_vf_k141_264 | Not-determined |
| NC3_vf_k141_1990 | Not-determined |
| NC3_vf_k141_389 | Not-determined |
| NC3_vf_k141_1432 | Not-determined |
| NC3_vf_k141_1006 | Not-determined |
| NC3_vf_k141_1274 | Not-determined |
| NC3_vf_k141_1722 | Not-determined |
| NC3_vf_k141_892 | Not-determined |
| NC3_vf_k141_1991 | Not-determined |
| NC3_vf_k141_390 | Not-determined |
| NC3_vf_k141_1591 | Not-determined |
| NC3_vf_k141_754 | Not-determined |
| NC3_vf_k141_654 | Low-quality |
| NC3_vf_k141_265 | Not-determined |
| NC3_vf_k141_1433 | Not-determined |
| NC3_vf_k141_1592 | Not-determined |
| NC3_vf_k141_1992 | Not-determined |
| NC3_vf_k141_1723 | Not-determined |
| NC3_vf_k141_2106 | Not-determined |
| NC3_vf_k141_893 | Not-determined |
| NC3_vf_k141_655 | Not-determined |
| NC3_vf_k141_1724 | Not-determined |
| NC3_vf_k141_755 | Not-determined |
| NC3_vf_k141_391 | Not-determined |
| NC3_vf_k141_1434 | Not-determined |
| NC3_vf_k141_1275 | Not-determined |
| NC3_vf_k141_1993 | Not-determined |
| NC3_vf_k141_492 | Not-determined |
| NC3_vf_k141_2107 | Not-determined |
| NC3_vf_k141_656 | Not-determined |
| NC3_vf_k141_392 | Not-determined |
| NC3_vf_k141_1593 | Not-determined |
| NC3_vf_k141_894 | Not-determined |
| NC3_vf_k141_2108 | Not-determined |
| NC3_vf_k141_1725 | Not-determined |
| NC3_vf_k141_1994 | Not-determined |
| NC3_vf_k141_1276 | Not-determined |
| NC3_vf_k141_657 | Low-quality |
| NC3_vf_k141_1726 | Low-quality |
| NC3_vf_k141_1435 | Not-determined |
| NC3_vf_k141_895 | Not-determined |
| NC3_vf_k141_493 | Not-determined |
| NC3_vf_k141_393 | Not-determined |
| NC3_vf_k141_1277 | Not-determined |
| NC3_vf_k141_1007 | Low-quality |
| NC3_vf_k141_1594 | Not-determined |
| NC3_vf_k141_658 | Not-determined |
| NC3_vf_k141_1436 | Not-determined |
| NC3_vf_k141_394 | Not-determined |
| NC3_vf_k141_1595 | Not-determined |
| NC3_vf_k141_1437 | Not-determined |
| NC3_vf_k141_896 | Not-determined |
| NC3_vf_k141_395 | Not-determined |
| NC3_vf_k141_1727 | Low-quality |
| NC3_vf_k141_396 | Low-quality |
| NC3_vf_k141_1190 | Not-determined |
| NC3_vf_k141_1278 | Not-determined |
| NC3_vf_k141_397 | Not-determined |
| NC3_vf_k141_1008 | Not-determined |
| NC3_vf_k141_897 | Not-determined |
| NC3_vf_k141_1728 | Not-determined |
| NC3_vf_k141_756 | Not-determined |
| NC3_vf_k141_1438 | Not-determined |
| NC3_vf_k141_1191 | Not-determined |
| NC3_vf_k141_898 | Not-determined |
| NC3_vf_k141_398 | Low-quality |
| NC3_vf_k141_1279 | Not-determined |
| NC3_vf_k141_757 | Not-determined |
| NC3_vf_k141_1192 | Not-determined |
| NC3_vf_k141_899 | Not-determined |
| NC3_vf_k141_1439 | Not-determined |
| NC3_vf_k141_1280 | Not-determined |
| NC3_vf_k141_1009 | Not-determined |
| NC3_vf_k141_758 | Not-determined |
| NC3_vf_k141_900 | Not-determined |
| NC3_vf_k141_1281 | Not-determined |
| NC3_vf_k141_1010 | Not-determined |
| NC3_vf_k141_759 | Not-determined |
| NC3_vf_k141_1440 | Not-determined |
| NC3_vf_k141_901 | Not-determined |
| NC3_vf_k141_494 | Not-determined |
| NC3_vf_k141_1193 | Not-determined |
| NC3_vf_k141_902 | Not-determined |
| NC3_vf_k141_659 | Not-determined |
| NC3_vf_k141_760 | Not-determined |
| NC3_vf_k141_1441 | Not-determined |
| NC3_vf_k141_495 | Not-determined |
| NC3_vf_k141_1282 | Not-determined |
| NC3_vf_k141_660 | Low-quality |
| NC3_vf_k141_1194 | Not-determined |
| NC3_vf_k141_761 | Not-determined |
| NC3_vf_k141_1442 | Not-determined |
| NC3_vf_k141_1195 | Not-determined |
| NC3_vf_k141_496 | Not-determined |
| NC3_vf_k141_1011 | Not-determined |
| NC3_vf_k141_661 | Low-quality |
| NC3_vf_k141_1443 | Not-determined |
| NC3_vf_k141_1283 | Not-determined |
| NC3_vf_k141_762 | Not-determined |
| NC3_vf_k141_662 | Not-determined |
| NC3_vf_k141_497 | Low-quality |
| NC3_vf_k141_903 | Not-determined |
| NC3_vf_k141_663 | Not-determined |
| NC3_vf_k141_1444 | Not-determined |
| NC3_vf_k141_763 | Not-determined |
| NC3_vf_k141_1284 | Not-determined |
| NC3_vf_k141_664 | Not-determined |
| NC3_vf_k141_498 | Not-determined |
| NC3_vf_k141_1012 | Low-quality |
| NC3_vf_k141_764 | Not-determined |
| NC3_vf_k141_1285 | Not-determined |
| NC3_vf_k141_499 | Not-determined |
| NC3_vf_k141_904 | Low-quality |
| NC3_vf_k141_1286 | Not-determined |
| NC3_vf_k141_500 | Not-determined |
| NC3_vf_k141_1445 | Low-quality |
| NC3_vf_k141_1013 | Low-quality |
| NC3_vf_k141_1287 | Not-determined |
| NC3_vf_k141_765 | Not-determined |
| NC3_vf_k141_1446 | Not-determined |
| NC3_vf_k141_1014 | Not-determined |
| NC3_vf_k141_1288 | Not-determined |
| NC3_vf_k141_1447 | Not-determined |
| NC3_vf_k141_1015 | Not-determined |
| NC3_vf_k141_766 | Not-determined |
| NC3_vf_k141_1196 | Not-determined |
| NC3_vf_k141_501 | Low-quality |
| NC3_vf_k141_1289 | Not-determined |
| NC3_vf_k141_2109 | Not-determined |
| NC3_vf_k141_502 | Not-determined |
| NC3_vf_k141_1448 | Low-quality |
| NC3_vf_k141_2110 | Not-determined |
| NC3_vf_k141_767 | Not-determined |
| NC3_vf_k141_503 | Not-determined |
| NC3_vf_k141_1016 | Not-determined |
| NC3_vf_k141_1449 | Not-determined |
| NC3_vf_k141_905 | Low-quality |
| NC3_vf_k141_2111 | Low-quality |
| NC3_vf_k141_1290 | Low-quality |
| NC3_vf_k141_1017 | Not-determined |
| NC3_vf_k141_1450 | Not-determined |
| NC3_vf_k141_2112 | Not-determined |
| NC3_vf_k141_2113 | Not-determined |
| NC3_vf_k141_1018 | Not-determined |
| NC3_vf_k141_2114 | Not-determined |
| NC3_vf_k141_1291 | Not-determined |
| NC3_vf_k141_1451 | Not-determined |
| NC3_vf_k141_504 | Not-determined |
| NC3_vf_k141_2115 | Not-determined |
| NC3_vf_k141_1452 | Not-determined |
| NC3_vf_k141_1292 | Not-determined |
| NC3_vf_k141_2116 | Not-determined |
| NC3_vf_k141_1453 | Not-determined |
| NC3_vf_k141_2117 | Not-determined |
| NC3_vf_k141_1293 | Low-quality |
| NC3_vf_k141_1454 | Not-determined |
| NC3_vf_k141_2118 | Not-determined |
| NC3_vf_k141_768 | Not-determined |
| NC3_vf_k141_1294 | Not-determined |
| NC3_vf_k141_906 | Not-determined |
| NC3_vf_k141_2119 | Not-determined |
| NC3_vf_k141_1455 | Not-determined |
| NC3_vf_k141_2120 | Not-determined |
| NC3_vf_k141_769 | Not-determined |
| NC3_vf_k141_907 | Not-determined |
| NC3_vf_k141_2121 | Not-determined |
| NC3_vf_k141_1295 | Not-determined |
| NC3_vf_k141_1456 | Not-determined |
| NC3_vf_k141_1019 | Low-quality |
| NC3_vf_k141_770 | Not-determined |
| NC3_vf_k141_908 | Low-quality |
| NC3_vf_k141_909 | Not-determined |
| NC3_vf_k141_1457 | Not-determined |
| NC3_vf_k141_771 | Not-determined |
| NC3_vf_k141_1020 | Not-determined |
| NC3_vf_k141_1458 | Not-determined |
| NC3_vf_k141_772 | Not-determined |
| NC3_vf_k141_1459 | Not-determined |
| NC3_vf_k141_1021 | Low-quality |
| NC3_vf_k141_505 | Low-quality |
| NC3_vf_k141_773 | Not-determined |
| NC3_vf_k141_1460 | Not-determined |
| NC3_vf_k141_506 | Not-determined |
| NC3_vf_k141_774 | Not-determined |
| NC3_vf_k141_1022 | Not-determined |
| NC3_vf_k141_1296 | Not-determined |
| NC3_vf_k141_1023 | Not-determined |
| NC3_vf_k141_775 | Not-determined |
| NC3_vf_k141_1461 | Low-quality |
| NC3_vf_k141_1024 | Low-quality |
| NC3_vf_k141_507 | Not-determined |
| NC3_vf_k141_1297 | Not-determined |
| NC3_vf_k141_1462 | Not-determined |
| NC3_vf_k141_1298 | Not-determined |
| NC3_vf_k141_1025 | Not-determined |
| NC3_vf_k141_508 | Not-determined |
| NC3_vf_k141_910 | Not-determined |
| NC3_vf_k141_776 | Not-determined |
| NC3_vf_k141_509 | Not-determined |
| NC3_vf_k141_1299 | Not-determined |
| NC3_vf_k141_777 | Low-quality |
| NC3_vf_k141_1026 | Not-determined |
| NC3_vf_k141_510 | Not-determined |
| NC3_vf_k141_1300 | Not-determined |
| NC3_vf_k141_1027 | Not-determined |
| NC3_vf_k141_911 | Not-determined |
| NC3_vf_k141_1028 | Not-determined |
| NC3_vf_k141_1301 | Not-determined |
| NC3_vf_k141_511 | Low-quality |
| NC3_vf_k141_778 | Not-determined |
| NC3_vf_k141_912 | Not-determined |
| NC3_vf_k141_1302 | Not-determined |
| NC3_vf_k141_1029 | Low-quality |
| NC3_vf_k141_779 | Not-determined |
| NC3_vf_k141_780 | Not-determined |
| NC3_vf_k141_1030 | Not-determined |
| NC3_vf_k141_512 | Not-determined |
| NC3_vf_k141_1303 | Not-determined |
| NC3_vf_k141_781 | Not-determined |
| NC3_vf_k141_1304 | Not-determined |
| NC3_vf_k141_782 | Not-determined |
| NC3_vf_k141_1031 | Not-determined |
| NC3_vf_k141_913 | Low-quality |
| NC3_vf_k141_1305 | Low-quality |
| NC3_vf_k141_1032 | Not-determined |
| NC3_vf_k141_783 | Not-determined |
| NC3_vf_k141_513 | Not-determined |
| NC3_vf_k141_914 | Not-determined |
| NC3_vf_k141_1306 | Not-determined |
| NC3_vf_k141_784 | Not-determined |
| NC3_vf_k141_2122 | Low-quality |
| NC3_vf_k141_915 | Not-determined |
| NC3_vf_k141_514 | Not-determined |
| NC3_vf_k141_1307 | Not-determined |
| NC3_vf_k141_1033 | Low-quality |
| NC3_vf_k141_785 | Not-determined |
| NC3_vf_k141_2123 | Not-determined |
| NC3_vf_k141_1308 | Not-determined |
| NC3_vf_k141_1034 | Not-determined |
| NC3_vf_k141_2124 | Not-determined |
| NC3_vf_k141_786 | Not-determined |
| NC3_vf_k141_1309 | Not-determined |
| NC3_vf_k141_916 | Low-quality |
| NC3_vf_k141_2125 | Low-quality |
| NC3_vf_k141_2126 | Not-determined |
| NC3_vf_k141_787 | Low-quality |
| NC3_vf_k141_2127 | Not-determined |
| NC3_vf_k141_515 | Low-quality |
| NC3_vf_k141_1310 | Not-determined |
| NC3_vf_k141_917 | Not-determined |
| NC3_vf_k141_788 | Not-determined |
| NC3_vf_k141_789 | Not-determined |
| NC3_vf_k141_918 | Not-determined |
| NC3_vf_k141_1035 | Low-quality |
| NC3_vf_k141_1311 | Not-determined |
| NC3_vf_k141_516 | Not-determined |
| NC3_vf_k141_790 | Not-determined |
| NC3_vf_k141_1036 | Not-determined |
| NC3_vf_k141_517 | Not-determined |
| NC3_vf_k141_1312 | Not-determined |
| NC3_vf_k141_919 | Not-determined |
| NC3_vf_k141_791 | Not-determined |
| NC3_vf_k141_518 | Not-determined |
| NC3_vf_k141_920 | Not-determined |
| NC3_vf_k141_1037 | Not-determined |
| NC3_vf_k141_792 | Not-determined |
| NC3_vf_k141_519 | Not-determined |
| NC3_vf_k141_921 | Not-determined |
| NC3_vf_k141_1038 | Not-determined |
| NC3_vf_k141_520 | Low-quality |
| NC3_vf_k141_1313 | Not-determined |
| NC3_vf_k141_793 | Not-determined |
| NC3_vf_k141_521 | Not-determined |
| NC3_vf_k141_922 | Low-quality |
| NC3_vf_k141_1314 | Not-determined |
| NC3_vf_k141_923 | Not-determined |
| NC3_vf_k141_794 | Not-determined |
| NC3_vf_k141_924 | Not-determined |
| NC3_vf_k141_795 | Not-determined |
| NC3_vf_k141_522 | Low-quality |
| NC3_vf_k141_1315 | Not-determined |
| NC3_vf_k141_796 | Not-determined |
| NC3_vf_k141_523 | Not-determined |
| NC3_vf_k141_925 | Not-determined |
| NC3_vf_k141_1316 | Not-determined |
| NC3_vf_k141_797 | Not-determined |
| NC3_vf_k141_524 | Not-determined |
| NC3_vf_k141_926 | Not-determined |
| NC3_vf_k141_1317 | Not-determined |
| NC3_vf_k141_1039 | Not-determined |
| NC3_vf_k141_525 | Not-determined |
| NC3_vf_k141_1318 | Not-determined |
| NC3_vf_k141_526 | Not-determined |
| NC3_vf_k141_1040 | Not-determined |
| NC3_vf_k141_1319 | Not-determined |
| NC3_vf_k141_927 | Not-determined |
| NC3_vf_k141_527 | Not-determined |
| NC3_vf_k141_1041 | Not-determined |
| NC3_vf_k141_1320 | Not-determined |
| NC3_vf_k141_1042 | Low-quality |
| NC3_vf_k141_928 | Not-determined |
| NC3_vf_k141_528 | Not-determined |
| NC3_vf_k141_1321 | Not-determined |
| NC3_vf_k141_1043 | Not-determined |
| NC3_vf_k141_1322 | Not-determined |
| NC3_vf_k141_929 | Not-determined |
| NC3_vf_k141_1323 | Not-determined |
| NC3_vf_k141_529 | Not-determined |
| NC3_vf_k141_930 | Low-quality |
| NC3_vf_k141_1044 | Not-determined |
| NC3_vf_k141_530 | Not-determined |
| NC3_vf_k141_1324 | Not-determined |
| NC3_vf_k141_1045 | Not-determined |
| NC3_vf_k141_1046 | Not-determined |
| NC3_vf_k141_531 | Not-determined |
| NC3_vf_k141_1325 | Not-determined |
| NC3_vf_k141_1326 | Not-determined |
| NC3_vf_k141_1047 | Not-determined |
| NC3_vf_k141_1327 | Low-quality |
| NC3_vf_k141_1048 | Not-determined |
| NC3_vf_k141_1328 | Not-determined |
| NC3_vf_k141_1049 | Not-determined |
| NC3_vf_k141_1050 | Not-determined |
| NC3_vf_k141_1329 | Not-determined |
| NC3_vf_k141_1051 | Low-quality |
| NC3_vf_k141_1052 | Low-quality |
| NC3_vf_k141_1053 | Not-determined |
| NC3_vf_k141_1054 | Not-determined |
| NC3_vf_k141_1055 | Not-determined |
| NC3_vf_k141_1056 | Not-determined |
| NC3_vf_k141_1057 | Low-quality |
| NC3_vf_k141_1058 | Not-determined |
| NC3_vf_k141_1059 | Not-determined |
| NC3_vf_k141_1060 | Not-determined |
| NC3_vf_k141_1061 | Not-determined |
| NC3_vf_k141_1062 | Not-determined |
| NC3_vf_k141_1063 | Not-determined |
| NC3_vs_k141_1866\|\|full | Not-determined |
| NC3_vs_k141_1200\|\|full | Low-quality |
| NC3_vs_k141_136\|\|full | Low-quality |
| NC3_vs_k141_1999\|\|full | Low-quality |
| NC3_vs_k141_275\|\|full | Low-quality |
| NC3_vs_k141_939\|\|full | Low-quality |
| NC3_vs_k141_280\|\|full | Low-quality |
| NC3_vs_k141_281\|\|full | Low-quality |
| NC3_vs_k141_284\|\|full | Low-quality |
| NC3_vs_k141_814\|\|full | Not-determined |
| NC3_vs_k141_1206\|\|full | Low-quality |
| NC3_vs_k141_563\|\|full | Low-quality |
| NC3_vs_k141_1496\|\|full | Low-quality |
| NC3_vs_k141_1210\|\|full | Low-quality |
| NC3_vs_k141_956\|\|full | Low-quality |
| NC3_vs_k141_1633\|\|full | Low-quality |
| NC3_vs_k141_957\|\|full | Low-quality |
| NC3_vs_k141_1507\|\|full | Low-quality |
| NC3_vs_k141_1106\|\|full | Low-quality |
| NC3_vs_k141_181\|\|full | Not-determined |
| NC3_vs_k141_1779\|\|full | Not-determined |
| NC3_vs_k141_582\|\|full | Low-quality |
| NC3_vs_k141_707\|\|full | Low-quality |
| NC3_vs_k141_1113\|\|full | Low-quality |
| NC3_vs_k141_843\|\|full | Low-quality |
| NC3_vs_k141_443\|\|full | Low-quality |
| NC3_vs_k141_1794\|\|full | Low-quality |
| NC3_vs_k141_195\|\|full | Low-quality |
| NC3_vs_k141_719\|\|full | Low-quality |
| NC3_vs_k141_71\|\|full | Low-quality |
| NC3_vs_k141_449\|\|full | Low-quality |
| NC3_vs_k141_1236\|\|full | Low-quality |
| NC3_vs_k141_598\|\|full | Low-quality |
| NC3_vs_k141_1934\|\|full | Low-quality |
| NC3_vs_k141_328\|\|full | Low-quality |
| NC3_vs_k141_334\|\|full | Low-quality |
| NC3_vs_k141_1149\|\|full | Low-quality |
| NC3_vs_k141_1154\|\|full | Low-quality |
| NC3_vs_k141_986\|\|full | Low-quality |
| NC3_vs_k141_1954\|\|full | Low-quality |
| NC3_vs_k141_347\|\|full | Low-quality |
| NC3_vs_k141_2073\|\|full | Not-determined |
| NC3_vs_k141_1965\|\|full | Low-quality |
| NC3_vs_k141_1168\|\|full | Low-quality |
| NC3_vs_k141_1967\|\|full | Low-quality |
| NC3_vs_k141_1174\|\|full | Low-quality |
| NC3_vs_k141_245\|\|full | Low-quality |
| NC3_vs_k141_997\|\|full | Low-quality |
| NC3_vs_k141_1575\|\|full | Low-quality |
| NC3_vs_k141_1268\|\|full | Not-determined |
| NC3_vs_k141_1858\|\|full | Low-quality |
| NC3_vs_k141_254\|\|full | Low-quality |
| NC3_vs_k141_380\|\|full | Low-quality |
| NC3_vs_k141_1713\|\|full | Low-quality |
| NC3_vs_k141_381\|\|full | Not-determined |
| NC3_vs_k141_131\|\|full | Low-quality |
| NC3_vs_k141_753\|\|full | Low-quality |
| NC3_vs_k141_1007\|\|full | Low-quality |
| NC3_vs_k141_1594\|\|full | Not-determined |
| NC3_vs_k141_1012\|\|full | Low-quality |
| NC3_vs_k141_501\|\|full | Low-quality |
| NC3_vs_k141_1019\|\|full | Low-quality |
| NC3_vs_k141_1296\|\|full | Not-determined |
| NC3_vs_k141_2122\|\|full | Low-quality |
| NC3_vs_k141_515\|\|full | Low-quality |
| NC3_vs_k141_1051\|\|full | Low-quality |
| NC3_vs_k141_266\|\|lt2gene | Not-determined |
| NC3_vs_k141_536\|\|lt2gene | Not-determined |
| NC3_vs_k141_1066\|\|lt2gene | Low-quality |

| | |
|---|---|
| NC3_vs_k141_936\|\|lt2gene | Not-determined |
| NC3_vs_k141_1604\|\|lt2gene | Low-quality |
| NC3_vs_k141_18\|\|lt2gene | Not-determined |
| NC3_vs_k141_678\|\|lt2gene | Not-determined |
| NC3_vs_k141_288\|\|lt2gene | Low-quality |
| NC3_vs_k141_1350\|\|lt2gene | Not-determined |
| NC3_vs_k141_2023\|\|lt2gene | Not-determined |
| NC3_vs_k141_567\|\|lt2gene | Not-determined |
| NC3_vs_k141_1631\|\|lt2gene | Not-determined |
| NC3_vs_k141_1903\|\|lt2gene | Low-quality |
| NC3_vs_k141_421\|\|lt2gene | Low-quality |
| NC3_vs_k141_575\|\|lt2gene | Low-quality |
| NC3_vs_k141_303\|\|lt2gene | Low-quality |
| NC3_vs_k141_49\|\|lt2gene | Low-quality |
| NC3_vs_k141_1224\|\|lt2gene | Not-determined |
| NC3_vs_k141_841\|\|lt2gene | Not-determined |
| NC3_vs_k141_586\|\|lt2gene | Not-determined |
| NC3_vs_k141_310\|\|lt2gene | Not-determined |
| NC3_vs_k141_1118\|\|lt2gene | Not-determined |
| NC3_vs_k141_1790\|\|lt2gene | Not-determined |
| NC3_vs_k141_318\|\|lt2gene | Not-determined |
| NC3_vs_k141_1921\|\|lt2gene | Not-determined |
| NC3_vs_k141_1229\|\|lt2gene | Not-determined |
| NC3_vs_k141_850\|\|lt2gene | Not-determined |
| NC3_vs_k141_206\|\|lt2gene | Not-determined |
| NC3_vs_k141_1241\|\|lt2gene | Low-quality |
| NC3_vs_k141_1669\|\|lt2gene | Not-determined |
| NC3_vs_k141_2055\|\|lt2gene | Not-determined |
| NC3_vs_k141_95\|\|lt2gene | Low-quality |
| NC3_vs_k141_983\|\|lt2gene | Low-quality |
| NC3_vs_k141_2064\|\|lt2gene | Not-determined |
| NC3_vs_k141_351\|\|lt2gene | Low-quality |
| NC3_vs_k141_1957\|\|lt2gene | Low-quality |
| NC3_vs_k141_1687\|\|lt2gene | Low-quality |
| NC3_vs_k141_1169\|\|lt2gene | Not-determined |
| NC3_vs_k141_1565\|\|lt2gene | Not-determined |
| NC3_vs_k141_1692\|\|lt2gene | Not-determined |
| NC3_vs_k141_1416\|\|lt2gene | Low-quality |
| NC3_vs_k141_127\|\|lt2gene | Low-quality |
| NC3_vs_k141_1711\|\|lt2gene | Not-determined |
| NC3_vs_k141_489\|\|lt2gene | Not-determined |
| NC3_vs_k141_650\|\|lt2gene | Low-quality |
| NC3_vs_k141_1273\|\|lt2gene | Not-determined |
| NC3_vs_k141_1435\|\|lt2gene | Not-determined |
| NC3_vs_k141_1727\|\|lt2gene | Low-quality |
| NC3_vs_k141_396\|\|lt2gene | Low-quality |
| NC3_vs_k141_660\|\|lt2gene | Low-quality |
| NC3_vs_k141_783\|\|lt2gene | Not-determined |
| NC3_vs_k141_787\|\|lt2gene | Low-quality |
| NC3_vs_k141_1035\|\|lt2gene | Low-quality |
| NC3_vs_k141_522\|\|lt2gene | Low-quality |
| NC3_vs_k141_928\|\|lt2gene | Not-determined |
| S16_extracted_1 | Not-determined |
| S16_extracted_2 | Not-determined |
| S16_extracted_13 | Not-determined |
| S16_extracted_14 | Low-quality |
| S16_extracted_30 | Not-determined |
| S16_extracted_50 | Not-determined |
| S16_extracted_63 | Not-determined |
| S16_extracted_77 | Not-determined |
| S16_extracted_94 | Not-determined |
| S16_extracted_96 | Not-determined |
| S16_extracted_115 | Not-determined |
| S16_extracted_121 | Not-determined |
| S16_extracted_129 | Not-determined |
| S16_extracted_144 | Not-determined |
| S16_extracted_155 | Low-quality |
| S16_extracted_167 | Not-determined |
| S16_extracted_177 | Not-determined |
| S16_extracted_184 | Not-determined |
| S16_extracted_187 | Not-determined |
| S16_extracted_210 | Low-quality |
| S16_extracted_215 | Not-determined |
| S16_extracted_229 | Low-quality |
| S16_extracted_237 | Not-determined |
| S16_extracted_247 | Not-determined |
| S16_extracted_249 | Not-determined |
| S16_extracted_259 | Not-determined |
| S16_extracted_265 | Not-determined |
| S16_extracted_302 | Not-determined |
| S16_extracted_326 | Not-determined |
| S16_extracted_333 | Not-determined |
| S16_extracted_354 | Not-determined |
| S16_extracted_356 | Not-determined |
| S16_extracted_369 | Not-determined |
| S16_extracted_378 | Not-determined |
| S16_extracted_395 | Low-quality |
| S16_extracted_410 | Not-determined |
| S16_extracted_411 | Not-determined |
| S16_extracted_416 | Not-determined |
| S16_extracted_420 | Not-determined |
| S16_extracted_424 | Not-determined |
| S16_extracted_429 | Not-determined |
| S16_extracted_455 | Not-determined |
| S16_extracted_458 | Not-determined |
| S16_extracted_465 | Not-determined |
| S16_extracted_467 | Not-determined |
| S16_extracted_483 | Not-determined |
| S16_extracted_487 | Not-determined |
| S16_extracted_498 | Low-quality |
| S16_extracted_548 | Not-determined |
| S16_extracted_552 | Low-quality |
| S16_extracted_560 | Not-determined |
| S16_extracted_566 | Not-determined |
| S16_extracted_575 | Not-determined |
| S16_extracted_609 | Not-determined |
| S16_extracted_626 | Not-determined |
| S16_extracted_643 | Not-determined |
| S16_extracted_660 | Not-determined |
| S16_extracted_683 | Not-determined |
| S16_extracted_686 | Not-determined |
| S16_extracted_688 | Low-quality |
| S16_extracted_695 | Not-determined |
| S16_extracted_712 | Not-determined |
| S16_extracted_721 | Not-determined |
| S16_extracted_731 | Not-determined |
| S16_extracted_733 | Not-determined |
| S16_extracted_743 | Not-determined |
| S16_extracted_749 | Not-determined |
| S16_extracted_750 | Not-determined |
| S16_extracted_762 | Not-determined |
| S16_extracted_764 | Not-determined |
| S16_extracted_806 | Not-determined |
| S16_extracted_809 | Not-determined |
| S16_extracted_825 | Low-quality |
| S16_extracted_827 | Not-determined |
| S16_extracted_830 | Low-quality |
| S16_extracted_839 | Not-determined |
| S16_extracted_848 | Not-determined |
| S16_extracted_872 | Low-quality |
| S16_extracted_875 | Not-determined |
| S16_extracted_884 | Not-determined |
| S16_extracted_907 | Low-quality |
| S16_extracted_912 | Not-determined |
| S16_extracted_914 | Not-determined |
| S16_extracted_917 | Not-determined |
| S16_extracted_942 | Not-determined |
| S16_extracted_945 | Not-determined |
| S16_extracted_966 | Not-determined |
| S16_extracted_971 | Not-determined |
| S16_extracted_973 | Not-determined |
| S16_extracted_986 | Not-determined |
| S16_extracted_995 | Not-determined |
| S16_extracted_1005 | Low-quality |
| S16_extracted_1009 | Low-quality |
| S16_extracted_1011 | Not-determined |
| S16_extracted_1018 | Not-determined |
| S16_extracted_1030 | Not-determined |
| S16_vs_10622\|\|full | Medium-quality |
| S16_extracted_1039 | Low-quality |
| S16_extracted_1043 | Not-determined |
| S16_extracted_1044 | Low-quality |
| S16_extracted_1064 | Not-determined |
| S16_extracted_1065 | Not-determined |
| S16_extracted_1067 | Not-determined |
| S16_extracted_1070 | Not-determined |
| S16_extracted_1080 | Not-determined |
| S16_extracted_1090 | Not-determined |
| S16_extracted_1093 | Not-determined |
| S16_extracted_1101 | Not-determined |
| S16_extracted_1103 | Not-determined |
| S16_extracted_1130 | Not-determined |
| S16_extracted_1169 | Not-determined |
| S16_extracted_1179 | Not-determined |
| S16_extracted_1192 | Not-determined |
| S16_extracted_1204 | Low-quality |
| S16_extracted_1218 | Not-determined |
| S16_extracted_1222 | Not-determined |
| S16_extracted_1237 | Not-determined |
| S16_extracted_1269 | Not-determined |
| S16_extracted_1278 | Not-determined |
| S16_extracted_1289 | Not-determined |
| S16_extracted_1314 | Not-determined |
| S16_extracted_1318 | Not-determined |
| S16_extracted_1329 | Not-determined |
| S16_extracted_1338 | Not-determined |
| S16_extracted_1344 | Low-quality |
| S16_extracted_1365 | Low-quality |
| S16_extracted_1367 | Not-determined |
| S16_extracted_1374 | Not-determined |
| S16_extracted_1389 | Not-determined |
| S16_extracted_1390 | Low-quality |
| S16_extracted_1393 | Low-quality |
| S16_extracted_1408 | Low-quality |
| S16_extracted_1414 | Not-determined |
| S16_extracted_1420 | Not-determined |
| S16_extracted_1438 | Not-determined |
| S16_extracted_1449 | Not-determined |
| S16_extracted_1452 | Low-quality |
| S16_extracted_1457 | Not-determined |
| S16_extracted_1460 | Not-determined |
| S16_extracted_1463 | Not-determined |
| S16_extracted_1479 | Not-determined |
| S16_extracted_1485 | Not-determined |
| S16_extracted_1489 | Not-determined |
| S16_extracted_1490 | Not-determined |
| S16_extracted_1491 | Not-determined |
| S16_extracted_1492 | Not-determined |
| S16_extracted_1506 | Low-quality |
| S16_extracted_1514 | Not-determined |
| S16_extracted_1530 | Not-determined |
| S16_extracted_1551 | Not-determined |
| S16_extracted_1557 | Not-determined |
| S16_extracted_1571 | Not-determined |
| S16_extracted_1572 | Not-determined |
| S16_extracted_1574 | Low-quality |
| S16_extracted_1584 | Not-determined |
| S16_extracted_1586 | Not-determined |
| S16_extracted_1588 | Not-determined |
| S16_extracted_1591 | Not-determined |
| S16_extracted_1601 | Not-determined |
| S16_extracted_1605 | Not-determined |
| S16_extracted_1615 | Not-determined |
| S16_extracted_1622 | Not-determined |
| S16_extracted_1630 | Not-determined |
| S16_extracted_1635 | Not-determined |
| S16_extracted_1646 | Not-determined |
| S16_extracted_1654 | Not-determined |
| S16_extracted_1692 | Not-determined |
| S16_extracted_1702 | Low-quality |
| S16_extracted_1703 | Not-determined |
| S16_extracted_1707 | Not-determined |
| S16_extracted_1708 | Not-determined |
| S16_extracted_1712 | Not-determined |
| S16_extracted_1715 | Low-quality |
| S16_extracted_1720 | Not-determined |
| S16_extracted_1722 | Not-determined |
| S16_extracted_1726 | Not-determined |
| S16_extracted_1729 | Not-determined |
| S16_extracted_1741 | Not-determined |
| S16_extracted_1742 | Not-determined |
| S16_extracted_1747 | Not-determined |
| S16_extracted_1752 | Not-determined |
| S16_extracted_1763 | Not-determined |
| S16_extracted_1765 | Not-determined |
| S16_extracted_1770 | Low-quality |
| S16_extracted_1798 | Not-determined |
| S16_extracted_1828 | Not-determined |
| S16_extracted_1841 | Low-quality |
| S16_extracted_1844 | Low-quality |
| S16_extracted_1850 | Not-determined |
| S16_extracted_1877 | Not-determined |
| S16_extracted_1879 | Not-determined |
| S16_extracted_1881 | Not-determined |
| S16_extracted_1924 | Not-determined |
| S16_extracted_1927 | Not-determined |
| S16_extracted_1929 | Not-determined |
| S16_extracted_1959 | Not-determined |
| S16_extracted_1960 | Not-determined |
| S16_extracted_1969 | Not-determined |
| S16_extracted_2013 | Not-determined |
| S16_extracted_2019 | Low-quality |
| S16_extracted_2024 | Not-determined |
| S16_extracted_2039 | Not-determined |
| S16_extracted_2048 | Low-quality |
| S16_extracted_2050 | Not-determined |
| S16_extracted_2059 | Not-determined |
| S16_extracted_2100 | Low-quality |
| S16_extracted_2109 | Not-determined |
| S16_extracted_2114 | Not-determined |
| S16_extracted_2165 | Not-determined |
| S16_extracted_2173 | Not-determined |
| S16_extracted_2180 | Low-quality |
| S16_extracted_2184 | Not-determined |
| S16_extracted_2216 | Not-determined |
| S16_extracted_2244 | Not-determined |
| S16_extracted_2247 | Not-determined |
| S16_extracted_2260 | Not-determined |
| S16_extracted_2282 | Not-determined |
| S16_extracted_2294 | Not-determined |
| S16_extracted_2301 | Not-determined |
| S16_extracted_2319 | Not-determined |
| S16_extracted_2323 | Low-quality |
| S16_extracted_2329 | Not-determined |
| S16_extracted_2333 | Low-quality |
| S16_extracted_2337 | Not-determined |
| S16_extracted_2344 | Not-determined |
| S16_extracted_2347 | Not-determined |
| S16_extracted_2362 | Not-determined |
| S16_extracted_2364 | Not-determined |
| S16_extracted_2391 | Low-quality |
| S16_extracted_2415 | Not-determined |
| S16_extracted_2445 | Not-determined |
| S16_extracted_2449 | Low-quality |
| S16_extracted_2452 | Not-determined |
| S16_extracted_2453 | Not-determined |
| S16_extracted_2463 | Not-determined |
| S16_extracted_2477 | Not-determined |
| S16_extracted_2482 | Low-quality |
| S16_extracted_2494 | Not-determined |
| S16_extracted_2496 | Not-determined |
| S16_extracted_2525 | Not-determined |
| S16_extracted_2590 | Low-quality |
| S16_extracted_2591 | Not-determined |
| S16_extracted_2593 | Not-determined |
| S16_extracted_2610 | Not-determined |
| S16_extracted_2612 | Not-determined |
| S16_extracted_2614 | Not-determined |
| S16_extracted_2620 | Not-determined |
| S16_extracted_2624 | Not-determined |
| S16_extracted_2628 | Low-quality |
| S16_extracted_2633 | Not-determined |
| S16_extracted_2635 | Not-determined |
| S16_extracted_2637 | Not-determined |
| S16_extracted_2644 | Not-determined |
| S16_extracted_2646 | Not-determined |
| S16_extracted_2655 | Not-determined |
| S16_extracted_2669 | Low-quality |
| S16_extracted_2672 | Low-quality |
| S16_extracted_2678 | Not-determined |
| S16_extracted_2704 | Low-quality |
| S16_extracted_2709 | Low-quality |
| S16_extracted_2713 | Not-determined |
| S16_extracted_2719 | Low-quality |
| S16_extracted_2727 | Not-determined |
| S16_extracted_2735 | Low-quality |
| S16_extracted_2736 | Low-quality |
| S16_extracted_2750 | Not-determined |
| S16_extracted_2761 | Not-determined |
| S16_extracted_2766 | Not-determined |
| S16_extracted_2767 | Not-determined |
| S16_extracted_2768 | Not-determined |
| S16_extracted_2782 | Not-determined |
| S16_extracted_2788 | Low-quality |
| S16_extracted_2812 | Not-determined |
| S16_extracted_2825 | Not-determined |
| S16_extracted_2855 | Not-determined |
| S16_extracted_2858 | Not-determined |
| S16_extracted_2860 | Low-quality |
| S16_extracted_2862 | Not-determined |
| S16_extracted_2865 | Not-determined |
| S16_extracted_2883 | Not-determined |
| S16_extracted_2897 | Not-determined |
| S16_extracted_2899 | Low-quality |
| S16_extracted_2900 | Not-determined |
| S16_extracted_2913 | Not-determined |
| S16_extracted_2928 | Not-determined |
| S16_extracted_2966 | Not-determined |
| S16_extracted_2969 | Not-determined |
| S16_extracted_2973 | Not-determined |
| S16_extracted_2976 | Not-determined |
| S16_extracted_2981 | Not-determined |
| S16_extracted_2994 | Not-determined |
| S16_extracted_2997 | Low-quality |
| S16_extracted_3003 | Not-determined |
| S16_extracted_3010 | Not-determined |
| S16_extracted_3026 | Not-determined |
| S16_extracted_3034 | Not-determined |
| S16_extracted_3052 | Not-determined |
| S16_extracted_3072 | Not-determined |
| S16_extracted_3077 | Not-determined |
| S16_extracted_3085 | Not-determined |
| S16_extracted_3093 | Not-determined |
| S16_extracted_3098 | Not-determined |
| S16_extracted_3115 | Not-determined |
| S16_extracted_3119 | Not-determined |
| S16_extracted_3122 | Not-determined |
| S16_extracted_3124 | Not-determined |
| S16_extracted_3127 | Not-determined |
| S16_extracted_3151 | Not-determined |
| S16_extracted_3155 | Not-determined |
| S16_extracted_3162 | Not-determined |
| S16_extracted_3164 | Not-determined |
| S16_extracted_3170 | Not-determined |
| S16_extracted_3175 | Not-determined |
| S16_extracted_3207 | Not-determined |
| S16_extracted_3210 | Not-determined |
| S16_extracted_3232 | Not-determined |
| S16_extracted_3254 | Not-determined |
| S16_extracted_3263 | Low-quality |
| S16_extracted_3283 | Low-quality |
| S16_extracted_3291 | Not-determined |
| S16_extracted_3315 | Not-determined |
| S16_extracted_3317 | Not-determined |
| S16_extracted_3318 | Not-determined |
| S16_extracted_3326 | Low-quality |
| S16_extracted_3328 | Not-determined |
| S16_extracted_3331 | Not-determined |
| S16_extracted_3332 | Not-determined |
| S16_extracted_3334 | Not-determined |
| S16_extracted_3336 | Not-determined |
| S16_extracted_3337 | Not-determined |
| S16_extracted_3347 | Not-determined |
| S16_extracted_3348 | Low-quality |
| S16_extracted_3381 | Not-determined |
| S16_extracted_3390 | Not-determined |
| S16_extracted_3425 | Not-determined |
| S16_extracted_3429 | Not-determined |
| S16_extracted_3433 | Low-quality |
| S16_extracted_3450 | Not-determined |
| S16_extracted_3452 | Not-determined |
| S16_extracted_3461 | Low-quality |
| S16_extracted_3479 | Not-determined |
| S16_extracted_3505 | Not-determined |
| S16_extracted_3519 | Not-determined |
| S16_extracted_3520 | Not-determined |
| S16_extracted_3543 | Not-determined |
| S16_extracted_3554 | Not-determined |
| S16_extracted_3571 | Not-determined |
| S16_extracted_3582 | Not-determined |
| S16_extracted_3583 | Not-determined |
| S16_extracted_3590 | Not-determined |
| S16_extracted_3604 | Low-quality |
| S16_extracted_3611 | Not-determined |
| S16_extracted_3618 | Not-determined |
| S16_extracted_3634 | Not-determined |
| S16_extracted_3637 | Not-determined |
| S16_extracted_3639 | Not-determined |
| S16_extracted_3641 | Not-determined |
| S16_extracted_3647 | Not-determined |
| S16_extracted_3652 | Not-determined |
| S16_extracted_3657 | Not-determined |
| S16_extracted_3664 | Not-determined |
| S16_extracted_3665 | Not-determined |
| S16_extracted_3680 | Not-determined |
| S16_extracted_3684 | Low-quality |
| S16_extracted_3691 | Not-determined |
| S16_extracted_3692 | Not-determined |
| S16_extracted_3700 | Not-determined |
| S16_extracted_3704 | Not-determined |
| S16_extracted_3722 | Not-determined |
| S16_extracted_3726 | Not-determined |
| S16_extracted_3743 | Not-determined |
| S16_extracted_3744 | Not-determined |
| S16_extracted_3757 | Not-determined |
| S16_extracted_3760 | Not-determined |
| S16_extracted_3762 | Not-determined |
| S16_extracted_3769 | Not-determined |
| S16_extracted_3771 | Not-determined |
| S16_extracted_3790 | Not-determined |
| S16_extracted_3796 | Low-quality |
| S16_extracted_3797 | Not-determined |
| S16_extracted_3808 | Not-determined |
| S16_extracted_3812 | Low-quality |
| S16_extracted_3823 | Not-determined |
| S16_extracted_3844 | Not-determined |
| S16_extracted_3852 | Not-determined |
| S16_extracted_3878 | Not-determined |
| S16_extracted_3895 | Not-determined |
| S16_extracted_3897 | Low-quality |
| S16_extracted_3924 | Not-determined |
| S16_extracted_3935 | Not-determined |
| S16_extracted_3938 | Not-determined |
| S16_extracted_3940 | Not-determined |
| S16_extracted_3942 | Low-quality |
| S16_extracted_3943 | Not-determined |
| S16_extracted_3944 | Not-determined |
| S16_extracted_3957 | Not-determined |
| S16_extracted_3979 | Not-determined |
| S16_extracted_3997 | Not-determined |
| S16_extracted_4003 | Low-quality |
| S16_extracted_4024 | Low-quality |
| S16_extracted_4033 | Low-quality |
| S16_extracted_4037 | Low-quality |
| S16_extracted_4041 | Not-determined |
| S16_extracted_4059 | Low-quality |
| S16_extracted_4061 | Not-determined |
| S16_extracted_4081 | Not-determined |
| S16_extracted_4082 | Low-quality |
| S16_extracted_4105 | Not-determined |
| S16_extracted_4127 | Not-determined |
| S16_extracted_4131 | Low-quality |
| S16_extracted_4134 | Low-quality |
| S16_extracted_4145 | Low-quality |
| S16_extracted_4150 | Not-determined |
| S16_extracted_4153 | Not-determined |
| S16_extracted_4161 | Low-quality |
| S16_extracted_4176 | Low-quality |
| S16_extracted_4205 | Not-determined |
| S16_extracted_4211 | Not-determined |
| S16_extracted_4263 | Not-determined |
| S16_extracted_4266 | Not-determined |
| S16_extracted_4268 | Not-determined |
| S16_extracted_4292 | Not-determined |
| S16_extracted_4305 | Not-determined |
| S16_extracted_4318 | Not-determined |
| S16_extracted_4321 | Not-determined |
| S16_extracted_4323 | Not-determined |
| S16_extracted_4332 | Not-determined |
| S16_extracted_4341 | Not-determined |
| S16_extracted_4375 | Not-determined |
| S16_extracted_4387 | Not-determined |
| S16_extracted_4409 | Not-determined |
| S16_extracted_4418 | Not-determined |
| S16_extracted_4419 | Not-determined |
| S16_extracted_4421 | Not-determined |
| S16_extracted_4424 | Not-determined |
| S16_extracted_4428 | Not-determined |
| S16_extracted_4474 | Not-determined |
| S16_extracted_4486 | Not-determined |
| S16_extracted_4489 | Not-determined |
| S16_extracted_4503 | Not-determined |
| S16_extracted_4508 | Not-determined |
| S16_extracted_4510 | Not-determined |
| S16_extracted_4513 | Not-determined |
| S16_extracted_4519 | Not-determined |
| S16_extracted_4531 | Not-determined |
| S16_extracted_4533 | Not-determined |
| S16_extracted_4549 | Not-determined |
| S16_extracted_4554 | Not-determined |
| S16_extracted_4567 | Not-determined |
| S16_extracted_4571 | Not-determined |
| S16_extracted_4581 | Low-quality |
| S16_extracted_4589 | Low-quality |
| S16_extracted_4591 | Not-determined |
| S16_extracted_4593 | Not-determined |
| S16_extracted_4595 | Not-determined |
| S16_extracted_4601 | Not-determined |
| S16_extracted_4615 | Not-determined |
| S16_extracted_4618 | Not-determined |
| S16_extracted_4625 | Not-determined |
| S16_extracted_4643 | Not-determined |
| S16_extracted_4654 | Low-quality |
| S16_extracted_4656 | Not-determined |
| S16_extracted_4664 | Not-determined |
| S16_extracted_4667 | Not-determined |
| S16_extracted_4680 | Low-quality |
| S16_extracted_4685 | Low-quality |
| S16_extracted_4698 | Not-determined |
| S16_extracted_4702 | Low-quality |
| S16_extracted_4705 | Not-determined |
| S16_extracted_4706 | Not-determined |
| S16_extracted_4731 | Not-determined |
| S16_extracted_4743 | Not-determined |
| S16_extracted_4751 | Not-determined |
| S16_extracted_4759 | Not-determined |
| S16_extracted_4761 | Not-determined |
| S16_extracted_4762 | Not-determined |
| S16_extracted_4763 | Not-determined |
| S16_extracted_4773 | Not-determined |
| S16_extracted_4779 | Not-determined |
| S16_extracted_4788 | Low-quality |
| S16_extracted_4796 | Not-determined |
| S16_extracted_4807 | Not-determined |
| S16_extracted_4814 | Not-determined |
| S16_extracted_4816 | Not-determined |
| S16_extracted_4819 | Not-determined |
| S16_extracted_4840 | Not-determined |
| S16_extracted_4875 | Not-determined |
| S16_extracted_4876 | Not-determined |
| S16_extracted_4885 | Not-determined |
| S16_extracted_4915 | Not-determined |
| S16_extracted_4937 | Not-determined |
| S16_extracted_4956 | Not-determined |
| S16_extracted_4958 | Not-determined |
| S16_extracted_4977 | Not-determined |
| S16_extracted_4992 | Not-determined |
| S16_extracted_4994 | Not-determined |
| S16_extracted_4999 | Not-determined |
| S16_extracted_5018 | Not-determined |
| S16_extracted_5031 | Not-determined |
| S16_extracted_5045 | Not-determined |
| S16_extracted_5047 | Not-determined |
| S16_extracted_5049 | Not-determined |
| S16_extracted_5051 | Not-determined |
| S16_extracted_5054 | Not-determined |
| S16_extracted_5063 | Low-quality |
| S16_extracted_5069 | Not-determined |
| S16_extracted_5071 | Not-determined |
| S16_extracted_5082 | Not-determined |
| S16_extracted_5084 | Not-determined |
| S16_extracted_5090 | Not-determined |
| S16_extracted_5099 | Not-determined |
| S16_extracted_5106 | Not-determined |
| S16_extracted_5109 | Not-determined |
| S16_extracted_5113 | Not-determined |
| S16_extracted_5120 | Not-determined |
| S16_extracted_5121 | Not-determined |
| S16_extracted_5122 | Not-determined |
| S16_extracted_5140 | Low-quality |
| S16_extracted_5153 | Not-determined |
| S16_extracted_5166 | Not-determined |
| S16_extracted_5170 | Not-determined |
| S16_extracted_5205 | Low-quality |
| S16_extracted_5207 | Not-determined |
| S16_extracted_5209 | Not-determined |
| S16_extracted_5229 | Not-determined |
| S16_extracted_5234 | Not-determined |
| S16_extracted_5240 | Not-determined |
| S16_extracted_5250 | Low-quality |
| S16_extracted_5251 | Not-determined |
| S16_extracted_5255 | Low-quality |
| S16_extracted_5257 | Not-determined |
| S16_extracted_5263 | Not-determined |
| S16_extracted_5268 | Not-determined |
| S14_vs_k141_117161\|\|full | High-quality |
| S16_extracted_5273 | Not-determined |
| S16_extracted_5274 | Low-quality |
| S16_extracted_5275 | Not-determined |

| | |
|---|---|
| S16_extracted_5289 | Not-determined |
| S16_extracted_5301 | Not-determined |
| S16_extracted_5304 | Not-determined |
| S16_extracted_5322 | Low-quality |
| S16_extracted_5327 | Not-determined |
| S16_extracted_5332 | Low-quality |
| S16_extracted_5337 | Not-determined |
| S16_extracted_5355 | Not-determined |
| S16_extracted_5368 | Not-determined |
| S16_extracted_5373 | Not-determined |
| S16_extracted_5396 | Not-determined |
| S16_extracted_5403 | Not-determined |
| S16_extracted_5405 | Not-determined |
| S16_extracted_5408 | Not-determined |
| S16_extracted_5412 | Not-determined |
| S16_extracted_5413 | Not-determined |
| S16_extracted_5416 | Not-determined |
| S16_extracted_5441 | Low-quality |
| S16_extracted_5442 | Not-determined |
| S16_extracted_5447 | Not-determined |
| S16_extracted_5466 | Not-determined |
| S16_extracted_5483 | Not-determined |
| S16_extracted_5537 | Not-determined |
| S16_extracted_5540 | Not-determined |
| S16_extracted_5545 | Not-determined |
| S16_extracted_5546 | Not-determined |
| S16_extracted_5561 | Not-determined |
| S16_extracted_5563 | Not-determined |
| S16_extracted_5566 | Low-quality |
| S16_extracted_5567 | Not-determined |
| S16_extracted_5586 | Not-determined |
| S16_extracted_5608 | Not-determined |
| S16_extracted_5621 | Not-determined |
| S16_extracted_5630 | Not-determined |
| S16_extracted_5650 | Not-determined |
| S16_extracted_5669 | Not-determined |
| S16_extracted_5672 | Not-determined |
| S16_extracted_5674 | Not-determined |
| S16_extracted_5676 | Not-determined |
| S16_extracted_5677 | Not-determined |
| S16_extracted_5697 | Not-determined |
| S16_extracted_5716 | Low-quality |
| S16_extracted_5737 | Low-quality |
| S16_extracted_5741 | Not-determined |
| S16_extracted_5754 | Not-determined |
| S16_extracted_5761 | Not-determined |
| S16_extracted_5775 | Not-determined |
| S16_extracted_5786 | Low-quality |
| S16_extracted_5792 | Not-determined |
| S16_extracted_5801 | Not-determined |
| S16_extracted_5804 | Not-determined |
| S13_extracted_147396 | Complete |
| S16_extracted_5810 | Low-quality |
| S16_extracted_5820 | Not-determined |
| S16_extracted_5821 | Not-determined |
| S16_extracted_5835 | Not-determined |
| S16_extracted_5836 | Low-quality |
| S16_extracted_5859 | Not-determined |
| S16_extracted_5862 | Not-determined |
| S16_extracted_5896 | Not-determined |
| S16_extracted_5898 | Not-determined |
| S16_extracted_5906 | Not-determined |
| S16_extracted_5911 | Low-quality |
| S16_extracted_5924 | Low-quality |
| S16_extracted_5952 | Low-quality |
| S16_extracted_5954 | Not-determined |
| S16_extracted_5960 | Not-determined |
| S16_extracted_5962 | Low-quality |
| S16_extracted_5964 | Not-determined |
| S16_extracted_5965 | Not-determined |
| S16_extracted_5982 | Not-determined |
| S16_extracted_5987 | Not-determined |
| S16_extracted_6003 | Not-determined |
| S16_extracted_6011 | Not-determined |
| S16_extracted_6012 | Not-determined |
| S16_extracted_6024 | Not-determined |
| S16_extracted_6040 | Not-determined |
| S16_extracted_6047 | Not-determined |
| S16_extracted_6075 | Not-determined |
| S16_extracted_6077 | Not-determined |
| S16_extracted_6080 | Not-determined |
| S16_extracted_6092 | Not-determined |
| S5_vs_k141_113268\|\|full | Medium-quality |
| S16_extracted_6109 | Not-determined |
| S16_extracted_6115 | Not-determined |
| S16_extracted_6130 | Not-determined |
| S16_extracted_6131 | Not-determined |
| S16_extracted_6147 | Not-determined |
| S16_extracted_6175 | Not-determined |
| S16_extracted_6177 | Not-determined |
| S16_extracted_6195 | Low-quality |
| S16_extracted_6197 | Not-determined |
| S16_extracted_6198 | Not-determined |
| S16_extracted_6210 | Low-quality |
| S16_extracted_6213 | Not-determined |
| S16_extracted_6241 | Not-determined |
| S16_extracted_6242 | Not-determined |
| S16_extracted_6246 | Not-determined |
| S16_extracted_6248 | Not-determined |
| S16_extracted_6254 | Low-quality |
| S16_extracted_6293 | Not-determined |
| S16_extracted_6296 | Not-determined |
| S16_extracted_6308 | Low-quality |
| S16_extracted_6314 | Not-determined |
| S16_extracted_6324 | Not-determined |
| S16_extracted_6327 | Low-quality |
| S16_extracted_6330 | Not-determined |
| S16_extracted_6332 | Not-determined |
| S16_extracted_6355 | Not-determined |
| S16_extracted_6359 | Not-determined |
| S16_extracted_6372 | Low-quality |
| S16_extracted_6374 | Not-determined |
| S16_extracted_6383 | Not-determined |
| S16_extracted_6398 | Not-determined |
| S16_extracted_6412 | Not-determined |
| S16_extracted_6437 | Not-determined |
| S16_extracted_6441 | Not-determined |
| S16_extracted_6444 | Not-determined |
| S16_extracted_6494 | Not-determined |
| S16_extracted_6509 | Not-determined |
| S16_extracted_6521 | Not-determined |
| S16_extracted_6528 | Not-determined |
| S16_extracted_6542 | Low-quality |
| S16_extracted_6552 | Not-determined |
| S16_extracted_6566 | Not-determined |
| S16_extracted_6579 | Not-determined |
| S16_extracted_6649 | Not-determined |
| S16_extracted_6654 | Low-quality |
| S16_extracted_6663 | Not-determined |
| S16_extracted_6686 | Low-quality |
| S16_extracted_6711 | Not-determined |
| S16_extracted_6713 | Not-determined |
| S16_extracted_6721 | Not-determined |
| S16_extracted_6723 | Not-determined |
| S16_extracted_6730 | Not-determined |
| S16_extracted_6745 | Not-determined |
| S16_extracted_6762 | Low-quality |
| S16_extracted_6771 | Not-determined |
| S16_extracted_6776 | Not-determined |
| S16_extracted_6779 | Not-determined |
| S16_extracted_6799 | Not-determined |
| S16_extracted_6814 | Not-determined |
| S16_extracted_6822 | Not-determined |
| S16_extracted_6823 | Not-determined |
| S16_extracted_6833 | Not-determined |
| S16_extracted_6838 | Not-determined |
| S16_extracted_6846 | Not-determined |
| S16_extracted_6855 | Not-determined |
| S16_extracted_6860 | Not-determined |
| S16_extracted_6871 | Not-determined |
| S16_extracted_6873 | Not-determined |
| S16_extracted_6891 | Not-determined |
| S16_extracted_6892 | Not-determined |
| S16_extracted_6915 | Not-determined |
| S16_extracted_6916 | Not-determined |
| S16_extracted_6920 | Not-determined |
| S16_extracted_6926 | Not-determined |
| S16_extracted_6942 | Not-determined |
| S16_extracted_6977 | Not-determined |
| S16_extracted_6978 | Not-determined |
| S16_extracted_6980 | Low-quality |
| S16_extracted_6982 | Not-determined |
| S16_extracted_6985 | Not-determined |
| S16_extracted_6986 | Not-determined |
| S16_extracted_6990 | Low-quality |
| S16_extracted_6993 | Not-determined |
| S16_extracted_7020 | Not-determined |
| S16_extracted_7021 | Not-determined |
| S16_extracted_7025 | Not-determined |
| S16_extracted_7038 | Not-determined |
| S16_extracted_7056 | Low-quality |
| S16_extracted_7071 | Not-determined |
| S16_extracted_7084 | Low-quality |
| S16_extracted_7093 | Not-determined |
| S16_extracted_7105 | Low-quality |
| S16_extracted_7108 | Not-determined |
| S16_extracted_7139 | Not-determined |
| S16_extracted_7143 | Not-determined |
| S16_extracted_7147 | Low-quality |
| S16_extracted_7164 | Not-determined |
| S16_extracted_7179 | Not-determined |
| S16_extracted_7219 | Not-determined |
| S16_extracted_7226 | Not-determined |
| S16_extracted_7247 | Not-determined |
| S16_extracted_7255 | Low-quality |
| S16_extracted_7268 | Not-determined |
| S16_extracted_7282 | Low-quality |
| S16_extracted_7293 | Not-determined |
| S16_extracted_7298 | Not-determined |
| S16_extracted_7314 | Not-determined |
| S16_extracted_7319 | Not-determined |
| S16_extracted_7324 | Not-determined |
| S16_extracted_7329 | Not-determined |
| S16_extracted_7335 | Not-determined |
| S16_extracted_7349 | Not-determined |
| S16_extracted_7372 | Not-determined |
| S16_extracted_7374 | Not-determined |
| S16_extracted_7387 | Not-determined |
| S16_extracted_7395 | Not-determined |
| S16_extracted_7402 | Not-determined |
| S16_extracted_7414 | Not-determined |
| S16_extracted_7429 | Not-determined |
| S16_extracted_7431 | Not-determined |
| S16_extracted_7444 | Not-determined |
| S16_extracted_7448 | Not-determined |
| S16_extracted_7451 | Not-determined |
| S16_extracted_7467 | Not-determined |
| S16_extracted_7486 | Low-quality |
| S16_extracted_7504 | Low-quality |
| S16_extracted_7548 | Not-determined |
| S16_extracted_7550 | Not-determined |
| S16_extracted_7552 | Not-determined |
| S16_extracted_7559 | Not-determined |
| S16_extracted_7561 | Not-determined |
| S16_extracted_7562 | Not-determined |
| S16_extracted_7570 | Not-determined |
| S16_extracted_7575 | Low-quality |
| S16_extracted_7576 | Not-determined |
| S16_extracted_7580 | Not-determined |
| S16_extracted_7610 | Not-determined |
| S16_extracted_7624 | Not-determined |
| S16_extracted_7633 | Not-determined |
| S16_extracted_7634 | Not-determined |
| S16_extracted_7637 | Not-determined |
| S16_extracted_7646 | Low-quality |
| S16_extracted_7653 | Not-determined |
| S16_extracted_7658 | Not-determined |
| S16_extracted_7664 | Not-determined |
| S16_extracted_7699 | Not-determined |
| S16_extracted_7732 | Not-determined |
| S16_extracted_7734 | Not-determined |
| S16_extracted_7747 | Not-determined |
| S16_extracted_7768 | Not-determined |
| S16_extracted_7773 | Not-determined |
| S16_extracted_7789 | Low-quality |
| S16_extracted_7794 | Low-quality |
| S16_extracted_7798 | Low-quality |
| S16_extracted_7800 | Not-determined |
| S16_extracted_7801 | Not-determined |
| S16_extracted_7806 | Not-determined |
| S16_extracted_7810 | Not-determined |
| S16_extracted_7815 | Not-determined |
| S16_extracted_7846 | Not-determined |
| S16_extracted_7857 | Low-quality |
| S16_extracted_7865 | Low-quality |
| S16_extracted_7875 | Not-determined |
| S16_extracted_7876 | Not-determined |
| S16_extracted_7882 | Not-determined |
| S16_extracted_7894 | Not-determined |
| S16_extracted_7900 | Not-determined |
| S16_extracted_7911 | Low-quality |
| S16_extracted_7922 | Not-determined |
| S16_extracted_7927 | Not-determined |
| S16_extracted_7931 | Low-quality |
| S16_extracted_7936 | Not-determined |
| S16_extracted_7944 | Low-quality |
| S16_extracted_7949 | Not-determined |
| S16_extracted_7977 | Not-determined |
| S16_extracted_8001 | Not-determined |
| S16_extracted_8003 | Not-determined |
| S16_extracted_8006 | Not-determined |
| S16_extracted_8010 | Not-determined |
| S16_extracted_8013 | Not-determined |
| S16_extracted_8024 | Low-quality |
| S16_extracted_8025 | Not-determined |
| S16_extracted_8047 | Not-determined |
| S16_extracted_8049 | Not-determined |
| S16_extracted_8062 | Low-quality |
| S16_extracted_8071 | Low-quality |
| S16_extracted_8078 | Not-determined |
| S16_extracted_8084 | Not-determined |
| S16_extracted_8090 | Not-determined |
| S16_extracted_8092 | Not-determined |
| S16_extracted_8102 | Not-determined |
| S16_extracted_8105 | Not-determined |
| S16_extracted_8107 | Not-determined |
| S16_extracted_8136 | Not-determined |
| S16_extracted_8157 | Not-determined |
| S16_extracted_8161 | Not-determined |
| S16_extracted_8164 | Not-determined |
| S16_extracted_8172 | Not-determined |
| S16_extracted_8180 | Not-determined |
| S16_extracted_8181 | Not-determined |
| S16_extracted_8202 | Not-determined |
| S16_extracted_8212 | Not-determined |
| S16_extracted_8243 | Not-determined |
| S16_extracted_8260 | Not-determined |
| S16_extracted_8261 | Not-determined |
| S16_extracted_8289 | Not-determined |
| S16_extracted_8304 | Low-quality |
| S16_extracted_8314 | Low-quality |
| S16_extracted_8331 | Not-determined |
| S16_extracted_8340 | Not-determined |
| S16_extracted_8358 | Not-determined |
| S16_extracted_8360 | Not-determined |
| S16_extracted_8366 | Not-determined |
| S16_extracted_8370 | Low-quality |
| S16_extracted_8373 | Not-determined |
| S16_extracted_8378 | Not-determined |
| S16_extracted_8379 | Not-determined |
| S16_extracted_8385 | Low-quality |
| S16_extracted_8390 | Low-quality |
| S16_extracted_8391 | Not-determined |
| S16_extracted_8406 | Low-quality |
| S16_extracted_8409 | Not-determined |
| S16_extracted_8419 | Not-determined |
| S16_extracted_8430 | Not-determined |
| S16_extracted_8432 | Not-determined |
| S16_extracted_8437 | Low-quality |
| S16_extracted_8446 | Not-determined |
| S16_extracted_8467 | Not-determined |
| S16_extracted_8481 | Not-determined |
| S16_extracted_8484 | Not-determined |
| S16_extracted_8486 | Not-determined |
| S16_extracted_8489 | Not-determined |
| S10_vs_120570\|\|full | High-quality |
| S16_extracted_8498 | Not-determined |
| S16_extracted_8505 | Low-quality |
| S16_extracted_8507 | Not-determined |
| S16_extracted_8520 | Not-determined |
| S16_extracted_8523 | Not-determined |
| S16_extracted_8525 | Low-quality |
| S16_extracted_8531 | Not-determined |
| S16_extracted_8545 | Low-quality |
| S16_extracted_8553 | Low-quality |
| S16_extracted_8563 | Not-determined |
| S16_extracted_8567 | Low-quality |
| S16_extracted_8582 | Not-determined |
| S16_extracted_8583 | Not-determined |
| S16_extracted_8598 | Not-determined |
| S16_extracted_8614 | Not-determined |
| S16_extracted_8634 | Not-determined |
| S16_extracted_8643 | Not-determined |
| S16_extracted_8647 | Low-quality |
| S16_extracted_8648 | Low-quality |
| S16_extracted_8654 | Not-determined |
| S16_extracted_8657 | Not-determined |
| S16_extracted_8660 | Not-determined |
| S16_extracted_8663 | Low-quality |
| S16_extracted_8668 | Not-determined |
| S16_extracted_8680 | Not-determined |
| S16_extracted_8683 | Not-determined |
| S16_extracted_8693 | Not-determined |
| S16_extracted_8695 | Not-determined |
| S16_extracted_8696 | Not-determined |
| S16_extracted_8714 | Not-determined |
| S16_extracted_8720 | Not-determined |
| S16_extracted_8749 | Not-determined |
| S16_extracted_8750 | Not-determined |
| S16_extracted_8761 | Low-quality |
| S16_extracted_8773 | Not-determined |
| S16_extracted_8787 | Not-determined |
| S16_extracted_8789 | Not-determined |
| S16_extracted_8792 | Not-determined |
| S16_extracted_8798 | Not-determined |
| S16_extracted_8807 | Not-determined |
| S16_extracted_8810 | Not-determined |
| S16_extracted_8825 | Not-determined |
| S16_extracted_8832 | Not-determined |
| S16_extracted_8845 | Low-quality |
| S16_extracted_8847 | Not-determined |
| S16_extracted_8851 | Not-determined |
| S16_extracted_8861 | Not-determined |
| S16_extracted_8862 | Low-quality |
| S16_extracted_8893 | Not-determined |
| S16_extracted_8902 | Not-determined |
| S16_extracted_8903 | Not-determined |
| S16_extracted_8907 | Not-determined |
| S16_extracted_8909 | Not-determined |
| S16_extracted_8925 | Not-determined |
| S16_extracted_8934 | Not-determined |
| S16_extracted_8956 | Not-determined |
| S16_extracted_8960 | Not-determined |
| S16_extracted_8965 | Low-quality |
| S16_extracted_8979 | Not-determined |
| S16_extracted_8998 | Not-determined |
| S16_extracted_9003 | Not-determined |
| S16_extracted_9022 | Not-determined |
| S16_extracted_9045 | Not-determined |
| S16_extracted_9055 | Low-quality |
| S16_extracted_9057 | Low-quality |
| S16_extracted_9088 | Not-determined |
| S16_extracted_9113 | Not-determined |
| S16_extracted_9116 | Not-determined |
| S16_extracted_9119 | Not-determined |
| S16_extracted_9128 | Not-determined |
| S16_extracted_9134 | Low-quality |
| S16_extracted_9136 | Not-determined |
| S16_extracted_9142 | Not-determined |
| S16_extracted_9143 | Not-determined |
| S16_extracted_9147 | Not-determined |
| S16_extracted_9151 | Not-determined |
| S16_extracted_9152 | Not-determined |
| S16_extracted_9163 | Not-determined |
| S16_extracted_9177 | Low-quality |
| S16_extracted_9192 | Not-determined |
| S16_extracted_9201 | Not-determined |
| S16_extracted_9202 | Not-determined |
| S16_extracted_9205 | Not-determined |
| S16_extracted_9208 | Not-determined |
| S16_extracted_9216 | Low-quality |
| S16_extracted_9241 | Not-determined |
| S16_extracted_9244 | Not-determined |
| S16_extracted_9253 | Not-determined |
| S16_extracted_9260 | Not-determined |
| S7_vs_134784\|\|full | High-quality |
| S16_extracted_9274 | Not-determined |
| S16_extracted_9294 | Not-determined |
| S16_extracted_9295 | Not-determined |
| S16_extracted_9296 | Not-determined |
| S16_extracted_9297 | Not-determined |
| S16_extracted_9314 | Not-determined |
| S16_extracted_9336 | Not-determined |
| S16_extracted_9338 | Not-determined |
| S16_extracted_9352 | Not-determined |
| S16_extracted_9359 | Not-determined |
| S16_extracted_9367 | Not-determined |
| S16_extracted_9374 | Not-determined |
| S16_extracted_9378 | Not-determined |
| S16_extracted_9385 | Low-quality |
| S16_extracted_9398 | Not-determined |
| S16_extracted_9401 | Not-determined |
| S16_extracted_9411 | Not-determined |
| S16_extracted_9421 | Not-determined |
| S16_extracted_9425 | Not-determined |
| S16_extracted_9432 | Not-determined |
| S16_extracted_9443 | Not-determined |
| S16_extracted_9456 | Not-determined |
| S16_extracted_9466 | Low-quality |
| S16_extracted_9471 | Low-quality |
| S16_extracted_9473 | Not-determined |
| S16_extracted_9476 | Not-determined |
| S16_extracted_9478 | Not-determined |
| S16_extracted_9497 | Not-determined |
| S16_extracted_9500 | Not-determined |
| S16_extracted_9537 | Low-quality |
| S16_extracted_9538 | Low-quality |
| S16_extracted_9568 | Not-determined |
| S16_extracted_9617 | Not-determined |
| S16_extracted_9621 | Not-determined |
| S16_extracted_9627 | Not-determined |
| S16_extracted_9631 | Not-determined |
| S16_extracted_9636 | Not-determined |
| S16_extracted_9668 | Not-determined |
| S16_extracted_9670 | Not-determined |
| S16_extracted_9676 | Not-determined |
| S16_extracted_9687 | Not-determined |
| S16_extracted_9697 | Not-determined |
| S16_extracted_9699 | Not-determined |
| S16_extracted_9734 | Low-quality |
| S16_extracted_9740 | Not-determined |
| S16_extracted_9762 | Not-determined |
| S16_extracted_9766 | Not-determined |
| S16_extracted_9768 | Not-determined |
| S16_extracted_9777 | Not-determined |
| S16_extracted_9786 | Not-determined |
| S16_extracted_9792 | Not-determined |
| S16_extracted_9809 | Not-determined |
| S16_extracted_9851 | Not-determined |
| S16_extracted_9864 | Low-quality |
| S16_extracted_9866 | Low-quality |
| S16_extracted_9871 | Not-determined |
| S16_extracted_9896 | Not-determined |
| S16_extracted_9905 | Not-determined |
| S16_extracted_9910 | Not-determined |
| S16_extracted_9934 | Low-quality |
| S16_extracted_9950 | Not-determined |
| S16_extracted_9954 | Not-determined |
| S16_extracted_9956 | Low-quality |
| S16_extracted_9970 | Not-determined |
| S16_extracted_9976 | Not-determined |
| S16_extracted_10021 | Low-quality |
| S16_extracted_10025 | Low-quality |
| S16_extracted_10026 | Not-determined |
| S16_extracted_10031 | Not-determined |
| S16_extracted_10034 | Not-determined |
| S16_extracted_10036 | Low-quality |
| S16_extracted_10044 | Low-quality |
| S16_extracted_10051 | Not-determined |
| S16_extracted_10052 | Not-determined |
| S16_extracted_10060 | Not-determined |
| S16_extracted_10063 | Low-quality |
| S16_extracted_10073 | Not-determined |
| S16_extracted_10082 | Low-quality |
| S16_extracted_10086 | Not-determined |
| S16_extracted_10097 | Not-determined |
| S16_extracted_10142 | Not-determined |
| S16_extracted_10159 | Not-determined |
| S16_extracted_10183 | Not-determined |
| S16_extracted_10184 | Not-determined |
| S16_extracted_10185 | Not-determined |
| S16_extracted_10193 | Low-quality |
| S16_extracted_10195 | Not-determined |
| S16_extracted_10221 | Not-determined |
| S16_extracted_10241 | Low-quality |
| S16_extracted_10257 | Not-determined |
| S16_extracted_10258 | Not-determined |
| S16_extracted_10280 | Not-determined |
| S16_extracted_10287 | Not-determined |
| S16_extracted_10296 | Not-determined |
| S16_extracted_10299 | Not-determined |
| S16_extracted_10305 | Not-determined |
| S16_extracted_10307 | Low-quality |
| S16_extracted_10311 | Low-quality |
| S16_extracted_10314 | Not-determined |
| S16_extracted_10330 | Low-quality |
| S16_extracted_10339 | Not-determined |
| S16_extracted_10342 | Not-determined |
| S16_extracted_10370 | Not-determined |
| S16_extracted_10381 | Not-determined |
| S16_extracted_10388 | Not-determined |
| S16_extracted_10405 | Not-determined |
| S16_extracted_10428 | Not-determined |
| S16_extracted_10439 | Not-determined |
| S16_extracted_10441 | Not-determined |
| S16_extracted_10447 | Not-determined |
| S16_extracted_10449 | Not-determined |
| S16_extracted_10460 | Not-determined |
| S16_extracted_10465 | Low-quality |
| S16_extracted_10467 | Not-determined |
| S16_extracted_10482 | Not-determined |
| S16_extracted_10489 | Not-determined |
| S16_extracted_10504 | Not-determined |
| S16_extracted_10527 | Low-quality |
| S16_extracted_10542 | Not-determined |
| S16_extracted_10551 | Not-determined |
| S16_extracted_10567 | Not-determined |
| S16_extracted_10577 | Not-determined |
| S16_extracted_10579 | Not-determined |
| S16_extracted_10580 | Not-determined |
| S16_extracted_10592 | Not-determined |
| S16_extracted_10646 | Not-determined |
| S16_extracted_10649 | Not-determined |
| S16_extracted_10650 | Not-determined |
| S16_extracted_10659 | Low-quality |
| S16_extracted_10665 | Not-determined |
| S16_extracted_10705 | Not-determined |
| S16_extracted_10708 | Not-determined |
| S16_extracted_10714 | Not-determined |
| S16_extracted_10720 | Not-determined |
| S16_extracted_10730 | Not-determined |
| S16_extracted_10732 | Not-determined |
| S16_extracted_10740 | Not-determined |
| S16_extracted_10743 | Not-determined |
| S16_extracted_10745 | Not-determined |
| S16_extracted_10775 | Not-determined |
| S16_extracted_10778 | Not-determined |
| S16_extracted_10780 | Not-determined |
| S16_extracted_10805 | Not-determined |
| S16_extracted_10812 | Not-determined |
| S16_extracted_10818 | Not-determined |
| S16_extracted_10819 | Not-determined |
| S16_extracted_10827 | Not-determined |
| S16_extracted_10829 | Not-determined |
| S16_extracted_10831 | Not-determined |
| S16_extracted_10839 | Not-determined |
| S16_extracted_10841 | Low-quality |
| S16_extracted_10843 | Low-quality |
| S16_extracted_10846 | Not-determined |
| S16_extracted_10855 | Not-determined |
| S16_extracted_10876 | Low-quality |
| S16_extracted_10893 | Not-determined |
| S16_extracted_10894 | Not-determined |
| S16_extracted_10910 | Not-determined |
| S16_extracted_10917 | Low-quality |
| S16_extracted_10922 | Not-determined |
| S16_extracted_10923 | Not-determined |
| S16_extracted_10937 | Not-determined |
| S16_extracted_10938 | Low-quality |
| S16_extracted_10943 | Not-determined |
| S16_extracted_10973 | Not-determined |
| S16_extracted_10995 | Not-determined |
| S16_extracted_11006 | Not-determined |
| S16_extracted_11009 | Not-determined |
| S16_extracted_11012 | Not-determined |
| S16_extracted_11040 | Not-determined |
| S16_extracted_11041 | Not-determined |
| S16_extracted_11046 | Low-quality |
| S16_extracted_11050 | Not-determined |
| S16_extracted_11051 | Low-quality |
| S16_extracted_11058 | Low-quality |
| S16_extracted_11067 | Not-determined |
| S16_extracted_11071 | Not-determined |
| S16_extracted_11074 | Not-determined |
| S16_extracted_11083 | Not-determined |
| S16_extracted_11088 | Not-determined |
| S16_extracted_11093 | Not-determined |
| S16_extracted_11094 | Low-quality |
| S16_extracted_11104 | Not-determined |

| | |
|---|---|
| S16_extracted_11114 | Not-determined |
| S16_extracted_11141 | Low-quality |
| S16_extracted_11148 | Not-determined |
| S16_extracted_11151 | Not-determined |
| S16_extracted_11170 | Not-determined |
| S16_extracted_11195 | Not-determined |
| S16_extracted_11204 | Not-determined |
| S16_extracted_11214 | Not-determined |
| S16_extracted_11221 | Not-determined |
| S16_extracted_11246 | Not-determined |
| S16_extracted_11251 | Not-determined |
| S16_extracted_11259 | Low-quality |
| S16_extracted_11301 | Not-determined |
| S16_extracted_11324 | Not-determined |
| S16_extracted_11332 | Not-determined |
| S16_extracted_11334 | Not-determined |
| S16_extracted_11336 | Not-determined |
| S16_extracted_11343 | Not-determined |
| S16_extracted_11350 | Not-determined |
| S16_extracted_11371 | Not-determined |
| S16_extracted_11388 | Low-quality |
| S16_extracted_11391 | Not-determined |
| S16_extracted_11395 | Not-determined |
| S16_extracted_11402 | Not-determined |
| S16_extracted_11405 | Not-determined |
| S16_extracted_11406 | Not-determined |
| S16_extracted_11414 | Not-determined |
| S16_extracted_11422 | Not-determined |
| S16_extracted_11429 | Not-determined |
| S16_extracted_11446 | Not-determined |
| S16_extracted_11457 | Low-quality |
| S16_extracted_11470 | Low-quality |
| S16_extracted_11476 | Not-determined |
| S16_extracted_11484 | Not-determined |
| S16_extracted_11491 | Low-quality |
| S16_extracted_11495 | Not-determined |
| S16_extracted_11504 | Not-determined |
| S16_extracted_11520 | Not-determined |
| S16_extracted_11524 | Not-determined |
| S16_extracted_11530 | Not-determined |
| S16_extracted_11542 | Not-determined |
| S16_extracted_11573 | Low-quality |
| S16_extracted_11583 | Low-quality |
| S16_extracted_11584 | Not-determined |
| S16_extracted_11605 | Not-determined |
| S16_extracted_11611 | Not-determined |
| S16_extracted_11619 | Low-quality |
| S16_extracted_11629 | Not-determined |
| S16_extracted_11633 | Not-determined |
| S16_extracted_11635 | Not-determined |
| S16_extracted_11659 | Low-quality |
| S16_extracted_11664 | Not-determined |
| S16_extracted_11669 | Not-determined |
| S16_extracted_11671 | Not-determined |
| S16_extracted_11674 | Not-determined |
| S16_extracted_11675 | Not-determined |
| S16_extracted_11697 | Low-quality |
| S16_extracted_11759 | Low-quality |
| S16_extracted_11764 | Low-quality |
| S16_extracted_11766 | Not-determined |
| S16_extracted_11784 | Not-determined |
| S16_extracted_11801 | Low-quality |
| S16_extracted_11811 | Not-determined |
| S16_extracted_11812 | Not-determined |
| S16_extracted_11813 | Low-quality |
| S16_extracted_11815 | Not-determined |
| S16_extracted_11828 | Not-determined |
| S16_extracted_11845 | Not-determined |
| S16_extracted_11846 | Not-determined |
| S16_extracted_11848 | Not-determined |
| S16_extracted_11856 | Not-determined |
| S16_extracted_11869 | Low-quality |
| S16_extracted_11874 | Not-determined |
| S16_extracted_11896 | Not-determined |
| S16_extracted_11902 | Low-quality |
| S16_extracted_11905 | Not-determined |
| S16_extracted_11914 | Not-determined |
| S16_extracted_11938 | Not-determined |
| S16_extracted_11954 | Not-determined |
| S16_extracted_11962 | Not-determined |
| S16_extracted_11968 | Not-determined |
| S16_extracted_11974 | Not-determined |
| S16_extracted_12010 | Not-determined |
| S16_extracted_12026 | Not-determined |
| S16_extracted_12027 | Not-determined |
| S16_extracted_12046 | Not-determined |
| S16_extracted_12051 | Not-determined |
| S16_extracted_12074 | Not-determined |
| S16_extracted_12084 | Not-determined |
| S16_extracted_12090 | Not-determined |
| S16_extracted_12094 | Not-determined |
| S16_extracted_12116 | Not-determined |
| S16_extracted_12131 | Not-determined |
| S16_extracted_12132 | Not-determined |
| S16_extracted_12137 | Low-quality |
| S16_extracted_12144 | Not-determined |
| S16_extracted_12150 | Low-quality |
| S16_extracted_12151 | Not-determined |
| S16_extracted_12163 | Low-quality |
| S16_extracted_12165 | Low-quality |
| S16_extracted_12182 | Low-quality |
| S16_extracted_12186 | Not-determined |
| S16_extracted_12194 | Not-determined |
| S16_extracted_12199 | Low-quality |
| S16_extracted_12217 | Low-quality |
| S16_extracted_12235 | Low-quality |
| S16_extracted_12251 | Not-determined |
| S16_extracted_12266 | Not-determined |
| S16_extracted_12277 | Not-determined |
| S16_extracted_12281 | Low-quality |
| S16_extracted_12286 | Not-determined |
| S16_extracted_12290 | Not-determined |
| S16_extracted_12309 | Low-quality |
| S16_extracted_12324 | Not-determined |
| S16_extracted_12342 | Not-determined |
| S16_extracted_12345 | Low-quality |
| S16_extracted_12370 | Not-determined |
| S16_extracted_12374 | Not-determined |
| S16_extracted_12385 | Not-determined |
| S16_extracted_12390 | Low-quality |
| S16_extracted_12416 | Not-determined |
| S16_extracted_12418 | Low-quality |
| S16_extracted_12422 | Not-determined |
| S16_extracted_12427 | Low-quality |
| S16_extracted_12428 | Low-quality |
| S16_extracted_12434 | Not-determined |
| S16_extracted_12444 | Low-quality |
| S16_extracted_12464 | Not-determined |
| S16_extracted_12482 | Low-quality |
| S16_extracted_12488 | Not-determined |
| S16_extracted_12505 | Not-determined |
| S16_extracted_12508 | Not-determined |
| S16_extracted_12511 | Not-determined |
| S16_extracted_12519 | Not-determined |
| S9_vs_k141_48347\|\|full | High-quality |
| S16_extracted_12573 | Not-determined |
| S16_extracted_12580 | Not-determined |
| S16_extracted_12592 | Not-determined |
| S16_extracted_12599 | Not-determined |
| S16_extracted_12622 | Not-determined |
| S16_extracted_12630 | Not-determined |
| S16_extracted_12654 | Not-determined |
| S16_extracted_12689 | Not-determined |
| S16_extracted_12720 | Not-determined |
| S16_extracted_12725 | Not-determined |
| S16_extracted_12726 | Not-determined |
| S16_extracted_12739 | Not-determined |
| S16_extracted_12748 | Not-determined |
| S16_extracted_12751 | Not-determined |
| S16_extracted_12762 | Not-determined |
| S16_extracted_12764 | Not-determined |
| S16_extracted_12765 | Not-determined |
| S16_extracted_12776 | Low-quality |
| S16_extracted_12794 | Not-determined |
| S16_extracted_12819 | Low-quality |
| S16_extracted_12820 | Not-determined |
| S16_extracted_12838 | Not-determined |
| S16_extracted_12854 | Not-determined |
| S16_extracted_12856 | Not-determined |
| S16_extracted_12865 | Not-determined |
| S16_extracted_12874 | Not-determined |
| S16_extracted_12880 | Low-quality |
| S16_extracted_12881 | Not-determined |
| S16_extracted_12887 | Not-determined |
| S16_extracted_12893 | Low-quality |
| S16_extracted_12899 | Not-determined |
| S16_extracted_12903 | Not-determined |
| S16_extracted_12908 | Not-determined |
| S16_extracted_12911 | Not-determined |
| S16_extracted_12920 | Not-determined |
| S16_extracted_12936 | Low-quality |
| S16_extracted_12940 | Not-determined |
| S16_extracted_12944 | Not-determined |
| S16_extracted_12945 | Not-determined |
| S16_extracted_12954 | Not-determined |
| S16_extracted_12972 | Not-determined |
| S16_extracted_12976 | Not-determined |
| S16_extracted_12986 | Not-determined |
| S16_extracted_12988 | Not-determined |
| S16_extracted_12992 | Not-determined |
| S16_extracted_13004 | Not-determined |
| S16_extracted_13014 | Not-determined |
| S16_extracted_13018 | Not-determined |
| S16_extracted_13030 | Not-determined |
| S16_extracted_13038 | Not-determined |
| S16_extracted_13043 | Not-determined |
| S16_extracted_13046 | Low-quality |
| S16_extracted_13056 | Not-determined |
| S16_extracted_13068 | Not-determined |
| S16_extracted_13069 | Not-determined |
| S16_extracted_13072 | Low-quality |
| S16_extracted_13073 | Not-determined |
| S16_extracted_13087 | Not-determined |
| S16_extracted_13088 | Not-determined |
| S16_extracted_13094 | Not-determined |
| S16_extracted_13109 | Not-determined |
| S16_extracted_13136 | Not-determined |
| S16_extracted_13137 | Not-determined |
| S16_extracted_13140 | Not-determined |
| S16_extracted_13150 | Not-determined |
| S16_extracted_13154 | Not-determined |
| S16_extracted_13166 | Not-determined |
| S16_extracted_13179 | Not-determined |
| S16_extracted_13185 | Not-determined |
| S16_extracted_13190 | Not-determined |
| S16_extracted_13215 | Not-determined |
| S16_extracted_13235 | Not-determined |
| S16_extracted_13240 | Low-quality |
| S16_extracted_13252 | Not-determined |
| S16_extracted_13263 | Not-determined |
| S16_extracted_13269 | Not-determined |
| S16_extracted_13307 | Not-determined |
| S16_extracted_13314 | Not-determined |
| S16_extracted_13333 | Not-determined |
| S16_extracted_13334 | Not-determined |
| S16_extracted_13342 | Not-determined |
| S16_extracted_13354 | Not-determined |
| S16_extracted_13361 | Low-quality |
| S16_extracted_13384 | Not-determined |
| S16_extracted_13401 | Not-determined |
| S16_extracted_13412 | Not-determined |
| S16_extracted_13417 | Not-determined |
| S16_extracted_13418 | Low-quality |
| S16_extracted_13466 | Low-quality |
| S16_extracted_13478 | Not-determined |
| S16_extracted_13481 | Not-determined |
| S16_extracted_13485 | Low-quality |
| S16_extracted_13499 | Low-quality |
| S16_extracted_13503 | Not-determined |
| S16_extracted_13505 | Not-determined |
| S16_extracted_13508 | Not-determined |
| S16_extracted_13522 | Not-determined |
| S16_extracted_13535 | Not-determined |
| S16_extracted_13573 | Not-determined |
| S16_extracted_13574 | Not-determined |
| S16_extracted_13583 | Not-determined |
| S16_extracted_13584 | Not-determined |
| S16_extracted_13586 | Not-determined |
| S16_extracted_13623 | Not-determined |
| S16_extracted_13629 | Not-determined |
| S16_extracted_13636 | Not-determined |
| S16_extracted_13648 | Not-determined |
| S16_extracted_13658 | Not-determined |
| S16_extracted_13673 | Not-determined |
| S16_extracted_13679 | Not-determined |
| S16_extracted_13691 | Not-determined |
| S16_extracted_13696 | Not-determined |
| S16_extracted_13704 | Not-determined |
| S16_extracted_13706 | Not-determined |
| S16_extracted_13715 | Low-quality |
| S16_extracted_13730 | Not-determined |
| S16_extracted_13737 | Not-determined |
| S16_extracted_13751 | Not-determined |
| S16_extracted_13755 | Not-determined |
| S16_extracted_13761 | Low-quality |
| S16_extracted_13778 | Not-determined |
| S16_extracted_13817 | Not-determined |
| S16_extracted_13831 | Not-determined |
| S16_extracted_13856 | Not-determined |
| S16_extracted_13858 | Not-determined |
| S16_extracted_13900 | Not-determined |
| S16_extracted_13908 | Not-determined |
| S16_extracted_13912 | Low-quality |
| S16_extracted_13937 | Low-quality |
| S16_extracted_13938 | Not-determined |
| S16_extracted_13961 | Not-determined |
| S16_extracted_13962 | Low-quality |
| S16_extracted_13967 | Low-quality |
| S16_extracted_14007 | Not-determined |
| S16_extracted_14017 | Not-determined |
| S16_extracted_14025 | Not-determined |
| S16_extracted_14030 | Not-determined |
| S16_extracted_14034 | Not-determined |
| S16_extracted_14044 | Not-determined |
| S16_extracted_14048 | Not-determined |
| S16_extracted_14069 | Not-determined |
| S16_extracted_14080 | Not-determined |
| S16_extracted_14087 | Not-determined |
| S16_extracted_14089 | Not-determined |
| S16_extracted_14098 | Not-determined |
| S16_extracted_14106 | Not-determined |
| S16_extracted_14111 | Not-determined |
| S16_extracted_14123 | Not-determined |
| S16_extracted_14128 | Low-quality |
| S16_extracted_14154 | Low-quality |
| S16_extracted_14167 | Not-determined |
| S16_extracted_14171 | Not-determined |
| S16_extracted_14176 | Not-determined |
| S16_extracted_14182 | Low-quality |
| S16_extracted_14194 | Not-determined |
| S16_extracted_14201 | Low-quality |
| S16_extracted_14204 | Not-determined |
| S16_extracted_14206 | Not-determined |
| S16_extracted_14216 | Not-determined |
| S16_extracted_14217 | Not-determined |
| S16_extracted_14235 | Not-determined |
| S16_extracted_14248 | Not-determined |
| S16_extracted_14250 | Not-determined |
| S16_extracted_14255 | Not-determined |
| S16_extracted_14260 | Not-determined |
| S16_extracted_14263 | Not-determined |
| S16_extracted_14266 | Not-determined |
| S16_extracted_14269 | Not-determined |
| S16_extracted_14279 | Not-determined |
| S16_extracted_14301 | Low-quality |
| S16_extracted_14313 | Not-determined |
| S16_extracted_14321 | Not-determined |
| S16_extracted_14335 | Not-determined |
| S16_extracted_14348 | Not-determined |
| S16_extracted_14358 | Not-determined |
| S16_extracted_14368 | Not-determined |
| S16_extracted_14387 | Not-determined |
| S16_extracted_14392 | Not-determined |
| S16_extracted_14427 | Not-determined |
| S16_extracted_14434 | Not-determined |
| S16_extracted_14458 | Not-determined |
| S16_extracted_14468 | Not-determined |
| S16_extracted_14478 | Low-quality |
| S16_extracted_14485 | Not-determined |
| S16_extracted_14494 | Not-determined |
| S16_extracted_14498 | Low-quality |
| S16_extracted_14502 | Not-determined |
| S16_extracted_14579 | Not-determined |
| S16_extracted_14597 | Not-determined |
| S16_extracted_14607 | Not-determined |
| S16_extracted_14608 | Not-determined |
| S16_extracted_14633 | Not-determined |
| S16_extracted_14634 | Low-quality |
| S16_extracted_14642 | Not-determined |
| S16_extracted_14654 | Not-determined |
| S16_extracted_14659 | Not-determined |
| S16_extracted_14668 | Not-determined |
| S16_extracted_14670 | Not-determined |
| S16_extracted_14705 | Not-determined |
| S16_extracted_14728 | Not-determined |
| S16_extracted_14737 | Low-quality |
| S16_extracted_14745 | Not-determined |
| S16_extracted_14757 | Low-quality |
| S16_extracted_14776 | Not-determined |
| S16_extracted_14783 | Not-determined |
| S16_extracted_14793 | Not-determined |
| S16_extracted_14796 | Not-determined |
| S16_extracted_14798 | Not-determined |
| S16_extracted_14802 | Not-determined |
| S16_extracted_14803 | Not-determined |
| S16_extracted_14821 | Not-determined |
| S16_extracted_14854 | Low-quality |
| S16_extracted_14855 | Not-determined |
| S16_extracted_14869 | Not-determined |
| S16_extracted_14875 | Not-determined |
| S16_extracted_14879 | Not-determined |
| S16_extracted_14886 | Low-quality |
| S16_extracted_14887 | Not-determined |
| S16_extracted_14888 | Not-determined |
| S16_extracted_14909 | Not-determined |
| S16_extracted_14923 | Not-determined |
| S16_extracted_14933 | Not-determined |
| S16_extracted_14936 | Low-quality |
| S16_extracted_14953 | Not-determined |
| S16_extracted_14961 | Not-determined |
| S16_extracted_14967 | Not-determined |
| S16_extracted_14997 | Not-determined |
| S16_extracted_14998 | Not-determined |
| S16_extracted_15008 | Not-determined |
| S16_extracted_15022 | Not-determined |
| S16_extracted_15026 | Not-determined |
| S16_extracted_15045 | Not-determined |
| S16_extracted_15059 | Low-quality |
| S16_extracted_15078 | Not-determined |
| S16_extracted_15092 | Not-determined |
| S16_extracted_15111 | Not-determined |
| S16_extracted_15122 | Not-determined |
| S16_extracted_15142 | Not-determined |
| S16_extracted_15159 | Not-determined |
| S16_extracted_15167 | Not-determined |
| S16_extracted_15177 | Not-determined |
| S16_extracted_15220 | Not-determined |
| S16_extracted_15234 | Not-determined |
| S16_extracted_15240 | Not-determined |
| S16_extracted_15246 | Not-determined |
| S16_extracted_15250 | Not-determined |
| S16_extracted_15252 | Not-determined |
| S16_extracted_15278 | Not-determined |
| S16_extracted_15293 | Not-determined |
| S16_extracted_15317 | Not-determined |
| S16_extracted_15337 | Not-determined |
| S16_extracted_15355 | Low-quality |
| S16_extracted_15356 | Not-determined |
| S16_extracted_15359 | Not-determined |
| S16_extracted_15361 | Not-determined |
| S16_extracted_15375 | Not-determined |
| S16_extracted_15380 | Low-quality |
| S16_extracted_15391 | Not-determined |
| S16_extracted_15392 | Not-determined |
| S16_extracted_15399 | Not-determined |
| S16_extracted_15407 | Not-determined |
| S16_extracted_15410 | Not-determined |
| S16_extracted_15411 | Not-determined |
| S16_extracted_15422 | Not-determined |
| S16_extracted_15425 | Low-quality |
| S16_extracted_15441 | Not-determined |
| S16_extracted_15446 | Not-determined |
| S16_extracted_15452 | Not-determined |
| S16_extracted_15454 | Not-determined |
| S16_extracted_15455 | Not-determined |
| S16_extracted_15473 | Not-determined |
| S16_extracted_15489 | Not-determined |
| S16_extracted_15497 | Not-determined |
| S16_extracted_15505 | Low-quality |
| S16_extracted_15520 | Low-quality |
| S16_extracted_15521 | Not-determined |
| S16_extracted_15524 | Not-determined |
| S16_extracted_15526 | Not-determined |
| S16_extracted_15527 | Not-determined |
| S16_extracted_15546 | Not-determined |
| S16_extracted_15550 | Low-quality |
| S16_extracted_15552 | Not-determined |
| S16_extracted_15558 | Not-determined |
| S16_extracted_15565 | Low-quality |
| S16_extracted_15566 | Low-quality |
| S16_extracted_15570 | Not-determined |
| S16_extracted_15588 | Not-determined |
| S16_extracted_15590 | Not-determined |
| S16_extracted_15600 | Not-determined |
| S16_extracted_15607 | Not-determined |
| S16_extracted_15614 | Not-determined |
| S16_extracted_15636 | Low-quality |
| S16_extracted_15642 | Not-determined |
| S16_extracted_15656 | Not-determined |
| S16_extracted_15665 | Not-determined |
| S16_extracted_15666 | Not-determined |
| S16_extracted_15669 | Not-determined |
| S16_extracted_15680 | Not-determined |
| S16_extracted_15711 | Not-determined |
| S16_extracted_15712 | Not-determined |
| S16_extracted_15714 | Not-determined |
| S16_extracted_15716 | Not-determined |
| S16_extracted_15739 | Not-determined |
| S16_extracted_15762 | Not-determined |
| S16_extracted_15767 | Not-determined |
| S16_extracted_15779 | Not-determined |
| S16_extracted_15780 | Not-determined |
| S16_extracted_15783 | Not-determined |
| S16_extracted_15793 | Not-determined |
| S16_extracted_15805 | Not-determined |
| S16_extracted_15806 | Not-determined |
| S16_extracted_15830 | Not-determined |
| S16_extracted_15843 | Not-determined |
| S16_extracted_15847 | Low-quality |
| S16_extracted_15852 | Not-determined |
| S16_extracted_15861 | Not-determined |
| S16_extracted_15873 | Not-determined |
| S16_extracted_15881 | Not-determined |
| S16_extracted_15910 | Not-determined |
| S16_extracted_15920 | Not-determined |
| S16_extracted_15932 | Not-determined |
| S16_extracted_15951 | Not-determined |
| S16_extracted_15956 | Not-determined |
| S16_extracted_15960 | Not-determined |
| S16_extracted_15973 | Not-determined |
| S16_extracted_15978 | Not-determined |
| S16_extracted_15980 | Not-determined |
| S16_extracted_16006 | Low-quality |
| S16_extracted_16019 | Low-quality |
| S16_extracted_16020 | Not-determined |
| S16_extracted_16025 | Low-quality |
| S16_extracted_16026 | Not-determined |
| S16_extracted_16043 | Not-determined |
| S16_extracted_16046 | Not-determined |
| S16_extracted_16052 | Not-determined |
| S16_extracted_16094 | Low-quality |
| S16_extracted_16097 | Not-determined |
| S16_extracted_16100 | Low-quality |
| S16_extracted_16104 | Not-determined |
| S16_extracted_16105 | Not-determined |
| S16_extracted_16116 | Not-determined |
| S16_extracted_16118 | Not-determined |
| S16_extracted_16120 | Not-determined |
| S16_extracted_16121 | Not-determined |
| S16_extracted_16127 | Low-quality |
| S16_extracted_16128 | Not-determined |
| S16_extracted_16138 | Low-quality |
| S16_extracted_16144 | Not-determined |
| S16_extracted_16156 | Low-quality |
| S16_extracted_16157 | Not-determined |
| S16_extracted_16159 | Not-determined |
| S16_extracted_16167 | Not-determined |
| S16_extracted_16170 | Low-quality |
| S16_extracted_16174 | Not-determined |
| S16_extracted_16176 | Not-determined |
| S16_extracted_16180 | Not-determined |
| S13_vs_147396\|\|full | High-quality |
| S16_extracted_16205 | Not-determined |
| S16_extracted_16218 | Not-determined |
| S16_extracted_16221 | Not-determined |
| S16_extracted_16238 | Not-determined |
| S16_extracted_16240 | Not-determined |
| S16_extracted_16268 | Not-determined |
| S16_extracted_16281 | Low-quality |
| S16_extracted_16315 | Not-determined |
| S16_extracted_16346 | Not-determined |
| S16_extracted_16351 | Not-determined |
| S16_extracted_16355 | Not-determined |
| S16_extracted_16357 | Not-determined |
| S16_extracted_16362 | Not-determined |
| S16_extracted_16372 | Not-determined |
| S16_extracted_16377 | Not-determined |
| S16_extracted_16387 | Not-determined |
| S16_extracted_16402 | Not-determined |
| S16_extracted_16403 | Not-determined |
| S16_extracted_16405 | Low-quality |
| S16_extracted_16427 | Not-determined |
| S16_extracted_16428 | Low-quality |
| S16_extracted_16430 | Not-determined |
| S16_extracted_16453 | Not-determined |
| S16_extracted_16457 | Low-quality |
| S16_extracted_16462 | Not-determined |
| S16_extracted_16468 | Not-determined |
| S16_extracted_16486 | Not-determined |
| S16_extracted_16511 | Low-quality |
| S16_extracted_16523 | Not-determined |
| S16_extracted_16527 | Not-determined |
| S16_extracted_16549 | Not-determined |
| S16_extracted_16565 | Not-determined |
| S16_extracted_16570 | Not-determined |
| S16_extracted_16571 | Low-quality |
| S16_extracted_16576 | Not-determined |
| S16_extracted_16579 | Not-determined |
| S16_extracted_16583 | Low-quality |
| S16_extracted_16588 | Not-determined |
| S16_extracted_16594 | Not-determined |
| S16_extracted_16595 | Not-determined |
| S16_extracted_16598 | Not-determined |
| S16_extracted_16602 | Not-determined |
| S16_extracted_16615 | Not-determined |
| S16_extracted_16620 | Not-determined |
| S16_extracted_16627 | Low-quality |
| S16_extracted_16635 | Not-determined |
| S16_extracted_16639 | Not-determined |
| S16_extracted_16649 | Low-quality |
| S16_extracted_16657 | Low-quality |
| S16_extracted_16659 | Not-determined |
| S16_extracted_16667 | Low-quality |
| S16_extracted_16678 | Not-determined |
| S16_extracted_16689 | Not-determined |
| S16_extracted_16699 | Not-determined |
| S16_extracted_16715 | Not-determined |
| S16_extracted_16725 | Not-determined |
| S16_extracted_16730 | Not-determined |
| S16_extracted_16745 | Not-determined |
| S16_extracted_16753 | Not-determined |
| S16_extracted_16800 | Not-determined |
| S16_extracted_16801 | Not-determined |
| S16_extracted_16822 | Low-quality |
| S16_extracted_16828 | Not-determined |
| S16_extracted_16841 | Low-quality |
| S16_extracted_16846 | Not-determined |
| S16_extracted_16856 | Not-determined |
| S16_extracted_16860 | Not-determined |
| S16_extracted_16861 | Not-determined |
| S16_extracted_16862 | Not-determined |
| S16_extracted_16866 | Low-quality |
| S16_extracted_16868 | Not-determined |
| S16_extracted_16869 | Not-determined |
| S16_extracted_16873 | Not-determined |
| S16_extracted_16880 | Low-quality |
| S16_extracted_16881 | Not-determined |
| S16_extracted_16890 | Not-determined |
| S16_extracted_16893 | Low-quality |
| S16_extracted_16894 | Not-determined |
| S16_extracted_16899 | Not-determined |
| S16_extracted_16900 | Low-quality |
| S16_extracted_16902 | Not-determined |
| S16_extracted_16905 | Not-determined |
| S16_extracted_16910 | Not-determined |
| S16_extracted_16913 | Not-determined |
| S16_extracted_16920 | Not-determined |
| S16_extracted_16937 | Not-determined |
| S16_extracted_16967 | Not-determined |
| S16_extracted_16972 | Not-determined |
| S16_extracted_16975 | Not-determined |
| S16_extracted_16994 | Not-determined |
| S16_extracted_16995 | Not-determined |
| S16_extracted_17022 | Not-determined |
| S16_extracted_17027 | Not-determined |
| S16_extracted_17028 | Not-determined |
| S16_extracted_17038 | Not-determined |
| S16_extracted_17053 | Not-determined |
| S16_extracted_17078 | Not-determined |

| | |
|---|---|
| S16_extracted_17107 | Not-determined |
| S16_extracted_17113 | Not-determined |
| S16_extracted_17128 | Not-determined |
| S16_extracted_17131 | Not-determined |
| S16_extracted_17146 | Not-determined |
| S16_extracted_17166 | Low-quality |
| S16_extracted_17185 | Not-determined |
| S16_extracted_17186 | Low-quality |
| S16_extracted_17190 | Not-determined |
| S16_extracted_17192 | Not-determined |
| S16_extracted_17194 | Low-quality |
| S16_extracted_17200 | Not-determined |
| S16_extracted_17213 | Not-determined |
| S16_extracted_17223 | Not-determined |
| S16_extracted_17231 | Low-quality |
| S16_extracted_17236 | Not-determined |
| S16_extracted_17245 | Not-determined |
| S16_extracted_17256 | Not-determined |
| S16_extracted_17260 | Not-determined |
| S16_extracted_17263 | Low-quality |
| S16_extracted_17270 | Low-quality |
| S16_extracted_17272 | Not-determined |
| S16_extracted_17276 | Not-determined |
| S16_extracted_17279 | Not-determined |
| S16_extracted_17286 | Not-determined |
| S16_extracted_17289 | Not-determined |
| S16_extracted_17290 | Not-determined |
| S16_extracted_17296 | Not-determined |
| S16_extracted_17305 | Not-determined |
| S16_extracted_17317 | Low-quality |
| S16_extracted_17331 | Not-determined |
| S16_extracted_17333 | Low-quality |
| S16_extracted_17339 | Not-determined |
| S16_extracted_17341 | Not-determined |
| S16_extracted_17345 | Not-determined |
| S16_extracted_17346 | Not-determined |
| S16_extracted_17356 | Not-determined |
| S16_extracted_17366 | Not-determined |
| S16_extracted_17367 | Not-determined |
| S16_extracted_17372 | Not-determined |
| S16_extracted_17373 | Not-determined |
| S16_extracted_17393 | Not-determined |
| S16_extracted_17404 | Not-determined |
| S16_extracted_17410 | Not-determined |
| S16_extracted_17421 | Not-determined |
| S16_extracted_17451 | Not-determined |
| S16_extracted_17459 | Not-determined |
| S16_extracted_17476 | Not-determined |
| S16_extracted_17477 | Low-quality |
| S16_extracted_17484 | Not-determined |
| S16_extracted_17485 | Not-determined |
| S16_extracted_17487 | Not-determined |
| S16_extracted_17490 | Not-determined |
| S16_extracted_17506 | Low-quality |
| S16_extracted_17515 | Not-determined |
| S16_extracted_17518 | Not-determined |
| S16_extracted_17533 | Not-determined |
| S16_extracted_17552 | Not-determined |
| S16_extracted_17557 | Not-determined |
| S16_extracted_17574 | Not-determined |
| S16_extracted_17606 | Not-determined |
| S16_extracted_17619 | Not-determined |
| S16_extracted_17638 | Not-determined |
| S16_extracted_17641 | Not-determined |
| S16_extracted_17645 | Not-determined |
| S16_extracted_17650 | Not-determined |
| S16_extracted_17651 | Not-determined |
| S16_extracted_17653 | Not-determined |
| S16_extracted_17655 | Not-determined |
| S16_extracted_17658 | Low-quality |
| S16_extracted_17670 | Low-quality |
| S16_extracted_17679 | Low-quality |
| S16_extracted_17683 | Not-determined |
| S16_extracted_17685 | Not-determined |
| S16_extracted_17696 | Not-determined |
| S16_extracted_17713 | Not-determined |
| S16_extracted_17719 | Not-determined |
| S16_extracted_17729 | Not-determined |
| S16_extracted_17735 | Low-quality |
| S16_extracted_17747 | Not-determined |
| S16_extracted_17754 | Not-determined |
| S16_extracted_17767 | Not-determined |
| S16_extracted_17801 | Not-determined |
| S16_extracted_17807 | Not-determined |
| S16_extracted_17814 | Not-determined |
| S16_extracted_17823 | Not-determined |
| S16_extracted_17827 | Not-determined |
| S16_extracted_17841 | Not-determined |
| S16_extracted_17865 | Not-determined |
| S16_extracted_17888 | Not-determined |
| S16_extracted_17896 | Not-determined |
| S16_extracted_17902 | Low-quality |
| S16_extracted_17904 | Not-determined |
| S16_extracted_17906 | Not-determined |
| S16_extracted_17916 | Not-determined |
| S16_extracted_17921 | Not-determined |
| S16_extracted_17923 | Not-determined |
| S16_extracted_17929 | Not-determined |
| S16_extracted_17938 | Not-determined |
| S16_extracted_17939 | Not-determined |
| S16_extracted_17941 | Not-determined |
| S16_extracted_17944 | Not-determined |
| S16_extracted_17947 | Not-determined |
| S16_extracted_17952 | Not-determined |
| S16_extracted_17961 | Not-determined |
| S16_extracted_17974 | Not-determined |
| S16_extracted_17986 | Not-determined |
| S16_extracted_17996 | Not-determined |
| S16_extracted_18015 | Not-determined |
| S16_extracted_18019 | Not-determined |
| S16_extracted_18025 | Not-determined |
| S16_extracted_18036 | Not-determined |
| S16_extracted_18041 | Not-determined |
| S16_extracted_18048 | Not-determined |
| S16_extracted_18059 | Not-determined |
| S16_extracted_18087 | Not-determined |
| S16_extracted_18092 | Not-determined |
| S16_extracted_18093 | Not-determined |
| S16_extracted_18106 | Not-determined |
| S16_extracted_18107 | Not-determined |
| S16_extracted_18138 | Not-determined |
| S16_extracted_18142 | Low-quality |
| S16_extracted_18147 | Not-determined |
| S16_extracted_18151 | Low-quality |
| S16_extracted_18154 | Not-determined |
| S16_extracted_18157 | Not-determined |
| S16_extracted_18161 | Not-determined |
| S16_extracted_18162 | Not-determined |
| S16_extracted_18166 | Low-quality |
| S16_extracted_18169 | Not-determined |
| S16_extracted_18183 | Not-determined |
| S16_extracted_18184 | Not-determined |
| S16_extracted_18185 | Not-determined |
| S16_extracted_18194 | Not-determined |
| S16_extracted_18205 | Low-quality |
| S16_extracted_18217 | Not-determined |
| S16_extracted_18218 | Not-determined |
| S16_extracted_18230 | Not-determined |
| S16_extracted_18245 | Low-quality |
| S16_extracted_18257 | Not-determined |
| S16_extracted_18270 | Not-determined |
| S16_extracted_18273 | Not-determined |
| S16_extracted_18280 | Not-determined |
| S16_extracted_18302 | Not-determined |
| S16_extracted_18310 | Not-determined |
| S16_extracted_18313 | Not-determined |
| S16_extracted_18317 | Not-determined |
| S16_extracted_18329 | Not-determined |
| S16_extracted_18336 | Low-quality |
| S16_extracted_18352 | Low-quality |
| S16_extracted_18359 | Low-quality |
| S16_extracted_18376 | Not-determined |
| S16_extracted_18382 | Low-quality |
| S16_extracted_18393 | Not-determined |
| S16_extracted_18413 | Low-quality |
| S16_extracted_18416 | Not-determined |
| S16_extracted_18428 | Low-quality |
| S16_extracted_18453 | Not-determined |
| S16_extracted_18457 | Not-determined |
| S16_extracted_18482 | Not-determined |
| S16_extracted_18485 | Low-quality |
| S16_extracted_18487 | Not-determined |
| S16_extracted_18490 | Not-determined |
| S16_extracted_18493 | Low-quality |
| S16_extracted_18499 | Not-determined |
| S16_extracted_18508 | Not-determined |
| S16_extracted_18509 | Not-determined |
| S16_extracted_18513 | Not-determined |
| S16_extracted_18516 | Not-determined |
| S16_extracted_18517 | Not-determined |
| S16_extracted_18522 | Not-determined |
| S16_extracted_18532 | Low-quality |
| S16_extracted_18536 | Not-determined |
| S16_extracted_18560 | Not-determined |
| S16_extracted_18570 | Not-determined |
| S16_extracted_18573 | Not-determined |
| S16_extracted_18601 | Not-determined |
| S16_extracted_18605 | Not-determined |
| S16_extracted_18635 | Low-quality |
| S16_extracted_18654 | Not-determined |
| S16_extracted_18655 | Not-determined |
| S16_extracted_18661 | Not-determined |
| S16_extracted_18665 | Low-quality |
| S16_extracted_18678 | Low-quality |
| S16_extracted_18681 | Not-determined |
| S16_extracted_18688 | Not-determined |
| S16_extracted_18692 | Not-determined |
| S16_extracted_18718 | Low-quality |
| S16_extracted_18731 | Not-determined |
| S16_extracted_18740 | Not-determined |
| S16_extracted_18742 | Low-quality |
| S16_extracted_18752 | Not-determined |
| S16_extracted_18778 | Not-determined |
| S16_extracted_18780 | Not-determined |
| S16_extracted_18842 | Not-determined |
| S16_extracted_18852 | Not-determined |
| S16_extracted_18855 | Not-determined |
| S16_extracted_18860 | Not-determined |
| S16_extracted_18864 | Not-determined |
| S16_extracted_18865 | Low-quality |
| S16_extracted_18907 | Low-quality |
| S16_extracted_18911 | Not-determined |
| S16_extracted_18919 | Low-quality |
| S16_extracted_18926 | Low-quality |
| S16_extracted_18954 | Not-determined |
| S16_extracted_18964 | Not-determined |
| S16_extracted_18967 | Not-determined |
| S16_extracted_18987 | Not-determined |
| S16_extracted_18988 | Low-quality |
| S16_extracted_18995 | Not-determined |
| S16_extracted_19000 | Not-determined |
| S16_extracted_19012 | Not-determined |
| S16_extracted_19017 | Not-determined |
| S16_extracted_19035 | Low-quality |
| S16_extracted_19037 | Low-quality |
| S16_extracted_19039 | Low-quality |
| S16_extracted_19040 | Not-determined |
| S16_extracted_19052 | Not-determined |
| S16_extracted_19055 | Not-determined |
| S16_extracted_19063 | Not-determined |
| S16_extracted_19082 | Not-determined |
| S16_extracted_19086 | Not-determined |
| S16_extracted_19088 | Low-quality |
| S16_extracted_19089 | Not-determined |
| S16_extracted_19104 | Not-determined |
| S16_extracted_19117 | Not-determined |
| S16_extracted_19126 | Low-quality |
| S16_extracted_19135 | Not-determined |
| S16_extracted_19140 | Not-determined |
| S16_extracted_19158 | Not-determined |
| S16_extracted_19161 | Not-determined |
| S16_extracted_19162 | Not-determined |
| S16_extracted_19166 | Not-determined |
| S16_extracted_19175 | Not-determined |
| S16_extracted_19180 | Not-determined |
| S16_extracted_19185 | Not-determined |
| S16_extracted_19188 | Low-quality |
| S16_extracted_19191 | Not-determined |
| S16_extracted_19196 | Not-determined |
| S16_extracted_19198 | Low-quality |
| S16_extracted_19218 | Not-determined |
| S16_extracted_19229 | Not-determined |
| S16_extracted_19233 | Not-determined |
| S16_extracted_19235 | Not-determined |
| S16_extracted_19243 | Low-quality |
| S16_extracted_19249 | Not-determined |
| S16_extracted_19271 | Not-determined |
| S16_extracted_19281 | Low-quality |
| S16_extracted_19338 | Low-quality |
| S16_extracted_19347 | Not-determined |
| S16_extracted_19351 | Not-determined |
| S16_extracted_19363 | Not-determined |
| S16_extracted_19385 | Not-determined |
| S16_extracted_19411 | Not-determined |
| S16_extracted_19424 | Not-determined |
| S16_extracted_19425 | Not-determined |
| S16_extracted_19461 | Not-determined |
| S16_extracted_19478 | Not-determined |
| S16_extracted_19485 | Not-determined |
| S16_extracted_19494 | Not-determined |
| S16_extracted_19499 | Not-determined |
| S16_extracted_19500 | Not-determined |
| S16_extracted_19507 | Not-determined |
| S16_extracted_19509 | Not-determined |
| S16_extracted_19510 | Not-determined |
| S16_extracted_19515 | Low-quality |
| S16_extracted_19516 | Not-determined |
| S16_extracted_19540 | Not-determined |
| S16_extracted_19565 | Not-determined |
| S16_extracted_19609 | Not-determined |
| S16_extracted_19611 | Not-determined |
| S16_extracted_19613 | Not-determined |
| S16_extracted_19689 | Not-determined |
| S16_extracted_19723 | Low-quality |
| S16_extracted_19731 | Not-determined |
| S16_extracted_19736 | Not-determined |
| S16_extracted_19742 | Not-determined |
| S16_extracted_19757 | Not-determined |
| S16_extracted_19773 | Not-determined |
| S16_extracted_19779 | Not-determined |
| S16_extracted_19786 | Not-determined |
| S16_extracted_19789 | Not-determined |
| S16_extracted_19790 | Not-determined |
| S16_extracted_19791 | Not-determined |
| S16_extracted_19797 | Not-determined |
| S16_extracted_19800 | Not-determined |
| S16_extracted_19812 | Not-determined |
| S16_extracted_19823 | Not-determined |
| S16_extracted_19825 | Low-quality |
| S16_extracted_19837 | Not-determined |
| S16_extracted_19838 | Not-determined |
| S16_extracted_19839 | Not-determined |
| S16_extracted_19840 | Not-determined |
| S16_extracted_19846 | Low-quality |
| S16_extracted_19849 | Not-determined |
| S16_extracted_19854 | Not-determined |
| S16_extracted_19867 | Not-determined |
| S16_extracted_19868 | Not-determined |
| S16_extracted_19899 | Low-quality |
| S16_extracted_19912 | Not-determined |
| S16_extracted_19915 | Not-determined |
| S16_extracted_19951 | Not-determined |
| S16_extracted_19953 | Not-determined |
| S16_extracted_19970 | Not-determined |
| S16_extracted_19987 | Not-determined |
| S16_extracted_20013 | Not-determined |
| S16_extracted_20014 | Not-determined |
| S16_extracted_20029 | Low-quality |
| S16_extracted_20033 | Not-determined |
| S16_extracted_20049 | Not-determined |
| S16_extracted_20056 | Low-quality |
| S16_extracted_20071 | Low-quality |
| S16_extracted_20082 | Not-determined |
| S16_extracted_20090 | Not-determined |
| S16_extracted_20091 | Not-determined |
| S16_extracted_20093 | Low-quality |
| S16_extracted_20096 | Not-determined |
| S16_extracted_20104 | Not-determined |
| S16_extracted_20108 | Not-determined |
| S16_extracted_20115 | Not-determined |
| S16_extracted_20119 | Not-determined |
| S16_extracted_20173 | Not-determined |
| S16_extracted_20175 | Not-determined |
| S16_extracted_20180 | Low-quality |
| S16_extracted_20186 | Not-determined |
| S16_extracted_20189 | Not-determined |
| S16_extracted_20204 | Not-determined |
| S16_extracted_20209 | Not-determined |
| S16_extracted_20237 | Low-quality |
| S16_extracted_20244 | Not-determined |
| S16_extracted_20264 | Not-determined |
| S16_extracted_20277 | Not-determined |
| S16_extracted_20285 | Low-quality |
| S16_extracted_20287 | Not-determined |
| S16_extracted_20289 | Not-determined |
| S16_extracted_20310 | Not-determined |
| S16_extracted_20316 | Not-determined |
| S16_extracted_20319 | Not-determined |
| S16_extracted_20328 | Not-determined |
| S16_extracted_20340 | Not-determined |
| S16_extracted_20341 | Not-determined |
| S16_extracted_20348 | Not-determined |
| S16_extracted_20357 | Not-determined |
| S16_extracted_20359 | Not-determined |
| S16_extracted_20366 | Low-quality |
| S16_extracted_20378 | Not-determined |
| S16_extracted_20391 | Not-determined |
| S16_extracted_20397 | Not-determined |
| S16_extracted_20419 | Not-determined |
| S16_extracted_20425 | Not-determined |
| S16_extracted_20441 | Not-determined |
| S16_extracted_20458 | Not-determined |
| S16_extracted_20478 | Not-determined |
| S16_extracted_20483 | Not-determined |
| S16_extracted_20487 | Low-quality |
| S16_extracted_20506 | Not-determined |
| S16_extracted_20514 | Not-determined |
| S16_extracted_20519 | Not-determined |
| S16_extracted_20524 | Not-determined |
| S16_extracted_20528 | Not-determined |
| S16_extracted_20531 | Low-quality |
| S16_extracted_20534 | Not-determined |
| S16_extracted_20550 | Not-determined |
| S16_extracted_20557 | Not-determined |
| S16_extracted_20595 | Not-determined |
| S16_extracted_20606 | Not-determined |
| S16_extracted_20627 | Not-determined |
| S16_extracted_20632 | Not-determined |
| S16_extracted_20633 | Not-determined |
| S16_extracted_20640 | Not-determined |
| S16_extracted_20661 | Low-quality |
| S16_extracted_20664 | Not-determined |
| S16_extracted_20665 | Low-quality |
| S16_extracted_20668 | Not-determined |
| S16_extracted_20696 | Not-determined |
| S16_extracted_20720 | Low-quality |
| S16_extracted_20722 | Low-quality |
| S16_extracted_20725 | Low-quality |
| S16_extracted_20732 | Not-determined |
| S16_extracted_20776 | Low-quality |
| S16_extracted_20791 | Low-quality |
| S16_extracted_20793 | Not-determined |
| S16_extracted_20794 | Not-determined |
| S16_extracted_20811 | Not-determined |
| S16_extracted_20817 | Not-determined |
| S16_extracted_20822 | Not-determined |
| S16_extracted_20823 | Not-determined |
| S16_extracted_20843 | Not-determined |
| S16_extracted_20848 | Not-determined |
| S16_extracted_20868 | Low-quality |
| S16_extracted_20892 | Not-determined |
| S16_extracted_20901 | Not-determined |
| S16_extracted_20902 | Not-determined |
| S16_extracted_20918 | Not-determined |
| S16_extracted_20920 | Not-determined |
| S16_extracted_20927 | Not-determined |
| S16_extracted_20935 | Not-determined |
| S16_extracted_20936 | Not-determined |
| S16_extracted_20956 | Low-quality |
| S16_extracted_20959 | Not-determined |
| S16_extracted_20965 | Not-determined |
| S16_extracted_20966 | Not-determined |
| S16_extracted_20984 | Not-determined |
| S16_extracted_20996 | Low-quality |
| S16_extracted_21002 | Not-determined |
| S16_extracted_21006 | Low-quality |
| S16_extracted_21011 | Low-quality |
| S16_extracted_21014 | Not-determined |
| S16_extracted_21015 | Not-determined |
| S16_extracted_21019 | Not-determined |
| S16_extracted_21042 | Not-determined |
| S16_extracted_21049 | Not-determined |
| S16_extracted_21051 | Not-determined |
| S16_extracted_21064 | Not-determined |
| S16_extracted_21076 | Not-determined |
| S16_extracted_21084 | Low-quality |
| S16_extracted_21119 | Not-determined |
| S16_extracted_21121 | Not-determined |
| S16_extracted_21136 | Not-determined |
| S16_extracted_21137 | Not-determined |
| S16_extracted_21159 | Low-quality |
| S16_extracted_21163 | Not-determined |
| S16_extracted_21172 | Not-determined |
| S16_extracted_21176 | Not-determined |
| S16_extracted_21192 | Not-determined |
| S16_extracted_21199 | Not-determined |
| S16_extracted_21222 | Low-quality |
| S16_extracted_21257 | Not-determined |
| S16_extracted_21266 | Not-determined |
| S16_extracted_21286 | Not-determined |
| S16_extracted_21295 | Not-determined |
| S16_extracted_21306 | Not-determined |
| S16_extracted_21333 | Not-determined |
| S16_extracted_21354 | Not-determined |
| S16_extracted_21383 | Not-determined |
| S16_extracted_21406 | Low-quality |
| S16_extracted_21409 | Not-determined |
| S16_extracted_21417 | Not-determined |
| S16_extracted_21426 | Not-determined |
| S16_extracted_21429 | Not-determined |
| S16_extracted_21432 | Not-determined |
| S16_extracted_21441 | Not-determined |
| S16_extracted_21451 | Not-determined |
| S16_extracted_21453 | Low-quality |
| S16_extracted_21465 | Not-determined |
| S16_extracted_21473 | Not-determined |
| S16_extracted_21489 | Low-quality |
| S16_extracted_21496 | Low-quality |
| S16_extracted_21508 | Not-determined |
| S16_extracted_21521 | Not-determined |
| S16_extracted_21567 | Not-determined |
| S16_extracted_21568 | Not-determined |
| S16_extracted_21576 | Not-determined |
| S16_extracted_21578 | Not-determined |
| S16_extracted_21584 | Not-determined |
| S16_extracted_21594 | Not-determined |
| S16_extracted_21601 | Not-determined |
| S16_extracted_21607 | Not-determined |
| S16_extracted_21613 | Not-determined |
| S16_extracted_21614 | Not-determined |
| S16_extracted_21626 | Not-determined |
| S16_extracted_21629 | Not-determined |
| S16_extracted_21636 | Not-determined |
| S16_extracted_21651 | Low-quality |
| S16_extracted_21660 | Not-determined |
| S16_extracted_21667 | Not-determined |
| S16_extracted_21681 | Not-determined |
| S16_extracted_21684 | Not-determined |
| S16_extracted_21694 | Not-determined |
| S16_extracted_21695 | Not-determined |
| S16_extracted_21705 | Not-determined |
| S16_extracted_21718 | Not-determined |
| S16_extracted_21719 | Not-determined |
| S16_extracted_21732 | Not-determined |
| S16_extracted_21734 | Not-determined |
| S16_extracted_21737 | Not-determined |
| S16_extracted_21746 | Not-determined |
| S16_extracted_21751 | Not-determined |
| S16_extracted_21754 | Not-determined |
| S16_extracted_21757 | Not-determined |
| S16_extracted_21758 | Low-quality |
| S16_extracted_21772 | Low-quality |
| S16_extracted_21778 | Not-determined |
| S16_extracted_21787 | Not-determined |
| S16_extracted_21791 | Not-determined |
| S16_extracted_21807 | Not-determined |
| S16_extracted_21809 | Not-determined |
| S16_extracted_21813 | Low-quality |
| S16_extracted_21817 | Not-determined |
| S16_extracted_21818 | Low-quality |
| S16_extracted_21829 | Not-determined |
| S16_extracted_21830 | Not-determined |
| S16_extracted_21835 | Not-determined |
| S16_extracted_21846 | Not-determined |
| S16_extracted_21848 | Not-determined |
| S16_extracted_21851 | Not-determined |
| S16_extracted_21860 | Not-determined |
| S16_extracted_21883 | Not-determined |
| S16_extracted_21886 | Not-determined |
| S16_extracted_21890 | Not-determined |
| S16_extracted_21891 | Not-determined |
| S16_extracted_21910 | Not-determined |
| S16_extracted_21958 | Not-determined |
| S16_extracted_21962 | Low-quality |
| S16_extracted_21973 | Not-determined |
| S16_extracted_21974 | Not-determined |
| S16_extracted_21981 | Not-determined |
| S16_extracted_21982 | Not-determined |
| S16_extracted_21988 | Not-determined |
| S16_extracted_21995 | Not-determined |
| S16_extracted_22013 | Not-determined |
| S16_extracted_22019 | Not-determined |
| S16_extracted_22024 | Not-determined |
| S16_extracted_22039 | Not-determined |
| S16_extracted_22047 | Not-determined |
| S16_extracted_22052 | Not-determined |
| S16_extracted_22059 | Not-determined |
| S16_extracted_22064 | Not-determined |
| S16_extracted_22086 | Not-determined |
| S16_extracted_22094 | Not-determined |
| S16_extracted_22110 | Not-determined |
| S16_extracted_22111 | Not-determined |
| S16_extracted_22122 | Not-determined |
| S16_extracted_22136 | Low-quality |
| S16_extracted_22142 | Low-quality |
| S16_extracted_22163 | Not-determined |
| S16_extracted_22175 | Not-determined |
| S16_extracted_22198 | Not-determined |
| S16_extracted_22200 | Low-quality |
| S16_extracted_22208 | Low-quality |
| S16_extracted_22213 | Not-determined |
| S16_extracted_22217 | Low-quality |
| S16_extracted_22244 | Not-determined |
| S16_extracted_22262 | Not-determined |
| S16_extracted_22285 | Not-determined |
| S16_extracted_22287 | Not-determined |
| S16_extracted_22297 | Not-determined |
| S16_extracted_22325 | Low-quality |
| S16_extracted_22327 | Not-determined |
| S16_extracted_22336 | Not-determined |
| S16_extracted_22337 | Not-determined |
| S16_extracted_22341 | Low-quality |
| S16_extracted_22347 | Not-determined |
| S16_extracted_22352 | Not-determined |
| S16_extracted_22369 | Not-determined |
| S16_extracted_22385 | Not-determined |
| S16_extracted_22391 | Not-determined |
| S16_extracted_22395 | Not-determined |
| S16_extracted_22399 | Not-determined |
| S16_extracted_22401 | Not-determined |
| S16_extracted_22405 | Not-determined |
| S16_extracted_22415 | Not-determined |
| S16_extracted_22418 | Not-determined |
| S16_extracted_22426 | Not-determined |
| S16_extracted_22429 | Not-determined |
| S16_extracted_22433 | Not-determined |
| S16_extracted_22434 | Low-quality |
| S16_extracted_22438 | Not-determined |
| S16_extracted_22459 | Not-determined |
| S16_extracted_22473 | Not-determined |
| S16_extracted_22478 | Not-determined |
| S16_extracted_22479 | Not-determined |
| S16_extracted_22482 | Not-determined |
| S16_extracted_22487 | Low-quality |
| S16_extracted_22496 | Not-determined |
| S16_extracted_22497 | Low-quality |
| S16_extracted_22506 | Not-determined |
| S16_extracted_22507 | Low-quality |
| S16_extracted_22509 | Low-quality |
| S16_extracted_22519 | Not-determined |
| S16_extracted_22534 | Not-determined |
| S16_extracted_22541 | Not-determined |
| S16_extracted_22546 | Not-determined |
| S16_extracted_22554 | Not-determined |
| S16_extracted_22556 | Not-determined |
| S16_extracted_22577 | Not-determined |
| S16_extracted_22581 | Not-determined |
| S16_extracted_22584 | Not-determined |
| S16_extracted_22593 | Not-determined |
| S16_extracted_22623 | Not-determined |
| S16_extracted_22630 | Low-quality |
| S16_extracted_22632 | Low-quality |
| S16_extracted_22649 | Not-determined |
| S16_extracted_22659 | Low-quality |
| S16_extracted_22679 | Not-determined |
| S16_extracted_22712 | Not-determined |
| S16_extracted_22714 | Not-determined |
| S16_extracted_22732 | Low-quality |
| S16_extracted_22739 | Not-determined |

| | |
|---|---|
| S16_extracted_22753 | Not-determined |
| S16_extracted_22771 | Not-determined |
| S16_extracted_22780 | Not-determined |
| S16_extracted_22789 | Not-determined |
| S16_extracted_22828 | Not-determined |
| S16_extracted_22830 | Not-determined |
| S16_extracted_22836 | Low-quality |
| S16_extracted_22847 | Not-determined |
| S16_extracted_22849 | Not-determined |
| S16_extracted_22877 | Not-determined |
| S16_extracted_22880 | Low-quality |
| S16_extracted_22903 | Not-determined |
| S16_extracted_22925 | Low-quality |
| S16_extracted_22926 | Low-quality |
| S16_extracted_22933 | Not-determined |
| S16_extracted_22972 | Not-determined |
| S16_extracted_22975 | Not-determined |
| S16_extracted_22996 | Low-quality |
| S16_extracted_23011 | Not-determined |
| S16_extracted_23015 | Not-determined |
| S16_extracted_23019 | Not-determined |
| S16_extracted_23023 | Not-determined |
| S16_extracted_23039 | Low-quality |
| S16_extracted_23045 | Not-determined |
| S16_extracted_23052 | Not-determined |
| S16_extracted_23066 | Not-determined |
| S16_extracted_23081 | Not-determined |
| S16_extracted_23086 | Not-determined |
| S16_extracted_23090 | Not-determined |
| S16_extracted_23092 | Not-determined |
| S16_extracted_23120 | Low-quality |
| S16_extracted_23128 | Not-determined |
| S16_extracted_23146 | Not-determined |
| S16_extracted_23156 | Not-determined |
| S16_extracted_23160 | Not-determined |
| S16_extracted_23172 | Not-determined |
| S16_extracted_23174 | Low-quality |
| S16_extracted_23181 | Not-determined |
| S16_extracted_23192 | Not-determined |
| S16_extracted_23195 | Not-determined |
| S16_extracted_23221 | Not-determined |
| S16_extracted_23240 | Not-determined |
| S16_extracted_23244 | Not-determined |
| S16_extracted_23275 | Not-determined |
| S16_extracted_23282 | Not-determined |
| S16_extracted_23300 | Not-determined |
| S16_extracted_23304 | Not-determined |
| S16_extracted_23336 | Not-determined |
| S16_extracted_23346 | Not-determined |
| S16_extracted_23351 | Not-determined |
| S16_extracted_23352 | Not-determined |
| S16_extracted_23363 | Not-determined |
| S16_extracted_23368 | Not-determined |
| S16_extracted_23383 | Not-determined |
| S16_extracted_23397 | Low-quality |
| S16_extracted_23413 | Not-determined |
| S16_extracted_23424 | Not-determined |
| S16_extracted_23428 | Not-determined |
| S16_extracted_23469 | Low-quality |
| S16_extracted_23471 | Not-determined |
| S16_extracted_23479 | Not-determined |
| S16_extracted_23496 | Not-determined |
| S16_extracted_23502 | Not-determined |
| S16_extracted_23508 | Not-determined |
| S16_extracted_23513 | Not-determined |
| S16_extracted_23519 | Not-determined |
| S16_extracted_23530 | Not-determined |
| S16_extracted_23542 | Not-determined |
| S16_extracted_23546 | Not-determined |
| S16_extracted_23559 | Not-determined |
| S16_extracted_23574 | Not-determined |
| S16_extracted_23575 | Not-determined |
| S16_extracted_23602 | Low-quality |
| S16_extracted_23603 | Not-determined |
| S16_extracted_23618 | Not-determined |
| S16_extracted_23646 | Not-determined |
| S16_extracted_23647 | Not-determined |
| S16_extracted_23653 | Not-determined |
| S16_extracted_23674 | Not-determined |
| S16_extracted_23682 | Not-determined |
| S16_extracted_23710 | Low-quality |
| S16_extracted_23715 | Not-determined |
| S16_extracted_23730 | Not-determined |
| S16_extracted_23737 | Not-determined |
| S16_extracted_23747 | Not-determined |
| S16_extracted_23748 | Not-determined |
| S16_extracted_23755 | Not-determined |
| S16_extracted_23756 | Not-determined |
| S16_extracted_23758 | Low-quality |
| S16_extracted_23762 | Low-quality |
| S16_extracted_23765 | Not-determined |
| S16_extracted_23768 | Not-determined |
| S16_extracted_23785 | Not-determined |
| S16_extracted_23787 | Not-determined |
| S16_extracted_23842 | Low-quality |
| S16_extracted_23846 | Not-determined |
| S16_extracted_23852 | Not-determined |
| S16_extracted_23856 | Not-determined |
| S16_extracted_23873 | Not-determined |
| S16_extracted_23881 | Not-determined |
| S16_extracted_23891 | Low-quality |
| S16_extracted_23906 | Not-determined |
| S16_extracted_23911 | Not-determined |
| S16_extracted_23912 | Not-determined |
| S16_extracted_23926 | Low-quality |
| S16_extracted_23937 | Not-determined |
| S16_extracted_23958 | Not-determined |
| S16_extracted_24002 | Low-quality |
| S16_extracted_24014 | Not-determined |
| S16_extracted_24047 | Not-determined |
| S16_extracted_24052 | Not-determined |
| S16_extracted_24056 | Not-determined |
| S16_extracted_24076 | Low-quality |
| S16_extracted_24090 | Not-determined |
| S16_extracted_24107 | Not-determined |
| S16_extracted_24108 | Not-determined |
| S16_extracted_24118 | Not-determined |
| S16_extracted_24125 | Not-determined |
| S16_extracted_24126 | Not-determined |
| S16_extracted_24141 | Not-determined |
| S16_extracted_24176 | Not-determined |
| S16_extracted_24198 | Low-quality |
| S16_extracted_24219 | Not-determined |
| S16_extracted_24224 | Low-quality |
| S16_extracted_24237 | Not-determined |
| S16_extracted_24267 | Not-determined |
| S16_extracted_24269 | Low-quality |
| S16_extracted_24274 | Not-determined |
| S16_extracted_24279 | Low-quality |
| S16_extracted_24285 | Not-determined |
| S16_extracted_24295 | Low-quality |
| S16_extracted_24308 | Not-determined |
| S16_extracted_24309 | Not-determined |
| S16_extracted_24315 | Not-determined |
| S16_extracted_24327 | Not-determined |
| S16_extracted_24343 | Not-determined |
| S16_extracted_24345 | Not-determined |
| S16_extracted_24347 | Not-determined |
| S16_extracted_24358 | Not-determined |
| S16_extracted_24363 | Not-determined |
| S16_extracted_24365 | Low-quality |
| S16_extracted_24369 | Not-determined |
| S16_extracted_24375 | Not-determined |
| S16_extracted_24380 | Not-determined |
| S16_extracted_24385 | Not-determined |
| S16_extracted_24391 | Not-determined |
| S16_extracted_24398 | Not-determined |
| S16_extracted_24399 | Low-quality |
| S16_extracted_24410 | Not-determined |
| S16_extracted_24412 | Not-determined |
| S16_extracted_24419 | Not-determined |
| S16_extracted_24424 | Not-determined |
| S16_extracted_24443 | Not-determined |
| S16_extracted_24460 | Not-determined |
| S16_extracted_24470 | Not-determined |
| S16_extracted_24472 | Not-determined |
| S16_extracted_24490 | Not-determined |
| S16_extracted_24503 | Not-determined |
| S16_extracted_24508 | Not-determined |
| S16_extracted_24511 | Not-determined |
| S16_extracted_24515 | Not-determined |
| S16_extracted_24526 | Not-determined |
| S16_extracted_24534 | Not-determined |
| S16_extracted_24541 | Low-quality |
| S16_extracted_24546 | Low-quality |
| S16_extracted_24555 | Not-determined |
| S16_extracted_24568 | Not-determined |
| S16_extracted_24570 | Not-determined |
| S16_extracted_24578 | Low-quality |
| S16_extracted_24590 | Not-determined |
| S16_extracted_24594 | Not-determined |
| S16_extracted_24607 | Not-determined |
| S16_extracted_24609 | Not-determined |
| S16_extracted_24610 | Not-determined |
| S16_extracted_24629 | Low-quality |
| S16_extracted_24646 | Not-determined |
| S16_extracted_24647 | Not-determined |
| S16_extracted_24649 | Not-determined |
| S16_extracted_24661 | Low-quality |
| S16_extracted_24663 | Not-determined |
| S16_extracted_24673 | Not-determined |
| S16_extracted_24697 | Not-determined |
| S16_extracted_24701 | Not-determined |
| S16_extracted_24712 | Not-determined |
| S16_extracted_24721 | Not-determined |
| S16_extracted_24723 | Not-determined |
| S16_extracted_24727 | Low-quality |
| S16_extracted_24770 | Not-determined |
| S16_extracted_24774 | Not-determined |
| S16_extracted_24780 | Not-determined |
| S16_extracted_24798 | Not-determined |
| S16_extracted_24801 | Not-determined |
| S16_extracted_24814 | Not-determined |
| S16_extracted_24816 | Low-quality |
| S16_extracted_24817 | Not-determined |
| S16_extracted_24821 | Not-determined |
| S16_extracted_24827 | Not-determined |
| S16_extracted_24834 | Not-determined |
| S16_extracted_24835 | Not-determined |
| S16_extracted_24842 | Low-quality |
| S16_extracted_24844 | Low-quality |
| S16_extracted_24856 | Not-determined |
| S16_extracted_24866 | Low-quality |
| S16_extracted_24890 | Not-determined |
| S16_extracted_24903 | Not-determined |
| S16_extracted_24906 | Not-determined |
| S16_extracted_24908 | Not-determined |
| S16_extracted_24931 | Low-quality |
| S16_extracted_24934 | Low-quality |
| S16_extracted_24946 | Not-determined |
| S16_extracted_24948 | Low-quality |
| S16_extracted_24957 | Not-determined |
| S16_extracted_24958 | Not-determined |
| S16_extracted_24960 | Low-quality |
| S16_extracted_24974 | Not-determined |
| S16_extracted_24980 | Not-determined |
| S16_extracted_24981 | Not-determined |
| S16_extracted_24985 | Low-quality |
| S16_extracted_24996 | Not-determined |
| S16_extracted_25014 | Not-determined |
| S16_extracted_25019 | Not-determined |
| S16_extracted_25026 | Not-determined |
| S16_extracted_25032 | Not-determined |
| S16_extracted_25077 | Not-determined |
| S16_extracted_25089 | Low-quality |
| S16_extracted_25115 | Not-determined |
| S16_extracted_25128 | Not-determined |
| S16_extracted_25133 | Not-determined |
| S16_extracted_25140 | Not-determined |
| S16_extracted_25146 | Not-determined |
| S16_extracted_25158 | Not-determined |
| S16_extracted_25175 | Not-determined |
| S16_extracted_25178 | Not-determined |
| S16_extracted_25184 | Not-determined |
| S16_extracted_25192 | Not-determined |
| S16_extracted_25193 | Not-determined |
| S16_extracted_25206 | Not-determined |
| S16_extracted_25220 | Not-determined |
| S16_extracted_25229 | Not-determined |
| S16_extracted_25230 | Not-determined |
| S16_extracted_25236 | Not-determined |
| S16_extracted_25243 | Not-determined |
| S16_extracted_25246 | Not-determined |
| S16_extracted_25247 | Not-determined |
| S16_extracted_25250 | Not-determined |
| S16_extracted_25256 | Not-determined |
| S16_extracted_25259 | Not-determined |
| S16_extracted_25261 | Not-determined |
| S16_extracted_25264 | Not-determined |
| S16_extracted_25283 | Not-determined |
| S16_extracted_25288 | Low-quality |
| S16_extracted_25292 | Not-determined |
| S16_extracted_25297 | Not-determined |
| S16_extracted_25306 | Not-determined |
| S16_extracted_25321 | Not-determined |
| S16_extracted_25328 | Not-determined |
| S16_extracted_25329 | Low-quality |
| S16_extracted_25375 | Low-quality |
| S16_extracted_25390 | Not-determined |
| S16_extracted_25421 | Not-determined |
| S16_extracted_25435 | Low-quality |
| S16_extracted_25438 | Not-determined |
| S16_extracted_25482 | Low-quality |
| S16_extracted_25484 | Not-determined |
| S16_extracted_25495 | Not-determined |
| S16_extracted_25509 | Not-determined |
| S16_extracted_25513 | Not-determined |
| S16_extracted_25530 | Not-determined |
| S16_extracted_25558 | Low-quality |
| S16_extracted_25564 | Not-determined |
| S16_extracted_25585 | Not-determined |
| S16_extracted_25589 | Low-quality |
| S16_extracted_25592 | Not-determined |
| S16_extracted_25609 | Not-determined |
| S16_extracted_25614 | Not-determined |
| S16_extracted_25617 | Not-determined |
| S16_extracted_25644 | Not-determined |
| S16_extracted_25647 | Not-determined |
| S16_extracted_25652 | Not-determined |
| S16_extracted_25656 | Not-determined |
| S16_extracted_25679 | Not-determined |
| S16_extracted_25696 | Not-determined |
| S16_extracted_25706 | Not-determined |
| S16_extracted_25726 | Not-determined |
| S16_extracted_25730 | Not-determined |
| S16_extracted_25732 | Low-quality |
| S16_extracted_25745 | Not-determined |
| S16_extracted_25749 | Not-determined |
| S16_extracted_25783 | Not-determined |
| S16_extracted_25789 | Not-determined |
| S16_extracted_25790 | Not-determined |
| S16_extracted_25799 | Not-determined |
| S16_extracted_25801 | Not-determined |
| S16_extracted_25809 | Not-determined |
| S16_extracted_25826 | Not-determined |
| S16_extracted_25845 | Not-determined |
| S16_extracted_25846 | Not-determined |
| S16_extracted_25847 | Not-determined |
| S16_extracted_25863 | Not-determined |
| S16_extracted_25869 | Not-determined |
| S16_extracted_25885 | Not-determined |
| S16_extracted_25891 | Not-determined |
| S16_extracted_25903 | Low-quality |
| S16_extracted_25906 | Low-quality |
| S16_extracted_25907 | Not-determined |
| S16_extracted_25916 | Not-determined |
| S16_extracted_25926 | Not-determined |
| S16_extracted_25936 | Not-determined |
| S16_extracted_25947 | Not-determined |
| S16_extracted_25957 | Low-quality |
| S16_extracted_25987 | Not-determined |
| S16_extracted_26005 | Low-quality |
| S16_extracted_26011 | Not-determined |
| S16_extracted_26021 | Not-determined |
| S16_extracted_26035 | Not-determined |
| S16_extracted_26042 | Not-determined |
| S16_extracted_26055 | Not-determined |
| S16_extracted_26061 | Low-quality |
| S16_extracted_26063 | Not-determined |
| S16_extracted_26068 | Not-determined |
| S16_extracted_26102 | Not-determined |
| S16_extracted_26105 | Not-determined |
| S16_extracted_26111 | Not-determined |
| S16_extracted_26118 | Not-determined |
| S16_extracted_26119 | Not-determined |
| S16_extracted_26150 | Not-determined |
| S16_extracted_26161 | Not-determined |
| S16_extracted_26181 | Not-determined |
| S16_extracted_26189 | Not-determined |
| S16_extracted_26210 | Not-determined |
| S16_extracted_26220 | Not-determined |
| S16_extracted_26222 | Not-determined |
| S16_extracted_26238 | Not-determined |
| S16_extracted_26260 | Low-quality |
| S16_extracted_26284 | Low-quality |
| S16_extracted_26293 | Not-determined |
| S16_extracted_26296 | Not-determined |
| S16_extracted_26297 | Not-determined |
| S16_extracted_26303 | Not-determined |
| S16_extracted_26308 | Not-determined |
| S16_extracted_26310 | Not-determined |
| S16_extracted_26321 | Not-determined |
| S16_extracted_26344 | Not-determined |
| S16_extracted_26347 | Low-quality |
| S16_extracted_26351 | Not-determined |
| S16_extracted_26359 | Not-determined |
| S16_extracted_26368 | Not-determined |
| S16_extracted_26378 | Not-determined |
| S16_extracted_26380 | Not-determined |
| S16_extracted_26385 | Low-quality |
| S16_extracted_26392 | Not-determined |
| S16_extracted_26393 | Not-determined |
| S16_extracted_26420 | Not-determined |
| S16_extracted_26426 | Not-determined |
| S16_extracted_26447 | Not-determined |
| S16_extracted_26453 | Not-determined |
| S16_extracted_26465 | Not-determined |
| S16_extracted_26485 | Not-determined |
| S16_extracted_26494 | Not-determined |
| S16_extracted_26514 | Not-determined |
| S16_extracted_26527 | Not-determined |
| S16_extracted_26532 | Low-quality |
| S16_extracted_26535 | Not-determined |
| S16_extracted_26548 | Not-determined |
| S16_extracted_26553 | Not-determined |
| S16_extracted_26593 | Not-determined |
| S16_extracted_26601 | Not-determined |
| S16_extracted_26614 | Not-determined |
| S16_extracted_26621 | Not-determined |
| S16_extracted_26624 | Low-quality |
| S16_extracted_26627 | Not-determined |
| S16_extracted_26636 | Not-determined |
| S16_extracted_26640 | Not-determined |
| S16_extracted_26646 | Not-determined |
| S16_extracted_26673 | Low-quality |
| S16_extracted_26680 | Not-determined |
| S16_extracted_26683 | Not-determined |
| S16_extracted_26690 | Not-determined |
| S16_extracted_26700 | Not-determined |
| S16_extracted_26702 | Not-determined |
| S16_extracted_26706 | Not-determined |
| S16_extracted_26707 | Not-determined |
| S16_extracted_26712 | Not-determined |
| S16_extracted_26714 | Not-determined |
| S16_extracted_26715 | Low-quality |
| S16_extracted_26721 | Low-quality |
| S16_extracted_26723 | Not-determined |
| S16_extracted_26725 | Not-determined |
| S16_extracted_26742 | Not-determined |
| S16_extracted_26748 | Not-determined |
| S16_extracted_26755 | Low-quality |
| S16_extracted_26762 | Not-determined |
| S16_extracted_26778 | Low-quality |
| S16_extracted_26781 | Not-determined |
| S16_extracted_26783 | Not-determined |
| S16_extracted_26808 | Not-determined |
| S16_extracted_26811 | Not-determined |
| S16_extracted_26816 | Not-determined |
| S16_extracted_26817 | Not-determined |
| S16_extracted_26820 | Not-determined |
| S16_extracted_26826 | Not-determined |
| S16_extracted_26827 | Not-determined |
| S16_extracted_26847 | Not-determined |
| S16_extracted_26849 | Not-determined |
| S16_extracted_26859 | Not-determined |
| S16_extracted_26861 | Not-determined |
| S16_extracted_26886 | Low-quality |
| S16_extracted_26907 | Not-determined |
| S16_extracted_26930 | Not-determined |
| S16_extracted_26934 | Not-determined |
| S16_extracted_26962 | Not-determined |
| S16_extracted_26985 | Low-quality |
| S16_extracted_26998 | Not-determined |
| S16_extracted_27004 | Low-quality |
| S16_extracted_27020 | Not-determined |
| S16_extracted_27069 | Not-determined |
| S16_extracted_27070 | Low-quality |
| S16_extracted_27072 | Not-determined |
| S16_extracted_27078 | Low-quality |
| S16_extracted_27080 | Not-determined |
| S16_extracted_27097 | Not-determined |
| S16_extracted_27099 | Not-determined |
| S16_extracted_27121 | Not-determined |
| S16_extracted_27160 | Not-determined |
| S16_extracted_27194 | Not-determined |
| S16_extracted_27224 | Low-quality |
| S16_extracted_27265 | Not-determined |
| S16_extracted_27272 | Not-determined |
| S16_extracted_27316 | Not-determined |
| S16_extracted_27330 | Low-quality |
| S16_extracted_27363 | Not-determined |
| S16_extracted_27364 | Low-quality |
| S16_extracted_27368 | Not-determined |
| S16_extracted_27369 | Not-determined |
| S16_extracted_27372 | Not-determined |
| S16_extracted_27386 | Low-quality |
| S16_extracted_27422 | Not-determined |
| S16_extracted_27432 | Not-determined |
| S16_extracted_27440 | Not-determined |
| S16_extracted_27448 | Not-determined |
| S16_extracted_27449 | Not-determined |
| S16_extracted_27455 | Not-determined |
| S16_extracted_27466 | Low-quality |
| S16_extracted_27481 | Not-determined |
| S16_extracted_27484 | Not-determined |
| S16_extracted_27485 | Not-determined |
| S16_extracted_27499 | Not-determined |
| S16_extracted_27505 | Low-quality |
| S16_extracted_27524 | Not-determined |
| S16_extracted_27530 | Low-quality |
| S16_extracted_27559 | Not-determined |
| S16_extracted_27566 | Not-determined |
| S16_extracted_27567 | Low-quality |
| S16_extracted_27574 | Not-determined |
| S16_extracted_27633 | Not-determined |
| S16_extracted_27648 | Low-quality |
| S16_extracted_27656 | Not-determined |
| S16_extracted_27675 | Not-determined |
| S16_extracted_27685 | Not-determined |
| S16_extracted_27703 | Not-determined |
| S16_extracted_27708 | Not-determined |
| S16_extracted_27711 | Not-determined |
| S16_extracted_27717 | Not-determined |
| S16_extracted_27754 | Not-determined |
| S16_extracted_27764 | Not-determined |
| S16_extracted_27766 | Not-determined |
| S16_extracted_27781 | Not-determined |
| S16_extracted_27789 | Low-quality |
| S16_extracted_27791 | Not-determined |
| S16_extracted_27797 | Not-determined |
| S16_extracted_27801 | Not-determined |
| S16_extracted_27803 | Low-quality |
| S16_extracted_27809 | Not-determined |
| S16_extracted_27811 | Not-determined |
| S16_extracted_27824 | Not-determined |
| S16_extracted_27828 | Not-determined |
| S16_extracted_27832 | Not-determined |
| S16_extracted_27836 | Not-determined |
| S16_extracted_27844 | Not-determined |
| S16_extracted_27848 | Not-determined |
| S16_extracted_27857 | Low-quality |
| S16_extracted_27861 | Low-quality |
| S16_extracted_27862 | Not-determined |
| S16_extracted_27880 | Not-determined |
| S16_extracted_27881 | Not-determined |
| S16_extracted_27884 | Not-determined |
| S16_extracted_27885 | Not-determined |
| S16_extracted_27890 | Low-quality |
| S16_extracted_27891 | Not-determined |
| S16_extracted_27896 | Not-determined |
| S16_extracted_27898 | Not-determined |
| S16_extracted_27902 | Not-determined |
| S16_extracted_27907 | Not-determined |
| S16_extracted_27919 | Not-determined |
| S16_extracted_27929 | Not-determined |
| S16_extracted_27945 | Not-determined |
| S16_extracted_27954 | Not-determined |
| S16_extracted_27964 | Not-determined |
| S16_extracted_27979 | Not-determined |
| S16_extracted_27985 | Not-determined |
| S16_extracted_28013 | Not-determined |
| S16_extracted_28041 | Not-determined |
| S16_extracted_28055 | Not-determined |
| S16_extracted_28083 | Low-quality |
| S16_extracted_28095 | Not-determined |
| S16_extracted_28098 | Not-determined |
| S16_extracted_28109 | Not-determined |
| S16_extracted_28117 | Not-determined |
| S16_extracted_28118 | Not-determined |
| S16_extracted_28125 | Not-determined |
| S16_extracted_28136 | Not-determined |
| S16_extracted_28141 | Not-determined |
| S16_extracted_28149 | Not-determined |
| S16_extracted_28162 | Not-determined |
| S16_extracted_28174 | Low-quality |
| S16_extracted_28186 | Not-determined |
| S16_extracted_28199 | Not-determined |
| S16_extracted_28203 | Not-determined |
| S16_extracted_28204 | Low-quality |
| S16_extracted_28206 | Not-determined |
| S16_extracted_28207 | Not-determined |
| S16_extracted_28214 | Not-determined |
| S16_extracted_28215 | Not-determined |
| S16_extracted_28218 | Not-determined |
| S16_extracted_28225 | Not-determined |
| S16_extracted_28246 | Not-determined |
| S16_extracted_28253 | Not-determined |
| S16_extracted_28255 | Not-determined |
| S16_extracted_28258 | Not-determined |
| S16_extracted_28272 | Low-quality |
| S16_extracted_28277 | Not-determined |
| S16_extracted_28292 | Not-determined |
| S16_extracted_28294 | Not-determined |
| S16_extracted_28310 | Not-determined |
| S16_extracted_28324 | Not-determined |
| S16_extracted_28336 | Not-determined |
| S16_extracted_28355 | Not-determined |
| S16_extracted_28356 | Not-determined |
| S16_extracted_28375 | Not-determined |
| S16_extracted_28383 | Not-determined |
| S16_extracted_28407 | Not-determined |
| S16_extracted_28410 | Not-determined |
| S16_extracted_28413 | Not-determined |
| S16_extracted_28422 | Not-determined |
| S16_extracted_28426 | Not-determined |
| S16_extracted_28429 | Not-determined |
| S16_extracted_28465 | Not-determined |
| S16_extracted_28467 | Not-determined |
| S16_extracted_28488 | Not-determined |
| S16_extracted_28499 | Not-determined |
| S16_extracted_28525 | Not-determined |
| S16_extracted_28530 | Low-quality |
| S16_extracted_28547 | Not-determined |
| S16_extracted_28551 | Low-quality |
| S16_extracted_28563 | Not-determined |
| S16_extracted_28580 | Not-determined |
| S16_extracted_28593 | Not-determined |
| S16_extracted_28623 | Not-determined |
| S16_extracted_28629 | Not-determined |
| S16_extracted_28639 | Not-determined |
| S16_extracted_28643 | Not-determined |
| S16_extracted_28644 | Not-determined |
| S16_extracted_28668 | Not-determined |
| S16_extracted_28688 | Not-determined |
| S16_extracted_28703 | Not-determined |
| S16_extracted_28730 | Low-quality |
| S16_extracted_28734 | Low-quality |
| S16_extracted_28738 | Low-quality |
| S16_extracted_28742 | Low-quality |
| S16_extracted_28744 | Low-quality |
| S16_extracted_28745 | Not-determined |
| S16_extracted_28756 | Not-determined |
| S16_extracted_28765 | Not-determined |
| S16_extracted_28798 | Low-quality |
| S16_extracted_28801 | Not-determined |
| S16_extracted_28813 | Not-determined |
| S16_extracted_28842 | Low-quality |
| S16_extracted_28844 | Not-determined |
| S16_extracted_28860 | Low-quality |
| S16_extracted_28861 | Not-determined |
| S16_extracted_28890 | Not-determined |
| S16_extracted_28891 | Not-determined |
| S16_extracted_28895 | Not-determined |
| S16_extracted_28900 | Low-quality |
| S16_extracted_28921 | Not-determined |
| S16_extracted_28946 | Low-quality |
| S16_extracted_28957 | Not-determined |
| S16_extracted_28971 | Not-determined |
| S16_extracted_28997 | Low-quality |

| | |
|---|---|
| S16_extracted_29004 | Not-determined |
| S16_extracted_29011 | Not-determined |
| S16_extracted_29030 | Not-determined |
| S16_extracted_29042 | Not-determined |
| S16_extracted_29049 | Not-determined |
| S16_extracted_29061 | Not-determined |
| S16_extracted_29088 | Not-determined |
| S16_extracted_29093 | Not-determined |
| S16_extracted_29112 | Not-determined |
| S16_extracted_29130 | Not-determined |
| S16_extracted_29134 | Not-determined |
| S16_extracted_29147 | Not-determined |
| S16_extracted_29154 | Not-determined |
| S16_extracted_29155 | Not-determined |
| S16_extracted_29157 | Not-determined |
| S16_extracted_29159 | Not-determined |
| S16_extracted_29163 | Not-determined |
| S16_extracted_29169 | Not-determined |
| S16_extracted_29170 | Not-determined |
| S16_extracted_29174 | Not-determined |
| S16_extracted_29179 | Not-determined |
| S16_extracted_29209 | Not-determined |
| S16_extracted_29213 | Not-determined |
| S16_extracted_29221 | Not-determined |
| S16_extracted_29234 | Not-determined |
| S16_extracted_29271 | Not-determined |
| S16_extracted_29282 | Not-determined |
| S16_extracted_29289 | Not-determined |
| S16_extracted_29296 | Not-determined |
| S16_extracted_29310 | Low-quality |
| S16_extracted_29325 | Not-determined |
| S16_extracted_29342 | Not-determined |
| S16_extracted_29344 | Not-determined |
| S16_extracted_29351 | Not-determined |
| S16_extracted_29352 | Not-determined |
| S16_extracted_29358 | Not-determined |
| S16_extracted_29361 | Not-determined |
| S16_extracted_29382 | Not-determined |
| S16_extracted_29392 | Not-determined |
| S16_extracted_29393 | Not-determined |
| S16_extracted_29413 | Not-determined |
| S16_extracted_29416 | Not-determined |
| S16_extracted_29418 | Not-determined |
| S16_extracted_29429 | Not-determined |
| S16_extracted_29435 | Not-determined |
| S16_extracted_29483 | Not-determined |
| S16_extracted_29488 | Not-determined |
| S16_extracted_29490 | Low-quality |
| S16_extracted_29500 | Not-determined |
| S16_extracted_29512 | Not-determined |
| S16_extracted_29525 | Not-determined |
| S16_extracted_29542 | Not-determined |
| S16_extracted_29545 | Not-determined |
| S16_extracted_29546 | Not-determined |
| S16_extracted_29569 | Not-determined |
| S16_extracted_29597 | Not-determined |
| S16_extracted_29604 | Not-determined |
| S16_extracted_29610 | Not-determined |
| S16_extracted_29611 | Not-determined |
| S16_extracted_29631 | Not-determined |
| S16_extracted_29634 | Low-quality |
| S16_extracted_29645 | Not-determined |
| S16_extracted_29646 | Low-quality |
| S16_extracted_29665 | Not-determined |
| S16_extracted_29666 | Not-determined |
| S16_extracted_29669 | Not-determined |
| S16_extracted_29680 | Not-determined |
| S16_extracted_29690 | Not-determined |
| S16_extracted_29726 | Not-determined |
| S16_extracted_29736 | Not-determined |
| S16_extracted_29741 | Not-determined |
| S16_extracted_29742 | Not-determined |
| S16_extracted_29759 | Low-quality |
| S16_extracted_29761 | Not-determined |
| S16_extracted_29776 | Not-determined |
| S16_extracted_29804 | Not-determined |
| S16_extracted_29805 | Not-determined |
| S16_extracted_29806 | Not-determined |
| S16_extracted_29846 | Not-determined |
| S16_extracted_29858 | Not-determined |
| S16_extracted_29859 | Not-determined |
| S16_extracted_29871 | Not-determined |
| S16_extracted_29874 | Not-determined |
| S16_extracted_29876 | Low-quality |
| S16_extracted_29879 | Not-determined |
| S16_extracted_29899 | Not-determined |
| S16_extracted_29903 | Not-determined |
| S16_extracted_29923 | Low-quality |
| S16_extracted_29940 | Not-determined |
| S16_extracted_29969 | Not-determined |
| S16_extracted_29974 | Not-determined |
| S16_extracted_29979 | Low-quality |
| S16_extracted_29992 | Low-quality |
| S16_extracted_29998 | Not-determined |
| S16_extracted_30019 | Not-determined |
| S16_extracted_30033 | Not-determined |
| S16_extracted_30043 | Not-determined |
| S16_extracted_30052 | Low-quality |
| S16_extracted_30062 | Not-determined |
| S16_extracted_30064 | Not-determined |
| S16_extracted_30075 | Not-determined |
| S16_extracted_30088 | Not-determined |
| S16_extracted_30093 | Not-determined |
| S16_extracted_30114 | Not-determined |
| S16_extracted_30124 | Not-determined |
| S16_extracted_30145 | Low-quality |
| S16_extracted_30158 | Not-determined |
| S16_extracted_30159 | Not-determined |
| S16_extracted_30173 | Not-determined |
| S16_extracted_30178 | Not-determined |
| S16_extracted_30194 | Not-determined |
| S16_extracted_30215 | Not-determined |
| S16_extracted_30238 | Not-determined |
| S16_extracted_30248 | Not-determined |
| S16_extracted_30261 | Not-determined |
| S16_extracted_30262 | Low-quality |
| S16_extracted_30264 | Not-determined |
| S16_extracted_30268 | Not-determined |
| S16_extracted_30279 | Not-determined |
| S16_extracted_30285 | Not-determined |
| S16_extracted_30299 | Not-determined |
| S16_extracted_30302 | Not-determined |
| S16_extracted_30311 | Not-determined |
| S16_extracted_30312 | Not-determined |
| S16_extracted_30313 | Low-quality |
| S16_extracted_30314 | Not-determined |
| S16_extracted_30318 | Not-determined |
| S16_extracted_30320 | Not-determined |
| S16_extracted_30322 | Not-determined |
| S16_extracted_30329 | Not-determined |
| S16_extracted_30341 | Not-determined |
| S16_extracted_30345 | Not-determined |
| S16_extracted_30348 | Not-determined |
| S16_extracted_30349 | Not-determined |
| S16_extracted_30356 | Not-determined |
| S16_extracted_30363 | Not-determined |
| S16_extracted_30370 | Not-determined |
| S16_extracted_30373 | Not-determined |
| S16_extracted_30395 | Not-determined |
| S16_extracted_30432 | Not-determined |
| S16_extracted_30435 | Not-determined |
| S16_extracted_30456 | Not-determined |
| S16_extracted_30457 | Low-quality |
| S16_extracted_30465 | Not-determined |
| S16_extracted_30467 | Not-determined |
| S16_extracted_30470 | Not-determined |
| S16_extracted_30498 | Not-determined |
| S16_extracted_30510 | Not-determined |
| S16_extracted_30535 | Not-determined |
| S16_extracted_30566 | Not-determined |
| S16_extracted_30583 | Not-determined |
| S16_extracted_30606 | Not-determined |
| S16_extracted_30636 | Not-determined |
| S16_extracted_30652 | Not-determined |
| S16_extracted_30654 | Low-quality |
| S16_extracted_30655 | Low-quality |
| S16_extracted_30691 | Not-determined |
| S16_extracted_30712 | Low-quality |
| S16_extracted_30722 | Not-determined |
| S16_extracted_30723 | Not-determined |
| S16_extracted_30735 | Not-determined |
| S16_extracted_30737 | Not-determined |
| S16_extracted_30738 | Not-determined |
| S16_extracted_30739 | Not-determined |
| S16_extracted_30749 | Not-determined |
| S16_extracted_30750 | Not-determined |
| S16_extracted_30751 | Not-determined |
| S16_extracted_30752 | Not-determined |
| S16_extracted_30756 | Not-determined |
| S16_extracted_30775 | Not-determined |
| S16_extracted_30780 | Not-determined |
| S16_extracted_30784 | Not-determined |
| S16_extracted_30786 | Low-quality |
| S16_extracted_30813 | Not-determined |
| S16_extracted_30818 | Not-determined |
| S16_extracted_30820 | Not-determined |
| S16_extracted_30827 | Not-determined |
| S16_extracted_30828 | Not-determined |
| S16_extracted_30850 | Not-determined |
| S16_extracted_30854 | Not-determined |
| S16_extracted_30858 | Not-determined |
| S16_extracted_30868 | Not-determined |
| S16_extracted_30869 | Not-determined |
| S16_extracted_30871 | Not-determined |
| S16_extracted_30903 | Not-determined |
| S16_extracted_30908 | Not-determined |
| S16_extracted_30919 | Not-determined |
| S16_extracted_30925 | Not-determined |
| S16_extracted_30926 | Low-quality |
| S16_extracted_30951 | Not-determined |
| S16_extracted_30967 | Not-determined |
| S16_extracted_30974 | Not-determined |
| S16_extracted_30994 | Not-determined |
| S16_extracted_30996 | Not-determined |
| S16_extracted_31002 | Not-determined |
| S16_extracted_31023 | Not-determined |
| S16_extracted_31034 | Not-determined |
| S16_extracted_31060 | Not-determined |
| S16_extracted_31076 | Not-determined |
| S16_extracted_31078 | Not-determined |
| S16_extracted_31107 | Not-determined |
| S16_extracted_31111 | Not-determined |
| S16_extracted_31124 | Not-determined |
| S16_extracted_31126 | Not-determined |
| S16_extracted_31149 | Not-determined |
| S16_extracted_31160 | Not-determined |
| S16_extracted_31173 | Not-determined |
| S16_extracted_31192 | Not-determined |
| S16_extracted_31219 | Not-determined |
| S16_extracted_31240 | Not-determined |
| S16_extracted_31248 | Low-quality |
| S16_extracted_31265 | Not-determined |
| S16_extracted_31266 | Not-determined |
| S16_extracted_31270 | Not-determined |
| S16_extracted_31272 | Not-determined |
| S16_extracted_31286 | Not-determined |
| S16_extracted_31288 | Low-quality |
| S16_extracted_31296 | Not-determined |
| S16_extracted_31299 | Not-determined |
| S16_extracted_31314 | Low-quality |
| S16_extracted_31326 | Low-quality |
| S16_extracted_31329 | Low-quality |
| S16_extracted_31355 | Not-determined |
| S16_extracted_31366 | Not-determined |
| S16_extracted_31367 | Not-determined |
| S16_extracted_31390 | Not-determined |
| S16_extracted_31392 | Not-determined |
| S16_extracted_31401 | Low-quality |
| S16_extracted_31409 | Not-determined |
| S16_extracted_31413 | Not-determined |
| S16_extracted_31417 | Not-determined |
| S16_extracted_31427 | Not-determined |
| S16_extracted_31429 | Not-determined |
| S16_extracted_31435 | Not-determined |
| S16_extracted_31439 | Not-determined |
| S16_extracted_31443 | Not-determined |
| S16_extracted_31444 | Low-quality |
| S16_extracted_31457 | Not-determined |
| S16_extracted_31471 | Not-determined |
| S16_extracted_31487 | Not-determined |
| S16_extracted_31493 | Not-determined |
| S16_extracted_31517 | Not-determined |
| S16_extracted_31521 | Not-determined |
| S16_extracted_31522 | Not-determined |
| S16_extracted_31525 | Not-determined |
| S16_extracted_31528 | Low-quality |
| S16_extracted_31535 | Not-determined |
| S16_extracted_31545 | Not-determined |
| S16_extracted_31549 | Not-determined |
| S16_extracted_31592 | Not-determined |
| S16_extracted_31602 | Not-determined |
| S16_extracted_31608 | Not-determined |
| S16_extracted_31625 | Not-determined |
| S16_extracted_31640 | Not-determined |
| S16_extracted_31642 | Not-determined |
| S16_extracted_31661 | Not-determined |
| S16_extracted_31680 | Not-determined |
| S16_extracted_31686 | Not-determined |
| S16_extracted_31689 | Not-determined |
| S16_extracted_31702 | Not-determined |
| S16_extracted_31717 | Not-determined |
| S16_extracted_31725 | Not-determined |
| S16_extracted_31737 | Low-quality |
| S16_extracted_31741 | Not-determined |
| S16_extracted_31743 | Not-determined |
| S16_extracted_31762 | Not-determined |
| S16_extracted_31782 | Low-quality |
| S16_extracted_31791 | Not-determined |
| S16_extracted_31797 | Not-determined |
| S16_extracted_31813 | Not-determined |
| S16_extracted_31848 | Not-determined |
| S16_extracted_31857 | Not-determined |
| S16_extracted_31860 | Not-determined |
| S16_extracted_31863 | Not-determined |
| S16_extracted_31893 | Not-determined |
| S16_extracted_31903 | Not-determined |
| S16_extracted_31908 | Not-determined |
| S16_extracted_31910 | Not-determined |
| S16_extracted_31913 | Not-determined |
| S16_extracted_31932 | Not-determined |
| S16_extracted_31933 | Not-determined |
| S16_extracted_31943 | Not-determined |
| S16_extracted_31944 | Not-determined |
| S16_extracted_31953 | Not-determined |
| S16_extracted_31955 | Low-quality |
| S16_extracted_31956 | Not-determined |
| S16_extracted_31991 | Low-quality |
| S16_extracted_32005 | Not-determined |
| S16_extracted_32010 | Not-determined |
| S16_extracted_32020 | Not-determined |
| S16_extracted_32030 | Not-determined |
| S16_extracted_32032 | Not-determined |
| S16_extracted_32045 | Not-determined |
| S16_extracted_32051 | Not-determined |
| S16_extracted_32055 | Low-quality |
| S16_extracted_32101 | Not-determined |
| S16_extracted_32114 | Not-determined |
| S16_extracted_32119 | Not-determined |
| S16_extracted_32135 | Not-determined |
| S16_extracted_32140 | Not-determined |
| S16_extracted_32200 | Not-determined |
| S16_extracted_32205 | Not-determined |
| S16_extracted_32208 | Not-determined |
| S16_extracted_32220 | Not-determined |
| S16_extracted_32235 | Not-determined |
| S16_extracted_32241 | Low-quality |
| S16_extracted_32245 | Not-determined |
| S16_extracted_32255 | Not-determined |
| S16_extracted_32258 | Not-determined |
| S16_extracted_32260 | Low-quality |
| S16_extracted_32282 | Not-determined |
| S16_extracted_32289 | Not-determined |
| S16_extracted_32301 | Not-determined |
| S16_extracted_32329 | Not-determined |
| S16_extracted_32336 | Not-determined |
| S8_vs_k141_204260\|\|full | Complete |
| S16_extracted_32377 | Not-determined |
| S16_extracted_32378 | Not-determined |
| S16_extracted_32379 | Not-determined |
| S16_extracted_32380 | Not-determined |
| S16_extracted_32381 | Not-determined |
| S16_extracted_32393 | Not-determined |
| S16_extracted_32430 | Not-determined |
| S16_extracted_32432 | Not-determined |
| S16_extracted_32441 | Low-quality |
| S16_extracted_32464 | Low-quality |
| S16_extracted_32466 | Low-quality |
| S16_extracted_32472 | Not-determined |
| S16_extracted_32511 | Not-determined |
| S16_extracted_32515 | Not-determined |
| S16_extracted_32517 | Not-determined |
| S16_extracted_32531 | Not-determined |
| S16_extracted_32536 | Low-quality |
| S16_extracted_32553 | Not-determined |
| S16_extracted_32554 | Not-determined |
| S16_extracted_32558 | Not-determined |
| S16_extracted_32565 | Not-determined |
| S16_extracted_32583 | Not-determined |
| S16_extracted_32589 | Not-determined |
| S16_extracted_32592 | Not-determined |
| S16_extracted_32593 | Not-determined |
| S16_extracted_32618 | Not-determined |
| S16_extracted_32621 | Not-determined |
| S16_extracted_32633 | Not-determined |
| S16_extracted_32644 | Not-determined |
| S16_extracted_32650 | Not-determined |
| S16_extracted_32680 | Not-determined |
| S16_extracted_32686 | Not-determined |
| S16_extracted_32687 | Low-quality |
| S16_extracted_32695 | Not-determined |
| S16_extracted_32719 | Not-determined |
| S16_extracted_32737 | Not-determined |
| S16_extracted_32740 | Not-determined |
| S16_extracted_32749 | Not-determined |
| S16_extracted_32763 | Not-determined |
| S16_extracted_32772 | Low-quality |
| S16_extracted_32773 | Low-quality |
| S16_extracted_32786 | Not-determined |
| S16_extracted_32788 | Not-determined |
| S16_extracted_32790 | Low-quality |
| S16_extracted_32799 | Not-determined |
| S16_extracted_32802 | Not-determined |
| S16_extracted_32804 | Not-determined |
| S16_extracted_32811 | Not-determined |
| S16_extracted_32823 | Low-quality |
| S16_extracted_32825 | Not-determined |
| S16_extracted_32826 | Not-determined |
| S16_extracted_32828 | Not-determined |
| S16_extracted_32834 | Not-determined |
| S16_extracted_32846 | Not-determined |
| S16_extracted_32858 | Not-determined |
| S16_extracted_32872 | Not-determined |
| S16_extracted_32883 | Low-quality |
| S16_extracted_32886 | Not-determined |
| S16_extracted_32895 | Not-determined |
| S16_extracted_32897 | Not-determined |
| S16_extracted_32905 | Not-determined |
| S16_extracted_32911 | Not-determined |
| S16_extracted_32919 | Not-determined |
| S16_extracted_32921 | Not-determined |
| S16_extracted_32929 | Not-determined |
| S16_extracted_32933 | Not-determined |
| S16_extracted_32941 | Not-determined |
| S16_extracted_32952 | Not-determined |
| S16_extracted_32954 | Not-determined |
| S16_extracted_32957 | Not-determined |
| S16_extracted_32965 | Not-determined |
| S16_extracted_32994 | Not-determined |
| S16_extracted_32995 | Not-determined |
| S16_extracted_33002 | Not-determined |
| S16_extracted_33026 | Not-determined |
| S16_extracted_33027 | Low-quality |
| S16_extracted_33031 | Not-determined |
| S16_extracted_33032 | Not-determined |
| S16_extracted_33033 | Not-determined |
| S16_extracted_33040 | Not-determined |
| S16_extracted_33059 | Not-determined |
| S16_extracted_33066 | Not-determined |
| S16_extracted_33116 | Low-quality |
| S16_extracted_33118 | Not-determined |
| S16_extracted_33135 | Not-determined |
| S16_extracted_33165 | Not-determined |
| S16_extracted_33172 | Not-determined |
| S16_extracted_33192 | Not-determined |
| S16_extracted_33196 | Low-quality |
| S16_extracted_33197 | Not-determined |
| S8_vs_k141_110896\|\|full | Medium-quality |
| S16_extracted_33229 | Not-determined |
| S16_extracted_33295 | Not-determined |
| S16_extracted_33297 | Not-determined |
| S16_extracted_33317 | Not-determined |
| S16_extracted_33318 | Not-determined |
| S16_extracted_33331 | Low-quality |
| S16_extracted_33332 | Not-determined |
| S16_extracted_33333 | Low-quality |
| S16_extracted_33335 | Not-determined |
| S16_extracted_33345 | Low-quality |
| S16_extracted_33355 | Not-determined |
| S16_extracted_33381 | Not-determined |
| S16_extracted_33383 | Not-determined |
| S16_extracted_33404 | Not-determined |
| S16_extracted_33416 | Not-determined |
| S16_extracted_33419 | Low-quality |
| S16_extracted_33421 | Not-determined |
| S16_extracted_33441 | Not-determined |
| S16_extracted_33444 | Not-determined |
| S16_extracted_33449 | Not-determined |
| S16_extracted_33454 | Not-determined |
| S16_extracted_33456 | Not-determined |
| S16_extracted_33472 | Not-determined |
| S16_extracted_33474 | Not-determined |
| S16_extracted_33480 | Not-determined |
| S16_extracted_33485 | Not-determined |
| S16_extracted_33488 | Not-determined |
| S16_extracted_33492 | Not-determined |
| S16_extracted_33534 | Not-determined |
| S16_extracted_33537 | Not-determined |
| S16_extracted_33550 | Not-determined |
| S16_extracted_33552 | Not-determined |
| S16_extracted_33554 | Not-determined |
| S16_extracted_33559 | Not-determined |
| S16_extracted_33572 | Not-determined |
| S16_extracted_33623 | Not-determined |
| S16_extracted_33624 | Low-quality |
| S16_extracted_33626 | Not-determined |
| S16_extracted_33629 | Not-determined |
| S16_extracted_33653 | Low-quality |
| S16_extracted_33654 | Not-determined |
| S16_extracted_33676 | Low-quality |
| S16_extracted_33679 | Not-determined |
| S16_extracted_33684 | Low-quality |
| S16_extracted_33698 | Not-determined |
| S16_extracted_33702 | Not-determined |
| S16_extracted_33705 | Not-determined |
| S16_extracted_33707 | Not-determined |
| S16_extracted_33719 | Not-determined |
| S16_extracted_33725 | Low-quality |
| S16_extracted_33729 | Not-determined |
| S16_extracted_33732 | Not-determined |
| S16_extracted_33735 | Not-determined |
| S16_extracted_33751 | Low-quality |
| S16_extracted_33759 | Not-determined |
| S16_extracted_33779 | Not-determined |
| S16_extracted_33783 | Not-determined |
| S16_extracted_33786 | Not-determined |
| S16_extracted_33794 | Not-determined |
| S16_extracted_33801 | Not-determined |
| S16_extracted_33805 | Not-determined |
| S16_extracted_33808 | Not-determined |
| S16_extracted_33815 | Low-quality |
| S16_extracted_33834 | Not-determined |
| S16_extracted_33856 | Not-determined |
| S16_extracted_33858 | Not-determined |
| S16_extracted_33870 | Not-determined |
| S16_extracted_33880 | Not-determined |
| S16_extracted_33881 | Not-determined |
| S16_extracted_33885 | Not-determined |
| S16_extracted_33888 | Not-determined |
| S16_extracted_33891 | Not-determined |
| S16_extracted_33892 | Not-determined |
| S16_extracted_33895 | Not-determined |
| S16_extracted_33905 | Not-determined |
| S16_extracted_33918 | Not-determined |
| S16_extracted_33922 | Not-determined |
| S16_extracted_33950 | Not-determined |
| S16_extracted_33953 | Not-determined |
| S16_extracted_33962 | Not-determined |
| S16_extracted_34016 | Not-determined |
| S16_extracted_34061 | Not-determined |
| S16_extracted_34062 | Not-determined |
| S16_extracted_34066 | Low-quality |
| S16_extracted_34074 | Not-determined |
| S16_extracted_34076 | Not-determined |
| S16_extracted_34086 | Not-determined |
| S16_extracted_34092 | Not-determined |
| S16_extracted_34097 | Not-determined |
| S16_extracted_34111 | Not-determined |
| S16_extracted_34115 | Not-determined |
| S16_extracted_34118 | Not-determined |
| S16_extracted_34128 | Not-determined |
| S16_extracted_34163 | Not-determined |
| S16_extracted_34176 | Not-determined |
| S16_extracted_34186 | Not-determined |
| S16_extracted_34198 | Low-quality |
| S16_extracted_34200 | Not-determined |
| S16_extracted_34234 | Not-determined |
| S16_extracted_34293 | Not-determined |
| S16_extracted_34295 | Not-determined |
| S16_extracted_34311 | Low-quality |
| S16_extracted_34323 | Not-determined |
| S16_extracted_34335 | Low-quality |
| S16_extracted_34338 | Low-quality |
| S16_extracted_34342 | Not-determined |
| S16_extracted_34344 | Not-determined |
| S16_extracted_34399 | Not-determined |
| S16_extracted_34405 | Not-determined |
| S16_extracted_34409 | Not-determined |
| S16_extracted_34410 | Not-determined |
| S16_extracted_34419 | Not-determined |
| S16_extracted_34423 | Not-determined |
| S16_extracted_34425 | Not-determined |
| S16_extracted_34431 | Not-determined |
| S16_extracted_34442 | Low-quality |
| S16_extracted_34443 | Not-determined |
| S16_extracted_34451 | Not-determined |
| S16_extracted_34461 | Not-determined |
| S16_extracted_34467 | Not-determined |
| S16_extracted_34469 | Low-quality |
| S16_extracted_34470 | Not-determined |
| S16_extracted_34475 | Not-determined |
| S16_extracted_34505 | Not-determined |
| S16_extracted_34523 | Low-quality |
| S16_extracted_34526 | Not-determined |
| S16_extracted_34527 | Not-determined |
| S16_extracted_34528 | Not-determined |
| S16_extracted_34554 | Not-determined |
| S16_extracted_34559 | Not-determined |
| S16_extracted_34589 | Not-determined |
| S16_extracted_34590 | Not-determined |
| S16_extracted_34593 | Low-quality |
| S16_extracted_34603 | Not-determined |
| S16_extracted_34611 | Not-determined |
| S16_extracted_34624 | Not-determined |
| S16_extracted_34627 | Not-determined |
| S16_extracted_34634 | Not-determined |
| S16_extracted_34643 | Low-quality |
| S16_extracted_34682 | Not-determined |
| S16_extracted_34692 | Not-determined |
| S16_extracted_34702 | Low-quality |
| S16_extracted_34726 | Not-determined |
| S16_extracted_34728 | Not-determined |
| S16_extracted_34729 | Not-determined |
| S16_extracted_34739 | Not-determined |
| S16_extracted_34745 | Not-determined |
| S16_extracted_34765 | Not-determined |
| S16_extracted_34778 | Low-quality |
| S16_extracted_34787 | Not-determined |
| S16_extracted_34821 | Low-quality |
| S16_extracted_34834 | Low-quality |
| S16_extracted_34846 | Low-quality |
| S16_extracted_34856 | Not-determined |
| S16_extracted_34883 | Not-determined |
| S16_extracted_34887 | Low-quality |
| S16_extracted_34889 | Not-determined |
| S16_extracted_34892 | Not-determined |
| S16_extracted_34897 | Not-determined |
| S16_extracted_34900 | Not-determined |
| S16_extracted_34902 | Not-determined |
| S16_extracted_34904 | Not-determined |
| S16_extracted_34921 | Not-determined |
| S16_extracted_34930 | Low-quality |
| S16_extracted_34938 | Not-determined |
| S16_extracted_34945 | Not-determined |
| S16_extracted_34960 | Not-determined |
| S16_extracted_34966 | Not-determined |
| S16_extracted_34974 | Not-determined |
| S16_extracted_34986 | Low-quality |
| S16_extracted_35017 | Not-determined |
| S16_extracted_35024 | Low-quality |
| S16_extracted_35027 | Not-determined |
| S16_extracted_35032 | Not-determined |
| S16_extracted_35034 | Not-determined |
| S16_extracted_35045 | Not-determined |
| S16_extracted_35055 | Not-determined |
| S16_extracted_35059 | Not-determined |
| S16_extracted_35086 | Not-determined |
| S16_extracted_35103 | Not-determined |
| S16_extracted_35104 | Not-determined |
| S16_extracted_35108 | Not-determined |

| | |
|---|---|
| S16_extracted_35124 | Not-determined |
| S16_extracted_35130 | Not-determined |
| S16_extracted_35132 | Not-determined |
| S16_extracted_35133 | Not-determined |
| S16_extracted_35146 | Not-determined |
| S16_extracted_35153 | Not-determined |
| S16_extracted_35165 | Not-determined |
| S16_extracted_35175 | Not-determined |
| S16_extracted_35180 | Not-determined |
| S16_extracted_35181 | Not-determined |
| S16_extracted_35182 | Not-determined |
| S16_extracted_35204 | Low-quality |
| S16_extracted_35205 | Not-determined |
| S16_extracted_35216 | Low-quality |
| S16_extracted_35217 | Not-determined |
| S16_extracted_35218 | Not-determined |
| S16_extracted_35227 | Not-determined |
| S16_extracted_35234 | Not-determined |
| S16_extracted_35254 | Not-determined |
| S16_extracted_35261 | Not-determined |
| S16_extracted_35271 | Not-determined |
| S16_extracted_35273 | Not-determined |
| S16_extracted_35274 | Not-determined |
| S16_extracted_35280 | Low-quality |
| S16_extracted_35283 | Not-determined |
| S16_extracted_35293 | Low-quality |
| S16_extracted_35296 | Not-determined |
| S16_extracted_35298 | Not-determined |
| S16_extracted_35299 | Not-determined |
| S16_extracted_35300 | Not-determined |
| S16_extracted_35301 | Not-determined |
| S16_extracted_35324 | Low-quality |
| S16_extracted_35330 | Not-determined |
| S16_extracted_35332 | Not-determined |
| S16_extracted_35343 | Low-quality |
| S16_extracted_35384 | Not-determined |
| S16_extracted_35388 | Not-determined |
| S16_extracted_35389 | Not-determined |
| S16_extracted_35390 | Not-determined |
| S16_extracted_35396 | Not-determined |
| S16_extracted_35414 | Not-determined |
| S16_extracted_35417 | Not-determined |
| S16_extracted_35423 | Not-determined |
| S16_extracted_35424 | Low-quality |
| S16_extracted_35466 | Not-determined |
| S16_extracted_35468 | Low-quality |
| S16_extracted_35480 | Low-quality |
| S16_extracted_35486 | Low-quality |
| S16_extracted_35493 | Not-determined |
| S16_extracted_35494 | Not-determined |
| S16_extracted_35505 | Low-quality |
| S16_extracted_35531 | Not-determined |
| S16_extracted_35539 | Low-quality |
| S16_extracted_35551 | Not-determined |
| S16_extracted_35553 | Not-determined |
| S16_extracted_35556 | Not-determined |
| S16_extracted_35566 | Not-determined |
| S16_extracted_35569 | Not-determined |
| S16_extracted_35582 | Not-determined |
| S16_extracted_35584 | Low-quality |
| S16_extracted_35599 | Not-determined |
| S16_extracted_35601 | Not-determined |
| S16_extracted_35608 | Not-determined |
| S16_extracted_35611 | Not-determined |
| S16_extracted_35618 | Not-determined |
| S16_extracted_35634 | Not-determined |
| S16_extracted_35646 | Not-determined |
| S16_extracted_35647 | Low-quality |
| S16_extracted_35649 | Low-quality |
| S16_extracted_35652 | Not-determined |
| S16_extracted_35665 | Not-determined |
| S16_extracted_35684 | Not-determined |
| S16_extracted_35710 | Not-determined |
| S16_extracted_35711 | Low-quality |
| S16_extracted_35712 | Low-quality |
| S16_extracted_35751 | Not-determined |
| S16_extracted_35768 | Not-determined |
| S16_extracted_35769 | Not-determined |
| S16_extracted_35791 | Low-quality |
| S16_extracted_35793 | Not-determined |
| S16_extracted_35813 | Low-quality |
| S16_extracted_35820 | Low-quality |
| S16_extracted_35829 | Not-determined |
| S16_extracted_35839 | Not-determined |
| S16_extracted_35887 | Low-quality |
| S16_extracted_35893 | Not-determined |
| S16_extracted_35897 | Low-quality |
| S16_extracted_35901 | Not-determined |
| S16_extracted_35906 | Not-determined |
| S16_extracted_35907 | Not-determined |
| S16_extracted_35912 | Not-determined |
| S16_extracted_35916 | Not-determined |
| S16_extracted_35931 | Low-quality |
| S16_extracted_35955 | Not-determined |
| S16_extracted_35962 | Not-determined |
| S16_extracted_35963 | Not-determined |
| S16_extracted_35977 | Low-quality |
| S16_extracted_35981 | Not-determined |
| S16_extracted_36005 | Not-determined |
| S16_extracted_36011 | Low-quality |
| S16_extracted_36017 | Not-determined |
| S16_extracted_36018 | Not-determined |
| S16_extracted_36032 | Not-determined |
| S16_extracted_36037 | Not-determined |
| S16_extracted_36045 | Not-determined |
| S16_extracted_36046 | Not-determined |
| S16_extracted_36050 | Not-determined |
| S16_extracted_36051 | Not-determined |
| S16_extracted_36055 | Low-quality |
| S16_extracted_36068 | Low-quality |
| S16_extracted_36075 | Not-determined |
| S16_extracted_36088 | Low-quality |
| S16_extracted_36089 | Low-quality |
| S16_extracted_36090 | Not-determined |
| S16_extracted_36095 | Not-determined |
| S16_extracted_36102 | Low-quality |
| S16_extracted_36111 | Not-determined |
| S16_extracted_36130 | Not-determined |
| S16_extracted_36137 | Low-quality |
| S16_extracted_36141 | Not-determined |
| S16_extracted_36142 | Not-determined |
| S16_extracted_36144 | Not-determined |
| S7_vs_138096\|\|full | High-quality |
| S16_extracted_36153 | Not-determined |
| S16_extracted_36167 | Not-determined |
| S16_extracted_36177 | Low-quality |
| S16_extracted_36195 | Not-determined |
| S16_extracted_36221 | Not-determined |
| S16_extracted_36224 | Not-determined |
| S16_extracted_36232 | Not-determined |
| S16_extracted_36264 | Not-determined |
| S16_extracted_36285 | Not-determined |
| S16_extracted_36286 | Low-quality |
| S16_extracted_36303 | Not-determined |
| S16_extracted_36322 | Low-quality |
| S16_extracted_36327 | Not-determined |
| S16_extracted_36330 | Not-determined |
| S16_extracted_36365 | Not-determined |
| S16_extracted_36371 | Not-determined |
| S16_extracted_36404 | Not-determined |
| S16_extracted_36407 | Not-determined |
| S16_extracted_36409 | Not-determined |
| S16_extracted_36426 | Not-determined |
| S16_extracted_36427 | Not-determined |
| S16_extracted_36452 | Not-determined |
| S16_extracted_36462 | Not-determined |
| S16_extracted_36463 | Not-determined |
| S16_extracted_36472 | Low-quality |
| S16_extracted_36476 | Low-quality |
| S16_extracted_36486 | Not-determined |
| S16_extracted_36487 | Not-determined |
| S16_extracted_36508 | Not-determined |
| S16_extracted_36513 | Low-quality |
| S16_extracted_36540 | Not-determined |
| S16_extracted_36548 | Not-determined |
| S16_extracted_36570 | Not-determined |
| S16_extracted_36574 | Not-determined |
| S16_extracted_36585 | Not-determined |
| S16_extracted_36617 | Not-determined |
| S16_extracted_36618 | Not-determined |
| S16_extracted_36625 | Not-determined |
| S16_extracted_36629 | Not-determined |
| S16_extracted_36630 | Not-determined |
| S16_extracted_36631 | Not-determined |
| S16_extracted_36642 | Not-determined |
| S16_extracted_36644 | Low-quality |
| S16_extracted_36645 | Not-determined |
| S16_extracted_36657 | Not-determined |
| S16_extracted_36662 | Not-determined |
| S16_extracted_36666 | Not-determined |
| S16_extracted_36676 | Not-determined |
| S16_extracted_36683 | Not-determined |
| S16_extracted_36692 | Low-quality |
| S16_extracted_36720 | Not-determined |
| S16_extracted_36736 | Not-determined |
| S16_extracted_36740 | Not-determined |
| S16_extracted_36749 | Not-determined |
| S16_extracted_36753 | Not-determined |
| S16_extracted_36756 | Not-determined |
| S16_extracted_36757 | Low-quality |
| S16_extracted_36762 | Not-determined |
| S16_extracted_36765 | Low-quality |
| S16_extracted_36781 | Not-determined |
| S16_extracted_36788 | Not-determined |
| S16_extracted_36795 | Not-determined |
| S16_extracted_36827 | Not-determined |
| S16_extracted_36832 | Low-quality |
| S16_extracted_36840 | Low-quality |
| S16_extracted_36843 | Low-quality |
| S16_extracted_36846 | Not-determined |
| S16_extracted_36854 | Not-determined |
| S16_extracted_36860 | Not-determined |
| S16_extracted_36863 | Not-determined |
| S16_extracted_36867 | Not-determined |
| S16_extracted_36899 | Low-quality |
| S16_extracted_36905 | Not-determined |
| S16_extracted_36922 | Not-determined |
| S16_extracted_36944 | Not-determined |
| S16_extracted_36958 | Not-determined |
| S16_extracted_36964 | Not-determined |
| S16_extracted_36994 | Not-determined |
| S16_extracted_37017 | Not-determined |
| S16_extracted_37018 | Not-determined |
| S16_extracted_37042 | Not-determined |
| S16_extracted_37073 | Not-determined |
| S16_extracted_37084 | Not-determined |
| S16_extracted_37089 | Not-determined |
| S16_extracted_37105 | Not-determined |
| S16_extracted_37109 | Not-determined |
| S16_extracted_37114 | Not-determined |
| S16_extracted_37126 | Not-determined |
| S16_extracted_37141 | Not-determined |
| S16_extracted_37143 | Not-determined |
| S16_extracted_37144 | Not-determined |
| S16_extracted_37150 | Not-determined |
| S16_extracted_37160 | Not-determined |
| S16_extracted_37183 | Low-quality |
| S16_extracted_37199 | Not-determined |
| S16_extracted_37218 | Not-determined |
| S16_extracted_37225 | Not-determined |
| S16_extracted_37237 | Low-quality |
| S16_extracted_37252 | Not-determined |
| S16_extracted_37256 | Not-determined |
| S16_extracted_37261 | Not-determined |
| S16_extracted_37275 | Not-determined |
| S16_extracted_37278 | Not-determined |
| S16_extracted_37316 | Not-determined |
| S16_extracted_37329 | Not-determined |
| S16_extracted_37353 | Not-determined |
| S16_extracted_37357 | Not-determined |
| S16_extracted_37373 | Not-determined |
| S16_extracted_37389 | Not-determined |
| S16_extracted_37392 | Not-determined |
| S16_extracted_37400 | Not-determined |
| S16_extracted_37401 | Not-determined |
| S16_extracted_37408 | Not-determined |
| S16_extracted_37475 | Low-quality |
| S16_extracted_37484 | Not-determined |
| S16_extracted_37503 | Not-determined |
| S16_extracted_37511 | Low-quality |
| S16_extracted_37522 | Not-determined |
| S16_extracted_37528 | Low-quality |
| S16_extracted_37529 | Not-determined |
| S16_extracted_37537 | Low-quality |
| S16_extracted_37544 | Not-determined |
| S16_extracted_37555 | Not-determined |
| S16_extracted_37556 | Not-determined |
| S16_extracted_37557 | Low-quality |
| S16_extracted_37569 | Not-determined |
| S16_extracted_37570 | Not-determined |
| S16_extracted_37572 | Not-determined |
| S16_extracted_37575 | Not-determined |
| S16_extracted_37592 | Not-determined |
| S16_extracted_37629 | Low-quality |
| S16_extracted_37635 | Not-determined |
| S16_extracted_37637 | Low-quality |
| S16_extracted_37638 | Not-determined |
| S16_extracted_37658 | Low-quality |
| S16_extracted_37671 | Not-determined |
| S16_extracted_37674 | Not-determined |
| S16_extracted_37675 | Not-determined |
| S16_extracted_37677 | Not-determined |
| S16_extracted_37683 | Not-determined |
| S16_extracted_37701 | Not-determined |
| S16_extracted_37709 | Low-quality |
| S16_extracted_37715 | Not-determined |
| S16_extracted_37739 | Not-determined |
| S16_extracted_37743 | Not-determined |
| S16_extracted_37748 | Not-determined |
| S16_extracted_37750 | Not-determined |
| S16_extracted_37753 | Not-determined |
| S16_extracted_37754 | Not-determined |
| S16_extracted_37778 | Low-quality |
| S16_extracted_37783 | Not-determined |
| S16_extracted_37796 | Not-determined |
| S16_extracted_37802 | Not-determined |
| S16_extracted_37812 | Not-determined |
| S16_extracted_37813 | Not-determined |
| S16_extracted_37828 | Not-determined |
| S16_extracted_37836 | Not-determined |
| S16_extracted_37838 | Not-determined |
| S16_extracted_37856 | Not-determined |
| S16_extracted_37859 | Not-determined |
| S16_extracted_37875 | Not-determined |
| S16_extracted_37880 | Not-determined |
| S16_extracted_37889 | Not-determined |
| S16_extracted_37898 | Not-determined |
| S16_extracted_37907 | Not-determined |
| S16_extracted_37912 | Not-determined |
| S16_extracted_37917 | Not-determined |
| S16_extracted_37928 | Not-determined |
| S16_extracted_37931 | Low-quality |
| S16_extracted_37933 | Not-determined |
| S16_extracted_37937 | Low-quality |
| S16_extracted_37938 | Not-determined |
| S16_extracted_37942 | Not-determined |
| S16_extracted_37947 | Not-determined |
| S16_extracted_37956 | Not-determined |
| S16_extracted_37960 | Low-quality |
| S16_extracted_37976 | Not-determined |
| S16_extracted_38000 | Not-determined |
| S16_extracted_38006 | Not-determined |
| S16_extracted_38017 | Low-quality |
| S16_extracted_38032 | Low-quality |
| S16_extracted_38034 | Not-determined |
| S16_extracted_38035 | Low-quality |
| S16_extracted_38041 | Not-determined |
| S16_extracted_38067 | Not-determined |
| S16_extracted_38069 | Not-determined |
| S16_extracted_38072 | Not-determined |
| S16_extracted_38076 | Not-determined |
| S16_extracted_38106 | Not-determined |
| S16_extracted_38123 | Not-determined |
| S16_extracted_38131 | Not-determined |
| S16_extracted_38149 | Not-determined |
| S16_extracted_38150 | Not-determined |
| S16_extracted_38159 | Low-quality |
| S16_extracted_38176 | Not-determined |
| S16_extracted_38179 | Not-determined |
| S16_extracted_38181 | Not-determined |
| S16_extracted_38206 | Not-determined |
| S16_extracted_38208 | Not-determined |
| S16_extracted_38213 | Not-determined |
| S16_extracted_38223 | Low-quality |
| S16_extracted_38232 | Low-quality |
| S16_extracted_38246 | Not-determined |
| S16_extracted_38248 | Not-determined |
| S16_extracted_38257 | Not-determined |
| S16_extracted_38276 | Not-determined |
| S16_extracted_38282 | Not-determined |
| S16_extracted_38285 | Not-determined |
| S16_extracted_38290 | Not-determined |
| S16_extracted_38297 | Not-determined |
| S16_extracted_38315 | Not-determined |
| S16_extracted_38318 | Not-determined |
| S16_extracted_38339 | Not-determined |
| S16_extracted_38341 | Not-determined |
| S16_extracted_38368 | Not-determined |
| S16_extracted_38389 | Low-quality |
| S16_extracted_38392 | Not-determined |
| S16_extracted_38395 | Not-determined |
| S16_extracted_38414 | Low-quality |
| S16_extracted_38430 | Not-determined |
| S16_extracted_38449 | Not-determined |
| S16_extracted_38454 | Not-determined |
| S16_extracted_38459 | Not-determined |
| S16_extracted_38471 | Not-determined |
| S16_extracted_38475 | Not-determined |
| S16_extracted_38518 | Low-quality |
| S16_extracted_38521 | Low-quality |
| S16_extracted_38540 | Not-determined |
| S16_extracted_38543 | Low-quality |
| S16_extracted_38563 | Not-determined |
| S16_extracted_38567 | Not-determined |
| S16_extracted_38573 | Low-quality |
| S16_extracted_38579 | Low-quality |
| S16_extracted_38593 | Not-determined |
| S16_extracted_38626 | Not-determined |
| S16_extracted_38628 | Not-determined |
| S16_extracted_38641 | Not-determined |
| S16_extracted_38644 | Not-determined |
| S16_extracted_38649 | Not-determined |
| S16_extracted_38667 | Not-determined |
| S16_extracted_38691 | Not-determined |
| S16_extracted_38697 | Not-determined |
| S16_extracted_38702 | Not-determined |
| S16_extracted_38719 | Not-determined |
| S16_extracted_38728 | Not-determined |
| S16_extracted_38747 | Not-determined |
| S16_extracted_38750 | Not-determined |
| S16_extracted_38762 | Not-determined |
| S16_extracted_38769 | Not-determined |
| S16_extracted_38786 | Not-determined |
| S16_extracted_38793 | Not-determined |
| S16_extracted_38799 | Not-determined |
| S16_extracted_38809 | Not-determined |
| S16_extracted_38817 | Not-determined |
| S16_extracted_38821 | Not-determined |
| S16_extracted_38844 | Low-quality |
| S16_extracted_38861 | Not-determined |
| S16_extracted_38868 | Not-determined |
| S16_extracted_38880 | Not-determined |
| S16_extracted_38902 | Not-determined |
| S16_extracted_38910 | Not-determined |
| S16_extracted_38911 | Not-determined |
| S16_extracted_38932 | Not-determined |
| S16_extracted_38935 | Not-determined |
| S16_extracted_38949 | Not-determined |
| S16_extracted_38959 | Low-quality |
| S16_extracted_38964 | Not-determined |
| S16_extracted_38965 | Not-determined |
| S16_extracted_38967 | Not-determined |
| S16_extracted_38983 | Not-determined |
| S16_extracted_38996 | Not-determined |
| S16_extracted_39032 | Not-determined |
| S16_extracted_39041 | Not-determined |
| S16_extracted_39045 | Not-determined |
| S16_extracted_39047 | Not-determined |
| S16_extracted_39053 | Low-quality |
| S16_extracted_39062 | Not-determined |
| S16_extracted_39067 | Not-determined |
| S16_extracted_39094 | Not-determined |
| S16_extracted_39100 | Not-determined |
| S16_extracted_39103 | Not-determined |
| S16_extracted_39123 | Not-determined |
| S16_extracted_39135 | Not-determined |
| S16_extracted_39145 | Not-determined |
| S16_extracted_39164 | Not-determined |
| S16_extracted_39167 | Not-determined |
| S16_extracted_39170 | Not-determined |
| S16_extracted_39189 | Not-determined |
| S16_extracted_39208 | Not-determined |
| S16_extracted_39211 | Not-determined |
| S16_extracted_39213 | Low-quality |
| S16_extracted_39236 | Not-determined |
| S16_extracted_39258 | Not-determined |
| S16_extracted_39265 | Not-determined |
| S16_extracted_39270 | Not-determined |
| S16_extracted_39280 | Low-quality |
| S16_extracted_39284 | Low-quality |
| S16_extracted_39305 | Not-determined |
| S16_extracted_39307 | Not-determined |
| S16_extracted_39319 | Not-determined |
| S16_extracted_39344 | Not-determined |
| S16_extracted_39356 | Not-determined |
| S16_extracted_39367 | Not-determined |
| S16_extracted_39387 | Not-determined |
| S16_extracted_39401 | Not-determined |
| S16_extracted_39426 | Not-determined |
| S16_extracted_39442 | Not-determined |
| S16_extracted_39447 | Not-determined |
| S16_extracted_39456 | Not-determined |
| S16_extracted_39457 | Not-determined |
| S16_extracted_39459 | Low-quality |
| S16_extracted_39462 | Not-determined |
| S16_extracted_39477 | Not-determined |
| S16_extracted_39478 | Not-determined |
| S16_extracted_39491 | Low-quality |
| S16_extracted_39496 | Not-determined |
| S16_extracted_39497 | Not-determined |
| S16_extracted_39508 | Not-determined |
| S16_extracted_39515 | Not-determined |
| S16_extracted_39517 | Not-determined |
| S16_extracted_39522 | Not-determined |
| S16_extracted_39561 | Not-determined |
| S16_extracted_39586 | Low-quality |
| S16_extracted_39594 | Not-determined |
| S16_extracted_39596 | Not-determined |
| S16_extracted_39610 | Not-determined |
| S16_extracted_39616 | Not-determined |
| S16_extracted_39636 | Not-determined |
| S16_extracted_39639 | Not-determined |
| S16_extracted_39644 | Not-determined |
| S16_extracted_39661 | Low-quality |
| S16_extracted_39669 | Not-determined |
| S16_extracted_39701 | Not-determined |
| S16_extracted_39702 | Not-determined |
| S16_extracted_39708 | Not-determined |
| S16_extracted_39709 | Not-determined |
| S16_extracted_39728 | Not-determined |
| S16_extracted_39734 | Not-determined |
| S16_extracted_39737 | Low-quality |
| S16_extracted_39741 | Not-determined |
| S16_extracted_39752 | Not-determined |
| S16_extracted_39759 | Not-determined |
| S16_extracted_39767 | Not-determined |
| S16_extracted_39772 | Low-quality |
| S16_extracted_39777 | Not-determined |
| S16_extracted_39813 | Not-determined |
| S16_extracted_39817 | Not-determined |
| S16_extracted_39826 | Low-quality |
| S16_extracted_39831 | Not-determined |
| S16_extracted_39852 | Not-determined |
| S16_extracted_39867 | Low-quality |
| S16_extracted_39890 | Not-determined |
| S16_extracted_39902 | Low-quality |
| S16_extracted_39909 | Not-determined |
| S16_extracted_39921 | Not-determined |
| S16_extracted_39947 | Not-determined |
| S16_extracted_39952 | Not-determined |
| S16_extracted_39964 | Low-quality |
| S16_extracted_39965 | Not-determined |
| S16_extracted_39993 | Low-quality |
| S16_extracted_40006 | Not-determined |
| S16_extracted_40019 | Not-determined |
| S16_extracted_40025 | Not-determined |
| S16_extracted_40034 | Not-determined |
| S16_extracted_40035 | Not-determined |
| S16_extracted_40043 | Not-determined |
| S16_extracted_40047 | Low-quality |
| S16_extracted_40051 | Not-determined |
| S16_extracted_40061 | Low-quality |
| S16_extracted_40062 | Not-determined |
| S16_extracted_40064 | Not-determined |
| S16_extracted_40071 | Low-quality |
| S16_extracted_40090 | Low-quality |
| S16_extracted_40099 | Not-determined |
| S16_extracted_40102 | Not-determined |
| S16_extracted_40122 | Not-determined |
| S16_extracted_40130 | Not-determined |
| S16_extracted_40147 | Low-quality |
| S16_extracted_40151 | Not-determined |
| S16_extracted_40165 | Not-determined |
| S16_extracted_40178 | Not-determined |
| S16_extracted_40183 | Not-determined |
| S16_extracted_40191 | Not-determined |
| S16_extracted_40198 | Not-determined |
| S16_extracted_40204 | Not-determined |
| S16_extracted_40206 | Not-determined |
| S16_extracted_40209 | Not-determined |
| S16_extracted_40221 | Not-determined |
| S16_extracted_40243 | Not-determined |
| S16_extracted_40246 | Not-determined |
| S16_extracted_40247 | Not-determined |
| S16_extracted_40252 | Not-determined |
| S16_extracted_40257 | Low-quality |
| S16_extracted_40279 | Not-determined |
| S16_extracted_40281 | Not-determined |
| S16_extracted_40301 | Not-determined |
| S16_extracted_40313 | Low-quality |
| S16_extracted_40319 | Not-determined |
| S16_extracted_40320 | Not-determined |
| S16_extracted_40324 | Not-determined |
| S16_extracted_40328 | Not-determined |
| S16_extracted_40329 | Not-determined |
| S16_extracted_40336 | Not-determined |
| S16_extracted_40340 | Not-determined |
| S16_extracted_40346 | Not-determined |
| S16_extracted_40353 | Not-determined |
| S16_extracted_40354 | Not-determined |
| S16_extracted_40389 | Not-determined |
| S16_extracted_40391 | Not-determined |
| S16_extracted_40402 | Not-determined |
| S16_extracted_40413 | Not-determined |
| S16_extracted_40417 | Not-determined |
| S16_extracted_40428 | Not-determined |
| S16_extracted_40437 | Not-determined |
| S16_extracted_40468 | Not-determined |
| S16_extracted_40471 | Not-determined |
| S16_extracted_40473 | Not-determined |
| S16_extracted_40479 | Not-determined |
| S16_extracted_40485 | Not-determined |
| S16_extracted_40499 | Not-determined |
| S16_extracted_40515 | Not-determined |
| S16_extracted_40519 | Not-determined |
| S16_extracted_40520 | Not-determined |
| S16_extracted_40568 | Not-determined |
| S16_extracted_40575 | Not-determined |
| S16_extracted_40587 | Not-determined |
| S16_extracted_40591 | Not-determined |
| S16_extracted_40602 | Not-determined |
| S16_extracted_40603 | Low-quality |
| S16_extracted_40637 | Not-determined |
| S16_extracted_40651 | Low-quality |
| S16_extracted_40656 | Not-determined |
| S16_extracted_40657 | Not-determined |
| S16_extracted_40660 | Low-quality |
| S16_extracted_40666 | Not-determined |
| S16_extracted_40667 | Not-determined |
| S16_extracted_40675 | Not-determined |
| S16_extracted_40699 | Not-determined |
| S16_extracted_40703 | Low-quality |
| S16_extracted_40707 | Low-quality |
| S16_extracted_40727 | Not-determined |
| S16_extracted_40743 | Not-determined |
| S16_extracted_40754 | Not-determined |
| S16_extracted_40756 | Not-determined |
| S16_extracted_40767 | Not-determined |
| S16_extracted_40770 | Not-determined |
| S16_extracted_40785 | Not-determined |
| S16_extracted_40787 | Not-determined |
| S16_extracted_40790 | Not-determined |
| S16_extracted_40794 | Not-determined |
| S16_extracted_40802 | Not-determined |
| S16_extracted_40821 | Not-determined |
| S16_extracted_40829 | Not-determined |
| S16_extracted_40841 | Low-quality |
| S16_extracted_40856 | Low-quality |
| S16_extracted_40875 | Not-determined |
| S16_extracted_40890 | Not-determined |
| S16_extracted_40896 | Not-determined |
| S16_extracted_40905 | Low-quality |
| S16_extracted_40908 | Not-determined |
| S16_extracted_40915 | Low-quality |
| S16_extracted_40917 | Not-determined |
| S16_extracted_40956 | Not-determined |
| S16_extracted_40963 | Not-determined |
| S16_extracted_40972 | Not-determined |
| S16_extracted_41007 | Not-determined |

| | |
|---|---|
| S16_extracted_41010 | Not-determined |
| S16_extracted_41013 | Low-quality |
| S16_extracted_41017 | Not-determined |
| S16_extracted_41027 | Not-determined |
| S16_extracted_41041 | Low-quality |
| S16_extracted_41081 | Not-determined |
| S16_extracted_41091 | Not-determined |
| S16_extracted_41106 | Not-determined |
| S16_extracted_41118 | Not-determined |
| S16_extracted_41127 | Not-determined |
| S16_extracted_41128 | Not-determined |
| S16_extracted_41139 | Not-determined |
| S16_extracted_41149 | Not-determined |
| S16_extracted_41153 | Not-determined |
| S16_extracted_41170 | Not-determined |
| S16_extracted_41171 | Low-quality |
| S16_extracted_41184 | Not-determined |
| S16_extracted_41185 | Not-determined |
| S16_extracted_41191 | Not-determined |
| S16_extracted_41211 | Not-determined |
| S16_extracted_41213 | Not-determined |
| S16_extracted_41243 | Low-quality |
| S16_extracted_41247 | Not-determined |
| S16_extracted_41250 | Not-determined |
| S16_extracted_41251 | Not-determined |
| S16_extracted_41259 | Not-determined |
| S16_extracted_41261 | Low-quality |
| S16_extracted_41266 | Not-determined |
| S16_extracted_41280 | Not-determined |
| S16_extracted_41294 | Not-determined |
| S16_extracted_41301 | Not-determined |
| S16_extracted_41302 | Not-determined |
| S16_extracted_41316 | Not-determined |
| S16_extracted_41321 | Not-determined |
| S16_extracted_41355 | Not-determined |
| S16_extracted_41363 | Low-quality |
| S16_extracted_41366 | Not-determined |
| S16_extracted_41368 | Low-quality |
| S16_extracted_41370 | Not-determined |
| S16_extracted_41388 | Not-determined |
| S16_extracted_41394 | Not-determined |
| S16_extracted_41399 | Not-determined |
| S16_extracted_41408 | Not-determined |
| S16_extracted_41415 | Not-determined |
| S16_extracted_41422 | Not-determined |
| S16_extracted_41434 | Not-determined |
| S16_extracted_41443 | Low-quality |
| S16_extracted_41471 | Not-determined |
| S16_extracted_41482 | Low-quality |
| S16_extracted_41484 | Low-quality |
| S16_extracted_41498 | Not-determined |
| S16_extracted_41540 | Not-determined |
| S16_extracted_41541 | Not-determined |
| S16_extracted_41589 | Low-quality |
| S16_extracted_41591 | Low-quality |
| S16_extracted_41594 | Not-determined |
| S16_extracted_41598 | Not-determined |
| S16_extracted_41627 | Not-determined |
| S16_extracted_41637 | Not-determined |
| S16_extracted_41648 | Not-determined |
| S16_extracted_41653 | Not-determined |
| S16_extracted_41659 | Not-determined |
| S16_extracted_41661 | Low-quality |
| S16_extracted_41663 | Low-quality |
| S16_extracted_41664 | Low-quality |
| S16_extracted_41668 | Not-determined |
| S16_extracted_41677 | Not-determined |
| S16_extracted_41684 | Not-determined |
| S16_extracted_41686 | Not-determined |
| S16_extracted_41694 | Not-determined |
| S16_extracted_41707 | Not-determined |
| S16_extracted_41712 | Not-determined |
| S16_extracted_41720 | Low-quality |
| S16_extracted_41737 | Low-quality |
| S16_extracted_41748 | Not-determined |
| S16_extracted_41762 | Not-determined |
| S16_extracted_41770 | Not-determined |
| S16_extracted_41783 | Not-determined |
| S16_extracted_41785 | Not-determined |
| S16_extracted_41795 | Not-determined |
| S16_extracted_41800 | Not-determined |
| S16_extracted_41802 | Not-determined |
| S16_extracted_41817 | Not-determined |
| S16_extracted_41819 | Not-determined |
| S16_extracted_41822 | Not-determined |
| S16_extracted_41827 | Low-quality |
| S16_extracted_41848 | Low-quality |
| S16_extracted_41849 | Low-quality |
| S16_extracted_41854 | Not-determined |
| S16_extracted_41862 | Not-determined |
| S16_extracted_41864 | Not-determined |
| S16_extracted_41874 | Not-determined |
| S16_extracted_41902 | Low-quality |
| S16_extracted_41939 | Not-determined |
| S16_extracted_41946 | Not-determined |
| S16_extracted_41957 | Not-determined |
| S16_extracted_41960 | Not-determined |
| S16_extracted_41966 | Not-determined |
| S16_extracted_41976 | Not-determined |
| S16_extracted_41988 | Not-determined |
| S16_extracted_41990 | Low-quality |
| S16_extracted_41996 | Low-quality |
| S16_extracted_42001 | Low-quality |
| S16_extracted_42008 | Not-determined |
| S16_extracted_42013 | Not-determined |
| S16_extracted_42015 | Not-determined |
| S16_extracted_42020 | Not-determined |
| S16_extracted_42038 | Not-determined |
| S16_extracted_42053 | Not-determined |
| S16_extracted_42055 | Not-determined |
| S16_extracted_42060 | Not-determined |
| S16_extracted_42069 | Low-quality |
| S16_extracted_42071 | Not-determined |
| S16_extracted_42087 | Not-determined |
| S16_extracted_42089 | Not-determined |
| S16_extracted_42100 | Low-quality |
| S16_extracted_42101 | Not-determined |
| S16_extracted_42114 | Not-determined |
| S16_extracted_42126 | Not-determined |
| S16_extracted_42134 | Not-determined |
| S16_extracted_42137 | Not-determined |
| S16_extracted_42174 | Low-quality |
| S16_extracted_42178 | Low-quality |
| S16_extracted_42183 | Not-determined |
| S16_extracted_42186 | Not-determined |
| S16_extracted_42209 | Not-determined |
| S16_extracted_42244 | Low-quality |
| S16_extracted_42264 | Low-quality |
| S16_extracted_42276 | Not-determined |
| S16_extracted_42280 | Not-determined |
| S16_extracted_42295 | Not-determined |
| S16_extracted_42296 | Not-determined |
| S16_extracted_42305 | Not-determined |
| S16_extracted_42307 | Not-determined |
| S16_extracted_42318 | Low-quality |
| S16_extracted_42322 | Not-determined |
| S16_extracted_42331 | Not-determined |
| S16_extracted_42371 | Not-determined |
| S16_extracted_42405 | Not-determined |
| S16_extracted_42421 | Not-determined |
| S16_extracted_42428 | Not-determined |
| S16_extracted_42431 | Not-determined |
| S16_extracted_42443 | Not-determined |
| S16_extracted_42449 | Not-determined |
| S16_extracted_42455 | Not-determined |
| S16_extracted_42466 | Not-determined |
| S16_extracted_42474 | Not-determined |
| S16_extracted_42481 | Not-determined |
| S16_extracted_42491 | Not-determined |
| S16_extracted_42509 | Not-determined |
| S16_extracted_42531 | Not-determined |
| S16_extracted_42534 | Not-determined |
| S16_extracted_42546 | Low-quality |
| S16_extracted_42557 | Not-determined |
| S16_extracted_42578 | Not-determined |
| S16_extracted_42590 | Not-determined |
| S16_extracted_42591 | Not-determined |
| S16_extracted_42592 | Not-determined |
| S16_extracted_42601 | Not-determined |
| S16_extracted_42603 | Low-quality |
| S16_extracted_42605 | Low-quality |
| S16_extracted_42640 | Not-determined |
| S16_extracted_42652 | Not-determined |
| S16_extracted_42659 | Not-determined |
| S16_extracted_42672 | Not-determined |
| S16_extracted_42675 | Low-quality |
| S16_extracted_42692 | Not-determined |
| S16_extracted_42696 | Not-determined |
| S16_extracted_42699 | Not-determined |
| S14_vs_k141_30566\|\|full | Medium-quality |
| S16_extracted_42708 | Not-determined |
| S16_extracted_42734 | Not-determined |
| S16_extracted_42758 | Not-determined |
| S16_extracted_42765 | Not-determined |
| S16_extracted_42780 | Not-determined |
| S16_extracted_42784 | Not-determined |
| S16_extracted_42820 | Not-determined |
| S16_extracted_42823 | Not-determined |
| S16_extracted_42825 | Not-determined |
| S16_extracted_42827 | Not-determined |
| S16_extracted_42836 | Not-determined |
| S16_extracted_42839 | Not-determined |
| S16_extracted_42844 | Not-determined |
| S16_extracted_42853 | Not-determined |
| S16_extracted_42855 | Not-determined |
| S16_extracted_42866 | Not-determined |
| S16_extracted_42873 | Not-determined |
| S16_extracted_42875 | Not-determined |
| S16_extracted_42890 | Not-determined |
| S16_extracted_42892 | Not-determined |
| S16_extracted_42896 | Not-determined |
| S16_extracted_42901 | Low-quality |
| S16_extracted_42930 | Not-determined |
| S16_extracted_42942 | Not-determined |
| S16_extracted_42949 | Not-determined |
| S16_extracted_42955 | Low-quality |
| S16_extracted_42969 | Not-determined |
| S16_extracted_42980 | Not-determined |
| S16_extracted_42981 | Not-determined |
| S16_extracted_42998 | Not-determined |
| S16_extracted_43003 | Low-quality |
| S16_extracted_43031 | Low-quality |
| S16_extracted_43039 | Not-determined |
| S16_extracted_43044 | Not-determined |
| S16_extracted_43067 | Low-quality |
| S16_extracted_43072 | Low-quality |
| S16_extracted_43095 | Not-determined |
| S16_extracted_43102 | Not-determined |
| S16_extracted_43119 | Not-determined |
| S16_extracted_43126 | Not-determined |
| S16_extracted_43136 | Not-determined |
| S16_extracted_43142 | Low-quality |
| S16_extracted_43144 | Low-quality |
| S16_extracted_43145 | Not-determined |
| S16_extracted_43146 | Not-determined |
| S16_extracted_43147 | Not-determined |
| S16_extracted_43151 | Not-determined |
| S16_extracted_43153 | Not-determined |
| S16_extracted_43160 | Not-determined |
| S16_extracted_43200 | Not-determined |
| S16_extracted_43201 | Not-determined |
| S16_extracted_43203 | Not-determined |
| S16_extracted_43208 | Not-determined |
| S16_extracted_43211 | Low-quality |
| S16_extracted_43219 | Low-quality |
| S16_extracted_43220 | Not-determined |
| S16_extracted_43226 | Not-determined |
| S16_extracted_43238 | Not-determined |
| S16_extracted_43239 | Low-quality |
| S16_extracted_43242 | Not-determined |
| S16_extracted_43246 | Low-quality |
| S16_extracted_43254 | Not-determined |
| S16_extracted_43270 | Not-determined |
| S16_extracted_43288 | Not-determined |
| S16_extracted_43292 | Not-determined |
| S16_extracted_43293 | Not-determined |
| S16_extracted_43297 | Not-determined |
| S16_extracted_43343 | Not-determined |
| S16_extracted_43349 | Not-determined |
| S16_extracted_43350 | Low-quality |
| S16_extracted_43362 | Not-determined |
| S16_extracted_43363 | Not-determined |
| S16_extracted_43383 | Not-determined |
| S16_extracted_43387 | Not-determined |
| S16_extracted_43409 | Not-determined |
| S16_extracted_43411 | Not-determined |
| S16_extracted_43420 | Not-determined |
| S16_extracted_43441 | Not-determined |
| S16_extracted_43443 | Not-determined |
| S16_extracted_43446 | Low-quality |
| S16_extracted_43447 | Not-determined |
| S16_extracted_43466 | Not-determined |
| S16_extracted_43487 | Not-determined |
| S16_extracted_43489 | Not-determined |
| S16_extracted_43496 | Not-determined |
| S16_extracted_43519 | Not-determined |
| S16_extracted_43540 | Not-determined |
| S16_extracted_43546 | Not-determined |
| S16_extracted_43547 | Not-determined |
| S16_extracted_43569 | Not-determined |
| S16_extracted_43576 | Not-determined |
| S16_extracted_43581 | Low-quality |
| S16_extracted_43582 | Not-determined |
| S16_extracted_43610 | Low-quality |
| S16_extracted_43612 | Low-quality |
| S16_extracted_43624 | Low-quality |
| S16_extracted_43642 | Not-determined |
| S16_extracted_43671 | Not-determined |
| S16_extracted_43688 | Not-determined |
| S16_extracted_43723 | Low-quality |
| S16_extracted_43726 | Low-quality |
| S16_extracted_43763 | Not-determined |
| S16_extracted_43770 | Not-determined |
| S16_extracted_43773 | Not-determined |
| S16_extracted_43779 | Not-determined |
| S16_extracted_43780 | Not-determined |
| S16_extracted_43810 | Low-quality |
| S16_extracted_43823 | Not-determined |
| S16_extracted_43827 | Low-quality |
| S16_extracted_43831 | Not-determined |
| S16_extracted_43839 | Low-quality |
| S16_extracted_43860 | Low-quality |
| S16_extracted_43874 | Not-determined |
| S16_extracted_43886 | Not-determined |
| S16_extracted_43906 | Not-determined |
| S16_extracted_43911 | Not-determined |
| S16_extracted_43924 | Low-quality |
| S16_extracted_43928 | Not-determined |
| S16_extracted_43933 | Not-determined |
| S16_extracted_43939 | Not-determined |
| S16_extracted_43960 | Not-determined |
| S16_extracted_43961 | Not-determined |
| S16_extracted_43974 | Not-determined |
| S16_extracted_43976 | Not-determined |
| S16_extracted_43980 | Not-determined |
| S16_extracted_43986 | Low-quality |
| S16_extracted_43997 | Not-determined |
| S16_extracted_44031 | Not-determined |
| S16_extracted_44034 | Not-determined |
| S16_extracted_44061 | Not-determined |
| S16_extracted_44062 | Not-determined |
| S16_extracted_44071 | Not-determined |
| S16_extracted_44073 | Not-determined |
| S16_extracted_44103 | Low-quality |
| S16_extracted_44123 | Not-determined |
| S16_extracted_44127 | Not-determined |
| S16_extracted_44135 | Not-determined |
| S16_extracted_44140 | Not-determined |
| S16_extracted_44163 | Not-determined |
| S16_extracted_44168 | Low-quality |
| S16_extracted_44180 | Not-determined |
| S16_extracted_44199 | Not-determined |
| S16_extracted_44201 | Not-determined |
| S16_extracted_44203 | Not-determined |
| S16_extracted_44204 | Not-determined |
| S16_extracted_44222 | Not-determined |
| S16_extracted_44232 | Not-determined |
| S16_extracted_44259 | Not-determined |
| S16_extracted_44264 | Low-quality |
| S16_extracted_44272 | Not-determined |
| S16_extracted_44282 | Not-determined |
| S16_extracted_44301 | Not-determined |
| S16_extracted_44302 | Not-determined |
| S16_extracted_44317 | Not-determined |
| S16_extracted_44321 | Not-determined |
| S16_extracted_44329 | Not-determined |
| S16_extracted_44335 | Not-determined |
| S16_extracted_44352 | Not-determined |
| S16_extracted_44357 | Not-determined |
| S16_extracted_44359 | Not-determined |
| S16_extracted_44370 | Not-determined |
| S16_extracted_44372 | Not-determined |
| S16_extracted_44373 | Not-determined |
| S16_extracted_44378 | Low-quality |
| S16_extracted_44383 | Not-determined |
| S16_extracted_44393 | Not-determined |
| S16_extracted_44404 | Not-determined |
| S16_extracted_44407 | Not-determined |
| S16_extracted_44418 | Not-determined |
| S16_extracted_44459 | Not-determined |
| S16_extracted_44474 | Not-determined |
| S16_extracted_44485 | Not-determined |
| S16_extracted_44487 | Not-determined |
| S16_extracted_44492 | Not-determined |
| S16_extracted_44500 | Low-quality |
| S16_extracted_44501 | Not-determined |
| S16_extracted_44503 | Not-determined |
| S16_extracted_44507 | Not-determined |
| S16_extracted_44510 | Not-determined |
| S16_extracted_44511 | Not-determined |
| S16_extracted_44538 | Not-determined |
| S16_extracted_44547 | Low-quality |
| S16_extracted_44561 | Not-determined |
| S16_extracted_44569 | Not-determined |
| S16_extracted_44572 | Not-determined |
| S16_extracted_44591 | Not-determined |
| S16_extracted_44603 | Not-determined |
| S16_extracted_44615 | Low-quality |
| S16_extracted_44636 | Not-determined |
| S16_extracted_44656 | Not-determined |
| S16_extracted_44659 | Not-determined |
| S16_extracted_44665 | Not-determined |
| S16_extracted_44689 | Low-quality |
| S16_extracted_44695 | Not-determined |
| S16_extracted_44697 | Not-determined |
| S16_extracted_44704 | Not-determined |
| S16_extracted_44708 | Not-determined |
| S16_extracted_44734 | Not-determined |
| S16_extracted_44770 | Not-determined |
| S16_extracted_44771 | Not-determined |
| S16_extracted_44775 | Not-determined |
| S16_extracted_44788 | Not-determined |
| S16_extracted_44790 | Not-determined |
| S16_extracted_44794 | Not-determined |
| S16_extracted_44796 | Not-determined |
| S16_extracted_44815 | Not-determined |
| S16_extracted_44837 | Not-determined |
| S16_extracted_44849 | Not-determined |
| S16_extracted_44864 | Not-determined |
| S16_extracted_44869 | Not-determined |
| S16_extracted_44872 | Not-determined |
| S16_extracted_44875 | Not-determined |
| S16_extracted_44876 | Not-determined |
| S16_extracted_44879 | Not-determined |
| S16_extracted_44884 | Low-quality |
| S16_extracted_44898 | Low-quality |
| S16_extracted_44916 | Not-determined |
| S16_extracted_44925 | Not-determined |
| S16_extracted_44937 | Not-determined |
| S16_extracted_44944 | Not-determined |
| S16_extracted_44957 | Low-quality |
| S16_extracted_44982 | Not-determined |
| S16_extracted_44988 | Not-determined |
| S16_extracted_44992 | Not-determined |
| S16_extracted_45000 | Low-quality |
| S16_extracted_45015 | Not-determined |
| S16_extracted_45019 | Not-determined |
| S16_extracted_45034 | Not-determined |
| S16_extracted_45060 | Not-determined |
| S16_extracted_45061 | Not-determined |
| S16_extracted_45070 | Not-determined |
| S16_extracted_45088 | Not-determined |
| S16_extracted_45096 | Low-quality |
| S16_extracted_45105 | Not-determined |
| S16_extracted_45115 | Not-determined |
| S16_extracted_45118 | Not-determined |
| S16_extracted_45122 | Not-determined |
| S16_extracted_45127 | Not-determined |
| S16_extracted_45131 | Not-determined |
| S16_extracted_45132 | Not-determined |
| S16_extracted_45142 | Not-determined |
| S16_extracted_45155 | Not-determined |
| S16_extracted_45172 | Low-quality |
| S16_extracted_45174 | Low-quality |
| S16_extracted_45184 | Not-determined |
| S16_extracted_45185 | Not-determined |
| S16_extracted_45186 | Low-quality |
| S16_extracted_45191 | Not-determined |
| S16_extracted_45203 | Not-determined |
| S16_extracted_45214 | Not-determined |
| S16_extracted_45220 | Not-determined |
| S16_extracted_45267 | Low-quality |
| S16_extracted_45271 | Not-determined |
| S16_extracted_45283 | Not-determined |
| S16_extracted_45305 | Not-determined |
| S16_extracted_45342 | Not-determined |
| S16_extracted_45346 | Low-quality |
| S16_extracted_45371 | Not-determined |
| S16_extracted_45379 | Low-quality |
| S16_extracted_45383 | Not-determined |
| S16_extracted_45384 | Not-determined |
| S16_extracted_45390 | Not-determined |
| S16_extracted_45398 | Not-determined |
| S16_extracted_45411 | Not-determined |
| S16_extracted_45412 | Not-determined |
| S16_extracted_45418 | Low-quality |
| S16_extracted_45424 | Not-determined |
| S16_extracted_45433 | Low-quality |
| S16_extracted_45436 | Not-determined |
| S16_extracted_45450 | Not-determined |
| S16_extracted_45454 | Low-quality |
| S16_extracted_45523 | Not-determined |
| S16_extracted_45537 | Low-quality |
| S16_extracted_45539 | Low-quality |
| S16_extracted_45546 | Not-determined |
| S16_extracted_45551 | Not-determined |
| S16_extracted_45556 | Not-determined |
| S16_extracted_45570 | Not-determined |
| S16_extracted_45597 | Not-determined |
| S16_extracted_45599 | Not-determined |
| S16_extracted_45616 | Not-determined |
| S16_extracted_45620 | Not-determined |
| S16_extracted_45621 | Not-determined |
| S16_extracted_45650 | Not-determined |
| S16_extracted_45652 | Low-quality |
| S16_extracted_45671 | Not-determined |
| S16_extracted_45676 | Not-determined |
| S16_extracted_45696 | Not-determined |
| S16_extracted_45703 | Not-determined |
| S16_extracted_45704 | Low-quality |
| S16_extracted_45711 | Low-quality |
| S16_extracted_45719 | Not-determined |
| S16_extracted_45723 | Not-determined |
| S16_extracted_45736 | Not-determined |
| S16_extracted_45751 | Low-quality |
| S16_extracted_45763 | Not-determined |
| S16_extracted_45766 | Low-quality |
| S16_extracted_45776 | Not-determined |
| S16_extracted_45778 | Low-quality |
| S16_extracted_45786 | Not-determined |
| S16_extracted_45788 | Low-quality |
| S16_extracted_45789 | Not-determined |
| S16_extracted_45795 | Not-determined |
| S16_extracted_45797 | Not-determined |
| S16_extracted_45799 | Not-determined |
| S16_extracted_45801 | Not-determined |
| S16_extracted_45808 | Not-determined |
| S16_extracted_45835 | Low-quality |
| S16_extracted_45841 | Low-quality |
| S16_extracted_45843 | Not-determined |
| S16_extracted_45850 | Low-quality |
| S16_extracted_45852 | Low-quality |
| S16_extracted_45859 | Not-determined |
| S16_extracted_45874 | Not-determined |
| S16_extracted_45877 | Not-determined |
| S16_extracted_45884 | Not-determined |
| S16_extracted_45893 | Not-determined |
| S16_extracted_45896 | Low-quality |
| S16_extracted_45898 | Not-determined |
| S16_extracted_45899 | Not-determined |
| S16_extracted_45908 | Not-determined |
| S16_extracted_45930 | Not-determined |
| S16_extracted_45937 | Not-determined |
| S16_extracted_45955 | Not-determined |
| S16_extracted_45959 | Not-determined |
| S16_extracted_45965 | Low-quality |
| S16_extracted_45966 | Not-determined |
| S16_extracted_45974 | Not-determined |
| S16_extracted_45978 | Low-quality |
| S16_extracted_45979 | Not-determined |
| S16_extracted_45982 | Not-determined |
| S16_extracted_46008 | Not-determined |
| S16_extracted_46016 | Low-quality |
| S16_extracted_46020 | Not-determined |
| S16_extracted_46025 | Not-determined |
| S16_extracted_46030 | Not-determined |
| S16_extracted_46040 | Not-determined |
| S16_extracted_46065 | Not-determined |
| S16_extracted_46067 | Not-determined |
| S16_extracted_46070 | Low-quality |
| S16_extracted_46078 | Not-determined |
| S16_extracted_46080 | Not-determined |
| S16_extracted_46083 | Not-determined |
| S16_extracted_46088 | Not-determined |
| S16_extracted_46125 | Not-determined |
| S16_extracted_46128 | Low-quality |
| S16_extracted_46158 | Not-determined |
| S16_extracted_46172 | Not-determined |
| S16_extracted_46176 | Not-determined |
| S16_extracted_46179 | Not-determined |
| S16_extracted_46183 | Not-determined |
| S16_extracted_46192 | Not-determined |
| S16_extracted_46193 | Not-determined |
| S16_extracted_46198 | Not-determined |
| S16_extracted_46206 | Not-determined |
| S16_extracted_46213 | Not-determined |
| S16_extracted_46217 | Not-determined |
| S16_extracted_46228 | Not-determined |
| S16_extracted_46230 | Not-determined |
| S16_extracted_46256 | Not-determined |
| S16_extracted_46266 | Not-determined |
| S16_extracted_46282 | Not-determined |
| S16_extracted_46285 | Not-determined |
| S16_extracted_46297 | Low-quality |
| S16_extracted_46299 | Low-quality |
| S16_extracted_46307 | Not-determined |
| S16_extracted_46310 | Not-determined |
| S16_extracted_46314 | Not-determined |
| S16_extracted_46349 | Not-determined |
| S16_extracted_46360 | Not-determined |
| S16_extracted_46370 | Low-quality |
| S16_extracted_46378 | Not-determined |
| S16_extracted_46386 | Not-determined |
| S16_extracted_46423 | Not-determined |
| S16_extracted_46425 | Not-determined |
| S16_extracted_46429 | Not-determined |
| S16_extracted_46438 | Not-determined |
| S16_extracted_46456 | Not-determined |
| S16_extracted_46458 | Not-determined |
| S16_extracted_46475 | Not-determined |
| S16_extracted_46491 | Low-quality |
| S16_extracted_46509 | Not-determined |
| S16_extracted_46523 | Not-determined |
| S16_extracted_46529 | Not-determined |
| S16_extracted_46533 | Low-quality |
| S16_extracted_46540 | Not-determined |
| S16_extracted_46541 | Not-determined |
| S16_extracted_46554 | Not-determined |
| S16_extracted_46585 | Low-quality |
| S16_extracted_46638 | Not-determined |
| S16_extracted_46643 | Low-quality |
| S16_extracted_46650 | Not-determined |
| S16_extracted_46653 | Not-determined |
| S16_extracted_46659 | Not-determined |
| S16_extracted_46665 | Not-determined |
| S16_extracted_46668 | Not-determined |
| S16_extracted_46680 | Not-determined |
| S16_extracted_46683 | Low-quality |
| S16_extracted_46691 | Not-determined |
| S16_extracted_46702 | Not-determined |
| S16_extracted_46713 | Not-determined |
| S16_extracted_46737 | Not-determined |
| S16_extracted_46757 | Low-quality |
| S16_extracted_46761 | Not-determined |
| S16_extracted_46779 | Not-determined |
| S16_extracted_46780 | Not-determined |
| S16_extracted_46800 | Not-determined |
| S16_extracted_46839 | Not-determined |
| S16_extracted_46847 | Low-quality |
| S16_extracted_46857 | Not-determined |
| S16_extracted_46861 | Not-determined |
| S16_extracted_46868 | Not-determined |
| S16_extracted_46888 | Not-determined |
| S16_extracted_46894 | Low-quality |
| S16_extracted_46895 | Not-determined |
| S16_extracted_46900 | Not-determined |
| S16_extracted_46917 | Low-quality |
| S16_extracted_46921 | Not-determined |
| S16_extracted_46925 | Not-determined |

| | |
|---|---|
| S16_extracted_46940 | Not-determined |
| S16_extracted_46942 | Not-determined |
| S16_extracted_46953 | Not-determined |
| S16_extracted_46961 | Low-quality |
| S16_extracted_46985 | Not-determined |
| S16_extracted_46987 | Not-determined |
| S16_extracted_47005 | Not-determined |
| S16_extracted_47010 | Not-determined |
| S16_extracted_47013 | Not-determined |
| S16_extracted_47034 | Not-determined |
| S16_extracted_47037 | Not-determined |
| S16_extracted_47038 | Low-quality |
| S16_extracted_47041 | Not-determined |
| S16_extracted_47045 | Not-determined |
| S16_extracted_47048 | Not-determined |
| S16_extracted_47058 | Not-determined |
| S16_extracted_47066 | Low-quality |
| S16_extracted_47069 | Not-determined |
| S16_extracted_47081 | Not-determined |
| S16_extracted_47082 | Not-determined |
| S16_extracted_47089 | Not-determined |
| S16_extracted_47093 | Not-determined |
| S16_extracted_47097 | Not-determined |
| S16_extracted_47104 | Not-determined |
| S16_extracted_47114 | Low-quality |
| S16_extracted_47135 | Not-determined |
| S16_extracted_47159 | Low-quality |
| S16_extracted_47179 | Not-determined |
| S16_extracted_47181 | Not-determined |
| S16_extracted_47192 | Low-quality |
| S16_extracted_47210 | Not-determined |
| S16_extracted_47229 | Not-determined |
| S16_extracted_47270 | Not-determined |
| S16_extracted_47271 | Not-determined |
| S16_extracted_47272 | Not-determined |
| S16_extracted_47277 | Not-determined |
| S16_extracted_47300 | Not-determined |
| S16_extracted_47314 | Not-determined |
| S16_extracted_47322 | Low-quality |
| S16_extracted_47323 | Not-determined |
| S16_extracted_47338 | Not-determined |
| S16_extracted_47346 | Low-quality |
| S16_extracted_47352 | Not-determined |
| S16_extracted_47357 | Not-determined |
| S16_extracted_47371 | Not-determined |
| S16_extracted_47381 | Low-quality |
| S16_extracted_47400 | Not-determined |
| S16_extracted_47409 | Low-quality |
| S16_extracted_47412 | Not-determined |
| S16_extracted_47419 | Low-quality |
| S16_extracted_47431 | Not-determined |
| S16_extracted_47446 | Not-determined |
| S16_extracted_47450 | Not-determined |
| S16_extracted_47455 | Not-determined |
| S16_extracted_47462 | Not-determined |
| S16_extracted_47484 | Not-determined |
| S16_extracted_47504 | Not-determined |
| S16_extracted_47516 | Not-determined |
| S16_extracted_47532 | Not-determined |
| S16_extracted_47548 | Not-determined |
| S7_extracted_124517 | Medium-quality |
| S16_extracted_47576 | Not-determined |
| S16_extracted_47604 | Not-determined |
| S16_extracted_47606 | Not-determined |
| S16_extracted_47607 | Not-determined |
| S16_extracted_47620 | Not-determined |
| S16_extracted_47623 | Not-determined |
| S16_extracted_47629 | Not-determined |
| S16_extracted_47635 | Not-determined |
| S16_extracted_47639 | Not-determined |
| S16_extracted_47650 | Not-determined |
| S16_extracted_47656 | Not-determined |
| S16_extracted_47674 | Low-quality |
| S16_extracted_47684 | Low-quality |
| S16_extracted_47689 | Not-determined |
| S16_extracted_47704 | Not-determined |
| S16_extracted_47746 | Not-determined |
| S16_extracted_47757 | Not-determined |
| S16_extracted_47763 | Low-quality |
| S16_extracted_47767 | Not-determined |
| S16_extracted_47770 | Low-quality |
| S16_extracted_47776 | Not-determined |
| S16_extracted_47804 | Not-determined |
| S16_extracted_47809 | Low-quality |
| S16_extracted_47819 | Not-determined |
| S16_extracted_47828 | Not-determined |
| S16_extracted_47851 | Not-determined |
| S16_extracted_47866 | Not-determined |
| S16_extracted_47880 | Not-determined |
| S16_extracted_47894 | Not-determined |
| S16_extracted_47899 | Not-determined |
| S16_extracted_47907 | Not-determined |
| S16_extracted_47949 | Not-determined |
| S16_extracted_47960 | Low-quality |
| S16_extracted_47967 | Not-determined |
| S16_extracted_47976 | Not-determined |
| S16_extracted_47980 | Not-determined |
| S16_extracted_47983 | Not-determined |
| S16_extracted_47990 | Low-quality |
| S16_extracted_48007 | Not-determined |
| S16_extracted_48016 | Not-determined |
| S16_extracted_48038 | Not-determined |
| S16_extracted_48073 | Not-determined |
| S16_extracted_48093 | Low-quality |
| S16_extracted_48096 | Not-determined |
| S16_extracted_48102 | Not-determined |
| S16_extracted_48118 | Not-determined |
| S7_vs_124517\|\|full | Medium-quality |
| S16_extracted_48141 | Not-determined |
| S16_extracted_48152 | Not-determined |
| S16_extracted_48182 | Low-quality |
| S16_extracted_48209 | Not-determined |
| S16_extracted_48214 | Low-quality |
| S16_extracted_48218 | Not-determined |
| S16_extracted_48233 | Not-determined |
| S16_extracted_48240 | Low-quality |
| S16_extracted_48247 | Not-determined |
| S16_extracted_48269 | Not-determined |
| S16_extracted_48271 | Not-determined |
| S16_extracted_48281 | Not-determined |
| S16_extracted_48282 | Low-quality |
| S16_extracted_48299 | Not-determined |
| S16_extracted_48309 | Not-determined |
| S16_extracted_48319 | Not-determined |
| S16_extracted_48328 | Not-determined |
| S16_extracted_48330 | Low-quality |
| S16_extracted_48336 | Not-determined |
| S16_extracted_48340 | Not-determined |
| S16_extracted_48343 | Not-determined |
| S16_extracted_48344 | Not-determined |
| S16_extracted_48362 | Not-determined |
| S16_extracted_48372 | Low-quality |
| S16_extracted_48379 | Not-determined |
| S16_extracted_48382 | Not-determined |
| S16_extracted_48384 | Low-quality |
| S16_extracted_48399 | Not-determined |
| S16_extracted_48401 | Not-determined |
| S16_extracted_48415 | Low-quality |
| S16_extracted_48429 | Not-determined |
| S16_extracted_48452 | Low-quality |
| S16_extracted_48453 | Not-determined |
| S16_extracted_48462 | Not-determined |
| S16_extracted_48471 | Not-determined |
| S16_extracted_48487 | Low-quality |
| S16_extracted_48508 | Not-determined |
| S16_extracted_48511 | Not-determined |
| S16_extracted_48518 | Not-determined |
| S16_extracted_48521 | Not-determined |
| S16_extracted_48525 | Low-quality |
| S16_extracted_48536 | Not-determined |
| S16_extracted_48545 | Low-quality |
| S16_extracted_48574 | Not-determined |
| S16_extracted_48607 | Low-quality |
| S16_extracted_48629 | Low-quality |
| S16_extracted_48641 | Low-quality |
| S16_extracted_48647 | Low-quality |
| S16_extracted_48650 | Not-determined |
| S16_extracted_48653 | Not-determined |
| S16_extracted_48669 | Not-determined |
| S16_extracted_48679 | Not-determined |
| S16_extracted_48685 | Not-determined |
| S16_extracted_48687 | Not-determined |
| S16_extracted_48693 | Not-determined |
| S16_extracted_48696 | Not-determined |
| S16_extracted_48697 | Not-determined |
| S16_extracted_48718 | Not-determined |
| S16_extracted_48728 | Not-determined |
| S16_extracted_48731 | Not-determined |
| S16_extracted_48786 | Not-determined |
| S16_extracted_48792 | Not-determined |
| S16_extracted_48809 | Not-determined |
| S16_extracted_48817 | Not-determined |
| S16_extracted_48872 | Not-determined |
| S16_extracted_48875 | Low-quality |
| S16_extracted_48881 | Not-determined |
| S16_extracted_48891 | Not-determined |
| S16_extracted_48893 | Not-determined |
| S16_extracted_48905 | Not-determined |
| S16_extracted_48909 | Not-determined |
| S16_extracted_48923 | Not-determined |
| S16_extracted_48934 | Not-determined |
| S16_extracted_48955 | Not-determined |
| S16_extracted_48961 | Not-determined |
| S16_extracted_48977 | Not-determined |
| S16_extracted_48979 | Not-determined |
| S16_extracted_48980 | Not-determined |
| S16_extracted_48998 | Low-quality |
| S16_extracted_48999 | Not-determined |
| S16_extracted_49006 | Not-determined |
| S16_extracted_49012 | Not-determined |
| S16_extracted_49016 | Not-determined |
| S16_extracted_49072 | Not-determined |
| S16_extracted_49074 | Not-determined |
| S16_extracted_49086 | Not-determined |
| S16_extracted_49090 | Not-determined |
| S16_extracted_49096 | Low-quality |
| S16_extracted_49108 | Not-determined |
| S16_extracted_49115 | Low-quality |
| S16_extracted_49138 | Not-determined |
| S16_extracted_49146 | Low-quality |
| S16_extracted_49153 | Not-determined |
| S16_extracted_49156 | Not-determined |
| S16_extracted_49158 | Not-determined |
| S16_extracted_49167 | Not-determined |
| S16_extracted_49181 | Low-quality |
| S16_extracted_49190 | Not-determined |
| S16_extracted_49196 | Not-determined |
| S16_extracted_49211 | Not-determined |
| S16_extracted_49214 | Low-quality |
| S16_extracted_49217 | Not-determined |
| S16_extracted_49221 | Low-quality |
| S16_extracted_49227 | Not-determined |
| S16_extracted_49228 | Low-quality |
| S16_extracted_49230 | Not-determined |
| S16_extracted_49264 | Not-determined |
| S16_extracted_49284 | Not-determined |
| S16_extracted_49288 | Not-determined |
| S16_extracted_49290 | Not-determined |
| S16_extracted_49293 | Not-determined |
| S16_extracted_49308 | Not-determined |
| S16_extracted_49323 | Low-quality |
| S16_extracted_49339 | Not-determined |
| S16_extracted_49347 | Not-determined |
| S16_extracted_49360 | Not-determined |
| S16_extracted_49371 | Not-determined |
| S16_extracted_49376 | Not-determined |
| S16_extracted_49384 | Not-determined |
| S16_extracted_49392 | Low-quality |
| S16_extracted_49396 | Not-determined |
| S16_extracted_49406 | Not-determined |
| S16_extracted_49409 | Not-determined |
| S16_extracted_49410 | Not-determined |
| S16_extracted_49413 | Not-determined |
| S16_extracted_49421 | Not-determined |
| S16_extracted_49422 | Not-determined |
| S16_extracted_49436 | Low-quality |
| S16_extracted_49441 | Not-determined |
| S16_extracted_49446 | Not-determined |
| S16_extracted_49459 | Not-determined |
| S16_extracted_49461 | Not-determined |
| S16_extracted_49476 | Not-determined |
| S16_extracted_49478 | Not-determined |
| S16_extracted_49492 | Not-determined |
| S16_extracted_49495 | Low-quality |
| S16_extracted_49510 | Not-determined |
| S16_extracted_49513 | Not-determined |
| S16_extracted_49516 | Not-determined |
| S16_extracted_49528 | Not-determined |
| S16_extracted_49534 | Not-determined |
| S16_extracted_49537 | Not-determined |
| S16_extracted_49539 | Low-quality |
| S16_extracted_49541 | Not-determined |
| S16_extracted_49543 | Low-quality |
| S16_extracted_49557 | Not-determined |
| S16_extracted_49571 | Not-determined |
| S16_extracted_49573 | Low-quality |
| S16_extracted_49578 | Not-determined |
| S16_extracted_49597 | Not-determined |
| S16_extracted_49599 | Low-quality |
| S16_extracted_49601 | Not-determined |
| S16_extracted_49608 | Not-determined |
| S16_extracted_49615 | Not-determined |
| S16_extracted_49618 | Not-determined |
| S12_vs_k141_188828\|\|full | Medium-quality |
| S16_extracted_49629 | Not-determined |
| S16_extracted_49644 | Low-quality |
| S16_extracted_49651 | Not-determined |
| S16_extracted_49652 | Not-determined |
| S16_extracted_49657 | Not-determined |
| S16_extracted_49658 | Not-determined |
| S16_extracted_49670 | Not-determined |
| S16_extracted_49671 | Not-determined |
| S16_extracted_49676 | Not-determined |
| S16_extracted_49692 | Low-quality |
| S16_extracted_49700 | Low-quality |
| S16_extracted_49720 | Not-determined |
| S16_extracted_49723 | Not-determined |
| S16_extracted_49742 | Not-determined |
| S16_extracted_49744 | Not-determined |
| S16_extracted_49747 | Not-determined |
| S16_extracted_49771 | Not-determined |
| S16_extracted_49774 | Low-quality |
| S16_extracted_49781 | Not-determined |
| S16_extracted_49783 | Not-determined |
| S16_extracted_49789 | Not-determined |
| S16_extracted_49815 | Not-determined |
| S16_extracted_49832 | Not-determined |
| S16_extracted_49839 | Not-determined |
| S16_extracted_49856 | Not-determined |
| S16_extracted_49877 | Not-determined |
| S16_extracted_49879 | Not-determined |
| S16_extracted_49888 | Not-determined |
| S16_extracted_49889 | Not-determined |
| S16_extracted_49893 | Low-quality |
| S16_extracted_49894 | Not-determined |
| S16_extracted_49898 | Not-determined |
| S16_extracted_49925 | Not-determined |
| S16_extracted_49929 | Low-quality |
| S16_extracted_49939 | Not-determined |
| S16_extracted_49952 | Not-determined |
| S16_extracted_49966 | Not-determined |
| S16_extracted_49972 | Low-quality |
| S16_extracted_49978 | Not-determined |
| S16_extracted_49983 | Not-determined |
| S16_extracted_50006 | Low-quality |
| S16_extracted_50011 | Not-determined |
| S16_extracted_50032 | Not-determined |
| S16_extracted_50034 | Not-determined |
| S16_extracted_50061 | Not-determined |
| S16_extracted_50075 | Not-determined |
| S16_extracted_50080 | Not-determined |
| S16_extracted_50092 | Not-determined |
| S16_extracted_50114 | Not-determined |
| S16_extracted_50125 | Not-determined |
| S16_extracted_50133 | Not-determined |
| S16_extracted_50140 | Low-quality |
| S16_extracted_50162 | Not-determined |
| S16_extracted_50166 | Not-determined |
| S16_extracted_50176 | Low-quality |
| S16_extracted_50194 | Not-determined |
| S16_extracted_50205 | Not-determined |
| S16_extracted_50221 | Not-determined |
| S16_extracted_50223 | Not-determined |
| S16_extracted_50244 | Not-determined |
| S16_extracted_50249 | Not-determined |
| S16_extracted_50277 | Not-determined |
| S16_extracted_50280 | Not-determined |
| S16_extracted_50289 | Not-determined |
| S16_extracted_50290 | Low-quality |
| S16_extracted_50295 | Not-determined |
| S16_extracted_50299 | Not-determined |
| S16_extracted_50305 | Low-quality |
| S16_extracted_50309 | Not-determined |
| S16_extracted_50328 | Not-determined |
| S16_extracted_50329 | Not-determined |
| S16_extracted_50350 | Not-determined |
| S16_extracted_50354 | Not-determined |
| S16_extracted_50360 | Not-determined |
| S16_extracted_50364 | Low-quality |
| S16_extracted_50373 | Not-determined |
| S16_extracted_50405 | Not-determined |
| S16_extracted_50453 | Not-determined |
| S16_extracted_50470 | Not-determined |
| S16_extracted_50488 | Not-determined |
| S16_extracted_50491 | Not-determined |
| S16_extracted_50497 | Not-determined |
| S16_extracted_50503 | Not-determined |
| S16_extracted_50506 | Not-determined |
| S16_extracted_50512 | Not-determined |
| S16_extracted_50513 | Low-quality |
| S16_extracted_50543 | Not-determined |
| S16_extracted_50555 | Low-quality |
| S16_extracted_50570 | Not-determined |
| S16_extracted_50577 | Not-determined |
| S16_extracted_50597 | Not-determined |
| S16_extracted_50607 | Not-determined |
| S16_extracted_50611 | Not-determined |
| S16_extracted_50620 | Not-determined |
| S16_extracted_50624 | Not-determined |
| S16_extracted_50632 | Low-quality |
| S16_extracted_50635 | Not-determined |
| S16_extracted_50641 | Not-determined |
| S16_extracted_50646 | Not-determined |
| S16_extracted_50653 | Not-determined |
| S16_extracted_50668 | Not-determined |
| S16_extracted_50669 | Not-determined |
| S16_extracted_50671 | Not-determined |
| S16_extracted_50677 | Not-determined |
| S16_extracted_50681 | Not-determined |
| S16_extracted_50689 | Not-determined |
| S16_extracted_50699 | Not-determined |
| S16_extracted_50708 | Low-quality |
| S16_extracted_50714 | Low-quality |
| S16_extracted_50721 | Not-determined |
| S16_extracted_50730 | Low-quality |
| S16_extracted_50742 | Low-quality |
| S16_extracted_50743 | Low-quality |
| S16_extracted_50759 | Not-determined |
| S16_extracted_50767 | Not-determined |
| S16_extracted_50768 | Not-determined |
| S16_extracted_50777 | Low-quality |
| S16_extracted_50778 | Not-determined |
| S16_extracted_50780 | Not-determined |
| S16_extracted_50781 | Not-determined |
| S16_extracted_50791 | Not-determined |
| S16_extracted_50801 | Not-determined |
| S16_extracted_50813 | Low-quality |
| S16_extracted_50815 | Not-determined |
| S16_extracted_50817 | Not-determined |
| S16_extracted_50822 | Not-determined |
| S16_extracted_50846 | Not-determined |
| S16_extracted_50853 | Not-determined |
| S16_extracted_50857 | Not-determined |
| S16_extracted_50864 | Not-determined |
| S16_extracted_50866 | Not-determined |
| S16_extracted_50870 | Not-determined |
| S16_extracted_50897 | Not-determined |
| S16_extracted_50911 | Not-determined |
| S16_extracted_50917 | Low-quality |
| S16_extracted_50924 | Not-determined |
| S16_extracted_50929 | Not-determined |
| S16_extracted_50943 | Not-determined |
| S16_extracted_50945 | Not-determined |
| S16_extracted_50958 | Not-determined |
| S16_extracted_50965 | Not-determined |
| S16_extracted_50969 | Low-quality |
| S16_extracted_50976 | Not-determined |
| S16_extracted_50977 | Not-determined |
| S16_extracted_50987 | Not-determined |
| S16_extracted_50989 | Not-determined |
| S16_extracted_50990 | Low-quality |
| S16_extracted_50991 | Not-determined |
| S16_extracted_50995 | Not-determined |
| S16_extracted_51010 | Not-determined |
| S16_extracted_51027 | Not-determined |
| S16_extracted_51030 | Not-determined |
| S16_extracted_51031 | Not-determined |
| S16_extracted_51034 | Low-quality |
| S16_extracted_51039 | Low-quality |
| S16_extracted_51054 | Not-determined |
| S16_extracted_51056 | Not-determined |
| S16_extracted_51057 | Not-determined |
| S16_extracted_51060 | Not-determined |
| S16_extracted_51064 | Not-determined |
| S16_extracted_51090 | Not-determined |
| S16_extracted_51101 | Not-determined |
| S16_extracted_51109 | Low-quality |
| S16_extracted_51134 | Not-determined |
| S16_extracted_51137 | Not-determined |
| S16_extracted_51152 | Not-determined |
| S16_extracted_51157 | Not-determined |
| S16_extracted_51179 | Not-determined |
| S16_extracted_51187 | Not-determined |
| S16_extracted_51219 | Not-determined |
| S16_extracted_51225 | Not-determined |
| S16_extracted_51238 | Not-determined |
| S16_extracted_51242 | Not-determined |
| S16_extracted_51261 | Low-quality |
| S16_extracted_51268 | Not-determined |
| S16_extracted_51278 | Not-determined |
| S16_extracted_51293 | Not-determined |
| S16_extracted_51295 | Not-determined |
| S16_extracted_51302 | Not-determined |
| S16_extracted_51305 | Not-determined |
| S16_extracted_51307 | Not-determined |
| S16_extracted_51310 | Not-determined |
| S16_extracted_51312 | Not-determined |
| S16_extracted_51338 | Not-determined |
| S16_extracted_51346 | Low-quality |
| S16_extracted_51353 | Not-determined |
| S16_extracted_51355 | Not-determined |
| S16_extracted_51363 | Not-determined |
| S16_extracted_51367 | Not-determined |
| S16_extracted_51379 | Not-determined |
| S16_extracted_51382 | Not-determined |
| S16_extracted_51391 | Not-determined |
| S16_extracted_51406 | Not-determined |
| S16_extracted_51427 | Low-quality |
| S16_extracted_51443 | Not-determined |
| S16_extracted_51451 | Low-quality |
| S16_extracted_51452 | Low-quality |
| S16_extracted_51466 | Not-determined |
| S16_extracted_51473 | Not-determined |
| S16_extracted_51478 | Not-determined |
| S16_extracted_51484 | Not-determined |
| S16_extracted_51531 | Not-determined |
| S16_extracted_51538 | Not-determined |
| S16_extracted_51543 | Not-determined |
| S16_extracted_51550 | Not-determined |
| S16_extracted_51552 | Not-determined |
| S16_extracted_51553 | Not-determined |
| S16_extracted_51554 | Not-determined |
| S16_extracted_51577 | Not-determined |
| S16_extracted_51582 | Not-determined |
| S16_extracted_51588 | Not-determined |
| S16_extracted_51603 | Not-determined |
| S16_extracted_51605 | Not-determined |
| S16_extracted_51609 | Not-determined |
| S16_extracted_51616 | Low-quality |
| S16_extracted_51617 | Not-determined |
| S16_extracted_51627 | Not-determined |
| S16_extracted_51646 | Not-determined |
| S16_extracted_51648 | Not-determined |
| S16_extracted_51657 | Low-quality |
| S16_extracted_51673 | Not-determined |
| S16_extracted_51711 | Low-quality |
| S16_extracted_51720 | Not-determined |
| S16_extracted_51745 | Not-determined |
| S16_extracted_51750 | Not-determined |
| S16_extracted_51755 | Not-determined |
| S16_extracted_51768 | Not-determined |
| S16_extracted_51772 | Not-determined |
| S16_extracted_51774 | Not-determined |
| S16_extracted_51792 | Not-determined |
| S16_extracted_51803 | Not-determined |
| S16_extracted_51804 | Not-determined |
| S16_extracted_51805 | Not-determined |
| S16_extracted_51811 | Low-quality |
| S16_extracted_51817 | Not-determined |
| S16_extracted_51823 | Not-determined |
| S16_extracted_51845 | Not-determined |
| S16_extracted_51865 | Not-determined |
| S16_extracted_51871 | Not-determined |
| S16_extracted_51887 | Low-quality |
| S16_extracted_51890 | Not-determined |
| S16_extracted_51892 | Not-determined |
| S16_extracted_51902 | Not-determined |
| S16_extracted_51906 | Not-determined |
| S16_extracted_51912 | Not-determined |
| S16_extracted_51913 | Not-determined |
| S16_extracted_51924 | Not-determined |
| S16_extracted_51925 | Not-determined |
| S16_extracted_51946 | Not-determined |
| S16_extracted_51947 | Not-determined |
| S16_extracted_51957 | Not-determined |
| S16_extracted_51958 | Not-determined |
| S16_extracted_51965 | Low-quality |
| S16_extracted_51966 | Not-determined |
| S16_extracted_51980 | Not-determined |
| S16_extracted_51988 | Not-determined |
| S16_extracted_51996 | Not-determined |
| S16_extracted_52017 | Low-quality |
| S16_extracted_52037 | Not-determined |
| S16_extracted_52052 | Not-determined |
| S16_extracted_52054 | Not-determined |
| S16_extracted_52058 | Not-determined |
| S16_extracted_52083 | Not-determined |
| S16_extracted_52085 | Not-determined |
| S16_extracted_52088 | Not-determined |
| S16_extracted_52092 | Not-determined |
| S16_extracted_52101 | Not-determined |
| S16_extracted_52104 | Not-determined |
| S16_extracted_52109 | Not-determined |
| S16_extracted_52110 | Not-determined |
| S16_extracted_52111 | Low-quality |
| S16_extracted_52124 | Not-determined |
| S16_extracted_52125 | Not-determined |
| S16_extracted_52141 | Not-determined |
| S16_extracted_52142 | Not-determined |
| S16_extracted_52144 | Not-determined |
| S16_extracted_52145 | Low-quality |
| S16_extracted_52158 | Not-determined |
| S16_extracted_52163 | Not-determined |
| S16_extracted_52172 | Not-determined |
| S16_extracted_52182 | Not-determined |
| S16_extracted_52186 | Not-determined |
| S16_extracted_52195 | Not-determined |
| S16_extracted_52197 | Not-determined |
| S16_extracted_52201 | Not-determined |
| S16_extracted_52212 | Not-determined |
| S16_extracted_52218 | Not-determined |
| S16_extracted_52226 | Not-determined |
| S16_extracted_52234 | Not-determined |
| S16_extracted_52235 | Not-determined |
| S16_extracted_52242 | Not-determined |
| S16_extracted_52245 | Not-determined |
| S16_extracted_52250 | Not-determined |
| S16_extracted_52259 | Low-quality |
| S16_extracted_52269 | Not-determined |
| S16_extracted_52279 | Not-determined |
| S16_extracted_52285 | Not-determined |
| S16_extracted_52300 | Not-determined |
| S16_extracted_52308 | Not-determined |
| S16_extracted_52318 | Low-quality |
| S16_extracted_52322 | Not-determined |
| S16_extracted_52352 | Not-determined |
| S16_extracted_52355 | Not-determined |
| S16_extracted_52370 | Not-determined |
| S16_extracted_52382 | Not-determined |
| S16_extracted_52391 | Not-determined |
| S16_extracted_52403 | Low-quality |
| S16_extracted_52417 | Not-determined |
| S16_extracted_52418 | Not-determined |
| S16_extracted_52421 | Not-determined |
| S16_extracted_52423 | Not-determined |
| S16_extracted_52426 | Not-determined |
| S16_extracted_52438 | Not-determined |
| S16_extracted_52442 | Not-determined |
| S16_extracted_52452 | Not-determined |
| S16_extracted_52483 | Not-determined |
| S16_extracted_52487 | Not-determined |
| S16_extracted_52493 | Not-determined |
| S16_extracted_52494 | Not-determined |
| S3_extracted_k141_56363 | Medium-quality |
| S16_extracted_52515 | Not-determined |
| S16_extracted_52523 | Not-determined |
| S16_extracted_52529 | Not-determined |
| S16_extracted_52534 | Not-determined |
| S16_extracted_52550 | Not-determined |

| | |
|---|---|
| S16_extracted_S2561 | Not-determined |
| S16_extracted_S2564 | Not-determined |
| S16_extracted_S2569 | Not-determined |
| S16_extracted_S2575 | Not-determined |
| S16_extracted_S2589 | Not-determined |
| S16_extracted_S2612 | Not-determined |
| S16_extracted_S2614 | Not-determined |
| S16_extracted_S2622 | Not-determined |
| S16_extracted_S2624 | Not-determined |
| S16_extracted_S2638 | Low-quality |
| S16_extracted_S2639 | Not-determined |
| S16_extracted_S2663 | Not-determined |
| S16_extracted_S2692 | Low-quality |
| S16_extracted_S2723 | Not-determined |
| S16_extracted_S2725 | Low-quality |
| S16_extracted_S2727 | Not-determined |
| S16_extracted_S2729 | Low-quality |
| S16_extracted_S2733 | Not-determined |
| S16_extracted_S2736 | Low-quality |
| S16_extracted_S2740 | Not-determined |
| S16_extracted_S2754 | Not-determined |
| S16_extracted_S2768 | Not-determined |
| S16_extracted_S2773 | Not-determined |
| S16_extracted_S2779 | Not-determined |
| S16_extracted_S2814 | Not-determined |
| S16_extracted_S2815 | Not-determined |
| S16_extracted_S2820 | Not-determined |
| S16_extracted_S2821 | Not-determined |
| S16_extracted_S2836 | Not-determined |
| S16_extracted_S2842 | Not-determined |
| S16_extracted_S2846 | Not-determined |
| S16_extracted_S2861 | Not-determined |
| S16_extracted_S2871 | Not-determined |
| S16_extracted_S2873 | Not-determined |
| S16_extracted_S2882 | Not-determined |
| S16_extracted_S2891 | Low-quality |
| S16_extracted_S2894 | Low-quality |
| S16_extracted_S2904 | Not-determined |
| S16_extracted_S2908 | Not-determined |
| S16_extracted_S2910 | Not-determined |
| S16_extracted_S2926 | Not-determined |
| S16_extracted_S2930 | Not-determined |
| S16_extracted_S2985 | Not-determined |
| S16_extracted_S2994 | Low-quality |
| S16_extracted_S3015 | Not-determined |
| S16_extracted_S3018 | Low-quality |
| S16_extracted_S3019 | Not-determined |
| S16_extracted_S3043 | Not-determined |
| S16_extracted_S3058 | Not-determined |
| S16_extracted_S3065 | Not-determined |
| S16_extracted_S3066 | Low-quality |
| S16_extracted_S3075 | Not-determined |
| S16_extracted_S3080 | Not-determined |
| S16_extracted_S3097 | Not-determined |
| S16_extracted_S3112 | Low-quality |
| S16_extracted_S3128 | Not-determined |
| S16_extracted_S3162 | Low-quality |
| S16_extracted_S3222 | Not-determined |
| S16_extracted_S3228 | Low-quality |
| S16_extracted_S3230 | Not-determined |
| S16_extracted_S3239 | Not-determined |
| S16_extracted_S3243 | Not-determined |
| S16_extracted_S3274 | Not-determined |
| S16_extracted_S3278 | Not-determined |
| S16_extracted_S3283 | Not-determined |
| S16_extracted_S3285 | Not-determined |
| S16_extracted_S3299 | Not-determined |
| S16_extracted_S3301 | Not-determined |
| S16_extracted_S3314 | Not-determined |
| S16_extracted_S3334 | Not-determined |
| S16_extracted_S3336 | Not-determined |
| S16_extracted_S3351 | Low-quality |
| S16_extracted_S3358 | Not-determined |
| S16_extracted_S3366 | Not-determined |
| S16_extracted_S3371 | Not-determined |
| S16_extracted_S3376 | Not-determined |
| S16_extracted_S3391 | Not-determined |
| S16_extracted_S3417 | Not-determined |
| S16_extracted_S3430 | Low-quality |
| S16_extracted_S3438 | Not-determined |
| S16_extracted_S3447 | Not-determined |
| S16_extracted_S3449 | Not-determined |
| S16_extracted_S3454 | Not-determined |
| S16_extracted_S3495 | Not-determined |
| S16_extracted_S3505 | Low-quality |
| S16_extracted_S3507 | Not-determined |
| S16_extracted_S3513 | Not-determined |
| S16_extracted_S3519 | Not-determined |
| S16_extracted_S3523 | Not-determined |
| S16_extracted_S3525 | Not-determined |
| S16_extracted_S3532 | Not-determined |
| S16_extracted_S3535 | Not-determined |
| S16_extracted_S3537 | Not-determined |
| S16_extracted_S3540 | Low-quality |
| S16_extracted_S3541 | Not-determined |
| S16_extracted_S3542 | Not-determined |
| S16_extracted_S3543 | Low-quality |
| S16_extracted_S3547 | Not-determined |
| S16_extracted_S3550 | Not-determined |
| S16_extracted_S3555 | Not-determined |
| S16_extracted_S3567 | Not-determined |
| S16_extracted_S3573 | Low-quality |
| S16_extracted_S3581 | Not-determined |
| S16_extracted_S3601 | Not-determined |
| S16_extracted_S3627 | Not-determined |
| S16_extracted_S3638 | Not-determined |
| S16_extracted_S3642 | Not-determined |
| S16_extracted_S3649 | Not-determined |
| S16_extracted_S3659 | Not-determined |
| S16_extracted_S3670 | Low-quality |
| S16_extracted_S3673 | Not-determined |
| S16_extracted_S3689 | Not-determined |
| S16_extracted_S3694 | Not-determined |
| S16_extracted_S3701 | Not-determined |
| S16_extracted_S3709 | Not-determined |
| S16_extracted_S3723 | Not-determined |
| S16_extracted_S3729 | Not-determined |
| S16_extracted_S3733 | Not-determined |
| S16_extracted_S3736 | Not-determined |
| S16_extracted_S3741 | Not-determined |
| S16_extracted_S3758 | Not-determined |
| S16_extracted_S3760 | Not-determined |
| S16_extracted_S3764 | Not-determined |
| S16_extracted_S3766 | Not-determined |
| S16_extracted_S3795 | Not-determined |
| S16_extracted_S3820 | Not-determined |
| S16_extracted_S3829 | Not-determined |
| S16_extracted_S3832 | Not-determined |
| S16_extracted_S3834 | Not-determined |
| S16_extracted_S3842 | Not-determined |
| S16_extracted_S3854 | Not-determined |
| S16_extracted_S3855 | Not-determined |
| S16_extracted_S3873 | Not-determined |
| S16_extracted_S3886 | Not-determined |
| S16_extracted_S3897 | Not-determined |
| S16_extracted_S3933 | Low-quality |
| S16_extracted_S3951 | Not-determined |
| S16_extracted_S3958 | Low-quality |
| S16_extracted_S3961 | Not-determined |
| S16_extracted_S3974 | Low-quality |
| S16_extracted_S3982 | Not-determined |
| S16_extracted_S4012 | Not-determined |
| S16_extracted_S4017 | Not-determined |
| S16_extracted_S4052 | Not-determined |
| S16_extracted_S4059 | Not-determined |
| S16_extracted_S4065 | Low-quality |
| S16_extracted_S4074 | Not-determined |
| S16_extracted_S4075 | Not-determined |
| S16_extracted_S4084 | Low-quality |
| S16_extracted_S4089 | Not-determined |
| S16_extracted_S4096 | Low-quality |
| S16_extracted_S4097 | Not-determined |
| S16_extracted_S4104 | Not-determined |
| S16_extracted_S4113 | Not-determined |
| S16_extracted_S4114 | Not-determined |
| S16_extracted_S4115 | Not-determined |
| S16_extracted_S4126 | Not-determined |
| S16_extracted_S4156 | Not-determined |
| S16_extracted_S4163 | Not-determined |
| S16_extracted_S4174 | Not-determined |
| S16_extracted_S4183 | Low-quality |
| S16_extracted_S4191 | Not-determined |
| S16_extracted_S4201 | Low-quality |
| S16_extracted_S4218 | Not-determined |
| S16_extracted_S4219 | Not-determined |
| S16_extracted_S4256 | Low-quality |
| S16_extracted_S4263 | Not-determined |
| S16_extracted_S4279 | Low-quality |
| S16_extracted_S4282 | Not-determined |
| S16_extracted_S4292 | Not-determined |
| S16_extracted_S4305 | Not-determined |
| S16_extracted_S4314 | Not-determined |
| S16_extracted_S4315 | Not-determined |
| S16_extracted_S4326 | Not-determined |
| S16_extracted_S4329 | Not-determined |
| S16_extracted_S4335 | Not-determined |
| S16_extracted_S4349 | Not-determined |
| S16_extracted_S4360 | Not-determined |
| S16_extracted_S4362 | Low-quality |
| S16_extracted_S4363 | Not-determined |
| S16_extracted_S4404 | Not-determined |
| S16_extracted_S4410 | Not-determined |
| S16_extracted_S4417 | Not-determined |
| S16_extracted_S4448 | Not-determined |
| S16_extracted_S4453 | Not-determined |
| S16_extracted_S4473 | Not-determined |
| S16_extracted_S4475 | Not-determined |
| S16_extracted_S4476 | Not-determined |
| S16_extracted_S4481 | Low-quality |
| S16_extracted_S4499 | Not-determined |
| S16_extracted_S4507 | Not-determined |
| S16_extracted_S4514 | Not-determined |
| S16_extracted_S4519 | Not-determined |
| S16_extracted_S4529 | Not-determined |
| S16_extracted_S4542 | Not-determined |
| S16_extracted_S4544 | Not-determined |
| S16_extracted_S4546 | Not-determined |
| S16_extracted_S4549 | Not-determined |
| S16_extracted_S4560 | Not-determined |
| S16_extracted_S4573 | Not-determined |
| S16_extracted_S4576 | Not-determined |
| S16_extracted_S4589 | Not-determined |
| S16_extracted_S4595 | Not-determined |
| S16_extracted_S4598 | Not-determined |
| S16_extracted_S4600 | Not-determined |
| S16_extracted_S4608 | Not-determined |
| S16_extracted_S4615 | Not-determined |
| S16_extracted_S4637 | Not-determined |
| S16_extracted_S4668 | Not-determined |
| S16_extracted_S4675 | Not-determined |
| S16_extracted_S4677 | Low-quality |
| S16_extracted_S4685 | Not-determined |
| S16_extracted_S4693 | Not-determined |
| S16_extracted_S4708 | Not-determined |
| S16_extracted_S4711 | Not-determined |
| S16_extracted_S4718 | Not-determined |
| S16_extracted_S4727 | Not-determined |
| S16_extracted_S4732 | Not-determined |
| S16_extracted_S4752 | Not-determined |
| S16_extracted_S4757 | Not-determined |
| S16_extracted_S4758 | Low-quality |
| S16_extracted_S4766 | Low-quality |
| S16_extracted_S4773 | Not-determined |
| S16_extracted_S4793 | Not-determined |
| S16_extracted_S4800 | Not-determined |
| S16_extracted_S4809 | Not-determined |
| S16_extracted_S4812 | Low-quality |
| S16_extracted_S4814 | Not-determined |
| S16_extracted_S4827 | Not-determined |
| S16_extracted_S4850 | Not-determined |
| S16_extracted_S4851 | Not-determined |
| S16_extracted_S4864 | Not-determined |
| S16_extracted_S4872 | Not-determined |
| S16_extracted_S4876 | Low-quality |
| S16_extracted_S4884 | Low-quality |
| S16_extracted_S4898 | Not-determined |
| S16_extracted_S4908 | Not-determined |
| S16_extracted_S4912 | Low-quality |
| S16_extracted_S4924 | Not-determined |
| S16_extracted_S4930 | Not-determined |
| S16_extracted_S4944 | Not-determined |
| S16_extracted_S4956 | Not-determined |
| S16_extracted_S4967 | Low-quality |
| S16_extracted_S4980 | Not-determined |
| S16_extracted_S4985 | Not-determined |
| S16_extracted_S4991 | Low-quality |
| S16_extracted_S5025 | Not-determined |
| S16_extracted_S5038 | Not-determined |
| S16_extracted_S5076 | Not-determined |
| S16_extracted_S5081 | Not-determined |
| S16_extracted_S5087 | Not-determined |
| S16_extracted_S5103 | Not-determined |
| S16_extracted_S5107 | Not-determined |
| S16_extracted_S5109 | Low-quality |
| S16_extracted_S5120 | Not-determined |
| S16_extracted_S5124 | Low-quality |
| S16_extracted_S5127 | Not-determined |
| S16_extracted_S5129 | Not-determined |
| S16_extracted_S5132 | Not-determined |
| S16_extracted_S5150 | Low-quality |
| S16_extracted_S5159 | Not-determined |
| S16_extracted_S5161 | Not-determined |
| S16_extracted_S5167 | Not-determined |
| S16_extracted_S5186 | Not-determined |
| S16_extracted_S5193 | Not-determined |
| S16_extracted_S5202 | Low-quality |
| S16_extracted_S5220 | Low-quality |
| S16_extracted_S5228 | Not-determined |
| S16_extracted_S5233 | Not-determined |
| S16_extracted_S5244 | Not-determined |
| S16_extracted_S5254 | Not-determined |
| S16_extracted_S5263 | Not-determined |
| S16_extracted_S5271 | Low-quality |
| S16_extracted_S5273 | Not-determined |
| S16_extracted_S5283 | Not-determined |
| S16_extracted_S5292 | Low-quality |
| S16_extracted_S5298 | Not-determined |
| S16_extracted_S5305 | Not-determined |
| S16_extracted_S5309 | Not-determined |
| S16_extracted_S5339 | Low-quality |
| S16_extracted_S5346 | Low-quality |
| S16_extracted_S5363 | Not-determined |
| S16_extracted_S5389 | Not-determined |
| S16_extracted_S5390 | Not-determined |
| S16_extracted_S5393 | Not-determined |
| S16_extracted_S5426 | Not-determined |
| S16_extracted_S5459 | Not-determined |
| S16_extracted_S5472 | Not-determined |
| S16_extracted_S5476 | Not-determined |
| S16_extracted_S5503 | Not-determined |
| S16_extracted_S5518 | Not-determined |
| S16_extracted_S5529 | Not-determined |
| S16_extracted_S5537 | Not-determined |
| S16_extracted_S5541 | Not-determined |
| S16_extracted_S5551 | Low-quality |
| S16_extracted_S5564 | Not-determined |
| S16_extracted_S5569 | Not-determined |
| S16_extracted_S5576 | Not-determined |
| S16_extracted_S5591 | Not-determined |
| S16_extracted_S5628 | Not-determined |
| S16_extracted_S5651 | Not-determined |
| S16_extracted_S5657 | Not-determined |
| S16_extracted_S5671 | Not-determined |
| S16_extracted_S5682 | Low-quality |
| S16_extracted_S5684 | Low-quality |
| S16_extracted_S5696 | Not-determined |
| S16_extracted_S5698 | Not-determined |
| S16_extracted_S5701 | Not-determined |
| S16_extracted_S5712 | Not-determined |
| S16_extracted_S5714 | Low-quality |
| S16_extracted_S5724 | Not-determined |
| S16_extracted_S5737 | Not-determined |
| S16_extracted_S5739 | Not-determined |
| S16_extracted_S5740 | Not-determined |
| S16_extracted_S5752 | Not-determined |
| S16_extracted_S5754 | Not-determined |
| S16_extracted_S5760 | Not-determined |
| S16_extracted_S5764 | Not-determined |
| S16_extracted_S5775 | Not-determined |
| S16_extracted_S5777 | Not-determined |
| S16_extracted_S5779 | Not-determined |
| S16_extracted_S5798 | Not-determined |
| S16_extracted_S5806 | Not-determined |
| S16_extracted_S5820 | Not-determined |
| S16_extracted_S5822 | Not-determined |
| S16_extracted_S5825 | Not-determined |
| S16_extracted_S5826 | Not-determined |
| S16_extracted_S5828 | Low-quality |
| S16_extracted_S5860 | Not-determined |
| S16_extracted_S5882 | Low-quality |
| S16_extracted_S5902 | Not-determined |
| S16_extracted_S5908 | Not-determined |
| S16_extracted_S5928 | Not-determined |
| S16_extracted_S5934 | Low-quality |
| S16_extracted_S5944 | Not-determined |
| S16_extracted_S5986 | Low-quality |
| S16_extracted_S5988 | Not-determined |
| S16_extracted_S5990 | Not-determined |
| S16_extracted_S5998 | Not-determined |
| S16_extracted_S6005 | Not-determined |
| S16_extracted_S6012 | Not-determined |
| S16_extracted_S6025 | Low-quality |
| S16_extracted_S6028 | Not-determined |
| S16_extracted_S6035 | Not-determined |
| S16_extracted_S6040 | Not-determined |
| S16_extracted_S6044 | Not-determined |
| S16_extracted_S6063 | Low-quality |
| S16_extracted_S6066 | Not-determined |
| S16_extracted_S6072 | Low-quality |
| S16_extracted_S6073 | Not-determined |
| S16_extracted_S6105 | Not-determined |
| S16_extracted_S6111 | Not-determined |
| S16_extracted_S6114 | Not-determined |
| S16_extracted_S6132 | Not-determined |
| S16_extracted_S6156 | Not-determined |
| S16_extracted_S6168 | Not-determined |
| S16_extracted_S6175 | Not-determined |
| S16_extracted_S6177 | Not-determined |
| S16_extracted_S6182 | Not-determined |
| S16_extracted_S6194 | Not-determined |
| S16_extracted_S6196 | Low-quality |
| S16_extracted_S6213 | Not-determined |
| S16_extracted_S6236 | Not-determined |
| S16_extracted_S6279 | Low-quality |
| S16_extracted_S6285 | Not-determined |
| S16_extracted_S6303 | Low-quality |
| S16_extracted_S6311 | Not-determined |
| S16_extracted_S6316 | Not-determined |
| S16_extracted_S6340 | Not-determined |
| S16_extracted_S6351 | Not-determined |
| S16_extracted_S6354 | Not-determined |
| S16_extracted_S6384 | Not-determined |
| S16_extracted_S6397 | Not-determined |
| S16_extracted_S6400 | Low-quality |
| S16_extracted_S6402 | Low-quality |
| S16_extracted_S6403 | Not-determined |
| S16_extracted_S6404 | Not-determined |
| S16_extracted_S6416 | Low-quality |
| S16_extracted_S6440 | Not-determined |
| S16_extracted_S6444 | Not-determined |
| S16_extracted_S6450 | Low-quality |
| S16_extracted_S6458 | Not-determined |
| S16_extracted_S6467 | Low-quality |
| S16_extracted_S6490 | Not-determined |
| S16_extracted_S6491 | Low-quality |
| S16_extracted_S6498 | Not-determined |
| S16_extracted_S6503 | Low-quality |
| S16_extracted_S6512 | Not-determined |
| S16_extracted_S6513 | Not-determined |
| S16_extracted_S6531 | Not-determined |
| S16_extracted_S6535 | Not-determined |
| S16_extracted_S6537 | Not-determined |
| S16_extracted_S6542 | Not-determined |
| S16_extracted_S6550 | Not-determined |
| S16_extracted_S6551 | Not-determined |
| S16_extracted_S6555 | Not-determined |
| S16_extracted_S6556 | Low-quality |
| S16_extracted_S6566 | Not-determined |
| S16_extracted_S6567 | Not-determined |
| S16_extracted_S6580 | Not-determined |
| S16_extracted_S6590 | Not-determined |
| S16_extracted_S6599 | Not-determined |
| S16_extracted_S6616 | Not-determined |
| S16_extracted_S6661 | Not-determined |
| S16_extracted_S6677 | Not-determined |
| S16_extracted_S6694 | Not-determined |
| S16_extracted_S6703 | Not-determined |
| S16_extracted_S6724 | Not-determined |
| S16_extracted_S6730 | Not-determined |
| S16_extracted_S6732 | Not-determined |
| S16_extracted_S6736 | Not-determined |
| S16_extracted_S6746 | Not-determined |
| S16_extracted_S6754 | Not-determined |
| S16_extracted_S6774 | Not-determined |
| S16_extracted_S6787 | Not-determined |
| S16_extracted_S6788 | Not-determined |
| S16_extracted_S6794 | Not-determined |
| S16_extracted_S6795 | Not-determined |
| S16_extracted_S6810 | Not-determined |
| S16_extracted_S6814 | Not-determined |
| S16_extracted_S6822 | Not-determined |
| S16_extracted_S6824 | Not-determined |
| S16_extracted_S6837 | Not-determined |
| S16_extracted_S6853 | Low-quality |
| S16_extracted_S6856 | Not-determined |
| S16_extracted_S6869 | Not-determined |
| S16_extracted_S6881 | Low-quality |
| S16_extracted_S6891 | Not-determined |
| S16_extracted_S6900 | Not-determined |
| S16_extracted_S6910 | Not-determined |
| S16_extracted_S6913 | Not-determined |
| S16_extracted_S6924 | Not-determined |
| S16_extracted_S6928 | Not-determined |
| S16_extracted_S6931 | Not-determined |
| S16_extracted_S6944 | Low-quality |
| S16_extracted_S6958 | Not-determined |
| S16_extracted_S6973 | Not-determined |
| S16_extracted_S6984 | Not-determined |
| S16_extracted_S6993 | Not-determined |
| S16_extracted_S6997 | Not-determined |
| S16_extracted_S7013 | Low-quality |
| S16_extracted_S7025 | Not-determined |
| S16_extracted_S7026 | Not-determined |
| S16_extracted_S7038 | Not-determined |
| S16_extracted_S7054 | Not-determined |
| S16_extracted_S7067 | Not-determined |
| S16_extracted_S7099 | Low-quality |
| S16_extracted_S7102 | Not-determined |
| S16_extracted_S7104 | Not-determined |
| S16_extracted_S7107 | Not-determined |
| S16_extracted_S7133 | Not-determined |
| S16_extracted_S7136 | Not-determined |
| S16_extracted_S7154 | Not-determined |
| S16_extracted_S7162 | Not-determined |
| S16_extracted_S7171 | Not-determined |
| S16_extracted_S7179 | Not-determined |
| S16_extracted_S7191 | Not-determined |
| S16_extracted_S7200 | Not-determined |
| S16_extracted_S7215 | Not-determined |
| S16_extracted_S7217 | Not-determined |
| S16_extracted_S7224 | Not-determined |
| S16_extracted_S7225 | Not-determined |
| S16_extracted_S7229 | Not-determined |
| S16_extracted_S7232 | Not-determined |
| S16_extracted_S7240 | Not-determined |
| S16_extracted_S7241 | Not-determined |
| S16_extracted_S7265 | Low-quality |
| S16_extracted_S7276 | Low-quality |
| S16_extracted_S7283 | Not-determined |
| S16_extracted_S7295 | Not-determined |
| S16_extracted_S7297 | Low-quality |
| S3_vs_k141_56363\|\|full | Medium-quality |
| S16_extracted_S7315 | Not-determined |
| S16_extracted_S7322 | Not-determined |
| S16_extracted_S7339 | Not-determined |
| S16_extracted_S7352 | Not-determined |
| S16_extracted_S7369 | Not-determined |
| S16_extracted_S7377 | Not-determined |
| S16_extracted_S7382 | Not-determined |
| S16_extracted_S7386 | Not-determined |
| S16_extracted_S7402 | Not-determined |
| S16_extracted_S7403 | Not-determined |
| S16_extracted_S7413 | Not-determined |
| S16_extracted_S7426 | Not-determined |
| S16_extracted_S7434 | Not-determined |
| S16_extracted_S7437 | Not-determined |
| S16_extracted_S7449 | Low-quality |
| S16_extracted_S7450 | Not-determined |
| S16_extracted_S7458 | Not-determined |
| S16_extracted_S7467 | Not-determined |
| S16_extracted_S7468 | Not-determined |
| S16_extracted_S7469 | Not-determined |
| S16_extracted_S7471 | Low-quality |
| S16_extracted_S7474 | Not-determined |
| S16_extracted_S7494 | Low-quality |
| S16_extracted_S7497 | Not-determined |
| S16_extracted_S7505 | Low-quality |
| S16_extracted_S7509 | Not-determined |
| S16_extracted_S7523 | Low-quality |
| S16_extracted_S7530 | Not-determined |
| S16_extracted_S7539 | Not-determined |
| S16_extracted_S7540 | Not-determined |
| S16_extracted_S7542 | Low-quality |
| S16_extracted_S7550 | Not-determined |
| S16_extracted_S7555 | Not-determined |
| S16_extracted_S7565 | Low-quality |
| S16_extracted_S7567 | Not-determined |
| S16_extracted_S7575 | Not-determined |
| S16_extracted_S7599 | Not-determined |
| S16_extracted_S7610 | Not-determined |
| S16_extracted_S7620 | Not-determined |
| S16_extracted_S7625 | Low-quality |
| S16_extracted_S7648 | Not-determined |
| S16_extracted_S7651 | Not-determined |
| S16_extracted_S7686 | Not-determined |
| S16_extracted_S7690 | Not-determined |
| S16_extracted_S7698 | Low-quality |
| S16_extracted_S7702 | Not-determined |
| S16_extracted_S7711 | Not-determined |
| S16_extracted_S7719 | Not-determined |
| S16_extracted_S7723 | Not-determined |
| S16_extracted_S7774 | Low-quality |
| S16_extracted_S7785 | Not-determined |
| S16_extracted_S7790 | Not-determined |
| S16_extracted_S7792 | Not-determined |
| S16_extracted_S7808 | Not-determined |
| S16_extracted_S7814 | Not-determined |
| S16_extracted_S7824 | Not-determined |
| S16_extracted_S7861 | Not-determined |
| S16_extracted_S7872 | Not-determined |
| S16_extracted_S7878 | Not-determined |
| S16_extracted_S7882 | Not-determined |
| S16_extracted_S7885 | Not-determined |
| S16_extracted_S7906 | Not-determined |
| S16_extracted_S7908 | Low-quality |
| S16_extracted_S7921 | Not-determined |
| S16_extracted_S7943 | Not-determined |
| S16_extracted_S7952 | Not-determined |
| S16_extracted_S7961 | Not-determined |
| S16_extracted_S7962 | Not-determined |
| S16_extracted_S7969 | Not-determined |
| S16_extracted_S7977 | Not-determined |
| S16_extracted_S8011 | Not-determined |
| S16_extracted_S8016 | Not-determined |
| S16_extracted_S8019 | Not-determined |
| S16_extracted_S8022 | Not-determined |
| S16_extracted_S8038 | Low-quality |
| S16_extracted_S8039 | Low-quality |
| S16_extracted_S8043 | Not-determined |
| S16_extracted_S8049 | Low-quality |
| S16_extracted_S8082 | Not-determined |
| S16_extracted_S8093 | Not-determined |
| S16_extracted_S8096 | Not-determined |
| S16_extracted_S8113 | Not-determined |
| S16_extracted_S8151 | Not-determined |
| S16_extracted_S8163 | Not-determined |
| S16_extracted_S8177 | Not-determined |
| S16_extracted_S8190 | Not-determined |
| S16_extracted_S8215 | Not-determined |
| S16_extracted_S8237 | Not-determined |
| S16_extracted_S8251 | Not-determined |
| S16_extracted_S8275 | Not-determined |
| S16_extracted_S8282 | Not-determined |
| S16_extracted_S8286 | Not-determined |
| S16_extracted_S8305 | Not-determined |
| S16_extracted_S8328 | Not-determined |
| S16_extracted_S8338 | Not-determined |
| S16_extracted_S8360 | Not-determined |
| S16_extracted_S8361 | Not-determined |
| S16_extracted_S8363 | Not-determined |
| S16_extracted_S8368 | Low-quality |
| S16_extracted_S8371 | Not-determined |
| S16_extracted_S8374 | Not-determined |
| S16_extracted_S8394 | Not-determined |
| S16_extracted_S8426 | Not-determined |
| S16_extracted_S8433 | Not-determined |
| S16_extracted_S8434 | Not-determined |
| S16_extracted_S8439 | Not-determined |
| S16_extracted_S8447 | Not-determined |
| S16_extracted_S8472 | Not-determined |
| S16_extracted_S8484 | Not-determined |
| S16_extracted_S8490 | Low-quality |
| S16_extracted_S8510 | Not-determined |
| S16_extracted_S8512 | Low-quality |
| S16_extracted_S8521 | Not-determined |
| S16_extracted_S8533 | Not-determined |
| S16_extracted_S8551 | Not-determined |
| S16_extracted_S8564 | Not-determined |
| S16_extracted_S8573 | Low-quality |

| | |
|---|---|
| S16_extracted_58592 | Low-quality |
| S16_extracted_58593 | Not-determined |
| S16_extracted_58601 | Not-determined |
| S16_extracted_58602 | Not-determined |
| S16_extracted_58606 | Not-determined |
| S16_extracted_58615 | Not-determined |
| S16_extracted_58668 | Not-determined |
| S16_extracted_58675 | Not-determined |
| S16_extracted_58684 | Not-determined |
| S16_extracted_58709 | Low-quality |
| S16_extracted_58724 | Not-determined |
| S16_extracted_58727 | Low-quality |
| S16_extracted_58730 | Not-determined |
| S16_extracted_58779 | Not-determined |
| S16_extracted_58791 | Low-quality |
| S16_extracted_58795 | Not-determined |
| S16_extracted_58801 | Not-determined |
| S16_extracted_58802 | Not-determined |
| S16_extracted_58812 | Not-determined |
| S16_extracted_58813 | Not-determined |
| S16_extracted_58822 | Not-determined |
| S16_extracted_58847 | Low-quality |
| S16_extracted_58850 | Not-determined |
| S16_extracted_58851 | Not-determined |
| S16_extracted_58863 | Not-determined |
| S16_extracted_58866 | Not-determined |
| S16_extracted_58871 | Not-determined |
| S16_extracted_58873 | Not-determined |
| S16_extracted_58877 | Not-determined |
| S16_extracted_58890 | Not-determined |
| S16_extracted_58895 | Not-determined |
| S16_extracted_58913 | Not-determined |
| S16_extracted_58917 | Not-determined |
| S16_extracted_58925 | Not-determined |
| S16_extracted_58969 | Not-determined |
| S16_extracted_58984 | Not-determined |
| S16_extracted_59014 | Low-quality |
| S16_extracted_59023 | Not-determined |
| S16_extracted_59048 | Not-determined |
| S16_extracted_59073 | Not-determined |
| S16_extracted_59083 | Not-determined |
| S16_extracted_59085 | Not-determined |
| S16_extracted_59094 | Not-determined |
| S16_extracted_59098 | Not-determined |
| S16_extracted_59110 | Not-determined |
| S16_extracted_59116 | Not-determined |
| S16_extracted_59126 | Not-determined |
| S16_extracted_59137 | Not-determined |
| S16_extracted_59140 | Low-quality |
| S16_extracted_59155 | Not-determined |
| S16_extracted_59167 | Low-quality |
| S16_extracted_59168 | Not-determined |
| S16_extracted_59172 | Not-determined |
| S16_extracted_59180 | Not-determined |
| S16_extracted_59188 | Not-determined |
| S16_extracted_59193 | Low-quality |
| S16_extracted_59224 | Low-quality |
| S16_extracted_59252 | Not-determined |
| S16_extracted_59285 | Not-determined |
| S16_extracted_59297 | Not-determined |
| S16_extracted_59317 | Not-determined |
| S16_extracted_59322 | Not-determined |
| S16_extracted_59345 | Not-determined |
| S16_extracted_59353 | Not-determined |
| S16_extracted_59366 | Not-determined |
| S16_extracted_59379 | Not-determined |
| S16_extracted_59382 | Not-determined |
| S16_extracted_59443 | Not-determined |
| S16_extracted_59445 | Not-determined |
| S16_extracted_59463 | Not-determined |
| S16_extracted_59466 | Not-determined |
| S16_extracted_59485 | Not-determined |
| S16_extracted_59487 | Not-determined |
| S16_extracted_59493 | Not-determined |
| S16_extracted_59511 | Not-determined |
| S16_extracted_59513 | Not-determined |
| S16_extracted_59522 | Not-determined |
| S16_extracted_59541 | Low-quality |
| S16_extracted_59546 | Low-quality |
| S16_extracted_59550 | Not-determined |
| S16_extracted_59556 | Not-determined |
| S16_extracted_59563 | Low-quality |
| S16_extracted_59575 | Not-determined |
| S16_extracted_59586 | Not-determined |
| S16_extracted_59631 | Not-determined |
| S16_extracted_59639 | Not-determined |
| S16_extracted_59640 | Not-determined |
| S16_extracted_59656 | Not-determined |
| S16_extracted_59664 | Not-determined |
| S16_extracted_59665 | Low-quality |
| S16_extracted_59680 | Not-determined |
| S16_extracted_59694 | Low-quality |
| S16_extracted_59699 | Not-determined |
| S16_extracted_59716 | Not-determined |
| S16_extracted_59737 | Not-determined |
| S16_extracted_59756 | Not-determined |
| S16_extracted_59769 | Not-determined |
| S16_extracted_59785 | Not-determined |
| S16_extracted_59809 | Not-determined |
| S16_extracted_59813 | Not-determined |
| S16_extracted_59816 | Not-determined |
| S16_extracted_59818 | Low-quality |
| S16_extracted_59826 | Not-determined |
| S16_extracted_59832 | Not-determined |
| S16_extracted_59854 | Not-determined |
| S16_extracted_59860 | Not-determined |
| S16_extracted_59871 | Not-determined |
| S16_extracted_59902 | Not-determined |
| S16_extracted_59908 | Not-determined |
| S16_extracted_59913 | Not-determined |
| S16_extracted_59919 | Not-determined |
| S16_extracted_59934 | Not-determined |
| S16_extracted_59938 | Not-determined |
| S16_extracted_59939 | Not-determined |
| S16_extracted_59940 | Not-determined |
| S16_extracted_59952 | Not-determined |
| S16_extracted_59957 | Not-determined |
| S16_extracted_59965 | Not-determined |
| S16_extracted_59978 | Not-determined |
| S16_extracted_59984 | Not-determined |
| S16_extracted_60001 | Not-determined |
| S16_extracted_60004 | Not-determined |
| S16_extracted_60005 | Not-determined |
| S16_extracted_60007 | Not-determined |
| S16_extracted_60025 | Not-determined |
| S16_extracted_60032 | Not-determined |
| S16_extracted_60049 | Not-determined |
| S16_extracted_60058 | Not-determined |
| S16_extracted_60067 | Not-determined |
| S16_extracted_60069 | Not-determined |
| S16_extracted_60070 | Not-determined |
| S16_extracted_60074 | Not-determined |
| S16_extracted_60076 | Not-determined |
| S16_extracted_60077 | Not-determined |
| S16_extracted_60080 | Not-determined |
| S16_extracted_60090 | Low-quality |
| S16_extracted_60101 | Not-determined |
| S16_extracted_60106 | Not-determined |
| S16_extracted_60129 | Not-determined |
| S16_extracted_60140 | Not-determined |
| S16_extracted_60171 | Not-determined |
| S16_extracted_60174 | Not-determined |
| S16_extracted_60193 | Not-determined |
| S16_extracted_60197 | Low-quality |
| S16_extracted_60206 | Low-quality |
| S16_extracted_60234 | Not-determined |
| S16_extracted_60238 | Not-determined |
| S16_extracted_60247 | Not-determined |
| S16_extracted_60250 | Not-determined |
| S16_extracted_60259 | Not-determined |
| S16_extracted_60264 | Not-determined |
| S16_extracted_60270 | Low-quality |
| S16_extracted_60277 | Low-quality |
| S16_extracted_60301 | Not-determined |
| S16_extracted_60320 | Not-determined |
| S16_extracted_60332 | Not-determined |
| S16_extracted_60337 | Low-quality |
| S16_extracted_60338 | Not-determined |
| S16_extracted_60350 | Not-determined |
| S16_extracted_60352 | Not-determined |
| S16_extracted_60382 | Not-determined |
| S16_extracted_60415 | Not-determined |
| S16_extracted_60422 | Low-quality |
| S16_extracted_60424 | Not-determined |
| S16_extracted_60428 | Not-determined |
| S16_extracted_60433 | Not-determined |
| S16_extracted_60441 | Low-quality |
| S16_extracted_60471 | Not-determined |
| S16_extracted_60481 | Not-determined |
| S16_extracted_60483 | Not-determined |
| S16_extracted_60525 | Not-determined |
| S16_extracted_60530 | Low-quality |
| S16_extracted_60539 | Not-determined |
| S16_extracted_60540 | Not-determined |
| S16_extracted_60545 | Low-quality |
| S16_extracted_60558 | Not-determined |
| S16_extracted_60561 | Not-determined |
| S16_extracted_60562 | Not-determined |
| S16_extracted_60566 | Low-quality |
| S16_extracted_60602 | Low-quality |
| S16_extracted_60605 | Low-quality |
| S16_extracted_60609 | Not-determined |
| S16_extracted_60616 | Not-determined |
| S16_extracted_60618 | Not-determined |
| S16_extracted_60633 | Not-determined |
| S16_extracted_60638 | Not-determined |
| S16_extracted_60642 | Not-determined |
| S16_extracted_60643 | Not-determined |
| S16_extracted_60647 | Not-determined |
| S16_extracted_60651 | Low-quality |
| S16_extracted_60657 | Not-determined |
| S16_extracted_60660 | Not-determined |
| S16_extracted_60661 | Not-determined |
| S16_extracted_60674 | Not-determined |
| S16_extracted_60680 | Not-determined |
| S16_extracted_60703 | Low-quality |
| S16_extracted_60713 | Low-quality |
| S16_extracted_60715 | Low-quality |
| S16_extracted_60716 | Not-determined |
| S16_extracted_60721 | Low-quality |
| S16_extracted_60724 | Not-determined |
| S16_extracted_60741 | Not-determined |
| S16_extracted_60750 | Low-quality |
| S16_extracted_60766 | Not-determined |
| S16_extracted_60770 | Low-quality |
| S16_extracted_60778 | Not-determined |
| S16_extracted_60781 | Not-determined |
| S16_extracted_60788 | Not-determined |
| S16_extracted_60812 | Not-determined |
| S16_extracted_60824 | Not-determined |
| S16_extracted_60835 | Not-determined |
| S16_extracted_60838 | Low-quality |
| S16_extracted_60841 | Not-determined |
| S16_extracted_60888 | Not-determined |
| S16_extracted_60899 | Not-determined |
| S16_extracted_60921 | Not-determined |
| S16_extracted_60930 | Not-determined |
| S16_extracted_60931 | Not-determined |
| S16_extracted_60932 | Low-quality |
| S16_extracted_60939 | Not-determined |
| S16_extracted_60951 | Not-determined |
| S16_extracted_60953 | Not-determined |
| S16_extracted_60954 | Not-determined |
| S16_extracted_60966 | Not-determined |
| S16_extracted_60973 | Not-determined |
| S16_extracted_60980 | Not-determined |
| S16_extracted_61004 | Not-determined |
| S16_extracted_61019 | Not-determined |
| S16_extracted_61025 | Not-determined |
| S16_extracted_61027 | Not-determined |
| S16_extracted_61034 | Not-determined |
| S16_extracted_61035 | Not-determined |
| S16_extracted_61037 | Not-determined |
| S16_extracted_61058 | Not-determined |
| S16_extracted_61063 | Not-determined |
| S16_extracted_61080 | Not-determined |
| S16_extracted_61097 | Not-determined |
| S16_extracted_61103 | Low-quality |
| S16_extracted_61105 | Low-quality |
| S16_extracted_61122 | Not-determined |
| S16_extracted_61125 | Not-determined |
| S16_extracted_61145 | Not-determined |
| S16_extracted_61160 | Low-quality |
| S16_extracted_61165 | Not-determined |
| S16_extracted_61179 | Not-determined |
| S16_extracted_61185 | Not-determined |
| S16_extracted_61208 | Not-determined |
| S16_extracted_61229 | Not-determined |
| S16_extracted_61242 | Not-determined |
| S16_extracted_61284 | Not-determined |
| S16_extracted_61293 | Low-quality |
| S16_extracted_61304 | Not-determined |
| S16_extracted_61309 | Not-determined |
| S16_extracted_61322 | Not-determined |
| S16_extracted_61323 | Not-determined |
| S16_extracted_61330 | Not-determined |
| S16_extracted_61361 | Not-determined |
| S16_extracted_61363 | Not-determined |
| S16_extracted_61369 | Not-determined |
| S16_extracted_61370 | Not-determined |
| S16_extracted_61410 | Not-determined |
| S16_extracted_61438 | Not-determined |
| S16_extracted_61440 | Not-determined |
| S16_extracted_61477 | Not-determined |
| S16_extracted_61478 | Not-determined |
| S16_extracted_61489 | Not-determined |
| S16_extracted_61495 | Not-determined |
| S16_extracted_61496 | Not-determined |
| S16_extracted_61497 | Not-determined |
| S16_extracted_61500 | Not-determined |
| S16_extracted_61514 | Not-determined |
| S16_extracted_61520 | Low-quality |
| S16_extracted_61538 | Not-determined |
| S16_extracted_61544 | Low-quality |
| S16_extracted_61554 | Low-quality |
| S16_extracted_61597 | Not-determined |
| S16_extracted_61603 | Not-determined |
| S16_extracted_61635 | Low-quality |
| S16_extracted_61643 | Not-determined |
| S16_extracted_61646 | Low-quality |
| S16_extracted_61650 | Not-determined |
| S16_extracted_61660 | Not-determined |
| S16_extracted_61665 | Low-quality |
| S16_extracted_61667 | Not-determined |
| S16_extracted_61675 | Not-determined |
| S16_extracted_61728 | Not-determined |
| S16_extracted_61738 | Not-determined |
| S16_extracted_61781 | Not-determined |
| S16_extracted_61821 | Not-determined |
| S16_extracted_61828 | Not-determined |
| S16_extracted_61830 | Not-determined |
| S16_extracted_61843 | Not-determined |
| S16_extracted_61855 | Not-determined |
| S16_extracted_61864 | Not-determined |
| S16_extracted_61866 | Not-determined |
| S16_extracted_61876 | Not-determined |
| S16_extracted_61877 | Not-determined |
| S16_extracted_61880 | Not-determined |
| S16_extracted_61881 | Not-determined |
| S16_extracted_61903 | Not-determined |
| S16_extracted_61917 | Not-determined |
| S16_extracted_61943 | Low-quality |
| S16_extracted_61945 | Not-determined |
| S16_extracted_61947 | Not-determined |
| S16_extracted_61960 | Low-quality |
| S16_extracted_61970 | Not-determined |
| S16_extracted_61991 | Not-determined |
| S16_extracted_61994 | Not-determined |
| S16_extracted_61996 | Not-determined |
| S16_extracted_62000 | Not-determined |
| S16_extracted_62001 | Not-determined |
| S16_extracted_62012 | Not-determined |
| S16_extracted_62033 | Not-determined |
| S16_extracted_62038 | Not-determined |
| S16_extracted_62044 | Not-determined |
| S16_extracted_62055 | Low-quality |
| S16_extracted_62093 | Not-determined |
| S16_extracted_62095 | Not-determined |
| S16_extracted_62096 | Low-quality |
| S16_extracted_62102 | Not-determined |
| S16_extracted_62109 | Not-determined |
| S16_extracted_62143 | Not-determined |
| S16_extracted_62160 | Not-determined |
| S16_extracted_62162 | Not-determined |
| S16_extracted_62167 | Low-quality |
| S16_extracted_62170 | Not-determined |
| S16_extracted_62171 | Low-quality |
| S16_extracted_62181 | Low-quality |
| S16_extracted_62189 | Not-determined |
| S16_extracted_62190 | Not-determined |
| S16_extracted_62195 | Not-determined |
| S16_extracted_62203 | Not-determined |
| S16_extracted_62222 | Not-determined |
| S16_extracted_62224 | Not-determined |
| S16_extracted_62243 | Not-determined |
| S16_extracted_62264 | Not-determined |
| S16_extracted_62295 | Not-determined |
| S16_extracted_62302 | Not-determined |
| S16_extracted_62313 | Not-determined |
| S16_extracted_62357 | Not-determined |
| S16_extracted_62371 | Not-determined |
| S16_extracted_62374 | Low-quality |
| S16_extracted_62376 | Not-determined |
| S16_extracted_62380 | Not-determined |
| S16_extracted_62381 | Not-determined |
| S16_extracted_62385 | Low-quality |
| S16_extracted_62401 | Not-determined |
| S16_extracted_62433 | Not-determined |
| S16_extracted_62456 | Not-determined |
| S16_extracted_62462 | Not-determined |
| S16_extracted_62519 | Not-determined |
| S16_extracted_62532 | Not-determined |
| S16_extracted_62547 | Not-determined |
| S16_extracted_62550 | Not-determined |
| S16_extracted_62565 | Not-determined |
| S16_extracted_62569 | Not-determined |
| S16_extracted_62575 | Not-determined |
| S16_extracted_62595 | Not-determined |
| S16_extracted_62618 | Not-determined |
| S16_extracted_62621 | Not-determined |
| S16_extracted_62669 | Not-determined |
| S16_extracted_62672 | Not-determined |
| S16_extracted_62675 | Low-quality |
| S16_extracted_62683 | Not-determined |
| S16_extracted_62691 | Not-determined |
| S16_extracted_62698 | Not-determined |
| S16_extracted_62701 | Not-determined |
| S16_extracted_62706 | Low-quality |
| S16_extracted_62707 | Not-determined |
| S16_extracted_62708 | Not-determined |
| S16_extracted_62712 | Not-determined |
| S16_extracted_62713 | Low-quality |
| S16_extracted_62725 | Not-determined |
| S16_extracted_62744 | Not-determined |
| S16_extracted_62783 | Not-determined |
| S16_extracted_62790 | Not-determined |
| S16_extracted_62810 | Not-determined |
| S16_extracted_62834 | Not-determined |
| S16_extracted_62835 | Not-determined |
| S16_extracted_62837 | Not-determined |
| S16_extracted_62842 | Not-determined |
| S16_extracted_62857 | Not-determined |
| S16_extracted_62858 | Not-determined |
| S16_extracted_62874 | Not-determined |
| S16_extracted_62876 | Not-determined |
| S16_extracted_62891 | Not-determined |
| S16_extracted_62902 | Not-determined |
| S16_extracted_62906 | Low-quality |
| S16_extracted_62912 | Not-determined |
| S16_extracted_62917 | Not-determined |
| S16_extracted_62923 | Not-determined |
| S16_extracted_62928 | Low-quality |
| S16_extracted_62978 | Not-determined |
| S16_extracted_62979 | Not-determined |
| S16_extracted_62981 | Not-determined |
| S16_extracted_62993 | Not-determined |
| S16_extracted_63003 | Not-determined |
| S16_extracted_63007 | Not-determined |
| S16_extracted_63025 | Not-determined |
| S16_extracted_63028 | Not-determined |
| S16_extracted_63040 | Not-determined |
| S16_extracted_63062 | Not-determined |
| S16_extracted_63064 | Not-determined |
| S16_extracted_63084 | Not-determined |
| S16_extracted_63090 | Not-determined |
| S16_extracted_63096 | Not-determined |
| S16_extracted_63128 | Not-determined |
| S16_extracted_63137 | Not-determined |
| S16_extracted_63146 | Not-determined |
| S16_extracted_63168 | Low-quality |
| S16_extracted_63195 | Not-determined |
| S16_extracted_63200 | Not-determined |
| S16_extracted_63232 | Not-determined |
| S16_extracted_63240 | Not-determined |
| S16_extracted_63269 | Not-determined |
| S16_extracted_63276 | Not-determined |
| S16_extracted_63280 | Low-quality |
| S16_extracted_63281 | Not-determined |
| S16_extracted_63300 | Not-determined |
| S16_extracted_63302 | Low-quality |
| S16_extracted_63307 | Not-determined |
| S16_extracted_63315 | Not-determined |
| S16_extracted_63316 | Not-determined |
| S16_extracted_63336 | Not-determined |
| S16_extracted_63358 | Not-determined |
| S16_extracted_63365 | Low-quality |
| S16_extracted_63376 | Not-determined |
| S16_extracted_63378 | Low-quality |
| S16_extracted_63385 | Not-determined |
| S16_extracted_63393 | Low-quality |
| S16_extracted_63406 | Low-quality |
| S16_extracted_63410 | Low-quality |
| S16_extracted_63414 | Not-determined |
| S16_extracted_63427 | Not-determined |
| S16_extracted_63429 | Low-quality |
| S16_extracted_63440 | Not-determined |
| S16_extracted_63446 | Low-quality |
| S16_extracted_63451 | Not-determined |
| S16_extracted_63456 | Not-determined |
| S16_extracted_63460 | Not-determined |
| S16_extracted_63461 | Not-determined |
| S16_extracted_63462 | Not-determined |
| S16_extracted_63469 | Not-determined |
| S16_extracted_63479 | Not-determined |
| S16_extracted_63489 | Low-quality |
| S16_extracted_63493 | Not-determined |
| S16_extracted_63494 | Not-determined |
| S16_extracted_63502 | Not-determined |
| S16_extracted_63523 | Not-determined |
| S16_extracted_63540 | Not-determined |
| S16_extracted_63541 | Not-determined |
| S16_extracted_63551 | Not-determined |
| S16_extracted_63557 | Not-determined |
| S16_extracted_63559 | Not-determined |
| S16_extracted_63582 | Low-quality |
| S16_extracted_63583 | Not-determined |
| S16_extracted_63599 | Not-determined |
| S16_extracted_63605 | Not-determined |
| S16_extracted_63609 | Not-determined |
| S16_extracted_63637 | Not-determined |
| S16_extracted_63641 | Low-quality |
| S16_extracted_63660 | Not-determined |
| S16_extracted_63675 | Not-determined |
| S16_extracted_63723 | Not-determined |
| S16_extracted_63734 | Not-determined |
| S16_extracted_63736 | Not-determined |
| S16_extracted_63743 | Not-determined |
| S16_extracted_63778 | Not-determined |
| S16_extracted_63784 | Not-determined |
| S16_extracted_63785 | Not-determined |
| S16_extracted_63797 | Low-quality |
| S16_extracted_63798 | Not-determined |
| S16_extracted_63806 | Not-determined |
| S16_extracted_63817 | Low-quality |
| S16_extracted_63828 | Not-determined |
| S16_extracted_63832 | Not-determined |
| S16_extracted_63833 | Not-determined |
| S16_extracted_63843 | Not-determined |
| S16_extracted_63853 | Not-determined |
| S16_extracted_63860 | Not-determined |
| S16_extracted_63864 | Not-determined |
| S16_extracted_63865 | Not-determined |
| S16_extracted_63876 | Not-determined |
| S16_extracted_63880 | Not-determined |
| S16_extracted_63903 | Not-determined |
| S16_extracted_63904 | Not-determined |
| S16_extracted_63910 | Not-determined |
| S16_extracted_63927 | Not-determined |
| S16_extracted_63932 | Not-determined |
| S16_extracted_63955 | Not-determined |
| S16_extracted_63968 | Not-determined |
| S16_extracted_63970 | Not-determined |
| S16_extracted_63973 | Not-determined |
| S16_extracted_63975 | Not-determined |
| S16_extracted_64006 | Not-determined |
| S16_extracted_64017 | Not-determined |
| S16_extracted_64019 | Low-quality |
| S16_extracted_64024 | Not-determined |
| S16_extracted_64036 | Low-quality |
| S16_extracted_64041 | Not-determined |
| S16_extracted_64050 | Not-determined |
| S16_extracted_64067 | Low-quality |
| S16_extracted_64069 | Not-determined |
| S16_extracted_64072 | Not-determined |
| S16_extracted_64075 | Not-determined |
| S16_extracted_64081 | Not-determined |
| S16_extracted_64084 | Not-determined |
| S16_extracted_64086 | Low-quality |
| S16_extracted_64097 | Not-determined |
| S16_extracted_64098 | Not-determined |
| S16_extracted_64100 | Not-determined |
| S16_extracted_64130 | Not-determined |
| S16_extracted_64136 | Not-determined |
| S16_extracted_64141 | Not-determined |
| S16_extracted_64142 | Not-determined |
| S16_extracted_64148 | Not-determined |
| S16_extracted_64158 | Low-quality |
| S16_extracted_64162 | Not-determined |
| S16_extracted_64163 | Not-determined |
| S16_extracted_64186 | Not-determined |
| S16_extracted_64200 | Not-determined |
| S16_extracted_64204 | Low-quality |
| S16_extracted_64228 | Not-determined |
| S16_extracted_64234 | Not-determined |
| S16_extracted_64276 | Not-determined |
| S16_extracted_64281 | Not-determined |
| S16_extracted_64291 | Low-quality |
| S16_extracted_64316 | Not-determined |
| S16_extracted_64323 | Not-determined |
| S16_extracted_64333 | Not-determined |
| S16_extracted_64334 | Low-quality |
| S16_extracted_64338 | Not-determined |
| S16_extracted_64341 | Not-determined |
| S16_extracted_64349 | Not-determined |
| S16_extracted_64372 | Not-determined |
| S16_extracted_64384 | Not-determined |
| S16_extracted_64393 | Not-determined |
| S16_extracted_64394 | Low-quality |
| S16_extracted_64399 | Not-determined |
| S16_extracted_64408 | Not-determined |
| S16_extracted_64421 | Not-determined |
| S16_extracted_64424 | Not-determined |
| S16_extracted_64427 | Low-quality |
| S16_extracted_64452 | Not-determined |
| S16_extracted_64454 | Low-quality |
| S16_extracted_64464 | Not-determined |
| S16_extracted_64489 | Not-determined |
| S16_extracted_64498 | Not-determined |
| S16_extracted_64506 | Low-quality |
| S16_extracted_64507 | Not-determined |
| S16_extracted_64518 | Not-determined |
| S16_extracted_64540 | Not-determined |
| S16_extracted_64554 | Not-determined |
| S16_extracted_64562 | Not-determined |
| S16_extracted_64566 | Not-determined |
| S16_extracted_64567 | Not-determined |
| S16_extracted_64573 | Not-determined |
| S16_extracted_64589 | Low-quality |
| S16_extracted_64599 | Not-determined |
| S16_extracted_64606 | Low-quality |
| S16_extracted_64609 | Low-quality |
| S16_extracted_64610 | Not-determined |
| S16_extracted_64619 | Not-determined |
| S16_extracted_64622 | Not-determined |
| S16_extracted_64624 | Low-quality |
| S16_extracted_64680 | Low-quality |
| S16_extracted_64686 | Low-quality |
| S16_extracted_64690 | Low-quality |
| S16_extracted_64700 | Not-determined |
| S16_extracted_64725 | Not-determined |
| S16_extracted_64728 | Not-determined |
| S16_extracted_64729 | Not-determined |
| S16_extracted_64739 | Not-determined |
| S16_extracted_64741 | Not-determined |
| S16_extracted_64766 | Not-determined |
| S16_extracted_64774 | Not-determined |
| S16_extracted_64781 | Not-determined |
| S16_extracted_64814 | Not-determined |
| S16_extracted_64815 | Not-determined |
| S16_extracted_64824 | Not-determined |
| S16_extracted_64826 | Not-determined |
| S16_extracted_64835 | Low-quality |
| S16_extracted_64842 | Not-determined |
| S16_extracted_64847 | Not-determined |
| S16_extracted_64854 | Not-determined |
| S16_extracted_64865 | Not-determined |
| S16_extracted_64876 | Low-quality |
| S16_extracted_64881 | Not-determined |
| S16_extracted_64885 | Low-quality |

| | |
|---|---|
| S16_extracted_64888 | Not-determined |
| S16_extracted_64905 | Not-determined |
| S16_extracted_64920 | Not-determined |
| S16_extracted_64927 | Not-determined |
| S16_extracted_64934 | Low-quality |
| S16_extracted_64950 | Not-determined |
| S16_extracted_64954 | Low-quality |
| S16_extracted_64980 | Not-determined |
| S16_extracted_65019 | Not-determined |
| S16_extracted_65022 | Not-determined |
| S16_extracted_65036 | Not-determined |
| S16_extracted_65041 | Low-quality |
| S16_extracted_65052 | Not-determined |
| S16_extracted_65056 | Not-determined |
| S16_extracted_65059 | Not-determined |
| S16_extracted_65070 | Not-determined |
| S16_extracted_65107 | Not-determined |
| S16_extracted_65111 | Not-determined |
| S16_extracted_65153 | Not-determined |
| S16_extracted_65173 | Not-determined |
| S16_extracted_65180 | Not-determined |
| S16_extracted_65185 | Not-determined |
| S16_extracted_65214 | Low-quality |
| S16_extracted_65217 | Not-determined |
| S16_extracted_65247 | Not-determined |
| S16_extracted_65251 | Not-determined |
| S16_extracted_65258 | Low-quality |
| S16_extracted_65267 | Not-determined |
| S16_extracted_65270 | Not-determined |
| S16_extracted_65278 | Not-determined |
| S16_extracted_65279 | Not-determined |
| S16_extracted_65299 | Not-determined |
| S16_extracted_65309 | Not-determined |
| S16_extracted_65310 | Not-determined |
| S16_extracted_65314 | Low-quality |
| S16_extracted_65318 | Not-determined |
| S16_extracted_65334 | Not-determined |
| S16_extracted_65342 | Not-determined |
| S16_extracted_65360 | Not-determined |
| S16_extracted_65364 | Not-determined |
| S16_extracted_65367 | Not-determined |
| S16_extracted_65370 | Low-quality |
| S16_extracted_65374 | Not-determined |
| S16_extracted_65380 | Not-determined |
| S16_extracted_65401 | Not-determined |
| S16_extracted_65402 | Not-determined |
| S16_extracted_65410 | Not-determined |
| S16_extracted_65416 | Not-determined |
| S16_extracted_65420 | Not-determined |
| S16_extracted_65438 | Not-determined |
| S16_extracted_65461 | Not-determined |
| S16_extracted_65463 | Low-quality |
| S16_extracted_65464 | Not-determined |
| S16_extracted_65473 | Not-determined |
| S16_extracted_65481 | Not-determined |
| S16_extracted_65497 | Not-determined |
| S16_extracted_65499 | Not-determined |
| S16_extracted_65507 | Not-determined |
| S16_extracted_65517 | Not-determined |
| S16_extracted_65523 | Not-determined |
| S16_extracted_65538 | Not-determined |
| S16_extracted_65542 | Not-determined |
| S16_extracted_65554 | Not-determined |
| S16_extracted_65586 | Not-determined |
| S16_extracted_65589 | Not-determined |
| S16_extracted_65590 | Low-quality |
| S16_extracted_65592 | Not-determined |
| S16_extracted_65596 | Not-determined |
| S16_extracted_65599 | Not-determined |
| S16_extracted_65601 | Low-quality |
| S16_extracted_65607 | Not-determined |
| S16_extracted_65622 | Low-quality |
| S16_extracted_65655 | Not-determined |
| S16_extracted_65679 | Not-determined |
| S16_extracted_65718 | Not-determined |
| S16_extracted_65744 | Not-determined |
| S16_extracted_65757 | Low-quality |
| S16_extracted_65776 | Not-determined |
| S16_extracted_65779 | Not-determined |
| S16_extracted_65782 | Low-quality |
| S16_extracted_65801 | Not-determined |
| S16_extracted_65802 | Not-determined |
| S16_extracted_65805 | Not-determined |
| S16_extracted_65837 | Low-quality |
| S16_extracted_65853 | Not-determined |
| S16_extracted_65862 | Not-determined |
| S16_extracted_65877 | Not-determined |
| S16_extracted_65901 | Not-determined |
| S16_extracted_65913 | Not-determined |
| S16_extracted_65914 | Low-quality |
| S16_extracted_65929 | Not-determined |
| S16_extracted_65931 | Not-determined |
| S16_extracted_65936 | Not-determined |
| S16_extracted_65938 | Low-quality |
| S16_extracted_65952 | Not-determined |
| S16_extracted_65953 | Not-determined |
| S16_extracted_65957 | Not-determined |
| S16_extracted_65965 | Not-determined |
| S16_extracted_65972 | Not-determined |
| S16_extracted_65973 | Not-determined |
| S16_extracted_65988 | Not-determined |
| S16_extracted_66009 | Not-determined |
| S16_extracted_66021 | Not-determined |
| S16_extracted_66022 | Low-quality |
| S16_extracted_66029 | Not-determined |
| S16_extracted_66043 | Not-determined |
| S16_extracted_66061 | Not-determined |
| S16_extracted_66069 | Not-determined |
| S16_extracted_66078 | Not-determined |
| S16_extracted_66080 | Not-determined |
| S16_extracted_66082 | Not-determined |
| S16_extracted_66098 | Not-determined |
| S16_extracted_66109 | Not-determined |
| S16_extracted_66115 | Not-determined |
| S16_extracted_66132 | Not-determined |
| S16_extracted_66133 | Not-determined |
| S16_extracted_66136 | Low-quality |
| S16_extracted_66157 | Not-determined |
| S16_extracted_66166 | Not-determined |
| S16_extracted_66171 | Not-determined |
| S16_extracted_66174 | Not-determined |
| S16_extracted_66176 | Low-quality |
| S16_extracted_66190 | Not-determined |
| S16_extracted_66193 | Not-determined |
| S16_extracted_66194 | Not-determined |
| S16_extracted_66207 | Not-determined |
| S16_extracted_66212 | Low-quality |
| S16_extracted_66218 | Not-determined |
| S16_extracted_66220 | Not-determined |
| S16_extracted_66236 | Not-determined |
| S16_extracted_66238 | Not-determined |
| S16_extracted_66250 | Not-determined |
| S16_extracted_66254 | Not-determined |
| S16_extracted_66266 | Not-determined |
| S16_extracted_66274 | Not-determined |
| S16_extracted_66281 | Low-quality |
| S16_extracted_66288 | Low-quality |
| S16_extracted_66299 | Low-quality |
| S16_extracted_66303 | Not-determined |
| S16_extracted_66304 | Not-determined |
| S16_extracted_66305 | Not-determined |
| S16_extracted_66318 | Low-quality |
| S16_extracted_66358 | Not-determined |
| S16_extracted_66365 | Not-determined |
| S16_extracted_66391 | Low-quality |
| S16_extracted_66406 | Not-determined |
| S16_extracted_66407 | Not-determined |
| S16_extracted_66410 | Not-determined |
| S16_extracted_66421 | Low-quality |
| S16_extracted_66422 | Not-determined |
| S16_extracted_66454 | Not-determined |
| S16_extracted_66493 | Not-determined |
| S16_extracted_66499 | Not-determined |
| S16_extracted_66501 | Not-determined |
| S16_extracted_66516 | Not-determined |
| S16_extracted_66522 | Low-quality |
| S16_extracted_66528 | Low-quality |
| S16_extracted_66537 | Not-determined |
| S16_extracted_66542 | Not-determined |
| S16_extracted_66574 | Not-determined |
| S16_extracted_66594 | Not-determined |
| S16_extracted_66600 | Not-determined |
| S16_extracted_66609 | Low-quality |
| S16_extracted_66619 | Not-determined |
| S16_extracted_66622 | Not-determined |
| S16_extracted_66623 | Not-determined |
| S16_extracted_66628 | Not-determined |
| S16_extracted_66631 | Not-determined |
| S16_extracted_66636 | Not-determined |
| S16_extracted_66661 | Not-determined |
| S16_extracted_66669 | Not-determined |
| S16_extracted_66670 | Low-quality |
| S16_extracted_66680 | Not-determined |
| S16_extracted_66686 | Not-determined |
| S16_extracted_66688 | Not-determined |
| S16_extracted_66693 | Not-determined |
| S16_extracted_66702 | Not-determined |
| S16_extracted_66708 | Not-determined |
| S16_extracted_66723 | Not-determined |
| S16_extracted_66736 | Not-determined |
| S16_extracted_66757 | Not-determined |
| S16_extracted_66794 | Not-determined |
| S16_extracted_66802 | Low-quality |
| S16_extracted_66813 | Not-determined |
| S16_extracted_66816 | Not-determined |
| S16_extracted_66829 | Not-determined |
| S16_extracted_66833 | Not-determined |
| S16_extracted_66877 | Not-determined |
| S16_extracted_66878 | Not-determined |
| S16_extracted_66894 | Not-determined |
| S16_extracted_66901 | Not-determined |
| S16_extracted_66903 | Low-quality |
| S16_extracted_66904 | Not-determined |
| S16_extracted_66911 | Not-determined |
| S16_extracted_66930 | Not-determined |
| S16_extracted_66932 | Not-determined |
| S16_extracted_66941 | Low-quality |
| S16_extracted_66944 | Not-determined |
| S16_extracted_66955 | Not-determined |
| S16_extracted_66961 | Not-determined |
| S16_extracted_66973 | Not-determined |
| S16_extracted_66974 | Not-determined |
| S16_extracted_66983 | Not-determined |
| S16_extracted_66995 | Not-determined |
| S16_extracted_67007 | Not-determined |
| S16_extracted_67028 | Not-determined |
| S16_extracted_67034 | Not-determined |
| S16_extracted_67041 | Low-quality |
| S16_extracted_67068 | Not-determined |
| S16_extracted_67078 | Not-determined |
| S16_extracted_67091 | Not-determined |
| S16_extracted_67099 | Not-determined |
| S16_extracted_67125 | Not-determined |
| S16_extracted_67126 | Not-determined |
| S16_extracted_67127 | Not-determined |
| S16_extracted_67148 | Not-determined |
| S16_extracted_67155 | Not-determined |
| S16_extracted_67158 | Not-determined |
| S16_extracted_67184 | Not-determined |
| S16_extracted_67185 | Not-determined |
| S16_extracted_67186 | Not-determined |
| S16_extracted_67195 | Low-quality |
| S16_extracted_67197 | Not-determined |
| S16_extracted_67213 | Not-determined |
| S16_extracted_67214 | Not-determined |
| S16_extracted_67230 | Not-determined |
| S16_extracted_67249 | Not-determined |
| S16_extracted_67253 | Not-determined |
| S16_extracted_67260 | Not-determined |
| S16_extracted_67262 | Not-determined |
| S16_extracted_67315 | Not-determined |
| S16_extracted_67316 | Not-determined |
| S16_extracted_67317 | Low-quality |
| S16_extracted_67327 | Low-quality |
| S16_extracted_67328 | Not-determined |
| S16_extracted_67333 | Not-determined |
| S16_extracted_67340 | Low-quality |
| S16_extracted_67351 | Not-determined |
| S16_extracted_67373 | Not-determined |
| S16_extracted_67379 | Not-determined |
| S16_extracted_67401 | Not-determined |
| S16_extracted_67416 | Not-determined |
| S16_extracted_67428 | Not-determined |
| S16_extracted_67461 | Not-determined |
| S16_extracted_67479 | Not-determined |
| S16_extracted_67497 | Not-determined |
| S16_extracted_67499 | Not-determined |
| S16_extracted_67506 | Not-determined |
| S16_extracted_67518 | Not-determined |
| S16_extracted_67559 | Low-quality |
| S16_extracted_67584 | Not-determined |
| S16_extracted_67593 | Not-determined |
| S16_extracted_67596 | Not-determined |
| S16_extracted_67599 | Not-determined |
| S16_extracted_67602 | Not-determined |
| S16_extracted_67613 | Not-determined |
| S16_extracted_67626 | Not-determined |
| S16_extracted_67630 | Not-determined |
| S16_extracted_67632 | Not-determined |
| S16_extracted_67633 | Low-quality |
| S16_extracted_67634 | Low-quality |
| S16_extracted_67639 | Not-determined |
| S16_extracted_67655 | Not-determined |
| S16_extracted_67657 | Not-determined |
| S16_extracted_67675 | Low-quality |
| S16_extracted_67692 | Not-determined |
| S16_extracted_67697 | Not-determined |
| S16_extracted_67706 | Not-determined |
| S16_extracted_67713 | Not-determined |
| S16_extracted_67718 | Not-determined |
| S16_extracted_67720 | Not-determined |
| S16_extracted_67733 | Not-determined |
| S16_extracted_67743 | Not-determined |
| S16_extracted_67745 | Not-determined |
| S16_extracted_67751 | Not-determined |
| S16_extracted_67766 | Not-determined |
| S16_extracted_67791 | Not-determined |
| S16_extracted_67794 | Not-determined |
| S16_extracted_67800 | Not-determined |
| S16_extracted_67831 | Not-determined |
| S16_extracted_67840 | Not-determined |
| S16_extracted_67845 | Not-determined |
| S16_extracted_67848 | Low-quality |
| S16_extracted_67852 | Not-determined |
| S16_extracted_67858 | Not-determined |
| S16_extracted_67880 | Not-determined |
| S16_extracted_67895 | Not-determined |
| S16_extracted_67906 | Low-quality |
| S16_extracted_67909 | Not-determined |
| S16_extracted_67917 | Not-determined |
| S16_extracted_67930 | Not-determined |
| S16_extracted_67932 | Not-determined |
| S16_extracted_67943 | Not-determined |
| S16_extracted_67944 | Low-quality |
| S16_extracted_67950 | Not-determined |
| S16_extracted_67961 | Low-quality |
| S16_extracted_67971 | Not-determined |
| S16_extracted_67974 | Low-quality |
| S16_extracted_67975 | Not-determined |
| S16_extracted_67978 | Not-determined |
| S16_extracted_67981 | Not-determined |
| S16_extracted_68001 | Not-determined |
| S16_extracted_68010 | Not-determined |
| S16_extracted_68024 | Low-quality |
| S16_extracted_68037 | Not-determined |
| S16_extracted_68047 | Low-quality |
| S16_extracted_68071 | Not-determined |
| S16_extracted_68072 | Not-determined |
| S16_extracted_68081 | Not-determined |
| S16_extracted_68083 | Not-determined |
| S16_extracted_68088 | Not-determined |
| S16_extracted_68109 | Not-determined |
| S16_extracted_68114 | Not-determined |
| S16_extracted_68121 | Not-determined |
| S16_extracted_68132 | Not-determined |
| S16_extracted_68137 | Not-determined |
| S16_extracted_68139 | Not-determined |
| S16_extracted_68142 | Not-determined |
| S16_extracted_68161 | Not-determined |
| S16_extracted_68164 | Not-determined |
| S16_extracted_68167 | Not-determined |
| S16_extracted_68194 | Not-determined |
| S16_extracted_68199 | Not-determined |
| S16_extracted_68216 | Not-determined |
| S16_extracted_68222 | Not-determined |
| S16_extracted_68235 | Low-quality |
| S16_extracted_68244 | Not-determined |
| S16_extracted_68267 | Not-determined |
| S16_extracted_68269 | Not-determined |
| S16_extracted_68280 | Not-determined |
| S16_extracted_68286 | Not-determined |
| S16_extracted_68301 | Not-determined |
| S16_extracted_68323 | Not-determined |
| S16_extracted_68361 | Not-determined |
| S16_extracted_68364 | Not-determined |
| S16_extracted_68367 | Not-determined |
| S16_extracted_68369 | Not-determined |
| S16_extracted_68372 | Not-determined |
| S16_extracted_68373 | Not-determined |
| S16_extracted_68386 | Not-determined |
| S16_extracted_68409 | Not-determined |
| S16_extracted_68419 | Not-determined |
| S16_extracted_68424 | Not-determined |
| S16_extracted_68425 | Not-determined |
| S16_extracted_68428 | Not-determined |
| S16_extracted_68438 | Not-determined |
| S16_extracted_68460 | Not-determined |
| S16_extracted_68469 | Not-determined |
| S16_extracted_68485 | Not-determined |
| S16_extracted_68489 | Not-determined |
| S16_extracted_68497 | Not-determined |
| S16_extracted_68503 | Not-determined |
| S16_extracted_68506 | Not-determined |
| S16_extracted_68507 | Low-quality |
| S16_extracted_68530 | Not-determined |
| S16_extracted_68532 | Not-determined |
| S16_extracted_68542 | Not-determined |
| S16_extracted_68544 | Not-determined |
| S16_extracted_68550 | Not-determined |
| S16_extracted_68558 | Not-determined |
| S16_extracted_68570 | Not-determined |
| S16_extracted_68614 | Low-quality |
| S16_extracted_68621 | Low-quality |
| S16_extracted_68636 | Not-determined |
| S16_extracted_68655 | Low-quality |
| S16_extracted_68656 | Not-determined |
| S16_extracted_68662 | Not-determined |
| S16_extracted_68671 | Low-quality |
| S16_extracted_68689 | Low-quality |
| S16_extracted_68699 | Not-determined |
| S16_extracted_68700 | Not-determined |
| S16_extracted_68714 | Low-quality |
| S16_extracted_68721 | Not-determined |
| S16_extracted_68727 | Not-determined |
| S16_extracted_68750 | Not-determined |
| S16_extracted_68751 | Not-determined |
| S16_extracted_68754 | Not-determined |
| S16_extracted_68780 | Not-determined |
| S16_extracted_68781 | Not-determined |
| S16_extracted_68785 | Not-determined |
| S16_extracted_68788 | Not-determined |
| S16_extracted_68806 | Not-determined |
| S16_extracted_68834 | Not-determined |
| S16_extracted_68865 | Low-quality |
| S16_extracted_68877 | Low-quality |
| S16_extracted_68895 | Not-determined |
| S16_extracted_68896 | Not-determined |
| S16_extracted_68902 | Not-determined |
| S16_extracted_68918 | Not-determined |
| S16_extracted_68932 | Low-quality |
| S16_extracted_68936 | Low-quality |
| S16_extracted_68943 | Not-determined |
| S16_extracted_68952 | Not-determined |
| S16_extracted_68955 | Not-determined |
| S16_extracted_68972 | Not-determined |
| S16_extracted_68977 | Not-determined |
| S16_extracted_68979 | Not-determined |
| S16_extracted_68981 | Not-determined |
| S16_extracted_68988 | Not-determined |
| S16_extracted_68996 | Not-determined |
| S16_extracted_69008 | Not-determined |
| S16_extracted_69016 | Low-quality |
| S16_extracted_69017 | Not-determined |
| S16_extracted_69019 | Not-determined |
| S16_extracted_69022 | Not-determined |
| S16_extracted_69030 | Low-quality |
| S16_extracted_69059 | Not-determined |
| S16_extracted_69075 | Not-determined |
| S16_extracted_69077 | Not-determined |
| S16_extracted_69097 | Not-determined |
| S16_extracted_69098 | Low-quality |
| S16_extracted_69130 | Not-determined |
| S16_extracted_69142 | Not-determined |
| S16_extracted_69145 | Not-determined |
| S16_extracted_69154 | Not-determined |
| S16_extracted_69163 | Not-determined |
| S16_extracted_69166 | Not-determined |
| S16_extracted_69177 | Low-quality |
| S16_extracted_69187 | Low-quality |
| S16_extracted_69205 | Not-determined |
| S16_extracted_69224 | Low-quality |
| S16_extracted_69232 | Not-determined |
| S16_extracted_69233 | Not-determined |
| S16_extracted_69237 | Not-determined |
| S16_extracted_69245 | Not-determined |
| S16_extracted_69250 | Not-determined |
| S16_extracted_69251 | Not-determined |
| S16_extracted_69252 | Low-quality |
| S16_extracted_69255 | Not-determined |
| S16_extracted_69256 | Low-quality |
| S16_extracted_69261 | Low-quality |
| S16_extracted_69288 | Not-determined |
| S16_extracted_69301 | Not-determined |
| S16_extracted_69330 | Low-quality |
| S16_extracted_69353 | Not-determined |
| S16_extracted_69364 | Not-determined |
| S16_extracted_69367 | Not-determined |
| S16_extracted_69369 | Not-determined |
| S16_extracted_69370 | Not-determined |
| S16_extracted_69371 | Not-determined |
| S16_extracted_69386 | Not-determined |
| S16_extracted_69388 | Not-determined |
| S16_extracted_69391 | Low-quality |
| S16_extracted_69399 | Not-determined |
| S16_extracted_69417 | Not-determined |
| S16_extracted_69426 | Not-determined |
| S16_extracted_69428 | Not-determined |
| S16_extracted_69433 | Not-determined |
| S16_extracted_69452 | Not-determined |
| S16_extracted_69509 | Not-determined |
| S16_extracted_69525 | Low-quality |
| S16_extracted_69533 | Not-determined |
| S16_extracted_69547 | Not-determined |
| S16_extracted_69554 | Not-determined |
| S16_extracted_69559 | Not-determined |
| S16_extracted_69563 | Not-determined |
| S16_extracted_69577 | Not-determined |
| S16_extracted_69580 | Not-determined |
| S16_extracted_69638 | Low-quality |
| S16_extracted_69639 | Not-determined |
| S16_extracted_69642 | Not-determined |
| S16_extracted_69646 | Not-determined |
| S16_extracted_69661 | Not-determined |
| S16_extracted_69675 | Not-determined |
| S16_extracted_69691 | Not-determined |
| S16_extracted_69703 | Not-determined |
| S16_extracted_69714 | Not-determined |
| S16_extracted_69745 | Not-determined |
| S16_extracted_69762 | Not-determined |
| S16_extracted_69769 | Not-determined |
| S16_extracted_69775 | Not-determined |
| S16_extracted_69780 | Low-quality |
| S16_extracted_69785 | Not-determined |
| S16_extracted_69792 | Not-determined |
| S16_extracted_69804 | Not-determined |
| S16_extracted_69805 | Not-determined |
| S16_extracted_69811 | Not-determined |
| S16_extracted_69815 | Not-determined |
| S16_extracted_69818 | Not-determined |
| S16_extracted_69826 | Not-determined |
| S16_extracted_69834 | Not-determined |
| S16_extracted_69870 | Not-determined |
| S16_extracted_69884 | Not-determined |
| S16_extracted_69885 | Not-determined |
| S16_extracted_69894 | Low-quality |
| S16_extracted_69914 | Not-determined |
| S16_extracted_69925 | Not-determined |
| S16_extracted_69937 | Not-determined |
| S16_extracted_69938 | Not-determined |
| S16_extracted_69946 | Not-determined |
| S16_extracted_69953 | Not-determined |
| S16_extracted_69967 | Not-determined |
| S16_extracted_69973 | Not-determined |
| S16_extracted_70002 | Not-determined |
| S16_extracted_70004 | Not-determined |
| S16_extracted_70013 | Not-determined |
| S16_extracted_70020 | Not-determined |
| S16_extracted_70024 | Not-determined |
| S16_extracted_70030 | Not-determined |
| S16_extracted_70031 | Not-determined |
| S16_extracted_70041 | Not-determined |
| S16_extracted_70057 | Not-determined |
| S16_extracted_70058 | Not-determined |
| S16_extracted_70080 | Not-determined |
| S16_extracted_70085 | Low-quality |
| S16_extracted_70088 | Not-determined |
| S16_extracted_70097 | Not-determined |
| S16_extracted_70111 | Not-determined |
| S16_extracted_70112 | Low-quality |
| S16_extracted_70116 | Not-determined |
| S16_extracted_70133 | Low-quality |
| S16_extracted_70140 | Low-quality |
| S16_extracted_70148 | Not-determined |
| S16_extracted_70151 | Not-determined |
| S16_extracted_70166 | Low-quality |
| S16_extracted_70181 | Not-determined |
| S16_extracted_70184 | Not-determined |
| S16_extracted_70194 | Not-determined |
| S16_extracted_70197 | Not-determined |
| S16_extracted_70200 | Low-quality |
| S16_extracted_70207 | Low-quality |
| S16_extracted_70212 | Not-determined |
| S16_extracted_70266 | Not-determined |
| S16_extracted_70289 | Not-determined |
| S16_extracted_70293 | Not-determined |
| S16_extracted_70294 | Not-determined |
| S16_extracted_70295 | Not-determined |
| S16_extracted_70307 | Low-quality |
| S16_extracted_70314 | Not-determined |
| S16_extracted_70336 | Not-determined |
| S16_extracted_70338 | Not-determined |
| S16_extracted_70342 | Not-determined |
| S16_extracted_70343 | Low-quality |
| S16_extracted_70347 | Not-determined |
| S16_extracted_70363 | Not-determined |
| S16_extracted_70364 | Low-quality |
| S16_extracted_70387 | Low-quality |
| S16_extracted_70393 | Not-determined |
| S16_extracted_70396 | Not-determined |
| S16_extracted_70424 | Not-determined |
| S16_extracted_70425 | Not-determined |
| S16_extracted_70434 | Not-determined |
| S16_extracted_70435 | Not-determined |
| S16_extracted_70458 | Not-determined |
| S16_extracted_70477 | Not-determined |
| S16_extracted_70481 | Not-determined |
| S16_extracted_70487 | Not-determined |
| S16_extracted_70493 | Not-determined |
| S16_extracted_70497 | Not-determined |
| S16_extracted_70517 | Not-determined |
| S16_extracted_70541 | Not-determined |
| S16_extracted_70562 | Low-quality |
| S16_extracted_70579 | Not-determined |
| S16_extracted_70611 | Low-quality |
| S16_extracted_70612 | Not-determined |
| S16_extracted_70626 | Not-determined |
| S16_extracted_70686 | Not-determined |
| S16_extracted_70692 | Not-determined |
| S16_extracted_70711 | Not-determined |
| S16_extracted_70723 | Not-determined |
| S16_extracted_70724 | Not-determined |
| S16_extracted_70745 | Not-determined |
| S16_extracted_70752 | Not-determined |
| S16_extracted_70780 | Not-determined |
| S16_extracted_70782 | Low-quality |
| S16_extracted_70784 | Not-determined |
| S16_extracted_70796 | Not-determined |
| S16_extracted_70814 | Not-determined |
| S16_extracted_70816 | Low-quality |
| S16_extracted_70817 | Not-determined |
| S16_extracted_70825 | Not-determined |
| S16_extracted_70836 | Low-quality |
| S16_extracted_70848 | Not-determined |
| S16_extracted_70874 | Not-determined |
| S16_extracted_70885 | Low-quality |
| S16_extracted_70891 | Low-quality |
| S16_extracted_70893 | Not-determined |
| S16_extracted_70898 | Not-determined |
| S16_extracted_70904 | Not-determined |
| S16_extracted_70909 | Not-determined |
| S16_extracted_70921 | Not-determined |
| S16_extracted_70932 | Not-determined |
| S16_extracted_70943 | Not-determined |
| S16_extracted_70964 | Not-determined |
| S16_extracted_70969 | Not-determined |
| S16_extracted_70970 | Not-determined |
| S16_extracted_70976 | Not-determined |
| S16_extracted_70977 | Not-determined |
| S16_extracted_70987 | Not-determined |

| | |
|---|---|
| S16_extracted_70988 | Not-determined |
| S16_extracted_70989 | Not-determined |
| S16_extracted_70998 | Not-determined |
| S16_extracted_71025 | Not-determined |
| S16_extracted_71027 | Not-determined |
| S16_extracted_71030 | Not-determined |
| S16_extracted_71033 | Not-determined |
| S16_extracted_71034 | Not-determined |
| S16_extracted_71037 | Not-determined |
| S16_extracted_71043 | Not-determined |
| S16_extracted_71045 | Not-determined |
| S16_extracted_71048 | Not-determined |
| S16_extracted_71053 | Not-determined |
| S16_extracted_71061 | Not-determined |
| S16_extracted_71080 | Not-determined |
| S16_extracted_71103 | Not-determined |
| S16_extracted_71116 | Low-quality |
| S16_extracted_71122 | Not-determined |
| S16_extracted_71131 | Not-determined |
| S16_extracted_71140 | Not-determined |
| S16_extracted_71142 | Not-determined |
| S16_extracted_71198 | Not-determined |
| S16_extracted_71213 | Not-determined |
| S16_extracted_71218 | Not-determined |
| S16_extracted_71220 | Not-determined |
| S16_extracted_71227 | Not-determined |
| S16_extracted_71234 | Not-determined |
| S16_extracted_71235 | Not-determined |
| S16_extracted_71245 | Not-determined |
| S16_extracted_71254 | Not-determined |
| S16_extracted_71260 | Low-quality |
| S16_extracted_71265 | Not-determined |
| S16_extracted_71267 | Not-determined |
| S16_extracted_71269 | Low-quality |
| S16_extracted_71270 | Not-determined |
| S16_extracted_71274 | Not-determined |
| S16_extracted_71292 | Not-determined |
| S16_extracted_71310 | Not-determined |
| S16_extracted_71314 | Not-determined |
| S16_extracted_71322 | Low-quality |
| S16_extracted_71333 | Not-determined |
| S16_extracted_71346 | Low-quality |
| S16_extracted_71350 | Not-determined |
| S16_extracted_71358 | Not-determined |
| S16_extracted_71359 | Not-determined |
| S16_extracted_71364 | Not-determined |
| S16_extracted_71366 | Not-determined |
| S16_extracted_71367 | Not-determined |
| S16_extracted_71378 | Not-determined |
| S16_extracted_71398 | Not-determined |
| S16_extracted_71420 | Low-quality |
| S16_extracted_71425 | Not-determined |
| S16_extracted_71426 | Not-determined |
| S16_extracted_71466 | Not-determined |
| S16_extracted_71467 | Not-determined |
| S16_extracted_71478 | Not-determined |
| S16_extracted_71495 | Not-determined |
| S16_extracted_71501 | Not-determined |
| S16_extracted_71502 | Not-determined |
| S16_extracted_71503 | Not-determined |
| S16_extracted_71510 | Not-determined |
| S16_extracted_71520 | Not-determined |
| S16_extracted_71563 | Not-determined |
| S16_extracted_71568 | Not-determined |
| S16_extracted_71582 | Not-determined |
| S16_extracted_71587 | Not-determined |
| S16_extracted_71588 | Low-quality |
| S16_extracted_71601 | Low-quality |
| S16_extracted_71633 | Not-determined |
| S16_extracted_71646 | Not-determined |
| S16_extracted_71651 | Not-determined |
| S16_extracted_71658 | Not-determined |
| S16_extracted_71664 | Not-determined |
| S16_extracted_71665 | Not-determined |
| S16_extracted_71684 | Not-determined |
| S16_extracted_71704 | Low-quality |
| S16_extracted_71725 | Low-quality |
| S16_extracted_71731 | Not-determined |
| S16_extracted_71753 | Not-determined |
| S16_extracted_71758 | Not-determined |
| S16_extracted_71766 | Not-determined |
| S16_extracted_71778 | Not-determined |
| S16_extracted_71794 | Not-determined |
| S16_extracted_71810 | Not-determined |
| S16_extracted_71861 | Not-determined |
| S16_extracted_71880 | Not-determined |
| S16_extracted_71893 | Not-determined |
| S16_extracted_71923 | Not-determined |
| S16_extracted_71925 | Not-determined |
| S16_extracted_71937 | Not-determined |
| S16_extracted_71943 | Not-determined |
| S16_extracted_71952 | Not-determined |
| S16_extracted_71979 | Not-determined |
| S16_extracted_71984 | Not-determined |
| S16_extracted_71997 | Not-determined |
| S16_extracted_71998 | Not-determined |
| S16_extracted_72001 | Not-determined |
| S16_extracted_72004 | Low-quality |
| S16_extracted_72021 | Not-determined |
| S16_extracted_72024 | Not-determined |
| S16_extracted_72025 | Low-quality |
| S16_extracted_72029 | Not-determined |
| S16_extracted_72032 | Not-determined |
| S16_extracted_72035 | Low-quality |
| S16_extracted_72062 | Low-quality |
| S16_extracted_72063 | Not-determined |
| S16_extracted_72064 | Not-determined |
| S16_extracted_72074 | Not-determined |
| S16_extracted_72095 | Not-determined |
| S16_extracted_72121 | Not-determined |
| S16_extracted_72134 | Not-determined |
| S16_extracted_72146 | Not-determined |
| S16_extracted_72205 | Not-determined |
| S16_extracted_72220 | Not-determined |
| S16_extracted_72222 | Low-quality |
| S16_extracted_72225 | Not-determined |
| S16_extracted_72238 | Not-determined |
| S16_extracted_72261 | Low-quality |
| S16_extracted_72267 | Low-quality |
| S16_extracted_72277 | Not-determined |
| S16_extracted_72283 | Low-quality |
| S16_extracted_72286 | Low-quality |
| S16_extracted_72300 | Not-determined |
| S16_extracted_72308 | Low-quality |
| S16_extracted_72312 | Not-determined |
| S16_extracted_72318 | Low-quality |
| S16_extracted_72325 | Not-determined |
| S16_extracted_72341 | Not-determined |
| S16_extracted_72363 | Not-determined |
| S16_extracted_72386 | Not-determined |
| S16_extracted_72391 | Not-determined |
| S16_extracted_72433 | Low-quality |
| S16_extracted_72440 | Not-determined |
| S16_extracted_72450 | Not-determined |
| S16_extracted_72461 | Not-determined |
| S16_extracted_72465 | Not-determined |
| S16_extracted_72487 | Not-determined |
| S16_extracted_72493 | Not-determined |
| S16_extracted_72497 | Not-determined |
| S16_extracted_72521 | Not-determined |
| S16_extracted_72538 | Low-quality |
| S16_extracted_72547 | Not-determined |
| S16_extracted_72563 | Not-determined |
| S16_extracted_72576 | Not-determined |
| S16_extracted_72582 | Not-determined |
| S16_extracted_72611 | Not-determined |
| S16_extracted_72665 | Not-determined |
| S16_extracted_72667 | Low-quality |
| S16_extracted_72678 | Not-determined |
| S16_extracted_72691 | Low-quality |
| S16_extracted_72696 | Not-determined |
| S16_extracted_72701 | Not-determined |
| S16_extracted_72709 | Not-determined |
| S16_extracted_72712 | Not-determined |
| S16_extracted_72721 | Not-determined |
| S16_extracted_72728 | Not-determined |
| S16_extracted_72740 | Not-determined |
| S16_extracted_72743 | Not-determined |
| S16_extracted_72744 | Not-determined |
| S16_extracted_72770 | Not-determined |
| S16_extracted_72771 | Not-determined |
| S16_extracted_72796 | Not-determined |
| S16_extracted_72801 | Not-determined |
| S16_extracted_72812 | Not-determined |
| S16_extracted_72818 | Not-determined |
| S16_extracted_72822 | Not-determined |
| S16_extracted_72834 | Not-determined |
| S16_extracted_72848 | Not-determined |
| S16_extracted_72849 | Not-determined |
| S16_extracted_72889 | Not-determined |
| S16_extracted_72891 | Not-determined |
| S16_extracted_72920 | Not-determined |
| S16_extracted_72925 | Low-quality |
| S16_extracted_72940 | Low-quality |
| S16_extracted_72943 | Low-quality |
| S16_extracted_72951 | Low-quality |
| S16_extracted_72952 | Not-determined |
| S16_extracted_72956 | Not-determined |
| S16_extracted_72969 | Not-determined |
| S16_extracted_72971 | Not-determined |
| S16_extracted_72986 | Not-determined |
| S16_extracted_72987 | Low-quality |
| S16_extracted_73014 | Not-determined |
| S16_extracted_73022 | Low-quality |
| S16_extracted_73030 | Not-determined |
| S16_extracted_73046 | Not-determined |
| S16_extracted_73058 | Low-quality |
| S16_extracted_73062 | Not-determined |
| S16_extracted_73070 | Not-determined |
| S16_extracted_73098 | Not-determined |
| S16_extracted_73116 | Not-determined |
| S16_extracted_73151 | Not-determined |
| S16_extracted_73155 | Not-determined |
| S16_extracted_73174 | Not-determined |
| S16_extracted_73179 | Not-determined |
| S16_extracted_73188 | Not-determined |
| S16_extracted_73198 | Not-determined |
| S16_extracted_73205 | Not-determined |
| S16_extracted_73209 | Not-determined |
| S16_extracted_73214 | Not-determined |
| S16_extracted_73217 | Not-determined |
| S16_extracted_73224 | Not-determined |
| S16_extracted_73228 | Not-determined |
| S16_extracted_73232 | Not-determined |
| S16_extracted_73244 | Not-determined |
| S16_extracted_73247 | Not-determined |
| S16_extracted_73251 | Not-determined |
| S16_extracted_73252 | Not-determined |
| S16_extracted_73258 | Not-determined |
| S16_extracted_73264 | Not-determined |
| S16_extracted_73274 | Low-quality |
| S16_extracted_73277 | Not-determined |
| S16_extracted_73280 | Not-determined |
| S16_extracted_73290 | Not-determined |
| S16_extracted_73300 | Not-determined |
| S16_extracted_73307 | Not-determined |
| S16_extracted_73318 | Not-determined |
| S16_extracted_73329 | Not-determined |
| S16_extracted_73333 | Not-determined |
| S16_extracted_73385 | Not-determined |
| S16_extracted_73387 | Not-determined |
| S16_extracted_73392 | Low-quality |
| S16_extracted_73395 | Not-determined |
| S16_extracted_73397 | Not-determined |
| S16_extracted_73414 | Low-quality |
| S16_extracted_73429 | Not-determined |
| S16_extracted_73433 | Not-determined |
| S16_extracted_73458 | Not-determined |
| S16_extracted_73462 | Not-determined |
| S16_extracted_73470 | Not-determined |
| S16_extracted_73486 | Not-determined |
| S16_extracted_73495 | Not-determined |
| S16_extracted_73497 | Not-determined |
| S16_extracted_73508 | Not-determined |
| S16_extracted_73518 | Not-determined |
| S16_extracted_73522 | Not-determined |
| S16_extracted_73526 | Not-determined |
| S16_extracted_73538 | Not-determined |
| S16_extracted_73545 | Not-determined |
| S16_extracted_73546 | Not-determined |
| S16_extracted_73550 | Low-quality |
| S16_extracted_73555 | Not-determined |
| S16_extracted_73558 | Low-quality |
| S16_extracted_73566 | Not-determined |
| S16_extracted_73597 | Not-determined |
| S16_extracted_73618 | Low-quality |
| S16_extracted_73651 | Not-determined |
| S16_extracted_73654 | Not-determined |
| S16_extracted_73656 | Not-determined |
| S16_extracted_73674 | Not-determined |
| S16_extracted_73689 | Not-determined |
| S16_extracted_73694 | Not-determined |
| S16_extracted_73695 | Not-determined |
| S16_extracted_73714 | Not-determined |
| S16_extracted_73719 | Not-determined |
| S16_extracted_73731 | Not-determined |
| S16_extracted_73734 | Not-determined |
| S16_extracted_73739 | Not-determined |
| S16_extracted_73743 | Not-determined |
| S16_extracted_73747 | Not-determined |
| S16_extracted_73754 | Not-determined |
| S16_extracted_73756 | Not-determined |
| S16_extracted_73763 | Low-quality |
| S16_extracted_73774 | Not-determined |
| S16_extracted_73779 | Not-determined |
| S16_extracted_73781 | Not-determined |
| S16_extracted_73797 | Not-determined |
| S16_extracted_73805 | Not-determined |
| S16_extracted_73817 | Low-quality |
| S16_extracted_73854 | Not-determined |
| S16_extracted_73867 | Not-determined |
| S16_extracted_73884 | Not-determined |
| S16_extracted_73910 | Not-determined |
| S16_extracted_73918 | Not-determined |
| S16_extracted_73924 | Not-determined |
| S16_extracted_73944 | Not-determined |
| S16_extracted_73955 | Low-quality |
| S16_extracted_73956 | Not-determined |
| S16_extracted_73970 | Not-determined |
| S16_extracted_73996 | Low-quality |
| S16_extracted_74001 | Not-determined |
| S16_extracted_74011 | Low-quality |
| S16_extracted_74014 | Low-quality |
| S16_extracted_74019 | Not-determined |
| S16_extracted_74045 | Not-determined |
| S16_extracted_74071 | Not-determined |
| S16_extracted_74078 | Not-determined |
| S16_extracted_74085 | Not-determined |
| S16_extracted_74095 | Low-quality |
| S16_extracted_74097 | Not-determined |
| S16_extracted_74104 | Low-quality |
| S16_extracted_74105 | Not-determined |
| S16_extracted_74112 | Not-determined |
| S16_extracted_74118 | Not-determined |
| S16_extracted_74173 | Not-determined |
| S16_extracted_74176 | Low-quality |
| S16_extracted_74181 | Low-quality |
| S16_extracted_74190 | Not-determined |
| S16_extracted_74220 | Not-determined |
| S16_extracted_74225 | Not-determined |
| S16_extracted_74227 | Not-determined |
| S16_extracted_74228 | Not-determined |
| S16_extracted_74249 | Not-determined |
| S16_extracted_74266 | Not-determined |
| S16_extracted_74272 | Not-determined |
| S16_extracted_74276 | Not-determined |
| S16_extracted_74277 | Low-quality |
| S16_extracted_74280 | Not-determined |
| S16_extracted_74310 | Low-quality |
| S16_extracted_74324 | Not-determined |
| S16_extracted_74340 | Not-determined |
| S16_extracted_74342 | Not-determined |
| S16_extracted_74343 | Not-determined |
| S16_extracted_74352 | Not-determined |
| S16_extracted_74357 | Not-determined |
| S16_extracted_74363 | Not-determined |
| S16_extracted_74375 | Not-determined |
| S16_extracted_74376 | Not-determined |
| S16_extracted_74377 | Not-determined |
| S16_extracted_74383 | Not-determined |
| S16_extracted_74388 | Not-determined |
| S16_extracted_74398 | Not-determined |
| S16_extracted_74404 | Low-quality |
| S16_extracted_74406 | Low-quality |
| S16_extracted_74410 | Not-determined |
| S16_extracted_74432 | Low-quality |
| S16_extracted_74437 | Not-determined |
| S16_extracted_74442 | Not-determined |
| S16_extracted_74461 | Not-determined |
| S16_extracted_74464 | Not-determined |
| S16_extracted_74496 | Not-determined |
| S16_extracted_74501 | Not-determined |
| S16_extracted_74512 | Not-determined |
| S16_extracted_74533 | Not-determined |
| S16_extracted_74542 | Not-determined |
| S16_extracted_74556 | Not-determined |
| S16_extracted_74579 | Not-determined |
| S16_extracted_74600 | Low-quality |
| S16_extracted_74608 | Not-determined |
| S16_extracted_74615 | Not-determined |
| S16_extracted_74622 | Not-determined |
| S16_extracted_74624 | Low-quality |
| S16_extracted_74627 | Not-determined |
| S16_extracted_74635 | Not-determined |
| S16_extracted_74638 | Not-determined |
| S16_extracted_74665 | Not-determined |
| S16_extracted_74691 | Not-determined |
| S16_extracted_74716 | Not-determined |
| S16_extracted_74723 | Not-determined |
| S16_extracted_74728 | Not-determined |
| S16_extracted_74752 | Not-determined |
| S16_extracted_74762 | Not-determined |
| S16_extracted_74767 | Not-determined |
| S16_extracted_74771 | Low-quality |
| S16_extracted_74811 | Not-determined |
| S16_extracted_74818 | Not-determined |
| S16_extracted_74821 | Low-quality |
| S16_extracted_74828 | Not-determined |
| S16_extracted_74836 | Low-quality |
| S16_extracted_74852 | Not-determined |
| S16_extracted_74863 | Not-determined |
| S16_extracted_74866 | Not-determined |
| S16_extracted_74887 | Not-determined |
| S16_extracted_74891 | Not-determined |
| S16_extracted_74897 | Not-determined |
| S16_extracted_74904 | Not-determined |
| S16_extracted_74921 | Not-determined |
| S16_extracted_74929 | Not-determined |
| S16_extracted_74936 | Not-determined |
| S16_extracted_74937 | Not-determined |
| S16_extracted_74938 | Not-determined |
| S16_extracted_74954 | Not-determined |
| S16_extracted_74972 | Low-quality |
| S16_extracted_74977 | Not-determined |
| S16_extracted_74997 | Not-determined |
| S16_extracted_74999 | Not-determined |
| S16_extracted_75003 | Not-determined |
| S16_extracted_75011 | Not-determined |
| S16_extracted_75023 | Not-determined |
| S16_extracted_75026 | Not-determined |
| S16_extracted_75043 | Not-determined |
| S16_extracted_75057 | Not-determined |
| S16_extracted_75098 | Not-determined |
| S16_extracted_75101 | Low-quality |
| S16_extracted_75102 | Not-determined |
| S16_extracted_75117 | Not-determined |
| S16_extracted_75159 | Not-determined |
| S16_extracted_75160 | Not-determined |
| S16_extracted_75161 | Not-determined |
| S16_extracted_75188 | Not-determined |
| S16_extracted_75190 | Not-determined |
| S16_extracted_75195 | Low-quality |
| S16_extracted_75196 | Not-determined |
| S16_extracted_75197 | Not-determined |
| S16_extracted_75208 | Not-determined |
| S16_extracted_75240 | Not-determined |
| S16_extracted_75243 | Not-determined |
| S16_extracted_75253 | Not-determined |
| S16_extracted_75290 | Not-determined |
| S16_extracted_75297 | Not-determined |
| S16_extracted_75302 | Not-determined |
| S16_extracted_75303 | Low-quality |
| S16_extracted_75305 | Low-quality |
| S16_extracted_75318 | Not-determined |
| S16_extracted_75320 | Not-determined |
| S16_extracted_75323 | Low-quality |
| S16_extracted_75334 | Not-determined |
| S16_extracted_75335 | Low-quality |
| S16_extracted_75336 | Low-quality |
| S16_extracted_75355 | Not-determined |
| S16_extracted_75364 | Low-quality |
| S16_extracted_75377 | Not-determined |
| S16_extracted_75382 | Not-determined |
| S16_extracted_75387 | Not-determined |
| S16_extracted_75390 | Not-determined |
| S16_extracted_75393 | Not-determined |
| S16_extracted_75398 | Not-determined |
| S16_extracted_75400 | Not-determined |
| S16_extracted_75409 | Not-determined |
| S16_extracted_75438 | Not-determined |
| S16_extracted_75464 | Low-quality |
| S16_extracted_75475 | Not-determined |
| S16_extracted_75481 | Not-determined |
| S16_extracted_75482 | Not-determined |
| S16_extracted_75488 | Low-quality |
| S16_extracted_75496 | Not-determined |
| S16_extracted_75499 | Not-determined |
| S16_extracted_75505 | Not-determined |
| S16_extracted_75531 | Not-determined |
| S16_extracted_75572 | Not-determined |
| S16_extracted_75574 | Not-determined |
| S16_extracted_75591 | Not-determined |
| S16_extracted_75622 | Not-determined |
| S16_extracted_75629 | Not-determined |
| S16_extracted_75654 | Not-determined |
| S16_extracted_75665 | Not-determined |
| S16_extracted_75675 | Not-determined |
| S16_extracted_75679 | Not-determined |
| S16_extracted_75697 | Not-determined |
| S16_extracted_75700 | Not-determined |
| S16_extracted_75701 | Not-determined |
| S16_extracted_75705 | Not-determined |
| S16_extracted_75723 | Not-determined |
| S16_extracted_75734 | Not-determined |
| S16_extracted_75737 | Low-quality |
| S16_extracted_75750 | Not-determined |
| S16_extracted_75752 | Not-determined |
| S16_extracted_75756 | Low-quality |
| S16_extracted_75790 | Low-quality |
| S16_extracted_75797 | Not-determined |
| S16_extracted_75799 | Low-quality |
| S16_extracted_75806 | Not-determined |
| S16_extracted_75828 | Not-determined |
| S16_extracted_75833 | Not-determined |
| S16_extracted_75836 | Not-determined |
| S16_extracted_75846 | Not-determined |
| S16_extracted_75863 | Not-determined |
| S16_extracted_75874 | Not-determined |
| S16_extracted_75885 | Not-determined |
| S16_extracted_75894 | Not-determined |
| S16_extracted_75900 | Not-determined |
| S16_extracted_75910 | Low-quality |
| S16_extracted_75936 | Not-determined |
| S16_extracted_75956 | Not-determined |
| S16_extracted_75977 | Not-determined |
| S16_extracted_75998 | Not-determined |
| S16_extracted_76005 | Not-determined |
| S16_extracted_76006 | Not-determined |
| S16_extracted_76025 | Low-quality |
| S16_extracted_76027 | Not-determined |
| S16_extracted_76038 | Not-determined |
| S16_extracted_76057 | Not-determined |
| S16_extracted_76065 | Not-determined |
| S16_extracted_76089 | Not-determined |
| S16_extracted_76092 | Not-determined |
| S16_extracted_76102 | Not-determined |
| S16_extracted_76107 | Not-determined |
| S16_extracted_76112 | Not-determined |
| S16_extracted_76115 | Not-determined |
| S16_extracted_76137 | Not-determined |
| S16_extracted_76147 | Not-determined |
| S16_extracted_76158 | Low-quality |
| S16_extracted_76164 | Not-determined |
| S16_extracted_76172 | Not-determined |
| S16_extracted_76176 | Not-determined |
| S16_extracted_76193 | Not-determined |
| S16_extracted_76205 | Low-quality |
| S16_extracted_76218 | Not-determined |
| S16_extracted_76226 | Not-determined |
| S16_extracted_76242 | Not-determined |
| S16_extracted_76266 | Not-determined |
| S16_extracted_76274 | Not-determined |
| S16_extracted_76284 | Not-determined |
| S16_extracted_76287 | Not-determined |
| S16_extracted_76292 | Low-quality |
| S16_extracted_76293 | Not-determined |
| S16_extracted_76308 | Low-quality |
| S16_extracted_76309 | Not-determined |
| S16_extracted_76315 | Not-determined |
| S16_extracted_76352 | Not-determined |
| S16_extracted_76366 | Not-determined |
| S16_extracted_76371 | Low-quality |
| S16_extracted_76393 | Not-determined |
| S16_extracted_76402 | Not-determined |
| S16_extracted_76411 | Not-determined |
| S16_extracted_76420 | Not-determined |
| S16_extracted_76425 | Low-quality |
| S16_extracted_76429 | Not-determined |
| S16_extracted_76436 | Low-quality |
| S16_extracted_76450 | Not-determined |
| S16_extracted_76465 | Not-determined |
| S16_extracted_76469 | Not-determined |
| S16_extracted_76484 | Not-determined |
| S16_extracted_76496 | Not-determined |
| S16_extracted_76508 | Not-determined |
| S16_extracted_76512 | Not-determined |
| S16_extracted_76525 | Not-determined |
| S16_extracted_76534 | Not-determined |
| S16_extracted_76546 | Not-determined |
| S16_extracted_76549 | Not-determined |
| S16_extracted_76553 | Not-determined |
| S16_extracted_76581 | Not-determined |
| S16_extracted_76588 | Low-quality |
| S16_extracted_76598 | Not-determined |
| S16_extracted_76628 | Not-determined |
| S16_extracted_76637 | Not-determined |
| S16_extracted_76660 | Not-determined |
| S16_extracted_76706 | Not-determined |
| S16_extracted_76711 | Not-determined |
| S16_extracted_76744 | Low-quality |
| S16_extracted_76757 | Not-determined |
| S16_extracted_76771 | Low-quality |
| S16_extracted_76773 | Not-determined |
| S16_extracted_76779 | Not-determined |
| S16_extracted_76805 | Not-determined |
| S16_extracted_76811 | Not-determined |
| S16_extracted_76816 | Not-determined |
| S16_extracted_76824 | Not-determined |
| S16_extracted_76837 | Not-determined |
| S16_extracted_76848 | Low-quality |
| S16_extracted_76861 | Not-determined |
| S16_extracted_76880 | Not-determined |
| S16_extracted_76890 | Not-determined |
| S16_extracted_76906 | Not-determined |
| S16_extracted_76909 | Not-determined |
| S16_extracted_76922 | Not-determined |
| S16_extracted_76925 | Not-determined |
| S16_extracted_76935 | Not-determined |
| S16_extracted_76940 | Not-determined |
| S16_extracted_76952 | Not-determined |
| S16_extracted_76956 | Not-determined |
| S16_extracted_76960 | Not-determined |
| S16_extracted_76968 | Low-quality |
| S16_extracted_76986 | Not-determined |
| S16_extracted_77002 | Not-determined |
| S16_extracted_77012 | Not-determined |
| S16_extracted_77016 | Not-determined |
| S16_extracted_77019 | Not-determined |
| S16_extracted_77038 | Not-determined |
| S16_extracted_77042 | Not-determined |
| S16_extracted_77049 | Not-determined |
| S16_extracted_77081 | Not-determined |
| S16_extracted_77087 | Not-determined |
| S16_extracted_77093 | Low-quality |
| S16_extracted_77099 | Not-determined |
| S16_extracted_77109 | Not-determined |
| S16_extracted_77110 | Not-determined |
| S16_extracted_77138 | Not-determined |
| S16_extracted_77143 | Not-determined |
| S16_extracted_77160 | Not-determined |
| S16_extracted_77161 | Not-determined |
| S16_extracted_77166 | Not-determined |
| S16_extracted_77168 | Low-quality |
| S16_extracted_77177 | Low-quality |
| S16_extracted_77182 | Not-determined |
| S16_extracted_77192 | Not-determined |
| S16_extracted_77204 | Not-determined |
| S16_extracted_77208 | Not-determined |
| S16_extracted_77212 | Not-determined |
| S16_extracted_77217 | Low-quality |
| S16_extracted_77223 | Not-determined |
| S16_extracted_77226 | Not-determined |
| S16_extracted_77262 | Not-determined |
| S16_extracted_77267 | Not-determined |
| S16_extracted_77268 | Low-quality |
| S16_extracted_77269 | Not-determined |
| S16_extracted_77289 | Not-determined |
| S16_extracted_77298 | Not-determined |
| S16_extracted_77327 | Not-determined |

| | |
|---|---|
| S16_extracted_77329 | Not-determined |
| S16_extracted_77337 | Not-determined |
| S16_extracted_77347 | Not-determined |
| S16_extracted_77349 | Not-determined |
| S16_extracted_77355 | Low-quality |
| S16_extracted_77362 | Not-determined |
| S16_extracted_77368 | Not-determined |
| S16_extracted_77370 | Not-determined |
| S16_extracted_77376 | Not-determined |
| S16_extracted_77398 | Not-determined |
| S16_extracted_77435 | Not-determined |
| S16_extracted_77442 | Not-determined |
| S16_extracted_77447 | Not-determined |
| S16_extracted_77448 | Low-quality |
| S16_extracted_77453 | Not-determined |
| S16_extracted_77482 | Not-determined |
| S16_extracted_77496 | Not-determined |
| S16_extracted_77525 | Not-determined |
| S16_extracted_77528 | Not-determined |
| S16_extracted_77541 | Not-determined |
| S16_extracted_77542 | Not-determined |
| S16_extracted_77555 | Not-determined |
| S16_extracted_77581 | Not-determined |
| S16_extracted_77589 | Not-determined |
| S16_extracted_77593 | Not-determined |
| S16_extracted_77600 | Not-determined |
| S16_extracted_77614 | Not-determined |
| S16_extracted_77624 | Not-determined |
| S16_extracted_77626 | Not-determined |
| S16_extracted_77629 | Not-determined |
| S16_extracted_77637 | Not-determined |
| S16_extracted_77638 | Not-determined |
| S16_extracted_77646 | Low-quality |
| S16_extracted_77647 | Not-determined |
| S16_extracted_77658 | Not-determined |
| S16_extracted_77661 | Not-determined |
| S16_extracted_77673 | Not-determined |
| S16_extracted_77678 | Not-determined |
| S16_extracted_77680 | Not-determined |
| S16_extracted_77703 | Low-quality |
| S16_extracted_77713 | Not-determined |
| S16_extracted_77716 | Low-quality |
| S16_extracted_77721 | Not-determined |
| S16_extracted_77724 | Not-determined |
| S16_extracted_77751 | Not-determined |
| S16_extracted_77755 | Not-determined |
| S16_extracted_77778 | Not-determined |
| S16_extracted_77794 | Not-determined |
| S16_extracted_77803 | Not-determined |
| S16_extracted_77810 | Not-determined |
| S16_extracted_77815 | Not-determined |
| S16_extracted_77830 | Not-determined |
| S16_extracted_77833 | Not-determined |
| S16_extracted_77835 | Not-determined |
| S16_extracted_77840 | Not-determined |
| S16_extracted_77848 | Low-quality |
| S16_extracted_77856 | Not-determined |
| S16_extracted_77863 | Low-quality |
| S16_extracted_77864 | Low-quality |
| S16_extracted_77867 | Not-determined |
| S16_extracted_77896 | Low-quality |
| S16_extracted_77900 | Not-determined |
| S16_extracted_77909 | Not-determined |
| S16_extracted_77914 | Low-quality |
| S16_extracted_77929 | Not-determined |
| S16_extracted_77936 | Not-determined |
| S16_extracted_77939 | Not-determined |
| S16_extracted_77960 | Not-determined |
| S16_extracted_77966 | Not-determined |
| S16_extracted_77967 | Not-determined |
| S16_extracted_77974 | Not-determined |
| S16_extracted_77980 | Not-determined |
| S16_extracted_77981 | Not-determined |
| S16_extracted_77987 | Low-quality |
| S16_extracted_77989 | Not-determined |
| S16_extracted_77994 | Not-determined |
| S16_extracted_78006 | Not-determined |
| S16_extracted_78013 | Not-determined |
| S16_extracted_78021 | Not-determined |
| S16_extracted_78037 | Not-determined |
| S16_extracted_78039 | Not-determined |
| S16_extracted_78085 | Not-determined |
| S16_extracted_78119 | Not-determined |
| S16_extracted_78130 | Not-determined |
| S16_extracted_78138 | Not-determined |
| S16_extracted_78143 | Not-determined |
| S16_extracted_78157 | Not-determined |
| S16_extracted_78172 | Not-determined |
| S16_extracted_78217 | Not-determined |
| S16_extracted_78219 | Not-determined |
| S16_extracted_78230 | Not-determined |
| S16_extracted_78239 | Not-determined |
| S16_extracted_78240 | Not-determined |
| S16_extracted_78250 | Not-determined |
| S16_extracted_78264 | Not-determined |
| S16_extracted_78285 | Low-quality |
| S16_extracted_78288 | Not-determined |
| S16_extracted_78307 | Not-determined |
| S16_extracted_78318 | Not-determined |
| S16_extracted_78323 | Not-determined |
| S16_extracted_78327 | Low-quality |
| S16_extracted_78333 | Not-determined |
| S16_extracted_78339 | Low-quality |
| S16_extracted_78359 | Not-determined |
| S16_extracted_78379 | Not-determined |
| S16_extracted_78381 | Not-determined |
| S16_extracted_78386 | Not-determined |
| S16_extracted_78389 | Low-quality |
| S16_extracted_78391 | Not-determined |
| S16_extracted_78394 | Not-determined |
| S16_extracted_78402 | Not-determined |
| S16_extracted_78413 | Low-quality |
| S16_extracted_78425 | Not-determined |
| S16_extracted_78442 | Low-quality |
| S16_extracted_78443 | Not-determined |
| S16_extracted_78451 | Not-determined |
| S16_extracted_78467 | Not-determined |
| S16_extracted_78476 | Not-determined |
| S16_extracted_78480 | Not-determined |
| S16_extracted_78483 | Not-determined |
| S16_extracted_78501 | Not-determined |
| S16_extracted_78502 | Not-determined |
| S16_extracted_78507 | Not-determined |
| S16_extracted_78509 | Not-determined |
| S16_extracted_78513 | Not-determined |
| S16_extracted_78515 | Not-determined |
| S16_extracted_78520 | Not-determined |
| S16_extracted_78521 | Not-determined |
| S16_extracted_78530 | Not-determined |
| S16_extracted_78532 | Not-determined |
| S16_extracted_78535 | Not-determined |
| S16_extracted_78552 | Not-determined |
| S16_extracted_78556 | Not-determined |
| S16_extracted_78574 | Low-quality |
| S16_extracted_78600 | Not-determined |
| S16_extracted_78625 | Not-determined |
| S16_extracted_78638 | Not-determined |
| S16_extracted_78645 | Low-quality |
| S16_extracted_78691 | Not-determined |
| S16_extracted_78696 | Not-determined |
| S16_extracted_78724 | Not-determined |
| S16_extracted_78727 | Not-determined |
| S16_extracted_78752 | Not-determined |
| S16_extracted_78755 | Not-determined |
| S16_extracted_78772 | Not-determined |
| S16_extracted_78776 | Not-determined |
| S16_extracted_78785 | Not-determined |
| S16_extracted_78790 | Not-determined |
| S16_extracted_78795 | Not-determined |
| S16_extracted_78804 | Not-determined |
| S16_extracted_78818 | Not-determined |
| S16_extracted_78821 | Not-determined |
| S16_extracted_78827 | Not-determined |
| S16_extracted_78844 | Not-determined |
| S16_extracted_78848 | Not-determined |
| S16_extracted_78864 | Low-quality |
| S16_extracted_78867 | Not-determined |
| S16_extracted_78869 | Not-determined |
| S16_extracted_78870 | Not-determined |
| S16_extracted_78886 | Not-determined |
| S16_extracted_78890 | Not-determined |
| S16_extracted_78910 | Not-determined |
| S16_extracted_78933 | Not-determined |
| S16_extracted_78947 | Not-determined |
| S16_extracted_78956 | Not-determined |
| S16_extracted_78960 | Not-determined |
| S16_extracted_78967 | Low-quality |
| S16_extracted_78968 | Not-determined |
| S16_extracted_78977 | Not-determined |
| S16_extracted_78987 | Not-determined |
| S16_extracted_78998 | Not-determined |
| S16_extracted_79026 | Not-determined |
| S16_extracted_79054 | Not-determined |
| S16_extracted_79069 | Not-determined |
| S16_extracted_79085 | Low-quality |
| S16_extracted_79086 | Not-determined |
| S16_extracted_79099 | Low-quality |
| S16_extracted_79133 | Low-quality |
| S16_extracted_79155 | Not-determined |
| S16_extracted_79171 | Not-determined |
| S16_extracted_79189 | Not-determined |
| S16_extracted_79197 | Not-determined |
| S16_extracted_79207 | Not-determined |
| S16_extracted_79229 | Low-quality |
| S16_extracted_79255 | Not-determined |
| S16_extracted_79265 | Not-determined |
| S16_extracted_79275 | Low-quality |
| S16_extracted_79281 | Not-determined |
| S16_extracted_79282 | Not-determined |
| S16_extracted_79305 | Not-determined |
| S16_extracted_79313 | Not-determined |
| S16_extracted_79347 | Low-quality |
| S16_extracted_79358 | Not-determined |
| S16_extracted_79373 | Not-determined |
| S16_extracted_79376 | Not-determined |
| S16_extracted_79380 | Low-quality |
| S16_extracted_79390 | Low-quality |
| S16_extracted_79415 | Not-determined |
| S16_extracted_79418 | Not-determined |
| S16_extracted_79422 | Not-determined |
| S16_extracted_79426 | Not-determined |
| S16_extracted_79436 | Not-determined |
| S16_extracted_79446 | Not-determined |
| S16_extracted_79491 | Not-determined |
| S16_extracted_79498 | Not-determined |
| S16_extracted_79511 | Low-quality |
| S16_extracted_79514 | Not-determined |
| S16_extracted_79515 | Low-quality |
| S16_extracted_79569 | Not-determined |
| S16_extracted_79581 | Not-determined |
| S16_extracted_79598 | Not-determined |
| S16_extracted_79612 | Not-determined |
| S16_extracted_79615 | Not-determined |
| S16_extracted_79618 | Not-determined |
| S16_extracted_79625 | Low-quality |
| S16_extracted_79628 | Not-determined |
| S16_extracted_79654 | Not-determined |
| S16_extracted_79661 | Not-determined |
| S16_extracted_79663 | Low-quality |
| S16_extracted_79684 | Not-determined |
| S16_extracted_79694 | Not-determined |
| S16_extracted_79724 | Not-determined |
| S16_extracted_79728 | Not-determined |
| S16_extracted_79733 | Not-determined |
| S16_extracted_79738 | Not-determined |
| S16_extracted_79751 | Not-determined |
| S16_extracted_79770 | Not-determined |
| S16_extracted_79774 | Low-quality |
| S16_extracted_79776 | Not-determined |
| S16_extracted_79778 | Low-quality |
| S16_extracted_79802 | Low-quality |
| S16_extracted_79809 | Not-determined |
| S16_extracted_79826 | Low-quality |
| S16_extracted_79835 | Low-quality |
| S16_extracted_79836 | Not-determined |
| S16_extracted_79839 | Not-determined |
| S16_extracted_79853 | Low-quality |
| S16_extracted_79858 | Low-quality |
| S16_extracted_79873 | Not-determined |
| S16_extracted_79878 | Not-determined |
| S16_extracted_79889 | Not-determined |
| S16_extracted_79894 | Low-quality |
| S16_extracted_79895 | Not-determined |
| S16_extracted_79945 | Low-quality |
| S16_extracted_79948 | Not-determined |
| S16_extracted_79950 | Not-determined |
| S16_extracted_79955 | Not-determined |
| S16_extracted_79970 | Not-determined |
| S16_extracted_79994 | Low-quality |
| S16_extracted_80006 | Low-quality |
| S16_extracted_80016 | Not-determined |
| S16_extracted_80019 | Not-determined |
| S16_extracted_80027 | Not-determined |
| S16_extracted_80103 | Not-determined |
| S16_extracted_80123 | Not-determined |
| S16_extracted_80130 | Not-determined |
| S16_extracted_80132 | Not-determined |
| S16_extracted_80134 | Not-determined |
| S16_extracted_80152 | Not-determined |
| S16_extracted_80161 | Not-determined |
| S16_extracted_80163 | Not-determined |
| S16_extracted_80169 | Not-determined |
| S16_extracted_80171 | Not-determined |
| S16_extracted_80195 | Not-determined |
| S16_extracted_80207 | Not-determined |
| S16_extracted_80221 | Not-determined |
| S16_extracted_80222 | Not-determined |
| S16_extracted_80223 | Low-quality |
| S16_extracted_80242 | Not-determined |
| S16_extracted_80251 | Not-determined |
| S16_extracted_80254 | Low-quality |
| S16_extracted_80261 | Not-determined |
| S16_extracted_80276 | Not-determined |
| S16_extracted_80280 | Not-determined |
| S16_extracted_80299 | Not-determined |
| S16_extracted_80300 | Not-determined |
| S16_extracted_80302 | Low-quality |
| S16_extracted_80321 | Not-determined |
| S16_extracted_80322 | Not-determined |
| S16_extracted_80329 | Not-determined |
| S16_extracted_80330 | Not-determined |
| S16_extracted_80341 | Not-determined |
| S16_extracted_80353 | Not-determined |
| S16_extracted_80354 | Not-determined |
| S16_extracted_80369 | Not-determined |
| S16_extracted_80370 | Low-quality |
| S16_extracted_80371 | Not-determined |
| S16_extracted_80385 | Not-determined |
| S16_extracted_80386 | Not-determined |
| S16_extracted_80391 | Not-determined |
| S16_extracted_80394 | Not-determined |
| S16_extracted_80399 | Not-determined |
| S16_extracted_80428 | Not-determined |
| S16_extracted_80465 | Not-determined |
| S16_extracted_80466 | Not-determined |
| S16_extracted_80484 | Not-determined |
| S16_extracted_80487 | Not-determined |
| S16_extracted_80498 | Not-determined |
| S16_extracted_80503 | Not-determined |
| S16_extracted_80510 | Low-quality |
| S16_extracted_80512 | Low-quality |
| S16_extracted_80513 | Not-determined |
| S16_extracted_80519 | Not-determined |
| S16_extracted_80523 | Not-determined |
| S16_extracted_80526 | Low-quality |
| S16_extracted_80542 | Not-determined |
| S16_extracted_80547 | Not-determined |
| S16_extracted_80571 | Not-determined |
| S16_extracted_80573 | Not-determined |
| S16_extracted_80586 | Low-quality |
| S16_extracted_80595 | Not-determined |
| S16_extracted_80607 | Not-determined |
| S16_extracted_80613 | Not-determined |
| S16_extracted_80628 | Not-determined |
| S16_extracted_80629 | Not-determined |
| S16_extracted_80631 | Not-determined |
| S16_extracted_80662 | Low-quality |
| S16_extracted_80678 | Not-determined |
| S16_extracted_80680 | Not-determined |
| S16_extracted_80699 | Not-determined |
| S16_extracted_80711 | Not-determined |
| S16_extracted_80712 | Not-determined |
| S16_extracted_80719 | Low-quality |
| S16_extracted_80747 | Not-determined |
| S16_extracted_80754 | Not-determined |
| S16_extracted_80771 | Not-determined |
| S16_extracted_80775 | Not-determined |
| S16_extracted_80789 | Not-determined |
| S16_extracted_80802 | Not-determined |
| S16_extracted_80803 | Not-determined |
| S16_extracted_80815 | Not-determined |
| S16_extracted_80816 | Low-quality |
| S16_extracted_80844 | Not-determined |
| S16_extracted_80849 | Not-determined |
| S16_extracted_80858 | Low-quality |
| S16_extracted_80861 | Not-determined |
| S16_extracted_80896 | Not-determined |
| S16_extracted_80914 | Not-determined |
| S16_extracted_80926 | Not-determined |
| S16_extracted_80952 | Low-quality |
| S16_extracted_80954 | Not-determined |
| S16_extracted_80963 | Not-determined |
| S16_extracted_80974 | Not-determined |
| S16_extracted_80977 | Not-determined |
| S16_extracted_81009 | Not-determined |
| S16_extracted_81020 | Not-determined |
| S16_extracted_81037 | Not-determined |
| S16_extracted_81072 | Low-quality |
| S16_extracted_81075 | Low-quality |
| S16_extracted_81079 | Low-quality |
| S16_extracted_81080 | Low-quality |
| S16_extracted_81110 | Low-quality |
| S16_extracted_81126 | Not-determined |
| S16_extracted_81136 | Not-determined |
| S16_extracted_81142 | Not-determined |
| S16_extracted_81157 | Not-determined |
| S16_extracted_81167 | Not-determined |
| S16_extracted_81180 | Low-quality |
| S16_extracted_81181 | Not-determined |
| S16_extracted_81210 | Not-determined |
| S16_extracted_81217 | Not-determined |
| S16_extracted_81223 | Not-determined |
| S16_extracted_81225 | Not-determined |
| S16_extracted_81227 | Low-quality |
| S16_extracted_81228 | Not-determined |
| S16_extracted_81234 | Not-determined |
| S16_extracted_81250 | Not-determined |
| S16_extracted_81260 | Not-determined |
| S16_extracted_81270 | Not-determined |
| S16_extracted_81290 | Not-determined |
| S16_extracted_81301 | Not-determined |
| S16_extracted_81307 | Not-determined |
| S16_extracted_81313 | Not-determined |
| S16_extracted_81336 | Not-determined |
| S16_extracted_81341 | Not-determined |
| S16_extracted_81348 | Low-quality |
| S16_extracted_81357 | Not-determined |
| S16_extracted_81363 | Not-determined |
| S16_extracted_81366 | Not-determined |
| S16_extracted_81390 | Not-determined |
| S16_extracted_81393 | Not-determined |
| S16_extracted_81397 | Not-determined |
| S16_extracted_81418 | Not-determined |
| S16_extracted_81433 | Not-determined |
| S16_extracted_81448 | Not-determined |
| S16_extracted_81469 | Not-determined |
| S16_extracted_81472 | Not-determined |
| S16_extracted_81485 | Not-determined |
| S16_extracted_81494 | Not-determined |
| S16_extracted_81497 | Not-determined |
| S16_extracted_81513 | Not-determined |
| S16_extracted_81519 | Not-determined |
| S16_extracted_81523 | Not-determined |
| S16_extracted_81532 | Not-determined |
| S16_extracted_81535 | Not-determined |
| S16_extracted_81537 | Not-determined |
| S16_extracted_81547 | Not-determined |
| S16_extracted_81548 | Not-determined |
| S16_extracted_81551 | Low-quality |
| S16_extracted_81570 | Not-determined |
| S16_extracted_81580 | Not-determined |
| S16_extracted_81590 | Not-determined |
| S16_extracted_81596 | Not-determined |
| S16_extracted_81601 | Not-determined |
| S16_extracted_81606 | Not-determined |
| S16_extracted_81643 | Not-determined |
| S16_extracted_81651 | Not-determined |
| S16_extracted_81657 | Not-determined |
| S16_extracted_81676 | Not-determined |
| S16_extracted_81695 | Not-determined |
| S16_extracted_81696 | Not-determined |
| S16_extracted_81706 | Not-determined |
| S16_extracted_81715 | Not-determined |
| S16_extracted_81717 | Low-quality |
| S16_extracted_81718 | Not-determined |
| S16_extracted_81719 | Not-determined |
| S16_extracted_81720 | Not-determined |
| S16_extracted_81732 | Not-determined |
| S16_extracted_81738 | Low-quality |
| S16_extracted_81744 | Not-determined |
| S16_extracted_81748 | Not-determined |
| S16_extracted_81750 | Not-determined |
| S16_extracted_81758 | Not-determined |
| S16_extracted_81762 | Not-determined |
| S16_extracted_81764 | Not-determined |
| S16_extracted_81772 | Not-determined |
| S16_extracted_81775 | Not-determined |
| S16_extracted_81781 | Not-determined |
| S16_extracted_81794 | Not-determined |
| S16_extracted_81823 | Not-determined |
| S16_extracted_81846 | Not-determined |
| S16_extracted_81882 | Not-determined |
| S16_extracted_81883 | Low-quality |
| S16_extracted_81885 | Not-determined |
| S16_extracted_81886 | Not-determined |
| S16_extracted_81921 | Not-determined |
| S16_extracted_81934 | Not-determined |
| S16_extracted_81948 | Not-determined |
| S16_extracted_81953 | Not-determined |
| S16_extracted_81955 | Not-determined |
| S16_extracted_81968 | Not-determined |
| S16_extracted_81969 | Low-quality |
| S16_extracted_81984 | Not-determined |
| S16_extracted_81989 | Not-determined |
| S16_extracted_81999 | Not-determined |
| S16_extracted_82003 | Not-determined |
| S16_extracted_82010 | Not-determined |
| S16_extracted_82028 | Low-quality |
| S16_extracted_82034 | Not-determined |
| S16_extracted_82047 | Not-determined |
| S16_extracted_82050 | Not-determined |
| S16_extracted_82053 | Not-determined |
| S16_extracted_82079 | Low-quality |
| S16_extracted_82099 | Not-determined |
| S16_extracted_82103 | Not-determined |
| S16_extracted_82106 | Not-determined |
| S16_extracted_82110 | Not-determined |
| S16_extracted_82111 | Not-determined |
| S16_extracted_82120 | Not-determined |
| S16_extracted_82125 | Not-determined |
| S16_extracted_82130 | Not-determined |
| S16_extracted_82131 | Not-determined |
| S16_extracted_82135 | Not-determined |
| S16_extracted_82158 | Not-determined |
| S16_extracted_82161 | Not-determined |
| S16_extracted_82180 | Low-quality |
| S16_extracted_82181 | Not-determined |
| S16_extracted_82199 | Not-determined |
| S16_extracted_82201 | Low-quality |
| S16_extracted_82207 | Not-determined |
| S16_extracted_82214 | Not-determined |
| S16_extracted_82218 | Not-determined |
| S16_extracted_82219 | Not-determined |
| S16_extracted_82232 | Not-determined |
| S16_extracted_82233 | Low-quality |
| S16_extracted_82237 | Not-determined |
| S16_extracted_82245 | Not-determined |
| S16_extracted_82246 | Not-determined |
| S16_extracted_82280 | Not-determined |
| S16_extracted_82283 | Not-determined |
| S16_extracted_82296 | Not-determined |
| S16_extracted_82308 | Not-determined |
| S16_extracted_82311 | Not-determined |
| S16_extracted_82336 | Not-determined |
| S16_extracted_82338 | Not-determined |
| S16_extracted_82345 | Not-determined |
| S16_extracted_82365 | Low-quality |
| S16_extracted_82372 | Not-determined |
| S16_extracted_82379 | Not-determined |
| S16_extracted_82385 | Not-determined |
| S16_extracted_82393 | Low-quality |
| S16_extracted_82395 | Not-determined |
| S16_extracted_82396 | Not-determined |
| S16_extracted_82412 | Not-determined |
| S16_extracted_82420 | Low-quality |
| S16_extracted_82450 | Not-determined |
| S16_extracted_82469 | Not-determined |
| S16_extracted_82473 | Not-determined |
| S16_extracted_82479 | Not-determined |
| S16_extracted_82493 | Not-determined |
| S16_extracted_82496 | Not-determined |
| S16_extracted_82497 | Not-determined |
| S16_extracted_82572 | Not-determined |
| S16_extracted_82575 | Not-determined |
| S16_extracted_82583 | Not-determined |
| S16_extracted_82585 | Not-determined |
| S16_extracted_82593 | Not-determined |
| S16_extracted_82608 | Not-determined |
| S16_extracted_82612 | Not-determined |
| S16_extracted_82619 | Not-determined |
| S16_extracted_82629 | Not-determined |
| S16_extracted_82636 | Low-quality |
| S16_extracted_82652 | Not-determined |
| S16_extracted_82657 | Not-determined |
| S16_extracted_82668 | Not-determined |
| S16_extracted_82678 | Not-determined |
| S16_extracted_82695 | Not-determined |
| S16_extracted_82700 | Not-determined |
| S16_extracted_82702 | Not-determined |
| S16_extracted_82705 | Not-determined |
| S16_extracted_82730 | Not-determined |
| S16_extracted_82731 | Not-determined |
| S16_extracted_82757 | Not-determined |
| S16_extracted_82770 | Not-determined |
| S16_extracted_82781 | Not-determined |
| S16_extracted_82785 | Not-determined |
| S16_extracted_82804 | Not-determined |
| S16_extracted_82812 | Not-determined |
| S16_extracted_82813 | Not-determined |
| S16_extracted_82817 | Not-determined |
| S16_extracted_82833 | Not-determined |
| S16_extracted_82843 | Not-determined |
| S16_extracted_82862 | Low-quality |
| S16_extracted_82873 | Not-determined |
| S16_extracted_82879 | Not-determined |
| S16_extracted_82893 | Not-determined |
| S16_extracted_82897 | Not-determined |
| S16_extracted_82903 | Not-determined |
| S16_extracted_82932 | Low-quality |
| S16_extracted_82933 | Not-determined |
| S16_extracted_82936 | Not-determined |
| S16_extracted_82943 | Not-determined |
| S16_extracted_82950 | Not-determined |
| S16_extracted_82955 | Not-determined |
| S16_extracted_82961 | Not-determined |
| S16_extracted_82976 | Not-determined |
| S16_extracted_82977 | Not-determined |
| S16_extracted_82978 | Not-determined |
| S16_extracted_82980 | Not-determined |
| S16_extracted_82990 | Not-determined |
| S16_extracted_82991 | Not-determined |
| S16_extracted_83006 | Not-determined |
| S16_extracted_83013 | Not-determined |
| S16_extracted_83020 | Not-determined |
| S16_extracted_83023 | Not-determined |
| S16_extracted_83024 | Low-quality |
| S16_extracted_83060 | Not-determined |
| S16_extracted_83062 | Not-determined |
| S16_extracted_83067 | Not-determined |
| S16_extracted_83087 | Not-determined |
| S16_extracted_83104 | Not-determined |
| S16_extracted_83118 | Not-determined |
| S16_extracted_83139 | Not-determined |
| S16_extracted_83144 | Low-quality |
| S16_extracted_83163 | Not-determined |
| S16_extracted_83187 | Not-determined |
| S16_extracted_83206 | Low-quality |
| S16_extracted_83214 | Not-determined |
| S16_extracted_83222 | Not-determined |
| S16_extracted_83227 | Not-determined |
| S16_extracted_83243 | Not-determined |
| S16_extracted_83244 | Not-determined |
| S16_extracted_83250 | Not-determined |
| S16_extracted_83257 | Not-determined |
| S16_extracted_83285 | Not-determined |
| S16_extracted_83307 | Not-determined |
| S16_extracted_83341 | Not-determined |
| S16_extracted_83354 | Not-determined |
| S16_extracted_83368 | Not-determined |
| S16_extracted_83371 | Not-determined |
| S16_extracted_83379 | Not-determined |
| S16_extracted_83401 | Low-quality |
| S16_extracted_83416 | Not-determined |
| S16_extracted_83426 | Not-determined |
| S16_extracted_83429 | Not-determined |
| S16_extracted_83432 | Not-determined |
| S16_extracted_83444 | Not-determined |
| S16_extracted_83478 | Not-determined |
| S16_extracted_83489 | Not-determined |

| | |
|---|---|
| S16_extracted_83494 | Not-determined |
| S16_extracted_83503 | Low-quality |
| S16_extracted_83505 | Not-determined |
| S16_extracted_83521 | Not-determined |
| S16_extracted_83524 | Not-determined |
| S16_extracted_83525 | Not-determined |
| S16_extracted_83535 | Not-determined |
| S16_extracted_83547 | Not-determined |
| S16_extracted_83550 | Not-determined |
| S16_extracted_83552 | Not-determined |
| S16_extracted_83557 | Not-determined |
| S16_extracted_83568 | Not-determined |
| S16_extracted_83580 | Not-determined |
| S16_extracted_83591 | Not-determined |
| S16_extracted_83594 | Not-determined |
| S16_extracted_83606 | Not-determined |
| S16_extracted_83624 | Low-quality |
| S16_extracted_83659 | Not-determined |
| S16_extracted_83663 | Low-quality |
| S16_extracted_83669 | Not-determined |
| S16_extracted_83675 | Not-determined |
| S16_extracted_83691 | Not-determined |
| S16_extracted_83709 | Not-determined |
| S16_extracted_83718 | Low-quality |
| S16_extracted_83719 | Not-determined |
| S16_extracted_83760 | Not-determined |
| S16_extracted_83761 | Not-determined |
| S16_extracted_83765 | Not-determined |
| S16_extracted_83769 | Not-determined |
| S16_extracted_83789 | Not-determined |
| S16_extracted_83797 | Not-determined |
| S16_extracted_83809 | Not-determined |
| S16_extracted_83812 | Not-determined |
| S16_extracted_83834 | Not-determined |
| S16_extracted_83852 | Not-determined |
| S16_extracted_83855 | Not-determined |
| S16_extracted_83857 | Not-determined |
| S16_extracted_83865 | Not-determined |
| S16_extracted_83869 | Low-quality |
| S16_extracted_83872 | Not-determined |
| S16_extracted_83874 | Low-quality |
| S16_extracted_83915 | Not-determined |
| S16_extracted_83918 | Not-determined |
| S16_extracted_83927 | Not-determined |
| S16_extracted_83929 | Not-determined |
| S16_extracted_83938 | Low-quality |
| S16_extracted_83949 | Not-determined |
| S16_extracted_83952 | Low-quality |
| S16_extracted_83970 | Not-determined |
| S16_extracted_83974 | Not-determined |
| S16_extracted_83975 | Low-quality |
| S16_extracted_83983 | Not-determined |
| S16_extracted_83985 | Not-determined |
| S16_extracted_83997 | Not-determined |
| S16_extracted_84001 | Not-determined |
| S16_extracted_84008 | Not-determined |
| S16_extracted_84023 | Low-quality |
| S16_extracted_84036 | Not-determined |
| S16_extracted_84053 | Not-determined |
| S16_extracted_84064 | Low-quality |
| S16_extracted_84075 | Not-determined |
| S16_extracted_84078 | Not-determined |
| S16_extracted_84083 | Not-determined |
| S16_extracted_84086 | Not-determined |
| S16_extracted_84090 | Not-determined |
| S16_extracted_84095 | Not-determined |
| S16_extracted_84098 | Not-determined |
| S16_extracted_84105 | Not-determined |
| S16_extracted_84114 | Not-determined |
| S16_extracted_84154 | Not-determined |
| S16_extracted_84175 | Not-determined |
| S16_extracted_84179 | Not-determined |
| S16_extracted_84194 | Low-quality |
| S16_extracted_84216 | Low-quality |
| S16_extracted_84229 | Not-determined |
| S16_extracted_84257 | Not-determined |
| S16_extracted_84265 | Not-determined |
| S16_extracted_84268 | Low-quality |
| S16_extracted_84270 | Not-determined |
| S16_extracted_84275 | Not-determined |
| S16_extracted_84283 | Not-determined |
| S16_extracted_84286 | Not-determined |
| S16_extracted_84292 | Not-determined |
| S16_extracted_84298 | Not-determined |
| S16_extracted_84311 | Not-determined |
| S16_extracted_84312 | Not-determined |
| S16_extracted_84323 | Low-quality |
| S16_extracted_84335 | Not-determined |
| S16_extracted_84340 | Not-determined |
| S16_extracted_84358 | Not-determined |
| S16_extracted_84359 | Low-quality |
| S16_extracted_84363 | Not-determined |
| S16_extracted_84364 | Not-determined |
| S16_extracted_84367 | Not-determined |
| S16_extracted_84369 | Not-determined |
| S16_extracted_84371 | Not-determined |
| S16_extracted_84375 | Not-determined |
| S16_extracted_84409 | Low-quality |
| S16_extracted_84432 | Not-determined |
| S16_extracted_84451 | Not-determined |
| S16_extracted_84457 | Not-determined |
| S3_vs_k141_127410\|\|full | High-quality |
| S16_extracted_84468 | Not-determined |
| S16_extracted_84472 | Low-quality |
| S16_extracted_84477 | Not-determined |
| S16_extracted_84490 | Not-determined |
| S16_extracted_84499 | Not-determined |
| S16_extracted_84507 | Not-determined |
| S16_extracted_84522 | Not-determined |
| S16_extracted_84528 | Not-determined |
| S16_extracted_84554 | Not-determined |
| S16_extracted_84564 | Low-quality |
| S16_extracted_84566 | Not-determined |
| S16_extracted_84583 | Not-determined |
| S16_extracted_84601 | Low-quality |
| S16_extracted_84613 | Not-determined |
| S16_extracted_84624 | Low-quality |
| S16_extracted_84629 | Not-determined |
| S16_extracted_84638 | Not-determined |
| S16_extracted_84660 | Not-determined |
| S16_extracted_84662 | Not-determined |
| S16_extracted_84664 | Not-determined |
| S16_extracted_84667 | Not-determined |
| S16_extracted_84671 | Not-determined |
| S16_extracted_84690 | Not-determined |
| S16_extracted_84692 | Not-determined |
| S16_extracted_84708 | Not-determined |
| S16_extracted_84724 | Not-determined |
| S16_extracted_84731 | Not-determined |
| S16_extracted_84733 | Not-determined |
| S16_extracted_84734 | Not-determined |
| S16_extracted_84746 | Not-determined |
| S16_extracted_84779 | Not-determined |
| S16_extracted_84786 | Not-determined |
| S16_extracted_84807 | Not-determined |
| S16_extracted_84813 | Low-quality |
| S16_extracted_84814 | Not-determined |
| S16_extracted_84835 | Not-determined |
| S16_extracted_84836 | Not-determined |
| S16_extracted_84863 | Not-determined |
| S16_extracted_84878 | Not-determined |
| S16_extracted_84886 | Not-determined |
| S16_extracted_84896 | Not-determined |
| S16_extracted_84914 | Low-quality |
| S16_extracted_84916 | Not-determined |
| S16_extracted_84933 | Not-determined |
| S16_extracted_84959 | Not-determined |
| S16_extracted_84993 | Not-determined |
| S16_extracted_84994 | Not-determined |
| S16_extracted_85009 | Not-determined |
| S16_extracted_85024 | Not-determined |
| S16_extracted_85032 | Not-determined |
| S16_extracted_85038 | Not-determined |
| S16_extracted_85059 | Not-determined |
| S16_extracted_85061 | Not-determined |
| S16_extracted_85066 | Not-determined |
| S16_extracted_85068 | Not-determined |
| S16_extracted_85080 | Not-determined |
| S16_extracted_85094 | Not-determined |
| S16_extracted_85105 | Low-quality |
| S16_extracted_85116 | Not-determined |
| S16_extracted_85147 | Not-determined |
| S16_extracted_85148 | Not-determined |
| S16_extracted_85150 | Not-determined |
| S16_extracted_85162 | Not-determined |
| S16_extracted_85165 | Not-determined |
| S16_extracted_85174 | Not-determined |
| S16_extracted_85188 | Not-determined |
| S16_extracted_85203 | Not-determined |
| S16_extracted_85214 | Not-determined |
| S16_extracted_85229 | Not-determined |
| S16_extracted_85231 | Low-quality |
| S16_extracted_85244 | Not-determined |
| S16_extracted_85253 | Not-determined |
| S16_extracted_85257 | Not-determined |
| S16_extracted_85262 | Low-quality |
| S16_extracted_85272 | Not-determined |
| S16_extracted_85278 | Not-determined |
| S16_extracted_85281 | Not-determined |
| S16_extracted_85288 | Not-determined |
| S16_extracted_85289 | Not-determined |
| S16_extracted_85297 | Not-determined |
| S16_extracted_85298 | Not-determined |
| S16_extracted_85302 | Not-determined |
| S16_extracted_85313 | Not-determined |
| S16_extracted_85325 | Not-determined |
| S16_extracted_85349 | Not-determined |
| S16_extracted_85354 | Not-determined |
| S16_extracted_85355 | Low-quality |
| S16_extracted_85397 | Not-determined |
| S16_extracted_85412 | Not-determined |
| S16_extracted_85425 | Not-determined |
| S16_extracted_85428 | Not-determined |
| S16_extracted_85431 | Not-determined |
| S16_extracted_85445 | Low-quality |
| S16_extracted_85452 | Not-determined |
| S16_extracted_85460 | Not-determined |
| S16_extracted_85462 | Not-determined |
| S16_extracted_85464 | Not-determined |
| S16_extracted_85465 | Not-determined |
| S16_extracted_85469 | Not-determined |
| S16_extracted_85491 | Not-determined |
| S16_extracted_85494 | Not-determined |
| S16_extracted_85507 | Not-determined |
| S16_extracted_85512 | Not-determined |
| S16_extracted_85527 | Low-quality |
| S16_extracted_85547 | Low-quality |
| S16_extracted_85573 | Not-determined |
| S16_extracted_85579 | Not-determined |
| S16_extracted_85580 | Not-determined |
| S16_extracted_85587 | Not-determined |
| S16_extracted_85594 | Low-quality |
| S16_extracted_85595 | Low-quality |
| S16_extracted_85609 | Not-determined |
| S16_extracted_85631 | Not-determined |
| S16_extracted_85636 | Not-determined |
| S16_extracted_85638 | Not-determined |
| S16_extracted_85648 | Not-determined |
| S16_extracted_85666 | Not-determined |
| S16_extracted_85696 | Not-determined |
| S16_extracted_85720 | Not-determined |
| S16_extracted_85722 | Not-determined |
| S16_extracted_85735 | Not-determined |
| S16_extracted_85738 | Not-determined |
| S16_extracted_85748 | Not-determined |
| S16_extracted_85760 | Not-determined |
| S16_extracted_85765 | Not-determined |
| S16_extracted_85771 | Not-determined |
| S16_extracted_85777 | Low-quality |
| S16_extracted_85802 | Not-determined |
| S16_extracted_85804 | Low-quality |
| S16_extracted_85814 | Not-determined |
| S16_extracted_85815 | Not-determined |
| S16_extracted_85816 | Low-quality |
| S16_extracted_85818 | Not-determined |
| S16_extracted_85822 | Not-determined |
| S16_extracted_85825 | Not-determined |
| S16_extracted_85827 | Not-determined |
| S16_extracted_85854 | Not-determined |
| S16_extracted_85857 | Not-determined |
| S16_extracted_85864 | Not-determined |
| S16_extracted_85870 | Low-quality |
| S16_extracted_85892 | Not-determined |
| S16_extracted_85935 | Not-determined |
| S16_extracted_85943 | Not-determined |
| S16_extracted_85947 | Not-determined |
| S16_extracted_85955 | Not-determined |
| S16_extracted_85965 | Low-quality |
| S16_extracted_85971 | Not-determined |
| S16_extracted_85975 | Not-determined |
| S16_extracted_85982 | Not-determined |
| S16_extracted_85985 | Not-determined |
| S16_extracted_86035 | Not-determined |
| S16_extracted_86047 | Not-determined |
| S16_extracted_86065 | Low-quality |
| S16_extracted_86079 | Low-quality |
| S16_extracted_86083 | Not-determined |
| S16_extracted_86108 | Not-determined |
| S16_extracted_86128 | Not-determined |
| S16_extracted_86129 | Not-determined |
| S16_extracted_86135 | Not-determined |
| S16_extracted_86141 | Not-determined |
| S16_extracted_86145 | Not-determined |
| S16_extracted_86146 | Not-determined |
| S16_extracted_86187 | Not-determined |
| S16_extracted_86194 | Not-determined |
| S16_extracted_86202 | Not-determined |
| S16_extracted_86206 | Not-determined |
| S16_extracted_86211 | Not-determined |
| S16_extracted_86213 | Not-determined |
| S16_extracted_86219 | Low-quality |
| S16_extracted_86227 | Not-determined |
| S16_extracted_86233 | Not-determined |
| S16_extracted_86242 | Not-determined |
| S16_extracted_86248 | Not-determined |
| S16_extracted_86261 | Not-determined |
| S16_extracted_86272 | Not-determined |
| S16_extracted_86276 | Not-determined |
| S16_extracted_86294 | Low-quality |
| S16_extracted_86296 | Not-determined |
| S16_extracted_86306 | Not-determined |
| S16_extracted_86315 | Not-determined |
| S16_extracted_86320 | Not-determined |
| S16_extracted_86349 | Not-determined |
| S16_extracted_86354 | Not-determined |
| S16_extracted_86358 | Not-determined |
| S16_extracted_86360 | Not-determined |
| S16_extracted_86367 | Not-determined |
| S16_extracted_86376 | Not-determined |
| S16_extracted_86379 | Not-determined |
| S16_extracted_86381 | Not-determined |
| S16_extracted_86384 | Not-determined |
| S16_extracted_86394 | Not-determined |
| S16_extracted_86406 | Not-determined |
| S16_extracted_86425 | Not-determined |
| S16_extracted_86428 | Not-determined |
| S16_extracted_86436 | Not-determined |
| S16_extracted_86438 | Not-determined |
| S16_extracted_86446 | Not-determined |
| S16_extracted_86474 | Not-determined |
| S16_extracted_86478 | Not-determined |
| S16_extracted_86490 | Not-determined |
| S16_extracted_86493 | Not-determined |
| S16_extracted_86503 | Low-quality |
| S16_extracted_86510 | Not-determined |
| S16_extracted_86512 | Low-quality |
| S16_extracted_86515 | Not-determined |
| S16_extracted_86517 | Not-determined |
| S16_extracted_86537 | Not-determined |
| S16_extracted_86554 | Not-determined |
| S16_extracted_86560 | Not-determined |
| S16_extracted_86570 | Not-determined |
| S16_extracted_86572 | Not-determined |
| S16_extracted_86592 | Not-determined |
| S16_extracted_86606 | Not-determined |
| S16_extracted_86615 | Not-determined |
| S16_extracted_86625 | Not-determined |
| S16_extracted_86636 | Not-determined |
| S16_extracted_86640 | Not-determined |
| S16_extracted_86645 | Not-determined |
| S16_extracted_86646 | Not-determined |
| S16_extracted_86649 | Not-determined |
| S16_extracted_86655 | Not-determined |
| S16_extracted_86657 | Not-determined |
| S16_extracted_86680 | Not-determined |
| S16_extracted_86684 | Not-determined |
| S16_extracted_86687 | Not-determined |
| S16_extracted_86702 | Not-determined |
| S16_extracted_86718 | Not-determined |
| S16_extracted_86728 | Not-determined |
| S16_extracted_86730 | Not-determined |
| S16_extracted_86740 | Low-quality |
| S16_extracted_86744 | Not-determined |
| S16_extracted_86759 | Low-quality |
| S16_extracted_86774 | Not-determined |
| S16_extracted_86775 | Low-quality |
| S16_extracted_86797 | Not-determined |
| S16_extracted_86809 | Not-determined |
| S16_extracted_86811 | Low-quality |
| S16_extracted_86826 | Not-determined |
| S16_extracted_86828 | Low-quality |
| S16_extracted_86830 | Not-determined |
| S16_extracted_86862 | Low-quality |
| S16_extracted_86882 | Not-determined |
| S16_extracted_86891 | Not-determined |
| S16_extracted_86898 | Not-determined |
| S16_extracted_86908 | Not-determined |
| S16_extracted_86911 | Low-quality |
| S16_extracted_86913 | Not-determined |
| S16_extracted_86920 | Not-determined |
| S16_extracted_86928 | Not-determined |
| S16_extracted_86931 | Not-determined |
| S16_extracted_86935 | Low-quality |
| S16_extracted_86953 | Not-determined |
| S16_extracted_86969 | Not-determined |
| S16_extracted_86975 | Low-quality |
| S16_extracted_86977 | Not-determined |
| S16_extracted_86986 | Not-determined |
| S16_extracted_86993 | Not-determined |
| S16_extracted_86998 | Low-quality |
| S16_extracted_87002 | Low-quality |
| S16_extracted_87009 | Not-determined |
| S16_extracted_87012 | Not-determined |
| S16_extracted_87038 | Not-determined |
| S16_extracted_87045 | Low-quality |
| S16_extracted_87068 | Not-determined |
| S16_extracted_87076 | Not-determined |
| S16_extracted_87077 | Not-determined |
| S16_extracted_87087 | Low-quality |
| S16_extracted_87090 | Low-quality |
| S16_extracted_87094 | Not-determined |
| S16_extracted_87099 | Not-determined |
| S16_extracted_87112 | Not-determined |
| S16_extracted_87121 | Not-determined |
| S16_extracted_87125 | Not-determined |
| S16_extracted_87141 | Not-determined |
| S16_extracted_87147 | Low-quality |
| S16_extracted_87150 | Not-determined |
| S16_extracted_87158 | Not-determined |
| S16_extracted_87165 | Not-determined |
| S16_extracted_87167 | Not-determined |
| S16_extracted_87183 | Not-determined |
| S16_extracted_87189 | Not-determined |
| S7_vs_109519\|\|full | High-quality |
| S16_extracted_87210 | Not-determined |
| S16_extracted_87218 | Not-determined |
| S16_extracted_87220 | Not-determined |
| S16_extracted_87224 | Not-determined |
| S16_extracted_87232 | Not-determined |
| S16_extracted_87254 | Not-determined |
| S16_extracted_87259 | Not-determined |
| S16_extracted_87261 | Not-determined |
| S16_extracted_87277 | Low-quality |
| S16_extracted_87281 | Not-determined |
| S16_extracted_87284 | Not-determined |
| S16_extracted_87287 | Not-determined |
| S16_extracted_87300 | Not-determined |
| S16_extracted_87313 | Not-determined |
| S16_extracted_87321 | Not-determined |
| S16_extracted_87325 | Not-determined |
| S16_extracted_87341 | Not-determined |
| S16_extracted_87344 | Not-determined |
| S16_extracted_87349 | Not-determined |
| S16_extracted_87357 | Not-determined |
| S16_extracted_87430 | Not-determined |
| S16_extracted_87442 | Low-quality |
| S16_extracted_87444 | Not-determined |
| S16_extracted_87465 | Not-determined |
| S16_extracted_87469 | Not-determined |
| S16_extracted_87477 | Not-determined |
| S16_extracted_87487 | Not-determined |
| S16_extracted_87492 | Not-determined |
| S16_extracted_87510 | Not-determined |
| S16_extracted_87529 | Not-determined |
| S16_extracted_87534 | Not-determined |
| S16_extracted_87558 | Not-determined |
| S16_extracted_87565 | Low-quality |
| S16_extracted_87573 | Not-determined |
| S16_extracted_87580 | Not-determined |
| S16_extracted_87586 | Not-determined |
| S16_extracted_87631 | Not-determined |
| S16_extracted_87651 | Not-determined |
| S16_extracted_87665 | Not-determined |
| S16_extracted_87669 | Not-determined |
| S16_extracted_87673 | Not-determined |
| S16_extracted_87675 | Not-determined |
| S16_extracted_87704 | Not-determined |
| S16_extracted_87713 | Low-quality |
| S16_extracted_87719 | Not-determined |
| S16_extracted_87738 | Not-determined |
| S16_extracted_87744 | Not-determined |
| S16_extracted_87745 | Low-quality |
| S16_extracted_87754 | Not-determined |
| S16_extracted_87755 | Not-determined |
| S16_extracted_87760 | Not-determined |
| S16_extracted_87795 | Not-determined |
| S16_extracted_87800 | Not-determined |
| S16_extracted_87803 | Not-determined |
| S16_extracted_87813 | Not-determined |
| S16_extracted_87838 | Not-determined |
| S16_extracted_87842 | Not-determined |
| S16_extracted_87865 | Not-determined |
| S16_extracted_87910 | Not-determined |
| S16_extracted_87913 | Not-determined |
| S16_extracted_87922 | Low-quality |
| S16_extracted_87935 | Not-determined |
| S16_extracted_87957 | Not-determined |
| S16_extracted_87969 | Not-determined |
| S16_extracted_87970 | Not-determined |
| S16_extracted_87975 | Low-quality |
| S16_extracted_87979 | Not-determined |
| S16_extracted_88011 | Not-determined |
| S16_extracted_88017 | Low-quality |
| S16_extracted_88018 | Not-determined |
| S16_extracted_88029 | Not-determined |
| S16_extracted_88050 | Not-determined |
| S16_extracted_88058 | Not-determined |
| S16_extracted_88062 | Not-determined |
| S16_extracted_88075 | Low-quality |
| S16_extracted_88081 | Not-determined |
| S16_extracted_88084 | Not-determined |
| S16_extracted_88090 | Not-determined |
| S16_extracted_88104 | Low-quality |
| S16_extracted_88117 | Not-determined |
| S16_extracted_88130 | Not-determined |
| S16_extracted_88133 | Not-determined |
| S16_extracted_88134 | Not-determined |
| S16_extracted_88135 | Low-quality |
| S16_extracted_88170 | Not-determined |
| S16_extracted_88187 | Not-determined |
| S16_extracted_88214 | Not-determined |
| S16_extracted_88224 | Not-determined |
| S16_extracted_88243 | Low-quality |
| S16_extracted_88251 | Not-determined |
| S16_extracted_88260 | Not-determined |
| S16_extracted_88262 | Not-determined |
| S16_extracted_88263 | Not-determined |
| S16_extracted_88270 | Not-determined |
| S16_extracted_88272 | Not-determined |
| S16_extracted_88281 | Not-determined |
| S16_extracted_88303 | Not-determined |
| S16_extracted_88324 | Not-determined |
| S16_extracted_88325 | Not-determined |
| S16_extracted_88329 | Not-determined |
| S16_extracted_88337 | Not-determined |
| S16_extracted_88365 | Not-determined |
| S16_extracted_88367 | Low-quality |
| S16_extracted_88369 | Not-determined |
| S16_extracted_88390 | Not-determined |
| S16_extracted_88409 | Low-quality |
| S16_extracted_88427 | Not-determined |
| S16_extracted_88429 | Not-determined |
| S16_extracted_88435 | Not-determined |
| S16_extracted_88445 | Not-determined |
| S16_extracted_88447 | Not-determined |
| S16_extracted_88457 | Low-quality |
| S16_extracted_88478 | Not-determined |
| S16_extracted_88485 | Not-determined |
| S16_extracted_88494 | Not-determined |
| S16_extracted_88506 | Not-determined |
| S16_extracted_88510 | Not-determined |
| S16_extracted_88524 | Not-determined |
| S16_extracted_88528 | Not-determined |
| S16_extracted_88532 | Not-determined |
| S16_extracted_88541 | Not-determined |
| S16_extracted_88547 | Not-determined |
| S16_extracted_88549 | Not-determined |
| S16_extracted_88550 | Not-determined |
| S16_extracted_88568 | Not-determined |
| S16_extracted_88572 | Low-quality |
| S16_extracted_88581 | Not-determined |
| S16_extracted_88594 | Not-determined |
| S16_extracted_88603 | Not-determined |
| S16_extracted_88616 | Not-determined |
| S16_extracted_88621 | Not-determined |
| S16_extracted_88625 | Not-determined |
| S16_extracted_88657 | Not-determined |
| S16_extracted_88674 | Not-determined |
| S16_extracted_88678 | Not-determined |
| S16_extracted_88683 | Not-determined |
| S16_extracted_88705 | Not-determined |
| S16_extracted_88721 | Not-determined |
| S16_extracted_88751 | Not-determined |
| S16_extracted_88761 | Not-determined |
| S16_extracted_88784 | Not-determined |
| S16_extracted_88806 | Not-determined |
| S16_extracted_88808 | Not-determined |
| S16_extracted_88815 | Not-determined |
| S16_extracted_88827 | Not-determined |
| S16_extracted_88840 | Not-determined |
| S16_extracted_88849 | Not-determined |
| S16_extracted_88851 | Not-determined |
| S16_extracted_88853 | Not-determined |
| S16_extracted_88857 | Not-determined |
| S16_extracted_88868 | Not-determined |
| S16_extracted_88876 | Not-determined |
| S16_extracted_88905 | Low-quality |
| S16_extracted_88915 | Low-quality |
| S16_extracted_88933 | Not-determined |
| S16_extracted_88952 | Not-determined |
| S16_extracted_88954 | Not-determined |
| S16_extracted_88973 | Not-determined |
| S16_extracted_89005 | Not-determined |
| S16_extracted_89006 | Not-determined |
| S16_extracted_89014 | Not-determined |
| S16_extracted_89021 | Not-determined |
| S16_extracted_89058 | Not-determined |
| S16_extracted_89065 | Not-determined |
| S16_extracted_89067 | Low-quality |
| S16_extracted_89087 | Not-determined |
| S16_extracted_89094 | Not-determined |
| S16_extracted_89104 | Not-determined |
| S16_extracted_89117 | Not-determined |
| S16_extracted_89119 | Not-determined |
| S16_extracted_89129 | Low-quality |
| S16_extracted_89146 | Not-determined |
| S16_extracted_89150 | Low-quality |
| S16_extracted_89161 | Not-determined |
| S16_extracted_89173 | Low-quality |
| S16_extracted_89176 | Not-determined |
| S16_extracted_89194 | Low-quality |
| S16_extracted_89239 | Low-quality |
| S16_extracted_89260 | Not-determined |
| S16_extracted_89261 | Not-determined |
| S16_extracted_89265 | Not-determined |
| S16_extracted_89280 | Low-quality |
| S16_extracted_89287 | Not-determined |
| S16_extracted_89290 | Not-determined |
| S16_extracted_89293 | Not-determined |
| S16_extracted_89318 | Not-determined |
| S16_extracted_89340 | Not-determined |
| S16_extracted_89345 | Not-determined |
| S16_extracted_89358 | Not-determined |
| S16_extracted_89393 | Not-determined |
| S16_extracted_89396 | Not-determined |
| S16_extracted_89398 | Not-determined |
| S16_extracted_89405 | Not-determined |
| S16_extracted_89435 | Not-determined |
| S16_extracted_89444 | Low-quality |
| S16_extracted_89450 | Not-determined |
| S16_extracted_89452 | Not-determined |
| S16_extracted_89456 | Low-quality |
| S16_extracted_89465 | Not-determined |
| S16_extracted_89470 | Low-quality |
| S16_extracted_89471 | Not-determined |
| S16_extracted_89478 | Not-determined |
| S16_extracted_89479 | Low-quality |
| S16_extracted_89480 | Not-determined |
| S16_extracted_89509 | Not-determined |

| | |
|---|---|
| S16_extracted_89512 | Not-determined |
| S16_extracted_89525 | Not-determined |
| S16_extracted_89526 | Not-determined |
| S16_extracted_89541 | Low-quality |
| S16_extracted_89547 | Low-quality |
| S16_extracted_89557 | Not-determined |
| S16_extracted_89558 | Not-determined |
| S16_extracted_89576 | Not-determined |
| S16_extracted_89589 | Low-quality |
| S16_extracted_89593 | Low-quality |
| S16_extracted_89620 | Not-determined |
| S16_extracted_89623 | Not-determined |
| S16_extracted_89636 | Not-determined |
| S16_extracted_89662 | Low-quality |
| S16_extracted_89665 | Low-quality |
| S16_extracted_89674 | Not-determined |
| S16_extracted_89686 | Not-determined |
| S16_extracted_89690 | Low-quality |
| S16_extracted_89696 | Not-determined |
| S16_extracted_89699 | Not-determined |
| S16_extracted_89702 | Not-determined |
| S16_extracted_89715 | Not-determined |
| S16_extracted_89733 | Not-determined |
| S16_extracted_89738 | Not-determined |
| S16_extracted_89744 | Low-quality |
| S16_extracted_89753 | Not-determined |
| S16_extracted_89755 | Not-determined |
| S16_extracted_89766 | Not-determined |
| S16_extracted_89768 | Low-quality |
| S16_extracted_89775 | Not-determined |
| S16_extracted_89778 | Not-determined |
| S16_extracted_89802 | Not-determined |
| S16_extracted_89805 | Low-quality |
| S16_extracted_89806 | Low-quality |
| S16_extracted_89847 | Not-determined |
| S16_extracted_89853 | Not-determined |
| S16_extracted_89858 | Low-quality |
| S16_extracted_89880 | Not-determined |
| S16_extracted_89902 | Not-determined |
| S16_extracted_89912 | Not-determined |
| S16_extracted_89914 | Not-determined |
| S16_extracted_89916 | Not-determined |
| S16_extracted_89933 | Not-determined |
| S16_extracted_89943 | Low-quality |
| S16_extracted_89961 | Not-determined |
| S16_extracted_89993 | Not-determined |
| S16_extracted_90004 | Low-quality |
| S16_extracted_90011 | Low-quality |
| S16_extracted_90014 | Not-determined |
| S16_extracted_90020 | Not-determined |
| S16_extracted_90022 | Not-determined |
| S16_extracted_90043 | Not-determined |
| S16_extracted_90045 | Not-determined |
| S16_extracted_90046 | Not-determined |
| S11_extracted_k141_104062 | Complete |
| S16_extracted_90061 | Not-determined |
| S16_extracted_90067 | Not-determined |
| S16_extracted_90072 | Low-quality |
| S16_extracted_90077 | Not-determined |
| S16_extracted_90082 | Low-quality |
| S16_extracted_90090 | Not-determined |
| S16_extracted_90093 | Low-quality |
| S16_extracted_90100 | Not-determined |
| S16_extracted_90113 | Not-determined |
| S16_extracted_90118 | Not-determined |
| S16_extracted_90123 | Not-determined |
| S16_extracted_90137 | Not-determined |
| S16_extracted_90156 | Not-determined |
| S16_extracted_90160 | Not-determined |
| S16_extracted_90179 | Not-determined |
| S16_extracted_90183 | Not-determined |
| S16_extracted_90185 | Not-determined |
| S16_extracted_90187 | Low-quality |
| S16_extracted_90190 | Not-determined |
| S16_extracted_90199 | Not-determined |
| S16_extracted_90202 | Not-determined |
| S16_extracted_90208 | Not-determined |
| S16_extracted_90211 | Low-quality |
| S16_extracted_90214 | Not-determined |
| S16_extracted_90235 | Not-determined |
| S16_extracted_90243 | Not-determined |
| S16_extracted_90267 | Not-determined |
| S16_extracted_90271 | Not-determined |
| S16_extracted_90273 | Not-determined |
| S16_extracted_90289 | Not-determined |
| S16_extracted_90290 | Not-determined |
| S3_vs_k141_127407\|\|full | High-quality |
| S16_extracted_90330 | Not-determined |
| S16_extracted_90343 | Low-quality |
| S16_extracted_90344 | Not-determined |
| S16_extracted_90352 | Not-determined |
| S16_extracted_90378 | Low-quality |
| S16_extracted_90380 | Not-determined |
| S16_extracted_90388 | Low-quality |
| S16_extracted_90401 | Low-quality |
| S16_extracted_90407 | Not-determined |
| S16_extracted_90435 | Not-determined |
| S16_extracted_90445 | Not-determined |
| S16_extracted_90449 | Not-determined |
| S16_extracted_90463 | Not-determined |
| S16_extracted_90470 | Not-determined |
| S16_extracted_90472 | Not-determined |
| S16_extracted_90483 | Not-determined |
| S16_extracted_90486 | Not-determined |
| S16_extracted_90493 | Not-determined |
| S16_extracted_90508 | Not-determined |
| S16_extracted_90517 | Not-determined |
| S16_extracted_90521 | Not-determined |
| S16_extracted_90540 | Low-quality |
| S16_extracted_90542 | Not-determined |
| S16_extracted_90559 | Not-determined |
| S16_extracted_90565 | Low-quality |
| S16_extracted_90574 | Not-determined |
| S16_extracted_90579 | Not-determined |
| S16_extracted_90582 | Low-quality |
| S16_extracted_90598 | Not-determined |
| S16_extracted_90603 | Not-determined |
| S16_extracted_90604 | Low-quality |
| S16_extracted_90613 | Not-determined |
| S16_extracted_90617 | Not-determined |
| S16_extracted_90631 | Not-determined |
| S16_extracted_90634 | Not-determined |
| S16_extracted_90640 | Not-determined |
| S16_extracted_90671 | Low-quality |
| S16_extracted_90676 | Not-determined |
| S16_extracted_90681 | Low-quality |
| S16_extracted_90686 | Not-determined |
| S16_extracted_90695 | Low-quality |
| S16_extracted_90699 | Not-determined |
| S16_extracted_90717 | Low-quality |
| S16_extracted_90721 | Not-determined |
| S16_extracted_90723 | Not-determined |
| S16_extracted_90728 | Not-determined |
| S16_extracted_90734 | Not-determined |
| S16_extracted_90739 | Low-quality |
| S16_extracted_90740 | Not-determined |
| S16_extracted_90774 | Not-determined |
| S16_extracted_90776 | Not-determined |
| S16_extracted_90781 | Not-determined |
| S16_extracted_90794 | Not-determined |
| S16_extracted_90806 | Not-determined |
| S16_extracted_90814 | Not-determined |
| S16_extracted_90815 | Not-determined |
| S16_extracted_90824 | Not-determined |
| S16_extracted_90833 | Not-determined |
| S16_extracted_90836 | Not-determined |
| S16_extracted_90848 | Not-determined |
| S16_extracted_90849 | Not-determined |
| S16_extracted_90850 | Not-determined |
| S16_extracted_90859 | Not-determined |
| S16_extracted_90890 | Not-determined |
| S16_extracted_90893 | Not-determined |
| S16_extracted_90897 | Not-determined |
| S16_extracted_90898 | Not-determined |
| S16_extracted_90901 | Low-quality |
| S16_extracted_90903 | Not-determined |
| S16_extracted_90904 | Not-determined |
| S16_extracted_90949 | Not-determined |
| S16_extracted_90952 | Not-determined |
| S16_extracted_90953 | Not-determined |
| S16_extracted_90965 | Low-quality |
| S16_extracted_90968 | Not-determined |
| S16_extracted_91006 | Not-determined |
| S16_extracted_91017 | Not-determined |
| S16_extracted_91022 | Not-determined |
| S16_extracted_91039 | Not-determined |
| S16_extracted_91047 | Not-determined |
| S16_extracted_91061 | Not-determined |
| S16_extracted_91116 | Not-determined |
| S16_extracted_91122 | Not-determined |
| S16_extracted_91140 | Not-determined |
| S16_extracted_91144 | Not-determined |
| S16_extracted_91159 | Not-determined |
| S16_extracted_91175 | Not-determined |
| S16_extracted_91194 | Not-determined |
| S16_extracted_91196 | Not-determined |
| S16_extracted_91220 | Not-determined |
| S16_extracted_91226 | Not-determined |
| S16_extracted_91227 | Not-determined |
| S16_extracted_91238 | Not-determined |
| S16_extracted_91241 | Not-determined |
| S16_extracted_91264 | Not-determined |
| S16_extracted_91269 | Not-determined |
| S16_extracted_91280 | Not-determined |
| S16_extracted_91281 | Not-determined |
| S16_extracted_91283 | Low-quality |
| S16_extracted_91293 | Not-determined |
| S16_extracted_91304 | Not-determined |
| S16_extracted_91307 | Not-determined |
| S16_extracted_91310 | Not-determined |
| S16_extracted_91312 | Not-determined |
| S16_extracted_91314 | Not-determined |
| S16_extracted_91319 | Not-determined |
| S16_extracted_91332 | Not-determined |
| S16_extracted_91343 | Low-quality |
| S16_extracted_91345 | Not-determined |
| S16_extracted_91348 | Not-determined |
| S16_extracted_91349 | Not-determined |
| S16_extracted_91355 | Not-determined |
| S16_extracted_91370 | Not-determined |
| S16_extracted_91377 | Not-determined |
| S16_extracted_91380 | Not-determined |
| S16_extracted_91382 | Low-quality |
| S16_extracted_91394 | Not-determined |
| S16_extracted_91402 | Not-determined |
| S16_extracted_91410 | Low-quality |
| S16_extracted_91413 | Not-determined |
| S16_extracted_91425 | Not-determined |
| S16_extracted_91429 | Not-determined |
| S16_extracted_91438 | Low-quality |
| S16_extracted_91442 | Not-determined |
| S16_extracted_91443 | Not-determined |
| S16_extracted_91457 | Not-determined |
| S16_extracted_91458 | Not-determined |
| S16_extracted_91460 | Not-determined |
| S16_extracted_91500 | Not-determined |
| S16_extracted_91501 | Not-determined |
| S16_extracted_91505 | Low-quality |
| S16_extracted_91507 | Not-determined |
| S16_extracted_91517 | Not-determined |
| S16_extracted_91519 | Not-determined |
| S16_extracted_91553 | Not-determined |
| S16_extracted_91559 | Not-determined |
| S16_extracted_91560 | Not-determined |
| S16_extracted_91561 | Low-quality |
| S16_extracted_91571 | Not-determined |
| S16_extracted_91581 | Low-quality |
| S16_extracted_91593 | Low-quality |
| S16_extracted_91604 | Not-determined |
| S16_extracted_91605 | Not-determined |
| S16_extracted_91606 | Not-determined |
| S16_extracted_91616 | Not-determined |
| S16_extracted_91650 | Not-determined |
| S16_extracted_91656 | Low-quality |
| S16_extracted_91662 | Not-determined |
| S16_extracted_91685 | Not-determined |
| S16_extracted_91716 | Not-determined |
| S16_extracted_91725 | Not-determined |
| S16_extracted_91727 | Not-determined |
| S16_extracted_91732 | Low-quality |
| S16_extracted_91733 | Not-determined |
| S16_extracted_91737 | Not-determined |
| S16_extracted_91754 | Not-determined |
| S16_extracted_91772 | Not-determined |
| S16_extracted_91786 | Not-determined |
| S16_extracted_91788 | Not-determined |
| S16_extracted_91798 | Not-determined |
| S16_extracted_91809 | Not-determined |
| S16_extracted_91813 | Not-determined |
| S16_extracted_91830 | Not-determined |
| S16_extracted_91831 | Not-determined |
| S16_extracted_91866 | Not-determined |
| S16_extracted_91869 | Not-determined |
| S16_extracted_91874 | Low-quality |
| S16_extracted_91895 | Low-quality |
| S16_extracted_91899 | Not-determined |
| S16_extracted_91900 | Not-determined |
| S16_extracted_91901 | Not-determined |
| S16_extracted_91925 | Low-quality |
| S16_extracted_91936 | Not-determined |
| S16_extracted_91938 | Not-determined |
| S16_extracted_91940 | Not-determined |
| S16_extracted_91943 | Not-determined |
| S16_extracted_91975 | Not-determined |
| S16_extracted_91979 | Not-determined |
| S16_extracted_91987 | Not-determined |
| S16_extracted_92000 | Not-determined |
| S16_extracted_92035 | Not-determined |
| S16_extracted_92038 | Low-quality |
| S16_extracted_92050 | Low-quality |
| S16_extracted_92054 | Not-determined |
| S16_extracted_92060 | Low-quality |
| S16_extracted_92066 | Low-quality |
| S16_extracted_92079 | Not-determined |
| S16_extracted_92103 | Not-determined |
| S16_extracted_92115 | Not-determined |
| S16_extracted_92129 | Not-determined |
| S16_extracted_92133 | Not-determined |
| S16_extracted_92151 | Not-determined |
| S16_extracted_92153 | Not-determined |
| S16_extracted_92163 | Not-determined |
| S16_extracted_92167 | Not-determined |
| S16_extracted_92172 | Not-determined |
| S16_extracted_92176 | Not-determined |
| S16_extracted_92196 | Not-determined |
| S16_extracted_92199 | Not-determined |
| S16_extracted_92200 | Not-determined |
| S16_extracted_92204 | Not-determined |
| S16_extracted_92207 | Low-quality |
| S16_extracted_92221 | Not-determined |
| S16_extracted_92234 | Not-determined |
| S16_extracted_92241 | Not-determined |
| S16_extracted_92258 | Not-determined |
| S16_extracted_92260 | Low-quality |
| S16_extracted_92263 | Not-determined |
| S16_extracted_92266 | Not-determined |
| S16_extracted_92270 | Not-determined |
| S16_extracted_92291 | Low-quality |
| S16_extracted_92313 | Not-determined |
| S16_extracted_92327 | Low-quality |
| S16_extracted_92335 | Low-quality |
| S16_extracted_92341 | Not-determined |
| S16_extracted_92342 | Not-determined |
| S16_extracted_92358 | Not-determined |
| S16_extracted_92362 | Not-determined |
| S16_extracted_92364 | Not-determined |
| S16_extracted_92368 | Not-determined |
| S16_extracted_92388 | Low-quality |
| S16_extracted_92394 | Not-determined |
| S16_extracted_92396 | Not-determined |
| S16_extracted_92414 | Not-determined |
| S16_extracted_92432 | Not-determined |
| S16_extracted_92435 | Not-determined |
| S16_extracted_92439 | Not-determined |
| S16_extracted_92462 | Not-determined |
| S16_extracted_92486 | Low-quality |
| S16_extracted_92488 | Not-determined |
| S16_extracted_92499 | Low-quality |
| S16_extracted_92506 | Not-determined |
| S16_extracted_92513 | Not-determined |
| S16_extracted_92521 | Not-determined |
| S16_extracted_92536 | Not-determined |
| S16_extracted_92537 | Not-determined |
| S16_extracted_92538 | Not-determined |
| S16_extracted_92560 | Not-determined |
| S16_extracted_92566 | Not-determined |
| S16_extracted_92573 | Not-determined |
| S16_extracted_92576 | Not-determined |
| S16_extracted_92580 | Low-quality |
| S16_extracted_92620 | Low-quality |
| S16_extracted_92627 | Low-quality |
| S16_extracted_92640 | Not-determined |
| S16_extracted_92669 | Not-determined |
| S16_extracted_92685 | Not-determined |
| S16_extracted_92689 | Not-determined |
| S16_extracted_92694 | Not-determined |
| S16_extracted_92706 | Not-determined |
| S16_extracted_92708 | Low-quality |
| S16_extracted_92709 | Not-determined |
| S16_extracted_92718 | Not-determined |
| S16_extracted_92754 | Not-determined |
| S16_extracted_92758 | Not-determined |
| S16_extracted_92774 | Low-quality |
| S16_extracted_92777 | Not-determined |
| S16_extracted_92781 | Not-determined |
| S16_extracted_92784 | Low-quality |
| S16_extracted_92792 | Low-quality |
| S16_extracted_92809 | Low-quality |
| S16_extracted_92827 | Not-determined |
| S16_extracted_92832 | Not-determined |
| S16_extracted_92839 | Low-quality |
| S16_extracted_92851 | Not-determined |
| S16_extracted_92853 | Not-determined |
| S16_extracted_92857 | Low-quality |
| S16_extracted_92864 | Low-quality |
| S16_extracted_92865 | Not-determined |
| S16_extracted_92868 | Not-determined |
| S16_extracted_92908 | Not-determined |
| S16_extracted_92909 | Not-determined |
| S16_extracted_92914 | Not-determined |
| S16_extracted_92925 | Not-determined |
| S16_extracted_92942 | Not-determined |
| S16_extracted_92947 | Not-determined |
| S16_extracted_92952 | Not-determined |
| S16_extracted_92974 | Not-determined |
| S16_extracted_92975 | Not-determined |
| S16_extracted_92978 | Low-quality |
| S16_extracted_92983 | Not-determined |
| S16_extracted_93003 | Low-quality |
| S16_extracted_93026 | Not-determined |
| S16_extracted_93031 | Not-determined |
| S16_extracted_93041 | Not-determined |
| S16_extracted_93048 | Low-quality |
| S16_extracted_93049 | Low-quality |
| S16_extracted_93052 | Not-determined |
| S16_extracted_93057 | Low-quality |
| S16_extracted_93073 | Not-determined |
| S16_extracted_93103 | Not-determined |
| S16_extracted_93104 | Not-determined |
| S16_extracted_93110 | Not-determined |
| S16_extracted_93118 | Low-quality |
| S16_extracted_93140 | Not-determined |
| S16_extracted_93143 | Not-determined |
| S16_extracted_93145 | Not-determined |
| S16_extracted_93159 | Not-determined |
| S16_extracted_93162 | Not-determined |
| S16_extracted_93193 | Low-quality |
| S16_extracted_93197 | Not-determined |
| S16_extracted_93199 | Low-quality |
| S16_extracted_93244 | Not-determined |
| S16_extracted_93254 | Low-quality |
| S16_extracted_93266 | Not-determined |
| S16_extracted_93271 | Not-determined |
| S16_extracted_93284 | Not-determined |
| S16_extracted_93286 | Not-determined |
| S16_extracted_93289 | Not-determined |
| S16_extracted_93305 | Low-quality |
| S16_extracted_93317 | Not-determined |
| S16_extracted_93321 | Not-determined |
| S16_extracted_93353 | Low-quality |
| S16_extracted_93356 | Not-determined |
| S16_extracted_93357 | Not-determined |
| S16_extracted_93390 | Not-determined |
| S16_extracted_93399 | Not-determined |
| S16_extracted_93404 | Not-determined |
| S16_extracted_93412 | Not-determined |
| S16_extracted_93414 | Not-determined |
| S16_extracted_93420 | Not-determined |
| S16_extracted_93439 | Not-determined |
| S16_extracted_93447 | Low-quality |
| S16_extracted_93486 | Not-determined |
| S16_extracted_93513 | Not-determined |
| S16_extracted_93514 | Not-determined |
| S16_extracted_93526 | Not-determined |
| S16_extracted_93538 | Not-determined |
| S16_extracted_93560 | Not-determined |
| S16_extracted_93562 | Not-determined |
| S16_extracted_93571 | Not-determined |
| S16_extracted_93601 | Low-quality |
| S16_extracted_93631 | Not-determined |
| S16_extracted_93632 | Not-determined |
| S16_extracted_93636 | Not-determined |
| S16_extracted_93646 | Not-determined |
| S16_extracted_93651 | Not-determined |
| S16_extracted_93653 | Low-quality |
| S16_extracted_93661 | Not-determined |
| S16_extracted_93669 | Not-determined |
| S16_extracted_93671 | Not-determined |
| S16_extracted_93678 | Not-determined |
| S16_extracted_93704 | Not-determined |
| S16_extracted_93732 | Low-quality |
| S16_extracted_93768 | Not-determined |
| S16_extracted_93769 | Not-determined |
| S16_extracted_93772 | Not-determined |
| S16_extracted_93790 | Not-determined |
| S16_extracted_93802 | Not-determined |
| S16_extracted_93803 | Not-determined |
| S16_extracted_93817 | Not-determined |
| S16_extracted_93831 | Not-determined |
| S16_extracted_93857 | Not-determined |
| S16_extracted_93877 | Not-determined |
| S16_extracted_93886 | Not-determined |
| S16_extracted_93892 | Not-determined |
| S16_extracted_93897 | Not-determined |
| S16_extracted_93898 | Not-determined |
| S16_extracted_93903 | Not-determined |
| S16_extracted_93912 | Not-determined |
| S16_extracted_93917 | Not-determined |
| S16_extracted_93935 | Not-determined |
| S16_extracted_93939 | Not-determined |
| S16_extracted_93945 | Not-determined |
| S16_extracted_93947 | Low-quality |
| S16_extracted_93952 | Not-determined |
| S16_extracted_93954 | Not-determined |
| S16_extracted_93990 | Low-quality |
| S16_extracted_94000 | Low-quality |
| S16_extracted_94006 | Not-determined |
| S16_extracted_94030 | Not-determined |
| S16_extracted_94034 | Not-determined |
| S16_extracted_94046 | Not-determined |
| S16_extracted_94048 | Not-determined |
| S16_extracted_94050 | Not-determined |
| S16_extracted_94052 | Not-determined |
| S16_extracted_94062 | Not-determined |
| S16_extracted_94064 | Low-quality |
| S16_extracted_94067 | Not-determined |
| S16_extracted_94086 | Not-determined |
| S16_extracted_94118 | Not-determined |
| S16_extracted_94122 | Not-determined |
| S16_extracted_94149 | Low-quality |
| S16_extracted_94183 | Not-determined |
| S16_extracted_94186 | Low-quality |
| S16_extracted_94189 | Not-determined |
| S16_extracted_94212 | Not-determined |
| S16_extracted_94213 | Low-quality |
| S16_extracted_94217 | Not-determined |
| S16_extracted_94218 | Not-determined |
| S16_extracted_94225 | Not-determined |
| S16_extracted_94226 | Not-determined |
| S16_extracted_94239 | Not-determined |
| S16_extracted_94267 | Not-determined |
| S16_extracted_94269 | Low-quality |
| S16_extracted_94273 | Not-determined |
| S16_extracted_94285 | Low-quality |
| S16_extracted_94287 | Not-determined |
| S16_extracted_94291 | Not-determined |
| S16_extracted_94298 | Not-determined |
| S16_extracted_94300 | Low-quality |
| S16_extracted_94307 | Not-determined |
| S16_extracted_94335 | Not-determined |
| S16_extracted_94337 | Not-determined |
| S16_extracted_94338 | Low-quality |
| S16_extracted_94349 | Not-determined |
| S16_extracted_94352 | Not-determined |
| S16_extracted_94373 | Not-determined |
| S16_extracted_94384 | Not-determined |
| S16_extracted_94396 | Low-quality |
| S16_extracted_94422 | Low-quality |
| S16_extracted_94426 | Not-determined |
| S16_extracted_94432 | Not-determined |
| S16_extracted_94436 | Not-determined |
| S16_extracted_94441 | Not-determined |
| S16_extracted_94461 | Not-determined |
| S16_extracted_94466 | Low-quality |
| S16_extracted_94475 | Not-determined |
| S16_extracted_94492 | Not-determined |
| S16_extracted_94496 | Not-determined |
| S16_extracted_94502 | Not-determined |
| S16_extracted_94524 | Not-determined |
| S16_extracted_94539 | Not-determined |
| S16_extracted_94554 | Not-determined |
| S16_extracted_94559 | Not-determined |
| S16_extracted_94615 | Not-determined |
| S16_extracted_94624 | Not-determined |
| S16_extracted_94675 | Low-quality |
| S16_extracted_94679 | Not-determined |
| S16_extracted_94683 | Not-determined |
| S16_extracted_94684 | Not-determined |
| S16_extracted_94685 | Not-determined |
| S16_extracted_94695 | Not-determined |
| S16_extracted_94710 | Not-determined |
| S16_extracted_94711 | Low-quality |
| S16_extracted_94712 | Low-quality |
| S16_extracted_94762 | Not-determined |
| S16_extracted_94771 | Not-determined |
| S16_extracted_94802 | Low-quality |
| S16_extracted_94817 | Low-quality |
| S16_extracted_94823 | Not-determined |
| S16_extracted_94832 | Not-determined |
| S16_extracted_94836 | Low-quality |
| S7_extracted_13614 | Complete |
| S16_extracted_94871 | Not-determined |
| S16_extracted_94873 | Low-quality |
| S16_extracted_94883 | Not-determined |
| S16_extracted_94891 | Not-determined |
| S16_extracted_94896 | Low-quality |
| S16_extracted_94912 | Not-determined |
| S16_extracted_94931 | Not-determined |
| S16_extracted_94938 | Low-quality |
| S16_extracted_94987 | Not-determined |
| S16_extracted_94989 | Not-determined |
| S16_extracted_94994 | Not-determined |
| S16_extracted_94999 | Not-determined |
| S16_extracted_95002 | Not-determined |
| S16_extracted_95005 | Not-determined |
| S16_extracted_95019 | Low-quality |
| S16_extracted_95022 | Not-determined |
| S16_extracted_95025 | Low-quality |
| S16_extracted_95030 | Not-determined |
| S16_extracted_95035 | Not-determined |
| S16_extracted_95053 | Not-determined |
| S16_extracted_95066 | Not-determined |
| S16_extracted_95083 | Not-determined |
| S16_extracted_95088 | Not-determined |
| S16_extracted_95112 | Not-determined |
| S16_extracted_95118 | Not-determined |
| S16_extracted_95123 | Not-determined |
| S16_extracted_95129 | Not-determined |
| S16_extracted_95162 | Not-determined |
| S16_extracted_95163 | Not-determined |
| S16_extracted_95170 | Not-determined |
| S16_extracted_95181 | Not-determined |
| S16_extracted_95184 | Not-determined |
| S16_extracted_95185 | Not-determined |
| S16_extracted_95222 | Not-determined |
| S16_extracted_95237 | Not-determined |
| S16_extracted_95246 | Low-quality |
| S16_extracted_95268 | Not-determined |
| S16_extracted_95271 | Not-determined |
| S16_extracted_95278 | Not-determined |
| S16_extracted_95283 | Not-determined |
| S16_extracted_95310 | Not-determined |
| S16_extracted_95320 | Not-determined |
| S16_extracted_95323 | Not-determined |
| S16_extracted_95333 | Not-determined |
| S16_extracted_95344 | Not-determined |
| S16_extracted_95348 | Not-determined |
| S16_extracted_95358 | Not-determined |
| S16_extracted_95382 | Low-quality |
| S16_extracted_95394 | Not-determined |
| S16_extracted_95400 | Not-determined |
| S16_extracted_95406 | Not-determined |
| S16_extracted_95414 | Not-determined |
| S16_extracted_95423 | Not-determined |
| S16_extracted_95424 | Not-determined |
| S16_extracted_95435 | Not-determined |
| S16_extracted_95456 | Not-determined |
| S16_extracted_95461 | Not-determined |
| S16_extracted_95466 | Low-quality |

| | |
|---|---|
| S16_extracted_95470 | Not-determined |
| S16_extracted_95472 | Not-determined |
| S16_extracted_95474 | Not-determined |
| S16_extracted_95478 | Not-determined |
| S16_extracted_95482 | Not-determined |
| S16_extracted_95493 | Not-determined |
| S16_extracted_95495 | Not-determined |
| S16_extracted_95508 | Low-quality |
| S16_extracted_95510 | Not-determined |
| S16_extracted_95513 | Not-determined |
| S16_extracted_95547 | Not-determined |
| S16_extracted_95561 | Not-determined |
| S16_extracted_95576 | Not-determined |
| S16_extracted_95600 | Not-determined |
| S16_extracted_95602 | Low-quality |
| S16_extracted_95620 | Not-determined |
| S16_extracted_95625 | Not-determined |
| S16_extracted_95628 | Not-determined |
| S16_extracted_95641 | Not-determined |
| S16_extracted_95653 | Not-determined |
| S16_extracted_95661 | Not-determined |
| S16_extracted_95665 | Not-determined |
| S16_extracted_95677 | Not-determined |
| S16_extracted_95679 | Not-determined |
| S16_extracted_95690 | Not-determined |
| S16_extracted_95703 | Not-determined |
| S16_extracted_95704 | Not-determined |
| S16_extracted_95753 | Not-determined |
| S16_extracted_95759 | Not-determined |
| S16_extracted_95763 | Not-determined |
| S16_extracted_95768 | Low-quality |
| S16_extracted_95772 | Not-determined |
| S16_extracted_95787 | Not-determined |
| S16_extracted_95788 | Not-determined |
| S16_extracted_95789 | Not-determined |
| S16_extracted_95818 | Not-determined |
| S16_extracted_95825 | Low-quality |
| S16_extracted_95831 | Not-determined |
| S16_extracted_95833 | Not-determined |
| S16_extracted_95835 | Not-determined |
| S16_extracted_95840 | Low-quality |
| S16_extracted_95842 | Not-determined |
| S16_extracted_95848 | Not-determined |
| S16_extracted_95851 | Not-determined |
| S16_extracted_95852 | Not-determined |
| S16_extracted_95865 | Not-determined |
| S16_extracted_95913 | Not-determined |
| S16_extracted_95919 | Not-determined |
| S16_extracted_95921 | Not-determined |
| S16_extracted_95924 | Not-determined |
| S16_extracted_95942 | Not-determined |
| S16_extracted_95956 | Not-determined |
| S16_extracted_95960 | Not-determined |
| S16_extracted_95966 | Low-quality |
| S16_extracted_95983 | Not-determined |
| S16_extracted_95993 | Not-determined |
| S16_extracted_96004 | Not-determined |
| S16_extracted_96006 | Not-determined |
| S16_extracted_96009 | Not-determined |
| S16_extracted_96015 | Not-determined |
| S16_extracted_96021 | Not-determined |
| S16_extracted_96026 | Not-determined |
| S16_extracted_96029 | Not-determined |
| S16_extracted_96038 | Not-determined |
| S16_extracted_96049 | Not-determined |
| S16_extracted_96052 | Not-determined |
| S16_extracted_96056 | Not-determined |
| S16_extracted_96073 | Not-determined |
| S16_extracted_96076 | Not-determined |
| S16_extracted_96089 | Not-determined |
| S16_extracted_96093 | Not-determined |
| S16_extracted_96097 | Not-determined |
| S16_extracted_96102 | Not-determined |
| S16_extracted_96107 | Low-quality |
| S16_extracted_96117 | Not-determined |
| S16_extracted_96143 | Low-quality |
| S16_extracted_96159 | Low-quality |
| S16_extracted_96161 | Not-determined |
| S16_extracted_96165 | Low-quality |
| S16_extracted_96169 | Not-determined |
| S16_extracted_96182 | Not-determined |
| S16_extracted_96194 | Not-determined |
| S16_extracted_96201 | Low-quality |
| S16_extracted_96223 | Not-determined |
| S16_extracted_96226 | Not-determined |
| S16_extracted_96238 | Not-determined |
| S16_extracted_96247 | Not-determined |
| S16_extracted_96251 | Not-determined |
| S16_extracted_96261 | Not-determined |
| S16_extracted_96267 | Not-determined |
| S16_extracted_96279 | Not-determined |
| S16_extracted_96283 | Low-quality |
| S16_extracted_96301 | Not-determined |
| S16_extracted_96315 | Not-determined |
| S16_extracted_96334 | Not-determined |
| S16_extracted_96339 | Not-determined |
| S16_extracted_96341 | Not-determined |
| S16_extracted_96344 | Not-determined |
| S16_extracted_96345 | Low-quality |
| S16_extracted_96354 | Not-determined |
| S16_extracted_96360 | Not-determined |
| S16_extracted_96370 | Not-determined |
| S16_extracted_96378 | Not-determined |
| S16_extracted_96394 | Not-determined |
| S16_extracted_96405 | Low-quality |
| S16_extracted_96415 | Not-determined |
| S16_extracted_96424 | Not-determined |
| S16_extracted_96427 | Not-determined |
| S16_extracted_96434 | Not-determined |
| S16_extracted_96436 | Not-determined |
| S16_extracted_96437 | Not-determined |
| S16_extracted_96442 | Not-determined |
| S16_extracted_96444 | Not-determined |
| S16_extracted_96453 | Not-determined |
| S16_extracted_96457 | Not-determined |
| S16_extracted_96467 | Not-determined |
| S16_extracted_96473 | Not-determined |
| S16_extracted_96476 | Not-determined |
| S16_extracted_96480 | Low-quality |
| S16_vs_38863\|\|full | Medium-quality |
| S14_vs_k141_117160\|\|full | Medium-quality |
| S16_extracted_96487 | Not-determined |
| S16_extracted_96496 | Not-determined |
| S16_extracted_96513 | Low-quality |
| S16_extracted_96517 | Not-determined |
| S16_extracted_96526 | Not-determined |
| S16_extracted_96530 | Not-determined |
| S16_extracted_96552 | Not-determined |
| S16_extracted_96562 | Not-determined |
| S16_extracted_96564 | Not-determined |
| S16_extracted_96594 | Not-determined |
| S16_extracted_96596 | Not-determined |
| S16_extracted_96620 | Not-determined |
| S16_extracted_96621 | Not-determined |
| S16_extracted_96625 | Not-determined |
| S16_extracted_96638 | Not-determined |
| S16_extracted_96653 | Not-determined |
| S16_extracted_96674 | Not-determined |
| S16_extracted_96682 | Not-determined |
| S16_extracted_96684 | Not-determined |
| S16_extracted_96701 | Not-determined |
| S16_extracted_96705 | Low-quality |
| S16_extracted_96707 | Low-quality |
| S16_extracted_96722 | Not-determined |
| S16_extracted_96734 | Not-determined |
| S16_extracted_96737 | Not-determined |
| S16_extracted_96741 | Not-determined |
| S16_extracted_96748 | Not-determined |
| S16_extracted_96762 | Not-determined |
| S16_extracted_96763 | Not-determined |
| S16_extracted_96779 | Not-determined |
| S16_extracted_96784 | Not-determined |
| S16_extracted_96796 | Not-determined |
| S16_extracted_96838 | Not-determined |
| S16_extracted_96840 | Not-determined |
| S16_extracted_96857 | Not-determined |
| S16_extracted_96876 | Not-determined |
| S16_extracted_96897 | Not-determined |
| S16_extracted_96899 | Not-determined |
| S16_extracted_96901 | Low-quality |
| S16_extracted_96910 | Not-determined |
| S16_extracted_96933 | Not-determined |
| S16_extracted_96938 | Not-determined |
| S16_extracted_96941 | Not-determined |
| S16_extracted_96947 | Not-determined |
| S16_extracted_96949 | Not-determined |
| S16_extracted_96953 | Low-quality |
| S16_extracted_96971 | Not-determined |
| S16_extracted_96982 | Not-determined |
| S16_extracted_96983 | Not-determined |
| S16_extracted_96989 | Not-determined |
| S16_extracted_97015 | Not-determined |
| S16_extracted_97038 | Not-determined |
| S16_extracted_97047 | Not-determined |
| S16_extracted_97054 | Low-quality |
| S16_extracted_97068 | Not-determined |
| S16_extracted_97075 | Not-determined |
| S16_extracted_97094 | Not-determined |
| S16_extracted_97097 | Not-determined |
| S16_extracted_97098 | Not-determined |
| S16_extracted_97101 | Not-determined |
| S16_extracted_97107 | Not-determined |
| S16_extracted_97111 | Not-determined |
| S16_extracted_97143 | Low-quality |
| S16_extracted_97160 | Not-determined |
| S16_extracted_97161 | Not-determined |
| S16_extracted_97179 | Low-quality |
| S16_extracted_97187 | Low-quality |
| S16_extracted_97193 | Not-determined |
| S16_extracted_97206 | Not-determined |
| S16_extracted_97207 | Not-determined |
| S16_extracted_97211 | Not-determined |
| S16_extracted_97218 | Not-determined |
| S16_extracted_97222 | Not-determined |
| S16_extracted_97224 | Not-determined |
| S16_extracted_97230 | Not-determined |
| S16_extracted_97242 | Not-determined |
| S16_extracted_97244 | Not-determined |
| S16_extracted_97249 | Not-determined |
| S16_extracted_97254 | Low-quality |
| S16_extracted_97270 | Not-determined |
| S5_vs_k141_118168\|\|full | Medium-quality |
| S16_extracted_97296 | Not-determined |
| S16_extracted_97298 | Not-determined |
| S16_extracted_97301 | Not-determined |
| S16_extracted_97308 | Not-determined |
| S16_extracted_97314 | Not-determined |
| S16_extracted_97337 | Low-quality |
| S16_extracted_97346 | Not-determined |
| S16_extracted_97365 | Not-determined |
| S16_extracted_97389 | Not-determined |
| S16_extracted_97390 | Not-determined |
| S16_extracted_97391 | Not-determined |
| S16_extracted_97413 | Not-determined |
| S16_extracted_97421 | Not-determined |
| S16_extracted_97431 | Not-determined |
| S16_extracted_97449 | Not-determined |
| S16_extracted_97452 | Not-determined |
| S16_extracted_97457 | Not-determined |
| S16_extracted_97476 | Not-determined |
| S16_extracted_97495 | Not-determined |
| S16_extracted_97501 | Low-quality |
| S16_extracted_97506 | Not-determined |
| S16_extracted_97507 | Not-determined |
| S16_extracted_97511 | Not-determined |
| S16_extracted_97514 | Not-determined |
| S16_extracted_97519 | Not-determined |
| S16_extracted_97520 | Not-determined |
| S16_extracted_97555 | Not-determined |
| S16_extracted_97562 | Low-quality |
| S16_extracted_97571 | Not-determined |
| S16_extracted_97573 | Not-determined |
| S16_extracted_97595 | Not-determined |
| S16_extracted_97619 | Low-quality |
| S16_extracted_97634 | Low-quality |
| S16_extracted_97639 | Not-determined |
| S16_extracted_97641 | Not-determined |
| S16_extracted_97648 | Not-determined |
| S16_extracted_97675 | Not-determined |
| S16_extracted_97677 | Not-determined |
| S16_extracted_97679 | Low-quality |
| S16_extracted_97693 | Low-quality |
| S16_extracted_97694 | Not-determined |
| S16_extracted_97695 | Not-determined |
| S16_extracted_97699 | Not-determined |
| S16_extracted_97703 | Not-determined |
| S16_extracted_97707 | Not-determined |
| S16_extracted_97712 | Not-determined |
| S16_extracted_97721 | Not-determined |
| S16_extracted_97731 | Not-determined |
| S16_extracted_97749 | Not-determined |
| S16_extracted_97788 | Not-determined |
| S16_extracted_97793 | Not-determined |
| S16_extracted_97800 | Not-determined |
| S16_extracted_97803 | Not-determined |
| S16_extracted_97817 | Not-determined |
| S16_extracted_97819 | Not-determined |
| S16_extracted_97822 | Low-quality |
| S16_extracted_97846 | Not-determined |
| S16_extracted_97847 | Not-determined |
| S16_extracted_97849 | Not-determined |
| S16_extracted_97851 | Not-determined |
| S16_extracted_97880 | Not-determined |
| S16_extracted_97881 | Low-quality |
| S16_extracted_97882 | Not-determined |
| S16_extracted_97885 | Not-determined |
| S16_extracted_97888 | Low-quality |
| S16_extracted_97903 | Not-determined |
| S16_extracted_97906 | Not-determined |
| S16_extracted_97908 | Not-determined |
| S16_extracted_97924 | Not-determined |
| S16_extracted_97929 | Not-determined |
| S16_extracted_97952 | Not-determined |
| S16_extracted_97954 | Not-determined |
| S16_extracted_97956 | Not-determined |
| S16_extracted_97965 | Not-determined |
| S16_extracted_97986 | Not-determined |
| S16_extracted_97991 | Not-determined |
| S16_extracted_97996 | Low-quality |
| S16_extracted_98001 | Not-determined |
| S16_extracted_98005 | Not-determined |
| S16_extracted_98013 | Not-determined |
| S16_extracted_98021 | Not-determined |
| S16_extracted_98029 | Not-determined |
| S16_extracted_98031 | Not-determined |
| S16_extracted_98043 | Not-determined |
| S16_extracted_98048 | Not-determined |
| S16_extracted_98052 | Not-determined |
| S16_extracted_98075 | Not-determined |
| S16_extracted_98081 | Low-quality |
| S16_extracted_98088 | Not-determined |
| S16_extracted_98111 | Not-determined |
| S16_extracted_98113 | Not-determined |
| S16_extracted_98115 | Not-determined |
| S16_extracted_98120 | Not-determined |
| S16_extracted_98126 | Not-determined |
| S16_extracted_98147 | Not-determined |
| S16_extracted_98153 | Not-determined |
| S16_extracted_98161 | Not-determined |
| S16_extracted_98167 | Not-determined |
| S16_extracted_98183 | Not-determined |
| S16_extracted_98186 | Not-determined |
| S16_extracted_98189 | Not-determined |
| S16_extracted_98210 | Not-determined |
| S16_extracted_98238 | Not-determined |
| S16_extracted_98241 | Not-determined |
| S16_extracted_98251 | Not-determined |
| S16_extracted_98255 | Not-determined |
| S16_extracted_98262 | Not-determined |
| S16_extracted_98279 | Not-determined |
| S16_extracted_98290 | Not-determined |
| S16_extracted_98303 | Low-quality |
| S16_extracted_98305 | Not-determined |
| S16_extracted_98317 | Not-determined |
| S16_extracted_98324 | Not-determined |
| S16_extracted_98341 | Not-determined |
| S16_extracted_98342 | Low-quality |
| S16_extracted_98343 | Not-determined |
| S16_extracted_98354 | Low-quality |
| S16_extracted_98366 | Not-determined |
| S16_extracted_98376 | Not-determined |
| S16_extracted_98395 | Low-quality |
| S16_extracted_98402 | Not-determined |
| S16_extracted_98422 | Not-determined |
| S16_extracted_98427 | Not-determined |
| S16_extracted_98432 | Not-determined |
| S16_extracted_98436 | Not-determined |
| S16_extracted_98478 | Not-determined |
| S16_extracted_98483 | Not-determined |
| S16_extracted_98488 | Not-determined |
| S16_extracted_98496 | Not-determined |
| S16_extracted_98502 | Not-determined |
| S16_extracted_98504 | Not-determined |
| S16_extracted_98508 | Not-determined |
| S16_extracted_98515 | Low-quality |
| S16_extracted_98528 | Not-determined |
| S16_extracted_98530 | Not-determined |
| S16_extracted_98532 | Not-determined |
| S16_extracted_98539 | Not-determined |
| S16_extracted_98548 | Not-determined |
| S16_extracted_98573 | Not-determined |
| S16_extracted_98578 | Not-determined |
| S16_extracted_98589 | Not-determined |
| S16_extracted_98614 | Not-determined |
| S16_extracted_98619 | Not-determined |
| S16_extracted_98626 | Not-determined |
| S16_extracted_98633 | Low-quality |
| S16_extracted_98637 | Not-determined |
| S16_extracted_98641 | Low-quality |
| S16_extracted_98648 | Not-determined |
| S16_extracted_98653 | Not-determined |
| S16_extracted_98655 | Low-quality |
| S16_extracted_98669 | Not-determined |
| S16_extracted_98676 | Not-determined |
| S16_extracted_98689 | Low-quality |
| S16_extracted_98709 | Not-determined |
| S16_extracted_98712 | Not-determined |
| S16_extracted_98714 | Not-determined |
| S16_extracted_98717 | Not-determined |
| S16_extracted_98723 | Not-determined |
| S16_extracted_98736 | Not-determined |
| S16_extracted_98753 | Not-determined |
| S16_extracted_98759 | Not-determined |
| S16_extracted_98760 | Not-determined |
| S16_extracted_98761 | Not-determined |
| S16_extracted_98788 | Not-determined |
| S16_extracted_98796 | Low-quality |
| S16_extracted_98805 | Not-determined |
| S16_extracted_98844 | Not-determined |
| S16_extracted_98848 | Not-determined |
| S16_extracted_98869 | Not-determined |
| S16_extracted_98870 | Not-determined |
| S16_extracted_98913 | Not-determined |
| S16_extracted_98916 | Not-determined |
| S16_extracted_98933 | Not-determined |
| S16_extracted_98942 | Not-determined |
| S16_extracted_98968 | Low-quality |
| S16_extracted_98979 | Not-determined |
| S16_extracted_98985 | Not-determined |
| S16_extracted_98989 | Low-quality |
| S16_extracted_98991 | Not-determined |
| S16_extracted_99004 | Low-quality |
| S16_extracted_99014 | Not-determined |
| S16_extracted_99015 | Low-quality |
| S16_extracted_99017 | Not-determined |
| S16_extracted_99019 | Not-determined |
| S16_extracted_99025 | Not-determined |
| S16_extracted_99050 | Not-determined |
| S16_extracted_99056 | Not-determined |
| S16_extracted_99063 | Low-quality |
| S16_extracted_99066 | Not-determined |
| S16_extracted_99070 | Not-determined |
| S16_extracted_99071 | Not-determined |
| S16_extracted_99076 | Not-determined |
| S16_extracted_99094 | Not-determined |
| S16_extracted_99102 | Not-determined |
| S16_extracted_99106 | Not-determined |
| S16_extracted_99112 | Not-determined |
| S16_extracted_99120 | Not-determined |
| S16_extracted_99156 | Not-determined |
| S16_extracted_99166 | Not-determined |
| S16_extracted_99169 | Not-determined |
| S16_extracted_99182 | Not-determined |
| S16_extracted_99183 | Low-quality |
| S16_extracted_99192 | Not-determined |
| S16_extracted_99195 | Not-determined |
| S16_extracted_99211 | Low-quality |
| S16_extracted_99215 | Not-determined |
| S16_extracted_99227 | Not-determined |
| S16_extracted_99236 | Not-determined |
| S16_extracted_99241 | Not-determined |
| S16_extracted_99244 | Not-determined |
| S16_extracted_99252 | Not-determined |
| S16_extracted_99263 | Not-determined |
| S16_extracted_99281 | Not-determined |
| S16_extracted_99331 | Low-quality |
| S16_extracted_99332 | Not-determined |
| S16_extracted_99337 | Low-quality |
| S16_extracted_99374 | Not-determined |
| S16_extracted_99375 | Not-determined |
| S16_extracted_99382 | Not-determined |
| S16_extracted_99402 | Not-determined |
| S16_extracted_99403 | Not-determined |
| S16_extracted_99408 | Not-determined |
| S16_extracted_99409 | Not-determined |
| S16_extracted_99421 | Not-determined |
| S16_extracted_99422 | Low-quality |
| S16_extracted_99437 | Not-determined |
| S16_extracted_99442 | Not-determined |
| S16_extracted_99447 | Not-determined |
| S16_extracted_99492 | Low-quality |
| S16_extracted_99493 | Not-determined |
| S16_extracted_99503 | Not-determined |
| S16_extracted_99517 | Not-determined |
| S16_extracted_99524 | Low-quality |
| S16_extracted_99532 | Not-determined |
| S16_extracted_99553 | Not-determined |
| S16_extracted_99567 | Low-quality |
| S16_extracted_99581 | Not-determined |
| S16_extracted_99582 | Not-determined |
| S16_extracted_99600 | Not-determined |
| S16_extracted_99607 | Not-determined |
| S16_extracted_99608 | Not-determined |
| S16_extracted_99611 | Not-determined |
| S16_extracted_99621 | Not-determined |
| S16_extracted_99622 | Not-determined |
| S16_extracted_99652 | Not-determined |
| S16_extracted_99654 | Not-determined |
| S16_extracted_99658 | Not-determined |
| S16_extracted_99672 | Low-quality |
| S16_extracted_99684 | Low-quality |
| S16_extracted_99692 | Not-determined |
| S16_extracted_99714 | Not-determined |
| S16_extracted_99731 | Not-determined |
| S16_extracted_99732 | Not-determined |
| S16_extracted_99737 | Not-determined |
| S16_extracted_99743 | Not-determined |
| S16_extracted_99745 | Low-quality |
| S16_extracted_99752 | Low-quality |
| S16_extracted_99759 | Not-determined |
| S16_extracted_99760 | Low-quality |
| S16_extracted_99778 | Not-determined |
| S16_extracted_99807 | Not-determined |
| S16_extracted_99814 | Not-determined |
| S16_extracted_99827 | Not-determined |
| S16_extracted_99829 | Low-quality |
| S16_extracted_99838 | Not-determined |
| S16_extracted_99839 | Low-quality |
| S16_extracted_99856 | Not-determined |
| S16_extracted_99864 | Not-determined |
| S16_extracted_99865 | Not-determined |
| S16_extracted_99868 | Low-quality |
| S16_extracted_99876 | Low-quality |
| S16_extracted_99878 | Not-determined |
| S16_extracted_99880 | Low-quality |
| S16_extracted_99881 | Not-determined |
| S16_extracted_99890 | Low-quality |
| S16_extracted_99909 | Not-determined |
| S16_extracted_99918 | Not-determined |
| S16_extracted_99931 | Not-determined |
| S16_extracted_99939 | Not-determined |
| S16_extracted_99950 | Low-quality |
| S16_extracted_99951 | Not-determined |
| S16_extracted_99985 | Not-determined |
| S16_extracted_99986 | Not-determined |
| S16_extracted_99993 | Not-determined |
| S16_extracted_100001 | Not-determined |
| S16_extracted_100019 | Not-determined |
| S16_extracted_100034 | Not-determined |
| S16_extracted_100044 | Not-determined |
| S16_extracted_100056 | Not-determined |
| S16_extracted_100070 | Not-determined |
| S16_extracted_100088 | Not-determined |
| S16_extracted_100090 | Not-determined |
| S16_extracted_100095 | Low-quality |
| S16_extracted_100105 | Not-determined |
| S16_extracted_100107 | Low-quality |
| S16_extracted_100128 | Not-determined |
| S16_extracted_100147 | Low-quality |
| S16_extracted_100166 | Not-determined |
| S16_extracted_100213 | Not-determined |
| S16_extracted_100220 | Not-determined |
| S16_extracted_100224 | Low-quality |
| S16_extracted_100229 | Not-determined |
| S16_extracted_100230 | Not-determined |
| S16_extracted_100234 | Low-quality |
| S7_vs_53264\|\|full | Medium-quality |
| S16_extracted_100237 | Not-determined |
| S16_extracted_100250 | Not-determined |
| S16_extracted_100259 | Low-quality |
| S16_extracted_100269 | Not-determined |
| S16_extracted_100281 | Not-determined |
| S16_extracted_100283 | Not-determined |
| S16_extracted_100285 | Not-determined |
| S16_extracted_100287 | Not-determined |
| S16_extracted_100293 | Not-determined |
| S16_extracted_100302 | Not-determined |
| S16_extracted_100303 | Not-determined |
| S16_extracted_100311 | Not-determined |
| S16_extracted_100312 | Not-determined |
| S16_extracted_100320 | Not-determined |
| S16_extracted_100324 | Not-determined |
| S16_extracted_100336 | Not-determined |
| S16_extracted_100338 | Not-determined |
| S16_extracted_100339 | Not-determined |
| S16_extracted_100366 | Not-determined |
| S7_vs_13614\|\|full | High-quality |
| S16_extracted_100379 | Low-quality |
| S16_extracted_100392 | Not-determined |
| S16_extracted_100404 | Not-determined |
| S16_extracted_100412 | Not-determined |
| S16_extracted_100430 | Not-determined |
| S16_extracted_100443 | Not-determined |
| S16_extracted_100444 | Low-quality |
| S16_extracted_100450 | Low-quality |
| S16_extracted_100451 | Low-quality |
| S16_extracted_100458 | Low-quality |
| S16_extracted_100472 | Not-determined |
| S7_vs_136164\|\|full | High-quality |
| S16_extracted_100484 | Not-determined |
| S16_extracted_100490 | Not-determined |
| S16_extracted_100491 | Not-determined |
| S16_extracted_100506 | Low-quality |
| S16_extracted_100514 | Not-determined |
| S16_extracted_100546 | Not-determined |
| S16_extracted_100553 | Not-determined |
| S16_extracted_100589 | Not-determined |
| S16_extracted_100592 | Not-determined |
| S16_extracted_100599 | Not-determined |
| S16_extracted_100606 | Not-determined |
| S16_extracted_100609 | Not-determined |
| S16_extracted_100637 | Not-determined |
| S16_extracted_100654 | Not-determined |
| S16_extracted_100665 | Not-determined |
| S16_extracted_100666 | Not-determined |
| S16_extracted_100688 | Not-determined |
| S16_extracted_100695 | Not-determined |
| S16_extracted_100697 | Not-determined |
| S16_extracted_100709 | Not-determined |
| S16_extracted_100717 | Not-determined |
| S16_extracted_100759 | Not-determined |
| S16_extracted_100765 | Low-quality |
| S16_extracted_100777 | Not-determined |
| S16_extracted_100790 | Not-determined |
| S16_extracted_100792 | Not-determined |
| S16_extracted_100800 | Not-determined |
| S16_extracted_100803 | Not-determined |
| S16_extracted_100831 | Not-determined |
| S16_extracted_100838 | Not-determined |
| S16_extracted_100864 | Not-determined |
| S16_extracted_100880 | Not-determined |
| S16_extracted_100882 | Not-determined |
| S16_extracted_100884 | Not-determined |
| S16_extracted_100890 | Not-determined |
| S16_extracted_100934 | Not-determined |
| S16_extracted_100944 | Not-determined |
| S16_extracted_100958 | Not-determined |
| S16_extracted_100959 | Not-determined |
| S16_extracted_100980 | Not-determined |
| S16_extracted_100981 | Not-determined |
| S16_extracted_100984 | Not-determined |
| S16_extracted_101016 | Low-quality |
| S16_extracted_101022 | Not-determined |
| S16_extracted_101033 | Not-determined |
| S16_extracted_101050 | Low-quality |
| S16_extracted_101059 | Low-quality |
| S16_extracted_101061 | Not-determined |
| S16_extracted_101063 | Not-determined |
| S16_extracted_101066 | Not-determined |

| | |
|---|---|
| S16_extracted_101086 | Low-quality |
| S16_extracted_101116 | Not-determined |
| S16_extracted_101154 | Not-determined |
| S16_extracted_101168 | Not-determined |
| S16_extracted_101174 | Not-determined |
| S16_extracted_101183 | Not-determined |
| S16_extracted_101195 | Low-quality |
| S16_extracted_101202 | Low-quality |
| S16_extracted_101206 | Low-quality |
| S16_extracted_101210 | Not-determined |
| S16_extracted_101221 | Low-quality |
| S16_extracted_101234 | Not-determined |
| S16_extracted_101237 | Low-quality |
| S16_extracted_101245 | Not-determined |
| S16_extracted_101248 | Low-quality |
| S16_extracted_101257 | Not-determined |
| S16_extracted_101258 | Not-determined |
| S16_extracted_101263 | Not-determined |
| S16_extracted_101264 | Low-quality |
| S16_extracted_101286 | Low-quality |
| S16_extracted_101308 | Not-determined |
| S16_extracted_101320 | Not-determined |
| S16_extracted_101321 | Not-determined |
| S16_extracted_101334 | Not-determined |
| S16_extracted_101335 | Low-quality |
| S16_extracted_101336 | Not-determined |
| S16_extracted_101339 | Low-quality |
| S16_extracted_101340 | Not-determined |
| S16_extracted_101342 | Not-determined |
| S16_extracted_101343 | Low-quality |
| S16_extracted_101348 | Not-determined |
| S16_extracted_101352 | Not-determined |
| S16_extracted_101358 | Not-determined |
| S16_extracted_101360 | Not-determined |
| S16_extracted_101376 | Not-determined |
| S16_extracted_101399 | Not-determined |
| S16_extracted_101402 | Not-determined |
| S16_extracted_101404 | Not-determined |
| S16_extracted_101426 | Not-determined |
| S16_extracted_101431 | Not-determined |
| S16_extracted_101434 | Not-determined |
| S16_extracted_101437 | Low-quality |
| S16_extracted_101443 | Not-determined |
| S16_extracted_101453 | Not-determined |
| S16_extracted_101454 | Not-determined |
| S16_extracted_101458 | Not-determined |
| S16_extracted_101481 | Not-determined |
| S16_extracted_101490 | Not-determined |
| S16_extracted_101495 | Not-determined |
| S16_extracted_101506 | Not-determined |
| S16_extracted_101512 | Not-determined |
| S16_extracted_101521 | Not-determined |
| S16_extracted_101540 | Not-determined |
| S16_extracted_101544 | Not-determined |
| S16_extracted_101553 | Not-determined |
| S16_extracted_101554 | Low-quality |
| S16_extracted_101555 | Not-determined |
| S16_extracted_101569 | Not-determined |
| S16_extracted_101578 | Not-determined |
| S16_extracted_101586 | Not-determined |
| S16_extracted_101590 | Not-determined |
| S16_extracted_101594 | Not-determined |
| S16_extracted_101596 | Not-determined |
| S16_extracted_101602 | Not-determined |
| S16_extracted_101607 | Not-determined |
| S16_extracted_101620 | Not-determined |
| S16_extracted_101639 | Not-determined |
| S16_extracted_101640 | Low-quality |
| S16_extracted_101647 | Low-quality |
| S16_extracted_101655 | Not-determined |
| S16_extracted_101660 | Not-determined |
| S16_extracted_101672 | Not-determined |
| S16_extracted_101673 | Not-determined |
| S16_extracted_101678 | Not-determined |
| S16_extracted_101693 | Not-determined |
| S16_extracted_101700 | Not-determined |
| S16_extracted_101705 | Not-determined |
| S16_extracted_101706 | Not-determined |
| S16_extracted_101724 | Not-determined |
| S16_extracted_101726 | Not-determined |
| S16_extracted_101729 | Not-determined |
| S16_extracted_101736 | Not-determined |
| S16_extracted_101745 | Not-determined |
| S16_extracted_101758 | Not-determined |
| S16_extracted_101766 | Not-determined |
| S16_extracted_101767 | Not-determined |
| S16_extracted_101792 | Not-determined |
| S16_extracted_101795 | Not-determined |
| S16_extracted_101814 | Not-determined |
| S16_extracted_101815 | Not-determined |
| S16_extracted_101826 | Low-quality |
| S16_extracted_101849 | Not-determined |
| S16_extracted_101857 | Low-quality |
| S16_extracted_101866 | Not-determined |
| S16_extracted_101879 | Not-determined |
| S16_extracted_101895 | Not-determined |
| S16_extracted_101898 | Not-determined |
| S16_extracted_101905 | Not-determined |
| S16_extracted_101910 | Not-determined |
| S16_extracted_101914 | Not-determined |
| S16_extracted_101931 | Not-determined |
| S16_extracted_101951 | Not-determined |
| S16_extracted_101954 | Not-determined |
| S16_extracted_101971 | Not-determined |
| S16_extracted_101985 | Not-determined |
| S16_extracted_101987 | Not-determined |
| S16_extracted_101994 | Not-determined |
| S16_extracted_102000 | Not-determined |
| S16_extracted_102002 | Not-determined |
| S16_extracted_102006 | Not-determined |
| S16_extracted_102009 | Not-determined |
| S16_extracted_102012 | Not-determined |
| S16_extracted_102022 | Not-determined |
| S16_extracted_102040 | Not-determined |
| S16_extracted_102066 | Not-determined |
| S16_extracted_102070 | Not-determined |
| S16_extracted_102096 | Not-determined |
| S16_extracted_102109 | Not-determined |
| S16_extracted_102111 | Not-determined |
| S16_extracted_102120 | Not-determined |
| S16_extracted_102162 | Not-determined |
| S16_extracted_102165 | Low-quality |
| S16_extracted_102170 | Not-determined |
| S16_extracted_102178 | Not-determined |
| S16_extracted_102191 | Not-determined |
| S16_extracted_102196 | Not-determined |
| S16_extracted_102203 | Not-determined |
| S16_extracted_102212 | Not-determined |
| S16_extracted_102234 | Not-determined |
| S16_extracted_102237 | Low-quality |
| S16_extracted_102268 | Not-determined |
| S16_extracted_102282 | Low-quality |
| S16_extracted_102284 | Not-determined |
| S16_extracted_102288 | Not-determined |
| S16_extracted_102289 | Not-determined |
| S16_extracted_102308 | Not-determined |
| S16_extracted_102314 | Not-determined |
| S16_extracted_102319 | Not-determined |
| S16_extracted_102321 | Not-determined |
| S16_extracted_102324 | Low-quality |
| S16_extracted_102336 | Not-determined |
| S16_extracted_102346 | Not-determined |
| S16_extracted_102347 | Low-quality |
| S16_extracted_102348 | Not-determined |
| S16_extracted_102349 | Low-quality |
| S16_extracted_102350 | Not-determined |
| S16_extracted_102351 | Not-determined |
| S16_extracted_102355 | Not-determined |
| S16_extracted_102364 | Not-determined |
| S16_extracted_102365 | Not-determined |
| S16_extracted_102373 | Not-determined |
| S16_extracted_102406 | Not-determined |
| S16_extracted_102421 | Not-determined |
| S16_extracted_102425 | Not-determined |
| S16_extracted_102426 | Not-determined |
| S16_extracted_102434 | Not-determined |
| S16_extracted_102439 | Not-determined |
| S16_extracted_102463 | Not-determined |
| S16_extracted_102466 | Not-determined |
| S16_extracted_102468 | Not-determined |
| S16_extracted_102478 | Not-determined |
| S16_extracted_102487 | Not-determined |
| S16_extracted_102503 | Not-determined |
| S16_extracted_102531 | Not-determined |
| S16_extracted_102534 | Low-quality |
| S16_extracted_102535 | Low-quality |
| S16_extracted_102542 | Not-determined |
| S16_extracted_102543 | Low-quality |
| S16_extracted_102545 | Low-quality |
| S16_extracted_102547 | Not-determined |
| S16_extracted_102593 | Low-quality |
| S16_extracted_102596 | Low-quality |
| S16_extracted_102599 | Not-determined |
| S16_extracted_102622 | Not-determined |
| S16_extracted_102637 | Not-determined |
| S16_extracted_102642 | Not-determined |
| S16_extracted_102653 | Not-determined |
| S16_extracted_102656 | Not-determined |
| S16_extracted_102667 | Low-quality |
| S16_extracted_102689 | Not-determined |
| S16_extracted_102690 | Not-determined |
| S16_extracted_102698 | Low-quality |
| S16_extracted_102704 | Low-quality |
| S16_extracted_102711 | Not-determined |
| S16_extracted_102721 | Not-determined |
| S16_extracted_102737 | Not-determined |
| S16_extracted_102748 | Not-determined |
| S16_extracted_102751 | Not-determined |
| S16_extracted_102774 | Not-determined |
| S16_extracted_102796 | Not-determined |
| S16_extracted_102800 | Not-determined |
| S16_extracted_102805 | Not-determined |
| S16_extracted_102815 | Not-determined |
| S16_extracted_102826 | Not-determined |
| S16_extracted_102833 | Not-determined |
| S16_extracted_102859 | Not-determined |
| S16_extracted_102868 | Not-determined |
| S16_extracted_102895 | Low-quality |
| S16_extracted_102911 | Not-determined |
| S16_extracted_102946 | Not-determined |
| S16_extracted_102959 | Not-determined |
| S16_extracted_102974 | Not-determined |
| S16_extracted_102975 | Not-determined |
| S16_extracted_102983 | Not-determined |
| S16_extracted_102996 | Not-determined |
| S16_extracted_102998 | Low-quality |
| S16_extracted_103004 | Not-determined |
| S16_extracted_103014 | Not-determined |
| S16_extracted_103016 | Low-quality |
| S16_extracted_103050 | Not-determined |
| S16_extracted_103053 | Not-determined |
| S16_extracted_103069 | Not-determined |
| S16_extracted_103109 | Not-determined |
| S16_extracted_103118 | Not-determined |
| S16_extracted_103121 | Not-determined |
| S16_extracted_103124 | Not-determined |
| S16_extracted_103125 | Not-determined |
| S16_extracted_103142 | Low-quality |
| S16_extracted_103156 | Low-quality |
| S16_extracted_103185 | Not-determined |
| S16_extracted_103202 | Not-determined |
| S16_extracted_103213 | Not-determined |
| S16_extracted_103230 | Low-quality |
| S16_extracted_103231 | Not-determined |
| S16_extracted_103243 | Not-determined |
| S16_extracted_103256 | Not-determined |
| S16_extracted_103287 | Low-quality |
| S16_extracted_103291 | Not-determined |
| S16_extracted_103306 | Low-quality |
| S16_extracted_103307 | Not-determined |
| S16_extracted_103309 | Not-determined |
| S16_extracted_103311 | Not-determined |
| S16_extracted_103340 | Not-determined |
| S16_extracted_103341 | Not-determined |
| S16_extracted_103360 | Not-determined |
| S16_extracted_103367 | Not-determined |
| S16_extracted_103375 | Not-determined |
| S16_extracted_103379 | Not-determined |
| S16_extracted_103389 | Not-determined |
| S16_extracted_103400 | Not-determined |
| S16_extracted_103406 | Not-determined |
| S16_extracted_103418 | Not-determined |
| S16_extracted_103426 | Not-determined |
| S16_extracted_103428 | Not-determined |
| S16_extracted_103432 | Not-determined |
| S16_extracted_103453 | Not-determined |
| S16_extracted_103455 | Not-determined |
| S16_extracted_103459 | Not-determined |
| S16_extracted_103461 | Low-quality |
| S16_extracted_103466 | Not-determined |
| S16_extracted_103473 | Not-determined |
| S16_extracted_103479 | Not-determined |
| S16_extracted_103503 | Not-determined |
| S16_extracted_103505 | Not-determined |
| S16_extracted_103509 | Not-determined |
| S16_extracted_103512 | Not-determined |
| S16_extracted_103530 | Not-determined |
| S16_extracted_103531 | Not-determined |
| S16_extracted_103544 | Not-determined |
| S16_extracted_103558 | Low-quality |
| S16_extracted_103569 | Not-determined |
| S16_extracted_103629 | Not-determined |
| S16_extracted_103648 | Not-determined |
| S16_extracted_103655 | Not-determined |
| S16_extracted_103668 | Not-determined |
| S16_extracted_103683 | Not-determined |
| S16_extracted_103694 | Not-determined |
| S16_extracted_103706 | Not-determined |
| S16_extracted_103730 | Not-determined |
| S16_extracted_103734 | Not-determined |
| S16_extracted_103737 | Low-quality |
| S16_extracted_103750 | Not-determined |
| S16_extracted_103763 | Not-determined |
| S16_extracted_103764 | Not-determined |
| S16_extracted_103765 | Not-determined |
| S16_extracted_103768 | Not-determined |
| S16_extracted_103774 | Low-quality |
| S16_extracted_103785 | Low-quality |
| S16_extracted_103791 | Low-quality |
| S16_extracted_103801 | Low-quality |
| S16_extracted_103804 | Not-determined |
| S16_extracted_103810 | Not-determined |
| S16_extracted_103815 | Not-determined |
| S16_extracted_103821 | Not-determined |
| S16_extracted_103824 | Low-quality |
| S16_extracted_103829 | Not-determined |
| S16_extracted_103835 | Not-determined |
| S16_extracted_103858 | Not-determined |
| S16_extracted_103859 | Not-determined |
| S16_extracted_103872 | Not-determined |
| S16_extracted_103903 | Not-determined |
| S16_extracted_103920 | Not-determined |
| S16_extracted_103927 | Low-quality |
| S11_vs_k141_76784\|\|full | High-quality |
| S16_extracted_103952 | Not-determined |
| S16_extracted_103968 | Not-determined |
| S16_extracted_103973 | Not-determined |
| S16_extracted_103978 | Not-determined |
| S16_extracted_103983 | Low-quality |
| S16_extracted_104002 | Not-determined |
| S16_extracted_104019 | Not-determined |
| S16_extracted_104028 | Not-determined |
| S16_extracted_104029 | Low-quality |
| S16_extracted_104047 | Not-determined |
| S16_extracted_104060 | Not-determined |
| S16_extracted_104094 | Not-determined |
| S16_extracted_104096 | Not-determined |
| S16_extracted_104099 | Low-quality |
| S16_extracted_104103 | Not-determined |
| S16_extracted_104112 | Not-determined |
| S16_extracted_104115 | Not-determined |
| S16_extracted_104121 | Low-quality |
| S16_extracted_104125 | Not-determined |
| S16_extracted_104131 | Not-determined |
| S16_extracted_104132 | Not-determined |
| S16_extracted_104151 | Low-quality |
| S16_extracted_104157 | Not-determined |
| S16_extracted_104163 | Not-determined |
| S16_extracted_104164 | Not-determined |
| S16_extracted_104169 | Low-quality |
| S16_extracted_104180 | Not-determined |
| S16_extracted_104181 | Not-determined |
| S16_extracted_104200 | Not-determined |
| S16_extracted_104202 | Not-determined |
| S16_extracted_104223 | Not-determined |
| S16_extracted_104231 | Not-determined |
| S16_extracted_104233 | Not-determined |
| S16_extracted_104238 | Not-determined |
| S16_extracted_104241 | Not-determined |
| S16_extracted_104248 | Not-determined |
| S16_extracted_104263 | Low-quality |
| S16_extracted_104275 | Low-quality |
| S16_extracted_104281 | Not-determined |
| S16_extracted_104285 | Not-determined |
| S16_extracted_104299 | Not-determined |
| S16_extracted_104301 | Low-quality |
| S16_extracted_104304 | Low-quality |
| S16_extracted_104330 | Not-determined |
| S16_extracted_104341 | Not-determined |
| S16_extracted_104343 | Not-determined |
| S16_extracted_104356 | Not-determined |
| S16_extracted_104357 | Not-determined |
| S16_extracted_104359 | Low-quality |
| S16_extracted_104362 | Not-determined |
| S16_extracted_104366 | Low-quality |
| S16_extracted_104377 | Not-determined |
| S16_extracted_104380 | Not-determined |
| S16_extracted_104402 | Not-determined |
| S16_extracted_104407 | Not-determined |
| S16_extracted_104409 | Not-determined |
| S16_extracted_104415 | Not-determined |
| S16_extracted_104425 | Low-quality |
| S16_extracted_104431 | Not-determined |
| S16_extracted_104432 | Not-determined |
| S16_extracted_104439 | Not-determined |
| S16_extracted_104449 | Not-determined |
| S16_extracted_104457 | Not-determined |
| S16_extracted_104459 | Not-determined |
| S16_extracted_104463 | Low-quality |
| S16_extracted_104468 | Not-determined |
| S16_extracted_104469 | Not-determined |
| S16_extracted_104479 | Low-quality |
| S16_extracted_104485 | Not-determined |
| S16_extracted_104489 | Not-determined |
| S16_extracted_104495 | Not-determined |
| S16_extracted_104501 | Not-determined |
| S16_extracted_104506 | Not-determined |
| S16_extracted_104509 | Low-quality |
| S16_extracted_104511 | Not-determined |
| S16_extracted_104512 | Not-determined |
| S16_extracted_104518 | Not-determined |
| S16_extracted_104525 | Not-determined |
| S16_extracted_104533 | Not-determined |
| S16_extracted_104555 | Not-determined |
| S16_extracted_104581 | Not-determined |
| S16_extracted_104583 | Not-determined |
| S16_extracted_104603 | Not-determined |
| S16_extracted_104608 | Not-determined |
| S16_extracted_104618 | Not-determined |
| S16_extracted_104622 | Not-determined |
| S16_extracted_104646 | Not-determined |
| S16_extracted_104664 | Not-determined |
| S16_extracted_104686 | Not-determined |
| S16_extracted_104688 | Not-determined |
| S16_extracted_104690 | Low-quality |
| S16_extracted_104692 | Not-determined |
| S16_extracted_104704 | Not-determined |
| S16_extracted_104713 | Not-determined |
| S16_extracted_104732 | Not-determined |
| S16_extracted_104749 | Low-quality |
| S16_extracted_104768 | Not-determined |
| S16_extracted_104782 | Not-determined |
| S16_extracted_104802 | Not-determined |
| S16_extracted_104806 | Not-determined |
| S16_extracted_104813 | Not-determined |
| S16_extracted_104815 | Not-determined |
| S16_extracted_104820 | Low-quality |
| S16_extracted_104821 | Not-determined |
| S16_extracted_104827 | Not-determined |
| S16_extracted_104870 | Not-determined |
| S16_extracted_104871 | Not-determined |
| S16_extracted_104895 | Low-quality |
| S16_extracted_104900 | Not-determined |
| S16_extracted_104904 | Not-determined |
| S16_extracted_104922 | Not-determined |
| S16_extracted_104939 | Not-determined |
| S16_extracted_104942 | Not-determined |
| S16_extracted_104946 | Not-determined |
| S16_extracted_104961 | Not-determined |
| S16_extracted_104977 | Not-determined |
| S16_extracted_104982 | Not-determined |
| S16_extracted_105000 | Not-determined |
| S16_extracted_105003 | Not-determined |
| S16_extracted_105019 | Not-determined |
| S16_extracted_105039 | Not-determined |
| S16_extracted_105041 | Not-determined |
| S16_extracted_105042 | Not-determined |
| S16_extracted_105051 | Not-determined |
| S16_extracted_105058 | Not-determined |
| S16_extracted_105060 | Not-determined |
| S16_extracted_105103 | Not-determined |
| S16_extracted_105135 | Not-determined |
| S16_extracted_105136 | Low-quality |
| S16_extracted_105144 | Not-determined |
| S16_extracted_105164 | Low-quality |
| S16_extracted_105173 | Not-determined |
| S16_extracted_105175 | Not-determined |
| S16_extracted_105176 | Not-determined |
| S16_extracted_105179 | Low-quality |
| S16_extracted_105180 | Low-quality |
| S16_extracted_105198 | Not-determined |
| S16_extracted_105206 | Not-determined |
| S16_extracted_105212 | Not-determined |
| S16_extracted_105213 | Low-quality |
| S16_extracted_105228 | Not-determined |
| S16_extracted_105231 | Low-quality |
| S16_extracted_105232 | Not-determined |
| S16_extracted_105246 | Not-determined |
| S16_extracted_105255 | Not-determined |
| S16_extracted_105259 | Not-determined |
| S16_extracted_105260 | Not-determined |
| S16_extracted_105272 | Not-determined |
| S16_extracted_105276 | Low-quality |
| S16_extracted_105278 | Not-determined |
| S16_extracted_105283 | Not-determined |
| S16_extracted_105285 | Not-determined |
| S16_extracted_105320 | Not-determined |
| S16_extracted_105340 | Not-determined |
| S16_extracted_105346 | Not-determined |
| S16_extracted_105363 | Not-determined |
| S16_extracted_105372 | Not-determined |
| S16_extracted_105386 | Not-determined |
| S16_extracted_105387 | Not-determined |
| S16_extracted_105389 | Not-determined |
| S16_extracted_105393 | Low-quality |
| S16_extracted_105400 | Not-determined |
| S16_extracted_105409 | Not-determined |
| S16_extracted_105421 | Not-determined |
| S16_extracted_105429 | Not-determined |
| S16_extracted_105442 | Not-determined |
| S16_extracted_105449 | Not-determined |
| S16_extracted_105461 | Not-determined |
| S16_extracted_105473 | Not-determined |
| S16_extracted_105475 | Not-determined |
| S16_extracted_105478 | Low-quality |
| S16_extracted_105480 | Not-determined |
| S16_extracted_105482 | Not-determined |
| S16_extracted_105487 | Low-quality |
| S16_extracted_105496 | Not-determined |
| S16_extracted_105504 | Not-determined |
| S16_extracted_105513 | Not-determined |
| S16_extracted_105520 | Not-determined |
| S16_extracted_105527 | Not-determined |
| S16_extracted_105553 | Not-determined |
| S16_extracted_105554 | Not-determined |
| S16_extracted_105558 | Not-determined |
| S16_extracted_105563 | Not-determined |
| S16_extracted_105574 | Not-determined |
| S16_extracted_105577 | Not-determined |
| S16_extracted_105607 | Not-determined |
| S16_extracted_105619 | Not-determined |
| S16_extracted_105622 | Not-determined |
| S16_extracted_105625 | Not-determined |
| S16_extracted_105635 | Low-quality |
| S16_extracted_105642 | Not-determined |
| S16_extracted_105649 | Not-determined |
| S16_extracted_105653 | Not-determined |
| S16_extracted_105657 | Low-quality |
| S16_extracted_105659 | Not-determined |
| S16_extracted_105674 | Not-determined |
| S16_extracted_105682 | Not-determined |
| S16_extracted_105698 | Not-determined |
| S16_extracted_105734 | Not-determined |
| S16_extracted_105747 | Low-quality |
| S16_extracted_105751 | Not-determined |
| S16_extracted_105761 | Not-determined |
| S16_extracted_105762 | Not-determined |
| S16_extracted_105765 | Not-determined |
| S16_extracted_105799 | Not-determined |
| S16_extracted_105800 | Not-determined |
| S16_extracted_105804 | Not-determined |
| S16_extracted_105808 | Not-determined |
| S16_extracted_105817 | Not-determined |
| S16_extracted_105838 | Not-determined |
| S16_extracted_105853 | Not-determined |
| S16_extracted_105856 | Not-determined |
| S16_extracted_105857 | Not-determined |
| S16_extracted_105868 | Not-determined |
| S16_extracted_105876 | Not-determined |
| S16_extracted_105880 | Not-determined |
| S16_extracted_105887 | Not-determined |
| S16_extracted_105888 | Not-determined |
| S16_extracted_105897 | Not-determined |
| S16_extracted_105907 | Not-determined |
| S16_extracted_105926 | Not-determined |
| S16_extracted_105936 | Not-determined |
| S16_extracted_105937 | Not-determined |
| S16_extracted_105941 | Not-determined |
| S16_extracted_105949 | Not-determined |
| S16_extracted_105975 | Low-quality |
| S16_extracted_105991 | Low-quality |
| S16_extracted_105999 | Low-quality |
| S16_extracted_106009 | Not-determined |
| S16_extracted_106010 | Not-determined |
| S16_extracted_106038 | Low-quality |
| S16_extracted_106044 | Not-determined |
| S16_extracted_106064 | Not-determined |
| S16_extracted_106076 | Not-determined |
| S16_extracted_106089 | Not-determined |
| S16_extracted_106091 | Low-quality |
| S16_extracted_106115 | Not-determined |
| S16_extracted_106124 | Not-determined |
| S16_extracted_106156 | Low-quality |
| S16_extracted_106159 | Not-determined |
| S16_extracted_106166 | Not-determined |
| S16_extracted_106171 | Not-determined |
| S16_extracted_106199 | Low-quality |
| S16_extracted_106206 | Not-determined |
| S16_extracted_106218 | Not-determined |
| S16_extracted_106221 | Not-determined |
| S16_extracted_106224 | Not-determined |
| S16_extracted_106301 | Not-determined |
| S16_extracted_106369 | Not-determined |
| S16_extracted_106370 | Not-determined |
| S16_extracted_106376 | Not-determined |
| S16_extracted_106401 | Not-determined |
| S16_extracted_106424 | Not-determined |
| S16_extracted_106429 | Not-determined |
| S16_extracted_106436 | Not-determined |
| S16_extracted_106445 | Not-determined |
| S16_extracted_106448 | Not-determined |
| S16_extracted_106470 | Not-determined |
| S16_extracted_106489 | Not-determined |
| S16_extracted_106496 | Not-determined |
| S16_extracted_106518 | Not-determined |
| S16_extracted_106524 | Not-determined |
| S16_extracted_106527 | Not-determined |
| S16_extracted_106535 | Not-determined |
| S16_extracted_106541 | Low-quality |
| S16_extracted_106559 | Not-determined |
| S16_extracted_106583 | Not-determined |
| S16_extracted_106595 | Not-determined |
| S16_extracted_106601 | Not-determined |
| S16_extracted_106604 | Not-determined |
| S16_extracted_106608 | Not-determined |
| S16_extracted_106609 | Not-determined |
| S16_extracted_106618 | Not-determined |
| S16_extracted_106630 | Not-determined |
| S16_extracted_106631 | Not-determined |
| S16_extracted_106659 | Low-quality |
| S16_extracted_106663 | Not-determined |
| S16_extracted_106675 | Not-determined |
| S16_extracted_106683 | Not-determined |
| S16_extracted_106690 | Not-determined |
| S16_extracted_106696 | Not-determined |
| S16_extracted_106706 | Not-determined |
| S16_extracted_106708 | Not-determined |
| S16_extracted_106721 | Not-determined |
| S16_extracted_106739 | Low-quality |
| S16_extracted_106742 | Not-determined |
| S16_extracted_106750 | Not-determined |
| S16_extracted_106762 | Not-determined |
| S16_extracted_106763 | Not-determined |
| S16_extracted_106767 | Not-determined |
| S16_extracted_106790 | Not-determined |
| S16_extracted_106797 | Not-determined |
| S16_extracted_106801 | Not-determined |
| S16_extracted_106802 | Not-determined |
| S16_extracted_106854 | Not-determined |

| | |
|---|---|
| S16_extracted_106865 | Not-determined |
| S16_extracted_106871 | Not-determined |
| S16_extracted_106873 | Not-determined |
| S16_extracted_106875 | Not-determined |
| S16_extracted_106897 | Not-determined |
| S16_extracted_106914 | Not-determined |
| S16_extracted_106919 | Not-determined |
| S16_extracted_106921 | Not-determined |
| S16_extracted_106925 | Low-quality |
| S16_extracted_106926 | Not-determined |
| S16_extracted_106933 | Not-determined |
| S16_extracted_106949 | Not-determined |
| S16_extracted_106951 | Not-determined |
| S16_extracted_106982 | Not-determined |
| S16_extracted_106985 | Not-determined |
| S16_extracted_106999 | Not-determined |
| S16_extracted_107001 | Not-determined |
| S16_extracted_107003 | Not-determined |
| S16_extracted_107013 | Not-determined |
| S16_extracted_107014 | Not-determined |
| S16_extracted_107025 | Low-quality |
| S16_extracted_107026 | Not-determined |
| S16_extracted_107036 | Low-quality |
| S16_extracted_107051 | Not-determined |
| S16_extracted_107065 | Not-determined |
| S16_extracted_107080 | Not-determined |
| S16_extracted_107087 | Not-determined |
| S16_extracted_107105 | Not-determined |
| S16_extracted_107126 | Not-determined |
| S16_extracted_107128 | Not-determined |
| S16_extracted_107130 | Not-determined |
| S16_extracted_107140 | Not-determined |
| S16_extracted_107147 | Not-determined |
| S16_extracted_107148 | Not-determined |
| S16_extracted_107150 | Not-determined |
| S16_extracted_107158 | Not-determined |
| S16_extracted_107164 | Not-determined |
| S16_extracted_107167 | Not-determined |
| S16_extracted_107171 | Not-determined |
| S16_extracted_107177 | Not-determined |
| S16_extracted_107213 | Not-determined |
| S16_extracted_107223 | Not-determined |
| S16_extracted_107226 | Not-determined |
| S16_extracted_107234 | Not-determined |
| S16_extracted_107241 | Low-quality |
| S16_extracted_107251 | Low-quality |
| S16_extracted_107255 | Not-determined |
| S16_extracted_107257 | Not-determined |
| S16_extracted_107270 | Not-determined |
| S16_extracted_107284 | Not-determined |
| S16_extracted_107308 | Not-determined |
| S16_extracted_107342 | Not-determined |
| S16_extracted_107362 | Not-determined |
| S16_extracted_107366 | Not-determined |
| S16_extracted_107396 | Not-determined |
| S16_extracted_107398 | Not-determined |
| S16_extracted_107419 | Not-determined |
| S16_extracted_107428 | Not-determined |
| S16_extracted_107433 | Not-determined |
| S16_extracted_107439 | Not-determined |
| S16_extracted_107457 | Not-determined |
| S16_extracted_107460 | Not-determined |
| S16_extracted_107461 | Not-determined |
| S16_extracted_107480 | Not-determined |
| S16_extracted_107494 | Not-determined |
| S16_extracted_107503 | Not-determined |
| S16_extracted_107537 | Not-determined |
| S16_extracted_107539 | Not-determined |
| S16_extracted_107544 | Not-determined |
| S16_extracted_107551 | Low-quality |
| S16_extracted_107559 | Not-determined |
| S16_extracted_107561 | Not-determined |
| S16_extracted_107578 | Not-determined |
| S16_extracted_107579 | Low-quality |
| S16_extracted_107586 | Not-determined |
| S16_extracted_107594 | Not-determined |
| S16_extracted_107607 | Not-determined |
| S16_extracted_107612 | Not-determined |
| S16_extracted_107616 | Low-quality |
| S16_extracted_107624 | Not-determined |
| S16_extracted_107625 | Low-quality |
| S16_extracted_107639 | Low-quality |
| S16_extracted_107643 | Not-determined |
| S16_extracted_107644 | Not-determined |
| S16_extracted_107653 | Not-determined |
| S16_extracted_107656 | Low-quality |
| S16_extracted_107657 | Not-determined |
| S16_extracted_107668 | Not-determined |
| S16_extracted_107681 | Not-determined |
| S16_extracted_107697 | Not-determined |
| S16_extracted_107711 | Low-quality |
| S16_extracted_107722 | Not-determined |
| S16_extracted_107723 | Not-determined |
| S16_extracted_107731 | Not-determined |
| S16_extracted_107737 | Not-determined |
| S16_extracted_107741 | Not-determined |
| S16_extracted_107747 | Not-determined |
| S16_extracted_107751 | Low-quality |
| S16_extracted_107752 | Not-determined |
| S16_extracted_107757 | Not-determined |
| S16_extracted_107759 | Not-determined |
| S16_extracted_107765 | Low-quality |
| S16_extracted_107772 | Not-determined |
| S16_extracted_107782 | Low-quality |
| S16_extracted_107783 | Not-determined |
| S16_extracted_107787 | Not-determined |
| S16_extracted_107795 | Not-determined |
| S16_extracted_107835 | Not-determined |
| S16_extracted_107839 | Not-determined |
| S16_extracted_107850 | Not-determined |
| S16_extracted_107858 | Not-determined |
| S16_extracted_107870 | Not-determined |
| S16_extracted_107897 | Not-determined |
| S16_extracted_107923 | Not-determined |
| S16_extracted_107938 | Not-determined |
| S16_extracted_107945 | Low-quality |
| S16_extracted_107954 | Not-determined |
| S16_extracted_107955 | Not-determined |
| S16_extracted_107975 | Low-quality |
| S16_extracted_107977 | Not-determined |
| S16_extracted_107983 | Not-determined |
| S16_extracted_107991 | Not-determined |
| S16_extracted_107993 | Not-determined |
| S16_extracted_107996 | Not-determined |
| S16_extracted_108020 | Not-determined |
| S16_extracted_108030 | Not-determined |
| S16_extracted_108040 | Not-determined |
| S16_extracted_108049 | Not-determined |
| S16_extracted_108055 | Not-determined |
| S16_extracted_108062 | Not-determined |
| S16_extracted_108071 | Not-determined |
| S16_extracted_108072 | Not-determined |
| S16_extracted_108105 | Not-determined |
| S16_extracted_108110 | Not-determined |
| S16_extracted_108124 | Not-determined |
| S16_extracted_108145 | Not-determined |
| S16_extracted_108150 | Not-determined |
| S16_extracted_108154 | Not-determined |
| S16_extracted_108163 | Not-determined |
| S16_extracted_108164 | Not-determined |
| S16_extracted_108169 | Not-determined |
| S16_extracted_108172 | Not-determined |
| S16_extracted_108177 | Not-determined |
| S16_extracted_108182 | Low-quality |
| S16_extracted_108199 | Not-determined |
| S16_extracted_108202 | Low-quality |
| S16_extracted_108219 | Not-determined |
| S16_extracted_108232 | Not-determined |
| S16_extracted_108233 | Low-quality |
| S16_extracted_108236 | Not-determined |
| S16_extracted_108238 | Not-determined |
| S16_extracted_108260 | Not-determined |
| S16_extracted_108262 | Not-determined |
| S16_extracted_108271 | Not-determined |
| S16_extracted_108277 | Not-determined |
| S16_extracted_108291 | Not-determined |
| S16_extracted_108296 | Low-quality |
| S16_extracted_108314 | Not-determined |
| S16_extracted_108320 | Not-determined |
| S16_extracted_108332 | Not-determined |
| S16_extracted_108342 | Not-determined |
| S16_extracted_108344 | Not-determined |
| S16_extracted_108387 | Not-determined |
| S16_extracted_108396 | Not-determined |
| S16_extracted_108403 | Not-determined |
| S16_extracted_108409 | Not-determined |
| S16_extracted_108415 | Not-determined |
| S16_extracted_108420 | Not-determined |
| S16_extracted_108423 | Not-determined |
| S16_extracted_108463 | Not-determined |
| S16_extracted_108479 | Not-determined |
| S16_extracted_108481 | Not-determined |
| S16_extracted_108508 | Not-determined |
| S16_extracted_108521 | Not-determined |
| S16_extracted_108528 | Not-determined |
| S16_extracted_108533 | Not-determined |
| S16_extracted_108540 | Not-determined |
| S16_extracted_108543 | Not-determined |
| S16_extracted_108546 | Low-quality |
| S16_extracted_108547 | Not-determined |
| S16_extracted_108549 | Not-determined |
| S13_vs_114213\|\|full | Medium-quality |
| S16_extracted_108566 | Not-determined |
| S16_extracted_108569 | Not-determined |
| S16_extracted_108582 | Not-determined |
| S16_extracted_108610 | Not-determined |
| S16_extracted_108611 | Not-determined |
| S16_extracted_108614 | Not-determined |
| S16_extracted_108628 | Low-quality |
| S16_extracted_108635 | Not-determined |
| S16_extracted_108645 | Not-determined |
| S16_extracted_108654 | Not-determined |
| S16_extracted_108676 | Not-determined |
| S16_extracted_108706 | Not-determined |
| S16_extracted_108711 | Not-determined |
| S16_extracted_108714 | Not-determined |
| S16_extracted_108715 | Not-determined |
| S16_extracted_108717 | Not-determined |
| S16_extracted_108722 | Not-determined |
| S16_extracted_108724 | Not-determined |
| S16_extracted_108727 | Not-determined |
| S16_extracted_108744 | Not-determined |
| S16_extracted_108747 | Not-determined |
| S16_extracted_108749 | Not-determined |
| S16_extracted_108762 | Not-determined |
| S16_extracted_108768 | Low-quality |
| S16_extracted_108811 | Low-quality |
| S16_extracted_108822 | Not-determined |
| S16_extracted_108825 | Not-determined |
| S16_extracted_108859 | Not-determined |
| S16_extracted_108860 | Low-quality |
| S16_extracted_108873 | Not-determined |
| S16_extracted_108890 | Not-determined |
| S16_extracted_108891 | Not-determined |
| S16_extracted_108916 | Not-determined |
| S16_extracted_108921 | Not-determined |
| S16_extracted_108922 | Not-determined |
| S16_extracted_108934 | Not-determined |
| S16_extracted_108975 | Not-determined |
| S16_extracted_108978 | Not-determined |
| S16_extracted_109008 | Low-quality |
| S16_extracted_109017 | Low-quality |
| S16_extracted_109062 | Not-determined |
| S16_extracted_109072 | Not-determined |
| S16_extracted_109084 | Not-determined |
| S16_extracted_109113 | Not-determined |
| S16_extracted_109118 | Not-determined |
| S16_extracted_109128 | Not-determined |
| S16_extracted_109140 | Low-quality |
| S16_extracted_109145 | Not-determined |
| S16_extracted_109158 | Low-quality |
| S16_extracted_109169 | Not-determined |
| S16_extracted_109180 | Not-determined |
| S16_extracted_109232 | Not-determined |
| S16_extracted_109253 | Low-quality |
| S16_extracted_109263 | Not-determined |
| S16_extracted_109276 | Not-determined |
| S16_extracted_109282 | Not-determined |
| S16_extracted_109292 | Not-determined |
| S16_extracted_109323 | Not-determined |
| S16_extracted_109325 | Not-determined |
| S16_extracted_109332 | Not-determined |
| S16_extracted_109347 | Not-determined |
| S16_extracted_109357 | Not-determined |
| S16_extracted_109361 | Low-quality |
| S16_extracted_109363 | Not-determined |
| S16_extracted_109368 | Not-determined |
| S16_extracted_109379 | Low-quality |
| S16_extracted_109399 | Not-determined |
| S16_extracted_109411 | Not-determined |
| S16_extracted_109415 | Not-determined |
| S16_extracted_109420 | Low-quality |
| S16_extracted_109439 | Not-determined |
| S16_extracted_109440 | Low-quality |
| S16_extracted_109468 | Low-quality |
| S16_extracted_109474 | Low-quality |
| S16_extracted_109477 | Not-determined |
| S16_extracted_109489 | Not-determined |
| S16_extracted_109495 | Not-determined |
| S16_extracted_109499 | Not-determined |
| S16_extracted_109500 | Not-determined |
| S16_extracted_109501 | Not-determined |
| S16_extracted_109516 | Not-determined |
| S16_extracted_109538 | Not-determined |
| S16_extracted_109552 | Low-quality |
| S16_extracted_109571 | Not-determined |
| S16_extracted_109573 | Not-determined |
| S16_extracted_109574 | Not-determined |
| S16_extracted_109581 | Not-determined |
| S16_extracted_109582 | Not-determined |
| S16_extracted_109603 | Not-determined |
| S16_extracted_109605 | Not-determined |
| S16_extracted_109621 | Not-determined |
| S16_extracted_109639 | Not-determined |
| S16_extracted_109659 | Low-quality |
| S16_extracted_109664 | Not-determined |
| S16_extracted_109667 | Not-determined |
| S16_extracted_109676 | Low-quality |
| S16_extracted_109688 | Not-determined |
| S16_extracted_109691 | Not-determined |
| S16_extracted_109699 | Not-determined |
| S16_extracted_109722 | Not-determined |
| S16_extracted_109726 | Not-determined |
| S16_extracted_109731 | Not-determined |
| S16_extracted_109733 | Not-determined |
| S16_extracted_109734 | Not-determined |
| S16_extracted_109755 | Not-determined |
| S16_extracted_109776 | Not-determined |
| S16_extracted_109795 | Not-determined |
| S16_extracted_109797 | Not-determined |
| S16_extracted_109807 | Not-determined |
| S16_extracted_109816 | Not-determined |
| S16_extracted_109824 | Not-determined |
| S16_extracted_109825 | Not-determined |
| S16_extracted_109840 | Not-determined |
| S16_extracted_109846 | Not-determined |
| S16_extracted_109852 | Not-determined |
| S16_extracted_109858 | Not-determined |
| S16_extracted_109877 | Low-quality |
| S16_extracted_109881 | Not-determined |
| S16_extracted_109882 | Not-determined |
| S16_extracted_109920 | Not-determined |
| S16_extracted_109928 | Low-quality |
| S16_extracted_109936 | Not-determined |
| S16_extracted_109949 | Not-determined |
| S16_extracted_109959 | Not-determined |
| S16_extracted_109962 | Not-determined |
| S16_extracted_109965 | Not-determined |
| S16_extracted_109976 | Not-determined |
| S16_extracted_110006 | Low-quality |
| S16_extracted_110040 | Not-determined |
| S16_extracted_110068 | Not-determined |
| S16_extracted_110093 | Not-determined |
| S16_extracted_110095 | Not-determined |
| S16_extracted_110104 | Not-determined |
| S16_extracted_110108 | Low-quality |
| S16_extracted_110115 | Not-determined |
| S16_extracted_110117 | Not-determined |
| S16_extracted_110124 | Not-determined |
| S16_extracted_110132 | Low-quality |
| S16_extracted_110134 | Low-quality |
| S16_extracted_110141 | Not-determined |
| S16_extracted_110149 | Not-determined |
| S16_extracted_110157 | Not-determined |
| S16_extracted_110168 | Not-determined |
| S16_extracted_110174 | Not-determined |
| S16_extracted_110181 | Not-determined |
| S16_extracted_110185 | Not-determined |
| S16_extracted_110187 | Not-determined |
| S16_extracted_110191 | Not-determined |
| S16_extracted_110192 | Not-determined |
| S16_extracted_110197 | Not-determined |
| S16_extracted_110216 | Not-determined |
| S16_extracted_110225 | Low-quality |
| S16_extracted_110228 | Not-determined |
| S16_extracted_110230 | Not-determined |
| S16_extracted_110231 | Not-determined |
| S16_extracted_110248 | Not-determined |
| S16_extracted_110250 | Not-determined |
| S16_extracted_110274 | Not-determined |
| S16_extracted_110281 | Not-determined |
| S16_extracted_110287 | Low-quality |
| S16_extracted_110311 | Not-determined |
| S16_extracted_110324 | Low-quality |
| S16_extracted_110377 | Not-determined |
| S16_extracted_110401 | Not-determined |
| S16_extracted_110429 | Not-determined |
| S16_extracted_110440 | Low-quality |
| S16_extracted_110465 | Low-quality |
| S16_extracted_110468 | Not-determined |
| S16_extracted_110472 | Not-determined |
| S16_extracted_110478 | Not-determined |
| S16_extracted_110485 | Not-determined |
| S16_extracted_110498 | Not-determined |
| S16_extracted_110523 | Not-determined |
| S16_extracted_110530 | Not-determined |
| S16_extracted_110537 | Not-determined |
| S16_extracted_110542 | Low-quality |
| S16_extracted_110549 | Not-determined |
| S16_extracted_110558 | Not-determined |
| S16_extracted_110563 | Not-determined |
| S16_extracted_110568 | Not-determined |
| S16_extracted_110576 | Low-quality |
| S16_extracted_110582 | Not-determined |
| S16_extracted_110585 | Not-determined |
| S16_extracted_110587 | Not-determined |
| S16_extracted_110604 | Not-determined |
| S16_extracted_110609 | Not-determined |
| S16_extracted_110623 | Not-determined |
| S16_extracted_110641 | Not-determined |
| S16_extracted_110652 | Not-determined |
| S16_extracted_110662 | Not-determined |
| S16_extracted_110671 | Not-determined |
| S16_extracted_110677 | Not-determined |
| S16_extracted_110678 | Low-quality |
| S16_extracted_110687 | Not-determined |
| S16_extracted_110700 | Not-determined |
| S16_extracted_110711 | Not-determined |
| S16_extracted_110713 | Not-determined |
| S16_extracted_110723 | Low-quality |
| S16_extracted_110724 | Not-determined |
| S16_extracted_110735 | Not-determined |
| S16_extracted_110737 | Not-determined |
| S16_extracted_110757 | Not-determined |
| S16_extracted_110771 | Not-determined |
| S16_extracted_110775 | Not-determined |
| S16_extracted_110788 | Not-determined |
| S16_extracted_110800 | Not-determined |
| S16_extracted_110812 | Not-determined |
| S16_extracted_110837 | Not-determined |
| S16_extracted_110840 | Not-determined |
| S16_extracted_110851 | Not-determined |
| S16_extracted_110859 | Not-determined |
| S16_extracted_110860 | Not-determined |
| S16_extracted_110863 | Not-determined |
| S16_extracted_110865 | Low-quality |
| S16_extracted_110874 | Low-quality |
| S16_extracted_110877 | Not-determined |
| S16_extracted_110886 | Not-determined |
| S16_extracted_110889 | Not-determined |
| S16_extracted_110892 | Low-quality |
| S16_extracted_110896 | Not-determined |
| S16_extracted_110897 | Not-determined |
| S16_extracted_110901 | Not-determined |
| S16_extracted_110903 | Low-quality |
| S16_extracted_110905 | Not-determined |
| S16_extracted_110912 | Not-determined |
| S16_extracted_110919 | Low-quality |
| S16_extracted_110920 | Not-determined |
| S16_extracted_110931 | Not-determined |
| S16_extracted_110948 | Not-determined |
| S16_extracted_110950 | Low-quality |
| S16_extracted_110965 | Not-determined |
| S16_extracted_110979 | Not-determined |
| S16_extracted_110986 | Low-quality |
| S16_extracted_110990 | Not-determined |
| S16_extracted_110993 | Not-determined |
| S16_extracted_110997 | Not-determined |
| S16_extracted_111027 | Not-determined |
| S16_extracted_111036 | Not-determined |
| S16_extracted_111040 | Not-determined |
| S16_extracted_111050 | Not-determined |
| S16_extracted_111057 | Not-determined |
| S16_extracted_111062 | Not-determined |
| S16_extracted_111074 | Low-quality |
| S16_extracted_111077 | Not-determined |
| S16_extracted_111079 | Not-determined |
| S16_extracted_111083 | Not-determined |
| S16_extracted_111097 | Low-quality |
| S16_extracted_111122 | Not-determined |
| S16_extracted_111125 | Not-determined |
| S16_extracted_111127 | Not-determined |
| S16_extracted_111141 | Not-determined |
| S16_extracted_111144 | Low-quality |
| S16_extracted_111149 | Low-quality |
| S16_extracted_111150 | Low-quality |
| S16_extracted_111158 | Not-determined |
| S16_extracted_111181 | Not-determined |
| S16_extracted_111186 | Not-determined |
| S16_extracted_111192 | Not-determined |
| S16_extracted_111226 | Not-determined |
| S16_extracted_111230 | Not-determined |
| S16_extracted_111255 | Not-determined |
| S16_extracted_111264 | Low-quality |
| S16_extracted_111277 | Not-determined |
| S16_extracted_111280 | Not-determined |
| S16_extracted_111287 | Not-determined |
| S16_extracted_111290 | Not-determined |
| S16_extracted_111307 | Not-determined |
| S16_extracted_111340 | Not-determined |
| S16_extracted_111349 | Not-determined |
| S16_extracted_111355 | Not-determined |
| S16_extracted_111358 | Not-determined |
| S16_extracted_111362 | Not-determined |
| S16_extracted_111366 | Low-quality |
| S16_extracted_111382 | Not-determined |
| S16_extracted_111383 | Not-determined |
| S16_extracted_111388 | Low-quality |
| S16_extracted_111398 | Not-determined |
| S16_extracted_111417 | Not-determined |
| S16_extracted_111424 | Not-determined |
| S16_extracted_111435 | Not-determined |
| S16_extracted_111438 | Not-determined |
| S16_extracted_111441 | Not-determined |
| S16_extracted_111462 | Not-determined |
| S16_extracted_111464 | Not-determined |
| S16_extracted_111468 | Not-determined |
| S16_extracted_111470 | Not-determined |
| S16_extracted_111512 | Low-quality |
| S16_extracted_111518 | Low-quality |
| S16_extracted_111531 | Not-determined |
| S16_extracted_111540 | Low-quality |
| S16_extracted_111550 | Not-determined |
| S16_extracted_111551 | Not-determined |
| S16_extracted_111561 | Not-determined |
| S16_extracted_111568 | Low-quality |
| S16_extracted_111607 | Not-determined |
| S16_extracted_111609 | Not-determined |
| S16_extracted_111623 | Not-determined |
| S16_extracted_111633 | Low-quality |
| S16_extracted_111635 | Not-determined |
| S16_extracted_111656 | Not-determined |
| S16_extracted_111660 | Not-determined |
| S16_extracted_111664 | Low-quality |
| S16_extracted_111671 | Low-quality |
| S16_extracted_111680 | Not-determined |
| S16_extracted_111694 | Not-determined |
| S16_extracted_111702 | Low-quality |
| S16_extracted_111708 | Not-determined |
| S16_extracted_111718 | Not-determined |
| S16_extracted_111725 | Low-quality |
| S16_extracted_111733 | Not-determined |
| S16_extracted_111743 | Low-quality |
| S16_extracted_111755 | Low-quality |
| S16_extracted_111783 | Not-determined |
| S16_extracted_111786 | Low-quality |
| S16_extracted_111787 | Not-determined |
| S16_extracted_111792 | Not-determined |
| S16_extracted_111796 | Not-determined |
| S16_extracted_111804 | Not-determined |
| S16_extracted_111823 | Not-determined |
| S16_extracted_111824 | Not-determined |
| S16_extracted_111837 | Not-determined |
| S16_extracted_111842 | Not-determined |
| S16_extracted_111863 | Low-quality |
| S16_extracted_111873 | Not-determined |
| S16_extracted_111882 | Not-determined |
| S16_extracted_111894 | Not-determined |
| S16_extracted_111907 | Not-determined |
| S16_extracted_111925 | Low-quality |
| S16_extracted_111936 | Low-quality |
| S16_extracted_111961 | Not-determined |
| S16_extracted_111980 | Not-determined |
| S16_extracted_111982 | Not-determined |
| S16_extracted_111991 | Not-determined |
| S16_extracted_112003 | Low-quality |
| S16_extracted_112005 | Not-determined |
| S16_extracted_112007 | Not-determined |
| S16_extracted_112028 | Not-determined |
| S16_extracted_112041 | Low-quality |
| S16_extracted_112043 | Not-determined |
| S16_extracted_112051 | Not-determined |
| S16_extracted_112057 | Low-quality |
| S16_extracted_112062 | Not-determined |
| S16_extracted_112063 | Not-determined |
| S16_extracted_112067 | Not-determined |
| S16_extracted_112085 | Not-determined |
| S16_extracted_112092 | Not-determined |
| S16_extracted_112093 | Not-determined |
| S16_extracted_112102 | Low-quality |
| S16_extracted_112104 | Not-determined |
| S16_extracted_112112 | Not-determined |
| S16_extracted_112115 | Not-determined |
| S16_extracted_112122 | Low-quality |
| S16_extracted_112123 | Not-determined |
| S16_extracted_112139 | Not-determined |
| S16_extracted_112155 | Not-determined |
| S16_extracted_112186 | Not-determined |
| S16_extracted_112196 | Not-determined |
| S16_extracted_112198 | Low-quality |
| S16_extracted_112199 | Not-determined |
| S16_extracted_112202 | Not-determined |
| S16_extracted_112205 | Not-determined |
| S16_extracted_112227 | Not-determined |
| S16_extracted_112230 | Low-quality |
| S16_extracted_112239 | Not-determined |
| S16_extracted_112253 | Not-determined |
| S16_extracted_112257 | Not-determined |
| S16_extracted_112266 | Not-determined |
| S16_extracted_112278 | Not-determined |
| S16_extracted_112287 | Low-quality |
| S16_extracted_112304 | Low-quality |
| S16_extracted_112314 | Not-determined |
| S16_extracted_112326 | Not-determined |
| S16_extracted_112377 | Not-determined |
| S16_extracted_112380 | Not-determined |
| S16_extracted_112385 | Not-determined |
| S16_extracted_112402 | Low-quality |
| S16_extracted_112407 | Not-determined |
| S16_extracted_112409 | Not-determined |
| S16_extracted_112456 | Not-determined |
| S16_extracted_112496 | Not-determined |
| S16_extracted_112503 | Not-determined |
| S16_extracted_112521 | Low-quality |
| S16_extracted_112529 | Not-determined |
| S16_extracted_112544 | Low-quality |
| S16_extracted_112548 | Not-determined |
| S16_extracted_112568 | Not-determined |
| S16_extracted_112570 | Not-determined |
| S16_extracted_112574 | Not-determined |
| S16_extracted_112578 | Not-determined |
| S16_extracted_112579 | Not-determined |
| S16_extracted_112590 | Not-determined |
| S16_extracted_112606 | Not-determined |
| S16_extracted_112617 | Not-determined |
| S16_extracted_112631 | Not-determined |
| S16_extracted_112633 | Not-determined |
| S16_extracted_112638 | Not-determined |
| S16_extracted_112643 | Not-determined |
| S16_extracted_112675 | Not-determined |
| S16_extracted_112680 | Low-quality |
| S16_extracted_112681 | Not-determined |
| S16_extracted_112692 | Not-determined |
| S16_extracted_112701 | Not-determined |
| S16_extracted_112737 | Not-determined |
| S16_extracted_112739 | Not-determined |
| S16_extracted_112761 | Low-quality |
| S16_extracted_112776 | Not-determined |
| S16_extracted_112781 | Not-determined |

| | |
|---|---|
| S16_extracted_112788 | Not-determined |
| S16_extracted_112802 | Not-determined |
| S16_extracted_112844 | Not-determined |
| S16_extracted_112846 | Low-quality |
| S16_extracted_112874 | Not-determined |
| S16_extracted_112880 | Not-determined |
| S16_extracted_112896 | Low-quality |
| S16_extracted_112920 | Low-quality |
| S16_extracted_112922 | Not-determined |
| S16_extracted_112927 | Not-determined |
| S16_extracted_112937 | Not-determined |
| S16_extracted_112967 | Low-quality |
| S16_extracted_112975 | Not-determined |
| S16_extracted_112983 | Not-determined |
| S16_extracted_112996 | Low-quality |
| S16_extracted_113028 | Not-determined |
| S16_extracted_113047 | Not-determined |
| S16_extracted_113059 | Low-quality |
| S16_extracted_113071 | Not-determined |
| S16_extracted_113074 | Not-determined |
| S16_extracted_113089 | Not-determined |
| S16_extracted_113100 | Low-quality |
| S16_extracted_113105 | Not-determined |
| S16_extracted_113109 | Not-determined |
| S16_extracted_113130 | Not-determined |
| S16_extracted_113147 | Not-determined |
| S16_extracted_113154 | Not-determined |
| S16_extracted_113161 | Not-determined |
| S16_extracted_113179 | Not-determined |
| S16_extracted_113180 | Not-determined |
| S16_extracted_113188 | Not-determined |
| S16_extracted_113206 | Not-determined |
| S16_extracted_113207 | Not-determined |
| S16_extracted_113218 | Not-determined |
| S16_extracted_113219 | Not-determined |
| S16_extracted_113220 | Not-determined |
| S16_extracted_113229 | Not-determined |
| S16_extracted_113236 | Not-determined |
| S16_extracted_113244 | Not-determined |
| S16_extracted_113246 | Low-quality |
| S16_extracted_113273 | Not-determined |
| S16_extracted_113274 | Not-determined |
| S16_extracted_113280 | Not-determined |
| S16_extracted_113300 | Not-determined |
| S16_extracted_113309 | Not-determined |
| S16_extracted_113322 | Low-quality |
| S16_extracted_113324 | Low-quality |
| S16_extracted_113337 | Not-determined |
| S16_extracted_113356 | Not-determined |
| S16_extracted_113364 | Not-determined |
| S16_extracted_113393 | Not-determined |
| S16_extracted_113405 | Low-quality |
| S16_extracted_113407 | Not-determined |
| S16_extracted_113434 | Not-determined |
| S16_extracted_113437 | Not-determined |
| S16_extracted_113440 | Not-determined |
| S16_extracted_113444 | Low-quality |
| S16_extracted_113453 | Not-determined |
| S16_extracted_113454 | Not-determined |
| S16_extracted_113455 | Not-determined |
| S16_extracted_113468 | Not-determined |
| S16_extracted_113484 | Not-determined |
| S16_extracted_113491 | Not-determined |
| S16_extracted_113519 | Not-determined |
| S16_extracted_113528 | Not-determined |
| S16_extracted_113536 | Not-determined |
| S16_extracted_113543 | Low-quality |
| S16_extracted_113546 | Not-determined |
| S16_extracted_113555 | Not-determined |
| S16_extracted_113561 | Not-determined |
| S16_extracted_113562 | Not-determined |
| S16_extracted_113585 | Not-determined |
| S16_extracted_113611 | Not-determined |
| S16_extracted_113620 | Not-determined |
| S16_extracted_113624 | Low-quality |
| S16_extracted_113632 | Not-determined |
| S16_extracted_113654 | Not-determined |
| S16_extracted_113671 | Not-determined |
| S16_extracted_113682 | Not-determined |
| S16_extracted_113683 | Not-determined |
| S16_extracted_113693 | Not-determined |
| S16_extracted_113694 | Not-determined |
| S16_extracted_113696 | Not-determined |
| S16_extracted_113698 | Not-determined |
| S16_extracted_113700 | Low-quality |
| S16_extracted_113705 | Not-determined |
| S16_extracted_113707 | Not-determined |
| S16_extracted_113715 | Not-determined |
| S16_extracted_113731 | Not-determined |
| S16_extracted_113738 | Not-determined |
| S16_extracted_113741 | Not-determined |
| S16_extracted_113749 | Low-quality |
| S16_extracted_113773 | Not-determined |
| S16_extracted_113782 | Low-quality |
| S16_extracted_113793 | Not-determined |
| S16_extracted_113794 | Not-determined |
| S16_extracted_113798 | Low-quality |
| S16_extracted_113807 | Not-determined |
| S16_extracted_113812 | Not-determined |
| S16_extracted_113813 | Not-determined |
| S16_extracted_113829 | Not-determined |
| S16_extracted_113833 | Low-quality |
| S16_extracted_113836 | Not-determined |
| S16_extracted_113843 | Not-determined |
| S16_extracted_113870 | Not-determined |
| S16_extracted_113874 | Not-determined |
| S16_extracted_113881 | Not-determined |
| S16_extracted_113913 | Low-quality |
| S16_extracted_113917 | Not-determined |
| S16_extracted_113919 | Not-determined |
| S16_extracted_113920 | Not-determined |
| S16_extracted_113924 | Not-determined |
| S16_extracted_113926 | Not-determined |
| S16_extracted_113929 | Not-determined |
| S16_extracted_113931 | Not-determined |
| S16_extracted_113933 | Not-determined |
| S16_extracted_113943 | Not-determined |
| S16_extracted_113945 | Not-determined |
| S16_extracted_113947 | Not-determined |
| S16_extracted_113953 | Not-determined |
| S16_extracted_113958 | Low-quality |
| S16_extracted_113964 | Not-determined |
| S16_extracted_113984 | Not-determined |
| S16_extracted_114010 | Low-quality |
| S16_extracted_114021 | Low-quality |
| S16_extracted_114030 | Not-determined |
| S16_extracted_114039 | Low-quality |
| S16_extracted_114041 | Not-determined |
| S16_extracted_114042 | Not-determined |
| S16_extracted_114045 | Not-determined |
| S16_extracted_114047 | Low-quality |
| S16_extracted_114059 | Not-determined |
| S16_extracted_114061 | Low-quality |
| S16_extracted_114063 | Not-determined |
| S16_extracted_114064 | Low-quality |
| S16_extracted_114066 | Not-determined |
| S16_extracted_114069 | Not-determined |
| S16_extracted_114071 | Not-determined |
| S16_extracted_114082 | Not-determined |
| S16_extracted_114100 | Not-determined |
| S16_extracted_114112 | Low-quality |
| S16_extracted_114126 | Not-determined |
| S16_extracted_114129 | Not-determined |
| S16_extracted_114132 | Not-determined |
| S16_extracted_114153 | Low-quality |
| S16_extracted_114165 | Not-determined |
| S16_extracted_114182 | Not-determined |
| S16_extracted_114189 | Not-determined |
| S16_extracted_114210 | Not-determined |
| S16_extracted_114213 | Not-determined |
| S16_extracted_114215 | Low-quality |
| S16_extracted_114216 | Not-determined |
| S16_extracted_114219 | Not-determined |
| S16_extracted_114222 | Not-determined |
| S16_extracted_114224 | Low-quality |
| S16_extracted_114231 | Not-determined |
| S16_extracted_114233 | Not-determined |
| S16_extracted_114236 | Not-determined |
| S16_extracted_114265 | Not-determined |
| S16_extracted_114271 | Low-quality |
| S16_extracted_114285 | Not-determined |
| S16_extracted_114286 | Not-determined |
| S16_extracted_114295 | Not-determined |
| S16_extracted_114310 | Not-determined |
| S16_extracted_114313 | Not-determined |
| S16_extracted_114332 | Low-quality |
| S16_extracted_114334 | Not-determined |
| S16_extracted_114345 | Not-determined |
| S16_extracted_114346 | Not-determined |
| S16_extracted_114367 | Not-determined |
| S16_extracted_114396 | Not-determined |
| S16_extracted_114416 | Low-quality |
| S16_extracted_114438 | Not-determined |
| S16_extracted_114443 | Not-determined |
| S16_extracted_114468 | Not-determined |
| S16_extracted_114469 | Not-determined |
| S16_extracted_114470 | Not-determined |
| S16_extracted_114473 | Low-quality |
| S16_extracted_114475 | Not-determined |
| S16_extracted_114488 | Not-determined |
| S16_extracted_114502 | Not-determined |
| S16_extracted_114510 | Low-quality |
| S16_extracted_114511 | Not-determined |
| S16_extracted_114519 | Not-determined |
| S16_extracted_114526 | Not-determined |
| S16_extracted_114531 | Not-determined |
| S16_extracted_114534 | Not-determined |
| S16_extracted_114544 | Not-determined |
| S16_extracted_114555 | Not-determined |
| S16_extracted_114558 | Not-determined |
| S16_extracted_114560 | Not-determined |
| S16_extracted_114578 | Not-determined |
| S16_extracted_114581 | Not-determined |
| S16_extracted_114583 | Not-determined |
| S16_extracted_114598 | Not-determined |
| S16_extracted_114612 | Not-determined |
| S16_extracted_114614 | Not-determined |
| S16_extracted_114627 | Not-determined |
| S16_extracted_114635 | Low-quality |
| S16_extracted_114653 | Not-determined |
| S16_extracted_114659 | Low-quality |
| S16_extracted_114662 | Not-determined |
| S16_extracted_114667 | Not-determined |
| S16_extracted_114668 | Not-determined |
| S16_extracted_114686 | Not-determined |
| S16_extracted_114713 | Not-determined |
| S16_extracted_114736 | Not-determined |
| S16_extracted_114746 | Not-determined |
| S16_extracted_114787 | Not-determined |
| S16_extracted_114791 | Not-determined |
| S16_extracted_114796 | Not-determined |
| S16_extracted_114798 | Not-determined |
| S16_extracted_114800 | Not-determined |
| S16_extracted_114814 | Not-determined |
| S16_extracted_114843 | Not-determined |
| S16_extracted_114845 | Not-determined |
| S16_extracted_114875 | Not-determined |
| S16_extracted_114911 | Not-determined |
| S16_extracted_114924 | Not-determined |
| S16_extracted_114925 | Not-determined |
| S16_extracted_114944 | Not-determined |
| S16_extracted_114946 | Low-quality |
| S16_extracted_114963 | Not-determined |
| S16_extracted_114971 | Low-quality |
| S16_extracted_114974 | Not-determined |
| S16_extracted_114992 | Not-determined |
| S16_extracted_114995 | Not-determined |
| S16_extracted_114999 | Not-determined |
| S16_extracted_115012 | Low-quality |
| S16_extracted_115015 | Not-determined |
| S16_extracted_115021 | Not-determined |
| S16_extracted_115030 | Not-determined |
| S16_extracted_115035 | Not-determined |
| S16_extracted_115036 | Not-determined |
| S16_extracted_115061 | Low-quality |
| S16_extracted_115066 | Not-determined |
| S16_extracted_115106 | Low-quality |
| S16_extracted_115113 | Not-determined |
| S16_extracted_115114 | Not-determined |
| S16_extracted_115125 | Not-determined |
| S16_extracted_115130 | Not-determined |
| S16_extracted_115136 | Not-determined |
| S16_extracted_115145 | Not-determined |
| S16_extracted_115146 | Not-determined |
| S16_extracted_115149 | Not-determined |
| S16_extracted_115150 | Not-determined |
| S16_extracted_115178 | Low-quality |
| S16_extracted_115188 | Low-quality |
| S16_extracted_115204 | Not-determined |
| S16_extracted_115209 | Not-determined |
| S16_extracted_115217 | Not-determined |
| S16_extracted_115222 | Low-quality |
| S16_extracted_115229 | Not-determined |
| S16_extracted_115239 | Not-determined |
| S16_extracted_115247 | Not-determined |
| S16_extracted_115256 | Not-determined |
| S16_extracted_115275 | Not-determined |
| S16_extracted_115277 | Not-determined |
| S16_extracted_115292 | Low-quality |
| S16_extracted_115302 | Not-determined |
| S16_extracted_115305 | Not-determined |
| S16_extracted_115306 | Low-quality |
| S16_extracted_115337 | Not-determined |
| S16_extracted_115345 | Not-determined |
| S16_extracted_115347 | Not-determined |
| S16_extracted_115350 | Not-determined |
| S16_extracted_115357 | Not-determined |
| S16_extracted_115362 | Not-determined |
| S16_extracted_115364 | Not-determined |
| S16_extracted_115375 | Not-determined |
| S16_extracted_115387 | Not-determined |
| S16_extracted_115401 | Not-determined |
| S16_extracted_115424 | Not-determined |
| S16_extracted_115483 | Not-determined |
| S16_extracted_115517 | Not-determined |
| S16_extracted_115521 | Not-determined |
| S16_extracted_115540 | Not-determined |
| S16_extracted_115544 | Not-determined |
| S16_extracted_115545 | Not-determined |
| S16_extracted_115547 | Not-determined |
| S16_extracted_115548 | Not-determined |
| S16_extracted_115551 | Not-determined |
| S16_extracted_115552 | Not-determined |
| S16_extracted_115560 | Not-determined |
| S16_extracted_115565 | Not-determined |
| S16_extracted_115589 | Not-determined |
| S16_extracted_115600 | Not-determined |
| S16_extracted_115610 | Not-determined |
| S16_extracted_115613 | Low-quality |
| S16_extracted_115617 | Not-determined |
| S16_extracted_115632 | Not-determined |
| S16_extracted_115639 | Low-quality |
| S16_extracted_115640 | Not-determined |
| S16_extracted_115641 | Not-determined |
| S16_extracted_115642 | Not-determined |
| S16_extracted_115647 | Not-determined |
| S16_extracted_115662 | Not-determined |
| S16_extracted_115679 | Not-determined |
| S16_extracted_115683 | Not-determined |
| S16_extracted_115692 | Not-determined |
| S16_extracted_115696 | Not-determined |
| S16_extracted_115720 | Not-determined |
| S16_extracted_115724 | Not-determined |
| S16_extracted_115728 | Low-quality |
| S16_extracted_115734 | Not-determined |
| S16_extracted_115735 | Not-determined |
| S16_extracted_115754 | Not-determined |
| S16_extracted_115793 | Not-determined |
| S16_extracted_115799 | Not-determined |
| S16_extracted_115802 | Not-determined |
| S16_extracted_115809 | Not-determined |
| S16_extracted_115817 | Not-determined |
| S16_extracted_115865 | Low-quality |
| S16_extracted_115872 | Not-determined |
| S16_extracted_115879 | Not-determined |
| S16_extracted_115891 | Not-determined |
| S16_extracted_115896 | Not-determined |
| S16_extracted_115899 | Not-determined |
| S16_extracted_115901 | Not-determined |
| S16_extracted_115911 | Low-quality |
| S16_extracted_115912 | Not-determined |
| S16_extracted_115913 | Low-quality |
| S16_extracted_115922 | Not-determined |
| S16_extracted_115930 | Not-determined |
| S16_extracted_115934 | Not-determined |
| S16_extracted_115942 | Not-determined |
| S16_extracted_115949 | Not-determined |
| S16_extracted_115950 | Not-determined |
| S16_extracted_115959 | Not-determined |
| S16_extracted_115967 | Not-determined |
| S16_extracted_115969 | Not-determined |
| S16_extracted_116006 | Not-determined |
| S16_extracted_116010 | Low-quality |
| S16_extracted_116035 | Not-determined |
| S16_extracted_116042 | Not-determined |
| S16_extracted_116072 | Not-determined |
| S16_extracted_116092 | Not-determined |
| S16_extracted_116102 | Not-determined |
| S16_extracted_116103 | Not-determined |
| S16_extracted_116106 | Not-determined |
| S16_extracted_116123 | Not-determined |
| S16_extracted_116135 | Not-determined |
| S16_extracted_116142 | Not-determined |
| S16_extracted_116145 | Not-determined |
| S16_extracted_116163 | Not-determined |
| S16_extracted_116183 | Low-quality |
| S16_extracted_116191 | Not-determined |
| S16_extracted_116198 | Not-determined |
| S16_extracted_116215 | Not-determined |
| S16_extracted_116222 | Not-determined |
| S16_extracted_116223 | Not-determined |
| S16_extracted_116244 | Not-determined |
| S16_extracted_116276 | Not-determined |
| S16_extracted_116296 | Not-determined |
| S16_extracted_116300 | Not-determined |
| S16_extracted_116326 | Not-determined |
| S16_extracted_116347 | Not-determined |
| S16_extracted_116355 | Not-determined |
| S16_extracted_116356 | Not-determined |
| S16_extracted_116369 | Not-determined |
| S16_extracted_116372 | Not-determined |
| S16_extracted_116385 | Not-determined |
| S16_extracted_116394 | Not-determined |
| S16_extracted_116398 | Not-determined |
| S16_extracted_116405 | Not-determined |
| S16_extracted_116406 | Not-determined |
| S16_extracted_116410 | Low-quality |
| S16_extracted_116429 | Not-determined |
| S16_extracted_116433 | Not-determined |
| S16_extracted_116443 | Not-determined |
| S16_extracted_116445 | Low-quality |
| S16_extracted_116454 | Not-determined |
| S16_extracted_116485 | Not-determined |
| S16_extracted_116492 | Not-determined |
| S16_extracted_116493 | Not-determined |
| S16_extracted_116511 | Not-determined |
| S16_extracted_116515 | Not-determined |
| S16_extracted_116517 | Low-quality |
| S16_extracted_116523 | Not-determined |
| S16_extracted_116539 | Not-determined |
| S16_extracted_116541 | Not-determined |
| S16_extracted_116542 | Not-determined |
| S16_extracted_116543 | Not-determined |
| S16_extracted_116545 | Low-quality |
| S16_extracted_116551 | Not-determined |
| S16_extracted_116570 | Not-determined |
| S16_extracted_116574 | Not-determined |
| S16_extracted_116579 | Low-quality |
| S16_extracted_116580 | Low-quality |
| S16_extracted_116586 | Not-determined |
| S16_extracted_116599 | Not-determined |
| S16_extracted_116611 | Not-determined |
| S16_extracted_116613 | Not-determined |
| S16_extracted_116618 | Not-determined |
| S16_extracted_116639 | Not-determined |
| S16_extracted_116665 | Not-determined |
| S16_extracted_116668 | Not-determined |
| S16_extracted_116671 | Not-determined |
| S16_extracted_116673 | Not-determined |
| S16_extracted_116688 | Not-determined |
| S16_extracted_116706 | Not-determined |
| S16_extracted_116746 | Low-quality |
| S16_extracted_116749 | Not-determined |
| S16_extracted_116751 | Not-determined |
| S16_extracted_116784 | Not-determined |
| S16_extracted_116800 | Not-determined |
| S16_extracted_116803 | Not-determined |
| S16_extracted_116855 | Not-determined |
| S16_extracted_116860 | Not-determined |
| S16_extracted_116865 | Not-determined |
| S16_extracted_116881 | Low-quality |
| S16_extracted_116884 | Not-determined |
| S16_extracted_116901 | Not-determined |
| S16_extracted_116922 | Not-determined |
| S16_extracted_116926 | Not-determined |
| S16_extracted_116938 | Not-determined |
| S16_extracted_116945 | Not-determined |
| S16_extracted_116965 | Not-determined |
| S16_extracted_116970 | Not-determined |
| S16_extracted_116974 | Low-quality |
| S16_extracted_116997 | Not-determined |
| S16_extracted_117002 | Not-determined |
| S16_extracted_117020 | Not-determined |
| S16_extracted_117031 | Not-determined |
| S16_extracted_117032 | Not-determined |
| S16_extracted_117045 | Not-determined |
| S16_extracted_117050 | Not-determined |
| S16_extracted_117064 | Low-quality |
| S16_extracted_117094 | Not-determined |
| S16_extracted_117097 | Not-determined |
| S16_extracted_117100 | Not-determined |
| S16_extracted_117118 | Low-quality |
| S16_extracted_117186 | Low-quality |
| S16_extracted_117193 | Low-quality |
| S16_extracted_117196 | Not-determined |
| S16_extracted_117240 | Not-determined |
| S16_extracted_117246 | Not-determined |
| S16_extracted_117267 | Low-quality |
| S16_extracted_117301 | Not-determined |
| S16_extracted_117304 | Not-determined |
| S16_extracted_117335 | Not-determined |
| S16_extracted_117355 | Low-quality |
| S16_extracted_117358 | Not-determined |
| S16_extracted_117361 | Not-determined |
| S16_extracted_117372 | Not-determined |
| S16_extracted_117380 | Not-determined |
| S16_extracted_117382 | Not-determined |
| S16_extracted_117383 | Not-determined |
| S16_extracted_117394 | Low-quality |
| S16_extracted_117412 | Not-determined |
| S16_extracted_117435 | Not-determined |
| S16_extracted_117438 | Not-determined |
| S16_extracted_117449 | Not-determined |
| S16_extracted_117462 | Not-determined |
| S16_extracted_117485 | Not-determined |
| S16_extracted_117489 | Not-determined |
| S16_extracted_117493 | Not-determined |
| S16_extracted_117500 | Not-determined |
| S16_extracted_117503 | Low-quality |
| S16_extracted_117504 | Not-determined |
| S16_extracted_117515 | Not-determined |
| S16_extracted_117519 | Low-quality |
| S16_extracted_117522 | Low-quality |
| S16_extracted_117542 | Not-determined |
| S16_extracted_117543 | Low-quality |
| S16_extracted_117545 | Not-determined |
| S16_extracted_117564 | Not-determined |
| S16_extracted_117595 | Low-quality |
| S16_extracted_117607 | Not-determined |
| S16_extracted_117609 | Not-determined |
| S16_extracted_117625 | Not-determined |
| S16_extracted_117641 | Not-determined |
| S16_extracted_117656 | Not-determined |
| S16_extracted_117666 | Not-determined |
| S16_extracted_117669 | Not-determined |
| S16_extracted_117675 | Not-determined |
| S16_extracted_117682 | Not-determined |
| S16_extracted_117683 | Not-determined |
| S16_extracted_117690 | Low-quality |
| S16_extracted_117695 | Not-determined |
| S16_extracted_117722 | Not-determined |
| S16_extracted_117750 | Low-quality |
| S16_extracted_117752 | Not-determined |
| S16_extracted_117764 | Not-determined |
| S16_extracted_117769 | Not-determined |
| S16_extracted_117786 | Not-determined |
| S16_extracted_117796 | Not-determined |
| S16_extracted_117802 | Not-determined |
| S16_extracted_117818 | Not-determined |
| S16_extracted_117830 | Not-determined |
| S16_extracted_117841 | Low-quality |
| S16_extracted_117852 | Not-determined |
| S16_extracted_117859 | Not-determined |
| S16_extracted_117865 | Not-determined |
| S16_extracted_117879 | Not-determined |
| S16_extracted_117886 | Not-determined |
| S16_extracted_117887 | Not-determined |
| S16_extracted_117904 | Low-quality |
| S16_extracted_117907 | Not-determined |
| S16_extracted_117930 | Not-determined |
| S16_extracted_117940 | Not-determined |
| S16_extracted_117947 | Not-determined |
| S16_extracted_117949 | Not-determined |
| S16_extracted_117955 | Not-determined |
| S16_extracted_117963 | Not-determined |
| S16_extracted_117975 | Not-determined |
| S16_extracted_117977 | Not-determined |
| S16_extracted_117986 | Not-determined |
| S16_extracted_117989 | Not-determined |
| S16_extracted_117991 | Not-determined |
| S16_extracted_118001 | Not-determined |
| S16_extracted_118013 | Not-determined |
| S16_extracted_118016 | Not-determined |
| S16_extracted_118027 | Not-determined |
| S16_extracted_118033 | Not-determined |
| S16_extracted_118038 | Not-determined |
| S16_extracted_118049 | Low-quality |
| S16_extracted_118054 | Not-determined |
| S16_extracted_118055 | Not-determined |
| S16_extracted_118061 | Not-determined |
| S16_extracted_118069 | Not-determined |
| S16_extracted_118086 | Not-determined |
| S16_extracted_118089 | Not-determined |
| S16_extracted_118097 | Not-determined |
| S16_extracted_118099 | Not-determined |
| S16_extracted_118107 | Not-determined |
| S16_extracted_118109 | Low-quality |
| S16_extracted_118110 | Not-determined |
| S16_extracted_118114 | Not-determined |
| S16_extracted_118128 | Not-determined |
| S16_extracted_118130 | Not-determined |
| S16_extracted_118150 | Low-quality |
| S16_extracted_118155 | Not-determined |
| S16_extracted_118168 | Low-quality |
| S16_extracted_118169 | Not-determined |
| S16_extracted_118173 | Not-determined |
| S16_extracted_118181 | Not-determined |
| S16_extracted_118183 | Not-determined |
| S16_extracted_118189 | Not-determined |
| S16_extracted_118193 | Not-determined |
| S16_extracted_118198 | Not-determined |
| S16_extracted_118207 | Low-quality |
| S16_extracted_118209 | Low-quality |
| S16_extracted_118211 | Not-determined |
| S16_extracted_118214 | Low-quality |
| S16_extracted_118219 | Not-determined |
| S16_extracted_118223 | Low-quality |
| S16_extracted_118232 | Not-determined |
| S16_extracted_118250 | Low-quality |
| S16_extracted_118282 | Not-determined |
| S16_extracted_118325 | Not-determined |
| S16_extracted_118329 | Not-determined |
| S16_extracted_118347 | Not-determined |
| S16_extracted_118351 | Not-determined |
| S16_extracted_118360 | Not-determined |
| S16_extracted_118363 | Not-determined |
| S16_extracted_118376 | Not-determined |
| S16_extracted_118381 | Not-determined |
| S16_extracted_118386 | Not-determined |
| S16_extracted_118392 | Low-quality |
| S16_extracted_118418 | Not-determined |
| S16_extracted_118427 | Not-determined |
| S16_extracted_118428 | Low-quality |
| S16_extracted_118434 | Not-determined |
| S16_extracted_118438 | Not-determined |
| S16_extracted_118452 | Not-determined |
| S16_extracted_118476 | Not-determined |
| S16_extracted_118482 | Not-determined |
| S16_extracted_118492 | Not-determined |
| S16_extracted_118496 | Not-determined |
| S16_extracted_118506 | Low-quality |
| S16_extracted_118554 | Not-determined |
| S16_extracted_118574 | Not-determined |
| S16_extracted_118600 | Not-determined |
| S16_extracted_118610 | Low-quality |
| S16_extracted_118620 | Not-determined |
| S16_extracted_118634 | Low-quality |
| S16_extracted_118650 | Not-determined |
| S16_extracted_118651 | Not-determined |
| S16_extracted_118657 | Not-determined |
| S16_extracted_118660 | Not-determined |
| S16_extracted_118678 | Low-quality |
| S16_extracted_118680 | Low-quality |
| S16_extracted_118683 | Low-quality |
| S16_extracted_118690 | Not-determined |
| S16_extracted_118692 | Low-quality |
| S16_extracted_118708 | Not-determined |
| S16_extracted_118709 | Not-determined |
| S16_extracted_118710 | Not-determined |

| | |
|---|---|
| S16_extracted_118711 | Not-determined |
| S16_extracted_118713 | Low-quality |
| S16_extracted_118739 | Low-quality |
| S16_extracted_118751 | Not-determined |
| S16_extracted_118768 | Not-determined |
| S16_extracted_118777 | Not-determined |
| S16_extracted_118783 | Not-determined |
| S16_extracted_118789 | Low-quality |
| S16_extracted_118798 | Not-determined |
| S16_extracted_118803 | Low-quality |
| S16_extracted_118832 | Low-quality |
| S16_extracted_118835 | Not-determined |
| S16_extracted_118841 | Not-determined |
| S16_extracted_118852 | Not-determined |
| S16_extracted_118856 | Not-determined |
| S16_extracted_118859 | Not-determined |
| S16_extracted_118862 | Not-determined |
| S16_extracted_118890 | Low-quality |
| S16_extracted_118909 | Low-quality |
| S16_extracted_118910 | Not-determined |
| S16_extracted_118914 | Not-determined |
| S16_extracted_118916 | Not-determined |
| S16_extracted_118921 | Low-quality |
| S16_extracted_118925 | Not-determined |
| S16_extracted_118931 | Not-determined |
| S16_extracted_118937 | Not-determined |
| S16_extracted_118943 | Not-determined |
| S16_extracted_118945 | Not-determined |
| S16_extracted_118948 | Not-determined |
| S16_extracted_118960 | Not-determined |
| S16_extracted_119002 | Not-determined |
| S16_extracted_119019 | Not-determined |
| S16_extracted_119050 | Not-determined |
| S16_extracted_119055 | Not-determined |
| S16_extracted_119056 | Low-quality |
| S16_extracted_119061 | Low-quality |
| S16_extracted_119077 | Low-quality |
| S16_extracted_119107 | Not-determined |
| S16_extracted_119124 | Not-determined |
| S16_extracted_119133 | Not-determined |
| S16_extracted_119157 | Not-determined |
| S16_extracted_119160 | Not-determined |
| S16_extracted_119163 | Low-quality |
| S16_extracted_119165 | Low-quality |
| S16_extracted_119169 | Not-determined |
| S16_extracted_119170 | Not-determined |
| S16_extracted_119179 | Not-determined |
| S16_extracted_119182 | Low-quality |
| S16_extracted_119190 | Low-quality |
| S16_extracted_119210 | Not-determined |
| S16_extracted_119213 | Not-determined |
| S16_extracted_119252 | Not-determined |
| S16_extracted_119276 | Not-determined |
| S16_extracted_119295 | Not-determined |
| S16_extracted_119330 | Low-quality |
| S16_extracted_119347 | Not-determined |
| S16_extracted_119355 | Not-determined |
| S16_extracted_119364 | Not-determined |
| S16_extracted_119374 | Not-determined |
| S16_extracted_119408 | Not-determined |
| S16_extracted_119419 | Not-determined |
| S16_extracted_119423 | Low-quality |
| S16_extracted_119427 | Not-determined |
| S16_extracted_119428 | Not-determined |
| S16_extracted_119445 | Not-determined |
| S16_extracted_119452 | Not-determined |
| S16_extracted_119460 | Low-quality |
| S16_extracted_119471 | Low-quality |
| S16_extracted_119479 | Not-determined |
| S16_extracted_119489 | Not-determined |
| S16_extracted_119503 | Low-quality |
| S16_extracted_119534 | Not-determined |
| S16_extracted_119541 | Not-determined |
| S16_extracted_119546 | Not-determined |
| S16_extracted_119569 | Not-determined |
| S16_extracted_119578 | Not-determined |
| S16_extracted_119589 | Not-determined |
| S16_extracted_119590 | Not-determined |
| S16_extracted_119598 | Not-determined |
| S16_extracted_119602 | Not-determined |
| S16_extracted_119614 | Not-determined |
| S16_extracted_119629 | Not-determined |
| S16_extracted_119637 | Low-quality |
| S16_extracted_119639 | Not-determined |
| S16_extracted_119641 | Not-determined |
| S16_extracted_119645 | Not-determined |
| S16_extracted_119667 | Low-quality |
| S16_extracted_119672 | Low-quality |
| S16_extracted_119675 | Not-determined |
| S16_extracted_119685 | Low-quality |
| S16_extracted_119707 | Not-determined |
| S16_extracted_119710 | Not-determined |
| S16_extracted_119711 | Not-determined |
| S16_extracted_119713 | Not-determined |
| S16_extracted_119724 | Low-quality |
| S16_extracted_119729 | Not-determined |
| S16_extracted_119730 | Low-quality |
| S16_extracted_119736 | Not-determined |
| S16_extracted_119738 | Not-determined |
| S16_extracted_119757 | Not-determined |
| S16_extracted_119777 | Not-determined |
| S16_extracted_119783 | Not-determined |
| S16_extracted_119787 | Not-determined |
| S16_extracted_119792 | Not-determined |
| S16_extracted_119802 | Not-determined |
| S16_extracted_119811 | Not-determined |
| S16_extracted_119823 | Not-determined |
| S11_vs_k141_104062\|\|full | High-quality |
| S16_extracted_119873 | Not-determined |
| S16_extracted_119882 | Low-quality |
| S16_extracted_119893 | Not-determined |
| S16_extracted_119911 | Not-determined |
| S16_extracted_119916 | Low-quality |
| S16_extracted_119919 | Not-determined |
| S16_extracted_119928 | Not-determined |
| S16_extracted_119940 | Not-determined |
| S16_extracted_119947 | Not-determined |
| S16_extracted_119969 | Not-determined |
| S16_extracted_119980 | Not-determined |
| S16_extracted_119982 | Not-determined |
| S16_extracted_119984 | Not-determined |
| S16_extracted_119999 | Not-determined |
| S16_extracted_120016 | Low-quality |
| S16_extracted_120018 | Not-determined |
| S16_extracted_120036 | Not-determined |
| S16_extracted_120045 | Not-determined |
| S16_extracted_120061 | Not-determined |
| S16_extracted_120082 | Not-determined |
| S16_extracted_120086 | Not-determined |
| S16_extracted_120095 | Not-determined |
| S16_extracted_120105 | Not-determined |
| S16_extracted_120108 | Not-determined |
| S16_extracted_120137 | Not-determined |
| S16_extracted_120138 | Low-quality |
| S16_extracted_120139 | Not-determined |
| S16_extracted_120141 | Not-determined |
| S16_extracted_120165 | Low-quality |
| S16_extracted_120169 | Low-quality |
| S16_extracted_120180 | Not-determined |
| S16_extracted_120184 | Not-determined |
| S16_extracted_120206 | Not-determined |
| S16_extracted_120233 | Not-determined |
| S16_extracted_120256 | Low-quality |
| S16_extracted_120260 | Not-determined |
| S16_extracted_120267 | Not-determined |
| S16_extracted_120269 | Not-determined |
| S16_extracted_120270 | Not-determined |
| S16_extracted_120273 | Not-determined |
| S16_extracted_120288 | Not-determined |
| S16_extracted_120299 | Not-determined |
| S16_extracted_120314 | Not-determined |
| S16_extracted_120319 | Not-determined |
| S16_extracted_120331 | Not-determined |
| S16_extracted_120338 | Not-determined |
| S16_extracted_120343 | Not-determined |
| S16_extracted_120356 | Not-determined |
| S16_extracted_120359 | Low-quality |
| S16_extracted_120369 | Not-determined |
| S16_extracted_120375 | Low-quality |
| S16_extracted_120391 | Not-determined |
| S16_extracted_120397 | Not-determined |
| S16_extracted_120401 | Not-determined |
| S16_extracted_120404 | Not-determined |
| S16_extracted_120411 | Not-determined |
| S16_extracted_120417 | Not-determined |
| S16_extracted_120447 | Low-quality |
| S16_extracted_120451 | Not-determined |
| S16_extracted_120474 | Not-determined |
| S16_extracted_120480 | Not-determined |
| S16_extracted_120485 | Not-determined |
| S16_extracted_120492 | Not-determined |
| S16_extracted_120503 | Not-determined |
| S16_extracted_120507 | Not-determined |
| S16_extracted_120520 | Not-determined |
| S16_extracted_120540 | Not-determined |
| S16_extracted_120544 | Not-determined |
| S16_extracted_120555 | Not-determined |
| S16_extracted_120575 | Not-determined |
| S16_extracted_120581 | Not-determined |
| S16_extracted_120584 | Not-determined |
| S16_extracted_120585 | Low-quality |
| S16_extracted_120588 | Not-determined |
| S16_extracted_120620 | Not-determined |
| S16_extracted_120634 | Not-determined |
| S16_extracted_120658 | Not-determined |
| S16_extracted_120665 | Not-determined |
| S16_extracted_120674 | Not-determined |
| S16_extracted_120677 | Not-determined |
| S16_extracted_120680 | Not-determined |
| S16_extracted_120687 | Not-determined |
| S16_extracted_120696 | Not-determined |
| S16_extracted_120703 | Not-determined |
| S16_extracted_120732 | Low-quality |
| S16_extracted_120738 | Not-determined |
| S16_extracted_120755 | Not-determined |
| S16_extracted_120758 | Not-determined |
| S16_extracted_120760 | Not-determined |
| S16_extracted_120762 | Not-determined |
| S16_extracted_120778 | Not-determined |
| S16_extracted_120779 | Not-determined |
| S16_extracted_120783 | Not-determined |
| S16_extracted_120791 | Not-determined |
| S16_extracted_120802 | Not-determined |
| S16_extracted_120804 | Not-determined |
| S16_extracted_120835 | Not-determined |
| S16_extracted_120838 | Not-determined |
| S16_extracted_120839 | Not-determined |
| S16_extracted_120861 | Not-determined |
| S16_extracted_120865 | Low-quality |
| S16_extracted_120878 | Not-determined |
| S16_extracted_120881 | Not-determined |
| S16_extracted_120889 | Low-quality |
| S16_extracted_120912 | Not-determined |
| S16_extracted_120922 | Not-determined |
| S16_extracted_120923 | Not-determined |
| S16_extracted_120938 | Low-quality |
| S16_extracted_120944 | Not-determined |
| S16_extracted_120951 | Low-quality |
| S16_extracted_120963 | Not-determined |
| S16_extracted_120965 | Not-determined |
| S16_extracted_120968 | Not-determined |
| S16_extracted_120970 | Not-determined |
| S16_extracted_120978 | Not-determined |
| S16_extracted_120979 | Not-determined |
| S16_extracted_120982 | Low-quality |
| S16_extracted_121013 | Not-determined |
| S16_extracted_121016 | Not-determined |
| S16_extracted_121044 | Not-determined |
| S16_extracted_121051 | Not-determined |
| S16_extracted_121078 | Not-determined |
| S16_extracted_121096 | Low-quality |
| S16_extracted_121123 | Not-determined |
| S16_extracted_121126 | Not-determined |
| S16_extracted_121137 | Not-determined |
| S16_extracted_121145 | Not-determined |
| S16_extracted_121168 | Not-determined |
| S16_extracted_121173 | Not-determined |
| S16_extracted_121176 | Low-quality |
| S16_extracted_121179 | Not-determined |
| S16_extracted_121181 | Low-quality |
| S16_extracted_121187 | Not-determined |
| S16_extracted_121213 | Not-determined |
| S16_extracted_121222 | Not-determined |
| S16_extracted_121234 | Not-determined |
| S16_extracted_121241 | Not-determined |
| S16_extracted_121244 | Not-determined |
| S16_extracted_121252 | Low-quality |
| S16_extracted_121257 | Not-determined |
| S16_extracted_121258 | Low-quality |
| S16_extracted_121259 | Not-determined |
| S16_extracted_121275 | Not-determined |
| S16_extracted_121278 | Not-determined |
| S16_extracted_121281 | Not-determined |
| S16_extracted_121292 | Low-quality |
| S16_extracted_121295 | Not-determined |
| S16_extracted_121299 | Low-quality |
| S16_extracted_121303 | Not-determined |
| S16_extracted_121306 | Not-determined |
| S16_extracted_121319 | Not-determined |
| S16_extracted_121329 | Low-quality |
| S16_extracted_121336 | Low-quality |
| S16_extracted_121344 | Not-determined |
| S16_extracted_121366 | Not-determined |
| S16_extracted_121375 | Not-determined |
| S16_extracted_121381 | Not-determined |
| S16_extracted_121385 | Not-determined |
| S16_extracted_121400 | Low-quality |
| S16_extracted_121409 | Not-determined |
| S16_extracted_121429 | Not-determined |
| S16_extracted_121430 | Not-determined |
| S16_extracted_121434 | Not-determined |
| S16_extracted_121437 | Not-determined |
| S16_extracted_121463 | Not-determined |
| S16_extracted_121477 | Not-determined |
| S16_extracted_121490 | Not-determined |
| S16_extracted_121502 | Low-quality |
| S16_extracted_121504 | Not-determined |
| S16_extracted_121524 | Not-determined |
| S16_extracted_121533 | Not-determined |
| S16_extracted_121540 | Not-determined |
| S16_extracted_121543 | Not-determined |
| S16_extracted_121549 | Not-determined |
| S16_extracted_121553 | Not-determined |
| S16_extracted_121562 | Not-determined |
| S16_extracted_121567 | Not-determined |
| S16_extracted_121594 | Low-quality |
| S16_extracted_121602 | Not-determined |
| S16_extracted_121609 | Low-quality |
| S16_extracted_121623 | Not-determined |
| S16_extracted_121627 | Not-determined |
| S16_extracted_121629 | Low-quality |
| S16_extracted_121632 | Not-determined |
| S16_extracted_121645 | Not-determined |
| S16_extracted_121667 | Not-determined |
| S16_extracted_121683 | Not-determined |
| S16_extracted_121700 | Not-determined |
| S16_extracted_121724 | Not-determined |
| S16_extracted_121751 | Not-determined |
| S16_extracted_121753 | Not-determined |
| S16_extracted_121755 | Not-determined |
| S16_extracted_121764 | Not-determined |
| S16_extracted_121778 | Not-determined |
| S16_extracted_121783 | Low-quality |
| S16_extracted_121799 | Not-determined |
| S16_extracted_121808 | Not-determined |
| S16_extracted_121816 | Not-determined |
| S16_extracted_121817 | Low-quality |
| S16_extracted_121822 | Not-determined |
| S16_extracted_121849 | Not-determined |
| S16_extracted_121862 | Not-determined |
| S16_extracted_121883 | Not-determined |
| S16_extracted_121887 | Not-determined |
| S16_extracted_121897 | Not-determined |
| S16_extracted_121908 | Not-determined |
| S16_extracted_121918 | Not-determined |
| S16_extracted_121924 | Not-determined |
| S16_extracted_121956 | Not-determined |
| S16_extracted_122003 | Not-determined |
| S16_extracted_122020 | Not-determined |
| S16_extracted_122042 | Low-quality |
| S16_extracted_122052 | Not-determined |
| S16_extracted_122053 | Not-determined |
| S16_extracted_122061 | Not-determined |
| S16_extracted_122064 | Low-quality |
| S16_extracted_122090 | Not-determined |
| S16_extracted_122091 | Not-determined |
| S16_extracted_122093 | Not-determined |
| S16_extracted_122125 | Not-determined |
| S16_extracted_122126 | Low-quality |
| S16_extracted_122131 | Not-determined |
| S16_extracted_122132 | Not-determined |
| S16_extracted_122137 | Not-determined |
| S16_extracted_122143 | Not-determined |
| S16_extracted_122159 | Not-determined |
| S16_extracted_122181 | Not-determined |
| S16_extracted_122195 | Not-determined |
| S16_extracted_122212 | Not-determined |
| S16_extracted_122215 | Not-determined |
| S16_extracted_122230 | Low-quality |
| S16_extracted_122234 | Not-determined |
| S16_extracted_122244 | Not-determined |
| S16_extracted_122255 | Not-determined |
| S16_extracted_122263 | Not-determined |
| S16_extracted_122276 | Not-determined |
| S16_extracted_122294 | Not-determined |
| S16_extracted_122295 | Not-determined |
| S16_extracted_122306 | Not-determined |
| S16_extracted_122327 | Not-determined |
| S16_extracted_122337 | Low-quality |
| S16_extracted_122351 | Low-quality |
| S16_extracted_122354 | Not-determined |
| S16_extracted_122362 | Not-determined |
| S16_extracted_122366 | Low-quality |
| S16_extracted_122373 | Not-determined |
| S16_extracted_122389 | Not-determined |
| S16_extracted_122390 | Not-determined |
| S16_extracted_122393 | Not-determined |
| S16_extracted_122401 | Not-determined |
| S16_extracted_122408 | Not-determined |
| S16_extracted_122426 | Not-determined |
| S16_extracted_122430 | Not-determined |
| S16_extracted_122444 | Not-determined |
| S16_extracted_122453 | Not-determined |
| S16_extracted_122480 | Not-determined |
| S16_extracted_122485 | Not-determined |
| S16_extracted_122488 | Not-determined |
| S16_extracted_122493 | Not-determined |
| S16_extracted_122494 | Not-determined |
| S16_extracted_122502 | Not-determined |
| S16_extracted_122508 | Not-determined |
| S16_extracted_122513 | Not-determined |
| S16_extracted_122517 | Not-determined |
| S16_extracted_122550 | Not-determined |
| S16_extracted_122581 | Not-determined |
| S16_extracted_122600 | Low-quality |
| S16_extracted_122605 | Not-determined |
| S16_extracted_122606 | Not-determined |
| S16_extracted_122610 | Low-quality |
| S16_extracted_122621 | Not-determined |
| S16_extracted_122634 | Not-determined |
| S16_extracted_122642 | Not-determined |
| S16_extracted_122644 | Not-determined |
| S16_extracted_122646 | Not-determined |
| S16_extracted_122663 | Not-determined |
| S16_extracted_122668 | Not-determined |
| S16_extracted_122675 | Not-determined |
| S16_extracted_122678 | Not-determined |
| S16_extracted_122680 | Not-determined |
| S16_extracted_122694 | Not-determined |
| S16_extracted_122698 | Not-determined |
| S16_extracted_122703 | Not-determined |
| S16_extracted_122713 | Low-quality |
| S16_extracted_122723 | Not-determined |
| S16_extracted_122724 | Not-determined |
| S16_extracted_122728 | Not-determined |
| S16_extracted_122732 | Low-quality |
| S16_extracted_122739 | Not-determined |
| S16_extracted_122742 | Not-determined |
| S16_extracted_122753 | Not-determined |
| S16_extracted_122771 | Low-quality |
| S16_extracted_122778 | Low-quality |
| S16_extracted_122781 | Not-determined |
| S16_extracted_122782 | Not-determined |
| S16_extracted_122793 | Not-determined |
| S16_extracted_122808 | Not-determined |
| S16_extracted_122811 | Not-determined |
| S16_extracted_122812 | Not-determined |
| S16_extracted_122824 | Not-determined |
| S16_extracted_122859 | Low-quality |
| S16_extracted_122862 | Not-determined |
| S16_extracted_122863 | Low-quality |
| S16_extracted_122865 | Not-determined |
| S16_extracted_122867 | Not-determined |
| S16_extracted_122897 | Not-determined |
| S16_extracted_122906 | Not-determined |
| S16_extracted_122911 | Not-determined |
| S16_extracted_122920 | Not-determined |
| S16_extracted_122937 | Not-determined |
| S16_extracted_122960 | Not-determined |
| S16_extracted_122975 | Not-determined |
| S16_extracted_122980 | Not-determined |
| S16_extracted_122988 | Not-determined |
| S16_extracted_123001 | Not-determined |
| S16_extracted_123060 | Low-quality |
| S16_extracted_123084 | Not-determined |
| S16_extracted_123125 | Low-quality |
| S16_extracted_123139 | Not-determined |
| S16_extracted_123149 | Not-determined |
| S16_extracted_123152 | Not-determined |
| S16_extracted_123154 | Not-determined |
| S16_extracted_123156 | Not-determined |
| S16_extracted_123160 | Not-determined |
| S16_extracted_123168 | Not-determined |
| S16_extracted_123171 | Not-determined |
| S16_extracted_123181 | Not-determined |
| S16_extracted_123184 | Not-determined |
| S16_extracted_123186 | Not-determined |
| S16_extracted_123203 | Not-determined |
| S16_extracted_123206 | Not-determined |
| S16_extracted_123246 | Not-determined |
| S16_extracted_123255 | Low-quality |
| S16_extracted_123259 | Not-determined |
| S16_extracted_123261 | Not-determined |
| S16_extracted_123277 | Not-determined |
| S16_extracted_123287 | Not-determined |
| S16_extracted_123292 | Not-determined |
| S16_extracted_123307 | Not-determined |
| S16_extracted_123311 | Not-determined |
| S16_extracted_123318 | Not-determined |
| S16_extracted_123319 | Low-quality |
| S16_extracted_123353 | Low-quality |
| S16_extracted_123361 | Not-determined |
| S16_extracted_123368 | Not-determined |
| S16_extracted_123377 | Not-determined |
| S16_extracted_123384 | Not-determined |
| S16_extracted_123395 | Low-quality |
| S16_extracted_123403 | Not-determined |
| S16_extracted_123410 | Not-determined |
| S16_extracted_123423 | Not-determined |
| S16_extracted_123425 | Not-determined |
| S16_extracted_123429 | Not-determined |
| S16_extracted_123432 | Not-determined |
| S16_extracted_123468 | Not-determined |
| S16_extracted_123501 | Not-determined |
| S16_extracted_123509 | Not-determined |
| S16_extracted_123519 | Not-determined |
| S16_extracted_123540 | Not-determined |
| S16_extracted_123543 | Not-determined |
| S16_extracted_123547 | Not-determined |
| S16_extracted_123548 | Not-determined |
| S16_extracted_123553 | Low-quality |
| S16_extracted_123555 | Not-determined |
| S16_extracted_123565 | Not-determined |
| S16_extracted_123579 | Not-determined |
| S16_extracted_123607 | Not-determined |
| S16_extracted_123609 | Not-determined |
| S16_extracted_123610 | Not-determined |
| S16_extracted_123627 | Low-quality |
| S16_extracted_123636 | Not-determined |
| S16_extracted_123645 | Not-determined |
| S16_extracted_123649 | Not-determined |
| S16_extracted_123668 | Not-determined |
| S16_extracted_123676 | Not-determined |
| S16_extracted_123685 | Not-determined |
| S16_extracted_123694 | Not-determined |
| S16_extracted_123705 | Low-quality |
| S16_extracted_123720 | Not-determined |
| S16_extracted_123723 | Not-determined |
| S16_extracted_123727 | Not-determined |
| S16_extracted_123734 | Not-determined |
| S16_extracted_123736 | Not-determined |
| S16_extracted_123761 | Not-determined |
| S16_extracted_123764 | Not-determined |
| S16_extracted_123766 | Not-determined |
| S16_extracted_123775 | Not-determined |
| S16_extracted_123778 | Low-quality |
| S16_extracted_123787 | Not-determined |
| S16_extracted_123799 | Not-determined |
| S16_extracted_123828 | Not-determined |
| S16_extracted_123833 | Not-determined |
| S16_extracted_123838 | Not-determined |
| S16_extracted_123843 | Not-determined |
| S16_extracted_123857 | Not-determined |
| S16_extracted_123866 | Not-determined |
| S16_extracted_123868 | Low-quality |
| S16_extracted_123877 | Not-determined |
| S16_extracted_123892 | Not-determined |
| S16_extracted_123900 | Low-quality |
| S16_extracted_123915 | Not-determined |
| S16_extracted_123919 | Not-determined |
| S16_extracted_123920 | Not-determined |
| S16_extracted_123929 | Not-determined |
| S16_extracted_123940 | Not-determined |
| S16_extracted_123943 | Not-determined |
| S16_extracted_123947 | Not-determined |
| S16_extracted_123960 | Not-determined |
| S16_extracted_123969 | Not-determined |
| S16_extracted_123971 | Not-determined |
| S16_extracted_123987 | Low-quality |
| S16_extracted_124011 | Not-determined |
| S16_extracted_124016 | Not-determined |
| S16_extracted_124029 | Not-determined |
| S16_extracted_124030 | Not-determined |
| S16_extracted_124035 | Not-determined |
| S16_extracted_124039 | Not-determined |
| S16_extracted_124048 | Not-determined |
| S16_extracted_124054 | Low-quality |
| S16_extracted_124074 | Not-determined |
| S16_extracted_124077 | Not-determined |
| S16_extracted_124099 | Not-determined |
| S16_extracted_124107 | Not-determined |
| S16_extracted_124114 | Not-determined |
| S16_extracted_124115 | Not-determined |
| S16_extracted_124123 | Low-quality |
| S16_extracted_124140 | Not-determined |
| S16_extracted_124145 | Not-determined |
| S16_extracted_124151 | Not-determined |
| S16_extracted_124156 | Not-determined |
| S16_extracted_124171 | Not-determined |
| S16_extracted_124174 | Not-determined |
| S16_extracted_124184 | Not-determined |
| S16_extracted_124185 | Not-determined |
| S16_extracted_124213 | Not-determined |
| S16_extracted_124226 | Not-determined |
| S16_extracted_124232 | Not-determined |
| S16_extracted_124238 | Not-determined |
| S16_extracted_124249 | Not-determined |
| S16_extracted_124262 | Not-determined |
| S16_extracted_124276 | Not-determined |
| S16_extracted_124277 | Not-determined |
| S16_extracted_124300 | Not-determined |
| S16_extracted_124301 | Not-determined |
| S16_extracted_124304 | Not-determined |
| S16_extracted_124310 | Not-determined |
| S16_extracted_124321 | Not-determined |
| S16_extracted_124327 | Not-determined |
| S16_extracted_124332 | Not-determined |
| S16_extracted_124334 | Not-determined |
| S16_extracted_124353 | Low-quality |
| S16_extracted_124358 | Not-determined |
| S16_extracted_124364 | Not-determined |
| S16_extracted_124370 | Not-determined |
| S16_extracted_124376 | Not-determined |
| S16_extracted_124379 | Not-determined |
| S16_extracted_124384 | Not-determined |
| S16_extracted_124394 | Not-determined |
| S16_extracted_124402 | Not-determined |
| S16_extracted_124404 | Not-determined |
| S16_extracted_124419 | Not-determined |
| S16_extracted_124433 | Not-determined |
| S16_extracted_124438 | Low-quality |
| S16_extracted_124445 | Not-determined |
| S16_extracted_124462 | Not-determined |
| S16_extracted_124467 | Not-determined |
| S16_extracted_124468 | Not-determined |
| S16_extracted_124486 | Not-determined |
| S16_extracted_124501 | Not-determined |
| S16_extracted_124509 | Not-determined |
| S16_extracted_124519 | Low-quality |
| S16_extracted_124534 | Not-determined |
| S16_extracted_124535 | Not-determined |
| S16_extracted_124536 | Not-determined |
| S16_extracted_124537 | Not-determined |
| S16_extracted_124556 | Not-determined |
| S16_extracted_124565 | Not-determined |
| S16_extracted_124569 | Not-determined |
| S16_extracted_124570 | Not-determined |
| S16_extracted_124573 | Not-determined |
| S16_extracted_124577 | Not-determined |

| | |
|---|---|
| S16_extracted_124594 | Not-determined |
| S16_extracted_124612 | Not-determined |
| S16_extracted_124614 | Not-determined |
| S16_extracted_124625 | Low-quality |
| S16_extracted_124644 | Not-determined |
| S16_extracted_124651 | Not-determined |
| S16_extracted_124661 | Not-determined |
| S10_vs_46819\|\|full | High-quality |
| S16_extracted_124687 | Not-determined |
| S16_extracted_124688 | Not-determined |
| S16_extracted_124727 | Not-determined |
| S16_extracted_124729 | Not-determined |
| S16_extracted_124740 | Not-determined |
| S16_extracted_124744 | Not-determined |
| S16_extracted_124749 | Not-determined |
| S16_extracted_124750 | Not-determined |
| S16_extracted_124755 | Not-determined |
| S16_extracted_124756 | Low-quality |
| S16_extracted_124758 | Low-quality |
| S16_extracted_124763 | Not-determined |
| S16_extracted_124787 | Not-determined |
| S16_extracted_124789 | Not-determined |
| S16_extracted_124804 | Not-determined |
| S16_extracted_124815 | Not-determined |
| S16_extracted_124825 | Not-determined |
| S16_extracted_124829 | Not-determined |
| S16_extracted_124830 | Not-determined |
| S16_extracted_124832 | Not-determined |
| S16_extracted_124837 | Not-determined |
| S16_extracted_124849 | Not-determined |
| S16_extracted_124852 | Not-determined |
| S16_extracted_124857 | Not-determined |
| S16_extracted_124860 | Not-determined |
| S16_extracted_124864 | Not-determined |
| S16_extracted_124866 | Not-determined |
| S16_extracted_124902 | Not-determined |
| S16_extracted_124929 | Not-determined |
| S16_extracted_124933 | Low-quality |
| S16_extracted_124943 | Not-determined |
| S16_extracted_124962 | Not-determined |
| S16_extracted_125003 | Not-determined |
| S16_extracted_125009 | Not-determined |
| S16_extracted_125042 | Not-determined |
| S16_extracted_125055 | Low-quality |
| S16_extracted_125071 | Low-quality |
| S16_extracted_125075 | Not-determined |
| S16_extracted_125080 | Not-determined |
| S16_extracted_125083 | Not-determined |
| S16_extracted_125091 | Not-determined |
| S16_extracted_125130 | Low-quality |
| S16_extracted_125135 | Not-determined |
| S16_extracted_125153 | Not-determined |
| S16_extracted_125168 | Low-quality |
| S16_extracted_125174 | Not-determined |
| S16_extracted_125183 | Not-determined |
| S16_extracted_125200 | Not-determined |
| S16_extracted_125205 | Not-determined |
| S16_extracted_125219 | Not-determined |
| S16_extracted_125240 | Not-determined |
| S16_extracted_125244 | Not-determined |
| S16_extracted_125247 | Not-determined |
| S16_extracted_125248 | Low-quality |
| S16_extracted_125266 | Not-determined |
| S16_extracted_125280 | Not-determined |
| S16_extracted_125287 | Not-determined |
| S16_extracted_125303 | Not-determined |
| S16_extracted_125305 | Not-determined |
| S16_extracted_125316 | Not-determined |
| S16_extracted_125319 | Not-determined |
| S16_extracted_125334 | Not-determined |
| S16_extracted_125340 | Not-determined |
| S16_extracted_125343 | Not-determined |
| S16_extracted_125362 | Low-quality |
| S16_extracted_125368 | Not-determined |
| S16_extracted_125375 | Not-determined |
| S16_extracted_125378 | Not-determined |
| S16_extracted_125389 | Low-quality |
| S16_extracted_125395 | Not-determined |
| S16_extracted_125396 | Low-quality |
| S16_extracted_125430 | Not-determined |
| S16_extracted_125439 | Not-determined |
| S16_extracted_125445 | Not-determined |
| S16_extracted_125448 | Not-determined |
| S16_extracted_125456 | Not-determined |
| S16_extracted_125465 | Not-determined |
| S16_extracted_125470 | Not-determined |
| S16_extracted_125475 | Not-determined |
| S16_extracted_125478 | Not-determined |
| S16_extracted_125520 | Not-determined |
| S16_extracted_125521 | Not-determined |
| S16_extracted_125527 | Low-quality |
| S16_extracted_125537 | Not-determined |
| S16_extracted_125542 | Low-quality |
| S16_extracted_125545 | Not-determined |
| S16_extracted_125549 | Not-determined |
| S16_extracted_125552 | Not-determined |
| S16_extracted_125554 | Not-determined |
| S16_extracted_125557 | Low-quality |
| S16_extracted_125561 | Low-quality |
| S16_extracted_125568 | Not-determined |
| S16_extracted_125584 | Low-quality |
| S16_extracted_125585 | Not-determined |
| S16_extracted_125591 | Low-quality |
| S16_extracted_125609 | Not-determined |
| S16_extracted_125616 | Not-determined |
| S16_extracted_125623 | Not-determined |
| S16_extracted_125629 | Not-determined |
| S16_extracted_125655 | Not-determined |
| S16_extracted_125664 | Low-quality |
| S16_extracted_125696 | Not-determined |
| S16_extracted_125753 | Not-determined |
| S16_extracted_125755 | Not-determined |
| S16_extracted_125789 | Low-quality |
| S16_extracted_125800 | Not-determined |
| S16_extracted_125807 | Low-quality |
| S16_extracted_125810 | Low-quality |
| S16_extracted_125812 | Not-determined |
| S16_extracted_125837 | Not-determined |
| S16_extracted_125840 | Low-quality |
| S16_extracted_125841 | Not-determined |
| S16_extracted_125850 | Not-determined |
| S16_extracted_125868 | Not-determined |
| S16_extracted_125873 | Not-determined |
| S16_extracted_125897 | Not-determined |
| S16_extracted_125908 | Not-determined |
| S16_extracted_125916 | Not-determined |
| S16_extracted_125951 | Not-determined |
| S16_extracted_125975 | Not-determined |
| S16_extracted_125978 | Not-determined |
| S16_extracted_125987 | Not-determined |
| S16_extracted_125994 | Not-determined |
| S16_extracted_125998 | Not-determined |
| S16_extracted_126020 | Not-determined |
| S16_extracted_126024 | Not-determined |
| S16_extracted_126029 | Not-determined |
| S16_extracted_126030 | Not-determined |
| S16_extracted_126050 | Not-determined |
| S16_extracted_126060 | Not-determined |
| S16_extracted_126061 | Not-determined |
| S16_extracted_126065 | Not-determined |
| S16_extracted_126093 | Not-determined |
| S16_extracted_126094 | Low-quality |
| S16_extracted_126098 | Not-determined |
| S16_extracted_126109 | Not-determined |
| S16_extracted_126132 | Not-determined |
| S16_extracted_126163 | Not-determined |
| S16_extracted_126166 | Not-determined |
| S16_extracted_126167 | Not-determined |
| S16_extracted_126170 | Not-determined |
| S16_extracted_126204 | Not-determined |
| S16_extracted_126210 | Not-determined |
| S16_extracted_126221 | Not-determined |
| S16_extracted_126229 | Not-determined |
| S16_extracted_126255 | Low-quality |
| S16_extracted_126299 | Not-determined |
| S16_extracted_126305 | Not-determined |
| S16_extracted_126318 | Not-determined |
| S16_extracted_126332 | Not-determined |
| S16_extracted_126338 | Not-determined |
| S16_extracted_126352 | Not-determined |
| S16_extracted_126354 | Not-determined |
| S16_extracted_126370 | Not-determined |
| S16_extracted_126376 | Not-determined |
| S16_extracted_126388 | Low-quality |
| S16_extracted_126407 | Not-determined |
| S16_extracted_126417 | Not-determined |
| S16_extracted_126431 | Not-determined |
| S16_extracted_126442 | Not-determined |
| S16_extracted_126449 | Not-determined |
| S16_extracted_126469 | Not-determined |
| S16_extracted_126478 | Not-determined |
| S16_extracted_126487 | Not-determined |
| S16_extracted_126496 | Not-determined |
| S16_extracted_126497 | Not-determined |
| S16_extracted_126509 | Not-determined |
| S16_extracted_126521 | Not-determined |
| S16_extracted_126530 | Not-determined |
| S16_extracted_126533 | Low-quality |
| S16_extracted_126537 | Not-determined |
| S16_extracted_126538 | Low-quality |
| S16_extracted_126545 | Low-quality |
| S16_extracted_126554 | Not-determined |
| S16_extracted_126569 | Not-determined |
| S16_extracted_126575 | Not-determined |
| S16_extracted_126576 | Not-determined |
| S16_extracted_126577 | Not-determined |
| S16_extracted_126603 | Not-determined |
| S16_extracted_126619 | Not-determined |
| S16_extracted_126626 | Not-determined |
| S16_extracted_126633 | Not-determined |
| S16_extracted_126635 | Not-determined |
| S16_extracted_126640 | Not-determined |
| S16_extracted_126660 | Not-determined |
| S16_extracted_126672 | Not-determined |
| S16_extracted_126683 | Not-determined |
| S16_extracted_126691 | Not-determined |
| S16_extracted_126694 | Not-determined |
| S16_extracted_126707 | Not-determined |
| S16_extracted_126709 | Not-determined |
| S16_extracted_126714 | Not-determined |
| S16_extracted_126726 | Not-determined |
| S16_extracted_126728 | Not-determined |
| S16_extracted_126738 | Not-determined |
| S16_extracted_126744 | Not-determined |
| S16_extracted_126748 | Not-determined |
| S16_extracted_126755 | Low-quality |
| S16_extracted_126791 | Low-quality |
| S16_extracted_126794 | Not-determined |
| S16_extracted_126795 | Not-determined |
| S16_extracted_126796 | Not-determined |
| S16_extracted_126797 | Not-determined |
| S16_extracted_126814 | Not-determined |
| S16_extracted_126820 | Not-determined |
| S16_extracted_126822 | Not-determined |
| S16_extracted_126825 | Low-quality |
| S16_extracted_126831 | Not-determined |
| S16_extracted_126842 | Low-quality |
| S16_extracted_126857 | Not-determined |
| S16_extracted_126864 | Low-quality |
| S16_extracted_126874 | Not-determined |
| S16_extracted_126888 | Not-determined |
| S16_extracted_126890 | Not-determined |
| S16_extracted_126899 | Not-determined |
| S16_extracted_126905 | Not-determined |
| S16_extracted_126927 | Not-determined |
| S16_extracted_126939 | Not-determined |
| S16_extracted_126945 | Not-determined |
| S16_extracted_126949 | Not-determined |
| S16_extracted_126958 | Not-determined |
| S16_extracted_126967 | Not-determined |
| S16_extracted_126983 | Not-determined |
| S16_extracted_126995 | Not-determined |
| S16_extracted_126997 | Not-determined |
| S16_extracted_127002 | Not-determined |
| S16_extracted_127014 | Not-determined |
| S16_extracted_127017 | Not-determined |
| S16_extracted_127027 | Not-determined |
| S16_extracted_127036 | Not-determined |
| S16_extracted_127049 | Not-determined |
| S16_extracted_127074 | Low-quality |
| S16_extracted_127098 | Not-determined |
| S16_extracted_127102 | Not-determined |
| S16_extracted_127103 | Not-determined |
| S16_extracted_127108 | Not-determined |
| S16_extracted_127114 | Not-determined |
| S16_extracted_127121 | Not-determined |
| S16_extracted_127123 | Not-determined |
| S16_extracted_127147 | Not-determined |
| S16_extracted_127168 | Not-determined |
| S16_extracted_127169 | Not-determined |
| S16_extracted_127177 | Not-determined |
| S16_extracted_127186 | Not-determined |
| S16_extracted_127187 | Low-quality |
| S16_extracted_127196 | Not-determined |
| S16_extracted_127219 | Not-determined |
| S16_extracted_127242 | Not-determined |
| S16_extracted_127255 | Not-determined |
| S16_extracted_127274 | Not-determined |
| S16_extracted_127275 | Not-determined |
| S16_extracted_127286 | Not-determined |
| S16_extracted_127291 | Low-quality |
| S16_extracted_127310 | Not-determined |
| S16_extracted_127311 | Not-determined |
| S16_extracted_127315 | Not-determined |
| S16_extracted_127320 | Not-determined |
| S16_extracted_127327 | Not-determined |
| S16_extracted_127352 | Not-determined |
| S16_extracted_127372 | Not-determined |
| S16_extracted_127374 | Low-quality |
| S16_extracted_127384 | Not-determined |
| S16_extracted_127386 | Not-determined |
| S16_extracted_127399 | Not-determined |
| S16_extracted_127402 | Not-determined |
| S16_extracted_127410 | Not-determined |
| S16_extracted_127411 | Not-determined |
| S16_extracted_127414 | Not-determined |
| S16_extracted_127418 | Not-determined |
| S16_extracted_127437 | Not-determined |
| S16_extracted_127449 | Low-quality |
| S16_extracted_127455 | Low-quality |
| S16_extracted_127465 | Not-determined |
| S16_extracted_127479 | Not-determined |
| S16_extracted_127483 | Not-determined |
| S16_extracted_127493 | Not-determined |
| S16_extracted_127495 | Low-quality |
| S16_extracted_127538 | Not-determined |
| S16_extracted_127551 | Not-determined |
| S16_extracted_127552 | Not-determined |
| S16_extracted_127560 | Low-quality |
| S16_extracted_127565 | Not-determined |
| S16_extracted_127610 | Not-determined |
| S16_extracted_127612 | Not-determined |
| S16_extracted_127613 | Not-determined |
| S16_extracted_127630 | Low-quality |
| S16_extracted_127632 | Not-determined |
| S16_extracted_127634 | Not-determined |
| S16_extracted_127635 | Not-determined |
| S16_extracted_127642 | Not-determined |
| S16_extracted_127648 | Not-determined |
| S16_extracted_127658 | Not-determined |
| S16_extracted_127667 | Low-quality |
| S16_extracted_127673 | Not-determined |
| S16_extracted_127702 | Not-determined |
| S16_extracted_127730 | Not-determined |
| S16_extracted_127744 | Not-determined |
| S16_extracted_127762 | Not-determined |
| S16_extracted_127768 | Low-quality |
| S16_extracted_127772 | Low-quality |
| S16_extracted_127788 | Not-determined |
| S16_extracted_127802 | Low-quality |
| S16_extracted_127803 | Not-determined |
| S16_extracted_127806 | Not-determined |
| S16_extracted_127844 | Not-determined |
| S16_extracted_127886 | Not-determined |
| S16_extracted_127909 | Not-determined |
| S16_extracted_127912 | Not-determined |
| S16_extracted_127919 | Low-quality |
| S16_extracted_127932 | Not-determined |
| S16_extracted_127935 | Not-determined |
| S16_extracted_127963 | Not-determined |
| S16_extracted_127981 | Not-determined |
| S16_extracted_127994 | Not-determined |
| S16_extracted_127996 | Not-determined |
| S16_extracted_128010 | Not-determined |
| S16_extracted_128028 | Not-determined |
| S16_extracted_128037 | Not-determined |
| S16_extracted_128038 | Not-determined |
| S16_extracted_128052 | Not-determined |
| S16_extracted_128082 | Not-determined |
| S16_extracted_128112 | Not-determined |
| S16_extracted_128120 | Not-determined |
| S16_extracted_128131 | Low-quality |
| S16_extracted_128133 | Not-determined |
| S16_extracted_128141 | Not-determined |
| S16_extracted_128144 | Low-quality |
| S16_extracted_128146 | Not-determined |
| S16_extracted_128161 | Not-determined |
| S16_extracted_128163 | Not-determined |
| S16_extracted_128218 | Low-quality |
| S16_extracted_128230 | Not-determined |
| S16_extracted_128242 | Not-determined |
| S16_extracted_128260 | Not-determined |
| S16_extracted_128270 | Not-determined |
| S16_extracted_128293 | Not-determined |
| S16_extracted_128311 | Low-quality |
| S16_extracted_128323 | Not-determined |
| S16_extracted_128324 | Not-determined |
| S16_extracted_128333 | Not-determined |
| S16_extracted_128335 | Not-determined |
| S16_extracted_128336 | Not-determined |
| S16_extracted_128337 | Not-determined |
| S16_extracted_128352 | Not-determined |
| S16_extracted_128362 | Not-determined |
| S16_extracted_128372 | Not-determined |
| S16_extracted_128380 | Not-determined |
| S16_extracted_128384 | Not-determined |
| S16_extracted_128401 | Not-determined |
| S16_extracted_128413 | Not-determined |
| S16_extracted_128419 | Not-determined |
| S16_extracted_128427 | Low-quality |
| S16_extracted_128429 | Not-determined |
| S16_extracted_128442 | Low-quality |
| S16_extracted_128443 | Not-determined |
| S16_extracted_128446 | Low-quality |
| S16_extracted_128448 | Not-determined |
| S16_extracted_128467 | Not-determined |
| S16_extracted_128472 | Not-determined |
| S16_extracted_128479 | Not-determined |
| S16_extracted_128494 | Low-quality |
| S16_extracted_128499 | Not-determined |
| S16_extracted_128501 | Not-determined |
| S16_extracted_128503 | Not-determined |
| S16_extracted_128508 | Not-determined |
| S16_extracted_128513 | Not-determined |
| S16_extracted_128515 | Not-determined |
| S16_extracted_128524 | Not-determined |
| S16_extracted_128525 | Not-determined |
| S16_extracted_128530 | Not-determined |
| S16_extracted_128538 | Not-determined |
| S16_extracted_128544 | Not-determined |
| S16_extracted_128568 | Not-determined |
| S16_extracted_128581 | Not-determined |
| S16_extracted_128585 | Not-determined |
| S16_extracted_128588 | Not-determined |
| S16_extracted_128598 | Not-determined |
| S16_extracted_128607 | Not-determined |
| S16_extracted_128623 | Not-determined |
| S16_extracted_128657 | Not-determined |
| S16_extracted_128684 | Not-determined |
| S16_extracted_128709 | Not-determined |
| S16_extracted_128768 | Not-determined |
| S16_extracted_128769 | Not-determined |
| S16_extracted_128803 | Not-determined |
| S16_extracted_128810 | Not-determined |
| S16_extracted_128814 | Not-determined |
| S16_extracted_128816 | Not-determined |
| S16_extracted_128818 | Not-determined |
| S16_extracted_128829 | Not-determined |
| S16_extracted_128833 | Not-determined |
| S16_extracted_128834 | Not-determined |
| S16_extracted_128894 | Not-determined |
| S16_extracted_128916 | Not-determined |
| S16_extracted_128920 | Not-determined |
| S16_extracted_128922 | Not-determined |
| S16_extracted_128923 | Not-determined |
| S16_extracted_128930 | Not-determined |
| S16_extracted_128931 | Not-determined |
| S16_extracted_128935 | Not-determined |
| S16_extracted_128946 | Not-determined |
| S16_extracted_128965 | Low-quality |
| S16_extracted_128967 | Not-determined |
| S16_extracted_128970 | Not-determined |
| S16_extracted_128986 | Not-determined |
| S16_extracted_128990 | Not-determined |
| S16_extracted_129006 | Low-quality |
| S16_extracted_129015 | Not-determined |
| S16_extracted_129018 | Not-determined |
| S16_extracted_129029 | Not-determined |
| S16_extracted_129051 | Not-determined |
| S16_extracted_129084 | Not-determined |
| S16_extracted_129102 | Not-determined |
| S16_extracted_129106 | Not-determined |
| S16_extracted_129121 | Not-determined |
| S16_extracted_129133 | Not-determined |
| S16_extracted_129151 | Low-quality |
| S16_extracted_129157 | Not-determined |
| S16_extracted_129173 | Low-quality |
| S16_extracted_129175 | Not-determined |
| S16_extracted_129184 | Not-determined |
| S16_extracted_129206 | Not-determined |
| S16_extracted_129221 | Not-determined |
| S16_extracted_129233 | Not-determined |
| S16_extracted_129239 | Not-determined |
| S16_extracted_129245 | Not-determined |
| S16_extracted_129258 | Low-quality |
| S16_extracted_129268 | Not-determined |
| S16_extracted_129284 | Not-determined |
| S16_extracted_129288 | Low-quality |
| S16_extracted_129306 | Low-quality |
| S16_extracted_129330 | Not-determined |
| S16_extracted_129331 | Not-determined |
| S16_extracted_129332 | Not-determined |
| S16_extracted_129338 | Not-determined |
| S16_extracted_129348 | Not-determined |
| S16_extracted_129363 | Low-quality |
| S16_extracted_129368 | Not-determined |
| S16_extracted_129369 | Not-determined |
| S16_extracted_129370 | Not-determined |
| S16_extracted_129374 | Not-determined |
| S16_extracted_129376 | Not-determined |
| S16_extracted_129387 | Not-determined |
| S16_extracted_129394 | Low-quality |
| S16_extracted_129404 | Not-determined |
| S16_extracted_129415 | Not-determined |
| S16_extracted_129420 | Not-determined |
| S16_extracted_129422 | Low-quality |
| S16_extracted_129423 | Not-determined |
| S16_extracted_129426 | Not-determined |
| S16_extracted_129429 | Low-quality |
| S16_extracted_129435 | Not-determined |
| S16_extracted_129445 | Not-determined |
| S16_extracted_129463 | Not-determined |
| S16_extracted_129465 | Not-determined |
| S16_extracted_129486 | Low-quality |
| S16_extracted_129511 | Not-determined |
| S16_extracted_129524 | Not-determined |
| S16_extracted_129525 | Low-quality |
| S16_extracted_129528 | Not-determined |
| S16_extracted_129532 | Not-determined |
| S16_extracted_129562 | Not-determined |
| S16_extracted_129580 | Not-determined |
| S16_extracted_129590 | Not-determined |
| S16_extracted_129603 | Not-determined |
| S16_extracted_129604 | Not-determined |
| S16_extracted_129617 | Not-determined |
| S16_extracted_129629 | Not-determined |
| S16_extracted_129632 | Low-quality |
| S16_extracted_129636 | Not-determined |
| S16_extracted_129640 | Low-quality |
| S16_extracted_129641 | Not-determined |
| S16_extracted_129656 | Low-quality |
| S16_extracted_129657 | Not-determined |
| S16_extracted_129664 | Not-determined |
| S16_extracted_129666 | Not-determined |
| S16_extracted_129672 | Not-determined |
| S16_extracted_129675 | Not-determined |
| S16_extracted_129679 | Not-determined |
| S16_extracted_129682 | Not-determined |
| S16_extracted_129690 | Not-determined |
| S16_extracted_129704 | Not-determined |
| S16_extracted_129705 | Not-determined |
| S16_extracted_129711 | Low-quality |
| S16_extracted_129715 | Not-determined |
| S16_extracted_129728 | Not-determined |
| S16_extracted_129743 | Not-determined |
| S16_extracted_129753 | Not-determined |
| S16_extracted_129757 | Not-determined |
| S16_extracted_129764 | Not-determined |
| S16_extracted_129768 | Low-quality |
| S16_extracted_129769 | Not-determined |
| S16_extracted_129777 | Not-determined |
| S16_extracted_129786 | Not-determined |
| S16_extracted_129787 | Not-determined |
| S16_extracted_129802 | Not-determined |
| S16_extracted_129809 | Not-determined |
| S16_extracted_129835 | Low-quality |
| S16_extracted_129837 | Not-determined |
| S16_extracted_129839 | Not-determined |
| S16_extracted_129862 | Low-quality |
| S16_extracted_129865 | Not-determined |
| S16_extracted_129872 | Not-determined |
| S16_extracted_129877 | Not-determined |
| S16_extracted_129889 | Not-determined |
| S16_extracted_129892 | Not-determined |
| S16_extracted_129917 | Not-determined |
| S16_extracted_129922 | Not-determined |
| S16_extracted_129939 | Not-determined |
| S16_extracted_129941 | Not-determined |
| S16_extracted_129951 | Not-determined |
| S16_extracted_129955 | Not-determined |
| S16_extracted_129959 | Not-determined |
| S16_extracted_129960 | Not-determined |
| S16_extracted_129961 | Not-determined |
| S16_extracted_129966 | Not-determined |
| S16_extracted_129969 | Not-determined |
| S16_extracted_129972 | Not-determined |
| S16_extracted_129992 | Not-determined |
| S16_extracted_130003 | Not-determined |
| S16_extracted_130007 | Low-quality |
| S16_extracted_130011 | Low-quality |
| S16_extracted_130048 | Not-determined |
| S16_extracted_130052 | Not-determined |
| S16_extracted_130067 | Not-determined |
| S16_extracted_130081 | Not-determined |
| S16_extracted_130104 | Not-determined |
| S16_extracted_130110 | Not-determined |
| S16_extracted_130112 | Not-determined |
| S16_extracted_130126 | Not-determined |
| S16_extracted_130147 | Not-determined |
| S16_extracted_130154 | Not-determined |
| S16_extracted_130169 | Not-determined |
| S16_extracted_130177 | Not-determined |
| S16_extracted_130189 | Low-quality |
| S16_extracted_130194 | Not-determined |
| S16_extracted_130203 | Not-determined |
| S16_extracted_130223 | Not-determined |
| S16_extracted_130232 | Not-determined |
| S16_extracted_130244 | Not-determined |
| S16_extracted_130255 | Not-determined |
| S16_extracted_130276 | Not-determined |
| S16_extracted_130280 | Not-determined |
| S16_extracted_130300 | Not-determined |
| S16_extracted_130303 | Not-determined |
| S16_extracted_130333 | Low-quality |
| S16_extracted_130345 | Low-quality |
| S16_extracted_130347 | Low-quality |
| S16_extracted_130363 | Not-determined |
| S16_extracted_130389 | Not-determined |
| S16_extracted_130390 | Low-quality |
| S16_extracted_130400 | Not-determined |
| S16_extracted_130405 | Low-quality |
| S16_extracted_130412 | Low-quality |
| S16_extracted_130428 | Not-determined |
| S16_extracted_130461 | Not-determined |
| S16_extracted_130481 | Not-determined |
| S16_extracted_130489 | Not-determined |
| S16_extracted_130500 | Not-determined |
| S16_extracted_130501 | Not-determined |
| S16_extracted_130558 | Not-determined |
| S16_extracted_130559 | Not-determined |
| S16_extracted_130566 | Low-quality |
| S16_extracted_130569 | Not-determined |
| S16_extracted_130576 | Not-determined |
| S16_extracted_130578 | Low-quality |
| S16_extracted_130590 | Not-determined |
| S16_extracted_130595 | Not-determined |
| S16_extracted_130602 | Not-determined |
| S16_extracted_130605 | Not-determined |
| S16_extracted_130617 | Low-quality |
| S16_extracted_130618 | Not-determined |
| S16_extracted_130619 | Not-determined |
| S16_extracted_130621 | Not-determined |
| S16_extracted_130627 | Not-determined |
| S16_extracted_130636 | Not-determined |
| S16_extracted_130641 | Not-determined |
| S16_extracted_130656 | Not-determined |
| S16_extracted_130663 | Not-determined |
| S16_extracted_130666 | Not-determined |
| S16_extracted_130667 | Not-determined |

| | |
|---|---|
| S16_extracted_130686 | Low-quality |
| S16_extracted_130708 | Not-determined |
| S16_extracted_130729 | Not-determined |
| S16_extracted_130731 | Not-determined |
| S16_extracted_130732 | Not-determined |
| S16_extracted_130739 | Not-determined |
| S16_extracted_130746 | Not-determined |
| S16_extracted_130748 | Not-determined |
| S16_extracted_130783 | Not-determined |
| S16_extracted_130797 | Not-determined |
| S16_extracted_130799 | Not-determined |
| S16_extracted_130801 | Not-determined |
| S16_extracted_130850 | Not-determined |
| S16_extracted_130854 | Not-determined |
| S16_extracted_130858 | Not-determined |
| S16_extracted_130867 | Low-quality |
| S16_extracted_130875 | Not-determined |
| S16_extracted_130881 | Not-determined |
| S16_extracted_130884 | Not-determined |
| S16_extracted_130894 | Not-determined |
| S16_extracted_130906 | Not-determined |
| S16_extracted_130920 | Not-determined |
| S16_extracted_130923 | Not-determined |
| S16_extracted_130944 | Not-determined |
| S16_extracted_130957 | Not-determined |
| S16_extracted_130965 | Not-determined |
| S16_extracted_130972 | Not-determined |
| S16_extracted_130978 | Not-determined |
| S16_extracted_130996 | Not-determined |
| S16_extracted_131001 | Not-determined |
| S16_extracted_131022 | Low-quality |
| S16_extracted_131026 | Not-determined |
| S16_extracted_131037 | Not-determined |
| S16_extracted_131042 | Not-determined |
| S16_extracted_131043 | Not-determined |
| S16_extracted_131054 | Not-determined |
| S16_extracted_131060 | Not-determined |
| S16_extracted_131066 | Not-determined |
| S16_extracted_131067 | Not-determined |
| S16_extracted_131083 | Not-determined |
| S16_extracted_131092 | Not-determined |
| S16_extracted_131116 | Low-quality |
| S16_extracted_131121 | Not-determined |
| S16_extracted_131124 | Not-determined |
| S16_extracted_131125 | Not-determined |
| S16_extracted_131136 | Not-determined |
| S16_extracted_131137 | Not-determined |
| S16_extracted_131146 | Not-determined |
| S16_extracted_131156 | Not-determined |
| S16_extracted_131175 | Not-determined |
| S16_extracted_131188 | Not-determined |
| S16_extracted_131200 | Not-determined |
| S16_extracted_131218 | Not-determined |
| S16_extracted_131233 | Not-determined |
| S16_extracted_131242 | Low-quality |
| S16_extracted_131252 | Not-determined |
| S16_extracted_131285 | Not-determined |
| S16_extracted_131304 | Not-determined |
| S16_extracted_131317 | Not-determined |
| S16_extracted_131326 | Not-determined |
| S16_extracted_131338 | Not-determined |
| S16_extracted_131358 | Not-determined |
| S16_extracted_131363 | Not-determined |
| S16_extracted_131367 | Not-determined |
| S16_extracted_131371 | Low-quality |
| S16_extracted_131376 | Not-determined |
| S16_extracted_131377 | Not-determined |
| S16_extracted_131403 | Not-determined |
| S16_extracted_131412 | Not-determined |
| S16_extracted_131422 | Not-determined |
| S16_extracted_131438 | Not-determined |
| S16_extracted_131439 | Low-quality |
| S16_extracted_131450 | Low-quality |
| S16_extracted_131455 | Not-determined |
| S16_extracted_131460 | Not-determined |
| S16_extracted_131476 | Not-determined |
| S16_extracted_131478 | Not-determined |
| S16_extracted_131482 | Not-determined |
| S16_extracted_131529 | Not-determined |
| S16_extracted_131546 | Not-determined |
| S16_extracted_131559 | Not-determined |
| S16_extracted_131568 | Not-determined |
| S16_extracted_131584 | Not-determined |
| S16_extracted_131589 | Not-determined |
| S16_extracted_131593 | Not-determined |
| S16_extracted_131617 | Not-determined |
| S16_extracted_131622 | Low-quality |
| S16_extracted_131624 | Not-determined |
| S16_extracted_131660 | Not-determined |
| S16_extracted_131667 | Not-determined |
| S16_extracted_131688 | Not-determined |
| S16_extracted_131689 | Not-determined |
| S16_extracted_131691 | Not-determined |
| S16_extracted_131698 | Not-determined |
| S16_extracted_131714 | Not-determined |
| S16_extracted_131735 | Low-quality |
| S16_extracted_131739 | Not-determined |
| S16_extracted_131749 | Not-determined |
| S16_extracted_131751 | Not-determined |
| S16_extracted_131762 | Low-quality |
| S16_extracted_131766 | Low-quality |
| S16_extracted_131775 | Not-determined |
| S16_extracted_131791 | Low-quality |
| S16_extracted_131796 | Not-determined |
| S16_extracted_131804 | Not-determined |
| S16_extracted_131807 | Not-determined |
| S16_extracted_131819 | Not-determined |
| S16_extracted_131828 | Not-determined |
| S16_extracted_131851 | Not-determined |
| S16_extracted_131856 | Not-determined |
| S16_extracted_131857 | Low-quality |
| S16_extracted_131860 | Not-determined |
| S16_extracted_131871 | Not-determined |
| S16_extracted_131878 | Low-quality |
| S16_extracted_131880 | Not-determined |
| S16_extracted_131881 | Not-determined |
| S16_extracted_131903 | Low-quality |
| S16_extracted_131905 | Not-determined |
| S16_extracted_131909 | Not-determined |
| S16_extracted_131910 | Not-determined |
| S16_extracted_131917 | Not-determined |
| S16_extracted_131919 | Low-quality |
| S16_extracted_131920 | Not-determined |
| S16_extracted_131938 | Not-determined |
| S16_extracted_131949 | Low-quality |
| S16_extracted_131969 | Not-determined |
| S16_extracted_131975 | Not-determined |
| S16_extracted_131985 | Not-determined |
| S16_extracted_131996 | Low-quality |
| S16_extracted_132056 | Not-determined |
| S16_extracted_132089 | Not-determined |
| S16_extracted_132093 | Low-quality |
| S16_extracted_132122 | Not-determined |
| S16_extracted_132124 | Not-determined |
| S16_extracted_132136 | Not-determined |
| S16_extracted_132144 | Not-determined |
| S16_extracted_132156 | Not-determined |
| S16_extracted_132159 | Not-determined |
| S16_extracted_132175 | Low-quality |
| S16_extracted_132181 | Not-determined |
| S16_extracted_132197 | Low-quality |
| S16_extracted_132224 | Not-determined |
| S16_extracted_132237 | Not-determined |
| S16_extracted_132248 | Not-determined |
| S16_extracted_132258 | Not-determined |
| S16_extracted_132270 | Not-determined |
| S16_extracted_132274 | Not-determined |
| S16_extracted_132279 | Not-determined |
| S16_extracted_132295 | Low-quality |
| S16_extracted_132311 | Not-determined |
| S16_extracted_132315 | Not-determined |
| S16_extracted_132320 | Not-determined |
| S16_extracted_132321 | Not-determined |
| S16_extracted_132336 | Not-determined |
| S16_extracted_132350 | Not-determined |
| S16_extracted_132356 | Not-determined |
| S16_extracted_132363 | Low-quality |
| S16_extracted_132380 | Low-quality |
| S16_extracted_132382 | Not-determined |
| S16_extracted_132414 | Not-determined |
| S16_extracted_132419 | Not-determined |
| S16_extracted_132429 | Not-determined |
| S16_extracted_132431 | Not-determined |
| S16_extracted_132440 | Low-quality |
| S16_extracted_132451 | Not-determined |
| S16_extracted_132471 | Not-determined |
| S16_extracted_132476 | Not-determined |
| S16_extracted_132487 | Not-determined |
| S16_extracted_132491 | Not-determined |
| S16_extracted_132500 | Low-quality |
| S16_extracted_132516 | Not-determined |
| S16_extracted_132535 | Not-determined |
| S16_extracted_132537 | Not-determined |
| S16_extracted_132543 | Not-determined |
| S16_extracted_132546 | Not-determined |
| S16_extracted_132554 | Not-determined |
| S16_extracted_132558 | Not-determined |
| S16_extracted_132571 | Not-determined |
| S16_extracted_132582 | Not-determined |
| S16_extracted_132586 | Low-quality |
| S16_extracted_132598 | Low-quality |
| S16_extracted_132615 | Not-determined |
| S16_extracted_132617 | Not-determined |
| S16_extracted_132620 | Not-determined |
| S16_extracted_132621 | Not-determined |
| S16_extracted_132622 | Low-quality |
| S16_extracted_132624 | Not-determined |
| S16_extracted_132626 | Low-quality |
| S16_extracted_132628 | Not-determined |
| S16_extracted_132630 | Not-determined |
| S16_extracted_132632 | Not-determined |
| S16_extracted_132634 | Not-determined |
| S16_extracted_132648 | Not-determined |
| S16_extracted_132654 | Not-determined |
| S16_extracted_132656 | Not-determined |
| S16_extracted_132659 | Low-quality |
| S16_extracted_132673 | Not-determined |
| S16_extracted_132674 | Low-quality |
| S16_extracted_132706 | Not-determined |
| S16_extracted_132710 | Low-quality |
| S16_extracted_132733 | Not-determined |
| S16_extracted_132747 | Not-determined |
| S16_extracted_132782 | Not-determined |
| S16_extracted_132787 | Not-determined |
| S16_extracted_132796 | Not-determined |
| S16_extracted_132844 | Not-determined |
| S16_extracted_132847 | Not-determined |
| S16_extracted_132856 | Not-determined |
| S16_extracted_132857 | Not-determined |
| S16_extracted_132863 | Not-determined |
| S16_extracted_132892 | Not-determined |
| S16_extracted_132898 | Not-determined |
| S16_extracted_132899 | Not-determined |
| S16_extracted_132919 | Not-determined |
| S16_extracted_132921 | Not-determined |
| S16_extracted_132932 | Not-determined |
| S16_extracted_132936 | Not-determined |
| S16_extracted_132958 | Low-quality |
| S16_extracted_132968 | Low-quality |
| S16_extracted_132989 | Not-determined |
| S16_extracted_132990 | Not-determined |
| S16_extracted_132999 | Not-determined |
| S16_extracted_133000 | Not-determined |
| S16_extracted_133007 | Not-determined |
| S16_extracted_133014 | Not-determined |
| S16_extracted_133017 | Not-determined |
| S16_extracted_133021 | Not-determined |
| S16_extracted_133022 | Not-determined |
| S16_extracted_133024 | Not-determined |
| S16_extracted_133045 | Not-determined |
| S16_extracted_133066 | Not-determined |
| S16_extracted_133068 | Not-determined |
| S16_extracted_133078 | Not-determined |
| S16_extracted_133121 | Not-determined |
| S16_extracted_133123 | Not-determined |
| S16_extracted_133128 | Not-determined |
| S16_extracted_133134 | Not-determined |
| S16_extracted_133135 | Low-quality |
| S16_extracted_133146 | Low-quality |
| S16_extracted_133151 | Not-determined |
| S16_extracted_133155 | Not-determined |
| S16_extracted_133177 | Not-determined |
| S16_extracted_133178 | Not-determined |
| S16_extracted_133180 | Not-determined |
| S16_extracted_133230 | Not-determined |
| S16_extracted_133238 | Low-quality |
| S16_extracted_133253 | Not-determined |
| S16_extracted_133265 | Low-quality |
| S16_extracted_133289 | Not-determined |
| S16_extracted_133291 | Not-determined |
| S16_extracted_133309 | Not-determined |
| S16_extracted_133321 | Not-determined |
| S16_extracted_133342 | Not-determined |
| S16_extracted_133347 | Not-determined |
| S16_extracted_133376 | Low-quality |
| S16_extracted_133377 | Not-determined |
| S16_extracted_133378 | Not-determined |
| S16_extracted_133390 | Not-determined |
| S16_extracted_133394 | Not-determined |
| S16_extracted_133398 | Not-determined |
| S16_extracted_133405 | Not-determined |
| S16_extracted_133432 | Not-determined |
| S16_extracted_133448 | Not-determined |
| S16_extracted_133458 | Not-determined |
| S16_extracted_133483 | Not-determined |
| S16_extracted_133498 | Not-determined |
| S16_extracted_133499 | Not-determined |
| S16_extracted_133508 | Low-quality |
| S16_extracted_133523 | Not-determined |
| S16_extracted_133573 | Not-determined |
| S16_extracted_133576 | Not-determined |
| S16_extracted_133588 | Not-determined |
| S16_extracted_133592 | Low-quality |
| S16_extracted_133622 | Low-quality |
| S16_extracted_133623 | Not-determined |
| S16_extracted_133624 | Not-determined |
| S16_extracted_133632 | Low-quality |
| S16_extracted_133650 | Low-quality |
| S16_extracted_133664 | Not-determined |
| S16_extracted_133671 | Not-determined |
| S16_extracted_133680 | Not-determined |
| S16_extracted_133688 | Not-determined |
| S16_extracted_133692 | Not-determined |
| S16_extracted_133696 | Not-determined |
| S16_extracted_133703 | Not-determined |
| S16_extracted_133715 | Not-determined |
| S16_extracted_133720 | Low-quality |
| S16_extracted_133755 | Not-determined |
| S16_extracted_133775 | Not-determined |
| S16_extracted_133788 | Low-quality |
| S16_extracted_133792 | Not-determined |
| S16_extracted_133801 | Not-determined |
| S16_extracted_133819 | Not-determined |
| S16_extracted_133848 | Low-quality |
| S16_extracted_133855 | Not-determined |
| S16_extracted_133862 | Not-determined |
| S16_extracted_133863 | Not-determined |
| S16_extracted_133880 | Not-determined |
| S16_extracted_133881 | Not-determined |
| S16_extracted_134003 | Not-determined |
| S16_extracted_134004 | Not-determined |
| S16_extracted_134033 | Not-determined |
| S16_extracted_134040 | Not-determined |
| S16_extracted_134082 | Not-determined |
| S16_extracted_134089 | Not-determined |
| S16_extracted_134093 | Not-determined |
| S16_extracted_134094 | Not-determined |
| S16_extracted_134119 | Not-determined |
| S16_extracted_134128 | Low-quality |
| S16_extracted_134143 | Not-determined |
| S16_extracted_134158 | Not-determined |
| S16_extracted_134167 | Low-quality |
| S16_extracted_134180 | Low-quality |
| S16_extracted_134181 | Not-determined |
| S16_extracted_134197 | Not-determined |
| S16_extracted_134221 | Not-determined |
| S16_extracted_134226 | Not-determined |
| S16_extracted_134229 | Low-quality |
| S16_extracted_134235 | Not-determined |
| S16_extracted_134243 | Not-determined |
| S16_extracted_134271 | Not-determined |
| S16_extracted_134282 | Not-determined |
| S16_extracted_134294 | Not-determined |
| S16_extracted_134299 | Not-determined |
| S16_extracted_134343 | Not-determined |
| S16_extracted_134360 | Not-determined |
| S16_extracted_134380 | Not-determined |
| S16_extracted_134381 | Not-determined |
| S16_extracted_134403 | Not-determined |
| S16_extracted_134422 | Not-determined |
| S16_extracted_134451 | Low-quality |
| S16_extracted_134474 | Low-quality |
| S16_extracted_134477 | Not-determined |
| S16_extracted_134485 | Low-quality |
| S16_extracted_134502 | Not-determined |
| S16_extracted_134507 | Not-determined |
| S16_extracted_134508 | Not-determined |
| S16_extracted_134531 | Not-determined |
| S16_extracted_134532 | Low-quality |
| S16_extracted_134547 | Not-determined |
| S16_extracted_134565 | Not-determined |
| S16_extracted_134569 | Not-determined |
| S16_extracted_134577 | Low-quality |
| S16_extracted_134587 | Not-determined |
| S16_extracted_134592 | Not-determined |
| S16_extracted_134600 | Low-quality |
| S16_extracted_134612 | Low-quality |
| S16_extracted_134614 | Not-determined |
| S16_extracted_134616 | Not-determined |
| S16_extracted_134621 | Not-determined |
| S16_extracted_134622 | Not-determined |
| S16_extracted_134638 | Not-determined |
| S16_extracted_134648 | Not-determined |
| S16_extracted_134660 | Not-determined |
| S16_extracted_134673 | Not-determined |
| S16_extracted_134676 | Not-determined |
| S16_extracted_134695 | Low-quality |
| S16_extracted_134700 | Not-determined |
| S8_extracted_k141_147338 | Medium-quality |
| S16_extracted_134709 | Not-determined |
| S16_extracted_134721 | Not-determined |
| S16_extracted_134724 | Not-determined |
| S16_extracted_134728 | Not-determined |
| S16_extracted_134737 | Not-determined |
| S16_extracted_134741 | Not-determined |
| S16_extracted_134774 | Not-determined |
| S16_extracted_134775 | Low-quality |
| S16_extracted_134804 | Not-determined |
| S16_extracted_134808 | Not-determined |
| S16_extracted_134843 | Not-determined |
| S16_extracted_134848 | Low-quality |
| S16_extracted_134851 | Not-determined |
| S16_extracted_134853 | Not-determined |
| S16_extracted_134886 | Not-determined |
| S16_extracted_134899 | Low-quality |
| S16_extracted_134925 | Not-determined |
| S16_extracted_134927 | Low-quality |
| S16_extracted_134937 | Low-quality |
| S16_extracted_134953 | Not-determined |
| S16_extracted_134967 | Not-determined |
| S16_extracted_134975 | Not-determined |
| S16_extracted_134982 | Not-determined |
| S16_extracted_134999 | Low-quality |
| S16_extracted_135006 | Not-determined |
| S16_extracted_135034 | Not-determined |
| S16_extracted_135042 | Low-quality |
| S16_extracted_135070 | Not-determined |
| S16_extracted_135074 | Not-determined |
| S16_extracted_135111 | Not-determined |
| S16_extracted_135122 | Not-determined |
| S16_extracted_135130 | Not-determined |
| S16_extracted_135144 | Not-determined |
| S16_extracted_135153 | Not-determined |
| S16_extracted_135155 | Not-determined |
| S16_extracted_135161 | Not-determined |
| S16_extracted_135166 | Not-determined |
| S16_extracted_135171 | Not-determined |
| S16_extracted_135182 | Not-determined |
| S16_extracted_135183 | Not-determined |
| S16_extracted_135208 | Not-determined |
| S16_extracted_135214 | Not-determined |
| S16_extracted_135247 | Not-determined |
| S16_extracted_135268 | Not-determined |
| S16_extracted_135273 | Not-determined |
| S16_extracted_135282 | Not-determined |
| S16_extracted_135284 | Not-determined |
| S16_extracted_135324 | Not-determined |
| S16_extracted_135355 | Not-determined |
| S16_extracted_135357 | Not-determined |
| S16_extracted_135365 | Low-quality |
| S16_extracted_135367 | Not-determined |
| S16_extracted_135410 | Not-determined |
| S16_extracted_135414 | Not-determined |
| S16_extracted_135427 | Not-determined |
| S16_extracted_135431 | Not-determined |
| S16_extracted_135432 | Not-determined |
| S16_extracted_135438 | Low-quality |
| S16_extracted_135448 | Not-determined |
| S16_extracted_135469 | Low-quality |
| S16_extracted_135477 | Not-determined |
| S16_extracted_135479 | Not-determined |
| S16_extracted_135483 | Not-determined |
| S16_extracted_135487 | Low-quality |
| S16_extracted_135494 | Not-determined |
| S16_extracted_135502 | Not-determined |
| S16_extracted_135520 | Not-determined |
| S16_extracted_135543 | Not-determined |
| S16_extracted_135557 | Not-determined |
| S16_extracted_135586 | Not-determined |
| S16_extracted_135603 | Low-quality |
| S16_extracted_135609 | Not-determined |
| S16_extracted_135614 | Not-determined |
| S16_extracted_135619 | Low-quality |
| S16_extracted_135622 | Not-determined |
| S16_extracted_135627 | Low-quality |
| S16_extracted_135629 | Not-determined |
| S16_extracted_135633 | Not-determined |
| S16_extracted_135637 | Low-quality |
| S16_extracted_135657 | Not-determined |
| S16_extracted_135659 | Not-determined |
| S16_extracted_135667 | Not-determined |
| S16_extracted_135675 | Low-quality |
| S16_extracted_135679 | Low-quality |
| S16_extracted_135682 | Not-determined |
| S16_extracted_135684 | Low-quality |
| S16_extracted_135686 | Not-determined |
| S16_extracted_135689 | Not-determined |
| S16_extracted_135691 | Not-determined |
| S16_extracted_135695 | Not-determined |
| S16_extracted_135699 | Not-determined |
| S16_extracted_135722 | Not-determined |
| S16_extracted_135727 | Not-determined |
| S16_extracted_135735 | Not-determined |
| S16_extracted_135737 | Low-quality |
| S16_extracted_135777 | Not-determined |
| S16_extracted_135791 | Not-determined |
| S16_extracted_135792 | Low-quality |
| S16_extracted_135796 | Not-determined |
| S16_extracted_135828 | Not-determined |
| S16_extracted_135836 | Not-determined |
| S16_extracted_135838 | Not-determined |
| S16_extracted_135845 | Not-determined |
| S16_extracted_135846 | Not-determined |
| S16_extracted_135852 | Not-determined |
| S16_extracted_135853 | Not-determined |
| S16_extracted_135862 | Not-determined |
| S16_extracted_135863 | Not-determined |
| S16_extracted_135872 | Low-quality |
| S16_extracted_135873 | Not-determined |
| S16_extracted_135894 | Not-determined |
| S16_extracted_135908 | Low-quality |
| S16_extracted_135915 | Not-determined |
| S16_extracted_135928 | Not-determined |
| S16_extracted_135930 | Not-determined |
| S16_extracted_135931 | Not-determined |
| S16_extracted_135954 | Not-determined |
| S16_extracted_135955 | Low-quality |
| S16_extracted_135960 | Not-determined |
| S16_extracted_135964 | Not-determined |
| S16_extracted_135969 | Not-determined |
| S16_extracted_135979 | Low-quality |
| S16_extracted_135985 | Not-determined |
| S16_extracted_135996 | Not-determined |
| S16_extracted_136023 | Not-determined |
| S16_extracted_136036 | Not-determined |
| S16_extracted_136056 | Not-determined |
| S16_extracted_136066 | Not-determined |
| S16_extracted_136081 | Not-determined |
| S16_extracted_136086 | Not-determined |
| S16_extracted_136114 | Not-determined |
| S16_extracted_136125 | Not-determined |
| S16_extracted_136138 | Not-determined |
| S16_extracted_136147 | Low-quality |
| S16_extracted_136151 | Not-determined |
| S16_extracted_136152 | Not-determined |
| S16_extracted_136158 | Not-determined |
| S16_extracted_136179 | Not-determined |
| S16_extracted_136209 | Not-determined |
| S16_extracted_136218 | Not-determined |
| S16_extracted_136225 | Not-determined |
| S16_extracted_136232 | Not-determined |
| S16_extracted_136248 | Not-determined |
| S16_extracted_136251 | Not-determined |
| S16_extracted_136269 | Not-determined |
| S16_extracted_136276 | Not-determined |
| S16_extracted_136286 | Not-determined |
| S16_extracted_136290 | Not-determined |
| S16_extracted_136291 | Not-determined |
| S16_extracted_136293 | Not-determined |
| S16_extracted_136294 | Not-determined |
| S16_extracted_136305 | Not-determined |
| S16_extracted_136314 | Not-determined |
| S16_extracted_136315 | Not-determined |
| S16_extracted_136318 | Not-determined |
| S16_extracted_136346 | Not-determined |
| S16_extracted_136351 | Not-determined |
| S16_extracted_136358 | Not-determined |
| S16_extracted_136397 | Not-determined |
| S16_extracted_136401 | Low-quality |
| S16_extracted_136406 | Not-determined |
| S16_extracted_136429 | Not-determined |
| S16_extracted_136434 | Not-determined |
| S16_extracted_136437 | Low-quality |
| S16_extracted_136441 | Not-determined |
| S16_extracted_136453 | Not-determined |
| S16_extracted_136486 | Not-determined |
| S16_extracted_136490 | Low-quality |
| S16_extracted_136500 | Not-determined |
| S16_extracted_136506 | Not-determined |
| S16_extracted_136515 | Not-determined |
| S16_extracted_136517 | Not-determined |
| S16_extracted_136529 | Not-determined |
| S16_extracted_136533 | Not-determined |
| S16_extracted_136540 | Not-determined |
| S16_extracted_136544 | Not-determined |
| S16_extracted_136547 | Low-quality |
| S16_extracted_136553 | Not-determined |
| S16_extracted_136555 | Low-quality |
| S16_extracted_136577 | Not-determined |
| S16_extracted_136578 | Low-quality |
| S16_extracted_136605 | Not-determined |
| S16_extracted_136611 | Low-quality |
| S16_extracted_136616 | Not-determined |
| S16_extracted_136630 | Not-determined |
| S16_extracted_136635 | Low-quality |
| S16_extracted_136639 | Low-quality |
| S16_extracted_136656 | Not-determined |
| S16_extracted_136664 | Low-quality |
| S16_extracted_136675 | Not-determined |
| S16_extracted_136680 | Not-determined |
| S16_extracted_136704 | Not-determined |
| S16_extracted_136734 | Not-determined |
| S16_extracted_136742 | Low-quality |
| S16_extracted_136750 | Not-determined |
| S16_extracted_136751 | Not-determined |
| S16_extracted_136752 | Low-quality |
| S16_extracted_136755 | Not-determined |
| S16_extracted_136756 | Not-determined |
| S16_extracted_136758 | Not-determined |
| S16_extracted_136770 | Not-determined |
| S16_extracted_136778 | Low-quality |
| S16_extracted_136791 | Not-determined |
| S16_extracted_136797 | Not-determined |
| S16_extracted_136810 | Not-determined |
| S16_extracted_136811 | Not-determined |
| S16_extracted_136823 | Not-determined |
| S16_extracted_136851 | Low-quality |
| S16_extracted_136858 | Not-determined |
| S16_extracted_136861 | Not-determined |
| S16_extracted_136877 | Not-determined |
| S16_extracted_136879 | Not-determined |
| S16_extracted_136892 | Not-determined |
| S16_extracted_136905 | Not-determined |
| S16_extracted_136913 | Not-determined |
| S16_extracted_136955 | Not-determined |
| S16_extracted_136965 | Not-determined |
| S16_extracted_136977 | Not-determined |
| S16_extracted_136998 | Not-determined |
| S16_extracted_137000 | Low-quality |
| S16_extracted_137021 | Low-quality |
| S16_extracted_137033 | Not-determined |
| S16_extracted_137047 | Low-quality |
| S16_extracted_137048 | Not-determined |
| S16_extracted_137079 | Not-determined |
| S16_extracted_137088 | Not-determined |
| S16_extracted_137101 | Not-determined |
| S16_extracted_137102 | Low-quality |
| S16_extracted_137113 | Not-determined |
| S16_extracted_137123 | Not-determined |

| | |
|---|---|
| S16_extracted_137147 | Not-determined |
| S16_extracted_137154 | Not-determined |
| S16_extracted_137156 | Not-determined |
| S16_extracted_137164 | Not-determined |
| S16_extracted_137165 | Low-quality |
| S16_extracted_137168 | Low-quality |
| S16_extracted_137170 | Not-determined |
| S16_extracted_137178 | Not-determined |
| S16_extracted_137179 | Not-determined |
| S16_extracted_137185 | Not-determined |
| S16_extracted_137195 | Not-determined |
| S16_extracted_137207 | Not-determined |
| S16_extracted_137214 | Not-determined |
| S16_extracted_137236 | Not-determined |
| S16_extracted_137249 | Not-determined |
| S16_extracted_137260 | Not-determined |
| S16_extracted_137318 | Not-determined |
| S16_extracted_137329 | Not-determined |
| S16_extracted_137341 | Not-determined |
| S16_extracted_137361 | Not-determined |
| S16_extracted_137365 | Not-determined |
| S16_extracted_137367 | Not-determined |
| S16_extracted_137370 | Not-determined |
| S16_extracted_137374 | Not-determined |
| S16_extracted_137382 | Not-determined |
| S16_extracted_137392 | Not-determined |
| S16_extracted_137417 | Not-determined |
| S16_extracted_137434 | Not-determined |
| S16_extracted_137436 | Not-determined |
| S16_extracted_137445 | Not-determined |
| S16_extracted_137458 | Low-quality |
| S16_extracted_137462 | Not-determined |
| S16_extracted_137473 | Not-determined |
| S16_extracted_137477 | Not-determined |
| S16_extracted_137489 | Not-determined |
| S16_extracted_137501 | Not-determined |
| S16_extracted_137505 | Not-determined |
| S16_extracted_137519 | Not-determined |
| S16_extracted_137562 | Low-quality |
| S16_extracted_137563 | Not-determined |
| S16_extracted_137567 | Not-determined |
| S16_extracted_137577 | Not-determined |
| S16_extracted_137593 | Not-determined |
| S16_extracted_137605 | Not-determined |
| S16_extracted_137606 | Not-determined |
| S16_extracted_137608 | Not-determined |
| S16_extracted_137636 | Not-determined |
| S16_extracted_137647 | Not-determined |
| S16_extracted_137657 | Not-determined |
| S16_extracted_137676 | Low-quality |
| S16_extracted_137679 | Not-determined |
| S16_extracted_137682 | Not-determined |
| S16_extracted_137684 | Not-determined |
| S16_extracted_137687 | Low-quality |
| S16_extracted_137694 | Not-determined |
| S16_extracted_137696 | Not-determined |
| S16_extracted_137716 | Not-determined |
| S16_extracted_137729 | Not-determined |
| S16_extracted_137730 | Not-determined |
| S16_extracted_137739 | Not-determined |
| S16_extracted_137756 | Not-determined |
| S16_extracted_137765 | Not-determined |
| S16_extracted_137771 | Not-determined |
| S16_extracted_137785 | Not-determined |
| S16_extracted_137787 | Not-determined |
| S16_extracted_137797 | Not-determined |
| S16_extracted_137806 | Not-determined |
| S16_extracted_137809 | Low-quality |
| S16_extracted_137823 | Not-determined |
| S16_extracted_137829 | Not-determined |
| S16_extracted_137837 | Not-determined |
| S16_extracted_137857 | Not-determined |
| S16_extracted_137866 | Low-quality |
| S16_extracted_137881 | Not-determined |
| S16_extracted_137895 | Low-quality |
| S16_extracted_137899 | Not-determined |
| S16_extracted_137907 | Not-determined |
| S16_extracted_137927 | Not-determined |
| S16_extracted_137936 | Not-determined |
| S16_extracted_137950 | Not-determined |
| S16_extracted_137954 | Not-determined |
| S16_extracted_137956 | Not-determined |
| S16_extracted_137973 | Not-determined |
| S16_extracted_137976 | Not-determined |
| S16_extracted_137983 | Low-quality |
| S16_extracted_137999 | Low-quality |
| S16_extracted_138006 | Not-determined |
| S16_extracted_138013 | Not-determined |
| S16_extracted_138028 | Low-quality |
| S16_extracted_138029 | Not-determined |
| S16_extracted_138040 | Low-quality |
| S16_extracted_138053 | Low-quality |
| S16_extracted_138064 | Not-determined |
| S16_extracted_138065 | Not-determined |
| S16_extracted_138072 | Not-determined |
| S16_extracted_138103 | Low-quality |
| S16_extracted_138104 | Not-determined |
| S16_extracted_138111 | Not-determined |
| S16_extracted_138140 | Not-determined |
| S16_extracted_138150 | Not-determined |
| S16_extracted_138167 | Low-quality |
| S16_extracted_138185 | Not-determined |
| S16_extracted_138218 | Not-determined |
| S16_extracted_138225 | Low-quality |
| S16_extracted_138235 | Not-determined |
| S16_extracted_138237 | Not-determined |
| S16_extracted_138260 | Not-determined |
| S16_extracted_138275 | Not-determined |
| S16_extracted_138283 | Not-determined |
| S16_extracted_138305 | Not-determined |
| S16_extracted_138309 | Not-determined |
| S16_extracted_138331 | Not-determined |
| S16_extracted_138334 | Not-determined |
| S16_extracted_138338 | Not-determined |
| S16_extracted_138350 | Not-determined |
| S16_extracted_138363 | Not-determined |
| S16_extracted_138387 | Not-determined |
| S16_extracted_138396 | Not-determined |
| S16_extracted_138406 | Low-quality |
| S16_extracted_138426 | Not-determined |
| S16_extracted_138437 | Low-quality |
| S16_extracted_138443 | Not-determined |
| S16_extracted_138447 | Not-determined |
| S16_extracted_138452 | Not-determined |
| S16_extracted_138468 | Not-determined |
| S16_extracted_138477 | Not-determined |
| S16_extracted_138487 | Not-determined |
| S16_extracted_138493 | Not-determined |
| S16_extracted_138519 | Not-determined |
| S16_extracted_138528 | Not-determined |
| S16_extracted_138532 | Not-determined |
| S16_extracted_138536 | Low-quality |
| S16_extracted_138551 | Not-determined |
| S16_extracted_138553 | Not-determined |
| S16_extracted_138557 | Not-determined |
| S16_extracted_138564 | Not-determined |
| S16_extracted_138569 | Not-determined |
| S16_extracted_138581 | Not-determined |
| S16_extracted_138590 | Low-quality |
| S16_extracted_138596 | Not-determined |
| S16_extracted_138610 | Not-determined |
| S16_extracted_138612 | Not-determined |
| S16_extracted_138619 | Not-determined |
| S16_extracted_138620 | Not-determined |
| S16_extracted_138629 | Not-determined |
| S16_extracted_138633 | Not-determined |
| S16_extracted_138642 | Not-determined |
| S16_extracted_138652 | Not-determined |
| S16_extracted_138663 | Not-determined |
| S16_extracted_138671 | Not-determined |
| S16_extracted_138698 | Not-determined |
| S16_extracted_138704 | Low-quality |
| S16_extracted_138706 | Not-determined |
| S16_extracted_138717 | Not-determined |
| S16_extracted_138728 | Not-determined |
| S16_extracted_138733 | Low-quality |
| S16_extracted_138749 | Not-determined |
| S16_extracted_138764 | Low-quality |
| S16_extracted_138773 | Not-determined |
| S16_extracted_138795 | Not-determined |
| S16_extracted_138812 | Not-determined |
| S16_extracted_138817 | Low-quality |
| S16_extracted_138835 | Low-quality |
| S16_extracted_138838 | Low-quality |
| S16_extracted_138846 | Not-determined |
| S16_extracted_138847 | Not-determined |
| S16_extracted_138848 | Not-determined |
| S16_extracted_138854 | Not-determined |
| S16_extracted_138868 | Not-determined |
| S16_extracted_138880 | Not-determined |
| S16_extracted_138882 | Low-quality |
| S16_extracted_138883 | Not-determined |
| S16_extracted_138902 | Not-determined |
| S16_extracted_138906 | Not-determined |
| S16_extracted_138925 | Not-determined |
| S16_extracted_138954 | Not-determined |
| S16_extracted_138955 | Not-determined |
| S16_extracted_138996 | Not-determined |
| S16_extracted_139004 | Not-determined |
| S16_extracted_139008 | Not-determined |
| S16_extracted_139032 | Low-quality |
| S16_extracted_139042 | Not-determined |
| S16_extracted_139073 | Not-determined |
| S16_extracted_139080 | Not-determined |
| S16_extracted_139096 | Not-determined |
| S16_extracted_139105 | Not-determined |
| S16_extracted_139108 | Low-quality |
| S16_extracted_139120 | Not-determined |
| S16_extracted_139121 | Low-quality |
| S16_extracted_139124 | Not-determined |
| S16_extracted_139129 | Not-determined |
| S16_extracted_139136 | Low-quality |
| S16_extracted_139141 | Not-determined |
| S16_extracted_139146 | Not-determined |
| S16_extracted_139147 | Low-quality |
| S16_extracted_139148 | Not-determined |
| S16_extracted_139150 | Not-determined |
| S16_extracted_139155 | Low-quality |
| S16_extracted_139162 | Not-determined |
| S16_extracted_139178 | Low-quality |
| S16_extracted_139181 | Not-determined |
| S16_extracted_139183 | Not-determined |
| S16_extracted_139185 | Not-determined |
| S16_extracted_139188 | Not-determined |
| S16_extracted_139189 | Not-determined |
| S16_extracted_139196 | Not-determined |
| S16_extracted_139213 | Low-quality |
| S16_extracted_139214 | Not-determined |
| S16_extracted_139220 | Not-determined |
| S16_extracted_139224 | Low-quality |
| S16_extracted_139231 | Low-quality |
| S16_extracted_139232 | Not-determined |
| S16_extracted_139256 | Not-determined |
| S16_extracted_139260 | Not-determined |
| S16_extracted_139261 | Not-determined |
| S16_extracted_139262 | Not-determined |
| S16_extracted_139264 | Low-quality |
| S16_extracted_139289 | Not-determined |
| S16_extracted_139313 | Not-determined |
| S16_extracted_139349 | Not-determined |
| S16_extracted_139353 | Not-determined |
| S16_extracted_139357 | Low-quality |
| S16_extracted_139363 | Not-determined |
| S16_extracted_139382 | Not-determined |
| S16_extracted_139387 | Low-quality |
| S16_extracted_139393 | Low-quality |
| S16_extracted_139400 | Not-determined |
| S16_extracted_139402 | Low-quality |
| S16_extracted_139404 | Not-determined |
| S16_extracted_139429 | Not-determined |
| S16_extracted_139433 | Not-determined |
| S16_extracted_139434 | Low-quality |
| S16_extracted_139435 | Not-determined |
| S16_extracted_139436 | Not-determined |
| S16_extracted_139441 | Not-determined |
| S16_extracted_139453 | Not-determined |
| S16_extracted_139455 | Low-quality |
| S16_extracted_139469 | Low-quality |
| S16_extracted_139472 | Not-determined |
| S16_extracted_139488 | Not-determined |
| S16_extracted_139496 | Not-determined |
| S16_extracted_139504 | Not-determined |
| S16_extracted_139526 | Not-determined |
| S16_extracted_139527 | Not-determined |
| S16_extracted_139529 | Not-determined |
| S16_extracted_139530 | Not-determined |
| S16_extracted_139531 | Not-determined |
| S16_extracted_139537 | Not-determined |
| S16_extracted_139547 | Not-determined |
| S16_extracted_139554 | Not-determined |
| S16_extracted_139560 | Not-determined |
| S16_extracted_139564 | Not-determined |
| S16_extracted_139601 | Not-determined |
| S16_extracted_139605 | Not-determined |
| S16_extracted_139613 | Not-determined |
| S16_extracted_139615 | Not-determined |
| S16_extracted_139620 | Not-determined |
| S16_extracted_139644 | Not-determined |
| S16_extracted_139676 | Low-quality |
| S16_extracted_139695 | Not-determined |
| S16_extracted_139698 | Low-quality |
| S16_extracted_139712 | Not-determined |
| S16_extracted_139720 | Not-determined |
| S16_extracted_139730 | Not-determined |
| S16_extracted_139739 | Not-determined |
| S16_extracted_139763 | Not-determined |
| S16_extracted_139769 | Not-determined |
| S16_extracted_139775 | Low-quality |
| S16_extracted_139783 | Not-determined |
| S16_extracted_139792 | Not-determined |
| S16_extracted_139798 | Not-determined |
| S16_extracted_139806 | Not-determined |
| S16_extracted_139809 | Not-determined |
| S16_extracted_139810 | Not-determined |
| S16_extracted_139813 | Not-determined |
| S16_extracted_139816 | Not-determined |
| S16_extracted_139820 | Not-determined |
| S16_extracted_139827 | Not-determined |
| S16_extracted_139859 | Not-determined |
| S16_extracted_139873 | Not-determined |
| S16_extracted_139875 | Low-quality |
| S16_extracted_139876 | Low-quality |
| S16_extracted_139885 | Not-determined |
| S16_extracted_139890 | Low-quality |
| S16_extracted_139897 | Not-determined |
| S16_extracted_139913 | Not-determined |
| S16_extracted_139919 | Low-quality |
| S16_extracted_139920 | Not-determined |
| S16_extracted_139944 | Not-determined |
| S16_extracted_139959 | Not-determined |
| S16_extracted_139971 | Not-determined |
| S16_extracted_139976 | Not-determined |
| S16_extracted_139978 | Low-quality |
| S16_extracted_139985 | Not-determined |
| S16_extracted_140003 | Not-determined |
| S16_extracted_140024 | Not-determined |
| S16_extracted_140029 | Not-determined |
| S16_extracted_140032 | Low-quality |
| S16_extracted_140035 | Not-determined |
| S16_extracted_140051 | Not-determined |
| S16_extracted_140079 | Low-quality |
| S16_extracted_140082 | Not-determined |
| S16_extracted_140086 | Low-quality |
| S16_extracted_140087 | Not-determined |
| S16_extracted_140095 | Not-determined |
| S16_extracted_140097 | Not-determined |
| S16_extracted_140105 | Low-quality |
| S16_extracted_140113 | Not-determined |
| S16_extracted_140118 | Not-determined |
| S16_extracted_140132 | Not-determined |
| S16_extracted_140151 | Low-quality |
| S16_extracted_140163 | Not-determined |
| S16_extracted_140164 | Not-determined |
| S16_extracted_140166 | Not-determined |
| S16_extracted_140169 | Not-determined |
| S16_extracted_140195 | Not-determined |
| S16_extracted_140215 | Not-determined |
| S16_extracted_140224 | Not-determined |
| S16_extracted_140239 | Not-determined |
| S16_extracted_140240 | Not-determined |
| S16_extracted_140266 | Low-quality |
| S16_extracted_140272 | Not-determined |
| S16_extracted_140299 | Not-determined |
| S16_extracted_140310 | Not-determined |
| S16_extracted_140315 | Not-determined |
| S16_extracted_140318 | Low-quality |
| S16_extracted_140332 | Not-determined |
| S16_extracted_140344 | Not-determined |
| S16_extracted_140346 | Not-determined |
| S16_extracted_140387 | Low-quality |
| S16_extracted_140398 | Not-determined |
| S16_extracted_140419 | Not-determined |
| S16_extracted_140440 | Not-determined |
| S16_extracted_140447 | Low-quality |
| S16_extracted_140455 | Not-determined |
| S16_extracted_140467 | Not-determined |
| S16_extracted_140469 | Not-determined |
| S16_extracted_140471 | Not-determined |
| S16_extracted_140502 | Not-determined |
| S16_extracted_140504 | Not-determined |
| S16_extracted_140512 | Not-determined |
| S16_extracted_140520 | Not-determined |
| S16_extracted_140526 | Not-determined |
| S16_extracted_140531 | Not-determined |
| S16_extracted_140540 | Not-determined |
| S16_extracted_140543 | Not-determined |
| S16_extracted_140580 | Low-quality |
| S16_extracted_140583 | Not-determined |
| S16_extracted_140584 | Low-quality |
| S16_extracted_140590 | Not-determined |
| S16_extracted_140604 | Not-determined |
| S16_extracted_140607 | Not-determined |
| S16_extracted_140614 | Not-determined |
| S16_extracted_140628 | Not-determined |
| S16_extracted_140641 | Not-determined |
| S16_extracted_140662 | Low-quality |
| S16_extracted_140665 | Not-determined |
| S16_extracted_140676 | Not-determined |
| S16_extracted_140685 | Not-determined |
| S16_extracted_140686 | Not-determined |
| S16_extracted_140708 | Not-determined |
| S16_extracted_140725 | Low-quality |
| S16_extracted_140727 | Not-determined |
| S16_extracted_140728 | Not-determined |
| S16_extracted_140730 | Not-determined |
| S16_extracted_140733 | Not-determined |
| S16_extracted_140743 | Not-determined |
| S16_extracted_140745 | Not-determined |
| S16_extracted_140757 | Not-determined |
| S16_extracted_140760 | Not-determined |
| S16_extracted_140769 | Not-determined |
| S16_extracted_140774 | Not-determined |
| S16_extracted_140778 | Not-determined |
| S16_extracted_140780 | Not-determined |
| S16_extracted_140790 | Not-determined |
| S16_extracted_140791 | Not-determined |
| S16_extracted_140793 | Not-determined |
| S16_extracted_140806 | Not-determined |
| S16_extracted_140811 | Not-determined |
| S16_extracted_140828 | Not-determined |
| S16_extracted_140832 | Not-determined |
| S16_extracted_140838 | Not-determined |
| S16_extracted_140844 | Not-determined |
| S16_extracted_140849 | Not-determined |
| S16_extracted_140850 | Not-determined |
| S16_extracted_140851 | Not-determined |
| S16_extracted_140866 | Not-determined |
| S16_extracted_140873 | Not-determined |
| S16_extracted_140905 | Not-determined |
| S16_extracted_140907 | Not-determined |
| S16_extracted_140910 | Not-determined |
| S16_extracted_140959 | Not-determined |
| S16_extracted_140965 | Not-determined |
| S16_extracted_140986 | Not-determined |
| S16_extracted_141007 | Not-determined |
| S16_extracted_141013 | Low-quality |
| S16_extracted_141023 | Low-quality |
| S16_extracted_141040 | Not-determined |
| S16_extracted_141041 | Not-determined |
| S16_extracted_141050 | Not-determined |
| S16_extracted_141053 | Not-determined |
| S16_extracted_141086 | Not-determined |
| S16_extracted_141105 | Not-determined |
| S16_extracted_141111 | Low-quality |
| S16_extracted_141115 | Not-determined |
| S16_extracted_141116 | Not-determined |
| S16_extracted_141135 | Not-determined |
| S16_extracted_141137 | Not-determined |
| S16_extracted_141147 | Not-determined |
| S16_extracted_141169 | Not-determined |
| S16_extracted_141178 | Not-determined |
| S16_extracted_141198 | Not-determined |
| S16_extracted_141220 | Not-determined |
| S16_extracted_141224 | Not-determined |
| S16_extracted_141230 | Not-determined |
| S16_extracted_141238 | Low-quality |
| S16_extracted_141241 | Not-determined |
| S16_extracted_141242 | Not-determined |
| S16_extracted_141243 | Low-quality |
| S16_extracted_141264 | Low-quality |
| S16_extracted_141265 | Not-determined |
| S16_extracted_141278 | Not-determined |
| S16_extracted_141287 | Not-determined |
| S16_extracted_141297 | Not-determined |
| S16_extracted_141308 | Not-determined |
| S16_extracted_141328 | Low-quality |
| S16_extracted_141351 | Low-quality |
| S16_extracted_141352 | Not-determined |
| S16_extracted_141367 | Not-determined |
| S16_extracted_141371 | Low-quality |
| S16_extracted_141391 | Low-quality |
| S16_extracted_141394 | Low-quality |
| S16_extracted_141412 | Low-quality |
| S16_extracted_141425 | Not-determined |
| S16_extracted_141480 | Not-determined |
| S16_extracted_141489 | Not-determined |
| S16_extracted_141510 | Not-determined |
| S16_extracted_141514 | Not-determined |
| S16_extracted_141527 | Not-determined |
| S16_extracted_141531 | Not-determined |
| S16_extracted_141535 | Not-determined |
| S16_extracted_141567 | Not-determined |
| S16_extracted_141575 | Not-determined |
| S16_extracted_141608 | Not-determined |
| S16_extracted_141626 | Not-determined |
| S16_extracted_141632 | Not-determined |
| S16_extracted_141635 | Not-determined |
| S16_extracted_141638 | Not-determined |
| S16_extracted_141648 | Low-quality |
| S16_extracted_141653 | Low-quality |
| S16_extracted_141659 | Not-determined |
| S16_extracted_141677 | Low-quality |
| S16_extracted_141679 | Low-quality |
| S16_extracted_141688 | Low-quality |
| S16_extracted_141695 | Not-determined |
| S16_extracted_141704 | Not-determined |
| S16_extracted_141708 | Not-determined |
| S16_extracted_141714 | Not-determined |
| S16_extracted_141717 | Not-determined |
| S16_extracted_141720 | Not-determined |
| S16_extracted_141724 | Low-quality |
| S16_extracted_141749 | Not-determined |
| S16_extracted_141766 | Not-determined |
| S16_extracted_141767 | Not-determined |
| S16_extracted_141776 | Not-determined |
| S16_extracted_141780 | Low-quality |
| S16_extracted_141785 | Not-determined |
| S16_extracted_141796 | Not-determined |
| S16_extracted_141832 | Not-determined |
| S16_extracted_141837 | Not-determined |
| S16_extracted_141847 | Not-determined |
| S16_extracted_141877 | Not-determined |
| S16_extracted_141888 | Not-determined |
| S16_extracted_141889 | Not-determined |
| S16_extracted_141896 | Not-determined |
| S16_extracted_141912 | Not-determined |
| S16_extracted_141920 | Not-determined |
| S16_extracted_141926 | Not-determined |
| S16_extracted_141935 | Not-determined |
| S16_extracted_141948 | Not-determined |
| S16_extracted_141958 | Not-determined |
| S16_extracted_141963 | Not-determined |
| S16_extracted_141977 | Not-determined |
| S16_extracted_141983 | Not-determined |
| S16_extracted_141999 | Not-determined |
| S16_extracted_142003 | Not-determined |
| S16_extracted_142010 | Low-quality |
| S16_extracted_142019 | Low-quality |
| S16_extracted_142025 | Not-determined |
| S16_extracted_142030 | Not-determined |
| S16_extracted_142037 | Low-quality |
| S16_extracted_142044 | Not-determined |
| S16_extracted_142047 | Not-determined |
| S16_extracted_142057 | Not-determined |
| S16_extracted_142058 | Not-determined |
| S16_extracted_142073 | Low-quality |
| S16_extracted_142075 | Not-determined |
| S16_extracted_142076 | Not-determined |
| S16_extracted_142103 | Not-determined |
| S16_extracted_142117 | Not-determined |
| S16_extracted_142140 | Not-determined |
| S16_extracted_142146 | Low-quality |
| S16_extracted_142150 | Not-determined |
| S16_extracted_142174 | Not-determined |
| S16_extracted_142176 | Not-determined |
| S16_extracted_142208 | Not-determined |
| S16_extracted_142214 | Not-determined |
| S16_extracted_142216 | Not-determined |
| S16_extracted_142217 | Not-determined |
| S16_extracted_142223 | Not-determined |
| S16_extracted_142229 | Low-quality |
| S16_extracted_142239 | Not-determined |
| S16_extracted_142245 | Not-determined |
| S16_extracted_142248 | Not-determined |
| S16_extracted_142266 | Not-determined |
| S16_extracted_142272 | Not-determined |
| S16_extracted_142274 | Low-quality |
| S16_extracted_142280 | Not-determined |
| S16_extracted_142299 | Not-determined |
| S16_extracted_142306 | Not-determined |
| S16_extracted_142309 | Not-determined |
| S16_extracted_142314 | Not-determined |
| S16_extracted_142320 | Not-determined |
| S16_extracted_142324 | Not-determined |
| S16_extracted_142342 | Not-determined |
| S16_extracted_142352 | Not-determined |
| S16_extracted_142355 | Low-quality |
| S16_extracted_142376 | Not-determined |
| S16_extracted_142384 | Not-determined |
| S16_extracted_142390 | Low-quality |
| S16_extracted_142393 | Low-quality |
| S16_extracted_142416 | Low-quality |
| S16_extracted_142435 | Not-determined |
| S16_extracted_142457 | Not-determined |
| S16_extracted_142460 | Not-determined |
| S16_extracted_142463 | Not-determined |
| S16_extracted_142466 | Not-determined |
| S16_extracted_142471 | Not-determined |
| S16_extracted_142476 | Low-quality |
| S16_extracted_142483 | Not-determined |
| S16_extracted_142485 | Not-determined |
| S16_extracted_142486 | Not-determined |
| S16_extracted_142496 | Not-determined |
| S16_extracted_142498 | Low-quality |
| S16_extracted_142536 | Low-quality |
| S16_extracted_142550 | Not-determined |
| S16_extracted_142558 | Not-determined |
| S16_extracted_142559 | Low-quality |
| S16_extracted_142580 | Not-determined |
| S16_extracted_142589 | Low-quality |
| S16_extracted_142596 | Not-determined |
| S16_extracted_142600 | Not-determined |
| S16_extracted_142620 | Not-determined |
| S16_extracted_142624 | Not-determined |
| S16_extracted_142640 | Not-determined |
| S16_extracted_142643 | Not-determined |
| S16_extracted_142646 | Not-determined |
| S16_extracted_142662 | Low-quality |
| S16_extracted_142682 | Not-determined |
| S16_extracted_142689 | Not-determined |
| S16_extracted_142691 | Not-determined |
| S16_extracted_142697 | Not-determined |
| S16_extracted_142698 | Low-quality |
| S16_extracted_142711 | Not-determined |
| S16_extracted_142717 | Low-quality |
| S16_extracted_142720 | Low-quality |
| S16_extracted_142729 | Not-determined |
| S16_extracted_142733 | Not-determined |
| S16_extracted_142735 | Not-determined |
| S16_extracted_142741 | Not-determined |
| S16_extracted_142745 | Not-determined |
| S16_extracted_142749 | Not-determined |
| S16_extracted_142750 | Not-determined |
| S16_extracted_142752 | Not-determined |
| S16_extracted_142756 | Not-determined |
| S16_extracted_142769 | Not-determined |
| S16_extracted_142771 | Not-determined |
| S16_extracted_142773 | Not-determined |
| S16_extracted_142807 | Not-determined |
| S16_extracted_142823 | Not-determined |
| S16_extracted_142841 | Not-determined |
| S16_extracted_142847 | Not-determined |
| S16_extracted_142850 | Not-determined |
| S16_extracted_142856 | Not-determined |
| S16_extracted_142863 | Not-determined |
| S16_extracted_142865 | Not-determined |
| S16_extracted_142880 | Not-determined |
| S16_extracted_142906 | Not-determined |
| S16_extracted_142915 | Low-quality |

| | |
|---|---|
| S16_extracted_142919 | Not-determined |
| S16_extracted_142927 | Low-quality |
| S16_extracted_142933 | Not-determined |
| S16_extracted_142936 | Not-determined |
| S16_extracted_142942 | Not-determined |
| S16_extracted_142954 | Not-determined |
| S16_extracted_142960 | Not-determined |
| S16_extracted_142961 | Not-determined |
| S16_extracted_142966 | Not-determined |
| S16_extracted_142967 | Not-determined |
| S16_extracted_143000 | Not-determined |
| S16_extracted_143006 | Not-determined |
| S16_extracted_143009 | Not-determined |
| S16_extracted_143010 | Not-determined |
| S16_extracted_143013 | Not-determined |
| S16_extracted_143017 | Not-determined |
| S16_extracted_143023 | Not-determined |
| S16_extracted_143033 | Not-determined |
| S16_extracted_143042 | Low-quality |
| S16_extracted_143044 | Not-determined |
| S16_extracted_143058 | Low-quality |
| S16_extracted_143079 | Not-determined |
| S16_extracted_143091 | Not-determined |
| S16_extracted_143122 | Not-determined |
| S16_extracted_143124 | Not-determined |
| S16_extracted_143130 | Not-determined |
| S16_extracted_143133 | Low-quality |
| S16_extracted_143181 | Not-determined |
| S16_extracted_143189 | Not-determined |
| S16_extracted_143210 | Not-determined |
| S16_extracted_143212 | Not-determined |
| S16_extracted_143221 | Not-determined |
| S16_extracted_143228 | Not-determined |
| S16_extracted_143259 | Not-determined |
| S16_extracted_143260 | Not-determined |
| S16_extracted_143263 | Not-determined |
| S16_extracted_143265 | Not-determined |
| S16_extracted_143274 | Not-determined |
| S16_extracted_143275 | Not-determined |
| S16_extracted_143282 | Low-quality |
| S16_extracted_143286 | Not-determined |
| S16_extracted_143308 | Not-determined |
| S16_extracted_143316 | Not-determined |
| S16_extracted_143324 | Not-determined |
| S16_extracted_143346 | Not-determined |
| S16_extracted_143367 | Low-quality |
| S16_extracted_143407 | Low-quality |
| S16_extracted_143408 | Not-determined |
| S16_extracted_143410 | Not-determined |
| S16_extracted_143426 | Not-determined |
| S16_extracted_143427 | Not-determined |
| S16_extracted_143428 | Low-quality |
| S16_extracted_143432 | Low-quality |
| S16_extracted_143435 | Low-quality |
| S16_extracted_143448 | Not-determined |
| S16_extracted_143457 | Not-determined |
| S16_extracted_143475 | Not-determined |
| S16_extracted_143490 | Not-determined |
| S16_extracted_143496 | Low-quality |
| S16_extracted_143508 | Not-determined |
| S16_extracted_143520 | Not-determined |
| S16_extracted_143526 | Not-determined |
| S16_extracted_143531 | Not-determined |
| S16_extracted_143534 | Not-determined |
| S16_extracted_143538 | Low-quality |
| S16_extracted_143539 | Not-determined |
| S16_extracted_143548 | Not-determined |
| S16_extracted_143564 | Low-quality |
| S16_extracted_143578 | Low-quality |
| S16_extracted_143581 | Not-determined |
| S16_extracted_143584 | Not-determined |
| S16_extracted_143586 | Not-determined |
| S16_extracted_143591 | Not-determined |
| S16_extracted_143639 | Not-determined |
| S16_extracted_143643 | Low-quality |
| S16_extracted_143645 | Low-quality |
| S16_extracted_143655 | Not-determined |
| S16_extracted_143658 | Not-determined |
| S16_extracted_143667 | Not-determined |
| S16_extracted_143679 | Low-quality |
| S16_extracted_143681 | Not-determined |
| S16_extracted_143699 | Low-quality |
| S16_extracted_143714 | Not-determined |
| S16_extracted_143717 | Not-determined |
| S16_extracted_143731 | Not-determined |
| S16_extracted_143733 | Not-determined |
| S16_extracted_143746 | Not-determined |
| S16_extracted_143751 | Not-determined |
| S16_extracted_143755 | Not-determined |
| S16_extracted_143756 | Not-determined |
| S16_extracted_143767 | Not-determined |
| S16_extracted_143768 | Not-determined |
| S16_extracted_143770 | Not-determined |
| S16_extracted_143776 | Not-determined |
| S16_extracted_143784 | Not-determined |
| S16_extracted_143795 | Not-determined |
| S16_extracted_143797 | Not-determined |
| S16_extracted_143803 | Not-determined |
| S16_extracted_143807 | Low-quality |
| S16_extracted_143814 | Not-determined |
| S16_extracted_143816 | Not-determined |
| S16_extracted_143827 | Low-quality |
| S16_extracted_143837 | Not-determined |
| S16_extracted_143854 | Low-quality |
| S16_extracted_143858 | Not-determined |
| S16_extracted_143879 | Low-quality |
| S16_extracted_143889 | Not-determined |
| S16_extracted_143898 | Not-determined |
| S16_extracted_143900 | Not-determined |
| S16_extracted_143935 | Not-determined |
| S16_extracted_143952 | Not-determined |
| S16_extracted_143963 | Not-determined |
| S16_extracted_143970 | Not-determined |
| S16_extracted_143973 | Not-determined |
| S16_extracted_143983 | Not-determined |
| S16_extracted_143984 | Not-determined |
| S16_extracted_144015 | Not-determined |
| S16_extracted_144029 | Not-determined |
| S16_extracted_144046 | Not-determined |
| S16_extracted_144058 | Not-determined |
| S16_extracted_144073 | Not-determined |
| S16_extracted_144086 | Not-determined |
| S16_extracted_144112 | Not-determined |
| S16_extracted_144122 | Low-quality |
| S16_extracted_144129 | Not-determined |
| S16_extracted_144131 | Not-determined |
| S16_extracted_144145 | Not-determined |
| S16_extracted_144160 | Not-determined |
| S16_extracted_144169 | Not-determined |
| S16_extracted_144180 | Not-determined |
| S16_extracted_144183 | Low-quality |
| S16_extracted_144195 | Not-determined |
| S16_extracted_144213 | Not-determined |
| S16_extracted_144220 | Not-determined |
| S16_extracted_144231 | Not-determined |
| S16_extracted_144234 | Not-determined |
| S16_extracted_144237 | Not-determined |
| S16_extracted_144247 | Not-determined |
| S16_extracted_144251 | Low-quality |
| S16_extracted_144274 | Not-determined |
| S16_extracted_144287 | Low-quality |
| S16_extracted_144324 | Not-determined |
| S16_extracted_144336 | Low-quality |
| S16_extracted_144340 | Low-quality |
| S16_extracted_144344 | Low-quality |
| S16_extracted_144346 | Not-determined |
| S16_extracted_144352 | Not-determined |
| S16_extracted_144354 | Not-determined |
| S16_extracted_144359 | Not-determined |
| S16_extracted_144374 | Not-determined |
| S16_extracted_144385 | Low-quality |
| S16_extracted_144395 | Not-determined |
| S16_extracted_144406 | Not-determined |
| S16_extracted_144415 | Not-determined |
| S16_extracted_144440 | Not-determined |
| S16_extracted_144444 | Not-determined |
| S16_extracted_144447 | Not-determined |
| S16_extracted_144468 | Not-determined |
| S16_extracted_144475 | Not-determined |
| S16_extracted_144481 | Not-determined |
| S16_extracted_144489 | Low-quality |
| S16_extracted_144498 | Not-determined |
| S16_extracted_144557 | Not-determined |
| S16_extracted_144562 | Not-determined |
| S16_extracted_144565 | Not-determined |
| S16_extracted_144590 | Not-determined |
| S16_extracted_144596 | Low-quality |
| S16_extracted_144597 | Not-determined |
| S16_extracted_144599 | Not-determined |
| S16_extracted_144607 | Not-determined |
| S16_extracted_144646 | Low-quality |
| S16_extracted_144673 | Low-quality |
| S16_extracted_144679 | Not-determined |
| S16_extracted_144686 | Not-determined |
| S16_extracted_144688 | Low-quality |
| S16_extracted_144695 | Low-quality |
| S16_extracted_144746 | Not-determined |
| S16_extracted_144748 | Not-determined |
| S16_extracted_144763 | Not-determined |
| S16_extracted_144768 | Low-quality |
| S16_extracted_144781 | Low-quality |
| S16_extracted_144800 | Not-determined |
| S16_extracted_144801 | Not-determined |
| S16_extracted_144804 | Not-determined |
| S16_extracted_144818 | Not-determined |
| S16_extracted_144822 | Not-determined |
| S16_extracted_144841 | Not-determined |
| S16_extracted_144848 | Not-determined |
| S16_extracted_144860 | Not-determined |
| S16_extracted_144862 | Not-determined |
| S16_extracted_144864 | Low-quality |
| S16_extracted_144865 | Not-determined |
| S16_extracted_144874 | Low-quality |
| S16_extracted_144888 | Not-determined |
| S16_extracted_144893 | Not-determined |
| S16_extracted_144897 | Low-quality |
| S16_extracted_144911 | Not-determined |
| S16_extracted_144920 | Not-determined |
| S16_extracted_144937 | Low-quality |
| S16_extracted_144966 | Not-determined |
| S16_extracted_144968 | Low-quality |
| S16_extracted_144983 | Not-determined |
| S16_extracted_144991 | Not-determined |
| S16_extracted_144992 | Not-determined |
| S16_extracted_145012 | Not-determined |
| S16_extracted_145030 | Not-determined |
| S16_extracted_145037 | Not-determined |
| S16_extracted_145050 | Low-quality |
| S16_extracted_145075 | Not-determined |
| S16_extracted_145105 | Not-determined |
| S16_extracted_145119 | Not-determined |
| S16_extracted_145127 | Not-determined |
| S16_extracted_145143 | Not-determined |
| S16_extracted_145148 | Not-determined |
| S16_extracted_145153 | Not-determined |
| S16_extracted_145160 | Low-quality |
| S16_extracted_145165 | Not-determined |
| S16_extracted_145167 | Not-determined |
| S16_extracted_145173 | Not-determined |
| S16_extracted_145174 | Not-determined |
| S16_extracted_145184 | Not-determined |
| S16_extracted_145186 | Not-determined |
| S16_extracted_145198 | Not-determined |
| S16_extracted_145200 | Not-determined |
| S16_extracted_145203 | Not-determined |
| S16_extracted_145205 | Not-determined |
| S16_extracted_145208 | Not-determined |
| S16_extracted_145225 | Low-quality |
| S16_extracted_145243 | Not-determined |
| S16_extracted_145251 | Not-determined |
| S16_extracted_145261 | Not-determined |
| S16_extracted_145267 | Low-quality |
| S16_extracted_145273 | Not-determined |
| S16_extracted_145309 | Not-determined |
| S16_extracted_145326 | Not-determined |
| S16_extracted_145336 | Not-determined |
| S16_extracted_145339 | Not-determined |
| S16_extracted_145341 | Not-determined |
| S16_extracted_145347 | Not-determined |
| S16_extracted_145356 | Low-quality |
| S16_extracted_145367 | Not-determined |
| S16_extracted_145381 | Not-determined |
| S16_extracted_145386 | Not-determined |
| S16_extracted_145388 | Not-determined |
| S16_extracted_145396 | Not-determined |
| S16_extracted_145397 | Not-determined |
| S16_extracted_145402 | Low-quality |
| S16_extracted_145421 | Not-determined |
| S16_extracted_145433 | Not-determined |
| S16_extracted_145446 | Not-determined |
| S16_extracted_145463 | Not-determined |
| S16_extracted_145467 | Not-determined |
| S16_extracted_145475 | Low-quality |
| S16_extracted_145478 | Not-determined |
| S16_extracted_145479 | Not-determined |
| S16_extracted_145481 | Low-quality |
| S16_extracted_145491 | Not-determined |
| S16_extracted_145502 | Not-determined |
| S16_extracted_145514 | Not-determined |
| S16_extracted_145525 | Not-determined |
| S16_extracted_145526 | Not-determined |
| S16_extracted_145536 | Not-determined |
| S16_extracted_145544 | Not-determined |
| S16_extracted_145552 | Low-quality |
| S16_extracted_145568 | Not-determined |
| S16_extracted_145570 | Not-determined |
| S16_extracted_145572 | Low-quality |
| S16_extracted_145577 | Not-determined |
| S16_extracted_145581 | Not-determined |
| S16_extracted_145585 | Not-determined |
| S16_extracted_145587 | Not-determined |
| S16_extracted_145611 | Low-quality |
| S16_extracted_145615 | Not-determined |
| S16_extracted_145619 | Not-determined |
| S16_extracted_145628 | Not-determined |
| S16_extracted_145635 | Not-determined |
| S16_extracted_145653 | Low-quality |
| S16_extracted_145657 | Not-determined |
| S16_extracted_145659 | Not-determined |
| S16_extracted_145675 | Not-determined |
| S16_extracted_145695 | Not-determined |
| S16_extracted_145700 | Not-determined |
| S16_extracted_145705 | Not-determined |
| S16_extracted_145721 | Low-quality |
| S16_extracted_145740 | Not-determined |
| S16_extracted_145768 | Not-determined |
| S16_extracted_145772 | Not-determined |
| S16_extracted_145776 | Not-determined |
| S16_extracted_145780 | Not-determined |
| S16_extracted_145789 | Low-quality |
| S16_extracted_145802 | Low-quality |
| S16_extracted_145805 | Not-determined |
| S16_extracted_145820 | Not-determined |
| S16_extracted_145825 | Low-quality |
| S16_extracted_145835 | Not-determined |
| S16_extracted_145838 | Not-determined |
| S16_extracted_145841 | Not-determined |
| S16_extracted_145858 | Not-determined |
| S16_extracted_145864 | Not-determined |
| S16_extracted_145867 | Not-determined |
| S16_extracted_145871 | Low-quality |
| S16_extracted_145886 | Not-determined |
| S16_extracted_145905 | Not-determined |
| S16_extracted_145925 | Not-determined |
| S16_extracted_145956 | Not-determined |
| S16_extracted_145965 | Not-determined |
| S16_extracted_145982 | Not-determined |
| S16_extracted_145984 | Not-determined |
| S16_extracted_145986 | Low-quality |
| S16_extracted_145989 | Not-determined |
| S16_extracted_145995 | Low-quality |
| S16_extracted_146005 | Low-quality |
| S16_extracted_146024 | Not-determined |
| S16_extracted_146031 | Low-quality |
| S16_extracted_146035 | Low-quality |
| S16_extracted_146039 | Not-determined |
| S16_extracted_146044 | Not-determined |
| S16_extracted_146047 | Not-determined |
| S16_extracted_146053 | Not-determined |
| S16_extracted_146078 | Low-quality |
| S16_extracted_146081 | Not-determined |
| S16_extracted_146083 | Not-determined |
| S16_extracted_146085 | Not-determined |
| S16_extracted_146092 | Not-determined |
| S16_extracted_146096 | Not-determined |
| S16_extracted_146101 | Low-quality |
| S16_extracted_146108 | Not-determined |
| S16_extracted_146110 | Not-determined |
| S16_extracted_146117 | Not-determined |
| S16_extracted_146127 | Low-quality |
| S16_extracted_146131 | Not-determined |
| S16_extracted_146134 | Not-determined |
| S16_extracted_146139 | Not-determined |
| S16_extracted_146141 | Not-determined |
| S16_extracted_146148 | Not-determined |
| S16_extracted_146162 | Not-determined |
| S16_extracted_146169 | Not-determined |
| S16_extracted_146175 | Not-determined |
| S16_extracted_146181 | Not-determined |
| S16_extracted_146187 | Not-determined |
| S16_extracted_146188 | Not-determined |
| S16_extracted_146189 | Not-determined |
| S16_extracted_146214 | Not-determined |
| S16_extracted_146230 | Not-determined |
| S16_extracted_146244 | Not-determined |
| S16_extracted_146245 | Not-determined |
| S16_extracted_146253 | Not-determined |
| S16_extracted_146264 | Not-determined |
| S16_extracted_146271 | Not-determined |
| S16_extracted_146280 | Not-determined |
| S16_extracted_146292 | Not-determined |
| S16_extracted_146305 | Low-quality |
| S16_extracted_146306 | Not-determined |
| S16_extracted_146330 | Not-determined |
| S16_extracted_146345 | Not-determined |
| S16_extracted_146347 | Not-determined |
| S16_extracted_146357 | Not-determined |
| S16_extracted_146360 | Not-determined |
| S16_extracted_146365 | Not-determined |
| S16_extracted_146369 | Low-quality |
| S16_extracted_146388 | Low-quality |
| S16_extracted_146389 | Not-determined |
| S16_extracted_146390 | Low-quality |
| S16_extracted_146400 | Low-quality |
| S16_extracted_146405 | Not-determined |
| S16_extracted_146416 | Not-determined |
| S16_extracted_146438 | Not-determined |
| S16_extracted_146450 | Not-determined |
| S16_extracted_146455 | Not-determined |
| S16_extracted_146471 | Not-determined |
| S16_extracted_146472 | Not-determined |
| S16_extracted_146500 | Not-determined |
| S16_extracted_146505 | Not-determined |
| S16_extracted_146538 | Not-determined |
| S16_extracted_146551 | Not-determined |
| S16_extracted_146561 | Not-determined |
| S16_extracted_146565 | Not-determined |
| S16_extracted_146579 | Not-determined |
| S16_extracted_146584 | Low-quality |
| S16_extracted_146589 | Not-determined |
| S16_extracted_146595 | Not-determined |
| S16_extracted_146598 | Not-determined |
| S16_extracted_146600 | Not-determined |
| S16_extracted_146606 | Not-determined |
| S16_extracted_146619 | Not-determined |
| S16_extracted_146631 | Not-determined |
| S16_extracted_146635 | Not-determined |
| S16_extracted_146653 | Not-determined |
| S16_extracted_146691 | Not-determined |
| S16_extracted_146703 | Not-determined |
| S16_extracted_146704 | Not-determined |
| S16_extracted_146720 | Not-determined |
| S16_extracted_146729 | Not-determined |
| S16_extracted_146733 | Not-determined |
| S16_extracted_146750 | Not-determined |
| S16_extracted_146754 | Not-determined |
| S16_extracted_146755 | Low-quality |
| S16_extracted_146792 | Not-determined |
| S16_extracted_146793 | Low-quality |
| S16_extracted_146805 | Not-determined |
| S16_extracted_146814 | Not-determined |
| S16_extracted_146836 | Not-determined |
| S16_extracted_146843 | Not-determined |
| S16_extracted_146862 | Not-determined |
| S16_extracted_146879 | Not-determined |
| S16_extracted_146889 | Not-determined |
| S16_extracted_146894 | Not-determined |
| S16_extracted_146907 | Not-determined |
| S16_extracted_146913 | Not-determined |
| S16_extracted_146922 | Not-determined |
| S16_extracted_146925 | Not-determined |
| S16_extracted_146928 | Not-determined |
| S16_extracted_146947 | Not-determined |
| S16_extracted_146958 | Not-determined |
| S16_extracted_146964 | Not-determined |
| S16_extracted_146965 | Not-determined |
| S16_extracted_146968 | Low-quality |
| S16_extracted_146975 | Not-determined |
| S16_extracted_146985 | Not-determined |
| S16_extracted_146989 | Not-determined |
| S16_extracted_146993 | Not-determined |
| S16_extracted_146995 | Not-determined |
| S16_extracted_146996 | Low-quality |
| S16_extracted_147002 | Not-determined |
| S16_extracted_147007 | Not-determined |
| S16_extracted_147008 | Not-determined |
| S16_extracted_147010 | Not-determined |
| S16_extracted_147047 | Not-determined |
| S16_extracted_147051 | Low-quality |
| S16_extracted_147062 | Not-determined |
| S16_extracted_147067 | Not-determined |
| S16_extracted_147073 | Not-determined |
| S16_extracted_147076 | Not-determined |
| S16_extracted_147080 | Not-determined |
| S16_extracted_147087 | Low-quality |
| S16_extracted_147095 | Low-quality |
| S16_extracted_147104 | Not-determined |
| S16_extracted_147112 | Not-determined |
| S16_extracted_147116 | Not-determined |
| S16_extracted_147129 | Not-determined |
| S16_extracted_147132 | Not-determined |
| S16_extracted_147135 | Not-determined |
| S16_extracted_147158 | Not-determined |
| S16_extracted_147164 | Not-determined |
| S16_extracted_147173 | Not-determined |
| S16_extracted_147174 | Not-determined |
| S16_extracted_147183 | Not-determined |
| S16_extracted_147191 | Not-determined |
| S16_extracted_147198 | Not-determined |
| S16_extracted_147204 | Not-determined |
| S16_extracted_147218 | Not-determined |
| S16_extracted_147226 | Not-determined |
| S16_extracted_147235 | Not-determined |
| S16_extracted_147254 | Not-determined |
| S16_extracted_147260 | Not-determined |
| S16_extracted_147267 | Not-determined |
| S16_extracted_147288 | Not-determined |
| S16_extracted_147317 | Not-determined |
| S16_extracted_147322 | Not-determined |
| S3_extracted_k141_111893 | Complete |
| S16_extracted_147336 | Not-determined |
| S16_extracted_147340 | Low-quality |
| S16_extracted_147349 | Not-determined |
| S16_extracted_147350 | Not-determined |
| S16_extracted_147369 | Not-determined |
| S16_extracted_147372 | Not-determined |
| S16_extracted_147376 | Not-determined |
| S16_extracted_147384 | Not-determined |
| S16_extracted_147406 | Not-determined |
| S16_extracted_147419 | Not-determined |
| S16_extracted_147428 | Not-determined |
| S16_extracted_147459 | Not-determined |
| S16_extracted_147466 | Not-determined |
| S16_extracted_147472 | Not-determined |
| S16_extracted_147480 | Not-determined |
| S16_extracted_147488 | Not-determined |
| S16_extracted_147493 | Not-determined |
| S16_extracted_147511 | Not-determined |
| S16_extracted_147528 | Low-quality |
| S16_extracted_147538 | Not-determined |
| S16_extracted_147553 | Not-determined |
| S16_extracted_147555 | Not-determined |
| S16_extracted_147569 | Not-determined |
| S16_extracted_147595 | Not-determined |
| S16_extracted_147598 | Not-determined |
| S16_extracted_147618 | Low-quality |
| S16_extracted_147635 | Not-determined |
| S16_extracted_147650 | Not-determined |
| S16_extracted_147664 | Not-determined |
| S16_extracted_147671 | Not-determined |
| S16_extracted_147673 | Not-determined |
| S16_extracted_147679 | Not-determined |
| S16_extracted_147680 | Not-determined |
| S16_extracted_147697 | Not-determined |
| S16_extracted_147713 | Not-determined |
| S16_extracted_147714 | Not-determined |
| S16_extracted_147715 | Not-determined |
| S16_extracted_147718 | Not-determined |
| S16_extracted_147723 | Not-determined |
| S16_extracted_147733 | Not-determined |
| S16_extracted_147736 | Low-quality |
| S16_extracted_147743 | Not-determined |
| S16_extracted_147795 | Low-quality |
| S16_extracted_147820 | Not-determined |
| S16_extracted_147843 | Low-quality |
| S16_extracted_147844 | Not-determined |
| S16_extracted_147845 | Not-determined |
| S16_extracted_147852 | Not-determined |
| S16_extracted_147866 | Not-determined |
| S16_extracted_147896 | Not-determined |
| S16_extracted_147920 | Not-determined |
| S16_extracted_147926 | Not-determined |
| S16_extracted_147935 | Not-determined |
| S16_extracted_147945 | Not-determined |
| S16_extracted_147946 | Not-determined |
| S16_extracted_147959 | Not-determined |
| S16_extracted_147961 | Not-determined |
| S16_extracted_147972 | Not-determined |
| S16_extracted_147975 | Not-determined |
| S16_extracted_147987 | Low-quality |
| S16_extracted_147994 | Not-determined |
| S16_extracted_148005 | Not-determined |
| S16_extracted_148007 | Not-determined |
| S16_extracted_148025 | Not-determined |
| S16_extracted_148027 | Low-quality |
| S16_extracted_148031 | Not-determined |
| S16_extracted_148033 | Not-determined |
| S16_extracted_148046 | Not-determined |
| S16_extracted_148048 | Not-determined |
| S16_extracted_148053 | Not-determined |
| S16_extracted_148058 | Not-determined |
| S16_extracted_148082 | Not-determined |
| S16_extracted_148117 | Not-determined |
| S16_extracted_148119 | Low-quality |
| S16_extracted_148120 | Not-determined |
| S16_extracted_148132 | Not-determined |
| S16_extracted_148135 | Not-determined |
| S16_extracted_148178 | Not-determined |
| S16_extracted_148186 | Not-determined |
| S16_extracted_148187 | Not-determined |
| S16_extracted_148195 | Not-determined |
| S16_extracted_148201 | Not-determined |
| S16_extracted_148205 | Not-determined |
| S16_extracted_148237 | Not-determined |
| S16_extracted_148260 | Not-determined |
| S16_extracted_148264 | Not-determined |
| S16_extracted_148267 | Not-determined |
| S16_extracted_148274 | Not-determined |
| S16_extracted_148277 | Not-determined |
| S16_extracted_148284 | Not-determined |
| S16_extracted_148307 | Low-quality |
| S16_extracted_148314 | Not-determined |
| S16_extracted_148338 | Not-determined |
| S16_extracted_148340 | Not-determined |
| S16_extracted_148343 | Not-determined |
| S16_extracted_148345 | Low-quality |
| S16_extracted_148361 | Not-determined |
| S16_extracted_148372 | Not-determined |
| S16_extracted_148389 | Low-quality |
| S16_extracted_148395 | Not-determined |
| S16_extracted_148409 | Not-determined |
| S16_extracted_148416 | Not-determined |
| S16_extracted_148417 | Not-determined |
| S16_extracted_148418 | Low-quality |
| S16_extracted_148421 | Not-determined |
| S16_extracted_148464 | Not-determined |
| S16_extracted_148466 | Not-determined |
| S16_extracted_148490 | Not-determined |
| S16_extracted_148505 | Not-determined |
| S16_extracted_148520 | Not-determined |
| S16_extracted_148536 | Not-determined |
| S16_extracted_148556 | Not-determined |
| S16_extracted_148557 | Not-determined |
| S16_extracted_148585 | Not-determined |
| S16_extracted_148604 | Not-determined |
| S16_extracted_148607 | Not-determined |
| S16_extracted_148620 | Not-determined |
| S16_extracted_148632 | Not-determined |
| S16_extracted_148640 | Not-determined |
| S16_extracted_148647 | Not-determined |
| S16_extracted_148659 | Low-quality |
| S16_extracted_148672 | Not-determined |
| S16_extracted_148692 | Not-determined |
| S16_extracted_148700 | Not-determined |
| S16_extracted_148701 | Not-determined |
| S16_extracted_148708 | Low-quality |
| S16_extracted_148709 | Not-determined |
| S16_extracted_148720 | Not-determined |
| S16_extracted_148722 | Not-determined |

| | |
|---|---|
| S16_extracted_148732 | Not-determined |
| S16_extracted_148740 | Not-determined |
| S16_extracted_148741 | Not-determined |
| S16_extracted_148750 | Not-determined |
| S16_extracted_148753 | Not-determined |
| S16_extracted_148756 | Not-determined |
| S16_extracted_148757 | Not-determined |
| S16_extracted_148769 | Not-determined |
| S16_extracted_148774 | Not-determined |
| S16_extracted_148803 | Not-determined |
| S16_extracted_148815 | Not-determined |
| S16_extracted_148825 | Low-quality |
| S16_extracted_148829 | Not-determined |
| S16_extracted_148833 | Not-determined |
| S16_extracted_148835 | Not-determined |
| S16_extracted_148840 | Not-determined |
| S16_extracted_148851 | Not-determined |
| S16_extracted_148858 | Not-determined |
| S16_extracted_148876 | Not-determined |
| S16_extracted_148882 | Not-determined |
| S16_extracted_148906 | Not-determined |
| S16_extracted_148920 | Not-determined |
| S16_extracted_148936 | Not-determined |
| S16_extracted_148946 | Not-determined |
| S16_extracted_148958 | Not-determined |
| S16_extracted_148976 | Low-quality |
| S16_extracted_148987 | Not-determined |
| S16_extracted_149006 | Not-determined |
| S16_extracted_149009 | Not-determined |
| S16_extracted_149025 | Not-determined |
| S16_extracted_149026 | Not-determined |
| S16_extracted_149027 | Not-determined |
| S16_extracted_149036 | Low-quality |
| S16_extracted_149039 | Not-determined |
| S16_extracted_149040 | Not-determined |
| S16_extracted_149047 | Not-determined |
| S16_extracted_149089 | Not-determined |
| S16_extracted_149092 | Low-quality |
| S16_extracted_149110 | Not-determined |
| S16_extracted_149111 | Not-determined |
| S16_extracted_149126 | Not-determined |
| S16_extracted_149151 | Not-determined |
| S16_extracted_149171 | Not-determined |
| S16_extracted_149175 | Not-determined |
| S16_extracted_149182 | Not-determined |
| S16_extracted_149185 | Not-determined |
| S16_extracted_149187 | Not-determined |
| S16_extracted_149228 | Not-determined |
| S16_extracted_149231 | Low-quality |
| S16_extracted_149239 | Not-determined |
| S16_extracted_149251 | Not-determined |
| S16_extracted_149264 | Not-determined |
| S16_extracted_149267 | Low-quality |
| S16_extracted_149286 | Not-determined |
| S16_extracted_149287 | Not-determined |
| S16_extracted_149299 | Low-quality |
| S16_extracted_149307 | Not-determined |
| S16_extracted_149315 | Not-determined |
| S16_extracted_149343 | Not-determined |
| S16_extracted_149350 | Not-determined |
| S16_extracted_149368 | Low-quality |
| S16_extracted_149393 | Not-determined |
| S16_extracted_149394 | Low-quality |
| S16_extracted_149398 | Not-determined |
| S16_extracted_149403 | Not-determined |
| S16_extracted_149406 | Not-determined |
| S16_extracted_149413 | Not-determined |
| S16_extracted_149446 | Not-determined |
| S16_extracted_149461 | Not-determined |
| S16_extracted_149462 | Not-determined |
| S16_extracted_149495 | Not-determined |
| S16_extracted_149503 | Low-quality |
| S16_extracted_149518 | Not-determined |
| S16_extracted_149527 | Not-determined |
| S16_extracted_149543 | Low-quality |
| S16_extracted_149546 | Not-determined |
| S16_extracted_149553 | Not-determined |
| S16_extracted_149558 | Not-determined |
| S16_extracted_149563 | Not-determined |
| S16_extracted_149575 | Not-determined |
| S16_extracted_149579 | Low-quality |
| S16_extracted_149600 | Not-determined |
| S16_extracted_149604 | Not-determined |
| S16_extracted_149617 | Not-determined |
| S16_extracted_149643 | Not-determined |
| S16_extracted_149645 | Not-determined |
| S16_extracted_149648 | Not-determined |
| S16_extracted_149665 | Low-quality |
| S16_extracted_149684 | Not-determined |
| S16_extracted_149694 | Not-determined |
| S16_extracted_149704 | Not-determined |
| S16_extracted_149714 | Not-determined |
| S16_extracted_149717 | Not-determined |
| S16_extracted_149727 | Low-quality |
| S16_extracted_149732 | Low-quality |
| S16_extracted_149733 | Not-determined |
| S16_extracted_149748 | Not-determined |
| S16_extracted_149764 | Not-determined |
| S16_extracted_149784 | Not-determined |
| S16_extracted_149788 | Not-determined |
| S16_extracted_149792 | Not-determined |
| S16_extracted_149793 | Low-quality |
| S16_extracted_149814 | Not-determined |
| S16_extracted_149817 | Not-determined |
| S16_extracted_149823 | Low-quality |
| S16_extracted_149912 | Not-determined |
| S16_extracted_149916 | Low-quality |
| S16_extracted_149934 | Low-quality |
| S16_extracted_149936 | Not-determined |
| S16_extracted_149937 | Not-determined |
| S16_extracted_149940 | Not-determined |
| S16_extracted_149942 | Low-quality |
| S16_extracted_149949 | Not-determined |
| S16_extracted_149950 | Not-determined |
| S16_extracted_149953 | Not-determined |
| S16_extracted_149959 | Not-determined |
| S16_extracted_149965 | Not-determined |
| S16_extracted_149968 | Not-determined |
| S16_extracted_149978 | Not-determined |
| S16_extracted_149989 | Not-determined |
| S16_extracted_150009 | Not-determined |
| S16_extracted_150035 | Low-quality |
| S16_extracted_150045 | Not-determined |
| S16_extracted_150046 | Not-determined |
| S16_extracted_150059 | Not-determined |
| S16_extracted_150072 | Not-determined |
| S16_extracted_150077 | Not-determined |
| S16_extracted_150081 | Not-determined |
| S16_extracted_150092 | Not-determined |
| S16_extracted_150097 | Not-determined |
| S16_extracted_150104 | Low-quality |
| S16_extracted_150113 | Not-determined |
| S16_extracted_150126 | Not-determined |
| S16_extracted_150150 | Not-determined |
| S16_extracted_150151 | Not-determined |
| S16_extracted_150171 | Not-determined |
| S16_extracted_150172 | Not-determined |
| S16_extracted_150190 | Not-determined |
| S16_extracted_150192 | Low-quality |
| S16_extracted_150206 | Not-determined |
| S16_extracted_150211 | Not-determined |
| S16_extracted_150220 | Not-determined |
| S16_extracted_150228 | Not-determined |
| S16_extracted_150234 | Not-determined |
| S16_extracted_150236 | Not-determined |
| S16_extracted_150239 | Not-determined |
| S16_extracted_150240 | Not-determined |
| S16_extracted_150272 | Not-determined |
| S16_extracted_150298 | Not-determined |
| S16_extracted_150310 | Not-determined |
| S16_extracted_150312 | Not-determined |
| S16_extracted_150328 | Low-quality |
| S16_extracted_150347 | Not-determined |
| S16_extracted_150369 | Not-determined |
| S16_extracted_150373 | Not-determined |
| S16_extracted_150394 | Not-determined |
| S16_extracted_150422 | Not-determined |
| S16_extracted_150425 | Not-determined |
| S16_extracted_150427 | Not-determined |
| S16_extracted_150428 | Low-quality |
| S16_extracted_150436 | Not-determined |
| S16_extracted_150457 | Not-determined |
| S16_extracted_150470 | Not-determined |
| S16_extracted_150478 | Not-determined |
| S16_extracted_150481 | Not-determined |
| S16_extracted_150482 | Not-determined |
| S16_extracted_150484 | Not-determined |
| S16_extracted_150485 | Not-determined |
| S16_extracted_150511 | Low-quality |
| S16_extracted_150515 | Not-determined |
| S16_extracted_150523 | Not-determined |
| S16_extracted_150539 | Not-determined |
| S16_extracted_150597 | Not-determined |
| S16_extracted_150607 | Not-determined |
| S16_extracted_150613 | Not-determined |
| S16_extracted_150618 | Not-determined |
| S16_extracted_150627 | Low-quality |
| S16_extracted_150629 | Low-quality |
| S16_extracted_150666 | Not-determined |
| S16_extracted_150667 | Not-determined |
| S16_extracted_150676 | Not-determined |
| S16_extracted_150715 | Low-quality |
| S16_extracted_150727 | Not-determined |
| S16_extracted_150768 | Not-determined |
| S16_extracted_150770 | Low-quality |
| S16_extracted_150792 | Low-quality |
| S16_extracted_150799 | Low-quality |
| S16_extracted_150802 | Not-determined |
| S16_extracted_150810 | Not-determined |
| S16_extracted_150811 | Not-determined |
| S16_extracted_150814 | Not-determined |
| S16_extracted_150815 | Low-quality |
| S16_extracted_150830 | Not-determined |
| S16_extracted_150832 | Not-determined |
| S16_extracted_150836 | Not-determined |
| S16_extracted_150849 | Not-determined |
| S16_extracted_150873 | Not-determined |
| S16_extracted_150889 | Not-determined |
| S16_extracted_150891 | Not-determined |
| S16_extracted_150894 | Not-determined |
| S16_extracted_150905 | Not-determined |
| S16_extracted_150907 | Not-determined |
| S16_extracted_150913 | Not-determined |
| S16_extracted_150915 | Not-determined |
| S16_extracted_150921 | Not-determined |
| S16_extracted_150932 | Not-determined |
| S16_extracted_150936 | Not-determined |
| S16_extracted_150968 | Not-determined |
| S16_extracted_150988 | Not-determined |
| S16_extracted_150995 | Low-quality |
| S16_extracted_151002 | Not-determined |
| S16_extracted_151010 | Not-determined |
| S16_extracted_151021 | Not-determined |
| S16_extracted_151030 | Not-determined |
| S16_extracted_151033 | Not-determined |
| S16_extracted_151049 | Not-determined |
| S16_extracted_151050 | Not-determined |
| S16_extracted_151052 | Not-determined |
| S16_extracted_151069 | Not-determined |
| S16_extracted_151099 | Not-determined |
| S16_extracted_151114 | Not-determined |
| S16_extracted_151122 | Not-determined |
| S16_extracted_151133 | Not-determined |
| S16_extracted_151136 | Not-determined |
| S16_extracted_151143 | Not-determined |
| S16_extracted_151169 | Not-determined |
| S16_extracted_151171 | Not-determined |
| S16_extracted_151180 | Not-determined |
| S16_extracted_151184 | Not-determined |
| S16_extracted_151187 | Not-determined |
| S16_extracted_151219 | Not-determined |
| S16_extracted_151228 | Low-quality |
| S16_extracted_151233 | Not-determined |
| S16_extracted_151236 | Not-determined |
| S16_extracted_151239 | Low-quality |
| S16_extracted_151244 | Not-determined |
| S16_extracted_151245 | Low-quality |
| S16_extracted_151253 | Not-determined |
| S16_extracted_151264 | Not-determined |
| S16_extracted_151267 | Not-determined |
| S16_extracted_151271 | Not-determined |
| S16_extracted_151275 | Not-determined |
| S16_extracted_151276 | Not-determined |
| S16_extracted_151308 | Not-determined |
| S16_extracted_151309 | Not-determined |
| S16_extracted_151315 | Not-determined |
| S16_extracted_151331 | Not-determined |
| S16_extracted_151333 | Low-quality |
| S16_extracted_151339 | Not-determined |
| S16_extracted_151353 | Low-quality |
| S16_extracted_151366 | Not-determined |
| S16_extracted_151370 | Not-determined |
| S16_extracted_151396 | Low-quality |
| S16_extracted_151397 | Not-determined |
| S16_extracted_151398 | Low-quality |
| S16_extracted_151416 | Not-determined |
| S16_extracted_151432 | Not-determined |
| S16_extracted_151447 | Not-determined |
| S12_extracted_k141_73926 | Complete |
| S16_extracted_151452 | Not-determined |
| S16_extracted_151460 | Low-quality |
| S16_extracted_151474 | Low-quality |
| S16_extracted_151482 | Not-determined |
| S16_extracted_151484 | Not-determined |
| S16_extracted_151488 | Not-determined |
| S16_extracted_151505 | Not-determined |
| S16_extracted_151506 | Not-determined |
| S16_extracted_151528 | Not-determined |
| S16_extracted_151554 | Low-quality |
| S16_extracted_151556 | Not-determined |
| S16_extracted_151567 | Not-determined |
| S16_extracted_151592 | Not-determined |
| S16_extracted_151598 | Not-determined |
| S16_extracted_151604 | Low-quality |
| S16_extracted_151607 | Not-determined |
| S16_extracted_151609 | Not-determined |
| S16_extracted_151625 | Low-quality |
| S16_extracted_151637 | Not-determined |
| S16_extracted_151663 | Not-determined |
| S16_extracted_151668 | Low-quality |
| S16_extracted_151672 | Not-determined |
| S16_extracted_151673 | Not-determined |
| S16_extracted_151681 | Low-quality |
| S16_extracted_151684 | Not-determined |
| S16_extracted_151685 | Not-determined |
| S16_extracted_151688 | Not-determined |
| S16_extracted_151690 | Not-determined |
| S16_extracted_151698 | Not-determined |
| S16_extracted_151704 | Not-determined |
| S16_extracted_151725 | Not-determined |
| S16_extracted_151733 | Low-quality |
| S16_extracted_151743 | Not-determined |
| S16_extracted_151745 | Not-determined |
| S16_extracted_151753 | Not-determined |
| S16_extracted_151754 | Not-determined |
| S16_extracted_151764 | Not-determined |
| S16_extracted_151765 | Not-determined |
| S16_extracted_151776 | Not-determined |
| S16_extracted_151796 | Not-determined |
| S16_extracted_151821 | Not-determined |
| S16_extracted_151822 | Low-quality |
| S16_extracted_151840 | Not-determined |
| S16_extracted_151846 | Not-determined |
| S16_extracted_151854 | Not-determined |
| S16_extracted_151863 | Not-determined |
| S16_extracted_151865 | Not-determined |
| S16_extracted_151888 | Not-determined |
| S16_extracted_151891 | Not-determined |
| S16_extracted_151906 | Not-determined |
| S16_extracted_151959 | Not-determined |
| S16_extracted_151974 | Not-determined |
| S16_extracted_151982 | Not-determined |
| S16_extracted_151991 | Not-determined |
| S16_extracted_152018 | Not-determined |
| S16_extracted_152027 | Not-determined |
| S16_extracted_152036 | Low-quality |
| S16_extracted_152052 | Not-determined |
| S16_extracted_152061 | Not-determined |
| S16_extracted_152064 | Not-determined |
| S16_extracted_152082 | Low-quality |
| S16_extracted_152089 | Not-determined |
| S16_extracted_152091 | Not-determined |
| S16_extracted_152102 | Not-determined |
| S16_extracted_152113 | Not-determined |
| S16_extracted_152114 | Not-determined |
| S16_extracted_152119 | Not-determined |
| S16_extracted_152128 | Not-determined |
| S16_extracted_152137 | Not-determined |
| S16_extracted_152141 | Not-determined |
| S16_extracted_152156 | Not-determined |
| S16_extracted_152160 | Not-determined |
| S16_extracted_152161 | Not-determined |
| S16_extracted_152190 | Low-quality |
| S16_extracted_152191 | Not-determined |
| S16_extracted_152192 | Not-determined |
| S16_extracted_152200 | Low-quality |
| S16_extracted_152213 | Low-quality |
| S16_extracted_152221 | Not-determined |
| S16_extracted_152236 | Not-determined |
| S16_extracted_152240 | Not-determined |
| S16_extracted_152254 | Low-quality |
| S16_extracted_152261 | Not-determined |
| S16_extracted_152262 | Not-determined |
| S16_extracted_152276 | Not-determined |
| S16_extracted_152297 | Not-determined |
| S16_extracted_152325 | Low-quality |
| S16_extracted_152332 | Not-determined |
| S16_extracted_152345 | Not-determined |
| S16_extracted_152346 | Low-quality |
| S16_extracted_152355 | Not-determined |
| S16_extracted_152360 | Not-determined |
| S16_extracted_152366 | Not-determined |
| S16_extracted_152368 | Not-determined |
| S16_extracted_152377 | Not-determined |
| S16_extracted_152416 | Not-determined |
| S16_extracted_152422 | Low-quality |
| S16_extracted_152430 | Not-determined |
| S16_extracted_152433 | Not-determined |
| S16_extracted_152436 | Not-determined |
| S16_extracted_152454 | Not-determined |
| S16_extracted_152469 | Not-determined |
| S16_extracted_152488 | Low-quality |
| S16_extracted_152495 | Not-determined |
| S16_extracted_152505 | Not-determined |
| S16_extracted_152506 | Not-determined |
| S16_extracted_152508 | Not-determined |
| S16_extracted_152512 | Not-determined |
| S16_extracted_152529 | Not-determined |
| S16_extracted_152534 | Not-determined |
| S16_extracted_152552 | Not-determined |
| S16_extracted_152556 | Not-determined |
| S16_extracted_152569 | Not-determined |
| S16_extracted_152590 | Not-determined |
| S16_extracted_152591 | Not-determined |
| S16_extracted_152595 | Not-determined |
| S16_extracted_152612 | Not-determined |
| S16_extracted_152613 | Not-determined |
| S16_extracted_152620 | Not-determined |
| S16_extracted_152625 | Low-quality |
| S16_extracted_152663 | Not-determined |
| S16_extracted_152674 | Not-determined |
| S16_extracted_152677 | Not-determined |
| S16_extracted_152690 | Not-determined |
| S16_extracted_152692 | Not-determined |
| S16_extracted_152694 | Not-determined |
| S16_extracted_152695 | Low-quality |
| S16_extracted_152700 | Not-determined |
| S16_extracted_152738 | Not-determined |
| S16_extracted_152755 | Not-determined |
| S16_extracted_152773 | Low-quality |
| S16_extracted_152775 | Low-quality |
| S16_extracted_152781 | Low-quality |
| S16_extracted_152788 | Not-determined |
| S16_extracted_152790 | Not-determined |
| S16_extracted_152807 | Low-quality |
| S16_extracted_152808 | Not-determined |
| S16_extracted_152817 | Not-determined |
| S16_extracted_152824 | Not-determined |
| S16_extracted_152832 | Not-determined |
| S16_extracted_152834 | Not-determined |
| S16_extracted_152843 | Not-determined |
| S16_extracted_152848 | Not-determined |
| S16_extracted_152852 | Not-determined |
| S16_extracted_152859 | Not-determined |
| S16_extracted_152864 | Not-determined |
| S16_extracted_152892 | Not-determined |
| S16_extracted_152893 | Not-determined |
| S16_extracted_152903 | Low-quality |
| S16_extracted_152911 | Not-determined |
| S16_extracted_152916 | Not-determined |
| S16_extracted_152922 | Not-determined |
| S16_extracted_152924 | Not-determined |
| S16_extracted_152939 | Not-determined |
| S16_extracted_152957 | Not-determined |
| S16_extracted_152966 | Not-determined |
| S16_extracted_152972 | Not-determined |
| S16_extracted_152977 | Not-determined |
| S16_extracted_152985 | Not-determined |
| S16_extracted_152993 | Not-determined |
| S16_extracted_152995 | Low-quality |
| S16_extracted_153014 | Not-determined |
| S16_extracted_153017 | Not-determined |
| S16_extracted_153020 | Not-determined |
| S16_extracted_153022 | Low-quality |
| S16_extracted_153026 | Not-determined |
| S16_extracted_153028 | Not-determined |
| S16_extracted_153029 | Not-determined |
| S16_extracted_153035 | Not-determined |
| S16_extracted_153047 | Not-determined |
| S16_extracted_153048 | Low-quality |
| S16_extracted_153049 | Low-quality |
| S16_extracted_153061 | Not-determined |
| S16_extracted_153063 | Not-determined |
| S16_extracted_153073 | Not-determined |
| S16_extracted_153087 | Not-determined |
| S16_extracted_153094 | Not-determined |
| S16_extracted_153100 | Not-determined |
| S16_extracted_153109 | Not-determined |
| S16_extracted_153132 | Not-determined |
| S16_extracted_153147 | Not-determined |
| S16_extracted_153158 | Not-determined |
| S16_extracted_153181 | Low-quality |
| S16_extracted_153189 | Low-quality |
| S16_extracted_153190 | Not-determined |
| S16_extracted_153198 | Low-quality |
| S16_extracted_153199 | Not-determined |
| S16_extracted_153213 | Not-determined |
| S16_extracted_153222 | Low-quality |
| S16_extracted_153225 | Not-determined |
| S16_extracted_153226 | Not-determined |
| S16_extracted_153230 | Not-determined |
| S16_extracted_153243 | Not-determined |
| S16_extracted_153257 | Not-determined |
| S16_extracted_153270 | Not-determined |
| S16_extracted_153280 | Not-determined |
| S16_extracted_153285 | Not-determined |
| S16_extracted_153289 | Not-determined |
| S16_extracted_153341 | Not-determined |
| S16_extracted_153363 | Low-quality |
| S16_extracted_153378 | Not-determined |
| S16_extracted_153393 | Not-determined |
| S16_extracted_153398 | Low-quality |
| S16_extracted_153401 | Low-quality |
| S16_extracted_153405 | Low-quality |
| S16_extracted_153419 | Not-determined |
| S16_extracted_153423 | Low-quality |
| S16_extracted_153430 | Not-determined |
| S16_extracted_153433 | Not-determined |
| S16_extracted_153459 | Not-determined |
| S16_extracted_153461 | Not-determined |
| S16_extracted_153465 | Low-quality |
| S16_extracted_153468 | Low-quality |
| S16_extracted_153484 | Not-determined |
| S16_extracted_153489 | Not-determined |
| S16_extracted_153492 | Not-determined |
| S16_extracted_153502 | Not-determined |
| S16_extracted_153514 | Not-determined |
| S16_extracted_153535 | Not-determined |
| S16_extracted_153542 | Low-quality |
| S16_extracted_153547 | Not-determined |
| S16_extracted_153552 | Not-determined |
| S16_extracted_153554 | Not-determined |
| S16_extracted_153555 | Not-determined |
| S16_extracted_153562 | Not-determined |
| S16_extracted_153578 | Not-determined |
| S16_extracted_153608 | Low-quality |
| S16_extracted_153623 | Not-determined |
| S16_extracted_153642 | Low-quality |
| S16_extracted_153656 | Not-determined |
| S16_extracted_153667 | Not-determined |
| S16_extracted_153668 | Not-determined |
| S16_extracted_153678 | Not-determined |
| S16_extracted_153695 | Not-determined |
| S16_extracted_153699 | Not-determined |
| S16_extracted_153706 | Not-determined |
| S16_extracted_153713 | Not-determined |
| S16_extracted_153727 | Not-determined |
| S16_extracted_153730 | Not-determined |
| S16_extracted_153769 | Not-determined |
| S16_extracted_153771 | Low-quality |
| S16_extracted_153781 | Not-determined |
| S16_extracted_153801 | Not-determined |
| S16_extracted_153814 | Not-determined |
| S16_extracted_153844 | Not-determined |
| S16_extracted_153851 | Not-determined |
| S16_extracted_153876 | Not-determined |
| S16_extracted_153940 | Not-determined |
| S16_extracted_153943 | Not-determined |
| S16_extracted_153952 | Not-determined |
| S16_extracted_153964 | Not-determined |
| S16_extracted_153976 | Not-determined |
| S16_extracted_154001 | Not-determined |
| S16_extracted_154002 | Not-determined |
| S16_extracted_154023 | Not-determined |
| S16_extracted_154038 | Not-determined |
| S16_extracted_154054 | Not-determined |
| S16_extracted_154078 | Not-determined |
| S16_extracted_154096 | Not-determined |
| S16_extracted_154104 | Low-quality |
| S16_extracted_154105 | Not-determined |
| S16_extracted_154136 | Not-determined |
| S16_extracted_154138 | Not-determined |
| S16_extracted_154144 | Not-determined |
| S16_extracted_154153 | Not-determined |
| S16_extracted_154170 | Not-determined |
| S16_extracted_154177 | Not-determined |
| S16_extracted_154183 | Not-determined |
| S16_extracted_154198 | Low-quality |
| S16_extracted_154199 | Not-determined |
| S16_extracted_154219 | Low-quality |
| S16_extracted_154222 | Low-quality |
| S16_extracted_154225 | Low-quality |
| S16_extracted_154254 | Not-determined |
| S16_extracted_154257 | Not-determined |
| S16_extracted_154265 | Not-determined |
| S16_extracted_154269 | Not-determined |
| S16_extracted_154286 | Low-quality |
| S16_extracted_154292 | Not-determined |
| S16_extracted_154311 | Low-quality |
| S16_extracted_154320 | Low-quality |
| S16_extracted_154323 | Not-determined |
| S16_extracted_154331 | Not-determined |
| S16_extracted_154336 | Not-determined |
| S16_extracted_154341 | Not-determined |
| S16_extracted_154367 | Not-determined |
| S16_extracted_154381 | Not-determined |
| S16_extracted_154384 | Not-determined |
| S16_extracted_154393 | Not-determined |
| S16_extracted_154403 | Not-determined |
| S16_extracted_154407 | Not-determined |
| S16_extracted_154415 | Low-quality |
| S16_extracted_154430 | Not-determined |
| S16_extracted_154435 | Not-determined |
| S16_extracted_154442 | Not-determined |
| S16_extracted_154471 | Not-determined |
| S16_extracted_154480 | Low-quality |
| S16_extracted_154493 | Not-determined |
| S16_extracted_154507 | Not-determined |
| S16_extracted_154516 | Not-determined |
| S16_extracted_154548 | Not-determined |
| S16_extracted_154558 | Not-determined |
| S16_extracted_154560 | Not-determined |
| S16_extracted_154566 | Not-determined |
| S16_extracted_154575 | Not-determined |
| S16_extracted_154577 | Not-determined |
| S16_extracted_154604 | Not-determined |
| S16_extracted_154621 | Not-determined |
| S16_extracted_154622 | Not-determined |
| S16_extracted_154641 | Not-determined |
| S16_extracted_154644 | Not-determined |
| S16_extracted_154645 | Low-quality |
| S16_extracted_154658 | Not-determined |
| S16_extracted_154664 | Low-quality |
| S16_extracted_154668 | Not-determined |
| S16_extracted_154703 | Low-quality |
| S16_extracted_154714 | Not-determined |
| S16_extracted_154730 | Not-determined |
| S16_extracted_154731 | Not-determined |
| S16_extracted_154742 | Low-quality |
| S16_extracted_154744 | Not-determined |
| S16_extracted_154753 | Not-determined |
| S16_extracted_154763 | Not-determined |
| S16_extracted_154791 | Low-quality |
| S16_extracted_154794 | Not-determined |
| S16_extracted_154795 | Not-determined |
| S16_extracted_154800 | Not-determined |
| S16_extracted_154804 | Not-determined |
| S16_extracted_154805 | Not-determined |
| S16_extracted_154831 | Low-quality |

| | |
|---|---|
| S16_extracted_154848 | Not-determined |
| S16_extracted_154851 | Not-determined |
| S16_extracted_154875 | Low-quality |
| S16_extracted_154882 | Not-determined |
| S16_extracted_154889 | Not-determined |
| S16_extracted_154891 | Not-determined |
| S16_extracted_154915 | Not-determined |
| S16_extracted_154931 | Not-determined |
| S16_extracted_154935 | Not-determined |
| S16_extracted_154940 | Low-quality |
| S16_extracted_154946 | Not-determined |
| S16_extracted_154948 | Not-determined |
| S16_extracted_154954 | Not-determined |
| S16_extracted_154970 | Not-determined |
| S16_extracted_154974 | Not-determined |
| S16_extracted_154978 | Low-quality |
| S16_extracted_155043 | Not-determined |
| S16_extracted_155046 | Not-determined |
| S16_extracted_155063 | Not-determined |
| S16_extracted_155071 | Not-determined |
| S16_extracted_155073 | Not-determined |
| S16_extracted_155079 | Not-determined |
| S16_extracted_155080 | Not-determined |
| S16_extracted_155081 | Not-determined |
| S16_extracted_155083 | Low-quality |
| S16_extracted_155092 | Low-quality |
| S16_extracted_155100 | Not-determined |
| S16_extracted_155107 | Not-determined |
| S16_extracted_155112 | Low-quality |
| S16_extracted_155122 | Not-determined |
| S16_extracted_155130 | Not-determined |
| S16_extracted_155149 | Not-determined |
| S16_extracted_155150 | Low-quality |
| S16_extracted_155176 | Low-quality |
| S16_extracted_155211 | Not-determined |
| S16_extracted_155224 | Not-determined |
| S16_extracted_155232 | Low-quality |
| S16_extracted_155234 | Not-determined |
| S16_extracted_155235 | Not-determined |
| S16_extracted_155244 | Not-determined |
| S16_extracted_155261 | Not-determined |
| S16_extracted_155274 | Not-determined |
| S16_extracted_155286 | Not-determined |
| S16_extracted_155287 | Not-determined |
| S16_extracted_155303 | Not-determined |
| S16_extracted_155339 | Not-determined |
| S16_extracted_155340 | Not-determined |
| S16_extracted_155354 | Not-determined |
| S16_extracted_155357 | Not-determined |
| S16_extracted_155364 | Not-determined |
| S16_extracted_155384 | Not-determined |
| S16_extracted_155390 | Not-determined |
| S16_extracted_155394 | Not-determined |
| S16_extracted_155402 | Low-quality |
| S16_extracted_155410 | Not-determined |
| S16_extracted_155439 | Low-quality |
| S16_extracted_155446 | Not-determined |
| S16_extracted_155451 | Not-determined |
| S16_extracted_155456 | Not-determined |
| S16_extracted_155457 | Not-determined |
| S16_extracted_155459 | Not-determined |
| S16_extracted_155461 | Not-determined |
| S16_extracted_155474 | Not-determined |
| S16_extracted_155481 | Low-quality |
| S16_extracted_155491 | Not-determined |
| S16_extracted_155494 | Low-quality |
| S16_extracted_155499 | Not-determined |
| S16_extracted_155502 | Not-determined |
| S16_extracted_155508 | Not-determined |
| S16_extracted_155536 | Not-determined |
| S16_extracted_155549 | Not-determined |
| S16_extracted_155551 | Not-determined |
| S16_extracted_155569 | Not-determined |
| S16_extracted_155576 | Not-determined |
| S16_extracted_155584 | Not-determined |
| S16_extracted_155589 | Low-quality |
| S16_extracted_155591 | Not-determined |
| S16_extracted_155596 | Not-determined |
| S16_extracted_155610 | Not-determined |
| S16_extracted_155624 | Not-determined |
| S16_extracted_155637 | Not-determined |
| S16_extracted_155643 | Not-determined |
| S16_extracted_155644 | Low-quality |
| S16_extracted_155650 | Low-quality |
| S16_extracted_155662 | Not-determined |
| S16_extracted_155668 | Low-quality |
| S16_extracted_155669 | Not-determined |
| S16_extracted_155672 | Not-determined |
| S16_extracted_155673 | Not-determined |
| S16_extracted_155676 | Not-determined |
| S16_extracted_155685 | Not-determined |
| S16_extracted_155696 | Not-determined |
| S16_extracted_155709 | Not-determined |
| S16_extracted_155726 | Not-determined |
| S16_extracted_155730 | Not-determined |
| S16_extracted_155738 | Not-determined |
| S16_extracted_155739 | Not-determined |
| S16_extracted_155741 | Not-determined |
| S16_extracted_155746 | Low-quality |
| S16_extracted_155747 | Not-determined |
| S16_extracted_155751 | Not-determined |
| S16_extracted_155752 | Not-determined |
| S16_extracted_155757 | Not-determined |
| S16_extracted_155773 | Not-determined |
| S16_extracted_155783 | Not-determined |
| S16_extracted_155790 | Not-determined |
| S16_extracted_155797 | Not-determined |
| S16_extracted_155811 | Low-quality |
| S16_extracted_155826 | Low-quality |
| S16_extracted_155831 | Not-determined |
| S16_extracted_155849 | Not-determined |
| S16_extracted_155859 | Not-determined |
| S16_extracted_155874 | Low-quality |
| S16_extracted_155875 | Low-quality |
| S16_extracted_155890 | Not-determined |
| S16_extracted_155891 | Not-determined |
| S16_extracted_155905 | Not-determined |
| S16_extracted_155912 | Not-determined |
| S16_extracted_155914 | Low-quality |
| S16_extracted_155919 | Not-determined |
| S16_extracted_155933 | Not-determined |
| S16_extracted_155940 | Not-determined |
| S16_extracted_155943 | Not-determined |
| S16_extracted_156009 | Not-determined |
| S16_extracted_156018 | Not-determined |
| S16_extracted_156024 | Not-determined |
| S16_extracted_156029 | Not-determined |
| S16_extracted_156044 | Not-determined |
| S16_extracted_156066 | Not-determined |
| S16_extracted_156086 | Not-determined |
| S16_extracted_156105 | Not-determined |
| S16_extracted_156126 | Not-determined |
| S16_extracted_156137 | Not-determined |
| S16_extracted_156140 | Not-determined |
| S16_extracted_156148 | Not-determined |
| S16_extracted_156156 | Not-determined |
| S16_extracted_156157 | Not-determined |
| S16_extracted_156158 | Low-quality |
| S16_extracted_156161 | Not-determined |
| S16_extracted_156171 | Not-determined |
| S16_extracted_156181 | Not-determined |
| S16_extracted_156193 | Not-determined |
| S16_extracted_156214 | Low-quality |
| S16_extracted_156224 | Not-determined |
| S16_extracted_156233 | Low-quality |
| S16_extracted_156250 | Not-determined |
| S16_extracted_156255 | Not-determined |
| S16_extracted_156259 | Low-quality |
| S16_extracted_156267 | Low-quality |
| S16_extracted_156268 | Not-determined |
| S16_extracted_156271 | Not-determined |
| S16_extracted_156272 | Not-determined |
| S16_extracted_156278 | Low-quality |
| S16_extracted_156287 | Not-determined |
| S16_extracted_156290 | Not-determined |
| S16_extracted_156314 | Not-determined |
| S16_extracted_156319 | Not-determined |
| S16_extracted_156343 | Not-determined |
| S16_extracted_156358 | Not-determined |
| S16_extracted_156365 | Not-determined |
| S16_extracted_156369 | Low-quality |
| S16_extracted_156381 | Not-determined |
| S16_extracted_156391 | Not-determined |
| S16_extracted_156395 | Not-determined |
| S16_extracted_156400 | Not-determined |
| S16_extracted_156406 | Not-determined |
| S16_extracted_156420 | Not-determined |
| S16_extracted_156434 | Not-determined |
| S16_extracted_156435 | Low-quality |
| S16_extracted_156451 | Low-quality |
| S16_extracted_156465 | Not-determined |
| S16_extracted_156475 | Low-quality |
| S16_extracted_156501 | Low-quality |
| S16_extracted_156503 | Not-determined |
| S16_extracted_156511 | Not-determined |
| S16_extracted_156512 | Low-quality |
| S16_extracted_156529 | Not-determined |
| S16_extracted_156531 | Low-quality |
| S16_extracted_156551 | Not-determined |
| S16_extracted_156567 | Not-determined |
| S16_extracted_156569 | Not-determined |
| S16_extracted_156613 | Not-determined |
| S16_extracted_156614 | Low-quality |
| S16_extracted_156630 | Low-quality |
| S16_extracted_156653 | Not-determined |
| S16_extracted_156658 | Not-determined |
| S16_extracted_156682 | Not-determined |
| S16_extracted_156690 | Not-determined |
| S16_extracted_156694 | Not-determined |
| S16_extracted_156698 | Not-determined |
| S16_extracted_156722 | Not-determined |
| S16_extracted_156729 | Not-determined |
| S16_extracted_156733 | Not-determined |
| S16_extracted_156742 | Not-determined |
| S16_extracted_156749 | Not-determined |
| S16_extracted_156762 | Not-determined |
| S16_extracted_156773 | Not-determined |
| S16_extracted_156780 | Not-determined |
| S16_extracted_156788 | Low-quality |
| S16_extracted_156790 | Not-determined |
| S16_extracted_156848 | Not-determined |
| S16_extracted_156851 | Not-determined |
| S16_extracted_156861 | Not-determined |
| S16_extracted_156863 | Not-determined |
| S16_extracted_156874 | Not-determined |
| S16_extracted_156876 | Not-determined |
| S16_extracted_156887 | Not-determined |
| S16_extracted_156891 | Not-determined |
| S16_extracted_156907 | Not-determined |
| S16_extracted_156911 | Not-determined |
| S16_extracted_156922 | Low-quality |
| S16_extracted_156923 | Not-determined |
| S16_extracted_156941 | Not-determined |
| S16_extracted_156948 | Not-determined |
| S16_extracted_156951 | Not-determined |
| S16_extracted_156963 | Low-quality |
| S16_extracted_156994 | Not-determined |
| S16_extracted_156995 | Not-determined |
| S16_extracted_156997 | Not-determined |
| S16_extracted_157007 | Not-determined |
| S16_extracted_157014 | Not-determined |
| S16_extracted_157021 | Not-determined |
| S16_extracted_157022 | Not-determined |
| S16_extracted_157023 | Not-determined |
| S16_extracted_157026 | Not-determined |
| S16_extracted_157032 | Not-determined |
| S16_extracted_157053 | Not-determined |
| S16_extracted_157078 | Not-determined |
| S16_extracted_157080 | Not-determined |
| S16_extracted_157098 | Not-determined |
| S16_extracted_157101 | Low-quality |
| S16_extracted_157103 | Not-determined |
| S16_extracted_157112 | Not-determined |
| S16_extracted_157114 | Not-determined |
| S16_extracted_157138 | Not-determined |
| S16_extracted_157147 | Not-determined |
| S16_extracted_157154 | Not-determined |
| S16_extracted_157155 | Not-determined |
| S16_extracted_157160 | Not-determined |
| S16_extracted_157170 | Not-determined |
| S16_extracted_157191 | Not-determined |
| S16_extracted_157205 | Not-determined |
| S16_extracted_157217 | Not-determined |
| S16_extracted_157218 | Not-determined |
| S16_extracted_157228 | Not-determined |
| S16_extracted_157238 | Not-determined |
| S16_extracted_157244 | Not-determined |
| S16_extracted_157246 | Not-determined |
| S16_extracted_157247 | Not-determined |
| S16_extracted_157251 | Not-determined |
| S16_extracted_157264 | Not-determined |
| S16_extracted_157268 | Not-determined |
| S16_extracted_157274 | Not-determined |
| S16_extracted_157282 | Not-determined |
| S16_extracted_157283 | Not-determined |
| S16_extracted_157286 | Not-determined |
| S16_extracted_157291 | Not-determined |
| S16_extracted_157303 | Low-quality |
| S16_extracted_157304 | Not-determined |
| S16_extracted_157321 | Not-determined |
| S16_extracted_157334 | Not-determined |
| S16_extracted_157346 | Not-determined |
| S16_extracted_157353 | Not-determined |
| S16_extracted_157355 | Not-determined |
| S16_extracted_157370 | Low-quality |
| S16_extracted_157378 | Not-determined |
| S16_extracted_157387 | Not-determined |
| S16_extracted_157392 | Not-determined |
| S16_extracted_157396 | Not-determined |
| S16_extracted_157397 | Not-determined |
| S16_extracted_157399 | Not-determined |
| S16_extracted_157404 | Low-quality |
| S16_extracted_157408 | Not-determined |
| S16_extracted_157419 | Not-determined |
| S16_extracted_157427 | Not-determined |
| S16_extracted_157429 | Not-determined |
| S16_extracted_157434 | Not-determined |
| S16_extracted_157444 | Not-determined |
| S16_extracted_157456 | Not-determined |
| S16_extracted_157457 | Not-determined |
| S16_extracted_157465 | Not-determined |
| S16_extracted_157476 | Not-determined |
| S16_extracted_157483 | Low-quality |
| S16_extracted_157506 | Not-determined |
| S16_extracted_157508 | Not-determined |
| S16_extracted_157514 | Not-determined |
| S16_extracted_157523 | Not-determined |
| S16_extracted_157524 | Not-determined |
| S16_extracted_157527 | Low-quality |
| S16_extracted_157531 | Not-determined |
| S16_extracted_157551 | Not-determined |
| S16_extracted_157556 | Not-determined |
| S16_extracted_157559 | Not-determined |
| S16_extracted_157572 | Not-determined |
| S16_extracted_157588 | Not-determined |
| S16_extracted_157592 | Not-determined |
| S16_extracted_157621 | Not-determined |
| S16_extracted_157625 | Low-quality |
| S16_extracted_157638 | Not-determined |
| S16_extracted_157667 | Not-determined |
| S16_extracted_157671 | Not-determined |
| S16_extracted_157681 | Low-quality |
| S16_extracted_157685 | Not-determined |
| S16_extracted_157686 | Not-determined |
| S16_extracted_157689 | Not-determined |
| S16_extracted_157701 | Not-determined |
| S16_extracted_157715 | Low-quality |
| S16_extracted_157719 | Not-determined |
| S16_extracted_157731 | Not-determined |
| S16_extracted_157746 | Not-determined |
| S16_extracted_157751 | Not-determined |
| S16_extracted_157781 | Low-quality |
| S16_extracted_157795 | Not-determined |
| S16_extracted_157804 | Not-determined |
| S16_extracted_157831 | Not-determined |
| S16_extracted_157852 | Not-determined |
| S16_extracted_157860 | Not-determined |
| S16_extracted_157872 | Not-determined |
| S16_extracted_157879 | Not-determined |
| S16_extracted_157903 | Low-quality |
| S16_extracted_157920 | Not-determined |
| S16_extracted_157931 | Not-determined |
| S16_extracted_157951 | Not-determined |
| S16_extracted_157955 | Not-determined |
| S16_extracted_157965 | Low-quality |
| S16_extracted_157968 | Not-determined |
| S16_extracted_157974 | Not-determined |
| S16_extracted_157978 | Not-determined |
| S16_extracted_157980 | Not-determined |
| S16_extracted_158002 | Not-determined |
| S16_extracted_158034 | Not-determined |
| S16_extracted_158035 | Not-determined |
| S16_extracted_158044 | Not-determined |
| S16_extracted_158055 | Not-determined |
| S16_extracted_158056 | Not-determined |
| S16_extracted_158060 | Not-determined |
| S16_extracted_158074 | Low-quality |
| S16_extracted_158090 | Not-determined |
| S16_extracted_158097 | Not-determined |
| S16_extracted_158100 | Not-determined |
| S16_extracted_158106 | Low-quality |
| S16_extracted_158125 | Not-determined |
| S16_extracted_158130 | Not-determined |
| S16_extracted_158133 | Not-determined |
| S16_extracted_158145 | Not-determined |
| S16_extracted_158160 | Not-determined |
| S16_extracted_158162 | Not-determined |
| S16_extracted_158192 | Not-determined |
| S16_extracted_158202 | Not-determined |
| S16_extracted_158219 | Not-determined |
| S16_extracted_158227 | Not-determined |
| S16_extracted_158233 | Not-determined |
| S16_extracted_158236 | Low-quality |
| S16_extracted_158254 | Not-determined |
| S16_extracted_158261 | Not-determined |
| S16_extracted_158289 | Not-determined |
| S16_extracted_158292 | Not-determined |
| S16_extracted_158303 | Low-quality |
| S16_extracted_158311 | Low-quality |
| S16_extracted_158324 | Not-determined |
| S16_extracted_158350 | Not-determined |
| S16_extracted_158368 | Not-determined |
| S16_extracted_158383 | Not-determined |
| S16_extracted_158384 | Not-determined |
| S16_extracted_158394 | Not-determined |
| S16_extracted_158405 | Not-determined |
| S16_extracted_158406 | Not-determined |
| S16_extracted_158413 | Not-determined |
| S16_extracted_158418 | Not-determined |
| S16_extracted_158421 | Not-determined |
| S16_extracted_158430 | Not-determined |
| S16_extracted_158434 | Not-determined |
| S16_extracted_158444 | Not-determined |
| S16_extracted_158462 | Not-determined |
| S16_extracted_158505 | Not-determined |
| S16_extracted_158506 | Not-determined |
| S16_extracted_158514 | Not-determined |
| S16_extracted_158536 | Not-determined |
| S16_extracted_158538 | Low-quality |
| S16_extracted_158539 | Not-determined |
| S16_extracted_158564 | Not-determined |
| S16_extracted_158596 | Not-determined |
| S16_extracted_158601 | Not-determined |
| S16_extracted_158630 | Not-determined |
| S16_extracted_158662 | Not-determined |
| S16_extracted_158673 | Not-determined |
| S16_extracted_158683 | Not-determined |
| S16_extracted_158722 | Low-quality |
| S16_extracted_158728 | Not-determined |
| S16_extracted_158733 | Low-quality |
| S16_extracted_158752 | Not-determined |
| S16_extracted_158770 | Low-quality |
| S16_extracted_158789 | Not-determined |
| S16_extracted_158794 | Low-quality |
| S16_extracted_158815 | Not-determined |
| S16_extracted_158823 | Not-determined |
| S16_extracted_158827 | Not-determined |
| S16_extracted_158837 | Not-determined |
| S16_extracted_158870 | Not-determined |
| S16_extracted_158875 | Not-determined |
| S16_extracted_158877 | Not-determined |
| S16_extracted_158887 | Not-determined |
| S16_extracted_158896 | Not-determined |
| S16_extracted_158905 | Not-determined |
| S16_extracted_158911 | Low-quality |
| S16_extracted_158920 | Low-quality |
| S16_extracted_158958 | Low-quality |
| S16_extracted_158961 | Low-quality |
| S16_extracted_158975 | Not-determined |
| S16_extracted_158980 | Not-determined |
| S16_extracted_158984 | Not-determined |
| S16_extracted_158995 | Not-determined |
| S16_extracted_159003 | Not-determined |
| S16_extracted_159004 | Not-determined |
| S16_extracted_159006 | Not-determined |
| S16_extracted_159008 | Low-quality |
| S16_extracted_159012 | Not-determined |
| S16_extracted_159020 | Not-determined |
| S16_extracted_159031 | Not-determined |
| S16_extracted_159065 | Low-quality |
| S16_extracted_159094 | Not-determined |
| S16_extracted_159102 | Not-determined |
| S16_extracted_159104 | Not-determined |
| S16_extracted_159165 | Not-determined |
| S16_extracted_159183 | Not-determined |
| S16_extracted_159213 | Not-determined |
| S16_extracted_159224 | Not-determined |
| S16_extracted_159243 | Low-quality |
| S16_extracted_159246 | Low-quality |
| S16_extracted_159260 | Not-determined |
| S16_extracted_159271 | Not-determined |
| S16_extracted_159279 | Not-determined |
| S16_extracted_159282 | Not-determined |
| S16_extracted_159301 | Not-determined |
| S16_extracted_159310 | Not-determined |
| S16_extracted_159314 | Not-determined |
| S16_extracted_159318 | Not-determined |
| S16_extracted_159324 | Not-determined |
| S16_extracted_159334 | Not-determined |
| S16_extracted_159354 | Not-determined |
| S16_extracted_159367 | Low-quality |
| S16_extracted_159381 | Low-quality |
| S16_extracted_159387 | Low-quality |
| S16_extracted_159388 | Low-quality |
| S16_extracted_159403 | Not-determined |
| S16_extracted_159412 | Low-quality |
| S16_extracted_159446 | Not-determined |
| S16_extracted_159450 | Not-determined |
| S16_extracted_159452 | Not-determined |
| S16_extracted_159458 | Not-determined |
| S16_extracted_159459 | Not-determined |
| S16_extracted_159468 | Not-determined |
| S16_extracted_159479 | Not-determined |
| S16_extracted_159494 | Low-quality |
| S16_extracted_159498 | Not-determined |
| S16_extracted_159513 | Not-determined |
| S16_extracted_159521 | Not-determined |
| S16_extracted_159522 | Not-determined |
| S16_extracted_159537 | Not-determined |
| S16_extracted_159544 | Not-determined |
| S16_extracted_159545 | Not-determined |
| S16_extracted_159585 | Not-determined |
| S16_extracted_159589 | Not-determined |
| S16_extracted_159591 | Not-determined |
| S16_extracted_159592 | Not-determined |
| S16_extracted_159593 | Not-determined |
| S16_extracted_159594 | Low-quality |
| S16_extracted_159595 | Not-determined |
| S16_extracted_159600 | Not-determined |
| S16_extracted_159602 | Not-determined |
| S16_extracted_159613 | Not-determined |
| S16_extracted_159651 | Not-determined |
| S16_extracted_159672 | Not-determined |
| S16_extracted_159684 | Low-quality |
| S16_extracted_159696 | Not-determined |
| S16_extracted_159712 | Not-determined |
| S16_extracted_159718 | Not-determined |
| S16_extracted_159730 | Not-determined |
| S16_extracted_159737 | Not-determined |
| S16_extracted_159740 | Low-quality |
| S16_extracted_159752 | Low-quality |
| S16_extracted_159753 | Not-determined |
| S16_extracted_159755 | Not-determined |
| S16_extracted_159762 | Not-determined |
| S16_extracted_159777 | Not-determined |
| S16_extracted_159780 | Low-quality |
| S16_extracted_159795 | Low-quality |
| S16_extracted_159797 | Not-determined |
| S16_extracted_159802 | Not-determined |
| S16_extracted_159814 | Not-determined |
| S16_extracted_159836 | Not-determined |
| S16_extracted_159838 | Not-determined |
| S16_extracted_159849 | Low-quality |
| S16_extracted_159876 | Not-determined |
| S16_extracted_159878 | Not-determined |
| S16_extracted_159884 | Not-determined |
| S16_extracted_159899 | Not-determined |
| S16_extracted_159927 | Not-determined |
| S16_extracted_159932 | Not-determined |
| S16_extracted_159936 | Low-quality |
| S16_extracted_159937 | Not-determined |
| S16_extracted_159943 | Not-determined |
| S16_extracted_159958 | Low-quality |
| S16_extracted_159961 | Not-determined |
| S16_extracted_159974 | Not-determined |
| S16_extracted_159988 | Not-determined |
| S16_extracted_159990 | Not-determined |
| S16_extracted_160000 | Not-determined |
| S16_extracted_160016 | Not-determined |
| S16_extracted_160026 | Low-quality |
| S16_extracted_160036 | Low-quality |
| S16_extracted_160065 | Not-determined |
| S16_extracted_160066 | Low-quality |
| S16_extracted_160075 | Low-quality |
| S16_extracted_160076 | Not-determined |
| S16_extracted_160096 | Low-quality |
| S16_extracted_160098 | Low-quality |
| S16_extracted_160132 | Not-determined |
| S16_extracted_160147 | Not-determined |
| S16_extracted_160154 | Not-determined |
| S16_extracted_160155 | Not-determined |
| S16_extracted_160171 | Not-determined |
| S16_extracted_160177 | Not-determined |
| S16_extracted_160192 | Not-determined |
| S16_extracted_160193 | Not-determined |
| S16_extracted_160199 | Not-determined |
| S16_extracted_160207 | Not-determined |
| S16_extracted_160233 | Not-determined |
| S16_extracted_160234 | Not-determined |
| S16_extracted_160254 | Not-determined |
| S16_extracted_160260 | Low-quality |
| S16_extracted_160272 | Not-determined |
| S16_extracted_160280 | Not-determined |
| S16_extracted_160285 | Low-quality |
| S16_extracted_160301 | Not-determined |
| S16_extracted_160302 | Not-determined |
| S16_extracted_160312 | Not-determined |
| S16_extracted_160315 | Not-determined |
| S16_extracted_160321 | Not-determined |
| S16_extracted_160326 | Not-determined |
| S16_extracted_160331 | Not-determined |
| S16_extracted_160336 | Not-determined |
| S16_extracted_160345 | Not-determined |
| S16_extracted_160360 | Not-determined |
| S16_extracted_160365 | Not-determined |
| S16_extracted_160377 | Not-determined |
| S16_extracted_160389 | Not-determined |
| S16_extracted_160394 | Low-quality |
| S16_extracted_160400 | Not-determined |
| S16_extracted_160405 | Low-quality |
| S16_extracted_160419 | Not-determined |
| S16_extracted_160433 | Not-determined |
| S16_extracted_160434 | Not-determined |
| S16_extracted_160440 | Not-determined |
| S16_extracted_160451 | Not-determined |
| S16_extracted_160454 | Not-determined |
| S16_extracted_160464 | Not-determined |
| S16_extracted_160480 | Not-determined |
| S16_extracted_160484 | Not-determined |
| S16_extracted_160488 | Not-determined |
| S16_extracted_160490 | Not-determined |
| S16_extracted_160497 | Not-determined |
| S16_extracted_160500 | Not-determined |
| S16_extracted_160528 | Low-quality |
| S16_extracted_160530 | Not-determined |
| S16_extracted_160551 | Not-determined |
| S16_extracted_160573 | Not-determined |
| S16_extracted_160581 | Not-determined |
| S16_extracted_160584 | Not-determined |
| S16_extracted_160585 | Low-quality |
| S16_extracted_160587 | Not-determined |
| S16_extracted_160625 | Not-determined |
| S16_extracted_160631 | Low-quality |
| S16_extracted_160639 | Not-determined |
| S16_extracted_160644 | Not-determined |
| S16_extracted_160659 | Not-determined |
| S16_extracted_160668 | Not-determined |
| S16_extracted_160673 | Not-determined |
| S16_extracted_160721 | Low-quality |
| S16_extracted_160726 | Low-quality |
| S16_extracted_160730 | Not-determined |
| S16_extracted_160751 | Not-determined |
| S16_extracted_160753 | Low-quality |
| S16_extracted_160762 | Low-quality |
| S16_extracted_160765 | Not-determined |
| S16_extracted_160770 | Not-determined |
| S16_extracted_160791 | Not-determined |
| S16_extracted_160795 | Not-determined |
| S16_extracted_160798 | Not-determined |

| | |
|---|---|
| S16_extracted_160807 | Not-determined |
| S16_extracted_160840 | Not-determined |
| S16_extracted_160852 | Not-determined |
| S16_extracted_160863 | Not-determined |
| S16_extracted_160869 | Not-determined |
| S16_extracted_160881 | Not-determined |
| S16_extracted_160888 | Low-quality |
| S16_extracted_160900 | Not-determined |
| S16_extracted_160904 | Low-quality |
| S16_extracted_160931 | Not-determined |
| S16_extracted_160942 | Not-determined |
| S16_extracted_160954 | Low-quality |
| S16_extracted_160974 | Not-determined |
| S16_extracted_160978 | Not-determined |
| S16_extracted_160996 | Not-determined |
| S16_extracted_161008 | Not-determined |
| S16_extracted_161010 | Not-determined |
| S16_extracted_161021 | Not-determined |
| S16_extracted_161039 | Not-determined |
| S16_extracted_161056 | Not-determined |
| S16_extracted_161076 | Not-determined |
| S16_extracted_161080 | Not-determined |
| S16_extracted_161090 | Not-determined |
| S16_extracted_161111 | Not-determined |
| S16_extracted_161156 | Not-determined |
| S16_extracted_161158 | Not-determined |
| S16_extracted_161163 | Not-determined |
| S16_extracted_161165 | Not-determined |
| S16_extracted_161180 | Not-determined |
| S16_extracted_161212 | Not-determined |
| S16_extracted_161221 | Not-determined |
| S16_extracted_161226 | Not-determined |
| S16_extracted_161253 | Not-determined |
| S16_extracted_161268 | Not-determined |
| S16_extracted_161281 | Low-quality |
| S16_extracted_161283 | Low-quality |
| S16_extracted_161285 | Not-determined |
| S16_extracted_161295 | Not-determined |
| S16_extracted_161321 | Not-determined |
| S16_extracted_161347 | Not-determined |
| S16_extracted_161358 | Not-determined |
| S16_extracted_161360 | Not-determined |
| S16_extracted_161361 | Not-determined |
| S16_extracted_161365 | Not-determined |
| S16_extracted_161373 | Not-determined |
| S16_extracted_161398 | Not-determined |
| S16_extracted_161401 | Not-determined |
| S16_extracted_161404 | Not-determined |
| S16_extracted_161422 | Not-determined |
| S16_extracted_161452 | Not-determined |
| S16_extracted_161459 | Not-determined |
| S16_extracted_161470 | Not-determined |
| S16_extracted_161489 | Not-determined |
| S16_extracted_161490 | Not-determined |
| S16_extracted_161551 | Not-determined |
| S16_extracted_161554 | Not-determined |
| S16_extracted_161573 | Not-determined |
| S16_extracted_161593 | Not-determined |
| S16_extracted_161614 | Not-determined |
| S16_extracted_161616 | Not-determined |
| S16_extracted_161621 | Not-determined |
| S16_extracted_161632 | Not-determined |
| S16_extracted_161644 | Not-determined |
| S16_extracted_161645 | Not-determined |
| S16_extracted_161656 | Not-determined |
| S14_extracted_k141_57689 | Medium-quality |
| S16_extracted_161675 | Low-quality |
| S16_extracted_161676 | Not-determined |
| S16_extracted_161679 | Not-determined |
| S16_extracted_161702 | Not-determined |
| S16_extracted_161708 | Not-determined |
| S16_extracted_161709 | Not-determined |
| S16_extracted_161717 | Not-determined |
| S16_extracted_161726 | Low-quality |
| S16_extracted_161727 | Low-quality |
| S16_extracted_161730 | Not-determined |
| S16_extracted_161733 | Not-determined |
| S16_extracted_161734 | Not-determined |
| S16_extracted_161739 | Low-quality |
| S16_extracted_161741 | Not-determined |
| S16_extracted_161759 | Not-determined |
| S16_extracted_161777 | Not-determined |
| S16_extracted_161794 | Not-determined |
| S16_extracted_161799 | Not-determined |
| S16_extracted_161809 | Not-determined |
| S16_extracted_161812 | Low-quality |
| S16_extracted_161819 | Not-determined |
| S16_extracted_161820 | Not-determined |
| S16_extracted_161836 | Not-determined |
| S16_extracted_161852 | Not-determined |
| S16_extracted_161875 | Not-determined |
| S16_extracted_161878 | Not-determined |
| S16_extracted_161894 | Not-determined |
| S16_extracted_161906 | Low-quality |
| S16_extracted_161914 | Not-determined |
| S16_extracted_161916 | Not-determined |
| S16_extracted_161938 | Not-determined |
| S16_extracted_161951 | Not-determined |
| S16_extracted_161965 | Low-quality |
| S16_extracted_161966 | Not-determined |
| S16_extracted_161977 | Not-determined |
| S16_extracted_161996 | Not-determined |
| S16_extracted_162003 | Not-determined |
| S16_extracted_162027 | Not-determined |
| S16_extracted_162030 | Not-determined |
| S16_extracted_162036 | Not-determined |
| S16_extracted_162058 | Not-determined |
| S16_extracted_162063 | Not-determined |
| S16_extracted_162086 | Not-determined |
| S16_extracted_162090 | Not-determined |
| S16_extracted_162094 | Not-determined |
| S16_extracted_162109 | Low-quality |
| S16_extracted_162118 | Not-determined |
| S16_extracted_162122 | Not-determined |
| S16_extracted_162136 | Low-quality |
| S16_extracted_162137 | Not-determined |
| S16_extracted_162149 | Not-determined |
| S16_extracted_162158 | Not-determined |
| S16_extracted_162164 | Not-determined |
| S16_extracted_162169 | Not-determined |
| S16_extracted_162191 | Not-determined |
| S16_extracted_162217 | Not-determined |
| S16_extracted_162226 | Low-quality |
| S16_extracted_162237 | Not-determined |
| S16_extracted_162247 | Not-determined |
| S16_extracted_162255 | Not-determined |
| S16_extracted_162261 | Not-determined |
| S16_extracted_162263 | Not-determined |
| S16_extracted_162268 | Not-determined |
| S16_extracted_162271 | Not-determined |
| S16_extracted_162296 | Not-determined |
| S16_extracted_162308 | Low-quality |
| S16_extracted_162317 | Not-determined |
| S16_extracted_162333 | Not-determined |
| S16_extracted_162335 | Low-quality |
| S16_extracted_162340 | Not-determined |
| S16_extracted_162344 | Not-determined |
| S16_extracted_162353 | Not-determined |
| S16_extracted_162354 | Not-determined |
| S16_extracted_162359 | Not-determined |
| S16_extracted_162378 | Not-determined |
| S16_extracted_162381 | Low-quality |
| S16_extracted_162389 | Not-determined |
| S16_extracted_162394 | Not-determined |
| S16_extracted_162395 | Low-quality |
| S16_extracted_162408 | Not-determined |
| S16_extracted_162417 | Not-determined |
| S16_extracted_162424 | Not-determined |
| S16_extracted_162449 | Not-determined |
| S16_extracted_162451 | Not-determined |
| S16_extracted_162457 | Not-determined |
| S16_extracted_162474 | Not-determined |
| S16_extracted_162477 | Not-determined |
| S16_extracted_162486 | Low-quality |
| S16_extracted_162491 | Not-determined |
| S16_extracted_162494 | Not-determined |
| S16_extracted_162501 | Not-determined |
| S16_extracted_162504 | Not-determined |
| S16_extracted_162512 | Not-determined |
| S16_extracted_162519 | Not-determined |
| S16_extracted_162535 | Low-quality |
| S16_extracted_162541 | Not-determined |
| S16_extracted_162550 | Not-determined |
| S16_extracted_162565 | Low-quality |
| S16_extracted_162574 | Low-quality |
| S16_extracted_162579 | Not-determined |
| S16_extracted_162583 | Not-determined |
| S16_extracted_162621 | Not-determined |
| S16_extracted_162625 | Not-determined |
| S16_extracted_162634 | Not-determined |
| S16_extracted_162641 | Low-quality |
| S16_extracted_162645 | Low-quality |
| S16_extracted_162661 | Not-determined |
| S16_extracted_162665 | Not-determined |
| S16_extracted_162669 | Not-determined |
| S16_extracted_162679 | Low-quality |
| S16_extracted_162695 | Not-determined |
| S16_extracted_162698 | Not-determined |
| S16_extracted_162708 | Not-determined |
| S16_extracted_162723 | Not-determined |
| S16_extracted_162732 | Not-determined |
| S16_extracted_162747 | Low-quality |
| S16_extracted_162748 | Not-determined |
| S16_extracted_162752 | Low-quality |
| S16_extracted_162756 | Low-quality |
| S16_extracted_162799 | Not-determined |
| S16_extracted_162830 | Low-quality |
| S16_extracted_162832 | Low-quality |
| S16_extracted_162833 | Not-determined |
| S16_extracted_162835 | Not-determined |
| S16_extracted_162837 | Not-determined |
| S16_extracted_162840 | Not-determined |
| S16_extracted_162852 | Not-determined |
| S16_extracted_162857 | Not-determined |
| S16_extracted_162858 | Not-determined |
| S16_extracted_162875 | Not-determined |
| S16_extracted_162877 | Not-determined |
| S16_extracted_162879 | Not-determined |
| S16_extracted_162889 | Not-determined |
| S16_extracted_162892 | Low-quality |
| S16_extracted_162917 | Not-determined |
| S16_extracted_162941 | Low-quality |
| S16_extracted_162944 | Low-quality |
| S16_extracted_162946 | Low-quality |
| S16_extracted_162963 | Not-determined |
| S16_extracted_162984 | Not-determined |
| S16_extracted_162998 | Low-quality |
| S16_extracted_163004 | Low-quality |
| S16_extracted_163005 | Not-determined |
| S16_extracted_163008 | Not-determined |
| S16_extracted_163016 | Not-determined |
| S16_extracted_163019 | Not-determined |
| S16_extracted_163034 | Low-quality |
| S16_extracted_163037 | Not-determined |
| S16_extracted_163072 | Not-determined |
| S16_extracted_163097 | Not-determined |
| S16_extracted_163111 | Not-determined |
| S16_extracted_163112 | Low-quality |
| S16_extracted_163118 | Not-determined |
| S16_extracted_163137 | Low-quality |
| S16_extracted_163144 | Not-determined |
| S16_extracted_163151 | Low-quality |
| S16_extracted_163165 | Not-determined |
| S16_extracted_163171 | Not-determined |
| S16_extracted_163180 | Not-determined |
| S16_extracted_163197 | Not-determined |
| S16_extracted_163208 | Low-quality |
| S16_extracted_163210 | Low-quality |
| S16_extracted_163211 | Not-determined |
| S16_extracted_163221 | Not-determined |
| S16_extracted_163246 | Low-quality |
| S16_extracted_163262 | Not-determined |
| S16_extracted_163269 | Not-determined |
| S16_extracted_163273 | Not-determined |
| S16_extracted_163305 | Low-quality |
| S16_extracted_163337 | Low-quality |
| S16_extracted_163362 | Not-determined |
| S16_extracted_163368 | Not-determined |
| S16_extracted_163369 | Not-determined |
| S16_extracted_163376 | Not-determined |
| S16_extracted_163377 | Not-determined |
| S16_extracted_163384 | Not-determined |
| S16_extracted_163387 | Not-determined |
| S16_extracted_163388 | Low-quality |
| S16_extracted_163394 | Not-determined |
| S16_extracted_163419 | Not-determined |
| S16_extracted_163455 | Not-determined |
| S16_extracted_163459 | Not-determined |
| S16_extracted_163463 | Not-determined |
| S16_extracted_163484 | Not-determined |
| S16_extracted_163493 | Not-determined |
| S16_extracted_163497 | Not-determined |
| S16_extracted_163504 | Not-determined |
| S16_extracted_163507 | Not-determined |
| S16_extracted_163513 | Not-determined |
| S16_extracted_163545 | Not-determined |
| S16_extracted_163548 | Not-determined |
| S16_extracted_163551 | Low-quality |
| S16_extracted_163556 | Low-quality |
| S16_extracted_163558 | Not-determined |
| S16_extracted_163560 | Not-determined |
| S16_extracted_163562 | Not-determined |
| S16_extracted_163567 | Not-determined |
| S16_extracted_163570 | Low-quality |
| S16_extracted_163580 | Not-determined |
| S16_extracted_163587 | Not-determined |
| S16_extracted_163590 | Not-determined |
| S16_extracted_163592 | Not-determined |
| S16_extracted_163602 | Not-determined |
| S16_extracted_163603 | Not-determined |
| S16_extracted_163605 | Not-determined |
| S16_extracted_163619 | Not-determined |
| S16_extracted_163628 | Not-determined |
| S16_extracted_163654 | Not-determined |
| S16_extracted_163674 | Not-determined |
| S16_extracted_163702 | Not-determined |
| S16_extracted_163704 | Not-determined |
| S16_extracted_163712 | Not-determined |
| S16_extracted_163721 | Low-quality |
| S16_extracted_163723 | Not-determined |
| S16_extracted_163729 | Not-determined |
| S16_extracted_163757 | Not-determined |
| S16_extracted_163765 | Not-determined |
| S16_extracted_163775 | Not-determined |
| S16_extracted_163785 | Not-determined |
| S16_extracted_163791 | Not-determined |
| S16_extracted_163797 | Not-determined |
| S16_extracted_163799 | Not-determined |
| S16_extracted_163820 | Not-determined |
| S16_extracted_163823 | Not-determined |
| S16_extracted_163830 | Not-determined |
| S16_extracted_163831 | Not-determined |
| S16_extracted_163835 | Not-determined |
| S16_extracted_163847 | Not-determined |
| S16_extracted_163855 | Not-determined |
| S16_extracted_163866 | Not-determined |
| S16_extracted_163902 | Not-determined |
| S16_extracted_163916 | Low-quality |
| S16_extracted_163920 | Low-quality |
| S16_extracted_163921 | Not-determined |
| S16_extracted_163934 | Not-determined |
| S16_extracted_163948 | Not-determined |
| S16_extracted_163951 | Not-determined |
| S16_extracted_163963 | Not-determined |
| S16_extracted_164015 | Not-determined |
| S16_extracted_164032 | Not-determined |
| S16_extracted_164037 | Not-determined |
| S16_extracted_164045 | Not-determined |
| S16_extracted_164068 | Not-determined |
| S16_extracted_164069 | Low-quality |
| S16_extracted_164087 | Not-determined |
| S16_extracted_164091 | Not-determined |
| S16_extracted_164099 | Not-determined |
| S16_extracted_164117 | Not-determined |
| S16_extracted_164146 | Not-determined |
| S16_extracted_164162 | Not-determined |
| S16_extracted_164167 | Not-determined |
| S16_extracted_164168 | Not-determined |
| S16_extracted_164174 | Not-determined |
| S16_extracted_164178 | Not-determined |
| S16_extracted_164182 | Not-determined |
| S16_extracted_164220 | Not-determined |
| S16_extracted_164236 | Not-determined |
| S16_extracted_164252 | Not-determined |
| S16_extracted_164260 | Not-determined |
| S16_extracted_164261 | Not-determined |
| S16_extracted_164303 | Not-determined |
| S16_extracted_164305 | Not-determined |
| S16_extracted_164316 | Not-determined |
| S16_extracted_164320 | Not-determined |
| S16_extracted_164321 | Not-determined |
| S16_extracted_164337 | Not-determined |
| S16_extracted_164347 | Not-determined |
| S16_extracted_164353 | Not-determined |
| S16_extracted_164391 | Not-determined |
| S16_extracted_164396 | Not-determined |
| S16_extracted_164409 | Not-determined |
| S16_extracted_164420 | Not-determined |
| S16_extracted_164437 | Not-determined |
| S16_extracted_164439 | Not-determined |
| S16_extracted_164450 | Not-determined |
| S16_extracted_164451 | Not-determined |
| S16_extracted_164459 | Low-quality |
| S16_extracted_164467 | Not-determined |
| S16_extracted_164495 | Not-determined |
| S16_extracted_164509 | Low-quality |
| S16_extracted_164514 | Not-determined |
| S16_extracted_164534 | Not-determined |
| S16_extracted_164553 | Not-determined |
| S16_extracted_164564 | Not-determined |
| S16_extracted_164572 | Low-quality |
| S16_extracted_164575 | Not-determined |
| S16_extracted_164579 | Low-quality |
| S16_extracted_164591 | Not-determined |
| S16_extracted_164610 | Not-determined |
| S16_extracted_164639 | Low-quality |
| S16_extracted_164641 | Not-determined |
| S16_extracted_164647 | Not-determined |
| S16_extracted_164667 | Not-determined |
| S16_extracted_164675 | Not-determined |
| S16_extracted_164689 | Not-determined |
| S16_extracted_164704 | Not-determined |
| S16_extracted_164730 | Not-determined |
| S16_extracted_164732 | Not-determined |
| S16_extracted_164750 | Not-determined |
| S16_extracted_164757 | Not-determined |
| S16_extracted_164773 | Not-determined |
| S16_extracted_164776 | Not-determined |
| S16_extracted_164778 | Not-determined |
| S16_extracted_164791 | Not-determined |
| S16_extracted_164794 | Not-determined |
| S16_extracted_164797 | Low-quality |
| S16_extracted_164799 | Not-determined |
| S16_extracted_164814 | Not-determined |
| S16_extracted_164823 | Not-determined |
| S16_extracted_164851 | Not-determined |
| S16_extracted_164887 | Not-determined |
| S16_extracted_164893 | Not-determined |
| S16_extracted_164912 | Not-determined |
| S3_vs_k141_72118\|\|full | Medium-quality |
| S16_extracted_164919 | Not-determined |
| S16_extracted_164928 | Not-determined |
| S16_extracted_164940 | Not-determined |
| S16_extracted_164941 | Not-determined |
| S16_extracted_164952 | Not-determined |
| S16_extracted_164960 | Low-quality |
| S16_extracted_164974 | Low-quality |
| S16_extracted_164988 | Not-determined |
| S16_extracted_164990 | Not-determined |
| S16_extracted_164994 | Not-determined |
| S16_extracted_164998 | Low-quality |
| S16_extracted_165020 | Not-determined |
| S16_extracted_165043 | Low-quality |
| S16_extracted_165068 | Not-determined |
| S16_extracted_165087 | Not-determined |
| S16_extracted_165090 | Not-determined |
| S16_extracted_165094 | Not-determined |
| S16_extracted_165101 | Not-determined |
| S16_extracted_165113 | Not-determined |
| S16_extracted_165126 | Low-quality |
| S16_extracted_165129 | Not-determined |
| S16_extracted_165146 | Not-determined |
| S16_extracted_165148 | Not-determined |
| S16_extracted_165170 | Not-determined |
| S16_extracted_165171 | Low-quality |
| S16_extracted_165179 | Not-determined |
| S16_extracted_165197 | Not-determined |
| S16_extracted_165210 | Not-determined |
| S16_extracted_165238 | Not-determined |
| S16_extracted_165262 | Not-determined |
| S16_extracted_165275 | Not-determined |
| S16_extracted_165276 | Not-determined |
| S16_extracted_165305 | Not-determined |
| S16_extracted_165311 | Low-quality |
| S16_extracted_165313 | Not-determined |
| S16_extracted_165322 | Not-determined |
| S16_extracted_165327 | Not-determined |
| S16_extracted_165332 | Not-determined |
| S16_extracted_165373 | Low-quality |
| S16_extracted_165389 | Not-determined |
| S16_extracted_165403 | Low-quality |
| S16_extracted_165411 | Not-determined |
| S16_extracted_165423 | Not-determined |
| S16_extracted_165431 | Low-quality |
| S16_extracted_165439 | Not-determined |
| S16_extracted_165440 | Low-quality |
| S16_extracted_165444 | Not-determined |
| S16_extracted_165452 | Not-determined |
| S16_extracted_165455 | Not-determined |
| S16_extracted_165456 | Not-determined |
| S16_extracted_165461 | Not-determined |
| S16_extracted_165463 | Not-determined |
| S16_extracted_165481 | Not-determined |
| S16_extracted_165504 | Not-determined |
| S16_extracted_165507 | Not-determined |
| S16_extracted_165517 | Low-quality |
| S16_extracted_165521 | Not-determined |
| S16_extracted_165523 | Not-determined |
| S16_extracted_165528 | Low-quality |
| S16_extracted_165545 | Not-determined |
| S16_extracted_165591 | Not-determined |
| S16_extracted_165601 | Not-determined |
| S16_extracted_165604 | Not-determined |
| S16_extracted_165607 | Not-determined |
| S16_extracted_165618 | Not-determined |
| S16_extracted_165627 | Not-determined |
| S16_extracted_165644 | Not-determined |
| S16_extracted_165651 | Low-quality |
| S16_extracted_165660 | Not-determined |
| S16_extracted_165696 | Not-determined |
| S16_extracted_165700 | Not-determined |
| S16_extracted_165705 | Not-determined |
| S16_extracted_165707 | Not-determined |
| S16_extracted_165719 | Low-quality |
| S16_extracted_165739 | Low-quality |
| S16_extracted_165751 | Low-quality |
| S16_extracted_165759 | Not-determined |
| S16_extracted_165765 | Not-determined |
| S16_extracted_165795 | Not-determined |
| S16_extracted_165810 | Not-determined |
| S16_extracted_165817 | Not-determined |
| S16_extracted_165820 | Not-determined |
| S16_extracted_165826 | Not-determined |
| S16_extracted_165827 | Not-determined |
| S16_extracted_165830 | Low-quality |
| S16_extracted_165836 | Not-determined |
| S16_extracted_165869 | Not-determined |
| S16_extracted_165870 | Not-determined |
| S16_extracted_165871 | Not-determined |
| S16_extracted_165876 | Not-determined |
| S16_extracted_165882 | Low-quality |
| S16_extracted_165883 | Not-determined |
| S16_extracted_165886 | Low-quality |
| S16_extracted_165908 | Not-determined |
| S16_extracted_165913 | Not-determined |
| S16_extracted_165934 | Not-determined |
| S16_extracted_165963 | Not-determined |
| S16_extracted_165974 | Not-determined |
| S16_extracted_165975 | Not-determined |
| S16_extracted_165992 | Not-determined |
| S16_extracted_165993 | Not-determined |
| S16_extracted_166006 | Not-determined |
| S16_extracted_166008 | Low-quality |
| S16_extracted_166021 | Not-determined |
| S16_extracted_166027 | Not-determined |
| S16_extracted_166033 | Not-determined |
| S16_extracted_166046 | Not-determined |
| S16_extracted_166055 | Not-determined |
| S16_extracted_166056 | Low-quality |
| S16_extracted_166057 | Not-determined |
| S16_extracted_166066 | Not-determined |
| S16_extracted_166092 | Not-determined |
| S16_extracted_166099 | Not-determined |
| S16_extracted_166125 | Not-determined |
| S16_extracted_166133 | Not-determined |
| S16_extracted_166134 | Low-quality |
| S16_extracted_166137 | Not-determined |
| S16_extracted_166142 | Not-determined |
| S16_extracted_166145 | Not-determined |
| S16_extracted_166154 | Not-determined |
| S16_extracted_166171 | Not-determined |
| S16_extracted_166174 | Not-determined |
| S16_extracted_166175 | Low-quality |
| S16_extracted_166180 | Not-determined |
| S16_extracted_166205 | Not-determined |
| S16_extracted_166218 | Not-determined |
| S16_extracted_166228 | Not-determined |
| S16_extracted_166234 | Not-determined |
| S16_extracted_166243 | Not-determined |
| S16_extracted_166255 | Low-quality |
| S16_extracted_166258 | Not-determined |
| S16_extracted_166259 | Low-quality |
| S16_extracted_166272 | Low-quality |
| S16_extracted_166275 | Not-determined |
| S16_extracted_166287 | Not-determined |
| S16_extracted_166289 | Not-determined |
| S16_extracted_166293 | Not-determined |
| S16_extracted_166304 | Low-quality |
| S16_extracted_166313 | Not-determined |
| S16_extracted_166316 | Not-determined |
| S16_extracted_166335 | Low-quality |
| S16_extracted_166336 | Not-determined |
| S16_extracted_166339 | Not-determined |
| S16_extracted_166348 | Low-quality |
| S16_extracted_166372 | Low-quality |
| S16_extracted_166379 | Not-determined |
| S16_extracted_166387 | Not-determined |
| S16_extracted_166390 | Not-determined |
| S16_extracted_166403 | Not-determined |
| S16_extracted_166433 | Not-determined |
| S16_extracted_166434 | Not-determined |
| S16_extracted_166438 | Not-determined |
| S16_extracted_166463 | Not-determined |
| S16_extracted_166486 | Not-determined |
| S16_extracted_166491 | Not-determined |
| S16_extracted_166493 | Not-determined |
| S16_extracted_166503 | Not-determined |
| S16_extracted_166511 | Not-determined |
| S16_extracted_166516 | Low-quality |
| S16_extracted_166522 | Not-determined |
| S16_extracted_166528 | Not-determined |
| S16_extracted_166529 | Not-determined |
| S16_extracted_166539 | Not-determined |
| S16_extracted_166549 | Low-quality |
| S16_extracted_166565 | Not-determined |
| S16_extracted_166572 | Not-determined |
| S16_extracted_166576 | Not-determined |
| S16_extracted_166579 | Not-determined |
| S16_extracted_166591 | Low-quality |
| S16_extracted_166613 | Not-determined |
| S16_extracted_166617 | Not-determined |
| S16_extracted_166623 | Not-determined |
| S16_extracted_166626 | Not-determined |
| S16_extracted_166652 | Not-determined |
| S16_extracted_166667 | Not-determined |
| S16_extracted_166677 | Not-determined |
| S16_extracted_166688 | Low-quality |
| S16_extracted_166693 | Not-determined |
| S16_extracted_166700 | Not-determined |
| S16_extracted_166701 | Low-quality |
| S16_extracted_166719 | Low-quality |
| S16_extracted_166748 | Not-determined |
| S16_extracted_166751 | Not-determined |
| S16_extracted_166755 | Not-determined |
| S16_extracted_166771 | Low-quality |
| S16_extracted_166802 | Not-determined |
| S16_extracted_166840 | Not-determined |
| S16_extracted_166842 | Not-determined |
| S16_extracted_166845 | Not-determined |
| S16_extracted_166847 | Not-determined |
| S16_extracted_166856 | Not-determined |
| S16_extracted_166863 | Not-determined |
| S16_extracted_166864 | Not-determined |
| S16_extracted_166870 | Not-determined |
| S16_extracted_166904 | Not-determined |
| S16_extracted_166905 | Not-determined |
| S16_extracted_166912 | Not-determined |
| S16_extracted_166918 | Low-quality |
| S16_extracted_166920 | Not-determined |
| S16_extracted_166923 | Not-determined |
| S16_extracted_166929 | Not-determined |
| S16_extracted_166940 | Not-determined |
| S16_extracted_166945 | Not-determined |
| S16_extracted_166967 | Low-quality |
| S16_extracted_166973 | Not-determined |
| S16_extracted_166974 | Not-determined |
| S16_extracted_167012 | Not-determined |
| S16_extracted_167027 | Not-determined |
| S16_extracted_167036 | Not-determined |

| | |
|---|---|
| S16_extracted_167044 | Not-determined |
| S16_extracted_167046 | Not-determined |
| S16_extracted_167048 | Not-determined |
| S16_extracted_167059 | Not-determined |
| S16_extracted_167062 | Not-determined |
| S16_extracted_167067 | Not-determined |
| S16_extracted_167108 | Not-determined |
| S16_extracted_167130 | Low-quality |
| S16_extracted_167132 | Not-determined |
| S16_extracted_167135 | Not-determined |
| S16_extracted_167140 | Not-determined |
| S16_extracted_167144 | Not-determined |
| S16_extracted_167158 | Not-determined |
| S16_extracted_167173 | Not-determined |
| S16_extracted_167186 | Low-quality |
| S16_extracted_167220 | Not-determined |
| S16_extracted_167226 | Not-determined |
| S16_extracted_167250 | Low-quality |
| S16_extracted_167257 | Not-determined |
| S16_extracted_167259 | Not-determined |
| S16_extracted_167270 | Low-quality |
| S16_extracted_167276 | Not-determined |
| S16_extracted_167288 | Not-determined |
| S16_extracted_167298 | Not-determined |
| S16_extracted_167304 | Not-determined |
| S16_extracted_167312 | Not-determined |
| S16_extracted_167315 | Not-determined |
| S16_extracted_167317 | Not-determined |
| S16_extracted_167321 | Not-determined |
| S16_extracted_167337 | Not-determined |
| S16_extracted_167340 | Not-determined |
| S16_extracted_167380 | Low-quality |
| S16_extracted_167411 | Not-determined |
| S16_extracted_167425 | Low-quality |
| S16_extracted_167429 | Not-determined |
| S16_extracted_167441 | Not-determined |
| S16_extracted_167450 | Not-determined |
| S16_extracted_167454 | Not-determined |
| S16_extracted_167468 | Not-determined |
| S16_extracted_167484 | Not-determined |
| S16_extracted_167494 | Not-determined |
| S16_extracted_167505 | Not-determined |
| S16_extracted_167525 | Not-determined |
| S16_extracted_167528 | Not-determined |
| S16_extracted_167529 | Not-determined |
| S16_extracted_167537 | Low-quality |
| S16_extracted_167541 | Not-determined |
| S16_extracted_167565 | Not-determined |
| S16_extracted_167576 | Not-determined |
| S16_extracted_167580 | Not-determined |
| S16_extracted_167590 | Low-quality |
| S16_extracted_167593 | Not-determined |
| S16_extracted_167624 | Not-determined |
| S16_extracted_167639 | Not-determined |
| S16_extracted_167642 | Not-determined |
| S16_extracted_167653 | Not-determined |
| S16_extracted_167666 | Not-determined |
| S16_extracted_167674 | Not-determined |
| S16_extracted_167685 | Not-determined |
| S16_extracted_167706 | Not-determined |
| S16_extracted_167715 | Not-determined |
| S16_extracted_167724 | Not-determined |
| S16_extracted_167732 | Not-determined |
| S16_extracted_167742 | Low-quality |
| S16_extracted_167750 | Low-quality |
| S16_extracted_167751 | Low-quality |
| S16_extracted_167755 | Not-determined |
| S16_extracted_167766 | Not-determined |
| S16_extracted_167782 | Not-determined |
| S16_extracted_167800 | Not-determined |
| S16_extracted_167824 | Not-determined |
| S16_extracted_167830 | Not-determined |
| S16_extracted_167832 | Not-determined |
| S16_extracted_167848 | Not-determined |
| S16_extracted_167852 | Low-quality |
| S16_extracted_167861 | Not-determined |
| S16_extracted_167887 | Low-quality |
| S16_extracted_167921 | Not-determined |
| S16_extracted_167926 | Not-determined |
| S16_extracted_167944 | Not-determined |
| S16_extracted_167964 | Low-quality |
| S16_extracted_167983 | Not-determined |
| S16_extracted_167986 | Not-determined |
| S16_extracted_167988 | Not-determined |
| S16_extracted_167992 | Not-determined |
| S16_extracted_168042 | Not-determined |
| S16_extracted_168048 | Low-quality |
| S16_extracted_168049 | Not-determined |
| S16_extracted_168092 | Not-determined |
| S16_extracted_168093 | Not-determined |
| S16_extracted_168095 | Not-determined |
| S16_extracted_168110 | Low-quality |
| S16_extracted_168116 | Not-determined |
| S16_extracted_168117 | Not-determined |
| S16_extracted_168124 | Low-quality |
| S16_extracted_168148 | Not-determined |
| S16_extracted_168157 | Not-determined |
| S16_extracted_168167 | Not-determined |
| S16_extracted_168173 | Low-quality |
| S16_extracted_168175 | Not-determined |
| S16_extracted_168191 | Not-determined |
| S16_extracted_168194 | Not-determined |
| S16_extracted_168196 | Not-determined |
| S16_extracted_168213 | Not-determined |
| S16_extracted_168216 | Not-determined |
| S16_extracted_168221 | Not-determined |
| S16_extracted_168222 | Not-determined |
| S16_extracted_168241 | Not-determined |
| S16_extracted_168243 | Not-determined |
| S16_extracted_168246 | Not-determined |
| S16_extracted_168257 | Not-determined |
| S16_extracted_168260 | Not-determined |
| S16_extracted_168270 | Not-determined |
| S16_extracted_168276 | Not-determined |
| S16_extracted_168283 | Not-determined |
| S16_extracted_168292 | Not-determined |
| S16_extracted_168302 | Not-determined |
| S16_extracted_168305 | Not-determined |
| S16_extracted_168308 | Not-determined |
| S16_extracted_168336 | Not-determined |
| S16_extracted_168346 | Not-determined |
| S16_extracted_168357 | Not-determined |
| S16_extracted_168360 | Not-determined |
| S16_extracted_168369 | Not-determined |
| S16_extracted_168379 | Not-determined |
| S16_extracted_168381 | Not-determined |
| S16_extracted_168390 | Not-determined |
| S16_extracted_168404 | Not-determined |
| S16_extracted_168418 | Not-determined |
| S16_extracted_168420 | Not-determined |
| S16_extracted_168428 | Not-determined |
| S16_extracted_168430 | Not-determined |
| S16_extracted_168439 | Low-quality |
| S16_extracted_168464 | Not-determined |
| S16_extracted_168492 | Low-quality |
| S16_extracted_168494 | Low-quality |
| S16_extracted_168527 | Not-determined |
| S16_extracted_168532 | Not-determined |
| S16_extracted_168556 | Not-determined |
| S16_extracted_168568 | Not-determined |
| S16_extracted_168605 | Not-determined |
| S16_extracted_168629 | Not-determined |
| S16_extracted_168634 | Not-determined |
| S16_extracted_168697 | Not-determined |
| S16_extracted_168698 | Not-determined |
| S16_extracted_168702 | Not-determined |
| S16_extracted_168704 | Not-determined |
| S16_extracted_168736 | Low-quality |
| S16_extracted_168754 | Low-quality |
| S16_extracted_168755 | Low-quality |
| S16_extracted_168771 | Low-quality |
| S16_extracted_168774 | Not-determined |
| S16_extracted_168814 | Low-quality |
| S16_extracted_168816 | Not-determined |
| S16_extracted_168836 | Not-determined |
| S16_extracted_168837 | Not-determined |
| S16_extracted_168864 | Not-determined |
| S16_extracted_168867 | Not-determined |
| S16_extracted_168883 | Not-determined |
| S16_extracted_168886 | Not-determined |
| S16_extracted_168893 | Low-quality |
| S16_extracted_168906 | Low-quality |
| S16_extracted_168933 | Not-determined |
| S16_extracted_168937 | Not-determined |
| S16_extracted_168955 | Low-quality |
| S16_extracted_168967 | Not-determined |
| S16_extracted_168976 | Not-determined |
| S16_extracted_168978 | Not-determined |
| S16_extracted_168982 | Low-quality |
| S16_extracted_169009 | Not-determined |
| S16_extracted_169015 | Not-determined |
| S16_extracted_169022 | Not-determined |
| S16_extracted_169027 | Not-determined |
| S16_extracted_169032 | Not-determined |
| S16_extracted_169038 | Not-determined |
| S16_extracted_169053 | Not-determined |
| S16_extracted_169054 | Not-determined |
| S16_extracted_169065 | Low-quality |
| S16_extracted_169077 | Not-determined |
| S16_extracted_169093 | Not-determined |
| S16_extracted_169094 | Not-determined |
| S16_extracted_169095 | Low-quality |
| S16_extracted_169104 | Low-quality |
| S16_extracted_169119 | Not-determined |
| S16_extracted_169134 | Not-determined |
| S16_extracted_169139 | Not-determined |
| S16_extracted_169151 | Not-determined |
| S16_extracted_169176 | Low-quality |
| S16_extracted_169180 | Not-determined |
| S16_extracted_169181 | Not-determined |
| S16_extracted_169190 | Not-determined |
| S16_extracted_169199 | Not-determined |
| S16_extracted_169204 | Not-determined |
| S16_extracted_169206 | Not-determined |
| S16_extracted_169223 | Not-determined |
| S16_extracted_169238 | Not-determined |
| S16_extracted_169241 | Not-determined |
| S16_extracted_169245 | Not-determined |
| S16_extracted_169252 | Not-determined |
| S16_extracted_169254 | Not-determined |
| S16_extracted_169259 | Not-determined |
| S16_extracted_169289 | Low-quality |
| S16_extracted_169293 | Not-determined |
| S16_extracted_169297 | Not-determined |
| S16_extracted_169309 | Not-determined |
| S16_extracted_169310 | Not-determined |
| S16_extracted_169336 | Not-determined |
| S16_extracted_169365 | Not-determined |
| S16_extracted_169375 | Not-determined |
| S16_extracted_169377 | Not-determined |
| S16_extracted_169397 | Not-determined |
| S16_extracted_169433 | Not-determined |
| S16_extracted_169435 | Not-determined |
| S16_extracted_169445 | Low-quality |
| S16_extracted_169449 | Not-determined |
| S16_extracted_169460 | Not-determined |
| S16_extracted_169467 | Not-determined |
| S16_extracted_169475 | Not-determined |
| S16_extracted_169476 | Not-determined |
| S16_extracted_169487 | Not-determined |
| S16_extracted_169502 | Not-determined |
| S16_extracted_169509 | Not-determined |
| S16_extracted_169524 | Low-quality |
| S16_extracted_169525 | Not-determined |
| S16_extracted_169526 | Low-quality |
| S16_extracted_169542 | Not-determined |
| S16_extracted_169543 | Not-determined |
| S16_extracted_169545 | Not-determined |
| S16_extracted_169555 | Not-determined |
| S16_extracted_169558 | Not-determined |
| S16_extracted_169567 | Not-determined |
| S16_extracted_169573 | Not-determined |
| S16_extracted_169596 | Not-determined |
| S16_extracted_169610 | Not-determined |
| S16_extracted_169611 | Not-determined |
| S16_extracted_169614 | Not-determined |
| S16_extracted_169623 | Not-determined |
| S16_extracted_169628 | Not-determined |
| S16_extracted_169638 | Low-quality |
| S16_extracted_169653 | Not-determined |
| S16_extracted_169683 | Not-determined |
| S16_extracted_169687 | Not-determined |
| S16_extracted_169691 | Not-determined |
| S16_extracted_169694 | Not-determined |
| S16_extracted_169725 | Not-determined |
| S16_extracted_169730 | Not-determined |
| S16_extracted_169752 | Not-determined |
| S16_extracted_169759 | Low-quality |
| S16_extracted_169768 | Not-determined |
| S16_extracted_169769 | Not-determined |
| S16_extracted_169776 | Not-determined |
| S16_extracted_169788 | Not-determined |
| S16_extracted_169789 | Not-determined |
| S16_extracted_169797 | Not-determined |
| S16_vs_173096\|\|full | High-quality |
| S16_extracted_169827 | Not-determined |
| S16_extracted_169828 | Not-determined |
| S16_extracted_169837 | Not-determined |
| S16_extracted_169857 | Not-determined |
| S16_extracted_169864 | Not-determined |
| S16_extracted_169885 | Not-determined |
| S16_extracted_169888 | Not-determined |
| S3_vs_k141_111893\|\|full | High-quality |
| S16_extracted_169899 | Low-quality |
| S16_extracted_169901 | Not-determined |
| S16_extracted_169904 | Not-determined |
| S16_extracted_169906 | Not-determined |
| S16_extracted_169907 | Low-quality |
| S16_extracted_169913 | Not-determined |
| S16_extracted_169914 | Not-determined |
| S16_extracted_169921 | Not-determined |
| S16_extracted_169929 | Not-determined |
| S16_extracted_169947 | Not-determined |
| S16_extracted_169948 | Not-determined |
| S16_extracted_169949 | Not-determined |
| S16_extracted_169961 | Not-determined |
| S16_extracted_169970 | Not-determined |
| S16_extracted_169971 | Not-determined |
| S16_extracted_169978 | Not-determined |
| S16_extracted_169980 | Not-determined |
| S16_extracted_169983 | Not-determined |
| S12_vs_k141_73926\|\|full | High-quality |
| S16_extracted_169993 | Not-determined |
| S16_extracted_170014 | Not-determined |
| S16_extracted_170016 | Low-quality |
| S16_extracted_170024 | Not-determined |
| S16_extracted_170030 | Not-determined |
| S16_extracted_170032 | Not-determined |
| S16_extracted_170044 | Not-determined |
| S16_extracted_170045 | Not-determined |
| S16_extracted_170054 | Not-determined |
| S16_extracted_170055 | Not-determined |
| S16_extracted_170060 | Not-determined |
| S16_extracted_170062 | Not-determined |
| S16_extracted_170077 | Low-quality |
| S16_extracted_170088 | Not-determined |
| S16_extracted_170092 | Not-determined |
| S16_extracted_170105 | Not-determined |
| S16_extracted_170112 | Not-determined |
| S16_extracted_170116 | Low-quality |
| S16_extracted_170141 | Not-determined |
| S16_extracted_170152 | Not-determined |
| S16_extracted_170153 | Not-determined |
| S16_extracted_170177 | Not-determined |
| S16_extracted_170185 | Not-determined |
| S16_extracted_170196 | Low-quality |
| S16_extracted_170215 | Not-determined |
| S16_extracted_170227 | Not-determined |
| S16_extracted_170264 | Not-determined |
| S16_extracted_170268 | Not-determined |
| S16_extracted_170284 | Low-quality |
| S16_extracted_170302 | Not-determined |
| S16_extracted_170305 | Not-determined |
| S16_extracted_170311 | Not-determined |
| S16_extracted_170337 | Not-determined |
| S16_extracted_170344 | Low-quality |
| S16_extracted_170347 | Not-determined |
| S16_extracted_170362 | Low-quality |
| S16_extracted_170374 | Not-determined |
| S16_extracted_170382 | Not-determined |
| S16_extracted_170395 | Not-determined |
| S16_extracted_170396 | Not-determined |
| S16_extracted_170428 | Not-determined |
| S16_extracted_170438 | Not-determined |
| S16_extracted_170443 | Not-determined |
| S16_extracted_170447 | Not-determined |
| S16_extracted_170459 | Not-determined |
| S16_extracted_170463 | Not-determined |
| S16_extracted_170477 | Not-determined |
| S16_extracted_170488 | Not-determined |
| S16_extracted_170508 | Not-determined |
| S16_extracted_170512 | Not-determined |
| S16_extracted_170519 | Not-determined |
| S16_extracted_170528 | Not-determined |
| S16_extracted_170543 | Not-determined |
| S16_extracted_170548 | Not-determined |
| S16_extracted_170556 | Not-determined |
| S16_extracted_170562 | Not-determined |
| S16_extracted_170563 | Not-determined |
| S16_extracted_170576 | Low-quality |
| S16_extracted_170592 | Not-determined |
| S16_extracted_170599 | Low-quality |
| S16_extracted_170614 | Not-determined |
| S16_extracted_170628 | Low-quality |
| S16_extracted_170641 | Not-determined |
| S16_extracted_170654 | Not-determined |
| S16_extracted_170655 | Not-determined |
| S16_extracted_170663 | Not-determined |
| S16_extracted_170669 | Not-determined |
| S16_extracted_170671 | Not-determined |
| S16_extracted_170690 | Not-determined |
| S16_extracted_170692 | Not-determined |
| S16_extracted_170709 | Not-determined |
| S16_extracted_170746 | Not-determined |
| S16_extracted_170757 | Not-determined |
| S16_extracted_170761 | Not-determined |
| S16_extracted_170772 | Not-determined |
| S16_extracted_170781 | Not-determined |
| S16_extracted_170786 | Not-determined |
| S16_extracted_170799 | Not-determined |
| S16_extracted_170803 | Low-quality |
| S16_extracted_170815 | Not-determined |
| S16_extracted_170818 | Low-quality |
| S16_extracted_170822 | Low-quality |
| S16_extracted_170827 | Not-determined |
| S16_extracted_170828 | Not-determined |
| S16_extracted_170829 | Not-determined |
| S16_extracted_170834 | Not-determined |
| S16_extracted_170846 | Not-determined |
| S16_extracted_170859 | Not-determined |
| S16_extracted_170860 | Not-determined |
| S16_extracted_170872 | Not-determined |
| S16_extracted_170882 | Low-quality |
| S16_extracted_170894 | Not-determined |
| S16_extracted_170896 | Not-determined |
| S16_extracted_170909 | Low-quality |
| S16_extracted_170922 | Not-determined |
| S16_extracted_170924 | Not-determined |
| S16_extracted_170930 | Not-determined |
| S16_extracted_170944 | Not-determined |
| S16_extracted_170956 | Not-determined |
| S16_extracted_170958 | Not-determined |
| S16_extracted_170961 | Not-determined |
| S16_extracted_170973 | Not-determined |
| S16_extracted_170974 | Low-quality |
| S16_extracted_170985 | Low-quality |
| S16_extracted_170995 | Not-determined |
| S16_extracted_170998 | Not-determined |
| S16_extracted_171001 | Not-determined |
| S16_extracted_171015 | Not-determined |
| S16_extracted_171048 | Not-determined |
| S16_extracted_171053 | Not-determined |
| S16_extracted_171061 | Not-determined |
| S16_extracted_171065 | Low-quality |
| S16_extracted_171092 | Not-determined |
| S16_extracted_171102 | Low-quality |
| S16_extracted_171127 | Not-determined |
| S16_extracted_171135 | Low-quality |
| S16_extracted_171137 | Low-quality |
| S16_extracted_171150 | Not-determined |
| S16_extracted_171176 | Not-determined |
| S16_extracted_171196 | Not-determined |
| S16_extracted_171205 | Not-determined |
| S16_extracted_171213 | Not-determined |
| S16_extracted_171238 | Not-determined |
| S16_extracted_171244 | Not-determined |
| S16_extracted_171316 | Not-determined |
| S16_extracted_171322 | Not-determined |
| S16_extracted_171348 | Not-determined |
| S16_extracted_171355 | Not-determined |
| S16_extracted_171377 | Not-determined |
| S16_extracted_171380 | Not-determined |
| S16_extracted_171388 | Not-determined |
| S16_extracted_171409 | Not-determined |
| S16_extracted_171437 | Low-quality |
| S16_extracted_171441 | Low-quality |
| S16_extracted_171443 | Not-determined |
| S16_extracted_171444 | Not-determined |
| S16_extracted_171447 | Not-determined |
| S16_extracted_171454 | Low-quality |
| S16_extracted_171460 | Not-determined |
| S16_extracted_171478 | Not-determined |
| S16_extracted_171481 | Not-determined |
| S16_extracted_171485 | Not-determined |
| S16_extracted_171489 | Not-determined |
| S16_extracted_171497 | Low-quality |
| S16_extracted_171505 | Not-determined |
| S16_extracted_171517 | Not-determined |
| S16_extracted_171520 | Not-determined |
| S16_extracted_171523 | Low-quality |
| S16_extracted_171529 | Not-determined |
| S16_extracted_171531 | Not-determined |
| S16_extracted_171535 | Not-determined |
| S16_extracted_171537 | Not-determined |
| S16_extracted_171561 | Low-quality |
| S16_extracted_171580 | Not-determined |
| S16_extracted_171583 | Not-determined |
| S16_extracted_171586 | Not-determined |
| S16_extracted_171588 | Not-determined |
| S16_extracted_171589 | Not-determined |
| S16_extracted_171610 | Not-determined |
| S16_extracted_171624 | Not-determined |
| S16_extracted_171625 | Not-determined |
| S16_extracted_171627 | Not-determined |
| S16_extracted_171635 | Not-determined |
| S16_extracted_171649 | Not-determined |
| S16_extracted_171650 | Not-determined |
| S16_extracted_171651 | Not-determined |
| S16_extracted_171654 | Not-determined |
| S16_extracted_171661 | Low-quality |
| S16_extracted_171662 | Not-determined |
| S16_extracted_171674 | Low-quality |
| S16_extracted_171677 | Not-determined |
| S16_extracted_171678 | Not-determined |
| S16_extracted_171703 | Not-determined |
| S16_extracted_171733 | Not-determined |
| S16_extracted_171735 | Not-determined |
| S16_extracted_171748 | Not-determined |
| S16_extracted_171755 | Not-determined |
| S16_extracted_171757 | Not-determined |
| S16_extracted_171760 | Not-determined |
| S16_extracted_171764 | Not-determined |
| S16_extracted_171766 | Low-quality |
| S16_extracted_171785 | Not-determined |
| S16_extracted_171787 | Not-determined |
| S16_extracted_171806 | Not-determined |
| S16_extracted_171814 | Not-determined |
| S16_extracted_171816 | Not-determined |
| S16_extracted_171829 | Not-determined |
| S16_extracted_171830 | Not-determined |
| S16_extracted_171833 | Not-determined |
| S16_extracted_171840 | Not-determined |
| S16_extracted_171845 | Not-determined |
| S16_extracted_171847 | Not-determined |
| S16_extracted_171856 | Not-determined |
| S16_extracted_171873 | Not-determined |
| S16_extracted_171902 | Not-determined |
| S16_extracted_171926 | Not-determined |
| S16_extracted_171930 | Not-determined |
| S16_extracted_171934 | Not-determined |
| S16_extracted_171936 | Not-determined |
| S16_extracted_171947 | Not-determined |
| S16_extracted_171953 | Low-quality |
| S16_extracted_171961 | Not-determined |
| S16_extracted_171977 | Not-determined |
| S16_extracted_171980 | Not-determined |
| S16_extracted_171984 | Not-determined |
| S16_extracted_172017 | Low-quality |
| S16_extracted_172018 | Not-determined |
| S16_extracted_172037 | Not-determined |
| S16_extracted_172042 | Not-determined |
| S16_extracted_172045 | Low-quality |
| S16_extracted_172049 | Not-determined |
| S16_extracted_172063 | Not-determined |
| S16_extracted_172069 | Not-determined |
| S16_extracted_172081 | Not-determined |
| S16_extracted_172086 | Not-determined |
| S16_extracted_172087 | Not-determined |
| S16_extracted_172116 | Not-determined |
| S16_extracted_172118 | Not-determined |
| S16_extracted_172133 | Not-determined |
| S16_extracted_172136 | Low-quality |
| S16_extracted_172151 | Not-determined |
| S16_extracted_172176 | Not-determined |
| S16_extracted_172181 | Not-determined |
| S16_extracted_172210 | Low-quality |
| S16_extracted_172214 | Not-determined |
| S16_extracted_172216 | Low-quality |
| S16_extracted_172218 | Not-determined |
| S16_extracted_172223 | Not-determined |
| S16_extracted_172231 | Not-determined |
| S16_extracted_172237 | Not-determined |
| S16_extracted_172247 | Not-determined |
| S16_extracted_172260 | Low-quality |
| S16_extracted_172266 | Not-determined |
| S16_extracted_172273 | Not-determined |
| S16_extracted_172294 | Not-determined |
| S16_extracted_172297 | Not-determined |
| S16_extracted_172312 | Not-determined |
| S16_extracted_172314 | Low-quality |
| S16_extracted_172322 | Not-determined |
| S16_extracted_172330 | Low-quality |
| S16_extracted_172335 | Low-quality |
| S16_extracted_172338 | Not-determined |
| S16_extracted_172340 | Not-determined |
| S16_extracted_172356 | Not-determined |
| S16_extracted_172368 | Low-quality |
| S16_extracted_172369 | Not-determined |
| S16_extracted_172400 | Not-determined |
| S16_extracted_172413 | Not-determined |
| S16_extracted_172415 | Not-determined |
| S16_extracted_172417 | Low-quality |
| S16_extracted_172419 | Not-determined |
| S16_extracted_172445 | Not-determined |
| S16_extracted_172465 | Not-determined |
| S16_extracted_172470 | Not-determined |
| S16_extracted_172486 | Not-determined |
| S16_extracted_172487 | Not-determined |
| S16_extracted_172489 | Not-determined |
| S16_extracted_172497 | Not-determined |
| S16_extracted_172506 | Not-determined |
| S16_extracted_172509 | Low-quality |
| S16_extracted_172515 | Not-determined |
| S16_extracted_172522 | Not-determined |
| S16_extracted_172544 | Not-determined |
| S16_extracted_172545 | Low-quality |
| S16_extracted_172546 | Not-determined |
| S16_extracted_172550 | Not-determined |
| S16_extracted_172552 | Not-determined |
| S16_extracted_172560 | Not-determined |
| S16_extracted_172568 | Not-determined |
| S16_extracted_172570 | Not-determined |
| S16_extracted_172571 | Not-determined |
| S16_extracted_172577 | Not-determined |
| S16_extracted_172583 | Not-determined |
| S16_extracted_172588 | Not-determined |
| S16_extracted_172599 | Not-determined |
| S16_extracted_172612 | Not-determined |
| S16_extracted_172618 | Not-determined |
| S16_extracted_172624 | Not-determined |
| S16_extracted_172625 | Low-quality |
| S16_extracted_172633 | Not-determined |
| S16_extracted_172634 | Low-quality |
| S16_extracted_172644 | Not-determined |
| S16_extracted_172645 | Not-determined |
| S16_extracted_172649 | Not-determined |
| S16_extracted_172650 | Not-determined |
| S16_extracted_172653 | Not-determined |
| S16_extracted_172657 | Not-determined |
| S16_extracted_172696 | Not-determined |
| S16_extracted_172705 | Not-determined |
| S16_extracted_172715 | Not-determined |
| S16_extracted_172729 | Not-determined |
| S16_extracted_172735 | Not-determined |
| S16_extracted_172737 | Low-quality |
| S16_extracted_172748 | Not-determined |
| S16_extracted_172768 | Not-determined |
| S16_extracted_172776 | Not-determined |
| S16_extracted_172780 | Not-determined |
| S16_extracted_172782 | Not-determined |
| S16_extracted_172785 | Not-determined |
| S16_extracted_172791 | Not-determined |
| S16_extracted_172816 | Low-quality |
| S16_extracted_172823 | Low-quality |
| S16_extracted_172825 | Not-determined |
| S16_extracted_172830 | Not-determined |
| S16_extracted_172836 | Low-quality |
| S16_extracted_172846 | Not-determined |
| S16_extracted_172870 | Not-determined |

| | |
|---|---|
| S16_extracted_172877 | Not-determined |
| S16_extracted_172891 | Not-determined |
| S16_extracted_172892 | Not-determined |
| S16_extracted_172895 | Low-quality |
| S17_vs_k141_121041\|\|full | Medium-quality |
| S16_extracted_172902 | Low-quality |
| S16_extracted_172925 | Not-determined |
| S16_extracted_172927 | Not-determined |
| S8_vs_k141_147338\|\|full | Medium-quality |
| S16_extracted_172967 | Not-determined |
| S16_extracted_172971 | Not-determined |
| S16_extracted_172973 | Not-determined |
| S16_extracted_172975 | Not-determined |
| S16_extracted_172981 | Not-determined |
| S16_extracted_172997 | Not-determined |
| S16_extracted_172999 | Low-quality |
| S16_extracted_173000 | Not-determined |
| S16_extracted_173005 | Low-quality |
| S16_extracted_173012 | Not-determined |
| S16_extracted_173026 | Not-determined |
| S16_extracted_173051 | Not-determined |
| S16_extracted_173061 | Low-quality |
| S16_extracted_173074 | Not-determined |
| S16_extracted_173086 | Not-determined |
| S16_extracted_173092 | Low-quality |
| S11_vs_k141_104038\|\|full | Medium-quality |
| S14_extracted_k141_92240 | High-quality |
| S16_extracted_173101 | Not-determined |
| S16_extracted_173103 | Not-determined |
| S14_vs_k141_92240\|\|full | High-quality |
| S16_extracted_173107 | Not-determined |
| S16_extracted_173108 | Low-quality |
| S9_vs_k141_5676\|\|full | Complete |
| S16_extracted_173114 | Not-determined |
| S14_vs_k141_3251\|\|full | High-quality |
| S16_extracted_173139 | Not-determined |
| S16_extracted_173142 | Not-determined |
| S16_extracted_173150 | Not-determined |
| S16_extracted_173155 | Not-determined |
| S16_extracted_173162 | Not-determined |
| S16_extracted_173163 | Not-determined |
| S16_extracted_173164 | Not-determined |
| S16_extracted_173174 | Not-determined |
| S16_extracted_173177 | Low-quality |
| S16_extracted_173183 | Not-determined |
| S16_extracted_173184 | Low-quality |
| S16_extracted_173187 | Not-determined |
| S16_extracted_173191 | Not-determined |
| S16_extracted_173196 | Not-determined |
| S16_extracted_173197 | Not-determined |
| S16_extracted_173199 | Not-determined |
| S16_extracted_173201 | Not-determined |
| S16_extracted_173205 | Not-determined |
| S16_extracted_173206 | Not-determined |
| S16_extracted_173209 | Not-determined |
| S16_extracted_173210 | Not-determined |
| S16_extracted_173212 | Not-determined |
| S16_extracted_173213 | Not-determined |
| S16_extracted_173218 | Not-determined |
| S16_extracted_173220 | Not-determined |
| S16_extracted_173225 | Not-determined |
| S16_extracted_173228 | Not-determined |
| S16_extracted_173229 | Not-determined |
| S16_extracted_173235 | Not-determined |
| S16_extracted_173237 | Not-determined |
| S7_extracted_27309 | Complete |
| S16_vs_2044\|\|full | Not-determined |
| S3_vs_k141_98675\|\|full | High-quality |
| S16_vs_8916\|\|full | Not-determined |
| S16_vs_13362\|\|full | Not-determined |
| S5_vs_k141_124251\|\|full | High-quality |
| S11_vs_k141_98193\|\|full | Medium-quality |
| S13_vs_13129\|\|full | High-quality |
| S16_vs_25826\|\|full | Not-determined |
| S16_vs_26408\|\|full | Low-quality |
| S1_vs_k141_73320\|\|full | High-quality |
| S9_extracted_k141_79919 | Complete |
| S7_vs_111831\|\|full | High-quality |
| S7_vs_43529\|\|full | Medium-quality |
| S14_vs_k141_30439\|\|full | High-quality |
| S3_vs_k141_28402\|\|full | High-quality |
| S10_vs_120489\|\|full | High-quality |
| S7_vs_27309\|\|full | High-quality |
| S16_vs_103726\|\|full | High-quality |
| S9_vs_k141_79919\|\|full | High-quality |
| S16_vs_128557\|\|full | Not-determined |
| S16_vs_139034\|\|full | Not-determined |
| S7_vs_122151\|\|full | High-quality |
| S8_vs_k141_154231\|\|full | High-quality |
| S8_vs_k141_40014\|\|full | High-quality |
| S7_vs_86377\|\|full | High-quality |
| S4_vs_VIRSorter_16803-cat_2 | High-quality |
| S9_vs_k141_48240\|\|full | Medium-quality |
| S1_vs_k141_117317\|\|full | Medium-quality |
| S7_extracted_67910 | High-quality |
| S6_vs_k141_56498\|\|full | High-quality |
| S14_vs_k141_110290\|\|full | High-quality |
| S3_vs_k141_90341\|\|full | High-quality |
| S7_vs_67910\|\|full | High-quality |
| S16_vs_116956\|\|full | Medium-quality |
| S3_vs_k141_86593\|\|full | Medium-quality |
| S5_vs_k141_67246\|\|full | High-quality |
| S16_vs_173113\|\|full | Not-determined |
| S16_vs_173114\|\|full | Not-determined |
| S16_vs_173117\|\|full | Not-determined |
| S16_vs_173119\|\|full | Low-quality |
| S16_vs_173128\|\|full | Not-determined |
| S7_vs_72737\|\|0_partial | High-quality |
| S16_vs_173136\|\|full | Low-quality |
| S5_vs_k141_111650\|\|full | High-quality |
| S9_extracted_k141_4115 | Medium-quality |
| S16_vs_173146\|\|full | Not-determined |
| S16_vs_68\|\|full | Low-quality |
| S16_vs_97\|\|full | Not-determined |
| S16_vs_103\|\|full | Low-quality |
| S16_vs_155\|\|full | Low-quality |
| S16_vs_178\|\|full | Low-quality |
| S16_vs_210\|\|full | Low-quality |
| S16_vs_241\|\|full | Not-determined |
| S16_vs_250\|\|full | Low-quality |
| S16_vs_256\|\|full | Low-quality |
| S16_vs_276\|\|full | Not-determined |
| S16_vs_312\|\|full | Not-determined |
| S16_vs_434\|\|full | Not-determined |
| S16_vs_438\|\|full | Not-determined |
| S16_vs_440\|\|full | Low-quality |
| S16_vs_464\|\|full | Low-quality |
| S16_vs_489\|\|full | Low-quality |
| S16_vs_497\|\|full | Not-determined |
| S16_vs_506\|\|full | Low-quality |
| S16_vs_507\|\|full | Low-quality |
| S16_vs_528\|\|full | Low-quality |
| S16_vs_583\|\|full | Low-quality |
| S16_vs_588\|\|full | Not-determined |
| S16_vs_591\|\|full | Low-quality |
| S16_vs_607\|\|full | Low-quality |
| S16_vs_611\|\|full | Not-determined |
| S16_vs_629\|\|full | Not-determined |
| S16_vs_640\|\|full | Low-quality |
| S16_vs_680\|\|full | Low-quality |
| S16_vs_694\|\|full | Low-quality |
| S16_vs_706\|\|full | Not-determined |
| S16_vs_726\|\|full | Low-quality |
| S16_vs_748\|\|full | Not-determined |
| S16_vs_754\|\|full | Not-determined |
| S16_vs_780\|\|full | Not-determined |
| S16_vs_786\|\|full | Not-determined |
| S16_vs_798\|\|full | Low-quality |
| S16_vs_825\|\|full | Low-quality |
| S16_vs_838\|\|full | Not-determined |
| S16_vs_846\|\|full | Not-determined |
| S16_vs_847\|\|full | Not-determined |
| S16_vs_877\|\|full | Not-determined |
| S16_vs_924\|\|full | Low-quality |
| S16_vs_959\|\|full | Low-quality |
| S16_vs_988\|\|full | Not-determined |
| S16_vs_990\|\|full | Not-determined |
| S16_vs_1025\|\|full | Not-determined |
| S16_vs_1032\|\|full | Not-determined |
| S16_vs_1044\|\|full | Low-quality |
| S16_vs_1064\|\|full | Not-determined |
| S16_vs_1087\|\|full | Low-quality |
| S16_vs_1094\|\|full | Low-quality |
| S16_vs_1100\|\|full | Not-determined |
| S16_vs_1105\|\|full | Low-quality |
| S16_vs_1124\|\|full | Low-quality |
| S16_vs_1134\|\|full | Low-quality |
| S16_vs_1203\|\|full | Not-determined |
| S16_vs_1260\|\|full | Not-determined |
| S16_vs_1321\|\|full | Not-determined |
| S16_vs_1328\|\|full | Low-quality |
| S16_vs_1355\|\|full | Not-determined |
| S16_vs_1365\|\|full | Low-quality |
| S16_vs_1368\|\|full | Not-determined |
| S16_vs_1370\|\|full | Low-quality |
| S16_vs_1386\|\|full | Not-determined |
| S16_vs_1402\|\|full | Not-determined |
| S16_vs_1407\|\|full | Not-determined |
| S16_vs_1434\|\|full | Not-determined |
| S16_vs_1452\|\|full | Low-quality |
| S16_vs_1473\|\|full | Not-determined |
| S16_vs_1476\|\|full | Low-quality |
| S16_vs_1496\|\|full | Not-determined |
| S16_vs_1506\|\|full | Low-quality |
| S16_vs_1527\|\|full | Low-quality |
| S16_vs_1544\|\|full | Low-quality |
| S16_vs_1554\|\|full | Not-determined |
| S16_vs_1574\|\|full | Low-quality |
| S16_vs_1603\|\|full | Not-determined |
| S16_vs_1626\|\|full | Not-determined |
| S16_vs_1634\|\|full | Not-determined |
| S16_vs_1715\|\|full | Low-quality |
| S16_vs_1738\|\|full | Low-quality |
| S16_vs_1763\|\|full | Not-determined |
| S16_vs_1773\|\|full | Low-quality |
| S16_vs_1780\|\|full | Low-quality |
| S16_vs_1801\|\|full | Low-quality |
| S16_vs_1844\|\|full | Low-quality |
| S16_vs_1853\|\|full | Low-quality |
| S16_vs_1858\|\|full | Low-quality |
| S16_vs_1926\|\|full | Low-quality |
| S16_vs_1954\|\|full | Not-determined |
| S16_vs_1993\|\|full | Low-quality |
| S16_vs_1994\|\|full | Low-quality |
| S16_vs_2017\|\|full | Low-quality |
| S16_vs_2031\|\|full | Not-determined |
| S16_vs_2049\|\|full | Low-quality |
| S16_vs_2073\|\|full | Low-quality |
| S16_vs_2107\|\|full | Not-determined |
| S16_vs_2121\|\|full | Low-quality |
| S16_vs_2148\|\|full | Not-determined |
| S16_vs_2205\|\|full | Not-determined |
| S16_vs_2220\|\|full | Not-determined |
| S16_vs_2248\|\|full | Low-quality |
| S16_vs_2273\|\|full | Low-quality |
| S16_vs_2308\|\|full | Not-determined |
| S16_vs_2323\|\|full | Low-quality |
| S16_vs_2336\|\|full | Low-quality |
| S16_vs_2359\|\|full | Low-quality |
| S16_vs_2449\|\|full | Low-quality |
| S16_vs_2465\|\|full | Low-quality |
| S16_vs_2488\|\|full | Not-determined |
| S16_vs_2499\|\|full | Low-quality |
| S16_vs_2521\|\|full | Not-determined |
| S16_vs_2540\|\|full | Low-quality |
| S16_vs_2564\|\|full | Not-determined |
| S16_vs_2578\|\|full | Not-determined |
| S16_vs_2588\|\|full | Not-determined |
| S16_vs_2645\|\|full | Low-quality |
| S16_vs_2647\|\|full | Low-quality |
| S16_vs_2669\|\|full | Low-quality |
| S16_vs_2719\|\|full | Low-quality |
| S16_vs_2736\|\|full | Low-quality |
| S16_vs_2759\|\|full | Low-quality |
| S16_vs_2763\|\|full | Low-quality |
| S16_vs_2770\|\|full | Not-determined |
| S16_vs_2788\|\|full | Low-quality |
| S16_vs_2800\|\|full | Not-determined |
| S16_vs_2811\|\|full | Not-determined |
| S16_vs_2844\|\|full | Low-quality |
| S16_vs_2864\|\|full | Low-quality |
| S16_vs_2899\|\|full | Low-quality |
| S16_vs_2929\|\|full | Not-determined |
| S16_vs_2967\|\|full | Not-determined |
| S16_vs_2997\|\|full | Low-quality |
| S16_vs_3040\|\|full | Low-quality |
| S16_vs_3041\|\|full | Low-quality |
| S16_vs_3046\|\|full | Not-determined |
| S16_vs_3058\|\|full | Not-determined |
| S16_vs_3091\|\|full | Not-determined |
| S16_vs_3111\|\|full | Low-quality |
| S16_vs_3138\|\|full | Not-determined |
| S16_vs_3193\|\|full | Low-quality |
| S16_vs_3253\|\|full | Low-quality |
| S16_vs_3330\|\|full | Not-determined |
| S16_vs_3342\|\|full | Low-quality |
| S16_vs_3348\|\|full | Low-quality |
| S16_vs_3383\|\|full | Low-quality |
| S16_vs_3386\|\|full | Low-quality |
| S16_vs_3436\|\|full | Low-quality |
| S16_vs_3440\|\|full | Low-quality |
| S16_vs_3638\|\|full | Low-quality |
| S16_vs_3677\|\|full | Low-quality |
| S16_vs_3706\|\|full | Low-quality |
| S16_vs_3734\|\|full | Low-quality |
| S16_vs_3777\|\|full | Low-quality |
| S16_vs_3806\|\|full | Low-quality |
| S16_vs_3841\|\|full | Low-quality |
| S16_vs_3856\|\|full | Not-determined |
| S16_vs_3872\|\|full | Not-determined |
| S16_vs_3942\|\|full | Low-quality |
| S16_vs_3965\|\|full | Not-determined |
| S16_vs_4004\|\|full | Not-determined |
| S16_vs_4049\|\|full | Low-quality |
| S16_vs_4059\|\|full | Low-quality |
| S16_vs_4147\|\|full | Not-determined |
| S16_vs_4161\|\|full | Low-quality |
| S16_vs_4176\|\|full | Low-quality |
| S16_vs_4203\|\|full | Low-quality |
| S16_vs_4218\|\|full | Low-quality |
| S16_vs_4225\|\|full | Low-quality |
| S16_vs_4229\|\|full | Not-determined |
| S16_vs_4247\|\|full | Low-quality |
| S16_vs_4261\|\|full | Not-determined |
| S16_vs_4282\|\|full | Not-determined |
| S16_vs_4284\|\|full | Low-quality |
| S16_vs_4310\|\|full | Low-quality |
| S16_vs_4461\|\|full | Not-determined |
| S16_vs_4477\|\|full | Low-quality |
| S16_vs_4504\|\|full | Low-quality |
| S16_vs_4548\|\|full | Not-determined |
| S16_vs_4558\|\|full | Not-determined |
| S16_vs_4588\|\|full | Not-determined |
| S16_vs_4604\|\|full | Not-determined |
| S7_vs_28150\|\|full | High-quality |
| S16_vs_4685\|\|full | Low-quality |
| S16_vs_4733\|\|full | Not-determined |
| S16_vs_4791\|\|full | Low-quality |
| S16_vs_4801\|\|full | Not-determined |
| S16_vs_4853\|\|full | Not-determined |
| S16_vs_4860\|\|full | Not-determined |
| S16_vs_4895\|\|full | Low-quality |
| S16_vs_4976\|\|full | Not-determined |
| S16_vs_5012\|\|full | Not-determined |
| S16_vs_5036\|\|full | Low-quality |
| S16_vs_5067\|\|full | Not-determined |
| S16_vs_5076\|\|full | Low-quality |
| S16_vs_5087\|\|full | Not-determined |
| S16_vs_5088\|\|full | Not-determined |
| S16_vs_5204\|\|full | Low-quality |
| S16_vs_5250\|\|full | Low-quality |
| S16_vs_5258\|\|full | Not-determined |
| S16_vs_5261\|\|full | Not-determined |
| S16_vs_5266\|\|full | Low-quality |
| S16_vs_124703\|\|full | Medium-quality |
| S16_vs_5293\|\|full | Low-quality |
| S16_vs_5353\|\|full | Low-quality |
| S16_vs_5379\|\|full | Low-quality |
| S16_vs_5411\|\|full | Low-quality |
| S16_vs_5441\|\|full | Low-quality |
| S16_vs_5444\|\|full | Not-determined |
| S16_vs_5476\|\|full | Not-determined |
| S16_vs_5524\|\|full | Low-quality |
| S16_vs_5544\|\|full | Not-determined |
| S16_vs_5716\|\|full | Low-quality |
| S16_vs_5726\|\|full | Low-quality |
| S16_vs_5737\|\|full | Low-quality |
| S16_vs_5784\|\|full | Low-quality |
| S16_vs_5786\|\|full | Low-quality |
| S16_vs_5804\|\|full | Not-determined |
| S16_vs_5808\|\|full | Not-determined |
| S3_vs_k141_114043\|\|full | High-quality |
| S16_vs_5836\|\|full | Low-quality |
| S16_vs_5849\|\|full | Not-determined |
| S16_vs_5854\|\|full | Low-quality |
| S16_vs_5925\|\|full | Not-determined |
| S16_vs_5952\|\|full | Low-quality |
| S16_vs_5956\|\|full | Low-quality |
| S16_vs_6050\|\|full | Not-determined |
| S11_vs_k141_104047\|\|full | Medium-quality |
| S16_vs_6144\|\|full | Low-quality |
| S16_vs_6151\|\|full | Low-quality |
| S16_vs_6158\|\|full | Not-determined |
| S16_vs_6181\|\|full | Low-quality |
| S16_vs_6222\|\|full | Not-determined |
| S16_vs_6261\|\|full | Low-quality |
| S16_vs_6352\|\|full | Low-quality |
| S16_vs_6393\|\|full | Not-determined |
| S16_vs_6415\|\|full | Low-quality |
| S16_vs_6438\|\|full | Low-quality |
| S16_vs_6442\|\|full | Low-quality |
| S16_vs_6459\|\|full | Low-quality |
| S16_vs_6475\|\|full | Low-quality |
| S16_vs_6514\|\|full | Low-quality |
| S16_vs_6526\|\|full | Not-determined |
| S16_vs_6539\|\|full | Low-quality |
| S16_vs_6545\|\|full | Low-quality |
| S16_vs_6556\|\|full | Low-quality |
| S16_vs_6585\|\|full | Not-determined |
| S16_vs_6601\|\|full | Not-determined |
| S16_vs_6605\|\|full | Not-determined |
| S16_vs_6608\|\|full | Not-determined |
| S16_vs_6618\|\|full | Not-determined |
| S16_vs_6657\|\|full | Low-quality |
| S16_vs_6659\|\|full | Low-quality |
| S16_vs_6675\|\|full | Low-quality |
| S16_vs_6768\|\|full | Not-determined |
| S16_vs_6792\|\|full | Low-quality |
| S16_vs_6882\|\|full | Not-determined |
| S16_vs_6889\|\|full | Not-determined |
| S16_vs_6898\|\|full | Not-determined |
| S16_vs_6905\|\|full | Not-determined |
| S16_vs_6913\|\|full | Not-determined |
| S16_vs_6953\|\|full | Low-quality |
| S16_vs_6965\|\|full | Not-determined |
| S16_vs_6977\|\|full | Not-determined |
| S16_vs_6990\|\|full | Low-quality |
| S16_vs_7040\|\|full | Not-determined |
| S16_vs_7050\|\|full | Not-determined |
| S16_vs_7056\|\|full | Low-quality |
| S16_vs_7084\|\|full | Low-quality |
| S16_vs_7091\|\|full | Not-determined |
| S16_vs_7094\|\|full | Not-determined |
| S16_vs_7203\|\|full | Low-quality |
| S16_vs_7257\|\|full | Low-quality |
| S16_vs_7271\|\|full | Low-quality |
| S16_vs_7379\|\|full | Not-determined |
| S16_vs_7501\|\|full | Not-determined |
| S16_vs_7566\|\|full | Not-determined |
| S16_vs_7569\|\|full | Not-determined |
| S16_vs_7582\|\|full | Not-determined |
| S16_vs_7583\|\|full | Not-determined |
| S14_extracted_k141_51134 | Complete |
| S16_vs_7656\|\|full | Not-determined |
| S16_vs_7750\|\|full | Low-quality |
| S16_vs_7756\|\|full | Not-determined |
| S16_vs_7759\|\|full | Low-quality |
| S16_vs_7763\|\|full | Not-determined |
| S16_vs_7813\|\|full | Low-quality |
| S16_vs_7817\|\|full | Low-quality |
| S16_vs_7832\|\|full | Low-quality |
| S16_vs_7865\|\|full | Low-quality |
| S16_vs_7871\|\|full | Not-determined |
| S16_vs_7873\|\|full | Low-quality |
| S16_vs_7893\|\|full | Low-quality |
| S16_vs_7944\|\|full | Low-quality |
| S16_vs_7965\|\|full | Not-determined |
| S16_vs_7987\|\|full | Not-determined |
| S16_vs_8018\|\|full | Low-quality |
| S16_vs_8048\|\|full | Low-quality |
| S16_vs_8061\|\|full | Low-quality |
| S16_vs_8107\|\|full | Not-determined |
| S16_vs_8144\|\|full | Low-quality |
| S16_vs_8146\|\|full | Low-quality |
| S16_vs_8173\|\|full | Low-quality |
| S16_vs_8209\|\|full | Low-quality |
| S16_vs_8265\|\|full | Low-quality |
| S16_vs_8304\|\|full | Low-quality |
| S16_vs_8372\|\|full | Not-determined |
| S16_vs_8406\|\|full | Low-quality |
| S16_vs_8412\|\|full | Not-determined |
| S16_vs_8413\|\|full | Not-determined |
| S16_vs_8417\|\|full | Not-determined |
| S16_vs_8431\|\|full | Low-quality |
| S16_vs_8436\|\|full | Not-determined |
| S16_vs_8470\|\|full | Low-quality |
| S16_vs_8485\|\|full | Low-quality |
| S9_vs_k141_88847\|\|full | Medium-quality |
| S16_vs_8497\|\|full | Low-quality |
| S16_vs_8502\|\|full | Low-quality |
| S16_vs_8505\|\|full | Low-quality |
| S16_vs_8515\|\|full | Low-quality |
| S16_vs_8545\|\|full | Low-quality |
| S16_vs_8585\|\|full | Not-determined |
| S16_vs_8589\|\|full | Low-quality |
| S16_vs_8596\|\|full | Low-quality |
| S16_vs_8610\|\|full | Low-quality |
| S16_vs_8619\|\|full | Low-quality |
| S16_vs_8647\|\|full | Low-quality |
| S16_vs_8659\|\|full | Low-quality |
| S16_vs_8663\|\|full | Low-quality |
| S16_vs_8685\|\|full | Not-determined |
| S16_vs_8701\|\|full | Not-determined |
| S16_vs_8727\|\|full | Low-quality |
| S16_vs_8775\|\|full | Not-determined |
| S16_vs_8799\|\|full | Low-quality |
| S16_vs_8833\|\|full | Not-determined |
| S16_vs_8845\|\|full | Low-quality |
| S16_vs_8929\|\|full | Low-quality |
| S16_vs_9027\|\|full | Low-quality |
| S16_vs_9052\|\|full | Not-determined |
| S16_vs_9092\|\|full | Low-quality |
| S16_vs_9128\|\|full | Not-determined |
| S16_vs_9176\|\|full | Low-quality |
| S16_vs_9193\|\|full | Not-determined |
| S16_vs_9209\|\|full | Low-quality |
| S16_vs_9215\|\|full | Low-quality |
| S16_vs_9218\|\|full | Low-quality |
| S16_vs_9219\|\|full | Low-quality |
| S16_vs_9234\|\|full | Low-quality |
| S16_vs_9248\|\|full | Low-quality |
| S16_vs_9258\|\|full | Low-quality |
| S16_vs_9269\|\|full | Not-determined |
| S3_vs_k141_73361\|\|full | High-quality |
| S16_vs_9321\|\|full | Not-determined |
| S16_vs_9347\|\|full | Low-quality |
| S16_vs_9385\|\|full | Low-quality |
| S16_vs_9429\|\|full | Not-determined |
| S16_vs_9525\|\|full | Low-quality |
| S16_vs_9530\|\|full | Not-determined |
| S16_vs_9560\|\|full | Low-quality |
| S16_vs_9606\|\|full | Low-quality |
| S16_vs_9703\|\|full | Not-determined |
| S16_vs_9722\|\|full | Not-determined |
| S16_vs_9727\|\|full | Low-quality |
| S16_vs_9774\|\|full | Not-determined |
| S16_vs_9864\|\|full | Low-quality |
| S16_vs_9866\|\|full | Low-quality |
| S16_vs_9869\|\|full | Not-determined |
| S16_vs_9906\|\|full | Not-determined |
| S16_vs_9913\|\|full | Not-determined |
| S16_vs_9917\|\|full | Low-quality |
| S16_vs_9918\|\|full | Not-determined |
| S16_vs_9925\|\|full | Low-quality |
| S16_vs_9936\|\|full | Low-quality |
| S16_vs_9981\|\|full | Not-determined |
| S16_vs_10002\|\|full | Low-quality |
| S16_vs_10021\|\|full | Low-quality |
| S16_vs_10025\|\|full | Low-quality |
| S16_vs_10028\|\|full | Low-quality |
| S16_vs_10036\|\|full | Low-quality |
| S16_vs_10082\|\|full | Low-quality |
| S16_vs_10095\|\|full | Low-quality |
| S16_vs_10123\|\|full | Not-determined |
| S16_vs_10146\|\|full | Not-determined |
| S16_vs_10147\|\|full | Not-determined |
| S16_vs_10205\|\|full | Low-quality |
| S16_vs_10249\|\|full | Low-quality |
| S16_vs_10252\|\|full | Not-determined |
| S16_vs_10277\|\|full | Not-determined |
| S16_vs_10307\|\|full | Low-quality |
| S16_vs_10311\|\|full | Low-quality |
| S16_vs_10318\|\|full | Low-quality |
| S16_vs_10330\|\|full | Low-quality |
| S16_vs_10333\|\|full | Not-determined |
| S16_vs_10432\|\|full | Not-determined |
| S16_vs_10492\|\|full | Not-determined |
| S16_vs_10496\|\|full | Low-quality |
| S16_vs_10506\|\|full | Not-determined |
| S16_vs_10527\|\|full | Low-quality |
| S16_vs_10556\|\|full | Low-quality |
| S16_vs_10569\|\|full | Not-determined |
| S5_vs_k141_49011\|\|full | High-quality |
| S16_vs_10626\|\|full | Not-determined |
| S16_vs_10634\|\|full | Low-quality |
| S16_vs_10688\|\|full | Not-determined |
| S16_vs_10748\|\|full | Low-quality |
| S16_vs_10810\|\|full | Low-quality |
| S16_vs_10830\|\|full | Not-determined |
| S16_vs_10833\|\|full | Low-quality |
| S16_vs_10885\|\|full | Low-quality |
| S16_vs_10896\|\|full | Low-quality |
| S16_vs_10924\|\|full | Low-quality |
| S16_vs_10925\|\|full | Low-quality |
| S16_vs_10938\|\|full | Low-quality |
| S16_vs_10984\|\|full | Low-quality |
| S16_vs_11017\|\|full | Low-quality |
| S16_vs_11023\|\|full | Not-determined |
| S16_vs_11043\|\|full | Low-quality |
| S16_vs_11046\|\|full | Low-quality |
| S16_vs_11051\|\|full | Low-quality |
| S16_vs_11075\|\|full | Not-determined |
| S16_vs_11111\|\|full | Not-determined |
| S16_vs_11114\|\|full | Not-determined |
| S16_vs_11165\|\|full | Not-determined |
| S16_vs_11222\|\|full | Low-quality |
| S16_vs_11225\|\|full | Low-quality |
| S16_vs_11244\|\|full | Not-determined |
| S16_vs_11259\|\|full | Low-quality |
| S16_vs_11294\|\|full | Not-determined |
| S16_vs_11338\|\|full | Low-quality |
| S16_vs_11342\|\|full | Low-quality |
| S16_vs_11388\|\|full | Low-quality |
| S16_vs_11401\|\|full | Low-quality |
| S16_vs_11457\|\|full | Low-quality |
| S16_vs_11470\|\|full | Low-quality |
| S16_vs_11514\|\|full | Not-determined |
| S16_vs_11557\|\|full | Not-determined |
| S16_vs_11571\|\|full | Low-quality |
| S16_vs_11583\|\|full | Low-quality |
| S16_vs_11619\|\|full | Low-quality |
| S16_vs_11647\|\|full | Low-quality |
| S5_vs_k141_57511\|\|full | Medium-quality |
| S16_vs_11662\|\|full | Not-determined |
| S16_vs_11667\|\|full | Low-quality |
| S16_vs_11668\|\|full | Low-quality |
| S16_vs_11702\|\|full | Low-quality |
| S16_vs_11747\|\|full | Not-determined |
| S16_vs_11769\|\|full | Low-quality |
| S16_vs_11791\|\|full | Low-quality |
| S16_vs_11869\|\|full | Low-quality |
| S16_vs_11885\|\|full | Not-determined |
| S16_vs_11899\|\|full | Low-quality |
| S16_vs_11902\|\|full | Low-quality |
| S16_vs_11996\|\|full | Low-quality |
| S16_vs_12005\|\|full | Not-determined |
| S16_vs_12037\|\|full | Not-determined |
| S16_vs_12080\|\|full | Low-quality |
| S16_vs_12136\|\|full | Not-determined |
| S16_vs_12141\|\|full | Not-determined |
| S16_vs_12145\|\|full | Not-determined |
| S16_vs_12150\|\|full | Low-quality |
| S16_vs_12165\|\|full | Low-quality |
| S16_vs_12167\|\|full | Not-determined |
| S16_vs_12175\|\|full | Not-determined |
| S16_vs_12199\|\|full | Low-quality |
| S16_vs_12228\|\|full | Not-determined |
| S16_vs_12232\|\|full | Not-determined |
| S16_vs_12247\|\|full | Low-quality |
| S16_vs_12254\|\|full | Low-quality |
| S16_vs_12331\|\|full | Low-quality |
| S16_vs_12353\|\|full | Low-quality |
| S16_vs_12412\|\|full | Low-quality |
| S16_vs_12415\|\|full | Low-quality |
| S16_vs_12472\|\|full | Not-determined |
| S16_vs_12477\|\|full | Not-determined |
| S16_vs_12536\|\|full | Low-quality |
| S6_vs_k141_5141\|\|full | Medium-quality |
| S16_vs_12615\|\|full | Low-quality |

| | |
|---|---|
| S16_vs_12620\|\|full | Not-determined |
| S16_vs_12638\|\|full | Low-quality |
| S16_vs_12669\|\|full | Low-quality |
| S16_vs_12716\|\|full | Not-determined |
| S16_vs_12730\|\|full | Low-quality |
| S16_vs_12736\|\|full | Not-determined |
| S16_vs_12741\|\|full | Not-determined |
| S16_vs_12768\|\|full | Low-quality |
| S16_vs_12793\|\|full | Not-determined |
| S16_vs_12805\|\|full | Not-determined |
| S16_vs_12812\|\|full | Low-quality |
| S16_vs_12863\|\|full | Not-determined |
| S16_vs_12881\|\|full | Not-determined |
| S16_vs_12884\|\|full | Not-determined |
| S16_vs_12929\|\|full | Not-determined |
| S16_vs_12939\|\|full | Low-quality |
| S16_vs_12949\|\|full | Not-determined |
| S16_vs_12953\|\|full | Not-determined |
| S16_vs_12990\|\|full | Not-determined |
| S16_vs_13032\|\|full | Low-quality |
| S16_vs_13039\|\|full | Not-determined |
| S16_vs_13146\|\|full | Not-determined |
| S16_vs_13161\|\|full | Low-quality |
| S16_vs_13191\|\|full | Low-quality |
| S16_vs_13242\|\|full | Not-determined |
| S16_vs_13243\|\|full | Low-quality |
| S16_vs_13310\|\|full | Not-determined |
| S16_vs_13320\|\|full | Not-determined |
| S16_vs_13360\|\|full | Low-quality |
| S16_vs_13370\|\|full | Not-determined |
| S16_vs_13393\|\|full | Low-quality |
| S16_vs_13411\|\|full | Not-determined |
| S16_vs_13416\|\|full | Not-determined |
| S16_vs_13418\|\|full | Low-quality |
| S16_vs_13438\|\|full | Not-determined |
| S16_vs_13441\|\|full | Not-determined |
| S16_vs_13456\|\|full | Low-quality |
| S16_vs_13458\|\|full | Low-quality |
| S16_vs_13486\|\|full | Low-quality |
| S16_vs_13489\|\|full | Not-determined |
| S16_vs_13491\|\|full | Low-quality |
| S16_vs_13499\|\|full | Low-quality |
| S16_vs_13505\|\|full | Not-determined |
| S16_vs_13511\|\|full | Low-quality |
| S16_vs_13649\|\|full | Low-quality |
| S16_vs_13677\|\|full | Not-determined |
| S16_vs_13702\|\|full | Low-quality |
| S16_vs_13708\|\|full | Not-determined |
| S16_vs_13710\|\|full | Low-quality |
| S16_vs_13746\|\|full | Low-quality |
| S16_vs_13777\|\|full | Low-quality |
| S16_vs_13782\|\|full | Not-determined |
| S16_vs_13815\|\|full | Not-determined |
| S16_vs_13824\|\|full | Low-quality |
| S16_vs_13839\|\|full | Low-quality |
| [illegible] | [illegible] |

| | |
|---|---|
| S16_vs_27364\|\|full | Low-quality |
| S16_vs_27374\|\|full | Low-quality |
| S16_vs_27387\|\|full | Not-determined |
| S16_vs_27411\|\|full | Low-quality |
| S16_vs_27420\|\|full | Not-determined |
| S16_vs_27466\|\|full | Low-quality |
| S16_vs_27545\|\|full | Not-determined |
| S16_vs_27554\|\|full | Low-quality |
| S16_vs_27598\|\|full | Not-determined |
| S16_vs_27638\|\|full | Low-quality |
| S16_vs_27679\|\|full | Low-quality |
| S16_vs_27690\|\|full | Low-quality |
| S16_vs_27698\|\|full | Not-determined |
| S16_vs_27729\|\|full | Low-quality |
| S16_vs_27741\|\|full | Not-determined |
| S16_vs_27759\|\|full | Low-quality |
| S16_vs_27777\|\|full | Not-determined |
| S16_vs_27789\|\|full | Low-quality |
| S16_vs_27798\|\|full | Not-determined |
| S16_vs_27812\|\|full | Low-quality |
| S16_vs_27829\|\|full | Not-determined |
| S16_vs_27853\|\|full | Not-determined |
| S16_vs_27890\|\|full | Low-quality |
| S16_vs_27899\|\|full | Low-quality |
| S16_vs_27980\|\|full | Not-determined |
| S16_vs_27996\|\|full | Low-quality |
| S16_vs_28035\|\|full | Low-quality |
| S16_vs_28039\|\|full | Low-quality |
| S16_vs_28086\|\|full | Low-quality |
| S16_vs_28088\|\|full | Low-quality |
| S16_vs_28106\|\|full | Not-determined |
| S16_vs_28121\|\|full | Low-quality |
| S16_vs_28165\|\|full | Low-quality |
| S16_vs_28174\|\|full | Low-quality |
| S16_vs_28175\|\|full | Not-determined |
| S16_vs_28204\|\|full | Low-quality |
| S16_vs_28224\|\|full | Low-quality |
| S16_vs_28237\|\|full | Not-determined |
| S16_vs_28240\|\|full | Low-quality |
| S16_vs_28247\|\|full | Low-quality |
| S16_vs_28249\|\|full | Not-determined |
| S16_vs_28256\|\|full | Low-quality |
| S16_vs_28257\|\|full | Not-determined |
| S16_vs_28272\|\|full | Low-quality |
| S16_vs_28295\|\|full | Not-determined |
| S16_vs_28348\|\|full | Low-quality |
| S16_vs_28368\|\|full | Not-determined |
| S16_vs_28403\|\|full | Not-determined |
| S16_vs_28409\|\|full | Not-determined |
| S16_vs_28416\|\|full | Not-determined |
| S16_vs_28530\|\|full | Low-quality |
| S16_vs_28540\|\|full | Not-determined |
| S16_vs_28551\|\|full | Low-quality |
| S16_vs_28580\|\|full | Not-determined |
| S16_vs_28616\|\|full | Low-quality |
| S16_vs_28723\|\|full | Not-determined |
| S16_vs_28728\|\|full | Low-quality |
| S16_vs_28798\|\|full | Low-quality |
| S16_vs_28804\|\|full | Low-quality |
| S16_vs_28848\|\|full | Low-quality |
| S16_vs_28860\|\|full | Low-quality |
| S16_vs_28879\|\|full | Low-quality |
| S16_vs_28882\|\|full | Not-determined |
| S16_vs_28900\|\|full | Low-quality |
| S16_vs_28928\|\|full | Low-quality |
| S16_vs_28941\|\|full | Not-determined |
| S16_vs_28952\|\|full | Low-quality |
| S16_vs_28960\|\|full | Low-quality |
| S16_vs_28971\|\|full | Not-determined |
| S16_vs_28997\|\|full | Low-quality |
| S16_vs_29023\|\|full | Not-determined |
| S16_vs_29126\|\|full | Low-quality |
| S16_vs_29137\|\|full | Not-determined |
| S16_vs_29172\|\|full | Not-determined |
| S16_vs_29184\|\|full | Not-determined |
| S16_vs_29206\|\|full | Low-quality |
| S16_vs_29224\|\|full | Low-quality |
| S16_vs_29230\|\|full | Low-quality |
| S16_vs_29235\|\|full | Low-quality |
| S16_vs_29298\|\|full | Low-quality |
| S16_vs_29353\|\|full | Low-quality |
| S16_vs_29374\|\|full | Not-determined |
| S16_vs_29387\|\|full | Low-quality |
| S16_vs_29497\|\|full | Not-determined |
| S16_vs_29506\|\|full | Low-quality |
| S16_vs_29507\|\|full | Low-quality |
| S16_vs_29521\|\|full | Not-determined |
| S16_vs_29552\|\|full | Not-determined |
| S16_vs_29553\|\|full | Low-quality |
| S16_vs_29560\|\|full | Low-quality |
| S16_vs_29585\|\|full | Low-quality |
| S16_vs_29587\|\|full | Not-determined |
| S16_vs_29606\|\|full | Not-determined |
| S16_vs_29646\|\|full | Low-quality |
| S16_vs_29698\|\|full | Not-determined |
| S16_vs_29714\|\|full | Low-quality |
| S16_vs_29716\|\|full | Not-determined |
| S16_vs_29720\|\|full | Not-determined |
| S16_vs_29737\|\|full | Not-determined |
| S16_vs_29739\|\|full | Low-quality |
| S16_vs_29743\|\|full | Not-determined |
| S16_vs_29811\|\|full | Not-determined |
| S16_vs_29840\|\|full | Low-quality |
| S16_vs_29959\|\|full | Low-quality |
| S16_vs_29977\|\|full | Low-quality |
| S16_vs_29992\|\|full | Low-quality |
| S16_vs_30029\|\|full | Not-determined |
| S16_vs_30034\|\|full | Not-determined |
| S16_vs_30043\|\|full | Not-determined |
| S16_vs_30067\|\|full | Low-quality |
| S16_vs_30110\|\|full | Not-determined |
| S16_vs_30111\|\|full | Low-quality |
| S16_vs_30116\|\|full | Low-quality |
| S16_vs_30128\|\|full | Low-quality |
| S16_vs_30143\|\|full | Low-quality |
| S16_vs_30145\|\|full | Low-quality |
| S16_vs_30174\|\|full | Not-determined |
| S16_vs_30253\|\|full | Low-quality |
| S16_vs_30274\|\|full | Not-determined |
| S16_vs_30293\|\|full | Low-quality |
| S16_vs_30313\|\|full | Low-quality |
| S16_vs_30329\|\|full | Not-determined |
| S16_vs_30378\|\|full | Low-quality |
| S16_vs_30441\|\|full | Low-quality |
| S16_vs_30446\|\|full | Low-quality |
| S16_vs_30449\|\|full | Low-quality |
| S16_vs_30547\|\|full | Low-quality |
| S16_vs_30548\|\|full | Low-quality |
| S16_vs_30551\|\|full | Low-quality |
| S16_vs_30586\|\|full | Low-quality |
| S16_vs_30597\|\|full | Not-determined |
| S16_vs_30654\|\|full | Low-quality |
| S16_vs_30655\|\|full | Low-quality |
| S16_vs_30700\|\|full | Not-determined |
| S16_vs_30809\|\|full | Not-determined |
| S16_vs_30888\|\|full | Low-quality |
| S16_vs_30926\|\|full | Low-quality |
| S16_vs_30929\|\|full | Not-determined |
| S16_vs_30959\|\|full | Not-determined |
| S16_vs_30992\|\|full | Low-quality |
| S16_vs_31017\|\|full | Not-determined |
| S16_vs_31109\|\|full | Low-quality |
| S16_vs_31155\|\|full | Not-determined |
| S16_vs_31213\|\|full | Low-quality |
| S16_vs_31259\|\|full | Low-quality |
| S16_vs_31282\|\|full | Low-quality |
| S16_vs_31285\|\|full | Low-quality |
| S16_vs_31326\|\|full | Low-quality |
| S16_vs_31347\|\|full | Not-determined |
| S16_vs_31350\|\|full | Low-quality |
| S16_vs_31375\|\|full | Not-determined |
| S16_vs_31383\|\|full | Not-determined |
| S16_vs_31404\|\|full | Low-quality |
| S16_vs_31456\|\|full | Not-determined |
| S16_vs_31507\|\|full | Not-determined |
| S16_vs_31530\|\|full | Low-quality |
| S16_vs_31542\|\|full | Low-quality |
| S16_vs_31644\|\|full | Low-quality |
| S16_vs_31786\|\|full | Not-determined |
| S16_vs_31832\|\|full | Not-determined |
| S16_vs_31852\|\|full | Low-quality |
| S16_vs_31934\|\|full | Not-determined |
| S16_vs_31948\|\|full | Not-determined |
| S16_vs_31970\|\|full | Not-determined |
| S16_vs_32022\|\|full | Low-quality |
| S16_vs_32040\|\|full | Low-quality |
| S16_vs_32055\|\|full | Not-determined |
| S16_vs_32086\|\|full | Low-quality |
| S16_vs_32099\|\|full | Not-determined |
| S16_vs_32101\|\|full | Not-determined |
| S16_vs_32110\|\|full | Not-determined |
| S16_vs_32133\|\|full | Low-quality |
| S16_vs_32137\|\|full | Low-quality |
| S16_vs_32141\|\|full | Low-quality |
| S16_vs_32151\|\|full | Not-determined |
| S16_vs_32153\|\|full | Low-quality |
| S16_vs_32157\|\|full | Not-determined |
| S16_vs_32196\|\|full | Not-determined |
| S16_vs_32306\|\|full | Low-quality |
| S16_vs_32311\|\|full | Low-quality |
| S16_vs_32314\|\|full | Low-quality |
| S6_vs_k141_22217\|\|full | Medium-quality |
| S16_vs_32536\|\|full | Low-quality |
| S16_vs_32564\|\|full | Not-determined |
| S16_vs_32575\|\|full | Not-determined |
| S16_vs_32606\|\|full | Low-quality |
| S16_vs_32613\|\|full | Not-determined |
| S16_vs_32640\|\|full | Not-determined |
| S16_vs_32646\|\|full | Low-quality |
| S16_vs_32689\|\|full | Low-quality |
| S16_vs_32716\|\|full | Low-quality |
| S16_vs_32729\|\|full | Not-determined |
| S16_vs_32761\|\|full | Not-determined |
| S9_vs_k141_4115\|\|full | Medium-quality |
| S16_vs_32772\|\|full | Low-quality |
| S16_vs_32823\|\|full | Low-quality |
| S16_vs_32836\|\|full | Low-quality |
| S16_vs_32861\|\|full | Not-determined |
| S16_vs_32866\|\|full | Low-quality |
| S16_vs_32868\|\|full | Low-quality |
| S16_vs_32871\|\|full | Not-determined |
| S16_vs_32886\|\|full | Not-determined |
| S16_vs_32889\|\|full | Not-determined |
| S16_vs_32904\|\|full | Low-quality |
| S16_vs_32918\|\|full | Low-quality |
| S16_vs_32928\|\|full | Low-quality |
| S16_vs_32989\|\|full | Low-quality |
| S16_vs_32998\|\|full | Low-quality |
| S16_vs_33009\|\|full | Not-determined |
| S16_vs_33023\|\|full | Low-quality |
| S16_vs_33041\|\|full | Low-quality |
| S16_vs_33072\|\|full | Low-quality |
| S16_vs_33109\|\|full | Not-determined |
| S16_vs_33149\|\|full | Not-determined |
| S16_vs_33180\|\|full | Not-determined |
| S16_vs_33198\|\|full | Not-determined |
| S16_vs_33211\|\|full | Low-quality |
| S4_extracted_31982 | Complete |
| S16_vs_33270\|\|full | Not-determined |
| S8_vs_k141_72117\|\|full | High-quality |
| S16_vs_33282\|\|full | Not-determined |
| S16_vs_33341\|\|full | Not-determined |
| S16_vs_33377\|\|full | Low-quality |
| S16_vs_33393\|\|full | Not-determined |
| S16_vs_33476\|\|full | Not-determined |
| S16_vs_33483\|\|full | Low-quality |
| S16_vs_33524\|\|full | Not-determined |
| S16_vs_33563\|\|full | Low-quality |
| S16_vs_33582\|\|full | Not-determined |
| S16_vs_33607\|\|full | Not-determined |
| S16_vs_33610\|\|full | Low-quality |
| S16_vs_33624\|\|full | Low-quality |
| S16_vs_33645\|\|full | Low-quality |
| S16_vs_33661\|\|full | Low-quality |
| S16_vs_33663\|\|full | Low-quality |
| S16_vs_33665\|\|full | Low-quality |
| S16_vs_33709\|\|full | Low-quality |
| S16_vs_33722\|\|full | Not-determined |
| S16_vs_33726\|\|full | Low-quality |
| S16_vs_33745\|\|full | Low-quality |
| S16_vs_33751\|\|full | Low-quality |
| S16_vs_33753\|\|full | Not-determined |
| S16_vs_33766\|\|full | Not-determined |
| S16_vs_33789\|\|full | Low-quality |
| S16_vs_33793\|\|full | Not-determined |
| S16_vs_33797\|\|full | Low-quality |
| S16_vs_33815\|\|full | Low-quality |
| S16_vs_33819\|\|full | Low-quality |
| S16_vs_33875\|\|full | Low-quality |
| S16_vs_33904\|\|full | Low-quality |
| S16_vs_34019\|\|full | Not-determined |
| S4_vs_VIRSorter_31982-circular-cat | High-quality |
| S16_vs_34066\|\|full | Low-quality |
| S16_vs_34072\|\|full | Low-quality |
| S16_vs_34131\|\|full | Not-determined |
| S16_vs_34165\|\|full | Not-determined |
| S16_vs_34167\|\|full | Low-quality |
| S16_vs_34198\|\|full | Low-quality |
| S16_vs_34236\|\|full | Not-determined |
| S16_vs_34241\|\|full | Not-determined |
| S16_vs_34264\|\|full | Not-determined |
| S16_vs_34408\|\|full | Low-quality |
| S16_vs_34465\|\|full | Low-quality |
| S16_vs_34469\|\|full | Low-quality |
| S16_vs_34616\|\|full | Low-quality |
| S16_vs_34621\|\|full | Not-determined |
| S16_vs_34680\|\|full | Low-quality |
| S16_vs_34685\|\|full | Low-quality |
| S16_vs_34690\|\|full | Low-quality |
| S14_extracted_k141_85122 | Medium-quality |
| S16_vs_34766\|\|full | Low-quality |
| S16_vs_34778\|\|full | Low-quality |
| S16_vs_34799\|\|full | Low-quality |
| S16_vs_34801\|\|full | Low-quality |
| S16_vs_34821\|\|full | Low-quality |
| S16_vs_34834\|\|full | Low-quality |
| S16_vs_34846\|\|full | Low-quality |
| S16_vs_34862\|\|full | Low-quality |
| S16_vs_34870\|\|full | Low-quality |
| S16_vs_34885\|\|full | Not-determined |
| S16_vs_34942\|\|full | Low-quality |
| S16_vs_34966\|\|full | Not-determined |
| S14_vs_k141_85122\|\|full | Medium-quality |
| S16_vs_35145\|\|full | Not-determined |
| S16_vs_35204\|\|full | Low-quality |
| S16_vs_35211\|\|full | Not-determined |
| S16_vs_35243\|\|full | Not-determined |
| S16_vs_35255\|\|full | Low-quality |
| S16_vs_35280\|\|full | Low-quality |
| S16_vs_35299\|\|full | Not-determined |
| S16_vs_35320\|\|full | Not-determined |
| S16_vs_35340\|\|full | Not-determined |
| S16_vs_35343\|\|full | Low-quality |
| S16_vs_35379\|\|full | Not-determined |
| S16_vs_35486\|\|full | Low-quality |
| S16_vs_35520\|\|full | Low-quality |
| S16_vs_35530\|\|full | Not-determined |
| S16_vs_35609\|\|full | Not-determined |
| S16_vs_35640\|\|full | Low-quality |
| S16_vs_35671\|\|full | Not-determined |
| S16_vs_35709\|\|full | Not-determined |
| S16_vs_35732\|\|full | Not-determined |
| S16_vs_35740\|\|full | Not-determined |
| S16_vs_35758\|\|full | Low-quality |
| S16_vs_35813\|\|full | Low-quality |
| S16_vs_35820\|\|full | Low-quality |
| S16_vs_35842\|\|full | Low-quality |
| S16_vs_35889\|\|full | Not-determined |
| S16_vs_35894\|\|full | Low-quality |
| S16_vs_35897\|\|full | Low-quality |
| S16_vs_35919\|\|full | Not-determined |
| S16_vs_35977\|\|full | Low-quality |
| S16_vs_36016\|\|full | Not-determined |
| S16_vs_36028\|\|full | Not-determined |
| S16_vs_36048\|\|full | Low-quality |
| S16_vs_36055\|\|full | Low-quality |
| S16_vs_36064\|\|full | Not-determined |
| S16_vs_36092\|\|full | Not-determined |
| S16_vs_36102\|\|full | Low-quality |
| S16_vs_36113\|\|full | Not-determined |
| S16_vs_36133\|\|full | Not-determined |
| S16_vs_36137\|\|full | Low-quality |
| S15_vs_39804\|\|full | Medium-quality |
| S16_vs_36197\|\|full | Not-determined |
| S16_vs_36260\|\|full | Low-quality |
| S16_vs_36287\|\|full | Not-determined |
| S16_vs_36292\|\|full | Low-quality |
| S16_vs_36294\|\|full | Not-determined |
| S16_vs_36316\|\|full | Low-quality |
| S16_vs_36322\|\|full | Low-quality |
| S16_vs_36334\|\|full | Low-quality |
| S16_vs_36339\|\|full | Not-determined |
| S16_vs_36394\|\|full | Low-quality |
| S16_vs_36449\|\|full | Not-determined |
| S16_vs_36469\|\|full | Not-determined |
| S16_vs_36474\|\|full | Low-quality |
| S16_vs_36481\|\|full | Not-determined |
| S16_vs_36500\|\|full | Not-determined |
| S16_vs_36506\|\|full | Not-determined |
| S16_vs_36527\|\|full | Low-quality |
| S16_vs_36546\|\|full | Low-quality |
| S16_vs_36575\|\|full | Low-quality |
| S16_vs_36599\|\|full | Low-quality |
| S16_vs_36644\|\|full | Low-quality |
| S16_vs_36658\|\|full | Low-quality |
| S16_vs_36663\|\|full | Low-quality |
| S16_vs_36664\|\|full | Not-determined |
| S16_vs_36703\|\|full | Low-quality |
| S16_vs_36708\|\|full | Low-quality |
| S16_vs_36765\|\|full | Low-quality |
| S16_vs_36771\|\|full | Low-quality |
| S16_vs_36799\|\|full | Not-determined |
| S16_vs_36831\|\|full | Low-quality |
| S16_vs_36853\|\|full | Low-quality |
| S16_vs_36866\|\|full | Low-quality |
| S16_vs_36873\|\|full | Low-quality |
| S16_vs_36902\|\|full | Low-quality |
| S16_vs_36914\|\|full | Not-determined |
| S16_vs_36930\|\|full | Low-quality |
| S16_vs_36942\|\|full | Not-determined |
| S16_vs_37062\|\|full | Low-quality |
| S16_vs_37159\|\|full | Not-determined |
| S16_vs_37180\|\|full | Low-quality |
| S16_vs_37183\|\|full | Low-quality |
| S16_vs_37194\|\|full | Not-determined |
| S16_vs_37207\|\|full | Not-determined |
| S16_vs_37218\|\|full | Not-determined |
| S16_vs_37233\|\|full | Not-determined |
| S16_vs_37247\|\|full | Low-quality |
| S16_vs_37276\|\|full | Not-determined |
| S16_vs_37300\|\|full | Not-determined |
| S16_vs_37307\|\|full | Not-determined |
| S16_vs_37343\|\|full | Low-quality |
| S16_vs_37361\|\|full | Not-determined |
| S16_vs_37423\|\|full | Low-quality |
| S16_vs_37448\|\|full | Not-determined |
| S16_vs_37475\|\|full | Low-quality |
| S16_vs_37481\|\|full | Not-determined |
| S16_vs_37549\|\|full | Low-quality |
| S16_vs_37568\|\|full | Low-quality |
| S16_vs_37599\|\|full | Not-determined |
| S16_vs_37613\|\|full | Low-quality |
| S16_vs_37651\|\|full | Low-quality |
| S16_vs_37685\|\|full | Not-determined |
| S16_vs_37694\|\|full | Low-quality |
| S16_vs_37697\|\|full | Not-determined |
| S16_vs_37709\|\|full | Low-quality |
| S16_vs_37766\|\|full | Not-determined |
| S16_vs_37780\|\|full | Not-determined |
| S16_vs_37872\|\|full | Low-quality |
| S16_vs_37912\|\|full | Not-determined |
| S16_vs_37919\|\|full | Not-determined |
| S16_vs_37935\|\|full | Not-determined |
| S16_vs_38017\|\|full | Low-quality |
| S16_vs_38105\|\|full | Not-determined |
| S16_vs_38174\|\|full | Low-quality |
| S16_vs_38183\|\|full | Not-determined |
| S16_vs_38191\|\|full | Low-quality |
| S16_vs_38235\|\|full | Low-quality |
| S16_vs_38261\|\|full | Low-quality |
| S16_vs_38262\|\|full | Low-quality |
| S16_vs_38264\|\|full | Low-quality |
| S16_vs_38272\|\|full | Low-quality |
| S16_vs_38313\|\|full | Low-quality |
| S16_vs_38373\|\|full | Low-quality |
| S16_vs_38393\|\|full | Low-quality |
| S16_vs_38404\|\|full | Not-determined |
| S16_vs_38415\|\|full | Low-quality |
| S16_vs_38436\|\|full | Low-quality |
| S16_vs_38466\|\|full | Not-determined |
| S16_vs_38503\|\|full | Not-determined |
| S16_vs_38528\|\|full | Low-quality |
| S16_vs_38533\|\|full | Not-determined |
| S16_vs_38536\|\|full | Not-determined |
| S16_vs_38561\|\|full | Not-determined |
| S16_vs_38577\|\|full | Not-determined |
| S16_vs_38649\|\|full | Not-determined |
| S16_vs_38663\|\|full | Low-quality |
| S16_vs_38664\|\|full | Not-determined |
| S16_vs_38696\|\|full | Low-quality |
| S16_vs_38698\|\|full | Low-quality |
| S16_vs_38711\|\|full | Not-determined |
| S16_vs_38734\|\|full | Not-determined |
| S16_vs_38777\|\|full | Not-determined |
| S16_vs_38779\|\|full | Low-quality |
| S16_vs_38804\|\|full | Low-quality |
| S3_vs_k141_127397\|\|full | High-quality |
| S16_vs_38912\|\|full | Low-quality |
| S16_vs_38951\|\|full | Low-quality |
| S16_vs_39022\|\|full | Low-quality |
| S16_vs_39028\|\|full | Not-determined |
| S16_vs_39082\|\|full | Low-quality |
| S16_vs_39175\|\|full | Low-quality |
| S16_vs_39179\|\|full | Low-quality |
| S16_vs_39244\|\|full | Not-determined |
| S16_vs_39284\|\|full | Low-quality |
| S16_vs_39300\|\|full | Not-determined |
| S16_vs_39321\|\|full | Low-quality |
| S16_vs_39354\|\|full | Low-quality |
| S16_vs_39417\|\|full | Low-quality |
| S16_vs_39419\|\|full | Not-determined |
| S16_vs_39452\|\|full | Low-quality |
| S16_vs_39459\|\|full | Low-quality |
| S16_vs_39518\|\|full | Low-quality |
| S16_vs_39531\|\|full | Low-quality |
| S16_vs_39544\|\|full | Not-determined |
| S16_vs_39547\|\|full | Low-quality |
| S16_vs_39550\|\|full | Not-determined |
| S16_vs_39552\|\|full | Not-determined |
| S16_vs_39567\|\|full | Low-quality |
| S16_vs_39597\|\|full | Low-quality |
| S16_vs_39668\|\|full | Low-quality |
| S16_vs_39736\|\|full | Low-quality |
| S16_vs_39756\|\|full | Low-quality |
| S16_vs_39803\|\|full | Low-quality |
| S16_vs_39827\|\|full | Not-determined |
| S16_vs_39828\|\|full | Low-quality |
| S16_vs_39858\|\|full | Not-determined |
| S16_vs_39865\|\|full | Not-determined |
| S16_vs_39867\|\|full | Low-quality |
| S16_vs_39957\|\|full | Low-quality |
| S16_vs_39964\|\|full | Low-quality |
| S16_vs_40057\|\|full | Low-quality |
| S16_vs_40071\|\|full | Low-quality |
| S16_vs_40103\|\|full | Not-determined |
| S16_vs_40111\|\|full | Low-quality |
| S16_vs_40137\|\|full | Not-determined |
| S16_vs_40144\|\|full | Not-determined |
| S16_vs_40164\|\|full | Low-quality |
| S16_vs_40185\|\|full | Not-determined |
| S16_vs_40282\|\|full | Low-quality |
| S16_vs_40286\|\|full | Low-quality |
| S16_vs_40398\|\|full | Not-determined |
| S16_vs_40425\|\|full | Low-quality |
| S16_vs_40455\|\|full | Low-quality |
| S16_vs_40469\|\|full | Not-determined |
| S16_vs_40481\|\|full | Low-quality |
| S16_vs_40508\|\|full | Low-quality |
| S16_vs_40541\|\|full | Not-determined |
| S16_vs_40603\|\|full | Low-quality |
| S16_vs_40645\|\|full | Low-quality |
| S16_vs_40660\|\|full | Low-quality |
| S16_vs_40708\|\|full | Low-quality |
| S16_vs_40788\|\|full | Not-determined |
| S16_vs_40791\|\|full | Not-determined |
| S16_vs_40809\|\|full | Not-determined |
| S16_vs_40853\|\|full | Not-determined |
| S16_vs_40915\|\|full | Low-quality |
| S16_vs_40944\|\|full | Low-quality |
| S16_vs_40968\|\|full | Low-quality |
| S16_vs_40980\|\|full | Low-quality |
| S16_vs_41037\|\|full | Not-determined |
| S16_vs_41073\|\|full | Low-quality |
| S16_vs_41158\|\|full | Not-determined |
| S16_vs_41164\|\|full | Low-quality |
| S16_vs_41166\|\|full | Not-determined |
| S16_vs_41171\|\|full | Low-quality |
| S16_vs_41175\|\|full | Not-determined |
| S16_vs_41181\|\|full | Not-determined |
| S16_vs_41195\|\|full | Not-determined |
| S16_vs_41212\|\|full | Not-determined |
| S16_vs_41220\|\|full | Low-quality |
| S16_vs_41306\|\|full | Not-determined |
| S16_vs_41316\|\|full | Not-determined |
| S16_vs_41334\|\|full | Low-quality |
| S16_vs_41363\|\|full | Low-quality |
| S16_vs_41403\|\|full | Low-quality |
| S16_vs_41427\|\|full | Not-determined |
| S16_vs_41428\|\|full | Not-determined |
| S16_vs_41437\|\|full | Not-determined |
| S16_vs_41439\|\|full | Low-quality |
| S16_vs_41482\|\|full | Low-quality |
| S16_vs_41486\|\|full | Low-quality |
| S16_vs_41529\|\|full | Not-determined |
| S16_vs_41545\|\|full | Low-quality |
| S16_vs_41589\|\|full | Low-quality |
| S16_vs_41591\|\|full | Low-quality |
| S16_vs_41612\|\|full | Low-quality |
| S16_vs_41637\|\|full | Not-determined |
| S16_vs_41661\|\|full | Low-quality |
| S16_vs_41713\|\|full | Not-determined |
| S16_vs_41720\|\|full | Low-quality |
| S16_vs_41721\|\|full | Not-determined |
| S16_vs_41722\|\|full | Low-quality |
| S16_vs_41773\|\|full | Low-quality |
| S16_vs_41810\|\|full | Not-determined |
| S16_vs_41954\|\|full | Not-determined |
| S16_vs_42065\|\|full | Low-quality |
| S16_vs_42072\|\|full | Low-quality |
| S16_vs_42081\|\|full | Not-determined |
| S16_vs_42102\|\|full | Not-determined |
| S16_vs_42141\|\|full | Low-quality |
| S16_vs_42143\|\|full | Low-quality |
| S16_vs_42144\|\|full | Low-quality |
| S16_vs_42206\|\|full | Not-determined |
| S16_vs_42222\|\|full | Low-quality |
| S16_vs_42228\|\|full | Not-determined |
| S16_vs_42250\|\|full | Not-determined |
| S16_vs_42264\|\|full | Low-quality |
| S16_vs_42319\|\|full | Low-quality |
| S16_vs_42329\|\|full | Not-determined |
| S16_vs_42365\|\|full | Low-quality |
| S16_vs_42410\|\|full | Low-quality |
| S16_vs_42428\|\|full | Not-determined |
| S16_vs_42505\|\|full | Low-quality |
| S16_vs_42605\|\|full | Low-quality |
| S16_vs_42643\|\|full | Low-quality |
| S16_vs_42669\|\|full | Not-determined |
| S16_vs_42677\|\|full | Not-determined |
| S16_vs_42695\|\|full | Not-determined |
| S17_vs_k141_5656\|\|full | High-quality |
| S16_vs_42710\|\|full | Not-determined |
| S16_vs_42742\|\|full | Not-determined |
| S16_vs_42769\|\|full | Not-determined |
| S16_vs_42801\|\|full | Low-quality |
| S16_vs_42824\|\|full | Low-quality |
| S16_vs_42836\|\|full | Not-determined |
| S16_vs_42839\|\|full | Not-determined |
| S16_vs_42901\|\|full | Low-quality |
| S16_vs_42948\|\|full | Low-quality |
| S16_vs_42955\|\|full | Low-quality |
| S16_vs_42990\|\|full | Not-determined |
| S16_vs_43012\|\|full | Low-quality |
| S16_vs_43015\|\|full | Low-quality |
| S16_vs_43019\|\|full | Not-determined |
| S16_vs_43055\|\|full | Not-determined |
| S16_vs_43067\|\|full | Low-quality |
| S16_vs_43072\|\|full | Low-quality |
| S16_vs_43115\|\|full | Low-quality |
| S16_vs_43141\|\|full | Low-quality |
| S16_vs_43159\|\|full | Low-quality |
| S16_vs_43204\|\|full | Not-determined |
| S16_vs_43227\|\|full | Low-quality |
| S16_vs_43241\|\|full | Not-determined |
| S16_vs_43246\|\|full | Low-quality |
| S16_vs_43249\|\|full | Low-quality |
| S16_vs_43280\|\|full | Low-quality |
| S16_vs_43305\|\|full | Not-determined |
| S16_vs_43334\|\|full | Low-quality |
| S16_vs_43350\|\|full | Low-quality |
| S16_vs_43472\|\|full | Not-determined |
| S16_vs_43581\|\|full | Low-quality |
| S16_vs_43599\|\|full | Not-determined |
| S16_vs_43638\|\|full | Not-determined |
| S16_vs_43651\|\|full | Not-determined |
| S16_vs_43673\|\|full | Not-determined |
| S16_vs_43689\|\|full | Not-determined |
| S16_vs_43690\|\|full | Not-determined |
| S16_vs_43703\|\|full | Low-quality |

S16_vs_43728||full Not-determined
S16_vs_43755||full Low-quality
S16_vs_43764||full Low-quality
S16_vs_43774||full Low-quality
S16_vs_43779||full Not-determined
S16_vs_43780||full Not-determined
S16_vs_43805||full Low-quality
S16_vs_43808||full Not-determined
S16_vs_43810||full Low-quality
S16_vs_43827||full Low-quality
S16_vs_43849||full Low-quality
S16_vs_43917||full Low-quality
S16_vs_43925||full Not-determined
S16_vs_43952||full Not-determined
S16_vs_43957||full Low-quality
S16_vs_44005||full Low-quality
S16_vs_44064||full Not-determined
S16_vs_44114||full Low-quality
S16_vs_44172||full Low-quality
S16_vs_44207||full Low-quality
S16_vs_44217||full Low-quality
S16_vs_44219||full Low-quality
S16_vs_44243||full Not-determined
S16_vs_44259||full Not-determined
S16_vs_44264||full Low-quality
S16_vs_44268||full Low-quality
S16_vs_44330||full Not-determined
S16_vs_44335||full Not-determined
S16_vs_44343||full Low-quality
S16_vs_44363||full Low-quality
S16_vs_44425||full Low-quality
S16_vs_44483||full Not-determined
S16_vs_44491||full Not-determined
S16_vs_44514||full Not-determined
S16_vs_44530||full Low-quality
S16_vs_44537||full Low-quality
S16_vs_44579||full Low-quality
S16_vs_44619||full Not-determined
S16_vs_44657||full Low-quality
S16_vs_44679||full Low-quality
S16_vs_44687||full Not-determined
S16_vs_44736||full Low-quality
S16_vs_44766||full Low-quality
S16_vs_44797||full Low-quality
S16_vs_44802||full Not-determined
S16_vs_44833||full Low-quality
S16_vs_44862||full Not-determined
S16_vs_44868||full Not-determined
S16_vs_44927||full Low-quality
S16_vs_44929||full Not-determined
S16_vs_44938||full Low-quality
S16_vs_44978||full Not-determined
S16_vs_44990||full Low-quality
S16_vs_45001||full Low-quality
S16_vs_45027||full Low-quality
S16_vs_45059||full Not-determined
S16_vs_45096||full Low-quality
S16_vs_45098||full Not-determined
S16_vs_45186||full Low-quality
S16_vs_45193||full Low-quality
S16_vs_45200||full Low-quality
S16_vs_45229||full Not-determined
S16_vs_45232||full Low-quality
S16_vs_45264||full Low-quality
S16_vs_45270||full Low-quality
S16_vs_45279||full Low-quality
S16_vs_45294||full Low-quality
S16_vs_45346||full Low-quality
S16_vs_45379||full Low-quality
S16_vs_45403||full Not-determined
S16_vs_45413||full Low-quality
S16_vs_45418||full Low-quality
S16_vs_45425||full Low-quality
S16_vs_45443||full Not-determined
S16_vs_45493||full Not-determined
S16_vs_45499||full Not-determined
S16_vs_45517||full Low-quality
S16_vs_45528||full Low-quality
S16_vs_45537||full Low-quality
S16_vs_45684||full Not-determined
S16_vs_45685||full Not-determined
S16_vs_45695||full Low-quality
S16_vs_45697||full Not-determined
S16_vs_45711||full Low-quality
S16_vs_45720||full Low-quality
S16_vs_45824||full Low-quality
S16_vs_45850||full Low-quality
S16_vs_45851||full Low-quality
S16_vs_45928||full Not-determined
S16_vs_45951||full Not-determined
S16_vs_45955||full Not-determined
S16_vs_45965||full Low-quality
S16_vs_45973||full Low-quality
S16_vs_45995||full Not-determined
S16_vs_46056||full Low-quality
S16_vs_46139||full Not-determined
S16_vs_46147||full Low-quality
S16_vs_46149||full Low-quality
S16_vs_46151||full Low-quality
S16_vs_46186||full Low-quality
S16_vs_46191||full Not-determined
S16_vs_46202||full Not-determined
S16_vs_46303||full Not-determined
S16_vs_46344||full Low-quality
S16_vs_46359||full Low-quality
S16_vs_46365||full Not-determined
S16_vs_46370||full Low-quality
S16_vs_46371||full Low-quality
S16_vs_46384||full Not-determined
S16_vs_46391||full Not-determined
S16_vs_46404||full Low-quality
S16_vs_46405||full Not-determined
S16_vs_46407||full Not-determined
S16_vs_46409||full Not-determined
S16_vs_46414||full Not-determined
S16_vs_46491||full Low-quality
S16_vs_46507||full Low-quality
S16_vs_46579||full Low-quality
S16_vs_46616||full Low-quality
S16_vs_46672||full Low-quality
S16_vs_46704||full Not-determined
S16_vs_46825||full Not-determined
S16_vs_46830||full Not-determined
S16_vs_46870||full Not-determined
S16_vs_46894||full Low-quality
S16_vs_46917||full Low-quality
S16_vs_46982||full Not-determined
S16_vs_47045||full Not-determined
S16_vs_47053||full Not-determined
S16_vs_47111||full Not-determined
S16_vs_47114||full Low-quality
S16_vs_47117||full Not-determined
S16_vs_47177||full Not-determined
S16_vs_47205||full Low-quality
S16_vs_47209||full Not-determined
S16_vs_47226||full Low-quality
S16_vs_47250||full Not-determined
S16_vs_47287||full Not-determined
S16_vs_47325||full Low-quality
S16_vs_47332||full Not-determined
S16_vs_47346||full Low-quality
S16_vs_47381||full Low-quality
S16_vs_47415||full Low-quality
S16_vs_47419||full Low-quality
S16_vs_47440||full Low-quality
S16_vs_47549||full Not-determined
S16_vs_47596||full Not-determined
S16_vs_47622||full Not-determined
S16_vs_47631||full Low-quality
S16_vs_47661||full Low-quality
S16_vs_47675||full Low-quality
S16_vs_47712||full Not-determined
S16_vs_47730||full Low-quality
S16_vs_47759||full Low-quality
S16_vs_47763||full Low-quality
S16_vs_47770||full Low-quality
S16_vs_47795||full Not-determined
S16_vs_47809||full Low-quality
S16_vs_47814||full Low-quality
S16_vs_47863||full Low-quality
S16_vs_47892||full Low-quality
S16_vs_47897||full Low-quality
S16_vs_47926||full Not-determined
S16_vs_47990||full Low-quality
S16_vs_48005||full Low-quality
S16_vs_48042||full Not-determined
S16_vs_48046||full Not-determined
S16_vs_48070||full Not-determined
S16_vs_48075||full Not-determined
S16_vs_48149||full Low-quality
S16_vs_48169||full Not-determined
S16_vs_48215||full Low-quality
S16_vs_48275||full Not-determined
S16_vs_48369||full Low-quality
S16_vs_48371||full Low-quality
S16_vs_48425||full Not-determined
S16_vs_48441||full Low-quality
S16_vs_48456||full Low-quality
S16_vs_48478||full Low-quality
S16_vs_48483||full Low-quality
S16_vs_48525||full Low-quality
S16_vs_48530||full Not-determined
S16_vs_48559||full Not-determined
S16_vs_48614||full Low-quality
S16_vs_48620||full Low-quality
S16_vs_48622||full Low-quality
S16_vs_48637||full Not-determined
S16_vs_48702||full Low-quality
S1_extracted_k141_57540 Medium-quality
S16_vs_48725||full Low-quality
S16_vs_48805||full Low-quality
S16_vs_48838||full Low-quality
S16_vs_48950||full Low-quality
S16_vs_48988||full Low-quality
S16_vs_49043||full Low-quality
S16_vs_49133||full Low-quality
S16_vs_49137||full Low-quality
S16_vs_49139||full Not-determined
S16_vs_49142||full Not-determined
S16_vs_49190||full Not-determined
S16_vs_49198||full Low-quality
S16_vs_49200||full Low-quality
S16_vs_49204||full Low-quality
S16_vs_49214||full Not-determined
S16_vs_49221||full Low-quality
S16_vs_49239||full Not-determined
S16_vs_49252||full Low-quality
S16_vs_49278||full Not-determined
S16_vs_49310||full Low-quality
S16_vs_49323||full Low-quality
S16_vs_49324||full Low-quality
S16_vs_49349||full Not-determined
S16_vs_49373||full Low-quality
S16_vs_49377||full Low-quality
S16_vs_49395||full Not-determined
S16_vs_49401||full Low-quality
S16_vs_49436||full Low-quality
S16_vs_49456||full Low-quality
S16_vs_49495||full Low-quality
S16_vs_49506||full Low-quality
S16_vs_49539||full Low-quality
S16_vs_49579||full Low-quality
S16_vs_49599||full Low-quality
S16_vs_49620||full Low-quality
S10_vs_17919||full Medium-quality
S16_vs_49660||full Not-determined
S16_vs_49701||full Low-quality
S16_vs_49759||full Low-quality
S16_vs_49801||full Low-quality
S16_vs_49848||full Not-determined
S16_vs_49862||full Not-determined
S16_vs_49875||full Low-quality
S16_vs_49884||full Low-quality
S16_vs_49919||full Low-quality
S16_vs_49947||full Low-quality
S16_vs_49960||full Not-determined
S16_vs_50027||full Low-quality
S16_vs_50055||full Low-quality
S16_vs_50059||full Low-quality
S16_vs_50063||full Not-determined
S16_vs_50070||full Low-quality
S16_vs_50084||full Low-quality
S16_vs_50088||full Low-quality
S16_vs_50135||full Low-quality
S16_vs_50235||full Not-determined
S16_vs_50242||full Low-quality
S16_vs_50317||full Low-quality
S16_vs_50322||full Low-quality
S16_vs_50354||full Not-determined
S16_vs_50356||full Not-determined
S16_vs_50358||full Not-determined
S16_vs_50368||full Not-determined
S16_vs_50372||full Not-determined
S16_vs_50425||full Low-quality
S16_vs_50434||full Not-determined
S16_vs_50452||full Low-quality
S16_vs_50465||full Low-quality
S16_vs_50507||full Not-determined
S16_vs_50530||full Low-quality
S16_vs_50553||full Low-quality
S16_vs_50555||full Low-quality
S16_vs_50569||full Not-determined
S16_vs_50583||full Low-quality
S16_vs_50594||full Not-determined
S16_vs_50632||full Low-quality
S16_vs_50657||full Low-quality
S16_vs_50666||full Not-determined
S16_vs_50684||full Low-quality
S16_vs_50708||full Low-quality
S16_vs_50714||full Low-quality
S16_vs_50739||full Not-determined
S16_vs_50741||full Low-quality
S16_vs_50742||full Low-quality
S16_vs_50748||full Not-determined
S16_vs_50761||full Low-quality
S16_vs_50780||full Not-determined
S16_vs_50805||full Low-quality
S16_vs_50813||full Low-quality
S16_vs_50890||full Low-quality
S16_vs_50945||full Not-determined
S16_vs_51005||full Not-determined
S16_vs_51006||full Not-determined
S16_vs_51025||full Not-determined
S16_vs_51039||full Low-quality
S16_vs_51057||full Not-determined
S16_vs_51264||full Not-determined
S16_vs_51314||full Low-quality
S16_vs_51330||full Not-determined
S16_vs_51346||full Low-quality
S16_vs_51356||full Low-quality
S16_vs_51425||full Low-quality
S16_vs_51439||full Low-quality
S16_vs_51452||full Low-quality
S16_vs_51474||full Not-determined
S16_vs_51486||full Not-determined
S16_vs_51492||full Low-quality
S16_vs_51512||full Not-determined
S16_vs_51586||full Not-determined
S16_vs_51616||full Low-quality
S16_vs_51655||full Low-quality
S16_vs_51659||full Low-quality
S16_vs_51692||full Not-determined
S16_vs_51713||full Not-determined
S16_vs_51727||full Not-determined
S16_vs_51757||full Low-quality
S16_vs_51771||full Not-determined
S16_vs_51807||full Low-quality
S16_vs_51870||full Low-quality
S12_vs_k141_48045||full Medium-quality
S16_vs_51908||full Not-determined
S16_vs_51926||full Low-quality
S16_vs_51965||full Low-quality
S16_vs_52005||full Low-quality
S16_vs_52010||full Low-quality
S16_vs_52012||full Not-determined
S16_vs_52016||full Low-quality
S16_vs_52017||full Low-quality
S16_vs_52045||full Not-determined
S16_vs_52068||full Low-quality
S16_vs_52087||full Low-quality
S16_vs_52090||full Not-determined
S16_vs_52177||full Not-determined
S16_vs_52187||full Low-quality
S16_vs_52209||full Not-determined
S16_vs_52214||full Not-determined
S16_vs_52237||full Not-determined
S16_vs_52278||full Not-determined
S16_vs_52318||full Low-quality
S16_vs_52345||full Low-quality
S16_vs_52351||full Not-determined
S16_vs_52379||full Not-determined
S16_vs_52403||full Low-quality
S16_vs_52417||full Not-determined
S16_vs_52457||full Not-determined
S16_vs_52479||full Not-determined
S16_vs_52508||full Not-determined
S1_vs_k141_21439||full Medium-quality
S16_vs_52567||full Not-determined
S16_vs_52600||full Low-quality
S16_vs_52604||full Not-determined
S16_vs_52609||full Low-quality
S16_vs_52610||full Not-determined
S16_vs_52638||full Low-quality
S16_vs_52643||full Low-quality
S16_vs_52668||full Low-quality
S16_vs_52685||full Low-quality
S16_vs_52692||full Low-quality
S16_vs_52724||full Low-quality
S16_vs_52772||full Low-quality
S16_vs_52778||full Not-determined
S16_vs_52821||full Not-determined
S16_vs_52829||full Low-quality
S16_vs_52841||full Low-quality
S16_vs_52865||full Not-determined
S16_vs_52914||full Not-determined
S16_vs_52917||full Not-determined
S16_vs_52923||full Low-quality
S16_vs_52956||full Low-quality
S16_vs_52962||full Low-quality
S16_vs_52980||full Not-determined
S16_vs_53013||full Low-quality
S16_vs_53061||full Not-determined
S16_vs_53109||full Not-determined
S16_vs_53120||full Low-quality
S16_vs_53174||full Not-determined
S16_vs_53197||full Low-quality
S16_vs_53203||full Low-quality
S16_vs_53210||full Not-determined
S16_vs_53255||full Low-quality
S16_vs_53269||full Low-quality
S16_vs_53276||full Not-determined
S16_vs_53300||full Not-determined
S16_vs_53303||full Low-quality
S16_vs_53340||full Not-determined
S16_vs_53411||full Low-quality
S16_vs_53452||full Not-determined
S16_vs_53463||full Low-quality
S16_vs_53483||full Not-determined
S16_vs_53505||full Low-quality
S16_vs_53527||full Low-quality
S16_vs_53578||full Not-determined
S16_vs_53620||full Low-quality
S16_vs_53623||full Not-determined
S16_vs_53637||full Low-quality
S16_vs_53645||full Low-quality
S16_vs_53670||full Low-quality
S16_vs_53671||full Low-quality
S16_vs_53753||full Low-quality
S16_vs_53775||full Low-quality
S16_vs_53800||full Low-quality
S16_vs_53927||full Not-determined
S16_vs_53938||full Not-determined
S16_vs_53955||full Low-quality
S16_vs_53974||full Low-quality
S16_vs_53992||full Low-quality
S16_vs_54005||full Low-quality
S16_vs_54056||full Low-quality
S16_vs_54065||full Low-quality
S16_vs_54071||full Low-quality
S16_vs_54084||full Low-quality
S16_vs_54099||full Low-quality
S16_vs_54175||full Not-determined
S16_vs_54183||full Low-quality
S16_vs_54201||full Low-quality
S16_vs_54239||full Low-quality
S16_vs_54240||full Not-determined
S4_extracted_11308 High-quality
S16_vs_54256||full Low-quality
S16_vs_54257||full Low-quality
S16_vs_54277||full Low-quality
S16_vs_54279||full Low-quality
S16_vs_54280||full Not-determined
S16_vs_54342||full Not-determined
S16_vs_54378||full Not-determined
S16_vs_54395||full Not-determined
S16_vs_54432||full Low-quality
S16_vs_54462||full Low-quality
S16_vs_54469||full Low-quality
S16_vs_54472||full Low-quality
S16_vs_54481||full Low-quality
S16_vs_54482||full Not-determined
S16_vs_54518||full Low-quality
S16_vs_54557||full Low-quality
S4_vs_VIRSorter_11308-cat_2 High-quality
S16_vs_54658||full Low-quality
S16_vs_54669||full Not-determined
S16_vs_54681||full Low-quality
S16_vs_54708||full Not-determined
S16_vs_54734||full Low-quality
S16_vs_54812||full Low-quality
S16_vs_54820||full Not-determined
S16_vs_54822||full Low-quality
S16_vs_54971||full Low-quality
S16_vs_55071||full Low-quality
S16_vs_55104||full Not-determined
S16_vs_55124||full Low-quality
S16_vs_55131||full Not-determined
S16_vs_55133||full Not-determined
S16_vs_55164||full Not-determined
S16_vs_55223||full Not-determined
S16_vs_55275||full Not-determined
S16_vs_55306||full Not-determined
S16_vs_55339||full Low-quality
S16_vs_55360||full Not-determined
S16_vs_55380||full Not-determined
S16_vs_55382||full Low-quality
S16_vs_55397||full Low-quality
S16_vs_55429||full Low-quality
S16_vs_55458||full Low-quality
S16_vs_55481||full Low-quality
S16_vs_55487||full Low-quality
S16_vs_55500||full Not-determined
S16_vs_55521||full Low-quality
S16_vs_55527||full Not-determined
S16_vs_55600||full Not-determined
S16_vs_55603||full Low-quality
S16_vs_55604||full Not-determined
S16_vs_55608||full Low-quality
S16_vs_55610||full Not-determined
S16_vs_55615||full Not-determined
S16_vs_55675||full Low-quality
S16_vs_55677||full Low-quality
S16_vs_55714||full Low-quality
S16_vs_55718||full Low-quality
S16_vs_55725||full Low-quality
S16_vs_55775||full Not-determined
S16_vs_55817||full Low-quality
S16_vs_55837||full Low-quality
S16_vs_55882||full Low-quality
S16_vs_55892||full Not-determined
S16_vs_55899||full Low-quality
S16_vs_55922||full Low-quality
S16_vs_55934||full Low-quality
S16_vs_55937||full Low-quality
S16_vs_55948||full Not-determined
S16_vs_55970||full Low-quality
S16_vs_55996||full Not-determined
S16_vs_56004||full Not-determined
S16_vs_56016||full Low-quality
S16_vs_56025||full Low-quality
S16_vs_56039||full Low-quality
S16_vs_56042||full Not-determined
S16_vs_56079||full Not-determined
S16_vs_56089||full Not-determined
S16_vs_56095||full Not-determined
S16_vs_56164||full Low-quality
S16_vs_56169||full Low-quality
S16_vs_56239||full Not-determined
S16_vs_56244||full Not-determined
S16_vs_56279||full Low-quality
S16_vs_56301||full Low-quality
S16_vs_56313||full Not-determined
S16_vs_56368||full Not-determined
S16_vs_56402||full Low-quality
S16_vs_56428||full Low-quality
S16_vs_56467||full Low-quality
S16_vs_56491||full Low-quality
S16_vs_56546||full Low-quality
S16_vs_56579||full Not-determined
S16_vs_56674||full Low-quality
S16_vs_56685||full Low-quality
S16_vs_56700||full Low-quality
S16_vs_56719||full Not-determined
S16_vs_56738||full Not-determined
S16_vs_56792||full Not-determined
S16_vs_56808||full Not-determined
S16_vs_56843||full Low-quality
S16_vs_56858||full Not-determined
S16_vs_56859||full Low-quality
S16_vs_56871||full Low-quality
S16_vs_56899||full Not-determined
S16_vs_56933||full Low-quality
S16_vs_56940||full Not-determined
S16_vs_56950||full Not-determined
S16_vs_56957||full Not-determined
S16_vs_56969||full Not-determined
S16_vs_57064||full Low-quality
S16_vs_57162||full Not-determined
S16_vs_57170||full Not-determined
S16_vs_57192||full Low-quality
S12_extracted_k141_67576 High-quality
S16_vs_57345||full Low-quality
S16_vs_57351||full Low-quality
S16_vs_57366||full Low-quality
S16_vs_57429||full Not-determined
S16_vs_57483||full Low-quality
S16_vs_57497||full Not-determined
S16_vs_57523||full Low-quality
S16_vs_57553||full Low-quality
S16_vs_57609||full Not-determined
S16_vs_57662||full Low-quality
S16_vs_57672||full Not-determined
S16_vs_57683||full Not-determined
S16_vs_57735||full Not-determined
S16_vs_57802||full Low-quality
S16_vs_57803||full Low-quality
S16_vs_57805||full Low-quality
S16_vs_57818||full Low-quality
S16_vs_57838||full Not-determined
S16_vs_57874||full Not-determined
S16_vs_57923||full Not-determined
S16_vs_57940||full Low-quality
S16_vs_57944||full Not-determined
S16_vs_57963||full Not-determined
S16_vs_57971||full Low-quality
S16_vs_57984||full Not-determined
S16_vs_58039||full Low-quality
S16_vs_58067||full Not-determined
S16_vs_58126||full Low-quality
S16_vs_58144||full Not-determined
S16_vs_58182||full Not-determined
S16_vs_58186||full Not-determined
S16_vs_58327||full Not-determined
S16_vs_58354||full Low-quality
S16_vs_58359||full Not-determined
S16_vs_58368||full Low-quality
S16_vs_58402||full Not-determined
S16_vs_58403||full Not-determined
S16_vs_58431||full Low-quality
S16_vs_58490||full Low-quality
S16_vs_58507||full Not-determined
S16_vs_58512||full Low-quality
S16_vs_58525||full Low-quality
S16_vs_58549||full Not-determined
S16_vs_58592||full Low-quality
S16_vs_58623||full Not-determined
S16_vs_58698||full Low-quality
S16_vs_58727||full Low-quality
S16_vs_58738||full Low-quality
S16_vs_58753||full Low-quality
S16_vs_58837||full Low-quality
S16_vs_58847||full Low-quality
S16_vs_58876||full Not-determined
S16_vs_58887||full Low-quality
S16_vs_58942||full Not-determined
S16_vs_58992||full Not-determined
S16_vs_59032||full Low-quality
S16_vs_59042||full Low-quality
S16_vs_59076||full Not-determined
S16_vs_59084||full Low-quality
S16_vs_59131||full Low-quality
S16_vs_59132||full Low-quality
S16_vs_59151||full Not-determined
S16_vs_59235||full Not-determined
S16_vs_59259||full Not-determined
S16_vs_59286||full Not-determined
S16_vs_59306||full Not-determined
S16_vs_59307||full Low-quality
S16_vs_59340||full Not-determined

| | |
|---|---|
| S16_vs_59347\|\|full | Low-quality |
| S16_vs_59360\|\|full | Not-determined |
| S16_vs_59381\|\|full | Not-determined |
| S16_vs_59395\|\|full | Not-determined |
| S16_vs_59448\|\|full | Not-determined |
| S16_vs_59450\|\|full | Low-quality |
| S16_vs_59500\|\|full | Low-quality |
| S16_vs_59521\|\|full | Low-quality |
| S16_vs_59541\|\|full | Low-quality |
| S16_vs_59552\|\|full | Low-quality |
| S16_vs_59593\|\|full | Not-determined |
| S16_vs_59636\|\|full | Low-quality |
| S16_vs_59654\|\|full | Low-quality |
| S16_vs_59680\|\|full | Not-determined |
| S16_vs_59687\|\|full | Not-determined |
| S16_vs_59750\|\|full | Low-quality |
| S16_vs_59764\|\|full | Low-quality |
| S16_vs_59776\|\|full | Not-determined |
| S16_vs_59787\|\|full | Not-determined |
| S16_vs_59796\|\|full | Low-quality |
| S16_vs_59856\|\|full | Not-determined |
| S16_vs_59857\|\|full | Low-quality |
| S16_vs_59858\|\|full | Low-quality |
| S16_vs_59861\|\|full | Low-quality |
| S16_vs_59890\|\|full | Not-determined |
| S16_vs_59946\|\|full | Not-determined |
| S16_vs_59992\|\|full | Low-quality |
| S16_vs_60019\|\|full | Not-determined |
| S16_vs_60039\|\|full | Not-determined |
| S16_vs_60042\|\|full | Low-quality |
| S16_vs_60142\|\|full | Not-determined |
| S16_vs_60186\|\|full | Not-determined |
| S16_vs_60274\|\|full | Low-quality |
| S16_vs_60320\|\|full | Not-determined |
| S16_vs_60337\|\|full | Low-quality |
| S16_vs_60359\|\|full | Not-determined |
| S16_vs_60396\|\|full | Low-quality |
| S16_vs_60397\|\|full | Low-quality |
| S16_vs_60401\|\|full | Not-determined |
| S16_vs_60425\|\|full | Low-quality |
| S16_vs_60468\|\|full | Low-quality |
| S16_vs_60475\|\|full | Not-determined |
| S16_vs_60511\|\|full | Low-quality |
| S16_vs_60522\|\|full | Low-quality |
| S16_vs_60544\|\|full | Not-determined |
| S16_vs_60570\|\|full | Low-quality |
| S16_vs_60581\|\|full | Low-quality |
| S16_vs_60588\|\|full | Not-determined |
| S16_vs_60589\|\|full | Low-quality |
| S16_vs_60605\|\|full | Low-quality |
| S16_vs_60608\|\|full | Low-quality |
| S16_vs_60661\|\|full | Not-determined |
| S16_vs_60663\|\|full | Low-quality |
| S16_vs_60737\|\|full | Low-quality |
| S16_vs_60742\|\|full | Not-determined |
| S16_vs_60747\|\|full | Low-quality |
| S16_vs_60768\|\|full | Not-determined |
| S16_vs_60817\|\|full | Not-determined |
| S16_vs_60838\|\|full | Low-quality |
| S16_vs_60875\|\|full | Not-determined |
| S16_vs_60877\|\|full | Not-determined |
| S16_vs_60882\|\|full | Low-quality |
| S16_vs_60953\|\|full | Not-determined |
| S16_vs_60977\|\|full | Not-determined |
| S16_vs_61042\|\|full | Not-determined |
| S16_vs_61081\|\|full | Low-quality |
| S16_vs_61095\|\|full | Low-quality |
| S16_vs_61103\|\|full | Low-quality |
| S16_vs_61112\|\|full | Low-quality |
| S16_vs_61133\|\|full | Low-quality |
| S16_vs_61187\|\|full | Low-quality |
| S16_vs_61219\|\|full | Not-determined |
| S16_vs_61229\|\|full | Not-determined |
| S16_vs_61250\|\|full | Not-determined |
| S16_vs_61264\|\|full | Low-quality |
| S16_vs_61311\|\|full | Not-determined |
| S16_vs_61402\|\|full | Low-quality |
| S16_vs_61425\|\|full | Not-determined |
| S16_vs_61498\|\|full | Low-quality |
| S16_vs_61511\|\|full | Not-determined |
| S16_vs_61567\|\|full | Low-quality |
| S16_vs_61613\|\|full | Low-quality |
| S16_vs_61617\|\|full | Not-determined |
| S16_vs_61619\|\|full | Not-determined |
| S16_vs_61646\|\|full | Low-quality |
| S16_vs_61648\|\|full | Not-determined |
| S16_vs_61696\|\|full | Not-determined |
| S16_vs_61732\|\|full | Not-determined |
| S16_vs_61753\|\|full | Not-determined |
| S16_vs_61794\|\|full | Not-determined |
| S16_vs_61800\|\|full | Not-determined |
| S16_vs_61831\|\|full | Low-quality |
| S16_vs_61834\|\|full | Not-determined |
| S16_vs_61889\|\|full | Low-quality |
| S16_vs_61928\|\|full | Not-determined |
| S16_vs_61951\|\|full | Not-determined |
| S16_vs_62003\|\|full | Low-quality |
| S16_vs_62064\|\|full | Not-determined |
| S16_vs_62081\|\|full | Low-quality |
| S16_vs_62086\|\|full | Low-quality |
| S16_vs_62089\|\|full | Not-determined |
| S16_vs_62097\|\|full | Not-determined |
| S16_vs_62112\|\|full | Not-determined |
| S16_vs_62132\|\|full | Low-quality |
| S16_vs_62150\|\|full | Low-quality |
| S16_vs_62226\|\|full | Low-quality |
| S16_vs_62229\|\|full | Not-determined |
| S16_vs_62235\|\|full | Low-quality |
| S16_vs_62239\|\|full | Not-determined |
| S16_vs_62255\|\|full | Low-quality |
| S16_vs_62257\|\|full | Low-quality |
| S16_vs_62273\|\|full | Not-determined |
| S16_vs_62316\|\|full | Not-determined |
| S16_vs_62327\|\|full | Not-determined |
| S16_vs_62446\|\|full | Not-determined |
| S16_vs_62450\|\|full | Not-determined |
| S16_vs_62495\|\|full | Not-determined |
| S16_vs_62521\|\|full | Low-quality |
| S16_vs_62537\|\|full | Not-determined |
| S16_vs_62565\|\|full | Not-determined |
| S16_vs_62580\|\|full | Not-determined |
| S16_vs_62597\|\|full | Not-determined |
| S16_vs_62599\|\|full | Not-determined |
| S16_vs_62644\|\|full | Low-quality |
| S16_vs_62671\|\|full | Low-quality |
| S16_vs_62680\|\|full | Low-quality |
| S16_vs_62713\|\|full | Low-quality |
| S16_vs_62761\|\|full | Low-quality |
| S16_vs_62763\|\|full | Low-quality |
| S16_vs_62797\|\|full | Not-determined |
| S16_vs_62843\|\|full | Low-quality |
| S16_vs_62871\|\|full | Low-quality |
| S16_vs_62875\|\|full | Low-quality |
| S16_vs_62880\|\|full | Low-quality |
| S16_vs_62892\|\|full | Not-determined |
| S16_vs_62906\|\|full | Low-quality |
| S16_vs_62907\|\|full | Low-quality |
| S16_vs_62928\|\|full | Low-quality |
| S16_vs_62934\|\|full | Not-determined |
| S16_vs_62989\|\|full | Not-determined |
| S16_vs_63008\|\|full | Low-quality |
| S16_vs_63012\|\|full | Not-determined |
| S16_vs_63020\|\|full | Low-quality |
| S16_vs_63154\|\|full | Low-quality |
| S16_vs_63164\|\|full | Not-determined |
| S16_vs_63184\|\|full | Not-determined |
| S16_vs_63209\|\|full | Low-quality |
| S16_vs_63302\|\|full | Low-quality |
| S16_vs_63318\|\|full | Not-determined |
| S16_vs_63345\|\|full | Not-determined |
| S16_vs_63365\|\|full | Low-quality |
| S16_vs_63426\|\|full | Not-determined |
| S16_vs_63429\|\|full | Low-quality |
| S16_vs_63446\|\|full | Low-quality |
| S16_vs_63468\|\|full | Not-determined |
| S16_vs_63499\|\|full | Low-quality |
| S16_vs_63585\|\|full | Not-determined |
| S16_vs_63635\|\|full | Low-quality |
| S16_vs_63674\|\|full | Not-determined |
| S16_vs_63690\|\|full | Not-determined |
| S16_vs_63699\|\|full | Low-quality |
| S16_vs_63717\|\|full | Low-quality |
| S16_vs_63762\|\|full | Low-quality |
| S16_vs_63782\|\|full | Not-determined |
| S16_vs_63809\|\|full | Low-quality |
| S16_vs_63816\|\|full | Not-determined |
| S16_vs_63817\|\|full | Low-quality |
| S16_vs_63863\|\|full | Not-determined |
| S16_vs_63874\|\|full | Not-determined |
| S16_vs_63890\|\|full | Low-quality |
| S16_vs_63986\|\|full | Not-determined |
| S16_vs_64015\|\|full | Low-quality |
| S16_vs_64018\|\|full | Low-quality |
| S16_vs_64062\|\|full | Not-determined |
| S16_vs_64067\|\|full | Low-quality |
| S16_vs_64092\|\|full | Low-quality |
| S16_vs_64143\|\|full | Low-quality |
| S16_vs_64161\|\|full | Low-quality |
| S16_vs_64177\|\|full | Not-determined |
| S16_vs_64183\|\|full | Not-determined |
| S16_vs_64184\|\|full | Not-determined |
| S16_vs_64204\|\|full | Low-quality |
| S16_vs_64217\|\|full | Not-determined |
| S16_vs_64273\|\|full | Not-determined |
| S16_vs_64289\|\|full | Not-determined |
| S16_vs_64308\|\|full | Not-determined |
| S16_vs_64393\|\|full | Not-determined |
| S16_vs_64440\|\|full | Low-quality |
| S16_vs_64454\|\|full | Low-quality |
| S16_vs_64478\|\|full | Not-determined |
| S16_vs_64491\|\|full | Low-quality |
| S16_vs_64502\|\|full | Not-determined |
| S16_vs_64530\|\|full | Low-quality |
| S16_vs_64541\|\|full | Low-quality |
| S16_vs_64560\|\|full | Not-determined |
| S9_vs_k141_75050\|\|full | High-quality |
| S16_vs_64589\|\|full | Low-quality |
| S16_vs_64603\|\|full | Low-quality |
| S16_vs_64613\|\|full | Low-quality |
| S16_vs_64663\|\|full | Low-quality |
| S16_vs_64680\|\|full | Low-quality |
| S16_vs_64686\|\|full | Low-quality |
| S16_vs_64701\|\|full | Not-determined |
| S16_vs_64751\|\|full | Low-quality |
| S16_vs_64774\|\|full | Not-determined |
| S16_vs_64798\|\|full | Low-quality |
| S16_vs_64835\|\|full | Low-quality |
| S16_vs_65002\|\|full | Not-determined |
| S16_vs_65033\|\|full | Not-determined |
| S16_vs_65041\|\|full | Low-quality |
| S16_vs_65080\|\|full | Low-quality |
| S16_vs_65187\|\|full | Low-quality |
| S16_vs_65219\|\|full | Low-quality |
| S16_vs_65222\|\|full | Low-quality |
| S16_vs_65239\|\|full | Low-quality |
| S16_vs_65280\|\|full | Low-quality |
| S16_vs_65341\|\|full | Low-quality |
| S16_vs_65350\|\|full | Low-quality |
| S16_vs_65403\|\|full | Not-determined |
| S16_vs_65518\|\|full | Low-quality |
| S16_vs_65530\|\|full | Not-determined |
| S16_vs_65579\|\|full | Low-quality |
| S16_vs_65590\|\|full | Low-quality |
| S16_vs_65631\|\|full | Low-quality |
| S16_vs_65731\|\|full | Low-quality |
| S16_vs_65755\|\|full | Low-quality |
| S16_vs_65814\|\|full | Low-quality |
| S16_vs_65888\|\|full | Low-quality |
| S16_vs_65900\|\|full | Low-quality |
| S16_vs_65924\|\|full | Not-determined |
| S16_vs_65957\|\|full | Not-determined |
| S16_vs_65962\|\|full | Low-quality |
| S16_vs_65964\|\|full | Not-determined |
| S16_vs_65968\|\|full | Not-determined |
| S16_vs_66036\|\|full | Low-quality |
| S16_vs_66051\|\|full | Low-quality |
| S16_vs_66083\|\|full | Not-determined |
| S16_vs_66097\|\|full | Not-determined |
| S16_vs_66116\|\|full | Low-quality |
| S16_vs_66159\|\|full | Low-quality |
| S16_vs_66202\|\|full | Low-quality |
| S16_vs_66234\|\|full | Low-quality |
| S16_vs_66279\|\|full | Low-quality |
| S16_vs_66295\|\|full | Low-quality |
| S16_vs_66299\|\|full | Low-quality |
| S16_vs_66402\|\|full | Low-quality |
| S16_vs_66421\|\|full | Low-quality |
| S16_vs_66450\|\|full | Not-determined |
| S16_vs_66459\|\|full | Low-quality |
| S16_vs_66528\|\|full | Low-quality |
| S16_vs_66534\|\|full | Low-quality |
| S16_vs_66543\|\|full | Not-determined |
| S16_vs_66575\|\|full | Low-quality |
| S16_vs_66577\|\|full | Not-determined |
| S16_vs_66592\|\|full | Not-determined |
| S16_vs_66621\|\|full | Low-quality |
| S16_vs_66631\|\|full | Not-determined |
| S16_vs_66634\|\|full | Not-determined |
| S16_vs_66644\|\|full | Low-quality |
| S16_vs_66649\|\|full | Low-quality |
| S16_vs_66659\|\|full | Low-quality |
| S16_vs_66700\|\|full | Not-determined |
| S16_vs_66753\|\|full | Low-quality |
| S16_vs_66766\|\|full | Not-determined |
| S16_vs_66841\|\|full | Not-determined |
| S16_vs_66850\|\|full | Low-quality |
| S16_vs_66903\|\|full | Low-quality |
| S16_vs_66916\|\|full | Low-quality |
| S16_vs_66918\|\|full | Not-determined |
| S16_vs_66949\|\|full | Not-determined |
| S16_vs_66963\|\|full | Not-determined |
| S16_vs_67002\|\|full | Low-quality |
| S16_vs_67030\|\|full | Low-quality |
| S16_vs_67073\|\|full | Not-determined |
| S16_vs_67081\|\|full | Not-determined |
| S16_vs_67232\|\|full | Not-determined |
| S16_vs_67271\|\|full | Low-quality |
| S16_vs_67292\|\|full | Low-quality |
| S16_vs_67294\|\|full | Not-determined |
| S16_vs_67337\|\|full | Low-quality |
| S16_vs_67352\|\|full | Low-quality |
| S16_vs_67366\|\|full | Low-quality |
| S16_vs_67373\|\|full | Not-determined |
| S16_vs_67417\|\|full | Not-determined |
| S16_vs_67559\|\|full | Low-quality |
| S16_vs_67587\|\|full | Low-quality |
| S16_vs_67634\|\|full | Low-quality |
| S16_vs_67638\|\|full | Not-determined |
| S16_vs_67646\|\|full | Low-quality |
| S16_vs_67684\|\|full | Low-quality |
| S16_vs_67713\|\|full | Not-determined |
| S16_vs_67726\|\|full | Not-determined |
| S16_vs_67735\|\|full | Low-quality |
| S16_vs_67740\|\|full | Not-determined |
| S16_vs_67848\|\|full | Low-quality |
| S16_vs_67864\|\|full | Low-quality |
| S16_vs_67867\|\|full | Low-quality |
| S16_vs_67896\|\|full | Not-determined |
| S16_vs_67908\|\|full | Low-quality |
| S16_vs_67970\|\|full | Low-quality |
| S16_vs_67974\|\|full | Low-quality |
| S16_vs_67982\|\|full | Not-determined |
| S16_vs_67990\|\|full | Not-determined |
| S16_vs_68008\|\|full | Low-quality |
| S16_vs_68017\|\|full | Low-quality |
| S16_vs_68036\|\|full | Not-determined |
| S16_vs_68044\|\|full | Not-determined |
| S16_vs_68060\|\|full | Low-quality |
| S16_vs_68117\|\|full | Not-determined |
| S16_vs_68118\|\|full | Low-quality |
| S16_vs_68148\|\|full | Low-quality |
| S16_vs_68160\|\|full | Not-determined |
| S16_vs_68179\|\|full | Low-quality |
| S16_vs_68226\|\|full | Not-determined |
| S16_vs_68235\|\|full | Low-quality |
| S16_vs_68254\|\|full | Not-determined |
| S16_vs_68257\|\|full | Low-quality |
| S16_vs_68324\|\|full | Not-determined |
| S16_vs_68327\|\|full | Not-determined |
| S16_vs_68331\|\|full | Not-determined |
| S16_vs_68350\|\|full | Not-determined |
| S16_vs_68370\|\|full | Low-quality |
| S16_vs_68388\|\|full | Low-quality |
| S16_vs_68507\|\|full | Low-quality |
| S16_vs_68511\|\|full | Not-determined |
| S16_vs_68535\|\|full | Low-quality |
| S16_vs_68539\|\|full | Low-quality |
| S16_vs_68619\|\|full | Low-quality |
| S16_vs_68655\|\|full | Low-quality |
| S16_vs_68659\|\|full | Low-quality |
| S16_vs_68689\|\|full | Low-quality |
| S16_vs_68696\|\|full | Not-determined |
| S16_vs_68715\|\|full | Not-determined |
| S16_vs_68739\|\|full | Low-quality |
| S16_vs_68764\|\|full | Not-determined |
| S16_vs_68774\|\|full | Not-determined |
| S16_vs_68806\|\|full | Not-determined |
| S16_vs_68845\|\|full | Low-quality |
| S16_vs_68877\|\|full | Low-quality |
| S16_vs_68884\|\|full | Low-quality |
| S16_vs_68899\|\|full | Not-determined |
| S16_vs_68913\|\|full | Low-quality |
| S16_vs_68931\|\|full | Not-determined |
| S16_vs_68934\|\|full | Not-determined |
| S16_vs_69013\|\|full | Not-determined |
| S16_vs_69016\|\|full | Low-quality |
| S16_vs_69030\|\|full | Low-quality |
| S16_vs_69038\|\|full | Low-quality |
| S16_vs_69098\|\|full | Low-quality |
| S16_vs_69129\|\|full | Not-determined |
| S16_vs_69141\|\|full | Low-quality |
| S16_vs_69182\|\|full | Low-quality |
| S16_vs_69219\|\|full | Low-quality |
| S16_vs_69230\|\|full | Not-determined |
| S16_vs_69256\|\|full | Low-quality |
| S16_vs_69336\|\|full | Not-determined |
| S16_vs_69391\|\|full | Low-quality |
| S16_vs_69402\|\|full | Not-determined |
| S16_vs_69434\|\|full | Low-quality |
| S16_vs_69518\|\|full | Not-determined |
| S16_vs_69520\|\|full | Not-determined |
| S16_vs_69549\|\|full | Low-quality |
| S16_vs_69647\|\|full | Low-quality |
| S16_vs_69660\|\|full | Not-determined |
| S16_vs_69693\|\|full | Not-determined |
| S16_vs_69730\|\|full | Low-quality |
| S16_vs_69737\|\|full | Low-quality |
| S16_vs_69763\|\|full | Low-quality |
| S16_vs_69772\|\|full | Low-quality |
| S16_vs_69780\|\|full | Low-quality |
| S16_vs_69807\|\|full | Not-determined |
| S16_vs_69838\|\|full | Not-determined |
| S16_vs_69858\|\|full | Not-determined |
| S16_vs_69929\|\|full | Not-determined |
| S16_vs_69945\|\|full | Low-quality |
| S16_vs_70076\|\|full | Low-quality |
| S16_vs_70085\|\|full | Low-quality |
| S16_vs_70140\|\|full | Low-quality |
| S16_vs_70156\|\|full | Low-quality |
| S16_vs_70162\|\|full | Low-quality |
| S16_vs_70166\|\|full | Low-quality |
| S16_vs_70195\|\|full | Low-quality |
| S16_vs_70196\|\|full | Not-determined |
| S16_vs_70201\|\|full | Low-quality |
| S16_vs_70205\|\|full | Not-determined |
| S16_vs_70206\|\|full | Low-quality |
| S16_vs_70245\|\|full | Not-determined |
| S16_vs_70251\|\|full | Not-determined |
| S16_vs_70256\|\|full | Not-determined |
| S16_vs_70295\|\|full | Not-determined |
| S16_vs_70296\|\|full | Low-quality |
| S16_vs_70343\|\|full | Low-quality |
| S16_vs_70377\|\|full | Not-determined |
| S16_vs_70381\|\|full | Not-determined |
| S16_vs_70414\|\|full | Low-quality |
| S16_vs_70423\|\|full | Low-quality |
| S16_vs_70436\|\|full | Not-determined |
| S16_vs_70465\|\|full | Low-quality |
| S16_vs_70504\|\|full | Not-determined |
| S16_vs_70548\|\|full | Low-quality |
| S16_vs_70549\|\|full | Low-quality |
| S16_vs_70579\|\|full | Not-determined |
| S16_vs_70585\|\|full | Low-quality |
| S16_vs_70599\|\|full | Not-determined |
| S16_vs_70624\|\|full | Not-determined |
| S16_vs_70662\|\|full | Low-quality |
| S16_vs_70672\|\|full | Low-quality |
| S16_vs_70687\|\|full | Low-quality |
| S16_vs_70696\|\|full | Not-determined |
| S16_vs_70709\|\|full | Not-determined |
| S16_vs_70753\|\|full | Low-quality |
| S16_vs_70782\|\|full | Low-quality |
| S16_vs_70816\|\|full | Low-quality |
| S16_vs_70844\|\|full | Not-determined |
| S16_vs_70867\|\|full | Low-quality |
| S16_vs_70933\|\|full | Not-determined |
| S16_vs_71000\|\|full | Not-determined |
| S16_vs_71011\|\|full | Not-determined |
| S16_vs_71024\|\|full | Low-quality |
| S16_vs_71041\|\|full | Not-determined |
| S16_vs_71046\|\|full | Not-determined |
| S16_vs_71071\|\|full | Low-quality |
| S16_vs_71115\|\|full | Not-determined |
| S16_vs_71116\|\|full | Low-quality |
| S16_vs_71136\|\|full | Not-determined |
| S16_vs_71154\|\|full | Not-determined |
| S16_vs_71172\|\|full | Not-determined |
| S16_vs_71180\|\|full | Low-quality |
| S16_vs_71187\|\|full | Not-determined |
| S16_vs_71213\|\|full | Not-determined |
| S16_vs_71259\|\|full | Low-quality |
| S16_vs_71260\|\|full | Low-quality |
| S16_vs_71269\|\|full | Low-quality |
| S16_vs_71316\|\|full | Not-determined |
| S16_vs_71322\|\|full | Low-quality |
| S16_vs_71346\|\|full | Low-quality |
| S16_vs_71386\|\|full | Not-determined |
| S16_vs_71418\|\|full | Not-determined |
| S16_vs_71434\|\|full | Low-quality |
| S16_vs_71451\|\|full | Not-determined |
| S16_vs_71457\|\|full | Low-quality |
| S16_vs_71473\|\|full | Not-determined |
| S16_vs_71544\|\|full | Not-determined |
| S16_vs_71567\|\|full | Not-determined |
| S16_vs_71598\|\|full | Not-determined |
| S16_vs_71601\|\|full | Low-quality |
| S16_vs_71618\|\|full | Low-quality |
| S16_vs_71626\|\|full | Low-quality |
| S16_vs_71631\|\|full | Low-quality |
| S16_vs_71663\|\|full | Not-determined |
| S16_vs_71704\|\|full | Low-quality |
| S16_vs_71762\|\|full | Low-quality |
| S16_vs_71806\|\|full | Low-quality |
| S16_vs_71832\|\|full | Low-quality |
| S16_vs_71835\|\|full | Not-determined |
| S16_vs_71856\|\|full | Not-determined |
| S16_vs_71872\|\|full | Low-quality |
| S16_vs_71924\|\|full | Not-determined |
| S16_vs_71973\|\|full | Not-determined |
| S16_vs_71975\|\|full | Not-determined |
| S16_vs_72057\|\|full | Low-quality |
| S16_vs_72062\|\|full | Low-quality |
| S16_vs_72073\|\|full | Not-determined |
| S16_vs_72085\|\|full | Low-quality |
| S16_vs_72111\|\|full | Not-determined |
| S16_vs_72170\|\|full | Low-quality |
| S16_vs_72248\|\|full | Low-quality |
| S16_vs_72261\|\|full | Low-quality |
| S16_vs_72283\|\|full | Low-quality |
| S16_vs_72285\|\|full | Not-determined |
| S16_vs_72346\|\|full | Low-quality |
| S16_vs_72387\|\|full | Low-quality |
| S16_vs_72395\|\|full | Not-determined |
| S16_vs_72405\|\|full | Not-determined |
| S16_vs_72425\|\|full | Low-quality |
| S16_vs_72465\|\|full | Not-determined |
| S16_vs_72517\|\|full | Not-determined |
| S16_vs_72519\|\|full | Low-quality |
| S16_vs_72567\|\|full | Low-quality |
| S16_vs_72631\|\|full | Not-determined |
| S16_vs_72655\|\|full | Low-quality |
| S16_vs_72686\|\|full | Low-quality |
| S16_vs_72689\|\|full | Not-determined |
| S16_vs_72714\|\|full | Low-quality |
| S16_vs_72766\|\|full | Not-determined |
| S16_vs_72815\|\|full | Low-quality |
| S16_vs_72819\|\|full | Not-determined |
| S16_vs_72827\|\|full | Not-determined |
| S16_vs_72853\|\|full | Not-determined |
| S16_vs_72916\|\|full | Not-determined |
| S16_vs_72921\|\|full | Not-determined |
| S16_vs_72966\|\|full | Low-quality |
| S16_vs_72974\|\|full | Not-determined |
| S16_vs_72982\|\|full | Low-quality |
| S16_vs_73012\|\|full | Low-quality |
| S16_vs_73059\|\|full | Low-quality |
| S16_vs_73068\|\|full | Low-quality |
| S16_vs_73083\|\|full | Not-determined |
| S16_vs_73085\|\|full | Low-quality |
| S16_vs_73128\|\|full | Not-determined |
| S16_vs_73135\|\|full | Low-quality |
| S16_vs_73349\|\|full | Not-determined |
| S16_vs_73360\|\|full | Not-determined |
| S16_vs_73394\|\|full | Not-determined |
| S16_vs_73427\|\|full | Not-determined |
| S16_vs_73503\|\|full | Low-quality |
| S16_vs_73516\|\|full | Not-determined |
| S16_vs_73565\|\|full | Low-quality |
| S16_vs_73593\|\|full | Low-quality |
| S16_vs_73684\|\|full | Low-quality |
| S16_vs_73808\|\|full | Not-determined |
| S16_vs_73851\|\|full | Not-determined |
| S16_vs_73889\|\|full | Not-determined |
| S16_vs_73906\|\|full | Low-quality |
| S16_vs_73955\|\|full | Low-quality |
| S16_vs_73963\|\|full | Low-quality |
| S16_vs_73984\|\|full | Not-determined |
| S16_vs_74011\|\|full | Low-quality |
| S16_vs_74047\|\|full | Not-determined |
| S16_vs_74062\|\|full | Low-quality |
| S16_vs_74097\|\|full | Not-determined |
| S16_vs_74111\|\|full | Low-quality |
| S16_vs_74175\|\|full | Low-quality |
| S16_vs_74182\|\|full | Low-quality |
| S16_vs_74218\|\|full | Low-quality |
| S16_vs_74256\|\|full | Not-determined |
| S16_vs_74267\|\|full | Low-quality |
| S16_vs_74277\|\|full | Low-quality |
| S16_vs_74430\|\|full | Low-quality |
| S16_vs_74445\|\|full | Low-quality |
| S16_vs_74516\|\|full | Low-quality |
| S16_vs_74555\|\|full | Low-quality |
| S16_vs_74556\|\|full | Not-determined |
| S16_vs_74579\|\|full | Not-determined |
| S16_vs_74582\|\|full | Not-determined |
| S16_vs_74592\|\|full | Low-quality |
| S16_vs_74620\|\|full | Low-quality |
| S16_vs_74668\|\|full | Low-quality |
| S16_vs_74675\|\|full | Low-quality |
| S16_vs_74764\|\|full | Low-quality |
| S16_vs_74781\|\|full | Not-determined |
| S16_vs_74799\|\|full | Low-quality |
| S16_vs_74800\|\|full | Low-quality |
| S16_vs_74805\|\|full | Low-quality |
| S16_vs_74815\|\|full | Low-quality |
| S16_vs_74821\|\|full | Low-quality |
| S16_vs_74836\|\|full | Low-quality |
| S16_vs_74859\|\|full | Not-determined |
| S16_vs_74919\|\|full | Low-quality |
| S16_vs_74925\|\|full | Low-quality |
| S16_vs_74934\|\|full | Not-determined |
| S16_vs_74935\|\|full | Not-determined |
| S16_vs_74986\|\|full | Low-quality |
| S16_vs_75079\|\|full | Not-determined |
| S16_vs_75080\|\|full | Not-determined |
| S16_vs_75084\|\|full | Not-determined |
| S16_vs_75150\|\|full | Low-quality |
| S16_vs_75220\|\|full | Not-determined |
| S16_vs_75224\|\|full | Not-determined |
| S16_vs_75251\|\|full | Low-quality |
| S16_vs_75306\|\|full | Low-quality |
| S16_vs_75335\|\|full | Not-determined |
| S16_vs_75404\|\|full | Low-quality |
| S16_vs_75405\|\|full | Not-determined |
| S16_vs_75430\|\|full | Not-determined |
| S16_vs_75436\|\|full | Low-quality |
| S16_vs_75445\|\|full | Low-quality |
| S16_vs_75496\|\|full | Not-determined |
| S16_vs_75497\|\|full | Low-quality |
| S16_vs_75517\|\|full | Low-quality |
| S16_vs_75540\|\|full | Not-determined |
| S16_vs_75547\|\|full | Low-quality |
| S16_vs_75549\|\|full | Not-determined |
| S16_vs_75595\|\|full | Low-quality |
| S16_vs_75603\|\|full | Low-quality |
| S16_vs_75616\|\|full | Low-quality |
| S16_vs_75654\|\|full | Not-determined |
| S16_vs_75737\|\|full | Low-quality |
| S16_vs_75747\|\|full | Not-determined |
| S16_vs_75797\|\|full | Not-determined |
| S16_vs_75800\|\|full | Low-quality |
| S16_vs_75839\|\|full | Not-determined |
| S16_vs_75849\|\|full | Not-determined |
| S16_vs_75863\|\|full | Not-determined |
| S16_vs_75870\|\|full | Not-determined |
| S16_vs_75876\|\|full | Low-quality |
| S16_vs_75901\|\|full | Low-quality |
| S16_vs_75950\|\|full | Not-determined |
| S16_vs_75987\|\|full | Not-determined |

| | |
|---|---|
| S16_vs_76001\|\|full | Low-quality |
| S16_vs_76010\|\|full | Low-quality |
| S16_vs_76025\|\|full | Low-quality |
| S16_vs_76074\|\|full | Low-quality |
| S16_vs_76089\|\|full | Not-determined |
| S16_vs_76092\|\|full | Not-determined |
| S16_vs_76120\|\|full | Low-quality |
| S16_vs_76158\|\|full | Low-quality |
| S16_vs_76192\|\|full | Not-determined |
| S16_vs_76194\|\|full | Low-quality |
| S16_vs_76219\|\|full | Low-quality |
| S16_vs_76308\|\|full | Low-quality |
| S16_vs_76327\|\|full | Not-determined |
| S16_vs_76333\|\|full | Not-determined |
| S16_vs_76336\|\|full | Low-quality |
| S16_vs_76362\|\|full | Not-determined |
| S16_vs_76370\|\|full | Low-quality |
| S16_vs_76422\|\|full | Not-determined |
| S16_vs_76540\|\|full | Low-quality |
| S16_vs_76587\|\|full | Not-determined |
| S16_vs_76591\|\|full | Not-determined |
| S16_vs_76599\|\|full | Low-quality |
| S16_vs_76605\|\|full | Low-quality |
| S16_vs_76631\|\|full | Low-quality |
| S16_vs_76636\|\|full | Low-quality |
| S16_vs_76645\|\|full | Low-quality |
| S16_vs_76756\|\|full | Low-quality |
| S16_vs_76771\|\|full | Low-quality |
| S16_vs_76778\|\|full | Not-determined |
| S16_vs_76849\|\|full | Low-quality |
| S16_vs_76906\|\|full | Not-determined |
| S16_vs_76914\|\|full | Not-determined |
| S16_vs_76928\|\|full | Low-quality |
| S16_vs_76934\|\|full | Low-quality |
| S16_vs_76992\|\|full | Low-quality |
| S16_vs_77020\|\|full | Not-determined |
| S16_vs_77044\|\|full | Not-determined |
| S16_vs_77061\|\|full | Not-determined |
| S16_vs_77072\|\|full | Low-quality |
| S16_vs_77073\|\|full | Not-determined |
| S16_vs_77135\|\|full | Not-determined |
| S16_vs_77177\|\|full | Low-quality |
| S16_vs_77186\|\|full | Low-quality |
| S16_vs_77212\|\|full | Not-determined |
| S16_vs_77217\|\|full | Low-quality |
| S16_vs_77219\|\|full | Not-determined |
| S16_vs_77252\|\|full | Low-quality |
| S16_vs_77258\|\|full | Not-determined |
| S16_vs_77385\|\|full | Low-quality |
| S16_vs_77396\|\|full | Low-quality |
| S16_vs_77436\|\|full | Not-determined |
| S16_vs_77448\|\|full | Low-quality |
| S16_vs_77457\|\|full | Not-determined |
| S16_vs_77471\|\|full | Not-determined |
| S16_vs_77473\|\|full | Low-quality |
| S16_vs_77484\|\|full | Low-quality |
| S16_vs_77497\|\|full | Low-quality |
| S16_vs_77501\|\|full | Not-determined |
| S16_vs_77506\|\|full | Low-quality |
| S16_vs_77511\|\|full | Not-determined |
| S16_vs_77554\|\|full | Low-quality |
| S16_vs_77656\|\|full | Not-determined |
| S16_vs_77692\|\|full | Not-determined |
| S16_vs_77703\|\|full | Low-quality |
| S16_vs_77704\|\|full | Not-determined |
| S16_vs_77761\|\|full | Not-determined |
| S16_vs_77803\|\|full | Not-determined |
| S16_vs_77808\|\|full | Not-determined |
| S16_vs_77812\|\|full | Not-determined |
| S16_vs_77848\|\|full | Low-quality |
| S16_vs_77900\|\|full | Low-quality |
| S16_vs_77914\|\|full | Low-quality |
| S16_vs_77919\|\|full | Low-quality |
| S16_vs_77931\|\|full | Not-determined |
| S16_vs_77940\|\|full | Low-quality |
| S16_vs_78011\|\|full | Not-determined |
| [illegible] | [illegible] |

| | |
|---|---|
| S16_vs_92554\|\|full | Low-quality |
| S16_vs_92555\|\|full | Not-determined |
| S16_vs_92644\|\|full | Low-quality |
| S16_vs_92650\|\|full | Not-determined |
| S16_vs_92723\|\|full | Low-quality |
| S16_vs_92728\|\|full | Low-quality |
| S16_vs_92784\|\|full | Low-quality |
| S16_vs_92843\|\|full | Low-quality |
| S16_vs_92870\|\|full | Not-determined |
| S16_vs_92910\|\|full | Not-determined |
| S16_vs_92975\|\|full | Not-determined |
| S16_vs_92978\|\|full | Low-quality |
| S16_vs_92990\|\|full | Low-quality |
| S16_vs_93040\|\|full | Low-quality |
| S16_vs_93048\|\|full | Low-quality |
| S16_vs_93049\|\|full | Low-quality |
| S16_vs_93103\|\|full | Not-determined |
| S16_vs_93112\|\|full | Low-quality |
| S16_vs_93131\|\|full | Low-quality |
| S16_vs_93161\|\|full | Low-quality |
| S16_vs_93167\|\|full | Not-determined |
| S16_vs_93186\|\|full | Low-quality |
| S16_vs_93195\|\|full | Low-quality |
| S16_vs_93201\|\|full | Low-quality |
| S16_vs_93206\|\|full | Not-determined |
| S16_vs_93216\|\|full | Not-determined |
| S16_vs_93228\|\|full | Low-quality |
| S16_vs_93234\|\|full | Low-quality |
| S16_vs_93254\|\|full | Low-quality |
| S16_vs_93295\|\|full | Low-quality |
| S16_vs_93305\|\|full | Low-quality |
| S16_vs_93314\|\|full | Low-quality |
| S16_vs_93380\|\|full | Not-determined |
| S16_vs_93396\|\|full | Not-determined |
| S16_vs_93409\|\|full | Low-quality |
| S16_vs_93484\|\|full | Not-determined |
| S16_vs_93491\|\|full | Low-quality |
| S16_vs_93494\|\|full | Low-quality |
| S16_vs_93544\|\|full | Not-determined |
| S16_vs_93675\|\|full | Not-determined |
| S16_vs_93697\|\|full | Low-quality |
| S16_vs_93726\|\|full | Low-quality |
| S16_vs_93732\|\|full | Low-quality |
| S16_vs_93776\|\|full | Not-determined |
| S16_vs_93811\|\|full | Low-quality |
| S16_vs_93816\|\|full | Low-quality |
| S16_vs_93915\|\|full | Low-quality |
| S16_vs_93925\|\|full | Low-quality |
| S16_vs_93949\|\|full | Not-determined |
| S16_vs_93993\|\|full | Low-quality |
| S16_vs_93995\|\|full | Not-determined |
| S16_vs_94000\|\|full | Low-quality |
| S16_vs_94002\|\|full | Low-quality |
| S16_vs_94037\|\|full | Not-determined |
| S16_vs_94075\|\|full | Not-determined |
| S16_vs_94098\|\|full | Low-quality |
| S16_vs_94112\|\|full | Not-determined |
| S16_vs_94166\|\|full | Low-quality |
| S16_vs_94176\|\|full | Not-determined |
| S16_vs_94186\|\|full | Low-quality |
| S16_vs_94259\|\|full | Low-quality |
| S16_vs_94284\|\|full | Low-quality |
| S16_vs_94300\|\|full | Low-quality |
| S16_vs_94319\|\|full | Low-quality |
| S16_vs_94340\|\|full | Not-determined |
| S16_vs_94346\|\|full | Low-quality |
| S16_vs_94357\|\|full | Not-determined |
| S16_vs_94379\|\|full | Low-quality |
| S16_vs_94389\|\|full | Low-quality |
| S16_vs_94396\|\|full | Low-quality |
| S16_vs_94400\|\|full | Not-determined |
| S16_vs_94421\|\|full | Low-quality |
| S16_vs_94422\|\|full | Low-quality |
| S16_vs_94431\|\|full | Not-determined |
| S16_vs_94488\|\|full | Low-quality |
| S16_vs_94519\|\|full | Low-quality |
| S16_vs_94531\|\|full | Not-determined |
| S16_vs_94556\|\|full | Not-determined |
| S16_vs_94619\|\|full | Low-quality |
| S16_vs_94625\|\|full | Low-quality |
| S16_vs_94687\|\|full | Low-quality |
| S16_vs_94701\|\|full | Low-quality |
| S16_vs_94712\|\|full | Low-quality |
| S16_vs_94741\|\|full | Low-quality |
| S16_vs_94793\|\|full | Low-quality |
| S16_vs_94801\|\|full | Low-quality |
| S16_vs_94802\|\|full | Not-determined |
| S16_vs_94841\|\|full | Not-determined |
| S16_vs_94850\|\|full | Low-quality |
| S16_vs_81805\|\|full | Medium-quality |
| S16_vs_94911\|\|full | Low-quality |
| S16_vs_94938\|\|full | Low-quality |
| S16_vs_94948\|\|full | Not-determined |
| S16_vs_94972\|\|full | Low-quality |
| S16_vs_94990\|\|full | Low-quality |
| S16_vs_95019\|\|full | Low-quality |
| S16_vs_95043\|\|full | Not-determined |
| S16_vs_95063\|\|full | Not-determined |
| S16_vs_95076\|\|full | Low-quality |
| S16_vs_95077\|\|full | Low-quality |
| S16_vs_95164\|\|full | Low-quality |
| S16_vs_95175\|\|full | Low-quality |
| S16_vs_95178\|\|full | Low-quality |
| S16_vs_95182\|\|full | Not-determined |
| S16_vs_95188\|\|full | Low-quality |
| S16_vs_95190\|\|full | Low-quality |
| S16_vs_95209\|\|full | Not-determined |
| S16_vs_95246\|\|full | Low-quality |
| S16_vs_95272\|\|full | Not-determined |
| S16_vs_95285\|\|full | Low-quality |
| S16_vs_95309\|\|full | Not-determined |
| S16_vs_95382\|\|full | Low-quality |
| S16_vs_95410\|\|full | Low-quality |
| S16_vs_95463\|\|full | Low-quality |
| S16_vs_95508\|\|full | Low-quality |
| S16_vs_95575\|\|full | Low-quality |
| S16_vs_95582\|\|full | Not-determined |
| S16_vs_95602\|\|full | Low-quality |
| S16_vs_95604\|\|full | Low-quality |
| S16_vs_95632\|\|full | Low-quality |
| S16_vs_95670\|\|full | Low-quality |
| S16_vs_95674\|\|full | Low-quality |
| S16_vs_95680\|\|full | Low-quality |
| S16_vs_95689\|\|full | Low-quality |
| S16_vs_95692\|\|full | Not-determined |
| S16_vs_95717\|\|full | Not-determined |
| S16_vs_95721\|\|full | Low-quality |
| S16_vs_95742\|\|full | Not-determined |
| S16_vs_95746\|\|full | Not-determined |
| S16_vs_95790\|\|full | Low-quality |
| S16_vs_95806\|\|full | Low-quality |
| S16_vs_95823\|\|full | Low-quality |
| S16_vs_95850\|\|full | Low-quality |
| S16_vs_95854\|\|full | Not-determined |
| S16_vs_95879\|\|full | Low-quality |
| S16_vs_95892\|\|full | Low-quality |
| S16_vs_95901\|\|full | Low-quality |
| S16_vs_95909\|\|full | Not-determined |
| S16_vs_95910\|\|full | Not-determined |
| S16_vs_95915\|\|full | Not-determined |
| S16_vs_95931\|\|full | Low-quality |
| S16_vs_95937\|\|full | Not-determined |
| S16_vs_95940\|\|full | Low-quality |
| S16_vs_95962\|\|full | Low-quality |
| S16_vs_95968\|\|full | Not-determined |
| S16_vs_95991\|\|full | Not-determined |
| S16_vs_96050\|\|full | Not-determined |
| S16_vs_96051\|\|full | Not-determined |
| S16_vs_96072\|\|full | Not-determined |
| S16_vs_96124\|\|full | Not-determined |
| S16_vs_96171\|\|full | Not-determined |
| S16_vs_96263\|\|full | Not-determined |
| S16_vs_96268\|\|full | Low-quality |
| S16_vs_96278\|\|full | Not-determined |
| S16_vs_96307\|\|full | Low-quality |
| S16_vs_96340\|\|full | Not-determined |
| S16_vs_96341\|\|full | Not-determined |
| S16_vs_96393\|\|full | Low-quality |
| S16_vs_96400\|\|full | Not-determined |
| S16_vs_96470\|\|full | Low-quality |
| S16_vs_96480\|\|full | Low-quality |
| S11_vs_k141_47632\|\|full | High-quality |
| S1_vs_k141_57546\|\|full | Medium-quality |
| S16_vs_96531\|\|full | Low-quality |
| S16_vs_96545\|\|full | Low-quality |
| S16_vs_96561\|\|full | Not-determined |
| S16_vs_96575\|\|full | Low-quality |
| S16_vs_96584\|\|full | Not-determined |
| S16_vs_96607\|\|full | Not-determined |
| S16_vs_96630\|\|full | Low-quality |
| S16_vs_96671\|\|full | Not-determined |
| S16_vs_96687\|\|full | Not-determined |
| S16_vs_96721\|\|full | Not-determined |
| S16_vs_96732\|\|full | Low-quality |
| S16_vs_96822\|\|full | Not-determined |
| S16_vs_96844\|\|full | Not-determined |
| S14_vs_k141_62178\|\|full | High-quality |
| S16_vs_96884\|\|full | Not-determined |
| S16_vs_96906\|\|full | Low-quality |
| S16_vs_96947\|\|full | Not-determined |
| S16_vs_96975\|\|full | Not-determined |
| S16_vs_97117\|\|full | Not-determined |
| S16_vs_97121\|\|full | Not-determined |
| S16_vs_97139\|\|full | Low-quality |
| S16_vs_97177\|\|full | Low-quality |
| S16_vs_97214\|\|full | Not-determined |
| S16_vs_97258\|\|full | Not-determined |
| S16_vs_97273\|\|full | Not-determined |
| S4_vs_VIRSorter_13488-cat_2 | High-quality |
| S16_vs_97287\|\|full | Low-quality |
| S16_vs_97306\|\|full | Low-quality |
| S16_vs_97308\|\|full | Not-determined |
| S16_vs_97312\|\|full | Not-determined |
| S16_vs_97324\|\|full | Not-determined |
| S16_vs_97337\|\|full | Low-quality |
| S16_vs_97409\|\|full | Not-determined |
| S16_vs_97442\|\|full | Not-determined |
| S16_vs_97447\|\|full | Low-quality |
| S16_vs_97459\|\|full | Low-quality |
| S16_vs_97466\|\|full | Low-quality |
| S16_vs_97470\|\|full | Low-quality |
| S16_vs_97485\|\|full | Low-quality |
| S16_vs_97501\|\|full | Low-quality |
| S11_vs_k141_3006\|\|full | Medium-quality |
| S16_vs_97514\|\|full | Not-determined |
| S16_vs_97552\|\|full | Low-quality |
| S16_vs_97578\|\|full | Low-quality |
| S16_vs_97608\|\|full | Low-quality |
| S16_vs_97611\|\|full | Low-quality |
| S16_vs_97619\|\|full | Low-quality |
| S16_vs_97634\|\|full | Low-quality |
| S16_vs_97655\|\|full | Low-quality |
| S16_vs_97670\|\|full | Low-quality |
| S16_vs_97679\|\|full | Low-quality |
| S16_vs_97751\|\|full | Not-determined |
| S16_vs_97840\|\|full | Low-quality |
| S16_vs_97893\|\|full | Not-determined |
| S16_vs_97959\|\|full | Low-quality |
| S16_vs_98051\|\|full | Low-quality |
| S16_vs_98065\|\|full | Low-quality |
| S16_vs_98081\|\|full | Low-quality |
| S16_vs_98140\|\|full | Low-quality |
| S16_vs_98148\|\|full | Low-quality |
| S16_vs_98174\|\|full | Not-determined |
| S16_vs_98178\|\|full | Low-quality |
| S16_vs_98184\|\|full | Not-determined |
| S16_vs_98242\|\|full | Low-quality |
| S16_vs_98257\|\|full | Low-quality |
| S16_vs_98284\|\|full | Low-quality |
| S16_vs_98347\|\|full | Low-quality |
| S16_vs_98354\|\|full | Low-quality |
| S16_vs_98437\|\|full | Low-quality |
| S16_vs_98449\|\|full | Low-quality |
| S16_vs_98473\|\|full | Low-quality |
| S16_vs_98492\|\|full | Low-quality |
| S16_vs_98507\|\|full | Low-quality |
| S16_vs_98547\|\|full | Low-quality |
| S16_vs_98626\|\|full | Not-determined |
| S16_vs_98641\|\|full | Low-quality |
| S16_vs_98646\|\|full | Low-quality |
| S16_vs_98647\|\|full | Low-quality |
| S16_vs_98743\|\|full | Low-quality |
| S16_vs_98750\|\|full | Not-determined |
| S16_vs_98757\|\|full | Low-quality |
| S16_vs_98809\|\|full | Not-determined |
| S16_vs_98901\|\|full | Not-determined |
| S16_vs_99010\|\|full | Not-determined |
| S16_vs_99082\|\|full | Low-quality |
| S16_vs_99124\|\|full | Not-determined |
| S16_vs_99139\|\|full | Low-quality |
| S16_vs_99172\|\|full | Not-determined |
| S16_vs_99183\|\|full | Low-quality |
| S16_vs_99248\|\|full | Not-determined |
| S16_vs_99288\|\|full | Not-determined |
| S16_vs_99350\|\|full | Not-determined |
| S16_vs_99368\|\|full | Low-quality |
| S16_vs_99422\|\|full | Low-quality |
| S16_vs_99438\|\|full | Not-determined |
| S16_vs_99444\|\|full | Low-quality |
| S16_vs_99460\|\|full | Not-determined |
| S16_vs_99476\|\|full | Low-quality |
| S16_vs_99492\|\|full | Low-quality |
| S16_vs_99540\|\|full | Low-quality |
| S16_vs_99620\|\|full | Low-quality |
| S16_vs_99689\|\|full | Low-quality |
| S16_vs_99701\|\|full | Not-determined |
| S16_vs_99712\|\|full | Low-quality |
| S16_vs_99715\|\|full | Low-quality |
| S16_vs_99718\|\|full | Not-determined |
| S16_vs_99773\|\|full | Not-determined |
| S16_vs_99797\|\|full | Not-determined |
| S16_vs_99826\|\|full | Low-quality |
| S16_vs_99835\|\|full | Low-quality |
| S16_vs_99870\|\|full | Low-quality |
| S16_vs_99886\|\|full | Low-quality |
| S16_vs_99916\|\|full | Low-quality |
| S16_vs_100035\|\|full | Low-quality |
| S16_vs_100064\|\|full | Not-determined |
| S16_vs_100088\|\|full | Not-determined |
| S16_vs_100107\|\|full | Low-quality |
| S16_vs_100113\|\|full | Low-quality |
| S16_vs_100160\|\|full | Low-quality |
| S16_vs_100170\|\|full | Low-quality |
| S16_vs_100214\|\|full | Not-determined |
| S16_vs_100221\|\|full | Low-quality |
| S16_vs_100224\|\|full | Low-quality |
| S16_vs_100225\|\|full | Not-determined |
| S16_vs_100234\|\|full | Low-quality |
| S7_extracted_68546 | Medium-quality |
| S16_vs_100294\|\|full | Low-quality |
| S16_vs_100337\|\|full | Not-determined |
| S16_vs_100358\|\|full | Low-quality |
| S16_vs_100359\|\|full | Low-quality |
| S17_extracted_k141_21718 | High-quality |
| S16_vs_100381\|\|full | Not-determined |
| S16_vs_100405\|\|full | Not-determined |
| S5_vs_k141_145597\|\|full | High-quality |
| S16_vs_100505\|\|full | Low-quality |
| S16_vs_100506\|\|full | Low-quality |
| S16_vs_100513\|\|full | Low-quality |
| S16_vs_100525\|\|full | Not-determined |
| S16_vs_100596\|\|full | Not-determined |
| S16_vs_100684\|\|full | Not-determined |
| S16_vs_100703\|\|full | Not-determined |
| S16_vs_100770\|\|full | Low-quality |
| S16_vs_100787\|\|full | Low-quality |
| S16_vs_100825\|\|full | Not-determined |
| S16_vs_100886\|\|full | Low-quality |
| S16_vs_100887\|\|full | Low-quality |
| S16_vs_100929\|\|full | Low-quality |
| S16_vs_100932\|\|full | Low-quality |
| S16_vs_100935\|\|full | Not-determined |
| S16_vs_100944\|\|full | Not-determined |
| S16_vs_100947\|\|full | Not-determined |
| S16_vs_100971\|\|full | Low-quality |
| S16_vs_101028\|\|full | Not-determined |
| S16_vs_101029\|\|full | Low-quality |
| S16_vs_101034\|\|full | Low-quality |
| S16_vs_101037\|\|full | Not-determined |
| S16_vs_101059\|\|full | Low-quality |
| S12_vs_k141_32781\|\|full | High-quality |
| S16_vs_101086\|\|full | Low-quality |
| S16_vs_101101\|\|full | Not-determined |
| S16_vs_101102\|\|full | Not-determined |
| S16_vs_101126\|\|full | Low-quality |
| S16_vs_101151\|\|full | Low-quality |
| S16_vs_101188\|\|full | Not-determined |
| S16_vs_101193\|\|full | Low-quality |
| S16_vs_101203\|\|full | Low-quality |
| S16_vs_101221\|\|full | Low-quality |
| S16_vs_101262\|\|full | Not-determined |
| S16_vs_101264\|\|full | Low-quality |
| S16_vs_101300\|\|full | Not-determined |
| S16_vs_101323\|\|full | Low-quality |
| S16_vs_101328\|\|full | Low-quality |
| S16_vs_101370\|\|full | Low-quality |
| S16_vs_101407\|\|full | Low-quality |
| S16_vs_101437\|\|full | Low-quality |
| S16_vs_101491\|\|full | Low-quality |
| S16_vs_101494\|\|full | Not-determined |
| S16_vs_101510\|\|full | Not-determined |
| S16_vs_101517\|\|full | Not-determined |
| S16_vs_101542\|\|full | Not-determined |
| S16_vs_101575\|\|full | Low-quality |
| S16_vs_101592\|\|full | Low-quality |
| S16_vs_101612\|\|full | Low-quality |
| S16_vs_101614\|\|full | Not-determined |
| S16_vs_101615\|\|full | Not-determined |
| S1_vs_k141_57540\|\|full | Medium-quality |
| S16_vs_101681\|\|full | Low-quality |
| S16_vs_101799\|\|full | Not-determined |
| S16_vs_101805\|\|full | Low-quality |
| S16_vs_101826\|\|full | Low-quality |
| S16_vs_101876\|\|full | Low-quality |
| S16_vs_101886\|\|full | Not-determined |
| S16_vs_102019\|\|full | Not-determined |
| S16_vs_102052\|\|full | Not-determined |
| S16_vs_102069\|\|full | Not-determined |
| S16_vs_102081\|\|full | Not-determined |
| S16_vs_102148\|\|full | Low-quality |
| S16_vs_102178\|\|full | Not-determined |
| S16_vs_102186\|\|full | Low-quality |
| S16_vs_102206\|\|full | Low-quality |
| S16_vs_102334\|\|full | Not-determined |
| S16_vs_102369\|\|full | Not-determined |
| S16_vs_102384\|\|full | Not-determined |
| S16_vs_102432\|\|full | Not-determined |
| S16_vs_102474\|\|full | Not-determined |
| S16_vs_102498\|\|full | Low-quality |
| S16_vs_102506\|\|full | Not-determined |
| S16_vs_102534\|\|full | Low-quality |
| S16_vs_102535\|\|full | Low-quality |
| S16_vs_102538\|\|full | Low-quality |
| S16_vs_102575\|\|full | Low-quality |
| S16_vs_102593\|\|full | Low-quality |
| S16_vs_102596\|\|full | Low-quality |
| S16_vs_102597\|\|full | Low-quality |
| S16_vs_102627\|\|full | Low-quality |
| S16_vs_102665\|\|full | Low-quality |
| S16_vs_102667\|\|full | Low-quality |
| S16_vs_102683\|\|full | Low-quality |
| S16_vs_102692\|\|full | Not-determined |
| S16_vs_102769\|\|full | Low-quality |
| S16_vs_102782\|\|full | Low-quality |
| S16_vs_102792\|\|full | Low-quality |
| S16_vs_102835\|\|full | Not-determined |
| S16_vs_102847\|\|full | Not-determined |
| S16_vs_102895\|\|full | Low-quality |
| S16_vs_102929\|\|full | Not-determined |
| S16_vs_102964\|\|full | Low-quality |
| S16_vs_102985\|\|full | Low-quality |
| S16_vs_103105\|\|full | Not-determined |
| S16_vs_103149\|\|full | Low-quality |
| S16_vs_103179\|\|full | Low-quality |
| S16_vs_103197\|\|full | Not-determined |
| S16_vs_103213\|\|full | Not-determined |
| S16_vs_103230\|\|full | Low-quality |
| S16_vs_103240\|\|full | Low-quality |
| S16_vs_103252\|\|full | Low-quality |
| S16_vs_103287\|\|full | Low-quality |
| S16_vs_103288\|\|full | Not-determined |
| S16_vs_103295\|\|full | Not-determined |
| S16_vs_103311\|\|full | Not-determined |
| S16_vs_103315\|\|full | Not-determined |
| S16_vs_103404\|\|full | Not-determined |
| S16_vs_103442\|\|full | Not-determined |
| S12_extracted_k141_69136 | Medium-quality |
| S16_vs_103461\|\|full | Low-quality |
| S16_vs_103477\|\|full | Not-determined |
| S16_vs_103485\|\|full | Low-quality |
| S16_vs_103570\|\|full | Low-quality |
| S16_vs_103579\|\|full | Low-quality |
| S16_vs_103619\|\|full | Low-quality |
| S16_vs_103663\|\|full | Not-determined |
| S7_vs_68546\|\|full | Medium-quality |
| S16_vs_103769\|\|full | Low-quality |
| S16_vs_103824\|\|full | Low-quality |
| S16_vs_103881\|\|full | Low-quality |
| S16_vs_103887\|\|full | Low-quality |
| S16_vs_103900\|\|full | Not-determined |
| S13_vs_9438\|\|full | Medium-quality |
| S16_vs_103983\|\|full | Low-quality |
| S16_vs_104015\|\|full | Not-determined |
| S16_vs_104052\|\|full | Low-quality |
| S16_vs_104058\|\|full | Not-determined |
| S16_vs_104089\|\|full | Low-quality |
| S16_vs_104130\|\|full | Low-quality |
| S16_vs_104148\|\|full | Low-quality |
| S16_vs_104261\|\|full | Low-quality |
| S16_vs_104304\|\|full | Low-quality |
| S16_vs_104356\|\|full | Not-determined |
| S16_vs_104419\|\|full | Not-determined |
| S16_vs_104423\|\|full | Low-quality |
| S16_vs_104509\|\|full | Low-quality |
| S16_vs_104690\|\|full | Low-quality |
| S16_vs_104693\|\|full | Not-determined |
| S16_vs_104716\|\|full | Low-quality |
| S16_vs_104749\|\|full | Low-quality |
| S16_vs_104811\|\|full | Not-determined |
| S16_vs_104817\|\|full | Low-quality |
| S16_vs_104860\|\|full | Low-quality |
| S16_vs_104895\|\|full | Low-quality |
| S16_vs_104931\|\|full | Low-quality |
| S16_vs_104974\|\|full | Low-quality |
| S16_vs_104991\|\|full | Low-quality |
| S16_vs_105035\|\|full | Not-determined |
| S16_vs_105064\|\|full | Not-determined |
| S16_vs_105065\|\|full | Not-determined |
| S16_vs_105082\|\|full | Not-determined |
| S16_vs_105096\|\|full | Not-determined |
| S16_vs_105115\|\|full | Low-quality |
| S16_vs_105213\|\|full | Low-quality |
| S16_vs_105227\|\|full | Not-determined |
| S16_vs_105229\|\|full | Low-quality |
| S16_vs_105256\|\|full | Not-determined |
| S16_vs_105314\|\|full | Not-determined |
| S16_vs_105323\|\|full | Low-quality |
| S16_vs_105401\|\|full | Low-quality |
| S16_vs_105406\|\|full | Low-quality |
| S16_vs_105470\|\|full | Low-quality |
| S16_vs_105546\|\|full | Not-determined |
| S16_vs_105559\|\|full | Low-quality |
| S16_vs_105588\|\|full | Not-determined |
| S16_vs_105609\|\|full | Low-quality |
| S16_vs_105612\|\|full | Low-quality |
| S16_vs_105624\|\|full | Low-quality |
| S16_vs_105639\|\|full | Not-determined |
| S16_vs_105657\|\|full | Low-quality |
| S16_vs_105672\|\|full | Not-determined |
| S16_vs_105705\|\|full | Not-determined |
| S16_vs_105718\|\|full | Low-quality |
| S16_vs_105724\|\|full | Low-quality |
| S16_vs_105735\|\|full | Low-quality |
| S16_vs_105775\|\|full | Not-determined |
| S16_vs_105783\|\|full | Low-quality |
| S16_vs_105803\|\|full | Not-determined |
| S16_vs_105828\|\|full | Not-determined |
| S16_vs_105842\|\|full | Low-quality |
| S16_vs_105884\|\|full | Low-quality |
| S16_vs_105910\|\|full | Not-determined |
| S16_vs_105929\|\|full | Low-quality |
| S16_vs_105931\|\|full | Low-quality |
| S16_vs_105984\|\|full | Low-quality |
| S16_vs_105991\|\|full | Low-quality |
| S16_vs_106028\|\|full | Not-determined |
| S16_vs_106061\|\|full | Low-quality |
| S16_vs_106076\|\|full | Not-determined |
| S16_vs_106188\|\|full | Not-determined |
| S16_vs_106247\|\|full | Not-determined |
| S16_vs_106250\|\|full | Low-quality |
| S16_vs_106298\|\|full | Low-quality |
| S16_vs_106316\|\|full | Low-quality |
| S16_vs_106348\|\|full | Not-determined |
| S16_vs_106355\|\|full | Low-quality |
| S16_vs_106366\|\|full | Not-determined |
| S16_vs_106367\|\|full | Low-quality |
| S16_vs_106368\|\|full | Low-quality |
| S16_vs_106372\|\|full | Low-quality |
| S16_vs_106388\|\|full | Not-determined |
| S16_vs_106394\|\|full | Not-determined |
| S16_vs_106427\|\|full | Not-determined |
| S16_vs_106466\|\|full | Not-determined |
| S16_vs_106480\|\|full | Low-quality |
| S16_vs_106495\|\|full | Not-determined |
| S16_vs_106508\|\|full | Not-determined |
| S16_vs_106512\|\|full | Low-quality |
| S16_vs_106541\|\|full | Low-quality |
| S16_vs_106548\|\|full | Low-quality |
| S16_vs_106561\|\|full | Low-quality |
| S16_vs_106588\|\|full | Not-determined |
| S16_vs_106621\|\|full | Not-determined |
| S16_vs_106644\|\|full | Not-determined |
| S16_vs_106652\|\|full | Not-determined |
| S16_vs_106773\|\|full | Not-determined |
| S16_vs_106816\|\|full | Low-quality |
| S16_vs_106828\|\|full | Low-quality |
| S16_vs_106836\|\|full | Not-determined |
| S16_vs_106843\|\|full | Not-determined |
| S16_vs_106856\|\|full | Low-quality |
| S16_vs_106897\|\|full | Not-determined |
| S16_vs_106899\|\|full | Low-quality |
| S16_vs_106927\|\|full | Not-determined |
| S16_vs_106942\|\|full | Not-determined |
| S16_vs_106966\|\|full | Low-quality |
| S16_vs_107022\|\|full | Not-determined |
| S16_vs_107025\|\|full | Low-quality |
| S16_vs_107033\|\|full | Not-determined |
| S16_vs_107035\|\|full | Not-determined |
| S16_vs_107047\|\|full | Low-quality |
| S16_vs_107060\|\|full | Not-determined |
| S16_vs_107189\|\|full | Not-determined |
| S16_vs_107209\|\|full | Low-quality |
| S7_vs_56529\|\|full | High-quality |
| S16_vs_107274\|\|full | Not-determined |
| S16_vs_107335\|\|full | Not-determined |
| S16_vs_107347\|\|full | Not-determined |
| S16_vs_107358\|\|full | Not-determined |
| S16_vs_107363\|\|full | Not-determined |
| S16_vs_107415\|\|full | Low-quality |
| S16_vs_107426\|\|full | Low-quality |
| S16_vs_107481\|\|full | Low-quality |
| S16_vs_107518\|\|full | Not-determined |
| S16_vs_107557\|\|full | Not-determined |
| S16_vs_107563\|\|full | Low-quality |
| S16_vs_107579\|\|full | Low-quality |
| S16_vs_107616\|\|full | Low-quality |
| S16_vs_107621\|\|full | Low-quality |
| S16_vs_107628\|\|full | Not-determined |
| S16_vs_107633\|\|full | Not-determined |
| S16_vs_107654\|\|full | Low-quality |
| S16_vs_107684\|\|full | Not-determined |
| S16_vs_107771\|\|full | Not-determined |
| S16_vs_107782\|\|full | Low-quality |
| S16_vs_107790\|\|full | Not-determined |
| S16_vs_107804\|\|full | Not-determined |
| S16_vs_107837\|\|full | Low-quality |
| S16_vs_107853\|\|full | Not-determined |
| S16_vs_107866\|\|full | Low-quality |
| S16_vs_107916\|\|full | Low-quality |
| S16_vs_107945\|\|full | Low-quality |
| S16_vs_107979\|\|full | Not-determined |
| S16_vs_108009\|\|full | Not-determined |
| S16_vs_108129\|\|full | Low-quality |
| S16_vs_108153\|\|full | Low-quality |
| S16_vs_108177\|\|full | Not-determined |
| S16_vs_108334\|\|full | Low-quality |
| S16_vs_108337\|\|full | Low-quality |
| S16_vs_108356\|\|full | Not-determined |
| S16_vs_108365\|\|full | Low-quality |
| S16_vs_108386\|\|full | Low-quality |
| S16_vs_108405\|\|full | Low-quality |
| S16_vs_108460\|\|full | Low-quality |
| S16_vs_108462\|\|full | Not-determined |
| S16_vs_108467\|\|full | Low-quality |
| S16_vs_108471\|\|full | Not-determined |
| S16_vs_108551\|\|full | Not-determined |
| S8_vs_k141_208117\|\|full | High-quality |
| S16_vs_108588\|\|full | Low-quality |
| S16_vs_108596\|\|full | Not-determined |
| S16_vs_108743\|\|full | Not-determined |
| S16_vs_108766\|\|full | Not-determined |
| S16_vs_108767\|\|full | Not-determined |
| S16_vs_108778\|\|full | Not-determined |
| S16_vs_108796\|\|full | Low-quality |
| S16_vs_108828\|\|full | Not-determined |
| S16_vs_108851\|\|full | Low-quality |
| S16_vs_108860\|\|full | Low-quality |
| S16_vs_108947\|\|full | Low-quality |
| S16_vs_108949\|\|full | Not-determined |

| | |
|---|---|
| S16_vs_108983||full | Not-determined |
| S16_vs_109089||full | Low-quality |
| S16_vs_109093||full | Not-determined |
| S16_vs_109096||full | Low-quality |
| S16_vs_109101||full | Not-determined |
| S16_vs_109115||full | Low-quality |
| S16_vs_109135||full | Low-quality |
| S16_vs_109140||full | Low-quality |
| S16_vs_109161||full | Not-determined |
| S16_vs_109211||full | Low-quality |
| S16_vs_109236||full | Low-quality |
| S16_vs_109253||full | Low-quality |
| S16_vs_109271||full | Not-determined |
| S16_vs_109299||full | Low-quality |
| S16_vs_109309||full | Low-quality |
| S16_vs_109420||full | Low-quality |
| S16_vs_109445||full | Low-quality |
| S16_vs_109465||full | Not-determined |
| S16_vs_109480||full | Low-quality |
| S16_vs_109540||full | Low-quality |
| S16_vs_109568||full | Low-quality |
| S16_vs_109576||full | Not-determined |
| S16_vs_109617||full | Not-determined |
| S16_vs_109630||full | Not-determined |
| S16_vs_109633||full | Not-determined |
| S16_vs_109646||full | Low-quality |
| S16_vs_109668||full | Not-determined |
| S16_vs_109670||full | Not-determined |
| S16_vs_109716||full | Low-quality |
| S16_vs_109815||full | Not-determined |
| S16_vs_109842||full | Not-determined |
| S16_vs_109843||full | Low-quality |
| S16_vs_109874||full | Low-quality |
| S16_vs_109896||full | Not-determined |
| S16_vs_109980||full | Not-determined |
| S16_vs_110035||full | Not-determined |
| S16_vs_110082||full | Not-determined |
| S16_vs_110107||full | Low-quality |
| S16_vs_110136||full | Not-determined |
| S16_vs_110143||full | Low-quality |
| S16_vs_110171||full | Not-determined |
| S16_vs_110218||full | Low-quality |
| S16_vs_110229||full | Low-quality |
| S16_vs_110252||full | Low-quality |
| S16_vs_110268||full | Not-determined |
| S16_vs_110282||full | Low-quality |
| S16_vs_110287||full | Low-quality |
| S16_vs_110324||full | Low-quality |
| S16_vs_110369||full | Not-determined |
| S16_vs_110371||full | Low-quality |
| S16_vs_110403||full | Low-quality |
| S16_vs_110440||full | Low-quality |
| S16_vs_110465||full | Low-quality |
| S16_vs_110480||full | Low-quality |
| S16_vs_110499||full | Low-quality |
| S16_vs_110508||full | Low-quality |
| S16_vs_110573||full | Not-determined |
| S16_vs_110605||full | Low-quality |
| S16_vs_110612||full | Low-quality |
| S16_vs_110637||full | Low-quality |
| S16_vs_110655||full | Low-quality |
| S16_vs_110682||full | Not-determined |
| S16_vs_110746||full | Not-determined |
| S16_vs_110754||full | Not-determined |
| S16_vs_110836||full | Low-quality |
| S16_vs_110846||full | Low-quality |
| S16_vs_110898||full | Low-quality |
| S16_vs_110905||full | Not-determined |
| S16_vs_110949||full | Low-quality |
| S16_vs_110986||full | Low-quality |
| S16_vs_111062||full | Not-determined |
| S16_vs_111097||full | Low-quality |
| S16_vs_111136||full | Not-determined |
| S16_vs_111247||full | Low-quality |
| S16_vs_111254||full | Low-quality |
| S16_vs_111276||full | Low-quality |
| S16_vs_111301||full | Low-quality |
| S16_vs_111335||full | Not-determined |
| S16_vs_111351||full | Not-determined |
| S16_vs_111365||full | Not-determined |
| S16_vs_111413||full | Low-quality |
| S16_vs_111433||full | Not-determined |
| S16_vs_111459||full | Low-quality |
| S16_vs_111474||full | Low-quality |
| S16_vs_111513||full | Low-quality |
| S16_vs_111582||full | Low-quality |
| S16_vs_111597||full | Not-determined |
| S16_vs_111606||full | Low-quality |
| S16_vs_111664||full | Low-quality |
| S16_vs_111702||full | Low-quality |
| S16_vs_111743||full | Low-quality |
| S16_vs_111755||full | Low-quality |
| S16_vs_111826||full | Low-quality |
| S16_vs_111836||full | Low-quality |
| S16_vs_111888||full | Not-determined |
| S16_vs_111896||full | Not-determined |
| S16_vs_111919||full | Not-determined |
| S16_vs_111925||full | Low-quality |
| S16_vs_111936||full | Low-quality |
| S16_vs_111940||full | Low-quality |
| S16_vs_111967||full | Not-determined |
| S16_vs_111992||full | Low-quality |
| S16_vs_112078||full | Low-quality |
| S16_vs_112163||full | Low-quality |
| S16_vs_112170||full | Not-determined |
| S16_vs_112181||full | Low-quality |
| S16_vs_112193||full | Not-determined |
| S16_vs_112197||full | Low-quality |
| S16_vs_112204||full | Not-determined |
| S16_vs_112306||full | Low-quality |
| S16_vs_112375||full | Low-quality |
| S16_vs_112386||full | Low-quality |
| S16_vs_112395||full | Not-determined |
| S16_vs_112453||full | Low-quality |
| S16_vs_112484||full | Low-quality |
| S16_vs_112508||full | Low-quality |
| S16_vs_112513||full | Not-determined |
| S16_vs_112526||full | Low-quality |
| S16_vs_112562||full | Low-quality |
| S16_vs_112601||full | Low-quality |
| S16_vs_112603||full | Not-determined |
| S16_vs_112646||full | Not-determined |
| S16_vs_112666||full | Not-determined |
| S16_vs_112672||full | Not-determined |
| S16_vs_112696||full | Low-quality |
| S16_vs_112707||full | Not-determined |
| S16_vs_112711||full | Not-determined |
| S16_vs_112756||full | Low-quality |
| S16_vs_112765||full | Not-determined |
| S16_vs_112790||full | Not-determined |
| S16_vs_112830||full | Not-determined |
| S16_vs_112846||full | Low-quality |
| S16_vs_112895||full | Low-quality |
| S16_vs_112920||full | Low-quality |
| S16_vs_112933||full | Low-quality |
| S16_vs_112982||full | Low-quality |
| S16_vs_112991||full | Not-determined |
| S16_vs_112996||full | Low-quality |
| S16_vs_113002||full | Not-determined |
| S16_vs_113019||full | Not-determined |
| S16_vs_113037||full | Low-quality |
| S16_vs_113100||full | Low-quality |
| S16_vs_113101||full | Not-determined |
| S16_vs_113108||full | Low-quality |
| S16_vs_113153||full | Low-quality |
| S16_vs_113174||full | Low-quality |
| S16_vs_113216||full | Not-determined |
| S16_vs_113234||full | Not-determined |
| S16_vs_113246||full | Low-quality |
| S16_vs_113249||full | Not-determined |
| S16_vs_113282||full | Low-quality |
| S16_vs_113283||full | Low-quality |
| S16_vs_113324||full | Low-quality |
| S16_vs_113442||full | Not-determined |
| S16_vs_113444||full | Low-quality |
| S16_vs_113462||full | Not-determined |
| S16_vs_113493||full | Not-determined |
| S16_vs_113528||full | Not-determined |
| S16_vs_113535||full | Not-determined |
| S16_vs_113563||full | Not-determined |
| S16_vs_113585||full | Not-determined |
| S16_vs_113598||full | Low-quality |
| S16_vs_113652||full | Low-quality |
| S16_vs_113668||full | Low-quality |
| S16_vs_113682||full | Not-determined |
| S16_vs_113683||full | Not-determined |
| S16_vs_113700||full | Low-quality |
| S16_vs_113727||full | Not-determined |
| S16_vs_113778||full | Low-quality |
| S16_vs_113784||full | Low-quality |
| S16_vs_113789||full | Not-determined |
| S16_vs_113809||full | Not-determined |
| S16_vs_113823||full | Not-determined |
| S16_vs_113841||full | Low-quality |
| S16_vs_113851||full | Low-quality |
| S16_vs_113853||full | Low-quality |
| S16_vs_113875||full | Low-quality |
| S16_vs_113911||full | Not-determined |
| S16_vs_113986||full | Low-quality |
| S16_vs_114001||full | Not-determined |
| S16_vs_114014||full | Not-determined |
| S16_vs_114025||full | Low-quality |
| S16_vs_114031||full | Low-quality |
| S16_vs_114034||full | Low-quality |
| S16_vs_114044||full | Not-determined |
| S16_vs_114061||full | Low-quality |
| S16_vs_114076||full | Low-quality |
| S16_vs_114152||full | Low-quality |
| S16_vs_114187||full | Low-quality |
| S16_vs_114192||full | Not-determined |
| S16_vs_114206||full | Not-determined |
| S16_vs_114224||full | Low-quality |
| S16_vs_114227||full | Low-quality |
| S16_vs_114271||full | Low-quality |
| S16_vs_114333||full | Not-determined |
| S16_vs_114372||full | Low-quality |
| S16_vs_114375||full | Not-determined |
| S16_vs_114395||full | Low-quality |
| S16_vs_114416||full | Low-quality |
| S16_vs_114444||full | Not-determined |
| S16_vs_114510||full | Low-quality |
| S16_vs_114513||full | Low-quality |
| S16_vs_114530||full | Low-quality |
| S16_vs_114568||full | Not-determined |
| S16_vs_114604||full | Low-quality |
| S16_vs_114605||full | Low-quality |
| S16_vs_114618||full | Not-determined |
| S16_vs_114625||full | Low-quality |
| S16_vs_114653||full | Not-determined |
| S16_vs_114659||full | Low-quality |
| S16_vs_114676||full | Not-determined |
| S16_vs_114736||full | Not-determined |
| S16_vs_114778||full | Not-determined |
| S16_vs_114785||full | Not-determined |
| S16_vs_114793||full | Low-quality |
| S16_vs_114814||full | Not-determined |
| S16_vs_114850||full | Not-determined |
| S16_vs_114901||full | Low-quality |
| S16_vs_114933||full | Low-quality |
| S16_vs_114935||full | Low-quality |
| S16_vs_114943||full | Not-determined |
| S16_vs_114946||full | Low-quality |
| S16_vs_114976||full | Low-quality |
| S16_vs_114983||full | Not-determined |
| S16_vs_115061||full | Low-quality |
| S16_vs_115067||full | Low-quality |
| S16_vs_115068||full | Not-determined |
| S16_vs_115104||full | Not-determined |
| S16_vs_115135||full | Low-quality |
| S16_vs_115159||full | Not-determined |
| S16_vs_115188||full | Low-quality |
| S16_vs_115193||full | Low-quality |
| S16_vs_115292||full | Low-quality |
| S16_vs_115306||full | Low-quality |
| S16_vs_115312||full | Not-determined |
| S16_vs_115313||full | Low-quality |
| S16_vs_115314||full | Low-quality |
| S16_vs_115322||full | Low-quality |
| S16_vs_115374||full | Not-determined |
| S16_vs_115441||full | Low-quality |
| S16_vs_115477||full | Low-quality |
| S16_vs_115478||full | Not-determined |
| S16_vs_115488||full | Not-determined |
| S16_vs_115489||full | Low-quality |
| S16_vs_115501||full | Not-determined |
| S16_vs_115576||full | Not-determined |
| S16_vs_115588||full | Low-quality |
| S16_vs_115591||full | Low-quality |
| S16_vs_115606||full | Not-determined |
| S16_vs_115609||full | Not-determined |
| S16_vs_115644||full | Not-determined |
| S16_vs_115664||full | Low-quality |
| S16_vs_115698||full | Not-determined |
| S16_vs_115712||full | Not-determined |
| S16_vs_115731||full | Not-determined |
| S16_vs_115743||full | Not-determined |
| S16_vs_115750||full | Low-quality |
| S16_vs_115763||full | Low-quality |
| S16_vs_115769||full | Low-quality |
| S16_vs_115791||full | Not-determined |
| S16_vs_115795||full | Not-determined |
| S16_vs_115799||full | Not-determined |
| S16_vs_115848||full | Not-determined |
| S16_vs_115856||full | Not-determined |
| S16_vs_115879||full | Not-determined |
| S16_vs_115898||full | Not-determined |
| S16_vs_115933||full | Not-determined |
| S16_vs_115968||full | Low-quality |
| S16_vs_115969||full | Not-determined |
| S16_vs_116023||full | Not-determined |
| S16_vs_116049||full | Low-quality |
| S16_vs_116073||full | Low-quality |
| S16_vs_116091||full | Low-quality |
| S16_vs_116095||full | Low-quality |
| S16_vs_116128||full | Not-determined |
| S16_vs_116161||full | Not-determined |
| S16_vs_116198||full | Not-determined |
| S16_vs_116225||full | Low-quality |
| S16_vs_116233||full | Low-quality |
| S16_vs_116248||full | Not-determined |
| S16_vs_116364||full | Not-determined |
| S16_vs_116393||full | Not-determined |
| S16_vs_116417||full | Low-quality |
| S16_vs_116448||full | Not-determined |
| S16_vs_116468||full | Not-determined |
| S16_vs_116486||full | Low-quality |
| S16_vs_116507||full | Low-quality |
| S16_vs_116524||full | Not-determined |
| S16_vs_116545||full | Low-quality |
| S16_vs_116564||full | Not-determined |
| S16_vs_116576||full | Not-determined |
| S16_vs_116577||full | Low-quality |
| S16_vs_116580||full | Low-quality |
| S16_vs_116604||full | Not-determined |
| S16_vs_116615||full | Low-quality |
| S16_vs_116702||full | Low-quality |
| S16_vs_116757||full | Low-quality |
| S16_vs_116789||full | Not-determined |
| S16_vs_116834||full | Not-determined |
| S16_vs_116844||full | Low-quality |
| S16_vs_116854||full | Low-quality |
| S16_vs_116881||full | Low-quality |
| S16_vs_116900||full | Low-quality |
| S16_vs_116913||full | Low-quality |
| S16_vs_116915||full | Low-quality |
| S16_vs_116932||full | Not-determined |
| S16_vs_116936||full | Not-determined |
| S8_extracted_k141_101283 | Medium-quality |
| S16_vs_116978||full | Low-quality |
| S16_vs_116996||full | Not-determined |
| S16_vs_117030||full | Not-determined |
| S16_vs_117035||full | Not-determined |
| S16_vs_117053||full | Low-quality |
| S16_vs_117074||full | Not-determined |
| S16_vs_117093||full | Not-determined |
| S16_vs_117110||full | Low-quality |
| S16_vs_117133||full | Low-quality |
| S16_vs_117145||full | Low-quality |
| S16_vs_117191||full | Not-determined |
| S16_vs_117239||full | Low-quality |
| S16_vs_117299||full | Low-quality |
| S16_vs_117328||full | Low-quality |
| S16_vs_117353||full | Not-determined |
| S16_vs_117355||full | Low-quality |
| S16_vs_117356||full | Low-quality |
| S16_vs_117379||full | Not-determined |
| S16_vs_117394||full | Low-quality |
| S16_vs_117434||full | Not-determined |
| S16_vs_117437||full | Low-quality |
| S16_vs_117463||full | Not-determined |
| S16_vs_117501||full | Low-quality |
| S16_vs_117519||full | Low-quality |
| S16_vs_117536||full | Low-quality |
| S16_vs_117568||full | Not-determined |
| S16_vs_117592||full | Not-determined |
| S16_vs_117649||full | Not-determined |
| S16_vs_117654||full | Low-quality |
| S16_vs_117681||full | Low-quality |
| S16_vs_117695||full | Not-determined |
| S16_vs_117736||full | Not-determined |
| S16_vs_117738||full | Not-determined |
| S16_vs_117750||full | Low-quality |
| S16_vs_117760||full | Low-quality |
| S16_vs_117904||full | Low-quality |
| S16_vs_117914||full | Not-determined |
| S16_vs_117939||full | Not-determined |
| S16_vs_117993||full | Not-determined |
| S16_vs_117996||full | Low-quality |
| S16_vs_118024||full | Low-quality |
| S16_vs_103457||full | Medium-quality |
| S16_vs_118070||full | Not-determined |
| S16_vs_118096||full | Low-quality |
| S16_vs_118150||full | Low-quality |
| S16_vs_118170||full | Low-quality |
| S16_vs_118257||full | Not-determined |
| S16_vs_118320||full | Not-determined |
| S16_vs_118343||full | Low-quality |
| S16_vs_118349||full | Low-quality |
| S16_vs_118372||full | Not-determined |
| S16_vs_118406||full | Low-quality |
| S16_vs_118424||full | Not-determined |
| S16_vs_118433||full | Low-quality |
| S16_vs_118435||full | Low-quality |
| S16_vs_118440||full | Low-quality |
| S16_vs_118469||full | Low-quality |
| S16_vs_118495||full | Low-quality |
| S16_vs_118550||full | Not-determined |
| S16_vs_118558||full | Low-quality |
| S16_vs_118590||full | Low-quality |
| S16_vs_118604||full | Not-determined |
| S16_vs_118610||full | Low-quality |
| S16_vs_118618||full | Not-determined |
| S16_vs_118634||full | Low-quality |
| S16_vs_118637||full | Not-determined |
| S16_vs_118663||full | Not-determined |
| S16_vs_118669||full | Not-determined |
| S16_vs_118677||full | Not-determined |
| S16_vs_118678||full | Low-quality |
| S16_vs_118681||full | Low-quality |
| S16_vs_118683||full | Low-quality |
| S16_vs_118684||full | Not-determined |
| S16_vs_118713||full | Low-quality |
| S16_vs_118736||full | Not-determined |
| S16_vs_118811||full | Not-determined |
| S16_vs_118883||full | Not-determined |
| S16_vs_118909||full | Low-quality |
| S16_vs_118921||full | Low-quality |
| S16_vs_118929||full | Not-determined |
| S16_vs_119029||full | Low-quality |
| S16_vs_119037||full | Not-determined |
| S16_vs_119039||full | Low-quality |
| S16_vs_119059||full | Low-quality |
| S16_vs_119061||full | Low-quality |
| S16_vs_119074||full | Not-determined |
| S16_vs_119077||full | Low-quality |
| S16_vs_119084||full | Low-quality |
| S16_vs_119113||full | Low-quality |
| S16_vs_119128||full | Low-quality |
| S16_vs_119130||full | Not-determined |
| S16_vs_119152||full | Not-determined |
| S16_vs_119230||full | Low-quality |
| S16_vs_119232||full | Low-quality |
| S16_vs_119250||full | Not-determined |
| S16_vs_119293||full | Low-quality |
| S16_vs_119305||full | Not-determined |
| S16_vs_119322||full | Not-determined |
| S16_vs_119323||full | Not-determined |
| S16_vs_119362||full | Low-quality |
| S16_vs_119370||full | Low-quality |
| S16_vs_119381||full | Low-quality |
| S16_vs_119428||full | Not-determined |
| S16_vs_119454||full | Low-quality |
| S16_vs_119460||full | Low-quality |
| S16_vs_119470||full | Low-quality |
| S16_vs_119473||full | Low-quality |
| S16_vs_119503||full | Low-quality |
| S16_vs_119505||full | Low-quality |
| S16_vs_119572||full | Low-quality |
| S16_vs_119597||full | Low-quality |
| S16_vs_119634||full | Low-quality |
| S16_vs_119730||full | Low-quality |
| S16_vs_119747||full | Not-determined |
| S16_vs_119752||full | Low-quality |
| S16_vs_119788||full | Low-quality |
| S16_vs_119819||full | Low-quality |
| S10_vs_2538||full | High-quality |
| S16_vs_119841||full | Low-quality |
| S16_vs_119842||full | Not-determined |
| S16_vs_119846||full | Low-quality |
| S16_vs_119859||full | Not-determined |
| S16_vs_119882||full | Low-quality |
| S16_vs_119956||full | Low-quality |
| S16_vs_119992||full | Low-quality |
| S16_vs_120016||full | Low-quality |
| S16_vs_120116||full | Not-determined |
| S6_vs_k141_37812||full | Medium-quality |
| S16_vs_120165||full | Low-quality |
| S16_vs_120234||full | Low-quality |
| S16_vs_120256||full | Low-quality |
| S16_vs_120293||full | Not-determined |
| S16_vs_120363||full | Low-quality |
| S16_vs_120371||full | Low-quality |
| S16_vs_120388||full | Low-quality |
| S16_vs_120392||full | Low-quality |
| S16_vs_120409||full | Not-determined |
| S16_vs_120434||full | Low-quality |
| S16_vs_120508||full | Not-determined |
| S16_vs_120512||full | Low-quality |
| S16_vs_120572||full | Not-determined |
| S16_vs_120574||full | Not-determined |
| S16_vs_120585||full | Low-quality |
| S12_vs_k141_188833||full | High-quality |
| S5_extracted_k141_11450 | Complete |
| S16_vs_120635||full | Not-determined |
| S16_vs_120788||full | Not-determined |
| S16_vs_120817||full | Not-determined |
| S16_vs_120846||full | Low-quality |
| S16_vs_120865||full | Low-quality |
| S16_vs_120889||full | Low-quality |
| S16_vs_120895||full | Not-determined |
| S16_vs_120907||full | Low-quality |
| S16_vs_120909||full | Not-determined |
| S16_vs_120914||full | Low-quality |
| S16_vs_120916||full | Not-determined |
| S16_vs_120938||full | Low-quality |
| S16_vs_120951||full | Low-quality |
| S16_vs_120953||full | Not-determined |
| S16_vs_120968||full | Not-determined |
| S16_vs_120971||full | Low-quality |
| S16_vs_120976||full | Low-quality |
| S16_vs_121030||full | Low-quality |
| S16_vs_121037||full | Not-determined |
| S16_vs_121040||full | Low-quality |
| S16_vs_121046||full | Not-determined |
| S16_vs_121088||full | Low-quality |
| S16_vs_121096||full | Low-quality |
| S16_vs_121129||full | Low-quality |
| S16_vs_121137||full | Not-determined |
| S16_vs_121161||full | Low-quality |
| S16_vs_121181||full | Low-quality |
| S16_vs_121222||full | Not-determined |
| S16_vs_121253||full | Not-determined |
| S16_vs_121278||full | Not-determined |
| S16_vs_121285||full | Low-quality |
| S16_vs_121290||full | Not-determined |
| S16_vs_121299||full | Low-quality |
| S16_vs_121315||full | Low-quality |
| S16_vs_121336||full | Low-quality |
| S16_vs_121365||full | Not-determined |
| S16_vs_121439||full | Not-determined |
| S16_vs_121461||full | Low-quality |
| S16_vs_121485||full | Low-quality |
| S16_vs_121515||full | Low-quality |
| S16_vs_121584||full | Low-quality |
| S16_vs_121594||full | Low-quality |
| S16_vs_121652||full | Low-quality |
| S16_vs_121678||full | Low-quality |
| S16_vs_121697||full | Not-determined |
| S16_vs_121710||full | Low-quality |
| S16_vs_121730||full | Not-determined |
| S16_vs_121748||full | Low-quality |
| S16_vs_121784||full | Low-quality |
| S16_vs_121802||full | Not-determined |
| S16_vs_121814||full | Not-determined |
| S16_vs_121843||full | Not-determined |
| S16_vs_121844||full | Low-quality |
| S16_vs_121885||full | Low-quality |
| S16_vs_121956||full | Not-determined |
| S16_vs_121967||full | Not-determined |
| S16_vs_121972||full | Low-quality |
| S16_vs_121979||full | Not-determined |
| S16_vs_122064||full | Low-quality |
| S16_vs_122126||full | Low-quality |
| S16_vs_122157||full | Not-determined |
| S16_vs_122163||full | Not-determined |
| S16_vs_122171||full | Low-quality |
| S16_vs_122173||full | Not-determined |
| S16_vs_122230||full | Low-quality |
| S16_vs_122308||full | Low-quality |
| S16_vs_122311||full | Not-determined |
| S16_vs_122329||full | Not-determined |
| S16_vs_122366||full | Low-quality |
| S16_vs_122382||full | Low-quality |
| S16_vs_122421||full | Low-quality |
| S16_vs_122429||full | Low-quality |
| S16_vs_122458||full | Not-determined |
| S16_vs_122488||full | Not-determined |
| S16_vs_122502||full | Not-determined |
| S16_vs_122521||full | Low-quality |
| S16_vs_122537||full | Low-quality |
| S16_vs_122600||full | Low-quality |
| S16_vs_122604||full | Not-determined |
| S16_vs_122611||full | Not-determined |
| S16_vs_122639||full | Not-determined |
| S16_vs_122720||full | Low-quality |
| S16_vs_122732||full | Low-quality |
| S16_vs_122746||full | Low-quality |
| S16_vs_122762||full | Not-determined |
| S16_vs_122778||full | Low-quality |
| S16_vs_122799||full | Low-quality |
| S16_vs_122804||full | Not-determined |
| S16_vs_122863||full | Low-quality |
| S16_vs_122866||full | Low-quality |
| S16_vs_122877||full | Low-quality |
| S16_vs_122887||full | Low-quality |
| S16_vs_122917||full | Not-determined |
| S16_vs_122928||full | Low-quality |
| S16_vs_122977||full | Low-quality |
| S16_vs_123060||full | Low-quality |
| S16_vs_123067||full | Not-determined |
| S16_vs_123071||full | Low-quality |
| S16_vs_123087||full | Low-quality |
| S16_vs_123114||full | Not-determined |
| S16_vs_123117||full | Not-determined |
| S16_vs_123123||full | Not-determined |
| S16_vs_123125||full | Low-quality |
| S16_vs_123128||full | Not-determined |
| S16_vs_123133||full | Not-determined |
| S16_vs_123147||full | Low-quality |
| S16_vs_123183||full | Not-determined |
| S16_vs_123223||full | Not-determined |
| S16_vs_123229||full | Not-determined |
| S16_vs_123231||full | Not-determined |
| S16_vs_123255||full | Low-quality |
| S16_vs_123264||full | Low-quality |
| S16_vs_123317||full | Low-quality |
| S16_vs_123319||full | Low-quality |
| S16_vs_123342||full | Not-determined |
| S16_vs_123353||full | Low-quality |
| S16_vs_123360||full | Low-quality |
| S16_vs_123435||full | Not-determined |
| S16_vs_123439||full | Not-determined |
| S16_vs_123450||full | Not-determined |
| S16_vs_123453||full | Low-quality |
| S16_vs_123484||full | Low-quality |
| S16_vs_123548||full | Not-determined |
| S16_vs_123550||full | Low-quality |
| S16_vs_123551||full | Low-quality |
| S16_vs_123553||full | Low-quality |
| S16_vs_123619||full | Low-quality |
| S16_vs_123627||full | Low-quality |
| S16_vs_123691||full | Low-quality |
| S16_vs_123695||full | Low-quality |
| S16_vs_123704||full | Low-quality |
| S16_vs_123706||full | Low-quality |
| S16_vs_123804||full | Low-quality |
| S16_vs_123820||full | Low-quality |
| S16_vs_123825||full | Not-determined |
| S16_vs_123842||full | Low-quality |

| | |
|---|---|
| S16_vs_123967\|\|full | Not-determined |
| S16_vs_123983\|\|full | Not-determined |
| S16_vs_123991\|\|full | Low-quality |
| S16_vs_124001\|\|full | Not-determined |
| S16_vs_124006\|\|full | Low-quality |
| S16_vs_124012\|\|full | Low-quality |
| S16_vs_124050\|\|full | Low-quality |
| S16_vs_124062\|\|full | Low-quality |
| S16_vs_124075\|\|full | Low-quality |
| S16_vs_124120\|\|full | Not-determined |
| S16_vs_124138\|\|full | Low-quality |
| S16_vs_124143\|\|full | Low-quality |
| S16_vs_124146\|\|full | Not-determined |
| S16_vs_124148\|\|full | Low-quality |
| S16_vs_124164\|\|full | Not-determined |
| S16_vs_124213\|\|full | Not-determined |
| S16_vs_124293\|\|full | Not-determined |
| S16_vs_124313\|\|full | Low-quality |
| S16_vs_124314\|\|full | Not-determined |
| S16_vs_124316\|\|full | Not-determined |
| S16_vs_124331\|\|full | Low-quality |
| S16_vs_124349\|\|full | Not-determined |
| S16_vs_124355\|\|full | Not-determined |
| S16_vs_124373\|\|full | Not-determined |
| S16_vs_124401\|\|full | Not-determined |
| S16_vs_124438\|\|full | Low-quality |
| S16_vs_124474\|\|full | Not-determined |
| S16_vs_124486\|\|full | Not-determined |
| S16_vs_124497\|\|full | Low-quality |
| S16_vs_124511\|\|full | Not-determined |
| S16_vs_124513\|\|full | Not-determined |
| S16_vs_124530\|\|full | Not-determined |
| S16_vs_124535\|\|full | Not-determined |
| S16_vs_124552\|\|full | Not-determined |
| S16_vs_124634\|\|full | Not-determined |
| S16_vs_124650\|\|full | Not-determined |
| S9_extracted_k141_15012 | High-quality |
| S16_vs_124674\|\|full | Low-quality |
| S16_vs_124679\|\|full | Not-determined |
| S16_vs_124681\|\|full | Not-determined |
| S16_vs_124686\|\|full | Not-determined |
| S9_vs_k141_15012\|\|full | High-quality |
| S16_vs_124708\|\|full | Low-quality |
| S16_vs_124729\|\|full | Not-determined |
| S16_vs_124756\|\|full | Low-quality |
| S16_vs_124760\|\|full | Not-determined |
| S16_vs_124876\|\|full | Low-quality |
| S16_vs_124879\|\|full | Low-quality |
| S16_vs_124934\|\|full | Low-quality |
| S16_vs_124936\|\|full | Not-determined |
| S16_vs_124941\|\|full | Not-determined |
| S16_vs_124951\|\|full | Not-determined |
| S16_vs_124981\|\|full | Low-quality |
| S16_vs_125068\|\|full | Low-quality |
| S16_vs_125069\|\|full | Low-quality |
| S16_vs_125130\|\|full | Low-quality |
| S16_vs_125136\|\|full | Low-quality |
| S16_vs_125137\|\|full | Low-quality |
| S16_vs_125227\|\|full | Low-quality |
| S16_vs_125228\|\|full | Not-determined |
| S16_vs_125248\|\|full | Low-quality |
| S16_vs_125272\|\|full | Not-determined |
| S16_vs_125296\|\|full | Low-quality |
| S16_vs_125298\|\|full | Low-quality |
| S16_vs_125300\|\|full | Low-quality |
| S16_vs_125309\|\|full | Not-determined |
| S16_vs_125320\|\|full | Low-quality |
| S16_vs_125437\|\|full | Low-quality |
| S16_vs_125451\|\|full | Not-determined |
| S16_vs_125462\|\|full | Low-quality |
| S16_vs_125463\|\|full | Low-quality |
| S16_vs_125472\|\|full | Not-determined |
| S16_vs_125484\|\|full | Low-quality |
| S16_vs_125494\|\|full | Low-quality |
| S16_vs_125513\|\|full | Low-quality |
| S16_vs_125541\|\|full | Low-quality |
| S16_vs_125542\|\|full | Low-quality |
| S16_vs_125546\|\|full | Not-determined |
| S16_vs_125580\|\|full | Low-quality |
| S16_vs_125584\|\|full | Low-quality |
| S16_vs_125620\|\|full | Not-determined |
| S16_vs_125716\|\|full | Low-quality |
| S16_vs_125766\|\|full | Low-quality |
| S16_vs_125793\|\|full | Low-quality |
| S16_vs_125802\|\|full | Not-determined |
| S16_vs_125816\|\|full | Low-quality |
| S16_vs_125860\|\|full | Low-quality |
| S16_vs_125892\|\|full | Low-quality |
| S16_vs_125911\|\|full | Not-determined |
| S16_vs_125912\|\|full | Not-determined |
| S16_vs_125938\|\|full | Low-quality |
| S16_vs_125996\|\|full | Low-quality |
| S16_vs_126000\|\|full | Low-quality |
| S16_vs_126094\|\|full | Low-quality |
| S16_vs_126149\|\|full | Not-determined |
| S16_vs_126196\|\|full | Low-quality |
| S16_vs_126212\|\|full | Not-determined |
| S16_vs_126216\|\|full | Not-determined |
| S16_vs_126243\|\|full | Not-determined |
| S16_vs_126244\|\|full | Low-quality |
| S16_vs_126247\|\|full | Not-determined |
| S16_vs_126310\|\|full | Low-quality |
| S16_vs_126358\|\|full | Low-quality |
| S16_vs_126368\|\|full | Not-determined |
| S16_vs_126373\|\|full | Not-determined |
| S16_vs_126388\|\|full | Low-quality |
| S16_vs_126471\|\|full | Low-quality |
| S16_vs_126545\|\|full | Low-quality |
| S16_vs_126555\|\|full | Low-quality |
| S16_vs_126563\|\|full | Not-determined |
| S16_vs_126613\|\|full | Not-determined |
| S16_vs_126644\|\|full | Not-determined |
| S16_vs_126701\|\|full | Low-quality |
| S16_vs_126704\|\|full | Not-determined |
| S16_vs_126755\|\|full | Low-quality |
| S16_vs_126813\|\|full | Low-quality |
| S16_vs_126823\|\|full | Low-quality |
| S16_vs_126824\|\|full | Not-determined |
| S16_vs_126956\|\|full | Low-quality |
| S16_vs_126965\|\|full | Not-determined |
| S16_vs_126981\|\|full | Low-quality |
| S16_vs_126993\|\|full | Low-quality |
| S16_vs_127056\|\|full | Not-determined |
| S16_vs_127077\|\|full | Low-quality |
| S16_vs_127117\|\|full | Low-quality |
| S16_vs_127145\|\|full | Low-quality |
| S16_vs_127170\|\|full | Low-quality |
| S16_vs_127174\|\|full | Not-determined |
| S16_vs_127179\|\|full | Not-determined |
| S16_vs_127245\|\|full | Low-quality |
| S16_vs_127263\|\|full | Low-quality |
| S16_vs_127357\|\|full | Not-determined |
| S16_vs_127360\|\|full | Not-determined |
| S16_vs_127406\|\|full | Not-determined |
| S16_vs_127447\|\|full | Not-determined |
| S16_vs_127449\|\|full | Low-quality |
| S16_vs_127455\|\|full | Low-quality |
| S16_vs_127459\|\|full | Not-determined |
| S16_vs_127505\|\|full | Not-determined |
| S16_vs_127530\|\|full | Low-quality |
| S16_vs_127560\|\|full | Low-quality |
| S16_vs_127579\|\|full | Low-quality |
| S16_vs_127594\|\|full | Not-determined |
| S16_vs_127611\|\|full | Low-quality |
| S16_vs_127637\|\|full | Not-determined |
| S16_vs_127661\|\|full | Low-quality |
| S16_vs_127663\|\|full | Not-determined |
| S5_vs_k141_13090\|\|full | High-quality |
| S16_vs_127670\|\|full | Not-determined |
| S16_vs_127674\|\|full | Not-determined |
| S16_vs_127684\|\|full | Low-quality |
| S16_vs_127690\|\|full | Low-quality |
| S16_vs_127713\|\|full | Low-quality |
| S16_vs_127718\|\|full | Low-quality |
| S16_vs_127772\|\|full | Low-quality |
| S16_vs_127802\|\|full | Low-quality |
| S16_vs_127819\|\|full | Low-quality |
| S16_vs_127920\|\|full | Low-quality |
| S16_vs_127936\|\|full | Low-quality |
| S16_vs_127937\|\|full | Low-quality |
| S16_vs_127939\|\|full | Low-quality |
| S16_vs_127945\|\|full | Low-quality |
| S16_vs_127963\|\|full | Not-determined |
| S16_vs_127971\|\|full | Not-determined |
| S16_vs_128031\|\|full | Not-determined |
| S16_vs_128068\|\|full | Not-determined |
| S16_vs_128076\|\|full | Low-quality |
| S16_vs_128080\|\|full | Not-determined |
| S16_vs_128129\|\|full | Not-determined |
| S16_vs_128131\|\|full | Low-quality |
| S16_vs_128144\|\|full | Low-quality |
| S16_vs_128152\|\|full | Low-quality |
| S16_vs_128207\|\|full | Low-quality |
| S16_vs_128226\|\|full | Not-determined |
| S16_vs_128270\|\|full | Not-determined |
| S16_vs_128283\|\|full | Not-determined |
| S16_vs_128333\|\|full | Not-determined |
| S16_vs_128336\|\|full | Not-determined |
| S16_vs_128358\|\|full | Low-quality |
| S16_vs_128414\|\|full | Low-quality |
| S16_vs_128427\|\|full | Low-quality |
| S16_vs_128449\|\|full | Low-quality |
| S16_vs_128463\|\|full | Not-determined |
| S16_vs_128465\|\|full | Low-quality |
| S16_vs_128475\|\|full | Low-quality |
| S16_vs_128490\|\|full | Not-determined |
| S16_vs_128494\|\|full | Low-quality |
| S16_vs_128517\|\|full | Not-determined |
| S16_vs_128543\|\|full | Low-quality |
| S16_vs_128563\|\|full | Not-determined |
| S16_vs_128572\|\|full | Not-determined |
| S16_vs_128573\|\|full | Low-quality |
| S16_vs_128616\|\|full | Not-determined |
| S16_vs_128627\|\|full | Low-quality |
| S16_vs_128694\|\|full | Not-determined |
| S16_vs_128706\|\|full | Not-determined |
| S16_vs_128754\|\|full | Low-quality |
| S16_vs_128813\|\|full | Not-determined |
| S16_vs_128849\|\|full | Not-determined |
| S16_vs_128966\|\|full | Low-quality |
| S16_vs_129049\|\|full | Not-determined |
| S16_vs_129057\|\|full | Not-determined |
| S16_vs_129119\|\|full | Low-quality |
| S16_vs_129145\|\|full | Low-quality |
| S16_vs_129215\|\|full | Low-quality |
| S16_vs_129249\|\|full | Not-determined |
| S16_vs_129288\|\|full | Low-quality |
| S16_vs_129311\|\|full | Low-quality |
| S16_vs_129315\|\|full | Low-quality |
| S16_vs_129326\|\|full | Low-quality |
| S16_vs_129357\|\|full | Not-determined |
| S16_vs_129364\|\|full | Low-quality |
| S16_vs_129366\|\|full | Low-quality |
| S16_vs_129382\|\|full | Low-quality |
| S16_vs_129388\|\|full | Low-quality |
| S16_vs_129412\|\|full | Not-determined |
| S16_vs_129422\|\|full | Low-quality |
| S16_vs_129442\|\|full | Low-quality |
| S16_vs_129451\|\|full | Low-quality |
| S16_vs_129452\|\|full | Not-determined |
| S16_vs_129478\|\|full | Low-quality |
| S16_vs_129485\|\|full | Not-determined |
| S16_vs_129540\|\|full | Low-quality |
| S16_vs_129551\|\|full | Not-determined |
| S16_vs_129644\|\|full | Low-quality |
| S16_vs_129656\|\|full | Low-quality |
| S16_vs_129665\|\|full | Low-quality |
| S16_vs_129734\|\|full | Low-quality |
| S16_vs_129747\|\|full | Not-determined |
| S16_vs_129768\|\|full | Low-quality |
| S16_vs_129772\|\|full | Low-quality |
| S16_vs_129779\|\|full | Not-determined |
| S16_vs_129870\|\|full | Low-quality |
| S16_vs_129890\|\|full | Not-determined |
| S16_vs_129925\|\|full | Not-determined |
| S16_vs_129938\|\|full | Low-quality |
| S16_vs_129975\|\|full | Low-quality |
| S16_vs_129983\|\|full | Low-quality |
| S16_vs_129996\|\|full | Low-quality |
| S16_vs_130024\|\|full | Not-determined |
| S16_vs_130033\|\|full | Low-quality |
| S16_vs_130037\|\|full | Not-determined |
| S16_vs_130106\|\|full | Low-quality |
| S16_vs_130129\|\|full | Low-quality |
| S16_vs_130135\|\|full | Not-determined |
| S16_vs_130141\|\|full | Not-determined |
| S16_vs_130143\|\|full | Low-quality |
| S16_vs_130238\|\|full | Low-quality |
| S16_vs_130283\|\|full | Low-quality |
| S16_vs_130332\|\|full | Not-determined |
| S16_vs_130333\|\|full | Low-quality |
| S16_vs_130369\|\|full | Not-determined |
| S16_vs_130379\|\|full | Low-quality |
| S16_vs_130397\|\|full | Not-determined |
| S16_vs_130408\|\|full | Not-determined |
| S16_vs_130471\|\|full | Low-quality |
| S16_vs_130496\|\|full | Not-determined |
| S16_vs_130513\|\|full | Low-quality |
| S16_vs_130524\|\|full | Not-determined |
| S16_vs_130529\|\|full | Not-determined |
| S16_vs_130550\|\|full | Low-quality |
| S16_vs_130566\|\|full | Low-quality |
| S16_vs_130588\|\|full | Low-quality |
| S16_vs_130610\|\|full | Low-quality |
| S16_vs_130617\|\|full | Low-quality |
| S16_vs_130649\|\|full | Low-quality |
| S16_vs_130686\|\|full | Low-quality |
| S16_vs_130711\|\|full | Not-determined |
| S16_vs_130817\|\|full | Not-determined |
| S16_vs_130843\|\|full | Not-determined |
| S16_vs_130860\|\|full | Not-determined |
| S16_vs_130867\|\|full | Low-quality |
| S16_vs_130887\|\|full | Not-determined |
| S16_vs_130896\|\|full | Not-determined |
| S16_vs_130900\|\|full | Low-quality |
| S16_vs_130906\|\|full | Not-determined |
| S16_vs_130959\|\|full | Low-quality |
| S16_vs_130969\|\|full | Not-determined |
| S16_vs_130987\|\|full | Not-determined |
| S16_vs_130994\|\|full | Low-quality |
| S16_vs_131023\|\|full | Not-determined |
| S16_vs_131036\|\|full | Not-determined |
| S16_vs_131115\|\|full | Low-quality |
| S16_vs_131116\|\|full | Low-quality |
| S16_vs_131117\|\|full | Low-quality |
| S16_vs_131118\|\|full | Not-determined |
| S16_vs_131126\|\|full | Low-quality |
| S16_vs_131144\|\|full | Not-determined |
| S16_vs_131200\|\|full | Not-determined |
| S16_vs_131275\|\|full | Not-determined |
| S16_vs_131284\|\|full | Not-determined |
| S16_vs_131334\|\|full | Low-quality |
| S16_vs_131369\|\|full | Low-quality |
| S16_vs_131447\|\|full | Low-quality |
| S16_vs_131450\|\|full | Low-quality |
| S16_vs_131472\|\|full | Not-determined |
| S16_vs_131477\|\|full | Low-quality |
| S16_vs_131502\|\|full | Not-determined |
| S16_vs_131508\|\|full | Low-quality |
| S16_vs_131516\|\|full | Low-quality |
| S16_vs_131521\|\|full | Low-quality |
| S16_vs_131524\|\|full | Not-determined |
| S16_vs_131542\|\|full | Not-determined |
| S16_vs_131552\|\|full | Low-quality |
| S16_vs_131604\|\|full | Low-quality |
| S16_vs_131621\|\|full | Not-determined |
| S16_vs_131665\|\|full | Low-quality |
| S16_vs_131666\|\|full | Not-determined |
| S16_vs_131669\|\|full | Low-quality |
| S16_vs_131713\|\|full | Not-determined |
| S16_vs_131717\|\|full | Low-quality |
| S16_vs_131720\|\|full | Not-determined |
| S16_vs_131749\|\|full | Not-determined |
| S16_vs_131797\|\|full | Not-determined |
| S16_vs_131810\|\|full | Not-determined |
| S16_vs_131857\|\|full | Low-quality |
| S16_vs_131919\|\|full | Low-quality |
| S16_vs_131949\|\|full | Low-quality |
| S16_vs_131978\|\|full | Low-quality |
| S16_vs_131986\|\|full | Not-determined |
| S16_vs_132038\|\|full | Low-quality |
| S16_vs_132040\|\|full | Low-quality |
| S16_vs_132052\|\|full | Low-quality |
| S16_vs_132069\|\|full | Not-determined |
| S16_vs_132073\|\|full | Not-determined |
| S16_vs_132201\|\|full | Low-quality |
| S16_vs_132235\|\|full | Low-quality |
| S16_vs_132258\|\|full | Not-determined |
| S16_vs_132273\|\|full | Not-determined |
| S16_vs_132289\|\|full | Low-quality |
| S16_vs_132294\|\|full | Low-quality |
| S16_vs_132343\|\|full | Not-determined |
| S16_vs_132345\|\|full | Not-determined |
| S16_vs_132410\|\|full | Low-quality |
| S16_vs_132412\|\|full | Not-determined |
| S16_vs_132417\|\|full | Low-quality |
| S16_vs_132440\|\|full | Low-quality |
| S16_vs_132449\|\|full | Not-determined |
| S16_vs_132539\|\|full | Not-determined |
| S16_vs_132547\|\|full | Not-determined |
| S16_vs_132619\|\|full | Not-determined |
| S16_vs_132657\|\|full | Low-quality |
| S16_vs_132659\|\|full | Low-quality |
| S16_vs_132661\|\|full | Not-determined |
| S16_vs_132705\|\|full | Not-determined |
| S16_vs_132710\|\|full | Low-quality |
| S16_vs_132716\|\|full | Not-determined |
| S16_vs_132721\|\|full | Not-determined |
| S16_vs_132754\|\|full | Not-determined |
| S16_vs_132758\|\|full | Low-quality |
| S16_vs_132793\|\|full | Not-determined |
| S16_vs_132904\|\|full | Not-determined |
| S16_vs_132938\|\|full | Low-quality |
| S16_vs_132940\|\|full | Not-determined |
| S16_vs_132943\|\|full | Low-quality |
| S5_vs_k141_56083\|\|full | High-quality |
| S16_vs_133019\|\|full | Low-quality |
| S16_vs_133065\|\|full | Low-quality |
| S16_vs_133069\|\|full | Low-quality |
| S16_vs_133100\|\|full | Low-quality |
| S16_vs_133152\|\|full | Low-quality |
| S16_vs_133166\|\|full | Low-quality |
| S16_vs_133180\|\|full | Not-determined |
| S16_vs_133208\|\|full | Not-determined |
| S16_vs_133238\|\|full | Low-quality |
| S16_vs_133269\|\|full | Low-quality |
| S16_vs_133282\|\|full | Low-quality |
| S16_vs_133382\|\|full | Low-quality |
| S16_vs_133392\|\|full | Not-determined |
| S16_vs_133406\|\|full | Not-determined |
| S16_vs_133428\|\|full | Low-quality |
| S16_vs_133429\|\|full | Low-quality |
| S16_vs_133437\|\|full | Low-quality |
| S16_vs_133472\|\|full | Low-quality |
| S16_vs_133504\|\|full | Not-determined |
| S16_vs_133519\|\|full | Not-determined |
| S16_vs_133575\|\|full | Not-determined |
| S16_vs_133579\|\|full | Not-determined |
| S16_vs_133598\|\|full | Not-determined |
| S16_vs_133662\|\|full | Low-quality |
| S16_vs_133688\|\|full | Not-determined |
| S16_vs_133720\|\|full | Low-quality |
| S16_vs_133725\|\|full | Low-quality |
| S16_vs_133736\|\|full | Low-quality |
| S16_vs_133752\|\|full | Low-quality |
| S16_vs_133794\|\|full | Not-determined |
| S16_vs_133838\|\|full | Low-quality |
| S16_vs_133863\|\|full | Not-determined |
| S16_vs_133929\|\|full | Not-determined |
| S16_vs_133934\|\|full | Not-determined |
| S16_vs_133986\|\|full | Low-quality |
| S16_vs_133993\|\|full | Low-quality |
| S16_vs_134070\|\|full | Low-quality |
| S16_vs_134077\|\|full | Low-quality |
| S16_vs_134105\|\|full | Low-quality |
| S16_vs_134123\|\|full | Low-quality |
| S16_vs_134144\|\|full | Not-determined |
| S16_vs_134229\|\|full | Low-quality |
| S16_vs_134320\|\|full | Not-determined |
| S16_vs_134359\|\|full | Not-determined |
| S16_vs_134363\|\|full | Low-quality |
| S16_vs_134374\|\|full | Not-determined |
| S16_vs_134387\|\|full | Low-quality |
| S16_vs_134421\|\|full | Not-determined |
| S16_vs_134429\|\|full | Not-determined |
| S16_vs_134475\|\|full | Low-quality |
| S16_vs_134480\|\|full | Low-quality |
| S16_vs_134499\|\|full | Low-quality |
| S16_vs_134504\|\|full | Low-quality |
| S16_vs_134509\|\|full | Low-quality |
| S16_vs_134550\|\|full | Not-determined |
| S16_vs_134556\|\|full | Low-quality |
| S16_vs_134570\|\|full | Low-quality |
| S16_vs_134601\|\|full | Low-quality |
| S16_vs_134621\|\|full | Not-determined |
| S16_vs_134649\|\|full | Not-determined |
| S5_vs_k141_11450\|\|full | High-quality |
| S16_vs_134707\|\|full | Low-quality |
| S16_vs_134708\|\|full | Low-quality |
| S16_vs_134766\|\|full | Not-determined |
| S16_vs_134773\|\|full | Low-quality |
| S16_vs_134775\|\|full | Low-quality |
| S16_vs_134788\|\|full | Low-quality |
| S16_vs_134814\|\|full | Not-determined |
| S16_vs_134834\|\|full | Low-quality |
| S16_vs_134840\|\|full | Not-determined |
| S16_vs_134896\|\|full | Low-quality |
| S16_vs_134923\|\|full | Not-determined |
| S16_vs_134927\|\|full | Low-quality |
| S16_vs_134942\|\|full | Not-determined |
| S16_vs_134951\|\|full | Low-quality |
| S16_vs_134968\|\|full | Low-quality |
| S16_vs_134998\|\|full | Not-determined |
| S16_vs_134999\|\|full | Low-quality |
| S16_vs_135031\|\|full | Not-determined |
| S16_vs_135033\|\|full | Low-quality |
| S16_vs_135193\|\|full | Not-determined |
| S16_vs_135218\|\|full | Not-determined |
| S16_vs_135255\|\|full | Low-quality |
| S16_vs_135280\|\|full | Low-quality |
| S16_vs_135297\|\|full | Low-quality |
| S16_vs_135303\|\|full | Low-quality |
| S16_vs_135455\|\|full | Not-determined |
| S16_vs_135475\|\|full | Low-quality |
| S16_vs_135501\|\|full | Not-determined |
| S16_vs_135517\|\|full | Not-determined |
| S16_vs_135600\|\|full | Low-quality |
| S16_vs_135637\|\|full | Low-quality |
| S16_vs_135679\|\|full | Low-quality |
| S16_vs_135682\|\|full | Not-determined |
| S16_vs_135708\|\|full | Low-quality |
| S16_vs_135782\|\|full | Not-determined |
| S16_vs_135874\|\|full | Not-determined |
| S16_vs_135877\|\|full | Low-quality |
| S16_vs_135940\|\|full | Not-determined |
| S16_vs_136002\|\|full | Low-quality |
| S16_vs_136062\|\|full | Not-determined |
| S16_vs_136108\|\|full | Not-determined |
| S16_vs_136117\|\|full | Not-determined |
| S16_vs_136127\|\|full | Low-quality |
| S16_vs_136147\|\|full | Low-quality |
| S16_vs_136150\|\|full | Low-quality |
| S16_vs_136203\|\|full | Not-determined |
| S16_vs_136208\|\|full | Low-quality |
| S16_vs_136220\|\|full | Low-quality |
| S16_vs_136280\|\|full | Low-quality |
| S16_vs_136312\|\|full | Low-quality |
| S16_vs_136333\|\|full | Not-determined |
| S16_vs_136361\|\|full | Low-quality |
| S16_vs_136385\|\|full | Low-quality |
| S16_vs_136405\|\|full | Low-quality |
| S16_vs_136451\|\|full | Low-quality |
| S16_vs_136465\|\|full | Not-determined |
| S16_vs_136479\|\|full | Not-determined |
| S16_vs_136490\|\|full | Low-quality |
| S16_vs_136513\|\|full | Low-quality |
| S16_vs_136559\|\|full | Not-determined |
| S16_vs_136581\|\|full | Low-quality |
| S16_vs_136596\|\|full | Low-quality |
| S16_vs_136700\|\|full | Not-determined |
| S16_vs_136724\|\|full | Low-quality |
| S16_vs_136747\|\|full | Low-quality |
| S16_vs_136809\|\|full | Low-quality |
| S16_vs_136810\|\|full | Not-determined |
| S16_vs_136910\|\|full | Not-determined |
| S16_vs_136918\|\|full | Low-quality |
| S16_vs_136928\|\|full | Low-quality |
| S12_vs_k141_67576\|\|full | Medium-quality |
| S16_vs_136980\|\|full | Not-determined |
| S16_vs_136989\|\|full | Low-quality |
| S16_vs_136990\|\|full | Low-quality |
| S16_vs_137094\|\|full | Low-quality |
| S16_vs_137194\|\|full | Low-quality |
| S14_vs_k141_95416\|\|full | Medium-quality |
| S16_vs_137223\|\|full | Not-determined |
| S16_vs_137261\|\|full | Low-quality |
| S16_vs_137288\|\|full | Low-quality |
| S16_vs_137298\|\|full | Low-quality |
| S16_vs_137317\|\|full | Low-quality |
| S16_vs_137321\|\|full | Not-determined |
| S16_vs_137322\|\|full | Low-quality |
| S16_vs_137391\|\|full | Low-quality |
| S16_vs_137450\|\|full | Not-determined |
| S16_vs_137562\|\|full | Low-quality |
| S16_vs_137583\|\|full | Low-quality |
| S16_vs_137676\|\|full | Low-quality |
| S16_vs_137687\|\|full | Low-quality |
| S16_vs_137734\|\|full | Low-quality |
| S16_vs_137785\|\|full | Not-determined |
| S16_vs_137809\|\|full | Low-quality |
| S16_vs_137810\|\|full | Low-quality |
| S16_vs_137828\|\|full | Not-determined |
| S16_vs_137880\|\|full | Not-determined |
| S16_vs_137889\|\|full | Not-determined |
| S16_vs_137960\|\|full | Not-determined |
| S16_vs_137980\|\|full | Low-quality |
| S16_vs_137983\|\|full | Low-quality |
| S16_vs_137999\|\|full | Low-quality |
| S16_vs_138032\|\|full | Low-quality |
| S16_vs_138062\|\|full | Not-determined |
| S16_vs_138077\|\|full | Not-determined |
| S16_vs_138167\|\|full | Low-quality |
| S16_vs_138234\|\|full | Low-quality |
| S16_vs_138279\|\|full | Low-quality |
| S16_vs_138284\|\|full | Low-quality |
| S16_vs_138297\|\|full | Low-quality |
| S16_vs_138311\|\|full | Low-quality |
| S16_vs_138323\|\|full | Not-determined |
| S16_vs_138340\|\|full | Not-determined |
| S16_vs_138346\|\|full | Not-determined |
| S16_vs_138373\|\|full | Not-determined |
| S16_vs_138395\|\|full | Not-determined |
| S16_vs_138406\|\|full | Low-quality |
| S16_vs_138407\|\|full | Low-quality |
| S16_vs_138410\|\|full | Not-determined |
| S16_vs_138419\|\|full | Not-determined |
| S16_vs_138438\|\|full | Low-quality |
| S16_vs_138537\|\|full | Low-quality |
| S16_vs_138561\|\|full | Not-determined |
| S16_vs_138702\|\|full | Low-quality |
| S16_vs_138704\|\|full | Low-quality |
| S16_vs_138721\|\|full | Low-quality |
| S16_vs_138731\|\|full | Not-determined |
| S16_vs_138733\|\|full | Low-quality |
| S16_vs_138765\|\|full | Low-quality |
| S16_vs_138801\|\|full | Not-determined |
| S16_vs_138838\|\|full | Low-quality |
| S16_vs_138844\|\|full | Low-quality |
| S16_vs_138861\|\|full | Not-determined |
| S16_vs_138950\|\|full | Low-quality |
| S16_vs_138980\|\|full | Low-quality |
| S16_vs_139108\|\|full | Low-quality |
| S16_vs_139172\|\|full | Low-quality |
| S16_vs_139176\|\|full | Not-determined |
| S16_vs_139186\|\|full | Low-quality |
| S16_vs_139231\|\|full | Low-quality |
| S16_vs_139242\|\|full | Not-determined |
| S16_vs_139302\|\|full | Low-quality |
| S16_vs_139321\|\|full | Not-determined |
| S16_vs_139362\|\|full | Low-quality |
| S16_vs_139375\|\|full | Not-determined |
| S16_vs_139378\|\|full | Not-determined |
| S16_vs_139405\|\|full | Low-quality |
| S16_vs_139455\|\|full | Low-quality |
| S16_vs_139469\|\|full | Low-quality |
| S16_vs_139475\|\|full | Not-determined |
| S16_vs_139476\|\|full | Not-determined |
| S16_vs_139510\|\|full | Not-determined |
| S16_vs_139557\|\|full | Low-quality |
| S16_vs_139642\|\|full | Not-determined |
| S16_vs_139676\|\|full | Low-quality |
| S16_vs_139678\|\|full | Low-quality |
| S16_vs_139751\|\|full | Low-quality |
| S16_vs_139762\|\|full | Low-quality |
| S16_vs_139805\|\|full | Low-quality |
| S16_vs_139807\|\|full | Low-quality |
| S16_vs_139864\|\|full | Low-quality |
| S16_vs_139868\|\|full | Not-determined |
| S16_vs_139877\|\|full | Low-quality |
| S16_vs_139890\|\|full | Low-quality |
| S16_vs_139891\|\|full | Low-quality |
| S16_vs_139921\|\|full | Low-quality |
| S16_vs_139922\|\|full | Not-determined |
| S16_vs_139934\|\|full | Low-quality |
| S16_vs_140005\|\|full | Low-quality |
| S16_vs_140006\|\|full | Not-determined |
| S16_vs_140027\|\|full | Not-determined |
| S16_vs_140032\|\|full | Low-quality |
| S16_vs_140116\|\|full | Not-determined |

| | |
|---|---|
| S16_vs_140119\|\|full | Not-determined |
| S16_vs_140128\|\|full | Not-determined |
| S16_vs_140160\|\|full | Low-quality |
| S16_vs_140162\|\|full | Not-determined |
| S16_vs_140187\|\|full | Not-determined |
| S16_vs_140237\|\|full | Low-quality |
| S16_vs_140283\|\|full | Low-quality |
| S16_vs_140332\|\|full | Not-determined |
| S16_vs_140343\|\|full | Not-determined |
| S16_vs_140370\|\|full | Low-quality |
| S16_vs_140372\|\|full | Not-determined |
| S16_vs_140379\|\|full | Low-quality |
| S16_vs_140387\|\|full | Low-quality |
| S16_vs_140413\|\|full | Low-quality |
| S16_vs_140422\|\|full | Not-determined |
| S16_vs_140448\|\|full | Not-determined |
| S16_vs_140462\|\|full | Not-determined |
| S12_vs_k141_69136\|\|full | Medium-quality |
| S16_vs_140522\|\|full | Low-quality |
| S16_vs_140580\|\|full | Low-quality |
| S16_vs_140581\|\|full | Not-determined |
| S16_vs_140584\|\|full | Low-quality |
| S16_vs_140653\|\|full | Not-determined |
| S16_vs_140670\|\|full | Not-determined |
| S4_vs_VIRSorter_15112-cat_2 | Medium-quality |
| S16_vs_140721\|\|full | Not-determined |
| S16_vs_140736\|\|full | Not-determined |
| S16_vs_140738\|\|full | Not-determined |
| S16_vs_140822\|\|full | Not-determined |
| S16_vs_140843\|\|full | Low-quality |
| S16_vs_140885\|\|full | Low-quality |
| S16_vs_140889\|\|full | Low-quality |
| S16_vs_140934\|\|full | Not-determined |
| S16_vs_140940\|\|full | Low-quality |
| S16_vs_140952\|\|full | Not-determined |
| S16_vs_140970\|\|full | Not-determined |
| S16_vs_141058\|\|full | Not-determined |
| S16_vs_141099\|\|full | Not-determined |
| S16_vs_141123\|\|full | Low-quality |
| S16_vs_141177\|\|full | Low-quality |
| S16_vs_141184\|\|full | Not-determined |
| S17_extracted_k141_111210 | Medium-quality |
| S16_vs_141238\|\|full | Low-quality |
| S16_vs_141291\|\|full | Not-determined |
| S16_vs_141299\|\|full | Low-quality |
| S16_vs_141334\|\|full | Low-quality |
| S16_vs_141352\|\|full | Not-determined |
| S16_vs_141368\|\|full | Not-determined |
| S16_vs_141371\|\|full | Low-quality |
| S16_vs_141419\|\|full | Low-quality |
| S16_vs_141428\|\|full | Not-determined |
| S16_vs_141472\|\|full | Low-quality |
| S16_vs_141518\|\|full | Low-quality |
| S16_vs_141535\|\|full | Not-determined |
| S16_vs_141648\|\|full | Low-quality |
| S16_vs_141653\|\|full | Low-quality |
| S16_vs_141664\|\|full | Not-determined |
| S16_vs_141755\|\|full | Not-determined |
| S16_vs_141786\|\|full | Low-quality |
| S16_vs_141914\|\|full | Low-quality |
| S16_vs_141919\|\|full | Not-determined |
| S16_vs_141988\|\|full | Low-quality |
| S16_vs_141994\|\|full | Low-quality |
| S16_vs_142010\|\|full | Low-quality |
| S16_vs_142012\|\|full | Not-determined |
| S16_vs_142038\|\|full | Not-determined |
| S16_vs_142090\|\|full | Not-determined |
| S16_vs_142140\|\|full | Not-determined |
| S16_vs_142146\|\|full | Low-quality |
| S16_vs_142202\|\|full | Low-quality |
| S16_vs_142234\|\|full | Low-quality |
| S16_vs_142271\|\|full | Not-determined |
| S16_vs_142334\|\|full | Low-quality |
| S16_vs_142354\|\|full | Low-quality |
| S16_vs_142364\|\|full | Low-quality |
| S16_vs_142390\|\|full | Low-quality |
| S16_vs_142393\|\|full | Low-quality |
| S16_vs_142414\|\|full | Not-determined |
| S16_vs_142416\|\|full | Low-quality |
| S16_vs_142498\|\|full | Low-quality |
| S16_vs_142513\|\|full | Low-quality |
| S16_vs_142545\|\|full | Not-determined |
| S16_vs_142554\|\|full | Low-quality |
| S5_vs_k141_13254\|\|full | Medium-quality |
| S16_vs_142672\|\|full | Low-quality |
| S16_vs_142715\|\|full | Not-determined |
| S16_vs_142717\|\|full | Low-quality |
| S16_vs_142802\|\|full | Not-determined |
| S16_vs_142809\|\|full | Not-determined |
| S16_vs_142850\|\|full | Not-determined |
| S16_vs_142917\|\|full | Not-determined |
| S16_vs_142922\|\|full | Low-quality |
| S16_vs_142924\|\|full | Not-determined |
| S16_vs_142925\|\|full | Not-determined |
| S16_vs_142931\|\|full | Not-determined |
| S16_vs_142951\|\|full | Low-quality |
| S16_vs_142980\|\|full | Not-determined |
| S16_vs_142994\|\|full | Low-quality |
| S16_vs_143001\|\|full | Low-quality |
| S16_vs_143020\|\|full | Low-quality |
| S16_vs_143106\|\|full | Not-determined |
| S16_vs_143123\|\|full | Not-determined |
| S16_vs_143140\|\|full | Low-quality |
| S16_vs_143147\|\|full | Not-determined |
| S16_vs_143173\|\|full | Not-determined |
| S16_vs_143197\|\|full | Low-quality |
| S16_vs_143252\|\|full | Low-quality |
| S16_vs_143262\|\|full | Low-quality |
| S16_vs_143269\|\|full | Not-determined |
| S16_vs_143270\|\|full | Low-quality |
| S16_vs_143328\|\|full | Low-quality |
| S16_vs_143329\|\|full | Low-quality |
| S16_vs_143361\|\|full | Not-determined |
| S16_vs_143422\|\|full | Low-quality |
| S16_vs_143435\|\|full | Low-quality |
| S16_vs_143446\|\|full | Not-determined |
| S16_vs_143448\|\|full | Not-determined |
| S16_vs_143482\|\|full | Not-determined |
| S16_vs_143518\|\|full | Not-determined |
| S16_vs_143538\|\|full | Low-quality |
| S16_vs_143564\|\|full | Low-quality |
| S16_vs_143585\|\|full | Not-determined |
| S16_vs_143611\|\|full | Not-determined |
| S16_vs_143621\|\|full | Low-quality |
| S16_vs_143641\|\|full | Not-determined |
| S16_vs_143645\|\|full | Low-quality |
| S16_vs_143679\|\|full | Low-quality |
| S16_vs_143688\|\|full | Not-determined |
| S16_vs_143703\|\|full | Low-quality |
| S16_vs_143708\|\|full | Low-quality |
| S16_vs_143713\|\|full | Not-determined |
| S16_vs_143722\|\|full | Low-quality |
| S16_vs_143849\|\|full | Low-quality |
| S16_vs_143856\|\|full | Not-determined |
| S16_vs_143867\|\|full | Low-quality |
| S16_vs_143871\|\|full | Not-determined |
| S16_vs_143942\|\|full | Not-determined |
| S16_vs_143944\|\|full | Low-quality |
| S16_vs_143955\|\|full | Not-determined |
| S16_vs_143986\|\|full | Not-determined |
| S16_vs_144031\|\|full | Low-quality |
| S16_vs_144051\|\|full | Low-quality |
| S16_vs_144129\|\|full | Not-determined |
| S16_vs_144132\|\|full | Low-quality |
| S16_vs_144183\|\|full | Low-quality |
| S16_vs_144198\|\|full | Low-quality |
| S16_vs_144201\|\|full | Low-quality |
| S16_vs_144209\|\|full | Low-quality |
| S16_vs_144218\|\|full | Not-determined |
| S16_vs_144239\|\|full | Not-determined |
| S16_vs_144240\|\|full | Low-quality |
| S16_vs_144302\|\|full | Low-quality |
| S16_vs_144303\|\|full | Low-quality |
| S16_vs_144307\|\|full | Low-quality |
| S16_vs_144335\|\|full | Low-quality |
| S16_vs_144340\|\|full | Low-quality |
| S16_vs_144376\|\|full | Low-quality |
| S16_vs_144378\|\|full | Not-determined |
| S16_vs_144384\|\|full | Low-quality |
| S16_vs_144392\|\|full | Not-determined |
| S16_vs_144438\|\|full | Low-quality |
| S16_vs_144477\|\|full | Not-determined |
| S16_vs_144492\|\|full | Not-determined |
| S16_vs_144518\|\|full | Low-quality |
| S16_vs_144539\|\|full | Not-determined |
| S16_vs_144561\|\|full | Low-quality |
| S16_vs_144588\|\|full | Not-determined |
| S16_vs_144596\|\|full | Low-quality |
| S16_vs_144673\|\|full | Low-quality |
| S16_vs_144680\|\|full | Not-determined |
| S16_vs_144728\|\|full | Low-quality |
| S16_vs_144731\|\|full | Low-quality |
| S16_vs_144749\|\|full | Not-determined |
| S16_vs_144786\|\|full | Low-quality |
| S16_vs_144798\|\|full | Low-quality |
| S16_vs_144825\|\|full | Low-quality |
| S16_vs_144847\|\|full | Not-determined |
| S16_vs_144882\|\|full | Low-quality |
| S16_vs_144896\|\|full | Low-quality |
| S16_vs_144897\|\|full | Low-quality |
| S16_vs_144985\|\|full | Not-determined |
| S16_vs_144989\|\|full | Not-determined |
| S16_vs_145080\|\|full | Low-quality |
| S16_vs_145160\|\|full | Low-quality |
| S16_vs_145245\|\|full | Low-quality |
| S16_vs_145262\|\|full | Low-quality |
| S16_vs_145283\|\|full | Low-quality |
| S16_vs_145295\|\|full | Low-quality |
| S16_vs_145296\|\|full | Low-quality |
| S16_vs_145356\|\|full | Low-quality |
| S16_vs_145382\|\|full | Low-quality |
| S16_vs_145383\|\|full | Not-determined |
| S16_vs_145436\|\|full | Low-quality |
| S16_vs_145461\|\|full | Not-determined |
| S16_vs_145475\|\|full | Low-quality |
| S16_vs_145481\|\|full | Low-quality |
| S16_vs_145504\|\|full | Low-quality |
| S16_vs_145523\|\|full | Low-quality |
| S16_vs_145532\|\|full | Low-quality |
| S16_vs_145564\|\|full | Not-determined |
| S16_vs_145567\|\|full | Not-determined |
| S16_vs_145575\|\|full | Low-quality |
| S16_vs_145611\|\|full | Low-quality |
| S16_vs_145621\|\|full | Low-quality |
| S16_vs_145698\|\|full | Low-quality |
| S16_vs_145724\|\|full | Low-quality |
| S16_vs_145733\|\|full | Not-determined |
| S16_vs_145745\|\|full | Low-quality |
| S16_vs_145758\|\|full | Low-quality |
| S16_vs_145759\|\|full | Low-quality |
| S16_vs_145787\|\|full | Low-quality |
| S16_vs_145793\|\|full | Not-determined |
| S16_vs_145796\|\|full | Not-determined |
| S16_vs_145819\|\|full | Not-determined |
| S16_vs_145871\|\|full | Low-quality |
| S16_vs_145907\|\|full | Not-determined |
| S16_vs_145928\|\|full | Low-quality |
| S16_vs_145934\|\|full | Low-quality |
| S16_vs_145962\|\|full | Not-determined |
| S16_vs_146035\|\|full | Low-quality |
| S16_vs_146080\|\|full | Low-quality |
| S16_vs_146127\|\|full | Low-quality |
| S16_vs_146221\|\|full | Not-determined |
| S16_vs_146223\|\|full | Low-quality |
| S16_vs_146267\|\|full | Not-determined |
| S16_vs_146287\|\|full | Not-determined |
| S16_vs_146301\|\|full | Low-quality |
| S16_vs_146317\|\|full | Low-quality |
| S16_vs_146327\|\|full | Low-quality |
| S16_vs_146331\|\|full | Low-quality |
| S16_vs_146364\|\|full | Low-quality |
| S16_vs_146369\|\|full | Low-quality |
| S16_vs_146390\|\|full | Low-quality |
| S16_vs_146400\|\|full | Low-quality |
| S16_vs_146432\|\|full | Low-quality |
| S16_vs_146454\|\|full | Not-determined |
| S16_vs_146459\|\|full | Low-quality |
| S16_vs_146532\|\|full | Low-quality |
| S16_vs_146544\|\|full | Low-quality |
| S16_vs_146610\|\|full | Low-quality |
| S16_vs_146611\|\|full | Not-determined |
| S16_vs_146612\|\|full | Not-determined |
| S16_vs_146646\|\|full | Not-determined |
| S16_vs_146647\|\|full | Not-determined |
| S16_vs_146657\|\|full | Not-determined |
| S16_vs_146705\|\|full | Low-quality |
| S16_vs_146716\|\|full | Not-determined |
| S16_vs_146720\|\|full | Not-determined |
| S16_vs_146749\|\|full | Low-quality |
| S12_vs_k141_71525\|\|full | Medium-quality |
| S16_vs_146828\|\|full | Low-quality |
| S16_vs_146840\|\|full | Not-determined |
| S16_vs_146867\|\|full | Low-quality |
| S16_vs_146873\|\|full | Not-determined |
| S16_vs_146901\|\|full | Not-determined |
| S16_vs_146941\|\|full | Not-determined |
| S16_vs_146952\|\|full | Low-quality |
| S16_vs_147015\|\|full | Low-quality |
| S16_vs_147023\|\|full | Not-determined |
| S16_vs_147087\|\|full | Low-quality |
| S16_vs_147213\|\|full | Not-determined |
| S16_vs_147230\|\|full | Not-determined |
| S16_vs_147326\|\|full | Low-quality |
| S16_vs_147330\|\|full | Not-determined |
| S16_vs_147340\|\|full | Low-quality |
| S16_vs_147366\|\|full | Not-determined |
| S16_vs_147388\|\|full | Not-determined |
| S16_vs_147405\|\|full | Low-quality |
| S16_vs_147492\|\|full | Not-determined |
| S16_vs_147503\|\|full | Not-determined |
| S16_vs_147528\|\|full | Low-quality |
| S16_vs_147573\|\|full | Low-quality |
| S16_vs_147601\|\|full | Low-quality |
| S16_vs_147603\|\|full | Not-determined |
| S16_vs_147618\|\|full | Low-quality |
| S16_vs_147641\|\|full | Low-quality |
| S16_vs_147705\|\|full | Low-quality |
| S16_vs_147798\|\|full | Low-quality |
| S16_vs_147808\|\|full | Low-quality |
| S16_vs_147843\|\|full | Low-quality |
| S16_vs_147868\|\|full | Not-determined |
| S16_vs_147882\|\|full | Not-determined |
| S16_vs_147891\|\|full | Not-determined |
| S16_vs_147934\|\|full | Low-quality |
| S16_vs_147941\|\|full | Low-quality |
| S16_vs_147957\|\|full | Not-determined |
| S16_vs_148019\|\|full | Low-quality |
| S16_vs_148065\|\|full | Not-determined |
| S16_vs_148101\|\|full | Low-quality |
| S16_vs_148119\|\|full | Low-quality |
| S16_vs_148210\|\|full | Low-quality |
| S16_vs_148411\|\|full | Not-determined |
| S16_vs_148451\|\|full | Low-quality |
| S16_vs_148517\|\|full | Low-quality |
| S16_vs_148529\|\|full | Low-quality |
| S16_vs_148537\|\|full | Not-determined |
| S16_vs_148554\|\|full | Low-quality |
| S16_vs_148567\|\|full | Not-determined |
| S16_vs_148580\|\|full | Low-quality |
| S16_vs_148599\|\|full | Not-determined |
| S16_vs_148659\|\|full | Low-quality |
| S16_vs_148675\|\|full | Low-quality |
| S16_vs_148745\|\|full | Low-quality |
| S16_vs_148748\|\|full | Not-determined |
| S16_vs_148792\|\|full | Not-determined |
| S16_vs_148817\|\|full | Low-quality |
| S16_vs_148823\|\|full | Not-determined |
| S16_vs_148826\|\|full | Low-quality |
| S16_vs_148868\|\|full | Not-determined |
| S16_vs_148885\|\|full | Low-quality |
| S16_vs_148886\|\|full | Not-determined |
| S16_vs_148888\|\|full | Low-quality |
| S16_vs_148976\|\|full | Low-quality |
| S16_vs_149036\|\|full | Low-quality |
| S16_vs_149059\|\|full | Not-determined |
| S16_vs_149070\|\|full | Not-determined |
| S16_vs_149084\|\|full | Not-determined |
| S16_vs_149086\|\|full | Low-quality |
| S16_vs_149152\|\|full | Not-determined |
| S16_vs_149187\|\|full | Not-determined |
| S16_vs_149213\|\|full | Not-determined |
| S16_vs_149234\|\|full | Not-determined |
| S16_vs_149244\|\|full | Not-determined |
| S16_vs_149255\|\|full | Not-determined |
| S16_vs_149270\|\|full | Not-determined |
| S16_vs_149365\|\|full | Low-quality |
| S16_vs_149379\|\|full | Low-quality |
| S16_vs_149381\|\|full | Not-determined |
| S16_vs_149394\|\|full | Low-quality |
| S16_vs_149421\|\|full | Low-quality |
| S16_vs_149454\|\|full | Not-determined |
| S16_vs_149470\|\|full | Low-quality |
| S16_vs_149487\|\|full | Not-determined |
| S16_vs_149503\|\|full | Low-quality |
| S16_vs_149520\|\|full | Low-quality |
| S16_vs_149534\|\|full | Low-quality |
| S16_vs_149557\|\|full | Low-quality |
| S16_vs_149579\|\|full | Low-quality |
| S16_vs_149719\|\|full | Not-determined |
| S16_vs_149727\|\|full | Low-quality |
| S16_vs_149750\|\|full | Not-determined |
| S16_vs_149790\|\|full | Low-quality |
| S16_vs_149812\|\|full | Not-determined |
| S16_vs_149813\|\|full | Low-quality |
| S16_vs_150035\|\|full | Low-quality |
| S16_vs_150058\|\|full | Low-quality |
| S16_vs_150135\|\|full | Low-quality |
| S16_vs_150155\|\|full | Not-determined |
| S16_vs_150229\|\|full | Not-determined |
| S16_vs_150239\|\|full | Not-determined |
| S16_vs_150247\|\|full | Not-determined |
| S16_vs_150252\|\|full | Low-quality |
| S16_vs_150286\|\|full | Not-determined |
| S16_vs_150288\|\|full | Not-determined |
| S16_vs_150295\|\|full | Not-determined |
| S16_vs_150391\|\|full | Low-quality |
| S16_vs_150433\|\|full | Not-determined |
| S16_vs_150465\|\|full | Low-quality |
| S16_vs_150474\|\|full | Low-quality |
| S16_vs_150500\|\|full | Low-quality |
| S16_vs_150547\|\|full | Not-determined |
| S16_vs_150569\|\|full | Low-quality |
| S16_vs_150629\|\|full | Low-quality |
| S16_vs_150709\|\|full | Not-determined |
| S16_vs_150763\|\|full | Not-determined |
| S16_vs_150782\|\|full | Low-quality |
| S16_vs_150827\|\|full | Not-determined |
| S16_vs_150853\|\|full | Low-quality |
| S16_vs_150908\|\|full | Low-quality |
| S16_vs_150919\|\|full | Not-determined |
| S16_vs_150920\|\|full | Low-quality |
| S16_vs_150929\|\|full | Low-quality |
| S16_vs_150937\|\|full | Low-quality |
| S16_vs_150956\|\|full | Low-quality |
| S16_vs_150995\|\|full | Low-quality |
| S16_vs_151028\|\|full | Not-determined |
| S16_vs_151035\|\|full | Low-quality |
| S16_vs_151043\|\|full | Low-quality |
| S16_vs_151134\|\|full | Low-quality |
| S16_vs_151135\|\|full | Not-determined |
| S16_vs_151199\|\|full | Low-quality |
| S16_vs_151228\|\|full | Low-quality |
| S16_vs_151237\|\|full | Low-quality |
| S16_vs_151270\|\|full | Low-quality |
| S16_vs_151291\|\|full | Not-determined |
| S16_vs_151341\|\|full | Low-quality |
| S16_vs_151351\|\|full | Low-quality |
| S16_vs_151353\|\|full | Low-quality |
| S16_vs_151398\|\|full | Low-quality |
| S16_vs_151426\|\|full | Low-quality |
| S16_vs_151457\|\|full | Low-quality |
| S16_vs_151531\|\|full | Low-quality |
| S16_vs_151538\|\|full | Not-determined |
| S16_vs_151588\|\|full | Not-determined |
| S16_vs_151593\|\|full | Low-quality |
| S16_vs_151604\|\|full | Low-quality |
| S16_vs_151625\|\|full | Low-quality |
| S16_vs_151638\|\|full | Not-determined |
| S16_vs_151668\|\|full | Low-quality |
| S16_vs_151733\|\|full | Low-quality |
| S16_vs_151788\|\|full | Low-quality |
| S16_vs_151825\|\|full | Low-quality |
| S16_vs_151857\|\|full | Not-determined |
| S16_vs_151912\|\|full | Low-quality |
| S16_vs_152020\|\|full | Low-quality |
| S16_vs_152034\|\|full | Not-determined |
| S16_vs_152043\|\|full | Low-quality |
| S16_vs_152059\|\|full | Low-quality |
| S16_vs_152145\|\|full | Not-determined |
| S16_vs_152168\|\|full | Low-quality |
| S16_vs_152190\|\|full | Low-quality |
| S16_vs_152200\|\|full | Low-quality |
| S16_vs_152210\|\|full | Low-quality |
| S16_vs_152254\|\|full | Low-quality |
| S16_vs_152268\|\|full | Low-quality |
| S16_vs_152296\|\|full | Low-quality |
| S16_vs_152323\|\|full | Not-determined |
| S16_vs_152325\|\|full | Low-quality |
| S16_vs_152400\|\|full | Not-determined |
| S16_vs_152414\|\|full | Not-determined |
| S16_vs_152420\|\|full | Low-quality |
| S16_vs_152459\|\|full | Low-quality |
| S16_vs_152487\|\|full | Not-determined |
| S16_vs_152488\|\|full | Low-quality |
| S16_vs_152543\|\|full | Not-determined |
| S16_vs_152565\|\|full | Low-quality |
| S16_vs_152570\|\|full | Low-quality |
| S16_vs_152577\|\|full | Low-quality |
| S16_vs_152590\|\|full | Not-determined |
| S16_vs_152644\|\|full | Low-quality |
| S16_vs_152680\|\|full | Low-quality |
| S16_vs_152689\|\|full | Not-determined |
| S16_vs_152695\|\|full | Low-quality |
| S16_vs_152728\|\|full | Not-determined |
| S16_vs_152749\|\|full | Low-quality |
| S16_vs_152773\|\|full | Low-quality |
| S16_vs_152807\|\|full | Low-quality |
| S16_vs_152833\|\|full | Not-determined |
| S16_vs_152865\|\|full | Low-quality |
| S16_vs_152872\|\|full | Low-quality |
| S16_vs_152892\|\|full | Not-determined |
| S16_vs_152902\|\|full | Low-quality |
| S16_vs_153022\|\|full | Low-quality |
| S16_vs_153074\|\|full | Not-determined |
| S16_vs_153087\|\|full | Not-determined |
| S16_vs_153118\|\|full | Low-quality |
| S16_vs_153162\|\|full | Not-determined |
| S16_vs_153181\|\|full | Low-quality |
| S16_vs_153198\|\|full | Low-quality |
| S16_vs_153256\|\|full | Low-quality |
| S16_vs_153257\|\|full | Not-determined |
| S16_vs_153260\|\|full | Not-determined |
| S16_vs_153336\|\|full | Low-quality |
| S16_vs_153373\|\|full | Not-determined |
| S16_vs_153413\|\|full | Low-quality |
| S16_vs_153465\|\|full | Low-quality |
| S16_vs_153494\|\|full | Not-determined |
| S16_vs_153513\|\|full | Not-determined |
| S16_vs_153524\|\|full | Low-quality |
| S16_vs_153537\|\|full | Not-determined |
| S16_vs_153545\|\|full | Not-determined |
| S16_vs_153564\|\|full | Not-determined |
| S16_vs_153622\|\|full | Not-determined |
| S16_vs_153636\|\|full | Not-determined |
| S16_vs_153642\|\|full | Low-quality |
| S16_vs_153654\|\|full | Not-determined |
| S16_vs_153696\|\|full | Low-quality |
| S16_vs_153794\|\|full | Not-determined |
| S16_vs_153817\|\|full | Not-determined |
| S16_vs_153824\|\|full | Low-quality |
| S16_vs_153861\|\|full | Low-quality |
| S16_vs_153946\|\|full | Low-quality |
| S16_vs_153968\|\|full | Low-quality |
| S16_vs_153993\|\|full | Low-quality |
| S16_vs_154045\|\|full | Not-determined |
| S16_vs_154070\|\|full | Low-quality |
| S16_vs_154179\|\|full | Not-determined |
| S16_vs_154222\|\|full | Low-quality |
| S16_vs_154225\|\|full | Low-quality |
| S16_vs_154246\|\|full | Low-quality |
| S16_vs_154311\|\|full | Low-quality |
| S16_vs_154361\|\|full | Low-quality |
| S16_vs_154427\|\|full | Low-quality |
| S16_vs_154450\|\|full | Not-determined |
| S16_vs_154460\|\|full | Low-quality |
| S16_vs_154544\|\|full | Not-determined |
| S16_vs_154549\|\|full | Not-determined |
| S16_vs_154556\|\|full | Not-determined |
| S16_vs_154584\|\|full | Not-determined |
| S16_vs_154693\|\|full | Not-determined |
| S16_vs_154698\|\|full | Not-determined |
| S16_vs_154773\|\|full | Not-determined |
| S16_vs_154796\|\|full | Low-quality |
| S16_vs_154803\|\|full | Low-quality |
| S16_vs_154841\|\|full | Low-quality |
| S16_vs_154881\|\|full | Low-quality |
| S16_vs_154982\|\|full | Low-quality |
| S16_vs_154988\|\|full | Low-quality |
| S16_vs_155042\|\|full | Not-determined |
| S16_vs_155112\|\|full | Low-quality |
| S16_vs_155114\|\|full | Not-determined |
| S16_vs_155121\|\|full | Low-quality |
| S16_vs_155149\|\|full | Not-determined |
| S16_vs_155161\|\|full | Not-determined |
| S16_vs_155171\|\|full | Not-determined |
| S16_vs_155176\|\|full | Low-quality |
| S16_vs_155178\|\|full | Not-determined |
| S16_vs_155195\|\|full | Low-quality |
| S16_vs_155240\|\|full | Low-quality |
| S16_vs_155250\|\|full | Low-quality |
| S16_vs_155259\|\|full | Low-quality |
| S16_vs_155316\|\|full | Not-determined |
| S16_vs_155317\|\|full | Not-determined |
| S16_vs_155321\|\|full | Low-quality |
| S16_vs_155324\|\|full | Low-quality |
| S16_vs_155426\|\|full | Not-determined |
| S16_vs_155485\|\|full | Not-determined |
| S16_vs_155505\|\|full | Not-determined |
| S16_vs_155520\|\|full | Not-determined |
| S16_vs_155547\|\|full | Not-determined |
| S16_vs_155644\|\|full | Low-quality |
| S16_vs_155695\|\|full | Not-determined |
| S16_vs_155727\|\|full | Low-quality |
| S16_vs_155810\|\|full | Not-determined |
| S16_vs_155847\|\|full | Low-quality |
| S16_vs_155864\|\|full | Low-quality |
| S16_vs_155875\|\|full | Low-quality |
| S16_vs_155880\|\|full | Low-quality |
| S16_vs_155914\|\|full | Low-quality |
| S16_vs_155956\|\|full | Low-quality |
| S16_vs_155965\|\|full | Not-determined |
| S16_vs_156030\|\|full | Low-quality |
| S16_vs_156087\|\|full | Low-quality |
| S16_vs_156089\|\|full | Low-quality |
| S16_vs_156096\|\|full | Not-determined |
| S16_vs_156134\|\|full | Not-determined |
| S16_vs_156148\|\|full | Not-determined |
| S16_vs_156187\|\|full | Low-quality |
| S16_vs_156196\|\|full | Low-quality |
| S16_vs_156199\|\|full | Not-determined |
| S16_vs_156221\|\|full | Low-quality |
| S16_vs_156248\|\|full | Low-quality |
| S16_vs_156266\|\|full | Not-determined |
| S16_vs_156315\|\|full | Not-determined |
| S16_vs_156377\|\|full | Not-determined |
| S16_vs_156391\|\|full | Not-determined |
| S16_vs_156393\|\|full | Low-quality |
| S16_vs_156424\|\|full | Low-quality |
| S16_vs_156438\|\|full | Low-quality |
| S16_vs_156442\|\|full | Not-determined |
| S16_vs_156461\|\|full | Not-determined |
| S16_vs_156466\|\|full | Low-quality |
| S16_vs_156482\|\|full | Low-quality |
| S16_vs_156501\|\|full | Low-quality |
| S16_vs_156512\|\|full | Low-quality |
| S16_vs_156549\|\|full | Not-determined |
| S16_vs_156568\|\|full | Not-determined |
| S16_vs_156573\|\|full | Not-determined |
| S16_vs_156584\|\|full | Not-determined |
| S16_vs_156592\|\|full | Not-determined |
| S16_vs_156623\|\|full | Low-quality |
| S16_vs_156641\|\|full | Low-quality |
| S16_vs_156650\|\|full | Low-quality |
| S16_vs_156660\|\|full | Not-determined |
| S16_vs_156687\|\|full | Not-determined |
| S16_vs_156690\|\|full | Not-determined |
| S16_vs_156704\|\|full | Low-quality |
| S16_vs_156750\|\|full | Not-determined |
| S16_vs_156754\|\|full | Not-determined |
| S16_vs_156831\|\|full | Low-quality |
| S16_vs_156844\|\|full | Not-determined |
| S16_vs_156845\|\|full | Not-determined |
| S16_vs_156864\|\|full | Low-quality |
| S16_vs_156865\|\|full | Not-determined |
| S16_vs_156916\|\|full | Not-determined |
| S16_vs_156921\|\|full | Not-determined |
| S16_vs_157089\|\|full | Low-quality |
| S16_vs_157107\|\|full | Not-determined |
| S16_vs_157123\|\|full | Low-quality |
| S16_vs_157191\|\|full | Not-determined |
| S16_vs_157193\|\|full | Not-determined |
| S16_vs_157204\|\|full | Not-determined |
| S16_vs_157303\|\|full | Low-quality |
| S16_vs_157310\|\|full | Low-quality |
| S16_vs_157324\|\|full | Not-determined |
| S16_vs_157365\|\|full | Not-determined |
| S16_vs_157373\|\|full | Low-quality |
| S16_vs_157380\|\|full | Not-determined |
| S16_vs_157481\|\|full | Not-determined |

| | |
|---|---|
| S16_vs_157483\|\|full | Low-quality |
| S16_vs_157485\|\|full | Low-quality |
| S16_vs_157549\|\|full | Low-quality |
| S16_vs_157568\|\|full | Not-determined |
| S16_vs_157640\|\|full | Low-quality |
| S16_vs_157681\|\|full | Low-quality |
| S16_vs_157684\|\|full | Not-determined |
| S16_vs_157694\|\|full | Low-quality |
| S16_vs_157702\|\|full | Low-quality |
| S16_vs_157708\|\|full | Not-determined |
| S16_vs_157747\|\|full | Not-determined |
| S16_vs_157764\|\|full | Not-determined |
| S16_vs_157781\|\|full | Low-quality |
| S16_vs_157797\|\|full | Not-determined |
| S16_vs_157800\|\|full | Low-quality |
| S16_vs_157891\|\|full | Low-quality |
| S16_vs_157908\|\|full | Low-quality |
| S16_vs_157930\|\|full | Low-quality |
| S16_vs_158000\|\|full | Low-quality |
| S16_vs_158028\|\|full | Not-determined |
| S16_vs_158057\|\|full | Low-quality |
| S16_vs_158080\|\|full | Not-determined |
| S16_vs_158098\|\|full | Not-determined |
| S16_vs_158127\|\|full | Low-quality |
| S16_vs_158220\|\|full | Not-determined |
| S16_vs_158224\|\|full | Low-quality |
| S16_vs_158279\|\|full | Low-quality |
| S16_vs_158285\|\|full | Not-determined |
| S16_vs_158416\|\|full | Low-quality |
| S16_vs_158420\|\|full | Not-determined |
| S16_vs_158446\|\|full | Not-determined |
| S16_vs_158565\|\|full | Low-quality |
| S16_vs_158572\|\|full | Low-quality |
| S16_vs_158574\|\|full | Low-quality |
| S16_vs_158713\|\|full | Low-quality |
| S16_vs_158718\|\|full | Not-determined |
| S16_vs_158722\|\|full | Low-quality |
| S16_vs_158726\|\|full | Low-quality |
| S16_vs_158794\|\|full | Low-quality |
| S16_vs_158806\|\|full | Low-quality |
| S16_vs_158818\|\|full | Low-quality |
| S16_vs_158820\|\|full | Low-quality |
| S16_vs_158850\|\|full | Not-determined |
| S16_vs_158921\|\|full | Low-quality |
| S16_vs_158938\|\|full | Not-determined |
| S16_vs_158974\|\|full | Low-quality |
| S16_vs_158978\|\|full | Not-determined |
| S16_vs_159003\|\|full | Not-determined |
| S16_vs_159019\|\|full | Not-determined |
| S16_vs_159084\|\|full | Low-quality |
| S16_vs_159089\|\|full | Not-determined |
| S16_vs_159096\|\|full | Low-quality |
| S16_vs_159104\|\|full | Not-determined |
| S16_vs_159111\|\|full | Not-determined |
| S16_vs_159113\|\|full | Low-quality |
| S16_vs_159133\|\|full | Not-determined |
| S16_vs_159137\|\|full | Low-quality |
| S16_vs_159181\|\|full | Low-quality |
| S16_vs_159186\|\|full | Low-quality |
| S16_vs_159193\|\|full | Not-determined |
| S16_vs_159210\|\|full | Low-quality |
| S16_vs_159236\|\|full | Not-determined |
| S16_vs_159243\|\|full | Low-quality |
| S16_vs_159246\|\|full | Low-quality |
| S16_vs_159264\|\|full | Not-determined |
| S16_vs_159276\|\|full | Not-determined |
| S16_vs_159297\|\|full | Low-quality |
| S16_vs_159338\|\|full | Low-quality |
| S16_vs_159355\|\|full | Not-determined |
| S16_vs_159387\|\|full | Low-quality |
| S16_vs_159388\|\|full | Low-quality |
| S16_vs_159413\|\|full | Low-quality |
| S16_vs_159439\|\|full | Not-determined |
| S16_vs_159506\|\|full | Not-determined |
| S16_vs_159618\|\|full | Low-quality |
| S16_vs_159623\|\|full | Low-quality |
| S16_vs_159656\|\|full | Not-determined |
| S16_vs_159724\|\|full | Low-quality |
| S16_vs_159756\|\|full | Not-determined |
| S16_vs_159757\|\|full | Low-quality |
| S16_vs_159772\|\|full | Not-determined |
| S16_vs_159779\|\|full | Low-quality |
| S16_vs_159780\|\|full | Low-quality |
| S16_vs_159832\|\|full | Not-determined |
| S16_vs_159873\|\|full | Low-quality |
| S16_vs_159890\|\|full | Low-quality |
| S16_vs_159936\|\|full | Low-quality |
| S16_vs_159971\|\|full | Not-determined |
| S16_vs_160053\|\|full | Low-quality |
| S16_vs_160066\|\|full | Low-quality |
| S16_vs_160082\|\|full | Not-determined |
| S16_vs_160096\|\|full | Low-quality |
| S16_vs_160109\|\|full | Not-determined |
| S16_vs_160126\|\|full | Low-quality |
| S16_vs_160203\|\|full | Not-determined |
| S16_vs_160207\|\|full | Not-determined |
| S16_vs_160232\|\|full | Low-quality |
| S16_vs_160243\|\|full | Not-determined |
| S16_vs_160289\|\|full | Low-quality |
| S16_vs_160350\|\|full | Not-determined |
| S16_vs_160405\|\|full | Low-quality |
| S16_vs_160407\|\|full | Low-quality |
| S16_vs_160413\|\|full | Not-determined |
| S16_vs_160446\|\|full | Low-quality |
| S16_vs_160527\|\|full | Low-quality |
| S16_vs_160577\|\|full | Not-determined |
| S16_vs_160650\|\|full | Not-determined |
| S16_vs_160653\|\|full | Not-determined |
| S16_vs_160690\|\|full | Low-quality |
| S16_vs_160696\|\|full | Low-quality |
| S16_vs_160716\|\|full | Not-determined |
| S16_vs_160724\|\|full | Not-determined |
| S16_vs_160726\|\|full | Low-quality |
| S16_vs_160733\|\|full | Low-quality |
| S16_vs_160748\|\|full | Low-quality |
| S16_vs_160753\|\|full | Low-quality |
| S16_vs_160776\|\|full | Not-determined |
| S16_vs_160789\|\|full | Not-determined |
| S16_vs_160831\|\|full | Not-determined |
| S16_vs_160838\|\|full | Not-determined |
| S16_vs_160860\|\|full | Not-determined |
| S16_vs_160861\|\|full | Low-quality |
| S16_vs_160888\|\|full | Low-quality |
| S16_vs_160893\|\|full | Not-determined |
| S16_vs_160899\|\|full | Low-quality |
| S16_vs_160928\|\|full | Low-quality |
| S16_vs_160974\|\|full | Low-quality |
| S16_vs_160975\|\|full | Low-quality |
| S16_vs_161010\|\|full | Not-determined |
| S16_vs_161018\|\|full | Not-determined |
| S16_vs_161056\|\|full | Not-determined |
| S16_vs_161146\|\|full | Low-quality |
| S16_vs_161152\|\|full | Low-quality |
| S16_vs_161186\|\|full | Low-quality |
| S16_vs_161195\|\|full | Not-determined |
| S16_vs_161247\|\|full | Not-determined |
| S16_vs_161285\|\|full | Not-determined |
| S16_vs_161310\|\|full | Not-determined |
| S16_vs_161315\|\|full | Low-quality |
| S16_vs_161425\|\|full | Not-determined |
| S16_vs_161446\|\|full | Low-quality |
| S16_vs_161531\|\|full | Low-quality |
| S16_vs_161587\|\|full | Not-determined |
| S16_vs_161606\|\|full | Low-quality |
| S16_vs_161610\|\|full | Low-quality |
| S17_vs_k141_127519\|\|full | Medium-quality |
| S16_vs_161726\|\|full | Low-quality |
| S16_vs_161763\|\|full | Low-quality |
| S16_vs_161792\|\|full | Low-quality |
| S16_vs_161856\|\|full | Low-quality |
| S16_vs_161896\|\|full | Low-quality |
| S16_vs_161906\|\|full | Low-quality |
| S16_vs_161921\|\|full | Low-quality |
| S16_vs_161955\|\|full | Low-quality |
| S16_vs_161965\|\|full | Low-quality |
| S16_vs_161982\|\|full | Not-determined |
| S16_vs_162034\|\|full | Not-determined |
| S16_vs_162040\|\|full | Low-quality |
| S16_vs_162109\|\|full | Low-quality |
| S16_vs_162112\|\|full | Low-quality |
| S16_vs_162166\|\|full | Low-quality |
| S8_vs_k141_101283\|\|full | Medium-quality |
| S16_vs_162227\|\|full | Low-quality |
| S16_vs_162256\|\|full | Not-determined |
| S16_vs_162270\|\|full | Low-quality |
| S16_vs_162316\|\|full | Low-quality |
| S16_vs_162328\|\|full | Not-determined |
| S16_vs_162336\|\|full | Low-quality |
| S16_vs_162337\|\|full | Not-determined |
| S16_vs_162410\|\|full | Low-quality |
| S16_vs_162450\|\|full | Not-determined |
| S16_vs_162466\|\|full | Not-determined |
| S16_vs_162483\|\|full | Low-quality |
| S16_vs_162486\|\|full | Low-quality |
| S16_vs_162488\|\|full | Low-quality |
| S16_vs_162527\|\|full | Low-quality |
| S16_vs_162565\|\|full | Low-quality |
| S16_vs_162586\|\|full | Not-determined |
| S16_vs_162622\|\|full | Low-quality |
| S16_vs_162641\|\|full | Low-quality |
| S16_vs_162644\|\|full | Not-determined |
| S16_vs_162645\|\|full | Low-quality |
| S16_vs_162665\|\|full | Not-determined |
| S16_vs_162687\|\|full | Low-quality |
| S16_vs_162749\|\|full | Not-determined |
| S16_vs_162752\|\|full | Low-quality |
| S16_vs_162767\|\|full | Low-quality |
| S16_vs_162768\|\|full | Not-determined |
| S16_vs_162809\|\|full | Not-determined |
| S16_vs_162844\|\|full | Low-quality |
| S16_vs_162883\|\|full | Low-quality |
| S16_vs_162916\|\|full | Not-determined |
| S16_vs_162928\|\|full | Low-quality |
| S16_vs_162933\|\|full | Not-determined |
| S16_vs_162935\|\|full | Low-quality |
| S16_vs_162944\|\|full | Low-quality |
| S16_vs_162977\|\|full | Low-quality |
| S16_vs_163024\|\|full | Low-quality |
| S16_vs_163041\|\|full | Not-determined |
| S16_vs_163079\|\|full | Low-quality |
| S16_vs_163112\|\|full | Low-quality |
| S16_vs_163123\|\|full | Low-quality |
| S16_vs_163138\|\|full | Low-quality |
| S16_vs_163163\|\|full | Not-determined |
| S16_vs_163210\|\|full | Low-quality |
| S16_vs_163243\|\|full | Not-determined |
| S16_vs_163297\|\|full | Low-quality |
| S16_vs_163305\|\|full | Low-quality |
| S16_vs_163339\|\|full | Not-determined |
| S16_vs_163397\|\|full | Low-quality |
| S16_vs_163404\|\|full | Not-determined |
| S16_vs_163505\|\|full | Not-determined |
| S16_vs_163537\|\|full | Low-quality |
| S16_vs_163551\|\|full | Low-quality |
| S16_vs_163586\|\|full | Not-determined |
| S16_vs_163663\|\|full | Not-determined |
| S16_vs_163672\|\|full | Not-determined |
| S16_vs_163674\|\|full | Not-determined |
| S16_vs_163742\|\|full | Not-determined |
| S16_vs_163744\|\|full | Low-quality |
| S16_vs_163753\|\|full | Not-determined |
| S16_vs_163808\|\|full | Low-quality |
| S16_vs_163844\|\|full | Not-determined |
| S16_vs_163906\|\|full | Low-quality |
| S16_vs_163916\|\|full | Low-quality |
| S16_vs_163921\|\|full | Not-determined |
| S16_vs_163935\|\|full | Not-determined |
| S16_vs_164037\|\|full | Not-determined |
| S16_vs_164060\|\|full | Low-quality |
| S16_vs_164083\|\|full | Not-determined |
| S16_vs_164131\|\|full | Not-determined |
| S16_vs_164175\|\|full | Not-determined |
| S16_vs_164192\|\|full | Not-determined |
| S16_vs_164218\|\|full | Low-quality |
| S16_vs_164221\|\|full | Low-quality |
| S16_vs_164252\|\|full | Not-determined |
| S16_vs_164339\|\|full | Not-determined |
| S16_vs_164342\|\|full | Low-quality |
| S16_vs_164370\|\|full | Low-quality |
| S16_vs_164392\|\|full | Not-determined |
| S16_vs_164461\|\|full | Low-quality |
| S16_vs_164497\|\|full | Low-quality |
| S16_vs_164498\|\|full | Not-determined |
| S16_vs_164521\|\|full | Not-determined |
| S16_vs_164572\|\|full | Low-quality |
| S16_vs_164579\|\|full | Low-quality |
| S16_vs_164612\|\|full | Low-quality |
| S16_vs_164617\|\|full | Low-quality |
| S16_vs_164656\|\|full | Low-quality |
| S16_vs_164732\|\|full | Not-determined |
| S16_vs_164747\|\|full | Low-quality |
| S16_vs_164770\|\|full | Low-quality |
| S16_vs_164792\|\|full | Not-determined |
| S16_vs_164797\|\|full | Low-quality |
| S16_vs_164798\|\|full | Not-determined |
| S16_vs_164805\|\|full | Low-quality |
| S16_vs_164858\|\|full | Not-determined |
| S16_vs_164904\|\|full | Low-quality |
| S13_vs_49889\|\|full | Medium-quality |
| S16_vs_164974\|\|full | Low-quality |
| S16_vs_164975\|\|full | Low-quality |
| S16_vs_164981\|\|full | Not-determined |
| S16_vs_164998\|\|full | Low-quality |
| S16_vs_165020\|\|full | Low-quality |
| S16_vs_165124\|\|full | Low-quality |
| S16_vs_165156\|\|full | Not-determined |
| S16_vs_165192\|\|full | Not-determined |
| S16_vs_165198\|\|full | Not-determined |
| S16_vs_165200\|\|full | Not-determined |
| S16_vs_165240\|\|full | Low-quality |
| S16_vs_165283\|\|full | Not-determined |
| S16_vs_165326\|\|full | Not-determined |
| S16_vs_165363\|\|full | Low-quality |
| S16_vs_165365\|\|full | Low-quality |
| S16_vs_165404\|\|full | Low-quality |
| S16_vs_165446\|\|full | Not-determined |
| S16_vs_165454\|\|full | Low-quality |
| S16_vs_165480\|\|full | Low-quality |
| S16_vs_165495\|\|full | Not-determined |
| S16_vs_165499\|\|full | Low-quality |
| S16_vs_165517\|\|full | Not-determined |
| S16_vs_165571\|\|full | Not-determined |
| S16_vs_165599\|\|full | Low-quality |
| S16_vs_165618\|\|full | Not-determined |
| S16_vs_165630\|\|full | Low-quality |
| S16_vs_165655\|\|full | Not-determined |
| S11_vs_k141_52577\|\|full | Medium-quality |
| S16_vs_165667\|\|full | Low-quality |
| S16_vs_165674\|\|full | Low-quality |
| S16_vs_165687\|\|full | Low-quality |
| S16_vs_165688\|\|full | Not-determined |
| S16_vs_165719\|\|full | Low-quality |
| S16_vs_165751\|\|full | Low-quality |
| S16_vs_165755\|\|full | Low-quality |
| S16_vs_165769\|\|full | Low-quality |
| S16_vs_165781\|\|full | Not-determined |
| S16_vs_165789\|\|full | Not-determined |
| S16_vs_165851\|\|full | Low-quality |
| S16_vs_165874\|\|full | Not-determined |
| S16_vs_166014\|\|full | Low-quality |
| S16_vs_166056\|\|full | Low-quality |
| S16_vs_166105\|\|full | Low-quality |
| S16_vs_166106\|\|full | Not-determined |
| S16_vs_166114\|\|full | Not-determined |
| S16_vs_166165\|\|full | Low-quality |
| S16_vs_166186\|\|full | Low-quality |
| S16_vs_166188\|\|full | Low-quality |
| S16_vs_166190\|\|full | Low-quality |
| S16_vs_166208\|\|full | Low-quality |
| S16_vs_166242\|\|full | Not-determined |
| S16_vs_166246\|\|full | Not-determined |
| S16_vs_166272\|\|full | Low-quality |
| S16_vs_166309\|\|full | Not-determined |
| S16_vs_166345\|\|full | Low-quality |
| S16_vs_166367\|\|full | Not-determined |
| S16_vs_166433\|\|full | Not-determined |
| S16_vs_166471\|\|full | Low-quality |
| S16_vs_166540\|\|full | Not-determined |
| S16_vs_166545\|\|full | Not-determined |
| S16_vs_166570\|\|full | Low-quality |
| S16_vs_166604\|\|full | Not-determined |
| S16_vs_166642\|\|full | Low-quality |
| S16_vs_166677\|\|full | Not-determined |
| S16_vs_166719\|\|full | Low-quality |
| S16_vs_166731\|\|full | Low-quality |
| S16_vs_166852\|\|full | Not-determined |
| S16_vs_166938\|\|full | Not-determined |
| S16_vs_166972\|\|full | Not-determined |
| S16_vs_166989\|\|full | Not-determined |
| S16_vs_166997\|\|full | Not-determined |
| S16_vs_166998\|\|full | Low-quality |
| S16_vs_167035\|\|full | Low-quality |
| S16_vs_167097\|\|full | Low-quality |
| S16_vs_167130\|\|full | Low-quality |
| S16_vs_167159\|\|full | Low-quality |
| S16_vs_167172\|\|full | Low-quality |
| S16_vs_167206\|\|full | Not-determined |
| S16_vs_167234\|\|full | Not-determined |
| S16_vs_167235\|\|full | Not-determined |
| S16_vs_167261\|\|full | Low-quality |
| S16_vs_167265\|\|full | Low-quality |
| S16_vs_167270\|\|full | Low-quality |
| S16_vs_167278\|\|full | Low-quality |
| S16_vs_167285\|\|full | Not-determined |
| S16_vs_167287\|\|full | Low-quality |
| S16_vs_167324\|\|full | Not-determined |
| S16_vs_167327\|\|full | Low-quality |
| S16_vs_167354\|\|full | Low-quality |
| S16_vs_167380\|\|full | Low-quality |
| S16_vs_167414\|\|full | Not-determined |
| S16_vs_167439\|\|full | Not-determined |
| S16_vs_167464\|\|full | Low-quality |
| S16_vs_167532\|\|full | Not-determined |
| S16_vs_167537\|\|full | Low-quality |
| S16_vs_167553\|\|full | Low-quality |
| S16_vs_167562\|\|full | Low-quality |
| S16_vs_167579\|\|full | Low-quality |
| S16_vs_167605\|\|full | Not-determined |
| S16_vs_167607\|\|full | Not-determined |
| S16_vs_167613\|\|full | Low-quality |
| S16_vs_167641\|\|full | Not-determined |
| S16_vs_167709\|\|full | Low-quality |
| S16_vs_167710\|\|full | Low-quality |
| S16_vs_167718\|\|full | Not-determined |
| S16_vs_167751\|\|full | Low-quality |
| S16_vs_167762\|\|full | Not-determined |
| S16_vs_167791\|\|full | Not-determined |
| S16_vs_167836\|\|full | Not-determined |
| S16_vs_167852\|\|full | Low-quality |
| S16_vs_167867\|\|full | Not-determined |
| S16_vs_167894\|\|full | Not-determined |
| S16_vs_167905\|\|full | Not-determined |
| S16_vs_167984\|\|full | Low-quality |
| S16_vs_167986\|\|full | Not-determined |
| S16_vs_168040\|\|full | Not-determined |
| S16_vs_168068\|\|full | Not-determined |
| S16_vs_168087\|\|full | Not-determined |
| S16_vs_168110\|\|full | Low-quality |
| S16_vs_168114\|\|full | Not-determined |
| S16_vs_168124\|\|full | Low-quality |
| S16_vs_168173\|\|full | Low-quality |
| S16_vs_168256\|\|full | Not-determined |
| S16_vs_168287\|\|full | Not-determined |
| S16_vs_168377\|\|full | Not-determined |
| S16_vs_168494\|\|full | Low-quality |
| S16_vs_168572\|\|full | Low-quality |
| S16_vs_168621\|\|full | Low-quality |
| S16_vs_168649\|\|full | Not-determined |
| S16_vs_168662\|\|full | Not-determined |
| S16_vs_168682\|\|full | Not-determined |
| S16_vs_168730\|\|full | Low-quality |
| S16_vs_168736\|\|full | Low-quality |
| S16_vs_168746\|\|full | Low-quality |
| S16_vs_168751\|\|full | Low-quality |
| S16_vs_168757\|\|full | Not-determined |
| S16_vs_168784\|\|full | Not-determined |
| S16_vs_168795\|\|full | Low-quality |
| S16_vs_168827\|\|full | Low-quality |
| S16_vs_168829\|\|full | Not-determined |
| S16_vs_168877\|\|full | Not-determined |
| S16_vs_168934\|\|full | Low-quality |
| S16_vs_169113\|\|full | Not-determined |
| S16_vs_169121\|\|full | Not-determined |
| S16_vs_169142\|\|full | Low-quality |
| S16_vs_169202\|\|full | Not-determined |
| S16_vs_169212\|\|full | Not-determined |
| S16_vs_169228\|\|full | Not-determined |
| S16_vs_169261\|\|full | Low-quality |
| S16_vs_169273\|\|full | Not-determined |
| S16_vs_169320\|\|full | Not-determined |
| S16_vs_169361\|\|full | Not-determined |
| S16_vs_169368\|\|full | Low-quality |
| S16_vs_169377\|\|full | Not-determined |
| S16_vs_169426\|\|full | Low-quality |
| S16_vs_169500\|\|full | Low-quality |
| S16_vs_169526\|\|full | Low-quality |
| S16_vs_169539\|\|full | Low-quality |
| S16_vs_169560\|\|full | Low-quality |
| S16_vs_169591\|\|full | Not-determined |
| S16_vs_169650\|\|full | Not-determined |
| S16_vs_169690\|\|full | Low-quality |
| S16_vs_169759\|\|full | Low-quality |
| S16_vs_169763\|\|full | Low-quality |
| S9_vs_k141_34898\|\|full | Medium-quality |
| S16_vs_169807\|\|full | Low-quality |
| S16_vs_169854\|\|full | Low-quality |
| S16_vs_169856\|\|full | Low-quality |
| S16_vs_169872\|\|full | Low-quality |
| S14_extracted_k141_106156 | Medium-quality |
| S16_vs_169899\|\|full | Low-quality |
| S16_vs_169915\|\|full | Low-quality |
| S16_vs_169934\|\|full | Low-quality |
| S16_vs_169944\|\|full | Low-quality |
| S16_vs_170009\|\|full | Not-determined |
| S16_vs_170013\|\|full | Low-quality |
| S16_vs_170033\|\|full | Low-quality |
| S16_vs_170040\|\|full | Not-determined |
| S16_vs_170041\|\|full | Low-quality |
| S16_vs_170074\|\|full | Low-quality |
| S16_vs_170076\|\|full | Low-quality |
| S16_vs_170123\|\|full | Low-quality |
| S16_vs_170187\|\|full | Low-quality |
| S16_vs_170196\|\|full | Low-quality |
| S16_vs_170208\|\|full | Low-quality |
| S16_vs_170213\|\|full | Low-quality |
| S16_vs_170216\|\|full | Not-determined |
| S16_vs_170254\|\|full | Not-determined |
| S16_vs_170265\|\|full | Low-quality |
| S16_vs_170284\|\|full | Low-quality |
| S16_vs_170351\|\|full | Low-quality |
| S16_vs_170356\|\|full | Not-determined |
| S16_vs_170425\|\|full | Not-determined |
| S16_vs_170430\|\|full | Low-quality |
| S16_vs_170504\|\|full | Not-determined |
| S16_vs_170516\|\|full | Not-determined |
| S16_vs_170520\|\|full | Not-determined |
| S16_vs_170524\|\|full | Low-quality |
| S16_vs_170533\|\|full | Not-determined |
| S16_vs_170537\|\|full | Not-determined |
| S16_vs_170570\|\|full | Low-quality |
| S16_vs_170576\|\|full | Low-quality |
| S16_vs_170594\|\|full | Low-quality |
| S16_vs_170619\|\|full | Not-determined |
| S16_vs_170648\|\|full | Low-quality |
| S16_vs_170696\|\|full | Low-quality |
| S16_vs_170711\|\|full | Low-quality |
| S16_vs_170737\|\|full | Not-determined |
| S16_vs_170738\|\|full | Not-determined |
| S16_vs_170800\|\|full | Not-determined |
| S16_vs_170805\|\|full | Low-quality |
| S16_vs_170824\|\|full | Low-quality |
| S16_vs_170856\|\|full | Not-determined |
| S16_vs_170900\|\|full | Not-determined |
| S16_vs_170909\|\|full | Low-quality |
| S16_vs_170969\|\|full | Low-quality |
| S16_vs_170971\|\|full | Low-quality |
| S16_vs_170972\|\|full | Low-quality |
| S16_vs_170974\|\|full | Low-quality |
| S16_vs_170985\|\|full | Low-quality |
| S16_vs_170988\|\|full | Not-determined |
| S16_vs_171000\|\|full | Low-quality |
| S16_vs_171006\|\|full | Low-quality |
| S16_vs_171042\|\|full | Low-quality |
| S16_vs_171129\|\|full | Not-determined |
| S16_vs_171137\|\|full | Low-quality |
| S16_vs_171155\|\|full | Low-quality |
| S16_vs_171170\|\|full | Low-quality |
| S16_vs_171192\|\|full | Not-determined |
| S16_vs_171219\|\|full | Low-quality |
| S16_vs_171236\|\|full | Low-quality |
| S16_vs_171243\|\|full | Not-determined |
| S16_vs_171249\|\|full | Not-determined |
| S16_vs_171270\|\|full | Low-quality |
| S16_vs_171295\|\|full | Low-quality |
| S16_vs_171423\|\|full | Low-quality |
| S16_vs_171431\|\|full | Not-determined |
| S16_vs_171441\|\|full | Low-quality |
| S16_vs_171455\|\|full | Not-determined |
| S16_vs_171478\|\|full | Not-determined |
| S16_vs_171493\|\|full | Low-quality |
| S16_vs_171598\|\|full | Not-determined |
| S16_vs_171601\|\|full | Low-quality |
| S16_vs_171664\|\|full | Not-determined |
| S16_vs_171674\|\|full | Low-quality |
| S16_vs_171681\|\|full | Low-quality |
| S16_vs_171707\|\|full | Low-quality |
| S16_vs_171726\|\|full | Not-determined |
| S16_vs_171745\|\|full | Not-determined |
| S16_vs_171770\|\|full | Low-quality |
| S16_vs_171820\|\|full | Not-determined |
| S16_vs_171863\|\|full | Low-quality |
| S16_vs_171887\|\|full | Low-quality |
| S16_vs_171940\|\|full | Low-quality |
| S16_vs_171944\|\|full | Not-determined |
| S16_vs_171945\|\|full | Not-determined |
| S16_vs_171982\|\|full | Low-quality |
| S16_vs_172000\|\|full | Low-quality |
| S16_vs_172017\|\|full | Low-quality |
| S16_vs_172045\|\|full | Low-quality |
| S16_vs_172135\|\|full | Low-quality |
| S16_vs_172136\|\|full | Low-quality |
| S16_vs_172167\|\|full | Not-determined |
| S16_vs_172195\|\|full | Low-quality |
| S16_vs_172203\|\|full | Low-quality |
| S16_vs_172242\|\|full | Not-determined |
| S16_vs_172262\|\|full | Not-determined |
| S16_vs_172299\|\|full | Low-quality |
| S16_vs_172320\|\|full | Low-quality |
| S16_vs_172330\|\|full | Low-quality |
| S16_vs_172335\|\|full | Low-quality |
| S16_vs_172343\|\|full | Low-quality |
| S16_vs_172367\|\|full | Low-quality |
| S16_vs_172373\|\|full | Low-quality |
| S16_vs_172379\|\|full | Low-quality |
| S16_vs_172412\|\|full | Not-determined |
| S16_vs_172417\|\|full | Low-quality |
| S16_vs_172431\|\|full | Low-quality |
| S16_vs_172443\|\|full | Not-determined |
| S16_vs_172509\|\|full | Low-quality |
| S16_vs_172573\|\|full | Not-determined |
| S16_vs_172609\|\|full | Not-determined |
| S16_vs_172642\|\|full | Low-quality |
| S16_vs_172677\|\|full | Not-determined |
| S16_vs_172721\|\|full | Low-quality |
| S16_vs_172730\|\|full | Not-determined |
| S16_vs_172744\|\|full | Low-quality |
| S16_vs_172769\|\|full | Not-determined |
| S16_vs_172785\|\|full | Not-determined |
| S16_vs_172796\|\|full | Not-determined |
| S16_vs_172821\|\|full | Not-determined |
| S16_vs_172862\|\|full | Not-determined |
| S16_vs_172884\|\|full | Not-determined |
| S16_vs_172885\|\|full | Not-determined |
| S16_vs_172949\|\|full | Not-determined |
| S16_vs_172968\|\|full | Not-determined |
| S16_vs_172990\|\|full | Low-quality |
| S16_vs_172993\|\|full | Low-quality |
| S16_vs_173004\|\|full | Low-quality |
| S16_vs_173044\|\|full | Low-quality |
| S16_vs_173061\|\|full | Low-quality |
| S16_vs_173080\|\|full | Not-determined |
| S16_vs_7947\|\|0_partial | Low-quality |
| S16_vs_13598\|\|0_partial | Low-quality |
| S16_vs_14307\|\|0_partial | Not-determined |
| S16_vs_14307\|\|1_partial | Low-quality |
| S16_vs_15775\|\|0_partial | Not-determined |
| S16_vs_29367\|\|0_partial | Low-quality |
| S16_vs_31632\|\|0_partial | Not-determined |
| S16_vs_32077\|\|0_partial | Not-determined |
| S16_vs_34591\|\|0_partial | Not-determined |
| S16_vs_36988\|\|0_partial | Not-determined |
| S16_vs_60695\|\|0_partial | Not-determined |
| S16_vs_64695\|\|0_partial | Low-quality |
| S16_vs_88202\|\|0_partial | Not-determined |
| S16_vs_88268\|\|0_partial | Not-determined |
| S16_vs_92916\|\|0_partial | Not-determined |
| S16_vs_102633\|\|0_partial | Low-quality |
| S16_vs_105031\|\|0_partial | Low-quality |
| S16_vs_113020\|\|0_partial | Low-quality |
| S16_vs_116081\|\|0_partial | Not-determined |
| S16_vs_121387\|\|0_partial | Not-determined |
| S16_vs_135204\|\|0_partial | Not-determined |
| S16_vs_140776\|\|0_partial | Not-determined |
| S16_vs_143298\|\|0_partial | Low-quality |
| S16_vs_147914\|\|0_partial | Not-determined |
| S16_vs_152197\|\|0_partial | Not-determined |
| S16_vs_165292\|\|0_partial | Low-quality |
| S16_vs_165292\|\|1_partial | Low-quality |
| S16_vs_171302\|\|0_partial | Low-quality |

| | |
|---|---|
| S16_vs_16\|\|lt2gene | Low-quality |
| S16_vs_79\|\|lt2gene | Low-quality |
| S16_vs_99\|\|lt2gene | Not-determined |
| S16_vs_400\|\|lt2gene | Low-quality |
| S16_vs_552\|\|lt2gene | Low-quality |
| S16_vs_638\|\|lt2gene | Low-quality |
| S16_vs_644\|\|lt2gene | Not-determined |
| S16_vs_701\|\|lt2gene | Low-quality |
| S16_vs_809\|\|lt2gene | Not-determined |
| S16_vs_868\|\|lt2gene | Low-quality |
| S16_vs_869\|\|lt2gene | Low-quality |
| S16_vs_871\|\|lt2gene | Low-quality |
| S16_vs_894\|\|lt2gene | Not-determined |
| S16_vs_1005\|\|lt2gene | Low-quality |
| S16_vs_1204\|\|lt2gene | Low-quality |
| S16_vs_1393\|\|lt2gene | Low-quality |
| S16_vs_1447\|\|lt2gene | Low-quality |
| S16_vs_1543\|\|lt2gene | Low-quality |
| S16_vs_1589\|\|lt2gene | Low-quality |
| S16_vs_1768\|\|lt2gene | Low-quality |
| S16_vs_1799\|\|lt2gene | Low-quality |
| S16_vs_1957\|\|lt2gene | Low-quality |
| S16_vs_1980\|\|lt2gene | Low-quality |
| S16_vs_2048\|\|lt2gene | Low-quality |
| S16_vs_2062\|\|lt2gene | Not-determined |
| S16_vs_2152\|\|lt2gene | Not-determined |
| S16_vs_2166\|\|lt2gene | Low-quality |
| S16_vs_2236\|\|lt2gene | Low-quality |
| S16_vs_2387\|\|lt2gene | Low-quality |
| S16_vs_2452\|\|lt2gene | Not-determined |
| S16_vs_2460\|\|lt2gene | Low-quality |
| S16_vs_2569\|\|lt2gene | Not-determined |
| S16_vs_2628\|\|lt2gene | Low-quality |
| S16_vs_2672\|\|lt2gene | Low-quality |
| S16_vs_2735\|\|lt2gene | Low-quality |
| S16_vs_2857\|\|lt2gene | Low-quality |
| S16_vs_2940\|\|lt2gene | Not-determined |
| S16_vs_3036\|\|lt2gene | Low-quality |
| S16_vs_3089\|\|lt2gene | Low-quality |
| S16_vs_3129\|\|lt2gene | Low-quality |
| S16_vs_3285\|\|lt2gene | Low-quality |
| S16_vs_3353\|\|lt2gene | Not-determined |
| S16_vs_3395\|\|lt2gene | Not-determined |
| S16_vs_3447\|\|lt2gene | Low-quality |
| S16_vs_3468\|\|lt2gene | Low-quality |
| S16_vs_3500\|\|lt2gene | Low-quality |
| S16_vs_3554\|\|lt2gene | Not-determined |
| S16_vs_3657\|\|lt2gene | Not-determined |
| S16_vs_3730\|\|lt2gene | Low-quality |
| S16_vs_3796\|\|lt2gene | Low-quality |
| S16_vs_3909\|\|lt2gene | Low-quality |
| S16_vs_4024\|\|lt2gene | Low-quality |
| S16_vs_4145\|\|lt2gene | Low-quality |
| S16_vs_4216\|\|lt2gene | Not-determined |
| S16_vs_4269\|\|lt2gene | Low-quality |
| S16_vs_4287\|\|lt2gene | Low-quality |
| S16_vs_4385\|\|lt2gene | Not-determined |
| S16_vs_4414\|\|lt2gene | Low-quality |
| S16_vs_4466\|\|lt2gene | Not-determined |
| S16_vs_4468\|\|lt2gene | Low-quality |
| S16_vs_4696\|\|lt2gene | Low-quality |
| S16_vs_4741\|\|lt2gene | Low-quality |
| S16_vs_4867\|\|lt2gene | Not-determined |
| S16_vs_5246\|\|lt2gene | Not-determined |
| S16_vs_5325\|\|lt2gene | Low-quality |
| S16_vs_5332\|\|lt2gene | Low-quality |
| S16_vs_5356\|\|lt2gene | Not-determined |
| S16_vs_5417\|\|lt2gene | Low-quality |
| S16_vs_5458\|\|lt2gene | Low-quality |
| S16_vs_5525\|\|lt2gene | Low-quality |
| S16_vs_5757\|\|lt2gene | Not-determined |
| S16_vs_5858\|\|lt2gene | Low-quality |
| S16_vs_5962\|\|lt2gene | Low-quality |
| S16_vs_5977\|\|lt2gene | Low-quality |
| S16_vs_6095\|\|lt2gene | Low-quality |
| S16_vs_6195\|\|lt2gene | Low-quality |
| S16_vs_6204\|\|lt2gene | Low-quality |
| S16_vs_6209\|\|lt2gene | Low-quality |
| S16_vs_6246\|\|lt2gene | Not-determined |
| S16_vs_6269\|\|lt2gene | Not-determined |
| S16_vs_6301\|\|lt2gene | Low-quality |
| S16_vs_6313\|\|lt2gene | Low-quality |
| S16_vs_6322\|\|lt2gene | Low-quality |
| S16_vs_6484\|\|lt2gene | Low-quality |
| S16_vs_6542\|\|lt2gene | Low-quality |
| S16_vs_6598\|\|lt2gene | Low-quality |
| S16_vs_6812\|\|lt2gene | Not-determined |
| S16_vs_6845\|\|lt2gene | Low-quality |
| S16_vs_6899\|\|lt2gene | Not-determined |
| S16_vs_7304\|\|lt2gene | Not-determined |
| S16_vs_7437\|\|lt2gene | Low-quality |
| S16_vs_7523\|\|lt2gene | Low-quality |
| S16_vs_7663\|\|lt2gene | Low-quality |
| S16_vs_7692\|\|lt2gene | Not-determined |
| S16_vs_7696\|\|lt2gene | Not-determined |
| S16_vs_7704\|\|lt2gene | Low-quality |
| S16_vs_7730\|\|lt2gene | Low-quality |
| S16_vs_7732\|\|lt2gene | Not-determined |
| S16_vs_7766\|\|lt2gene | Low-quality |
| S16_vs_7798\|\|lt2gene | Low-quality |
| S16_vs_8027\|\|lt2gene | Low-quality |
| S16_vs_8041\|\|lt2gene | Low-quality |
| S16_vs_8062\|\|lt2gene | Low-quality |
| S16_vs_8071\|\|lt2gene | Low-quality |
| S16_vs_8130\|\|lt2gene | Low-quality |
| S16_vs_8255\|\|lt2gene | Not-determined |
| S16_vs_8314\|\|lt2gene | Low-quality |
| S16_vs_8370\|\|lt2gene | Low-quality |
| S16_vs_8441\|\|lt2gene | Not-determined |
| S16_vs_8609\|\|lt2gene | Low-quality |
| S16_vs_8634\|\|lt2gene | Not-determined |
| S16_vs_8651\|\|lt2gene | Low-quality |
| S16_vs_8810\|\|lt2gene | Not-determined |
| S16_vs_8872\|\|lt2gene | Low-quality |
| S16_vs_9017\|\|lt2gene | Low-quality |
| S16_vs_9175\|\|lt2gene | Low-quality |
| S16_vs_9466\|\|lt2gene | Low-quality |
| S16_vs_9511\|\|lt2gene | Low-quality |
| S16_vs_9517\|\|lt2gene | Not-determined |
| S16_vs_9532\|\|lt2gene | Low-quality |
| S16_vs_9734\|\|lt2gene | Low-quality |
| S16_vs_9893\|\|lt2gene | Low-quality |
| S16_vs_9934\|\|lt2gene | Low-quality |
| S16_vs_9982\|\|lt2gene | Low-quality |
| S16_vs_10111\|\|lt2gene | Low-quality |
| S16_vs_10272\|\|lt2gene | Low-quality |
| S16_vs_10276\|\|lt2gene | Low-quality |
| S16_vs_10484\|\|lt2gene | Low-quality |
| S16_vs_10581\|\|lt2gene | Low-quality |
| S16_vs_10639\|\|lt2gene | Not-determined |
| S16_vs_10674\|\|lt2gene | Low-quality |
| S16_vs_10841\|\|lt2gene | Low-quality |
| S16_vs_11053\|\|lt2gene | Low-quality |
| S16_vs_11134\|\|lt2gene | Low-quality |
| S16_vs_11257\|\|lt2gene | Low-quality |
| S16_vs_11270\|\|lt2gene | Not-determined |
| S16_vs_11278\|\|lt2gene | Not-determined |
| S16_vs_11351\|\|lt2gene | Low-quality |
| S16_vs_11529\|\|lt2gene | Not-determined |
| S16_vs_11716\|\|lt2gene | Low-quality |
| S16_vs_11880\|\|lt2gene | Low-quality |
| S16_vs_11924\|\|lt2gene | Low-quality |
| S16_vs_12197\|\|lt2gene | Low-quality |
| S16_vs_12217\|\|lt2gene | Low-quality |
| S16_vs_12227\|\|lt2gene | Low-quality |
| S16_vs_12326\|\|lt2gene | Low-quality |
| S16_vs_12371\|\|lt2gene | Low-quality |
| S16_vs_12390\|\|lt2gene | Low-quality |
| S16_vs_12451\|\|lt2gene | Not-determined |
| S16_vs_12680\|\|lt2gene | Low-quality |
| S16_vs_12819\|\|lt2gene | Low-quality |
| S16_vs_12825\|\|lt2gene | Low-quality |
| S16_vs_12862\|\|lt2gene | Low-quality |
| S16_vs_12936\|\|lt2gene | Low-quality |
| S16_vs_13046\|\|lt2gene | Low-quality |
| S16_vs_13154\|\|lt2gene | Not-determined |
| S16_vs_13170\|\|lt2gene | Low-quality |
| S16_vs_13213\|\|lt2gene | Not-determined |
| S16_vs_13216\|\|lt2gene | Low-quality |
| S16_vs_13230\|\|lt2gene | Not-determined |
| S16_vs_13308\|\|lt2gene | Not-determined |
| S16_vs_13323\|\|lt2gene | Low-quality |
| S16_vs_13415\|\|lt2gene | Low-quality |
| S16_vs_13531\|\|lt2gene | Low-quality |
| S16_vs_13739\|\|lt2gene | Low-quality |
| S16_vs_13748\|\|lt2gene | Low-quality |
| S16_vs_13753\|\|lt2gene | Not-determined |
| S16_vs_13804\|\|lt2gene | Low-quality |
| S16_vs_14059\|\|lt2gene | Not-determined |
| S16_vs_14138\|\|lt2gene | Not-determined |
| S16_vs_14169\|\|lt2gene | Not-determined |
| S16_vs_14182\|\|lt2gene | Low-quality |
| S16_vs_14189\|\|lt2gene | Not-determined |
| S16_vs_14282\|\|lt2gene | Low-quality |
| S16_vs_14360\|\|lt2gene | Low-quality |
| S16_vs_14451\|\|lt2gene | Low-quality |
| S16_vs_14452\|\|lt2gene | Low-quality |
| S16_vs_14569\|\|lt2gene | Low-quality |
| S16_vs_14573\|\|lt2gene | Low-quality |
| S16_vs_14611\|\|lt2gene | Low-quality |
| S16_vs_14622\|\|lt2gene | Low-quality |
| S16_vs_14711\|\|lt2gene | Low-quality |
| S16_vs_14820\|\|lt2gene | Low-quality |
| S16_vs_14844\|\|lt2gene | Low-quality |
| S16_vs_14936\|\|lt2gene | Low-quality |
| S16_vs_15253\|\|lt2gene | Low-quality |
| S16_vs_15657\|\|lt2gene | Low-quality |
| S16_vs_15701\|\|lt2gene | Low-quality |
| S16_vs_15702\|\|lt2gene | Low-quality |
| S16_vs_15797\|\|lt2gene | Low-quality |
| S16_vs_15869\|\|lt2gene | Not-determined |
| S16_vs_15886\|\|lt2gene | Not-determined |
| S16_vs_16030\|\|lt2gene | Low-quality |
| S16_vs_16056\|\|lt2gene | Low-quality |
| S16_vs_16170\|\|lt2gene | Low-quality |
| S16_vs_16219\|\|lt2gene | Low-quality |
| S16_vs_16431\|\|lt2gene | Low-quality |
| S16_vs_16880\|\|lt2gene | Low-quality |
| S16_vs_17004\|\|lt2gene | Not-determined |
| S16_vs_17047\|\|lt2gene | Low-quality |
| S16_vs_17174\|\|lt2gene | Low-quality |
| S16_vs_17181\|\|lt2gene | Low-quality |
| S16_vs_17269\|\|lt2gene | Low-quality |
| S16_vs_17310\|\|lt2gene | Not-determined |
| S16_vs_17333\|\|lt2gene | Low-quality |
| S16_vs_17631\|\|lt2gene | Low-quality |
| S16_vs_17637\|\|lt2gene | Low-quality |
| S16_vs_17686\|\|lt2gene | Low-quality |
| S16_vs_18245\|\|lt2gene | Low-quality |
| S16_vs_18313\|\|lt2gene | Not-determined |
| S16_vs_18428\|\|lt2gene | Low-quality |
| S16_vs_18468\|\|lt2gene | Low-quality |
| S16_vs_18493\|\|lt2gene | Low-quality |
| S16_vs_18658\|\|lt2gene | Low-quality |
| S16_vs_18678\|\|lt2gene | Low-quality |
| S16_vs_18742\|\|lt2gene | Low-quality |
| S16_vs_18865\|\|lt2gene | Low-quality |
| S16_vs_19037\|\|lt2gene | Low-quality |
| S16_vs_19087\|\|lt2gene | Not-determined |
| S16_vs_19211\|\|lt2gene | Low-quality |
| S16_vs_19228\|\|lt2gene | Not-determined |
| S16_vs_19230\|\|lt2gene | Low-quality |
| S16_vs_19450\|\|lt2gene | Low-quality |
| S16_vs_19511\|\|lt2gene | Low-quality |
| S16_vs_19693\|\|lt2gene | Low-quality |
| S16_vs_19706\|\|lt2gene | Low-quality |
| S16_vs_19734\|\|lt2gene | Not-determined |
| S16_vs_19832\|\|lt2gene | Low-quality |
| S16_vs_20009\|\|lt2gene | Not-determined |
| S16_vs_20119\|\|lt2gene | Not-determined |
| S16_vs_20478\|\|lt2gene | Not-determined |
| S16_vs_20480\|\|lt2gene | Not-determined |
| S16_vs_20573\|\|lt2gene | Low-quality |
| S16_vs_20585\|\|lt2gene | Not-determined |
| S16_vs_20722\|\|lt2gene | Low-quality |
| S16_vs_20742\|\|lt2gene | Low-quality |
| S16_vs_20776\|\|lt2gene | Low-quality |
| S16_vs_20800\|\|lt2gene | Low-quality |
| S16_vs_20808\|\|lt2gene | Low-quality |
| S16_vs_20861\|\|lt2gene | Not-determined |
| S16_vs_20864\|\|lt2gene | Low-quality |
| S16_vs_20968\|\|lt2gene | Low-quality |
| S16_vs_20996\|\|lt2gene | Low-quality |
| S16_vs_21092\|\|lt2gene | Low-quality |
| S16_vs_21187\|\|lt2gene | Low-quality |
| S16_vs_21238\|\|lt2gene | Low-quality |
| S16_vs_21321\|\|lt2gene | Low-quality |
| S16_vs_21517\|\|lt2gene | Low-quality |
| S16_vs_21537\|\|lt2gene | Not-determined |
| S16_vs_21758\|\|lt2gene | Low-quality |
| S16_vs_21841\|\|lt2gene | Low-quality |
| S16_vs_21875\|\|lt2gene | Not-determined |
| S16_vs_21965\|\|lt2gene | Low-quality |
| S16_vs_21971\|\|lt2gene | Low-quality |
| S16_vs_22168\|\|lt2gene | Low-quality |
| S16_vs_22208\|\|lt2gene | Low-quality |
| S16_vs_22265\|\|lt2gene | Low-quality |
| S16_vs_22341\|\|lt2gene | Low-quality |
| S16_vs_22358\|\|lt2gene | Low-quality |
| S16_vs_22401\|\|lt2gene | Not-determined |
| S16_vs_22563\|\|lt2gene | Not-determined |
| S16_vs_22587\|\|lt2gene | Low-quality |
| S16_vs_22850\|\|lt2gene | Not-determined |
| S16_vs_22856\|\|lt2gene | Low-quality |
| S16_vs_22866\|\|lt2gene | Not-determined |
| S16_vs_22876\|\|lt2gene | Not-determined |
| S16_vs_22905\|\|lt2gene | Low-quality |
| S16_vs_22913\|\|lt2gene | Low-quality |
| S16_vs_23039\|\|lt2gene | Low-quality |
| S16_vs_23055\|\|lt2gene | Low-quality |
| S16_vs_23070\|\|lt2gene | Low-quality |
| S16_vs_23137\|\|lt2gene | Low-quality |
| S16_vs_23304\|\|lt2gene | Not-determined |
| S16_vs_23426\|\|lt2gene | Not-determined |
| S16_vs_23518\|\|lt2gene | Low-quality |
| S16_vs_23571\|\|lt2gene | Low-quality |
| S16_vs_23576\|\|lt2gene | Low-quality |
| S16_vs_23641\|\|lt2gene | Not-determined |
| S16_vs_23703\|\|lt2gene | Not-determined |
| S16_vs_23718\|\|lt2gene | Low-quality |
| S16_vs_23767\|\|lt2gene | Low-quality |
| S16_vs_23960\|\|lt2gene | Not-determined |
| S16_vs_24211\|\|lt2gene | Low-quality |
| S16_vs_24226\|\|lt2gene | Low-quality |
| S16_vs_24362\|\|lt2gene | Low-quality |
| S16_vs_24490\|\|lt2gene | Not-determined |
| S16_vs_24552\|\|lt2gene | Low-quality |
| S16_vs_24558\|\|lt2gene | Not-determined |
| S16_vs_24674\|\|lt2gene | Low-quality |
| S16_vs_24793\|\|lt2gene | Low-quality |
| S16_vs_24948\|\|lt2gene | Low-quality |
| S16_vs_25023\|\|lt2gene | Low-quality |
| S16_vs_25089\|\|lt2gene | Low-quality |
| S16_vs_25127\|\|lt2gene | Not-determined |
| S16_vs_25186\|\|lt2gene | Low-quality |
| S16_vs_25266\|\|lt2gene | Low-quality |
| S16_vs_25329\|\|lt2gene | Low-quality |
| S16_vs_25386\|\|lt2gene | Low-quality |
| S16_vs_25547\|\|lt2gene | Low-quality |
| S16_vs_25570\|\|lt2gene | Low-quality |
| S16_vs_25635\|\|lt2gene | Low-quality |
| S16_vs_25732\|\|lt2gene | Low-quality |
| S16_vs_25850\|\|lt2gene | Not-determined |
| S16_vs_25948\|\|lt2gene | Low-quality |
| S16_vs_26304\|\|lt2gene | Not-determined |
| S16_vs_26307\|\|lt2gene | Not-determined |
| S16_vs_26385\|\|lt2gene | Low-quality |
| S16_vs_26561\|\|lt2gene | Low-quality |
| S16_vs_26631\|\|lt2gene | Low-quality |
| S16_vs_26659\|\|lt2gene | Not-determined |
| S16_vs_26670\|\|lt2gene | Low-quality |
| S16_vs_26755\|\|lt2gene | Low-quality |
| S16_vs_26761\|\|lt2gene | Not-determined |
| S16_vs_26965\|\|lt2gene | Low-quality |
| S16_vs_26982\|\|lt2gene | Not-determined |
| S16_vs_27293\|\|lt2gene | Low-quality |
| S16_vs_27331\|\|lt2gene | Low-quality |
| S16_vs_27417\|\|lt2gene | Not-determined |
| S16_vs_27490\|\|lt2gene | Low-quality |
| S16_vs_27607\|\|lt2gene | Low-quality |
| S16_vs_27938\|\|lt2gene | Not-determined |
| S16_vs_28043\|\|lt2gene | Low-quality |
| S16_vs_28168\|\|lt2gene | Not-determined |
| S16_vs_28294\|\|lt2gene | Not-determined |
| S16_vs_28296\|\|lt2gene | Not-determined |
| S16_vs_28362\|\|lt2gene | Low-quality |
| S16_vs_28407\|\|lt2gene | Not-determined |
| S16_vs_28507\|\|lt2gene | Low-quality |
| S16_vs_28688\|\|lt2gene | Not-determined |
| S16_vs_28712\|\|lt2gene | Not-determined |
| S16_vs_28730\|\|lt2gene | Low-quality |
| S16_vs_28896\|\|lt2gene | Low-quality |
| S16_vs_29080\|\|lt2gene | Low-quality |
| S16_vs_29093\|\|lt2gene | Not-determined |
| S16_vs_29176\|\|lt2gene | Low-quality |
| S16_vs_29310\|\|lt2gene | Low-quality |
| S16_vs_29438\|\|lt2gene | Low-quality |
| S16_vs_29452\|\|lt2gene | Not-determined |
| S16_vs_29514\|\|lt2gene | Low-quality |
| S16_vs_29730\|\|lt2gene | Low-quality |
| S16_vs_29876\|\|lt2gene | Low-quality |
| S16_vs_30017\|\|lt2gene | Low-quality |
| S16_vs_30040\|\|lt2gene | Low-quality |
| S16_vs_30052\|\|lt2gene | Low-quality |
| S16_vs_30074\|\|lt2gene | Low-quality |
| S16_vs_30122\|\|lt2gene | Low-quality |
| S16_vs_30148\|\|lt2gene | Low-quality |
| S16_vs_30161\|\|lt2gene | Not-determined |
| S16_vs_30234\|\|lt2gene | Not-determined |
| S16_vs_30236\|\|lt2gene | Not-determined |
| S16_vs_30261\|\|lt2gene | Not-determined |
| S16_vs_30288\|\|lt2gene | Low-quality |
| S16_vs_30326\|\|lt2gene | Low-quality |
| S16_vs_30362\|\|lt2gene | Low-quality |
| S16_vs_30365\|\|lt2gene | Not-determined |
| S16_vs_30389\|\|lt2gene | Not-determined |
| S16_vs_30420\|\|lt2gene | Not-determined |
| S16_vs_30482\|\|lt2gene | Low-quality |
| S16_vs_30491\|\|lt2gene | Not-determined |
| S16_vs_30660\|\|lt2gene | Not-determined |
| S16_vs_30711\|\|lt2gene | Low-quality |
| S16_vs_30820\|\|lt2gene | Not-determined |
| S16_vs_30842\|\|lt2gene | Low-quality |
| S16_vs_30852\|\|lt2gene | Low-quality |
| S16_vs_31082\|\|lt2gene | Low-quality |
| S16_vs_31108\|\|lt2gene | Low-quality |
| S16_vs_31121\|\|lt2gene | Low-quality |
| S16_vs_31248\|\|lt2gene | Low-quality |
| S16_vs_31322\|\|lt2gene | Low-quality |
| S16_vs_31508\|\|lt2gene | Low-quality |
| S16_vs_31819\|\|lt2gene | Low-quality |
| S16_vs_32070\|\|lt2gene | Low-quality |
| S16_vs_32122\|\|lt2gene | Low-quality |
| S16_vs_32277\|\|lt2gene | Low-quality |
| S16_vs_32464\|\|lt2gene | Low-quality |
| S16_vs_32534\|\|lt2gene | Low-quality |
| S16_vs_32572\|\|lt2gene | Low-quality |
| S16_vs_32595\|\|lt2gene | Not-determined |
| S16_vs_32629\|\|lt2gene | Low-quality |
| S16_vs_32685\|\|lt2gene | Not-determined |
| S16_vs_32751\|\|lt2gene | Low-quality |
| S16_vs_32803\|\|lt2gene | Low-quality |
| S16_vs_32851\|\|lt2gene | Not-determined |
| S16_vs_32917\|\|lt2gene | Low-quality |
| S16_vs_32926\|\|lt2gene | Low-quality |
| S16_vs_33011\|\|lt2gene | Low-quality |
| S16_vs_33027\|\|lt2gene | Low-quality |
| S16_vs_33047\|\|lt2gene | Low-quality |
| S16_vs_33048\|\|lt2gene | Low-quality |
| S16_vs_33088\|\|lt2gene | Not-determined |
| S16_vs_33415\|\|lt2gene | Not-determined |
| S16_vs_33536\|\|lt2gene | Not-determined |
| S16_vs_33676\|\|lt2gene | Low-quality |
| S16_vs_33699\|\|lt2gene | Low-quality |
| S16_vs_33725\|\|lt2gene | Low-quality |
| S16_vs_33729\|\|lt2gene | Not-determined |
| S16_vs_33848\|\|lt2gene | Not-determined |
| S16_vs_33979\|\|lt2gene | Low-quality |
| S16_vs_33995\|\|lt2gene | Low-quality |
| S16_vs_34121\|\|lt2gene | Low-quality |
| S16_vs_34186\|\|lt2gene | Not-determined |
| S16_vs_34338\|\|lt2gene | Low-quality |
| S16_vs_34348\|\|lt2gene | Low-quality |
| S16_vs_34366\|\|lt2gene | Not-determined |
| S16_vs_34477\|\|lt2gene | Not-determined |
| S16_vs_34643\|\|lt2gene | Low-quality |
| S16_vs_34713\|\|lt2gene | Low-quality |
| S16_vs_34717\|\|lt2gene | Not-determined |
| S16_vs_34732\|\|lt2gene | Low-quality |
| S16_vs_34756\|\|lt2gene | Low-quality |
| S16_vs_34761\|\|lt2gene | Low-quality |
| S16_vs_34975\|\|lt2gene | Low-quality |
| S16_vs_35025\|\|lt2gene | Not-determined |
| S16_vs_35216\|\|lt2gene | Low-quality |
| S16_vs_35282\|\|lt2gene | Low-quality |
| S16_vs_35457\|\|lt2gene | Low-quality |
| S16_vs_35647\|\|lt2gene | Low-quality |
| S16_vs_35649\|\|lt2gene | Low-quality |
| S16_vs_35711\|\|lt2gene | Low-quality |
| S16_vs_35741\|\|lt2gene | Low-quality |
| S16_vs_35747\|\|lt2gene | Low-quality |
| S16_vs_35774\|\|lt2gene | Low-quality |
| S16_vs_35803\|\|lt2gene | Low-quality |
| S16_vs_35931\|\|lt2gene | Low-quality |
| S16_vs_35947\|\|lt2gene | Low-quality |
| S16_vs_36003\|\|lt2gene | Low-quality |
| S16_vs_36068\|\|lt2gene | Low-quality |
| S16_vs_36177\|\|lt2gene | Low-quality |
| S16_vs_36244\|\|lt2gene | Low-quality |
| S16_vs_36351\|\|lt2gene | Low-quality |
| S16_vs_36434\|\|lt2gene | Low-quality |
| S16_vs_36536\|\|lt2gene | Low-quality |
| S16_vs_36609\|\|lt2gene | Low-quality |
| S16_vs_36732\|\|lt2gene | Not-determined |
| S16_vs_36757\|\|lt2gene | Low-quality |
| S16_vs_36762\|\|lt2gene | Not-determined |
| S16_vs_36790\|\|lt2gene | Low-quality |
| S16_vs_36888\|\|lt2gene | Low-quality |
| S16_vs_36899\|\|lt2gene | Low-quality |
| S16_vs_36934\|\|lt2gene | Low-quality |
| S16_vs_36983\|\|lt2gene | Low-quality |
| S16_vs_37016\|\|lt2gene | Low-quality |
| S16_vs_37070\|\|lt2gene | Not-determined |
| S16_vs_37219\|\|lt2gene | Low-quality |
| S16_vs_37396\|\|lt2gene | Low-quality |
| S16_vs_37492\|\|lt2gene | Not-determined |
| S16_vs_37528\|\|lt2gene | Low-quality |
| S16_vs_37638\|\|lt2gene | Not-determined |
| S16_vs_37691\|\|lt2gene | Not-determined |
| S16_vs_37740\|\|lt2gene | Not-determined |
| S16_vs_37907\|\|lt2gene | Not-determined |
| S16_vs_38029\|\|lt2gene | Not-determined |
| S16_vs_38271\|\|lt2gene | Low-quality |
| S16_vs_38521\|\|lt2gene | Low-quality |
| S16_vs_38638\|\|lt2gene | Low-quality |
| S16_vs_38688\|\|lt2gene | Low-quality |
| S16_vs_38745\|\|lt2gene | Low-quality |
| S16_vs_39015\|\|lt2gene | Low-quality |
| S16_vs_39286\|\|lt2gene | Low-quality |
| S16_vs_39462\|\|lt2gene | Not-determined |
| S16_vs_39477\|\|lt2gene | Not-determined |
| S16_vs_39700\|\|lt2gene | Low-quality |
| S16_vs_39737\|\|lt2gene | Low-quality |
| S16_vs_39826\|\|lt2gene | Low-quality |
| S16_vs_40050\|\|lt2gene | Low-quality |
| S16_vs_40051\|\|lt2gene | Not-determined |
| S16_vs_40216\|\|lt2gene | Not-determined |
| S16_vs_40245\|\|lt2gene | Low-quality |
| S16_vs_40254\|\|lt2gene | Not-determined |
| S16_vs_40279\|\|lt2gene | Not-determined |
| S16_vs_40519\|\|lt2gene | Not-determined |
| S16_vs_40552\|\|lt2gene | Low-quality |
| S16_vs_40623\|\|lt2gene | Low-quality |
| S16_vs_40663\|\|lt2gene | Low-quality |
| S16_vs_40671\|\|lt2gene | Low-quality |
| S16_vs_40703\|\|lt2gene | Low-quality |
| S16_vs_40747\|\|lt2gene | Low-quality |
| S16_vs_40945\|\|lt2gene | Low-quality |
| S16_vs_41013\|\|lt2gene | Low-quality |
| S16_vs_41041\|\|lt2gene | Low-quality |
| S16_vs_41068\|\|lt2gene | Low-quality |
| S16_vs_41125\|\|lt2gene | Low-quality |
| S16_vs_41197\|\|lt2gene | Low-quality |
| S16_vs_41296\|\|lt2gene | Not-determined |
| S16_vs_41350\|\|lt2gene | Not-determined |
| S16_vs_41358\|\|lt2gene | Not-determined |
| S16_vs_41664\|\|lt2gene | Low-quality |
| S16_vs_41870\|\|lt2gene | Not-determined |
| S16_vs_41990\|\|lt2gene | Low-quality |
| S16_vs_41999\|\|lt2gene | Low-quality |
| S16_vs_42163\|\|lt2gene | Low-quality |
| S16_vs_42175\|\|lt2gene | Not-determined |
| S16_vs_42244\|\|lt2gene | Low-quality |
| S16_vs_42310\|\|lt2gene | Low-quality |
| S16_vs_42355\|\|lt2gene | Low-quality |
| S16_vs_42401\|\|lt2gene | Not-determined |
| S16_vs_42427\|\|lt2gene | Low-quality |
| S16_vs_42589\|\|lt2gene | Low-quality |
| S16_vs_42675\|\|lt2gene | Low-quality |
| S16_vs_42987\|\|lt2gene | Not-determined |
| S16_vs_43031\|\|lt2gene | Low-quality |
| S16_vs_43142\|\|lt2gene | Low-quality |
| S16_vs_43154\|\|lt2gene | Low-quality |
| S16_vs_43211\|\|lt2gene | Low-quality |
| S16_vs_43380\|\|lt2gene | Low-quality |
| S16_vs_43907\|\|lt2gene | Low-quality |
| S16_vs_43913\|\|lt2gene | Low-quality |
| S16_vs_44067\|\|lt2gene | Low-quality |
| S16_vs_44089\|\|lt2gene | Low-quality |
| S16_vs_44250\|\|lt2gene | Low-quality |
| S16_vs_44314\|\|lt2gene | Low-quality |
| S16_vs_44318\|\|lt2gene | Not-determined |
| S16_vs_44539\|\|lt2gene | Not-determined |
| S16_vs_44718\|\|lt2gene | Low-quality |
| S16_vs_44778\|\|lt2gene | Not-determined |
| S16_vs_44817\|\|lt2gene | Low-quality |
| S16_vs_44835\|\|lt2gene | Low-quality |
| S16_vs_44935\|\|lt2gene | Low-quality |
| S16_vs_45005\|\|lt2gene | Low-quality |
| S16_vs_45067\|\|lt2gene | Low-quality |
| S16_vs_45068\|\|lt2gene | Low-quality |
| S16_vs_45092\|\|lt2gene | Low-quality |
| S16_vs_45100\|\|lt2gene | Low-quality |
| S16_vs_45204\|\|lt2gene | Low-quality |
| S16_vs_45420\|\|lt2gene | Low-quality |
| S16_vs_45769\|\|lt2gene | Low-quality |
| S16_vs_45780\|\|lt2gene | Low-quality |
| S16_vs_45817\|\|lt2gene | Low-quality |
| S16_vs_45821\|\|lt2gene | Not-determined |
| S16_vs_45835\|\|lt2gene | Low-quality |
| S16_vs_45956\|\|lt2gene | Not-determined |
| S16_vs_46016\|\|lt2gene | Low-quality |
| S16_vs_46224\|\|lt2gene | Low-quality |
| S16_vs_46229\|\|lt2gene | Low-quality |
| S16_vs_46297\|\|lt2gene | Low-quality |
| S16_vs_46330\|\|lt2gene | Low-quality |
| S16_vs_46589\|\|lt2gene | Not-determined |
| S16_vs_46609\|\|lt2gene | Low-quality |
| S16_vs_46619\|\|lt2gene | Low-quality |
| S16_vs_46643\|\|lt2gene | Low-quality |
| S16_vs_46771\|\|lt2gene | Low-quality |
| S16_vs_46799\|\|lt2gene | Low-quality |
| S16_vs_46810\|\|lt2gene | Low-quality |
| S16_vs_46816\|\|lt2gene | Low-quality |
| S16_vs_46969\|\|lt2gene | Low-quality |
| S16_vs_47309\|\|lt2gene | Low-quality |
| S16_vs_47340\|\|lt2gene | Low-quality |
| S16_vs_47409\|\|lt2gene | Low-quality |
| S16_vs_47411\|\|lt2gene | Not-determined |
| S16_vs_47433\|\|lt2gene | Low-quality |
| S16_vs_47461\|\|lt2gene | Low-quality |
| S16_vs_47527\|\|lt2gene | Low-quality |
| S16_vs_47637\|\|lt2gene | Low-quality |
| S16_vs_47670\|\|lt2gene | Low-quality |
| S16_vs_47684\|\|lt2gene | Low-quality |
| S16_vs_47960\|\|lt2gene | Low-quality |
| S16_vs_48137\|\|lt2gene | Low-quality |
| S16_vs_48156\|\|lt2gene | Low-quality |
| S16_vs_48184\|\|lt2gene | Low-quality |
| S16_vs_48192\|\|lt2gene | Not-determined |
| S16_vs_48240\|\|lt2gene | Low-quality |
| S16_vs_48256\|\|lt2gene | Not-determined |
| S16_vs_48282\|\|lt2gene | Low-quality |
| S16_vs_48303\|\|lt2gene | Low-quality |
| S16_vs_48372\|\|lt2gene | Low-quality |
| S16_vs_48403\|\|lt2gene | Low-quality |
| S16_vs_48452\|\|lt2gene | Low-quality |
| S16_vs_48487\|\|lt2gene | Low-quality |
| S16_vs_48519\|\|lt2gene | Low-quality |
| S16_vs_48545\|\|lt2gene | Low-quality |
| S16_vs_48561\|\|lt2gene | Low-quality |
| S16_vs_48785\|\|lt2gene | Low-quality |
| S16_vs_48995\|\|lt2gene | Not-determined |
| S16_vs_49019\|\|lt2gene | Low-quality |
| S16_vs_49071\|\|lt2gene | Low-quality |
| S16_vs_49412\|\|lt2gene | Low-quality |
| S16_vs_49546\|\|lt2gene | Low-quality |
| S16_vs_49557\|\|lt2gene | Not-determined |
| S16_vs_49625\|\|lt2gene | Low-quality |
| S16_vs_49641\|\|lt2gene | Low-quality |
| S16_vs_49664\|\|lt2gene | Not-determined |
| S16_vs_49716\|\|lt2gene | Low-quality |
| S16_vs_49779\|\|lt2gene | Not-determined |
| S16_vs_49815\|\|lt2gene | Not-determined |
| S16_vs_49854\|\|lt2gene | Low-quality |
| S16_vs_49869\|\|lt2gene | Low-quality |
| S16_vs_49877\|\|lt2gene | Not-determined |
| S16_vs_50189\|\|lt2gene | Low-quality |
| S16_vs_50208\|\|lt2gene | Low-quality |

S16_vs_50226||lt2gene Low-quality
S16_vs_50391||lt2gene Not-determined
S16_vs_50757||lt2gene Not-determined
S16_vs_50969||lt2gene Low-quality
S16_vs_50980||lt2gene Low-quality
S16_vs_51034||lt2gene Low-quality
S16_vs_51170||lt2gene Not-determined
S16_vs_51203||lt2gene Not-determined
S16_vs_51261||lt2gene Low-quality
S16_vs_51300||lt2gene Not-determined
S16_vs_51421||lt2gene Low-quality
S16_vs_51427||lt2gene Low-quality
S16_vs_51516||lt2gene Low-quality
S16_vs_51630||lt2gene Low-quality
S16_vs_51666||lt2gene Not-determined
S16_vs_51731||lt2gene Low-quality
S16_vs_51811||lt2gene Low-quality
S16_vs_51874||lt2gene Low-quality
S16_vs_51895||lt2gene Low-quality
S16_vs_51942||lt2gene Low-quality
S16_vs_51955||lt2gene Low-quality
S16_vs_52002||lt2gene Low-quality
S16_vs_52259||lt2gene Low-quality
S16_vs_52284||lt2gene Low-quality
S16_vs_52364||lt2gene Low-quality
S16_vs_52392||lt2gene Low-quality
S16_vs_52739||lt2gene Low-quality
S16_vs_52891||lt2gene Low-quality
S16_vs_52894||lt2gene Low-quality
S16_vs_53066||lt2gene Low-quality
S16_vs_53146||lt2gene Not-determined
S16_vs_53321||lt2gene Low-quality
S16_vs_53430||lt2gene Low-quality
S16_vs_53485||lt2gene Low-quality
S16_vs_53780||lt2gene Low-quality
S16_vs_53798||lt2gene Not-determined
S16_vs_53933||lt2gene Low-quality
S16_vs_53939||lt2gene Low-quality
S16_vs_53941||lt2gene Not-determined
S16_vs_53950||lt2gene Low-quality
S16_vs_53972||lt2gene Low-quality
S16_vs_54184||lt2gene Low-quality
S16_vs_54188||lt2gene Low-quality
S16_vs_54200||lt2gene Not-determined
S16_vs_54357||lt2gene Low-quality
S16_vs_54375||lt2gene Not-determined
S16_vs_54526||lt2gene Low-quality
S16_vs_54532||lt2gene Low-quality
S16_vs_54553||lt2gene Low-quality
S16_vs_54618||lt2gene Not-determined
S16_vs_54630||lt2gene Not-determined
S16_vs_54700||lt2gene Low-quality
S16_vs_54742||lt2gene Low-quality
S16_vs_54743||lt2gene Low-quality
S16_vs_54885||lt2gene Low-quality
S16_vs_54912||lt2gene Low-quality
S16_vs_54952||lt2gene Low-quality
S16_vs_55008||lt2gene Low-quality
S16_vs_55109||lt2gene Low-quality
S16_vs_55177||lt2gene Low-quality
S16_vs_55226||lt2gene Low-quality
S16_vs_55304||lt2gene Not-determined
S16_vs_55395||lt2gene Low-quality
S16_vs_55437||lt2gene Low-quality
S16_vs_55442||lt2gene Low-quality
S16_vs_55598||lt2gene Not-determined
S16_vs_55667||lt2gene Low-quality
S16_vs_55763||lt2gene Not-determined
S16_vs_55819||lt2gene Low-quality
S16_vs_55829||lt2gene Low-quality
S16_vs_55966||lt2gene Low-quality
S16_vs_56023||lt2gene Low-quality
S16_vs_56147||lt2gene Not-determined
S16_vs_56157||lt2gene Not-determined
S16_vs_56416||lt2gene Low-quality
S16_vs_56427||lt2gene Low-quality
S16_vs_56458||lt2gene Not-determined
S16_vs_56556||lt2gene Low-quality
S16_vs_56604||lt2gene Low-quality
S16_vs_56633||lt2gene Low-quality
S16_vs_56648||lt2gene Low-quality
S16_vs_56707||lt2gene Low-quality
S16_vs_56805||lt2gene Low-quality
S16_vs_56854||lt2gene Low-quality
S16_vs_56863||lt2gene Low-quality
S16_vs_56888||lt2gene Low-quality
S16_vs_57010||lt2gene Low-quality
S16_vs_57103||lt2gene Low-quality
S16_vs_57117||lt2gene Not-determined
S16_vs_57239||lt2gene Not-determined
S16_vs_57297||lt2gene Low-quality
S16_vs_57324||lt2gene Not-determined
S16_vs_57329||lt2gene Low-quality
S16_vs_57449||lt2gene Low-quality
S16_vs_57505||lt2gene Low-quality
S16_vs_57512||lt2gene Low-quality
S16_vs_57594||lt2gene Low-quality
S16_vs_57602||lt2gene Low-quality
S16_vs_57698||lt2gene Low-quality
S16_vs_57908||lt2gene Low-quality
S16_vs_57929||lt2gene Low-quality
S16_vs_58038||lt2gene Low-quality
S16_vs_58340||lt2gene Not-determined
S16_vs_58363||lt2gene Not-determined
S16_vs_58474||lt2gene Not-determined
S16_vs_58508||lt2gene Low-quality
S16_vs_58518||lt2gene Low-quality
S16_vs_58605||lt2gene Not-determined
S16_vs_58752||lt2gene Low-quality
S16_vs_58802||lt2gene Not-determined
S16_vs_58856||lt2gene Low-quality
S16_vs_58857||lt2gene Low-quality
S16_vs_58864||lt2gene Low-quality
S16_vs_58882||lt2gene Low-quality
S16_vs_58917||lt2gene Not-determined
S16_vs_58920||lt2gene Low-quality
S16_vs_58938||lt2gene Low-quality
S16_vs_59446||lt2gene Not-determined
S16_vs_59480||lt2gene Low-quality
S16_vs_59489||lt2gene Low-quality
S16_vs_59494||lt2gene Low-quality
S16_vs_59531||lt2gene Low-quality
S16_vs_59536||lt2gene Not-determined
S16_vs_59658||lt2gene Low-quality
S16_vs_59793||lt2gene Low-quality
S16_vs_59808||lt2gene Low-quality
S16_vs_59811||lt2gene Not-determined
S16_vs_59951||lt2gene Not-determined
S16_vs_60160||lt2gene Low-quality
S16_vs_60226||lt2gene Not-determined
S16_vs_60262||lt2gene Low-quality
S16_vs_60277||lt2gene Low-quality
S16_vs_60566||lt2gene Low-quality
S16_vs_60713||lt2gene Low-quality
S16_vs_60721||lt2gene Low-quality
S16_vs_60741||lt2gene Not-determined
S16_vs_60750||lt2gene Low-quality
S16_vs_60944||lt2gene Low-quality
S16_vs_61072||lt2gene Low-quality
S16_vs_61135||lt2gene Low-quality
S16_vs_61160||lt2gene Low-quality
S16_vs_61360||lt2gene Low-quality
S16_vs_61367||lt2gene Not-determined
S16_vs_61398||lt2gene Low-quality
S16_vs_61401||lt2gene Low-quality
S16_vs_61408||lt2gene Not-determined
S16_vs_61515||lt2gene Low-quality
S16_vs_61635||lt2gene Low-quality
S16_vs_61674||lt2gene Not-determined
S16_vs_61736||lt2gene Low-quality
S16_vs_61943||lt2gene Low-quality
S16_vs_61946||lt2gene Low-quality
S16_vs_62167||lt2gene Low-quality
S16_vs_62305||lt2gene Low-quality
S16_vs_62310||lt2gene Low-quality
S16_vs_62335||lt2gene Not-determined
S16_vs_62482||lt2gene Low-quality
S16_vs_62655||lt2gene Not-determined
S16_vs_62664||lt2gene Not-determined
S16_vs_62675||lt2gene Low-quality
S16_vs_62706||lt2gene Low-quality
S16_vs_62784||lt2gene Low-quality
S16_vs_62876||lt2gene Not-determined
S16_vs_62924||lt2gene Low-quality
S16_vs_62999||lt2gene Low-quality
S16_vs_63033||lt2gene Low-quality
S16_vs_63331||lt2gene Low-quality
S16_vs_63369||lt2gene Low-quality
S16_vs_63393||lt2gene Low-quality
S16_vs_63406||lt2gene Low-quality
S16_vs_63410||lt2gene Low-quality
S16_vs_63493||lt2gene Not-determined
S16_vs_63600||lt2gene Low-quality
S16_vs_63697||lt2gene Low-quality
S16_vs_63926||lt2gene Not-determined
S16_vs_64036||lt2gene Low-quality
S16_vs_64100||lt2gene Not-determined
S16_vs_64334||lt2gene Low-quality
S16_vs_64470||lt2gene Low-quality
S16_vs_64606||lt2gene Low-quality
S16_vs_64779||lt2gene Low-quality
S16_vs_64859||lt2gene Low-quality
S16_vs_64866||lt2gene Not-determined
S16_vs_65005||lt2gene Not-determined
S16_vs_65129||lt2gene Low-quality
S16_vs_65288||lt2gene Not-determined
S16_vs_65314||lt2gene Low-quality
S16_vs_65324||lt2gene Low-quality
S16_vs_65442||lt2gene Low-quality
S16_vs_65463||lt2gene Low-quality
S16_vs_65493||lt2gene Low-quality
S16_vs_65766||lt2gene Low-quality
S16_vs_65850||lt2gene Low-quality
S16_vs_65870||lt2gene Not-determined
S16_vs_65938||lt2gene Low-quality
S16_vs_66022||lt2gene Low-quality
S16_vs_66029||lt2gene Not-determined
S16_vs_66212||lt2gene Low-quality
S16_vs_66391||lt2gene Low-quality
S16_vs_66678||lt2gene Not-determined
S16_vs_66698||lt2gene Low-quality
S16_vs_66750||lt2gene Low-quality
S16_vs_66802||lt2gene Low-quality
S16_vs_66942||lt2gene Low-quality
S16_vs_67021||lt2gene Low-quality
S16_vs_67212||lt2gene Low-quality
S16_vs_67317||lt2gene Low-quality
S16_vs_67340||lt2gene Low-quality
S16_vs_67712||lt2gene Not-determined
S16_vs_67787||lt2gene Low-quality
S16_vs_67832||lt2gene Low-quality
S16_vs_67870||lt2gene Low-quality
S16_vs_68172||lt2gene Not-determined
S16_vs_68299||lt2gene Low-quality
S16_vs_68432||lt2gene Low-quality
S16_vs_68464||lt2gene Not-determined
S16_vs_68513||lt2gene Low-quality
S16_vs_68932||lt2gene Low-quality
S16_vs_68965||lt2gene Not-determined
S16_vs_68982||lt2gene Not-determined
S16_vs_69011||lt2gene Low-quality
S16_vs_69018||lt2gene Low-quality
S16_vs_69185||lt2gene Low-quality
S16_vs_69269||lt2gene Low-quality
S16_vs_69338||lt2gene Low-quality
S16_vs_69468||lt2gene Not-determined
S16_vs_69634||lt2gene Low-quality
S16_vs_69662||lt2gene Not-determined
S16_vs_69694||lt2gene Not-determined
S16_vs_69710||lt2gene Low-quality
S16_vs_69908||lt2gene Low-quality
S16_vs_70110||lt2gene Low-quality
S16_vs_70200||lt2gene Low-quality
S16_vs_70246||lt2gene Not-determined
S16_vs_70283||lt2gene Not-determined
S16_vs_70523||lt2gene Low-quality
S16_vs_70637||lt2gene Low-quality
S16_vs_70937||lt2gene Not-determined
S16_vs_71397||lt2gene Low-quality
S16_vs_71461||lt2gene Not-determined
S16_vs_71578||lt2gene Not-determined
S16_vs_72157||lt2gene Low-quality
S16_vs_72286||lt2gene Low-quality
S16_vs_72359||lt2gene Not-determined
S16_vs_72399||lt2gene Not-determined
S16_vs_72448||lt2gene Low-quality
S16_vs_72538||lt2gene Low-quality
S16_vs_72738||lt2gene Low-quality
S16_vs_72778||lt2gene Low-quality
S16_vs_72806||lt2gene Low-quality
S16_vs_72875||lt2gene Low-quality
S16_vs_72922||lt2gene Not-determined
S16_vs_72939||lt2gene Low-quality
S16_vs_72979||lt2gene Not-determined
S16_vs_73201||lt2gene Low-quality
S16_vs_73331||lt2gene Low-quality
S16_vs_73353||lt2gene Not-determined
S16_vs_73404||lt2gene Not-determined
S16_vs_73524||lt2gene Not-determined
S16_vs_73558||lt2gene Low-quality
S16_vs_73618||lt2gene Low-quality
S16_vs_73763||lt2gene Low-quality
S16_vs_73775||lt2gene Not-determined
S16_vs_73783||lt2gene Low-quality
S16_vs_73857||lt2gene Not-determined
S16_vs_73868||lt2gene Low-quality
S16_vs_73976||lt2gene Low-quality
S16_vs_73982||lt2gene Low-quality
S16_vs_73996||lt2gene Low-quality
S16_vs_74088||lt2gene Low-quality
S16_vs_74095||lt2gene Low-quality
S16_vs_74249||lt2gene Not-determined
S16_vs_74365||lt2gene Not-determined
S16_vs_74406||lt2gene Low-quality
S16_vs_74436||lt2gene Not-determined
S16_vs_74438||lt2gene Low-quality
S16_vs_74491||lt2gene Low-quality
S16_vs_74591||lt2gene Low-quality
S16_vs_74768||lt2gene Low-quality
S16_vs_74975||lt2gene Not-determined
S16_vs_75057||lt2gene Not-determined
S16_vs_75101||lt2gene Low-quality
S16_vs_75130||lt2gene Not-determined
S16_vs_75137||lt2gene Low-quality
S16_vs_75235||lt2gene Not-determined
S16_vs_75263||lt2gene Low-quality
S16_vs_75321||lt2gene Low-quality
S16_vs_75336||lt2gene Low-quality
S16_vs_75338||lt2gene Low-quality
S16_vs_75373||lt2gene Low-quality
S16_vs_75409||lt2gene Not-determined
S16_vs_75414||lt2gene Low-quality
S16_vs_75464||lt2gene Low-quality
S16_vs_75489||lt2gene Low-quality
S16_vs_75570||lt2gene Not-determined
S16_vs_75732||lt2gene Low-quality
S16_vs_75799||lt2gene Low-quality
S16_vs_75848||lt2gene Low-quality
S16_vs_75910||lt2gene Low-quality
S16_vs_76077||lt2gene Not-determined
S16_vs_76140||lt2gene Not-determined
S16_vs_76248||lt2gene Not-determined
S16_vs_76371||lt2gene Low-quality
S16_vs_76425||lt2gene Low-quality
S16_vs_76483||lt2gene Low-quality
S16_vs_76535||lt2gene Low-quality
S16_vs_76666||lt2gene Low-quality
S16_vs_76780||lt2gene Low-quality
S16_vs_76848||lt2gene Low-quality
S16_vs_76998||lt2gene Low-quality
S16_vs_77268||lt2gene Low-quality
S16_vs_77270||lt2gene Low-quality
S16_vs_77356||lt2gene Low-quality
S16_vs_77365||lt2gene Low-quality
S16_vs_77536||lt2gene Low-quality
S16_vs_77558||lt2gene Low-quality
S16_vs_77640||lt2gene Low-quality
S16_vs_77646||lt2gene Low-quality
S16_vs_77711||lt2gene Low-quality
S16_vs_77826||lt2gene Low-quality
S16_vs_77954||lt2gene Low-quality
S16_vs_77966||lt2gene Not-determined
S16_vs_77967||lt2gene Not-determined
S16_vs_77974||lt2gene Not-determined
S16_vs_77978||lt2gene Not-determined
S16_vs_78285||lt2gene Low-quality
S16_vs_78286||lt2gene Low-quality
S16_vs_78302||lt2gene Not-determined
S16_vs_78327||lt2gene Low-quality
S16_vs_78374||lt2gene Low-quality
S16_vs_78386||lt2gene Not-determined
S16_vs_78466||lt2gene Low-quality
S16_vs_78596||lt2gene Low-quality
S16_vs_78711||lt2gene Low-quality
S16_vs_78826||lt2gene Not-determined
S16_vs_78890||lt2gene Not-determined
S16_vs_78899||lt2gene Low-quality
S16_vs_79007||lt2gene Low-quality
S16_vs_79241||lt2gene Not-determined
S16_vs_79411||lt2gene Low-quality
S16_vs_79584||lt2gene Low-quality
S16_vs_79680||lt2gene Not-determined
S16_vs_79860||lt2gene Low-quality
S16_vs_80039||lt2gene Low-quality
S16_vs_80165||lt2gene Low-quality
S16_vs_80268||lt2gene Low-quality
S16_vs_80322||lt2gene Not-determined
S16_vs_80373||lt2gene Not-determined
S16_vs_80526||lt2gene Low-quality
S16_vs_80692||lt2gene Low-quality
S16_vs_80727||lt2gene Low-quality
S16_vs_80876||lt2gene Low-quality
S16_vs_80906||lt2gene Low-quality
S16_vs_80941||lt2gene Low-quality
S16_vs_81072||lt2gene Low-quality
S16_vs_81081||lt2gene Low-quality
S16_vs_81180||lt2gene Low-quality
S16_vs_81255||lt2gene Low-quality
S16_vs_81281||lt2gene Low-quality
S16_vs_81444||lt2gene Low-quality
S16_vs_81524||lt2gene Not-determined
S16_vs_81646||lt2gene Not-determined
S16_vs_81711||lt2gene Low-quality
S16_vs_81801||lt2gene Low-quality
S16_vs_82254||lt2gene Not-determined
S16_vs_82273||lt2gene Low-quality
S16_vs_82404||lt2gene Low-quality
S16_vs_82522||lt2gene Low-quality
S16_vs_82550||lt2gene Low-quality
S16_vs_82608||lt2gene Not-determined
S16_vs_82622||lt2gene Low-quality
S16_vs_82636||lt2gene Low-quality
S16_vs_83159||lt2gene Low-quality
S16_vs_83179||lt2gene Low-quality
S16_vs_83194||lt2gene Low-quality
S16_vs_83198||lt2gene Low-quality
S16_vs_83237||lt2gene Low-quality
S16_vs_83252||lt2gene Not-determined
S16_vs_83401||lt2gene Low-quality
S16_vs_83522||lt2gene Not-determined
S16_vs_83535||lt2gene Not-determined
S16_vs_83615||lt2gene Low-quality
S16_vs_83669||lt2gene Not-determined
S16_vs_83805||lt2gene Low-quality
S16_vs_83938||lt2gene Low-quality
S16_vs_84209||lt2gene Low-quality
S16_vs_84216||lt2gene Low-quality
S16_vs_84296||lt2gene Low-quality
S16_vs_84345||lt2gene Low-quality
S16_vs_84366||lt2gene Low-quality
S16_vs_84410||lt2gene Low-quality
S16_vs_84433||lt2gene Not-determined
S16_vs_84624||lt2gene Low-quality
S16_vs_84657||lt2gene Not-determined
S16_vs_84710||lt2gene Low-quality
S16_vs_84914||lt2gene Low-quality
S16_vs_85034||lt2gene Low-quality
S16_vs_85094||lt2gene Not-determined
S16_vs_85175||lt2gene Not-determined
S16_vs_85336||lt2gene Low-quality
S16_vs_85341||lt2gene Low-quality
S16_vs_85400||lt2gene Not-determined
S16_vs_85527||lt2gene Low-quality
S16_vs_85593||lt2gene Low-quality
S16_vs_85651||lt2gene Low-quality
S16_vs_85852||lt2gene Low-quality
S16_vs_85934||lt2gene Low-quality
S16_vs_85954||lt2gene Not-determined
S16_vs_86013||lt2gene Low-quality
S16_vs_86170||lt2gene Low-quality
S16_vs_86226||lt2gene Low-quality
S16_vs_86359||lt2gene Not-determined
S16_vs_86503||lt2gene Low-quality
S16_vs_86512||lt2gene Low-quality
S16_vs_86635||lt2gene Low-quality
S16_vs_86679||lt2gene Low-quality
S16_vs_86963||lt2gene Low-quality
S16_vs_87087||lt2gene Low-quality
S16_vs_87131||lt2gene Not-determined
S16_vs_87277||lt2gene Low-quality
S16_vs_87330||lt2gene Not-determined
S16_vs_87343||lt2gene Low-quality
S16_vs_87351||lt2gene Not-determined
S16_vs_87372||lt2gene Not-determined
S16_vs_87381||lt2gene Low-quality
S16_vs_87418||lt2gene Not-determined
S16_vs_87442||lt2gene Low-quality
S16_vs_87580||lt2gene Not-determined
S16_vs_87605||lt2gene Not-determined
S16_vs_87684||lt2gene Low-quality
S16_vs_87702||lt2gene Low-quality
S16_vs_87713||lt2gene Low-quality
S16_vs_87825||lt2gene Low-quality
S16_vs_87871||lt2gene Low-quality
S16_vs_87975||lt2gene Low-quality
S16_vs_87993||lt2gene Not-determined
S16_vs_88135||lt2gene Low-quality
S16_vs_88150||lt2gene Low-quality
S16_vs_88312||lt2gene Not-determined
S16_vs_88334||lt2gene Low-quality
S16_vs_88376||lt2gene Low-quality
S16_vs_88705||lt2gene Not-determined
S16_vs_88848||lt2gene Low-quality
S16_vs_88936||lt2gene Low-quality
S16_vs_88951||lt2gene Low-quality
S16_vs_89110||lt2gene Not-determined
S16_vs_89112||lt2gene Not-determined
S16_vs_89252||lt2gene Low-quality
S16_vs_89444||lt2gene Low-quality
S16_vs_89479||lt2gene Low-quality
S16_vs_89529||lt2gene Not-determined
S16_vs_89558||lt2gene Not-determined
S16_vs_89770||lt2gene Low-quality
S16_vs_89791||lt2gene Low-quality
S16_vs_89806||lt2gene Low-quality
S16_vs_89807||lt2gene Not-determined
S16_vs_89883||lt2gene Low-quality
S16_vs_89950||lt2gene Low-quality
S16_vs_90254||lt2gene Low-quality
S16_vs_90267||lt2gene Not-determined
S16_vs_90343||lt2gene Low-quality
S16_vs_90393||lt2gene Not-determined
S16_vs_90507||lt2gene Low-quality
S16_vs_90509||lt2gene Low-quality
S16_vs_90681||lt2gene Low-quality
S16_vs_90717||lt2gene Low-quality
S16_vs_90721||lt2gene Not-determined
S16_vs_90803||lt2gene Low-quality
S16_vs_90845||lt2gene Low-quality
S16_vs_90965||lt2gene Low-quality
S16_vs_91186||lt2gene Low-quality
S16_vs_91363||lt2gene Not-determined
S16_vs_91438||lt2gene Low-quality
S16_vs_91498||lt2gene Low-quality
S16_vs_91593||lt2gene Low-quality
S16_vs_91708||lt2gene Not-determined
S16_vs_91724||lt2gene Low-quality
S16_vs_91743||lt2gene Low-quality
S16_vs_91880||lt2gene Low-quality
S16_vs_91933||lt2gene Low-quality
S16_vs_92028||lt2gene Not-determined
S16_vs_92060||lt2gene Low-quality
S16_vs_92066||lt2gene Low-quality
S16_vs_92282||lt2gene Low-quality
S16_vs_92283||lt2gene Not-determined
S16_vs_92293||lt2gene Not-determined
S16_vs_92327||lt2gene Low-quality
S16_vs_92349||lt2gene Low-quality
S16_vs_92372||lt2gene Low-quality
S16_vs_92374||lt2gene Not-determined
S16_vs_92536||lt2gene Not-determined
S16_vs_92606||lt2gene Low-quality
S16_vs_92627||lt2gene Low-quality
S16_vs_92677||lt2gene Low-quality
S16_vs_92708||lt2gene Low-quality
S16_vs_92809||lt2gene Low-quality
S16_vs_92874||lt2gene Low-quality
S16_vs_92983||lt2gene Not-determined
S16_vs_93085||lt2gene Not-determined
S16_vs_93170||lt2gene Not-determined
S16_vs_93193||lt2gene Low-quality
S16_vs_93199||lt2gene Low-quality
S16_vs_93358||lt2gene Low-quality
S16_vs_93442||lt2gene Low-quality
S16_vs_93496||lt2gene Low-quality
S16_vs_93571||lt2gene Not-determined
S16_vs_93572||lt2gene Not-determined
S16_vs_93821||lt2gene Low-quality
S16_vs_93951||lt2gene Not-determined
S16_vs_93990||lt2gene Low-quality
S16_vs_94152||lt2gene Low-quality
S16_vs_94213||lt2gene Low-quality
S16_vs_94404||lt2gene Low-quality
S16_vs_94526||lt2gene Low-quality
S16_vs_94739||lt2gene Low-quality
S16_vs_94947||lt2gene Low-quality
S16_vs_95053||lt2gene Not-determined
S16_vs_95168||lt2gene Low-quality
S16_vs_95194||lt2gene Low-quality
S16_vs_95466||lt2gene Low-quality
S16_vs_95527||lt2gene Low-quality
S16_vs_95541||lt2gene Low-quality
S16_vs_95761||lt2gene Low-quality
S16_vs_95766||lt2gene Low-quality
S16_vs_95816||lt2gene Low-quality
S16_vs_95893||lt2gene Not-determined
S16_vs_96013||lt2gene Not-determined
S16_vs_96201||lt2gene Low-quality
S16_vs_96318||lt2gene Low-quality
S16_vs_96351||lt2gene Not-determined
S16_vs_96430||lt2gene Low-quality
S16_vs_96705||lt2gene Low-quality
S16_vs_96828||lt2gene Low-quality
S16_vs_96934||lt2gene Low-quality
S16_vs_96964||lt2gene Not-determined
S16_vs_97050||lt2gene Low-quality
S16_vs_97076||lt2gene Low-quality
S16_vs_97272||lt2gene Low-quality
S16_vs_97276||lt2gene Low-quality
S16_vs_97384||lt2gene Low-quality
S16_vs_97454||lt2gene Low-quality
S16_vs_97498||lt2gene Low-quality
S16_vs_97516||lt2gene Low-quality
S16_vs_97560||lt2gene Low-quality
S16_vs_97562||lt2gene Low-quality
S16_vs_97590||lt2gene Low-quality
S16_vs_97720||lt2gene Low-quality
S16_vs_97800||lt2gene Not-determined
S16_vs_97822||lt2gene Low-quality
S16_vs_98146||lt2gene Low-quality
S16_vs_98224||lt2gene Low-quality
S16_vs_98275||lt2gene Low-quality
S16_vs_98422||lt2gene Not-determined
S16_vs_98433||lt2gene Low-quality
S16_vs_98476||lt2gene Not-determined
S16_vs_98557||lt2gene Low-quality
S16_vs_98689||lt2gene Low-quality
S16_vs_98865||lt2gene Low-quality
S16_vs_98999||lt2gene Low-quality
S16_vs_99046||lt2gene Low-quality
S16_vs_99116||lt2gene Low-quality
S16_vs_99345||lt2gene Low-quality
S16_vs_99363||lt2gene Low-quality
S16_vs_99408||lt2gene Not-determined
S16_vs_99524||lt2gene Low-quality
S16_vs_99539||lt2gene Low-quality
S16_vs_99684||lt2gene Low-quality
S16_vs_99745||lt2gene Low-quality
S16_vs_99836||lt2gene Low-quality
S16_vs_99841||lt2gene Low-quality
S16_vs_99843||lt2gene Low-quality
S16_vs_99847||lt2gene Low-quality
S16_vs_99872||lt2gene Not-determined
S16_vs_99880||lt2gene Low-quality
S16_vs_99947||lt2gene Low-quality
S16_vs_99950||lt2gene Low-quality
S16_vs_100284||lt2gene Not-determined
S16_vs_100356||lt2gene Low-quality
S16_vs_100379||lt2gene Low-quality
S16_vs_100458||lt2gene Low-quality
S16_vs_100528||lt2gene Low-quality
S16_vs_100606||lt2gene Not-determined
S16_vs_100710||lt2gene Not-determined
S16_vs_100781||lt2gene Low-quality
S16_vs_100854||lt2gene Low-quality
S16_vs_100998||lt2gene Low-quality
S16_vs_101055||lt2gene Low-quality
S16_vs_101096||lt2gene Low-quality
S16_vs_101165||lt2gene Low-quality

S16_vs_101178||lt2gene Low-quality
S16_vs_101195||lt2gene Low-quality
S16_vs_101245||lt2gene Not-determined
S16_vs_101270||lt2gene Low-quality
S16_vs_101286||lt2gene Low-quality
S16_vs_101343||lt2gene Low-quality
S16_vs_101775||lt2gene Low-quality
S16_vs_101801||lt2gene Low-quality
S16_vs_101878||lt2gene Low-quality
S16_vs_102036||lt2gene Low-quality
S16_vs_102076||lt2gene Low-quality
S16_vs_102131||lt2gene Low-quality
S16_vs_102165||lt2gene Low-quality
S16_vs_102172||lt2gene Low-quality
S16_vs_102185||lt2gene Low-quality
S16_vs_102195||lt2gene Not-determined
S16_vs_102294||lt2gene Not-determined
S16_vs_102324||lt2gene Low-quality
S16_vs_102543||lt2gene Low-quality
S16_vs_102704||lt2gene Low-quality
S16_vs_102731||lt2gene Not-determined
S16_vs_102819||lt2gene Low-quality
S16_vs_102877||lt2gene Not-determined
S16_vs_102934||lt2gene Not-determined
S16_vs_103007||lt2gene Not-determined
S16_vs_103020||lt2gene Low-quality
S16_vs_103193||lt2gene Not-determined
S16_vs_103225||lt2gene Low-quality
S16_vs_103306||lt2gene Low-quality
S16_vs_103376||lt2gene Low-quality
S16_vs_103578||lt2gene Low-quality
S16_vs_103582||lt2gene Low-quality
S16_vs_103691||lt2gene Low-quality
S16_vs_103927||lt2gene Low-quality
S16_vs_104076||lt2gene Low-quality
S16_vs_104099||lt2gene Low-quality
S16_vs_104121||lt2gene Low-quality
S16_vs_104167||lt2gene Not-determined
S16_vs_104245||lt2gene Low-quality
S16_vs_104269||lt2gene Low-quality
S16_vs_104275||lt2gene Low-quality
S16_vs_104366||lt2gene Low-quality
S16_vs_104502||lt2gene Low-quality
S16_vs_104563||lt2gene Low-quality
S16_vs_104650||lt2gene Low-quality
S16_vs_105128||lt2gene Low-quality
S16_vs_105196||lt2gene Not-determined
S16_vs_105241||lt2gene Low-quality
S16_vs_105453||lt2gene Low-quality
S16_vs_105854||lt2gene Low-quality
S16_vs_106139||lt2gene Low-quality
S16_vs_106175||lt2gene Low-quality
S16_vs_106199||lt2gene Low-quality
S16_vs_106236||lt2gene Low-quality
S16_vs_106633||lt2gene Not-determined
S16_vs_106670||lt2gene Not-determined
S16_vs_106737||lt2gene Not-determined
S16_vs_106739||lt2gene Low-quality
S16_vs_106825||lt2gene Low-quality
S16_vs_106889||lt2gene Low-quality
S16_vs_106913||lt2gene Low-quality
S16_vs_107151||lt2gene Not-determined
S16_vs_107178||lt2gene Low-quality
S16_vs_107333||lt2gene Low-quality
S16_vs_107740||lt2gene Low-quality
S16_vs_107912||lt2gene Low-quality
S16_vs_107938||lt2gene Not-determined
S16_vs_107950||lt2gene Low-quality
S16_vs_107952||lt2gene Low-quality
S16_vs_108128||lt2gene Not-determined
S16_vs_108233||lt2gene Low-quality
S16_vs_108378||lt2gene Not-determined
S16_vs_108430||lt2gene Not-determined
S16_vs_108668||lt2gene Low-quality
S16_vs_108706||lt2gene Not-determined
S16_vs_108730||lt2gene Low-quality
S16_vs_108895||lt2gene Low-quality
S16_vs_108955||lt2gene Low-quality
S16_vs_108981||lt2gene Not-determined
S16_vs_109017||lt2gene Low-quality
S16_vs_109167||lt2gene Low-quality
S16_vs_109298||lt2gene Not-determined
S16_vs_109376||lt2gene Low-quality
S16_vs_109402||lt2gene Low-quality
S16_vs_109457||lt2gene Low-quality
S16_vs_109502||lt2gene Low-quality
S16_vs_109645||lt2gene Low-quality
S16_vs_109688||lt2gene Not-determined
S16_vs_109708||lt2gene Low-quality
S16_vs_109796||lt2gene Not-determined
S16_vs_109877||lt2gene Low-quality
S16_vs_109965||lt2gene Not-determined
S16_vs_109997||lt2gene Low-quality
S16_vs_110006||lt2gene Low-quality
S16_vs_110028||lt2gene Not-determined
S16_vs_110032||lt2gene Low-quality
S16_vs_110045||lt2gene Low-quality
S16_vs_110052||lt2gene Low-quality
S16_vs_110132||lt2gene Low-quality
S16_vs_110305||lt2gene Low-quality
S16_vs_110354||lt2gene Low-quality
S16_vs_110661||lt2gene Low-quality
S16_vs_110689||lt2gene Not-determined
S16_vs_110843||lt2gene Low-quality
S16_vs_110919||lt2gene Low-quality
S16_vs_110959||lt2gene Low-quality
S16_vs_111030||lt2gene Low-quality
S16_vs_111093||lt2gene Low-quality
S16_vs_111237||lt2gene Low-quality
S16_vs_111388||lt2gene Low-quality
S16_vs_111512||lt2gene Low-quality
S16_vs_111633||lt2gene Low-quality
S16_vs_111671||lt2gene Low-quality
S16_vs_111725||lt2gene Low-quality
S16_vs_111790||lt2gene Low-quality
S16_vs_111946||lt2gene Low-quality
S16_vs_112003||lt2gene Low-quality
S16_vs_112045||lt2gene Low-quality
S16_vs_112102||lt2gene Low-quality
S16_vs_112135||lt2gene Low-quality
S16_vs_112177||lt2gene Not-determined
S16_vs_112493||lt2gene Not-determined
S16_vs_112503||lt2gene Not-determined
S16_vs_112869||lt2gene Not-determined
S16_vs_112893||lt2gene Not-determined
S16_vs_112967||lt2gene Low-quality
S16_vs_112971||lt2gene Not-determined
S16_vs_113016||lt2gene Not-determined
S16_vs_113018||lt2gene Low-quality
S16_vs_113059||lt2gene Low-quality
S16_vs_113140||lt2gene Low-quality
S16_vs_113221||lt2gene Low-quality
S16_vs_113543||lt2gene Low-quality
S16_vs_113562||lt2gene Not-determined
S16_vs_113676||lt2gene Low-quality
S16_vs_113798||lt2gene Low-quality
S16_vs_113871||lt2gene Low-quality
S16_vs_113922||lt2gene Not-determined
S16_vs_114010||lt2gene Low-quality
S16_vs_114047||lt2gene Low-quality
S16_vs_114074||lt2gene Low-quality
S16_vs_114120||lt2gene Low-quality
S16_vs_114143||lt2gene Low-quality
S16_vs_114291||lt2gene Not-determined
S16_vs_114298||lt2gene Not-determined
S16_vs_114402||lt2gene Low-quality
S16_vs_114590||lt2gene Not-determined
S16_vs_114629||lt2gene Low-quality
S16_vs_114665||lt2gene Low-quality
S16_vs_114670||lt2gene Low-quality
S16_vs_114752||lt2gene Not-determined
S16_vs_114821||lt2gene Low-quality
S16_vs_114919||lt2gene Low-quality
S16_vs_114931||lt2gene Low-quality
S16_vs_114963||lt2gene Not-determined
S16_vs_114989||lt2gene Not-determined
S16_vs_115023||lt2gene Low-quality
S16_vs_115178||lt2gene Low-quality
S16_vs_115222||lt2gene Low-quality
S16_vs_115247||lt2gene Not-determined
S16_vs_115273||lt2gene Low-quality
S16_vs_115341||lt2gene Not-determined
S16_vs_115396||lt2gene Low-quality
S16_vs_115461||lt2gene Low-quality
S16_vs_115710||lt2gene Low-quality
S16_vs_115756||lt2gene Low-quality
S16_vs_115907||lt2gene Low-quality
S16_vs_115930||lt2gene Not-determined
S16_vs_115993||lt2gene Low-quality
S16_vs_116262||lt2gene Not-determined
S16_vs_116263||lt2gene Not-determined
S16_vs_116273||lt2gene Low-quality
S16_vs_116314||lt2gene Low-quality
S16_vs_116318||lt2gene Not-determined
S16_vs_116325||lt2gene Not-determined
S16_vs_116395||lt2gene Low-quality
S16_vs_116467||lt2gene Low-quality
S16_vs_116508||lt2gene Low-quality
S16_vs_116517||lt2gene Low-quality
S16_vs_116639||lt2gene Not-determined
S16_vs_117118||lt2gene Low-quality
S16_vs_117160||lt2gene Not-determined
S16_vs_117162||lt2gene Low-quality
S16_vs_117543||lt2gene Low-quality
S16_vs_117547||lt2gene Low-quality
S16_vs_117581||lt2gene Not-determined
S16_vs_117596||lt2gene Low-quality
S16_vs_118054||lt2gene Not-determined
S16_vs_118109||lt2gene Low-quality
S16_vs_118198||lt2gene Not-determined
S16_vs_118207||lt2gene Low-quality
S16_vs_118214||lt2gene Low-quality
S16_vs_118222||lt2gene Low-quality
S16_vs_118337||lt2gene Not-determined
S16_vs_118603||lt2gene Low-quality
S16_vs_118666||lt2gene Low-quality
S16_vs_118885||lt2gene Not-determined
S16_vs_118905||lt2gene Not-determined
S16_vs_119279||lt2gene Low-quality
S16_vs_119347||lt2gene Not-determined
S16_vs_119419||lt2gene Not-determined
S16_vs_119423||lt2gene Low-quality
S16_vs_119471||lt2gene Low-quality
S16_vs_119623||lt2gene Low-quality
S16_vs_119930||lt2gene Not-determined
S16_vs_119945||lt2gene Low-quality
S16_vs_119950||lt2gene Low-quality
S16_vs_120037||lt2gene Low-quality
S16_vs_120203||lt2gene Low-quality
S16_vs_120255||lt2gene Low-quality
S16_vs_120284||lt2gene Not-determined
S16_vs_120323||lt2gene Not-determined
S16_vs_120355||lt2gene Low-quality
S16_vs_120359||lt2gene Low-quality
S16_vs_120361||lt2gene Not-determined
S16_vs_120511||lt2gene Low-quality
S16_vs_120651||lt2gene Low-quality
S16_vs_120699||lt2gene Low-quality
S16_vs_120819||lt2gene Low-quality
S16_vs_121402||lt2gene Not-determined
S16_vs_121419||lt2gene Not-determined
S16_vs_121526||lt2gene Not-determined
S16_vs_121643||lt2gene Low-quality
S16_vs_121729||lt2gene Not-determined
S16_vs_121758||lt2gene Low-quality
S16_vs_121940||lt2gene Low-quality
S16_vs_122337||lt2gene Low-quality
S16_vs_122610||lt2gene Low-quality
S16_vs_122617||lt2gene Low-quality
S16_vs_122714||lt2gene Low-quality
S16_vs_122734||lt2gene Low-quality
S16_vs_122758||lt2gene Low-quality
S16_vs_122763||lt2gene Low-quality
S16_vs_122890||lt2gene Not-determined
S16_vs_123110||lt2gene Low-quality
S16_vs_123142||lt2gene Low-quality
S16_vs_123189||lt2gene Low-quality
S16_vs_123286||lt2gene Low-quality
S16_vs_123417||lt2gene Not-determined
S16_vs_123502||lt2gene Low-quality
S16_vs_123569||lt2gene Not-determined
S16_vs_123683||lt2gene Not-determined
S16_vs_123696||lt2gene Low-quality
S16_vs_123778||lt2gene Low-quality
S16_vs_123793||lt2gene Low-quality
S16_vs_124069||lt2gene Low-quality
S16_vs_124488||lt2gene Not-determined
S16_vs_124499||lt2gene Low-quality
S16_vs_124558||lt2gene Low-quality
S16_vs_124604||lt2gene Low-quality
S16_vs_124608||lt2gene Low-quality
S16_vs_124661||lt2gene Not-determined
S16_vs_124672||lt2gene Low-quality
S16_vs_124690||lt2gene Not-determined
S16_vs_124728||lt2gene Low-quality
S16_vs_124911||lt2gene Low-quality
S16_vs_124918||lt2gene Low-quality
S16_vs_124965||lt2gene Low-quality
S16_vs_125028||lt2gene Low-quality
S16_vs_125071||lt2gene Low-quality
S16_vs_125221||lt2gene Low-quality
S16_vs_125263||lt2gene Low-quality
S16_vs_125279||lt2gene Not-determined
S16_vs_125394||lt2gene Low-quality
S16_vs_125454||lt2gene Low-quality
S16_vs_125557||lt2gene Low-quality
S16_vs_125561||lt2gene Low-quality
S16_vs_125568||lt2gene Not-determined
S16_vs_125650||lt2gene Not-determined
S16_vs_126201||lt2gene Low-quality
S16_vs_126255||lt2gene Low-quality
S16_vs_126276||lt2gene Low-quality
S16_vs_126303||lt2gene Not-determined
S16_vs_126378||lt2gene Not-determined
S16_vs_126382||lt2gene Low-quality
S16_vs_126477||lt2gene Low-quality
S16_vs_126533||lt2gene Low-quality
S16_vs_126612||lt2gene Not-determined
S16_vs_126760||lt2gene Low-quality
S16_vs_126791||lt2gene Low-quality
S16_vs_126835||lt2gene Low-quality
S16_vs_126842||lt2gene Low-quality
S16_vs_126869||lt2gene Not-determined
S16_vs_127046||lt2gene Low-quality
S16_vs_127074||lt2gene Low-quality
S16_vs_127094||lt2gene Low-quality
S16_vs_127214||lt2gene Low-quality
S16_vs_127247||lt2gene Low-quality
S16_vs_127370||lt2gene Low-quality
S16_vs_127441||lt2gene Low-quality
S16_vs_127899||lt2gene Low-quality
S16_vs_128157||lt2gene Not-determined
S16_vs_128159||lt2gene Not-determined
S16_vs_128312||lt2gene Low-quality
S16_vs_128317||lt2gene Low-quality
S16_vs_128388||lt2gene Low-quality
S16_vs_128439||lt2gene Low-quality
S16_vs_128447||lt2gene Low-quality
S16_vs_128652||lt2gene Low-quality
S16_vs_128663||lt2gene Low-quality
S16_vs_128721||lt2gene Low-quality
S16_vs_128733||lt2gene Not-determined
S16_vs_128780||lt2gene Low-quality
S16_vs_128797||lt2gene Low-quality
S16_vs_128817||lt2gene Low-quality
S16_vs_129047||lt2gene Low-quality
S16_vs_129059||lt2gene Low-quality
S16_vs_129068||lt2gene Not-determined
S16_vs_129157||lt2gene Not-determined
S16_vs_129304||lt2gene Low-quality
S16_vs_129363||lt2gene Low-quality
S16_vs_129380||lt2gene Not-determined
S16_vs_129429||lt2gene Low-quality
S16_vs_129525||lt2gene Low-quality
S16_vs_129681||lt2gene Low-quality
S16_vs_129757||lt2gene Not-determined
S16_vs_129778||lt2gene Low-quality
S16_vs_129828||lt2gene Not-determined
S16_vs_129889||lt2gene Not-determined
S16_vs_129972||lt2gene Not-determined
S16_vs_129987||lt2gene Low-quality
S16_vs_130189||lt2gene Low-quality
S16_vs_130223||lt2gene Not-determined
S16_vs_130520||lt2gene Low-quality
S16_vs_130652||lt2gene Low-quality
S16_vs_130658||lt2gene Low-quality
S16_vs_130670||lt2gene Low-quality
S16_vs_130700||lt2gene Low-quality
S16_vs_130742||lt2gene Low-quality
S16_vs_130927||lt2gene Low-quality
S16_vs_130943||lt2gene Low-quality
S16_vs_130962||lt2gene Low-quality
S16_vs_131016||lt2gene Low-quality
S16_vs_131075||lt2gene Low-quality
S16_vs_131135||lt2gene Low-quality
S16_vs_131160||lt2gene Low-quality
S16_vs_131302||lt2gene Low-quality
S16_vs_131371||lt2gene Low-quality
S16_vs_131480||lt2gene Low-quality
S16_vs_131535||lt2gene Low-quality
S16_vs_131576||lt2gene Low-quality
S16_vs_131622||lt2gene Low-quality
S16_vs_131759||lt2gene Low-quality
S16_vs_131766||lt2gene Low-quality
S16_vs_131868||lt2gene Low-quality
S16_vs_131903||lt2gene Low-quality
S16_vs_132039||lt2gene Low-quality
S16_vs_132175||lt2gene Low-quality
S16_vs_132207||lt2gene Not-determined
S16_vs_132266||lt2gene Low-quality
S16_vs_132308||lt2gene Low-quality
S16_vs_132380||lt2gene Low-quality
S16_vs_132432||lt2gene Not-determined
S16_vs_132476||lt2gene Not-determined
S16_vs_132500||lt2gene Low-quality
S16_vs_132580||lt2gene Low-quality
S16_vs_132618||lt2gene Low-quality
S16_vs_132628||lt2gene Not-determined
S16_vs_132662||lt2gene Not-determined
S16_vs_132672||lt2gene Low-quality
S16_vs_132673||lt2gene Not-determined
S16_vs_132826||lt2gene Not-determined
S16_vs_132958||lt2gene Low-quality
S16_vs_133030||lt2gene Low-quality
S16_vs_133302||lt2gene Low-quality
S16_vs_133444||lt2gene Low-quality
S16_vs_133460||lt2gene Low-quality
S16_vs_133650||lt2gene Low-quality
S16_vs_133943||lt2gene Low-quality
S16_vs_133951||lt2gene Not-determined
S16_vs_134011||lt2gene Not-determined
S16_vs_134033||lt2gene Not-determined
S16_vs_134176||lt2gene Low-quality
S16_vs_134293||lt2gene Low-quality
S16_vs_134369||lt2gene Low-quality
S16_vs_134507||lt2gene Not-determined
S16_vs_134565||lt2gene Not-determined
S16_vs_134806||lt2gene Low-quality
S16_vs_134894||lt2gene Not-determined
S16_vs_134943||lt2gene Low-quality
S16_vs_134958||lt2gene Not-determined
S16_vs_134996||lt2gene Not-determined
S16_vs_135020||lt2gene Low-quality
S16_vs_135222||lt2gene Low-quality
S16_vs_135388||lt2gene Low-quality
S16_vs_135423||lt2gene Low-quality
S16_vs_135453||lt2gene Low-quality
S16_vs_135480||lt2gene Not-determined
S16_vs_135487||lt2gene Low-quality
S16_vs_135507||lt2gene Not-determined
S16_vs_135564||lt2gene Low-quality
S16_vs_135859||lt2gene Low-quality
S16_vs_135908||lt2gene Low-quality
S16_vs_136015||lt2gene Not-determined
S16_vs_136042||lt2gene Low-quality
S16_vs_136316||lt2gene Low-quality
S16_vs_136537||lt2gene Low-quality
S16_vs_136557||lt2gene Low-quality
S16_vs_136573||lt2gene Low-quality
S16_vs_136838||lt2gene Not-determined
S16_vs_136839||lt2gene Low-quality
S16_vs_136888||lt2gene Not-determined
S16_vs_136934||lt2gene Low-quality
S16_vs_137021||lt2gene Low-quality
S16_vs_137047||lt2gene Low-quality
S16_vs_137081||lt2gene Not-determined
S16_vs_137161||lt2gene Low-quality
S16_vs_137204||lt2gene Low-quality
S16_vs_137251||lt2gene Low-quality
S16_vs_137453||lt2gene Not-determined
S16_vs_137454||lt2gene Low-quality
S16_vs_137467||lt2gene Not-determined
S16_vs_137597||lt2gene Not-determined
S16_vs_137602||lt2gene Low-quality
S16_vs_137641||lt2gene Low-quality
S16_vs_137783||lt2gene Not-determined
S16_vs_137822||lt2gene Low-quality
S16_vs_138018||lt2gene Not-determined
S16_vs_138041||lt2gene Low-quality
S16_vs_138103||lt2gene Low-quality
S16_vs_138316||lt2gene Low-quality
S16_vs_138358||lt2gene Low-quality
S16_vs_138443||lt2gene Not-determined
S16_vs_138461||lt2gene Low-quality
S16_vs_138498||lt2gene Low-quality
S16_vs_138499||lt2gene Not-determined
S16_vs_138541||lt2gene Not-determined
S16_vs_138573||lt2gene Low-quality
S16_vs_138590||lt2gene Low-quality
S16_vs_138764||lt2gene Low-quality
S16_vs_138817||lt2gene Low-quality
S16_vs_138882||lt2gene Low-quality
S16_vs_138929||lt2gene Low-quality
S16_vs_138978||lt2gene Low-quality
S16_vs_139027||lt2gene Low-quality
S16_vs_139035||lt2gene Low-quality
S16_vs_139264||lt2gene Low-quality
S16_vs_139275||lt2gene Low-quality
S16_vs_139300||lt2gene Low-quality
S16_vs_139341||lt2gene Low-quality
S16_vs_139357||lt2gene Low-quality
S16_vs_139388||lt2gene Low-quality
S16_vs_139473||lt2gene Low-quality
S16_vs_139501||lt2gene Low-quality
S16_vs_139611||lt2gene Low-quality
S16_vs_139664||lt2gene Low-quality
S16_vs_139847||lt2gene Low-quality
S16_vs_139896||lt2gene Low-quality
S16_vs_139910||lt2gene Not-determined
S16_vs_139978||lt2gene Low-quality
S16_vs_140043||lt2gene Low-quality
S16_vs_140079||lt2gene Low-quality
S16_vs_140151||lt2gene Low-quality
S16_vs_140303||lt2gene Low-quality
S16_vs_140355||lt2gene Not-determined
S16_vs_140399||lt2gene Not-determined
S16_vs_140406||lt2gene Low-quality
S16_vs_140407||lt2gene Not-determined
S16_vs_140443||lt2gene Low-quality
S16_vs_140626||lt2gene Low-quality
S16_vs_140922||lt2gene Not-determined
S16_vs_140963||lt2gene Low-quality
S16_vs_141063||lt2gene Not-determined
S16_vs_141169||lt2gene Not-determined
S16_vs_141221||lt2gene Low-quality
S16_vs_141264||lt2gene Low-quality
S16_vs_141282||lt2gene Low-quality
S16_vs_141335||lt2gene Not-determined
S16_vs_141338||lt2gene Low-quality
S16_vs_141347||lt2gene Not-determined
S16_vs_141412||lt2gene Low-quality
S16_vs_141421||lt2gene Low-quality
S16_vs_141445||lt2gene Not-determined
S16_vs_141460||lt2gene Low-quality
S16_vs_141605||lt2gene Low-quality
S16_vs_141616||lt2gene Low-quality
S16_vs_141638||lt2gene Not-determined
S16_vs_141672||lt2gene Low-quality
S16_vs_141780||lt2gene Low-quality
S16_vs_141796||lt2gene Not-determined
S16_vs_141871||lt2gene Low-quality
S16_vs_141910||lt2gene Low-quality
S16_vs_141984||lt2gene Low-quality
S16_vs_142002||lt2gene Low-quality
S16_vs_142037||lt2gene Low-quality
S16_vs_142073||lt2gene Low-quality
S16_vs_142156||lt2gene Low-quality
S16_vs_142274||lt2gene Low-quality
S16_vs_142355||lt2gene Low-quality
S16_vs_142362||lt2gene Low-quality
S16_vs_142559||lt2gene Low-quality
S16_vs_142589||lt2gene Low-quality
S16_vs_142642||lt2gene Low-quality
S16_vs_142668||lt2gene Low-quality
S16_vs_142720||lt2gene Low-quality
S16_vs_142738||lt2gene Low-quality
S16_vs_142779||lt2gene Not-determined
S16_vs_142848||lt2gene Low-quality
S16_vs_142909||lt2gene Low-quality
S16_vs_142932||lt2gene Low-quality
S16_vs_143031||lt2gene Low-quality
S16_vs_143069||lt2gene Low-quality
S16_vs_143169||lt2gene Not-determined
S16_vs_143282||lt2gene Low-quality
S16_vs_143285||lt2gene Low-quality
S16_vs_143339||lt2gene Low-quality
S16_vs_143390||lt2gene Low-quality
S16_vs_143407||lt2gene Low-quality
S16_vs_143428||lt2gene Low-quality
S16_vs_143565||lt2gene Low-quality
S16_vs_143625||lt2gene Low-quality
S16_vs_143661||lt2gene Low-quality
S16_vs_143827||lt2gene Low-quality
S16_vs_143879||lt2gene Low-quality
S16_vs_144083||lt2gene Not-determined
S16_vs_144100||lt2gene Not-determined
S16_vs_144136||lt2gene Low-quality
S16_vs_144202||lt2gene Low-quality
S16_vs_144246||lt2gene Low-quality
S16_vs_144251||lt2gene Low-quality
S16_vs_144268||lt2gene Not-determined
S16_vs_144336||lt2gene Low-quality
S16_vs_144420||lt2gene Low-quality
S16_vs_144431||lt2gene Not-determined
S16_vs_144489||lt2gene Low-quality
S16_vs_144689||lt2gene Low-quality
S16_vs_144803||lt2gene Low-quality
S16_vs_144830||lt2gene Low-quality
S16_vs_144931||lt2gene Low-quality
S16_vs_144951||lt2gene Not-determined
S16_vs_145049||lt2gene Not-determined
S16_vs_145189||lt2gene Not-determined
S16_vs_145204||lt2gene Not-determined
S16_vs_145226||lt2gene Not-determined
S16_vs_145233||lt2gene Low-quality
S16_vs_145596||lt2gene Not-determined
S16_vs_145653||lt2gene Low-quality
S16_vs_145701||lt2gene Low-quality
S16_vs_145765||lt2gene Low-quality
S16_vs_145789||lt2gene Low-quality
S16_vs_145980||lt2gene Not-determined
S16_vs_146070||lt2gene Low-quality
S16_vs_146116||lt2gene Low-quality
S16_vs_146213||lt2gene Low-quality
S16_vs_146283||lt2gene Not-determined
S16_vs_146355||lt2gene Not-determined
S16_vs_146376||lt2gene Not-determined
S16_vs_146534||lt2gene Not-determined
S16_vs_146667||lt2gene Low-quality
S16_vs_146782||lt2gene Not-determined
S16_vs_146850||lt2gene Low-quality
S16_vs_146877||lt2gene Low-quality
S16_vs_147046||lt2gene Not-determined
S16_vs_147069||lt2gene Low-quality
S16_vs_147235||lt2gene Not-determined
S16_vs_147414||lt2gene Low-quality
S16_vs_147454||lt2gene Low-quality
S16_vs_147563||lt2gene Low-quality
S16_vs_147606||lt2gene Low-quality
S16_vs_147637||lt2gene Not-determined
S16_vs_147669||lt2gene Low-quality
S16_vs_147688||lt2gene Not-determined
S16_vs_147736||lt2gene Low-quality
S16_vs_147738||lt2gene Low-quality
S16_vs_147866||lt2gene Not-determined
S16_vs_147887||lt2gene Low-quality
S16_vs_147932||lt2gene Low-quality
S16_vs_147979||lt2gene Not-determined
S16_vs_148008||lt2gene Low-quality
S16_vs_148021||lt2gene Low-quality
S16_vs_148056||lt2gene Low-quality
S16_vs_148074||lt2gene Low-quality
S16_vs_148109||lt2gene Low-quality
S16_vs_148146||lt2gene Low-quality
S16_vs_148374||lt2gene Low-quality
S16_vs_148389||lt2gene Low-quality
S16_vs_148696||lt2gene Low-quality
S16_vs_148794||lt2gene Not-determined
S16_vs_148963||lt2gene Low-quality
S16_vs_149018||lt2gene Low-quality
S16_vs_149055||lt2gene Low-quality
S16_vs_149195||lt2gene Low-quality
S16_vs_149215||lt2gene Low-quality
S16_vs_149231||lt2gene Low-quality
S16_vs_149238||lt2gene Low-quality
S16_vs_149299||lt2gene Low-quality
S16_vs_149341||lt2gene Not-determined
S16_vs_149406||lt2gene Not-determined
S16_vs_149413||lt2gene Not-determined

S16_vs_149427||lt2gene Low-quality
S16_vs_149467||lt2gene Low-quality
S16_vs_149517||lt2gene Low-quality
S16_vs_149665||lt2gene Low-quality
S16_vs_149704||lt2gene Not-determined
S16_vs_149706||lt2gene Low-quality
S16_vs_149714||lt2gene Not-determined
S16_vs_149801||lt2gene Low-quality
S16_vs_149828||lt2gene Low-quality
S16_vs_149847||lt2gene Low-quality
S16_vs_149885||lt2gene Low-quality
S16_vs_149934||lt2gene Low-quality
S16_vs_149953||lt2gene Not-determined
S16_vs_149966||lt2gene Not-determined
S16_vs_149992||lt2gene Low-quality
S16_vs_150010||lt2gene Not-determined
S16_vs_150061||lt2gene Low-quality
S16_vs_150104||lt2gene Low-quality
S16_vs_150129||lt2gene Low-quality
S16_vs_150194||lt2gene Low-quality
S16_vs_150293||lt2gene Low-quality
S16_vs_150480||lt2gene Not-determined
S16_vs_150580||lt2gene Low-quality
S16_vs_150621||lt2gene Low-quality
S16_vs_150627||lt2gene Low-quality
S16_vs_150715||lt2gene Low-quality
S16_vs_150730||lt2gene Not-determined
S16_vs_150815||lt2gene Low-quality
S16_vs_150832||lt2gene Not-determined
S16_vs_150976||lt2gene Low-quality
S16_vs_151088||lt2gene Low-quality
S16_vs_151113||lt2gene Low-quality
S16_vs_151210||lt2gene Not-determined
S16_vs_151235||lt2gene Low-quality
S16_vs_151271||lt2gene Not-determined
S16_vs_151370||lt2gene Not-determined
S16_vs_151424||lt2gene Low-quality
S16_vs_151480||lt2gene Not-determined
S16_vs_151554||lt2gene Low-quality
S16_vs_151680||lt2gene Low-quality
S16_vs_151692||lt2gene Not-determined
S16_vs_151726||lt2gene Low-quality
S16_vs_151729||lt2gene Not-determined
S16_vs_151773||lt2gene Low-quality
S16_vs_151845||lt2gene Low-quality
S16_vs_151898||lt2gene Low-quality
S16_vs_152058||lt2gene Low-quality
S16_vs_152137||lt2gene Not-determined
S16_vs_152444||lt2gene Low-quality
S16_vs_152550||lt2gene Not-determined
S16_vs_152634||lt2gene Low-quality
S16_vs_152686||lt2gene Low-quality
S16_vs_152692||lt2gene Not-determined
S16_vs_152936||lt2gene Low-quality
S16_vs_152975||lt2gene Low-quality
S16_vs_153009||lt2gene Low-quality
S16_vs_153049||lt2gene Low-quality
S16_vs_153139||lt2gene Low-quality
S16_vs_153164||lt2gene Low-quality
S16_vs_153166||lt2gene Low-quality
S16_vs_153179||lt2gene Low-quality
S16_vs_153185||lt2gene Low-quality
S16_vs_153189||lt2gene Low-quality
S16_vs_153234||lt2gene Low-quality
S16_vs_153243||lt2gene Not-determined
S16_vs_153398||lt2gene Low-quality
S16_vs_153530||lt2gene Low-quality
S16_vs_153601||lt2gene Low-quality
S16_vs_153688||lt2gene Low-quality
S16_vs_153941||lt2gene Low-quality
S16_vs_154049||lt2gene Low-quality
S16_vs_154103||lt2gene Not-determined
S16_vs_154122||lt2gene Low-quality
S16_vs_154219||lt2gene Low-quality
S16_vs_154245||lt2gene Low-quality
S16_vs_154277||lt2gene Low-quality
S16_vs_154343||lt2gene Low-quality
S16_vs_154687||lt2gene Low-quality
S16_vs_154750||lt2gene Not-determined
S16_vs_154760||lt2gene Low-quality
S16_vs_154837||lt2gene Low-quality
S16_vs_154928||lt2gene Low-quality
S16_vs_154940||lt2gene Low-quality
S16_vs_154944||lt2gene Low-quality
S16_vs_154950||lt2gene Low-quality
S16_vs_154966||lt2gene Not-determined
S16_vs_154995||lt2gene Low-quality
S16_vs_155053||lt2gene Low-quality
S16_vs_155083||lt2gene Low-quality
S16_vs_155092||lt2gene Low-quality
S16_vs_155109||lt2gene Low-quality
S16_vs_155138||lt2gene Low-quality
S16_vs_155580||lt2gene Low-quality
S16_vs_155622||lt2gene Not-determined
S16_vs_155771||lt2gene Not-determined
S16_vs_155849||lt2gene Not-determined
S16_vs_155860||lt2gene Low-quality
S16_vs_155902||lt2gene Low-quality
S16_vs_155918||lt2gene Low-quality
S16_vs_155921||lt2gene Not-determined
S16_vs_155931||lt2gene Low-quality
S16_vs_156002||lt2gene Low-quality
S16_vs_156091||lt2gene Low-quality
S16_vs_156167||lt2gene Low-quality
S16_vs_156312||lt2gene Not-determined
S16_vs_156329||lt2gene Low-quality
S16_vs_156364||lt2gene Not-determined
S16_vs_156369||lt2gene Low-quality
S16_vs_156548||lt2gene Low-quality
S16_vs_156625||lt2gene Low-quality
S16_vs_156640||lt2gene Low-quality
S16_vs_156671||lt2gene Low-quality
S16_vs_156748||lt2gene Low-quality
S16_vs_156812||lt2gene Low-quality
S16_vs_156892||lt2gene Low-quality
S16_vs_156963||lt2gene Low-quality
S16_vs_157004||lt2gene Not-determined
S16_vs_157011||lt2gene Low-quality
S16_vs_157101||lt2gene Low-quality
S16_vs_157108||lt2gene Low-quality
S16_vs_157229||lt2gene Not-determined
S16_vs_157240||lt2gene Low-quality
S16_vs_157522||lt2gene Low-quality
S16_vs_157545||lt2gene Low-quality
S16_vs_157620||lt2gene Low-quality
S16_vs_157641||lt2gene Low-quality
S16_vs_157731||lt2gene Not-determined
S16_vs_157880||lt2gene Not-determined
S16_vs_157945||lt2gene Low-quality
S16_vs_157965||lt2gene Low-quality
S16_vs_157991||lt2gene Not-determined
S16_vs_158116||lt2gene Low-quality
S16_vs_158191||lt2gene Not-determined
S16_vs_158287||lt2gene Low-quality
S16_vs_158396||lt2gene Low-quality
S16_vs_158480||lt2gene Not-determined
S16_vs_158920||lt2gene Low-quality
S16_vs_158961||lt2gene Low-quality
S16_vs_158965||lt2gene Low-quality
S16_vs_158993||lt2gene Low-quality
S16_vs_159011||lt2gene Low-quality
S16_vs_159035||lt2gene Low-quality
S16_vs_159173||lt2gene Low-quality
S16_vs_159229||lt2gene Low-quality
S16_vs_159250||lt2gene Low-quality
S16_vs_159306||lt2gene Not-determined
S16_vs_159334||lt2gene Not-determined
S16_vs_159381||lt2gene Low-quality
S16_vs_159684||lt2gene Low-quality
S16_vs_159702||lt2gene Not-determined
S16_vs_159713||lt2gene Low-quality
S16_vs_159740||lt2gene Low-quality
S16_vs_159752||lt2gene Low-quality
S16_vs_159828||lt2gene Low-quality
S16_vs_159923||lt2gene Not-determined
S16_vs_159927||lt2gene Not-determined
S16_vs_160027||lt2gene Low-quality
S16_vs_160079||lt2gene Low-quality
S16_vs_160224||lt2gene Not-determined
S16_vs_160260||lt2gene Low-quality
S16_vs_160279||lt2gene Low-quality
S16_vs_160483||lt2gene Not-determined
S16_vs_160530||lt2gene Not-determined
S16_vs_160585||lt2gene Low-quality
S16_vs_160739||lt2gene Not-determined
S16_vs_160769||lt2gene Low-quality
S16_vs_160842||lt2gene Not-determined
S16_vs_160963||lt2gene Low-quality
S16_vs_161273||lt2gene Low-quality
S16_vs_161281||lt2gene Low-quality
S16_vs_161511||lt2gene Not-determined
S16_vs_161648||lt2gene Not-determined
S16_vs_161675||lt2gene Low-quality
S16_vs_161731||lt2gene Low-quality
S16_vs_161848||lt2gene Low-quality
S16_vs_161914||lt2gene Not-determined
S16_vs_162171||lt2gene Low-quality
S16_vs_162226||lt2gene Low-quality
S16_vs_162276||lt2gene Low-quality
S16_vs_162430||lt2gene Low-quality
S16_vs_162490||lt2gene Low-quality
S16_vs_162518||lt2gene Low-quality
S16_vs_162626||lt2gene Not-determined
S16_vs_162642||lt2gene Not-determined
S16_vs_162717||lt2gene Low-quality
S16_vs_162742||lt2gene Low-quality
S16_vs_162747||lt2gene Low-quality
S16_vs_162776||lt2gene Low-quality
S16_vs_162803||lt2gene Low-quality
S16_vs_162854||lt2gene Low-quality
S16_vs_162892||lt2gene Low-quality
S16_vs_163058||lt2gene Low-quality
S16_vs_163137||lt2gene Low-quality
S16_vs_163151||lt2gene Low-quality
S16_vs_163321||lt2gene Low-quality
S16_vs_163407||lt2gene Low-quality
S16_vs_163416||lt2gene Not-determined
S16_vs_163489||lt2gene Low-quality
S16_vs_163843||lt2gene Not-determined
S16_vs_163858||lt2gene Not-determined
S16_vs_163927||lt2gene Not-determined
S16_vs_164069||lt2gene Low-quality
S16_vs_164264||lt2gene Not-determined
S16_vs_164299||lt2gene Low-quality
S16_vs_164494||lt2gene Low-quality
S16_vs_164524||lt2gene Low-quality
S16_vs_164528||lt2gene Low-quality
S16_vs_164564||lt2gene Not-determined
S16_vs_164588||lt2gene Low-quality
S16_vs_164639||lt2gene Low-quality
S16_vs_164701||lt2gene Low-quality
S16_vs_164751||lt2gene Low-quality
S16_vs_164949||lt2gene Low-quality
S16_vs_164988||lt2gene Not-determined
S16_vs_165110||lt2gene Not-determined
S16_vs_165186||lt2gene Not-determined
S16_vs_165251||lt2gene Low-quality
S16_vs_165312||lt2gene Low-quality
S16_vs_165373||lt2gene Low-quality
S16_vs_165375||lt2gene Not-determined
S16_vs_165471||lt2gene Not-determined
S16_vs_165475||lt2gene Low-quality
S16_vs_165519||lt2gene Not-determined
S16_vs_165521||lt2gene Not-determined
S16_vs_165676||lt2gene Low-quality
S16_vs_166048||lt2gene Not-determined
S16_vs_166076||lt2gene Not-determined
S16_vs_166095||lt2gene Low-quality
S16_vs_166134||lt2gene Low-quality
S16_vs_166149||lt2gene Low-quality
S16_vs_166206||lt2gene Low-quality
S16_vs_166390||lt2gene Not-determined
S16_vs_166447||lt2gene Low-quality
S16_vs_166461||lt2gene Not-determined
S16_vs_166463||lt2gene Not-determined
S16_vs_166577||lt2gene Low-quality
S16_vs_166591||lt2gene Low-quality
S16_vs_166682||lt2gene Low-quality
S16_vs_166707||lt2gene Low-quality
S16_vs_166914||lt2gene Low-quality
S16_vs_166923||lt2gene Not-determined
S16_vs_166967||lt2gene Low-quality
S16_vs_166991||lt2gene Not-determined
S16_vs_167066||lt2gene Low-quality
S16_vs_167116||lt2gene Not-determined
S16_vs_167216||lt2gene Low-quality
S16_vs_167317||lt2gene Not-determined
S16_vs_167601||lt2gene Low-quality
S16_vs_167742||lt2gene Low-quality
S16_vs_167750||lt2gene Low-quality
S16_vs_167798||lt2gene Low-quality
S16_vs_167924||lt2gene Low-quality
S16_vs_167948||lt2gene Low-quality
S16_vs_167964||lt2gene Low-quality
S16_vs_167965||lt2gene Low-quality
S16_vs_167970||lt2gene Not-determined
S16_vs_168157||lt2gene Not-determined
S16_vs_168338||lt2gene Low-quality
S16_vs_168384||lt2gene Low-quality
S16_vs_168501||lt2gene Low-quality
S16_vs_168841||lt2gene Low-quality
S16_vs_168893||lt2gene Low-quality
S16_vs_169171||lt2gene Low-quality
S16_vs_169436||lt2gene Low-quality
S16_vs_169488||lt2gene Not-determined
S16_vs_169516||lt2gene Low-quality
S16_vs_169554||lt2gene Low-quality
S16_vs_169594||lt2gene Low-quality
S16_vs_169633||lt2gene Low-quality
S16_vs_169686||lt2gene Low-quality
S16_vs_169941||lt2gene Low-quality
S16_vs_170211||lt2gene Not-determined
S16_vs_170242||lt2gene Low-quality
S16_vs_170563||lt2gene Not-determined
S16_vs_170758||lt2gene Low-quality
S16_vs_170859||lt2gene Not-determined
S16_vs_171046||lt2gene Not-determined
S16_vs_171216||lt2gene Low-quality
S16_vs_171312||lt2gene Not-determined
S16_vs_171497||lt2gene Low-quality
S16_vs_171851||lt2gene Not-determined
S16_vs_171866||lt2gene Not-determined
S16_vs_171941||lt2gene Not-determined
S16_vs_172168||lt2gene Low-quality
S16_vs_172183||lt2gene Low-quality
S16_vs_172216||lt2gene Low-quality
S16_vs_172258||lt2gene Low-quality
S16_vs_172278||lt2gene Low-quality
S16_vs_172287||lt2gene Low-quality
S16_vs_172485||lt2gene Low-quality
S16_vs_172778||lt2gene Low-quality
S16_vs_172806||lt2gene Not-determined
S16_vs_172816||lt2gene Low-quality
S16_vs_172823||lt2gene Low-quality
S16_vs_172902||lt2gene Low-quality
S16_vs_173059||lt2gene Low-quality
S10_extracted_1 Not-determined
S10_extracted_11 Low-quality
S10_extracted_12 Not-determined
S10_extracted_15 Not-determined
S10_extracted_20 Not-determined
S10_extracted_28 Not-determined
S10_extracted_41 Not-determined
S10_extracted_49 Not-determined
S10_extracted_64 Not-determined
S10_extracted_74 Not-determined
S10_extracted_82 Not-determined
S10_extracted_83 Not-determined
S10_extracted_88 Not-determined
S10_extracted_101 Not-determined
S10_extracted_114 Low-quality
S10_extracted_143 Not-determined
S10_extracted_174 Low-quality
S10_extracted_182 Low-quality
S10_extracted_183 Not-determined
S10_extracted_189 Low-quality
S10_extracted_196 Not-determined
S10_extracted_211 Not-determined
S10_extracted_219 Low-quality
S10_extracted_228 Not-determined
S10_extracted_242 Not-determined
S10_extracted_250 Not-determined
S10_extracted_251 Not-determined
S10_extracted_279 Not-determined
S10_extracted_289 Not-determined
S10_extracted_295 Not-determined
S10_extracted_307 Not-determined
S10_extracted_312 Not-determined
S10_extracted_313 Low-quality
S10_extracted_315 Not-determined
S10_extracted_324 Not-determined
S10_extracted_344 Not-determined
S10_extracted_353 Not-determined
S10_extracted_365 Not-determined
S10_extracted_370 Not-determined
S10_extracted_373 Not-determined
S10_extracted_376 Not-determined
S10_extracted_377 Not-determined
S10_extracted_388 Not-determined
S10_extracted_399 Not-determined
S10_extracted_428 Not-determined
S10_extracted_432 Not-determined
S10_extracted_453 Not-determined
S10_extracted_491 Not-determined
S10_extracted_506 Low-quality
S10_extracted_529 Not-determined
S10_extracted_538 Not-determined
S10_extracted_564 Not-determined
S10_extracted_572 Not-determined
S10_extracted_595 Not-determined
S10_extracted_604 Not-determined
S10_extracted_613 Not-determined
S10_extracted_614 Not-determined
S10_extracted_622 Low-quality
S10_extracted_629 Not-determined
S10_extracted_650 Not-determined
S10_extracted_658 Not-determined
S10_extracted_659 Not-determined
S10_extracted_661 Not-determined
S10_extracted_678 Low-quality
S10_extracted_682 Not-determined
S10_extracted_703 Not-determined
S10_extracted_706 Low-quality
S10_extracted_727 Not-determined
S10_extracted_737 Low-quality
S10_extracted_744 Low-quality
S10_extracted_755 Not-determined
S10_extracted_780 Not-determined
S10_extracted_807 Not-determined
S10_extracted_816 Not-determined
S10_extracted_818 Not-determined
S10_extracted_821 Not-determined
S10_extracted_836 Not-determined
S10_extracted_874 Not-determined
S10_extracted_878 Low-quality
S10_extracted_890 Not-determined
S10_extracted_895 Not-determined
S10_extracted_900 Not-determined
S10_extracted_911 Not-determined
S10_extracted_919 Low-quality
S10_extracted_930 Not-determined
S10_extracted_932 Not-determined
S10_extracted_941 Not-determined
S10_extracted_945 Not-determined
S10_extracted_962 Not-determined
S10_extracted_996 Not-determined
S10_extracted_1004 Not-determined
S10_extracted_1012 Low-quality
S10_extracted_1014 Low-quality
S10_extracted_1019 Low-quality
S10_extracted_1020 Low-quality
S10_extracted_1034 Not-determined
S10_extracted_1038 Low-quality
S10_extracted_1043 Not-determined
S10_extracted_1044 Low-quality
S10_extracted_1060 Not-determined
S10_extracted_1067 Not-determined
S10_extracted_1068 Not-determined
S10_extracted_1078 Not-determined
S10_extracted_1081 Not-determined
S10_extracted_1102 Not-determined
S10_extracted_1109 Not-determined
S10_extracted_1127 Not-determined
S10_extracted_1138 Not-determined
S10_extracted_1149 Not-determined
S10_extracted_1160 Low-quality
S10_extracted_1167 Not-determined
S10_extracted_1171 Not-determined
S10_extracted_1176 Low-quality
S10_extracted_1178 Not-determined
S10_extracted_1191 Not-determined
S10_extracted_1196 Not-determined
S10_extracted_1198 Not-determined
S10_extracted_1207 Not-determined
S10_extracted_1219 Not-determined
S10_extracted_1220 Not-determined
S10_extracted_1235 Low-quality
S10_extracted_1244 Not-determined
S10_extracted_1248 Not-determined
S10_extracted_1253 Not-determined
S10_extracted_1262 Not-determined
S10_extracted_1275 Low-quality
S10_extracted_1276 Not-determined
S10_extracted_1292 Not-determined
S10_extracted_1299 Low-quality
S10_extracted_1305 Not-determined
S10_extracted_1316 Not-determined
S10_extracted_1321 Not-determined
S10_extracted_1346 Low-quality
S10_extracted_1347 Not-determined
S10_extracted_1355 Not-determined
S10_extracted_1359 Low-quality
S10_extracted_1374 Not-determined
S10_extracted_1375 Not-determined
S10_extracted_1382 Not-determined
S10_extracted_1384 Not-determined
S10_extracted_1392 Not-determined
S10_extracted_1401 Not-determined
S10_extracted_1405 Not-determined
S10_extracted_1407 Low-quality
S10_extracted_1412 Not-determined
S10_extracted_1415 Low-quality
S10_extracted_1423 Not-determined
S10_extracted_1427 Not-determined
S10_extracted_1440 Low-quality
S10_extracted_1443 Not-determined
S10_extracted_1452 Not-determined
S10_extracted_1461 Not-determined
S10_extracted_1484 Not-determined
S10_extracted_1486 Not-determined
S10_extracted_1518 Not-determined
S10_extracted_1519 Not-determined
S10_extracted_1520 Not-determined
S10_extracted_1523 Not-determined
S10_extracted_1532 Not-determined
S10_extracted_1537 Not-determined
S10_extracted_1542 Low-quality
S10_extracted_1547 Not-determined
S10_extracted_1553 Not-determined
S10_extracted_1575 Not-determined
S10_extracted_1590 Not-determined
S10_extracted_1619 Not-determined
S10_extracted_1632 Not-determined
S10_extracted_1643 Not-determined
S10_extracted_1649 Not-determined
S10_extracted_1661 Not-determined
S10_extracted_1670 Not-determined
S10_extracted_1685 Not-determined
S10_extracted_1688 Not-determined
S10_extracted_1696 Not-determined
S10_extracted_1704 Not-determined
S10_extracted_1722 Not-determined
S10_extracted_1726 Not-determined
S10_extracted_1729 Not-determined
S10_extracted_1740 Not-determined
S10_extracted_1752 Not-determined
S10_extracted_1754 Low-quality
S10_extracted_1758 Not-determined
S10_extracted_1768 Not-determined
S10_extracted_1780 Not-determined
S10_extracted_1791 Not-determined
S10_extracted_1806 Not-determined
S10_extracted_1808 Not-determined
S10_extracted_1840 Not-determined
S10_extracted_1867 Low-quality
S10_extracted_1878 Not-determined
S10_extracted_1892 Not-determined
S10_extracted_1905 Not-determined
S10_extracted_1906 Not-determined
S10_extracted_1923 Low-quality
S10_extracted_1928 Not-determined
S10_extracted_1944 Not-determined
S10_extracted_1958 Not-determined
S10_extracted_1977 Not-determined
S10_extracted_2002 Low-quality
S10_extracted_2010 Not-determined
S10_extracted_2016 Not-determined
S10_extracted_2038 Not-determined
S10_extracted_2042 Low-quality
S10_extracted_2045 Not-determined
S10_extracted_2048 Not-determined
S10_extracted_2068 Not-determined
S10_extracted_2086 Not-determined
S10_extracted_2088 Not-determined
S10_extracted_2112 Not-determined
S10_extracted_2121 Not-determined
S10_extracted_2125 Not-determined
S10_extracted_2141 Not-determined
S10_extracted_2157 Not-determined
S10_extracted_2163 Low-quality
S10_extracted_2168 Not-determined
S10_extracted_2169 Not-determined
S10_extracted_2172 Low-quality
S10_extracted_2174 Not-determined
S10_extracted_2180 Not-determined
S10_extracted_2188 Not-determined
S10_extracted_2223 Not-determined
S10_extracted_2248 Not-determined
S10_extracted_2259 Not-determined
S10_extracted_2269 Low-quality
S10_extracted_2275 Not-determined
S10_extracted_2314 Low-quality
S10_extracted_2329 Not-determined
S10_extracted_2360 Not-determined
S10_extracted_2368 Not-determined
S10_extracted_2373 Not-determined
S10_extracted_2384 Not-determined
S10_extracted_2390 Low-quality
S10_extracted_2404 Low-quality
S10_extracted_2420 Not-determined
S10_extracted_2423 Not-determined
S10_extracted_2443 Not-determined
S10_extracted_2451 Low-quality
S10_extracted_2457 Not-determined
S10_extracted_2489 Low-quality
S10_extracted_2501 Not-determined
S10_extracted_2519 Not-determined
S10_extracted_2523 Not-determined
S10_extracted_2543 Not-determined
S10_extracted_2557 Not-determined
S10_extracted_2579 Not-determined
S10_extracted_2588 Low-quality
S10_extracted_2589 Not-determined
S10_extracted_2593 Not-determined
S10_extracted_2597 Not-determined
S10_extracted_2598 Not-determined
S10_extracted_2601 Not-determined
S10_extracted_2609 Low-quality
S10_extracted_2626 Not-determined
S10_extracted_2631 Not-determined
S10_extracted_2632 Not-determined
S10_extracted_2633 Not-determined
S10_extracted_2634 Not-determined
S10_extracted_2644 Low-quality
S10_extracted_2665 Not-determined
S10_extracted_2669 Not-determined
S10_extracted_2684 Low-quality
S10_extracted_2693 Not-determined
S10_extracted_2694 Not-determined
S10_extracted_2710 Not-determined
S10_extracted_2730 Low-quality
S10_extracted_2747 Not-determined
S10_extracted_2755 Low-quality
S10_extracted_2760 Not-determined
S10_extracted_2768 Not-determined
S10_extracted_2791 Not-determined
S10_extracted_2792 Not-determined
S10_extracted_2794 Low-quality
S10_extracted_2820 Low-quality
S10_extracted_2849 Not-determined
S10_extracted_2854 Not-determined
S10_extracted_2873 Not-determined
S10_extracted_2892 Not-determined
S10_extracted_2926 Not-determined
S10_extracted_2933 Not-determined
S10_extracted_2942 Not-determined
S10_extracted_2970 Low-quality
S10_extracted_2972 Not-determined
S10_extracted_2973 Not-determined
S10_extracted_2980 Not-determined
S10_extracted_2983 Not-determined
S10_extracted_3009 Low-quality
S10_extracted_3010 Low-quality
S10_extracted_3016 Low-quality
S10_extracted_3018 Not-determined
S10_extracted_3042 Not-determined
S10_extracted_3055 Not-determined
S10_extracted_3056 Not-determined
S10_extracted_3061 Not-determined

| | |
|---|---|
| S10_extracted_3065 | Not-determined |
| S10_extracted_3084 | Not-determined |
| S10_extracted_3086 | Not-determined |
| S10_extracted_3093 | Not-determined |
| S10_extracted_3094 | Not-determined |
| S10_extracted_3121 | Not-determined |
| S10_extracted_3131 | Not-determined |
| S10_extracted_3140 | Low-quality |
| S10_extracted_3144 | Not-determined |
| S10_extracted_3146 | Not-determined |
| S10_extracted_3150 | Not-determined |
| S10_extracted_3158 | Not-determined |
| S10_extracted_3172 | Not-determined |
| S10_extracted_3178 | Not-determined |
| S10_extracted_3187 | Not-determined |
| S10_extracted_3191 | Not-determined |
| S10_extracted_3192 | Low-quality |
| S10_extracted_3198 | Not-determined |
| S10_extracted_3218 | Not-determined |
| S10_extracted_3219 | Not-determined |
| S10_extracted_3230 | Not-determined |
| S10_extracted_3233 | Not-determined |
| S10_extracted_3237 | Low-quality |
| S10_extracted_3240 | Not-determined |
| S10_extracted_3242 | Not-determined |
| S10_extracted_3251 | Not-determined |
| S10_extracted_3253 | Not-determined |
| S10_extracted_3255 | Low-quality |
| S10_extracted_3256 | Not-determined |
| S10_extracted_3267 | Low-quality |
| S10_extracted_3286 | Low-quality |
| S10_extracted_3287 | Not-determined |
| S10_extracted_3306 | Low-quality |
| S10_extracted_3309 | Not-determined |
| S10_extracted_3336 | Not-determined |
| S10_extracted_3338 | Not-determined |
| S10_extracted_3361 | Low-quality |
| S10_extracted_3369 | Not-determined |
| S10_extracted_3374 | Not-determined |
| S10_extracted_3377 | Not-determined |
| S10_extracted_3384 | Not-determined |
| S10_extracted_3387 | Not-determined |
| S10_extracted_3399 | Not-determined |
| S10_extracted_3402 | Not-determined |
| S10_extracted_3434 | Not-determined |
| S10_extracted_3435 | Not-determined |
| S10_extracted_3439 | Not-determined |
| S10_extracted_3446 | Low-quality |
| S10_extracted_3457 | Not-determined |
| S10_extracted_3465 | Not-determined |
| S10_extracted_3472 | Not-determined |
| S10_extracted_3481 | Low-quality |
| S10_extracted_3493 | Low-quality |
| S10_extracted_3549 | Low-quality |
| S10_extracted_3560 | Not-determined |
| S10_extracted_3561 | Not-determined |
| S10_extracted_3575 | Not-determined |
| S10_extracted_3588 | Not-determined |
| S10_extracted_3590 | Not-determined |
| S10_extracted_3600 | Not-determined |
| S10_extracted_3622 | Not-determined |
| S11_vs_k141_19892\|\|full | Medium-quality |
| S10_extracted_3672 | Not-determined |
| S10_extracted_3686 | Low-quality |
| S10_extracted_3695 | Not-determined |
| S10_extracted_3699 | Not-determined |
| S10_extracted_3700 | Not-determined |
| S10_extracted_3703 | Not-determined |
| S10_extracted_3712 | Not-determined |
| S10_extracted_3724 | Not-determined |
| S10_extracted_3738 | Not-determined |
| S10_extracted_3747 | Not-determined |
| S10_extracted_3784 | Not-determined |
| S10_extracted_3786 | Not-determined |
| S10_extracted_3811 | Not-determined |
| S10_extracted_3813 | Not-determined |
| S10_extracted_3824 | Low-quality |
| S10_extracted_3826 | Not-determined |
| S10_extracted_3868 | Not-determined |
| S10_extracted_3869 | Not-determined |
| S10_extracted_3895 | Not-determined |
| S10_extracted_3902 | Not-determined |
| S10_extracted_3910 | Not-determined |
| S10_extracted_3911 | Not-determined |
| S10_extracted_3914 | Not-determined |
| S10_extracted_3916 | Low-quality |
| S10_extracted_3918 | Not-determined |
| S10_extracted_3928 | Not-determined |
| S10_extracted_3932 | Not-determined |
| S10_extracted_3958 | Not-determined |
| S10_extracted_3973 | Not-determined |
| S10_extracted_3984 | Not-determined |
| S10_extracted_3986 | Not-determined |
| S10_extracted_3994 | Not-determined |
| S10_extracted_3995 | Not-determined |
| S10_extracted_4023 | Not-determined |
| S10_extracted_4048 | Not-determined |
| S10_extracted_4049 | Not-determined |
| S10_extracted_4054 | Not-determined |
| S10_extracted_4055 | Not-determined |
| S10_extracted_4062 | Low-quality |
| S10_extracted_4070 | Not-determined |
| S10_extracted_4075 | Not-determined |
| S10_extracted_4079 | Not-determined |
| S10_extracted_4081 | Not-determined |
| S10_extracted_4087 | Not-determined |
| S10_extracted_4088 | Not-determined |
| S10_extracted_4106 | Low-quality |
| S10_extracted_4111 | Not-determined |
| S10_extracted_4124 | Not-determined |
| S10_extracted_4127 | Not-determined |
| S10_extracted_4130 | Not-determined |
| S10_extracted_4143 | Not-determined |
| S10_extracted_4155 | Not-determined |
| S10_extracted_4179 | Low-quality |
| S10_extracted_4183 | Not-determined |
| S10_extracted_4188 | Not-determined |
| S10_extracted_4198 | Low-quality |
| S10_extracted_4212 | Not-determined |
| S10_extracted_4218 | Not-determined |
| S10_extracted_4229 | Not-determined |
| S10_extracted_4238 | Not-determined |
| S10_extracted_4252 | Not-determined |
| S10_extracted_4271 | Not-determined |
| S10_extracted_4273 | Not-determined |
| S10_extracted_4283 | Not-determined |
| S10_extracted_4297 | Not-determined |
| S10_extracted_4319 | Not-determined |
| S10_extracted_4323 | Not-determined |
| S10_extracted_4341 | Not-determined |
| S10_extracted_4345 | Not-determined |
| S10_extracted_4357 | Low-quality |
| S10_extracted_4364 | Low-quality |
| S10_extracted_4373 | Not-determined |
| S10_extracted_4381 | Not-determined |
| S10_extracted_4386 | Not-determined |
| S10_extracted_4387 | Low-quality |
| S10_extracted_4391 | Not-determined |
| S10_extracted_4393 | Not-determined |
| S10_extracted_4396 | Low-quality |
| S10_extracted_4420 | Not-determined |
| S10_extracted_4440 | Not-determined |
| S10_extracted_4462 | Low-quality |
| S10_extracted_4464 | Not-determined |
| S10_extracted_4471 | Not-determined |
| S10_extracted_4473 | Not-determined |
| S10_extracted_4494 | Not-determined |
| S10_extracted_4496 | Not-determined |
| S10_extracted_4514 | Not-determined |
| S10_extracted_4541 | Not-determined |
| S10_extracted_4567 | Not-determined |
| S10_extracted_4568 | Not-determined |
| S10_extracted_4573 | Not-determined |
| S10_extracted_4592 | Low-quality |
| S10_extracted_4593 | Not-determined |
| S10_extracted_4608 | Not-determined |
| S10_extracted_4616 | Not-determined |
| S10_extracted_4621 | Not-determined |
| S10_extracted_4638 | Not-determined |
| S10_extracted_4639 | Not-determined |
| S10_extracted_4659 | Not-determined |
| S10_extracted_4661 | Not-determined |
| S10_extracted_4665 | Not-determined |
| S10_extracted_4704 | Low-quality |
| S10_extracted_4706 | Low-quality |
| S10_extracted_4711 | Not-determined |
| S10_extracted_4712 | Not-determined |
| S10_extracted_4724 | Not-determined |
| S10_extracted_4726 | Not-determined |
| S10_extracted_4756 | Not-determined |
| S10_extracted_4757 | Not-determined |
| S10_extracted_4765 | Not-determined |
| S10_extracted_4772 | Not-determined |
| S10_extracted_4782 | Not-determined |
| S10_extracted_4798 | Not-determined |
| S10_extracted_4807 | Low-quality |
| S10_extracted_4815 | Not-determined |
| S10_extracted_4835 | Not-determined |
| S10_extracted_4841 | Low-quality |
| S10_extracted_4845 | Not-determined |
| S10_extracted_4855 | Not-determined |
| S10_extracted_4870 | Low-quality |
| S10_extracted_4874 | Not-determined |
| S10_extracted_4878 | Low-quality |
| S10_extracted_4879 | Not-determined |
| S10_extracted_4893 | Not-determined |
| S10_extracted_4896 | Not-determined |
| S10_extracted_4940 | Not-determined |
| S10_extracted_4967 | Not-determined |
| S10_extracted_4969 | Not-determined |
| S10_extracted_4974 | Not-determined |
| S10_extracted_4992 | Not-determined |
| S10_extracted_4998 | Not-determined |
| S10_extracted_5005 | Not-determined |
| S10_extracted_5017 | Not-determined |
| S10_extracted_5018 | Not-determined |
| S10_extracted_5021 | Not-determined |
| S10_extracted_5023 | Not-determined |
| S10_extracted_5032 | Not-determined |
| S10_extracted_5036 | Not-determined |
| S10_extracted_5039 | Not-determined |
| S10_extracted_5040 | Not-determined |
| S10_extracted_5053 | Not-determined |
| S10_extracted_5056 | Not-determined |
| S10_extracted_5066 | Not-determined |
| S10_extracted_5068 | Not-determined |
| S10_extracted_5071 | Not-determined |
| S10_extracted_5122 | Not-determined |
| S10_extracted_5129 | Not-determined |
| S10_extracted_5132 | Not-determined |
| S10_extracted_5134 | Not-determined |
| S10_extracted_5139 | Not-determined |
| S10_extracted_5147 | Not-determined |
| S10_extracted_5148 | Not-determined |
| S10_extracted_5149 | Not-determined |
| S10_extracted_5159 | Not-determined |
| S10_extracted_5165 | Not-determined |
| S10_extracted_5175 | Not-determined |
| S10_extracted_5181 | Not-determined |
| S10_extracted_5183 | Not-determined |
| S10_extracted_5199 | Not-determined |
| S10_extracted_5207 | Not-determined |
| S10_extracted_5208 | Not-determined |
| S10_extracted_5209 | Not-determined |
| S10_extracted_5213 | Not-determined |
| S10_extracted_5222 | Not-determined |
| S10_extracted_5227 | Not-determined |
| S10_extracted_5228 | Not-determined |
| S10_extracted_5237 | Not-determined |
| S10_extracted_5238 | Not-determined |
| S10_extracted_5255 | Not-determined |
| S10_extracted_5274 | Not-determined |
| S10_extracted_5303 | Not-determined |
| S10_extracted_5308 | Low-quality |
| S10_extracted_5316 | Not-determined |
| S10_extracted_5352 | Not-determined |
| S10_extracted_5373 | Not-determined |
| S10_extracted_5374 | Not-determined |
| S10_extracted_5392 | Not-determined |
| S10_extracted_5394 | Not-determined |
| S10_extracted_5396 | Not-determined |
| S10_extracted_5401 | Not-determined |
| S10_extracted_5403 | Not-determined |
| S10_extracted_5411 | Not-determined |
| S10_extracted_5422 | Low-quality |
| S10_extracted_5440 | Not-determined |
| S10_extracted_5445 | Not-determined |
| S10_extracted_5463 | Not-determined |
| S10_extracted_5467 | Not-determined |
| S10_extracted_5480 | Not-determined |
| S10_extracted_5486 | Not-determined |
| S10_extracted_5490 | Not-determined |
| S10_extracted_5501 | Low-quality |
| S10_extracted_5506 | Not-determined |
| S10_extracted_5508 | Not-determined |
| S10_extracted_5514 | Not-determined |
| S10_extracted_5515 | Not-determined |
| S10_extracted_5522 | Not-determined |
| S10_extracted_5534 | Low-quality |
| S10_extracted_5542 | Not-determined |
| S10_extracted_5550 | Low-quality |
| S10_extracted_5566 | Not-determined |
| S10_extracted_5574 | Not-determined |
| S10_extracted_5586 | Not-determined |
| S10_extracted_5590 | Not-determined |
| S10_extracted_5592 | Low-quality |
| S10_extracted_5609 | Low-quality |
| S10_extracted_5639 | Low-quality |
| S10_extracted_5647 | Not-determined |
| S10_extracted_5651 | Not-determined |
| S10_extracted_5652 | Not-determined |
| S10_extracted_5660 | Low-quality |
| S10_extracted_5667 | Not-determined |
| S10_extracted_5677 | Not-determined |
| S10_extracted_5691 | Not-determined |
| S10_extracted_5695 | Not-determined |
| S10_extracted_5700 | Not-determined |
| S10_extracted_5706 | Low-quality |
| S10_extracted_5717 | Not-determined |
| S10_extracted_5723 | Not-determined |
| S10_extracted_5727 | Not-determined |
| S10_extracted_5736 | Not-determined |
| S10_extracted_5752 | Low-quality |
| S10_extracted_5756 | Low-quality |
| S10_extracted_5759 | Not-determined |
| S10_extracted_5762 | Not-determined |
| S10_extracted_5772 | Not-determined |
| S10_extracted_5776 | Not-determined |
| S10_extracted_5777 | Not-determined |
| S10_extracted_5779 | Not-determined |
| S10_extracted_5781 | Not-determined |
| S10_extracted_5783 | Not-determined |
| S10_extracted_5784 | Not-determined |
| S10_extracted_5786 | Not-determined |
| S10_extracted_5797 | Not-determined |
| S10_extracted_5805 | Not-determined |
| S10_extracted_5829 | Not-determined |
| S10_extracted_5830 | Not-determined |
| S10_extracted_5840 | Not-determined |
| S10_extracted_5843 | Not-determined |
| S10_extracted_5856 | Not-determined |
| S10_extracted_5866 | Not-determined |
| S10_extracted_5877 | Not-determined |
| S10_extracted_5901 | Not-determined |
| S10_extracted_5907 | Not-determined |
| S10_extracted_5910 | Not-determined |
| S10_extracted_5920 | Not-determined |
| S10_extracted_5937 | Low-quality |
| S10_extracted_5938 | Not-determined |
| S10_extracted_5953 | Not-determined |
| S10_extracted_5970 | Not-determined |
| S10_extracted_6002 | Not-determined |
| S10_extracted_6006 | Not-determined |
| S10_extracted_6019 | Not-determined |
| S10_extracted_6050 | Not-determined |
| S10_extracted_6056 | Not-determined |
| S10_extracted_6074 | Not-determined |
| S10_extracted_6083 | Not-determined |
| S10_extracted_6091 | Not-determined |
| S10_extracted_6111 | Not-determined |
| S10_extracted_6138 | Not-determined |
| S10_extracted_6191 | Not-determined |
| S10_extracted_6193 | Not-determined |
| S10_extracted_6211 | Not-determined |
| S10_extracted_6231 | Not-determined |
| S10_extracted_6240 | Not-determined |
| S10_extracted_6242 | Not-determined |
| S10_extracted_6251 | Low-quality |
| S10_extracted_6263 | Not-determined |
| S10_extracted_6265 | Not-determined |
| S10_extracted_6273 | Low-quality |
| S10_extracted_6280 | Not-determined |
| S10_extracted_6282 | Not-determined |
| S10_extracted_6298 | Low-quality |
| S10_extracted_6301 | Not-determined |
| S10_extracted_6320 | Not-determined |
| S10_extracted_6331 | Low-quality |
| S10_extracted_6343 | Not-determined |
| S10_extracted_6344 | Low-quality |
| S10_extracted_6351 | Not-determined |
| S10_extracted_6363 | Low-quality |
| S10_extracted_6385 | Not-determined |
| S10_extracted_6395 | Low-quality |
| S10_extracted_6400 | Not-determined |
| S10_extracted_6414 | Not-determined |
| S10_extracted_6419 | Not-determined |
| S10_extracted_6427 | Not-determined |
| S10_extracted_6430 | Not-determined |
| S10_extracted_6432 | Low-quality |
| S10_extracted_6439 | Not-determined |
| S10_extracted_6449 | Low-quality |
| S10_extracted_6459 | Not-determined |
| S10_extracted_6491 | Not-determined |
| S10_extracted_6498 | Not-determined |
| S10_extracted_6516 | Not-determined |
| S10_extracted_6520 | Not-determined |
| S10_extracted_6528 | Not-determined |
| S10_extracted_6603 | Not-determined |
| S10_extracted_6648 | Not-determined |
| S10_extracted_6662 | Not-determined |
| S10_extracted_6665 | Low-quality |
| S10_extracted_6669 | Not-determined |
| S10_extracted_6673 | Not-determined |
| S10_extracted_6681 | Low-quality |
| S10_extracted_6687 | Not-determined |
| S10_extracted_6695 | Low-quality |
| S10_extracted_6700 | Low-quality |
| S10_extracted_6706 | Not-determined |
| S10_extracted_6727 | Not-determined |
| S10_extracted_6740 | Low-quality |
| S10_extracted_6758 | Not-determined |
| S10_extracted_6765 | Not-determined |
| S10_extracted_6767 | Not-determined |
| S10_extracted_6775 | Not-determined |
| S10_extracted_6783 | Low-quality |
| S10_extracted_6790 | Not-determined |
| S10_extracted_6794 | Not-determined |
| S10_extracted_6801 | Not-determined |
| S10_extracted_6806 | Not-determined |
| S10_extracted_6810 | Not-determined |
| S10_extracted_6818 | Not-determined |
| S10_extracted_6825 | Not-determined |
| S10_extracted_6834 | Not-determined |
| S10_extracted_6837 | Low-quality |
| S10_extracted_6838 | Low-quality |
| S10_extracted_6841 | Not-determined |
| S10_extracted_6851 | Not-determined |
| S10_extracted_6878 | Not-determined |
| S10_extracted_6880 | Not-determined |
| S10_extracted_6886 | Not-determined |
| S10_extracted_6897 | Not-determined |
| S10_extracted_6917 | Not-determined |
| S10_extracted_6953 | Low-quality |
| S10_extracted_6961 | Not-determined |
| S10_extracted_6968 | Not-determined |
| S10_extracted_7001 | Not-determined |
| S10_extracted_7024 | Not-determined |
| S10_extracted_7056 | Not-determined |
| S10_extracted_7074 | Not-determined |
| S10_extracted_7083 | Low-quality |
| S10_extracted_7091 | Not-determined |
| S10_extracted_7111 | Not-determined |
| S10_extracted_7125 | Not-determined |
| S10_extracted_7154 | Not-determined |
| S10_extracted_7155 | Not-determined |
| S10_extracted_7159 | Not-determined |
| S10_extracted_7168 | Low-quality |
| S10_extracted_7169 | Not-determined |
| S10_extracted_7178 | Not-determined |
| S10_extracted_7201 | Not-determined |
| S10_extracted_7224 | Low-quality |
| S10_extracted_7239 | Not-determined |
| S10_extracted_7255 | Not-determined |
| S10_extracted_7268 | Low-quality |
| S10_extracted_7269 | Not-determined |
| S10_extracted_7271 | Not-determined |
| S10_extracted_7282 | Not-determined |
| S10_extracted_7301 | Not-determined |
| S10_extracted_7302 | Not-determined |
| S10_extracted_7318 | Not-determined |
| S10_extracted_7319 | Not-determined |
| S10_extracted_7325 | Not-determined |
| S10_extracted_7329 | Not-determined |
| S10_extracted_7336 | Not-determined |
| S10_extracted_7339 | Low-quality |
| S10_extracted_7366 | Not-determined |
| S10_extracted_7370 | Not-determined |
| S10_extracted_7371 | Low-quality |
| S10_extracted_7395 | Not-determined |
| S10_extracted_7419 | Low-quality |
| S10_extracted_7431 | Not-determined |
| S10_extracted_7459 | Not-determined |
| S10_extracted_7467 | Not-determined |
| S10_extracted_7476 | Not-determined |
| S10_extracted_7479 | Not-determined |
| S10_extracted_7499 | Low-quality |
| S10_extracted_7509 | Not-determined |
| S10_extracted_7514 | Low-quality |
| S10_extracted_7535 | Not-determined |
| S10_extracted_7542 | Not-determined |
| S10_extracted_7555 | Not-determined |
| S10_extracted_7562 | Not-determined |
| S10_extracted_7566 | Not-determined |
| S10_extracted_7585 | Low-quality |
| S10_extracted_7594 | Not-determined |
| S10_extracted_7602 | Not-determined |
| S10_extracted_7618 | Not-determined |
| S10_extracted_7639 | Not-determined |
| S10_extracted_7644 | Not-determined |
| S10_extracted_7653 | Not-determined |
| S10_extracted_7662 | Not-determined |
| S10_extracted_7665 | Not-determined |
| S10_extracted_7667 | Not-determined |
| S10_extracted_7676 | Not-determined |
| S10_extracted_7679 | Not-determined |
| S10_extracted_7695 | Not-determined |
| S10_extracted_7730 | Not-determined |
| S10_extracted_7734 | Not-determined |
| S10_extracted_7753 | Not-determined |
| S10_extracted_7760 | Not-determined |
| S10_extracted_7763 | Not-determined |
| S10_extracted_7766 | Not-determined |
| S10_extracted_7778 | Not-determined |
| S10_extracted_7782 | Not-determined |
| S10_extracted_7789 | Not-determined |
| S10_extracted_7792 | Not-determined |
| S10_extracted_7797 | Low-quality |
| S10_extracted_7809 | Not-determined |
| S10_extracted_7822 | Not-determined |
| S10_extracted_7825 | Not-determined |
| S10_extracted_7840 | Not-determined |
| S10_extracted_7848 | Not-determined |
| S10_extracted_7859 | Not-determined |
| S10_extracted_7874 | Not-determined |
| S10_extracted_7910 | Not-determined |
| S10_extracted_7913 | Not-determined |
| S10_extracted_7914 | Not-determined |
| S10_extracted_7923 | Not-determined |
| S10_extracted_7924 | Not-determined |
| S10_extracted_7929 | Not-determined |
| S10_extracted_7931 | Not-determined |
| S10_extracted_7936 | Not-determined |
| S10_extracted_7940 | Not-determined |
| S10_extracted_7945 | Low-quality |
| S10_extracted_7950 | Low-quality |
| S10_extracted_7952 | Not-determined |
| S10_extracted_7970 | Not-determined |
| S10_extracted_7971 | Not-determined |
| S10_extracted_7976 | Not-determined |
| S10_extracted_8000 | Not-determined |
| S10_extracted_8008 | Not-determined |
| S10_extracted_8024 | Not-determined |
| S10_extracted_8027 | Low-quality |
| S10_extracted_8028 | Not-determined |
| S10_extracted_8036 | Low-quality |
| S10_extracted_8042 | Not-determined |
| S10_extracted_8060 | Not-determined |
| S10_extracted_8089 | Low-quality |
| S10_extracted_8091 | Not-determined |
| S10_extracted_8093 | Not-determined |
| S10_extracted_8098 | Not-determined |
| S10_extracted_8102 | Not-determined |
| S10_extracted_8103 | Not-determined |
| S10_extracted_8118 | Not-determined |
| S10_extracted_8133 | Not-determined |
| S10_extracted_8140 | Not-determined |
| S10_extracted_8150 | Not-determined |
| S10_extracted_8167 | Not-determined |
| S10_extracted_8179 | Not-determined |
| S10_extracted_8180 | Not-determined |
| S10_extracted_8184 | Not-determined |
| S10_extracted_8195 | Low-quality |
| S10_extracted_8218 | Not-determined |
| S10_extracted_8223 | Not-determined |
| S10_extracted_8231 | Not-determined |
| S10_extracted_8256 | Low-quality |
| S10_extracted_8258 | Low-quality |
| S10_extracted_8262 | Not-determined |
| S10_extracted_8318 | Not-determined |
| S10_extracted_8331 | Not-determined |
| S10_extracted_8338 | Not-determined |
| S10_extracted_8374 | Not-determined |
| S10_extracted_8389 | Not-determined |
| S10_extracted_8415 | Not-determined |
| S10_extracted_8440 | Not-determined |
| S10_extracted_8447 | Not-determined |
| S10_extracted_8448 | Not-determined |
| S10_extracted_8455 | Not-determined |
| S10_extracted_8469 | Not-determined |
| S10_extracted_8473 | Not-determined |
| S10_extracted_8505 | Not-determined |
| S10_extracted_8507 | Not-determined |
| S10_extracted_8512 | Low-quality |
| S10_extracted_8525 | Not-determined |
| S10_extracted_8528 | Not-determined |
| S10_extracted_8537 | Not-determined |
| S10_extracted_8541 | Not-determined |
| S10_extracted_8544 | Low-quality |
| S10_extracted_8552 | Not-determined |
| S10_extracted_8554 | Low-quality |
| S10_extracted_8555 | Not-determined |
| S10_extracted_8576 | Not-determined |
| S10_extracted_8579 | Not-determined |
| S10_extracted_8592 | Not-determined |
| S10_extracted_8603 | Not-determined |
| S10_extracted_8605 | Not-determined |
| S10_extracted_8611 | Low-quality |
| S10_extracted_8612 | Not-determined |
| S10_extracted_8628 | Not-determined |
| S10_extracted_8642 | Not-determined |
| S10_extracted_8650 | Not-determined |
| S10_extracted_8654 | Not-determined |
| S10_extracted_8691 | Not-determined |
| S10_extracted_8693 | Low-quality |
| S10_extracted_8700 | Not-determined |
| S10_extracted_8701 | Not-determined |
| S10_extracted_8717 | Not-determined |
| S10_extracted_8723 | Not-determined |
| S10_extracted_8728 | Not-determined |
| S10_extracted_8731 | Not-determined |
| S10_extracted_8752 | Not-determined |
| S10_extracted_8764 | Not-determined |
| S10_extracted_8792 | Not-determined |
| S10_extracted_8798 | Not-determined |
| S10_extracted_8799 | Not-determined |
| S10_extracted_8808 | Not-determined |
| S10_extracted_8811 | Not-determined |
| S10_extracted_8833 | Not-determined |
| S10_extracted_8837 | Low-quality |
| S10_extracted_8860 | Not-determined |
| S10_extracted_8862 | Low-quality |
| S10_extracted_8864 | Not-determined |
| S10_extracted_8869 | Low-quality |
| S10_extracted_8876 | Not-determined |
| S10_extracted_8883 | Not-determined |
| S10_extracted_8886 | Not-determined |
| S10_extracted_8897 | Low-quality |
| S10_extracted_8902 | Not-determined |
| S10_extracted_8909 | Not-determined |
| S10_extracted_8910 | Not-determined |
| S10_extracted_8919 | Not-determined |
| S10_extracted_8927 | Not-determined |
| S10_extracted_8935 | Not-determined |
| S10_extracted_8942 | Not-determined |
| S10_extracted_8943 | Not-determined |
| S10_extracted_8947 | Not-determined |
| S10_extracted_8948 | Not-determined |
| S10_extracted_8952 | Low-quality |
| S10_extracted_8977 | Not-determined |

| | |
|---|---|
| S10_extracted_8993 | Not-determined |
| S10_extracted_8995 | Not-determined |
| S10_extracted_8996 | Low-quality |
| S10_extracted_9011 | Not-determined |
| S10_extracted_9019 | Not-determined |
| S10_extracted_9040 | Not-determined |
| S10_extracted_9046 | Not-determined |
| S10_extracted_9060 | Not-determined |
| S10_extracted_9061 | Not-determined |
| S10_extracted_9067 | Not-determined |
| S10_extracted_9084 | Not-determined |
| S10_extracted_9105 | Not-determined |
| S10_extracted_9124 | Not-determined |
| S10_extracted_9144 | Not-determined |
| S10_extracted_9164 | Not-determined |
| S10_extracted_9165 | Not-determined |
| S10_extracted_9167 | Not-determined |
| S10_extracted_9178 | Not-determined |
| S10_extracted_9204 | Not-determined |
| S10_extracted_9206 | Not-determined |
| S10_extracted_9233 | Not-determined |
| S10_extracted_9236 | Not-determined |
| S10_extracted_9243 | Not-determined |
| S10_extracted_9246 | Low-quality |
| S10_extracted_9247 | Not-determined |
| S10_extracted_9258 | Not-determined |
| S10_extracted_9293 | Low-quality |
| S10_extracted_9294 | Not-determined |
| S10_extracted_9320 | Not-determined |
| S10_extracted_9331 | Not-determined |
| S10_extracted_9337 | Not-determined |
| S10_extracted_9361 | Not-determined |
| S10_extracted_9369 | Not-determined |
| S10_extracted_9373 | Not-determined |
| S10_extracted_9398 | Not-determined |
| S10_extracted_9404 | Not-determined |
| S10_extracted_9414 | Not-determined |
| S10_extracted_9418 | Not-determined |
| S10_extracted_9420 | Low-quality |
| S10_extracted_9431 | Low-quality |
| S10_extracted_9440 | Not-determined |
| S10_extracted_9452 | Not-determined |
| S10_extracted_9464 | Not-determined |
| S10_extracted_9468 | Low-quality |
| S10_extracted_9469 | Not-determined |
| S10_extracted_9520 | Not-determined |
| S10_extracted_9550 | Not-determined |
| S10_extracted_9567 | Low-quality |
| S10_extracted_9574 | Not-determined |
| S10_extracted_9580 | Not-determined |
| S10_extracted_9591 | Low-quality |
| S10_extracted_9592 | Not-determined |
| S10_extracted_9595 | Not-determined |
| S10_extracted_9604 | Not-determined |
| S10_extracted_9613 | Not-determined |
| S10_extracted_9619 | Not-determined |
| S10_extracted_9639 | Not-determined |
| S10_extracted_9659 | Not-determined |
| S10_extracted_9666 | Not-determined |
| S10_extracted_9668 | Low-quality |
| S10_extracted_9670 | Not-determined |
| S10_extracted_9680 | Not-determined |
| S10_extracted_9692 | Not-determined |
| S10_extracted_9703 | Not-determined |
| S10_extracted_9709 | Not-determined |
| S10_extracted_9718 | Not-determined |
| S10_extracted_9723 | Not-determined |
| S10_extracted_9726 | Not-determined |
| S10_extracted_9736 | Not-determined |
| S10_extracted_9739 | Low-quality |
| S10_extracted_9753 | Not-determined |
| S10_extracted_9771 | Not-determined |
| S10_extracted_9776 | Not-determined |
| S10_extracted_9780 | Not-determined |
| S10_extracted_9796 | Low-quality |
| S10_extracted_9812 | Low-quality |
| S10_extracted_9834 | Not-determined |
| S10_extracted_9847 | Not-determined |
| S10_extracted_9851 | Not-determined |
| S10_extracted_9857 | Not-determined |
| S10_extracted_9873 | Not-determined |
| S10_extracted_9879 | Not-determined |
| S10_extracted_9882 | Not-determined |
| S10_extracted_9898 | Not-determined |
| S10_extracted_9911 | Not-determined |
| S10_extracted_9914 | Low-quality |
| S10_extracted_9915 | Not-determined |
| S10_extracted_9923 | Not-determined |
| S10_extracted_9927 | Not-determined |
| S10_extracted_9952 | Not-determined |
| S10_extracted_9970 | Not-determined |
| S10_extracted_9978 | Low-quality |
| S10_extracted_9987 | Not-determined |
| S10_extracted_9994 | Not-determined |
| S10_extracted_9995 | Not-determined |
| S10_extracted_10015 | Not-determined |
| S10_extracted_10030 | Not-determined |
| S10_extracted_10031 | Not-determined |
| S10_extracted_10050 | Not-determined |
| S10_extracted_10052 | Low-quality |
| S10_extracted_10070 | Not-determined |
| S10_extracted_10079 | Low-quality |
| S10_extracted_10085 | Not-determined |
| S10_extracted_10114 | Not-determined |
| S10_extracted_10129 | Not-determined |
| S10_extracted_10138 | Not-determined |
| S10_extracted_10151 | Not-determined |
| S10_extracted_10167 | Not-determined |
| S10_extracted_10197 | Not-determined |
| S10_extracted_10198 | Low-quality |
| S10_extracted_10217 | Not-determined |
| S10_extracted_10231 | Not-determined |
| S10_extracted_10243 | Not-determined |
| S10_extracted_10281 | Not-determined |
| S10_extracted_10282 | Not-determined |
| S10_extracted_10284 | Not-determined |
| S10_extracted_10286 | Not-determined |
| S10_extracted_10299 | Not-determined |
| S10_extracted_10300 | Not-determined |
| S10_extracted_10305 | Not-determined |
| S10_extracted_10310 | Not-determined |
| S10_extracted_10321 | Low-quality |
| S10_extracted_10322 | Not-determined |
| S10_extracted_10330 | Not-determined |
| S10_extracted_10332 | Not-determined |
| S10_extracted_10340 | Not-determined |
| S10_extracted_10342 | Not-determined |
| S10_extracted_10381 | Not-determined |
| S10_extracted_10382 | Not-determined |
| S10_extracted_10398 | Not-determined |
| S10_extracted_10401 | Not-determined |
| S10_extracted_10402 | Not-determined |
| S10_extracted_10408 | Not-determined |
| S10_extracted_10410 | Not-determined |
| S10_extracted_10417 | Not-determined |
| S10_extracted_10419 | Not-determined |
| S10_extracted_10456 | Not-determined |
| S10_extracted_10460 | Not-determined |
| S10_extracted_10500 | Not-determined |
| S10_extracted_10503 | Not-determined |
| S10_extracted_10518 | Not-determined |
| S10_extracted_10521 | Not-determined |
| S10_extracted_10527 | Low-quality |
| S10_extracted_10531 | Not-determined |
| S10_extracted_10551 | Not-determined |
| S10_extracted_10556 | Not-determined |
| S10_extracted_10566 | Not-determined |
| S10_extracted_10590 | Not-determined |
| S10_extracted_10599 | Not-determined |
| S10_extracted_10607 | Not-determined |
| S10_extracted_10647 | Not-determined |
| S10_extracted_10656 | Not-determined |
| S10_extracted_10676 | Not-determined |
| S10_extracted_10690 | Not-determined |
| S10_extracted_10691 | Not-determined |
| S10_extracted_10734 | Low-quality |
| S10_extracted_10735 | Not-determined |
| S10_extracted_10743 | Low-quality |
| S10_extracted_10747 | Not-determined |
| S10_extracted_10754 | Not-determined |
| S10_extracted_10768 | Not-determined |
| S10_extracted_10787 | Not-determined |
| S10_extracted_10789 | Not-determined |
| S10_extracted_10796 | Not-determined |
| S10_extracted_10824 | Not-determined |
| S10_extracted_10849 | Not-determined |
| S10_extracted_10852 | Not-determined |
| S10_extracted_10854 | Low-quality |
| S10_extracted_10857 | Low-quality |
| S10_extracted_10867 | Not-determined |
| S10_extracted_10869 | Low-quality |
| S10_extracted_10878 | Not-determined |
| S10_extracted_10879 | Not-determined |
| S10_extracted_10886 | Not-determined |
| S10_extracted_10891 | Not-determined |
| S10_extracted_10895 | Not-determined |
| S10_extracted_10904 | Not-determined |
| S10_extracted_10916 | Not-determined |
| S10_extracted_10950 | Not-determined |
| S10_extracted_10955 | Not-determined |
| S10_extracted_10957 | Not-determined |
| S10_extracted_10963 | Not-determined |
| S10_extracted_10972 | Not-determined |
| S10_extracted_10982 | Not-determined |
| S10_extracted_11006 | Not-determined |
| S10_extracted_11012 | Low-quality |
| S10_extracted_11033 | Not-determined |
| S10_extracted_11044 | Not-determined |
| S10_extracted_11049 | Not-determined |
| S10_extracted_11071 | Not-determined |
| S10_extracted_11072 | Not-determined |
| S10_extracted_11076 | Not-determined |
| S10_extracted_11082 | Not-determined |
| S10_extracted_11089 | Low-quality |
| S10_extracted_11092 | Low-quality |
| S10_extracted_11110 | Not-determined |
| S10_extracted_11121 | Not-determined |
| S10_extracted_11128 | Not-determined |
| S10_extracted_11130 | Not-determined |
| S10_extracted_11132 | Not-determined |
| S10_extracted_11141 | Low-quality |
| S10_extracted_11145 | Not-determined |
| S10_extracted_11156 | Not-determined |
| S10_extracted_11169 | Not-determined |
| S10_extracted_11191 | Not-determined |
| S10_extracted_11198 | Not-determined |
| S10_extracted_11201 | Not-determined |
| S10_extracted_11217 | Not-determined |
| S10_extracted_11218 | Not-determined |
| S10_extracted_11225 | Not-determined |
| S10_extracted_11232 | Not-determined |
| S10_extracted_11233 | Not-determined |
| S10_extracted_11237 | Not-determined |
| S10_extracted_11257 | Not-determined |
| S10_extracted_11270 | Not-determined |
| S10_extracted_11273 | Not-determined |
| S10_extracted_11283 | Not-determined |
| S10_extracted_11296 | Not-determined |
| S10_extracted_11304 | Low-quality |
| S10_extracted_11310 | Not-determined |
| S10_extracted_11342 | Not-determined |
| S10_extracted_11357 | Not-determined |
| S10_extracted_11362 | Not-determined |
| S10_extracted_11369 | Not-determined |
| S10_extracted_11372 | Not-determined |
| S10_extracted_11373 | Not-determined |
| S10_extracted_11375 | Not-determined |
| S10_extracted_11401 | Not-determined |
| S10_extracted_11413 | Not-determined |
| S10_extracted_11425 | Not-determined |
| S10_extracted_11438 | Low-quality |
| S10_extracted_11441 | Not-determined |
| S10_extracted_11442 | Not-determined |
| S10_extracted_11457 | Low-quality |
| S10_extracted_11472 | Not-determined |
| S10_extracted_11488 | Low-quality |
| S10_extracted_11498 | Not-determined |
| S10_extracted_11501 | Not-determined |
| S10_extracted_11530 | Not-determined |
| S10_extracted_11536 | Low-quality |
| S10_extracted_11550 | Not-determined |
| S10_extracted_11559 | Not-determined |
| S10_extracted_11575 | Not-determined |
| S10_extracted_11616 | Not-determined |
| S10_extracted_11619 | Not-determined |
| S10_extracted_11620 | Not-determined |
| S10_extracted_11627 | Not-determined |
| S10_extracted_11644 | Not-determined |
| S10_extracted_11659 | Not-determined |
| S10_extracted_11676 | Not-determined |
| S10_extracted_11683 | Not-determined |
| S10_extracted_11689 | Not-determined |
| S10_extracted_11703 | Not-determined |
| S10_extracted_11715 | Not-determined |
| S10_extracted_11730 | Low-quality |
| S10_extracted_11736 | Low-quality |
| S10_extracted_11741 | Not-determined |
| S10_extracted_11748 | Not-determined |
| S10_extracted_11757 | Not-determined |
| S10_extracted_11758 | Not-determined |
| S10_extracted_11768 | Not-determined |
| S10_extracted_11794 | Not-determined |
| S10_extracted_11802 | Not-determined |
| S10_extracted_11803 | Not-determined |
| S10_extracted_11815 | Not-determined |
| S10_extracted_11856 | Not-determined |
| S10_extracted_11859 | Not-determined |
| S10_extracted_11864 | Not-determined |
| S10_extracted_11875 | Not-determined |
| S10_extracted_11901 | Not-determined |
| S10_extracted_11903 | Not-determined |
| S10_extracted_11916 | Not-determined |
| S10_extracted_11924 | Not-determined |
| S10_extracted_11925 | Not-determined |
| S10_extracted_11942 | Not-determined |
| S10_extracted_11953 | Low-quality |
| S10_extracted_11971 | Low-quality |
| S10_extracted_11972 | Low-quality |
| S10_extracted_11978 | Not-determined |
| S10_extracted_11988 | Not-determined |
| S10_extracted_11990 | Not-determined |
| S10_extracted_11996 | Not-determined |
| S10_extracted_12002 | Not-determined |
| S10_extracted_12005 | Not-determined |
| S10_extracted_12007 | Not-determined |
| S10_extracted_12008 | Not-determined |
| S10_extracted_12014 | Not-determined |
| S10_extracted_12029 | Not-determined |
| S10_extracted_12032 | Not-determined |
| S10_extracted_12038 | Not-determined |
| S10_extracted_12043 | Not-determined |
| S10_extracted_12047 | Not-determined |
| S10_extracted_12056 | Not-determined |
| S10_extracted_12086 | Not-determined |
| S10_extracted_12088 | Not-determined |
| S10_extracted_12092 | Not-determined |
| S10_extracted_12096 | Low-quality |
| S10_extracted_12119 | Low-quality |
| S10_extracted_12136 | Not-determined |
| S10_extracted_12142 | Not-determined |
| S10_extracted_12143 | Not-determined |
| S10_extracted_12157 | Not-determined |
| S10_extracted_12181 | Not-determined |
| S10_extracted_12203 | Not-determined |
| S10_extracted_12211 | Low-quality |
| S10_extracted_12213 | Not-determined |
| S10_extracted_12217 | Not-determined |
| S10_extracted_12224 | Not-determined |
| S10_extracted_12226 | Not-determined |
| S10_extracted_12258 | Low-quality |
| S10_extracted_12308 | Not-determined |
| S10_extracted_12339 | Not-determined |
| S10_extracted_12345 | Not-determined |
| S10_extracted_12348 | Not-determined |
| S10_extracted_12351 | Not-determined |
| S10_extracted_12356 | Not-determined |
| S10_extracted_12363 | Low-quality |
| S10_extracted_12390 | Low-quality |
| S10_extracted_12392 | Not-determined |
| S10_extracted_12401 | Not-determined |
| S10_extracted_12418 | Not-determined |
| S10_extracted_12443 | Not-determined |
| S10_extracted_12449 | Not-determined |
| S10_extracted_12455 | Not-determined |
| S10_extracted_12456 | Low-quality |
| S10_extracted_12462 | Not-determined |
| S10_extracted_12466 | Not-determined |
| S10_extracted_12473 | Not-determined |
| S10_extracted_12476 | Not-determined |
| S10_extracted_12487 | Not-determined |
| S10_extracted_12509 | Not-determined |
| S10_extracted_12511 | Low-quality |
| S10_extracted_12513 | Not-determined |
| S10_extracted_12518 | Not-determined |
| S10_extracted_12520 | Not-determined |
| S10_extracted_12526 | Not-determined |
| S10_extracted_12536 | Not-determined |
| S10_extracted_12542 | Low-quality |
| S10_extracted_12557 | Not-determined |
| S10_extracted_12559 | Low-quality |
| S10_extracted_12577 | Not-determined |
| S10_extracted_12605 | Not-determined |
| S10_extracted_12621 | Not-determined |
| S10_extracted_12627 | Low-quality |
| S10_extracted_12649 | Not-determined |
| S10_extracted_12650 | Not-determined |
| S10_extracted_12670 | Not-determined |
| S10_extracted_12677 | Not-determined |
| S10_extracted_12682 | Not-determined |
| S10_extracted_12686 | Not-determined |
| S10_extracted_12697 | Not-determined |
| S10_extracted_12710 | Not-determined |
| S10_extracted_12715 | Not-determined |
| S10_extracted_12729 | Not-determined |
| S10_extracted_12738 | Not-determined |
| S10_extracted_12739 | Not-determined |
| S10_extracted_12776 | Low-quality |
| S10_extracted_12822 | Not-determined |
| S10_extracted_12825 | Not-determined |
| S10_extracted_12833 | Not-determined |
| S10_extracted_12834 | Not-determined |
| S10_extracted_12854 | Not-determined |
| S10_extracted_12856 | Not-determined |
| S10_extracted_12863 | Low-quality |
| S10_extracted_12866 | Low-quality |
| S10_extracted_12870 | Not-determined |
| S10_extracted_12878 | Low-quality |
| S10_extracted_12887 | Low-quality |
| S10_extracted_12890 | Not-determined |
| S10_extracted_12903 | Not-determined |
| S10_extracted_12905 | Not-determined |
| S10_extracted_12906 | Not-determined |
| S10_extracted_12937 | Not-determined |
| S10_extracted_12952 | Not-determined |
| S10_extracted_12954 | Not-determined |
| S10_extracted_12959 | Not-determined |
| S10_extracted_12972 | Not-determined |
| S10_extracted_13009 | Not-determined |
| S10_extracted_13023 | Not-determined |
| S10_extracted_13037 | Not-determined |
| S10_extracted_13043 | Not-determined |
| S10_extracted_13044 | Not-determined |
| S10_extracted_13049 | Low-quality |
| S10_extracted_13053 | Not-determined |
| S10_extracted_13057 | Low-quality |
| S10_extracted_13065 | Low-quality |
| S10_extracted_13066 | Not-determined |
| S10_extracted_13075 | Not-determined |
| S10_extracted_13081 | Low-quality |
| S10_extracted_13084 | Not-determined |
| S10_extracted_13098 | Not-determined |
| S10_extracted_13109 | Not-determined |
| S10_extracted_13110 | Not-determined |
| S10_extracted_13113 | Not-determined |
| S10_extracted_13124 | Not-determined |
| S10_extracted_13131 | Not-determined |
| S10_extracted_13134 | Low-quality |
| S10_extracted_13136 | Not-determined |
| S10_extracted_13139 | Not-determined |
| S10_extracted_13147 | Not-determined |
| S10_extracted_13155 | Not-determined |
| S10_extracted_13164 | Low-quality |
| S10_extracted_13166 | Not-determined |
| S10_extracted_13176 | Not-determined |
| S10_extracted_13179 | Low-quality |
| S10_extracted_13184 | Low-quality |
| S10_extracted_13187 | Not-determined |
| S10_extracted_13190 | Low-quality |
| S10_extracted_13192 | Low-quality |
| S10_extracted_13198 | Not-determined |
| S10_extracted_13199 | Not-determined |
| S10_extracted_13211 | Not-determined |
| S10_extracted_13213 | Low-quality |
| S10_extracted_13226 | Not-determined |
| S10_extracted_13229 | Not-determined |
| S10_extracted_13233 | Not-determined |
| S10_extracted_13237 | Not-determined |
| S10_extracted_13244 | Not-determined |
| S10_extracted_13258 | Not-determined |
| S10_extracted_13260 | Not-determined |
| S10_extracted_13264 | Not-determined |
| S10_extracted_13279 | Not-determined |
| S10_extracted_13282 | Not-determined |
| S10_extracted_13284 | Not-determined |
| S10_extracted_13294 | Not-determined |
| S10_extracted_13299 | Not-determined |
| S10_extracted_13328 | Not-determined |
| S10_extracted_13386 | Low-quality |
| S10_extracted_13412 | Not-determined |
| S10_extracted_13431 | Not-determined |
| S10_extracted_13434 | Not-determined |
| S10_extracted_13439 | Low-quality |
| S10_extracted_13444 | Not-determined |
| S10_extracted_13448 | Not-determined |
| S10_extracted_13461 | Not-determined |
| S10_extracted_13466 | Not-determined |
| S10_extracted_13477 | Not-determined |
| S10_extracted_13482 | Not-determined |
| S10_extracted_13488 | Low-quality |
| S10_extracted_13492 | Not-determined |
| S10_extracted_13515 | Not-determined |
| S10_extracted_13516 | Not-determined |
| S10_extracted_13518 | Not-determined |
| S10_extracted_13532 | Not-determined |
| S10_extracted_13533 | Low-quality |
| S10_extracted_13541 | Not-determined |
| S10_extracted_13543 | Not-determined |
| S10_extracted_13555 | Not-determined |
| S10_extracted_13573 | Not-determined |
| S10_extracted_13578 | Not-determined |
| S10_extracted_13581 | Not-determined |
| S10_extracted_13600 | Not-determined |
| S10_extracted_13601 | Not-determined |
| S10_extracted_13609 | Not-determined |
| S10_extracted_13618 | Not-determined |
| S10_extracted_13649 | Not-determined |
| S10_extracted_13657 | Not-determined |
| S10_extracted_13660 | Low-quality |
| S10_extracted_13677 | Not-determined |
| S10_extracted_13678 | Not-determined |
| S10_extracted_13679 | Not-determined |
| S10_extracted_13711 | Not-determined |
| S10_extracted_13716 | Not-determined |
| S10_extracted_13719 | Not-determined |
| S10_extracted_13724 | Not-determined |
| S10_extracted_13752 | Not-determined |
| S10_extracted_13760 | Not-determined |
| S10_extracted_13761 | Not-determined |
| S10_extracted_13766 | Not-determined |
| S10_extracted_13769 | Not-determined |
| S10_extracted_13783 | Not-determined |
| S10_extracted_13784 | Not-determined |
| S10_extracted_13791 | Not-determined |
| S10_extracted_13794 | Not-determined |
| S10_extracted_13796 | Not-determined |
| S10_extracted_13802 | Not-determined |
| S10_extracted_13812 | Not-determined |
| S10_extracted_13820 | Not-determined |
| S10_extracted_13826 | Not-determined |
| S10_extracted_13834 | Low-quality |
| S10_extracted_13852 | Not-determined |
| S10_extracted_13873 | Not-determined |
| S10_extracted_13891 | Not-determined |
| S10_extracted_13903 | Low-quality |
| S10_extracted_13912 | Not-determined |
| S10_extracted_13913 | Not-determined |
| S10_extracted_13937 | Low-quality |
| S10_extracted_13943 | Low-quality |
| S10_extracted_13947 | Not-determined |
| S10_extracted_13950 | Not-determined |
| S10_extracted_13966 | Not-determined |
| S10_extracted_13994 | Not-determined |
| S10_extracted_14006 | Not-determined |
| S10_extracted_14008 | Not-determined |
| S10_extracted_14012 | Not-determined |
| S10_extracted_14017 | Low-quality |
| S10_extracted_14021 | Not-determined |
| S10_extracted_14031 | Not-determined |
| S10_extracted_14049 | Not-determined |
| S10_extracted_14052 | Not-determined |
| S10_extracted_14064 | Not-determined |
| S10_extracted_14067 | Not-determined |
| S10_extracted_14072 | Not-determined |
| S10_extracted_14083 | Not-determined |
| S10_extracted_14089 | Not-determined |
| S10_extracted_14094 | Not-determined |
| S10_extracted_14112 | Low-quality |
| S10_extracted_14130 | Not-determined |
| S10_extracted_14150 | Not-determined |
| S10_extracted_14165 | Not-determined |
| S10_extracted_14174 | Low-quality |
| S10_extracted_14175 | Not-determined |
| S10_extracted_14193 | Not-determined |
| S10_extracted_14210 | Not-determined |
| S10_extracted_14263 | Not-determined |
| S10_extracted_14264 | Low-quality |
| S10_extracted_14276 | Not-determined |
| S10_extracted_14277 | Not-determined |
| S10_extracted_14316 | Not-determined |
| S10_extracted_14321 | Not-determined |
| S10_extracted_14330 | Not-determined |
| S10_extracted_14351 | Low-quality |
| S10_extracted_14359 | Not-determined |
| S10_extracted_14380 | Not-determined |
| S10_extracted_14388 | Not-determined |
| S10_extracted_14389 | Not-determined |
| S10_extracted_14394 | Not-determined |
| S10_extracted_14398 | Not-determined |
| S10_extracted_14411 | Not-determined |
| S10_extracted_14415 | Low-quality |
| S10_extracted_14417 | Low-quality |
| S10_extracted_14419 | Not-determined |
| S10_extracted_14425 | Not-determined |
| S10_extracted_14445 | Low-quality |
| S10_extracted_14464 | Not-determined |
| S10_extracted_14468 | Not-determined |
| S10_extracted_14471 | Low-quality |
| S10_extracted_14481 | Not-determined |
| S10_extracted_14492 | Not-determined |
| S10_extracted_14495 | Not-determined |
| S10_extracted_14496 | Not-determined |
| S10_extracted_14502 | Not-determined |
| S10_extracted_14513 | Not-determined |
| S10_extracted_14516 | Not-determined |
| S10_extracted_14520 | Not-determined |
| S10_extracted_14528 | Not-determined |
| S10_extracted_14532 | Low-quality |
| S10_extracted_14543 | Low-quality |
| S10_extracted_14566 | Not-determined |
| S10_extracted_14568 | Not-determined |
| S10_extracted_14574 | Not-determined |
| S10_extracted_14577 | Not-determined |
| S10_extracted_14588 | Not-determined |
| S10_extracted_14591 | Not-determined |
| S10_extracted_14593 | Not-determined |
| S10_extracted_14614 | Not-determined |
| S10_extracted_14619 | Not-determined |
| S10_extracted_14625 | Not-determined |
| S10_extracted_14633 | Not-determined |
| S10_extracted_14651 | Not-determined |
| S10_extracted_14654 | Not-determined |
| S10_extracted_14658 | Not-determined |
| S10_extracted_14665 | Not-determined |
| S10_extracted_14666 | Not-determined |
| S10_extracted_14670 | Not-determined |
| S10_extracted_14675 | Not-determined |
| S10_extracted_14676 | Low-quality |
| S10_extracted_14685 | Not-determined |
| S10_extracted_14700 | Low-quality |
| S10_extracted_14704 | Not-determined |
| S10_extracted_14726 | Not-determined |
| S10_extracted_14739 | Low-quality |
| S10_extracted_14742 | Not-determined |
| S10_extracted_14765 | Not-determined |
| S10_extracted_14771 | Not-determined |
| S10_extracted_14773 | Low-quality |
| S10_extracted_14775 | Not-determined |
| S10_extracted_14779 | Not-determined |
| S10_extracted_14809 | Low-quality |
| S10_extracted_14812 | Not-determined |
| S10_extracted_14815 | Not-determined |
| S10_extracted_14820 | Not-determined |
| S10_extracted_14830 | Low-quality |
| S10_extracted_14836 | Low-quality |
| S10_extracted_14837 | Not-determined |
| S10_extracted_14840 | Not-determined |
| S10_extracted_14845 | Not-determined |
| S10_extracted_14849 | Not-determined |
| S10_extracted_14864 | Low-quality |

| | |
|---|---|
| S10_extracted_14873 | Not-determined |
| S10_extracted_14881 | Not-determined |
| S10_extracted_14882 | Not-determined |
| S10_extracted_14953 | Low-quality |
| S10_extracted_14955 | Not-determined |
| S10_extracted_14960 | Not-determined |
| S10_extracted_14965 | Low-quality |
| S10_extracted_14994 | Not-determined |
| S10_extracted_14998 | Not-determined |
| S10_extracted_15029 | Not-determined |
| S10_extracted_15046 | Not-determined |
| S10_extracted_15050 | Not-determined |
| S10_extracted_15069 | Not-determined |
| S10_extracted_15101 | Low-quality |
| S10_extracted_15102 | Not-determined |
| S10_extracted_15117 | Not-determined |
| S10_extracted_15126 | Not-determined |
| S10_extracted_15129 | Not-determined |
| S10_extracted_15143 | Not-determined |
| S10_extracted_15147 | Not-determined |
| S10_extracted_15166 | Not-determined |
| S10_extracted_15167 | Not-determined |
| S10_extracted_15174 | Not-determined |
| S10_extracted_15183 | Not-determined |
| S10_extracted_15198 | Not-determined |
| S10_extracted_15200 | Not-determined |
| S10_extracted_15205 | Low-quality |
| S10_extracted_15215 | Not-determined |
| S10_extracted_15217 | Not-determined |
| S10_extracted_15218 | Not-determined |
| S10_extracted_15252 | Not-determined |
| S10_extracted_15296 | Not-determined |
| S10_extracted_15297 | Not-determined |
| S10_extracted_15304 | Not-determined |
| S10_extracted_15309 | Not-determined |
| S10_extracted_15314 | Low-quality |
| S10_extracted_15317 | Low-quality |
| S10_extracted_15333 | Not-determined |
| S10_extracted_15354 | Not-determined |
| S10_extracted_15358 | Low-quality |
| S10_extracted_15412 | Not-determined |
| S10_extracted_15435 | Not-determined |
| S10_extracted_15445 | Not-determined |
| S10_extracted_15457 | Not-determined |
| S10_extracted_15500 | Not-determined |
| S10_extracted_15510 | Not-determined |
| S10_extracted_15518 | Not-determined |
| S10_extracted_15520 | Low-quality |
| S10_extracted_15524 | Not-determined |
| S10_extracted_15525 | Not-determined |
| S10_extracted_15527 | Not-determined |
| S10_extracted_15531 | Not-determined |
| S10_extracted_15546 | Not-determined |
| S10_extracted_15548 | Not-determined |
| S10_extracted_15558 | Not-determined |
| S7_vs_13041\|\|full | High-quality |
| S10_extracted_15571 | Not-determined |
| S10_extracted_15573 | Not-determined |
| S10_extracted_15597 | Low-quality |
| S10_extracted_15611 | Not-determined |
| S10_extracted_15632 | Not-determined |
| S10_extracted_15636 | Not-determined |
| S10_extracted_15656 | Not-determined |
| S10_extracted_15657 | Not-determined |
| S10_extracted_15665 | Not-determined |
| S10_extracted_15695 | Not-determined |
| S10_extracted_15699 | Not-determined |
| S10_extracted_15705 | Not-determined |
| S10_extracted_15733 | Not-determined |
| S10_extracted_15757 | Not-determined |
| S10_extracted_15758 | Not-determined |
| S10_extracted_15767 | Not-determined |
| S10_extracted_15778 | Not-determined |
| S10_extracted_15785 | Low-quality |
| S10_extracted_15805 | Low-quality |
| S10_extracted_15813 | Not-determined |
| S10_extracted_15829 | Not-determined |
| S10_extracted_15849 | Not-determined |
| S10_extracted_15854 | Low-quality |
| S10_extracted_15857 | Not-determined |
| S10_extracted_15876 | Not-determined |
| S10_extracted_15877 | Not-determined |
| S10_extracted_15880 | Not-determined |
| S10_extracted_15882 | Not-determined |
| S10_extracted_15887 | Not-determined |
| S10_extracted_15889 | Low-quality |
| S10_extracted_15906 | Not-determined |
| S10_extracted_15910 | Not-determined |
| S10_extracted_15915 | Low-quality |
| S10_extracted_15917 | Low-quality |
| S10_extracted_15922 | Not-determined |
| S10_extracted_15930 | Low-quality |
| S10_extracted_15944 | Not-determined |
| S10_extracted_15955 | Not-determined |
| S10_extracted_15957 | Not-determined |
| S10_extracted_15958 | Not-determined |
| S10_extracted_15997 | Not-determined |
| S10_extracted_16008 | Not-determined |
| S10_extracted_16024 | Not-determined |
| S10_extracted_16032 | Not-determined |
| S10_extracted_16033 | Low-quality |
| S10_extracted_16035 | Not-determined |
| S10_extracted_16038 | Low-quality |
| S10_extracted_16047 | Not-determined |
| S10_extracted_16067 | Not-determined |
| S10_extracted_16094 | Not-determined |
| S10_extracted_16095 | Not-determined |
| S10_extracted_16114 | Not-determined |
| S10_extracted_16136 | Not-determined |
| S10_extracted_16140 | Not-determined |
| S10_extracted_16154 | Low-quality |
| S10_extracted_16164 | Not-determined |
| S10_extracted_16191 | Low-quality |
| S10_extracted_16194 | Not-determined |
| S10_extracted_16201 | Not-determined |
| S10_extracted_16202 | Not-determined |
| S10_extracted_16205 | Not-determined |
| S10_extracted_16230 | Not-determined |
| S10_extracted_16235 | Not-determined |
| S10_extracted_16250 | Not-determined |
| S10_extracted_16276 | Not-determined |
| S10_extracted_16280 | Not-determined |
| S10_extracted_16282 | Not-determined |
| S10_extracted_16284 | Low-quality |
| S10_extracted_16314 | Not-determined |
| S10_extracted_16315 | Not-determined |
| S10_extracted_16343 | Not-determined |
| S10_extracted_16360 | Not-determined |
| S10_extracted_16372 | Not-determined |
| S10_extracted_16393 | Not-determined |
| S10_extracted_16396 | Not-determined |
| S10_extracted_16409 | Not-determined |
| S10_extracted_16411 | Low-quality |
| S10_extracted_16416 | Not-determined |
| S10_extracted_16421 | Not-determined |
| S10_extracted_16424 | Not-determined |
| S10_extracted_16435 | Not-determined |
| S10_extracted_16442 | Not-determined |
| S10_extracted_16449 | Not-determined |
| S10_extracted_16453 | Not-determined |
| S10_extracted_16458 | Not-determined |
| S10_extracted_16471 | Not-determined |
| S10_extracted_16489 | Not-determined |
| S10_extracted_16506 | Not-determined |
| S10_extracted_16514 | Not-determined |
| S10_extracted_16518 | Low-quality |
| S10_extracted_16520 | Not-determined |
| S10_extracted_16522 | Not-determined |
| S10_extracted_16526 | Low-quality |
| S10_extracted_16562 | Not-determined |
| S10_extracted_16567 | Low-quality |
| S10_extracted_16570 | Not-determined |
| S10_extracted_16582 | Not-determined |
| S10_extracted_16597 | Not-determined |
| S10_extracted_16617 | Not-determined |
| S10_extracted_16621 | Not-determined |
| S10_extracted_16652 | Not-determined |
| S10_extracted_16674 | Not-determined |
| S10_extracted_16711 | Not-determined |
| S10_extracted_16717 | Not-determined |
| S10_extracted_16727 | Low-quality |
| S10_extracted_16733 | Not-determined |
| S10_extracted_16764 | Not-determined |
| S10_extracted_16769 | Not-determined |
| S10_extracted_16777 | Not-determined |
| S10_extracted_16782 | Not-determined |
| S10_extracted_16786 | Low-quality |
| S10_extracted_16810 | Not-determined |
| S10_extracted_16833 | Low-quality |
| S10_extracted_16848 | Not-determined |
| S10_extracted_16854 | Low-quality |
| S10_extracted_16858 | Not-determined |
| S10_extracted_16871 | Not-determined |
| S10_extracted_16874 | Not-determined |
| S10_extracted_16876 | Not-determined |
| S10_extracted_16878 | Not-determined |
| S10_extracted_16881 | Not-determined |
| S10_extracted_16884 | Not-determined |
| S10_extracted_16894 | Not-determined |
| S10_extracted_16897 | Low-quality |
| S10_extracted_16898 | Not-determined |
| S10_extracted_16903 | Not-determined |
| S10_extracted_16908 | Not-determined |
| S10_extracted_16917 | Not-determined |
| S10_extracted_16924 | Not-determined |
| S10_extracted_16932 | Not-determined |
| S10_extracted_16938 | Low-quality |
| S10_extracted_16945 | Low-quality |
| S10_extracted_16953 | Low-quality |
| S10_extracted_16956 | Low-quality |
| S10_extracted_16969 | Not-determined |
| S10_extracted_16971 | Not-determined |
| S10_extracted_16985 | Low-quality |
| S10_extracted_17020 | Not-determined |
| S10_extracted_17063 | Not-determined |
| S10_extracted_17070 | Not-determined |
| S10_extracted_17073 | Not-determined |
| S10_extracted_17081 | Low-quality |
| S10_extracted_17084 | Not-determined |
| S10_extracted_17098 | Not-determined |
| S10_extracted_17108 | Not-determined |
| S10_extracted_17122 | Not-determined |
| S10_extracted_17134 | Not-determined |
| S10_extracted_17145 | Low-quality |
| S10_extracted_17146 | Not-determined |
| S10_extracted_17151 | Not-determined |
| S10_extracted_17153 | Not-determined |
| S10_extracted_17160 | Not-determined |
| S10_extracted_17167 | Not-determined |
| S10_extracted_17170 | Not-determined |
| S10_extracted_17185 | Low-quality |
| S10_extracted_17188 | Low-quality |
| S10_extracted_17193 | Not-determined |
| S10_extracted_17196 | Not-determined |
| S10_extracted_17208 | Not-determined |
| S10_extracted_17231 | Not-determined |
| S10_extracted_17232 | Not-determined |
| S10_extracted_17233 | Not-determined |
| S10_extracted_17251 | Not-determined |
| S10_extracted_17253 | Low-quality |
| S10_extracted_17256 | Low-quality |
| S10_extracted_17260 | Not-determined |
| S10_extracted_17274 | Not-determined |
| S10_extracted_17277 | Not-determined |
| S10_extracted_17281 | Not-determined |
| S10_extracted_17284 | Not-determined |
| S10_extracted_17291 | Not-determined |
| S10_extracted_17305 | Not-determined |
| S10_extracted_17318 | Not-determined |
| S10_extracted_17332 | Not-determined |
| S10_extracted_17357 | Not-determined |
| S10_extracted_17397 | Not-determined |
| S10_extracted_17406 | Not-determined |
| S10_extracted_17422 | Not-determined |
| S10_extracted_17427 | Not-determined |
| S10_extracted_17465 | Not-determined |
| S10_extracted_17493 | Not-determined |
| S10_extracted_17498 | Not-determined |
| S10_extracted_17505 | Not-determined |
| S10_extracted_17543 | Not-determined |
| S10_extracted_17548 | Not-determined |
| S10_extracted_17565 | Not-determined |
| S10_extracted_17571 | Low-quality |
| S10_extracted_17605 | Not-determined |
| S10_extracted_17629 | Not-determined |
| S10_extracted_17634 | Not-determined |
| S10_extracted_17657 | Low-quality |
| S10_extracted_17663 | Not-determined |
| S10_extracted_17665 | Not-determined |
| S10_extracted_17711 | Not-determined |
| S10_extracted_17720 | Low-quality |
| S10_extracted_17722 | Not-determined |
| S10_extracted_17728 | Not-determined |
| S10_extracted_17739 | Not-determined |
| S10_extracted_17740 | Not-determined |
| S10_extracted_17756 | Not-determined |
| S10_extracted_17760 | Not-determined |
| S10_extracted_17762 | Not-determined |
| S10_extracted_17775 | Low-quality |
| S10_extracted_17776 | Not-determined |
| S10_extracted_17814 | Low-quality |
| S10_extracted_17817 | Not-determined |
| S10_extracted_17837 | Not-determined |
| S10_extracted_17839 | Not-determined |
| S10_extracted_17844 | Not-determined |
| S10_extracted_17845 | Not-determined |
| S10_extracted_17858 | Not-determined |
| S10_extracted_17861 | Not-determined |
| S10_extracted_17867 | Not-determined |
| S10_extracted_17875 | Not-determined |
| S10_extracted_17880 | Not-determined |
| S10_extracted_17902 | Low-quality |
| S10_extracted_17905 | Not-determined |
| S10_extracted_17906 | Not-determined |
| S10_extracted_17926 | Not-determined |
| S10_extracted_17937 | Low-quality |
| S10_extracted_17939 | Not-determined |
| S10_extracted_17940 | Low-quality |
| S10_extracted_17941 | Not-determined |
| S10_extracted_17944 | Not-determined |
| S10_extracted_17949 | Not-determined |
| S10_extracted_17971 | Not-determined |
| S10_extracted_17991 | Not-determined |
| S10_extracted_18015 | Low-quality |
| S10_extracted_18016 | Low-quality |
| S10_extracted_18024 | Not-determined |
| S10_extracted_18032 | Not-determined |
| S10_extracted_18039 | Not-determined |
| S10_extracted_18052 | Not-determined |
| S10_extracted_18055 | Low-quality |
| S10_extracted_18057 | Not-determined |
| S10_extracted_18061 | Not-determined |
| S10_extracted_18074 | Not-determined |
| S10_extracted_18141 | Not-determined |
| S10_extracted_18156 | Not-determined |
| S10_extracted_18168 | Not-determined |
| S10_extracted_18178 | Not-determined |
| S10_extracted_18179 | Low-quality |
| S10_extracted_18192 | Not-determined |
| S10_extracted_18193 | Not-determined |
| S10_extracted_18237 | Not-determined |
| S10_extracted_18250 | Not-determined |
| S10_extracted_18255 | Not-determined |
| S10_extracted_18264 | Not-determined |
| S10_extracted_18272 | Not-determined |
| S10_extracted_18309 | Not-determined |
| S10_extracted_18312 | Not-determined |
| S10_extracted_18314 | Not-determined |
| S10_extracted_18325 | Not-determined |
| S10_extracted_18332 | Not-determined |
| S10_extracted_18334 | Not-determined |
| S10_extracted_18359 | Not-determined |
| S10_extracted_18374 | Not-determined |
| S10_extracted_18385 | Low-quality |
| S10_extracted_18390 | Not-determined |
| S10_extracted_18394 | Not-determined |
| S10_extracted_18418 | Not-determined |
| S10_extracted_18422 | Not-determined |
| S10_extracted_18440 | Not-determined |
| S10_extracted_18453 | Not-determined |
| S10_extracted_18455 | Not-determined |
| S10_extracted_18464 | Low-quality |
| S10_extracted_18465 | Low-quality |
| S10_extracted_18472 | Not-determined |
| S10_extracted_18480 | Not-determined |
| S10_extracted_18492 | Not-determined |
| S10_extracted_18502 | Not-determined |
| S10_extracted_18506 | Not-determined |
| S10_extracted_18513 | Not-determined |
| S10_extracted_18528 | Low-quality |
| S10_extracted_18530 | Not-determined |
| S10_extracted_18556 | Low-quality |
| S10_extracted_18559 | Not-determined |
| S10_extracted_18573 | Low-quality |
| S10_extracted_18579 | Not-determined |
| S10_extracted_18584 | Not-determined |
| S10_extracted_18588 | Not-determined |
| S10_extracted_18594 | Low-quality |
| S10_extracted_18597 | Not-determined |
| S10_extracted_18602 | Low-quality |
| S10_extracted_18616 | Not-determined |
| S10_extracted_18621 | Not-determined |
| S10_extracted_18628 | Not-determined |
| S10_extracted_18634 | Not-determined |
| S10_extracted_18639 | Not-determined |
| S10_extracted_18661 | Not-determined |
| S14_vs_k141_106156\|\|full | Medium-quality |
| S10_extracted_18682 | Low-quality |
| S10_extracted_18692 | Not-determined |
| S10_extracted_18700 | Not-determined |
| S10_extracted_18703 | Not-determined |
| S10_extracted_18708 | Not-determined |
| S10_extracted_18710 | Not-determined |
| S10_extracted_18713 | Not-determined |
| S10_extracted_18719 | Low-quality |
| S10_extracted_18725 | Not-determined |
| S10_extracted_18727 | Low-quality |
| S10_extracted_18728 | Not-determined |
| S10_extracted_18730 | Not-determined |
| S10_extracted_18747 | Not-determined |
| S10_extracted_18756 | Not-determined |
| S10_extracted_18763 | Not-determined |
| S10_extracted_18767 | Not-determined |
| S10_extracted_18780 | Not-determined |
| S10_extracted_18782 | Not-determined |
| S10_extracted_18787 | Not-determined |
| S10_extracted_18792 | Not-determined |
| S10_extracted_18800 | Not-determined |
| S10_extracted_18810 | Not-determined |
| S10_extracted_18820 | Low-quality |
| S10_extracted_18854 | Not-determined |
| S10_extracted_18859 | Not-determined |
| S10_extracted_18889 | Not-determined |
| S10_extracted_18899 | Not-determined |
| S10_extracted_18909 | Not-determined |
| S10_extracted_18910 | Not-determined |
| S10_extracted_18912 | Low-quality |
| S10_extracted_18916 | Low-quality |
| S10_extracted_18932 | Not-determined |
| S10_extracted_18945 | Not-determined |
| S10_extracted_18947 | Low-quality |
| S10_extracted_18954 | Not-determined |
| S10_extracted_18956 | Not-determined |
| S10_extracted_18977 | Not-determined |
| S10_extracted_18980 | Not-determined |
| S10_extracted_18989 | Not-determined |
| S10_extracted_18993 | Not-determined |
| S10_extracted_19007 | Not-determined |
| S10_extracted_19010 | Not-determined |
| S10_extracted_19039 | Not-determined |
| S10_extracted_19060 | Not-determined |
| S10_extracted_19062 | Not-determined |
| S10_extracted_19082 | Not-determined |
| S10_extracted_19169 | Not-determined |
| S10_extracted_19174 | Low-quality |
| S10_extracted_19175 | Not-determined |
| S10_extracted_19178 | Not-determined |
| S10_extracted_19201 | Not-determined |
| S10_extracted_19208 | Not-determined |
| S10_extracted_19225 | Not-determined |
| S10_extracted_19228 | Not-determined |
| S10_extracted_19253 | Not-determined |
| S10_extracted_19259 | Low-quality |
| S10_extracted_19270 | Not-determined |
| S10_extracted_19272 | Not-determined |
| S10_extracted_19284 | Not-determined |
| S10_extracted_19298 | Not-determined |
| S10_extracted_19299 | Not-determined |
| S10_extracted_19319 | Not-determined |
| S10_extracted_19324 | Not-determined |
| S10_extracted_19355 | Not-determined |
| S10_extracted_19379 | Not-determined |
| S10_extracted_19391 | Not-determined |
| S10_extracted_19405 | Not-determined |
| S10_extracted_19419 | Not-determined |
| S10_extracted_19447 | Not-determined |
| S10_extracted_19455 | Not-determined |
| S10_extracted_19459 | Not-determined |
| S10_extracted_19463 | Not-determined |
| S10_extracted_19467 | Not-determined |
| S10_extracted_19495 | Not-determined |
| S10_extracted_19499 | Low-quality |
| S10_extracted_19514 | Not-determined |
| S10_extracted_19554 | Low-quality |
| S10_extracted_19563 | Not-determined |
| S10_extracted_19564 | Not-determined |
| S10_extracted_19571 | Not-determined |
| S10_extracted_19604 | Not-determined |
| S10_extracted_19619 | Low-quality |
| S10_extracted_19623 | Not-determined |
| S10_extracted_19636 | Not-determined |
| S10_extracted_19641 | Not-determined |
| S10_extracted_19651 | Low-quality |
| S10_extracted_19659 | Not-determined |
| S10_extracted_19662 | Not-determined |
| S10_extracted_19663 | Not-determined |
| S10_extracted_19671 | Not-determined |
| S10_extracted_19698 | Not-determined |
| S10_extracted_19699 | Not-determined |
| S10_extracted_19704 | Not-determined |
| S10_extracted_19707 | Not-determined |
| S10_extracted_19714 | Not-determined |
| S10_extracted_19716 | Not-determined |
| S10_extracted_19720 | Not-determined |
| S10_extracted_19722 | Not-determined |
| S10_extracted_19723 | Not-determined |
| S10_extracted_19724 | Not-determined |
| S10_extracted_19727 | Not-determined |
| S10_extracted_19740 | Not-determined |
| S10_extracted_19752 | Not-determined |
| S10_extracted_19762 | Not-determined |
| S10_extracted_19764 | Not-determined |
| S10_extracted_19770 | Not-determined |
| S10_extracted_19778 | Not-determined |
| S10_extracted_19784 | Low-quality |
| S10_extracted_19787 | Not-determined |
| S10_extracted_19793 | Not-determined |
| S10_extracted_19795 | Not-determined |
| S10_extracted_19797 | Not-determined |
| S10_extracted_19812 | Not-determined |
| S10_extracted_19819 | Low-quality |
| S10_extracted_19820 | Low-quality |
| S10_extracted_19824 | Not-determined |
| S10_extracted_19827 | Not-determined |
| S10_extracted_19830 | Low-quality |
| S10_extracted_19831 | Not-determined |
| S10_extracted_19843 | Low-quality |
| S10_extracted_19859 | Not-determined |
| S10_extracted_19870 | Not-determined |
| S10_extracted_19873 | Not-determined |
| S10_extracted_19882 | Not-determined |
| S10_extracted_19896 | Not-determined |
| S10_extracted_19900 | Low-quality |
| S10_extracted_19910 | Not-determined |
| S10_extracted_19931 | Not-determined |
| S10_extracted_19934 | Low-quality |
| S10_extracted_19952 | Not-determined |
| S10_extracted_19979 | Low-quality |
| S10_extracted_20028 | Low-quality |
| S10_extracted_20042 | Not-determined |
| S10_extracted_20043 | Not-determined |
| S10_extracted_20052 | Not-determined |
| S10_extracted_20067 | Not-determined |
| S10_extracted_20074 | Not-determined |
| S10_extracted_20082 | Not-determined |
| S10_extracted_20095 | Not-determined |
| S10_extracted_20105 | Not-determined |
| S10_extracted_20109 | Not-determined |
| S10_extracted_20110 | Low-quality |
| S10_extracted_20128 | Not-determined |
| S10_extracted_20136 | Not-determined |
| S10_extracted_20142 | Not-determined |
| S10_extracted_20145 | Not-determined |
| S10_extracted_20156 | Not-determined |
| S10_extracted_20163 | Not-determined |
| S10_extracted_20169 | Not-determined |
| S10_extracted_20191 | Not-determined |
| S10_extracted_20198 | Low-quality |
| S10_extracted_20203 | Not-determined |
| S10_extracted_20211 | Not-determined |
| S10_extracted_20214 | Not-determined |
| S10_extracted_20222 | Not-determined |
| S10_extracted_20223 | Not-determined |
| S10_extracted_20236 | Not-determined |
| S10_extracted_20239 | Not-determined |
| S10_extracted_20242 | Not-determined |
| S10_extracted_20246 | Not-determined |
| S10_extracted_20247 | Not-determined |
| S10_extracted_20257 | Not-determined |
| S10_extracted_20297 | Low-quality |
| S10_extracted_20319 | Not-determined |
| S10_extracted_20321 | Not-determined |
| S10_extracted_20330 | Not-determined |
| S10_extracted_20332 | Low-quality |
| S10_extracted_20333 | Not-determined |
| S10_extracted_20336 | Not-determined |
| S10_extracted_20343 | Not-determined |
| S10_extracted_20347 | Not-determined |
| S10_extracted_20353 | Not-determined |
| S10_extracted_20367 | Not-determined |
| S10_extracted_20374 | Not-determined |
| S10_extracted_20405 | Not-determined |
| S10_extracted_20414 | Not-determined |
| S10_extracted_20422 | Not-determined |
| S10_extracted_20446 | Not-determined |
| S10_extracted_20462 | Not-determined |
| S10_extracted_20471 | Not-determined |
| S10_extracted_20477 | Not-determined |
| S10_extracted_20480 | Not-determined |
| S10_extracted_20482 | Not-determined |
| S10_extracted_20497 | Not-determined |
| S10_extracted_20501 | Not-determined |
| S10_extracted_20524 | Low-quality |
| S10_extracted_20526 | Not-determined |
| S10_extracted_20533 | Not-determined |
| S10_extracted_20548 | Not-determined |
| S10_extracted_20555 | Not-determined |
| S10_extracted_20563 | Not-determined |
| S10_extracted_20576 | Not-determined |
| S10_extracted_20582 | Not-determined |
| S10_extracted_20598 | Low-quality |
| S10_extracted_20599 | Not-determined |
| S10_extracted_20602 | Not-determined |
| S10_extracted_20611 | Not-determined |
| S10_extracted_20617 | Not-determined |
| S10_extracted_20622 | Not-determined |
| S10_extracted_20624 | Not-determined |
| S10_extracted_20640 | Not-determined |
| S10_extracted_20645 | Not-determined |
| S10_extracted_20654 | Not-determined |
| S10_extracted_20657 | Low-quality |
| S10_extracted_20671 | Not-determined |
| S10_extracted_20675 | Not-determined |
| S10_extracted_20686 | Not-determined |
| S10_extracted_20693 | Not-determined |
| S10_extracted_20701 | Not-determined |
| S10_extracted_20714 | Not-determined |
| S10_extracted_20722 | Not-determined |
| S10_extracted_20745 | Not-determined |
| S10_extracted_20755 | Not-determined |
| S10_extracted_20761 | Not-determined |
| S10_extracted_20776 | Not-determined |
| S10_extracted_20792 | Not-determined |
| S10_extracted_20795 | Not-determined |
| S10_extracted_20799 | Not-determined |
| S10_extracted_20803 | Not-determined |
| S7_vs_55409\|\|full | Medium-quality |
| S10_extracted_20828 | Low-quality |
| S10_extracted_20847 | Not-determined |
| S10_extracted_20854 | Not-determined |
| S10_extracted_20858 | Not-determined |
| S10_extracted_20876 | Not-determined |
| S10_extracted_20880 | Not-determined |
| S10_extracted_20887 | Not-determined |
| S10_extracted_20895 | Not-determined |
| S10_extracted_20902 | Low-quality |
| S10_extracted_20905 | Not-determined |
| S10_extracted_20908 | Not-determined |
| S10_extracted_20914 | Not-determined |
| S10_extracted_20917 | Not-determined |
| S10_extracted_20919 | Not-determined |
| S10_extracted_20920 | Not-determined |
| S10_extracted_20921 | Not-determined |
| S10_extracted_20935 | Low-quality |
| S10_extracted_20969 | Not-determined |
| S10_extracted_20983 | Not-determined |
| S10_extracted_21004 | Not-determined |
| S10_extracted_21010 | Not-determined |
| S10_extracted_21012 | Not-determined |

| | |
|---|---|
| S10_extracted_21013 | Low-quality |
| S10_extracted_21026 | Not-determined |
| S10_extracted_21031 | Not-determined |
| S10_extracted_21040 | Low-quality |
| S10_extracted_21042 | Low-quality |
| S10_extracted_21067 | Not-determined |
| S10_extracted_21069 | Not-determined |
| S10_extracted_21086 | Not-determined |
| S10_extracted_21093 | Low-quality |
| S10_extracted_21096 | Not-determined |
| S10_extracted_21097 | Not-determined |
| S10_extracted_21100 | Not-determined |
| S10_extracted_21104 | Not-determined |
| S10_extracted_21110 | Not-determined |
| S10_extracted_21127 | Not-determined |
| S10_extracted_21135 | Not-determined |
| S10_extracted_21144 | Low-quality |
| S10_extracted_21146 | Not-determined |
| S10_extracted_21156 | Not-determined |
| S10_extracted_21160 | Not-determined |
| S10_extracted_21166 | Not-determined |
| S10_extracted_21179 | Not-determined |
| S10_extracted_21185 | Not-determined |
| S10_extracted_21188 | Not-determined |
| S10_extracted_21206 | Not-determined |
| S10_extracted_21215 | Not-determined |
| S10_extracted_21216 | Not-determined |
| S10_extracted_21234 | Not-determined |
| S10_extracted_21243 | Not-determined |
| S10_extracted_21267 | Low-quality |
| S10_extracted_21271 | Low-quality |
| S10_extracted_21272 | Not-determined |
| S10_extracted_21287 | Low-quality |
| S10_extracted_21294 | Not-determined |
| S10_extracted_21312 | Not-determined |
| S10_extracted_21342 | Not-determined |
| S10_extracted_21352 | Not-determined |
| S10_extracted_21376 | Low-quality |
| S10_extracted_21399 | Not-determined |
| S10_extracted_21403 | Not-determined |
| S10_extracted_21407 | Not-determined |
| S10_extracted_21410 | Not-determined |
| S10_extracted_21432 | Not-determined |
| S10_extracted_21460 | Not-determined |
| S10_extracted_21475 | Not-determined |
| S10_extracted_21492 | Not-determined |
| S10_extracted_21511 | Not-determined |
| S10_extracted_21560 | Not-determined |
| S10_extracted_21573 | Low-quality |
| S10_extracted_21594 | Not-determined |
| S10_extracted_21599 | Not-determined |
| S10_extracted_21606 | Not-determined |
| S10_extracted_21609 | Low-quality |
| S10_extracted_21632 | Not-determined |
| S10_extracted_21633 | Not-determined |
| S10_extracted_21640 | Not-determined |
| S10_extracted_21642 | Not-determined |
| S10_extracted_21654 | Not-determined |
| S10_extracted_21661 | Not-determined |
| S10_extracted_21666 | Low-quality |
| S10_extracted_21669 | Not-determined |
| S10_extracted_21698 | Not-determined |
| S10_extracted_21700 | Not-determined |
| S10_extracted_21738 | Not-determined |
| S10_extracted_21744 | Not-determined |
| S10_extracted_21761 | Low-quality |
| S10_extracted_21770 | Low-quality |
| S10_extracted_21791 | Not-determined |
| S10_extracted_21800 | Not-determined |
| S10_extracted_21809 | Not-determined |
| S10_extracted_21811 | Low-quality |
| S10_extracted_21845 | Not-determined |
| S10_extracted_21851 | Low-quality |
| S10_extracted_21852 | Not-determined |
| S10_extracted_21853 | Low-quality |
| S10_extracted_21857 | Not-determined |
| S10_extracted_21865 | Low-quality |
| S10_extracted_21870 | Not-determined |
| S10_extracted_21872 | Not-determined |
| S10_extracted_21883 | Not-determined |
| S10_extracted_21884 | Low-quality |
| S10_extracted_21903 | Not-determined |
| S10_extracted_21913 | Not-determined |
| S10_extracted_21917 | Not-determined |
| S10_extracted_21921 | Not-determined |
| S10_extracted_21925 | Not-determined |
| S10_extracted_21931 | Not-determined |
| S10_extracted_21950 | Not-determined |
| S10_extracted_21957 | Not-determined |
| S10_extracted_21969 | Not-determined |
| S10_extracted_21984 | Not-determined |
| S10_extracted_21991 | Not-determined |
| S10_extracted_22007 | Not-determined |
| S10_extracted_22008 | Not-determined |
| S10_extracted_22020 | Not-determined |
| S10_extracted_22037 | Not-determined |
| S10_extracted_22039 | Not-determined |
| S10_extracted_22054 | Low-quality |
| S10_extracted_22058 | Not-determined |
| S10_extracted_22059 | Not-determined |
| S10_extracted_22066 | Not-determined |
| S10_extracted_22067 | Not-determined |
| S10_extracted_22072 | Low-quality |
| S10_extracted_22083 | Low-quality |
| S10_extracted_22102 | Not-determined |
| S10_extracted_22107 | Low-quality |
| S10_extracted_22132 | Low-quality |
| S10_extracted_22138 | Low-quality |
| S10_extracted_22146 | Not-determined |
| S10_extracted_22159 | Not-determined |
| S10_extracted_22164 | Not-determined |
| S10_extracted_22187 | Low-quality |
| S10_extracted_22189 | Low-quality |
| S10_extracted_22190 | Not-determined |
| S10_extracted_22191 | Low-quality |
| S10_extracted_22201 | Not-determined |
| S10_extracted_22215 | Not-determined |
| S10_extracted_22223 | Low-quality |
| S10_extracted_22224 | Low-quality |
| S10_extracted_22229 | Not-determined |
| S10_extracted_22231 | Not-determined |
| S10_extracted_22235 | Low-quality |
| S10_extracted_22238 | Not-determined |
| S10_extracted_22239 | Not-determined |
| S10_extracted_22240 | Not-determined |
| S10_extracted_22266 | Low-quality |
| S10_extracted_22280 | Not-determined |
| S10_extracted_22296 | Low-quality |
| S10_extracted_22299 | Not-determined |
| S10_extracted_22317 | Not-determined |
| S10_extracted_22321 | Low-quality |
| S10_extracted_22329 | Not-determined |
| S10_extracted_22345 | Not-determined |
| S10_extracted_22353 | Not-determined |
| S10_extracted_22375 | Not-determined |
| S10_extracted_22377 | Not-determined |
| S10_extracted_22386 | Not-determined |
| S10_extracted_22387 | Not-determined |
| S10_extracted_22391 | Not-determined |
| S10_extracted_22405 | Not-determined |
| S10_extracted_22410 | Not-determined |
| S10_extracted_22422 | Not-determined |
| S10_extracted_22442 | Not-determined |
| S10_extracted_22460 | Not-determined |
| S10_extracted_22468 | Low-quality |
| S10_extracted_22476 | Not-determined |
| S10_extracted_22489 | Not-determined |
| S10_extracted_22501 | Not-determined |
| S10_extracted_22503 | Not-determined |
| S10_extracted_22508 | Not-determined |
| S10_extracted_22518 | Low-quality |
| S10_extracted_22519 | Not-determined |
| S10_extracted_22521 | Not-determined |
| S10_extracted_22544 | Not-determined |
| S10_extracted_22545 | Low-quality |
| S10_extracted_22548 | Not-determined |
| S10_extracted_22565 | Not-determined |
| S10_extracted_22570 | Not-determined |
| S10_extracted_22576 | Not-determined |
| S10_extracted_22583 | Not-determined |
| S10_extracted_22590 | Not-determined |
| S10_extracted_22600 | Not-determined |
| S10_extracted_22605 | Not-determined |
| S10_extracted_22609 | Low-quality |
| S10_extracted_22615 | Not-determined |
| S10_extracted_22619 | Low-quality |
| S10_extracted_22621 | Not-determined |
| S10_extracted_22622 | Low-quality |
| S10_extracted_22623 | Not-determined |
| S10_extracted_22633 | Not-determined |
| S10_extracted_22640 | Not-determined |
| S10_extracted_22641 | Not-determined |
| S10_extracted_22650 | Not-determined |
| S10_extracted_22653 | Not-determined |
| S10_extracted_22657 | Not-determined |
| S10_extracted_22662 | Not-determined |
| S10_extracted_22672 | Not-determined |
| S10_extracted_22706 | Low-quality |
| S10_extracted_22719 | Not-determined |
| S10_extracted_22722 | Not-determined |
| S10_extracted_22737 | Not-determined |
| S10_extracted_22740 | Not-determined |
| S10_extracted_22775 | Not-determined |
| S10_extracted_22776 | Not-determined |
| S10_extracted_22807 | Not-determined |
| S10_extracted_22830 | Not-determined |
| S10_extracted_22837 | Not-determined |
| S10_extracted_22871 | Not-determined |
| S10_extracted_22875 | Low-quality |
| S10_extracted_22901 | Not-determined |
| S10_extracted_22906 | Not-determined |
| S10_extracted_22918 | Not-determined |
| S10_extracted_22950 | Not-determined |
| S10_extracted_22952 | Not-determined |
| S10_extracted_22953 | Not-determined |
| S10_extracted_22958 | Not-determined |
| S10_extracted_22974 | Not-determined |
| S10_extracted_22985 | Not-determined |
| S10_extracted_22995 | Low-quality |
| S10_extracted_23001 | Not-determined |
| S10_extracted_23012 | Not-determined |
| S10_extracted_23017 | Low-quality |
| S10_extracted_23019 | Not-determined |
| S10_extracted_23023 | Not-determined |
| S10_extracted_23024 | Low-quality |
| S10_extracted_23044 | Low-quality |
| S10_extracted_23053 | Not-determined |
| S10_extracted_23054 | Not-determined |
| S10_extracted_23061 | Low-quality |
| S10_extracted_23072 | Not-determined |
| S10_extracted_23075 | Not-determined |
| S10_extracted_23082 | Not-determined |
| S10_extracted_23102 | Low-quality |
| S10_extracted_23116 | Not-determined |
| S10_extracted_23131 | Not-determined |
| S10_extracted_23141 | Not-determined |
| S10_extracted_23159 | Not-determined |
| S10_extracted_23167 | Not-determined |
| S10_extracted_23175 | Not-determined |
| S10_extracted_23208 | Not-determined |
| S10_extracted_23212 | Not-determined |
| S10_extracted_23220 | Not-determined |
| S10_extracted_23225 | Not-determined |
| S10_extracted_23229 | Not-determined |
| S10_extracted_23232 | Not-determined |
| S10_extracted_23256 | Not-determined |
| S10_extracted_23266 | Low-quality |
| S10_extracted_23274 | Not-determined |
| S10_extracted_23283 | Not-determined |
| S10_extracted_23305 | Low-quality |
| S10_extracted_23307 | Not-determined |
| S10_extracted_23323 | Not-determined |
| S10_extracted_23347 | Low-quality |
| S10_extracted_23350 | Not-determined |
| S10_extracted_23352 | Not-determined |
| S10_extracted_23372 | Not-determined |
| S10_extracted_23383 | Not-determined |
| S10_extracted_23384 | Not-determined |
| S10_extracted_23422 | Not-determined |
| S10_extracted_23442 | Not-determined |
| S10_extracted_23449 | Not-determined |
| S10_extracted_23495 | Low-quality |
| S10_extracted_23499 | Low-quality |
| S10_extracted_23508 | Not-determined |
| S10_extracted_23515 | Not-determined |
| S10_extracted_23523 | Not-determined |
| S10_extracted_23556 | Not-determined |
| S10_extracted_23574 | Not-determined |
| S10_extracted_23575 | Not-determined |
| S10_extracted_23581 | Not-determined |
| S10_extracted_23586 | Not-determined |
| S10_extracted_23587 | Not-determined |
| S10_extracted_23588 | Not-determined |
| S10_extracted_23595 | Not-determined |
| S10_extracted_23598 | Not-determined |
| S10_extracted_23603 | Not-determined |
| S10_extracted_23605 | Not-determined |
| S10_extracted_23665 | Low-quality |
| S10_extracted_23668 | Not-determined |
| S10_extracted_23685 | Low-quality |
| S10_extracted_23690 | Not-determined |
| S10_extracted_23704 | Not-determined |
| S10_extracted_23731 | Not-determined |
| S10_extracted_23735 | Low-quality |
| S10_extracted_23741 | Not-determined |
| S10_extracted_23745 | Low-quality |
| S10_extracted_23753 | Not-determined |
| S10_extracted_23756 | Not-determined |
| S10_extracted_23761 | Not-determined |
| S10_extracted_23775 | Not-determined |
| S10_extracted_23779 | Low-quality |
| S10_extracted_23781 | Not-determined |
| S10_extracted_23802 | Not-determined |
| S10_extracted_23809 | Not-determined |
| S10_extracted_23837 | Low-quality |
| S10_extracted_23873 | Not-determined |
| S10_extracted_23878 | Not-determined |
| S10_extracted_23879 | Not-determined |
| S10_extracted_23889 | Not-determined |
| S10_extracted_23909 | Not-determined |
| S10_extracted_23919 | Low-quality |
| S10_extracted_23920 | Not-determined |
| S10_extracted_23927 | Not-determined |
| S10_extracted_23928 | Low-quality |
| S10_extracted_23932 | Not-determined |
| S10_extracted_23937 | Not-determined |
| S10_extracted_23940 | Not-determined |
| S10_extracted_23947 | Not-determined |
| S10_extracted_23949 | Not-determined |
| S10_extracted_23957 | Not-determined |
| S10_extracted_23978 | Not-determined |
| S10_extracted_23987 | Not-determined |
| S10_extracted_23989 | Not-determined |
| S10_extracted_24002 | Not-determined |
| S10_extracted_24019 | Not-determined |
| S10_extracted_24022 | Not-determined |
| S10_extracted_24024 | Not-determined |
| S10_extracted_24045 | Not-determined |
| S10_extracted_24048 | Not-determined |
| S10_extracted_24055 | Not-determined |
| S10_extracted_24057 | Not-determined |
| S10_extracted_24059 | Not-determined |
| S10_extracted_24061 | Not-determined |
| S10_extracted_24067 | Not-determined |
| S10_extracted_24070 | Not-determined |
| S10_extracted_24083 | Not-determined |
| S10_extracted_24093 | Not-determined |
| S10_extracted_24098 | Not-determined |
| S10_extracted_24106 | Not-determined |
| S10_extracted_24107 | Not-determined |
| S10_extracted_24112 | Not-determined |
| S10_extracted_24116 | Not-determined |
| S10_extracted_24143 | Not-determined |
| S10_extracted_24147 | Not-determined |
| S10_extracted_24148 | Not-determined |
| S10_extracted_24149 | Not-determined |
| S10_extracted_24164 | Not-determined |
| S10_extracted_24178 | Not-determined |
| S10_extracted_24191 | Not-determined |
| S10_extracted_24192 | Not-determined |
| S10_extracted_24198 | Not-determined |
| S10_extracted_24199 | Not-determined |
| S10_extracted_24208 | Not-determined |
| S10_extracted_24210 | Not-determined |
| S10_extracted_24216 | Not-determined |
| S10_extracted_24217 | Not-determined |
| S10_extracted_24234 | Not-determined |
| S10_extracted_24237 | Low-quality |
| S10_extracted_24240 | Not-determined |
| S10_extracted_24243 | Not-determined |
| S10_extracted_24261 | Not-determined |
| S10_extracted_24267 | Low-quality |
| S10_extracted_24269 | Low-quality |
| S10_extracted_24290 | Not-determined |
| S10_extracted_24315 | Not-determined |
| S10_extracted_24318 | Not-determined |
| S10_extracted_24321 | Not-determined |
| S10_extracted_24345 | Not-determined |
| S10_extracted_24356 | Not-determined |
| S10_extracted_24359 | Not-determined |
| S10_extracted_24373 | Not-determined |
| S10_extracted_24383 | Not-determined |
| S10_extracted_24396 | Low-quality |
| S10_extracted_24408 | Not-determined |
| S10_extracted_24410 | Not-determined |
| S10_extracted_24424 | Not-determined |
| S10_extracted_24435 | Not-determined |
| S10_extracted_24439 | Not-determined |
| S10_extracted_24445 | Not-determined |
| S10_extracted_24455 | Not-determined |
| S10_extracted_24465 | Not-determined |
| S10_extracted_24467 | Low-quality |
| S10_extracted_24474 | Not-determined |
| S10_extracted_24485 | Not-determined |
| S10_extracted_24489 | Not-determined |
| S10_extracted_24505 | Not-determined |
| S10_extracted_24519 | Not-determined |
| S10_extracted_24536 | Not-determined |
| S10_extracted_24541 | Not-determined |
| S10_extracted_24558 | Low-quality |
| S10_extracted_24571 | Not-determined |
| S10_extracted_24573 | Not-determined |
| S10_extracted_24574 | Low-quality |
| S10_extracted_24575 | Not-determined |
| S10_extracted_24582 | Not-determined |
| S10_extracted_24616 | Not-determined |
| S10_extracted_24619 | Low-quality |
| S10_extracted_24632 | Not-determined |
| S10_extracted_24648 | Not-determined |
| S10_extracted_24668 | Not-determined |
| S10_extracted_24691 | Not-determined |
| S10_extracted_24716 | Low-quality |
| S10_extracted_24722 | Not-determined |
| S10_extracted_24751 | Not-determined |
| S10_extracted_24752 | Not-determined |
| S10_extracted_24758 | Not-determined |
| S10_extracted_24765 | Not-determined |
| S10_extracted_24775 | Not-determined |
| S10_extracted_24790 | Not-determined |
| S7_extracted_8332 | Medium-quality |
| S10_extracted_24805 | Low-quality |
| S10_extracted_24808 | Not-determined |
| S10_extracted_24816 | Not-determined |
| S10_extracted_24828 | Not-determined |
| S10_extracted_24834 | Not-determined |
| S10_extracted_24839 | Not-determined |
| S10_extracted_24840 | Not-determined |
| S10_extracted_24845 | Not-determined |
| S10_extracted_24854 | Not-determined |
| S10_extracted_24858 | Not-determined |
| S10_extracted_24882 | Not-determined |
| S10_extracted_24889 | Not-determined |
| S10_extracted_24897 | Not-determined |
| S10_extracted_24904 | Not-determined |
| S10_extracted_24917 | Not-determined |
| S10_extracted_24928 | Not-determined |
| S10_extracted_24930 | Low-quality |
| S10_extracted_24953 | Not-determined |
| S10_extracted_24975 | Not-determined |
| S10_extracted_24979 | Not-determined |
| S10_extracted_24991 | Not-determined |
| S10_extracted_24994 | Not-determined |
| S10_extracted_25004 | Not-determined |
| S10_extracted_25006 | Not-determined |
| S10_extracted_25008 | Not-determined |
| S10_extracted_25015 | Not-determined |
| S10_extracted_25024 | Not-determined |
| S10_extracted_25039 | Not-determined |
| S10_extracted_25075 | Not-determined |
| S10_extracted_25082 | Not-determined |
| S10_extracted_25088 | Not-determined |
| S10_extracted_25098 | Not-determined |
| S10_extracted_25101 | Not-determined |
| S10_extracted_25105 | Not-determined |
| S10_extracted_25110 | Not-determined |
| S10_extracted_25126 | Not-determined |
| S10_extracted_25127 | Not-determined |
| S10_extracted_25129 | Not-determined |
| S10_extracted_25142 | Not-determined |
| S10_extracted_25145 | Not-determined |
| S10_extracted_25155 | Not-determined |
| S10_extracted_25158 | Not-determined |
| S10_extracted_25184 | Not-determined |
| S10_extracted_25185 | Low-quality |
| S10_extracted_25186 | Not-determined |
| S10_extracted_25194 | Not-determined |
| S10_extracted_25209 | Low-quality |
| S10_extracted_25219 | Not-determined |
| S10_extracted_25233 | Not-determined |
| S16_vs_101072\|\|full | Medium-quality |
| S10_extracted_25243 | Not-determined |
| S10_extracted_25276 | Not-determined |
| S10_extracted_25291 | Not-determined |
| S10_extracted_25294 | Low-quality |
| S10_extracted_25303 | Low-quality |
| S10_extracted_25305 | Not-determined |
| S10_extracted_25308 | Low-quality |
| S10_extracted_25316 | Low-quality |
| S10_extracted_25331 | Not-determined |
| S10_extracted_25361 | Low-quality |
| S10_extracted_25364 | Not-determined |
| S10_extracted_25391 | Not-determined |
| S10_extracted_25409 | Not-determined |
| S10_extracted_25415 | Low-quality |
| S10_extracted_25420 | Low-quality |
| S10_extracted_25424 | Not-determined |
| S10_extracted_25432 | Not-determined |
| S10_extracted_25455 | Not-determined |
| S10_extracted_25457 | Low-quality |
| S10_extracted_25492 | Not-determined |
| S10_extracted_25494 | Not-determined |
| S10_extracted_25501 | Not-determined |
| S10_extracted_25511 | Not-determined |
| S10_extracted_25524 | Not-determined |
| S10_extracted_25527 | Not-determined |
| S10_extracted_25530 | Low-quality |
| S10_extracted_25540 | Not-determined |
| S10_extracted_25542 | Not-determined |
| S10_extracted_25552 | Low-quality |
| S10_extracted_25563 | Not-determined |
| S10_extracted_25565 | Not-determined |
| S10_extracted_25575 | Not-determined |
| S10_extracted_25578 | Not-determined |
| S10_extracted_25585 | Not-determined |
| S10_extracted_25596 | Not-determined |
| S10_extracted_25624 | Not-determined |
| S10_extracted_25625 | Low-quality |
| S10_extracted_25632 | Not-determined |
| S10_extracted_25634 | Not-determined |
| S10_extracted_25639 | Not-determined |
| S10_extracted_25643 | Not-determined |
| S10_extracted_25646 | Not-determined |
| S10_extracted_25652 | Low-quality |
| S10_extracted_25655 | Not-determined |
| S10_extracted_25665 | Not-determined |
| S10_extracted_25674 | Not-determined |
| S10_extracted_25676 | Not-determined |
| S10_extracted_25697 | Low-quality |
| S10_extracted_25722 | Not-determined |
| S10_extracted_25728 | Not-determined |
| S10_extracted_25739 | Low-quality |
| S10_extracted_25751 | Not-determined |
| S10_extracted_25752 | Not-determined |
| S10_extracted_25753 | Not-determined |
| S10_extracted_25761 | Not-determined |
| S10_extracted_25790 | Not-determined |
| S10_extracted_25821 | Low-quality |
| S10_extracted_25823 | Not-determined |
| S10_extracted_25855 | Low-quality |
| S10_extracted_25875 | Not-determined |
| S10_extracted_25884 | Not-determined |
| S10_extracted_25889 | Not-determined |
| S10_extracted_25904 | Not-determined |
| S10_extracted_25917 | Low-quality |
| S10_extracted_25951 | Low-quality |
| S10_extracted_25956 | Low-quality |
| S10_extracted_25958 | Low-quality |
| S10_extracted_25967 | Not-determined |
| S10_extracted_25972 | Low-quality |
| S10_extracted_25976 | Not-determined |
| S10_extracted_25978 | Not-determined |
| S10_extracted_25997 | Low-quality |
| S10_extracted_26001 | Low-quality |
| S10_extracted_26010 | Not-determined |
| S10_extracted_26011 | Not-determined |
| S10_extracted_26014 | Not-determined |
| S10_extracted_26016 | Not-determined |
| S10_extracted_26022 | Not-determined |
| S10_extracted_26028 | Not-determined |
| S10_extracted_26038 | Not-determined |
| S10_extracted_26039 | Not-determined |
| S10_extracted_26042 | Not-determined |
| S10_extracted_26051 | Not-determined |
| S10_extracted_26079 | Not-determined |
| S10_extracted_26082 | Not-determined |
| S10_extracted_26086 | Low-quality |
| S10_extracted_26088 | Not-determined |
| S10_extracted_26095 | Not-determined |
| S10_extracted_26096 | Low-quality |
| S10_extracted_26105 | Not-determined |
| S10_extracted_26110 | Not-determined |
| S10_extracted_26112 | Not-determined |
| S10_extracted_26125 | Not-determined |
| S10_extracted_26149 | Low-quality |
| S10_extracted_26176 | Not-determined |
| S10_extracted_26177 | Not-determined |
| S10_extracted_26185 | Not-determined |
| S10_extracted_26188 | Not-determined |
| S10_extracted_26194 | Low-quality |
| S10_extracted_26195 | Not-determined |
| S10_extracted_26200 | Not-determined |
| S10_extracted_26202 | Low-quality |
| S10_extracted_26222 | Not-determined |
| S10_extracted_26223 | Not-determined |
| S10_extracted_26224 | Low-quality |
| S10_extracted_26225 | Not-determined |
| S10_extracted_26247 | Not-determined |
| S10_extracted_26253 | Not-determined |
| S10_extracted_26280 | Not-determined |
| S10_extracted_26281 | Not-determined |
| S10_extracted_26292 | Not-determined |
| S10_extracted_26299 | Not-determined |
| S10_extracted_26306 | Not-determined |
| S10_extracted_26307 | Not-determined |
| S10_extracted_26315 | Not-determined |
| S10_extracted_26318 | Not-determined |
| S10_extracted_26319 | Low-quality |
| S10_extracted_26323 | Not-determined |
| S10_extracted_26325 | Not-determined |
| S10_extracted_26332 | Not-determined |
| S10_extracted_26345 | Not-determined |
| S10_extracted_26353 | Not-determined |
| S10_extracted_26390 | Not-determined |
| S10_extracted_26392 | Not-determined |
| S10_extracted_26413 | Not-determined |
| S10_extracted_26437 | Not-determined |
| S10_extracted_26441 | Low-quality |
| S10_extracted_26450 | Low-quality |
| S10_extracted_26451 | Not-determined |
| S10_extracted_26453 | Not-determined |
| S10_extracted_26457 | Not-determined |
| S10_extracted_26467 | Not-determined |
| S10_extracted_26474 | Not-determined |
| S10_extracted_26485 | Not-determined |
| S10_extracted_26500 | Not-determined |
| S10_extracted_26501 | Not-determined |
| S10_extracted_26511 | Low-quality |
| S10_extracted_26519 | Not-determined |
| S10_extracted_26522 | Not-determined |
| S10_extracted_26525 | Low-quality |
| S10_extracted_26540 | Not-determined |
| S10_extracted_26542 | Not-determined |
| S10_extracted_26547 | Not-determined |
| S10_extracted_26563 | Not-determined |
| S10_extracted_26568 | Not-determined |
| S10_extracted_26573 | Not-determined |
| S10_extracted_26578 | Not-determined |
| S10_extracted_26590 | Not-determined |
| S10_extracted_26603 | Not-determined |
| S10_extracted_26635 | Not-determined |
| S10_extracted_26638 | Not-determined |
| S10_extracted_26640 | Low-quality |
| S10_extracted_26660 | Not-determined |
| S10_extracted_26685 | Low-quality |
| S10_extracted_26688 | Not-determined |
| S10_extracted_26709 | Not-determined |
| S10_extracted_26711 | Not-determined |
| S10_extracted_26720 | Low-quality |
| S10_extracted_26727 | Low-quality |

| | |
|---|---|
| S10_extracted_26733 | Not-determined |
| S10_extracted_26736 | Not-determined |
| S10_extracted_26754 | Low-quality |
| S10_extracted_26757 | Low-quality |
| S10_extracted_26795 | Not-determined |
| S10_extracted_26796 | Low-quality |
| S10_extracted_26808 | Not-determined |
| S10_extracted_26815 | Not-determined |
| S10_extracted_26816 | Not-determined |
| S10_extracted_26826 | Low-quality |
| S10_extracted_26832 | Not-determined |
| S10_extracted_26843 | Not-determined |
| S10_extracted_26854 | Not-determined |
| S10_extracted_26855 | Not-determined |
| S10_extracted_26859 | Not-determined |
| S10_extracted_26862 | Not-determined |
| S10_extracted_26868 | Not-determined |
| S10_extracted_26873 | Not-determined |
| S10_extracted_26879 | Not-determined |
| S10_extracted_26887 | Not-determined |
| S10_extracted_26908 | Not-determined |
| S10_extracted_26912 | Not-determined |
| S10_extracted_26921 | Not-determined |
| S10_extracted_26942 | Not-determined |
| S10_extracted_26964 | Not-determined |
| S10_extracted_26967 | Low-quality |
| S10_extracted_26968 | Not-determined |
| S10_extracted_26986 | Not-determined |
| S10_extracted_27007 | Not-determined |
| S10_extracted_27010 | Low-quality |
| S10_extracted_27011 | Not-determined |
| S10_extracted_27021 | Low-quality |
| S10_extracted_27056 | Not-determined |
| S10_extracted_27065 | Not-determined |
| S10_extracted_27076 | Not-determined |
| S10_extracted_27082 | Low-quality |
| S10_extracted_27090 | Not-determined |
| S10_extracted_27127 | Not-determined |
| S10_extracted_27138 | Not-determined |
| S10_extracted_27149 | Low-quality |
| S10_extracted_27154 | Not-determined |
| S10_extracted_27155 | Not-determined |
| S10_extracted_27157 | Not-determined |
| S10_extracted_27165 | Not-determined |
| S10_extracted_27178 | Not-determined |
| S10_extracted_27187 | Not-determined |
| S10_extracted_27192 | Not-determined |
| S10_extracted_27204 | Not-determined |
| S10_extracted_27229 | Not-determined |
| S10_extracted_27239 | Low-quality |
| S10_extracted_27244 | Not-determined |
| S10_extracted_27251 | Not-determined |
| S10_extracted_27256 | Not-determined |
| S10_extracted_27262 | Not-determined |
| S10_extracted_27272 | Not-determined |
| S10_extracted_27297 | Not-determined |
| S10_extracted_27308 | Low-quality |
| S10_extracted_27309 | Not-determined |
| S10_extracted_27356 | Not-determined |
| S10_extracted_27357 | Low-quality |
| S10_extracted_27358 | Not-determined |
| S10_extracted_27387 | Not-determined |
| S10_extracted_27396 | Low-quality |
| S10_extracted_27398 | Not-determined |
| S10_extracted_27409 | Not-determined |
| S10_extracted_27422 | Not-determined |
| S10_extracted_27432 | Low-quality |
| S10_extracted_27435 | Not-determined |
| S10_extracted_27461 | Not-determined |
| S10_extracted_27496 | Not-determined |
| S10_extracted_27505 | Not-determined |
| S10_extracted_27542 | Not-determined |
| S10_extracted_27578 | Not-determined |
| S10_extracted_27611 | Not-determined |
| S10_extracted_27632 | Not-determined |
| S10_extracted_27639 | Not-determined |
| S10_extracted_27645 | Low-quality |
| S10_extracted_27648 | Not-determined |
| S10_extracted_27664 | Not-determined |
| S10_extracted_27670 | Not-determined |
| S10_extracted_27678 | Not-determined |
| S10_extracted_27679 | Not-determined |
| S10_extracted_27685 | Not-determined |
| S10_extracted_27687 | Not-determined |
| S10_extracted_27704 | Not-determined |
| S10_extracted_27717 | Not-determined |
| S10_extracted_27731 | Not-determined |
| S10_extracted_27737 | Not-determined |
| S10_extracted_27739 | Low-quality |
| S10_extracted_27742 | Not-determined |
| S10_extracted_27745 | Not-determined |
| S10_extracted_27756 | Not-determined |
| S10_extracted_27769 | Not-determined |
| S10_extracted_27790 | Not-determined |
| S10_extracted_27792 | Not-determined |
| S10_extracted_27811 | Not-determined |
| S10_extracted_27828 | Low-quality |
| S10_extracted_27833 | Not-determined |
| S10_extracted_27861 | Not-determined |
| S10_extracted_27866 | Not-determined |
| S10_extracted_27893 | Not-determined |
| S10_extracted_27926 | Low-quality |
| S10_extracted_27928 | Not-determined |
| S10_extracted_27942 | Not-determined |
| S10_extracted_27952 | Not-determined |
| S10_extracted_27963 | Not-determined |
| S10_extracted_27972 | Not-determined |
| S10_extracted_27975 | Not-determined |
| S10_extracted_27982 | Not-determined |
| S10_extracted_27983 | Not-determined |
| S10_extracted_27987 | Not-determined |
| S10_extracted_27993 | Not-determined |
| S10_extracted_28000 | Not-determined |
| S10_extracted_28007 | Not-determined |
| S10_extracted_28008 | Not-determined |
| S10_extracted_28027 | Not-determined |
| S10_extracted_28033 | Not-determined |
| S10_extracted_28042 | Not-determined |
| S10_extracted_28046 | Not-determined |
| S10_extracted_28059 | Not-determined |
| S10_extracted_28070 | Not-determined |
| S10_extracted_28072 | Not-determined |
| S10_extracted_28074 | Not-determined |
| S10_extracted_28114 | Not-determined |
| S10_extracted_28130 | Not-determined |
| S10_extracted_28135 | Not-determined |
| S10_extracted_28137 | Not-determined |
| S10_extracted_28149 | Not-determined |
| S10_extracted_28168 | Not-determined |
| S10_extracted_28172 | Not-determined |
| S10_extracted_28207 | Not-determined |
| S10_extracted_28211 | Not-determined |
| S10_extracted_28214 | Not-determined |
| S10_extracted_28217 | Not-determined |
| S10_extracted_28220 | Not-determined |
| S10_extracted_28228 | Not-determined |
| S10_extracted_28238 | Not-determined |
| S10_extracted_28278 | Not-determined |
| S10_extracted_28281 | Not-determined |
| S10_extracted_28282 | Not-determined |
| S10_extracted_28303 | Not-determined |
| S10_extracted_28315 | Not-determined |
| S10_extracted_28327 | Not-determined |
| S10_extracted_28328 | Not-determined |
| S10_extracted_28352 | Low-quality |
| S10_extracted_28364 | Not-determined |
| S10_extracted_28373 | Not-determined |
| S10_extracted_28375 | Not-determined |
| S10_extracted_28379 | Low-quality |
| S10_extracted_28396 | Not-determined |
| S10_extracted_28398 | Not-determined |
| S10_extracted_28400 | Not-determined |
| S10_extracted_28435 | Not-determined |
| S10_extracted_28436 | Not-determined |
| S10_extracted_28441 | Not-determined |
| S10_extracted_28447 | Not-determined |
| S10_extracted_28460 | Not-determined |
| S10_extracted_28479 | Not-determined |
| S10_extracted_28486 | Not-determined |
| S10_extracted_28490 | Not-determined |
| S10_extracted_28506 | Not-determined |
| S10_extracted_28527 | Not-determined |
| S10_extracted_28539 | Not-determined |
| S10_extracted_28555 | Not-determined |
| S10_extracted_28558 | Not-determined |
| S10_extracted_28569 | Not-determined |
| S10_extracted_28577 | Not-determined |
| S10_extracted_28581 | Not-determined |
| S10_extracted_28592 | Not-determined |
| S10_extracted_28612 | Not-determined |
| S10_extracted_28617 | Low-quality |
| S10_extracted_28620 | Not-determined |
| S10_extracted_28623 | Not-determined |
| S10_extracted_28633 | Not-determined |
| S10_extracted_28641 | Not-determined |
| S10_extracted_28665 | Not-determined |
| S10_extracted_28669 | Not-determined |
| S10_extracted_28676 | Not-determined |
| S10_extracted_28687 | Not-determined |
| S10_extracted_28694 | Not-determined |
| S10_extracted_28714 | Not-determined |
| S10_extracted_28715 | Low-quality |
| S10_extracted_28719 | Not-determined |
| S10_extracted_28722 | Not-determined |
| S10_extracted_28727 | Not-determined |
| S10_extracted_28735 | Low-quality |
| S10_extracted_28761 | Not-determined |
| S10_extracted_28766 | Low-quality |
| S10_extracted_28795 | Not-determined |
| S10_extracted_28797 | Not-determined |
| S10_extracted_28802 | Not-determined |
| S10_extracted_28808 | Low-quality |
| S10_extracted_28819 | Not-determined |
| S10_extracted_28840 | Not-determined |
| S10_extracted_28850 | Not-determined |
| S10_extracted_28866 | Not-determined |
| S10_extracted_28894 | Not-determined |
| S10_extracted_28899 | Not-determined |
| S10_extracted_28930 | Not-determined |
| S10_extracted_28934 | Not-determined |
| S10_extracted_28951 | Not-determined |
| S10_extracted_28953 | Not-determined |
| S10_extracted_28970 | Not-determined |
| S10_extracted_28972 | Not-determined |
| S10_extracted_28985 | Not-determined |
| S10_extracted_29020 | Not-determined |
| S10_extracted_29037 | Not-determined |
| S10_extracted_29039 | Not-determined |
| S10_extracted_29043 | Not-determined |
| S10_extracted_29078 | Not-determined |
| S10_extracted_29082 | Not-determined |
| S10_extracted_29102 | Not-determined |
| S10_extracted_29106 | Not-determined |
| S10_extracted_29107 | Not-determined |
| S10_extracted_29112 | Not-determined |
| S10_extracted_29115 | Not-determined |
| S10_extracted_29129 | Not-determined |
| S10_extracted_29141 | Not-determined |
| S10_extracted_29172 | Low-quality |
| S10_extracted_29174 | Not-determined |
| S10_extracted_29191 | Not-determined |
| S10_extracted_29205 | Not-determined |
| S10_extracted_29209 | Not-determined |
| S10_extracted_29216 | Not-determined |
| S10_extracted_29220 | Not-determined |
| S10_extracted_29225 | Not-determined |
| S10_extracted_29229 | Not-determined |
| S10_extracted_29237 | Not-determined |
| S10_extracted_29263 | Not-determined |
| S17_vs_k141_111210\|\|full | Medium-quality |
| S10_extracted_29271 | Not-determined |
| S10_extracted_29284 | Not-determined |
| S10_extracted_29293 | Low-quality |
| S10_extracted_29304 | Not-determined |
| S10_extracted_29307 | Not-determined |
| S10_extracted_29310 | Not-determined |
| S10_extracted_29328 | Low-quality |
| S10_extracted_29329 | Low-quality |
| S10_extracted_29337 | Not-determined |
| S10_extracted_29347 | Not-determined |
| S10_extracted_29348 | Not-determined |
| S10_extracted_29376 | Not-determined |
| S10_extracted_29405 | Not-determined |
| S10_extracted_29419 | Not-determined |
| S10_extracted_29425 | Not-determined |
| S10_extracted_29453 | Low-quality |
| S10_extracted_29493 | Not-determined |
| S10_extracted_29499 | Not-determined |
| S10_extracted_29507 | Low-quality |
| S10_extracted_29521 | Not-determined |
| S10_extracted_29541 | Low-quality |
| S10_extracted_29579 | Not-determined |
| S10_extracted_29609 | Low-quality |
| S10_extracted_29611 | Not-determined |
| S10_extracted_29629 | Not-determined |
| S10_extracted_29632 | Not-determined |
| S10_extracted_29679 | Not-determined |
| S10_extracted_29689 | Not-determined |
| S10_extracted_29691 | Not-determined |
| S10_extracted_29705 | Low-quality |
| S10_extracted_29715 | Not-determined |
| S10_extracted_29724 | Not-determined |
| S10_extracted_29749 | Not-determined |
| S10_extracted_29754 | Not-determined |
| S10_extracted_29773 | Low-quality |
| S10_extracted_29786 | Not-determined |
| S10_extracted_29788 | Low-quality |
| S10_extracted_29797 | Low-quality |
| S10_extracted_29800 | Not-determined |
| S10_extracted_29804 | Not-determined |
| S10_extracted_29805 | Not-determined |
| S10_extracted_29814 | Not-determined |
| S10_extracted_29828 | Not-determined |
| S10_extracted_29838 | Not-determined |
| S10_extracted_29840 | Not-determined |
| S10_extracted_29846 | Not-determined |
| S10_extracted_29849 | Not-determined |
| S10_extracted_29851 | Not-determined |
| S10_extracted_29866 | Not-determined |
| S10_extracted_29871 | Not-determined |
| S10_extracted_29874 | Not-determined |
| S10_extracted_29891 | Not-determined |
| S10_extracted_29898 | Not-determined |
| S10_extracted_29914 | Not-determined |
| S10_extracted_29916 | Not-determined |
| S10_extracted_29926 | Not-determined |
| S10_extracted_29928 | Not-determined |
| S10_extracted_29933 | Not-determined |
| S10_extracted_29936 | Not-determined |
| S10_extracted_29940 | Not-determined |
| S10_extracted_29963 | Not-determined |
| S10_extracted_29971 | Not-determined |
| S10_extracted_29993 | Not-determined |
| S10_extracted_30009 | Not-determined |
| S10_extracted_30010 | Not-determined |
| S10_extracted_30017 | Low-quality |
| S10_extracted_30033 | Not-determined |
| S10_extracted_30038 | Not-determined |
| S10_extracted_30045 | Not-determined |
| S10_extracted_30047 | Not-determined |
| S10_extracted_30057 | Not-determined |
| S10_extracted_30059 | Not-determined |
| S10_extracted_30071 | Not-determined |
| S10_extracted_30072 | Not-determined |
| S10_extracted_30112 | Not-determined |
| S10_extracted_30113 | Not-determined |
| S10_extracted_30115 | Not-determined |
| S10_extracted_30132 | Not-determined |
| S10_extracted_30133 | Not-determined |
| S10_extracted_30164 | Not-determined |
| S10_extracted_30165 | Not-determined |
| S10_extracted_30173 | Low-quality |
| S10_extracted_30198 | Not-determined |
| S10_extracted_30201 | Not-determined |
| S10_extracted_30205 | Low-quality |
| S10_extracted_30207 | Not-determined |
| S10_extracted_30216 | Not-determined |
| S10_extracted_30232 | Not-determined |
| S10_extracted_30238 | Low-quality |
| S10_extracted_30253 | Low-quality |
| S10_extracted_30262 | Not-determined |
| S10_extracted_30270 | Not-determined |
| S10_extracted_30280 | Not-determined |
| S10_extracted_30295 | Low-quality |
| S10_extracted_30300 | Low-quality |
| S10_extracted_30307 | Not-determined |
| S10_extracted_30318 | Low-quality |
| S10_extracted_30328 | Low-quality |
| S10_extracted_30332 | Low-quality |
| S10_extracted_30351 | Not-determined |
| S10_extracted_30370 | Not-determined |
| S10_extracted_30376 | Low-quality |
| S10_extracted_30383 | Low-quality |
| S10_extracted_30396 | Not-determined |
| S10_extracted_30404 | Not-determined |
| S10_extracted_30428 | Not-determined |
| S10_extracted_30434 | Low-quality |
| S10_extracted_30445 | Not-determined |
| S10_extracted_30470 | Not-determined |
| S10_extracted_30473 | Not-determined |
| S10_extracted_30481 | Not-determined |
| S10_extracted_30484 | Not-determined |
| S10_extracted_30493 | Low-quality |
| S10_extracted_30498 | Not-determined |
| S10_extracted_30502 | Not-determined |
| S10_extracted_30506 | Not-determined |
| S10_extracted_30511 | Not-determined |
| S10_extracted_30512 | Low-quality |
| S10_extracted_30521 | Not-determined |
| S10_extracted_30527 | Not-determined |
| S10_extracted_30530 | Not-determined |
| S10_extracted_30531 | Not-determined |
| S10_extracted_30532 | Not-determined |
| S10_extracted_30535 | Not-determined |
| S10_extracted_30545 | Not-determined |
| S10_extracted_30552 | Not-determined |
| S10_extracted_30554 | Not-determined |
| S10_extracted_30555 | Not-determined |
| S10_extracted_30559 | Not-determined |
| S10_extracted_30563 | Not-determined |
| S10_extracted_30573 | Not-determined |
| S10_extracted_30581 | Not-determined |
| S10_extracted_30630 | Not-determined |
| S10_extracted_30640 | Not-determined |
| S10_extracted_30644 | Not-determined |
| S10_extracted_30645 | Not-determined |
| S10_extracted_30647 | Not-determined |
| S10_extracted_30650 | Not-determined |
| S10_extracted_30653 | Not-determined |
| S10_extracted_30682 | Not-determined |
| S10_extracted_30687 | Not-determined |
| S10_extracted_30691 | Not-determined |
| S10_extracted_30692 | Not-determined |
| S10_extracted_30694 | Not-determined |
| S10_extracted_30713 | Not-determined |
| S10_extracted_30723 | Not-determined |
| S10_extracted_30733 | Not-determined |
| S10_extracted_30738 | Not-determined |
| S10_extracted_30740 | Not-determined |
| S10_extracted_30742 | Low-quality |
| S10_extracted_30748 | Not-determined |
| S10_extracted_30761 | Not-determined |
| S10_extracted_30786 | Not-determined |
| S10_extracted_30798 | Not-determined |
| S10_extracted_30815 | Not-determined |
| S10_extracted_30821 | Not-determined |
| S10_extracted_30838 | Not-determined |
| S10_extracted_30841 | Not-determined |
| S10_extracted_30845 | Not-determined |
| S10_extracted_30854 | Low-quality |
| S10_extracted_30877 | Low-quality |
| S10_extracted_30879 | Not-determined |
| S10_extracted_30883 | Not-determined |
| S10_extracted_30885 | Not-determined |
| S10_extracted_30887 | Not-determined |
| S10_extracted_30892 | Not-determined |
| S10_extracted_30895 | Low-quality |
| S10_extracted_30908 | Not-determined |
| S10_extracted_30910 | Low-quality |
| S10_extracted_30923 | Not-determined |
| S10_extracted_30924 | Low-quality |
| S10_extracted_30926 | Not-determined |
| S10_extracted_30928 | Not-determined |
| S10_extracted_30952 | Not-determined |
| S10_extracted_30957 | Not-determined |
| S10_extracted_30971 | Not-determined |
| S10_extracted_30985 | Not-determined |
| S10_extracted_30988 | Not-determined |
| S10_extracted_31003 | Not-determined |
| S10_extracted_31014 | Not-determined |
| S10_extracted_31020 | Not-determined |
| S10_extracted_31031 | Not-determined |
| S10_extracted_31047 | Not-determined |
| S10_extracted_31054 | Low-quality |
| S10_extracted_31056 | Not-determined |
| S10_extracted_31060 | Not-determined |
| S10_extracted_31061 | Not-determined |
| S10_extracted_31104 | Not-determined |
| S10_extracted_31132 | Not-determined |
| S10_extracted_31136 | Low-quality |
| S10_extracted_31141 | Not-determined |
| S10_extracted_31150 | Not-determined |
| S10_extracted_31154 | Not-determined |
| S10_extracted_31160 | Not-determined |
| S10_extracted_31161 | Not-determined |
| S10_extracted_31173 | Not-determined |
| S10_extracted_31190 | Not-determined |
| S10_extracted_31204 | Low-quality |
| S10_extracted_31207 | Not-determined |
| S10_extracted_31225 | Not-determined |
| S10_extracted_31253 | Not-determined |
| S10_extracted_31257 | Not-determined |
| S10_extracted_31278 | Not-determined |
| S10_extracted_31287 | Not-determined |
| S10_extracted_31295 | Low-quality |
| S10_extracted_31302 | Low-quality |
| S10_extracted_31304 | Not-determined |
| S10_extracted_31307 | Low-quality |
| S10_extracted_31317 | Not-determined |
| S10_extracted_31322 | Not-determined |
| S10_extracted_31339 | Not-determined |
| S10_extracted_31340 | Low-quality |
| S10_extracted_31359 | Not-determined |
| S10_extracted_31362 | Low-quality |
| S10_extracted_31364 | Not-determined |
| S10_extracted_31380 | Not-determined |
| S10_extracted_31415 | Not-determined |
| S10_extracted_31427 | Not-determined |
| S10_extracted_31429 | Not-determined |
| S10_extracted_31435 | Not-determined |
| S10_extracted_31439 | Not-determined |
| S10_extracted_31444 | Low-quality |
| S10_extracted_31448 | Not-determined |
| S10_extracted_31466 | Not-determined |
| S10_extracted_31475 | Not-determined |
| S10_extracted_31487 | Not-determined |
| S10_extracted_31491 | Not-determined |
| S10_extracted_31497 | Not-determined |
| S10_extracted_31525 | Not-determined |
| S10_extracted_31531 | Low-quality |
| S10_extracted_31548 | Not-determined |
| S10_extracted_31557 | Not-determined |
| S10_extracted_31559 | Not-determined |
| S10_extracted_31562 | Not-determined |
| S10_extracted_31581 | Not-determined |
| S10_extracted_31592 | Not-determined |
| S10_extracted_31603 | Not-determined |
| S10_extracted_31606 | Not-determined |
| S10_extracted_31619 | Not-determined |
| S10_extracted_31625 | Low-quality |
| S10_extracted_31633 | Low-quality |
| S10_extracted_31642 | Not-determined |
| S10_extracted_31646 | Not-determined |
| S10_extracted_31665 | Low-quality |
| S10_extracted_31667 | Not-determined |
| S10_extracted_31672 | Not-determined |
| S10_extracted_31675 | Not-determined |
| S10_extracted_31688 | Not-determined |
| S10_extracted_31728 | Not-determined |
| S10_extracted_31729 | Not-determined |
| S10_extracted_31734 | Not-determined |
| S10_extracted_31775 | Not-determined |
| S10_extracted_31790 | Not-determined |
| S10_extracted_31812 | Not-determined |
| S10_extracted_31814 | Not-determined |
| S10_extracted_31818 | Not-determined |
| S10_extracted_31819 | Not-determined |
| S10_extracted_31826 | Not-determined |
| S10_extracted_31827 | Not-determined |
| S10_extracted_31829 | Low-quality |
| S10_extracted_31831 | Not-determined |
| S10_extracted_31843 | Not-determined |
| S10_extracted_31849 | Not-determined |
| S10_extracted_31851 | Low-quality |
| S10_extracted_31860 | Not-determined |
| S10_extracted_31869 | Not-determined |
| S10_extracted_31872 | Not-determined |
| S10_extracted_31884 | Low-quality |
| S10_extracted_31910 | Not-determined |
| S10_extracted_31930 | Low-quality |
| S10_extracted_31944 | Not-determined |
| S10_extracted_31946 | Not-determined |
| S10_extracted_31971 | Not-determined |
| S10_extracted_32003 | Not-determined |
| S10_extracted_32036 | Not-determined |
| S10_extracted_32040 | Not-determined |
| S10_extracted_32048 | Not-determined |
| S10_extracted_32062 | Not-determined |
| S10_extracted_32075 | Not-determined |
| S10_extracted_32099 | Not-determined |
| S10_extracted_32106 | Low-quality |
| S10_extracted_32114 | Not-determined |
| S10_extracted_32116 | Not-determined |
| S10_extracted_32117 | Not-determined |
| S10_extracted_32119 | Not-determined |
| S10_extracted_32122 | Not-determined |
| S10_extracted_32127 | Not-determined |
| S10_extracted_32152 | Not-determined |
| S10_extracted_32153 | Not-determined |
| S10_extracted_32166 | Not-determined |
| S10_extracted_32199 | Not-determined |
| S10_extracted_32201 | Not-determined |
| S10_extracted_32207 | Not-determined |
| S10_extracted_32224 | Not-determined |
| S10_extracted_32236 | Not-determined |
| S10_extracted_32264 | Not-determined |
| S10_extracted_32265 | Not-determined |
| S10_extracted_32302 | Not-determined |
| S10_extracted_32306 | Not-determined |
| S10_extracted_32317 | Not-determined |
| S10_extracted_32325 | Low-quality |
| S10_extracted_32338 | Not-determined |
| S10_extracted_32365 | Not-determined |
| S10_extracted_32388 | Low-quality |
| S10_extracted_32389 | Not-determined |
| S10_extracted_32390 | Low-quality |
| S10_extracted_32394 | Not-determined |
| S10_extracted_32418 | Low-quality |
| S10_extracted_32455 | Not-determined |
| S10_extracted_32469 | Not-determined |
| S10_extracted_32470 | Not-determined |
| S10_extracted_32487 | Low-quality |
| S10_extracted_32490 | Low-quality |
| S10_extracted_32506 | Not-determined |
| S10_extracted_32517 | Not-determined |
| S10_extracted_32523 | Not-determined |
| S10_extracted_32545 | Not-determined |
| S10_extracted_32546 | Not-determined |
| S10_extracted_32549 | Not-determined |
| S10_extracted_32557 | Not-determined |
| S10_extracted_32558 | Not-determined |
| S10_extracted_32563 | Low-quality |
| S10_extracted_32568 | Not-determined |
| S10_extracted_32575 | Not-determined |
| S10_extracted_32603 | Not-determined |
| S10_extracted_32607 | Not-determined |
| S10_extracted_32630 | Not-determined |
| S10_extracted_32659 | Not-determined |
| S10_extracted_32660 | Not-determined |
| S10_extracted_32662 | Not-determined |
| S10_extracted_32663 | Low-quality |
| S10_extracted_32664 | Not-determined |
| S10_extracted_32693 | Not-determined |
| S10_extracted_32695 | Not-determined |
| S10_extracted_32702 | Not-determined |
| S10_extracted_32703 | Not-determined |
| S10_extracted_32704 | Not-determined |
| S10_extracted_32725 | Not-determined |
| S10_extracted_32729 | Low-quality |
| S10_extracted_32733 | Not-determined |
| S10_extracted_32779 | Not-determined |
| S10_extracted_32784 | Not-determined |
| S10_extracted_32795 | Low-quality |
| S10_extracted_32804 | Not-determined |
| S10_extracted_32826 | Low-quality |
| S10_extracted_32829 | Not-determined |
| S10_extracted_32834 | Not-determined |
| S10_extracted_32852 | Not-determined |
| S10_extracted_32855 | Not-determined |
| S10_extracted_32857 | Low-quality |
| S10_extracted_32859 | Not-determined |
| S10_extracted_32872 | Low-quality |
| S10_extracted_32874 | Not-determined |
| S10_extracted_32892 | Not-determined |
| S10_extracted_32897 | Not-determined |
| S10_extracted_32937 | Low-quality |
| S10_extracted_32944 | Not-determined |
| S10_extracted_32953 | Not-determined |
| S10_extracted_32964 | Not-determined |

| | |
|---|---|
| S10_extracted_32969 | Not-determined |
| S10_extracted_32974 | Not-determined |
| S10_extracted_32999 | Not-determined |
| S10_extracted_33007 | Not-determined |
| S10_extracted_33064 | Low-quality |
| S10_extracted_33072 | Not-determined |
| S10_extracted_33074 | Low-quality |
| S10_extracted_33075 | Not-determined |
| S10_extracted_33100 | Not-determined |
| S10_extracted_33109 | Not-determined |
| S10_extracted_33117 | Not-determined |
| S10_extracted_33135 | Low-quality |
| S10_extracted_33150 | Not-determined |
| S10_extracted_33155 | Not-determined |
| S10_extracted_33170 | Not-determined |
| S10_extracted_33186 | Not-determined |
| S10_extracted_33191 | Not-determined |
| S10_extracted_33213 | Not-determined |
| S10_extracted_33261 | Low-quality |
| S10_extracted_33263 | Not-determined |
| S10_extracted_33272 | Not-determined |
| S10_extracted_33294 | Low-quality |
| S10_extracted_33307 | Not-determined |
| S10_extracted_33312 | Not-determined |
| S10_extracted_33323 | Low-quality |
| S10_extracted_33335 | Not-determined |
| S10_extracted_33342 | Not-determined |
| S10_extracted_33361 | Not-determined |
| S10_extracted_33367 | Not-determined |
| S10_extracted_33375 | Not-determined |
| S10_extracted_33386 | Not-determined |
| S10_extracted_33418 | Not-determined |
| S10_extracted_33432 | Not-determined |
| S10_extracted_33437 | Not-determined |
| S10_extracted_33451 | Not-determined |
| S10_extracted_33456 | Not-determined |
| S10_extracted_33459 | Not-determined |
| S10_extracted_33469 | Not-determined |
| S10_extracted_33471 | Not-determined |
| S10_extracted_33472 | Not-determined |
| S10_extracted_33484 | Not-determined |
| S10_extracted_33490 | Not-determined |
| S10_extracted_33500 | Not-determined |
| S10_extracted_33501 | Not-determined |
| S10_extracted_33503 | Not-determined |
| S10_extracted_33514 | Not-determined |
| S10_extracted_33517 | Not-determined |
| S10_extracted_33540 | Low-quality |
| S10_extracted_33547 | Not-determined |
| S10_extracted_33555 | Not-determined |
| S10_extracted_33576 | Not-determined |
| S10_extracted_33578 | Low-quality |
| S10_extracted_33580 | Not-determined |
| S10_extracted_33584 | Not-determined |
| S10_extracted_33589 | Not-determined |
| S10_extracted_33590 | Not-determined |
| S10_extracted_33591 | Not-determined |
| S10_extracted_33612 | Not-determined |
| S10_extracted_33629 | Not-determined |
| S10_extracted_33637 | Not-determined |
| S10_extracted_33649 | Not-determined |
| S10_extracted_33676 | Not-determined |
| S10_extracted_33683 | Not-determined |
| S10_extracted_33685 | Low-quality |
| S10_extracted_33692 | Not-determined |
| S10_extracted_33726 | Not-determined |
| S10_extracted_33740 | Not-determined |
| S10_extracted_33749 | Low-quality |
| S10_extracted_33759 | Not-determined |
| S10_extracted_33778 | Not-determined |
| S10_extracted_33781 | Low-quality |
| S10_extracted_33786 | Not-determined |
| S10_extracted_33787 | Not-determined |
| S10_extracted_33809 | Not-determined |
| S10_extracted_33839 | Low-quality |
| S10_extracted_33849 | Not-determined |
| S10_extracted_33855 | Not-determined |
| S10_extracted_33856 | Not-determined |
| S10_extracted_33857 | Not-determined |
| S10_extracted_33870 | Not-determined |
| S10_extracted_33881 | Not-determined |
| S10_extracted_33911 | Not-determined |
| S10_extracted_33913 | Not-determined |
| S10_extracted_33927 | Not-determined |
| S10_extracted_33947 | Low-quality |
| S10_extracted_33969 | Not-determined |
| S10_extracted_33970 | Not-determined |
| S10_extracted_33975 | Not-determined |
| S10_extracted_34002 | Low-quality |
| S10_extracted_34006 | Not-determined |
| S10_extracted_34021 | Not-determined |
| S10_extracted_34024 | Low-quality |
| S10_extracted_34034 | Not-determined |
| S10_extracted_34035 | Not-determined |
| S10_extracted_34038 | Not-determined |
| S10_extracted_34039 | Not-determined |
| S10_extracted_34048 | Not-determined |
| S10_extracted_34050 | Low-quality |
| S10_extracted_34051 | Not-determined |
| S10_extracted_34055 | Not-determined |
| S10_extracted_34056 | Not-determined |
| S10_extracted_34079 | Not-determined |
| S10_extracted_34080 | Not-determined |
| S10_extracted_34081 | Not-determined |
| S10_extracted_34083 | Low-quality |
| S10_extracted_34088 | Not-determined |
| S10_extracted_34104 | Not-determined |
| S10_extracted_34110 | Not-determined |
| S10_extracted_34122 | Not-determined |
| S10_extracted_34128 | Low-quality |
| S10_extracted_34138 | Not-determined |
| S10_extracted_34159 | Not-determined |
| S10_extracted_34168 | Not-determined |
| S10_extracted_34182 | Not-determined |
| S10_extracted_34214 | Not-determined |
| S10_extracted_34216 | Not-determined |
| S10_extracted_34233 | Not-determined |
| S10_extracted_34254 | Not-determined |
| S10_extracted_34255 | Not-determined |
| S10_extracted_34285 | Not-determined |
| S10_extracted_34298 | Low-quality |
| S10_extracted_34322 | Low-quality |
| S10_extracted_34330 | Not-determined |
| S10_extracted_34331 | Not-determined |
| S10_extracted_34338 | Not-determined |
| S10_extracted_34339 | Not-determined |
| S10_extracted_34351 | Not-determined |
| S10_extracted_34354 | Not-determined |
| S10_extracted_34363 | Not-determined |
| S10_extracted_34395 | Not-determined |
| S10_extracted_34407 | Not-determined |
| S10_extracted_34414 | Not-determined |
| S10_extracted_34431 | Low-quality |
| S10_extracted_34453 | Not-determined |
| S10_extracted_34478 | Not-determined |
| S10_extracted_34491 | Not-determined |
| S10_extracted_34536 | Low-quality |
| S10_extracted_34540 | Not-determined |
| S10_extracted_34541 | Not-determined |
| S10_extracted_34556 | Not-determined |
| S10_extracted_34558 | Not-determined |
| S10_extracted_34567 | Not-determined |
| S10_extracted_34570 | Not-determined |
| S10_extracted_34574 | Not-determined |
| S10_extracted_34578 | Not-determined |
| S10_extracted_34583 | Not-determined |
| S10_extracted_34585 | Not-determined |
| S10_extracted_34590 | Not-determined |
| S10_extracted_34596 | Not-determined |
| S10_extracted_34597 | Not-determined |
| S10_extracted_34601 | Not-determined |
| S10_extracted_34614 | Not-determined |
| S10_extracted_34617 | Not-determined |
| S10_extracted_34639 | Not-determined |
| S10_extracted_34642 | Not-determined |
| S10_extracted_34646 | Not-determined |
| S10_extracted_34663 | Not-determined |
| S10_extracted_34672 | Not-determined |
| S10_extracted_34682 | Not-determined |
| S10_extracted_34686 | Low-quality |
| S10_extracted_34701 | Not-determined |
| S10_extracted_34709 | Not-determined |
| S10_extracted_34710 | Not-determined |
| S10_extracted_34719 | Not-determined |
| S10_extracted_34741 | Not-determined |
| S10_extracted_34760 | Not-determined |
| S10_extracted_34762 | Not-determined |
| S10_extracted_34763 | Low-quality |
| S10_extracted_34768 | Not-determined |
| S10_extracted_34772 | Low-quality |
| S10_extracted_34797 | Not-determined |
| S10_extracted_34816 | Not-determined |
| S10_extracted_34817 | Not-determined |
| S10_extracted_34818 | Low-quality |
| S10_extracted_34834 | Not-determined |
| S10_extracted_34858 | Not-determined |
| S10_extracted_34863 | Not-determined |
| S10_extracted_34872 | Low-quality |
| S10_extracted_34897 | Not-determined |
| S10_extracted_34909 | Low-quality |
| S10_extracted_34925 | Not-determined |
| S10_extracted_34936 | Not-determined |
| S10_extracted_34939 | Low-quality |
| S10_extracted_34948 | Not-determined |
| S10_extracted_34949 | Not-determined |
| S10_extracted_34960 | Not-determined |
| S10_extracted_34966 | Not-determined |
| S10_extracted_34970 | Not-determined |
| S10_extracted_34984 | Not-determined |
| S10_extracted_34990 | Not-determined |
| S10_extracted_34994 | Low-quality |
| S10_extracted_34995 | Not-determined |
| S10_extracted_35019 | Not-determined |
| S10_extracted_35042 | Low-quality |
| S10_extracted_35093 | Not-determined |
| S10_extracted_35096 | Not-determined |
| S10_extracted_35100 | Not-determined |
| S10_extracted_35106 | Not-determined |
| S10_extracted_35118 | Not-determined |
| S10_extracted_35157 | Not-determined |
| S10_extracted_35158 | Not-determined |
| S10_extracted_35166 | Low-quality |
| S10_extracted_35186 | Not-determined |
| S10_extracted_35200 | Low-quality |
| S10_extracted_35205 | Not-determined |
| S10_extracted_35226 | Not-determined |
| S10_extracted_35230 | Low-quality |
| S10_extracted_35246 | Not-determined |
| S10_extracted_35266 | Not-determined |
| S10_extracted_35271 | Not-determined |
| S10_extracted_35297 | Not-determined |
| S10_extracted_35304 | Not-determined |
| S10_extracted_35309 | Not-determined |
| S10_extracted_35311 | Not-determined |
| S10_extracted_35321 | Not-determined |
| S10_extracted_35322 | Not-determined |
| S10_extracted_35329 | Not-determined |
| S10_extracted_35349 | Low-quality |
| S10_extracted_35350 | Not-determined |
| S10_extracted_35367 | Low-quality |
| S10_extracted_35381 | Not-determined |
| S10_extracted_35402 | Not-determined |
| S10_extracted_35406 | Not-determined |
| S10_extracted_35407 | Not-determined |
| S10_extracted_35418 | Not-determined |
| S10_extracted_35430 | Low-quality |
| S10_extracted_35459 | Not-determined |
| S10_extracted_35460 | Not-determined |
| S10_extracted_35464 | Not-determined |
| S10_extracted_35472 | Not-determined |
| S10_extracted_35478 | Not-determined |
| S10_extracted_35487 | Not-determined |
| S10_extracted_35495 | Not-determined |
| S10_extracted_35508 | Not-determined |
| S10_extracted_35511 | Not-determined |
| S10_extracted_35528 | Not-determined |
| S10_extracted_35566 | Not-determined |
| S10_extracted_35583 | Not-determined |
| S10_extracted_35584 | Not-determined |
| S10_extracted_35586 | Not-determined |
| S10_extracted_35598 | Not-determined |
| S10_extracted_35603 | Not-determined |
| S10_extracted_35606 | Not-determined |
| S10_extracted_35610 | Not-determined |
| S10_extracted_35613 | Not-determined |
| S10_extracted_35632 | Low-quality |
| S10_extracted_35654 | Not-determined |
| S10_extracted_35658 | Not-determined |
| S10_extracted_35663 | Low-quality |
| S10_extracted_35665 | Not-determined |
| S10_extracted_35669 | Not-determined |
| S10_extracted_35681 | Not-determined |
| S10_extracted_35682 | Not-determined |
| S10_extracted_35684 | Not-determined |
| S10_extracted_35687 | Not-determined |
| S10_extracted_35693 | Not-determined |
| S10_extracted_35706 | Not-determined |
| S10_extracted_35718 | Not-determined |
| S10_extracted_35740 | Not-determined |
| S10_extracted_35754 | Not-determined |
| S10_extracted_35787 | Not-determined |
| S10_extracted_35799 | Not-determined |
| S10_extracted_35803 | Not-determined |
| S10_extracted_35811 | Not-determined |
| S10_extracted_35815 | Not-determined |
| S10_extracted_35844 | Not-determined |
| S10_extracted_35857 | Not-determined |
| S10_extracted_35864 | Not-determined |
| S10_extracted_35871 | Not-determined |
| S10_extracted_35888 | Low-quality |
| S10_extracted_35911 | Not-determined |
| S10_extracted_35938 | Not-determined |
| S10_extracted_35992 | Low-quality |
| S10_extracted_36010 | Not-determined |
| S10_extracted_36011 | Not-determined |
| S10_extracted_36012 | Not-determined |
| S10_extracted_36020 | Not-determined |
| S10_extracted_36024 | Not-determined |
| S10_extracted_36035 | Not-determined |
| S10_extracted_36044 | Not-determined |
| S10_extracted_36071 | Not-determined |
| S10_extracted_36079 | Not-determined |
| S10_extracted_36084 | Not-determined |
| S10_extracted_36086 | Not-determined |
| S10_extracted_36102 | Not-determined |
| S10_extracted_36122 | Not-determined |
| S10_extracted_36123 | Low-quality |
| S10_extracted_36157 | Not-determined |
| S10_extracted_36158 | Not-determined |
| S10_extracted_36178 | Not-determined |
| S10_extracted_36179 | Not-determined |
| S10_extracted_36195 | Low-quality |
| S10_extracted_36210 | Not-determined |
| S10_extracted_36211 | Low-quality |
| S10_extracted_36247 | Not-determined |
| S10_extracted_36248 | Not-determined |
| S10_extracted_36249 | Not-determined |
| S10_extracted_36257 | Not-determined |
| S10_extracted_36259 | Not-determined |
| S10_extracted_36273 | Not-determined |
| S10_extracted_36281 | Not-determined |
| S10_extracted_36290 | Not-determined |
| S10_extracted_36333 | Not-determined |
| S10_extracted_36389 | Not-determined |
| S10_extracted_36393 | Not-determined |
| S10_extracted_36403 | Not-determined |
| S10_extracted_36406 | Not-determined |
| S10_extracted_36429 | Not-determined |
| S10_extracted_36431 | Not-determined |
| S10_extracted_36455 | Low-quality |
| S10_extracted_36458 | Not-determined |
| S10_extracted_36466 | Not-determined |
| S10_extracted_36468 | Not-determined |
| S10_extracted_36475 | Low-quality |
| S10_extracted_36479 | Not-determined |
| S10_extracted_36491 | Low-quality |
| S10_extracted_36515 | Low-quality |
| S10_extracted_36537 | Not-determined |
| S10_extracted_36543 | Not-determined |
| S10_extracted_36546 | Not-determined |
| S10_extracted_36551 | Not-determined |
| S10_extracted_36553 | Not-determined |
| S10_extracted_36564 | Not-determined |
| S10_extracted_36572 | Not-determined |
| S10_extracted_36586 | Low-quality |
| S10_extracted_36590 | Not-determined |
| S10_extracted_36606 | Not-determined |
| S10_extracted_36608 | Not-determined |
| S10_extracted_36617 | Not-determined |
| S10_extracted_36621 | Low-quality |
| S10_extracted_36642 | Not-determined |
| S10_extracted_36663 | Not-determined |
| S10_extracted_36666 | Not-determined |
| S10_extracted_36671 | Not-determined |
| S10_extracted_36677 | Low-quality |
| S10_extracted_36685 | Not-determined |
| S10_extracted_36695 | Not-determined |
| S10_extracted_36706 | Not-determined |
| S10_extracted_36729 | Not-determined |
| S10_extracted_36733 | Not-determined |
| S10_extracted_36742 | Not-determined |
| S10_extracted_36744 | Not-determined |
| S10_extracted_36748 | Not-determined |
| S10_extracted_36772 | Not-determined |
| S10_extracted_36834 | Not-determined |
| S10_extracted_36836 | Not-determined |
| S10_extracted_36850 | Not-determined |
| S10_extracted_36858 | Not-determined |
| S10_extracted_36876 | Not-determined |
| S3_extracted_k141_127450 | Complete |
| S10_extracted_36882 | Not-determined |
| S10_extracted_36885 | Not-determined |
| S10_extracted_36889 | Not-determined |
| S10_extracted_36892 | Not-determined |
| S10_extracted_36894 | Not-determined |
| S10_extracted_36899 | Not-determined |
| S10_extracted_36914 | Not-determined |
| S10_extracted_36920 | Not-determined |
| S10_extracted_36931 | Low-quality |
| S10_extracted_36935 | Not-determined |
| S10_extracted_36965 | Not-determined |
| S10_extracted_36978 | Low-quality |
| S10_extracted_36998 | Not-determined |
| S10_extracted_37001 | Not-determined |
| S10_extracted_37003 | Not-determined |
| S10_extracted_37005 | Not-determined |
| S10_extracted_37013 | Low-quality |
| S10_extracted_37021 | Not-determined |
| S10_extracted_37029 | Not-determined |
| S10_extracted_37052 | Not-determined |
| S10_extracted_37062 | Not-determined |
| S10_extracted_37070 | Not-determined |
| S10_extracted_37075 | Not-determined |
| S10_extracted_37077 | Not-determined |
| S10_extracted_37079 | Low-quality |
| S10_extracted_37087 | Not-determined |
| S10_extracted_37104 | Not-determined |
| S10_extracted_37114 | Not-determined |
| S10_extracted_37129 | Not-determined |
| S10_extracted_37137 | Not-determined |
| S10_extracted_37145 | Low-quality |
| S10_extracted_37157 | Low-quality |
| S10_extracted_37161 | Not-determined |
| S10_extracted_37180 | Not-determined |
| S10_extracted_37182 | Not-determined |
| S10_extracted_37198 | Not-determined |
| S10_extracted_37207 | Not-determined |
| S10_extracted_37211 | Not-determined |
| S10_extracted_37217 | Low-quality |
| S10_extracted_37238 | Not-determined |
| S10_extracted_37240 | Not-determined |
| S10_extracted_37244 | Not-determined |
| S10_extracted_37255 | Not-determined |
| S10_extracted_37271 | Not-determined |
| S10_extracted_37284 | Not-determined |
| S10_extracted_37285 | Low-quality |
| S10_extracted_37292 | Not-determined |
| S10_extracted_37296 | Not-determined |
| S10_extracted_37341 | Not-determined |
| S10_extracted_37358 | Not-determined |
| S10_extracted_37375 | Not-determined |
| S10_extracted_37376 | Not-determined |
| S10_extracted_37389 | Not-determined |
| S10_extracted_37412 | Low-quality |
| S10_extracted_37421 | Not-determined |
| S10_extracted_37425 | Not-determined |
| S10_extracted_37445 | Not-determined |
| S10_extracted_37462 | Not-determined |
| S10_extracted_37479 | Not-determined |
| S10_extracted_37520 | Low-quality |
| S10_extracted_37521 | Not-determined |
| S10_extracted_37543 | Not-determined |
| S10_extracted_37544 | Low-quality |
| S10_extracted_37552 | Not-determined |
| S10_extracted_37574 | Not-determined |
| S10_extracted_37586 | Not-determined |
| S10_extracted_37639 | Not-determined |
| S10_extracted_37649 | Not-determined |
| S10_extracted_37654 | Not-determined |
| S10_extracted_37671 | Not-determined |
| S10_extracted_37681 | Not-determined |
| S10_extracted_37696 | Not-determined |
| S10_extracted_37700 | Not-determined |
| S10_extracted_37701 | Not-determined |
| S10_extracted_37714 | Not-determined |
| S10_extracted_37716 | Not-determined |
| S10_extracted_37739 | Not-determined |
| S10_extracted_37750 | Not-determined |
| S10_extracted_37762 | Not-determined |
| S10_extracted_37799 | Not-determined |
| S10_extracted_37811 | Low-quality |
| S10_extracted_37826 | Not-determined |
| S10_extracted_37850 | Not-determined |
| S10_extracted_37873 | Not-determined |
| S10_extracted_37875 | Not-determined |
| S10_extracted_37882 | Not-determined |
| S10_extracted_37896 | Low-quality |
| S10_extracted_37904 | Not-determined |
| S10_extracted_37917 | Not-determined |
| S10_extracted_37922 | Not-determined |
| S10_extracted_37927 | Not-determined |
| S10_extracted_37934 | Low-quality |
| S10_extracted_37968 | Low-quality |
| S10_extracted_37969 | Low-quality |
| S10_extracted_37978 | Not-determined |
| S10_extracted_37999 | Low-quality |
| S10_extracted_38001 | Not-determined |
| S10_extracted_38020 | Low-quality |
| S10_extracted_38032 | Not-determined |
| S10_extracted_38034 | Not-determined |
| S10_extracted_38057 | Not-determined |
| S10_extracted_38060 | Not-determined |
| S10_extracted_38069 | Not-determined |
| S10_extracted_38097 | Low-quality |
| S10_extracted_38113 | Not-determined |
| S10_extracted_38121 | Not-determined |
| S10_extracted_38149 | Not-determined |
| S10_extracted_38150 | Not-determined |
| S10_extracted_38157 | Not-determined |
| S10_extracted_38165 | Not-determined |
| S10_extracted_38166 | Not-determined |
| S10_extracted_38178 | Not-determined |
| S10_extracted_38180 | Not-determined |
| S10_extracted_38184 | Low-quality |
| S10_extracted_38190 | Not-determined |
| S10_extracted_38201 | Not-determined |
| S10_extracted_38217 | Low-quality |
| S10_extracted_38252 | Not-determined |
| S10_extracted_38259 | Low-quality |
| S10_extracted_38260 | Not-determined |
| S10_extracted_38266 | Not-determined |
| S10_extracted_38273 | Not-determined |
| S10_extracted_38294 | Not-determined |
| S10_extracted_38303 | Low-quality |
| S10_extracted_38304 | Not-determined |
| S10_extracted_38327 | Not-determined |
| S10_extracted_38358 | Not-determined |
| S10_extracted_38377 | Not-determined |
| S10_extracted_38382 | Not-determined |
| S10_extracted_38388 | Not-determined |
| S10_extracted_38389 | Not-determined |
| S10_extracted_38390 | Not-determined |
| S10_extracted_38411 | Low-quality |
| S10_extracted_38422 | Not-determined |
| S10_extracted_38436 | Not-determined |
| S10_extracted_38446 | Not-determined |
| S10_extracted_38452 | Not-determined |
| S10_extracted_38465 | Not-determined |
| S10_extracted_38482 | Not-determined |
| S10_extracted_38483 | Not-determined |
| S10_extracted_38486 | Not-determined |
| S10_extracted_38489 | Not-determined |
| S10_extracted_38502 | Not-determined |
| S10_extracted_38510 | Low-quality |
| S10_extracted_38529 | Low-quality |
| S10_extracted_38539 | Not-determined |
| S10_extracted_38541 | Not-determined |
| S10_extracted_38625 | Not-determined |
| S10_extracted_38637 | Not-determined |
| S10_extracted_38641 | Not-determined |
| S10_extracted_38642 | Low-quality |
| S10_extracted_38643 | Not-determined |
| S10_extracted_38649 | Not-determined |
| S10_extracted_38660 | Not-determined |
| S10_extracted_38666 | Low-quality |
| S10_extracted_38674 | Low-quality |
| S10_extracted_38693 | Not-determined |
| S10_extracted_38701 | Low-quality |
| S10_extracted_38702 | Not-determined |
| S10_extracted_38714 | Not-determined |
| S10_extracted_38715 | Not-determined |
| S10_extracted_38720 | Not-determined |
| S10_extracted_38722 | Low-quality |
| S10_extracted_38732 | Not-determined |
| S10_extracted_38745 | Not-determined |
| S10_extracted_38749 | Not-determined |
| S10_extracted_38753 | Not-determined |
| S10_extracted_38758 | Not-determined |
| S10_extracted_38767 | Not-determined |
| S10_extracted_38775 | Not-determined |
| S10_extracted_38777 | Not-determined |
| S10_extracted_38785 | Not-determined |
| S10_extracted_38786 | Low-quality |
| S10_extracted_38793 | Not-determined |
| S10_extracted_38796 | Low-quality |
| S10_extracted_38801 | Not-determined |
| S10_extracted_38826 | Low-quality |
| S10_extracted_38829 | Not-determined |
| S10_extracted_38832 | Not-determined |
| S10_extracted_38842 | Not-determined |
| S10_extracted_38904 | Not-determined |
| S10_extracted_38913 | Low-quality |
| S10_extracted_38934 | Not-determined |
| S10_extracted_38937 | Low-quality |
| S10_extracted_38938 | Not-determined |
| S10_extracted_38950 | Not-determined |
| S10_extracted_38951 | Not-determined |
| S10_extracted_38952 | Not-determined |
| S10_extracted_38958 | Not-determined |
| S10_extracted_38974 | Not-determined |
| S10_extracted_38975 | Not-determined |
| S10_extracted_38989 | Not-determined |
| S10_extracted_39006 | Not-determined |
| S10_extracted_39010 | Not-determined |
| S10_extracted_39015 | Not-determined |
| S10_extracted_39054 | Not-determined |
| S10_extracted_39056 | Not-determined |
| S10_extracted_39059 | Not-determined |
| S10_extracted_39066 | Not-determined |
| S10_extracted_39075 | Not-determined |
| S10_extracted_39080 | Not-determined |
| S10_extracted_39105 | Not-determined |
| S10_extracted_39109 | Not-determined |
| S10_extracted_39139 | Low-quality |
| S10_extracted_39143 | Not-determined |
| S10_extracted_39144 | Not-determined |
| S10_extracted_39145 | Low-quality |
| S10_extracted_39147 | Not-determined |
| S10_extracted_39156 | Not-determined |
| S10_extracted_39169 | Not-determined |
| S10_extracted_39195 | Not-determined |
| S10_extracted_39214 | Not-determined |
| S10_extracted_39215 | Not-determined |
| S10_extracted_39250 | Not-determined |
| S10_extracted_39265 | Not-determined |
| S10_extracted_39267 | Not-determined |
| S10_extracted_39268 | Low-quality |
| S10_extracted_39280 | Not-determined |
| S10_extracted_39284 | Not-determined |
| S10_extracted_39307 | Not-determined |
| S10_extracted_39318 | Not-determined |
| S10_extracted_39324 | Not-determined |
| S10_extracted_39341 | Not-determined |
| S10_extracted_39363 | Not-determined |
| S10_extracted_39410 | Not-determined |
| S10_extracted_39413 | Not-determined |
| S10_extracted_39424 | Not-determined |
| S10_extracted_39433 | Not-determined |
| S10_extracted_39439 | Not-determined |
| S10_extracted_39447 | Not-determined |
| S10_extracted_39453 | Not-determined |
| S10_extracted_39456 | Not-determined |
| S10_extracted_39467 | Not-determined |
| S10_extracted_39469 | Low-quality |
| S10_extracted_39478 | Low-quality |
| S10_extracted_39486 | Not-determined |
| S10_extracted_39507 | Not-determined |
| S10_extracted_39527 | Not-determined |
| S10_extracted_39543 | Not-determined |

| | |
|---|---|
| S10_extracted_39549 | Not-determined |
| S10_extracted_39554 | Not-determined |
| S10_extracted_39555 | Not-determined |
| S10_extracted_39559 | Not-determined |
| S10_extracted_39562 | Not-determined |
| S10_extracted_39576 | Not-determined |
| S10_extracted_39579 | Not-determined |
| S10_extracted_39583 | Low-quality |
| S10_extracted_39596 | Low-quality |
| S10_extracted_39601 | Low-quality |
| S10_extracted_39609 | Low-quality |
| S10_extracted_39610 | Not-determined |
| S10_extracted_39613 | Not-determined |
| S10_extracted_39619 | Not-determined |
| S10_extracted_39625 | Not-determined |
| S10_extracted_39634 | Not-determined |
| S10_extracted_39651 | Not-determined |
| S10_extracted_39660 | Low-quality |
| S10_extracted_39661 | Not-determined |
| S10_extracted_39689 | Not-determined |
| S10_extracted_39695 | Not-determined |
| S10_extracted_39706 | Not-determined |
| S10_extracted_39707 | Not-determined |
| S10_extracted_39718 | Not-determined |
| S10_extracted_39724 | Not-determined |
| S10_extracted_39732 | Not-determined |
| S10_extracted_39734 | Not-determined |
| S10_extracted_39745 | Not-determined |
| S10_extracted_39748 | Low-quality |
| S10_extracted_39753 | Not-determined |
| S10_extracted_39755 | Not-determined |
| S10_extracted_39757 | Not-determined |
| S10_extracted_39779 | Not-determined |
| S10_extracted_39792 | Not-determined |
| S10_extracted_39847 | Not-determined |
| S10_extracted_39848 | Not-determined |
| S10_extracted_39863 | Not-determined |
| S10_extracted_39886 | Low-quality |
| S10_extracted_39926 | Not-determined |
| S10_extracted_39929 | Not-determined |
| S10_extracted_39935 | Not-determined |
| S10_extracted_39948 | Not-determined |
| S10_extracted_39972 | Not-determined |
| S10_extracted_39975 | Not-determined |
| S10_extracted_39981 | Low-quality |
| S10_extracted_39988 | Not-determined |
| S10_extracted_39994 | Not-determined |
| S10_extracted_40047 | Not-determined |
| S10_extracted_40052 | Not-determined |
| S10_extracted_40053 | Not-determined |
| S10_extracted_40060 | Low-quality |
| S10_extracted_40069 | Low-quality |
| S10_extracted_40073 | Not-determined |
| S10_extracted_40085 | Not-determined |
| S10_extracted_40093 | Low-quality |
| S10_extracted_40106 | Not-determined |
| S10_extracted_40112 | Not-determined |
| S10_extracted_40117 | Not-determined |
| S10_extracted_40125 | Low-quality |
| S10_extracted_40130 | Not-determined |
| S10_extracted_40134 | Not-determined |
| S10_extracted_40141 | Not-determined |
| S10_extracted_40153 | Not-determined |
| S10_extracted_40163 | Not-determined |
| S10_extracted_40174 | Not-determined |
| S10_extracted_40196 | Low-quality |
| S10_extracted_40200 | Not-determined |
| S10_extracted_40210 | Not-determined |
| S10_extracted_40211 | Low-quality |
| S10_extracted_40213 | Low-quality |
| S10_extracted_40251 | Not-determined |
| S10_extracted_40280 | Not-determined |
| S10_extracted_40313 | Not-determined |
| S10_extracted_40320 | Not-determined |
| S10_extracted_40324 | Not-determined |
| S10_extracted_40325 | Low-quality |
| S10_extracted_40328 | Low-quality |
| S10_extracted_40353 | Not-determined |
| S10_extracted_40360 | Not-determined |
| S10_extracted_40369 | Not-determined |
| S10_extracted_40383 | Not-determined |
| S10_extracted_40388 | Low-quality |
| S10_extracted_40403 | Not-determined |
| S10_extracted_40419 | Not-determined |
| S10_extracted_40422 | Not-determined |
| S10_extracted_40424 | Not-determined |
| S10_extracted_40425 | Not-determined |
| S10_extracted_40433 | Not-determined |
| S10_extracted_40457 | Not-determined |
| S10_extracted_40462 | Not-determined |
| S10_extracted_40470 | Not-determined |
| S10_extracted_40485 | Not-determined |
| S10_extracted_40490 | Not-determined |
| S10_extracted_40523 | Not-determined |
| S10_extracted_40534 | Low-quality |
| S10_extracted_40540 | Not-determined |
| S10_extracted_40547 | Not-determined |
| S10_extracted_40549 | Not-determined |
| S10_extracted_40555 | Not-determined |
| S10_extracted_40558 | Low-quality |
| S10_extracted_40561 | Not-determined |
| S10_extracted_40612 | Not-determined |
| S10_extracted_40619 | Low-quality |
| S10_extracted_40628 | Not-determined |
| S10_extracted_40641 | Not-determined |
| S10_extracted_40651 | Not-determined |
| S10_extracted_40660 | Not-determined |
| S10_extracted_40666 | Not-determined |
| S10_extracted_40668 | Not-determined |
| S10_extracted_40669 | Not-determined |
| S10_extracted_40679 | Not-determined |
| S10_extracted_40686 | Not-determined |
| S10_extracted_40689 | Not-determined |
| S10_extracted_40716 | Not-determined |
| S10_extracted_40721 | Not-determined |
| S10_extracted_40726 | Low-quality |
| S10_extracted_40731 | Not-determined |
| S10_extracted_40752 | Not-determined |
| S10_extracted_40778 | Not-determined |
| S10_extracted_40780 | Not-determined |
| S10_extracted_40799 | Not-determined |
| S10_extracted_40801 | Not-determined |
| S10_extracted_40814 | Not-determined |
| S10_extracted_40816 | Not-determined |
| S10_extracted_40828 | Not-determined |
| S10_extracted_40830 | Not-determined |
| S10_extracted_40831 | Not-determined |
| S10_extracted_40836 | Not-determined |
| S10_extracted_40842 | Not-determined |
| S10_extracted_40848 | Not-determined |
| S10_extracted_40865 | Not-determined |
| S10_extracted_40866 | Not-determined |
| S10_extracted_40869 | Not-determined |
| S10_extracted_40886 | Not-determined |
| S10_extracted_40887 | Low-quality |
| S10_extracted_40917 | Not-determined |
| S10_extracted_40932 | Not-determined |
| S10_extracted_40942 | Not-determined |
| S10_extracted_40956 | Not-determined |
| S10_extracted_40957 | Not-determined |
| S10_extracted_40970 | Low-quality |
| S10_extracted_40975 | Not-determined |
| S10_extracted_40979 | Not-determined |
| S10_extracted_40994 | Not-determined |
| S10_extracted_41008 | Not-determined |
| S10_extracted_41013 | Not-determined |
| S10_extracted_41026 | Not-determined |
| S10_extracted_41051 | Not-determined |
| S10_extracted_41052 | Not-determined |
| S10_extracted_41058 | Low-quality |
| S10_extracted_41067 | Not-determined |
| S10_extracted_41076 | Not-determined |
| S10_extracted_41122 | Not-determined |
| S10_extracted_41128 | Not-determined |
| S10_extracted_41132 | Not-determined |
| S10_extracted_41140 | Not-determined |
| S10_extracted_41163 | Not-determined |
| S10_extracted_41164 | Not-determined |
| S10_extracted_41180 | Low-quality |
| S10_extracted_41185 | Not-determined |
| S10_extracted_41218 | Not-determined |
| S10_extracted_41219 | Not-determined |
| S10_extracted_41226 | Low-quality |
| S10_extracted_41228 | Not-determined |
| S10_extracted_41250 | Not-determined |
| S10_extracted_41253 | Not-determined |
| S10_extracted_41258 | Not-determined |
| S10_extracted_41274 | Not-determined |
| S10_extracted_41284 | Not-determined |
| S10_extracted_41285 | Not-determined |
| S10_extracted_41289 | Not-determined |
| S10_extracted_41291 | Low-quality |
| S10_extracted_41292 | Not-determined |
| S10_extracted_41297 | Low-quality |
| S10_extracted_41305 | Not-determined |
| S10_extracted_41318 | Not-determined |
| S10_extracted_41344 | Not-determined |
| S10_extracted_41348 | Not-determined |
| S10_extracted_41356 | Not-determined |
| S10_extracted_41372 | Not-determined |
| S10_extracted_41373 | Not-determined |
| S10_extracted_41380 | Not-determined |
| S10_extracted_41381 | Not-determined |
| S10_extracted_41384 | Not-determined |
| S10_extracted_41396 | Not-determined |
| S10_extracted_41427 | Not-determined |
| S10_extracted_41430 | Not-determined |
| S10_extracted_41446 | Not-determined |
| S10_extracted_41447 | Not-determined |
| S10_extracted_41452 | Low-quality |
| S10_extracted_41453 | Not-determined |
| S10_extracted_41456 | Not-determined |
| S10_extracted_41458 | Not-determined |
| S10_extracted_41481 | Not-determined |
| S10_extracted_41508 | Low-quality |
| S10_extracted_41509 | Low-quality |
| S10_extracted_41520 | Not-determined |
| S10_extracted_41527 | Not-determined |
| S10_extracted_41543 | Not-determined |
| S10_extracted_41545 | Not-determined |
| S10_extracted_41561 | Not-determined |
| S10_extracted_41562 | Not-determined |
| S10_extracted_41564 | Low-quality |
| S10_extracted_41597 | Not-determined |
| S10_extracted_41606 | Low-quality |
| S10_extracted_41611 | Not-determined |
| S10_extracted_41613 | Low-quality |
| S10_extracted_41632 | Not-determined |
| S10_extracted_41633 | Not-determined |
| S10_extracted_41661 | Not-determined |
| S10_extracted_41664 | Not-determined |
| S10_extracted_41672 | Not-determined |
| S10_extracted_41676 | Not-determined |
| S10_extracted_41682 | Not-determined |
| S10_extracted_41686 | Low-quality |
| S10_extracted_41710 | Not-determined |
| S10_extracted_41714 | Low-quality |
| S10_extracted_41724 | Not-determined |
| S10_extracted_41759 | Not-determined |
| S10_extracted_41760 | Not-determined |
| S10_extracted_41795 | Not-determined |
| S10_extracted_41802 | Low-quality |
| S10_extracted_41807 | Not-determined |
| S10_extracted_41817 | Not-determined |
| S10_extracted_41837 | Not-determined |
| S10_extracted_41843 | Not-determined |
| S10_extracted_41855 | Not-determined |
| S10_extracted_41862 | Low-quality |
| S10_extracted_41863 | Not-determined |
| S10_extracted_41880 | Not-determined |
| S10_extracted_41889 | Not-determined |
| S10_extracted_41922 | Not-determined |
| S10_extracted_41934 | Not-determined |
| S10_extracted_41944 | Not-determined |
| S10_extracted_41950 | Not-determined |
| S10_extracted_41953 | Not-determined |
| S10_extracted_41954 | Not-determined |
| S10_extracted_41957 | Not-determined |
| S10_extracted_41959 | Not-determined |
| S10_extracted_41962 | Not-determined |
| S10_extracted_41965 | Not-determined |
| S10_extracted_41969 | Not-determined |
| S10_extracted_41978 | Not-determined |
| S10_extracted_41979 | Low-quality |
| S10_extracted_41991 | Low-quality |
| S10_extracted_41998 | Not-determined |
| S10_extracted_42007 | Low-quality |
| S10_extracted_42009 | Not-determined |
| S10_extracted_42013 | Not-determined |
| S10_extracted_42017 | Not-determined |
| S10_extracted_42020 | Not-determined |
| S10_extracted_42022 | Not-determined |
| S10_extracted_42059 | Not-determined |
| S10_extracted_42060 | Not-determined |
| S10_extracted_42062 | Not-determined |
| S10_extracted_42070 | Not-determined |
| S10_extracted_42097 | Not-determined |
| S10_extracted_42115 | Low-quality |
| S10_extracted_42118 | Not-determined |
| S10_extracted_42129 | Not-determined |
| S10_extracted_42137 | Not-determined |
| S10_extracted_42144 | Not-determined |
| S10_extracted_42147 | Not-determined |
| S10_extracted_42149 | Not-determined |
| S10_extracted_42153 | Low-quality |
| S10_extracted_42160 | Not-determined |
| S10_extracted_42172 | Not-determined |
| S10_extracted_42180 | Not-determined |
| S10_extracted_42239 | Not-determined |
| S10_extracted_42246 | Not-determined |
| S10_extracted_42251 | Not-determined |
| S10_extracted_42276 | Low-quality |
| S10_extracted_42282 | Not-determined |
| S10_extracted_42283 | Not-determined |
| S10_extracted_42285 | Not-determined |
| S10_extracted_42289 | Not-determined |
| S10_extracted_42292 | Not-determined |
| S10_extracted_42315 | Not-determined |
| S10_extracted_42331 | Low-quality |
| S10_extracted_42344 | Not-determined |
| S10_extracted_42353 | Low-quality |
| S10_extracted_42356 | Not-determined |
| S10_extracted_42372 | Not-determined |
| S10_extracted_42375 | Low-quality |
| S10_extracted_42378 | Not-determined |
| S10_extracted_42394 | Not-determined |
| S10_extracted_42395 | Low-quality |
| S10_extracted_42398 | Not-determined |
| S10_extracted_42409 | Not-determined |
| S10_extracted_42420 | Not-determined |
| S10_extracted_42424 | Not-determined |
| S10_extracted_42432 | Not-determined |
| S10_extracted_42435 | Not-determined |
| S10_extracted_42442 | Not-determined |
| S10_extracted_42455 | Not-determined |
| S10_extracted_42460 | Not-determined |
| S10_extracted_42464 | Not-determined |
| S10_extracted_42480 | Not-determined |
| S10_extracted_42483 | Low-quality |
| S10_extracted_42495 | Not-determined |
| S10_extracted_42498 | Not-determined |
| S10_extracted_42504 | Not-determined |
| S10_extracted_42505 | Not-determined |
| S10_extracted_42509 | Not-determined |
| S10_extracted_42512 | Not-determined |
| S10_extracted_42533 | Low-quality |
| S10_extracted_42544 | Low-quality |
| S10_extracted_42564 | Not-determined |
| S10_extracted_42601 | Not-determined |
| S10_extracted_42623 | Low-quality |
| S10_extracted_42637 | Not-determined |
| S10_extracted_42657 | Not-determined |
| S10_extracted_42664 | Low-quality |
| S10_extracted_42675 | Not-determined |
| S10_extracted_42687 | Not-determined |
| S10_extracted_42689 | Not-determined |
| S10_extracted_42706 | Low-quality |
| S10_extracted_42709 | Not-determined |
| S10_extracted_42710 | Not-determined |
| S10_extracted_42713 | Not-determined |
| S10_extracted_42717 | Not-determined |
| S10_extracted_42724 | Low-quality |
| S10_extracted_42747 | Low-quality |
| S10_extracted_42751 | Not-determined |
| S10_extracted_42753 | Not-determined |
| S10_extracted_42762 | Not-determined |
| S10_extracted_42777 | Not-determined |
| S10_extracted_42789 | Not-determined |
| S10_extracted_42790 | Not-determined |
| S10_extracted_42794 | Low-quality |
| S10_extracted_42796 | Not-determined |
| S10_extracted_42827 | Not-determined |
| S10_extracted_42828 | Not-determined |
| S10_extracted_42836 | Low-quality |
| S10_extracted_42843 | Not-determined |
| S10_extracted_42844 | Not-determined |
| S10_extracted_42866 | Not-determined |
| S10_extracted_42892 | Not-determined |
| S10_extracted_42893 | Not-determined |
| S10_extracted_42897 | Not-determined |
| S10_extracted_42902 | Not-determined |
| S10_extracted_42907 | Low-quality |
| S10_extracted_42912 | Not-determined |
| S10_extracted_42922 | Not-determined |
| S10_extracted_42924 | Not-determined |
| S10_extracted_42967 | Not-determined |
| S10_extracted_42974 | Not-determined |
| S10_extracted_42975 | Not-determined |
| S10_extracted_42978 | Not-determined |
| S10_extracted_42982 | Not-determined |
| S10_extracted_42986 | Low-quality |
| S10_extracted_42988 | Not-determined |
| S10_extracted_42995 | Not-determined |
| S10_extracted_43006 | Not-determined |
| S10_extracted_43012 | Not-determined |
| S10_extracted_43016 | Not-determined |
| S10_extracted_43032 | Not-determined |
| S10_extracted_43042 | Low-quality |
| S10_extracted_43093 | Not-determined |
| S10_extracted_43103 | Low-quality |
| S10_extracted_43122 | Not-determined |
| S10_extracted_43138 | Not-determined |
| S10_extracted_43143 | Not-determined |
| S10_extracted_43144 | Not-determined |
| S10_extracted_43156 | Not-determined |
| S10_extracted_43159 | Low-quality |
| S10_extracted_43162 | Not-determined |
| S10_extracted_43172 | Not-determined |
| S10_extracted_43182 | Not-determined |
| S10_extracted_43186 | Not-determined |
| S10_extracted_43193 | Low-quality |
| S10_extracted_43219 | Low-quality |
| S10_extracted_43237 | Not-determined |
| S10_extracted_43244 | Not-determined |
| S10_extracted_43252 | Not-determined |
| S10_extracted_43253 | Not-determined |
| S10_extracted_43260 | Low-quality |
| S10_extracted_43279 | Not-determined |
| S10_extracted_43296 | Not-determined |
| S10_extracted_43345 | Low-quality |
| S10_extracted_43346 | Not-determined |
| S10_extracted_43353 | Not-determined |
| S10_extracted_43385 | Not-determined |
| S10_extracted_43387 | Not-determined |
| S10_extracted_43388 | Not-determined |
| S10_extracted_43401 | Not-determined |
| S10_extracted_43407 | Not-determined |
| S10_extracted_43418 | Not-determined |
| S10_extracted_43421 | Not-determined |
| S10_extracted_43433 | Not-determined |
| S10_extracted_43440 | Not-determined |
| S10_extracted_43474 | Not-determined |
| S10_extracted_43493 | Low-quality |
| S10_extracted_43512 | Not-determined |
| S10_extracted_43525 | Not-determined |
| S10_extracted_43527 | Low-quality |
| S10_extracted_43545 | Not-determined |
| S10_extracted_43561 | Not-determined |
| S10_extracted_43569 | Not-determined |
| S10_extracted_43571 | Not-determined |
| S10_extracted_43597 | Not-determined |
| S10_extracted_43604 | Not-determined |
| S10_extracted_43614 | Not-determined |
| S10_extracted_43616 | Not-determined |
| S10_extracted_43617 | Not-determined |
| S10_extracted_43623 | Not-determined |
| S10_extracted_43624 | Not-determined |
| S10_extracted_43628 | Not-determined |
| S10_extracted_43635 | Not-determined |
| S10_extracted_43636 | Not-determined |
| S10_extracted_43639 | Not-determined |
| S10_extracted_43666 | Not-determined |
| S10_extracted_43678 | Not-determined |
| S10_extracted_43726 | Not-determined |
| S10_extracted_43727 | Not-determined |
| S10_extracted_43728 | Not-determined |
| S10_extracted_43733 | Not-determined |
| S10_extracted_43763 | Not-determined |
| S10_extracted_43764 | Not-determined |
| S10_extracted_43770 | Not-determined |
| S10_extracted_43773 | Not-determined |
| S10_extracted_43786 | Not-determined |
| S10_extracted_43799 | Not-determined |
| S10_extracted_43805 | Low-quality |
| S10_extracted_43810 | Not-determined |
| S10_extracted_43824 | Low-quality |
| S10_extracted_43827 | Not-determined |
| S10_extracted_43830 | Not-determined |
| S10_extracted_43840 | Not-determined |
| S10_extracted_43867 | Not-determined |
| S10_extracted_43876 | Not-determined |
| S10_extracted_43880 | Not-determined |
| S10_extracted_43893 | Not-determined |
| S10_extracted_43903 | Not-determined |
| S10_extracted_43912 | Not-determined |
| S10_extracted_43913 | Not-determined |
| S10_extracted_43934 | Not-determined |
| S10_extracted_43935 | Not-determined |
| S10_extracted_43963 | Not-determined |
| S3_extracted_k141_100440 | Medium-quality |
| S10_extracted_43991 | Not-determined |
| S10_extracted_43997 | Low-quality |
| S10_extracted_43999 | Not-determined |
| S10_extracted_44005 | Not-determined |
| S10_extracted_44008 | Low-quality |
| S10_extracted_44080 | Not-determined |
| S10_extracted_44082 | Low-quality |
| S10_extracted_44102 | Not-determined |
| S10_extracted_44117 | Low-quality |
| S10_extracted_44120 | Not-determined |
| S10_extracted_44123 | Not-determined |
| S10_extracted_44140 | Not-determined |
| S10_extracted_44149 | Not-determined |
| S10_extracted_44152 | Not-determined |
| S10_extracted_44160 | Not-determined |
| S10_extracted_44161 | Not-determined |
| S10_extracted_44217 | Low-quality |
| S10_extracted_44246 | Not-determined |
| S10_extracted_44252 | Not-determined |
| S10_extracted_44262 | Not-determined |
| S10_extracted_44264 | Not-determined |
| S10_extracted_44286 | Not-determined |
| S10_extracted_44290 | Not-determined |
| S10_extracted_44316 | Not-determined |
| S10_extracted_44321 | Not-determined |
| S10_extracted_44322 | Not-determined |
| S10_extracted_44327 | Not-determined |
| S10_extracted_44332 | Not-determined |
| S10_extracted_44367 | Not-determined |
| S10_extracted_44375 | Not-determined |
| S10_extracted_44379 | Not-determined |
| S10_extracted_44393 | Not-determined |
| S10_extracted_44398 | Low-quality |
| S10_extracted_44411 | Not-determined |
| S10_extracted_44414 | Not-determined |
| S10_extracted_44432 | Low-quality |
| S10_extracted_44447 | Not-determined |
| S10_extracted_44448 | Not-determined |
| S10_extracted_44450 | Not-determined |
| S10_extracted_44460 | Not-determined |
| S10_extracted_44461 | Not-determined |
| S10_extracted_44474 | Not-determined |
| S10_extracted_44476 | Not-determined |
| S10_extracted_44482 | Low-quality |
| S10_extracted_44500 | Low-quality |
| S10_extracted_44501 | Not-determined |
| S10_extracted_44503 | Not-determined |
| S10_extracted_44505 | Low-quality |
| S10_extracted_44509 | Not-determined |
| S10_extracted_44517 | Not-determined |
| S10_extracted_44525 | Not-determined |
| S10_extracted_44526 | Not-determined |
| S10_extracted_44540 | Low-quality |
| S10_extracted_44560 | Low-quality |
| S10_extracted_44562 | Not-determined |
| S10_extracted_44568 | Not-determined |
| S10_extracted_44583 | Not-determined |
| S10_extracted_44588 | Not-determined |
| S10_extracted_44617 | Low-quality |
| S10_extracted_44619 | Not-determined |
| S10_extracted_44625 | Low-quality |
| S10_extracted_44632 | Not-determined |
| S10_extracted_44637 | Not-determined |
| S10_extracted_44646 | Low-quality |
| S10_extracted_44653 | Not-determined |
| S10_extracted_44654 | Not-determined |
| S10_extracted_44700 | Not-determined |
| S10_extracted_44704 | Not-determined |
| S10_extracted_44720 | Not-determined |
| S10_extracted_44723 | Not-determined |
| S10_extracted_44724 | Low-quality |
| S10_extracted_44736 | Not-determined |
| S10_extracted_44737 | Not-determined |
| S10_extracted_44738 | Not-determined |
| S10_extracted_44750 | Not-determined |
| S10_extracted_44756 | Not-determined |
| S10_extracted_44770 | Not-determined |
| S10_extracted_44781 | Not-determined |
| S10_extracted_44792 | Not-determined |
| S10_extracted_44807 | Not-determined |
| S10_extracted_44814 | Low-quality |
| S10_extracted_44817 | Not-determined |
| S10_extracted_44818 | Not-determined |
| S10_extracted_44821 | Not-determined |
| S10_extracted_44826 | Not-determined |
| S10_extracted_44827 | Not-determined |
| S10_extracted_44836 | Not-determined |
| S10_extracted_44858 | Low-quality |
| S10_extracted_44859 | Not-determined |
| S10_extracted_44860 | Not-determined |
| S10_extracted_44877 | Not-determined |
| S10_extracted_44884 | Not-determined |
| S10_extracted_44895 | Not-determined |
| S10_extracted_44900 | Not-determined |
| S10_extracted_44928 | Not-determined |
| S10_extracted_44931 | Low-quality |
| S10_extracted_44949 | Not-determined |
| S10_extracted_44950 | Not-determined |
| S10_extracted_44957 | Not-determined |
| S10_extracted_44993 | Not-determined |
| S10_extracted_45003 | Not-determined |
| S10_extracted_45004 | Not-determined |
| S10_extracted_45038 | Low-quality |
| S10_extracted_45058 | Not-determined |
| S10_extracted_45068 | Not-determined |
| S10_extracted_45114 | Not-determined |
| S10_extracted_45117 | Not-determined |
| S10_extracted_45119 | Not-determined |
| S10_extracted_45124 | Not-determined |
| S10_extracted_45140 | Not-determined |
| S10_extracted_45152 | Not-determined |
| S10_extracted_45157 | Not-determined |
| S10_extracted_45161 | Low-quality |
| S10_extracted_45174 | Not-determined |
| S10_extracted_45186 | Low-quality |
| S10_extracted_45199 | Not-determined |
| S10_extracted_45219 | Not-determined |
| S10_extracted_45235 | Low-quality |
| S10_extracted_45238 | Not-determined |
| S10_extracted_45272 | Not-determined |
| S10_extracted_45280 | Not-determined |
| S10_extracted_45297 | Not-determined |
| S10_extracted_45312 | Not-determined |
| S10_extracted_45320 | Not-determined |
| S10_extracted_45352 | Not-determined |
| S10_extracted_45362 | Not-determined |
| S10_extracted_45370 | Low-quality |
| S10_extracted_45373 | Not-determined |
| S10_extracted_45388 | Not-determined |
| S10_extracted_45400 | Not-determined |
| S10_extracted_45405 | Not-determined |
| S10_extracted_45408 | Not-determined |
| S10_extracted_45421 | Not-determined |
| S10_extracted_45462 | Low-quality |
| S10_extracted_45466 | Not-determined |
| S10_extracted_45473 | Low-quality |
| S10_extracted_45480 | Not-determined |
| S10_extracted_45495 | Not-determined |
| S10_extracted_45499 | Not-determined |
| S10_extracted_45501 | Not-determined |
| S10_extracted_45528 | Not-determined |
| S10_extracted_45529 | Not-determined |
| S10_extracted_45538 | Not-determined |
| S10_extracted_45542 | Low-quality |
| S10_extracted_45559 | Not-determined |
| S10_extracted_45569 | Not-determined |
| S10_extracted_45570 | Not-determined |
| S10_extracted_45580 | Not-determined |
| S10_extracted_45617 | Not-determined |

| | |
|---|---|
| S10_extracted_45638 | Not-determined |
| S10_extracted_45641 | Not-determined |
| S10_extracted_45666 | Not-determined |
| S10_extracted_45668 | Not-determined |
| S10_extracted_45677 | Low-quality |
| S10_extracted_45686 | Not-determined |
| S10_extracted_45689 | Not-determined |
| S10_extracted_45691 | Not-determined |
| S10_extracted_45704 | Not-determined |
| S10_extracted_45714 | Not-determined |
| S10_extracted_45738 | Not-determined |
| S10_extracted_45744 | Not-determined |
| S10_extracted_45750 | Not-determined |
| S10_extracted_45756 | Not-determined |
| S10_extracted_45758 | Not-determined |
| S10_extracted_45771 | Low-quality |
| S10_extracted_45776 | Not-determined |
| S10_extracted_45777 | Not-determined |
| S10_extracted_45781 | Not-determined |
| S10_extracted_45782 | Not-determined |
| S10_extracted_45791 | Not-determined |
| S10_extracted_45792 | Not-determined |
| S10_extracted_45801 | Not-determined |
| S10_extracted_45819 | Low-quality |
| S10_extracted_45834 | Not-determined |
| S10_extracted_45845 | Not-determined |
| S10_extracted_45851 | Not-determined |
| S10_extracted_45857 | Not-determined |
| S10_extracted_45866 | Not-determined |
| S10_extracted_45868 | Not-determined |
| S10_extracted_45900 | Not-determined |
| S10_extracted_45919 | Not-determined |
| S10_extracted_45920 | Not-determined |
| S10_extracted_45948 | Not-determined |
| S10_extracted_45956 | Not-determined |
| S10_extracted_45958 | Not-determined |
| S10_extracted_45959 | Not-determined |
| S10_extracted_45975 | Not-determined |
| S10_extracted_45980 | Not-determined |
| S10_extracted_46005 | Low-quality |
| S10_extracted_46021 | Not-determined |
| S10_extracted_46023 | Low-quality |
| S10_extracted_46031 | Not-determined |
| S10_extracted_46034 | Not-determined |
| S10_extracted_46049 | Not-determined |
| S10_extracted_46069 | Not-determined |
| S10_extracted_46078 | Low-quality |
| S10_extracted_46087 | Not-determined |
| S10_extracted_46090 | Not-determined |
| S10_extracted_46095 | Not-determined |
| S10_extracted_46106 | Not-determined |
| S10_extracted_46127 | Low-quality |
| S10_extracted_46130 | Not-determined |
| S10_extracted_46139 | Not-determined |
| S10_extracted_46147 | Low-quality |
| S10_extracted_46148 | Low-quality |
| S10_extracted_46149 | Not-determined |
| S10_extracted_46152 | Not-determined |
| S10_extracted_46153 | Not-determined |
| S10_extracted_46157 | Not-determined |
| S10_extracted_46171 | Low-quality |
| S10_extracted_46172 | Not-determined |
| S10_extracted_46177 | Low-quality |
| S10_extracted_46201 | Not-determined |
| S10_extracted_46241 | Not-determined |
| S10_extracted_46251 | Not-determined |
| S10_extracted_46255 | Not-determined |
| S10_extracted_46257 | Not-determined |
| S10_extracted_46281 | Not-determined |
| S7_vs_8332\|\|full | Medium-quality |
| S10_extracted_46285 | Not-determined |
| S10_extracted_46287 | Not-determined |
| S10_extracted_46292 | Not-determined |
| S10_extracted_46302 | Not-determined |
| S10_extracted_46307 | Not-determined |
| S10_extracted_46310 | Not-determined |
| S10_extracted_46326 | Low-quality |
| S10_extracted_46327 | Low-quality |
| S3_vs_k141_100440\|\|full | Medium-quality |
| S10_extracted_46356 | Low-quality |
| S10_extracted_46365 | Not-determined |
| S10_extracted_46376 | Not-determined |
| S10_extracted_46377 | Not-determined |
| S10_extracted_46379 | Not-determined |
| S10_extracted_46386 | Not-determined |
| S10_extracted_46387 | Not-determined |
| S10_extracted_46410 | Not-determined |
| S10_extracted_46417 | Not-determined |
| S10_extracted_46444 | Not-determined |
| S10_extracted_46460 | Not-determined |
| S10_extracted_46462 | Not-determined |
| S10_extracted_46483 | Not-determined |
| S10_extracted_46486 | Not-determined |
| S10_extracted_46505 | Not-determined |
| S10_extracted_46522 | Not-determined |
| S10_extracted_46526 | Not-determined |
| S10_extracted_46537 | Not-determined |
| S10_extracted_46538 | Not-determined |
| S10_extracted_46555 | Not-determined |
| S10_extracted_46563 | Not-determined |
| S10_extracted_46568 | Not-determined |
| S10_extracted_46584 | Low-quality |
| S10_extracted_46591 | Low-quality |
| S10_extracted_46602 | Not-determined |
| S10_extracted_46609 | Not-determined |
| S10_extracted_46626 | Not-determined |
| S10_extracted_46638 | Not-determined |
| S10_extracted_46644 | Not-determined |
| S10_extracted_46675 | Low-quality |
| S10_extracted_46676 | Not-determined |
| S10_extracted_46686 | Not-determined |
| S10_extracted_46711 | Not-determined |
| S10_extracted_46732 | Not-determined |
| S10_extracted_46753 | Not-determined |
| S10_extracted_46760 | Not-determined |
| S10_extracted_46766 | Low-quality |
| S10_extracted_46770 | Not-determined |
| S10_extracted_46792 | Not-determined |
| S10_extracted_46807 | Low-quality |
| S10_extracted_46824 | Not-determined |
| S10_extracted_46828 | Not-determined |
| S10_extracted_46857 | Not-determined |
| S10_extracted_46862 | Not-determined |
| S10_extracted_46871 | Not-determined |
| S10_extracted_46880 | Low-quality |
| S10_extracted_46885 | Low-quality |
| S10_extracted_46886 | Low-quality |
| S10_extracted_46890 | Not-determined |
| S10_extracted_46900 | Not-determined |
| S10_extracted_46911 | Not-determined |
| S10_extracted_46916 | Not-determined |
| S10_extracted_46954 | Not-determined |
| S10_extracted_46963 | Not-determined |
| S10_extracted_46964 | Not-determined |
| S10_extracted_46968 | Low-quality |
| S10_extracted_46984 | Not-determined |
| S10_extracted_46992 | Not-determined |
| S10_extracted_47005 | Low-quality |
| S10_extracted_47026 | Not-determined |
| S10_extracted_47042 | Not-determined |
| S10_extracted_47057 | Not-determined |
| S10_extracted_47066 | Not-determined |
| S10_extracted_47069 | Not-determined |
| S10_extracted_47079 | Not-determined |
| S10_extracted_47089 | Low-quality |
| S10_extracted_47106 | Low-quality |
| S10_extracted_47126 | Not-determined |
| S10_extracted_47130 | Low-quality |
| S10_extracted_47145 | Not-determined |
| S10_extracted_47146 | Not-determined |
| S10_extracted_47157 | Not-determined |
| S10_extracted_47163 | Not-determined |
| S3_vs_k141_127450\|\|full | High-quality |
| S10_extracted_47177 | Not-determined |
| S10_extracted_47190 | Not-determined |
| S10_extracted_47194 | Low-quality |
| S10_extracted_47196 | Not-determined |
| S10_extracted_47204 | Not-determined |
| S10_extracted_47212 | Not-determined |
| S10_extracted_47225 | Not-determined |
| S10_extracted_47232 | Not-determined |
| S10_extracted_47239 | Not-determined |
| S10_extracted_47242 | Not-determined |
| S10_extracted_47251 | Not-determined |
| S10_extracted_47286 | Not-determined |
| S10_extracted_47296 | Not-determined |
| S10_extracted_47301 | Not-determined |
| S10_extracted_47308 | Not-determined |
| S10_extracted_47330 | Not-determined |
| S10_extracted_47337 | Not-determined |
| S10_extracted_47339 | Not-determined |
| S10_extracted_47364 | Not-determined |
| S10_extracted_47371 | Not-determined |
| S10_extracted_47373 | Not-determined |
| S10_extracted_47419 | Not-determined |
| S10_extracted_47430 | Low-quality |
| S10_extracted_47442 | Not-determined |
| S10_extracted_47444 | Not-determined |
| S10_extracted_47448 | Not-determined |
| S10_extracted_47451 | Not-determined |
| S10_extracted_47456 | Not-determined |
| S10_extracted_47463 | Not-determined |
| S10_extracted_47470 | Low-quality |
| S10_extracted_47476 | Not-determined |
| S10_extracted_47477 | Not-determined |
| S10_extracted_47481 | Not-determined |
| S10_extracted_47491 | Not-determined |
| S10_extracted_47493 | Not-determined |
| S10_extracted_47508 | Not-determined |
| S10_extracted_47511 | Not-determined |
| S10_extracted_47513 | Low-quality |
| S10_extracted_47515 | Not-determined |
| S10_extracted_47525 | Not-determined |
| S10_extracted_47537 | Not-determined |
| S10_extracted_47538 | Not-determined |
| S10_extracted_47539 | Not-determined |
| S10_extracted_47561 | Not-determined |
| S10_extracted_47563 | Not-determined |
| S10_extracted_47567 | Not-determined |
| S10_extracted_47573 | Not-determined |
| S10_extracted_47587 | Not-determined |
| S10_extracted_47600 | Not-determined |
| S10_extracted_47615 | Not-determined |
| S10_extracted_47621 | Not-determined |
| S10_extracted_47660 | Not-determined |
| S10_extracted_47665 | Not-determined |
| S10_extracted_47679 | Not-determined |
| S10_extracted_47717 | Not-determined |
| S10_extracted_47721 | Low-quality |
| S10_extracted_47725 | Not-determined |
| S10_extracted_47731 | Not-determined |
| S10_extracted_47738 | Not-determined |
| S10_extracted_47743 | Not-determined |
| S10_extracted_47747 | Not-determined |
| S10_extracted_47757 | Not-determined |
| S10_extracted_47758 | Not-determined |
| S10_extracted_47771 | Not-determined |
| S10_extracted_47792 | Not-determined |
| S10_extracted_47804 | Not-determined |
| S10_extracted_47812 | Low-quality |
| S10_extracted_47814 | Not-determined |
| S10_extracted_47816 | Not-determined |
| S10_extracted_47817 | Not-determined |
| S10_extracted_47827 | Not-determined |
| S10_extracted_47842 | Not-determined |
| S10_extracted_47850 | Low-quality |
| S10_extracted_47859 | Not-determined |
| S10_extracted_47864 | Not-determined |
| S10_extracted_47872 | Not-determined |
| S10_extracted_47873 | Not-determined |
| S10_extracted_47877 | Not-determined |
| S10_extracted_47878 | Low-quality |
| S10_extracted_47896 | Not-determined |
| S10_extracted_47912 | Not-determined |
| S10_extracted_47913 | Not-determined |
| S10_extracted_47923 | Not-determined |
| S10_extracted_47926 | Not-determined |
| S10_extracted_47941 | Not-determined |
| S10_extracted_47948 | Not-determined |
| S10_extracted_47949 | Not-determined |
| S10_extracted_47951 | Not-determined |
| S10_extracted_47958 | Low-quality |
| S10_extracted_47962 | Not-determined |
| S10_extracted_47968 | Not-determined |
| S10_extracted_47970 | Not-determined |
| S10_extracted_47971 | Not-determined |
| S10_extracted_47983 | Not-determined |
| S10_extracted_47992 | Not-determined |
| S10_extracted_47994 | Low-quality |
| S10_extracted_48012 | Not-determined |
| S10_extracted_48051 | Not-determined |
| S10_extracted_48064 | Low-quality |
| S10_extracted_48065 | Not-determined |
| S10_extracted_48070 | Not-determined |
| S10_extracted_48071 | Not-determined |
| S10_extracted_48085 | Low-quality |
| S10_extracted_48086 | Not-determined |
| S10_extracted_48089 | Not-determined |
| S10_extracted_48100 | Not-determined |
| S10_extracted_48109 | Not-determined |
| S10_extracted_48112 | Not-determined |
| S10_extracted_48115 | Not-determined |
| S10_extracted_48119 | Not-determined |
| S10_extracted_48132 | Not-determined |
| S10_extracted_48137 | Not-determined |
| S10_extracted_48154 | Not-determined |
| S10_extracted_48157 | Not-determined |
| S10_extracted_48163 | Not-determined |
| S10_extracted_48194 | Not-determined |
| S10_extracted_48206 | Low-quality |
| S10_extracted_48228 | Not-determined |
| S10_extracted_48230 | Not-determined |
| S10_extracted_48239 | Not-determined |
| S10_extracted_48244 | Not-determined |
| S10_extracted_48256 | Not-determined |
| S10_extracted_48266 | Not-determined |
| S10_extracted_48275 | Not-determined |
| S10_extracted_48282 | Not-determined |
| S10_extracted_48287 | Not-determined |
| S10_extracted_48292 | Low-quality |
| S10_extracted_48316 | Low-quality |
| S10_extracted_48317 | Not-determined |
| S10_extracted_48326 | Not-determined |
| S10_extracted_48329 | Not-determined |
| S10_extracted_48332 | Not-determined |
| S10_extracted_48341 | Not-determined |
| S10_extracted_48350 | Not-determined |
| S10_extracted_48366 | Not-determined |
| S10_extracted_48398 | Not-determined |
| S10_extracted_48407 | Not-determined |
| S10_extracted_48425 | Not-determined |
| S10_extracted_48430 | Low-quality |
| S10_extracted_48436 | Low-quality |
| S10_extracted_48452 | Low-quality |
| S10_extracted_48461 | Not-determined |
| S10_extracted_48506 | Not-determined |
| S10_extracted_48515 | Not-determined |
| S10_extracted_48552 | Low-quality |
| S10_extracted_48553 | Not-determined |
| S10_extracted_48563 | Not-determined |
| S10_extracted_48570 | Not-determined |
| S10_extracted_48583 | Not-determined |
| S10_extracted_48591 | Not-determined |
| S10_extracted_48606 | Not-determined |
| S10_extracted_48628 | Not-determined |
| S10_extracted_48653 | Not-determined |
| S10_extracted_48658 | Not-determined |
| S10_extracted_48663 | Not-determined |
| S10_extracted_48665 | Not-determined |
| S10_extracted_48672 | Low-quality |
| S10_extracted_48682 | Not-determined |
| S10_extracted_48690 | Not-determined |
| S10_extracted_48691 | Not-determined |
| S10_extracted_48696 | Low-quality |
| S10_extracted_48709 | Not-determined |
| S10_extracted_48727 | Not-determined |
| S10_extracted_48729 | Not-determined |
| S10_extracted_48738 | Not-determined |
| S10_extracted_48751 | Not-determined |
| S10_extracted_48764 | Not-determined |
| S10_extracted_48765 | Low-quality |
| S10_extracted_48797 | Not-determined |
| S10_extracted_48829 | Not-determined |
| S10_extracted_48832 | Not-determined |
| S10_extracted_48834 | Not-determined |
| S10_extracted_48840 | Not-determined |
| S10_extracted_48853 | Low-quality |
| S10_extracted_48861 | Not-determined |
| S10_extracted_48870 | Low-quality |
| S10_extracted_48879 | Not-determined |
| S10_extracted_48889 | Not-determined |
| S10_extracted_48894 | Not-determined |
| S10_extracted_48901 | Not-determined |
| S10_extracted_48908 | Not-determined |
| S10_extracted_48938 | Not-determined |
| S10_extracted_48946 | Not-determined |
| S10_extracted_48948 | Not-determined |
| S10_extracted_48956 | Not-determined |
| S10_extracted_48965 | Not-determined |
| S10_extracted_48978 | Low-quality |
| S10_extracted_48981 | Not-determined |
| S10_extracted_48998 | Low-quality |
| S10_extracted_49044 | Not-determined |
| S10_extracted_49056 | Not-determined |
| S10_extracted_49061 | Not-determined |
| S10_extracted_49066 | Not-determined |
| S10_extracted_49069 | Low-quality |
| S10_extracted_49080 | Not-determined |
| S10_extracted_49084 | Not-determined |
| S10_extracted_49087 | Not-determined |
| S10_extracted_49091 | Not-determined |
| S10_extracted_49108 | Not-determined |
| S10_extracted_49112 | Not-determined |
| S10_extracted_49120 | Not-determined |
| S10_extracted_49133 | Not-determined |
| S10_extracted_49142 | Not-determined |
| S10_extracted_49150 | Not-determined |
| S10_extracted_49157 | Not-determined |
| S10_extracted_49165 | Not-determined |
| S10_extracted_49180 | Not-determined |
| S10_extracted_49181 | Low-quality |
| S10_extracted_49200 | Not-determined |
| S10_extracted_49246 | Low-quality |
| S10_extracted_49249 | Low-quality |
| S10_extracted_49254 | Low-quality |
| S10_extracted_49257 | Low-quality |
| S10_extracted_49258 | Low-quality |
| S10_extracted_49259 | Not-determined |
| S10_extracted_49284 | Not-determined |
| S10_extracted_49294 | Not-determined |
| S10_extracted_49313 | Not-determined |
| S10_extracted_49322 | Not-determined |
| S10_extracted_49324 | Not-determined |
| S10_extracted_49325 | Not-determined |
| S10_extracted_49331 | Not-determined |
| S10_extracted_49356 | Low-quality |
| S10_extracted_49397 | Not-determined |
| S10_extracted_49401 | Not-determined |
| S10_extracted_49408 | Low-quality |
| S10_extracted_49409 | Not-determined |
| S10_extracted_49413 | Not-determined |
| S10_extracted_49414 | Not-determined |
| S10_extracted_49416 | Not-determined |
| S10_extracted_49420 | Not-determined |
| S10_extracted_49422 | Not-determined |
| S10_extracted_49425 | Not-determined |
| S10_extracted_49428 | Not-determined |
| S10_extracted_49446 | Not-determined |
| S10_extracted_49450 | Not-determined |
| S10_extracted_49454 | Not-determined |
| S10_extracted_49467 | Not-determined |
| S10_extracted_49468 | Not-determined |
| S10_extracted_49480 | Not-determined |
| S10_extracted_49486 | Not-determined |
| S10_extracted_49487 | Low-quality |
| S10_extracted_49492 | Not-determined |
| S10_extracted_49514 | Low-quality |
| S10_extracted_49519 | Not-determined |
| S10_extracted_49531 | Not-determined |
| S10_extracted_49532 | Not-determined |
| S10_extracted_49551 | Not-determined |
| S10_extracted_49552 | Not-determined |
| S10_extracted_49557 | Not-determined |
| S10_extracted_49563 | Not-determined |
| S10_extracted_49590 | Not-determined |
| S10_extracted_49603 | Low-quality |
| S10_extracted_49613 | Not-determined |
| S10_extracted_49647 | Not-determined |
| S10_extracted_49657 | Not-determined |
| S10_extracted_49667 | Not-determined |
| S10_extracted_49682 | Not-determined |
| S10_extracted_49686 | Not-determined |
| S10_extracted_49687 | Low-quality |
| S10_extracted_49692 | Not-determined |
| S10_extracted_49693 | Low-quality |
| S10_extracted_49703 | Not-determined |
| S10_extracted_49749 | Low-quality |
| S10_extracted_49755 | Not-determined |
| S10_extracted_49757 | Not-determined |
| S10_extracted_49767 | Not-determined |
| S10_extracted_49777 | Not-determined |
| S10_extracted_49781 | Not-determined |
| S10_extracted_49782 | Not-determined |
| S10_extracted_49793 | Not-determined |
| S10_extracted_49825 | Not-determined |
| S10_extracted_49829 | Not-determined |
| S10_extracted_49837 | Not-determined |
| S10_extracted_49849 | Not-determined |
| S10_extracted_49851 | Not-determined |
| S10_extracted_49885 | Not-determined |
| S10_extracted_49913 | Not-determined |
| S10_extracted_49914 | Not-determined |
| S10_extracted_49922 | Not-determined |
| S10_extracted_49924 | Not-determined |
| S10_extracted_49929 | Not-determined |
| S10_extracted_49938 | Low-quality |
| S10_extracted_49947 | Not-determined |
| S10_extracted_49977 | Low-quality |
| S10_extracted_49994 | Not-determined |
| S10_extracted_49996 | Not-determined |
| S10_extracted_50001 | Not-determined |
| S10_extracted_50010 | Not-determined |
| S10_extracted_50014 | Not-determined |
| S10_extracted_50017 | Not-determined |
| S10_extracted_50031 | Not-determined |
| S10_extracted_50042 | Not-determined |
| S10_extracted_50078 | Low-quality |
| S10_extracted_50100 | Not-determined |
| S10_extracted_50108 | Low-quality |
| S10_extracted_50121 | Not-determined |
| S10_extracted_50126 | Not-determined |
| S10_extracted_50135 | Not-determined |
| S10_extracted_50139 | Low-quality |
| S10_extracted_50149 | Not-determined |
| S10_extracted_50180 | Not-determined |
| S10_extracted_50199 | Not-determined |
| S10_extracted_50204 | Not-determined |
| S10_extracted_50209 | Not-determined |
| S10_extracted_50215 | Not-determined |
| S10_extracted_50239 | Not-determined |
| S10_extracted_50246 | Not-determined |
| S10_extracted_50250 | Not-determined |
| S10_extracted_50251 | Not-determined |
| S10_extracted_50265 | Not-determined |
| S10_extracted_50272 | Not-determined |
| S10_extracted_50293 | Low-quality |
| S10_extracted_50302 | Not-determined |
| S10_extracted_50303 | Not-determined |
| S10_extracted_50307 | Not-determined |
| S10_extracted_50315 | Not-determined |
| S10_extracted_50317 | Low-quality |
| S10_extracted_50335 | Not-determined |
| S10_extracted_50344 | Not-determined |
| S10_extracted_50347 | Not-determined |
| S10_extracted_50363 | Not-determined |
| S10_extracted_50382 | Not-determined |
| S10_extracted_50384 | Not-determined |
| S10_extracted_50387 | Not-determined |
| S10_extracted_50389 | Not-determined |
| S10_extracted_50397 | Not-determined |
| S10_extracted_50406 | Not-determined |
| S10_extracted_50408 | Not-determined |
| S10_extracted_50409 | Not-determined |
| S10_extracted_50421 | Not-determined |
| S10_extracted_50422 | Low-quality |
| S10_extracted_50437 | Not-determined |
| S10_extracted_50440 | Not-determined |
| S10_extracted_50447 | Low-quality |
| S10_extracted_50452 | Not-determined |
| S10_extracted_50457 | Not-determined |
| S10_extracted_50487 | Not-determined |
| S10_extracted_50490 | Not-determined |
| S10_extracted_50510 | Not-determined |
| S10_extracted_50513 | Not-determined |
| S10_extracted_50515 | Not-determined |
| S10_extracted_50518 | Low-quality |
| S10_extracted_50520 | Not-determined |
| S10_extracted_50522 | Low-quality |
| S10_extracted_50526 | Not-determined |
| S10_extracted_50545 | Not-determined |
| S10_extracted_50547 | Not-determined |
| S10_extracted_50550 | Not-determined |
| S10_extracted_50553 | Not-determined |
| S10_extracted_50599 | Not-determined |
| S10_extracted_50608 | Not-determined |
| S10_extracted_50635 | Not-determined |
| S10_extracted_50652 | Not-determined |
| S10_extracted_50657 | Not-determined |
| S10_extracted_50681 | Not-determined |
| S10_extracted_50698 | Not-determined |
| S10_extracted_50699 | Not-determined |
| S10_extracted_50703 | Not-determined |
| S10_extracted_50706 | Not-determined |
| S10_extracted_50718 | Not-determined |
| S10_extracted_50722 | Not-determined |
| S10_extracted_50727 | Not-determined |
| S10_extracted_50735 | Not-determined |
| S10_extracted_50746 | Not-determined |
| S10_extracted_50763 | Not-determined |
| S10_extracted_50767 | Not-determined |
| S10_extracted_50797 | Not-determined |
| S10_extracted_50801 | Low-quality |
| S10_extracted_50805 | Not-determined |
| S10_extracted_50806 | Low-quality |
| S10_extracted_50810 | Not-determined |
| S10_extracted_50844 | Not-determined |
| S10_extracted_50850 | Not-determined |
| S10_extracted_50856 | Not-determined |
| S10_extracted_50864 | Not-determined |
| S10_extracted_50871 | Not-determined |
| S10_extracted_50877 | Not-determined |
| S10_extracted_50890 | Not-determined |
| S10_extracted_50891 | Not-determined |
| S10_extracted_50936 | Not-determined |
| S10_extracted_50937 | Low-quality |
| S10_extracted_50938 | Not-determined |
| S10_extracted_50941 | Low-quality |
| S10_extracted_50951 | Not-determined |
| S10_extracted_50963 | Not-determined |
| S10_extracted_50984 | Not-determined |
| S10_extracted_50986 | Not-determined |
| S10_extracted_50995 | Not-determined |
| S10_extracted_50998 | Not-determined |
| S10_extracted_50999 | Not-determined |
| S10_extracted_51014 | Not-determined |
| S10_extracted_51030 | Low-quality |
| S10_extracted_51031 | Not-determined |
| S10_extracted_51037 | Not-determined |
| S10_extracted_51051 | Not-determined |
| S10_extracted_51054 | Low-quality |
| S10_extracted_51059 | Not-determined |
| S10_extracted_51063 | Low-quality |
| S10_extracted_51096 | Not-determined |
| S10_extracted_51103 | Not-determined |
| S10_extracted_51116 | Not-determined |
| S10_extracted_51119 | Not-determined |
| S10_extracted_51127 | Low-quality |
| S10_extracted_51160 | Low-quality |
| S10_extracted_51167 | Not-determined |
| S10_extracted_51179 | Not-determined |
| S10_extracted_51210 | Not-determined |
| S10_extracted_51226 | Not-determined |
| S10_extracted_51227 | Not-determined |
| S10_extracted_51262 | Not-determined |
| S10_extracted_51278 | Not-determined |
| S10_extracted_51279 | Not-determined |
| S10_extracted_51280 | Low-quality |
| S10_extracted_51283 | Not-determined |
| S10_extracted_51286 | Low-quality |
| S10_extracted_51287 | Not-determined |
| S10_extracted_51288 | Not-determined |
| S10_extracted_51295 | Not-determined |
| S10_extracted_51311 | Not-determined |
| S10_extracted_51317 | Not-determined |
| S10_extracted_51322 | Not-determined |
| S10_extracted_51325 | Not-determined |
| S10_extracted_51346 | Not-determined |
| S10_extracted_51347 | Not-determined |
| S10_extracted_51354 | Not-determined |
| S10_extracted_51355 | Not-determined |
| S10_extracted_51357 | Low-quality |
| S10_extracted_51367 | Not-determined |
| S10_extracted_51368 | Not-determined |
| S10_extracted_51379 | Not-determined |
| S10_extracted_51418 | Not-determined |
| S10_extracted_51430 | Not-determined |
| S10_extracted_51434 | Not-determined |
| S10_extracted_51447 | Not-determined |
| S10_extracted_51451 | Low-quality |

| | |
|---|---|
| S10_extracted_S1460 | Low-quality |
| S10_extracted_S1463 | Low-quality |
| S10_extracted_S1476 | Not-determined |
| S10_extracted_S1486 | Not-determined |
| S10_extracted_S1494 | Not-determined |
| S10_extracted_S1495 | Not-determined |
| S10_extracted_S1496 | Not-determined |
| S10_extracted_S1526 | Not-determined |
| S10_extracted_S1529 | Low-quality |
| S10_extracted_S1543 | Not-determined |
| S10_extracted_S1573 | Not-determined |
| S10_extracted_S1574 | Low-quality |
| S10_extracted_S1587 | Not-determined |
| S10_extracted_S1589 | Not-determined |
| S10_extracted_S1599 | Not-determined |
| S10_extracted_S1612 | Not-determined |
| S10_extracted_S1619 | Not-determined |
| S10_extracted_S1622 | Not-determined |
| S10_extracted_S1635 | Not-determined |
| S10_extracted_S1646 | Not-determined |
| S10_extracted_S1665 | Not-determined |
| S10_extracted_S1672 | Not-determined |
| S10_extracted_S1682 | Not-determined |
| S10_extracted_S1687 | Not-determined |
| S10_extracted_S1707 | Not-determined |
| S10_extracted_S1715 | Low-quality |
| S10_extracted_S1720 | Not-determined |
| S10_extracted_S1731 | Not-determined |
| S10_extracted_S1742 | Not-determined |
| S10_extracted_S1792 | Not-determined |
| S10_extracted_S1800 | Not-determined |
| S10_extracted_S1810 | Not-determined |
| S10_extracted_S1812 | Not-determined |
| S10_extracted_S1820 | Not-determined |
| S10_extracted_S1836 | Not-determined |
| S10_extracted_S1854 | Low-quality |
| S10_extracted_S1860 | Not-determined |
| S10_extracted_S1863 | Not-determined |
| S10_extracted_S1876 | Low-quality |
| S10_extracted_S1887 | Not-determined |
| S10_extracted_S1901 | Not-determined |
| S10_extracted_S1904 | Not-determined |
| S10_extracted_S1911 | Not-determined |
| S10_extracted_S1914 | Not-determined |
| S10_extracted_S1921 | Not-determined |
| S10_extracted_S1923 | Not-determined |
| S10_extracted_S1927 | Not-determined |
| S10_extracted_S1928 | Not-determined |
| S10_extracted_S1932 | Low-quality |
| S10_extracted_S1935 | Not-determined |
| S10_extracted_S1952 | Not-determined |
| S10_extracted_S1965 | Not-determined |
| S10_extracted_S1998 | Not-determined |
| S10_extracted_S2000 | Not-determined |
| S10_extracted_S2007 | Not-determined |
| S10_extracted_S2015 | Low-quality |
| S10_extracted_S2018 | Not-determined |
| S10_extracted_S2021 | Not-determined |
| S10_extracted_S2027 | Not-determined |
| S10_extracted_S2060 | Not-determined |
| S10_extracted_S2061 | Not-determined |
| S10_extracted_S2063 | Not-determined |
| S10_extracted_S2094 | Not-determined |
| S10_extracted_S2101 | Not-determined |
| S10_extracted_S2113 | Not-determined |
| S10_extracted_S2119 | Not-determined |
| S10_extracted_S2136 | Not-determined |
| S10_extracted_S2154 | Not-determined |
| S10_extracted_S2156 | Not-determined |
| S10_extracted_S2160 | Low-quality |
| S10_extracted_S2169 | Not-determined |
| S10_extracted_S2188 | Low-quality |
| S10_extracted_S2196 | Low-quality |
| S10_extracted_S2199 | Not-determined |
| S10_extracted_S2233 | Not-determined |
| S10_extracted_S2242 | Not-determined |
| S10_extracted_S2247 | Not-determined |
| S10_extracted_S2253 | Not-determined |
| S10_extracted_S2255 | Not-determined |
| S10_extracted_S2262 | Not-determined |
| S10_extracted_S2265 | Not-determined |
| S10_extracted_S2305 | Not-determined |
| S10_extracted_S2320 | Not-determined |
| S10_extracted_S2321 | Not-determined |
| S10_extracted_S2335 | Not-determined |
| S10_extracted_S2346 | Not-determined |
| S10_extracted_S2355 | Not-determined |
| S10_extracted_S2365 | Low-quality |
| S10_extracted_S2374 | Not-determined |
| S10_extracted_S2379 | Low-quality |
| S10_extracted_S2382 | Not-determined |
| S10_extracted_S2390 | Not-determined |
| S10_extracted_S2392 | Not-determined |
| S10_extracted_S2399 | Not-determined |
| S10_extracted_S2400 | Not-determined |
| S10_extracted_S2417 | Not-determined |
| S10_extracted_S2430 | Not-determined |
| S10_extracted_S2437 | Not-determined |
| S10_extracted_S2461 | Low-quality |
| S10_extracted_S2471 | Not-determined |
| S10_extracted_S2473 | Low-quality |
| S10_extracted_S2475 | Not-determined |
| S10_extracted_S2483 | Not-determined |
| S10_extracted_S2486 | Not-determined |
| S10_extracted_S2490 | Not-determined |
| S10_extracted_S2499 | Not-determined |
| S10_extracted_S2501 | Not-determined |
| S10_extracted_S2504 | Not-determined |
| S10_extracted_S2507 | Not-determined |
| S10_extracted_S2510 | Not-determined |
| S10_extracted_S2511 | Not-determined |
| S10_extracted_S2516 | Not-determined |
| S10_extracted_S2539 | Not-determined |
| S10_extracted_S2541 | Not-determined |
| S10_extracted_S2553 | Not-determined |
| S10_extracted_S2559 | Not-determined |
| S10_extracted_S2577 | Not-determined |
| S10_extracted_S2578 | Not-determined |
| S10_extracted_S2584 | Not-determined |
| S10_extracted_S2587 | Not-determined |
| S10_extracted_S2614 | Not-determined |
| S10_extracted_S2618 | Not-determined |
| S10_extracted_S2625 | Not-determined |
| S10_extracted_S2628 | Not-determined |
| S10_extracted_S2629 | Not-determined |
| S10_extracted_S2634 | Not-determined |
| S10_extracted_S2637 | Not-determined |
| S10_extracted_S2645 | Not-determined |
| S10_extracted_S2651 | Not-determined |
| S10_extracted_S2655 | Not-determined |
| S10_extracted_S2666 | Not-determined |
| S10_extracted_S2674 | Not-determined |
| S10_extracted_S2682 | Not-determined |
| S10_extracted_S2685 | Not-determined |
| S10_extracted_S2704 | Low-quality |
| S10_extracted_S2713 | Not-determined |
| S10_extracted_S2724 | Not-determined |
| S10_extracted_S2742 | Not-determined |
| S10_extracted_S2754 | Not-determined |
| S10_extracted_S2802 | Not-determined |
| S10_extracted_S2807 | Not-determined |
| S10_extracted_S2810 | Not-determined |
| S10_extracted_S2811 | Not-determined |
| S10_extracted_S2825 | Low-quality |
| S10_extracted_S2837 | Not-determined |
| S10_extracted_S2841 | Low-quality |
| S10_extracted_S2845 | Low-quality |
| S10_extracted_S2852 | Low-quality |
| S10_extracted_S2858 | Not-determined |
| S10_extracted_S2861 | Not-determined |
| S10_extracted_S2882 | Not-determined |
| S10_extracted_S2893 | Not-determined |
| S10_extracted_S2903 | Not-determined |
| S10_extracted_S2904 | Not-determined |
| S10_extracted_S2916 | Not-determined |
| S10_extracted_S2921 | Low-quality |
| S10_extracted_S2927 | Not-determined |
| S10_extracted_S2928 | Low-quality |
| S10_extracted_S2932 | Not-determined |
| S10_extracted_S2936 | Not-determined |
| S10_extracted_S2953 | Not-determined |
| S10_extracted_S2970 | Not-determined |
| S10_extracted_S2979 | Not-determined |
| S10_extracted_S2984 | Low-quality |
| S10_extracted_S2989 | Low-quality |
| S10_extracted_S3018 | Not-determined |
| S10_extracted_S3025 | Not-determined |
| S10_extracted_S3034 | Not-determined |
| S10_extracted_S3044 | Not-determined |
| S10_extracted_S3045 | Not-determined |
| S10_extracted_S3051 | Not-determined |
| S10_extracted_S3052 | Not-determined |
| S10_extracted_S3063 | Not-determined |
| S10_extracted_S3064 | Not-determined |
| S10_extracted_S3069 | Not-determined |
| S10_extracted_S3083 | Low-quality |
| S10_extracted_S3094 | Not-determined |
| S10_extracted_S3099 | Not-determined |
| S10_extracted_S3101 | Not-determined |
| S10_extracted_S3128 | Not-determined |
| S10_extracted_S3137 | Not-determined |
| S10_extracted_S3141 | Not-determined |
| S10_extracted_S3144 | Not-determined |
| S10_extracted_S3150 | Not-determined |
| S10_extracted_S3157 | Not-determined |
| S10_extracted_S3162 | Not-determined |
| S10_extracted_S3169 | Not-determined |
| S10_extracted_S3177 | Not-determined |
| S10_extracted_S3193 | Low-quality |
| S10_extracted_S3216 | Not-determined |
| S10_extracted_S3254 | Not-determined |
| S10_extracted_S3266 | Not-determined |
| S10_extracted_S3270 | Not-determined |
| S10_extracted_S3288 | Not-determined |
| S10_extracted_S3299 | Not-determined |
| S10_extracted_S3308 | Not-determined |
| S10_extracted_S3309 | Not-determined |
| S10_extracted_S3312 | Not-determined |
| S10_extracted_S3315 | Not-determined |
| S10_extracted_S3330 | Not-determined |
| S10_extracted_S3333 | Low-quality |
| S10_extracted_S3338 | Not-determined |
| S10_extracted_S3373 | Low-quality |
| S10_extracted_S3392 | Not-determined |
| S10_extracted_S3394 | Not-determined |
| S10_extracted_S3401 | Not-determined |
| S10_extracted_S3406 | Not-determined |
| S10_extracted_S3422 | Not-determined |
| S10_extracted_S3424 | Low-quality |
| S10_extracted_S3426 | Not-determined |
| S10_extracted_S3431 | Not-determined |
| S10_extracted_S3437 | Not-determined |
| S10_extracted_S3439 | Not-determined |
| S10_extracted_S3440 | Low-quality |
| S10_extracted_S3446 | Not-determined |
| S10_extracted_S3447 | Not-determined |
| S10_extracted_S3448 | Not-determined |
| S10_extracted_S3449 | Not-determined |
| S10_extracted_S3454 | Low-quality |
| S10_extracted_S3462 | Low-quality |
| S10_extracted_S3477 | Not-determined |
| S10_extracted_S3492 | Not-determined |
| S10_extracted_S3527 | Not-determined |
| S10_extracted_S3530 | Low-quality |
| S10_extracted_S3542 | Not-determined |
| S10_extracted_S3543 | Not-determined |
| S10_extracted_S3566 | Not-determined |
| S10_extracted_S3576 | Not-determined |
| S10_extracted_S3590 | Not-determined |
| S10_extracted_S3591 | Not-determined |
| S10_extracted_S3598 | Not-determined |
| S10_extracted_S3604 | Not-determined |
| S10_extracted_S3609 | Not-determined |
| S10_extracted_S3636 | Not-determined |
| S10_extracted_S3637 | Not-determined |
| S10_extracted_S3640 | Not-determined |
| S10_extracted_S3657 | Not-determined |
| S10_extracted_S3665 | Not-determined |
| S10_extracted_S3675 | Not-determined |
| S10_extracted_S3676 | Not-determined |
| S10_extracted_S3678 | Not-determined |
| S10_extracted_S3680 | Not-determined |
| S10_extracted_S3684 | Not-determined |
| S10_extracted_S3709 | Not-determined |
| S10_extracted_S3712 | Not-determined |
| S10_extracted_S3719 | Not-determined |
| S10_extracted_S3723 | Not-determined |
| S10_extracted_S3724 | Not-determined |
| S10_extracted_S3727 | Low-quality |
| S10_extracted_S3728 | Not-determined |
| S10_extracted_S3748 | Not-determined |
| S10_extracted_S3753 | Low-quality |
| S10_extracted_S3756 | Not-determined |
| S10_extracted_S3758 | Low-quality |
| S10_extracted_S3770 | Low-quality |
| S10_extracted_S3778 | Not-determined |
| S10_extracted_S3784 | Not-determined |
| S10_extracted_S3793 | Not-determined |
| S10_extracted_S3794 | Not-determined |
| S10_extracted_S3803 | Not-determined |
| S10_extracted_S3809 | Not-determined |
| S10_extracted_S3810 | Not-determined |
| S10_extracted_S3811 | Not-determined |
| S10_extracted_S3812 | Not-determined |
| S10_extracted_S3816 | Not-determined |
| S10_extracted_S3827 | Not-determined |
| S10_extracted_S3832 | Not-determined |
| S10_extracted_S3841 | Low-quality |
| S10_extracted_S3852 | Low-quality |
| S10_extracted_S3884 | Not-determined |
| S10_extracted_S3904 | Not-determined |
| S10_extracted_S3927 | Not-determined |
| S10_extracted_S3933 | Not-determined |
| S10_extracted_S3936 | Not-determined |
| S10_extracted_S3943 | Not-determined |
| S10_extracted_S3974 | Not-determined |
| S10_extracted_S3976 | Not-determined |
| S10_extracted_S3995 | Low-quality |
| S10_extracted_S4014 | Not-determined |
| S10_extracted_S4021 | Low-quality |
| S10_extracted_S4031 | Low-quality |
| S10_extracted_S4035 | Low-quality |
| S10_extracted_S4040 | Not-determined |
| S10_extracted_S4043 | Not-determined |
| S10_extracted_S4044 | Not-determined |
| S10_extracted_S4057 | Not-determined |
| S10_extracted_S4067 | Not-determined |
| S10_extracted_S4095 | Not-determined |
| S10_extracted_S4096 | Not-determined |
| S10_extracted_S4126 | Not-determined |
| S10_extracted_S4131 | Not-determined |
| S10_extracted_S4137 | Not-determined |
| S10_extracted_S4150 | Not-determined |
| S10_extracted_S4154 | Not-determined |
| S10_extracted_S4166 | Not-determined |
| S10_extracted_S4169 | Not-determined |
| S10_extracted_S4195 | Not-determined |
| S10_extracted_S4199 | Not-determined |
| S10_extracted_S4204 | Not-determined |
| S10_extracted_S4236 | Low-quality |
| S10_extracted_S4248 | Not-determined |
| S10_extracted_S4250 | Low-quality |
| S10_extracted_S4273 | Not-determined |
| S10_extracted_S4274 | Low-quality |
| S10_extracted_S4280 | Not-determined |
| S10_extracted_S4284 | Not-determined |
| S10_extracted_S4286 | Not-determined |
| S10_extracted_S4290 | Not-determined |
| S10_extracted_S4301 | Low-quality |
| S10_extracted_S4305 | Not-determined |
| S10_extracted_S4326 | Low-quality |
| S10_extracted_S4341 | Not-determined |
| S10_extracted_S4342 | Low-quality |
| S10_extracted_S4345 | Not-determined |
| S10_extracted_S4348 | Not-determined |
| S10_extracted_S4359 | Not-determined |
| S10_extracted_S4363 | Not-determined |
| S10_extracted_S4367 | Not-determined |
| S10_extracted_S4386 | Not-determined |
| S10_extracted_S4389 | Not-determined |
| S10_extracted_S4390 | Not-determined |
| S10_extracted_S4402 | Not-determined |
| S10_extracted_S4404 | Not-determined |
| S10_extracted_S4411 | Not-determined |
| S10_extracted_S4434 | Not-determined |
| S10_extracted_S4440 | Low-quality |
| S10_extracted_S4460 | Not-determined |
| S10_extracted_S4461 | Not-determined |
| S10_extracted_S4481 | Not-determined |
| S10_extracted_S4483 | Not-determined |
| S10_extracted_S4489 | Not-determined |
| S10_extracted_S4515 | Not-determined |
| S10_extracted_S4525 | Not-determined |
| S10_extracted_S4539 | Not-determined |
| S10_extracted_S4545 | Low-quality |
| S10_extracted_S4550 | Not-determined |
| S10_extracted_S4559 | Not-determined |
| S10_extracted_S4591 | Not-determined |
| S10_extracted_S4593 | Not-determined |
| S10_extracted_S4600 | Not-determined |
| S10_extracted_S4601 | Not-determined |
| S10_extracted_S4602 | Not-determined |
| S10_extracted_S4608 | Not-determined |
| S10_extracted_S4619 | Not-determined |
| S10_extracted_S4623 | Not-determined |
| S10_extracted_S4634 | Not-determined |
| S10_extracted_S4639 | Not-determined |
| S10_extracted_S4661 | Low-quality |
| S10_extracted_S4707 | Not-determined |
| S10_extracted_S4708 | Not-determined |
| S10_extracted_S4720 | Low-quality |
| S10_extracted_S4732 | Not-determined |
| S10_extracted_S4740 | Not-determined |
| S10_extracted_S4743 | Not-determined |
| S10_extracted_S4750 | Not-determined |
| S10_extracted_S4757 | Not-determined |
| S10_extracted_S4759 | Not-determined |
| S10_extracted_S4761 | Not-determined |
| S10_extracted_S4762 | Not-determined |
| S10_extracted_S4763 | Not-determined |
| S10_extracted_S4796 | Not-determined |
| S10_extracted_S4836 | Not-determined |
| S10_extracted_S4842 | Low-quality |
| S10_extracted_S4850 | Not-determined |
| S10_extracted_S4869 | Not-determined |
| S10_extracted_S4870 | Not-determined |
| S10_extracted_S4894 | Not-determined |
| S10_extracted_S4897 | Not-determined |
| S10_extracted_S4899 | Not-determined |
| S10_extracted_S4901 | Not-determined |
| S10_extracted_S4904 | Not-determined |
| S10_extracted_S4933 | Not-determined |
| S10_extracted_S4954 | Low-quality |
| S10_extracted_S4969 | Not-determined |
| S10_extracted_S4974 | Not-determined |
| S10_extracted_S4981 | Not-determined |
| S10_extracted_S4991 | Not-determined |
| S10_extracted_S4999 | Not-determined |
| S10_extracted_S5011 | Not-determined |
| S10_extracted_S5038 | Not-determined |
| S10_extracted_S5080 | Not-determined |
| S10_extracted_S5083 | Not-determined |
| S10_extracted_S5104 | Not-determined |
| S10_extracted_S5115 | Not-determined |
| S10_extracted_S5135 | Not-determined |
| S10_extracted_S5139 | Not-determined |
| S10_extracted_S5169 | Low-quality |
| S10_extracted_S5171 | Low-quality |
| S10_extracted_S5180 | Not-determined |
| S10_extracted_S5184 | Not-determined |
| S10_extracted_S5186 | Not-determined |
| S10_extracted_S5208 | Not-determined |
| S10_extracted_S5232 | Not-determined |
| S10_extracted_S5249 | Not-determined |
| S10_extracted_S5251 | Not-determined |
| S10_extracted_S5258 | Not-determined |
| S10_extracted_S5268 | Low-quality |
| S10_extracted_S5284 | Not-determined |
| S10_extracted_S5290 | Low-quality |
| S10_extracted_S5295 | Not-determined |
| S10_extracted_S5302 | Not-determined |
| S10_extracted_S5307 | Not-determined |
| S10_extracted_S5308 | Not-determined |
| S10_extracted_S5314 | Not-determined |
| S10_extracted_S5390 | Not-determined |
| S10_extracted_S5397 | Not-determined |
| S10_extracted_S5409 | Not-determined |
| S10_extracted_S5435 | Not-determined |
| S10_extracted_S5443 | Not-determined |
| S10_extracted_S5445 | Low-quality |
| S10_extracted_S5453 | Not-determined |
| S10_extracted_S5468 | Not-determined |
| S10_extracted_S5474 | Not-determined |
| S10_extracted_S5480 | Not-determined |
| S10_extracted_S5483 | Not-determined |
| S10_extracted_S5485 | Not-determined |
| S10_extracted_S5496 | Not-determined |
| S10_extracted_S5512 | Low-quality |
| S10_extracted_S5526 | Low-quality |
| S10_extracted_S5529 | Not-determined |
| S10_extracted_S5532 | Not-determined |
| S10_extracted_S5534 | Not-determined |
| S10_extracted_S5547 | Not-determined |
| S10_extracted_S5550 | Not-determined |
| S10_extracted_S5551 | Not-determined |
| S10_extracted_S5556 | Not-determined |
| S10_extracted_S5624 | Low-quality |
| S10_extracted_S5643 | Not-determined |
| S10_extracted_S5681 | Not-determined |
| S10_extracted_S5682 | Not-determined |
| S10_extracted_S5687 | Not-determined |
| S10_extracted_S5688 | Low-quality |
| S10_extracted_S5715 | Low-quality |
| S10_extracted_S5717 | Not-determined |
| S10_extracted_S5735 | Low-quality |
| S10_extracted_S5745 | Not-determined |
| S10_extracted_S5760 | Not-determined |
| S10_extracted_S5777 | Not-determined |
| S10_extracted_S5779 | Not-determined |
| S10_extracted_S5818 | Not-determined |
| S10_extracted_S5831 | Not-determined |
| S10_extracted_S5834 | Not-determined |
| S10_extracted_S5835 | Not-determined |
| S10_extracted_S5838 | Not-determined |
| S10_extracted_S5844 | Not-determined |
| S10_extracted_S5848 | Not-determined |
| S10_extracted_S5854 | Not-determined |
| S10_extracted_S5868 | Not-determined |
| S10_extracted_S5869 | Low-quality |
| S10_extracted_S5872 | Not-determined |
| S10_extracted_S5880 | Not-determined |
| S10_extracted_S5884 | Not-determined |
| S10_extracted_S5909 | Not-determined |
| S10_extracted_S5913 | Not-determined |
| S10_extracted_S5920 | Low-quality |
| S10_extracted_S5945 | Not-determined |
| S10_extracted_S5955 | Low-quality |
| S10_extracted_S5957 | Not-determined |
| S10_extracted_S5977 | Not-determined |
| S10_extracted_S5984 | Not-determined |
| S10_extracted_S5997 | Not-determined |
| S10_extracted_S6006 | Not-determined |
| S10_extracted_S6021 | Low-quality |
| S10_extracted_S6024 | Not-determined |
| S10_extracted_S6030 | Not-determined |
| S10_extracted_S6056 | Not-determined |
| S10_extracted_S6062 | Not-determined |
| S10_extracted_S6081 | Not-determined |
| S10_extracted_S6084 | Not-determined |
| S10_extracted_S6099 | Not-determined |
| S10_extracted_S6110 | Not-determined |
| S10_extracted_S6111 | Not-determined |
| S10_extracted_S6123 | Low-quality |
| S10_extracted_S6133 | Not-determined |
| S10_extracted_S6145 | Not-determined |
| S10_extracted_S6153 | Not-determined |
| S10_extracted_S6154 | Not-determined |
| S10_extracted_S6156 | Not-determined |
| S10_extracted_S6170 | Not-determined |
| S10_extracted_S6173 | Not-determined |
| S10_extracted_S6176 | Not-determined |
| S10_extracted_S6199 | Not-determined |
| S10_extracted_S6213 | Not-determined |
| S10_extracted_S6214 | Not-determined |
| S10_extracted_S6221 | Not-determined |
| S10_extracted_S6225 | Not-determined |
| S10_extracted_S6248 | Not-determined |
| S10_extracted_S6258 | Not-determined |
| S10_extracted_S6281 | Not-determined |
| S10_extracted_S6292 | Not-determined |
| S10_extracted_S6294 | Not-determined |
| S10_extracted_S6301 | Low-quality |
| S10_extracted_S6309 | Low-quality |
| S10_extracted_S6324 | Not-determined |
| S10_extracted_S6347 | Not-determined |
| S10_extracted_S6358 | Not-determined |
| S10_extracted_S6359 | Not-determined |
| S10_extracted_S6373 | Not-determined |
| S10_extracted_S6397 | Not-determined |
| S10_extracted_S6398 | Not-determined |
| S10_extracted_S6414 | Not-determined |
| S10_extracted_S6419 | Not-determined |
| S10_extracted_S6427 | Not-determined |
| S10_extracted_S6433 | Low-quality |
| S10_extracted_S6448 | Not-determined |
| S10_extracted_S6501 | Low-quality |
| S10_extracted_S6511 | Not-determined |
| S10_extracted_S6514 | Low-quality |
| S10_extracted_S6518 | Not-determined |
| S10_extracted_S6527 | Not-determined |
| S10_extracted_S6540 | Not-determined |
| S10_extracted_S6542 | Not-determined |
| S10_extracted_S6559 | Not-determined |
| S10_extracted_S6564 | Not-determined |
| S10_extracted_S6569 | Not-determined |
| S10_extracted_S6570 | Not-determined |
| S10_extracted_S6585 | Not-determined |
| S10_extracted_S6595 | Not-determined |
| S10_extracted_S6601 | Not-determined |
| S10_extracted_S6604 | Not-determined |
| S10_extracted_S6613 | Not-determined |
| S10_extracted_S6614 | Not-determined |
| S10_extracted_S6624 | Not-determined |
| S10_extracted_S6663 | Not-determined |
| S10_extracted_S6678 | Low-quality |
| S10_extracted_S6682 | Not-determined |
| S10_extracted_S6699 | Not-determined |
| S10_extracted_S6706 | Not-determined |
| S10_extracted_S6709 | Not-determined |
| S10_extracted_S6710 | Not-determined |
| S10_extracted_S6712 | Not-determined |
| S10_extracted_S6739 | Not-determined |
| S10_extracted_S6762 | Not-determined |
| S10_extracted_S6768 | Not-determined |
| S10_extracted_S6781 | Not-determined |
| S10_extracted_S6788 | Not-determined |
| S10_extracted_S6791 | Not-determined |
| S10_extracted_S6792 | Not-determined |
| S10_extracted_S6794 | Not-determined |
| S10_extracted_S6802 | Not-determined |
| S10_extracted_S6808 | Not-determined |
| S10_extracted_S6836 | Not-determined |
| S10_extracted_S6841 | Not-determined |
| S10_extracted_S6861 | Not-determined |
| S10_extracted_S6867 | Not-determined |
| S10_extracted_S6868 | Not-determined |
| S10_extracted_S6874 | Not-determined |
| S10_extracted_S6890 | Not-determined |
| S10_extracted_S6901 | Not-determined |
| S10_extracted_S6908 | Not-determined |
| S10_extracted_S6909 | Not-determined |
| S10_extracted_S6911 | Not-determined |
| S10_extracted_S6914 | Not-determined |
| S10_extracted_S6940 | Low-quality |
| S10_extracted_S6943 | Not-determined |
| S10_extracted_S6958 | Not-determined |
| S10_extracted_S6983 | Not-determined |
| S10_extracted_S6985 | Low-quality |
| S10_extracted_S7007 | Not-determined |
| S10_extracted_S7022 | Not-determined |
| S10_extracted_S7036 | Not-determined |
| S10_extracted_S7038 | Not-determined |
| S10_extracted_S7040 | Not-determined |
| S10_extracted_S7053 | Low-quality |
| S10_extracted_S7063 | Low-quality |
| S10_extracted_S7068 | Not-determined |
| S10_extracted_S7075 | Low-quality |
| S10_extracted_S7100 | Not-determined |
| S10_extracted_S7106 | Not-determined |
| S10_extracted_S7109 | Not-determined |
| S10_extracted_S7111 | Low-quality |
| S10_extracted_S7122 | Low-quality |
| S10_extracted_S7140 | Not-determined |
| S10_extracted_S7144 | Not-determined |
| S10_extracted_S7146 | Not-determined |
| S10_extracted_S7163 | Low-quality |
| S10_extracted_S7185 | Not-determined |
| S10_extracted_S7192 | Not-determined |
| S10_extracted_S7207 | Not-determined |
| S10_extracted_S7214 | Not-determined |
| S10_extracted_S7228 | Not-determined |
| S10_extracted_S7230 | Not-determined |
| S10_extracted_S7232 | Not-determined |
| S10_extracted_S7233 | Not-determined |
| S10_extracted_S7238 | Not-determined |
| S10_extracted_S7242 | Not-determined |
| S10_extracted_S7245 | Low-quality |
| S10_extracted_S7250 | Not-determined |
| S10_extracted_S7251 | Not-determined |

| | |
|---|---|
| S10_extracted_57263 | Low-quality |
| S10_extracted_57264 | Not-determined |
| S10_extracted_57268 | Not-determined |
| S10_extracted_57269 | Not-determined |
| S10_extracted_57271 | Not-determined |
| S10_extracted_57304 | Not-determined |
| S10_extracted_57307 | Not-determined |
| S10_extracted_57308 | Low-quality |
| S10_extracted_57314 | Not-determined |
| S10_extracted_57348 | Not-determined |
| S10_extracted_57350 | Not-determined |
| S10_extracted_57358 | Not-determined |
| S10_extracted_57359 | Low-quality |
| S10_extracted_57364 | Not-determined |
| S10_extracted_57370 | Low-quality |
| S10_extracted_57399 | Not-determined |
| S10_extracted_57405 | Not-determined |
| S10_extracted_57413 | Not-determined |
| S10_extracted_57430 | Not-determined |
| S10_extracted_57437 | Not-determined |
| S10_extracted_57447 | Not-determined |
| S10_extracted_57455 | Not-determined |
| S10_extracted_57472 | Low-quality |
| S10_extracted_57475 | Not-determined |
| S10_extracted_57480 | Low-quality |
| S10_extracted_57483 | Not-determined |
| S10_extracted_57484 | Not-determined |
| S10_extracted_57499 | Not-determined |
| S10_extracted_57525 | Not-determined |
| S10_extracted_57535 | Not-determined |
| S10_extracted_57542 | Not-determined |
| S10_extracted_57552 | Low-quality |
| S10_extracted_57555 | Not-determined |
| S10_extracted_57572 | Not-determined |
| S10_extracted_57591 | Not-determined |
| S10_extracted_57593 | Not-determined |
| S10_extracted_57596 | Not-determined |
| S10_extracted_57609 | Not-determined |
| S10_extracted_57616 | Low-quality |
| S10_extracted_57627 | Not-determined |
| S10_extracted_57638 | Not-determined |
| S10_extracted_57642 | Not-determined |
| S10_extracted_57644 | Not-determined |
| S10_extracted_57688 | Low-quality |
| S10_extracted_57701 | Not-determined |
| S8_vs_k141_152553\|\|full | Medium-quality |
| S10_extracted_57712 | Low-quality |
| S10_extracted_57728 | Not-determined |
| S10_extracted_57736 | Not-determined |
| S10_extracted_57766 | Not-determined |
| S10_extracted_57769 | Not-determined |
| S10_extracted_57770 | Not-determined |
| S10_extracted_57776 | Not-determined |
| S10_extracted_57780 | Not-determined |
| S10_extracted_57799 | Not-determined |
| S10_extracted_57800 | Low-quality |
| S10_extracted_57809 | Not-determined |
| S10_extracted_57820 | Not-determined |
| S10_extracted_57823 | Not-determined |
| S10_extracted_57825 | Not-determined |
| S10_extracted_57837 | Not-determined |
| S10_extracted_57839 | Not-determined |
| S10_extracted_57843 | Not-determined |
| S10_extracted_57858 | Not-determined |
| S10_extracted_57880 | Not-determined |
| S10_extracted_57881 | Not-determined |
| S10_extracted_57884 | Not-determined |
| S10_extracted_57891 | Not-determined |
| S10_extracted_57901 | Not-determined |
| S10_extracted_57903 | Not-determined |
| S10_extracted_57930 | Not-determined |
| S10_extracted_57935 | Not-determined |
| S10_extracted_57942 | Not-determined |
| S10_extracted_57953 | Low-quality |
| S10_extracted_57956 | Not-determined |
| S10_extracted_57957 | Low-quality |
| S10_extracted_57959 | Not-determined |
| S10_extracted_57967 | Not-determined |
| S10_extracted_57976 | Not-determined |
| S10_extracted_58010 | Not-determined |
| S10_extracted_58016 | Not-determined |
| S10_extracted_58018 | Not-determined |
| S10_extracted_58038 | Not-determined |
| S10_extracted_58046 | Not-determined |
| S10_extracted_58058 | Low-quality |
| S10_extracted_58062 | Not-determined |
| S10_extracted_58069 | Low-quality |
| S10_extracted_58074 | Not-determined |
| S10_extracted_58078 | Low-quality |
| S10_extracted_58088 | Low-quality |
| S10_extracted_58089 | Low-quality |
| S10_extracted_58101 | Not-determined |
| S10_extracted_58123 | Not-determined |
| S10_extracted_58125 | Not-determined |
| S10_extracted_58127 | Not-determined |
| S10_extracted_58168 | Not-determined |
| S10_extracted_58197 | Not-determined |
| S10_extracted_58205 | Not-determined |
| S10_extracted_58211 | Not-determined |
| S10_extracted_58213 | Not-determined |
| S10_extracted_58253 | Not-determined |
| S10_extracted_58254 | Not-determined |
| S10_extracted_58271 | Not-determined |
| S10_extracted_58277 | Low-quality |
| S10_extracted_58300 | Low-quality |
| S10_extracted_58301 | Not-determined |
| S10_extracted_58303 | Not-determined |
| S10_extracted_58307 | Not-determined |
| S10_extracted_58340 | Not-determined |
| S10_extracted_58341 | Not-determined |
| S10_extracted_58343 | Not-determined |
| S10_extracted_58354 | Low-quality |
| S10_extracted_58355 | Not-determined |
| S10_extracted_58359 | Not-determined |
| S10_extracted_58364 | Low-quality |
| S10_extracted_58380 | Not-determined |
| S10_extracted_58387 | Not-determined |
| S10_extracted_58388 | Not-determined |
| S10_extracted_58389 | Not-determined |
| S10_extracted_58396 | Not-determined |
| S10_extracted_58407 | Not-determined |
| S10_extracted_58408 | Not-determined |
| S10_extracted_58465 | Not-determined |
| S10_extracted_58521 | Not-determined |
| S10_extracted_58524 | Not-determined |
| S10_extracted_58528 | Not-determined |
| S10_extracted_58538 | Not-determined |
| S10_extracted_58540 | Not-determined |
| S10_extracted_58574 | Low-quality |
| S10_extracted_58577 | Not-determined |
| S10_extracted_58616 | Not-determined |
| S10_extracted_58621 | Low-quality |
| S10_extracted_58642 | Low-quality |
| S10_extracted_58644 | Not-determined |
| S10_extracted_58648 | Not-determined |
| S10_extracted_58650 | Low-quality |
| S10_extracted_58651 | Not-determined |
| S10_extracted_58655 | Not-determined |
| S10_extracted_58661 | Not-determined |
| S10_extracted_58665 | Not-determined |
| S10_extracted_58673 | Not-determined |
| S10_extracted_58675 | Not-determined |
| S10_extracted_58679 | Not-determined |
| S10_extracted_58687 | Not-determined |
| S10_extracted_58697 | Not-determined |
| S10_extracted_58699 | Not-determined |
| S10_extracted_58707 | Not-determined |
| S10_extracted_58709 | Not-determined |
| S10_extracted_58713 | Not-determined |
| S10_extracted_58741 | Low-quality |
| S10_extracted_58762 | Low-quality |
| S10_extracted_58765 | Not-determined |
| S10_extracted_58766 | Not-determined |
| S10_extracted_58775 | Not-determined |
| S10_extracted_58790 | Low-quality |
| S10_extracted_58799 | Low-quality |
| S10_extracted_58800 | Low-quality |
| S10_extracted_58802 | Not-determined |
| S10_extracted_58825 | Not-determined |
| S10_extracted_58834 | Low-quality |
| S10_extracted_58835 | Not-determined |
| S10_extracted_58842 | Not-determined |
| S10_extracted_58845 | Not-determined |
| S10_extracted_58849 | Not-determined |
| S10_extracted_58874 | Not-determined |
| S10_extracted_58879 | Low-quality |
| S10_extracted_58915 | Not-determined |
| S10_extracted_58929 | Not-determined |
| S10_extracted_58934 | Not-determined |
| S10_extracted_58940 | Not-determined |
| S10_extracted_58954 | Not-determined |
| S10_extracted_58962 | Not-determined |
| S10_extracted_58978 | Not-determined |
| S10_extracted_58979 | Not-determined |
| S10_extracted_58984 | Not-determined |
| S10_extracted_58998 | Not-determined |
| S10_extracted_59011 | Not-determined |
| S10_extracted_59014 | Low-quality |
| S10_extracted_59015 | Not-determined |
| S10_extracted_59019 | Not-determined |
| S10_extracted_59020 | Not-determined |
| S10_extracted_59028 | Not-determined |
| S10_extracted_59047 | Not-determined |
| S10_extracted_59068 | Low-quality |
| S10_extracted_59073 | Not-determined |
| S10_extracted_59085 | Not-determined |
| S10_extracted_59090 | Not-determined |
| S10_extracted_59099 | Not-determined |
| S10_extracted_59113 | Not-determined |
| S10_extracted_59150 | Not-determined |
| S10_extracted_59178 | Not-determined |
| S10_extracted_59182 | Not-determined |
| S10_extracted_59206 | Low-quality |
| S10_extracted_59209 | Low-quality |
| S10_extracted_59232 | Not-determined |
| S10_extracted_59244 | Not-determined |
| S10_extracted_59245 | Not-determined |
| S10_extracted_59246 | Not-determined |
| S10_extracted_59257 | Not-determined |
| S10_extracted_59260 | Not-determined |
| S10_extracted_59270 | Not-determined |
| S10_extracted_59283 | Not-determined |
| S10_extracted_59284 | Low-quality |
| S10_extracted_59291 | Not-determined |
| S10_extracted_59293 | Not-determined |
| S10_extracted_59297 | Not-determined |
| S10_extracted_59331 | Not-determined |
| S10_extracted_59338 | Not-determined |
| S10_extracted_59369 | Not-determined |
| S10_extracted_59382 | Not-determined |
| S10_extracted_59414 | Not-determined |
| S10_extracted_59422 | Low-quality |
| S10_extracted_59428 | Not-determined |
| S10_extracted_59430 | Not-determined |
| S10_extracted_59435 | Low-quality |
| S10_extracted_59447 | Not-determined |
| S10_extracted_59449 | Not-determined |
| S10_extracted_59460 | Not-determined |
| S10_extracted_59461 | Not-determined |
| S10_extracted_59474 | Not-determined |
| S10_extracted_59537 | Not-determined |
| S10_extracted_59538 | Not-determined |
| S10_extracted_59558 | Not-determined |
| S10_extracted_59570 | Not-determined |
| S10_extracted_59575 | Not-determined |
| S10_extracted_59579 | Not-determined |
| S10_extracted_59582 | Not-determined |
| S10_extracted_59607 | Not-determined |
| S10_extracted_59617 | Not-determined |
| S10_extracted_59623 | Not-determined |
| S10_extracted_59643 | Low-quality |
| S10_extracted_59644 | Not-determined |
| S10_extracted_59651 | Not-determined |
| S10_extracted_59671 | Not-determined |
| S10_extracted_59689 | Not-determined |
| S10_extracted_59712 | Not-determined |
| S10_extracted_59714 | Low-quality |
| S10_extracted_59718 | Not-determined |
| S10_extracted_59720 | Not-determined |
| S10_extracted_59722 | Not-determined |
| S10_extracted_59732 | Not-determined |
| S10_extracted_59737 | Not-determined |
| S10_extracted_59742 | Not-determined |
| S10_extracted_59761 | Not-determined |
| S10_extracted_59763 | Not-determined |
| S10_extracted_59770 | Not-determined |
| S10_extracted_59772 | Not-determined |
| S10_extracted_59785 | Not-determined |
| S10_extracted_59800 | Not-determined |
| S10_extracted_59810 | Not-determined |
| S10_extracted_59821 | Not-determined |
| S10_extracted_59827 | Not-determined |
| S10_extracted_59839 | Not-determined |
| S10_extracted_59844 | Not-determined |
| S10_extracted_59890 | Not-determined |
| S10_extracted_59906 | Not-determined |
| S10_extracted_59920 | Not-determined |
| S10_extracted_59921 | Not-determined |
| S10_extracted_59939 | Not-determined |
| S10_extracted_59942 | Low-quality |
| S10_extracted_59951 | Not-determined |
| S10_extracted_59952 | Low-quality |
| S10_extracted_59964 | Not-determined |
| S10_extracted_59968 | Not-determined |
| S10_extracted_59972 | Not-determined |
| S10_extracted_59974 | Not-determined |
| S10_extracted_60003 | Not-determined |
| S10_extracted_60007 | Not-determined |
| S10_extracted_60012 | Not-determined |
| S10_extracted_60042 | Not-determined |
| S10_extracted_60050 | Low-quality |
| S10_extracted_60060 | Not-determined |
| S10_extracted_60063 | Low-quality |
| S10_extracted_60075 | Not-determined |
| S10_extracted_60084 | Not-determined |
| S10_extracted_60095 | Not-determined |
| S10_extracted_60096 | Not-determined |
| S10_extracted_60107 | Not-determined |
| S10_extracted_60108 | Not-determined |
| S10_extracted_60111 | Low-quality |
| S10_extracted_60122 | Not-determined |
| S10_extracted_60155 | Not-determined |
| S10_extracted_60170 | Not-determined |
| S10_extracted_60173 | Not-determined |
| S10_extracted_60192 | Not-determined |
| S10_extracted_60205 | Not-determined |
| S10_extracted_60208 | Not-determined |
| S10_extracted_60220 | Low-quality |
| S10_extracted_60225 | Low-quality |
| S10_extracted_60227 | Not-determined |
| S10_extracted_60237 | Not-determined |
| S10_extracted_60264 | Not-determined |
| S10_extracted_60280 | Not-determined |
| S10_extracted_60289 | Not-determined |
| S10_extracted_60303 | Not-determined |
| S10_extracted_60305 | Not-determined |
| S10_extracted_60350 | Not-determined |
| S10_extracted_60351 | Not-determined |
| S10_extracted_60355 | Not-determined |
| S10_extracted_60370 | Not-determined |
| S10_extracted_60375 | Not-determined |
| S10_extracted_60379 | Not-determined |
| S10_extracted_60390 | Low-quality |
| S10_extracted_60392 | Not-determined |
| S10_extracted_60396 | Not-determined |
| S10_extracted_60398 | Not-determined |
| S10_extracted_60429 | Not-determined |
| S10_extracted_60436 | Not-determined |
| S10_extracted_60440 | Not-determined |
| S10_extracted_60443 | Not-determined |
| S10_extracted_60444 | Not-determined |
| S10_extracted_60447 | Not-determined |
| S10_extracted_60449 | Not-determined |
| S10_extracted_60458 | Not-determined |
| S10_extracted_60460 | Not-determined |
| S10_extracted_60465 | Low-quality |
| S10_extracted_60466 | Not-determined |
| S10_extracted_60474 | Not-determined |
| S10_extracted_60485 | Not-determined |
| S10_extracted_60488 | Not-determined |
| S10_extracted_60492 | Low-quality |
| S10_extracted_60496 | Not-determined |
| S10_extracted_60522 | Not-determined |
| S10_extracted_60523 | Not-determined |
| S10_extracted_60529 | Not-determined |
| S10_extracted_60541 | Not-determined |
| S10_extracted_60545 | Not-determined |
| S10_extracted_60548 | Not-determined |
| S10_extracted_60559 | Not-determined |
| S10_extracted_60575 | Not-determined |
| S10_extracted_60582 | Not-determined |
| S10_extracted_60583 | Not-determined |
| S10_extracted_60586 | Low-quality |
| S10_extracted_60587 | Not-determined |
| S10_extracted_60610 | Not-determined |
| S10_extracted_60611 | Not-determined |
| S10_extracted_60618 | Low-quality |
| S10_extracted_60620 | Not-determined |
| S10_extracted_60635 | Not-determined |
| S10_extracted_60662 | Not-determined |
| S10_extracted_60663 | Not-determined |
| S10_extracted_60691 | Not-determined |
| S10_extracted_60692 | Not-determined |
| S10_extracted_60698 | Not-determined |
| S10_extracted_60708 | Not-determined |
| S10_extracted_60712 | Not-determined |
| S10_extracted_60714 | Not-determined |
| S10_extracted_60715 | Low-quality |
| S10_extracted_60754 | Not-determined |
| S10_extracted_60756 | Not-determined |
| S10_extracted_60762 | Not-determined |
| S10_extracted_60763 | Not-determined |
| S10_extracted_60772 | Not-determined |
| S10_extracted_60774 | Not-determined |
| S10_extracted_60775 | Not-determined |
| S10_extracted_60793 | Not-determined |
| S10_extracted_60815 | Not-determined |
| S10_extracted_60818 | Not-determined |
| S10_extracted_60828 | Low-quality |
| S10_extracted_60847 | Low-quality |
| S10_extracted_60851 | Not-determined |
| S10_extracted_60868 | Not-determined |
| S10_extracted_60871 | Low-quality |
| S10_extracted_60889 | Not-determined |
| S10_extracted_60890 | Not-determined |
| S10_extracted_60891 | Not-determined |
| S10_extracted_60897 | Not-determined |
| S10_extracted_60938 | Not-determined |
| S10_extracted_60953 | Not-determined |
| S10_extracted_60960 | Not-determined |
| S10_extracted_60966 | Not-determined |
| S10_extracted_60971 | Not-determined |
| S10_extracted_60993 | Not-determined |
| S10_extracted_60998 | Not-determined |
| S10_extracted_61001 | Low-quality |
| S10_extracted_61031 | Not-determined |
| S10_extracted_61048 | Not-determined |
| S10_extracted_61051 | Low-quality |
| S10_extracted_61057 | Not-determined |
| S10_extracted_61070 | Not-determined |
| S10_extracted_61074 | Not-determined |
| S10_extracted_61082 | Not-determined |
| S10_extracted_61091 | Not-determined |
| S10_extracted_61125 | Not-determined |
| S10_extracted_61131 | Low-quality |
| S10_extracted_61136 | Low-quality |
| S10_extracted_61154 | Not-determined |
| S10_extracted_61163 | Not-determined |
| S10_extracted_61190 | Low-quality |
| S10_extracted_61198 | Not-determined |
| S10_extracted_61218 | Not-determined |
| S10_extracted_61236 | Not-determined |
| S10_extracted_61244 | Not-determined |
| S10_extracted_61264 | Not-determined |
| S10_extracted_61283 | Not-determined |
| S10_extracted_61330 | Not-determined |
| S10_extracted_61337 | Low-quality |
| S10_extracted_61345 | Not-determined |
| S10_extracted_61352 | Not-determined |
| S10_extracted_61363 | Not-determined |
| S10_extracted_61382 | Low-quality |
| S10_extracted_61391 | Low-quality |
| S10_extracted_61392 | Not-determined |
| S10_extracted_61396 | Not-determined |
| S10_extracted_61407 | Not-determined |
| S10_extracted_61417 | Not-determined |
| S10_extracted_61428 | Not-determined |
| S10_extracted_61437 | Low-quality |
| S10_extracted_61443 | Low-quality |
| S10_extracted_61449 | Not-determined |
| S10_extracted_61462 | Not-determined |
| S10_extracted_61469 | Not-determined |
| S10_extracted_61480 | Not-determined |
| S10_extracted_61490 | Low-quality |
| S10_extracted_61492 | Not-determined |
| S10_extracted_61493 | Not-determined |
| S10_extracted_61494 | Not-determined |
| S10_extracted_61496 | Not-determined |
| S10_extracted_61503 | Not-determined |
| S10_extracted_61505 | Not-determined |
| S10_extracted_61515 | Not-determined |
| S10_extracted_61516 | Not-determined |
| S10_extracted_61540 | Not-determined |
| S10_extracted_61547 | Not-determined |
| S10_extracted_61548 | Not-determined |
| S10_extracted_61549 | Not-determined |
| S10_extracted_61553 | Not-determined |
| S10_extracted_61564 | Not-determined |
| S10_extracted_61593 | Not-determined |
| S10_extracted_61599 | Not-determined |
| S10_extracted_61604 | Low-quality |
| S10_extracted_61611 | Not-determined |
| S10_extracted_61624 | Not-determined |
| S10_extracted_61631 | Not-determined |
| S10_extracted_61678 | Not-determined |
| S5_vs_k141_8847\|\|full | Medium-quality |
| S10_extracted_61684 | Not-determined |
| S10_extracted_61685 | Not-determined |
| S10_extracted_61699 | Not-determined |
| S10_extracted_61700 | Not-determined |
| S10_extracted_61714 | Low-quality |
| S10_extracted_61716 | Not-determined |
| S10_extracted_61748 | Not-determined |
| S10_extracted_61750 | Not-determined |
| S10_extracted_61765 | Not-determined |
| S10_extracted_61770 | Not-determined |
| S10_extracted_61771 | Not-determined |
| S10_extracted_61786 | Not-determined |
| S10_extracted_61792 | Not-determined |
| S10_extracted_61797 | Low-quality |
| S10_extracted_61809 | Not-determined |
| S10_extracted_61824 | Not-determined |
| S10_extracted_61830 | Not-determined |
| S10_extracted_61831 | Low-quality |
| S10_extracted_61849 | Not-determined |
| S10_extracted_61853 | Not-determined |
| S10_extracted_61854 | Low-quality |
| S10_extracted_61873 | Not-determined |
| S10_extracted_61893 | Not-determined |
| S10_extracted_61896 | Not-determined |
| S10_extracted_61900 | Not-determined |
| S10_extracted_61905 | Not-determined |
| S10_extracted_61942 | Not-determined |
| S10_extracted_61946 | Not-determined |
| S10_extracted_61950 | Not-determined |
| S10_extracted_61974 | Low-quality |
| S10_extracted_61977 | Low-quality |
| S10_extracted_61979 | Not-determined |
| S10_extracted_61990 | Not-determined |
| S10_extracted_62022 | Not-determined |
| S10_extracted_62040 | Low-quality |
| S10_extracted_62042 | Not-determined |
| S10_extracted_62044 | Not-determined |
| S10_extracted_62047 | Not-determined |
| S10_extracted_62051 | Not-determined |
| S10_extracted_62065 | Not-determined |
| S10_extracted_62075 | Not-determined |
| S10_extracted_62076 | Low-quality |
| S10_extracted_62080 | Low-quality |
| S10_extracted_62082 | Not-determined |
| S10_extracted_62090 | Not-determined |
| S10_extracted_62098 | Not-determined |
| S10_extracted_62134 | Low-quality |
| S10_extracted_62137 | Not-determined |
| S10_extracted_62146 | Not-determined |
| S10_extracted_62152 | Not-determined |
| S10_extracted_62172 | Not-determined |
| S10_extracted_62182 | Not-determined |
| S10_extracted_62198 | Not-determined |
| S10_extracted_62208 | Not-determined |
| S10_extracted_62210 | Not-determined |
| S10_extracted_62211 | Not-determined |
| S10_extracted_62221 | Not-determined |
| S14_extracted_k141_44452 | High-quality |
| S10_extracted_62242 | Not-determined |
| S10_extracted_62255 | Not-determined |
| S10_extracted_62257 | Not-determined |
| S10_extracted_62264 | Not-determined |
| S10_extracted_62270 | Not-determined |
| S10_extracted_62273 | Not-determined |
| S10_extracted_62282 | Not-determined |
| S10_extracted_62301 | Not-determined |
| S10_extracted_62313 | Not-determined |
| S10_extracted_62317 | Not-determined |
| S10_extracted_62320 | Low-quality |
| S10_extracted_62352 | Not-determined |
| S10_extracted_62358 | Not-determined |
| S10_extracted_62362 | Not-determined |
| S10_extracted_62364 | Not-determined |
| S10_extracted_62365 | Not-determined |
| S10_extracted_62375 | Not-determined |
| S10_extracted_62388 | Not-determined |
| S10_extracted_62390 | Not-determined |
| S10_extracted_62423 | Not-determined |
| S10_extracted_62425 | Not-determined |
| S10_extracted_62426 | Not-determined |
| S10_extracted_62436 | Low-quality |
| S10_extracted_62439 | Not-determined |
| S10_extracted_62442 | Not-determined |
| S10_extracted_62457 | Not-determined |
| S10_extracted_62470 | Not-determined |
| S10_extracted_62475 | Not-determined |
| S10_extracted_62480 | Not-determined |
| S10_extracted_62495 | Not-determined |
| S10_extracted_62504 | Not-determined |
| S10_extracted_62508 | Not-determined |
| S10_extracted_62509 | Low-quality |
| S10_extracted_62520 | Not-determined |
| S10_extracted_62536 | Not-determined |
| S10_extracted_62557 | Low-quality |
| S10_extracted_62566 | Not-determined |
| S10_extracted_62569 | Not-determined |
| S10_extracted_62582 | Not-determined |
| S10_extracted_62587 | Low-quality |
| S10_extracted_62588 | Not-determined |
| S10_extracted_62596 | Not-determined |
| S10_extracted_62597 | Not-determined |
| S10_extracted_62608 | Not-determined |
| S10_extracted_62637 | Not-determined |
| S10_extracted_62639 | Not-determined |
| S10_extracted_62644 | Not-determined |
| S10_extracted_62659 | Not-determined |
| S10_extracted_62660 | Not-determined |
| S10_extracted_62675 | Not-determined |
| S10_extracted_62683 | Not-determined |
| S10_extracted_62694 | Low-quality |
| S10_extracted_62718 | Not-determined |
| S10_extracted_62727 | Not-determined |
| S10_extracted_62728 | Not-determined |
| S10_extracted_62732 | Not-determined |
| S10_extracted_62744 | Not-determined |
| S10_extracted_62768 | Not-determined |
| S10_extracted_62790 | Not-determined |
| S10_extracted_62791 | Not-determined |
| S10_extracted_62802 | Not-determined |
| S10_extracted_62808 | Not-determined |
| S10_extracted_62825 | Not-determined |
| S10_extracted_62827 | Not-determined |
| S10_extracted_62829 | Not-determined |
| S10_extracted_62844 | Not-determined |
| S10_extracted_62847 | Not-determined |
| S10_extracted_62873 | Not-determined |
| S10_extracted_62881 | Not-determined |
| S10_extracted_62901 | Not-determined |
| S10_extracted_62904 | Not-determined |
| S10_extracted_62905 | Low-quality |
| S10_extracted_62908 | Not-determined |
| S10_extracted_62910 | Not-determined |
| S10_extracted_62922 | Not-determined |
| S10_extracted_62948 | Not-determined |
| S10_extracted_62966 | Not-determined |
| S10_extracted_62968 | Not-determined |
| S10_extracted_62972 | Low-quality |
| S10_extracted_62975 | Not-determined |
| S10_extracted_62986 | Not-determined |
| S10_extracted_62992 | Not-determined |
| S10_extracted_63024 | Not-determined |
| S10_extracted_63029 | Not-determined |
| S10_extracted_63060 | Not-determined |
| S10_extracted_63087 | Not-determined |
| S10_extracted_63089 | Not-determined |
| S10_extracted_63096 | Not-determined |
| S10_extracted_63105 | Not-determined |
| S10_extracted_63111 | Not-determined |
| S10_extracted_63127 | Not-determined |
| S10_extracted_63131 | Not-determined |
| S10_extracted_63153 | Low-quality |
| S10_extracted_63159 | Not-determined |
| S10_extracted_63169 | Not-determined |

| | |
|---|---|
| S10_extracted_63175 | Not-determined |
| S10_extracted_63177 | Not-determined |
| S10_extracted_63180 | Low-quality |
| S10_extracted_63185 | Not-determined |
| S10_extracted_63195 | Not-determined |
| S10_extracted_63209 | Not-determined |
| S10_extracted_63211 | Not-determined |
| S10_extracted_63217 | Not-determined |
| S10_extracted_63218 | Not-determined |
| S10_extracted_63221 | Not-determined |
| S10_extracted_63227 | Not-determined |
| S10_extracted_63228 | Not-determined |
| S10_extracted_63233 | Low-quality |
| S10_extracted_63234 | Not-determined |
| S10_extracted_63271 | Not-determined |
| S10_extracted_63273 | Not-determined |
| S10_extracted_63275 | Not-determined |
| S10_extracted_63301 | Not-determined |
| S10_extracted_63304 | Not-determined |
| S10_extracted_63323 | Low-quality |
| S10_extracted_63325 | Not-determined |
| S10_extracted_63327 | Low-quality |
| S10_extracted_63338 | Not-determined |
| S10_extracted_63345 | Not-determined |
| S10_extracted_63356 | Not-determined |
| S10_extracted_63363 | Not-determined |
| S10_extracted_63376 | Not-determined |
| S10_extracted_63399 | Low-quality |
| S10_extracted_63401 | Not-determined |
| S10_extracted_63405 | Low-quality |
| S10_extracted_63412 | Not-determined |
| S10_extracted_63438 | Not-determined |
| S10_extracted_63461 | Not-determined |
| S10_extracted_63462 | Not-determined |
| S10_extracted_63473 | Not-determined |
| S10_extracted_63481 | Not-determined |
| S10_extracted_63483 | Not-determined |
| S10_extracted_63484 | Not-determined |
| S10_extracted_63488 | Not-determined |
| S10_extracted_63497 | Low-quality |
| S10_extracted_63511 | Low-quality |
| S10_extracted_63512 | Low-quality |
| S10_extracted_63515 | Low-quality |
| S10_extracted_63526 | Not-determined |
| S10_extracted_63558 | Not-determined |
| S10_extracted_63562 | Low-quality |
| S10_extracted_63567 | Not-determined |
| S10_extracted_63580 | Not-determined |
| S10_extracted_63588 | Not-determined |
| S10_extracted_63593 | Not-determined |
| S10_extracted_63601 | Not-determined |
| S10_extracted_63615 | Not-determined |
| S10_extracted_63633 | Not-determined |
| S10_extracted_63653 | Not-determined |
| S10_extracted_63658 | Not-determined |
| S10_extracted_63667 | Not-determined |
| S10_extracted_63690 | Not-determined |
| S10_extracted_63697 | Low-quality |
| S10_extracted_63707 | Not-determined |
| S10_extracted_63715 | Not-determined |
| S10_extracted_63722 | Not-determined |
| S10_extracted_63724 | Low-quality |
| S10_extracted_63737 | Not-determined |
| S10_extracted_63756 | Not-determined |
| S10_extracted_63758 | Not-determined |
| S10_extracted_63762 | Low-quality |
| S10_extracted_63766 | Not-determined |
| S10_extracted_63797 | Not-determined |
| S10_extracted_63810 | Not-determined |
| S10_extracted_63815 | Low-quality |
| S10_extracted_63828 | Not-determined |
| S10_extracted_63831 | Not-determined |
| S10_extracted_63847 | Not-determined |
| S10_extracted_63852 | Low-quality |
| S10_extracted_63893 | Low-quality |
| S10_extracted_63900 | Not-determined |
| S10_extracted_63914 | Not-determined |
| S10_extracted_63922 | Not-determined |
| S10_extracted_63926 | Not-determined |
| S10_extracted_63934 | Not-determined |
| S10_extracted_63935 | Not-determined |
| S10_extracted_63936 | Low-quality |
| S10_extracted_63937 | Not-determined |
| S10_extracted_63940 | Not-determined |
| S10_extracted_63945 | Low-quality |
| S10_extracted_63946 | Not-determined |
| S10_extracted_63952 | Not-determined |
| S10_extracted_63976 | Not-determined |
| S10_extracted_64003 | Not-determined |
| S10_extracted_64006 | Not-determined |
| S10_extracted_64012 | Not-determined |
| S10_extracted_64020 | Not-determined |
| S10_extracted_64029 | Not-determined |
| S10_extracted_64037 | Not-determined |
| S10_extracted_64041 | Not-determined |
| S10_extracted_64058 | Not-determined |
| S10_extracted_64060 | Not-determined |
| S10_extracted_64062 | Not-determined |
| S10_extracted_64066 | Not-determined |
| S10_extracted_64074 | Low-quality |
| S10_extracted_64080 | Low-quality |
| S10_extracted_64081 | Not-determined |
| S10_extracted_64090 | Not-determined |
| S10_extracted_64108 | Not-determined |
| S10_extracted_64114 | Not-determined |
| S10_extracted_64123 | Not-determined |
| S10_extracted_64131 | Not-determined |
| S10_extracted_64136 | Low-quality |
| S10_extracted_64164 | Not-determined |
| S10_extracted_64181 | Not-determined |
| S10_extracted_64195 | Not-determined |
| S10_extracted_64200 | Not-determined |
| S10_extracted_64202 | Not-determined |
| S10_extracted_64224 | Not-determined |
| S10_extracted_64226 | Not-determined |
| S10_extracted_64230 | Not-determined |
| S10_extracted_64231 | Not-determined |
| S10_extracted_64234 | Not-determined |
| S10_extracted_64237 | Not-determined |
| S10_extracted_64243 | Not-determined |
| S10_extracted_64247 | Not-determined |
| S10_extracted_64253 | Not-determined |
| S10_extracted_64264 | Not-determined |
| S10_extracted_64265 | Low-quality |
| S10_extracted_64273 | Not-determined |
| S10_extracted_64275 | Low-quality |
| S10_extracted_64278 | Not-determined |
| S10_extracted_64287 | Not-determined |
| S10_extracted_64300 | Not-determined |
| S10_extracted_64308 | Not-determined |
| S10_extracted_64317 | Not-determined |
| S10_extracted_64323 | Not-determined |
| S10_extracted_64336 | Not-determined |
| S10_extracted_64339 | Not-determined |
| S10_extracted_64355 | Low-quality |
| S10_extracted_64356 | Low-quality |
| S5_extracted_k141_6032 | High-quality |
| S10_extracted_64375 | Low-quality |
| S10_extracted_64378 | Not-determined |
| S10_extracted_64383 | Not-determined |
| S10_extracted_64389 | Not-determined |
| S10_extracted_64406 | Not-determined |
| S10_extracted_64422 | Not-determined |
| S10_extracted_64448 | Not-determined |
| S10_extracted_64449 | Not-determined |
| S10_extracted_64453 | Not-determined |
| S10_extracted_64458 | Not-determined |
| S10_extracted_64461 | Not-determined |
| S10_extracted_64466 | Not-determined |
| S10_extracted_64478 | Not-determined |
| S10_extracted_64479 | Not-determined |
| S10_extracted_64488 | Not-determined |
| S10_extracted_64490 | Not-determined |
| S10_extracted_64496 | Not-determined |
| S10_extracted_64504 | Low-quality |
| S10_extracted_64508 | Not-determined |
| S10_extracted_64522 | Not-determined |
| S10_extracted_64529 | Not-determined |
| S10_extracted_64549 | Not-determined |
| S10_extracted_64558 | Not-determined |
| S10_extracted_64564 | Not-determined |
| S10_extracted_64576 | Not-determined |
| S10_extracted_64615 | Not-determined |
| S10_extracted_64659 | Not-determined |
| S10_extracted_64662 | Not-determined |
| S10_extracted_64667 | Not-determined |
| S10_extracted_64669 | Low-quality |
| S10_extracted_64671 | Low-quality |
| S10_extracted_64696 | Not-determined |
| S10_extracted_64701 | Not-determined |
| S10_extracted_64719 | Not-determined |
| S10_extracted_64734 | Not-determined |
| S10_extracted_64745 | Not-determined |
| S10_extracted_64763 | Not-determined |
| S10_extracted_64767 | Low-quality |
| S10_extracted_64775 | Not-determined |
| S10_extracted_64776 | Not-determined |
| S10_extracted_64783 | Low-quality |
| S10_extracted_64796 | Not-determined |
| S10_extracted_64797 | Not-determined |
| S10_extracted_64802 | Not-determined |
| S10_extracted_64811 | Not-determined |
| S10_extracted_64815 | Not-determined |
| S10_extracted_64844 | Not-determined |
| S10_extracted_64849 | Not-determined |
| S10_extracted_64858 | Not-determined |
| S10_extracted_64864 | Not-determined |
| S10_extracted_64867 | Not-determined |
| S10_extracted_64872 | Low-quality |
| S10_extracted_64884 | Not-determined |
| S10_extracted_64899 | Not-determined |
| S10_extracted_64917 | Not-determined |
| S10_extracted_64930 | Not-determined |
| S10_extracted_64932 | Not-determined |
| S10_extracted_64933 | Not-determined |
| S10_extracted_64935 | Low-quality |
| S10_extracted_64941 | Not-determined |
| S10_extracted_64953 | Low-quality |
| S10_extracted_64993 | Low-quality |
| S10_extracted_65002 | Not-determined |
| S10_extracted_65013 | Not-determined |
| S10_extracted_65014 | Not-determined |
| S10_extracted_65015 | Not-determined |
| S10_extracted_65021 | Not-determined |
| S10_extracted_65033 | Not-determined |
| S10_extracted_65035 | Low-quality |
| S10_extracted_65058 | Not-determined |
| S10_extracted_65088 | Low-quality |
| S10_extracted_65090 | Not-determined |
| S10_extracted_65093 | Not-determined |
| S10_extracted_65097 | Not-determined |
| S10_extracted_65113 | Not-determined |
| S10_extracted_65116 | Not-determined |
| S10_extracted_65124 | Not-determined |
| S10_extracted_65139 | Not-determined |
| S10_extracted_65142 | Not-determined |
| S10_extracted_65146 | Not-determined |
| S10_extracted_65179 | Not-determined |
| S10_extracted_65207 | Not-determined |
| S10_extracted_65209 | Low-quality |
| S10_extracted_65216 | Not-determined |
| S10_extracted_65228 | Not-determined |
| S10_extracted_65232 | Not-determined |
| S10_extracted_65238 | Not-determined |
| S10_extracted_65260 | Not-determined |
| S10_extracted_65262 | Not-determined |
| S10_extracted_65266 | Not-determined |
| S10_extracted_65269 | Low-quality |
| S10_extracted_65292 | Not-determined |
| S10_extracted_65305 | Not-determined |
| S10_extracted_65311 | Not-determined |
| S10_extracted_65317 | Not-determined |
| S10_extracted_65333 | Not-determined |
| S10_extracted_65347 | Low-quality |
| S10_extracted_65377 | Low-quality |
| S10_extracted_65388 | Not-determined |
| S10_extracted_65389 | Not-determined |
| S10_extracted_65395 | Not-determined |
| S10_extracted_65399 | Not-determined |
| S10_extracted_65400 | Low-quality |
| S10_extracted_65408 | Not-determined |
| S10_extracted_65410 | Not-determined |
| S10_extracted_65419 | Not-determined |
| S10_extracted_65425 | Not-determined |
| S10_extracted_65434 | Low-quality |
| S10_extracted_65440 | Not-determined |
| S10_extracted_65445 | Not-determined |
| S10_extracted_65450 | Not-determined |
| S10_extracted_65455 | Not-determined |
| S10_extracted_65498 | Not-determined |
| S10_extracted_65499 | Not-determined |
| S10_extracted_65508 | Not-determined |
| S10_extracted_65528 | Not-determined |
| S10_extracted_65546 | Low-quality |
| S10_extracted_65557 | Low-quality |
| S10_extracted_65586 | Not-determined |
| S10_extracted_65589 | Not-determined |
| S10_extracted_65594 | Not-determined |
| S10_extracted_65598 | Low-quality |
| S10_extracted_65609 | Not-determined |
| S10_extracted_65611 | Not-determined |
| S10_extracted_65616 | Not-determined |
| S10_extracted_65621 | Not-determined |
| S10_extracted_65675 | Not-determined |
| S10_extracted_65698 | Not-determined |
| S10_extracted_65702 | Not-determined |
| S10_extracted_65746 | Not-determined |
| S10_extracted_65749 | Not-determined |
| S10_extracted_65750 | Not-determined |
| S10_extracted_65753 | Not-determined |
| S10_extracted_65765 | Not-determined |
| S10_extracted_65769 | Not-determined |
| S10_extracted_65801 | Not-determined |
| S10_extracted_65829 | Not-determined |
| S10_extracted_65832 | Not-determined |
| S10_extracted_65834 | Not-determined |
| S10_extracted_65842 | Not-determined |
| S10_extracted_65844 | Low-quality |
| S10_extracted_65854 | Not-determined |
| S10_extracted_65860 | Not-determined |
| S10_extracted_65900 | Not-determined |
| S10_extracted_65906 | Not-determined |
| S10_extracted_65920 | Not-determined |
| S10_extracted_65921 | Not-determined |
| S10_extracted_65958 | Not-determined |
| S10_extracted_65966 | Not-determined |
| S10_extracted_65970 | Not-determined |
| S10_extracted_65971 | Low-quality |
| S10_extracted_66007 | Not-determined |
| S10_extracted_66010 | Not-determined |
| S10_extracted_66028 | Not-determined |
| S10_extracted_66030 | Not-determined |
| S10_extracted_66049 | Not-determined |
| S10_extracted_66059 | Not-determined |
| S10_extracted_66067 | Not-determined |
| S10_extracted_66079 | Not-determined |
| S10_extracted_66082 | Not-determined |
| S10_extracted_66084 | Low-quality |
| S10_extracted_66089 | Not-determined |
| S10_extracted_66102 | Not-determined |
| S10_extracted_66115 | Not-determined |
| S10_extracted_66137 | Not-determined |
| S10_extracted_66162 | Not-determined |
| S10_extracted_66163 | Not-determined |
| S10_extracted_66169 | Not-determined |
| S10_extracted_66183 | Not-determined |
| S10_extracted_66190 | Not-determined |
| S10_extracted_66197 | Not-determined |
| S10_extracted_66203 | Low-quality |
| S10_extracted_66206 | Not-determined |
| S10_extracted_66215 | Not-determined |
| S10_extracted_66231 | Not-determined |
| S10_extracted_66240 | Not-determined |
| S10_extracted_66242 | Not-determined |
| S10_extracted_66247 | Not-determined |
| S10_extracted_66249 | Not-determined |
| S10_extracted_66254 | Not-determined |
| S10_extracted_66257 | Not-determined |
| S10_extracted_66260 | Not-determined |
| S10_extracted_66263 | Not-determined |
| S10_extracted_66265 | Not-determined |
| S10_extracted_66268 | Not-determined |
| S10_extracted_66313 | Not-determined |
| S10_extracted_66338 | Low-quality |
| S10_extracted_66345 | Low-quality |
| S10_extracted_66350 | Low-quality |
| S10_extracted_66359 | Not-determined |
| S10_extracted_66360 | Not-determined |
| S10_extracted_66363 | Not-determined |
| S10_extracted_66374 | Low-quality |
| S10_extracted_66381 | Low-quality |
| S10_extracted_66410 | Low-quality |
| S10_extracted_66445 | Not-determined |
| S10_extracted_66465 | Not-determined |
| S10_extracted_66491 | Not-determined |
| S10_extracted_66511 | Not-determined |
| S10_extracted_66520 | Not-determined |
| S10_extracted_66525 | Not-determined |
| S10_extracted_66526 | Not-determined |
| S10_extracted_66527 | Low-quality |
| S10_extracted_66528 | Not-determined |
| S10_extracted_66553 | Not-determined |
| S10_extracted_66591 | Low-quality |
| S10_extracted_66595 | Low-quality |
| S10_extracted_66597 | Low-quality |
| S10_extracted_66599 | Not-determined |
| S10_extracted_66622 | Not-determined |
| S10_extracted_66623 | Low-quality |
| S10_extracted_66633 | Not-determined |
| S10_extracted_66635 | Not-determined |
| S10_extracted_66643 | Not-determined |
| S10_extracted_66647 | Not-determined |
| S10_extracted_66648 | Not-determined |
| S10_extracted_66660 | Not-determined |
| S10_extracted_66673 | Not-determined |
| S10_extracted_66694 | Low-quality |
| S10_extracted_66717 | Low-quality |
| S10_extracted_66721 | Low-quality |
| S10_extracted_66745 | Not-determined |
| S10_extracted_66751 | Not-determined |
| S10_extracted_66753 | Not-determined |
| S10_extracted_66755 | Not-determined |
| S10_extracted_66757 | Not-determined |
| S10_extracted_66760 | Low-quality |
| S10_extracted_66775 | Low-quality |
| S10_extracted_66778 | Not-determined |
| S10_extracted_66793 | Not-determined |
| S10_extracted_66794 | Low-quality |
| S10_extracted_66827 | Not-determined |
| S10_extracted_66831 | Not-determined |
| S10_extracted_66854 | Not-determined |
| S10_extracted_66866 | Not-determined |
| S10_extracted_66875 | Not-determined |
| S10_extracted_66891 | Low-quality |
| S10_extracted_66900 | Low-quality |
| S10_extracted_66910 | Not-determined |
| S10_extracted_66926 | Not-determined |
| S10_extracted_66938 | Not-determined |
| S10_extracted_66945 | Not-determined |
| S10_extracted_66946 | Low-quality |
| S10_extracted_66961 | Not-determined |
| S10_extracted_66962 | Not-determined |
| S10_extracted_66967 | Not-determined |
| S10_extracted_66968 | Not-determined |
| S10_extracted_66971 | Not-determined |
| S10_extracted_66975 | Not-determined |
| S10_extracted_66978 | Not-determined |
| S10_extracted_66985 | Not-determined |
| S10_extracted_66997 | Not-determined |
| S10_extracted_67002 | Low-quality |
| S10_extracted_67009 | Not-determined |
| S10_extracted_67024 | Low-quality |
| S10_extracted_67030 | Not-determined |
| S10_extracted_67031 | Not-determined |
| S10_extracted_67042 | Not-determined |
| S10_extracted_67050 | Not-determined |
| S10_extracted_67058 | Not-determined |
| S10_extracted_67062 | Not-determined |
| S10_extracted_67078 | Not-determined |
| S10_extracted_67085 | Not-determined |
| S10_extracted_67119 | Not-determined |
| S10_extracted_67154 | Not-determined |
| S10_extracted_67158 | Not-determined |
| S10_extracted_67161 | Not-determined |
| S10_extracted_67163 | Not-determined |
| S10_extracted_67169 | Not-determined |
| S10_extracted_67171 | Not-determined |
| S10_extracted_67178 | Not-determined |
| S10_extracted_67184 | Not-determined |
| S10_extracted_67189 | Not-determined |
| S10_extracted_67196 | Not-determined |
| S10_extracted_67198 | Not-determined |
| S10_extracted_67220 | Low-quality |
| S10_extracted_67228 | Not-determined |
| S10_extracted_67236 | Not-determined |
| S10_extracted_67249 | Low-quality |
| S10_extracted_67259 | Not-determined |
| S10_extracted_67261 | Low-quality |
| S10_extracted_67268 | Not-determined |
| S10_extracted_67285 | Not-determined |
| S10_extracted_67295 | Low-quality |
| S10_extracted_67304 | Not-determined |
| S10_extracted_67334 | Not-determined |
| S10_extracted_67338 | Not-determined |
| S10_extracted_67365 | Not-determined |
| S10_extracted_67381 | Not-determined |
| S10_extracted_67394 | Low-quality |
| S10_extracted_67402 | Not-determined |
| S10_extracted_67419 | Not-determined |
| S10_extracted_67420 | Not-determined |
| S10_extracted_67427 | Not-determined |
| S10_extracted_67434 | Not-determined |
| S10_extracted_67444 | Not-determined |
| S10_extracted_67459 | Not-determined |
| S10_extracted_67468 | Not-determined |
| S10_extracted_67484 | Not-determined |
| S10_extracted_67490 | Not-determined |
| S10_extracted_67492 | Not-determined |
| S10_extracted_67500 | Not-determined |
| S10_extracted_67550 | Not-determined |
| S10_extracted_67564 | Not-determined |
| S10_extracted_67569 | Low-quality |
| S10_extracted_67580 | Not-determined |
| S10_extracted_67581 | Low-quality |
| S10_extracted_67586 | Not-determined |
| S10_extracted_67629 | Not-determined |
| S10_extracted_67634 | Not-determined |
| S10_extracted_67635 | Not-determined |
| S10_extracted_67646 | Not-determined |
| S10_extracted_67647 | Not-determined |
| S10_extracted_67649 | Not-determined |
| S10_extracted_67678 | Not-determined |
| S10_extracted_67679 | Not-determined |
| S10_extracted_67685 | Not-determined |
| S10_extracted_67691 | Low-quality |
| S10_extracted_67701 | Not-determined |
| S10_extracted_67710 | Not-determined |
| S10_extracted_67715 | Not-determined |
| S10_extracted_67717 | Not-determined |
| S10_extracted_67718 | Not-determined |
| S10_extracted_67751 | Not-determined |
| S10_extracted_67752 | Not-determined |
| S10_extracted_67775 | Not-determined |
| S10_extracted_67778 | Not-determined |
| S10_extracted_67788 | Not-determined |
| S10_extracted_67793 | Not-determined |
| S10_extracted_67794 | Not-determined |
| S10_extracted_67814 | Low-quality |
| S10_extracted_67817 | Not-determined |
| S10_extracted_67871 | Not-determined |
| S10_extracted_67873 | Not-determined |
| S10_extracted_67878 | Not-determined |
| S10_extracted_67893 | Not-determined |
| S10_extracted_67904 | Not-determined |
| S10_extracted_67910 | Not-determined |
| S10_extracted_67914 | Not-determined |
| S10_extracted_67928 | Not-determined |
| S10_extracted_67943 | Not-determined |
| S10_extracted_67964 | Not-determined |
| S10_extracted_67965 | Not-determined |
| S10_extracted_67979 | Not-determined |
| S10_extracted_67993 | Not-determined |
| S10_extracted_67995 | Not-determined |
| S10_extracted_67997 | Not-determined |
| S10_extracted_68003 | Not-determined |
| S10_extracted_68004 | Not-determined |
| S10_extracted_68011 | Not-determined |
| S10_extracted_68045 | Not-determined |
| S10_extracted_68097 | Not-determined |
| S10_extracted_68107 | Not-determined |
| S10_extracted_68109 | Not-determined |
| S10_extracted_68115 | Not-determined |
| S10_extracted_68121 | Not-determined |
| S10_extracted_68129 | Not-determined |
| S10_extracted_68141 | Not-determined |
| S10_extracted_68142 | Not-determined |
| S10_extracted_68144 | Not-determined |
| S10_extracted_68150 | Not-determined |
| S10_extracted_68152 | Not-determined |
| S10_extracted_68153 | Not-determined |
| S10_extracted_68160 | Not-determined |
| S10_extracted_68161 | Not-determined |
| S10_extracted_68167 | Low-quality |
| S10_extracted_68178 | Not-determined |
| S10_extracted_68196 | Low-quality |
| S10_extracted_68198 | Not-determined |
| S10_extracted_68201 | Not-determined |
| S10_extracted_68204 | Low-quality |
| S10_extracted_68205 | Low-quality |
| S10_extracted_68209 | Not-determined |
| S10_extracted_68223 | Not-determined |
| S10_extracted_68233 | Not-determined |
| S10_extracted_68238 | Not-determined |
| S10_extracted_68239 | Not-determined |
| S10_extracted_68251 | Not-determined |
| S10_extracted_68278 | Not-determined |
| S10_extracted_68281 | Not-determined |
| S10_extracted_68318 | Not-determined |
| S10_extracted_68319 | Not-determined |
| S10_extracted_68321 | Not-determined |
| S10_extracted_68338 | Not-determined |
| S10_extracted_68346 | Not-determined |
| S10_extracted_68360 | Not-determined |
| S10_extracted_68367 | Not-determined |
| S10_extracted_68375 | Not-determined |
| S10_extracted_68376 | Not-determined |
| S10_extracted_68393 | Not-determined |
| S10_extracted_68397 | Not-determined |
| S10_extracted_68404 | Not-determined |
| S10_extracted_68407 | Not-determined |
| S10_extracted_68408 | Not-determined |
| S10_extracted_68409 | Not-determined |
| S10_extracted_68415 | Not-determined |
| S10_extracted_68430 | Not-determined |
| S10_extracted_68432 | Not-determined |
| S10_extracted_68437 | Low-quality |
| S10_extracted_68459 | Low-quality |
| S10_extracted_68469 | Not-determined |
| S10_extracted_68471 | Not-determined |
| S10_extracted_68501 | Not-determined |
| S10_extracted_68510 | Low-quality |
| S10_extracted_68517 | Low-quality |
| S10_extracted_68526 | Not-determined |
| S10_extracted_68537 | Not-determined |
| S10_extracted_68539 | Not-determined |
| S10_extracted_68545 | Not-determined |
| S10_extracted_68562 | Not-determined |
| S10_extracted_68567 | Not-determined |
| S10_extracted_68571 | Not-determined |
| S10_extracted_68589 | Not-determined |
| S10_extracted_68592 | Low-quality |
| S10_extracted_68597 | Not-determined |
| S10_extracted_68602 | Not-determined |
| S10_extracted_68615 | Low-quality |
| S10_extracted_68622 | Not-determined |
| S10_extracted_68632 | Not-determined |
| S10_extracted_68633 | Not-determined |
| S10_extracted_68638 | Not-determined |
| S10_extracted_68641 | Not-determined |
| S10_extracted_68649 | Not-determined |
| S10_extracted_68650 | Not-determined |
| S10_extracted_68678 | Low-quality |
| S10_extracted_68684 | Not-determined |
| S10_extracted_68690 | Not-determined |
| S10_extracted_68696 | Not-determined |
| S10_extracted_68706 | Not-determined |
| S10_extracted_68718 | Not-determined |
| S10_extracted_68726 | Not-determined |
| S10_extracted_68728 | Not-determined |
| S10_extracted_68736 | Not-determined |
| S10_extracted_68738 | Not-determined |
| S10_extracted_68749 | Not-determined |
| S10_extracted_68757 | Not-determined |
| S10_extracted_68769 | Not-determined |
| S10_extracted_68775 | Not-determined |
| S10_extracted_68794 | Low-quality |
| S10_extracted_68818 | Not-determined |
| S10_extracted_68830 | Not-determined |
| S10_extracted_68840 | Not-determined |
| S10_extracted_68845 | Not-determined |
| S10_extracted_68848 | Not-determined |
| S10_extracted_68860 | Not-determined |
| S10_extracted_68871 | Not-determined |
| S10_extracted_68877 | Not-determined |
| S10_extracted_68881 | Not-determined |
| S10_extracted_68894 | Not-determined |
| S10_extracted_68926 | Low-quality |
| S10_extracted_68933 | Not-determined |
| S10_extracted_68936 | Not-determined |

| | |
|---|---|
| S10_extracted_68999 | Not-determined |
| S10_extracted_69001 | Low-quality |
| S10_extracted_69028 | Not-determined |
| S10_extracted_69030 | Low-quality |
| S10_extracted_69038 | Not-determined |
| S10_extracted_69045 | Not-determined |
| S10_extracted_69050 | Low-quality |
| S10_extracted_69053 | Not-determined |
| S10_extracted_69077 | Not-determined |
| S10_extracted_69092 | Not-determined |
| S10_extracted_69096 | Not-determined |
| S10_extracted_69112 | Low-quality |
| S10_extracted_69156 | Not-determined |
| S10_extracted_69179 | Not-determined |
| S10_extracted_69210 | Not-determined |
| S10_extracted_69219 | Not-determined |
| S10_extracted_69228 | Low-quality |
| S10_extracted_69256 | Not-determined |
| S10_extracted_69270 | Not-determined |
| S10_extracted_69279 | Not-determined |
| S10_extracted_69301 | Not-determined |
| S10_extracted_69306 | Not-determined |
| S10_extracted_69315 | Not-determined |
| S10_extracted_69321 | Not-determined |
| S10_extracted_69325 | Not-determined |
| S10_extracted_69334 | Not-determined |
| S10_extracted_69338 | Not-determined |
| S10_extracted_69339 | Not-determined |
| S10_extracted_69340 | Not-determined |
| S10_extracted_69342 | Not-determined |
| S10_extracted_69344 | Low-quality |
| S10_extracted_69370 | Not-determined |
| S10_extracted_69372 | Not-determined |
| S10_extracted_69379 | Not-determined |
| S10_extracted_69383 | Low-quality |
| S10_extracted_69406 | Low-quality |
| S10_extracted_69407 | Not-determined |
| S10_extracted_69415 | Not-determined |
| S10_extracted_69416 | Not-determined |
| S10_extracted_69446 | Not-determined |
| S10_extracted_69452 | Not-determined |
| S10_extracted_69467 | Not-determined |
| S10_extracted_69478 | Not-determined |
| S10_extracted_69483 | Not-determined |
| S10_extracted_69500 | Not-determined |
| S10_extracted_69507 | Not-determined |
| S10_extracted_69517 | Not-determined |
| S10_extracted_69524 | Not-determined |
| S10_extracted_69529 | Not-determined |
| S10_extracted_69541 | Not-determined |
| S10_extracted_69558 | Not-determined |
| S10_extracted_69569 | Not-determined |
| S10_extracted_69576 | Not-determined |
| S10_extracted_69615 | Not-determined |
| S10_extracted_69629 | Not-determined |
| [illegible] | [illegible] |

| | |
|---|---|
| S10_extracted_75174 | Not-determined |
| S10_extracted_75183 | Not-determined |
| S10_extracted_75218 | Not-determined |
| S10_extracted_75222 | Not-determined |
| S10_extracted_75225 | Not-determined |
| S10_extracted_75228 | Low-quality |
| S10_extracted_75231 | Not-determined |
| S10_extracted_75234 | Not-determined |
| S10_extracted_75243 | Not-determined |
| S10_extracted_75251 | Not-determined |
| S10_extracted_75276 | Not-determined |
| S10_extracted_75286 | Not-determined |
| S10_extracted_75290 | Not-determined |
| S10_extracted_75302 | Not-determined |
| S10_extracted_75308 | Not-determined |
| S10_extracted_75339 | Not-determined |
| S10_extracted_75341 | Not-determined |
| S10_extracted_75370 | Not-determined |
| S10_extracted_75390 | Not-determined |
| S10_extracted_75398 | Not-determined |
| S10_extracted_75415 | Not-determined |
| S10_extracted_75424 | Low-quality |
| S10_extracted_75431 | Not-determined |
| S10_extracted_75432 | Not-determined |
| S10_extracted_75437 | Not-determined |
| S10_extracted_75451 | Not-determined |
| S10_extracted_75455 | Not-determined |
| S10_extracted_75461 | Not-determined |
| S10_extracted_75487 | Not-determined |
| S10_extracted_75497 | Not-determined |
| S10_extracted_75499 | Not-determined |
| S10_extracted_75503 | Not-determined |
| S10_extracted_75506 | Not-determined |
| S10_extracted_75507 | Not-determined |
| S10_extracted_75527 | Not-determined |
| S10_extracted_75530 | Not-determined |
| S10_extracted_75537 | Low-quality |
| S10_extracted_75542 | Low-quality |
| S10_extracted_75543 | Low-quality |
| S10_extracted_75549 | Not-determined |
| S10_extracted_75560 | Not-determined |
| S10_extracted_75569 | Not-determined |
| S10_extracted_75571 | Not-determined |
| S10_extracted_75583 | Not-determined |
| S10_extracted_75588 | Not-determined |
| S10_extracted_75614 | Not-determined |
| S10_extracted_75636 | Not-determined |
| S10_extracted_75662 | Not-determined |
| S10_extracted_75692 | Not-determined |
| S10_extracted_75734 | Not-determined |
| S10_extracted_75743 | Not-determined |
| S10_extracted_75745 | Not-determined |
| S10_extracted_75752 | Not-determined |
| S10_extracted_75760 | Not-determined |
| S10_extracted_75800 | Not-determined |
| S10_extracted_75803 | Not-determined |
| S10_extracted_75825 | Low-quality |
| S10_extracted_75829 | Not-determined |
| S10_extracted_75834 | Not-determined |
| S10_extracted_75844 | Not-determined |
| S10_extracted_75895 | Not-determined |
| S10_extracted_75904 | Low-quality |
| S10_extracted_75909 | Not-determined |
| S10_extracted_75917 | Not-determined |
| S10_extracted_75919 | Not-determined |
| S10_extracted_75920 | Not-determined |
| S10_extracted_75937 | Not-determined |
| S10_extracted_75942 | Low-quality |
| S10_extracted_75950 | Not-determined |
| S10_extracted_75954 | Not-determined |
| S10_extracted_75976 | Not-determined |
| S10_extracted_75982 | Not-determined |
| S10_extracted_75985 | Not-determined |
| S10_extracted_76003 | Not-determined |
| S10_extracted_76056 | Not-determined |
| S10_extracted_76079 | Low-quality |
| S10_extracted_76084 | Low-quality |
| S10_extracted_76087 | Not-determined |
| S10_extracted_76102 | Not-determined |
| S10_extracted_76104 | Not-determined |
| S10_extracted_76110 | Not-determined |
| S10_extracted_76112 | Not-determined |
| S10_extracted_76132 | Not-determined |
| S10_extracted_76140 | Not-determined |
| S10_extracted_76148 | Not-determined |
| S10_extracted_76187 | Not-determined |
| S10_extracted_76198 | Low-quality |
| S10_extracted_76212 | Not-determined |
| S10_extracted_76221 | Not-determined |
| S10_extracted_76224 | Not-determined |
| S10_extracted_76231 | Not-determined |
| S10_extracted_76240 | Not-determined |
| S10_extracted_76245 | Not-determined |
| S10_extracted_76259 | Not-determined |
| S10_extracted_76265 | Low-quality |
| S10_extracted_76276 | Not-determined |
| S10_extracted_76286 | Not-determined |
| S10_extracted_76299 | Not-determined |
| S10_extracted_76305 | Low-quality |
| S10_extracted_76307 | Not-determined |
| S10_extracted_76316 | Not-determined |
| S10_extracted_76326 | Not-determined |
| S10_extracted_76330 | Not-determined |
| S10_extracted_76336 | Low-quality |
| S10_extracted_76338 | Not-determined |
| S10_extracted_76352 | Low-quality |
| S10_extracted_76363 | Not-determined |
| S10_extracted_76373 | Low-quality |
| S10_extracted_76388 | Not-determined |
| S10_extracted_76389 | Not-determined |
| S10_extracted_76424 | Not-determined |
| S10_extracted_76454 | Not-determined |
| S10_extracted_76459 | Not-determined |
| S10_extracted_76460 | Low-quality |
| S10_extracted_76471 | Not-determined |
| S10_extracted_76498 | Not-determined |
| S10_extracted_76502 | Not-determined |
| S10_extracted_76519 | Not-determined |
| S10_extracted_76531 | Not-determined |
| S10_extracted_76548 | Not-determined |
| S10_extracted_76565 | Not-determined |
| S10_extracted_76582 | Not-determined |
| S10_extracted_76606 | Low-quality |
| S10_extracted_76608 | Not-determined |
| S10_extracted_76618 | Not-determined |
| S10_extracted_76625 | Not-determined |
| S10_extracted_76629 | Not-determined |
| S10_extracted_76643 | Not-determined |
| S10_extracted_76647 | Not-determined |
| S10_extracted_76650 | Low-quality |
| S10_extracted_76679 | Not-determined |
| S10_extracted_76696 | Not-determined |
| S10_extracted_76705 | Not-determined |
| S10_extracted_76716 | Not-determined |
| S10_extracted_76720 | Not-determined |
| S10_extracted_76721 | Not-determined |
| S10_extracted_76727 | Not-determined |
| S10_extracted_76730 | Not-determined |
| S10_extracted_76731 | Not-determined |
| S10_extracted_76746 | Low-quality |
| S10_extracted_76754 | Not-determined |
| S10_extracted_76764 | Not-determined |
| S10_extracted_76766 | Not-determined |
| S10_extracted_76790 | Not-determined |
| S10_extracted_76801 | Not-determined |
| S10_extracted_76819 | Not-determined |
| S10_extracted_76820 | Not-determined |
| S10_extracted_76831 | Low-quality |
| S10_extracted_76833 | Not-determined |
| S10_extracted_76859 | Not-determined |
| S10_extracted_76869 | Not-determined |
| S10_extracted_76897 | Low-quality |
| S10_extracted_76901 | Not-determined |
| S10_extracted_76911 | Low-quality |
| S10_extracted_76913 | Not-determined |
| S10_extracted_76918 | Not-determined |
| S10_extracted_76927 | Not-determined |
| S10_extracted_76938 | Low-quality |
| S10_extracted_76945 | Not-determined |
| S10_extracted_76951 | Not-determined |
| S10_extracted_76960 | Not-determined |
| S10_extracted_76963 | Low-quality |
| S10_extracted_76970 | Not-determined |
| S10_extracted_76990 | Not-determined |
| S10_extracted_77008 | Not-determined |
| S10_extracted_77012 | Not-determined |
| S10_extracted_77027 | Not-determined |
| S10_extracted_77036 | Not-determined |
| S10_extracted_77051 | Not-determined |
| S10_extracted_77064 | Low-quality |
| S10_extracted_77065 | Not-determined |
| S10_extracted_77073 | Low-quality |
| S10_extracted_77075 | Low-quality |
| S10_extracted_77083 | Not-determined |
| S10_extracted_77084 | Not-determined |
| S10_extracted_77088 | Not-determined |
| S10_extracted_77090 | Not-determined |
| S10_extracted_77092 | Not-determined |
| S10_extracted_77096 | Not-determined |
| S10_extracted_77111 | Low-quality |
| S10_extracted_77118 | Low-quality |
| S10_extracted_77119 | Not-determined |
| S10_extracted_77126 | Not-determined |
| S10_extracted_77132 | Not-determined |
| S10_extracted_77147 | Not-determined |
| S10_extracted_77169 | Not-determined |
| S10_extracted_77194 | Not-determined |
| S10_extracted_77202 | Not-determined |
| S10_extracted_77225 | Not-determined |
| S10_extracted_77230 | Low-quality |
| S10_extracted_77241 | Not-determined |
| S10_extracted_77266 | Not-determined |
| S10_extracted_77274 | Not-determined |
| S10_extracted_77284 | Low-quality |
| S10_extracted_77286 | Not-determined |
| S10_extracted_77301 | Not-determined |
| S10_extracted_77305 | Not-determined |
| S10_extracted_77308 | Not-determined |
| S10_extracted_77323 | Not-determined |
| S10_extracted_77346 | Not-determined |
| S10_extracted_77351 | Not-determined |
| S10_extracted_77367 | Not-determined |
| S10_extracted_77371 | Not-determined |
| S10_extracted_77374 | Not-determined |
| S10_extracted_77381 | Not-determined |
| S10_extracted_77388 | Not-determined |
| S10_extracted_77390 | Not-determined |
| S10_extracted_77400 | Low-quality |
| S10_extracted_77402 | Not-determined |
| S10_extracted_77406 | Not-determined |
| S10_extracted_77427 | Low-quality |
| S10_extracted_77431 | Not-determined |
| S10_extracted_77432 | Low-quality |
| S10_extracted_77440 | Not-determined |
| S10_extracted_77442 | Not-determined |
| S10_extracted_77447 | Not-determined |
| S10_extracted_77452 | Not-determined |
| S10_extracted_77466 | Not-determined |
| S10_extracted_77490 | Not-determined |
| S10_extracted_77499 | Not-determined |
| S10_extracted_77504 | Not-determined |
| S10_extracted_77515 | Not-determined |
| S10_extracted_77521 | Not-determined |
| S10_extracted_77535 | Not-determined |
| S10_extracted_77567 | Not-determined |
| S10_extracted_77572 | Not-determined |
| S10_extracted_77576 | Not-determined |
| S10_extracted_77583 | Not-determined |
| S10_extracted_77591 | Not-determined |
| S10_extracted_77601 | Not-determined |
| S10_extracted_77604 | Not-determined |
| S10_extracted_77607 | Low-quality |
| S10_extracted_77620 | Not-determined |
| S10_extracted_77641 | Not-determined |
| S10_extracted_77643 | Not-determined |
| S10_extracted_77648 | Not-determined |
| S10_extracted_77649 | Not-determined |
| S10_extracted_77654 | Not-determined |
| S10_extracted_77658 | Not-determined |
| S10_extracted_77668 | Not-determined |
| S10_extracted_77676 | Low-quality |
| S10_extracted_77682 | Not-determined |
| S10_extracted_77685 | Not-determined |
| S10_extracted_77696 | Not-determined |
| S10_extracted_77706 | Not-determined |
| S10_extracted_77709 | Not-determined |
| S10_extracted_77712 | Not-determined |
| S10_extracted_77722 | Not-determined |
| S10_extracted_77762 | Not-determined |
| S10_extracted_77763 | Not-determined |
| S10_extracted_77765 | Not-determined |
| S10_extracted_77775 | Not-determined |
| S10_extracted_77779 | Not-determined |
| S10_extracted_77784 | Not-determined |
| S10_extracted_77785 | Not-determined |
| S10_extracted_77799 | Not-determined |
| S10_extracted_77820 | Not-determined |
| S10_extracted_77832 | Low-quality |
| S10_extracted_77842 | Not-determined |
| S10_extracted_77862 | Not-determined |
| S10_extracted_77869 | Not-determined |
| S10_extracted_77880 | Not-determined |
| S10_extracted_77887 | Not-determined |
| S10_extracted_77901 | Not-determined |
| S10_extracted_77907 | Not-determined |
| S10_extracted_77910 | Not-determined |
| S10_extracted_77932 | Low-quality |
| S10_extracted_77939 | Not-determined |
| S10_extracted_77972 | Not-determined |
| S10_extracted_77977 | Not-determined |
| S10_extracted_77991 | Not-determined |
| S10_extracted_78000 | Not-determined |
| S10_extracted_78004 | Not-determined |
| S10_extracted_78009 | Not-determined |
| S10_extracted_78020 | Not-determined |
| S10_extracted_78025 | Not-determined |
| S10_extracted_78040 | Not-determined |
| S10_extracted_78047 | Not-determined |
| S10_extracted_78054 | Not-determined |
| S10_extracted_78055 | Not-determined |
| S10_extracted_78060 | Not-determined |
| S10_extracted_78061 | Low-quality |
| S10_extracted_78063 | Not-determined |
| S10_extracted_78064 | Not-determined |
| S10_extracted_78069 | Not-determined |
| S10_extracted_78114 | Not-determined |
| S10_extracted_78117 | Not-determined |
| S10_extracted_78124 | Not-determined |
| S10_extracted_78125 | Not-determined |
| S10_extracted_78149 | Not-determined |
| S10_extracted_78161 | Not-determined |
| S10_extracted_78183 | Not-determined |
| S10_extracted_78207 | Not-determined |
| S10_extracted_78231 | Low-quality |
| S10_extracted_78250 | Not-determined |
| S10_extracted_78261 | Not-determined |
| S10_extracted_78265 | Low-quality |
| S10_extracted_78274 | Not-determined |
| S10_extracted_78281 | Not-determined |
| S10_extracted_78289 | Not-determined |
| S10_extracted_78303 | Not-determined |
| S10_extracted_78308 | Low-quality |
| S10_extracted_78347 | Not-determined |
| S10_extracted_78368 | Low-quality |
| S10_extracted_78375 | Not-determined |
| S10_extracted_78399 | Not-determined |
| S10_extracted_78400 | Not-determined |
| S10_extracted_78405 | Not-determined |
| S10_extracted_78407 | Not-determined |
| S10_extracted_78415 | Not-determined |
| S10_extracted_78419 | Low-quality |
| S10_extracted_78426 | Not-determined |
| S10_extracted_78427 | Low-quality |
| S10_extracted_78429 | Not-determined |
| S10_extracted_78431 | Not-determined |
| S10_extracted_78442 | Not-determined |
| S10_extracted_78447 | Low-quality |
| S10_extracted_78458 | Not-determined |
| S10_extracted_78475 | Not-determined |
| S10_extracted_78483 | Low-quality |
| S10_extracted_78484 | Not-determined |
| S10_extracted_78510 | Low-quality |
| S10_extracted_78518 | Not-determined |
| S10_extracted_78521 | Not-determined |
| S10_extracted_78526 | Not-determined |
| S10_extracted_78527 | Low-quality |
| S10_extracted_78568 | Not-determined |
| S10_extracted_78569 | Not-determined |
| S10_extracted_78578 | Not-determined |
| S10_extracted_78586 | Not-determined |
| S10_extracted_78593 | Not-determined |
| S10_extracted_78595 | Not-determined |
| S10_extracted_78605 | Not-determined |
| S10_extracted_78614 | Not-determined |
| S10_extracted_78620 | Not-determined |
| S10_extracted_78636 | Not-determined |
| S10_extracted_78637 | Not-determined |
| S10_extracted_78653 | Low-quality |
| S10_extracted_78654 | Not-determined |
| S10_extracted_78689 | Low-quality |
| S10_extracted_78693 | Low-quality |
| S10_extracted_78700 | Not-determined |
| S10_extracted_78703 | Not-determined |
| S10_extracted_78722 | Not-determined |
| S10_extracted_78730 | Not-determined |
| S10_extracted_78739 | Low-quality |
| S10_extracted_78766 | Not-determined |
| S10_extracted_78780 | Not-determined |
| S10_extracted_78813 | Not-determined |
| S10_extracted_78816 | Not-determined |
| S10_extracted_78818 | Low-quality |
| S10_extracted_78857 | Not-determined |
| S10_extracted_78859 | Not-determined |
| S10_extracted_78871 | Not-determined |
| S10_extracted_78875 | Not-determined |
| S10_extracted_78877 | Not-determined |
| S10_extracted_78881 | Not-determined |
| S10_extracted_78894 | Not-determined |
| S10_extracted_78896 | Not-determined |
| S10_extracted_78907 | Low-quality |
| S10_extracted_78908 | Not-determined |
| S10_extracted_78912 | Not-determined |
| S10_extracted_78918 | Not-determined |
| S10_extracted_78919 | Low-quality |
| S10_extracted_78924 | Not-determined |
| S10_extracted_78928 | Not-determined |
| S10_extracted_78929 | Not-determined |
| S10_extracted_78931 | Not-determined |
| S10_extracted_78943 | Not-determined |
| S10_extracted_78952 | Low-quality |
| S10_extracted_78957 | Not-determined |
| S10_extracted_78997 | Not-determined |
| S10_extracted_79000 | Low-quality |
| S10_extracted_79062 | Low-quality |
| S10_extracted_79065 | Not-determined |
| S10_extracted_79082 | Not-determined |
| S10_extracted_79108 | Not-determined |
| S10_extracted_79115 | Low-quality |
| S10_extracted_79116 | Not-determined |
| S10_extracted_79164 | Not-determined |
| S10_extracted_79174 | Not-determined |
| S10_extracted_79197 | Not-determined |
| S10_extracted_79204 | Not-determined |
| S10_extracted_79212 | Not-determined |
| S10_extracted_79216 | Not-determined |
| S10_extracted_79222 | Not-determined |
| S10_extracted_79233 | Not-determined |
| S10_extracted_79244 | Not-determined |
| S10_extracted_79250 | Not-determined |
| S10_extracted_79257 | Not-determined |
| S10_extracted_79285 | Not-determined |
| S10_extracted_79301 | Not-determined |
| S10_extracted_79307 | Not-determined |
| S10_extracted_79311 | Low-quality |
| S10_extracted_79313 | Low-quality |
| S10_extracted_79321 | Not-determined |
| S10_extracted_79333 | Not-determined |
| S10_extracted_79337 | Not-determined |
| S10_extracted_79339 | Not-determined |
| S10_extracted_79351 | Not-determined |
| S10_extracted_79366 | Low-quality |
| S10_extracted_79383 | Not-determined |
| S10_extracted_79384 | Not-determined |
| S10_extracted_79389 | Not-determined |
| S10_extracted_79406 | Not-determined |
| S10_extracted_79409 | Not-determined |
| S10_extracted_79410 | Not-determined |
| S10_extracted_79416 | Not-determined |
| S10_extracted_79417 | Not-determined |
| S10_extracted_79425 | Not-determined |
| S10_extracted_79428 | Not-determined |
| S10_extracted_79443 | Not-determined |
| S10_extracted_79450 | Not-determined |
| S10_extracted_79453 | Not-determined |
| S10_extracted_79482 | Not-determined |
| S10_extracted_79485 | Not-determined |
| S10_extracted_79490 | Not-determined |
| S10_extracted_79492 | Low-quality |
| S10_extracted_79495 | Not-determined |
| S10_extracted_79503 | Not-determined |
| S10_extracted_79509 | Not-determined |
| S10_extracted_79512 | Not-determined |
| S10_extracted_79529 | Not-determined |
| S10_extracted_79531 | Not-determined |
| S10_extracted_79532 | Not-determined |
| S10_extracted_79534 | Not-determined |
| S10_extracted_79540 | Not-determined |
| S10_extracted_79552 | Not-determined |
| S10_extracted_79583 | Not-determined |
| S10_extracted_79584 | Not-determined |
| S10_extracted_79586 | Not-determined |
| S10_extracted_79590 | Not-determined |
| S10_extracted_79591 | Not-determined |
| S10_extracted_79600 | Not-determined |
| S10_extracted_79626 | Not-determined |
| S10_extracted_79641 | Not-determined |
| S10_extracted_79651 | Not-determined |
| S10_extracted_79659 | Not-determined |
| S10_extracted_79670 | Not-determined |
| S10_extracted_79678 | Not-determined |
| S10_extracted_79694 | Low-quality |
| S10_extracted_79699 | Not-determined |
| S10_extracted_79717 | Not-determined |
| S10_extracted_79718 | Low-quality |
| S10_extracted_79721 | Not-determined |
| S10_extracted_79730 | Not-determined |
| S10_extracted_79737 | Not-determined |
| S10_extracted_79742 | Not-determined |
| S10_extracted_79743 | Not-determined |
| S10_extracted_79751 | Not-determined |
| S10_extracted_79754 | Not-determined |
| S10_extracted_79800 | Not-determined |
| S10_extracted_79820 | Not-determined |
| S10_extracted_79836 | Not-determined |
| S10_extracted_79838 | Not-determined |
| S10_extracted_79848 | Not-determined |
| S10_extracted_79858 | Not-determined |
| S10_extracted_79875 | Not-determined |
| S10_extracted_79882 | Not-determined |
| S10_extracted_79886 | Not-determined |
| S10_extracted_79889 | Not-determined |
| S10_extracted_79903 | Not-determined |
| S10_extracted_79912 | Low-quality |
| S10_extracted_79917 | Not-determined |
| S10_extracted_79935 | Not-determined |
| S10_extracted_79953 | Not-determined |
| S10_extracted_79955 | Not-determined |
| S10_extracted_79965 | Not-determined |
| S10_extracted_79981 | Not-determined |
| S10_extracted_79984 | Not-determined |
| S10_extracted_80002 | Low-quality |
| S10_extracted_80022 | Not-determined |
| S10_extracted_80042 | Low-quality |
| S10_extracted_80064 | Not-determined |
| S10_extracted_80076 | Not-determined |
| S10_extracted_80078 | Not-determined |
| S10_extracted_80082 | Not-determined |
| S10_extracted_80088 | Not-determined |
| S10_extracted_80096 | Not-determined |
| S10_extracted_80097 | Not-determined |
| S10_extracted_80103 | Not-determined |
| S10_extracted_80120 | Not-determined |
| S10_extracted_80128 | Low-quality |
| S10_extracted_80158 | Not-determined |
| S10_extracted_80169 | Not-determined |
| S10_extracted_80184 | Not-determined |
| S10_extracted_80189 | Not-determined |
| S10_extracted_80209 | Not-determined |
| S10_extracted_80214 | Not-determined |
| S10_extracted_80227 | Not-determined |
| S10_extracted_80228 | Not-determined |
| S10_extracted_80231 | Not-determined |
| S10_extracted_80240 | Not-determined |
| S10_extracted_80251 | Not-determined |
| S10_extracted_80259 | Not-determined |
| S10_extracted_80266 | Not-determined |
| S10_extracted_80274 | Low-quality |
| S10_extracted_80304 | Low-quality |
| S10_extracted_80308 | Not-determined |
| S10_extracted_80317 | Not-determined |
| S10_extracted_80319 | Not-determined |
| S10_extracted_80351 | Not-determined |
| S10_extracted_80373 | Not-determined |
| S10_extracted_80396 | Not-determined |
| S10_extracted_80410 | Not-determined |
| S10_extracted_80414 | Not-determined |
| S10_extracted_80418 | Not-determined |
| S10_extracted_80428 | Not-determined |
| S10_extracted_80429 | Not-determined |
| S10_extracted_80433 | Not-determined |
| S10_extracted_80437 | Not-determined |
| S10_extracted_80490 | Not-determined |
| S10_extracted_80498 | Not-determined |
| S10_extracted_80504 | Not-determined |
| S10_extracted_80511 | Not-determined |
| S10_extracted_80516 | Not-determined |
| S10_extracted_80517 | Not-determined |
| S10_extracted_80521 | Not-determined |
| S10_extracted_80526 | Not-determined |
| S10_extracted_80527 | Low-quality |
| S10_extracted_80538 | Not-determined |
| S10_extracted_80541 | Not-determined |
| S10_extracted_80549 | Not-determined |
| S10_extracted_80572 | Not-determined |
| S10_extracted_80592 | Not-determined |
| S10_extracted_80603 | Not-determined |
| S10_extracted_80613 | Low-quality |
| S10_extracted_80653 | Low-quality |
| S10_extracted_80671 | Not-determined |
| S7_vs_136782\|\|full | Medium-quality |
| S10_extracted_80716 | Not-determined |
| S10_extracted_80734 | Not-determined |
| S10_extracted_80753 | Not-determined |
| S10_extracted_80760 | Not-determined |
| S10_extracted_80772 | Not-determined |
| S10_extracted_80778 | Not-determined |
| S10_extracted_80783 | Not-determined |
| S10_extracted_80788 | Not-determined |
| S10_extracted_80799 | Not-determined |
| S10_extracted_80800 | Not-determined |
| S10_extracted_80812 | Low-quality |
| S10_extracted_80837 | Not-determined |
| S10_extracted_80838 | Not-determined |
| S10_extracted_80852 | Not-determined |
| S10_extracted_80853 | Not-determined |
| S10_extracted_80860 | Not-determined |
| S10_extracted_80868 | Not-determined |
| S10_extracted_80876 | Not-determined |
| S10_extracted_80880 | Not-determined |
| S10_extracted_80898 | Not-determined |
| S10_extracted_80899 | Not-determined |
| S10_extracted_80929 | Not-determined |
| S10_extracted_80960 | Low-quality |
| S10_extracted_80964 | Not-determined |
| S10_extracted_80969 | Not-determined |
| S10_extracted_80970 | Not-determined |
| S10_extracted_80986 | Not-determined |
| S10_extracted_81003 | Low-quality |
| S10_extracted_81007 | Low-quality |
| S10_extracted_81021 | Not-determined |
| S10_extracted_81030 | Not-determined |
| S10_extracted_81036 | Not-determined |
| S10_extracted_81078 | Not-determined |
| S10_extracted_81081 | Not-determined |
| S10_extracted_81088 | Not-determined |
| S10_extracted_81100 | Not-determined |
| S10_extracted_81101 | Not-determined |
| S10_extracted_81110 | Not-determined |
| S10_extracted_81118 | Not-determined |
| S10_extracted_81122 | Not-determined |
| S10_extracted_81136 | Not-determined |
| S10_extracted_81141 | Low-quality |
| S10_extracted_81155 | Not-determined |
| S10_extracted_81166 | Not-determined |
| S10_extracted_81197 | Not-determined |
| S10_extracted_81204 | Not-determined |
| S10_extracted_81208 | Not-determined |
| S10_extracted_81214 | Not-determined |
| S10_extracted_81230 | Not-determined |
| S10_extracted_81238 | Not-determined |
| S10_extracted_81244 | Not-determined |
| S10_extracted_81248 | Not-determined |
| S10_extracted_81252 | Not-determined |
| S10_extracted_81253 | Not-determined |
| S10_extracted_81262 | Not-determined |
| S10_extracted_81275 | Not-determined |
| S10_extracted_81278 | Not-determined |
| S10_extracted_81302 | Not-determined |
| S10_extracted_81322 | Not-determined |

| | |
|---|---|
| S10_extracted_B1330 | Not-determined |
| S10_extracted_B1333 | Not-determined |
| S10_extracted_B1353 | Not-determined |
| S10_extracted_B1368 | Not-determined |
| S10_extracted_B1369 | Not-determined |
| S10_extracted_B1380 | Not-determined |
| S10_extracted_B1394 | Not-determined |
| S10_extracted_B1397 | Not-determined |
| S10_extracted_B1402 | Not-determined |
| S10_extracted_B1409 | Not-determined |
| S10_extracted_B1412 | Not-determined |
| S10_extracted_B1425 | Low-quality |
| S10_extracted_B1451 | Not-determined |
| S10_extracted_B1460 | Low-quality |
| S10_extracted_B1468 | Low-quality |
| S10_extracted_B1494 | Low-quality |
| S10_extracted_B1504 | Not-determined |
| S10_extracted_B1508 | Not-determined |
| S10_extracted_B1512 | Not-determined |
| S10_extracted_B1524 | Not-determined |
| S10_extracted_B1542 | Not-determined |
| S10_extracted_B1552 | Not-determined |
| S10_extracted_B1553 | Not-determined |
| S10_extracted_B1563 | Not-determined |
| S10_extracted_B1570 | Low-quality |
| S10_extracted_B1571 | Not-determined |
| S10_extracted_B1596 | Not-determined |
| S10_extracted_B1598 | Not-determined |
| S10_extracted_B1601 | Not-determined |
| S10_extracted_B1605 | Not-determined |
| S10_extracted_B1619 | Not-determined |
| S10_extracted_B1624 | Not-determined |
| S10_extracted_B1625 | Not-determined |
| S10_extracted_B1636 | Not-determined |
| S10_extracted_B1645 | Not-determined |
| S10_extracted_B1647 | Not-determined |
| S10_extracted_B1648 | Low-quality |
| S10_extracted_B1653 | Not-determined |
| S10_extracted_B1668 | Low-quality |
| S10_extracted_B1673 | Not-determined |
| S10_extracted_B1674 | Not-determined |
| S10_extracted_B1686 | Not-determined |
| S10_extracted_B1694 | Low-quality |
| S10_extracted_B1697 | Not-determined |
| S10_extracted_B1699 | Not-determined |
| S10_extracted_B1703 | Not-determined |
| S10_extracted_B1709 | Not-determined |
| S10_extracted_B1716 | Not-determined |
| S10_extracted_B1719 | Not-determined |
| S10_extracted_B1744 | Not-determined |
| S10_extracted_B1745 | Not-determined |
| S10_extracted_B1748 | Low-quality |
| S10_extracted_B1757 | Low-quality |
| S10_extracted_B1762 | Not-determined |
| S10_extracted_B1782 | Not-determined |
| S10_extracted_B1785 | Low-quality |
| S10_extracted_B1790 | Not-determined |
| S10_extracted_B1801 | Not-determined |
| S10_extracted_B1802 | Not-determined |
| S10_extracted_B1805 | Low-quality |
| S10_extracted_B1812 | Low-quality |
| S10_extracted_B1814 | Not-determined |
| S10_extracted_B1841 | Not-determined |
| S10_extracted_B1842 | Not-determined |
| S10_extracted_B1849 | Not-determined |
| S10_extracted_B1857 | Not-determined |
| S10_extracted_B1866 | Not-determined |
| S10_extracted_B1867 | Not-determined |
| S10_extracted_B1873 | Not-determined |
| S10_extracted_B1876 | Not-determined |
| S10_extracted_B1880 | Not-determined |
| S10_extracted_B1900 | Low-quality |
| S10_extracted_B1904 | Not-determined |
| S10_extracted_B1905 | Not-determined |
| S10_extracted_B1909 | Not-determined |
| S10_extracted_B1935 | Low-quality |
| S10_extracted_B1955 | Not-determined |
| S10_extracted_B1956 | Not-determined |
| S10_extracted_B1957 | Not-determined |
| S10_extracted_B1963 | Not-determined |
| S10_extracted_B1967 | Not-determined |
| S10_extracted_B1974 | Not-determined |
| S10_extracted_B1977 | Not-determined |
| S10_extracted_B2002 | Low-quality |
| S10_extracted_B2016 | Not-determined |
| S10_extracted_B2023 | Low-quality |
| S10_extracted_B2047 | Not-determined |
| S10_extracted_B2075 | Not-determined |
| S10_extracted_B2078 | Not-determined |
| S10_extracted_B2089 | Not-determined |
| S10_extracted_B2106 | Not-determined |
| S10_extracted_B2124 | Not-determined |
| S10_extracted_B2125 | Not-determined |
| S10_extracted_B2131 | Not-determined |
| S10_extracted_B2133 | Low-quality |
| S10_extracted_B2140 | Not-determined |
| S10_extracted_B2145 | Not-determined |
| S10_extracted_B2153 | Low-quality |
| S10_extracted_B2169 | Not-determined |
| S10_extracted_B2171 | Not-determined |
| S10_extracted_B2179 | Not-determined |
| S10_extracted_B2181 | Low-quality |
| S10_extracted_B2206 | Low-quality |
| S10_extracted_B2211 | Not-determined |
| S10_extracted_B2212 | Not-determined |
| S10_extracted_B2218 | Not-determined |
| S10_extracted_B2233 | Low-quality |
| S10_extracted_B2251 | Not-determined |
| S10_extracted_B2253 | Not-determined |
| S10_extracted_B2299 | Not-determined |
| S10_extracted_B2315 | Not-determined |
| S10_extracted_B2316 | Not-determined |
| S10_extracted_B2319 | Not-determined |
| S10_extracted_B2320 | Not-determined |
| S10_extracted_B2323 | Not-determined |
| S10_extracted_B2326 | Not-determined |
| S10_extracted_B2362 | Not-determined |
| S10_extracted_B2364 | Not-determined |
| S10_extracted_B2371 | Not-determined |
| S10_extracted_B2379 | Not-determined |
| S10_extracted_B2388 | Low-quality |
| S10_extracted_B2389 | Not-determined |
| S10_extracted_B2391 | Not-determined |
| S10_extracted_B2393 | Not-determined |
| S10_extracted_B2405 | Not-determined |
| S10_extracted_B2408 | Not-determined |
| S10_extracted_B2413 | Not-determined |
| S10_extracted_B2427 | Not-determined |
| S10_extracted_B2433 | Not-determined |
| S10_extracted_B2450 | Low-quality |
| S10_extracted_B2484 | Not-determined |
| S10_extracted_B2489 | Not-determined |
| S10_extracted_B2491 | Not-determined |
| S10_extracted_B2493 | Low-quality |
| S10_extracted_B2502 | Not-determined |
| S10_extracted_B2509 | Low-quality |
| S10_extracted_B2529 | Not-determined |
| S10_extracted_B2533 | Not-determined |
| S10_extracted_B2536 | Not-determined |
| S10_extracted_B2543 | Low-quality |
| S10_extracted_B2546 | Not-determined |
| S10_extracted_B2561 | Low-quality |
| S10_extracted_B2574 | Not-determined |
| S10_extracted_B2598 | Not-determined |
| S10_extracted_B2603 | Not-determined |
| S10_extracted_B2618 | Not-determined |
| S10_extracted_B2623 | Not-determined |
| S10_extracted_B2624 | Not-determined |
| S10_extracted_B2630 | Not-determined |
| S10_extracted_B2634 | Not-determined |
| S10_extracted_B2654 | Not-determined |
| S10_extracted_B2661 | Not-determined |
| S10_extracted_B2662 | Not-determined |
| S10_extracted_B2669 | Not-determined |
| S10_extracted_B2680 | Not-determined |
| S10_extracted_B2692 | Not-determined |
| S10_extracted_B2693 | Low-quality |
| S10_extracted_B2698 | Not-determined |
| S10_extracted_B2711 | Not-determined |
| S10_extracted_B2722 | Not-determined |
| S10_extracted_B2725 | Not-determined |
| S10_extracted_B2752 | Not-determined |
| S10_extracted_B2760 | Not-determined |
| S10_extracted_B2762 | Low-quality |
| S10_extracted_B2771 | Not-determined |
| S10_extracted_B2772 | Low-quality |
| S10_extracted_B2789 | Not-determined |
| S10_extracted_B2794 | Not-determined |
| S10_extracted_B2804 | Not-determined |
| S10_extracted_B2809 | Not-determined |
| S10_extracted_B2821 | Not-determined |
| S10_extracted_B2845 | Not-determined |
| S10_extracted_B2846 | Not-determined |
| S10_extracted_B2859 | Not-determined |
| S10_extracted_B2872 | Not-determined |
| S10_extracted_B2875 | Not-determined |
| S10_extracted_B2876 | Not-determined |
| S10_extracted_B2878 | Not-determined |
| S10_extracted_B2896 | Not-determined |
| S10_extracted_B2899 | Low-quality |
| S10_extracted_B2928 | Not-determined |
| S10_extracted_B2938 | Not-determined |
| S10_extracted_B2942 | Not-determined |
| S10_extracted_B2955 | Low-quality |
| S10_extracted_B2986 | Not-determined |
| S10_extracted_B3013 | Not-determined |
| S10_extracted_B3015 | Not-determined |
| S10_extracted_B3028 | Not-determined |
| S10_extracted_B3043 | Low-quality |
| S10_extracted_B3050 | Not-determined |
| S10_extracted_B3051 | Not-determined |
| S10_extracted_B3077 | Not-determined |
| S10_extracted_B3083 | Low-quality |
| S10_extracted_B3088 | Not-determined |
| S10_extracted_B3098 | Not-determined |
| S10_extracted_B3100 | Low-quality |
| S10_extracted_B3104 | Not-determined |
| S10_extracted_B3122 | Not-determined |
| S10_extracted_B3126 | Low-quality |
| S10_extracted_B3134 | Not-determined |
| S10_extracted_B3168 | Not-determined |
| S10_extracted_B3169 | Not-determined |
| S10_extracted_B3172 | Not-determined |
| S10_extracted_B3179 | Not-determined |
| S10_extracted_B3181 | Not-determined |
| S10_extracted_B3204 | Low-quality |
| S10_extracted_B3205 | Low-quality |
| S10_extracted_B3206 | Not-determined |
| S10_extracted_B3208 | Not-determined |
| S10_extracted_B3217 | Low-quality |
| S10_extracted_B3218 | Not-determined |
| S10_extracted_B3219 | Not-determined |
| S10_extracted_B3223 | Low-quality |
| S10_extracted_B3227 | Not-determined |
| S10_extracted_B3231 | Not-determined |
| S10_extracted_B3241 | Not-determined |
| S10_extracted_B3242 | Low-quality |
| S10_extracted_B3244 | Not-determined |
| S10_extracted_B3245 | Not-determined |
| S10_extracted_B3252 | Not-determined |
| S10_extracted_B3277 | Low-quality |
| S10_extracted_B3301 | Not-determined |
| S10_extracted_B3319 | Not-determined |
| S10_extracted_B3343 | Not-determined |
| S10_extracted_B3348 | Not-determined |
| S10_extracted_B3357 | Not-determined |
| S10_extracted_B3364 | Not-determined |
| S10_extracted_B3381 | Not-determined |
| S10_extracted_B3385 | Not-determined |
| S10_extracted_B3392 | Low-quality |
| S10_extracted_B3404 | Not-determined |
| S10_extracted_B3416 | Not-determined |
| S10_extracted_B3423 | Not-determined |
| S10_extracted_B3428 | Not-determined |
| S10_extracted_B3434 | Not-determined |
| S10_extracted_B3482 | Not-determined |
| S10_extracted_B3497 | Not-determined |
| S10_extracted_B3503 | Not-determined |
| S10_extracted_B3524 | Low-quality |
| S10_extracted_B3526 | Not-determined |
| S10_extracted_B3544 | Not-determined |
| S10_extracted_B3545 | Not-determined |
| S10_extracted_B3551 | Low-quality |
| S10_extracted_B3567 | Not-determined |
| S10_extracted_B3586 | Low-quality |
| S10_extracted_B3592 | Not-determined |
| S10_extracted_B3604 | Not-determined |
| S10_extracted_B3619 | Not-determined |
| S10_extracted_B3622 | Not-determined |
| S10_extracted_B3623 | Not-determined |
| S10_extracted_B3635 | Not-determined |
| S10_extracted_B3666 | Not-determined |
| S10_extracted_B3667 | Not-determined |
| S10_extracted_B3677 | Not-determined |
| S10_extracted_B3680 | Not-determined |
| S10_extracted_B3709 | Not-determined |
| S10_extracted_B3729 | Not-determined |
| S10_extracted_B3737 | Not-determined |
| S10_extracted_B3739 | Not-determined |
| S10_extracted_B3744 | Not-determined |
| S10_extracted_B3745 | Not-determined |
| S10_extracted_B3753 | Low-quality |
| S10_extracted_B3754 | Not-determined |
| S10_extracted_B3757 | Not-determined |
| S10_extracted_B3787 | Low-quality |
| S10_extracted_B3814 | Not-determined |
| S10_extracted_B3829 | Not-determined |
| S10_extracted_B3840 | Not-determined |
| S10_extracted_B3846 | Not-determined |
| S10_extracted_B3879 | Not-determined |
| S10_extracted_B3911 | Low-quality |
| S10_extracted_B3936 | Not-determined |
| S10_extracted_B3941 | Not-determined |
| S10_extracted_B3943 | Not-determined |
| S10_extracted_B3954 | Not-determined |
| S10_extracted_B3964 | Low-quality |
| S10_extracted_B3982 | Low-quality |
| S10_extracted_B4003 | Not-determined |
| S10_extracted_B4006 | Not-determined |
| S10_extracted_B4009 | Not-determined |
| S10_extracted_B4020 | Not-determined |
| S10_extracted_B4030 | Not-determined |
| S10_extracted_B4043 | Not-determined |
| S10_extracted_B4064 | Not-determined |
| S10_extracted_B4070 | Not-determined |
| S10_extracted_B4096 | Not-determined |
| S10_extracted_B4098 | Not-determined |
| S10_extracted_B4121 | Not-determined |
| S10_extracted_B4133 | Not-determined |
| S10_extracted_B4156 | Not-determined |
| S10_extracted_B4160 | Not-determined |
| S10_extracted_B4163 | Low-quality |
| S10_extracted_B4183 | Low-quality |
| S10_extracted_B4205 | Not-determined |
| S10_extracted_B4207 | Not-determined |
| S10_extracted_B4215 | Not-determined |
| S10_extracted_B4250 | Not-determined |
| S10_extracted_B4256 | Not-determined |
| S10_extracted_B4259 | Not-determined |
| S10_extracted_B4261 | Not-determined |
| S10_extracted_B4276 | Not-determined |
| S10_extracted_B4285 | Not-determined |
| S10_extracted_B4299 | Not-determined |
| S10_extracted_B4310 | Not-determined |
| S10_extracted_B4314 | Not-determined |
| S10_extracted_B4320 | Not-determined |
| S10_extracted_B4342 | Not-determined |
| S10_extracted_B4347 | Not-determined |
| S10_extracted_B4360 | Not-determined |
| S10_extracted_B4384 | Not-determined |
| S10_extracted_B4401 | Not-determined |
| S10_extracted_B4406 | Not-determined |
| S10_extracted_B4412 | Not-determined |
| S10_extracted_B4422 | Not-determined |
| S10_extracted_B4434 | Low-quality |
| S10_extracted_B4442 | Not-determined |
| S10_extracted_B4451 | Not-determined |
| S10_extracted_B4453 | Not-determined |
| S10_extracted_B4493 | Low-quality |
| S10_extracted_B4510 | Not-determined |
| S10_extracted_B4517 | Not-determined |
| S10_extracted_B4540 | Not-determined |
| S10_extracted_B4541 | Not-determined |
| S10_extracted_B4551 | Not-determined |
| S10_extracted_B4565 | Not-determined |
| S10_extracted_B4577 | Not-determined |
| S10_extracted_B4591 | Not-determined |
| S10_extracted_B4597 | Not-determined |
| S10_extracted_B4601 | Not-determined |
| S10_extracted_B4615 | Not-determined |
| S10_extracted_B4638 | Not-determined |
| S10_extracted_B4657 | Not-determined |
| S10_extracted_B4664 | Not-determined |
| S10_extracted_B4679 | Not-determined |
| S10_extracted_B4695 | Not-determined |
| S10_extracted_B4721 | Not-determined |
| S10_extracted_B4730 | Not-determined |
| S10_extracted_B4756 | Not-determined |
| S10_extracted_B4758 | Not-determined |
| S10_extracted_B4773 | Not-determined |
| S10_extracted_B4775 | Low-quality |
| S10_extracted_B4793 | Not-determined |
| S10_extracted_B4797 | Not-determined |
| S10_extracted_B4806 | Not-determined |
| S10_extracted_B4814 | Not-determined |
| S10_extracted_B4815 | Not-determined |
| S10_extracted_B4831 | Not-determined |
| S10_extracted_B4844 | Not-determined |
| S10_extracted_B4848 | Not-determined |
| S10_extracted_B4851 | Low-quality |
| S10_extracted_B4857 | Not-determined |
| S10_extracted_B4863 | Not-determined |
| S10_extracted_B4870 | Not-determined |
| S10_extracted_B4874 | Not-determined |
| S10_extracted_B4875 | Not-determined |
| S10_extracted_B4886 | Not-determined |
| S10_extracted_B4891 | Low-quality |
| S10_extracted_B4893 | Low-quality |
| S10_extracted_B4898 | Not-determined |
| S10_extracted_B4903 | Not-determined |
| S10_extracted_B4912 | Low-quality |
| S10_extracted_B4924 | Not-determined |
| S10_extracted_B4940 | Not-determined |
| S10_extracted_B4945 | Not-determined |
| S10_extracted_B4947 | Not-determined |
| S10_extracted_B4960 | Low-quality |
| S10_extracted_B4961 | Not-determined |
| S10_extracted_B4970 | Low-quality |
| S10_extracted_B4975 | Not-determined |
| S10_extracted_B4980 | Not-determined |
| S10_extracted_B4985 | Not-determined |
| S10_extracted_B5023 | Low-quality |
| S10_extracted_B5025 | Not-determined |
| S10_extracted_B5039 | Not-determined |
| S10_extracted_B5042 | Not-determined |
| S10_extracted_B5046 | Not-determined |
| S10_extracted_B5059 | Not-determined |
| S10_extracted_B5082 | Not-determined |
| S10_extracted_B5084 | Low-quality |
| S10_extracted_B5111 | Not-determined |
| S10_extracted_B5118 | Low-quality |
| S10_extracted_B5121 | Not-determined |
| S10_extracted_B5122 | Not-determined |
| S10_extracted_B5124 | Low-quality |
| S10_extracted_B5140 | Not-determined |
| S10_extracted_B5145 | Not-determined |
| S10_extracted_B5155 | Not-determined |
| S10_extracted_B5166 | Not-determined |
| S10_extracted_B5172 | Not-determined |
| S10_extracted_B5178 | Not-determined |
| S10_extracted_B5196 | Not-determined |
| S10_extracted_B5198 | Not-determined |
| S10_extracted_B5225 | Not-determined |
| S10_extracted_B5243 | Not-determined |
| S10_extracted_B5253 | Not-determined |
| S10_extracted_B5268 | Not-determined |
| S10_extracted_B5269 | Not-determined |
| S10_extracted_B5275 | Not-determined |
| S10_extracted_B5295 | Not-determined |
| S10_extracted_B5308 | Not-determined |
| S10_extracted_B5312 | Not-determined |
| S10_extracted_B5334 | Not-determined |
| S10_extracted_B5362 | Low-quality |
| S10_extracted_B5372 | Not-determined |
| S10_extracted_B5374 | Not-determined |
| S10_extracted_B5384 | Low-quality |
| S10_extracted_B5403 | Not-determined |
| S10_extracted_B5411 | Not-determined |
| S10_extracted_B5427 | Not-determined |
| S10_extracted_B5431 | Not-determined |
| S10_extracted_B5433 | Not-determined |
| S10_extracted_B5443 | Not-determined |
| S10_extracted_B5446 | Not-determined |
| S10_extracted_B5449 | Not-determined |
| S10_extracted_B5457 | Not-determined |
| S10_extracted_B5458 | Not-determined |
| S10_extracted_B5459 | Not-determined |
| S10_extracted_B5460 | Not-determined |
| S10_extracted_B5462 | Not-determined |
| S10_extracted_B5477 | Not-determined |
| S10_extracted_B5479 | Not-determined |
| S10_extracted_B5490 | Not-determined |
| S10_extracted_B5503 | Not-determined |
| S10_extracted_B5505 | Not-determined |
| S10_extracted_B5510 | Not-determined |
| S10_extracted_B5511 | Not-determined |
| S10_extracted_B5513 | Low-quality |
| S10_extracted_B5520 | Not-determined |
| S10_extracted_B5521 | Not-determined |
| S10_extracted_B5524 | Not-determined |
| S10_extracted_B5533 | Not-determined |
| S10_extracted_B5566 | Not-determined |
| S10_extracted_B5569 | Low-quality |
| S10_extracted_B5570 | Not-determined |
| S10_extracted_B5574 | Not-determined |
| S10_extracted_B5578 | Not-determined |
| S10_extracted_B5582 | Not-determined |
| S10_extracted_B5584 | Not-determined |
| S10_extracted_B5594 | Low-quality |
| S10_extracted_B5605 | Not-determined |
| S10_extracted_B5617 | Not-determined |
| S10_extracted_B5642 | Low-quality |
| S10_extracted_B5643 | Not-determined |
| S10_extracted_B5668 | Not-determined |
| S10_extracted_B5675 | Low-quality |
| S10_extracted_B5686 | Not-determined |
| S10_extracted_B5697 | Not-determined |
| S10_extracted_B5727 | Not-determined |
| S10_extracted_B5743 | Not-determined |
| S10_extracted_B5752 | Not-determined |
| S10_extracted_B5760 | Not-determined |
| S10_extracted_B5778 | Not-determined |
| S10_extracted_B5797 | Not-determined |
| S10_extracted_B5816 | Not-determined |
| S10_extracted_B5821 | Not-determined |
| S10_extracted_B5849 | Not-determined |
| S10_extracted_B5865 | Not-determined |
| S10_extracted_B5872 | Low-quality |
| S10_extracted_B5875 | Not-determined |
| S10_extracted_B5891 | Not-determined |
| S10_extracted_B5893 | Not-determined |
| S10_extracted_B5913 | Not-determined |
| S10_extracted_B5921 | Low-quality |
| S10_extracted_B5949 | Not-determined |
| S10_extracted_B5955 | Not-determined |
| S10_extracted_B5985 | Not-determined |
| S10_extracted_B6000 | Not-determined |
| S10_extracted_B6009 | Not-determined |
| S10_extracted_B6011 | Not-determined |
| S10_extracted_B6014 | Low-quality |
| S10_extracted_B6016 | Not-determined |
| S10_extracted_B6020 | Not-determined |
| S10_extracted_B6025 | Not-determined |
| S10_extracted_B6063 | Not-determined |
| S10_extracted_B6064 | Not-determined |
| S10_extracted_B6078 | Not-determined |
| S10_extracted_B6085 | Low-quality |
| S10_extracted_B6088 | Not-determined |
| S10_extracted_B6101 | Low-quality |
| S10_extracted_B6102 | Not-determined |
| S10_extracted_B6108 | Not-determined |
| S10_extracted_B6121 | Not-determined |
| S10_extracted_B6129 | Not-determined |
| S10_extracted_B6135 | Not-determined |
| S10_extracted_B6163 | Not-determined |
| S10_extracted_B6171 | Not-determined |
| S10_extracted_B6178 | Not-determined |
| S10_extracted_B6183 | Not-determined |
| S10_extracted_B6196 | Not-determined |
| S10_extracted_B6197 | Not-determined |
| S10_extracted_B6204 | Not-determined |
| S10_extracted_B6205 | Not-determined |
| S10_extracted_B6207 | Not-determined |
| S10_extracted_B6211 | Low-quality |
| S10_extracted_B6219 | Not-determined |
| S10_extracted_B6227 | Not-determined |
| S10_extracted_B6234 | Not-determined |
| S10_extracted_B6245 | Not-determined |
| S10_extracted_B6255 | Not-determined |
| S10_extracted_B6260 | Not-determined |
| S10_extracted_B6279 | Not-determined |
| S10_extracted_B6283 | Not-determined |
| S10_extracted_B6321 | Not-determined |
| S10_extracted_B6334 | Not-determined |
| S10_extracted_B6344 | Not-determined |
| S10_extracted_B6346 | Low-quality |
| S10_extracted_B6351 | Not-determined |
| S10_extracted_B6367 | Not-determined |
| S10_extracted_B6371 | Not-determined |
| S10_extracted_B6379 | Not-determined |
| S10_extracted_B6392 | Not-determined |
| S10_extracted_B6398 | Not-determined |
| S10_extracted_B6404 | Not-determined |
| S10_extracted_B6405 | Not-determined |
| S10_extracted_B6421 | Not-determined |
| S10_extracted_B6428 | Not-determined |
| S10_extracted_B6439 | Not-determined |
| S10_extracted_B6451 | Not-determined |
| S10_extracted_B6452 | Not-determined |
| S10_extracted_B6454 | Low-quality |
| S10_extracted_B6455 | Not-determined |
| S10_extracted_B6460 | Not-determined |
| S10_extracted_B6464 | Low-quality |
| S10_extracted_B6468 | Not-determined |
| S10_extracted_B6472 | Low-quality |
| S10_extracted_B6478 | Low-quality |
| S10_extracted_B6485 | Not-determined |
| S10_extracted_B6486 | Not-determined |
| S10_extracted_B6489 | Not-determined |
| S10_extracted_B6493 | Not-determined |
| S10_extracted_B6517 | Not-determined |
| S10_extracted_B6537 | Not-determined |
| S10_extracted_B6539 | Low-quality |
| S10_extracted_B6566 | Not-determined |
| S10_extracted_B6570 | Not-determined |
| S10_extracted_B6576 | Not-determined |
| S10_extracted_B6590 | Not-determined |
| S10_extracted_B6637 | Not-determined |
| S10_extracted_B6653 | Not-determined |
| S10_extracted_B6655 | Not-determined |
| S10_extracted_B6684 | Not-determined |
| S10_extracted_B6686 | Low-quality |
| S10_extracted_B6694 | Not-determined |
| S10_extracted_B6723 | Not-determined |
| S10_extracted_B6734 | Low-quality |
| S10_extracted_B6751 | Not-determined |
| S10_extracted_B6755 | Not-determined |
| S10_extracted_B6756 | Not-determined |
| S10_extracted_B6776 | Low-quality |
| S10_extracted_B6781 | Low-quality |
| S10_extracted_B6784 | Not-determined |
| S10_extracted_B6796 | Low-quality |
| S10_extracted_B6800 | Not-determined |
| S10_extracted_B6808 | Not-determined |
| S10_extracted_B6819 | Not-determined |
| S10_extracted_B6825 | Low-quality |
| S10_extracted_B6828 | Low-quality |
| S10_extracted_B6842 | Low-quality |
| S10_extracted_B6850 | Not-determined |
| S10_extracted_B6862 | Not-determined |
| S10_extracted_B6878 | Not-determined |
| S10_extracted_B6880 | Not-determined |
| S10_extracted_B6909 | Not-determined |
| S10_extracted_B6911 | Not-determined |
| S10_extracted_B6929 | Low-quality |
| S10_extracted_B6930 | Not-determined |
| S10_extracted_B6931 | Not-determined |
| S10_extracted_B6951 | Not-determined |
| S10_extracted_B6962 | Not-determined |
| S10_extracted_B6964 | Not-determined |
| S10_extracted_B6987 | Low-quality |
| S10_extracted_B6999 | Not-determined |
| S10_extracted_B7000 | Not-determined |
| S10_extracted_B7013 | Not-determined |
| S10_extracted_B7015 | Not-determined |
| S10_extracted_B7016 | Not-determined |
| S10_extracted_B7035 | Not-determined |
| S10_extracted_B7041 | Not-determined |
| S10_extracted_B7066 | Not-determined |
| S10_extracted_B7072 | Not-determined |
| S10_extracted_B7093 | Not-determined |
| S10_extracted_B7096 | Not-determined |
| S10_extracted_B7112 | Not-determined |
| S10_extracted_B7114 | Not-determined |
| S10_extracted_B7138 | Not-determined |
| S10_extracted_B7142 | Not-determined |
| S10_extracted_B7144 | Not-determined |
| S10_extracted_B7151 | Not-determined |
| S10_extracted_B7159 | Not-determined |

| | |
|---|---|
| S10_extracted_87160 | Low-quality |
| S10_extracted_87172 | Not-determined |
| S10_extracted_87177 | Low-quality |
| S10_extracted_87186 | Low-quality |
| S10_extracted_87211 | Not-determined |
| S10_extracted_87223 | Not-determined |
| S10_extracted_87231 | Not-determined |
| S10_extracted_87236 | Not-determined |
| S10_extracted_87244 | Not-determined |
| S10_extracted_87246 | Not-determined |
| S10_extracted_87266 | Not-determined |
| S10_extracted_87267 | Not-determined |
| S10_extracted_87276 | Not-determined |
| S10_extracted_87280 | Not-determined |
| S10_extracted_87283 | Low-quality |
| S10_extracted_87297 | Not-determined |
| S10_extracted_87308 | Not-determined |
| S10_extracted_87319 | Not-determined |
| S10_extracted_87329 | Not-determined |
| S10_extracted_87331 | Not-determined |
| S10_extracted_87340 | Not-determined |
| S10_extracted_87354 | Not-determined |
| S10_extracted_87355 | Not-determined |
| S10_extracted_87366 | Low-quality |
| S10_extracted_87376 | Not-determined |
| S10_extracted_87377 | Not-determined |
| S10_extracted_87387 | Low-quality |
| S10_extracted_87411 | Not-determined |
| S10_extracted_87419 | Not-determined |
| S10_extracted_87435 | Not-determined |
| S10_extracted_87437 | Not-determined |
| S10_extracted_87443 | Not-determined |
| S10_extracted_87467 | Low-quality |
| S10_extracted_87471 | Not-determined |
| S10_extracted_87475 | Not-determined |
| S10_extracted_87482 | Low-quality |
| S10_extracted_87500 | Not-determined |
| S10_extracted_87508 | Not-determined |
| S10_extracted_87518 | Not-determined |
| S10_extracted_87534 | Not-determined |
| S10_extracted_87558 | Not-determined |
| S10_extracted_87563 | Not-determined |
| S10_extracted_87566 | Not-determined |
| S10_extracted_87581 | Not-determined |
| S10_extracted_87583 | Not-determined |
| S10_extracted_87597 | Not-determined |
| S10_extracted_87599 | Not-determined |
| S10_extracted_87613 | Not-determined |
| S10_extracted_87622 | Not-determined |
| S10_extracted_87641 | Not-determined |
| S10_extracted_87644 | Not-determined |
| S10_extracted_87651 | Not-determined |
| S10_extracted_87658 | Not-determined |
| S10_extracted_87669 | Not-determined |
| S10_extracted_87681 | Not-determined |
| S10_extracted_87684 | Not-determined |
| S10_extracted_87685 | Not-determined |
| S10_extracted_87686 | Low-quality |
| S10_extracted_87690 | Not-determined |
| S10_extracted_87701 | Not-determined |
| S10_extracted_87704 | Low-quality |
| S10_extracted_87724 | Not-determined |
| S10_extracted_87726 | Not-determined |
| S10_extracted_87730 | Not-determined |
| S10_extracted_87738 | Low-quality |
| S10_extracted_87741 | Not-determined |
| S10_extracted_87746 | Not-determined |
| S10_extracted_87753 | Not-determined |
| S10_extracted_87785 | Not-determined |
| S10_extracted_87792 | Not-determined |
| S10_extracted_87796 | Not-determined |
| S10_extracted_87800 | Not-determined |
| S10_extracted_87813 | Not-determined |
| S10_extracted_87827 | Not-determined |
| S10_extracted_87858 | Not-determined |
| S10_extracted_87862 | Not-determined |
| S10_extracted_87864 | Not-determined |
| S10_extracted_87865 | Not-determined |
| S10_extracted_87869 | Not-determined |
| S10_extracted_87871 | Not-determined |
| S10_extracted_87895 | Not-determined |
| S10_extracted_87901 | Not-determined |
| S10_extracted_87903 | Not-determined |
| S10_extracted_87909 | Not-determined |
| S10_extracted_87910 | Not-determined |
| S10_extracted_87911 | Not-determined |
| S10_extracted_87926 | Not-determined |
| S10_extracted_87927 | Not-determined |
| S10_extracted_87929 | Not-determined |
| S10_extracted_87936 | Low-quality |
| S10_extracted_87946 | Not-determined |
| S10_extracted_87974 | Not-determined |
| S10_extracted_87977 | Not-determined |
| S10_extracted_87990 | Not-determined |
| S10_extracted_88003 | Not-determined |
| S10_extracted_88008 | Not-determined |
| S10_extracted_88009 | Not-determined |
| S10_extracted_88053 | Not-determined |
| S10_extracted_88075 | Not-determined |
| S10_extracted_88076 | Not-determined |
| S10_extracted_88077 | Low-quality |
| S10_extracted_88080 | Not-determined |
| S10_extracted_88100 | Not-determined |
| S10_extracted_88104 | Not-determined |
| S10_extracted_88127 | Low-quality |
| S10_extracted_88133 | Not-determined |
| S10_extracted_88140 | Not-determined |
| S10_extracted_88150 | Not-determined |
| S10_extracted_88166 | Not-determined |
| S10_extracted_88167 | Not-determined |
| S10_extracted_88168 | Not-determined |
| S10_extracted_88172 | Not-determined |
| S10_extracted_88202 | Low-quality |
| S10_extracted_88208 | Not-determined |
| S10_extracted_88211 | Not-determined |
| S10_extracted_88229 | Not-determined |
| S10_extracted_88237 | Not-determined |
| S10_extracted_88250 | Not-determined |
| S10_extracted_88268 | Not-determined |
| S10_extracted_88281 | Not-determined |
| S10_extracted_88285 | Not-determined |
| S10_extracted_88293 | Not-determined |
| S10_extracted_88308 | Not-determined |
| S10_extracted_88321 | Not-determined |
| S10_extracted_88329 | Low-quality |
| S10_extracted_88332 | Not-determined |
| S10_extracted_88333 | Not-determined |
| S10_extracted_88371 | Not-determined |
| S10_extracted_88374 | Low-quality |
| S10_extracted_88375 | Not-determined |
| S10_extracted_88376 | Not-determined |
| S10_extracted_88387 | Not-determined |
| S10_extracted_88419 | Low-quality |
| S10_extracted_88425 | Not-determined |
| S10_extracted_88457 | Not-determined |
| S10_extracted_88464 | Not-determined |
| S10_extracted_88480 | Low-quality |
| S10_extracted_88485 | Not-determined |
| S10_extracted_88496 | Not-determined |
| S10_extracted_88499 | Not-determined |
| S10_extracted_88571 | Low-quality |
| S10_extracted_88582 | Not-determined |
| S10_extracted_88589 | Not-determined |
| S10_extracted_88591 | Not-determined |
| S10_extracted_88595 | Not-determined |
| S10_extracted_88608 | Not-determined |
| S10_extracted_88609 | Not-determined |
| S10_extracted_88619 | Not-determined |
| S10_extracted_88636 | Not-determined |
| S10_extracted_88648 | Not-determined |
| S10_extracted_88687 | Not-determined |
| S10_extracted_88695 | Not-determined |
| S10_extracted_88700 | Not-determined |
| S10_extracted_88706 | Not-determined |
| S10_extracted_88711 | Low-quality |
| S10_extracted_88715 | Not-determined |
| S10_extracted_88725 | Not-determined |
| S10_extracted_88726 | Not-determined |
| S10_extracted_88733 | Not-determined |
| S10_extracted_88739 | Low-quality |
| S10_extracted_88743 | Not-determined |
| S10_extracted_88751 | Not-determined |
| S10_extracted_88763 | Not-determined |
| S10_extracted_88800 | Not-determined |
| S10_extracted_88836 | Not-determined |
| S10_extracted_88845 | Not-determined |
| S10_extracted_88856 | Not-determined |
| S10_extracted_88871 | Not-determined |
| S10_extracted_88878 | Not-determined |
| S10_extracted_88935 | Not-determined |
| S10_extracted_88954 | Not-determined |
| S10_extracted_88967 | Not-determined |
| S10_extracted_88970 | Not-determined |
| S10_extracted_88973 | Not-determined |
| S10_extracted_88985 | Not-determined |
| S10_extracted_88995 | Not-determined |
| S10_extracted_89005 | Not-determined |
| S10_extracted_89007 | Not-determined |
| S10_extracted_89009 | Not-determined |
| S10_extracted_89021 | Not-determined |
| S10_extracted_89025 | Not-determined |
| S10_extracted_89033 | Not-determined |
| S10_extracted_89052 | Not-determined |
| S10_extracted_89062 | Low-quality |
| S10_extracted_89070 | Low-quality |
| S10_extracted_89085 | Not-determined |
| S10_extracted_89087 | Not-determined |
| S10_extracted_89112 | Not-determined |
| S10_extracted_89114 | Not-determined |
| S10_extracted_89120 | Not-determined |
| S10_extracted_89121 | Low-quality |
| S10_extracted_89123 | Not-determined |
| S10_extracted_89129 | Low-quality |
| S10_extracted_89136 | Not-determined |
| S10_extracted_89152 | Not-determined |
| S10_extracted_89169 | Not-determined |
| S10_extracted_89179 | Not-determined |
| S10_extracted_89186 | Not-determined |
| S10_extracted_89187 | Not-determined |
| S10_extracted_89193 | Not-determined |
| S10_extracted_89211 | Not-determined |
| S10_extracted_89217 | Not-determined |
| S10_extracted_89224 | Low-quality |
| S10_extracted_89236 | Low-quality |
| S10_extracted_89246 | Not-determined |
| S10_extracted_89248 | Not-determined |
| S10_extracted_89258 | Not-determined |
| S10_extracted_89285 | Not-determined |
| S10_extracted_89290 | Not-determined |
| S10_extracted_89292 | Not-determined |
| S10_extracted_89295 | Not-determined |
| S10_extracted_89323 | Not-determined |
| S10_extracted_89327 | Not-determined |
| S10_extracted_89332 | Not-determined |
| S10_extracted_89367 | Not-determined |
| S10_extracted_89388 | Not-determined |
| S10_extracted_89402 | Not-determined |
| S10_extracted_89404 | Low-quality |
| S10_extracted_89407 | Not-determined |
| S10_extracted_89425 | Not-determined |
| S10_extracted_89431 | Not-determined |
| S10_extracted_89436 | Low-quality |
| S10_extracted_89450 | Not-determined |
| S10_extracted_89514 | Not-determined |
| S10_extracted_89515 | Not-determined |
| S10_extracted_89540 | Not-determined |
| S10_extracted_89552 | Not-determined |
| S10_extracted_89580 | Not-determined |
| S10_extracted_89591 | Not-determined |
| S10_extracted_89603 | Not-determined |
| S10_extracted_89604 | Not-determined |
| S10_extracted_89641 | Not-determined |
| S10_extracted_89670 | Not-determined |
| S10_extracted_89683 | Not-determined |
| S10_extracted_89685 | Low-quality |
| S10_extracted_89691 | Not-determined |
| S10_extracted_89708 | Not-determined |
| S10_extracted_89713 | Not-determined |
| S10_extracted_89724 | Not-determined |
| S10_extracted_89735 | Low-quality |
| S10_extracted_89740 | Low-quality |
| S10_extracted_89742 | Not-determined |
| S10_extracted_89747 | Not-determined |
| S10_extracted_89754 | Low-quality |
| S10_extracted_89793 | Not-determined |
| S10_extracted_89823 | Not-determined |
| S10_extracted_89849 | Not-determined |
| S10_extracted_89876 | Not-determined |
| S10_extracted_89920 | Not-determined |
| S10_extracted_89934 | Low-quality |
| S10_extracted_89976 | Not-determined |
| S10_extracted_89979 | Not-determined |
| S10_extracted_89980 | Not-determined |
| S10_extracted_89990 | Not-determined |
| S10_extracted_90021 | Not-determined |
| S10_extracted_90046 | Not-determined |
| S10_extracted_90047 | Not-determined |
| S10_extracted_90050 | Not-determined |
| S10_extracted_90053 | Not-determined |
| S10_extracted_90054 | Not-determined |
| S10_extracted_90059 | Low-quality |
| S10_extracted_90075 | Not-determined |
| S10_extracted_90086 | Low-quality |
| S10_extracted_90106 | Not-determined |
| S10_extracted_90133 | Not-determined |
| S10_extracted_90159 | Not-determined |
| S10_extracted_90163 | Not-determined |
| S10_extracted_90167 | Not-determined |
| S10_extracted_90170 | Not-determined |
| S10_extracted_90175 | Not-determined |
| S10_extracted_90184 | Not-determined |
| S10_extracted_90190 | Low-quality |
| S10_extracted_90206 | Not-determined |
| S10_extracted_90211 | Low-quality |
| S10_extracted_90215 | Not-determined |
| S10_extracted_90225 | Not-determined |
| S10_extracted_90231 | Not-determined |
| S10_extracted_90233 | Low-quality |
| S10_extracted_90246 | Not-determined |
| S10_extracted_90248 | Not-determined |
| S10_extracted_90252 | Not-determined |
| S10_extracted_90271 | Not-determined |
| S10_extracted_90286 | Not-determined |
| S10_extracted_90289 | Not-determined |
| S10_extracted_90293 | Not-determined |
| S10_extracted_90308 | Low-quality |
| S10_extracted_90318 | Not-determined |
| S10_extracted_90331 | Not-determined |
| S10_extracted_90351 | Not-determined |
| S10_extracted_90381 | Low-quality |
| S10_extracted_90384 | Not-determined |
| S10_extracted_90387 | Not-determined |
| S10_extracted_90391 | Not-determined |
| S10_extracted_90410 | Not-determined |
| S10_extracted_90411 | Not-determined |
| S10_extracted_90424 | Low-quality |
| S10_extracted_90430 | Not-determined |
| S10_extracted_90458 | Not-determined |
| S10_extracted_90462 | Low-quality |
| S10_extracted_90485 | Not-determined |
| S10_extracted_90489 | Not-determined |
| S10_extracted_90520 | Low-quality |
| S10_extracted_90521 | Not-determined |
| S10_extracted_90530 | Not-determined |
| S10_extracted_90558 | Not-determined |
| S10_extracted_90564 | Low-quality |
| S10_extracted_90565 | Not-determined |
| S10_extracted_90576 | Not-determined |
| S10_extracted_90602 | Not-determined |
| S10_extracted_90618 | Low-quality |
| S10_extracted_90628 | Not-determined |
| S10_extracted_90647 | Not-determined |
| S10_extracted_90649 | Not-determined |
| S10_extracted_90660 | Low-quality |
| S10_extracted_90662 | Not-determined |
| S10_extracted_90665 | Not-determined |
| S10_extracted_90669 | Not-determined |
| S10_extracted_90671 | Not-determined |
| S10_extracted_90685 | Low-quality |
| S10_extracted_90698 | Not-determined |
| S10_extracted_90701 | Not-determined |
| S10_extracted_90703 | Not-determined |
| S10_extracted_90723 | Not-determined |
| S10_extracted_90753 | Not-determined |
| S10_extracted_90758 | Low-quality |
| S10_extracted_90775 | Low-quality |
| S10_extracted_90781 | Not-determined |
| S10_extracted_90792 | Not-determined |
| S10_extracted_90808 | Not-determined |
| S10_extracted_90829 | Not-determined |
| S10_extracted_90830 | Not-determined |
| S10_extracted_90832 | Not-determined |
| S10_extracted_90838 | Not-determined |
| S10_extracted_90846 | Not-determined |
| S10_extracted_90855 | Not-determined |
| S10_extracted_90862 | Not-determined |
| S10_extracted_90878 | Not-determined |
| S10_extracted_90888 | Not-determined |
| S10_extracted_90919 | Not-determined |
| S10_extracted_90924 | Not-determined |
| S10_extracted_90942 | Not-determined |
| S10_extracted_90944 | Not-determined |
| S10_extracted_90950 | Not-determined |
| S10_extracted_90957 | Not-determined |
| S10_extracted_90958 | Not-determined |
| S10_extracted_90963 | Not-determined |
| S10_extracted_90964 | Not-determined |
| S10_extracted_90967 | Not-determined |
| S10_extracted_90992 | Not-determined |
| S10_extracted_91002 | Not-determined |
| S10_extracted_91011 | Not-determined |
| S10_extracted_91023 | Not-determined |
| S10_extracted_91030 | Not-determined |
| S10_extracted_91036 | Not-determined |
| S10_extracted_91051 | Low-quality |
| S10_extracted_91053 | Not-determined |
| S10_extracted_91060 | Not-determined |
| S10_extracted_91075 | Low-quality |
| S10_extracted_91080 | Not-determined |
| S10_extracted_91084 | Low-quality |
| S10_extracted_91086 | Not-determined |
| S10_extracted_91103 | Not-determined |
| S10_extracted_91111 | Not-determined |
| S10_extracted_91113 | Low-quality |
| S10_extracted_91117 | Not-determined |
| S10_extracted_91127 | Not-determined |
| S10_extracted_91151 | Not-determined |
| S10_extracted_91157 | Low-quality |
| S10_extracted_91162 | Not-determined |
| S10_extracted_91179 | Low-quality |
| S10_extracted_91182 | Not-determined |
| S10_extracted_91197 | Not-determined |
| S10_extracted_91210 | Not-determined |
| S10_extracted_91211 | Not-determined |
| S10_extracted_91242 | Not-determined |
| S10_extracted_91250 | Not-determined |
| S10_extracted_91258 | Not-determined |
| S10_extracted_91270 | Not-determined |
| S10_extracted_91275 | Not-determined |
| S10_extracted_91282 | Not-determined |
| S10_extracted_91287 | Not-determined |
| S10_extracted_91300 | Not-determined |
| S10_extracted_91310 | Not-determined |
| S10_extracted_91323 | Not-determined |
| S10_extracted_91324 | Not-determined |
| S10_extracted_91331 | Not-determined |
| S10_extracted_91335 | Not-determined |
| S10_extracted_91347 | Not-determined |
| S10_extracted_91349 | Not-determined |
| S10_extracted_91355 | Low-quality |
| S10_extracted_91403 | Not-determined |
| S10_extracted_91423 | Not-determined |
| S10_extracted_91466 | Not-determined |
| S10_extracted_91498 | Not-determined |
| S10_extracted_91509 | Not-determined |
| S10_extracted_91527 | Not-determined |
| S10_extracted_91544 | Not-determined |
| S10_extracted_91555 | Not-determined |
| S10_extracted_91556 | Not-determined |
| S10_extracted_91557 | Low-quality |
| S10_extracted_91569 | Not-determined |
| S10_extracted_91587 | Not-determined |
| S10_extracted_91602 | Not-determined |
| S10_extracted_91614 | Not-determined |
| S10_extracted_91625 | Not-determined |
| S10_extracted_91635 | Not-determined |
| S10_extracted_91639 | Not-determined |
| S10_extracted_91650 | Not-determined |
| S10_extracted_91651 | Not-determined |
| S10_extracted_91661 | Not-determined |
| S10_extracted_91671 | Not-determined |
| S10_extracted_91677 | Low-quality |
| S10_extracted_91694 | Not-determined |
| S10_extracted_91723 | Not-determined |
| S10_extracted_91728 | Low-quality |
| S10_extracted_91732 | Not-determined |
| S10_extracted_91764 | Not-determined |
| S10_extracted_91767 | Not-determined |
| S10_extracted_91776 | Not-determined |
| S10_extracted_91786 | Not-determined |
| S10_extracted_91789 | Not-determined |
| S10_extracted_91798 | Not-determined |
| S10_extracted_91812 | Not-determined |
| S10_extracted_91823 | Not-determined |
| S10_extracted_91862 | Not-determined |
| S10_extracted_91863 | Low-quality |
| S10_extracted_91869 | Not-determined |
| S10_extracted_91871 | Low-quality |
| S10_extracted_91882 | Low-quality |
| S10_extracted_91909 | Low-quality |
| S10_extracted_91911 | Not-determined |
| S10_extracted_91912 | Not-determined |
| S10_extracted_91922 | Not-determined |
| S10_extracted_91927 | Low-quality |
| S10_extracted_91931 | Not-determined |
| S10_extracted_91940 | Low-quality |
| S10_extracted_91943 | Not-determined |
| S10_extracted_91981 | Low-quality |
| S10_extracted_92003 | Low-quality |
| S10_extracted_92009 | Not-determined |
| S10_extracted_92011 | Not-determined |
| S10_extracted_92014 | Not-determined |
| S10_extracted_92025 | Not-determined |
| S10_extracted_92042 | Not-determined |
| S10_extracted_92060 | Not-determined |
| S10_extracted_92063 | Not-determined |
| S10_extracted_92064 | Not-determined |
| S10_extracted_92066 | Not-determined |
| S10_extracted_92075 | Not-determined |
| S10_extracted_92104 | Not-determined |
| S10_extracted_92107 | Not-determined |
| S10_extracted_92108 | Not-determined |
| S10_extracted_92172 | Not-determined |
| S10_extracted_92177 | Not-determined |
| S10_extracted_92181 | Not-determined |
| S10_extracted_92184 | Not-determined |
| S10_extracted_92192 | Not-determined |
| S10_extracted_92247 | Low-quality |
| S10_extracted_92253 | Not-determined |
| S10_extracted_92255 | Not-determined |
| S10_extracted_92285 | Not-determined |
| S10_extracted_92301 | Not-determined |
| S10_extracted_92305 | Not-determined |
| S10_extracted_92328 | Not-determined |
| S10_extracted_92347 | Not-determined |
| S10_extracted_92376 | Not-determined |
| S10_extracted_92393 | Not-determined |
| S10_extracted_92400 | Not-determined |
| S10_extracted_92412 | Low-quality |
| S10_extracted_92418 | Not-determined |
| S10_extracted_92430 | Low-quality |
| S10_extracted_92451 | Not-determined |
| S10_extracted_92459 | Not-determined |
| S10_extracted_92470 | Not-determined |
| S10_extracted_92479 | Not-determined |
| S10_extracted_92492 | Low-quality |
| S10_extracted_92495 | Not-determined |
| S10_extracted_92516 | Not-determined |
| S10_extracted_92531 | Not-determined |
| S10_extracted_92532 | Low-quality |
| S10_extracted_92547 | Low-quality |
| S10_extracted_92550 | Not-determined |
| S10_extracted_92554 | Not-determined |
| S10_extracted_92563 | Not-determined |
| S10_extracted_92579 | Not-determined |
| S10_extracted_92580 | Not-determined |
| S10_extracted_92585 | Not-determined |
| S10_extracted_92590 | Not-determined |
| S10_extracted_92592 | Not-determined |
| S10_extracted_92596 | Not-determined |
| S10_extracted_92597 | Not-determined |
| S10_extracted_92607 | Not-determined |
| S10_extracted_92615 | Not-determined |
| S10_extracted_92631 | Not-determined |
| S10_extracted_92650 | Low-quality |
| S10_extracted_92653 | Not-determined |
| S10_extracted_92673 | Not-determined |
| S10_extracted_92677 | Not-determined |
| S10_extracted_92711 | Not-determined |
| S10_extracted_92714 | Not-determined |
| S10_extracted_92716 | Not-determined |
| S10_extracted_92727 | Not-determined |
| S10_extracted_92741 | Not-determined |
| S10_extracted_92748 | Low-quality |
| S10_extracted_92759 | Low-quality |
| S10_extracted_92804 | Not-determined |
| S10_extracted_92834 | Not-determined |
| S10_extracted_92836 | Low-quality |
| S10_extracted_92838 | Not-determined |
| S10_extracted_92862 | Not-determined |
| S10_extracted_92864 | Not-determined |
| S10_extracted_92869 | Low-quality |
| S10_extracted_92886 | Not-determined |
| S10_extracted_92915 | Not-determined |
| S10_extracted_92918 | Not-determined |
| S10_extracted_92930 | Not-determined |
| S10_extracted_92931 | Not-determined |
| S10_extracted_92980 | Not-determined |
| S10_extracted_92982 | Not-determined |
| S10_extracted_92983 | Not-determined |
| S10_extracted_92986 | Not-determined |
| S10_extracted_92990 | Not-determined |
| S10_extracted_93013 | Not-determined |
| S10_extracted_93029 | Not-determined |
| S10_extracted_93032 | Not-determined |
| S10_extracted_93047 | Not-determined |
| S10_extracted_93055 | Not-determined |
| S10_extracted_93061 | Not-determined |
| S10_extracted_93064 | Not-determined |
| S10_extracted_93070 | Not-determined |
| S10_extracted_93076 | Not-determined |
| S10_extracted_93097 | Not-determined |
| S10_extracted_93110 | Not-determined |
| S10_extracted_93122 | Low-quality |
| S10_extracted_93127 | Not-determined |
| S10_extracted_93142 | Not-determined |
| S10_extracted_93144 | Not-determined |
| S10_extracted_93165 | Not-determined |
| S10_extracted_93166 | Not-determined |
| S10_extracted_93170 | Not-determined |
| S10_extracted_93178 | Not-determined |
| S10_extracted_93194 | Not-determined |
| S10_extracted_93198 | Not-determined |
| S10_extracted_93213 | Not-determined |
| S10_extracted_93215 | Low-quality |
| S10_extracted_93218 | Not-determined |
| S10_extracted_93228 | Not-determined |
| S10_extracted_93233 | Not-determined |
| S10_extracted_93244 | Not-determined |
| S10_extracted_93255 | Not-determined |
| S10_extracted_93258 | Not-determined |
| S10_extracted_93266 | Not-determined |
| S10_extracted_93268 | Not-determined |
| S10_extracted_93286 | Not-determined |
| S10_extracted_93293 | Not-determined |
| S10_extracted_93294 | Not-determined |
| S10_extracted_93295 | Not-determined |
| S10_extracted_93301 | Not-determined |
| S10_extracted_93314 | Not-determined |
| S10_extracted_93323 | Not-determined |
| S10_extracted_93359 | Not-determined |
| S10_extracted_93365 | Not-determined |
| S10_extracted_93375 | Not-determined |
| S10_extracted_93378 | Not-determined |
| S10_extracted_93380 | Not-determined |
| S10_extracted_93387 | Not-determined |
| S10_extracted_93389 | Not-determined |
| S10_extracted_93412 | Not-determined |
| S10_extracted_93445 | Not-determined |
| S10_extracted_93447 | Not-determined |
| S10_extracted_93459 | Not-determined |
| S10_extracted_93494 | Not-determined |
| S10_extracted_93500 | Not-determined |
| S10_extracted_93521 | Not-determined |
| S10_extracted_93534 | Not-determined |
| S10_extracted_93554 | Not-determined |
| S10_extracted_93576 | Not-determined |
| S10_extracted_93593 | Not-determined |
| S10_extracted_93599 | Not-determined |
| S10_extracted_93621 | Not-determined |
| S10_extracted_93648 | Not-determined |
| S10_extracted_93649 | Not-determined |
| S10_extracted_93671 | Not-determined |
| S10_extracted_93695 | Not-determined |
| S10_extracted_93699 | Not-determined |
| S10_extracted_93702 | Not-determined |
| S10_extracted_93703 | Not-determined |
| S10_extracted_93708 | Not-determined |
| S10_extracted_93712 | Not-determined |

| | |
|---|---|
| S10_extracted_93716 | Low-quality |
| S10_extracted_93723 | Not-determined |
| S10_extracted_93733 | Not-determined |
| S10_extracted_93749 | Not-determined |
| S10_extracted_93750 | Not-determined |
| S10_extracted_93761 | Not-determined |
| S10_extracted_93764 | Not-determined |
| S10_extracted_93776 | Low-quality |
| S10_extracted_93777 | Low-quality |
| S10_extracted_93783 | Not-determined |
| S10_extracted_93813 | Not-determined |
| S10_extracted_93831 | Not-determined |
| S10_extracted_93833 | Not-determined |
| S10_extracted_93834 | Low-quality |
| S10_extracted_93836 | Not-determined |
| S10_extracted_93842 | Low-quality |
| S10_extracted_93848 | Not-determined |
| S10_extracted_93871 | Not-determined |
| S10_extracted_93894 | Low-quality |
| S10_extracted_93895 | Not-determined |
| S10_extracted_93900 | Not-determined |
| S10_extracted_93901 | Not-determined |
| S10_extracted_93907 | Not-determined |
| S10_extracted_93912 | Not-determined |
| S10_extracted_93915 | Low-quality |
| S10_extracted_93935 | Low-quality |
| S10_extracted_93955 | Not-determined |
| S10_extracted_93958 | Not-determined |
| S10_extracted_93989 | Not-determined |
| S10_extracted_94029 | Not-determined |
| S10_extracted_94033 | Low-quality |
| S10_extracted_94045 | Not-determined |
| S10_extracted_94055 | Low-quality |
| S10_extracted_94092 | Not-determined |
| S10_extracted_94106 | Not-determined |
| S10_extracted_94108 | Not-determined |
| S10_extracted_94126 | Not-determined |
| S10_extracted_94127 | Not-determined |
| S10_extracted_94180 | Not-determined |
| S10_extracted_94189 | Not-determined |
| S10_extracted_94198 | Not-determined |
| S10_extracted_94205 | Not-determined |
| S10_extracted_94215 | Not-determined |
| S10_extracted_94227 | Not-determined |
| S10_extracted_94242 | Low-quality |
| S10_extracted_94252 | Not-determined |
| S10_extracted_94259 | Not-determined |
| S10_extracted_94276 | Not-determined |
| S10_extracted_94283 | Not-determined |
| S10_extracted_94299 | Not-determined |
| S10_extracted_94327 | Low-quality |
| S10_extracted_94333 | Not-determined |
| S10_extracted_94339 | Not-determined |
| S10_extracted_94357 | Not-determined |
| S10_extracted_94365 | Not-determined |
| S10_extracted_94370 | Not-determined |
| S10_extracted_94387 | Not-determined |
| S10_extracted_94388 | Not-determined |
| S10_extracted_94395 | Low-quality |
| S10_extracted_94399 | Not-determined |
| S10_extracted_94406 | Not-determined |
| S10_extracted_94419 | Not-determined |
| S10_extracted_94429 | Not-determined |
| S10_extracted_94442 | Not-determined |
| S10_extracted_94456 | Not-determined |
| S10_extracted_94464 | Not-determined |
| S10_extracted_94465 | Not-determined |
| S10_extracted_94474 | Not-determined |
| S10_extracted_94475 | Low-quality |
| S10_extracted_94476 | Not-determined |
| S10_extracted_94479 | Not-determined |
| S10_extracted_94485 | Low-quality |
| S10_extracted_94488 | Not-determined |
| S10_extracted_94489 | Low-quality |
| S10_extracted_94495 | Low-quality |
| S10_extracted_94520 | Not-determined |
| S10_extracted_94526 | Not-determined |
| S10_extracted_94535 | Not-determined |
| S10_extracted_94538 | Not-determined |
| S10_extracted_94543 | Not-determined |
| S10_extracted_94547 | Not-determined |
| S10_extracted_94552 | Not-determined |
| S10_extracted_94555 | Low-quality |
| S10_extracted_94592 | Low-quality |
| S10_extracted_94596 | Not-determined |
| S10_extracted_94603 | Not-determined |
| S10_extracted_94619 | Not-determined |
| S10_extracted_94625 | Low-quality |
| S10_extracted_94632 | Not-determined |
| S10_extracted_94636 | Low-quality |
| S10_extracted_94638 | Not-determined |
| S10_extracted_94646 | Not-determined |
| S10_extracted_94660 | Not-determined |
| S10_extracted_94663 | Not-determined |
| S10_extracted_94665 | Not-determined |
| S10_extracted_94666 | Not-determined |
| S10_extracted_94691 | Not-determined |
| S10_extracted_94696 | Not-determined |
| S10_extracted_94703 | Not-determined |
| S10_extracted_94704 | Not-determined |
| S10_extracted_94750 | Not-determined |
| S10_extracted_94780 | Not-determined |
| S10_extracted_94791 | Low-quality |
| S10_extracted_94796 | Not-determined |
| S10_extracted_94797 | Not-determined |
| S10_extracted_94799 | Not-determined |
| S10_extracted_94802 | Not-determined |
| S10_extracted_94803 | Not-determined |
| S10_extracted_94832 | Not-determined |
| S10_extracted_94839 | Low-quality |
| S10_extracted_94840 | Low-quality |
| S10_extracted_94855 | Not-determined |
| S10_extracted_94867 | Not-determined |
| S10_extracted_94873 | Not-determined |
| S10_extracted_94886 | Low-quality |
| S10_extracted_94902 | Low-quality |
| S10_extracted_94924 | Not-determined |
| S10_extracted_94933 | Low-quality |
| S10_extracted_94934 | Not-determined |
| S10_extracted_94945 | Not-determined |
| S10_extracted_94947 | Low-quality |
| S10_extracted_94956 | Not-determined |
| S10_extracted_94965 | Not-determined |
| S10_extracted_94973 | Low-quality |
| S10_extracted_94974 | Not-determined |
| S10_extracted_94990 | Not-determined |
| S10_extracted_95012 | Not-determined |
| S10_extracted_95018 | Not-determined |
| S10_extracted_95022 | Low-quality |
| S10_extracted_95027 | Not-determined |
| S10_extracted_95032 | Not-determined |
| S10_extracted_95042 | Not-determined |
| S10_extracted_95049 | Not-determined |
| S10_extracted_95067 | Low-quality |
| S10_extracted_95084 | Low-quality |
| S10_extracted_95093 | Not-determined |
| S10_extracted_95099 | Low-quality |
| S10_extracted_95112 | Low-quality |
| S10_extracted_95116 | Not-determined |
| S10_extracted_95124 | Not-determined |
| S10_extracted_95131 | Not-determined |
| S10_extracted_95135 | Not-determined |
| S10_extracted_95142 | Not-determined |
| S10_extracted_95159 | Not-determined |
| S10_extracted_95193 | Not-determined |
| S10_extracted_95197 | Not-determined |
| S10_extracted_95217 | Not-determined |
| S10_extracted_95220 | Not-determined |
| S10_extracted_95231 | Not-determined |
| S10_extracted_95257 | Low-quality |
| S10_extracted_95268 | Not-determined |
| S10_extracted_95272 | Not-determined |
| S10_extracted_95275 | Not-determined |
| S10_extracted_95295 | Not-determined |
| S10_extracted_95296 | Not-determined |
| S10_extracted_95308 | Not-determined |
| S10_extracted_95309 | Low-quality |
| S10_extracted_95316 | Low-quality |
| S10_extracted_95323 | Not-determined |
| S10_extracted_95352 | Not-determined |
| S10_extracted_95366 | Not-determined |
| S10_extracted_95374 | Low-quality |
| S10_extracted_95395 | Not-determined |
| S10_extracted_95400 | Not-determined |
| S10_extracted_95401 | Not-determined |
| S10_extracted_95408 | Not-determined |
| S10_extracted_95460 | Low-quality |
| S10_extracted_95488 | Not-determined |
| S10_extracted_95493 | Not-determined |
| S10_extracted_95510 | Not-determined |
| S10_extracted_95519 | Not-determined |
| S10_extracted_95546 | Not-determined |
| S10_extracted_95552 | Not-determined |
| S10_extracted_95567 | Not-determined |
| S10_extracted_95589 | Not-determined |
| S10_extracted_95627 | Not-determined |
| S10_extracted_95641 | Not-determined |
| S10_extracted_95679 | Low-quality |
| S10_extracted_95694 | Not-determined |
| S10_extracted_95705 | Not-determined |
| S10_extracted_95732 | Low-quality |
| S10_extracted_95755 | Not-determined |
| S10_extracted_95766 | Not-determined |
| S10_extracted_95768 | Not-determined |
| S10_extracted_95774 | Low-quality |
| S10_extracted_95796 | Not-determined |
| S10_extracted_95803 | Not-determined |
| S10_extracted_95808 | Not-determined |
| S10_extracted_95823 | Not-determined |
| S10_extracted_95833 | Not-determined |
| S10_extracted_95842 | Not-determined |
| S10_extracted_95875 | Not-determined |
| S10_extracted_95886 | Not-determined |
| S10_extracted_95941 | Not-determined |
| S10_extracted_95951 | Not-determined |
| S10_extracted_95963 | Not-determined |
| S10_extracted_95969 | Not-determined |
| S10_extracted_95973 | Not-determined |
| S10_extracted_96008 | Not-determined |
| S10_extracted_96010 | Not-determined |
| S10_extracted_96013 | Not-determined |
| S10_extracted_96020 | Not-determined |
| S10_extracted_96038 | Not-determined |
| S10_extracted_96040 | Not-determined |
| S10_extracted_96049 | Not-determined |
| S10_extracted_96056 | Not-determined |
| S10_extracted_96060 | Not-determined |
| S10_extracted_96076 | Not-determined |
| S10_extracted_96077 | Not-determined |
| S10_extracted_96112 | Not-determined |
| S10_extracted_96120 | Not-determined |
| S10_extracted_96124 | Not-determined |
| S10_extracted_96125 | Not-determined |
| S10_extracted_96166 | Not-determined |
| S10_extracted_96178 | Low-quality |
| S10_extracted_96179 | Not-determined |
| S10_extracted_96193 | Not-determined |
| S10_extracted_96196 | Not-determined |
| S10_extracted_96209 | Not-determined |
| S10_extracted_96220 | Not-determined |
| S10_extracted_96228 | Not-determined |
| S10_extracted_96229 | Not-determined |
| S10_extracted_96233 | Not-determined |
| S10_extracted_96256 | Not-determined |
| S10_extracted_96262 | Not-determined |
| S10_extracted_96284 | Not-determined |
| S10_extracted_96289 | Not-determined |
| S10_extracted_96296 | Not-determined |
| S10_extracted_96299 | Not-determined |
| S10_extracted_96308 | Not-determined |
| S10_extracted_96313 | Low-quality |
| S10_extracted_96334 | Not-determined |
| S10_extracted_96373 | Not-determined |
| S10_extracted_96376 | Not-determined |
| S10_extracted_96397 | Not-determined |
| S10_extracted_96400 | Not-determined |
| S10_extracted_96414 | Not-determined |
| S10_extracted_96416 | Not-determined |
| S10_extracted_96432 | Low-quality |
| S10_extracted_96434 | Not-determined |
| S10_extracted_96445 | Not-determined |
| S10_extracted_96459 | Not-determined |
| S10_extracted_96473 | Not-determined |
| S10_extracted_96474 | Not-determined |
| S10_extracted_96480 | Not-determined |
| S10_extracted_96489 | Not-determined |
| S10_extracted_96491 | Not-determined |
| S10_extracted_96517 | Not-determined |
| S10_extracted_96518 | Not-determined |
| S10_extracted_96529 | Not-determined |
| S10_extracted_96531 | Low-quality |
| S10_extracted_96560 | Not-determined |
| S10_extracted_96564 | Not-determined |
| S10_extracted_96579 | Low-quality |
| S10_extracted_96582 | Not-determined |
| S10_extracted_96594 | Not-determined |
| S10_extracted_96604 | Not-determined |
| S10_extracted_96606 | Not-determined |
| S10_extracted_96619 | Not-determined |
| S10_extracted_96628 | Not-determined |
| S10_extracted_96630 | Not-determined |
| S10_extracted_96637 | Not-determined |
| S10_extracted_96642 | Not-determined |
| S10_extracted_96653 | Not-determined |
| S10_extracted_96659 | Not-determined |
| S10_extracted_96671 | Not-determined |
| S10_extracted_96683 | Not-determined |
| S10_extracted_96690 | Not-determined |
| S10_extracted_96695 | Not-determined |
| S10_extracted_96700 | Low-quality |
| S10_extracted_96737 | Not-determined |
| S10_extracted_96758 | Not-determined |
| S10_extracted_96769 | Not-determined |
| S10_extracted_96772 | Low-quality |
| S10_extracted_96785 | Not-determined |
| S10_extracted_96790 | Not-determined |
| S10_extracted_96792 | Not-determined |
| S10_extracted_96794 | Not-determined |
| S10_extracted_96801 | Not-determined |
| S10_extracted_96805 | Not-determined |
| S10_extracted_96827 | Low-quality |
| S10_extracted_96831 | Low-quality |
| S10_extracted_96837 | Low-quality |
| S10_extracted_96865 | Low-quality |
| S10_extracted_96877 | Not-determined |
| S10_extracted_96895 | Not-determined |
| S10_extracted_96898 | Not-determined |
| S10_extracted_96908 | Not-determined |
| S10_extracted_96919 | Not-determined |
| S10_extracted_96930 | Low-quality |
| S10_extracted_96933 | Not-determined |
| S10_extracted_96944 | Not-determined |
| S10_extracted_96945 | Not-determined |
| S10_extracted_96954 | Not-determined |
| S10_extracted_96968 | Not-determined |
| S10_extracted_96969 | Not-determined |
| S10_extracted_96976 | Not-determined |
| S10_extracted_96985 | Not-determined |
| S10_extracted_96999 | Low-quality |
| S10_extracted_97003 | Not-determined |
| S10_extracted_97005 | Not-determined |
| S10_extracted_97016 | Not-determined |
| S10_extracted_97038 | Low-quality |
| S10_extracted_97042 | Low-quality |
| S10_extracted_97055 | Not-determined |
| S10_extracted_97070 | Not-determined |
| S10_extracted_97088 | Not-determined |
| S10_extracted_97130 | Not-determined |
| S10_extracted_97163 | Not-determined |
| S10_extracted_97170 | Not-determined |
| S10_extracted_97201 | Not-determined |
| S10_extracted_97222 | Not-determined |
| S10_extracted_97235 | Not-determined |
| S10_extracted_97242 | Not-determined |
| S10_extracted_97259 | Not-determined |
| S10_extracted_97263 | Low-quality |
| S10_extracted_97268 | Not-determined |
| S10_extracted_97280 | Low-quality |
| S10_extracted_97305 | Not-determined |
| S10_extracted_97309 | Not-determined |
| S10_extracted_97311 | Not-determined |
| S10_extracted_97314 | Not-determined |
| S10_extracted_97321 | Low-quality |
| S10_extracted_97333 | Not-determined |
| S10_extracted_97340 | Not-determined |
| S10_extracted_97354 | Not-determined |
| S10_extracted_97360 | Low-quality |
| S10_extracted_97370 | Not-determined |
| S10_extracted_97387 | Not-determined |
| S10_extracted_97403 | Low-quality |
| S10_extracted_97406 | Not-determined |
| S10_extracted_97429 | Low-quality |
| S10_extracted_97434 | Not-determined |
| S10_extracted_97443 | Not-determined |
| S10_extracted_97444 | Not-determined |
| S10_extracted_97448 | Not-determined |
| S10_extracted_97465 | Not-determined |
| S10_extracted_97482 | Not-determined |
| S10_extracted_97490 | Not-determined |
| S10_extracted_97492 | Low-quality |
| S10_extracted_97493 | Not-determined |
| S10_extracted_97499 | Low-quality |
| S10_extracted_97522 | Not-determined |
| S10_extracted_97524 | Low-quality |
| S10_extracted_97542 | Not-determined |
| S10_extracted_97549 | Not-determined |
| S10_extracted_97555 | Low-quality |
| S10_extracted_97560 | Not-determined |
| S10_extracted_97568 | Low-quality |
| S10_extracted_97611 | Low-quality |
| S10_extracted_97613 | Not-determined |
| S10_extracted_97624 | Not-determined |
| S10_extracted_97625 | Low-quality |
| S10_extracted_97628 | Not-determined |
| S10_extracted_97631 | Not-determined |
| S10_extracted_97676 | Not-determined |
| S10_extracted_97679 | Not-determined |
| S10_extracted_97682 | Not-determined |
| S10_extracted_97700 | Not-determined |
| S10_extracted_97706 | Not-determined |
| S10_extracted_97728 | Not-determined |
| S10_extracted_97741 | Not-determined |
| S10_extracted_97745 | Not-determined |
| S10_extracted_97748 | Not-determined |
| S10_extracted_97751 | Not-determined |
| S10_extracted_97757 | Not-determined |
| S10_extracted_97762 | Low-quality |
| S10_extracted_97775 | Low-quality |
| S10_extracted_97778 | Not-determined |
| S10_extracted_97783 | Not-determined |
| S10_extracted_97795 | Not-determined |
| S10_extracted_97802 | Not-determined |
| S10_extracted_97805 | Not-determined |
| S10_extracted_97813 | Not-determined |
| S10_extracted_97820 | Not-determined |
| S10_extracted_97829 | Not-determined |
| S10_extracted_97853 | Not-determined |
| S10_extracted_97857 | Not-determined |
| S10_extracted_97862 | Low-quality |
| S10_extracted_97864 | Not-determined |
| S10_extracted_97865 | Low-quality |
| S10_extracted_97867 | Not-determined |
| S10_extracted_97925 | Not-determined |
| S10_extracted_97931 | Not-determined |
| S10_extracted_97938 | Not-determined |
| S10_extracted_97948 | Low-quality |
| S10_extracted_97960 | Low-quality |
| S10_extracted_97969 | Not-determined |
| S10_extracted_97972 | Not-determined |
| S10_extracted_97986 | Not-determined |
| S10_extracted_97992 | Not-determined |
| S10_extracted_97993 | Not-determined |
| S10_extracted_98029 | Not-determined |
| S10_extracted_98054 | Not-determined |
| S10_extracted_98061 | Low-quality |
| S10_extracted_98062 | Not-determined |
| S10_extracted_98069 | Not-determined |
| S10_extracted_98070 | Low-quality |
| S10_extracted_98075 | Not-determined |
| S10_extracted_98099 | Low-quality |
| S10_extracted_98104 | Not-determined |
| S10_extracted_98136 | Not-determined |
| S10_extracted_98164 | Not-determined |
| S10_extracted_98182 | Not-determined |
| S10_extracted_98187 | Not-determined |
| S10_extracted_98216 | Not-determined |
| S10_extracted_98244 | Not-determined |
| S10_extracted_98251 | Not-determined |
| S10_extracted_98257 | Low-quality |
| S10_extracted_98264 | Not-determined |
| S10_extracted_98275 | Not-determined |
| S10_extracted_98295 | Low-quality |
| S10_extracted_98308 | Low-quality |
| S10_extracted_98311 | Not-determined |
| S10_extracted_98313 | Not-determined |
| S10_extracted_98333 | Low-quality |
| S10_extracted_98334 | Low-quality |
| S10_extracted_98337 | Not-determined |
| S10_extracted_98339 | Low-quality |
| S10_extracted_98345 | Not-determined |
| S10_extracted_98364 | Not-determined |
| S10_extracted_98370 | Low-quality |
| S10_extracted_98392 | Not-determined |
| S10_extracted_98393 | Not-determined |
| S10_extracted_98397 | Not-determined |
| S10_extracted_98420 | Not-determined |
| S10_extracted_98432 | Not-determined |
| S10_extracted_98441 | Not-determined |
| S10_extracted_98461 | Not-determined |
| S10_extracted_98462 | Not-determined |
| S10_extracted_98466 | Not-determined |
| S10_extracted_98467 | Not-determined |
| S10_extracted_98469 | Low-quality |
| S10_extracted_98500 | Not-determined |
| S10_extracted_98516 | Not-determined |
| S10_extracted_98539 | Not-determined |
| S10_extracted_98542 | Not-determined |
| S10_extracted_98555 | Not-determined |
| S10_extracted_98556 | Not-determined |
| S10_extracted_98568 | Not-determined |
| S10_extracted_98579 | Not-determined |
| S10_extracted_98582 | Not-determined |
| S10_extracted_98594 | Low-quality |
| S10_extracted_98608 | Not-determined |
| S10_extracted_98615 | Not-determined |
| S10_extracted_98621 | Not-determined |
| S10_extracted_98630 | Not-determined |
| S10_extracted_98649 | Not-determined |
| S10_extracted_98651 | Not-determined |
| S10_extracted_98653 | Not-determined |
| S10_extracted_98654 | Not-determined |
| S10_extracted_98662 | Not-determined |
| S10_extracted_98663 | Not-determined |
| S10_extracted_98666 | Not-determined |
| S10_extracted_98673 | Not-determined |
| S10_extracted_98675 | Low-quality |
| S10_extracted_98687 | Low-quality |
| S10_extracted_98696 | Not-determined |
| S10_extracted_98712 | Not-determined |
| S10_extracted_98723 | Not-determined |
| S10_extracted_98725 | Not-determined |
| S10_extracted_98737 | Not-determined |
| S10_extracted_98745 | Not-determined |
| S10_extracted_98750 | Not-determined |
| S10_extracted_98810 | Not-determined |
| S10_extracted_98815 | Low-quality |
| S10_extracted_98818 | Not-determined |
| S10_extracted_98819 | Not-determined |
| S10_extracted_98830 | Not-determined |
| S10_extracted_98840 | Not-determined |
| S10_extracted_98843 | Not-determined |
| S10_extracted_98845 | Not-determined |
| S10_extracted_98846 | Low-quality |
| S10_extracted_98867 | Not-determined |
| S10_extracted_98887 | Not-determined |
| S10_extracted_98890 | Not-determined |
| S10_extracted_98900 | Not-determined |
| S10_extracted_98907 | Low-quality |
| S10_extracted_98910 | Not-determined |
| S10_extracted_98926 | Low-quality |
| S10_extracted_98962 | Not-determined |
| S10_extracted_98978 | Not-determined |
| S10_extracted_98981 | Not-determined |
| S10_extracted_98986 | Not-determined |
| S10_extracted_99008 | Low-quality |
| S10_extracted_99038 | Not-determined |
| S10_extracted_99042 | Not-determined |
| S10_extracted_99055 | Not-determined |
| S10_extracted_99067 | Not-determined |
| S10_extracted_99082 | Low-quality |
| S10_extracted_99083 | Low-quality |
| S10_extracted_99113 | Not-determined |
| S10_extracted_99116 | Not-determined |
| S10_extracted_99160 | Not-determined |
| S10_extracted_99172 | Not-determined |
| S10_extracted_99180 | Low-quality |
| S10_extracted_99190 | Not-determined |
| S10_extracted_99199 | Not-determined |
| S10_extracted_99214 | Not-determined |
| S10_extracted_99240 | Not-determined |
| S10_extracted_99256 | Not-determined |
| S10_extracted_99263 | Low-quality |
| S10_extracted_99290 | Not-determined |
| S10_extracted_99293 | Not-determined |
| S10_extracted_99308 | Not-determined |
| S10_extracted_99324 | Low-quality |
| S10_extracted_99334 | Not-determined |
| S10_extracted_99350 | Not-determined |
| S10_extracted_99383 | Not-determined |
| S10_extracted_99391 | Not-determined |
| S10_extracted_99398 | Not-determined |
| S10_extracted_99430 | Not-determined |
| S10_extracted_99463 | Not-determined |
| S10_extracted_99475 | Low-quality |
| S10_extracted_99477 | Low-quality |
| S10_extracted_99487 | Low-quality |
| S10_extracted_99488 | Not-determined |
| S10_extracted_99495 | Not-determined |
| S10_extracted_99501 | Not-determined |
| S10_extracted_99513 | Not-determined |
| S10_extracted_99534 | Low-quality |
| S10_extracted_99548 | Not-determined |
| S10_extracted_99549 | Not-determined |
| S10_extracted_99551 | Low-quality |
| S10_extracted_99567 | Not-determined |
| S10_extracted_99580 | Not-determined |
| S10_extracted_99604 | Not-determined |
| S10_extracted_99607 | Not-determined |
| S10_extracted_99619 | Not-determined |
| S10_extracted_99632 | Not-determined |
| S10_extracted_99647 | Not-determined |
| S10_extracted_99651 | Not-determined |
| S10_extracted_99671 | Not-determined |
| S10_extracted_99672 | Low-quality |
| S10_extracted_99675 | Not-determined |
| S10_extracted_99697 | Not-determined |
| S10_extracted_99702 | Not-determined |
| S10_extracted_99703 | Not-determined |
| S10_extracted_99741 | Not-determined |
| S10_extracted_99753 | Not-determined |
| S10_extracted_99756 | Not-determined |
| S10_extracted_99758 | Not-determined |
| S10_extracted_99781 | Not-determined |
| S10_extracted_99783 | Not-determined |
| S10_extracted_99793 | Not-determined |
| S10_extracted_99817 | Not-determined |
| S10_extracted_99827 | Not-determined |
| S10_extracted_99828 | Not-determined |
| S10_extracted_99840 | Not-determined |
| S10_extracted_99843 | Low-quality |
| S10_extracted_99845 | Not-determined |
| S10_extracted_99853 | Not-determined |
| S10_extracted_99877 | Not-determined |
| S10_extracted_99879 | Low-quality |
| S10_extracted_99896 | Not-determined |
| S10_extracted_99900 | Not-determined |
| S10_extracted_99908 | Not-determined |
| S10_extracted_99913 | Not-determined |
| S10_extracted_99915 | Not-determined |
| S10_extracted_99926 | Not-determined |
| S10_extracted_99934 | Not-determined |
| S10_extracted_99948 | Not-determined |
| S10_extracted_99955 | Low-quality |
| S10_extracted_99971 | Low-quality |
| S10_extracted_99979 | Not-determined |
| S10_extracted_99980 | Not-determined |
| S10_extracted_100003 | Not-determined |
| S10_extracted_100027 | Not-determined |
| S10_extracted_100028 | Not-determined |
| S10_extracted_100033 | Not-determined |
| S10_extracted_100055 | Not-determined |
| S10_extracted_100078 | Not-determined |
| S10_extracted_100088 | Low-quality |
| S10_extracted_100100 | Not-determined |
| S10_extracted_100127 | Not-determined |
| S10_extracted_100162 | Not-determined |
| S10_extracted_100197 | Not-determined |
| S10_extracted_100199 | Not-determined |
| S10_extracted_100206 | Not-determined |
| S10_extracted_100211 | Not-determined |
| S10_extracted_100236 | Not-determined |
| S10_extracted_100237 | Low-quality |
| S10_extracted_100245 | Not-determined |
| S10_extracted_100256 | Not-determined |
| S10_extracted_100275 | Not-determined |
| S10_extracted_100295 | Not-determined |
| S10_extracted_100308 | Not-determined |
| S10_extracted_100311 | Not-determined |
| S10_extracted_100336 | Not-determined |

| | |
|---|---|
| S10_extracted_100338 | Not-determined |
| S10_extracted_100339 | Not-determined |
| S10_extracted_100341 | Not-determined |
| S10_extracted_100342 | Not-determined |
| S10_extracted_100358 | Not-determined |
| S10_extracted_100361 | Not-determined |
| S10_extracted_100392 | Not-determined |
| S10_extracted_100426 | Not-determined |
| S10_extracted_100427 | Low-quality |
| S10_extracted_100442 | Not-determined |
| S10_extracted_100445 | Not-determined |
| S10_extracted_100453 | Not-determined |
| S10_extracted_100467 | Not-determined |
| S10_extracted_100480 | Not-determined |
| S10_extracted_100511 | Not-determined |
| S10_extracted_100526 | Not-determined |
| S10_extracted_100535 | Not-determined |
| S10_extracted_100560 | Not-determined |
| S10_extracted_100565 | Not-determined |
| S10_extracted_100572 | Not-determined |
| S10_extracted_100580 | Not-determined |
| S10_extracted_100593 | Not-determined |
| S10_extracted_100598 | Not-determined |
| S10_extracted_100599 | Not-determined |
| S10_extracted_100617 | Not-determined |
| S10_extracted_100620 | Low-quality |
| S10_extracted_100640 | Not-determined |
| S10_extracted_100642 | Not-determined |
| S10_extracted_100672 | Not-determined |
| S10_extracted_100679 | Not-determined |
| S10_extracted_100689 | Low-quality |
| S10_extracted_100702 | Low-quality |
| S10_extracted_100707 | Low-quality |
| S10_extracted_100715 | Not-determined |
| S10_extracted_100732 | Low-quality |
| S10_extracted_100740 | Not-determined |
| S10_extracted_100755 | Not-determined |
| S10_extracted_100759 | Low-quality |
| S10_extracted_100761 | Not-determined |
| S10_extracted_100780 | Not-determined |
| S10_extracted_100802 | Not-determined |
| S10_extracted_100803 | Not-determined |
| S10_extracted_100804 | Not-determined |
| S10_extracted_100811 | Not-determined |
| S10_extracted_100818 | Not-determined |
| S10_extracted_100821 | Not-determined |
| S10_extracted_100823 | Not-determined |
| S10_extracted_100830 | Not-determined |
| S10_extracted_100839 | Not-determined |
| S10_extracted_100846 | Not-determined |
| S10_extracted_100850 | Not-determined |
| S10_extracted_100872 | Not-determined |
| S10_extracted_100889 | Low-quality |
| S10_extracted_100899 | Not-determined |
| S10_extracted_100902 | Not-determined |
| S10_extracted_100911 | Not-determined |
| S10_extracted_100922 | Not-determined |
| S10_extracted_100941 | Not-determined |
| S10_extracted_100945 | Not-determined |
| S10_extracted_100951 | Not-determined |
| S10_extracted_100956 | Not-determined |
| S10_extracted_100957 | Not-determined |
| S10_extracted_100959 | Not-determined |
| S10_extracted_100967 | Not-determined |
| S10_extracted_100976 | Low-quality |
| S10_extracted_100978 | Not-determined |
| S10_extracted_100982 | Not-determined |
| S10_extracted_100984 | Not-determined |
| S10_extracted_100986 | Not-determined |
| S10_extracted_100993 | Not-determined |
| S10_extracted_101000 | Low-quality |
| S10_extracted_101007 | Not-determined |
| S10_extracted_101020 | Low-quality |
| S10_extracted_101022 | Not-determined |
| S10_extracted_101023 | Not-determined |
| S10_extracted_101037 | Not-determined |
| S10_extracted_101058 | Not-determined |
| S10_extracted_101059 | Not-determined |
| S10_extracted_101071 | Not-determined |
| S10_extracted_101072 | Not-determined |
| S10_extracted_101096 | Not-determined |
| S10_extracted_101103 | Not-determined |
| S10_extracted_101108 | Not-determined |
| S10_extracted_101119 | Not-determined |
| S10_extracted_101129 | Not-determined |
| S10_extracted_101144 | Not-determined |
| S10_extracted_101169 | Low-quality |
| S10_extracted_101190 | Not-determined |
| S10_extracted_101197 | Not-determined |
| S10_extracted_101212 | Not-determined |
| S10_extracted_101213 | Not-determined |
| S10_extracted_101235 | Not-determined |
| S10_extracted_101236 | Not-determined |
| S10_extracted_101244 | Not-determined |
| S10_extracted_101247 | Not-determined |
| S10_extracted_101254 | Not-determined |
| S10_extracted_101273 | Not-determined |
| S10_extracted_101299 | Not-determined |
| S10_extracted_101306 | Low-quality |
| S10_extracted_101322 | Not-determined |
| S10_extracted_101328 | Not-determined |
| S10_extracted_101338 | Not-determined |
| S10_extracted_101342 | Not-determined |
| S10_extracted_101350 | Not-determined |
| S10_extracted_101357 | Not-determined |
| S10_extracted_101380 | Low-quality |
| S10_extracted_101396 | Not-determined |
| S10_extracted_101418 | Not-determined |
| S10_extracted_101421 | Not-determined |
| S10_extracted_101427 | Not-determined |
| S10_extracted_101436 | Not-determined |
| S10_extracted_101456 | Not-determined |
| S10_extracted_101461 | Not-determined |
| S10_extracted_101465 | Not-determined |
| S10_extracted_101518 | Not-determined |
| S10_extracted_101520 | Not-determined |
| S10_extracted_101525 | Not-determined |
| S10_extracted_101527 | Not-determined |
| S10_extracted_101529 | Not-determined |
| S10_extracted_101533 | Not-determined |
| S10_extracted_101551 | Not-determined |
| S10_extracted_101557 | Not-determined |
| S10_extracted_101559 | Not-determined |
| S10_extracted_101562 | Not-determined |
| S10_extracted_101567 | Not-determined |
| S10_extracted_101573 | Not-determined |
| S10_extracted_101580 | Not-determined |
| S10_extracted_101601 | Not-determined |
| S10_extracted_101606 | Not-determined |
| S10_extracted_101614 | Not-determined |
| S10_extracted_101620 | Not-determined |
| S10_extracted_101650 | Low-quality |
| S10_extracted_101651 | Not-determined |
| S10_extracted_101655 | Not-determined |
| S10_extracted_101665 | Not-determined |
| S10_extracted_101670 | Not-determined |
| S10_extracted_101718 | Not-determined |
| S10_extracted_101729 | Not-determined |
| S10_extracted_101738 | Low-quality |
| S10_extracted_101741 | Not-determined |
| S10_extracted_101745 | Low-quality |
| S10_extracted_101746 | Not-determined |
| S10_extracted_101753 | Not-determined |
| S10_extracted_101765 | Not-determined |
| S10_extracted_101780 | Not-determined |
| S10_extracted_101796 | Low-quality |
| S10_extracted_101803 | Not-determined |
| S10_extracted_101813 | Not-determined |
| S10_extracted_101818 | Not-determined |
| S10_extracted_101850 | Not-determined |
| S10_extracted_101853 | Not-determined |
| S10_extracted_101856 | Low-quality |
| S10_extracted_101867 | Not-determined |
| S10_extracted_101872 | Not-determined |
| S10_extracted_101892 | Not-determined |
| S10_extracted_101893 | Not-determined |
| S10_extracted_101896 | Low-quality |
| S10_extracted_101919 | Not-determined |
| S10_extracted_101928 | Not-determined |
| S10_extracted_101929 | Low-quality |
| S10_extracted_101942 | Low-quality |
| S10_extracted_101943 | Not-determined |
| S10_extracted_101963 | Not-determined |
| S10_extracted_101965 | Not-determined |
| S10_extracted_101973 | Not-determined |
| S10_extracted_102011 | Not-determined |
| S10_extracted_102015 | Not-determined |
| S10_extracted_102019 | Not-determined |
| S10_extracted_102027 | Not-determined |
| S10_extracted_102040 | Not-determined |
| S10_extracted_102048 | Not-determined |
| S10_extracted_102058 | Not-determined |
| S10_extracted_102098 | Not-determined |
| S10_extracted_102115 | Not-determined |
| S10_extracted_102184 | Not-determined |
| S10_extracted_102187 | Not-determined |
| S10_extracted_102194 | Not-determined |
| S10_extracted_102199 | Not-determined |
| S10_extracted_102201 | Low-quality |
| S10_extracted_102204 | Not-determined |
| S10_extracted_102213 | Not-determined |
| S10_extracted_102235 | Low-quality |
| S10_extracted_102237 | Not-determined |
| S10_extracted_102245 | Not-determined |
| S10_extracted_102246 | Not-determined |
| S10_extracted_102262 | Not-determined |
| S10_extracted_102276 | Low-quality |
| S10_extracted_102285 | Not-determined |
| S10_extracted_102319 | Not-determined |
| S10_extracted_102332 | Not-determined |
| S10_extracted_102338 | Not-determined |
| S10_extracted_102353 | Not-determined |
| S10_extracted_102381 | Not-determined |
| S10_extracted_102384 | Low-quality |
| S10_extracted_102388 | Low-quality |
| S10_extracted_102403 | Low-quality |
| S10_extracted_102415 | Not-determined |
| S10_extracted_102435 | Not-determined |
| S10_extracted_102454 | Low-quality |
| S10_extracted_102480 | Not-determined |
| S10_extracted_102502 | Low-quality |
| S10_extracted_102507 | Not-determined |
| S10_extracted_102512 | Not-determined |
| S10_extracted_102523 | Not-determined |
| S10_extracted_102525 | Not-determined |
| S10_extracted_102543 | Not-determined |
| S10_extracted_102547 | Not-determined |
| S10_extracted_102558 | Not-determined |
| S10_extracted_102573 | Not-determined |
| S10_extracted_102593 | Not-determined |
| S10_extracted_102596 | Not-determined |
| S10_extracted_102615 | Not-determined |
| S10_extracted_102625 | Low-quality |
| S10_extracted_102634 | Low-quality |
| S10_extracted_102639 | Not-determined |
| S10_extracted_102648 | Not-determined |
| S10_extracted_102651 | Not-determined |
| S10_extracted_102662 | Not-determined |
| S10_extracted_102688 | Not-determined |
| S10_extracted_102695 | Not-determined |
| S10_extracted_102704 | Not-determined |
| S10_extracted_102716 | Not-determined |
| S10_extracted_102719 | Low-quality |
| S10_extracted_102725 | Not-determined |
| S10_extracted_102730 | Low-quality |
| S10_extracted_102731 | Not-determined |
| S10_extracted_102739 | Not-determined |
| S10_extracted_102747 | Not-determined |
| S10_extracted_102755 | Not-determined |
| S10_extracted_102756 | Not-determined |
| S10_extracted_102757 | Not-determined |
| S10_extracted_102761 | Not-determined |
| S10_extracted_102765 | Not-determined |
| S10_extracted_102767 | Not-determined |
| S10_extracted_102776 | Not-determined |
| S10_extracted_102782 | Not-determined |
| S10_extracted_102797 | Not-determined |
| S10_extracted_102823 | Not-determined |
| S10_extracted_102824 | Not-determined |
| S10_extracted_102851 | Not-determined |
| S10_extracted_102853 | Not-determined |
| S10_extracted_102884 | Low-quality |
| S10_extracted_102887 | Not-determined |
| S10_extracted_102895 | Not-determined |
| S10_extracted_102929 | Not-determined |
| S10_extracted_102932 | Not-determined |
| S10_extracted_102936 | Not-determined |
| S10_extracted_102941 | Low-quality |
| S10_extracted_102944 | Not-determined |
| S10_extracted_102963 | Low-quality |
| S10_extracted_102986 | Not-determined |
| S10_extracted_102995 | Not-determined |
| S10_extracted_102996 | Not-determined |
| S10_extracted_103006 | Low-quality |
| S10_extracted_103018 | Not-determined |
| S10_extracted_103027 | Not-determined |
| S10_extracted_103048 | Not-determined |
| S10_extracted_103055 | Not-determined |
| S10_extracted_103075 | Not-determined |
| S10_extracted_103106 | Not-determined |
| S10_extracted_103124 | Not-determined |
| S10_extracted_103132 | Not-determined |
| S10_extracted_103154 | Not-determined |
| S10_extracted_103170 | Low-quality |
| S10_extracted_103171 | Not-determined |
| S10_extracted_103172 | Not-determined |
| S10_extracted_103202 | Not-determined |
| S10_extracted_103206 | Not-determined |
| S10_extracted_103226 | Not-determined |
| S10_extracted_103228 | Not-determined |
| S10_extracted_103238 | Not-determined |
| S10_extracted_103242 | Not-determined |
| S10_extracted_103243 | Not-determined |
| S10_extracted_103247 | Not-determined |
| S10_extracted_103307 | Not-determined |
| S10_extracted_103308 | Not-determined |
| S10_extracted_103314 | Not-determined |
| S10_extracted_103318 | Not-determined |
| S10_extracted_103325 | Not-determined |
| S10_extracted_103334 | Not-determined |
| S10_extracted_103335 | Not-determined |
| S10_extracted_103340 | Not-determined |
| S10_extracted_103345 | Not-determined |
| S10_extracted_103347 | Not-determined |
| S10_extracted_103359 | Not-determined |
| S10_extracted_103382 | Not-determined |
| S10_extracted_103385 | Not-determined |
| S10_extracted_103386 | Not-determined |
| S10_extracted_103391 | Not-determined |
| S10_extracted_103400 | Not-determined |
| S10_extracted_103428 | Not-determined |
| S10_extracted_103438 | Not-determined |
| S10_extracted_103439 | Not-determined |
| S10_extracted_103452 | Not-determined |
| S10_extracted_103456 | Not-determined |
| S10_extracted_103466 | Not-determined |
| S10_extracted_103472 | Not-determined |
| S10_extracted_103478 | Not-determined |
| S10_extracted_103482 | Low-quality |
| S10_extracted_103520 | Not-determined |
| S10_extracted_103522 | Not-determined |
| S10_extracted_103523 | Not-determined |
| S10_extracted_103525 | Not-determined |
| S10_extracted_103526 | Low-quality |
| S10_extracted_103533 | Not-determined |
| S10_extracted_103540 | Not-determined |
| S10_extracted_103542 | Not-determined |
| S10_extracted_103556 | Not-determined |
| S10_extracted_103558 | Not-determined |
| S10_extracted_103579 | Not-determined |
| S10_extracted_103588 | Not-determined |
| S10_extracted_103595 | Low-quality |
| S10_extracted_103610 | Not-determined |
| S10_extracted_103616 | Not-determined |
| S10_extracted_103617 | Not-determined |
| S10_extracted_103630 | Not-determined |
| S10_extracted_103644 | Not-determined |
| S10_extracted_103662 | Not-determined |
| S10_extracted_103673 | Not-determined |
| S10_extracted_103678 | Not-determined |
| S10_extracted_103681 | Not-determined |
| S10_extracted_103693 | Not-determined |
| S10_extracted_103708 | Not-determined |
| S10_extracted_103716 | Low-quality |
| S10_extracted_103720 | Not-determined |
| S10_extracted_103742 | Not-determined |
| S10_extracted_103754 | Not-determined |
| S10_extracted_103760 | Low-quality |
| S10_extracted_103767 | Not-determined |
| S10_extracted_103795 | Not-determined |
| S10_extracted_103796 | Not-determined |
| S10_extracted_103811 | Not-determined |
| S10_extracted_103814 | Not-determined |
| S10_extracted_103823 | Not-determined |
| S10_extracted_103837 | Not-determined |
| S10_extracted_103841 | Not-determined |
| S10_extracted_103850 | Not-determined |
| S10_extracted_103856 | Not-determined |
| S10_extracted_103872 | Low-quality |
| S10_extracted_103873 | Not-determined |
| S10_extracted_103888 | Low-quality |
| S10_extracted_103910 | Not-determined |
| S10_extracted_103925 | Not-determined |
| S10_extracted_103929 | Not-determined |
| S10_extracted_103938 | Not-determined |
| S10_extracted_103943 | Not-determined |
| S10_extracted_103947 | Low-quality |
| S10_extracted_103953 | Low-quality |
| S10_extracted_103956 | Not-determined |
| S10_extracted_103965 | Not-determined |
| S10_extracted_103966 | Not-determined |
| S10_extracted_103967 | Not-determined |
| S10_extracted_103968 | Not-determined |
| S10_extracted_103976 | Not-determined |
| S10_extracted_103983 | Not-determined |
| S10_extracted_103994 | Not-determined |
| S10_extracted_104006 | Not-determined |
| S10_extracted_104013 | Not-determined |
| S10_extracted_104014 | Low-quality |
| S10_extracted_104038 | Not-determined |
| S10_extracted_104055 | Low-quality |
| S10_extracted_104057 | Not-determined |
| S10_extracted_104072 | Low-quality |
| S10_extracted_104083 | Not-determined |
| S10_extracted_104091 | Not-determined |
| S10_extracted_104093 | Not-determined |
| S10_extracted_104095 | Not-determined |
| S10_extracted_104096 | Not-determined |
| S10_extracted_104103 | Not-determined |
| S10_extracted_104120 | Not-determined |
| S10_extracted_104168 | Not-determined |
| S10_extracted_104178 | Not-determined |
| S10_extracted_104190 | Not-determined |
| S10_extracted_104191 | Not-determined |
| S10_extracted_104192 | Low-quality |
| S10_extracted_104213 | Not-determined |
| S10_extracted_104224 | Not-determined |
| S10_extracted_104230 | Not-determined |
| S10_extracted_104254 | Not-determined |
| S10_extracted_104265 | Low-quality |
| S10_extracted_104269 | Not-determined |
| S10_extracted_104276 | Not-determined |
| S10_extracted_104279 | Low-quality |
| S10_extracted_104290 | Not-determined |
| S10_extracted_104291 | Not-determined |
| S10_extracted_104311 | Not-determined |
| S10_extracted_104320 | Not-determined |
| S10_extracted_104325 | Low-quality |
| S10_extracted_104327 | Not-determined |
| S10_extracted_104330 | Not-determined |
| S10_extracted_104347 | Not-determined |
| S10_extracted_104385 | Low-quality |
| S10_extracted_104390 | Not-determined |
| S10_extracted_104417 | Not-determined |
| S10_extracted_104419 | Not-determined |
| S10_extracted_104438 | Not-determined |
| S10_extracted_104451 | Not-determined |
| S10_extracted_104457 | Not-determined |
| S10_extracted_104479 | Not-determined |
| S10_extracted_104484 | Not-determined |
| S10_extracted_104494 | Not-determined |
| S10_extracted_104506 | Not-determined |
| S10_extracted_104522 | Not-determined |
| S10_extracted_104536 | Not-determined |
| S10_extracted_104548 | Not-determined |
| S10_extracted_104555 | Not-determined |
| S10_extracted_104558 | Not-determined |
| S10_extracted_104560 | Not-determined |
| S10_extracted_104606 | Not-determined |
| S10_extracted_104609 | Not-determined |
| S10_extracted_104611 | Not-determined |
| S10_extracted_104622 | Not-determined |
| S10_extracted_104625 | Not-determined |
| S10_extracted_104635 | Low-quality |
| S10_extracted_104637 | Not-determined |
| S10_extracted_104654 | Low-quality |
| S10_extracted_104661 | Low-quality |
| S10_extracted_104662 | Not-determined |
| S10_extracted_104668 | Not-determined |
| S10_extracted_104670 | Not-determined |
| S10_extracted_104678 | Not-determined |
| S10_extracted_104686 | Not-determined |
| S10_extracted_104697 | Not-determined |
| S10_extracted_104701 | Not-determined |
| S10_extracted_104715 | Not-determined |
| S10_extracted_104717 | Not-determined |
| S10_extracted_104718 | Not-determined |
| S10_extracted_104720 | Not-determined |
| S10_extracted_104731 | Low-quality |
| S10_extracted_104734 | Not-determined |
| S10_extracted_104735 | Not-determined |
| S10_extracted_104742 | Not-determined |
| S10_extracted_104745 | Not-determined |
| S10_extracted_104765 | Not-determined |
| S10_extracted_104780 | Not-determined |
| S10_extracted_104795 | Not-determined |
| S10_extracted_104796 | Low-quality |
| S10_extracted_104802 | Not-determined |
| S10_extracted_104813 | Not-determined |
| S10_extracted_104838 | Not-determined |
| S10_extracted_104848 | Not-determined |
| S10_extracted_104868 | Not-determined |
| S10_extracted_104869 | Not-determined |
| S10_extracted_104876 | Not-determined |
| S10_extracted_104879 | Not-determined |
| S10_extracted_104900 | Low-quality |
| S10_extracted_104912 | Not-determined |
| S10_extracted_104921 | Not-determined |
| S10_extracted_104931 | Not-determined |
| S10_extracted_104933 | Not-determined |
| S10_extracted_104942 | Low-quality |
| S10_extracted_104965 | Not-determined |
| S10_extracted_104966 | Low-quality |
| S10_extracted_104969 | Not-determined |
| S10_extracted_104982 | Not-determined |
| S10_extracted_104989 | Low-quality |
| S10_extracted_104994 | Not-determined |
| S10_extracted_105030 | Not-determined |
| S10_extracted_105037 | Not-determined |
| S10_extracted_105045 | Not-determined |
| S10_extracted_105055 | Not-determined |
| S10_extracted_105083 | Not-determined |
| S10_extracted_105084 | Low-quality |
| S10_extracted_105099 | Not-determined |
| S10_extracted_105107 | Not-determined |
| S10_extracted_105108 | Not-determined |
| S10_extracted_105109 | Not-determined |
| S10_extracted_105111 | Not-determined |
| S10_extracted_105112 | Not-determined |
| S10_extracted_105123 | Not-determined |
| S10_extracted_105149 | Not-determined |
| S10_extracted_105150 | Not-determined |
| S10_extracted_105174 | Not-determined |
| S10_extracted_105181 | Not-determined |
| S10_extracted_105203 | Not-determined |
| S10_extracted_105206 | Low-quality |
| S10_extracted_105207 | Not-determined |
| S10_extracted_105209 | Low-quality |
| S10_extracted_105215 | Not-determined |
| S10_extracted_105218 | Not-determined |
| S10_extracted_105226 | Not-determined |
| S10_extracted_105246 | Not-determined |
| S10_extracted_105249 | Low-quality |
| S10_extracted_105251 | Low-quality |
| S10_extracted_105261 | Not-determined |
| S10_extracted_105329 | Not-determined |
| S10_extracted_105331 | Low-quality |
| S10_extracted_105332 | Not-determined |
| S10_extracted_105335 | Not-determined |
| S10_extracted_105336 | Not-determined |
| S10_extracted_105337 | Not-determined |
| S10_extracted_105345 | Not-determined |
| S10_extracted_105355 | Not-determined |
| S10_extracted_105358 | Not-determined |
| S10_extracted_105375 | Not-determined |
| S10_extracted_105406 | Low-quality |
| S10_extracted_105430 | Not-determined |
| S10_extracted_105434 | Not-determined |
| S10_extracted_105436 | Not-determined |
| S10_extracted_105445 | Not-determined |
| S10_extracted_105475 | Not-determined |
| S10_extracted_105498 | Not-determined |
| S10_extracted_105509 | Low-quality |
| S10_extracted_105519 | Not-determined |
| S10_extracted_105525 | Not-determined |
| S10_extracted_105526 | Not-determined |
| S10_extracted_105532 | Not-determined |
| S10_extracted_105544 | Not-determined |
| S10_extracted_105551 | Low-quality |
| S10_extracted_105553 | Not-determined |
| S10_extracted_105560 | Not-determined |
| S10_extracted_105561 | Not-determined |
| S10_extracted_105578 | Not-determined |
| S10_extracted_105601 | Not-determined |
| S10_extracted_105603 | Not-determined |
| S10_extracted_105631 | Not-determined |
| S10_extracted_105641 | Not-determined |
| S10_extracted_105652 | Not-determined |
| S10_extracted_105676 | Not-determined |
| S10_extracted_105683 | Not-determined |
| S10_extracted_105689 | Not-determined |
| S10_extracted_105696 | Not-determined |
| S10_extracted_105718 | Not-determined |
| S10_extracted_105736 | Not-determined |
| S10_extracted_105740 | Not-determined |
| S10_extracted_105745 | Not-determined |
| S10_extracted_105761 | Not-determined |
| S10_extracted_105768 | Not-determined |
| S10_extracted_105769 | Not-determined |
| S10_extracted_105770 | Not-determined |
| S10_extracted_105799 | Not-determined |
| S10_extracted_105805 | Not-determined |
| S10_extracted_105807 | Not-determined |
| S10_extracted_105808 | Not-determined |
| S10_extracted_105810 | Not-determined |
| S10_extracted_105813 | Not-determined |
| S10_extracted_105831 | Not-determined |
| S10_extracted_105841 | Not-determined |
| S10_extracted_105873 | Not-determined |
| S10_extracted_105883 | Not-determined |
| S10_extracted_105884 | Not-determined |
| S10_extracted_105886 | Not-determined |
| S10_extracted_105927 | Low-quality |
| S10_extracted_105938 | Not-determined |
| S10_extracted_105973 | Not-determined |
| S10_extracted_105987 | Not-determined |
| S10_extracted_105993 | Not-determined |
| S10_extracted_105994 | Not-determined |
| S10_extracted_105998 | Not-determined |
| S10_extracted_106000 | Not-determined |
| S10_extracted_106002 | Not-determined |
| S10_extracted_106005 | Not-determined |
| S10_extracted_106014 | Not-determined |
| S10_extracted_106035 | Not-determined |
| S10_extracted_106046 | Not-determined |
| S10_extracted_106053 | Not-determined |
| S10_extracted_106057 | Not-determined |
| S10_extracted_106072 | Not-determined |
| S10_extracted_106073 | Not-determined |
| S10_extracted_106074 | Not-determined |
| S10_extracted_106086 | Not-determined |
| S10_extracted_106104 | Not-determined |
| S10_extracted_106110 | Not-determined |
| S10_extracted_106116 | Not-determined |
| S10_extracted_106123 | Not-determined |
| S10_extracted_106127 | Not-determined |
| S10_extracted_106135 | Not-determined |
| S10_extracted_106138 | Not-determined |
| S10_extracted_106166 | Not-determined |
| S10_extracted_106184 | Low-quality |
| S10_extracted_106193 | Low-quality |
| S10_extracted_106195 | Not-determined |
| S10_extracted_106209 | Low-quality |
| S10_extracted_106222 | Low-quality |
| S10_extracted_106250 | Not-determined |
| S10_extracted_106260 | Not-determined |
| S10_extracted_106270 | Not-determined |
| S10_extracted_106271 | Low-quality |
| S10_extracted_106275 | Not-determined |
| S10_extracted_106280 | Not-determined |
| S10_extracted_106296 | Not-determined |
| S10_extracted_106309 | Not-determined |
| S10_extracted_106329 | Not-determined |
| S10_extracted_106335 | Not-determined |
| S10_extracted_106337 | Not-determined |
| S10_extracted_106341 | Not-determined |
| S10_extracted_106342 | Not-determined |
| S10_extracted_106346 | Not-determined |
| S10_extracted_106352 | Not-determined |

| | |
|---|---|
| S10_extracted_106370 | Not-determined |
| S10_extracted_106390 | Not-determined |
| S10_extracted_106402 | Not-determined |
| S10_extracted_106408 | Not-determined |
| S10_extracted_106420 | Not-determined |
| S10_extracted_106435 | Not-determined |
| S10_extracted_106448 | Not-determined |
| S10_extracted_106466 | Not-determined |
| S10_extracted_106506 | Not-determined |
| S10_extracted_106543 | Not-determined |
| S10_extracted_106555 | Not-determined |
| S10_extracted_106561 | Not-determined |
| S10_extracted_106563 | Not-determined |
| S10_extracted_106566 | Not-determined |
| S10_extracted_106573 | Not-determined |
| S10_extracted_106576 | Not-determined |
| S10_extracted_106590 | Not-determined |
| S10_extracted_106594 | Low-quality |
| S10_extracted_106597 | Not-determined |
| S10_extracted_106600 | Not-determined |
| S10_extracted_106605 | Not-determined |
| S10_extracted_106606 | Not-determined |
| S10_extracted_106611 | Not-determined |
| S10_extracted_106619 | Not-determined |
| S10_extracted_106622 | Not-determined |
| S10_extracted_106636 | Low-quality |
| S10_extracted_106637 | Not-determined |
| S10_extracted_106657 | Not-determined |
| S10_extracted_106665 | Not-determined |
| S10_extracted_106667 | Not-determined |
| S10_extracted_106668 | Not-determined |
| S10_extracted_106685 | Not-determined |
| S10_extracted_106717 | Not-determined |
| S10_extracted_106742 | Not-determined |
| S10_extracted_106750 | Not-determined |
| S10_extracted_106751 | Low-quality |
| S10_extracted_106774 | Not-determined |
| S10_extracted_106775 | Not-determined |
| S10_extracted_106791 | Not-determined |
| S10_extracted_106796 | Not-determined |
| S10_extracted_106797 | Not-determined |
| S10_extracted_106808 | Not-determined |
| S10_extracted_106810 | Not-determined |
| S10_extracted_106813 | Not-determined |
| S10_extracted_106837 | Low-quality |
| S10_extracted_106856 | Low-quality |
| S10_extracted_106861 | Not-determined |
| S10_extracted_106868 | Not-determined |
| S10_extracted_106869 | Not-determined |
| S10_extracted_106877 | Low-quality |
| S10_extracted_106884 | Not-determined |
| S10_extracted_106888 | Not-determined |
| S10_extracted_106889 | Not-determined |
| S10_extracted_106891 | Low-quality |
| S10_extracted_106897 | Not-determined |
| S10_extracted_106910 | Not-determined |
| S10_extracted_106915 | Not-determined |
| S10_extracted_106923 | Not-determined |
| S10_extracted_106930 | Not-determined |
| S10_extracted_106945 | Not-determined |
| S10_extracted_106949 | Not-determined |
| S10_extracted_106957 | Not-determined |
| S10_extracted_106962 | Not-determined |
| S10_extracted_106966 | Not-determined |
| S10_extracted_106968 | Not-determined |
| S10_extracted_106970 | Not-determined |
| S10_extracted_106983 | Not-determined |
| S10_extracted_106986 | Not-determined |
| S10_extracted_107008 | Not-determined |
| S10_extracted_107012 | Low-quality |
| S10_extracted_107020 | Not-determined |
| S10_extracted_107021 | Not-determined |
| S10_extracted_107036 | Not-determined |
| S10_extracted_107041 | Low-quality |
| S10_extracted_107046 | Not-determined |
| S10_extracted_107059 | Not-determined |
| S10_extracted_107061 | Not-determined |
| S10_extracted_107072 | Not-determined |
| S10_extracted_107078 | Low-quality |
| S10_extracted_107081 | Not-determined |
| S10_extracted_107082 | Not-determined |
| S10_extracted_107092 | Not-determined |
| S10_extracted_107095 | Low-quality |
| S10_extracted_107125 | Not-determined |
| S10_extracted_107128 | Not-determined |
| S10_extracted_107138 | Low-quality |
| S10_extracted_107155 | Not-determined |
| S10_extracted_107174 | Not-determined |
| S10_extracted_107177 | Not-determined |
| S10_extracted_107198 | Not-determined |
| S10_extracted_107227 | Not-determined |
| S10_extracted_107231 | Not-determined |
| S10_extracted_107235 | Not-determined |
| S10_extracted_107237 | Not-determined |
| S10_extracted_107241 | Not-determined |
| S10_extracted_107253 | Not-determined |
| S10_extracted_107254 | Low-quality |
| S10_extracted_107263 | Low-quality |
| S10_extracted_107274 | Not-determined |
| S10_extracted_107277 | Low-quality |
| S10_extracted_107290 | Not-determined |
| S10_extracted_107326 | Not-determined |
| S10_extracted_107342 | Low-quality |
| S10_extracted_107356 | Not-determined |
| S10_extracted_107361 | Not-determined |
| S10_extracted_107364 | Low-quality |
| S10_extracted_107383 | Not-determined |
| S10_extracted_107407 | Not-determined |
| S10_extracted_107422 | Not-determined |
| S10_extracted_107426 | Not-determined |
| S10_extracted_107431 | Not-determined |
| S10_extracted_107439 | Not-determined |
| S10_extracted_107452 | Not-determined |
| S10_extracted_107470 | Low-quality |
| S10_extracted_107494 | Not-determined |
| S10_extracted_107500 | Not-determined |
| S10_extracted_107501 | Not-determined |
| S10_extracted_107505 | Not-determined |
| S10_extracted_107517 | Not-determined |
| S10_extracted_107538 | Not-determined |
| S10_extracted_107575 | Not-determined |
| S10_extracted_107594 | Not-determined |
| S10_extracted_107598 | Not-determined |
| S10_extracted_107621 | Not-determined |
| S10_extracted_107624 | Not-determined |
| S10_extracted_107629 | Not-determined |
| S10_extracted_107631 | Not-determined |
| S10_extracted_107680 | Not-determined |
| S10_extracted_107682 | Low-quality |
| S10_extracted_107690 | Not-determined |
| S10_extracted_107692 | Low-quality |
| S10_extracted_107699 | Not-determined |
| S10_extracted_107713 | Not-determined |
| S10_extracted_107722 | Low-quality |
| S10_extracted_107770 | Not-determined |
| S10_extracted_107772 | Not-determined |
| S10_extracted_107780 | Low-quality |
| S10_extracted_107783 | Not-determined |
| S10_extracted_107812 | Not-determined |
| S10_extracted_107824 | Not-determined |
| S10_extracted_107843 | Not-determined |
| S10_extracted_107848 | Not-determined |
| S10_extracted_107892 | Not-determined |
| S10_extracted_107895 | Not-determined |
| S10_extracted_107898 | Not-determined |
| S10_extracted_107911 | Not-determined |
| S10_extracted_107915 | Not-determined |
| S10_extracted_107928 | Not-determined |
| S10_extracted_107937 | Not-determined |
| S10_extracted_107945 | Not-determined |
| S10_extracted_107958 | Not-determined |
| S10_extracted_107972 | Not-determined |
| S10_extracted_107975 | Not-determined |
| S10_extracted_107977 | Not-determined |
| S10_extracted_107981 | Not-determined |
| S10_extracted_107982 | Not-determined |
| S10_extracted_107991 | Low-quality |
| S10_extracted_107994 | Not-determined |
| S10_extracted_107997 | Not-determined |
| S10_extracted_108001 | Not-determined |
| S10_extracted_108010 | Not-determined |
| S10_extracted_108011 | Not-determined |
| S10_extracted_108019 | Not-determined |
| S10_extracted_108024 | Not-determined |
| S10_extracted_108054 | Not-determined |
| S10_extracted_108057 | Not-determined |
| S10_extracted_108060 | Not-determined |
| S10_extracted_108077 | Low-quality |
| S10_extracted_108080 | Not-determined |
| S10_extracted_108093 | Not-determined |
| S10_extracted_108101 | Low-quality |
| S10_extracted_108115 | Not-determined |
| S10_extracted_108118 | Not-determined |
| S10_extracted_108162 | Low-quality |
| S10_extracted_108196 | Low-quality |
| S10_extracted_108203 | Not-determined |
| S10_extracted_108215 | Not-determined |
| S10_extracted_108233 | Not-determined |
| S10_extracted_108248 | Not-determined |
| S10_extracted_108255 | Not-determined |
| S10_extracted_108259 | Not-determined |
| S10_extracted_108268 | Not-determined |
| S10_extracted_108271 | Low-quality |
| S10_extracted_108273 | Not-determined |
| S10_extracted_108284 | Not-determined |
| S10_extracted_108287 | Not-determined |
| S10_extracted_108297 | Not-determined |
| S10_extracted_108312 | Not-determined |
| S10_extracted_108316 | Not-determined |
| S10_extracted_108323 | Not-determined |
| S10_extracted_108336 | Not-determined |
| S10_extracted_108337 | Not-determined |
| S10_extracted_108351 | Not-determined |
| S10_extracted_108358 | Not-determined |
| S10_extracted_108383 | Not-determined |
| S10_extracted_108397 | Low-quality |
| S10_extracted_108410 | Not-determined |
| S10_extracted_108414 | Not-determined |
| S10_extracted_108416 | Not-determined |
| S10_extracted_108459 | Not-determined |
| S10_extracted_108471 | Not-determined |
| S10_extracted_108476 | Not-determined |
| S10_extracted_108484 | Not-determined |
| S10_extracted_108492 | Not-determined |
| S10_extracted_108503 | Not-determined |
| S10_extracted_108506 | Not-determined |
| S10_extracted_108512 | Not-determined |
| S10_extracted_108515 | Not-determined |
| S10_extracted_108517 | Not-determined |
| S10_extracted_108525 | Not-determined |
| S10_extracted_108527 | Low-quality |
| S10_extracted_108530 | Not-determined |
| S10_extracted_108548 | Not-determined |
| S10_extracted_108568 | Not-determined |
| S10_extracted_108572 | Not-determined |
| S10_extracted_108576 | Not-determined |
| S10_extracted_108577 | Not-determined |
| S10_extracted_108601 | Not-determined |
| S10_extracted_108609 | Not-determined |
| S10_extracted_108628 | Not-determined |
| S10_extracted_108634 | Not-determined |
| S10_extracted_108642 | Not-determined |
| S10_extracted_108647 | Not-determined |
| S10_extracted_108651 | Not-determined |
| S10_extracted_108661 | Not-determined |
| S10_extracted_108669 | Not-determined |
| S10_extracted_108671 | Not-determined |
| S10_extracted_108686 | Not-determined |
| S10_extracted_108704 | Low-quality |
| S10_extracted_108718 | Not-determined |
| S10_extracted_108721 | Low-quality |
| S10_extracted_108736 | Not-determined |
| S10_extracted_108740 | Not-determined |
| S10_extracted_108742 | Not-determined |
| S10_extracted_108753 | Not-determined |
| S10_extracted_108769 | Not-determined |
| S10_extracted_108785 | Low-quality |
| S10_extracted_108794 | Not-determined |
| S10_extracted_108799 | Not-determined |
| S10_extracted_108801 | Not-determined |
| S10_extracted_108805 | Not-determined |
| S10_extracted_108812 | Low-quality |
| S10_extracted_108823 | Not-determined |
| S10_extracted_108831 | Not-determined |
| S10_extracted_108842 | Not-determined |
| S10_extracted_108848 | Low-quality |
| S10_extracted_108850 | Not-determined |
| S10_extracted_108864 | Not-determined |
| S10_extracted_108874 | Not-determined |
| S10_extracted_108875 | Not-determined |
| S10_extracted_108888 | Not-determined |
| S10_extracted_108898 | Low-quality |
| S10_extracted_108910 | Not-determined |
| S10_extracted_108924 | Not-determined |
| S10_extracted_108932 | Not-determined |
| S10_extracted_108933 | Not-determined |
| S10_extracted_108937 | Not-determined |
| S10_extracted_108957 | Not-determined |
| S10_extracted_108981 | Not-determined |
| S10_extracted_108982 | Not-determined |
| S10_extracted_108987 | Not-determined |
| S10_extracted_109000 | Low-quality |
| S10_extracted_109019 | Not-determined |
| S10_extracted_109023 | Not-determined |
| S10_extracted_109028 | Not-determined |
| S10_extracted_109035 | Not-determined |
| S10_extracted_109036 | Not-determined |
| S10_extracted_109049 | Low-quality |
| S10_extracted_109064 | Low-quality |
| S10_extracted_109066 | Not-determined |
| S10_extracted_109074 | Not-determined |
| S10_extracted_109089 | Not-determined |
| S10_extracted_109091 | Not-determined |
| S10_extracted_109095 | Not-determined |
| S10_extracted_109097 | Not-determined |
| S10_extracted_109100 | Not-determined |
| S10_extracted_109109 | Not-determined |
| S10_extracted_109115 | Low-quality |
| S10_extracted_109128 | Not-determined |
| S10_extracted_109147 | Not-determined |
| S10_extracted_109151 | Low-quality |
| S10_extracted_109158 | Not-determined |
| S10_extracted_109161 | Not-determined |
| S10_extracted_109164 | Not-determined |
| S10_extracted_109171 | Not-determined |
| S10_extracted_109173 | Not-determined |
| S10_extracted_109174 | Low-quality |
| S10_extracted_109180 | Not-determined |
| S10_extracted_109195 | Low-quality |
| S10_extracted_109204 | Not-determined |
| S10_extracted_109214 | Not-determined |
| S10_extracted_109229 | Not-determined |
| S10_extracted_109244 | Not-determined |
| S10_extracted_109246 | Not-determined |
| S10_extracted_109247 | Not-determined |
| S10_extracted_109253 | Low-quality |
| S10_extracted_109273 | Not-determined |
| S10_extracted_109274 | Not-determined |
| S10_extracted_109301 | Not-determined |
| S10_extracted_109314 | Not-determined |
| S10_extracted_109319 | Not-determined |
| S10_extracted_109323 | Not-determined |
| S10_extracted_109333 | Low-quality |
| S10_extracted_109337 | Not-determined |
| S10_extracted_109352 | Not-determined |
| S10_extracted_109364 | Not-determined |
| S10_extracted_109377 | Not-determined |
| S10_extracted_109408 | Not-determined |
| S10_extracted_109433 | Not-determined |
| S10_extracted_109438 | Not-determined |
| S10_extracted_109439 | Low-quality |
| S10_extracted_109443 | Not-determined |
| S10_extracted_109453 | Not-determined |
| S10_extracted_109460 | Not-determined |
| S10_extracted_109474 | Not-determined |
| S10_extracted_109483 | Not-determined |
| S10_extracted_109489 | Not-determined |
| S10_extracted_109490 | Low-quality |
| S10_extracted_109491 | Not-determined |
| S10_extracted_109503 | Not-determined |
| S10_extracted_109516 | Low-quality |
| S10_extracted_109553 | Not-determined |
| S10_extracted_109556 | Not-determined |
| S10_extracted_109558 | Not-determined |
| S10_extracted_109577 | Not-determined |
| S10_extracted_109594 | Low-quality |
| S10_extracted_109619 | Low-quality |
| S10_extracted_109639 | Not-determined |
| S10_extracted_109642 | Not-determined |
| S10_extracted_109645 | Not-determined |
| S10_extracted_109657 | Not-determined |
| S10_extracted_109701 | Not-determined |
| S10_extracted_109702 | Not-determined |
| S10_extracted_109708 | Not-determined |
| S10_extracted_109718 | Not-determined |
| S10_extracted_109721 | Low-quality |
| S10_extracted_109728 | Low-quality |
| S10_extracted_109734 | Not-determined |
| S10_extracted_109742 | Not-determined |
| S10_extracted_109752 | Not-determined |
| S10_extracted_109767 | Not-determined |
| S10_extracted_109770 | Not-determined |
| S10_extracted_109787 | Low-quality |
| S10_extracted_109788 | Low-quality |
| S10_extracted_109800 | Not-determined |
| S10_extracted_109802 | Not-determined |
| S10_extracted_109819 | Not-determined |
| S10_extracted_109832 | Not-determined |
| S10_extracted_109839 | Not-determined |
| S10_extracted_109869 | Low-quality |
| S10_extracted_109885 | Not-determined |
| S10_extracted_109893 | Not-determined |
| S10_extracted_109913 | Not-determined |
| S10_extracted_109928 | Not-determined |
| S10_extracted_109957 | Not-determined |
| S10_extracted_109958 | Not-determined |
| S10_extracted_109967 | Not-determined |
| S10_extracted_109991 | Not-determined |
| S10_extracted_110004 | Not-determined |
| S10_extracted_110005 | Not-determined |
| S10_extracted_110017 | Low-quality |
| S10_extracted_110023 | Low-quality |
| S10_extracted_110047 | Not-determined |
| S10_extracted_110068 | Not-determined |
| S10_extracted_110079 | Not-determined |
| S10_extracted_110094 | Low-quality |
| S10_extracted_110107 | Not-determined |
| S10_extracted_110121 | Not-determined |
| S10_extracted_110165 | Not-determined |
| S10_extracted_110179 | Not-determined |
| S10_extracted_110180 | Not-determined |
| S10_extracted_110183 | Not-determined |
| S10_extracted_110184 | Low-quality |
| S10_extracted_110186 | Not-determined |
| S10_extracted_110187 | Not-determined |
| S10_extracted_110198 | Not-determined |
| S10_extracted_110205 | Low-quality |
| S10_extracted_110208 | Low-quality |
| S10_extracted_110217 | Not-determined |
| S10_extracted_110237 | Not-determined |
| S10_extracted_110247 | Not-determined |
| S10_extracted_110256 | Not-determined |
| S10_extracted_110258 | Not-determined |
| S10_extracted_110286 | Not-determined |
| S10_extracted_110289 | Not-determined |
| S10_extracted_110295 | Not-determined |
| S10_extracted_110296 | Not-determined |
| S10_extracted_110309 | Low-quality |
| S10_extracted_110334 | Not-determined |
| S10_extracted_110335 | Not-determined |
| S10_extracted_110347 | Not-determined |
| S10_extracted_110366 | Not-determined |
| S10_extracted_110388 | Not-determined |
| S10_extracted_110399 | Not-determined |
| S10_extracted_110402 | Not-determined |
| S10_extracted_110423 | Not-determined |
| S10_extracted_110447 | Not-determined |
| S10_extracted_110448 | Not-determined |
| S10_extracted_110449 | Not-determined |
| S10_extracted_110457 | Not-determined |
| S10_extracted_110461 | Not-determined |
| S10_extracted_110464 | Not-determined |
| S10_extracted_110467 | Not-determined |
| S10_extracted_110478 | Not-determined |
| S10_extracted_110481 | Not-determined |
| S10_extracted_110497 | Not-determined |
| S10_extracted_110500 | Not-determined |
| S10_extracted_110508 | Not-determined |
| S10_extracted_110514 | Not-determined |
| S10_extracted_110517 | Not-determined |
| S10_extracted_110521 | Not-determined |
| S10_extracted_110530 | Not-determined |
| S10_extracted_110533 | Not-determined |
| S10_extracted_110537 | Not-determined |
| S10_extracted_110543 | Not-determined |
| S10_extracted_110545 | Not-determined |
| S10_extracted_110549 | Not-determined |
| S10_extracted_110566 | Low-quality |
| S10_extracted_110572 | Not-determined |
| S10_extracted_110579 | Not-determined |
| S10_extracted_110583 | Not-determined |
| S10_extracted_110586 | Not-determined |
| S10_extracted_110589 | Low-quality |
| S10_extracted_110592 | Not-determined |
| S10_extracted_110598 | Not-determined |
| S10_extracted_110614 | Not-determined |
| S10_extracted_110617 | Not-determined |
| S10_extracted_110635 | Not-determined |
| S10_extracted_110653 | Not-determined |
| S10_extracted_110661 | Not-determined |
| S10_extracted_110666 | Not-determined |
| S10_extracted_110680 | Low-quality |
| S10_extracted_110694 | Not-determined |
| S10_extracted_110696 | Not-determined |
| S10_extracted_110699 | Not-determined |
| S10_extracted_110702 | Not-determined |
| S10_extracted_110703 | Not-determined |
| S10_extracted_110710 | Not-determined |
| S10_extracted_110715 | Not-determined |
| S10_extracted_110728 | Low-quality |
| S10_extracted_110734 | Not-determined |
| S10_extracted_110738 | Not-determined |
| S10_extracted_110755 | Not-determined |
| S10_extracted_110759 | Not-determined |
| S10_extracted_110783 | Low-quality |
| S10_extracted_110804 | Not-determined |
| S10_extracted_110805 | Not-determined |
| S10_extracted_110812 | Low-quality |
| S10_extracted_110813 | Low-quality |
| S10_extracted_110815 | Not-determined |
| S10_extracted_110820 | Not-determined |
| S10_extracted_110828 | Not-determined |
| S10_extracted_110831 | Not-determined |
| S10_extracted_110832 | Low-quality |
| S10_extracted_110834 | Not-determined |
| S10_extracted_110837 | Low-quality |
| S10_extracted_110840 | Not-determined |
| S10_extracted_110841 | Not-determined |
| S10_extracted_110856 | Not-determined |
| S10_extracted_110857 | Not-determined |
| S10_extracted_110859 | Not-determined |
| S10_extracted_110862 | Not-determined |
| S10_extracted_110866 | Not-determined |
| S10_extracted_110893 | Not-determined |
| S10_extracted_110899 | Not-determined |
| S10_extracted_110909 | Not-determined |
| S10_extracted_110912 | Not-determined |
| S10_extracted_110918 | Not-determined |
| S10_extracted_110921 | Not-determined |
| S10_extracted_110943 | Low-quality |
| S10_extracted_110946 | Not-determined |
| S10_extracted_110982 | Not-determined |
| S10_extracted_111004 | Low-quality |
| S10_extracted_111023 | Not-determined |
| S10_extracted_111026 | Not-determined |
| S10_extracted_111027 | Not-determined |
| S10_extracted_111038 | Not-determined |
| S10_extracted_111045 | Low-quality |
| S10_extracted_111046 | Low-quality |
| S10_extracted_111063 | Not-determined |
| S10_extracted_111072 | Low-quality |
| S10_extracted_111075 | Not-determined |
| S10_extracted_111089 | Not-determined |
| S10_extracted_111093 | Not-determined |
| S10_extracted_111100 | Not-determined |
| S10_extracted_111104 | Not-determined |
| S10_extracted_111105 | Not-determined |
| S10_extracted_111107 | Not-determined |
| S10_extracted_111143 | Low-quality |
| S10_extracted_111150 | Not-determined |
| S10_extracted_111154 | Not-determined |
| S10_extracted_111171 | Not-determined |
| S10_extracted_111175 | Not-determined |
| S10_extracted_111225 | Not-determined |
| S10_extracted_111250 | Not-determined |
| S10_extracted_111254 | Not-determined |
| S10_extracted_111260 | Not-determined |
| S10_extracted_111261 | Not-determined |
| S10_extracted_111276 | Not-determined |
| S10_extracted_111291 | Not-determined |
| S10_extracted_111296 | Not-determined |
| S10_extracted_111325 | Not-determined |
| S10_extracted_111341 | Not-determined |
| S10_extracted_111350 | Not-determined |
| S10_extracted_111357 | Low-quality |
| S10_extracted_111367 | Not-determined |
| S10_extracted_111369 | Not-determined |
| S10_extracted_111380 | Not-determined |
| S10_extracted_111384 | Not-determined |
| S10_extracted_111394 | Not-determined |
| S10_extracted_111400 | Not-determined |
| S10_extracted_111409 | Not-determined |
| S10_extracted_111417 | Not-determined |
| S10_extracted_111432 | Not-determined |
| S10_extracted_111435 | Not-determined |
| S10_extracted_111441 | Low-quality |
| S10_extracted_111446 | Not-determined |
| S10_extracted_111462 | Not-determined |
| S10_extracted_111464 | Not-determined |
| S10_extracted_111469 | Not-determined |
| S10_extracted_111489 | Not-determined |
| S10_extracted_111491 | Not-determined |
| S10_extracted_111495 | Not-determined |
| S10_extracted_111513 | Not-determined |
| S10_extracted_111516 | Not-determined |
| S10_extracted_111526 | Not-determined |
| S10_extracted_111535 | Not-determined |
| S10_extracted_111537 | Not-determined |
| S10_extracted_111543 | Not-determined |
| S10_extracted_111556 | Not-determined |
| S10_extracted_111561 | Not-determined |
| S10_extracted_111598 | Low-quality |
| S10_extracted_111621 | Not-determined |
| S10_extracted_111626 | Low-quality |
| S10_extracted_111639 | Not-determined |
| S10_extracted_111667 | Not-determined |
| S10_extracted_111668 | Not-determined |
| S10_extracted_111679 | Not-determined |
| S10_extracted_111690 | Not-determined |
| S10_extracted_111711 | Not-determined |
| S10_extracted_111716 | Not-determined |
| S10_extracted_111719 | Low-quality |
| S10_extracted_111731 | Low-quality |
| S10_extracted_111768 | Not-determined |
| S10_extracted_111771 | Not-determined |
| S10_extracted_111781 | Low-quality |
| S10_extracted_111785 | Not-determined |
| S10_extracted_111798 | Not-determined |
| S10_extracted_111799 | Not-determined |
| S10_extracted_111805 | Not-determined |
| S10_extracted_111814 | Not-determined |
| S10_extracted_111820 | Not-determined |
| S10_extracted_111821 | Not-determined |
| S10_extracted_111873 | Not-determined |
| S10_extracted_111898 | Not-determined |
| S10_extracted_111907 | Not-determined |
| S10_extracted_111919 | Not-determined |
| S10_extracted_111930 | Not-determined |
| S10_extracted_111932 | Not-determined |
| S10_extracted_111933 | Not-determined |
| S10_extracted_111936 | Not-determined |
| S10_extracted_111938 | Not-determined |
| S10_extracted_111959 | Not-determined |
| S10_extracted_111967 | Not-determined |
| S10_extracted_111969 | Not-determined |
| S10_extracted_111980 | Not-determined |
| S10_extracted_111988 | Not-determined |
| S10_extracted_111996 | Not-determined |
| S10_extracted_112003 | Low-quality |
| S10_extracted_112005 | Not-determined |
| S10_extracted_112016 | Not-determined |
| S10_extracted_112018 | Not-determined |
| S10_extracted_112020 | Not-determined |
| S10_extracted_112021 | Low-quality |
| S10_extracted_112037 | Not-determined |
| S10_extracted_112060 | Not-determined |
| S10_extracted_112068 | Not-determined |
| S10_extracted_112073 | Not-determined |
| S10_extracted_112096 | Not-determined |
| S10_extracted_112108 | Not-determined |
| S10_extracted_112113 | Not-determined |
| S10_extracted_112140 | Not-determined |
| S10_extracted_112142 | Not-determined |
| S10_extracted_112147 | Not-determined |
| S10_extracted_112149 | Not-determined |
| S10_extracted_112151 | Not-determined |
| S10_extracted_112158 | Not-determined |
| S10_extracted_112165 | Not-determined |
| S10_extracted_112171 | Not-determined |

| | |
|---|---|
| S10_extracted_112178 | Low-quality |
| S10_extracted_112186 | Not-determined |
| S10_extracted_112208 | Not-determined |
| S10_extracted_112213 | Not-determined |
| S10_extracted_112219 | Not-determined |
| S10_extracted_112231 | Not-determined |
| S10_extracted_112236 | Not-determined |
| S10_extracted_112243 | Not-determined |
| S10_extracted_112245 | Not-determined |
| S10_extracted_112257 | Low-quality |
| S10_extracted_112275 | Not-determined |
| S10_extracted_112280 | Not-determined |
| S10_extracted_112282 | Not-determined |
| S10_extracted_112288 | Not-determined |
| S10_extracted_112302 | Not-determined |
| S10_extracted_112313 | Not-determined |
| S10_extracted_112314 | Not-determined |
| S10_extracted_112316 | Low-quality |
| S10_extracted_112317 | Low-quality |
| S10_extracted_112326 | Not-determined |
| S10_extracted_112333 | Not-determined |
| S10_extracted_112348 | Not-determined |
| S10_extracted_112349 | Not-determined |
| S10_extracted_112366 | Not-determined |
| S10_extracted_112382 | Not-determined |
| S10_extracted_112391 | Not-determined |
| S10_extracted_112402 | Not-determined |
| S10_extracted_112405 | Not-determined |
| S10_extracted_112406 | Not-determined |
| S10_extracted_112420 | Not-determined |
| S10_extracted_112474 | Not-determined |
| S10_extracted_112490 | Not-determined |
| S10_extracted_112501 | Not-determined |
| S10_extracted_112518 | Not-determined |
| S10_extracted_112526 | Low-quality |
| S10_extracted_112529 | Not-determined |
| S10_extracted_112537 | Not-determined |
| S10_extracted_112550 | Not-determined |
| S10_extracted_112563 | Not-determined |
| S10_extracted_112586 | Not-determined |
| S10_extracted_112592 | Not-determined |
| S10_extracted_112596 | Low-quality |
| S10_extracted_112597 | Not-determined |
| S10_extracted_112600 | Not-determined |
| S10_extracted_112615 | Not-determined |
| S10_extracted_112640 | Not-determined |
| S10_extracted_112642 | Not-determined |
| S10_extracted_112651 | Not-determined |
| S10_extracted_112658 | Not-determined |
| S10_extracted_112681 | Not-determined |
| S10_extracted_112688 | Not-determined |
| S10_extracted_112710 | Not-determined |
| S10_extracted_112729 | Low-quality |
| S10_extracted_112735 | Not-determined |
| S10_extracted_112773 | Not-determined |
| S10_extracted_112774 | Not-determined |
| S10_extracted_112795 | Not-determined |
| S10_extracted_112804 | Low-quality |
| S10_extracted_112816 | Not-determined |
| S10_extracted_112817 | Not-determined |
| S10_extracted_112831 | Not-determined |
| S10_extracted_112845 | Not-determined |
| S10_extracted_112848 | Low-quality |
| S10_extracted_112865 | Not-determined |
| S10_extracted_112872 | Low-quality |
| S10_extracted_112886 | Not-determined |
| S10_extracted_112919 | Low-quality |
| S10_extracted_112928 | Low-quality |
| S10_extracted_112932 | Low-quality |
| S10_extracted_112934 | Low-quality |
| S10_extracted_112941 | Not-determined |
| S10_extracted_112947 | Not-determined |
| S10_extracted_112957 | Not-determined |
| S10_extracted_112958 | Not-determined |
| S10_extracted_112962 | Not-determined |
| S10_extracted_112994 | Low-quality |
| S10_extracted_112999 | Low-quality |
| S10_extracted_113003 | Not-determined |
| S10_extracted_113008 | Not-determined |
| S10_extracted_113045 | Not-determined |
| S10_extracted_113059 | Not-determined |
| S10_extracted_113060 | Not-determined |
| S10_extracted_113061 | Low-quality |
| S10_extracted_113067 | Not-determined |
| S10_extracted_113073 | Not-determined |
| S10_extracted_113079 | Not-determined |
| S10_extracted_113105 | Not-determined |
| S10_extracted_113129 | Not-determined |
| S10_extracted_113133 | Not-determined |
| S10_extracted_113134 | Low-quality |
| S10_extracted_113158 | Not-determined |
| S10_extracted_113170 | Not-determined |
| S10_extracted_113171 | Not-determined |
| S10_extracted_113180 | Not-determined |
| S10_extracted_113181 | Not-determined |
| S10_extracted_113182 | Low-quality |
| S10_extracted_113185 | Not-determined |
| S10_extracted_113195 | Not-determined |
| S10_extracted_113208 | Not-determined |
| S10_extracted_113222 | Not-determined |
| S10_extracted_113259 | Not-determined |
| S10_extracted_113264 | Not-determined |
| S10_extracted_113266 | Not-determined |
| S10_extracted_113278 | Not-determined |
| S10_extracted_113283 | Low-quality |
| S10_extracted_113284 | Low-quality |
| S10_extracted_113305 | Not-determined |
| S10_extracted_113312 | Not-determined |
| S10_extracted_113342 | Not-determined |
| S10_extracted_113346 | Not-determined |
| S10_extracted_113352 | Not-determined |
| S10_extracted_113356 | Not-determined |
| S10_extracted_113366 | Not-determined |
| S10_extracted_113372 | Not-determined |
| S10_extracted_113373 | Not-determined |
| S10_extracted_113384 | Not-determined |
| S10_extracted_113400 | Not-determined |
| S10_extracted_113410 | Not-determined |
| S10_extracted_113437 | Not-determined |
| S10_extracted_113440 | Not-determined |
| S10_extracted_113442 | Not-determined |
| S10_extracted_113450 | Not-determined |
| S10_extracted_113465 | Not-determined |
| S1_extracted_k141_136526 | Medium-quality |
| S10_extracted_113508 | Not-determined |
| S10_extracted_113510 | Not-determined |
| S10_extracted_113528 | Not-determined |
| S10_extracted_113537 | Low-quality |
| S10_extracted_113543 | Not-determined |
| S10_extracted_113563 | Not-determined |
| S10_extracted_113569 | Not-determined |
| S10_extracted_113589 | Not-determined |
| S10_extracted_113616 | Not-determined |
| S10_extracted_113643 | Not-determined |
| S10_extracted_113645 | Not-determined |
| S10_extracted_113661 | Not-determined |
| S10_extracted_113665 | Not-determined |
| S10_extracted_113678 | Not-determined |
| S10_extracted_113719 | Low-quality |
| S10_extracted_113754 | Not-determined |
| S10_extracted_113785 | Not-determined |
| S10_extracted_113791 | Low-quality |
| S10_extracted_113794 | Not-determined |
| S10_extracted_113806 | Not-determined |
| S10_extracted_113835 | Not-determined |
| S10_extracted_113840 | Not-determined |
| S10_extracted_113850 | Not-determined |
| S10_extracted_113854 | Not-determined |
| S10_extracted_113856 | Low-quality |
| S10_extracted_113860 | Not-determined |
| S10_extracted_113865 | Not-determined |
| S10_extracted_113874 | Not-determined |
| S10_extracted_113876 | Not-determined |
| S10_extracted_113887 | Not-determined |
| S10_extracted_113898 | Not-determined |
| S10_extracted_113914 | Low-quality |
| S10_extracted_113918 | Not-determined |
| S10_extracted_113924 | Not-determined |
| S10_extracted_113935 | Low-quality |
| S10_extracted_113936 | Not-determined |
| S10_extracted_113937 | Not-determined |
| S10_extracted_113955 | Not-determined |
| S10_extracted_113987 | Not-determined |
| S10_extracted_113995 | Not-determined |
| S10_extracted_114003 | Not-determined |
| S10_extracted_114019 | Not-determined |
| S10_extracted_114038 | Not-determined |
| S10_extracted_114064 | Not-determined |
| S10_extracted_114077 | Not-determined |
| S10_extracted_114109 | Low-quality |
| S10_extracted_114113 | Not-determined |
| S10_extracted_114122 | Not-determined |
| S10_extracted_114133 | Not-determined |
| S10_extracted_114134 | Not-determined |
| S10_extracted_114151 | Not-determined |
| S10_extracted_114153 | Low-quality |
| S10_extracted_114161 | Not-determined |
| S10_extracted_114172 | Not-determined |
| S10_extracted_114186 | Not-determined |
| S10_extracted_114201 | Low-quality |
| S10_extracted_114204 | Not-determined |
| S10_extracted_114210 | Not-determined |
| S10_extracted_114219 | Low-quality |
| S10_extracted_114224 | Not-determined |
| S10_extracted_114235 | Not-determined |
| S10_extracted_114260 | Not-determined |
| S10_extracted_114262 | Not-determined |
| S10_extracted_114268 | Not-determined |
| S10_extracted_114289 | Not-determined |
| S10_extracted_114309 | Low-quality |
| S10_extracted_114325 | Not-determined |
| S10_extracted_114330 | Not-determined |
| S10_extracted_114338 | Not-determined |
| S10_extracted_114344 | Not-determined |
| S10_extracted_114347 | Not-determined |
| S10_extracted_114348 | Not-determined |
| S10_extracted_114349 | Low-quality |
| S10_extracted_114363 | Not-determined |
| S10_extracted_114375 | Low-quality |
| S10_extracted_114396 | Not-determined |
| S10_extracted_114397 | Not-determined |
| S10_extracted_114401 | Not-determined |
| S10_extracted_114402 | Not-determined |
| S10_extracted_114413 | Not-determined |
| S10_extracted_114427 | Low-quality |
| S10_extracted_114433 | Not-determined |
| S10_extracted_114434 | Not-determined |
| S10_extracted_114437 | Low-quality |
| S10_extracted_114438 | Not-determined |
| S10_extracted_114439 | Not-determined |
| S10_extracted_114443 | Not-determined |
| S10_extracted_114477 | Low-quality |
| S10_extracted_114488 | Not-determined |
| S10_extracted_114515 | Not-determined |
| S10_extracted_114517 | Not-determined |
| S10_extracted_114518 | Not-determined |
| S10_extracted_114528 | Not-determined |
| S10_extracted_114529 | Not-determined |
| S10_extracted_114532 | Not-determined |
| S10_extracted_114536 | Not-determined |
| S10_extracted_114547 | Not-determined |
| S10_extracted_114569 | Not-determined |
| S10_extracted_114582 | Low-quality |
| S10_extracted_114591 | Not-determined |
| S10_extracted_114593 | Not-determined |
| S10_extracted_114596 | Not-determined |
| S10_extracted_114600 | Not-determined |
| S10_extracted_114604 | Not-determined |
| S10_extracted_114605 | Not-determined |
| S10_extracted_114610 | Not-determined |
| S10_extracted_114622 | Not-determined |
| S10_extracted_114626 | Low-quality |
| S10_extracted_114632 | Not-determined |
| S10_extracted_114654 | Low-quality |
| S10_extracted_114656 | Not-determined |
| S10_extracted_114658 | Not-determined |
| S10_extracted_114665 | Low-quality |
| S10_extracted_114671 | Not-determined |
| S10_extracted_114675 | Not-determined |
| S10_extracted_114679 | Not-determined |
| S10_extracted_114680 | Not-determined |
| S10_extracted_114710 | Not-determined |
| S10_extracted_114728 | Not-determined |
| S10_extracted_114729 | Not-determined |
| S10_extracted_114736 | Not-determined |
| S10_extracted_114751 | Not-determined |
| S10_extracted_114760 | Not-determined |
| S10_extracted_114770 | Not-determined |
| S10_extracted_114784 | Not-determined |
| S10_extracted_114787 | Not-determined |
| S10_extracted_114790 | Not-determined |
| S10_extracted_114802 | Not-determined |
| S10_extracted_114804 | Not-determined |
| S10_extracted_114807 | Not-determined |
| S10_extracted_114815 | Not-determined |
| S10_extracted_114817 | Not-determined |
| S10_extracted_114818 | Not-determined |
| S10_extracted_114823 | Not-determined |
| S10_extracted_114832 | Not-determined |
| S10_extracted_114836 | Not-determined |
| S10_extracted_114856 | Not-determined |
| S10_extracted_114863 | Not-determined |
| S10_extracted_114867 | Low-quality |
| S10_extracted_114870 | Not-determined |
| S10_extracted_114874 | Not-determined |
| S10_extracted_114885 | Not-determined |
| S10_extracted_114886 | Not-determined |
| S10_extracted_114918 | Not-determined |
| S10_extracted_114919 | Not-determined |
| S10_extracted_114923 | Not-determined |
| S10_extracted_114924 | Not-determined |
| S10_extracted_114930 | Not-determined |
| S10_extracted_114933 | Not-determined |
| S10_extracted_114945 | Not-determined |
| S10_extracted_114993 | Not-determined |
| S10_extracted_114995 | Not-determined |
| S10_extracted_114996 | Not-determined |
| S10_extracted_115003 | Not-determined |
| S10_extracted_115019 | Not-determined |
| S10_extracted_115026 | Not-determined |
| S10_extracted_115028 | Not-determined |
| S10_extracted_115029 | Not-determined |
| S10_extracted_115035 | Not-determined |
| S10_extracted_115047 | Low-quality |
| S10_extracted_115077 | Not-determined |
| S10_extracted_115082 | Not-determined |
| S10_extracted_115091 | Not-determined |
| S10_extracted_115133 | Low-quality |
| S10_extracted_115143 | Not-determined |
| S10_extracted_115156 | Not-determined |
| S10_extracted_115174 | Not-determined |
| S10_extracted_115184 | Not-determined |
| S10_extracted_115185 | Not-determined |
| S10_extracted_115186 | Not-determined |
| S10_extracted_115190 | Not-determined |
| S10_extracted_115201 | Not-determined |
| S10_extracted_115209 | Not-determined |
| S10_extracted_115231 | Not-determined |
| S10_extracted_115257 | Not-determined |
| S10_extracted_115265 | Not-determined |
| S10_extracted_115274 | Not-determined |
| S10_extracted_115276 | Not-determined |
| S10_extracted_115282 | Not-determined |
| S10_extracted_115295 | Low-quality |
| S10_extracted_115297 | Low-quality |
| S10_extracted_115299 | Not-determined |
| S10_extracted_115304 | Not-determined |
| S10_extracted_115306 | Not-determined |
| S10_extracted_115322 | Not-determined |
| S10_extracted_115326 | Not-determined |
| S10_extracted_115340 | Not-determined |
| S10_extracted_115341 | Not-determined |
| S10_extracted_115349 | Not-determined |
| S10_extracted_115352 | Not-determined |
| S10_extracted_115355 | Not-determined |
| S10_extracted_115374 | Low-quality |
| S10_extracted_115379 | Not-determined |
| S10_extracted_115387 | Not-determined |
| S10_extracted_115390 | Not-determined |
| S10_extracted_115397 | Not-determined |
| S10_extracted_115402 | Not-determined |
| S10_extracted_115405 | Not-determined |
| S10_extracted_115407 | Not-determined |
| S10_extracted_115408 | Not-determined |
| S10_extracted_115410 | Low-quality |
| S10_extracted_115422 | Not-determined |
| S10_extracted_115441 | Not-determined |
| S10_extracted_115448 | Low-quality |
| S10_extracted_115451 | Not-determined |
| S10_extracted_115464 | Not-determined |
| S10_extracted_115467 | Not-determined |
| S10_extracted_115471 | Not-determined |
| S10_extracted_115474 | Low-quality |
| S10_extracted_115481 | Not-determined |
| S10_extracted_115488 | Not-determined |
| S10_extracted_115509 | Not-determined |
| S10_extracted_115517 | Not-determined |
| S10_extracted_115521 | Not-determined |
| S10_extracted_115535 | Not-determined |
| S10_extracted_115537 | Not-determined |
| S10_extracted_115544 | Not-determined |
| S10_extracted_115545 | Not-determined |
| S10_extracted_115563 | Not-determined |
| S10_extracted_115577 | Low-quality |
| S10_extracted_115578 | Not-determined |
| S10_extracted_115588 | Not-determined |
| S10_extracted_115590 | Not-determined |
| S10_extracted_115595 | Not-determined |
| S10_extracted_115599 | Not-determined |
| S10_extracted_115600 | Not-determined |
| S10_extracted_115606 | Low-quality |
| S10_extracted_115612 | Not-determined |
| S10_extracted_115623 | Not-determined |
| S10_extracted_115634 | Not-determined |
| S10_extracted_115645 | Not-determined |
| S10_extracted_115647 | Not-determined |
| S10_extracted_115651 | Not-determined |
| S10_extracted_115652 | Not-determined |
| S10_extracted_115654 | Not-determined |
| S10_extracted_115658 | Not-determined |
| S10_extracted_115661 | Not-determined |
| S10_extracted_115668 | Not-determined |
| S10_extracted_115674 | Not-determined |
| S10_extracted_115682 | Not-determined |
| S10_extracted_115689 | Not-determined |
| S10_extracted_115691 | Low-quality |
| S10_extracted_115717 | Not-determined |
| S10_extracted_115729 | Not-determined |
| S10_extracted_115736 | Not-determined |
| S10_extracted_115737 | Low-quality |
| S10_extracted_115738 | Not-determined |
| S10_extracted_115741 | Not-determined |
| S10_extracted_115747 | Not-determined |
| S10_extracted_115749 | Not-determined |
| S10_extracted_115751 | Not-determined |
| S10_extracted_115755 | Not-determined |
| S10_extracted_115772 | Not-determined |
| S10_extracted_115794 | Not-determined |
| S10_extracted_115830 | Not-determined |
| S10_extracted_115852 | Low-quality |
| S10_extracted_115855 | Not-determined |
| S10_extracted_115862 | Not-determined |
| S10_extracted_115911 | Low-quality |
| S10_extracted_115927 | Not-determined |
| S10_extracted_115942 | Not-determined |
| S10_extracted_115955 | Not-determined |
| S10_extracted_115963 | Low-quality |
| S10_extracted_115974 | Not-determined |
| S10_extracted_115978 | Not-determined |
| S10_extracted_115984 | Not-determined |
| S10_extracted_116013 | Not-determined |
| S10_extracted_116017 | Not-determined |
| S10_extracted_116018 | Low-quality |
| S10_extracted_116023 | Not-determined |
| S10_extracted_116031 | Low-quality |
| S10_extracted_116043 | Not-determined |
| S10_extracted_116051 | Not-determined |
| S10_extracted_116072 | Not-determined |
| S10_extracted_116079 | Not-determined |
| S10_extracted_116092 | Not-determined |
| S10_extracted_116115 | Low-quality |
| S10_extracted_116118 | Not-determined |
| S10_extracted_116234 | Low-quality |
| S10_extracted_116241 | Not-determined |
| S10_extracted_116242 | Not-determined |
| S10_extracted_116254 | Not-determined |
| S10_extracted_116263 | Not-determined |
| S10_extracted_116267 | Not-determined |
| S10_extracted_116271 | Not-determined |
| S10_extracted_116272 | Not-determined |
| S10_extracted_116301 | Not-determined |
| S10_extracted_116302 | Not-determined |
| S10_extracted_116320 | Not-determined |
| S10_extracted_116332 | Low-quality |
| S10_extracted_116345 | Not-determined |
| S10_extracted_116346 | Not-determined |
| S10_extracted_116350 | Not-determined |
| S10_extracted_116352 | Low-quality |
| S10_extracted_116354 | Not-determined |
| S10_extracted_116385 | Not-determined |
| S10_extracted_116398 | Low-quality |
| S10_extracted_116402 | Not-determined |
| S10_extracted_116405 | Not-determined |
| S10_extracted_116409 | Not-determined |
| S10_extracted_116415 | Not-determined |
| S10_extracted_116428 | Not-determined |
| S10_extracted_116431 | Not-determined |
| S10_extracted_116447 | Not-determined |
| S10_extracted_116448 | Not-determined |
| S10_extracted_116461 | Not-determined |
| S10_extracted_116462 | Not-determined |
| S10_extracted_116482 | Not-determined |
| S10_extracted_116496 | Not-determined |
| S10_extracted_116509 | Not-determined |
| S10_extracted_116516 | Not-determined |
| S10_extracted_116526 | Low-quality |
| S10_extracted_116534 | Not-determined |
| S10_extracted_116551 | Not-determined |
| S10_extracted_116553 | Not-determined |
| S10_extracted_116562 | Not-determined |
| S10_extracted_116584 | Not-determined |
| S10_extracted_116590 | Not-determined |
| S10_extracted_116603 | Not-determined |
| S10_extracted_116615 | Not-determined |
| S10_extracted_116623 | Not-determined |
| S10_extracted_116653 | Not-determined |
| S10_extracted_116664 | Not-determined |
| S10_extracted_116673 | Low-quality |
| S10_extracted_116688 | Not-determined |
| S10_extracted_116705 | Not-determined |
| S10_extracted_116706 | Not-determined |
| S10_extracted_116709 | Not-determined |
| S10_extracted_116713 | Not-determined |
| S10_extracted_116715 | Not-determined |
| S10_extracted_116716 | Not-determined |
| S10_extracted_116718 | Low-quality |
| S10_extracted_116721 | Low-quality |
| S10_extracted_116730 | Not-determined |
| S10_extracted_116734 | Not-determined |
| S10_extracted_116743 | Not-determined |
| S10_extracted_116755 | Low-quality |
| S10_extracted_116762 | Not-determined |
| S10_extracted_116771 | Not-determined |
| S10_extracted_116791 | Not-determined |
| S10_extracted_116796 | Not-determined |
| S10_extracted_116803 | Not-determined |
| S10_extracted_116809 | Low-quality |
| S10_extracted_116815 | Not-determined |
| S10_extracted_116818 | Low-quality |
| S10_extracted_116821 | Low-quality |
| S10_extracted_116835 | Low-quality |
| S10_extracted_116838 | Not-determined |
| S10_extracted_116842 | Not-determined |
| S10_extracted_116875 | Not-determined |
| S10_extracted_116877 | Not-determined |
| S10_extracted_116882 | Not-determined |
| S10_extracted_116883 | Low-quality |
| S10_extracted_116890 | Not-determined |
| S10_extracted_116891 | Not-determined |
| S10_extracted_116900 | Not-determined |
| S10_extracted_116948 | Not-determined |
| S10_extracted_116949 | Low-quality |
| S10_extracted_116953 | Not-determined |
| S10_extracted_116954 | Not-determined |
| S10_extracted_116962 | Not-determined |
| S10_extracted_116992 | Not-determined |
| S10_extracted_116996 | Not-determined |
| S10_extracted_116997 | Not-determined |
| S10_extracted_117034 | Low-quality |
| S10_extracted_117053 | Not-determined |
| S10_extracted_117068 | Not-determined |
| S10_extracted_117079 | Not-determined |
| S10_extracted_117083 | Not-determined |
| S10_extracted_117084 | Not-determined |
| S10_extracted_117086 | Not-determined |
| S10_extracted_117091 | Not-determined |
| S10_extracted_117092 | Not-determined |
| S10_extracted_117099 | Not-determined |
| S10_extracted_117126 | Not-determined |
| S10_extracted_117162 | Not-determined |
| S10_extracted_117170 | Not-determined |
| S10_extracted_117186 | Not-determined |
| S10_extracted_117188 | Not-determined |
| S10_extracted_117194 | Not-determined |
| S10_extracted_117206 | Low-quality |
| S10_extracted_117212 | Not-determined |
| S10_extracted_117214 | Not-determined |
| S10_extracted_117244 | Not-determined |
| S10_extracted_117268 | Not-determined |
| S10_extracted_117280 | Not-determined |
| S10_extracted_117282 | Not-determined |
| S10_extracted_117295 | Not-determined |
| S10_extracted_117298 | Not-determined |
| S10_extracted_117302 | Not-determined |
| S10_extracted_117303 | Not-determined |
| S10_extracted_117307 | Not-determined |
| S10_extracted_117308 | Low-quality |
| S10_extracted_117312 | Not-determined |
| S10_extracted_117361 | Not-determined |
| S10_extracted_117388 | Not-determined |
| S10_extracted_117389 | Not-determined |
| S10_extracted_117394 | Not-determined |
| S10_extracted_117399 | Not-determined |
| S10_extracted_117400 | Not-determined |
| S10_extracted_117409 | Not-determined |
| S10_extracted_117432 | Not-determined |
| S10_extracted_117454 | Low-quality |
| S10_extracted_117477 | Not-determined |
| S10_extracted_117490 | Low-quality |
| S10_extracted_117500 | Not-determined |
| S10_extracted_117503 | Not-determined |
| S10_extracted_117514 | Not-determined |
| S10_extracted_117519 | Not-determined |
| S10_extracted_117534 | Not-determined |
| S10_extracted_117542 | Not-determined |
| S10_extracted_117553 | Not-determined |
| S10_extracted_117556 | Not-determined |
| S10_extracted_117570 | Not-determined |
| S10_extracted_117571 | Low-quality |
| S10_extracted_117582 | Not-determined |
| S10_extracted_117593 | Not-determined |
| S10_extracted_117604 | Not-determined |
| S10_extracted_117625 | Not-determined |
| S10_extracted_117638 | Not-determined |
| S10_extracted_117661 | Not-determined |
| S10_extracted_117680 | Low-quality |
| S10_extracted_117694 | Not-determined |
| S10_extracted_117696 | Not-determined |
| S10_extracted_117708 | Not-determined |
| S10_extracted_117727 | Not-determined |
| S10_extracted_117734 | Not-determined |
| S10_extracted_117735 | Not-determined |
| S10_extracted_117738 | Not-determined |
| S10_extracted_117739 | Low-quality |
| S10_extracted_117788 | Not-determined |
| S10_extracted_117807 | Not-determined |
| S10_extracted_117814 | Low-quality |
| S10_extracted_117823 | Low-quality |
| S10_extracted_117825 | Not-determined |
| S10_extracted_117826 | Low-quality |
| S10_extracted_117828 | Not-determined |
| S10_extracted_117829 | Not-determined |
| S10_extracted_117831 | Not-determined |
| S10_extracted_117838 | Not-determined |
| S10_extracted_117844 | Not-determined |
| S10_extracted_117847 | Not-determined |
| S10_extracted_117854 | Low-quality |
| S10_extracted_117866 | Not-determined |
| S10_extracted_117867 | Not-determined |
| S10_extracted_117883 | Not-determined |
| S10_extracted_117893 | Not-determined |
| S10_extracted_117904 | Not-determined |
| S10_extracted_117905 | Not-determined |
| S10_extracted_117919 | Not-determined |
| S10_extracted_117930 | Not-determined |
| S10_extracted_117931 | Not-determined |
| S10_extracted_117944 | Not-determined |
| S10_extracted_117948 | Low-quality |
| S10_extracted_117954 | Not-determined |
| S10_extracted_117966 | Not-determined |
| S10_extracted_117994 | Not-determined |
| S10_extracted_118018 | Not-determined |
| S10_extracted_118025 | Not-determined |
| S10_extracted_118038 | Not-determined |
| S10_extracted_118054 | Not-determined |
| S10_extracted_118063 | Not-determined |
| S10_extracted_118064 | Not-determined |
| S10_extracted_118072 | Not-determined |
| S10_extracted_118080 | Low-quality |

S10_extracted_118082 Not-determined
S10_extracted_118092 Low-quality
S10_extracted_118097 Not-determined
S10_extracted_118145 Not-determined
S10_extracted_118156 Not-determined
S10_extracted_118167 Not-determined
S10_extracted_118186 Not-determined
S10_extracted_118191 Not-determined
S10_extracted_118195 Not-determined
S10_extracted_118203 Not-determined
S10_extracted_118215 Not-determined
S10_extracted_118218 Not-determined
S10_extracted_118245 Low-quality
S10_extracted_118253 Low-quality
S10_extracted_118260 Not-determined
S10_extracted_118262 Not-determined
S10_extracted_118279 Low-quality
S10_extracted_118280 Low-quality
S10_extracted_118283 Not-determined
S10_extracted_118317 Not-determined
S10_extracted_118338 Not-determined
S10_extracted_118342 Not-determined
S10_extracted_118352 Low-quality
S10_extracted_118359 Not-determined
S10_extracted_118363 Not-determined
S10_extracted_118366 Not-determined
S10_extracted_118393 Not-determined
S10_extracted_118395 Not-determined
S10_extracted_118417 Not-determined
S10_extracted_118419 Not-determined
S10_extracted_118425 Not-determined
S10_extracted_118428 Not-determined
S10_extracted_118432 Not-determined
S10_extracted_118455 Not-determined
S10_extracted_118465 Low-quality
S10_extracted_118481 Not-determined
S10_extracted_118496 Not-determined
S10_extracted_118505 Not-determined
S10_extracted_118518 Not-determined
S10_extracted_118523 Not-determined
S10_extracted_118528 Not-determined
S10_extracted_118542 Low-quality
S10_extracted_118555 Not-determined
S10_extracted_118558 Not-determined
S10_extracted_118588 Not-determined
S10_extracted_118606 Not-determined
S10_extracted_118612 Not-determined
S10_extracted_118613 Not-determined
S10_extracted_118624 Not-determined
S10_extracted_118629 Not-determined
S10_extracted_118630 Not-determined
S10_extracted_118632 Not-determined
S10_extracted_118635 Not-determined
S10_extracted_118639 Low-quality
S10_extracted_118648 Not-determined
S10_extracted_118658 Not-determined
S10_extracted_118663 Not-determined
S10_extracted_118674 Not-determined
S10_extracted_118685 Not-determined
S10_extracted_118690 Not-determined
S10_extracted_118726 Not-determined
S10_extracted_118744 Not-determined
S10_extracted_118746 Low-quality
S10_extracted_118755 Not-determined
S10_extracted_118756 Not-determined
S10_extracted_118772 Not-determined
S10_extracted_118779 Not-determined
S10_extracted_118791 Not-determined
S10_extracted_118801 Not-determined
S10_extracted_118807 Not-determined
S10_extracted_118812 Not-determined
S10_extracted_118836 Not-determined
S10_extracted_118846 Low-quality
S10_extracted_118847 Not-determined
S10_extracted_118857 Not-determined
S10_extracted_118860 Not-determined
S10_extracted_118872 Not-determined
S10_extracted_118878 Not-determined
S10_extracted_118892 Not-determined
S10_extracted_118899 Not-determined
S10_extracted_118917 Not-determined
S10_extracted_118936 Not-determined
S10_extracted_118947 Not-determined
S10_extracted_118980 Low-quality
S10_extracted_118993 Not-determined
S10_extracted_119008 Low-quality
S10_extracted_119012 Not-determined
S10_extracted_119019 Low-quality
S10_extracted_119059 Not-determined
S10_extracted_119078 Not-determined
S10_extracted_119098 Not-determined
S10_extracted_119118 Not-determined
S10_extracted_119119 Not-determined
S10_extracted_119122 Not-determined
S10_extracted_119139 Not-determined
S10_extracted_119140 Not-determined
S10_extracted_119146 Not-determined
S10_extracted_119155 Not-determined
S10_extracted_119164 Not-determined
S10_extracted_119181 Not-determined
S10_extracted_119193 Not-determined
S10_extracted_119202 Not-determined
S10_extracted_119210 Low-quality
S10_extracted_119214 Not-determined
S10_extracted_119222 Not-determined
S10_extracted_119243 Not-determined
S10_extracted_119254 Not-determined
S10_extracted_119266 Not-determined
S10_extracted_119293 Not-determined
S10_extracted_119312 Not-determined
S10_extracted_119316 Low-quality
S10_extracted_119319 Not-determined
S10_extracted_119348 Not-determined
S10_extracted_119349 Not-determined
S10_extracted_119353 Not-determined
S10_extracted_119357 Not-determined
S10_extracted_119372 Not-determined
S10_extracted_119382 Not-determined
S10_extracted_119394 Not-determined
S10_extracted_119399 Not-determined
S10_extracted_119407 Not-determined
S10_extracted_119408 Not-determined
S10_extracted_119413 Not-determined
S10_extracted_119430 Not-determined
S10_extracted_119435 Low-quality
S10_extracted_119441 Low-quality
S10_extracted_119459 Not-determined
S10_extracted_119466 Not-determined
S10_extracted_119490 Not-determined
S10_extracted_119507 Not-determined
S10_extracted_119518 Not-determined
S10_extracted_119528 Not-determined
S10_extracted_119529 Not-determined
S10_extracted_119552 Not-determined
S10_extracted_119559 Low-quality
S10_extracted_119585 Not-determined
S10_extracted_119587 Not-determined
S10_extracted_119591 Not-determined
S10_extracted_119606 Low-quality
S10_extracted_119607 Not-determined
S10_extracted_119611 Not-determined
S10_extracted_119612 Not-determined
S10_extracted_119617 Not-determined
S10_extracted_119619 Low-quality
S10_extracted_119640 Low-quality
S10_extracted_119664 Not-determined
S10_extracted_119674 Not-determined
S10_extracted_119677 Not-determined
S10_extracted_119698 Not-determined
S10_extracted_119706 Not-determined
S10_extracted_119713 Not-determined
S10_extracted_119722 Not-determined
S10_extracted_119727 Low-quality
S10_extracted_119760 Low-quality
S10_extracted_119773 Not-determined
S10_extracted_119792 Low-quality
S10_extracted_119795 Low-quality
S10_extracted_119808 Not-determined
S10_extracted_119842 Not-determined
S10_extracted_119857 Not-determined
S10_extracted_119873 Not-determined
S10_extracted_119875 Not-determined
S10_extracted_119908 Not-determined
S10_extracted_119919 Not-determined
S10_extracted_119924 Not-determined
S10_extracted_119943 Not-determined
S10_extracted_119950 Not-determined
S10_extracted_119957 Not-determined
S10_extracted_119964 Not-determined
S10_extracted_119972 Not-determined
S10_extracted_119981 Low-quality
S10_extracted_120002 Not-determined
S10_extracted_120006 Not-determined
S10_extracted_120007 Not-determined
S10_extracted_120016 Not-determined
S10_extracted_120041 Not-determined
S10_extracted_120052 Not-determined
S10_extracted_120055 Not-determined
S10_extracted_120070 Not-determined
S10_extracted_120074 Not-determined
S10_extracted_120076 Not-determined
S10_extracted_120105 Not-determined
S10_extracted_120106 Not-determined
S10_extracted_120108 Not-determined
S10_extracted_120109 Not-determined
S10_extracted_120113 Not-determined
S10_extracted_120122 Not-determined
S10_extracted_120126 Low-quality
S10_extracted_120135 Not-determined
S10_extracted_120170 Not-determined
S10_extracted_120186 Not-determined
S10_extracted_120192 Not-determined
S10_extracted_120197 Not-determined
S10_extracted_120207 Low-quality
S10_extracted_120217 Not-determined
S10_extracted_120239 Not-determined
S10_extracted_120254 Not-determined
S10_extracted_120259 Not-determined
S10_extracted_120269 Low-quality
S10_extracted_120273 Not-determined
S10_extracted_120276 Not-determined
S10_extracted_120317 Not-determined
S10_extracted_120339 Not-determined
S10_extracted_120354 Not-determined
S10_extracted_120362 Not-determined
S10_extracted_120371 Not-determined
S10_extracted_120379 Not-determined
S10_extracted_120388 Not-determined
S10_extracted_120394 Not-determined
S10_extracted_120398 Not-determined
S10_extracted_120404 Not-determined
S10_extracted_120408 Not-determined
S10_extracted_120419 Not-determined
S10_extracted_120424 Not-determined
S10_extracted_120427 Not-determined
S10_extracted_120429 Not-determined
S10_extracted_120454 Not-determined
S10_extracted_120457 Not-determined
S10_extracted_120460 Not-determined
S10_extracted_120464 Not-determined
S10_extracted_120495 Not-determined
S10_extracted_120522 Not-determined
S10_extracted_120546 Not-determined
S10_extracted_120551 Not-determined
S10_extracted_120558 Not-determined
S10_extracted_120566 Not-determined
S10_extracted_120594 Not-determined
S10_extracted_120611 Low-quality
S10_extracted_120613 Not-determined
S10_extracted_120620 Not-determined
S10_extracted_120679 Not-determined
S10_extracted_120683 Not-determined
S10_extracted_120691 Not-determined
S10_extracted_120692 Low-quality
S10_extracted_120695 Not-determined
S10_extracted_120702 Not-determined
S10_extracted_120703 Not-determined
S10_extracted_120714 Not-determined
S10_extracted_120727 Not-determined
S1_vs_k141_136526||full Medium-quality
S10_extracted_120740 Not-determined
S10_extracted_120753 Not-determined
S10_extracted_120762 Not-determined
S10_extracted_120763 Not-determined
S10_extracted_120794 Not-determined
S10_extracted_120800 Not-determined
S10_extracted_120808 Not-determined
S10_extracted_120812 Not-determined
S10_extracted_120818 Not-determined
S10_extracted_120823 Not-determined
S10_extracted_120824 Not-determined
S10_extracted_120832 Not-determined
S10_extracted_120833 Not-determined
S10_extracted_120835 Not-determined
S10_extracted_120840 Low-quality
S10_extracted_120848 Not-determined
S10_extracted_120857 Not-determined
S10_extracted_120858 Not-determined
S10_extracted_120871 Not-determined
S10_extracted_120875 Not-determined
S10_extracted_120885 Not-determined
S10_extracted_120886 Not-determined
S10_extracted_120889 Not-determined
S10_extracted_120892 Not-determined
S10_extracted_120899 Not-determined
S10_extracted_120903 Not-determined
S10_extracted_120910 Not-determined
S10_extracted_120913 Not-determined
S10_extracted_120918 Not-determined
S10_extracted_120923 Not-determined
S10_extracted_120934 Low-quality
S10_extracted_120944 Not-determined
S10_extracted_120958 Not-determined
S10_extracted_120963 Not-determined
S10_extracted_120971 Not-determined
S10_extracted_120981 Not-determined
S10_extracted_121001 Not-determined
S10_extracted_121009 Low-quality
S10_extracted_121015 Not-determined
S10_extracted_121047 Not-determined
S10_extracted_121050 Not-determined
S10_extracted_121054 Not-determined
S10_extracted_121059 Not-determined
S10_extracted_121064 Not-determined
S10_extracted_121082 Not-determined
S10_extracted_121095 Not-determined
S10_extracted_121103 Not-determined
S10_extracted_121117 Not-determined
S10_extracted_121120 Not-determined
S10_extracted_121123 Not-determined
S10_extracted_121131 Not-determined
S10_extracted_121142 Not-determined
S10_extracted_121167 Low-quality
S10_extracted_121202 Not-determined
S10_extracted_121210 Not-determined
S10_extracted_121223 Low-quality
S10_extracted_121227 Not-determined
S10_extracted_121228 Low-quality
S10_extracted_121244 Not-determined
S10_extracted_121258 Low-quality
S10_extracted_121259 Not-determined
S10_extracted_121260 Not-determined
S10_vs_1702||full Not-determined
S10_vs_8605||full Not-determined
S11_vs_k141_5077||full High-quality
S16_vs_132979||full Medium-quality
S10_vs_23835||full Not-determined
S14_vs_k141_56251||full Medium-quality
S16_vs_18293||full Medium-quality
S16_extracted_100378 Medium-quality
S16_vs_100378||full Medium-quality
S10_vs_58432||full Low-quality
S16_vs_35031||full Medium-quality
S7_vs_13370||full Medium-quality
S10_vs_92936||full Not-determined
S10_vs_106834||full Medium-quality
S14_vs_k141_96286||full High-quality
S3_extracted_k141_70746 Medium-quality
S5_vs_k141_6032||full High-quality
S10_vs_120745||full Low-quality
S10_vs_120762||full Not-determined
S10_vs_120773||full Low-quality
S10_vs_120815||full Not-determined
S10_vs_120862||full Low-quality
S10_vs_51||full Low-quality
S10_vs_55||full Not-determined
S10_vs_72||full Not-determined
S10_vs_102||full Not-determined
S10_vs_145||full Not-determined
S10_vs_171||full Not-determined
S10_vs_179||full Not-determined
S10_vs_193||full Low-quality
S10_vs_206||full Not-determined
S10_vs_231||full Not-determined
S10_vs_313||full Low-quality
S10_vs_349||full Not-determined
S10_vs_358||full Not-determined
S10_vs_379||full Not-determined
S10_vs_408||full Not-determined
S10_vs_473||full Not-determined
S10_vs_499||full Not-determined
S10_vs_523||full Low-quality
S10_vs_635||full Low-quality
S10_vs_802||full Low-quality
S10_vs_947||full Low-quality
S10_vs_967||full Not-determined
S10_vs_1012||full Low-quality
S10_vs_1014||full Low-quality
S10_vs_1019||full Low-quality
S10_vs_1089||full Not-determined
S10_vs_1126||full Not-determined
S10_vs_1160||full Low-quality
S10_vs_1259||full Low-quality
S10_vs_1265||full Low-quality
S10_vs_1280||full Low-quality
S10_vs_1299||full Low-quality
S10_vs_1307||full Low-quality
S10_vs_1324||full Low-quality
S10_vs_1341||full Low-quality
S10_vs_1359||full Low-quality
S10_vs_1432||full Low-quality
S10_vs_1441||full Not-determined
S10_vs_1468||full Not-determined
S10_vs_1476||full Not-determined
S10_vs_1493||full Not-determined
S10_vs_1496||full Not-determined
S10_vs_1567||full Not-determined
S10_vs_1583||full Not-determined
S10_vs_1701||full Not-determined
S10_vs_1754||full Low-quality
S10_vs_1759||full Low-quality
S10_vs_1884||full Low-quality
S10_vs_1890||full Low-quality
S10_vs_1915||full Low-quality
S10_vs_1963||full Not-determined
S10_vs_1976||full Low-quality
S10_vs_1980||full Low-quality
S10_vs_2042||full Low-quality
S10_vs_2071||full Not-determined
S10_vs_2091||full Low-quality
S10_vs_2095||full Low-quality
S10_vs_2115||full Not-determined
S10_vs_2163||full Low-quality
S10_vs_2230||full Low-quality
S10_vs_2314||full Low-quality
S10_vs_2318||full Low-quality
S10_vs_2359||full Low-quality
S10_vs_2434||full Not-determined
S10_vs_2435||full Low-quality
S10_vs_2499||full Not-determined
S12_vs_k141_22853||full Medium-quality
S10_vs_2595||full Low-quality
S10_vs_2639||full Not-determined
S10_vs_2656||full Low-quality
S10_vs_2672||full Low-quality
S10_vs_2684||full Low-quality
S10_vs_2726||full Not-determined
S10_vs_2780||full Low-quality
S10_vs_2781||full Low-quality
S10_vs_2794||full Low-quality
S5_vs_k141_49480||full High-quality
S10_vs_2904||full Low-quality
S10_vs_2961||full Not-determined
S10_vs_2970||full Low-quality
S10_vs_3009||full Low-quality
S10_vs_3011||full Not-determined
S10_vs_3128||full Low-quality
S10_vs_3137||full Low-quality
S10_vs_3169||full Not-determined
S10_vs_3223||full Low-quality
S10_vs_3247||full Low-quality
S10_vs_3304||full Low-quality
S10_vs_3328||full Low-quality
S10_vs_3398||full Not-determined
S10_vs_3447||full Not-determined
S10_vs_3456||full Not-determined
S10_vs_3473||full Not-determined
S10_vs_3481||full Low-quality
S10_vs_3493||full Low-quality
S10_vs_3596||full Low-quality
S10_vs_3629||full Not-determined
S7_vs_26235||full Medium-quality
S10_vs_3666||full Not-determined
S10_vs_3691||full Not-determined
S10_vs_3702||full Not-determined
S10_vs_3709||full Low-quality
S10_vs_3730||full Low-quality
S10_vs_3751||full Not-determined
S10_vs_3778||full Low-quality
S10_vs_3836||full Low-quality
S10_vs_3849||full Low-quality
S10_vs_3891||full Not-determined
S10_vs_3934||full Low-quality
S10_vs_3974||full Low-quality
S10_vs_4027||full Low-quality
S10_vs_4114||full Not-determined
S10_vs_4123||full Not-determined
S10_vs_4214||full Low-quality
S10_vs_4229||full Not-determined
S10_vs_4343||full Low-quality
S10_vs_4364||full Low-quality
S10_vs_4387||full Low-quality
S10_vs_4417||full Not-determined
S10_vs_4456||full Low-quality
S10_vs_4462||full Low-quality
S10_vs_4470||full Low-quality
S10_vs_4519||full Low-quality
S10_vs_4550||full Low-quality
S10_vs_4592||full Low-quality
S10_vs_4624||full Not-determined
S10_vs_4627||full Not-determined
S10_vs_4640||full Low-quality
S10_vs_4673||full Low-quality
S10_vs_4693||full Low-quality
S10_vs_4710||full Low-quality
S10_vs_4714||full Low-quality
S10_vs_4750||full Low-quality
S10_vs_4790||full Not-determined
S10_vs_4812||full Not-determined
S10_vs_4841||full Low-quality
S10_vs_4870||full Low-quality
S10_vs_4874||full Not-determined
S10_vs_4878||full Low-quality
S10_vs_4933||full Not-determined
S10_vs_5005||full Not-determined
S10_vs_5124||full Not-determined
S10_vs_5163||full Low-quality
S10_vs_5173||full Not-determined
S10_vs_5177||full Not-determined
S10_vs_5214||full Low-quality
S10_vs_5252||full Low-quality
S10_vs_5254||full Low-quality
S10_vs_5288||full Low-quality
S10_vs_5384||full Low-quality
S10_vs_5400||full Low-quality
S10_vs_5422||full Low-quality
S10_vs_5540||full Not-determined
S10_vs_5550||full Low-quality
S10_vs_5588||full Low-quality
S10_vs_5592||full Low-quality
S10_vs_5608||full Not-determined
S10_vs_5632||full Not-determined
S10_vs_5707||full Not-determined
S10_vs_5775||full Low-quality
S10_vs_5825||full Not-determined
S10_vs_5877||full Not-determined
S10_vs_5924||full Not-determined
S10_vs_5938||full Not-determined
S10_vs_5982||full Not-determined
S10_vs_6009||full Not-determined
S10_vs_6013||full Low-quality
S10_vs_6015||full Not-determined
S10_vs_6251||full Low-quality
S10_vs_6273||full Low-quality
S10_vs_6344||full Low-quality
S10_vs_6345||full Low-quality
S10_vs_6393||full Low-quality
S10_vs_6408||full Low-quality
S10_vs_6499||full Low-quality
S10_vs_6557||full Not-determined
S10_vs_6587||full Low-quality
S10_vs_6600||full Low-quality
S10_vs_6613||full Not-determined
S10_vs_6660||full Not-determined
S10_vs_6705||full Low-quality
S10_vs_6745||full Not-determined
S10_vs_6774||full Low-quality
S10_vs_6796||full Low-quality
S10_vs_6838||full Low-quality
S10_vs_6848||full Not-determined
S10_vs_6867||full Not-determined
S10_vs_6949||full Low-quality
S10_vs_6980||full Not-determined
S10_vs_7008||full Low-quality
S10_vs_7020||full Not-determined
S10_vs_7023||full Low-quality
S10_vs_7049||full Low-quality
S10_vs_7050||full Not-determined
S10_vs_7083||full Low-quality
S10_vs_7173||full Not-determined
S10_vs_7218||full Not-determined
S10_vs_7250||full Low-quality
S10_vs_7302||full Not-determined
S10_vs_7321||full Low-quality
S10_vs_7326||full Not-determined
S10_vs_7367||full Not-determined
S10_vs_7393||full Not-determined
S10_vs_7442||full Not-determined
S10_vs_7499||full Low-quality
S10_vs_7527||full Low-quality
S10_vs_7558||full Low-quality
S10_vs_7579||full Not-determined
S10_vs_7587||full Low-quality
S10_vs_7647||full Low-quality
S10_vs_7651||full Low-quality
S10_vs_7665||full Not-determined
S10_vs_7669||full Low-quality
S10_vs_7686||full Low-quality
S10_vs_7700||full Low-quality
S10_vs_7709||full Low-quality
S10_vs_7850||full Not-determined
S10_vs_7854||full Not-determined
S10_vs_7874||full Not-determined
S10_vs_7901||full Not-determined
S10_vs_7905||full Not-determined
S10_vs_7945||full Low-quality
S10_vs_7968||full Low-quality
S10_vs_7992||full Low-quality
S10_vs_8046||full Not-determined
S10_vs_8076||full Low-quality
S10_vs_8087||full Low-quality
S10_vs_8106||full Not-determined
S10_vs_8127||full Not-determined
S10_vs_8136||full Low-quality
S10_vs_8195||full Low-quality
S10_vs_8199||full Not-determined
S10_vs_8236||full Low-quality
S10_vs_8251||full Low-quality
S10_vs_8258||full Low-quality
S10_vs_8270||full Not-determined
S10_vs_8317||full Not-determined
S10_vs_8512||full Low-quality
S10_vs_8532||full Low-quality
S10_vs_8554||full Low-quality
S10_vs_8592||full Not-determined
S10_vs_8611||full Low-quality
S10_vs_8616||full Not-determined
S10_vs_8722||full Not-determined
S10_vs_8750||full Low-quality
S10_vs_8751||full Low-quality
S10_vs_8778||full Low-quality
S10_vs_8833||full Not-determined
S10_vs_8836||full Low-quality
S10_vs_8837||full Low-quality
S1_vs_k141_155183||full Medium-quality
S10_vs_8855||full Not-determined
S10_vs_8936||full Low-quality
S10_vs_8952||full Low-quality
S10_vs_9087||full Low-quality
S10_vs_9159||full Not-determined
S10_vs_9313||full Not-determined
S10_vs_9330||full Low-quality
S10_vs_9337||full Not-determined
S10_vs_9376||full Low-quality
S10_vs_9420||full Low-quality
S10_vs_9432||full Not-determined
S10_vs_9451||full Low-quality
S10_vs_9467||full Not-determined
S10_vs_9484||full Low-quality

| | |
|---|---|
| S10_vs_9508\|\|full | Low-quality |
| S10_vs_9720\|\|full | Not-determined |
| S10_vs_9723\|\|full | Not-determined |
| S10_vs_9728\|\|full | Not-determined |
| S10_vs_9780\|\|full | Not-determined |
| S10_vs_9829\|\|full | Low-quality |
| S10_vs_9841\|\|full | Not-determined |
| S10_vs_9875\|\|full | Not-determined |
| S10_vs_10042\|\|full | Not-determined |
| S10_vs_10044\|\|full | Not-determined |
| S10_vs_10062\|\|full | Low-quality |
| S10_vs_10068\|\|full | Low-quality |
| S10_vs_10188\|\|full | Low-quality |
| S10_vs_10268\|\|full | Low-quality |
| S10_vs_10326\|\|full | Low-quality |
| S10_vs_10366\|\|full | Not-determined |
| S10_vs_10373\|\|full | Not-determined |
| S10_vs_10488\|\|full | Low-quality |
| S10_vs_10528\|\|full | Low-quality |
| S10_vs_10721\|\|full | Not-determined |
| S10_vs_10776\|\|full | Low-quality |
| S10_vs_10778\|\|full | Not-determined |
| S10_vs_10806\|\|full | Not-determined |
| S10_vs_10832\|\|full | Low-quality |
| S10_vs_10839\|\|full | Low-quality |
| S10_vs_10854\|\|full | Low-quality |
| S10_vs_10869\|\|full | Low-quality |
| S10_vs_10883\|\|full | Not-determined |
| S10_vs_10887\|\|full | Not-determined |
| S10_vs_10901\|\|full | Low-quality |
| S10_vs_10935\|\|full | Low-quality |
| S10_vs_11005\|\|full | Not-determined |
| S10_vs_11028\|\|full | Not-determined |
| S10_vs_11101\|\|full | Low-quality |
| S10_vs_11143\|\|full | Not-determined |
| S10_vs_11166\|\|full | Low-quality |
| S10_vs_11223\|\|full | Not-determined |
| S10_vs_11289\|\|full | Not-determined |
| S10_vs_11304\|\|full | Low-quality |
| S10_vs_11310\|\|full | Not-determined |
| S10_vs_11356\|\|full | Low-quality |
| S10_vs_11420\|\|full | Low-quality |
| S10_vs_11457\|\|full | Low-quality |
| S10_vs_11488\|\|full | Low-quality |
| S10_vs_11637\|\|full | Low-quality |
| S10_vs_11730\|\|full | Low-quality |
| S10_vs_11809\|\|full | Not-determined |
| S10_vs_11837\|\|full | Low-quality |
| S10_vs_12075\|\|full | Low-quality |
| S10_vs_12229\|\|full | Low-quality |
| S10_vs_12255\|\|full | Not-determined |
| S10_vs_12299\|\|full | Not-determined |
| S10_vs_12363\|\|full | Low-quality |
| S10_vs_12460\|\|full | Not-determined |
| S10_vs_12495\|\|full | Not-determined |
| S10_vs_12515\|\|full | Not-determined |
| S10_vs_12552\|\|full | Low-quality |
| S10_vs_12559\|\|full | Not-determined |
| S10_vs_12619\|\|full | Low-quality |
| S10_vs_12626\|\|full | Low-quality |
| S10_vs_12645\|\|full | Low-quality |
| S10_vs_12776\|\|full | Low-quality |
| S10_vs_12795\|\|full | Not-determined |
| S10_vs_12804\|\|full | Low-quality |
| S10_vs_12810\|\|full | Low-quality |
| S10_vs_12830\|\|full | Not-determined |
| S10_vs_12866\|\|full | Low-quality |
| S10_vs_12868\|\|full | Not-determined |
| S10_vs_12882\|\|full | Low-quality |
| S10_vs_12887\|\|full | Low-quality |
| S10_vs_12980\|\|full | Not-determined |
| S10_vs_12988\|\|full | Low-quality |
| S10_vs_13014\|\|full | Low-quality |
| S10_vs_13049\|\|full | Low-quality |
| S10_vs_13065\|\|full | Low-quality |
| S10_vs_13080\|\|full | Not-determined |
| S10_vs_13081\|\|full | Low-quality |
| S10_vs_13090\|\|full | Not-determined |
| S10_vs_13112\|\|full | Low-quality |
| S10_vs_13119\|\|full | Not-determined |
| S10_vs_13131\|\|full | Not-determined |
| S10_vs_13140\|\|full | Not-determined |
| S10_vs_13145\|\|full | Low-quality |
| S10_vs_13213\|\|full | Low-quality |
| S10_vs_13251\|\|full | Not-determined |
| S10_vs_13315\|\|full | Low-quality |
| S10_vs_13436\|\|full | Low-quality |
| S10_vs_13439\|\|full | Low-quality |
| S10_vs_13486\|\|full | Not-determined |
| S10_vs_13511\|\|full | Not-determined |
| S10_vs_13533\|\|full | Low-quality |
| S10_vs_13536\|\|full | Low-quality |
| S10_vs_13567\|\|full | Not-determined |
| S10_vs_13587\|\|full | Low-quality |
| S10_vs_13617\|\|full | Low-quality |
| S10_vs_13639\|\|full | Low-quality |
| S10_vs_13705\|\|full | Not-determined |
| S10_vs_13744\|\|full | Not-determined |
| S10_vs_13792\|\|full | Low-quality |
| S10_vs_13807\|\|full | Low-quality |
| S10_vs_13828\|\|full | Not-determined |
| S10_vs_13868\|\|full | Not-determined |
| S10_vs_13933\|\|full | Low-quality |
| S10_vs_13983\|\|full | Not-determined |
| S10_vs_13996\|\|full | Low-quality |
| S10_vs_14001\|\|full | Not-determined |
| S10_vs_14042\|\|full | Not-determined |
| S10_vs_14075\|\|full | Low-quality |
| S10_vs_14332\|\|full | Not-determined |
| S10_vs_14381\|\|full | Low-quality |
| S10_vs_14460\|\|full | Low-quality |
| S10_vs_14471\|\|full | Low-quality |
| S10_vs_14475\|\|full | Not-determined |
| S10_vs_14486\|\|full | Not-determined |
| S3_vs_k141_70746\|\|full | Medium-quality |
| S10_vs_14595\|\|full | Not-determined |
| S10_vs_14667\|\|full | Low-quality |
| S10_vs_14700\|\|full | Low-quality |
| S10_vs_14739\|\|full | Low-quality |
| S10_vs_14777\|\|full | Low-quality |
| S10_vs_14796\|\|full | Low-quality |
| S10_vs_14817\|\|full | Low-quality |
| S10_vs_14830\|\|full | Low-quality |
| S10_vs_14839\|\|full | Low-quality |
| S10_vs_14888\|\|full | Low-quality |
| S10_vs_14903\|\|full | Not-determined |
| S10_vs_14940\|\|full | Low-quality |
| S10_vs_14949\|\|full | Not-determined |
| S10_vs_14988\|\|full | Not-determined |
| S10_vs_14991\|\|full | Not-determined |
| S10_vs_15002\|\|full | Not-determined |
| S10_vs_15103\|\|full | Low-quality |
| S10_vs_15161\|\|full | Low-quality |
| S10_vs_15207\|\|full | Low-quality |
| S10_vs_15216\|\|full | Low-quality |
| S10_vs_15248\|\|full | Not-determined |
| S10_vs_15287\|\|full | Low-quality |
| S10_vs_15314\|\|full | Low-quality |
| S10_vs_15349\|\|full | Not-determined |
| S10_vs_15359\|\|full | Low-quality |
| S10_vs_15365\|\|full | Not-determined |
| S10_vs_15433\|\|full | Not-determined |
| S10_vs_15437\|\|full | Low-quality |
| S10_vs_15497\|\|full | Not-determined |
| S10_vs_15520\|\|full | Low-quality |
| S10_vs_15542\|\|full | Not-determined |
| S6_vs_k141_46091\|\|full | High-quality |
| S10_vs_15567\|\|full | Low-quality |
| S10_vs_15568\|\|full | Low-quality |
| S10_vs_15612\|\|full | Low-quality |
| S10_vs_15639\|\|full | Not-determined |
| S10_vs_15641\|\|full | Not-determined |
| S10_vs_15642\|\|full | Not-determined |
| S10_vs_15696\|\|full | Low-quality |
| S10_vs_15728\|\|full | Low-quality |
| S10_vs_15763\|\|full | Low-quality |
| S10_vs_15764\|\|full | Not-determined |
| S10_vs_15805\|\|full | Low-quality |
| S10_vs_15827\|\|full | Not-determined |
| S10_vs_15854\|\|full | Low-quality |
| S10_vs_15894\|\|full | Low-quality |
| S10_vs_15945\|\|full | Low-quality |
| S10_vs_15960\|\|full | Not-determined |
| S10_vs_16005\|\|full | Low-quality |
| S10_vs_16023\|\|full | Not-determined |
| S10_vs_16038\|\|full | Low-quality |
| S10_vs_16154\|\|full | Low-quality |
| S10_vs_16177\|\|full | Not-determined |
| S10_vs_16182\|\|full | Not-determined |
| S10_vs_16185\|\|full | Not-determined |
| S10_vs_16225\|\|full | Not-determined |
| S10_vs_16253\|\|full | Low-quality |
| S10_vs_16263\|\|full | Not-determined |
| S10_vs_16269\|\|full | Low-quality |
| S10_vs_16333\|\|full | Not-determined |
| S10_vs_16355\|\|full | Not-determined |
| S10_vs_16358\|\|full | Low-quality |
| S10_vs_16368\|\|full | Not-determined |
| S10_vs_16372\|\|full | Not-determined |
| S10_vs_16380\|\|full | Not-determined |
| S10_vs_16429\|\|full | Low-quality |
| S10_vs_16519\|\|full | Not-determined |
| S10_vs_16567\|\|full | Low-quality |
| S10_vs_16572\|\|full | Not-determined |
| S10_vs_16592\|\|full | Low-quality |
| S10_vs_16632\|\|full | Not-determined |
| S10_vs_16658\|\|full | Not-determined |
| S10_vs_16833\|\|full | Low-quality |
| S10_vs_16938\|\|full | Low-quality |
| S10_vs_16947\|\|full | Low-quality |
| S10_vs_16972\|\|full | Not-determined |
| S10_vs_17005\|\|full | Low-quality |
| S10_vs_17019\|\|full | Not-determined |
| S10_vs_17068\|\|full | Not-determined |
| S10_vs_17091\|\|full | Not-determined |
| S10_vs_17188\|\|full | Low-quality |
| S10_vs_17200\|\|full | Not-determined |
| S10_vs_17272\|\|full | Low-quality |
| S10_vs_17287\|\|full | Not-determined |
| S10_vs_17292\|\|full | Low-quality |
| S10_vs_17333\|\|full | Low-quality |
| S10_vs_17363\|\|full | Not-determined |
| S10_vs_17395\|\|full | Not-determined |
| S10_vs_17402\|\|full | Not-determined |
| S10_vs_17465\|\|full | Low-quality |
| S10_vs_17497\|\|full | Not-determined |
| S10_vs_17525\|\|full | Not-determined |
| S10_vs_17561\|\|full | Low-quality |
| S10_vs_17569\|\|full | Low-quality |
| S10_vs_17571\|\|full | Low-quality |
| S10_vs_17661\|\|full | Low-quality |
| S10_vs_17702\|\|full | Low-quality |
| S10_vs_17731\|\|full | Not-determined |
| S10_vs_17746\|\|full | Low-quality |
| S10_vs_17750\|\|full | Not-determined |
| S10_vs_17773\|\|full | Not-determined |
| S10_vs_17775\|\|full | Low-quality |
| S10_vs_17882\|\|full | Low-quality |
| S16_vs_17493\|\|full | Medium-quality |
| S10_vs_17940\|\|full | Low-quality |
| S10_vs_17944\|\|full | Not-determined |
| S10_vs_18007\|\|full | Low-quality |
| S10_vs_18009\|\|full | Low-quality |
| S10_vs_18019\|\|full | Not-determined |
| S10_vs_18131\|\|full | Not-determined |
| S10_vs_18157\|\|full | Not-determined |
| S10_vs_18179\|\|full | Low-quality |
| S10_vs_18238\|\|full | Not-determined |
| S10_vs_18297\|\|full | Low-quality |
| S10_vs_18366\|\|full | Low-quality |
| S10_vs_18385\|\|full | Low-quality |
| S10_vs_18423\|\|full | Low-quality |
| S10_vs_18443\|\|full | Not-determined |
| S10_vs_18488\|\|full | Not-determined |
| S10_vs_18495\|\|full | Not-determined |
| S10_vs_18537\|\|full | Not-determined |
| S10_vs_18553\|\|full | Low-quality |
| S10_vs_18570\|\|full | Low-quality |
| S10_vs_18579\|\|full | Not-determined |
| S10_vs_18623\|\|full | Not-determined |
| S10_vs_18727\|\|full | Low-quality |
| S10_vs_18745\|\|full | Low-quality |
| S10_vs_18779\|\|full | Low-quality |
| S10_vs_18810\|\|full | Not-determined |
| S10_vs_18820\|\|full | Low-quality |
| S10_vs_18845\|\|full | Low-quality |
| S10_vs_18864\|\|full | Not-determined |
| S5_vs_k141_93839\|\|full | Medium-quality |
| S10_vs_18931\|\|full | Not-determined |
| S10_vs_18988\|\|full | Not-determined |
| S10_vs_19036\|\|full | Not-determined |
| S10_vs_19049\|\|full | Not-determined |
| S10_vs_19052\|\|full | Low-quality |
| S10_vs_19074\|\|full | Low-quality |
| S10_vs_19079\|\|full | Not-determined |
| S10_vs_19131\|\|full | Not-determined |
| S10_vs_19146\|\|full | Low-quality |
| S10_vs_19259\|\|full | Low-quality |
| S10_vs_19260\|\|full | Low-quality |
| S10_vs_19284\|\|full | Not-determined |
| S10_vs_19332\|\|full | Not-determined |
| S10_vs_19337\|\|full | Low-quality |
| S10_vs_19339\|\|full | Not-determined |
| S10_vs_19435\|\|full | Not-determined |
| S10_vs_19448\|\|full | Not-determined |
| S10_vs_19451\|\|full | Low-quality |
| S10_vs_19466\|\|full | Not-determined |
| S10_vs_19554\|\|full | Low-quality |
| S10_vs_19597\|\|full | Not-determined |
| S10_vs_19691\|\|full | Not-determined |
| S10_vs_19698\|\|full | Not-determined |
| S10_vs_19713\|\|full | Not-determined |
| S10_vs_19734\|\|full | Not-determined |
| S10_vs_19787\|\|full | Not-determined |
| S10_vs_19788\|\|full | Low-quality |
| S10_vs_19810\|\|full | Not-determined |
| S10_vs_19819\|\|full | Low-quality |
| S10_vs_19821\|\|full | Low-quality |
| S10_vs_19843\|\|full | Low-quality |
| S10_vs_19878\|\|full | Not-determined |
| S10_vs_19900\|\|full | Low-quality |
| S10_vs_19955\|\|full | Low-quality |
| S10_vs_19976\|\|full | Not-determined |
| S10_vs_20000\|\|full | Not-determined |
| S10_vs_20057\|\|full | Not-determined |
| S10_vs_20087\|\|full | Low-quality |
| S10_vs_20110\|\|full | Low-quality |
| S10_vs_20140\|\|full | Not-determined |
| S10_vs_20158\|\|full | Not-determined |
| S10_vs_20219\|\|full | Low-quality |
| S10_vs_20268\|\|full | Low-quality |
| S10_vs_20338\|\|full | Not-determined |
| S10_vs_20372\|\|full | Low-quality |
| S4_vs_VIRSorter_31171-circular-cat | Medium-quality |
| S10_vs_20402\|\|full | Low-quality |
| S10_vs_20419\|\|full | Low-quality |
| S10_vs_20452\|\|full | Not-determined |
| S10_vs_20464\|\|full | Not-determined |
| S10_vs_20496\|\|full | Not-determined |
| S10_vs_20525\|\|full | Not-determined |
| S10_vs_20536\|\|full | Low-quality |
| S10_vs_20544\|\|full | Low-quality |
| S10_vs_20665\|\|full | Low-quality |
| S10_vs_20668\|\|full | Not-determined |
| S10_vs_20747\|\|full | Not-determined |
| S10_vs_20768\|\|full | Not-determined |
| S10_vs_20789\|\|full | Low-quality |
| S10_vs_20802\|\|full | Not-determined |
| S10_vs_20902\|\|full | Low-quality |
| S10_vs_20915\|\|full | Not-determined |
| S10_vs_20930\|\|full | Not-determined |
| S10_vs_20946\|\|full | Not-determined |
| S10_vs_20993\|\|full | Not-determined |
| S10_vs_21014\|\|full | Not-determined |
| S10_vs_21040\|\|full | Low-quality |
| S10_vs_21046\|\|full | Low-quality |
| S10_vs_21058\|\|full | Low-quality |
| S10_vs_21154\|\|full | Low-quality |
| S10_vs_21189\|\|full | Not-determined |
| S10_vs_21207\|\|full | Low-quality |
| S10_vs_21267\|\|full | Low-quality |
| S10_vs_21324\|\|full | Low-quality |
| S10_vs_21329\|\|full | Low-quality |
| S10_vs_21370\|\|full | Not-determined |
| S10_vs_21389\|\|full | Low-quality |
| S10_vs_21396\|\|full | Low-quality |
| S10_vs_21459\|\|full | Low-quality |
| S10_vs_21613\|\|full | Not-determined |
| S10_vs_21643\|\|full | Not-determined |
| S10_vs_21668\|\|full | Low-quality |
| S10_vs_21671\|\|full | Not-determined |
| S10_vs_21718\|\|full | Not-determined |
| S10_vs_21761\|\|full | Low-quality |
| S10_vs_21858\|\|full | Not-determined |
| S10_vs_21946\|\|full | Not-determined |
| S10_vs_22040\|\|full | Low-quality |
| S10_vs_22072\|\|full | Low-quality |
| S10_vs_22116\|\|full | Low-quality |
| S10_vs_22138\|\|full | Low-quality |
| S10_vs_22189\|\|full | Low-quality |
| S10_vs_22223\|\|full | Low-quality |
| S10_vs_22238\|\|full | Not-determined |
| S10_vs_22321\|\|full | Low-quality |
| S10_vs_22365\|\|full | Not-determined |
| S10_vs_22372\|\|full | Not-determined |
| S10_vs_22396\|\|full | Low-quality |
| S10_vs_22502\|\|full | Not-determined |
| S10_vs_22599\|\|full | Low-quality |
| S10_vs_22607\|\|full | Not-determined |
| S10_vs_22626\|\|full | Not-determined |
| S10_vs_22676\|\|full | Not-determined |
| S10_vs_22681\|\|full | Not-determined |
| S10_vs_22728\|\|full | Not-determined |
| S10_vs_22798\|\|full | Low-quality |
| S10_vs_22821\|\|full | Not-determined |
| S10_vs_22842\|\|full | Not-determined |
| S10_vs_22856\|\|full | Low-quality |
| S10_vs_22903\|\|full | Low-quality |
| S10_vs_22995\|\|full | Low-quality |
| S10_vs_23059\|\|full | Not-determined |
| S10_vs_23061\|\|full | Low-quality |
| S10_vs_23073\|\|full | Low-quality |
| S10_vs_23088\|\|full | Not-determined |
| S10_vs_23120\|\|full | Not-determined |
| S10_vs_23207\|\|full | Not-determined |
| S10_vs_23260\|\|full | Not-determined |
| S10_vs_23266\|\|full | Low-quality |
| S10_vs_23344\|\|full | Not-determined |
| S10_vs_23347\|\|full | Low-quality |
| S10_vs_23360\|\|full | Not-determined |
| S10_vs_23361\|\|full | Not-determined |
| S10_vs_23378\|\|full | Not-determined |
| S10_vs_23387\|\|full | Low-quality |
| S10_vs_23423\|\|full | Not-determined |
| S10_vs_23468\|\|full | Low-quality |
| S10_vs_23470\|\|full | Not-determined |
| S10_vs_23474\|\|full | Low-quality |
| S10_vs_23497\|\|full | Not-determined |
| S10_vs_23593\|\|full | Not-determined |
| S10_vs_23601\|\|full | Low-quality |
| S10_vs_23666\|\|full | Low-quality |
| S10_vs_23697\|\|full | Not-determined |
| S10_vs_23702\|\|full | Low-quality |
| S10_vs_23735\|\|full | Low-quality |
| S10_vs_23771\|\|full | Not-determined |
| S10_vs_23837\|\|full | Low-quality |
| S10_vs_23851\|\|full | Low-quality |
| S10_vs_23853\|\|full | Not-determined |
| S10_vs_23883\|\|full | Low-quality |
| S10_vs_23900\|\|full | Not-determined |
| S10_vs_23919\|\|full | Low-quality |
| S10_vs_23928\|\|full | Low-quality |
| S10_vs_23950\|\|full | Not-determined |
| S10_vs_24022\|\|full | Not-determined |
| S10_vs_24051\|\|full | Low-quality |
| S10_vs_24098\|\|full | Not-determined |
| S10_vs_24102\|\|full | Low-quality |
| S10_vs_24159\|\|full | Not-determined |
| S10_vs_24170\|\|full | Low-quality |
| S10_vs_24227\|\|full | Low-quality |
| S10_vs_24229\|\|full | Not-determined |
| S10_vs_24272\|\|full | Low-quality |
| S10_vs_24295\|\|full | Not-determined |
| S10_vs_24298\|\|full | Not-determined |
| S10_vs_24313\|\|full | Low-quality |
| S10_vs_24333\|\|full | Low-quality |
| S10_vs_24341\|\|full | Not-determined |
| S10_vs_24357\|\|full | Low-quality |
| S10_vs_24379\|\|full | Not-determined |
| S10_vs_24382\|\|full | Low-quality |
| S10_vs_24396\|\|full | Low-quality |
| S10_vs_24507\|\|full | Low-quality |
| S10_vs_24543\|\|full | Not-determined |
| S10_vs_24668\|\|full | Not-determined |
| S10_vs_24673\|\|full | Low-quality |
| S10_vs_24687\|\|full | Low-quality |
| S10_vs_24700\|\|full | Low-quality |
| S10_vs_24716\|\|full | Low-quality |
| S10_vs_24751\|\|full | Not-determined |
| S10_vs_24781\|\|full | Not-determined |
| S7_vs_30640\|\|full | Medium-quality |
| S10_vs_24826\|\|full | Low-quality |
| S10_vs_24863\|\|full | Not-determined |
| S10_vs_24902\|\|full | Low-quality |
| S10_vs_25102\|\|full | Not-determined |
| S10_vs_25164\|\|full | Not-determined |
| S10_vs_25172\|\|full | Low-quality |
| S10_vs_25200\|\|full | Low-quality |
| S12_extracted_k141_173666 | High-quality |
| S10_vs_25316\|\|full | Low-quality |
| S10_vs_25368\|\|full | Low-quality |
| S10_vs_25371\|\|full | Low-quality |
| S10_vs_25388\|\|full | Not-determined |
| S10_vs_25390\|\|full | Not-determined |
| S10_vs_25414\|\|full | Low-quality |
| S10_vs_25420\|\|full | Low-quality |
| S10_vs_25453\|\|full | Low-quality |
| S10_vs_25468\|\|full | Not-determined |
| S10_vs_25477\|\|full | Low-quality |
| S10_vs_25512\|\|full | Low-quality |
| S10_vs_25566\|\|full | Not-determined |
| S10_vs_25580\|\|full | Not-determined |
| S10_vs_25620\|\|full | Not-determined |
| S10_vs_25731\|\|full | Low-quality |
| S10_vs_25912\|\|full | Low-quality |
| S10_vs_25958\|\|full | Low-quality |
| S10_vs_25963\|\|full | Not-determined |
| S10_vs_25966\|\|full | Low-quality |
| S10_vs_25971\|\|full | Not-determined |
| S10_vs_25972\|\|full | Low-quality |
| S10_vs_25992\|\|full | Not-determined |
| S10_vs_26086\|\|full | Low-quality |
| S10_vs_26093\|\|full | Not-determined |
| S10_vs_26131\|\|full | Not-determined |
| S10_vs_26149\|\|full | Low-quality |
| S10_vs_26164\|\|full | Low-quality |
| S10_vs_26185\|\|full | Not-determined |
| S10_vs_26188\|\|full | Not-determined |
| S10_vs_26194\|\|full | Low-quality |
| S10_vs_26251\|\|full | Not-determined |
| S10_vs_26265\|\|full | Low-quality |
| S10_vs_26319\|\|full | Low-quality |
| S10_vs_26343\|\|full | Not-determined |
| S14_vs_k141_44452\|\|full | High-quality |
| S10_vs_26402\|\|full | Not-determined |
| S10_vs_26450\|\|full | Low-quality |
| S10_vs_26458\|\|full | Low-quality |
| S10_vs_26484\|\|full | Low-quality |
| S10_vs_26525\|\|full | Low-quality |
| S10_vs_26573\|\|full | Not-determined |
| S10_vs_26583\|\|full | Not-determined |
| S10_vs_26620\|\|full | Low-quality |
| S10_vs_26638\|\|full | Not-determined |
| S10_vs_26675\|\|full | Not-determined |
| S10_vs_26707\|\|full | Not-determined |
| S10_vs_26720\|\|full | Low-quality |
| S10_vs_26727\|\|full | Low-quality |
| S10_vs_26754\|\|full | Low-quality |
| S10_vs_26766\|\|full | Low-quality |
| S10_vs_26768\|\|full | Low-quality |
| S10_vs_26793\|\|full | Not-determined |
| S10_vs_26903\|\|full | Low-quality |
| S10_vs_26915\|\|full | Low-quality |
| S10_vs_26967\|\|full | Low-quality |
| S10_vs_26977\|\|full | Low-quality |
| S10_vs_27021\|\|full | Low-quality |
| S10_vs_27023\|\|full | Not-determined |
| S10_vs_27043\|\|full | Low-quality |
| S10_vs_27053\|\|full | Low-quality |
| S10_vs_27082\|\|full | Low-quality |
| S10_vs_27083\|\|full | Low-quality |
| S10_vs_27093\|\|full | Not-determined |
| S10_vs_27116\|\|full | Not-determined |
| S10_vs_27125\|\|full | Low-quality |
| S10_vs_27149\|\|full | Low-quality |
| S10_vs_27150\|\|full | Low-quality |
| S10_vs_27151\|\|full | Low-quality |
| S10_vs_27190\|\|full | Low-quality |
| S10_vs_27223\|\|full | Not-determined |
| S10_vs_27239\|\|full | Low-quality |
| S10_vs_27308\|\|full | Low-quality |
| S10_vs_27335\|\|full | Not-determined |
| S10_vs_27357\|\|full | Low-quality |
| S10_vs_27361\|\|full | Low-quality |
| S10_vs_27401\|\|full | Not-determined |
| S10_vs_27500\|\|full | Low-quality |
| S10_vs_27548\|\|full | Low-quality |
| S10_vs_27586\|\|full | Not-determined |
| S10_vs_27615\|\|full | Not-determined |
| S10_vs_27626\|\|full | Low-quality |
| S10_vs_27704\|\|full | Not-determined |
| S10_vs_27772\|\|full | Not-determined |
| S10_vs_27783\|\|full | Low-quality |
| S10_vs_27785\|\|full | Low-quality |
| S10_vs_27786\|\|full | Low-quality |
| S10_vs_27802\|\|full | Not-determined |
| S10_vs_27805\|\|full | Low-quality |
| S10_vs_27834\|\|full | Not-determined |
| S10_vs_27865\|\|full | Not-determined |
| S10_vs_27874\|\|full | Low-quality |
| S10_vs_27901\|\|full | Low-quality |
| S10_vs_27922\|\|full | Low-quality |
| S10_vs_27926\|\|full | Low-quality |
| S10_vs_28041\|\|full | Low-quality |
| S10_vs_28066\|\|full | Not-determined |
| S10_vs_28108\|\|full | Not-determined |
| S10_vs_28166\|\|full | Low-quality |
| S10_vs_28299\|\|full | Low-quality |
| S10_vs_28382\|\|full | Not-determined |
| S10_vs_28484\|\|full | Not-determined |
| S10_vs_28596\|\|full | Not-determined |
| S10_vs_28607\|\|full | Low-quality |
| S10_vs_28646\|\|full | Not-determined |
| S10_vs_28658\|\|full | Not-determined |
| S10_vs_28659\|\|full | Low-quality |
| S10_vs_28742\|\|full | Low-quality |
| S10_vs_28824\|\|full | Not-determined |
| S10_vs_28841\|\|full | Low-quality |
| S10_vs_28883\|\|full | Low-quality |
| S10_vs_28916\|\|full | Not-determined |
| S10_vs_29013\|\|full | Not-determined |
| S10_vs_29023\|\|full | Not-determined |
| S10_vs_29024\|\|full | Not-determined |
| S10_vs_29103\|\|full | Not-determined |
| S10_vs_29200\|\|full | Not-determined |
| S10_vs_29259\|\|full | Not-determined |
| S12_vs_k141_173666\|\|full | High-quality |
| S10_vs_29280\|\|full | Not-determined |
| S10_vs_29329\|\|full | Low-quality |
| S10_vs_29334\|\|full | Not-determined |
| S10_vs_29395\|\|full | Not-determined |
| S10_vs_29396\|\|full | Low-quality |
| S10_vs_29426\|\|full | Low-quality |
| S10_vs_29450\|\|full | Not-determined |
| S10_vs_29560\|\|full | Not-determined |
| S10_vs_29609\|\|full | Low-quality |
| S10_vs_29645\|\|full | Low-quality |
| S10_vs_29651\|\|full | Not-determined |
| S10_vs_29704\|\|full | Not-determined |
| S10_vs_29737\|\|full | Low-quality |
| S10_vs_29788\|\|full | Low-quality |
| S10_vs_29793\|\|full | Not-determined |
| S10_vs_29794\|\|full | Low-quality |
| S10_vs_29797\|\|full | Low-quality |
| S10_vs_29865\|\|full | Low-quality |
| S10_vs_29888\|\|full | Low-quality |
| S10_vs_29910\|\|full | Low-quality |
| S10_vs_29922\|\|full | Low-quality |
| S10_vs_29923\|\|full | Not-determined |
| S10_vs_29983\|\|full | Not-determined |
| S10_vs_30001\|\|full | Low-quality |
| S10_vs_30031\|\|full | Not-determined |
| S10_vs_30148\|\|full | Not-determined |
| S10_vs_30202\|\|full | Low-quality |
| S10_vs_30208\|\|full | Low-quality |
| S10_vs_30270\|\|full | Not-determined |
| S10_vs_30295\|\|full | Low-quality |
| S10_vs_30296\|\|full | Low-quality |
| S10_vs_30328\|\|full | Low-quality |
| S10_vs_30355\|\|full | Not-determined |
| S10_vs_30356\|\|full | Low-quality |
| S10_vs_30383\|\|full | Low-quality |
| S10_vs_30468\|\|full | Not-determined |
| S10_vs_30546\|\|full | Low-quality |
| S10_vs_30560\|\|full | Low-quality |
| S10_vs_30570\|\|full | Low-quality |
| S10_vs_30669\|\|full | Low-quality |
| S10_vs_30742\|\|full | Low-quality |
| S10_vs_30801\|\|full | Not-determined |

S10_vs_30836||full Low-quality
S10_vs_30877||full Low-quality
S10_vs_30931||full Not-determined
S10_vs_31030||full Not-determined
S10_vs_31048||full Not-determined
S10_vs_31055||full Not-determined
S10_vs_31086||full Low-quality
S10_vs_31236||full Not-determined
S10_vs_31238||full Not-determined
S10_vs_31302||full Low-quality
S10_vs_31445||full Not-determined
S10_vs_31633||full Low-quality
S10_vs_31670||full Low-quality
S10_vs_31720||full Low-quality
S10_vs_31731||full Not-determined
S10_vs_31792||full Low-quality
S10_vs_31829||full Low-quality
S10_vs_31855||full Not-determined
S10_vs_31861||full Low-quality
S10_vs_31883||full Low-quality
S10_vs_31895||full Not-determined
S10_vs_31955||full Not-determined
S10_vs_31974||full Not-determined
S10_vs_32008||full Not-determined
S10_vs_32076||full Not-determined
S10_vs_32106||full Low-quality
S10_vs_32301||full Low-quality
S10_vs_32315||full Not-determined
S10_vs_32330||full Low-quality
S10_vs_32390||full Low-quality
S10_vs_32418||full Low-quality
S10_vs_32445||full Low-quality
S10_vs_32553||full Low-quality
S10_vs_32643||full Not-determined
S10_vs_32658||full Low-quality
S10_vs_32678||full Low-quality
S10_vs_32687||full Low-quality
S10_vs_32729||full Low-quality
S10_vs_32747||full Not-determined
S10_vs_32774||full Not-determined
S10_vs_32789||full Low-quality
S10_vs_32796||full Not-determined
S10_vs_32938||full Not-determined
S10_vs_32950||full Low-quality
S10_vs_32955||full Low-quality
S10_vs_32967||full Not-determined
S10_vs_32972||full Low-quality
S10_vs_33006||full Low-quality
S10_vs_33009||full Not-determined
S10_vs_33035||full Low-quality
S10_vs_33115||full Low-quality
S10_vs_33122||full Not-determined
S10_vs_33129||full Not-determined
S10_vs_33135||full Low-quality
S10_vs_33189||full Not-determined
S10_vs_33215||full Low-quality
S10_vs_33270||full Not-determined
S10_vs_33293||full Low-quality
S10_vs_33354||full Low-quality
S10_vs_33366||full Not-determined
S10_vs_33457||full Low-quality
S10_vs_33463||full Low-quality
S10_vs_33467||full Low-quality
S10_vs_33473||full Not-determined
S10_vs_33565||full Not-determined
S10_vs_33592||full Not-determined
S10_vs_33670||full Not-determined
S10_vs_33749||full Low-quality
S10_vs_33781||full Low-quality
S10_vs_33835||full Low-quality
S10_vs_33897||full Low-quality
S10_vs_33898||full Not-determined
S10_vs_33902||full Low-quality
S10_vs_33943||full Low-quality
S10_vs_33950||full Not-determined
S10_vs_33980||full Low-quality
S10_vs_33984||full Low-quality
S10_vs_34000||full Not-determined
S10_vs_34002||full Low-quality
S10_vs_34024||full Low-quality
S10_vs_34031||full Low-quality
S10_vs_34039||full Not-determined
S10_vs_34060||full Low-quality
S10_vs_34117||full Not-determined
S10_vs_34126||full Low-quality
S10_vs_34205||full Low-quality
S10_vs_34208||full Low-quality
S10_vs_34228||full Low-quality
S10_vs_34248||full Low-quality
S10_vs_34260||full Low-quality
S10_vs_34322||full Low-quality
S10_vs_34362||full Low-quality
S10_vs_34431||full Low-quality
S10_vs_34438||full Low-quality
S10_vs_34443||full Not-determined
S10_vs_34536||full Low-quality
S10_vs_34543||full Low-quality
S10_vs_34574||full Not-determined
S10_vs_34624||full Not-determined
S10_vs_34627||full Not-determined
S10_vs_34715||full Low-quality
S10_vs_34717||full Low-quality
S10_vs_34730||full Low-quality
S10_vs_34763||full Low-quality
S10_vs_34782||full Not-determined
S10_vs_34833||full Not-determined
S10_vs_34843||full Not-determined
S10_vs_34872||full Low-quality
S10_vs_34889||full Low-quality
S10_vs_34923||full Not-determined
S10_vs_34954||full Low-quality
S10_vs_34981||full Low-quality
S10_vs_35001||full Low-quality
S10_vs_35068||full Not-determined
S10_vs_35079||full Not-determined
S10_vs_35113||full Not-determined
S10_vs_35118||full Not-determined
S10_vs_35122||full Not-determined
S10_vs_35128||full Low-quality
S10_vs_35219||full Low-quality
S10_vs_35308||full Not-determined
S10_vs_35382||full Low-quality
S10_vs_35399||full Low-quality
S10_vs_35401||full Not-determined
S10_vs_35430||full Low-quality
S10_vs_35431||full Low-quality
S10_vs_35465||full Not-determined
S10_vs_35468||full Low-quality
S10_vs_35479||full Not-determined
S10_vs_35547||full Not-determined
S10_vs_35553||full Not-determined
S10_vs_35639||full Low-quality
S10_vs_35646||full Not-determined
S10_vs_35663||full Low-quality
S10_vs_35828||full Not-determined
S10_vs_35850||full Low-quality
S5_vs_k141_74199||0_partial Medium-quality
S10_vs_35857||full Not-determined
S10_vs_35875||full Low-quality
S10_vs_35893||full Not-determined
S10_vs_35904||full Low-quality
S10_vs_35919||full Low-quality
S10_vs_35960||full Low-quality
S10_vs_36039||full Not-determined
S10_vs_36066||full Not-determined
S10_vs_36081||full Not-determined
S10_vs_36169||full Low-quality
S10_vs_36174||full Low-quality
S10_vs_36195||full Low-quality
S10_vs_36276||full Low-quality
S10_vs_36280||full Not-determined
S10_vs_36296||full Not-determined
S10_vs_36344||full Low-quality
S10_vs_36347||full Not-determined
S6_vs_k141_13043||full Medium-quality
S10_vs_36429||full Not-determined
S10_vs_36457||full Not-determined
S10_vs_36473||full Not-determined
S10_vs_36477||full Low-quality
S10_vs_36489||full Not-determined
S10_vs_36497||full Not-determined
S10_vs_36501||full Not-determined
S10_vs_36515||full Low-quality
S16_vs_80124||full Medium-quality
S10_vs_36623||full Low-quality
S10_vs_36677||full Low-quality
S10_vs_36678||full Low-quality
S10_vs_36718||full Low-quality
S10_vs_36728||full Not-determined
S10_vs_36729||full Not-determined
S10_vs_36734||full Low-quality
S10_vs_36747||full Low-quality
S10_vs_36790||full Low-quality
S10_vs_36858||full Not-determined
S10_vs_36861||full Not-determined
S10_vs_36954||full Low-quality
S10_vs_37120||full Low-quality
S10_vs_37121||full Low-quality
S10_vs_37155||full Not-determined
S10_vs_37159||full Not-determined
S10_vs_37169||full Low-quality
S10_vs_37217||full Low-quality
S10_vs_37280||full Low-quality
S10_vs_37335||full Not-determined
S10_vs_37337||full Not-determined
S16_extracted_169806 Medium-quality
S10_vs_37393||full Not-determined
S10_vs_37395||full Low-quality
S10_vs_37410||full Not-determined
S10_vs_37537||full Not-determined
S10_vs_37539||full Not-determined
S10_vs_37543||full Not-determined
S10_vs_37664||full Not-determined
S16_vs_169806||full Medium-quality
S10_vs_37701||full Low-quality
S10_vs_37707||full Low-quality
S10_vs_37752||full Low-quality
S10_vs_37757||full Low-quality
S10_vs_37766||full Not-determined
S10_vs_37796||full Not-determined
S10_vs_37845||full Not-determined
S10_vs_37896||full Low-quality
S10_vs_37917||full Not-determined
S10_vs_37933||full Not-determined
S10_vs_37968||full Low-quality
S10_vs_38020||full Low-quality
S10_vs_38063||full Low-quality
S10_vs_38074||full Low-quality
S10_vs_38110||full Not-determined
S10_vs_38112||full Low-quality
S10_vs_38179||full Low-quality
S10_vs_38203||full Not-determined
S10_vs_38228||full Low-quality
S10_vs_38278||full Low-quality
S10_vs_38280||full Low-quality
S10_vs_38297||full Not-determined
S10_vs_38407||full Low-quality
S10_vs_38427||full Not-determined
S10_vs_38434||full Low-quality
S10_vs_38476||full Not-determined
S10_vs_38500||full Low-quality
S10_vs_38501||full Low-quality
S10_vs_38529||full Low-quality
S10_vs_38554||full Not-determined
S10_vs_38567||full Not-determined
S10_vs_38674||full Low-quality
S10_vs_38694||full Low-quality
S10_vs_38772||full Not-determined
S10_vs_38796||full Low-quality
S10_vs_38826||full Low-quality
S10_vs_38913||full Low-quality
S10_vs_38948||full Low-quality
S10_vs_38957||full Not-determined
S10_vs_39026||full Not-determined
S10_vs_39047||full Not-determined
S10_vs_39050||full Low-quality
S10_vs_39182||full Low-quality
S10_vs_39274||full Not-determined
S10_vs_39279||full Low-quality
S10_vs_39329||full Not-determined
S10_vs_39359||full Low-quality
S10_vs_39400||full Not-determined
S10_vs_39401||full Not-determined
S10_vs_39407||full Not-determined
S10_vs_39409||full Low-quality
S10_vs_39493||full Low-quality
S10_vs_39496||full Low-quality
S10_vs_39583||full Low-quality
S10_vs_39603||full Low-quality
S10_vs_39609||full Low-quality
S10_vs_39612||full Low-quality
S10_vs_39617||full Not-determined
S10_vs_39644||full Low-quality
S10_vs_39660||full Low-quality
S10_vs_39674||full Low-quality
S10_vs_39683||full Not-determined
S10_vs_39702||full Low-quality
S10_vs_39717||full Not-determined
S10_vs_39730||full Low-quality
S10_vs_39754||full Not-determined
S10_vs_39796||full Not-determined
S12_vs_k141_118154||full High-quality
S10_vs_39886||full Low-quality
S10_vs_39919||full Not-determined
S10_vs_39961||full Not-determined
S10_vs_40126||full Low-quality
S10_vs_40150||full Low-quality
S10_vs_40315||full Low-quality
S10_vs_40328||full Low-quality
S10_vs_40380||full Low-quality
S10_vs_40381||full Not-determined
S10_vs_40388||full Low-quality
S10_vs_40507||full Not-determined
S10_vs_40557||full Not-determined
S10_vs_40619||full Low-quality
S10_vs_40642||full Low-quality
S10_vs_40661||full Not-determined
S10_vs_40676||full Not-determined
S10_vs_40746||full Low-quality
S10_vs_40791||full Low-quality
S10_vs_40792||full Not-determined
S10_vs_40890||full Not-determined
S10_vs_40909||full Not-determined
S10_vs_40985||full Low-quality
S10_vs_41003||full Not-determined
S10_vs_41044||full Not-determined
S10_vs_41049||full Not-determined
S10_vs_41055||full Not-determined
S10_vs_41105||full Not-determined
S10_vs_41112||full Low-quality
S10_vs_41130||full Low-quality
S10_vs_41138||full Low-quality
S10_vs_41166||full Low-quality
S10_vs_41243||full Not-determined
S10_vs_41287||full Low-quality
S10_vs_41304||full Low-quality
S10_vs_41305||full Not-determined
S10_vs_41310||full Not-determined
S10_vs_41326||full Low-quality
S10_vs_41346||full Not-determined
S10_vs_41406||full Not-determined
S10_vs_41422||full Not-determined
S10_vs_41564||full Low-quality
S10_vs_41590||full Low-quality
S10_vs_41650||full Low-quality
S10_vs_41706||full Not-determined
S10_vs_41714||full Low-quality
S10_vs_41745||full Low-quality
S10_vs_41831||full Low-quality
S10_vs_41887||full Low-quality
S10_vs_42000||full Low-quality
S10_vs_42089||full Not-determined
S10_vs_42153||full Low-quality
S10_vs_42159||full Low-quality
S10_vs_42214||full Low-quality
S10_vs_42300||full Not-determined
S10_vs_42331||full Low-quality
S10_vs_42350||full Not-determined
S10_vs_42373||full Low-quality
S10_vs_42387||full Low-quality
S10_vs_42392||full Not-determined
S10_vs_42413||full Not-determined
S10_vs_42417||full Not-determined
S10_vs_42460||full Not-determined
S10_vs_42463||full Not-determined
S10_vs_42477||full Not-determined
S10_vs_42483||full Low-quality
S10_vs_42487||full Not-determined
S10_vs_42578||full Not-determined
S10_vs_42592||full Low-quality
S10_vs_42626||full Low-quality
S10_vs_42649||full Low-quality
S10_vs_42747||full Low-quality
S10_vs_42752||full Low-quality
S10_vs_42794||full Low-quality
S10_vs_42914||full Low-quality
S10_vs_42922||full Not-determined
S10_vs_42933||full Low-quality
S10_vs_42978||full Not-determined
S10_vs_43001||full Low-quality
S10_vs_43009||full Not-determined
S10_vs_43025||full Not-determined
S10_vs_43042||full Low-quality
S10_vs_43089||full Not-determined
S10_vs_43115||full Not-determined
S10_vs_43165||full Low-quality
S14_vs_k141_61530||full Medium-quality
S10_vs_43219||full Low-quality
S10_vs_43284||full Low-quality
S10_vs_43319||full Low-quality
S10_vs_43377||full Not-determined
S10_vs_43402||full Not-determined
S10_vs_43403||full Not-determined
S10_vs_43435||full Not-determined
S10_vs_43450||full Low-quality
S10_vs_43493||full Low-quality
S10_vs_43527||full Low-quality
S10_vs_43569||full Not-determined
S10_vs_43682||full Not-determined
S10_vs_43707||full Low-quality
S10_vs_43715||full Low-quality
S10_vs_43723||full Low-quality
S10_vs_43771||full Not-determined
S10_vs_43775||full Not-determined
S10_vs_43817||full Low-quality
S10_vs_43847||full Low-quality
S10_vs_43882||full Low-quality
S10_vs_43904||full Not-determined
S10_vs_43951||full Low-quality
S17_vs_k141_13596||full Medium-quality
S10_vs_43966||full Low-quality
S10_vs_44029||full Not-determined
S10_vs_44037||full Low-quality
S10_vs_44076||full Not-determined
S10_vs_44128||full Not-determined
S10_vs_44217||full Low-quality
S10_vs_44253||full Not-determined
S10_vs_44319||full Not-determined
S10_vs_44412||full Not-determined
S10_vs_44425||full Low-quality
S10_vs_44453||full Low-quality
S10_vs_44482||full Low-quality
S10_vs_44498||full Low-quality
S10_vs_44500||full Low-quality
S10_vs_44551||full Not-determined
S10_vs_44560||full Low-quality
S10_vs_44573||full Not-determined
S10_vs_44590||full Low-quality
S10_vs_44649||full Not-determined
S10_vs_44681||full Low-quality
S10_vs_44684||full Not-determined
S10_vs_44724||full Low-quality
S10_vs_44739||full Not-determined
S10_vs_44784||full Low-quality
S10_vs_44833||full Not-determined
S10_vs_44847||full Not-determined
S10_vs_44879||full Not-determined
S10_vs_44891||full Not-determined
S10_vs_44931||full Low-quality
S10_vs_44963||full Not-determined
S10_vs_45001||full Low-quality
S10_vs_45039||full Not-determined
S10_vs_45083||full Low-quality
S10_vs_45193||full Low-quality
S10_vs_45221||full Not-determined
S10_vs_45314||full Low-quality
S10_vs_45345||full Not-determined
S10_vs_45422||full Low-quality
S10_vs_45424||full Not-determined
S10_vs_45431||full Not-determined
S10_vs_45609||full Low-quality
S10_vs_45624||full Low-quality
S10_vs_45629||full Not-determined
S10_vs_45670||full Low-quality
S10_vs_45710||full Not-determined
S10_vs_45720||full Not-determined
S10_vs_45733||full Low-quality
S10_vs_45754||full Low-quality
S10_vs_45805||full Low-quality
S10_vs_45832||full Not-determined
S10_vs_45978||full Low-quality
S10_vs_46005||full Low-quality
S10_vs_46008||full Low-quality
S10_vs_46014||full Not-determined
S10_vs_46096||full Not-determined
S10_vs_46133||full Not-determined
S10_vs_46170||full Low-quality
S10_vs_46186||full Low-quality
S10_vs_46195||full Low-quality
S10_vs_46228||full Not-determined
S10_vs_46263||full Low-quality
S10_vs_46265||full Not-determined
S10_vs_46279||full Low-quality
S17_extracted_k141_140653 High-quality
S17_vs_k141_140653||full High-quality
S10_vs_46333||full Low-quality
S10_vs_46480||full Low-quality
S10_vs_46487||full Not-determined
S10_vs_46508||full Not-determined
S10_vs_46533||full Low-quality
S10_vs_46587||full Low-quality
S10_vs_46593||full Low-quality
S10_vs_46632||full Not-determined
S10_vs_46663||full Not-determined
S10_vs_46681||full Not-determined
S10_vs_46722||full Low-quality
S10_vs_46807||full Low-quality
S14_vs_k141_73301||full Medium-quality
S10_vs_46880||full Low-quality
S10_vs_46884||full Not-determined
S10_vs_46886||full Low-quality
S10_vs_46939||full Low-quality
S10_vs_46952||full Not-determined
S10_vs_47089||full Low-quality
S10_vs_47106||full Low-quality
S10_vs_47170||full Low-quality
S10_vs_47194||full Low-quality
S10_vs_47199||full Not-determined
S10_vs_47206||full Low-quality
S10_vs_47235||full Not-determined
S10_vs_47243||full Not-determined
S10_vs_47389||full Not-determined
S10_vs_47430||full Low-quality
S10_vs_47432||full Not-determined
S10_vs_47513||full Low-quality
S10_vs_47615||full Not-determined
S10_vs_47640||full Not-determined
S10_vs_47641||full Low-quality
S10_vs_47670||full Low-quality
S10_vs_47672||full Not-determined
S10_vs_47722||full Not-determined
S10_vs_47750||full Low-quality
S10_vs_47850||full Low-quality
S10_vs_47865||full Low-quality
S10_vs_47882||full Low-quality
S10_vs_47957||full Not-determined
S10_vs_48024||full Not-determined
S10_vs_48098||full Low-quality
S10_vs_48131||full Low-quality
S10_vs_48135||full Low-quality
S10_vs_48141||full Not-determined
S10_vs_48234||full Low-quality
S10_vs_48269||full Low-quality
S10_vs_48271||full Low-quality
S10_vs_48301||full Low-quality
S10_vs_48333||full Not-determined
S10_vs_48335||full Low-quality
S10_vs_48360||full Not-determined
S10_vs_48389||full Low-quality
S10_vs_48393||full Not-determined
S10_vs_48438||full Low-quality
S10_vs_48452||full Low-quality
S10_vs_48527||full Low-quality
S10_vs_48576||full Not-determined
S10_vs_48618||full Not-determined
S10_vs_48654||full Not-determined
S10_vs_48672||full Low-quality
S10_vs_48684||full Low-quality
S10_vs_48765||full Low-quality
S10_vs_48809||full Low-quality
S10_vs_48870||full Low-quality
S10_vs_48928||full Low-quality
S10_vs_48960||full Low-quality
S10_vs_48978||full Low-quality
S10_vs_48993||full Not-determined
S10_vs_49001||full Not-determined
S10_vs_49075||full Low-quality
S10_vs_49124||full Not-determined
S10_vs_49136||full Not-determined
S10_vs_49148||full Not-determined
S10_vs_49181||full Low-quality
S10_vs_49241||full Low-quality
S10_vs_49243||full Not-determined
S10_vs_49246||full Low-quality
S10_vs_49258||full Low-quality
S10_vs_49267||full Low-quality
S10_vs_49290||full Not-determined
S10_vs_49293||full Low-quality
S10_vs_49298||full Low-quality
S10_vs_49307||full Low-quality
S10_vs_49323||full Low-quality
S10_vs_49336||full Low-quality
S10_vs_49394||full Not-determined
S10_vs_49429||full Not-determined
S10_vs_49431||full Low-quality
S10_vs_49445||full Low-quality
S10_vs_49473||full Not-determined
S10_vs_49487||full Low-quality
S10_vs_49540||full Not-determined
S10_vs_49599||full Not-determined
S10_vs_49610||full Low-quality
S10_vs_49618||full Not-determined
S10_vs_49627||full Not-determined
S10_vs_49694||full Not-determined
S10_vs_49716||full Not-determined
S10_vs_49726||full Not-determined
S10_vs_49751||full Not-determined
S10_vs_49918||full Not-determined
S10_vs_49985||full Not-determined
S10_vs_49996||full Not-determined
S10_vs_50036||full Low-quality
S10_vs_50078||full Low-quality
S10_vs_50093||full Low-quality
S10_vs_50108||full Low-quality
S10_vs_50130||full Not-determined
S10_vs_50139||full Low-quality
S10_vs_50274||full Low-quality
S10_vs_50293||full Low-quality
S10_vs_50314||full Low-quality
S10_vs_50319||full Not-determined
S10_vs_50350||full Not-determined
S10_vs_50362||full Low-quality
S10_vs_50459||full Not-determined
S10_vs_50469||full Low-quality
S10_vs_50518||full Low-quality
S10_vs_50532||full Low-quality
S10_vs_50568||full Not-determined
S10_vs_50624||full Low-quality
S10_vs_50646||full Low-quality
S10_vs_50717||full Low-quality
S10_vs_50738||full Low-quality
S10_vs_50753||full Low-quality
S10_vs_50756||full Low-quality
S10_vs_50796||full Not-determined
S10_vs_50801||full Low-quality
S10_vs_50856||full Not-determined
S10_vs_50884||full Low-quality
S10_vs_50937||full Low-quality
S10_vs_51001||full Not-determined
S10_vs_51114||full Low-quality
S10_vs_51142||full Low-quality
S10_vs_51300||full Low-quality
S10_vs_51319||full Low-quality
S10_vs_51321||full Not-determined
S10_vs_51357||full Low-quality
S10_vs_51363||full Low-quality
S10_vs_51371||full Not-determined
S10_vs_51391||full Low-quality
S10_vs_51393||full Low-quality
S10_vs_51407||full Not-determined
S10_vs_51412||full Not-determined
S10_vs_51415||full Low-quality
S10_vs_51418||full Not-determined
S10_vs_51435||full Low-quality
S10_vs_51490||full Not-determined
S10_vs_51514||full Not-determined
S10_vs_51515||full Not-determined
S10_vs_51537||full Not-determined
S10_vs_51574||full Low-quality

| | |
|---|---|
| S10_vs_S1578\|\|full | Low-quality |
| S10_vs_S1581\|\|full | Low-quality |
| S10_vs_S1588\|\|full | Not-determined |
| S10_vs_S1593\|\|full | Low-quality |
| S10_vs_S1602\|\|full | Not-determined |
| S10_vs_S1623\|\|full | Low-quality |
| S10_vs_S1649\|\|full | Low-quality |
| S10_vs_S1653\|\|full | Not-determined |
| S10_vs_S1675\|\|full | Low-quality |
| S10_vs_S1691\|\|full | Not-determined |
| S10_vs_S1706\|\|full | Low-quality |
| S10_vs_S1715\|\|full | Low-quality |
| S10_vs_S1743\|\|full | Low-quality |
| S10_vs_S1752\|\|full | Not-determined |
| S10_vs_S1754\|\|full | Not-determined |
| S10_vs_S1779\|\|full | Not-determined |
| S10_vs_S1880\|\|full | Low-quality |
| S10_vs_S1888\|\|full | Low-quality |
| S10_vs_S1917\|\|full | Low-quality |
| S10_vs_S1946\|\|full | Low-quality |
| S10_vs_S1962\|\|full | Not-determined |
| S10_vs_S2115\|\|full | Not-determined |
| S10_vs_S2161\|\|full | Low-quality |
| S10_vs_S2359\|\|full | Low-quality |
| S10_vs_S2394\|\|full | Not-determined |
| S10_vs_S2404\|\|full | Low-quality |
| S10_vs_S2426\|\|full | Low-quality |
| S10_vs_S2443\|\|full | Low-quality |
| S10_vs_S2461\|\|full | Low-quality |
| S10_vs_S2536\|\|full | Low-quality |
| S10_vs_S2573\|\|full | Low-quality |
| S10_vs_S2647\|\|full | Not-determined |
| S10_vs_S2662\|\|full | Low-quality |
| S10_vs_S2692\|\|full | Not-determined |
| S10_vs_S2814\|\|full | Not-determined |
| S10_vs_S2918\|\|full | Low-quality |
| S10_vs_S2930\|\|full | Low-quality |
| S10_vs_S2990\|\|full | Not-determined |
| S10_vs_S3006\|\|full | Low-quality |
| S10_vs_S3108\|\|full | Low-quality |
| S10_vs_S3249\|\|full | Low-quality |
| S10_vs_S3281\|\|full | Low-quality |
| S10_vs_S3289\|\|full | Not-determined |
| S10_vs_S3399\|\|full | Low-quality |
| S10_vs_S3462\|\|full | Low-quality |
| S10_vs_S3479\|\|full | Not-determined |
| S10_vs_S3485\|\|full | Low-quality |
| S10_vs_S3547\|\|full | Not-determined |
| S10_vs_S3552\|\|full | Not-determined |
| S10_vs_S3553\|\|full | Not-determined |
| S10_vs_S3581\|\|full | Low-quality |
| S10_vs_S3687\|\|full | Not-determined |
| S10_vs_S3715\|\|full | Not-determined |
| S10_vs_S3727\|\|full | Low-quality |
| S10_vs_S3758\|\|full | Low-quality |
| S10_vs_S3781\|\|full | Not-determined |
| S10_vs_S3816\|\|full | Not-determined |
| S10_vs_S3845\|\|full | Low-quality |
| S10_vs_S3852\|\|full | Low-quality |
| S10_vs_S3909\|\|full | Not-determined |
| S10_vs_S3995\|\|full | Low-quality |
| S10_vs_S3999\|\|full | Low-quality |
| S10_vs_S4021\|\|full | Low-quality |
| S10_vs_S4035\|\|full | Low-quality |
| S10_vs_S4100\|\|full | Low-quality |
| S10_vs_S4116\|\|full | Low-quality |
| S10_vs_S4117\|\|full | Low-quality |
| S10_vs_S4164\|\|full | Not-determined |
| S10_vs_S4202\|\|full | Not-determined |
| S10_vs_S4211\|\|full | Not-determined |
| S10_vs_S4237\|\|full | Low-quality |
| S10_vs_S4261\|\|full | Low-quality |
| S10_vs_S4268\|\|full | Low-quality |
| S10_vs_S4301\|\|full | Low-quality |
| S10_vs_S4333\|\|full | Not-determined |
| S10_vs_S4341\|\|full | Not-determined |
| S10_vs_S4342\|\|full | Low-quality |
| S10_vs_S4408\|\|full | Not-determined |
| S10_vs_S4410\|\|full | Not-determined |
| S10_vs_S4440\|\|full | Low-quality |
| S10_vs_S4478\|\|full | Low-quality |
| S10_vs_S4538\|\|full | Low-quality |
| S10_vs_S4546\|\|full | Low-quality |
| S10_vs_S4627\|\|full | Not-determined |
| S10_vs_S4640\|\|full | Not-determined |
| S10_vs_S4659\|\|full | Not-determined |
| S10_vs_S4670\|\|full | Not-determined |
| S10_vs_S4706\|\|full | Low-quality |
| S10_vs_S4717\|\|full | Not-determined |
| S10_vs_S4733\|\|full | Low-quality |
| S10_vs_S4815\|\|full | Low-quality |
| S10_vs_S4827\|\|full | Not-determined |
| S10_vs_S4861\|\|full | Not-determined |
| S10_vs_S4875\|\|full | Low-quality |
| S10_vs_S5034\|\|full | Low-quality |
| S10_vs_S5043\|\|full | Not-determined |
| S10_vs_S5098\|\|full | Low-quality |
| S10_vs_S5112\|\|full | Not-determined |
| S10_vs_S5138\|\|full | Low-quality |
| S10_vs_S5214\|\|full | Low-quality |
| S10_vs_S5276\|\|full | Not-determined |
| S10_vs_S5290\|\|full | Low-quality |
| S10_vs_S5361\|\|full | Not-determined |
| S10_vs_S5465\|\|full | Low-quality |
| S10_vs_S5466\|\|full | Not-determined |
| S10_vs_S5497\|\|full | Not-determined |
| S10_vs_S5520\|\|full | Low-quality |
| S10_vs_S5624\|\|full | Low-quality |
| S10_vs_S5696\|\|full | Low-quality |
| S10_vs_S5724\|\|full | Not-determined |
| S10_vs_S5725\|\|full | Not-determined |
| S10_vs_S5735\|\|full | Low-quality |
| S10_vs_S5742\|\|full | Low-quality |
| S10_vs_S5748\|\|full | Low-quality |
| S10_vs_S5783\|\|full | Not-determined |
| S10_vs_S5790\|\|full | Not-determined |
| S10_vs_S5799\|\|full | Not-determined |
| S10_vs_S5809\|\|full | Low-quality |
| S10_vs_S5810\|\|full | Low-quality |
| S10_vs_S5869\|\|full | Low-quality |
| S10_vs_S5877\|\|full | Not-determined |
| S10_vs_S5895\|\|full | Not-determined |
| S10_vs_S5898\|\|full | Low-quality |
| S10_vs_S5925\|\|full | Not-determined |
| S10_vs_S5967\|\|full | Low-quality |
| S10_vs_S5983\|\|full | Low-quality |
| S10_vs_S6000\|\|full | Not-determined |
| S10_vs_S6007\|\|full | Not-determined |
| S10_vs_S6012\|\|full | Low-quality |
| S10_vs_S6015\|\|full | Low-quality |
| S10_vs_S6023\|\|full | Low-quality |
| S10_vs_S6043\|\|full | Low-quality |
| S10_vs_S6081\|\|full | Not-determined |
| S10_vs_S6118\|\|full | Low-quality |
| S10_vs_S6123\|\|full | Low-quality |
| S10_vs_S6147\|\|full | Not-determined |
| S10_vs_S6161\|\|full | Not-determined |
| S10_vs_S6165\|\|full | Low-quality |
| S10_vs_S6167\|\|full | Low-quality |
| S10_vs_S6199\|\|full | Not-determined |
| S10_vs_S6306\|\|full | Not-determined |
| S10_vs_S6319\|\|full | Low-quality |
| S10_vs_S6344\|\|full | Not-determined |
| S10_vs_S6387\|\|full | Not-determined |
| S10_vs_S6413\|\|full | Low-quality |
| S10_vs_S6420\|\|full | Not-determined |
| S10_vs_S6421\|\|full | Not-determined |
| S10_vs_S6436\|\|full | Low-quality |
| S10_vs_S6444\|\|full | Low-quality |
| S10_vs_S6498\|\|full | Low-quality |
| S10_vs_S6500\|\|full | Low-quality |
| S10_vs_S6573\|\|full | Low-quality |
| S10_vs_S6614\|\|full | Not-determined |
| S10_vs_S6724\|\|full | Low-quality |
| S10_vs_S6737\|\|full | Low-quality |
| S10_vs_S6760\|\|full | Low-quality |
| S10_vs_S6780\|\|full | Not-determined |
| S10_vs_S6834\|\|full | Not-determined |
| S10_vs_S6855\|\|full | Not-determined |
| S10_vs_S6940\|\|full | Low-quality |
| S10_vs_S7007\|\|full | Not-determined |
| S10_vs_S7053\|\|full | Low-quality |
| S10_vs_S7054\|\|full | Low-quality |
| S10_vs_S7099\|\|full | Not-determined |
| S10_vs_S7101\|\|full | Not-determined |
| S10_vs_S7111\|\|full | Low-quality |
| S10_vs_S7120\|\|full | Not-determined |
| S10_vs_S7263\|\|full | Low-quality |
| S10_vs_S7300\|\|full | Low-quality |
| S10_vs_S7366\|\|full | Low-quality |
| S10_vs_S7385\|\|full | Low-quality |
| S10_vs_S7389\|\|full | Low-quality |
| S10_vs_S7393\|\|full | Low-quality |
| S10_vs_S7413\|\|full | Not-determined |
| S10_vs_S7416\|\|full | Low-quality |
| S10_vs_S7421\|\|full | Low-quality |
| S10_vs_S7436\|\|full | Not-determined |
| S10_vs_S7439\|\|full | Not-determined |
| S10_vs_S7442\|\|full | Low-quality |
| S10_vs_S7486\|\|full | Not-determined |
| S10_vs_S7517\|\|full | Not-determined |
| S10_vs_S7536\|\|full | Low-quality |
| S10_vs_S7552\|\|full | Low-quality |
| S10_vs_S7584\|\|full | Not-determined |
| S10_vs_S7643\|\|full | Low-quality |
| S10_vs_S7653\|\|full | Not-determined |
| S10_vs_S7686\|\|full | Not-determined |
| S16_vs_92150\|\|full | Medium-quality |
| S10_vs_S7711\|\|full | Not-determined |
| S10_vs_S7821\|\|full | Low-quality |
| S10_vs_S7846\|\|full | Low-quality |
| S10_vs_S7848\|\|full | Not-determined |
| S10_vs_S7957\|\|full | Low-quality |
| S10_vs_S8009\|\|full | Not-determined |
| S11_vs_k141_104054\|\|full | Medium-quality |
| S10_vs_S8048\|\|full | Not-determined |
| S10_vs_S8058\|\|full | Low-quality |
| S10_vs_S8078\|\|full | Low-quality |
| S10_vs_S8081\|\|full | Low-quality |
| S10_vs_S8129\|\|full | Low-quality |
| S10_vs_S8204\|\|full | Not-determined |
| S10_vs_S8223\|\|full | Not-determined |
| S10_vs_S8232\|\|full | Low-quality |
| S10_vs_S8277\|\|full | Low-quality |
| S10_vs_S8406\|\|full | Low-quality |
| S10_vs_S8478\|\|full | Low-quality |
| S10_vs_S8482\|\|full | Low-quality |
| S10_vs_S8503\|\|full | Low-quality |
| S10_vs_S8505\|\|full | Not-determined |
| S10_vs_S8567\|\|full | Not-determined |
| S10_vs_S8574\|\|full | Low-quality |
| S10_vs_S8615\|\|full | Low-quality |
| S10_vs_S8722\|\|full | Not-determined |
| S10_vs_S8741\|\|full | Low-quality |
| S10_vs_S8745\|\|full | Not-determined |
| S10_vs_S8799\|\|full | Low-quality |
| S10_vs_S8800\|\|full | Low-quality |
| S10_vs_S8802\|\|full | Not-determined |
| S10_vs_S8843\|\|full | Not-determined |
| S10_vs_S8865\|\|full | Low-quality |
| S10_vs_S8891\|\|full | Not-determined |
| S10_vs_S8897\|\|full | Low-quality |
| S10_vs_S8908\|\|full | Low-quality |
| S10_vs_S8976\|\|full | Low-quality |
| S10_vs_S9004\|\|full | Not-determined |
| S10_vs_S9025\|\|full | Low-quality |
| S10_vs_S9107\|\|full | Low-quality |
| S10_vs_S9177\|\|full | Not-determined |
| S10_vs_S9188\|\|full | Not-determined |
| S10_vs_S9206\|\|full | Low-quality |
| S9_vs_k141_84046\|\|full | High-quality |
| S10_vs_S9230\|\|full | Not-determined |
| S10_vs_S9256\|\|full | Not-determined |
| S10_vs_S9270\|\|full | Not-determined |
| S10_vs_S9284\|\|full | Low-quality |
| S10_vs_S9300\|\|full | Low-quality |
| S10_vs_S9302\|\|full | Not-determined |
| S10_vs_S9352\|\|full | Not-determined |
| S10_vs_S9414\|\|full | Not-determined |
| S10_vs_S9487\|\|full | Not-determined |
| S10_vs_S9566\|\|full | Low-quality |
| S10_vs_S9697\|\|full | Not-determined |
| S10_vs_S9767\|\|full | Low-quality |
| S10_vs_S9886\|\|full | Low-quality |
| S10_vs_S9887\|\|full | Low-quality |
| S10_vs_S9891\|\|full | Low-quality |
| S10_vs_S9917\|\|full | Low-quality |
| S10_vs_S9927\|\|full | Low-quality |
| S10_vs_S9929\|\|full | Not-determined |
| S10_vs_S60063\|\|full | Low-quality |
| S10_vs_S60092\|\|full | Low-quality |
| S10_vs_S60112\|\|full | Not-determined |
| S10_vs_S60135\|\|full | Low-quality |
| S10_vs_S60240\|\|full | Not-determined |
| S10_vs_S60276\|\|full | Not-determined |
| S10_vs_S60294\|\|full | Not-determined |
| S10_vs_S60316\|\|full | Not-determined |
| S10_vs_S60353\|\|full | Low-quality |
| S10_vs_S60364\|\|full | Low-quality |
| S10_vs_S60404\|\|full | Not-determined |
| S10_vs_S60564\|\|full | Low-quality |
| S10_vs_S60578\|\|full | Low-quality |
| S10_vs_S60579\|\|full | Low-quality |
| S10_vs_S60618\|\|full | Low-quality |
| S10_vs_S60621\|\|full | Not-determined |
| S10_vs_S60637\|\|full | Low-quality |
| S10_vs_S60645\|\|full | Low-quality |
| S10_vs_S60675\|\|full | Not-determined |
| S10_vs_S60677\|\|full | Not-determined |
| S10_vs_S60684\|\|full | Not-determined |
| S10_vs_S60704\|\|full | Low-quality |
| S10_vs_S60726\|\|full | Low-quality |
| S10_vs_S60742\|\|full | Not-determined |
| S10_vs_S60781\|\|full | Low-quality |
| S10_vs_S60810\|\|full | Not-determined |
| S10_vs_S60937\|\|full | Not-determined |
| S10_vs_S61009\|\|full | Low-quality |
| S10_vs_S61048\|\|full | Not-determined |
| S10_vs_S61189\|\|full | Low-quality |
| S10_vs_S61191\|\|full | Low-quality |
| S10_vs_S61200\|\|full | Not-determined |
| S10_vs_S61203\|\|full | Not-determined |
| S10_vs_S61265\|\|full | Low-quality |
| S10_vs_S61337\|\|full | Low-quality |
| S10_vs_S61347\|\|full | Low-quality |
| S10_vs_S61490\|\|full | Low-quality |
| S10_vs_S61511\|\|full | Low-quality |
| S10_vs_S61580\|\|full | Low-quality |
| S10_vs_S61582\|\|full | Low-quality |
| S10_vs_S61583\|\|full | Not-determined |
| S10_vs_S61603\|\|full | Low-quality |
| S10_vs_S61658\|\|full | Not-determined |
| S10_vs_S61831\|\|full | Low-quality |
| S10_vs_S61854\|\|full | Low-quality |
| S10_vs_S61861\|\|full | Low-quality |
| S10_vs_S61869\|\|full | Not-determined |
| S10_vs_S61888\|\|full | Not-determined |
| S10_vs_S61922\|\|full | Low-quality |
| S10_vs_S61964\|\|full | Not-determined |
| S10_vs_S62003\|\|full | Not-determined |
| S10_vs_S62040\|\|full | Low-quality |
| S10_vs_S62076\|\|full | Low-quality |
| S10_vs_S62084\|\|full | Low-quality |
| S10_vs_S62141\|\|full | Low-quality |
| S10_vs_S62142\|\|full | Not-determined |
| S10_vs_S62150\|\|full | Low-quality |
| S10_vs_S62158\|\|full | Low-quality |
| S6_vs_k141_22637\|\|full | Medium-quality |
| S10_vs_S62236\|\|full | Not-determined |
| S10_vs_S62248\|\|full | Low-quality |
| S10_vs_S62296\|\|full | Not-determined |
| S10_vs_S62320\|\|full | Low-quality |
| S10_vs_S62338\|\|full | Low-quality |
| S10_vs_S62341\|\|full | Not-determined |
| S10_vs_S62384\|\|full | Not-determined |
| S10_vs_S62408\|\|full | Low-quality |
| S10_vs_S62438\|\|full | Not-determined |
| S10_vs_S62496\|\|full | Not-determined |
| S10_vs_S62567\|\|full | Low-quality |
| S10_vs_S62629\|\|full | Not-determined |
| S10_vs_S62678\|\|full | Low-quality |
| S10_vs_36598\|\|full | Medium-quality |
| S10_vs_S62777\|\|full | Not-determined |
| S10_vs_S62866\|\|full | Not-determined |
| S10_vs_S62892\|\|full | Not-determined |
| S10_vs_S63001\|\|full | Not-determined |
| S10_vs_S63017\|\|full | Low-quality |
| S10_vs_S63030\|\|full | Low-quality |
| S10_vs_S63045\|\|full | Not-determined |
| S10_vs_S63105\|\|full | Not-determined |
| S10_vs_S63211\|\|full | Not-determined |
| S10_vs_S63246\|\|full | Not-determined |
| S10_vs_S63272\|\|full | Not-determined |
| S10_vs_S63281\|\|full | Low-quality |
| S10_vs_S63296\|\|full | Low-quality |
| S10_vs_S63318\|\|full | Low-quality |
| S10_vs_S63340\|\|full | Low-quality |
| S10_vs_S63344\|\|full | Not-determined |
| S10_vs_S63346\|\|full | Not-determined |
| S10_vs_S63373\|\|full | Not-determined |
| S10_vs_S63389\|\|full | Not-determined |
| S10_vs_S63418\|\|full | Not-determined |
| S10_vs_S63478\|\|full | Low-quality |
| S10_vs_S63479\|\|full | Low-quality |
| S10_vs_S63489\|\|full | Not-determined |
| S10_vs_S63515\|\|full | Low-quality |
| S10_vs_S63546\|\|full | Not-determined |
| S10_vs_S63561\|\|full | Low-quality |
| S10_vs_S63587\|\|full | Low-quality |
| S10_vs_S63632\|\|full | Not-determined |
| S10_vs_S63668\|\|full | Not-determined |
| S10_vs_S63681\|\|full | Not-determined |
| S10_vs_S63704\|\|full | Not-determined |
| S10_vs_S63724\|\|full | Low-quality |
| S10_vs_S63798\|\|full | Low-quality |
| S10_vs_S63894\|\|full | Not-determined |
| S10_vs_S63901\|\|full | Low-quality |
| S10_vs_S63919\|\|full | Not-determined |
| S10_vs_S63936\|\|full | Low-quality |
| S10_vs_S63979\|\|full | Not-determined |
| S10_vs_S63995\|\|full | Not-determined |
| S10_vs_S64037\|\|full | Not-determined |
| S10_vs_S64065\|\|full | Not-determined |
| S10_vs_S64099\|\|full | Low-quality |
| S10_vs_S64149\|\|full | Low-quality |
| S10_vs_S64204\|\|full | Not-determined |
| S10_vs_S64239\|\|full | Not-determined |
| S10_vs_S64262\|\|full | Low-quality |
| S10_vs_S64265\|\|full | Low-quality |
| S10_vs_S64335\|\|full | Low-quality |
| S10_vs_S64349\|\|full | Low-quality |
| S10_vs_S64376\|\|full | Low-quality |
| S10_vs_S64492\|\|full | Not-determined |
| S10_vs_S64499\|\|full | Low-quality |
| S10_vs_S64510\|\|full | Not-determined |
| S10_vs_S64527\|\|full | Not-determined |
| S10_vs_S64544\|\|full | Not-determined |
| S10_vs_S64669\|\|full | Low-quality |
| S10_vs_S64688\|\|full | Low-quality |
| S10_vs_S64737\|\|full | Not-determined |
| S10_vs_S64739\|\|full | Low-quality |
| S10_vs_S64741\|\|full | Not-determined |
| S10_vs_S64832\|\|full | Low-quality |
| S10_vs_S64847\|\|full | Not-determined |
| S10_vs_S64872\|\|full | Low-quality |
| S10_vs_S64937\|\|full | Low-quality |
| S10_vs_S64951\|\|full | Low-quality |
| S10_vs_S64953\|\|full | Low-quality |
| S10_vs_S65011\|\|full | Not-determined |
| S10_vs_S65050\|\|full | Low-quality |
| S10_vs_S65147\|\|full | Low-quality |
| S10_vs_S65196\|\|full | Not-determined |
| S10_vs_S65257\|\|full | Low-quality |
| S10_vs_S65278\|\|full | Not-determined |
| S10_vs_S65281\|\|full | Not-determined |
| S10_vs_S65313\|\|full | Not-determined |
| S10_vs_S65523\|\|full | Low-quality |
| S10_vs_S65536\|\|full | Low-quality |
| S10_vs_S65546\|\|full | Low-quality |
| S10_vs_S65718\|\|full | Not-determined |
| S10_vs_S65844\|\|full | Low-quality |
| S10_vs_S65858\|\|full | Low-quality |
| S10_vs_S65887\|\|full | Low-quality |
| S10_vs_S65915\|\|full | Not-determined |
| S10_vs_S65976\|\|full | Not-determined |
| S10_vs_S65979\|\|full | Not-determined |
| S10_vs_S65988\|\|full | Low-quality |
| S10_vs_S66111\|\|full | Low-quality |
| S10_vs_S66153\|\|full | Not-determined |
| S10_vs_S66208\|\|full | Not-determined |
| S10_vs_S66275\|\|full | Low-quality |
| S10_vs_S66338\|\|full | Low-quality |
| S10_vs_S66374\|\|full | Low-quality |
| S10_vs_S66379\|\|full | Low-quality |
| S10_vs_S66385\|\|full | Low-quality |
| S10_vs_S66404\|\|full | Not-determined |
| S10_vs_S66415\|\|full | Low-quality |
| S10_vs_S66421\|\|full | Low-quality |
| S10_vs_S66452\|\|full | Not-determined |
| S10_vs_S66472\|\|full | Low-quality |
| S10_vs_S66591\|\|full | Low-quality |
| S10_vs_S66618\|\|full | Low-quality |
| S10_vs_S66649\|\|full | Low-quality |
| S10_vs_S66700\|\|full | Low-quality |
| S10_vs_S66702\|\|full | Low-quality |
| S10_vs_S66709\|\|full | Not-determined |
| S10_vs_S66717\|\|full | Low-quality |
| S10_vs_S66728\|\|full | Not-determined |
| S10_vs_S66775\|\|full | Low-quality |
| S10_vs_S66783\|\|full | Not-determined |
| S10_vs_S66784\|\|full | Low-quality |
| S10_vs_S66791\|\|full | Low-quality |
| S10_vs_S66836\|\|full | Low-quality |
| S10_vs_S66847\|\|full | Low-quality |
| S10_vs_S66926\|\|full | Not-determined |
| S10_vs_S66962\|\|full | Not-determined |
| S10_vs_S67025\|\|full | Low-quality |
| S10_vs_S67058\|\|full | Not-determined |
| S10_vs_S67069\|\|full | Not-determined |
| S10_vs_S67070\|\|full | Low-quality |
| S10_vs_S67094\|\|full | Not-determined |
| S10_vs_S67130\|\|full | Low-quality |
| S10_vs_S67135\|\|full | Not-determined |
| S10_vs_S67195\|\|full | Low-quality |
| S10_vs_S67247\|\|full | Not-determined |
| S10_vs_S67249\|\|full | Low-quality |
| S10_vs_S67258\|\|full | Low-quality |
| S10_vs_S67295\|\|full | Low-quality |
| S10_vs_S67319\|\|full | Not-determined |
| S10_vs_S67322\|\|full | Low-quality |
| S10_vs_S67343\|\|full | Not-determined |
| S10_vs_S67382\|\|full | Not-determined |
| S10_vs_S67429\|\|full | Not-determined |
| S10_vs_S67436\|\|full | Low-quality |
| S5_extracted_k141_18461 | Medium-quality |
| S10_vs_S67518\|\|full | Not-determined |
| S10_vs_S67549\|\|full | Low-quality |
| S10_vs_S67588\|\|full | Not-determined |
| S10_vs_S67607\|\|full | Not-determined |
| S10_vs_S67611\|\|full | Low-quality |
| S10_vs_S67624\|\|full | Not-determined |
| S10_vs_S67625\|\|full | Low-quality |
| S10_vs_S67626\|\|full | Not-determined |
| S10_vs_S67643\|\|full | Not-determined |
| S10_vs_S67664\|\|full | Not-determined |
| S10_vs_S67667\|\|full | Low-quality |
| S10_vs_S67682\|\|full | Not-determined |
| S10_vs_S67691\|\|full | Low-quality |
| S10_vs_S67758\|\|full | Low-quality |
| S10_vs_S67770\|\|full | Not-determined |
| S10_vs_S67772\|\|full | Not-determined |
| S10_vs_S67781\|\|full | Low-quality |
| S10_vs_S67795\|\|full | Not-determined |
| S10_vs_S67858\|\|full | Not-determined |
| S10_vs_S67958\|\|full | Not-determined |
| S10_vs_S67983\|\|full | Not-determined |
| S10_vs_S68027\|\|full | Low-quality |
| S10_vs_S68036\|\|full | Not-determined |
| S10_vs_S68060\|\|full | Not-determined |
| S10_vs_S68062\|\|full | Low-quality |
| S10_vs_S68167\|\|full | Low-quality |
| S10_vs_S68211\|\|full | Low-quality |
| S10_vs_S68267\|\|full | Low-quality |
| S10_vs_S68299\|\|full | Low-quality |
| S10_vs_S68303\|\|full | Low-quality |
| S10_vs_S68378\|\|full | Low-quality |
| S10_vs_S68405\|\|full | Low-quality |
| S10_vs_S68442\|\|full | Low-quality |
| S10_vs_S68477\|\|full | Low-quality |
| S10_vs_S68510\|\|full | Low-quality |
| S10_vs_S68515\|\|full | Not-determined |
| S10_vs_S68653\|\|full | Low-quality |
| S10_vs_S68658\|\|full | Not-determined |
| S10_vs_S68674\|\|full | Low-quality |
| S10_vs_S68706\|\|full | Not-determined |
| S10_vs_S68746\|\|full | Not-determined |
| S10_vs_S68794\|\|full | Low-quality |
| S10_vs_S68799\|\|full | Not-determined |
| S10_vs_S68814\|\|full | Low-quality |
| S10_vs_S68898\|\|full | Low-quality |
| S10_vs_S68920\|\|full | Low-quality |
| S10_vs_S68967\|\|full | Low-quality |
| S10_vs_S68985\|\|full | Low-quality |
| S10_vs_S68997\|\|full | Low-quality |
| S10_vs_S69001\|\|full | Low-quality |
| S10_vs_S69030\|\|full | Low-quality |
| S10_vs_S69098\|\|full | Not-determined |
| S10_vs_S69112\|\|full | Low-quality |
| S10_vs_S69176\|\|full | Low-quality |
| S10_vs_S69189\|\|full | Low-quality |
| S10_vs_S69228\|\|full | Low-quality |
| S10_vs_S69242\|\|full | Low-quality |
| S10_vs_S69281\|\|full | Low-quality |
| S10_vs_S69294\|\|full | Not-determined |
| S10_vs_S69358\|\|full | Low-quality |
| S10_vs_S69415\|\|full | Not-determined |
| S10_vs_S69439\|\|full | Not-determined |
| S10_vs_S69520\|\|full | Low-quality |
| S10_vs_S69561\|\|full | Not-determined |
| S10_vs_S69596\|\|full | Not-determined |
| S10_vs_S69638\|\|full | Low-quality |
| S10_vs_S69671\|\|full | Not-determined |
| S10_vs_S69675\|\|full | Not-determined |
| S10_vs_S69703\|\|full | Low-quality |
| S10_vs_S69801\|\|full | Not-determined |
| S10_vs_S69845\|\|full | Not-determined |
| S10_vs_S69918\|\|full | Low-quality |
| S10_vs_S69933\|\|full | Low-quality |
| S10_vs_S70040\|\|full | Low-quality |
| S10_vs_S70082\|\|full | Not-determined |
| S10_vs_S70151\|\|full | Not-determined |
| S10_vs_S70155\|\|full | Not-determined |
| S10_vs_S70156\|\|full | Not-determined |
| S10_vs_S70182\|\|full | Low-quality |
| S10_vs_S70190\|\|full | Low-quality |
| S10_vs_S70194\|\|full | Not-determined |
| S10_vs_S70198\|\|full | Low-quality |
| S10_vs_S70222\|\|full | Not-determined |
| S10_vs_S70242\|\|full | Low-quality |
| S10_vs_S70247\|\|full | Not-determined |
| S10_vs_S70275\|\|full | Low-quality |
| S10_vs_S70386\|\|full | Low-quality |
| S10_vs_S70402\|\|full | Low-quality |
| S10_vs_S70411\|\|full | Low-quality |
| S10_vs_S70464\|\|full | Low-quality |
| S10_vs_S70521\|\|full | Not-determined |
| S10_vs_S70563\|\|full | Low-quality |
| S10_vs_S70606\|\|full | Low-quality |
| S10_vs_S70610\|\|full | Low-quality |
| S10_vs_S70704\|\|full | Low-quality |
| S10_vs_S70707\|\|full | Not-determined |
| S10_vs_S70720\|\|full | Low-quality |
| S10_vs_S70727\|\|full | Low-quality |
| S10_vs_S70845\|\|full | Not-determined |
| S10_vs_S71002\|\|full | Low-quality |
| S10_vs_S71003\|\|full | Not-determined |
| S10_vs_S71023\|\|full | Low-quality |
| S10_vs_S71029\|\|full | Low-quality |
| S10_vs_S71030\|\|full | Low-quality |
| S10_vs_S71091\|\|full | Not-determined |
| S10_vs_S71229\|\|full | Low-quality |
| S10_vs_S71236\|\|full | Low-quality |
| S10_vs_S71315\|\|full | Not-determined |
| S10_vs_S71396\|\|full | Not-determined |
| S10_vs_S71405\|\|full | Not-determined |
| S10_vs_S71470\|\|full | Low-quality |
| S10_vs_S71572\|\|full | Not-determined |
| S10_vs_S71584\|\|full | Not-determined |
| S10_vs_S71642\|\|full | Low-quality |
| S10_vs_S71659\|\|full | Low-quality |
| S10_vs_S71708\|\|full | Low-quality |
| S10_vs_S71731\|\|full | Not-determined |
| S10_vs_S71736\|\|full | Low-quality |
| S10_vs_S71739\|\|full | Not-determined |
| S10_vs_S71823\|\|full | Low-quality |
| S10_vs_S71863\|\|full | Low-quality |
| S10_vs_S71882\|\|full | Low-quality |
| S12_vs_k141_26847\|\|full | Medium-quality |
| S10_vs_S71911\|\|full | Not-determined |
| S10_vs_S71918\|\|full | Not-determined |
| S10_vs_S71985\|\|full | Low-quality |
| S10_vs_S72004\|\|full | Not-determined |
| S10_vs_S72019\|\|full | Low-quality |
| S10_vs_S72061\|\|full | Not-determined |
| S10_vs_S72075\|\|full | Not-determined |
| S10_vs_S72101\|\|full | Not-determined |
| S10_vs_S72116\|\|full | Not-determined |
| S10_vs_S72134\|\|full | Low-quality |
| S10_vs_S72166\|\|full | Low-quality |

| | |
|---|---|
| S10_vs_72177\|\|full | Low-quality |
| S10_vs_72217\|\|full | Not-determined |
| S10_vs_72224\|\|full | Low-quality |
| S10_vs_72239\|\|full | Low-quality |
| S10_vs_72263\|\|full | Not-determined |
| S10_vs_72395\|\|full | Low-quality |
| S10_vs_72432\|\|full | Not-determined |
| S10_vs_72483\|\|full | Not-determined |
| S10_vs_72495\|\|full | Low-quality |
| S10_vs_72525\|\|full | Low-quality |
| S10_vs_72574\|\|full | Not-determined |
| S10_vs_72590\|\|full | Low-quality |
| S10_vs_72632\|\|full | Low-quality |
| S10_vs_72667\|\|full | Low-quality |
| S10_vs_72713\|\|full | Low-quality |
| S10_vs_72768\|\|full | Not-determined |
| S10_vs_72801\|\|full | Not-determined |
| S10_vs_72879\|\|full | Low-quality |
| S10_vs_72953\|\|full | Low-quality |
| S10_vs_72962\|\|full | Low-quality |
| S10_vs_72981\|\|full | Not-determined |
| S10_vs_73001\|\|full | Not-determined |
| S10_vs_73039\|\|full | Not-determined |
| S10_vs_73129\|\|full | Low-quality |
| S10_vs_73144\|\|full | Low-quality |
| S10_vs_73191\|\|full | Low-quality |
| S10_vs_73218\|\|full | Low-quality |
| S10_vs_73229\|\|full | Low-quality |
| S10_vs_73238\|\|full | Not-determined |
| S10_vs_73292\|\|full | Low-quality |
| S10_vs_73341\|\|full | Low-quality |
| S10_vs_73354\|\|full | Low-quality |
| S10_vs_73370\|\|full | Low-quality |
| S10_vs_73379\|\|full | Low-quality |
| S10_vs_73397\|\|full | Low-quality |
| S10_vs_73482\|\|full | Low-quality |
| S10_vs_73521\|\|full | Not-determined |
| S10_vs_73543\|\|full | Low-quality |
| S10_vs_73566\|\|full | Low-quality |
| S10_vs_73637\|\|full | Low-quality |
| S10_vs_73688\|\|full | Not-determined |
| S10_vs_73745\|\|full | Low-quality |
| S10_vs_73781\|\|full | Low-quality |
| S10_vs_73794\|\|full | Not-determined |
| S10_vs_73878\|\|full | Not-determined |
| S10_vs_73979\|\|full | Not-determined |
| S10_vs_74019\|\|full | Not-determined |
| S10_vs_74028\|\|full | Not-determined |
| S10_vs_74087\|\|full | Low-quality |
| S10_vs_74095\|\|full | Low-quality |
| S10_vs_74110\|\|full | Not-determined |
| S10_vs_74121\|\|full | Not-determined |
| S10_vs_74216\|\|full | Not-determined |
| S10_vs_74263\|\|full | Low-quality |
| S10_vs_74332\|\|full | Low-quality |
| S10_vs_74415\|\|full | Low-quality |
| S10_vs_74460\|\|full | Low-quality |
| S10_vs_74463\|\|full | Not-determined |
| S10_vs_74543\|\|full | Low-quality |
| S10_vs_74563\|\|full | Not-determined |
| S10_vs_74701\|\|full | Low-quality |
| S10_vs_74752\|\|full | Low-quality |
| S10_vs_74792\|\|full | Low-quality |
| S10_vs_74897\|\|full | Low-quality |
| S10_vs_74899\|\|full | Not-determined |
| S10_vs_75035\|\|full | Not-determined |
| S10_vs_75036\|\|full | Low-quality |
| S10_vs_75163\|\|full | Not-determined |
| S10_vs_75187\|\|full | Not-determined |
| S10_vs_75194\|\|full | Low-quality |
| S10_vs_75228\|\|full | Low-quality |
| S10_vs_75238\|\|full | Low-quality |
| S10_vs_75252\|\|full | Low-quality |
| S10_vs_75305\|\|full | Low-quality |
| S10_vs_75336\|\|full | Low-quality |
| S10_vs_75338\|\|full | Not-determined |
| S10_vs_75365\|\|full | Low-quality |
| S10_vs_75424\|\|full | Low-quality |
| S10_vs_75439\|\|full | Low-quality |
| S10_vs_75543\|\|full | Low-quality |
| S10_vs_75590\|\|full | Not-determined |
| S10_vs_75630\|\|full | Not-determined |
| S10_vs_75682\|\|full | Not-determined |
| S10_vs_75757\|\|full | Not-determined |
| S10_vs_75786\|\|full | Low-quality |
| S10_vs_75810\|\|full | Low-quality |
| S10_vs_75847\|\|full | Low-quality |
| S10_vs_75931\|\|full | Not-determined |
| S10_vs_75935\|\|full | Not-determined |
| S10_vs_75942\|\|full | Low-quality |
| S10_vs_75957\|\|full | Not-determined |
| S10_vs_75970\|\|full | Not-determined |
| S10_vs_75977\|\|full | Low-quality |
| S10_vs_75981\|\|full | Low-quality |
| S10_vs_75992\|\|full | Low-quality |
| S10_vs_75993\|\|full | Low-quality |
| S10_vs_76048\|\|full | Not-determined |
| S10_vs_76139\|\|full | Not-determined |
| S10_vs_76166\|\|full | Not-determined |
| S10_vs_76213\|\|full | Not-determined |
| S10_vs_76241\|\|full | Not-determined |
| S10_vs_76265\|\|full | Low-quality |
| S10_vs_76272\|\|full | Not-determined |
| S10_vs_76281\|\|full | Not-determined |
| S10_vs_76289\|\|full | Low-quality |
| S10_vs_76305\|\|full | Low-quality |
| S10_vs_76332\|\|full | Low-quality |
| S10_vs_76336\|\|full | Low-quality |
| S10_vs_76398\|\|full | Not-determined |
| S10_vs_76464\|\|full | Low-quality |
| S10_vs_76491\|\|full | Low-quality |
| S10_vs_76493\|\|full | Low-quality |
| S10_vs_76504\|\|full | Low-quality |
| S10_vs_76509\|\|full | Not-determined |
| S10_vs_76545\|\|full | Low-quality |
| S10_vs_76647\|\|full | Not-determined |
| S10_vs_76660\|\|full | Not-determined |
| S10_vs_76664\|\|full | Not-determined |
| S10_vs_76746\|\|full | Low-quality |
| S10_vs_76787\|\|full | Low-quality |
| S10_vs_76831\|\|full | Low-quality |
| S10_vs_76854\|\|full | Not-determined |
| S10_vs_76911\|\|full | Low-quality |
| S10_vs_76928\|\|full | Low-quality |
| S10_vs_76938\|\|full | Low-quality |
| S10_vs_76963\|\|full | Low-quality |
| S9_vs_k141_44331\|\|full | Medium-quality |
| S10_vs_77029\|\|full | Low-quality |
| S10_vs_77228\|\|full | Not-determined |
| S10_vs_77235\|\|full | Low-quality |
| S10_vs_77269\|\|full | Low-quality |
| S10_vs_77274\|\|full | Not-determined |
| S10_vs_77284\|\|full | Low-quality |
| S10_vs_77325\|\|full | Low-quality |
| S10_vs_77363\|\|full | Not-determined |
| S10_vs_77401\|\|full | Not-determined |
| S10_vs_77403\|\|full | Not-determined |
| S10_vs_77429\|\|full | Low-quality |
| S10_vs_77432\|\|full | Low-quality |
| S10_vs_77460\|\|full | Low-quality |
| S10_vs_77554\|\|full | Not-determined |
| S10_vs_77656\|\|full | Not-determined |
| S10_vs_77663\|\|full | Low-quality |
| S10_vs_77672\|\|full | Low-quality |
| S10_vs_77680\|\|full | Low-quality |
| S10_vs_77726\|\|full | Low-quality |
| S10_vs_77760\|\|full | Low-quality |
| S10_vs_77839\|\|full | Not-determined |
| S10_vs_77848\|\|full | Low-quality |
| S10_vs_77929\|\|full | Low-quality |
| S10_vs_77969\|\|full | Not-determined |
| S10_vs_78080\|\|full | Not-determined |
| S10_vs_78098\|\|full | Not-determined |
| S10_vs_78104\|\|full | Low-quality |
| S10_vs_78176\|\|full | Not-determined |
| S10_vs_78203\|\|full | Not-determined |
| S10_vs_78217\|\|full | Not-determined |
| S10_vs_78267\|\|full | Low-quality |
| S10_vs_78281\|\|full | Not-determined |
| S10_vs_78312\|\|full | Low-quality |
| S10_vs_78483\|\|full | Low-quality |
| S10_vs_78490\|\|full | Not-determined |
| S10_vs_78510\|\|full | Low-quality |
| S10_vs_78527\|\|full | Low-quality |
| S10_vs_78539\|\|full | Not-determined |
| S10_vs_78555\|\|full | Low-quality |
| S10_vs_78557\|\|full | Low-quality |
| S10_vs_78565\|\|full | Not-determined |
| S10_vs_78603\|\|full | Low-quality |
| S10_vs_78653\|\|full | Low-quality |
| S10_vs_78657\|\|full | Not-determined |
| S10_vs_78666\|\|full | Low-quality |
| S10_vs_78689\|\|full | Low-quality |
| S10_vs_78693\|\|full | Low-quality |
| S10_vs_78704\|\|full | Not-determined |
| S10_vs_78745\|\|full | Low-quality |
| S10_vs_78757\|\|full | Low-quality |
| S10_vs_78818\|\|full | Low-quality |
| S10_vs_78830\|\|full | Low-quality |
| S10_vs_78872\|\|full | Not-determined |
| S10_vs_78919\|\|full | Low-quality |
| S10_vs_78956\|\|full | Not-determined |
| S10_vs_78983\|\|full | Low-quality |
| S10_vs_78988\|\|full | Not-determined |
| S10_vs_79013\|\|full | Low-quality |
| S10_vs_79053\|\|full | Low-quality |
| S10_vs_79062\|\|full | Low-quality |
| S10_vs_79064\|\|full | Low-quality |
| S10_vs_79191\|\|full | Not-determined |
| S10_vs_79237\|\|full | Not-determined |
| S10_vs_79283\|\|full | Low-quality |
| S10_vs_79412\|\|full | Low-quality |
| S10_vs_79418\|\|full | Not-determined |
| S10_vs_79429\|\|full | Not-determined |
| S10_vs_79492\|\|full | Low-quality |
| S10_vs_79518\|\|full | Not-determined |
| S10_vs_79521\|\|full | Not-determined |
| S10_vs_79534\|\|full | Not-determined |
| S10_vs_79541\|\|full | Not-determined |
| S10_vs_79584\|\|full | Not-determined |
| S10_vs_79667\|\|full | Low-quality |
| S10_vs_79688\|\|full | Low-quality |
| S10_vs_79705\|\|full | Low-quality |
| S10_vs_79718\|\|full | Low-quality |
| S10_vs_79721\|\|full | Not-determined |
| S10_vs_79763\|\|full | Not-determined |
| S10_vs_79807\|\|full | Low-quality |
| S10_vs_79814\|\|full | Low-quality |
| S10_vs_79821\|\|full | Low-quality |
| S10_vs_79844\|\|full | Low-quality |
| S10_vs_79867\|\|full | Low-quality |
| S10_vs_79912\|\|full | Low-quality |
| S10_vs_79968\|\|full | Low-quality |
| S10_vs_80002\|\|full | Low-quality |
| S10_vs_80009\|\|full | Low-quality |
| S10_vs_80011\|\|full | Low-quality |
| S10_vs_80026\|\|full | Not-determined |
| S10_vs_80035\|\|full | Low-quality |
| S10_vs_80063\|\|full | Low-quality |
| S10_vs_80106\|\|full | Low-quality |
| S10_vs_80107\|\|full | Low-quality |
| S10_vs_80128\|\|full | Low-quality |
| S10_vs_80202\|\|full | Low-quality |
| S10_vs_80213\|\|full | Low-quality |
| S10_vs_80216\|\|full | Not-determined |
| S10_vs_80306\|\|full | Low-quality |
| S10_vs_80314\|\|full | Not-determined |
| S10_vs_80342\|\|full | Low-quality |
| S10_vs_80355\|\|full | Low-quality |
| S10_vs_80381\|\|full | Not-determined |
| S10_vs_80409\|\|full | Not-determined |
| S10_vs_80411\|\|full | Low-quality |
| S10_vs_80427\|\|full | Not-determined |
| S10_vs_80447\|\|full | Low-quality |
| S10_vs_80452\|\|full | Low-quality |
| S10_vs_80498\|\|full | Not-determined |
| S10_vs_80512\|\|full | Not-determined |
| S10_vs_80527\|\|full | Low-quality |
| S10_vs_80547\|\|full | Low-quality |
| S10_vs_80556\|\|full | Not-determined |
| S10_vs_80559\|\|full | Not-determined |
| S10_vs_80580\|\|full | Not-determined |
| S10_vs_80589\|\|full | Low-quality |
| S10_vs_80595\|\|full | Not-determined |
| S10_vs_80628\|\|full | Low-quality |
| S10_vs_80645\|\|full | Low-quality |
| S10_vs_80687\|\|full | Low-quality |
| S5_vs_k141_18461\|\|full | Medium-quality |
| S10_vs_80717\|\|full | Low-quality |
| S10_vs_80923\|\|full | Not-determined |
| S10_vs_80933\|\|full | Low-quality |
| S10_vs_80967\|\|full | Not-determined |
| S10_vs_80995\|\|full | Low-quality |
| S10_vs_80999\|\|full | Low-quality |
| S10_vs_81007\|\|full | Low-quality |
| S10_vs_81025\|\|full | Low-quality |
| S10_vs_81126\|\|full | Low-quality |
| S10_vs_81137\|\|full | Low-quality |
| S10_vs_81214\|\|full | Not-determined |
| S10_vs_81293\|\|full | Low-quality |
| S10_vs_81305\|\|full | Low-quality |
| S10_vs_81340\|\|full | Not-determined |
| S10_vs_81360\|\|full | Not-determined |
| S10_vs_81393\|\|full | Not-determined |
| S10_vs_81417\|\|full | Not-determined |
| S10_vs_81456\|\|full | Not-determined |
| S10_vs_81457\|\|full | Not-determined |
| S10_vs_81521\|\|full | Low-quality |
| S10_vs_81562\|\|full | Not-determined |
| S10_vs_81570\|\|full | Low-quality |
| S10_vs_81581\|\|full | Low-quality |
| S10_vs_81614\|\|full | Not-determined |
| S10_vs_81618\|\|full | Low-quality |
| S10_vs_81743\|\|full | Not-determined |
| S10_vs_81757\|\|full | Low-quality |
| S10_vs_81791\|\|full | Low-quality |
| S10_vs_81829\|\|full | Not-determined |
| S10_vs_81869\|\|full | Low-quality |
| S10_vs_81933\|\|full | Not-determined |
| S10_vs_81935\|\|full | Low-quality |
| S10_vs_81943\|\|full | Not-determined |
| S10_vs_81988\|\|full | Not-determined |
| S10_vs_82011\|\|full | Not-determined |
| S10_vs_82029\|\|full | Low-quality |
| S10_vs_82069\|\|full | Low-quality |
| S10_vs_82090\|\|full | Low-quality |
| S10_vs_82119\|\|full | Not-determined |
| S10_vs_82166\|\|full | Low-quality |
| S10_vs_82208\|\|full | Not-determined |
| S10_vs_82232\|\|full | Not-determined |
| S10_vs_82234\|\|full | Low-quality |
| S10_vs_82248\|\|full | Low-quality |
| S10_vs_82285\|\|full | Not-determined |
| S10_vs_82286\|\|full | Low-quality |
| S10_vs_82295\|\|full | Not-determined |
| S10_vs_82334\|\|full | Low-quality |
| S10_vs_82343\|\|full | Low-quality |
| S10_vs_82418\|\|full | Not-determined |
| S10_vs_82435\|\|full | Not-determined |
| S10_vs_82437\|\|full | Low-quality |
| S10_vs_82450\|\|full | Low-quality |
| S10_vs_82503\|\|full | Not-determined |
| S10_vs_82512\|\|full | Not-determined |
| S10_vs_82560\|\|full | Low-quality |
| S10_vs_82571\|\|full | Not-determined |
| S10_vs_82633\|\|full | Not-determined |
| S10_vs_82667\|\|full | Not-determined |
| S10_vs_82754\|\|full | Not-determined |
| S10_vs_82760\|\|full | Not-determined |
| S10_vs_82796\|\|full | Not-determined |
| S10_vs_82851\|\|full | Not-determined |
| S10_vs_82887\|\|full | Not-determined |
| S10_vs_82921\|\|full | Not-determined |
| S10_vs_82963\|\|full | Not-determined |
| S10_vs_82978\|\|full | Low-quality |
| S10_vs_82996\|\|full | Low-quality |
| S10_vs_83040\|\|full | Not-determined |
| S10_vs_83049\|\|full | Low-quality |
| S10_vs_83052\|\|full | Not-determined |
| S10_vs_83057\|\|full | Low-quality |
| S10_vs_83083\|\|full | Low-quality |
| S10_vs_83217\|\|full | Low-quality |
| S10_vs_83223\|\|full | Low-quality |
| S10_vs_83265\|\|full | Low-quality |
| S10_vs_83289\|\|full | Not-determined |
| S10_vs_83342\|\|full | Not-determined |
| S10_vs_83366\|\|full | Not-determined |
| S10_vs_83400\|\|full | Low-quality |
| S10_vs_83423\|\|full | Not-determined |
| S10_vs_83462\|\|full | Not-determined |
| S10_vs_83538\|\|full | Low-quality |
| S10_vs_83548\|\|full | Not-determined |
| S10_vs_83585\|\|full | Not-determined |
| S10_vs_83619\|\|full | Not-determined |
| S10_vs_83642\|\|full | Low-quality |
| S10_vs_83674\|\|full | Low-quality |
| S10_vs_83731\|\|full | Low-quality |
| S10_vs_83748\|\|full | Not-determined |
| S10_vs_83787\|\|full | Low-quality |
| S10_vs_83796\|\|full | Not-determined |
| S10_vs_83898\|\|full | Low-quality |
| S10_vs_83907\|\|full | Low-quality |
| S10_vs_83918\|\|full | Not-determined |
| S10_vs_83920\|\|full | Not-determined |
| S10_vs_83963\|\|full | Low-quality |
| S10_vs_84001\|\|full | Low-quality |
| S10_vs_84051\|\|full | Not-determined |
| S10_vs_84082\|\|full | Not-determined |
| S10_vs_84102\|\|full | Low-quality |
| S10_vs_84124\|\|full | Not-determined |
| S10_vs_84149\|\|full | Low-quality |
| S10_vs_84150\|\|full | Not-determined |
| S10_vs_84183\|\|full | Low-quality |
| S10_vs_84224\|\|full | Low-quality |
| S10_vs_84271\|\|full | Low-quality |
| S10_vs_84284\|\|full | Low-quality |
| S10_vs_84288\|\|full | Low-quality |
| S10_vs_84336\|\|full | Not-determined |
| S10_vs_84366\|\|full | Not-determined |
| S10_vs_84429\|\|full | Low-quality |
| S10_vs_84431\|\|full | Not-determined |
| S10_vs_84686\|\|full | Not-determined |
| S10_vs_84692\|\|full | Low-quality |
| S10_vs_84694\|\|full | Not-determined |
| S10_vs_84713\|\|full | Not-determined |
| S10_vs_84760\|\|full | Not-determined |
| S10_vs_84820\|\|full | Low-quality |
| S10_vs_84822\|\|full | Not-determined |
| S10_vs_84880\|\|full | Not-determined |
| S10_vs_84891\|\|full | Low-quality |
| S10_vs_84893\|\|full | Low-quality |
| S10_vs_84897\|\|full | Low-quality |
| S10_vs_84953\|\|full | Low-quality |
| S10_vs_84964\|\|full | Low-quality |
| S10_vs_85023\|\|full | Low-quality |
| S10_vs_85061\|\|full | Low-quality |
| S10_vs_85069\|\|full | Low-quality |
| S10_vs_85083\|\|full | Not-determined |
| S10_vs_85084\|\|full | Low-quality |
| S10_vs_85085\|\|full | Not-determined |
| S10_vs_85090\|\|full | Not-determined |
| S10_vs_85091\|\|full | Not-determined |
| S10_vs_85092\|\|full | Not-determined |
| S10_vs_85118\|\|full | Low-quality |
| S10_vs_85124\|\|full | Low-quality |
| S10_vs_85156\|\|full | Not-determined |
| S10_vs_85235\|\|full | Not-determined |
| S10_vs_85347\|\|full | Not-determined |
| S10_vs_85350\|\|full | Not-determined |
| S10_vs_85362\|\|full | Low-quality |
| S10_vs_85380\|\|full | Low-quality |
| S10_vs_85384\|\|full | Low-quality |
| S10_vs_85387\|\|full | Not-determined |
| S10_vs_85569\|\|full | Low-quality |
| S10_vs_85685\|\|full | Low-quality |
| S10_vs_85687\|\|full | Low-quality |
| S10_vs_85757\|\|full | Low-quality |
| S10_vs_85768\|\|full | Not-determined |
| S10_vs_85782\|\|full | Not-determined |
| S10_vs_85791\|\|full | Low-quality |
| S10_vs_85820\|\|full | Low-quality |
| S10_vs_85834\|\|full | Not-determined |
| S10_vs_85836\|\|full | Low-quality |
| S10_vs_85852\|\|full | Low-quality |
| S10_vs_85896\|\|full | Low-quality |
| S10_vs_85920\|\|full | Low-quality |
| S10_vs_86014\|\|full | Low-quality |
| S10_vs_86036\|\|full | Low-quality |
| S10_vs_86052\|\|full | Low-quality |
| S10_vs_86082\|\|full | Low-quality |
| S10_vs_86085\|\|full | Low-quality |
| S10_vs_86092\|\|full | Low-quality |
| S10_vs_86197\|\|full | Not-determined |
| S10_vs_86211\|\|full | Low-quality |
| S10_vs_86255\|\|full | Not-determined |
| S10_vs_86294\|\|full | Not-determined |
| S10_vs_86300\|\|full | Not-determined |
| S10_vs_86333\|\|full | Low-quality |
| S10_vs_86346\|\|full | Low-quality |
| S10_vs_86403\|\|full | Low-quality |
| S10_vs_86458\|\|full | Not-determined |
| S10_vs_86480\|\|full | Low-quality |
| S10_vs_86604\|\|full | Not-determined |
| S10_vs_86673\|\|full | Not-determined |
| S10_vs_86711\|\|full | Not-determined |
| S10_vs_86716\|\|full | Low-quality |
| S10_vs_86740\|\|full | Low-quality |
| S10_vs_86745\|\|full | Not-determined |
| S10_vs_86796\|\|full | Low-quality |
| S10_vs_86840\|\|full | Low-quality |
| S10_vs_86852\|\|full | Not-determined |
| S10_vs_86864\|\|full | Not-determined |
| S10_vs_86883\|\|full | Low-quality |
| S10_vs_86963\|\|full | Low-quality |
| S10_vs_86987\|\|full | Low-quality |
| S10_vs_87009\|\|full | Not-determined |
| S10_vs_87051\|\|full | Not-determined |
| S10_vs_87074\|\|full | Low-quality |
| S10_vs_87160\|\|full | Low-quality |
| S10_vs_87162\|\|full | Not-determined |
| S10_vs_87178\|\|full | Low-quality |
| S10_vs_87250\|\|full | Not-determined |
| S10_vs_87292\|\|full | Low-quality |
| S10_vs_87378\|\|full | Not-determined |
| S5_vs_k141_28263\|\|full | Medium-quality |
| S10_vs_87467\|\|full | Low-quality |
| S10_vs_87555\|\|full | Not-determined |
| S10_vs_87561\|\|full | Not-determined |
| S10_vs_87600\|\|full | Not-determined |
| S10_vs_87613\|\|full | Not-determined |
| S10_vs_87667\|\|full | Low-quality |
| S10_vs_87738\|\|full | Low-quality |
| S10_vs_87748\|\|full | Not-determined |
| S10_vs_87763\|\|full | Low-quality |
| S10_vs_87837\|\|full | Not-determined |
| S10_vs_87897\|\|full | Low-quality |
| S10_vs_87933\|\|full | Not-determined |
| S10_vs_88022\|\|full | Low-quality |
| S10_vs_88034\|\|full | Not-determined |
| S10_vs_88078\|\|full | Low-quality |
| S10_vs_88082\|\|full | Low-quality |
| S10_vs_88098\|\|full | Low-quality |
| S10_vs_88110\|\|full | Not-determined |
| S10_vs_88113\|\|full | Low-quality |
| S10_vs_88119\|\|full | Low-quality |
| S10_vs_88120\|\|full | Not-determined |
| S10_vs_88137\|\|full | Not-determined |
| S10_vs_88194\|\|full | Low-quality |
| S10_vs_88202\|\|full | Low-quality |
| S10_vs_88215\|\|full | Low-quality |
| S10_vs_88264\|\|full | Low-quality |
| S10_vs_88266\|\|full | Not-determined |
| S10_vs_88294\|\|full | Not-determined |
| S10_vs_88354\|\|full | Low-quality |
| S10_vs_88374\|\|full | Low-quality |
| S10_vs_88397\|\|full | Not-determined |
| S10_vs_88419\|\|full | Low-quality |
| S10_vs_88429\|\|full | Low-quality |
| S10_vs_88484\|\|full | Not-determined |
| S10_vs_88519\|\|full | Low-quality |
| S10_vs_88546\|\|full | Not-determined |
| S10_vs_88576\|\|full | Low-quality |
| S10_vs_88716\|\|full | Not-determined |
| S10_vs_88740\|\|full | Not-determined |
| S10_vs_88794\|\|full | Not-determined |
| S10_vs_88872\|\|full | Not-determined |
| S10_vs_88896\|\|full | Low-quality |
| S10_vs_88942\|\|full | Not-determined |
| S3_extracted_k141_127408 | Complete |
| S10_vs_89108\|\|full | Not-determined |
| S10_vs_89129\|\|full | Low-quality |
| S10_vs_89160\|\|full | Low-quality |
| S10_vs_89255\|\|full | Not-determined |
| S10_vs_89371\|\|full | Low-quality |
| S10_vs_89396\|\|full | Low-quality |
| S10_vs_89399\|\|full | Not-determined |
| S10_vs_89427\|\|full | Not-determined |
| S10_vs_89462\|\|full | Not-determined |
| S10_vs_89464\|\|full | Low-quality |
| S10_vs_89468\|\|full | Low-quality |
| S10_vs_89521\|\|full | Low-quality |
| S10_vs_89553\|\|full | Low-quality |
| S10_vs_89590\|\|full | Not-determined |
| S10_vs_89595\|\|full | Low-quality |
| S10_vs_89667\|\|full | Not-determined |
| S10_vs_89690\|\|full | Not-determined |
| S10_vs_89710\|\|full | Low-quality |
| S10_vs_89729\|\|full | Low-quality |
| S10_vs_89760\|\|full | Low-quality |
| S10_vs_89767\|\|full | Not-determined |
| S10_vs_89819\|\|full | Low-quality |
| S10_vs_89828\|\|full | Low-quality |
| S10_vs_89834\|\|full | Low-quality |
| S10_vs_89977\|\|full | Low-quality |
| S10_vs_90069\|\|full | Low-quality |
| S10_vs_90084\|\|full | Not-determined |
| S10_vs_90086\|\|full | Low-quality |
| S10_vs_90132\|\|full | Not-determined |
| S10_vs_90209\|\|full | Not-determined |
| S10_vs_90211\|\|full | Low-quality |
| S10_vs_90257\|\|full | Low-quality |
| S10_vs_90264\|\|full | Not-determined |
| S10_vs_90284\|\|full | Low-quality |
| S10_vs_90363\|\|full | Low-quality |
| S10_vs_90365\|\|full | Not-determined |
| S10_vs_90408\|\|full | Not-determined |
| S10_vs_90410\|\|full | Not-determined |
| S10_vs_90452\|\|full | Not-determined |
| S10_vs_90462\|\|full | Low-quality |
| S10_vs_90497\|\|full | Not-determined |
| S10_vs_90579\|\|full | Low-quality |
| S10_vs_90660\|\|full | Low-quality |
| S10_vs_90661\|\|full | Low-quality |
| S10_vs_90702\|\|full | Not-determined |
| S10_vs_90704\|\|full | Not-determined |
| S10_vs_90872\|\|full | Not-determined |
| S10_vs_90917\|\|full | Low-quality |
| S10_vs_91051\|\|full | Low-quality |
| S10_vs_91075\|\|full | Low-quality |
| S10_vs_91082\|\|full | Not-determined |
| S10_vs_91084\|\|full | Low-quality |
| S10_vs_91091\|\|full | Not-determined |
| S10_vs_91228\|\|full | Low-quality |
| S10_vs_91346\|\|full | Not-determined |
| S10_vs_91354\|\|full | Not-determined |
| S10_vs_91365\|\|full | Low-quality |
| S10_vs_91561\|\|full | Low-quality |
| S10_vs_91566\|\|full | Low-quality |
| S10_vs_91677\|\|full | Low-quality |
| S10_vs_91730\|\|full | Low-quality |
| S10_vs_91758\|\|full | Low-quality |
| S10_vs_91839\|\|full | Low-quality |
| S10_vs_91843\|\|full | Low-quality |
| S10_vs_91867\|\|full | Low-quality |
| S10_vs_91871\|\|full | Low-quality |
| S10_vs_91917\|\|full | Low-quality |
| S10_vs_91927\|\|full | Low-quality |
| S10_vs_92005\|\|full | Low-quality |
| S10_vs_92079\|\|full | Low-quality |
| S10_vs_92084\|\|full | Low-quality |
| S10_vs_92088\|\|full | Low-quality |
| S10_vs_92184\|\|full | Not-determined |
| S10_vs_92185\|\|full | Low-quality |
| S10_vs_92351\|\|full | Not-determined |
| S10_vs_92412\|\|full | Low-quality |
| S10_vs_92430\|\|full | Low-quality |
| S10_vs_92467\|\|full | Low-quality |
| S10_vs_92487\|\|full | Low-quality |
| S10_vs_92492\|\|full | Low-quality |
| S10_vs_92493\|\|full | Low-quality |
| S10_vs_92511\|\|full | Low-quality |
| S10_vs_92518\|\|full | Low-quality |
| S10_vs_92534\|\|full | Low-quality |
| S10_vs_92547\|\|full | Low-quality |
| S10_vs_92564\|\|full | Low-quality |
| S10_vs_92637\|\|full | Not-determined |
| S10_vs_92650\|\|full | Low-quality |
| S10_vs_92720\|\|full | Not-determined |
| S10_vs_92721\|\|full | Not-determined |
| S10_vs_92748\|\|full | Low-quality |
| S10_vs_92752\|\|full | Low-quality |
| S10_vs_92770\|\|full | Not-determined |
| S10_vs_92798\|\|full | Low-quality |
| S10_vs_92804\|\|full | Not-determined |
| S10_vs_92814\|\|full | Low-quality |
| S10_vs_92888\|\|full | Not-determined |
| S10_vs_92896\|\|full | Low-quality |
| S10_vs_92928\|\|full | Not-determined |
| S10_vs_93022\|\|full | Low-quality |

| | |
|---|---|
| S10_vs_93044\|\|full | Low-quality |
| S10_vs_93084\|\|full | Not-determined |
| S10_vs_93126\|\|full | Low-quality |
| S10_vs_93391\|\|full | Not-determined |
| S10_vs_93431\|\|full | Not-determined |
| S10_vs_93479\|\|full | Not-determined |
| S10_vs_93551\|\|full | Low-quality |
| S10_vs_93578\|\|full | Not-determined |
| S10_vs_93603\|\|full | Not-determined |
| S10_vs_93636\|\|full | Low-quality |
| S10_vs_93697\|\|full | Not-determined |
| S10_vs_93700\|\|full | Low-quality |
| S10_vs_93763\|\|full | Not-determined |
| S10_vs_93837\|\|full | Low-quality |
| S10_vs_93879\|\|full | Low-quality |
| S10_vs_93894\|\|full | Low-quality |
| S10_vs_93910\|\|full | Low-quality |
| S10_vs_93912\|\|full | Not-determined |
| S10_vs_93951\|\|full | Low-quality |
| S10_vs_93959\|\|full | Not-determined |
| S10_vs_93976\|\|full | Low-quality |
| S10_vs_94007\|\|full | Not-determined |
| S10_vs_94138\|\|full | Not-determined |
| S10_vs_94170\|\|full | Not-determined |
| S10_vs_94317\|\|full | Not-determined |
| S10_vs_94363\|\|full | Not-determined |
| S10_vs_94395\|\|full | Low-quality |
| S10_vs_94475\|\|full | Low-quality |
| S10_vs_94482\|\|full | Low-quality |
| S10_vs_94489\|\|full | Low-quality |
| S10_vs_94555\|\|full | Low-quality |
| S10_vs_94592\|\|full | Low-quality |
| S10_vs_94600\|\|full | Low-quality |
| S10_vs_94607\|\|full | Low-quality |
| S10_vs_94625\|\|full | Low-quality |
| S10_vs_94644\|\|full | Not-determined |
| S10_vs_94746\|\|full | Low-quality |
| S10_vs_94797\|\|full | Not-determined |
| S10_vs_94839\|\|full | Low-quality |
| S10_vs_94851\|\|full | Low-quality |
| S10_vs_94873\|\|full | Not-determined |
| S10_vs_94973\|\|full | Low-quality |
| S10_vs_95098\|\|full | Not-determined |
| S10_vs_95236\|\|full | Not-determined |
| S10_vs_95263\|\|full | Not-determined |
| S10_vs_95312\|\|full | Low-quality |
| S10_vs_95357\|\|full | Not-determined |
| S10_vs_95358\|\|full | Not-determined |
| S10_vs_95374\|\|full | Low-quality |
| S10_vs_95402\|\|full | Low-quality |
| S10_vs_95452\|\|full | Not-determined |
| S10_vs_95459\|\|full | Low-quality |
| S10_vs_95472\|\|full | Low-quality |
| S10_vs_95495\|\|full | Not-determined |
| S10_vs_95609\|\|full | Not-determined |
| S10_vs_95651\|\|full | Low-quality |
| S10_vs_95731\|\|full | Low-quality |
| S10_vs_95732\|\|full | Low-quality |
| S10_vs_95773\|\|full | Not-determined |
| S10_vs_95831\|\|full | Not-determined |
| S10_vs_95893\|\|full | Low-quality |
| S10_vs_95954\|\|full | Not-determined |
| S10_vs_96018\|\|full | Not-determined |
| S10_vs_96081\|\|full | Not-determined |
| S10_vs_96118\|\|full | Low-quality |
| S10_vs_96121\|\|full | Low-quality |
| S10_vs_96174\|\|full | Low-quality |
| S10_vs_96230\|\|full | Not-determined |
| S10_vs_96277\|\|full | Not-determined |
| S10_vs_96340\|\|full | Not-determined |
| S10_vs_96347\|\|full | Low-quality |
| S10_vs_96422\|\|full | Not-determined |
| S10_vs_96437\|\|full | Not-determined |
| S10_vs_96462\|\|full | Not-determined |
| S10_vs_96505\|\|full | Low-quality |
| S10_vs_96517\|\|full | Not-determined |
| S10_vs_96631\|\|full | Low-quality |
| S10_vs_96678\|\|full | Not-determined |
| S10_vs_96688\|\|full | Not-determined |
| S10_vs_96771\|\|full | Low-quality |
| S10_vs_96772\|\|full | Low-quality |
| S10_vs_96773\|\|full | Not-determined |
| S10_vs_96780\|\|full | Low-quality |
| S10_vs_96824\|\|full | Not-determined |
| S10_vs_96835\|\|full | Low-quality |
| S10_vs_96947\|\|full | Low-quality |
| S10_vs_97032\|\|full | Low-quality |
| S10_vs_97038\|\|full | Not-determined |
| S10_vs_97143\|\|full | Not-determined |
| S10_vs_97158\|\|full | Low-quality |
| S10_vs_97208\|\|full | Not-determined |
| S10_vs_97221\|\|full | Not-determined |
| S10_vs_97316\|\|full | Low-quality |
| S10_vs_97320\|\|full | Not-determined |
| S10_vs_97352\|\|full | Not-determined |
| S10_vs_97360\|\|full | Low-quality |
| S10_vs_97366\|\|full | Not-determined |
| S10_vs_97375\|\|full | Low-quality |
| S10_vs_97402\|\|full | Low-quality |
| S10_vs_97420\|\|full | Not-determined |
| S10_vs_97425\|\|full | Not-determined |
| S10_vs_97441\|\|full | Low-quality |
| S10_vs_97524\|\|full | Low-quality |
| S10_vs_97554\|\|full | Low-quality |
| S10_vs_97559\|\|full | Low-quality |
| S10_vs_97568\|\|full | Low-quality |
| S10_vs_97593\|\|full | Not-determined |
| S10_vs_97611\|\|full | Low-quality |
| S10_vs_97625\|\|full | Low-quality |
| S10_vs_97629\|\|full | Not-determined |
| S10_vs_97680\|\|full | Not-determined |
| S10_vs_97712\|\|full | Not-determined |
| S10_vs_97719\|\|full | Low-quality |
| S10_vs_97778\|\|full | Not-determined |
| S10_vs_97822\|\|full | Not-determined |
| S10_vs_97823\|\|full | Not-determined |
| S10_vs_97862\|\|full | Low-quality |
| S10_vs_97865\|\|full | Low-quality |
| S10_vs_97915\|\|full | Low-quality |
| S10_vs_97947\|\|full | Not-determined |
| S10_vs_97996\|\|full | Not-determined |
| S10_vs_98006\|\|full | Not-determined |
| S10_vs_98099\|\|full | Low-quality |
| S10_vs_98153\|\|full | Not-determined |
| S10_vs_98169\|\|full | Low-quality |
| S10_vs_98293\|\|full | Low-quality |
| S10_vs_98317\|\|full | Not-determined |
| S10_vs_98318\|\|full | Low-quality |
| S10_vs_98325\|\|full | Not-determined |
| S10_vs_98339\|\|full | Low-quality |
| S10_vs_98340\|\|full | Not-determined |
| S10_vs_98359\|\|full | Low-quality |
| S10_vs_98407\|\|full | Not-determined |
| S10_vs_98456\|\|full | Low-quality |
| S10_vs_98469\|\|full | Low-quality |
| S10_vs_98527\|\|full | Not-determined |
| S10_vs_98573\|\|full | Low-quality |
| S10_vs_98662\|\|full | Not-determined |
| S10_vs_98675\|\|full | Low-quality |
| S10_vs_98692\|\|full | Low-quality |
| S10_vs_98846\|\|full | Low-quality |
| S10_vs_98853\|\|full | Low-quality |
| S10_vs_98883\|\|full | Low-quality |
| S10_vs_98904\|\|full | Low-quality |
| S10_vs_98905\|\|full | Not-determined |
| S10_vs_98909\|\|full | Not-determined |
| S10_vs_98926\|\|full | Low-quality |
| S10_vs_98957\|\|full | Low-quality |
| S10_vs_98959\|\|full | Not-determined |
| S10_vs_99008\|\|full | Low-quality |
| S10_vs_99083\|\|full | Low-quality |
| S10_vs_99092\|\|full | Low-quality |
| S10_vs_99163\|\|full | Not-determined |
| S10_vs_99193\|\|full | Not-determined |
| S10_vs_99207\|\|full | Low-quality |
| S10_vs_99208\|\|full | Not-determined |
| S10_vs_99216\|\|full | Low-quality |
| S10_vs_99250\|\|full | Not-determined |
| S10_vs_99263\|\|full | Low-quality |
| S10_vs_99267\|\|full | Not-determined |
| S10_vs_99307\|\|full | Not-determined |
| S10_vs_99407\|\|full | Low-quality |
| S10_vs_99459\|\|full | Low-quality |
| S10_vs_99482\|\|full | Not-determined |
| S10_vs_99521\|\|full | Not-determined |
| S10_vs_99530\|\|full | Low-quality |
| S10_vs_99562\|\|full | Not-determined |
| S10_vs_99582\|\|full | Not-determined |
| S10_vs_99609\|\|full | Not-determined |
| S10_vs_99672\|\|full | Low-quality |
| S10_vs_99694\|\|full | Low-quality |
| S10_vs_99730\|\|full | Low-quality |
| S10_vs_99822\|\|full | Not-determined |
| S10_vs_99852\|\|full | Low-quality |
| S10_vs_99919\|\|full | Not-determined |
| S10_vs_99922\|\|full | Low-quality |
| S10_vs_99933\|\|full | Low-quality |
| S10_vs_99943\|\|full | Low-quality |
| S10_vs_99944\|\|full | Low-quality |
| S10_vs_100019\|\|full | Low-quality |
| S10_vs_100068\|\|full | Not-determined |
| S10_vs_100090\|\|full | Not-determined |
| S10_vs_100103\|\|full | Not-determined |
| S10_vs_100131\|\|full | Not-determined |
| S10_vs_100313\|\|full | Not-determined |
| S10_vs_100391\|\|full | Not-determined |
| S10_vs_100440\|\|full | Low-quality |
| S10_vs_100742\|\|full | Not-determined |
| S10_vs_100837\|\|full | Low-quality |
| S10_vs_100868\|\|full | Not-determined |
| S10_vs_100929\|\|full | Low-quality |
| S10_vs_100937\|\|full | Low-quality |
| S10_vs_100972\|\|full | Not-determined |
| S10_vs_100986\|\|full | Not-determined |
| S10_vs_100998\|\|full | Low-quality |
| S10_vs_101000\|\|full | Low-quality |
| S10_vs_101066\|\|full | Low-quality |
| S10_vs_101095\|\|full | Not-determined |
| S10_vs_101146\|\|full | Low-quality |
| S10_vs_101150\|\|full | Low-quality |
| S10_vs_101170\|\|full | Not-determined |
| S10_vs_101201\|\|full | Not-determined |
| S10_vs_101293\|\|full | Low-quality |
| S10_vs_101298\|\|full | Not-determined |
| S10_vs_101376\|\|full | Low-quality |
| S10_vs_101445\|\|full | Low-quality |
| S10_vs_101479\|\|full | Not-determined |
| S10_vs_101522\|\|full | Low-quality |
| S10_vs_101535\|\|full | Not-determined |
| S10_vs_101612\|\|full | Not-determined |
| S10_vs_101681\|\|full | Low-quality |
| S10_vs_101684\|\|full | Low-quality |
| S10_vs_101693\|\|full | Low-quality |
| S10_vs_101697\|\|full | Not-determined |
| S10_vs_101702\|\|full | Not-determined |
| S10_vs_101738\|\|full | Low-quality |
| S10_vs_101747\|\|full | Low-quality |
| S10_vs_101749\|\|full | Not-determined |
| S10_vs_101843\|\|full | Not-determined |
| S10_vs_101856\|\|full | Low-quality |
| S10_vs_101858\|\|full | Low-quality |
| S10_vs_101903\|\|full | Not-determined |
| S10_vs_101913\|\|full | Low-quality |
| S10_vs_101942\|\|full | Not-determined |
| S10_vs_101949\|\|full | Low-quality |
| S10_vs_101957\|\|full | Not-determined |
| S10_vs_101981\|\|full | Low-quality |
| S10_vs_102086\|\|full | Low-quality |
| S10_vs_102127\|\|full | Not-determined |
| S10_vs_102178\|\|full | Not-determined |
| S10_vs_102216\|\|full | Low-quality |
| S10_vs_102253\|\|full | Not-determined |
| S10_vs_102292\|\|full | Low-quality |
| S10_vs_102315\|\|full | Low-quality |
| S10_vs_102339\|\|full | Not-determined |
| S10_vs_102355\|\|full | Not-determined |
| S10_vs_102368\|\|full | Not-determined |
| S10_vs_102378\|\|full | Low-quality |
| S10_vs_102388\|\|full | Low-quality |
| S10_vs_102389\|\|full | Not-determined |
| S10_vs_102416\|\|full | Not-determined |
| S10_vs_102447\|\|full | Not-determined |
| S10_vs_102454\|\|full | Low-quality |
| S10_vs_102460\|\|full | Low-quality |
| S10_vs_102488\|\|full | Low-quality |
| S10_vs_102502\|\|full | Low-quality |
| S10_vs_102625\|\|full | Low-quality |
| S10_vs_102630\|\|full | Low-quality |
| S10_vs_102633\|\|full | Not-determined |
| S10_vs_102644\|\|full | Low-quality |
| S10_vs_102688\|\|full | Not-determined |
| S10_vs_102707\|\|full | Low-quality |
| S10_vs_102745\|\|full | Not-determined |
| S10_vs_102884\|\|full | Low-quality |
| S10_vs_102891\|\|full | Low-quality |
| S10_vs_102965\|\|full | Not-determined |
| S10_vs_102982\|\|full | Low-quality |
| S10_vs_103046\|\|full | Low-quality |
| S10_vs_103085\|\|full | Not-determined |
| S10_vs_103190\|\|full | Low-quality |
| S10_vs_103266\|\|full | Not-determined |
| S10_vs_103267\|\|full | Not-determined |
| S10_vs_103280\|\|full | Low-quality |
| S10_vs_103285\|\|full | Low-quality |
| S10_vs_103404\|\|full | Low-quality |
| S10_vs_103420\|\|full | Low-quality |
| S10_vs_103421\|\|full | Low-quality |
| S10_vs_103423\|\|full | Low-quality |
| S10_vs_103462\|\|full | Low-quality |
| S10_vs_103550\|\|full | Low-quality |
| S10_vs_103557\|\|full | Low-quality |
| S10_vs_103563\|\|full | Not-determined |
| S10_vs_103581\|\|full | Not-determined |
| S10_vs_103709\|\|full | Not-determined |
| S10_vs_103716\|\|full | Low-quality |
| S10_vs_103735\|\|full | Low-quality |
| S10_vs_103738\|\|full | Low-quality |
| S10_vs_103740\|\|full | Low-quality |
| S10_vs_103760\|\|full | Low-quality |
| S10_vs_103769\|\|full | Low-quality |
| S10_vs_103827\|\|full | Low-quality |
| S10_vs_103886\|\|full | Low-quality |
| S10_vs_103901\|\|full | Low-quality |
| S10_vs_103945\|\|full | Low-quality |
| S10_vs_103958\|\|full | Low-quality |
| S10_vs_103973\|\|full | Low-quality |
| S10_vs_104037\|\|full | Low-quality |
| S10_vs_104054\|\|full | Low-quality |
| S10_vs_104069\|\|full | Low-quality |
| S10_vs_104113\|\|full | Not-determined |
| S10_vs_104152\|\|full | Low-quality |
| S10_vs_104165\|\|full | Low-quality |
| S10_vs_104188\|\|full | Low-quality |
| S10_vs_104198\|\|full | Low-quality |
| S10_vs_104244\|\|full | Low-quality |
| S10_vs_104245\|\|full | Not-determined |
| S10_vs_104279\|\|full | Low-quality |
| S10_vs_104304\|\|full | Not-determined |
| S10_vs_104342\|\|full | Not-determined |
| S10_vs_104380\|\|full | Low-quality |
| S10_vs_104455\|\|full | Not-determined |
| S10_vs_104463\|\|full | Not-determined |
| S10_vs_104517\|\|full | Low-quality |
| S10_vs_104536\|\|full | Not-determined |
| S10_vs_104537\|\|full | Not-determined |
| S10_vs_104550\|\|full | Not-determined |
| S10_vs_104554\|\|full | Low-quality |
| S10_vs_104571\|\|full | Not-determined |
| S10_vs_104589\|\|full | Low-quality |
| S10_vs_104628\|\|full | Not-determined |
| S10_vs_104644\|\|full | Not-determined |
| S10_vs_104654\|\|full | Low-quality |
| S10_vs_104660\|\|full | Low-quality |
| S10_vs_104671\|\|full | Not-determined |
| S10_vs_104775\|\|full | Low-quality |
| S10_vs_104844\|\|full | Low-quality |
| S10_vs_104909\|\|full | Not-determined |
| S10_vs_104922\|\|full | Not-determined |
| S10_vs_104990\|\|full | Not-determined |
| S10_vs_105067\|\|full | Not-determined |
| S10_vs_105068\|\|full | Not-determined |
| S10_vs_105091\|\|full | Low-quality |
| S10_vs_105117\|\|full | Low-quality |
| S10_vs_105137\|\|full | Low-quality |
| S10_vs_105157\|\|full | Low-quality |
| S10_vs_105164\|\|full | Not-determined |
| S10_vs_105183\|\|full | Not-determined |
| S10_vs_105200\|\|full | Low-quality |
| S10_vs_105202\|\|full | Low-quality |
| S10_vs_105226\|\|full | Not-determined |
| S10_vs_105239\|\|full | Low-quality |
| S10_vs_105251\|\|full | Low-quality |
| S10_vs_105331\|\|full | Low-quality |
| S10_vs_105399\|\|full | Not-determined |
| S10_vs_105415\|\|full | Not-determined |
| S10_vs_105429\|\|full | Not-determined |
| S10_vs_105451\|\|full | Low-quality |
| S10_vs_105458\|\|full | Not-determined |
| S10_vs_105547\|\|full | Not-determined |
| S10_vs_105597\|\|full | Not-determined |
| S10_vs_105689\|\|full | Not-determined |
| S10_vs_105832\|\|full | Not-determined |
| S10_vs_105862\|\|full | Low-quality |
| S10_vs_105873\|\|full | Not-determined |
| S10_vs_106024\|\|full | Not-determined |
| S10_vs_106047\|\|full | Low-quality |
| S10_vs_106078\|\|full | Low-quality |
| S10_vs_106086\|\|full | Not-determined |
| S10_vs_106090\|\|full | Not-determined |
| S10_vs_106107\|\|full | Not-determined |
| S10_vs_106193\|\|full | Low-quality |
| S10_vs_106197\|\|full | Not-determined |
| S10_vs_106209\|\|full | Low-quality |
| S10_vs_106229\|\|full | Low-quality |
| S10_vs_106371\|\|full | Low-quality |
| S10_vs_106373\|\|full | Not-determined |
| S10_vs_106453\|\|full | Low-quality |
| S10_vs_106525\|\|full | Low-quality |
| S10_vs_106537\|\|full | Low-quality |
| S10_vs_106591\|\|full | Not-determined |
| S10_vs_106625\|\|full | Not-determined |
| S10_vs_106627\|\|full | Low-quality |
| S10_vs_106631\|\|full | Not-determined |
| S10_vs_106645\|\|full | Low-quality |
| S10_vs_106690\|\|full | Not-determined |
| S10_vs_106735\|\|full | Low-quality |
| S10_vs_106736\|\|full | Low-quality |
| S10_vs_106737\|\|full | Low-quality |
| S10_vs_106751\|\|full | Low-quality |
| S10_vs_106787\|\|full | Not-determined |
| S10_vs_106825\|\|full | Not-determined |
| S3_vs_k141_3335\|\|full | Medium-quality |
| S10_vs_106838\|\|full | Low-quality |
| S10_vs_106971\|\|full | Low-quality |
| S10_vs_107012\|\|full | Low-quality |
| S10_vs_107054\|\|full | Not-determined |
| S10_vs_107093\|\|full | Not-determined |
| S10_vs_107131\|\|full | Not-determined |
| S10_vs_107173\|\|full | Low-quality |
| S10_vs_107190\|\|full | Low-quality |
| S10_vs_107201\|\|full | Not-determined |
| S10_vs_107213\|\|full | Low-quality |
| S10_vs_107254\|\|full | Low-quality |
| S10_vs_107260\|\|full | Low-quality |
| S10_vs_107307\|\|full | Not-determined |
| S10_vs_107312\|\|full | Low-quality |
| S10_vs_107320\|\|full | Low-quality |
| S10_vs_107335\|\|full | Low-quality |
| S10_vs_107369\|\|full | Not-determined |
| S10_vs_107402\|\|full | Not-determined |
| S10_vs_107423\|\|full | Not-determined |
| S10_vs_107424\|\|full | Not-determined |
| S10_vs_107441\|\|full | Not-determined |
| S10_vs_107465\|\|full | Low-quality |
| S10_vs_107470\|\|full | Low-quality |
| S10_vs_107477\|\|full | Not-determined |
| S10_vs_107517\|\|full | Not-determined |
| S10_vs_107526\|\|full | Low-quality |
| S10_vs_107576\|\|full | Low-quality |
| S10_vs_107577\|\|full | Not-determined |
| S10_vs_107588\|\|full | Low-quality |
| S10_vs_107693\|\|full | Low-quality |
| S10_vs_107823\|\|full | Low-quality |
| S10_vs_107853\|\|full | Low-quality |
| S10_vs_107890\|\|full | Not-determined |
| S10_vs_107991\|\|full | Low-quality |
| S10_vs_108016\|\|full | Low-quality |
| S10_vs_108044\|\|full | Low-quality |
| S10_vs_108048\|\|full | Not-determined |
| S10_vs_108072\|\|full | Low-quality |
| S10_vs_108096\|\|full | Low-quality |
| S10_vs_108137\|\|full | Not-determined |
| S10_vs_108208\|\|full | Not-determined |
| S10_vs_108285\|\|full | Not-determined |
| S10_vs_108363\|\|full | Low-quality |
| S10_vs_108372\|\|full | Not-determined |
| S10_vs_108376\|\|full | Low-quality |
| S10_vs_108435\|\|full | Not-determined |
| S10_vs_108439\|\|full | Not-determined |
| S10_vs_108477\|\|full | Low-quality |
| S10_vs_108489\|\|full | Not-determined |
| S10_vs_108525\|\|full | Not-determined |
| S10_vs_108532\|\|full | Not-determined |
| S10_vs_108565\|\|full | Low-quality |
| S10_vs_108652\|\|full | Not-determined |
| S10_vs_108670\|\|full | Not-determined |
| S10_vs_108708\|\|full | Not-determined |
| S10_vs_108735\|\|full | Low-quality |
| S10_vs_108825\|\|full | Not-determined |
| S10_vs_108840\|\|full | Not-determined |
| S10_vs_108870\|\|full | Low-quality |
| S10_vs_108884\|\|full | Not-determined |
| S10_vs_108886\|\|full | Low-quality |
| S10_vs_108927\|\|full | Low-quality |
| S10_vs_108991\|\|full | Low-quality |
| S10_vs_109000\|\|full | Low-quality |
| S10_vs_109056\|\|full | Low-quality |
| S10_vs_109090\|\|full | Low-quality |
| S10_vs_109119\|\|full | Not-determined |
| S10_vs_109134\|\|full | Low-quality |
| S10_vs_109162\|\|full | Low-quality |
| S10_vs_109169\|\|full | Not-determined |
| S10_vs_109209\|\|full | Not-determined |
| S10_vs_109253\|\|full | Low-quality |
| S10_vs_109342\|\|full | Low-quality |
| S10_vs_109377\|\|full | Not-determined |
| S10_vs_109385\|\|full | Not-determined |
| S10_vs_109398\|\|full | Not-determined |
| S10_vs_109406\|\|full | Low-quality |
| S10_vs_109454\|\|full | Low-quality |
| S10_vs_109468\|\|full | Not-determined |
| S10_vs_109470\|\|full | Low-quality |
| S10_vs_109490\|\|full | Low-quality |
| S10_vs_109569\|\|full | Low-quality |
| S10_vs_109594\|\|full | Low-quality |
| S10_vs_109686\|\|full | Not-determined |
| S10_vs_109731\|\|full | Not-determined |
| S10_vs_109744\|\|full | Low-quality |
| S10_vs_109773\|\|full | Not-determined |
| S10_vs_109788\|\|full | Low-quality |
| S10_vs_109821\|\|full | Low-quality |
| S10_vs_109867\|\|full | Not-determined |
| S10_vs_109884\|\|full | Low-quality |
| S10_vs_110070\|\|full | Low-quality |
| S10_vs_110074\|\|full | Low-quality |
| S10_vs_110099\|\|full | Not-determined |
| S10_vs_110106\|\|full | Low-quality |
| S10_vs_110111\|\|full | Not-determined |
| S10_vs_110138\|\|full | Low-quality |
| S10_vs_110152\|\|full | Low-quality |
| S10_vs_110167\|\|full | Not-determined |
| S10_vs_110184\|\|full | Low-quality |
| S10_vs_110195\|\|full | Low-quality |
| S10_vs_110202\|\|full | Low-quality |
| S10_vs_110208\|\|full | Low-quality |
| S10_vs_110305\|\|full | Not-determined |
| S10_vs_110361\|\|full | Low-quality |
| S10_vs_110390\|\|full | Low-quality |
| S10_vs_110437\|\|full | Not-determined |
| S10_vs_110456\|\|full | Low-quality |
| S10_vs_110502\|\|full | Low-quality |
| S10_vs_110520\|\|full | Not-determined |
| S10_vs_110542\|\|full | Low-quality |
| S10_vs_110553\|\|full | Not-determined |
| S10_vs_110569\|\|full | Not-determined |
| S10_vs_110658\|\|full | Not-determined |
| S10_vs_110697\|\|full | Low-quality |
| S10_vs_110709\|\|full | Low-quality |
| S10_vs_110762\|\|full | Not-determined |
| S10_vs_110796\|\|full | Not-determined |
| S10_vs_110812\|\|full | Low-quality |
| S10_vs_110835\|\|full | Not-determined |
| S10_vs_110851\|\|full | Low-quality |
| S10_vs_110892\|\|full | Low-quality |
| S10_vs_110943\|\|full | Low-quality |
| S10_vs_110997\|\|full | Low-quality |
| S10_vs_111041\|\|full | Low-quality |
| S10_vs_111067\|\|full | Low-quality |
| S10_vs_111143\|\|full | Low-quality |
| S10_vs_111170\|\|full | Low-quality |
| S10_vs_111232\|\|full | Low-quality |
| S10_vs_111340\|\|full | Low-quality |
| S10_vs_111371\|\|full | Not-determined |
| S10_vs_111439\|\|full | Low-quality |
| S10_vs_111557\|\|full | Not-determined |
| S10_vs_111564\|\|full | Not-determined |
| S10_vs_111586\|\|full | Low-quality |
| S10_vs_111603\|\|full | Not-determined |
| S10_vs_111703\|\|full | Low-quality |
| S10_vs_111719\|\|full | Low-quality |
| S10_vs_111768\|\|full | Not-determined |
| S10_vs_112023\|\|full | Not-determined |
| S10_vs_112125\|\|full | Low-quality |
| S10_vs_112152\|\|full | Low-quality |
| S10_vs_112224\|\|full | Not-determined |
| S10_vs_112285\|\|full | Low-quality |
| S10_vs_112309\|\|full | Low-quality |
| S10_vs_112316\|\|full | Low-quality |
| S10_vs_112317\|\|full | Low-quality |
| S10_vs_112378\|\|full | Not-determined |
| S10_vs_112484\|\|full | Low-quality |
| S10_vs_112496\|\|full | Not-determined |
| S10_vs_112497\|\|full | Not-determined |
| S10_vs_112524\|\|full | Not-determined |
| S10_vs_112525\|\|full | Not-determined |
| S10_vs_112570\|\|full | Low-quality |
| S10_vs_112580\|\|full | Low-quality |
| S10_vs_112614\|\|full | Low-quality |
| S10_vs_112631\|\|full | Low-quality |
| S10_vs_112725\|\|full | Low-quality |
| S10_vs_112729\|\|full | Low-quality |
| S10_vs_112732\|\|full | Low-quality |
| S10_vs_112848\|\|full | Low-quality |
| S10_vs_112861\|\|full | Not-determined |
| S10_vs_112942\|\|full | Low-quality |
| S10_vs_112964\|\|full | Not-determined |
| S10_vs_112999\|\|full | Low-quality |
| S10_vs_113021\|\|full | Low-quality |
| S10_vs_113072\|\|full | Low-quality |
| S10_vs_113182\|\|full | Low-quality |
| S10_vs_113218\|\|full | Not-determined |
| S10_vs_113241\|\|full | Not-determined |
| S10_vs_113297\|\|full | Low-quality |
| S10_vs_113330\|\|full | Low-quality |
| S10_vs_113364\|\|full | Not-determined |
| S10_vs_113371\|\|full | Low-quality |
| S10_vs_113405\|\|full | Not-determined |
| S3_vs_k141_127408\|\|full | High-quality |
| S10_vs_113620\|\|full | Low-quality |
| S10_vs_113717\|\|full | Low-quality |
| S10_vs_113719\|\|full | Low-quality |
| S10_vs_113730\|\|full | Not-determined |
| S10_vs_113732\|\|full | Not-determined |
| S10_vs_113791\|\|full | Low-quality |
| S10_vs_113842\|\|full | Low-quality |
| S10_vs_113884\|\|full | Low-quality |
| S10_vs_113914\|\|full | Low-quality |
| S10_vs_113955\|\|full | Not-determined |
| S10_vs_113962\|\|full | Not-determined |
| S10_vs_113999\|\|full | Low-quality |
| S10_vs_114047\|\|full | Not-determined |
| S10_vs_114061\|\|full | Low-quality |
| S10_vs_114072\|\|full | Low-quality |
| S10_vs_114073\|\|full | Low-quality |
| S10_vs_114077\|\|full | Not-determined |
| S10_vs_114093\|\|full | Low-quality |
| S10_vs_114144\|\|full | Low-quality |
| S10_vs_114153\|\|full | Low-quality |
| S10_vs_114188\|\|full | Low-quality |
| S10_vs_114200\|\|full | Low-quality |
| S10_vs_114217\|\|full | Not-determined |
| S10_vs_114219\|\|full | Low-quality |
| S10_vs_114234\|\|full | Low-quality |
| S10_vs_114248\|\|full | Low-quality |
| S10_vs_114388\|\|full | Not-determined |
| S10_vs_114397\|\|full | Not-determined |
| S10_vs_114405\|\|full | Not-determined |
| S10_vs_114417\|\|full | Low-quality |
| S10_vs_114418\|\|full | Not-determined |
| S10_vs_114479\|\|full | Low-quality |
| S10_vs_114527\|\|full | Not-determined |
| S10_vs_114583\|\|full | Not-determined |
| S10_vs_114606\|\|full | Not-determined |
| S10_vs_114618\|\|full | Low-quality |
| S10_vs_114656\|\|full | Not-determined |
| S10_vs_114673\|\|full | Not-determined |
| S10_vs_114791\|\|full | Low-quality |
| S10_vs_114820\|\|full | Low-quality |
| S10_vs_114860\|\|full | Low-quality |

S10_vs_114966||full Not-determined
S10_vs_115003||full Not-determined
S10_vs_115015||full Not-determined
S10_vs_115033||full Not-determined
S10_vs_115054||full Low-quality
S10_vs_115060||full Not-determined
S10_vs_115133||full Low-quality
S10_vs_115155||full Not-determined
S10_vs_115210||full Low-quality
S10_vs_115231||full Not-determined
S10_vs_115253||full Not-determined
S10_vs_115295||full Low-quality
S10_vs_115314||full Not-determined
S10_vs_115321||full Low-quality
S10_vs_115361||full Not-determined
S10_vs_115374||full Low-quality
S10_vs_115452||full Low-quality
S10_vs_115501||full Not-determined
S10_vs_115537||full Not-determined
S10_vs_115545||full Not-determined
S10_vs_115577||full Low-quality
S10_vs_115610||full Not-determined
S10_vs_115722||full Not-determined
S10_vs_115737||full Low-quality
S10_vs_115787||full Low-quality
S10_vs_115798||full Not-determined
S10_vs_115833||full Low-quality
S10_vs_115859||full Low-quality
S10_vs_115877||full Not-determined
S10_vs_115903||full Not-determined
S10_vs_115935||full Low-quality
S10_vs_116031||full Low-quality
S10_vs_116047||full Not-determined
S10_vs_116092||full Not-determined
S10_vs_116127||full Not-determined
S10_vs_116137||full Not-determined
S10_vs_116151||full Low-quality
S10_vs_116167||full Not-determined
S10_vs_116200||full Low-quality
S10_vs_116234||full Low-quality
S10_vs_116237||full Low-quality
S10_vs_116257||full Low-quality
S10_vs_116287||full Not-determined
S10_vs_116297||full Not-determined
S10_vs_116352||full Low-quality
S10_vs_116365||full Not-determined
S10_vs_116426||full Not-determined
S10_vs_116474||full Not-determined
S10_vs_116551||full Not-determined
S10_vs_116576||full Low-quality
S10_vs_116578||full Not-determined
S10_vs_116625||full Low-quality
S10_vs_116636||full Not-determined
S10_vs_116670||full Not-determined
S10_vs_116681||full Not-determined
S10_vs_116684||full Low-quality
S10_vs_116718||full Low-quality
S10_vs_116759||full Low-quality
S10_vs_116779||full Not-determined
S10_vs_116800||full Low-quality
S10_vs_116925||full Low-quality
S10_vs_116946||full Low-quality
S10_vs_116949||full Low-quality
S10_vs_116966||full Not-determined
S10_vs_116968||full Low-quality
S10_vs_116993||full Low-quality
S10_vs_116997||full Not-determined
S10_vs_117019||full Not-determined
S10_vs_117022||full Low-quality
S10_vs_117048||full Low-quality
S10_vs_117123||full Low-quality
S10_vs_117150||full Not-determined
S10_vs_117165||full Low-quality
S10_vs_117206||full Low-quality
S10_vs_117254||full Low-quality
S10_vs_117351||full Low-quality
S10_vs_117383||full Not-determined
S10_vs_117411||full Low-quality
S10_vs_117414||full Not-determined
S10_vs_117447||full Low-quality
S10_vs_117496||full Low-quality
S10_vs_117499||full Low-quality
S10_vs_117571||full Low-quality
S10_vs_117586||full Low-quality
S10_vs_117600||full Low-quality
S10_vs_117641||full Low-quality
S10_vs_117699||full Not-determined
S10_vs_117836||full Not-determined
S10_vs_117854||full Low-quality
S10_vs_117921||full Not-determined
S10_vs_117922||full Low-quality
S10_vs_117945||full Not-determined
S10_vs_117969||full Low-quality
S10_vs_118090||full Not-determined
S10_vs_118148||full Low-quality
S10_vs_118179||full Low-quality
S10_vs_118184||full Not-determined
S10_vs_118189||full Not-determined
S10_vs_118236||full Low-quality
S10_vs_118253||full Low-quality
S10_vs_118254||full Not-determined
S10_vs_118320||full Not-determined
S10_vs_118352||full Low-quality
S10_vs_118365||full Low-quality
S10_vs_118435||full Low-quality
S10_vs_118446||full Not-determined
S10_vs_118493||full Not-determined
S10_vs_118542||full Low-quality
S10_vs_118575||full Not-determined
S10_vs_118580||full Low-quality
S10_vs_118639||full Low-quality
S10_vs_118641||full Low-quality
S10_vs_118666||full Low-quality
S10_vs_118724||full Low-quality
S10_vs_118746||full Low-quality
S10_vs_118770||full Low-quality
S10_vs_118776||full Not-determined
S10_vs_118923||full Low-quality
S10_vs_118957||full Low-quality
S10_vs_118980||full Low-quality
S10_vs_119001||full Low-quality
S10_vs_119008||full Low-quality
S10_vs_119022||full Not-determined
S10_vs_119035||full Not-determined
S10_vs_119084||full Not-determined
S10_vs_119100||full Low-quality
S10_vs_119118||full Not-determined
S10_vs_119161||full Not-determined
S10_vs_119168||full Low-quality
S10_vs_119220||full Not-determined
S10_vs_119245||full Not-determined
S10_vs_119250||full Low-quality
S10_vs_119265||full Low-quality
S14_vs_k141_96111||full Medium-quality
S10_vs_119308||full Low-quality
S10_vs_119317||full Not-determined
S10_vs_119376||full Not-determined
S10_vs_119389||full Not-determined
S10_vs_119435||full Low-quality
S10_vs_119492||full Low-quality
S10_vs_119523||full Not-determined
S10_vs_119559||full Low-quality
S10_vs_119575||full Low-quality
S10_vs_119628||full Not-determined
S10_vs_119640||full Low-quality
S10_vs_119715||full Low-quality
S10_vs_119727||full Low-quality
S10_vs_119797||full Low-quality
S10_vs_119812||full Low-quality
S10_vs_119822||full Not-determined
S10_vs_119833||full Not-determined
S10_vs_119874||full Low-quality
S10_vs_119900||full Low-quality
S10_vs_119947||full Not-determined
S10_vs_119953||full Not-determined
S10_vs_120079||full Low-quality
S10_vs_120114||full Not-determined
S10_vs_120115||full Not-determined
S10_vs_120118||full Low-quality
S10_vs_120193||full Not-determined
S10_vs_120212||full Not-determined
S10_vs_120214||full Not-determined
S10_vs_120250||full Low-quality
S10_vs_120255||full Low-quality
S10_vs_120366||full Not-determined
S10_vs_120367||full Not-determined
S10_vs_120410||full Not-determined
S10_vs_120421||full Low-quality
S10_vs_120431||full Not-determined
S10_vs_120490||full Low-quality
S10_vs_120517||full Low-quality
S10_vs_120542||full Low-quality
S10_vs_120576||full Low-quality
S10_vs_120611||full Low-quality
S10_vs_120624||full Not-determined
S10_vs_120734||full Not-determined
S10_vs_120749||full Not-determined
S10_vs_120755||full Not-determined
S10_vs_120767||full Low-quality
S10_vs_120799||full Not-determined
S10_vs_120955||full Low-quality
S10_vs_120959||full Low-quality
S10_vs_121014||full Not-determined
S10_vs_121040||full Low-quality
S10_vs_121042||full Not-determined
S10_vs_121078||full Low-quality
S10_vs_121091||full Low-quality
S10_vs_121194||full Not-determined
S10_vs_121252||full Low-quality
S10_vs_12211||0_partial Low-quality
S10_vs_13905||0_partial Not-determined
S10_vs_17793||0_partial Low-quality
S10_vs_20642||0_partial Not-determined
S10_vs_21758||0_partial Not-determined
S10_vs_33805||0_partial Not-determined
S10_vs_42162||0_partial Not-determined
S10_vs_52220||0_partial Low-quality
S10_vs_62668||0_partial Low-quality
S10_vs_62668||1_partial Not-determined
S10_vs_68606||0_partial Low-quality
S10_vs_70031||0_partial Not-determined
S10_vs_83125||0_partial Not-determined
S10_vs_87403||0_partial Low-quality
S10_vs_93079||0_partial Low-quality
S10_vs_98758||0_partial Low-quality
S10_vs_99476||0_partial Low-quality
S10_vs_1||lt2gene Not-determined
S10_vs_82||lt2gene Not-determined
S10_vs_282||lt2gene Low-quality
S10_vs_623||lt2gene Low-quality
S10_vs_633||lt2gene Low-quality
S10_vs_662||lt2gene Low-quality
S10_vs_676||lt2gene Low-quality
S10_vs_788||lt2gene Low-quality
S10_vs_1147||lt2gene Low-quality
S10_vs_1275||lt2gene Low-quality
S10_vs_1314||lt2gene Low-quality
S10_vs_1440||lt2gene Low-quality
S10_vs_1471||lt2gene Low-quality
S10_vs_1542||lt2gene Low-quality
S10_vs_1559||lt2gene Low-quality
S10_vs_1867||lt2gene Low-quality
S10_vs_1878||lt2gene Not-determined
S10_vs_1897||lt2gene Low-quality
S10_vs_1924||lt2gene Low-quality
S10_vs_2171||lt2gene Low-quality
S10_vs_2205||lt2gene Not-determined
S10_vs_2383||lt2gene Low-quality
S10_vs_2730||lt2gene Low-quality
S10_vs_2818||lt2gene Not-determined
S10_vs_2847||lt2gene Low-quality
S10_vs_2872||lt2gene Low-quality
S10_vs_3052||lt2gene Low-quality
S10_vs_3249||lt2gene Low-quality
S10_vs_3340||lt2gene Low-quality
S10_vs_3486||lt2gene Low-quality
S10_vs_3554||lt2gene Low-quality
S10_vs_3591||lt2gene Low-quality
S10_vs_3606||lt2gene Low-quality
S10_vs_3800||lt2gene Not-determined
S10_vs_3871||lt2gene Not-determined
S10_vs_3916||lt2gene Low-quality
S10_vs_4128||lt2gene Not-determined
S10_vs_4235||lt2gene Not-determined
S10_vs_4331||lt2gene Low-quality
S10_vs_4354||lt2gene Low-quality
S10_vs_4774||lt2gene Low-quality
S10_vs_4849||lt2gene Low-quality
S10_vs_4851||lt2gene Low-quality
S10_vs_4902||lt2gene Not-determined
S10_vs_5049||lt2gene Low-quality
S10_vs_5088||lt2gene Not-determined
S10_vs_5282||lt2gene Low-quality
S10_vs_5284||lt2gene Low-quality
S10_vs_5308||lt2gene Low-quality
S10_vs_5332||lt2gene Low-quality
S10_vs_5360||lt2gene Low-quality
S10_vs_5474||lt2gene Not-determined
S10_vs_5579||lt2gene Low-quality
S10_vs_5623||lt2gene Low-quality
S10_vs_5670||lt2gene Not-determined
S10_vs_5745||lt2gene Low-quality
S10_vs_5756||lt2gene Low-quality
S10_vs_5937||lt2gene Low-quality
S10_vs_6010||lt2gene Low-quality
S10_vs_6011||lt2gene Low-quality
S10_vs_6255||lt2gene Low-quality
S10_vs_6331||lt2gene Low-quality
S10_vs_6338||lt2gene Low-quality
S10_vs_6363||lt2gene Low-quality
S10_vs_6410||lt2gene Not-determined
S10_vs_6420||lt2gene Low-quality
S10_vs_6446||lt2gene Low-quality
S10_vs_6449||lt2gene Low-quality
S10_vs_6569||lt2gene Low-quality
S10_vs_6595||lt2gene Low-quality
S10_vs_6681||lt2gene Low-quality
S10_vs_6736||lt2gene Not-determined
S10_vs_6806||lt2gene Not-determined
S10_vs_6808||lt2gene Low-quality
S10_vs_6837||lt2gene Low-quality
S10_vs_6887||lt2gene Low-quality
S10_vs_6922||lt2gene Low-quality
S10_vs_7054||lt2gene Low-quality
S10_vs_7165||lt2gene Low-quality
S10_vs_7255||lt2gene Not-determined
S10_vs_7399||lt2gene Low-quality
S10_vs_7407||lt2gene Low-quality
S10_vs_7419||lt2gene Low-quality
S10_vs_7430||lt2gene Low-quality
S10_vs_7583||lt2gene Low-quality
S10_vs_7942||lt2gene Low-quality
S10_vs_8036||lt2gene Low-quality
S10_vs_8245||lt2gene Not-determined
S10_vs_8263||lt2gene Low-quality
S10_vs_8299||lt2gene Not-determined
S10_vs_8313||lt2gene Low-quality
S10_vs_8473||lt2gene Not-determined
S10_vs_8523||lt2gene Low-quality
S10_vs_8607||lt2gene Low-quality
S10_vs_8862||lt2gene Low-quality
S10_vs_8897||lt2gene Low-quality
S10_vs_9026||lt2gene Not-determined
S10_vs_9132||lt2gene Not-determined
S10_vs_9157||lt2gene Low-quality
S10_vs_9240||lt2gene Not-determined
S10_vs_9252||lt2gene Low-quality
S10_vs_9317||lt2gene Not-determined
S10_vs_9342||lt2gene Not-determined
S10_vs_9362||lt2gene Low-quality
S10_vs_9380||lt2gene Low-quality
S10_vs_9431||lt2gene Low-quality
S10_vs_9442||lt2gene Low-quality
S10_vs_9444||lt2gene Low-quality
S10_vs_9602||lt2gene Low-quality
S10_vs_9796||lt2gene Low-quality
S10_vs_9844||lt2gene Low-quality
S10_vs_10125||lt2gene Not-determined
S10_vs_10198||lt2gene Low-quality
S10_vs_10348||lt2gene Not-determined
S10_vs_10690||lt2gene Not-determined
S10_vs_10727||lt2gene Low-quality
S10_vs_10837||lt2gene Low-quality
S10_vs_10905||lt2gene Low-quality
S10_vs_10930||lt2gene Not-determined
S10_vs_11349||lt2gene Low-quality
S10_vs_11384||lt2gene Not-determined
S10_vs_11438||lt2gene Low-quality
S10_vs_11449||lt2gene Low-quality
S10_vs_11732||lt2gene Low-quality
S10_vs_11736||lt2gene Low-quality
S10_vs_11969||lt2gene Low-quality
S10_vs_11971||lt2gene Low-quality
S10_vs_11972||lt2gene Low-quality
S10_vs_12005||lt2gene Not-determined
S10_vs_12086||lt2gene Not-determined
S10_vs_12091||lt2gene Low-quality
S10_vs_12125||lt2gene Low-quality
S10_vs_12193||lt2gene Not-determined
S10_vs_12258||lt2gene Low-quality
S10_vs_12316||lt2gene Low-quality
S10_vs_12400||lt2gene Low-quality
S10_vs_12421||lt2gene Low-quality
S10_vs_12476||lt2gene Not-determined
S10_vs_12485||lt2gene Low-quality
S10_vs_12494||lt2gene Low-quality
S10_vs_12523||lt2gene Low-quality
S10_vs_12892||lt2gene Low-quality
S10_vs_13039||lt2gene Low-quality
S10_vs_13180||lt2gene Low-quality
S10_vs_13379||lt2gene Low-quality
S10_vs_13443||lt2gene Low-quality
S10_vs_13488||lt2gene Low-quality
S10_vs_13859||lt2gene Low-quality
S10_vs_14036||lt2gene Low-quality
S10_vs_14161||lt2gene Low-quality
S10_vs_14182||lt2gene Not-determined
S10_vs_14385||lt2gene Not-determined
S10_vs_14415||lt2gene Low-quality
S10_vs_14445||lt2gene Low-quality
S10_vs_14477||lt2gene Low-quality
S10_vs_14680||lt2gene Not-determined
S10_vs_14755||lt2gene Low-quality
S10_vs_14773||lt2gene Low-quality
S10_vs_14836||lt2gene Low-quality
S10_vs_14864||lt2gene Low-quality
S10_vs_15091||lt2gene Low-quality
S10_vs_15101||lt2gene Low-quality
S10_vs_15205||lt2gene Low-quality
S10_vs_15712||lt2gene Low-quality
S10_vs_15741||lt2gene Low-quality
S10_vs_15768||lt2gene Low-quality
S10_vs_16261||lt2gene Low-quality
S10_vs_16713||lt2gene Low-quality
S10_vs_16766||lt2gene Not-determined
S10_vs_16865||lt2gene Low-quality
S10_vs_17081||lt2gene Low-quality
S10_vs_17126||lt2gene Low-quality
S10_vs_17176||lt2gene Low-quality
S10_vs_17195||lt2gene Not-determined
S10_vs_17303||lt2gene Not-determined
S10_vs_17450||lt2gene Low-quality
S10_vs_17664||lt2gene Not-determined
S10_vs_17727||lt2gene Low-quality
S10_vs_17759||lt2gene Low-quality
S10_vs_17867||lt2gene Not-determined
S10_vs_17937||lt2gene Low-quality
S10_vs_18023||lt2gene Not-determined
S10_vs_18465||lt2gene Low-quality
S10_vs_18541||lt2gene Low-quality
S10_vs_18690||lt2gene Low-quality
S10_vs_18696||lt2gene Low-quality
S10_vs_18790||lt2gene Low-quality
S10_vs_18856||lt2gene Low-quality
S10_vs_18886||lt2gene Low-quality
S10_vs_18888||lt2gene Low-quality
S10_vs_19048||lt2gene Low-quality
S10_vs_19056||lt2gene Low-quality
S10_vs_19275||lt2gene Low-quality
S10_vs_19377||lt2gene Low-quality
S10_vs_19436||lt2gene Low-quality
S10_vs_19490||lt2gene Low-quality
S10_vs_19653||lt2gene Not-determined
S10_vs_19674||lt2gene Low-quality
S10_vs_19744||lt2gene Low-quality
S10_vs_19754||lt2gene Low-quality
S10_vs_19802||lt2gene Low-quality
S10_vs_19816||lt2gene Low-quality
S10_vs_19864||lt2gene Low-quality
S10_vs_19992||lt2gene Low-quality
S10_vs_20055||lt2gene Not-determined
S10_vs_20070||lt2gene Not-determined
S10_vs_20130||lt2gene Low-quality
S10_vs_20197||lt2gene Low-quality
S10_vs_20230||lt2gene Low-quality
S10_vs_20349||lt2gene Not-determined
S10_vs_20363||lt2gene Low-quality
S10_vs_20393||lt2gene Low-quality
S10_vs_20466||lt2gene Low-quality
S10_vs_20502||lt2gene Low-quality
S10_vs_20528||lt2gene Low-quality
S10_vs_20529||lt2gene Low-quality
S10_vs_20664||lt2gene Not-determined
S10_vs_20675||lt2gene Not-determined
S10_vs_20752||lt2gene Low-quality
S10_vs_20803||lt2gene Not-determined
S10_vs_20983||lt2gene Not-determined
S10_vs_21028||lt2gene Low-quality
S10_vs_21105||lt2gene Not-determined
S10_vs_21166||lt2gene Not-determined
S10_vs_21287||lt2gene Low-quality
S10_vs_21450||lt2gene Low-quality
S10_vs_21492||lt2gene Not-determined
S10_vs_21590||lt2gene Low-quality
S10_vs_21609||lt2gene Low-quality
S10_vs_21793||lt2gene Low-quality
S10_vs_21811||lt2gene Low-quality
S10_vs_21872||lt2gene Not-determined
S10_vs_22006||lt2gene Low-quality
S10_vs_22159||lt2gene Not-determined
S10_vs_22187||lt2gene Low-quality
S10_vs_22278||lt2gene Low-quality
S10_vs_22401||lt2gene Low-quality
S10_vs_22518||lt2gene Low-quality
S10_vs_22617||lt2gene Low-quality
S10_vs_22702||lt2gene Low-quality
S10_vs_22797||lt2gene Not-determined
S10_vs_22875||lt2gene Low-quality
S10_vs_22879||lt2gene Low-quality
S10_vs_23057||lt2gene Not-determined
S10_vs_23102||lt2gene Low-quality
S10_vs_23287||lt2gene Low-quality
S10_vs_23476||lt2gene Low-quality
S10_vs_23707||lt2gene Not-determined
S10_vs_23815||lt2gene Low-quality
S10_vs_23976||lt2gene Not-determined
S10_vs_23980||lt2gene Low-quality
S10_vs_24212||lt2gene Low-quality
S10_vs_24267||lt2gene Low-quality
S10_vs_24321||lt2gene Not-determined
S10_vs_24434||lt2gene Low-quality
S10_vs_24438||lt2gene Low-quality
S10_vs_24768||lt2gene Low-quality
S10_vs_24916||lt2gene Not-determined
S10_vs_24950||lt2gene Not-determined
S10_vs_25039||lt2gene Not-determined
S10_vs_25062||lt2gene Not-determined
S10_vs_25209||lt2gene Low-quality
S10_vs_25415||lt2gene Low-quality
S10_vs_25530||lt2gene Low-quality
S10_vs_25533||lt2gene Low-quality
S10_vs_25552||lt2gene Low-quality
S10_vs_25691||lt2gene Low-quality
S10_vs_25715||lt2gene Not-determined
S10_vs_25778||lt2gene Low-quality
S10_vs_25855||lt2gene Low-quality
S10_vs_25876||lt2gene Low-quality
S10_vs_25993||lt2gene Low-quality
S10_vs_26000||lt2gene Low-quality
S10_vs_26096||lt2gene Low-quality
S10_vs_26559||lt2gene Not-determined
S10_vs_26633||lt2gene Not-determined
S10_vs_26756||lt2gene Low-quality
S10_vs_26895||lt2gene Not-determined
S10_vs_27005||lt2gene Not-determined
S10_vs_27161||lt2gene Low-quality
S10_vs_27168||lt2gene Low-quality
S10_vs_27376||lt2gene Low-quality
S10_vs_27420||lt2gene Low-quality
S10_vs_27454||lt2gene Low-quality
S10_vs_27751||lt2gene Not-determined
S10_vs_27794||lt2gene Low-quality
S10_vs_27949||lt2gene Low-quality
S10_vs_27959||lt2gene Low-quality
S10_vs_28099||lt2gene Not-determined
S10_vs_28150||lt2gene Low-quality
S10_vs_28177||lt2gene Low-quality
S10_vs_28261||lt2gene Low-quality
S10_vs_28273||lt2gene Low-quality
S10_vs_28352||lt2gene Low-quality
S10_vs_28379||lt2gene Low-quality
S10_vs_28457||lt2gene Low-quality
S10_vs_28501||lt2gene Low-quality
S10_vs_28531||lt2gene Low-quality
S10_vs_28643||lt2gene Low-quality
S10_vs_28772||lt2gene Low-quality
S10_vs_28781||lt2gene Low-quality
S10_vs_28795||lt2gene Not-determined
S10_vs_28808||lt2gene Low-quality
S10_vs_28964||lt2gene Low-quality
S10_vs_29127||lt2gene Low-quality
S10_vs_29148||lt2gene Low-quality
S10_vs_29222||lt2gene Low-quality
S10_vs_29357||lt2gene Low-quality
S10_vs_29705||lt2gene Low-quality
S10_vs_29773||lt2gene Low-quality
S10_vs_29844||lt2gene Low-quality
S10_vs_29895||lt2gene Not-determined
S10_vs_29941||lt2gene Low-quality
S10_vs_30174||lt2gene Low-quality
S10_vs_30318||lt2gene Low-quality
S10_vs_30492||lt2gene Not-determined
S10_vs_30832||lt2gene Low-quality
S10_vs_30854||lt2gene Low-quality
S10_vs_30895||lt2gene Low-quality
S10_vs_30902||lt2gene Not-determined
S10_vs_30924||lt2gene Low-quality
S10_vs_31054||lt2gene Low-quality
S10_vs_31258||lt2gene Low-quality
S10_vs_31344||lt2gene Low-quality
S10_vs_31400||lt2gene Low-quality
S10_vs_31450||lt2gene Low-quality
S10_vs_31572||lt2gene Not-determined
S10_vs_31574||lt2gene Low-quality
S10_vs_31597||lt2gene Not-determined
S10_vs_31625||lt2gene Low-quality
S10_vs_31884||lt2gene Low-quality
S10_vs_31896||lt2gene Low-quality
S10_vs_31938||lt2gene Low-quality
S10_vs_31947||lt2gene Low-quality
S10_vs_31970||lt2gene Not-determined
S10_vs_32047||lt2gene Not-determined
S10_vs_32203||lt2gene Not-determined
S10_vs_32354||lt2gene Low-quality
S10_vs_32517||lt2gene Not-determined
S10_vs_32599||lt2gene Low-quality
S10_vs_32602||lt2gene Low-quality
S10_vs_32695||lt2gene Not-determined
S10_vs_33031||lt2gene Not-determined
S10_vs_33044||lt2gene Low-quality
S10_vs_33064||lt2gene Low-quality
S10_vs_33074||lt2gene Low-quality
S10_vs_33091||lt2gene Low-quality
S10_vs_33154||lt2gene Low-quality
S10_vs_33291||lt2gene Low-quality
S10_vs_33294||lt2gene Low-quality
S10_vs_33323||lt2gene Low-quality
S10_vs_33358||lt2gene Low-quality
S10_vs_33560||lt2gene Low-quality
S10_vs_33579||lt2gene Low-quality
S10_vs_33651||lt2gene Not-determined
S10_vs_33817||lt2gene Low-quality
S10_vs_33888||lt2gene Not-determined
S10_vs_33923||lt2gene Not-determined
S10_vs_33930||lt2gene Not-determined
S10_vs_34084||lt2gene Low-quality
S10_vs_34170||lt2gene Not-determined
S10_vs_34215||lt2gene Low-quality
S10_vs_34298||lt2gene Low-quality
S10_vs_34309||lt2gene Low-quality
S10_vs_34399||lt2gene Not-determined
S10_vs_34403||lt2gene Not-determined
S10_vs_34415||lt2gene Not-determined
S10_vs_34428||lt2gene Not-determined
S10_vs_34560||lt2gene Low-quality
S10_vs_34755||lt2gene Low-quality
S10_vs_34761||lt2gene Low-quality
S10_vs_34787||lt2gene Low-quality
S10_vs_34832||lt2gene Low-quality
S10_vs_34868||lt2gene Not-determined
S10_vs_34909||lt2gene Low-quality
S10_vs_34916||lt2gene Not-determined
S10_vs_35009||lt2gene Low-quality
S10_vs_35042||lt2gene Low-quality
S10_vs_35092||lt2gene Low-quality
S10_vs_35200||lt2gene Low-quality
S10_vs_35208||lt2gene Not-determined
S10_vs_35264||lt2gene Low-quality
S10_vs_35266||lt2gene Not-determined

| | |
|---|---|
| S10_vs_35349\|\|lt2gene | Low-quality |
| S10_vs_35496\|\|lt2gene | Low-quality |
| S10_vs_35524\|\|lt2gene | Low-quality |
| S10_vs_35771\|\|lt2gene | Low-quality |
| S10_vs_35992\|\|lt2gene | Low-quality |
| S10_vs_36227\|\|lt2gene | Not-determined |
| S10_vs_36320\|\|lt2gene | Low-quality |
| S10_vs_36416\|\|lt2gene | Not-determined |
| S10_vs_36447\|\|lt2gene | Not-determined |
| S10_vs_36458\|\|lt2gene | Not-determined |
| S10_vs_36712\|\|lt2gene | Not-determined |
| S10_vs_36785\|\|lt2gene | Low-quality |
| S10_vs_36839\|\|lt2gene | Low-quality |
| S10_vs_36844\|\|lt2gene | Low-quality |
| S10_vs_36872\|\|lt2gene | Not-determined |
| S10_vs_37055\|\|lt2gene | Low-quality |
| S10_vs_37069\|\|lt2gene | Low-quality |
| S10_vs_37110\|\|lt2gene | Low-quality |
| S10_vs_37157\|\|lt2gene | Low-quality |
| S10_vs_37239\|\|lt2gene | Low-quality |
| S10_vs_37263\|\|lt2gene | Low-quality |
| S10_vs_37285\|\|lt2gene | Low-quality |
| S10_vs_37561\|\|lt2gene | Low-quality |
| S10_vs_37644\|\|lt2gene | Low-quality |
| S10_vs_37673\|\|lt2gene | Low-quality |
| S10_vs_37729\|\|lt2gene | Low-quality |
| S10_vs_37843\|\|lt2gene | Low-quality |
| S10_vs_37969\|\|lt2gene | Low-quality |
| S10_vs_38223\|\|lt2gene | Low-quality |
| S10_vs_38303\|\|lt2gene | Low-quality |
| S10_vs_38339\|\|lt2gene | Low-quality |
| S10_vs_38358\|\|lt2gene | Not-determined |
| S10_vs_38426\|\|lt2gene | Low-quality |
| S10_vs_38622\|\|lt2gene | Low-quality |
| S10_vs_38680\|\|lt2gene | Not-determined |
| S10_vs_38691\|\|lt2gene | Low-quality |
| S10_vs_38787\|\|lt2gene | Low-quality |
| S10_vs_38892\|\|lt2gene | Low-quality |
| S10_vs_38911\|\|lt2gene | Low-quality |
| S10_vs_39121\|\|lt2gene | Not-determined |
| S10_vs_39248\|\|lt2gene | Low-quality |
| S10_vs_39275\|\|lt2gene | Low-quality |
| S10_vs_39282\|\|lt2gene | Not-determined |
| S10_vs_39348\|\|lt2gene | Not-determined |
| S10_vs_39523\|\|lt2gene | Low-quality |
| S10_vs_39594\|\|lt2gene | Low-quality |
| S10_vs_40048\|\|lt2gene | Low-quality |
| S10_vs_40083\|\|lt2gene | Not-determined |
| S10_vs_40115\|\|lt2gene | Not-determined |
| S10_vs_40352\|\|lt2gene | Low-quality |
| S10_vs_40379\|\|lt2gene | Low-quality |
| S10_vs_40476\|\|lt2gene | Low-quality |
| S10_vs_40527\|\|lt2gene | Not-determined |
| S10_vs_40726\|\|lt2gene | Low-quality |
| S10_vs_40831\|\|lt2gene | Not-determined |
| S10_vs_40849\|\|lt2gene | Low-quality |
| S10_vs_40860\|\|lt2gene | Not-determined |
| S10_vs_40962\|\|lt2gene | Low-quality |
| S10_vs_40987\|\|lt2gene | Low-quality |
| S10_vs_41081\|\|lt2gene | Low-quality |
| S10_vs_41226\|\|lt2gene | Low-quality |
| S10_vs_41271\|\|lt2gene | Low-quality |
| S10_vs_41418\|\|lt2gene | Low-quality |
| S10_vs_41466\|\|lt2gene | Low-quality |
| S10_vs_41475\|\|lt2gene | Low-quality |
| S10_vs_41508\|\|lt2gene | Low-quality |
| S10_vs_41606\|\|lt2gene | Low-quality |
| S10_vs_41920\|\|lt2gene | Low-quality |
| S10_vs_41979\|\|lt2gene | Low-quality |
| S10_vs_42011\|\|lt2gene | Low-quality |
| S10_vs_42076\|\|lt2gene | Low-quality |
| S10_vs_42188\|\|lt2gene | Low-quality |
| S10_vs_42217\|\|lt2gene | Low-quality |
| S10_vs_42456\|\|lt2gene | Low-quality |
| S10_vs_42533\|\|lt2gene | Low-quality |
| S10_vs_42969\|\|lt2gene | Low-quality |
| S10_vs_43268\|\|lt2gene | Low-quality |
| S10_vs_43372\|\|lt2gene | Not-determined |
| S10_vs_43458\|\|lt2gene | Not-determined |
| S10_vs_43551\|\|lt2gene | Low-quality |
| S10_vs_43671\|\|lt2gene | Low-quality |
| S10_vs_43864\|\|lt2gene | Not-determined |
| S10_vs_43979\|\|lt2gene | Not-determined |
| S10_vs_44126\|\|lt2gene | Low-quality |
| S10_vs_44145\|\|lt2gene | Not-determined |
| S10_vs_44169\|\|lt2gene | Low-quality |
| S10_vs_44180\|\|lt2gene | Low-quality |
| S10_vs_44521\|\|lt2gene | Low-quality |
| S10_vs_44526\|\|lt2gene | Not-determined |
| S10_vs_44571\|\|lt2gene | Low-quality |
| S10_vs_44610\|\|lt2gene | Not-determined |
| S10_vs_44823\|\|lt2gene | Low-quality |
| S10_vs_44884\|\|lt2gene | Not-determined |
| S10_vs_44890\|\|lt2gene | Low-quality |
| S10_vs_45162\|\|lt2gene | Low-quality |
| S10_vs_45215\|\|lt2gene | Low-quality |
| S10_vs_45235\|\|lt2gene | Low-quality |
| S10_vs_45370\|\|lt2gene | Low-quality |
| S10_vs_45555\|\|lt2gene | Low-quality |
| S10_vs_45577\|\|lt2gene | Not-determined |
| S10_vs_45677\|\|lt2gene | Low-quality |
| S10_vs_45679\|\|lt2gene | Low-quality |
| S10_vs_46222\|\|lt2gene | Not-determined |
| S10_vs_46326\|\|lt2gene | Low-quality |
| S10_vs_46327\|\|lt2gene | Low-quality |
| S10_vs_46356\|\|lt2gene | Low-quality |
| S10_vs_46512\|\|lt2gene | Not-determined |
| S10_vs_46546\|\|lt2gene | Low-quality |
| S10_vs_46571\|\|lt2gene | Low-quality |
| S10_vs_46891\|\|lt2gene | Low-quality |
| S10_vs_46945\|\|lt2gene | Not-determined |
| S10_vs_46968\|\|lt2gene | Low-quality |
| S10_vs_47326\|\|lt2gene | Low-quality |
| S10_vs_47365\|\|lt2gene | Low-quality |
| S10_vs_47566\|\|lt2gene | Not-determined |
| S10_vs_47653\|\|lt2gene | Not-determined |
| S10_vs_47958\|\|lt2gene | Low-quality |
| S10_vs_47976\|\|lt2gene | Low-quality |
| S10_vs_47994\|\|lt2gene | Low-quality |
| S10_vs_48167\|\|lt2gene | Low-quality |
| S10_vs_48195\|\|lt2gene | Low-quality |
| S10_vs_48277\|\|lt2gene | Low-quality |
| S10_vs_48355\|\|lt2gene | Low-quality |
| S10_vs_48656\|\|lt2gene | Low-quality |
| S10_vs_48764\|\|lt2gene | Not-determined |
| S10_vs_48976\|\|lt2gene | Not-determined |
| S10_vs_49029\|\|lt2gene | Low-quality |
| S10_vs_49069\|\|lt2gene | Low-quality |
| S10_vs_49247\|\|lt2gene | Not-determined |
| S10_vs_49257\|\|lt2gene | Low-quality |
| S10_vs_49299\|\|lt2gene | Low-quality |
| S10_vs_49319\|\|lt2gene | Low-quality |
| S10_vs_49878\|\|lt2gene | Low-quality |
| S10_vs_49904\|\|lt2gene | Low-quality |
| S10_vs_49938\|\|lt2gene | Low-quality |
| S10_vs_50099\|\|lt2gene | Not-determined |
| S10_vs_50254\|\|lt2gene | Not-determined |
| S10_vs_50317\|\|lt2gene | Low-quality |
| S10_vs_50422\|\|lt2gene | Low-quality |
| S10_vs_50541\|\|lt2gene | Low-quality |
| S10_vs_50806\|\|lt2gene | Low-quality |
| S10_vs_50829\|\|lt2gene | Not-determined |
| S10_vs_51028\|\|lt2gene | Not-determined |
| S10_vs_51199\|\|lt2gene | Not-determined |
| S10_vs_51250\|\|lt2gene | Not-determined |
| S10_vs_51280\|\|lt2gene | Low-quality |
| S10_vs_51287\|\|lt2gene | Not-determined |
| S10_vs_51294\|\|lt2gene | Low-quality |
| S10_vs_51422\|\|lt2gene | Low-quality |
| S10_vs_51460\|\|lt2gene | Low-quality |
| S10_vs_51476\|\|lt2gene | Not-determined |
| S10_vs_51529\|\|lt2gene | Low-quality |
| S10_vs_51851\|\|lt2gene | Low-quality |
| S10_vs_51862\|\|lt2gene | Not-determined |
| S10_vs_51955\|\|lt2gene | Low-quality |
| S10_vs_52052\|\|lt2gene | Low-quality |
| S10_vs_52280\|\|lt2gene | Low-quality |
| S10_vs_52422\|\|lt2gene | Low-quality |
| S10_vs_52731\|\|lt2gene | Low-quality |
| S10_vs_52740\|\|lt2gene | Low-quality |
| S10_vs_52934\|\|lt2gene | Low-quality |
| S10_vs_53117\|\|lt2gene | Low-quality |
| S10_vs_53245\|\|lt2gene | Low-quality |
| S10_vs_53251\|\|lt2gene | Low-quality |
| S10_vs_53381\|\|lt2gene | Low-quality |
| S10_vs_53454\|\|lt2gene | Low-quality |
| S10_vs_53457\|\|lt2gene | Low-quality |
| S10_vs_53618\|\|lt2gene | Not-determined |
| S10_vs_53630\|\|lt2gene | Low-quality |
| S10_vs_53679\|\|lt2gene | Low-quality |
| S10_vs_53741\|\|lt2gene | Not-determined |
| S10_vs_53787\|\|lt2gene | Low-quality |
| S10_vs_53949\|\|lt2gene | Not-determined |
| S10_vs_54132\|\|lt2gene | Low-quality |
| S10_vs_54162\|\|lt2gene | Low-quality |
| S10_vs_54323\|\|lt2gene | Not-determined |
| S10_vs_54375\|\|lt2gene | Low-quality |
| S10_vs_54443\|\|lt2gene | Low-quality |
| S10_vs_54720\|\|lt2gene | Low-quality |
| S10_vs_54996\|\|lt2gene | Low-quality |
| S10_vs_55268\|\|lt2gene | Low-quality |
| S10_vs_55314\|\|lt2gene | Not-determined |
| S10_vs_55327\|\|lt2gene | Not-determined |
| S10_vs_55385\|\|lt2gene | Low-quality |
| S10_vs_55475\|\|lt2gene | Low-quality |
| S10_vs_55635\|\|lt2gene | Low-quality |
| S10_vs_55670\|\|lt2gene | Not-determined |
| S10_vs_55743\|\|lt2gene | Not-determined |
| S10_vs_56148\|\|lt2gene | Low-quality |
| S10_vs_56201\|\|lt2gene | Not-determined |
| S10_vs_56301\|\|lt2gene | Low-quality |
| S10_vs_56309\|\|lt2gene | Low-quality |
| S10_vs_56357\|\|lt2gene | Not-determined |
| S10_vs_56362\|\|lt2gene | Low-quality |
| S10_vs_56475\|\|lt2gene | Not-determined |
| S10_vs_56530\|\|lt2gene | Not-determined |
| S10_vs_56697\|\|lt2gene | Low-quality |
| S10_vs_57040\|\|lt2gene | Not-determined |
| S10_vs_57163\|\|lt2gene | Low-quality |
| S10_vs_57197\|\|lt2gene | Low-quality |
| S10_vs_57227\|\|lt2gene | Low-quality |
| S10_vs_57462\|\|lt2gene | Not-determined |
| S10_vs_57541\|\|lt2gene | Low-quality |
| S10_vs_57570\|\|lt2gene | Low-quality |
| S10_vs_57579\|\|lt2gene | Not-determined |
| S10_vs_57582\|\|lt2gene | Low-quality |
| S10_vs_57591\|\|lt2gene | Not-determined |
| S10_vs_57616\|\|lt2gene | Low-quality |
| S10_vs_57706\|\|lt2gene | Low-quality |
| S10_vs_57753\|\|lt2gene | Low-quality |
| S10_vs_57800\|\|lt2gene | Low-quality |
| S10_vs_57833\|\|lt2gene | Low-quality |
| S10_vs_57953\|\|lt2gene | Low-quality |
| S10_vs_58105\|\|lt2gene | Not-determined |
| S10_vs_58197\|\|lt2gene | Not-determined |
| S10_vs_58449\|\|lt2gene | Not-determined |
| S10_vs_58589\|\|lt2gene | Low-quality |
| S10_vs_58646\|\|lt2gene | Not-determined |
| S10_vs_58727\|\|lt2gene | Low-quality |
| S10_vs_58893\|\|lt2gene | Low-quality |
| S10_vs_58974\|\|lt2gene | Low-quality |
| S10_vs_59209\|\|lt2gene | Low-quality |
| S10_vs_59238\|\|lt2gene | Low-quality |
| S10_vs_59296\|\|lt2gene | Low-quality |
| S10_vs_59476\|\|lt2gene | Low-quality |
| S10_vs_59643\|\|lt2gene | Low-quality |
| S10_vs_59776\|\|lt2gene | Low-quality |
| S10_vs_59783\|\|lt2gene | Not-determined |
| S10_vs_60002\|\|lt2gene | Low-quality |
| S10_vs_60053\|\|lt2gene | Low-quality |
| S10_vs_60145\|\|lt2gene | Not-determined |
| S10_vs_60222\|\|lt2gene | Not-determined |
| S10_vs_60241\|\|lt2gene | Low-quality |
| S10_vs_60245\|\|lt2gene | Low-quality |
| S10_vs_60252\|\|lt2gene | Low-quality |
| S10_vs_60551\|\|lt2gene | Not-determined |
| S10_vs_60581\|\|lt2gene | Low-quality |
| S10_vs_60586\|\|lt2gene | Low-quality |
| S10_vs_60758\|\|lt2gene | Low-quality |
| S10_vs_60847\|\|lt2gene | Low-quality |
| S10_vs_60901\|\|lt2gene | Low-quality |
| S10_vs_60938\|\|lt2gene | Not-determined |
| S10_vs_61254\|\|lt2gene | Low-quality |
| S10_vs_61283\|\|lt2gene | Not-determined |
| S10_vs_61286\|\|lt2gene | Low-quality |
| S10_vs_61384\|\|lt2gene | Low-quality |
| S10_vs_61462\|\|lt2gene | Not-determined |
| S10_vs_61472\|\|lt2gene | Low-quality |
| S10_vs_61645\|\|lt2gene | Low-quality |
| S10_vs_62015\|\|lt2gene | Low-quality |
| S10_vs_62049\|\|lt2gene | Not-determined |
| S10_vs_62069\|\|lt2gene | Low-quality |
| S10_vs_62254\|\|lt2gene | Not-determined |
| S10_vs_62396\|\|lt2gene | Not-determined |
| S10_vs_62436\|\|lt2gene | Low-quality |
| S10_vs_62509\|\|lt2gene | Low-quality |
| S10_vs_62638\|\|lt2gene | Low-quality |
| S10_vs_62714\|\|lt2gene | Low-quality |
| S10_vs_62825\|\|lt2gene | Not-determined |
| S10_vs_62937\|\|lt2gene | Low-quality |
| S10_vs_63006\|\|lt2gene | Not-determined |
| S10_vs_63009\|\|lt2gene | Low-quality |
| S10_vs_63070\|\|lt2gene | Low-quality |
| S10_vs_63233\|\|lt2gene | Low-quality |
| S10_vs_63387\|\|lt2gene | Low-quality |
| S10_vs_63436\|\|lt2gene | Low-quality |
| S10_vs_63488\|\|lt2gene | Not-determined |
| S10_vs_63639\|\|lt2gene | Low-quality |
| S10_vs_63855\|\|lt2gene | Low-quality |
| S10_vs_64136\|\|lt2gene | Low-quality |
| S10_vs_64144\|\|lt2gene | Low-quality |
| S10_vs_64364\|\|lt2gene | Low-quality |
| S10_vs_64445\|\|lt2gene | Low-quality |
| S10_vs_64689\|\|lt2gene | Not-determined |
| S10_vs_64692\|\|lt2gene | Low-quality |
| S10_vs_64854\|\|lt2gene | Low-quality |
| S10_vs_64912\|\|lt2gene | Not-determined |
| S10_vs_64982\|\|lt2gene | Low-quality |
| S10_vs_64993\|\|lt2gene | Low-quality |
| S10_vs_65035\|\|lt2gene | Low-quality |
| S10_vs_65114\|\|lt2gene | Low-quality |
| S10_vs_65248\|\|lt2gene | Not-determined |
| S10_vs_65473\|\|lt2gene | Not-determined |
| S10_vs_65485\|\|lt2gene | Low-quality |
| S10_vs_65514\|\|lt2gene | Low-quality |
| S10_vs_65722\|\|lt2gene | Not-determined |
| S10_vs_65862\|\|lt2gene | Not-determined |
| S10_vs_66016\|\|lt2gene | Not-determined |
| S10_vs_66363\|\|lt2gene | Not-determined |
| S10_vs_66381\|\|lt2gene | Low-quality |
| S10_vs_66439\|\|lt2gene | Low-quality |
| S10_vs_66542\|\|lt2gene | Low-quality |
| S10_vs_66595\|\|lt2gene | Low-quality |
| S10_vs_66667\|\|lt2gene | Not-determined |
| S10_vs_66830\|\|lt2gene | Low-quality |
| S10_vs_66832\|\|lt2gene | Not-determined |
| S10_vs_66852\|\|lt2gene | Low-quality |
| S10_vs_67221\|\|lt2gene | Low-quality |
| S10_vs_67393\|\|lt2gene | Not-determined |
| S10_vs_67477\|\|lt2gene | Not-determined |
| S10_vs_67488\|\|lt2gene | Low-quality |
| S10_vs_67583\|\|lt2gene | Low-quality |
| S10_vs_67733\|\|lt2gene | Low-quality |
| S10_vs_67738\|\|lt2gene | Not-determined |
| S10_vs_67901\|\|lt2gene | Not-determined |
| S10_vs_67937\|\|lt2gene | Low-quality |
| S10_vs_67944\|\|lt2gene | Low-quality |
| S10_vs_68263\|\|lt2gene | Low-quality |
| S10_vs_68327\|\|lt2gene | Low-quality |
| S10_vs_68672\|\|lt2gene | Not-determined |
| S10_vs_68702\|\|lt2gene | Low-quality |
| S10_vs_68767\|\|lt2gene | Low-quality |
| S10_vs_68786\|\|lt2gene | Low-quality |
| S10_vs_68912\|\|lt2gene | Low-quality |
| S10_vs_68928\|\|lt2gene | Not-determined |
| S10_vs_68935\|\|lt2gene | Not-determined |
| S10_vs_69120\|\|lt2gene | Low-quality |
| S10_vs_69130\|\|lt2gene | Low-quality |
| S10_vs_69306\|\|lt2gene | Not-determined |
| S10_vs_69317\|\|lt2gene | Not-determined |
| S10_vs_69541\|\|lt2gene | Not-determined |
| S10_vs_69553\|\|lt2gene | Not-determined |
| S10_vs_69580\|\|lt2gene | Low-quality |
| S10_vs_69828\|\|lt2gene | Low-quality |
| S10_vs_69971\|\|lt2gene | Not-determined |
| S10_vs_70021\|\|lt2gene | Not-determined |
| S10_vs_70022\|\|lt2gene | Low-quality |
| S10_vs_70117\|\|lt2gene | Low-quality |
| S10_vs_70188\|\|lt2gene | Not-determined |
| S10_vs_70202\|\|lt2gene | Low-quality |
| S10_vs_70255\|\|lt2gene | Low-quality |
| S10_vs_70306\|\|lt2gene | Low-quality |
| S10_vs_70319\|\|lt2gene | Low-quality |
| S10_vs_70371\|\|lt2gene | Low-quality |
| S10_vs_70405\|\|lt2gene | Low-quality |
| S10_vs_70539\|\|lt2gene | Low-quality |
| S10_vs_70564\|\|lt2gene | Low-quality |
| S10_vs_70592\|\|lt2gene | Low-quality |
| S10_vs_70693\|\|lt2gene | Low-quality |
| S10_vs_70735\|\|lt2gene | Low-quality |
| S10_vs_70803\|\|lt2gene | Low-quality |
| S10_vs_70998\|\|lt2gene | Not-determined |
| S10_vs_71103\|\|lt2gene | Low-quality |
| S10_vs_71143\|\|lt2gene | Not-determined |
| S10_vs_71225\|\|lt2gene | Not-determined |
| S10_vs_71272\|\|lt2gene | Not-determined |
| S10_vs_71287\|\|lt2gene | Low-quality |
| S10_vs_71378\|\|lt2gene | Low-quality |
| S10_vs_71474\|\|lt2gene | Not-determined |
| S10_vs_71552\|\|lt2gene | Low-quality |
| S10_vs_71655\|\|lt2gene | Low-quality |
| S10_vs_71839\|\|lt2gene | Low-quality |
| S10_vs_71903\|\|lt2gene | Low-quality |
| S10_vs_71955\|\|lt2gene | Not-determined |
| S10_vs_72175\|\|lt2gene | Low-quality |
| S10_vs_72212\|\|lt2gene | Low-quality |
| S10_vs_72215\|\|lt2gene | Low-quality |
| S10_vs_72311\|\|lt2gene | Low-quality |
| S10_vs_72349\|\|lt2gene | Low-quality |
| S10_vs_72445\|\|lt2gene | Not-determined |
| S10_vs_72583\|\|lt2gene | Low-quality |
| S10_vs_72752\|\|lt2gene | Low-quality |
| S10_vs_73044\|\|lt2gene | Not-determined |
| S10_vs_73394\|\|lt2gene | Not-determined |
| S10_vs_73479\|\|lt2gene | Low-quality |
| S10_vs_73550\|\|lt2gene | Low-quality |
| S10_vs_73690\|\|lt2gene | Low-quality |
| S10_vs_73708\|\|lt2gene | Low-quality |
| S10_vs_73798\|\|lt2gene | Not-determined |
| S10_vs_74038\|\|lt2gene | Low-quality |
| S10_vs_74125\|\|lt2gene | Low-quality |
| S10_vs_74402\|\|lt2gene | Low-quality |
| S10_vs_74508\|\|lt2gene | Low-quality |
| S10_vs_74694\|\|lt2gene | Not-determined |
| S10_vs_74794\|\|lt2gene | Low-quality |
| S10_vs_74859\|\|lt2gene | Low-quality |
| S10_vs_74934\|\|lt2gene | Low-quality |
| S10_vs_74939\|\|lt2gene | Low-quality |
| S10_vs_75394\|\|lt2gene | Low-quality |
| S10_vs_75578\|\|lt2gene | Low-quality |
| S10_vs_75595\|\|lt2gene | Not-determined |
| S10_vs_75924\|\|lt2gene | Low-quality |
| S10_vs_75932\|\|lt2gene | Low-quality |
| S10_vs_76059\|\|lt2gene | Not-determined |
| S10_vs_76084\|\|lt2gene | Low-quality |
| S10_vs_76159\|\|lt2gene | Low-quality |
| S10_vs_76411\|\|lt2gene | Not-determined |
| S10_vs_76606\|\|lt2gene | Low-quality |
| S10_vs_76620\|\|lt2gene | Not-determined |
| S10_vs_76638\|\|lt2gene | Low-quality |
| S10_vs_76869\|\|lt2gene | Not-determined |
| S10_vs_77056\|\|lt2gene | Low-quality |
| S10_vs_77064\|\|lt2gene | Low-quality |
| S10_vs_77204\|\|lt2gene | Low-quality |
| S10_vs_77300\|\|lt2gene | Low-quality |
| S10_vs_77336\|\|lt2gene | Low-quality |
| S10_vs_77400\|\|lt2gene | Low-quality |
| S10_vs_77478\|\|lt2gene | Low-quality |
| S10_vs_77501\|\|lt2gene | Not-determined |
| S10_vs_77518\|\|lt2gene | Low-quality |
| S10_vs_77523\|\|lt2gene | Low-quality |
| S10_vs_77725\|\|lt2gene | Low-quality |
| S10_vs_77734\|\|lt2gene | Low-quality |
| S10_vs_77840\|\|lt2gene | Low-quality |
| S10_vs_78075\|\|lt2gene | Low-quality |
| S10_vs_78118\|\|lt2gene | Low-quality |
| S10_vs_78136\|\|lt2gene | Not-determined |
| S10_vs_78305\|\|lt2gene | Low-quality |
| S10_vs_78721\|\|lt2gene | Not-determined |
| S10_vs_78776\|\|lt2gene | Low-quality |
| S10_vs_78800\|\|lt2gene | Low-quality |
| S10_vs_78844\|\|lt2gene | Low-quality |
| S10_vs_78897\|\|lt2gene | Low-quality |
| S10_vs_79052\|\|lt2gene | Low-quality |
| S10_vs_79192\|\|lt2gene | Low-quality |
| S10_vs_79266\|\|lt2gene | Low-quality |
| S10_vs_79549\|\|lt2gene | Low-quality |
| S10_vs_79574\|\|lt2gene | Low-quality |
| S10_vs_79613\|\|lt2gene | Not-determined |
| S10_vs_79701\|\|lt2gene | Low-quality |
| S10_vs_79708\|\|lt2gene | Not-determined |
| S10_vs_79851\|\|lt2gene | Low-quality |
| S10_vs_80490\|\|lt2gene | Not-determined |
| S10_vs_80613\|\|lt2gene | Low-quality |
| S10_vs_80637\|\|lt2gene | Not-determined |
| S10_vs_80653\|\|lt2gene | Low-quality |
| S10_vs_80862\|\|lt2gene | Low-quality |
| S10_vs_80918\|\|lt2gene | Not-determined |
| S10_vs_81003\|\|lt2gene | Low-quality |
| S10_vs_81050\|\|lt2gene | Low-quality |
| S10_vs_81081\|\|lt2gene | Not-determined |
| S10_vs_81261\|\|lt2gene | Low-quality |
| S10_vs_81268\|\|lt2gene | Low-quality |
| S10_vs_81460\|\|lt2gene | Low-quality |
| S10_vs_81642\|\|lt2gene | Not-determined |
| S10_vs_81720\|\|lt2gene | Low-quality |
| S10_vs_81781\|\|lt2gene | Low-quality |
| S10_vs_81879\|\|lt2gene | Not-determined |
| S10_vs_81980\|\|lt2gene | Low-quality |
| S10_vs_82060\|\|lt2gene | Low-quality |
| S10_vs_82063\|\|lt2gene | Low-quality |
| S10_vs_82074\|\|lt2gene | Low-quality |
| S10_vs_82133\|\|lt2gene | Low-quality |
| S10_vs_82184\|\|lt2gene | Low-quality |
| S10_vs_82235\|\|lt2gene | Low-quality |
| S10_vs_82251\|\|lt2gene | Not-determined |
| S10_vs_82348\|\|lt2gene | Low-quality |
| S10_vs_82391\|\|lt2gene | Not-determined |
| S10_vs_82407\|\|lt2gene | Low-quality |
| S10_vs_82543\|\|lt2gene | Low-quality |
| S10_vs_82674\|\|lt2gene | Low-quality |
| S10_vs_82693\|\|lt2gene | Low-quality |
| S10_vs_82769\|\|lt2gene | Low-quality |
| S10_vs_82772\|\|lt2gene | Low-quality |
| S10_vs_82803\|\|lt2gene | Low-quality |
| S10_vs_83043\|\|lt2gene | Low-quality |
| S10_vs_83104\|\|lt2gene | Not-determined |
| S10_vs_83156\|\|lt2gene | Low-quality |
| S10_vs_83246\|\|lt2gene | Not-determined |
| S10_vs_83387\|\|lt2gene | Low-quality |
| S10_vs_83391\|\|lt2gene | Not-determined |
| S10_vs_83430\|\|lt2gene | Low-quality |
| S10_vs_83551\|\|lt2gene | Low-quality |
| S10_vs_83553\|\|lt2gene | Not-determined |
| S10_vs_83586\|\|lt2gene | Low-quality |
| S10_vs_83634\|\|lt2gene | Not-determined |
| S10_vs_83670\|\|lt2gene | Low-quality |
| S10_vs_83703\|\|lt2gene | Low-quality |
| S10_vs_83753\|\|lt2gene | Low-quality |
| S10_vs_83854\|\|lt2gene | Not-determined |
| S10_vs_83862\|\|lt2gene | Not-determined |
| S10_vs_83881\|\|lt2gene | Low-quality |
| S10_vs_83911\|\|lt2gene | Low-quality |
| S10_vs_83960\|\|lt2gene | Not-determined |
| S10_vs_83964\|\|lt2gene | Low-quality |
| S10_vs_83985\|\|lt2gene | Not-determined |
| S10_vs_84057\|\|lt2gene | Low-quality |
| S10_vs_84210\|\|lt2gene | Low-quality |
| S10_vs_84220\|\|lt2gene | Low-quality |
| S10_vs_84339\|\|lt2gene | Low-quality |
| S10_vs_84385\|\|lt2gene | Low-quality |
| S10_vs_84434\|\|lt2gene | Low-quality |
| S10_vs_84496\|\|lt2gene | Not-determined |
| S10_vs_84557\|\|lt2gene | Not-determined |
| S10_vs_84678\|\|lt2gene | Low-quality |
| S10_vs_84767\|\|lt2gene | Low-quality |
| S10_vs_84776\|\|lt2gene | Low-quality |
| S10_vs_84813\|\|lt2gene | Not-determined |
| S10_vs_84816\|\|lt2gene | Low-quality |
| S10_vs_84912\|\|lt2gene | Low-quality |
| S10_vs_84954\|\|lt2gene | Low-quality |
| S10_vs_85065\|\|lt2gene | Low-quality |
| S10_vs_85077\|\|lt2gene | Low-quality |
| S10_vs_85195\|\|lt2gene | Low-quality |
| S10_vs_85210\|\|lt2gene | Low-quality |
| S10_vs_85275\|\|lt2gene | Not-determined |
| S10_vs_85351\|\|lt2gene | Low-quality |
| S10_vs_85566\|\|lt2gene | Not-determined |
| S10_vs_85594\|\|lt2gene | Low-quality |
| S10_vs_85629\|\|lt2gene | Low-quality |
| S10_vs_85651\|\|lt2gene | Low-quality |
| S10_vs_85670\|\|lt2gene | Low-quality |
| S10_vs_85684\|\|lt2gene | Low-quality |
| S10_vs_85903\|\|lt2gene | Low-quality |
| S10_vs_85939\|\|lt2gene | Low-quality |
| S10_vs_85947\|\|lt2gene | Not-determined |
| S10_vs_86013\|\|lt2gene | Low-quality |
| S10_vs_86060\|\|lt2gene | Low-quality |
| S10_vs_86101\|\|lt2gene | Low-quality |
| S10_vs_86472\|\|lt2gene | Low-quality |
| S10_vs_86632\|\|lt2gene | Low-quality |
| S10_vs_86686\|\|lt2gene | Low-quality |
| S10_vs_86736\|\|lt2gene | Not-determined |
| S10_vs_87061\|\|lt2gene | Low-quality |
| S10_vs_87177\|\|lt2gene | Low-quality |
| S10_vs_87366\|\|lt2gene | Low-quality |
| S10_vs_87408\|\|lt2gene | Low-quality |
| S10_vs_87428\|\|lt2gene | Low-quality |
| S10_vs_87466\|\|lt2gene | Low-quality |
| S10_vs_87486\|\|lt2gene | Low-quality |
| S10_vs_87653\|\|lt2gene | Low-quality |
| S10_vs_87658\|\|lt2gene | Not-determined |
| S10_vs_87822\|\|lt2gene | Low-quality |
| S10_vs_87996\|\|lt2gene | Low-quality |
| S10_vs_88123\|\|lt2gene | Low-quality |
| S10_vs_88127\|\|lt2gene | Low-quality |
| S10_vs_88171\|\|lt2gene | Low-quality |
| S10_vs_88526\|\|lt2gene | Low-quality |
| S10_vs_88529\|\|lt2gene | Low-quality |
| S10_vs_88581\|\|lt2gene | Low-quality |
| S10_vs_88637\|\|lt2gene | Low-quality |
| S10_vs_88824\|\|lt2gene | Low-quality |
| S10_vs_88825\|\|lt2gene | Low-quality |
| S10_vs_88949\|\|lt2gene | Low-quality |
| S10_vs_89016\|\|lt2gene | Low-quality |
| S10_vs_89019\|\|lt2gene | Low-quality |
| S10_vs_89070\|\|lt2gene | Low-quality |
| S10_vs_89087\|\|lt2gene | Not-determined |
| S10_vs_89128\|\|lt2gene | Not-determined |
| S10_vs_89236\|\|lt2gene | Low-quality |
| S10_vs_89491\|\|lt2gene | Low-quality |
| S10_vs_89515\|\|lt2gene | Not-determined |
| S10_vs_89567\|\|lt2gene | Not-determined |
| S10_vs_89570\|\|lt2gene | Low-quality |
| S10_vs_89649\|\|lt2gene | Not-determined |
| S10_vs_89736\|\|lt2gene | Not-determined |
| S10_vs_89974\|\|lt2gene | Not-determined |
| S10_vs_90150\|\|lt2gene | Low-quality |
| S10_vs_90414\|\|lt2gene | Low-quality |
| S10_vs_90514\|\|lt2gene | Low-quality |
| S10_vs_90582\|\|lt2gene | Not-determined |
| S10_vs_90610\|\|lt2gene | Low-quality |
| S10_vs_90802\|\|lt2gene | Not-determined |
| S10_vs_90804\|\|lt2gene | Not-determined |
| S10_vs_90808\|\|lt2gene | Not-determined |
| S10_vs_90902\|\|lt2gene | Low-quality |
| S10_vs_90918\|\|lt2gene | Not-determined |
| S10_vs_91031\|\|lt2gene | Low-quality |
| S10_vs_91061\|\|lt2gene | Not-determined |
| S10_vs_91098\|\|lt2gene | Low-quality |
| S10_vs_91157\|\|lt2gene | Low-quality |
| S10_vs_91310\|\|lt2gene | Not-determined |
| S10_vs_91349\|\|lt2gene | Not-determined |
| S10_vs_91374\|\|lt2gene | Low-quality |
| S10_vs_91423\|\|lt2gene | Not-determined |
| S10_vs_91520\|\|lt2gene | Low-quality |
| S10_vs_91542\|\|lt2gene | Low-quality |
| S10_vs_91584\|\|lt2gene | Low-quality |
| S10_vs_91688\|\|lt2gene | Low-quality |
| S10_vs_91754\|\|lt2gene | Not-determined |
| S10_vs_91858\|\|lt2gene | Low-quality |
| S10_vs_91861\|\|lt2gene | Low-quality |
| S10_vs_92029\|\|lt2gene | Low-quality |
| S10_vs_92042\|\|lt2gene | Not-determined |
| S10_vs_92133\|\|lt2gene | Low-quality |
| S10_vs_92244\|\|lt2gene | Low-quality |
| S10_vs_92247\|\|lt2gene | Low-quality |
| S10_vs_92501\|\|lt2gene | Not-determined |
| S10_vs_92509\|\|lt2gene | Not-determined |
| S10_vs_92541\|\|lt2gene | Low-quality |
| S10_vs_92609\|\|lt2gene | Not-determined |
| S10_vs_92759\|\|lt2gene | Low-quality |
| S10_vs_92849\|\|lt2gene | Low-quality |
| S10_vs_93365\|\|lt2gene | Not-determined |
| S10_vs_93520\|\|lt2gene | Low-quality |
| S10_vs_93589\|\|lt2gene | Low-quality |
| S10_vs_93628\|\|lt2gene | Low-quality |
| S10_vs_93703\|\|lt2gene | Not-determined |
| S10_vs_93804\|\|lt2gene | Not-determined |

| | |
|---|---|
| S10_vs_93915\|\|lt2gene | Low-quality |
| S10_vs_94033\|\|lt2gene | Low-quality |
| S10_vs_94416\|\|lt2gene | Low-quality |
| S10_vs_94791\|\|lt2gene | Low-quality |
| S10_vs_94878\|\|lt2gene | Low-quality |
| S10_vs_94880\|\|lt2gene | Low-quality |
| S10_vs_94947\|\|lt2gene | Low-quality |
| S10_vs_94970\|\|lt2gene | Low-quality |
| S10_vs_95067\|\|lt2gene | Low-quality |
| S10_vs_95348\|\|lt2gene | Not-determined |
| S10_vs_95461\|\|lt2gene | Low-quality |
| S10_vs_95656\|\|lt2gene | Not-determined |
| S10_vs_95674\|\|lt2gene | Low-quality |
| S10_vs_95774\|\|lt2gene | Low-quality |
| S10_vs_95932\|\|lt2gene | Low-quality |
| S10_vs_96162\|\|lt2gene | Low-quality |
| S10_vs_96397\|\|lt2gene | Not-determined |
| S10_vs_96479\|\|lt2gene | Low-quality |
| S10_vs_96496\|\|lt2gene | Low-quality |
| S10_vs_96595\|\|lt2gene | Not-determined |
| S10_vs_96618\|\|lt2gene | Low-quality |
| S10_vs_96700\|\|lt2gene | Low-quality |
| S10_vs_96705\|\|lt2gene | Low-quality |
| S10_vs_96714\|\|lt2gene | Not-determined |
| S10_vs_96717\|\|lt2gene | Not-determined |
| S10_vs_96770\|\|lt2gene | Not-determined |
| S10_vs_96775\|\|lt2gene | Low-quality |
| S10_vs_96826\|\|lt2gene | Low-quality |
| S10_vs_96900\|\|lt2gene | Not-determined |
| S10_vs_96977\|\|lt2gene | Not-determined |
| S10_vs_96987\|\|lt2gene | Low-quality |
| S10_vs_96999\|\|lt2gene | Low-quality |
| S10_vs_97007\|\|lt2gene | Low-quality |
| S10_vs_97012\|\|lt2gene | Low-quality |
| S10_vs_97280\|\|lt2gene | Low-quality |
| S10_vs_97385\|\|lt2gene | Low-quality |
| S10_vs_97428\|\|lt2gene | Low-quality |
| S10_vs_97429\|\|lt2gene | Low-quality |
| S10_vs_97452\|\|lt2gene | Low-quality |
| S10_vs_97883\|\|lt2gene | Low-quality |
| S10_vs_97960\|\|lt2gene | Low-quality |
| S10_vs_98266\|\|lt2gene | Low-quality |
| S10_vs_98343\|\|lt2gene | Not-determined |
| S10_vs_98480\|\|lt2gene | Not-determined |
| S10_vs_98627\|\|lt2gene | Not-determined |
| S10_vs_98767\|\|lt2gene | Not-determined |
| S10_vs_98907\|\|lt2gene | Low-quality |
| S10_vs_99035\|\|lt2gene | Low-quality |
| S10_vs_99140\|\|lt2gene | Not-determined |
| S10_vs_99180\|\|lt2gene | Low-quality |
| S10_vs_99455\|\|lt2gene | Low-quality |
| S10_vs_99855\|\|lt2gene | Low-quality |
| S10_vs_99860\|\|lt2gene | Low-quality |
| S10_vs_99995\|\|lt2gene | Not-determined |
| S10_vs_100059\|\|lt2gene | Low-quality |
| S10_vs_100079\|\|lt2gene | Low-quality |
| S10_vs_100137\|\|lt2gene | Low-quality |
| S10_vs_100172\|\|lt2gene | Low-quality |
| S10_vs_100282\|\|lt2gene | Not-determined |
| S10_vs_100307\|\|lt2gene | Not-determined |
| S10_vs_100558\|\|lt2gene | Not-determined |
| S10_vs_100561\|\|lt2gene | Low-quality |
| S10_vs_100620\|\|lt2gene | Low-quality |
| S10_vs_100707\|\|lt2gene | Low-quality |
| S10_vs_100789\|\|lt2gene | Low-quality |
| S10_vs_101020\|\|lt2gene | Low-quality |
| S10_vs_101297\|\|lt2gene | Low-quality |
| S10_vs_101476\|\|lt2gene | Low-quality |
| S10_vs_101625\|\|lt2gene | Low-quality |
| S10_vs_101902\|\|lt2gene | Not-determined |
| S10_vs_101936\|\|lt2gene | Low-quality |
| S10_vs_101980\|\|lt2gene | Low-quality |
| S10_vs_102041\|\|lt2gene | Low-quality |
| S10_vs_102052\|\|lt2gene | Low-quality |
| S10_vs_102113\|\|lt2gene | Low-quality |
| S10_vs_102744\|\|lt2gene | Low-quality |
| S10_vs_102988\|\|lt2gene | Low-quality |
| S10_vs_103077\|\|lt2gene | Not-determined |
| S10_vs_103115\|\|lt2gene | Low-quality |
| S10_vs_103142\|\|lt2gene | Low-quality |
| S10_vs_103157\|\|lt2gene | Low-quality |
| S10_vs_103526\|\|lt2gene | Low-quality |
| S10_vs_103582\|\|lt2gene | Low-quality |
| S10_vs_103835\|\|lt2gene | Low-quality |
| S10_vs_104055\|\|lt2gene | Low-quality |
| S10_vs_104072\|\|lt2gene | Low-quality |
| S10_vs_104168\|\|lt2gene | Not-determined |
| S10_vs_104219\|\|lt2gene | Not-determined |
| S10_vs_104334\|\|lt2gene | Low-quality |
| S10_vs_104374\|\|lt2gene | Low-quality |
| S10_vs_104421\|\|lt2gene | Low-quality |
| S10_vs_104666\|\|lt2gene | Not-determined |
| S10_vs_104876\|\|lt2gene | Not-determined |
| S10_vs_104996\|\|lt2gene | Low-quality |
| S10_vs_105010\|\|lt2gene | Low-quality |
| S10_vs_105084\|\|lt2gene | Low-quality |
| S10_vs_105268\|\|lt2gene | Low-quality |
| S10_vs_105377\|\|lt2gene | Low-quality |
| S10_vs_105509\|\|lt2gene | Low-quality |
| S10_vs_105521\|\|lt2gene | Low-quality |
| S10_vs_105628\|\|lt2gene | Not-determined |
| S10_vs_105691\|\|lt2gene | Low-quality |
| S10_vs_105692\|\|lt2gene | Low-quality |
| S10_vs_106026\|\|lt2gene | Low-quality |
| S10_vs_106196\|\|lt2gene | Low-quality |
| S10_vs_106202\|\|lt2gene | Low-quality |
| S10_vs_106360\|\|lt2gene | Not-determined |
| S10_vs_106374\|\|lt2gene | Low-quality |
| S10_vs_106377\|\|lt2gene | Low-quality |
| S10_vs_106594\|\|lt2gene | Low-quality |
| S10_vs_106676\|\|lt2gene | Low-quality |
| S10_vs_106750\|\|lt2gene | Not-determined |
| S10_vs_106805\|\|lt2gene | Not-determined |
| S10_vs_106837\|\|lt2gene | Low-quality |
| S10_vs_106877\|\|lt2gene | Low-quality |
| S10_vs_106983\|\|lt2gene | Not-determined |
| S10_vs_107095\|\|lt2gene | Low-quality |
| S10_vs_107364\|\|lt2gene | Low-quality |
| S10_vs_107520\|\|lt2gene | Low-quality |
| S10_vs_107623\|\|lt2gene | Low-quality |
| S10_vs_107668\|\|lt2gene | Low-quality |
| S10_vs_107679\|\|lt2gene | Low-quality |
| S10_vs_107691\|\|lt2gene | Low-quality |
| S10_vs_107780\|\|lt2gene | Low-quality |
| S10_vs_107974\|\|lt2gene | Low-quality |
| S10_vs_108162\|\|lt2gene | Low-quality |
| S10_vs_108196\|\|lt2gene | Low-quality |
| S10_vs_108217\|\|lt2gene | Not-determined |
| S10_vs_108236\|\|lt2gene | Not-determined |
| S10_vs_108331\|\|lt2gene | Low-quality |
| S10_vs_108458\|\|lt2gene | Low-quality |
| S10_vs_108480\|\|lt2gene | Low-quality |
| S10_vs_108498\|\|lt2gene | Low-quality |
| S10_vs_108527\|\|lt2gene | Low-quality |
| S10_vs_108534\|\|lt2gene | Low-quality |
| S10_vs_108586\|\|lt2gene | Not-determined |
| S10_vs_108734\|\|lt2gene | Low-quality |
| S10_vs_108818\|\|lt2gene | Low-quality |
| S10_vs_108895\|\|lt2gene | Low-quality |
| S10_vs_109021\|\|lt2gene | Low-quality |
| S10_vs_109106\|\|lt2gene | Low-quality |
| S10_vs_109151\|\|lt2gene | Low-quality |
| S10_vs_109250\|\|lt2gene | Not-determined |
| S10_vs_109321\|\|lt2gene | Low-quality |
| S10_vs_109330\|\|lt2gene | Not-determined |
| S10_vs_109333\|\|lt2gene | Low-quality |
| S10_vs_109384\|\|lt2gene | Low-quality |
| S10_vs_109479\|\|lt2gene | Low-quality |
| S10_vs_109547\|\|lt2gene | Low-quality |
| S10_vs_109627\|\|lt2gene | Low-quality |
| S10_vs_109715\|\|lt2gene | Low-quality |
| S10_vs_109787\|\|lt2gene | Low-quality |
| S10_vs_109804\|\|lt2gene | Low-quality |
| S10_vs_109819\|\|lt2gene | Not-determined |
| S10_vs_109956\|\|lt2gene | Not-determined |
| S10_vs_110092\|\|lt2gene | Low-quality |
| S10_vs_110094\|\|lt2gene | Low-quality |
| S10_vs_110205\|\|lt2gene | Low-quality |
| S10_vs_110267\|\|lt2gene | Not-determined |
| S10_vs_110303\|\|lt2gene | Low-quality |
| S10_vs_110434\|\|lt2gene | Low-quality |
| S10_vs_110554\|\|lt2gene | Low-quality |
| S10_vs_110579\|\|lt2gene | Not-determined |
| S10_vs_110715\|\|lt2gene | Not-determined |
| S10_vs_110734\|\|lt2gene | Not-determined |
| S10_vs_110863\|\|lt2gene | Low-quality |
| S10_vs_110941\|\|lt2gene | Low-quality |
| S10_vs_111002\|\|lt2gene | Low-quality |
| S10_vs_111085\|\|lt2gene | Low-quality |
| S10_vs_111131\|\|lt2gene | Low-quality |
| S10_vs_111357\|\|lt2gene | Low-quality |
| S10_vs_111555\|\|lt2gene | Not-determined |
| S10_vs_111626\|\|lt2gene | Low-quality |
| S10_vs_111781\|\|lt2gene | Low-quality |
| S10_vs_111864\|\|lt2gene | Low-quality |
| S10_vs_111901\|\|lt2gene | Not-determined |
| S10_vs_111903\|\|lt2gene | Not-determined |
| S10_vs_111948\|\|lt2gene | Low-quality |
| S10_vs_111998\|\|lt2gene | Not-determined |
| S10_vs_112243\|\|lt2gene | Not-determined |
| S10_vs_112365\|\|lt2gene | Low-quality |
| S10_vs_112872\|\|lt2gene | Low-quality |
| S10_vs_113010\|\|lt2gene | Not-determined |
| S10_vs_113027\|\|lt2gene | Low-quality |
| S10_vs_113156\|\|lt2gene | Low-quality |
| S10_vs_113178\|\|lt2gene | Low-quality |
| S10_vs_113438\|\|lt2gene | Low-quality |
| S10_vs_113495\|\|lt2gene | Not-determined |
| S10_vs_113737\|\|lt2gene | Low-quality |
| S10_vs_113854\|\|lt2gene | Not-determined |
| S10_vs_113866\|\|lt2gene | Not-determined |
| S10_vs_113912\|\|lt2gene | Low-quality |
| S10_vs_113915\|\|lt2gene | Low-quality |
| S10_vs_113929\|\|lt2gene | Low-quality |
| S10_vs_114033\|\|lt2gene | Not-determined |
| S10_vs_114070\|\|lt2gene | Low-quality |
| S10_vs_114138\|\|lt2gene | Low-quality |
| S10_vs_114140\|\|lt2gene | Low-quality |
| S10_vs_114193\|\|lt2gene | Not-determined |
| S10_vs_114239\|\|lt2gene | Low-quality |
| S10_vs_114324\|\|lt2gene | Low-quality |
| S10_vs_114427\|\|lt2gene | Low-quality |
| S10_vs_114437\|\|lt2gene | Low-quality |
| S10_vs_114515\|\|lt2gene | Not-determined |
| S10_vs_114797\|\|lt2gene | Low-quality |
| S10_vs_114901\|\|lt2gene | Not-determined |
| S10_vs_114902\|\|lt2gene | Low-quality |
| S10_vs_115088\|\|lt2gene | Low-quality |
| S10_vs_115160\|\|lt2gene | Low-quality |
| S10_vs_115339\|\|lt2gene | Low-quality |
| S10_vs_115410\|\|lt2gene | Low-quality |
| S10_vs_115551\|\|lt2gene | Not-determined |
| S10_vs_115598\|\|lt2gene | Not-determined |
| S10_vs_115732\|\|lt2gene | Not-determined |
| S10_vs_115825\|\|lt2gene | Low-quality |
| S10_vs_115871\|\|lt2gene | Low-quality |
| S10_vs_116018\|\|lt2gene | Low-quality |
| S10_vs_116415\|\|lt2gene | Not-determined |
| S10_vs_116486\|\|lt2gene | Low-quality |
| S10_vs_116593\|\|lt2gene | Not-determined |
| S10_vs_116715\|\|lt2gene | Not-determined |
| S10_vs_116771\|\|lt2gene | Not-determined |
| S10_vs_116821\|\|lt2gene | Low-quality |
| S10_vs_116827\|\|lt2gene | Low-quality |
| S10_vs_116832\|\|lt2gene | Low-quality |
| S10_vs_116834\|\|lt2gene | Not-determined |
| S10_vs_116923\|\|lt2gene | Low-quality |
| S10_vs_116962\|\|lt2gene | Not-determined |
| S10_vs_117053\|\|lt2gene | Not-determined |
| S10_vs_117148\|\|lt2gene | Not-determined |
| S10_vs_117541\|\|lt2gene | Low-quality |
| S10_vs_117568\|\|lt2gene | Not-determined |
| S10_vs_117594\|\|lt2gene | Low-quality |
| S10_vs_117637\|\|lt2gene | Low-quality |
| S10_vs_117693\|\|lt2gene | Low-quality |
| S10_vs_117724\|\|lt2gene | Not-determined |
| S10_vs_117768\|\|lt2gene | Low-quality |
| S10_vs_117794\|\|lt2gene | Low-quality |
| S10_vs_117824\|\|lt2gene | Low-quality |
| S10_vs_117883\|\|lt2gene | Not-determined |
| S10_vs_117973\|\|lt2gene | Low-quality |
| S10_vs_117994\|\|lt2gene | Not-determined |
| S10_vs_118071\|\|lt2gene | Low-quality |
| S10_vs_118734\|\|lt2gene | Low-quality |
| S10_vs_118871\|\|lt2gene | Not-determined |
| S10_vs_118937\|\|lt2gene | Low-quality |
| S10_vs_119167\|\|lt2gene | Low-quality |
| S10_vs_119173\|\|lt2gene | Low-quality |
| S10_vs_119215\|\|lt2gene | Not-determined |
| S10_vs_119323\|\|lt2gene | Not-determined |
| S10_vs_119326\|\|lt2gene | Low-quality |
| S10_vs_119377\|\|lt2gene | Low-quality |
| S10_vs_119431\|\|lt2gene | Low-quality |
| S10_vs_119441\|\|lt2gene | Low-quality |
| S10_vs_119507\|\|lt2gene | Not-determined |
| S10_vs_119619\|\|lt2gene | Low-quality |
| S10_vs_119860\|\|lt2gene | Low-quality |
| S10_vs_119981\|\|lt2gene | Low-quality |
| S10_vs_120113\|\|lt2gene | Not-determined |
| S10_vs_120135\|\|lt2gene | Not-determined |
| S10_vs_120662\|\|lt2gene | Low-quality |
| S10_vs_121122\|\|lt2gene | Low-quality |
| S10_vs_121228\|\|lt2gene | Low-quality |
| S12_extracted_k141_86614 | Not-determined |
| S12_extracted_k141_110236 | Not-determined |
| S12_extracted_k141_149602 | Not-determined |
| S12_extracted_k141_47245 | Not-determined |
| S12_extracted_k141_141731 | Not-determined |
| S12_extracted_k141_62995 | Not-determined |
| S12_extracted_k141_181096 | Not-determined |
| S12_extracted_k141_181097 | Not-determined |
| S12_extracted_k141_94490 | Not-determined |
| S12_extracted_k141_55120 | Not-determined |
| S12_extracted_k141_110241 | Not-determined |
| S12_extracted_k141_55121 | Not-determined |
| S12_extracted_k141_7 | Not-determined |
| S12_extracted_k141_181099 | Not-determined |
| S12_extracted_k141_47250 | Low-quality |
| S12_extracted_k141_165354 | Not-determined |
| S12_extracted_k141_8 | Low-quality |
| S12_extracted_k141_133862 | Not-determined |
| S12_extracted_k141_10 | Not-determined |
| S12_extracted_k141_125990 | Not-determined |
| S12_extracted_k141_157478 | Not-determined |
| S12_extracted_k141_63001 | Low-quality |
| S12_extracted_k141_110246 | Not-determined |
| S12_extracted_k141_110247 | Not-determined |
| S12_extracted_k141_181104 | Low-quality |
| S12_extracted_k141_55124 | Not-determined |
| S12_extracted_k141_15755 | Not-determined |
| S12_extracted_k141_102371 | Not-determined |
| S12_extracted_k141_133871 | Not-determined |
| S12_extracted_k141_78749 | Not-determined |
| S12_extracted_k141_55127 | Not-determined |
| S12_extracted_k141_19 | Not-determined |
| S12_extracted_k141_181111 | Not-determined |
| S12_extracted_k141_55128 | Not-determined |
| S12_extracted_k141_133873 | Not-determined |
| S12_extracted_k141_23 | Not-determined |
| S12_extracted_k141_63009 | Not-determined |
| S12_extracted_k141_63012 | Not-determined |
| S12_extracted_k141_125996 | Not-determined |
| S12_extracted_k141_86631 | Not-determined |
| S12_extracted_k141_23640 | Not-determined |
| S12_extracted_k141_63014 | Not-determined |
| S12_extracted_k141_7882 | Not-determined |
| S12_extracted_k141_63015 | Not-determined |
| S12_extracted_k141_181113 | Not-determined |
| S12_extracted_k141_125999 | Low-quality |
| S12_extracted_k141_86634 | Not-determined |
| S12_extracted_k141_15766 | Not-determined |
| S12_extracted_k141_55135 | Not-determined |
| S12_extracted_k141_157491 | Not-determined |
| S12_extracted_k141_157493 | Not-determined |
| S12_extracted_k141_141741 | Low-quality |
| S12_extracted_k141_102376 | Not-determined |
| S12_extracted_k141_141743 | Not-determined |
| S12_extracted_k141_86637 | Not-determined |
| S12_extracted_k141_15769 | Not-determined |
| S12_extracted_k141_32 | Not-determined |
| S12_extracted_k141_23650 | Not-determined |
| S12_extracted_k141_31510 | Not-determined |
| S12_extracted_k141_7894 | Not-determined |
| S12_extracted_k141_149619 | Not-determined |
| S12_extracted_k141_15776 | Not-determined |
| S12_extracted_k141_86641 | Not-determined |
| S12_extracted_k141_23651 | Not-determined |
| S12_extracted_k141_31513 | Not-determined |
| S12_extracted_k141_78770 | Not-determined |
| S12_extracted_k141_126011 | Not-determined |
| S12_extracted_k141_102383 | Not-determined |
| S12_extracted_k141_181123 | Not-determined |
| S12_extracted_k141_165383 | Not-determined |
| S12_extracted_k141_102385 | Not-determined |
| S12_extracted_k141_133887 | Not-determined |
| S12_extracted_k141_102386 | Not-determined |
| S12_extracted_k141_181126 | Not-determined |
| S12_extracted_k141_165385 | Not-determined |
| S12_extracted_k141_78773 | Not-determined |
| S12_extracted_k141_165386 | Not-determined |
| S12_extracted_k141_86647 | Low-quality |
| S12_extracted_k141_165388 | Not-determined |
| S12_extracted_k141_102391 | Not-determined |
| S12_extracted_k141_149627 | Not-determined |
| S12_extracted_k141_23659 | Not-determined |
| S12_extracted_k141_86649 | Not-determined |
| S12_extracted_k141_94518 | Not-determined |
| S12_extracted_k141_86651 | Not-determined |
| S12_extracted_k141_149631 | Not-determined |
| S12_extracted_k141_157512 | Not-determined |
| S12_extracted_k141_31523 | Not-determined |
| S12_extracted_k141_126018 | Not-determined |
| S12_extracted_k141_86655 | Not-determined |
| S12_extracted_k141_173250 | Not-determined |
| S12_extracted_k141_47 | Not-determined |
| S12_extracted_k141_94524 | Not-determined |
| S12_extracted_k141_133896 | Not-determined |
| S12_extracted_k141_94530 | Not-determined |
| S12_extracted_k141_94533 | Not-determined |
| S12_extracted_k141_126028 | Not-determined |
| S12_extracted_k141_70897 | Not-determined |
| S12_extracted_k141_118131 | Not-determined |
| S12_extracted_k141_126029 | Not-determined |
| S12_extracted_k141_118132 | Not-determined |
| S12_extracted_k141_94534 | Not-determined |
| S12_extracted_k141_157521 | Not-determined |
| S12_extracted_k141_133899 | Not-determined |
| S12_extracted_k141_133903 | Not-determined |
| S12_extracted_k141_133904 | Not-determined |
| S12_extracted_k141_23671 | Low-quality |
| S12_extracted_k141_47269 | Not-determined |
| S12_extracted_k141_133907 | Not-determined |
| S12_extracted_k141_94542 | Not-determined |
| S12_extracted_k141_31542 | Not-determined |
| S12_extracted_k141_110267 | Not-determined |
| S12_extracted_k141_31546 | Not-determined |
| S12_extracted_k141_23677 | Not-determined |
| S12_extracted_k141_133915 | Not-determined |
| S12_extracted_k141_110269 | Not-determined |
| S12_extracted_k141_31552 | Not-determined |
| S12_extracted_k141_86689 | Not-determined |
| S12_extracted_k141_165413 | Not-determined |
| S12_extracted_k141_55145 | Not-determined |
| S12_extracted_k141_110272 | Not-determined |
| S12_extracted_k141_70913 | Low-quality |
| S12_extracted_k141_126046 | Not-determined |
| S12_extracted_k141_110274 | Not-determined |
| S12_extracted_k141_157534 | Not-determined |
| S12_extracted_k141_118148 | Not-determined |
| S12_extracted_k141_157539 | Not-determined |
| S12_extracted_k141_110278 | Not-determined |
| S12_extracted_k141_70916 | Low-quality |
| S12_extracted_k141_94549 | Not-determined |
| S12_extracted_k141_149666 | Not-determined |
| S12_extracted_k141_165420 | Low-quality |
| S12_extracted_k141_165421 | Not-determined |
| S12_extracted_k141_149669 | Not-determined |
| S12_extracted_k141_7910 | Not-determined |
| S12_extracted_k141_63044 | Not-determined |
| S12_extracted_k141_78783 | Not-determined |
| S12_extracted_k141_126060 | Not-determined |
| S12_extracted_k141_86707 | Not-determined |
| S12_extracted_k141_102431 | Low-quality |
| S12_extracted_k141_23686 | Not-determined |
| S12_extracted_k141_181146 | Low-quality |
| S12_extracted_k141_126068 | Not-determined |
| S12_extracted_k141_181147 | Not-determined |
| S12_extracted_k141_102434 | Not-determined |
| S12_extracted_k141_126070 | Not-determined |
| S12_extracted_k141_70931 | Not-determined |
| S12_extracted_k141_78789 | Low-quality |
| S12_extracted_k141_78790 | Not-determined |
| S12_extracted_k141_157549 | Not-determined |
| S12_extracted_k141_102437 | Not-determined |
| S12_extracted_k141_63055 | Not-determined |
| S12_extracted_k141_110294 | Low-quality |
| S12_extracted_k141_173292 | Not-determined |
| S12_extracted_k141_90 | Not-determined |
| S12_extracted_k141_86712 | Not-determined |
| S12_extracted_k141_7928 | Not-determined |
| S12_extracted_k141_86713 | Not-determined |
| S12_extracted_k141_7932 | Not-determined |
| S12_extracted_k141_181153 | Low-quality |
| S12_extracted_k141_110299 | Not-determined |
| S12_extracted_k141_149682 | Not-determined |
| S12_extracted_k141_181157 | Not-determined |
| S12_extracted_k141_70942 | Not-determined |
| S12_extracted_k141_7937 | Not-determined |
| S12_extracted_k141_110303 | Not-determined |
| S12_extracted_k141_78809 | Low-quality |
| S12_extracted_k141_78810 | Not-determined |
| S12_extracted_k141_31564 | Low-quality |
| S12_extracted_k141_165440 | Not-determined |
| S12_extracted_k141_31569 | Not-determined |
| S12_extracted_k141_157562 | Not-determined |
| S12_extracted_k141_31571 | Not-determined |
| S12_extracted_k141_7943 | Not-determined |
| S12_extracted_k141_110306 | Not-determined |
| S12_extracted_k141_102450 | Not-determined |
| S12_extracted_k141_181161 | Not-determined |
| S12_extracted_k141_173301 | Not-determined |
| S12_extracted_k141_157565 | Not-determined |
| S12_extracted_k141_78818 | Not-determined |
| S12_extracted_k141_102452 | Not-determined |
| S12_extracted_k141_23695 | Not-determined |
| S12_extracted_k141_78820 | Not-determined |
| S12_extracted_k141_70950 | Not-determined |
| S12_extracted_k141_173303 | Low-quality |
| S12_extracted_k141_47273 | Not-determined |
| S12_extracted_k141_165448 | Low-quality |
| S12_extracted_k141_120 | Not-determined |
| S12_extracted_k141_126090 | Not-determined |
| S12_extracted_k141_39413 | Not-determined |
| S12_extracted_k141_7950 | Low-quality |
| S12_extracted_k141_181169 | Not-determined |
| S12_extracted_k141_173306 | Not-determined |
| S12_extracted_k141_78829 | Low-quality |
| S12_extracted_k141_157574 | Not-determined |
| S12_extracted_k141_55165 | Not-determined |
| S12_extracted_k141_173311 | Not-determined |
| S12_extracted_k141_181176 | Low-quality |
| S12_extracted_k141_173312 | Low-quality |
| S12_extracted_k141_129 | Not-determined |
| S12_extracted_k141_55169 | Not-determined |
| S12_extracted_k141_23704 | Not-determined |
| S12_extracted_k141_165461 | Not-determined |
| S12_extracted_k141_47284 | Not-determined |
| S12_extracted_k141_165463 | Not-determined |
| S12_extracted_k141_118161 | Not-determined |
| S12_extracted_k141_23707 | Not-determined |
| S12_extracted_k141_15800 | Not-determined |
| S12_extracted_k141_149688 | Low-quality |
| S12_extracted_k141_181182 | Not-determined |
| S12_extracted_k141_126110 | Not-determined |
| S12_extracted_k141_126111 | Low-quality |
| S12_extracted_k141_137 | Not-determined |
| S12_extracted_k141_173325 | Not-determined |
| S12_extracted_k141_70967 | Not-determined |
| S12_extracted_k141_173328 | Not-determined |
| S12_extracted_k141_126113 | Not-determined |
| S12_extracted_k141_165473 | Not-determined |
| S12_extracted_k141_15807 | Not-determined |
| S12_extracted_k141_15809 | Not-determined |
| S12_extracted_k141_149694 | Not-determined |
| S12_extracted_k141_7977 | Not-determined |
| S12_extracted_k141_149697 | Not-determined |
| S12_extracted_k141_23717 | Not-determined |
| S12_extracted_k141_149698 | Not-determined |
| S12_extracted_k141_15815 | Not-determined |
| S12_extracted_k141_118179 | Not-determined |
| S12_extracted_k141_141763 | Not-determined |
| S12_extracted_k141_165480 | Not-determined |
| S12_extracted_k141_23721 | Not-determined |
| S12_extracted_k141_7980 | Not-determined |
| S12_extracted_k141_141 | Not-determined |
| S12_extracted_k141_126123 | Not-determined |
| S12_extracted_k141_23726 | Not-determined |
| S12_extracted_k141_126125 | Not-determined |
| S12_extracted_k141_144 | Not-determined |
| S12_extracted_k141_110320 | Not-determined |
| S12_extracted_k141_118187 | Not-determined |
| S12_extracted_k141_149711 | Not-determined |
| S12_extracted_k141_23734 | Not-determined |
| S12_extracted_k141_110330 | Not-determined |
| S12_extracted_k141_173340 | Not-determined |
| S12_extracted_k141_63075 | Not-determined |
| S12_extracted_k141_110332 | Not-determined |
| S12_extracted_k141_63076 | Not-determined |
| S12_extracted_k141_94581 | Not-determined |
| S12_extracted_k141_70980 | Not-determined |
| S12_extracted_k141_63080 | Not-determined |
| S12_extracted_k141_70983 | Not-determined |
| S12_extracted_k141_15819 | Not-determined |
| S12_extracted_k141_118200 | Not-determined |
| S12_extracted_k141_63082 | Not-determined |
| S12_extracted_k141_70984 | Not-determined |
| S12_extracted_k141_70985 | Not-determined |
| S12_extracted_k141_133937 | Low-quality |
| S12_extracted_k141_15823 | Not-determined |
| S12_extracted_k141_86737 | Low-quality |
| S12_extracted_k141_157587 | Low-quality |
| S12_extracted_k141_126144 | Not-determined |
| S12_extracted_k141_165493 | Not-determined |
| S12_extracted_k141_141783 | Not-determined |
| S12_extracted_k141_157590 | Not-determined |
| S12_extracted_k141_173346 | Low-quality |
| S12_extracted_k141_181193 | Not-determined |
| S12_extracted_k141_110338 | Low-quality |
| S12_extracted_k141_71000 | Not-determined |
| S12_extracted_k141_8007 | Not-determined |
| S12_extracted_k141_149725 | Not-determined |
| S12_extracted_k141_15833 | Not-determined |
| S12_extracted_k141_39446 | Not-determined |
| S12_extracted_k141_71004 | Low-quality |
| S12_extracted_k141_71005 | Not-determined |
| S12_extracted_k141_94599 | Not-determined |
| S12_extracted_k141_15835 | Not-determined |
| S12_extracted_k141_94600 | Not-determined |
| S12_extracted_k141_165495 | Not-determined |
| S12_extracted_k141_110343 | Not-determined |
| S12_extracted_k141_71007 | Not-determined |
| S12_extracted_k141_173355 | Not-determined |
| S12_extracted_k141_173356 | Low-quality |
| S12_extracted_k141_94604 | Not-determined |
| S12_extracted_k141_94606 | Not-determined |
| S12_extracted_k141_110346 | Low-quality |
| S12_extracted_k141_181205 | Not-determined |
| S12_extracted_k141_133953 | Not-determined |
| S12_extracted_k141_71012 | Not-determined |
| S12_extracted_k141_71015 | Not-determined |
| S12_extracted_k141_15837 | Low-quality |
| S12_extracted_k141_102488 | Not-determined |
| S12_extracted_k141_133958 | Low-quality |
| S12_extracted_k141_126149 | Not-determined |
| S12_extracted_k141_78866 | Not-determined |
| S12_extracted_k141_39462 | Not-determined |
| S12_extracted_k141_118214 | Low-quality |
| S12_extracted_k141_178 | Not-determined |
| S12_extracted_k141_181212 | Low-quality |
| S12_extracted_k141_180 | Not-determined |
| S12_extracted_k141_63090 | Not-determined |
| S12_extracted_k141_78869 | Not-determined |
| S12_extracted_k141_181214 | Not-determined |
| S12_extracted_k141_78870 | Not-determined |
| S12_extracted_k141_118218 | Not-determined |
| S12_extracted_k141_86762 | Not-determined |
| S14_vs_k141_38905\|\|full | Medium-quality |
| S12_extracted_k141_133968 | Not-determined |
| S12_extracted_k141_181218 | Not-determined |
| S12_extracted_k141_94610 | Not-determined |
| S12_extracted_k141_141813 | Not-determined |
| S12_extracted_k141_63103 | Not-determined |
| S12_extracted_k141_23773 | Not-determined |
| S12_extracted_k141_110352 | Not-determined |
| S12_extracted_k141_133974 | Not-determined |
| S12_extracted_k141_8027 | Not-determined |
| S12_extracted_k141_181225 | Not-determined |
| S12_extracted_k141_165504 | Not-determined |
| S12_extracted_k141_94619 | Not-determined |
| S12_extracted_k141_141817 | Not-determined |
| S12_extracted_k141_141821 | Not-determined |
| S12_extracted_k141_141825 | Not-determined |
| S12_extracted_k141_110358 | Not-determined |
| S12_extracted_k141_86777 | Not-determined |
| S12_extracted_k141_94623 | Not-determined |
| S12_extracted_k141_39481 | Not-determined |
| S12_extracted_k141_86785 | Low-quality |
| S12_extracted_k141_198 | Not-determined |
| S12_extracted_k141_39488 | Not-determined |

| | |
|---|---|
| S12_extracted_k141_63116 | Not-determined |
| S12_extracted_k141_102500 | Low-quality |
| S12_extracted_k141_126164 | Low-quality |
| S12_extracted_k141_63119 | Not-determined |
| S12_extracted_k141_86791 | Low-quality |
| S12_extracted_k141_118241 | Not-determined |
| S12_extracted_k141_55195 | Not-determined |
| S12_extracted_k141_63120 | Not-determined |
| S12_extracted_k141_8047 | Not-determined |
| S12_extracted_k141_181239 | Not-determined |
| S12_extracted_k141_165527 | Low-quality |
| S12_extracted_k141_165528 | Not-determined |
| S12_extracted_k141_141833 | Not-determined |
| S12_extracted_k141_86799 | Not-determined |
| S12_extracted_k141_110370 | Not-determined |
| S12_extracted_k141_47325 | Low-quality |
| S12_extracted_k141_173374 | Not-determined |
| S12_extracted_k141_47327 | Not-determined |
| S12_extracted_k141_71024 | Not-determined |
| S12_extracted_k141_141839 | Not-determined |
| S12_extracted_k141_110376 | Not-determined |
| S12_extracted_k141_165536 | Not-determined |
| S12_extracted_k141_55200 | Not-determined |
| S12_extracted_k141_118253 | Not-determined |
| S12_extracted_k141_47333 | Not-determined |
| S12_extracted_k141_47337 | Not-determined |
| S12_extracted_k141_94638 | Not-determined |
| S12_extracted_k141_86807 | Not-determined |
| S12_extracted_k141_71034 | Not-determined |
| S12_extracted_k141_118255 | Low-quality |
| S12_extracted_k141_221 | Not-determined |
| S12_extracted_k141_118258 | Not-determined |
| S12_extracted_k141_15843 | Not-determined |
| S12_extracted_k141_173387 | Not-determined |
| S12_extracted_k141_94643 | Not-determined |
| S12_extracted_k141_133996 | Not-determined |
| S12_extracted_k141_173391 | Not-determined |
| S12_extracted_k141_149748 | Not-determined |
| S12_extracted_k141_86815 | Low-quality |
| S12_extracted_k141_47340 | Not-determined |
| S12_extracted_k141_71043 | Not-determined |
| S12_extracted_k141_55207 | Not-determined |
| S12_extracted_k141_165546 | Not-determined |
| S12_extracted_k141_63132 | Not-determined |
| S12_extracted_k141_165547 | Not-determined |
| S12_extracted_k141_173395 | Not-determined |
| S12_extracted_k141_229 | Not-determined |
| S12_extracted_k141_102512 | Not-determined |
| S12_extracted_k141_118270 | Not-determined |
| S12_extracted_k141_94646 | Low-quality |
| S12_extracted_k141_181262 | Not-determined |
| S12_extracted_k141_157598 | Not-determined |
| S12_extracted_k141_181266 | Not-determined |
| S12_extracted_k141_234 | Not-determined |
| S12_extracted_k141_102521 | Not-determined |
| S12_extracted_k141_23791 | Not-determined |
| S12_extracted_k141_86826 | Not-determined |
| S12_extracted_k141_47346 | Not-determined |
| S12_extracted_k141_47347 | Low-quality |
| S12_extracted_k141_71053 | Not-determined |
| S12_extracted_k141_165552 | Not-determined |
| S12_extracted_k141_181273 | Not-determined |
| S12_extracted_k141_134008 | Not-determined |
| S12_extracted_k141_165556 | Not-determined |
| S12_extracted_k141_47352 | Not-determined |
| S12_extracted_k141_134009 | Not-determined |
| S12_extracted_k141_102529 | Not-determined |
| S12_extracted_k141_78890 | Not-determined |
| S12_extracted_k141_47363 | Not-determined |
| S12_extracted_k141_165571 | Low-quality |
| S12_extracted_k141_31627 | Not-determined |
| S12_extracted_k141_141872 | Not-determined |
| S12_extracted_k141_94674 | Not-determined |
| S12_extracted_k141_86847 | Not-determined |
| S12_extracted_k141_71066 | Not-determined |
| S12_extracted_k141_157623 | Not-determined |
| S12_extracted_k141_47368 | Low-quality |
| S12_extracted_k141_254 | Not-determined |
| S12_extracted_k141_47370 | Not-determined |
| S12_extracted_k141_165580 | Not-determined |
| S12_extracted_k141_78907 | Not-determined |
| S12_extracted_k141_102544 | Not-determined |
| S12_extracted_k141_257 | Low-quality |
| S12_extracted_k141_173402 | Low-quality |
| S12_extracted_k141_55221 | Low-quality |
| S12_extracted_k141_173404 | Not-determined |
| S12_extracted_k141_118289 | Low-quality |
| S12_extracted_k141_165587 | Not-determined |
| S12_extracted_k141_47376 | Not-determined |
| S12_extracted_k141_173409 | Not-determined |
| S12_extracted_k141_157630 | Not-determined |
| S12_extracted_k141_55229 | Not-determined |
| S12_extracted_k141_134025 | Not-determined |
| S12_extracted_k141_110400 | Not-determined |
| S12_extracted_k141_55234 | Not-determined |
| S12_extracted_k141_47384 | Not-determined |
| S12_extracted_k141_181292 | Not-determined |
| S12_extracted_k141_181293 | Not-determined |
| S12_extracted_k141_264 | Not-determined |
| S12_extracted_k141_102561 | Not-determined |
| S12_extracted_k141_173423 | Not-determined |
| S12_extracted_k141_165601 | Not-determined |
| S12_extracted_k141_110406 | Not-determined |
| S12_extracted_k141_118304 | Not-determined |
| S12_extracted_k141_102565 | Not-determined |
| S12_extracted_k141_8053 | Not-determined |
| S12_extracted_k141_94691 | Not-determined |
| S12_extracted_k141_157643 | Not-determined |
| S12_extracted_k141_110409 | Not-determined |
| S12_extracted_k141_173426 | Low-quality |
| S12_extracted_k141_118312 | Not-determined |
| S12_extracted_k141_102570 | Low-quality |
| S12_extracted_k141_86875 | Not-determined |
| S12_extracted_k141_118314 | Not-determined |
| S12_extracted_k141_110410 | Not-determined |
| S12_extracted_k141_270 | Not-determined |
| S12_extracted_k141_118316 | Not-determined |
| S12_extracted_k141_94695 | Not-determined |
| S12_extracted_k141_94697 | Not-determined |
| S12_extracted_k141_55243 | Not-determined |
| S12_extracted_k141_94698 | Not-determined |
| S12_extracted_k141_134046 | Not-determined |
| S12_extracted_k141_86878 | Low-quality |
| S12_extracted_k141_165604 | Low-quality |
| S12_extracted_k141_118319 | Not-determined |
| S12_extracted_k141_94704 | Not-determined |
| S12_extracted_k141_165606 | Not-determined |
| S12_extracted_k141_15853 | Not-determined |
| S12_extracted_k141_63169 | Not-determined |
| S12_extracted_k141_165610 | Not-determined |
| S12_extracted_k141_157655 | Not-determined |
| S12_extracted_k141_134049 | Not-determined |
| S12_extracted_k141_86886 | Not-determined |
| S12_extracted_k141_165613 | Not-determined |
| S12_extracted_k141_141890 | Not-determined |
| S12_extracted_k141_157659 | Not-determined |
| S12_extracted_k141_102586 | Not-determined |
| S12_extracted_k141_94714 | Low-quality |
| S12_extracted_k141_63177 | Not-determined |
| S12_extracted_k141_165617 | Not-determined |
| S12_extracted_k141_110420 | Low-quality |
| S12_extracted_k141_63179 | Not-determined |
| S12_extracted_k141_63180 | Low-quality |
| S12_extracted_k141_71077 | Not-determined |
| S12_extracted_k141_94716 | Low-quality |
| S12_extracted_k141_78946 | Low-quality |
| S12_extracted_k141_94720 | Not-determined |
| S12_extracted_k141_8073 | Not-determined |
| S12_extracted_k141_181298 | Low-quality |
| S12_extracted_k141_102593 | Low-quality |
| S12_extracted_k141_173445 | Not-determined |
| S12_extracted_k141_110424 | Not-determined |
| S12_extracted_k141_118340 | Not-determined |
| S12_extracted_k141_8082 | Not-determined |
| S12_extracted_k141_94734 | Not-determined |
| S12_extracted_k141_8086 | Not-determined |
| S12_extracted_k141_110428 | Not-determined |
| S12_extracted_k141_78956 | Not-determined |
| S12_extracted_k141_141898 | Not-determined |
| S12_extracted_k141_149779 | Not-determined |
| S12_extracted_k141_55248 | Not-determined |
| S12_extracted_k141_55251 | Not-determined |
| S12_extracted_k141_39508 | Not-determined |
| S12_extracted_k141_71089 | Not-determined |
| S12_extracted_k141_181315 | Not-determined |
| S12_extracted_k141_134058 | Not-determined |
| S12_extracted_k141_63205 | Not-determined |
| S12_extracted_k141_181319 | Not-determined |
| S12_extracted_k141_110447 | Not-determined |
| S12_extracted_k141_181320 | Not-determined |
| S12_extracted_k141_126202 | Not-determined |
| S12_extracted_k141_86918 | Not-determined |
| S12_extracted_k141_296 | Low-quality |
| S12_extracted_k141_15867 | Not-determined |
| S12_extracted_k141_94748 | Not-determined |
| S12_extracted_k141_86923 | Not-determined |
| S12_extracted_k141_149793 | Not-determined |
| S12_extracted_k141_299 | Low-quality |
| S12_extracted_k141_110453 | Not-determined |
| S12_extracted_k141_301 | Not-determined |
| S12_extracted_k141_302 | Not-determined |
| S12_extracted_k141_78982 | Not-determined |
| S12_extracted_k141_39522 | Not-determined |
| S12_extracted_k141_55255 | Low-quality |
| S12_extracted_k141_134064 | Not-determined |
| S12_extracted_k141_15870 | Not-determined |
| S12_extracted_k141_141914 | Not-determined |
| S12_extracted_k141_126211 | Not-determined |
| S12_extracted_k141_110457 | Not-determined |
| S12_extracted_k141_63217 | Not-determined |
| S12_extracted_k141_39525 | Not-determined |
| S12_extracted_k141_110459 | Not-determined |
| S12_extracted_k141_173476 | Low-quality |
| S12_extracted_k141_71105 | Not-determined |
| S12_extracted_k141_310 | Not-determined |
| S12_extracted_k141_78988 | Low-quality |
| S12_extracted_k141_94756 | Not-determined |
| S12_extracted_k141_110462 | Not-determined |
| S12_extracted_k141_55258 | Not-determined |
| S12_extracted_k141_149805 | Not-determined |
| S12_extracted_k141_134068 | Not-determined |
| S12_extracted_k141_94759 | Not-determined |
| S12_extracted_k141_157694 | Not-determined |
| S12_extracted_k141_8099 | Not-determined |
| S12_extracted_k141_55263 | Not-determined |
| S12_extracted_k141_94766 | Not-determined |
| S12_extracted_k141_110470 | Not-determined |
| S12_extracted_k141_102624 | Not-determined |
| S12_extracted_k141_78999 | Not-determined |
| S12_extracted_k141_79000 | Not-determined |
| S12_extracted_k141_157700 | Not-determined |
| S12_extracted_k141_181331 | Not-determined |
| S12_extracted_k141_181332 | Not-determined |
| S12_extracted_k141_94768 | Not-determined |
| S12_extracted_k141_327 | Not-determined |
| S12_extracted_k141_173490 | Not-determined |
| S12_extracted_k141_173491 | Not-determined |
| S12_extracted_k141_126233 | Not-determined |
| S12_extracted_k141_157709 | Not-determined |
| S12_extracted_k141_126235 | Not-determined |
| S12_extracted_k141_173493 | Not-determined |
| S12_extracted_k141_149821 | Not-determined |
| S12_extracted_k141_149823 | Not-determined |
| S12_extracted_k141_149825 | Not-determined |
| S12_extracted_k141_126242 | Not-determined |
| S12_extracted_k141_110477 | Not-determined |
| S12_extracted_k141_39548 | Not-determined |
| S12_extracted_k141_332 | Not-determined |
| S12_extracted_k141_39549 | Not-determined |
| S12_extracted_k141_126243 | Not-determined |
| S12_extracted_k141_181345 | Not-determined |
| S12_extracted_k141_134087 | Low-quality |
| S12_extracted_k141_141922 | Not-determined |
| S12_extracted_k141_126246 | Not-determined |
| S12_extracted_k141_39552 | Not-determined |
| S12_extracted_k141_71119 | Not-determined |
| S12_extracted_k141_173502 | Not-determined |
| S12_extracted_k141_141924 | Not-determined |
| S12_extracted_k141_110478 | Not-determined |
| S12_extracted_k141_8119 | Not-determined |
| S12_extracted_k141_39558 | Not-determined |
| S12_extracted_k141_102643 | Not-determined |
| S12_extracted_k141_8125 | Not-determined |
| S12_extracted_k141_338 | Not-determined |
| S12_extracted_k141_110482 | Not-determined |
| S12_extracted_k141_15885 | Low-quality |
| S12_extracted_k141_63246 | Not-determined |
| S12_extracted_k141_8127 | Not-determined |
| S12_extracted_k141_8131 | Not-determined |
| S12_extracted_k141_94792 | Not-determined |
| S12_extracted_k141_102650 | Not-determined |
| S12_extracted_k141_63249 | Not-determined |
| S12_extracted_k141_55279 | Not-determined |
| S12_extracted_k141_343 | Not-determined |
| S12_extracted_k141_126258 | Not-determined |
| S12_extracted_k141_94794 | Not-determined |
| S12_extracted_k141_94795 | Not-determined |
| S12_extracted_k141_165653 | Not-determined |
| S12_extracted_k141_141935 | Not-determined |
| S12_extracted_k141_149840 | Not-determined |
| S12_extracted_k141_102657 | Not-determined |
| S12_extracted_k141_71142 | Low-quality |
| S12_extracted_k141_55282 | Not-determined |
| S12_extracted_k141_39578 | Not-determined |
| S12_extracted_k141_102665 | Not-determined |
| S12_extracted_k141_141939 | Not-determined |
| S12_extracted_k141_39587 | Not-determined |
| S12_extracted_k141_39591 | Not-determined |
| S12_extracted_k141_55290 | Not-determined |
| S12_extracted_k141_63253 | Low-quality |
| S12_extracted_k141_55294 | Not-determined |
| S12_extracted_k141_47410 | Low-quality |
| S12_extracted_k141_126271 | Not-determined |
| S12_extracted_k141_369 | Low-quality |
| S12_extracted_k141_165659 | Not-determined |
| S12_extracted_k141_55299 | Low-quality |
| S12_extracted_k141_181386 | Not-determined |
| S12_extracted_k141_39602 | Not-determined |
| S12_extracted_k141_15896 | Not-determined |
| S12_extracted_k141_94806 | Not-determined |
| S12_extracted_k141_15897 | Not-determined |
| S12_extracted_k141_39607 | Not-determined |
| S12_extracted_k141_39609 | Low-quality |
| S12_extracted_k141_15901 | Not-determined |
| S12_extracted_k141_63267 | Not-determined |
| S12_extracted_k141_110515 | Not-determined |
| S12_extracted_k141_94811 | Not-determined |
| S12_extracted_k141_71166 | Not-determined |
| S12_extracted_k141_63276 | Not-determined |
| S12_extracted_k141_8143 | Not-determined |
| S12_extracted_k141_102695 | Not-determined |
| S12_extracted_k141_8145 | Not-determined |
| S12_extracted_k141_141961 | Low-quality |
| S12_extracted_k141_8147 | Not-determined |
| S12_extracted_k141_55322 | Not-determined |
| S12_extracted_k141_134131 | Not-determined |
| S12_extracted_k141_110525 | Not-determined |
| S12_extracted_k141_157721 | Not-determined |
| S12_extracted_k141_110527 | Low-quality |
| S12_extracted_k141_134134 | Low-quality |
| S12_extracted_k141_141969 | Low-quality |
| S12_extracted_k141_47428 | Not-determined |
| S3_vs_k141_119771\|\|full | Medium-quality |
| S12_extracted_k141_110532 | Not-determined |
| S12_extracted_k141_47431 | Not-determined |
| S12_extracted_k141_47433 | Not-determined |
| S16_vs_19320\|\|full | Medium-quality |
| S12_extracted_k141_15922 | Not-determined |
| S12_extracted_k141_55336 | Low-quality |
| S12_extracted_k141_79008 | Low-quality |
| S12_extracted_k141_134142 | Not-determined |
| S12_extracted_k141_15927 | Low-quality |
| S12_extracted_k141_384 | Not-determined |
| S12_extracted_k141_385 | Not-determined |
| S12_extracted_k141_134147 | Not-determined |
| S12_extracted_k141_141987 | Not-determined |
| S12_extracted_k141_388 | Not-determined |
| S12_extracted_k141_126303 | Not-determined |
| S12_extracted_k141_393 | Not-determined |
| S12_extracted_k141_141993 | Not-determined |
| S12_extracted_k141_134156 | Not-determined |
| S12_extracted_k141_71198 | Not-determined |
| S12_extracted_k141_79024 | Low-quality |
| S12_extracted_k141_86960 | Not-determined |
| S12_extracted_k141_110550 | Not-determined |
| S12_extracted_k141_126311 | Not-determined |
| S12_extracted_k141_47462 | Not-determined |
| S12_extracted_k141_39642 | Not-determined |
| S12_extracted_k141_126313 | Not-determined |
| S12_extracted_k141_142001 | Not-determined |
| S12_extracted_k141_86970 | Not-determined |
| S13_extracted_38708 | Medium-quality |
| S12_extracted_k141_39652 | Not-determined |
| S12_extracted_k141_8181 | Not-determined |
| S12_extracted_k141_157729 | Not-determined |
| S12_extracted_k141_8183 | Not-determined |
| S12_extracted_k141_157730 | Not-determined |
| S12_extracted_k141_8184 | Not-determined |
| S12_extracted_k141_157731 | Not-determined |
| S12_extracted_k141_118357 | Not-determined |
| S12_extracted_k141_79044 | Low-quality |
| S12_extracted_k141_15948 | Not-determined |
| S12_extracted_k141_157742 | Not-determined |
| S12_extracted_k141_86989 | Not-determined |
| S12_extracted_k141_427 | Not-determined |
| S12_extracted_k141_63309 | Not-determined |
| S12_extracted_k141_79049 | Low-quality |
| S12_extracted_k141_110583 | Not-determined |
| S12_extracted_k141_118373 | Not-determined |
| S12_extracted_k141_157748 | Not-determined |
| S12_extracted_k141_149859 | Not-determined |
| S12_extracted_k141_142035 | Not-determined |
| S12_extracted_k141_55356 | Not-determined |
| S12_extracted_k141_149862 | Not-determined |
| S12_extracted_k141_126338 | Not-determined |
| S12_extracted_k141_431 | Not-determined |
| S12_extracted_k141_55363 | Not-determined |
| S12_extracted_k141_142041 | Not-determined |
| S12_extracted_k141_165678 | Not-determined |
| S12_extracted_k141_118386 | Not-determined |
| S12_extracted_k141_126348 | Not-determined |
| S12_extracted_k141_126349 | Not-determined |
| S12_extracted_k141_86996 | Not-determined |
| S12_extracted_k141_63319 | Not-determined |
| S12_extracted_k141_63323 | Not-determined |
| S12_extracted_k141_126351 | Not-determined |
| S12_extracted_k141_63327 | Not-determined |
| S12_extracted_k141_126356 | Not-determined |
| S12_extracted_k141_142049 | Not-determined |
| S12_extracted_k141_63329 | Not-determined |
| S12_extracted_k141_87011 | Not-determined |
| S12_extracted_k141_63334 | Not-determined |
| S12_extracted_k141_15959 | Not-determined |
| S12_extracted_k141_87014 | Not-determined |
| S12_extracted_k141_118399 | Low-quality |
| S12_extracted_k141_87015 | Not-determined |
| S12_extracted_k141_126361 | Low-quality |
| S12_extracted_k141_142055 | Not-determined |
| S12_extracted_k141_126363 | Not-determined |
| S12_extracted_k141_55382 | Not-determined |
| S12_extracted_k141_102704 | Not-determined |
| S12_extracted_k141_39672 | Not-determined |
| S12_extracted_k141_31643 | Low-quality |
| S12_extracted_k141_126369 | Low-quality |
| S12_extracted_k141_39677 | Not-determined |
| S12_extracted_k141_63341 | Not-determined |
| S12_extracted_k141_118411 | Not-determined |
| S12_extracted_k141_15978 | Not-determined |
| S12_extracted_k141_15980 | Not-determined |
| S12_extracted_k141_31651 | Low-quality |
| S12_extracted_k141_118416 | Not-determined |
| S12_extracted_k141_39687 | Not-determined |
| S12_extracted_k141_118419 | Not-determined |
| S12_extracted_k141_126379 | Not-determined |
| S12_extracted_k141_435 | Not-determined |
| S12_extracted_k141_436 | Not-determined |
| S12_extracted_k141_55390 | Not-determined |
| S12_extracted_k141_126383 | Not-determined |
| S12_extracted_k141_39695 | Not-determined |
| S12_extracted_k141_149871 | Low-quality |
| S12_extracted_k141_118420 | Low-quality |
| S12_extracted_k141_39699 | Not-determined |
| S12_extracted_k141_31654 | Not-determined |
| S12_extracted_k141_157776 | Not-determined |
| S12_extracted_k141_31656 | Not-determined |
| S12_extracted_k141_39701 | Not-determined |
| S12_extracted_k141_31657 | Not-determined |
| S12_extracted_k141_31661 | Not-determined |
| S12_extracted_k141_63359 | Not-determined |
| S12_extracted_k141_440 | Not-determined |
| S12_extracted_k141_157784 | Not-determined |
| S12_extracted_k141_31671 | Not-determined |
| S12_extracted_k141_39709 | Not-determined |
| S12_extracted_k141_87038 | Not-determined |
| S12_extracted_k141_142066 | Not-determined |
| S12_extracted_k141_126392 | Not-determined |
| S12_extracted_k141_87042 | Not-determined |
| S12_extracted_k141_39715 | Not-determined |
| S12_extracted_k141_79081 | Not-determined |
| S12_extracted_k141_55404 | Not-determined |
| S12_extracted_k141_39720 | Not-determined |
| S12_extracted_k141_142072 | Not-determined |
| S12_extracted_k141_39721 | Not-determined |
| S12_extracted_k141_157790 | Low-quality |
| S12_extracted_k141_94822 | Not-determined |
| S12_extracted_k141_79091 | Low-quality |
| S12_extracted_k141_165693 | Low-quality |
| S12_extracted_k141_79096 | Not-determined |
| S12_extracted_k141_87055 | Not-determined |
| S12_extracted_k141_455 | Not-determined |
| S12_extracted_k141_55418 | Not-determined |
| S12_extracted_k141_142080 | Not-determined |
| S12_extracted_k141_94837 | Not-determined |
| S12_extracted_k141_126405 | Low-quality |
| S12_extracted_k141_465 | Not-determined |
| S12_extracted_k141_126409 | Not-determined |
| S12_extracted_k141_142082 | Not-determined |
| S12_extracted_k141_31687 | Not-determined |
| S12_extracted_k141_470 | Not-determined |
| S12_extracted_k141_173529 | Not-determined |
| S12_extracted_k141_39725 | Not-determined |
| S12_extracted_k141_63391 | Not-determined |
| S12_extracted_k141_79112 | Not-determined |
| S12_extracted_k141_39729 | Not-determined |
| S12_extracted_k141_165706 | Low-quality |
| S12_extracted_k141_63395 | Low-quality |
| S12_extracted_k141_173540 | Not-determined |
| S12_extracted_k141_142101 | Not-determined |
| S12_extracted_k141_39731 | Low-quality |
| S12_extracted_k141_31704 | Not-determined |
| S12_extracted_k141_87058 | Not-determined |
| S12_extracted_k141_157801 | Low-quality |
| S12_extracted_k141_31710 | Not-determined |
| S12_extracted_k141_31713 | Low-quality |
| S12_extracted_k141_149892 | Not-determined |
| S12_extracted_k141_39743 | Not-determined |
| S12_extracted_k141_118447 | Not-determined |
| S12_extracted_k141_94875 | Not-determined |
| S12_extracted_k141_493 | Not-determined |
| S12_extracted_k141_94877 | Not-determined |
| S12_extracted_k141_31725 | Not-determined |
| S12_extracted_k141_157810 | Not-determined |
| S12_extracted_k141_110591 | Not-determined |
| S12_extracted_k141_142122 | Not-determined |
| S12_extracted_k141_94881 | Not-determined |
| S12_extracted_k141_499 | Not-determined |
| S12_extracted_k141_94887 | Not-determined |
| S12_extracted_k141_157817 | Not-determined |
| S12_extracted_k141_39751 | Not-determined |
| S12_extracted_k141_31738 | Not-determined |
| S12_extracted_k141_15984 | Not-determined |
| S12_extracted_k141_15985 | Not-determined |
| S12_extracted_k141_55434 | Low-quality |
| S12_extracted_k141_149912 | Not-determined |
| S12_extracted_k141_87079 | Not-determined |
| S12_extracted_k141_94893 | Not-determined |
| S12_extracted_k141_55438 | Not-determined |
| S12_extracted_k141_110600 | Not-determined |
| S12_extracted_k141_181407 | Not-determined |
| S12_extracted_k141_110601 | Not-determined |
| S12_extracted_k141_55440 | Not-determined |
| S12_extracted_k141_165725 | Low-quality |
| S12_extracted_k141_39754 | Not-determined |
| S12_extracted_k141_181409 | Not-determined |
| S12_extracted_k141_142134 | Not-determined |
| S12_extracted_k141_55442 | Not-determined |
| S12_extracted_k141_87081 | Low-quality |
| S12_extracted_k141_31745 | Not-determined |
| S12_extracted_k141_173552 | Not-determined |
| S12_extracted_k141_79130 | Not-determined |
| S12_extracted_k141_94904 | Not-determined |
| S12_extracted_k141_509 | Not-determined |
| S12_extracted_k141_173554 | Low-quality |
| S12_extracted_k141_79131 | Not-determined |
| S12_extracted_k141_79133 | Not-determined |
| S12_extracted_k141_173560 | Not-determined |
| S12_extracted_k141_94915 | Not-determined |
| S12_extracted_k141_523 | Not-determined |
| S12_extracted_k141_79138 | Not-determined |
| S12_extracted_k141_181422 | Not-determined |
| S12_extracted_k141_94919 | Not-determined |
| S12_extracted_k141_71227 | Not-determined |
| S12_extracted_k141_39761 | Not-determined |
| S12_extracted_k141_94920 | Not-determined |
| S12_extracted_k141_31766 | Not-determined |
| S12_extracted_k141_31767 | Not-determined |
| S12_extracted_k141_157841 | Not-determined |
| S12_extracted_k141_94924 | Low-quality |
| S12_extracted_k141_142154 | Not-determined |
| S12_extracted_k141_181430 | Not-determined |
| S12_extracted_k141_126471 | Not-determined |
| S12_extracted_k141_181433 | Not-determined |
| S12_extracted_k141_157845 | Not-determined |
| S12_extracted_k141_16003 | Not-determined |
| S12_extracted_k141_79152 | Not-determined |
| S12_extracted_k141_540 | Not-determined |
| S12_extracted_k141_157851 | Not-determined |
| S12_extracted_k141_126478 | Not-determined |
| S12_extracted_k141_23809 | Not-determined |
| S12_extracted_k141_142170 | Not-determined |
| S12_extracted_k141_181444 | Not-determined |
| S12_extracted_k141_16013 | Not-determined |
| S12_extracted_k141_31792 | Not-determined |
| S12_extracted_k141_181448 | Not-determined |
| S12_extracted_k141_23813 | Not-determined |
| S12_extracted_k141_16016 | Not-determined |
| S12_extracted_k141_79165 | Low-quality |
| S12_extracted_k141_63412 | Not-determined |
| S12_extracted_k141_16021 | Not-determined |
| S12_extracted_k141_71249 | Not-determined |
| S12_extracted_k141_142175 | Not-determined |
| S12_extracted_k141_126485 | Not-determined |
| S12_extracted_k141_63420 | Not-determined |
| S12_extracted_k141_71256 | Not-determined |
| S12_extracted_k141_181462 | Not-determined |
| S12_extracted_k141_142181 | Not-determined |
| S12_extracted_k141_157862 | Low-quality |
| S12_extracted_k141_23827 | Not-determined |
| S12_extracted_k141_31809 | Low-quality |
| S12_extracted_k141_157866 | Not-determined |
| S12_extracted_k141_142183 | Not-determined |
| S12_extracted_k141_31813 | Not-determined |
| S12_extracted_k141_23833 | Not-determined |
| S12_extracted_k141_16033 | Not-determined |
| S12_extracted_k141_31815 | Not-determined |
| S12_extracted_k141_16034 | Low-quality |
| S12_extracted_k141_63430 | Not-determined |
| S12_extracted_k141_23839 | Not-determined |
| S12_extracted_k141_63431 | Not-determined |
| S12_extracted_k141_23840 | Not-determined |
| S12_extracted_k141_118509 | Low-quality |
| S12_extracted_k141_181468 | Not-determined |
| S12_extracted_k141_79184 | Not-determined |
| S12_extracted_k141_31821 | Low-quality |
| S12_extracted_k141_87092 | Not-determined |
| S12_extracted_k141_87094 | Not-determined |
| S12_extracted_k141_142192 | Not-determined |
| S12_extracted_k141_157881 | Not-determined |
| S12_extracted_k141_87097 | Not-determined |
| S12_extracted_k141_87100 | Not-determined |
| S12_extracted_k141_79189 | Not-determined |
| S12_extracted_k141_55466 | Not-determined |
| S12_extracted_k141_142195 | Not-determined |
| S12_extracted_k141_560 | Low-quality |
| S12_extracted_k141_564 | Not-determined |
| S12_extracted_k141_126490 | Low-quality |
| S12_extracted_k141_63445 | Not-determined |
| S12_extracted_k141_16046 | Not-determined |
| S12_extracted_k141_157895 | Low-quality |
| S12_extracted_k141_23863 | Not-determined |
| S12_extracted_k141_118530 | Not-determined |
| S12_extracted_k141_149927 | Not-determined |
| S12_extracted_k141_149928 | Not-determined |
| S12_extracted_k141_149931 | Not-determined |
| S12_extracted_k141_572 | Not-determined |
| S12_extracted_k141_63453 | Not-determined |
| S12_extracted_k141_63454 | Not-determined |
| S12_extracted_k141_142215 | Not-determined |
| S12_extracted_k141_63455 | Not-determined |
| S12_extracted_k141_63457 | Low-quality |
| S12_extracted_k141_579 | Not-determined |
| S12_extracted_k141_16049 | Not-determined |
| S12_extracted_k141_581 | Not-determined |
| S12_extracted_k141_142216 | Low-quality |
| S12_extracted_k141_16052 | Not-determined |
| S12_extracted_k141_583 | Not-determined |
| S12_extracted_k141_157902 | Not-determined |
| S12_extracted_k141_157904 | Not-determined |
| S12_extracted_k141_149945 | Not-determined |

| | |
|---|---|
| S12_extracted_k141_149946 | Not-determined |
| S12_extracted_k141_586 | Not-determined |
| S12_extracted_k141_589 | Not-determined |
| S12_extracted_k141_126512 | Not-determined |
| S12_extracted_k141_79201 | Not-determined |
| S12_extracted_k141_118546 | Not-determined |
| S12_extracted_k141_142217 | Not-determined |
| S12_extracted_k141_591 | Not-determined |
| S12_extracted_k141_23874 | Not-determined |
| S12_extracted_k141_592 | Not-determined |
| S12_extracted_k141_63470 | Not-determined |
| S12_extracted_k141_23876 | Not-determined |
| S12_extracted_k141_142219 | Not-determined |
| S12_extracted_k141_157907 | Not-determined |
| S12_extracted_k141_63474 | Not-determined |
| S12_extracted_k141_63475 | Not-determined |
| S12_extracted_k141_134163 | Not-determined |
| S12_extracted_k141_596 | Not-determined |
| S12_extracted_k141_165752 | Not-determined |
| S12_extracted_k141_118551 | Not-determined |
| S12_extracted_k141_79211 | Not-determined |
| S12_extracted_k141_31846 | Not-determined |
| S12_extracted_k141_126521 | Not-determined |
| S12_extracted_k141_23887 | Not-determined |
| S12_extracted_k141_126525 | Not-determined |
| S12_extracted_k141_602 | Low-quality |
| S12_extracted_k141_55501 | Not-determined |
| S12_extracted_k141_134165 | Not-determined |
| S12_extracted_k141_165760 | Not-determined |
| S12_extracted_k141_126528 | Not-determined |
| S12_extracted_k141_16064 | Not-determined |
| S12_extracted_k141_47482 | Not-determined |
| S12_extracted_k141_79218 | Not-determined |
| S12_extracted_k141_63484 | Not-determined |
| S12_extracted_k141_8206 | Not-determined |
| S12_extracted_k141_165767 | Not-determined |
| S12_extracted_k141_63487 | Not-determined |
| S12_extracted_k141_149959 | Not-determined |
| S12_extracted_k141_134174 | Not-determined |
| S12_extracted_k141_118560 | Not-determined |
| S12_extracted_k141_87123 | Not-determined |
| S12_extracted_k141_55510 | Not-determined |
| S12_extracted_k141_612 | Not-determined |
| S12_extracted_k141_55511 | Not-determined |
| S12_extracted_k141_87125 | Not-determined |
| S12_extracted_k141_87127 | Low-quality |
| S12_extracted_k141_79224 | Not-determined |
| S12_extracted_k141_39790 | Not-determined |
| S12_extracted_k141_23905 | Not-determined |
| S12_extracted_k141_63498 | Not-determined |
| S12_extracted_k141_87131 | Not-determined |
| S12_extracted_k141_149968 | Not-determined |
| S12_extracted_k141_31867 | Low-quality |
| S12_extracted_k141_165772 | Not-determined |
| S12_extracted_k141_55517 | Not-determined |
| S12_extracted_k141_181496 | Not-determined |
| S12_extracted_k141_47494 | Not-determined |
| S12_extracted_k141_39795 | Not-determined |
| S12_extracted_k141_39796 | Not-determined |
| S12_extracted_k141_126532 | Low-quality |
| S12_extracted_k141_55520 | Not-determined |
| S12_extracted_k141_134183 | Not-determined |
| S12_extracted_k141_31868 | Not-determined |
| S12_extracted_k141_149973 | Not-determined |
| S12_extracted_k141_55526 | Not-determined |
| S12_extracted_k141_16073 | Not-determined |
| S12_extracted_k141_134188 | Not-determined |
| S12_extracted_k141_31874 | Not-determined |
| S12_extracted_k141_165777 | Not-determined |
| S12_extracted_k141_102746 | Not-determined |
| S12_extracted_k141_142254 | Low-quality |
| S12_extracted_k141_102749 | Not-determined |
| S12_extracted_k141_23919 | Not-determined |
| S12_extracted_k141_63512 | Not-determined |
| S12_extracted_k141_102753 | Not-determined |
| S12_extracted_k141_165785 | Not-determined |
| S12_extracted_k141_87141 | Not-determined |
| S12_extracted_k141_55535 | Not-determined |
| S12_extracted_k141_623 | Not-determined |
| S12_extracted_k141_157930 | Not-determined |
| S12_extracted_k141_16075 | Not-determined |
| S12_extracted_k141_165795 | Not-determined |
| S12_extracted_k141_87147 | Not-determined |
| S12_extracted_k141_628 | Low-quality |
| S12_extracted_k141_16080 | Not-determined |
| S12_extracted_k141_134198 | Not-determined |
| S12_extracted_k141_39810 | Not-determined |
| S12_extracted_k141_16084 | Not-determined |
| S12_extracted_k141_87154 | Not-determined |
| S12_extracted_k141_8218 | Not-determined |
| S12_extracted_k141_110627 | Not-determined |
| S12_extracted_k141_165807 | Low-quality |
| S12_extracted_k141_39813 | Not-determined |
| S12_extracted_k141_181516 | Not-determined |
| S12_extracted_k141_165809 | Not-determined |
| S12_extracted_k141_8227 | Not-determined |
| S12_extracted_k141_181522 | Not-determined |
| S12_extracted_k141_110639 | Not-determined |
| S12_extracted_k141_23939 | Not-determined |
| S12_extracted_k141_149990 | Low-quality |
| S12_extracted_k141_638 | Not-determined |
| S12_extracted_k141_102762 | Not-determined |
| S12_extracted_k141_102763 | Not-determined |
| S12_extracted_k141_149996 | Not-determined |
| S12_extracted_k141_47511 | Not-determined |
| S12_extracted_k141_149997 | Low-quality |
| S12_extracted_k141_118583 | Not-determined |
| S12_extracted_k141_8236 | Low-quality |
| S12_extracted_k141_650 | Not-determined |
| S12_extracted_k141_118584 | Not-determined |
| S12_extracted_k141_102768 | Not-determined |
| S12_extracted_k141_142288 | Not-determined |
| S12_extracted_k141_79255 | Not-determined |
| S12_extracted_k141_157956 | Not-determined |
| S12_extracted_k141_102771 | Not-determined |
| S12_extracted_k141_8243 | Not-determined |
| S12_extracted_k141_8244 | Not-determined |
| S12_extracted_k141_39822 | Not-determined |
| S12_extracted_k141_655 | Not-determined |
| S12_extracted_k141_657 | Not-determined |
| S12_extracted_k141_157959 | Not-determined |
| S12_extracted_k141_87165 | Not-determined |
| S12_extracted_k141_150001 | Low-quality |
| S12_extracted_k141_142290 | Not-determined |
| S12_extracted_k141_118592 | Low-quality |
| S12_extracted_k141_79270 | Not-determined |
| S12_extracted_k141_659 | Not-determined |
| S12_extracted_k141_94937 | Not-determined |
| S12_extracted_k141_150007 | Not-determined |
| S12_extracted_k141_662 | Not-determined |
| S12_extracted_k141_181533 | Not-determined |
| S12_extracted_k141_55556 | Not-determined |
| S12_extracted_k141_63520 | Low-quality |
| S12_extracted_k141_23963 | Not-determined |
| S12_extracted_k141_23964 | Not-determined |
| S12_extracted_k141_181534 | Low-quality |
| S12_extracted_k141_165833 | Not-determined |
| S12_extracted_k141_8249 | Low-quality |
| S12_extracted_k141_87173 | Not-determined |
| S12_extracted_k141_71303 | Not-determined |
| S12_extracted_k141_118602 | Not-determined |
| S12_extracted_k141_47531 | Low-quality |
| S12_extracted_k141_47532 | Not-determined |
| S12_extracted_k141_670 | Not-determined |
| S12_extracted_k141_79276 | Not-determined |
| S12_extracted_k141_87174 | Not-determined |
| S12_extracted_k141_118606 | Not-determined |
| S12_extracted_k141_47534 | Not-determined |
| S12_extracted_k141_181540 | Not-determined |
| S12_extracted_k141_181541 | Not-determined |
| S12_extracted_k141_118608 | Not-determined |
| S12_extracted_k141_672 | Not-determined |
| S12_extracted_k141_8257 | Low-quality |
| S12_extracted_k141_673 | Not-determined |
| S12_extracted_k141_150016 | Low-quality |
| S12_extracted_k141_165840 | Not-determined |
| S12_extracted_k141_47538 | Not-determined |
| S12_extracted_k141_181544 | Not-determined |
| S12_extracted_k141_165843 | Not-determined |
| S12_extracted_k141_157988 | Not-determined |
| S12_extracted_k141_165844 | Low-quality |
| S12_extracted_k141_126557 | Not-determined |
| S12_extracted_k141_181548 | Not-determined |
| S12_extracted_k141_87180 | Not-determined |
| S12_extracted_k141_71306 | Not-determined |
| S12_extracted_k141_87187 | Not-determined |
| S12_extracted_k141_71310 | Not-determined |
| S12_extracted_k141_150029 | Not-determined |
| S12_extracted_k141_150032 | Not-determined |
| S12_extracted_k141_118610 | Low-quality |
| S12_extracted_k141_118612 | Not-determined |
| S12_extracted_k141_126564 | Not-determined |
| S12_extracted_k141_150038 | Not-determined |
| S12_extracted_k141_23983 | Not-determined |
| S12_extracted_k141_142312 | Not-determined |
| S12_extracted_k141_126567 | Not-determined |
| S12_extracted_k141_23984 | Not-determined |
| S12_extracted_k141_142321 | Not-determined |
| S12_extracted_k141_695 | Low-quality |
| S12_extracted_k141_8281 | Not-determined |
| S12_extracted_k141_39848 | Not-determined |
| S12_extracted_k141_8282 | Not-determined |
| S12_extracted_k141_697 | Not-determined |
| S12_extracted_k141_8284 | Not-determined |
| S12_extracted_k141_79284 | Not-determined |
| S12_extracted_k141_703 | Low-quality |
| S12_extracted_k141_31890 | Low-quality |
| S12_extracted_k141_55584 | Not-determined |
| S12_extracted_k141_150054 | Low-quality |
| S12_extracted_k141_126570 | Low-quality |
| S12_extracted_k141_150055 | Not-determined |
| S12_extracted_k141_79290 | Not-determined |
| S12_extracted_k141_181571 | Not-determined |
| S12_extracted_k141_150059 | Not-determined |
| S12_extracted_k141_8291 | Not-determined |
| S12_extracted_k141_118634 | Not-determined |
| S12_extracted_k141_118635 | Not-determined |
| S12_extracted_k141_150063 | Not-determined |
| S12_extracted_k141_711 | Not-determined |
| S12_extracted_k141_126578 | Not-determined |
| S12_extracted_k141_158005 | Not-determined |
| S12_extracted_k141_165861 | Not-determined |
| S12_extracted_k141_142342 | Not-determined |
| S12_extracted_k141_71340 | Low-quality |
| S12_extracted_k141_63554 | Not-determined |
| S12_extracted_k141_134204 | Not-determined |
| S12_extracted_k141_71342 | Not-determined |
| S12_extracted_k141_150068 | Not-determined |
| S12_extracted_k141_118642 | Not-determined |
| S12_extracted_k141_158007 | Not-determined |
| S12_extracted_k141_126586 | Not-determined |
| S12_extracted_k141_102790 | Not-determined |
| S12_extracted_k141_71350 | Not-determined |
| S12_extracted_k141_142345 | Not-determined |
| S12_extracted_k141_102796 | Low-quality |
| S12_extracted_k141_126591 | Not-determined |
| S12_extracted_k141_118649 | Low-quality |
| S12_extracted_k141_102797 | Not-determined |
| S12_extracted_k141_55594 | Not-determined |
| S12_extracted_k141_31898 | Not-determined |
| S12_extracted_k141_134219 | Not-determined |
| S12_extracted_k141_94983 | Not-determined |
| S12_extracted_k141_31901 | Low-quality |
| S12_extracted_k141_55597 | Not-determined |
| S12_extracted_k141_126592 | Not-determined |
| S12_extracted_k141_126593 | Low-quality |
| S12_extracted_k141_94989 | Not-determined |
| S12_extracted_k141_110672 | Not-determined |
| S12_extracted_k141_31904 | Not-determined |
| S12_extracted_k141_118659 | Not-determined |
| S12_extracted_k141_79313 | Low-quality |
| S12_extracted_k141_31907 | Not-determined |
| S12_extracted_k141_110679 | Not-determined |
| S12_extracted_k141_71364 | Not-determined |
| S12_extracted_k141_94996 | Not-determined |
| S12_extracted_k141_31909 | Not-determined |
| S12_extracted_k141_181605 | Not-determined |
| S12_extracted_k141_63578 | Not-determined |
| S12_extracted_k141_95000 | Not-determined |
| S12_extracted_k141_118665 | Not-determined |
| S12_extracted_k141_55605 | Not-determined |
| S12_extracted_k141_79323 | Not-determined |
| S12_extracted_k141_134239 | Not-determined |
| S12_extracted_k141_55606 | Not-determined |
| S12_extracted_k141_134240 | Not-determined |
| S12_extracted_k141_31917 | Not-determined |
| S12_extracted_k141_142348 | Not-determined |
| S12_extracted_k141_31920 | Not-determined |
| S12_extracted_k141_126612 | Low-quality |
| S12_extracted_k141_142353 | Low-quality |
| S12_extracted_k141_181618 | Not-determined |
| S12_extracted_k141_118677 | Not-determined |
| S12_extracted_k141_126620 | Low-quality |
| S12_extracted_k141_142356 | Not-determined |
| S12_extracted_k141_63594 | Not-determined |
| S12_extracted_k141_63595 | Low-quality |
| S12_extracted_k141_732 | Not-determined |
| S12_extracted_k141_39860 | Not-determined |
| S12_extracted_k141_126625 | Not-determined |
| S12_extracted_k141_134245 | Not-determined |
| S12_extracted_k141_150089 | Not-determined |
| S12_extracted_k141_39862 | Not-determined |
| S12_extracted_k141_24024 | Not-determined |
| S12_extracted_k141_110699 | Not-determined |
| S12_extracted_k141_95017 | Not-determined |
| S12_extracted_k141_126629 | Not-determined |
| S12_extracted_k141_102828 | Not-determined |
| S12_extracted_k141_735 | Low-quality |
| S12_extracted_k141_134250 | Not-determined |
| S12_extracted_k141_181630 | Not-determined |
| S12_extracted_k141_150099 | Not-determined |
| S12_extracted_k141_126633 | Not-determined |
| S12_extracted_k141_181633 | Not-determined |
| S12_extracted_k141_39869 | Not-determined |
| S12_extracted_k141_63601 | Not-determined |
| S12_extracted_k141_158027 | Not-determined |
| S12_extracted_k141_158029 | Not-determined |
| S12_extracted_k141_39871 | Not-determined |
| S13_vs_38708\|\|full | Medium-quality |
| S12_extracted_k141_142367 | Low-quality |
| S12_extracted_k141_173573 | Not-determined |
| S12_extracted_k141_118704 | Not-determined |
| S12_extracted_k141_173574 | Not-determined |
| S12_extracted_k141_181637 | Not-determined |
| S12_extracted_k141_142372 | Not-determined |
| S12_extracted_k141_142376 | Not-determined |
| S12_extracted_k141_134261 | Not-determined |
| S12_extracted_k141_95031 | Not-determined |
| S12_extracted_k141_181648 | Low-quality |
| S12_extracted_k141_8321 | Not-determined |
| S12_extracted_k141_55625 | Not-determined |
| S12_extracted_k141_55626 | Not-determined |
| S12_extracted_k141_747 | Not-determined |
| S12_extracted_k141_63620 | Low-quality |
| S12_extracted_k141_39885 | Not-determined |
| S12_extracted_k141_63624 | Not-determined |
| S12_extracted_k141_63625 | Not-determined |
| S12_extracted_k141_134271 | Not-determined |
| S12_extracted_k141_165906 | Not-determined |
| S12_extracted_k141_39887 | Not-determined |
| S12_extracted_k141_24045 | Not-determined |
| S12_extracted_k141_63627 | Low-quality |
| S12_extracted_k141_55632 | Not-determined |
| S12_extracted_k141_63631 | Not-determined |
| S12_extracted_k141_55634 | Not-determined |
| S12_extracted_k141_24054 | Not-determined |
| S12_extracted_k141_110721 | Not-determined |
| S12_extracted_k141_8332 | Not-determined |
| S12_extracted_k141_95043 | Not-determined |
| S12_extracted_k141_118719 | Not-determined |
| S12_extracted_k141_8334 | Not-determined |
| S12_extracted_k141_24058 | Not-determined |
| S12_extracted_k141_118721 | Not-determined |
| S12_extracted_k141_165918 | Not-determined |
| S12_extracted_k141_24064 | Not-determined |
| S12_extracted_k141_71407 | Not-determined |
| S12_extracted_k141_39892 | Not-determined |
| S12_extracted_k141_757 | Low-quality |
| S12_extracted_k141_173592 | Not-determined |
| S12_extracted_k141_31934 | Not-determined |
| S12_extracted_k141_55646 | Not-determined |
| S12_extracted_k141_8340 | Not-determined |
| S12_extracted_k141_63642 | Not-determined |
| S12_extracted_k141_8341 | Not-determined |
| S12_extracted_k141_110735 | Not-determined |
| S12_extracted_k141_158056 | Not-determined |
| S12_extracted_k141_31937 | Low-quality |
| S12_extracted_k141_150118 | Not-determined |
| S12_extracted_k141_102858 | Not-determined |
| S12_extracted_k141_16100 | Not-determined |
| S12_extracted_k141_181671 | Not-determined |
| S12_extracted_k141_24074 | Not-determined |
| S12_extracted_k141_150121 | Not-determined |
| S12_extracted_k141_110742 | Not-determined |
| S12_extracted_k141_150123 | Not-determined |
| S12_extracted_k141_173600 | Low-quality |
| S12_extracted_k141_102864 | Not-determined |
| S12_extracted_k141_63655 | Not-determined |
| S12_extracted_k141_8348 | Not-determined |
| S12_extracted_k141_142379 | Not-determined |
| S12_extracted_k141_150128 | Not-determined |
| S12_extracted_k141_31949 | Not-determined |
| S12_extracted_k141_95054 | Not-determined |
| S12_extracted_k141_63663 | Not-determined |
| S12_extracted_k141_142382 | Not-determined |
| S12_extracted_k141_150132 | Not-determined |
| S12_extracted_k141_150133 | Not-determined |
| S12_extracted_k141_8360 | Not-determined |
| S12_extracted_k141_173610 | Not-determined |
| S12_extracted_k141_181683 | Not-determined |
| S12_extracted_k141_8362 | Not-determined |
| S12_extracted_k141_102879 | Not-determined |
| S12_extracted_k141_63675 | Not-determined |
| S12_extracted_k141_63677 | Not-determined |
| S12_extracted_k141_55663 | Low-quality |
| S12_extracted_k141_150147 | Low-quality |
| S12_extracted_k141_785 | Not-determined |
| S12_extracted_k141_102884 | Not-determined |
| S12_extracted_k141_102885 | Not-determined |
| S12_extracted_k141_118744 | Not-determined |
| S12_extracted_k141_786 | Not-determined |
| S12_extracted_k141_787 | Not-determined |
| S12_extracted_k141_173628 | Not-determined |
| S12_extracted_k141_39912 | Low-quality |
| S12_extracted_k141_31961 | Not-determined |
| S12_extracted_k141_158062 | Not-determined |
| S12_extracted_k141_95061 | Not-determined |
| S12_extracted_k141_102895 | Not-determined |
| S12_extracted_k141_24088 | Not-determined |
| S12_extracted_k141_794 | Not-determined |
| S12_extracted_k141_150150 | Not-determined |
| S12_extracted_k141_79362 | Not-determined |
| S12_extracted_k141_71438 | Not-determined |
| S12_extracted_k141_158065 | Not-determined |
| S12_extracted_k141_118755 | Not-determined |
| S12_extracted_k141_118757 | Not-determined |
| S12_extracted_k141_142410 | Not-determined |
| S12_extracted_k141_24096 | Not-determined |
| S12_extracted_k141_102907 | Not-determined |
| S12_extracted_k141_39930 | Low-quality |
| S12_extracted_k141_110761 | Low-quality |
| S12_extracted_k141_8403 | Not-determined |
| S12_extracted_k141_158072 | Not-determined |
| S12_extracted_k141_71446 | Low-quality |
| S12_extracted_k141_173640 | Not-determined |
| S12_extracted_k141_134290 | Low-quality |
| S12_extracted_k141_118762 | Not-determined |
| S12_extracted_k141_102916 | Not-determined |
| S12_extracted_k141_95072 | Not-determined |
| S12_extracted_k141_8410 | Not-determined |
| S12_extracted_k141_150172 | Not-determined |
| S12_extracted_k141_181697 | Not-determined |
| S12_extracted_k141_39941 | Not-determined |
| S12_extracted_k141_181698 | Not-determined |
| S12_extracted_k141_173645 | Not-determined |
| S12_extracted_k141_71452 | Not-determined |
| S12_extracted_k141_39943 | Not-determined |
| S12_extracted_k141_142418 | Not-determined |
| S12_extracted_k141_71454 | Not-determined |
| S12_extracted_k141_181705 | Low-quality |
| S12_extracted_k141_158081 | Not-determined |
| S12_extracted_k141_142419 | Not-determined |
| S12_extracted_k141_87219 | Not-determined |
| S12_extracted_k141_71460 | Not-determined |
| S12_extracted_k141_173654 | Low-quality |
| S12_extracted_k141_142421 | Not-determined |
| S12_extracted_k141_134298 | Not-determined |
| S12_extracted_k141_150181 | Not-determined |
| S12_extracted_k141_158087 | Not-determined |
| S12_extracted_k141_181714 | Not-determined |
| S12_extracted_k141_181715 | Low-quality |
| S12_extracted_k141_55680 | Low-quality |
| S12_extracted_k141_32000 | Not-determined |
| S12_extracted_k141_16114 | Not-determined |
| S12_extracted_k141_71467 | Not-determined |
| S12_extracted_k141_150186 | Low-quality |
| S12_extracted_k141_16116 | Not-determined |
| S12_extracted_k141_16117 | Not-determined |
| S12_extracted_k141_47602 | Not-determined |
| S12_extracted_k141_71470 | Not-determined |
| S12_extracted_k141_16119 | Not-determined |
| S12_extracted_k141_63712 | Not-determined |
| S12_extracted_k141_87225 | Not-determined |
| S12_extracted_k141_158099 | Not-determined |
| S12_extracted_k141_110789 | Low-quality |
| S12_extracted_k141_71475 | Not-determined |
| S12_extracted_k141_102945 | Not-determined |
| S12_extracted_k141_47605 | Not-determined |
| S12_extracted_k141_142434 | Not-determined |
| S12_extracted_k141_8427 | Low-quality |
| S12_extracted_k141_818 | Not-determined |
| S12_extracted_k141_16124 | Not-determined |
| S12_extracted_k141_47610 | Not-determined |
| S12_extracted_k141_819 | Not-determined |
| S12_extracted_k141_134302 | Not-determined |
| S12_extracted_k141_165942 | Not-determined |
| S12_extracted_k141_95101 | Low-quality |
| S12_extracted_k141_71481 | Not-determined |
| S12_extracted_k141_32015 | Not-determined |
| S12_extracted_k141_8428 | Low-quality |
| S12_extracted_k141_102949 | Not-determined |
| S12_extracted_k141_102951 | Not-determined |
| S12_extracted_k141_102952 | Not-determined |
| S12_extracted_k141_173659 | Not-determined |
| S12_extracted_k141_47618 | Not-determined |
| S12_extracted_k141_173660 | Not-determined |
| S12_extracted_k141_118785 | Not-determined |
| S12_extracted_k141_79404 | Not-determined |
| S12_extracted_k141_87239 | Not-determined |
| S12_extracted_k141_24113 | Low-quality |
| S12_extracted_k141_95110 | Not-determined |
| S12_extracted_k141_165948 | Not-determined |
| S12_extracted_k141_55698 | Not-determined |
| S12_extracted_k141_95115 | Not-determined |
| S12_extracted_k141_165949 | Not-determined |
| S12_extracted_k141_55701 | Not-determined |
| S12_extracted_k141_32023 | Not-determined |
| S12_extracted_k141_118795 | Not-determined |
| S12_extracted_k141_39956 | Not-determined |
| S12_extracted_k141_71495 | Not-determined |
| S12_extracted_k141_24121 | Not-determined |
| S12_extracted_k141_16147 | Not-determined |
| S12_extracted_k141_8435 | Not-determined |
| S12_extracted_k141_87251 | Not-determined |
| S12_extracted_k141_87252 | Not-determined |
| S12_extracted_k141_134315 | Not-determined |
| S12_extracted_k141_142448 | Low-quality |
| S12_extracted_k141_16154 | Not-determined |
| S12_extracted_k141_95127 | Not-determined |
| S12_extracted_k141_71502 | Not-determined |
| S12_extracted_k141_71505 | Not-determined |
| S12_extracted_k141_165964 | Not-determined |
| S12_extracted_k141_102974 | Not-determined |
| S12_extracted_k141_87254 | Not-determined |
| S12_extracted_k141_118797 | Not-determined |
| S12_extracted_k141_87264 | Not-determined |
| S12_extracted_k141_63740 | Not-determined |
| S12_extracted_k141_32038 | Not-determined |
| S12_extracted_k141_165968 | Not-determined |
| S12_extracted_k141_87270 | Not-determined |
| S12_extracted_k141_79417 | Not-determined |
| S12_extracted_k141_79418 | Not-determined |
| S12_extracted_k141_32043 | Not-determined |
| S12_extracted_k141_134332 | Not-determined |
| S12_extracted_k141_142463 | Not-determined |
| S12_extracted_k141_47638 | Not-determined |
| S12_extracted_k141_142467 | Not-determined |
| S12_extracted_k141_24125 | Not-determined |
| S12_extracted_k141_87276 | Not-determined |
| S12_extracted_k141_181753 | Not-determined |
| S12_extracted_k141_39967 | Not-determined |
| S12_extracted_k141_118805 | Not-determined |
| S12_extracted_k141_16163 | Not-determined |
| S12_extracted_k141_102980 | Not-determined |
| S12_extracted_k141_134341 | Not-determined |
| S12_extracted_k141_118806 | Not-determined |
| S12_extracted_k141_165979 | Not-determined |
| S12_extracted_k141_39968 | Not-determined |
| S12_extracted_k141_102981 | Not-determined |
| S12_extracted_k141_24133 | Low-quality |
| S12_extracted_k141_102983 | Not-determined |
| S12_extracted_k141_134345 | Low-quality |
| S12_extracted_k141_79431 | Not-determined |
| S12_extracted_k141_95143 | Not-determined |
| S12_extracted_k141_134346 | Not-determined |
| S12_extracted_k141_142474 | Not-determined |
| S12_extracted_k141_165988 | Low-quality |
| S12_extracted_k141_102988 | Low-quality |
| S12_extracted_k141_142481 | Not-determined |
| S12_extracted_k141_150204 | Low-quality |
| S12_extracted_k141_79438 | Not-determined |
| S12_extracted_k141_95159 | Not-determined |
| S12_extracted_k141_32053 | Not-determined |
| S12_extracted_k141_87297 | Not-determined |
| S12_extracted_k141_79444 | Not-determined |
| S12_extracted_k141_142492 | Not-determined |
| S12_extracted_k141_79446 | Low-quality |
| S12_extracted_k141_95164 | Not-determined |
| S12_extracted_k141_142494 | Not-determined |
| S5_extracted_k141_25423 | High-quality |
| S12_extracted_k141_24157 | Not-determined |
| S12_extracted_k141_173667 | Low-quality |
| S12_extracted_k141_103002 | Not-determined |
| S12_extracted_k141_8451 | Low-quality |
| S12_extracted_k141_16180 | Not-determined |
| S12_extracted_k141_16182 | Low-quality |
| S12_extracted_k141_134362 | Not-determined |
| S12_extracted_k141_126647 | Not-determined |
| S12_extracted_k141_32056 | Not-determined |
| S12_extracted_k141_837 | Not-determined |
| S12_extracted_k141_838 | Not-determined |
| S12_extracted_k141_87311 | Not-determined |
| S12_extracted_k141_79460 | Low-quality |
| S12_extracted_k141_126652 | Not-determined |
| S12_extracted_k141_16194 | Not-determined |
| S12_extracted_k141_16195 | Not-determined |
| S12_extracted_k141_8460 | Not-determined |
| S12_extracted_k141_55709 | Not-determined |
| S12_extracted_k141_126656 | Not-determined |
| S12_extracted_k141_103016 | Not-determined |
| S12_extracted_k141_24171 | Not-determined |
| S12_extracted_k141_150226 | Not-determined |
| S12_extracted_k141_95180 | Not-determined |
| S12_extracted_k141_103021 | Not-determined |
| S12_extracted_k141_32061 | Not-determined |
| S12_extracted_k141_16205 | Not-determined |
| S12_extracted_k141_142519 | Not-determined |
| S12_extracted_k141_181765 | Not-determined |
| S12_extracted_k141_165993 | Not-determined |
| S12_extracted_k141_181767 | Not-determined |
| S12_extracted_k141_103024 | Not-determined |
| S12_extracted_k141_55713 | Not-determined |
| S12_extracted_k141_150230 | Not-determined |
| S12_extracted_k141_40009 | Not-determined |
| S12_extracted_k141_165995 | Low-quality |
| S12_extracted_k141_87325 | Not-determined |
| S12_extracted_k141_165997 | Not-determined |
| S12_extracted_k141_142527 | Not-determined |
| S12_extracted_k141_8470 | Not-determined |
| S12_extracted_k141_87331 | Not-determined |
| S12_extracted_k141_181774 | Not-determined |
| S12_extracted_k141_16211 | Low-quality |
| S12_extracted_k141_166001 | Not-determined |
| S12_extracted_k141_24185 | Not-determined |
| S12_extracted_k141_173675 | Not-determined |
| S12_extracted_k141_173676 | Not-determined |
| S12_extracted_k141_173677 | Not-determined |
| S12_extracted_k141_142535 | Not-determined |
| S12_extracted_k141_142536 | Low-quality |
| S12_extracted_k141_862 | Not-determined |
| S12_extracted_k141_181777 | Not-determined |
| S12_extracted_k141_166009 | Not-determined |
| S12_extracted_k141_126673 | Not-determined |
| S12_extracted_k141_40020 | Not-determined |
| S12_extracted_k141_865 | Not-determined |
| S12_extracted_k141_55723 | Not-determined |
| S12_extracted_k141_173685 | Not-determined |
| S12_extracted_k141_40024 | Not-determined |
| S12_extracted_k141_55728 | Not-determined |
| S12_extracted_k141_8481 | Low-quality |
| S12_extracted_k141_55730 | Not-determined |
| S12_extracted_k141_166019 | Not-determined |
| S12_extracted_k141_181782 | Not-determined |
| S12_extracted_k141_126683 | Low-quality |
| S12_extracted_k141_181786 | Not-determined |
| S12_extracted_k141_32087 | Not-determined |
| S12_extracted_k141_24201 | Low-quality |
| S12_extracted_k141_103047 | Low-quality |
| S12_extracted_k141_103048 | Not-determined |

| | |
|---|---|
| S12_extracted_k141_40032 | Not-determined |
| S12_extracted_k141_71527 | Not-determined |
| S12_extracted_k141_71528 | Not-determined |
| S12_extracted_k141_32093 | Not-determined |
| S12_extracted_k141_32096 | Not-determined |
| S12_extracted_k141_24209 | Not-determined |
| S12_extracted_k141_103053 | Not-determined |
| S12_extracted_k141_181789 | Low-quality |
| S12_extracted_k141_150253 | Not-determined |
| S12_extracted_k141_181791 | Low-quality |
| S12_extracted_k141_24211 | Not-determined |
| S12_extracted_k141_8487 | Not-determined |
| S12_extracted_k141_24212 | Not-determined |
| S12_extracted_k141_47644 | Not-determined |
| S12_extracted_k141_142553 | Not-determined |
| S12_extracted_k141_24215 | Not-determined |
| S12_extracted_k141_71539 | Not-determined |
| S12_extracted_k141_8491 | Not-determined |
| S12_extracted_k141_126691 | Low-quality |
| S12_extracted_k141_40053 | Not-determined |
| S12_extracted_k141_40054 | Not-determined |
| S12_extracted_k141_40055 | Low-quality |
| S12_extracted_k141_166042 | Not-determined |
| S12_extracted_k141_885 | Low-quality |
| S12_extracted_k141_24221 | Not-determined |
| S12_extracted_k141_24222 | Low-quality |
| S12_extracted_k141_887 | Not-determined |
| S12_extracted_k141_888 | Not-determined |
| S12_extracted_k141_24225 | Low-quality |
| S12_extracted_k141_32108 | Not-determined |
| S12_extracted_k141_16247 | Not-determined |
| S12_extracted_k141_142560 | Not-determined |
| S12_extracted_k141_16249 | Not-determined |
| S12_extracted_k141_142561 | Not-determined |
| S12_extracted_k141_24231 | Not-determined |
| S12_extracted_k141_87344 | Low-quality |
| S12_extracted_k141_103063 | Not-determined |
| S12_extracted_k141_55757 | Not-determined |
| S12_extracted_k141_103067 | Low-quality |
| S12_extracted_k141_16254 | Not-determined |
| S12_extracted_k141_181797 | Low-quality |
| S12_extracted_k141_87347 | Not-determined |
| S12_extracted_k141_126711 | Not-determined |
| S12_extracted_k141_103072 | Not-determined |
| S12_extracted_k141_158130 | Not-determined |
| S12_extracted_k141_8519 | Not-determined |
| S12_extracted_k141_8520 | Low-quality |
| S12_extracted_k141_103076 | Not-determined |
| S12_extracted_k141_24241 | Not-determined |
| S12_extracted_k141_103079 | Not-determined |
| S12_extracted_k141_166066 | Not-determined |
| S12_extracted_k141_87355 | Low-quality |
| S12_extracted_k141_8534 | Not-determined |
| S12_extracted_k141_181805 | Not-determined |
| S12_extracted_k141_55762 | Not-determined |
| S12_extracted_k141_55763 | Not-determined |
| S12_extracted_k141_902 | Not-determined |
| S12_extracted_k141_158144 | Not-determined |
| S12_extracted_k141_47651 | Not-determined |
| S12_extracted_k141_55765 | Not-determined |
| S12_extracted_k141_47652 | Not-determined |
| S12_extracted_k141_906 | Not-determined |
| S12_extracted_k141_907 | Not-determined |
| S12_extracted_k141_16270 | Not-determined |
| S12_extracted_k141_47656 | Not-determined |
| S12_extracted_k141_16274 | Not-determined |
| S12_extracted_k141_55773 | Low-quality |
| S12_extracted_k141_47661 | Not-determined |
| S12_extracted_k141_173721 | Not-determined |
| S12_extracted_k141_103097 | Not-determined |
| S12_extracted_k141_40072 | Not-determined |
| S12_extracted_k141_173724 | Not-determined |
| S12_extracted_k141_173725 | Not-determined |
| S12_extracted_k141_32142 | Not-determined |
| S12_extracted_k141_150281 | Not-determined |
| S12_extracted_k141_40075 | Not-determined |
| S12_extracted_k141_118855 | Not-determined |
| S12_extracted_k141_173728 | Not-determined |
| S12_extracted_k141_166085 | Not-determined |
| S12_extracted_k141_173730 | Not-determined |
| S12_extracted_k141_103103 | Not-determined |
| S12_extracted_k141_103104 | Low-quality |
| S12_extracted_k141_118858 | Not-determined |
| S12_extracted_k141_103106 | Not-determined |
| S12_extracted_k141_79466 | Not-determined |
| S12_extracted_k141_95200 | Low-quality |
| S12_extracted_k141_150289 | Not-determined |
| S12_extracted_k141_134419 | Low-quality |
| S12_extracted_k141_55789 | Not-determined |
| S12_extracted_k141_103111 | Not-determined |
| S12_extracted_k141_134420 | Not-determined |
| S12_extracted_k141_103112 | Not-determined |
| S12_extracted_k141_95203 | Not-determined |
| S12_extracted_k141_173748 | Not-determined |
| S12_extracted_k141_158163 | Not-determined |
| S12_extracted_k141_134424 | Not-determined |
| S12_extracted_k141_87365 | Not-determined |
| S12_extracted_k141_8552 | Not-determined |
| S12_extracted_k141_71565 | Low-quality |
| S12_extracted_k141_173755 | Not-determined |
| S12_extracted_k141_55800 | Not-determined |
| S12_extracted_k141_173760 | Not-determined |
| S12_extracted_k141_158176 | Not-determined |
| S12_extracted_k141_150299 | Not-determined |
| S12_extracted_k141_158177 | Not-determined |
| S12_extracted_k141_158178 | Not-determined |
| S12_extracted_k141_47671 | Not-determined |
| S12_extracted_k141_16280 | Not-determined |
| S12_extracted_k141_71572 | Not-determined |
| S12_extracted_k141_40100 | Not-determined |
| S12_extracted_k141_931 | Not-determined |
| S12_extracted_k141_150301 | Not-determined |
| S12_extracted_k141_173763 | Not-determined |
| S12_extracted_k141_173764 | Not-determined |
| S12_extracted_k141_150302 | Not-determined |
| S12_extracted_k141_47673 | Not-determined |
| S12_extracted_k141_103119 | Not-determined |
| S12_extracted_k141_16286 | Low-quality |
| S12_extracted_k141_150305 | Not-determined |
| S12_extracted_k141_142578 | Not-determined |
| S12_extracted_k141_142579 | Not-determined |
| S12_extracted_k141_16288 | Not-determined |
| S12_extracted_k141_150307 | Not-determined |
| S12_extracted_k141_150309 | Not-determined |
| S12_extracted_k141_16290 | Not-determined |
| S12_extracted_k141_181816 | Not-determined |
| S12_extracted_k141_142587 | Low-quality |
| S12_extracted_k141_173774 | Low-quality |
| S12_extracted_k141_103132 | Not-determined |
| S12_extracted_k141_142589 | Not-determined |
| S12_extracted_k141_142590 | Not-determined |
| S12_extracted_k141_181823 | Not-determined |
| S12_extracted_k141_8568 | Not-determined |
| S12_extracted_k141_32148 | Not-determined |
| S12_extracted_k141_24260 | Not-determined |
| S12_extracted_k141_55810 | Not-determined |
| S12_extracted_k141_47685 | Not-determined |
| S12_extracted_k141_24261 | Not-determined |
| S12_extracted_k141_40110 | Not-determined |
| S12_extracted_k141_63804 | Not-determined |
| S12_extracted_k141_158195 | Not-determined |
| S12_extracted_k141_79479 | Not-determined |
| S12_extracted_k141_181826 | Not-determined |
| S12_extracted_k141_24264 | Not-determined |
| S12_extracted_k141_142595 | Not-determined |
| S12_extracted_k141_158198 | Not-determined |
| S12_extracted_k141_24270 | Low-quality |
| S12_extracted_k141_40117 | Not-determined |
| S12_extracted_k141_40120 | Not-determined |
| S12_extracted_k141_142602 | Not-determined |
| S12_extracted_k141_940 | Not-determined |
| S12_extracted_k141_55816 | Not-determined |
| S12_extracted_k141_142604 | Not-determined |
| S12_extracted_k141_126717 | Low-quality |
| S12_extracted_k141_55818 | Not-determined |
| S12_extracted_k141_16307 | Not-determined |
| S12_extracted_k141_87394 | Not-determined |
| S12_extracted_k141_79497 | Low-quality |
| S12_extracted_k141_150321 | Not-determined |
| S12_extracted_k141_103143 | Not-determined |
| S12_extracted_k141_126723 | Not-determined |
| S12_extracted_k141_173796 | Not-determined |
| S12_extracted_k141_103145 | Not-determined |
| S12_extracted_k141_150328 | Not-determined |
| S12_extracted_k141_948 | Not-determined |
| S12_extracted_k141_32165 | Not-determined |
| S12_extracted_k141_951 | Not-determined |
| S12_extracted_k141_952 | Not-determined |
| S12_extracted_k141_71599 | Not-determined |
| S12_extracted_k141_142619 | Not-determined |
| S12_extracted_k141_32169 | Not-determined |
| S12_extracted_k141_103157 | Not-determined |
| S12_extracted_k141_32170 | Not-determined |
| S12_extracted_k141_71606 | Not-determined |
| S12_extracted_k141_150340 | Not-determined |
| S12_extracted_k141_32176 | Not-determined |
| S12_extracted_k141_181855 | Not-determined |
| S12_extracted_k141_71609 | Not-determined |
| S12_extracted_k141_118882 | Not-determined |
| S12_extracted_k141_40134 | Not-determined |
| S12_extracted_k141_103170 | Not-determined |
| S12_extracted_k141_166104 | Not-determined |
| S12_extracted_k141_79517 | Not-determined |
| S12_extracted_k141_16329 | Not-determined |
| S12_extracted_k141_959 | Low-quality |
| S12_extracted_k141_103172 | Not-determined |
| S12_extracted_k141_118884 | Not-determined |
| S12_extracted_k141_24309 | Not-determined |
| S12_extracted_k141_118887 | Not-determined |
| S12_extracted_k141_150345 | Not-determined |
| S12_extracted_k141_968 | Low-quality |
| S12_extracted_k141_166105 | Not-determined |
| S12_extracted_k141_173816 | Not-determined |
| S12_extracted_k141_118891 | Low-quality |
| S12_extracted_k141_55839 | Not-determined |
| S12_extracted_k141_55846 | Not-determined |
| S12_extracted_k141_103191 | Not-determined |
| S12_extracted_k141_63826 | Not-determined |
| S12_extracted_k141_166115 | Not-determined |
| S12_extracted_k141_79533 | Not-determined |
| S12_extracted_k141_79537 | Not-determined |
| S12_extracted_k141_79540 | Not-determined |
| S12_extracted_k141_55852 | Not-determined |
| S12_extracted_k141_983 | Not-determined |
| S12_extracted_k141_79544 | Not-determined |
| S12_extracted_k141_55856 | Not-determined |
| S12_extracted_k141_16363 | Not-determined |
| S12_extracted_k141_173819 | Not-determined |
| S12_extracted_k141_40145 | Not-determined |
| S12_extracted_k141_55858 | Not-determined |
| S12_extracted_k141_126733 | Not-determined |
| S12_extracted_k141_55862 | Not-determined |
| S12_extracted_k141_173827 | Not-determined |
| S12_extracted_k141_126736 | Low-quality |
| S12_extracted_k141_55870 | Not-determined |
| S12_extracted_k141_55871 | Not-determined |
| S12_extracted_k141_118902 | Not-determined |
| S12_extracted_k141_55874 | Not-determined |
| S12_extracted_k141_40148 | Not-determined |
| S12_extracted_k141_8613 | Not-determined |
| S12_extracted_k141_118904 | Not-determined |
| S12_extracted_k141_47705 | Not-determined |
| S12_extracted_k141_24339 | Not-determined |
| S12_extracted_k141_32219 | Not-determined |
| S12_extracted_k141_173832 | Not-determined |
| S12_extracted_k141_47708 | Not-determined |
| S12_extracted_k141_55877 | Not-determined |
| S12_extracted_k141_150358 | Not-determined |
| S12_extracted_k141_158246 | Low-quality |
| S12_extracted_k141_126743 | Not-determined |
| S12_extracted_k141_40154 | Not-determined |
| S12_extracted_k141_55881 | Not-determined |
| S12_extracted_k141_126746 | Low-quality |
| S12_extracted_k141_55885 | Not-determined |
| S12_extracted_k141_126747 | Not-determined |
| S12_extracted_k141_71649 | Not-determined |
| S12_extracted_k141_150366 | Not-determined |
| S12_extracted_k141_40162 | Not-determined |
| S12_extracted_k141_24349 | Low-quality |
| S12_extracted_k141_134447 | Not-determined |
| S12_extracted_k141_134448 | Not-determined |
| S12_extracted_k141_134450 | Not-determined |
| S12_extracted_k141_134452 | Not-determined |
| S12_extracted_k141_103204 | Low-quality |
| S12_extracted_k141_118920 | Not-determined |
| S12_extracted_k141_150379 | Not-determined |
| S12_extracted_k141_181883 | Not-determined |
| S12_extracted_k141_126757 | Not-determined |
| S12_extracted_k141_24355 | Not-determined |
| S12_extracted_k141_24357 | Low-quality |
| S12_extracted_k141_103212 | Not-determined |
| S12_extracted_k141_103214 | Not-determined |
| S12_extracted_k141_118926 | Low-quality |
| S12_extracted_k141_32231 | Not-determined |
| S12_extracted_k141_118934 | Not-determined |
| S12_extracted_k141_103226 | Not-determined |
| S12_extracted_k141_158281 | Not-determined |
| S12_extracted_k141_103229 | Not-determined |
| S12_extracted_k141_24372 | Not-determined |
| S12_extracted_k141_103230 | Not-determined |
| S12_extracted_k141_118942 | Not-determined |
| S12_extracted_k141_24376 | Not-determined |
| S12_extracted_k141_55905 | Not-determined |
| S12_extracted_k141_158287 | Low-quality |
| S12_extracted_k141_150389 | Not-determined |
| S12_extracted_k141_166141 | Not-determined |
| S12_extracted_k141_173869 | Not-determined |
| S12_extracted_k141_32240 | Not-determined |
| S12_extracted_k141_40179 | Not-determined |
| S12_extracted_k141_47740 | Not-determined |
| S12_extracted_k141_95256 | Not-determined |
| S12_extracted_k141_158293 | Not-determined |
| S12_extracted_k141_79568 | Not-determined |
| S12_extracted_k141_95258 | Not-determined |
| S12_extracted_k141_55910 | Not-determined |
| S12_extracted_k141_103242 | Not-determined |
| S12_extracted_k141_158300 | Not-determined |
| S12_extracted_k141_103246 | Not-determined |
| S12_extracted_k141_95264 | Not-determined |
| S12_extracted_k141_166147 | Not-determined |
| S12_extracted_k141_47746 | Not-determined |
| S12_extracted_k141_63862 | Not-determined |
| S12_extracted_k141_173876 | Low-quality |
| S12_extracted_k141_32256 | Not-determined |
| S12_extracted_k141_47749 | Not-determined |
| S12_extracted_k141_87404 | Not-determined |
| S12_extracted_k141_63864 | Not-determined |
| S12_extracted_k141_32258 | Not-determined |
| S12_extracted_k141_103252 | Not-determined |
| S12_extracted_k141_32259 | Low-quality |
| S12_extracted_k141_173881 | Not-determined |
| S12_extracted_k141_181890 | Low-quality |
| S12_extracted_k141_134476 | Not-determined |
| S12_extracted_k141_95273 | Not-determined |
| S12_extracted_k141_150411 | Not-determined |
| S12_extracted_k141_32270 | Low-quality |
| S12_extracted_k141_63872 | Not-determined |
| S12_extracted_k141_134484 | Low-quality |
| S12_extracted_k141_32271 | Not-determined |
| S12_extracted_k141_166166 | Not-determined |
| S12_extracted_k141_103257 | Not-determined |
| S12_extracted_k141_24395 | Not-determined |
| S12_extracted_k141_995 | Not-determined |
| S12_extracted_k141_32276 | Not-determined |
| S12_extracted_k141_63874 | Not-determined |
| S12_extracted_k141_95277 | Not-determined |
| S12_extracted_k141_134487 | Not-determined |
| S12_extracted_k141_47766 | Not-determined |
| S12_extracted_k141_134488 | Low-quality |
| S12_extracted_k141_126773 | Not-determined |
| S12_extracted_k141_24400 | Not-determined |
| S12_extracted_k141_47771 | Not-determined |
| S12_extracted_k141_40206 | Not-determined |
| S12_extracted_k141_103265 | Not-determined |
| S12_extracted_k141_40207 | Not-determined |
| S12_extracted_k141_24403 | Not-determined |
| S12_extracted_k141_142628 | Not-determined |
| S12_extracted_k141_40212 | Not-determined |
| S12_extracted_k141_87431 | Not-determined |
| S12_extracted_k141_87433 | Not-determined |
| S12_extracted_k141_126780 | Not-determined |
| S12_extracted_k141_95285 | Not-determined |
| S12_extracted_k141_63888 | Low-quality |
| S12_extracted_k141_71673 | Not-determined |
| S12_extracted_k141_71674 | Not-determined |
| S12_extracted_k141_173913 | Not-determined |
| S12_extracted_k141_158324 | Low-quality |
| S12_extracted_k141_95288 | Not-determined |
| S12_extracted_k141_63892 | Not-determined |
| S12_extracted_k141_24409 | Not-determined |
| S12_extracted_k141_55918 | Not-determined |
| S12_extracted_k141_166195 | Not-determined |
| S12_extracted_k141_166196 | Not-determined |
| S12_extracted_k141_32299 | Not-determined |
| S12_extracted_k141_32301 | Not-determined |
| S12_extracted_k141_181906 | Not-determined |
| S12_extracted_k141_32302 | Not-determined |
| S12_extracted_k141_40218 | Not-determined |
| S12_extracted_k141_63899 | Not-determined |
| S12_extracted_k141_40220 | Not-determined |
| S12_extracted_k141_103288 | Not-determined |
| S12_extracted_k141_32306 | Not-determined |
| S12_extracted_k141_166200 | Not-determined |
| S12_extracted_k141_55934 | Not-determined |
| S12_extracted_k141_47798 | Not-determined |
| S12_extracted_k141_103301 | Not-determined |
| S8_extracted_k141_65522 | Medium-quality |
| S12_extracted_k141_103302 | Not-determined |
| S12_extracted_k141_24424 | Not-determined |
| S12_extracted_k141_32311 | Not-determined |
| S12_extracted_k141_32312 | Not-determined |
| S12_extracted_k141_87454 | Not-determined |
| S12_extracted_k141_87457 | Not-determined |
| S12_extracted_k141_47809 | Not-determined |
| S12_extracted_k141_16380 | Not-determined |
| S12_extracted_k141_166218 | Not-determined |
| S12_extracted_k141_181918 | Not-determined |
| S12_extracted_k141_166219 | Not-determined |
| S12_extracted_k141_24432 | Not-determined |
| S12_extracted_k141_79601 | Not-determined |
| S12_extracted_k141_16386 | Not-determined |
| S12_extracted_k141_166225 | Low-quality |
| S12_extracted_k141_16387 | Low-quality |
| S12_extracted_k141_103313 | Not-determined |
| S12_extracted_k141_79603 | Not-determined |
| S12_extracted_k141_142651 | Not-determined |
| S12_extracted_k141_142654 | Not-determined |
| S12_extracted_k141_40240 | Not-determined |
| S12_extracted_k141_103318 | Not-determined |
| S12_extracted_k141_55956 | Not-determined |
| S12_extracted_k141_87468 | Not-determined |
| S12_extracted_k141_103320 | Not-determined |
| S12_extracted_k141_24448 | Not-determined |
| S12_extracted_k141_134529 | Not-determined |
| S12_extracted_k141_87477 | Not-determined |
| S12_extracted_k141_181933 | Not-determined |
| S12_extracted_k141_47820 | Not-determined |
| S12_extracted_k141_134535 | Not-determined |
| S12_extracted_k141_181936 | Not-determined |
| S12_extracted_k141_126791 | Not-determined |
| S12_extracted_k141_47824 | Not-determined |
| S12_extracted_k141_181938 | Not-determined |
| S12_extracted_k141_158343 | Not-determined |
| S12_extracted_k141_142659 | Not-determined |
| S12_extracted_k141_103325 | Not-determined |
| S12_extracted_k141_63914 | Not-determined |
| S12_extracted_k141_181942 | Not-determined |
| S12_extracted_k141_24451 | Not-determined |
| S12_extracted_k141_134544 | Not-determined |
| S12_extracted_k141_118978 | Not-determined |
| S12_extracted_k141_71695 | Not-determined |
| S12_extracted_k141_118984 | Not-determined |
| S12_extracted_k141_158352 | Not-determined |
| S12_extracted_k141_79621 | Not-determined |
| S12_extracted_k141_134551 | Not-determined |
| S12_extracted_k141_79624 | Not-determined |
| S12_extracted_k141_71696 | Not-determined |
| S12_extracted_k141_166233 | Not-determined |
| S12_extracted_k141_55982 | Not-determined |
| S12_extracted_k141_55983 | Not-determined |
| S12_extracted_k141_126805 | Not-determined |
| S12_extracted_k141_1041 | Not-determined |
| S12_extracted_k141_16398 | Not-determined |
| S12_extracted_k141_126813 | Not-determined |
| S12_extracted_k141_142668 | Not-determined |
| S12_extracted_k141_134561 | Not-determined |
| S12_extracted_k141_63942 | Not-determined |
| S12_extracted_k141_166251 | Not-determined |
| S12_extracted_k141_1052 | Not-determined |
| S12_extracted_k141_166255 | Low-quality |
| S12_extracted_k141_134570 | Not-determined |
| S12_extracted_k141_32329 | Not-determined |
| S12_extracted_k141_40255 | Not-determined |
| S12_extracted_k141_40257 | Not-determined |
| S12_extracted_k141_166262 | Not-determined |
| S12_extracted_k141_1056 | Not-determined |
| S12_extracted_k141_87496 | Not-determined |
| S12_extracted_k141_87498 | Not-determined |
| S12_extracted_k141_32341 | Not-determined |
| S12_extracted_k141_16409 | Not-determined |
| S12_extracted_k141_173935 | Not-determined |
| S12_extracted_k141_40265 | Not-determined |
| S12_extracted_k141_1070 | Not-determined |
| S12_extracted_k141_142684 | Not-determined |
| S12_extracted_k141_158381 | Low-quality |
| S12_extracted_k141_16416 | Not-determined |
| S12_extracted_k141_126818 | Low-quality |
| S12_extracted_k141_173937 | Not-determined |
| S12_extracted_k141_40270 | Not-determined |
| S12_extracted_k141_134588 | Not-determined |
| S12_extracted_k141_158383 | Not-determined |
| S12_extracted_k141_134592 | Not-determined |
| S12_extracted_k141_134593 | Not-determined |
| S12_extracted_k141_95326 | Not-determined |
| S12_extracted_k141_16421 | Not-determined |
| S12_extracted_k141_47878 | Not-determined |
| S12_extracted_k141_95328 | Not-determined |
| S12_extracted_k141_142698 | Not-determined |
| S12_extracted_k141_181973 | Not-determined |
| S12_extracted_k141_95336 | Not-determined |
| S12_extracted_k141_1072 | Not-determined |
| S12_extracted_k141_95341 | Not-determined |
| S12_extracted_k141_173940 | Not-determined |
| S12_extracted_k141_119007 | Not-determined |
| S12_extracted_k141_142711 | Not-determined |
| S12_extracted_k141_166281 | Not-determined |
| S12_extracted_k141_119012 | Not-determined |
| S12_extracted_k141_126836 | Not-determined |
| S12_extracted_k141_1090 | Not-determined |
| S12_extracted_k141_142717 | Low-quality |
| S12_extracted_k141_95349 | Not-determined |
| S12_extracted_k141_95351 | Not-determined |
| S12_extracted_k141_142722 | Not-determined |
| S12_extracted_k141_119020 | Not-determined |
| S12_extracted_k141_181997 | Not-determined |
| S12_extracted_k141_63983 | Not-determined |
| S12_extracted_k141_16431 | Not-determined |
| S12_extracted_k141_87517 | Not-determined |
| S12_extracted_k141_134603 | Not-determined |
| S12_extracted_k141_47887 | Not-determined |
| S12_extracted_k141_71705 | Low-quality |
| S12_extracted_k141_134608 | Not-determined |
| S12_extracted_k141_182004 | Low-quality |
| S12_extracted_k141_63990 | Not-determined |
| S12_extracted_k141_47892 | Not-determined |
| S12_extracted_k141_56019 | Low-quality |
| S12_extracted_k141_63993 | Not-determined |
| S12_extracted_k141_166287 | Not-determined |
| S12_extracted_k141_1096 | Not-determined |
| S12_extracted_k141_119027 | Not-determined |
| S12_extracted_k141_16436 | Not-determined |
| S12_extracted_k141_134613 | Not-determined |
| S12_extracted_k141_47895 | Not-determined |
| S12_extracted_k141_103346 | Not-determined |
| S12_extracted_k141_119030 | Not-determined |
| S12_extracted_k141_134615 | Not-determined |
| S12_extracted_k141_47896 | Low-quality |
| S12_extracted_k141_158400 | Not-determined |
| S12_extracted_k141_47898 | Not-determined |
| S12_extracted_k141_119036 | Not-determined |
| S12_extracted_k141_103357 | Not-determined |
| S12_extracted_k141_8628 | Not-determined |
| S12_extracted_k141_158406 | Not-determined |
| S12_extracted_k141_150453 | Not-determined |
| S12_extracted_k141_182012 | Not-determined |
| S12_extracted_k141_182013 | Not-determined |
| S12_extracted_k141_47909 | Not-determined |
| S12_extracted_k141_95369 | Not-determined |
| S12_extracted_k141_166303 | Not-determined |
| S12_extracted_k141_158412 | Not-determined |
| S12_extracted_k141_8636 | Not-determined |
| S12_extracted_k141_40287 | Not-determined |
| S12_extracted_k141_40288 | Not-determined |
| S12_extracted_k141_8640 | Not-determined |
| S12_extracted_k141_126856 | Not-determined |
| S12_extracted_k141_40291 | Not-determined |
| S12_extracted_k141_95371 | Not-determined |
| S12_extracted_k141_16459 | Not-determined |
| S12_extracted_k141_40292 | Not-determined |
| S12_extracted_k141_134640 | Not-determined |
| S12_extracted_k141_56030 | Not-determined |
| S12_extracted_k141_16462 | Low-quality |
| S12_extracted_k141_103384 | Not-determined |
| S12_extracted_k141_56034 | Not-determined |
| S12_extracted_k141_40299 | Low-quality |
| S12_extracted_k141_182030 | Not-determined |
| S12_extracted_k141_8647 | Not-determined |
| S12_extracted_k141_103390 | Not-determined |
| S12_extracted_k141_40305 | Not-determined |
| S12_extracted_k141_126872 | Not-determined |
| S12_extracted_k141_103393 | Low-quality |
| S12_extracted_k141_150471 | Not-determined |
| S12_extracted_k141_24488 | Not-determined |
| S12_extracted_k141_126875 | Low-quality |
| S12_extracted_k141_24490 | Not-determined |
| S12_extracted_k141_110811 | Not-determined |
| S12_extracted_k141_56061 | Not-determined |
| S12_extracted_k141_142735 | Not-determined |
| S12_extracted_k141_110812 | Not-determined |
| S12_extracted_k141_126884 | Not-determined |
| S12_extracted_k141_150477 | Not-determined |
| S12_extracted_k141_103400 | Not-determined |
| S12_extracted_k141_1101 | Not-determined |
| S12_extracted_k141_32349 | Not-determined |
| S12_extracted_k141_47919 | Not-determined |
| S12_extracted_k141_103405 | Not-determined |
| S12_extracted_k141_182053 | Low-quality |
| S12_extracted_k141_126891 | Not-determined |
| S12_extracted_k141_40324 | Not-determined |
| S12_extracted_k141_182057 | Not-determined |
| S12_extracted_k141_158427 | Not-determined |
| S12_extracted_k141_47929 | Not-determined |
| S12_extracted_k141_56077 | Not-determined |
| S12_extracted_k141_142751 | Not-determined |
| S12_extracted_k141_47931 | Low-quality |
| S12_extracted_k141_103421 | Not-determined |
| S12_extracted_k141_47933 | Not-determined |
| S12_extracted_k141_32360 | Low-quality |
| S12_extracted_k141_71715 | Not-determined |
| S12_extracted_k141_103422 | Not-determined |
| S12_extracted_k141_142756 | Not-determined |
| S12_extracted_k141_64017 | Not-determined |
| S12_extracted_k141_95397 | Not-determined |
| S12_extracted_k141_24495 | Not-determined |
| S12_extracted_k141_24496 | Not-determined |
| S12_extracted_k141_64021 | Not-determined |
| S12_extracted_k141_24497 | Not-determined |
| S12_extracted_k141_47939 | Not-determined |
| S12_extracted_k141_95399 | Not-determined |
| S12_extracted_k141_64022 | Low-quality |
| S12_extracted_k141_182061 | Low-quality |
| S12_extracted_k141_32368 | Not-determined |
| S12_extracted_k141_16489 | Not-determined |
| S12_extracted_k141_95402 | Not-determined |
| S12_extracted_k141_173953 | Not-determined |
| S12_extracted_k141_24502 | Not-determined |
| S12_extracted_k141_16491 | Not-determined |
| S12_extracted_k141_142767 | Low-quality |
| S12_extracted_k141_158447 | Not-determined |
| S12_extracted_k141_16496 | Low-quality |
| S12_extracted_k141_24508 | Low-quality |
| S12_extracted_k141_56082 | Not-determined |
| S12_extracted_k141_166328 | Low-quality |
| S12_extracted_k141_40337 | Not-determined |
| S12_extracted_k141_182079 | Not-determined |
| S12_extracted_k141_1136 | Not-determined |
| S12_extracted_k141_182082 | Not-determined |
| S12_extracted_k141_40344 | Not-determined |
| S12_extracted_k141_71734 | Not-determined |
| S12_extracted_k141_95420 | Not-determined |
| S12_extracted_k141_182086 | Low-quality |
| S12_extracted_k141_56096 | Low-quality |
| S12_extracted_k141_16507 | Not-determined |
| S12_extracted_k141_32385 | Not-determined |
| S12_extracted_k141_24524 | Not-determined |
| S12_extracted_k141_24525 | Not-determined |
| S12_extracted_k141_166342 | Not-determined |
| S12_extracted_k141_95425 | Not-determined |
| S12_extracted_k141_16511 | Not-determined |
| S12_extracted_k141_158462 | Not-determined |
| S12_extracted_k141_173969 | Low-quality |
| S12_extracted_k141_64042 | Not-determined |
| S12_extracted_k141_16515 | Not-determined |
| S12_extracted_k141_95430 | Not-determined |
| S12_extracted_k141_71744 | Not-determined |
| S12_extracted_k141_64046 | Not-determined |
| S12_extracted_k141_182101 | Not-determined |
| S12_extracted_k141_103433 | Not-determined |
| S12_extracted_k141_110845 | Not-determined |

| | |
|---|---|
| S12_extracted_k141_103436 | Not-determined |
| S12_extracted_k141_8693 | Low-quality |
| S12_extracted_k141_24541 | Not-determined |
| S12_extracted_k141_110850 | Not-determined |
| S12_extracted_k141_1150 | Not-determined |
| S12_extracted_k141_1152 | Not-determined |
| S12_extracted_k141_8697 | Low-quality |
| S12_extracted_k141_95444 | Not-determined |
| S12_extracted_k141_110856 | Not-determined |
| S12_extracted_k141_24548 | Not-determined |
| S12_extracted_k141_103444 | Not-determined |
| S12_extracted_k141_71758 | Not-determined |
| S12_extracted_k141_95448 | Not-determined |
| S12_extracted_k141_40349 | Not-determined |
| S12_extracted_k141_103450 | Not-determined |
| S12_extracted_k141_24553 | Low-quality |
| S12_extracted_k141_142782 | Not-determined |
| S12_extracted_k141_119070 | Not-determined |
| S12_extracted_k141_1169 | Not-determined |
| S12_extracted_k141_103453 | Not-determined |
| S12_extracted_k141_1170 | Low-quality |
| S12_extracted_k141_47979 | Low-quality |
| S12_extracted_k141_56112 | Not-determined |
| S12_extracted_k141_47980 | Not-determined |
| S12_extracted_k141_71762 | Not-determined |
| S12_extracted_k141_95459 | Not-determined |
| S12_extracted_k141_64068 | Low-quality |
| S12_extracted_k141_182116 | Not-determined |
| S12_extracted_k141_119077 | Not-determined |
| S12_extracted_k141_47986 | Not-determined |
| S12_extracted_k141_24562 | Not-determined |
| S12_extracted_k141_64077 | Not-determined |
| S12_extracted_k141_24566 | Not-determined |
| S12_extracted_k141_182124 | Not-determined |
| S12_extracted_k141_56122 | Not-determined |
| S12_extracted_k141_182126 | Not-determined |
| S12_extracted_k141_142796 | Not-determined |
| S12_extracted_k141_16525 | Not-determined |
| S12_extracted_k141_166366 | Not-determined |
| S12_extracted_k141_32408 | Not-determined |
| S12_extracted_k141_126931 | Not-determined |
| S12_extracted_k141_71771 | Not-determined |
| S12_extracted_k141_32410 | Not-determined |
| S12_extracted_k141_182128 | Low-quality |
| S12_extracted_k141_40371 | Low-quality |
| S12_extracted_k141_173983 | Not-determined |
| S12_extracted_k141_32412 | Not-determined |
| S12_extracted_k141_142806 | Not-determined |
| S12_extracted_k141_47993 | Not-determined |
| S12_extracted_k141_126938 | Not-determined |
| S12_extracted_k141_56129 | Not-determined |
| S12_extracted_k141_16530 | Not-determined |
| S12_extracted_k141_47994 | Not-determined |
| S12_extracted_k141_47997 | Not-determined |
| S12_extracted_k141_40376 | Not-determined |
| S12_extracted_k141_173995 | Not-determined |
| S12_extracted_k141_47999 | Not-determined |
| S12_extracted_k141_166378 | Not-determined |
| S12_extracted_k141_16533 | Not-determined |
| S12_extracted_k141_173998 | Not-determined |
| S12_extracted_k141_48001 | Not-determined |
| S12_extracted_k141_182140 | Not-determined |
| S12_extracted_k141_110883 | Not-determined |
| S12_extracted_k141_126945 | Not-determined |
| S12_extracted_k141_166383 | Not-determined |
| S12_extracted_k141_32423 | Low-quality |
| S12_extracted_k141_16538 | Not-determined |
| S12_extracted_k141_103462 | Not-determined |
| S12_extracted_k141_119086 | Not-determined |
| S12_extracted_k141_64094 | Not-determined |
| S12_extracted_k141_56147 | Not-determined |
| S12_extracted_k141_182151 | Not-determined |
| S12_extracted_k141_103467 | Low-quality |
| S12_extracted_k141_40386 | Not-determined |
| S12_extracted_k141_64100 | Not-determined |
| S12_extracted_k141_103469 | Not-determined |
| S12_extracted_k141_182158 | Not-determined |
| S12_extracted_k141_103471 | Not-determined |
| S12_extracted_k141_32435 | Not-determined |
| S12_extracted_k141_71794 | Not-determined |
| S12_extracted_k141_64105 | Not-determined |
| S12_extracted_k141_40388 | Not-determined |
| S12_extracted_k141_103474 | Not-determined |
| S12_extracted_k141_103477 | Not-determined |
| S12_extracted_k141_103478 | Not-determined |
| S12_extracted_k141_126975 | Not-determined |
| S12_extracted_k141_48026 | Not-determined |
| S12_extracted_k141_48027 | Not-determined |
| S12_extracted_k141_110911 | Not-determined |
| S12_extracted_k141_182173 | Not-determined |
| S12_extracted_k141_32446 | Not-determined |
| S12_extracted_k141_174022 | Not-determined |
| S12_extracted_k141_110916 | Not-determined |
| S12_extracted_k141_24597 | Low-quality |
| S12_extracted_k141_110917 | Not-determined |
| S12_extracted_k141_182182 | Not-determined |
| S12_extracted_k141_103488 | Not-determined |
| S12_extracted_k141_95497 | Low-quality |
| S12_extracted_k141_174031 | Not-determined |
| S12_extracted_k141_142815 | Low-quality |
| S12_extracted_k141_95502 | Not-determined |
| S12_extracted_k141_16559 | Low-quality |
| S12_extracted_k141_182188 | Not-determined |
| S12_extracted_k141_142819 | Not-determined |
| S12_extracted_k141_182191 | Not-determined |
| S12_extracted_k141_56169 | Not-determined |
| S12_extracted_k141_87583 | Not-determined |
| S12_extracted_k141_8749 | Low-quality |
| S12_extracted_k141_103494 | Not-determined |
| S12_extracted_k141_71814 | Not-determined |
| S12_extracted_k141_119095 | Not-determined |
| S12_extracted_k141_150552 | Not-determined |
| S12_extracted_k141_182204 | Not-determined |
| S12_extracted_k141_16576 | Not-determined |
| S12_extracted_k141_64131 | Not-determined |
| S12_extracted_k141_16577 | Not-determined |
| S12_extracted_k141_8762 | Low-quality |
| S12_extracted_k141_182207 | Not-determined |
| S12_extracted_k141_56177 | Not-determined |
| S12_extracted_k141_182210 | Not-determined |
| S12_extracted_k141_71819 | Not-determined |
| S12_extracted_k141_103498 | Not-determined |
| S12_extracted_k141_103500 | Not-determined |
| S12_extracted_k141_182216 | Not-determined |
| S12_extracted_k141_24621 | Not-determined |
| S12_extracted_k141_142839 | Not-determined |
| S12_extracted_k141_150569 | Not-determined |
| S12_extracted_k141_95518 | Not-determined |
| S12_extracted_k141_110943 | Not-determined |
| S12_extracted_k141_174052 | Not-determined |
| S12_extracted_k141_182222 | Not-determined |
| S12_extracted_k141_174057 | Not-determined |
| S12_extracted_k141_182227 | Not-determined |
| S12_extracted_k141_71827 | Not-determined |
| S12_extracted_k141_24629 | Not-determined |
| S12_extracted_k141_150578 | Not-determined |
| S12_extracted_k141_95526 | Not-determined |
| S12_extracted_k141_24631 | Not-determined |
| S12_extracted_k141_182232 | Not-determined |
| S12_extracted_k141_71831 | Not-determined |
| S12_extracted_k141_95530 | Not-determined |
| S12_extracted_k141_174065 | Not-determined |
| S12_extracted_k141_95532 | Not-determined |
| S12_extracted_k141_150584 | Not-determined |
| S12_extracted_k141_87607 | Low-quality |
| S12_extracted_k141_150587 | Not-determined |
| S12_extracted_k141_95537 | Not-determined |
| S8_vs_k141_65522\|\|full | Medium-quality |
| S12_extracted_k141_166438 | Not-determined |
| S12_extracted_k141_182240 | Not-determined |
| S12_extracted_k141_87614 | Not-determined |
| S12_extracted_k141_166441 | Not-determined |
| S12_extracted_k141_95543 | Not-determined |
| S12_extracted_k141_24640 | Not-determined |
| S12_extracted_k141_79650 | Not-determined |
| S12_extracted_k141_150594 | Not-determined |
| S12_extracted_k141_79651 | Low-quality |
| S12_extracted_k141_166443 | Not-determined |
| S12_extracted_k141_56214 | Not-determined |
| S12_extracted_k141_1209 | Not-determined |
| S12_extracted_k141_182242 | Not-determined |
| S12_extracted_k141_134672 | Not-determined |
| S12_extracted_k141_1211 | Not-determined |
| S12_extracted_k141_64143 | Not-determined |
| S12_extracted_k141_24647 | Low-quality |
| S12_extracted_k141_87618 | Not-determined |
| S12_extracted_k141_24649 | Not-determined |
| S12_extracted_k141_1215 | Not-determined |
| S12_extracted_k141_1217 | Not-determined |
| S12_extracted_k141_110955 | Not-determined |
| S12_extracted_k141_87623 | Not-determined |
| S12_extracted_k141_71835 | Not-determined |
| S12_extracted_k141_95553 | Not-determined |
| S12_extracted_k141_16606 | Not-determined |
| S12_extracted_k141_182248 | Not-determined |
| S12_extracted_k141_16610 | Low-quality |
| S12_extracted_k141_134674 | Not-determined |
| S12_extracted_k141_56227 | Not-determined |
| S12_extracted_k141_174083 | Low-quality |
| S12_extracted_k141_174087 | Not-determined |
| S12_extracted_k141_110962 | Low-quality |
| S12_extracted_k141_174088 | Not-determined |
| S12_extracted_k141_56232 | Not-determined |
| S12_extracted_k141_142852 | Not-determined |
| S12_extracted_k141_64153 | Not-determined |
| S12_extracted_k141_87634 | Not-determined |
| S12_extracted_k141_1221 | Not-determined |
| S12_extracted_k141_166472 | Not-determined |
| S12_extracted_k141_56240 | Not-determined |
| S12_extracted_k141_126990 | Low-quality |
| S12_extracted_k141_16621 | Low-quality |
| S12_extracted_k141_119123 | Not-determined |
| S12_extracted_k141_71852 | Not-determined |
| S12_extracted_k141_119125 | Not-determined |
| S12_extracted_k141_8781 | Not-determined |
| S12_extracted_k141_182258 | Not-determined |
| S12_extracted_k141_1230 | Not-determined |
| S12_extracted_k141_95561 | Not-determined |
| S12_extracted_k141_182260 | Not-determined |
| S12_extracted_k141_24673 | Not-determined |
| S12_extracted_k141_24674 | Not-determined |
| S12_extracted_k141_142860 | Not-determined |
| S12_extracted_k141_166482 | Not-determined |
| S12_extracted_k141_182261 | Low-quality |
| S12_extracted_k141_24677 | Not-determined |
| S12_extracted_k141_8783 | Low-quality |
| S12_extracted_k141_1235 | Not-determined |
| S12_extracted_k141_110977 | Not-determined |
| S12_extracted_k141_182268 | Not-determined |
| S12_extracted_k141_32454 | Low-quality |
| S12_extracted_k141_8787 | Not-determined |
| S12_extracted_k141_16636 | Not-determined |
| S12_extracted_k141_182271 | Not-determined |
| S12_extracted_k141_142868 | Not-determined |
| S12_extracted_k141_150615 | Low-quality |
| S12_extracted_k141_64156 | Not-determined |
| S12_extracted_k141_8788 | Not-determined |
| S12_extracted_k141_119146 | Not-determined |
| S12_extracted_k141_8792 | Not-determined |
| S12_extracted_k141_142872 | Not-determined |
| S12_extracted_k141_95580 | Not-determined |
| S12_extracted_k141_40455 | Not-determined |
| S12_extracted_k141_110991 | Not-determined |
| S12_extracted_k141_119151 | Not-determined |
| S12_extracted_k141_71877 | Low-quality |
| S12_extracted_k141_119154 | Low-quality |
| S12_extracted_k141_119155 | Not-determined |
| S12_extracted_k141_119156 | Not-determined |
| S12_extracted_k141_24697 | Low-quality |
| S12_extracted_k141_40472 | Not-determined |
| S12_extracted_k141_95588 | Not-determined |
| S12_extracted_k141_24701 | Low-quality |
| S12_extracted_k141_71887 | Not-determined |
| S12_extracted_k141_166506 | Not-determined |
| S12_extracted_k141_111003 | Not-determined |
| S12_extracted_k141_174101 | Not-determined |
| S12_extracted_k141_134681 | Not-determined |
| S12_extracted_k141_32462 | Not-determined |
| S12_extracted_k141_95595 | Not-determined |
| S12_extracted_k141_71894 | Not-determined |
| S12_extracted_k141_111011 | Not-determined |
| S12_extracted_k141_182278 | Low-quality |
| S12_extracted_k141_24709 | Not-determined |
| S12_extracted_k141_24717 | Not-determined |
| S12_extracted_k141_87658 | Not-determined |
| S12_extracted_k141_24720 | Not-determined |
| S12_extracted_k141_95602 | Not-determined |
| S12_extracted_k141_48076 | Not-determined |
| S12_extracted_k141_40481 | Not-determined |
| S12_extracted_k141_64163 | Not-determined |
| S12_extracted_k141_119178 | Not-determined |
| S12_extracted_k141_71902 | Not-determined |
| S12_extracted_k141_24723 | Not-determined |
| S12_extracted_k141_174120 | Not-determined |
| S12_extracted_k141_64173 | Not-determined |
| S12_extracted_k141_87661 | Not-determined |
| S12_extracted_k141_48083 | Not-determined |
| S12_extracted_k141_56253 | Not-determined |
| S12_extracted_k141_71911 | Not-determined |
| S12_extracted_k141_174123 | Not-determined |
| S12_extracted_k141_174124 | Low-quality |
| S12_extracted_k141_119186 | Not-determined |
| S12_extracted_k141_71912 | Not-determined |
| S12_extracted_k141_87664 | Not-determined |
| S12_extracted_k141_111019 | Not-determined |
| S12_extracted_k141_56260 | Not-determined |
| S12_extracted_k141_174126 | Not-determined |
| S12_extracted_k141_182296 | Not-determined |
| S12_extracted_k141_127011 | Not-determined |
| S12_extracted_k141_87668 | Not-determined |
| S12_extracted_k141_71920 | Not-determined |
| S12_extracted_k141_64191 | Not-determined |
| S12_extracted_k141_16668 | Not-determined |
| S12_extracted_k141_174137 | Not-determined |
| S12_extracted_k141_40506 | Not-determined |
| S12_extracted_k141_56267 | Not-determined |
| S12_extracted_k141_48105 | Not-determined |
| S5_vs_k141_112345\|\|full | Medium-quality |
| S12_extracted_k141_48110 | Not-determined |
| S12_extracted_k141_174143 | Not-determined |
| S12_extracted_k141_8810 | Low-quality |
| S12_extracted_k141_87682 | Not-determined |
| S12_extracted_k141_64206 | Not-determined |
| S12_extracted_k141_8811 | Low-quality |
| S12_extracted_k141_64211 | Not-determined |
| S12_extracted_k141_111038 | Not-determined |
| S12_extracted_k141_48119 | Not-determined |
| S12_extracted_k141_87684 | Not-determined |
| S12_extracted_k141_48120 | Not-determined |
| S12_extracted_k141_79676 | Not-determined |
| S12_extracted_k141_174153 | Not-determined |
| S12_extracted_k141_95637 | Not-determined |
| S12_extracted_k141_111044 | Low-quality |
| S12_extracted_k141_95639 | Not-determined |
| S12_extracted_k141_174157 | Not-determined |
| S12_extracted_k141_48128 | Not-determined |
| S12_extracted_k141_134695 | Not-determined |
| S12_extracted_k141_95640 | Not-determined |
| S12_extracted_k141_111046 | Not-determined |
| S12_extracted_k141_174160 | Not-determined |
| S12_extracted_k141_56291 | Not-determined |
| S12_extracted_k141_24749 | Not-determined |
| S12_extracted_k141_174163 | Not-determined |
| S12_extracted_k141_56295 | Not-determined |
| S12_extracted_k141_24751 | Not-determined |
| S12_extracted_k141_79678 | Low-quality |
| S12_extracted_k141_79680 | Not-determined |
| S12_extracted_k141_119217 | Not-determined |
| S12_extracted_k141_119221 | Not-determined |
| S12_extracted_k141_119223 | Not-determined |
| S12_extracted_k141_166519 | Low-quality |
| S12_extracted_k141_40528 | Not-determined |
| S12_extracted_k141_24760 | Not-determined |
| S12_extracted_k141_119227 | Not-determined |
| S12_extracted_k141_127025 | Not-determined |
| S12_extracted_k141_111062 | Not-determined |
| S12_extracted_k141_95647 | Not-determined |
| S12_extracted_k141_16672 | Low-quality |
| S12_extracted_k141_48146 | Not-determined |
| S12_extracted_k141_79695 | Not-determined |
| S12_extracted_k141_182312 | Not-determined |
| S12_extracted_k141_48150 | Not-determined |
| S12_extracted_k141_95651 | Not-determined |
| S12_extracted_k141_166527 | Not-determined |
| S12_extracted_k141_182315 | Not-determined |
| S12_extracted_k141_24766 | Not-determined |
| S12_extracted_k141_79704 | Not-determined |
| S12_extracted_k141_16681 | Not-determined |
| S12_extracted_k141_87690 | Not-determined |
| S12_extracted_k141_182328 | Low-quality |
| S12_extracted_k141_182329 | Not-determined |
| S12_extracted_k141_48158 | Not-determined |
| S12_extracted_k141_48162 | Not-determined |
| S12_extracted_k141_95662 | Not-determined |
| S12_extracted_k141_48165 | Low-quality |
| S12_extracted_k141_40549 | Not-determined |
| S12_extracted_k141_119240 | Not-determined |
| S12_extracted_k141_95669 | Not-determined |
| S12_extracted_k141_56321 | Not-determined |
| S12_extracted_k141_24788 | Not-determined |
| S12_extracted_k141_71954 | Not-determined |
| S12_extracted_k141_150634 | Not-determined |
| S12_extracted_k141_150635 | Not-determined |
| S12_extracted_k141_111078 | Not-determined |
| S12_extracted_k141_64229 | Not-determined |
| S12_extracted_k141_16685 | Low-quality |
| S12_extracted_k141_40560 | Not-determined |
| S12_extracted_k141_95683 | Not-determined |
| S12_extracted_k141_24797 | Not-determined |
| S12_extracted_k141_95684 | Not-determined |
| S12_extracted_k141_174171 | Not-determined |
| S12_extracted_k141_8822 | Not-determined |
| S12_extracted_k141_40561 | Not-determined |
| S12_extracted_k141_111082 | Not-determined |
| S12_extracted_k141_40562 | Not-determined |
| S12_extracted_k141_8824 | Not-determined |
| S12_extracted_k141_119248 | Not-determined |
| S12_extracted_k141_166559 | Low-quality |
| S12_extracted_k141_87716 | Not-determined |
| S12_extracted_k141_32475 | Not-determined |
| S12_extracted_k141_8830 | Not-determined |
| S12_extracted_k141_79717 | Not-determined |
| S12_extracted_k141_32478 | Not-determined |
| S12_extracted_k141_119259 | Not-determined |
| S12_extracted_k141_32481 | Not-determined |
| S12_extracted_k141_142884 | Not-determined |
| S12_extracted_k141_16697 | Not-determined |
| S12_extracted_k141_87726 | Low-quality |
| S12_extracted_k141_56344 | Not-determined |
| S12_extracted_k141_64256 | Low-quality |
| S12_extracted_k141_87731 | Low-quality |
| S12_extracted_k141_32488 | Not-determined |
| S12_extracted_k141_32489 | Not-determined |
| S12_extracted_k141_142890 | Not-determined |
| S12_extracted_k141_64266 | Not-determined |
| S12_extracted_k141_119265 | Low-quality |
| S12_extracted_k141_111110 | Not-determined |
| S12_extracted_k141_119267 | Not-determined |
| S12_extracted_k141_142891 | Not-determined |
| S12_extracted_k141_150655 | Not-determined |
| S12_extracted_k141_79726 | Not-determined |
| S12_extracted_k141_111114 | Not-determined |
| S12_extracted_k141_1252 | Not-determined |
| S12_extracted_k141_32497 | Not-determined |
| S12_extracted_k141_111121 | Not-determined |
| S12_extracted_k141_79736 | Not-determined |
| S12_extracted_k141_127062 | Not-determined |
| S12_extracted_k141_64278 | Not-determined |
| S12_extracted_k141_32501 | Not-determined |
| S12_extracted_k141_87746 | Not-determined |
| S12_extracted_k141_174198 | Not-determined |
| S12_extracted_k141_24802 | Not-determined |
| S12_extracted_k141_79738 | Not-determined |
| S12_extracted_k141_8837 | Not-determined |
| S12_extracted_k141_134712 | Not-determined |
| S12_extracted_k141_8839 | Not-determined |
| S12_extracted_k141_48172 | Not-determined |
| S12_extracted_k141_40580 | Not-determined |
| S12_extracted_k141_174203 | Not-determined |
| S12_extracted_k141_64290 | Not-determined |
| S12_extracted_k141_8846 | Not-determined |
| S12_extracted_k141_64291 | Not-determined |
| S12_extracted_k141_174205 | Not-determined |
| S12_extracted_k141_95721 | Not-determined |
| S12_extracted_k141_79739 | Not-determined |
| S12_extracted_k141_134720 | Not-determined |
| S12_extracted_k141_16706 | Not-determined |
| S12_extracted_k141_71960 | Not-determined |
| S12_extracted_k141_174210 | Not-determined |
| S12_extracted_k141_1265 | Low-quality |
| S12_extracted_k141_174211 | Not-determined |
| S12_extracted_k141_134722 | Not-determined |
| S12_extracted_k141_111130 | Not-determined |
| S12_extracted_k141_111132 | Not-determined |
| S12_extracted_k141_182345 | Not-determined |
| S12_extracted_k141_32511 | Not-determined |
| S12_extracted_k141_95728 | Low-quality |
| S12_extracted_k141_40597 | Not-determined |
| S12_extracted_k141_40598 | Not-determined |
| S12_extracted_k141_134734 | Not-determined |
| S12_extracted_k141_182356 | Not-determined |
| S12_extracted_k141_24819 | Not-determined |
| S12_extracted_k141_95732 | Low-quality |
| S12_extracted_k141_95733 | Not-determined |
| S12_extracted_k141_182361 | Not-determined |
| S12_extracted_k141_174216 | Not-determined |
| S12_extracted_k141_8861 | Not-determined |
| S12_extracted_k141_142916 | Not-determined |
| S12_extracted_k141_87759 | Not-determined |
| S12_extracted_k141_182369 | Not-determined |
| S12_extracted_k141_71975 | Not-determined |
| S12_extracted_k141_127076 | Not-determined |
| S12_extracted_k141_71977 | Not-determined |
| S12_extracted_k141_127079 | Not-determined |
| S12_extracted_k141_79760 | Not-determined |
| S12_extracted_k141_142926 | Not-determined |
| S12_extracted_k141_71983 | Not-determined |
| S12_extracted_k141_127086 | Not-determined |
| S12_extracted_k141_79763 | Not-determined |
| S12_extracted_k141_48189 | Not-determined |
| S12_extracted_k141_1284 | Not-determined |
| S12_extracted_k141_127089 | Not-determined |
| S12_extracted_k141_1285 | Low-quality |
| S12_extracted_k141_32523 | Not-determined |
| S12_extracted_k141_127093 | Not-determined |
| S12_extracted_k141_134752 | Not-determined |
| S12_extracted_k141_134753 | Low-quality |
| S12_extracted_k141_174220 | Not-determined |
| S12_extracted_k141_134754 | Not-determined |
| S12_extracted_k141_87774 | Not-determined |
| S12_extracted_k141_16725 | Not-determined |
| S12_extracted_k141_1288 | Not-determined |
| S12_extracted_k141_87775 | Not-determined |
| S12_extracted_k141_174222 | Not-determined |
| S12_extracted_k141_8876 | Not-determined |
| S12_extracted_k141_174224 | Low-quality |
| S12_extracted_k141_134757 | Not-determined |
| S12_extracted_k141_8881 | Not-determined |
| S12_extracted_k141_95744 | Low-quality |
| S12_extracted_k141_79772 | Not-determined |
| S12_extracted_k141_111138 | Low-quality |
| S12_extracted_k141_142938 | Low-quality |
| S12_extracted_k141_87782 | Not-determined |
| S12_extracted_k141_40607 | Not-determined |
| S12_extracted_k141_64312 | Not-determined |
| S12_extracted_k141_32536 | Not-determined |
| S12_extracted_k141_95748 | Not-determined |
| S12_extracted_k141_16734 | Not-determined |
| S12_extracted_k141_24825 | Not-determined |
| S12_extracted_k141_95754 | Not-determined |
| S12_extracted_k141_24826 | Not-determined |
| S12_extracted_k141_95757 | Low-quality |
| S12_extracted_k141_40616 | Low-quality |
| S12_extracted_k141_119291 | Not-determined |
| S12_extracted_k141_32544 | Not-determined |
| S12_extracted_k141_32550 | Low-quality |
| S12_extracted_k141_64316 | Not-determined |
| S12_extracted_k141_95758 | Not-determined |
| S12_extracted_k141_64317 | Not-determined |
| S12_extracted_k141_71995 | Not-determined |
| S12_extracted_k141_95759 | Not-determined |
| S12_extracted_k141_16745 | Not-determined |
| S12_extracted_k141_127107 | Low-quality |
| S12_extracted_k141_16747 | Not-determined |
| S12_extracted_k141_142958 | Not-determined |
| S12_extracted_k141_111153 | Not-determined |
| S12_extracted_k141_16750 | Not-determined |
| S12_extracted_k141_166561 | Not-determined |
| S12_extracted_k141_48213 | Not-determined |
| S12_extracted_k141_87805 | Not-determined |
| S12_extracted_k141_111159 | Not-determined |
| S12_extracted_k141_72004 | Not-determined |
| S12_extracted_k141_87811 | Not-determined |
| S12_extracted_k141_119294 | Not-determined |
| S12_extracted_k141_95768 | Not-determined |
| S12_extracted_k141_127112 | Not-determined |
| S12_extracted_k141_111166 | Not-determined |
| S12_extracted_k141_87814 | Not-determined |
| S12_extracted_k141_32561 | Not-determined |
| S12_extracted_k141_24841 | Not-determined |
| S12_extracted_k141_95770 | Not-determined |
| S12_extracted_k141_87820 | Low-quality |
| S12_extracted_k141_95775 | Not-determined |
| S12_extracted_k141_103534 | Not-determined |
| S12_extracted_k141_16770 | Not-determined |
| S12_extracted_k141_127116 | Not-determined |
| S12_extracted_k141_134766 | Low-quality |
| S12_extracted_k141_40637 | Not-determined |
| S12_extracted_k141_127119 | Not-determined |
| S12_extracted_k141_40639 | Not-determined |
| S12_extracted_k141_72018 | Not-determined |
| S12_extracted_k141_32573 | Not-determined |
| S12_extracted_k141_16775 | Not-determined |
| S12_extracted_k141_95786 | Not-determined |
| S12_extracted_k141_127125 | Not-determined |
| S12_extracted_k141_87835 | Not-determined |
| S12_extracted_k141_32582 | Not-determined |
| S12_extracted_k141_127131 | Not-determined |
| S12_extracted_k141_56390 | Not-determined |
| S12_extracted_k141_24855 | Not-determined |
| S12_extracted_k141_40649 | Not-determined |
| S12_extracted_k141_24857 | Not-determined |
| S12_extracted_k141_103553 | Not-determined |
| S12_extracted_k141_103554 | Not-determined |
| S12_extracted_k141_174255 | Not-determined |
| S12_extracted_k141_119323 | Not-determined |
| S12_extracted_k141_174257 | Not-determined |
| S12_extracted_k141_111191 | Not-determined |
| S12_extracted_k141_16790 | Not-determined |
| S12_extracted_k141_1317 | Not-determined |
| S12_extracted_k141_1318 | Not-determined |
| S12_extracted_k141_24863 | Not-determined |
| S12_extracted_k141_127139 | Not-determined |
| S12_extracted_k141_166592 | Not-determined |
| S12_extracted_k141_158486 | Not-determined |
| S12_extracted_k141_134777 | Not-determined |
| S12_extracted_k141_111196 | Not-determined |
| S12_extracted_k141_119330 | Not-determined |
| S12_extracted_k141_87851 | Not-determined |
| S12_extracted_k141_72044 | Low-quality |
| S12_extracted_k141_40657 | Not-determined |
| S12_extracted_k141_1337 | Low-quality |
| S12_extracted_k141_72048 | Not-determined |
| S12_extracted_k141_119347 | Not-determined |
| S12_extracted_k141_134787 | Not-determined |
| S12_extracted_k141_1339 | Not-determined |
| S12_extracted_k141_72050 | Not-determined |
| S12_extracted_k141_72051 | Not-determined |
| S12_extracted_k141_8929 | Not-determined |
| S12_extracted_k141_72054 | Not-determined |
| S12_extracted_k141_127157 | Not-determined |
| S12_extracted_k141_134795 | Not-determined |
| S12_extracted_k141_1346 | Not-determined |
| S12_extracted_k141_103568 | Not-determined |
| S12_extracted_k141_16795 | Not-determined |
| S12_extracted_k141_72061 | Not-determined |
| S12_extracted_k141_166616 | Not-determined |
| S12_extracted_k141_87873 | Not-determined |
| S12_extracted_k141_134803 | Not-determined |
| S12_extracted_k141_119354 | Low-quality |
| S12_extracted_k141_16800 | Not-determined |
| S12_extracted_k141_166619 | Not-determined |
| S12_extracted_k141_72067 | Not-determined |
| S12_extracted_k141_134807 | Not-determined |
| S12_extracted_k141_8945 | Not-determined |
| S12_extracted_k141_1354 | Not-determined |
| S12_extracted_k141_72070 | Not-determined |
| S12_extracted_k141_48266 | Not-determined |
| S12_extracted_k141_32597 | Not-determined |
| S12_extracted_k141_150667 | Not-determined |
| S12_extracted_k141_48268 | Not-determined |
| S12_extracted_k141_111226 | Not-determined |
| S12_extracted_k141_87882 | Not-determined |
| S12_extracted_k141_16812 | Not-determined |
| S12_extracted_k141_143026 | Not-determined |
| S12_extracted_k141_150671 | Not-determined |
| S12_extracted_k141_64340 | Not-determined |
| S12_extracted_k141_79803 | Not-determined |
| S12_extracted_k141_182380 | Not-determined |
| S12_extracted_k141_56430 | Low-quality |
| S12_extracted_k141_79804 | Not-determined |
| S12_extracted_k141_158513 | Not-determined |
| S12_extracted_k141_134823 | Not-determined |
| S12_extracted_k141_182383 | Not-determined |
| S12_extracted_k141_56435 | Not-determined |
| S12_extracted_k141_40674 | Not-determined |
| S12_extracted_k141_72082 | Not-determined |
| S12_extracted_k141_134826 | Not-determined |
| S12_extracted_k141_87889 | Low-quality |

| | |
|---|---|
| S12_extracted_k141_32604 | Not-determined |
| S12_extracted_k141_79814 | Not-determined |
| S12_extracted_k141_79815 | Not-determined |
| S12_extracted_k141_64357 | Low-quality |
| S12_extracted_k141_79822 | Low-quality |
| S12_extracted_k141_32607 | Not-determined |
| S12_extracted_k141_150690 | Not-determined |
| S12_extracted_k141_64361 | Not-determined |
| S12_extracted_k141_87900 | Not-determined |
| S12_extracted_k141_16829 | Low-quality |
| S12_extracted_k141_8967 | Not-determined |
| S12_extracted_k141_16830 | Not-determined |
| S12_extracted_k141_56443 | Not-determined |
| S12_extracted_k141_182394 | Low-quality |
| S12_extracted_k141_158521 | Not-determined |
| S12_extracted_k141_64363 | Not-determined |
| S12_extracted_k141_1363 | Not-determined |
| S12_extracted_k141_72090 | Low-quality |
| S12_extracted_k141_150692 | Low-quality |
| S12_extracted_k141_158523 | Not-determined |
| S12_extracted_k141_79829 | Not-determined |
| S12_extracted_k141_95805 | Low-quality |
| S12_extracted_k141_134832 | Not-determined |
| S12_extracted_k141_111240 | Low-quality |
| S12_extracted_k141_134833 | Not-determined |
| S12_extracted_k141_127165 | Not-determined |
| S12_extracted_k141_182400 | Not-determined |
| S12_extracted_k141_1370 | Not-determined |
| S12_extracted_k141_79835 | Not-determined |
| S12_extracted_k141_32613 | Low-quality |
| S12_extracted_k141_56448 | Not-determined |
| S12_extracted_k141_56449 | Not-determined |
| S12_extracted_k141_72101 | Not-determined |
| S12_extracted_k141_72103 | Not-determined |
| S12_extracted_k141_56453 | Not-determined |
| S12_extracted_k141_182409 | Not-determined |
| S12_extracted_k141_111244 | Not-determined |
| S12_extracted_k141_182410 | Not-determined |
| S12_extracted_k141_111246 | Not-determined |
| S12_extracted_k141_95825 | Low-quality |
| S12_extracted_k141_56458 | Not-determined |
| S12_extracted_k141_87913 | Not-determined |
| S12_extracted_k141_72111 | Not-determined |
| S12_extracted_k141_79838 | Not-determined |
| S12_extracted_k141_72112 | Not-determined |
| S12_extracted_k141_119370 | Low-quality |
| S12_extracted_k141_119372 | Not-determined |
| S12_extracted_k141_1390 | Not-determined |
| S12_extracted_k141_1392 | Not-determined |
| S12_extracted_k141_127191 | Not-determined |
| S12_extracted_k141_119377 | Not-determined |
| S12_extracted_k141_24903 | Low-quality |
| S12_extracted_k141_40685 | Not-determined |
| S12_extracted_k141_182428 | Low-quality |
| S12_extracted_k141_119383 | Not-determined |
| S12_extracted_k141_127201 | Not-determined |
| S12_extracted_k141_1398 | Not-determined |
| S12_extracted_k141_87925 | Not-determined |
| S12_extracted_k141_158533 | Not-determined |
| S12_extracted_k141_182433 | Low-quality |
| S12_extracted_k141_103585 | Not-determined |
| S12_extracted_k141_24908 | Not-determined |
| S12_extracted_k141_119385 | Not-determined |
| S12_extracted_k141_134855 | Not-determined |
| S12_extracted_k141_166658 | Not-determined |
| S12_extracted_k141_1408 | Not-determined |
| S12_extracted_k141_87935 | Low-quality |
| S12_extracted_k141_134868 | Not-determined |
| S12_extracted_k141_72139 | Not-determined |
| S12_extracted_k141_72140 | Not-determined |
| S12_extracted_k141_103605 | Not-determined |
| S12_extracted_k141_127211 | Low-quality |
| S12_extracted_k141_127212 | Not-determined |
| S6_vs_k141_53966\|\|full | Medium-quality |
| S12_extracted_k141_103612 | Not-determined |
| S12_extracted_k141_127219 | Not-determined |
| S12_extracted_k141_48286 | Not-determined |
| S12_extracted_k141_182446 | Not-determined |
| S12_extracted_k141_48288 | Low-quality |
| S12_extracted_k141_182449 | Not-determined |
| S12_extracted_k141_166679 | Not-determined |
| S12_extracted_k141_24941 | Not-determined |
| S12_extracted_k141_72160 | Not-determined |
| S12_extracted_k141_103630 | Not-determined |
| S12_extracted_k141_72166 | Not-determined |
| S12_extracted_k141_158569 | Not-determined |
| S12_extracted_k141_158570 | Low-quality |
| S12_extracted_k141_8986 | Not-determined |
| S12_extracted_k141_95865 | Not-determined |
| S12_extracted_k141_174268 | Not-determined |
| S12_extracted_k141_150701 | Low-quality |
| S12_extracted_k141_95868 | Not-determined |
| S12_extracted_k141_174271 | Not-determined |
| S12_extracted_k141_72178 | Not-determined |
| S12_extracted_k141_56473 | Not-determined |
| S12_extracted_k141_79851 | Not-determined |
| S12_extracted_k141_1444 | Not-determined |
| S12_extracted_k141_56474 | Not-determined |
| S12_extracted_k141_79852 | Not-determined |
| S12_extracted_k141_111274 | Not-determined |
| S12_extracted_k141_150708 | Not-determined |
| S12_extracted_k141_79857 | Low-quality |
| S12_extracted_k141_166709 | Not-determined |
| S12_extracted_k141_79860 | Not-determined |
| S12_extracted_k141_56479 | Not-determined |
| S12_extracted_k141_1455 | Not-determined |
| S12_extracted_k141_182468 | Not-determined |
| S12_extracted_k141_1457 | Not-determined |
| S12_extracted_k141_1458 | Not-determined |
| S12_extracted_k141_174291 | Not-determined |
| S12_extracted_k141_95879 | Not-determined |
| S12_extracted_k141_64389 | Not-determined |
| S12_extracted_k141_24951 | Low-quality |
| S12_extracted_k141_158585 | Not-determined |
| S12_extracted_k141_111282 | Not-determined |
| S12_extracted_k141_95881 | Not-determined |
| S12_extracted_k141_111284 | Not-determined |
| S12_extracted_k141_9015 | Not-determined |
| S12_extracted_k141_127257 | Not-determined |
| S12_extracted_k141_158590 | Not-determined |
| S12_extracted_k141_9018 | Not-determined |
| S12_extracted_k141_174299 | Not-determined |
| S12_extracted_k141_56491 | Not-determined |
| S12_extracted_k141_166731 | Not-determined |
| S12_extracted_k141_16905 | Not-determined |
| S12_extracted_k141_127263 | Not-determined |
| S12_extracted_k141_1467 | Not-determined |
| S12_extracted_k141_9022 | Not-determined |
| S12_extracted_k141_87991 | Not-determined |
| S12_extracted_k141_143030 | Not-determined |
| S12_extracted_k141_79863 | Not-determined |
| S12_extracted_k141_16908 | Low-quality |
| S12_extracted_k141_16912 | Not-determined |
| S12_extracted_k141_182476 | Not-determined |
| S12_extracted_k141_182477 | Not-determined |
| S12_extracted_k141_127269 | Not-determined |
| S12_extracted_k141_134904 | Not-determined |
| S12_extracted_k141_158593 | Low-quality |
| S12_extracted_k141_56508 | Not-determined |
| S12_extracted_k141_127272 | Not-determined |
| S12_extracted_k141_134905 | Not-determined |
| S12_extracted_k141_40727 | Not-determined |
| S12_extracted_k141_134910 | Not-determined |
| S12_extracted_k141_158605 | Not-determined |
| S12_extracted_k141_24973 | Not-determined |
| S12_extracted_k141_134913 | Not-determined |
| S12_extracted_k141_79868 | Not-determined |
| S12_extracted_k141_95892 | Not-determined |
| S12_extracted_k141_72194 | Not-determined |
| S12_extracted_k141_16922 | Not-determined |
| S12_extracted_k141_16924 | Not-determined |
| S12_extracted_k141_40738 | Not-determined |
| S12_extracted_k141_72198 | Not-determined |
| S12_extracted_k141_143039 | Not-determined |
| S12_extracted_k141_174321 | Not-determined |
| S12_extracted_k141_16928 | Not-determined |
| S12_extracted_k141_143044 | Not-determined |
| S12_extracted_k141_127288 | Not-determined |
| S12_extracted_k141_158619 | Not-determined |
| S12_extracted_k141_72204 | Not-determined |
| S12_extracted_k141_88007 | Not-determined |
| S12_extracted_k141_40741 | Not-determined |
| S12_extracted_k141_158621 | Not-determined |
| S12_extracted_k141_158622 | Not-determined |
| S12_extracted_k141_134932 | Not-determined |
| S12_extracted_k141_79873 | Not-determined |
| S12_extracted_k141_182498 | Not-determined |
| S12_extracted_k141_56525 | Not-determined |
| S12_extracted_k141_103643 | Not-determined |
| S12_extracted_k141_143055 | Not-determined |
| S12_extracted_k141_182502 | Not-determined |
| S12_extracted_k141_24983 | Not-determined |
| S12_extracted_k141_88017 | Not-determined |
| S12_extracted_k141_56528 | Not-determined |
| S12_extracted_k141_134940 | Not-determined |
| S12_extracted_k141_64428 | Not-determined |
| S12_extracted_k141_127302 | Not-determined |
| S12_extracted_k141_119399 | Not-determined |
| S12_extracted_k141_127303 | Not-determined |
| S12_extracted_k141_40744 | Low-quality |
| S12_extracted_k141_40745 | Not-determined |
| S12_extracted_k141_79887 | Not-determined |
| S12_extracted_k141_143058 | Low-quality |
| S12_extracted_k141_64434 | Not-determined |
| S12_extracted_k141_16939 | Low-quality |
| S12_extracted_k141_56536 | Not-determined |
| S12_extracted_k141_16940 | Not-determined |
| S12_extracted_k141_79891 | Not-determined |
| S12_extracted_k141_119410 | Not-determined |
| S12_extracted_k141_166755 | Not-determined |
| S12_extracted_k141_79892 | Not-determined |
| S12_extracted_k141_9066 | Not-determined |
| S12_extracted_k141_158641 | Not-determined |
| S12_extracted_k141_24995 | Not-determined |
| S12_extracted_k141_182518 | Not-determined |
| S12_extracted_k141_9068 | Not-determined |
| S12_extracted_k141_64447 | Low-quality |
| S12_extracted_k141_56545 | Not-determined |
| S12_extracted_k141_143074 | Low-quality |
| S12_extracted_k141_16946 | Low-quality |
| S12_extracted_k141_88025 | Not-determined |
| S12_extracted_k141_103671 | Not-determined |
| S12_extracted_k141_134958 | Not-determined |
| S12_extracted_k141_119422 | Low-quality |
| S12_extracted_k141_56549 | Not-determined |
| S12_extracted_k141_9078 | Not-determined |
| S12_extracted_k141_16951 | Not-determined |
| S12_extracted_k141_24998 | Not-determined |
| S12_extracted_k141_158655 | Not-determined |
| S12_extracted_k141_166764 | Not-determined |
| S12_extracted_k141_143084 | Not-determined |
| S12_extracted_k141_134969 | Not-determined |
| S12_extracted_k141_88037 | Not-determined |
| S12_extracted_k141_119433 | Not-determined |
| S12_extracted_k141_103681 | Not-determined |
| S12_extracted_k141_16958 | Not-determined |
| S12_extracted_k141_134972 | Not-determined |
| S12_extracted_k141_103685 | Not-determined |
| S12_extracted_k141_127328 | Not-determined |
| S12_extracted_k141_134976 | Not-determined |
| S12_extracted_k141_134979 | Not-determined |
| S12_extracted_k141_158663 | Not-determined |
| S12_extracted_k141_150719 | Not-determined |
| S12_extracted_k141_150720 | Not-determined |
| S12_extracted_k141_103695 | Not-determined |
| S12_extracted_k141_88049 | Not-determined |
| S12_extracted_k141_103702 | Not-determined |
| S12_extracted_k141_16975 | Not-determined |
| S12_extracted_k141_158669 | Low-quality |
| S12_extracted_k141_182533 | Not-determined |
| S12_extracted_k141_16978 | Low-quality |
| S12_extracted_k141_119451 | Not-determined |
| S12_extracted_k141_103707 | Not-determined |
| S12_extracted_k141_135004 | Not-determined |
| S12_extracted_k141_111287 | Not-determined |
| S12_extracted_k141_111289 | Not-determined |
| S12_extracted_k141_158680 | Not-determined |
| S12_extracted_k141_119456 | Not-determined |
| S12_extracted_k141_182543 | Not-determined |
| S12_extracted_k141_9099 | Not-determined |
| S12_extracted_k141_56576 | Not-determined |
| S12_extracted_k141_103718 | Not-determined |
| S12_extracted_k141_135017 | Not-determined |
| S12_extracted_k141_9103 | Not-determined |
| S12_extracted_k141_182545 | Not-determined |
| S12_extracted_k141_158686 | Not-determined |
| S12_extracted_k141_95925 | Not-determined |
| S12_extracted_k141_158688 | Not-determined |
| S12_extracted_k141_166783 | Not-determined |
| S12_extracted_k141_166784 | Not-determined |
| S12_extracted_k141_182550 | Not-determined |
| S12_extracted_k141_95930 | Low-quality |
| S12_extracted_k141_166787 | Not-determined |
| S12_extracted_k141_119466 | Not-determined |
| S12_extracted_k141_72212 | Not-determined |
| S12_extracted_k141_119469 | Not-determined |
| S12_extracted_k141_95933 | Low-quality |
| S12_extracted_k141_72222 | Not-determined |
| S12_extracted_k141_95938 | Not-determined |
| S12_extracted_k141_135038 | Not-determined |
| S12_extracted_k141_64462 | Not-determined |
| S12_extracted_k141_119478 | Not-determined |
| S12_extracted_k141_95940 | Not-determined |
| S12_extracted_k141_119484 | Not-determined |
| S12_extracted_k141_150744 | Not-determined |
| S12_extracted_k141_119486 | Not-determined |
| S12_extracted_k141_88073 | Not-determined |
| S12_extracted_k141_64472 | Not-determined |
| S12_extracted_k141_25032 | Not-determined |
| S12_extracted_k141_158709 | Not-determined |
| S12_extracted_k141_16989 | Not-determined |
| S12_extracted_k141_166793 | Not-determined |
| S12_extracted_k141_95950 | Low-quality |
| S12_extracted_k141_56599 | Not-determined |
| S12_extracted_k141_88081 | Not-determined |
| S12_extracted_k141_119499 | Not-determined |
| S12_extracted_k141_72240 | Not-determined |
| S12_extracted_k141_95958 | Not-determined |
| S12_extracted_k141_72243 | Not-determined |
| S12_extracted_k141_56607 | Not-determined |
| S12_extracted_k141_25043 | Not-determined |
| S12_extracted_k141_17001 | Not-determined |
| S12_extracted_k141_64475 | Not-determined |
| S12_extracted_k141_17003 | Not-determined |
| S12_extracted_k141_25046 | Not-determined |
| S12_extracted_k141_25048 | Not-determined |
| S12_extracted_k141_88083 | Not-determined |
| S12_extracted_k141_25049 | Not-determined |
| S12_extracted_k141_25050 | Not-determined |
| S12_extracted_k141_150765 | Not-determined |
| S12_extracted_k141_25054 | Not-determined |
| S12_extracted_k141_150767 | Not-determined |
| S12_extracted_k141_111334 | Not-determined |
| S12_extracted_k141_17008 | Not-determined |
| S12_extracted_k141_111335 | Not-determined |
| S12_extracted_k141_103747 | Not-determined |
| S12_extracted_k141_174326 | Not-determined |
| S12_extracted_k141_166795 | Not-determined |
| S12_extracted_k141_25061 | Not-determined |
| S12_extracted_k141_174329 | Not-determined |
| S12_extracted_k141_111341 | Not-determined |
| S12_extracted_k141_166797 | Not-determined |
| S12_extracted_k141_17013 | Not-determined |
| S12_extracted_k141_119512 | Not-determined |
| S12_extracted_k141_56612 | Low-quality |
| S12_extracted_k141_166801 | Not-determined |
| S10_vs_43210\|\|full | High-quality |
| S12_extracted_k141_103754 | Not-determined |
| S12_extracted_k141_95971 | Not-determined |
| S12_extracted_k141_135067 | Not-determined |
| S12_extracted_k141_95972 | Not-determined |
| S12_extracted_k141_79932 | Not-determined |
| S12_extracted_k141_135068 | Not-determined |
| S12_extracted_k141_166810 | Low-quality |
| S12_extracted_k141_32621 | Not-determined |
| S12_extracted_k141_103760 | Not-determined |
| S12_extracted_k141_72266 | Not-determined |
| S12_extracted_k141_135070 | Not-determined |
| S12_extracted_k141_32623 | Not-determined |
| S12_extracted_k141_25072 | Not-determined |
| S12_extracted_k141_32625 | Not-determined |
| S12_extracted_k141_150784 | Not-determined |
| S12_extracted_k141_150788 | Not-determined |
| S12_extracted_k141_25079 | Low-quality |
| S12_extracted_k141_135074 | Not-determined |
| S12_extracted_k141_174346 | Not-determined |
| S12_extracted_k141_72275 | Not-determined |
| S12_extracted_k141_143138 | Not-determined |
| S12_extracted_k141_103763 | Not-determined |
| S12_extracted_k141_182603 | Not-determined |
| S12_extracted_k141_95984 | Low-quality |
| S12_extracted_k141_1481 | Not-determined |
| S12_extracted_k141_9125 | Not-determined |
| S12_extracted_k141_143142 | Not-determined |
| S12_extracted_k141_150793 | Not-determined |
| S12_extracted_k141_119532 | Not-determined |
| S12_extracted_k141_119534 | Not-determined |
| S12_extracted_k141_111355 | Not-determined |
| S12_extracted_k141_79947 | Not-determined |
| S12_extracted_k141_25090 | Not-determined |
| S12_extracted_k141_166818 | Not-determined |
| S12_extracted_k141_135094 | Not-determined |
| S12_extracted_k141_166820 | Not-determined |
| S12_extracted_k141_25095 | Not-determined |
| S12_extracted_k141_135099 | Low-quality |
| S12_extracted_k141_25097 | Not-determined |
| S12_extracted_k141_72285 | Not-determined |
| S12_extracted_k141_135101 | Not-determined |
| S12_extracted_k141_135102 | Not-determined |
| S12_extracted_k141_174350 | Not-determined |
| S12_extracted_k141_182606 | Not-determined |
| S12_extracted_k141_56638 | Not-determined |
| S12_extracted_k141_119543 | Not-determined |
| S12_extracted_k141_1496 | Not-determined |
| S12_extracted_k141_150802 | Not-determined |
| S12_extracted_k141_182610 | Not-determined |
| S12_extracted_k141_166825 | Not-determined |
| S12_extracted_k141_25100 | Not-determined |
| S12_extracted_k141_72299 | Not-determined |
| S12_extracted_k141_166831 | Not-determined |
| S12_extracted_k141_25104 | Not-determined |
| S12_extracted_k141_135111 | Not-determined |
| S12_extracted_k141_9138 | Not-determined |
| S12_extracted_k141_166839 | Not-determined |
| S12_extracted_k141_1513 | Not-determined |
| S12_extracted_k141_9146 | Not-determined |
| S12_extracted_k141_9147 | Not-determined |
| S12_extracted_k141_174358 | Not-determined |
| S12_extracted_k141_72309 | Not-determined |
| S12_extracted_k141_1517 | Not-determined |
| S12_extracted_k141_174365 | Not-determined |
| S12_extracted_k141_9156 | Not-determined |
| S12_extracted_k141_72317 | Not-determined |
| S12_extracted_k141_72318 | Not-determined |
| S12_extracted_k141_119567 | Not-determined |
| S12_extracted_k141_72320 | Low-quality |
| S12_extracted_k141_72322 | Not-determined |
| S12_extracted_k141_56669 | Not-determined |
| S12_extracted_k141_135123 | Not-determined |
| S12_extracted_k141_119569 | Not-determined |
| S12_extracted_k141_64527 | Not-determined |
| S12_extracted_k141_166864 | Not-determined |
| S12_extracted_k141_96021 | Not-determined |
| S12_extracted_k141_64532 | Not-determined |
| S12_extracted_k141_103770 | Not-determined |
| S12_extracted_k141_103772 | Not-determined |
| S12_extracted_k141_64534 | Low-quality |
| S12_extracted_k141_25115 | Not-determined |
| S12_extracted_k141_9163 | Not-determined |
| S12_extracted_k141_72336 | Not-determined |
| S12_extracted_k141_9165 | Not-determined |
| S12_extracted_k141_9166 | Not-determined |
| S12_extracted_k141_166872 | Not-determined |
| S12_extracted_k141_166875 | Not-determined |
| S12_extracted_k141_119584 | Not-determined |
| S12_extracted_k141_17020 | Not-determined |
| S12_extracted_k141_32659 | Not-determined |
| S12_extracted_k141_25123 | Not-determined |
| S12_extracted_k141_79990 | Not-determined |
| S12_extracted_k141_17027 | Not-determined |
| S12_extracted_k141_17028 | Not-determined |
| S12_extracted_k141_88086 | Low-quality |
| S12_extracted_k141_17029 | Not-determined |
| S12_extracted_k141_1552 | Not-determined |
| S12_extracted_k141_72349 | Not-determined |
| S12_extracted_k141_96049 | Low-quality |
| S12_extracted_k141_143154 | Not-determined |
| S12_extracted_k141_88099 | Not-determined |
| S12_extracted_k141_25139 | Not-determined |
| S12_extracted_k141_25140 | Not-determined |
| S12_extracted_k141_96055 | Not-determined |
| S12_extracted_k141_64571 | Not-determined |
| S12_extracted_k141_72358 | Not-determined |
| S12_extracted_k141_9180 | Not-determined |
| S12_extracted_k141_56685 | Not-determined |
| S12_extracted_k141_56692 | Not-determined |
| S12_extracted_k141_64578 | Not-determined |
| S12_extracted_k141_88109 | Not-determined |
| S12_extracted_k141_17048 | Not-determined |
| S12_extracted_k141_72362 | Not-determined |
| S12_extracted_k141_9188 | Not-determined |
| S12_extracted_k141_88112 | Not-determined |
| S12_extracted_k141_25155 | Not-determined |
| S12_extracted_k141_56695 | Not-determined |
| S12_extracted_k141_25156 | Not-determined |
| S12_extracted_k141_119607 | Not-determined |
| S12_extracted_k141_56696 | Low-quality |
| S12_extracted_k141_119609 | Not-determined |
| S12_extracted_k141_143168 | Not-determined |
| S12_extracted_k141_166906 | Not-determined |
| S12_extracted_k141_103800 | Not-determined |
| S12_extracted_k141_119611 | Not-determined |
| S12_extracted_k141_64589 | Not-determined |
| S12_extracted_k141_150820 | Low-quality |
| S12_extracted_k141_72373 | Not-determined |
| S12_extracted_k141_135143 | Not-determined |
| S12_extracted_k141_56713 | Not-determined |
| S12_extracted_k141_182612 | Not-determined |
| S12_extracted_k141_143169 | Low-quality |
| S12_extracted_k141_72377 | Not-determined |
| S12_extracted_k141_103803 | Low-quality |
| S16_vs_4662\|\|full | Medium-quality |
| S12_extracted_k141_182614 | Not-determined |
| S12_extracted_k141_182616 | Not-determined |
| S12_extracted_k141_166916 | Not-determined |
| S12_extracted_k141_9201 | Not-determined |
| S12_extracted_k141_103817 | Not-determined |
| S12_extracted_k141_17061 | Low-quality |
| S12_extracted_k141_103819 | Not-determined |
| S12_extracted_k141_9206 | Not-determined |
| S12_extracted_k141_135159 | Not-determined |
| S12_extracted_k141_174385 | Not-determined |
| S12_extracted_k141_17064 | Not-determined |
| S12_extracted_k141_80016 | Low-quality |
| S12_extracted_k141_119629 | Not-determined |
| S12_extracted_k141_150840 | Not-determined |
| S12_extracted_k141_174390 | Not-determined |
| S12_extracted_k141_64601 | Not-determined |
| S12_extracted_k141_9210 | Not-determined |
| S12_extracted_k141_64604 | Not-determined |
| S12_extracted_k141_166923 | Not-determined |
| S12_extracted_k141_25179 | Not-determined |
| S12_extracted_k141_96066 | Not-determined |
| S12_extracted_k141_182626 | Low-quality |
| S12_extracted_k141_80029 | Not-determined |
| S12_extracted_k141_17076 | Low-quality |
| S12_extracted_k141_174402 | Not-determined |
| S12_extracted_k141_96072 | Not-determined |
| S12_extracted_k141_174403 | Not-determined |
| S12_extracted_k141_80035 | Not-determined |
| S12_extracted_k141_88136 | Not-determined |
| S12_extracted_k141_72407 | Not-determined |
| S12_extracted_k141_1598 | Low-quality |
| S12_extracted_k141_135185 | Not-determined |
| S12_extracted_k141_72411 | Not-determined |
| S12_extracted_k141_103830 | Not-determined |
| S12_extracted_k141_166936 | Not-determined |
| S12_extracted_k141_1600 | Not-determined |
| S12_extracted_k141_80041 | Not-determined |
| S12_extracted_k141_9218 | Low-quality |
| S12_extracted_k141_182632 | Not-determined |
| S12_extracted_k141_143211 | Not-determined |
| S12_extracted_k141_166944 | Not-determined |
| S12_extracted_k141_143212 | Not-determined |
| S12_extracted_k141_1607 | Not-determined |
| S12_extracted_k141_150863 | Not-determined |
| S12_extracted_k141_103833 | Not-determined |
| S12_extracted_k141_96085 | Not-determined |
| S12_extracted_k141_25188 | Not-determined |
| S12_extracted_k141_119662 | Not-determined |
| S12_extracted_k141_72422 | Not-determined |
| S12_extracted_k141_56741 | Not-determined |
| S12_extracted_k141_166952 | Not-determined |
| S12_extracted_k141_9230 | Not-determined |
| S12_extracted_k141_111367 | Not-determined |
| S12_extracted_k141_182634 | Not-determined |
| S12_extracted_k141_119673 | Low-quality |
| S12_extracted_k141_119674 | Not-determined |
| S12_extracted_k141_166956 | Not-determined |
| S12_extracted_k141_119675 | Not-determined |
| S12_extracted_k141_182638 | Not-determined |
| S12_extracted_k141_182639 | Not-determined |
| S12_extracted_k141_127388 | Not-determined |
| S12_extracted_k141_103839 | Not-determined |
| S12_extracted_k141_150875 | Not-determined |
| S12_extracted_k141_135199 | Not-determined |
| S12_extracted_k141_103841 | Not-determined |
| S12_extracted_k141_25203 | Not-determined |
| S12_extracted_k141_17101 | Low-quality |
| S12_extracted_k141_182651 | Not-determined |
| S12_extracted_k141_143219 | Not-determined |
| S12_extracted_k141_143221 | Not-determined |
| S12_extracted_k141_9241 | Not-determined |
| S12_extracted_k141_182659 | Not-determined |
| S12_extracted_k141_1631 | Low-quality |
| S12_extracted_k141_143225 | Not-determined |
| S12_extracted_k141_150882 | Not-determined |
| S12_extracted_k141_96098 | Not-determined |
| S12_extracted_k141_150883 | Not-determined |
| S12_extracted_k141_135217 | Not-determined |
| S12_extracted_k141_88159 | Not-determined |
| S12_extracted_k141_56756 | Low-quality |
| S12_extracted_k141_56757 | Not-determined |
| S12_extracted_k141_25222 | Not-determined |
| S12_extracted_k141_88165 | Not-determined |
| S12_extracted_k141_25223 | Not-determined |
| S12_extracted_k141_103868 | Not-determined |
| S12_extracted_k141_127407 | Not-determined |
| S12_extracted_k141_143231 | Not-determined |
| S12_extracted_k141_127410 | Not-determined |
| S12_extracted_k141_119705 | Not-determined |
| S12_extracted_k141_166967 | Not-determined |
| S12_extracted_k141_103870 | Not-determined |
| S12_extracted_k141_56763 | Not-determined |
| S12_extracted_k141_25230 | Low-quality |
| S12_extracted_k141_96106 | Not-determined |
| S12_extracted_k141_135224 | Not-determined |
| S12_extracted_k141_25234 | Not-determined |
| S12_extracted_k141_166973 | Not-determined |
| S12_extracted_k141_135228 | Not-determined |
| S12_extracted_k141_127417 | Not-determined |
| S12_extracted_k141_127419 | Not-determined |
| S12_extracted_k141_135230 | Not-determined |
| S12_extracted_k141_182681 | Low-quality |
| S12_extracted_k141_127424 | Not-determined |
| S12_extracted_k141_182685 | Not-determined |
| S12_extracted_k141_103882 | Not-determined |
| S12_extracted_k141_119726 | Not-determined |
| S12_extracted_k141_9249 | Not-determined |
| S12_extracted_k141_96112 | Not-determined |
| S12_extracted_k141_182688 | Not-determined |
| S12_extracted_k141_143245 | Not-determined |
| S12_extracted_k141_96113 | Low-quality |
| S12_extracted_k141_56768 | Not-determined |
| S12_extracted_k141_182694 | Not-determined |
| S12_extracted_k141_103889 | Not-determined |
| S12_extracted_k141_150905 | Not-determined |
| S12_extracted_k141_88195 | Not-determined |
| S12_extracted_k141_174452 | Not-determined |
| S12_extracted_k141_111397 | Not-determined |
| S12_extracted_k141_135246 | Not-determined |
| S12_extracted_k141_103894 | Not-determined |
| S12_extracted_k141_40753 | Low-quality |
| S12_extracted_k141_1652 | Low-quality |
| S12_extracted_k141_150914 | Not-determined |
| S12_extracted_k141_119740 | Not-determined |
| S12_extracted_k141_88200 | Not-determined |
| S12_extracted_k141_174468 | Not-determined |
| S12_extracted_k141_150917 | Not-determined |
| S12_extracted_k141_111408 | Low-quality |
| S8_extracted_k141_166403 | Medium-quality |
| S12_extracted_k141_127458 | Low-quality |
| S12_extracted_k141_48295 | Not-determined |
| S12_extracted_k141_111411 | Not-determined |
| S12_extracted_k141_174473 | Not-determined |
| S12_extracted_k141_119743 | Not-determined |
| S12_extracted_k141_1660 | Not-determined |
| S12_extracted_k141_96141 | Not-determined |
| S12_extracted_k141_96142 | Not-determined |
| S12_extracted_k141_143266 | Not-determined |
| S12_extracted_k141_182709 | Not-determined |
| S12_extracted_k141_182710 | Not-determined |
| S12_extracted_k141_40766 | Not-determined |
| S12_extracted_k141_40768 | Not-determined |
| S12_extracted_k141_119758 | Not-determined |
| S12_extracted_k141_40773 | Not-determined |
| S12_extracted_k141_40776 | Not-determined |
| S12_extracted_k141_9272 | Not-determined |
| S12_extracted_k141_127477 | Not-determined |

| | |
|---|---|
| S12_extracted_k141_40786 | Not-determined |
| S12_extracted_k141_182718 | Not-determined |
| S12_extracted_k141_127480 | Not-determined |
| S12_extracted_k141_9279 | Not-determined |
| S12_extracted_k141_119770 | Low-quality |
| S12_extracted_k141_127481 | Not-determined |
| S12_extracted_k141_64645 | Not-determined |
| S12_extracted_k141_119773 | Not-determined |
| S12_extracted_k141_1674 | Not-determined |
| S12_extracted_k141_9285 | Not-determined |
| S12_extracted_k141_56797 | Not-determined |
| S12_extracted_k141_119780 | Not-determined |
| S12_extracted_k141_182726 | Low-quality |
| S12_extracted_k141_9289 | Low-quality |
| S12_extracted_k141_150943 | Not-determined |
| S12_extracted_k141_96184 | Not-determined |
| S12_extracted_k141_96186 | Not-determined |
| S12_extracted_k141_96188 | Not-determined |
| S12_extracted_k141_40806 | Not-determined |
| S12_extracted_k141_64660 | Not-determined |
| S12_extracted_k141_56809 | Low-quality |
| S12_extracted_k141_111440 | Not-determined |
| S12_extracted_k141_48308 | Not-determined |
| S12_extracted_k141_9298 | Not-determined |
| S12_extracted_k141_127498 | Not-determined |
| S12_extracted_k141_1682 | Not-determined |
| S12_extracted_k141_96193 | Not-determined |
| S12_extracted_k141_119788 | Not-determined |
| S12_extracted_k141_17137 | Low-quality |
| S12_extracted_k141_127501 | Not-determined |
| S12_extracted_k141_1690 | Not-determined |
| S12_extracted_k141_48321 | Not-determined |
| S12_extracted_k141_127504 | Not-determined |
| S12_extracted_k141_1695 | Low-quality |
| S12_extracted_k141_111454 | Not-determined |
| S12_extracted_k141_64682 | Not-determined |
| S12_extracted_k141_17148 | Not-determined |
| S12_extracted_k141_111455 | Not-determined |
| S12_extracted_k141_96211 | Not-determined |
| S12_extracted_k141_48325 | Not-determined |
| S12_extracted_k141_64687 | Low-quality |
| S12_extracted_k141_111457 | Not-determined |
| S12_extracted_k141_119791 | Low-quality |
| S12_extracted_k141_127522 | Not-determined |
| S12_extracted_k141_96223 | Not-determined |
| S12_extracted_k141_56839 | Not-determined |
| S12_extracted_k141_64697 | Not-determined |
| S12_extracted_k141_17159 | Not-determined |
| S12_extracted_k141_167015 | Not-determined |
| S12_extracted_k141_9315 | Not-determined |
| S12_extracted_k141_182736 | Not-determined |
| S12_extracted_k141_135267 | Not-determined |
| S12_extracted_k141_135268 | Not-determined |
| S12_extracted_k141_96232 | Low-quality |
| S12_extracted_k141_56844 | Low-quality |
| S12_extracted_k141_182744 | Not-determined |
| S12_extracted_k141_127530 | Not-determined |
| S12_extracted_k141_174482 | Not-determined |
| S12_extracted_k141_111468 | Not-determined |
| S12_extracted_k141_150956 | Not-determined |
| S12_extracted_k141_40819 | Low-quality |
| S12_extracted_k141_143307 | Not-determined |
| S12_extracted_k141_127537 | Not-determined |
| S12_extracted_k141_56851 | Not-determined |
| S12_extracted_k141_143313 | Not-determined |
| S12_extracted_k141_96242 | Not-determined |
| S12_extracted_k141_56853 | Not-determined |
| S12_extracted_k141_72442 | Not-determined |
| S12_extracted_k141_167027 | Not-determined |
| S12_extracted_k141_72443 | Not-determined |
| S12_extracted_k141_143318 | Not-determined |
| S12_extracted_k141_96245 | Low-quality |
| S12_extracted_k141_143319 | Not-determined |
| S12_extracted_k141_127545 | Not-determined |
| S12_extracted_k141_103951 | Low-quality |
| S12_extracted_k141_25241 | Low-quality |
| S12_extracted_k141_72451 | Not-determined |
| S12_extracted_k141_119816 | Not-determined |
| S12_extracted_k141_25242 | Not-determined |
| S12_extracted_k141_103956 | Not-determined |
| S12_extracted_k141_25243 | Low-quality |
| S12_extracted_k141_48339 | Not-determined |
| S12_extracted_k141_64714 | Not-determined |
| S12_extracted_k141_103957 | Not-determined |
| S12_extracted_k141_72458 | Not-determined |
| S12_extracted_k141_25249 | Not-determined |
| S12_extracted_k141_72461 | Not-determined |
| S12_extracted_k141_25251 | Not-determined |
| S12_extracted_k141_167044 | Not-determined |
| S12_extracted_k141_143332 | Not-determined |
| S12_extracted_k141_143334 | Low-quality |
| S12_extracted_k141_119822 | Not-determined |
| S12_extracted_k141_103965 | Not-determined |
| S12_extracted_k141_167048 | Not-determined |
| S12_extracted_k141_111479 | Not-determined |
| S12_extracted_k141_111480 | Not-determined |
| S12_extracted_k141_127559 | Not-determined |
| S12_extracted_k141_103972 | Not-determined |
| S12_extracted_k141_127563 | Not-determined |
| S12_extracted_k141_25262 | Not-determined |
| S12_extracted_k141_119830 | Not-determined |
| S12_extracted_k141_25265 | Not-determined |
| S12_extracted_k141_1740 | Not-determined |
| S12_extracted_k141_72481 | Not-determined |
| S12_extracted_k141_25270 | Not-determined |
| S12_extracted_k141_64730 | Not-determined |
| S12_extracted_k141_40840 | Not-determined |
| S12_extracted_k141_96274 | Low-quality |
| S12_extracted_k141_48361 | Not-determined |
| S12_extracted_k141_25272 | Not-determined |
| S12_extracted_k141_127578 | Not-determined |
| S12_extracted_k141_119834 | Not-determined |
| S12_extracted_k141_143346 | Not-determined |
| S12_extracted_k141_25278 | Not-determined |
| S12_extracted_k141_96282 | Not-determined |
| S12_extracted_k141_143347 | Low-quality |
| S12_extracted_k141_96284 | Not-determined |
| S12_extracted_k141_56880 | Not-determined |
| S12_extracted_k141_17178 | Not-determined |
| S12_extracted_k141_40848 | Low-quality |
| S12_extracted_k141_25282 | Not-determined |
| S12_extracted_k141_119842 | Not-determined |
| S12_extracted_k141_17182 | Not-determined |
| S12_extracted_k141_40851 | Not-determined |
| S12_extracted_k141_17183 | Not-determined |
| S12_extracted_k141_1744 | Low-quality |
| S12_extracted_k141_64743 | Not-determined |
| S12_extracted_k141_40856 | Not-determined |
| S12_extracted_k141_1748 | Not-determined |
| S12_extracted_k141_111496 | Not-determined |
| S12_extracted_k141_103983 | Not-determined |
| S12_extracted_k141_1751 | Low-quality |
| S12_extracted_k141_111498 | Not-determined |
| S12_extracted_k141_25294 | Not-determined |
| S12_extracted_k141_96300 | Low-quality |
| S12_extracted_k141_56906 | Not-determined |
| S12_extracted_k141_56907 | Not-determined |
| S12_extracted_k141_40871 | Not-determined |
| S12_extracted_k141_40872 | Not-determined |
| S12_extracted_k141_17196 | Not-determined |
| S12_extracted_k141_127594 | Not-determined |
| S12_extracted_k141_72496 | Not-determined |
| S12_extracted_k141_72497 | Not-determined |
| S12_extracted_k141_72502 | Not-determined |
| S12_extracted_k141_72503 | Not-determined |
| S12_extracted_k141_127602 | Not-determined |
| S12_extracted_k141_48368 | Not-determined |
| S12_extracted_k141_48369 | Not-determined |
| S12_extracted_k141_96301 | Not-determined |
| S12_extracted_k141_135288 | Not-determined |
| S12_extracted_k141_127606 | Not-determined |
| S12_extracted_k141_104002 | Not-determined |
| S12_extracted_k141_40880 | Not-determined |
| S12_extracted_k141_1779 | Not-determined |
| S12_extracted_k141_48386 | Not-determined |
| S12_extracted_k141_96316 | Low-quality |
| S12_extracted_k141_96318 | Not-determined |
| S12_extracted_k141_167104 | Not-determined |
| S12_extracted_k141_96320 | Not-determined |
| S12_extracted_k141_111508 | Not-determined |
| S12_extracted_k141_64766 | Not-determined |
| S12_extracted_k141_40887 | Low-quality |
| S12_extracted_k141_96324 | Not-determined |
| S12_extracted_k141_111510 | Not-determined |
| S12_extracted_k141_104016 | Not-determined |
| S12_extracted_k141_127623 | Low-quality |
| S12_extracted_k141_64770 | Not-determined |
| S12_extracted_k141_104018 | Not-determined |
| S12_extracted_k141_1799 | Not-determined |
| S12_extracted_k141_127624 | Not-determined |
| S12_extracted_k141_72527 | Not-determined |
| S12_extracted_k141_96330 | Low-quality |
| S12_extracted_k141_96334 | Not-determined |
| S12_extracted_k141_119893 | Not-determined |
| S12_extracted_k141_182757 | Not-determined |
| S12_extracted_k141_127626 | Not-determined |
| S12_extracted_k141_56941 | Not-determined |
| S12_extracted_k141_56942 | Not-determined |
| S12_extracted_k141_111514 | Not-determined |
| S12_extracted_k141_48396 | Not-determined |
| S12_extracted_k141_104023 | Not-determined |
| S12_extracted_k141_119901 | Not-determined |
| S12_extracted_k141_143353 | Not-determined |
| S12_extracted_k141_127633 | Not-determined |
| S12_extracted_k141_1811 | Not-determined |
| S12_extracted_k141_111525 | Not-determined |
| S12_extracted_k141_72545 | Not-determined |
| S12_extracted_k141_119920 | Not-determined |
| S12_extracted_k141_143366 | Not-determined |
| S12_extracted_k141_143368 | Not-determined |
| S12_extracted_k141_1823 | Not-determined |
| S12_extracted_k141_17204 | Not-determined |
| S12_extracted_k141_72559 | Not-determined |
| S8_vs_k141_166403\|\|full | Medium-quality |
| S12_extracted_k141_48408 | Not-determined |
| S12_extracted_k141_40900 | Not-determined |
| S12_extracted_k141_25327 | Not-determined |
| S12_extracted_k141_96358 | Not-determined |
| S12_extracted_k141_1828 | Not-determined |
| S12_extracted_k141_96359 | Not-determined |
| S12_extracted_k141_48415 | Not-determined |
| S12_extracted_k141_25329 | Not-determined |
| S12_extracted_k141_40902 | Not-determined |
| S12_extracted_k141_119932 | Not-determined |
| S12_extracted_k141_17210 | Not-determined |
| S12_extracted_k141_40903 | Not-determined |
| S12_extracted_k141_96362 | Low-quality |
| S12_extracted_k141_40904 | Not-determined |
| S12_extracted_k141_96366 | Not-determined |
| S12_extracted_k141_25334 | Not-determined |
| S12_extracted_k141_119936 | Not-determined |
| S12_extracted_k141_96371 | Not-determined |
| S12_extracted_k141_1837 | Not-determined |
| S12_extracted_k141_56969 | Not-determined |
| S12_extracted_k141_119937 | Low-quality |
| S12_extracted_k141_88238 | Not-determined |
| S12_extracted_k141_119938 | Not-determined |
| S12_extracted_k141_40908 | Not-determined |
| S12_extracted_k141_56975 | Not-determined |
| S12_extracted_k141_56978 | Not-determined |
| S12_extracted_k141_17224 | Not-determined |
| S12_extracted_k141_25338 | Not-determined |
| S12_extracted_k141_167144 | Not-determined |
| S12_extracted_k141_119945 | Not-determined |
| S12_extracted_k141_174486 | Not-determined |
| S12_extracted_k141_119947 | Low-quality |
| S12_extracted_k141_25344 | Not-determined |
| S12_extracted_k141_143386 | Not-determined |
| S12_extracted_k141_182780 | Not-determined |
| S12_extracted_k141_143393 | Not-determined |
| S12_extracted_k141_9323 | Not-determined |
| S12_extracted_k141_119963 | Not-determined |
| S12_extracted_k141_111536 | Not-determined |
| S12_extracted_k141_119964 | Not-determined |
| S12_extracted_k141_25349 | Not-determined |
| S12_extracted_k141_143394 | Low-quality |
| S12_extracted_k141_111538 | Not-determined |
| S12_extracted_k141_143397 | Not-determined |
| S12_extracted_k141_17238 | Not-determined |
| S12_extracted_k141_96384 | Not-determined |
| S12_extracted_k141_1851 | Not-determined |
| S12_extracted_k141_104064 | Not-determined |
| S12_extracted_k141_9330 | Not-determined |
| S12_extracted_k141_96388 | Not-determined |
| S12_extracted_k141_88262 | Not-determined |
| S12_extracted_k141_104071 | Not-determined |
| S12_extracted_k141_56986 | Not-determined |
| S12_extracted_k141_135309 | Not-determined |
| S12_extracted_k141_135310 | Not-determined |
| S12_extracted_k141_111545 | Not-determined |
| S12_extracted_k141_9335 | Not-determined |
| S12_extracted_k141_9336 | Not-determined |
| S12_extracted_k141_48438 | Low-quality |
| S12_extracted_k141_151002 | Not-determined |
| S12_extracted_k141_32671 | Not-determined |
| S12_extracted_k141_151004 | Not-determined |
| S12_extracted_k141_88268 | Not-determined |
| S12_extracted_k141_135319 | Not-determined |
| S12_extracted_k141_143407 | Not-determined |
| S12_extracted_k141_88272 | Not-determined |
| S12_extracted_k141_119981 | Not-determined |
| S12_extracted_k141_25361 | Not-determined |
| S12_extracted_k141_48445 | Not-determined |
| S12_extracted_k141_9342 | Not-determined |
| S12_extracted_k141_111554 | Low-quality |
| S12_extracted_k141_17246 | Not-determined |
| S12_extracted_k141_96401 | Not-determined |
| S12_extracted_k141_48449 | Not-determined |
| S12_extracted_k141_40924 | Not-determined |
| S12_extracted_k141_96403 | Not-determined |
| S12_extracted_k141_48450 | Low-quality |
| S12_extracted_k141_40926 | Not-determined |
| S12_extracted_k141_48451 | Not-determined |
| S12_extracted_k141_32678 | Low-quality |
| S12_extracted_k141_57008 | Not-determined |
| S12_extracted_k141_32682 | Not-determined |
| S12_extracted_k141_40932 | Not-determined |
| S12_extracted_k141_104087 | Not-determined |
| S12_extracted_k141_96414 | Not-determined |
| S12_extracted_k141_25370 | Low-quality |
| S12_extracted_k141_40935 | Not-determined |
| S12_extracted_k141_40936 | Not-determined |
| S12_extracted_k141_32688 | Not-determined |
| S12_extracted_k141_135335 | Not-determined |
| S12_extracted_k141_32691 | Not-determined |
| S12_extracted_k141_96426 | Not-determined |
| S12_extracted_k141_48458 | Not-determined |
| S12_extracted_k141_17260 | Not-determined |
| S12_extracted_k141_32693 | Not-determined |
| S12_extracted_k141_104094 | Not-determined |
| S12_extracted_k141_96432 | Not-determined |
| S12_extracted_k141_96436 | Not-determined |
| S12_extracted_k141_57026 | Not-determined |
| S12_extracted_k141_32699 | Not-determined |
| S12_extracted_k141_104103 | Not-determined |
| S12_extracted_k141_32700 | Low-quality |
| S12_extracted_k141_88289 | Not-determined |
| S12_extracted_k141_25391 | Not-determined |
| S12_extracted_k141_72602 | Not-determined |
| S12_extracted_k141_80050 | Not-determined |
| S12_extracted_k141_72604 | Not-determined |
| S12_extracted_k141_111584 | Not-determined |
| S12_extracted_k141_32710 | Not-determined |
| S12_extracted_k141_32711 | Not-determined |
| S12_extracted_k141_80055 | Not-determined |
| S12_extracted_k141_25401 | Not-determined |
| S12_extracted_k141_111593 | Not-determined |
| S12_extracted_k141_80060 | Not-determined |
| S12_extracted_k141_111595 | Not-determined |
| S12_extracted_k141_111600 | Not-determined |
| S12_extracted_k141_104114 | Not-determined |
| S12_extracted_k141_88301 | Not-determined |
| S12_extracted_k141_40977 | Not-determined |
| S12_extracted_k141_104116 | Not-determined |
| S12_extracted_k141_80070 | Not-determined |
| S12_extracted_k141_104120 | Not-determined |
| S12_extracted_k141_111610 | Not-determined |
| S12_extracted_k141_182802 | Not-determined |
| S12_extracted_k141_57052 | Not-determined |
| S12_extracted_k141_104124 | Not-determined |
| S12_extracted_k141_120019 | Not-determined |
| S12_extracted_k141_80074 | Not-determined |
| S12_extracted_k141_96449 | Not-determined |
| S12_extracted_k141_120026 | Not-determined |
| S12_extracted_k141_182811 | Not-determined |
| S12_extracted_k141_96453 | Not-determined |
| S12_extracted_k141_120035 | Not-determined |
| S12_extracted_k141_143432 | Not-determined |
| S12_extracted_k141_182820 | Not-determined |
| S12_extracted_k141_182821 | Not-determined |
| S12_extracted_k141_40997 | Low-quality |
| S12_extracted_k141_40998 | Not-determined |
| S12_extracted_k141_88316 | Not-determined |
| S12_extracted_k141_17274 | Not-determined |
| S12_extracted_k141_17276 | Not-determined |
| S12_extracted_k141_96472 | Low-quality |
| S12_extracted_k141_1873 | Not-determined |
| S12_extracted_k141_88322 | Not-determined |
| S12_extracted_k141_48469 | Low-quality |
| S12_extracted_k141_182826 | Not-determined |
| S12_extracted_k141_182827 | Not-determined |
| S12_extracted_k141_41007 | Not-determined |
| S12_extracted_k141_25430 | Not-determined |
| S12_extracted_k141_25431 | Not-determined |
| S12_extracted_k141_9365 | Not-determined |
| S12_extracted_k141_57079 | Not-determined |
| S12_extracted_k141_96483 | Not-determined |
| S12_extracted_k141_80097 | Not-determined |
| S12_extracted_k141_41010 | Not-determined |
| S12_extracted_k141_104152 | Not-determined |
| S12_extracted_k141_120056 | Low-quality |
| S12_extracted_k141_25433 | Not-determined |
| S12_extracted_k141_80099 | Not-determined |
| S12_extracted_k141_111638 | Not-determined |
| S12_extracted_k141_104155 | Not-determined |
| S12_extracted_k141_64780 | Not-determined |
| S12_extracted_k141_127644 | Not-determined |
| S12_extracted_k141_151019 | Low-quality |
| S12_extracted_k141_41019 | Not-determined |
| S12_extracted_k141_151021 | Not-determined |
| S12_extracted_k141_182835 | Not-determined |
| S12_extracted_k141_9371 | Not-determined |
| S12_extracted_k141_1889 | Not-determined |
| S12_extracted_k141_111643 | Not-determined |
| S12_extracted_k141_104159 | Low-quality |
| S12_extracted_k141_143453 | Not-determined |
| S12_extracted_k141_48480 | Not-determined |
| S12_extracted_k141_80105 | Not-determined |
| S12_extracted_k141_96490 | Not-determined |
| S12_extracted_k141_182841 | Not-determined |
| S12_extracted_k141_48485 | Low-quality |
| S12_extracted_k141_64783 | Not-determined |
| S12_extracted_k141_9377 | Not-determined |
| S12_extracted_k141_127649 | Not-determined |
| S12_extracted_k141_25442 | Not-determined |
| S12_extracted_k141_120071 | Not-determined |
| S12_extracted_k141_25443 | Not-determined |
| S12_extracted_k141_143469 | Not-determined |
| S12_extracted_k141_64791 | Not-determined |
| S12_extracted_k141_127656 | Not-determined |
| S12_extracted_k141_32746 | Not-determined |
| S12_extracted_k141_72627 | Low-quality |
| S12_extracted_k141_104174 | Not-determined |
| S12_extracted_k141_9385 | Not-determined |
| S12_extracted_k141_72630 | Not-determined |
| S12_extracted_k141_143474 | Not-determined |
| S12_extracted_k141_1901 | Not-determined |
| S12_extracted_k141_25451 | Not-determined |
| S12_extracted_k141_143479 | Not-determined |
| S12_extracted_k141_104177 | Not-determined |
| S12_extracted_k141_127659 | Low-quality |
| S12_extracted_k141_25456 | Not-determined |
| S12_extracted_k141_41040 | Low-quality |
| S12_extracted_k141_151037 | Not-determined |
| S12_extracted_k141_111673 | Not-determined |
| S12_extracted_k141_104182 | Not-determined |
| S12_extracted_k141_41048 | Not-determined |
| S12_extracted_k141_151043 | Not-determined |
| S12_extracted_k141_143492 | Not-determined |
| S12_extracted_k141_104188 | Low-quality |
| S12_extracted_k141_88382 | Not-determined |
| S12_extracted_k141_111676 | Not-determined |
| S12_extracted_k141_104191 | Not-determined |
| S12_extracted_k141_111677 | Not-determined |
| S12_extracted_k141_143499 | Not-determined |
| S12_extracted_k141_17310 | Low-quality |
| S12_extracted_k141_96517 | Not-determined |
| S12_extracted_k141_96518 | Not-determined |
| S12_extracted_k141_104192 | Not-determined |
| S12_extracted_k141_96521 | Not-determined |
| S12_extracted_k141_143501 | Not-determined |
| S12_extracted_k141_143502 | Not-determined |
| S12_extracted_k141_1915 | Not-determined |
| S12_extracted_k141_25464 | Not-determined |
| S12_extracted_k141_41062 | Not-determined |
| S12_extracted_k141_80115 | Not-determined |
| S12_extracted_k141_1922 | Not-determined |
| S12_extracted_k141_48497 | Not-determined |
| S12_extracted_k141_57100 | Not-determined |
| S12_extracted_k141_158768 | Not-determined |
| S12_extracted_k141_41064 | Low-quality |
| S12_extracted_k141_48502 | Low-quality |
| S12_extracted_k141_17321 | Not-determined |
| S12_extracted_k141_104204 | Not-determined |
| S12_extracted_k141_17325 | Not-determined |
| S12_extracted_k141_32751 | Not-determined |
| S12_extracted_k141_104207 | Not-determined |
| S12_extracted_k141_182844 | Not-determined |
| S12_extracted_k141_151064 | Not-determined |
| S12_extracted_k141_104210 | Not-determined |
| S12_extracted_k141_135347 | Not-determined |
| S12_extracted_k141_96540 | Low-quality |
| S12_extracted_k141_32761 | Not-determined |
| S12_extracted_k141_17335 | Not-determined |
| S12_extracted_k141_135351 | Not-determined |
| S12_extracted_k141_96547 | Not-determined |
| S12_extracted_k141_57111 | Not-determined |
| S12_extracted_k141_111689 | Not-determined |
| S12_extracted_k141_17339 | Not-determined |
| S12_extracted_k141_1939 | Not-determined |
| S12_extracted_k141_41075 | Not-determined |
| S12_extracted_k141_41077 | Not-determined |
| S12_extracted_k141_151075 | Not-determined |
| S12_extracted_k141_135364 | Not-determined |
| S12_extracted_k141_88397 | Not-determined |
| S12_extracted_k141_182858 | Not-determined |
| S12_extracted_k141_64838 | Not-determined |
| S12_extracted_k141_120079 | Not-determined |
| S12_extracted_k141_9403 | Not-determined |
| S12_extracted_k141_158786 | Not-determined |
| S12_extracted_k141_1949 | Not-determined |
| S12_extracted_k141_9410 | Not-determined |
| S12_extracted_k141_111701 | Not-determined |
| S12_extracted_k141_104242 | Not-determined |
| S12_extracted_k141_143529 | Not-determined |
| S12_extracted_k141_1955 | Not-determined |
| S12_extracted_k141_1956 | Not-determined |
| S12_extracted_k141_41086 | Not-determined |
| S12_extracted_k141_17353 | Not-determined |
| S12_extracted_k141_64847 | Not-determined |
| S12_extracted_k141_158793 | Not-determined |
| S12_extracted_k141_25473 | Not-determined |
| S12_extracted_k141_41087 | Low-quality |
| S12_extracted_k141_143536 | Not-determined |
| S12_extracted_k141_25478 | Not-determined |
| S12_extracted_k141_32773 | Not-determined |
| S12_extracted_k141_41089 | Not-determined |
| S12_extracted_k141_64856 | Not-determined |
| S12_extracted_k141_96568 | Not-determined |
| S12_extracted_k141_135384 | Not-determined |
| S12_extracted_k141_48514 | Not-determined |
| S12_extracted_k141_96572 | Not-determined |
| S12_extracted_k141_57129 | Not-determined |
| S12_extracted_k141_9418 | Not-determined |
| S12_extracted_k141_88424 | Not-determined |
| S12_extracted_k141_111711 | Low-quality |
| S12_extracted_k141_127707 | Not-determined |
| S12_extracted_k141_1958 | Not-determined |
| S12_extracted_k141_17360 | Not-determined |
| S12_extracted_k141_9421 | Not-determined |
| S12_extracted_k141_25484 | Not-determined |
| S12_extracted_k141_151094 | Low-quality |
| S12_extracted_k141_158805 | Not-determined |
| S12_extracted_k141_143548 | Not-determined |
| S12_extracted_k141_135394 | Not-determined |
| S12_extracted_k141_104261 | Not-determined |
| S12_extracted_k141_48525 | Not-determined |
| S12_extracted_k141_158810 | Not-determined |
| S12_extracted_k141_64869 | Not-determined |
| S12_extracted_k141_88431 | Not-determined |
| S12_extracted_k141_158811 | Not-determined |
| S12_extracted_k141_151103 | Not-determined |
| S12_extracted_k141_151104 | Not-determined |
| S12_extracted_k141_88432 | Not-determined |
| S12_extracted_k141_17369 | Not-determined |
| S12_extracted_k141_167164 | Not-determined |
| S12_extracted_k141_57141 | Not-determined |
| S12_extracted_k141_41118 | Not-determined |
| S12_extracted_k141_158819 | Not-determined |
| S12_extracted_k141_9434 | Not-determined |
| S12_extracted_k141_104268 | Not-determined |
| S12_extracted_k141_17374 | Low-quality |
| S12_extracted_k141_151108 | Not-determined |
| S12_extracted_k141_64876 | Not-determined |
| S12_extracted_k141_88436 | Not-determined |
| S12_extracted_k141_127716 | Not-determined |
| S12_extracted_k141_57152 | Not-determined |
| S12_extracted_k141_104277 | Not-determined |
| S12_extracted_k141_135403 | Low-quality |
| S12_extracted_k141_127717 | Not-determined |
| S12_extracted_k141_25506 | Not-determined |
| S12_extracted_k141_9440 | Not-determined |
| S12_extracted_k141_9441 | Not-determined |
| S12_extracted_k141_64877 | Not-determined |
| S12_extracted_k141_104282 | Not-determined |
| S12_extracted_k141_104284 | Not-determined |
| S12_extracted_k141_9444 | Not-determined |
| S12_extracted_k141_64879 | Low-quality |
| S12_extracted_k141_158827 | Not-determined |
| S12_extracted_k141_57160 | Not-determined |
| S12_extracted_k141_143578 | Not-determined |
| S12_extracted_k141_57161 | Low-quality |
| S12_extracted_k141_167181 | Not-determined |
| S12_extracted_k141_17390 | Not-determined |
| S12_extracted_k141_104292 | Not-determined |
| S12_extracted_k141_17393 | Not-determined |
| S12_extracted_k141_17394 | Not-determined |
| S12_extracted_k141_167186 | Not-determined |
| S12_extracted_k141_64886 | Not-determined |
| S12_extracted_k141_158836 | Not-determined |
| S12_extracted_k141_158837 | Not-determined |
| S12_extracted_k141_9457 | Low-quality |
| S12_extracted_k141_111730 | Not-determined |
| S12_extracted_k141_104299 | Not-determined |
| S12_extracted_k141_111731 | Not-determined |
| S12_extracted_k141_88461 | Not-determined |
| S12_extracted_k141_96602 | Not-determined |
| S12_extracted_k141_17404 | Low-quality |
| S12_extracted_k141_127735 | Not-determined |
| S12_extracted_k141_174490 | Not-determined |
| S12_extracted_k141_96606 | Not-determined |
| S12_extracted_k141_17409 | Not-determined |
| S12_extracted_k141_111739 | Not-determined |
| S12_extracted_k141_120126 | Not-determined |
| S12_extracted_k141_1979 | Not-determined |
| S12_extracted_k141_120129 | Not-determined |
| S12_extracted_k141_158842 | Not-determined |
| S12_extracted_k141_17413 | Low-quality |
| S12_extracted_k141_17415 | Not-determined |
| S12_extracted_k141_88472 | Not-determined |
| S12_extracted_k141_88476 | Not-determined |
| S12_extracted_k141_17419 | Not-determined |
| S14_vs_k141_10656\|\|full | Medium-quality |
| S12_extracted_k141_143598 | Not-determined |
| S12_extracted_k141_96611 | Low-quality |
| S12_extracted_k141_120130 | Low-quality |
| S12_extracted_k141_80133 | Not-determined |
| S12_extracted_k141_88481 | Not-determined |
| S12_extracted_k141_120133 | Not-determined |
| S12_extracted_k141_120134 | Not-determined |
| S12_extracted_k141_182915 | Not-determined |
| S12_extracted_k141_182918 | Not-determined |
| S12_extracted_k141_41141 | Low-quality |
| S12_extracted_k141_174506 | Not-determined |
| S12_extracted_k141_151123 | Not-determined |
| S12_extracted_k141_88487 | Not-determined |
| S12_extracted_k141_120142 | Not-determined |
| S12_extracted_k141_88491 | Not-determined |
| S12_extracted_k141_158854 | Not-determined |
| S12_extracted_k141_182924 | Not-determined |
| S12_extracted_k141_120145 | Not-determined |
| S12_extracted_k141_158855 | Not-determined |
| S12_extracted_k141_182925 | Not-determined |
| S12_extracted_k141_25526 | Not-determined |
| S12_extracted_k141_17430 | Low-quality |
| S12_extracted_k141_135436 | Not-determined |
| S12_extracted_k141_151128 | Not-determined |
| S12_extracted_k141_41151 | Not-determined |
| S12_extracted_k141_41152 | Not-determined |
| S12_extracted_k141_96616 | Not-determined |
| S12_extracted_k141_96617 | Not-determined |
| S12_extracted_k141_151132 | Not-determined |
| S12_extracted_k141_96621 | Not-determined |
| S12_extracted_k141_88495 | Not-determined |
| S12_extracted_k141_120156 | Not-determined |
| S12_extracted_k141_88497 | Not-determined |
| S12_extracted_k141_88498 | Not-determined |
| S12_extracted_k141_182935 | Not-determined |
| S12_extracted_k141_143617 | Not-determined |

| | |
|---|---|
| S12_extracted_k141_151138 | Not-determined |
| S12_extracted_k141_48563 | Not-determined |
| S12_extracted_k141_96627 | Not-determined |
| S12_extracted_k141_151140 | Not-determined |
| S12_extracted_k141_151142 | Not-determined |
| S12_extracted_k141_96629 | Not-determined |
| S12_extracted_k141_9476 | Low-quality |
| S12_extracted_k141_174521 | Low-quality |
| S12_extracted_k141_57210 | Not-determined |
| S12_extracted_k141_158863 | Not-determined |
| S12_extracted_k141_96633 | Not-determined |
| S12_extracted_k141_9480 | Not-determined |
| S12_extracted_k141_167228 | Not-determined |
| S12_extracted_k141_25534 | Not-determined |
| S12_extracted_k141_48568 | Not-determined |
| S12_extracted_k141_17437 | Not-determined |
| S12_extracted_k141_64912 | Not-determined |
| S12_extracted_k141_80159 | Not-determined |
| S12_extracted_k141_167236 | Not-determined |
| S12_extracted_k141_167237 | Not-determined |
| S12_extracted_k141_64916 | Not-determined |
| S12_extracted_k141_135453 | Not-determined |
| S12_extracted_k141_96638 | Not-determined |
| S12_extracted_k141_104340 | Not-determined |
| S12_extracted_k141_151153 | Not-determined |
| S12_extracted_k141_64920 | Not-determined |
| S12_extracted_k141_17441 | Not-determined |
| S12_extracted_k141_96640 | Not-determined |
| S12_extracted_k141_17443 | Not-determined |
| S12_extracted_k141_174528 | Not-determined |
| S12_extracted_k141_9494 | Not-determined |
| S12_extracted_k141_96641 | Not-determined |
| S12_extracted_k141_80164 | Not-determined |
| S12_extracted_k141_127745 | Not-determined |
| S12_extracted_k141_48574 | Not-determined |
| S12_extracted_k141_135457 | Not-determined |
| S12_extracted_k141_143626 | Not-determined |
| S12_extracted_k141_88511 | Not-determined |
| S12_extracted_k141_127748 | Low-quality |
| S12_extracted_k141_32795 | Not-determined |
| S12_extracted_k141_127752 | Not-determined |
| S12_extracted_k141_41169 | Not-determined |
| S12_extracted_k141_135462 | Not-determined |
| S12_extracted_k141_174532 | Not-determined |
| S12_extracted_k141_25549 | Not-determined |
| S12_extracted_k141_48581 | Low-quality |
| S12_extracted_k141_174534 | Not-determined |
| S12_extracted_k141_41179 | Not-determined |
| S12_extracted_k141_120174 | Not-determined |
| S12_extracted_k141_25556 | Not-determined |
| S12_extracted_k141_41181 | Not-determined |
| S12_extracted_k141_17456 | Not-determined |
| S12_extracted_k141_25558 | Not-determined |
| S12_extracted_k141_174538 | Not-determined |
| S12_extracted_k141_174541 | Not-determined |
| S12_extracted_k141_25564 | Not-determined |
| S12_extracted_k141_143644 | Not-determined |
| S12_extracted_k141_25565 | Not-determined |
| S12_extracted_k141_120180 | Not-determined |
| S12_extracted_k141_143645 | Not-determined |
| S12_extracted_k141_25567 | Not-determined |
| S12_extracted_k141_25569 | Not-determined |
| S12_extracted_k141_48590 | Not-determined |
| S12_extracted_k141_41190 | Not-determined |
| S12_extracted_k141_174549 | Not-determined |
| S12_extracted_k141_135472 | Not-determined |
| S12_extracted_k141_41196 | Not-determined |
| S12_extracted_k141_1985 | Low-quality |
| S12_extracted_k141_1986 | Not-determined |
| S12_extracted_k141_25571 | Not-determined |
| S12_extracted_k141_48600 | Low-quality |
| S12_extracted_k141_41199 | Not-determined |
| S12_extracted_k141_17474 | Not-determined |
| S12_extracted_k141_41200 | Not-determined |
| S12_extracted_k141_25581 | Not-determined |
| S12_extracted_k141_17475 | Low-quality |
| S12_extracted_k141_174560 | Not-determined |
| S12_extracted_k141_17477 | Not-determined |
| S12_extracted_k141_17478 | Not-determined |
| S12_extracted_k141_1994 | Not-determined |
| S12_extracted_k141_25584 | Not-determined |
| S12_extracted_k141_120197 | Not-determined |
| S12_extracted_k141_41204 | Not-determined |
| S12_extracted_k141_120199 | Not-determined |
| S12_extracted_k141_57237 | Not-determined |
| S12_extracted_k141_143658 | Not-determined |
| S12_extracted_k141_120201 | Not-determined |
| S12_extracted_k141_48607 | Not-determined |
| S12_extracted_k141_80182 | Not-determined |
| S12_extracted_k141_48608 | Not-determined |
| S12_extracted_k141_174566 | Not-determined |
| S12_extracted_k141_96647 | Low-quality |
| S12_extracted_k141_48609 | Not-determined |
| S12_extracted_k141_127772 | Not-determined |
| S12_extracted_k141_57241 | Low-quality |
| S12_extracted_k141_2003 | Not-determined |
| S12_extracted_k141_64956 | Not-determined |
| S12_extracted_k141_96653 | Not-determined |
| S12_extracted_k141_41214 | Not-determined |
| S12_extracted_k141_88538 | Not-determined |
| S12_extracted_k141_2005 | Not-determined |
| S12_extracted_k141_182974 | Not-determined |
| S12_extracted_k141_182975 | Not-determined |
| S12_extracted_k141_96655 | Not-determined |
| S12_extracted_k141_158884 | Not-determined |
| S12_extracted_k141_48615 | Not-determined |
| S12_extracted_k141_120210 | Not-determined |
| S12_extracted_k141_25599 | Not-determined |
| S12_extracted_k141_158887 | Not-determined |
| S12_extracted_k141_88544 | Not-determined |
| S12_extracted_k141_88546 | Not-determined |
| S12_extracted_k141_48620 | Not-determined |
| S12_extracted_k141_120216 | Not-determined |
| S12_extracted_k141_167290 | Not-determined |
| S12_extracted_k141_48621 | Not-determined |
| S12_extracted_k141_88550 | Low-quality |
| S12_extracted_k141_9513 | Not-determined |
| S12_extracted_k141_158889 | Not-determined |
| S12_extracted_k141_158890 | Not-determined |
| S12_extracted_k141_48628 | Not-determined |
| S12_extracted_k141_25608 | Not-determined |
| S12_extracted_k141_32808 | Not-determined |
| S12_extracted_k141_9520 | Not-determined |
| S12_extracted_k141_2008 | Not-determined |
| S12_extracted_k141_80191 | Not-determined |
| S12_extracted_k141_96675 | Not-determined |
| S12_extracted_k141_167299 | Not-determined |
| S12_extracted_k141_41223 | Not-determined |
| S12_extracted_k141_32814 | Not-determined |
| S12_extracted_k141_41224 | Not-determined |
| S12_extracted_k141_48634 | Not-determined |
| S12_extracted_k141_80195 | Not-determined |
| S12_extracted_k141_48636 | Not-determined |
| S12_extracted_k141_41229 | Low-quality |
| S12_extracted_k141_88565 | Not-determined |
| S12_extracted_k141_41230 | Not-determined |
| S12_extracted_k141_32829 | Not-determined |
| S12_extracted_k141_2022 | Not-determined |
| S12_extracted_k141_32832 | Not-determined |
| S12_extracted_k141_57256 | Not-determined |
| S12_extracted_k141_2023 | Not-determined |
| S12_extracted_k141_88573 | Not-determined |
| S12_extracted_k141_143668 | Not-determined |
| S12_extracted_k141_25625 | Not-determined |
| S12_extracted_k141_64973 | Not-determined |
| S12_extracted_k141_80208 | Not-determined |
| S12_extracted_k141_64976 | Not-determined |
| S12_extracted_k141_25628 | Not-determined |
| S12_extracted_k141_41237 | Not-determined |
| S12_extracted_k141_64979 | Not-determined |
| S12_extracted_k141_41244 | Not-determined |
| S12_extracted_k141_104361 | Not-determined |
| S12_extracted_k141_182991 | Not-determined |
| S12_extracted_k141_9552 | Not-determined |
| S12_extracted_k141_158915 | Not-determined |
| S12_extracted_k141_182993 | Not-determined |
| S12_extracted_k141_9553 | Not-determined |
| S12_extracted_k141_41248 | Not-determined |
| S12_extracted_k141_104365 | Not-determined |
| S12_extracted_k141_17532 | Not-determined |
| S12_extracted_k141_158918 | Not-determined |
| S12_extracted_k141_80219 | Not-determined |
| S12_extracted_k141_41251 | Not-determined |
| S12_extracted_k141_158922 | Not-determined |
| S12_extracted_k141_143686 | Not-determined |
| S12_extracted_k141_143688 | Not-determined |
| S12_extracted_k141_2040 | Not-determined |
| S12_extracted_k141_41261 | Not-determined |
| S12_extracted_k141_57284 | Not-determined |
| S12_extracted_k141_88585 | Not-determined |
| S12_extracted_k141_57290 | Not-determined |
| S12_extracted_k141_143696 | Not-determined |
| S12_extracted_k141_135505 | Low-quality |
| S12_extracted_k141_88594 | Not-determined |
| S12_extracted_k141_64999 | Not-determined |
| S12_extracted_k141_174575 | Low-quality |
| S12_extracted_k141_41272 | Not-determined |
| S12_extracted_k141_174577 | Not-determined |
| S12_extracted_k141_41275 | Not-determined |
| S12_extracted_k141_158935 | Not-determined |
| S12_extracted_k141_9570 | Not-determined |
| S12_extracted_k141_174581 | Not-determined |
| S12_extracted_k141_183022 | Not-determined |
| S12_extracted_k141_88601 | Not-determined |
| S12_extracted_k141_135517 | Not-determined |
| S12_extracted_k141_127785 | Not-determined |
| S12_extracted_k141_57301 | Not-determined |
| S12_extracted_k141_135518 | Not-determined |
| S12_extracted_k141_32863 | Not-determined |
| S12_extracted_k141_80242 | Not-determined |
| S12_extracted_k141_48676 | Not-determined |
| S12_extracted_k141_174585 | Not-determined |
| S12_extracted_k141_17552 | Not-determined |
| S12_extracted_k141_25638 | Not-determined |
| S12_extracted_k141_135520 | Not-determined |
| S12_extracted_k141_96701 | Not-determined |
| S12_extracted_k141_88624 | Low-quality |
| S12_extracted_k141_88625 | Not-determined |
| S12_extracted_k141_127803 | Not-determined |
| S12_extracted_k141_41292 | Low-quality |
| S12_extracted_k141_158944 | Not-determined |
| S12_extracted_k141_32871 | Low-quality |
| S12_extracted_k141_96705 | Not-determined |
| S12_extracted_k141_17556 | Not-determined |
| S12_extracted_k141_9579 | Not-determined |
| S12_extracted_k141_41294 | Not-determined |
| S12_extracted_k141_9581 | Not-determined |
| S12_extracted_k141_120226 | Not-determined |
| S12_extracted_k141_32875 | Not-determined |
| S12_extracted_k141_57324 | Not-determined |
| S12_extracted_k141_120229 | Not-determined |
| S12_extracted_k141_88635 | Not-determined |
| S12_extracted_k141_41301 | Low-quality |
| S12_extracted_k141_174597 | Not-determined |
| S12_extracted_k141_183029 | Low-quality |
| S12_extracted_k141_96710 | Not-determined |
| S12_extracted_k141_41302 | Not-determined |
| S12_extracted_k141_183030 | Not-determined |
| S12_extracted_k141_183031 | Not-determined |
| S12_extracted_k141_135534 | Not-determined |
| S12_extracted_k141_120234 | Not-determined |
| S12_extracted_k141_158960 | Not-determined |
| S12_extracted_k141_9591 | Not-determined |
| S12_extracted_k141_127812 | Not-determined |
| S12_extracted_k141_167331 | Not-determined |
| S12_extracted_k141_127813 | Not-determined |
| S12_extracted_k141_158967 | Not-determined |
| S12_extracted_k141_143714 | Not-determined |
| S12_extracted_k141_158968 | Not-determined |
| S12_extracted_k141_167333 | Not-determined |
| S12_extracted_k141_143717 | Not-determined |
| S12_extracted_k141_143720 | Not-determined |
| S12_extracted_k141_25650 | Not-determined |
| S12_extracted_k141_96724 | Not-determined |
| S12_extracted_k141_41318 | Low-quality |
| S12_extracted_k141_80274 | Not-determined |
| S12_extracted_k141_183047 | Low-quality |
| S9_vs_k141_90860\|\|full | High-quality |
| S12_extracted_k141_9609 | Not-determined |
| S12_extracted_k141_96732 | Not-determined |
| S12_extracted_k141_48685 | Not-determined |
| S12_extracted_k141_183051 | Not-determined |
| S12_extracted_k141_80283 | Not-determined |
| S12_extracted_k141_25657 | Not-determined |
| S12_extracted_k141_143726 | Not-determined |
| S12_extracted_k141_65036 | Not-determined |
| S12_extracted_k141_32906 | Not-determined |
| S12_extracted_k141_80288 | Not-determined |
| S12_extracted_k141_127818 | Low-quality |
| S12_extracted_k141_25660 | Not-determined |
| S12_extracted_k141_158977 | Not-determined |
| S12_extracted_k141_127820 | Not-determined |
| S12_extracted_k141_96736 | Not-determined |
| S12_extracted_k141_120258 | Not-determined |
| S12_extracted_k141_135540 | Low-quality |
| S12_extracted_k141_57347 | Not-determined |
| S12_extracted_k141_41326 | Not-determined |
| S12_extracted_k141_48699 | Not-determined |
| S12_extracted_k141_25662 | Not-determined |
| S12_extracted_k141_48701 | Not-determined |
| S12_extracted_k141_135546 | Not-determined |
| S12_extracted_k141_135547 | Not-determined |
| S12_extracted_k141_65046 | Not-determined |
| S12_extracted_k141_80291 | Not-determined |
| S12_extracted_k141_80294 | Not-determined |
| S12_extracted_k141_32921 | Not-determined |
| S12_extracted_k141_25666 | Not-determined |
| S12_extracted_k141_80299 | Not-determined |
| S12_extracted_k141_41336 | Not-determined |
| S12_extracted_k141_41337 | Not-determined |
| S12_extracted_k141_158998 | Not-determined |
| S12_extracted_k141_9613 | Low-quality |
| S12_extracted_k141_183073 | Not-determined |
| S12_extracted_k141_127832 | Not-determined |
| S12_extracted_k141_80304 | Not-determined |
| S12_extracted_k141_9617 | Not-determined |
| S12_extracted_k141_32933 | Not-determined |
| S12_extracted_k141_17576 | Not-determined |
| S12_extracted_k141_17580 | Not-determined |
| S12_extracted_k141_143737 | Not-determined |
| S12_extracted_k141_2077 | Not-determined |
| S12_extracted_k141_135554 | Not-determined |
| S12_extracted_k141_48717 | Not-determined |
| S12_extracted_k141_159004 | Not-determined |
| S12_extracted_k141_120272 | Not-determined |
| S12_extracted_k141_143739 | Not-determined |
| S12_extracted_k141_104384 | Not-determined |
| S12_extracted_k141_104385 | Not-determined |
| S12_extracted_k141_2083 | Not-determined |
| S12_extracted_k141_159011 | Not-determined |
| S12_extracted_k141_17587 | Low-quality |
| S12_extracted_k141_48725 | Low-quality |
| S12_extracted_k141_159012 | Low-quality |
| S12_extracted_k141_80309 | Not-determined |
| S12_extracted_k141_143744 | Not-determined |
| S12_extracted_k141_159013 | Not-determined |
| S12_extracted_k141_17592 | Not-determined |
| S12_extracted_k141_120278 | Not-determined |
| S12_extracted_k141_143745 | Not-determined |
| S12_extracted_k141_80312 | Not-determined |
| S12_extracted_k141_143747 | Low-quality |
| S12_extracted_k141_48732 | Not-determined |
| S12_extracted_k141_104400 | Not-determined |
| S12_extracted_k141_159018 | Not-determined |
| S12_extracted_k141_183097 | Not-determined |
| S12_extracted_k141_80315 | Not-determined |
| S12_extracted_k141_17595 | Not-determined |
| S12_extracted_k141_48736 | Not-determined |
| S12_extracted_k141_174617 | Not-determined |
| S12_extracted_k141_143752 | Not-determined |
| S12_extracted_k141_143753 | Low-quality |
| S12_extracted_k141_2089 | Not-determined |
| S12_extracted_k141_159023 | Not-determined |
| S12_extracted_k141_80324 | Not-determined |
| S12_extracted_k141_80326 | Not-determined |
| S12_extracted_k141_65078 | Low-quality |
| S12_extracted_k141_41353 | Not-determined |
| S12_extracted_k141_159028 | Not-determined |
| S12_extracted_k141_120292 | Not-determined |
| S12_extracted_k141_48744 | Not-determined |
| S12_extracted_k141_143757 | Not-determined |
| S12_extracted_k141_80329 | Not-determined |
| S12_extracted_k141_80330 | Not-determined |
| S12_extracted_k141_17601 | Not-determined |
| S12_extracted_k141_41357 | Not-determined |
| S12_extracted_k141_104413 | Not-determined |
| S12_extracted_k141_174625 | Not-determined |
| S12_extracted_k141_159030 | Not-determined |
| S12_extracted_k141_120298 | Not-determined |
| S12_extracted_k141_96749 | Not-determined |
| S12_extracted_k141_174627 | Not-determined |
| S12_extracted_k141_80336 | Not-determined |
| S12_extracted_k141_57353 | Low-quality |
| S12_extracted_k141_96757 | Not-determined |
| S12_extracted_k141_48748 | Not-determined |
| S12_extracted_k141_32955 | Not-determined |
| S12_extracted_k141_80342 | Not-determined |
| S12_extracted_k141_120305 | Not-determined |
| S12_extracted_k141_32962 | Not-determined |
| S12_extracted_k141_17616 | Not-determined |
| S12_extracted_k141_32964 | Not-determined |
| S12_extracted_k141_120312 | Not-determined |
| S12_extracted_k141_96766 | Low-quality |
| S12_extracted_k141_127866 | Not-determined |
| S12_extracted_k141_96770 | Not-determined |
| S12_extracted_k141_32969 | Not-determined |
| S12_extracted_k141_167348 | Not-determined |
| S12_extracted_k141_104428 | Not-determined |
| S12_extracted_k141_127869 | Not-determined |
| S12_extracted_k141_104431 | Not-determined |
| S12_extracted_k141_183110 | Not-determined |
| S12_extracted_k141_167357 | Not-determined |
| S12_extracted_k141_174662 | Not-determined |
| S12_extracted_k141_174664 | Not-determined |
| S12_extracted_k141_96780 | Low-quality |
| S12_extracted_k141_25689 | Not-determined |
| S12_extracted_k141_127875 | Not-determined |
| S12_extracted_k141_25691 | Not-determined |
| S12_extracted_k141_167366 | Not-determined |
| S12_extracted_k141_32981 | Not-determined |
| S12_extracted_k141_143779 | Not-determined |
| S12_extracted_k141_96785 | Not-determined |
| S12_extracted_k141_32983 | Not-determined |
| S12_extracted_k141_25695 | Not-determined |
| S12_extracted_k141_143780 | Not-determined |
| S12_extracted_k141_104441 | Not-determined |
| S12_extracted_k141_183114 | Low-quality |
| S12_extracted_k141_9633 | Not-determined |
| S12_extracted_k141_88651 | Not-determined |
| S12_extracted_k141_32985 | Not-determined |
| S12_extracted_k141_135573 | Not-determined |
| S12_extracted_k141_9636 | Not-determined |
| S12_extracted_k141_167375 | Not-determined |
| S12_extracted_k141_174669 | Not-determined |
| S12_extracted_k141_159074 | Not-determined |
| S12_extracted_k141_104451 | Not-determined |
| S12_extracted_k141_159075 | Not-determined |
| S12_extracted_k141_183126 | Not-determined |
| S12_extracted_k141_80374 | Not-determined |
| S12_extracted_k141_41371 | Not-determined |
| S12_extracted_k141_80375 | Not-determined |
| S12_extracted_k141_65083 | Not-determined |
| S12_extracted_k141_65085 | Not-determined |
| S12_extracted_k141_135585 | Not-determined |
| S12_extracted_k141_104455 | Not-determined |
| S12_extracted_k141_25706 | Not-determined |
| S12_extracted_k141_80378 | Not-determined |
| S12_extracted_k141_9638 | Not-determined |
| S12_extracted_k141_57364 | Not-determined |
| S12_extracted_k141_41375 | Not-determined |
| S12_extracted_k141_9642 | Low-quality |
| S12_extracted_k141_25707 | Low-quality |
| S12_extracted_k141_167379 | Low-quality |
| S12_extracted_k141_167380 | Not-determined |
| S12_extracted_k141_88668 | Not-determined |
| S12_extracted_k141_65095 | Not-determined |
| S12_extracted_k141_183138 | Not-determined |
| S12_extracted_k141_104463 | Not-determined |
| S12_extracted_k141_151178 | Not-determined |
| S12_extracted_k141_104466 | Not-determined |
| S12_extracted_k141_104467 | Not-determined |
| S12_extracted_k141_2127 | Not-determined |
| S12_extracted_k141_151182 | Not-determined |
| S12_extracted_k141_135593 | Not-determined |
| S12_extracted_k141_57368 | Not-determined |
| S12_extracted_k141_41383 | Not-determined |
| S12_extracted_k141_33003 | Not-determined |
| S12_extracted_k141_96814 | Not-determined |
| S12_extracted_k141_104473 | Not-determined |
| S12_extracted_k141_174682 | Not-determined |
| S12_extracted_k141_41386 | Not-determined |
| S12_extracted_k141_143788 | Not-determined |
| S12_extracted_k141_80389 | Not-determined |
| S12_extracted_k141_151184 | Not-determined |
| S12_extracted_k141_96818 | Not-determined |
| S12_extracted_k141_80393 | Not-determined |
| S12_extracted_k141_25722 | Not-determined |
| S12_extracted_k141_9662 | Not-determined |
| S12_extracted_k141_143793 | Not-determined |
| S12_extracted_k141_88676 | Not-determined |
| S12_extracted_k141_174684 | Not-determined |
| S12_extracted_k141_2130 | Low-quality |
| S12_extracted_k141_135600 | Not-determined |
| S12_extracted_k141_104477 | Low-quality |
| S12_extracted_k141_151186 | Not-determined |
| S12_extracted_k141_25726 | Not-determined |
| S12_extracted_k141_33008 | Not-determined |
| S12_extracted_k141_143796 | Not-determined |
| S12_extracted_k141_17655 | Low-quality |
| S12_extracted_k141_9664 | Not-determined |
| S12_extracted_k141_151188 | Not-determined |
| S12_extracted_k141_135605 | Not-determined |
| S12_extracted_k141_120321 | Not-determined |
| S12_extracted_k141_80400 | Not-determined |
| S12_extracted_k141_65112 | Not-determined |
| S12_extracted_k141_174690 | Not-determined |
| S12_extracted_k141_17659 | Not-determined |
| S12_extracted_k141_33013 | Not-determined |
| S12_extracted_k141_104484 | Not-determined |
| S12_extracted_k141_104486 | Not-determined |
| S12_extracted_k141_80403 | Not-determined |
| S12_extracted_k141_135609 | Not-determined |
| S12_extracted_k141_33017 | Not-determined |
| S12_extracted_k141_9673 | Low-quality |
| S12_extracted_k141_120324 | Low-quality |
| S12_extracted_k141_57383 | Not-determined |
| S12_extracted_k141_80409 | Not-determined |
| S12_extracted_k141_9680 | Not-determined |
| S12_extracted_k141_151194 | Not-determined |
| S12_extracted_k141_48760 | Not-determined |
| S12_extracted_k141_57387 | Not-determined |
| S12_extracted_k141_183164 | Not-determined |
| S12_extracted_k141_57388 | Not-determined |
| S12_extracted_k141_2144 | Not-determined |
| S12_extracted_k141_88700 | Not-determined |
| S12_extracted_k141_104489 | Low-quality |
| S12_extracted_k141_88703 | Not-determined |
| S12_extracted_k141_104490 | Not-determined |
| S12_extracted_k141_143811 | Not-determined |
| S12_extracted_k141_41409 | Not-determined |
| S12_extracted_k141_88705 | Low-quality |
| S12_extracted_k141_48768 | Not-determined |
| S12_extracted_k141_174711 | Not-determined |
| S12_extracted_k141_127894 | Not-determined |
| S12_extracted_k141_127895 | Not-determined |
| S12_extracted_k141_17680 | Not-determined |
| S12_extracted_k141_135624 | Not-determined |
| S12_extracted_k141_183171 | Not-determined |
| S12_extracted_k141_174715 | Not-determined |
| S12_extracted_k141_127900 | Not-determined |
| S12_extracted_k141_33026 | Not-determined |
| S12_extracted_k141_17681 | Low-quality |
| S12_extracted_k141_25749 | Not-determined |
| S12_extracted_k141_135629 | Not-determined |
| S12_extracted_k141_96838 | Not-determined |
| S12_extracted_k141_25750 | Not-determined |
| S12_extracted_k141_151206 | Not-determined |
| S12_extracted_k141_111755 | Not-determined |
| S12_extracted_k141_183178 | Not-determined |
| S12_extracted_k141_167404 | Not-determined |
| S12_extracted_k141_159123 | Not-determined |
| S12_extracted_k141_183179 | Not-determined |
| S12_extracted_k141_151213 | Not-determined |
| S12_extracted_k141_159125 | Not-determined |
| S12_extracted_k141_135634 | Not-determined |
| S12_extracted_k141_174727 | Not-determined |
| S12_extracted_k141_2159 | Low-quality |
| S12_extracted_k141_17684 | Low-quality |
| S12_extracted_k141_174730 | Not-determined |
| S12_extracted_k141_48777 | Not-determined |
| S12_extracted_k141_41424 | Not-determined |
| S12_extracted_k141_135636 | Not-determined |
| S12_extracted_k141_174731 | Not-determined |
| S12_extracted_k141_48780 | Not-determined |
| S12_extracted_k141_174734 | Not-determined |
| S12_extracted_k141_48781 | Not-determined |
| S12_extracted_k141_65117 | Not-determined |
| S12_extracted_k141_167416 | Low-quality |
| S12_extracted_k141_65118 | Not-determined |
| S12_extracted_k141_174736 | Not-determined |
| S12_extracted_k141_151218 | Not-determined |
| S12_extracted_k141_135640 | Not-determined |
| S12_extracted_k141_151222 | Not-determined |
| S12_extracted_k141_17695 | Not-determined |
| S12_extracted_k141_111772 | Not-determined |
| S12_extracted_k141_151224 | Not-determined |
| S12_extracted_k141_80438 | Low-quality |
| S12_extracted_k141_151225 | Not-determined |
| S12_extracted_k141_174744 | Not-determined |
| S12_extracted_k141_183188 | Not-determined |
| S12_extracted_k141_2176 | Not-determined |
| S12_extracted_k141_41429 | Not-determined |
| S12_extracted_k141_80441 | Not-determined |
| S12_extracted_k141_111774 | Not-determined |
| S12_extracted_k141_25760 | Not-determined |
| S12_extracted_k141_183190 | Not-determined |
| S12_extracted_k141_2179 | Not-determined |
| S12_extracted_k141_111776 | Not-determined |
| S12_extracted_k141_120347 | Not-determined |
| S12_extracted_k141_167430 | Not-determined |
| S12_extracted_k141_33049 | Not-determined |
| S12_extracted_k141_9700 | Not-determined |
| S12_extracted_k141_88724 | Low-quality |
| S12_extracted_k141_174752 | Not-determined |
| S12_extracted_k141_9702 | Not-determined |
| S12_extracted_k141_17705 | Not-determined |
| S12_extracted_k141_48797 | Not-determined |
| S12_extracted_k141_9706 | Not-determined |
| S12_extracted_k141_17708 | Low-quality |
| S12_extracted_k141_88728 | Not-determined |
| S12_extracted_k141_183202 | Not-determined |
| S12_extracted_k141_17710 | Not-determined |
| S12_extracted_k141_151233 | Not-determined |
| S12_extracted_k141_135649 | Not-determined |
| S12_extracted_k141_48800 | Not-determined |
| S12_extracted_k141_41433 | Low-quality |
| S12_extracted_k141_17716 | Not-determined |
| S12_extracted_k141_183208 | Not-determined |
| S12_extracted_k141_88733 | Low-quality |
| S12_extracted_k141_159143 | Not-determined |
| S12_extracted_k141_183209 | Low-quality |
| S12_extracted_k141_111784 | Not-determined |
| S12_extracted_k141_159145 | Not-determined |
| S12_extracted_k141_2194 | Not-determined |
| S12_extracted_k141_9713 | Not-determined |
| S12_extracted_k141_17718 | Not-determined |
| S12_extracted_k141_2195 | Not-determined |
| S12_extracted_k141_33062 | Not-determined |
| S12_extracted_k141_25772 | Not-determined |
| S12_extracted_k141_41441 | Low-quality |
| S12_extracted_k141_159148 | Not-determined |
| S12_extracted_k141_25774 | Not-determined |
| S12_extracted_k141_159149 | Not-determined |
| S12_extracted_k141_57396 | Not-determined |
| S12_extracted_k141_159150 | Not-determined |
| S12_extracted_k141_2196 | Not-determined |
| S12_extracted_k141_57398 | Not-determined |
| S12_extracted_k141_33066 | Not-determined |
| S12_extracted_k141_120353 | Not-determined |
| S12_extracted_k141_9717 | Not-determined |
| S12_extracted_k141_88739 | Low-quality |
| S12_extracted_k141_96872 | Not-determined |
| S12_extracted_k141_104511 | Not-determined |
| S12_extracted_k141_65138 | Not-determined |
| S12_extracted_k141_120357 | Not-determined |
| S12_extracted_k141_104512 | Not-determined |
| S12_extracted_k141_80462 | Not-determined |
| S12_extracted_k141_96877 | Not-determined |
| S12_extracted_k141_17727 | Not-determined |
| S12_extracted_k141_104515 | Not-determined |
| S12_extracted_k141_111787 | Not-determined |
| S12_extracted_k141_159158 | Not-determined |
| S12_extracted_k141_17731 | Not-determined |
| S12_extracted_k141_174769 | Not-determined |
| S12_extracted_k141_167448 | Not-determined |
| S12_extracted_k141_65140 | Low-quality |
| S12_extracted_k141_120366 | Not-determined |
| S12_extracted_k141_25782 | Not-determined |
| S12_extracted_k141_33083 | Not-determined |
| S12_extracted_k141_167449 | Not-determined |
| S12_extracted_k141_151251 | Not-determined |
| S12_extracted_k141_159165 | Not-determined |
| S12_extracted_k141_111793 | Not-determined |
| S12_extracted_k141_80471 | Not-determined |
| S12_extracted_k141_33087 | Not-determined |
| S12_extracted_k141_135663 | Not-determined |
| S12_extracted_k141_48814 | Not-determined |
| S12_extracted_k141_111799 | Not-determined |
| S12_extracted_k141_174781 | Not-determined |

| | |
|---|---|
| S12_extracted_k141_151256 | Not-determined |
| S12_extracted_k141_33094 | Not-determined |
| S12_extracted_k141_174782 | Not-determined |
| S12_extracted_k141_88750 | Not-determined |
| S12_extracted_k141_65154 | Low-quality |
| S12_extracted_k141_88752 | Not-determined |
| S12_extracted_k141_135675 | Not-determined |
| S12_extracted_k141_183215 | Not-determined |
| S12_extracted_k141_120373 | Not-determined |
| S12_extracted_k141_174790 | Not-determined |
| S12_extracted_k141_120377 | Not-determined |
| S12_extracted_k141_127942 | Not-determined |
| S12_extracted_k141_88757 | Low-quality |
| S12_extracted_k141_25792 | Not-determined |
| S12_extracted_k141_65163 | Not-determined |
| S12_extracted_k141_143851 | Not-determined |
| S12_extracted_k141_96889 | Low-quality |
| S12_extracted_k141_48833 | Not-determined |
| S12_extracted_k141_57430 | Not-determined |
| S12_extracted_k141_65175 | Not-determined |
| S12_extracted_k141_33099 | Low-quality |
| S12_extracted_k141_2216 | Not-determined |
| S12_extracted_k141_151261 | Not-determined |
| S12_extracted_k141_2217 | Not-determined |
| S12_extracted_k141_41456 | Low-quality |
| S12_extracted_k141_48842 | Not-determined |
| S12_extracted_k141_183230 | Low-quality |
| S12_extracted_k141_57444 | Not-determined |
| S12_extracted_k141_183231 | Not-determined |
| S12_extracted_k141_57445 | Not-determined |
| S12_extracted_k141_151272 | Not-determined |
| S12_extracted_k141_80482 | Not-determined |
| S12_extracted_k141_96910 | Not-determined |
| S12_extracted_k141_151279 | Not-determined |
| S12_extracted_k141_48848 | Not-determined |
| S12_extracted_k141_48850 | Not-determined |
| S12_extracted_k141_80489 | Not-determined |
| S12_extracted_k141_174834 | Not-determined |
| S12_extracted_k141_80491 | Not-determined |
| S12_extracted_k141_151287 | Not-determined |
| S12_extracted_k141_120381 | Low-quality |
| S12_extracted_k141_96918 | Not-determined |
| S12_extracted_k141_135712 | Not-determined |
| S12_extracted_k141_80498 | Not-determined |
| S12_extracted_k141_96923 | Not-determined |
| S12_extracted_k141_41479 | Not-determined |
| S12_extracted_k141_80499 | Not-determined |
| S12_extracted_k141_33107 | Not-determined |
| S12_extracted_k141_135717 | Not-determined |
| S12_extracted_k141_41483 | Not-determined |
| S12_extracted_k141_33113 | Not-determined |
| S12_extracted_k141_135724 | Not-determined |
| S12_extracted_k141_88797 | Not-determined |
| S12_extracted_k141_120394 | Low-quality |
| S12_extracted_k141_96933 | Not-determined |
| S12_extracted_k141_41490 | Not-determined |
| S12_extracted_k141_96935 | Not-determined |
| S12_extracted_k141_33122 | Not-determined |
| S12_extracted_k141_120400 | Not-determined |
| S12_extracted_k141_120402 | Not-determined |
| S12_extracted_k141_143865 | Low-quality |
| S12_extracted_k141_88806 | Not-determined |
| S12_extracted_k141_174859 | Not-determined |
| S12_extracted_k141_25811 | Not-determined |
| S12_extracted_k141_120409 | Not-determined |
| S12_extracted_k141_120411 | Low-quality |
| S12_extracted_k141_174864 | Low-quality |
| S12_extracted_k141_135739 | Not-determined |
| S12_extracted_k141_88813 | Not-determined |
| S12_extracted_k141_33131 | Not-determined |
| S12_extracted_k141_120421 | Not-determined |
| S12_extracted_k141_174872 | Not-determined |
| S12_extracted_k141_127951 | Not-determined |
| S12_extracted_k141_159188 | Not-determined |
| S12_extracted_k141_25831 | Not-determined |
| S12_extracted_k141_143878 | Not-determined |
| S12_extracted_k141_159192 | Not-determined |
| S12_extracted_k141_80528 | Not-determined |
| S12_extracted_k141_159198 | Not-determined |
| S12_extracted_k141_120436 | Not-determined |
| S12_extracted_k141_120438 | Not-determined |
| S12_extracted_k141_127959 | Not-determined |
| S12_extracted_k141_57454 | Not-determined |
| S16_vs_162189\|\|full | Medium-quality |
| S12_extracted_k141_104518 | Not-determined |
| S12_extracted_k141_96940 | Low-quality |
| S12_extracted_k141_80537 | Not-determined |
| S12_extracted_k141_96942 | Not-determined |
| S12_extracted_k141_17753 | Not-determined |
| S12_extracted_k141_17754 | Not-determined |
| S12_extracted_k141_80541 | Not-determined |
| S12_extracted_k141_48872 | Not-determined |
| S12_extracted_k141_96945 | Not-determined |
| S12_extracted_k141_65195 | Not-determined |
| S12_extracted_k141_96949 | Not-determined |
| S12_extracted_k141_96957 | Not-determined |
| S12_extracted_k141_65201 | Not-determined |
| S12_extracted_k141_25843 | Not-determined |
| S12_extracted_k141_33157 | Not-determined |
| S12_extracted_k141_65204 | Not-determined |
| S12_extracted_k141_96959 | Not-determined |
| S12_extracted_k141_104525 | Not-determined |
| S12_extracted_k141_65206 | Not-determined |
| S12_extracted_k141_96960 | Not-determined |
| S12_extracted_k141_57475 | Not-determined |
| S12_extracted_k141_111822 | Low-quality |
| S12_extracted_k141_120454 | Low-quality |
| S12_extracted_k141_17767 | Not-determined |
| S12_extracted_k141_48894 | Low-quality |
| S12_extracted_k141_111829 | Low-quality |
| S12_extracted_k141_41532 | Not-determined |
| S12_extracted_k141_17771 | Not-determined |
| S12_extracted_k141_57482 | Not-determined |
| S12_extracted_k141_33170 | Not-determined |
| S12_extracted_k141_17774 | Low-quality |
| S12_extracted_k141_57485 | Not-determined |
| S12_extracted_k141_88839 | Not-determined |
| S12_extracted_k141_33173 | Not-determined |
| S12_extracted_k141_104532 | Not-determined |
| S12_extracted_k141_57487 | Not-determined |
| S12_extracted_k141_65219 | Not-determined |
| S12_extracted_k141_104539 | Not-determined |
| S12_extracted_k141_41546 | Not-determined |
| S12_extracted_k141_88857 | Not-determined |
| S12_extracted_k141_48899 | Not-determined |
| S12_extracted_k141_88861 | Not-determined |
| S12_extracted_k141_127971 | Not-determined |
| S12_extracted_k141_25850 | Not-determined |
| S12_extracted_k141_111840 | Not-determined |
| S12_extracted_k141_48908 | Not-determined |
| S12_extracted_k141_57496 | Not-determined |
| S12_extracted_k141_143940 | Not-determined |
| S12_extracted_k141_127973 | Not-determined |
| S12_extracted_k141_104550 | Not-determined |
| S12_extracted_k141_120488 | Not-determined |
| S12_extracted_k141_25856 | Not-determined |
| S12_extracted_k141_135744 | Not-determined |
| S12_extracted_k141_151293 | Not-determined |
| S12_extracted_k141_33190 | Not-determined |
| S12_extracted_k141_104559 | Not-determined |
| S7_vs_55526\|\|full | Medium-quality |
| S12_extracted_k141_120495 | Not-determined |
| S12_extracted_k141_120497 | Not-determined |
| S12_extracted_k141_88873 | Not-determined |
| S12_extracted_k141_88874 | Not-determined |
| S12_extracted_k141_80586 | Not-determined |
| S12_extracted_k141_33191 | Not-determined |
| S12_extracted_k141_120498 | Low-quality |
| S12_extracted_k141_135747 | Low-quality |
| S12_extracted_k141_135748 | Not-determined |
| S12_extracted_k141_48929 | Not-determined |
| S12_extracted_k141_65252 | Not-determined |
| S12_extracted_k141_48930 | Not-determined |
| S12_extracted_k141_65253 | Not-determined |
| S12_extracted_k141_80590 | Not-determined |
| S12_extracted_k141_48932 | Not-determined |
| S12_extracted_k141_80592 | Not-determined |
| S12_extracted_k141_104571 | Not-determined |
| S12_extracted_k141_127997 | Not-determined |
| S12_extracted_k141_80596 | Not-determined |
| S12_extracted_k141_80597 | Not-determined |
| S12_extracted_k141_65257 | Not-determined |
| S12_extracted_k141_80599 | Not-determined |
| S12_extracted_k141_120508 | Not-determined |
| S12_extracted_k141_2234 | Not-determined |
| S12_extracted_k141_104576 | Not-determined |
| S12_extracted_k141_48935 | Not-determined |
| S12_extracted_k141_135757 | Not-determined |
| S12_extracted_k141_65261 | Not-determined |
| S12_extracted_k141_80604 | Low-quality |
| S12_extracted_k141_41574 | Not-determined |
| S12_extracted_k141_17804 | Not-determined |
| S12_extracted_k141_183245 | Not-determined |
| S12_extracted_k141_2239 | Not-determined |
| S12_extracted_k141_135758 | Not-determined |
| S12_extracted_k141_183249 | Not-determined |
| S12_extracted_k141_41579 | Not-determined |
| S12_extracted_k141_183251 | Not-determined |
| S12_extracted_k141_17809 | Not-determined |
| S12_extracted_k141_183253 | Not-determined |
| S12_extracted_k141_97010 | Not-determined |
| S12_extracted_k141_57524 | Not-determined |
| S12_extracted_k141_88893 | Not-determined |
| S12_extracted_k141_143951 | Not-determined |
| S12_extracted_k141_97015 | Not-determined |
| S12_extracted_k141_57530 | Low-quality |
| S12_extracted_k141_57531 | Not-determined |
| S12_extracted_k141_183259 | Not-determined |
| S12_extracted_k141_97017 | Not-determined |
| S12_extracted_k141_104594 | Low-quality |
| S12_extracted_k141_88899 | Not-determined |
| S12_extracted_k141_41595 | Not-determined |
| S12_extracted_k141_128007 | Not-determined |
| S12_extracted_k141_104596 | Low-quality |
| S12_extracted_k141_151317 | Not-determined |
| S12_extracted_k141_104600 | Not-determined |
| S12_extracted_k141_97033 | Low-quality |
| S12_extracted_k141_25877 | Not-determined |
| S12_extracted_k141_88901 | Not-determined |
| S12_extracted_k141_17832 | Not-determined |
| S12_extracted_k141_97041 | Not-determined |
| S12_extracted_k141_183267 | Not-determined |
| S12_extracted_k141_17835 | Not-determined |
| S12_extracted_k141_174893 | Not-determined |
| S12_extracted_k141_57541 | Not-determined |
| S12_extracted_k141_88909 | Not-determined |
| S12_extracted_k141_57542 | Not-determined |
| S12_extracted_k141_135775 | Low-quality |
| S12_extracted_k141_57544 | Not-determined |
| S12_extracted_k141_128017 | Not-determined |
| S12_extracted_k141_57545 | Low-quality |
| S12_extracted_k141_128019 | Not-determined |
| S12_extracted_k141_97043 | Low-quality |
| S12_extracted_k141_135781 | Not-determined |
| S12_extracted_k141_97045 | Not-determined |
| S12_extracted_k141_135782 | Not-determined |
| S12_extracted_k141_97046 | Not-determined |
| S12_extracted_k141_174899 | Not-determined |
| S12_extracted_k141_97047 | Not-determined |
| S12_extracted_k141_135785 | Not-determined |
| S12_extracted_k141_41622 | Not-determined |
| S12_extracted_k141_135791 | Not-determined |
| S12_extracted_k141_88916 | Not-determined |
| S12_extracted_k141_57552 | Not-determined |
| S12_extracted_k141_88919 | Not-determined |
| S12_extracted_k141_174909 | Not-determined |
| S12_extracted_k141_80646 | Low-quality |
| S12_extracted_k141_174911 | Not-determined |
| S12_extracted_k141_65290 | Not-determined |
| S12_extracted_k141_159231 | Not-determined |
| S12_extracted_k141_128026 | Not-determined |
| S12_extracted_k141_151345 | Not-determined |
| S12_extracted_k141_128030 | Not-determined |
| S12_extracted_k141_159237 | Not-determined |
| S12_extracted_k141_135796 | Not-determined |
| S12_extracted_k141_174921 | Not-determined |
| S12_extracted_k141_41633 | Not-determined |
| S12_extracted_k141_159241 | Not-determined |
| S12_extracted_k141_174925 | Not-determined |
| S12_extracted_k141_151354 | Not-determined |
| S12_extracted_k141_128037 | Not-determined |
| S12_extracted_k141_41641 | Not-determined |
| S12_extracted_k141_128039 | Not-determined |
| S12_extracted_k141_41642 | Not-determined |
| S12_extracted_k141_135807 | Not-determined |
| S12_extracted_k141_135808 | Not-determined |
| S12_extracted_k141_135809 | Not-determined |
| S12_extracted_k141_2253 | Not-determined |
| S12_extracted_k141_174933 | Not-determined |
| S12_extracted_k141_174934 | Low-quality |
| S12_extracted_k141_17856 | Not-determined |
| S12_extracted_k141_174937 | Not-determined |
| S12_extracted_k141_80662 | Not-determined |
| S12_extracted_k141_41652 | Not-determined |
| S12_extracted_k141_104615 | Not-determined |
| S12_extracted_k141_159251 | Not-determined |
| S12_extracted_k141_80663 | Not-determined |
| S12_extracted_k141_2263 | Not-determined |
| S12_extracted_k141_104617 | Low-quality |
| S12_extracted_k141_65312 | Not-determined |
| S12_extracted_k141_128047 | Low-quality |
| S12_extracted_k141_25898 | Not-determined |
| S12_extracted_k141_159255 | Not-determined |
| S12_extracted_k141_17862 | Not-determined |
| S12_extracted_k141_48947 | Not-determined |
| S12_extracted_k141_48948 | Not-determined |
| S12_extracted_k141_17867 | Low-quality |
| S12_extracted_k141_80671 | Not-determined |
| S12_extracted_k141_57568 | Not-determined |
| S12_extracted_k141_41665 | Not-determined |
| S12_extracted_k141_33221 | Low-quality |
| S12_extracted_k141_120539 | Not-determined |
| S12_extracted_k141_41668 | Not-determined |
| S12_extracted_k141_33223 | Not-determined |
| S12_extracted_k141_2278 | Not-determined |
| S12_extracted_k141_104629 | Not-determined |
| S12_extracted_k141_33225 | Not-determined |
| S12_extracted_k141_159266 | Not-determined |
| S12_extracted_k141_159267 | Not-determined |
| S12_extracted_k141_143971 | Not-determined |
| S12_extracted_k141_120546 | Not-determined |
| S12_extracted_k141_80680 | Not-determined |
| S12_extracted_k141_2286 | Not-determined |
| S12_extracted_k141_159275 | Not-determined |
| S12_extracted_k141_104642 | Not-determined |
| S12_extracted_k141_183285 | Not-determined |
| S12_extracted_k141_159280 | Not-determined |
| S12_extracted_k141_33239 | Not-determined |
| S12_extracted_k141_57578 | Not-determined |
| S12_extracted_k141_57579 | Not-determined |
| S12_extracted_k141_159283 | Not-determined |
| S12_extracted_k141_104645 | Low-quality |
| S12_extracted_k141_135828 | Not-determined |
| S12_extracted_k141_104647 | Not-determined |
| S12_extracted_k141_135829 | Not-determined |
| S12_extracted_k141_48971 | Not-determined |
| S12_extracted_k141_159287 | Low-quality |
| S12_extracted_k141_97090 | Not-determined |
| S12_extracted_k141_48974 | Low-quality |
| S12_extracted_k141_97094 | Not-determined |
| S12_extracted_k141_80701 | Not-determined |
| S12_extracted_k141_120553 | Low-quality |
| S12_extracted_k141_111895 | Not-determined |
| S12_extracted_k141_88933 | Not-determined |
| S12_extracted_k141_151399 | Low-quality |
| S12_extracted_k141_183302 | Not-determined |
| S12_extracted_k141_80707 | Not-determined |
| S12_extracted_k141_135852 | Not-determined |
| S12_extracted_k141_111897 | Not-determined |
| S12_extracted_k141_88943 | Not-determined |
| S12_extracted_k141_88944 | Not-determined |
| S12_extracted_k141_2316 | Low-quality |
| S12_extracted_k141_128096 | Not-determined |
| S12_extracted_k141_104656 | Not-determined |
| S12_extracted_k141_41703 | Not-determined |
| S12_extracted_k141_174984 | Not-determined |
| S12_extracted_k141_104659 | Not-determined |
| S12_extracted_k141_17889 | Low-quality |
| S12_extracted_k141_17891 | Low-quality |
| S12_extracted_k141_80715 | Low-quality |
| S12_extracted_k141_120564 | Not-determined |
| S12_extracted_k141_80720 | Not-determined |
| S12_extracted_k141_57585 | Not-determined |
| S12_extracted_k141_33263 | Not-determined |
| S12_extracted_k141_175000 | Not-determined |
| S12_extracted_k141_80723 | Not-determined |
| S12_extracted_k141_128100 | Not-determined |
| S12_extracted_k141_57591 | Low-quality |
| S12_extracted_k141_88968 | Not-determined |
| S12_extracted_k141_135855 | Not-determined |
| S12_extracted_k141_135857 | Not-determined |
| S12_extracted_k141_97120 | Not-determined |
| S12_extracted_k141_97121 | Not-determined |
| S12_extracted_k141_183310 | Not-determined |
| S12_extracted_k141_57595 | Not-determined |
| S12_extracted_k141_57596 | Not-determined |
| S12_extracted_k141_33272 | Not-determined |
| S12_extracted_k141_88979 | Not-determined |
| S12_extracted_k141_80730 | Not-determined |
| S12_extracted_k141_49016 | Not-determined |
| S12_extracted_k141_80732 | Not-determined |
| S12_extracted_k141_88983 | Not-determined |
| S12_extracted_k141_120580 | Not-determined |
| S12_extracted_k141_151423 | Not-determined |
| S12_extracted_k141_49020 | Not-determined |
| S12_extracted_k141_25910 | Not-determined |
| S12_extracted_k141_143989 | Not-determined |
| S12_extracted_k141_175020 | Not-determined |
| S12_extracted_k141_111931 | Not-determined |
| S12_extracted_k141_65329 | Not-determined |
| S12_extracted_k141_135864 | Not-determined |
| S12_extracted_k141_151427 | Not-determined |
| S12_extracted_k141_33274 | Not-determined |
| S12_extracted_k141_143993 | Not-determined |
| S12_extracted_k141_33275 | Not-determined |
| S12_extracted_k141_65332 | Not-determined |
| S12_extracted_k141_88986 | Not-determined |
| S12_extracted_k141_183321 | Not-determined |
| S12_extracted_k141_135869 | Not-determined |
| S12_extracted_k141_25918 | Not-determined |
| S12_extracted_k141_41721 | Not-determined |
| S12_extracted_k141_2341 | Not-determined |
| S12_extracted_k141_17905 | Not-determined |
| S12_extracted_k141_144000 | Not-determined |
| S12_extracted_k141_183328 | Not-determined |
| S12_extracted_k141_144003 | Not-determined |
| S12_extracted_k141_183332 | Not-determined |
| S12_extracted_k141_135877 | Not-determined |
| S12_extracted_k141_120589 | Not-determined |
| S12_extracted_k141_65341 | Not-determined |
| S12_extracted_k141_2347 | Not-determined |
| S12_extracted_k141_25924 | Not-determined |
| S12_extracted_k141_135879 | Not-determined |
| S12_extracted_k141_135883 | Not-determined |
| S12_extracted_k141_25931 | Not-determined |
| S12_extracted_k141_151440 | Not-determined |
| S12_extracted_k141_135887 | Not-determined |
| S12_extracted_k141_104671 | Not-determined |
| S12_extracted_k141_183342 | Not-determined |
| S12_extracted_k141_183343 | Not-determined |
| S12_extracted_k141_135891 | Not-determined |
| S12_extracted_k141_151444 | Not-determined |
| S12_extracted_k141_183344 | Not-determined |
| S12_extracted_k141_183345 | Low-quality |
| S12_extracted_k141_183349 | Not-determined |
| S12_extracted_k141_65355 | Not-determined |
| S12_extracted_k141_135895 | Low-quality |
| S12_extracted_k141_111962 | Not-determined |
| S12_extracted_k141_88991 | Not-determined |
| S12_extracted_k141_135897 | Low-quality |
| S12_extracted_k141_65360 | Not-determined |
| S12_extracted_k141_144023 | Not-determined |
| S12_extracted_k141_104681 | Not-determined |
| S12_extracted_k141_65364 | Not-determined |
| S12_extracted_k141_128145 | Not-determined |
| S12_extracted_k141_65366 | Not-determined |
| S12_extracted_k141_128146 | Low-quality |
| S12_extracted_k141_111968 | Not-determined |
| S12_extracted_k141_65369 | Not-determined |
| S12_extracted_k141_49049 | Low-quality |
| S12_extracted_k141_111972 | Not-determined |
| S12_extracted_k141_104684 | Not-determined |
| S12_extracted_k141_135901 | Not-determined |
| S12_extracted_k141_33285 | Not-determined |
| S12_extracted_k141_25960 | Not-determined |
| S12_extracted_k141_89002 | Not-determined |
| S12_extracted_k141_97134 | Not-determined |
| S12_extracted_k141_25965 | Not-determined |
| S12_extracted_k141_104690 | Low-quality |
| S12_extracted_k141_128157 | Not-determined |
| S12_extracted_k141_25969 | Not-determined |
| S12_extracted_k141_97138 | Not-determined |
| S12_extracted_k141_25972 | Low-quality |
| S12_extracted_k141_25975 | Low-quality |
| S12_extracted_k141_41766 | Not-determined |
| S12_extracted_k141_97143 | Not-determined |
| S12_extracted_k141_17924 | Not-determined |
| S12_extracted_k141_128158 | Not-determined |
| S12_extracted_k141_183362 | Not-determined |
| S12_extracted_k141_144055 | Not-determined |
| S12_extracted_k141_25978 | Not-determined |
| S12_extracted_k141_144058 | Not-determined |
| S12_extracted_k141_2375 | Low-quality |
| S12_extracted_k141_183365 | Not-determined |
| S12_extracted_k141_17929 | Not-determined |
| S12_extracted_k141_41773 | Low-quality |
| S12_extracted_k141_2378 | Not-determined |
| S12_extracted_k141_104703 | Low-quality |
| S12_extracted_k141_89017 | Not-determined |
| S12_extracted_k141_111994 | Not-determined |
| S12_extracted_k141_128160 | Not-determined |
| S12_extracted_k141_89020 | Not-determined |
| S12_extracted_k141_135920 | Not-determined |
| S12_extracted_k141_25990 | Not-determined |
| S12_extracted_k141_17935 | Not-determined |
| S12_extracted_k141_97156 | Not-determined |
| S12_extracted_k141_65381 | Not-determined |
| S12_extracted_k141_2386 | Not-determined |
| S12_extracted_k141_183379 | Low-quality |
| S12_extracted_k141_183380 | Not-determined |
| S12_extracted_k141_183382 | Not-determined |
| S12_extracted_k141_183383 | Not-determined |
| S12_extracted_k141_183384 | Not-determined |
| S12_extracted_k141_41787 | Not-determined |
| S12_extracted_k141_183387 | Not-determined |
| S12_extracted_k141_151474 | Not-determined |
| S12_extracted_k141_183388 | Not-determined |
| S12_extracted_k141_57626 | Not-determined |
| S12_extracted_k141_159298 | Not-determined |
| S12_extracted_k141_17944 | Not-determined |
| S12_extracted_k141_57630 | Not-determined |
| S12_extracted_k141_151475 | Low-quality |
| S12_extracted_k141_65391 | Not-determined |
| S12_extracted_k141_80743 | Not-determined |
| S12_extracted_k141_41801 | Low-quality |
| S12_extracted_k141_89046 | Not-determined |
| S12_extracted_k141_80747 | Not-determined |
| S12_extracted_k141_80748 | Not-determined |
| S12_extracted_k141_144075 | Not-determined |
| S12_extracted_k141_80750 | Low-quality |
| S12_extracted_k141_41807 | Not-determined |
| S12_extracted_k141_144077 | Not-determined |
| S12_extracted_k141_159308 | Not-determined |
| S12_extracted_k141_65404 | Not-determined |
| S12_extracted_k141_159310 | Not-determined |
| S12_extracted_k141_151491 | Not-determined |
| S12_extracted_k141_89049 | Not-determined |
| S12_extracted_k141_128179 | Not-determined |
| S12_extracted_k141_128180 | Not-determined |
| S12_extracted_k141_89050 | Not-determined |
| S12_extracted_k141_151494 | Not-determined |
| S12_extracted_k141_151497 | Low-quality |
| S12_extracted_k141_33323 | Not-determined |
| S12_extracted_k141_80762 | Not-determined |
| S12_extracted_k141_159322 | Not-determined |
| S12_extracted_k141_112021 | Not-determined |
| S12_extracted_k141_159323 | Low-quality |
| S12_extracted_k141_26015 | Not-determined |
| S12_extracted_k141_104732 | Not-determined |
| S12_extracted_k141_128189 | Low-quality |
| S12_extracted_k141_80770 | Not-determined |
| S12_extracted_k141_104737 | Not-determined |
| S12_extracted_k141_57665 | Not-determined |
| S12_extracted_k141_128193 | Not-determined |
| S12_extracted_k141_159330 | Not-determined |
| S12_extracted_k141_57667 | Not-determined |
| S12_extracted_k141_112028 | Not-determined |
| S12_extracted_k141_112030 | Not-determined |
| S12_extracted_k141_72657 | Not-determined |
| S12_extracted_k141_72658 | Not-determined |
| S12_extracted_k141_49071 | Low-quality |
| S12_extracted_k141_144091 | Not-determined |
| S12_extracted_k141_97165 | Not-determined |
| S12_extracted_k141_159338 | Not-determined |
| S12_extracted_k141_97166 | Not-determined |
| S12_extracted_k141_159340 | Not-determined |
| S12_extracted_k141_151507 | Low-quality |
| S12_extracted_k141_175041 | Not-determined |
| S12_extracted_k141_112035 | Not-determined |
| S12_extracted_k141_104745 | Not-determined |
| S12_extracted_k141_65422 | Not-determined |
| S12_extracted_k141_151512 | Low-quality |
| S12_extracted_k141_128197 | Not-determined |
| S12_extracted_k141_65426 | Not-determined |
| S12_extracted_k141_80785 | Not-determined |
| S12_extracted_k141_151517 | Low-quality |
| S12_extracted_k141_112049 | Not-determined |
| S12_extracted_k141_97179 | Not-determined |
| S12_extracted_k141_65429 | Not-determined |
| S12_extracted_k141_183400 | Not-determined |
| S12_extracted_k141_33348 | Not-determined |
| S12_extracted_k141_49087 | Not-determined |
| S12_extracted_k141_183403 | Not-determined |
| S12_extracted_k141_33350 | Not-determined |
| S12_extracted_k141_104757 | Not-determined |
| S12_extracted_k141_65432 | Not-determined |
| S12_extracted_k141_80792 | Not-determined |
| S12_extracted_k141_183406 | Not-determined |
| S12_extracted_k141_65435 | Not-determined |
| S12_extracted_k141_128207 | Not-determined |
| S12_extracted_k141_97182 | Not-determined |
| S12_extracted_k141_104767 | Not-determined |
| S12_extracted_k141_33355 | Not-determined |
| S12_extracted_k141_49095 | Not-determined |
| S12_extracted_k141_183416 | Not-determined |
| S12_extracted_k141_183417 | Not-determined |
| S12_extracted_k141_128215 | Not-determined |
| S12_extracted_k141_135955 | Not-determined |
| S12_extracted_k141_97188 | Not-determined |
| S12_extracted_k141_33361 | Not-determined |
| S12_extracted_k141_41819 | Not-determined |
| S12_extracted_k141_175062 | Low-quality |
| S12_extracted_k141_49106 | Not-determined |
| S12_extracted_k141_104774 | Not-determined |
| S12_extracted_k141_41824 | Not-determined |
| S12_extracted_k141_120610 | Not-determined |
| S12_extracted_k141_104775 | Not-determined |
| S12_extracted_k141_175066 | Not-determined |
| S12_extracted_k141_72662 | Low-quality |
| S12_extracted_k141_120617 | Low-quality |
| S12_extracted_k141_41832 | Not-determined |
| S12_extracted_k141_128236 | Not-determined |
| S12_extracted_k141_80813 | Not-determined |
| S12_extracted_k141_57691 | Not-determined |
| S12_extracted_k141_80814 | Low-quality |
| S12_extracted_k141_183431 | Not-determined |
| S12_extracted_k141_80816 | Not-determined |
| S12_extracted_k141_72664 | Not-determined |
| S12_extracted_k141_120619 | Not-determined |
| S12_extracted_k141_41837 | Not-determined |
| S12_extracted_k141_72667 | Not-determined |
| S12_extracted_k141_97206 | Not-determined |
| S12_extracted_k141_135982 | Not-determined |
| S12_extracted_k141_144103 | Not-determined |
| S12_extracted_k141_97210 | Not-determined |
| S12_extracted_k141_89099 | Not-determined |
| S12_extracted_k141_72673 | Not-determined |
| S12_extracted_k141_97214 | Not-determined |
| S12_extracted_k141_120626 | Not-determined |
| S12_extracted_k141_135990 | Not-determined |
| S12_extracted_k141_57704 | Not-determined |
| S12_extracted_k141_135992 | Low-quality |
| S12_extracted_k141_97218 | Not-determined |
| S12_extracted_k141_183439 | Not-determined |
| S12_extracted_k141_17973 | Low-quality |
| S12_extracted_k141_41840 | Not-determined |
| S12_extracted_k141_57707 | Not-determined |
| S12_extracted_k141_144112 | Not-determined |
| S12_extracted_k141_17975 | Not-determined |
| S12_extracted_k141_144113 | Not-determined |
| S12_extracted_k141_104783 | Low-quality |
| S12_extracted_k141_72682 | Not-determined |
| S12_extracted_k141_135996 | Not-determined |
| S12_extracted_k141_120631 | Not-determined |
| S12_extracted_k141_151539 | Low-quality |
| S12_extracted_k141_57711 | Not-determined |
| S12_extracted_k141_151540 | Not-determined |
| S12_extracted_k141_41847 | Not-determined |
| S12_extracted_k141_104786 | Not-determined |
| S12_extracted_k141_136001 | Not-determined |
| S12_extracted_k141_57713 | Not-determined |
| S12_extracted_k141_151544 | Not-determined |
| S12_extracted_k141_49113 | Not-determined |
| S12_extracted_k141_17979 | Not-determined |
| S12_extracted_k141_151546 | Not-determined |
| S12_extracted_k141_26027 | Not-determined |
| S12_extracted_k141_151551 | Not-determined |
| S12_extracted_k141_120638 | Low-quality |
| S12_extracted_k141_89116 | Not-determined |

| | |
|---|---|
| S12_extracted_k141_136012 | Not-determined |
| S12_extracted_k141_57718 | Not-determined |
| S12_extracted_k141_183451 | Not-determined |
| S12_extracted_k141_144132 | Low-quality |
| S12_extracted_k141_136015 | Not-determined |
| S12_extracted_k141_89124 | Not-determined |
| S12_extracted_k141_72689 | Low-quality |
| S12_extracted_k141_97242 | Not-determined |
| S12_extracted_k141_136018 | Not-determined |
| S12_extracted_k141_151566 | Not-determined |
| S12_extracted_k141_41869 | Not-determined |
| S12_extracted_k141_104810 | Not-determined |
| S12_extracted_k141_175091 | Not-determined |
| S12_extracted_k141_151568 | Not-determined |
| S12_extracted_k141_80855 | Not-determined |
| S12_extracted_k141_89140 | Not-determined |
| S12_extracted_k141_80856 | Not-determined |
| S12_extracted_k141_65487 | Not-determined |
| S12_extracted_k141_97254 | Not-determined |
| S12_extracted_k141_151572 | Not-determined |
| S12_extracted_k141_2427 | Not-determined |
| S12_extracted_k141_183467 | Not-determined |
| S12_extracted_k141_183469 | Not-determined |
| S12_extracted_k141_97259 | Low-quality |
| S12_extracted_k141_120655 | Not-determined |
| S12_extracted_k141_80868 | Not-determined |
| S12_extracted_k141_120656 | Low-quality |
| S12_extracted_k141_120659 | Not-determined |
| S12_extracted_k141_18003 | Not-determined |
| S12_extracted_k141_112091 | Not-determined |
| S12_extracted_k141_89151 | Not-determined |
| S12_extracted_k141_65495 | Not-determined |
| S12_extracted_k141_136032 | Not-determined |
| S12_extracted_k141_41880 | Not-determined |
| S12_extracted_k141_26044 | Not-determined |
| S12_extracted_k141_2434 | Not-determined |
| S12_extracted_k141_151580 | Not-determined |
| S12_extracted_k141_167462 | Not-determined |
| S12_extracted_k141_151582 | Not-determined |
| S12_extracted_k141_136040 | Not-determined |
| S12_extracted_k141_136041 | Not-determined |
| S12_extracted_k141_120670 | Not-determined |
| S12_extracted_k141_112102 | Not-determined |
| S12_extracted_k141_151587 | Not-determined |
| S12_extracted_k141_80884 | Not-determined |
| S12_extracted_k141_120673 | Not-determined |
| S12_extracted_k141_80888 | Not-determined |
| S12_extracted_k141_80889 | Not-determined |
| S12_extracted_k141_167467 | Not-determined |
| S12_extracted_k141_49130 | Not-determined |
| S12_extracted_k141_144177 | Not-determined |
| S12_extracted_k141_26053 | Not-determined |
| S12_extracted_k141_89165 | Not-determined |
| S12_extracted_k141_49134 | Low-quality |
| S12_extracted_k141_136051 | Not-determined |
| S12_extracted_k141_9728 | Not-determined |
| S12_extracted_k141_33386 | Not-determined |
| S12_extracted_k141_80895 | Not-determined |
| S12_extracted_k141_26056 | Not-determined |
| S12_extracted_k141_112106 | Low-quality |
| S12_extracted_k141_159358 | Not-determined |
| S12_extracted_k141_18014 | Not-determined |
| S12_extracted_k141_112110 | Not-determined |
| S12_extracted_k141_26062 | Not-determined |
| S12_extracted_k141_89170 | Not-determined |
| S12_extracted_k141_151594 | Not-determined |
| S12_extracted_k141_151595 | Not-determined |
| S12_extracted_k141_41894 | Not-determined |
| S12_extracted_k141_167473 | Low-quality |
| S12_extracted_k141_49142 | Not-determined |
| S12_extracted_k141_104821 | Not-determined |
| S12_extracted_k141_80903 | Not-determined |
| S12_extracted_k141_144193 | Not-determined |
| S12_extracted_k141_104824 | Not-determined |
| S12_extracted_k141_89177 | Not-determined |
| S12_extracted_k141_136063 | Not-determined |
| S12_extracted_k141_104825 | Not-determined |
| S12_extracted_k141_104826 | Not-determined |
| S12_extracted_k141_65526 | Not-determined |
| S12_extracted_k141_104827 | Not-determined |
| S12_extracted_k141_9740 | Not-determined |
| S12_extracted_k141_151606 | Not-determined |
| S12_extracted_k141_65531 | Not-determined |
| S12_extracted_k141_80913 | Not-determined |
| S12_extracted_k141_136069 | Not-determined |
| S12_extracted_k141_49151 | Not-determined |
| S12_extracted_k141_72726 | Not-determined |
| S12_extracted_k141_112121 | Low-quality |
| S12_extracted_k141_57756 | Not-determined |
| S12_extracted_k141_72727 | Not-determined |
| S12_extracted_k141_112122 | Not-determined |
| S12_extracted_k141_183490 | Not-determined |
| S12_extracted_k141_57759 | Not-determined |
| S12_extracted_k141_9744 | Not-determined |
| S12_extracted_k141_112128 | Not-determined |
| S12_extracted_k141_2463 | Not-determined |
| S12_extracted_k141_49159 | Low-quality |
| S12_extracted_k141_41904 | Not-determined |
| S12_extracted_k141_65542 | Not-determined |
| S12_extracted_k141_97318 | Not-determined |
| S12_extracted_k141_2470 | Not-determined |
| S12_extracted_k141_136079 | Not-determined |
| S12_extracted_k141_26077 | Not-determined |
| S12_extracted_k141_80924 | Not-determined |
| S12_extracted_k141_41907 | Not-determined |
| S12_extracted_k141_159375 | Not-determined |
| S12_extracted_k141_183499 | Low-quality |
| S12_extracted_k141_2474 | Not-determined |
| S12_extracted_k141_183500 | Not-determined |
| S12_extracted_k141_80929 | Not-determined |
| S12_extracted_k141_49169 | Not-determined |
| S12_extracted_k141_9756 | Not-determined |
| S12_extracted_k141_89207 | Not-determined |
| S12_extracted_k141_41912 | Low-quality |
| S12_extracted_k141_65552 | Not-determined |
| S12_extracted_k141_41916 | Not-determined |
| S12_extracted_k141_112145 | Not-determined |
| S12_extracted_k141_41921 | Not-determined |
| S12_extracted_k141_89215 | Not-determined |
| S12_extracted_k141_89217 | Not-determined |
| S12_extracted_k141_159387 | Not-determined |
| S12_extracted_k141_18037 | Not-determined |
| S12_extracted_k141_183507 | Not-determined |
| S12_extracted_k141_112151 | Not-determined |
| S12_extracted_k141_72746 | Not-determined |
| S12_extracted_k141_159391 | Not-determined |
| S12_extracted_k141_41930 | Not-determined |
| S12_extracted_k141_41933 | Not-determined |
| S12_extracted_k141_9775 | Not-determined |
| S12_extracted_k141_41934 | Not-determined |
| S12_extracted_k141_72753 | Not-determined |
| S12_extracted_k141_89232 | Not-determined |
| S12_extracted_k141_18051 | Not-determined |
| S12_extracted_k141_89235 | Not-determined |
| S12_extracted_k141_18053 | Not-determined |
| S12_extracted_k141_159397 | Not-determined |
| S12_extracted_k141_167487 | Not-determined |
| S12_extracted_k141_57775 | Not-determined |
| S12_extracted_k141_72758 | Not-determined |
| S12_extracted_k141_2492 | Not-determined |
| S12_extracted_k141_57779 | Not-determined |
| S12_extracted_k141_97354 | Not-determined |
| S12_extracted_k141_33416 | Low-quality |
| S12_extracted_k141_183522 | Not-determined |
| S12_extracted_k141_167497 | Not-determined |
| S12_extracted_k141_57783 | Not-determined |
| S12_extracted_k141_159399 | Low-quality |
| S12_extracted_k141_57786 | Not-determined |
| S12_extracted_k141_33421 | Not-determined |
| S12_extracted_k141_89244 | Not-determined |
| S12_extracted_k141_97359 | Not-determined |
| S12_extracted_k141_41945 | Not-determined |
| S12_extracted_k141_183528 | Not-determined |
| S12_extracted_k141_183530 | Not-determined |
| S12_extracted_k141_112165 | Not-determined |
| S12_extracted_k141_57794 | Not-determined |
| S12_extracted_k141_65587 | Not-determined |
| S12_extracted_k141_65588 | Not-determined |
| S12_extracted_k141_128252 | Not-determined |
| S12_extracted_k141_65590 | Not-determined |
| S12_extracted_k141_2508 | Not-determined |
| S12_extracted_k141_159414 | Not-determined |
| S12_extracted_k141_144206 | Not-determined |
| S12_extracted_k141_183536 | Not-determined |
| S12_extracted_k141_2510 | Not-determined |
| S12_extracted_k141_89255 | Not-determined |
| S12_extracted_k141_41954 | Not-determined |
| S12_extracted_k141_167510 | Not-determined |
| S12_extracted_k141_80938 | Not-determined |
| S12_extracted_k141_167514 | Not-determined |
| S12_extracted_k141_128262 | Not-determined |
| S12_extracted_k141_120695 | Not-determined |
| S12_extracted_k141_80947 | Low-quality |
| S12_extracted_k141_65599 | Not-determined |
| S12_extracted_k141_167518 | Not-determined |
| S12_extracted_k141_41958 | Not-determined |
| S12_extracted_k141_97375 | Not-determined |
| S12_extracted_k141_167521 | Not-determined |
| S12_extracted_k141_159432 | Not-determined |
| S12_extracted_k141_80953 | Not-determined |
| S12_extracted_k141_136088 | Not-determined |
| S12_extracted_k141_18069 | Low-quality |
| S12_extracted_k141_18070 | Not-determined |
| S12_extracted_k141_18071 | Not-determined |
| S12_extracted_k141_144220 | Not-determined |
| S12_extracted_k141_2520 | Not-determined |
| S12_extracted_k141_89264 | Not-determined |
| S12_extracted_k141_128271 | Not-determined |
| S12_extracted_k141_65612 | Not-determined |
| S12_extracted_k141_97383 | Not-determined |
| S12_extracted_k141_18078 | Not-determined |
| S12_extracted_k141_89269 | Not-determined |
| S12_extracted_k141_26104 | Not-determined |
| S12_extracted_k141_33436 | Not-determined |
| S12_extracted_k141_18083 | Not-determined |
| S12_extracted_k141_159442 | Not-determined |
| S12_extracted_k141_65620 | Not-determined |
| S12_extracted_k141_33439 | Low-quality |
| S12_extracted_k141_144228 | Not-determined |
| S12_extracted_k141_175103 | Not-determined |
| S12_extracted_k141_33442 | Not-determined |
| S12_extracted_k141_104836 | Not-determined |
| S12_extracted_k141_136106 | Not-determined |
| S12_extracted_k141_159444 | Not-determined |
| S12_extracted_k141_97396 | Low-quality |
| S12_extracted_k141_144232 | Not-determined |
| S12_extracted_k141_136110 | Not-determined |
| S12_extracted_k141_97398 | Not-determined |
| S12_extracted_k141_175110 | Not-determined |
| S12_extracted_k141_183551 | Not-determined |
| S12_extracted_k141_72777 | Low-quality |
| S12_extracted_k141_33449 | Low-quality |
| S12_extracted_k141_183552 | Low-quality |
| S12_extracted_k141_183553 | Low-quality |
| S12_extracted_k141_33452 | Not-determined |
| S12_extracted_k141_159454 | Not-determined |
| S12_extracted_k141_175119 | Not-determined |
| S12_extracted_k141_41981 | Not-determined |
| S12_extracted_k141_18097 | Not-determined |
| S12_extracted_k141_33457 | Not-determined |
| S12_extracted_k141_33458 | Not-determined |
| S12_extracted_k141_183559 | Low-quality |
| S12_extracted_k141_183561 | Not-determined |
| S12_extracted_k141_175127 | Not-determined |
| S12_extracted_k141_2535 | Not-determined |
| S12_extracted_k141_175129 | Not-determined |
| S12_extracted_k141_18105 | Not-determined |
| S12_extracted_k141_33463 | Not-determined |
| S12_extracted_k141_159466 | Not-determined |
| S12_extracted_k141_18107 | Not-determined |
| S12_extracted_k141_89299 | Not-determined |
| S12_extracted_k141_18108 | Low-quality |
| S12_extracted_k141_80967 | Not-determined |
| S12_extracted_k141_41990 | Not-determined |
| S12_extracted_k141_183567 | Not-determined |
| S12_extracted_k141_72792 | Not-determined |
| S12_extracted_k141_80973 | Not-determined |
| S12_extracted_k141_112172 | Not-determined |
| S12_extracted_k141_175138 | Not-determined |
| S12_extracted_k141_175139 | Not-determined |
| S12_extracted_k141_112176 | Not-determined |
| S12_extracted_k141_41999 | Not-determined |
| S12_extracted_k141_42000 | Not-determined |
| S12_extracted_k141_18118 | Not-determined |
| S12_extracted_k141_128291 | Not-determined |
| S12_extracted_k141_72800 | Not-determined |
| S12_extracted_k141_159474 | Not-determined |
| S12_extracted_k141_128294 | Not-determined |
| S12_extracted_k141_175146 | Not-determined |
| S12_extracted_k141_128295 | Not-determined |
| S12_extracted_k141_136139 | Not-determined |
| S12_extracted_k141_128296 | Not-determined |
| S12_extracted_k141_42005 | Not-determined |
| S12_extracted_k141_136142 | Not-determined |
| S12_extracted_k141_42006 | Not-determined |
| S12_extracted_k141_18124 | Not-determined |
| S12_extracted_k141_136143 | Not-determined |
| S12_extracted_k141_167538 | Not-determined |
| S12_extracted_k141_42009 | Not-determined |
| S12_extracted_k141_175151 | Not-determined |
| S12_extracted_k141_9812 | Not-determined |
| S12_extracted_k141_42014 | Low-quality |
| S12_extracted_k141_104870 | Low-quality |
| S12_extracted_k141_175156 | Not-determined |
| S12_extracted_k141_72812 | Not-determined |
| S12_extracted_k141_175158 | Not-determined |
| S12_extracted_k141_97418 | Not-determined |
| S12_extracted_k141_97419 | Not-determined |
| S12_extracted_k141_72816 | Not-determined |
| S12_extracted_k141_112186 | Low-quality |
| S12_extracted_k141_167553 | Not-determined |
| S12_extracted_k141_72818 | Not-determined |
| S12_extracted_k141_80990 | Not-determined |
| S12_extracted_k141_183573 | Not-determined |
| S12_extracted_k141_128315 | Not-determined |
| S12_extracted_k141_97425 | Not-determined |
| S12_extracted_k141_57813 | Not-determined |
| S12_extracted_k141_144260 | Not-determined |
| S12_extracted_k141_18142 | Not-determined |
| S12_extracted_k141_144261 | Not-determined |
| S12_extracted_k141_89308 | Low-quality |
| S12_extracted_k141_57815 | Not-determined |
| S12_extracted_k141_42025 | Not-determined |
| S12_extracted_k141_144264 | Not-determined |
| S12_extracted_k141_167566 | Low-quality |
| S12_extracted_k141_167568 | Not-determined |
| S12_extracted_k141_33489 | Not-determined |
| S12_extracted_k141_167574 | Not-determined |
| S12_extracted_k141_175168 | Low-quality |
| S12_extracted_k141_33492 | Not-determined |
| S12_extracted_k141_167575 | Not-determined |
| S12_extracted_k141_97434 | Not-determined |
| S12_extracted_k141_42031 | Not-determined |
| S12_extracted_k141_144269 | Not-determined |
| S12_extracted_k141_9829 | Low-quality |
| S12_extracted_k141_183585 | Not-determined |
| S12_extracted_k141_80997 | Not-determined |
| S12_extracted_k141_18146 | Not-determined |
| S12_extracted_k141_2576 | Low-quality |
| S12_extracted_k141_65626 | Not-determined |
| S12_extracted_k141_144274 | Low-quality |
| S12_extracted_k141_57830 | Not-determined |
| S12_extracted_k141_151637 | Not-determined |
| S12_extracted_k141_175173 | Not-determined |
| S12_extracted_k141_2578 | Low-quality |
| S12_extracted_k141_57831 | Not-determined |
| S12_extracted_k141_144276 | Not-determined |
| S12_extracted_k141_26139 | Not-determined |
| S12_extracted_k141_33500 | Low-quality |
| S12_extracted_k141_183589 | Not-determined |
| S12_extracted_k141_183590 | Not-determined |
| S12_extracted_k141_2580 | Not-determined |
| S12_extracted_k141_26142 | Low-quality |
| S12_extracted_k141_26144 | Not-determined |
| S12_extracted_k141_2582 | Not-determined |
| S12_extracted_k141_167580 | Not-determined |
| S12_extracted_k141_57835 | Not-determined |
| S12_extracted_k141_65633 | Not-determined |
| S12_extracted_k141_81002 | Not-determined |
| S12_extracted_k141_72839 | Not-determined |
| S12_extracted_k141_183595 | Not-determined |
| S12_extracted_k141_42045 | Not-determined |
| S12_extracted_k141_26146 | Not-determined |
| S12_extracted_k141_57838 | Not-determined |
| S12_extracted_k141_42048 | Not-determined |
| S12_extracted_k141_167588 | Not-determined |
| S12_extracted_k141_57841 | Not-determined |
| S12_extracted_k141_42052 | Not-determined |
| S12_extracted_k141_26153 | Not-determined |
| S12_extracted_k141_97440 | Low-quality |
| S12_extracted_k141_18159 | Not-determined |
| S12_extracted_k141_81008 | Not-determined |
| S12_extracted_k141_57847 | Not-determined |
| S12_extracted_k141_65640 | Not-determined |
| S12_extracted_k141_104900 | Not-determined |
| S12_extracted_k141_175193 | Not-determined |
| S12_extracted_k141_26159 | Not-determined |
| S12_extracted_k141_144279 | Not-determined |
| S12_extracted_k141_33504 | Not-determined |
| S12_extracted_k141_183605 | Not-determined |
| S12_extracted_k141_104901 | Not-determined |
| S12_extracted_k141_136161 | Not-determined |
| S12_extracted_k141_65647 | Not-determined |
| S12_extracted_k141_89332 | Not-determined |
| S12_extracted_k141_136162 | Not-determined |
| S12_extracted_k141_183607 | Not-determined |
| S12_extracted_k141_81012 | Not-determined |
| S12_extracted_k141_175196 | Not-determined |
| S12_extracted_k141_136163 | Not-determined |
| S12_extracted_k141_89335 | Not-determined |
| S12_extracted_k141_183610 | Not-determined |
| S12_extracted_k141_65656 | Not-determined |
| S12_extracted_k141_18172 | Not-determined |
| S12_extracted_k141_175204 | Not-determined |
| S12_extracted_k141_159499 | Not-determined |
| S12_extracted_k141_33506 | Not-determined |
| S12_extracted_k141_2592 | Not-determined |
| S12_extracted_k141_81023 | Not-determined |
| S12_extracted_k141_136171 | Low-quality |
| S12_extracted_k141_65663 | Not-determined |
| S12_extracted_k141_136172 | Low-quality |
| S12_extracted_k141_175209 | Low-quality |
| S12_extracted_k141_81027 | Not-determined |
| S12_extracted_k141_144294 | Not-determined |
| S12_extracted_k141_57867 | Not-determined |
| S12_extracted_k141_18182 | Not-determined |
| S12_extracted_k141_57869 | Not-determined |
| S12_extracted_k141_136178 | Not-determined |
| S12_extracted_k141_57873 | Not-determined |
| S12_extracted_k141_2609 | Not-determined |
| S12_extracted_k141_2611 | Low-quality |
| S12_extracted_k141_72850 | Not-determined |
| S12_extracted_k141_26187 | Not-determined |
| S12_extracted_k141_97457 | Not-determined |
| S12_extracted_k141_57881 | Not-determined |
| S12_extracted_k141_104920 | Not-determined |
| S12_extracted_k141_72854 | Not-determined |
| S12_extracted_k141_89359 | Not-determined |
| S12_extracted_k141_2619 | Not-determined |
| S12_extracted_k141_57887 | Low-quality |
| S12_extracted_k141_33512 | Not-determined |
| S12_extracted_k141_175219 | Not-determined |
| S12_extracted_k141_89367 | Not-determined |
| S12_extracted_k141_2624 | Not-determined |
| S12_extracted_k141_33517 | Not-determined |
| S12_extracted_k141_2625 | Not-determined |
| S12_extracted_k141_175221 | Not-determined |
| S12_extracted_k141_2629 | Not-determined |
| S12_extracted_k141_136198 | Not-determined |
| S12_extracted_k141_33524 | Not-determined |
| S12_extracted_k141_104937 | Not-determined |
| S12_extracted_k141_175231 | Not-determined |
| S12_extracted_k141_159530 | Not-determined |
| S12_extracted_k141_57897 | Not-determined |
| S12_extracted_k141_167619 | Not-determined |
| S12_extracted_k141_81045 | Low-quality |
| S12_extracted_k141_57898 | Not-determined |
| S12_extracted_k141_42067 | Not-determined |
| S12_extracted_k141_151641 | Not-determined |
| S12_extracted_k141_65692 | Not-determined |
| S12_extracted_k141_72859 | Not-determined |
| S12_extracted_k141_151647 | Not-determined |
| S12_extracted_k141_136212 | Low-quality |
| S12_extracted_k141_2642 | Not-determined |
| S12_extracted_k141_72865 | Not-determined |
| S12_extracted_k141_151656 | Not-determined |
| S12_extracted_k141_9882 | Not-determined |
| S12_extracted_k141_151658 | Not-determined |
| S12_extracted_k141_151660 | Not-determined |
| S12_extracted_k141_104947 | Not-determined |
| S12_extracted_k141_89388 | Not-determined |
| S12_extracted_k141_42083 | Not-determined |
| S12_extracted_k141_57908 | Not-determined |
| S12_extracted_k141_104953 | Not-determined |
| S12_extracted_k141_9888 | Not-determined |
| S12_extracted_k141_57909 | Not-determined |
| S12_extracted_k141_167630 | Low-quality |
| S12_extracted_k141_81064 | Not-determined |
| S12_extracted_k141_167633 | Not-determined |
| S12_extracted_k141_167634 | Not-determined |
| S12_extracted_k141_151671 | Low-quality |
| S12_extracted_k141_49181 | Not-determined |
| S12_extracted_k141_104962 | Low-quality |
| S12_extracted_k141_167641 | Not-determined |
| S12_extracted_k141_136223 | Not-determined |
| S12_extracted_k141_144302 | Not-determined |
| S12_extracted_k141_2669 | Not-determined |
| S12_extracted_k141_144306 | Not-determined |
| S12_extracted_k141_81076 | Not-determined |
| S12_extracted_k141_144308 | Not-determined |
| S12_extracted_k141_57920 | Low-quality |
| S12_extracted_k141_183614 | Not-determined |
| S12_extracted_k141_49193 | Not-determined |
| S12_extracted_k141_42103 | Not-determined |
| S12_extracted_k141_167649 | Not-determined |
| S12_extracted_k141_49196 | Not-determined |
| S12_extracted_k141_33564 | Not-determined |
| S12_extracted_k141_175269 | Not-determined |
| S12_extracted_k141_33568 | Not-determined |
| S12_extracted_k141_104976 | Not-determined |
| S12_extracted_k141_144315 | Low-quality |
| S12_extracted_k141_42104 | Not-determined |
| S12_extracted_k141_42107 | Not-determined |
| S12_extracted_k141_144318 | Not-determined |
| S12_extracted_k141_42109 | Not-determined |
| S12_extracted_k141_2675 | Not-determined |
| S12_extracted_k141_144320 | Not-determined |
| S12_extracted_k141_65732 | Not-determined |
| S12_extracted_k141_175275 | Not-determined |
| S12_extracted_k141_159566 | Not-determined |
| S12_extracted_k141_167665 | Not-determined |
| S12_extracted_k141_97467 | Not-determined |
| S12_extracted_k141_49204 | Not-determined |
| S12_extracted_k141_57928 | Not-determined |
| S12_extracted_k141_49205 | Not-determined |
| S12_extracted_k141_112196 | Not-determined |
| S12_extracted_k141_183633 | Not-determined |
| S12_extracted_k141_57931 | Low-quality |
| S12_extracted_k141_89421 | Low-quality |
| S12_extracted_k141_49210 | Not-determined |
| S12_extracted_k141_57934 | Not-determined |
| S12_extracted_k141_9912 | Not-determined |
| S12_extracted_k141_144331 | Not-determined |
| S12_extracted_k141_57943 | Not-determined |
| S12_extracted_k141_104985 | Not-determined |
| S12_extracted_k141_175291 | Not-determined |
| S12_extracted_k141_112207 | Low-quality |
| S12_extracted_k141_18210 | Not-determined |
| S12_extracted_k141_57947 | Not-determined |
| S12_extracted_k141_9917 | Low-quality |
| S12_extracted_k141_144337 | Not-determined |
| S12_extracted_k141_57948 | Not-determined |
| S12_extracted_k141_167681 | Not-determined |
| S12_extracted_k141_18212 | Not-determined |
| S12_extracted_k141_183646 | Not-determined |
| S12_extracted_k141_183647 | Not-determined |
| S12_extracted_k141_9921 | Not-determined |
| S12_extracted_k141_9922 | Not-determined |
| S12_extracted_k141_9923 | Not-determined |
| S12_extracted_k141_167684 | Low-quality |
| S12_extracted_k141_42134 | Not-determined |
| S12_extracted_k141_81115 | Low-quality |
| S12_extracted_k141_81116 | Not-determined |
| S12_extracted_k141_9926 | Not-determined |
| S12_extracted_k141_33589 | Not-determined |
| S12_extracted_k141_144344 | Not-determined |
| S12_extracted_k141_144345 | Not-determined |
| S12_extracted_k141_49225 | Not-determined |
| S12_extracted_k141_81124 | Not-determined |
| S12_extracted_k141_49226 | Not-determined |
| S12_extracted_k141_112219 | Not-determined |
| S12_extracted_k141_81125 | Not-determined |
| S12_extracted_k141_112220 | Not-determined |
| S12_extracted_k141_136256 | Low-quality |
| S12_extracted_k141_9938 | Not-determined |
| S12_extracted_k141_105001 | Not-determined |
| S12_extracted_k141_112227 | Not-determined |
| S12_extracted_k141_33591 | Not-determined |
| S7_vs_28733\|\|full | High-quality |
| S12_extracted_k141_167688 | Low-quality |
| S12_extracted_k141_159576 | Not-determined |
| S12_extracted_k141_105007 | Not-determined |
| S12_extracted_k141_183666 | Not-determined |
| S12_extracted_k141_144364 | Not-determined |
| S12_extracted_k141_18240 | Not-determined |
| S12_extracted_k141_49229 | Not-determined |
| S12_extracted_k141_49230 | Not-determined |
| S12_extracted_k141_144372 | Not-determined |
| S12_extracted_k141_112245 | Low-quality |
| S12_extracted_k141_42165 | Not-determined |
| S12_extracted_k141_144374 | Not-determined |
| S12_extracted_k141_112247 | Not-determined |
| S12_extracted_k141_18250 | Not-determined |
| S12_extracted_k141_33614 | Not-determined |
| S12_extracted_k141_81144 | Not-determined |
| S12_extracted_k141_49234 | Not-determined |
| S12_extracted_k141_42175 | Not-determined |
| S12_extracted_k141_72888 | Not-determined |
| S12_extracted_k141_136279 | Not-determined |
| S12_extracted_k141_144376 | Low-quality |
| S12_extracted_k141_159581 | Not-determined |
| S12_extracted_k141_42178 | Low-quality |
| S12_extracted_k141_57953 | Not-determined |
| S12_extracted_k141_57954 | Not-determined |
| S12_extracted_k141_183693 | Not-determined |
| S12_extracted_k141_42184 | Not-determined |
| S12_extracted_k141_183697 | Not-determined |
| S12_extracted_k141_159587 | Not-determined |
| S12_extracted_k141_183698 | Not-determined |
| S12_extracted_k141_112261 | Low-quality |
| S12_extracted_k141_72895 | Not-determined |
| S12_extracted_k141_81153 | Not-determined |
| S12_extracted_k141_33628 | Not-determined |
| S12_extracted_k141_167707 | Not-determined |
| S12_extracted_k141_72899 | Low-quality |
| S12_extracted_k141_144384 | Not-determined |
| S12_extracted_k141_72900 | Not-determined |
| S12_extracted_k141_81161 | Not-determined |
| S12_extracted_k141_136287 | Not-determined |
| S12_extracted_k141_9954 | Not-determined |
| S12_extracted_k141_33632 | Not-determined |
| S12_extracted_k141_112263 | Low-quality |
| S12_extracted_k141_33638 | Not-determined |
| S12_extracted_k141_33642 | Not-determined |
| S12_extracted_k141_81168 | Not-determined |
| S12_extracted_k141_105011 | Not-determined |
| S12_extracted_k141_144393 | Not-determined |
| S12_extracted_k141_33644 | Low-quality |
| S12_extracted_k141_144394 | Not-determined |
| S12_extracted_k141_33645 | Not-determined |
| S12_extracted_k141_33649 | Not-determined |
| S12_extracted_k141_144397 | Not-determined |
| S12_extracted_k141_18258 | Low-quality |
| S12_extracted_k141_72921 | Not-determined |
| S12_extracted_k141_33652 | Low-quality |
| S12_extracted_k141_57966 | Not-determined |
| S12_extracted_k141_128349 | Not-determined |
| S12_extracted_k141_72927 | Low-quality |
| S12_extracted_k141_128350 | Low-quality |
| S12_extracted_k141_159617 | Not-determined |
| S12_extracted_k141_136298 | Not-determined |
| S12_extracted_k141_81184 | Not-determined |
| S12_extracted_k141_112288 | Not-determined |
| S12_extracted_k141_105035 | Not-determined |
| S12_extracted_k141_72928 | Not-determined |
| S12_extracted_k141_112291 | Not-determined |
| S12_extracted_k141_159624 | Low-quality |
| S12_extracted_k141_33663 | Not-determined |
| S12_extracted_k141_136304 | Low-quality |
| S12_extracted_k141_42191 | Not-determined |
| S12_extracted_k141_136305 | Not-determined |
| S12_extracted_k141_159628 | Not-determined |
| S12_extracted_k141_89463 | Not-determined |
| S12_extracted_k141_136306 | Not-determined |
| S12_extracted_k141_183715 | Not-determined |
| S12_extracted_k141_72933 | Not-determined |
| S12_extracted_k141_112298 | Not-determined |
| S12_extracted_k141_159635 | Not-determined |
| S12_extracted_k141_167726 | Not-determined |
| S12_extracted_k141_57984 | Not-determined |
| S12_extracted_k141_105040 | Not-determined |
| S12_extracted_k141_159636 | Low-quality |
| S12_extracted_k141_112305 | Not-determined |
| S12_extracted_k141_144422 | Not-determined |
| S12_extracted_k141_2700 | Not-determined |
| S12_extracted_k141_89476 | Not-determined |
| S12_extracted_k141_42206 | Not-determined |
| S12_extracted_k141_49259 | Not-determined |
| S12_extracted_k141_9977 | Not-determined |

| | |
|---|---|
| S12_extracted_k141_81206 | Not-determined |
| S12_extracted_k141_105054 | Not-determined |
| S12_extracted_k141_9981 | Low-quality |
| S12_extracted_k141_65771 | Not-determined |
| S12_extracted_k141_49265 | Not-determined |
| S12_extracted_k141_136324 | Not-determined |
| S12_extracted_k141_9985 | Not-determined |
| S12_extracted_k141_42213 | Not-determined |
| S12_extracted_k141_9988 | Not-determined |
| S12_extracted_k141_42215 | Not-determined |
| S12_extracted_k141_105056 | Not-determined |
| S12_extracted_k141_128373 | Not-determined |
| S12_extracted_k141_81211 | Not-determined |
| S12_extracted_k141_183728 | Not-determined |
| S12_extracted_k141_128375 | Not-determined |
| S12_extracted_k141_89481 | Not-determined |
| S12_extracted_k141_89482 | Not-determined |
| S12_extracted_k141_49273 | Not-determined |
| S12_extracted_k141_2709 | Not-determined |
| S12_extracted_k141_18294 | Not-determined |
| S12_extracted_k141_2710 | Not-determined |
| S12_extracted_k141_9995 | Not-determined |
| S12_extracted_k141_151742 | Not-determined |
| S12_extracted_k141_33697 | Not-determined |
| S12_extracted_k141_26241 | Not-determined |
| S12_extracted_k141_128380 | Not-determined |
| S12_extracted_k141_18297 | Not-determined |
| S12_extracted_k141_151744 | Not-determined |
| S12_extracted_k141_159642 | Low-quality |
| S12_extracted_k141_144425 | Not-determined |
| S12_extracted_k141_65786 | Not-determined |
| S12_extracted_k141_159644 | Low-quality |
| S12_extracted_k141_49280 | Not-determined |
| S12_extracted_k141_18299 | Not-determined |
| S12_extracted_k141_58001 | Not-determined |
| S12_extracted_k141_112327 | Not-determined |
| S12_extracted_k141_33706 | Not-determined |
| S12_extracted_k141_183744 | Not-determined |
| S12_extracted_k141_89491 | Low-quality |
| S12_extracted_k141_18303 | Not-determined |
| S12_extracted_k141_183745 | Not-determined |
| S12_extracted_k141_175323 | Not-determined |
| S12_extracted_k141_105071 | Not-determined |
| S12_extracted_k141_151751 | Low-quality |
| S12_extracted_k141_81220 | Not-determined |
| S12_extracted_k141_105077 | Not-determined |
| S12_extracted_k141_128390 | Not-determined |
| S12_extracted_k141_26254 | Not-determined |
| S12_extracted_k141_183748 | Not-determined |
| S12_extracted_k141_26257 | Not-determined |
| S12_extracted_k141_18312 | Not-determined |
| S12_extracted_k141_175330 | Not-determined |
| S12_extracted_k141_42227 | Not-determined |
| S12_extracted_k141_42234 | Not-determined |
| S12_extracted_k141_97520 | Not-determined |
| S12_extracted_k141_26263 | Not-determined |
| S12_extracted_k141_2717 | Low-quality |
| S12_extracted_k141_18314 | Not-determined |
| S12_extracted_k141_26269 | Not-determined |
| S12_extracted_k141_49292 | Low-quality |
| S12_extracted_k141_49293 | Not-determined |
| S12_extracted_k141_128396 | Not-determined |
| S12_extracted_k141_167741 | Not-determined |
| S12_extracted_k141_128398 | Not-determined |
| S12_extracted_k141_42237 | Not-determined |
| S12_extracted_k141_167743 | Not-determined |
| S12_extracted_k141_167744 | Not-determined |
| S12_extracted_k141_112350 | Not-determined |
| S12_extracted_k141_151770 | Not-determined |
| S12_extracted_k141_10008 | Not-determined |
| S12_extracted_k141_136328 | Not-determined |
| S12_extracted_k141_58026 | Low-quality |
| S12_extracted_k141_58027 | Not-determined |
| S12_extracted_k141_10011 | Not-determined |
| S12_extracted_k141_151777 | Not-determined |
| S12_extracted_k141_18325 | Not-determined |
| S12_extracted_k141_42241 | Not-determined |
| S12_extracted_k141_128403 | Not-determined |
| S12_extracted_k141_175348 | Not-determined |
| S12_extracted_k141_18327 | Not-determined |
| S12_extracted_k141_167751 | Not-determined |
| S12_extracted_k141_175349 | Not-determined |
| S12_extracted_k141_159668 | Not-determined |
| S12_extracted_k141_175351 | Not-determined |
| S12_extracted_k141_58035 | Low-quality |
| S12_extracted_k141_167755 | Not-determined |
| S12_extracted_k141_2734 | Not-determined |
| S12_extracted_k141_89517 | Not-determined |
| S12_extracted_k141_2735 | Not-determined |
| S12_extracted_k141_10018 | Not-determined |
| S12_extracted_k141_151783 | Not-determined |
| S12_extracted_k141_151785 | Not-determined |
| S12_extracted_k141_175358 | Not-determined |
| S12_extracted_k141_175359 | Not-determined |
| S12_extracted_k141_58041 | Not-determined |
| S12_extracted_k141_26292 | Not-determined |
| S12_extracted_k141_183758 | Not-determined |
| S12_extracted_k141_175366 | Not-determined |
| S12_extracted_k141_183760 | Low-quality |
| S12_extracted_k141_183761 | Not-determined |
| S12_extracted_k141_58044 | Not-determined |
| S12_extracted_k141_42262 | Not-determined |
| S12_extracted_k141_175372 | Not-determined |
| S12_extracted_k141_42264 | Not-determined |
| S12_extracted_k141_183767 | Not-determined |
| S12_extracted_k141_10024 | Not-determined |
| S12_extracted_k141_97535 | Not-determined |
| S12_extracted_k141_72947 | Low-quality |
| S12_extracted_k141_128422 | Low-quality |
| S12_extracted_k141_42274 | Not-determined |
| S12_extracted_k141_33745 | Not-determined |
| S12_extracted_k141_58051 | Not-determined |
| S12_extracted_k141_159681 | Not-determined |
| S12_extracted_k141_2761 | Not-determined |
| S12_extracted_k141_128431 | Not-determined |
| S12_extracted_k141_26302 | Not-determined |
| S12_extracted_k141_2766 | Not-determined |
| S12_extracted_k141_26303 | Not-determined |
| S12_extracted_k141_58055 | Not-determined |
| S12_extracted_k141_81260 | Not-determined |
| S12_extracted_k141_18355 | Not-determined |
| S12_extracted_k141_18357 | Not-determined |
| S12_extracted_k141_2778 | Not-determined |
| S12_extracted_k141_10037 | Not-determined |
| S12_extracted_k141_10039 | Low-quality |
| S12_extracted_k141_2780 | Not-determined |
| S12_extracted_k141_183780 | Not-determined |
| S12_extracted_k141_49321 | Not-determined |
| S12_extracted_k141_49322 | Not-determined |
| S12_extracted_k141_2783 | Not-determined |
| S12_extracted_k141_49324 | Not-determined |
| S12_extracted_k141_72967 | Low-quality |
| S12_extracted_k141_183786 | Not-determined |
| S12_extracted_k141_183788 | Not-determined |
| S12_extracted_k141_10045 | Low-quality |
| S12_extracted_k141_10046 | Not-determined |
| S12_extracted_k141_128437 | Low-quality |
| S12_extracted_k141_65799 | Low-quality |
| S12_extracted_k141_72973 | Not-determined |
| S12_extracted_k141_58070 | Not-determined |
| S12_extracted_k141_167773 | Not-determined |
| S12_extracted_k141_72979 | Not-determined |
| S12_extracted_k141_112368 | Not-determined |
| S12_extracted_k141_72982 | Not-determined |
| S12_extracted_k141_89548 | Not-determined |
| S12_extracted_k141_136339 | Low-quality |
| S12_extracted_k141_58078 | Not-determined |
| S12_extracted_k141_2797 | Not-determined |
| S12_extracted_k141_97546 | Not-determined |
| S12_extracted_k141_2799 | Not-determined |
| S12_extracted_k141_49341 | Not-determined |
| S12_extracted_k141_81280 | Not-determined |
| S12_extracted_k141_89550 | Not-determined |
| S12_extracted_k141_65812 | Not-determined |
| S12_extracted_k141_128442 | Not-determined |
| S12_extracted_k141_58079 | Not-determined |
| S12_extracted_k141_97550 | Low-quality |
| S12_extracted_k141_2800 | Not-determined |
| S12_extracted_k141_167785 | Not-determined |
| S12_extracted_k141_10055 | Low-quality |
| S12_extracted_k141_89557 | Not-determined |
| S12_extracted_k141_49349 | Not-determined |
| S12_extracted_k141_42293 | Not-determined |
| S12_extracted_k141_2806 | Not-determined |
| S12_extracted_k141_42294 | Not-determined |
| S12_extracted_k141_136353 | Not-determined |
| S12_extracted_k141_105107 | Not-determined |
| S12_extracted_k141_81297 | Not-determined |
| S12_extracted_k141_2808 | Not-determined |
| S12_extracted_k141_81298 | Not-determined |
| S12_extracted_k141_136355 | Not-determined |
| S12_extracted_k141_128453 | Not-determined |
| S12_extracted_k141_58091 | Not-determined |
| S12_extracted_k141_42301 | Not-determined |
| S12_extracted_k141_151796 | Not-determined |
| S12_extracted_k141_2811 | Not-determined |
| S12_extracted_k141_167800 | Not-determined |
| S12_extracted_k141_151798 | Not-determined |
| S12_extracted_k141_73000 | Not-determined |
| S12_extracted_k141_58095 | Not-determined |
| S12_extracted_k141_151799 | Not-determined |
| S12_extracted_k141_136361 | Not-determined |
| S12_extracted_k141_18368 | Not-determined |
| S12_extracted_k141_58098 | Low-quality |
| S12_extracted_k141_58099 | Not-determined |
| S12_extracted_k141_151804 | Not-determined |
| S12_extracted_k141_18373 | Not-determined |
| S12_extracted_k141_105119 | Not-determined |
| S12_extracted_k141_105124 | Not-determined |
| S12_extracted_k141_136372 | Not-determined |
| S12_extracted_k141_81305 | Not-determined |
| S12_extracted_k141_42314 | Not-determined |
| S12_extracted_k141_151811 | Not-determined |
| S12_extracted_k141_167821 | Not-determined |
| S12_extracted_k141_10076 | Not-determined |
| S12_extracted_k141_167824 | Not-determined |
| S12_extracted_k141_81307 | Low-quality |
| S12_extracted_k141_97581 | Not-determined |
| S12_extracted_k141_2819 | Not-determined |
| S12_extracted_k141_49369 | Low-quality |
| S12_extracted_k141_81308 | Low-quality |
| S12_extracted_k141_10079 | Not-determined |
| S12_extracted_k141_10081 | Not-determined |
| S12_extracted_k141_97586 | Not-determined |
| S12_extracted_k141_151821 | Not-determined |
| S12_extracted_k141_49377 | Low-quality |
| S12_extracted_k141_58108 | Not-determined |
| S12_extracted_k141_97592 | Not-determined |
| S12_extracted_k141_167831 | Not-determined |
| S12_extracted_k141_183828 | Not-determined |
| S12_extracted_k141_65855 | Low-quality |
| S12_extracted_k141_151829 | Not-determined |
| S12_extracted_k141_105137 | Not-determined |
| S12_extracted_k141_18395 | Not-determined |
| S12_extracted_k141_10099 | Low-quality |
| S12_extracted_k141_18396 | Not-determined |
| S12_extracted_k141_73029 | Not-determined |
| S12_extracted_k141_49383 | Not-determined |
| S12_extracted_k141_183837 | Not-determined |
| S12_extracted_k141_58118 | Not-determined |
| S12_extracted_k141_10102 | Not-determined |
| S12_extracted_k141_183838 | Not-determined |
| S12_extracted_k141_105146 | Not-determined |
| S12_extracted_k141_73031 | Not-determined |
| S12_extracted_k141_136393 | Not-determined |
| S12_extracted_k141_105149 | Not-determined |
| S12_extracted_k141_183839 | Low-quality |
| S12_extracted_k141_151833 | Not-determined |
| S12_extracted_k141_18402 | Not-determined |
| S12_extracted_k141_183841 | Not-determined |
| S12_extracted_k141_89572 | Low-quality |
| S12_extracted_k141_89573 | Not-determined |
| S12_extracted_k141_183846 | Not-determined |
| S12_extracted_k141_183847 | Not-determined |
| S12_extracted_k141_42344 | Not-determined |
| S12_extracted_k141_112410 | Not-determined |
| S12_extracted_k141_183849 | Low-quality |
| S12_extracted_k141_42346 | Not-determined |
| S12_extracted_k141_65866 | Low-quality |
| S12_extracted_k141_183850 | Low-quality |
| S12_extracted_k141_42349 | Not-determined |
| S12_extracted_k141_65868 | Not-determined |
| S12_extracted_k141_97613 | Not-determined |
| S12_extracted_k141_42353 | Not-determined |
| S12_extracted_k141_97614 | Not-determined |
| S12_extracted_k141_73055 | Not-determined |
| S12_extracted_k141_42358 | Not-determined |
| S12_extracted_k141_81340 | Not-determined |
| S12_extracted_k141_18421 | Not-determined |
| S12_extracted_k141_167845 | Not-determined |
| S12_extracted_k141_65880 | Low-quality |
| S12_extracted_k141_167847 | Not-determined |
| S12_extracted_k141_167849 | Not-determined |
| S12_extracted_k141_18425 | Not-determined |
| S12_extracted_k141_105160 | Low-quality |
| S12_extracted_k141_89593 | Low-quality |
| S12_extracted_k141_2841 | Not-determined |
| S12_extracted_k141_73061 | Not-determined |
| S12_extracted_k141_42365 | Not-determined |
| S12_extracted_k141_167857 | Not-determined |
| S12_extracted_k141_167859 | Not-determined |
| S12_extracted_k141_167861 | Not-determined |
| S12_extracted_k141_183864 | Low-quality |
| S12_extracted_k141_136418 | Low-quality |
| S12_extracted_k141_183866 | Low-quality |
| S12_extracted_k141_167869 | Not-determined |
| S12_extracted_k141_136428 | Not-determined |
| S12_extracted_k141_167880 | Not-determined |
| S12_extracted_k141_105185 | Not-determined |
| S12_extracted_k141_136434 | Not-determined |
| S12_extracted_k141_89607 | Not-determined |
| S12_extracted_k141_2853 | Not-determined |
| S12_extracted_k141_183876 | Not-determined |
| S12_extracted_k141_2855 | Not-determined |
| S12_extracted_k141_2856 | Not-determined |
| S12_extracted_k141_167883 | Not-determined |
| S12_extracted_k141_183878 | Not-determined |
| S12_extracted_k141_167884 | Not-determined |
| S12_extracted_k141_2860 | Not-determined |
| S12_extracted_k141_159709 | Not-determined |
| S12_extracted_k141_42385 | Not-determined |
| S12_extracted_k141_183885 | Low-quality |
| S12_extracted_k141_49398 | Low-quality |
| S12_extracted_k141_105199 | Not-determined |
| S12_extracted_k141_49399 | Not-determined |
| S12_extracted_k141_2865 | Not-determined |
| S12_extracted_k141_73072 | Not-determined |
| S12_extracted_k141_49410 | Not-determined |
| S12_extracted_k141_2875 | Low-quality |
| S12_extracted_k141_159722 | Not-determined |
| S12_extracted_k141_105208 | Not-determined |
| S12_extracted_k141_167899 | Not-determined |
| S12_extracted_k141_73077 | Not-determined |
| S12_extracted_k141_159727 | Not-determined |
| S12_extracted_k141_18441 | Not-determined |
| S12_extracted_k141_42395 | Not-determined |
| S12_extracted_k141_49417 | Not-determined |
| S12_extracted_k141_159728 | Not-determined |
| S12_extracted_k141_2887 | Not-determined |
| S12_extracted_k141_112461 | Not-determined |
| S12_extracted_k141_159733 | Not-determined |
| S12_extracted_k141_112463 | Not-determined |
| S12_extracted_k141_18451 | Not-determined |
| S12_extracted_k141_26342 | Not-determined |
| S12_extracted_k141_112467 | Not-determined |
| S12_extracted_k141_151842 | Not-determined |
| S12_extracted_k141_49432 | Not-determined |
| S12_extracted_k141_73091 | Not-determined |
| S12_extracted_k141_167923 | Not-determined |
| S12_extracted_k141_89635 | Not-determined |
| S12_extracted_k141_151851 | Not-determined |
| S12_extracted_k141_58158 | Not-determined |
| S12_extracted_k141_18462 | Not-determined |
| S12_extracted_k141_183904 | Not-determined |
| S12_extracted_k141_49440 | Not-determined |
| S12_extracted_k141_58160 | Not-determined |
| S12_extracted_k141_2902 | Not-determined |
| S12_extracted_k141_26354 | Not-determined |
| S12_extracted_k141_2907 | Not-determined |
| S12_extracted_k141_112487 | Not-determined |
| S12_extracted_k141_167928 | Not-determined |
| S12_extracted_k141_112488 | Not-determined |
| S12_extracted_k141_105237 | Not-determined |
| S12_extracted_k141_26364 | Not-determined |
| S12_extracted_k141_159755 | Not-determined |
| S12_extracted_k141_105238 | Not-determined |
| S12_extracted_k141_26367 | Low-quality |
| S12_extracted_k141_167931 | Not-determined |
| S12_extracted_k141_89656 | Not-determined |
| S12_extracted_k141_97650 | Not-determined |
| S12_extracted_k141_49461 | Not-determined |
| S12_extracted_k141_58166 | Not-determined |
| S12_extracted_k141_136445 | Not-determined |
| S12_extracted_k141_26373 | Not-determined |
| S12_extracted_k141_97654 | Not-determined |
| S12_extracted_k141_89664 | Not-determined |
| S12_extracted_k141_151860 | Low-quality |
| S12_extracted_k141_58175 | Low-quality |
| S12_extracted_k141_65897 | Not-determined |
| S12_extracted_k141_26378 | Not-determined |
| S12_extracted_k141_159766 | Not-determined |
| S12_extracted_k141_18483 | Not-determined |
| S12_extracted_k141_128458 | Low-quality |
| S12_extracted_k141_105245 | Not-determined |
| S12_extracted_k141_49468 | Not-determined |
| S12_extracted_k141_2909 | Not-determined |
| S12_extracted_k141_18487 | Not-determined |
| S12_extracted_k141_49470 | Not-determined |
| S12_extracted_k141_128460 | Not-determined |
| S12_extracted_k141_18488 | Not-determined |
| S12_extracted_k141_49473 | Not-determined |
| S12_extracted_k141_58182 | Not-determined |
| S12_extracted_k141_18493 | Not-determined |
| S12_extracted_k141_18495 | Not-determined |
| S12_extracted_k141_144431 | Not-determined |
| S12_extracted_k141_18499 | Not-determined |
| S12_extracted_k141_105254 | Not-determined |
| S12_extracted_k141_18502 | Not-determined |
| S12_extracted_k141_33753 | Not-determined |
| S12_extracted_k141_65904 | Not-determined |
| S12_extracted_k141_112503 | Not-determined |
| S12_extracted_k141_65907 | Not-determined |
| S12_extracted_k141_58192 | Not-determined |
| S12_extracted_k141_89679 | Not-determined |
| S12_extracted_k141_97666 | Not-determined |
| S12_extracted_k141_33755 | Not-determined |
| S12_extracted_k141_33760 | Not-determined |
| S12_extracted_k141_65914 | Not-determined |
| S12_extracted_k141_112513 | Not-determined |
| S12_extracted_k141_2928 | Not-determined |
| S12_extracted_k141_73136 | Not-determined |
| S12_extracted_k141_2931 | Not-determined |
| S12_extracted_k141_49491 | Not-determined |
| S12_extracted_k141_49492 | Not-determined |
| S12_extracted_k141_105271 | Not-determined |
| S12_extracted_k141_33767 | Not-determined |
| S12_extracted_k141_183924 | Not-determined |
| S12_extracted_k141_33769 | Low-quality |
| S12_extracted_k141_58196 | Not-determined |
| S12_extracted_k141_112524 | Not-determined |
| S12_extracted_k141_151867 | Not-determined |
| S9_vs_k141_19689\|\|full | Medium-quality |
| S12_extracted_k141_65929 | Not-determined |
| S12_extracted_k141_33775 | Not-determined |
| S12_extracted_k141_136467 | Not-determined |
| S12_extracted_k141_183928 | Not-determined |
| S12_extracted_k141_151874 | Not-determined |
| S12_extracted_k141_18531 | Not-determined |
| S12_extracted_k141_49504 | Not-determined |
| S12_extracted_k141_49506 | Not-determined |
| S12_extracted_k141_167955 | Not-determined |
| S12_extracted_k141_18533 | Not-determined |
| S12_extracted_k141_49508 | Not-determined |
| S12_extracted_k141_175399 | Not-determined |
| S12_extracted_k141_18535 | Not-determined |
| S12_extracted_k141_58203 | Not-determined |
| S12_extracted_k141_65939 | Not-determined |
| S12_extracted_k141_144456 | Not-determined |
| S12_extracted_k141_105280 | Not-determined |
| S12_extracted_k141_97680 | Not-determined |
| S12_extracted_k141_42412 | Not-determined |
| S12_extracted_k141_136480 | Low-quality |
| S12_extracted_k141_151884 | Not-determined |
| S12_extracted_k141_49511 | Not-determined |
| S12_extracted_k141_144463 | Not-determined |
| S12_extracted_k141_151888 | Not-determined |
| S12_extracted_k141_112541 | Not-determined |
| S12_extracted_k141_167967 | Not-determined |
| S12_extracted_k141_167970 | Not-determined |
| S12_extracted_k141_128489 | Not-determined |
| S12_extracted_k141_58213 | Not-determined |
| S12_extracted_k141_18548 | Not-determined |
| S12_extracted_k141_42418 | Not-determined |
| S12_extracted_k141_167975 | Not-determined |
| S12_extracted_k141_183944 | Not-determined |
| S12_extracted_k141_97695 | Not-determined |
| S12_extracted_k141_183946 | Not-determined |
| S12_extracted_k141_175411 | Not-determined |
| S12_extracted_k141_73155 | Not-determined |
| S12_extracted_k141_159777 | Not-determined |
| S12_extracted_k141_2955 | Not-determined |
| S12_extracted_k141_112551 | Not-determined |
| S12_extracted_k141_65956 | Not-determined |
| S12_extracted_k141_26405 | Not-determined |
| S12_extracted_k141_151904 | Low-quality |
| S12_extracted_k141_2959 | Not-determined |
| S12_extracted_k141_81383 | Not-determined |
| S12_extracted_k141_42424 | Not-determined |
| S12_extracted_k141_120710 | Not-determined |
| S12_extracted_k141_112555 | Not-determined |
| S12_extracted_k141_33803 | Not-determined |
| S12_extracted_k141_2963 | Not-determined |
| S12_extracted_k141_2964 | Not-determined |
| S12_extracted_k141_42428 | Not-determined |
| S12_extracted_k141_2965 | Not-determined |
| S12_extracted_k141_120714 | Low-quality |
| S12_extracted_k141_105303 | Not-determined |
| S12_extracted_k141_159781 | Not-determined |
| S12_extracted_k141_97713 | Not-determined |
| S12_extracted_k141_65963 | Not-determined |
| S12_extracted_k141_120717 | Not-determined |
| S12_extracted_k141_97715 | Not-determined |
| S12_extracted_k141_58226 | Not-determined |
| S12_extracted_k141_159785 | Low-quality |
| S12_extracted_k141_26415 | Low-quality |
| S12_extracted_k141_175418 | Not-determined |
| S12_extracted_k141_151917 | Not-determined |
| S12_extracted_k141_2968 | Not-determined |
| S12_extracted_k141_65969 | Not-determined |
| S12_extracted_k141_112563 | Not-determined |
| S12_extracted_k141_112565 | Not-determined |
| S12_extracted_k141_151921 | Not-determined |
| S12_extracted_k141_42440 | Not-determined |
| S12_extracted_k141_65975 | Not-determined |
| S12_extracted_k141_120724 | Not-determined |
| S12_extracted_k141_159796 | Not-determined |
| S12_extracted_k141_42445 | Not-determined |
| S12_extracted_k141_58236 | Not-determined |
| S12_extracted_k141_42449 | Not-determined |
| S12_extracted_k141_58237 | Not-determined |
| S12_extracted_k141_167998 | Not-determined |
| S12_extracted_k141_49522 | Not-determined |
| S12_extracted_k141_128512 | Not-determined |
| S12_extracted_k141_151925 | Not-determined |
| S12_extracted_k141_151927 | Not-determined |
| S12_extracted_k141_42459 | Not-determined |
| S12_extracted_k141_97734 | Not-determined |
| S12_extracted_k141_65984 | Not-determined |
| S12_extracted_k141_136519 | Not-determined |
| S12_extracted_k141_97736 | Not-determined |
| S12_extracted_k141_151932 | Not-determined |
| S12_extracted_k141_49528 | Not-determined |
| S12_extracted_k141_33821 | Not-determined |
| S12_extracted_k141_175436 | Not-determined |
| S12_extracted_k141_97739 | Not-determined |
| S12_extracted_k141_33823 | Not-determined |
| S12_extracted_k141_42471 | Not-determined |
| S12_extracted_k141_136527 | Not-determined |
| S12_extracted_k141_18590 | Not-determined |
| S12_extracted_k141_120731 | Not-determined |
| S12_extracted_k141_128524 | Not-determined |
| S12_extracted_k141_49533 | Not-determined |
| S12_extracted_k141_136536 | Not-determined |
| S12_extracted_k141_33828 | Not-determined |
| S12_extracted_k141_42474 | Not-determined |
| S12_extracted_k141_168014 | Not-determined |
| S12_extracted_k141_49541 | Not-determined |
| S12_extracted_k141_112583 | Not-determined |
| S12_extracted_k141_105311 | Low-quality |
| S12_extracted_k141_112584 | Not-determined |
| S12_extracted_k141_136541 | Low-quality |
| S12_extracted_k141_120736 | Not-determined |
| S12_extracted_k141_112587 | Not-determined |
| S12_extracted_k141_159827 | Not-determined |
| S12_extracted_k141_128536 | Not-determined |
| S12_extracted_k141_49544 | Not-determined |
| S12_extracted_k141_175441 | Not-determined |
| S12_extracted_k141_18602 | Not-determined |
| S12_extracted_k141_159837 | Not-determined |
| S12_extracted_k141_151951 | Not-determined |
| S12_extracted_k141_112595 | Not-determined |
| S12_extracted_k141_97751 | Not-determined |
| S12_extracted_k141_168017 | Low-quality |
| S12_extracted_k141_159840 | Not-determined |
| S12_extracted_k141_97754 | Not-determined |
| S12_extracted_k141_112598 | Low-quality |
| S12_extracted_k141_42492 | Not-determined |
| S12_extracted_k141_33839 | Low-quality |
| S12_extracted_k141_42493 | Not-determined |
| S12_extracted_k141_58248 | Not-determined |
| S12_extracted_k141_159844 | Not-determined |
| S12_extracted_k141_183956 | Not-determined |
| S12_extracted_k141_159846 | Not-determined |
| S12_extracted_k141_151962 | Not-determined |
| S12_extracted_k141_159852 | Not-determined |
| S12_extracted_k141_183964 | Not-determined |
| S12_extracted_k141_18617 | Not-determined |
| S12_extracted_k141_49557 | Not-determined |
| S12_extracted_k141_18619 | Not-determined |
| S12_extracted_k141_144472 | Not-determined |
| S12_extracted_k141_120748 | Not-determined |
| S12_extracted_k141_49561 | Not-determined |
| S12_extracted_k141_112612 | Not-determined |
| S12_extracted_k141_97774 | Not-determined |
| S12_extracted_k141_168024 | Not-determined |
| S12_extracted_k141_168027 | Not-determined |
| S12_extracted_k141_151970 | Not-determined |
| S12_extracted_k141_26421 | Not-determined |
| S12_extracted_k141_136546 | Not-determined |
| S12_extracted_k141_26423 | Not-determined |
| S12_extracted_k141_49569 | Not-determined |
| S12_extracted_k141_120758 | Not-determined |
| S12_extracted_k141_159868 | Not-determined |
| S12_extracted_k141_33851 | Not-determined |
| S12_extracted_k141_168030 | Not-determined |
| S12_extracted_k141_97779 | Not-determined |
| S12_extracted_k141_136554 | Not-determined |
| S12_extracted_k141_42508 | Not-determined |
| S12_extracted_k141_112618 | Not-determined |
| S12_extracted_k141_144485 | Not-determined |
| S12_extracted_k141_175465 | Not-determined |
| S12_extracted_k141_136556 | Not-determined |
| S12_extracted_k141_42513 | Low-quality |
| S12_extracted_k141_66000 | Not-determined |
| S12_extracted_k141_144486 | Not-determined |
| S12_extracted_k141_42515 | Not-determined |
| S12_extracted_k141_42516 | Not-determined |
| S12_extracted_k141_42517 | Not-determined |
| S12_extracted_k141_144488 | Not-determined |
| S12_extracted_k141_42519 | Not-determined |
| S12_extracted_k141_128554 | Not-determined |
| S12_extracted_k141_175469 | Not-determined |
| S12_extracted_k141_97784 | Not-determined |
| S12_extracted_k141_105339 | Not-determined |
| S12_extracted_k141_66005 | Low-quality |
| S12_extracted_k141_58271 | Not-determined |
| S12_extracted_k141_183970 | Not-determined |
| S12_extracted_k141_58272 | Low-quality |
| S12_extracted_k141_151975 | Not-determined |
| S12_extracted_k141_18645 | Not-determined |
| S12_extracted_k141_33864 | Not-determined |
| S12_extracted_k141_97790 | Not-determined |
| S12_extracted_k141_105342 | Not-determined |
| S12_extracted_k141_97795 | Not-determined |
| S12_extracted_k141_151979 | Not-determined |
| S12_extracted_k141_183976 | Not-determined |
| S12_extracted_k141_97798 | Not-determined |
| S12_extracted_k141_183977 | Not-determined |
| S12_extracted_k141_105346 | Not-determined |
| S12_extracted_k141_66007 | Not-determined |
| S12_extracted_k141_18655 | Not-determined |
| S12_extracted_k141_175489 | Not-determined |
| S12_extracted_k141_42529 | Not-determined |
| S12_extracted_k141_159886 | Low-quality |
| S12_extracted_k141_183993 | Not-determined |
| S12_extracted_k141_33874 | Low-quality |
| S12_extracted_k141_89702 | Not-determined |
| S12_extracted_k141_42535 | Not-determined |
| S12_extracted_k141_2988 | Not-determined |
| S12_extracted_k141_33879 | Not-determined |
| S12_extracted_k141_58283 | Not-determined |
| S12_extracted_k141_81392 | Not-determined |
| S12_extracted_k141_97816 | Not-determined |
| S12_extracted_k141_58286 | Not-determined |
| S12_extracted_k141_159893 | Not-determined |
| S12_extracted_k141_105355 | Not-determined |
| S12_extracted_k141_3000 | Not-determined |
| S12_extracted_k141_175501 | Not-determined |
| S12_extracted_k141_3002 | Not-determined |

| | |
|---|---|
| S12_extracted_k141_73163 | Not-determined |
| S12_extracted_k141_97825 | Not-determined |
| S12_extracted_k141_33888 | Not-determined |
| S12_extracted_k141_73165 | Not-determined |
| S12_extracted_k141_128572 | Low-quality |
| S12_extracted_k141_49591 | Not-determined |
| S12_extracted_k141_144500 | Not-determined |
| S12_extracted_k141_144502 | Not-determined |
| S12_extracted_k141_49592 | Not-determined |
| S12_extracted_k141_89720 | Not-determined |
| S12_extracted_k141_33896 | Not-determined |
| S12_extracted_k141_105364 | Not-determined |
| S12_extracted_k141_49595 | Not-determined |
| S12_extracted_k141_112623 | Not-determined |
| S12_extracted_k141_97834 | Not-determined |
| S12_extracted_k141_97837 | Not-determined |
| S12_extracted_k141_58303 | Low-quality |
| S12_extracted_k141_175515 | Not-determined |
| S12_extracted_k141_18665 | Not-determined |
| S12_extracted_k141_81413 | Not-determined |
| S12_extracted_k141_112624 | Not-determined |
| S12_extracted_k141_112625 | Low-quality |
| S12_extracted_k141_3014 | Low-quality |
| S12_extracted_k141_66017 | Not-determined |
| S12_extracted_k141_105370 | Not-determined |
| S12_extracted_k141_49605 | Not-determined |
| S12_extracted_k141_120769 | Not-determined |
| S12_extracted_k141_18679 | Not-determined |
| S12_extracted_k141_42545 | Not-determined |
| S12_extracted_k141_33910 | Not-determined |
| S12_extracted_k141_73187 | Not-determined |
| S12_extracted_k141_152014 | Not-determined |
| S12_extracted_k141_152015 | Low-quality |
| S12_extracted_k141_175522 | Not-determined |
| S12_extracted_k141_49608 | Not-determined |
| S12_extracted_k141_144519 | Not-determined |
| S12_extracted_k141_152017 | Low-quality |
| S12_extracted_k141_144521 | Not-determined |
| S12_extracted_k141_26434 | Not-determined |
| S12_extracted_k141_175529 | Not-determined |
| S12_extracted_k141_112629 | Not-determined |
| S12_extracted_k141_42551 | Not-determined |
| S12_extracted_k141_18684 | Not-determined |
| S12_extracted_k141_184005 | Not-determined |
| S12_extracted_k141_26437 | Not-determined |
| S12_extracted_k141_144526 | Not-determined |
| S12_extracted_k141_3019 | Low-quality |
| S12_extracted_k141_184007 | Not-determined |
| S12_extracted_k141_42555 | Low-quality |
| S12_extracted_k141_18691 | Not-determined |
| S12_extracted_k141_159916 | Not-determined |
| S12_extracted_k141_184009 | Low-quality |
| S12_extracted_k141_159918 | Not-determined |
| S12_extracted_k141_159920 | Not-determined |
| S12_extracted_k141_3029 | Not-determined |
| S12_extracted_k141_66033 | Not-determined |
| S12_extracted_k141_159923 | Not-determined |
| S12_extracted_k141_18698 | Not-determined |
| S12_extracted_k141_3031 | Low-quality |
| S12_extracted_k141_89747 | Not-determined |
| S12_extracted_k141_184012 | Not-determined |
| S12_extracted_k141_175541 | Not-determined |
| S12_extracted_k141_58317 | Not-determined |
| S12_extracted_k141_26449 | Not-determined |
| S12_extracted_k141_120779 | Not-determined |
| S12_extracted_k141_184015 | Not-determined |
| S12_extracted_k141_120784 | Not-determined |
| S12_extracted_k141_26453 | Not-determined |
| S12_extracted_k141_120786 | Low-quality |
| S12_extracted_k141_33934 | Not-determined |
| S12_extracted_k141_152025 | Not-determined |
| S12_extracted_k141_3039 | Not-determined |
| S12_extracted_k141_3041 | Not-determined |
| S12_extracted_k141_168082 | Not-determined |
| S12_extracted_k141_152027 | Not-determined |
| S12_extracted_k141_33938 | Not-determined |
| S12_extracted_k141_89759 | Low-quality |
| S12_extracted_k141_184017 | Not-determined |
| S12_extracted_k141_168084 | Not-determined |
| S12_extracted_k141_58326 | Not-determined |
| S12_extracted_k141_26467 | Low-quality |
| S12_extracted_k141_58330 | Not-determined |
| S12_extracted_k141_184020 | Not-determined |
| S12_extracted_k141_58332 | Not-determined |
| S12_extracted_k141_136589 | Not-determined |
| S12_extracted_k141_26471 | Not-determined |
| S12_extracted_k141_81424 | Not-determined |
| S12_extracted_k141_112661 | Not-determined |
| S12_extracted_k141_49616 | Not-determined |
| S12_extracted_k141_105412 | Not-determined |
| S12_extracted_k141_168091 | Not-determined |
| S12_extracted_k141_120805 | Not-determined |
| S12_extracted_k141_128576 | Not-determined |
| S12_extracted_k141_89774 | Not-determined |
| S12_extracted_k141_42573 | Not-determined |
| S12_extracted_k141_81425 | Low-quality |
| S12_extracted_k141_58335 | Not-determined |
| S12_extracted_k141_58337 | Not-determined |
| S12_extracted_k141_49623 | Low-quality |
| S12_extracted_k141_42580 | Not-determined |
| S12_extracted_k141_105421 | Not-determined |
| S12_extracted_k141_159937 | Not-determined |
| S12_extracted_k141_89776 | Not-determined |
| S12_extracted_k141_136599 | Not-determined |
| S12_extracted_k141_168100 | Not-determined |
| S12_extracted_k141_3053 | Not-determined |
| S12_extracted_k141_136602 | Not-determined |
| S12_extracted_k141_112679 | Not-determined |
| S12_extracted_k141_3059 | Not-determined |
| S12_extracted_k141_128583 | Not-determined |
| S12_extracted_k141_112681 | Not-determined |
| S12_extracted_k141_73216 | Not-determined |
| S12_extracted_k141_97876 | Not-determined |
| S12_extracted_k141_89786 | Not-determined |
| S12_extracted_k141_81432 | Not-determined |
| S12_extracted_k141_112684 | Not-determined |
| S12_extracted_k141_73222 | Not-determined |
| S12_extracted_k141_159955 | Not-determined |
| S12_extracted_k141_105437 | Not-determined |
| S12_extracted_k141_33967 | Not-determined |
| S12_extracted_k141_3064 | Low-quality |
| S12_extracted_k141_49640 | Not-determined |
| S12_extracted_k141_159957 | Not-determined |
| S12_extracted_k141_18732 | Not-determined |
| S12_extracted_k141_26500 | Not-determined |
| S12_extracted_k141_152055 | Not-determined |
| S12_extracted_k141_136614 | Not-determined |
| S12_extracted_k141_136616 | Not-determined |
| S12_extracted_k141_105447 | Not-determined |
| S12_extracted_k141_105448 | Not-determined |
| S12_extracted_k141_112695 | Not-determined |
| S12_extracted_k141_42591 | Low-quality |
| S12_extracted_k141_112698 | Not-determined |
| S12_extracted_k141_89801 | Not-determined |
| S12_extracted_k141_136620 | Not-determined |
| S12_extracted_k141_112702 | Low-quality |
| S12_extracted_k141_3076 | Not-determined |
| S12_extracted_k141_42596 | Not-determined |
| S12_extracted_k141_112704 | Not-determined |
| S12_extracted_k141_49658 | Not-determined |
| S12_extracted_k141_144561 | Not-determined |
| S12_extracted_k141_89808 | Not-determined |
| S12_extracted_k141_49664 | Not-determined |
| S12_extracted_k141_89811 | Not-determined |
| S12_extracted_k141_73242 | Not-determined |
| S12_extracted_k141_42606 | Not-determined |
| S12_extracted_k141_144570 | Not-determined |
| S12_extracted_k141_112710 | Not-determined |
| S12_extracted_k141_112711 | Not-determined |
| S12_extracted_k141_3089 | Not-determined |
| S12_extracted_k141_3091 | Not-determined |
| S12_extracted_k141_112715 | Not-determined |
| S12_extracted_k141_18743 | Not-determined |
| S12_extracted_k141_18744 | Not-determined |
| S12_extracted_k141_120848 | Not-determined |
| S12_extracted_k141_97890 | Not-determined |
| S12_extracted_k141_49680 | Not-determined |
| S12_extracted_k141_97891 | Not-determined |
| S12_extracted_k141_105464 | Not-determined |
| S12_extracted_k141_136632 | Not-determined |
| S12_extracted_k141_81463 | Low-quality |
| S12_extracted_k141_136635 | Not-determined |
| S12_extracted_k141_3106 | Not-determined |
| S12_extracted_k141_175586 | Not-determined |
| S12_extracted_k141_120859 | Not-determined |
| S12_extracted_k141_58353 | Not-determined |
| S12_extracted_k141_184033 | Not-determined |
| S12_extracted_k141_58360 | Not-determined |
| S12_extracted_k141_58362 | Not-determined |
| S12_extracted_k141_97898 | Not-determined |
| S12_extracted_k141_18757 | Not-determined |
| S12_extracted_k141_81471 | Not-determined |
| S12_extracted_k141_97899 | Not-determined |
| S12_extracted_k141_66046 | Not-determined |
| S12_extracted_k141_81472 | Not-determined |
| S12_extracted_k141_136650 | Low-quality |
| S12_extracted_k141_120872 | Not-determined |
| S12_extracted_k141_175595 | Not-determined |
| S12_extracted_k141_81477 | Not-determined |
| S12_extracted_k141_120873 | Not-determined |
| S12_extracted_k141_49699 | Not-determined |
| S12_extracted_k141_89827 | Not-determined |
| S12_extracted_k141_120877 | Not-determined |
| S12_extracted_k141_81478 | Not-determined |
| S12_extracted_k141_97903 | Not-determined |
| S12_extracted_k141_18766 | Not-determined |
| S12_extracted_k141_66063 | Low-quality |
| S12_extracted_k141_120881 | Not-determined |
| S12_extracted_k141_81482 | Low-quality |
| S12_extracted_k141_136656 | Low-quality |
| S12_extracted_k141_49712 | Not-determined |
| S12_extracted_k141_144596 | Not-determined |
| S12_extracted_k141_184055 | Not-determined |
| S12_extracted_k141_144599 | Not-determined |
| S12_extracted_k141_26525 | Not-determined |
| S12_extracted_k141_168109 | Not-determined |
| S12_extracted_k141_120891 | Not-determined |
| S12_extracted_k141_89838 | Not-determined |
| S12_extracted_k141_184063 | Low-quality |
| S12_extracted_k141_97913 | Low-quality |
| S12_extracted_k141_97914 | Not-determined |
| S12_extracted_k141_81500 | Not-determined |
| S12_extracted_k141_18776 | Low-quality |
| S12_extracted_k141_175619 | Not-determined |
| S12_extracted_k141_49728 | Not-determined |
| S12_extracted_k141_42639 | Low-quality |
| S12_extracted_k141_97923 | Not-determined |
| S12_extracted_k141_175624 | Not-determined |
| S12_extracted_k141_89850 | Not-determined |
| S12_extracted_k141_49735 | Not-determined |
| S12_extracted_k141_18781 | Not-determined |
| S12_extracted_k141_97928 | Not-determined |
| S12_extracted_k141_49736 | Low-quality |
| S12_extracted_k141_18782 | Not-determined |
| S12_extracted_k141_42643 | Not-determined |
| S12_extracted_k141_120906 | Not-determined |
| S12_extracted_k141_58400 | Not-determined |
| S12_extracted_k141_18788 | Not-determined |
| S12_extracted_k141_184079 | Low-quality |
| S12_extracted_k141_152075 | Not-determined |
| S12_extracted_k141_136691 | Not-determined |
| S12_extracted_k141_73287 | Not-determined |
| S12_extracted_k141_136697 | Not-determined |
| S12_extracted_k141_33976 | Not-determined |
| S12_extracted_k141_3127 | Not-determined |
| S12_extracted_k141_97940 | Not-determined |
| S12_extracted_k141_89866 | Not-determined |
| S12_extracted_k141_144623 | Not-determined |
| S12_extracted_k141_18803 | Not-determined |
| S12_extracted_k141_136705 | Not-determined |
| S12_extracted_k141_144624 | Not-determined |
| S12_extracted_k141_49743 | Low-quality |
| S12_extracted_k141_73294 | Not-determined |
| S12_extracted_k141_42654 | Not-determined |
| S12_extracted_k141_136708 | Not-determined |
| S12_extracted_k141_97946 | Not-determined |
| S12_extracted_k141_81523 | Not-determined |
| S12_extracted_k141_81524 | Not-determined |
| S12_extracted_k141_89874 | Not-determined |
| S12_extracted_k141_152092 | Not-determined |
| S12_extracted_k141_42660 | Not-determined |
| S12_extracted_k141_89875 | Not-determined |
| S12_extracted_k141_175645 | Not-determined |
| S12_extracted_k141_42664 | Not-determined |
| S12_extracted_k141_97951 | Not-determined |
| S12_extracted_k141_97952 | Not-determined |
| S12_extracted_k141_49749 | Not-determined |
| S12_extracted_k141_89882 | Not-determined |
| S12_extracted_k141_136718 | Low-quality |
| S12_extracted_k141_97960 | Not-determined |
| S12_extracted_k141_112754 | Not-determined |
| S12_extracted_k141_136722 | Not-determined |
| S12_extracted_k141_112756 | Not-determined |
| S12_extracted_k141_152101 | Not-determined |
| S12_extracted_k141_89886 | Not-determined |
| S12_extracted_k141_89892 | Not-determined |
| S12_extracted_k141_81530 | Not-determined |
| S12_extracted_k141_159963 | Not-determined |
| S12_extracted_k141_112768 | Not-determined |
| S12_extracted_k141_97973 | Low-quality |
| S12_extracted_k141_81534 | Not-determined |
| S12_extracted_k141_159965 | Not-determined |
| S12_extracted_k141_159966 | Not-determined |
| S12_extracted_k141_3153 | Not-determined |
| S12_extracted_k141_175667 | Not-determined |
| S12_extracted_k141_144641 | Not-determined |
| S12_extracted_k141_184093 | Not-determined |
| S12_extracted_k141_97974 | Not-determined |
| S12_extracted_k141_49769 | Not-determined |
| S12_extracted_k141_49770 | Not-determined |
| S12_extracted_k141_42685 | Not-determined |
| S12_extracted_k141_144649 | Not-determined |
| S12_extracted_k141_184099 | Not-determined |
| S12_extracted_k141_81545 | Not-determined |
| S12_extracted_k141_42692 | Not-determined |
| S12_extracted_k141_66076 | Not-determined |
| S12_extracted_k141_66077 | Low-quality |
| S12_extracted_k141_105487 | Not-determined |
| S12_extracted_k141_66080 | Not-determined |
| S12_extracted_k141_66082 | Not-determined |
| S12_extracted_k141_97987 | Low-quality |
| S12_extracted_k141_66083 | Not-determined |
| S12_extracted_k141_136731 | Not-determined |
| S12_extracted_k141_10136 | Low-quality |
| S12_extracted_k141_58439 | Not-determined |
| S12_extracted_k141_97992 | Not-determined |
| S12_extracted_k141_26557 | Not-determined |
| S12_extracted_k141_97996 | Not-determined |
| S12_extracted_k141_3175 | Not-determined |
| S12_extracted_k141_73314 | Low-quality |
| S12_extracted_k141_105496 | Low-quality |
| S12_extracted_k141_136744 | Not-determined |
| S12_extracted_k141_128589 | Not-determined |
| S12_extracted_k141_3177 | Not-determined |
| S12_extracted_k141_18829 | Not-determined |
| S12_extracted_k141_105507 | Not-determined |
| S12_extracted_k141_26566 | Low-quality |
| S12_extracted_k141_58454 | Not-determined |
| S12_extracted_k141_58459 | Not-determined |
| S12_extracted_k141_66090 | Not-determined |
| S12_extracted_k141_42712 | Not-determined |
| S12_extracted_k141_105517 | Not-determined |
| S12_extracted_k141_105520 | Not-determined |
| S12_extracted_k141_144653 | Low-quality |
| S12_extracted_k141_98011 | Low-quality |
| S12_extracted_k141_89919 | Not-determined |
| S12_extracted_k141_58464 | Low-quality |
| S12_extracted_k141_168154 | Low-quality |
| S12_extracted_k141_175685 | Not-determined |
| S12_extracted_k141_175687 | Not-determined |
| S12_extracted_k141_66093 | Not-determined |
| S12_extracted_k141_3197 | Not-determined |
| S12_extracted_k141_66100 | Not-determined |
| S12_extracted_k141_42724 | Not-determined |
| S12_extracted_k141_120961 | Not-determined |
| S12_extracted_k141_89925 | Not-determined |
| S12_extracted_k141_136763 | Not-determined |
| S12_extracted_k141_81561 | Not-determined |
| S12_extracted_k141_3201 | Not-determined |
| S12_extracted_k141_89931 | Not-determined |
| S12_extracted_k141_105524 | Low-quality |
| S12_extracted_k141_144664 | Not-determined |
| S12_extracted_k141_168167 | Not-determined |
| S12_extracted_k141_98029 | Low-quality |
| S12_extracted_k141_81566 | Not-determined |
| S12_extracted_k141_144665 | Not-determined |
| S12_extracted_k141_3207 | Low-quality |
| S12_extracted_k141_112782 | Not-determined |
| S12_extracted_k141_66110 | Not-determined |
| S12_extracted_k141_98030 | Not-determined |
| S12_extracted_k141_168169 | Not-determined |
| S12_extracted_k141_136768 | Not-determined |
| S12_extracted_k141_168170 | Low-quality |
| S12_extracted_k141_89938 | Not-determined |
| S12_extracted_k141_168171 | Low-quality |
| S12_extracted_k141_120967 | Not-determined |
| S12_extracted_k141_144666 | Not-determined |
| S12_extracted_k141_81567 | Low-quality |
| S12_extracted_k141_3215 | Not-determined |
| S12_extracted_k141_3216 | Not-determined |
| S12_extracted_k141_98034 | Not-determined |
| S12_extracted_k141_144669 | Not-determined |
| S12_extracted_k141_184119 | Not-determined |
| S12_extracted_k141_184120 | Not-determined |
| S12_extracted_k141_112790 | Low-quality |
| S12_extracted_k141_120974 | Not-determined |
| S12_extracted_k141_81571 | Not-determined |
| S12_extracted_k141_144675 | Not-determined |
| S12_extracted_k141_136779 | Not-determined |
| S12_extracted_k141_89952 | Not-determined |
| S12_extracted_k141_175716 | Not-determined |
| S12_extracted_k141_168187 | Low-quality |
| S12_extracted_k141_89957 | Not-determined |
| S12_extracted_k141_144679 | Not-determined |
| S12_extracted_k141_175719 | Not-determined |
| S12_extracted_k141_89958 | Not-determined |
| S12_extracted_k141_128600 | Not-determined |
| S12_extracted_k141_73343 | Low-quality |
| S12_extracted_k141_128601 | Not-determined |
| S12_extracted_k141_49792 | Not-determined |
| S12_extracted_k141_3232 | Not-determined |
| S12_extracted_k141_175721 | Not-determined |
| S12_extracted_k141_112810 | Not-determined |
| S12_extracted_k141_184137 | Not-determined |
| S12_extracted_k141_128606 | Not-determined |
| S12_extracted_k141_49797 | Not-determined |
| S12_extracted_k141_168191 | Not-determined |
| S12_extracted_k141_168192 | Not-determined |
| S12_extracted_k141_49800 | Not-determined |
| S12_extracted_k141_152147 | Not-determined |
| S12_extracted_k141_26595 | Not-determined |
| S12_extracted_k141_144685 | Not-determined |
| S12_extracted_k141_159998 | Not-determined |
| S12_extracted_k141_81577 | Not-determined |
| S12_extracted_k141_120988 | Not-determined |
| S12_extracted_k141_184151 | Not-determined |
| S12_extracted_k141_81579 | Not-determined |
| S12_extracted_k141_73345 | Not-determined |
| S12_extracted_k141_168200 | Not-determined |
| S12_extracted_k141_81580 | Not-determined |
| S12_extracted_k141_81582 | Not-determined |
| S12_extracted_k141_26603 | Not-determined |
| S12_extracted_k141_33992 | Low-quality |
| S12_extracted_k141_120996 | Not-determined |
| S12_extracted_k141_152156 | Not-determined |
| S12_extracted_k141_160005 | Not-determined |
| S12_extracted_k141_26607 | Not-determined |
| S12_extracted_k141_73350 | Not-determined |
| S12_extracted_k141_89975 | Not-determined |
| S12_extracted_k141_26614 | Not-determined |
| S12_extracted_k141_168210 | Not-determined |
| S12_extracted_k141_152160 | Not-determined |
| S12_extracted_k141_98063 | Not-determined |
| S12_extracted_k141_112830 | Not-determined |
| S12_extracted_k141_33999 | Not-determined |
| S12_extracted_k141_34000 | Not-determined |
| S12_extracted_k141_168216 | Not-determined |
| S12_extracted_k141_160011 | Not-determined |
| S12_extracted_k141_81593 | Not-determined |
| S12_extracted_k141_10191 | Not-determined |
| S12_extracted_k141_34007 | Not-determined |
| S12_extracted_k141_34008 | Not-determined |
| S12_extracted_k141_10194 | Not-determined |
| S12_extracted_k141_81598 | Not-determined |
| S12_extracted_k141_73362 | Not-determined |
| S12_extracted_k141_89986 | Not-determined |
| S12_extracted_k141_144714 | Not-determined |
| S12_extracted_k141_152165 | Not-determined |
| S12_extracted_k141_98074 | Not-determined |
| S12_extracted_k141_168224 | Not-determined |
| S12_extracted_k141_34013 | Not-determined |
| S12_extracted_k141_26627 | Not-determined |
| S12_extracted_k141_26629 | Not-determined |
| S12_extracted_k141_112839 | Not-determined |
| S12_extracted_k141_168227 | Not-determined |
| S12_extracted_k141_152175 | Low-quality |
| S12_extracted_k141_144719 | Not-determined |
| S12_extracted_k141_66118 | Not-determined |
| S12_extracted_k141_34018 | Not-determined |
| S12_extracted_k141_112842 | Low-quality |
| S12_extracted_k141_73372 | Not-determined |
| S12_extracted_k141_73373 | Low-quality |
| S12_extracted_k141_42752 | Not-determined |
| S12_extracted_k141_42754 | Not-determined |
| S12_extracted_k141_26639 | Low-quality |
| S12_extracted_k141_66121 | Not-determined |
| S12_extracted_k141_144727 | Not-determined |
| S12_extracted_k141_73382 | Not-determined |
| S12_extracted_k141_136817 | Not-determined |
| S12_extracted_k141_152187 | Not-determined |
| S12_extracted_k141_10200 | Not-determined |
| S12_extracted_k141_18858 | Not-determined |
| S12_extracted_k141_128629 | Not-determined |
| S12_extracted_k141_58503 | Low-quality |
| S12_extracted_k141_112855 | Not-determined |
| S12_extracted_k141_10206 | Not-determined |
| S12_extracted_k141_184166 | Not-determined |
| S12_extracted_k141_98082 | Not-determined |
| S12_extracted_k141_18864 | Not-determined |
| S12_extracted_k141_18865 | Not-determined |
| S12_extracted_k141_98084 | Low-quality |
| S12_extracted_k141_58507 | Not-determined |
| S12_extracted_k141_184170 | Low-quality |
| S12_extracted_k141_34025 | Low-quality |
| S12_extracted_k141_3263 | Not-determined |
| S12_extracted_k141_26659 | Not-determined |
| S12_extracted_k141_184175 | Not-determined |
| S12_extracted_k141_144734 | Not-determined |
| S12_extracted_k141_18871 | Not-determined |
| S12_extracted_k141_10216 | Not-determined |
| S12_extracted_k141_58512 | Not-determined |
| S14_vs_k141_30108\|\|full | Medium-quality |
| S12_extracted_k141_98089 | Not-determined |
| S12_extracted_k141_144739 | Not-determined |
| S12_extracted_k141_3266 | Not-determined |
| S12_extracted_k141_81629 | Not-determined |
| S12_extracted_k141_168245 | Not-determined |
| S12_extracted_k141_42772 | Not-determined |
| S12_extracted_k141_128640 | Not-determined |
| S12_extracted_k141_81632 | Not-determined |
| S12_extracted_k141_144744 | Not-determined |
| S12_extracted_k141_81633 | Not-determined |
| S12_extracted_k141_3273 | Low-quality |
| S12_extracted_k141_42774 | Low-quality |
| S12_extracted_k141_168252 | Not-determined |
| S12_extracted_k141_144749 | Not-determined |
| S12_extracted_k141_89998 | Not-determined |
| S12_extracted_k141_26669 | Not-determined |
| S12_extracted_k141_128646 | Not-determined |
| S12_extracted_k141_168255 | Low-quality |
| S12_extracted_k141_26672 | Not-determined |
| S12_extracted_k141_128647 | Not-determined |
| S12_extracted_k141_168261 | Not-determined |
| S12_extracted_k141_144757 | Not-determined |
| S12_extracted_k141_152212 | Not-determined |
| S12_extracted_k141_73409 | Low-quality |
| S12_extracted_k141_81647 | Not-determined |
| S12_extracted_k141_168264 | Not-determined |
| S12_extracted_k141_144766 | Not-determined |
| S12_extracted_k141_73414 | Not-determined |
| S12_extracted_k141_105550 | Not-determined |
| S12_extracted_k141_128658 | Not-determined |
| S12_extracted_k141_81649 | Not-determined |
| S12_extracted_k141_73416 | Not-determined |
| S12_extracted_k141_34046 | Not-determined |
| S12_extracted_k141_66145 | Not-determined |
| S12_extracted_k141_90013 | Low-quality |
| S12_extracted_k141_18892 | Not-determined |
| S12_extracted_k141_42786 | Low-quality |
| S12_extracted_k141_128666 | Not-determined |
| S12_extracted_k141_160069 | Not-determined |
| S12_extracted_k141_18897 | Low-quality |
| S12_extracted_k141_42791 | Low-quality |
| S12_extracted_k141_90015 | Low-quality |
| S12_extracted_k141_42796 | Not-determined |
| S12_extracted_k141_160073 | Not-determined |
| S12_extracted_k141_160074 | Not-determined |
| S12_extracted_k141_144782 | Not-determined |
| S12_extracted_k141_90021 | Not-determined |
| S12_extracted_k141_81660 | Low-quality |
| S12_extracted_k141_42802 | Not-determined |
| S12_extracted_k141_42804 | Not-determined |
| S12_extracted_k141_144789 | Not-determined |
| S12_extracted_k141_184204 | Not-determined |
| S12_extracted_k141_42807 | Not-determined |
| S12_extracted_k141_105569 | Not-determined |
| S12_extracted_k141_10252 | Not-determined |
| S12_extracted_k141_98115 | Not-determined |
| S12_extracted_k141_168291 | Not-determined |
| S12_extracted_k141_136845 | Not-determined |
| S12_extracted_k141_144793 | Low-quality |
| S12_extracted_k141_34056 | Not-determined |
| S12_extracted_k141_81674 | Not-determined |
| S12_extracted_k141_98125 | Not-determined |
| S12_extracted_k141_81678 | Not-determined |
| S12_extracted_k141_112875 | Not-determined |
| S12_extracted_k141_152229 | Not-determined |
| S12_extracted_k141_152230 | Low-quality |
| S12_extracted_k141_152232 | Not-determined |
| S12_extracted_k141_90038 | Not-determined |
| S12_extracted_k141_105592 | Not-determined |
| S12_extracted_k141_90040 | Not-determined |
| S12_extracted_k141_73424 | Low-quality |
| S12_extracted_k141_18900 | Low-quality |
| S12_extracted_k141_98132 | Low-quality |
| S12_extracted_k141_66164 | Not-determined |
| S12_extracted_k141_152237 | Not-determined |
| S12_extracted_k141_90048 | Not-determined |
| S12_extracted_k141_168315 | Not-determined |
| S12_extracted_k141_160090 | Not-determined |
| S12_extracted_k141_105598 | Not-determined |
| S12_extracted_k141_105599 | Not-determined |
| S12_extracted_k141_26684 | Not-determined |
| S12_extracted_k141_10261 | Not-determined |
| S12_extracted_k141_10264 | Not-determined |
| S12_extracted_k141_168324 | Not-determined |
| S12_extracted_k141_168325 | Not-determined |
| S12_extracted_k141_128677 | Low-quality |
| S12_extracted_k141_42837 | Not-determined |
| S12_extracted_k141_105604 | Low-quality |
| S12_extracted_k141_26692 | Not-determined |
| S12_extracted_k141_81702 | Not-determined |
| S12_extracted_k141_90066 | Not-determined |
| S12_extracted_k141_105606 | Low-quality |
| S12_extracted_k141_73438 | Not-determined |
| S12_extracted_k141_10275 | Low-quality |
| S12_extracted_k141_184234 | Not-determined |
| S12_extracted_k141_160104 | Low-quality |
| S12_extracted_k141_10278 | Not-determined |
| S12_extracted_k141_120999 | Not-determined |
| S12_extracted_k141_128684 | Not-determined |
| S12_extracted_k141_160108 | Not-determined |
| S12_extracted_k141_128686 | Not-determined |
| S12_extracted_k141_26703 | Not-determined |
| S12_extracted_k141_73445 | Not-determined |
| S12_extracted_k141_128692 | Not-determined |
| S12_extracted_k141_90077 | Not-determined |
| S12_extracted_k141_136863 | Not-determined |
| S12_extracted_k141_105619 | Not-determined |
| S12_extracted_k141_128696 | Not-determined |
| S12_extracted_k141_128699 | Not-determined |
| S12_extracted_k141_98143 | Not-determined |
| S12_extracted_k141_26711 | Not-determined |
| S12_extracted_k141_98144 | Not-determined |
| S12_extracted_k141_128701 | Not-determined |
| S12_extracted_k141_3306 | Not-determined |
| S12_extracted_k141_3307 | Low-quality |
| S12_extracted_k141_128704 | Low-quality |
| S12_extracted_k141_128705 | Low-quality |
| S12_extracted_k141_73454 | Not-determined |
| S12_extracted_k141_81724 | Not-determined |
| S12_extracted_k141_3310 | Not-determined |
| S12_extracted_k141_90088 | Not-determined |
| S12_extracted_k141_112904 | Not-determined |
| S12_extracted_k141_90090 | Not-determined |
| S12_extracted_k141_144829 | Not-determined |
| S12_extracted_k141_98150 | Not-determined |
| S12_extracted_k141_184248 | Not-determined |
| S12_extracted_k141_105627 | Not-determined |
| S12_extracted_k141_136870 | Not-determined |
| S12_extracted_k141_105628 | Not-determined |
| S12_extracted_k141_73462 | Not-determined |
| S12_extracted_k141_144836 | Low-quality |
| S12_extracted_k141_144837 | Not-determined |
| S12_extracted_k141_42860 | Not-determined |
| S12_extracted_k141_66206 | Not-determined |
| S12_extracted_k141_121017 | Low-quality |
| S12_extracted_k141_105631 | Not-determined |
| S12_extracted_k141_26719 | Not-determined |

| | |
|---|---|
| S12_extracted_k141_3317 | Low-quality |
| S12_extracted_k141_105635 | Not-determined |
| S12_extracted_k141_73468 | Not-determined |
| S12_extracted_k141_73470 | Not-determined |
| S12_extracted_k141_160137 | Not-determined |
| S12_extracted_k141_184259 | Not-determined |
| S12_extracted_k141_160138 | Not-determined |
| S12_extracted_k141_66217 | Not-determined |
| S12_extracted_k141_42865 | Low-quality |
| S12_extracted_k141_128710 | Low-quality |
| S12_extracted_k141_42866 | Low-quality |
| S12_extracted_k141_98157 | Not-determined |
| S12_extracted_k141_34076 | Not-determined |
| S12_extracted_k141_112925 | Not-determined |
| S12_extracted_k141_144850 | Not-determined |
| S12_extracted_k141_184262 | Not-determined |
| S12_extracted_k141_168345 | Not-determined |
| S12_extracted_k141_112927 | Not-determined |
| S12_extracted_k141_136886 | Not-determined |
| S12_extracted_k141_184265 | Not-determined |
| S12_extracted_k141_26733 | Not-determined |
| S12_extracted_k141_98162 | Not-determined |
| S12_extracted_k141_175739 | Not-determined |
| S12_extracted_k141_121043 | Not-determined |
| S12_extracted_k141_98170 | Not-determined |
| S12_extracted_k141_175742 | Not-determined |
| S12_extracted_k141_128722 | Not-determined |
| S12_extracted_k141_128726 | Not-determined |
| S12_extracted_k141_168365 | Low-quality |
| S12_extracted_k141_128728 | Not-determined |
| S12_extracted_k141_128729 | Not-determined |
| S12_extracted_k141_128730 | Not-determined |
| S12_extracted_k141_105661 | Not-determined |
| S12_extracted_k141_26746 | Not-determined |
| S12_extracted_k141_175748 | Low-quality |
| S12_extracted_k141_26748 | Not-determined |
| S12_extracted_k141_42892 | Not-determined |
| S12_extracted_k141_128739 | Not-determined |
| S12_extracted_k141_160161 | Not-determined |
| S12_extracted_k141_58558 | Low-quality |
| S12_extracted_k141_90116 | Not-determined |
| S12_extracted_k141_112944 | Low-quality |
| S12_extracted_k141_168367 | Low-quality |
| S12_extracted_k141_34088 | Low-quality |
| S12_extracted_k141_152257 | Low-quality |
| S12_extracted_k141_175759 | Not-determined |
| S12_extracted_k141_90125 | Low-quality |
| S12_extracted_k141_49849 | Not-determined |
| S12_extracted_k141_105677 | Not-determined |
| S12_extracted_k141_49852 | Not-determined |
| S12_extracted_k141_152264 | Not-determined |
| S12_extracted_k141_49853 | Not-determined |
| S12_extracted_k141_73504 | Not-determined |
| S12_extracted_k141_105678 | Not-determined |
| S12_extracted_k141_184277 | Low-quality |
| S12_extracted_k141_49856 | Low-quality |
| S12_extracted_k141_90131 | Not-determined |
| S12_extracted_k141_144882 | Not-determined |
| S12_extracted_k141_49862 | Not-determined |
| S12_extracted_k141_160182 | Not-determined |
| S12_extracted_k141_168377 | Not-determined |
| S12_extracted_k141_128762 | Not-determined |
| S12_extracted_k141_128763 | Not-determined |
| S12_extracted_k141_160187 | Not-determined |
| S12_extracted_k141_26756 | Not-determined |
| S12_extracted_k141_112961 | Not-determined |
| S12_extracted_k141_144887 | Not-determined |
| S12_extracted_k141_144888 | Not-determined |
| S12_extracted_k141_144890 | Not-determined |
| S12_extracted_k141_152276 | Not-determined |
| S12_extracted_k141_112966 | Not-determined |
| S12_extracted_k141_128770 | Not-determined |
| S12_extracted_k141_73520 | Not-determined |
| S12_extracted_k141_136897 | Not-determined |
| S12_extracted_k141_90134 | Not-determined |
| S12_extracted_k141_168388 | Not-determined |
| S12_extracted_k141_34093 | Not-determined |
| S12_extracted_k141_175776 | Not-determined |
| S12_extracted_k141_3368 | Not-determined |
| S12_extracted_k141_3370 | Not-determined |
| S12_extracted_k141_42918 | Not-determined |
| S12_extracted_k141_98179 | Low-quality |
| S12_extracted_k141_42919 | Not-determined |
| S12_extracted_k141_90138 | Not-determined |
| S12_extracted_k141_144904 | Not-determined |
| S12_extracted_k141_3375 | Low-quality |
| S12_extracted_k141_66246 | Not-determined |
| S12_extracted_k141_81744 | Not-determined |
| S12_extracted_k141_112981 | Not-determined |
| S12_extracted_k141_152289 | Low-quality |
| S12_extracted_k141_136913 | Not-determined |
| S12_extracted_k141_49889 | Not-determined |
| S12_extracted_k141_136914 | Not-determined |
| S12_extracted_k141_184294 | Low-quality |
| S12_extracted_k141_105707 | Not-determined |
| S12_extracted_k141_73524 | Not-determined |
| S12_extracted_k141_90159 | Not-determined |
| S12_extracted_k141_160195 | Not-determined |
| S12_extracted_k141_160199 | Not-determined |
| S12_extracted_k141_81762 | Not-determined |
| S12_extracted_k141_90167 | Not-determined |
| S12_extracted_k141_42935 | Not-determined |
| S12_extracted_k141_10294 | Not-determined |
| S12_extracted_k141_105715 | Not-determined |
| S12_extracted_k141_184311 | Not-determined |
| S12_extracted_k141_136931 | Not-determined |
| S12_extracted_k141_168407 | Not-determined |
| S12_extracted_k141_152304 | Not-determined |
| S12_extracted_k141_152307 | Not-determined |
| S12_extracted_k141_168409 | Low-quality |
| S12_extracted_k141_113004 | Low-quality |
| S12_extracted_k141_113005 | Not-determined |
| S12_extracted_k141_168412 | Not-determined |
| S12_extracted_k141_26777 | Not-determined |
| S12_extracted_k141_160215 | Not-determined |
| S12_extracted_k141_136935 | Not-determined |
| S12_extracted_k141_10305 | Not-determined |
| S12_extracted_k141_136936 | Not-determined |
| S12_extracted_k141_175806 | Not-determined |
| S12_extracted_k141_66263 | Not-determined |
| S12_extracted_k141_121045 | Low-quality |
| S12_extracted_k141_66267 | Not-determined |
| S12_extracted_k141_175812 | Not-determined |
| S12_extracted_k141_113009 | Low-quality |
| S12_extracted_k141_160230 | Not-determined |
| S12_extracted_k141_58561 | Not-determined |
| S12_extracted_k141_42946 | Not-determined |
| S12_extracted_k141_42947 | Low-quality |
| S12_extracted_k141_144927 | Not-determined |
| S12_extracted_k141_160234 | Not-determined |
| S12_extracted_k141_121053 | Not-determined |
| S12_extracted_k141_152325 | Not-determined |
| S12_extracted_k141_144930 | Not-determined |
| S12_extracted_k141_144932 | Not-determined |
| S12_extracted_k141_175819 | Not-determined |
| S12_extracted_k141_73545 | Not-determined |
| S12_extracted_k141_136955 | Not-determined |
| S12_extracted_k141_113016 | Not-determined |
| S12_extracted_k141_175821 | Low-quality |
| S12_extracted_k141_175822 | Not-determined |
| S12_extracted_k141_3415 | Not-determined |
| S12_extracted_k141_160240 | Not-determined |
| S12_extracted_k141_113021 | Not-determined |
| S12_extracted_k141_66284 | Low-quality |
| S12_extracted_k141_160242 | Not-determined |
| S12_extracted_k141_73551 | Not-determined |
| S12_extracted_k141_113024 | Not-determined |
| S12_extracted_k141_58576 | Not-determined |
| S12_extracted_k141_168435 | Not-determined |
| S12_extracted_k141_105741 | Not-determined |
| S12_extracted_k141_90201 | Not-determined |
| S12_extracted_k141_90205 | Not-determined |
| S12_extracted_k141_66294 | Not-determined |
| S12_extracted_k141_175829 | Not-determined |
| S12_extracted_k141_66298 | Not-determined |
| S12_extracted_k141_10336 | Not-determined |
| S12_extracted_k141_144944 | Not-determined |
| S12_extracted_k141_136963 | Not-determined |
| S12_extracted_k141_136964 | Not-determined |
| S12_extracted_k141_175830 | Not-determined |
| S12_extracted_k141_121072 | Not-determined |
| S12_extracted_k141_136967 | Not-determined |
| S12_extracted_k141_136968 | Low-quality |
| S12_extracted_k141_81777 | Not-determined |
| S14_vs_k141_23558\|\|full | Medium-quality |
| S12_extracted_k141_175840 | Not-determined |
| S12_extracted_k141_26781 | Not-determined |
| S12_extracted_k141_81780 | Not-determined |
| S12_extracted_k141_160258 | Not-determined |
| S12_extracted_k141_81781 | Not-determined |
| S12_extracted_k141_26783 | Not-determined |
| S12_extracted_k141_81782 | Not-determined |
| S12_extracted_k141_18907 | Not-determined |
| S12_extracted_k141_152328 | Low-quality |
| S12_extracted_k141_90221 | Not-determined |
| S12_extracted_k141_26785 | Not-determined |
| S12_extracted_k141_3444 | Not-determined |
| S12_extracted_k141_3446 | Not-determined |
| S12_extracted_k141_3447 | Not-determined |
| S12_extracted_k141_90225 | Not-determined |
| S12_extracted_k141_136979 | Not-determined |
| S12_extracted_k141_175851 | Not-determined |
| S12_extracted_k141_152332 | Not-determined |
| S12_extracted_k141_90226 | Not-determined |
| S12_extracted_k141_160263 | Not-determined |
| S12_extracted_k141_160265 | Low-quality |
| S12_extracted_k141_18916 | Not-determined |
| S12_extracted_k141_10356 | Not-determined |
| S12_extracted_k141_58595 | Not-determined |
| S12_extracted_k141_98222 | Low-quality |
| S12_extracted_k141_90229 | Not-determined |
| S12_extracted_k141_18918 | Not-determined |
| S12_extracted_k141_136985 | Not-determined |
| S12_extracted_k141_49926 | Low-quality |
| S12_extracted_k141_58599 | Not-determined |
| S12_extracted_k141_105770 | Not-determined |
| S12_extracted_k141_175860 | Low-quality |
| S12_extracted_k141_42975 | Not-determined |
| S12_extracted_k141_105772 | Not-determined |
| S12_extracted_k141_34154 | Not-determined |
| S12_extracted_k141_58603 | Not-determined |
| S12_extracted_k141_105773 | Not-determined |
| S12_extracted_k141_58605 | Not-determined |
| S12_extracted_k141_81800 | Not-determined |
| S12_extracted_k141_136994 | Not-determined |
| S12_extracted_k141_42976 | Not-determined |
| S12_extracted_k141_98229 | Not-determined |
| S12_extracted_k141_26799 | Not-determined |
| S1_extracted_k141_100150 | Medium-quality |
| S12_extracted_k141_144963 | Not-determined |
| S12_extracted_k141_113048 | Low-quality |
| S12_extracted_k141_90242 | Not-determined |
| S12_extracted_k141_144964 | Not-determined |
| S12_extracted_k141_42979 | Not-determined |
| S12_extracted_k141_136996 | Not-determined |
| S12_extracted_k141_49937 | Low-quality |
| S12_extracted_k141_42981 | Not-determined |
| S12_extracted_k141_18923 | Not-determined |
| S12_extracted_k141_26807 | Not-determined |
| S12_extracted_k141_168478 | Not-determined |
| S12_extracted_k141_168480 | Not-determined |
| S12_extracted_k141_160277 | Not-determined |
| S12_extracted_k141_90251 | Not-determined |
| S12_extracted_k141_73581 | Low-quality |
| S12_extracted_k141_42991 | Not-determined |
| S12_extracted_k141_42992 | Not-determined |
| S12_extracted_k141_42993 | Not-determined |
| S12_extracted_k141_144973 | Not-determined |
| S12_extracted_k141_152344 | Not-determined |
| S12_extracted_k141_168486 | Not-determined |
| S12_extracted_k141_128803 | Not-determined |
| S12_extracted_k141_137005 | Not-determined |
| S12_extracted_k141_90257 | Not-determined |
| S12_extracted_k141_42999 | Low-quality |
| S12_extracted_k141_26818 | Not-determined |
| S12_extracted_k141_58618 | Not-determined |
| S12_extracted_k141_128807 | Low-quality |
| S12_extracted_k141_105790 | Not-determined |
| S12_extracted_k141_43003 | Not-determined |
| S12_extracted_k141_152349 | Not-determined |
| S12_extracted_k141_168497 | Not-determined |
| S12_extracted_k141_43006 | Not-determined |
| S12_extracted_k141_128811 | Not-determined |
| S12_extracted_k141_58624 | Low-quality |
| S12_extracted_k141_128812 | Not-determined |
| S12_extracted_k141_105802 | Low-quality |
| S12_extracted_k141_90269 | Not-determined |
| S12_extracted_k141_137014 | Not-determined |
| S12_extracted_k141_34165 | Not-determined |
| S12_extracted_k141_168504 | Not-determined |
| S12_extracted_k141_144983 | Not-determined |
| S12_extracted_k141_81835 | Not-determined |
| S12_extracted_k141_26819 | Not-determined |
| S12_extracted_k141_152358 | Not-determined |
| S12_extracted_k141_175876 | Not-determined |
| S12_extracted_k141_98256 | Not-determined |
| S12_extracted_k141_160306 | Not-determined |
| S12_extracted_k141_81848 | Not-determined |
| S12_extracted_k141_58631 | Not-determined |
| S12_extracted_k141_175888 | Not-determined |
| S12_extracted_k141_34186 | Not-determined |
| S12_extracted_k141_144994 | Not-determined |
| S12_extracted_k141_105814 | Not-determined |
| S12_extracted_k141_160312 | Not-determined |
| S12_extracted_k141_105817 | Not-determined |
| S12_extracted_k141_34198 | Not-determined |
| S12_extracted_k141_90278 | Not-determined |
| S12_extracted_k141_26842 | Not-determined |
| S12_extracted_k141_34202 | Not-determined |
| S12_extracted_k141_137022 | Not-determined |
| S12_extracted_k141_137026 | Not-determined |
| S12_extracted_k141_113076 | Low-quality |
| S12_extracted_k141_98269 | Not-determined |
| S12_extracted_k141_34207 | Not-determined |
| S12_extracted_k141_3472 | Not-determined |
| S12_extracted_k141_105827 | Low-quality |
| S12_extracted_k141_58652 | Not-determined |
| S12_extracted_k141_90284 | Not-determined |
| S12_extracted_k141_152377 | Not-determined |
| S12_extracted_k141_152378 | Not-determined |
| S12_extracted_k141_58656 | Not-determined |
| S12_extracted_k141_58657 | Not-determined |
| S12_extracted_k141_34213 | Not-determined |
| S12_extracted_k141_128839 | Not-determined |
| S12_extracted_k141_184365 | Not-determined |
| S12_extracted_k141_105831 | Not-determined |
| S12_extracted_k141_160326 | Not-determined |
| S12_extracted_k141_105833 | Low-quality |
| S12_extracted_k141_58662 | Not-determined |
| S12_extracted_k141_73587 | Not-determined |
| S12_extracted_k141_152382 | Not-determined |
| S12_extracted_k141_58665 | Not-determined |
| S12_extracted_k141_98284 | Not-determined |
| S12_extracted_k141_175897 | Low-quality |
| S12_extracted_k141_168535 | Not-determined |
| S12_extracted_k141_58669 | Not-determined |
| S12_extracted_k141_105838 | Not-determined |
| S12_extracted_k141_43026 | Low-quality |
| S12_extracted_k141_175899 | Not-determined |
| S12_extracted_k141_152390 | Low-quality |
| S12_extracted_k141_73596 | Not-determined |
| S12_extracted_k141_145006 | Not-determined |
| S12_extracted_k141_152392 | Not-determined |
| S12_extracted_k141_128849 | Not-determined |
| S12_extracted_k141_73605 | Not-determined |
| S12_extracted_k141_18935 | Not-determined |
| S12_extracted_k141_137045 | Low-quality |
| S12_extracted_k141_10363 | Not-determined |
| S12_extracted_k141_81869 | Not-determined |
| S12_extracted_k141_128856 | Not-determined |
| S12_extracted_k141_43044 | Not-determined |
| S12_extracted_k141_66322 | Not-determined |
| S12_extracted_k141_168557 | Not-determined |
| S12_extracted_k141_184396 | Not-determined |
| S12_extracted_k141_66324 | Not-determined |
| S12_extracted_k141_184399 | Not-determined |
| S12_extracted_k141_137050 | Not-determined |
| S12_extracted_k141_43050 | Not-determined |
| S12_extracted_k141_105850 | Not-determined |
| S12_extracted_k141_168562 | Not-determined |
| S12_extracted_k141_168564 | Not-determined |
| S12_extracted_k141_66331 | Not-determined |
| S12_extracted_k141_81878 | Not-determined |
| S12_extracted_k141_58675 | Low-quality |
| S12_extracted_k141_137059 | Low-quality |
| S12_extracted_k141_34223 | Low-quality |
| S12_extracted_k141_10375 | Not-determined |
| S12_extracted_k141_18943 | Not-determined |
| S12_extracted_k141_34227 | Low-quality |
| S12_extracted_k141_128864 | Not-determined |
| S12_extracted_k141_160368 | Not-determined |
| S12_extracted_k141_168573 | Not-determined |
| S12_extracted_k141_160371 | Not-determined |
| S12_extracted_k141_10381 | Not-determined |
| S12_extracted_k141_160374 | Not-determined |
| S12_extracted_k141_90297 | Not-determined |
| S12_extracted_k141_152416 | Not-determined |
| S12_extracted_k141_184417 | Not-determined |
| S12_extracted_k141_58691 | Not-determined |
| S12_extracted_k141_90298 | Not-determined |
| S12_extracted_k141_58692 | Not-determined |
| S12_extracted_k141_128874 | Not-determined |
| S12_extracted_k141_34240 | Low-quality |
| S12_extracted_k141_152419 | Not-determined |
| S12_extracted_k141_98321 | Not-determined |
| S12_extracted_k141_152420 | Not-determined |
| S12_extracted_k141_34243 | Not-determined |
| S12_extracted_k141_10392 | Low-quality |
| S12_extracted_k141_18952 | Low-quality |
| S12_extracted_k141_49957 | Not-determined |
| S12_extracted_k141_105865 | Not-determined |
| S12_extracted_k141_98329 | Not-determined |
| S12_extracted_k141_113119 | Not-determined |
| S12_extracted_k141_3515 | Not-determined |
| S12_extracted_k141_26854 | Not-determined |
| S12_extracted_k141_18955 | Not-determined |
| S12_extracted_k141_34251 | Not-determined |
| S12_extracted_k141_137071 | Not-determined |
| S12_extracted_k141_152432 | Not-determined |
| S12_extracted_k141_160388 | Not-determined |
| S12_extracted_k141_105870 | Not-determined |
| S12_extracted_k141_66352 | Not-determined |
| S12_extracted_k141_49963 | Not-determined |
| S12_extracted_k141_18960 | Not-determined |
| S12_extracted_k141_34253 | Not-determined |
| S12_extracted_k141_168601 | Not-determined |
| S12_extracted_k141_152438 | Not-determined |
| S12_extracted_k141_34256 | Not-determined |
| S12_extracted_k141_137082 | Low-quality |
| S12_extracted_k141_73634 | Not-determined |
| S12_extracted_k141_81891 | Not-determined |
| S12_extracted_k141_10401 | Not-determined |
| S12_extracted_k141_160391 | Low-quality |
| S12_extracted_k141_128895 | Not-determined |
| S12_extracted_k141_98334 | Low-quality |
| S12_extracted_k141_66361 | Not-determined |
| S12_extracted_k141_160398 | Not-determined |
| S12_extracted_k141_175928 | Not-determined |
| S12_extracted_k141_73642 | Not-determined |
| S12_extracted_k141_184439 | Low-quality |
| S12_extracted_k141_168617 | Not-determined |
| S12_extracted_k141_43086 | Not-determined |
| S12_extracted_k141_18975 | Not-determined |
| S12_extracted_k141_73647 | Not-determined |
| S12_extracted_k141_34268 | Not-determined |
| S12_extracted_k141_3526 | Not-determined |
| S12_extracted_k141_73651 | Not-determined |
| S12_extracted_k141_49979 | Not-determined |
| S12_extracted_k141_98340 | Not-determined |
| S12_extracted_k141_34269 | Not-determined |
| S12_extracted_k141_128910 | Not-determined |
| S12_extracted_k141_184458 | Not-determined |
| S12_extracted_k141_184460 | Not-determined |
| S12_extracted_k141_49986 | Not-determined |
| S12_extracted_k141_49989 | Not-determined |
| S12_extracted_k141_10425 | Not-determined |
| S12_extracted_k141_81904 | Not-determined |
| S12_extracted_k141_66386 | Not-determined |
| S12_extracted_k141_98351 | Not-determined |
| S12_extracted_k141_128919 | Not-determined |
| S12_extracted_k141_43101 | Not-determined |
| S12_extracted_k141_98360 | Not-determined |
| S12_extracted_k141_43103 | Not-determined |
| S12_extracted_k141_81909 | Not-determined |
| S12_extracted_k141_43105 | Not-determined |
| S12_extracted_k141_128924 | Not-determined |
| S12_extracted_k141_43106 | Not-determined |
| S12_extracted_k141_98361 | Low-quality |
| S12_extracted_k141_3542 | Not-determined |
| S12_extracted_k141_128933 | Not-determined |
| S12_extracted_k141_98365 | Not-determined |
| S12_extracted_k141_128934 | Not-determined |
| S12_extracted_k141_66396 | Low-quality |
| S12_extracted_k141_105886 | Not-determined |
| S12_extracted_k141_98366 | Not-determined |
| S12_extracted_k141_10434 | Low-quality |
| S12_extracted_k141_18982 | Not-determined |
| S12_extracted_k141_184477 | Not-determined |
| S12_extracted_k141_3548 | Not-determined |
| S12_extracted_k141_50001 | Not-determined |
| S12_extracted_k141_34286 | Low-quality |
| S12_extracted_k141_81925 | Low-quality |
| S12_extracted_k141_10438 | Low-quality |
| S12_extracted_k141_73676 | Low-quality |
| S12_extracted_k141_10441 | Not-determined |
| S12_extracted_k141_66406 | Not-determined |
| S12_extracted_k141_50010 | Not-determined |
| S12_extracted_k141_50011 | Not-determined |
| S12_extracted_k141_128944 | Not-determined |
| S12_extracted_k141_81930 | Low-quality |
| S12_extracted_k141_3558 | Not-determined |
| S12_extracted_k141_3559 | Not-determined |
| S12_extracted_k141_26878 | Not-determined |
| S12_extracted_k141_152493 | Not-determined |
| S12_extracted_k141_66410 | Low-quality |
| S12_extracted_k141_128948 | Not-determined |
| S12_extracted_k141_43121 | Not-determined |
| S12_extracted_k141_152496 | Not-determined |
| S12_extracted_k141_113150 | Not-determined |
| S12_extracted_k141_90323 | Not-determined |
| S12_extracted_k141_73689 | Not-determined |
| S12_extracted_k141_66415 | Low-quality |
| S12_extracted_k141_81939 | Not-determined |
| S12_extracted_k141_3568 | Not-determined |
| S12_extracted_k141_43129 | Not-determined |
| S12_extracted_k141_113158 | Not-determined |
| S12_extracted_k141_113159 | Not-determined |
| S12_extracted_k141_34292 | Not-determined |
| S12_extracted_k141_175943 | Not-determined |
| S12_extracted_k141_81943 | Not-determined |
| S12_extracted_k141_184480 | Not-determined |
| S12_extracted_k141_152502 | Not-determined |
| S12_extracted_k141_113166 | Not-determined |
| S12_extracted_k141_73699 | Not-determined |
| S12_extracted_k141_3577 | Not-determined |
| S12_extracted_k141_175952 | Not-determined |
| S12_extracted_k141_184487 | Not-determined |
| S12_extracted_k141_10457 | Not-determined |
| S12_extracted_k141_105902 | Not-determined |
| S12_extracted_k141_105903 | Not-determined |
| S12_extracted_k141_81953 | Not-determined |
| S12_extracted_k141_98388 | Low-quality |
| S12_extracted_k141_90343 | Not-determined |
| S12_extracted_k141_34300 | Low-quality |
| S12_extracted_k141_81960 | Not-determined |
| S12_extracted_k141_50026 | Not-determined |
| S12_extracted_k141_81961 | Low-quality |
| S12_extracted_k141_81962 | Not-determined |
| S12_extracted_k141_184497 | Not-determined |
| S12_extracted_k141_73714 | Not-determined |
| S12_extracted_k141_184498 | Low-quality |
| S12_extracted_k141_66426 | Not-determined |
| S12_extracted_k141_66427 | Not-determined |
| S12_extracted_k141_81965 | Not-determined |
| S12_extracted_k141_34304 | Not-determined |
| S12_extracted_k141_98399 | Not-determined |
| S12_extracted_k141_81967 | Not-determined |
| S12_extracted_k141_175967 | Not-determined |
| S12_extracted_k141_98403 | Not-determined |
| S12_extracted_k141_98406 | Not-determined |
| S12_extracted_k141_3597 | Not-determined |
| S12_extracted_k141_3598 | Not-determined |
| S12_extracted_k141_3601 | Not-determined |
| S12_extracted_k141_152510 | Not-determined |
| S12_extracted_k141_152512 | Not-determined |
| S12_extracted_k141_10471 | Not-determined |
| S12_extracted_k141_73732 | Not-determined |
| S12_extracted_k141_10475 | Not-determined |
| S12_extracted_k141_3607 | Not-determined |
| S12_extracted_k141_81987 | Not-determined |
| S12_extracted_k141_175975 | Not-determined |
| S12_extracted_k141_121088 | Not-determined |
| S12_extracted_k141_184514 | Not-determined |
| S12_extracted_k141_81991 | Not-determined |
| S12_extracted_k141_160416 | Not-determined |
| S12_extracted_k141_50051 | Not-determined |
| S12_extracted_k141_34312 | Not-determined |
| S12_extracted_k141_66447 | Not-determined |
| S12_extracted_k141_152526 | Not-determined |
| S12_extracted_k141_19018 | Not-determined |
| S12_extracted_k141_82004 | Not-determined |
| S12_extracted_k141_90377 | Not-determined |
| S12_extracted_k141_90379 | Low-quality |
| S12_extracted_k141_34319 | Low-quality |
| S12_extracted_k141_43163 | Not-determined |
| S12_extracted_k141_152532 | Not-determined |
| S12_extracted_k141_98440 | Not-determined |
| S12_extracted_k141_43164 | Not-determined |
| S12_extracted_k141_152533 | Not-determined |
| S12_extracted_k141_82008 | Low-quality |
| S12_extracted_k141_98441 | Low-quality |
| S12_extracted_k141_50063 | Not-determined |
| S12_extracted_k141_34324 | Not-determined |
| S12_extracted_k141_66460 | Not-determined |
| S12_extracted_k141_168624 | Not-determined |
| S12_extracted_k141_73759 | Not-determined |
| S12_extracted_k141_98446 | Not-determined |
| S12_extracted_k141_3637 | Not-determined |
| S12_extracted_k141_137109 | Not-determined |
| S12_extracted_k141_3638 | Not-determined |
| S12_extracted_k141_50068 | Not-determined |
| S12_extracted_k141_10495 | Not-determined |
| S12_extracted_k141_73766 | Not-determined |
| S12_extracted_k141_19031 | Not-determined |
| S12_extracted_k141_3641 | Not-determined |
| S12_extracted_k141_43179 | Not-determined |
| S12_extracted_k141_34333 | Not-determined |
| S12_extracted_k141_176007 | Not-determined |
| S12_extracted_k141_3644 | Not-determined |
| S12_extracted_k141_184516 | Not-determined |
| S12_extracted_k141_184518 | Not-determined |
| S12_extracted_k141_137121 | Not-determined |
| S12_extracted_k141_50080 | Not-determined |
| S12_extracted_k141_121103 | Not-determined |
| S12_extracted_k141_66469 | Not-determined |
| S12_extracted_k141_176013 | Low-quality |
| S12_extracted_k141_160436 | Not-determined |
| S12_extracted_k141_34347 | Not-determined |
| S12_extracted_k141_50087 | Not-determined |
| S12_extracted_k141_50091 | Not-determined |
| S12_extracted_k141_43190 | Not-determined |
| S12_extracted_k141_26906 | Not-determined |
| S12_extracted_k141_121112 | Not-determined |
| S12_extracted_k141_98466 | Not-determined |
| S12_extracted_k141_184532 | Not-determined |
| S12_extracted_k141_168649 | Not-determined |
| S12_extracted_k141_176017 | Low-quality |
| S12_extracted_k141_105959 | Not-determined |
| S12_extracted_k141_90413 | Not-determined |
| S12_extracted_k141_10514 | Low-quality |
| S12_extracted_k141_168656 | Not-determined |
| S12_extracted_k141_90416 | Not-determined |
| S12_extracted_k141_105965 | Low-quality |
| S12_extracted_k141_82030 | Low-quality |
| S12_extracted_k141_34364 | Not-determined |
| S12_extracted_k141_82032 | Not-determined |
| S12_extracted_k141_34365 | Not-determined |
| S12_extracted_k141_82033 | Not-determined |
| S12_extracted_k141_3666 | Not-determined |
| S12_extracted_k141_50106 | Not-determined |
| S12_extracted_k141_160458 | Not-determined |
| S12_extracted_k141_10517 | Low-quality |
| S12_extracted_k141_50110 | Not-determined |
| S12_extracted_k141_90425 | Not-determined |
| S12_extracted_k141_160464 | Not-determined |
| S12_extracted_k141_137138 | Not-determined |
| S12_extracted_k141_105976 | Low-quality |
| S12_extracted_k141_43197 | Not-determined |
| S12_extracted_k141_184559 | Not-determined |
| S12_extracted_k141_73794 | Not-determined |
| S12_extracted_k141_26919 | Not-determined |
| S12_extracted_k141_160466 | Not-determined |
| S12_extracted_k141_50114 | Not-determined |
| S12_extracted_k141_10519 | Not-determined |
| S12_extracted_k141_160470 | Not-determined |
| S12_extracted_k141_43205 | Not-determined |
| S12_extracted_k141_26922 | Not-determined |
| S12_extracted_k141_98500 | Not-determined |
| S12_extracted_k141_160475 | Not-determined |
| S12_extracted_k141_34372 | Not-determined |
| S12_extracted_k141_3685 | Not-determined |
| S12_extracted_k141_3686 | Low-quality |
| S12_extracted_k141_10527 | Not-determined |
| S12_extracted_k141_3690 | Not-determined |
| S12_extracted_k141_10535 | Not-determined |
| S12_extracted_k141_113180 | Not-determined |
| S12_extracted_k141_73806 | Not-determined |
| S12_extracted_k141_3696 | Not-determined |
| S12_extracted_k141_3699 | Low-quality |

| | |
|---|---|
| S12_extracted_k141_176061 | Not-determined |
| S12_extracted_k141_137156 | Low-quality |
| S12_extracted_k141_98511 | Low-quality |
| S12_extracted_k141_106009 | Not-determined |
| S12_extracted_k141_90450 | Low-quality |
| S12_extracted_k141_10540 | Not-determined |
| S12_extracted_k141_90453 | Not-determined |
| S12_extracted_k141_113197 | Not-determined |
| S12_extracted_k141_43217 | Not-determined |
| S12_extracted_k141_26931 | Not-determined |
| S12_extracted_k141_121134 | Not-determined |
| S12_extracted_k141_160487 | Not-determined |
| S12_extracted_k141_10545 | Not-determined |
| S12_extracted_k141_106015 | Not-determined |
| S12_extracted_k141_184575 | Not-determined |
| S12_extracted_k141_98523 | Not-determined |
| S12_extracted_k141_168661 | Not-determined |
| S12_extracted_k141_184578 | Not-determined |
| S12_extracted_k141_113202 | Not-determined |
| S12_extracted_k141_176085 | Not-determined |
| S12_extracted_k141_43226 | Not-determined |
| S12_extracted_k141_98527 | Not-determined |
| S12_extracted_k141_106024 | Not-determined |
| S12_extracted_k141_113207 | Not-determined |
| S12_extracted_k141_113209 | Not-determined |
| S12_extracted_k141_152547 | Not-determined |
| S12_extracted_k141_137182 | Not-determined |
| S12_extracted_k141_168674 | Not-determined |
| S12_extracted_k141_137183 | Not-determined |
| S12_extracted_k141_43230 | Not-determined |
| S12_extracted_k141_3728 | Not-determined |
| S12_extracted_k141_176096 | Not-determined |
| S12_extracted_k141_168676 | Not-determined |
| S12_extracted_k141_106032 | Not-determined |
| S12_extracted_k141_98534 | Not-determined |
| S12_extracted_k141_113217 | Not-determined |
| S12_extracted_k141_184590 | Not-determined |
| S12_extracted_k141_82066 | Not-determined |
| S12_extracted_k141_152551 | Not-determined |
| S12_extracted_k141_152555 | Not-determined |
| S12_extracted_k141_184596 | Not-determined |
| S12_extracted_k141_26958 | Not-determined |
| S12_extracted_k141_50165 | Not-determined |
| S12_extracted_k141_3746 | Not-determined |
| S12_extracted_k141_43247 | Not-determined |
| S12_extracted_k141_50168 | Not-determined |
| S12_extracted_k141_66484 | Low-quality |
| S12_extracted_k141_66488 | Not-determined |
| S12_extracted_k141_128964 | Not-determined |
| S12_extracted_k141_160518 | Not-determined |
| S12_extracted_k141_137200 | Not-determined |
| S12_extracted_k141_128965 | Not-determined |
| S12_extracted_k141_90485 | Not-determined |
| S12_extracted_k141_82075 | Not-determined |
| S12_extracted_k141_184608 | Not-determined |
| S12_extracted_k141_137207 | Not-determined |
| S12_extracted_k141_128978 | Not-determined |
| S12_extracted_k141_160527 | Not-determined |
| S12_extracted_k141_128982 | Not-determined |
| S12_extracted_k141_160528 | Not-determined |
| S12_extracted_k141_160530 | Not-determined |
| S12_extracted_k141_106050 | Not-determined |
| S12_extracted_k141_82083 | Not-determined |
| S12_extracted_k141_184622 | Low-quality |
| S12_extracted_k141_106057 | Not-determined |
| S12_extracted_k141_43263 | Not-determined |
| S12_extracted_k141_160539 | Not-determined |
| S12_extracted_k141_184625 | Not-determined |
| S12_extracted_k141_113228 | Not-determined |
| S12_extracted_k141_106064 | Not-determined |
| S12_extracted_k141_43267 | Not-determined |
| S12_extracted_k141_26964 | Not-determined |
| S12_extracted_k141_184631 | Not-determined |
| S12_extracted_k141_3773 | Not-determined |
| S3_vs_k141_127387\|\|full | Medium-quality |
| S12_extracted_k141_82092 | Low-quality |
| S12_extracted_k141_145046 | Not-determined |
| S12_extracted_k141_50185 | Not-determined |
| S12_extracted_k141_10578 | Not-determined |
| S12_extracted_k141_145049 | Not-determined |
| S12_extracted_k141_26967 | Not-determined |
| S12_extracted_k141_50188 | Not-determined |
| S12_extracted_k141_106066 | Not-determined |
| S12_extracted_k141_82101 | Not-determined |
| S12_extracted_k141_106069 | Not-determined |
| S12_extracted_k141_82104 | Not-determined |
| S12_extracted_k141_152592 | Not-determined |
| S12_extracted_k141_160550 | Low-quality |
| S12_extracted_k141_26969 | Not-determined |
| S12_extracted_k141_73838 | Not-determined |
| S12_extracted_k141_3792 | Not-determined |
| S12_extracted_k141_43295 | Not-determined |
| S12_extracted_k141_82109 | Not-determined |
| S12_extracted_k141_113247 | Not-determined |
| S12_extracted_k141_50206 | Not-determined |
| S12_extracted_k141_160559 | Not-determined |
| S12_extracted_k141_66511 | Not-determined |
| S12_extracted_k141_19126 | Not-determined |
| S12_extracted_k141_121149 | Not-determined |
| S12_extracted_k141_106080 | Not-determined |
| S12_extracted_k141_98553 | Not-determined |
| S12_extracted_k141_43306 | Not-determined |
| S12_extracted_k141_73843 | Not-determined |
| S12_extracted_k141_73848 | Not-determined |
| S12_extracted_k141_50220 | Not-determined |
| S12_extracted_k141_90521 | Low-quality |
| S12_extracted_k141_106091 | Not-determined |
| S12_extracted_k141_113262 | Not-determined |
| S12_extracted_k141_176142 | Not-determined |
| S12_extracted_k141_121157 | Not-determined |
| S12_extracted_k141_184643 | Not-determined |
| S12_extracted_k141_145085 | Not-determined |
| S12_extracted_k141_184646 | Not-determined |
| S12_extracted_k141_43320 | Not-determined |
| S12_extracted_k141_50224 | Not-determined |
| S12_extracted_k141_73856 | Not-determined |
| S12_extracted_k141_43321 | Not-determined |
| S12_extracted_k141_10610 | Not-determined |
| S12_extracted_k141_50230 | Not-determined |
| S12_extracted_k141_98567 | Low-quality |
| S12_extracted_k141_90535 | Not-determined |
| S12_extracted_k141_90538 | Not-determined |
| S12_extracted_k141_121161 | Not-determined |
| S12_extracted_k141_43331 | Not-determined |
| S12_extracted_k141_10617 | Not-determined |
| S12_extracted_k141_128991 | Not-determined |
| S12_extracted_k141_160572 | Not-determined |
| S12_extracted_k141_121164 | Not-determined |
| S12_extracted_k141_128992 | Not-determined |
| S12_extracted_k141_90542 | Low-quality |
| S12_extracted_k141_176153 | Low-quality |
| S12_extracted_k141_137235 | Low-quality |
| S12_extracted_k141_73867 | Not-determined |
| S12_extracted_k141_121168 | Not-determined |
| S12_extracted_k141_121170 | Not-determined |
| S12_extracted_k141_90546 | Not-determined |
| S12_extracted_k141_121171 | Not-determined |
| S12_extracted_k141_128997 | Low-quality |
| S12_extracted_k141_152607 | Not-determined |
| S12_extracted_k141_43343 | Not-determined |
| S12_extracted_k141_3822 | Not-determined |
| S12_extracted_k141_160584 | Not-determined |
| S12_extracted_k141_19140 | Not-determined |
| S12_extracted_k141_19141 | Not-determined |
| S12_extracted_k141_121177 | Not-determined |
| S12_extracted_k141_121180 | Not-determined |
| S12_extracted_k141_176165 | Not-determined |
| S12_extracted_k141_3829 | Not-determined |
| S12_extracted_k141_160589 | Not-determined |
| S12_extracted_k141_10620 | Not-determined |
| S12_extracted_k141_106117 | Not-determined |
| S12_extracted_k141_137245 | Not-determined |
| S12_extracted_k141_184663 | Not-determined |
| S12_extracted_k141_176168 | Not-determined |
| S12_extracted_k141_121183 | Not-determined |
| S12_extracted_k141_176169 | Not-determined |
| S12_extracted_k141_58715 | Not-determined |
| S12_extracted_k141_152618 | Not-determined |
| S12_extracted_k141_58716 | Low-quality |
| S12_extracted_k141_50263 | Not-determined |
| S12_extracted_k141_50267 | Not-determined |
| S12_extracted_k141_129010 | Not-determined |
| S12_extracted_k141_90563 | Not-determined |
| S12_extracted_k141_160594 | Not-determined |
| S12_extracted_k141_90565 | Not-determined |
| S12_extracted_k141_50270 | Not-determined |
| S12_extracted_k141_160595 | Not-determined |
| S12_extracted_k141_90568 | Not-determined |
| S12_extracted_k141_160598 | Not-determined |
| S12_extracted_k141_145111 | Not-determined |
| S12_extracted_k141_121195 | Not-determined |
| S12_extracted_k141_3843 | Not-determined |
| S12_extracted_k141_90576 | Not-determined |
| S12_extracted_k141_90577 | Not-determined |
| S12_extracted_k141_176177 | Not-determined |
| S12_extracted_k141_137254 | Low-quality |
| S12_extracted_k141_184680 | Not-determined |
| S12_extracted_k141_10639 | Not-determined |
| S12_extracted_k141_3845 | Not-determined |
| S12_extracted_k141_98580 | Not-determined |
| S12_extracted_k141_10643 | Not-determined |
| S12_extracted_k141_121198 | Not-determined |
| S12_extracted_k141_106137 | Not-determined |
| S12_extracted_k141_160605 | Not-determined |
| S12_extracted_k141_176186 | Not-determined |
| S12_extracted_k141_98583 | Not-determined |
| S12_extracted_k141_58717 | Low-quality |
| S12_extracted_k141_106145 | Not-determined |
| S12_extracted_k141_82150 | Low-quality |
| S12_extracted_k141_3855 | Not-determined |
| S12_extracted_k141_145120 | Not-determined |
| S12_extracted_k141_145122 | Low-quality |
| S12_extracted_k141_58724 | Not-determined |
| S12_extracted_k141_137275 | Not-determined |
| S12_extracted_k141_137276 | Not-determined |
| S12_extracted_k141_121211 | Not-determined |
| S12_extracted_k141_121212 | Not-determined |
| S12_extracted_k141_176201 | Not-determined |
| S12_extracted_k141_98600 | Not-determined |
| S12_extracted_k141_137279 | Not-determined |
| S12_extracted_k141_160614 | Not-determined |
| S12_extracted_k141_137280 | Not-determined |
| S12_extracted_k141_98604 | Not-determined |
| S12_extracted_k141_184695 | Not-determined |
| S12_extracted_k141_137283 | Not-determined |
| S12_extracted_k141_137284 | Not-determined |
| S12_extracted_k141_26996 | Not-determined |
| S12_extracted_k141_90587 | Not-determined |
| S12_extracted_k141_58735 | Not-determined |
| S12_extracted_k141_98609 | Not-determined |
| S12_extracted_k141_98610 | Not-determined |
| S12_extracted_k141_58738 | Not-determined |
| S12_extracted_k141_145137 | Low-quality |
| S12_extracted_k141_90589 | Not-determined |
| S12_extracted_k141_137287 | Not-determined |
| S12_extracted_k141_98614 | Not-determined |
| S12_extracted_k141_176207 | Not-determined |
| S12_extracted_k141_73912 | Low-quality |
| S12_extracted_k141_90592 | Low-quality |
| S12_extracted_k141_82165 | Not-determined |
| S12_extracted_k141_90593 | Low-quality |
| S12_extracted_k141_19155 | Not-determined |
| S12_extracted_k141_90594 | Not-determined |
| S12_extracted_k141_176211 | Not-determined |
| S12_extracted_k141_137290 | Not-determined |
| S12_extracted_k141_73917 | Not-determined |
| S12_extracted_k141_137294 | Not-determined |
| S12_extracted_k141_98628 | Not-determined |
| S12_extracted_k141_10676 | Not-determined |
| S12_extracted_k141_27010 | Not-determined |
| S12_extracted_k141_27013 | Not-determined |
| S12_extracted_k141_43355 | Not-determined |
| S12_extracted_k141_145156 | Not-determined |
| S12_extracted_k141_27015 | Not-determined |
| S12_extracted_k141_121228 | Not-determined |
| S12_extracted_k141_3882 | Not-determined |
| S12_extracted_k141_43358 | Not-determined |
| S12_extracted_k141_43361 | Not-determined |
| S12_extracted_k141_121231 | Not-determined |
| S12_extracted_k141_10681 | Not-determined |
| S12_extracted_k141_58767 | Not-determined |
| S12_extracted_k141_145161 | Not-determined |
| S12_extracted_k141_10685 | Not-determined |
| S12_extracted_k141_184710 | Not-determined |
| S12_extracted_k141_27017 | Low-quality |
| S12_extracted_k141_3890 | Not-determined |
| S12_extracted_k141_98634 | Not-determined |
| S12_extracted_k141_137309 | Not-determined |
| S12_extracted_k141_27021 | Not-determined |
| S12_extracted_k141_34398 | Not-determined |
| S12_extracted_k141_10695 | Not-determined |
| S12_extracted_k141_152645 | Not-determined |
| S12_extracted_k141_137311 | Low-quality |
| S12_extracted_k141_34400 | Not-determined |
| S12_extracted_k141_98639 | Not-determined |
| S12_extracted_k141_160630 | Not-determined |
| S12_extracted_k141_160631 | Not-determined |
| S12_extracted_k141_145165 | Not-determined |
| S12_extracted_k141_106188 | Not-determined |
| S12_extracted_k141_106191 | Not-determined |
| S12_extracted_k141_129041 | Not-determined |
| S12_extracted_k141_34408 | Not-determined |
| S12_extracted_k141_184716 | Not-determined |
| S12_extracted_k141_50289 | Not-determined |
| S12_extracted_k141_137314 | Low-quality |
| S12_extracted_k141_50291 | Not-determined |
| S12_extracted_k141_137316 | Not-determined |
| S12_extracted_k141_137317 | Not-determined |
| S12_extracted_k141_129045 | Not-determined |
| S12_extracted_k141_98651 | Not-determined |
| S12_extracted_k141_113285 | Low-quality |
| S12_extracted_k141_27033 | Low-quality |
| S12_extracted_k141_113288 | Not-determined |
| S12_extracted_k141_129052 | Not-determined |
| S12_extracted_k141_106193 | Not-determined |
| S12_extracted_k141_152653 | Low-quality |
| S12_extracted_k141_106194 | Not-determined |
| S12_extracted_k141_3906 | Low-quality |
| S12_extracted_k141_129055 | Not-determined |
| S12_extracted_k141_34429 | Not-determined |
| S12_extracted_k141_10713 | Not-determined |
| S12_extracted_k141_152662 | Not-determined |
| S12_extracted_k141_137321 | Not-determined |
| S12_extracted_k141_137322 | Not-determined |
| S12_extracted_k141_98660 | Low-quality |
| S12_extracted_k141_176243 | Not-determined |
| S12_extracted_k141_129059 | Not-determined |
| S12_extracted_k141_58803 | Not-determined |
| S12_extracted_k141_129061 | Not-determined |
| S12_extracted_k141_129062 | Not-determined |
| S12_extracted_k141_98667 | Not-determined |
| S12_extracted_k141_129063 | Not-determined |
| S12_extracted_k141_50322 | Not-determined |
| S12_extracted_k141_152675 | Not-determined |
| S12_extracted_k141_184742 | Not-determined |
| S5_extracted_k141_106428 | Medium-quality |
| S12_extracted_k141_129066 | Low-quality |
| S12_extracted_k141_10729 | Not-determined |
| S12_extracted_k141_19184 | Not-determined |
| S12_extracted_k141_129068 | Not-determined |
| S12_extracted_k141_184747 | Not-determined |
| S12_extracted_k141_19185 | Not-determined |
| S12_extracted_k141_10732 | Not-determined |
| S12_extracted_k141_152687 | Not-determined |
| S12_extracted_k141_176254 | Not-determined |
| S12_extracted_k141_176256 | Not-determined |
| S12_extracted_k141_19190 | Not-determined |
| S12_extracted_k141_19192 | Not-determined |
| S12_extracted_k141_129079 | Not-determined |
| S12_extracted_k141_137350 | Low-quality |
| S12_extracted_k141_19196 | Not-determined |
| S12_extracted_k141_27053 | Not-determined |
| S12_extracted_k141_121249 | Not-determined |
| S12_extracted_k141_50335 | Not-determined |
| S12_extracted_k141_176260 | Not-determined |
| S12_extracted_k141_121251 | Low-quality |
| S12_extracted_k141_3921 | Not-determined |
| S12_extracted_k141_50338 | Not-determined |
| S12_extracted_k141_106197 | Low-quality |
| S12_extracted_k141_27059 | Not-determined |
| S12_extracted_k141_10744 | Not-determined |
| S12_extracted_k141_3928 | Not-determined |
| S12_extracted_k141_98680 | Low-quality |
| S12_extracted_k141_176261 | Not-determined |
| S12_extracted_k141_43395 | Not-determined |
| S12_extracted_k141_137359 | Not-determined |
| S12_extracted_k141_19210 | Not-determined |
| S12_extracted_k141_27070 | Not-determined |
| S12_extracted_k141_137360 | Not-determined |
| S12_extracted_k141_98687 | Not-determined |
| S12_extracted_k141_10753 | Not-determined |
| S12_extracted_k141_27071 | Not-determined |
| S12_extracted_k141_160645 | Not-determined |
| S12_extracted_k141_184770 | Not-determined |
| S12_extracted_k141_152695 | Not-determined |
| S12_extracted_k141_176271 | Not-determined |
| S12_extracted_k141_10758 | Low-quality |
| S12_extracted_k141_73953 | Not-determined |
| S12_extracted_k141_137367 | Not-determined |
| S12_extracted_k141_43404 | Not-determined |
| S12_extracted_k141_145176 | Not-determined |
| S12_extracted_k141_43405 | Not-determined |
| S12_extracted_k141_176273 | Not-determined |
| S12_extracted_k141_176275 | Not-determined |
| S12_extracted_k141_98701 | Not-determined |
| S12_extracted_k141_176279 | Not-determined |
| S12_extracted_k141_152702 | Not-determined |
| S12_extracted_k141_43410 | Not-determined |
| S12_extracted_k141_145181 | Not-determined |
| S12_extracted_k141_73965 | Not-determined |
| S12_extracted_k141_66521 | Not-determined |
| S12_extracted_k141_129103 | Not-determined |
| S12_extracted_k141_168688 | Not-determined |
| S12_extracted_k141_43415 | Not-determined |
| S12_extracted_k141_98710 | Not-determined |
| S12_extracted_k141_34473 | Not-determined |
| S12_extracted_k141_43418 | Not-determined |
| S12_extracted_k141_184785 | Not-determined |
| S12_extracted_k141_160657 | Not-determined |
| S12_extracted_k141_184787 | Not-determined |
| S12_extracted_k141_137379 | Not-determined |
| S12_extracted_k141_129106 | Not-determined |
| S12_extracted_k141_129107 | Not-determined |
| S12_extracted_k141_160660 | Not-determined |
| S12_extracted_k141_184788 | Not-determined |
| S12_extracted_k141_176286 | Not-determined |
| S12_extracted_k141_160662 | Not-determined |
| S12_extracted_k141_66527 | Not-determined |
| S12_extracted_k141_145191 | Not-determined |
| S12_extracted_k141_145192 | Not-determined |
| S12_extracted_k141_121261 | Not-determined |
| S12_extracted_k141_145195 | Not-determined |
| S12_extracted_k141_113335 | Not-determined |
| S12_extracted_k141_160671 | Not-determined |
| S12_extracted_k141_129113 | Not-determined |
| S12_extracted_k141_129114 | Not-determined |
| S12_extracted_k141_43439 | Low-quality |
| S12_extracted_k141_145207 | Low-quality |
| S12_extracted_k141_10770 | Not-determined |
| S12_extracted_k141_129119 | Not-determined |
| S12_extracted_k141_145208 | Not-determined |
| S12_extracted_k141_129120 | Not-determined |
| S12_extracted_k141_145209 | Not-determined |
| S12_extracted_k141_50355 | Not-determined |
| S12_extracted_k141_152727 | Not-determined |
| S12_extracted_k141_145215 | Low-quality |
| S12_extracted_k141_145216 | Not-determined |
| S12_extracted_k141_10779 | Not-determined |
| S12_extracted_k141_145217 | Not-determined |
| S12_extracted_k141_176307 | Not-determined |
| S12_extracted_k141_98747 | Not-determined |
| S12_extracted_k141_50361 | Not-determined |
| S12_extracted_k141_121280 | Not-determined |
| S12_extracted_k141_168712 | Not-determined |
| S12_extracted_k141_43445 | Not-determined |
| S12_extracted_k141_152736 | Not-determined |
| S12_extracted_k141_43446 | Not-determined |
| S12_extracted_k141_58820 | Not-determined |
| S12_extracted_k141_152740 | Not-determined |
| S12_extracted_k141_43450 | Not-determined |
| S12_extracted_k141_121284 | Not-determined |
| S12_extracted_k141_113351 | Not-determined |
| S12_extracted_k141_73976 | Not-determined |
| S12_extracted_k141_113352 | Not-determined |
| S12_extracted_k141_43452 | Low-quality |
| S12_extracted_k141_121288 | Not-determined |
| S12_extracted_k141_137389 | Not-determined |
| S12_extracted_k141_145236 | Not-determined |
| S12_extracted_k141_145237 | Not-determined |
| S12_extracted_k141_27101 | Not-determined |
| S12_extracted_k141_98761 | Not-determined |
| S12_extracted_k141_82189 | Not-determined |
| S12_extracted_k141_98763 | Low-quality |
| S12_extracted_k141_10799 | Not-determined |
| S12_extracted_k141_10800 | Not-determined |
| S12_extracted_k141_113363 | Not-determined |
| S12_extracted_k141_113364 | Not-determined |
| S12_extracted_k141_121293 | Not-determined |
| S12_extracted_k141_98765 | Not-determined |
| S12_extracted_k141_82197 | Not-determined |
| S12_extracted_k141_27108 | Not-determined |
| S12_extracted_k141_137396 | Not-determined |
| S12_extracted_k141_58833 | Low-quality |
| S12_extracted_k141_176337 | Low-quality |
| S12_extracted_k141_129132 | Low-quality |
| S12_extracted_k141_19246 | Not-determined |
| S12_extracted_k141_168739 | Not-determined |
| S12_extracted_k141_121294 | Not-determined |
| S12_extracted_k141_73983 | Not-determined |
| S12_extracted_k141_82199 | Not-determined |
| S12_extracted_k141_129141 | Not-determined |
| S12_extracted_k141_10815 | Not-determined |
| S12_extracted_k141_43463 | Not-determined |
| S12_extracted_k141_168745 | Low-quality |
| S12_extracted_k141_73986 | Not-determined |
| S12_extracted_k141_168747 | Not-determined |
| S12_extracted_k141_145257 | Not-determined |
| S12_extracted_k141_82208 | Not-determined |
| S12_extracted_k141_168748 | Not-determined |
| S12_extracted_k141_27124 | Not-determined |
| S12_extracted_k141_58845 | Not-determined |
| S12_extracted_k141_145261 | Not-determined |
| S12_extracted_k141_58846 | Not-determined |
| S12_extracted_k141_50382 | Not-determined |
| S12_extracted_k141_121303 | Not-determined |
| S12_extracted_k141_184817 | Not-determined |
| S12_extracted_k141_184821 | Not-determined |
| S12_extracted_k141_50388 | Not-determined |
| S12_extracted_k141_152774 | Not-determined |
| S12_extracted_k141_184824 | Not-determined |
| S12_extracted_k141_50389 | Low-quality |
| S12_extracted_k141_121311 | Not-determined |
| S12_extracted_k141_168758 | Not-determined |
| S12_extracted_k141_27138 | Not-determined |
| S12_extracted_k141_58864 | Not-determined |
| S12_extracted_k141_176359 | Not-determined |
| S12_extracted_k141_27141 | Not-determined |
| S12_extracted_k141_152778 | Low-quality |
| S12_extracted_k141_106206 | Not-determined |
| S12_extracted_k141_113403 | Not-determined |
| S12_extracted_k141_113405 | Low-quality |
| S12_extracted_k141_152783 | Not-determined |
| S12_extracted_k141_121320 | Not-determined |
| S12_extracted_k141_121323 | Not-determined |
| S12_extracted_k141_106213 | Not-determined |
| S12_extracted_k141_113409 | Not-determined |
| S12_extracted_k141_137420 | Not-determined |
| S12_extracted_k141_19251 | Not-determined |
| S12_extracted_k141_82218 | Not-determined |
| S12_extracted_k141_3940 | Not-determined |
| S12_extracted_k141_129166 | Not-determined |
| S12_extracted_k141_106219 | Low-quality |
| S12_extracted_k141_106220 | Not-determined |
| S12_extracted_k141_137426 | Not-determined |
| S12_extracted_k141_3946 | Not-determined |
| S12_extracted_k141_176373 | Low-quality |
| S12_extracted_k141_82223 | Not-determined |
| S12_extracted_k141_3948 | Not-determined |
| S12_extracted_k141_176376 | Not-determined |
| S12_extracted_k141_129172 | Not-determined |
| S12_extracted_k141_129173 | Not-determined |
| S12_extracted_k141_27172 | Not-determined |
| S12_extracted_k141_113422 | Not-determined |
| S12_extracted_k141_137430 | Not-determined |
| S12_extracted_k141_160679 | Not-determined |
| S12_extracted_k141_58884 | Not-determined |
| S12_extracted_k141_27174 | Not-determined |
| S12_extracted_k141_184831 | Not-determined |
| S12_extracted_k141_58885 | Low-quality |
| S12_extracted_k141_50404 | Not-determined |
| S12_extracted_k141_74020 | Not-determined |
| S12_extracted_k141_74024 | Not-determined |
| S12_extracted_k141_129188 | Low-quality |
| S12_extracted_k141_27180 | Not-determined |
| S12_extracted_k141_176378 | Not-determined |
| S12_extracted_k141_106236 | Low-quality |
| S12_extracted_k141_66549 | Not-determined |
| S12_extracted_k141_27183 | Not-determined |
| S12_extracted_k141_121347 | Not-determined |
| S12_extracted_k141_129193 | Not-determined |
| S12_extracted_k141_3958 | Not-determined |
| S12_extracted_k141_3959 | Not-determined |
| S12_extracted_k141_106241 | Not-determined |
| S12_extracted_k141_27187 | Not-determined |
| S12_extracted_k141_74026 | Not-determined |
| S12_extracted_k141_66553 | Not-determined |
| S12_extracted_k141_106242 | Not-determined |
| S12_extracted_k141_129198 | Not-determined |
| S12_extracted_k141_176383 | Not-determined |
| S12_extracted_k141_129199 | Low-quality |
| S12_extracted_k141_50413 | Not-determined |
| S12_extracted_k141_106246 | Not-determined |
| S12_extracted_k141_113435 | Not-determined |
| S12_extracted_k141_160695 | Not-determined |
| S12_extracted_k141_106248 | Not-determined |
| S12_extracted_k141_160696 | Not-determined |
| S12_extracted_k141_66557 | Not-determined |
| S12_extracted_k141_27191 | Not-determined |
| S12_extracted_k141_152796 | Not-determined |
| S12_extracted_k141_10824 | Not-determined |
| S12_extracted_k141_113440 | Not-determined |
| S12_extracted_k141_50421 | Not-determined |
| S12_extracted_k141_129206 | Not-determined |
| S12_extracted_k141_27195 | Not-determined |
| S12_extracted_k141_137450 | Not-determined |
| S12_extracted_k141_98786 | Not-determined |
| S12_extracted_k141_19271 | Not-determined |
| S12_extracted_k141_98789 | Not-determined |
| S12_extracted_k141_106260 | Not-determined |
| S12_extracted_k141_34477 | Not-determined |
| S12_extracted_k141_113445 | Not-determined |
| S12_extracted_k141_176394 | Not-determined |
| S12_extracted_k141_160710 | Not-determined |
| S12_extracted_k141_176396 | Not-determined |
| S12_extracted_k141_106267 | Not-determined |
| S12_extracted_k141_82247 | Not-determined |
| S12_extracted_k141_58903 | Not-determined |
| S12_extracted_k141_3981 | Not-determined |
| S12_extracted_k141_43486 | Not-determined |
| S12_extracted_k141_106270 | Not-determined |
| S12_extracted_k141_184840 | Not-determined |
| S12_extracted_k141_82249 | Not-determined |
| S12_extracted_k141_34485 | Not-determined |
| S12_extracted_k141_58909 | Not-determined |
| S12_extracted_k141_50428 | Not-determined |
| S12_extracted_k141_145274 | Not-determined |
| S12_extracted_k141_34486 | Not-determined |
| S12_extracted_k141_82253 | Not-determined |
| S12_extracted_k141_152805 | Not-determined |
| S12_extracted_k141_129226 | Not-determined |
| S12_extracted_k141_137460 | Not-determined |
| S12_extracted_k141_34489 | Not-determined |
| S12_extracted_k141_152807 | Not-determined |
| S12_extracted_k141_106273 | Not-determined |
| S12_extracted_k141_3991 | Not-determined |
| S12_extracted_k141_3992 | Not-determined |
| S12_extracted_k141_152810 | Not-determined |
| S12_extracted_k141_74033 | Not-determined |
| S12_extracted_k141_145285 | Not-determined |
| S12_extracted_k141_113461 | Not-determined |
| S12_extracted_k141_98816 | Not-determined |
| S12_extracted_k141_152816 | Low-quality |
| S12_extracted_k141_152817 | Not-determined |
| S12_extracted_k141_106283 | Not-determined |
| S12_extracted_k141_98819 | Low-quality |
| S12_extracted_k141_137479 | Not-determined |
| S12_extracted_k141_4001 | Not-determined |
| S12_extracted_k141_50455 | Not-determined |
| S12_extracted_k141_129253 | Not-determined |
| S12_extracted_k141_129254 | Not-determined |
| S12_extracted_k141_34507 | Not-determined |
| S12_extracted_k141_145296 | Not-determined |
| S12_extracted_k141_145297 | Not-determined |
| S12_extracted_k141_58940 | Not-determined |
| S12_extracted_k141_66575 | Not-determined |
| S12_extracted_k141_137493 | Not-determined |
| S12_extracted_k141_27222 | Not-determined |
| S12_extracted_k141_82260 | Not-determined |
| S12_extracted_k141_98830 | Not-determined |
| S12_extracted_k141_184858 | Not-determined |
| S12_extracted_k141_145300 | Not-determined |
| S12_extracted_k141_50464 | Not-determined |
| S12_extracted_k141_98831 | Low-quality |
| S12_extracted_k141_137497 | Low-quality |
| S12_extracted_k141_184859 | Not-determined |
| S12_extracted_k141_106296 | Not-determined |
| S12_extracted_k141_129262 | Not-determined |
| S12_extracted_k141_113478 | Not-determined |
| S12_extracted_k141_10857 | Not-determined |

| | |
|---|---|
| S12_extracted_k141_121356 | Not-determined |
| S12_extracted_k141_98834 | Not-determined |
| S12_extracted_k141_43506 | Not-determined |
| S12_extracted_k141_10862 | Not-determined |
| S12_extracted_k141_184866 | Low-quality |
| S12_extracted_k141_98836 | Not-determined |
| S12_extracted_k141_184867 | Not-determined |
| S12_extracted_k141_137508 | Not-determined |
| S12_extracted_k141_145317 | Not-determined |
| S12_extracted_k141_106307 | Not-determined |
| S12_extracted_k141_34519 | Not-determined |
| S12_extracted_k141_152840 | Not-determined |
| S12_extracted_k141_145320 | Not-determined |
| S12_extracted_k141_168768 | Not-determined |
| S12_extracted_k141_74037 | Not-determined |
| S12_extracted_k141_168771 | Not-determined |
| S12_extracted_k141_74040 | Not-determined |
| S12_extracted_k141_82271 | Not-determined |
| S12_extracted_k141_4016 | Not-determined |
| S12_extracted_k141_129280 | Not-determined |
| S12_extracted_k141_82274 | Not-determined |
| S12_extracted_k141_43513 | Not-determined |
| S12_extracted_k141_129281 | Not-determined |
| S12_extracted_k141_10874 | Not-determined |
| S12_extracted_k141_58953 | Not-determined |
| S12_extracted_k141_43517 | Not-determined |
| S12_extracted_k141_10881 | Not-determined |
| S12_extracted_k141_168780 | Not-determined |
| S12_extracted_k141_74052 | Not-determined |
| S12_extracted_k141_10882 | Not-determined |
| S12_extracted_k141_152846 | Not-determined |
| S12_extracted_k141_50477 | Not-determined |
| S12_extracted_k141_145329 | Not-determined |
| S12_extracted_k141_82277 | Not-determined |
| S12_extracted_k141_4025 | Not-determined |
| S12_extracted_k141_74059 | Not-determined |
| S12_extracted_k141_129298 | Not-determined |
| S12_extracted_k141_145338 | Low-quality |
| S12_extracted_k141_58965 | Not-determined |
| S12_extracted_k141_66597 | Not-determined |
| S12_extracted_k141_66598 | Not-determined |
| S12_extracted_k141_98849 | Low-quality |
| S12_extracted_k141_27257 | Not-determined |
| S12_extracted_k141_145348 | Not-determined |
| S12_extracted_k141_121366 | Not-determined |
| S12_extracted_k141_98852 | Not-determined |
| S12_extracted_k141_113503 | Not-determined |
| S12_extracted_k141_50496 | Not-determined |
| S12_extracted_k141_145352 | Not-determined |
| S12_extracted_k141_113506 | Not-determined |
| S12_extracted_k141_82289 | Not-determined |
| S12_extracted_k141_43541 | Not-determined |
| S12_extracted_k141_129311 | Not-determined |
| S12_extracted_k141_168805 | Not-determined |
| S12_extracted_k141_66606 | Not-determined |
| S12_extracted_k141_98856 | Not-determined |
| S12_extracted_k141_50499 | Not-determined |
| S12_extracted_k141_121374 | Low-quality |
| S12_extracted_k141_98859 | Not-determined |
| S12_extracted_k141_168808 | Low-quality |
| S12_extracted_k141_66614 | Not-determined |
| S12_extracted_k141_74077 | Not-determined |
| S12_extracted_k141_121377 | Low-quality |
| S12_extracted_k141_137545 | Not-determined |
| S12_extracted_k141_98864 | Not-determined |
| S12_extracted_k141_82297 | Not-determined |
| S12_extracted_k141_129327 | Not-determined |
| S12_extracted_k141_82303 | Not-determined |
| S12_extracted_k141_184892 | Not-determined |
| S12_extracted_k141_152866 | Not-determined |
| S12_extracted_k141_121386 | Not-determined |
| S12_extracted_k141_74085 | Not-determined |
| S12_extracted_k141_152867 | Not-determined |
| S12_extracted_k141_27277 | Not-determined |
| S12_extracted_k141_27278 | Not-determined |
| S12_extracted_k141_82311 | Not-determined |
| S12_extracted_k141_19320 | Not-determined |
| S12_extracted_k141_121391 | Not-determined |
| S12_extracted_k141_168810 | Not-determined |
| S12_extracted_k141_129333 | Low-quality |
| S12_extracted_k141_74095 | Not-determined |
| S12_extracted_k141_121396 | Not-determined |
| S12_extracted_k141_27289 | Low-quality |
| S12_extracted_k141_58974 | Not-determined |
| S12_extracted_k141_145383 | Not-determined |
| S12_extracted_k141_176402 | Low-quality |
| S12_extracted_k141_145384 | Not-determined |
| S12_extracted_k141_74099 | Not-determined |
| S12_extracted_k141_145385 | Low-quality |
| S12_extracted_k141_121403 | Not-determined |
| S12_extracted_k141_90642 | Not-determined |
| S12_extracted_k141_27295 | Not-determined |
| S12_extracted_k141_43553 | Not-determined |
| S12_extracted_k141_121405 | Not-determined |
| S12_extracted_k141_82322 | Not-determined |
| S12_extracted_k141_121407 | Not-determined |
| S12_extracted_k141_19333 | Not-determined |
| S12_extracted_k141_160722 | Not-determined |
| S12_extracted_k141_19337 | Not-determined |
| S12_extracted_k141_19338 | Not-determined |
| S12_extracted_k141_121409 | Not-determined |
| S12_extracted_k141_184907 | Not-determined |
| S12_extracted_k141_19339 | Not-determined |
| S12_extracted_k141_19340 | Not-determined |
| S12_extracted_k141_98898 | Not-determined |
| S12_extracted_k141_160725 | Not-determined |
| S12_extracted_k141_58984 | Not-determined |
| S12_extracted_k141_74110 | Not-determined |
| S12_extracted_k141_137575 | Not-determined |
| S12_extracted_k141_27305 | Not-determined |
| S12_extracted_k141_160732 | Not-determined |
| S12_extracted_k141_74115 | Not-determined |
| S12_extracted_k141_43561 | Not-determined |
| S12_extracted_k141_43563 | Not-determined |
| S12_extracted_k141_145388 | Low-quality |
| S12_extracted_k141_82324 | Not-determined |
| S12_extracted_k141_43567 | Not-determined |
| S12_extracted_k141_19347 | Not-determined |
| S12_extracted_k141_168839 | Not-determined |
| S12_extracted_k141_66629 | Not-determined |
| S12_extracted_k141_58992 | Not-determined |
| S12_extracted_k141_27317 | Not-determined |
| S12_extracted_k141_58995 | Not-determined |
| S12_extracted_k141_66633 | Not-determined |
| S12_extracted_k141_50551 | Not-determined |
| S12_extracted_k141_82335 | Not-determined |
| S12_extracted_k141_145398 | Not-determined |
| S12_extracted_k141_98921 | Low-quality |
| S12_extracted_k141_98925 | Not-determined |
| S12_extracted_k141_66644 | Not-determined |
| S12_extracted_k141_43586 | Not-determined |
| S12_extracted_k141_66645 | Not-determined |
| S12_extracted_k141_113525 | Not-determined |
| S12_extracted_k141_113526 | Not-determined |
| S12_extracted_k141_66647 | Not-determined |
| S12_extracted_k141_129346 | Not-determined |
| S12_extracted_k141_66649 | Not-determined |
| S12_extracted_k141_34587 | Not-determined |
| S12_extracted_k141_59004 | Low-quality |
| S12_extracted_k141_106336 | Low-quality |
| S12_extracted_k141_74144 | Not-determined |
| S12_extracted_k141_113534 | Not-determined |
| S12_extracted_k141_59005 | Not-determined |
| S12_extracted_k141_43599 | Not-determined |
| S12_extracted_k141_34593 | Not-determined |
| S12_extracted_k141_98941 | Not-determined |
| S12_extracted_k141_50566 | Not-determined |
| S12_extracted_k141_50567 | Not-determined |
| S12_extracted_k141_19368 | Low-quality |
| S12_extracted_k141_82354 | Not-determined |
| S12_extracted_k141_50568 | Not-determined |
| S12_extracted_k141_50569 | Not-determined |
| S12_extracted_k141_43607 | Not-determined |
| S12_extracted_k141_59013 | Not-determined |
| S12_extracted_k141_27346 | Not-determined |
| S12_extracted_k141_113545 | Not-determined |
| S12_extracted_k141_113546 | Low-quality |
| S12_extracted_k141_98943 | Low-quality |
| S1_vs_k141_100150\|\|full | Medium-quality |
| S12_extracted_k141_50575 | Not-determined |
| S12_extracted_k141_152881 | Not-determined |
| S12_extracted_k141_19371 | Not-determined |
| S12_extracted_k141_82362 | Not-determined |
| S12_extracted_k141_82363 | Not-determined |
| S12_extracted_k141_152885 | Low-quality |
| S12_extracted_k141_184931 | Not-determined |
| S12_extracted_k141_129367 | Not-determined |
| S12_extracted_k141_34619 | Not-determined |
| S12_extracted_k141_74160 | Not-determined |
| S12_extracted_k141_34620 | Not-determined |
| S12_extracted_k141_121439 | Not-determined |
| S12_extracted_k141_10895 | Not-determined |
| S12_extracted_k141_19376 | Not-determined |
| S12_extracted_k141_121440 | Not-determined |
| S12_extracted_k141_66676 | Not-determined |
| S12_extracted_k141_152894 | Not-determined |
| S12_extracted_k141_106345 | Not-determined |
| S12_extracted_k141_90660 | Not-determined |
| S12_extracted_k141_106349 | Not-determined |
| S12_extracted_k141_152897 | Not-determined |
| S12_extracted_k141_43629 | Not-determined |
| S12_extracted_k141_129373 | Not-determined |
| S12_extracted_k141_74169 | Not-determined |
| S12_extracted_k141_27368 | Not-determined |
| S12_extracted_k141_184947 | Not-determined |
| S12_extracted_k141_129374 | Not-determined |
| S12_extracted_k141_10900 | Not-determined |
| S12_extracted_k141_184950 | Not-determined |
| S12_extracted_k141_113559 | Not-determined |
| S12_extracted_k141_74177 | Not-determined |
| S12_extracted_k141_184951 | Not-determined |
| S12_extracted_k141_50589 | Not-determined |
| S12_extracted_k141_74178 | Not-determined |
| S12_extracted_k141_121446 | Not-determined |
| S12_extracted_k141_113562 | Not-determined |
| S12_extracted_k141_19389 | Not-determined |
| S12_extracted_k141_10901 | Low-quality |
| S12_extracted_k141_168863 | Not-determined |
| S12_extracted_k141_34633 | Not-determined |
| S12_extracted_k141_66681 | Not-determined |
| S12_extracted_k141_90673 | Not-determined |
| S12_extracted_k141_19394 | Not-determined |
| S12_extracted_k141_113570 | Not-determined |
| S12_extracted_k141_168866 | Not-determined |
| S12_extracted_k141_59034 | Not-determined |
| S12_extracted_k141_82389 | Not-determined |
| S12_extracted_k141_160747 | Low-quality |
| S12_extracted_k141_145445 | Not-determined |
| S12_extracted_k141_113577 | Not-determined |
| S12_extracted_k141_50594 | Low-quality |
| S12_extracted_k141_19411 | Not-determined |
| S12_extracted_k141_160751 | Not-determined |
| S12_extracted_k141_50598 | Not-determined |
| S12_extracted_k141_137605 | Not-determined |
| S12_extracted_k141_152912 | Low-quality |
| S12_extracted_k141_121459 | Not-determined |
| S12_extracted_k141_50602 | Not-determined |
| S12_extracted_k141_106362 | Not-determined |
| S12_extracted_k141_90683 | Not-determined |
| S12_extracted_k141_121463 | Not-determined |
| S12_extracted_k141_19421 | Not-determined |
| S12_extracted_k141_160755 | Not-determined |
| S12_extracted_k141_74191 | Low-quality |
| S12_extracted_k141_168879 | Not-determined |
| S12_extracted_k141_168881 | Low-quality |
| S12_extracted_k141_160758 | Not-determined |
| S12_extracted_k141_43665 | Not-determined |
| S12_extracted_k141_129394 | Not-determined |
| S12_extracted_k141_59050 | Not-determined |
| S7_vs_108680\|\|full | Medium-quality |
| S12_extracted_k141_50620 | Not-determined |
| S12_extracted_k141_59052 | Not-determined |
| S12_extracted_k141_19430 | Not-determined |
| S12_extracted_k141_66713 | Not-determined |
| S12_extracted_k141_27379 | Not-determined |
| S12_extracted_k141_129402 | Not-determined |
| S12_extracted_k141_10935 | Not-determined |
| S5_vs_k141_139401\|\|full | Medium-quality |
| S12_extracted_k141_74206 | Low-quality |
| S12_extracted_k141_34650 | Not-determined |
| S12_extracted_k141_152925 | Not-determined |
| S12_extracted_k141_4034 | Not-determined |
| S12_extracted_k141_168898 | Not-determined |
| S12_extracted_k141_129406 | Not-determined |
| S12_extracted_k141_66722 | Not-determined |
| S12_extracted_k141_98980 | Not-determined |
| S12_extracted_k141_98981 | Not-determined |
| S12_extracted_k141_160767 | Low-quality |
| S12_extracted_k141_160769 | Low-quality |
| S12_extracted_k141_160770 | Not-determined |
| S12_extracted_k141_50627 | Low-quality |
| S12_extracted_k141_152934 | Low-quality |
| S12_extracted_k141_160773 | Not-determined |
| S12_extracted_k141_27388 | Not-determined |
| S12_extracted_k141_82399 | Low-quality |
| S12_extracted_k141_184988 | Not-determined |
| S12_extracted_k141_160775 | Not-determined |
| S12_extracted_k141_160777 | Not-determined |
| S12_extracted_k141_129413 | Not-determined |
| S12_extracted_k141_137626 | Low-quality |
| S12_extracted_k141_82406 | Not-determined |
| S12_extracted_k141_27398 | Not-determined |
| S12_extracted_k141_27399 | Not-determined |
| S12_extracted_k141_152941 | Not-determined |
| S12_extracted_k141_82408 | Not-determined |
| S12_extracted_k141_176445 | Not-determined |
| S12_extracted_k141_176446 | Not-determined |
| S12_extracted_k141_152945 | Not-determined |
| S12_extracted_k141_90716 | Not-determined |
| S12_extracted_k141_176448 | Not-determined |
| S12_extracted_k141_184994 | Not-determined |
| S12_extracted_k141_184995 | Not-determined |
| S12_extracted_k141_152947 | Not-determined |
| S12_extracted_k141_176451 | Not-determined |
| S12_extracted_k141_66740 | Not-determined |
| S12_extracted_k141_10955 | Not-determined |
| S12_extracted_k141_10956 | Low-quality |
| S12_extracted_k141_74224 | Not-determined |
| S12_extracted_k141_10958 | Low-quality |
| S12_extracted_k141_145453 | Not-determined |
| S12_extracted_k141_34670 | Not-determined |
| S12_extracted_k141_90725 | Not-determined |
| S12_extracted_k141_152955 | Not-determined |
| S12_extracted_k141_160793 | Low-quality |
| S12_extracted_k141_10962 | Not-determined |
| S12_extracted_k141_82421 | Not-determined |
| S12_extracted_k141_27415 | Not-determined |
| S12_extracted_k141_152957 | Not-determined |
| S12_extracted_k141_90728 | Not-determined |
| S12_extracted_k141_160797 | Not-determined |
| S12_extracted_k141_121492 | Not-determined |
| S12_extracted_k141_99000 | Not-determined |
| S12_extracted_k141_99001 | Low-quality |
| S12_extracted_k141_34677 | Not-determined |
| S12_extracted_k141_185006 | Not-determined |
| S12_extracted_k141_145456 | Not-determined |
| S12_extracted_k141_185009 | Low-quality |
| S12_extracted_k141_99004 | Not-determined |
| S12_extracted_k141_27427 | Not-determined |
| S12_extracted_k141_185012 | Not-determined |
| S12_extracted_k141_137645 | Not-determined |
| S12_extracted_k141_160806 | Not-determined |
| S12_extracted_k141_34681 | Not-determined |
| S12_extracted_k141_160808 | Not-determined |
| S12_extracted_k141_185015 | Not-determined |
| S12_extracted_k141_82434 | Not-determined |
| S12_extracted_k141_145465 | Not-determined |
| S12_extracted_k141_185016 | Not-determined |
| S12_extracted_k141_59072 | Not-determined |
| S12_extracted_k141_34682 | Not-determined |
| S12_extracted_k141_185018 | Not-determined |
| S12_extracted_k141_121499 | Low-quality |
| S12_extracted_k141_43682 | Not-determined |
| S12_extracted_k141_19449 | Not-determined |
| S12_extracted_k141_34686 | Low-quality |
| S12_extracted_k141_43683 | Not-determined |
| S12_extracted_k141_34687 | Not-determined |
| S12_extracted_k141_121503 | Not-determined |
| S12_extracted_k141_27450 | Not-determined |
| S12_extracted_k141_185029 | Not-determined |
| S12_extracted_k141_34690 | Not-determined |
| S12_extracted_k141_185032 | Not-determined |
| S12_extracted_k141_10968 | Low-quality |
| S12_extracted_k141_145478 | Not-determined |
| S12_extracted_k141_106386 | Not-determined |
| S12_extracted_k141_50649 | Not-determined |
| S12_extracted_k141_106387 | Not-determined |
| S12_extracted_k141_82443 | Not-determined |
| S12_extracted_k141_145481 | Not-determined |
| S12_extracted_k141_121513 | Not-determined |
| S12_extracted_k141_152979 | Not-determined |
| S12_extracted_k141_27457 | Not-determined |
| S12_extracted_k141_4052 | Not-determined |
| S12_extracted_k141_121515 | Not-determined |
| S12_extracted_k141_168950 | Not-determined |
| S12_extracted_k141_152984 | Not-determined |
| S12_extracted_k141_137674 | Not-determined |
| S12_extracted_k141_185043 | Not-determined |
| S12_extracted_k141_74256 | Not-determined |
| S12_extracted_k141_43708 | Not-determined |
| S12_extracted_k141_152991 | Not-determined |
| S12_extracted_k141_50659 | Not-determined |
| S12_extracted_k141_19453 | Not-determined |
| S12_extracted_k141_152994 | Low-quality |
| S12_extracted_k141_43712 | Not-determined |
| S12_extracted_k141_168955 | Not-determined |
| S12_extracted_k141_4057 | Not-determined |
| S12_extracted_k141_59091 | Not-determined |
| S12_extracted_k141_185049 | Not-determined |
| S12_extracted_k141_106404 | Not-determined |
| S12_extracted_k141_34711 | Not-determined |
| S12_extracted_k141_74272 | Not-determined |
| S12_extracted_k141_121525 | Not-determined |
| S12_extracted_k141_145497 | Not-determined |
| S12_extracted_k141_152999 | Not-determined |
| S12_extracted_k141_90775 | Not-determined |
| S12_extracted_k141_106413 | Low-quality |
| S12_extracted_k141_50672 | Not-determined |
| S12_extracted_k141_153005 | Not-determined |
| S12_extracted_k141_34723 | Not-determined |
| S12_extracted_k141_153007 | Low-quality |
| S12_extracted_k141_145503 | Not-determined |
| S12_extracted_k141_50677 | Not-determined |
| S12_extracted_k141_4069 | Not-determined |
| S12_extracted_k141_10998 | Not-determined |
| S12_extracted_k141_160820 | Not-determined |
| S12_extracted_k141_34729 | Low-quality |
| S12_extracted_k141_11002 | Not-determined |
| S12_extracted_k141_50683 | Not-determined |
| S12_extracted_k141_43731 | Not-determined |
| S12_extracted_k141_90785 | Not-determined |
| S12_extracted_k141_27463 | Not-determined |
| S12_extracted_k141_59100 | Not-determined |
| S12_extracted_k141_106428 | Not-determined |
| S12_extracted_k141_168967 | Not-determined |
| S12_extracted_k141_121549 | Not-determined |
| S12_extracted_k141_113589 | Low-quality |
| S12_extracted_k141_74281 | Low-quality |
| S12_extracted_k141_50697 | Not-determined |
| S12_extracted_k141_99025 | Not-determined |
| S12_extracted_k141_137708 | Not-determined |
| S12_extracted_k141_160845 | Not-determined |
| S12_extracted_k141_66808 | Not-determined |
| S12_extracted_k141_19469 | Not-determined |
| S12_extracted_k141_106443 | Not-determined |
| S12_extracted_k141_106444 | Not-determined |
| S12_extracted_k141_34743 | Low-quality |
| S12_extracted_k141_113601 | Not-determined |
| S12_extracted_k141_19473 | Not-determined |
| S12_extracted_k141_43746 | Not-determined |
| S12_extracted_k141_185055 | Not-determined |
| S12_extracted_k141_160851 | Not-determined |
| S12_extracted_k141_113605 | Not-determined |
| S12_extracted_k141_160854 | Not-determined |
| S12_extracted_k141_113607 | Low-quality |
| S12_extracted_k141_145515 | Not-determined |
| S12_extracted_k141_4100 | Not-determined |
| S12_extracted_k141_160858 | Not-determined |
| S12_extracted_k141_50711 | Not-determined |
| S12_extracted_k141_4102 | Not-determined |
| S12_extracted_k141_168983 | Low-quality |
| S12_extracted_k141_66819 | Not-determined |
| S12_extracted_k141_153028 | Not-determined |
| S12_extracted_k141_137720 | Low-quality |
| S12_extracted_k141_137722 | Not-determined |
| S12_extracted_k141_74295 | Not-determined |
| S12_extracted_k141_34744 | Not-determined |
| S12_extracted_k141_4105 | Not-determined |
| S12_extracted_k141_34746 | Not-determined |
| S12_extracted_k141_160864 | Not-determined |
| S12_extracted_k141_145525 | Not-determined |
| S12_extracted_k141_106466 | Not-determined |
| S12_extracted_k141_113614 | Not-determined |
| S12_extracted_k141_113616 | Not-determined |
| S12_extracted_k141_145526 | Not-determined |
| S12_extracted_k141_90811 | Not-determined |
| S12_extracted_k141_27497 | Not-determined |
| S12_extracted_k141_121581 | Not-determined |
| S12_extracted_k141_106471 | Low-quality |
| S12_extracted_k141_27498 | Not-determined |
| S12_extracted_k141_153030 | Not-determined |
| S12_extracted_k141_90814 | Not-determined |
| S12_extracted_k141_66825 | Not-determined |
| S12_extracted_k141_129475 | Low-quality |
| S12_extracted_k141_121585 | Not-determined |
| S12_extracted_k141_27501 | Not-determined |
| S12_extracted_k141_4112 | Not-determined |
| S12_extracted_k141_90817 | Not-determined |
| S12_extracted_k141_185071 | Not-determined |
| S12_extracted_k141_185075 | Not-determined |
| S12_extracted_k141_4116 | Not-determined |
| S12_extracted_k141_145539 | Not-determined |
| S12_extracted_k141_90828 | Not-determined |
| S12_extracted_k141_19495 | Not-determined |
| S12_extracted_k141_66838 | Not-determined |
| S12_extracted_k141_106479 | Not-determined |
| S12_extracted_k141_169000 | Not-determined |
| S12_extracted_k141_113632 | Low-quality |
| S12_extracted_k141_4125 | Low-quality |
| S12_extracted_k141_59144 | Not-determined |
| S12_extracted_k141_121599 | Not-determined |
| S12_extracted_k141_34769 | Not-determined |
| S12_extracted_k141_153042 | Not-determined |
| S12_extracted_k141_66850 | Not-determined |
| S12_extracted_k141_99041 | Not-determined |
| S12_extracted_k141_145557 | Not-determined |
| S12_extracted_k141_185091 | Not-determined |
| S12_extracted_k141_27520 | Not-determined |
| S12_extracted_k141_50719 | Not-determined |
| S12_extracted_k141_121607 | Not-determined |
| S12_extracted_k141_121608 | Not-determined |
| S12_extracted_k141_66854 | Not-determined |
| S12_extracted_k141_169013 | Not-determined |
| S12_extracted_k141_43778 | Not-determined |
| S12_extracted_k141_43779 | Not-determined |
| S12_extracted_k141_185095 | Not-determined |
| S12_extracted_k141_185097 | Not-determined |
| S12_extracted_k141_19507 | Low-quality |
| S12_extracted_k141_90854 | Not-determined |
| S12_extracted_k141_43782 | Not-determined |
| S12_extracted_k141_99047 | Not-determined |
| S12_extracted_k141_121616 | Not-determined |
| S12_extracted_k141_113644 | Not-determined |
| S12_extracted_k141_50729 | Not-determined |
| S12_extracted_k141_169020 | Low-quality |
| S12_extracted_k141_145571 | Not-determined |
| S12_extracted_k141_153056 | Low-quality |
| S12_extracted_k141_59160 | Low-quality |
| S12_extracted_k141_137746 | Not-determined |
| S12_extracted_k141_169023 | Not-determined |
| S12_extracted_k141_99056 | Not-determined |
| S12_extracted_k141_185111 | Not-determined |
| S12_extracted_k141_27531 | Not-determined |
| S12_extracted_k141_59166 | Not-determined |
| S12_extracted_k141_137753 | Not-determined |
| S12_extracted_k141_27535 | Not-determined |
| S12_extracted_k141_185118 | Not-determined |
| S12_extracted_k141_145583 | Not-determined |
| S12_extracted_k141_27538 | Not-determined |
| S12_extracted_k141_59168 | Not-determined |
| S12_extracted_k141_121630 | Not-determined |
| S12_extracted_k141_106498 | Low-quality |
| S12_extracted_k141_137759 | Not-determined |
| S12_extracted_k141_106504 | Not-determined |
| S12_extracted_k141_90866 | Low-quality |
| S12_extracted_k141_137761 | Not-determined |
| S12_extracted_k141_99061 | Not-determined |
| S12_extracted_k141_160882 | Not-determined |
| S12_extracted_k141_82470 | Not-determined |
| S12_extracted_k141_4147 | Not-determined |
| S12_extracted_k141_106511 | Not-determined |
| S12_extracted_k141_113656 | Not-determined |
| S12_extracted_k141_27548 | Low-quality |
| S12_extracted_k141_106514 | Not-determined |
| S12_extracted_k141_160893 | Not-determined |
| S12_extracted_k141_185129 | Low-quality |
| S12_extracted_k141_59179 | Low-quality |
| S12_extracted_k141_169035 | Not-determined |
| S12_extracted_k141_160896 | Not-determined |
| S12_extracted_k141_19511 | Not-determined |
| S12_extracted_k141_82479 | Not-determined |
| S12_extracted_k141_59185 | Not-determined |
| S12_extracted_k141_11014 | Not-determined |
| S12_extracted_k141_19513 | Not-determined |
| S12_extracted_k141_106520 | Not-determined |
| S12_extracted_k141_145606 | Not-determined |
| S12_extracted_k141_137777 | Not-determined |
| S12_extracted_k141_145607 | Not-determined |
| S12_extracted_k141_82489 | Not-determined |
| S12_extracted_k141_59192 | Not-determined |
| S12_extracted_k141_160906 | Low-quality |
| S12_extracted_k141_137779 | Not-determined |
| S12_extracted_k141_160907 | Not-determined |
| S12_extracted_k141_19518 | Not-determined |
| S12_extracted_k141_160913 | Not-determined |
| S12_extracted_k141_50756 | Not-determined |
| S12_extracted_k141_99090 | Low-quality |
| S12_extracted_k141_59203 | Low-quality |
| S12_extracted_k141_50759 | Not-determined |
| S12_extracted_k141_59205 | Not-determined |
| S12_extracted_k141_106526 | Not-determined |
| S12_extracted_k141_50760 | Low-quality |
| S12_extracted_k141_160920 | Not-determined |
| S12_extracted_k141_185146 | Not-determined |
| S12_extracted_k141_34816 | Not-determined |
| S12_extracted_k141_153099 | Low-quality |
| S12_extracted_k141_129505 | Not-determined |
| S12_extracted_k141_113680 | Not-determined |
| S12_extracted_k141_169058 | Not-determined |
| S12_extracted_k141_59212 | Not-determined |
| S12_extracted_k141_113682 | Not-determined |
| S12_extracted_k141_106528 | Not-determined |
| S12_extracted_k141_169060 | Not-determined |
| S12_extracted_k141_27571 | Not-determined |
| S12_extracted_k141_129511 | Not-determined |
| S12_extracted_k141_50767 | Not-determined |
| S12_extracted_k141_106531 | Not-determined |
| S12_extracted_k141_90871 | Not-determined |
| S12_extracted_k141_90872 | Not-determined |
| S12_extracted_k141_11023 | Not-determined |
| S12_extracted_k141_160926 | Not-determined |
| S12_extracted_k141_90879 | Low-quality |
| S12_extracted_k141_43787 | Not-determined |
| S12_extracted_k141_19528 | Not-determined |
| S12_extracted_k141_4153 | Not-determined |
| S12_extracted_k141_106540 | Not-determined |
| S12_extracted_k141_185164 | Low-quality |
| S12_extracted_k141_34839 | Not-determined |
| S12_extracted_k141_4155 | Not-determined |
| S12_extracted_k141_153110 | Not-determined |
| S12_extracted_k141_153116 | Not-determined |
| S12_extracted_k141_185168 | Low-quality |
| S12_extracted_k141_129516 | Not-determined |
| S12_extracted_k141_121682 | Not-determined |
| S12_extracted_k141_185174 | Not-determined |
| S12_extracted_k141_137801 | Not-determined |
| S12_extracted_k141_185177 | Not-determined |
| S12_extracted_k141_169081 | Not-determined |
| S12_extracted_k141_129523 | Not-determined |
| S12_extracted_k141_50786 | Not-determined |
| S12_extracted_k141_82512 | Not-determined |
| S12_extracted_k141_137809 | Not-determined |
| S12_extracted_k141_185184 | Not-determined |
| S12_extracted_k141_153130 | Not-determined |
| S12_extracted_k141_169089 | Not-determined |
| S12_extracted_k141_129528 | Not-determined |
| S12_extracted_k141_90885 | Not-determined |
| S12_extracted_k141_106554 | Not-determined |
| S12_extracted_k141_169091 | Not-determined |
| S12_extracted_k141_19541 | Not-determined |
| S12_extracted_k141_82514 | Not-determined |
| S12_extracted_k141_90888 | Not-determined |
| S12_extracted_k141_19543 | Not-determined |
| S12_extracted_k141_160946 | Not-determined |
| S12_extracted_k141_145635 | Low-quality |
| S12_extracted_k141_11034 | Not-determined |
| S12_extracted_k141_185199 | Not-determined |
| S12_extracted_k141_169097 | Not-determined |
| S12_extracted_k141_169098 | Not-determined |
| S12_extracted_k141_153141 | Not-determined |
| S12_extracted_k141_50794 | Not-determined |
| S12_extracted_k141_145637 | Not-determined |
| S12_extracted_k141_169101 | Not-determined |
| S12_extracted_k141_153142 | Not-determined |
| S12_extracted_k141_82524 | Not-determined |
| S12_extracted_k141_11041 | Not-determined |
| S12_extracted_k141_169106 | Not-determined |
| S12_extracted_k141_153148 | Not-determined |
| S12_extracted_k141_34854 | Not-determined |

| | |
|---|---|
| S12_extracted_k141_82526 | Low-quality |
| S12_extracted_k141_43815 | Not-determined |
| S12_extracted_k141_4184 | Not-determined |
| S12_extracted_k141_145644 | Not-determined |
| S12_extracted_k141_160951 | Not-determined |
| S12_extracted_k141_82528 | Not-determined |
| S12_extracted_k141_11042 | Not-determined |
| S12_extracted_k141_27597 | Not-determined |
| S12_extracted_k141_145647 | Not-determined |
| S12_extracted_k141_19557 | Not-determined |
| S12_extracted_k141_185210 | Not-determined |
| S12_extracted_k141_90905 | Not-determined |
| S12_extracted_k141_11047 | Not-determined |
| S12_extracted_k141_160959 | Not-determined |
| S12_extracted_k141_27600 | Low-quality |
| S12_extracted_k141_99098 | Not-determined |
| S12_extracted_k141_185216 | Not-determined |
| S12_extracted_k141_90909 | Not-determined |
| S12_extracted_k141_19560 | Not-determined |
| S12_extracted_k141_19561 | Not-determined |
| S12_extracted_k141_145654 | Not-determined |
| S12_extracted_k141_11054 | Not-determined |
| S12_extracted_k141_90913 | Not-determined |
| S12_extracted_k141_160968 | Not-determined |
| S12_extracted_k141_145657 | Not-determined |
| S12_extracted_k141_50804 | Not-determined |
| S12_extracted_k141_43836 | Not-determined |
| S12_extracted_k141_145658 | Not-determined |
| S12_extracted_k141_50805 | Not-determined |
| S12_extracted_k141_59239 | Not-determined |
| S12_extracted_k141_160974 | Not-determined |
| S12_extracted_k141_185231 | Low-quality |
| S12_extracted_k141_19573 | Not-determined |
| S12_extracted_k141_74324 | Not-determined |
| S12_extracted_k141_121696 | Not-determined |
| S12_extracted_k141_50813 | Not-determined |
| S12_extracted_k141_99105 | Not-determined |
| S12_extracted_k141_121698 | Low-quality |
| S12_extracted_k141_160979 | Not-determined |
| S12_extracted_k141_59245 | Not-determined |
| S12_extracted_k141_19576 | Not-determined |
| S12_extracted_k141_66872 | Not-determined |
| S12_extracted_k141_99108 | Not-determined |
| S12_extracted_k141_145662 | Not-determined |
| S12_extracted_k141_90926 | Low-quality |
| S12_extracted_k141_145668 | Not-determined |
| S12_extracted_k141_90927 | Not-determined |
| S12_extracted_k141_90928 | Not-determined |
| S12_extracted_k141_145671 | Not-determined |
| S12_extracted_k141_90931 | Not-determined |
| S12_extracted_k141_185243 | Not-determined |
| S12_extracted_k141_27616 | Not-determined |
| S12_extracted_k141_137842 | Not-determined |
| S12_extracted_k141_137845 | Not-determined |
| S12_extracted_k141_185252 | Not-determined |
| S12_extracted_k141_137846 | Not-determined |
| S12_extracted_k141_106585 | Not-determined |
| S12_extracted_k141_169126 | Low-quality |
| S12_extracted_k141_27626 | Not-determined |
| S12_extracted_k141_34879 | Not-determined |
| S12_extracted_k141_43864 | Not-determined |
| S12_extracted_k141_121708 | Not-determined |
| S12_extracted_k141_99121 | Not-determined |
| S12_extracted_k141_27634 | Not-determined |
| S12_extracted_k141_121715 | Not-determined |
| S12_extracted_k141_59251 | Not-determined |
| S12_extracted_k141_169139 | Not-determined |
| S12_extracted_k141_59259 | Not-determined |
| S12_extracted_k141_153172 | Low-quality |
| S12_extracted_k141_59260 | Not-determined |
| S12_extracted_k141_11090 | Not-determined |
| S12_extracted_k141_74331 | Not-determined |
| S12_extracted_k141_11091 | Not-determined |
| S12_extracted_k141_34893 | Not-determined |
| S12_extracted_k141_153178 | Not-determined |
| S12_extracted_k141_74337 | Not-determined |
| S12_extracted_k141_82571 | Not-determined |
| S12_extracted_k141_106600 | Low-quality |
| S12_extracted_k141_153183 | Not-determined |
| S12_extracted_k141_99127 | Not-determined |
| S12_extracted_k141_153184 | Not-determined |
| S12_extracted_k141_4193 | Not-determined |
| S12_extracted_k141_99130 | Low-quality |
| S12_extracted_k141_153187 | Not-determined |
| S12_extracted_k141_4196 | Low-quality |
| S12_extracted_k141_74349 | Not-determined |
| S12_extracted_k141_169149 | Not-determined |
| S12_extracted_k141_74351 | Not-determined |
| S12_extracted_k141_153194 | Not-determined |
| S12_extracted_k141_4201 | Not-determined |
| S12_extracted_k141_34905 | Not-determined |
| S12_extracted_k141_11104 | Not-determined |
| S12_extracted_k141_99135 | Not-determined |
| S12_extracted_k141_137859 | Not-determined |
| S12_extracted_k141_34910 | Not-determined |
| S12_extracted_k141_4204 | Not-determined |
| S12_extracted_k141_34912 | Not-determined |
| S12_extracted_k141_106613 | Not-determined |
| S12_extracted_k141_145696 | Not-determined |
| S12_extracted_k141_74354 | Not-determined |
| S12_extracted_k141_137860 | Low-quality |
| S12_extracted_k141_27650 | Not-determined |
| S12_extracted_k141_59283 | Not-determined |
| S12_extracted_k141_74364 | Not-determined |
| S12_extracted_k141_4212 | Not-determined |
| S12_extracted_k141_153207 | Low-quality |
| S12_extracted_k141_113688 | Not-determined |
| S12_extracted_k141_113689 | Not-determined |
| S12_extracted_k141_169167 | Not-determined |
| S12_extracted_k141_121746 | Not-determined |
| S12_extracted_k141_121747 | Not-determined |
| S12_extracted_k141_113695 | Not-determined |
| S12_extracted_k141_129548 | Not-determined |
| S12_extracted_k141_99145 | Not-determined |
| S12_extracted_k141_161028 | Not-determined |
| S12_extracted_k141_74376 | Not-determined |
| S12_extracted_k141_34932 | Low-quality |
| S12_extracted_k141_99150 | Low-quality |
| S12_extracted_k141_82600 | Not-determined |
| S12_extracted_k141_99153 | Not-determined |
| S12_extracted_k141_169176 | Not-determined |
| S12_extracted_k141_11122 | Not-determined |
| S12_extracted_k141_11127 | Not-determined |
| S12_extracted_k141_4231 | Not-determined |
| S12_extracted_k141_82607 | Not-determined |
| S12_extracted_k141_82608 | Not-determined |
| S12_extracted_k141_11128 | Not-determined |
| S12_extracted_k141_137886 | Not-determined |
| S12_extracted_k141_129552 | Not-determined |
| S12_extracted_k141_50845 | Not-determined |
| S12_extracted_k141_82611 | Not-determined |
| S12_extracted_k141_169182 | Not-determined |
| S12_extracted_k141_129555 | Not-determined |
| S12_extracted_k141_82612 | Not-determined |
| S12_extracted_k141_161042 | Not-determined |
| S12_extracted_k141_137889 | Not-determined |
| S12_extracted_k141_74395 | Not-determined |
| S12_extracted_k141_43894 | Not-determined |
| S12_extracted_k141_185306 | Not-determined |
| S12_extracted_k141_27663 | Not-determined |
| S12_extracted_k141_185309 | Not-determined |
| S12_extracted_k141_137892 | Not-determined |
| S12_extracted_k141_137893 | Not-determined |
| S12_extracted_k141_59305 | Not-determined |
| S12_extracted_k141_161059 | Low-quality |
| S12_extracted_k141_137899 | Not-determined |
| S12_extracted_k141_99176 | Not-determined |
| S12_extracted_k141_161065 | Not-determined |
| S12_extracted_k141_27677 | Not-determined |
| S12_extracted_k141_50862 | Not-determined |
| S12_extracted_k141_27678 | Not-determined |
| S12_extracted_k141_106644 | Not-determined |
| S12_extracted_k141_4256 | Not-determined |
| S12_extracted_k141_27685 | Not-determined |
| S12_extracted_k141_27686 | Low-quality |
| S12_extracted_k141_161080 | Not-determined |
| S12_extracted_k141_137914 | Low-quality |
| S12_extracted_k141_153240 | Not-determined |
| S12_extracted_k141_106655 | Not-determined |
| S12_extracted_k141_11154 | Low-quality |
| S12_extracted_k141_90982 | Not-determined |
| S12_extracted_k141_66917 | Low-quality |
| S12_extracted_k141_106660 | Low-quality |
| S12_extracted_k141_90985 | Not-determined |
| S12_extracted_k141_59323 | Low-quality |
| S12_extracted_k141_74416 | Not-determined |
| S12_extracted_k141_66922 | Not-determined |
| S12_extracted_k141_145721 | Not-determined |
| S12_extracted_k141_153248 | Not-determined |
| S12_extracted_k141_66924 | Not-determined |
| S12_extracted_k141_153249 | Not-determined |
| S12_extracted_k141_66925 | Not-determined |
| S12_extracted_k141_27700 | Not-determined |
| S12_extracted_k141_90991 | Not-determined |
| S12_extracted_k141_145723 | Not-determined |
| S12_extracted_k141_106666 | Low-quality |
| S12_extracted_k141_50868 | Not-determined |
| S12_extracted_k141_145724 | Not-determined |
| S12_extracted_k141_66929 | Not-determined |
| S12_extracted_k141_90993 | Not-determined |
| S12_extracted_k141_153257 | Not-determined |
| S12_extracted_k141_27707 | Not-determined |
| S12_extracted_k141_66940 | Not-determined |
| S12_extracted_k141_43913 | Not-determined |
| S12_extracted_k141_74434 | Not-determined |
| S12_extracted_k141_145730 | Not-determined |
| S12_extracted_k141_4275 | Not-determined |
| S12_extracted_k141_34956 | Not-determined |
| S12_extracted_k141_74435 | Not-determined |
| S12_extracted_k141_66941 | Not-determined |
| S12_extracted_k141_59333 | Not-determined |
| S12_extracted_k141_153260 | Not-determined |
| S12_extracted_k141_34957 | Not-determined |
| S12_extracted_k141_43919 | Not-determined |
| S12_extracted_k141_74439 | Not-determined |
| S12_extracted_k141_82640 | Low-quality |
| S12_extracted_k141_106678 | Not-determined |
| S12_extracted_k141_106679 | Not-determined |
| S12_extracted_k141_82642 | Not-determined |
| S12_extracted_k141_66948 | Not-determined |
| S12_extracted_k141_34962 | Not-determined |
| S12_extracted_k141_34966 | Not-determined |
| S12_extracted_k141_59341 | Not-determined |
| S12_extracted_k141_106684 | Not-determined |
| S12_extracted_k141_99214 | Not-determined |
| S12_extracted_k141_43927 | Not-determined |
| S12_extracted_k141_137924 | Not-determined |
| S12_extracted_k141_99216 | Not-determined |
| S12_extracted_k141_106687 | Low-quality |
| S12_extracted_k141_27720 | Not-determined |
| S12_extracted_k141_169206 | Not-determined |
| S12_extracted_k141_43935 | Not-determined |
| S12_extracted_k141_11175 | Not-determined |
| S12_extracted_k141_66960 | Not-determined |
| S12_extracted_k141_27723 | Not-determined |
| S12_extracted_k141_27725 | Not-determined |
| S12_extracted_k141_50891 | Not-determined |
| S12_extracted_k141_106693 | Not-determined |
| S12_extracted_k141_27726 | Not-determined |
| S12_extracted_k141_82656 | Not-determined |
| S12_extracted_k141_4298 | Not-determined |
| S12_extracted_k141_66966 | Not-determined |
| S12_extracted_k141_169211 | Not-determined |
| S12_extracted_k141_113710 | Not-determined |
| S12_extracted_k141_11183 | Not-determined |
| S12_extracted_k141_50897 | Not-determined |
| S12_extracted_k141_153272 | Not-determined |
| S12_extracted_k141_113716 | Not-determined |
| S12_extracted_k141_161119 | Not-determined |
| S12_extracted_k141_11185 | Not-determined |
| S12_extracted_k141_50902 | Not-determined |
| S12_extracted_k141_11189 | Not-determined |
| S12_extracted_k141_145748 | Not-determined |
| S12_extracted_k141_82664 | Not-determined |
| S12_extracted_k141_145750 | Not-determined |
| S12_extracted_k141_66973 | Not-determined |
| S12_extracted_k141_66974 | Not-determined |
| S12_extracted_k141_66975 | Not-determined |
| S12_extracted_k141_137943 | Not-determined |
| S12_extracted_k141_4308 | Not-determined |
| S12_extracted_k141_59361 | Not-determined |
| S12_extracted_k141_59363 | Not-determined |
| S12_extracted_k141_169228 | Not-determined |
| S12_extracted_k141_11197 | Not-determined |
| S12_extracted_k141_66980 | Not-determined |
| S12_extracted_k141_153285 | Not-determined |
| S12_extracted_k141_137945 | Low-quality |
| S12_extracted_k141_11200 | Not-determined |
| S12_extracted_k141_161133 | Not-determined |
| S12_extracted_k141_161135 | Low-quality |
| S12_extracted_k141_27741 | Not-determined |
| S12_extracted_k141_137951 | Not-determined |
| S12_extracted_k141_50914 | Not-determined |
| S12_extracted_k141_4318 | Not-determined |
| S12_extracted_k141_4319 | Low-quality |
| S12_extracted_k141_169234 | Not-determined |
| S12_extracted_k141_145764 | Not-determined |
| S12_extracted_k141_161143 | Not-determined |
| S12_extracted_k141_161144 | Not-determined |
| S12_extracted_k141_50916 | Not-determined |
| S12_extracted_k141_169238 | Not-determined |
| S12_extracted_k141_43962 | Not-determined |
| S12_extracted_k141_59372 | Not-determined |
| S12_extracted_k141_59375 | Not-determined |
| S12_extracted_k141_43968 | Low-quality |
| S12_extracted_k141_43971 | Not-determined |
| S12_extracted_k141_43973 | Not-determined |
| S12_extracted_k141_43975 | Not-determined |
| S12_extracted_k141_137957 | Not-determined |
| S12_extracted_k141_137958 | Not-determined |
| S12_extracted_k141_153293 | Not-determined |
| S12_extracted_k141_82686 | Not-determined |
| S12_extracted_k141_27762 | Not-determined |
| S12_extracted_k141_4332 | Not-determined |
| S12_extracted_k141_161158 | Not-determined |
| S12_extracted_k141_137964 | Not-determined |
| S12_extracted_k141_4336 | Not-determined |
| S12_extracted_k141_137970 | Not-determined |
| S12_extracted_k141_11206 | Not-determined |
| S12_extracted_k141_137972 | Not-determined |
| S12_extracted_k141_4342 | Not-determined |
| S12_extracted_k141_161170 | Not-determined |
| S12_extracted_k141_82700 | Not-determined |
| S12_extracted_k141_137979 | Not-determined |
| S12_extracted_k141_145787 | Not-determined |
| S12_extracted_k141_161177 | Not-determined |
| S12_extracted_k141_161179 | Not-determined |
| S12_extracted_k141_113753 | Not-determined |
| S12_extracted_k141_185335 | Not-determined |
| S12_extracted_k141_113755 | Low-quality |
| S12_extracted_k141_82703 | Low-quality |
| S12_extracted_k141_27777 | Not-determined |
| S12_extracted_k141_82707 | Not-determined |
| S12_extracted_k141_161190 | Not-determined |
| S12_extracted_k141_161193 | Not-determined |
| S12_extracted_k141_91045 | Not-determined |
| S12_extracted_k141_137998 | Not-determined |
| S12_extracted_k141_50951 | Not-determined |
| S12_extracted_k141_82710 | Not-determined |
| S12_extracted_k141_185348 | Not-determined |
| S12_extracted_k141_185349 | Not-determined |
| S12_extracted_k141_82716 | Not-determined |
| S12_extracted_k141_161200 | Not-determined |
| S12_extracted_k141_50957 | Not-determined |
| S12_extracted_k141_59396 | Not-determined |
| S12_extracted_k141_74476 | Not-determined |
| S12_extracted_k141_59398 | Not-determined |
| S12_extracted_k141_91054 | Not-determined |
| S12_extracted_k141_50964 | Not-determined |
| S12_extracted_k141_66992 | Not-determined |
| S12_extracted_k141_161212 | Not-determined |
| S12_extracted_k141_91061 | Not-determined |
| S12_extracted_k141_161213 | Not-determined |
| S12_extracted_k141_138012 | Not-determined |
| S12_extracted_k141_35010 | Not-determined |
| S12_extracted_k141_82721 | Not-determined |
| S12_extracted_k141_27798 | Not-determined |
| S12_extracted_k141_11243 | Not-determined |
| S12_extracted_k141_4366 | Not-determined |
| S12_extracted_k141_44000 | Not-determined |
| S12_extracted_k141_4371 | Not-determined |
| S12_extracted_k141_161221 | Not-determined |
| S12_extracted_k141_161222 | Not-determined |
| S12_extracted_k141_35018 | Not-determined |
| S12_extracted_k141_91070 | Low-quality |
| S12_extracted_k141_161223 | Not-determined |
| S12_extracted_k141_91071 | Not-determined |
| S12_extracted_k141_91072 | Not-determined |
| S12_extracted_k141_11253 | Not-determined |
| S12_extracted_k141_67003 | Not-determined |
| S12_extracted_k141_35024 | Not-determined |
| S12_extracted_k141_138017 | Not-determined |
| S12_extracted_k141_138019 | Low-quality |
| S12_extracted_k141_185374 | Not-determined |
| S12_extracted_k141_161230 | Low-quality |
| S12_extracted_k141_82731 | Not-determined |
| S12_extracted_k141_153330 | Not-determined |
| S12_extracted_k141_82732 | Not-determined |
| S12_extracted_k141_74495 | Not-determined |
| S12_extracted_k141_145821 | Not-determined |
| S12_extracted_k141_4385 | Low-quality |
| S12_extracted_k141_153333 | Not-determined |
| S12_extracted_k141_4387 | Not-determined |
| S12_extracted_k141_161233 | Not-determined |
| S12_extracted_k141_44003 | Not-determined |
| S12_extracted_k141_59415 | Not-determined |
| S12_extracted_k141_161237 | Not-determined |
| S12_extracted_k141_185377 | Not-determined |
| S12_extracted_k141_44004 | Not-determined |
| S12_extracted_k141_59417 | Not-determined |
| S12_extracted_k141_44007 | Not-determined |
| S12_extracted_k141_153337 | Not-determined |
| S12_extracted_k141_35037 | Not-determined |
| S12_extracted_k141_35038 | Not-determined |
| S12_extracted_k141_82742 | Not-determined |
| S12_extracted_k141_91084 | Not-determined |
| S12_extracted_k141_44013 | Not-determined |
| S12_extracted_k141_153341 | Not-determined |
| S12_extracted_k141_74508 | Not-determined |
| S12_extracted_k141_161243 | Not-determined |
| S12_extracted_k141_44016 | Not-determined |
| S12_extracted_k141_35043 | Not-determined |
| S12_extracted_k141_50982 | Low-quality |
| S12_extracted_k141_145834 | Not-determined |
| S12_extracted_k141_145835 | Not-determined |
| S12_extracted_k141_82746 | Not-determined |
| S12_extracted_k141_169249 | Not-determined |
| S12_extracted_k141_161255 | Not-determined |
| S12_extracted_k141_50986 | Not-determined |
| S12_extracted_k141_82753 | Low-quality |
| S12_extracted_k141_113810 | Not-determined |
| S12_extracted_k141_113811 | Not-determined |
| S12_extracted_k141_82756 | Not-determined |
| S12_extracted_k141_91106 | Not-determined |
| S12_extracted_k141_35056 | Not-determined |
| S12_extracted_k141_74523 | Not-determined |
| S12_extracted_k141_50997 | Not-determined |
| S12_extracted_k141_44029 | Not-determined |
| S12_extracted_k141_74527 | Not-determined |
| S12_extracted_k141_35061 | Not-determined |
| S12_extracted_k141_185384 | Low-quality |
| S12_extracted_k141_169260 | Not-determined |
| S12_extracted_k141_91112 | Not-determined |
| S12_extracted_k141_185386 | Not-determined |
| S12_extracted_k141_113815 | Not-determined |
| S12_extracted_k141_185388 | Not-determined |
| S12_extracted_k141_4417 | Not-determined |
| S12_extracted_k141_44038 | Not-determined |
| S12_extracted_k141_67046 | Not-determined |
| S12_extracted_k141_161266 | Not-determined |
| S12_extracted_k141_129566 | Low-quality |
| S12_extracted_k141_51009 | Not-determined |
| S12_extracted_k141_74537 | Not-determined |
| S12_extracted_k141_74539 | Not-determined |
| S12_extracted_k141_106700 | Not-determined |
| S12_extracted_k141_44055 | Not-determined |
| S12_extracted_k141_106703 | Not-determined |
| S12_extracted_k141_106704 | Not-determined |
| S12_extracted_k141_35083 | Not-determined |
| S12_extracted_k141_106709 | Not-determined |
| S12_extracted_k141_67059 | Not-determined |
| S12_extracted_k141_106711 | Not-determined |
| S12_extracted_k141_185401 | Not-determined |
| S12_extracted_k141_35091 | Not-determined |
| S12_extracted_k141_35093 | Not-determined |
| S12_extracted_k141_44062 | Not-determined |
| S12_extracted_k141_145883 | Not-determined |
| S12_extracted_k141_138069 | Not-determined |
| S12_extracted_k141_145884 | Not-determined |
| S12_extracted_k141_185409 | Not-determined |
| S12_extracted_k141_91128 | Low-quality |
| S12_extracted_k141_129592 | Low-quality |
| S12_extracted_k141_145887 | Not-determined |
| S12_extracted_k141_145889 | Not-determined |
| S12_extracted_k141_44071 | Not-determined |
| S12_extracted_k141_91138 | Not-determined |
| S12_extracted_k141_74564 | Not-determined |
| S12_extracted_k141_44077 | Low-quality |
| S12_extracted_k141_67075 | Not-determined |
| S12_extracted_k141_67076 | Not-determined |
| S12_extracted_k141_44083 | Not-determined |
| S12_extracted_k141_74571 | Not-determined |
| S12_extracted_k141_74573 | Not-determined |
| S12_extracted_k141_51011 | Low-quality |
| S12_extracted_k141_67095 | Not-determined |
| S12_extracted_k141_74577 | Not-determined |
| S12_extracted_k141_44086 | Low-quality |
| S12_extracted_k141_51012 | Not-determined |
| S12_extracted_k141_74581 | Not-determined |
| S12_extracted_k141_67101 | Not-determined |
| S12_extracted_k141_145905 | Not-determined |
| S12_extracted_k141_19604 | Not-determined |
| S12_extracted_k141_138082 | Not-determined |
| S12_extracted_k141_19606 | Not-determined |
| S12_extracted_k141_44091 | Not-determined |
| S12_extracted_k141_82785 | Not-determined |
| S12_extracted_k141_161274 | Not-determined |
| S12_extracted_k141_35096 | Low-quality |
| S12_extracted_k141_185446 | Not-determined |
| S12_extracted_k141_185448 | Not-determined |
| S12_extracted_k141_169297 | Not-determined |
| S12_extracted_k141_67111 | Not-determined |
| S12_extracted_k141_113841 | Not-determined |
| S12_extracted_k141_169299 | Not-determined |
| S12_extracted_k141_113842 | Not-determined |
| S12_extracted_k141_19612 | Not-determined |
| S12_extracted_k141_59476 | Not-determined |
| S12_extracted_k141_176473 | Not-determined |
| S12_extracted_k141_44092 | Low-quality |
| S12_extracted_k141_44093 | Not-determined |
| S12_extracted_k141_91144 | Not-determined |
| S12_extracted_k141_138092 | Low-quality |
| S12_extracted_k141_106723 | Low-quality |
| S12_extracted_k141_169309 | Not-determined |
| S12_extracted_k141_106724 | Not-determined |
| S12_extracted_k141_169310 | Not-determined |
| S12_extracted_k141_67117 | Not-determined |
| S12_extracted_k141_35113 | Not-determined |
| S12_extracted_k141_82801 | Not-determined |
| S12_extracted_k141_35115 | Low-quality |
| S12_extracted_k141_51023 | Not-determined |
| S12_extracted_k141_91158 | Not-determined |
| S12_extracted_k141_106731 | Not-determined |
| S12_extracted_k141_74602 | Not-determined |
| S12_extracted_k141_185457 | Not-determined |
| S12_extracted_k141_67129 | Not-determined |
| S12_extracted_k141_67131 | Not-determined |
| S12_extracted_k141_35124 | Not-determined |
| S12_extracted_k141_51031 | Not-determined |
| S12_extracted_k141_145934 | Not-determined |
| S12_extracted_k141_161299 | Not-determined |
| S12_extracted_k141_176495 | Low-quality |
| S12_extracted_k141_91165 | Not-determined |
| S12_extracted_k141_145936 | Not-determined |
| S12_extracted_k141_161301 | Not-determined |
| S12_extracted_k141_161303 | Not-determined |
| S12_extracted_k141_67137 | Not-determined |
| S12_extracted_k141_129616 | Low-quality |
| S12_extracted_k141_91171 | Not-determined |
| S12_extracted_k141_113848 | Not-determined |
| S12_extracted_k141_161308 | Low-quality |
| S12_extracted_k141_145942 | Not-determined |
| S12_extracted_k141_51036 | Low-quality |
| S12_extracted_k141_176501 | Not-determined |
| S12_extracted_k141_51038 | Not-determined |
| S12_extracted_k141_145943 | Not-determined |
| S12_extracted_k141_169316 | Not-determined |
| S12_extracted_k141_106748 | Low-quality |
| S12_extracted_k141_185476 | Not-determined |
| S12_extracted_k141_145945 | Low-quality |
| S12_extracted_k141_161309 | Not-determined |
| S12_extracted_k141_153350 | Not-determined |
| S12_extracted_k141_185478 | Not-determined |
| S12_extracted_k141_113855 | Not-determined |
| S12_extracted_k141_161312 | Not-determined |
| S12_extracted_k141_27813 | Not-determined |
| S12_extracted_k141_99245 | Not-determined |
| S12_extracted_k141_91181 | Not-determined |
| S12_extracted_k141_161315 | Not-determined |
| S12_extracted_k141_4459 | Not-determined |
| S12_extracted_k141_145954 | Not-determined |
| S12_extracted_k141_145959 | Not-determined |
| S12_extracted_k141_185485 | Not-determined |
| S12_extracted_k141_99252 | Not-determined |
| S12_extracted_k141_27821 | Not-determined |
| S12_extracted_k141_169326 | Not-determined |
| S12_extracted_k141_176515 | Not-determined |
| S12_extracted_k141_176516 | Not-determined |
| S12_extracted_k141_161323 | Not-determined |
| S12_extracted_k141_113867 | Not-determined |
| S12_extracted_k141_74628 | Not-determined |
| S12_extracted_k141_153358 | Not-determined |
| S12_extracted_k141_74630 | Not-determined |
| S12_extracted_k141_4466 | Not-determined |
| S12_extracted_k141_74632 | Not-determined |
| S12_extracted_k141_145966 | Not-determined |
| S12_extracted_k141_27831 | Not-determined |
| S12_extracted_k141_99262 | Not-determined |
| S12_extracted_k141_51065 | Not-determined |
| S12_extracted_k141_74639 | Not-determined |
| S12_extracted_k141_51071 | Not-determined |
| S12_extracted_k141_153368 | Not-determined |
| S12_extracted_k141_129629 | Not-determined |
| S12_extracted_k141_74645 | Low-quality |
| S12_extracted_k141_145979 | Not-determined |
| S12_extracted_k141_129633 | Low-quality |
| S12_extracted_k141_113879 | Not-determined |
| S12_extracted_k141_91207 | Not-determined |
| S12_extracted_k141_161337 | Not-determined |
| S12_extracted_k141_91209 | Not-determined |
| S12_extracted_k141_99275 | Low-quality |
| S12_extracted_k141_4477 | Not-determined |
| S12_extracted_k141_113886 | Not-determined |
| S12_extracted_k141_51085 | Not-determined |
| S12_extracted_k141_106767 | Not-determined |
| S12_extracted_k141_35134 | Not-determined |
| S12_extracted_k141_145985 | Not-determined |
| S12_extracted_k141_113889 | Not-determined |
| S12_extracted_k141_129639 | Not-determined |
| S12_extracted_k141_4484 | Not-determined |
| S12_extracted_k141_106770 | Not-determined |
| S12_extracted_k141_161342 | Not-determined |
| S12_extracted_k141_153379 | Low-quality |
| S12_extracted_k141_176527 | Not-determined |
| S12_extracted_k141_113897 | Not-determined |
| S12_extracted_k141_129643 | Not-determined |
| S12_extracted_k141_4491 | Not-determined |
| S12_extracted_k141_59496 | Not-determined |
| S12_extracted_k141_74655 | Not-determined |
| S12_extracted_k141_161349 | Not-determined |
| S12_extracted_k141_27841 | Not-determined |
| S12_extracted_k141_145995 | Not-determined |
| S12_extracted_k141_176539 | Not-determined |
| S12_extracted_k141_113908 | Not-determined |
| S12_extracted_k141_169350 | Not-determined |
| S12_extracted_k141_153386 | Low-quality |
| S12_extracted_k141_51101 | Not-determined |
| S12_extracted_k141_99293 | Not-determined |
| S12_extracted_k141_153388 | Not-determined |
| S12_extracted_k141_185510 | Not-determined |
| S12_extracted_k141_113911 | Not-determined |
| S12_extracted_k141_145998 | Low-quality |
| S12_extracted_k141_44120 | Low-quality |
| S12_extracted_k141_82834 | Not-determined |
| S12_extracted_k141_113918 | Not-determined |
| S12_extracted_k141_44125 | Low-quality |
| S12_extracted_k141_185515 | Not-determined |
| S12_extracted_k141_113922 | Not-determined |
| S12_extracted_k141_59511 | Not-determined |
| S12_extracted_k141_176555 | Not-determined |
| S12_extracted_k141_113925 | Not-determined |
| S12_extracted_k141_44131 | Not-determined |
| S12_extracted_k141_161360 | Not-determined |
| S12_extracted_k141_161361 | Not-determined |
| S12_extracted_k141_11260 | Not-determined |
| S12_extracted_k141_161363 | Not-determined |
| S12_extracted_k141_146001 | Not-determined |
| S12_extracted_k141_74663 | Not-determined |
| S12_extracted_k141_161364 | Not-determined |
| S12_extracted_k141_44137 | Not-determined |
| S12_extracted_k141_74667 | Not-determined |
| S12_extracted_k141_138112 | Not-determined |
| S12_extracted_k141_35165 | Not-determined |
| S12_extracted_k141_91246 | Not-determined |
| S12_extracted_k141_11264 | Not-determined |
| S12_extracted_k141_44146 | Not-determined |
| S12_extracted_k141_35167 | Not-determined |
| S12_extracted_k141_161371 | Not-determined |
| S12_extracted_k141_11267 | Not-determined |

| | |
|---|---|
| S12_extracted_k141_82843 | Not-determined |
| S12_extracted_k141_19622 | Not-determined |
| S12_extracted_k141_27877 | Not-determined |
| S12_extracted_k141_138117 | Not-determined |
| S12_extracted_k141_27880 | Not-determined |
| S12_extracted_k141_59531 | Not-determined |
| S12_extracted_k141_11270 | Not-determined |
| S12_extracted_k141_44154 | Not-determined |
| S12_extracted_k141_113935 | Not-determined |
| S12_extracted_k141_161377 | Not-determined |
| S12_extracted_k141_153418 | Not-determined |
| S12_extracted_k141_59534 | Low-quality |
| S12_extracted_k141_153422 | Not-determined |
| S12_extracted_k141_153424 | Not-determined |
| S12_extracted_k141_35180 | Not-determined |
| S12_extracted_k141_4525 | Not-determined |
| S12_extracted_k141_44161 | Not-determined |
| S12_extracted_k141_169360 | Low-quality |
| S12_extracted_k141_4526 | Not-determined |
| S12_extracted_k141_19631 | Not-determined |
| S12_extracted_k141_19633 | Not-determined |
| S12_extracted_k141_35183 | Not-determined |
| S12_extracted_k141_153426 | Low-quality |
| S12_extracted_k141_169365 | Not-determined |
| S12_extracted_k141_113949 | Not-determined |
| S12_extracted_k141_35188 | Not-determined |
| S12_extracted_k141_129666 | Not-determined |
| S12_extracted_k141_169371 | Not-determined |
| S12_extracted_k141_11279 | Not-determined |
| S12_extracted_k141_185538 | Not-determined |
| S12_extracted_k141_185541 | Not-determined |
| S12_extracted_k141_74696 | Not-determined |
| S12_extracted_k141_138133 | Not-determined |
| S12_extracted_k141_153433 | Low-quality |
| S12_extracted_k141_106834 | Not-determined |
| S12_extracted_k141_106835 | Not-determined |
| S12_extracted_k141_169375 | Not-determined |
| S12_extracted_k141_91255 | Not-determined |
| S12_extracted_k141_4532 | Not-determined |
| S12_extracted_k141_169377 | Not-determined |
| S12_extracted_k141_185547 | Not-determined |
| S12_extracted_k141_44173 | Not-determined |
| S12_extracted_k141_4535 | Not-determined |
| S12_extracted_k141_35205 | Not-determined |
| S12_extracted_k141_138142 | Not-determined |
| S12_extracted_k141_106844 | Not-determined |
| S12_extracted_k141_176568 | Not-determined |
| S12_extracted_k141_146019 | Not-determined |
| S12_extracted_k141_153438 | Not-determined |
| S12_extracted_k141_44178 | Not-determined |
| S12_extracted_k141_35209 | Low-quality |
| S12_extracted_k141_146022 | Not-determined |
| S12_extracted_k141_146025 | Not-determined |
| S12_extracted_k141_44184 | Not-determined |
| S12_extracted_k141_153446 | Not-determined |
| S12_extracted_k141_4547 | Not-determined |
| S12_extracted_k141_106853 | Not-determined |
| S12_extracted_k141_59567 | Not-determined |
| S12_extracted_k141_161394 | Not-determined |
| S12_extracted_k141_129685 | Not-determined |
| S12_extracted_k141_176578 | Not-determined |
| S12_extracted_k141_169389 | Not-determined |
| S12_extracted_k141_185555 | Low-quality |
| S12_extracted_k141_106859 | Not-determined |
| S12_extracted_k141_44196 | Not-determined |
| S12_extracted_k141_91275 | Not-determined |
| S12_extracted_k141_91276 | Not-determined |
| S12_extracted_k141_19667 | Not-determined |
| S12_extracted_k141_169397 | Not-determined |
| S12_extracted_k141_106862 | Low-quality |
| S12_extracted_k141_106863 | Not-determined |
| S12_extracted_k141_138163 | Low-quality |
| S12_extracted_k141_59577 | Not-determined |
| S12_extracted_k141_138164 | Not-determined |
| S12_extracted_k141_153456 | Not-determined |
| S12_extracted_k141_146036 | Not-determined |
| S12_extracted_k141_185569 | Low-quality |
| S12_extracted_k141_153459 | Not-determined |
| S12_extracted_k141_106870 | Not-determined |
| S12_extracted_k141_161413 | Not-determined |
| S12_extracted_k141_185576 | Not-determined |
| S12_extracted_k141_27898 | Not-determined |
| S12_extracted_k141_169406 | Not-determined |
| S12_extracted_k141_27899 | Not-determined |
| S12_extracted_k141_106877 | Not-determined |
| S12_extracted_k141_138177 | Not-determined |
| S12_extracted_k141_106879 | Not-determined |
| S12_extracted_k141_129710 | Not-determined |
| S12_extracted_k141_185578 | Not-determined |
| S12_extracted_k141_153471 | Not-determined |
| S12_extracted_k141_146048 | Not-determined |
| S12_extracted_k141_138185 | Not-determined |
| S12_extracted_k141_161416 | Not-determined |
| S12_extracted_k141_106885 | Low-quality |
| S12_extracted_k141_113973 | Not-determined |
| S12_extracted_k141_113974 | Not-determined |
| S12_extracted_k141_161418 | Not-determined |
| S12_extracted_k141_113976 | Not-determined |
| S12_extracted_k141_146056 | Not-determined |
| S12_extracted_k141_59600 | Not-determined |
| S12_extracted_k141_121773 | Not-determined |
| S12_extracted_k141_121776 | Not-determined |
| S12_extracted_k141_153490 | Low-quality |
| S12_extracted_k141_113983 | Not-determined |
| S12_extracted_k141_67188 | Not-determined |
| S12_extracted_k141_11301 | Low-quality |
| S12_extracted_k141_11302 | Not-determined |
| S12_extracted_k141_121781 | Not-determined |
| S12_extracted_k141_11304 | Not-determined |
| S12_extracted_k141_11306 | Not-determined |
| S12_extracted_k141_146067 | Not-determined |
| S12_extracted_k141_82857 | Not-determined |
| S12_extracted_k141_91297 | Not-determined |
| S12_extracted_k141_138197 | Not-determined |
| S12_extracted_k141_91300 | Not-determined |
| S3_vs_k141_114271\|\|full | Medium-quality |
| S12_extracted_k141_185588 | Low-quality |
| S12_extracted_k141_74738 | Not-determined |
| S12_extracted_k141_153501 | Not-determined |
| S12_extracted_k141_67194 | Not-determined |
| S12_extracted_k141_121794 | Not-determined |
| S12_extracted_k141_106904 | Not-determined |
| S12_extracted_k141_27916 | Low-quality |
| S12_extracted_k141_82865 | Not-determined |
| S12_extracted_k141_185595 | Not-determined |
| S12_extracted_k141_27923 | Not-determined |
| S12_extracted_k141_138210 | Not-determined |
| S12_extracted_k141_59620 | Not-determined |
| S12_extracted_k141_67200 | Not-determined |
| S12_extracted_k141_161423 | Not-determined |
| S12_extracted_k141_121799 | Not-determined |
| S12_extracted_k141_99329 | Not-determined |
| S12_extracted_k141_185602 | Not-determined |
| S12_extracted_k141_74754 | Not-determined |
| S12_extracted_k141_161429 | Not-determined |
| S12_extracted_k141_99334 | Not-determined |
| S12_extracted_k141_185609 | Not-determined |
| S12_extracted_k141_74760 | Not-determined |
| S12_extracted_k141_138219 | Not-determined |
| S12_extracted_k141_176618 | Not-determined |
| S12_extracted_k141_82872 | Not-determined |
| S12_extracted_k141_146088 | Not-determined |
| S12_extracted_k141_185617 | Not-determined |
| S12_extracted_k141_121804 | Low-quality |
| S12_extracted_k141_114012 | Not-determined |
| S12_extracted_k141_82874 | Low-quality |
| S12_extracted_k141_138225 | Not-determined |
| S12_extracted_k141_176623 | Not-determined |
| S12_extracted_k141_82875 | Not-determined |
| S12_extracted_k141_67213 | Not-determined |
| S12_extracted_k141_74768 | Not-determined |
| S12_extracted_k141_82881 | Not-determined |
| S12_extracted_k141_4603 | Not-determined |
| S12_extracted_k141_161443 | Low-quality |
| S12_extracted_k141_74770 | Not-determined |
| S12_extracted_k141_67220 | Not-determined |
| S12_extracted_k141_99345 | Not-determined |
| S12_extracted_k141_161446 | Not-determined |
| S12_extracted_k141_153512 | Not-determined |
| S12_extracted_k141_153513 | Not-determined |
| S12_extracted_k141_176629 | Not-determined |
| S12_extracted_k141_176630 | Not-determined |
| S12_extracted_k141_146099 | Not-determined |
| S12_extracted_k141_153516 | Not-determined |
| S12_extracted_k141_67227 | Not-determined |
| S12_extracted_k141_19679 | Not-determined |
| S12_extracted_k141_153523 | Not-determined |
| S12_extracted_k141_91328 | Not-determined |
| S12_extracted_k141_114025 | Not-determined |
| S12_extracted_k141_99356 | Not-determined |
| S12_extracted_k141_114026 | Low-quality |
| S12_extracted_k141_106940 | Not-determined |
| S12_extracted_k141_19683 | Not-determined |
| S12_extracted_k141_4620 | Not-determined |
| S12_extracted_k141_121811 | Not-determined |
| S12_extracted_k141_106943 | Not-determined |
| S12_extracted_k141_185628 | Not-determined |
| S12_extracted_k141_27939 | Not-determined |
| S12_extracted_k141_114033 | Not-determined |
| S12_extracted_k141_82911 | Not-determined |
| S12_extracted_k141_153531 | Not-determined |
| S12_extracted_k141_67240 | Not-determined |
| S12_extracted_k141_114037 | Not-determined |
| S12_extracted_k141_185634 | Not-determined |
| S12_extracted_k141_59640 | Low-quality |
| S12_extracted_k141_44234 | Not-determined |
| S12_extracted_k141_82914 | Not-determined |
| S12_extracted_k141_59645 | Not-determined |
| S12_extracted_k141_121822 | Not-determined |
| S12_extracted_k141_19697 | Not-determined |
| S12_extracted_k141_176653 | Not-determined |
| S12_extracted_k141_114046 | Low-quality |
| S12_extracted_k141_161465 | Not-determined |
| S12_extracted_k141_138245 | Not-determined |
| S12_extracted_k141_27950 | Not-determined |
| S12_extracted_k141_35219 | Not-determined |
| S12_extracted_k141_59650 | Not-determined |
| S12_extracted_k141_129727 | Not-determined |
| S12_extracted_k141_35221 | Not-determined |
| S12_extracted_k141_129728 | Not-determined |
| S12_extracted_k141_67253 | Not-determined |
| S12_extracted_k141_35225 | Not-determined |
| S12_extracted_k141_129732 | Not-determined |
| S12_extracted_k141_11346 | Not-determined |
| S12_extracted_k141_114057 | Not-determined |
| S12_extracted_k141_59655 | Not-determined |
| S12_extracted_k141_114058 | Not-determined |
| S12_extracted_k141_185647 | Not-determined |
| S12_extracted_k141_91347 | Not-determined |
| S12_extracted_k141_27960 | Not-determined |
| S12_extracted_k141_114061 | Not-determined |
| S12_extracted_k141_67263 | Not-determined |
| S12_extracted_k141_138258 | Low-quality |
| S12_extracted_k141_82925 | Not-determined |
| S12_extracted_k141_35238 | Not-determined |
| S12_extracted_k141_161473 | Not-determined |
| S12_extracted_k141_35239 | Not-determined |
| S12_extracted_k141_114067 | Not-determined |
| S12_extracted_k141_106962 | Not-determined |
| S12_extracted_k141_67267 | Low-quality |
| S12_extracted_k141_99379 | Not-determined |
| S12_extracted_k141_161476 | Not-determined |
| S12_extracted_k141_153540 | Not-determined |
| S12_extracted_k141_4635 | Not-determined |
| S12_extracted_k141_82934 | Not-determined |
| S12_extracted_k141_114074 | Not-determined |
| S12_extracted_k141_161480 | Low-quality |
| S12_extracted_k141_153545 | Not-determined |
| S12_extracted_k141_44242 | Not-determined |
| S12_extracted_k141_35250 | Not-determined |
| S12_extracted_k141_19715 | Not-determined |
| S12_extracted_k141_153554 | Not-determined |
| S12_extracted_k141_74817 | Not-determined |
| S12_extracted_k141_74818 | Not-determined |
| S12_extracted_k141_74820 | Not-determined |
| S12_extracted_k141_153557 | Not-determined |
| S12_extracted_k141_106985 | Not-determined |
| S12_extracted_k141_67283 | Not-determined |
| S12_extracted_k141_121845 | Not-determined |
| S12_extracted_k141_4642 | Not-determined |
| S12_extracted_k141_99393 | Not-determined |
| S12_extracted_k141_185663 | Low-quality |
| S12_extracted_k141_44252 | Not-determined |
| S12_extracted_k141_27978 | Not-determined |
| S12_extracted_k141_129738 | Not-determined |
| S12_extracted_k141_121849 | Not-determined |
| S12_extracted_k141_44254 | Low-quality |
| S12_extracted_k141_185665 | Not-determined |
| S12_extracted_k141_121851 | Low-quality |
| S12_extracted_k141_4646 | Not-determined |
| S12_extracted_k141_114086 | Not-determined |
| S12_extracted_k141_74828 | Not-determined |
| S12_extracted_k141_106989 | Not-determined |
| S12_extracted_k141_185670 | Not-determined |
| S12_extracted_k141_169433 | Not-determined |
| S12_extracted_k141_176690 | Not-determined |
| S12_extracted_k141_185672 | Not-determined |
| S12_extracted_k141_82941 | Low-quality |
| S12_extracted_k141_129741 | Not-determined |
| S12_extracted_k141_185673 | Not-determined |
| S12_extracted_k141_67294 | Not-determined |
| S12_extracted_k141_114097 | Not-determined |
| S12_extracted_k141_27982 | Not-determined |
| S12_extracted_k141_44263 | Low-quality |
| S12_extracted_k141_74837 | Not-determined |
| S12_extracted_k141_176694 | Not-determined |
| S12_extracted_k141_169436 | Not-determined |
| S12_extracted_k141_106998 | Not-determined |
| S12_extracted_k141_35268 | Not-determined |
| S12_extracted_k141_153570 | Not-determined |
| S12_extracted_k141_121865 | Not-determined |
| S12_extracted_k141_11360 | Not-determined |
| S12_extracted_k141_161506 | Not-determined |
| S12_extracted_k141_4651 | Not-determined |
| S12_extracted_k141_129752 | Not-determined |
| S12_extracted_k141_74843 | Not-determined |
| S12_extracted_k141_91362 | Not-determined |
| S12_extracted_k141_67304 | Low-quality |
| S12_extracted_k141_27994 | Not-determined |
| S12_extracted_k141_51130 | Not-determined |
| S12_extracted_k141_91366 | Not-determined |
| S12_extracted_k141_114108 | Low-quality |
| S12_extracted_k141_51132 | Not-determined |
| S12_extracted_k141_51133 | Not-determined |
| S12_extracted_k141_51134 | Not-determined |
| S12_extracted_k141_35276 | Not-determined |
| S12_extracted_k141_153578 | Not-determined |
| S12_extracted_k141_67312 | Low-quality |
| S12_extracted_k141_107017 | Not-determined |
| S12_extracted_k141_153580 | Not-determined |
| S12_extracted_k141_153582 | Not-determined |
| S12_extracted_k141_121876 | Not-determined |
| S12_extracted_k141_107020 | Not-determined |
| S12_extracted_k141_4665 | Not-determined |
| S12_extracted_k141_99410 | Not-determined |
| S12_extracted_k141_169439 | Not-determined |
| S12_extracted_k141_153591 | Not-determined |
| S12_extracted_k141_161514 | Not-determined |
| S12_extracted_k141_67322 | Not-determined |
| S12_extracted_k141_146117 | Not-determined |
| S12_extracted_k141_129780 | Not-determined |
| S12_extracted_k141_114120 | Not-determined |
| S12_extracted_k141_185702 | Not-determined |
| S12_extracted_k141_67327 | Not-determined |
| S12_extracted_k141_153598 | Low-quality |
| S12_extracted_k141_146121 | Not-determined |
| S12_extracted_k141_51150 | Not-determined |
| S12_extracted_k141_121889 | Not-determined |
| S12_extracted_k141_129786 | Not-determined |
| S12_extracted_k141_185709 | Not-determined |
| S12_extracted_k141_107038 | Not-determined |
| S12_extracted_k141_91399 | Not-determined |
| S12_extracted_k141_153605 | Not-determined |
| S12_extracted_k141_74860 | Low-quality |
| S12_extracted_k141_146133 | Not-determined |
| S12_extracted_k141_59691 | Not-determined |
| S12_extracted_k141_44290 | Not-determined |
| S12_extracted_k141_107050 | Not-determined |
| S12_extracted_k141_91403 | Not-determined |
| S12_extracted_k141_11371 | Not-determined |
| S12_extracted_k141_185718 | Not-determined |
| S12_extracted_k141_11372 | Not-determined |
| S12_extracted_k141_161531 | Not-determined |
| S12_extracted_k141_99425 | Not-determined |
| S12_extracted_k141_114135 | Not-determined |
| S12_extracted_k141_114136 | Not-determined |
| S12_extracted_k141_107053 | Not-determined |
| S12_extracted_k141_129791 | Not-determined |
| S12_extracted_k141_161533 | Not-determined |
| S12_extracted_k141_185723 | Not-determined |
| S12_extracted_k141_185724 | Not-determined |
| S12_extracted_k141_35326 | Not-determined |
| S12_extracted_k141_185726 | Not-determined |
| S12_extracted_k141_146147 | Not-determined |
| S12_extracted_k141_169457 | Not-determined |
| S12_extracted_k141_35328 | Low-quality |
| S12_extracted_k141_11381 | Not-determined |
| S12_extracted_k141_161537 | Not-determined |
| S12_extracted_k141_59703 | Low-quality |
| S12_extracted_k141_99426 | Not-determined |
| S12_extracted_k141_169461 | Low-quality |
| S12_extracted_k141_161538 | Not-determined |
| S12_extracted_k141_114149 | Low-quality |
| S12_extracted_k141_129804 | Not-determined |
| S12_extracted_k141_91407 | Not-determined |
| S12_extracted_k141_59708 | Not-determined |
| S12_extracted_k141_161547 | Not-determined |
| S12_extracted_k141_153626 | Low-quality |
| S12_extracted_k141_129808 | Not-determined |
| S12_extracted_k141_153627 | Not-determined |
| S12_extracted_k141_146158 | Not-determined |
| S12_extracted_k141_91412 | Not-determined |
| S12_extracted_k141_19774 | Not-determined |
| S12_extracted_k141_19775 | Not-determined |
| S12_extracted_k141_169466 | Not-determined |
| S12_extracted_k141_114155 | Not-determined |
| S12_extracted_k141_161555 | Not-determined |
| S12_extracted_k141_91417 | Not-determined |
| S12_extracted_k141_169468 | Low-quality |
| S12_extracted_k141_138264 | Not-determined |
| S12_extracted_k141_169469 | Not-determined |
| S12_extracted_k141_161558 | Not-determined |
| S12_extracted_k141_129823 | Not-determined |
| S12_extracted_k141_169474 | Not-determined |
| S12_extracted_k141_169477 | Not-determined |
| S12_extracted_k141_44322 | Not-determined |
| S12_extracted_k141_107078 | Not-determined |
| S12_extracted_k141_153643 | Not-determined |
| S12_extracted_k141_153644 | Not-determined |
| S12_extracted_k141_44325 | Not-determined |
| S12_extracted_k141_28003 | Low-quality |
| S12_extracted_k141_67335 | Not-determined |
| S12_extracted_k141_153648 | Not-determined |
| S12_extracted_k141_107083 | Not-determined |
| S12_extracted_k141_11404 | Not-determined |
| S12_extracted_k141_129833 | Not-determined |
| S12_extracted_k141_176735 | Not-determined |
| S12_extracted_k141_107085 | Low-quality |
| S12_extracted_k141_114171 | Not-determined |
| S12_extracted_k141_153652 | Not-determined |
| S12_extracted_k141_121902 | Low-quality |
| S12_extracted_k141_114174 | Not-determined |
| S12_extracted_k141_107087 | Not-determined |
| S12_extracted_k141_121904 | Not-determined |
| S12_extracted_k141_28010 | Not-determined |
| S12_extracted_k141_91443 | Not-determined |
| S12_extracted_k141_153654 | Not-determined |
| S12_extracted_k141_28011 | Not-determined |
| S12_extracted_k141_91444 | Not-determined |
| S12_extracted_k141_4687 | Not-determined |
| S12_extracted_k141_153657 | Not-determined |
| S12_extracted_k141_44332 | Not-determined |
| S12_extracted_k141_138277 | Low-quality |
| S12_extracted_k141_28015 | Not-determined |
| S12_extracted_k141_44334 | Not-determined |
| S12_extracted_k141_44337 | Not-determined |
| S12_extracted_k141_107090 | Low-quality |
| S12_extracted_k141_4696 | Not-determined |
| S12_extracted_k141_99433 | Not-determined |
| S12_extracted_k141_99435 | Not-determined |
| S12_extracted_k141_107094 | Low-quality |
| S12_extracted_k141_146193 | Not-determined |
| S12_extracted_k141_91452 | Not-determined |
| S12_extracted_k141_169490 | Not-determined |
| S12_extracted_k141_107096 | Not-determined |
| S12_extracted_k141_185730 | Not-determined |
| S12_extracted_k141_146196 | Not-determined |
| S12_extracted_k141_74865 | Not-determined |
| S12_extracted_k141_51173 | Not-determined |
| S12_extracted_k141_28024 | Not-determined |
| S12_extracted_k141_161588 | Low-quality |
| S12_extracted_k141_82990 | Not-determined |
| S12_extracted_k141_161589 | Low-quality |
| S12_extracted_k141_74873 | Not-determined |
| S12_extracted_k141_114180 | Not-determined |
| S12_extracted_k141_99457 | Not-determined |
| S12_extracted_k141_11428 | Not-determined |
| S12_extracted_k141_44352 | Not-determined |
| S12_extracted_k141_99458 | Not-determined |
| S12_extracted_k141_19807 | Not-determined |
| S12_extracted_k141_19809 | Low-quality |
| S12_extracted_k141_99461 | Not-determined |
| S12_extracted_k141_11433 | Not-determined |
| S12_extracted_k141_51184 | Not-determined |
| S12_extracted_k141_44356 | Not-determined |
| S12_extracted_k141_185750 | Not-determined |
| S12_extracted_k141_176748 | Not-determined |
| S12_extracted_k141_59744 | Not-determined |
| S12_extracted_k141_99468 | Not-determined |
| S12_extracted_k141_107115 | Low-quality |
| S12_extracted_k141_44364 | Low-quality |
| S12_extracted_k141_114187 | Not-determined |
| S12_extracted_k141_44365 | Not-determined |
| S12_extracted_k141_51193 | Not-determined |
| S12_extracted_k141_185761 | Not-determined |
| S12_extracted_k141_51194 | Not-determined |
| S12_extracted_k141_176753 | Not-determined |
| S12_extracted_k141_51195 | Not-determined |
| S12_extracted_k141_44370 | Not-determined |
| S12_extracted_k141_138293 | Not-determined |
| S12_extracted_k141_44372 | Not-determined |
| S12_extracted_k141_146209 | Low-quality |
| S12_extracted_k141_169518 | Not-determined |
| S12_extracted_k141_161611 | Low-quality |
| S12_extracted_k141_138296 | Not-determined |
| S12_extracted_k141_19815 | Not-determined |
| S12_extracted_k141_169522 | Not-determined |
| S12_extracted_k141_19816 | Not-determined |
| S12_extracted_k141_114200 | Not-determined |
| S12_extracted_k141_59750 | Not-determined |
| S12_extracted_k141_185765 | Not-determined |
| S12_extracted_k141_59752 | Low-quality |
| S12_extracted_k141_4709 | Not-determined |
| S12_extracted_k141_19818 | Low-quality |
| S12_extracted_k141_19819 | Not-determined |
| S12_extracted_k141_59754 | Low-quality |
| S12_extracted_k141_51207 | Not-determined |
| S12_extracted_k141_99485 | Not-determined |
| S12_extracted_k141_74901 | Not-determined |
| S12_extracted_k141_51210 | Not-determined |
| S12_extracted_k141_11441 | Not-determined |
| S12_extracted_k141_176768 | Not-determined |
| S12_extracted_k141_185778 | Low-quality |
| S12_extracted_k141_161623 | Not-determined |
| S12_extracted_k141_4719 | Not-determined |
| S12_extracted_k141_176772 | Not-determined |
| S12_extracted_k141_44389 | Not-determined |
| S12_extracted_k141_161628 | Not-determined |
| S12_extracted_k141_4720 | Low-quality |
| S12_extracted_k141_161629 | Not-determined |
| S12_extracted_k141_28043 | Not-determined |
| S12_extracted_k141_4721 | Not-determined |
| S12_extracted_k141_28048 | Not-determined |
| S12_extracted_k141_28049 | Not-determined |
| S12_extracted_k141_176782 | Not-determined |
| S12_extracted_k141_11455 | Not-determined |
| S12_extracted_k141_114230 | Not-determined |
| S12_extracted_k141_161633 | Not-determined |
| S12_extracted_k141_44405 | Not-determined |
| S12_extracted_k141_161634 | Not-determined |
| S12_extracted_k141_67364 | Not-determined |
| S12_extracted_k141_74909 | Not-determined |
| S12_extracted_k141_28053 | Not-determined |
| S12_extracted_k141_74911 | Not-determined |
| S12_extracted_k141_161640 | Not-determined |
| S12_extracted_k141_51213 | Not-determined |
| S12_extracted_k141_99496 | Not-determined |
| S12_extracted_k141_28058 | Not-determined |
| S12_extracted_k141_67374 | Not-determined |
| S12_extracted_k141_146220 | Not-determined |
| S12_extracted_k141_28067 | Low-quality |
| S12_extracted_k141_59784 | Not-determined |
| S12_extracted_k141_161656 | Not-determined |
| S12_extracted_k141_169537 | Low-quality |
| S12_extracted_k141_83015 | Not-determined |
| S12_extracted_k141_83016 | Not-determined |
| S12_extracted_k141_169538 | Not-determined |
| S12_extracted_k141_11466 | Low-quality |
| S12_extracted_k141_19845 | Not-determined |
| S12_extracted_k141_99504 | Not-determined |
| S12_extracted_k141_114257 | Not-determined |
| S12_extracted_k141_185793 | Not-determined |
| S12_extracted_k141_59792 | Not-determined |
| S12_extracted_k141_59795 | Not-determined |
| S12_extracted_k141_59797 | Not-determined |
| S12_extracted_k141_146236 | Not-determined |
| S12_extracted_k141_67388 | Not-determined |
| S12_extracted_k141_153666 | Not-determined |
| S12_extracted_k141_146239 | Not-determined |
| S12_extracted_k141_146240 | Not-determined |
| S12_extracted_k141_11480 | Low-quality |
| S12_extracted_k141_169544 | Not-determined |
| S12_extracted_k141_91474 | Not-determined |
| S12_extracted_k141_161667 | Not-determined |
| S12_extracted_k141_59805 | Not-determined |
| S12_extracted_k141_138320 | Low-quality |
| S12_extracted_k141_99521 | Not-determined |
| S12_extracted_k141_138321 | Low-quality |
| S12_extracted_k141_51228 | Not-determined |
| S12_extracted_k141_59808 | Not-determined |
| S12_extracted_k141_107139 | Not-determined |
| S12_extracted_k141_138324 | Not-determined |
| S12_extracted_k141_28095 | Not-determined |
| S12_extracted_k141_4727 | Not-determined |
| S12_extracted_k141_59813 | Not-determined |
| S12_extracted_k141_28097 | Not-determined |
| S12_extracted_k141_44438 | Not-determined |
| S12_extracted_k141_28100 | Not-determined |
| S12_extracted_k141_67417 | Not-determined |
| S12_extracted_k141_4735 | Low-quality |
| S12_extracted_k141_83025 | Not-determined |
| S12_extracted_k141_138334 | Not-determined |
| S12_extracted_k141_107148 | Not-determined |
| S12_extracted_k141_138337 | Not-determined |
| S12_extracted_k141_138338 | Not-determined |
| S12_extracted_k141_11496 | Not-determined |
| S12_extracted_k141_185822 | Not-determined |
| S12_extracted_k141_28111 | Low-quality |
| S12_extracted_k141_169566 | Not-determined |
| S12_extracted_k141_161683 | Not-determined |
| S12_extracted_k141_11499 | Not-determined |
| S12_extracted_k141_28114 | Not-determined |
| S12_extracted_k141_185825 | Not-determined |
| S12_extracted_k141_91496 | Not-determined |
| S12_extracted_k141_169572 | Not-determined |
| S12_extracted_k141_185828 | Not-determined |
| S12_extracted_k141_153684 | Low-quality |
| S12_extracted_k141_176798 | Not-determined |
| S12_extracted_k141_129907 | Low-quality |
| S12_extracted_k141_146250 | Not-determined |
| S12_extracted_k141_4741 | Not-determined |
| S12_extracted_k141_185830 | Low-quality |
| S12_extracted_k141_35360 | Low-quality |
| S12_extracted_k141_114264 | Not-determined |
| S12_extracted_k141_146252 | Not-determined |
| S12_extracted_k141_28122 | Not-determined |
| S12_extracted_k141_161691 | Not-determined |
| S12_extracted_k141_129911 | Not-determined |
| S12_extracted_k141_169578 | Not-determined |
| S12_extracted_k141_51252 | Not-determined |
| S12_extracted_k141_153691 | Not-determined |
| S12_extracted_k141_51253 | Not-determined |
| S12_extracted_k141_74944 | Not-determined |
| S12_extracted_k141_107166 | Not-determined |
| S12_extracted_k141_107168 | Not-determined |
| S12_extracted_k141_185837 | Not-determined |
| S12_extracted_k141_67433 | Not-determined |
| S12_extracted_k141_67434 | Not-determined |
| S12_extracted_k141_161697 | Not-determined |
| S12_extracted_k141_67436 | Not-determined |
| S12_extracted_k141_129915 | Not-determined |
| S12_extracted_k141_4754 | Not-determined |
| S12_extracted_k141_185838 | Not-determined |
| S12_extracted_k141_74951 | Not-determined |
| S12_extracted_k141_99557 | Not-determined |
| S12_extracted_k141_146263 | Low-quality |
| S12_extracted_k141_67438 | Not-determined |
| S12_extracted_k141_129922 | Not-determined |
| S12_extracted_k141_59827 | Not-determined |
| S12_extracted_k141_185846 | Not-determined |
| S12_extracted_k141_153709 | Not-determined |
| S12_extracted_k141_129926 | Not-determined |
| S12_extracted_k141_91515 | Not-determined |
| S12_extracted_k141_185850 | Not-determined |
| S12_extracted_k141_161710 | Low-quality |
| S12_extracted_k141_59832 | Low-quality |
| S12_extracted_k141_91516 | Not-determined |
| S12_extracted_k141_59835 | Low-quality |
| S12_extracted_k141_138358 | Not-determined |

| | |
|---|---|
| S12_extracted_k141_107176 | Not-determined |
| S12_extracted_k141_67452 | Not-determined |
| S12_extracted_k141_129936 | Not-determined |
| S12_extracted_k141_4767 | Not-determined |
| S12_extracted_k141_161714 | Not-determined |
| S12_extracted_k141_129938 | Not-determined |
| S12_extracted_k141_138365 | Not-determined |
| S12_extracted_k141_176812 | Not-determined |
| S12_extracted_k141_35379 | Not-determined |
| S12_extracted_k141_67463 | Not-determined |
| S12_extracted_k141_107188 | Low-quality |
| S12_extracted_k141_138371 | Not-determined |
| S12_extracted_k141_59845 | Not-determined |
| S12_extracted_k141_138372 | Not-determined |
| S12_extracted_k141_35382 | Not-determined |
| S12_extracted_k141_176817 | Not-determined |
| S12_extracted_k141_74961 | Not-determined |
| S12_extracted_k141_67471 | Not-determined |
| S12_extracted_k141_107192 | Not-determined |
| S12_extracted_k141_138378 | Not-determined |
| S12_extracted_k141_83062 | Not-determined |
| S12_extracted_k141_35389 | Not-determined |
| S12_extracted_k141_4778 | Not-determined |
| S12_extracted_k141_35390 | Not-determined |
| S12_extracted_k141_83065 | Not-determined |
| S12_extracted_k141_67476 | Not-determined |
| S12_extracted_k141_146270 | Not-determined |
| S12_extracted_k141_91547 | Not-determined |
| S12_extracted_k141_67477 | Not-determined |
| S12_extracted_k141_4784 | Low-quality |
| S12_extracted_k141_176824 | Low-quality |
| S12_extracted_k141_146273 | Not-determined |
| S12_extracted_k141_35397 | Not-determined |
| S12_extracted_k141_107197 | Not-determined |
| S12_extracted_k141_11534 | Not-determined |
| S12_extracted_k141_138387 | Not-determined |
| S12_extracted_k141_107199 | Not-determined |
| S12_extracted_k141_35400 | Not-determined |
| S12_extracted_k141_44490 | Not-determined |
| S12_extracted_k141_169593 | Low-quality |
| S12_extracted_k141_169594 | Not-determined |
| S12_extracted_k141_169596 | Not-determined |
| S12_extracted_k141_176833 | Low-quality |
| S12_extracted_k141_59866 | Not-determined |
| S12_extracted_k141_35403 | Not-determined |
| S12_extracted_k141_153727 | Not-determined |
| S12_extracted_k141_67488 | Not-determined |
| S12_extracted_k141_19907 | Not-determined |
| S12_extracted_k141_107204 | Low-quality |
| S12_extracted_k141_91559 | Not-determined |
| S12_extracted_k141_28146 | Not-determined |
| S12_extracted_k141_138395 | Not-determined |
| S12_extracted_k141_153733 | Not-determined |
| S12_extracted_k141_107207 | Not-determined |
| S12_extracted_k141_44501 | Not-determined |
| S12_extracted_k141_107208 | Not-determined |
| S12_extracted_k141_107209 | Not-determined |
| S12_extracted_k141_107210 | Not-determined |
| S12_extracted_k141_161737 | Low-quality |
| S12_extracted_k141_51300 | Not-determined |
| S12_extracted_k141_19915 | Not-determined |
| S12_extracted_k141_44507 | Not-determined |
| S12_extracted_k141_51302 | Not-determined |
| S12_extracted_k141_176842 | Not-determined |
| S12_extracted_k141_83072 | Not-determined |
| S12_extracted_k141_99567 | Not-determined |
| S12_extracted_k141_51305 | Not-determined |
| S12_extracted_k141_67502 | Low-quality |
| S12_extracted_k141_176848 | Not-determined |
| S12_extracted_k141_146278 | Not-determined |
| S12_extracted_k141_146281 | Low-quality |
| S12_extracted_k141_107218 | Not-determined |
| S12_extracted_k141_114302 | Not-determined |
| S12_extracted_k141_67504 | Not-determined |
| S12_extracted_k141_146285 | Low-quality |
| S12_extracted_k141_146286 | Not-determined |
| S12_extracted_k141_4786 | Low-quality |
| S12_extracted_k141_153746 | Not-determined |
| S12_extracted_k141_169610 | Not-determined |
| S12_extracted_k141_19923 | Low-quality |
| S12_extracted_k141_169613 | Not-determined |
| S12_extracted_k141_67511 | Not-determined |
| S12_extracted_k141_107224 | Not-determined |
| S12_extracted_k141_4791 | Not-determined |
| S12_extracted_k141_44521 | Not-determined |
| S12_extracted_k141_74983 | Not-determined |
| S12_extracted_k141_146293 | Not-determined |
| S12_extracted_k141_4795 | Not-determined |
| S12_extracted_k141_44526 | Not-determined |
| S12_extracted_k141_153761 | Not-determined |
| S12_extracted_k141_146296 | Not-determined |
| S12_extracted_k141_11550 | Not-determined |
| S12_extracted_k141_67515 | Not-determined |
| S12_extracted_k141_67516 | Not-determined |
| S12_extracted_k141_4800 | Not-determined |
| S12_extracted_k141_67518 | Not-determined |
| S12_extracted_k141_11555 | Not-determined |
| S12_extracted_k141_169629 | Low-quality |
| S12_extracted_k141_146298 | Not-determined |
| S12_extracted_k141_161761 | Not-determined |
| S12_extracted_k141_146299 | Not-determined |
| S12_extracted_k141_114321 | Not-determined |
| S12_extracted_k141_114323 | Not-determined |
| S12_extracted_k141_4807 | Not-determined |
| S12_extracted_k141_153766 | Not-determined |
| S12_extracted_k141_138415 | Not-determined |
| S12_extracted_k141_91576 | Not-determined |
| S12_extracted_k141_83081 | Not-determined |
| S12_extracted_k141_129971 | Not-determined |
| S12_extracted_k141_99582 | Not-determined |
| S12_extracted_k141_161770 | Not-determined |
| S12_extracted_k141_4810 | Low-quality |
| S12_extracted_k141_176879 | Not-determined |
| S12_extracted_k141_91580 | Not-determined |
| S12_extracted_k141_28158 | Not-determined |
| S12_extracted_k141_4812 | Low-quality |
| S12_extracted_k141_83085 | Not-determined |
| S12_extracted_k141_4814 | Not-determined |
| S12_extracted_k141_83088 | Not-determined |
| S12_extracted_k141_129976 | Not-determined |
| S12_extracted_k141_4818 | Not-determined |
| S12_extracted_k141_169647 | Low-quality |
| S12_extracted_k141_185874 | Not-determined |
| S12_extracted_k141_153777 | Not-determined |
| S12_extracted_k141_185878 | Low-quality |
| S12_extracted_k141_161775 | Not-determined |
| S12_extracted_k141_176886 | Low-quality |
| S12_extracted_k141_83091 | Not-determined |
| S12_extracted_k141_129985 | Not-determined |
| S12_extracted_k141_176894 | Not-determined |
| S12_extracted_k141_138427 | Not-determined |
| S12_extracted_k141_19959 | Not-determined |
| S12_extracted_k141_59903 | Not-determined |
| S12_extracted_k141_138430 | Not-determined |
| S12_extracted_k141_107247 | Not-determined |
| S12_extracted_k141_185886 | Not-determined |
| S12_extracted_k141_75015 | Not-determined |
| S12_extracted_k141_59911 | Not-determined |
| S12_extracted_k141_91591 | Not-determined |
| S12_extracted_k141_121932 | Not-determined |
| S12_extracted_k141_28172 | Not-determined |
| S12_extracted_k141_19963 | Not-determined |
| S12_extracted_k141_121935 | Not-determined |
| S12_extracted_k141_91596 | Not-determined |
| S12_extracted_k141_51331 | Not-determined |
| S12_extracted_k141_121938 | Not-determined |
| S12_extracted_k141_114335 | Not-determined |
| S12_extracted_k141_35439 | Not-determined |
| S12_extracted_k141_146322 | Not-determined |
| S12_extracted_k141_35442 | Not-determined |
| S12_extracted_k141_138443 | Low-quality |
| S12_extracted_k141_176912 | Low-quality |
| S12_extracted_k141_130014 | Low-quality |
| S12_extracted_k141_35444 | Not-determined |
| S12_extracted_k141_59921 | Low-quality |
| S12_extracted_k141_83107 | Not-determined |
| S12_extracted_k141_176917 | Not-determined |
| S12_extracted_k141_185902 | Not-determined |
| S12_extracted_k141_161796 | Low-quality |
| S12_extracted_k141_121951 | Not-determined |
| S12_extracted_k141_114350 | Not-determined |
| S12_extracted_k141_121955 | Not-determined |
| S12_extracted_k141_114352 | Not-determined |
| S12_extracted_k141_51336 | Not-determined |
| S12_extracted_k141_59937 | Not-determined |
| S12_extracted_k141_121957 | Low-quality |
| S12_extracted_k141_130016 | Not-determined |
| S12_extracted_k141_35458 | Low-quality |
| S12_extracted_k141_107271 | Low-quality |
| S12_extracted_k141_114356 | Not-determined |
| S12_extracted_k141_138461 | Low-quality |
| S12_extracted_k141_44539 | Not-determined |
| S12_extracted_k141_4835 | Not-determined |
| S12_extracted_k141_161812 | Not-determined |
| S12_extracted_k141_161815 | Not-determined |
| S12_extracted_k141_35467 | Not-determined |
| S12_extracted_k141_99633 | Not-determined |
| S12_extracted_k141_44550 | Not-determined |
| S12_extracted_k141_107283 | Not-determined |
| S12_extracted_k141_114368 | Not-determined |
| S12_extracted_k141_59952 | Not-determined |
| S12_extracted_k141_35475 | Not-determined |
| S12_extracted_k141_4843 | Not-determined |
| S12_extracted_k141_185925 | Not-determined |
| S12_extracted_k141_161820 | Not-determined |
| S12_extracted_k141_169667 | Not-determined |
| S12_extracted_k141_91618 | Not-determined |
| S12_extracted_k141_121978 | Not-determined |
| S12_extracted_k141_138491 | Not-determined |
| S12_extracted_k141_99645 | Not-determined |
| S12_extracted_k141_83138 | Not-determined |
| S12_extracted_k141_107293 | Not-determined |
| S12_extracted_k141_91621 | Low-quality |
| S12_extracted_k141_130019 | Not-determined |
| S12_extracted_k141_91622 | Low-quality |
| S12_extracted_k141_176937 | Not-determined |
| S12_extracted_k141_114370 | Low-quality |
| S12_extracted_k141_130021 | Not-determined |
| S12_extracted_k141_114371 | Not-determined |
| S12_extracted_k141_107300 | Not-determined |
| S12_extracted_k141_121984 | Low-quality |
| S12_extracted_k141_35496 | Not-determined |
| S12_extracted_k141_67543 | Not-determined |
| S12_extracted_k141_176941 | Low-quality |
| S12_extracted_k141_169686 | Not-determined |
| S12_extracted_k141_19998 | Not-determined |
| S12_extracted_k141_28194 | Not-determined |
| S12_extracted_k141_161835 | Not-determined |
| S12_extracted_k141_185954 | Low-quality |
| S12_extracted_k141_114391 | Low-quality |
| S12_extracted_k141_67554 | Not-determined |
| S12_extracted_k141_83154 | Not-determined |
| S12_extracted_k141_51357 | Not-determined |
| S12_extracted_k141_83155 | Not-determined |
| S12_extracted_k141_114394 | Not-determined |
| S12_extracted_k141_44580 | Not-determined |
| S12_extracted_k141_75027 | Not-determined |
| S12_extracted_k141_44581 | Not-determined |
| S12_extracted_k141_67556 | Not-determined |
| S12_extracted_k141_28204 | Not-determined |
| S12_extracted_k141_169691 | Not-determined |
| S12_extracted_k141_107322 | Not-determined |
| S12_extracted_k141_99652 | Not-determined |
| S12_extracted_k141_161847 | Not-determined |
| S12_extracted_k141_67559 | Not-determined |
| S12_extracted_k141_67560 | Not-determined |
| S12_extracted_k141_169698 | Not-determined |
| S12_extracted_k141_51363 | Not-determined |
| S12_extracted_k141_44587 | Not-determined |
| S12_extracted_k141_121994 | Not-determined |
| S12_extracted_k141_20007 | Not-determined |
| S12_extracted_k141_75031 | Not-determined |
| S12_extracted_k141_91638 | Not-determined |
| S12_extracted_k141_107333 | Not-determined |
| S12_extracted_k141_161854 | Not-determined |
| S12_extracted_k141_44594 | Not-determined |
| S12_extracted_k141_169711 | Not-determined |
| S12_extracted_k141_185960 | Not-determined |
| S12_extracted_k141_122004 | Not-determined |
| S12_extracted_k141_122005 | Not-determined |
| S12_extracted_k141_138497 | Not-determined |
| S12_extracted_k141_99673 | Not-determined |
| S12_extracted_k141_20022 | Not-determined |
| S12_extracted_k141_138502 | Low-quality |
| S12_extracted_k141_138503 | Not-determined |
| S12_extracted_k141_107341 | Not-determined |
| S12_extracted_k141_99677 | Not-determined |
| S12_extracted_k141_185965 | Not-determined |
| S12_extracted_k141_107342 | Not-determined |
| S12_extracted_k141_107344 | Not-determined |
| S12_extracted_k141_44609 | Not-determined |
| S12_extracted_k141_169723 | Not-determined |
| S12_extracted_k141_20029 | Low-quality |
| S12_extracted_k141_176962 | Not-determined |
| S12_extracted_k141_44613 | Not-determined |
| S12_extracted_k141_169724 | Not-determined |
| S12_extracted_k141_75034 | Not-determined |
| S12_extracted_k141_107349 | Not-determined |
| S12_extracted_k141_161867 | Low-quality |
| S12_extracted_k141_107353 | Low-quality |
| S12_extracted_k141_114420 | Not-determined |
| S12_extracted_k141_138511 | Not-determined |
| S12_extracted_k141_185970 | Not-determined |
| S12_extracted_k141_114421 | Not-determined |
| S12_extracted_k141_122007 | Not-determined |
| S12_extracted_k141_28239 | Low-quality |
| S12_extracted_k141_114423 | Not-determined |
| S12_extracted_k141_122008 | Not-determined |
| S12_extracted_k141_185975 | Not-determined |
| S12_extracted_k141_185976 | Not-determined |
| S12_extracted_k141_153798 | Not-determined |
| S12_extracted_k141_107364 | Not-determined |
| S12_extracted_k141_67563 | Low-quality |
| S12_extracted_k141_83198 | Not-determined |
| S12_extracted_k141_83201 | Not-determined |
| S12_extracted_k141_28249 | Not-determined |
| S12_extracted_k141_185983 | Not-determined |
| S12_extracted_k141_138520 | Not-determined |
| S12_extracted_k141_153807 | Not-determined |
| S12_extracted_k141_51387 | Not-determined |
| S12_extracted_k141_185985 | Not-determined |
| S12_extracted_k141_185987 | Not-determined |
| S12_extracted_k141_44626 | Not-determined |
| S12_extracted_k141_83204 | Not-determined |
| S12_extracted_k141_20036 | Low-quality |
| S12_extracted_k141_161877 | Not-determined |
| S12_extracted_k141_169753 | Not-determined |
| S12_extracted_k141_153817 | Not-determined |
| S12_extracted_k141_169754 | Not-determined |
| S12_extracted_k141_114438 | Not-determined |
| S12_extracted_k141_91674 | Not-determined |
| S12_extracted_k141_153821 | Not-determined |
| S12_extracted_k141_44633 | Not-determined |
| S12_extracted_k141_185997 | Not-determined |
| S12_extracted_k141_130073 | Not-determined |
| S12_extracted_k141_169757 | Low-quality |
| S12_extracted_k141_75065 | Not-determined |
| S12_extracted_k141_186003 | Not-determined |
| S12_extracted_k141_4878 | Low-quality |
| S12_extracted_k141_186005 | Not-determined |
| S12_extracted_k141_186006 | Not-determined |
| S12_extracted_k141_130079 | Not-determined |
| S12_extracted_k141_4880 | Not-determined |
| S12_extracted_k141_186009 | Not-determined |
| S12_extracted_k141_91679 | Not-determined |
| S12_extracted_k141_130080 | Not-determined |
| S12_extracted_k141_161887 | Low-quality |
| S12_extracted_k141_138542 | Not-determined |
| S12_extracted_k141_4885 | Not-determined |
| S12_extracted_k141_75073 | Not-determined |
| S12_extracted_k141_138548 | Low-quality |
| S12_extracted_k141_99702 | Not-determined |
| S12_extracted_k141_130090 | Not-determined |
| S12_extracted_k141_75075 | Low-quality |
| S12_extracted_k141_4890 | Not-determined |
| S12_extracted_k141_75077 | Not-determined |
| S12_extracted_k141_28282 | Not-determined |
| S12_extracted_k141_186018 | Not-determined |
| S12_extracted_k141_28283 | Not-determined |
| S12_extracted_k141_161893 | Not-determined |
| S12_extracted_k141_99704 | Not-determined |
| S12_extracted_k141_99706 | Not-determined |
| S12_extracted_k141_20053 | Not-determined |
| S12_extracted_k141_186025 | Not-determined |
| S12_extracted_k141_186026 | Not-determined |
| S12_extracted_k141_44666 | Not-determined |
| S12_extracted_k141_138555 | Not-determined |
| S12_extracted_k141_75088 | Not-determined |
| S12_extracted_k141_99711 | Not-determined |
| S12_extracted_k141_51398 | Low-quality |
| S12_extracted_k141_28293 | Not-determined |
| S12_extracted_k141_138557 | Not-determined |
| S12_extracted_k141_35556 | Not-determined |
| S12_extracted_k141_169775 | Not-determined |
| S12_extracted_k141_153837 | Not-determined |
| S12_extracted_k141_138560 | Low-quality |
| S12_extracted_k141_51402 | Not-determined |
| S12_extracted_k141_51404 | Not-determined |
| S12_extracted_k141_161910 | Not-determined |
| S12_extracted_k141_35558 | Not-determined |
| S12_extracted_k141_44672 | Not-determined |
| S12_extracted_k141_186032 | Not-determined |
| S12_extracted_k141_35560 | Not-determined |
| S12_extracted_k141_138566 | Low-quality |
| S12_extracted_k141_114462 | Not-determined |
| S12_extracted_k141_153844 | Not-determined |
| S12_extracted_k141_153846 | Not-determined |
| S12_extracted_k141_28300 | Not-determined |
| S12_extracted_k141_99724 | Not-determined |
| S12_extracted_k141_176998 | Not-determined |
| S12_extracted_k141_177002 | Not-determined |
| S12_extracted_k141_107391 | Not-determined |
| S12_extracted_k141_153847 | Low-quality |
| S12_extracted_k141_20080 | Not-determined |
| S12_extracted_k141_35570 | Not-determined |
| S12_extracted_k141_107392 | Not-determined |
| S12_extracted_k141_107393 | Not-determined |
| S12_extracted_k141_107394 | Not-determined |
| S12_extracted_k141_51418 | Not-determined |
| S12_extracted_k141_153851 | Low-quality |
| S12_extracted_k141_153852 | Not-determined |
| S12_extracted_k141_4910 | Not-determined |
| S12_extracted_k141_99732 | Not-determined |
| S12_extracted_k141_177008 | Not-determined |
| S12_extracted_k141_130115 | Not-determined |
| S12_extracted_k141_20088 | Not-determined |
| S12_extracted_k141_186049 | Not-determined |
| S12_extracted_k141_186051 | Not-determined |
| S12_extracted_k141_177011 | Not-determined |
| S12_extracted_k141_28322 | Not-determined |
| S12_extracted_k141_20093 | Not-determined |
| S12_extracted_k141_177016 | Not-determined |
| S12_extracted_k141_186055 | Not-determined |
| S12_extracted_k141_83246 | Not-determined |
| S12_extracted_k141_177018 | Not-determined |
| S12_extracted_k141_146329 | Low-quality |
| S12_extracted_k141_146330 | Not-determined |
| S12_extracted_k141_169787 | Not-determined |
| S12_extracted_k141_130123 | Low-quality |
| S12_extracted_k141_161917 | Not-determined |
| S12_extracted_k141_51432 | Not-determined |
| S12_extracted_k141_130126 | Not-determined |
| S12_extracted_k141_130127 | Not-determined |
| S12_extracted_k141_20099 | Not-determined |
| S12_extracted_k141_130128 | Not-determined |
| S12_extracted_k141_169790 | Low-quality |
| S12_extracted_k141_107410 | Not-determined |
| S12_extracted_k141_51435 | Not-determined |
| S12_extracted_k141_153872 | Not-determined |
| S12_extracted_k141_186065 | Not-determined |
| S3_vs_k141_29077\|\|full | Medium-quality |
| S12_extracted_k141_107418 | Not-determined |
| S12_extracted_k141_153878 | Not-determined |
| S12_extracted_k141_130135 | Not-determined |
| S12_extracted_k141_107421 | Low-quality |
| S12_extracted_k141_107422 | Not-determined |
| S12_extracted_k141_44695 | Not-determined |
| S12_extracted_k141_44696 | Not-determined |
| S12_extracted_k141_44698 | Not-determined |
| S12_extracted_k141_146345 | Not-determined |
| S12_extracted_k141_138603 | Not-determined |
| S12_extracted_k141_99747 | Not-determined |
| S12_extracted_k141_146347 | Not-determined |
| S12_extracted_k141_44701 | Not-determined |
| S12_extracted_k141_44702 | Not-determined |
| S12_extracted_k141_138605 | Low-quality |
| S12_extracted_k141_186082 | Not-determined |
| S12_extracted_k141_51444 | Not-determined |
| S12_extracted_k141_130145 | Not-determined |
| S12_extracted_k141_28323 | Not-determined |
| S12_extracted_k141_146351 | Low-quality |
| S12_extracted_k141_67588 | Not-determined |
| S12_extracted_k141_146352 | Not-determined |
| S12_extracted_k141_161929 | Not-determined |
| S12_extracted_k141_122035 | Not-determined |
| S12_extracted_k141_130151 | Not-determined |
| S12_extracted_k141_44714 | Not-determined |
| S12_extracted_k141_51456 | Not-determined |
| S12_extracted_k141_99754 | Not-determined |
| S12_extracted_k141_91712 | Not-determined |
| S12_extracted_k141_44719 | Not-determined |
| S12_extracted_k141_161939 | Not-determined |
| S12_extracted_k141_122042 | Not-determined |
| S12_extracted_k141_161940 | Not-determined |
| S12_extracted_k141_91717 | Not-determined |
| S12_extracted_k141_122052 | Not-determined |
| S12_extracted_k141_67601 | Low-quality |
| S12_extracted_k141_186086 | Not-determined |
| S12_extracted_k141_44726 | Not-determined |
| S12_extracted_k141_4926 | Low-quality |
| S12_extracted_k141_153907 | Not-determined |
| S12_extracted_k141_107435 | Not-determined |
| S12_extracted_k141_51464 | Not-determined |
| S12_extracted_k141_99766 | Not-determined |
| S12_extracted_k141_28338 | Low-quality |
| S12_extracted_k141_153912 | Not-determined |
| S12_extracted_k141_35610 | Not-determined |
| S12_extracted_k141_99768 | Not-determined |
| S12_extracted_k141_177024 | Low-quality |
| S12_extracted_k141_186092 | Not-determined |
| S12_extracted_k141_186093 | Not-determined |
| S12_extracted_k141_114487 | Not-determined |
| S12_extracted_k141_51471 | Not-determined |
| S12_extracted_k141_114488 | Not-determined |
| S12_extracted_k141_35614 | Not-determined |
| S12_extracted_k141_28342 | Not-determined |
| S12_extracted_k141_20124 | Not-determined |
| S12_extracted_k141_177031 | Not-determined |
| S12_extracted_k141_138622 | Low-quality |
| S12_extracted_k141_28346 | Not-determined |
| S12_extracted_k141_177032 | Not-determined |
| S12_extracted_k141_51479 | Not-determined |
| S12_extracted_k141_20130 | Not-determined |
| S12_extracted_k141_99775 | Low-quality |
| S12_extracted_k141_83266 | Low-quality |
| S12_extracted_k141_177043 | Not-determined |
| S12_extracted_k141_75106 | Not-determined |
| S12_extracted_k141_130172 | Low-quality |
| S12_extracted_k141_130177 | Not-determined |
| S12_extracted_k141_138639 | Not-determined |
| S12_extracted_k141_146360 | Not-determined |
| S12_extracted_k141_186113 | Not-determined |
| S12_extracted_k141_75112 | Low-quality |
| S12_extracted_k141_130179 | Not-determined |
| S12_extracted_k141_67615 | Low-quality |
| S12_extracted_k141_107460 | Low-quality |
| S12_extracted_k141_161966 | Not-determined |
| S12_extracted_k141_161967 | Not-determined |
| S12_extracted_k141_83277 | Not-determined |
| S12_extracted_k141_114510 | Not-determined |
| S12_extracted_k141_161975 | Not-determined |
| S12_extracted_k141_107472 | Low-quality |
| S12_extracted_k141_177062 | Not-determined |
| S12_extracted_k141_138648 | Not-determined |
| S12_extracted_k141_122069 | Not-determined |
| S12_extracted_k141_153941 | Not-determined |
| S12_extracted_k141_177065 | Not-determined |
| S12_extracted_k141_67630 | Not-determined |
| S12_extracted_k141_177068 | Not-determined |
| S12_extracted_k141_122075 | Not-determined |
| S12_extracted_k141_20151 | Not-determined |
| S12_extracted_k141_107476 | Not-determined |
| S12_extracted_k141_44753 | Not-determined |
| S12_extracted_k141_153951 | Not-determined |
| S12_extracted_k141_91749 | Not-determined |
| S12_extracted_k141_4940 | Not-determined |
| S12_extracted_k141_99794 | Not-determined |
| S12_extracted_k141_28355 | Not-determined |
| S12_extracted_k141_177071 | Not-determined |
| S12_extracted_k141_177072 | Not-determined |
| S12_extracted_k141_67640 | Not-determined |
| S12_extracted_k141_122084 | Not-determined |
| S12_extracted_k141_114522 | Low-quality |
| S12_extracted_k141_67645 | Not-determined |
| S12_extracted_k141_186142 | Not-determined |
| S12_extracted_k141_138653 | Not-determined |
| S12_extracted_k141_153963 | Not-determined |
| S12_extracted_k141_20164 | Not-determined |
| S12_extracted_k141_107482 | Low-quality |
| S12_extracted_k141_186144 | Not-determined |
| S12_extracted_k141_122088 | Not-determined |
| S12_extracted_k141_11589 | Not-determined |
| S12_extracted_k141_99808 | Not-determined |
| S12_extracted_k141_44761 | Not-determined |
| S12_extracted_k141_11591 | Not-determined |
| S12_extracted_k141_28369 | Not-determined |
| S12_extracted_k141_99809 | Low-quality |
| S12_extracted_k141_91754 | Not-determined |
| S12_extracted_k141_44762 | Not-determined |
| S12_extracted_k141_20169 | Not-determined |
| S12_extracted_k141_130206 | Not-determined |
| S12_extracted_k141_91757 | Not-determined |
| S12_extracted_k141_177087 | Not-determined |
| S12_extracted_k141_162008 | Not-determined |
| S12_extracted_k141_130208 | Not-determined |
| S12_extracted_k141_177088 | Not-determined |
| S12_extracted_k141_130211 | Low-quality |
| S12_extracted_k141_51507 | Low-quality |
| S12_extracted_k141_186159 | Not-determined |
| S12_extracted_k141_177094 | Not-determined |
| S12_extracted_k141_28383 | Not-determined |
| S12_extracted_k141_28384 | Not-determined |
| S12_extracted_k141_138659 | Not-determined |
| S12_extracted_k141_138660 | Not-determined |
| S12_extracted_k141_130218 | Not-determined |
| S12_extracted_k141_177096 | Not-determined |
| S12_extracted_k141_91770 | Not-determined |
| S12_extracted_k141_186166 | Not-determined |
| S12_extracted_k141_44774 | Not-determined |
| S12_extracted_k141_11605 | Low-quality |
| S12_extracted_k141_44777 | Not-determined |
| S12_extracted_k141_130222 | Low-quality |
| S12_extracted_k141_51513 | Not-determined |
| S12_extracted_k141_107505 | Not-determined |
| S12_extracted_k141_99819 | Not-determined |
| S12_extracted_k141_162010 | Not-determined |
| S12_extracted_k141_162011 | Not-determined |
| S12_extracted_k141_138673 | Not-determined |
| S12_extracted_k141_107510 | Not-determined |
| S12_extracted_k141_4966 | Not-determined |
| S12_extracted_k141_186174 | Not-determined |
| S12_extracted_k141_177101 | Not-determined |
| S12_extracted_k141_28392 | Not-determined |
| S12_extracted_k141_44785 | Not-determined |
| S12_extracted_k141_51518 | Not-determined |
| S12_extracted_k141_67658 | Not-determined |
| S12_extracted_k141_11614 | Not-determined |
| S12_extracted_k141_130228 | Not-determined |
| S12_extracted_k141_177109 | Low-quality |
| S12_extracted_k141_186181 | Not-determined |
| S12_extracted_k141_138682 | Low-quality |
| S12_extracted_k141_186184 | Low-quality |
| S12_extracted_k141_75141 | Not-determined |
| S12_extracted_k141_114549 | Not-determined |
| S12_extracted_k141_186187 | Not-determined |
| S12_extracted_k141_122108 | Not-determined |
| S12_extracted_k141_130232 | Not-determined |
| S12_extracted_k141_130234 | Not-determined |
| S12_extracted_k141_122111 | Not-determined |
| S12_extracted_k141_130237 | Not-determined |
| S12_extracted_k141_4984 | Not-determined |
| S12_extracted_k141_146380 | Not-determined |
| S12_extracted_k141_35675 | Not-determined |
| S12_extracted_k141_51531 | Not-determined |
| S12_extracted_k141_75146 | Not-determined |
| S12_extracted_k141_122113 | Not-determined |
| S12_extracted_k141_51533 | Not-determined |
| S12_extracted_k141_153986 | Not-determined |
| S12_extracted_k141_75149 | Not-determined |
| S12_extracted_k141_114556 | Low-quality |
| S12_extracted_k141_44803 | Not-determined |
| S12_extracted_k141_107517 | Not-determined |
| S12_extracted_k141_11624 | Not-determined |
| S12_extracted_k141_138693 | Not-determined |
| S12_extracted_k141_83290 | Not-determined |
| S12_extracted_k141_186202 | Not-determined |
| S12_extracted_k141_20178 | Low-quality |
| S12_extracted_k141_99841 | Not-determined |
| S12_extracted_k141_35686 | Not-determined |
| S12_extracted_k141_67666 | Not-determined |
| S12_extracted_k141_35688 | Not-determined |
| S12_extracted_k141_186205 | Not-determined |
| S12_extracted_k141_107520 | Not-determined |
| S12_extracted_k141_107521 | Not-determined |
| S12_extracted_k141_114560 | Not-determined |
| S12_extracted_k141_44809 | Not-determined |
| S12_extracted_k141_114561 | Not-determined |
| S12_extracted_k141_91808 | Not-determined |
| S12_extracted_k141_67678 | Not-determined |

| | |
|---|---|
| S12_extracted_k141_11641 | Not-determined |
| S12_extracted_k141_107527 | Low-quality |
| S12_extracted_k141_130247 | Not-determined |
| S12_extracted_k141_11644 | Low-quality |
| S12_extracted_k141_107531 | Not-determined |
| S12_extracted_k141_162049 | Not-determined |
| S12_extracted_k141_186217 | Not-determined |
| S12_extracted_k141_130250 | Not-determined |
| S12_extracted_k141_162053 | Not-determined |
| S12_extracted_k141_75166 | Not-determined |
| S12_extracted_k141_75167 | Not-determined |
| S12_extracted_k141_130253 | Not-determined |
| S12_extracted_k141_114574 | Not-determined |
| S12_extracted_k141_162055 | Low-quality |
| S12_extracted_k141_154005 | Low-quality |
| S12_extracted_k141_162056 | Not-determined |
| S12_extracted_k141_154009 | Not-determined |
| S12_extracted_k141_107535 | Not-determined |
| S12_extracted_k141_51552 | Not-determined |
| S12_extracted_k141_75170 | Not-determined |
| S12_extracted_k141_35714 | Not-determined |
| S12_extracted_k141_114585 | Not-determined |
| S12_extracted_k141_146417 | Low-quality |
| S12_extracted_k141_51557 | Not-determined |
| S12_extracted_k141_75172 | Not-determined |
| S12_extracted_k141_20201 | Not-determined |
| S12_extracted_k141_99861 | Not-determined |
| S12_extracted_k141_146422 | Not-determined |
| S12_extracted_k141_20210 | Not-determined |
| S12_extracted_k141_146424 | Not-determined |
| S12_extracted_k141_146425 | Not-determined |
| S12_extracted_k141_51567 | Low-quality |
| S12_extracted_k141_114587 | Low-quality |
| S12_extracted_k141_67696 | Not-determined |
| S12_extracted_k141_186225 | Not-determined |
| S12_extracted_k141_67697 | Not-determined |
| S12_extracted_k141_186226 | Not-determined |
| S12_extracted_k141_107547 | Not-determined |
| S12_extracted_k141_75184 | Not-determined |
| S12_extracted_k141_11672 | Low-quality |
| S12_extracted_k141_130268 | Low-quality |
| S12_extracted_k141_162071 | Not-determined |
| S12_extracted_k141_5027 | Not-determined |
| S12_extracted_k141_177159 | Low-quality |
| S12_extracted_k141_20215 | Not-determined |
| S12_extracted_k141_44835 | Not-determined |
| S12_extracted_k141_186234 | Not-determined |
| S12_extracted_k141_146432 | Not-determined |
| S12_extracted_k141_162072 | Not-determined |
| S12_extracted_k141_186239 | Not-determined |
| S12_extracted_k141_107559 | Not-determined |
| S12_extracted_k141_162075 | Not-determined |
| S12_extracted_k141_20222 | Not-determined |
| S12_extracted_k141_177164 | Not-determined |
| S12_extracted_k141_114600 | Not-determined |
| S12_extracted_k141_154022 | Not-determined |
| S12_extracted_k141_114604 | Not-determined |
| S12_extracted_k141_28413 | Not-determined |
| S12_extracted_k141_107563 | Not-determined |
| S12_extracted_k141_114605 | Not-determined |
| S12_extracted_k141_138729 | Not-determined |
| S12_extracted_k141_75206 | Not-determined |
| S12_extracted_k141_186252 | Not-determined |
| S12_extracted_k141_51576 | Not-determined |
| S12_extracted_k141_99879 | Not-determined |
| S12_extracted_k141_177178 | Not-determined |
| S12_extracted_k141_44846 | Low-quality |
| S12_extracted_k141_162088 | Not-determined |
| S12_extracted_k141_186259 | Low-quality |
| S12_extracted_k141_51582 | Not-determined |
| S12_extracted_k141_75215 | Not-determined |
| S12_extracted_k141_122152 | Not-determined |
| S12_extracted_k141_107570 | Not-determined |
| S12_extracted_k141_51590 | Not-determined |
| S12_extracted_k141_91864 | Not-determined |
| S12_extracted_k141_11703 | Low-quality |
| S12_extracted_k141_83342 | Not-determined |
| S12_extracted_k141_186271 | Not-determined |
| S12_extracted_k141_67711 | Not-determined |
| S12_extracted_k141_91866 | Not-determined |
| S12_extracted_k141_162106 | Not-determined |
| S12_extracted_k141_44853 | Not-determined |
| S12_extracted_k141_122156 | Not-determined |
| S12_extracted_k141_11709 | Not-determined |
| S12_extracted_k141_186274 | Not-determined |
| S12_extracted_k141_28421 | Not-determined |
| S4_extracted_18803 | Medium-quality |
| S12_extracted_k141_138733 | Low-quality |
| S12_extracted_k141_162109 | Low-quality |
| S12_extracted_k141_177194 | Not-determined |
| S12_extracted_k141_51592 | Not-determined |
| S12_extracted_k141_138738 | Not-determined |
| S12_extracted_k141_146469 | Not-determined |
| S12_extracted_k141_51593 | Not-determined |
| S12_extracted_k141_67718 | Not-determined |
| S12_extracted_k141_51594 | Not-determined |
| S12_extracted_k141_177198 | Not-determined |
| S12_extracted_k141_162113 | Not-determined |
| S12_extracted_k141_67720 | Low-quality |
| S12_extracted_k141_186280 | Not-determined |
| S12_extracted_k141_177200 | Not-determined |
| S12_extracted_k141_91871 | Not-determined |
| S12_extracted_k141_91873 | Not-determined |
| S12_extracted_k141_114631 | Low-quality |
| S12_extracted_k141_51597 | Not-determined |
| S12_extracted_k141_67723 | Not-determined |
| S12_extracted_k141_177204 | Not-determined |
| S12_extracted_k141_44867 | Not-determined |
| S12_extracted_k141_11717 | Not-determined |
| S12_extracted_k141_138745 | Not-determined |
| S12_extracted_k141_91874 | Low-quality |
| S12_extracted_k141_146474 | Not-determined |
| S12_extracted_k141_138748 | Not-determined |
| S12_extracted_k141_186286 | Not-determined |
| S12_extracted_k141_75228 | Not-determined |
| S12_extracted_k141_138749 | Not-determined |
| S12_extracted_k141_169811 | Not-determined |
| S12_extracted_k141_122177 | Not-determined |
| S12_extracted_k141_51604 | Not-determined |
| S12_extracted_k141_162117 | Low-quality |
| S12_extracted_k141_5069 | Not-determined |
| S12_extracted_k141_177217 | Not-determined |
| S12_extracted_k141_91882 | Not-determined |
| S12_extracted_k141_44873 | Low-quality |
| S12_extracted_k141_162119 | Not-determined |
| S12_extracted_k141_51609 | Not-determined |
| S12_extracted_k141_35753 | Not-determined |
| S12_extracted_k141_186290 | Not-determined |
| S12_extracted_k141_11727 | Not-determined |
| S12_extracted_k141_146483 | Not-determined |
| S12_extracted_k141_11728 | Not-determined |
| S12_extracted_k141_114640 | Not-determined |
| S12_extracted_k141_75231 | Low-quality |
| S12_extracted_k141_122190 | Not-determined |
| S12_extracted_k141_130295 | Low-quality |
| S12_extracted_k141_162123 | Low-quality |
| S12_extracted_k141_35758 | Low-quality |
| S12_extracted_k141_51615 | Not-determined |
| S12_extracted_k141_91893 | Not-determined |
| S12_extracted_k141_107597 | Not-determined |
| S12_extracted_k141_20251 | Not-determined |
| S12_extracted_k141_186298 | Not-determined |
| S12_extracted_k141_122194 | Not-determined |
| S12_extracted_k141_99887 | Not-determined |
| S12_extracted_k141_75238 | Not-determined |
| S12_extracted_k141_122196 | Not-determined |
| S12_extracted_k141_11731 | Not-determined |
| S12_extracted_k141_177228 | Not-determined |
| S12_extracted_k141_11734 | Not-determined |
| S12_extracted_k141_138766 | Not-determined |
| S12_extracted_k141_122201 | Not-determined |
| S12_extracted_k141_107605 | Not-determined |
| S12_extracted_k141_122202 | Not-determined |
| S12_extracted_k141_114650 | Not-determined |
| S12_extracted_k141_130304 | Not-determined |
| S12_extracted_k141_138769 | Not-determined |
| S12_extracted_k141_28456 | Not-determined |
| S12_extracted_k141_67735 | Not-determined |
| S12_extracted_k141_138770 | Not-determined |
| S12_extracted_k141_67736 | Not-determined |
| S12_extracted_k141_51624 | Not-determined |
| S12_extracted_k141_177231 | Not-determined |
| S12_extracted_k141_5084 | Not-determined |
| S12_extracted_k141_28459 | Not-determined |
| S12_extracted_k141_5085 | Not-determined |
| S12_extracted_k141_20257 | Not-determined |
| S12_extracted_k141_5086 | Not-determined |
| S12_extracted_k141_44880 | Low-quality |
| S12_extracted_k141_75252 | Not-determined |
| S12_extracted_k141_122206 | Not-determined |
| S12_extracted_k141_28464 | Not-determined |
| S12_extracted_k141_177240 | Not-determined |
| S12_extracted_k141_107614 | Not-determined |
| S12_extracted_k141_169826 | Low-quality |
| S12_extracted_k141_20259 | Not-determined |
| S12_extracted_k141_28466 | Not-determined |
| S12_extracted_k141_5090 | Not-determined |
| S12_extracted_k141_122208 | Not-determined |
| S12_extracted_k141_177241 | Not-determined |
| S12_extracted_k141_35784 | Not-determined |
| S12_extracted_k141_186308 | Not-determined |
| S12_extracted_k141_20263 | Not-determined |
| S12_extracted_k141_177245 | Not-determined |
| S12_extracted_k141_114662 | Not-determined |
| S12_extracted_k141_186310 | Not-determined |
| S12_extracted_k141_186312 | Not-determined |
| S12_extracted_k141_5094 | Low-quality |
| S12_extracted_k141_44888 | Not-determined |
| S12_extracted_k141_5096 | Not-determined |
| S12_extracted_k141_51634 | Not-determined |
| S12_extracted_k141_138790 | Not-determined |
| S12_extracted_k141_130312 | Not-determined |
| S12_extracted_k141_99917 | Not-determined |
| S12_extracted_k141_122214 | Not-determined |
| S12_extracted_k141_107628 | Not-determined |
| S12_extracted_k141_130313 | Not-determined |
| S12_extracted_k141_99921 | Low-quality |
| S12_extracted_k141_75268 | Not-determined |
| S12_extracted_k141_99923 | Not-determined |
| S12_extracted_k141_146492 | Not-determined |
| S12_extracted_k141_138800 | Not-determined |
| S12_extracted_k141_138802 | Not-determined |
| S12_extracted_k141_130318 | Not-determined |
| S12_extracted_k141_146494 | Not-determined |
| S12_extracted_k141_122222 | Not-determined |
| S12_extracted_k141_35804 | Not-determined |
| S12_extracted_k141_5107 | Not-determined |
| S12_extracted_k141_177257 | Not-determined |
| S12_extracted_k141_130320 | Not-determined |
| S12_extracted_k141_177258 | Not-determined |
| S12_extracted_k141_122224 | Not-determined |
| S12_extracted_k141_75275 | Not-determined |
| S12_extracted_k141_20281 | Not-determined |
| S12_extracted_k141_107635 | Not-determined |
| S12_extracted_k141_146497 | Not-determined |
| S12_extracted_k141_35805 | Not-determined |
| S12_extracted_k141_169834 | Not-determined |
| S12_extracted_k141_11754 | Not-determined |
| S12_extracted_k141_146499 | Not-determined |
| S12_extracted_k141_107638 | Not-determined |
| S12_extracted_k141_138805 | Not-determined |
| S12_extracted_k141_122229 | Not-determined |
| S12_extracted_k141_11757 | Not-determined |
| S12_extracted_k141_83358 | Not-determined |
| S12_extracted_k141_138806 | Not-determined |
| S12_extracted_k141_130325 | Not-determined |
| S12_extracted_k141_28488 | Not-determined |
| S12_extracted_k141_83360 | Not-determined |
| S12_extracted_k141_20290 | Not-determined |
| S12_extracted_k141_138808 | Not-determined |
| S12_extracted_k141_28490 | Not-determined |
| S12_extracted_k141_20292 | Not-determined |
| S12_extracted_k141_169842 | Not-determined |
| S12_extracted_k141_107644 | Not-determined |
| S12_extracted_k141_138812 | Not-determined |
| S12_extracted_k141_146505 | Not-determined |
| S12_extracted_k141_28494 | Low-quality |
| S12_extracted_k141_28495 | Not-determined |
| S12_extracted_k141_83365 | Not-determined |
| S12_extracted_k141_83366 | Not-determined |
| S12_extracted_k141_146507 | Not-determined |
| S12_extracted_k141_75283 | Not-determined |
| S12_extracted_k141_177268 | Not-determined |
| S12_extracted_k141_169849 | Not-determined |
| S12_extracted_k141_122241 | Not-determined |
| S12_extracted_k141_114683 | Not-determined |
| S12_extracted_k141_122243 | Not-determined |
| S12_extracted_k141_59971 | Not-determined |
| S12_extracted_k141_59972 | Not-determined |
| S12_extracted_k141_146514 | Not-determined |
| S12_extracted_k141_122244 | Not-determined |
| S12_extracted_k141_75291 | Not-determined |
| S12_extracted_k141_59974 | Not-determined |
| S12_extracted_k141_154038 | Low-quality |
| S12_extracted_k141_28506 | Low-quality |
| S12_extracted_k141_75293 | Not-determined |
| S12_extracted_k141_67779 | Not-determined |
| S12_extracted_k141_169861 | Not-determined |
| S12_extracted_k141_83379 | Not-determined |
| S12_extracted_k141_154042 | Not-determined |
| S12_extracted_k141_67782 | Not-determined |
| S12_extracted_k141_59979 | Not-determined |
| S12_extracted_k141_154044 | Not-determined |
| S12_extracted_k141_59980 | Not-determined |
| S12_extracted_k141_177275 | Not-determined |
| S12_extracted_k141_75299 | Not-determined |
| S12_extracted_k141_154046 | Not-determined |
| S12_extracted_k141_114687 | Not-determined |
| S12_extracted_k141_51676 | Not-determined |
| S12_extracted_k141_107660 | Not-determined |
| S12_extracted_k141_75307 | Not-determined |
| S12_extracted_k141_107662 | Not-determined |
| S12_extracted_k141_28522 | Not-determined |
| S12_extracted_k141_59992 | Not-determined |
| S12_extracted_k141_107666 | Not-determined |
| S12_extracted_k141_99968 | Not-determined |
| S12_extracted_k141_75313 | Not-determined |
| S12_extracted_k141_114703 | Low-quality |
| S12_extracted_k141_122260 | Not-determined |
| S12_extracted_k141_44918 | Not-determined |
| S12_extracted_k141_177297 | Low-quality |
| S12_extracted_k141_177298 | Not-determined |
| S12_extracted_k141_138827 | Not-determined |
| S12_extracted_k141_44919 | Not-determined |
| S12_extracted_k141_177299 | Not-determined |
| S12_extracted_k141_107670 | Not-determined |
| S12_extracted_k141_107671 | Not-determined |
| S12_extracted_k141_177302 | Not-determined |
| S12_extracted_k141_114711 | Not-determined |
| S12_extracted_k141_186321 | Not-determined |
| S12_extracted_k141_35832 | Low-quality |
| S12_extracted_k141_11778 | Not-determined |
| S12_extracted_k141_138832 | Not-determined |
| S12_extracted_k141_20331 | Not-determined |
| S12_extracted_k141_177307 | Not-determined |
| S12_extracted_k141_177310 | Not-determined |
| S12_extracted_k141_114714 | Not-determined |
| S12_extracted_k141_186329 | Not-determined |
| S12_extracted_k141_75322 | Not-determined |
| S12_extracted_k141_60018 | Low-quality |
| S12_extracted_k141_75327 | Not-determined |
| S12_extracted_k141_28543 | Not-determined |
| S12_extracted_k141_44935 | Not-determined |
| S12_extracted_k141_186334 | Not-determined |
| S12_extracted_k141_91916 | Low-quality |
| S12_extracted_k141_91918 | Not-determined |
| S12_extracted_k141_83409 | Not-determined |
| S12_extracted_k141_28550 | Not-determined |
| S12_extracted_k141_44941 | Not-determined |
| S12_extracted_k141_91922 | Not-determined |
| S12_extracted_k141_107683 | Not-determined |
| S12_extracted_k141_91924 | Not-determined |
| S12_extracted_k141_28554 | Not-determined |
| S12_extracted_k141_138845 | Low-quality |
| S12_extracted_k141_138846 | Not-determined |
| S12_extracted_k141_28556 | Not-determined |
| S12_extracted_k141_28559 | Not-determined |
| S12_extracted_k141_35841 | Not-determined |
| S12_extracted_k141_20344 | Not-determined |
| S12_extracted_k141_99983 | Not-determined |
| S12_extracted_k141_83421 | Not-determined |
| S12_extracted_k141_177319 | Not-determined |
| S12_extracted_k141_177320 | Not-determined |
| S12_extracted_k141_138848 | Not-determined |
| S12_extracted_k141_138850 | Not-determined |
| S12_extracted_k141_91935 | Not-determined |
| S12_extracted_k141_107692 | Not-determined |
| S12_extracted_k141_138851 | Not-determined |
| S12_extracted_k141_162168 | Not-determined |
| S12_extracted_k141_162169 | Not-determined |
| S12_extracted_k141_99986 | Low-quality |
| S12_extracted_k141_107695 | Not-determined |
| S12_extracted_k141_177326 | Not-determined |
| S12_extracted_k141_99988 | Not-determined |
| S12_extracted_k141_28562 | Not-determined |
| S12_extracted_k141_11807 | Low-quality |
| S12_extracted_k141_146532 | Not-determined |
| S12_extracted_k141_35849 | Not-determined |
| S12_extracted_k141_28571 | Not-determined |
| S12_extracted_k141_154072 | Low-quality |
| S12_extracted_k141_154074 | Not-determined |
| S12_extracted_k141_186354 | Not-determined |
| S12_extracted_k141_162179 | Not-determined |
| S12_extracted_k141_83437 | Not-determined |
| S12_extracted_k141_146541 | Not-determined |
| S12_extracted_k141_5137 | Not-determined |
| S12_extracted_k141_162182 | Not-determined |
| S12_extracted_k141_130362 | Not-determined |
| S12_extracted_k141_162187 | Not-determined |
| S12_extracted_k141_138861 | Not-determined |
| S12_extracted_k141_114737 | Not-determined |
| S12_extracted_k141_107703 | Not-determined |
| S12_extracted_k141_186359 | Not-determined |
| S12_extracted_k141_154085 | Not-determined |
| S12_extracted_k141_114744 | Not-determined |
| S12_extracted_k141_138865 | Not-determined |
| S12_extracted_k141_122273 | Not-determined |
| S12_extracted_k141_138866 | Not-determined |
| S12_extracted_k141_162194 | Not-determined |
| S12_extracted_k141_11820 | Low-quality |
| S12_extracted_k141_186366 | Not-determined |
| S12_extracted_k141_146555 | Not-determined |
| S12_extracted_k141_122275 | Not-determined |
| S12_extracted_k141_169916 | Not-determined |
| S12_extracted_k141_11823 | Not-determined |
| S12_extracted_k141_169919 | Not-determined |
| S12_extracted_k141_186370 | Low-quality |
| S12_extracted_k141_169920 | Not-determined |
| S12_extracted_k141_114748 | Not-determined |
| S12_extracted_k141_20374 | Not-determined |
| S12_extracted_k141_11828 | Not-determined |
| S12_extracted_k141_60021 | Not-determined |
| S12_extracted_k141_122281 | Not-determined |
| S12_extracted_k141_20376 | Not-determined |
| S12_extracted_k141_146565 | Not-determined |
| S12_extracted_k141_20378 | Not-determined |
| S12_extracted_k141_122287 | Not-determined |
| S12_extracted_k141_162206 | Not-determined |
| S12_extracted_k141_100007 | Low-quality |
| S12_extracted_k141_100008 | Not-determined |
| S12_extracted_k141_100010 | Not-determined |
| S12_extracted_k141_83459 | Not-determined |
| S12_extracted_k141_114761 | Not-determined |
| S12_extracted_k141_146571 | Not-determined |
| S12_extracted_k141_162215 | Not-determined |
| S12_extracted_k141_75383 | Not-determined |
| S12_extracted_k141_138878 | Not-determined |
| S12_extracted_k141_162216 | Low-quality |
| S12_extracted_k141_146575 | Not-determined |
| S12_extracted_k141_75384 | Not-determined |
| S12_extracted_k141_138880 | Not-determined |
| S12_extracted_k141_75385 | Not-determined |
| S12_extracted_k141_83465 | Low-quality |
| S12_extracted_k141_162219 | Low-quality |
| S12_extracted_k141_162220 | Not-determined |
| S12_extracted_k141_75387 | Not-determined |
| S12_extracted_k141_169940 | Not-determined |
| S12_extracted_k141_154109 | Not-determined |
| S12_extracted_k141_154110 | Not-determined |
| S12_extracted_k141_5167 | Not-determined |
| S12_extracted_k141_146580 | Not-determined |
| S12_extracted_k141_20403 | Not-determined |
| S12_extracted_k141_100025 | Not-determined |
| S12_extracted_k141_138886 | Not-determined |
| S12_extracted_k141_114771 | Not-determined |
| S12_extracted_k141_11843 | Low-quality |
| S12_extracted_k141_91942 | Not-determined |
| S12_extracted_k141_11845 | Not-determined |
| S12_extracted_k141_83479 | Not-determined |
| S12_extracted_k141_154118 | Not-determined |
| S12_extracted_k141_138891 | Not-determined |
| S12_extracted_k141_100032 | Not-determined |
| S12_extracted_k141_75396 | Not-determined |
| S12_extracted_k141_75397 | Not-determined |
| S12_extracted_k141_51693 | Not-determined |
| S12_extracted_k141_51694 | Not-determined |
| S12_extracted_k141_154122 | Not-determined |
| S12_extracted_k141_107706 | Not-determined |
| S12_extracted_k141_100034 | Not-determined |
| S12_extracted_k141_114785 | Not-determined |
| S12_extracted_k141_107708 | Not-determined |
| S12_extracted_k141_28596 | Not-determined |
| S12_extracted_k141_5180 | Not-determined |
| S12_extracted_k141_146596 | Not-determined |
| S12_extracted_k141_122303 | Not-determined |
| S12_extracted_k141_28599 | Not-determined |
| S12_extracted_k141_122304 | Not-determined |
| S12_extracted_k141_91951 | Not-determined |
| S12_extracted_k141_186378 | Not-determined |
| S12_extracted_k141_35871 | Not-determined |
| S12_extracted_k141_146601 | Not-determined |
| S12_extracted_k141_122311 | Not-determined |
| S12_extracted_k141_122314 | Not-determined |
| S12_extracted_k141_169951 | Low-quality |
| S12_extracted_k141_154129 | Not-determined |
| S12_extracted_k141_20426 | Not-determined |
| S12_extracted_k141_169957 | Not-determined |
| S12_extracted_k141_146607 | Not-determined |
| S12_extracted_k141_122318 | Not-determined |
| S12_extracted_k141_146609 | Not-determined |
| S12_extracted_k141_91963 | Not-determined |
| S12_extracted_k141_154133 | Not-determined |
| S12_extracted_k141_114797 | Low-quality |
| S12_extracted_k141_114799 | Not-determined |
| S12_extracted_k141_20432 | Not-determined |
| S12_extracted_k141_91967 | Not-determined |
| S12_extracted_k141_11861 | Not-determined |
| S12_extracted_k141_100051 | Not-determined |
| S12_extracted_k141_162247 | Not-determined |
| S12_extracted_k141_75408 | Low-quality |
| S12_extracted_k141_44980 | Not-determined |
| S12_extracted_k141_162249 | Not-determined |
| S12_extracted_k141_169970 | Not-determined |
| S12_extracted_k141_11866 | Not-determined |
| S12_extracted_k141_51707 | Low-quality |
| S12_extracted_k141_44988 | Not-determined |
| S12_extracted_k141_100062 | Not-determined |
| S12_extracted_k141_130396 | Not-determined |
| S12_extracted_k141_122333 | Low-quality |
| S12_extracted_k141_100065 | Not-determined |
| S12_extracted_k141_44999 | Not-determined |
| S12_extracted_k141_154141 | Not-determined |
| S12_extracted_k141_169979 | Not-determined |
| S12_extracted_k141_138914 | Not-determined |
| S12_extracted_k141_169980 | Not-determined |
| S12_extracted_k141_162259 | Not-determined |
| S12_extracted_k141_169982 | Not-determined |
| S12_extracted_k141_154144 | Not-determined |
| S12_extracted_k141_169983 | Not-determined |
| S12_extracted_k141_162264 | Not-determined |
| S12_extracted_k141_130404 | Not-determined |
| S12_extracted_k141_138917 | Not-determined |
| S12_extracted_k141_28608 | Not-determined |
| S12_extracted_k141_138923 | Not-determined |
| S12_extracted_k141_114819 | Not-determined |
| S12_extracted_k141_138924 | Not-determined |
| S12_extracted_k141_146626 | Not-determined |
| S12_extracted_k141_146628 | Not-determined |
| S12_extracted_k141_75424 | Not-determined |
| S12_extracted_k141_122345 | Not-determined |
| S12_extracted_k141_114829 | Not-determined |
| S12_extracted_k141_28613 | Not-determined |
| S12_extracted_k141_138932 | Not-determined |
| S12_extracted_k141_138934 | Not-determined |
| S12_extracted_k141_114830 | Not-determined |
| S12_extracted_k141_5199 | Low-quality |
| S12_extracted_k141_51725 | Not-determined |
| S12_extracted_k141_75430 | Not-determined |
| S12_extracted_k141_51727 | Not-determined |
| S12_extracted_k141_91983 | Not-determined |
| S12_extracted_k141_107719 | Not-determined |
| S12_extracted_k141_138942 | Low-quality |
| S12_extracted_k141_107723 | Not-determined |
| S12_extracted_k141_75435 | Not-determined |
| S12_extracted_k141_5210 | Not-determined |
| S12_extracted_k141_28628 | Not-determined |
| S12_extracted_k141_75438 | Not-determined |
| S12_extracted_k141_100079 | Not-determined |
| S12_extracted_k141_100082 | Not-determined |
| S12_extracted_k141_186426 | Not-determined |
| S12_extracted_k141_146652 | Not-determined |
| S12_extracted_k141_170006 | Not-determined |
| S12_extracted_k141_28638 | Low-quality |
| S12_extracted_k141_35903 | Not-determined |
| S12_extracted_k141_92007 | Low-quality |
| S12_extracted_k141_186432 | Not-determined |
| S12_extracted_k141_75449 | Not-determined |
| S12_extracted_k141_130433 | Not-determined |
| S12_extracted_k141_60030 | Not-determined |
| S12_extracted_k141_92012 | Not-determined |
| S12_extracted_k141_154175 | Not-determined |
| S12_extracted_k141_130436 | Low-quality |
| S12_extracted_k141_20468 | Not-determined |
| S12_extracted_k141_75457 | Not-determined |
| S12_extracted_k141_186441 | Not-determined |
| S12_extracted_k141_20469 | Not-determined |
| S12_extracted_k141_122385 | Not-determined |
| S12_extracted_k141_162283 | Not-determined |
| S12_extracted_k141_20474 | Not-determined |
| S12_extracted_k141_154181 | Not-determined |
| S12_extracted_k141_67805 | Not-determined |
| S12_extracted_k141_20478 | Low-quality |
| S12_extracted_k141_186451 | Not-determined |
| S12_extracted_k141_20480 | Not-determined |
| S12_extracted_k141_60033 | Not-determined |
| S12_extracted_k141_35915 | Not-determined |
| S12_extracted_k141_67810 | Not-determined |
| S12_extracted_k141_5231 | Not-determined |
| S12_extracted_k141_162291 | Not-determined |
| S12_extracted_k141_51731 | Not-determined |
| S12_extracted_k141_186458 | Not-determined |
| S12_extracted_k141_75467 | Low-quality |
| S12_extracted_k141_130459 | Not-determined |
| S12_extracted_k141_11905 | Not-determined |
| S12_extracted_k141_138972 | Not-determined |
| S12_extracted_k141_92028 | Not-determined |
| S12_extracted_k141_162299 | Not-determined |
| S12_extracted_k141_100103 | Not-determined |
| S12_extracted_k141_186462 | Not-determined |
| S12_extracted_k141_170020 | Not-determined |
| S12_extracted_k141_83496 | Not-determined |
| S12_extracted_k141_138977 | Not-determined |
| S12_extracted_k141_67822 | Not-determined |
| S12_extracted_k141_162303 | Not-determined |
| S12_extracted_k141_20490 | Not-determined |
| S12_extracted_k141_146681 | Not-determined |
| S12_extracted_k141_83501 | Not-determined |
| S12_extracted_k141_186466 | Not-determined |
| S12_extracted_k141_170023 | Not-determined |
| S12_extracted_k141_138980 | Not-determined |
| S12_extracted_k141_100108 | Low-quality |
| S12_extracted_k141_114847 | Not-determined |
| S12_extracted_k141_162309 | Not-determined |
| S12_extracted_k141_60045 | Low-quality |
| S12_extracted_k141_51753 | Not-determined |
| S12_extracted_k141_114853 | Not-determined |
| S12_extracted_k141_75477 | Not-determined |
| S12_extracted_k141_114859 | Not-determined |
| S12_extracted_k141_122389 | Not-determined |
| S12_extracted_k141_11917 | Not-determined |
| S12_extracted_k141_11918 | Not-determined |
| S12_extracted_k141_75481 | Not-determined |
| S12_extracted_k141_67836 | Not-determined |
| S12_extracted_k141_75482 | Not-determined |
| S12_extracted_k141_170033 | Not-determined |
| S12_extracted_k141_130477 | Not-determined |
| S12_extracted_k141_35930 | Not-determined |
| S12_extracted_k141_114861 | Not-determined |
| S12_extracted_k141_92054 | Not-determined |
| S12_extracted_k141_114862 | Not-determined |
| S12_extracted_k141_122396 | Not-determined |
| S12_extracted_k141_35936 | Not-determined |
| S12_extracted_k141_92057 | Not-determined |
| S12_extracted_k141_35938 | Not-determined |
| S12_extracted_k141_170043 | Not-determined |
| S12_extracted_k141_146703 | Not-determined |
| S12_extracted_k141_162318 | Not-determined |
| S12_extracted_k141_20517 | Low-quality |
| S12_extracted_k141_100123 | Not-determined |
| S12_extracted_k141_75493 | Not-determined |
| S12_extracted_k141_186495 | Not-determined |
| S12_extracted_k141_67846 | Not-determined |
| S12_extracted_k141_60058 | Not-determined |
| S12_extracted_k141_5249 | Not-determined |
| S12_extracted_k141_51763 | Not-determined |
| S12_extracted_k141_114876 | Not-determined |
| S12_extracted_k141_60061 | Not-determined |
| S12_extracted_k141_75502 | Not-determined |
| S12_extracted_k141_162323 | Not-determined |

| | |
|---|---|
| S12_extracted_k141_20529 | Not-determined |
| S12_extracted_k141_114879 | Not-determined |
| S12_extracted_k141_130494 | Not-determined |
| S12_extracted_k141_92069 | Not-determined |
| S12_extracted_k141_60068 | Not-determined |
| S12_extracted_k141_60072 | Not-determined |
| S12_extracted_k141_114884 | Not-determined |
| S12_extracted_k141_139009 | Not-determined |
| S12_extracted_k141_100142 | Not-determined |
| S12_extracted_k141_5256 | Not-determined |
| S12_extracted_k141_51776 | Not-determined |
| S12_extracted_k141_83515 | Not-determined |
| S12_extracted_k141_122417 | Not-determined |
| S12_extracted_k141_83516 | Not-determined |
| S12_extracted_k141_60084 | Not-determined |
| S12_extracted_k141_67853 | Low-quality |
| S12_extracted_k141_60085 | Not-determined |
| S12_extracted_k141_45065 | Not-determined |
| S12_extracted_k141_20541 | Not-determined |
| S12_extracted_k141_162343 | Not-determined |
| S12_extracted_k141_122422 | Low-quality |
| S12_extracted_k141_92083 | Not-determined |
| S12_extracted_k141_20545 | Not-determined |
| S12_extracted_k141_162345 | Not-determined |
| S12_extracted_k141_130508 | Not-determined |
| S12_extracted_k141_60097 | Not-determined |
| S12_extracted_k141_5266 | Not-determined |
| S12_extracted_k141_20549 | Not-determined |
| S12_extracted_k141_114893 | Not-determined |
| S12_extracted_k141_67870 | Not-determined |
| S12_extracted_k141_107773 | Not-determined |
| S12_extracted_k141_92093 | Not-determined |
| S12_extracted_k141_154211 | Not-determined |
| S12_extracted_k141_20555 | Not-determined |
| S12_extracted_k141_170058 | Low-quality |
| S12_extracted_k141_130520 | Not-determined |
| S12_extracted_k141_100158 | Not-determined |
| S12_extracted_k141_162359 | Not-determined |
| S12_extracted_k141_92096 | Not-determined |
| S12_extracted_k141_20560 | Not-determined |
| S12_extracted_k141_186508 | Not-determined |
| S12_extracted_k141_146720 | Not-determined |
| S12_extracted_k141_92100 | Not-determined |
| S12_extracted_k141_122436 | Not-determined |
| S12_extracted_k141_11943 | Low-quality |
| S12_extracted_k141_186512 | Not-determined |
| S12_extracted_k141_11944 | Not-determined |
| S12_extracted_k141_83519 | Not-determined |
| S12_extracted_k141_83520 | Not-determined |
| S12_extracted_k141_162366 | Not-determined |
| S12_extracted_k141_5276 | Not-determined |
| S12_extracted_k141_67876 | Not-determined |
| S12_extracted_k141_154222 | Not-determined |
| S12_extracted_k141_100166 | Not-determined |
| S12_extracted_k141_186517 | Not-determined |
| S12_extracted_k141_92107 | Not-determined |
| S12_extracted_k141_162368 | Not-determined |
| S12_extracted_k141_67878 | Not-determined |
| S12_extracted_k141_146725 | Low-quality |
| S12_extracted_k141_122444 | Not-determined |
| S12_extracted_k141_20563 | Not-determined |
| S12_extracted_k141_139033 | Not-determined |
| S12_extracted_k141_154225 | Not-determined |
| S12_extracted_k141_122445 | Not-determined |
| S12_extracted_k141_139035 | Not-determined |
| S12_extracted_k141_11950 | Not-determined |
| S12_extracted_k141_122447 | Not-determined |
| S12_extracted_k141_146727 | Not-determined |
| S12_extracted_k141_83530 | Not-determined |
| S12_extracted_k141_5285 | Not-determined |
| S12_extracted_k141_92116 | Not-determined |
| S12_extracted_k141_67882 | Not-determined |
| S12_extracted_k141_170073 | Low-quality |
| S12_extracted_k141_130533 | Low-quality |
| S12_extracted_k141_154239 | Not-determined |
| S12_extracted_k141_92124 | Not-determined |
| S12_extracted_k141_60117 | Not-determined |
| S12_extracted_k141_83537 | Not-determined |
| S12_extracted_k141_83538 | Not-determined |
| S12_extracted_k141_162372 | Not-determined |
| S12_extracted_k141_83541 | Not-determined |
| S12_extracted_k141_114897 | Not-determined |
| S12_extracted_k141_154242 | Not-determined |
| S12_extracted_k141_83543 | Not-determined |
| S12_extracted_k141_75516 | Not-determined |
| S12_extracted_k141_92131 | Not-determined |
| S12_extracted_k141_114900 | Not-determined |
| S12_extracted_k141_5300 | Not-determined |
| S12_extracted_k141_139045 | Not-determined |
| S12_extracted_k141_130540 | Not-determined |
| S12_extracted_k141_67903 | Not-determined |
| S12_extracted_k141_130542 | Not-determined |
| S12_extracted_k141_139048 | Not-determined |
| S12_extracted_k141_162378 | Not-determined |
| S12_extracted_k141_67906 | Low-quality |
| S12_extracted_k141_130544 | Not-determined |
| S12_extracted_k141_75527 | Not-determined |
| S12_extracted_k141_20588 | Not-determined |
| S12_extracted_k141_83551 | Not-determined |
| S12_extracted_k141_154251 | Not-determined |
| S12_extracted_k141_154252 | Not-determined |
| S12_extracted_k141_60134 | Not-determined |
| S12_extracted_k141_20592 | Not-determined |
| S12_extracted_k141_130550 | Not-determined |
| S12_extracted_k141_162391 | Not-determined |
| S12_extracted_k141_107797 | Not-determined |
| S12_extracted_k141_162392 | Not-determined |
| S12_extracted_k141_177353 | Not-determined |
| S12_extracted_k141_51838 | Not-determined |
| S12_extracted_k141_5314 | Not-determined |
| S12_extracted_k141_51841 | Not-determined |
| S12_extracted_k141_177360 | Not-determined |
| S12_extracted_k141_51843 | Not-determined |
| S12_extracted_k141_5317 | Not-determined |
| S12_extracted_k141_162400 | Low-quality |
| S12_extracted_k141_139064 | Not-determined |
| S12_extracted_k141_75537 | Not-determined |
| S12_extracted_k141_162405 | Not-determined |
| S12_extracted_k141_51849 | Not-determined |
| S12_extracted_k141_5322 | Not-determined |
| S12_extracted_k141_60137 | Not-determined |
| S12_extracted_k141_139068 | Not-determined |
| S12_extracted_k141_130565 | Not-determined |
| S12_extracted_k141_177369 | Not-determined |
| S12_extracted_k141_75539 | Not-determined |
| S12_extracted_k141_146739 | Not-determined |
| S12_extracted_k141_122476 | Not-determined |
| S12_extracted_k141_177374 | Low-quality |
| S12_extracted_k141_139075 | Not-determined |
| S12_extracted_k141_5329 | Not-determined |
| S12_extracted_k141_114913 | Not-determined |
| S12_extracted_k141_139077 | Not-determined |
| S12_extracted_k141_130576 | Not-determined |
| S12_extracted_k141_146742 | Not-determined |
| S12_extracted_k141_162415 | Not-determined |
| S12_extracted_k141_139079 | Not-determined |
| S12_extracted_k141_130578 | Not-determined |
| S12_extracted_k141_75544 | Not-determined |
| S12_extracted_k141_162417 | Not-determined |
| S12_extracted_k141_75545 | Not-determined |
| S12_extracted_k141_177376 | Not-determined |
| S12_extracted_k141_139084 | Not-determined |
| S12_extracted_k141_122487 | Not-determined |
| S12_extracted_k141_146748 | Low-quality |
| S12_extracted_k141_35948 | Not-determined |
| S12_extracted_k141_75551 | Not-determined |
| S12_extracted_k141_35951 | Not-determined |
| S12_extracted_k141_5341 | Not-determined |
| S12_extracted_k141_146751 | Not-determined |
| S12_extracted_k141_100197 | Not-determined |
| S12_extracted_k141_92165 | Not-determined |
| S12_extracted_k141_177389 | Not-determined |
| S12_extracted_k141_35959 | Not-determined |
| S12_extracted_k141_60166 | Not-determined |
| S12_extracted_k141_75559 | Not-determined |
| S12_extracted_k141_60168 | Not-determined |
| S12_extracted_k141_122489 | Not-determined |
| S12_extracted_k141_122490 | Not-determined |
| S12_extracted_k141_162435 | Not-determined |
| S12_extracted_k141_170086 | Not-determined |
| S12_extracted_k141_75565 | Not-determined |
| S12_extracted_k141_100212 | Not-determined |
| S12_extracted_k141_139094 | Not-determined |
| S12_extracted_k141_100215 | Not-determined |
| S12_extracted_k141_162439 | Low-quality |
| S12_extracted_k141_162442 | Not-determined |
| S12_extracted_k141_11967 | Not-determined |
| S12_extracted_k141_11968 | Low-quality |
| S12_extracted_k141_177403 | Not-determined |
| S12_extracted_k141_177405 | Not-determined |
| S12_extracted_k141_177413 | Not-determined |
| S12_extracted_k141_122498 | Low-quality |
| S12_extracted_k141_170088 | Not-determined |
| S12_extracted_k141_130594 | Not-determined |
| S12_extracted_k141_130596 | Not-determined |
| S12_extracted_k141_170090 | Not-determined |
| S12_extracted_k141_146776 | Not-determined |
| S12_extracted_k141_122503 | Not-determined |
| S12_extracted_k141_162455 | Not-determined |
| S12_extracted_k141_11980 | Not-determined |
| S12_extracted_k141_20599 | Not-determined |
| S12_extracted_k141_139101 | Not-determined |
| S12_extracted_k141_139102 | Not-determined |
| S12_extracted_k141_162461 | Not-determined |
| S12_extracted_k141_177422 | Not-determined |
| S12_extracted_k141_20604 | Not-determined |
| S12_extracted_k141_100226 | Not-determined |
| S12_extracted_k141_139104 | Not-determined |
| S12_extracted_k141_100228 | Not-determined |
| S12_extracted_k141_60184 | Not-determined |
| S12_extracted_k141_100229 | Not-determined |
| S12_extracted_k141_146777 | Low-quality |
| S12_extracted_k141_122515 | Not-determined |
| S12_extracted_k141_20611 | Not-determined |
| S12_extracted_k141_107802 | Not-determined |
| S12_extracted_k141_60188 | Not-determined |
| S12_extracted_k141_75574 | Not-determined |
| S12_extracted_k141_60193 | Not-determined |
| S12_extracted_k141_139110 | Not-determined |
| S12_extracted_k141_177437 | Not-determined |
| S12_extracted_k141_60194 | Not-determined |
| S12_extracted_k141_75578 | Not-determined |
| S12_extracted_k141_130613 | Not-determined |
| S12_extracted_k141_162476 | Not-determined |
| S12_extracted_k141_177440 | Not-determined |
| S12_extracted_k141_130615 | Low-quality |
| S12_extracted_k141_107806 | Not-determined |
| S12_extracted_k141_162478 | Not-determined |
| S12_extracted_k141_100241 | Not-determined |
| S12_extracted_k141_11990 | Not-determined |
| S12_extracted_k141_5387 | Low-quality |
| S12_extracted_k141_100245 | Not-determined |
| S12_extracted_k141_75583 | Low-quality |
| S12_extracted_k141_162482 | Not-determined |
| S12_extracted_k141_170102 | Not-determined |
| S12_extracted_k141_154302 | Low-quality |
| S12_extracted_k141_154303 | Not-determined |
| S12_extracted_k141_170103 | Not-determined |
| S12_extracted_k141_170104 | Not-determined |
| S12_extracted_k141_5399 | Not-determined |
| S12_extracted_k141_170105 | Not-determined |
| S12_extracted_k141_154307 | Not-determined |
| S12_extracted_k141_162484 | Not-determined |
| S12_extracted_k141_170107 | Not-determined |
| S12_extracted_k141_20631 | Not-determined |
| S12_extracted_k141_51858 | Not-determined |
| S12_extracted_k141_75595 | Not-determined |
| S12_extracted_k141_35985 | Not-determined |
| S12_extracted_k141_146805 | Not-determined |
| S12_extracted_k141_12005 | Not-determined |
| S12_extracted_k141_139130 | Not-determined |
| S12_extracted_k141_146807 | Not-determined |
| S12_extracted_k141_12011 | Not-determined |
| S12_extracted_k141_162493 | Low-quality |
| S12_extracted_k141_51871 | Not-determined |
| S12_extracted_k141_146811 | Not-determined |
| S12_extracted_k141_162496 | Low-quality |
| S12_extracted_k141_122530 | Low-quality |
| S12_extracted_k141_107829 | Low-quality |
| S12_extracted_k141_5411 | Not-determined |
| S12_extracted_k141_75609 | Not-determined |
| S12_extracted_k141_5415 | Not-determined |
| S12_extracted_k141_162503 | Not-determined |
| S12_extracted_k141_122539 | Not-determined |
| S12_extracted_k141_146818 | Not-determined |
| S12_extracted_k141_75615 | Not-determined |
| S12_extracted_k141_20644 | Not-determined |
| S12_extracted_k141_146821 | Not-determined |
| S12_extracted_k141_177480 | Not-determined |
| S12_extracted_k141_162511 | Not-determined |
| S12_extracted_k141_5424 | Not-determined |
| S12_extracted_k141_100273 | Not-determined |
| S12_extracted_k141_122547 | Not-determined |
| S12_extracted_k141_122549 | Not-determined |
| S12_extracted_k141_162516 | Not-determined |
| S12_extracted_k141_186526 | Not-determined |
| S12_extracted_k141_186527 | Not-determined |
| S12_extracted_k141_122556 | Not-determined |
| S12_extracted_k141_122559 | Not-determined |
| S12_extracted_k141_5433 | Low-quality |
| S12_extracted_k141_83561 | Not-determined |
| S12_extracted_k141_114918 | Low-quality |
| S12_extracted_k141_100277 | Low-quality |
| S12_extracted_k141_122563 | Not-determined |
| S12_extracted_k141_75630 | Not-determined |
| S12_extracted_k141_114922 | Not-determined |
| S12_extracted_k141_75632 | Not-determined |
| S12_extracted_k141_122568 | Not-determined |
| S12_extracted_k141_177489 | Not-determined |
| S12_extracted_k141_114924 | Not-determined |
| S12_extracted_k141_12025 | Not-determined |
| S12_extracted_k141_170135 | Not-determined |
| S12_extracted_k141_170137 | Not-determined |
| S12_extracted_k141_186542 | Not-determined |
| S12_extracted_k141_186543 | Not-determined |
| S12_extracted_k141_20653 | Not-determined |
| S12_extracted_k141_83567 | Not-determined |
| S12_extracted_k141_83569 | Not-determined |
| S12_extracted_k141_51901 | Not-determined |
| S12_extracted_k141_139134 | Not-determined |
| S12_extracted_k141_20658 | Not-determined |
| S12_extracted_k141_139135 | Not-determined |
| S12_extracted_k141_177497 | Not-determined |
| S12_extracted_k141_170149 | Not-determined |
| S12_extracted_k141_154320 | Not-determined |
| S12_extracted_k141_20662 | Not-determined |
| S12_extracted_k141_170154 | Not-determined |
| S12_extracted_k141_12037 | Not-determined |
| S12_extracted_k141_177503 | Not-determined |
| S12_extracted_k141_146842 | Not-determined |
| S12_extracted_k141_170163 | Not-determined |
| S12_extracted_k141_12043 | Not-determined |
| S12_extracted_k141_51905 | Not-determined |
| S12_extracted_k141_100294 | Not-determined |
| S12_extracted_k141_36019 | Not-determined |
| S12_extracted_k141_139145 | Not-determined |
| S12_extracted_k141_51907 | Not-determined |
| S12_extracted_k141_51908 | Low-quality |
| S12_extracted_k141_100297 | Not-determined |
| S12_extracted_k141_83572 | Not-determined |
| S12_extracted_k141_36024 | Not-determined |
| S12_extracted_k141_83575 | Not-determined |
| S12_extracted_k141_75649 | Not-determined |
| S12_extracted_k141_170171 | Not-determined |
| S12_extracted_k141_5449 | Not-determined |
| S12_extracted_k141_51915 | Not-determined |
| S12_extracted_k141_100301 | Not-determined |
| S12_extracted_k141_177518 | Not-determined |
| S12_extracted_k141_114933 | Not-determined |
| S12_extracted_k141_75652 | Not-determined |
| S12_extracted_k141_170175 | Not-determined |
| S12_extracted_k141_139157 | Low-quality |
| S12_extracted_k141_75654 | Not-determined |
| S12_extracted_k141_100312 | Not-determined |
| S12_extracted_k141_114938 | Not-determined |
| S12_extracted_k141_162541 | Not-determined |
| S12_extracted_k141_122604 | Not-determined |
| S12_extracted_k141_122605 | Not-determined |
| S12_extracted_k141_45082 | Not-determined |
| S12_extracted_k141_36035 | Low-quality |
| S12_extracted_k141_122608 | Not-determined |
| S12_extracted_k141_139172 | Not-determined |
| S12_extracted_k141_122609 | Not-determined |
| S12_extracted_k141_122610 | Not-determined |
| S12_extracted_k141_186582 | Not-determined |
| S12_extracted_k141_45084 | Not-determined |
| S12_extracted_k141_83589 | Low-quality |
| S12_extracted_k141_100320 | Not-determined |
| S12_extracted_k141_100322 | Low-quality |
| S12_extracted_k141_36038 | Not-determined |
| S12_extracted_k141_36039 | Not-determined |
| S12_extracted_k141_122617 | Not-determined |
| S12_extracted_k141_114950 | Not-determined |
| S12_extracted_k141_170188 | Low-quality |
| S12_extracted_k141_100328 | Not-determined |
| S12_extracted_k141_5479 | Not-determined |
| S12_extracted_k141_114951 | Low-quality |
| S12_extracted_k141_177536 | Not-determined |
| S16_extracted_173109 | Complete |
| S12_extracted_k141_12051 | Not-determined |
| S12_extracted_k141_12052 | Low-quality |
| S12_extracted_k141_20701 | Not-determined |
| S12_extracted_k141_5484 | Not-determined |
| S12_extracted_k141_154342 | Low-quality |
| S12_extracted_k141_45092 | Not-determined |
| S12_extracted_k141_83601 | Not-determined |
| S12_extracted_k141_170201 | Low-quality |
| S12_extracted_k141_20705 | Not-determined |
| S12_extracted_k141_122626 | Low-quality |
| S16_extracted_36146 | Medium-quality |
| S12_extracted_k141_83608 | Not-determined |
| S12_extracted_k141_28686 | Not-determined |
| S12_extracted_k141_28687 | Not-determined |
| S12_extracted_k141_170205 | Not-determined |
| S12_extracted_k141_67918 | Not-determined |
| S12_extracted_k141_28688 | Not-determined |
| S12_extracted_k141_28689 | Not-determined |
| S12_extracted_k141_92200 | Not-determined |
| S12_extracted_k141_67920 | Not-determined |
| S12_extracted_k141_100341 | Not-determined |
| S12_extracted_k141_83614 | Not-determined |
| S12_extracted_k141_186593 | Not-determined |
| S12_extracted_k141_186594 | Not-determined |
| S12_extracted_k141_146877 | Not-determined |
| S12_extracted_k141_139199 | Not-determined |
| S12_extracted_k141_146883 | Not-determined |
| S12_extracted_k141_51938 | Not-determined |
| S12_extracted_k141_139205 | Not-determined |
| S12_extracted_k141_114962 | Not-determined |
| S12_extracted_k141_5504 | Not-determined |
| S12_extracted_k141_51943 | Not-determined |
| S12_extracted_k141_36052 | Low-quality |
| S12_extracted_k141_20716 | Not-determined |
| S12_extracted_k141_100362 | Not-determined |
| S12_extracted_k141_20721 | Not-determined |
| S12_extracted_k141_146892 | Not-determined |
| S12_extracted_k141_20722 | Not-determined |
| S12_extracted_k141_139212 | Not-determined |
| S12_extracted_k141_45118 | Not-determined |
| S12_extracted_k141_12081 | Not-determined |
| S12_extracted_k141_67938 | Low-quality |
| S12_extracted_k141_20723 | Not-determined |
| S12_extracted_k141_100364 | Not-determined |
| S12_extracted_k141_20724 | Not-determined |
| S12_extracted_k141_100365 | Not-determined |
| S12_extracted_k141_83628 | Not-determined |
| S12_extracted_k141_146898 | Low-quality |
| S12_extracted_k141_92206 | Not-determined |
| S12_extracted_k141_67947 | Not-determined |
| S12_extracted_k141_20726 | Not-determined |
| S12_extracted_k141_28707 | Low-quality |
| S12_extracted_k141_5517 | Not-determined |
| S12_extracted_k141_177542 | Not-determined |
| S12_extracted_k141_170228 | Not-determined |
| S12_extracted_k141_170234 | Not-determined |
| S12_extracted_k141_186613 | Not-determined |
| S12_extracted_k141_28710 | Low-quality |
| S12_extracted_k141_51960 | Low-quality |
| S12_extracted_k141_177557 | Low-quality |
| S12_extracted_k141_75665 | Not-determined |
| S12_extracted_k141_170241 | Not-determined |
| S12_extracted_k141_45132 | Not-determined |
| S12_extracted_k141_177559 | Not-determined |
| S12_extracted_k141_75669 | Not-determined |
| S12_extracted_k141_100378 | Not-determined |
| S12_extracted_k141_75670 | Not-determined |
| S12_extracted_k141_170245 | Not-determined |
| S12_extracted_k141_92228 | Not-determined |
| S12_extracted_k141_12099 | Not-determined |
| S12_extracted_k141_83648 | Not-determined |
| S12_extracted_k141_83649 | Not-determined |
| S12_extracted_k141_67957 | Not-determined |
| S12_extracted_k141_114985 | Not-determined |
| S12_extracted_k141_20732 | Low-quality |
| S12_extracted_k141_154364 | Low-quality |
| S12_extracted_k141_28720 | Not-determined |
| S12_extracted_k141_5521 | Not-determined |
| S12_extracted_k141_154365 | Not-determined |
| S12_extracted_k141_186622 | Not-determined |
| S12_extracted_k141_186624 | Not-determined |
| S12_extracted_k141_36087 | Not-determined |
| S12_extracted_k141_177572 | Not-determined |
| S12_extracted_k141_75692 | Not-determined |
| S12_extracted_k141_186630 | Not-determined |
| S12_extracted_k141_154371 | Low-quality |
| S12_extracted_k141_5527 | Not-determined |
| S12_extracted_k141_122642 | Not-determined |
| S12_extracted_k141_92235 | Not-determined |
| S12_extracted_k141_45143 | Not-determined |
| S12_extracted_k141_100392 | Not-determined |
| S12_extracted_k141_130641 | Not-determined |
| S12_extracted_k141_75695 | Not-determined |
| S12_extracted_k141_139226 | Not-determined |
| S12_extracted_k141_186636 | Not-determined |
| S12_extracted_k141_100397 | Not-determined |
| S12_extracted_k141_67972 | Not-determined |
| S12_extracted_k141_130651 | Not-determined |
| S12_extracted_k141_92239 | Not-determined |
| S12_extracted_k141_130653 | Not-determined |
| S12_extracted_k141_154380 | Not-determined |
| S12_extracted_k141_130654 | Not-determined |
| S12_extracted_k141_45154 | Not-determined |
| S12_extracted_k141_139239 | Not-determined |
| S12_extracted_k141_92249 | Not-determined |
| S12_extracted_k141_154391 | Low-quality |
| S12_extracted_k141_186648 | Not-determined |
| S12_extracted_k141_154392 | Not-determined |
| S12_extracted_k141_100402 | Low-quality |
| S12_extracted_k141_170270 | Low-quality |
| S12_extracted_k141_45160 | Not-determined |
| S12_extracted_k141_51996 | Not-determined |
| S12_extracted_k141_45161 | Not-determined |
| S12_extracted_k141_154397 | Not-determined |
| S12_extracted_k141_100405 | Not-determined |
| S12_extracted_k141_139249 | Not-determined |
| S12_extracted_k141_45163 | Not-determined |
| S12_extracted_k141_5547 | Not-determined |
| S12_extracted_k141_52003 | Not-determined |
| S12_extracted_k141_45168 | Low-quality |
| S12_extracted_k141_75706 | Not-determined |
| S12_extracted_k141_75707 | Not-determined |
| S12_extracted_k141_154404 | Not-determined |
| S12_extracted_k141_114995 | Not-determined |
| S12_extracted_k141_45170 | Not-determined |
| S12_extracted_k141_186655 | Not-determined |
| S12_extracted_k141_36098 | Not-determined |
| S12_extracted_k141_139260 | Not-determined |
| S12_extracted_k141_186657 | Not-determined |
| S12_extracted_k141_130675 | Low-quality |
| S12_extracted_k141_154409 | Not-determined |
| S12_extracted_k141_186663 | Not-determined |
| S12_extracted_k141_115007 | Not-determined |
| S12_extracted_k141_45174 | Not-determined |
| S12_extracted_k141_45175 | Not-determined |
| S12_extracted_k141_154414 | Not-determined |
| S12_extracted_k141_12113 | Not-determined |
| S12_extracted_k141_177592 | Not-determined |
| S12_extracted_k141_5565 | Not-determined |
| S12_extracted_k141_75723 | Low-quality |
| S12_extracted_k141_67999 | Not-determined |
| S12_extracted_k141_170289 | Not-determined |
| S12_extracted_k141_107845 | Not-determined |
| S12_extracted_k141_5567 | Not-determined |
| S12_extracted_k141_130682 | Not-determined |
| S12_extracted_k141_115013 | Not-determined |
| S12_extracted_k141_52024 | Not-determined |
| S12_extracted_k141_52025 | Not-determined |
| S12_extracted_k141_12117 | Not-determined |
| S12_extracted_k141_146931 | Not-determined |
| S12_extracted_k141_154423 | Not-determined |
| S12_extracted_k141_154424 | Not-determined |
| S12_extracted_k141_52026 | Not-determined |
| S12_extracted_k141_107855 | Not-determined |
| S12_extracted_k141_75732 | Not-determined |
| S12_extracted_k141_170295 | Low-quality |
| S12_extracted_k141_100433 | Not-determined |
| S12_extracted_k141_115023 | Not-determined |
| S12_extracted_k141_36112 | Not-determined |
| S12_extracted_k141_177616 | Not-determined |
| S12_extracted_k141_107865 | Not-determined |
| S12_extracted_k141_52031 | Not-determined |
| S12_extracted_k141_139265 | Not-determined |
| S12_extracted_k141_68014 | Not-determined |
| S12_extracted_k141_36114 | Not-determined |
| S12_extracted_k141_28784 | Not-determined |
| S12_extracted_k141_52035 | Not-determined |
| S12_extracted_k141_154449 | Not-determined |
| S12_extracted_k141_75739 | Low-quality |
| S12_extracted_k141_154450 | Not-determined |
| S12_extracted_k141_146933 | Not-determined |
| S12_extracted_k141_139269 | Not-determined |
| S12_extracted_k141_186674 | Not-determined |
| S12_extracted_k141_52040 | Not-determined |
| S12_extracted_k141_28788 | Not-determined |
| S12_extracted_k141_186682 | Not-determined |
| S12_extracted_k141_107877 | Not-determined |
| S12_extracted_k141_154458 | Not-determined |
| S12_extracted_k141_130708 | Not-determined |
| S12_extracted_k141_130710 | Low-quality |
| S12_extracted_k141_177636 | Not-determined |
| S12_extracted_k141_115034 | Not-determined |
| S12_extracted_k141_170308 | Not-determined |
| S12_extracted_k141_130712 | Low-quality |
| S12_extracted_k141_139281 | Low-quality |
| S12_extracted_k141_45201 | Low-quality |
| S12_extracted_k141_146946 | Low-quality |
| S12_extracted_k141_75754 | Not-determined |
| S12_extracted_k141_28798 | Not-determined |
| S12_extracted_k141_154468 | Not-determined |
| S12_extracted_k141_115044 | Not-determined |
| S12_extracted_k141_139286 | Not-determined |
| S12_extracted_k141_130719 | Not-determined |
| S12_extracted_k141_75757 | Not-determined |
| S12_extracted_k141_60228 | Not-determined |
| S12_extracted_k141_107884 | Not-determined |
| S12_extracted_k141_52065 | Not-determined |
| S12_extracted_k141_186691 | Not-determined |
| S12_extracted_k141_75765 | Not-determined |
| S12_extracted_k141_45207 | Not-determined |
| S12_extracted_k141_186692 | Not-determined |
| S12_extracted_k141_68030 | Low-quality |
| S12_extracted_k141_100445 | Not-determined |
| S12_extracted_k141_45208 | Not-determined |
| S12_extracted_k141_12143 | Not-determined |
| S12_extracted_k141_45210 | Not-determined |
| S12_extracted_k141_60239 | Not-determined |
| S12_extracted_k141_12148 | Low-quality |
| S12_extracted_k141_170321 | Not-determined |
| S12_extracted_k141_68034 | Not-determined |
| S12_extracted_k141_177649 | Not-determined |
| S12_extracted_k141_28800 | Low-quality |
| S12_extracted_k141_170322 | Not-determined |
| S12_extracted_k141_100451 | Not-determined |
| S12_extracted_k141_107891 | Not-determined |
| S12_extracted_k141_52074 | Not-determined |
| S12_extracted_k141_186699 | Low-quality |
| S12_extracted_k141_60240 | Not-determined |
| S12_extracted_k141_45217 | Not-determined |
| S12_extracted_k141_122659 | Low-quality |
| S12_extracted_k141_12157 | Not-determined |
| S12_extracted_k141_92277 | Not-determined |
| S12_extracted_k141_68041 | Not-determined |
| S12_extracted_k141_45223 | Not-determined |
| S12_extracted_k141_60246 | Not-determined |
| S12_extracted_k141_122665 | Not-determined |
| S12_extracted_k141_60247 | Not-determined |
| S12_extracted_k141_186711 | Not-determined |
| S12_extracted_k141_186714 | Not-determined |
| S12_extracted_k141_154497 | Low-quality |
| S12_extracted_k141_68052 | Not-determined |
| S12_extracted_k141_92286 | Not-determined |
| S12_extracted_k141_170334 | Not-determined |
| S12_extracted_k141_100462 | Not-determined |
| S12_extracted_k141_68056 | Low-quality |
| S12_extracted_k141_170338 | Not-determined |
| S12_extracted_k141_12168 | Not-determined |
| S12_extracted_k141_75790 | Not-determined |
| S12_extracted_k141_107902 | Not-determined |
| S12_extracted_k141_170346 | Not-determined |
| S12_extracted_k141_12174 | Low-quality |
| S12_extracted_k141_177665 | Not-determined |
| S12_extracted_k141_130739 | Not-determined |
| S12_extracted_k141_20737 | Low-quality |
| S12_extracted_k141_100464 | Low-quality |
| S12_extracted_k141_5600 | Not-determined |
| S12_extracted_k141_75801 | Not-determined |
| S12_extracted_k141_177669 | Low-quality |
| S12_extracted_k141_75802 | Not-determined |
| S12_extracted_k141_122679 | Not-determined |

| | |
|---|---|
| S12_extracted_k141_5603 | Not-determined |
| S12_extracted_k141_28824 | Not-determined |
| S12_extracted_k141_107910 | Not-determined |
| S12_extracted_k141_186732 | Not-determined |
| S12_extracted_k141_170351 | Not-determined |
| S12_extracted_k141_68071 | Low-quality |
| S12_extracted_k141_107912 | Not-determined |
| S12_extracted_k141_130751 | Not-determined |
| S12_extracted_k141_75806 | Not-determined |
| S12_extracted_k141_28826 | Low-quality |
| S12_extracted_k141_5606 | Not-determined |
| S12_extracted_k141_130753 | Not-determined |
| S12_extracted_k141_60263 | Not-determined |
| S12_extracted_k141_186737 | Not-determined |
| S12_extracted_k141_107916 | Not-determined |
| S12_extracted_k141_45237 | Not-determined |
| S12_extracted_k141_139326 | Not-determined |
| S12_extracted_k141_107918 | Not-determined |
| S12_extracted_k141_177681 | Not-determined |
| S12_extracted_k141_45239 | Not-determined |
| S12_extracted_k141_162596 | Low-quality |
| S12_extracted_k141_68074 | Not-determined |
| S12_extracted_k141_130758 | Not-determined |
| S12_extracted_k141_75814 | Not-determined |
| S12_extracted_k141_5614 | Not-determined |
| S12_extracted_k141_68077 | Not-determined |
| S12_extracted_k141_177691 | Not-determined |
| S12_extracted_k141_122695 | Not-determined |
| S12_extracted_k141_122696 | Not-determined |
| S12_extracted_k141_122697 | Not-determined |
| S12_extracted_k141_5616 | Not-determined |
| S12_extracted_k141_52099 | Not-determined |
| S12_extracted_k141_139335 | Not-determined |
| S12_extracted_k141_60285 | Not-determined |
| S12_extracted_k141_154528 | Low-quality |
| S12_extracted_k141_170365 | Not-determined |
| S12_extracted_k141_45249 | Not-determined |
| S12_extracted_k141_52103 | Not-determined |
| S12_extracted_k141_177703 | Low-quality |
| S12_extracted_k141_28843 | Not-determined |
| S12_extracted_k141_107943 | Not-determined |
| S12_extracted_k141_75821 | Low-quality |
| S12_extracted_k141_52118 | Not-determined |
| S12_extracted_k141_52119 | Not-determined |
| S12_extracted_k141_28852 | Not-determined |
| S12_extracted_k141_130774 | Not-determined |
| S12_extracted_k141_45257 | Not-determined |
| S12_extracted_k141_60300 | Not-determined |
| S12_extracted_k141_92317 | Not-determined |
| S12_extracted_k141_92318 | Low-quality |
| S12_extracted_k141_162620 | Not-determined |
| S12_extracted_k141_68087 | Low-quality |
| S12_extracted_k141_115058 | Low-quality |
| S12_extracted_k141_36118 | Not-determined |
| S12_extracted_k141_130782 | Not-determined |
| S12_extracted_k141_75840 | Not-determined |
| S12_extracted_k141_60313 | Not-determined |
| S12_extracted_k141_186773 | Not-determined |
| S12_extracted_k141_75842 | Not-determined |
| S12_extracted_k141_5638 | Not-determined |
| S12_extracted_k141_107953 | Low-quality |
| S12_extracted_k141_92332 | Not-determined |
| S12_extracted_k141_115064 | Low-quality |
| S12_extracted_k141_92334 | Not-determined |
| S12_extracted_k141_12200 | Not-determined |
| S12_extracted_k141_52139 | Not-determined |
| S12_extracted_k141_75852 | Not-determined |
| S12_extracted_k141_45269 | Not-determined |
| S12_extracted_k141_186779 | Low-quality |
| S12_extracted_k141_5643 | Not-determined |
| S12_extracted_k141_52140 | Not-determined |
| S12_extracted_k141_115067 | Not-determined |
| S12_extracted_k141_130797 | Not-determined |
| S12_extracted_k141_107956 | Low-quality |
| S12_extracted_k141_115070 | Not-determined |
| S12_extracted_k141_107957 | Not-determined |
| S12_extracted_k141_154546 | Not-determined |
| S12_extracted_k141_154548 | Not-determined |
| S12_extracted_k141_68105 | Not-determined |
| S12_extracted_k141_60317 | Not-determined |
| S12_extracted_k141_68106 | Not-determined |
| S12_extracted_k141_130800 | Not-determined |
| S12_extracted_k141_36124 | Not-determined |
| S12_extracted_k141_52150 | Not-determined |
| S12_extracted_k141_92342 | Not-determined |
| S12_extracted_k141_107966 | Not-determined |
| S12_extracted_k141_12210 | Not-determined |
| S12_extracted_k141_100499 | Not-determined |
| S12_extracted_k141_60319 | Not-determined |
| S12_extracted_k141_170393 | Not-determined |
| S12_extracted_k141_122725 | Not-determined |
| S12_extracted_k141_52154 | Not-determined |
| S12_extracted_k141_107971 | Not-determined |
| S12_extracted_k141_130804 | Not-determined |
| S12_extracted_k141_130805 | Not-determined |
| S12_extracted_k141_12214 | Not-determined |
| S12_extracted_k141_92351 | Not-determined |
| S12_extracted_k141_147037 | Not-determined |
| S12_extracted_k141_115078 | Not-determined |
| S12_extracted_k141_130808 | Not-determined |
| S12_extracted_k141_52158 | Not-determined |
| S12_extracted_k141_52160 | Not-determined |
| S12_extracted_k141_177709 | Not-determined |
| S12_extracted_k141_92358 | Not-determined |
| S12_extracted_k141_92359 | Not-determined |
| S12_extracted_k141_45282 | Not-determined |
| S12_extracted_k141_5665 | Not-determined |
| S12_extracted_k141_45283 | Not-determined |
| S12_extracted_k141_130815 | Not-determined |
| S12_extracted_k141_45284 | Not-determined |
| S12_extracted_k141_52170 | Not-determined |
| S12_extracted_k141_68109 | Not-determined |
| S12_extracted_k141_122737 | Not-determined |
| S12_extracted_k141_154555 | Not-determined |
| S12_extracted_k141_36141 | Low-quality |
| S12_extracted_k141_107981 | Low-quality |
| S12_extracted_k141_147047 | Not-determined |
| S12_extracted_k141_92369 | Not-determined |
| S12_extracted_k141_122742 | Not-determined |
| S12_extracted_k141_52174 | Low-quality |
| S12_extracted_k141_12231 | Not-determined |
| S12_extracted_k141_107984 | Not-determined |
| S12_extracted_k141_12232 | Not-determined |
| S12_extracted_k141_107987 | Not-determined |
| S12_extracted_k141_5677 | Not-determined |
| S12_extracted_k141_107989 | Not-determined |
| S12_extracted_k141_5678 | Not-determined |
| S12_extracted_k141_186811 | Not-determined |
| S12_extracted_k141_75885 | Not-determined |
| S12_extracted_k141_186813 | Not-determined |
| S12_extracted_k141_122752 | Not-determined |
| S12_extracted_k141_60334 | Not-determined |
| S12_extracted_k141_45297 | Not-determined |
| S12_extracted_k141_107993 | Not-determined |
| S12_extracted_k141_139367 | Not-determined |
| S12_extracted_k141_186819 | Not-determined |
| S12_extracted_k141_186820 | Not-determined |
| S12_extracted_k141_52188 | Not-determined |
| S12_extracted_k141_92377 | Not-determined |
| S12_extracted_k141_139370 | Not-determined |
| S12_extracted_k141_68115 | Not-determined |
| S12_extracted_k141_28887 | Not-determined |
| S12_extracted_k141_177717 | Not-determined |
| S12_extracted_k141_12248 | Not-determined |
| S12_extracted_k141_75895 | Low-quality |
| S12_extracted_k141_5693 | Not-determined |
| S12_extracted_k141_36171 | Not-determined |
| S12_extracted_k141_154578 | Not-determined |
| S12_extracted_k141_122764 | Not-determined |
| S12_extracted_k141_108006 | Not-determined |
| S12_extracted_k141_186832 | Not-determined |
| S12_extracted_k141_154584 | Not-determined |
| S12_extracted_k141_45313 | Not-determined |
| S12_extracted_k141_92388 | Not-determined |
| S12_extracted_k141_20769 | Not-determined |
| S12_extracted_k141_75910 | Not-determined |
| S12_extracted_k141_177727 | Not-determined |
| S12_extracted_k141_75911 | Not-determined |
| S12_extracted_k141_45318 | Not-determined |
| S12_extracted_k141_139387 | Not-determined |
| S12_extracted_k141_154593 | Not-determined |
| S12_extracted_k141_162680 | Not-determined |
| S12_extracted_k141_52192 | Low-quality |
| S12_extracted_k141_154596 | Not-determined |
| S12_extracted_k141_122771 | Not-determined |
| S12_extracted_k141_60356 | Low-quality |
| S12_extracted_k141_12261 | Not-determined |
| S12_extracted_k141_75919 | Not-determined |
| S12_extracted_k141_139391 | Not-determined |
| S12_extracted_k141_60359 | Not-determined |
| S12_extracted_k141_75924 | Not-determined |
| S12_extracted_k141_45328 | Not-determined |
| S12_extracted_k141_108015 | Not-determined |
| S12_extracted_k141_60362 | Not-determined |
| S12_extracted_k141_92412 | Not-determined |
| S12_extracted_k141_162689 | Not-determined |
| S12_extracted_k141_108018 | Not-determined |
| S12_extracted_k141_12270 | Not-determined |
| S12_extracted_k141_108022 | Low-quality |
| S12_extracted_k141_122781 | Not-determined |
| S12_extracted_k141_75932 | Not-determined |
| S12_extracted_k141_12275 | Not-determined |
| S12_extracted_k141_92422 | Low-quality |
| S12_extracted_k141_75934 | Not-determined |
| S12_extracted_k141_154615 | Not-determined |
| S12_extracted_k141_20779 | Not-determined |
| S12_extracted_k141_122785 | Not-determined |
| S12_extracted_k141_92425 | Not-determined |
| S12_extracted_k141_122786 | Not-determined |
| S12_extracted_k141_60377 | Not-determined |
| S12_extracted_k141_5724 | Not-determined |
| S12_extracted_k141_170419 | Low-quality |
| S12_extracted_k141_68150 | Low-quality |
| S12_extracted_k141_20785 | Not-determined |
| S12_extracted_k141_170421 | Not-determined |
| S12_extracted_k141_5728 | Not-determined |
| S12_extracted_k141_170424 | Not-determined |
| S12_extracted_k141_154621 | Not-determined |
| S12_extracted_k141_5730 | Not-determined |
| S12_extracted_k141_52215 | Not-determined |
| S12_extracted_k141_12277 | Not-determined |
| S12_extracted_k141_20786 | Not-determined |
| S12_extracted_k141_177745 | Not-determined |
| S12_extracted_k141_45353 | Not-determined |
| S12_extracted_k141_108037 | Not-determined |
| S12_extracted_k141_162696 | Not-determined |
| S12_extracted_k141_170427 | Not-determined |
| S12_extracted_k141_177751 | Not-determined |
| S12_extracted_k141_83659 | Not-determined |
| S12_extracted_k141_177752 | Not-determined |
| S12_extracted_k141_60385 | Not-determined |
| S12_extracted_k141_68153 | Not-determined |
| S12_extracted_k141_122800 | Not-determined |
| S12_extracted_k141_177755 | Not-determined |
| S12_extracted_k141_60386 | Not-determined |
| S12_extracted_k141_122805 | Not-determined |
| S12_extracted_k141_92441 | Not-determined |
| S12_extracted_k141_108046 | Not-determined |
| S12_extracted_k141_92442 | Not-determined |
| S12_extracted_k141_60387 | Not-determined |
| S12_extracted_k141_60388 | Not-determined |
| S12_extracted_k141_12292 | Not-determined |
| S12_extracted_k141_92448 | Not-determined |
| S12_extracted_k141_12294 | Not-determined |
| S12_extracted_k141_177760 | Low-quality |
| S12_extracted_k141_92451 | Not-determined |
| S12_extracted_k141_154629 | Not-determined |
| S12_extracted_k141_154631 | Low-quality |
| S12_extracted_k141_92453 | Not-determined |
| S12_extracted_k141_12301 | Not-determined |
| S12_extracted_k141_92455 | Not-determined |
| S12_extracted_k141_68172 | Not-determined |
| S12_extracted_k141_45367 | Not-determined |
| S12_extracted_k141_147053 | Not-determined |
| S12_extracted_k141_45371 | Not-determined |
| S12_extracted_k141_147055 | Not-determined |
| S12_extracted_k141_115094 | Not-determined |
| S12_extracted_k141_12306 | Not-determined |
| S12_extracted_k141_115095 | Not-determined |
| S12_extracted_k141_20806 | Low-quality |
| S12_extracted_k141_12307 | Not-determined |
| S12_extracted_k141_45375 | Not-determined |
| S12_extracted_k141_20808 | Not-determined |
| S12_extracted_k141_45377 | Not-determined |
| S12_extracted_k141_177776 | Not-determined |
| S12_extracted_k141_45378 | Not-determined |
| S12_extracted_k141_115098 | Not-determined |
| S12_extracted_k141_115099 | Not-determined |
| S12_extracted_k141_45382 | Not-determined |
| S12_extracted_k141_68185 | Not-determined |
| S12_extracted_k141_68186 | Not-determined |
| S12_extracted_k141_177785 | Not-determined |
| S12_extracted_k141_177787 | Not-determined |
| S12_extracted_k141_108063 | Not-determined |
| S12_extracted_k141_52249 | Not-determined |
| S12_extracted_k141_108067 | Not-determined |
| S12_extracted_k141_68191 | Not-determined |
| S12_extracted_k141_83694 | Not-determined |
| S12_extracted_k141_68194 | Not-determined |
| S12_extracted_k141_177794 | Not-determined |
| S12_extracted_k141_45387 | Not-determined |
| S12_extracted_k141_83695 | Not-determined |
| S12_extracted_k141_52257 | Not-determined |
| S12_extracted_k141_108070 | Not-determined |
| S12_extracted_k141_147070 | Not-determined |
| S12_extracted_k141_75979 | Low-quality |
| S12_extracted_k141_92465 | Not-determined |
| S12_extracted_k141_108076 | Not-determined |
| S12_extracted_k141_75985 | Not-determined |
| S12_extracted_k141_75986 | Not-determined |
| S12_extracted_k141_36213 | Not-determined |
| S12_extracted_k141_36214 | Not-determined |
| S12_extracted_k141_139416 | Not-determined |
| S12_extracted_k141_75989 | Not-determined |
| S12_extracted_k141_170432 | Not-determined |
| S12_extracted_k141_60412 | Not-determined |
| S12_extracted_k141_147079 | Not-determined |
| S12_extracted_k141_20828 | Not-determined |
| S12_extracted_k141_170439 | Low-quality |
| S12_extracted_k141_36217 | Not-determined |
| S12_extracted_k141_20831 | Not-determined |
| S12_extracted_k141_186906 | Low-quality |
| S12_extracted_k141_68200 | Not-determined |
| S12_extracted_k141_147091 | Not-determined |
| S12_extracted_k141_115120 | Not-determined |
| S12_extracted_k141_52269 | Not-determined |
| S12_extracted_k141_154639 | Not-determined |
| S12_extracted_k141_68204 | Not-determined |
| S12_extracted_k141_170444 | Not-determined |
| S12_extracted_k141_147094 | Not-determined |
| S12_extracted_k141_60416 | Not-determined |
| S12_extracted_k141_20843 | Not-determined |
| S12_extracted_k141_45405 | Not-determined |
| S12_extracted_k141_45406 | Not-determined |
| S12_extracted_k141_115125 | Not-determined |
| S12_extracted_k141_83704 | Not-determined |
| S12_extracted_k141_92481 | Not-determined |
| S12_extracted_k141_108085 | Not-determined |
| S12_extracted_k141_60420 | Not-determined |
| S12_extracted_k141_154645 | Not-determined |
| S12_extracted_k141_108088 | Not-determined |
| S12_extracted_k141_36238 | Not-determined |
| S12_extracted_k141_68215 | Not-determined |
| S12_extracted_k141_170451 | Not-determined |
| S12_extracted_k141_76005 | Not-determined |
| S12_extracted_k141_36239 | Not-determined |
| S12_extracted_k141_28921 | Low-quality |
| S12_extracted_k141_115134 | Not-determined |
| S12_extracted_k141_12337 | Low-quality |
| S12_extracted_k141_76011 | Not-determined |
| S12_extracted_k141_12339 | Not-determined |
| S12_extracted_k141_60423 | Not-determined |
| S12_extracted_k141_170460 | Not-determined |
| S12_extracted_k141_108092 | Low-quality |
| S12_extracted_k141_115142 | Not-determined |
| S12_extracted_k141_12340 | Not-determined |
| S12_extracted_k141_60425 | Not-determined |
| S12_extracted_k141_130883 | Not-determined |
| S12_extracted_k141_45416 | Low-quality |
| S12_extracted_k141_139421 | Not-determined |
| S12_extracted_k141_76020 | Not-determined |
| S12_extracted_k141_130889 | Not-determined |
| S12_extracted_k141_170469 | Not-determined |
| S12_extracted_k141_83715 | Not-determined |
| S12_extracted_k141_52298 | Low-quality |
| S12_extracted_k141_12349 | Not-determined |
| S12_extracted_k141_177826 | Not-determined |
| S12_extracted_k141_139430 | Not-determined |
| S12_extracted_k141_130893 | Not-determined |
| S12_extracted_k141_115148 | Not-determined |
| S12_extracted_k141_122815 | Not-determined |
| S12_extracted_k141_115150 | Not-determined |
| S12_extracted_k141_115151 | Not-determined |
| S12_extracted_k141_68234 | Not-determined |
| S12_extracted_k141_154660 | Not-determined |
| S12_extracted_k141_130897 | Not-determined |
| S12_extracted_k141_60439 | Low-quality |
| S12_extracted_k141_147114 | Not-determined |
| S12_extracted_k141_154666 | Not-determined |
| S12_extracted_k141_76028 | Not-determined |
| S12_extracted_k141_147117 | Not-determined |
| S12_extracted_k141_147118 | Not-determined |
| S12_extracted_k141_108110 | Not-determined |
| S12_extracted_k141_36261 | Not-determined |
| S12_extracted_k141_108111 | Not-determined |
| S12_extracted_k141_177839 | Not-determined |
| S12_extracted_k141_92498 | Not-determined |
| S12_extracted_k141_115154 | Not-determined |
| S12_extracted_k141_76032 | Low-quality |
| S12_extracted_k141_115155 | Not-determined |
| S12_extracted_k141_76033 | Not-determined |
| S12_extracted_k141_130903 | Low-quality |
| S12_extracted_k141_130904 | Not-determined |
| S12_extracted_k141_68244 | Not-determined |
| S12_extracted_k141_130906 | Not-determined |
| S12_extracted_k141_130907 | Not-determined |
| S12_extracted_k141_68246 | Not-determined |
| S12_extracted_k141_20864 | Not-determined |
| S12_extracted_k141_52318 | Not-determined |
| S12_extracted_k141_130910 | Not-determined |
| S12_extracted_k141_52319 | Not-determined |
| S12_extracted_k141_177854 | Not-determined |
| S12_extracted_k141_76042 | Not-determined |
| S12_extracted_k141_76043 | Not-determined |
| S12_extracted_k141_76044 | Not-determined |
| S12_extracted_k141_83732 | Low-quality |
| S12_extracted_k141_12365 | Not-determined |
| S12_extracted_k141_177861 | Not-determined |
| S12_extracted_k141_92509 | Not-determined |
| S12_extracted_k141_170493 | Low-quality |
| S12_extracted_k141_130913 | Not-determined |
| S12_extracted_k141_100583 | Not-determined |
| S12_extracted_k141_45446 | Not-determined |
| S12_extracted_k141_170495 | Not-determined |
| S12_extracted_k141_139457 | Not-determined |
| S12_extracted_k141_20881 | Not-determined |
| S12_extracted_k141_28956 | Not-determined |
| S12_extracted_k141_186950 | Not-determined |
| S12_extracted_k141_12371 | Not-determined |
| S12_extracted_k141_52337 | Not-determined |
| S12_extracted_k141_45460 | Not-determined |
| S12_extracted_k141_108128 | Low-quality |
| S12_extracted_k141_12374 | Not-determined |
| S12_extracted_k141_108129 | Not-determined |
| S12_extracted_k141_20883 | Not-determined |
| S12_extracted_k141_154685 | Not-determined |
| S12_extracted_k141_92519 | Not-determined |
| S12_extracted_k141_170497 | Not-determined |
| S12_extracted_k141_154688 | Not-determined |
| S12_extracted_k141_115187 | Not-determined |
| S12_extracted_k141_154689 | Not-determined |
| S12_extracted_k141_60467 | Not-determined |
| S12_extracted_k141_177876 | Low-quality |
| S12_extracted_k141_139471 | Not-determined |
| S12_extracted_k141_52342 | Not-determined |
| S12_extracted_k141_177877 | Not-determined |
| S12_extracted_k141_100595 | Not-determined |
| S12_extracted_k141_52345 | Not-determined |
| S12_extracted_k141_147146 | Not-determined |
| S12_extracted_k141_45466 | Not-determined |
| S12_extracted_k141_186962 | Not-determined |
| S12_extracted_k141_139476 | Not-determined |
| S12_extracted_k141_60474 | Not-determined |
| S12_extracted_k141_108137 | Not-determined |
| S12_extracted_k141_20888 | Low-quality |
| S12_extracted_k141_186965 | Low-quality |
| S12_extracted_k141_28966 | Not-determined |
| S12_extracted_k141_154696 | Not-determined |
| S12_extracted_k141_147153 | Not-determined |
| S12_extracted_k141_60478 | Not-determined |
| S12_extracted_k141_154698 | Not-determined |
| S12_extracted_k141_186966 | Not-determined |
| S12_extracted_k141_12383 | Not-determined |
| S12_extracted_k141_154699 | Not-determined |
| S12_extracted_k141_170508 | Not-determined |
| S12_extracted_k141_130944 | Low-quality |
| S12_extracted_k141_139482 | Not-determined |
| S12_extracted_k141_60482 | Not-determined |
| S12_extracted_k141_68262 | Not-determined |
| S12_extracted_k141_12387 | Not-determined |
| S12_extracted_k141_100606 | Not-determined |
| S12_extracted_k141_68265 | Not-determined |
| S12_extracted_k141_28975 | Not-determined |
| S12_extracted_k141_28976 | Not-determined |
| S12_extracted_k141_28977 | Not-determined |
| S12_extracted_k141_20900 | Not-determined |
| S12_extracted_k141_177898 | Not-determined |
| S12_extracted_k141_170514 | Not-determined |
| S12_extracted_k141_20903 | Not-determined |
| S12_extracted_k141_130950 | Not-determined |
| S12_extracted_k141_100613 | Not-determined |
| S12_extracted_k141_12394 | Low-quality |
| S12_extracted_k141_20905 | Not-determined |
| S12_extracted_k141_108153 | Not-determined |
| S12_extracted_k141_154708 | Low-quality |
| S12_extracted_k141_115192 | Not-determined |
| S12_extracted_k141_5822 | Not-determined |
| S12_extracted_k141_5823 | Not-determined |
| S12_extracted_k141_130955 | Not-determined |
| S12_extracted_k141_76048 | Not-determined |
| S12_extracted_k141_20911 | Not-determined |
| S12_extracted_k141_108158 | Not-determined |
| S12_extracted_k141_108161 | Not-determined |
| S12_extracted_k141_115200 | Not-determined |
| S12_extracted_k141_28989 | Not-determined |
| S12_extracted_k141_108162 | Not-determined |
| S12_extracted_k141_115201 | Not-determined |
| S12_extracted_k141_177903 | Not-determined |
| S12_extracted_k141_5829 | Low-quality |
| S12_extracted_k141_170524 | Not-determined |
| S12_extracted_k141_154714 | Not-determined |
| S12_extracted_k141_20917 | Not-determined |
| S12_extracted_k141_177907 | Not-determined |
| S12_extracted_k141_130962 | Not-determined |
| S12_extracted_k141_5834 | Not-determined |
| S12_extracted_k141_60500 | Not-determined |
| S12_extracted_k141_115208 | Not-determined |
| S12_extracted_k141_76051 | Not-determined |
| S12_extracted_k141_122856 | Low-quality |
| S12_extracted_k141_139498 | Not-determined |
| S12_extracted_k141_60501 | Low-quality |
| S12_extracted_k141_115212 | Not-determined |
| S12_extracted_k141_147166 | Not-determined |
| S12_extracted_k141_60502 | Not-determined |
| S12_extracted_k141_20924 | Not-determined |
| S12_extracted_k141_20925 | Not-determined |
| S12_extracted_k141_186986 | Not-determined |
| S12_extracted_k141_130967 | Not-determined |
| S12_extracted_k141_45486 | Not-determined |
| S12_extracted_k141_122862 | Not-determined |
| S12_extracted_k141_186989 | Not-determined |
| S12_extracted_k141_52372 | Not-determined |
| S12_extracted_k141_115220 | Not-determined |
| S12_extracted_k141_20929 | Low-quality |
| S12_extracted_k141_130975 | Not-determined |
| S12_extracted_k141_154730 | Low-quality |
| S12_extracted_k141_76060 | Low-quality |
| S12_extracted_k141_186999 | Not-determined |
| S12_extracted_k141_52380 | Not-determined |
| S12_extracted_k141_130984 | Not-determined |
| S12_extracted_k141_83762 | Not-determined |
| S12_extracted_k141_5848 | Not-determined |
| S12_extracted_k141_20943 | Not-determined |
| S12_extracted_k141_12427 | Not-determined |
| S12_extracted_k141_36290 | Not-determined |
| S12_extracted_k141_130987 | Not-determined |
| S12_extracted_k141_12429 | Not-determined |
| S12_extracted_k141_12433 | Low-quality |
| S12_extracted_k141_45493 | Not-determined |
| S12_extracted_k141_5854 | Not-determined |
| S12_extracted_k141_139520 | Low-quality |
| S12_extracted_k141_177934 | Low-quality |
| S12_extracted_k141_177935 | Low-quality |
| S12_extracted_k141_187015 | Not-determined |
| S12_extracted_k141_115241 | Not-determined |
| S12_extracted_k141_36298 | Not-determined |
| S12_extracted_k141_130991 | Not-determined |
| S12_extracted_k141_36299 | Not-determined |
| S12_extracted_k141_100654 | Not-determined |
| S12_extracted_k141_154742 | Not-determined |
| S12_extracted_k141_29011 | Not-determined |
| S12_extracted_k141_187017 | Not-determined |
| S12_extracted_k141_154748 | Not-determined |
| S12_extracted_k141_29015 | Not-determined |
| S12_extracted_k141_139524 | Not-determined |
| S12_extracted_k141_45506 | Not-determined |
| S12_extracted_k141_177945 | Not-determined |
| S12_extracted_k141_187023 | Low-quality |
| S12_extracted_k141_60509 | Not-determined |
| S12_extracted_k141_122877 | Not-determined |
| S12_extracted_k141_45511 | Not-determined |
| S12_extracted_k141_131007 | Low-quality |
| S12_extracted_k141_36312 | Low-quality |
| S12_extracted_k141_52398 | Low-quality |
| S12_extracted_k141_131011 | Low-quality |
| S12_extracted_k141_12446 | Not-determined |
| S12_extracted_k141_68286 | Not-determined |
| S12_extracted_k141_20965 | Not-determined |
| S12_extracted_k141_68289 | Not-determined |
| S12_extracted_k141_131015 | Not-determined |
| S12_extracted_k141_177953 | Not-determined |
| S12_extracted_k141_115253 | Not-determined |
| S12_extracted_k141_108169 | Not-determined |
| S12_extracted_k141_36315 | Low-quality |
| S12_extracted_k141_52402 | Not-determined |
| S12_extracted_k141_122889 | Not-determined |
| S12_extracted_k141_12449 | Not-determined |
| S12_extracted_k141_68295 | Not-determined |
| S12_extracted_k141_100672 | Low-quality |
| S12_extracted_k141_115260 | Not-determined |
| S12_extracted_k141_154769 | Not-determined |
| S12_extracted_k141_154772 | Not-determined |
| S12_extracted_k141_177969 | Not-determined |
| S12_extracted_k141_177970 | Low-quality |
| S12_extracted_k141_154774 | Not-determined |
| S12_extracted_k141_139540 | Low-quality |
| S12_extracted_k141_187039 | Not-determined |
| S12_extracted_k141_45520 | Not-determined |
| S12_extracted_k141_83773 | Not-determined |
| S12_extracted_k141_187040 | Not-determined |
| S12_extracted_k141_177972 | Not-determined |
| S12_extracted_k141_52418 | Not-determined |
| S12_extracted_k141_154779 | Not-determined |
| S12_extracted_k141_52420 | Not-determined |
| S12_extracted_k141_60531 | Not-determined |
| S12_extracted_k141_170553 | Not-determined |
| S12_extracted_k141_115265 | Not-determined |
| S12_extracted_k141_45523 | Not-determined |
| S12_extracted_k141_60533 | Not-determined |
| S12_extracted_k141_45524 | Not-determined |
| S12_extracted_k141_177974 | Not-determined |
| S12_extracted_k141_45525 | Not-determined |
| S12_extracted_k141_177976 | Not-determined |
| S12_extracted_k141_177977 | Not-determined |
| S12_extracted_k141_60537 | Low-quality |
| S12_extracted_k141_68300 | Not-determined |
| S12_extracted_k141_177982 | Not-determined |
| S12_extracted_k141_20975 | Not-determined |
| S12_extracted_k141_52423 | Not-determined |
| S12_extracted_k141_45530 | Not-determined |
| S12_extracted_k141_170559 | Not-determined |
| S12_extracted_k141_100682 | Not-determined |
| S12_extracted_k141_122917 | Not-determined |
| S12_extracted_k141_147172 | Not-determined |
| S12_extracted_k141_52427 | Not-determined |
| S12_extracted_k141_154794 | Not-determined |
| S12_extracted_k141_147173 | Not-determined |
| S12_extracted_k141_100684 | Low-quality |
| S12_extracted_k141_20978 | Not-determined |
| S12_extracted_k141_187053 | Not-determined |
| S12_extracted_k141_100686 | Not-determined |
| S12_extracted_k141_154797 | Low-quality |
| S12_extracted_k141_177988 | Not-determined |
| S12_extracted_k141_139544 | Not-determined |
| S12_extracted_k141_187056 | Not-determined |
| S12_extracted_k141_177991 | Not-determined |
| S12_extracted_k141_52433 | Not-determined |
| S12_extracted_k141_147188 | Not-determined |
| S12_extracted_k141_139550 | Not-determined |
| S12_extracted_k141_177994 | Low-quality |
| S12_extracted_k141_52439 | Not-determined |
| S12_extracted_k141_76082 | Not-determined |
| S12_extracted_k141_29049 | Not-determined |
| S12_extracted_k141_60558 | Not-determined |
| S12_extracted_k141_52440 | Not-determined |
| S12_extracted_k141_45553 | Not-determined |
| S12_extracted_k141_139552 | Low-quality |
| S12_extracted_k141_52442 | Low-quality |
| S12_extracted_k141_187062 | Not-determined |

| | |
|---|---|
| S12_extracted_k141_187064 | Low-quality |
| S12_extracted_k141_52448 | Not-determined |
| S12_extracted_k141_68312 | Not-determined |
| S12_extracted_k141_122925 | Not-determined |
| S12_extracted_k141_76096 | Not-determined |
| S12_extracted_k141_139561 | Not-determined |
| S12_extracted_k141_115298 | Not-determined |
| S12_extracted_k141_178005 | Not-determined |
| S12_extracted_k141_5876 | Not-determined |
| S12_extracted_k141_12477 | Not-determined |
| S12_extracted_k141_115301 | Not-determined |
| S12_extracted_k141_12480 | Not-determined |
| S12_extracted_k141_178008 | Not-determined |
| S12_extracted_k141_36326 | Not-determined |
| S12_extracted_k141_12482 | Not-determined |
| S12_extracted_k141_115304 | Not-determined |
| S12_extracted_k141_52454 | Not-determined |
| S12_extracted_k141_5881 | Not-determined |
| S12_extracted_k141_29059 | Not-determined |
| S12_extracted_k141_60578 | Not-determined |
| S12_extracted_k141_92532 | Not-determined |
| S12_extracted_k141_76105 | Not-determined |
| S12_extracted_k141_12487 | Not-determined |
| S12_extracted_k141_178013 | Low-quality |
| S12_extracted_k141_100695 | Not-determined |
| S12_extracted_k141_52457 | Low-quality |
| S12_extracted_k141_36331 | Not-determined |
| S12_extracted_k141_178017 | Not-determined |
| S12_extracted_k141_122938 | Not-determined |
| S12_extracted_k141_29069 | Not-determined |
| S12_extracted_k141_139579 | Not-determined |
| S12_extracted_k141_139580 | Not-determined |
| S12_extracted_k141_147208 | Not-determined |
| S12_extracted_k141_139581 | Not-determined |
| S12_extracted_k141_76110 | Not-determined |
| S12_extracted_k141_83802 | Not-determined |
| S12_extracted_k141_83803 | Not-determined |
| S12_extracted_k141_83804 | Not-determined |
| S12_extracted_k141_147211 | Not-determined |
| S12_extracted_k141_100701 | Low-quality |
| S12_extracted_k141_187089 | Not-determined |
| S12_extracted_k141_20994 | Not-determined |
| S12_extracted_k141_139589 | Not-determined |
| S12_extracted_k141_52468 | Not-determined |
| S12_extracted_k141_76118 | Not-determined |
| S12_extracted_k141_29080 | Low-quality |
| S12_extracted_k141_29081 | Not-determined |
| S12_extracted_k141_52470 | Not-determined |
| S12_extracted_k141_92545 | Not-determined |
| S12_extracted_k141_147218 | Not-determined |
| S12_extracted_k141_76123 | Not-determined |
| S12_extracted_k141_29084 | Not-determined |
| S12_extracted_k141_76124 | Not-determined |
| S12_extracted_k141_21000 | Not-determined |
| S12_extracted_k141_122953 | Not-determined |
| S12_extracted_k141_147222 | Not-determined |
| S12_extracted_k141_60597 | Not-determined |
| S12_extracted_k141_178033 | Low-quality |
| S12_extracted_k141_76130 | Not-determined |
| S12_extracted_k141_139600 | Not-determined |
| S12_extracted_k141_178036 | Not-determined |
| S12_extracted_k141_29092 | Not-determined |
| S12_extracted_k141_52474 | Not-determined |
| S12_extracted_k141_162712 | Not-determined |
| S12_extracted_k141_45578 | Not-determined |
| S12_extracted_k141_100715 | Not-determined |
| S12_extracted_k141_122962 | Low-quality |
| S12_extracted_k141_178041 | Not-determined |
| S12_extracted_k141_36349 | Not-determined |
| S12_extracted_k141_12509 | Not-determined |
| S12_extracted_k141_147233 | Not-determined |
| S12_extracted_k141_36351 | Low-quality |
| S12_extracted_k141_187095 | Low-quality |
| S12_extracted_k141_21013 | Not-determined |
| S12_extracted_k141_92559 | Not-determined |
| S12_extracted_k141_29104 | Not-determined |
| S12_extracted_k141_122971 | Not-determined |
| S12_extracted_k141_178049 | Not-determined |
| S12_extracted_k141_178050 | Not-determined |
| S12_extracted_k141_29107 | Not-determined |
| S12_extracted_k141_45591 | Not-determined |
| S12_extracted_k141_76147 | Not-determined |
| S12_extracted_k141_36354 | Not-determined |
| S12_extracted_k141_21018 | Low-quality |
| S12_extracted_k141_131074 | Low-quality |
| S12_extracted_k141_131075 | Not-determined |
| S12_extracted_k141_122977 | Not-determined |
| S12_extracted_k141_52487 | Not-determined |
| S12_extracted_k141_162717 | Not-determined |
| S12_extracted_k141_131078 | Not-determined |
| S12_extracted_k141_162718 | Not-determined |
| S12_extracted_k141_92561 | Not-determined |
| S12_extracted_k141_83819 | Not-determined |
| S12_extracted_k141_108204 | Not-determined |
| S12_extracted_k141_115312 | Low-quality |
| S12_extracted_k141_29116 | Low-quality |
| S12_extracted_k141_139632 | Not-determined |
| S12_extracted_k141_100738 | Not-determined |
| S12_extracted_k141_139636 | Not-determined |
| S12_extracted_k141_12523 | Not-determined |
| S12_extracted_k141_139638 | Not-determined |
| S12_extracted_k141_154818 | Not-determined |
| S12_extracted_k141_21030 | Not-determined |
| S12_extracted_k141_187116 | Not-determined |
| S12_extracted_k141_45606 | Not-determined |
| S12_extracted_k141_147248 | Not-determined |
| S12_extracted_k141_83827 | Not-determined |
| S12_extracted_k141_139642 | Not-determined |
| S12_extracted_k141_36374 | Low-quality |
| S12_extracted_k141_154824 | Not-determined |
| S12_extracted_k141_36376 | Not-determined |
| S12_extracted_k141_60603 | Not-determined |
| S12_extracted_k141_162730 | Not-determined |
| S12_extracted_k141_76169 | Not-determined |
| S12_extracted_k141_139652 | Not-determined |
| S12_extracted_k141_5906 | Not-determined |
| S12_extracted_k141_12534 | Low-quality |
| S12_extracted_k141_5907 | Not-determined |
| S12_extracted_k141_162734 | Not-determined |
| S12_extracted_k141_115325 | Not-determined |
| S12_extracted_k141_83835 | Not-determined |
| S12_extracted_k141_108222 | Not-determined |
| S12_extracted_k141_45615 | Not-determined |
| S12_extracted_k141_139653 | Low-quality |
| S12_extracted_k141_45618 | Not-determined |
| S12_extracted_k141_154832 | Not-determined |
| S12_extracted_k141_100753 | Not-determined |
| S12_extracted_k141_68336 | Not-determined |
| S12_extracted_k141_5919 | Not-determined |
| S12_extracted_k141_68341 | Not-determined |
| S12_extracted_k141_5921 | Not-determined |
| S12_extracted_k141_108233 | Low-quality |
| S12_extracted_k141_139670 | Not-determined |
| S12_extracted_k141_100763 | Not-determined |
| S12_extracted_k141_68344 | Not-determined |
| S12_extracted_k141_100765 | Not-determined |
| S12_extracted_k141_45626 | Not-determined |
| S12_extracted_k141_83841 | Not-determined |
| S12_extracted_k141_123006 | Not-determined |
| S12_extracted_k141_5932 | Not-determined |
| S12_extracted_k141_115343 | Not-determined |
| S12_extracted_k141_100767 | Not-determined |
| S12_extracted_k141_123009 | Not-determined |
| S12_extracted_k141_131091 | Not-determined |
| S12_extracted_k141_139674 | Not-determined |
| S12_extracted_k141_68353 | Not-determined |
| S12_extracted_k141_76197 | Low-quality |
| S12_extracted_k141_108244 | Not-determined |
| S12_extracted_k141_83849 | Not-determined |
| S12_extracted_k141_178088 | Low-quality |
| S12_extracted_k141_187143 | Not-determined |
| S12_extracted_k141_154846 | Not-determined |
| S12_extracted_k141_36388 | Low-quality |
| S12_extracted_k141_108248 | Not-determined |
| S12_extracted_k141_178094 | Not-determined |
| S12_extracted_k141_154852 | Low-quality |
| S12_extracted_k141_115354 | Not-determined |
| S12_extracted_k141_68362 | Low-quality |
| S12_extracted_k141_123022 | Not-determined |
| S12_extracted_k141_131097 | Not-determined |
| S12_extracted_k141_131098 | Not-determined |
| S12_extracted_k141_187148 | Not-determined |
| S12_extracted_k141_170581 | Not-determined |
| S12_extracted_k141_60628 | Not-determined |
| S12_extracted_k141_131101 | Not-determined |
| S12_extracted_k141_100783 | Not-determined |
| S12_extracted_k141_5946 | Not-determined |
| S12_extracted_k141_187153 | Not-determined |
| S12_extracted_k141_108255 | Not-determined |
| S12_extracted_k141_52512 | Low-quality |
| S12_extracted_k141_187155 | Not-determined |
| S12_extracted_k141_131107 | Not-determined |
| S12_extracted_k141_21055 | Low-quality |
| S12_extracted_k141_92587 | Not-determined |
| S12_extracted_k141_108259 | Not-determined |
| S12_extracted_k141_76210 | Not-determined |
| S12_extracted_k141_36401 | Not-determined |
| S12_extracted_k141_83863 | Low-quality |
| S12_extracted_k141_100790 | Not-determined |
| S12_extracted_k141_154872 | Not-determined |
| S12_extracted_k141_83865 | Not-determined |
| S12_extracted_k141_45637 | Not-determined |
| S12_extracted_k141_21059 | Not-determined |
| S12_extracted_k141_83866 | Not-determined |
| S12_extracted_k141_154876 | Not-determined |
| S12_extracted_k141_170594 | Not-determined |
| S12_extracted_k141_170595 | Not-determined |
| S12_extracted_k141_76215 | Not-determined |
| S12_extracted_k141_83869 | Not-determined |
| S12_extracted_k141_100799 | Not-determined |
| S12_extracted_k141_170596 | Not-determined |
| S12_extracted_k141_21067 | Low-quality |
| S12_extracted_k141_5959 | Low-quality |
| S12_extracted_k141_154883 | Not-determined |
| S12_extracted_k141_170597 | Not-determined |
| S12_extracted_k141_76222 | Not-determined |
| S12_extracted_k141_115357 | Not-determined |
| S12_extracted_k141_100804 | Not-determined |
| S12_extracted_k141_45653 | Not-determined |
| S12_extracted_k141_178108 | Low-quality |
| S12_extracted_k141_162771 | Not-determined |
| S12_extracted_k141_12565 | Not-determined |
| S12_extracted_k141_36416 | Not-determined |
| S12_extracted_k141_115364 | Not-determined |
| S12_extracted_k141_115365 | Not-determined |
| S12_extracted_k141_162775 | Not-determined |
| S12_extracted_k141_45657 | Not-determined |
| S12_extracted_k141_92602 | Low-quality |
| S12_extracted_k141_187170 | Not-determined |
| S12_extracted_k141_100811 | Not-determined |
| S12_extracted_k141_108273 | Not-determined |
| S12_extracted_k141_83879 | Not-determined |
| S12_extracted_k141_83880 | Not-determined |
| S12_extracted_k141_92607 | Not-determined |
| S12_extracted_k141_68380 | Not-determined |
| S12_extracted_k141_36428 | Not-determined |
| S12_extracted_k141_187177 | Not-determined |
| S12_extracted_k141_45662 | Low-quality |
| S12_extracted_k141_139686 | Not-determined |
| S12_extracted_k141_178121 | Low-quality |
| S12_extracted_k141_36431 | Not-determined |
| S12_extracted_k141_92617 | Not-determined |
| S12_extracted_k141_60655 | Not-determined |
| S12_extracted_k141_123041 | Not-determined |
| S12_extracted_k141_68390 | Not-determined |
| S12_extracted_k141_187182 | Not-determined |
| S12_extracted_k141_68393 | Not-determined |
| S12_extracted_k141_36436 | Not-determined |
| S12_extracted_k141_12576 | Low-quality |
| S12_extracted_k141_187185 | Not-determined |
| S12_extracted_k141_83890 | Not-determined |
| S12_extracted_k141_147265 | Not-determined |
| S12_extracted_k141_115389 | Not-determined |
| S12_extracted_k141_187186 | Not-determined |
| S12_extracted_k141_83892 | Not-determined |
| S12_extracted_k141_100836 | Not-determined |
| S12_extracted_k141_170613 | Not-determined |
| S12_extracted_k141_108289 | Not-determined |
| S12_extracted_k141_83895 | Low-quality |
| S12_extracted_k141_76235 | Not-determined |
| S12_extracted_k141_60660 | Low-quality |
| S12_extracted_k141_131110 | Low-quality |
| S12_extracted_k141_60662 | Not-determined |
| S12_extracted_k141_115397 | Not-determined |
| S12_extracted_k141_123055 | Not-determined |
| S12_extracted_k141_131115 | Not-determined |
| S12_extracted_k141_139708 | Not-determined |
| S12_extracted_k141_139709 | Not-determined |
| S12_extracted_k141_12585 | Not-determined |
| S12_extracted_k141_123061 | Not-determined |
| S12_extracted_k141_60665 | Not-determined |
| S12_extracted_k141_92640 | Not-determined |
| S12_extracted_k141_131122 | Not-determined |
| S12_extracted_k141_83905 | Not-determined |
| S12_extracted_k141_139714 | Not-determined |
| S12_extracted_k141_12591 | Not-determined |
| S12_extracted_k141_92643 | Not-determined |
| S12_extracted_k141_139717 | Not-determined |
| S12_extracted_k141_12594 | Not-determined |
| S12_extracted_k141_68411 | Not-determined |
| S12_extracted_k141_123065 | Not-determined |
| S12_extracted_k141_100860 | Not-determined |
| S12_extracted_k141_187200 | Not-determined |
| S12_extracted_k141_45676 | Not-determined |
| S12_extracted_k141_187201 | Not-determined |
| S12_extracted_k141_147298 | Not-determined |
| S16_vs_36146\|\|full | Medium-quality |
| S12_extracted_k141_170615 | Not-determined |
| S12_extracted_k141_29126 | Not-determined |
| S12_extracted_k141_45681 | Not-determined |
| S12_extracted_k141_45686 | Not-determined |
| S12_extracted_k141_147307 | Not-determined |
| S12_extracted_k141_115414 | Low-quality |
| S12_extracted_k141_123077 | Not-determined |
| S12_extracted_k141_154910 | Not-determined |
| S12_extracted_k141_5977 | Not-determined |
| S12_extracted_k141_139735 | Not-determined |
| S12_extracted_k141_68427 | Not-determined |
| S12_extracted_k141_29144 | Not-determined |
| S12_extracted_k141_12611 | Low-quality |
| S12_extracted_k141_29146 | Low-quality |
| S12_extracted_k141_170628 | Not-determined |
| S12_extracted_k141_154921 | Not-determined |
| S12_extracted_k141_147319 | Low-quality |
| S12_extracted_k141_162808 | Not-determined |
| S12_extracted_k141_12613 | Not-determined |
| S12_extracted_k141_131139 | Not-determined |
| S12_extracted_k141_29155 | Not-determined |
| S12_extracted_k141_108305 | Not-determined |
| S12_extracted_k141_187208 | Not-determined |
| S12_extracted_k141_12615 | Not-determined |
| S12_extracted_k141_21094 | Not-determined |
| S12_extracted_k141_187211 | Not-determined |
| S12_extracted_k141_139745 | Not-determined |
| S12_extracted_k141_21095 | Not-determined |
| S12_extracted_k141_147323 | Not-determined |
| S12_extracted_k141_21096 | Not-determined |
| S12_extracted_k141_147324 | Not-determined |
| S12_extracted_k141_162811 | Not-determined |
| S12_extracted_k141_123091 | Not-determined |
| S12_extracted_k141_115419 | Not-determined |
| S12_extracted_k141_29157 | Not-determined |
| S12_extracted_k141_115425 | Not-determined |
| S12_extracted_k141_12624 | Not-determined |
| S12_extracted_k141_162816 | Not-determined |
| S12_extracted_k141_29160 | Low-quality |
| S12_extracted_k141_29162 | Not-determined |
| S12_extracted_k141_154933 | Not-determined |
| S12_extracted_k141_29165 | Not-determined |
| S12_extracted_k141_178181 | Not-determined |
| S12_extracted_k141_115432 | Not-determined |
| S12_extracted_k141_108317 | Not-determined |
| S12_extracted_k141_147335 | Not-determined |
| S12_extracted_k141_21110 | Not-determined |
| S12_extracted_k141_100873 | Not-determined |
| S12_extracted_k141_92676 | Not-determined |
| S12_extracted_k141_139751 | Not-determined |
| S12_extracted_k141_21119 | Not-determined |
| S12_extracted_k141_123109 | Not-determined |
| S12_extracted_k141_162833 | Not-determined |
| S12_extracted_k141_83925 | Not-determined |
| S12_extracted_k141_108332 | Not-determined |
| S12_extracted_k141_147347 | Low-quality |
| S12_extracted_k141_162838 | Not-determined |
| S12_extracted_k141_108335 | Not-determined |
| S12_extracted_k141_154938 | Not-determined |
| S12_extracted_k141_21129 | Not-determined |
| S12_extracted_k141_170651 | Low-quality |
| S12_extracted_k141_115440 | Not-determined |
| S12_extracted_k141_139757 | Not-determined |
| S12_extracted_k141_123113 | Not-determined |
| S12_extracted_k141_83933 | Not-determined |
| S12_extracted_k141_131170 | Not-determined |
| S12_extracted_k141_68454 | Not-determined |
| S12_extracted_k141_83935 | Not-determined |
| S12_extracted_k141_131172 | Not-determined |
| S12_extracted_k141_45718 | Not-determined |
| S12_extracted_k141_115445 | Not-determined |
| S12_extracted_k141_147356 | Not-determined |
| S12_extracted_k141_92692 | Not-determined |
| S12_extracted_k141_178201 | Low-quality |
| S12_extracted_k141_68457 | Not-determined |
| S12_extracted_k141_115448 | Not-determined |
| S12_extracted_k141_147358 | Not-determined |
| S12_extracted_k141_108342 | Not-determined |
| S12_extracted_k141_12648 | Not-determined |
| S12_extracted_k141_83943 | Not-determined |
| S12_extracted_k141_131178 | Not-determined |
| S12_extracted_k141_187228 | Not-determined |
| S12_extracted_k141_108346 | Not-determined |
| S12_extracted_k141_6009 | Not-determined |
| S12_extracted_k141_92700 | Not-determined |
| S12_extracted_k141_92703 | Not-determined |
| S12_extracted_k141_139768 | Not-determined |
| S12_extracted_k141_170660 | Low-quality |
| S12_extracted_k141_68470 | Low-quality |
| S12_extracted_k141_170661 | Low-quality |
| S12_extracted_k141_6017 | Not-determined |
| S12_extracted_k141_68472 | Not-determined |
| S12_extracted_k141_12659 | Not-determined |
| S12_extracted_k141_36445 | Low-quality |
| S12_extracted_k141_108354 | Not-determined |
| S12_extracted_k141_108355 | Not-determined |
| S12_extracted_k141_115452 | Not-determined |
| S12_extracted_k141_12665 | Not-determined |
| S12_extracted_k141_68484 | Not-determined |
| S12_extracted_k141_162849 | Not-determined |
| S12_extracted_k141_139779 | Not-determined |
| S12_extracted_k141_12668 | Not-determined |
| S12_extracted_k141_187234 | Not-determined |
| S12_extracted_k141_147373 | Not-determined |
| S12_extracted_k141_162852 | Not-determined |
| S12_extracted_k141_29188 | Not-determined |
| S12_extracted_k141_162853 | Not-determined |
| S12_extracted_k141_108367 | Low-quality |
| S12_extracted_k141_100908 | Not-determined |
| S12_extracted_k141_108368 | Not-determined |
| S12_extracted_k141_123122 | Low-quality |
| S12_extracted_k141_21153 | Not-determined |
| S12_extracted_k141_100911 | Not-determined |
| S12_extracted_k141_123124 | Not-determined |
| S12_extracted_k141_12675 | Not-determined |
| S12_extracted_k141_115454 | Not-determined |
| S12_extracted_k141_123125 | Not-determined |
| S12_extracted_k141_12677 | Not-determined |
| S12_extracted_k141_36461 | Not-determined |
| S12_extracted_k141_123128 | Not-determined |
| S6_vs_k141_44671\|\|full | Medium-quality |
| S12_extracted_k141_21157 | Not-determined |
| S12_extracted_k141_170685 | Not-determined |
| S12_extracted_k141_187241 | Not-determined |
| S12_extracted_k141_6035 | Not-determined |
| S12_extracted_k141_147389 | Not-determined |
| S12_extracted_k141_108383 | Not-determined |
| S12_extracted_k141_60699 | Not-determined |
| S12_extracted_k141_108386 | Not-determined |
| S12_extracted_k141_100924 | Not-determined |
| S12_extracted_k141_21168 | Not-determined |
| S12_extracted_k141_83970 | Not-determined |
| S12_extracted_k141_21169 | Not-determined |
| S12_extracted_k141_187246 | Not-determined |
| S12_extracted_k141_178239 | Not-determined |
| S12_extracted_k141_83972 | Not-determined |
| S12_extracted_k141_187247 | Not-determined |
| S12_extracted_k141_147399 | Not-determined |
| S12_extracted_k141_187248 | Not-determined |
| S12_extracted_k141_21174 | Not-determined |
| S12_extracted_k141_162879 | Not-determined |
| S12_extracted_k141_60704 | Not-determined |
| S12_extracted_k141_162881 | Not-determined |
| S12_extracted_k141_162882 | Not-determined |
| S12_extracted_k141_92741 | Not-determined |
| S12_extracted_k141_147406 | Not-determined |
| S12_extracted_k141_108398 | Not-determined |
| S12_extracted_k141_21181 | Not-determined |
| S12_extracted_k141_170711 | Not-determined |
| S12_extracted_k141_68509 | Not-determined |
| S12_extracted_k141_162889 | Low-quality |
| S12_extracted_k141_187259 | Not-determined |
| S12_extracted_k141_115488 | Not-determined |
| S12_extracted_k141_178254 | Not-determined |
| S12_extracted_k141_92744 | Low-quality |
| S12_extracted_k141_12701 | Not-determined |
| S12_extracted_k141_21186 | Not-determined |
| S12_extracted_k141_100946 | Not-determined |
| S12_extracted_k141_12703 | Not-determined |
| S12_extracted_k141_76240 | Not-determined |
| S12_extracted_k141_6055 | Not-determined |
| S12_extracted_k141_162895 | Low-quality |
| S12_extracted_k141_178264 | Not-determined |
| S12_extracted_k141_108410 | Not-determined |
| S12_extracted_k141_162899 | Not-determined |
| S12_extracted_k141_92750 | Not-determined |
| S12_extracted_k141_147422 | Not-determined |
| S12_extracted_k141_147424 | Low-quality |
| S12_extracted_k141_68524 | Not-determined |
| S12_extracted_k141_108415 | Not-determined |
| S12_extracted_k141_76249 | Low-quality |
| S12_extracted_k141_100955 | Not-determined |
| S12_extracted_k141_100957 | Not-determined |
| S12_extracted_k141_12714 | Low-quality |
| S12_extracted_k141_92757 | Not-determined |
| S12_extracted_k141_92758 | Not-determined |
| S12_extracted_k141_60743 | Not-determined |
| S12_extracted_k141_108420 | Not-determined |
| S12_extracted_k141_36475 | Low-quality |
| S12_extracted_k141_115496 | Not-determined |
| S12_extracted_k141_108423 | Not-determined |
| S12_extracted_k141_60745 | Not-determined |
| S12_extracted_k141_6067 | Not-determined |
| S12_extracted_k141_6068 | Not-determined |
| S12_extracted_k141_6070 | Not-determined |
| S12_extracted_k141_60748 | Not-determined |
| S12_extracted_k141_68531 | Not-determined |
| S12_extracted_k141_12717 | Not-determined |
| S12_extracted_k141_21200 | Not-determined |
| S12_extracted_k141_139798 | Not-determined |
| S12_extracted_k141_115498 | Not-determined |
| S12_extracted_k141_6073 | Not-determined |
| S12_extracted_k141_12719 | Not-determined |
| S12_extracted_k141_123165 | Not-determined |
| S12_extracted_k141_108430 | Low-quality |
| S12_extracted_k141_60754 | Not-determined |
| S12_extracted_k141_21205 | Not-determined |
| S12_extracted_k141_187286 | Not-determined |
| S12_extracted_k141_178281 | Not-determined |
| S12_extracted_k141_139804 | Not-determined |
| S12_extracted_k141_178282 | Not-determined |
| S12_extracted_k141_36488 | Not-determined |
| S12_extracted_k141_154968 | Not-determined |
| S12_extracted_k141_60760 | Low-quality |
| S12_extracted_k141_68536 | Not-determined |
| S12_extracted_k141_36493 | Not-determined |
| S12_extracted_k141_68538 | Not-determined |
| S12_extracted_k141_68541 | Not-determined |
| S12_extracted_k141_154979 | Not-determined |
| S12_extracted_k141_60768 | Not-determined |
| S12_extracted_k141_178293 | Not-determined |
| S12_extracted_k141_178294 | Not-determined |
| S12_extracted_k141_115513 | Not-determined |
| S12_extracted_k141_139812 | Low-quality |
| S12_extracted_k141_147462 | Not-determined |
| S12_extracted_k141_123184 | Not-determined |
| S12_extracted_k141_139815 | Not-determined |
| S12_extracted_k141_123185 | Not-determined |
| S12_extracted_k141_170743 | Not-determined |
| S12_extracted_k141_170744 | Not-determined |
| S12_extracted_k141_115528 | Not-determined |
| S12_extracted_k141_154989 | Not-determined |
| S12_extracted_k141_52529 | Not-determined |
| S12_extracted_k141_92779 | Not-determined |
| S12_extracted_k141_147467 | Not-determined |
| S12_extracted_k141_21226 | Low-quality |
| S12_extracted_k141_147468 | Not-determined |
| S12_extracted_k141_6095 | Low-quality |
| S12_extracted_k141_108438 | Not-determined |
| S12_extracted_k141_68568 | Not-determined |
| S12_extracted_k141_92782 | Not-determined |
| S12_extracted_k141_108446 | Not-determined |
| S12_extracted_k141_52533 | Not-determined |
| S12_extracted_k141_92788 | Not-determined |
| S12_extracted_k141_68572 | Low-quality |
| S12_extracted_k141_76269 | Not-determined |
| S12_extracted_k141_76270 | Not-determined |
| S12_extracted_k141_115547 | Not-determined |
| S12_extracted_k141_154998 | Not-determined |
| S12_extracted_k141_123206 | Not-determined |
| S12_extracted_k141_131216 | Not-determined |
| S12_extracted_k141_68579 | Not-determined |
| S12_extracted_k141_139838 | Not-determined |
| S12_extracted_k141_162913 | Not-determined |
| S12_extracted_k141_21235 | Not-determined |
| S12_extracted_k141_162919 | Not-determined |
| S12_extracted_k141_100959 | Not-determined |
| S12_extracted_k141_68585 | Not-determined |
| S12_extracted_k141_131225 | Low-quality |
| S12_extracted_k141_92801 | Not-determined |
| S12_extracted_k141_170774 | Not-determined |
| S12_extracted_k141_84033 | Not-determined |
| S12_extracted_k141_92803 | Not-determined |
| S12_extracted_k141_155015 | Not-determined |
| S12_extracted_k141_155016 | Not-determined |
| S12_extracted_k141_170779 | Low-quality |
| S12_extracted_k141_108466 | Not-determined |
| S12_extracted_k141_92805 | Not-determined |
| S12_extracted_k141_92806 | Not-determined |
| S12_extracted_k141_139853 | Not-determined |
| S12_extracted_k141_155021 | Not-determined |
| S12_extracted_k141_21251 | Not-determined |
| S12_extracted_k141_162925 | Not-determined |
| S12_extracted_k141_139856 | Not-determined |
| S12_extracted_k141_108471 | Not-determined |
| S12_extracted_k141_170788 | Not-determined |
| S12_extracted_k141_131235 | Low-quality |
| S12_extracted_k141_92811 | Not-determined |
| S12_extracted_k141_52542 | Low-quality |
| S12_extracted_k141_52543 | Not-determined |
| S12_extracted_k141_92817 | Not-determined |
| S12_extracted_k141_115569 | Not-determined |
| S12_extracted_k141_12737 | Not-determined |
| S12_extracted_k141_131242 | Not-determined |
| S12_extracted_k141_131244 | Not-determined |
| S12_extracted_k141_155036 | Not-determined |
| S12_extracted_k141_123219 | Not-determined |
| S12_extracted_k141_155039 | Not-determined |
| S12_extracted_k141_131251 | Not-determined |
| S12_extracted_k141_45753 | Not-determined |
| S12_extracted_k141_178326 | Low-quality |
| S12_extracted_k141_84051 | Not-determined |
| S12_extracted_k141_155042 | Not-determined |
| S12_extracted_k141_155043 | Not-determined |
| S12_extracted_k141_115586 | Not-determined |
| S12_extracted_k141_21262 | Not-determined |
| S12_extracted_k141_60833 | Not-determined |
| S12_extracted_k141_115588 | Not-determined |
| S12_extracted_k141_21265 | Not-determined |
| S12_extracted_k141_84061 | Not-determined |
| S12_extracted_k141_92826 | Low-quality |
| S12_extracted_k141_92827 | Low-quality |
| S12_extracted_k141_155052 | Not-determined |
| S12_extracted_k141_45767 | Not-determined |
| S12_extracted_k141_147479 | Not-determined |
| S12_extracted_k141_170823 | Not-determined |
| S12_extracted_k141_162948 | Not-determined |
| S12_extracted_k141_6105 | Not-determined |
| S12_extracted_k141_139883 | Not-determined |
| S12_extracted_k141_187296 | Not-determined |
| S12_extracted_k141_147488 | Low-quality |
| S12_extracted_k141_123242 | Not-determined |
| S12_extracted_k141_52547 | Not-determined |
| S12_extracted_k141_147490 | Not-determined |
| S12_extracted_k141_170832 | Not-determined |
| S12_extracted_k141_92837 | Not-determined |
| S12_extracted_k141_52558 | Not-determined |
| S12_extracted_k141_21288 | Not-determined |
| S12_extracted_k141_123257 | Not-determined |
| S12_extracted_k141_6121 | Low-quality |
| S12_extracted_k141_21293 | Not-determined |
| S12_extracted_k141_147507 | Not-determined |
| S12_extracted_k141_147508 | Not-determined |
| S12_extracted_k141_21296 | Not-determined |
| S12_extracted_k141_29196 | Not-determined |
| S12_extracted_k141_21297 | Not-determined |
| S12_extracted_k141_84083 | Not-determined |
| S12_extracted_k141_6130 | Not-determined |
| S12_extracted_k141_6135 | Not-determined |
| S12_extracted_k141_76295 | Not-determined |
| S12_extracted_k141_36516 | Not-determined |
| S12_extracted_k141_178352 | Not-determined |

| | |
|---|---|
| S12_extracted_k141_139904 | Not-determined |
| S12_extracted_k141_92852 | Not-determined |
| S12_extracted_k141_45786 | Low-quality |
| S12_extracted_k141_92853 | Not-determined |
| S12_extracted_k141_147512 | Not-determined |
| S12_extracted_k141_45789 | Not-determined |
| S12_extracted_k141_123269 | Low-quality |
| S12_extracted_k141_178355 | Not-determined |
| S12_extracted_k141_6144 | Not-determined |
| S12_extracted_k141_147517 | Not-determined |
| S12_extracted_k141_21305 | Not-determined |
| S12_extracted_k141_29201 | Not-determined |
| S12_extracted_k141_29202 | Not-determined |
| S12_extracted_k141_92858 | Not-determined |
| S12_extracted_k141_162984 | Not-determined |
| S12_extracted_k141_178363 | Not-determined |
| S12_extracted_k141_92861 | Not-determined |
| S12_extracted_k141_92862 | Not-determined |
| S12_extracted_k141_21309 | Not-determined |
| S12_extracted_k141_139917 | Low-quality |
| S12_extracted_k141_36526 | Not-determined |
| S12_extracted_k141_147525 | Not-determined |
| S12_extracted_k141_139918 | Low-quality |
| S12_extracted_k141_52585 | Not-determined |
| S12_extracted_k141_52587 | Not-determined |
| S12_extracted_k141_187310 | Not-determined |
| S12_extracted_k141_45804 | Low-quality |
| S12_extracted_k141_101048 | Not-determined |
| S12_extracted_k141_36531 | Not-determined |
| S12_extracted_k141_76311 | Not-determined |
| S12_extracted_k141_12755 | Not-determined |
| S12_extracted_k141_29216 | Low-quality |
| S12_extracted_k141_178367 | Not-determined |
| S12_extracted_k141_178368 | Not-determined |
| S12_extracted_k141_12757 | Not-determined |
| S12_extracted_k141_29219 | Not-determined |
| S12_extracted_k141_101051 | Not-determined |
| S12_extracted_k141_36533 | Not-determined |
| S12_extracted_k141_36534 | Not-determined |
| S12_extracted_k141_29221 | Not-determined |
| S12_extracted_k141_60872 | Not-determined |
| S12_extracted_k141_6155 | Not-determined |
| S12_extracted_k141_52594 | Not-determined |
| S12_extracted_k141_123285 | Low-quality |
| S12_extracted_k141_178380 | Not-determined |
| S12_extracted_k141_29224 | Not-determined |
| S12_extracted_k141_45821 | Not-determined |
| S12_extracted_k141_29225 | Not-determined |
| S12_extracted_k141_36536 | Not-determined |
| S12_extracted_k141_36538 | Not-determined |
| S12_extracted_k141_178387 | Not-determined |
| S12_extracted_k141_131283 | Not-determined |
| S12_extracted_k141_21327 | Not-determined |
| S12_extracted_k141_12773 | Not-determined |
| S12_extracted_k141_131284 | Not-determined |
| S12_extracted_k141_60890 | Not-determined |
| S12_extracted_k141_76337 | Not-determined |
| S12_extracted_k141_163004 | Not-determined |
| S12_extracted_k141_29236 | Not-determined |
| S12_extracted_k141_29238 | Not-determined |
| S12_extracted_k141_187332 | Not-determined |
| S12_extracted_k141_12778 | Not-determined |
| S12_extracted_k141_12779 | Not-determined |
| S12_extracted_k141_131288 | Not-determined |
| S12_extracted_k141_76343 | Not-determined |
| S12_extracted_k141_163008 | Not-determined |
| S12_extracted_k141_36548 | Not-determined |
| S12_extracted_k141_68637 | Not-determined |
| S12_extracted_k141_101073 | Not-determined |
| S12_extracted_k141_36550 | Not-determined |
| S12_extracted_k141_36551 | Not-determined |
| S12_extracted_k141_170858 | Low-quality |
| S12_extracted_k141_123318 | Not-determined |
| S12_extracted_k141_101075 | Low-quality |
| S12_extracted_k141_139944 | Not-determined |
| S12_extracted_k141_6174 | Not-determined |
| S12_extracted_k141_21339 | Not-determined |
| S12_extracted_k141_123321 | Not-determined |
| S12_extracted_k141_45833 | Not-determined |
| S12_extracted_k141_45834 | Not-determined |
| S12_extracted_k141_21341 | Not-determined |
| S12_extracted_k141_123323 | Not-determined |
| S12_extracted_k141_6177 | Not-determined |
| S12_extracted_k141_170863 | Low-quality |
| S12_extracted_k141_60906 | Not-determined |
| S12_extracted_k141_45841 | Not-determined |
| S12_extracted_k141_123328 | Not-determined |
| S12_extracted_k141_123329 | Not-determined |
| S12_extracted_k141_21346 | Low-quality |
| S12_extracted_k141_178391 | Not-determined |
| S12_extracted_k141_60909 | Not-determined |
| S12_extracted_k141_101087 | Not-determined |
| S12_extracted_k141_187333 | Not-determined |
| S12_extracted_k141_147552 | Not-determined |
| S12_extracted_k141_131304 | Not-determined |
| S12_extracted_k141_36554 | Not-determined |
| S12_extracted_k141_52619 | Not-determined |
| S12_extracted_k141_76361 | Low-quality |
| S12_extracted_k141_36556 | Not-determined |
| S12_extracted_k141_76362 | Not-determined |
| S12_extracted_k141_101088 | Not-determined |
| S12_extracted_k141_170872 | Not-determined |
| S12_extracted_k141_21351 | Low-quality |
| S12_extracted_k141_29260 | Not-determined |
| S12_extracted_k141_163019 | Not-determined |
| S12_extracted_k141_60916 | Not-determined |
| S12_extracted_k141_170876 | Not-determined |
| S9_vs_k141_7881\|\|full | Medium-quality |
| S12_extracted_k141_21354 | Not-determined |
| S12_extracted_k141_163021 | Not-determined |
| S12_extracted_k141_178396 | Not-determined |
| S12_extracted_k141_163023 | Not-determined |
| S12_extracted_k141_52622 | Not-determined |
| S12_extracted_k141_131316 | Not-determined |
| S12_extracted_k141_170882 | Not-determined |
| S12_extracted_k141_60921 | Not-determined |
| S12_extracted_k141_101095 | Not-determined |
| S12_extracted_k141_178399 | Not-determined |
| S12_extracted_k141_139965 | Not-determined |
| S12_extracted_k141_178400 | Not-determined |
| S12_extracted_k141_178401 | Not-determined |
| S12_extracted_k141_52624 | Not-determined |
| S12_extracted_k141_101097 | Not-determined |
| S12_extracted_k141_123347 | Not-determined |
| S12_extracted_k141_6190 | Not-determined |
| S12_extracted_k141_101100 | Not-determined |
| S12_extracted_k141_147564 | Not-determined |
| S12_extracted_k141_139970 | Not-determined |
| S12_extracted_k141_108511 | Not-determined |
| S12_extracted_k141_139971 | Not-determined |
| S12_extracted_k141_29270 | Not-determined |
| S12_extracted_k141_101104 | Not-determined |
| S12_extracted_k141_60932 | Low-quality |
| S12_extracted_k141_6194 | Not-determined |
| S12_extracted_k141_12817 | Not-determined |
| S12_extracted_k141_76379 | Not-determined |
| S12_extracted_k141_12821 | Not-determined |
| S12_extracted_k141_131327 | Not-determined |
| S12_extracted_k141_108515 | Not-determined |
| S12_extracted_k141_12824 | Not-determined |
| S12_extracted_k141_123360 | Not-determined |
| S12_extracted_k141_60943 | Not-determined |
| S12_extracted_k141_12828 | Not-determined |
| S12_extracted_k141_163033 | Not-determined |
| S12_extracted_k141_163034 | Not-determined |
| S12_extracted_k141_131332 | Not-determined |
| S12_extracted_k141_108522 | Not-determined |
| S12_extracted_k141_36587 | Not-determined |
| S12_extracted_k141_6210 | Not-determined |
| S12_extracted_k141_108524 | Not-determined |
| S12_extracted_k141_76388 | Not-determined |
| S12_extracted_k141_155070 | Low-quality |
| S12_extracted_k141_147589 | Not-determined |
| S12_extracted_k141_155071 | Not-determined |
| S12_extracted_k141_6215 | Not-determined |
| S12_extracted_k141_84134 | Not-determined |
| S12_extracted_k141_147591 | Not-determined |
| S12_extracted_k141_155072 | Not-determined |
| S12_extracted_k141_12841 | Not-determined |
| S12_extracted_k141_52651 | Low-quality |
| S12_extracted_k141_123365 | Not-determined |
| S12_extracted_k141_178418 | Not-determined |
| S12_extracted_k141_52654 | Not-determined |
| S12_extracted_k141_170891 | Not-determined |
| S12_extracted_k141_101127 | Not-determined |
| S12_extracted_k141_163045 | Not-determined |
| S12_extracted_k141_147594 | Not-determined |
| S12_extracted_k141_84138 | Not-determined |
| S12_extracted_k141_6226 | Not-determined |
| S12_extracted_k141_139993 | Not-determined |
| S12_extracted_k141_36602 | Not-determined |
| S12_extracted_k141_139994 | Not-determined |
| S12_extracted_k141_60953 | Not-determined |
| S12_extracted_k141_76400 | Not-determined |
| S12_extracted_k141_131336 | Not-determined |
| S12_extracted_k141_155074 | Not-determined |
| S12_extracted_k141_108530 | Not-determined |
| S12_extracted_k141_139997 | Not-determined |
| S12_extracted_k141_68676 | Not-determined |
| S12_extracted_k141_123376 | Low-quality |
| S12_extracted_k141_108532 | Not-determined |
| S12_extracted_k141_108533 | Not-determined |
| S12_extracted_k141_163057 | Not-determined |
| S12_extracted_k141_178430 | Not-determined |
| S12_extracted_k141_123380 | Not-determined |
| S12_extracted_k141_163060 | Not-determined |
| S12_extracted_k141_187376 | Not-determined |
| S12_extracted_k141_123382 | Not-determined |
| S12_extracted_k141_21370 | Low-quality |
| S12_extracted_k141_84144 | Not-determined |
| S12_extracted_k141_68684 | Not-determined |
| S12_extracted_k141_187381 | Not-determined |
| S12_extracted_k141_68686 | Not-determined |
| S12_extracted_k141_101141 | Not-determined |
| S12_extracted_k141_187386 | Low-quality |
| S12_extracted_k141_163062 | Not-determined |
| S12_extracted_k141_21383 | Not-determined |
| S12_extracted_k141_123389 | Not-determined |
| S12_extracted_k141_84153 | Not-determined |
| S12_extracted_k141_84154 | Not-determined |
| S12_extracted_k141_21386 | Not-determined |
| S12_extracted_k141_68689 | Not-determined |
| S12_extracted_k141_60966 | Not-determined |
| S12_extracted_k141_123394 | Not-determined |
| S12_extracted_k141_29299 | Not-determined |
| S12_extracted_k141_108548 | Not-determined |
| S12_extracted_k141_163068 | Low-quality |
| S12_extracted_k141_163069 | Not-determined |
| S12_extracted_k141_84161 | Not-determined |
| S12_extracted_k141_123399 | Not-determined |
| S12_extracted_k141_163074 | Not-determined |
| S12_extracted_k141_163076 | Not-determined |
| S12_extracted_k141_52690 | Not-determined |
| S12_extracted_k141_155104 | Low-quality |
| S12_extracted_k141_68699 | Low-quality |
| S12_extracted_k141_45899 | Not-determined |
| S12_extracted_k141_155105 | Not-determined |
| S12_extracted_k141_68703 | Not-determined |
| S12_extracted_k141_108564 | Not-determined |
| S12_extracted_k141_84171 | Not-determined |
| S12_extracted_k141_178454 | Not-determined |
| S12_extracted_k141_178456 | Not-determined |
| S12_extracted_k141_60987 | Not-determined |
| S12_extracted_k141_140028 | Not-determined |
| S12_extracted_k141_68708 | Not-determined |
| S12_extracted_k141_45908 | Not-determined |
| S12_extracted_k141_170906 | Not-determined |
| S12_extracted_k141_170907 | Low-quality |
| S12_extracted_k141_108571 | Not-determined |
| S12_extracted_k141_84184 | Not-determined |
| S12_extracted_k141_123411 | Not-determined |
| S12_extracted_k141_140037 | Not-determined |
| S12_extracted_k141_155122 | Not-determined |
| S12_extracted_k141_45914 | Not-determined |
| S12_extracted_k141_178464 | Not-determined |
| S12_extracted_k141_36629 | Not-determined |
| S12_extracted_k141_29324 | Not-determined |
| S12_extracted_k141_131347 | Low-quality |
| S12_extracted_k141_52705 | Not-determined |
| S12_extracted_k141_163094 | Not-determined |
| S12_extracted_k141_29328 | Low-quality |
| S12_extracted_k141_140043 | Low-quality |
| S12_extracted_k141_187394 | Low-quality |
| S12_extracted_k141_76448 | Not-determined |
| S12_extracted_k141_36635 | Not-determined |
| S12_extracted_k141_84195 | Not-determined |
| S12_extracted_k141_170910 | Not-determined |
| S12_extracted_k141_140045 | Not-determined |
| S12_extracted_k141_92871 | Not-determined |
| S12_extracted_k141_108583 | Not-determined |
| S12_extracted_k141_52713 | Low-quality |
| S12_extracted_k141_108594 | Not-determined |
| S12_extracted_k141_76459 | Not-determined |
| S12_extracted_k141_76460 | Not-determined |
| S12_extracted_k141_45929 | Not-determined |
| S12_extracted_k141_29338 | Not-determined |
| S12_extracted_k141_76463 | Not-determined |
| S12_extracted_k141_108596 | Not-determined |
| S12_extracted_k141_115612 | Not-determined |
| S12_extracted_k141_84212 | Not-determined |
| S12_extracted_k141_52720 | Not-determined |
| S12_extracted_k141_52721 | Not-determined |
| S12_extracted_k141_131366 | Not-determined |
| S12_extracted_k141_21393 | Low-quality |
| S12_extracted_k141_187414 | Not-determined |
| S12_extracted_k141_187415 | Not-determined |
| S12_extracted_k141_108607 | Not-determined |
| S12_extracted_k141_21398 | Not-determined |
| S12_extracted_k141_108608 | Not-determined |
| S12_extracted_k141_140068 | Not-determined |
| S12_extracted_k141_163113 | Not-determined |
| S12_extracted_k141_178478 | Not-determined |
| S12_extracted_k141_108612 | Not-determined |
| S12_extracted_k141_6269 | Not-determined |
| S12_extracted_k141_108614 | Not-determined |
| S12_extracted_k141_163117 | Low-quality |
| S12_extracted_k141_178486 | Not-determined |
| S12_extracted_k141_163119 | Not-determined |
| S12_extracted_k141_178487 | Not-determined |
| S12_extracted_k141_131381 | Not-determined |
| S12_extracted_k141_123440 | Not-determined |
| S12_extracted_k141_108621 | Not-determined |
| S12_extracted_k141_187427 | Not-determined |
| S12_extracted_k141_178490 | Not-determined |
| S12_extracted_k141_147629 | Not-determined |
| S12_extracted_k141_131388 | Not-determined |
| S12_extracted_k141_6281 | Not-determined |
| S12_extracted_k141_163122 | Not-determined |
| S12_extracted_k141_187429 | Low-quality |
| S12_extracted_k141_68741 | Not-determined |
| S12_extracted_k141_123443 | Not-determined |
| S12_extracted_k141_178492 | Not-determined |
| S12_extracted_k141_108625 | Not-determined |
| S12_extracted_k141_36652 | Not-determined |
| S12_extracted_k141_12859 | Not-determined |
| S12_extracted_k141_123448 | Not-determined |
| S12_extracted_k141_21414 | Low-quality |
| S12_extracted_k141_76490 | Not-determined |
| S12_extracted_k141_178497 | Not-determined |
| S12_extracted_k141_131390 | Not-determined |
| S12_extracted_k141_68751 | Not-determined |
| S12_extracted_k141_29353 | Low-quality |
| S12_extracted_k141_131393 | Low-quality |
| S12_extracted_k141_68756 | Not-determined |
| S12_extracted_k141_84235 | Not-determined |
| S12_extracted_k141_178503 | Not-determined |
| S12_extracted_k141_76495 | Not-determined |
| S12_extracted_k141_6292 | Low-quality |
| S12_extracted_k141_6293 | Low-quality |
| S12_extracted_k141_101195 | Not-determined |
| S12_extracted_k141_29359 | Not-determined |
| S12_extracted_k141_12872 | Not-determined |
| S12_extracted_k141_76503 | Not-determined |
| S12_extracted_k141_101200 | Not-determined |
| S12_extracted_k141_147644 | Not-determined |
| S12_extracted_k141_178510 | Not-determined |
| S12_extracted_k141_163137 | Not-determined |
| S12_extracted_k141_29363 | Not-determined |
| S12_extracted_k141_84243 | Not-determined |
| S12_extracted_k141_187446 | Low-quality |
| S12_extracted_k141_101202 | Low-quality |
| S12_extracted_k141_178512 | Not-determined |
| S12_extracted_k141_123461 | Not-determined |
| S12_extracted_k141_101206 | Not-determined |
| S12_extracted_k141_45972 | Not-determined |
| S12_extracted_k141_29365 | Not-determined |
| S12_extracted_k141_29368 | Not-determined |
| S12_extracted_k141_163143 | Not-determined |
| S12_extracted_k141_108641 | Not-determined |
| S12_extracted_k141_12882 | Not-determined |
| S12_extracted_k141_163145 | Not-determined |
| S12_extracted_k141_147651 | Not-determined |
| S12_extracted_k141_101208 | Not-determined |
| S12_extracted_k141_178521 | Not-determined |
| S12_extracted_k141_147652 | Not-determined |
| S12_extracted_k141_131408 | Low-quality |
| S12_extracted_k141_140093 | Low-quality |
| S12_extracted_k141_68760 | Not-determined |
| S12_extracted_k141_178524 | Not-determined |
| S12_extracted_k141_108646 | Low-quality |
| S12_extracted_k141_131412 | Not-determined |
| S12_extracted_k141_123472 | Not-determined |
| S12_extracted_k141_21433 | Not-determined |
| S12_extracted_k141_140101 | Not-determined |
| S12_extracted_k141_76515 | Not-determined |
| S12_extracted_k141_108649 | Not-determined |
| S12_extracted_k141_147663 | Not-determined |
| S12_extracted_k141_131415 | Not-determined |
| S12_extracted_k141_45987 | Low-quality |
| S12_extracted_k141_52760 | Not-determined |
| S12_extracted_k141_108651 | Not-determined |
| S12_extracted_k141_61033 | Low-quality |
| S12_extracted_k141_178529 | Not-determined |
| S12_extracted_k141_163160 | Not-determined |
| S12_extracted_k141_68765 | Not-determined |
| S12_extracted_k141_76521 | Not-determined |
| S12_extracted_k141_36683 | Not-determined |
| S12_extracted_k141_61036 | Not-determined |
| S12_extracted_k141_108653 | Not-determined |
| S12_extracted_k141_68767 | Low-quality |
| S12_extracted_k141_21438 | Not-determined |
| S12_extracted_k141_147669 | Not-determined |
| S12_extracted_k141_76524 | Not-determined |
| S12_extracted_k141_61037 | Not-determined |
| S12_extracted_k141_155157 | Not-determined |
| S12_extracted_k141_45997 | Not-determined |
| S12_extracted_k141_108655 | Not-determined |
| S12_extracted_k141_84254 | Not-determined |
| S12_extracted_k141_52768 | Not-determined |
| S12_extracted_k141_108658 | Not-determined |
| S12_extracted_k141_36685 | Not-determined |
| S12_extracted_k141_123490 | Low-quality |
| S12_extracted_k141_155163 | Not-determined |
| S12_extracted_k141_115632 | Low-quality |
| S12_extracted_k141_52770 | Not-determined |
| S12_extracted_k141_140118 | Not-determined |
| S12_extracted_k141_101230 | Not-determined |
| S12_extracted_k141_178542 | Not-determined |
| S12_extracted_k141_163171 | Not-determined |
| S12_extracted_k141_115636 | Not-determined |
| S12_extracted_k141_123496 | Not-determined |
| S12_extracted_k141_115638 | Not-determined |
| S12_extracted_k141_68781 | Not-determined |
| S12_extracted_k141_84263 | Not-determined |
| S12_extracted_k141_84264 | Not-determined |
| S12_extracted_k141_84265 | Not-determined |
| S12_extracted_k141_101235 | Not-determined |
| S12_extracted_k141_140122 | Not-determined |
| S12_extracted_k141_115644 | Not-determined |
| S12_extracted_k141_155170 | Not-determined |
| S12_extracted_k141_140123 | Not-determined |
| S12_extracted_k141_21453 | Not-determined |
| S12_extracted_k141_21454 | Not-determined |
| S12_extracted_k141_131431 | Not-determined |
| S12_extracted_k141_101241 | Not-determined |
| S12_extracted_k141_101242 | Not-determined |
| S12_extracted_k141_147675 | Not-determined |
| S12_extracted_k141_76540 | Not-determined |
| S12_extracted_k141_52788 | Not-determined |
| S12_extracted_k141_178548 | Not-determined |
| S12_extracted_k141_101243 | Not-determined |
| S12_extracted_k141_170951 | Not-determined |
| S12_extracted_k141_21458 | Not-determined |
| S12_extracted_k141_61068 | Not-determined |
| S12_extracted_k141_101249 | Not-determined |
| S12_extracted_k141_115650 | Not-determined |
| S12_extracted_k141_147684 | Not-determined |
| S12_extracted_k141_163179 | Not-determined |
| S12_extracted_k141_155175 | Not-determined |
| S12_extracted_k141_131448 | Not-determined |
| S12_extracted_k141_115656 | Not-determined |
| S12_extracted_k141_155178 | Not-determined |
| S12_extracted_k141_108666 | Low-quality |
| S12_extracted_k141_108667 | Not-determined |
| S12_extracted_k141_140138 | Not-determined |
| S12_extracted_k141_140139 | Not-determined |
| S12_extracted_k141_36703 | Not-determined |
| S12_extracted_k141_170961 | Not-determined |
| S12_extracted_k141_108670 | Not-determined |
| S12_extracted_k141_170964 | Low-quality |
| S12_extracted_k141_170965 | Not-determined |
| S12_extracted_k141_108673 | Not-determined |
| S12_extracted_k141_84287 | Not-determined |
| S12_extracted_k141_36711 | Not-determined |
| S12_extracted_k141_12895 | Not-determined |
| S12_extracted_k141_36715 | Not-determined |
| S12_extracted_k141_36716 | Not-determined |
| S12_extracted_k141_108674 | Not-determined |
| S12_extracted_k141_147692 | Not-determined |
| S12_extracted_k141_84297 | Not-determined |
| S12_extracted_k141_147693 | Not-determined |
| S12_extracted_k141_108676 | Not-determined |
| S12_extracted_k141_12907 | Not-determined |
| S12_extracted_k141_108678 | Not-determined |
| S12_extracted_k141_21478 | Low-quality |
| S12_extracted_k141_115661 | Not-determined |
| S12_extracted_k141_76548 | Not-determined |
| S12_extracted_k141_115662 | Not-determined |
| S12_extracted_k141_123518 | Not-determined |
| S12_extracted_k141_46045 | Low-quality |
| S12_extracted_k141_131477 | Not-determined |
| S12_extracted_k141_170988 | Not-determined |
| S12_extracted_k141_163186 | Low-quality |
| S12_extracted_k141_140165 | Not-determined |
| S12_extracted_k141_140166 | Not-determined |
| S12_extracted_k141_187464 | Low-quality |
| S12_extracted_k141_84301 | Low-quality |
| S12_extracted_k141_155191 | Not-determined |
| S12_extracted_k141_68788 | Not-determined |
| S12_extracted_k141_21490 | Low-quality |
| S12_extracted_k141_155193 | Not-determined |
| S12_extracted_k141_108693 | Not-determined |
| S12_extracted_k141_21491 | Not-determined |
| S12_extracted_k141_187475 | Not-determined |
| S12_extracted_k141_68792 | Not-determined |
| S12_extracted_k141_21495 | Low-quality |
| S12_extracted_k141_52827 | Not-determined |
| S12_extracted_k141_29445 | Not-determined |
| S12_extracted_k141_29446 | Not-determined |
| S12_extracted_k141_36734 | Not-determined |
| S12_extracted_k141_171005 | Not-determined |
| S12_extracted_k141_155194 | Not-determined |
| S12_extracted_k141_108699 | Not-determined |
| S12_extracted_k141_101292 | Not-determined |
| S12_extracted_k141_140179 | Not-determined |
| S12_extracted_k141_131500 | Not-determined |
| S12_extracted_k141_12935 | Not-determined |
| S12_extracted_k141_101295 | Low-quality |
| S12_extracted_k141_101300 | Not-determined |
| S12_extracted_k141_178590 | Not-determined |
| S12_extracted_k141_115680 | Not-determined |
| S12_extracted_k141_84326 | Not-determined |
| S12_extracted_k141_155204 | Not-determined |
| S12_extracted_k141_36745 | Not-determined |
| S12_extracted_k141_36746 | Not-determined |
| S12_extracted_k141_46071 | Not-determined |
| S12_extracted_k141_36748 | Low-quality |
| S12_extracted_k141_163204 | Not-determined |
| S12_extracted_k141_46076 | Not-determined |
| S12_extracted_k141_46077 | Low-quality |
| S12_extracted_k141_155210 | Not-determined |
| S12_extracted_k141_101309 | Not-determined |
| S12_extracted_k141_36752 | Not-determined |
| S12_extracted_k141_163208 | Not-determined |
| S12_extracted_k141_36757 | Low-quality |
| S12_extracted_k141_101317 | Low-quality |
| S12_extracted_k141_108719 | Not-determined |
| S12_extracted_k141_68804 | Low-quality |
| S12_extracted_k141_123542 | Not-determined |
| S12_extracted_k141_76576 | Not-determined |
| S12_extracted_k141_155217 | Not-determined |
| S12_extracted_k141_101324 | Not-determined |
| S12_extracted_k141_163217 | Low-quality |
| S12_extracted_k141_155219 | Not-determined |
| S12_extracted_k141_171014 | Not-determined |
| S12_extracted_k141_147719 | Not-determined |
| S12_extracted_k141_163218 | Not-determined |
| S12_extracted_k141_76579 | Not-determined |
| S12_extracted_k141_131508 | Not-determined |
| S12_extracted_k141_6312 | Not-determined |
| S12_extracted_k141_147723 | Not-determined |
| S12_extracted_k141_36777 | Low-quality |
| S12_extracted_k141_123546 | Not-determined |
| S12_extracted_k141_76582 | Low-quality |
| S12_extracted_k141_101330 | Not-determined |
| S12_extracted_k141_68814 | Not-determined |
| S12_extracted_k141_187499 | Not-determined |
| S12_extracted_k141_123552 | Not-determined |
| S12_extracted_k141_84346 | Low-quality |
| S12_extracted_k141_12969 | Not-determined |
| S12_extracted_k141_84348 | Not-determined |
| S12_extracted_k141_12971 | Not-determined |
| S12_extracted_k141_76585 | Not-determined |
| S12_extracted_k141_140213 | Not-determined |
| S12_extracted_k141_84354 | Not-determined |
| S12_extracted_k141_147734 | Not-determined |
| S12_extracted_k141_131522 | Low-quality |
| S12_extracted_k141_147736 | Not-determined |
| S12_extracted_k141_84357 | Not-determined |
| S12_extracted_k141_61085 | Not-determined |
| S12_extracted_k141_68818 | Not-determined |
| S12_extracted_k141_171031 | Not-determined |
| S12_extracted_k141_163225 | Not-determined |
| S12_extracted_k141_52840 | Not-determined |
| S12_extracted_k141_6322 | Not-determined |
| S12_extracted_k141_21515 | Low-quality |
| S12_extracted_k141_29482 | Not-determined |
| S12_extracted_k141_46088 | Not-determined |
| S12_extracted_k141_140218 | Low-quality |
| S12_extracted_k141_187513 | Not-determined |
| S12_extracted_k141_29487 | Not-determined |
| S12_extracted_k141_163230 | Not-determined |
| S12_extracted_k141_101353 | Not-determined |
| S12_extracted_k141_21521 | Not-determined |
| S12_extracted_k141_163233 | Not-determined |
| S12_extracted_k141_6329 | Not-determined |
| S12_extracted_k141_163234 | Not-determined |
| S12_extracted_k141_52848 | Not-determined |
| S12_extracted_k141_29493 | Not-determined |
| S12_extracted_k141_163236 | Not-determined |
| S12_extracted_k141_131533 | Not-determined |
| S12_extracted_k141_163240 | Not-determined |
| S12_extracted_k141_76598 | Not-determined |
| S12_extracted_k141_187520 | Not-determined |
| S12_extracted_k141_147746 | Low-quality |
| S12_extracted_k141_187521 | Not-determined |
| S12_extracted_k141_29500 | Not-determined |
| S12_extracted_k141_52855 | Not-determined |
| S12_extracted_k141_52856 | Not-determined |
| S12_extracted_k141_92893 | Not-determined |
| S12_extracted_k141_108734 | Not-determined |
| S12_extracted_k141_140231 | Not-determined |
| S12_extracted_k141_115682 | Low-quality |
| S12_extracted_k141_115683 | Not-determined |
| S12_extracted_k141_6339 | Low-quality |
| S12_extracted_k141_101359 | Not-determined |
| S12_extracted_k141_147752 | Not-determined |
| S12_extracted_k141_147753 | Not-determined |
| S12_extracted_k141_131550 | Low-quality |
| S12_extracted_k141_163249 | Not-determined |
| S12_extracted_k141_140235 | Not-determined |
| S12_extracted_k141_52861 | Not-determined |
| S12_extracted_k141_92898 | Low-quality |
| S12_extracted_k141_21536 | Not-determined |
| S12_extracted_k141_140238 | Not-determined |
| S12_extracted_k141_76612 | Not-determined |
| S12_extracted_k141_178625 | Low-quality |
| S12_extracted_k141_140241 | Not-determined |
| S12_extracted_k141_76614 | Not-determined |
| S12_extracted_k141_61118 | Not-determined |
| S12_extracted_k141_12995 | Not-determined |
| S12_extracted_k141_76615 | Not-determined |
| S12_extracted_k141_163257 | Not-determined |
| S12_extracted_k141_52867 | Not-determined |
| S12_extracted_k141_46113 | Not-determined |
| S12_extracted_k141_52869 | Not-determined |
| S12_extracted_k141_46116 | Not-determined |
| S12_extracted_k141_155239 | Low-quality |
| S12_extracted_k141_187537 | Not-determined |
| S12_extracted_k141_61127 | Not-determined |
| S12_extracted_k141_187538 | Not-determined |
| S12_extracted_k141_52873 | Not-determined |
| S12_extracted_k141_147776 | Not-determined |
| S12_extracted_k141_140253 | Not-determined |
| S12_extracted_k141_52877 | Not-determined |
| S12_extracted_k141_84389 | Not-determined |
| S12_extracted_k141_68846 | Not-determined |
| S12_extracted_k141_46123 | Not-determined |
| S12_extracted_k141_108760 | Not-determined |

| | |
|---|---|
| S12_extracted_k141_163267 | Not-determined |
| S12_extracted_k141_101379 | Low-quality |
| S12_extracted_k141_131569 | Low-quality |
| S12_extracted_k141_76626 | Not-determined |
| S12_extracted_k141_61138 | Not-determined |
| S12_extracted_k141_68851 | Not-determined |
| S12_extracted_k141_6360 | Low-quality |
| S12_extracted_k141_108767 | Not-determined |
| S12_extracted_k141_101387 | Not-determined |
| S12_extracted_k141_155256 | Not-determined |
| S12_extracted_k141_147781 | Not-determined |
| S12_extracted_k141_46129 | Not-determined |
| S12_extracted_k141_115704 | Low-quality |
| S12_extracted_k141_76632 | Not-determined |
| S12_extracted_k141_6364 | Not-determined |
| S12_extracted_k141_36830 | Not-determined |
| S12_extracted_k141_140264 | Low-quality |
| S12_extracted_k141_131577 | Not-determined |
| S12_extracted_k141_13022 | Not-determined |
| S12_extracted_k141_171056 | Not-determined |
| S12_extracted_k141_147786 | Not-determined |
| S12_extracted_k141_140266 | Not-determined |
| S12_extracted_k141_101397 | Not-determined |
| S12_extracted_k141_115711 | Not-determined |
| S12_extracted_k141_101399 | Not-determined |
| S12_extracted_k141_68859 | Not-determined |
| S12_extracted_k141_76641 | Not-determined |
| S12_extracted_k141_178641 | Not-determined |
| S12_extracted_k141_36832 | Not-determined |
| S12_extracted_k141_6373 | Low-quality |
| S12_extracted_k141_140273 | Low-quality |
| S12_extracted_k141_101406 | Not-determined |
| S12_extracted_k141_123574 | Low-quality |
| S12_extracted_k141_178644 | Not-determined |
| S12_extracted_k141_46143 | Not-determined |
| S12_extracted_k141_123576 | Not-determined |
| S12_extracted_k141_108788 | Not-determined |
| S12_extracted_k141_123580 | Not-determined |
| S12_extracted_k141_108793 | Not-determined |
| S12_extracted_k141_46145 | Not-determined |
| S12_extracted_k141_61148 | Not-determined |
| S12_extracted_k141_115719 | Not-determined |
| S12_extracted_k141_36839 | Not-determined |
| S12_extracted_k141_92937 | Not-determined |
| S12_extracted_k141_187558 | Not-determined |
| S12_extracted_k141_101415 | Not-determined |
| S12_extracted_k141_140282 | Not-determined |
| S12_extracted_k141_36840 | Not-determined |
| S12_extracted_k141_131594 | Not-determined |
| S12_extracted_k141_108799 | Not-determined |
| S12_extracted_k141_36841 | Not-determined |
| S12_extracted_k141_92940 | Low-quality |
| S12_extracted_k141_68873 | Not-determined |
| S12_extracted_k141_171070 | Not-determined |
| S12_extracted_k141_108800 | Not-determined |
| S12_extracted_k141_108801 | Not-determined |
| S12_extracted_k141_68874 | Not-determined |
| S12_extracted_k141_36844 | Not-determined |
| S12_extracted_k141_92943 | Not-determined |
| S12_extracted_k141_187563 | Not-determined |
| S12_extracted_k141_61158 | Not-determined |
| S12_extracted_k141_187566 | Not-determined |
| S12_extracted_k141_108807 | Not-determined |
| S12_extracted_k141_140291 | Not-determined |
| S12_extracted_k141_187569 | Not-determined |
| S12_extracted_k141_68878 | Not-determined |
| S12_extracted_k141_68879 | Not-determined |
| S12_extracted_k141_171077 | Not-determined |
| S12_extracted_k141_92948 | Not-determined |
| S12_extracted_k141_115732 | Not-determined |
| S12_extracted_k141_36852 | Not-determined |
| S12_extracted_k141_108812 | Not-determined |
| S12_extracted_k141_155285 | Not-determined |
| S12_extracted_k141_101424 | Not-determined |
| S12_extracted_k141_68882 | Not-determined |
| S12_extracted_k141_171080 | Not-determined |
| S12_extracted_k141_131599 | Low-quality |
| S12_extracted_k141_46152 | Not-determined |
| S12_extracted_k141_101428 | Not-determined |
| S12_extracted_k141_171081 | Not-determined |
| S12_extracted_k141_123604 | Not-determined |
| S12_extracted_k141_68886 | Not-determined |
| S12_extracted_k141_131602 | Not-determined |
| S12_extracted_k141_13037 | Not-determined |
| S12_extracted_k141_178660 | Not-determined |
| S12_extracted_k141_13041 | Not-determined |
| S12_extracted_k141_187577 | Not-determined |
| S12_extracted_k141_108821 | Not-determined |
| S12_extracted_k141_123611 | Not-determined |
| S12_extracted_k141_108822 | Not-determined |
| S12_extracted_k141_155295 | Not-determined |
| S12_extracted_k141_92952 | Low-quality |
| S12_extracted_k141_187579 | Not-determined |
| S12_extracted_k141_46162 | Low-quality |
| S12_extracted_k141_187580 | Not-determined |
| S12_extracted_k141_101439 | Not-determined |
| S12_extracted_k141_187581 | Not-determined |
| S12_extracted_k141_29531 | Not-determined |
| S12_extracted_k141_92957 | Not-determined |
| S12_extracted_k141_13051 | Low-quality |
| S12_extracted_k141_163309 | Not-determined |
| S12_extracted_k141_36866 | Not-determined |
| S12_extracted_k141_178670 | Not-determined |
| S12_extracted_k141_13055 | Not-determined |
| S12_extracted_k141_140314 | Low-quality |
| S12_extracted_k141_36873 | Not-determined |
| S12_extracted_k141_140315 | Not-determined |
| S12_extracted_k141_21569 | Not-determined |
| S12_extracted_k141_36876 | Not-determined |
| S12_extracted_k141_36877 | Not-determined |
| S12_extracted_k141_163316 | Not-determined |
| S12_extracted_k141_29551 | Not-determined |
| S12_extracted_k141_36881 | Not-determined |
| S12_extracted_k141_115753 | Not-determined |
| S12_extracted_k141_36883 | Not-determined |
| S12_extracted_k141_187595 | Not-determined |
| S12_extracted_k141_147823 | Not-determined |
| S12_extracted_k141_101454 | Not-determined |
| S12_extracted_k141_178680 | Not-determined |
| S12_extracted_k141_108834 | Low-quality |
| S12_extracted_k141_101455 | Not-determined |
| S12_extracted_k141_178682 | Not-determined |
| S12_extracted_k141_155309 | Not-determined |
| S12_extracted_k141_29555 | Low-quality |
| S12_extracted_k141_92981 | Not-determined |
| S12_extracted_k141_13071 | Not-determined |
| S12_extracted_k141_115760 | Not-determined |
| S12_extracted_k141_84402 | Not-determined |
| S12_extracted_k141_140337 | Low-quality |
| S12_extracted_k141_171104 | Not-determined |
| S12_extracted_k141_84404 | Not-determined |
| S12_extracted_k141_36896 | Not-determined |
| S12_extracted_k141_131626 | Not-determined |
| S12_extracted_k141_147833 | Not-determined |
| S12_extracted_k141_115765 | Not-determined |
| S12_extracted_k141_171107 | Not-determined |
| S12_extracted_k141_36898 | Low-quality |
| S12_extracted_k141_187609 | Not-determined |
| S12_extracted_k141_155316 | Not-determined |
| S12_extracted_k141_68904 | Not-determined |
| S12_extracted_k141_29562 | Not-determined |
| S12_extracted_k141_46170 | Not-determined |
| S12_extracted_k141_76671 | Not-determined |
| S12_extracted_k141_171111 | Not-determined |
| S12_extracted_k141_52931 | Low-quality |
| S12_extracted_k141_13080 | Not-determined |
| S12_extracted_k141_46173 | Not-determined |
| S12_extracted_k141_13082 | Not-determined |
| S12_extracted_k141_115775 | Not-determined |
| S12_extracted_k141_140343 | Low-quality |
| S12_extracted_k141_29570 | Not-determined |
| S12_extracted_k141_171120 | Not-determined |
| S12_extracted_k141_187612 | Not-determined |
| S12_extracted_k141_46178 | Not-determined |
| S12_extracted_k141_29572 | Not-determined |
| S12_extracted_k141_108848 | Not-determined |
| S12_extracted_k141_92985 | Not-determined |
| S12_extracted_k141_52941 | Low-quality |
| S12_extracted_k141_61178 | Not-determined |
| S12_extracted_k141_187617 | Not-determined |
| S12_extracted_k141_84419 | Not-determined |
| S12_extracted_k141_171129 | Not-determined |
| S12_extracted_k141_140353 | Not-determined |
| S12_extracted_k141_101463 | Not-determined |
| S12_extracted_k141_171131 | Not-determined |
| S12_extracted_k141_155327 | Not-determined |
| S12_extracted_k141_84424 | Not-determined |
| S12_extracted_k141_140359 | Not-determined |
| S12_extracted_k141_29577 | Not-determined |
| S12_extracted_k141_29580 | Not-determined |
| S12_extracted_k141_76682 | Low-quality |
| S12_extracted_k141_108856 | Not-determined |
| S12_extracted_k141_115786 | Not-determined |
| S12_extracted_k141_178705 | Not-determined |
| S12_extracted_k141_178706 | Not-determined |
| S12_extracted_k141_108859 | Not-determined |
| S12_extracted_k141_108860 | Low-quality |
| S12_extracted_k141_84436 | Not-determined |
| S12_extracted_k141_29586 | Low-quality |
| S12_extracted_k141_115794 | Not-determined |
| S12_extracted_k141_61189 | Not-determined |
| S12_extracted_k141_36918 | Low-quality |
| S12_extracted_k141_187626 | Not-determined |
| S12_extracted_k141_187627 | Not-determined |
| S12_extracted_k141_178714 | Not-determined |
| S12_extracted_k141_178715 | Not-determined |
| S12_extracted_k141_108868 | Not-determined |
| S12_extracted_k141_140376 | Not-determined |
| S12_extracted_k141_187633 | Not-determined |
| S12_extracted_k141_68939 | Not-determined |
| S12_extracted_k141_178719 | Not-determined |
| S12_extracted_k141_163322 | Not-determined |
| S12_extracted_k141_171138 | Not-determined |
| S12_extracted_k141_61193 | Not-determined |
| S12_extracted_k141_163329 | Not-determined |
| S12_extracted_k141_29608 | Not-determined |
| S12_extracted_k141_187644 | Not-determined |
| S12_extracted_k141_178728 | Not-determined |
| S12_extracted_k141_147849 | Not-determined |
| S12_extracted_k141_163337 | Not-determined |
| S12_extracted_k141_93007 | Not-determined |
| S12_extracted_k141_101485 | Not-determined |
| S12_extracted_k141_13122 | Not-determined |
| S12_extracted_k141_187647 | Not-determined |
| S12_extracted_k141_61205 | Not-determined |
| S12_extracted_k141_101488 | Not-determined |
| S12_extracted_k141_171153 | Not-determined |
| S12_extracted_k141_68957 | Low-quality |
| S12_extracted_k141_178737 | Not-determined |
| S12_extracted_k141_178743 | Not-determined |
| S12_extracted_k141_115824 | Not-determined |
| S12_extracted_k141_46184 | Not-determined |
| S12_extracted_k141_68960 | Not-determined |
| S12_extracted_k141_46186 | Not-determined |
| S12_extracted_k141_68962 | Not-determined |
| S12_extracted_k141_76727 | Not-determined |
| S12_extracted_k141_76728 | Not-determined |
| S12_extracted_k141_101495 | Not-determined |
| S12_extracted_k141_187668 | Not-determined |
| S12_extracted_k141_178750 | Low-quality |
| S12_extracted_k141_101499 | Not-determined |
| S12_extracted_k141_21574 | Low-quality |
| S12_extracted_k141_115833 | Not-determined |
| S12_extracted_k141_140402 | Not-determined |
| S12_extracted_k141_147867 | Not-determined |
| S12_extracted_k141_61230 | Not-determined |
| S12_extracted_k141_46196 | Not-determined |
| S12_extracted_k141_76730 | Not-determined |
| S12_extracted_k141_115834 | Low-quality |
| S12_extracted_k141_93029 | Not-determined |
| S12_extracted_k141_163348 | Not-determined |
| S12_extracted_k141_140408 | Not-determined |
| S12_extracted_k141_131651 | Not-determined |
| S12_extracted_k141_13145 | Not-determined |
| S12_extracted_k141_140409 | Not-determined |
| S12_extracted_k141_76737 | Not-determined |
| S12_extracted_k141_140412 | Not-determined |
| S12_extracted_k141_68974 | Not-determined |
| S12_extracted_k141_140414 | Low-quality |
| S12_extracted_k141_131656 | Not-determined |
| S12_extracted_k141_46205 | Not-determined |
| S12_extracted_k141_131658 | Not-determined |
| S12_extracted_k141_101509 | Not-determined |
| S12_extracted_k141_187682 | Not-determined |
| S12_extracted_k141_131660 | Not-determined |
| S12_extracted_k141_163359 | Not-determined |
| S12_extracted_k141_171159 | Not-determined |
| S12_extracted_k141_93036 | Not-determined |
| S12_extracted_k141_93037 | Not-determined |
| S12_extracted_k141_13146 | Not-determined |
| S12_extracted_k141_36945 | Not-determined |
| S12_extracted_k141_178764 | Not-determined |
| S12_extracted_k141_21582 | Not-determined |
| S12_extracted_k141_131668 | Not-determined |
| S12_extracted_k141_52949 | Not-determined |
| S12_extracted_k141_163364 | Not-determined |
| S12_extracted_k141_101516 | Not-determined |
| S12_extracted_k141_131671 | Not-determined |
| S12_extracted_k141_163367 | Not-determined |
| S12_extracted_k141_13155 | Not-determined |
| S12_extracted_k141_29627 | Not-determined |
| S12_extracted_k141_68985 | Not-determined |
| S12_extracted_k141_93048 | Not-determined |
| S12_extracted_k141_101520 | Not-determined |
| S12_extracted_k141_171175 | Not-determined |
| S12_extracted_k141_29633 | Low-quality |
| S12_extracted_k141_115848 | Not-determined |
| S12_extracted_k141_131684 | Not-determined |
| S12_extracted_k141_52961 | Not-determined |
| S12_extracted_k141_93052 | Not-determined |
| S12_extracted_k141_163374 | Not-determined |
| S12_extracted_k141_76747 | Not-determined |
| S12_extracted_k141_93054 | Not-determined |
| S12_extracted_k141_93055 | Not-determined |
| S12_extracted_k141_93057 | Not-determined |
| S12_extracted_k141_147884 | Not-determined |
| S12_extracted_k141_171183 | Not-determined |
| S12_extracted_k141_13173 | Not-determined |
| S12_extracted_k141_52975 | Not-determined |
| S12_extracted_k141_52976 | Not-determined |
| S12_extracted_k141_140428 | Not-determined |
| S12_extracted_k141_93071 | Not-determined |
| S12_extracted_k141_52978 | Low-quality |
| S12_extracted_k141_93073 | Not-determined |
| S12_extracted_k141_101535 | Not-determined |
| S12_extracted_k141_147902 | Not-determined |
| S12_extracted_k141_76757 | Not-determined |
| S12_extracted_k141_171197 | Not-determined |
| S12_extracted_k141_69013 | Not-determined |
| S12_extracted_k141_76760 | Not-determined |
| S12_extracted_k141_187705 | Not-determined |
| S12_extracted_k141_69016 | Not-determined |
| S12_extracted_k141_115879 | Not-determined |
| S12_extracted_k141_61253 | Not-determined |
| S12_extracted_k141_13200 | Not-determined |
| S12_extracted_k141_171200 | Not-determined |
| S12_extracted_k141_163405 | Not-determined |
| S12_extracted_k141_13201 | Not-determined |
| S12_extracted_k141_171202 | Not-determined |
| S12_extracted_k141_131693 | Not-determined |
| S12_extracted_k141_13202 | Not-determined |
| S12_extracted_k141_61254 | Low-quality |
| S12_extracted_k141_171203 | Not-determined |
| S12_extracted_k141_171207 | Not-determined |
| S12_extracted_k141_36987 | Not-determined |
| S12_extracted_k141_140440 | Not-determined |
| S12_extracted_k141_13214 | Not-determined |
| S12_extracted_k141_101546 | Not-determined |
| S12_extracted_k141_93082 | Not-determined |
| S12_extracted_k141_36993 | Low-quality |
| S12_extracted_k141_36994 | Not-determined |
| S12_extracted_k141_171215 | Not-determined |
| S12_extracted_k141_115883 | Not-determined |
| S12_extracted_k141_171216 | Not-determined |
| S12_extracted_k141_171218 | Not-determined |
| S12_extracted_k141_147915 | Not-determined |
| S12_extracted_k141_131709 | Not-determined |
| S12_extracted_k141_37001 | Not-determined |
| S12_extracted_k141_53001 | Not-determined |
| S12_extracted_k141_37002 | Not-determined |
| S12_extracted_k141_140444 | Not-determined |
| S12_extracted_k141_140445 | Low-quality |
| S12_extracted_k141_147921 | Not-determined |
| S12_extracted_k141_61271 | Not-determined |
| S12_extracted_k141_13225 | Not-determined |
| S12_extracted_k141_6484 | Not-determined |
| S12_extracted_k141_46257 | Not-determined |
| S12_extracted_k141_6487 | Not-determined |
| S12_extracted_k141_131721 | Not-determined |
| S12_extracted_k141_187723 | Not-determined |
| S12_extracted_k141_37010 | Not-determined |
| S12_extracted_k141_13233 | Not-determined |
| S12_extracted_k141_187724 | Not-determined |
| S12_extracted_k141_37012 | Not-determined |
| S12_extracted_k141_171233 | Not-determined |
| S12_extracted_k141_171234 | Not-determined |
| S12_extracted_k141_171236 | Not-determined |
| S12_extracted_k141_147924 | Not-determined |
| S12_extracted_k141_61281 | Not-determined |
| S12_extracted_k141_108883 | Not-determined |
| S12_extracted_k141_76773 | Low-quality |
| S12_extracted_k141_76774 | Not-determined |
| S12_extracted_k141_69025 | Not-determined |
| S12_extracted_k141_69026 | Not-determined |
| S12_extracted_k141_131729 | Low-quality |
| S12_extracted_k141_187729 | Not-determined |
| S12_extracted_k141_69027 | Not-determined |
| S12_extracted_k141_108886 | Not-determined |
| S12_extracted_k141_61286 | Not-determined |
| S12_extracted_k141_61288 | Low-quality |
| S12_extracted_k141_140462 | Not-determined |
| S12_extracted_k141_187734 | Not-determined |
| S12_extracted_k141_131732 | Not-determined |
| S12_extracted_k141_101562 | Not-determined |
| S12_extracted_k141_123623 | Not-determined |
| S12_extracted_k141_46271 | Not-determined |
| S12_extracted_k141_13249 | Not-determined |
| S12_extracted_k141_76781 | Not-determined |
| S12_extracted_k141_115898 | Not-determined |
| S12_extracted_k141_13250 | Not-determined |
| S12_extracted_k141_187738 | Not-determined |
| S12_extracted_k141_101567 | Not-determined |
| S12_extracted_k141_37033 | Not-determined |
| S12_extracted_k141_37034 | Not-determined |
| S12_extracted_k141_76786 | Not-determined |
| S12_extracted_k141_178794 | Low-quality |
| S12_extracted_k141_76787 | Not-determined |
| S12_extracted_k141_53025 | Not-determined |
| S12_extracted_k141_69036 | Not-determined |
| S12_extracted_k141_123631 | Not-determined |
| S12_extracted_k141_93115 | Not-determined |
| S12_extracted_k141_53030 | Not-determined |
| S12_extracted_k141_29652 | Not-determined |
| S12_extracted_k141_93118 | Not-determined |
| S12_extracted_k141_131747 | Not-determined |
| S12_extracted_k141_163419 | Not-determined |
| S12_extracted_k141_147942 | Not-determined |
| S12_extracted_k141_108893 | Low-quality |
| S12_extracted_k141_101578 | Not-determined |
| S12_extracted_k141_29655 | Not-determined |
| S12_extracted_k141_6511 | Low-quality |
| S12_extracted_k141_131748 | Not-determined |
| S12_extracted_k141_101579 | Not-determined |
| S12_extracted_k141_13256 | Not-determined |
| S12_extracted_k141_108895 | Not-determined |
| S12_extracted_k141_155356 | Low-quality |
| S12_extracted_k141_53037 | Not-determined |
| S12_extracted_k141_29658 | Not-determined |
| S12_extracted_k141_53038 | Not-determined |
| S12_extracted_k141_131750 | Not-determined |
| S12_extracted_k141_53039 | Not-determined |
| S12_extracted_k141_6519 | Not-determined |
| S12_extracted_k141_140475 | Not-determined |
| S12_extracted_k141_115901 | Not-determined |
| S12_extracted_k141_163427 | Not-determined |
| S12_extracted_k141_108906 | Not-determined |
| S12_extracted_k141_123647 | Not-determined |
| S12_extracted_k141_93133 | Not-determined |
| S12_extracted_k141_21603 | Not-determined |
| S12_extracted_k141_53045 | Not-determined |
| S12_extracted_k141_29664 | Not-determined |
| S12_extracted_k141_147947 | Not-determined |
| S12_extracted_k141_115909 | Not-determined |
| S12_extracted_k141_131757 | Not-determined |
| S12_extracted_k141_140482 | Not-determined |
| S12_extracted_k141_147950 | Not-determined |
| S12_extracted_k141_6528 | Not-determined |
| S12_extracted_k141_163433 | Not-determined |
| S12_extracted_k141_115911 | Not-determined |
| S12_extracted_k141_123651 | Not-determined |
| S12_extracted_k141_53052 | Not-determined |
| S12_extracted_k141_147954 | Low-quality |
| S12_extracted_k141_29669 | Not-determined |
| S12_extracted_k141_21616 | Not-determined |
| S12_extracted_k141_93146 | Not-determined |
| S12_extracted_k141_147957 | Low-quality |
| S12_extracted_k141_13262 | Low-quality |
| S12_extracted_k141_13263 | Not-determined |
| S4_vs_VIRSorter_18803-circular-cat | Medium-quality |
| S12_extracted_k141_115915 | Not-determined |
| S12_extracted_k141_187761 | Not-determined |
| S12_extracted_k141_140486 | Not-determined |
| S12_extracted_k141_187762 | Not-determined |
| S12_extracted_k141_140488 | Not-determined |
| S12_extracted_k141_61304 | Not-determined |
| S12_extracted_k141_131771 | Not-determined |
| S12_extracted_k141_155379 | Not-determined |
| S12_extracted_k141_140489 | Not-determined |
| S12_extracted_k141_163441 | Not-determined |
| S12_extracted_k141_13270 | Not-determined |
| S12_extracted_k141_108917 | Not-determined |
| S12_extracted_k141_155381 | Not-determined |
| S12_extracted_k141_29679 | Not-determined |
| S12_extracted_k141_131772 | Not-determined |
| S12_extracted_k141_187766 | Not-determined |
| S12_extracted_k141_131774 | Not-determined |
| S12_extracted_k141_21624 | Not-determined |
| S12_extracted_k141_21629 | Not-determined |
| S12_extracted_k141_115924 | Not-determined |
| S12_extracted_k141_131778 | Not-determined |
| S12_extracted_k141_101600 | Low-quality |
| S12_extracted_k141_61315 | Not-determined |
| S12_extracted_k141_115927 | Not-determined |
| S12_extracted_k141_84500 | Not-determined |
| S12_extracted_k141_101602 | Low-quality |
| S12_extracted_k141_115930 | Low-quality |
| S12_extracted_k141_140498 | Not-determined |
| S12_extracted_k141_37053 | Not-determined |
| S12_extracted_k141_187770 | Not-determined |
| S12_extracted_k141_46285 | Not-determined |
| S12_extracted_k141_108927 | Not-determined |
| S12_extracted_k141_61319 | Not-determined |
| S12_extracted_k141_46287 | Not-determined |
| S12_extracted_k141_84511 | Low-quality |
| S12_extracted_k141_171264 | Low-quality |
| S12_extracted_k141_171265 | Not-determined |
| S12_extracted_k141_93160 | Not-determined |
| S12_extracted_k141_21637 | Not-determined |
| S12_extracted_k141_93161 | Not-determined |
| S12_extracted_k141_29698 | Not-determined |
| S12_extracted_k141_61325 | Not-determined |
| S12_extracted_k141_171269 | Not-determined |
| S12_extracted_k141_163457 | Not-determined |
| S12_extracted_k141_6532 | Low-quality |
| S12_extracted_k141_37068 | Low-quality |
| S12_extracted_k141_37069 | Not-determined |
| S12_extracted_k141_6533 | Not-determined |
| S12_extracted_k141_93169 | Not-determined |
| S12_extracted_k141_115942 | Not-determined |
| S12_extracted_k141_6537 | Not-determined |
| S12_extracted_k141_21648 | Not-determined |
| S12_extracted_k141_147964 | Low-quality |
| S12_extracted_k141_147965 | Not-determined |
| S12_extracted_k141_163466 | Not-determined |
| S12_extracted_k141_46305 | Not-determined |
| S12_extracted_k141_46306 | Not-determined |
| S12_extracted_k141_61331 | Not-determined |
| S12_extracted_k141_13278 | Not-determined |
| S12_extracted_k141_93177 | Not-determined |
| S12_extracted_k141_123677 | Not-determined |
| S12_extracted_k141_21658 | Not-determined |
| S12_extracted_k141_108937 | Not-determined |
| S12_extracted_k141_37082 | Not-determined |
| S12_extracted_k141_76829 | Not-determined |
| S12_extracted_k141_115948 | Not-determined |
| S12_extracted_k141_171285 | Not-determined |
| S12_extracted_k141_76832 | Not-determined |
| S12_extracted_k141_21665 | Not-determined |
| S12_extracted_k141_147976 | Not-determined |
| S12_extracted_k141_29735 | Not-determined |
| S12_extracted_k141_76835 | Not-determined |
| S12_extracted_k141_6558 | Not-determined |
| S12_extracted_k141_115957 | Not-determined |
| S12_extracted_k141_21671 | Not-determined |
| S12_extracted_k141_53076 | Not-determined |
| S12_extracted_k141_101622 | Low-quality |
| S12_extracted_k141_108953 | Not-determined |
| S12_extracted_k141_37095 | Not-determined |
| S12_extracted_k141_147985 | Not-determined |
| S12_extracted_k141_187777 | Not-determined |
| S12_extracted_k141_37096 | Not-determined |
| S12_extracted_k141_46318 | Not-determined |
| S12_extracted_k141_37097 | Not-determined |
| S12_extracted_k141_76840 | Not-determined |
| S12_extracted_k141_123701 | Not-determined |
| S12_extracted_k141_93200 | Not-determined |
| S12_extracted_k141_147991 | Not-determined |
| S12_extracted_k141_147992 | Not-determined |
| S12_extracted_k141_108960 | Not-determined |
| S12_extracted_k141_29746 | Not-determined |
| S12_extracted_k141_101630 | Not-determined |
| S12_extracted_k141_155444 | Not-determined |
| S12_extracted_k141_69069 | Not-determined |
| S12_extracted_k141_76848 | Not-determined |
| S12_extracted_k141_187785 | Not-determined |
| S12_extracted_k141_155445 | Not-determined |
| S12_extracted_k141_101634 | Not-determined |
| S12_extracted_k141_21683 | Not-determined |
| S12_extracted_k141_155446 | Not-determined |
| S12_extracted_k141_155447 | Not-determined |
| S12_extracted_k141_148004 | Not-determined |
| S12_extracted_k141_101640 | Not-determined |
| S12_extracted_k141_123711 | Not-determined |
| S12_extracted_k141_53086 | Not-determined |
| S12_extracted_k141_76853 | Not-determined |
| S12_extracted_k141_53091 | Not-determined |
| S12_extracted_k141_187794 | Not-determined |
| S12_extracted_k141_69081 | Not-determined |
| S12_extracted_k141_76859 | Not-determined |
| S12_extracted_k141_148012 | Not-determined |
| S12_extracted_k141_115969 | Not-determined |
| S12_extracted_k141_76860 | Not-determined |
| S12_extracted_k141_163473 | Not-determined |
| S12_extracted_k141_93220 | Not-determined |
| S12_extracted_k141_53096 | Not-determined |
| S12_extracted_k141_37103 | Not-determined |
| S12_extracted_k141_123718 | Not-determined |
| S12_extracted_k141_37104 | Not-determined |
| S12_extracted_k141_37105 | Not-determined |
| S12_extracted_k141_123720 | Not-determined |
| S12_extracted_k141_131810 | Not-determined |
| S12_extracted_k141_53102 | Not-determined |
| S12_extracted_k141_108981 | Not-determined |
| S12_extracted_k141_123726 | Not-determined |
| S12_extracted_k141_6565 | Not-determined |
| S12_extracted_k141_37113 | Not-determined |
| S12_extracted_k141_93224 | Not-determined |
| S12_extracted_k141_53110 | Not-determined |
| S12_extracted_k141_108986 | Not-determined |
| S12_extracted_k141_61369 | Not-determined |
| S12_extracted_k141_53111 | Not-determined |
| S12_extracted_k141_187805 | Not-determined |
| S12_extracted_k141_69102 | Not-determined |
| S12_extracted_k141_187806 | Not-determined |
| S12_extracted_k141_6572 | Not-determined |
| S12_extracted_k141_46327 | Not-determined |
| S12_extracted_k141_84553 | Not-determined |
| S12_extracted_k141_6576 | Not-determined |
| S12_extracted_k141_29783 | Not-determined |
| S12_extracted_k141_37120 | Not-determined |
| S12_extracted_k141_131827 | Not-determined |
| S12_extracted_k141_69109 | Not-determined |
| S12_extracted_k141_148024 | Not-determined |
| S12_extracted_k141_131835 | Not-determined |
| S12_extracted_k141_93236 | Not-determined |
| S12_extracted_k141_178806 | Not-determined |
| S12_extracted_k141_131838 | Not-determined |
| S12_extracted_k141_76886 | Not-determined |
| S12_extracted_k141_84562 | Not-determined |
| S12_extracted_k141_187829 | Not-determined |
| S12_extracted_k141_108999 | Not-determined |
| S12_extracted_k141_155477 | Low-quality |
| S12_extracted_k141_123744 | Not-determined |
| S12_extracted_k141_163488 | Not-determined |
| S12_extracted_k141_123746 | Not-determined |
| S12_extracted_k141_163489 | Low-quality |
| S12_extracted_k141_109002 | Not-determined |
| S12_extracted_k141_46345 | Not-determined |
| S12_extracted_k141_37124 | Not-determined |
| S12_extracted_k141_155487 | Not-determined |
| S12_extracted_k141_93253 | Not-determined |
| S12_extracted_k141_93256 | Not-determined |
| S12_extracted_k141_61382 | Not-determined |
| S12_extracted_k141_6589 | Not-determined |
| S12_extracted_k141_6591 | Not-determined |
| S12_extracted_k141_155494 | Low-quality |
| S12_extracted_k141_6593 | Not-determined |
| S12_extracted_k141_6594 | Not-determined |
| S12_extracted_k141_187842 | Not-determined |

| | |
|---|---|
| S12_extracted_k141_148032 | Low-quality |
| S12_extracted_k141_109014 | Not-determined |
| S12_extracted_k141_178822 | Not-determined |
| S12_extracted_k141_61388 | Not-determined |
| S12_extracted_k141_61391 | Not-determined |
| S12_extracted_k141_101682 | Not-determined |
| S12_extracted_k141_76898 | Low-quality |
| S12_extracted_k141_155504 | Not-determined |
| S12_extracted_k141_163502 | Not-determined |
| S12_extracted_k141_178828 | Not-determined |
| S12_extracted_k141_76900 | Low-quality |
| S12_extracted_k141_187847 | Not-determined |
| S12_extracted_k141_37132 | Not-determined |
| S12_extracted_k141_148036 | Not-determined |
| S12_extracted_k141_84579 | Not-determined |
| S12_extracted_k141_148037 | Not-determined |
| S12_extracted_k141_148038 | Low-quality |
| S12_extracted_k141_37135 | Not-determined |
| S12_extracted_k141_6605 | Not-determined |
| S12_extracted_k141_29817 | Not-determined |
| S12_extracted_k141_131856 | Not-determined |
| S12_extracted_k141_163509 | Not-determined |
| S12_extracted_k141_29818 | Not-determined |
| S12_extracted_k141_101688 | Not-determined |
| S12_extracted_k141_29820 | Not-determined |
| S12_extracted_k141_53126 | Low-quality |
| S12_extracted_k141_115981 | Not-determined |
| S12_extracted_k141_84584 | Not-determined |
| S12_extracted_k141_84585 | Low-quality |
| S12_extracted_k141_187859 | Low-quality |
| S12_extracted_k141_131859 | Not-determined |
| S12_extracted_k141_76908 | Not-determined |
| S12_extracted_k141_178833 | Not-determined |
| S12_extracted_k141_93268 | Not-determined |
| S12_extracted_k141_123768 | Not-determined |
| S12_extracted_k141_163520 | Not-determined |
| S12_extracted_k141_61403 | Not-determined |
| S12_extracted_k141_29827 | Not-determined |
| S12_extracted_k141_131864 | Not-determined |
| S12_extracted_k141_187867 | Not-determined |
| S12_extracted_k141_84590 | Not-determined |
| S12_extracted_k141_178838 | Not-determined |
| S12_extracted_k141_148056 | Not-determined |
| S12_extracted_k141_109041 | Not-determined |
| S12_extracted_k141_163523 | Not-determined |
| S12_extracted_k141_61407 | Not-determined |
| S12_extracted_k141_178843 | Low-quality |
| S12_extracted_k141_76912 | Not-determined |
| S12_extracted_k141_61408 | Not-determined |
| S12_extracted_k141_76913 | Not-determined |
| S12_extracted_k141_46356 | Not-determined |
| S12_extracted_k141_101704 | Low-quality |
| S12_extracted_k141_61416 | Not-determined |
| S12_extracted_k141_140544 | Not-determined |
| S12_extracted_k141_163532 | Not-determined |
| S12_extracted_k141_163533 | Low-quality |
| S12_extracted_k141_163537 | Not-determined |
| S12_extracted_k141_163538 | Not-determined |
| S12_extracted_k141_148062 | Not-determined |
| S12_extracted_k141_76923 | Not-determined |
| S12_extracted_k141_84606 | Not-determined |
| S12_extracted_k141_13311 | Not-determined |
| S12_extracted_k141_171343 | Low-quality |
| S12_extracted_k141_29840 | Not-determined |
| S12_extracted_k141_6624 | Not-determined |
| S12_extracted_k141_101715 | Not-determined |
| S12_extracted_k141_53149 | Not-determined |
| S12_extracted_k141_76933 | Not-determined |
| S12_extracted_k141_84616 | Not-determined |
| S12_extracted_k141_76934 | Not-determined |
| S12_extracted_k141_171351 | Not-determined |
| S12_extracted_k141_76937 | Not-determined |
| S12_extracted_k141_109044 | Not-determined |
| S12_extracted_k141_76940 | Not-determined |
| S12_extracted_k141_109045 | Not-determined |
| S12_extracted_k141_84623 | Not-determined |
| S12_extracted_k141_13325 | Not-determined |
| S12_extracted_k141_13326 | Not-determined |
| S12_extracted_k141_93276 | Low-quality |
| S12_extracted_k141_37148 | Not-determined |
| S12_extracted_k141_61427 | Not-determined |
| S12_extracted_k141_37149 | Not-determined |
| S12_extracted_k141_53157 | Low-quality |
| S12_extracted_k141_123786 | Not-determined |
| S12_extracted_k141_61432 | Not-determined |
| S12_extracted_k141_109050 | Low-quality |
| S12_extracted_k141_53164 | Not-determined |
| S7_extracted_29196 | Medium-quality |
| S12_extracted_k141_61439 | Not-determined |
| S12_extracted_k141_131897 | Not-determined |
| S12_extracted_k141_187883 | Low-quality |
| S12_extracted_k141_61441 | Not-determined |
| S12_extracted_k141_116019 | Not-determined |
| S12_extracted_k141_37159 | Not-determined |
| S12_extracted_k141_131903 | Not-determined |
| S12_extracted_k141_163572 | Not-determined |
| S12_extracted_k141_101719 | Not-determined |
| S12_extracted_k141_101720 | Not-determined |
| S12_extracted_k141_155507 | Not-determined |
| S7_vs_29196\|\|full | Medium-quality |
| S12_extracted_k141_37170 | Not-determined |
| S12_extracted_k141_155510 | Not-determined |
| S12_extracted_k141_61448 | Not-determined |
| S12_extracted_k141_123805 | Not-determined |
| S12_extracted_k141_140576 | Not-determined |
| S12_extracted_k141_101725 | Not-determined |
| S12_extracted_k141_6639 | Not-determined |
| S12_extracted_k141_76960 | Not-determined |
| S12_extracted_k141_123813 | Not-determined |
| S12_extracted_k141_187894 | Not-determined |
| S12_extracted_k141_6642 | Not-determined |
| S1_extracted_k141_28398 | Medium-quality |
| S12_extracted_k141_93306 | Low-quality |
| S12_extracted_k141_187900 | Low-quality |
| S12_extracted_k141_163582 | Not-determined |
| S12_extracted_k141_69159 | Not-determined |
| S12_extracted_k141_116022 | Not-determined |
| S12_extracted_k141_116023 | Not-determined |
| S12_extracted_k141_109079 | Not-determined |
| S12_extracted_k141_171357 | Not-determined |
| S12_extracted_k141_61467 | Low-quality |
| S12_extracted_k141_131918 | Not-determined |
| S12_extracted_k141_101739 | Not-determined |
| S12_extracted_k141_171363 | Not-determined |
| S12_extracted_k141_171364 | Not-determined |
| S12_extracted_k141_76980 | Not-determined |
| S12_extracted_k141_46362 | Not-determined |
| S12_extracted_k141_101743 | Not-determined |
| S12_extracted_k141_84636 | Not-determined |
| S12_extracted_k141_163592 | Not-determined |
| S12_extracted_k141_61474 | Not-determined |
| S12_extracted_k141_46365 | Not-determined |
| S12_extracted_k141_46366 | Not-determined |
| S12_extracted_k141_116037 | Not-determined |
| S12_extracted_k141_13364 | Low-quality |
| S12_extracted_k141_13365 | Not-determined |
| S12_extracted_k141_6664 | Not-determined |
| S12_extracted_k141_61477 | Not-determined |
| S12_extracted_k141_76989 | Not-determined |
| S12_extracted_k141_13366 | Not-determined |
| S12_extracted_k141_101751 | Not-determined |
| S12_extracted_k141_76991 | Not-determined |
| S12_extracted_k141_84641 | Not-determined |
| S12_extracted_k141_163599 | Not-determined |
| S12_extracted_k141_53204 | Low-quality |
| S12_extracted_k141_13370 | Not-determined |
| S12_extracted_k141_123821 | Not-determined |
| S12_extracted_k141_69175 | Not-determined |
| S12_extracted_k141_140595 | Not-determined |
| S12_extracted_k141_69178 | Not-determined |
| S12_extracted_k141_148065 | Low-quality |
| S12_extracted_k141_163609 | Not-determined |
| S12_extracted_k141_37190 | Not-determined |
| S12_extracted_k141_84647 | Not-determined |
| S12_extracted_k141_53210 | Not-determined |
| S12_extracted_k141_61497 | Not-determined |
| S12_extracted_k141_123827 | Not-determined |
| S12_extracted_k141_140600 | Not-determined |
| S12_extracted_k141_13379 | Low-quality |
| S12_extracted_k141_123830 | Not-determined |
| S12_extracted_k141_140601 | Not-determined |
| S12_extracted_k141_61502 | Not-determined |
| S12_extracted_k141_171381 | Not-determined |
| S12_extracted_k141_109100 | Low-quality |
| S12_extracted_k141_84652 | Not-determined |
| S12_extracted_k141_77009 | Not-determined |
| S12_extracted_k141_13382 | Not-determined |
| S12_extracted_k141_6685 | Not-determined |
| S12_extracted_k141_29867 | Low-quality |
| S12_extracted_k141_46384 | Not-determined |
| S12_extracted_k141_29868 | Not-determined |
| S12_extracted_k141_84654 | Not-determined |
| S12_extracted_k141_61509 | Not-determined |
| S12_extracted_k141_61510 | Not-determined |
| S12_extracted_k141_46388 | Not-determined |
| S12_extracted_k141_61511 | Not-determined |
| S12_extracted_k141_123841 | Not-determined |
| S12_extracted_k141_93333 | Not-determined |
| S12_extracted_k141_123844 | Not-determined |
| S12_extracted_k141_131943 | Not-determined |
| S12_extracted_k141_77019 | Not-determined |
| S12_extracted_k141_37208 | Not-determined |
| S12_extracted_k141_116071 | Not-determined |
| S12_extracted_k141_178861 | Not-determined |
| S12_extracted_k141_77020 | Not-determined |
| S12_extracted_k141_163637 | Not-determined |
| S12_extracted_k141_6688 | Not-determined |
| S12_extracted_k141_13391 | Not-determined |
| S12_extracted_k141_116072 | Not-determined |
| S12_extracted_k141_6689 | Not-determined |
| S12_extracted_k141_93337 | Not-determined |
| S12_extracted_k141_131949 | Not-determined |
| S12_extracted_k141_13394 | Not-determined |
| S12_extracted_k141_178866 | Not-determined |
| S12_extracted_k141_77026 | Not-determined |
| S12_extracted_k141_178867 | Not-determined |
| S12_extracted_k141_123847 | Low-quality |
| S12_extracted_k141_101763 | Not-determined |
| S12_extracted_k141_77029 | Not-determined |
| S12_extracted_k141_37213 | Not-determined |
| S12_extracted_k141_109109 | Not-determined |
| S12_extracted_k141_46399 | Not-determined |
| S12_extracted_k141_163643 | Not-determined |
| S12_extracted_k141_77031 | Not-determined |
| S12_extracted_k141_6695 | Not-determined |
| S12_extracted_k141_84675 | Not-determined |
| S12_extracted_k141_131954 | Not-determined |
| S12_extracted_k141_37221 | Not-determined |
| S12_extracted_k141_101767 | Not-determined |
| S12_extracted_k141_69201 | Not-determined |
| S12_extracted_k141_53231 | Not-determined |
| S12_extracted_k141_84678 | Not-determined |
| S12_extracted_k141_29881 | Not-determined |
| S12_extracted_k141_116089 | Not-determined |
| S12_extracted_k141_163652 | Not-determined |
| S12_extracted_k141_37229 | Not-determined |
| S12_extracted_k141_84684 | Not-determined |
| S12_extracted_k141_29886 | Not-determined |
| S12_extracted_k141_29888 | Not-determined |
| S12_extracted_k141_13415 | Not-determined |
| S12_extracted_k141_109128 | Low-quality |
| S12_extracted_k141_61532 | Not-determined |
| S12_extracted_k141_178881 | Not-determined |
| S12_extracted_k141_37233 | Not-determined |
| S12_extracted_k141_123868 | Not-determined |
| S12_extracted_k141_13420 | Not-determined |
| S12_extracted_k141_93354 | Not-determined |
| S12_extracted_k141_53248 | Not-determined |
| S12_extracted_k141_123871 | Not-determined |
| S12_extracted_k141_13423 | Not-determined |
| S12_extracted_k141_140609 | Not-determined |
| S12_extracted_k141_61542 | Not-determined |
| S12_extracted_k141_123878 | Not-determined |
| S12_extracted_k141_109140 | Not-determined |
| S12_extracted_k141_155527 | Not-determined |
| S12_extracted_k141_21692 | Not-determined |
| S12_extracted_k141_131984 | Not-determined |
| S12_extracted_k141_37248 | Not-determined |
| S12_extracted_k141_53257 | Not-determined |
| S12_extracted_k141_131986 | Not-determined |
| S12_extracted_k141_140621 | Low-quality |
| S12_extracted_k141_61556 | Low-quality |
| S12_extracted_k141_6708 | Not-determined |
| S12_extracted_k141_155531 | Not-determined |
| S12_extracted_k141_6709 | Not-determined |
| S12_extracted_k141_109150 | Not-determined |
| S12_extracted_k141_109152 | Not-determined |
| S12_extracted_k141_13438 | Not-determined |
| S12_extracted_k141_101782 | Not-determined |
| S12_extracted_k141_69224 | Not-determined |
| S12_extracted_k141_140629 | Not-determined |
| S12_extracted_k141_53269 | Not-determined |
| S12_extracted_k141_148104 | Not-determined |
| S12_extracted_k141_53273 | Low-quality |
| S12_extracted_k141_131997 | Not-determined |
| S12_extracted_k141_116116 | Not-determined |
| S12_extracted_k141_29910 | Not-determined |
| S12_extracted_k141_155538 | Not-determined |
| S12_extracted_k141_140636 | Not-determined |
| S12_extracted_k141_148109 | Not-determined |
| S12_extracted_k141_109162 | Not-determined |
| S12_extracted_k141_6727 | Not-determined |
| S12_extracted_k141_37258 | Not-determined |
| S12_extracted_k141_69240 | Not-determined |
| S12_extracted_k141_163674 | Not-determined |
| S12_extracted_k141_46424 | Not-determined |
| S12_extracted_k141_37264 | Not-determined |
| S12_extracted_k141_37265 | Low-quality |
| S12_extracted_k141_21709 | Not-determined |
| S12_extracted_k141_178906 | Low-quality |
| S12_extracted_k141_140647 | Not-determined |
| S12_extracted_k141_53287 | Not-determined |
| S12_extracted_k141_163678 | Not-determined |
| S12_extracted_k141_132006 | Not-determined |
| S12_extracted_k141_84709 | Not-determined |
| S12_extracted_k141_29920 | Not-determined |
| S12_extracted_k141_101788 | Not-determined |
| S12_extracted_k141_46434 | Not-determined |
| S12_extracted_k141_53290 | Not-determined |
| S12_extracted_k141_148120 | Not-determined |
| S12_extracted_k141_61575 | Not-determined |
| S12_extracted_k141_84711 | Not-determined |
| S12_extracted_k141_178912 | Not-determined |
| S12_extracted_k141_13452 | Not-determined |
| S12_extracted_k141_116127 | Not-determined |
| S12_extracted_k141_37270 | Not-determined |
| S12_extracted_k141_53295 | Not-determined |
| S12_extracted_k141_6735 | Not-determined |
| S12_extracted_k141_140655 | Not-determined |
| S12_extracted_k141_163686 | Not-determined |
| S12_extracted_k141_84717 | Not-determined |
| S12_extracted_k141_163688 | Not-determined |
| S12_extracted_k141_13456 | Low-quality |
| S12_extracted_k141_155553 | Not-determined |
| S12_extracted_k141_178918 | Not-determined |
| S12_extracted_k141_29927 | Not-determined |
| S12_extracted_k141_140662 | Not-determined |
| S12_extracted_k141_69255 | Low-quality |
| S12_extracted_k141_13463 | Not-determined |
| S12_extracted_k141_61582 | Not-determined |
| S12_extracted_k141_69257 | Not-determined |
| S12_extracted_k141_101798 | Not-determined |
| S12_extracted_k141_6739 | Not-determined |
| S12_extracted_k141_148133 | Not-determined |
| S12_extracted_k141_6741 | Not-determined |
| S12_extracted_k141_123896 | Not-determined |
| S12_extracted_k141_84728 | Not-determined |
| S12_extracted_k141_132016 | Not-determined |
| S12_extracted_k141_132020 | Low-quality |
| S12_extracted_k141_84734 | Not-determined |
| S12_extracted_k141_21724 | Not-determined |
| S12_extracted_k141_155571 | Not-determined |
| S12_extracted_k141_116145 | Not-determined |
| S12_extracted_k141_155578 | Not-determined |
| S12_extracted_k141_178929 | Not-determined |
| S12_extracted_k141_21729 | Not-determined |
| S12_extracted_k141_29945 | Not-determined |
| S12_extracted_k141_93361 | Not-determined |
| S12_extracted_k141_13473 | Not-determined |
| S12_extracted_k141_37272 | Not-determined |
| S12_extracted_k141_29946 | Not-determined |
| S12_extracted_k141_37274 | Low-quality |
| S12_extracted_k141_84739 | Not-determined |
| S12_extracted_k141_77039 | Low-quality |
| S12_extracted_k141_109188 | Not-determined |
| S12_extracted_k141_116154 | Not-determined |
| S6_vs_k141_49920\|\|full | Medium-quality |
| S12_extracted_k141_69262 | Not-determined |
| S12_extracted_k141_77043 | Not-determined |
| S12_extracted_k141_37280 | Not-determined |
| S12_extracted_k141_93364 | Not-determined |
| S12_extracted_k141_29950 | Not-determined |
| S12_extracted_k141_69265 | Not-determined |
| S12_extracted_k141_132024 | Not-determined |
| S12_extracted_k141_53324 | Not-determined |
| S12_extracted_k141_46465 | Low-quality |
| S12_extracted_k141_29955 | Not-determined |
| S12_extracted_k141_46466 | Not-determined |
| S12_extracted_k141_93370 | Low-quality |
| S12_extracted_k141_37285 | Not-determined |
| S12_extracted_k141_6763 | Low-quality |
| S12_extracted_k141_6765 | Not-determined |
| S12_extracted_k141_53327 | Not-determined |
| S12_extracted_k141_53328 | Not-determined |
| S12_extracted_k141_101829 | Not-determined |
| S12_extracted_k141_69276 | Not-determined |
| S12_extracted_k141_84743 | Not-determined |
| S12_extracted_k141_29963 | Not-determined |
| S12_extracted_k141_93376 | Not-determined |
| S12_extracted_k141_29965 | Not-determined |
| S12_extracted_k141_93377 | Not-determined |
| S12_extracted_k141_155584 | Low-quality |
| S12_extracted_k141_6769 | Not-determined |
| S12_extracted_k141_93378 | Not-determined |
| S12_extracted_k141_187950 | Not-determined |
| S12_extracted_k141_37290 | Not-determined |
| S12_extracted_k141_6771 | Not-determined |
| S12_extracted_k141_46481 | Low-quality |
| S12_extracted_k141_46483 | Not-determined |
| S12_extracted_k141_187952 | Not-determined |
| S12_extracted_k141_84747 | Not-determined |
| S12_extracted_k141_155594 | Not-determined |
| S12_extracted_k141_123917 | Not-determined |
| S12_extracted_k141_37296 | Not-determined |
| S12_extracted_k141_6775 | Low-quality |
| S12_extracted_k141_6776 | Low-quality |
| S12_extracted_k141_148150 | Not-determined |
| S12_extracted_k141_171397 | Not-determined |
| S12_extracted_k141_61616 | Not-determined |
| S12_extracted_k141_61618 | Not-determined |
| S12_extracted_k141_163712 | Not-determined |
| S12_extracted_k141_21734 | Not-determined |
| S12_extracted_k141_69289 | Not-determined |
| S12_extracted_k141_123920 | Not-determined |
| S12_extracted_k141_123924 | Not-determined |
| S12_extracted_k141_155605 | Not-determined |
| S12_extracted_k141_171409 | Not-determined |
| S12_extracted_k141_61621 | Low-quality |
| S12_extracted_k141_69294 | Not-determined |
| S12_extracted_k141_123926 | Not-determined |
| S12_extracted_k141_53343 | Not-determined |
| S12_extracted_k141_178951 | Not-determined |
| S12_extracted_k141_69299 | Not-determined |
| S12_extracted_k141_171420 | Not-determined |
| S12_extracted_k141_116166 | Not-determined |
| S12_extracted_k141_37305 | Not-determined |
| S12_extracted_k141_109207 | Not-determined |
| S12_extracted_k141_155614 | Not-determined |
| S12_extracted_k141_148165 | Not-determined |
| S12_extracted_k141_84769 | Not-determined |
| S12_extracted_k141_93397 | Low-quality |
| S12_extracted_k141_116170 | Not-determined |
| S12_extracted_k141_37309 | Not-determined |
| S12_extracted_k141_116171 | Not-determined |
| S12_extracted_k141_37311 | Not-determined |
| S12_extracted_k141_37312 | Not-determined |
| S12_extracted_k141_187986 | Not-determined |
| S12_extracted_k141_155617 | Not-determined |
| S12_extracted_k141_37314 | Not-determined |
| S12_extracted_k141_53360 | Not-determined |
| S12_extracted_k141_163733 | Not-determined |
| S12_extracted_k141_148175 | Not-determined |
| S12_extracted_k141_155624 | Not-determined |
| S12_extracted_k141_140710 | Not-determined |
| S12_extracted_k141_46510 | Not-determined |
| S12_extracted_k141_21749 | Not-determined |
| S12_extracted_k141_37318 | Low-quality |
| S12_extracted_k141_84779 | Not-determined |
| S12_extracted_k141_69302 | Not-determined |
| S12_extracted_k141_53369 | Not-determined |
| S12_extracted_k141_69303 | Not-determined |
| S12_extracted_k141_148183 | Not-determined |
| S12_extracted_k141_21755 | Not-determined |
| S12_extracted_k141_46517 | Not-determined |
| S12_extracted_k141_29998 | Not-determined |
| S12_extracted_k141_140717 | Not-determined |
| S12_extracted_k141_187995 | Not-determined |
| S12_extracted_k141_61654 | Not-determined |
| S12_extracted_k141_187998 | Not-determined |
| S12_extracted_k141_163745 | Not-determined |
| S12_extracted_k141_21765 | Not-determined |
| S12_extracted_k141_53375 | Low-quality |
| S12_extracted_k141_77102 | Not-determined |
| S12_extracted_k141_155631 | Not-determined |
| S12_extracted_k141_188000 | Low-quality |
| S12_extracted_k141_53381 | Not-determined |
| S12_extracted_k141_155635 | Not-determined |
| S12_extracted_k141_84792 | Not-determined |
| S12_extracted_k141_84793 | Not-determined |
| S12_extracted_k141_77110 | Not-determined |
| S12_extracted_k141_53382 | Not-determined |
| S12_extracted_k141_61659 | Not-determined |
| S12_extracted_k141_116190 | Not-determined |
| S12_extracted_k141_140731 | Not-determined |
| S12_extracted_k141_116192 | Not-determined |
| S12_extracted_k141_77114 | Not-determined |
| S12_extracted_k141_163752 | Not-determined |
| S12_extracted_k141_140736 | Not-determined |
| S12_extracted_k141_163754 | Not-determined |
| S12_extracted_k141_30014 | Not-determined |
| S12_extracted_k141_148201 | Not-determined |
| S12_extracted_k141_140739 | Not-determined |
| S12_extracted_k141_61666 | Not-determined |
| S12_extracted_k141_53388 | Not-determined |
| S12_extracted_k141_30017 | Not-determined |
| S12_extracted_k141_123955 | Not-determined |
| S12_extracted_k141_30018 | Not-determined |
| S12_extracted_k141_163756 | Not-determined |
| S12_extracted_k141_178963 | Not-determined |
| S12_extracted_k141_13537 | Not-determined |
| S12_extracted_k141_53393 | Not-determined |
| S12_extracted_k141_13538 | Low-quality |
| S12_extracted_k141_6782 | Not-determined |
| S12_extracted_k141_171451 | Not-determined |
| S12_extracted_k141_178967 | Not-determined |
| S12_extracted_k141_6784 | Not-determined |
| S12_extracted_k141_101848 | Not-determined |
| S12_extracted_k141_148213 | Not-determined |
| S12_extracted_k141_21776 | Not-determined |
| S12_extracted_k141_101851 | Not-determined |
| S12_extracted_k141_116204 | Not-determined |
| S12_extracted_k141_116205 | Low-quality |
| S12_extracted_k141_21779 | Low-quality |
| S12_extracted_k141_77130 | Not-determined |
| S12_extracted_k141_188016 | Not-determined |
| S12_extracted_k141_77132 | Not-determined |
| S12_extracted_k141_6794 | Not-determined |
| S12_extracted_k141_30030 | Not-determined |
| S12_extracted_k141_30031 | Not-determined |
| S12_extracted_k141_155643 | Not-determined |
| S12_extracted_k141_123976 | Not-determined |
| S12_extracted_k141_77141 | Not-determined |
| S12_extracted_k141_188023 | Not-determined |
| S12_extracted_k141_155650 | Low-quality |
| S12_extracted_k141_188024 | Not-determined |
| S12_extracted_k141_178985 | Not-determined |
| S12_extracted_k141_155652 | Not-determined |
| S12_extracted_k141_171470 | Not-determined |
| S12_extracted_k141_132053 | Not-determined |
| S12_extracted_k141_21796 | Not-determined |
| S12_extracted_k141_140763 | Not-determined |
| S12_extracted_k141_155660 | Not-determined |
| S12_extracted_k141_21798 | Not-determined |
| S12_extracted_k141_163767 | Not-determined |
| S12_extracted_k141_6804 | Not-determined |
| S12_extracted_k141_123983 | Not-determined |
| S12_extracted_k141_132057 | Not-determined |
| S12_extracted_k141_77150 | Not-determined |
| S12_extracted_k141_132058 | Not-determined |
| S12_extracted_k141_155664 | Not-determined |
| S12_extracted_k141_132059 | Low-quality |
| S12_extracted_k141_178997 | Not-determined |
| S12_extracted_k141_140766 | Low-quality |
| S12_extracted_k141_6810 | Not-determined |
| S12_extracted_k141_116208 | Not-determined |
| S12_extracted_k141_148231 | Not-determined |
| S12_extracted_k141_163777 | Not-determined |
| S12_extracted_k141_109244 | Not-determined |
| S12_extracted_k141_179002 | Not-determined |
| S12_extracted_k141_30055 | Low-quality |
| S12_extracted_k141_132065 | Not-determined |
| S12_extracted_k141_93433 | Not-determined |
| S12_extracted_k141_140777 | Not-determined |
| S12_extracted_k141_116214 | Not-determined |
| S12_extracted_k141_101862 | Not-determined |
| S12_extracted_k141_140778 | Not-determined |
| S12_extracted_k141_101864 | Not-determined |
| S12_extracted_k141_179004 | Not-determined |
| S12_extracted_k141_101865 | Low-quality |
| S12_extracted_k141_6822 | Not-determined |
| S12_extracted_k141_77163 | Not-determined |
| S12_extracted_k141_101866 | Not-determined |
| S12_extracted_k141_109250 | Low-quality |
| S12_extracted_k141_148241 | Low-quality |
| S12_extracted_k141_123988 | Not-determined |
| S12_extracted_k141_53424 | Low-quality |
| S12_extracted_k141_123989 | Not-determined |
| S12_extracted_k141_84812 | Not-determined |
| S12_extracted_k141_109251 | Not-determined |
| S12_extracted_k141_46554 | Not-determined |
| S12_extracted_k141_93446 | Low-quality |
| S12_extracted_k141_188050 | Not-determined |
| S12_extracted_k141_163782 | Low-quality |
| S12_extracted_k141_179012 | Low-quality |
| S12_extracted_k141_124002 | Not-determined |
| S12_extracted_k141_101872 | Low-quality |
| S12_extracted_k141_30071 | Not-determined |
| S12_extracted_k141_84821 | Low-quality |
| S12_extracted_k141_101876 | Low-quality |
| S12_extracted_k141_163789 | Not-determined |
| S12_extracted_k141_188059 | Not-determined |
| S12_extracted_k141_148248 | Not-determined |
| S12_extracted_k141_124014 | Not-determined |
| S12_extracted_k141_179017 | Not-determined |
| S12_extracted_k141_77173 | Not-determined |
| S12_extracted_k141_69350 | Not-determined |
| S12_extracted_k141_101886 | Not-determined |
| S12_extracted_k141_148255 | Not-determined |
| S12_extracted_k141_84828 | Not-determined |
| S12_extracted_k141_163795 | Not-determined |
| S12_extracted_k141_132091 | Not-determined |
| S12_extracted_k141_179024 | Not-determined |
| S12_extracted_k141_84833 | Not-determined |
| S12_extracted_k141_53446 | Not-determined |
| S12_extracted_k141_132093 | Not-determined |
| S12_extracted_k141_155676 | Low-quality |
| S12_extracted_k141_53447 | Not-determined |
| S12_extracted_k141_13576 | Not-determined |
| S12_extracted_k141_37363 | Not-determined |
| S12_extracted_k141_77191 | Low-quality |
| S12_extracted_k141_179026 | Not-determined |
| S12_extracted_k141_140804 | Not-determined |
| S12_extracted_k141_30096 | Not-determined |
| S12_extracted_k141_163808 | Not-determined |
| S12_extracted_k141_69366 | Not-determined |
| S12_extracted_k141_13589 | Not-determined |
| S12_extracted_k141_179029 | Not-determined |
| S12_extracted_k141_155679 | Not-determined |
| S12_extracted_k141_155680 | Not-determined |
| S12_extracted_k141_30102 | Not-determined |
| S12_extracted_k141_93475 | Not-determined |
| S12_extracted_k141_84841 | Not-determined |
| S12_extracted_k141_148268 | Not-determined |
| S12_extracted_k141_148269 | Not-determined |
| S12_extracted_k141_124039 | Not-determined |
| S12_extracted_k141_37376 | Not-determined |
| S12_extracted_k141_13598 | Not-determined |
| S12_extracted_k141_21839 | Not-determined |
| S12_extracted_k141_53454 | Low-quality |
| S12_extracted_k141_84848 | Not-determined |
| S12_extracted_k141_77203 | Not-determined |
| S12_extracted_k141_132103 | Not-determined |
| S12_extracted_k141_30108 | Not-determined |
| S12_extracted_k141_179043 | Not-determined |
| S12_extracted_k141_155687 | Not-determined |
| S12_extracted_k141_84851 | Not-determined |
| S12_extracted_k141_132104 | Not-determined |
| S12_extracted_k141_163818 | Not-determined |
| S12_extracted_k141_163820 | Not-determined |
| S12_extracted_k141_84855 | Not-determined |
| S12_extracted_k141_155692 | Not-determined |
| S12_extracted_k141_53458 | Not-determined |
| S12_extracted_k141_93501 | Not-determined |
| S12_extracted_k141_30117 | Not-determined |
| S12_extracted_k141_116243 | Not-determined |
| S12_extracted_k141_30121 | Low-quality |
| S12_extracted_k141_109273 | Not-determined |
| S12_extracted_k141_163831 | Not-determined |
| S12_extracted_k141_101935 | Low-quality |

| | |
|---|---|
| S12_extracted_k141_21852 | Low-quality |
| S12_extracted_k141_6839 | Not-determined |
| S12_extracted_k141_101941 | Not-determined |
| S12_extracted_k141_148276 | Low-quality |
| S12_extracted_k141_46613 | Not-determined |
| S12_extracted_k141_13625 | Not-determined |
| S12_extracted_k141_13626 | Not-determined |
| S12_extracted_k141_148277 | Not-determined |
| S12_extracted_k141_69398 | Not-determined |
| S12_extracted_k141_77223 | Not-determined |
| S12_extracted_k141_116250 | Not-determined |
| S12_extracted_k141_21855 | Not-determined |
| S12_extracted_k141_6850 | Not-determined |
| S12_extracted_k141_21856 | Not-determined |
| S12_extracted_k141_46617 | Not-determined |
| S12_extracted_k141_13629 | Not-determined |
| S12_extracted_k141_77229 | Not-determined |
| S12_extracted_k141_46620 | Not-determined |
| S12_extracted_k141_21857 | Not-determined |
| S12_extracted_k141_132115 | Not-determined |
| S12_extracted_k141_163842 | Not-determined |
| S12_extracted_k141_132116 | Not-determined |
| S12_extracted_k141_21860 | Low-quality |
| S12_extracted_k141_21863 | Not-determined |
| S12_extracted_k141_69408 | Not-determined |
| S12_extracted_k141_69410 | Low-quality |
| S12_extracted_k141_13635 | Not-determined |
| S12_extracted_k141_21865 | Not-determined |
| S12_extracted_k141_93521 | Low-quality |
| S12_extracted_k141_116266 | Not-determined |
| S12_extracted_k141_179056 | Low-quality |
| S12_extracted_k141_46632 | Not-determined |
| S12_extracted_k141_69415 | Not-determined |
| S12_extracted_k141_46636 | Not-determined |
| S12_extracted_k141_132121 | Not-determined |
| S12_extracted_k141_61680 | Not-determined |
| S12_extracted_k141_46638 | Not-determined |
| S12_extracted_k141_148289 | Not-determined |
| S12_extracted_k141_132123 | Not-determined |
| S12_extracted_k141_69422 | Not-determined |
| S12_extracted_k141_93529 | Not-determined |
| S12_extracted_k141_171513 | Not-determined |
| S12_extracted_k141_148296 | Not-determined |
| S12_extracted_k141_84865 | Not-determined |
| S12_extracted_k141_101953 | Not-determined |
| S12_extracted_k141_46644 | Not-determined |
| S12_extracted_k141_37399 | Not-determined |
| S12_extracted_k141_53506 | Not-determined |
| S12_extracted_k141_30151 | Not-determined |
| S12_extracted_k141_30155 | Not-determined |
| S12_extracted_k141_77251 | Not-determined |
| S12_extracted_k141_84872 | Not-determined |
| S12_extracted_k141_13642 | Not-determined |
| S12_extracted_k141_21878 | Not-determined |
| S12_extracted_k141_37409 | Not-determined |
| S12_extracted_k141_163857 | Not-determined |
| S12_extracted_k141_77256 | Not-determined |
| S12_extracted_k141_155701 | Not-determined |
| S12_extracted_k141_30164 | Low-quality |
| S12_extracted_k141_155703 | Not-determined |
| S12_extracted_k141_84885 | Low-quality |
| S12_extracted_k141_77262 | Not-determined |
| S12_extracted_k141_21888 | Not-determined |
| S12_extracted_k141_30172 | Not-determined |
| S12_extracted_k141_124045 | Not-determined |
| S12_extracted_k141_163874 | Not-determined |
| S12_extracted_k141_6869 | Not-determined |
| S12_extracted_k141_163875 | Not-determined |
| S12_extracted_k141_30180 | Not-determined |
| S12_extracted_k141_148319 | Not-determined |
| S12_extracted_k141_148320 | Not-determined |
| S12_extracted_k141_109307 | Not-determined |
| S12_extracted_k141_69454 | Not-determined |
| S12_extracted_k141_155712 | Not-determined |
| S12_extracted_k141_6874 | Not-determined |
| S12_extracted_k141_77272 | Not-determined |
| S12_extracted_k141_6875 | Not-determined |
| S12_extracted_k141_132156 | Not-determined |
| S12_extracted_k141_109313 | Not-determined |
| S12_extracted_k141_140817 | Not-determined |
| S12_extracted_k141_77278 | Not-determined |
| S12_extracted_k141_132160 | Not-determined |
| S12_extracted_k141_77280 | Not-determined |
| S12_extracted_k141_163889 | Low-quality |
| S12_extracted_k141_124058 | Not-determined |
| S12_extracted_k141_101965 | Not-determined |
| S12_extracted_k141_37432 | Not-determined |
| S12_extracted_k141_77283 | Not-determined |
| S12_extracted_k141_77284 | Not-determined |
| S12_extracted_k141_101968 | Not-determined |
| S12_extracted_k141_179079 | Not-determined |
| S12_extracted_k141_61689 | Not-determined |
| S12_extracted_k141_77287 | Not-determined |
| S12_extracted_k141_6886 | Low-quality |
| S12_extracted_k141_69474 | Not-determined |
| S12_extracted_k141_84911 | Not-determined |
| S12_extracted_k141_140828 | Low-quality |
| S12_extracted_k141_37439 | Not-determined |
| S12_extracted_k141_84916 | Not-determined |
| S12_extracted_k141_69477 | Low-quality |
| S12_extracted_k141_163901 | Not-determined |
| S12_extracted_k141_132170 | Not-determined |
| S12_extracted_k141_140830 | Not-determined |
| S12_extracted_k141_109331 | Not-determined |
| S12_extracted_k141_77293 | Not-determined |
| S12_extracted_k141_132175 | Not-determined |
| S12_extracted_k141_101979 | Not-determined |
| S12_extracted_k141_6893 | Not-determined |
| S12_extracted_k141_84923 | Not-determined |
| S12_extracted_k141_101983 | Not-determined |
| S12_extracted_k141_46674 | Not-determined |
| S12_extracted_k141_46676 | Not-determined |
| S12_extracted_k141_77301 | Not-determined |
| S12_extracted_k141_30198 | Not-determined |
| S12_extracted_k141_69491 | Not-determined |
| S12_extracted_k141_30199 | Low-quality |
| S12_extracted_k141_77305 | Not-determined |
| S12_extracted_k141_61711 | Not-determined |
| S12_extracted_k141_109346 | Not-determined |
| S12_extracted_k141_84933 | Not-determined |
| S12_extracted_k141_132182 | Not-determined |
| S12_extracted_k141_101998 | Not-determined |
| S12_extracted_k141_116322 | Not-determined |
| S12_extracted_k141_116326 | Not-determined |
| S12_extracted_k141_124080 | Not-determined |
| S12_extracted_k141_132194 | Not-determined |
| S12_extracted_k141_124081 | Not-determined |
| S12_extracted_k141_6901 | Not-determined |
| S12_extracted_k141_155755 | Not-determined |
| S12_extracted_k141_163915 | Not-determined |
| S12_extracted_k141_6902 | Not-determined |
| S12_extracted_k141_163918 | Low-quality |
| S12_extracted_k141_179096 | Not-determined |
| S12_extracted_k141_109364 | Not-determined |
| S12_extracted_k141_102006 | Not-determined |
| S12_extracted_k141_171539 | Not-determined |
| S12_extracted_k141_140858 | Not-determined |
| S12_extracted_k141_124086 | Not-determined |
| S12_extracted_k141_171541 | Not-determined |
| S12_extracted_k141_109369 | Low-quality |
| S12_extracted_k141_148353 | Not-determined |
| S12_extracted_k141_140865 | Low-quality |
| S12_extracted_k141_148354 | Not-determined |
| S12_extracted_k141_30218 | Not-determined |
| S12_extracted_k141_140866 | Not-determined |
| S12_extracted_k141_37462 | Not-determined |
| S12_extracted_k141_171549 | Not-determined |
| S12_extracted_k141_21898 | Not-determined |
| S12_extracted_k141_155774 | Not-determined |
| S12_extracted_k141_6916 | Not-determined |
| S12_extracted_k141_93572 | Not-determined |
| S12_extracted_k141_179107 | Not-determined |
| S12_extracted_k141_140872 | Not-determined |
| S12_extracted_k141_102013 | Not-determined |
| S12_extracted_k141_69509 | Low-quality |
| S12_extracted_k141_155779 | Not-determined |
| S12_extracted_k141_84966 | Not-determined |
| S12_extracted_k141_69513 | Low-quality |
| S12_extracted_k141_124096 | Not-determined |
| S12_extracted_k141_102016 | Not-determined |
| S12_extracted_k141_84967 | Not-determined |
| S12_extracted_k141_109381 | Not-determined |
| S12_extracted_k141_132208 | Not-determined |
| S12_extracted_k141_163944 | Not-determined |
| S12_extracted_k141_61746 | Not-determined |
| S12_extracted_k141_132210 | Not-determined |
| S12_extracted_k141_37474 | Not-determined |
| S12_extracted_k141_163949 | Not-determined |
| S12_extracted_k141_61749 | Not-determined |
| S12_extracted_k141_155786 | Not-determined |
| S12_extracted_k141_163952 | Not-determined |
| S12_extracted_k141_46690 | Not-determined |
| S12_extracted_k141_6927 | Not-determined |
| S12_extracted_k141_46691 | Not-determined |
| S12_extracted_k141_116334 | Not-determined |
| S12_extracted_k141_46692 | Not-determined |
| S12_extracted_k141_69525 | Not-determined |
| S12_extracted_k141_30235 | Not-determined |
| S12_extracted_k141_30236 | Not-determined |
| S12_extracted_k141_21915 | Not-determined |
| S12_extracted_k141_163958 | Not-determined |
| S12_extracted_k141_53572 | Not-determined |
| S12_extracted_k141_93586 | Not-determined |
| S12_extracted_k141_37486 | Not-determined |
| S12_extracted_k141_21917 | Low-quality |
| S12_extracted_k141_13660 | Not-determined |
| S12_extracted_k141_61759 | Not-determined |
| S12_extracted_k141_46700 | Not-determined |
| S12_extracted_k141_155803 | Not-determined |
| S12_extracted_k141_61763 | Not-determined |
| S12_extracted_k141_37488 | Not-determined |
| S12_extracted_k141_46703 | Low-quality |
| S12_extracted_k141_179133 | Not-determined |
| S12_extracted_k141_21924 | Not-determined |
| S12_extracted_k141_21925 | Not-determined |
| S12_extracted_k141_21927 | Not-determined |
| S12_extracted_k141_30241 | Low-quality |
| S12_extracted_k141_124113 | Not-determined |
| S12_extracted_k141_46707 | Not-determined |
| S12_extracted_k141_132227 | Not-determined |
| S12_extracted_k141_46708 | Low-quality |
| S12_extracted_k141_21931 | Not-determined |
| S12_extracted_k141_77310 | Not-determined |
| S12_extracted_k141_155807 | Not-determined |
| S12_extracted_k141_116342 | Not-determined |
| S12_extracted_k141_46713 | Not-determined |
| S12_extracted_k141_124118 | Not-determined |
| S12_extracted_k141_69542 | Not-determined |
| S12_extracted_k141_155811 | Not-determined |
| S12_extracted_k141_21937 | Not-determined |
| S12_extracted_k141_132229 | Not-determined |
| S12_extracted_k141_84986 | Not-determined |
| S12_extracted_k141_155817 | Not-determined |
| S12_extracted_k141_132230 | Not-determined |
| S12_extracted_k141_37499 | Low-quality |
| S12_extracted_k141_124121 | Not-determined |
| S12_extracted_k141_13680 | Not-determined |
| S12_extracted_k141_13682 | Not-determined |
| S12_extracted_k141_77321 | Not-determined |
| S12_extracted_k141_53592 | Not-determined |
| S12_extracted_k141_155822 | Not-determined |
| S12_extracted_k141_6967 | Not-determined |
| S12_extracted_k141_84994 | Not-determined |
| S12_extracted_k141_6969 | Not-determined |
| S12_extracted_k141_155824 | Not-determined |
| S12_extracted_k141_140880 | Not-determined |
| S12_extracted_k141_6970 | Not-determined |
| S12_extracted_k141_155826 | Not-determined |
| S12_extracted_k141_155827 | Not-determined |
| S12_extracted_k141_140882 | Not-determined |
| S12_extracted_k141_109413 | Not-determined |
| S12_extracted_k141_6974 | Not-determined |
| S12_extracted_k141_93619 | Not-determined |
| S12_extracted_k141_53602 | Not-determined |
| S12_extracted_k141_53606 | Not-determined |
| S12_extracted_k141_148393 | Not-determined |
| S12_extracted_k141_148394 | Not-determined |
| S12_extracted_k141_46729 | Not-determined |
| S12_extracted_k141_13700 | Not-determined |
| S12_extracted_k141_53610 | Not-determined |
| S12_extracted_k141_93623 | Not-determined |
| S12_extracted_k141_69554 | Not-determined |
| S12_extracted_k141_37511 | Not-determined |
| S12_extracted_k141_6979 | Not-determined |
| S12_extracted_k141_155837 | Not-determined |
| S12_extracted_k141_46736 | Not-determined |
| S12_extracted_k141_69561 | Not-determined |
| S12_extracted_k141_85004 | Not-determined |
| S12_extracted_k141_37514 | Not-determined |
| S12_extracted_k141_53614 | Not-determined |
| S12_extracted_k141_93631 | Not-determined |
| S12_extracted_k141_13707 | Not-determined |
| S12_extracted_k141_53615 | Not-determined |
| S12_extracted_k141_85009 | Not-determined |
| S12_extracted_k141_102031 | Not-determined |
| S12_extracted_k141_109422 | Not-determined |
| S12_extracted_k141_148406 | Not-determined |
| S12_extracted_k141_140898 | Not-determined |
| S12_extracted_k141_155844 | Low-quality |
| S12_extracted_k141_85017 | Not-determined |
| S12_extracted_k141_171564 | Not-determined |
| S12_extracted_k141_155846 | Low-quality |
| S12_extracted_k141_21969 | Not-determined |
| S12_extracted_k141_155847 | Not-determined |
| S12_extracted_k141_109427 | Not-determined |
| S12_extracted_k141_148408 | Low-quality |
| S12_extracted_k141_132265 | Not-determined |
| S12_extracted_k141_13719 | Not-determined |
| S12_extracted_k141_69579 | Not-determined |
| S12_extracted_k141_37536 | Not-determined |
| S12_extracted_k141_37537 | Not-determined |
| S12_extracted_k141_132273 | Not-determined |
| S12_extracted_k141_69581 | Not-determined |
| S12_extracted_k141_46758 | Not-determined |
| S12_extracted_k141_13724 | Not-determined |
| S12_extracted_k141_140910 | Not-determined |
| S12_extracted_k141_85022 | Not-determined |
| S12_extracted_k141_69586 | Not-determined |
| S12_extracted_k141_21987 | Not-determined |
| S12_extracted_k141_13728 | Not-determined |
| S12_extracted_k141_30249 | Not-determined |
| S12_extracted_k141_37543 | Not-determined |
| S12_extracted_k141_124129 | Not-determined |
| S12_extracted_k141_85024 | Not-determined |
| S12_extracted_k141_13730 | Low-quality |
| S12_extracted_k141_85025 | Not-determined |
| S12_extracted_k141_93649 | Not-determined |
| S12_extracted_k141_171581 | Not-determined |
| S12_extracted_k141_46762 | Low-quality |
| S12_extracted_k141_93650 | Not-determined |
| S12_extracted_k141_163982 | Not-determined |
| S12_extracted_k141_61774 | Not-determined |
| S12_extracted_k141_132275 | Low-quality |
| S12_extracted_k141_85029 | Not-determined |
| S12_extracted_k141_109447 | Not-determined |
| S12_extracted_k141_85031 | Not-determined |
| S12_extracted_k141_69594 | Low-quality |
| S12_extracted_k141_188090 | Not-determined |
| S12_extracted_k141_171586 | Not-determined |
| S12_extracted_k141_109449 | Not-determined |
| S12_extracted_k141_148427 | Not-determined |
| S12_extracted_k141_188091 | Not-determined |
| S12_extracted_k141_109451 | Not-determined |
| S12_extracted_k141_163989 | Low-quality |
| S12_extracted_k141_140923 | Not-determined |
| S12_extracted_k141_13736 | Not-determined |
| S12_extracted_k141_188092 | Not-determined |
| S12_extracted_k141_93661 | Not-determined |
| S12_extracted_k141_61779 | Not-determined |
| S12_extracted_k141_171593 | Not-determined |
| S12_extracted_k141_61780 | Not-determined |
| S12_extracted_k141_116367 | Low-quality |
| S12_extracted_k141_188096 | Low-quality |
| S12_extracted_k141_21998 | Not-determined |
| S12_extracted_k141_188097 | Low-quality |
| S12_extracted_k141_109458 | Not-determined |
| S12_extracted_k141_69604 | Not-determined |
| S12_extracted_k141_30266 | Not-determined |
| S12_extracted_k141_188099 | Low-quality |
| S12_extracted_k141_30268 | Not-determined |
| S12_extracted_k141_116375 | Not-determined |
| S12_extracted_k141_46778 | Not-determined |
| S12_extracted_k141_46781 | Not-determined |
| S12_extracted_k141_22006 | Not-determined |
| S12_extracted_k141_124146 | Not-determined |
| S12_extracted_k141_188106 | Not-determined |
| S12_extracted_k141_77346 | Not-determined |
| S12_extracted_k141_116384 | Not-determined |
| S12_extracted_k141_13744 | Not-determined |
| S12_extracted_k141_13747 | Not-determined |
| S12_extracted_k141_61789 | Not-determined |
| S12_extracted_k141_116386 | Not-determined |
| S12_extracted_k141_61790 | Not-determined |
| S12_extracted_k141_188111 | Not-determined |
| S12_extracted_k141_30278 | Low-quality |
| S12_extracted_k141_102057 | Not-determined |
| S12_extracted_k141_93674 | Not-determined |
| S12_extracted_k141_93677 | Not-determined |
| S12_extracted_k141_53666 | Not-determined |
| S12_extracted_k141_46787 | Not-determined |
| S12_extracted_k141_30284 | Not-determined |
| S12_extracted_k141_140937 | Not-determined |
| S12_extracted_k141_188119 | Not-determined |
| S12_extracted_k141_116388 | Not-determined |
| S12_extracted_k141_30288 | Not-determined |
| S12_extracted_k141_93679 | Not-determined |
| S12_extracted_k141_85067 | Not-determined |
| S12_extracted_k141_171614 | Not-determined |
| S12_extracted_k141_140941 | Not-determined |
| S12_extracted_k141_124154 | Not-determined |
| S12_extracted_k141_132292 | Low-quality |
| S12_extracted_k141_93682 | Not-determined |
| S12_extracted_k141_124157 | Not-determined |
| S12_extracted_k141_188124 | Not-determined |
| S12_extracted_k141_140946 | Not-determined |
| S12_extracted_k141_53675 | Not-determined |
| S12_extracted_k141_116397 | Not-determined |
| S12_extracted_k141_171622 | Not-determined |
| S12_extracted_k141_61816 | Not-determined |
| S12_extracted_k141_61818 | Low-quality |
| S12_extracted_k141_116405 | Not-determined |
| S12_extracted_k141_30294 | Not-determined |
| S12_extracted_k141_77376 | Not-determined |
| S12_extracted_k141_61821 | Not-determined |
| S12_extracted_k141_116411 | Not-determined |
| S12_extracted_k141_171635 | Not-determined |
| S12_extracted_k141_171636 | Not-determined |
| S12_extracted_k141_22018 | Not-determined |
| S12_extracted_k141_109488 | Not-determined |
| S12_extracted_k141_140957 | Not-determined |
| S12_extracted_k141_85081 | Not-determined |
| S12_extracted_k141_85083 | Low-quality |
| S12_extracted_k141_53688 | Not-determined |
| S12_extracted_k141_85086 | Low-quality |
| S12_extracted_k141_155855 | Low-quality |
| S12_extracted_k141_61834 | Not-determined |
| S12_extracted_k141_85089 | Not-determined |
| S12_extracted_k141_124179 | Not-determined |
| S12_extracted_k141_30300 | Not-determined |
| S12_extracted_k141_22023 | Not-determined |
| S12_extracted_k141_179168 | Not-determined |
| S12_extracted_k141_13793 | Low-quality |
| S12_extracted_k141_109496 | Low-quality |
| S12_extracted_k141_37583 | Not-determined |
| S12_extracted_k141_77393 | Not-determined |
| S12_extracted_k141_109497 | Not-determined |
| S12_extracted_k141_61838 | Not-determined |
| S12_extracted_k141_132308 | Low-quality |
| S12_extracted_k141_155858 | Not-determined |
| S12_extracted_k141_61839 | Not-determined |
| S12_extracted_k141_148461 | Not-determined |
| S12_extracted_k141_140966 | Not-determined |
| S12_extracted_k141_140968 | Not-determined |
| S12_extracted_k141_148462 | Low-quality |
| S12_extracted_k141_155864 | Not-determined |
| S12_extracted_k141_77400 | Not-determined |
| S12_extracted_k141_85099 | Low-quality |
| S12_extracted_k141_61846 | Not-determined |
| S12_extracted_k141_69647 | Not-determined |
| S12_extracted_k141_85102 | Not-determined |
| S12_extracted_k141_30310 | Not-determined |
| S12_extracted_k141_22034 | Not-determined |
| S12_extracted_k141_124192 | Not-determined |
| S12_extracted_k141_85105 | Not-determined |
| S12_extracted_k141_22035 | Not-determined |
| S12_extracted_k141_7019 | Not-determined |
| S12_extracted_k141_93707 | Not-determined |
| S12_extracted_k141_132322 | Not-determined |
| S12_extracted_k141_109508 | Not-determined |
| S12_extracted_k141_22037 | Not-determined |
| S12_extracted_k141_22040 | Not-determined |
| S12_extracted_k141_148464 | Low-quality |
| S12_extracted_k141_61855 | Not-determined |
| S12_extracted_k141_13799 | Low-quality |
| S12_extracted_k141_132329 | Not-determined |
| S12_extracted_k141_61859 | Not-determined |
| S12_extracted_k141_69659 | Not-determined |
| S12_extracted_k141_188157 | Not-determined |
| S12_extracted_k141_93719 | Not-determined |
| S12_extracted_k141_85113 | Not-determined |
| S12_extracted_k141_7027 | Not-determined |
| S12_extracted_k141_171650 | Not-determined |
| S12_extracted_k141_132339 | Not-determined |
| S12_extracted_k141_77418 | Not-determined |
| S12_extracted_k141_77422 | Not-determined |
| S12_extracted_k141_171655 | Not-determined |
| S12_extracted_k141_37605 | Low-quality |
| S12_extracted_k141_7035 | Not-determined |
| S12_extracted_k141_188161 | Low-quality |
| S12_extracted_k141_7036 | Not-determined |
| S12_extracted_k141_37606 | Not-determined |
| S12_extracted_k141_116416 | Low-quality |
| S12_extracted_k141_53713 | Not-determined |
| S12_extracted_k141_109532 | Low-quality |
| S12_extracted_k141_30325 | Not-determined |
| S12_extracted_k141_61873 | Low-quality |
| S12_extracted_k141_7044 | Not-determined |
| S12_extracted_k141_53715 | Not-determined |
| S12_extracted_k141_61875 | Not-determined |
| S12_extracted_k141_85128 | Not-determined |
| S12_extracted_k141_77431 | Not-determined |
| S12_extracted_k141_148479 | Not-determined |
| S12_extracted_k141_148480 | Not-determined |
| S12_extracted_k141_140987 | Not-determined |
| S12_extracted_k141_171665 | Not-determined |
| S12_extracted_k141_102089 | Not-determined |
| S12_extracted_k141_22063 | Not-determined |
| S12_extracted_k141_124203 | Not-determined |
| S12_extracted_k141_109539 | Not-determined |
| S12_extracted_k141_22064 | Not-determined |
| S12_extracted_k141_140990 | Not-determined |
| S12_extracted_k141_116431 | Not-determined |
| S12_extracted_k141_13816 | Not-determined |
| S12_extracted_k141_140995 | Not-determined |
| S12_extracted_k141_109543 | Not-determined |
| S12_extracted_k141_179181 | Not-determined |
| S12_extracted_k141_124205 | Low-quality |
| S12_extracted_k141_53721 | Not-determined |
| S12_extracted_k141_93737 | Not-determined |
| S12_extracted_k141_109544 | Not-determined |
| S12_extracted_k141_132357 | Low-quality |
| S12_extracted_k141_13818 | Not-determined |
| S12_extracted_k141_109545 | Not-determined |
| S12_extracted_k141_13819 | Low-quality |
| S12_extracted_k141_102095 | Not-determined |
| S12_extracted_k141_69679 | Not-determined |
| S12_extracted_k141_53733 | Not-determined |
| S12_extracted_k141_109548 | Not-determined |
| S12_extracted_k141_53734 | Not-determined |
| S12_extracted_k141_13823 | Not-determined |
| S12_extracted_k141_109550 | Not-determined |
| S12_extracted_k141_46807 | Not-determined |
| S12_extracted_k141_61898 | Not-determined |
| S12_extracted_k141_69681 | Not-determined |
| S12_extracted_k141_22081 | Not-determined |
| S12_extracted_k141_69682 | Not-determined |
| S12_extracted_k141_77446 | Not-determined |
| S12_extracted_k141_22083 | Not-determined |
| S12_extracted_k141_77447 | Not-determined |
| S12_extracted_k141_53742 | Not-determined |
| S12_extracted_k141_46810 | Not-determined |
| S12_extracted_k141_53743 | Not-determined |
| S12_extracted_k141_69685 | Not-determined |
| S12_extracted_k141_7057 | Not-determined |
| S12_extracted_k141_69686 | Not-determined |
| S12_extracted_k141_116445 | Not-determined |
| S12_extracted_k141_53747 | Not-determined |
| S12_extracted_k141_7059 | Not-determined |
| S12_extracted_k141_148497 | Not-determined |
| S12_extracted_k141_179195 | Not-determined |
| S12_extracted_k141_155906 | Not-determined |
| S12_extracted_k141_93752 | Not-determined |
| S12_extracted_k141_141016 | Low-quality |
| S12_extracted_k141_164003 | Not-determined |
| S12_extracted_k141_155908 | Not-determined |
| S12_extracted_k141_109556 | Low-quality |
| S12_extracted_k141_93756 | Low-quality |
| S12_extracted_k141_124226 | Not-determined |
| S12_extracted_k141_22095 | Not-determined |
| S12_extracted_k141_93758 | Not-determined |
| S12_extracted_k141_46816 | Low-quality |
| S12_extracted_k141_141019 | Not-determined |
| S12_extracted_k141_53755 | Not-determined |
| S12_extracted_k141_141021 | Not-determined |
| S12_extracted_k141_30342 | Not-determined |
| S12_extracted_k141_85150 | Not-determined |
| S12_extracted_k141_93765 | Not-determined |
| S12_extracted_k141_148504 | Not-determined |
| S12_extracted_k141_132373 | Not-determined |
| S12_extracted_k141_93766 | Not-determined |
| S12_extracted_k141_61922 | Not-determined |
| S12_extracted_k141_171685 | Low-quality |
| S12_extracted_k141_37641 | Low-quality |
| S12_extracted_k141_61925 | Not-determined |
| S12_extracted_k141_22106 | Not-determined |
| S12_extracted_k141_93770 | Not-determined |
| S12_extracted_k141_13845 | Low-quality |
| S12_extracted_k141_179209 | Low-quality |
| S12_extracted_k141_53762 | Low-quality |
| S12_extracted_k141_171690 | Low-quality |
| S12_extracted_k141_188179 | Not-determined |
| S12_extracted_k141_116468 | Not-determined |
| S12_extracted_k141_124230 | Not-determined |
| S12_extracted_k141_188180 | Not-determined |
| S12_extracted_k141_30358 | Not-determined |
| S12_extracted_k141_116470 | Not-determined |
| S12_extracted_k141_132383 | Low-quality |
| S12_extracted_k141_188183 | Not-determined |
| S12_extracted_k141_7071 | Not-determined |
| S12_extracted_k141_77485 | Not-determined |
| S12_extracted_k141_188186 | Not-determined |
| S12_extracted_k141_77486 | Not-determined |
| S12_extracted_k141_85167 | Not-determined |
| S12_extracted_k141_132391 | Not-determined |
| S12_extracted_k141_116471 | Low-quality |
| S12_extracted_k141_164025 | Not-determined |
| S12_extracted_k141_46833 | Not-determined |
| S12_extracted_k141_171700 | Not-determined |
| S12_extracted_k141_148516 | Not-determined |
| S12_extracted_k141_116474 | Not-determined |
| S12_extracted_k141_188189 | Not-determined |
| S12_extracted_k141_109579 | Not-determined |
| S12_extracted_k141_116477 | Low-quality |
| S12_extracted_k141_124236 | Low-quality |
| S12_extracted_k141_102142 | Not-determined |
| S12_extracted_k141_53773 | Not-determined |
| S12_extracted_k141_116481 | Not-determined |
| S12_extracted_k141_37652 | Low-quality |
| S12_extracted_k141_148524 | Not-determined |
| S12_extracted_k141_164030 | Low-quality |
| S12_extracted_k141_109589 | Not-determined |
| S12_extracted_k141_148529 | Not-determined |
| S12_extracted_k141_61928 | Not-determined |
| S12_extracted_k141_148531 | Not-determined |
| S12_extracted_k141_171710 | Low-quality |
| S12_extracted_k141_13864 | Not-determined |
| S12_extracted_k141_164034 | Not-determined |
| S12_extracted_k141_93803 | Not-determined |
| S12_extracted_k141_116488 | Not-determined |
| S12_extracted_k141_7085 | Not-determined |
| S12_extracted_k141_124244 | Not-determined |
| S12_extracted_k141_7087 | Not-determined |
| S12_extracted_k141_53793 | Not-determined |
| S12_extracted_k141_141040 | Not-determined |
| S12_extracted_k141_148536 | Not-determined |
| S12_extracted_k141_109602 | Not-determined |
| S12_extracted_k141_148538 | Not-determined |
| S12_extracted_k141_53800 | Not-determined |
| S12_extracted_k141_61937 | Not-determined |
| S12_extracted_k141_69738 | Not-determined |
| S12_extracted_k141_109606 | Not-determined |
| S12_extracted_k141_188207 | Not-determined |
| S12_extracted_k141_141048 | Not-determined |
| S12_extracted_k141_46842 | Not-determined |
| S12_extracted_k141_124254 | Not-determined |
| S12_extracted_k141_109608 | Not-determined |
| S12_extracted_k141_109609 | Not-determined |
| S12_extracted_k141_188211 | Not-determined |
| S12_extracted_k141_85182 | Not-determined |
| S12_extracted_k141_102148 | Low-quality |
| S12_extracted_k141_188214 | Not-determined |
| S12_extracted_k141_171726 | Not-determined |
| S12_extracted_k141_85185 | Not-determined |
| S12_extracted_k141_171729 | Not-determined |
| S12_extracted_k141_85190 | Not-determined |
| S12_extracted_k141_102157 | Not-determined |
| S12_extracted_k141_102159 | Not-determined |
| S12_extracted_k141_132403 | Not-determined |
| S12_extracted_k141_179242 | Not-determined |

| | |
|---|---|
| S12_extracted_k141_102162 | Not-determined |
| S12_extracted_k141_171733 | Not-determined |
| S12_extracted_k141_85196 | Not-determined |
| S12_extracted_k141_13878 | Not-determined |
| S12_extracted_k141_132405 | Not-determined |
| S12_extracted_k141_85198 | Not-determined |
| S12_extracted_k141_124260 | Not-determined |
| S12_extracted_k141_13882 | Not-determined |
| S12_extracted_k141_188225 | Not-determined |
| S12_extracted_k141_116499 | Not-determined |
| S12_extracted_k141_102168 | Not-determined |
| S12_extracted_k141_179255 | Not-determined |
| S12_extracted_k141_85203 | Not-determined |
| S12_extracted_k141_30390 | Not-determined |
| S12_extracted_k141_132412 | Not-determined |
| S12_extracted_k141_179257 | Not-determined |
| S12_extracted_k141_53807 | Not-determined |
| S12_extracted_k141_93831 | Not-determined |
| S12_extracted_k141_85204 | Not-determined |
| S12_extracted_k141_22160 | Not-determined |
| S12_extracted_k141_141061 | Not-determined |
| S12_extracted_k141_124270 | Not-determined |
| S12_extracted_k141_69768 | Not-determined |
| S12_extracted_k141_124272 | Not-determined |
| S12_extracted_k141_53813 | Low-quality |
| S12_extracted_k141_124274 | Not-determined |
| S12_extracted_k141_46864 | Not-determined |
| S12_extracted_k141_148570 | Not-determined |
| S12_extracted_k141_30398 | Not-determined |
| S12_extracted_k141_61955 | Not-determined |
| S12_extracted_k141_124279 | Not-determined |
| S12_extracted_k141_61956 | Not-determined |
| S12_extracted_k141_148574 | Not-determined |
| S12_extracted_k141_93839 | Not-determined |
| S12_extracted_k141_124280 | Not-determined |
| S12_extracted_k141_53819 | Low-quality |
| S12_extracted_k141_171743 | Not-determined |
| S12_extracted_k141_164059 | Not-determined |
| S12_extracted_k141_102188 | Not-determined |
| S12_extracted_k141_22171 | Not-determined |
| S12_extracted_k141_164065 | Not-determined |
| S12_extracted_k141_85222 | Not-determined |
| S12_extracted_k141_148580 | Low-quality |
| S12_extracted_k141_85223 | Not-determined |
| S12_extracted_k141_30414 | Not-determined |
| S12_extracted_k141_30415 | Not-determined |
| S12_extracted_k141_85227 | Not-determined |
| S12_extracted_k141_30421 | Not-determined |
| S12_extracted_k141_141084 | Low-quality |
| S12_extracted_k141_85236 | Not-determined |
| S12_extracted_k141_179261 | Not-determined |
| S12_extracted_k141_77538 | Not-determined |
| S12_extracted_k141_77540 | Not-determined |
| S12_extracted_k141_132435 | Not-determined |
| S12_extracted_k141_148593 | Not-determined |
| S12_extracted_k141_141088 | Not-determined |
| S12_extracted_k141_179266 | Not-determined |
| S12_extracted_k141_53831 | Not-determined |
| S12_extracted_k141_7135 | Not-determined |
| S12_extracted_k141_77545 | Not-determined |
| S12_extracted_k141_132439 | Not-determined |
| S12_extracted_k141_53834 | Not-determined |
| S12_extracted_k141_85244 | Not-determined |
| S12_extracted_k141_69791 | Not-determined |
| S12_extracted_k141_37679 | Not-determined |
| S12_extracted_k141_141089 | Not-determined |
| S12_extracted_k141_37681 | Not-determined |
| S12_extracted_k141_77552 | Not-determined |
| S12_extracted_k141_93868 | Not-determined |
| S12_extracted_k141_22200 | Not-determined |
| S12_extracted_k141_30442 | Not-determined |
| S12_extracted_k141_171771 | Not-determined |
| S12_extracted_k141_179269 | Not-determined |
| S12_extracted_k141_77559 | Not-determined |
| S12_extracted_k141_7149 | Not-determined |
| S12_extracted_k141_77561 | Low-quality |
| S12_extracted_k141_77562 | Not-determined |
| S12_extracted_k141_30449 | Not-determined |
| S12_extracted_k141_85264 | Not-determined |
| S12_extracted_k141_171778 | Not-determined |
| S12_extracted_k141_69798 | Not-determined |
| S12_extracted_k141_188237 | Not-determined |
| S12_extracted_k141_77569 | Not-determined |
| S12_extracted_k141_7164 | Not-determined |
| S12_extracted_k141_85270 | Not-determined |
| S12_extracted_k141_85273 | Not-determined |
| S12_extracted_k141_7166 | Not-determined |
| S12_extracted_k141_155955 | Not-determined |
| S12_extracted_k141_7167 | Not-determined |
| S12_extracted_k141_188238 | Low-quality |
| S12_extracted_k141_69805 | Not-determined |
| S12_extracted_k141_141110 | Not-determined |
| S12_extracted_k141_30464 | Not-determined |
| S12_extracted_k141_171791 | Not-determined |
| S12_extracted_k141_13901 | Not-determined |
| S12_extracted_k141_132470 | Not-determined |
| S12_extracted_k141_155957 | Low-quality |
| S12_extracted_k141_164102 | Low-quality |
| S12_extracted_k141_93873 | Not-determined |
| S12_extracted_k141_179278 | Not-determined |
| S12_extracted_k141_69807 | Not-determined |
| S12_extracted_k141_164105 | Not-determined |
| S12_extracted_k141_132473 | Not-determined |
| S12_extracted_k141_85282 | Low-quality |
| S12_extracted_k141_62006 | Not-determined |
| S12_extracted_k141_155958 | Not-determined |
| S12_extracted_k141_85284 | Not-determined |
| S12_extracted_k141_188243 | Not-determined |
| S12_extracted_k141_179280 | Not-determined |
| S12_extracted_k141_62008 | Not-determined |
| S12_extracted_k141_22227 | Not-determined |
| S12_extracted_k141_93877 | Not-determined |
| S12_extracted_k141_7174 | Not-determined |
| S12_extracted_k141_171796 | Not-determined |
| S12_extracted_k141_30469 | Not-determined |
| S12_extracted_k141_37706 | Low-quality |
| S12_extracted_k141_46896 | Not-determined |
| S12_extracted_k141_164116 | Not-determined |
| S12_extracted_k141_171799 | Not-determined |
| S12_extracted_k141_188254 | Not-determined |
| S12_extracted_k141_188255 | Low-quality |
| S12_extracted_k141_46902 | Low-quality |
| S12_extracted_k141_85297 | Not-determined |
| S12_extracted_k141_188256 | Not-determined |
| S12_extracted_k141_30472 | Low-quality |
| S12_extracted_k141_124315 | Not-determined |
| S12_extracted_k141_93889 | Not-determined |
| S12_extracted_k141_155964 | Not-determined |
| S12_extracted_k141_155965 | Not-determined |
| S12_extracted_k141_164129 | Not-determined |
| S12_extracted_k141_13918 | Not-determined |
| S12_extracted_k141_188266 | Not-determined |
| S12_extracted_k141_171804 | Not-determined |
| S12_extracted_k141_22242 | Not-determined |
| S12_extracted_k141_62016 | Not-determined |
| S12_extracted_k141_164135 | Not-determined |
| S12_extracted_k141_155969 | Low-quality |
| S12_extracted_k141_69829 | Not-determined |
| S12_extracted_k141_46916 | Not-determined |
| S12_extracted_k141_141132 | Not-determined |
| S12_extracted_k141_148652 | Not-determined |
| S12_extracted_k141_46917 | Not-determined |
| S12_extracted_k141_141135 | Not-determined |
| S12_extracted_k141_46918 | Not-determined |
| S12_extracted_k141_102199 | Not-determined |
| S12_extracted_k141_46921 | Low-quality |
| S12_extracted_k141_69833 | Not-determined |
| S12_extracted_k141_46922 | Low-quality |
| S12_extracted_k141_30488 | Not-determined |
| S12_extracted_k141_116538 | Not-determined |
| S12_extracted_k141_116539 | Not-determined |
| S12_extracted_k141_164149 | Not-determined |
| S12_extracted_k141_22257 | Not-determined |
| S12_extracted_k141_188272 | Not-determined |
| S12_extracted_k141_155980 | Not-determined |
| S12_extracted_k141_179302 | Not-determined |
| S12_extracted_k141_188273 | Not-determined |
| S12_extracted_k141_155981 | Not-determined |
| S12_extracted_k141_164151 | Not-determined |
| S12_extracted_k141_46925 | Not-determined |
| S12_extracted_k141_102204 | Not-determined |
| S12_extracted_k141_46926 | Not-determined |
| S12_extracted_k141_13937 | Low-quality |
| S12_extracted_k141_69839 | Low-quality |
| S12_extracted_k141_155983 | Low-quality |
| S12_extracted_k141_30496 | Not-determined |
| S12_extracted_k141_37722 | Not-determined |
| S12_extracted_k141_171808 | Not-determined |
| S12_extracted_k141_179310 | Not-determined |
| S12_extracted_k141_102206 | Not-determined |
| S12_extracted_k141_188281 | Not-determined |
| S12_extracted_k141_46935 | Not-determined |
| S12_extracted_k141_102208 | Not-determined |
| S12_extracted_k141_155987 | Not-determined |
| S12_extracted_k141_132513 | Not-determined |
| S12_extracted_k141_148669 | Not-determined |
| S12_extracted_k141_164160 | Not-determined |
| S12_extracted_k141_148671 | Not-determined |
| S12_extracted_k141_13941 | Low-quality |
| S12_extracted_k141_141155 | Not-determined |
| S12_extracted_k141_179316 | Not-determined |
| S12_extracted_k141_155993 | Not-determined |
| S12_extracted_k141_13943 | Not-determined |
| S12_extracted_k141_109633 | Low-quality |
| S12_extracted_k141_7201 | Not-determined |
| S12_extracted_k141_22268 | Not-determined |
| S12_extracted_k141_148677 | Not-determined |
| S12_extracted_k141_62043 | Not-determined |
| S12_extracted_k141_37731 | Not-determined |
| S12_extracted_k141_164167 | Not-determined |
| S12_extracted_k141_7205 | Not-determined |
| S12_extracted_k141_155997 | Not-determined |
| S12_extracted_k141_62045 | Not-determined |
| S12_extracted_k141_148689 | Not-determined |
| S12_extracted_k141_188295 | Not-determined |
| S12_extracted_k141_7211 | Not-determined |
| S12_extracted_k141_179322 | Not-determined |
| S12_extracted_k141_179324 | Not-determined |
| S12_extracted_k141_109639 | Not-determined |
| S12_extracted_k141_179326 | Not-determined |
| S12_extracted_k141_132532 | Not-determined |
| S12_extracted_k141_46947 | Not-determined |
| S12_extracted_k141_132533 | Not-determined |
| S12_extracted_k141_69869 | Not-determined |
| S12_extracted_k141_124353 | Not-determined |
| S12_extracted_k141_46950 | Low-quality |
| S12_extracted_k141_62060 | Not-determined |
| S12_extracted_k141_116564 | Not-determined |
| S12_extracted_k141_69872 | Not-determined |
| S12_extracted_k141_116567 | Not-determined |
| S12_extracted_k141_22289 | Not-determined |
| S12_extracted_k141_188306 | Not-determined |
| S12_extracted_k141_148703 | Not-determined |
| S12_extracted_k141_116574 | Not-determined |
| S12_extracted_k141_22290 | Not-determined |
| S12_extracted_k141_69877 | Not-determined |
| S12_extracted_k141_179330 | Not-determined |
| S12_extracted_k141_53857 | Low-quality |
| S12_extracted_k141_30529 | Not-determined |
| S12_extracted_k141_141166 | Not-determined |
| S12_extracted_k141_46958 | Not-determined |
| S12_extracted_k141_93915 | Not-determined |
| S12_extracted_k141_124362 | Not-determined |
| S12_extracted_k141_109661 | Not-determined |
| S12_extracted_k141_188317 | Not-determined |
| S12_extracted_k141_132539 | Not-determined |
| S12_extracted_k141_132541 | Not-determined |
| S12_extracted_k141_179338 | Not-determined |
| S12_extracted_k141_102230 | Not-determined |
| S12_extracted_k141_37754 | Not-determined |
| S12_extracted_k141_102231 | Not-determined |
| S12_extracted_k141_148716 | Not-determined |
| S12_extracted_k141_188325 | Not-determined |
| S12_extracted_k141_124370 | Not-determined |
| S12_extracted_k141_53863 | Not-determined |
| S12_extracted_k141_46969 | Not-determined |
| S12_extracted_k141_69888 | Not-determined |
| S12_extracted_k141_179342 | Not-determined |
| S12_extracted_k141_93927 | Low-quality |
| S12_extracted_k141_102236 | Not-determined |
| S12_extracted_k141_148719 | Not-determined |
| S12_extracted_k141_171831 | Not-determined |
| S12_extracted_k141_102239 | Not-determined |
| S12_extracted_k141_188331 | Not-determined |
| S12_extracted_k141_132551 | Not-determined |
| S12_extracted_k141_85305 | Not-determined |
| S12_extracted_k141_62085 | Not-determined |
| S12_extracted_k141_46970 | Low-quality |
| S12_extracted_k141_62086 | Not-determined |
| S12_extracted_k141_148726 | Not-determined |
| S12_extracted_k141_93935 | Not-determined |
| S12_extracted_k141_164207 | Not-determined |
| S12_extracted_k141_132557 | Not-determined |
| S12_extracted_k141_22297 | Not-determined |
| S12_extracted_k141_69895 | Not-determined |
| S12_extracted_k141_7238 | Not-determined |
| S12_extracted_k141_141189 | Low-quality |
| S12_extracted_k141_132560 | Not-determined |
| S12_extracted_k141_124379 | Not-determined |
| S12_extracted_k141_124381 | Not-determined |
| S12_extracted_k141_109665 | Low-quality |
| S12_extracted_k141_124383 | Not-determined |
| S12_extracted_k141_148739 | Low-quality |
| S12_extracted_k141_132572 | Not-determined |
| S12_extracted_k141_116604 | Not-determined |
| S12_extracted_k141_53881 | Not-determined |
| S12_extracted_k141_62098 | Not-determined |
| S12_extracted_k141_22308 | Low-quality |
| S12_extracted_k141_62099 | Not-determined |
| S12_extracted_k141_148742 | Not-determined |
| S12_extracted_k141_69910 | Not-determined |
| S12_extracted_k141_116611 | Low-quality |
| S12_extracted_k141_171846 | Not-determined |
| S12_extracted_k141_30539 | Not-determined |
| S12_extracted_k141_102251 | Not-determined |
| S12_extracted_k141_109672 | Not-determined |
| S12_extracted_k141_102252 | Not-determined |
| S12_extracted_k141_69914 | Not-determined |
| S12_extracted_k141_188352 | Not-determined |
| S12_extracted_k141_85317 | Not-determined |
| S12_extracted_k141_171850 | Not-determined |
| S12_extracted_k141_47003 | Not-determined |
| S12_extracted_k141_109674 | Not-determined |
| S12_extracted_k141_30542 | Not-determined |
| S12_extracted_k141_47005 | Not-determined |
| S12_extracted_k141_85322 | Not-determined |
| S12_extracted_k141_116619 | Low-quality |
| S12_extracted_k141_171854 | Not-determined |
| S12_extracted_k141_85325 | Not-determined |
| S12_extracted_k141_22323 | Not-determined |
| S12_extracted_k141_116622 | Not-determined |
| S12_extracted_k141_30546 | Not-determined |
| S12_extracted_k141_141205 | Not-determined |
| S12_extracted_k141_141206 | Not-determined |
| S12_extracted_k141_179369 | Not-determined |
| S12_extracted_k141_141207 | Not-determined |
| S12_extracted_k141_171859 | Not-determined |
| S12_extracted_k141_30550 | Not-determined |
| S12_extracted_k141_179370 | Not-determined |
| S12_extracted_k141_188364 | Not-determined |
| S12_extracted_k141_148752 | Not-determined |
| S12_extracted_k141_22332 | Not-determined |
| S12_extracted_k141_164231 | Not-determined |
| S12_extracted_k141_124401 | Not-determined |
| S12_extracted_k141_47014 | Low-quality |
| S12_extracted_k141_22336 | Not-determined |
| S12_extracted_k141_13971 | Not-determined |
| S12_extracted_k141_85337 | Not-determined |
| S12_extracted_k141_188371 | Not-determined |
| S12_extracted_k141_47018 | Low-quality |
| S12_extracted_k141_171872 | Not-determined |
| S12_extracted_k141_69925 | Not-determined |
| S12_extracted_k141_62126 | Not-determined |
| S12_extracted_k141_22341 | Not-determined |
| S12_extracted_k141_22345 | Not-determined |
| S12_extracted_k141_148762 | Not-determined |
| S12_extracted_k141_62127 | Low-quality |
| S12_extracted_k141_124412 | Not-determined |
| S12_extracted_k141_109687 | Not-determined |
| S12_extracted_k141_77598 | Not-determined |
| S12_extracted_k141_124413 | Not-determined |
| S12_extracted_k141_124414 | Not-determined |
| S12_extracted_k141_7268 | Not-determined |
| S12_extracted_k141_148767 | Not-determined |
| S12_extracted_k141_85346 | Low-quality |
| S12_extracted_k141_171886 | Not-determined |
| S12_extracted_k141_109690 | Not-determined |
| S12_extracted_k141_47034 | Not-determined |
| S12_extracted_k141_77607 | Not-determined |
| S12_extracted_k141_124421 | Not-determined |
| S12_extracted_k141_132600 | Low-quality |
| S12_extracted_k141_109698 | Not-determined |
| S12_extracted_k141_109704 | Not-determined |
| S12_extracted_k141_30555 | Not-determined |
| S12_extracted_k141_22361 | Low-quality |
| S12_extracted_k141_124431 | Low-quality |
| S12_extracted_k141_164258 | Not-determined |
| S12_extracted_k141_13987 | Not-determined |
| S12_extracted_k141_171903 | Not-determined |
| S12_extracted_k141_77622 | Not-determined |
| S12_extracted_k141_148777 | Not-determined |
| S12_extracted_k141_85370 | Low-quality |
| S12_extracted_k141_179385 | Not-determined |
| S12_extracted_k141_85373 | Not-determined |
| S12_extracted_k141_22370 | Not-determined |
| S12_extracted_k141_7278 | Low-quality |
| S12_extracted_k141_132616 | Not-determined |
| S12_extracted_k141_171909 | Not-determined |
| S12_extracted_k141_85379 | Low-quality |
| S12_extracted_k141_188396 | Not-determined |
| S12_extracted_k141_148782 | Not-determined |
| S12_extracted_k141_85384 | Not-determined |
| S12_extracted_k141_7288 | Not-determined |
| S12_extracted_k141_85385 | Low-quality |
| S12_extracted_k141_30568 | Not-determined |
| S12_extracted_k141_102293 | Not-determined |
| S12_extracted_k141_69961 | Not-determined |
| S12_extracted_k141_7292 | Not-determined |
| S12_extracted_k141_62164 | Not-determined |
| S12_extracted_k141_109721 | Not-determined |
| S12_extracted_k141_85390 | Not-determined |
| S12_extracted_k141_85391 | Not-determined |
| S12_extracted_k141_85392 | Not-determined |
| S12_extracted_k141_148791 | Not-determined |
| S12_extracted_k141_77634 | Not-determined |
| S12_extracted_k141_188403 | Not-determined |
| S12_extracted_k141_37780 | Not-determined |
| S12_extracted_k141_188407 | Not-determined |
| S12_extracted_k141_77636 | Not-determined |
| S12_extracted_k141_47063 | Not-determined |
| S12_extracted_k141_47064 | Not-determined |
| S12_extracted_k141_7302 | Not-determined |
| S12_extracted_k141_188411 | Not-determined |
| S12_extracted_k141_14001 | Not-determined |
| S12_extracted_k141_188417 | Not-determined |
| S12_extracted_k141_141246 | Not-determined |
| S12_extracted_k141_188420 | Not-determined |
| S12_extracted_k141_14005 | Not-determined |
| S12_extracted_k141_188421 | Not-determined |
| S12_extracted_k141_30579 | Not-determined |
| S12_extracted_k141_132637 | Not-determined |
| S12_extracted_k141_7310 | Not-determined |
| S12_extracted_k141_37794 | Low-quality |
| S12_extracted_k141_7316 | Not-determined |
| S12_extracted_k141_37796 | Not-determined |
| S12_extracted_k141_22391 | Not-determined |
| S12_extracted_k141_188428 | Not-determined |
| S12_extracted_k141_77650 | Not-determined |
| S12_extracted_k141_141254 | Not-determined |
| S12_extracted_k141_188429 | Not-determined |
| S12_extracted_k141_188432 | Not-determined |
| S12_extracted_k141_179402 | Low-quality |
| S12_extracted_k141_141259 | Not-determined |
| S12_extracted_k141_37808 | Not-determined |
| S12_extracted_k141_179406 | Low-quality |
| S12_extracted_k141_37809 | Not-determined |
| S12_extracted_k141_22401 | Not-determined |
| S12_extracted_k141_77657 | Not-determined |
| S12_extracted_k141_188440 | Not-determined |
| S12_extracted_k141_148806 | Not-determined |
| S12_extracted_k141_77660 | Not-determined |
| S12_extracted_k141_188442 | Not-determined |
| S12_extracted_k141_116647 | Not-determined |
| S12_extracted_k141_22406 | Not-determined |
| S12_extracted_k141_62183 | Low-quality |
| S12_extracted_k141_93953 | Not-determined |
| S12_extracted_k141_37814 | Not-determined |
| S12_extracted_k141_141270 | Not-determined |
| S12_extracted_k141_141271 | Not-determined |
| S12_extracted_k141_53939 | Not-determined |
| S12_extracted_k141_77661 | Low-quality |
| S12_extracted_k141_53944 | Not-determined |
| S12_extracted_k141_141280 | Not-determined |
| S12_extracted_k141_93960 | Not-determined |
| S12_extracted_k141_30597 | Not-determined |
| S12_extracted_k141_47097 | Low-quality |
| S12_extracted_k141_141283 | Not-determined |
| S12_extracted_k141_69985 | Low-quality |
| S12_extracted_k141_53952 | Not-determined |
| S12_extracted_k141_124441 | Low-quality |
| S12_extracted_k141_53955 | Not-determined |
| S12_extracted_k141_132670 | Not-determined |
| S12_extracted_k141_53957 | Not-determined |
| S12_extracted_k141_109751 | Low-quality |
| S12_extracted_k141_179426 | Not-determined |
| S12_extracted_k141_7344 | Not-determined |
| S12_extracted_k141_30606 | Not-determined |
| S12_extracted_k141_22418 | Not-determined |
| S12_extracted_k141_69990 | Not-determined |
| S12_extracted_k141_188448 | Not-determined |
| S12_extracted_k141_179428 | Not-determined |
| S12_extracted_k141_53960 | Not-determined |
| S12_extracted_k141_116660 | Low-quality |
| S12_extracted_k141_14046 | Not-determined |
| S12_extracted_k141_62204 | Low-quality |
| S12_extracted_k141_7352 | Not-determined |
| S12_extracted_k141_37829 | Not-determined |
| S12_extracted_k141_109757 | Not-determined |
| S12_extracted_k141_93968 | Not-determined |
| S12_extracted_k141_70003 | Not-determined |
| S12_extracted_k141_124456 | Low-quality |
| S12_extracted_k141_116668 | Not-determined |
| S12_extracted_k141_14053 | Not-determined |
| S12_extracted_k141_148839 | Not-determined |
| S12_extracted_k141_62210 | Not-determined |
| S12_extracted_k141_141300 | Not-determined |
| S12_extracted_k141_179444 | Not-determined |
| S12_extracted_k141_124457 | Not-determined |
| S12_extracted_k141_7361 | Not-determined |
| S12_extracted_k141_62213 | Not-determined |
| S12_extracted_k141_109764 | Not-determined |
| S12_extracted_k141_14057 | Not-determined |
| S12_extracted_k141_93975 | Not-determined |
| S12_extracted_k141_37839 | Not-determined |
| S12_extracted_k141_22433 | Not-determined |
| S12_extracted_k141_70015 | Not-determined |
| S12_extracted_k141_179450 | Not-determined |
| S12_extracted_k141_132681 | Not-determined |
| S12_extracted_k141_22436 | Not-determined |
| S12_extracted_k141_179451 | Not-determined |
| S12_extracted_k141_70020 | Low-quality |
| S12_extracted_k141_132685 | Not-determined |
| S12_extracted_k141_53974 | Not-determined |
| S12_extracted_k141_22438 | Low-quality |
| S12_extracted_k141_124461 | Not-determined |
| S12_extracted_k141_22441 | Not-determined |
| S12_extracted_k141_37847 | Low-quality |
| S12_extracted_k141_22442 | Not-determined |
| S12_extracted_k141_179457 | Not-determined |
| S12_extracted_k141_14068 | Not-determined |
| S12_extracted_k141_47115 | Low-quality |
| S12_extracted_k141_179459 | Not-determined |
| S12_extracted_k141_77683 | Not-determined |
| S12_extracted_k141_93988 | Low-quality |
| S12_extracted_k141_93990 | Not-determined |
| S12_extracted_k141_14073 | Not-determined |
| S12_extracted_k141_77685 | Not-determined |
| S12_extracted_k141_53988 | Not-determined |
| S12_extracted_k141_7376 | Not-determined |
| S12_extracted_k141_22447 | Not-determined |
| S12_extracted_k141_148854 | Not-determined |
| S12_extracted_k141_22449 | Not-determined |
| S12_extracted_k141_124476 | Not-determined |
| S12_extracted_k141_85455 | Not-determined |
| S12_extracted_k141_85456 | Not-determined |
| S12_extracted_k141_188471 | Not-determined |
| S12_extracted_k141_188473 | Not-determined |
| S12_extracted_k141_77693 | Not-determined |
| S12_extracted_k141_30628 | Not-determined |
| S12_extracted_k141_30629 | Not-determined |
| S12_extracted_k141_37863 | Low-quality |
| S12_extracted_k141_124482 | Low-quality |
| S12_extracted_k141_124483 | Not-determined |
| S12_extracted_k141_14079 | Not-determined |
| S12_extracted_k141_164279 | Not-determined |
| S12_extracted_k141_179479 | Not-determined |
| S12_extracted_k141_148861 | Not-determined |
| S12_extracted_k141_77698 | Not-determined |
| S12_extracted_k141_77700 | Not-determined |
| S12_extracted_k141_62222 | Low-quality |
| S12_extracted_k141_62223 | Not-determined |
| S12_extracted_k141_62224 | Not-determined |
| S12_extracted_k141_53996 | Not-determined |
| S12_extracted_k141_85467 | Low-quality |
| S12_extracted_k141_14087 | Not-determined |
| S12_extracted_k141_124488 | Not-determined |
| S12_extracted_k141_188483 | Not-determined |
| S12_extracted_k141_30636 | Not-determined |
| S12_extracted_k141_188486 | Not-determined |
| S12_extracted_k141_7390 | Not-determined |
| S12_extracted_k141_179489 | Not-determined |
| S12_extracted_k141_54007 | Not-determined |
| S12_extracted_k141_179492 | Not-determined |
| S12_extracted_k141_37872 | Not-determined |
| S12_extracted_k141_70049 | Not-determined |
| S12_extracted_k141_85472 | Low-quality |
| S12_extracted_k141_70051 | Not-determined |
| S12_extracted_k141_77708 | Not-determined |
| S12_extracted_k141_70053 | Not-determined |
| S12_extracted_k141_179496 | Not-determined |
| S12_extracted_k141_77709 | Not-determined |
| S12_extracted_k141_124503 | Not-determined |
| S12_extracted_k141_7396 | Not-determined |
| S12_extracted_k141_164294 | Not-determined |
| S12_extracted_k141_164297 | Not-determined |
| S12_extracted_k141_7403 | Not-determined |
| S12_extracted_k141_188498 | Not-determined |
| S12_extracted_k141_116708 | Not-determined |
| S12_extracted_k141_93994 | Not-determined |
| S12_extracted_k141_116710 | Not-determined |
| S12_extracted_k141_188501 | Not-determined |
| S12_extracted_k141_93996 | Not-determined |
| S12_extracted_k141_124513 | Not-determined |
| S12_extracted_k141_124515 | Not-determined |
| S12_extracted_k141_179509 | Not-determined |
| S12_extracted_k141_148877 | Not-determined |
| S12_extracted_k141_85484 | Not-determined |
| S12_extracted_k141_148879 | Not-determined |
| S12_extracted_k141_85486 | Not-determined |
| S12_extracted_k141_148880 | Low-quality |
| S12_extracted_k141_116717 | Not-determined |
| S12_extracted_k141_47140 | Not-determined |
| S12_extracted_k141_30657 | Not-determined |
| S12_extracted_k141_7410 | Not-determined |
| S12_extracted_k141_14110 | Not-determined |
| S12_extracted_k141_62257 | Not-determined |
| S12_extracted_k141_188504 | Not-determined |
| S12_extracted_k141_148883 | Not-determined |
| S12_extracted_k141_7414 | Not-determined |
| S12_extracted_k141_7415 | Not-determined |
| S12_extracted_k141_77721 | Not-determined |
| S12_extracted_k141_85490 | Not-determined |
| S12_extracted_k141_124520 | Not-determined |
| S12_extracted_k141_85492 | Not-determined |
| S12_extracted_k141_7417 | Not-determined |
| S12_extracted_k141_109795 | Not-determined |
| S12_extracted_k141_77727 | Not-determined |
| S12_extracted_k141_22490 | Not-determined |
| S12_extracted_k141_47143 | Not-determined |
| S12_extracted_k141_109796 | Not-determined |
| S12_extracted_k141_188512 | Not-determined |
| S12_extracted_k141_124529 | Low-quality |
| S12_extracted_k141_30674 | Not-determined |
| S12_extracted_k141_116724 | Not-determined |
| S12_extracted_k141_7424 | Not-determined |
| S12_extracted_k141_124530 | Not-determined |
| S12_extracted_k141_102343 | Not-determined |
| S12_extracted_k141_7426 | Not-determined |
| S12_extracted_k141_77734 | Not-determined |
| S12_extracted_k141_54030 | Not-determined |
| S12_extracted_k141_30678 | Not-determined |
| S12_extracted_k141_77735 | Not-determined |
| S12_extracted_k141_47148 | Not-determined |
| S12_extracted_k141_14124 | Low-quality |
| S12_extracted_k141_124533 | Not-determined |
| S12_extracted_k141_47149 | Not-determined |
| S12_extracted_k141_148891 | Not-determined |
| S12_extracted_k141_179535 | Not-determined |
| S12_extracted_k141_7434 | Not-determined |
| S12_extracted_k141_77743 | Not-determined |
| S12_extracted_k141_62275 | Not-determined |
| S12_extracted_k141_47151 | Not-determined |
| S12_extracted_k141_164304 | Not-determined |
| S12_extracted_k141_109816 | Not-determined |
| S12_extracted_k141_70059 | Not-determined |
| S12_extracted_k141_7441 | Low-quality |
| S12_extracted_k141_77754 | Not-determined |
| S12_extracted_k141_77755 | Not-determined |
| S12_extracted_k141_77757 | Not-determined |

| | |
|---|---|
| S12_extracted_k141_102355 | Not-determined |
| S12_extracted_k141_179547 | Not-determined |
| S12_extracted_k141_47157 | Not-determined |
| S12_extracted_k141_132703 | Not-determined |
| S12_extracted_k141_14136 | Not-determined |
| S12_extracted_k141_132706 | Not-determined |
| S12_extracted_k141_116742 | Low-quality |
| S12_extracted_k141_109817 | Low-quality |
| S12_extracted_k141_77763 | Not-determined |
| S12_extracted_k141_179552 | Not-determined |
| S12_extracted_k141_62292 | Not-determined |
| S12_extracted_k141_7451 | Not-determined |
| S12_extracted_k141_62294 | Low-quality |
| S12_extracted_k141_7452 | Not-determined |
| S12_extracted_k141_14143 | Not-determined |
| S12_extracted_k141_124546 | Not-determined |
| S12_extracted_k141_54048 | Not-determined |
| S12_extracted_k141_62296 | Low-quality |
| S12_extracted_k141_109820 | Not-determined |
| S12_extracted_k141_47162 | Not-determined |
| S12_extracted_k141_109821 | Not-determined |
| S12_extracted_k141_70074 | Not-determined |
| S12_extracted_k141_14149 | Not-determined |
| S12_extracted_k141_62297 | Not-determined |
| S12_extracted_k141_109827 | Not-determined |
| S12_extracted_k141_37892 | Not-determined |
| S12_extracted_k141_47170 | Not-determined |
| S12_extracted_k141_14155 | Not-determined |
| S12_extracted_k141_70083 | Not-determined |
| S12_extracted_k141_109836 | Not-determined |
| S12_extracted_k141_116754 | Not-determined |
| S12_extracted_k141_37900 | Not-determined |
| S12_extracted_k141_77774 | Not-determined |
| S12_extracted_k141_14157 | Not-determined |
| S12_extracted_k141_109839 | Not-determined |
| S12_extracted_k141_77775 | Not-determined |
| S12_extracted_k141_132725 | Not-determined |
| S12_extracted_k141_141343 | Not-determined |
| S12_extracted_k141_164328 | Not-determined |
| S12_extracted_k141_164329 | Not-determined |
| S12_extracted_k141_54063 | Low-quality |
| S12_extracted_k141_132728 | Low-quality |
| S12_extracted_k141_148928 | Not-determined |
| S12_extracted_k141_62308 | Not-determined |
| S12_extracted_k141_116763 | Not-determined |
| S12_extracted_k141_70088 | Not-determined |
| S12_extracted_k141_85506 | Not-determined |
| S12_extracted_k141_116765 | Not-determined |
| S12_extracted_k141_85507 | Not-determined |
| S12_extracted_k141_85509 | Not-determined |
| S12_extracted_k141_7457 | Not-determined |
| S12_extracted_k141_124564 | Not-determined |
| S12_extracted_k141_85511 | Not-determined |
| S12_extracted_k141_109850 | Not-determined |
| S12_extracted_k141_7458 | Not-determined |
| S12_extracted_k141_37907 | Not-determined |
| S12_extracted_k141_37908 | Not-determined |
| S12_extracted_k141_132730 | Not-determined |
| S12_extracted_k141_70094 | Not-determined |
| S12_extracted_k141_70095 | Not-determined |
| S12_extracted_k141_7461 | Not-determined |
| S12_extracted_k141_116768 | Not-determined |
| S12_extracted_k141_54068 | Not-determined |
| S12_extracted_k141_141361 | Not-determined |
| S12_extracted_k141_85515 | Not-determined |
| S12_extracted_k141_116777 | Not-determined |
| S12_extracted_k141_141366 | Not-determined |
| S12_extracted_k141_132741 | Low-quality |
| S12_extracted_k141_77792 | Not-determined |
| S12_extracted_k141_85519 | Not-determined |
| S12_extracted_k141_7471 | Not-determined |
| S12_extracted_k141_148941 | Not-determined |
| S12_extracted_k141_148943 | Not-determined |
| S12_extracted_k141_132750 | Not-determined |
| S12_extracted_k141_7476 | Not-determined |
| S12_extracted_k141_132751 | Low-quality |
| S12_extracted_k141_141373 | Not-determined |
| S12_extracted_k141_14171 | Not-determined |
| S12_extracted_k141_22522 | Not-determined |
| S12_extracted_k141_14172 | Not-determined |
| S12_extracted_k141_30685 | Not-determined |
| S12_extracted_k141_54089 | Not-determined |
| S12_extracted_k141_14178 | Not-determined |
| S12_extracted_k141_7481 | Not-determined |
| S12_extracted_k141_85531 | Not-determined |
| S12_extracted_k141_14181 | Low-quality |
| S12_extracted_k141_148950 | Not-determined |
| S12_extracted_k141_85534 | Not-determined |
| S12_extracted_k141_22530 | Not-determined |
| S12_extracted_k141_188524 | Not-determined |
| S12_extracted_k141_70115 | Not-determined |
| S12_extracted_k141_70116 | Not-determined |
| S12_extracted_k141_132756 | Not-determined |
| S12_extracted_k141_132757 | Not-determined |
| S12_extracted_k141_124583 | Not-determined |
| S12_extracted_k141_62321 | Not-determined |
| S12_extracted_k141_124585 | Not-determined |
| S12_extracted_k141_85544 | Not-determined |
| S12_extracted_k141_141392 | Not-determined |
| S12_extracted_k141_132763 | Not-determined |
| S12_extracted_k141_132764 | Not-determined |
| S12_extracted_k141_124591 | Not-determined |
| S12_extracted_k141_7501 | Not-determined |
| S12_extracted_k141_124594 | Not-determined |
| S12_extracted_k141_141398 | Not-determined |
| S12_extracted_k141_148957 | Not-determined |
| S12_extracted_k141_124598 | Not-determined |
| S12_extracted_k141_188537 | Not-determined |
| S12_extracted_k141_85555 | Not-determined |
| S12_extracted_k141_70128 | Not-determined |
| S12_extracted_k141_22541 | Not-determined |
| S12_extracted_k141_132773 | Not-determined |
| S12_extracted_k141_164345 | Not-determined |
| S12_extracted_k141_77813 | Not-determined |
| S12_extracted_k141_54109 | Not-determined |
| S12_extracted_k141_22544 | Not-determined |
| S12_extracted_k141_62332 | Not-determined |
| S12_extracted_k141_30691 | Low-quality |
| S12_extracted_k141_124606 | Low-quality |
| S12_extracted_k141_62337 | Low-quality |
| S12_extracted_k141_62338 | Not-determined |
| S12_extracted_k141_62340 | Not-determined |
| S12_extracted_k141_124612 | Not-determined |
| S12_extracted_k141_148977 | Not-determined |
| S12_extracted_k141_62343 | Not-determined |
| S12_extracted_k141_47204 | Not-determined |
| S12_extracted_k141_62345 | Not-determined |
| S12_extracted_k141_188555 | Not-determined |
| S12_extracted_k141_77833 | Not-determined |
| S12_extracted_k141_77839 | Not-determined |
| S12_extracted_k141_164362 | Low-quality |
| S12_extracted_k141_109890 | Not-determined |
| S12_extracted_k141_132783 | Not-determined |
| S12_extracted_k141_85576 | Not-determined |
| S12_extracted_k141_132784 | Low-quality |
| S12_extracted_k141_62359 | Not-determined |
| S12_extracted_k141_132787 | Low-quality |
| S12_extracted_k141_77845 | Low-quality |
| S12_extracted_k141_124632 | Not-determined |
| S12_extracted_k141_164368 | Not-determined |
| S12_extracted_k141_164370 | Not-determined |
| S12_extracted_k141_132791 | Not-determined |
| S12_extracted_k141_30705 | Low-quality |
| S12_extracted_k141_7521 | Not-determined |
| S12_extracted_k141_85580 | Not-determined |
| S12_extracted_k141_77849 | Not-determined |
| S12_extracted_k141_7524 | Not-determined |
| S12_extracted_k141_85583 | Not-determined |
| S12_extracted_k141_47212 | Low-quality |
| S12_extracted_k141_132799 | Not-determined |
| S12_extracted_k141_14211 | Not-determined |
| S12_extracted_k141_54138 | Low-quality |
| S12_extracted_k141_188565 | Not-determined |
| S12_extracted_k141_85592 | Low-quality |
| S12_extracted_k141_85593 | Not-determined |
| S12_extracted_k141_132811 | Not-determined |
| S12_extracted_k141_124645 | Not-determined |
| S12_extracted_k141_164382 | Not-determined |
| S12_extracted_k141_47216 | Not-determined |
| S12_extracted_k141_156021 | Not-determined |
| S12_extracted_k141_47220 | Not-determined |
| S12_extracted_k141_188578 | Not-determined |
| S12_extracted_k141_47221 | Not-determined |
| S12_extracted_k141_156025 | Not-determined |
| S12_extracted_k141_124648 | Low-quality |
| S12_extracted_k141_62369 | Not-determined |
| S12_extracted_k141_156028 | Not-determined |
| S12_extracted_k141_164388 | Low-quality |
| S12_extracted_k141_47224 | Not-determined |
| S12_extracted_k141_70164 | Not-determined |
| S12_extracted_k141_47230 | Not-determined |
| S12_extracted_k141_77869 | Not-determined |
| S12_extracted_k141_179637 | Not-determined |
| S12_extracted_k141_85604 | Not-determined |
| S12_extracted_k141_124655 | Low-quality |
| S12_extracted_k141_132820 | Not-determined |
| S12_extracted_k141_156038 | Not-determined |
| S12_extracted_k141_188588 | Not-determined |
| S12_extracted_k141_188589 | Not-determined |
| S12_extracted_k141_179641 | Not-determined |
| S12_extracted_k141_124660 | Low-quality |
| S12_extracted_k141_37960 | Not-determined |
| S12_extracted_k141_109905 | Not-determined |
| S12_extracted_k141_37962 | Not-determined |
| S12_extracted_k141_30714 | Not-determined |
| S12_extracted_k141_54167 | Not-determined |
| S12_extracted_k141_188593 | Not-determined |
| S12_extracted_k141_109909 | Not-determined |
| S12_extracted_k141_37964 | Not-determined |
| S12_extracted_k141_47240 | Not-determined |
| S12_extracted_k141_85609 | Low-quality |
| S12_extracted_k141_37966 | Not-determined |
| S12_extracted_k141_132831 | Not-determined |
| S12_extracted_k141_47243 | Not-determined |
| S12_extracted_k141_124668 | Not-determined |
| S12_extracted_k141_124669 | Low-quality |
| S12_extracted_k141_156047 | Not-determined |
| S12_extracted_k141_37970 | Low-quality |
| S12_extracted_k141_30725 | Not-determined |
| S12_extracted_k141_109918 | Low-quality |
| S12_extracted_k141_37972 | Not-determined |
| S12_extracted_k141_30727 | Not-determined |
| S12_extracted_k141_30728 | Not-determined |
| S12_extracted_k141_188606 | Not-determined |
| S12_extracted_k141_149014 | Not-determined |
| S12_extracted_k141_62391 | Not-determined |
| S12_extracted_k141_70175 | Low-quality |
| S12_extracted_k141_149018 | Not-determined |
| S12_extracted_k141_132842 | Not-determined |
| S12_extracted_k141_77903 | Not-determined |
| S12_extracted_k141_14247 | Not-determined |
| S12_extracted_k141_62398 | Not-determined |
| S12_extracted_k141_70180 | Not-determined |
| S12_extracted_k141_70181 | Not-determined |
| S12_extracted_k141_14251 | Not-determined |
| S12_extracted_k141_7531 | Low-quality |
| S12_extracted_k141_149023 | Not-determined |
| S12_extracted_k141_116846 | Not-determined |
| S12_extracted_k141_62402 | Not-determined |
| S12_extracted_k141_77908 | Not-determined |
| S12_extracted_k141_188618 | Not-determined |
| S12_extracted_k141_77909 | Not-determined |
| S12_extracted_k141_37980 | Low-quality |
| S12_extracted_k141_188619 | Not-determined |
| S12_extracted_k141_149029 | Not-determined |
| S12_extracted_k141_149030 | Not-determined |
| S12_extracted_k141_62407 | Not-determined |
| S12_extracted_k141_85620 | Not-determined |
| S12_extracted_k141_85621 | Low-quality |
| S12_extracted_k141_156066 | Not-determined |
| S12_extracted_k141_62409 | Not-determined |
| S12_extracted_k141_164413 | Not-determined |
| S12_extracted_k141_149036 | Low-quality |
| S12_extracted_k141_37985 | Not-determined |
| S12_extracted_k141_85623 | Not-determined |
| S12_extracted_k141_70186 | Not-determined |
| S12_extracted_k141_179654 | Low-quality |
| S12_extracted_k141_70189 | Not-determined |
| S12_extracted_k141_70190 | Not-determined |
| S12_extracted_k141_54184 | Low-quality |
| S12_extracted_k141_37990 | Not-determined |
| S12_extracted_k141_124675 | Not-determined |
| S12_extracted_k141_14267 | Not-determined |
| S12_extracted_k141_54189 | Low-quality |
| S12_extracted_k141_62419 | Not-determined |
| S12_extracted_k141_54191 | Low-quality |
| S12_extracted_k141_164431 | Not-determined |
| S12_extracted_k141_70197 | Not-determined |
| S12_extracted_k141_164432 | Low-quality |
| S12_extracted_k141_54194 | Not-determined |
| S12_extracted_k141_149056 | Not-determined |
| S12_extracted_k141_149057 | Not-determined |
| S12_extracted_k141_14273 | Low-quality |
| S12_extracted_k141_188633 | Not-determined |
| S12_extracted_k141_149062 | Not-determined |
| S12_extracted_k141_54201 | Not-determined |
| S12_extracted_k141_14279 | Not-determined |
| S12_extracted_k141_179672 | Not-determined |
| S12_extracted_k141_124680 | Not-determined |
| S12_extracted_k141_14283 | Not-determined |
| S12_extracted_k141_85636 | Not-determined |
| S12_extracted_k141_164441 | Not-determined |
| S12_extracted_k141_179675 | Not-determined |
| S12_extracted_k141_70213 | Not-determined |
| S12_extracted_k141_54208 | Not-determined |
| S12_extracted_k141_149074 | Not-determined |
| S12_extracted_k141_38002 | Not-determined |
| S12_extracted_k141_7540 | Not-determined |
| S12_extracted_k141_132873 | Not-determined |
| S12_extracted_k141_149075 | Not-determined |
| S12_extracted_k141_179684 | Not-determined |
| S12_extracted_k141_116855 | Not-determined |
| S12_extracted_k141_116856 | Not-determined |
| S12_extracted_k141_14290 | Not-determined |
| S12_extracted_k141_85646 | Not-determined |
| S12_extracted_k141_109960 | Not-determined |
| S12_extracted_k141_116857 | Not-determined |
| S12_extracted_k141_132878 | Not-determined |
| S12_extracted_k141_109964 | Not-determined |
| S12_extracted_k141_132880 | Not-determined |
| S12_extracted_k141_179694 | Not-determined |
| S12_extracted_k141_116864 | Not-determined |
| S12_extracted_k141_14302 | Low-quality |
| S12_extracted_k141_149087 | Not-determined |
| S12_extracted_k141_164457 | Not-determined |
| S12_extracted_k141_179698 | Not-determined |
| S12_extracted_k141_132893 | Not-determined |
| S12_extracted_k141_54218 | Not-determined |
| S12_extracted_k141_141408 | Not-determined |
| S12_extracted_k141_188650 | Not-determined |
| S12_extracted_k141_54224 | Low-quality |
| S12_extracted_k141_109981 | Not-determined |
| S12_extracted_k141_156087 | Low-quality |
| S12_extracted_k141_94021 | Not-determined |
| S12_extracted_k141_109983 | Low-quality |
| S12_extracted_k141_14320 | Not-determined |
| S12_extracted_k141_85652 | Not-determined |
| S12_extracted_k141_149100 | Not-determined |
| S12_extracted_k141_141418 | Not-determined |
| S12_extracted_k141_141420 | Not-determined |
| S12_extracted_k141_70232 | Low-quality |
| S12_extracted_k141_179709 | Not-determined |
| S12_extracted_k141_94029 | Not-determined |
| S12_extracted_k141_54232 | Not-determined |
| S12_extracted_k141_109993 | Not-determined |
| S12_extracted_k141_14327 | Not-determined |
| S12_extracted_k141_85657 | Not-determined |
| S12_extracted_k141_7547 | Not-determined |
| S12_extracted_k141_54237 | Not-determined |
| S12_extracted_k141_7548 | Low-quality |
| S12_extracted_k141_116881 | Not-determined |
| S12_extracted_k141_116883 | Not-determined |
| S12_extracted_k141_14331 | Not-determined |
| S12_extracted_k141_94038 | Not-determined |
| S12_extracted_k141_85659 | Not-determined |
| S12_extracted_k141_54240 | Not-determined |
| S12_extracted_k141_116884 | Not-determined |
| S12_extracted_k141_149109 | Low-quality |
| S12_extracted_k141_179716 | Not-determined |
| S12_extracted_k141_54245 | Not-determined |
| S12_extracted_k141_109998 | Not-determined |
| S12_extracted_k141_156097 | Not-determined |
| S12_extracted_k141_54252 | Not-determined |
| S12_extracted_k141_30775 | Not-determined |
| S12_extracted_k141_116890 | Not-determined |
| S12_extracted_k141_54253 | Not-determined |
| S12_extracted_k141_149116 | Not-determined |
| S12_extracted_k141_85664 | Not-determined |
| S12_extracted_k141_141432 | Not-determined |
| S12_extracted_k141_7563 | Not-determined |
| S12_extracted_k141_141433 | Not-determined |
| S12_extracted_k141_62479 | Not-determined |
| S12_extracted_k141_179729 | Low-quality |
| S12_extracted_k141_85672 | Not-determined |
| S12_extracted_k141_179730 | Not-determined |
| S12_extracted_k141_30783 | Not-determined |
| S12_extracted_k141_7570 | Not-determined |
| S12_extracted_k141_14343 | Not-determined |
| S12_extracted_k141_30786 | Not-determined |
| S12_extracted_k141_30794 | Not-determined |
| S12_extracted_k141_7576 | Not-determined |
| S12_extracted_k141_14348 | Not-determined |
| S12_extracted_k141_38027 | Not-determined |
| S12_extracted_k141_38028 | Low-quality |
| S12_extracted_k141_14350 | Not-determined |
| S12_extracted_k141_38035 | Not-determined |
| S12_extracted_k141_38036 | Not-determined |
| S12_extracted_k141_22560 | Not-determined |
| S12_extracted_k141_14363 | Low-quality |
| S12_extracted_k141_179739 | Not-determined |
| S12_extracted_k141_141455 | Not-determined |
| S12_extracted_k141_110018 | Not-determined |
| S12_extracted_k141_85691 | Not-determined |
| S12_extracted_k141_179742 | Low-quality |
| S12_extracted_k141_85696 | Not-determined |
| S12_extracted_k141_110025 | Not-determined |
| S12_extracted_k141_110027 | Not-determined |
| S12_extracted_k141_179749 | Not-determined |
| S12_extracted_k141_62493 | Low-quality |
| S12_extracted_k141_179755 | Not-determined |
| S12_extracted_k141_110037 | Not-determined |
| S12_extracted_k141_22587 | Not-determined |
| S12_extracted_k141_62496 | Not-determined |
| S12_extracted_k141_7592 | Not-determined |
| S12_extracted_k141_22590 | Not-determined |
| S12_extracted_k141_62499 | Low-quality |
| S12_extracted_k141_14371 | Not-determined |
| S12_extracted_k141_22594 | Not-determined |
| S12_extracted_k141_62503 | Low-quality |
| S12_extracted_k141_141473 | Low-quality |
| S12_extracted_k141_85714 | Low-quality |
| S12_extracted_k141_7604 | Not-determined |
| S12_extracted_k141_164514 | Low-quality |
| S12_extracted_k141_141478 | Not-determined |
| S12_extracted_k141_7608 | Not-determined |
| S12_extracted_k141_141480 | Not-determined |
| S12_extracted_k141_22605 | Not-determined |
| S12_extracted_k141_124703 | Low-quality |
| S12_extracted_k141_124704 | Not-determined |
| S12_extracted_k141_22607 | Not-determined |
| S12_extracted_k141_7614 | Not-determined |
| S12_extracted_k141_149156 | Not-determined |
| S12_extracted_k141_30819 | Not-determined |
| S12_extracted_k141_124708 | Not-determined |
| S12_extracted_k141_70297 | Not-determined |
| S12_extracted_k141_62517 | Not-determined |
| S12_extracted_k141_30834 | Low-quality |
| S12_extracted_k141_78011 | Not-determined |
| S12_extracted_k141_54276 | Not-determined |
| S12_extracted_k141_7628 | Not-determined |
| S12_extracted_k141_30836 | Not-determined |
| S12_extracted_k141_14383 | Not-determined |
| S12_extracted_k141_179784 | Not-determined |
| S12_extracted_k141_78014 | Not-determined |
| S12_extracted_k141_179785 | Not-determined |
| S12_extracted_k141_7632 | Not-determined |
| S12_extracted_k141_78015 | Low-quality |
| S12_extracted_k141_85728 | Not-determined |
| S12_extracted_k141_141506 | Not-determined |
| S12_extracted_k141_94063 | Not-determined |
| S12_extracted_k141_70305 | Low-quality |
| S12_extracted_k141_14388 | Not-determined |
| S12_extracted_k141_30838 | Not-determined |
| S12_extracted_k141_85731 | Not-determined |
| S12_extracted_k141_94064 | Not-determined |
| S12_extracted_k141_14389 | Not-determined |
| S12_extracted_k141_85732 | Low-quality |
| S12_extracted_k141_22619 | Not-determined |
| S12_extracted_k141_179792 | Low-quality |
| S12_extracted_k141_179793 | Not-determined |
| S12_extracted_k141_70311 | Not-determined |
| S12_extracted_k141_85734 | Not-determined |
| S12_extracted_k141_141511 | Low-quality |
| S12_extracted_k141_30847 | Not-determined |
| S12_extracted_k141_30848 | Not-determined |
| S12_extracted_k141_62535 | Not-determined |
| S12_extracted_k141_110055 | Not-determined |
| S12_extracted_k141_85744 | Not-determined |
| S12_extracted_k141_70320 | Not-determined |
| S12_extracted_k141_62538 | Not-determined |
| S12_extracted_k141_149170 | Not-determined |
| S12_extracted_k141_110059 | Not-determined |
| S12_extracted_k141_14406 | Not-determined |
| S12_extracted_k141_164549 | Not-determined |
| S12_extracted_k141_149171 | Not-determined |
| S12_extracted_k141_62540 | Not-determined |
| S12_extracted_k141_164553 | Not-determined |
| S12_extracted_k141_94079 | Not-determined |
| S12_extracted_k141_7642 | Not-determined |
| S12_extracted_k141_62542 | Not-determined |
| S12_extracted_k141_141516 | Low-quality |
| S12_extracted_k141_132963 | Not-determined |
| S12_extracted_k141_141519 | Not-determined |
| S12_extracted_k141_179806 | Low-quality |
| S12_extracted_k141_22636 | Not-determined |
| S12_extracted_k141_62544 | Not-determined |
| S12_extracted_k141_54302 | Not-determined |
| S12_extracted_k141_164558 | Not-determined |
| S12_extracted_k141_141520 | Not-determined |
| S12_extracted_k141_85748 | Not-determined |
| S12_extracted_k141_22639 | Not-determined |
| S12_extracted_k141_116925 | Not-determined |
| S12_extracted_k141_94090 | Not-determined |
| S12_extracted_k141_124737 | Not-determined |
| S12_extracted_k141_141524 | Not-determined |
| S12_extracted_k141_22641 | Not-determined |
| S12_extracted_k141_164564 | Not-determined |
| S12_extracted_k141_149185 | Not-determined |
| S12_extracted_k141_85756 | Not-determined |
| S12_extracted_k141_94094 | Not-determined |
| S12_extracted_k141_179816 | Not-determined |
| S12_extracted_k141_54310 | Low-quality |
| S12_extracted_k141_54315 | Not-determined |
| S12_extracted_k141_85762 | Not-determined |
| S12_extracted_k141_30853 | Not-determined |
| S12_extracted_k141_54319 | Not-determined |
| S12_extracted_k141_85767 | Not-determined |
| S12_extracted_k141_179828 | Not-determined |
| S12_extracted_k141_141544 | Not-determined |
| S12_extracted_k141_94099 | Not-determined |
| S12_extracted_k141_22663 | Not-determined |
| S12_extracted_k141_70346 | Not-determined |
| S12_extracted_k141_22664 | Low-quality |
| S12_extracted_k141_7669 | Low-quality |
| S12_extracted_k141_54321 | Low-quality |
| S12_extracted_k141_110096 | Not-determined |
| S12_extracted_k141_94113 | Not-determined |
| S12_extracted_k141_54325 | Not-determined |
| S12_extracted_k141_132987 | Not-determined |
| S12_extracted_k141_94115 | Not-determined |
| S12_extracted_k141_132988 | Not-determined |
| S12_extracted_k141_132989 | Not-determined |
| S12_extracted_k141_116945 | Low-quality |
| S12_extracted_k141_7680 | Not-determined |
| S12_extracted_k141_132993 | Not-determined |
| S12_extracted_k141_85787 | Low-quality |
| S12_extracted_k141_22676 | Not-determined |
| S12_extracted_k141_94118 | Not-determined |
| S12_extracted_k141_14446 | Not-determined |
| S12_extracted_k141_110105 | Not-determined |
| S12_extracted_k141_156101 | Low-quality |
| S12_extracted_k141_179852 | Not-determined |
| S12_extracted_k141_62568 | Not-determined |
| S12_extracted_k141_54340 | Not-determined |
| S12_extracted_k141_156108 | Not-determined |
| S12_extracted_k141_54344 | Not-determined |
| S12_extracted_k141_133004 | Not-determined |
| S12_extracted_k141_110111 | Not-determined |
| S12_extracted_k141_85792 | Not-determined |
| S12_extracted_k141_133007 | Not-determined |
| S12_extracted_k141_70374 | Not-determined |
| S12_extracted_k141_62573 | Not-determined |
| S12_extracted_k141_141562 | Not-determined |
| S12_extracted_k141_149214 | Not-determined |
| S12_extracted_k141_70378 | Not-determined |
| S12_extracted_k141_7692 | Not-determined |
| S12_extracted_k141_38068 | Not-determined |
| S12_extracted_k141_179858 | Not-determined |
| S12_extracted_k141_85794 | Low-quality |
| S12_extracted_k141_14459 | Low-quality |
| S12_extracted_k141_7698 | Not-determined |
| S12_extracted_k141_141567 | Not-determined |
| S12_extracted_k141_124744 | Not-determined |
| S12_extracted_k141_22695 | Not-determined |
| S12_extracted_k141_14463 | Not-determined |
| S12_extracted_k141_70382 | Not-determined |
| S12_extracted_k141_149221 | Not-determined |
| S12_extracted_k141_116958 | Not-determined |
| S12_extracted_k141_149222 | Not-determined |
| S12_extracted_k141_70384 | Not-determined |
| S12_extracted_k141_116962 | Not-determined |
| S12_extracted_k141_133018 | Not-determined |
| S12_extracted_k141_78059 | Not-determined |
| S12_extracted_k141_22706 | Not-determined |
| S12_extracted_k141_38082 | Not-determined |
| S12_extracted_k141_116968 | Not-determined |
| S12_extracted_k141_116970 | Not-determined |
| S12_extracted_k141_22708 | Not-determined |
| S12_extracted_k141_179862 | Low-quality |
| S12_extracted_k141_116974 | Not-determined |
| S12_extracted_k141_54362 | Low-quality |
| S12_extracted_k141_133025 | Not-determined |
| S12_extracted_k141_38084 | Not-determined |
| S12_extracted_k141_38085 | Low-quality |
| S12_extracted_k141_116975 | Not-determined |
| S12_extracted_k141_38087 | Not-determined |
| S12_extracted_k141_149225 | Not-determined |
| S12_extracted_k141_22720 | Not-determined |
| S12_extracted_k141_116978 | Not-determined |
| S12_extracted_k141_116979 | Not-determined |
| S12_extracted_k141_22723 | Not-determined |
| S12_extracted_k141_116983 | Not-determined |
| S12_extracted_k141_22725 | Not-determined |
| S12_extracted_k141_171924 | Not-determined |
| S12_extracted_k141_110142 | Not-determined |
| S12_extracted_k141_54381 | Not-determined |
| S12_extracted_k141_22735 | Not-determined |
| S12_extracted_k141_124759 | Not-determined |
| S12_extracted_k141_171935 | Not-determined |
| S12_extracted_k141_22741 | Not-determined |
| S12_extracted_k141_94128 | Not-determined |
| S12_extracted_k141_149242 | Not-determined |
| S12_extracted_k141_141604 | Low-quality |
| S12_extracted_k141_94130 | Not-determined |
| S12_extracted_k141_116996 | Not-determined |
| S12_extracted_k141_70412 | Not-determined |
| S12_extracted_k141_179886 | Low-quality |
| S12_extracted_k141_141606 | Not-determined |
| S12_extracted_k141_124765 | Not-determined |
| S12_extracted_k141_124766 | Not-determined |
| S12_extracted_k141_78088 | Not-determined |
| S12_extracted_k141_38098 | Not-determined |
| S12_extracted_k141_22747 | Low-quality |
| S12_extracted_k141_38099 | Not-determined |
| S12_extracted_k141_171945 | Not-determined |
| S12_extracted_k141_14473 | Low-quality |
| S12_extracted_k141_22751 | Not-determined |
| S12_extracted_k141_14475 | Not-determined |
| S12_extracted_k141_38107 | Not-determined |
| S12_extracted_k141_14477 | Not-determined |
| S12_extracted_k141_22757 | Not-determined |
| S12_extracted_k141_70425 | Not-determined |
| S12_extracted_k141_7712 | Not-determined |
| S12_extracted_k141_14479 | Low-quality |
| S12_extracted_k141_171953 | Not-determined |
| S12_extracted_k141_7714 | Not-determined |
| S12_extracted_k141_22761 | Not-determined |
| S12_extracted_k141_38114 | Not-determined |
| S12_extracted_k141_171957 | Not-determined |
| S12_extracted_k141_171958 | Not-determined |
| S12_extracted_k141_7716 | Not-determined |
| S12_extracted_k141_94145 | Not-determined |
| S12_extracted_k141_164602 | Not-determined |
| S12_extracted_k141_117013 | Not-determined |
| S12_extracted_k141_30942 | Not-determined |
| S12_extracted_k141_14494 | Not-determined |
| S12_extracted_k141_117015 | Not-determined |
| S12_extracted_k141_149265 | Not-determined |
| S12_extracted_k141_156125 | Low-quality |
| S12_extracted_k141_14498 | Not-determined |
| S12_extracted_k141_14499 | Not-determined |
| S12_extracted_k141_149271 | Not-determined |
| S12_extracted_k141_156130 | Not-determined |
| S12_extracted_k141_38127 | Not-determined |
| S12_extracted_k141_14501 | Not-determined |
| S12_extracted_k141_62605 | Not-determined |
| S12_extracted_k141_38128 | Not-determined |
| S12_extracted_k141_110170 | Not-determined |
| S12_extracted_k141_85804 | Not-determined |

| | |
|---|---|
| S12_extracted_k141_110171 | Not-determined |
| S12_extracted_k141_54395 | Not-determined |
| S12_extracted_k141_22777 | Low-quality |
| S12_extracted_k141_38131 | Not-determined |
| S12_extracted_k141_110172 | Not-determined |
| S12_extracted_k141_164609 | Not-determined |
| S12_extracted_k141_70441 | Not-determined |
| S12_extracted_k141_54397 | Low-quality |
| S12_extracted_k141_85807 | Low-quality |
| S12_extracted_k141_156136 | Not-determined |
| S12_extracted_k141_117018 | Low-quality |
| S12_extracted_k141_179910 | Not-determined |
| S12_extracted_k141_30959 | Not-determined |
| S12_extracted_k141_117019 | Low-quality |
| S12_extracted_k141_78102 | Not-determined |
| S12_extracted_k141_85813 | Not-determined |
| S12_extracted_k141_22785 | Not-determined |
| S12_extracted_k141_62612 | Not-determined |
| S12_extracted_k141_38135 | Not-determined |
| S12_extracted_k141_22786 | Not-determined |
| S12_extracted_k141_22788 | Not-determined |
| S12_extracted_k141_54403 | Not-determined |
| S12_extracted_k141_70448 | Not-determined |
| S12_extracted_k141_164618 | Not-determined |
| S12_extracted_k141_141649 | Not-determined |
| S12_extracted_k141_54408 | Not-determined |
| S12_extracted_k141_70452 | Not-determined |
| S12_extracted_k141_179914 | Not-determined |
| S12_extracted_k141_78112 | Not-determined |
| S12_extracted_k141_141652 | Not-determined |
| S12_extracted_k141_141653 | Not-determined |
| S12_extracted_k141_14520 | Not-determined |
| S12_extracted_k141_70458 | Not-determined |
| S12_extracted_k141_70462 | Not-determined |
| S12_extracted_k141_179922 | Not-determined |
| S12_extracted_k141_70464 | Not-determined |
| S12_extracted_k141_30968 | Not-determined |
| S12_extracted_k141_85834 | Not-determined |
| S12_extracted_k141_62629 | Not-determined |
| S12_extracted_k141_62631 | Not-determined |
| S12_extracted_k141_124777 | Not-determined |
| S12_extracted_k141_124778 | Not-determined |
| S12_extracted_k141_124780 | Not-determined |
| S12_extracted_k141_38155 | Not-determined |
| S12_extracted_k141_149287 | Low-quality |
| S12_extracted_k141_179936 | Not-determined |
| S12_extracted_k141_110207 | Low-quality |
| S12_extracted_k141_171988 | Not-determined |
| S12_extracted_k141_110208 | Low-quality |
| S12_extracted_k141_171992 | Not-determined |
| S12_extracted_k141_62645 | Not-determined |
| S12_extracted_k141_117024 | Not-determined |
| S12_extracted_k141_94153 | Not-determined |
| S12_extracted_k141_22810 | Low-quality |
| S12_extracted_k141_54420 | Not-determined |
| S12_extracted_k141_14525 | Low-quality |
| S12_extracted_k141_171998 | Not-determined |
| S12_extracted_k141_30990 | Not-determined |
| S12_extracted_k141_62650 | Not-determined |
| S12_extracted_k141_38162 | Not-determined |
| S12_extracted_k141_171999 | Not-determined |
| S12_extracted_k141_7741 | Low-quality |
| S12_extracted_k141_141692 | Not-determined |
| S12_extracted_k141_22814 | Not-determined |
| S12_extracted_k141_141693 | Not-determined |
| S12_extracted_k141_38164 | Not-determined |
| S12_extracted_k141_141694 | Not-determined |
| S12_extracted_k141_78146 | Low-quality |
| S12_extracted_k141_62656 | Not-determined |
| S12_extracted_k141_22818 | Not-determined |
| S12_extracted_k141_62657 | Not-determined |
| S12_extracted_k141_110215 | Not-determined |
| S12_extracted_k141_124797 | Not-determined |
| S12_extracted_k141_14527 | Not-determined |
| S12_extracted_k141_94173 | Low-quality |
| S12_extracted_k141_22823 | Not-determined |
| S12_extracted_k141_14529 | Not-determined |
| S12_extracted_k141_156148 | Not-determined |
| S12_extracted_k141_149302 | Not-determined |
| S12_extracted_k141_78152 | Not-determined |
| S12_extracted_k141_156149 | Not-determined |
| S12_extracted_k141_78153 | Not-determined |
| S12_extracted_k141_179954 | Not-determined |
| S12_extracted_k141_110226 | Not-determined |
| S12_extracted_k141_149309 | Not-determined |
| S12_extracted_k141_172018 | Not-determined |
| S12_extracted_k141_149310 | Not-determined |
| S12_extracted_k141_78155 | Not-determined |
| S12_extracted_k141_172019 | Not-determined |
| S12_extracted_k141_117027 | Not-determined |
| S12_extracted_k141_30996 | Not-determined |
| S12_extracted_k141_78158 | Not-determined |
| S12_extracted_k141_78159 | Not-determined |
| S12_extracted_k141_62676 | Not-determined |
| S12_extracted_k141_38194 | Not-determined |
| S12_extracted_k141_117030 | Low-quality |
| S12_extracted_k141_156159 | Not-determined |
| S12_extracted_k141_14541 | Not-determined |
| S12_extracted_k141_149320 | Not-determined |
| S12_extracted_k141_78163 | Not-determined |
| S12_extracted_k141_78167 | Not-determined |
| S12_extracted_k141_22833 | Not-determined |
| S12_extracted_k141_156169 | Not-determined |
| S12_extracted_k141_78168 | Not-determined |
| S12_extracted_k141_14547 | Not-determined |
| S12_extracted_k141_85843 | Not-determined |
| S12_extracted_k141_31005 | Not-determined |
| S12_extracted_k141_14551 | Not-determined |
| S12_extracted_k141_54422 | Not-determined |
| S12_extracted_k141_31007 | Not-determined |
| S12_extracted_k141_149329 | Not-determined |
| S12_extracted_k141_38210 | Not-determined |
| S12_extracted_k141_14555 | Not-determined |
| S12_extracted_k141_124817 | Not-determined |
| S12_extracted_k141_7769 | Not-determined |
| S12_extracted_k141_62694 | Not-determined |
| S12_extracted_k141_172045 | Not-determined |
| S12_extracted_k141_31014 | Not-determined |
| S12_extracted_k141_94190 | Not-determined |
| S12_extracted_k141_179974 | Not-determined |
| S12_extracted_k141_54432 | Not-determined |
| S12_extracted_k141_172048 | Not-determined |
| S12_extracted_k141_70472 | Not-determined |
| S12_extracted_k141_124827 | Not-determined |
| S12_extracted_k141_149341 | Not-determined |
| S12_extracted_k141_164654 | Low-quality |
| S12_extracted_k141_70479 | Not-determined |
| S12_extracted_k141_117041 | Not-determined |
| S12_extracted_k141_38214 | Not-determined |
| S12_extracted_k141_156183 | Not-determined |
| S12_extracted_k141_70483 | Not-determined |
| S12_extracted_k141_38218 | Not-determined |
| S12_extracted_k141_70485 | Not-determined |
| S12_extracted_k141_78186 | Not-determined |
| S12_extracted_k141_31017 | Not-determined |
| S12_extracted_k141_172055 | Not-determined |
| S12_extracted_k141_31019 | Not-determined |
| S12_extracted_k141_172057 | Not-determined |
| S12_extracted_k141_124836 | Not-determined |
| S12_extracted_k141_172065 | Not-determined |
| S12_extracted_k141_78194 | Not-determined |
| S12_extracted_k141_7785 | Not-determined |
| S12_extracted_k141_141705 | Not-determined |
| S12_extracted_k141_62718 | Low-quality |
| S12_extracted_k141_70491 | Not-determined |
| S12_extracted_k141_54454 | Not-determined |
| S12_extracted_k141_172070 | Not-determined |
| S12_extracted_k141_54456 | Not-determined |
| S12_extracted_k141_124841 | Not-determined |
| S12_extracted_k141_7793 | Not-determined |
| S12_extracted_k141_70495 | Not-determined |
| S12_extracted_k141_7794 | Not-determined |
| S12_extracted_k141_70496 | Low-quality |
| S12_extracted_k141_164666 | Not-determined |
| S12_extracted_k141_172072 | Not-determined |
| S12_extracted_k141_70503 | Not-determined |
| S12_extracted_k141_172080 | Not-determined |
| S12_extracted_k141_94214 | Not-determined |
| S12_extracted_k141_141719 | Not-determined |
| S12_extracted_k141_62720 | Not-determined |
| S12_extracted_k141_85877 | Low-quality |
| S12_extracted_k141_85879 | Not-determined |
| S12_extracted_k141_156207 | Not-determined |
| S12_extracted_k141_85881 | Low-quality |
| S12_extracted_k141_124861 | Not-determined |
| S12_extracted_k141_62728 | Not-determined |
| S12_extracted_k141_117071 | Not-determined |
| S12_extracted_k141_188657 | Not-determined |
| S12_extracted_k141_70513 | Not-determined |
| S12_extracted_k141_180003 | Not-determined |
| S12_extracted_k141_117072 | Not-determined |
| S12_extracted_k141_7815 | Low-quality |
| S12_extracted_k141_164681 | Not-determined |
| S12_extracted_k141_180007 | Not-determined |
| S12_extracted_k141_38231 | Not-determined |
| S12_extracted_k141_85887 | Not-determined |
| S12_extracted_k141_38232 | Not-determined |
| S12_extracted_k141_14602 | Low-quality |
| S12_extracted_k141_78200 | Not-determined |
| S12_extracted_k141_94221 | Not-determined |
| S12_extracted_k141_94225 | Not-determined |
| S12_extracted_k141_94228 | Low-quality |
| S12_extracted_k141_38242 | Not-determined |
| S12_extracted_k141_188669 | Not-determined |
| S12_extracted_k141_7821 | Not-determined |
| S12_extracted_k141_62738 | Not-determined |
| S12_extracted_k141_70517 | Low-quality |
| S12_extracted_k141_156218 | Not-determined |
| S12_extracted_k141_38244 | Not-determined |
| S12_extracted_k141_149375 | Not-determined |
| S12_extracted_k141_124884 | Not-determined |
| S12_extracted_k141_78208 | Low-quality |
| S12_extracted_k141_22856 | Not-determined |
| S12_extracted_k141_85896 | Not-determined |
| S12_extracted_k141_62752 | Low-quality |
| S12_extracted_k141_22868 | Not-determined |
| S12_extracted_k141_124894 | Not-determined |
| S12_extracted_k141_180018 | Not-determined |
| S12_extracted_k141_164698 | Not-determined |
| S12_extracted_k141_180019 | Not-determined |
| S12_extracted_k141_188675 | Not-determined |
| S12_extracted_k141_14626 | Not-determined |
| S12_extracted_k141_78232 | Not-determined |
| S12_extracted_k141_180021 | Not-determined |
| S12_extracted_k141_7831 | Not-determined |
| S12_extracted_k141_164699 | Not-determined |
| S12_extracted_k141_188681 | Not-determined |
| S12_extracted_k141_7833 | Not-determined |
| S12_extracted_k141_70521 | Not-determined |
| S12_extracted_k141_172127 | Not-determined |
| S12_extracted_k141_124899 | Not-determined |
| S12_extracted_k141_94245 | Low-quality |
| S12_extracted_k141_149387 | Not-determined |
| S12_extracted_k141_149388 | Not-determined |
| S12_extracted_k141_188685 | Not-determined |
| S12_extracted_k141_164707 | Not-determined |
| S12_extracted_k141_94247 | Low-quality |
| S12_extracted_k141_85905 | Not-determined |
| S12_extracted_k141_70529 | Low-quality |
| S12_extracted_k141_7839 | Low-quality |
| S12_extracted_k141_188692 | Not-determined |
| S12_extracted_k141_62764 | Not-determined |
| S12_extracted_k141_14631 | Not-determined |
| S12_extracted_k141_7842 | Not-determined |
| S12_extracted_k141_124909 | Not-determined |
| S12_extracted_k141_14638 | Not-determined |
| S12_extracted_k141_85911 | Low-quality |
| S12_extracted_k141_188699 | Not-determined |
| S12_extracted_k141_124912 | Not-determined |
| S12_extracted_k141_172133 | Not-determined |
| S12_extracted_k141_188709 | Not-determined |
| S12_extracted_k141_22885 | Low-quality |
| S12_extracted_k141_117104 | Not-determined |
| S12_extracted_k141_156227 | Not-determined |
| S12_extracted_k141_188714 | Not-determined |
| S12_extracted_k141_7857 | Low-quality |
| S12_extracted_k141_156232 | Not-determined |
| S12_extracted_k141_188717 | Not-determined |
| S12_extracted_k141_172135 | Low-quality |
| S12_extracted_k141_164727 | Not-determined |
| S12_extracted_k141_188719 | Not-determined |
| S12_extracted_k141_117114 | Not-determined |
| S12_extracted_k141_31025 | Not-determined |
| S12_extracted_k141_31027 | Not-determined |
| S12_extracted_k141_149396 | Not-determined |
| S12_extracted_k141_62791 | Not-determined |
| S12_extracted_k141_70537 | Not-determined |
| S12_extracted_k141_149402 | Not-determined |
| S12_extracted_k141_14647 | Not-determined |
| S12_extracted_k141_70546 | Not-determined |
| S12_extracted_k141_31039 | Not-determined |
| S12_extracted_k141_124936 | Low-quality |
| S12_extracted_k141_70550 | Not-determined |
| S12_extracted_k141_94270 | Not-determined |
| S12_extracted_k141_124942 | Not-determined |
| S12_extracted_k141_156247 | Not-determined |
| S12_extracted_k141_172146 | Not-determined |
| S12_extracted_k141_14654 | Not-determined |
| S12_extracted_k141_70552 | Not-determined |
| S12_extracted_k141_188732 | Not-determined |
| S12_extracted_k141_164743 | Not-determined |
| S12_extracted_k141_172150 | Not-determined |
| S12_extracted_k141_156251 | Not-determined |
| S12_extracted_k141_156252 | Not-determined |
| S12_extracted_k141_172153 | Not-determined |
| S12_extracted_k141_117134 | Not-determined |
| S12_extracted_k141_54526 | Not-determined |
| S12_extracted_k141_14660 | Not-determined |
| S12_extracted_k141_117136 | Low-quality |
| S12_extracted_k141_85934 | Not-determined |
| S12_extracted_k141_54528 | Not-determined |
| S12_extracted_k141_188739 | Not-determined |
| S12_extracted_k141_54529 | Not-determined |
| S12_extracted_k141_62811 | Not-determined |
| S12_extracted_k141_133083 | Not-determined |
| S12_extracted_k141_70560 | Not-determined |
| S12_extracted_k141_38264 | Not-determined |
| S12_extracted_k141_38265 | Not-determined |
| S12_extracted_k141_94282 | Not-determined |
| S12_extracted_k141_14665 | Not-determined |
| S12_extracted_k141_94283 | Not-determined |
| S12_extracted_k141_149414 | Not-determined |
| S12_extracted_k141_70562 | Not-determined |
| S12_extracted_k141_172160 | Low-quality |
| S12_extracted_k141_70565 | Not-determined |
| S12_extracted_k141_124954 | Not-determined |
| S12_extracted_k141_22932 | Not-determined |
| S12_extracted_k141_124957 | Not-determined |
| S12_extracted_k141_164753 | Not-determined |
| S12_extracted_k141_164755 | Not-determined |
| S12_extracted_k141_62816 | Not-determined |
| S12_extracted_k141_172165 | Not-determined |
| S12_extracted_k141_172167 | Not-determined |
| S12_extracted_k141_188745 | Not-determined |
| S12_extracted_k141_172169 | Not-determined |
| S12_extracted_k141_14674 | Not-determined |
| S12_extracted_k141_172171 | Not-determined |
| S12_extracted_k141_85951 | Not-determined |
| S12_extracted_k141_14677 | Not-determined |
| S12_extracted_k141_117152 | Not-determined |
| S12_extracted_k141_117153 | Not-determined |
| S12_extracted_k141_164759 | Not-determined |
| S12_extracted_k141_85954 | Not-determined |
| S12_extracted_k141_85955 | Not-determined |
| S12_extracted_k141_31068 | Not-determined |
| S12_extracted_k141_70582 | Not-determined |
| S12_extracted_k141_156271 | Not-determined |
| S12_extracted_k141_54543 | Not-determined |
| S12_extracted_k141_156272 | Not-determined |
| S12_extracted_k141_164762 | Not-determined |
| S12_extracted_k141_117157 | Not-determined |
| S12_extracted_k141_85964 | Low-quality |
| S12_extracted_k141_180056 | Not-determined |
| S12_extracted_k141_14691 | Not-determined |
| S12_extracted_k141_31081 | Not-determined |
| S12_extracted_k141_22957 | Not-determined |
| S12_extracted_k141_85970 | Not-determined |
| S12_extracted_k141_22958 | Not-determined |
| S12_extracted_k141_85973 | Not-determined |
| S12_extracted_k141_14698 | Not-determined |
| S12_extracted_k141_188766 | Not-determined |
| S12_extracted_k141_85976 | Not-determined |
| S12_extracted_k141_188768 | Not-determined |
| S12_extracted_k141_94305 | Not-determined |
| S12_extracted_k141_164779 | Low-quality |
| S12_extracted_k141_78284 | Not-determined |
| S12_extracted_k141_70591 | Not-determined |
| S12_extracted_k141_38280 | Low-quality |
| S12_extracted_k141_133113 | Not-determined |
| S12_extracted_k141_70595 | Not-determined |
| S12_extracted_k141_164782 | Not-determined |
| S12_extracted_k141_54558 | Not-determined |
| S12_extracted_k141_78289 | Not-determined |
| S12_extracted_k141_149435 | Not-determined |
| S12_extracted_k141_38282 | Low-quality |
| S12_extracted_k141_188779 | Not-determined |
| S12_extracted_k141_62842 | Not-determined |
| S12_extracted_k141_85991 | Not-determined |
| S12_extracted_k141_156296 | Not-determined |
| S12_extracted_k141_85993 | Not-determined |
| S12_extracted_k141_54562 | Not-determined |
| S12_extracted_k141_85995 | Not-determined |
| S12_extracted_k141_38286 | Not-determined |
| S12_extracted_k141_164790 | Not-determined |
| S12_extracted_k141_85997 | Not-determined |
| S12_extracted_k141_22968 | Not-determined |
| S12_extracted_k141_54566 | Not-determined |
| S12_extracted_k141_149439 | Not-determined |
| S12_extracted_k141_85999 | Not-determined |
| S12_extracted_k141_156304 | Not-determined |
| S12_extracted_k141_117182 | Not-determined |
| S12_extracted_k141_22972 | Not-determined |
| S12_extracted_k141_117183 | Low-quality |
| S12_extracted_k141_86000 | Not-determined |
| S12_extracted_k141_86001 | Not-determined |
| S12_extracted_k141_117188 | Not-determined |
| S12_extracted_k141_31104 | Not-determined |
| S12_extracted_k141_156311 | Not-determined |
| S12_extracted_k141_124997 | Not-determined |
| S12_extracted_k141_22985 | Not-determined |
| S12_extracted_k141_133130 | Not-determined |
| S12_extracted_k141_31108 | Not-determined |
| S12_extracted_k141_62851 | Not-determined |
| S12_extracted_k141_22989 | Not-determined |
| S12_extracted_k141_149454 | Not-determined |
| S12_extracted_k141_62856 | Not-determined |
| S12_extracted_k141_164802 | Not-determined |
| S12_extracted_k141_78320 | Not-determined |
| S12_extracted_k141_156321 | Not-determined |
| S12_extracted_k141_22991 | Not-determined |
| S12_extracted_k141_86008 | Not-determined |
| S12_extracted_k141_38300 | Not-determined |
| S12_extracted_k141_156323 | Not-determined |
| S12_extracted_k141_149458 | Not-determined |
| S12_extracted_k141_172186 | Not-determined |
| S12_extracted_k141_62859 | Not-determined |
| S12_extracted_k141_149461 | Not-determined |
| S12_extracted_k141_38306 | Not-determined |
| S12_extracted_k141_172190 | Not-determined |
| S12_extracted_k141_70624 | Not-determined |
| S12_extracted_k141_22993 | Not-determined |
| S12_extracted_k141_70626 | Not-determined |
| S12_extracted_k141_149466 | Not-determined |
| S12_extracted_k141_22996 | Not-determined |
| S12_extracted_k141_86012 | Not-determined |
| S12_extracted_k141_149470 | Not-determined |
| S12_extracted_k141_125014 | Not-determined |
| S12_extracted_k141_54583 | Not-determined |
| S12_extracted_k141_31120 | Not-determined |
| S12_extracted_k141_172193 | Not-determined |
| S12_extracted_k141_23000 | Not-determined |
| S12_extracted_k141_133144 | Not-determined |
| S12_extracted_k141_23004 | Not-determined |
| S12_extracted_k141_188814 | Not-determined |
| S12_extracted_k141_70629 | Not-determined |
| S12_extracted_k141_94327 | Not-determined |
| S12_extracted_k141_164808 | Not-determined |
| S12_extracted_k141_86019 | Not-determined |
| S12_extracted_k141_78329 | Not-determined |
| S12_extracted_k141_86020 | Low-quality |
| S12_extracted_k141_70631 | Not-determined |
| S12_extracted_k141_62874 | Not-determined |
| S12_extracted_k141_62881 | Not-determined |
| S12_extracted_k141_31131 | Not-determined |
| S12_extracted_k141_156340 | Not-determined |
| S12_extracted_k141_62883 | Not-determined |
| S12_extracted_k141_172204 | Not-determined |
| S12_extracted_k141_38328 | Not-determined |
| S12_extracted_k141_78339 | Not-determined |
| S12_extracted_k141_31135 | Not-determined |
| S12_extracted_k141_133158 | Not-determined |
| S12_extracted_k141_14743 | Not-determined |
| S12_extracted_k141_149497 | Not-determined |
| S12_extracted_k141_31143 | Not-determined |
| S12_extracted_k141_133167 | Not-determined |
| S12_extracted_k141_164820 | Not-determined |
| S12_extracted_k141_172215 | Not-determined |
| S12_extracted_k141_125042 | Low-quality |
| S12_extracted_k141_14754 | Not-determined |
| S12_extracted_k141_14755 | Low-quality |
| S12_extracted_k141_78344 | Not-determined |
| S12_extracted_k141_172220 | Low-quality |
| S12_extracted_k141_164823 | Not-determined |
| S12_extracted_k141_172222 | Not-determined |
| S12_extracted_k141_31158 | Not-determined |
| S12_extracted_k141_31160 | Not-determined |
| S12_extracted_k141_62898 | Not-determined |
| S12_extracted_k141_164829 | Not-determined |
| S12_extracted_k141_180070 | Not-determined |
| S12_extracted_k141_78355 | Not-determined |
| S12_extracted_k141_164831 | Not-determined |
| S12_extracted_k141_164832 | Not-determined |
| S12_extracted_k141_38336 | Not-determined |
| S12_extracted_k141_156368 | Low-quality |
| S12_extracted_k141_54614 | Low-quality |
| S12_extracted_k141_94340 | Not-determined |
| S12_extracted_k141_38342 | Not-determined |
| S12_extracted_k141_54619 | Not-determined |
| S12_extracted_k141_133178 | Not-determined |
| S12_extracted_k141_23023 | Not-determined |
| S12_extracted_k141_133181 | Not-determined |
| S12_extracted_k141_180076 | Not-determined |
| S12_extracted_k141_70657 | Not-determined |
| S12_extracted_k141_94350 | Not-determined |
| S12_extracted_k141_78370 | Not-determined |
| S12_extracted_k141_156377 | Not-determined |
| S12_extracted_k141_54621 | Not-determined |
| S12_extracted_k141_86058 | Not-determined |
| S12_extracted_k141_156380 | Not-determined |
| S12_extracted_k141_117235 | Not-determined |
| S12_extracted_k141_78375 | Not-determined |
| S12_extracted_k141_78376 | Not-determined |
| S12_extracted_k141_117241 | Not-determined |
| S12_extracted_k141_62915 | Not-determined |
| S12_extracted_k141_70673 | Not-determined |
| S12_extracted_k141_54635 | Not-determined |
| S12_extracted_k141_78386 | Not-determined |
| S12_extracted_k141_149512 | Not-determined |
| S12_extracted_k141_14772 | Low-quality |
| S12_extracted_k141_156391 | Low-quality |
| S12_extracted_k141_94371 | Not-determined |
| S12_extracted_k141_172240 | Not-determined |
| S12_extracted_k141_86071 | Not-determined |
| S12_extracted_k141_180089 | Not-determined |
| S12_extracted_k141_156394 | Not-determined |
| S12_extracted_k141_149517 | Not-determined |
| S12_extracted_k141_149518 | Not-determined |
| S12_extracted_k141_78401 | Not-determined |
| S12_extracted_k141_38354 | Not-determined |
| S12_extracted_k141_78402 | Not-determined |
| S12_extracted_k141_149523 | Not-determined |
| S12_extracted_k141_86081 | Not-determined |
| S12_extracted_k141_180096 | Not-determined |
| S12_extracted_k141_172254 | Not-determined |
| S12_extracted_k141_94376 | Not-determined |
| S12_extracted_k141_172255 | Low-quality |
| S12_extracted_k141_133210 | Low-quality |
| S12_extracted_k141_14781 | Not-determined |
| S12_extracted_k141_78410 | Not-determined |
| S12_extracted_k141_172259 | Not-determined |
| S12_extracted_k141_94384 | Not-determined |
| S12_extracted_k141_156414 | Not-determined |
| S12_extracted_k141_38366 | Not-determined |
| S12_extracted_k141_86088 | Not-determined |
| S12_extracted_k141_14786 | Not-determined |
| S12_extracted_k141_180106 | Not-determined |
| S12_extracted_k141_23055 | Not-determined |
| S12_extracted_k141_172263 | Not-determined |
| S12_extracted_k141_38371 | Not-determined |
| S12_extracted_k141_23058 | Low-quality |
| S12_extracted_k141_14791 | Not-determined |
| S12_extracted_k141_117259 | Not-determined |
| S12_extracted_k141_23063 | Low-quality |
| S12_extracted_k141_94389 | Not-determined |
| S12_extracted_k141_180117 | Not-determined |
| S12_extracted_k141_70688 | Not-determined |
| S12_extracted_k141_133225 | Not-determined |
| S12_extracted_k141_117265 | Not-determined |
| S12_extracted_k141_38380 | Not-determined |
| S12_extracted_k141_172274 | Not-determined |
| S12_extracted_k141_23069 | Not-determined |
| S12_extracted_k141_180120 | Not-determined |
| S12_extracted_k141_86103 | Not-determined |
| S12_extracted_k141_180121 | Low-quality |
| S12_extracted_k141_38382 | Not-determined |
| S12_extracted_k141_172277 | Not-determined |
| S12_extracted_k141_86107 | Not-determined |
| S12_extracted_k141_180126 | Low-quality |
| S12_extracted_k141_23076 | Not-determined |
| S12_extracted_k141_149549 | Not-determined |
| S12_extracted_k141_149552 | Not-determined |
| S12_extracted_k141_149553 | Not-determined |
| S12_extracted_k141_86111 | Not-determined |
| S12_extracted_k141_54651 | Not-determined |
| S12_extracted_k141_14808 | Not-determined |
| S12_extracted_k141_117274 | Not-determined |
| S12_extracted_k141_180133 | Not-determined |
| S12_extracted_k141_117276 | Not-determined |
| S12_extracted_k141_86115 | Not-determined |
| S12_extracted_k141_86119 | Not-determined |
| S12_extracted_k141_86120 | Low-quality |
| S12_extracted_k141_94412 | Not-determined |
| S12_extracted_k141_180137 | Not-determined |
| S12_extracted_k141_149570 | Not-determined |
| S12_extracted_k141_78439 | Not-determined |
| S12_extracted_k141_54664 | Not-determined |
| S12_extracted_k141_117281 | Low-quality |
| S12_extracted_k141_78449 | Not-determined |
| S12_extracted_k141_86131 | Not-determined |
| S12_extracted_k141_14816 | Not-determined |
| S12_extracted_k141_78452 | Not-determined |
| S12_extracted_k141_117290 | Not-determined |
| S12_extracted_k141_14820 | Not-determined |
| S12_extracted_k141_14822 | Not-determined |
| S12_extracted_k141_86136 | Not-determined |
| S12_extracted_k141_156431 | Not-determined |
| S12_extracted_k141_54674 | Not-determined |
| S12_extracted_k141_156435 | Low-quality |
| S12_extracted_k141_172298 | Not-determined |
| S12_extracted_k141_117295 | Not-determined |
| S12_extracted_k141_149575 | Not-determined |
| S12_extracted_k141_133232 | Not-determined |
| S12_extracted_k141_172300 | Not-determined |
| S12_extracted_k141_54681 | Not-determined |
| S12_extracted_k141_70706 | Not-determined |
| S12_extracted_k141_31180 | Not-determined |
| S12_extracted_k141_180157 | Not-determined |
| S12_extracted_k141_86148 | Not-determined |
| S12_extracted_k141_133238 | Not-determined |
| S12_extracted_k141_94437 | Not-determined |
| S12_extracted_k141_14832 | Not-determined |
| S12_extracted_k141_133239 | Not-determined |
| S12_extracted_k141_149583 | Not-determined |
| S12_extracted_k141_133242 | Not-determined |
| S12_extracted_k141_164837 | Not-determined |
| S12_extracted_k141_78471 | Not-determined |
| S12_extracted_k141_70718 | Not-determined |
| S12_extracted_k141_23104 | Not-determined |
| S12_extracted_k141_54690 | Low-quality |
| S12_extracted_k141_149592 | Not-determined |
| S12_extracted_k141_94446 | Not-determined |
| S12_extracted_k141_31192 | Not-determined |
| S12_extracted_k141_23110 | Not-determined |
| S12_extracted_k141_70725 | Low-quality |
| S12_extracted_k141_117314 | Not-determined |
| S12_extracted_k141_31194 | Not-determined |
| S12_extracted_k141_78479 | Not-determined |
| S12_extracted_k141_31197 | Not-determined |
| S12_extracted_k141_172321 | Not-determined |
| S12_extracted_k141_70730 | Not-determined |
| S12_extracted_k141_172323 | Not-determined |
| S12_extracted_k141_86170 | Low-quality |
| S12_extracted_k141_31200 | Not-determined |
| S12_extracted_k141_149601 | Not-determined |
| S12_extracted_k141_172327 | Not-determined |
| S12_extracted_k141_172328 | Not-determined |
| S12_extracted_k141_78485 | Not-determined |
| S12_extracted_k141_38435 | Not-determined |
| S12_extracted_k141_117320 | Low-quality |
| S12_extracted_k141_180164 | Not-determined |
| S12_extracted_k141_38436 | Not-determined |
| S12_extracted_k141_180165 | Not-determined |
| S12_extracted_k141_62949 | Not-determined |
| S12_extracted_k141_164863 | Not-determined |
| S12_extracted_k141_117332 | Not-determined |
| S12_extracted_k141_23112 | Not-determined |
| S12_extracted_k141_31211 | Low-quality |
| S12_extracted_k141_14853 | Not-determined |
| S12_extracted_k141_23113 | Not-determined |

| | |
|---|---|
| S12_extracted_k141_70745 | Not-determined |
| S12_extracted_k141_172344 | Not-determined |
| S12_extracted_k141_54711 | Not-determined |
| S12_extracted_k141_133254 | Low-quality |
| S12_extracted_k141_70746 | Low-quality |
| S12_extracted_k141_54713 | Low-quality |
| S12_extracted_k141_125071 | Not-determined |
| S12_extracted_k141_125074 | Not-determined |
| S12_extracted_k141_133260 | Not-determined |
| S12_extracted_k141_172351 | Not-determined |
| S12_extracted_k141_23127 | Not-determined |
| S12_extracted_k141_54717 | Low-quality |
| S12_extracted_k141_172353 | Not-determined |
| S12_extracted_k141_31227 | Not-determined |
| S12_extracted_k141_70755 | Low-quality |
| S12_extracted_k141_172356 | Not-determined |
| S12_extracted_k141_117343 | Low-quality |
| S12_extracted_k141_31231 | Not-determined |
| S12_extracted_k141_23134 | Not-determined |
| S12_extracted_k141_70758 | Low-quality |
| S12_extracted_k141_172358 | Not-determined |
| S12_extracted_k141_54723 | Not-determined |
| S12_extracted_k141_172359 | Not-determined |
| S12_extracted_k141_54725 | Not-determined |
| S12_extracted_k141_31236 | Not-determined |
| S12_extracted_k141_31237 | Not-determined |
| S12_extracted_k141_78509 | Not-determined |
| S12_extracted_k141_172362 | Not-determined |
| S12_extracted_k141_23143 | Not-determined |
| S12_extracted_k141_54730 | Not-determined |
| S12_extracted_k141_125102 | Not-determined |
| S12_extracted_k141_117354 | Not-determined |
| S12_extracted_k141_133272 | Not-determined |
| S12_extracted_k141_164896 | Not-determined |
| S12_extracted_k141_23153 | Not-determined |
| S12_extracted_k141_180170 | Not-determined |
| S12_extracted_k141_117361 | Not-determined |
| S12_extracted_k141_133274 | Low-quality |
| S12_extracted_k141_125109 | Not-determined |
| S12_extracted_k141_180181 | Not-determined |
| S12_extracted_k141_180182 | Not-determined |
| S12_extracted_k141_31256 | Not-determined |
| S12_extracted_k141_180185 | Not-determined |
| S12_extracted_k141_23161 | Not-determined |
| S12_extracted_k141_133280 | Not-determined |
| S12_extracted_k141_133283 | Not-determined |
| S12_extracted_k141_133288 | Not-determined |
| S12_extracted_k141_62974 | Not-determined |
| S12_extracted_k141_86219 | Not-determined |
| S12_extracted_k141_86220 | Not-determined |
| S12_extracted_k141_180193 | Not-determined |
| S12_extracted_k141_38485 | Not-determined |
| S12_extracted_k141_31268 | Not-determined |
| S12_extracted_k141_133294 | Not-determined |
| S12_extracted_k141_62979 | Not-determined |
| S12_extracted_k141_31271 | Not-determined |
| S12_extracted_k141_70775 | Not-determined |
| S12_extracted_k141_117373 | Not-determined |
| S12_extracted_k141_38491 | Not-determined |
| S12_extracted_k141_117374 | Not-determined |
| S12_extracted_k141_180199 | Not-determined |
| S12_extracted_k141_70777 | Not-determined |
| S12_extracted_k141_180203 | Not-determined |
| S12_extracted_k141_117380 | Not-determined |
| S12_extracted_k141_78540 | Not-determined |
| S12_extracted_k141_70783 | Not-determined |
| S12_extracted_k141_180208 | Not-determined |
| S12_extracted_k141_70786 | Not-determined |
| S12_extracted_k141_94470 | Not-determined |
| S12_extracted_k141_31286 | Not-determined |
| S12_extracted_k141_133300 | Not-determined |
| S12_extracted_k141_54733 | Low-quality |
| S12_extracted_k141_86235 | Not-determined |
| S12_extracted_k141_31289 | Not-determined |
| S12_extracted_k141_86236 | Not-determined |
| S12_extracted_k141_125111 | Not-determined |
| S12_extracted_k141_86238 | Not-determined |
| S12_extracted_k141_23171 | Not-determined |
| S12_extracted_k141_180222 | Not-determined |
| S12_extracted_k141_86243 | Low-quality |
| S12_extracted_k141_54741 | Not-determined |
| S12_extracted_k141_31297 | Not-determined |
| S12_extracted_k141_38513 | Not-determined |
| S12_extracted_k141_125123 | Not-determined |
| S12_extracted_k141_23177 | Not-determined |
| S12_extracted_k141_180227 | Not-determined |
| S12_extracted_k141_94485 | Not-determined |
| S12_extracted_k141_70804 | Low-quality |
| S12_extracted_k141_125132 | Not-determined |
| S12_extracted_k141_23184 | Not-determined |
| S12_extracted_k141_180233 | Not-determined |
| S12_extracted_k141_14860 | Not-determined |
| S12_extracted_k141_31310 | Not-determined |
| S12_extracted_k141_125136 | Not-determined |
| S12_extracted_k141_117390 | Not-determined |
| S12_extracted_k141_31313 | Not-determined |
| S12_extracted_k141_125141 | Not-determined |
| S12_extracted_k141_125143 | Not-determined |
| S12_extracted_k141_31316 | Not-determined |
| S12_extracted_k141_14869 | Not-determined |
| S12_extracted_k141_23191 | Low-quality |
| S12_extracted_k141_86272 | Not-determined |
| S12_extracted_k141_38539 | Not-determined |
| S12_extracted_k141_117397 | Not-determined |
| S12_extracted_k141_38543 | Not-determined |
| S12_extracted_k141_31326 | Not-determined |
| S12_extracted_k141_31329 | Not-determined |
| S12_extracted_k141_156472 | Not-determined |
| S12_extracted_k141_117403 | Not-determined |
| S12_extracted_k141_23203 | Not-determined |
| S12_extracted_k141_86289 | Not-determined |
| S12_extracted_k141_54784 | Not-determined |
| S12_extracted_k141_164946 | Not-determined |
| S12_extracted_k141_70825 | Not-determined |
| S12_extracted_k141_117408 | Not-determined |
| S12_extracted_k141_31335 | Not-determined |
| S12_extracted_k141_14882 | Not-determined |
| S12_extracted_k141_164947 | Not-determined |
| S12_extracted_k141_164949 | Not-determined |
| S12_extracted_k141_70835 | Not-determined |
| S12_extracted_k141_86297 | Not-determined |
| S12_extracted_k141_31346 | Not-determined |
| S12_extracted_k141_125171 | Not-determined |
| S12_extracted_k141_125172 | Not-determined |
| S12_extracted_k141_70839 | Not-determined |
| S12_extracted_k141_156494 | Low-quality |
| S12_extracted_k141_38549 | Not-determined |
| S12_extracted_k141_31351 | Not-determined |
| S12_extracted_k141_54797 | Not-determined |
| S12_extracted_k141_14893 | Not-determined |
| S12_extracted_k141_54799 | Not-determined |
| S12_extracted_k141_31355 | Not-determined |
| S12_extracted_k141_38556 | Not-determined |
| S12_extracted_k141_31360 | Not-determined |
| S12_extracted_k141_31361 | Not-determined |
| S12_extracted_k141_14902 | Not-determined |
| S12_extracted_k141_78608 | Not-determined |
| S12_extracted_k141_117432 | Not-determined |
| S12_extracted_k141_23214 | Not-determined |
| S12_extracted_k141_23215 | Not-determined |
| S12_extracted_k141_54806 | Not-determined |
| S12_extracted_k141_86311 | Not-determined |
| S12_extracted_k141_70847 | Not-determined |
| S12_extracted_k141_125194 | Not-determined |
| S12_extracted_k141_164979 | Not-determined |
| S12_extracted_k141_54816 | Not-determined |
| S12_extracted_k141_133346 | Not-determined |
| S12_extracted_k141_133348 | Not-determined |
| S12_extracted_k141_78627 | Low-quality |
| S16_extracted_32359 | Medium-quality |
| S12_extracted_k141_23231 | Not-determined |
| S12_extracted_k141_86326 | Low-quality |
| S12_extracted_k141_38581 | Not-determined |
| S12_extracted_k141_156524 | Not-determined |
| S12_extracted_k141_86328 | Not-determined |
| S12_extracted_k141_180260 | Low-quality |
| S12_extracted_k141_23234 | Not-determined |
| S12_extracted_k141_133351 | Low-quality |
| S12_extracted_k141_78636 | Not-determined |
| S12_extracted_k141_14918 | Not-determined |
| S12_extracted_k141_156532 | Not-determined |
| S12_extracted_k141_78645 | Not-determined |
| S12_extracted_k141_180276 | Low-quality |
| S12_extracted_k141_180277 | Not-determined |
| S12_extracted_k141_133358 | Not-determined |
| S12_extracted_k141_133362 | Not-determined |
| S12_extracted_k141_38596 | Not-determined |
| S12_extracted_k141_133363 | Not-determined |
| S12_extracted_k141_133364 | Not-determined |
| S12_extracted_k141_54827 | Not-determined |
| S12_extracted_k141_38600 | Not-determined |
| S12_extracted_k141_180284 | Not-determined |
| S12_extracted_k141_133370 | Not-determined |
| S12_extracted_k141_78657 | Low-quality |
| S12_extracted_k141_31397 | Not-determined |
| S12_extracted_k141_133372 | Not-determined |
| S12_extracted_k141_38605 | Not-determined |
| S12_extracted_k141_31398 | Not-determined |
| S12_extracted_k141_180292 | Not-determined |
| S12_extracted_k141_117448 | Low-quality |
| S12_extracted_k141_117452 | Not-determined |
| S12_extracted_k141_78667 | Not-determined |
| S12_extracted_k141_180297 | Low-quality |
| S12_extracted_k141_78670 | Not-determined |
| S12_extracted_k141_54842 | Not-determined |
| S12_extracted_k141_165031 | Not-determined |
| S12_extracted_k141_133382 | Not-determined |
| S12_extracted_k141_156560 | Not-determined |
| S12_extracted_k141_125256 | Not-determined |
| S12_extracted_k141_165037 | Not-determined |
| S12_extracted_k141_78675 | Not-determined |
| S12_extracted_k141_133387 | Not-determined |
| S12_extracted_k141_125258 | Not-determined |
| S12_extracted_k141_38620 | Low-quality |
| S12_extracted_k141_133390 | Not-determined |
| S12_extracted_k141_125259 | Not-determined |
| S12_extracted_k141_156570 | Not-determined |
| S12_extracted_k141_133398 | Not-determined |
| S12_extracted_k141_156575 | Not-determined |
| S12_extracted_k141_125262 | Low-quality |
| S12_extracted_k141_125264 | Not-determined |
| S12_extracted_k141_125265 | Not-determined |
| S12_extracted_k141_23272 | Not-determined |
| S12_extracted_k141_38633 | Not-determined |
| S12_extracted_k141_86365 | Not-determined |
| S12_extracted_k141_86366 | Not-determined |
| S12_extracted_k141_86370 | Not-determined |
| S12_extracted_k141_78692 | Low-quality |
| S12_extracted_k141_117465 | Not-determined |
| S12_extracted_k141_180306 | Not-determined |
| S12_extracted_k141_86373 | Low-quality |
| S12_extracted_k141_31432 | Not-determined |
| S12_extracted_k141_117468 | Not-determined |
| S12_extracted_k141_125274 | Not-determined |
| S12_extracted_k141_38642 | Low-quality |
| S12_extracted_k141_31437 | Not-determined |
| S12_extracted_k141_117472 | Not-determined |
| S12_extracted_k141_125282 | Not-determined |
| S12_extracted_k141_38651 | Not-determined |
| S12_extracted_k141_125283 | Not-determined |
| S12_extracted_k141_31441 | Not-determined |
| S12_extracted_k141_117482 | Not-determined |
| S12_extracted_k141_31442 | Not-determined |
| S12_extracted_k141_125285 | Not-determined |
| S12_extracted_k141_86384 | Not-determined |
| S12_extracted_k141_54877 | Low-quality |
| S12_extracted_k141_133424 | Low-quality |
| S12_extracted_k141_180315 | Not-determined |
| S12_extracted_k141_38659 | Not-determined |
| S12_extracted_k141_180319 | Not-determined |
| S12_extracted_k141_133429 | Not-determined |
| S12_extracted_k141_54891 | Low-quality |
| S12_extracted_k141_86400 | Not-determined |
| S12_extracted_k141_165053 | Not-determined |
| S12_extracted_k141_156594 | Not-determined |
| S12_extracted_k141_23300 | Not-determined |
| S12_extracted_k141_117496 | Low-quality |
| S12_extracted_k141_125292 | Not-determined |
| S12_extracted_k141_38676 | Not-determined |
| S12_extracted_k141_117502 | Not-determined |
| S12_extracted_k141_23306 | Not-determined |
| S12_extracted_k141_23307 | Not-determined |
| S12_extracted_k141_54904 | Not-determined |
| S12_extracted_k141_125299 | Not-determined |
| S12_extracted_k141_38682 | Not-determined |
| S12_extracted_k141_156605 | Not-determined |
| S12_extracted_k141_156609 | Not-determined |
| S12_extracted_k141_23316 | Not-determined |
| S12_extracted_k141_23317 | Not-determined |
| S12_extracted_k141_23318 | Not-determined |
| S12_extracted_k141_23319 | Not-determined |
| S12_extracted_k141_156612 | Low-quality |
| S12_extracted_k141_133460 | Not-determined |
| S12_extracted_k141_156615 | Not-determined |
| S12_extracted_k141_180353 | Not-determined |
| S12_extracted_k141_180354 | Not-determined |
| S12_extracted_k141_180355 | Not-determined |
| S12_extracted_k141_165064 | Not-determined |
| S12_extracted_k141_23331 | Not-determined |
| S12_extracted_k141_165069 | Not-determined |
| S12_extracted_k141_133464 | Not-determined |
| S12_extracted_k141_165074 | Not-determined |
| S12_extracted_k141_23342 | Not-determined |
| S12_extracted_k141_133473 | Not-determined |
| S12_extracted_k141_133474 | Not-determined |
| S12_extracted_k141_180369 | Not-determined |
| S12_extracted_k141_86447 | Not-determined |
| S12_extracted_k141_180370 | Not-determined |
| S12_extracted_k141_180375 | Not-determined |
| S12_extracted_k141_180376 | Not-determined |
| S12_extracted_k141_23347 | Not-determined |
| S12_extracted_k141_54923 | Low-quality |
| S12_extracted_k141_165096 | Not-determined |
| S12_extracted_k141_23352 | Low-quality |
| S12_extracted_k141_172392 | Not-determined |
| S12_extracted_k141_54929 | Not-determined |
| S12_extracted_k141_38699 | Not-determined |
| S12_extracted_k141_23356 | Not-determined |
| S12_extracted_k141_133486 | Not-determined |
| S12_extracted_k141_38710 | Not-determined |
| S12_extracted_k141_54933 | Not-determined |
| S12_extracted_k141_165107 | Low-quality |
| S12_extracted_k141_156635 | Not-determined |
| S12_extracted_k141_165115 | Not-determined |
| S12_extracted_k141_180389 | Not-determined |
| S12_extracted_k141_86487 | Not-determined |
| S12_extracted_k141_86488 | Not-determined |
| S12_extracted_k141_86489 | Not-determined |
| S12_extracted_k141_23370 | Not-determined |
| S12_extracted_k141_172414 | Low-quality |
| S12_extracted_k141_180393 | Not-determined |
| S12_extracted_k141_172419 | Not-determined |
| S12_extracted_k141_180394 | Not-determined |
| S12_extracted_k141_23376 | Not-determined |
| S12_extracted_k141_86494 | Not-determined |
| S12_extracted_k141_180397 | Low-quality |
| S12_extracted_k141_172423 | Not-determined |
| S12_extracted_k141_133504 | Not-determined |
| S12_extracted_k141_117514 | Not-determined |
| S12_extracted_k141_180401 | Not-determined |
| S12_extracted_k141_172430 | Not-determined |
| S12_extracted_k141_125323 | Not-determined |
| S12_extracted_k141_78717 | Not-determined |
| S12_extracted_k141_172441 | Not-determined |
| S12_extracted_k141_31451 | Not-determined |
| S12_extracted_k141_156653 | Not-determined |
| S12_extracted_k141_133517 | Not-determined |
| S12_extracted_k141_180418 | Not-determined |
| S12_extracted_k141_54978 | Not-determined |
| S12_extracted_k141_172453 | Not-determined |
| S12_extracted_k141_54982 | Not-determined |
| S12_extracted_k141_180426 | Not-determined |
| S12_extracted_k141_180427 | Not-determined |
| S12_extracted_k141_86510 | Not-determined |
| S16_vs_32359\|\|full | Medium-quality |
| S12_extracted_k141_86511 | Low-quality |
| S12_extracted_k141_14936 | Low-quality |
| S12_extracted_k141_172461 | Not-determined |
| S12_extracted_k141_156664 | Not-determined |
| S12_extracted_k141_54995 | Not-determined |
| S12_extracted_k141_86513 | Not-determined |
| S12_extracted_k141_172468 | Not-determined |
| S12_extracted_k141_55000 | Not-determined |
| S12_extracted_k141_38739 | Not-determined |
| S12_extracted_k141_125354 | Not-determined |
| S12_extracted_k141_86526 | Not-determined |
| S12_extracted_k141_172485 | Not-determined |
| S12_extracted_k141_165131 | Not-determined |
| S12_extracted_k141_165142 | Low-quality |
| S12_extracted_k141_172494 | Not-determined |
| S12_extracted_k141_156689 | Not-determined |
| S12_extracted_k141_55011 | Low-quality |
| S12_extracted_k141_38758 | Not-determined |
| S12_extracted_k141_133545 | Not-determined |
| S12_extracted_k141_165147 | Low-quality |
| S12_extracted_k141_133550 | Low-quality |
| S12_extracted_k141_165150 | Not-determined |
| S12_extracted_k141_180450 | Not-determined |
| S12_extracted_k141_125380 | Low-quality |
| S12_extracted_k141_55024 | Low-quality |
| S12_extracted_k141_23407 | Not-determined |
| S12_extracted_k141_165153 | Not-determined |
| S12_extracted_k141_125386 | Not-determined |
| S12_extracted_k141_172512 | Not-determined |
| S12_extracted_k141_23410 | Not-determined |
| S12_extracted_k141_125388 | Not-determined |
| S12_extracted_k141_172515 | Low-quality |
| S12_extracted_k141_172517 | Not-determined |
| S12_extracted_k141_156708 | Not-determined |
| S12_extracted_k141_23414 | Not-determined |
| S12_extracted_k141_117517 | Not-determined |
| S12_extracted_k141_55035 | Not-determined |
| S16_vs_173109\|\|full | High-quality |
| S12_extracted_k141_55037 | Not-determined |
| S12_extracted_k141_133572 | Not-determined |
| S12_extracted_k141_172523 | Low-quality |
| S12_extracted_k141_38775 | Not-determined |
| S12_extracted_k141_38776 | Not-determined |
| S12_extracted_k141_156717 | Not-determined |
| S12_extracted_k141_23430 | Not-determined |
| S12_extracted_k141_117520 | Low-quality |
| S12_extracted_k141_172534 | Not-determined |
| S12_extracted_k141_180481 | Not-determined |
| S12_extracted_k141_133587 | Not-determined |
| S12_extracted_k141_156726 | Not-determined |
| S12_extracted_k141_86529 | Not-determined |
| S12_extracted_k141_86532 | Not-determined |
| S12_extracted_k141_14983 | Not-determined |
| S12_extracted_k141_117534 | Not-determined |
| S12_extracted_k141_86534 | Not-determined |
| S12_extracted_k141_156729 | Low-quality |
| S12_extracted_k141_38793 | Not-determined |
| S12_extracted_k141_117536 | Not-determined |
| S12_extracted_k141_86535 | Not-determined |
| S12_extracted_k141_172546 | Not-determined |
| S12_extracted_k141_38795 | Not-determined |
| S12_extracted_k141_23446 | Not-determined |
| S12_extracted_k141_117539 | Not-determined |
| S12_extracted_k141_125394 | Low-quality |
| S12_extracted_k141_125397 | Not-determined |
| S12_extracted_k141_180497 | Not-determined |
| S12_extracted_k141_133612 | Not-determined |
| S12_extracted_k141_14987 | Not-determined |
| S12_extracted_k141_156733 | Not-determined |
| S12_extracted_k141_133613 | Not-determined |
| S12_extracted_k141_14997 | Low-quality |
| S12_extracted_k141_156742 | Not-determined |
| S12_extracted_k141_23463 | Not-determined |
| S12_extracted_k141_125405 | Not-determined |
| S12_extracted_k141_38824 | Not-determined |
| S12_extracted_k141_86555 | Not-determined |
| S12_extracted_k141_38827 | Not-determined |
| S12_extracted_k141_133636 | Not-determined |
| S12_extracted_k141_180519 | Low-quality |
| S12_extracted_k141_133639 | Not-determined |
| S12_extracted_k141_172573 | Not-determined |
| S12_extracted_k141_172577 | Not-determined |
| S12_extracted_k141_117557 | Not-determined |
| S12_extracted_k141_86563 | Not-determined |
| S12_extracted_k141_125420 | Low-quality |
| S12_extracted_k141_15022 | Not-determined |
| S12_extracted_k141_133644 | Not-determined |
| S12_extracted_k141_156748 | Not-determined |
| S12_extracted_k141_172587 | Not-determined |
| S12_extracted_k141_172590 | Low-quality |
| S12_extracted_k141_180533 | Not-determined |
| S12_extracted_k141_38842 | Not-determined |
| S12_extracted_k141_180535 | Not-determined |
| S12_extracted_k141_172594 | Not-determined |
| S12_extracted_k141_117566 | Not-determined |
| S12_extracted_k141_156758 | Not-determined |
| S12_extracted_k141_117569 | Not-determined |
| S12_extracted_k141_180541 | Not-determined |
| S12_extracted_k141_117570 | Not-determined |
| S12_extracted_k141_86577 | Not-determined |
| S12_extracted_k141_86579 | Not-determined |
| S12_extracted_k141_156764 | Not-determined |
| S12_extracted_k141_172607 | Not-determined |
| S12_extracted_k141_23479 | Not-determined |
| S12_extracted_k141_180545 | Not-determined |
| S12_extracted_k141_23480 | Low-quality |
| S12_extracted_k141_180546 | Not-determined |
| S12_extracted_k141_15037 | Not-determined |
| S12_extracted_k141_172612 | Not-determined |
| S12_extracted_k141_125451 | Not-determined |
| S12_extracted_k141_23488 | Not-determined |
| S12_extracted_k141_172619 | Not-determined |
| S12_extracted_k141_180557 | Low-quality |
| S12_extracted_k141_15049 | Not-determined |
| S12_extracted_k141_172626 | Low-quality |
| S12_extracted_k141_86586 | Not-determined |
| S12_extracted_k141_172627 | Not-determined |
| S12_extracted_k141_15054 | Not-determined |
| S12_extracted_k141_15055 | Not-determined |
| S12_extracted_k141_165229 | Not-determined |
| S12_extracted_k141_117594 | Not-determined |
| S12_extracted_k141_172635 | Not-determined |
| S12_extracted_k141_156769 | Not-determined |
| S12_extracted_k141_133655 | Not-determined |
| S12_extracted_k141_180569 | Low-quality |
| S12_extracted_k141_117601 | Not-determined |
| S12_extracted_k141_117604 | Not-determined |
| S12_extracted_k141_86606 | Not-determined |
| S12_extracted_k141_172643 | Not-determined |
| S12_extracted_k141_180579 | Not-determined |
| S12_extracted_k141_180580 | Not-determined |
| S12_extracted_k141_86608 | Not-determined |
| S12_extracted_k141_38879 | Not-determined |
| S12_extracted_k141_156793 | Not-determined |
| S12_extracted_k141_86612 | Not-determined |
| S12_extracted_k141_172650 | Low-quality |
| S12_extracted_k141_117627 | Not-determined |
| S12_extracted_k141_156796 | Not-determined |
| S12_extracted_k141_156797 | Low-quality |
| S12_extracted_k141_156798 | Not-determined |
| S12_extracted_k141_172655 | Low-quality |
| S12_extracted_k141_133677 | Not-determined |
| S12_extracted_k141_180588 | Not-determined |
| S12_extracted_k141_156802 | Not-determined |
| S12_extracted_k141_38891 | Not-determined |
| S12_extracted_k141_156805 | Not-determined |
| S12_extracted_k141_172660 | Not-determined |
| S12_extracted_k141_117637 | Not-determined |
| S12_extracted_k141_156807 | Not-determined |
| S12_extracted_k141_156808 | Not-determined |
| S12_extracted_k141_38901 | Not-determined |
| S12_extracted_k141_117647 | Not-determined |
| S12_extracted_k141_180600 | Not-determined |
| S12_extracted_k141_180605 | Not-determined |
| S12_extracted_k141_23528 | Not-determined |
| S12_extracted_k141_15101 | Not-determined |
| S12_extracted_k141_117659 | Low-quality |
| S12_extracted_k141_117660 | Not-determined |
| S12_extracted_k141_23532 | Not-determined |
| S12_extracted_k141_172684 | Not-determined |
| S12_extracted_k141_23535 | Not-determined |
| S12_extracted_k141_165245 | Not-determined |
| S12_extracted_k141_15107 | Not-determined |
| S12_extracted_k141_38915 | Not-determined |
| S12_extracted_k141_133709 | Not-determined |
| S12_extracted_k141_156825 | Not-determined |
| S12_extracted_k141_172688 | Not-determined |
| S12_extracted_k141_165250 | Low-quality |
| S12_extracted_k141_156827 | Not-determined |
| S12_extracted_k141_55080 | Not-determined |
| S12_extracted_k141_165256 | Not-determined |
| S12_extracted_k141_180617 | Not-determined |
| S12_extracted_k141_165257 | Not-determined |
| S12_extracted_k141_180621 | Not-determined |
| S12_extracted_k141_133730 | Not-determined |
| S12_extracted_k141_133735 | Not-determined |
| S12_extracted_k141_15123 | Not-determined |
| S12_extracted_k141_156844 | Not-determined |
| S12_extracted_k141_117676 | Not-determined |
| S12_extracted_k141_172703 | Not-determined |
| S12_extracted_k141_180625 | Not-determined |
| S12_extracted_k141_38924 | Not-determined |
| S12_extracted_k141_38927 | Not-determined |
| S12_extracted_k141_117687 | Not-determined |
| S5_vs_k141_25423\|\|full | High-quality |
| S12_extracted_k141_180627 | Not-determined |
| S12_extracted_k141_165271 | Not-determined |
| S12_extracted_k141_156850 | Not-determined |
| S12_extracted_k141_55097 | Not-determined |
| S12_extracted_k141_133748 | Not-determined |
| S12_extracted_k141_15142 | Not-determined |
| S12_extracted_k141_55099 | Not-determined |
| S12_extracted_k141_117695 | Not-determined |
| S12_extracted_k141_117697 | Not-determined |
| S12_extracted_k141_38943 | Low-quality |
| S12_extracted_k141_172729 | Not-determined |
| S12_extracted_k141_117719 | Low-quality |
| S12_extracted_k141_156880 | Low-quality |
| S12_extracted_k141_38950 | Not-determined |
| S12_extracted_k141_133760 | Not-determined |
| S12_extracted_k141_156885 | Not-determined |
| S12_extracted_k141_15160 | Not-determined |
| S12_extracted_k141_38957 | Not-determined |
| S12_extracted_k141_156891 | Not-determined |
| S12_extracted_k141_172744 | Not-determined |
| S12_extracted_k141_15162 | Not-determined |
| S12_extracted_k141_38962 | Not-determined |
| S12_extracted_k141_38966 | Not-determined |
| S12_extracted_k141_38968 | Low-quality |
| S12_extracted_k141_165289 | Not-determined |
| S12_extracted_k141_156898 | Not-determined |
| S12_extracted_k141_133764 | Not-determined |
| S12_extracted_k141_38973 | Not-determined |
| S12_extracted_k141_172756 | Not-determined |
| S12_extracted_k141_38982 | Not-determined |
| S12_extracted_k141_38989 | Low-quality |
| S12_extracted_k141_133782 | Not-determined |
| S12_extracted_k141_133783 | Not-determined |
| S12_extracted_k141_117735 | Not-determined |
| S12_extracted_k141_38990 | Low-quality |
| S12_extracted_k141_38994 | Not-determined |
| S12_extracted_k141_117739 | Not-determined |
| S12_extracted_k141_172791 | Not-determined |
| S12_extracted_k141_172792 | Not-determined |
| S12_extracted_k141_125459 | Not-determined |
| S12_extracted_k141_23579 | Not-determined |
| S12_extracted_k141_38999 | Low-quality |
| S12_extracted_k141_117745 | Not-determined |
| S12_extracted_k141_117747 | Not-determined |
| S12_extracted_k141_172797 | Not-determined |
| S12_extracted_k141_125464 | Not-determined |
| S12_extracted_k141_117750 | Not-determined |
| S12_extracted_k141_23583 | Not-determined |
| S12_extracted_k141_117751 | Low-quality |
| S12_extracted_k141_117753 | Not-determined |
| S12_extracted_k141_39006 | Not-determined |
| S12_extracted_k141_172804 | Not-determined |
| S12_extracted_k141_172808 | Low-quality |
| S12_extracted_k141_15176 | Low-quality |
| S12_extracted_k141_125467 | Not-determined |
| S12_extracted_k141_117765 | Not-determined |
| S12_extracted_k141_15185 | Not-determined |
| S12_extracted_k141_15188 | Not-determined |
| S12_extracted_k141_172821 | Not-determined |
| S12_extracted_k141_172822 | Not-determined |
| S12_extracted_k141_117772 | Not-determined |
| S12_extracted_k141_172826 | Not-determined |
| S12_extracted_k141_172828 | Not-determined |
| S12_extracted_k141_39012 | Low-quality |
| S12_extracted_k141_172829 | Not-determined |
| S12_extracted_k141_117783 | Not-determined |
| S12_extracted_k141_117787 | Not-determined |
| S12_extracted_k141_156931 | Not-determined |
| S12_extracted_k141_125486 | Not-determined |
| S12_extracted_k141_125489 | Not-determined |
| S12_extracted_k141_117795 | Not-determined |
| S12_extracted_k141_15211 | Not-determined |
| S5_vs_k141_106428\|\|full | Medium-quality |
| S12_extracted_k141_117801 | Not-determined |
| S12_extracted_k141_23592 | Not-determined |
| S12_extracted_k141_125501 | Not-determined |
| S12_extracted_k141_125502 | Low-quality |
| S11_vs_k141_67573\|\|full | Medium-quality |
| S12_extracted_k141_39040 | Not-determined |
| S12_extracted_k141_117806 | Low-quality |
| S12_extracted_k141_133832 | Not-determined |
| S12_extracted_k141_133834 | Not-determined |
| S12_extracted_k141_23598 | Not-determined |
| S12_extracted_k141_15237 | Not-determined |
| S12_extracted_k141_23601 | Not-determined |
| S12_extracted_k141_156943 | Not-determined |
| S12_extracted_k141_156944 | Not-determined |
| S12_extracted_k141_165308 | Not-determined |
| S12_extracted_k141_23607 | Not-determined |

| | |
|---|---|
| S12_extracted_k141_165309 | Not-determined |
| S12_extracted_k141_165310 | Not-determined |
| S12_extracted_k141_23608 | Not-determined |
| S12_extracted_k141_15252 | Not-determined |
| S12_extracted_k141_15254 | Not-determined |
| S12_extracted_k141_172883 | Not-determined |
| S12_extracted_k141_125523 | Not-determined |
| S12_extracted_k141_165341 | Not-determined |
| S12_extracted_k141_165343 | Not-determined |
| S12_extracted_k141_125528 | Not-determined |
| S12_extracted_k141_125529 | Not-determined |
| S12_extracted_k141_117841 | Not-determined |
| S12_extracted_k141_117844 | Not-determined |
| S12_extracted_k141_117846 | Not-determined |
| S12_extracted_k141_117852 | Not-determined |
| S12_extracted_k141_125550 | Not-determined |
| S12_extracted_k141_125553 | Not-determined |
| S12_extracted_k141_125554 | Not-determined |
| S12_extracted_k141_125558 | Not-determined |
| S12_extracted_k141_15271 | Not-determined |
| S12_extracted_k141_39057 | Low-quality |
| S12_extracted_k141_39060 | Not-determined |
| S12_extracted_k141_125568 | Not-determined |
| S12_extracted_k141_15290 | Not-determined |
| S12_extracted_k141_39075 | Not-determined |
| S12_extracted_k141_117877 | Low-quality |
| S12_extracted_k141_15306 | Not-determined |
| S12_extracted_k141_156981 | Not-determined |
| S12_extracted_k141_117885 | Not-determined |
| S12_extracted_k141_125580 | Not-determined |
| S12_extracted_k141_15310 | Not-determined |
| S12_extracted_k141_117890 | Not-determined |
| S12_extracted_k141_39091 | Low-quality |
| S12_extracted_k141_156997 | Not-determined |
| S12_extracted_k141_125600 | Not-determined |
| S12_extracted_k141_125602 | Not-determined |
| S12_extracted_k141_125603 | Not-determined |
| S12_extracted_k141_156999 | Not-determined |
| S12_extracted_k141_15323 | Not-determined |
| S12_extracted_k141_15335 | Not-determined |
| S12_extracted_k141_15336 | Not-determined |
| S12_extracted_k141_39096 | Not-determined |
| S12_extracted_k141_117907 | Not-determined |
| S12_extracted_k141_117909 | Not-determined |
| S12_extracted_k141_15343 | Not-determined |
| S12_extracted_k141_157023 | Low-quality |
| S12_extracted_k141_117912 | Not-determined |
| S12_extracted_k141_15346 | Not-determined |
| S12_extracted_k141_180634 | Not-determined |
| S12_extracted_k141_180636 | Low-quality |
| S12_extracted_k141_15353 | Not-determined |
| S12_extracted_k141_180639 | Not-determined |
| S12_extracted_k141_117919 | Not-determined |
| S12_extracted_k141_39107 | Not-determined |
| S12_extracted_k141_172943 | Not-determined |
| S12_extracted_k141_180645 | Not-determined |
| S12_extracted_k141_157035 | Not-determined |
| S12_extracted_k141_39118 | Not-determined |
| S12_extracted_k141_172950 | Not-determined |
| S12_extracted_k141_39119 | Not-determined |
| S12_extracted_k141_39120 | Not-determined |
| S12_extracted_k141_172952 | Not-determined |
| S12_extracted_k141_117929 | Not-determined |
| S12_extracted_k141_172953 | Not-determined |
| S12_extracted_k141_172954 | Low-quality |
| S12_extracted_k141_157043 | Not-determined |
| S12_extracted_k141_39123 | Not-determined |
| S12_extracted_k141_180657 | Not-determined |
| S9_vs_k141_28197\|\|full | Medium-quality |
| S12_extracted_k141_39125 | Not-determined |
| S12_extracted_k141_39126 | Not-determined |
| S12_extracted_k141_39130 | Not-determined |
| S12_extracted_k141_172971 | Not-determined |
| S12_extracted_k141_172972 | Not-determined |
| S12_extracted_k141_180671 | Low-quality |
| S12_extracted_k141_15387 | Not-determined |
| S12_extracted_k141_15392 | Not-determined |
| S12_extracted_k141_180679 | Not-determined |
| S12_extracted_k141_39141 | Not-determined |
| S12_extracted_k141_15400 | Not-determined |
| S12_extracted_k141_39147 | Not-determined |
| S12_extracted_k141_157067 | Not-determined |
| S12_extracted_k141_172990 | Not-determined |
| S12_extracted_k141_157075 | Low-quality |
| S12_extracted_k141_172992 | Not-determined |
| S12_extracted_k141_117956 | Not-determined |
| S12_extracted_k141_125648 | Not-determined |
| S12_extracted_k141_39158 | Not-determined |
| S12_extracted_k141_117961 | Low-quality |
| S12_extracted_k141_157083 | Low-quality |
| S12_extracted_k141_39160 | Not-determined |
| S12_extracted_k141_39163 | Not-determined |
| S12_extracted_k141_125655 | Low-quality |
| S12_extracted_k141_39166 | Not-determined |
| S12_extracted_k141_117970 | Not-determined |
| S12_extracted_k141_39168 | Not-determined |
| S12_extracted_k141_173009 | Not-determined |
| S12_extracted_k141_173012 | Not-determined |
| S12_extracted_k141_15432 | Not-determined |
| S12_extracted_k141_173014 | Not-determined |
| S12_extracted_k141_117978 | Low-quality |
| S12_extracted_k141_188835 | Low-quality |
| S12_extracted_k141_39181 | Low-quality |
| S12_extracted_k141_125673 | Not-determined |
| S12_extracted_k141_157111 | Not-determined |
| S12_extracted_k141_173039 | Not-determined |
| S12_extracted_k141_157115 | Not-determined |
| S12_extracted_k141_117990 | Not-determined |
| S10_vs_37694\|\|full | Medium-quality |
| S12_extracted_k141_39211 | Not-determined |
| S12_extracted_k141_125692 | Low-quality |
| S12_extracted_k141_180691 | Low-quality |
| S12_extracted_k141_157142 | Not-determined |
| S12_extracted_k141_173069 | Not-determined |
| S12_extracted_k141_39222 | Low-quality |
| S12_extracted_k141_173071 | Not-determined |
| S12_extracted_k141_188843 | Not-determined |
| S12_extracted_k141_39227 | Not-determined |
| S12_extracted_k141_173075 | Not-determined |
| S12_extracted_k141_188845 | Not-determined |
| S12_extracted_k141_125693 | Low-quality |
| S12_extracted_k141_157151 | Not-determined |
| S12_extracted_k141_173085 | Not-determined |
| S11_extracted_k141_66522 | High-quality |
| S12_extracted_k141_173089 | Not-determined |
| S12_extracted_k141_180702 | Low-quality |
| S12_extracted_k141_15476 | Not-determined |
| S12_extracted_k141_180715 | Low-quality |
| S12_extracted_k141_180719 | Not-determined |
| S12_extracted_k141_173101 | Not-determined |
| S12_extracted_k141_180721 | Not-determined |
| S12_extracted_k141_180723 | Not-determined |
| S12_extracted_k141_39249 | Low-quality |
| S12_extracted_k141_39250 | Not-determined |
| S12_extracted_k141_15493 | Not-determined |
| S12_extracted_k141_15498 | Not-determined |
| S12_extracted_k141_188865 | Not-determined |
| S12_extracted_k141_157178 | Not-determined |
| S12_extracted_k141_180735 | Low-quality |
| S12_extracted_k141_125712 | Not-determined |
| S12_extracted_k141_15506 | Not-determined |
| S12_extracted_k141_118009 | Not-determined |
| S12_extracted_k141_39263 | Not-determined |
| S12_extracted_k141_188874 | Not-determined |
| S12_extracted_k141_118011 | Not-determined |
| S12_extracted_k141_188876 | Not-determined |
| S12_extracted_k141_157184 | Not-determined |
| S12_extracted_k141_188878 | Not-determined |
| S12_extracted_k141_173130 | Not-determined |
| S12_extracted_k141_39272 | Not-determined |
| S12_extracted_k141_188880 | Not-determined |
| S12_extracted_k141_188881 | Not-determined |
| S12_extracted_k141_188882 | Not-determined |
| S12_extracted_k141_39275 | Not-determined |
| S12_extracted_k141_15520 | Not-determined |
| S12_extracted_k141_15521 | Not-determined |
| S12_extracted_k141_173141 | Low-quality |
| S12_extracted_k141_188900 | Not-determined |
| S12_extracted_k141_15529 | Not-determined |
| S12_extracted_k141_188902 | Not-determined |
| S12_extracted_k141_188905 | Not-determined |
| S12_extracted_k141_15535 | Not-determined |
| S12_extracted_k141_188907 | Not-determined |
| S12_extracted_k141_15536 | Not-determined |
| S12_extracted_k141_173144 | Not-determined |
| S12_extracted_k141_188909 | Not-determined |
| S12_extracted_k141_39286 | Not-determined |
| S12_extracted_k141_188913 | Not-determined |
| S12_extracted_k141_39291 | Not-determined |
| S12_extracted_k141_15544 | Not-determined |
| S12_extracted_k141_39292 | Not-determined |
| S12_extracted_k141_188917 | Not-determined |
| S12_extracted_k141_125730 | Low-quality |
| S12_extracted_k141_125731 | Low-quality |
| S12_extracted_k141_188919 | Not-determined |
| S12_extracted_k141_157204 | Not-determined |
| S12_extracted_k141_173151 | Not-determined |
| S12_extracted_k141_157205 | Not-determined |
| S12_extracted_k141_125738 | Not-determined |
| S12_extracted_k141_188929 | Not-determined |
| S12_extracted_k141_180763 | Not-determined |
| S12_extracted_k141_180764 | Not-determined |
| S12_extracted_k141_188932 | Not-determined |
| S12_extracted_k141_157209 | Not-determined |
| S12_extracted_k141_39300 | Not-determined |
| S12_extracted_k141_118026 | Not-determined |
| S12_extracted_k141_173164 | Not-determined |
| S12_extracted_k141_118030 | Not-determined |
| S12_extracted_k141_39303 | Not-determined |
| S12_extracted_k141_188946 | Not-determined |
| S12_extracted_k141_188948 | Not-determined |
| S12_extracted_k141_188949 | Not-determined |
| S12_extracted_k141_118037 | Not-determined |
| S12_extracted_k141_39308 | Not-determined |
| S12_extracted_k141_180769 | Not-determined |
| S12_extracted_k141_188958 | Not-determined |
| S12_extracted_k141_118043 | Not-determined |
| S12_extracted_k141_188961 | Not-determined |
| S12_extracted_k141_188962 | Not-determined |
| S12_extracted_k141_188963 | Not-determined |
| S12_extracted_k141_173169 | Not-determined |
| S12_extracted_k141_15568 | Not-determined |
| S12_extracted_k141_39320 | Not-determined |
| S12_extracted_k141_180784 | Not-determined |
| S12_extracted_k141_39322 | Not-determined |
| S12_extracted_k141_125747 | Not-determined |
| S12_extracted_k141_15576 | Not-determined |
| S12_extracted_k141_125749 | Not-determined |
| S12_extracted_k141_118062 | Not-determined |
| S12_extracted_k141_118065 | Not-determined |
| S12_extracted_k141_39329 | Not-determined |
| S12_extracted_k141_39331 | Not-determined |
| S12_extracted_k141_125751 | Not-determined |
| S12_extracted_k141_39334 | Not-determined |
| S12_extracted_k141_180791 | Not-determined |
| S12_extracted_k141_180793 | Not-determined |
| S12_extracted_k141_39338 | Not-determined |
| S12_extracted_k141_180794 | Not-determined |
| S12_extracted_k141_15590 | Not-determined |
| S12_extracted_k141_180797 | Not-determined |
| S12_extracted_k141_39350 | Not-determined |
| S12_extracted_k141_118080 | Not-determined |
| S12_extracted_k141_180799 | Not-determined |
| S12_extracted_k141_118086 | Not-determined |
| S12_extracted_k141_118093 | Not-determined |
| S12_extracted_k141_180811 | Not-determined |
| S12_extracted_k141_39365 | Not-determined |
| S12_extracted_k141_39367 | Not-determined |
| S12_extracted_k141_118100 | Not-determined |
| S12_extracted_k141_118101 | Low-quality |
| S12_extracted_k141_15607 | Not-determined |
| S12_extracted_k141_180827 | Not-determined |
| S12_extracted_k141_118103 | Not-determined |
| S12_extracted_k141_118108 | Not-determined |
| S12_extracted_k141_157221 | Low-quality |
| S12_extracted_k141_157225 | Not-determined |
| S12_extracted_k141_157234 | Not-determined |
| S12_extracted_k141_15630 | Not-determined |
| S12_extracted_k141_180843 | Not-determined |
| S12_extracted_k141_157247 | Not-determined |
| S12_extracted_k141_157253 | Not-determined |
| S12_extracted_k141_180855 | Not-determined |
| S12_extracted_k141_180856 | Not-determined |
| S12_extracted_k141_15650 | Not-determined |
| S12_extracted_k141_15654 | Not-determined |
| S12_extracted_k141_157270 | Not-determined |
| S12_extracted_k141_15656 | Not-determined |
| S12_extracted_k141_15662 | Not-determined |
| S12_extracted_k141_15667 | Low-quality |
| S12_extracted_k141_180872 | Not-determined |
| S12_extracted_k141_125811 | Not-determined |
| S12_extracted_k141_125817 | Not-determined |
| S12_extracted_k141_15684 | Not-determined |
| S12_extracted_k141_15685 | Not-determined |
| S12_extracted_k141_157302 | Not-determined |
| S12_extracted_k141_180880 | Not-determined |
| S12_extracted_k141_180882 | Not-determined |
| S12_extracted_k141_125820 | Low-quality |
| S12_extracted_k141_180886 | Not-determined |
| S12_extracted_k141_180893 | Not-determined |
| S12_extracted_k141_125835 | Not-determined |
| S12_extracted_k141_157328 | Not-determined |
| S12_extracted_k141_157331 | Not-determined |
| S12_extracted_k141_157332 | Low-quality |
| S12_extracted_k141_180901 | Not-determined |
| S12_extracted_k141_157350 | Not-determined |
| S12_extracted_k141_125849 | Not-determined |
| S12_extracted_k141_125850 | Not-determined |
| S12_extracted_k141_125854 | Not-determined |
| S12_extracted_k141_15697 | Not-determined |
| S12_extracted_k141_180917 | Not-determined |
| S12_extracted_k141_180923 | Not-determined |
| S12_extracted_k141_180933 | Not-determined |
| S12_extracted_k141_157379 | Not-determined |
| S12_extracted_k141_157380 | Not-determined |
| S12_extracted_k141_125877 | Not-determined |
| S12_extracted_k141_125878 | Not-determined |
| S12_extracted_k141_180940 | Not-determined |
| S12_extracted_k141_125881 | Not-determined |
| S12_extracted_k141_15721 | Not-determined |
| S12_extracted_k141_125895 | Not-determined |
| S12_extracted_k141_180956 | Not-determined |
| S12_extracted_k141_15740 | Not-determined |
| S8_vs_k141_169949\|\|full | Medium-quality |
| S12_extracted_k141_180963 | Not-determined |
| S12_extracted_k141_125914 | Not-determined |
| S12_extracted_k141_180994 | Low-quality |
| S12_extracted_k141_180995 | Not-determined |
| S12_extracted_k141_181002 | Not-determined |
| S12_extracted_k141_125938 | Not-determined |
| S12_extracted_k141_181008 | Not-determined |
| S12_extracted_k141_125949 | Not-determined |
| S12_extracted_k141_181017 | Not-determined |
| S12_extracted_k141_125971 | Not-determined |
| S12_extracted_k141_157412 | Not-determined |
| S12_extracted_k141_157427 | Not-determined |
| S12_extracted_k141_157437 | Not-determined |
| S12_extracted_k141_157459 | Not-determined |
| S12_extracted_k141_157470 | Not-determined |
| S11_vs_k141_56762\|\|full | Medium-quality |
| S12_extracted_k141_181038 | Not-determined |
| S12_extracted_k141_181049 | Low-quality |
| S12_extracted_k141_181054 | Low-quality |
| S12_extracted_k141_181065 | Not-determined |
| S12_extracted_k141_181069 | Not-determined |
| S13_vs_30025\|\|full | Medium-quality |
| S12_vs_k141_55201\|\|full | Not-determined |
| S11_extracted_k141_34008 | Medium-quality |
| S5_vs_k141_17520\|\|full | Medium-quality |
| S8_vs_k141_172088\|\|full | Medium-quality |
| S9_vs_k141_69322\|\|full | Medium-quality |
| S8_extracted_k141_129423 | High-quality |
| S12_vs_k141_28661\|\|full | Medium-quality |
| S8_vs_k141_4883\|\|full | Medium-quality |
| S12_vs_k141_174415\|\|full | Low-quality |
| S11_vs_k141_66522\|\|full | High-quality |
| S9_vs_k141_58086\|\|full | Medium-quality |
| S5_vs_k141_60507\|\|full | Medium-quality |
| S1_extracted_k141_123958 | Medium-quality |
| S10_extracted_113489 | Medium-quality |
| S10_vs_113489\|\|full | Medium-quality |
| S16_vs_88946\|\|full | Medium-quality |
| S12_vs_k141_105523\|\|full | Not-determined |
| S7_extracted_63894 | High-quality |
| S1_vs_k141_123958\|\|full | Medium-quality |
| S11_vs_k141_74968\|\|full | Medium-quality |
| S9_vs_k141_1966\|\|full | Medium-quality |
| S11_vs_k141_34008\|\|full | Medium-quality |
| S12_vs_k141_146000\|\|full | Low-quality |
| S3_vs_k141_7533\|\|full | Medium-quality |
| S5_extracted_k141_111742 | High-quality |
| S12_vs_k141_52623\|\|full | Low-quality |
| S7_vs_63894\|\|full | High-quality |
| S5_vs_k141_107105\|\|full | Medium-quality |
| S8_vs_k141_129423\|\|full | High-quality |
| S5_vs_k141_128881\|\|full | Medium-quality |
| S10_vs_71900\|\|full | Medium-quality |
| S8_extracted_k141_93976 | Medium-quality |
| S8_vs_k141_93976\|\|full | Medium-quality |
| S5_vs_k141_140946\|\|full | Medium-quality |
| S16_vs_64584\|\|full | Medium-quality |
| S16_extracted_90328 | Medium-quality |
| S12_vs_k141_188829\|\|full | Not-determined |
| S12_vs_k141_6836\|\|full | Medium-quality |
| S5_vs_k141_111742\|\|full | High-quality |
| S8_vs_k141_50431\|\|full | Medium-quality |
| S12_vs_k141_188834\|\|full | Not-determined |
| S12_vs_k141_188836\|\|full | Not-determined |
| S7_vs_118295\|\|full | Medium-quality |
| S12_vs_k141_188841\|\|full | Not-determined |
| S4_vs_VIRSorter_29959-cat_2 | Medium-quality |
| S12_vs_k141_188855\|\|full | Not-determined |
| S12_vs_k141_188863\|\|full | Not-determined |
| S12_vs_k141_188865\|\|full | Not-determined |
| S12_vs_k141_188873\|\|full | Low-quality |
| S12_vs_k141_188879\|\|full | Not-determined |
| S12_vs_k141_78741\|\|full | Low-quality |
| S12_vs_k141_47251\|\|full | Low-quality |
| S12_vs_k141_165365\|\|full | Not-determined |
| S12_vs_k141_133877\|\|full | Low-quality |
| S12_vs_k141_31501\|\|full | Low-quality |
| S12_vs_k141_141741\|\|full | Low-quality |
| S12_vs_k141_157497\|\|full | Low-quality |
| S12_vs_k141_86639\|\|full | Not-determined |
| S12_vs_k141_94505\|\|full | Not-determined |
| S12_vs_k141_110262\|\|full | Not-determined |
| S12_vs_k141_86641\|\|full | Not-determined |
| S12_vs_k141_181122\|\|full | Not-determined |
| S12_vs_k141_39391\|\|full | Low-quality |
| S12_vs_k141_7898\|\|full | Low-quality |
| S12_vs_k141_86647\|\|full | Low-quality |
| S12_vs_k141_23664\|\|full | Low-quality |
| S12_vs_k141_149639\|\|full | Low-quality |
| S12_vs_k141_102393\|\|full | Low-quality |
| S12_vs_k141_126025\|\|full | Not-determined |
| S12_vs_k141_53\|\|full | Not-determined |
| S12_vs_k141_23671\|\|full | Low-quality |
| S12_vs_k141_55\|\|full | Not-determined |
| S12_vs_k141_102403\|\|full | Not-determined |
| S12_vs_k141_65\|\|full | Not-determined |
| S12_vs_k141_157523\|\|full | Low-quality |
| S12_vs_k141_70916\|\|full | Low-quality |
| S12_vs_k141_165419\|\|full | Not-determined |
| S12_vs_k141_7908\|\|full | Not-determined |
| S12_vs_k141_94550\|\|full | Not-determined |
| S12_vs_k141_165420\|\|full | Low-quality |
| S12_vs_k141_181146\|\|full | Low-quality |
| S12_vs_k141_70934\|\|full | Not-determined |
| S12_vs_k141_110300\|\|full | Not-determined |
| S12_vs_k141_157557\|\|full | Not-determined |
| S12_vs_k141_78807\|\|full | Low-quality |
| S12_vs_k141_31564\|\|full | Low-quality |
| S12_vs_k141_70947\|\|full | Low-quality |
| S12_vs_k141_39403\|\|full | Not-determined |
| S12_vs_k141_181158\|\|full | Low-quality |
| S12_vs_k141_110305\|\|full | Low-quality |
| S12_vs_k141_165443\|\|full | Low-quality |
| S12_vs_k141_39408\|\|full | Low-quality |
| S12_vs_k141_70953\|\|full | Not-determined |
| S8_vs_k141_139391\|\|full | Medium-quality |
| S12_vs_k141_102461\|\|full | Low-quality |
| S12_vs_k141_157570\|\|full | Not-determined |
| S12_vs_k141_70957\|\|full | Not-determined |
| S12_vs_k141_173318\|\|full | Low-quality |
| S12_vs_k141_149688\|\|full | Low-quality |
| S12_vs_k141_126111\|\|full | Low-quality |
| S12_vs_k141_7973\|\|full | Low-quality |
| S12_vs_k141_70969\|\|full | Low-quality |
| S12_vs_k141_118171\|\|full | Not-determined |
| S12_vs_k141_149708\|\|full | Not-determined |
| S12_vs_k141_110322\|\|full | Low-quality |
| S12_vs_k141_126129\|\|full | Low-quality |
| S12_vs_k141_146\|\|full | Low-quality |
| S12_vs_k141_133937\|\|full | Low-quality |
| S12_vs_k141_149715\|\|full | Not-determined |
| S12_vs_k141_150\|\|full | Low-quality |
| S12_vs_k141_157587\|\|full | Low-quality |
| S12_vs_k141_181185\|\|full | Low-quality |
| S12_vs_k141_110338\|\|full | Low-quality |
| S12_vs_k141_102478\|\|full | Low-quality |
| S12_vs_k141_8011\|\|full | Low-quality |
| S12_vs_k141_173356\|\|full | Low-quality |
| S12_vs_k141_15836\|\|full | Low-quality |
| S12_vs_k141_86751\|\|full | Not-determined |
| S12_vs_k141_15837\|\|full | Low-quality |
| S12_vs_k141_39460\|\|full | Low-quality |
| S12_vs_k141_8022\|\|full | Low-quality |
| S12_vs_k141_110353\|\|full | Not-determined |
| S12_vs_k141_39477\|\|full | Not-determined |
| S12_vs_k141_78885\|\|full | Low-quality |
| S12_vs_k141_110359\|\|full | Low-quality |
| S12_vs_k141_86785\|\|full | Low-quality |
| S12_vs_k141_133979\|\|full | Not-determined |
| S12_vs_k141_86788\|\|full | Low-quality |
| S12_vs_k141_71020\|\|full | Low-quality |
| S12_vs_k141_63121\|\|full | Not-determined |
| S12_vs_k141_118245\|\|full | Not-determined |
| S12_vs_k141_204\|\|full | Not-determined |
| S12_vs_k141_118247\|\|full | Not-determined |
| S12_vs_k141_181245\|\|full | Low-quality |
| S12_vs_k141_165539\|\|full | Not-determined |
| S12_vs_k141_86806\|\|full | Low-quality |
| S12_vs_k141_165542\|\|full | Not-determined |
| S12_vs_k141_141848\|\|full | Not-determined |
| S12_vs_k141_63130\|\|full | Low-quality |
| S12_vs_k141_31598\|\|full | Low-quality |
| S12_vs_k141_63134\|\|full | Not-determined |
| S12_vs_k141_71046\|\|full | Low-quality |
| S12_vs_k141_78888\|\|full | Not-determined |
| S12_vs_k141_110381\|\|full | Low-quality |
| S12_vs_k141_157599\|\|full | Low-quality |
| S12_vs_k141_232\|\|full | Not-determined |
| S12_vs_k141_134002\|\|full | Not-determined |
| S12_vs_k141_118277\|\|full | Low-quality |
| S12_vs_k141_141856\|\|full | Low-quality |
| S12_vs_k141_238\|\|full | Low-quality |
| S12_vs_k141_63153\|\|full | Low-quality |
| S12_vs_k141_47368\|\|full | Low-quality |
| S12_vs_k141_102542\|\|full | Low-quality |
| S12_vs_k141_71069\|\|full | Low-quality |
| S12_vs_k141_86852\|\|full | Not-determined |
| S12_vs_k141_173402\|\|full | Low-quality |
| S12_vs_k141_165585\|\|full | Low-quality |
| S12_vs_k141_118289\|\|full | Low-quality |
| S12_vs_k141_78918\|\|full | Low-quality |
| S12_vs_k141_110403\|\|full | Low-quality |
| S12_vs_k141_173419\|\|full | Not-determined |
| S12_vs_k141_118308\|\|full | Low-quality |
| S12_vs_k141_110409\|\|full | Not-determined |
| S12_vs_k141_181297\|\|full | Not-determined |
| S12_vs_k141_63173\|\|full | Low-quality |
| S12_vs_k141_94714\|\|full | Low-quality |
| S12_vs_k141_102587\|\|full | Not-determined |
| S12_vs_k141_63178\|\|full | Not-determined |
| S12_vs_k141_110421\|\|full | Not-determined |
| S12_vs_k141_71075\|\|full | Low-quality |
| S12_vs_k141_86894\|\|full | Low-quality |
| S12_vs_k141_181298\|\|full | Low-quality |
| S12_vs_k141_181304\|\|full | Not-determined |
| S12_vs_k141_141900\|\|full | Low-quality |
| S12_vs_k141_94742\|\|full | Not-determined |
| S12_vs_k141_78966\|\|full | Not-determined |
| S12_vs_k141_55253\|\|full | Low-quality |
| S12_vs_k141_71090\|\|full | Low-quality |
| S12_vs_k141_149788\|\|full | Not-determined |
| S12_vs_k141_173460\|\|full | Low-quality |
| S12_vs_k141_299\|\|full | Low-quality |
| S12_vs_k141_126208\|\|full | Low-quality |
| S12_vs_k141_157689\|\|full | Low-quality |
| S12_vs_k141_110461\|\|full | Low-quality |
| S12_vs_k141_39535\|\|full | Low-quality |
| S12_vs_k141_327\|\|full | Not-determined |
| S12_vs_k141_94769\|\|full | Not-determined |
| S12_vs_k141_173493\|\|full | Not-determined |
| S12_vs_k141_8107\|\|full | Not-determined |
| S12_vs_k141_71116\|\|full | Not-determined |
| S12_vs_k141_71122\|\|full | Low-quality |
| S12_vs_k141_55273\|\|full | Not-determined |
| S12_vs_k141_94786\|\|full | Low-quality |
| S12_vs_k141_141929\|\|full | Low-quality |
| S12_vs_k141_134094\|\|full | Low-quality |
| S12_vs_k141_55277\|\|full | Not-determined |
| S12_vs_k141_39568\|\|full | Low-quality |
| S12_vs_k141_63249\|\|full | Not-determined |
| S12_vs_k141_102658\|\|full | Not-determined |
| S12_vs_k141_181367\|\|full | Not-determined |
| S12_vs_k141_110498\|\|full | Not-determined |
| S12_vs_k141_55290\|\|full | Not-determined |
| S12_vs_k141_47409\|\|full | Low-quality |
| S12_vs_k141_63253\|\|full | Low-quality |
| S12_vs_k141_102677\|\|full | Not-determined |
| S12_vs_k141_63255\|\|full | Low-quality |
| S12_vs_k141_94804\|\|full | Low-quality |
| S12_vs_k141_47414\|\|full | Not-determined |
| S12_vs_k141_181388\|\|full | Low-quality |
| S12_vs_k141_94805\|\|full | Not-determined |
| S12_vs_k141_86937\|\|full | Low-quality |
| S12_vs_k141_102691\|\|full | Low-quality |
| S12_vs_k141_102692\|\|full | Not-determined |
| S12_vs_k141_94813\|\|full | Not-determined |
| S12_vs_k141_8146\|\|full | Not-determined |
| S12_vs_k141_134134\|\|full | Low-quality |
| S12_vs_k141_141969\|\|full | Low-quality |
| S16_vs_90328\|\|full | Medium-quality |
| S12_vs_k141_71175\|\|full | Not-determined |
| S12_vs_k141_110538\|\|full | Low-quality |
| S12_vs_k141_86948\|\|full | Not-determined |
| S1_vs_k141_28398\|\|full | Medium-quality |
| S12_vs_k141_79008\|\|full | Low-quality |
| S12_vs_k141_79010\|\|full | Not-determined |
| S12_vs_k141_383\|\|full | Not-determined |
| S12_vs_k141_86956\|\|full | Not-determined |
| S12_vs_k141_15932\|\|full | Not-determined |
| S12_vs_k141_141992\|\|full | Low-quality |
| S12_vs_k141_71191\|\|full | Not-determined |
| S12_vs_k141_141993\|\|full | Not-determined |
| S12_vs_k141_141996\|\|full | Low-quality |
| S5_vs_k141_128398\|\|full | Medium-quality |
| S12_vs_k141_126316\|\|full | Low-quality |
| S12_vs_k141_63288\|\|full | Low-quality |
| S12_vs_k141_409\|\|full | Not-determined |
| S12_vs_k141_63289\|\|full | Low-quality |
| S12_vs_k141_110561\|\|full | Not-determined |
| S12_vs_k141_8187\|\|full | Low-quality |
| S12_vs_k141_79044\|\|full | Low-quality |
| S12_vs_k141_427\|\|full | Not-determined |
| S12_vs_k141_110578\|\|full | Low-quality |
| S12_vs_k141_79047\|\|full | Low-quality |
| S12_vs_k141_86992\|\|full | Low-quality |
| S12_vs_k141_157745\|\|full | Low-quality |
| S12_vs_k141_55350\|\|full | Not-determined |
| S12_vs_k141_79049\|\|full | Low-quality |
| S12_vs_k141_126336\|\|full | Not-determined |
| S12_vs_k141_79050\|\|full | Not-determined |
| S12_vs_k141_8194\|\|full | Low-quality |
| S12_vs_k141_86993\|\|full | Low-quality |
| S12_vs_k141_79056\|\|full | Low-quality |
| S12_vs_k141_8203\|\|full | Low-quality |
| S12_vs_k141_63327\|\|full | Not-determined |
| S12_vs_k141_118399\|\|full | Low-quality |
| S12_vs_k141_87024\|\|full | Not-determined |
| S12_vs_k141_126376\|\|full | Low-quality |
| S12_vs_k141_157771\|\|full | Low-quality |
| S12_vs_k141_63349\|\|full | Low-quality |
| S12_vs_k141_157772\|\|full | Not-determined |
| S12_vs_k141_55390\|\|full | Not-determined |
| S12_vs_k141_149871\|\|full | Low-quality |
| S12_vs_k141_79065\|\|full | Low-quality |
| S12_vs_k141_442\|\|full | Not-determined |
| S12_vs_k141_157790\|\|full | Low-quality |
| S12_vs_k141_447\|\|full | Low-quality |
| S12_vs_k141_165701\|\|full | Not-determined |
| S12_vs_k141_173535\|\|full | Low-quality |
| S12_vs_k141_149889\|\|full | Not-determined |
| S12_vs_k141_87057\|\|full | Not-determined |
| S12_vs_k141_149891\|\|full | Not-determined |
| S12_vs_k141_87065\|\|full | Not-determined |
| S12_vs_k141_126430\|\|full | Low-quality |
| S12_vs_k141_55429\|\|full | Not-determined |
| S12_vs_k141_173547\|\|full | Low-quality |
| S12_vs_k141_142138\|\|full | Low-quality |
| S12_vs_k141_31749\|\|full | Not-determined |
| S12_vs_k141_55445\|\|full | Low-quality |
| S12_vs_k141_79136\|\|full | Low-quality |
| S12_vs_k141_31762\|\|full | Low-quality |
| S12_vs_k141_157838\|\|full | Low-quality |
| S12_vs_k141_126460\|\|full | Low-quality |
| S12_vs_k141_118476\|\|full | Not-determined |
| S12_vs_k141_15994\|\|full | Not-determined |

| | |
|---|---|
| S12_vs_k141_94923\|\|full | Low-quality |
| S12_vs_k141_110617\|\|full | Not-determined |
| S12_vs_k141_71236\|\|full | Low-quality |
| S12_vs_k141_79153\|\|full | Low-quality |
| S12_vs_k141_31787\|\|full | Low-quality |
| S12_vs_k141_31801\|\|full | Not-determined |
| S12_vs_k141_157861\|\|full | Low-quality |
| S12_vs_k141_63426\|\|full | Low-quality |
| S12_vs_k141_39778\|\|full | Not-determined |
| S12_vs_k141_87093\|\|full | Low-quality |
| S12_vs_k141_118511\|\|full | Not-determined |
| S12_vs_k141_560\|\|full | Low-quality |
| S12_vs_k141_63442\|\|full | Not-determined |
| S12_vs_k141_142199\|\|full | Low-quality |
| S12_vs_k141_71293\|\|full | Not-determined |
| S12_vs_k141_23859\|\|full | Low-quality |
| S12_vs_k141_16045\|\|full | Not-determined |
| S12_vs_k141_157895\|\|full | Low-quality |
| S12_vs_k141_102720\|\|full | Low-quality |
| S12_vs_k141_149941\|\|full | Low-quality |
| S12_vs_k141_118540\|\|full | Not-determined |
| S12_vs_k141_126509\|\|full | Low-quality |
| S12_vs_k141_55487\|\|full | Low-quality |
| S12_vs_k141_165747\|\|full | Low-quality |
| S12_vs_k141_602\|\|full | Low-quality |
| S8_vs_k141_5211\|\|full | Medium-quality |
| S12_vs_k141_165760\|\|full | Not-determined |
| S12_vs_k141_63482\|\|full | Not-determined |
| S12_vs_k141_165762\|\|full | Low-quality |
| S12_vs_k141_118556\|\|full | Not-determined |
| S12_vs_k141_8205\|\|full | Low-quality |
| S12_vs_k141_149957\|\|full | Low-quality |
| S12_vs_k141_87122\|\|full | Not-determined |
| S12_vs_k141_149960\|\|full | Low-quality |
| S12_vs_k141_79219\|\|full | Low-quality |
| S12_vs_k141_63490\|\|full | Low-quality |
| S12_vs_k141_16067\|\|full | Low-quality |
| S12_vs_k141_79222\|\|full | Not-determined |
| S12_vs_k141_157916\|\|full | Not-determined |
| S12_vs_k141_126532\|\|full | Low-quality |
| S12_vs_k141_47495\|\|full | Not-determined |
| S12_vs_k141_165776\|\|full | Low-quality |
| S12_vs_k141_142252\|\|full | Low-quality |
| S12_vs_k141_39800\|\|full | Low-quality |
| S12_vs_k141_63513\|\|full | Low-quality |
| S12_vs_k141_181512\|\|full | Low-quality |
| S12_vs_k141_87144\|\|full | Low-quality |
| S12_vs_k141_47504\|\|full | Not-determined |
| S12_vs_k141_628\|\|full | Low-quality |
| S8_extracted_k141_104367 | Medium-quality |
| S12_vs_k141_118567\|\|full | Not-determined |
| S12_vs_k141_23923\|\|full | Not-determined |
| S12_vs_k141_118579\|\|full | Low-quality |
| S12_vs_k141_23944\|\|full | Low-quality |
| S12_vs_k141_640\|\|full | Low-quality |
| S12_vs_k141_181528\|\|full | Low-quality |
| S12_vs_k141_118581\|\|full | Not-determined |
| S12_vs_k141_8232\|\|full | Low-quality |
| S12_vs_k141_23947\|\|full | Low-quality |
| S12_vs_k141_102764\|\|full | Low-quality |
| S12_vs_k141_39820\|\|full | Low-quality |
| S12_vs_k141_110651\|\|full | Low-quality |
| S12_vs_k141_150001\|\|full | Low-quality |
| S12_vs_k141_150003\|\|full | Not-determined |
| S12_vs_k141_23960\|\|full | Not-determined |
| S12_vs_k141_63520\|\|full | Low-quality |
| S12_vs_k141_663\|\|full | Not-determined |
| S12_vs_k141_8249\|\|full | Low-quality |
| S12_vs_k141_142295\|\|full | Low-quality |
| S12_vs_k141_150016\|\|full | Low-quality |
| S12_vs_k141_181543\|\|full | Low-quality |
| S12_vs_k141_55559\|\|full | Not-determined |
| S12_vs_k141_87177\|\|full | Low-quality |
| S12_vs_k141_8262\|\|full | Low-quality |
| S12_vs_k141_87194\|\|full | Low-quality |
| S12_vs_k141_150035\|\|full | Low-quality |
| S12_vs_k141_55572\|\|full | Low-quality |
| S12_vs_k141_699\|\|full | Low-quality |
| S12_vs_k141_150053\|\|full | Low-quality |
| S12_vs_k141_142332\|\|full | Low-quality |
| S12_vs_k141_8287\|\|full | Low-quality |
| S12_vs_k141_110666\|\|full | Low-quality |
| S12_vs_k141_134200\|\|full | Not-determined |
| S12_vs_k141_713\|\|full | Not-determined |
| S12_vs_k141_150066\|\|full | Not-determined |
| S12_vs_k141_71340\|\|full | Low-quality |
| S12_vs_k141_150069\|\|full | Low-quality |
| S12_vs_k141_39854\|\|full | Low-quality |
| S12_vs_k141_158006\|\|full | Not-determined |
| S12_vs_k141_158009\|\|full | Low-quality |
| S12_vs_k141_63568\|\|full | Low-quality |
| S12_vs_k141_181590\|\|full | Not-determined |
| S12_vs_k141_134224\|\|full | Low-quality |
| S12_vs_k141_718\|\|full | Not-determined |
| S12_vs_k141_126592\|\|full | Not-determined |
| S12_vs_k141_150078\|\|full | Low-quality |
| S12_vs_k141_118659\|\|full | Not-determined |
| S12_vs_k141_79313\|\|full | Low-quality |
| S12_vs_k141_71360\|\|full | Low-quality |
| S12_vs_k141_126597\|\|full | Not-determined |
| S12_vs_k141_47540\|\|full | Low-quality |
| S12_vs_k141_142347\|\|full | Low-quality |
| S12_vs_k141_79326\|\|full | Not-determined |
| S12_vs_k141_71375\|\|full | Not-determined |
| S12_vs_k141_134245\|\|full | Not-determined |
| S12_vs_k141_8307\|\|full | Not-determined |
| S12_vs_k141_126627\|\|full | Low-quality |
| S12_vs_k141_735\|\|full | Low-quality |
| S12_vs_k141_63599\|\|full | Not-determined |
| S12_vs_k141_142366\|\|full | Not-determined |
| S12_vs_k141_745\|\|full | Not-determined |
| S12_vs_k141_71385\|\|full | Low-quality |
| S12_vs_k141_165903\|\|full | Low-quality |
| S12_vs_k141_150103\|\|full | Not-determined |
| S12_vs_k141_158040\|\|full | Low-quality |
| S12_vs_k141_63633\|\|full | Not-determined |
| S12_vs_k141_118718\|\|full | Low-quality |
| S12_vs_k141_63636\|\|full | Not-determined |
| S12_vs_k141_165917\|\|full | Low-quality |
| S12_vs_k141_71406\|\|full | Low-quality |
| S12_vs_k141_8347\|\|full | Not-determined |
| S12_vs_k141_95051\|\|full | Low-quality |
| S12_vs_k141_55651\|\|full | Low-quality |
| S12_vs_k141_71424\|\|full | Not-determined |
| S12_vs_k141_142393\|\|full | Low-quality |
| S12_vs_k141_71434\|\|full | Not-determined |
| S12_vs_k141_102905\|\|full | Low-quality |
| S12_vs_k141_16103\|\|full | Not-determined |
| S12_vs_k141_142412\|\|full | Low-quality |
| S12_vs_k141_110761\|\|full | Low-quality |
| S12_vs_k141_142415\|\|full | Not-determined |
| S12_vs_k141_110766\|\|full | Not-determined |
| S12_vs_k141_173641\|\|full | Low-quality |
| S12_vs_k141_102917\|\|full | Low-quality |
| S12_vs_k141_173647\|\|full | Low-quality |
| S12_vs_k141_16106\|\|full | Low-quality |
| S12_vs_k141_181707\|\|full | Not-determined |
| S12_vs_k141_142422\|\|full | Not-determined |
| S12_vs_k141_79378\|\|full | Not-determined |
| S12_vs_k141_8422\|\|full | Low-quality |
| S12_vs_k141_47603\|\|full | Low-quality |
| S12_vs_k141_39952\|\|full | Not-determined |
| S12_vs_k141_87226\|\|full | Not-determined |
| S12_vs_k141_142437\|\|full | Low-quality |
| S12_vs_k141_165940\|\|full | Low-quality |
| S12_vs_k141_95101\|\|full | Low-quality |
| S12_vs_k141_8428\|\|full | Low-quality |
| S12_vs_k141_118784\|\|full | Low-quality |
| S12_vs_k141_173660\|\|full | Not-determined |
| S12_vs_k141_24113\|\|full | Low-quality |
| S12_vs_k141_47622\|\|full | Low-quality |
| S12_vs_k141_55696\|\|full | Low-quality |
| S12_vs_k141_87242\|\|full | Not-determined |
| S12_vs_k141_24114\|\|full | Low-quality |
| S12_vs_k141_71488\|\|full | Low-quality |
| S12_vs_k141_142446\|\|full | Not-determined |
| S12_vs_k141_95118\|\|full | Not-determined |
| S12_vs_k141_16144\|\|full | Not-determined |
| S12_vs_k141_95123\|\|full | Not-determined |
| S12_vs_k141_165960\|\|full | Not-determined |
| S12_vs_k141_181741\|\|full | Low-quality |
| S12_vs_k141_71507\|\|full | Low-quality |
| S12_vs_k141_47637\|\|full | Low-quality |
| S12_vs_k141_32045\|\|full | Low-quality |
| S12_vs_k141_63747\|\|full | Low-quality |
| S12_vs_k141_142469\|\|full | Not-determined |
| S12_vs_k141_71513\|\|full | Not-determined |
| S12_vs_k141_87288\|\|full | Not-determined |
| S12_vs_k141_95152\|\|full | Low-quality |
| S12_vs_k141_24142\|\|full | Low-quality |
| S12_vs_k141_32053\|\|full | Not-determined |
| S12_vs_k141_830\|\|full | Not-determined |
| S8_vs_k141_104367\|\|full | Medium-quality |
| S12_vs_k141_833\|\|full | Not-determined |
| S12_vs_k141_8455\|\|full | Not-determined |
| S12_vs_k141_103006\|\|full | Not-determined |
| S12_vs_k141_142516\|\|full | Low-quality |
| S12_vs_k141_40002\|\|full | Low-quality |
| S12_vs_k141_150226\|\|full | Not-determined |
| S12_vs_k141_165991\|\|full | Low-quality |
| S12_vs_k141_126658\|\|full | Not-determined |
| S12_vs_k141_8463\|\|full | Low-quality |
| S12_vs_k141_165995\|\|full | Low-quality |
| S12_vs_k141_173674\|\|full | Low-quality |
| S12_vs_k141_181774\|\|full | Not-determined |
| S12_vs_k141_150236\|\|full | Low-quality |
| S12_vs_k141_24188\|\|full | Low-quality |
| S12_vs_k141_87334\|\|full | Low-quality |
| S12_vs_k141_181776\|\|full | Not-determined |
| S12_vs_k141_150241\|\|full | Low-quality |
| S12_vs_k141_16231\|\|full | Not-determined |
| S5_vs_k141_140710\|\|full | Medium-quality |
| S12_vs_k141_16234\|\|full | Low-quality |
| S12_vs_k141_40043\|\|full | Low-quality |
| S12_vs_k141_71535\|\|full | Low-quality |
| S12_vs_k141_150255\|\|full | Low-quality |
| S12_vs_k141_32097\|\|full | Not-determined |
| S12_vs_k141_150258\|\|full | Low-quality |
| S12_vs_k141_126691\|\|full | Low-quality |
| S12_vs_k141_166045\|\|full | Low-quality |
| S12_vs_k141_47647\|\|full | Not-determined |
| S12_vs_k141_55754\|\|full | Not-determined |
| S12_vs_k141_8507\|\|full | Not-determined |
| S12_vs_k141_87344\|\|full | Low-quality |
| S12_vs_k141_166048\|\|full | Not-determined |
| S12_vs_k141_181797\|\|full | Low-quality |
| S12_vs_k141_103068\|\|full | Not-determined |
| S12_vs_k141_24236\|\|full | Low-quality |
| S12_vs_k141_181802\|\|full | Low-quality |
| S12_vs_k141_55760\|\|full | Not-determined |
| S12_vs_k141_24242\|\|full | Not-determined |
| S12_vs_k141_901\|\|full | Low-quality |
| S12_vs_k141_16266\|\|full | Not-determined |
| S12_vs_k141_55764\|\|full | Not-determined |
| S12_vs_k141_166072\|\|full | Not-determined |
| S12_vs_k141_909\|\|full | Low-quality |
| S12_vs_k141_24251\|\|full | Not-determined |
| S12_vs_k141_8539\|\|full | Low-quality |
| S12_vs_k141_150279\|\|full | Not-determined |
| S12_vs_k141_40073\|\|full | Low-quality |
| S12_vs_k141_24252\|\|full | Not-determined |
| S12_vs_k141_158160\|\|full | Not-determined |
| S12_vs_k141_150286\|\|full | Low-quality |
| S12_vs_k141_79467\|\|full | Not-determined |
| S12_vs_k141_150294\|\|full | Not-determined |
| S12_vs_k141_917\|\|full | Low-quality |
| S12_vs_k141_71565\|\|full | Low-quality |
| S12_vs_k141_87373\|\|full | Not-determined |
| S12_vs_k141_47672\|\|full | Not-determined |
| S12_vs_k141_40101\|\|full | Not-determined |
| S12_vs_k141_8566\|\|full | Not-determined |
| S12_vs_k141_142587\|\|full | Low-quality |
| S12_vs_k141_173774\|\|full | Low-quality |
| S12_vs_k141_158192\|\|full | Low-quality |
| S12_vs_k141_79473\|\|full | Not-determined |
| S12_vs_k141_71583\|\|full | Not-determined |
| S12_vs_k141_142592\|\|full | Low-quality |
| S12_vs_k141_103140\|\|full | Low-quality |
| S12_vs_k141_142595\|\|full | Not-determined |
| S12_vs_k141_142601\|\|full | Low-quality |
| S12_vs_k141_24272\|\|full | Not-determined |
| S12_vs_k141_95221\|\|full | Low-quality |
| S12_vs_k141_79497\|\|full | Low-quality |
| S12_vs_k141_126719\|\|full | Not-determined |
| S12_vs_k141_95233\|\|full | Not-determined |
| S12_vs_k141_103144\|\|full | Low-quality |
| S12_vs_k141_71596\|\|full | Not-determined |
| S12_vs_k141_118875\|\|full | Low-quality |
| S12_vs_k141_95242\|\|full | Low-quality |
| S12_vs_k141_8577\|\|full | Low-quality |
| S12_vs_k141_150340\|\|full | Not-determined |
| S12_vs_k141_166097\|\|full | Low-quality |
| S12_vs_k141_150344\|\|full | Not-determined |
| S12_vs_k141_956\|\|full | Low-quality |
| S12_vs_k141_960\|\|full | Low-quality |
| S12_vs_k141_118891\|\|full | Low-quality |
| S12_vs_k141_181874\|\|full | Not-determined |
| S12_vs_k141_32186\|\|full | Low-quality |
| S12_vs_k141_63824\|\|full | Not-determined |
| S12_vs_k141_166114\|\|full | Low-quality |
| S12_vs_k141_977\|\|full | Not-determined |
| S12_vs_k141_24324\|\|full | Not-determined |
| S12_vs_k141_71627\|\|full | Low-quality |
| S12_vs_k141_40143\|\|full | Low-quality |
| S12_vs_k141_16368\|\|full | Not-determined |
| S12_vs_k141_173832\|\|full | Not-determined |
| S12_vs_k141_24348\|\|full | Low-quality |
| S12_vs_k141_24349\|\|full | Low-quality |
| S12_vs_k141_126751\|\|full | Not-determined |
| S12_vs_k141_40166\|\|full | Not-determined |
| S12_vs_k141_181881\|\|full | Low-quality |
| S12_vs_k141_103204\|\|full | Low-quality |
| S12_vs_k141_126755\|\|full | Not-determined |
| S12_vs_k141_150382\|\|full | Low-quality |
| S12_vs_k141_32226\|\|full | Not-determined |
| S12_vs_k141_79551\|\|full | Low-quality |
| S12_vs_k141_134462\|\|full | Low-quality |
| S12_vs_k141_32231\|\|full | Not-determined |
| S12_vs_k141_134465\|\|full | Not-determined |
| S12_vs_k141_87401\|\|full | Low-quality |
| S12_vs_k141_150389\|\|full | Not-determined |
| S12_vs_k141_95253\|\|full | Low-quality |
| S12_vs_k141_173870\|\|full | Low-quality |
| S12_vs_k141_24377\|\|full | Low-quality |
| S12_vs_k141_79569\|\|full | Not-determined |
| S12_vs_k141_181884\|\|full | Low-quality |
| S12_vs_k141_150403\|\|full | Not-determined |
| S12_vs_k141_181890\|\|full | Low-quality |
| S12_vs_k141_40190\|\|full | Not-determined |
| S12_vs_k141_40192\|\|full | Low-quality |
| S12_vs_k141_32270\|\|full | Low-quality |
| S12_vs_k141_134484\|\|full | Low-quality |
| S12_vs_k141_47759\|\|full | Low-quality |
| S12_vs_k141_150413\|\|full | Low-quality |
| S12_vs_k141_24396\|\|full | Low-quality |
| S17_extracted_k141_107742 | Medium-quality |
| S12_vs_k141_87420\|\|full | Not-determined |
| S12_vs_k141_32285\|\|full | Low-quality |
| S12_vs_k141_134493\|\|full | Not-determined |
| S12_vs_k141_63884\|\|full | Low-quality |
| S12_vs_k141_95283\|\|full | Not-determined |
| S12_vs_k141_142637\|\|full | Low-quality |
| S12_vs_k141_63894\|\|full | Not-determined |
| S12_vs_k141_158327\|\|full | Not-determined |
| S12_vs_k141_63897\|\|full | Low-quality |
| S12_vs_k141_40217\|\|full | Low-quality |
| S12_vs_k141_1008\|\|full | Not-determined |
| S12_vs_k141_24423\|\|full | Low-quality |
| S12_vs_k141_106329\|\|full | Medium-quality |
| S12_vs_k141_134519\|\|full | Low-quality |
| S12_vs_k141_79601\|\|full | Not-determined |
| S12_vs_k141_134525\|\|full | Not-determined |
| S12_vs_k141_181931\|\|full | Low-quality |
| S12_vs_k141_47819\|\|full | Not-determined |
| S12_vs_k141_95319\|\|full | Not-determined |
| S12_vs_k141_181942\|\|full | Not-determined |
| S12_vs_k141_142660\|\|full | Not-determined |
| S12_vs_k141_126798\|\|full | Low-quality |
| S12_vs_k141_40246\|\|full | Low-quality |
| S12_vs_k141_47827\|\|full | Low-quality |
| S12_vs_k141_71696\|\|full | Not-determined |
| S12_vs_k141_47834\|\|full | Low-quality |
| S12_vs_k141_79632\|\|full | Low-quality |
| S12_vs_k141_1041\|\|full | Not-determined |
| S12_vs_k141_63937\|\|full | Low-quality |
| S12_vs_k141_166250\|\|full | Not-determined |
| S12_vs_k141_1045\|\|full | Low-quality |
| S12_vs_k141_16406\|\|full | Not-determined |
| S12_vs_k141_55998\|\|full | Not-determined |
| S12_vs_k141_87499\|\|full | Low-quality |
| S12_vs_k141_119003\|\|full | Low-quality |
| S12_vs_k141_16412\|\|full | Not-determined |
| S12_vs_k141_158381\|\|full | Low-quality |
| S12_vs_k141_181967\|\|full | Not-determined |
| S12_vs_k141_126821\|\|full | Low-quality |
| S12_vs_k141_142707\|\|full | Low-quality |
| S12_vs_k141_181984\|\|full | Not-determined |
| S12_vs_k141_166283\|\|full | Low-quality |
| S12_vs_k141_1088\|\|full | Low-quality |
| S12_vs_k141_95347\|\|full | Low-quality |
| S12_vs_k141_142726\|\|full | Low-quality |
| S12_vs_k141_1096\|\|full | Not-determined |
| S12_vs_k141_142729\|\|full | Not-determined |
| S12_vs_k141_24466\|\|full | Not-determined |
| S12_vs_k141_63998\|\|full | Low-quality |
| S12_vs_k141_8633\|\|full | Not-determined |
| S12_vs_k141_16455\|\|full | Low-quality |
| S12_vs_k141_56035\|\|full | Not-determined |
| S12_vs_k141_150465\|\|full | Not-determined |
| S12_vs_k141_126867\|\|full | Not-determined |
| S12_vs_k141_8646\|\|full | Not-determined |
| S12_vs_k141_150468\|\|full | Not-determined |
| S12_vs_k141_24489\|\|full | Low-quality |
| S12_vs_k141_142732\|\|full | Low-quality |
| S12_vs_k141_79642\|\|full | Not-determined |
| S12_vs_k141_166321\|\|full | Low-quality |
| S12_vs_k141_95388\|\|full | Not-determined |
| S12_vs_k141_16483\|\|full | Not-determined |
| S12_vs_k141_150484\|\|full | Low-quality |
| S12_vs_k141_47931\|\|full | Low-quality |
| S12_vs_k141_150494\|\|full | Low-quality |
| S12_vs_k141_150498\|\|full | Low-quality |
| S12_vs_k141_24505\|\|full | Low-quality |
| S12_vs_k141_71723\|\|full | Not-determined |
| S12_vs_k141_1134\|\|full | Not-determined |
| S12_vs_k141_40333\|\|full | Low-quality |
| S12_vs_k141_40336\|\|full | Low-quality |
| S12_vs_k141_158456\|\|full | Low-quality |
| S12_vs_k141_1138\|\|full | Low-quality |
| S12_vs_k141_40346\|\|full | Low-quality |
| S12_vs_k141_173959\|\|full | Not-determined |
| S12_vs_k141_1140\|\|full | Not-determined |
| S12_vs_k141_126925\|\|full | Low-quality |
| S12_vs_k141_150508\|\|full | Not-determined |
| S12_vs_k141_173976\|\|full | Low-quality |
| S12_vs_k141_8693\|\|full | Low-quality |
| S12_vs_k141_150512\|\|full | Low-quality |
| S12_vs_k141_24543\|\|full | Low-quality |
| S12_vs_k141_142774\|\|full | Not-determined |
| S12_vs_k141_24553\|\|full | Low-quality |
| S12_vs_k141_47979\|\|full | Low-quality |
| S12_vs_k141_103454\|\|full | Not-determined |
| S12_vs_k141_119074\|\|full | Not-determined |
| S12_vs_k141_40358\|\|full | Low-quality |
| S12_vs_k141_95460\|\|full | Not-determined |
| S12_vs_k141_56117\|\|full | Not-determined |
| S12_vs_k141_16520\|\|full | Not-determined |
| S12_vs_k141_182122\|\|full | Low-quality |
| S12_vs_k141_119079\|\|full | Not-determined |
| S12_vs_k141_142803\|\|full | Low-quality |
| S12_vs_k141_142810\|\|full | Low-quality |
| S12_vs_k141_103456\|\|full | Not-determined |
| S12_vs_k141_64092\|\|full | Low-quality |
| S12_vs_k141_182150\|\|full | Low-quality |
| S12_vs_k141_64098\|\|full | Not-determined |
| S12_vs_k141_40386\|\|full | Not-determined |
| S12_vs_k141_95474\|\|full | Not-determined |
| S12_vs_k141_56149\|\|full | Low-quality |
| S12_vs_k141_71789\|\|full | Not-determined |
| S12_vs_k141_103473\|\|full | Low-quality |
| S12_vs_k141_126979\|\|full | Not-determined |
| S12_vs_k141_166406\|\|full | Not-determined |
| S12_vs_k141_95488\|\|full | Low-quality |
| S12_vs_k141_182174\|\|full | Low-quality |
| S12_vs_k141_174027\|\|full | Low-quality |
| S12_vs_k141_150542\|\|full | Not-determined |
| S12_vs_k141_56168\|\|full | Low-quality |
| S12_vs_k141_64127\|\|full | Low-quality |
| S12_vs_k141_24623\|\|full | Low-quality |
| S12_vs_k141_142844\|\|full | Low-quality |
| S12_vs_k141_95523\|\|full | Low-quality |
| S12_vs_k141_64139\|\|full | Low-quality |
| S12_vs_k141_24636\|\|full | Low-quality |
| S14_extracted_k141_109545 | Medium-quality |
| S12_vs_k141_150592\|\|full | Low-quality |
| S12_vs_k141_56209\|\|full | Not-determined |
| S12_vs_k141_24647\|\|full | Low-quality |
| S12_vs_k141_103529\|\|full | Not-determined |
| S12_vs_k141_87624\|\|full | Low-quality |
| S12_vs_k141_40425\|\|full | Not-determined |
| S12_vs_k141_40426\|\|full | Not-determined |
| S12_vs_k141_119118\|\|full | Not-determined |
| S12_vs_k141_56241\|\|full | Not-determined |
| S12_vs_k141_8776\|\|full | Low-quality |
| S12_vs_k141_8778\|\|full | Not-determined |
| S12_vs_k141_95558\|\|full | Low-quality |
| S3_extracted_k141_82571 | Medium-quality |
| S12_vs_k141_182261\|\|full | Low-quality |
| S12_vs_k141_8783\|\|full | Low-quality |
| S12_vs_k141_182268\|\|full | Not-determined |
| S12_vs_k141_32454\|\|full | Low-quality |
| S12_vs_k141_150614\|\|full | Low-quality |
| S12_vs_k141_16637\|\|full | Not-determined |
| S12_vs_k141_110978\|\|full | Low-quality |
| S12_vs_k141_142872\|\|full | Not-determined |
| S12_vs_k141_24692\|\|full | Low-quality |
| S12_vs_k141_95581\|\|full | Not-determined |
| S12_vs_k141_16647\|\|full | Not-determined |
| S12_vs_k141_142876\|\|full | Low-quality |
| S12_vs_k141_64160\|\|full | Not-determined |
| S12_vs_k141_119160\|\|full | Low-quality |
| S12_vs_k141_174100\|\|full | Not-determined |
| S12_vs_k141_182278\|\|full | Low-quality |
| S12_vs_k141_48069\|\|full | Low-quality |
| S12_vs_k141_174107\|\|full | Not-determined |
| S12_vs_k141_40479\|\|full | Low-quality |
| S12_vs_k141_119180\|\|full | Low-quality |
| S12_vs_k141_87660\|\|full | Not-determined |
| S12_vs_k141_24726\|\|full | Low-quality |
| S12_vs_k141_127010\|\|full | Not-determined |
| S12_vs_k141_48091\|\|full | Not-determined |
| S12_vs_k141_182298\|\|full | Low-quality |
| S12_vs_k141_174133\|\|full | Not-determined |
| S12_vs_k141_134689\|\|full | Not-determined |
| S12_vs_k141_64193\|\|full | Not-determined |
| S8_extracted_k141_111790 | Medium-quality |
| S12_vs_k141_79674\|\|full | Low-quality |
| S12_vs_k141_8811\|\|full | Low-quality |
| S12_vs_k141_79676\|\|full | Not-determined |
| S12_vs_k141_56285\|\|full | Low-quality |
| S12_vs_k141_174156\|\|full | Low-quality |
| S12_vs_k141_134695\|\|full | Not-determined |
| S12_vs_k141_24746\|\|full | Low-quality |
| S12_vs_k141_24750\|\|full | Not-determined |
| S12_vs_k141_79678\|\|full | Low-quality |
| S12_vs_k141_119220\|\|full | Low-quality |
| S12_vs_k141_40530\|\|full | Not-determined |
| S12_vs_k141_40531\|\|full | Low-quality |
| S12_vs_k141_79689\|\|full | Low-quality |
| S12_vs_k141_119229\|\|full | Not-determined |
| S12_vs_k141_16672\|\|full | Low-quality |
| S12_vs_k141_95655\|\|full | Low-quality |
| S12_vs_k141_166536\|\|full | Not-determined |
| S12_vs_k141_166537\|\|full | Low-quality |
| S12_vs_k141_119234\|\|full | Not-determined |
| S12_vs_k141_127035\|\|full | Not-determined |
| S12_vs_k141_127036\|\|full | Not-determined |
| S12_vs_k141_119240\|\|full | Not-determined |
| S12_vs_k141_24782\|\|full | Low-quality |
| S12_vs_k141_24787\|\|full | Not-determined |
| S12_vs_k141_40558\|\|full | Low-quality |
| S12_vs_k141_16685\|\|full | Low-quality |
| S12_vs_k141_111081\|\|full | Low-quality |
| S12_vs_k141_32473\|\|full | Low-quality |
| S12_vs_k141_119249\|\|full | Not-determined |
| S12_vs_k141_8829\|\|full | Not-determined |
| S12_vs_k141_150640\|\|full | Not-determined |
| S12_vs_k141_64248\|\|full | Low-quality |
| S12_vs_k141_150645\|\|full | Low-quality |
| S12_vs_k141_79725\|\|full | Low-quality |
| S12_vs_k141_32485\|\|full | Low-quality |
| S12_vs_k141_150649\|\|full | Not-determined |
| S12_vs_k141_119265\|\|full | Low-quality |
| S12_vs_k141_127051\|\|full | Low-quality |
| S12_vs_k141_127052\|\|full | Low-quality |
| S12_vs_k141_127056\|\|full | Low-quality |
| S12_vs_k141_142898\|\|full | Low-quality |
| S12_vs_k141_87745\|\|full | Low-quality |
| S12_vs_k141_142905\|\|full | Not-determined |
| S12_vs_k141_71960\|\|full | Not-determined |
| S12_vs_k141_40589\|\|full | Low-quality |
| S12_vs_k141_95725\|\|full | Low-quality |
| S12_vs_k141_8850\|\|full | Not-determined |
| S12_vs_k141_8851\|\|full | Low-quality |
| S12_vs_k141_142915\|\|full | Not-determined |
| S12_vs_k141_79749\|\|full | Low-quality |
| S12_vs_k141_1274\|\|full | Not-determined |
| S12_vs_k141_64305\|\|full | Not-determined |
| S12_vs_k141_1280\|\|full | Low-quality |
| S12_vs_k141_127084\|\|full | Low-quality |
| S12_vs_k141_16715\|\|full | Low-quality |
| S12_vs_k141_119282\|\|full | Low-quality |
| S12_vs_k141_174220\|\|full | Not-determined |
| S12_vs_k141_87776\|\|full | Low-quality |
| S12_vs_k141_119284\|\|full | Not-determined |
| S12_vs_k141_134758\|\|full | Low-quality |
| S12_vs_k141_16730\|\|full | Not-determined |
| S12_vs_k141_95752\|\|full | Not-determined |
| S12_vs_k141_182371\|\|full | Not-determined |
| S12_vs_k141_127100\|\|full | Not-determined |
| S12_vs_k141_111143\|\|full | Not-determined |
| S12_vs_k141_40616\|\|full | Low-quality |
| S12_vs_k141_142954\|\|full | Not-determined |
| S12_vs_k141_8888\|\|full | Not-determined |
| S12_vs_k141_127107\|\|full | Low-quality |
| S12_vs_k141_95760\|\|full | Low-quality |
| S12_vs_k141_119292\|\|full | Not-determined |
| S12_vs_k141_166562\|\|full | Not-determined |
| S12_vs_k141_142966\|\|full | Low-quality |
| S12_vs_k141_24837\|\|full | Not-determined |
| S12_vs_k141_111167\|\|full | Not-determined |
| S12_vs_k141_142971\|\|full | Not-determined |
| S12_vs_k141_64338\|\|full | Low-quality |
| S12_vs_k141_87824\|\|full | Low-quality |
| S12_vs_k141_16772\|\|full | Low-quality |
| S12_vs_k141_8894\|\|full | Low-quality |
| S12_vs_k141_8896\|\|full | Not-determined |
| S12_vs_k141_119310\|\|full | Not-determined |
| S12_vs_k141_142987\|\|full | Not-determined |
| S12_vs_k141_1311\|\|full | Not-determined |
| S12_vs_k141_127140\|\|full | Not-determined |
| S12_vs_k141_166606\|\|full | Low-quality |
| S12_vs_k141_87855\|\|full | Not-determined |
| S12_vs_k141_32589\|\|full | Low-quality |
| S12_vs_k141_134785\|\|full | Not-determined |
| S12_vs_k141_166611\|\|full | Low-quality |
| S12_vs_k141_56414\|\|full | Not-determined |
| S12_vs_k141_24886\|\|full | Not-determined |
| S12_vs_k141_119349\|\|full | Not-determined |
| S12_vs_k141_134802\|\|full | Not-determined |
| S12_vs_k141_72065\|\|full | Low-quality |
| S12_vs_k141_79797\|\|full | Low-quality |
| S12_vs_k141_134817\|\|full | Not-determined |
| S12_vs_k141_158512\|\|full | Not-determined |
| S12_vs_k141_150673\|\|full | Low-quality |
| S12_vs_k141_134829\|\|full | Low-quality |
| S12_vs_k141_182389\|\|full | Low-quality |
| S12_vs_k141_64357\|\|full | Low-quality |
| S12_vs_k141_16830\|\|full | Not-determined |
| S12_vs_k141_87901\|\|full | Not-determined |
| S12_vs_k141_134831\|\|full | Low-quality |
| S12_vs_k141_95805\|\|full | Low-quality |
| S12_vs_k141_111240\|\|full | Low-quality |
| S12_vs_k141_127165\|\|full | Not-determined |
| S12_vs_k141_79833\|\|full | Not-determined |
| S12_vs_k141_32613\|\|full | Low-quality |
| S12_vs_k141_182401\|\|full | Not-determined |
| S12_vs_k141_182402\|\|full | Not-determined |
| S12_vs_k141_56452\|\|full | Not-determined |
| S12_vs_k141_111244\|\|full | Not-determined |
| S12_vs_k141_95821\|\|full | Not-determined |
| S12_vs_k141_119370\|\|full | Low-quality |
| S12_vs_k141_87914\|\|full | Low-quality |
| S12_vs_k141_182423\|\|full | Low-quality |
| S12_vs_k141_1401\|\|full | Not-determined |
| S12_vs_k141_166657\|\|full | Not-determined |
| S12_vs_k141_24915\|\|full | Low-quality |
| S12_vs_k141_24929\|\|full | Low-quality |
| S12_vs_k141_48281\|\|full | Not-determined |
| S12_vs_k141_72153\|\|full | Low-quality |
| S12_vs_k141_182450\|\|full | Not-determined |
| S12_vs_k141_158568\|\|full | Not-determined |
| S12_vs_k141_158570\|\|full | Low-quality |
| S12_vs_k141_16879\|\|full | Low-quality |
| S12_vs_k141_64374\|\|full | Low-quality |
| S12_vs_k141_8983\|\|full | Not-determined |
| S12_vs_k141_8987\|\|full | Not-determined |
| S12_vs_k141_150701\|\|full | Low-quality |
| S12_vs_k141_56469\|\|full | Low-quality |

S12_vs_k141_40710||full Not-determined
S12_vs_k141_111272||full Low-quality
S12_vs_k141_150709||full Not-determined
S12_vs_k141_95872||full Low-quality
S12_vs_k141_79861||full Low-quality
S12_vs_k141_16893||full Not-determined
S12_vs_k141_72188||full Low-quality
S12_vs_k141_56490||full Low-quality
S12_vs_k141_134894||full Low-quality
S12_vs_k141_9022||full Not-determined
S12_vs_k141_1468||full Low-quality
S12_vs_k141_56504||full Low-quality
S12_vs_k141_158593||full Low-quality
S12_vs_k141_134909||full Not-determined
S12_vs_k141_24971||full Low-quality
S12_vs_k141_24974||full Not-determined
S12_vs_k141_79869||full Not-determined
S12_vs_k141_127287||full Not-determined
S12_vs_k141_16934||full Not-determined
S12_vs_k141_64424||full Not-determined
S12_vs_k141_134939||full Low-quality
S12_vs_k141_127298||full Low-quality
S12_vs_k141_158632||full Not-determined
S12_vs_k141_79885||full Low-quality
S12_vs_k141_24984||full Not-determined
S12_vs_k141_40744||full Low-quality
S12_vs_k141_127305||full Not-determined
S12_vs_k141_24985||full Not-determined
S12_vs_k141_79888||full Low-quality
S12_vs_k141_182512||full Not-determined
S12_vs_k141_9062||full Not-determined
S12_vs_k141_16942||full Low-quality
S12_vs_k141_182517||full Low-quality
S12_vs_k141_16946||full Low-quality
S12_vs_k141_119422||full Low-quality
S12_vs_k141_88026||full Not-determined
S12_vs_k141_24998||full Not-determined
S12_vs_k141_119431||full Not-determined
S12_vs_k141_119435||full Low-quality
S12_vs_k141_103699||full Not-determined
S12_vs_k141_150723||full Low-quality
S12_vs_k141_158669||full Low-quality
S12_vs_k141_16978||full Low-quality
S12_vs_k141_103704||full Not-determined
S12_vs_k141_25019||full Low-quality
S12_vs_k141_88058||full Not-determined
S8_vs_k141_111790||full Medium-quality
S12_vs_k141_158684||full Low-quality
S12_vs_k141_88061||full Not-determined
S12_vs_k141_143106||full Not-determined
S12_vs_k141_9115||full Low-quality
S12_vs_k141_95934||full Low-quality
S12_vs_k141_143109||full Low-quality
S12_vs_k141_158705||full Low-quality
S12_vs_k141_150744||full Not-determined
S12_vs_k141_166790||full Not-determined
S12_vs_k141_25031||full Low-quality
S12_vs_k141_88076||full Not-determined
S12_vs_k141_16995||full Not-determined
S12_vs_k141_135057||full Not-determined
S12_vs_k141_56610||full Not-determined
S12_vs_k141_25064||full Not-determined
S12_vs_k141_9123||full Not-determined
S12_vs_k141_79933||full Not-determined
S12_vs_k141_32622||full Not-determined
S12_vs_k141_143136||full Not-determined
S12_vs_k141_1479||full Low-quality
S12_vs_k141_79941||full Low-quality
S12_vs_k141_150794||full Not-determined
S12_vs_k141_72282||full Low-quality
S12_vs_k141_1492||full Not-determined
S12_vs_k141_64506||full Not-determined
S12_vs_k141_166853||full Not-determined
S12_vs_k141_79975||full Not-determined
S5_vs_k141_10089||full Medium-quality
S12_vs_k141_119568||full Low-quality
S12_vs_k141_25107||full Low-quality
S12_vs_k141_56669||full Not-determined
S12_vs_k141_25113||full Not-determined
S12_vs_k141_79983||full Not-determined
S12_vs_k141_166880||full Low-quality
S12_vs_k141_1548||full Low-quality
S12_vs_k141_103781||full Low-quality
S12_vs_k141_79993||full Low-quality
S12_vs_k141_88086||full Low-quality
S12_vs_k141_1552||full Not-determined
S12_vs_k141_96049||full Low-quality
S12_vs_k141_1557||full Low-quality
S12_vs_k141_9171||full Not-determined
S12_vs_k141_64563||full Low-quality
S12_vs_k141_96055||full Not-determined
S6_vs_k141_12176||full Medium-quality
S12_vs_k141_25148||full Not-determined
S12_vs_k141_72361||full Not-determined
S12_vs_k141_135135||full Not-determined
S12_vs_k141_135136||full Not-determined
S12_vs_k141_64581||full Low-quality
S12_vs_k141_72368||full Not-determined
S12_vs_k141_25157||full Not-determined
S12_vs_k141_143169||full Low-quality
S12_vs_k141_17054||full Not-determined
S12_vs_k141_103803||full Low-quality
S17_vs_k141_107742||full Medium-quality
S12_vs_k141_9198||full Not-determined
S12_vs_k141_174375||full Low-quality
S12_vs_k141_56720||full Low-quality
S12_vs_k141_56722||full Not-determined
S12_vs_k141_182624||full Not-determined
S12_vs_k141_80022||full Not-determined
S12_vs_k141_1590||full Not-determined
S12_vs_k141_174401||full Not-determined
S12_vs_k141_1592||full Low-quality
S12_vs_k141_72406||full Low-quality
S12_vs_k141_143202||full Not-determined
S12_vs_k141_103827||full Low-quality
S12_vs_k141_103832||full Low-quality
S12_vs_k141_1601||full Low-quality
S12_vs_k141_96080||full Not-determined
S12_vs_k141_56737||full Low-quality
S12_vs_k141_166953||full Low-quality
S12_vs_k141_182634||full Not-determined
S12_vs_k141_119681||full Low-quality
S12_vs_k141_1626||full Low-quality
S12_vs_k141_96092||full Not-determined
S12_vs_k141_17101||full Low-quality
S12_vs_k141_96093||full Low-quality
S12_vs_k141_174427||full Low-quality
S12_vs_k141_1631||full Low-quality
S12_vs_k141_17109||full Low-quality
S12_vs_k141_174429||full Low-quality
S12_vs_k141_135215||full Low-quality
S12_vs_k141_56756||full Low-quality
S12_vs_k141_119701||full Low-quality
S12_vs_k141_17125||full Not-determined
S12_vs_k141_119703||full Not-determined
S12_vs_k141_182671||full Not-determined
S12_vs_k141_150896||full Not-determined
S12_vs_k141_103877||full Low-quality
S12_vs_k141_174449||full Not-determined
S12_vs_k141_119718||full Not-determined
S12_vs_k141_96113||full Low-quality
S12_vs_k141_150904||full Low-quality
S12_vs_k141_150907||full Low-quality
S12_vs_k141_127443||full Not-determined
S12_vs_k141_56780||full Not-determined
S12_vs_k141_150918||full Not-determined
S12_vs_k141_166976||full Low-quality
S12_vs_k141_64638||full Not-determined
S12_vs_k141_40766||full Not-determined
S12_vs_k141_9266||full Low-quality
S12_vs_k141_1668||full Not-determined
S12_vs_k141_119760||full Not-determined
S12_vs_k141_166978||full Not-determined
S12_vs_k141_9276||full Not-determined
S12_vs_k141_64650||full Low-quality
S12_vs_k141_9289||full Low-quality
S12_vs_k141_64654||full Not-determined
S12_vs_k141_127493||full Low-quality
S12_vs_k141_111442||full Low-quality
S12_vs_k141_56815||full Low-quality
S12_vs_k141_96198||full Not-determined
S12_vs_k141_111446||full Not-determined
S12_vs_k141_1686||full Low-quality
S12_vs_k141_48323||full Not-determined
S12_vs_k141_40810||full Low-quality
S12_vs_k141_64687||full Low-quality
S12_vs_k141_9311||full Low-quality
S12_vs_k141_9312||full Low-quality
S12_vs_k141_17157||full Low-quality
S12_vs_k141_167013||full Not-determined
S12_vs_k141_111464||full Not-determined
S12_vs_k141_56847||full Low-quality
S12_vs_k141_40819||full Low-quality
S12_vs_k141_72440||full Low-quality
S12_vs_k141_64709||full Not-determined
S12_vs_k141_127541||full Not-determined
S12_vs_k141_143317||full Not-determined
S12_vs_k141_96245||full Low-quality
S12_vs_k141_103951||full Low-quality
S12_vs_k141_25243||full Low-quality
S12_vs_k141_96251||full Low-quality
S12_vs_k141_143334||full Low-quality
S12_vs_k141_48348||full Low-quality
S12_vs_k141_96254||full Low-quality
S12_vs_k141_111478||full Low-quality
S12_vs_k141_64716||full Low-quality
S12_vs_k141_143337||full Not-determined
S12_vs_k141_64723||full Not-determined
S12_vs_k141_96267||full Low-quality
S12_vs_k141_48358||full Not-determined
S12_vs_k141_56880||full Not-determined
S12_vs_k141_64742||full Not-determined
S12_vs_k141_25295||full Low-quality
S12_vs_k141_167085||full Low-quality
S12_vs_k141_103986||full Not-determined
S12_vs_k141_119859||full Low-quality
S12_vs_k141_56909||full Not-determined
S12_vs_k141_119863||full Not-determined
S12_vs_k141_1767||full Not-determined
S12_vs_k141_25299||full Not-determined
S12_vs_k141_104008||full Not-determined
S12_vs_k141_119888||full Low-quality
S12_vs_k141_127623||full Not-determined
S12_vs_k141_56934||full Not-determined
S12_vs_k141_56955||full Not-determined
S12_vs_k141_127635||full Low-quality
S12_vs_k141_25319||full Not-determined
S12_vs_k141_48402||full Low-quality
S3_vs_k141_82571||full Medium-quality
S12_vs_k141_56956||full Not-determined
S12_vs_k141_119927||full Not-determined
S12_vs_k141_25328||full Low-quality
S12_vs_k141_96362||full Low-quality
S12_vs_k141_25337||full Not-determined
S12_vs_k141_119937||full Low-quality
S12_vs_k141_167142||full Low-quality
S12_vs_k141_25338||full Not-determined
S12_vs_k141_143394||full Low-quality
S12_vs_k141_150997||full Not-determined
S12_vs_k141_143402||full Low-quality
S12_vs_k141_48438||full Low-quality
S12_vs_k141_135320||full Not-determined
S12_vs_k141_104082||full Not-determined
S12_vs_k141_40923||full Not-determined
S12_vs_k141_32678||full Low-quality
S12_vs_k141_48453||full Not-determined
S12_vs_k141_111563||full Not-determined
S12_vs_k141_104088||full Low-quality
S12_vs_k141_96424||full Low-quality
S12_vs_k141_104093||full Low-quality
S12_vs_k141_88274||full Low-quality
S12_vs_k141_32695||full Low-quality
S12_vs_k141_88282||full Not-determined
S12_vs_k141_80046||full Low-quality
S12_vs_k141_32702||full Low-quality
S12_vs_k141_40961||full Low-quality
S12_vs_k141_57039||full Low-quality
S12_vs_k141_57040||full Not-determined
S12_vs_k141_57041||full Low-quality
S12_vs_k141_72612||full Low-quality
S12_vs_k141_72619||full Low-quality
S12_vs_k141_182800||full Not-determined
S12_vs_k141_104137||full Not-determined
S12_vs_k141_48469||full Low-quality
S12_vs_k141_1878||full Not-determined
S12_vs_k141_151019||full Low-quality
S12_vs_k141_104156||full Not-determined
S12_vs_k141_80101||full Not-determined
S12_vs_k141_64781||full Not-determined
S12_vs_k141_88342||full Low-quality
S12_vs_k141_104163||full Not-determined
S12_vs_k141_32736||full Low-quality
S12_vs_k141_9379||full Not-determined
S12_vs_k141_143470||full Low-quality
S12_vs_k141_57081||full Low-quality
S12_vs_k141_64797||full Low-quality
S12_vs_k141_32748||full Not-determined
S12_vs_k141_127659||full Low-quality
S12_vs_k141_41040||full Low-quality
S12_vs_k141_143482||full Low-quality
S12_vs_k141_96506||full Not-determined
S12_vs_k141_151041||full Low-quality
S12_vs_k141_64812||full Not-determined
S12_vs_k141_17313||full Not-determined
S12_vs_k141_41064||full Low-quality
S12_vs_k141_151060||full Low-quality
S12_vs_k141_17317||full Not-determined
S12_vs_k141_104199||full Low-quality
S12_vs_k141_48502||full Low-quality
S12_vs_k141_41069||full Not-determined
S12_vs_k141_32750||full Not-determined
S12_vs_k141_1934||full Not-determined
S12_vs_k141_48507||full Not-determined
S12_vs_k141_127678||full Low-quality
S12_vs_k141_32770||full Not-determined
S12_vs_k141_158788||full Low-quality
S12_vs_k141_182862||full Not-determined
S12_vs_k141_64845||full Not-determined
S12_vs_k141_135376||full Not-determined
S12_vs_k141_96565||full Low-quality
S12_vs_k141_64854||full Low-quality
S12_vs_k141_127703||full Not-determined
S12_vs_k141_64857||full Low-quality
S12_vs_k141_1958||full Not-determined
S12_vs_k141_158801||full Low-quality
S12_vs_k141_135393||full Low-quality
S12_vs_k141_9431||full Low-quality
S12_vs_k141_48527||full Low-quality
S12_vs_k141_167163||full Low-quality
S12_vs_k141_17372||full Low-quality
S12_vs_k141_104268||full Not-determined
S12_vs_k141_1964||full Low-quality
S12_vs_k141_135400||full Low-quality
S12_vs_k141_96583||full Not-determined
S12_vs_k141_135403||full Low-quality
S12_vs_k141_143566||full Low-quality
S12_vs_k141_182900||full Not-determined
S12_vs_k141_111721||full Low-quality
S12_vs_k141_25514||full Not-determined
S12_vs_k141_167187||full Low-quality
S12_vs_k141_158844||full Not-determined
S12_vs_k141_9465||full Not-determined
S12_vs_k141_143597||full Low-quality
S12_vs_k141_96611||full Low-quality
S12_vs_k141_57182||full Low-quality
S12_vs_k141_41138||full Not-determined
S12_vs_k141_111742||full Low-quality
S12_vs_k141_88481||full Not-determined
S12_vs_k141_182913||full Low-quality
S12_vs_k141_135427||full Low-quality
S12_vs_k141_88485||full Not-determined
S12_vs_k141_174509||full Not-determined
S12_vs_k141_143613||full Low-quality
S12_vs_k141_167224||full Low-quality
S12_vs_k141_9476||full Low-quality
S12_vs_k141_174521||full Low-quality
S12_vs_k141_17432||full Not-determined
S12_vs_k141_158866||full Low-quality
S12_vs_k141_48575||full Not-determined
S12_vs_k141_158871||full Low-quality
S12_vs_k141_57221||full Not-determined
S12_vs_k141_143629||full Low-quality
S12_vs_k141_64935||full Low-quality
S12_vs_k141_167253||full Low-quality
S12_vs_k141_17457||full Not-determined
S12_vs_k141_120180||full Not-determined
S12_vs_k141_9502||full Low-quality
S12_vs_k141_143646||full Low-quality
S12_vs_k141_167265||full Not-determined
S12_vs_k141_17472||full Not-determined
S12_vs_k141_57232||full Not-determined
S12_vs_k141_41201||full Not-determined
S12_vs_k141_182969||full Low-quality
S12_vs_k141_32800||full Not-determined
S12_vs_k141_167286||full Not-determined
S12_vs_k141_25599||full Not-determined
S12_vs_k141_64964||full Not-determined
S12_vs_k141_135479||full Low-quality
S12_vs_k141_32809||full Not-determined
S12_vs_k141_80191||full Not-determined
S12_vs_k141_17503||full Low-quality
S12_vs_k141_135495||full Not-determined
S12_vs_k141_88573||full Not-determined
S12_vs_k141_41233||full Not-determined
S12_vs_k141_64975||full Low-quality
S12_vs_k141_57257||full Not-determined
S12_vs_k141_64978||full Low-quality
S12_vs_k141_17527||full Low-quality
S12_vs_k141_88577||full Not-determined
S12_vs_k141_32845||full Not-determined
S12_vs_k141_2029||full Low-quality
S12_vs_k141_48654||full Not-determined
S12_vs_k141_32849||full Low-quality
S12_vs_k141_104371||full Low-quality
S12_vs_k141_183002||full Not-determined
S12_vs_k141_32851||full Low-quality
S12_vs_k141_9557||full Low-quality
S12_vs_k141_104373||full Not-determined
S12_vs_k141_143694||full Low-quality
S12_vs_k141_135505||full Low-quality
S12_vs_k141_57292||full Not-determined
S12_vs_k141_143702||full Low-quality
S12_vs_k141_32857||full Low-quality
S12_vs_k141_143703||full Low-quality
S12_vs_k141_65000||full Not-determined
S12_vs_k141_174583||full Not-determined
S12_vs_k141_127787||full Low-quality
S12_vs_k141_17541||full Low-quality
S12_vs_k141_127794||full Low-quality
S12_vs_k141_48677||full Not-determined
S12_vs_k141_48680||full Not-determined
S12_vs_k141_88623||full Low-quality
S12_vs_k141_17553||full Not-determined
S12_vs_k141_96708||full Not-determined
S12_vs_k141_183029||full Low-quality
S12_vs_k141_80270||full Low-quality
S12_vs_k141_57340||full Low-quality
S12_vs_k141_80271||full Not-determined
S12_vs_k141_127816||full Not-determined
S12_extracted_k141_60694 Medium-quality
S12_vs_k141_48685||full Not-determined
S12_vs_k141_158970||full Not-determined
S12_vs_k141_41320||full Not-determined
S12_vs_k141_25658||full Low-quality
S12_vs_k141_57341||full Not-determined
S12_vs_k141_183054||full Not-determined
S12_vs_k141_2064||full Low-quality
S12_vs_k141_32914||full Not-determined
S12_vs_k141_65050||full Low-quality
S12_vs_k141_9612||full Low-quality
S12_vs_k141_48712||full Not-determined
S12_vs_k141_96741||full Not-determined
S12_vs_k141_41340||full Low-quality
S12_vs_k141_17581||full Low-quality
S12_vs_k141_41346||full Not-determined
S12_vs_k141_80307||full Not-determined
S12_vs_k141_17587||full Low-quality
S12_vs_k141_41349||full Not-determined
S12_vs_k141_57348||full Low-quality
S12_vs_k141_65068||full Low-quality
S12_vs_k141_127842||full Low-quality
S12_vs_k141_2086||full Not-determined
S12_vs_k141_32938||full Low-quality
S12_vs_k141_183104||full Not-determined
S12_vs_k141_167336||full Low-quality
S12_vs_k141_167340||full Not-determined
S12_vs_k141_57353||full Low-quality
S12_vs_k141_96755||full Not-determined
S12_vs_k141_17611||full Not-determined
S12_vs_k141_104423||full Low-quality
S12_vs_k141_25682||full Low-quality
S12_vs_k141_80353||full Low-quality
S12_vs_k141_96773||full Low-quality
S12_vs_k141_174658||full Not-determined
S12_vs_k141_127872||full Low-quality
S12_vs_k141_96780||full Low-quality
S12_vs_k141_25693||full Low-quality
S12_vs_k141_104444||full Low-quality
S12_vs_k141_80365||full Not-determined
S12_vs_k141_25697||full Not-determined
S12_vs_k141_17640||full Not-determined
S12_vs_k141_96800||full Not-determined
S12_vs_k141_2117||full Low-quality
S12_vs_k141_9642||full Low-quality
S12_vs_k141_88660||full Not-determined
S12_vs_k141_25707||full Low-quality
S12_vs_k141_167379||full Low-quality
S12_vs_k141_151177||full Not-determined
S12_vs_k141_183141||full Low-quality
S12_vs_k141_151184||full Not-determined
S12_vs_k141_65106||full Low-quality
S12_vs_k141_25721||full Not-determined
S12_vs_k141_88674||full Not-determined
S12_vs_k141_9663||full Not-determined
S12_vs_k141_2130||full Low-quality
S12_vs_k141_33007||full Not-determined
S12_vs_k141_96822||full Not-determined
S12_vs_k141_80404||full Low-quality
S12_vs_k141_183161||full Low-quality
S12_vs_k141_80412||full Low-quality
S12_vs_k141_96831||full Low-quality
S12_vs_k141_104490||full Not-determined
S12_vs_k141_17673||full Low-quality
S12_vs_k141_88705||full Low-quality
S12_vs_k141_25743||full Low-quality
S12_vs_k141_88709||full Not-determined
S12_vs_k141_17681||full Low-quality
S12_vs_k141_2154||full Not-determined
S12_vs_k141_25751||full Not-determined
S12_vs_k141_111757||full Low-quality
S12_vs_k141_17684||full Low-quality
S12_vs_k141_17687||full Low-quality
S12_vs_k141_41425||full Not-determined
S12_vs_k141_135639||full Low-quality
S12_vs_k141_9692||full Low-quality
S12_vs_k141_88719||full Low-quality
S12_vs_k141_135641||full Low-quality
S12_vs_k141_167428||full Low-quality
S12_vs_k141_135645||full Not-determined
S12_vs_k141_25761||full Low-quality
S12_vs_k141_135646||full Low-quality
S12_vs_k141_127909||full Not-determined
S12_vs_k141_159138||full Low-quality
S12_vs_k141_33055||full Not-determined
S12_vs_k141_25766||full Low-quality
S12_vs_k141_41441||full Low-quality
S12_vs_k141_167435||full Low-quality
S12_vs_k141_111787||full Not-determined
S12_vs_k141_167444||full Low-quality
S12_vs_k141_65140||full Low-quality
S12_vs_k141_111791||full Not-determined
S12_vs_k141_143839||full Not-determined
S12_vs_k141_48817||full Not-determined
S12_vs_k141_159175||full Low-quality
S12_vs_k141_96886||full Low-quality
S12_vs_k141_127944||full Low-quality
S12_vs_k141_183218||full Low-quality
S12_vs_k141_88763||full Low-quality
S12_vs_k141_57429||full Low-quality
S12_vs_k141_48834||full Low-quality
S12_vs_k141_135693||full Not-determined
S12_vs_k141_41456||full Low-quality
S12_vs_k141_48837||full Not-determined
S12_vs_k141_143860||full Low-quality
S12_vs_k141_174829||full Low-quality
S12_vs_k141_174832||full Low-quality
S12_vs_k141_48856||full Not-determined
S12_vs_k141_33106||full Low-quality
S12_vs_k141_174850||full Low-quality
S12_vs_k141_120398||full Not-determined
S12_vs_k141_88800||full Low-quality
S12_vs_k141_80517||full Low-quality
S12_vs_k141_41498||full Low-quality
S12_vs_k141_25832||full Low-quality
S12_vs_k141_174875||full Not-determined
S12_vs_k141_25833||full Low-quality
S12_vs_k141_33140||full Low-quality
S12_vs_k141_127958||full Not-determined
S12_vs_k141_33142||full Not-determined
S12_vs_k141_36322||full Medium-quality
S12_vs_k141_80542||full Low-quality
S12_vs_k141_57462||full Low-quality
S12_vs_k141_159213||full Not-determined
S12_vs_k141_57469||full Not-determined
S12_vs_k141_143909||full Low-quality
S12_vs_k141_111822||full Low-quality
S12_vs_k141_80553||full Not-determined
S12_vs_k141_33178||full Low-quality
S12_vs_k141_17789||full Not-determined
S12_vs_k141_120489||full Not-determined
S9_vs_k141_73764||full Medium-quality
S12_vs_k141_151296||full Low-quality
S12_vs_k141_33189||full Low-quality
S12_vs_k141_97001||full Low-quality
S14_vs_k141_109545||full Medium-quality
S12_vs_k141_135747||full Low-quality
S12_vs_k141_120502||full Low-quality
S12_vs_k141_127996||full Low-quality
S12_vs_k141_25861||full Low-quality
S12_vs_k141_104573||full Not-determined
S12_vs_k141_135756||full Not-determined
S12_vs_k141_2234||full Not-determined
S12_vs_k141_17809||full Not-determined
S12_vs_k141_143949||full Low-quality
S12_vs_k141_183254||full Not-determined
S12_vs_k141_25867||full Low-quality
S12_vs_k141_33193||full Low-quality
S12_vs_k141_104594||full Low-quality
S12_vs_k141_128004||full Low-quality
S12_vs_k141_17821||full Low-quality
S12_vs_k141_151316||full Low-quality
S12_vs_k141_104596||full Low-quality
S12_vs_k141_41604||full Not-determined
S12_vs_k141_120530||full Not-determined
S12_vs_k141_104610||full Not-determined
S12_vs_k141_88910||full Low-quality
S12_vs_k141_25892||full Low-quality
S12_vs_k141_159224||full Not-determined
S12_vs_k141_80646||full Low-quality
S12_vs_k141_65286||full Not-determined
S12_vs_k141_80648||full Not-determined
S12_vs_k141_57563||full Not-determined
S12_vs_k141_65297||full Low-quality
S12_vs_k141_151357||full Not-determined
S12_vs_k141_159248||full Not-determined
S12_vs_k141_80659||full Low-quality
S12_vs_k141_104614||full Low-quality
S12_vs_k141_65310||full Not-determined
S12_vs_k141_151368||full Low-quality
S12_vs_k141_128047||full Low-quality
S12_vs_k141_174947||full Low-quality
S12_vs_k141_33221||full Low-quality
S12_vs_k141_120544||full Not-determined
S12_vs_k141_104639||full Low-quality
S12_vs_k141_57575||full Low-quality
S12_vs_k141_183287||full Low-quality
S12_vs_k141_159284||full Not-determined
S12_vs_k141_41682||full Low-quality
S12_vs_k141_33247||full Low-quality
S12_vs_k141_80702||full Low-quality
S12_vs_k141_120553||full Low-quality
S12_vs_k141_151399||full Low-quality
S12_vs_k141_97096||full Not-determined
S12_vs_k141_41695||full Low-quality
S12_vs_k141_48988||full Not-determined
S12_vs_k141_104656||full Not-determined
S12_vs_k141_17889||full Low-quality
S12_vs_k141_17891||full Low-quality
S12_vs_k141_128099||full Low-quality
S12_vs_k141_120577||full Not-determined
S12_vs_k141_41710||full Low-quality
S12_vs_k141_57597||full Not-determined
S12_vs_k141_111923||full Not-determined
S12_vs_k141_183312||full Low-quality
S12_vs_k141_143990||full Not-determined
S12_vs_k141_17910||full Low-quality
S12_vs_k141_65345||full Not-determined
S12_vs_k141_57611||full Not-determined
S12_vs_k141_49045||full Low-quality
S12_vs_k141_33280||full Low-quality
S12_vs_k141_128147||full Not-determined
S12_vs_k141_41754||full Not-determined
S12_vs_k141_41760||full Low-quality
S12_vs_k141_111979||full Not-determined
S12_vs_k141_104690||full Low-quality
S12_vs_k141_2375||full Low-quality
S12_vs_k141_17931||full Not-determined
S12_vs_k141_17933||full Low-quality
S12_vs_k141_25992||full Not-determined

| | |
|---|---|
| S12_vs_k141_144066\|\|full | Not-determined |
| S12_vs_k141_65382\|\|full | Not-determined |
| S12_vs_k141_151470\|\|full | Low-quality |
| S12_vs_k141_128172\|\|full | Low-quality |
| S12_vs_k141_151472\|\|full | Not-determined |
| S12_vs_k141_57633\|\|full | Low-quality |
| S12_vs_k141_41804\|\|full | Not-determined |
| S12_vs_k141_17957\|\|full | Low-quality |
| S12_vs_k141_144076\|\|full | Low-quality |
| S12_vs_k141_183394\|\|full | Not-determined |
| S12_vs_k141_159321\|\|full | Low-quality |
| S12_vs_k141_72646\|\|full | Low-quality |
| S12_vs_k141_26017\|\|full | Not-determined |
| S12_vs_k141_144087\|\|full | Not-determined |
| S12_vs_k141_104741\|\|full | Low-quality |
| S12_vs_k141_151512\|\|full | Low-quality |
| S12_vs_k141_175045\|\|full | Not-determined |
| S12_vs_k141_49088\|\|full | Not-determined |
| S12_vs_k141_151523\|\|full | Not-determined |
| S12_vs_k141_112061\|\|full | Low-quality |
| S12_vs_k141_183420\|\|full | Low-quality |
| S12_vs_k141_175062\|\|full | Low-quality |
| S12_vs_k141_128224\|\|full | Not-determined |
| S12_vs_k141_112074\|\|full | Low-quality |
| S12_vs_k141_49108\|\|full | Not-determined |
| S12_vs_k141_112076\|\|full | Not-determined |
| S12_vs_k141_104778\|\|full | Not-determined |
| S12_vs_k141_72662\|\|full | Low-quality |
| S12_vs_k141_17963\|\|full | Not-determined |
| S12_vs_k141_144103\|\|full | Not-determined |
| S12_vs_k141_17968\|\|full | Low-quality |
| S12_vs_k141_144106\|\|full | Low-quality |
| S12_vs_k141_144114\|\|full | Not-determined |
| S12_vs_k141_65473\|\|full | Not-determined |
| S12_vs_k141_89108\|\|full | Low-quality |
| S12_vs_k141_89109\|\|full | Low-quality |
| S12_vs_k141_144121\|\|full | Low-quality |
| S12_vs_k141_49121\|\|full | Low-quality |
| S12_vs_k141_144136\|\|full | Not-determined |
| S12_vs_k141_175093\|\|full | Low-quality |
| S12_vs_k141_136027\|\|full | Not-determined |
| S12_vs_k141_26039\|\|full | Low-quality |
| S12_vs_k141_57724\|\|full | Low-quality |
| S12_vs_k141_18006\|\|full | Low-quality |
| S12_vs_k141_33384\|\|full | Low-quality |
| S12_vs_k141_65504\|\|full | Not-determined |
| S12_vs_k141_72714\|\|full | Not-determined |
| S12_vs_k141_72718\|\|full | Low-quality |
| S12_vs_k141_167473\|\|full | Low-quality |
| S12_vs_k141_80901\|\|full | Low-quality |
| S12_vs_k141_72726\|\|full | Not-determined |
| S12_vs_k141_112121\|\|full | Low-quality |
| S12_vs_k141_2460\|\|full | Low-quality |
| S12_vs_k141_49159\|\|full | Low-quality |
| S12_vs_k141_151609\|\|full | Not-determined |
| S12_vs_k141_159362\|\|full | Not-determined |
| S12_vs_k141_57763\|\|full | Not-determined |
| S12_vs_k141_33397\|\|full | Low-quality |
| S12_vs_k141_65548\|\|full | Not-determined |
| S12_vs_k141_80932\|\|full | Not-determined |
| S12_vs_k141_18027\|\|full | Not-determined |
| S12_vs_k141_33399\|\|full | Not-determined |
| S12_vs_k141_89217\|\|full | Not-determined |
| S12_vs_k141_72744\|\|full | Low-quality |
| S12_vs_k141_57765\|\|full | Low-quality |
| S12_vs_k141_112154\|\|full | Low-quality |
| S12_vs_k141_18046\|\|full | Not-determined |
| S12_vs_k141_167480\|\|full | Low-quality |
| S12_vs_k141_151619\|\|full | Low-quality |
| S12_vs_k141_72755\|\|full | Low-quality |
| S12_vs_k141_41936\|\|full | Low-quality |
| S12_vs_k141_57780\|\|full | Low-quality |
| S12_vs_k141_89237\|\|full | Not-determined |
| S12_vs_k141_33416\|\|full | Low-quality |
| S12_vs_k141_2500\|\|full | Low-quality |
| S12_vs_k141_65584\|\|full | Low-quality |
| S12_vs_k141_167504\|\|full | Not-determined |
| S12_vs_k141_120692\|\|full | Not-determined |
| S12_vs_k141_33431\|\|full | Low-quality |
| S12_vs_k141_41967\|\|full | Low-quality |
| S12_vs_k141_183539\|\|full | Not-determined |
| S12_vs_k141_2524\|\|full | Low-quality |
| S12_vs_k141_80956\|\|full | Low-quality |
| S12_vs_k141_72777\|\|full | Low-quality |
| S12_vs_k141_183552\|\|full | Low-quality |
| S12_vs_k141_80959\|\|full | Low-quality |
| S12_vs_k141_104847\|\|full | Not-determined |
| S12_vs_k141_97408\|\|full | Not-determined |
| S12_vs_k141_144247\|\|full | Low-quality |
| S12_vs_k141_97413\|\|full | Low-quality |
| S12_vs_k141_2541\|\|full | Not-determined |
| S12_vs_k141_112170\|\|full | Low-quality |
| S12_vs_k141_80974\|\|full | Not-determined |
| S12_vs_k141_144250\|\|full | Low-quality |
| S12_vs_k141_41995\|\|full | Not-determined |
| S12_vs_k141_80976\|\|full | Not-determined |
| S12_vs_k141_144251\|\|full | Not-determined |
| S12_vs_k141_104870\|\|full | Low-quality |
| S12_vs_k141_128307\|\|full | Low-quality |
| S12_vs_k141_26122\|\|full | Not-determined |
| S12_vs_k141_175162\|\|full | Low-quality |
| S12_vs_k141_89306\|\|full | Low-quality |
| S12_vs_k141_80992\|\|full | Not-determined |
| S12_vs_k141_26133\|\|full | Not-determined |
| S12_vs_k141_89309\|\|full | Not-determined |
| S12_vs_k141_151634\|\|full | Low-quality |
| S12_vs_k141_167576\|\|full | Low-quality |
| S12_vs_k141_104888\|\|full | Not-determined |
| S12_vs_k141_183588\|\|full | Not-determined |
| S12_vs_k141_33500\|\|full | Low-quality |
| S12_vs_k141_104889\|\|full | Low-quality |
| S12_vs_k141_2583\|\|full | Low-quality |
| S12_vs_k141_97440\|\|full | Low-quality |
| S12_vs_k141_89326\|\|full | Not-determined |
| S12_vs_k141_18170\|\|full | Low-quality |
| S12_vs_k141_167603\|\|full | Low-quality |
| S12_vs_k141_183609\|\|full | Not-determined |
| S12_vs_k141_89341\|\|full | Not-determined |
| S12_vs_k141_104906\|\|full | Not-determined |
| S12_vs_k141_65658\|\|full | Not-determined |
| S12_vs_k141_136172\|\|full | Low-quality |
| S12_vs_k141_175209\|\|full | Low-quality |
| S12_vs_k141_72840\|\|full | Low-quality |
| S12_vs_k141_89349\|\|full | Not-determined |
| S12_vs_k141_26187\|\|full | Not-determined |
| S12_vs_k141_42059\|\|full | Low-quality |
| S12_vs_k141_104921\|\|full | Low-quality |
| S12_vs_k141_136190\|\|full | Low-quality |
| S12_vs_k141_104931\|\|full | Not-determined |
| S12_vs_k141_89371\|\|full | Low-quality |
| S12_vs_k141_2628\|\|full | Not-determined |
| S12_vs_k141_26203\|\|full | Not-determined |
| S12_vs_k141_18190\|\|full | Not-determined |
| S12_vs_k141_136208\|\|full | Not-determined |
| S12_vs_k141_72865\|\|full | Not-determined |
| S12_vs_k141_72871\|\|full | Not-determined |
| S12_vs_k141_159545\|\|full | Not-determined |
| S12_vs_k141_42082\|\|full | Low-quality |
| S12_vs_k141_9887\|\|full | Not-determined |
| S12_vs_k141_2649\|\|full | Low-quality |
| S12_vs_k141_81062\|\|full | Not-determined |
| S12_vs_k141_42084\|\|full | Not-determined |
| S12_vs_k141_167630\|\|full | Low-quality |
| S12_vs_k141_159546\|\|full | Not-determined |
| S12_vs_k141_72878\|\|full | Low-quality |
| S12_vs_k141_65708\|\|full | Not-determined |
| S12_vs_k141_151673\|\|full | Low-quality |
| S12_vs_k141_104962\|\|full | Low-quality |
| S12_vs_k141_49187\|\|full | Low-quality |
| S12_vs_k141_183618\|\|full | Low-quality |
| S12_vs_k141_2673\|\|full | Not-determined |
| S12_vs_k141_144315\|\|full | Low-quality |
| S12_vs_k141_33582\|\|full | Low-quality |
| S12_vs_k141_167667\|\|full | Low-quality |
| S12_vs_k141_9917\|\|full | Low-quality |
| S12_vs_k141_112209\|\|full | Low-quality |
| S12_vs_k141_97489\|\|full | Low-quality |
| S12_vs_k141_136247\|\|full | Not-determined |
| S12_vs_k141_167685\|\|full | Low-quality |
| S12_vs_k141_175300\|\|full | Low-quality |
| S12_vs_k141_183653\|\|full | Not-determined |
| S12_vs_k141_18221\|\|full | Not-determined |
| S12_vs_k141_112223\|\|full | Low-quality |
| S12_vs_k141_105001\|\|full | Not-determined |
| S12_vs_k141_175309\|\|full | Not-determined |
| S12_vs_k141_167688\|\|full | Low-quality |
| S12_vs_k141_136269\|\|full | Not-determined |
| S12_vs_k141_81141\|\|full | Not-determined |
| S12_vs_k141_81145\|\|full | Low-quality |
| S12_vs_k141_183686\|\|full | Low-quality |
| S12_vs_k141_144376\|\|full | Low-quality |
| S12_vs_k141_89428\|\|full | Not-determined |
| S12_vs_k141_136286\|\|full | Low-quality |
| S12_vs_k141_57957\|\|full | Not-determined |
| S12_vs_k141_33631\|\|full | Low-quality |
| S12_vs_k141_112263\|\|full | Low-quality |
| S12_vs_k141_57959\|\|full | Low-quality |
| S12_vs_k141_81165\|\|full | Not-determined |
| S12_vs_k141_33644\|\|full | Low-quality |
| S12_vs_k141_18258\|\|full | Low-quality |
| S12_vs_k141_81174\|\|full | Low-quality |
| S12_vs_k141_128344\|\|full | Not-determined |
| S12_vs_k141_33652\|\|full | Low-quality |
| S12_vs_k141_81180\|\|full | Not-determined |
| S12_vs_k141_26215\|\|full | Not-determined |
| S12_vs_k141_159636\|\|full | Low-quality |
| S12_vs_k141_144421\|\|full | Low-quality |
| S12_vs_k141_183719\|\|full | Not-determined |
| S12_vs_k141_65767\|\|full | Not-determined |
| S12_vs_k141_2701\|\|full | Not-determined |
| S12_vs_k141_49267\|\|full | Not-determined |
| S12_vs_k141_105056\|\|full | Not-determined |
| S12_vs_k141_57999\|\|full | Not-determined |
| S12_vs_k141_9990\|\|full | Low-quality |
| S12_vs_k141_159642\|\|full | Low-quality |
| S12_vs_k141_183741\|\|full | Not-determined |
| S12_vs_k141_65791\|\|full | Low-quality |
| S12_vs_k141_97508\|\|full | Not-determined |
| S12_vs_k141_42225\|\|full | Low-quality |
| S12_vs_k141_81227\|\|full | Low-quality |
| S12_vs_k141_183751\|\|full | Not-determined |
| S12_vs_k141_151764\|\|full | Not-determined |
| S12_vs_k141_112344\|\|full | Not-determined |
| S12_vs_k141_81239\|\|full | Not-determined |
| S12_vs_k141_128399\|\|full | Not-determined |
| S12_vs_k141_112355\|\|full | Not-determined |
| S12_vs_k141_128406\|\|full | Not-determined |
| S12_vs_k141_128409\|\|full | Not-determined |
| S17_vs_k141_117507\|\|full | Medium-quality |
| S12_vs_k141_97523\|\|full | Low-quality |
| S12_vs_k141_33734\|\|full | Low-quality |
| S12_vs_k141_2749\|\|full | Low-quality |
| S12_vs_k141_10021\|\|full | Low-quality |
| S12_vs_k141_18339\|\|full | Low-quality |
| S12_vs_k141_81249\|\|full | Low-quality |
| S12_vs_k141_72947\|\|full | Low-quality |
| S12_vs_k141_10027\|\|full | Low-quality |
| S12_vs_k141_159680\|\|full | Low-quality |
| S12_vs_k141_26296\|\|full | Low-quality |
| S12_vs_k141_58056\|\|full | Low-quality |
| S12_vs_k141_72962\|\|full | Low-quality |
| S12_vs_k141_81263\|\|full | Not-determined |
| S12_vs_k141_128437\|\|full | Low-quality |
| S12_vs_k141_183796\|\|full | Not-determined |
| S12_vs_k141_26305\|\|full | Not-determined |
| S12_vs_k141_167780\|\|full | Low-quality |
| S12_vs_k141_26309\|\|full | Low-quality |
| S12_vs_k141_136350\|\|full | Low-quality |
| S12_vs_k141_97559\|\|full | Low-quality |
| S12_vs_k141_72992\|\|full | Low-quality |
| S12_vs_k141_183808\|\|full | Not-determined |
| S12_vs_k141_97571\|\|full | Not-determined |
| S12_vs_k141_42311\|\|full | Low-quality |
| S12_vs_k141_97572\|\|full | Not-determined |
| S12_vs_k141_73008\|\|full | Not-determined |
| S12_vs_k141_81307\|\|full | Low-quality |
| S12_vs_k141_105126\|\|full | Low-quality |
| S12_vs_k141_49369\|\|full | Low-quality |
| S12_vs_k141_81308\|\|full | Low-quality |
| S12_vs_k141_97582\|\|full | Low-quality |
| S12_vs_k141_42320\|\|full | Not-determined |
| S12_vs_k141_58107\|\|full | Not-determined |
| S12_vs_k141_136390\|\|full | Not-determined |
| S12_vs_k141_18392\|\|full | Not-determined |
| S12_vs_k141_10097\|\|full | Low-quality |
| S12_vs_k141_58116\|\|full | Low-quality |
| S12_vs_k141_183836\|\|full | Low-quality |
| S12_vs_k141_49387\|\|full | Not-determined |
| S12_vs_k141_136394\|\|full | Not-determined |
| S12_vs_k141_81327\|\|full | Not-determined |
| S12_vs_k141_89572\|\|full | Low-quality |
| S12_vs_k141_65866\|\|full | Low-quality |
| S12_vs_k141_183850\|\|full | Low-quality |
| S12_vs_k141_89575\|\|full | Low-quality |
| S12_vs_k141_112414\|\|full | Low-quality |
| S12_vs_k141_81336\|\|full | Low-quality |
| S12_vs_k141_97614\|\|full | Not-determined |
| S12_vs_k141_136412\|\|full | Not-determined |
| S12_vs_k141_136414\|\|full | Low-quality |
| S12_vs_k141_81347\|\|full | Not-determined |
| S12_vs_k141_2838\|\|full | Not-determined |
| S12_vs_k141_183852\|\|full | Not-determined |
| S12_vs_k141_167857\|\|full | Not-determined |
| S12_vs_k141_112426\|\|full | Low-quality |
| S12_vs_k141_136418\|\|full | Low-quality |
| S12_vs_k141_42373\|\|full | Low-quality |
| S12_vs_k141_167864\|\|full | Not-determined |
| S12_vs_k141_42379\|\|full | Not-determined |
| S12_vs_k141_183866\|\|full | Low-quality |
| S12_vs_k141_105184\|\|full | Not-determined |
| S12_vs_k141_175381\|\|full | Low-quality |
| S12_vs_k141_183873\|\|full | Low-quality |
| S12_vs_k141_112448\|\|full | Low-quality |
| S12_vs_k141_183885\|\|full | Low-quality |
| S12_vs_k141_49398\|\|full | Low-quality |
| S12_vs_k141_105197\|\|full | Not-determined |
| S12_vs_k141_159711\|\|full | Low-quality |
| S12_vs_k141_105202\|\|full | Low-quality |
| S12_vs_k141_49413\|\|full | Not-determined |
| S12_vs_k141_42393\|\|full | Not-determined |
| S12_vs_k141_2880\|\|full | Low-quality |
| S12_vs_k141_60694\|\|full | Medium-quality |
| S12_vs_k141_167907\|\|full | Low-quality |
| S12_vs_k141_159730\|\|full | Not-determined |
| S9_vs_k141_41371\|\|full | Medium-quality |
| S12_vs_k141_18450\|\|full | Not-determined |
| S12_vs_k141_18454\|\|full | Not-determined |
| S12_vs_k141_2892\|\|full | Not-determined |
| S12_vs_k141_58152\|\|full | Low-quality |
| S12_vs_k141_18455\|\|full | Low-quality |
| S12_vs_k141_18456\|\|full | Not-determined |
| S12_vs_k141_26343\|\|full | Not-determined |
| S12_vs_k141_49433\|\|full | Low-quality |
| S12_vs_k141_2897\|\|full | Not-determined |
| S12_vs_k141_167925\|\|full | Not-determined |
| S12_vs_k141_105230\|\|full | Low-quality |
| S12_vs_k141_151857\|\|full | Not-determined |
| S12_vs_k141_97639\|\|full | Not-determined |
| S12_vs_k141_49456\|\|full | Low-quality |
| S12_vs_k141_167929\|\|full | Low-quality |
| S12_vs_k141_18476\|\|full | Low-quality |
| S12_vs_k141_167933\|\|full | Not-determined |
| S12_vs_k141_151860\|\|full | Low-quality |
| S12_vs_k141_112497\|\|full | Not-determined |
| S12_vs_k141_105244\|\|full | Low-quality |
| S12_vs_k141_58178\|\|full | Low-quality |
| S12_vs_k141_65898\|\|full | Low-quality |
| S12_vs_k141_49476\|\|full | Not-determined |
| S12_vs_k141_89681\|\|full | Not-determined |
| S12_vs_k141_58193\|\|full | Not-determined |
| S12_vs_k141_151863\|\|full | Not-determined |
| S12_vs_k141_2927\|\|full | Not-determined |
| S12_vs_k141_89684\|\|full | Not-determined |
| S12_vs_k141_33763\|\|full | Low-quality |
| S12_vs_k141_2932\|\|full | Not-determined |
| S13_vs_2031\|\|full | Medium-quality |
| S12_vs_k141_89685\|\|full | Low-quality |
| S12_vs_k141_136466\|\|full | Not-determined |
| S12_vs_k141_18523\|\|full | Low-quality |
| S12_vs_k141_89689\|\|full | Low-quality |
| S12_vs_k141_33779\|\|full | Low-quality |
| S12_vs_k141_65936\|\|full | Low-quality |
| S12_vs_k141_33781\|\|full | Not-determined |
| S12_vs_k141_42411\|\|full | Not-determined |
| S12_vs_k141_33788\|\|full | Not-determined |
| S12_vs_k141_18552\|\|full | Not-determined |
| S12_vs_k141_2953\|\|full | Not-determined |
| S12_vs_k141_33793\|\|full | Not-determined |
| S12_vs_k141_112552\|\|full | Not-determined |
| S12_vs_k141_81382\|\|full | Not-determined |
| S12_vs_k141_120714\|\|full | Low-quality |
| S12_vs_k141_105303\|\|full | Not-determined |
| S12_vs_k141_97713\|\|full | Not-determined |
| S12_vs_k141_42432\|\|full | Low-quality |
| S12_vs_k141_18570\|\|full | Low-quality |
| S12_vs_k141_175421\|\|full | Low-quality |
| S12_vs_k141_18575\|\|full | Not-determined |
| S12_vs_k141_26417\|\|full | Not-determined |
| S12_vs_k141_58233\|\|full | Not-determined |
| S12_vs_k141_159792\|\|full | Low-quality |
| S12_vs_k141_136510\|\|full | Not-determined |
| S12_vs_k141_151923\|\|full | Low-quality |
| S12_vs_k141_18586\|\|full | Not-determined |
| S12_vs_k141_65991\|\|full | Low-quality |
| S12_vs_k141_136533\|\|full | Not-determined |
| S12_vs_k141_18597\|\|full | Not-determined |
| S12_vs_k141_168017\|\|full | Low-quality |
| S12_vs_k141_105322\|\|full | Low-quality |
| S12_vs_k141_112603\|\|full | Not-determined |
| S12_vs_k141_58254\|\|full | Low-quality |
| S12_vs_k141_168022\|\|full | Not-determined |
| S12_vs_k141_97773\|\|full | Not-determined |
| S12_vs_k141_159869\|\|full | Low-quality |
| S12_vs_k141_58264\|\|full | Not-determined |
| S12_vs_k141_97781\|\|full | Not-determined |
| S12_vs_k141_2982\|\|full | Not-determined |
| S12_vs_k141_183970\|\|full | Not-determined |
| S12_vs_k141_42524\|\|full | Not-determined |
| S12_vs_k141_159881\|\|full | Not-determined |
| S12_vs_k141_105345\|\|full | Low-quality |
| S12_vs_k141_18652\|\|full | Not-determined |
| S12_vs_k141_159883\|\|full | Not-determined |
| S12_vs_k141_183994\|\|full | Not-determined |
| S12_vs_k141_97814\|\|full | Not-determined |
| S12_vs_k141_89707\|\|full | Not-determined |
| S12_vs_k141_49586\|\|full | Not-determined |
| S12_vs_k141_89716\|\|full | Not-determined |
| S12_vs_k141_128572\|\|full | Low-quality |
| S12_vs_k141_97827\|\|full | Not-determined |
| S12_vs_k141_120765\|\|full | Not-determined |
| S12_vs_k141_89724\|\|full | Not-determined |
| S12_vs_k141_58303\|\|full | Low-quality |
| S12_vs_k141_112625\|\|full | Low-quality |
| S12_vs_k141_58308\|\|full | Not-determined |
| S12_vs_k141_184001\|\|full | Not-determined |
| S12_vs_k141_112628\|\|full | Low-quality |
| S12_vs_k141_97845\|\|full | Low-quality |
| S12_vs_k141_3021\|\|full | Low-quality |
| S12_vs_k141_42555\|\|full | Low-quality |
| S12_vs_k141_33923\|\|full | Low-quality |
| S12_vs_k141_89744\|\|full | Low-quality |
| S12_vs_k141_184011\|\|full | Not-determined |
| S12_vs_k141_184016\|\|full | Not-determined |
| S12_vs_k141_3037\|\|full | Low-quality |
| S12_vs_k141_144541\|\|full | Low-quality |
| S12_vs_k141_3042\|\|full | Not-determined |
| S12_vs_k141_81425\|\|full | Low-quality |
| S12_vs_k141_159933\|\|full | Low-quality |
| S12_vs_k141_184030\|\|full | Low-quality |
| S12_vs_k141_136598\|\|full | Not-determined |
| S12_vs_k141_168103\|\|full | Not-determined |
| S12_vs_k141_152043\|\|full | Not-determined |
| S12_vs_k141_26493\|\|full | Low-quality |
| S12_vs_k141_81435\|\|full | Not-determined |
| S12_vs_k141_112685\|\|full | Not-determined |
| S12_vs_k141_97878\|\|full | Not-determined |
| S12_vs_k141_112686\|\|full | Not-determined |
| S12_vs_k141_42585\|\|full | Low-quality |
| S12_vs_k141_159959\|\|full | Not-determined |
| S12_vs_k141_144559\|\|full | Not-determined |
| S12_vs_k141_105454\|\|full | Low-quality |
| S12_vs_k141_42599\|\|full | Low-quality |
| S12_vs_k141_42601\|\|full | Low-quality |
| S12_vs_k141_136621\|\|full | Not-determined |
| S12_vs_k141_18742\|\|full | Not-determined |
| S12_vs_k141_58341\|\|full | Low-quality |
| S12_vs_k141_105466\|\|full | Low-quality |
| S12_vs_k141_49689\|\|full | Low-quality |
| S12_vs_k141_175588\|\|full | Not-determined |
| S12_vs_k141_144586\|\|full | Not-determined |
| S12_vs_k141_3107\|\|full | Not-determined |
| S12_vs_k141_97901\|\|full | Low-quality |
| S12_vs_k141_58371\|\|full | Not-determined |
| S12_vs_k141_175597\|\|full | Not-determined |
| S12_vs_k141_81482\|\|full | Low-quality |
| S12_vs_k141_136656\|\|full | Low-quality |
| S12_vs_k141_120887\|\|full | Low-quality |
| S12_vs_k141_42630\|\|full | Not-determined |
| S12_vs_k141_144603\|\|full | Not-determined |
| S12_vs_k141_73279\|\|full | Not-determined |
| S12_vs_k141_184078\|\|full | Not-determined |
| S12_vs_k141_120910\|\|full | Not-determined |
| S12_vs_k141_136701\|\|full | Not-determined |
| S12_vs_k141_144623\|\|full | Not-determined |
| S12_vs_k141_175635\|\|full | Not-determined |
| S12_vs_k141_81519\|\|full | Low-quality |
| S12_vs_k141_112743\|\|full | Low-quality |
| S12_vs_k141_112746\|\|full | Low-quality |
| S12_vs_k141_3136\|\|full | Low-quality |
| S12_vs_k141_81527\|\|full | Not-determined |
| S12_vs_k141_97957\|\|full | Low-quality |
| S12_vs_k141_144629\|\|full | Low-quality |
| S12_vs_k141_97961\|\|full | Not-determined |
| S12_vs_k141_42674\|\|full | Low-quality |
| S12_vs_k141_168134\|\|full | Not-determined |
| S12_vs_k141_26541\|\|full | Not-determined |
| S12_vs_k141_89895\|\|full | Low-quality |
| S12_vs_k141_144641\|\|full | Not-determined |
| S12_vs_k141_26545\|\|full | Not-determined |
| S12_vs_k141_136727\|\|full | Not-determined |
| S12_vs_k141_105493\|\|full | Low-quality |
| S12_vs_k141_120932\|\|full | Not-determined |
| S12_vs_k141_136739\|\|full | Low-quality |
| S12_vs_k141_105496\|\|full | Low-quality |
| S12_vs_k141_168150\|\|full | Low-quality |
| S12_vs_k141_175672\|\|full | Low-quality |
| S12_vs_k141_168154\|\|full | Low-quality |
| S12_vs_k141_3197\|\|full | Not-determined |
| S12_vs_k141_58472\|\|full | Low-quality |
| S12_vs_k141_81552\|\|full | Not-determined |
| S12_vs_k141_168161\|\|full | Low-quality |
| S12_vs_k141_152120\|\|full | Low-quality |
| S12_vs_k141_66106\|\|full | Not-determined |
| S12_vs_k141_144666\|\|full | Not-determined |
| S12_vs_k141_152131\|\|full | Not-determined |
| S12_vs_k141_136770\|\|full | Low-quality |
| S12_vs_k141_112785\|\|full | Low-quality |
| S12_vs_k141_81569\|\|full | Low-quality |
| S12_vs_k141_184120\|\|full | Not-determined |
| S12_vs_k141_112790\|\|full | Low-quality |
| S12_vs_k141_136782\|\|full | Not-determined |
| S12_vs_k141_144680\|\|full | Not-determined |
| S12_vs_k141_112809\|\|full | Low-quality |
| S12_vs_k141_81574\|\|full | Low-quality |
| S12_vs_k141_98051\|\|full | Not-determined |
| S12_vs_k141_112821\|\|full | Not-determined |
| S12_vs_k141_136803\|\|full | Low-quality |
| S12_vs_k141_144705\|\|full | Low-quality |
| S12_vs_k141_160009\|\|full | Not-determined |
| S12_vs_k141_128614\|\|full | Not-determined |
| S12_vs_k141_160020\|\|full | Low-quality |
| S12_vs_k141_128619\|\|full | Low-quality |
| S12_vs_k141_58496\|\|full | Low-quality |
| S12_vs_k141_34023\|\|full | Not-determined |
| S12_vs_k141_136817\|\|full | Not-determined |
| S12_vs_k141_26645\|\|full | Low-quality |
| S12_vs_k141_26650\|\|full | Low-quality |
| S12_vs_k141_73392\|\|full | Not-determined |
| S12_vs_k141_152193\|\|full | Low-quality |
| S12_vs_k141_42759\|\|full | Low-quality |
| S12_vs_k141_66122\|\|full | Not-determined |
| S12_vs_k141_112861\|\|full | Low-quality |
| S12_vs_k141_168241\|\|full | Not-determined |
| S12_vs_k141_98088\|\|full | Not-determined |
| S12_vs_k141_93992\|\|full | Medium-quality |
| S12_vs_k141_34026\|\|full | Not-determined |
| S12_vs_k141_73398\|\|full | Not-determined |
| S12_vs_k141_73400\|\|full | Low-quality |
| S12_vs_k141_10222\|\|full | Not-determined |
| S12_vs_k141_10226\|\|full | Low-quality |
| S12_vs_k141_152205\|\|full | Not-determined |
| S12_vs_k141_128647\|\|full | Not-determined |
| S12_vs_k141_136840\|\|full | Low-quality |
| S12_vs_k141_152211\|\|full | Low-quality |
| S12_vs_k141_105548\|\|full | Low-quality |
| S12_vs_k141_81648\|\|full | Not-determined |
| S12_vs_k141_90010\|\|full | Low-quality |
| S12_vs_k141_42786\|\|full | Low-quality |
| S12_vs_k141_90015\|\|full | Low-quality |
| S12_vs_k141_168282\|\|full | Not-determined |
| S12_vs_k141_90023\|\|full | Not-determined |
| S12_vs_k141_73424\|\|full | Low-quality |
| S12_vs_k141_42828\|\|full | Not-determined |
| S12_vs_k141_105596\|\|full | Low-quality |
| S12_vs_k141_10253\|\|full | Low-quality |
| S12_vs_k141_66173\|\|full | Not-determined |
| S12_vs_k141_136855\|\|full | Low-quality |
| S12_vs_k141_90061\|\|full | Low-quality |
| S12_vs_k141_10268\|\|full | Low-quality |
| S12_vs_k141_112893\|\|full | Low-quality |
| S12_vs_k141_105604\|\|full | Low-quality |
| S12_vs_k141_105607\|\|full | Not-determined |
| S12_vs_k141_3290\|\|full | Not-determined |
| S12_vs_k141_112900\|\|full | Low-quality |
| S12_vs_k141_73442\|\|full | Low-quality |
| S12_vs_k141_121004\|\|full | Low-quality |
| S12_vs_k141_98138\|\|full | Low-quality |
| S12_vs_k141_66192\|\|full | Low-quality |
| S12_vs_k141_112901\|\|full | Not-determined |
| S12_vs_k141_98145\|\|full | Low-quality |
| S12_vs_k141_136864\|\|full | Not-determined |
| S12_vs_k141_128705\|\|full | Low-quality |
| S12_vs_k141_42854\|\|full | Not-determined |
| S12_vs_k141_98153\|\|full | Not-determined |
| S12_vs_k141_184249\|\|full | Not-determined |
| S12_vs_k141_112914\|\|full | Low-quality |
| S12_vs_k141_136873\|\|full | Low-quality |
| S12_vs_k141_42864\|\|full | Low-quality |
| S12_vs_k141_168339\|\|full | Not-determined |
| S12_vs_k141_184260\|\|full | Low-quality |
| S12_vs_k141_58538\|\|full | Low-quality |
| S12_vs_k141_121036\|\|full | Not-determined |
| S12_vs_k141_73477\|\|full | Low-quality |
| S12_vs_k141_128722\|\|full | Not-determined |
| S12_vs_k141_160150\|\|full | Not-determined |
| S12_vs_k141_168365\|\|full | Low-quality |
| S12_vs_k141_42889\|\|full | Low-quality |
| S12_vs_k141_175748\|\|full | Low-quality |
| S12_vs_k141_34084\|\|full | Low-quality |
| S12_vs_k141_58558\|\|full | Low-quality |
| S12_vs_k141_73500\|\|full | Low-quality |
| S12_vs_k141_42893\|\|full | Not-determined |
| S12_vs_k141_168367\|\|full | Low-quality |
| S12_vs_k141_175757\|\|full | Low-quality |
| S12_vs_k141_152257\|\|full | Low-quality |
| S12_vs_k141_160166\|\|full | Not-determined |
| S12_vs_k141_49850\|\|full | Low-quality |
| S12_vs_k141_152266\|\|full | Low-quality |
| S15_vs_32488\|\|full | Medium-quality |
| S12_vs_k141_160188\|\|full | Low-quality |
| S12_vs_k141_168382\|\|full | Not-determined |
| S12_vs_k141_3363\|\|full | Not-determined |
| S17_vs_k141_2079\|\|full | Medium-quality |
| S12_vs_k141_81739\|\|full | Not-determined |
| S12_vs_k141_168391\|\|full | Not-determined |
| S12_vs_k141_160192\|\|full | Not-determined |
| S12_vs_k141_168393\|\|full | Not-determined |
| S12_vs_k141_152289\|\|full | Low-quality |
| S12_vs_k141_90145\|\|full | Low-quality |
| S12_vs_k141_128778\|\|full | Low-quality |
| S12_vs_k141_112991\|\|full | Not-determined |
| S12_vs_k141_81761\|\|full | Not-determined |
| S12_vs_k141_112996\|\|full | Low-quality |
| S12_vs_k141_73528\|\|full | Not-determined |
| S12_vs_k141_26775\|\|full | Low-quality |
| S12_vs_k141_10300\|\|full | Low-quality |
| S12_vs_k141_66254\|\|full | Not-determined |
| S12_vs_k141_90175\|\|full | Not-determined |
| S12_vs_k141_113008\|\|full | Not-determined |
| S12_vs_k141_98207\|\|full | Low-quality |
| S4_vs_VIRSorter_9913-cat_1 | Medium-quality |
| S12_vs_k141_152317\|\|full | Not-determined |
| S12_vs_k141_73539\|\|full | Low-quality |
| S12_vs_k141_136948\|\|full | Low-quality |
| S12_vs_k141_160231\|\|full | Low-quality |
| S12_vs_k141_175821\|\|full | Low-quality |
| S12_vs_k141_175823\|\|full | Not-determined |
| S12_vs_k141_144936\|\|full | Not-determined |
| S12_vs_k141_168432\|\|full | Not-determined |
| S12_vs_k141_66292\|\|full | Not-determined |
| S12_vs_k141_3425\|\|full | Not-determined |
| S12_vs_k141_136959\|\|full | Low-quality |
| S12_vs_k141_144945\|\|full | Not-determined |
| S12_vs_k141_49905\|\|full | Low-quality |
| S12_vs_k141_66301\|\|full | Not-determined |
| S12_vs_k141_49908\|\|full | Low-quality |
| S12_vs_k141_113035\|\|full | Not-determined |
| S12_vs_k141_136972\|\|full | Not-determined |
| S12_vs_k141_81789\|\|full | Low-quality |

S12_vs_k141_160265||full Low-quality
S12_vs_k141_98222||full Low-quality
S12_vs_k141_98223||full Low-quality
S12_vs_k141_49926||full Low-quality
S12_vs_k141_144955||full Not-determined
S12_vs_k141_175865||full Not-determined
S12_vs_k141_152341||full Low-quality
S12_vs_k141_152342||full Low-quality
S12_vs_k141_98235||full Low-quality
S12_vs_k141_105785||full Low-quality
S12_vs_k141_144980||full Not-determined
S12_vs_k141_137004||full Not-determined
S12_vs_k141_168490||full Not-determined
S12_vs_k141_58624||full Low-quality
S12_vs_k141_168499||full Low-quality
S12_vs_k141_26819||full Not-determined
S12_vs_k141_34184||full Low-quality
S12_vs_k141_137018||full Not-determined
S12_vs_k141_34186||full Not-determined
S12_vs_k141_137019||full Not-determined
S12_vs_k141_90274||full Low-quality
S17_vs_k141_31654||full Medium-quality
S12_vs_k141_137023||full Low-quality
S12_vs_k141_43013||full Low-quality
S12_vs_k141_98273||full Not-determined
S12_vs_k141_43017||full Not-determined
S12_vs_k141_184362||full Low-quality
S12_vs_k141_105830||full Not-determined
S12_vs_k141_160325||full Low-quality
S12_vs_k141_184369||full Low-quality
S12_vs_k141_113080||full Low-quality
S12_vs_k141_26844||full Low-quality
S12_vs_k141_3483||full Not-determined
S12_vs_k141_137045||full Low-quality
S12_vs_k141_98292||full Not-determined
S12_vs_k141_81875||full Not-determined
S12_vs_k141_18942||full Not-determined
S12_vs_k141_152393||full Low-quality
S12_vs_k141_3498||full Not-determined
S12_vs_k141_58675||full Low-quality
S12_vs_k141_81884||full Low-quality
S12_vs_k141_3500||full Low-quality
S12_vs_k141_128863||full Low-quality
S12_vs_k141_160369||full Not-determined
S12_vs_k141_152407||full Low-quality
S12_vs_k141_73618||full Not-determined
S12_vs_k141_34234||full Low-quality
S12_vs_k141_105854||full Low-quality
S12_vs_k141_3510||full Not-determined
S12_vs_k141_184420||full Low-quality
S14_vs_k141_82685||full Medium-quality
S12_vs_k141_18949||full Low-quality
S12_vs_k141_145035||full Low-quality
S12_vs_k141_18952||full Low-quality
S12_vs_k141_98330||full Low-quality
S12_vs_k141_128884||full Not-determined
S12_vs_k141_73627||full Not-determined
S12_vs_k141_184434||full Not-determined
S12_vs_k141_168604||full Low-quality
S12_vs_k141_3521||full Not-determined
S12_vs_k141_73633||full Low-quality
S12_vs_k141_152443||full Low-quality
S12_vs_k141_160390||full Low-quality
S12_vs_k141_43087||full Low-quality
S12_vs_k141_128907||full Not-determined
S12_vs_k141_43091||full Low-quality
S12_vs_k141_43096||full Low-quality
S12_vs_k141_10419||full Low-quality
S12_vs_k141_43099||full Not-determined
S12_vs_k141_113123||full Not-determined
S12_vs_k141_34281||full Not-determined
S12_vs_k141_113131||full Not-determined
S12_vs_k141_73672||full Not-determined
S12_vs_k141_152482||full Not-determined
S12_vs_k141_66398||full Not-determined
S12_vs_k141_152483||full Low-quality
S12_vs_k141_34286||full Low-quality
S12_vs_k141_73676||full Low-quality
S12_vs_k141_3564||full Low-quality
S12_vs_k141_3565||full Low-quality
S12_vs_k141_66415||full Low-quality
S12_vs_k141_3566||full Low-quality
S12_vs_k141_113157||full Low-quality
S12_vs_k141_105898||full Not-determined
S12_vs_k141_105901||full Not-determined
S12_vs_k141_175954||full Low-quality
S12_vs_k141_90339||full Low-quality
S12_vs_k141_184491||full Low-quality
S12_vs_k141_73710||full Low-quality
S12_vs_k141_19011||full Low-quality
S12_vs_k141_90351||full Not-determined
S12_vs_k141_66440||full Not-determined
S12_vs_k141_43142||full Not-determined
S12_vs_k141_66441||full Not-determined
S12_vs_k141_81988||full Not-determined
S12_vs_k141_105933||full Not-determined
S12_vs_k141_34310||full Low-quality
S12_vs_k141_19014||full Low-quality
S12_vs_k141_43160||full Not-determined
S12_vs_k141_137101||full Low-quality
S12_vs_k141_82008||full Low-quality
S12_vs_k141_50061||full Not-determined
S12_vs_k141_34325||full Not-determined
S12_vs_k141_82010||full Not-determined
S12_vs_k141_168628||full Not-determined
S12_vs_k141_160426||full Low-quality
S12_vs_k141_3641||full Not-determined
S12_vs_k141_168638||full Low-quality
S12_vs_k141_98457||full Not-determined
S12_vs_k141_10498||full Not-determined
S12_vs_k141_176013||full Low-quality
S12_vs_k141_176014||full Low-quality
S12_vs_k141_137123||full Not-determined
S12_vs_k141_184529||full Low-quality
S12_vs_k141_50094||full Not-determined
S12_vs_k141_10514||full Low-quality
S12_vs_k141_137127||full Low-quality
S12_vs_k141_176030||full Not-determined
S12_vs_k141_137132||full Low-quality
S12_vs_k141_90420||full Low-quality
S12_vs_k141_10517||full Low-quality
S12_vs_k141_82040||full Not-determined
S12_vs_k141_176037||full Low-quality
S12_vs_k141_26914||full Not-determined
S12_vs_k141_105976||full Low-quality
S12_vs_k141_90431||full Low-quality
S12_vs_k141_26921||full Not-determined
S12_vs_k141_34370||full Low-quality
S12_vs_k141_3686||full Low-quality
S12_vs_k141_82054||full Not-determined
S12_vs_k141_66480||full Low-quality
S12_vs_k141_10531||full Not-determined
S12_vs_k141_73808||full Low-quality
S12_vs_k141_34383||full Not-determined
S12_vs_k141_34385||full Not-determined
S12_vs_k141_43216||full Not-determined
S12_vs_k141_26933||full Low-quality
S12_vs_k141_43222||full Low-quality
S12_vs_k141_50147||full Not-determined
S12_vs_k141_10551||full Low-quality
S12_vs_k141_90464||full Not-determined
S12_vs_k141_50154||full Low-quality
S12_vs_k141_19081||full Low-quality
S12_vs_k141_90470||full Not-determined
S12_vs_k141_152560||full Not-determined
S12_vs_k141_90478||full Not-determined
S12_vs_k141_3748||full Not-determined
S12_vs_k141_176097||full Low-quality
S12_vs_k141_3756||full Low-quality
S12_vs_k141_152579||full Not-determined
S12_vs_k141_82080||full Low-quality
S12_vs_k141_82084||full Low-quality
S7_extracted_114476 Medium-quality
S12_vs_k141_82092||full Low-quality
S12_vs_k141_26965||full Not-determined
S12_vs_k141_152583||full Low-quality
S12_vs_k141_160546||full Low-quality
S12_vs_k141_73837||full Not-determined
S12_vs_k141_106083||full Low-quality
S12_vs_k141_106089||full Low-quality
S12_vs_k141_90521||full Low-quality
S12_vs_k141_113261||full Low-quality
S12_vs_k141_90526||full Low-quality
S12_vs_k141_184648||full Not-determined
S12_vs_k141_82128||full Low-quality
S12_vs_k141_121160||full Not-determined
S12_vs_k141_50241||full Not-determined
S12_vs_k141_160583||full Not-determined
S12_vs_k141_121181||full Low-quality
S12_vs_k141_58716||full Low-quality
S12_vs_k141_73883||full Low-quality
S12_vs_k141_98571||full Low-quality
S12_vs_k141_145110||full Low-quality
S12_vs_k141_176170||full Low-quality
S12_vs_k141_152623||full Not-determined
S12_vs_k141_184681||full Low-quality
S12_vs_k141_98580||full Not-determined
S12_vs_k141_3846||full Not-determined
S12_vs_k141_137258||full Low-quality
S12_vs_k141_145115||full Low-quality
S12_vs_k141_90578||full Low-quality
S12_vs_k141_137265||full Low-quality
S12_vs_k141_58719||full Low-quality
S12_vs_k141_145118||full Not-determined
S12_vs_k141_90586||full Low-quality
S12_vs_k141_145131||full Not-determined
S12_vs_k141_121220||full Low-quality
S12_vs_k141_10668||full Not-determined
S12_vs_k141_184701||full Low-quality
S12_vs_k141_3872||full Not-determined
S12_vs_k141_73916||full Low-quality
S12_vs_k141_98623||full Not-determined
S12_vs_k141_90606||full Low-quality
S5_vs_k141_83886||full Medium-quality
S12_vs_k141_19164||full Low-quality
S12_vs_k141_129036||full Not-determined
S12_vs_k141_106180||full Low-quality
S12_vs_k141_152642||full Low-quality
S12_vs_k141_34398||full Not-determined
S12_vs_k141_152646||full Low-quality
S12_vs_k141_34409||full Low-quality
S12_vs_k141_184717||full Low-quality
S12_vs_k141_27033||full Low-quality
S12_vs_k141_98652||full Low-quality
S12_vs_k141_152653||full Low-quality
S12_vs_k141_98660||full Low-quality
S12_vs_k141_152663||full Not-determined
S12_vs_k141_3912||full Low-quality
S12_vs_k141_98669||full Low-quality
S12_vs_k141_121243||full Low-quality
S12_vs_k141_113295||full Not-determined
S12_vs_k141_19188||full Low-quality
S12_vs_k141_152690||full Not-determined
S12_vs_k141_3913||full Low-quality
S12_vs_k141_106197||full Low-quality
S12_vs_k141_98680||full Low-quality
S12_vs_k141_98683||full Low-quality
S12_vs_k141_10752||full Not-determined
S12_vs_k141_34450||full Low-quality
S12_vs_k141_152693||full Low-quality
S12_vs_k141_27075||full Not-determined
S12_vs_k141_129093||full Not-determined
S12_vs_k141_184774||full Not-determined
S12_vs_k141_27078||full Not-determined
S12_vs_k141_19217||full Not-determined
S12_vs_k141_176283||full Not-determined
S12_vs_k141_73967||full Low-quality
S12_vs_k141_98715||full Not-determined
S12_vs_k141_73970||full Low-quality
S12_vs_k141_66534||full Not-determined
S12_vs_k141_82177||full Low-quality
S12_vs_k141_58827||full Low-quality
S12_vs_k141_50362||full Not-determined
S12_vs_k141_129122||full Not-determined
S12_vs_k141_152749||full Not-determined
S12_vs_k141_145239||full Low-quality
S12_vs_k141_176337||full Low-quality
S12_vs_k141_73982||full Low-quality
S12_vs_k141_137405||full Not-determined
S12_vs_k141_27123||full Not-determined
S12_vs_k141_98769||full Low-quality
S12_vs_k141_73994||full Low-quality
S12_vs_k141_145269||full Not-determined
S12_vs_k141_27137||full Not-determined
S12_vs_k141_129156||full Low-quality
S12_vs_k141_113402||full Not-determined
S12_vs_k141_3939||full Low-quality
S12_vs_k141_106215||full Not-determined
S12_vs_k141_176370||full Not-determined
S12_vs_k141_3954||full Low-quality
S12_vs_k141_113421||full Low-quality
S12_vs_k141_27178||full Not-determined
S12_vs_k141_106235||full Not-determined
S12_vs_k141_82228||full Low-quality
S12_vs_k141_10824||full Not-determined
S12_vs_k141_129202||full Not-determined
S12_vs_k141_160698||full Not-determined
S12_vs_k141_58894||full Low-quality
S12_vs_k141_10831||full Not-determined
S12_vs_k141_27194||full Low-quality
S12_vs_k141_82242||full Not-determined
S12_vs_k141_176390||full Not-determined
S12_vs_k141_152798||full Low-quality
S12_vs_k141_98797||full Low-quality
S12_vs_k141_10843||full Not-determined
S12_vs_k141_34493||full Low-quality
S12_vs_k141_58916||full Not-determined
S12_vs_k141_145279||full Not-determined
S12_vs_k141_66565||full Not-determined
S12_vs_k141_137474||full Low-quality
S12_vs_k141_19292||full Not-determined
S12_vs_k141_106281||full Not-determined
S12_vs_k141_50448||full Not-determined
S12_vs_k141_184850||full Not-determined
S12_vs_k141_113472||full Not-determined
S12_vs_k141_66575||full Not-determined
S12_vs_k141_66576||full Low-quality
S12_vs_k141_4004||full Low-quality
S12_vs_k141_184860||full Not-determined
S12_vs_k141_113484||full Low-quality
S12_vs_k141_4007||full Not-determined
S12_vs_k141_152838||full Low-quality
S12_vs_k141_4008||full Low-quality
S12_vs_k141_98841||full Not-determined
S12_vs_k141_152840||full Not-determined
S12_vs_k141_82267||full Not-determined
S12_vs_k141_145328||full Not-determined
S12_vs_k141_82275||full Low-quality
S12_vs_k141_66579||full Low-quality
S12_vs_k141_184886||full Low-quality
S12_vs_k141_4023||full Not-determined
S12_vs_k141_90618||full Not-determined
S12_vs_k141_152852||full Low-quality
S12_vs_k141_113497||full Low-quality
S12_vs_k141_145343||full Not-determined
S12_vs_k141_74070||full Low-quality
S12_vs_k141_98849||full Low-quality
S12_vs_k141_82283||full Low-quality
S12_vs_k141_43533||full Not-determined
S12_vs_k141_129309||full Low-quality
S12_vs_k141_121367||full Low-quality
S12_vs_k141_168807||full Not-determined
S12_vs_k141_152860||full Low-quality
S12_vs_k141_43549||full Not-determined
S12_vs_k141_121377||full Low-quality
S12_vs_k141_152861||full Not-determined
S12_vs_k141_82305||full Low-quality
S12_vs_k141_121385||full Low-quality
S12_vs_k141_152868||full Not-determined
S12_vs_k141_137557||full Low-quality
S12_vs_k141_66620||full Not-determined
S12_vs_k141_129333||full Low-quality
S12_vs_k141_137559||full Low-quality
S12_vs_k141_184904||full Not-determined
S12_vs_k141_145385||full Low-quality
S12_vs_k141_50523||full Not-determined
S12_vs_k141_50526||full Low-quality
S12_vs_k141_145388||full Low-quality
S12_vs_k141_98905||full Not-determined
S12_vs_k141_27315||full Not-determined
S12_vs_k141_50539||full Low-quality
S12_vs_k141_34572||full Not-determined
S12_vs_k141_58997||full Not-determined
S11_vs_k141_26767||full Medium-quality
S12_vs_k141_34579||full Not-determined
S12_vs_k141_27326||full Low-quality
S12_vs_k141_50559||full Low-quality
S12_vs_k141_66652||full Not-determined
S12_vs_k141_82348||full Not-determined
S12_vs_k141_106336||full Low-quality
S12_vs_k141_82349||full Not-determined
S12_vs_k141_50565||full Low-quality
S12_vs_k141_74147||full Not-determined
S12_vs_k141_82360||full Not-determined
S12_vs_k141_98943||full Low-quality
S12_vs_k141_34612||full Low-quality
S12_vs_k141_43618||full Not-determined
S12_vs_k141_59018||full Low-quality
S12_vs_k141_184930||full Low-quality
S12_vs_k141_43620||full Not-determined
S12_vs_k141_27353||full Not-determined
S12_vs_k141_82365||full Low-quality
S12_vs_k141_129369||full Not-determined
S12_vs_k141_106343||full Low-quality
S12_vs_k141_176426||full Not-determined
S12_vs_k141_34627||full Low-quality
S12_vs_k141_19378||full Low-quality
S12_vs_k141_98946||full Not-determined
S12_vs_k141_176428||full Not-determined
S12_vs_k141_10901||full Low-quality
S12_vs_k141_34634||full Low-quality
S12_vs_k141_160747||full Low-quality
S12_vs_k141_129382||full Not-determined
S12_vs_k141_129383||full Not-determined
S12_vs_k141_66690||full Low-quality
S12_vs_k141_152912||full Low-quality
S12_vs_k141_34638||full Not-determined
S12_vs_k141_74191||full Low-quality
S12_vs_k141_129392||full Not-determined
S12_vs_k141_43668||full Not-determined
S12_extracted_k141_2062 Medium-quality
S12_vs_k141_2062||full Medium-quality
S12_vs_k141_74206||full Low-quality
S12_vs_k141_152926||full Low-quality
S12_vs_k141_129407||full Not-determined
S12_vs_k141_4039||full Not-determined
S12_vs_k141_152934||full Low-quality
S12_vs_k141_82399||full Low-quality
S12_vs_k141_137626||full Low-quality
S12_vs_k141_74216||full Not-determined
S12_vs_k141_66734||full Not-determined
S12_vs_k141_82409||full Low-quality
S12_vs_k141_50636||full Low-quality
S12_vs_k141_121486||full Not-determined
S12_vs_k141_168926||full Not-determined
S12_vs_k141_184996||full Low-quality
S12_vs_k141_82412||full Not-determined
S12_vs_k141_129427||full Low-quality
S12_vs_k141_27410||full Low-quality
S12_vs_k141_98994||full Low-quality
S12_vs_k141_137639||full Low-quality
S12_vs_k141_185004||full Low-quality
S12_vs_k141_152958||full Low-quality
S12_vs_k141_90736||full Low-quality
S12_vs_k141_66757||full Low-quality
S12_vs_k141_50639||full Not-determined
S12_vs_k141_66764||full Low-quality
S12_vs_k141_82435||full Low-quality
S12_vs_k141_43679||full Low-quality
S12_vs_k141_27444||full Low-quality
S12_vs_k141_168939||full Low-quality
S12_vs_k141_137656||full Low-quality
S12_vs_k141_185031||full Low-quality
S12_vs_k141_152971||full Not-determined
S12_vs_k141_59085||full Low-quality
S12_vs_k141_145481||full Not-determined
S12_vs_k141_185039||full Low-quality
S12_vs_k141_34703||full Low-quality
S12_vs_k141_10980||full Low-quality
S12_vs_k141_185040||full Not-determined
S12_vs_k141_90746||full Not-determined
S12_vs_k141_185041||full Low-quality
S12_vs_k141_168952||full Low-quality
S12_vs_k141_129444||full Low-quality
S12_vs_k141_50658||full Low-quality
S12_vs_k141_121521||full Low-quality
S12_vs_k141_99010||full Low-quality
S12_vs_k141_74274||full Low-quality
S12_vs_k141_145500||full Not-determined
S12_vs_k141_153007||full Low-quality
S12_vs_k141_82464||full Not-determined
S12_vs_k141_59096||full Low-quality
S12_vs_k141_168964||full Low-quality
S12_vs_k141_106429||full Not-determined
S12_vs_k141_99023||full Not-determined
S12_vs_k141_90793||full Not-determined
S10_extracted_15561 Medium-quality
S12_vs_k141_90794||full Low-quality
S12_vs_k141_50702||full Not-determined
S12_vs_k141_121566||full Low-quality
S12_vs_k141_4091||full Low-quality
S12_vs_k141_34743||full Low-quality
S12_vs_k141_4094||full Not-determined
S12_vs_k141_137711||full Not-determined
S12_vs_k141_66813||full Low-quality
S12_vs_k141_74291||full Not-determined
S12_vs_k141_168983||full Low-quality
S12_vs_k141_27490||full Low-quality
S12_vs_k141_129475||full Low-quality
S12_vs_k141_113626||full Low-quality
S12_vs_k141_153035||full Low-quality
S12_vs_k141_66844||full Low-quality
S12_vs_k141_4125||full Low-quality
S12_vs_k141_59144||full Not-determined
S12_vs_k141_27519||full Not-determined
S12_vs_k141_19504||full Low-quality
S12_vs_k141_34775||full Low-quality
S12_vs_k141_121614||full Not-determined
S12_vs_k141_34777||full Low-quality
S12_vs_k141_27528||full Low-quality
S12_vs_k141_121629||full Low-quality
S12_vs_k141_106498||full Low-quality
S12_vs_k141_50738||full Low-quality
S12_vs_k141_99059||full Not-determined
S12_vs_k141_121634||full Not-determined
S12_vs_k141_82471||full Not-determined
S12_vs_k141_160894||full Not-determined
S12_vs_k141_59182||full Low-quality
S12_vs_k141_59183||full Low-quality
S12_vs_k141_113662||full Not-determined
S12_vs_k141_99077||full Not-determined
S12_vs_k141_106518||full Not-determined
S12_vs_k141_129490||full Low-quality
S12_vs_k141_169047||full Not-determined
S12_vs_k141_129492||full Low-quality
S12_vs_k141_106521||full Not-determined
S12_vs_k141_59203||full Low-quality
S12_vs_k141_153097||full Low-quality
S12_vs_k141_82500||full Not-determined
S12_vs_k141_27571||full Not-determined
S12_vs_k141_50769||full Not-determined
S12_vs_k141_27579||full Low-quality
S12_vs_k141_121671||full Low-quality
S12_vs_k141_43795||full Not-determined
S12_vs_k141_153118||full Not-determined
S12_vs_k141_11026||full Not-determined
S12_vs_k141_82512||full Not-determined
S12_vs_k141_106556||full Low-quality
S12_vs_k141_11033||full Not-determined
S12_vs_k141_169096||full Low-quality
S12_vs_k141_19549||full Not-determined
S12_vs_k141_160949||full Low-quality
S10_vs_15561||full Medium-quality
S12_vs_k141_19554||full Low-quality
S12_vs_k141_74307||full Not-determined
S12_vs_k141_82534||full Not-determined
S12_vs_k141_59238||full Not-determined
S12_vs_k141_185231||full Low-quality
S12_vs_k141_27606||full Not-determined
S12_vs_k141_137839||full Low-quality
S12_vs_k141_11075||full Low-quality
S12_vs_k141_90939||full Low-quality
S12_vs_k141_169126||full Low-quality
S12_vs_k141_145672||full Not-determined
S12_vs_k141_27635||full Low-quality
S12_vs_k141_145681||full Not-determined
S12_vs_k141_34892||full Not-determined
S12_vs_k141_66887||full Not-determined
S12_vs_k141_59266||full Low-quality
S12_vs_k141_27640||full Not-determined
S12_vs_k141_160991||full Not-determined
S12_vs_k141_59271||full Not-determined
S12_vs_k141_145696||full Not-determined
S12_vs_k141_99141||full Low-quality
S12_vs_k141_137860||full Low-quality
S12_vs_k141_153207||full Low-quality
S12_vs_k141_27654||full Not-determined
S12_vs_k141_74370||full Not-determined
S12_vs_k141_99143||full Low-quality
S12_vs_k141_185295||full Low-quality
S12_vs_k141_106623||full Low-quality
S12_vs_k141_137879||full Low-quality
S12_vs_k141_34932||full Low-quality
S12_vs_k141_74378||full Low-quality
S12_vs_k141_50836||full Low-quality
S12_vs_k141_137880||full Low-quality
S12_vs_k141_50837||full Low-quality
S12_vs_k141_43884||full Not-determined
S12_vs_k141_129552||full Not-determined
S12_vs_k141_34943||full Low-quality
S12_vs_k141_59298||full Not-determined
S12_vs_k141_11131||full Low-quality
S12_vs_k141_99167||full Low-quality
S12_vs_k141_137893||full Not-determined
S12_vs_k141_50851||full Low-quality
S12_vs_k141_59306||full Low-quality
S12_vs_k141_43898||full Low-quality
S12_vs_k141_137900||full Low-quality
S12_vs_k141_11139||full Low-quality
S12_vs_k141_27676||full Not-determined
S12_vs_k141_27679||full Not-determined
S12_vs_k141_27683||full Not-determined
S12_vs_k141_99179||full Low-quality
S12_vs_k141_66917||full Low-quality
S12_vs_k141_59320||full Not-determined
S12_vs_k141_4267||full Low-quality
S12_vs_k141_74426||full Low-quality
S12_vs_k141_34952||full Not-determined
S12_vs_k141_145728||full Low-quality
S12_vs_k141_50874||full Not-determined
S12_vs_k141_99201||full Low-quality
S12_vs_k141_82633||full Not-determined
S12_vs_k141_50880||full Not-determined
S12_vs_k141_34957||full Not-determined
S12_vs_k141_185321||full Low-quality
S12_vs_k141_74441||full Not-determined
S12_vs_k141_185328||full Low-quality
S12_vs_k141_91000||full Not-determined
S12_vs_k141_137938||full Not-determined
S12_vs_k141_27728||full Not-determined
S12_vs_k141_34975||full Low-quality
S12_vs_k141_161121||full Low-quality
S12_vs_k141_59357||full Not-determined
S12_vs_k141_82666||full Not-determined
S12_vs_k141_43953||full Low-quality
S12_vs_k141_27754||full Low-quality
S12_vs_k141_66985||full Low-quality
S12_vs_k141_91009||full Low-quality
S12_vs_k141_161154||full Low-quality
S12_vs_k141_113737||full Low-quality
S12_vs_k141_4334||full Not-determined
S12_vs_k141_4354||full Low-quality
S12_vs_k141_50950||full Low-quality
S12_vs_k141_185343||full Not-determined
S12_vs_k141_59399||full Low-quality
S12_vs_k141_153323||full Low-quality
S12_vs_k141_82718||full Low-quality
S12_vs_k141_185361||full Not-determined
S12_vs_k141_91060||full Low-quality
S12_vs_k141_82719||full Not-determined
S12_vs_k141_145808||full Low-quality
S12_vs_k141_91070||full Low-quality
S7_vs_114476||full Medium-quality
S12_vs_k141_113789||full Not-determined
S12_vs_k141_185376||full Low-quality
S12_vs_k141_74496||full Not-determined
S12_vs_k141_74501||full Low-quality
S12_vs_k141_59420||full Not-determined
S12_vs_k141_99227||full Low-quality
S12_vs_k141_50982||full Low-quality
S12_vs_k141_113804||full Not-determined
S12_vs_k141_67027||full Not-determined
S12_vs_k141_138042||full Low-quality
S12_vs_k141_185384||full Low-quality
S12_vs_k141_161265||full Low-quality
S12_vs_k141_35073||full Not-determined
S12_vs_k141_4430||full Low-quality
S12_vs_k141_4435||full Low-quality
S12_vs_k141_129588||full Low-quality
S12_vs_k141_138066||full Low-quality
S12_vs_k141_91122||full Low-quality
S12_vs_k141_59459||full Low-quality
S12_vs_k141_74553||full Not-determined
S12_vs_k141_129592||full Low-quality
S12_vs_k141_67069||full Low-quality
S12_vs_k141_145891||full Not-determined
S12_vs_k141_169276||full Not-determined
S12_vs_k141_185423||full Not-determined
S12_vs_k141_51011||full Low-quality

| | |
|---|---|
| S12_vs_k141_44086\|\|full | Low-quality |
| S12_vs_k141_59462\|\|full | Not-determined |
| S12_vs_k141_19603\|\|full | Low-quality |
| S12_vs_k141_35096\|\|full | Low-quality |
| S12_vs_k141_51017\|\|full | Not-determined |
| S12_vs_k141_106722\|\|full | Low-quality |
| S12_vs_k141_106723\|\|full | Low-quality |
| S12_vs_k141_106727\|\|full | Not-determined |
| S12_vs_k141_51022\|\|full | Not-determined |
| S12_vs_k141_44101\|\|full | Not-determined |
| S12_vs_k141_161295\|\|full | Low-quality |
| S12_vs_k141_91161\|\|full | Not-determined |
| S12_vs_k141_82809\|\|full | Low-quality |
| S12_vs_k141_129616\|\|full | Low-quality |
| S12_vs_k141_51036\|\|full | Low-quality |
| S12_vs_k141_74619\|\|full | Not-determined |
| S12_vs_k141_185475\|\|full | Low-quality |
| S12_vs_k141_106748\|\|full | Low-quality |
| S12_vs_k141_113857\|\|full | Not-determined |
| S12_vs_k141_185480\|\|full | Low-quality |
| S12_vs_k141_4461\|\|full | Low-quality |
| S12_vs_k141_113864\|\|full | Not-determined |
| S12_vs_k141_161328\|\|full | Not-determined |
| S12_vs_k141_67169\|\|full | Not-determined |
| S12_vs_k141_74645\|\|full | Low-quality |
| S12_vs_k141_51079\|\|full | Not-determined |
| S12_vs_k141_4480\|\|full | Not-determined |
| S12_vs_k141_99278\|\|full | Low-quality |
| S12_vs_k141_138100\|\|full | Low-quality |
| S12_vs_k141_113890\|\|full | Low-quality |
| S12_vs_k141_99281\|\|full | Not-determined |
| S12_vs_k141_176525\|\|full | Not-determined |
| S12_vs_k141_27845\|\|full | Low-quality |
| S12_vs_k141_82831\|\|full | Low-quality |
| S12_vs_k141_106785\|\|full | Low-quality |
| S12_vs_k141_145998\|\|full | Low-quality |
| S12_vs_k141_59513\|\|full | Low-quality |
| S12_vs_k141_35158\|\|full | Not-determined |
| S12_vs_k141_91239\|\|full | Not-determined |
| S12_vs_k141_27863\|\|full | Low-quality |
| S12_vs_k141_99308\|\|full | Low-quality |
| S12_vs_k141_59519\|\|full | Not-determined |
| S12_vs_k141_169357\|\|full | Low-quality |
| S12_vs_k141_106815\|\|full | Low-quality |
| S12_vs_k141_113928\|\|full | Not-determined |
| S12_vs_k141_35171\|\|full | Low-quality |
| S12_vs_k141_11270\|\|full | Not-determined |
| S12_vs_k141_113935\|\|full | Not-determined |
| S12_vs_k141_59532\|\|full | Low-quality |
| S12_vs_k141_138125\|\|full | Not-determined |
| S12_vs_k141_59534\|\|full | Low-quality |
| S12_vs_k141_106826\|\|full | Not-determined |
| S12_vs_k141_11273\|\|full | Low-quality |
| S12_vs_k141_74689\|\|full | Not-determined |
| S12_vs_k141_35195\|\|full | Not-determined |
| S12_vs_k141_27884\|\|full | Low-quality |
| S12_vs_k141_11280\|\|full | Low-quality |
| S12_vs_k141_19655\|\|full | Low-quality |
| S12_vs_k141_169385\|\|full | Not-determined |
| S12_vs_k141_129684\|\|full | Low-quality |
| S12_vs_k141_161395\|\|full | Not-determined |
| S12_vs_k141_185553\|\|full | Low-quality |
| S12_vs_k141_146027\|\|full | Low-quality |
| S12_vs_k141_138156\|\|full | Low-quality |
| S12_vs_k141_146033\|\|full | Low-quality |
| S12_vs_k141_146034\|\|full | Not-determined |
| S12_vs_k141_185569\|\|full | Low-quality |
| S12_vs_k141_11290\|\|full | Low-quality |
| S12_vs_k141_113966\|\|full | Low-quality |
| S12_vs_k141_169406\|\|full | Not-determined |
| S12_vs_k141_91283\|\|full | Not-determined |
| S12_vs_k141_129707\|\|full | Low-quality |
| S12_vs_k141_106884\|\|full | Low-quality |
| S12_vs_k141_176594\|\|full | Not-determined |
| S12_vs_k141_113981\|\|full | Not-determined |
| S10_vs_36407\|\|full | Medium-quality |
| S12_vs_k141_185588\|\|full | Low-quality |
| S12_vs_k141_121792\|\|full | Not-determined |
| S12_vs_k141_176604\|\|full | Low-quality |
| S12_vs_k141_27916\|\|full | Low-quality |
| S12_vs_k141_138208\|\|full | Not-determined |
| S12_vs_k141_67197\|\|full | Not-determined |
| S12_vs_k141_99325\|\|full | Not-determined |
| S12_vs_k141_59622\|\|full | Low-quality |
| S12_vs_k141_91309\|\|full | Low-quality |
| S12_vs_k141_67205\|\|full | Not-determined |
| S12_vs_k141_99338\|\|full | Low-quality |
| S12_vs_k141_27934\|\|full | Low-quality |
| S12_vs_k141_121803\|\|full | Low-quality |
| S12_vs_k141_185618\|\|full | Not-determined |
| S12_vs_k141_153509\|\|full | Not-determined |
| S12_vs_k141_121807\|\|full | Low-quality |
| S12_vs_k141_114017\|\|full | Not-determined |
| S12_vs_k141_4617\|\|full | Not-determined |
| S12_vs_k141_121808\|\|full | Not-determined |
| S12_vs_k141_91329\|\|full | Low-quality |
| S12_vs_k141_106949\|\|full | Low-quality |
| S12_vs_k141_121816\|\|full | Low-quality |
| S12_vs_k141_185636\|\|full | Not-determined |
| S12_vs_k141_114046\|\|full | Low-quality |
| S12_vs_k141_99363\|\|full | Low-quality |
| S12_vs_k141_176655\|\|full | Low-quality |
| S12_vs_k141_27949\|\|full | Low-quality |
| S12_vs_k141_91335\|\|full | Low-quality |
| S12_vs_k141_35218\|\|full | Low-quality |
| S12_vs_k141_82922\|\|full | Not-determined |
| S12_vs_k141_114053\|\|full | Low-quality |
| S12_vs_k141_185648\|\|full | Low-quality |
| S12_vs_k141_138257\|\|full | Low-quality |
| S12_vs_k141_161472\|\|full | Low-quality |
| S12_vs_k141_35241\|\|full | Not-determined |
| S12_vs_k141_19710\|\|full | Low-quality |
| S12_vs_k141_4640\|\|full | Not-determined |
| S12_vs_k141_121834\|\|full | Low-quality |
| S12_vs_k141_67275\|\|full | Low-quality |
| S12_vs_k141_74814\|\|full | Not-determined |
| S12_vs_k141_19715\|\|full | Not-determined |
| S12_vs_k141_176684\|\|full | Low-quality |
| S12_vs_k141_35254\|\|full | Low-quality |
| S12_vs_k141_176687\|\|full | Not-determined |
| S12_vs_k141_74822\|\|full | Low-quality |
| S12_vs_k141_185664\|\|full | Not-determined |
| S12_vs_k141_99395\|\|full | Not-determined |
| S12_vs_k141_35259\|\|full | Not-determined |
| S12_vs_k141_35260\|\|full | Low-quality |
| S12_vs_k141_161495\|\|full | Not-determined |
| S12_vs_k141_4647\|\|full | Low-quality |
| S12_vs_k141_44263\|\|full | Low-quality |
| S12_vs_k141_11357\|\|full | Not-determined |
| S12_vs_k141_67301\|\|full | Not-determined |
| S12_vs_k141_4653\|\|full | Low-quality |
| S12_vs_k141_114108\|\|full | Low-quality |
| S12_vs_k141_121866\|\|full | Low-quality |
| S12_vs_k141_185689\|\|full | Not-determined |
| S12_vs_k141_176706\|\|full | Low-quality |
| S12_vs_k141_91371\|\|full | Not-determined |
| S12_vs_k141_44278\|\|full | Low-quality |
| S12_vs_k141_107027\|\|full | Not-determined |
| S12_vs_k141_99415\|\|full | Low-quality |
| S12_vs_k141_59687\|\|full | Not-determined |
| S12_vs_k141_44286\|\|full | Not-determined |
| S12_vs_k141_51157\|\|full | Low-quality |
| S12_vs_k141_185723\|\|full | Not-determined |
| S12_vs_k141_114142\|\|full | Low-quality |
| S12_vs_k141_176722\|\|full | Not-determined |
| S12_vs_k141_129795\|\|full | Low-quality |
| S12_vs_k141_169461\|\|full | Low-quality |
| S12_vs_k141_4673\|\|full | Not-determined |
| S11_vs_k141_66101\|\|full | Medium-quality |
| S12_vs_k141_161554\|\|full | Not-determined |
| S12_vs_k141_19778\|\|full | Low-quality |
| S12_vs_k141_169469\|\|full | Not-determined |
| S12_vs_k141_138267\|\|full | Low-quality |
| S12_vs_k141_121897\|\|full | Not-determined |
| S12_vs_k141_28003\|\|full | Low-quality |
| S12_vs_k141_107084\|\|full | Low-quality |
| S12_vs_k141_19799\|\|full | Low-quality |
| S12_vs_k141_138277\|\|full | Low-quality |
| S12_vs_k141_107093\|\|full | Low-quality |
| S12_vs_k141_185727\|\|full | Not-determined |
| S12_vs_k141_91452\|\|full | Not-determined |
| S12_vs_k141_4700\|\|full | Low-quality |
| S12_vs_k141_59726\|\|full | Low-quality |
| S12_vs_k141_4701\|\|full | Low-quality |
| S12_vs_k141_82989\|\|full | Not-determined |
| S12_vs_k141_185732\|\|full | Not-determined |
| S12_vs_k141_44348\|\|full | Not-determined |
| S12_vs_k141_114181\|\|full | Not-determined |
| S12_vs_k141_11433\|\|full | Not-determined |
| S12_vs_k141_185749\|\|full | Low-quality |
| S12_vs_k141_129855\|\|full | Low-quality |
| S12_vs_k141_176752\|\|full | Low-quality |
| S12_vs_k141_161611\|\|full | Low-quality |
| S12_vs_k141_114198\|\|full | Low-quality |
| S12_vs_k141_185763\|\|full | Low-quality |
| S12_vs_k141_19818\|\|full | Low-quality |
| S12_vs_k141_114207\|\|full | Not-determined |
| S12_vs_k141_99487\|\|full | Low-quality |
| S12_vs_k141_129872\|\|full | Not-determined |
| S12_vs_k141_44403\|\|full | Not-determined |
| S12_vs_k141_67373\|\|full | Low-quality |
| S12_vs_k141_11461\|\|full | Low-quality |
| S12_vs_k141_161648\|\|full | Low-quality |
| S12_vs_k141_51216\|\|full | Low-quality |
| S12_vs_k141_11464\|\|full | Low-quality |
| S12_vs_k141_169535\|\|full | Low-quality |
| S12_vs_k141_91465\|\|full | Low-quality |
| S12_vs_k141_67380\|\|full | Not-determined |
| S12_vs_k141_91467\|\|full | Low-quality |
| S12_vs_k141_44414\|\|full | Not-determined |
| S12_vs_k141_99507\|\|full | Low-quality |
| S12_vs_k141_35342\|\|full | Low-quality |
| S12_vs_k141_169548\|\|full | Not-determined |
| S12_vs_k141_138320\|\|full | Low-quality |
| S12_vs_k141_99524\|\|full | Low-quality |
| S12_vs_k141_169549\|\|full | Low-quality |
| S12_vs_k141_74927\|\|full | Not-determined |
| S12_vs_k141_59814\|\|full | Not-determined |
| S12_vs_k141_83019\|\|full | Not-determined |
| S12_vs_k141_59815\|\|full | Not-determined |
| S12_vs_k141_51237\|\|full | Not-determined |
| S12_vs_k141_185826\|\|full | Low-quality |
| S12_vs_k141_146257\|\|full | Low-quality |
| S12_vs_k141_28124\|\|full | Low-quality |
| S12_vs_k141_153697\|\|full | Low-quality |
| S12_vs_k141_176805\|\|full | Low-quality |
| S12_vs_k141_59826\|\|full | Low-quality |
| S12_vs_k141_51258\|\|full | Not-determined |
| S12_vs_k141_51264\|\|full | Low-quality |
| S12_vs_k141_59832\|\|full | Low-quality |
| S12_vs_k141_129931\|\|full | Not-determined |
| S12_vs_k141_4764\|\|full | Not-determined |
| S12_vs_k141_19881\|\|full | Not-determined |
| S12_vs_k141_99560\|\|full | Not-determined |
| S12_vs_k141_176810\|\|full | Not-determined |
| S12_vs_k141_138364\|\|full | Not-determined |
| S12_vs_k141_4771\|\|full | Low-quality |
| S12_vs_k141_19889\|\|full | Low-quality |
| S12_vs_k141_107188\|\|full | Low-quality |
| S12_vs_k141_51276\|\|full | Low-quality |
| S12_vs_k141_74958\|\|full | Not-determined |
| S12_vs_k141_44477\|\|full | Low-quality |
| S12_vs_k141_4775\|\|full | Low-quality |
| S12_vs_k141_107193\|\|full | Low-quality |
| S12_vs_k141_35388\|\|full | Low-quality |
| S12_vs_k141_51284\|\|full | Not-determined |
| S12_vs_k141_107194\|\|full | Not-determined |
| S12_vs_k141_35392\|\|full | Not-determined |
| S12_vs_k141_74969\|\|full | Low-quality |
| S12_vs_k141_107198\|\|full | Low-quality |
| S12_vs_k141_129953\|\|full | Not-determined |
| S12_vs_k141_153731\|\|full | Low-quality |
| S12_vs_k141_138393\|\|full | Not-determined |
| S12_vs_k141_176841\|\|full | Low-quality |
| S12_vs_k141_19914\|\|full | Low-quality |
| S12_vs_k141_35422\|\|full | Low-quality |
| S12_vs_k141_74982\|\|full | Low-quality |
| S12_vs_k141_51315\|\|full | Low-quality |
| S12_vs_k141_74985\|\|full | Low-quality |
| S12_vs_k141_138405\|\|full | Low-quality |
| S12_vs_k141_153765\|\|full | Not-determined |
| S7_vs_62347\|\|full | Medium-quality |
| S12_vs_k141_129971\|\|full | Not-determined |
| S12_vs_k141_4810\|\|full | Low-quality |
| S12_vs_k141_153776\|\|full | Not-determined |
| S12_vs_k141_176886\|\|full | Low-quality |
| S12_vs_k141_91581\|\|full | Not-determined |
| S12_vs_k141_91585\|\|full | Not-determined |
| S12_vs_k141_75009\|\|full | Low-quality |
| S12_vs_k141_185883\|\|full | Not-determined |
| S12_vs_k141_176899\|\|full | Not-determined |
| S12_vs_k141_59909\|\|full | Not-determined |
| S12_vs_k141_114326\|\|full | Not-determined |
| S12_vs_k141_28168\|\|full | Low-quality |
| S12_vs_k141_176904\|\|full | Not-determined |
| S12_vs_k141_130005\|\|full | Low-quality |
| S12_vs_k141_28176\|\|full | Low-quality |
| S12_vs_k141_130012\|\|full | Not-determined |
| S12_vs_k141_176911\|\|full | Low-quality |
| S12_vs_k141_19971\|\|full | Low-quality |
| S12_vs_k141_185901\|\|full | Not-determined |
| S12_vs_k141_138447\|\|full | Low-quality |
| S12_vs_k141_138448\|\|full | Low-quality |
| S12_vs_k141_176923\|\|full | Low-quality |
| S12_vs_k141_11563\|\|full | Low-quality |
| S12_vs_k141_59936\|\|full | Not-determined |
| S12_vs_k141_161809\|\|full | Low-quality |
| S12_vs_k141_161825\|\|full | Not-determined |
| S12_vs_k141_91621\|\|full | Low-quality |
| S12_vs_k141_114370\|\|full | Low-quality |
| S12_vs_k141_161827\|\|full | Low-quality |
| S12_vs_k141_121985\|\|full | Not-determined |
| S12_vs_k141_91627\|\|full | Not-determined |
| S12_vs_k141_35497\|\|full | Low-quality |
| S12_vs_k141_185943\|\|full | Not-determined |
| S12_vs_k141_130025\|\|full | Low-quality |
| S12_vs_k141_161838\|\|full | Low-quality |
| S12_vs_k141_67553\|\|full | Low-quality |
| S12_vs_k141_114391\|\|full | Low-quality |
| S12_vs_k141_121986\|\|full | Not-determined |
| S12_vs_k141_130029\|\|full | Not-determined |
| S12_vs_k141_35510\|\|full | Not-determined |
| S12_vs_k141_114398\|\|full | Low-quality |
| S12_vs_k141_51361\|\|full | Not-determined |
| S12_vs_k141_20005\|\|full | Low-quality |
| S12_vs_k141_83172\|\|full | Not-determined |
| S12_vs_k141_32669\|\|0_partial | Medium-quality |
| S12_vs_k141_4851\|\|full | Not-determined |
| S12_vs_k141_169721\|\|full | Not-determined |
| S12_vs_k141_161867\|\|full | Low-quality |
| S12_vs_k141_185967\|\|full | Not-determined |
| S12_vs_k141_4864\|\|full | Low-quality |
| S12_vs_k141_91659\|\|full | Not-determined |
| S12_vs_k141_107363\|\|full | Not-determined |
| S12_vs_k141_138513\|\|full | Low-quality |
| S12_vs_k141_51381\|\|full | Not-determined |
| S12_vs_k141_67563\|\|full | Low-quality |
| S12_vs_k141_44623\|\|full | Low-quality |
| S12_vs_k141_122016\|\|full | Low-quality |
| S12_vs_k141_4868\|\|full | Low-quality |
| S12_vs_k141_44626\|\|full | Not-determined |
| S12_vs_k141_153813\|\|full | Not-determined |
| S12_vs_k141_75054\|\|full | Low-quality |
| S12_vs_k141_28258\|\|full | Low-quality |
| S12_vs_k141_161882\|\|full | Low-quality |
| S12_vs_k141_4878\|\|full | Low-quality |
| S12_vs_k141_138538\|\|full | Low-quality |
| S12_vs_k141_91679\|\|full | Not-determined |
| S12_vs_k141_161887\|\|full | Low-quality |
| S12_vs_k141_51397\|\|full | Low-quality |
| S12_vs_k141_28278\|\|full | Low-quality |
| S12_vs_k141_91680\|\|full | Not-determined |
| S12_vs_k141_114444\|\|full | Not-determined |
| S12_vs_k141_75075\|\|full | Low-quality |
| S12_vs_k141_20050\|\|full | Not-determined |
| S12_vs_k141_153827\|\|full | Low-quality |
| S12_vs_k141_161896\|\|full | Low-quality |
| S12_vs_k141_75086\|\|full | Not-determined |
| S12_vs_k141_35552\|\|full | Low-quality |
| S12_vs_k141_51398\|\|full | Low-quality |
| S12_vs_k141_138564\|\|full | Not-determined |
| S12_vs_k141_161911\|\|full | Low-quality |
| S12_vs_k141_130106\|\|full | Not-determined |
| S12_vs_k141_28297\|\|full | Not-determined |
| S12_vs_k141_169783\|\|full | Low-quality |
| S12_vs_k141_35566\|\|full | Low-quality |
| S12_vs_k141_51419\|\|full | Not-determined |
| S12_vs_k141_146325\|\|full | Not-determined |
| S12_vs_k141_20089\|\|full | Not-determined |
| S12_vs_k141_146329\|\|full | Low-quality |
| S12_vs_k141_44680\|\|full | Not-determined |
| S12_vs_k141_169788\|\|full | Low-quality |
| S12_vs_k141_4925\|\|full | Not-determined |
| S14_vs_k141_115377\|\|full | Medium-quality |
| S12_vs_k141_107419\|\|full | Low-quality |
| S12_vs_k141_138600\|\|full | Not-determined |
| S12_vs_k141_130138\|\|full | Low-quality |
| S12_vs_k141_130151\|\|full | Not-determined |
| S12_vs_k141_44715\|\|full | Low-quality |
| S12_vs_k141_28329\|\|full | Low-quality |
| S12_vs_k141_67601\|\|full | Low-quality |
| S12_vs_k141_4926\|\|full | Low-quality |
| S12_vs_k141_122053\|\|full | Low-quality |
| S12_vs_k141_114475\|\|full | Low-quality |
| S12_vs_k141_67602\|\|full | Low-quality |
| S12_vs_k141_44730\|\|full | Low-quality |
| S12_vs_k141_28338\|\|full | Low-quality |
| S12_vs_k141_130159\|\|full | Not-determined |
| S12_vs_k141_107436\|\|full | Low-quality |
| S12_vs_k141_177024\|\|full | Low-quality |
| S12_vs_k141_138622\|\|full | Low-quality |
| S11_vs_k141_83666\|\|full | Medium-quality |
| S12_vs_k141_35618\|\|full | Low-quality |
| S12_vs_k141_83260\|\|full | Not-determined |
| S12_vs_k141_99775\|\|full | Low-quality |
| S12_vs_k141_161958\|\|full | Low-quality |
| S12_vs_k141_75112\|\|full | Low-quality |
| S12_vs_k141_177047\|\|full | Not-determined |
| S12_vs_k141_83270\|\|full | Low-quality |
| S12_vs_k141_114507\|\|full | Not-determined |
| S12_vs_k141_107472\|\|full | Low-quality |
| S12_vs_k141_122070\|\|full | Not-determined |
| S12_vs_k141_186130\|\|full | Low-quality |
| S12_vs_k141_67632\|\|full | Not-determined |
| S12_vs_k141_20163\|\|full | Low-quality |
| S12_vs_k141_130211\|\|full | Low-quality |
| S12_vs_k141_20171\|\|full | Low-quality |
| S12_vs_k141_122096\|\|full | Not-determined |
| S12_vs_k141_11605\|\|full | Low-quality |
| S12_vs_k141_130222\|\|full | Low-quality |
| S12_vs_k141_75135\|\|full | Low-quality |
| S12_vs_k141_130225\|\|full | Low-quality |
| S12_vs_k141_153982\|\|full | Low-quality |
| S12_vs_k141_28399\|\|full | Not-determined |
| S12_vs_k141_130229\|\|full | Low-quality |
| S12_vs_k141_91790\|\|full | Low-quality |
| S12_vs_k141_130239\|\|full | Not-determined |
| S12_vs_k141_114557\|\|full | Low-quality |
| S12_vs_k141_4999\|\|full | Not-determined |
| S12_vs_k141_83295\|\|full | Low-quality |
| S12_vs_k141_75166\|\|full | Not-determined |
| S12_vs_k141_177142\|\|full | Not-determined |
| S12_vs_k141_114573\|\|full | Low-quality |
| S12_vs_k141_162055\|\|full | Low-quality |
| S12_vs_k141_20197\|\|full | Not-determined |
| S12_vs_k141_138718\|\|full | Not-determined |
| S12_vs_k141_99859\|\|full | Low-quality |
| S12_vs_k141_44833\|\|full | Low-quality |
| S12_vs_k141_107550\|\|full | Low-quality |
| S12_vs_k141_177159\|\|full | Low-quality |
| S12_vs_k141_114590\|\|full | Low-quality |
| S12_vs_k141_138722\|\|full | Low-quality |
| S12_vs_k141_11689\|\|full | Low-quality |
| S12_vs_k141_75208\|\|full | Low-quality |
| S12_vs_k141_162088\|\|full | Not-determined |
| S12_vs_k141_146645\|\|full | Low-quality |
| S12_vs_k141_35745\|\|full | Low-quality |
| S12_vs_k141_122154\|\|full | Low-quality |
| S12_vs_k141_28418\|\|full | Low-quality |
| S12_vs_k141_20234\|\|full | Low-quality |
| S6_extracted_k141_40951 | Medium-quality |
| S12_vs_k141_91868\|\|full | Low-quality |
| S12_vs_k141_35747\|\|full | Not-determined |
| S12_vs_k141_138733\|\|full | Low-quality |
| S12_vs_k141_114630\|\|full | Low-quality |
| S12_vs_k141_51598\|\|full | Not-determined |
| S12_vs_k141_11717\|\|full | Not-determined |
| S12_vs_k141_186283\|\|full | Low-quality |
| S12_vs_k141_122170\|\|full | Not-determined |
| S12_vs_k141_162114\|\|full | Low-quality |
| S12_vs_k141_138750\|\|full | Not-determined |
| S12_vs_k141_162118\|\|full | Low-quality |
| S12_vs_k141_35753\|\|full | Not-determined |
| S12_vs_k141_51610\|\|full | Not-determined |
| S12_vs_k141_75231\|\|full | Low-quality |
| S12_vs_k141_162123\|\|full | Low-quality |
| S12_vs_k141_177220\|\|full | Not-determined |
| S12_vs_k141_67732\|\|full | Low-quality |
| S12_vs_k141_44879\|\|full | Not-determined |
| S12_vs_k141_169826\|\|full | Low-quality |
| S12_vs_k141_169828\|\|full | Low-quality |
| S12_vs_k141_107620\|\|full | Not-determined |
| S12_vs_k141_20267\|\|full | Not-determined |
| S12_vs_k141_114666\|\|full | Low-quality |
| S12_vs_k141_146489\|\|full | Low-quality |
| S12_vs_k141_130317\|\|full | Low-quality |
| S12_vs_k141_28486\|\|full | Not-determined |
| S12_vs_k141_35806\|\|full | Low-quality |
| S12_vs_k141_122230\|\|full | Low-quality |
| S12_vs_k141_146501\|\|full | Not-determined |
| S12_vs_k141_83362\|\|full | Low-quality |
| S12_vs_k141_75282\|\|full | Low-quality |
| S12_vs_k141_67768\|\|full | Not-determined |
| S12_vs_k141_59967\|\|full | Low-quality |
| S12_vs_k141_130335\|\|full | Not-determined |
| S12_vs_k141_177273\|\|full | Not-determined |
| S12_vs_k141_83379\|\|full | Not-determined |
| S12_vs_k141_162142\|\|full | Low-quality |
| S12_vs_k141_138819\|\|full | Not-determined |
| S12_vs_k141_138821\|\|full | Not-determined |
| S12_vs_k141_59986\|\|full | Not-determined |
| S12_vs_k141_67797\|\|full | Low-quality |
| S12_vs_k141_44909\|\|full | Not-determined |
| S12_vs_k141_177297\|\|full | Low-quality |
| S12_vs_k141_99975\|\|full | Not-determined |
| S12_vs_k141_162150\|\|full | Not-determined |
| S12_vs_k141_11785\|\|full | Low-quality |
| S12_vs_k141_28552\|\|full | Low-quality |
| S12_vs_k141_186344\|\|full | Not-determined |
| S12_vs_k141_83422\|\|full | Low-quality |
| S12_vs_k141_44952\|\|full | Not-determined |
| S12_vs_k141_83434\|\|full | Not-determined |
| S12_vs_k141_114726\|\|full | Not-determined |
| S12_vs_k141_35853\|\|full | Low-quality |
| S12_vs_k141_146545\|\|full | Low-quality |
| S12_vs_k141_83439\|\|full | Low-quality |
| S12_vs_k141_177345\|\|full | Not-determined |
| S12_vs_k141_146556\|\|full | Low-quality |
| S12_vs_k141_138867\|\|full | Low-quality |
| S12_vs_k141_11833\|\|full | Not-determined |
| S12_vs_k141_154112\|\|full | Low-quality |
| S12_vs_k141_83481\|\|full | Not-determined |
| S12_vs_k141_130378\|\|full | Not-determined |
| S12_vs_k141_5176\|\|full | Not-determined |
| S12_vs_k141_51695\|\|full | Low-quality |
| S12_vs_k141_130380\|\|full | Low-quality |
| S12_vs_k141_162230\|\|full | Not-determined |
| S12_vs_k141_28602\|\|full | Not-determined |
| S12_vs_k141_44965\|\|full | Not-determined |
| S12_vs_k141_75404\|\|full | Not-determined |
| S12_vs_k141_11859\|\|full | Not-determined |
| S12_vs_k141_114797\|\|full | Low-quality |
| S12_vs_k141_122319\|\|full | Not-determined |
| S12_vs_k141_83489\|\|full | Not-determined |
| S12_vs_k141_75408\|\|full | Low-quality |
| S12_vs_k141_130392\|\|full | Low-quality |
| S12_vs_k141_75412\|\|full | Low-quality |
| S12_vs_k141_122331\|\|full | Low-quality |
| S12_vs_k141_130395\|\|full | Low-quality |
| S12_vs_k141_154138\|\|full | Not-determined |
| S12_vs_k141_146614\|\|full | Not-determined |
| S12_vs_k141_186400\|\|full | Not-determined |
| S12_vs_k141_91977\|\|full | Low-quality |
| S12_vs_k141_186406\|\|full | Low-quality |
| S12_vs_k141_107716\|\|full | Not-determined |
| S12_vs_k141_5201\|\|full | Not-determined |
| S12_vs_k141_5205\|\|full | Not-determined |
| S12_vs_k141_169996\|\|full | Not-determined |
| S12_vs_k141_35894\|\|full | Low-quality |
| S12_vs_k141_130422\|\|full | Low-quality |
| S12_vs_k141_100080\|\|full | Low-quality |
| S12_vs_k141_35900\|\|full | Not-determined |
| S12_vs_k141_45029\|\|full | Low-quality |
| S12_vs_k141_107726\|\|full | Low-quality |
| S12_vs_k141_130430\|\|full | Not-determined |
| S12_vs_k141_75444\|\|full | Not-determined |
| S12_vs_k141_186430\|\|full | Low-quality |
| S12_vs_k141_146656\|\|full | Low-quality |
| S12_vs_k141_122373\|\|full | Not-determined |
| S12_vs_k141_162275\|\|full | Low-quality |
| S12_vs_k141_75461\|\|full | Not-determined |
| S12_vs_k141_45041\|\|full | Not-determined |
| S12_vs_k141_138964\|\|full | Not-determined |
| S12_vs_k141_186456\|\|full | Not-determined |
| S12_vs_k141_75467\|\|full | Low-quality |
| S12_vs_k141_51736\|\|full | Low-quality |
| S12_vs_k141_35916\|\|full | Low-quality |
| S12_vs_k141_5236\|\|full | Not-determined |
| S12_vs_k141_20488\|\|full | Not-determined |
| S12_vs_k141_83497\|\|full | Not-determined |
| S12_vs_k141_100108\|\|full | Low-quality |
| S12_vs_k141_45053\|\|full | Not-determined |
| S12_vs_k141_20501\|\|full | Not-determined |
| S12_vs_k141_67831\|\|full | Not-determined |
| S12_vs_k141_130475\|\|full | Not-determined |
| S12_vs_k141_162318\|\|full | Not-determined |
| S12_vs_k141_130485\|\|full | Low-quality |
| S12_vs_k141_51760\|\|full | Not-determined |
| S12_vs_k141_5248\|\|full | Not-determined |
| S12_vs_k141_83505\|\|full | Low-quality |
| S12_vs_k141_130491\|\|full | Low-quality |
| S12_vs_k141_11930\|\|full | Low-quality |
| S12_vs_k141_139004\|\|full | Not-determined |
| S6_vs_k141_40951\|\|full | Medium-quality |
| S12_vs_k141_83511\|\|full | Low-quality |
| S12_vs_k141_28663\|\|full | Not-determined |
| S12_vs_k141_51774\|\|full | Not-determined |
| S12_vs_k141_20539\|\|full | Low-quality |
| S12_vs_k141_20541\|\|full | Not-determined |
| S12_vs_k141_28671\|\|full | Low-quality |
| S12_vs_k141_122422\|\|full | Low-quality |
| S12_vs_k141_170049\|\|full | Low-quality |
| S12_vs_k141_107769\|\|full | Not-determined |
| S12_vs_k141_67857\|\|full | Not-determined |
| S12_vs_k141_107772\|\|full | Low-quality |
| S12_vs_k141_139017\|\|full | Not-determined |
| S12_vs_k141_51794\|\|full | Not-determined |
| S12_vs_k141_11934\|\|full | Not-determined |
| S12_vs_k141_170058\|\|full | Low-quality |
| S12_vs_k141_154215\|\|full | Not-determined |
| S12_vs_k141_130523\|\|full | Not-determined |
| S12_vs_k141_186514\|\|full | Low-quality |
| S12_vs_k141_146725\|\|full | Low-quality |
| S12_vs_k141_20563\|\|full | Not-determined |
| S12_vs_k141_92115\|\|full | Low-quality |
| S12_vs_k141_170074\|\|full | Not-determined |
| S12_vs_k141_75510\|\|full | Not-determined |
| S12_vs_k141_51820\|\|full | Not-determined |
| S12_vs_k141_92135\|\|full | Not-determined |
| S12_vs_k141_20584\|\|full | Not-determined |
| S12_vs_k141_20585\|\|full | Not-determined |
| S12_vs_k141_5303\|\|full | Not-determined |
| S12_vs_k141_5306\|\|full | Low-quality |
| S12_vs_k141_107799\|\|full | Low-quality |
| S12_vs_k141_177353\|\|full | Not-determined |
| S12_vs_k141_139061\|\|full | Low-quality |
| S12_vs_k141_146733\|\|full | Low-quality |
| S12_vs_k141_154277\|\|full | Low-quality |
| S12_vs_k141_146747\|\|full | Not-determined |
| S12_vs_k141_100196\|\|full | Not-determined |
| S12_vs_k141_100204\|\|full | Low-quality |
| S12_vs_k141_162428\|\|full | Not-determined |
| S12_vs_k141_35967\|\|full | Not-determined |
| S12_vs_k141_162439\|\|full | Low-quality |
| S12_vs_k141_5360\|\|full | Low-quality |
| S12_vs_k141_11974\|\|full | Low-quality |
| S12_vs_k141_130602\|\|full | Not-determined |
| S12_vs_k141_146777\|\|full | Low-quality |
| S12_vs_k141_5379\|\|full | Not-determined |
| S12_vs_k141_162475\|\|full | Low-quality |
| S12_vs_k141_170099\|\|full | Not-determined |
| S12_vs_k141_139111\|\|full | Low-quality |
| S12_vs_k141_177444\|\|full | Not-determined |
| S12_vs_k141_75583\|\|full | Low-quality |
| S12_vs_k141_75586\|\|full | Low-quality |
| S12_vs_k141_35977\|\|full | Not-determined |
| S12_vs_k141_100255\|\|full | Not-determined |
| S12_vs_k141_170114\|\|full | Low-quality |
| S12_vs_k141_177464\|\|full | Low-quality |
| S12_vs_k141_122530\|\|full | Low-quality |
| S12_vs_k141_20642\|\|full | Low-quality |
| S12_vs_k141_186523\|\|full | Low-quality |
| S12_vs_k141_20649\|\|full | Low-quality |
| S12_vs_k141_122557\|\|full | Low-quality |
| S12_vs_k141_5433\|\|full | Low-quality |
| S12_vs_k141_170128\|\|full | Low-quality |
| S12_vs_k141_114918\|\|full | Low-quality |
| S12_vs_k141_177488\|\|full | Not-determined |
| S12_vs_k141_162517\|\|full | Not-determined |

| | |
|---|---|
| S12_vs_k141_177496\|\|full | Low-quality |
| S12_vs_k141_139138\|\|full | Not-determined |
| S12_vs_k141_51903\|\|full | Low-quality |
| S12_vs_k141_170164\|\|full | Low-quality |
| S12_vs_k141_75643\|\|full | Low-quality |
| S12_vs_k141_154324\|\|full | Low-quality |
| S12_vs_k141_170168\|\|full | Low-quality |
| S12_vs_k141_92184\|\|full | Low-quality |
| S12_vs_k141_51916\|\|full | Low-quality |
| S12_vs_k141_139158\|\|full | Low-quality |
| S12_vs_k141_36035\|\|full | Low-quality |
| S12_vs_k141_186582\|\|full | Not-determined |
| S12_vs_k141_154330\|\|full | Not-determined |
| S12_vs_k141_139178\|\|full | Not-determined |
| S12_vs_k141_170184\|\|full | Low-quality |
| S12_vs_k141_162547\|\|full | Not-determined |
| S12_vs_k141_83591\|\|full | Not-determined |
| S12_vs_k141_83595\|\|full | Not-determined |
| S12_vs_k141_139184\|\|full | Low-quality |
| S12_vs_k141_20699\|\|full | Not-determined |
| S12_vs_k141_170201\|\|full | Low-quality |
| S12_vs_k141_12058\|\|full | Low-quality |
| S10_extracted_46328 | Medium-quality |
| S12_vs_k141_186590\|\|full | Low-quality |
| S12_vs_k141_114960\|\|full | Low-quality |
| S12_vs_k141_139194\|\|full | Not-determined |
| S12_vs_k141_92202\|\|full | Not-determined |
| S12_vs_k141_51935\|\|full | Low-quality |
| S12_vs_k141_114961\|\|full | Not-determined |
| S12_vs_k141_170214\|\|full | Low-quality |
| S12_vs_k141_83631\|\|full | Not-determined |
| S12_vs_k141_177540\|\|full | Low-quality |
| S12_vs_k141_20726\|\|full | Not-determined |
| S12_vs_k141_28707\|\|full | Low-quality |
| S12_vs_k141_186610\|\|full | Not-determined |
| S12_vs_k141_146903\|\|full | Low-quality |
| S12_vs_k141_114984\|\|full | Low-quality |
| S12_vs_k141_28713\|\|full | Not-determined |
| S12_vs_k141_51967\|\|full | Low-quality |
| S12_vs_k141_20732\|\|full | Low-quality |
| S12_vs_k141_186621\|\|full | Not-determined |
| S12_vs_k141_139215\|\|full | Low-quality |
| S12_vs_k141_177571\|\|full | Low-quality |
| S12_vs_k141_154371\|\|full | Low-quality |
| S12_vs_k141_36089\|\|full | Low-quality |
| S12_vs_k141_177577\|\|full | Not-determined |
| S12_vs_k141_45153\|\|full | Low-quality |
| S12_vs_k141_51985\|\|full | Low-quality |
| S12_vs_k141_154382\|\|full | Not-determined |
| S12_vs_k141_83654\|\|full | Not-determined |
| S12_vs_k141_28750\|\|full | Not-determined |
| S12_vs_k141_67976\|\|full | Not-determined |
| S12_vs_k141_100402\|\|full | Low-quality |
| S12_vs_k141_100408\|\|full | Not-determined |
| S12_vs_k141_154406\|\|full | Low-quality |
| S12_vs_k141_130677\|\|full | Not-determined |
| S12_vs_k141_115009\|\|full | Not-determined |
| S12_vs_k141_186664\|\|full | Not-determined |
| S12_vs_k141_107844\|\|full | Not-determined |
| S12_vs_k141_130679\|\|full | Low-quality |
| S12_vs_k141_68002\|\|full | Not-determined |
| S12_vs_k141_170295\|\|full | Low-quality |
| S12_vs_k141_36111\|\|full | Not-determined |
| S12_vs_k141_139264\|\|full | Low-quality |
| S12_vs_k141_68013\|\|full | Low-quality |
| S12_vs_k141_75739\|\|full | Low-quality |
| S12_vs_k141_177630\|\|full | Low-quality |
| S12_vs_k141_139278\|\|full | Not-determined |
| S12_vs_k141_146945\|\|full | Not-determined |
| S12_vs_k141_45198\|\|full | Not-determined |
| S12_vs_k141_28796\|\|full | Not-determined |
| S12_vs_k141_146946\|\|full | Low-quality |
| S12_vs_k141_12137\|\|full | Low-quality |
| S12_vs_k141_52064\|\|full | Low-quality |
| S12_vs_k141_60232\|\|full | Not-determined |
| S12_vs_k141_115052\|\|full | Not-determined |
| S12_vs_k141_170315\|\|full | Low-quality |
| S12_vs_k141_130723\|\|full | Low-quality |
| S12_vs_k141_186699\|\|full | Low-quality |
| S12_vs_k141_122659\|\|full | Low-quality |
| S12_vs_k141_154489\|\|full | Low-quality |
| S12_vs_k141_122661\|\|full | Low-quality |
| S12_vs_k141_122675\|\|full | Not-determined |
| S12_vs_k141_68061\|\|full | Low-quality |
| S12_vs_k141_107906\|\|full | Low-quality |
| S12_vs_k141_52078\|\|full | Not-determined |
| S12_vs_k141_130742\|\|full | Not-determined |
| S12_vs_k141_154508\|\|full | Low-quality |
| S12_vs_k141_68071\|\|full | Low-quality |
| S12_vs_k141_28826\|\|full | Low-quality |
| S12_vs_k141_28827\|\|full | Low-quality |
| S12_vs_k141_154512\|\|full | Low-quality |
| S12_vs_k141_162596\|\|full | Low-quality |
| S12_vs_k141_122693\|\|full | Low-quality |
| S12_vs_k141_60286\|\|full | Low-quality |
| S12_vs_k141_107939\|\|full | Not-determined |
| S12_vs_k141_170373\|\|full | Not-determined |
| S12_vs_k141_130770\|\|full | Low-quality |
| S12_vs_k141_75821\|\|full | Low-quality |
| S12_vs_k141_60305\|\|full | Low-quality |
| S12_vs_k141_122706\|\|full | Not-determined |
| S12_vs_k141_154534\|\|full | Low-quality |
| S12_vs_k141_186769\|\|full | Not-determined |
| S12_vs_k141_107952\|\|full | Low-quality |
| S12_vs_k141_154540\|\|full | Low-quality |
| S12_vs_k141_92330\|\|full | Not-determined |
| S12_vs_k141_75848\|\|full | Low-quality |
| S12_vs_k141_12199\|\|full | Not-determined |
| S12_vs_k141_130796\|\|full | Low-quality |
| S12_vs_k141_186779\|\|full | Low-quality |
| S12_vs_k141_147023\|\|full | Low-quality |
| S12_vs_k141_36122\|\|full | Not-determined |
| S12_vs_k141_60319\|\|full | Not-determined |
| S12_vs_k141_20746\|\|full | Not-determined |
| S12_vs_k141_5660\|\|full | Low-quality |
| S12_vs_k141_177709\|\|full | Not-determined |
| S12_vs_k141_92362\|\|full | Low-quality |
| S12_vs_k141_52171\|\|full | Low-quality |
| S12_vs_k141_154563\|\|full | Low-quality |
| S12_vs_k141_45292\|\|full | Not-determined |
| S12_vs_k141_130823\|\|full | Low-quality |
| S12_vs_k141_170396\|\|full | Low-quality |
| S12_vs_k141_75886\|\|full | Not-determined |
| S12_vs_k141_139368\|\|full | Low-quality |
| S12_vs_k141_75890\|\|full | Low-quality |
| S12_vs_k141_68114\|\|full | Not-determined |
| S12_vs_k141_5688\|\|full | Not-determined |
| S12_vs_k141_5689\|\|full | Not-determined |
| S12_vs_k141_75895\|\|full | Low-quality |
| S12_vs_k141_52191\|\|full | Not-determined |
| S12_vs_k141_92380\|\|full | Not-determined |
| S12_vs_k141_122762\|\|full | Not-determined |
| S12_vs_k141_20764\|\|full | Low-quality |
| S12_vs_k141_92382\|\|full | Not-determined |
| S12_vs_k141_68135\|\|full | Low-quality |
| S12_vs_k141_52192\|\|full | Low-quality |
| S12_vs_k141_52193\|\|full | Not-determined |
| S12_vs_k141_5711\|\|full | Low-quality |
| S12_vs_k141_68140\|\|full | Low-quality |
| S12_vs_k141_45331\|\|full | Low-quality |
| S12_vs_k141_45342\|\|full | Not-determined |
| S12_vs_k141_5721\|\|full | Not-determined |
| S12_vs_k141_177742\|\|full | Not-determined |
| S12_vs_k141_108038\|\|full | Low-quality |
| S12_vs_k141_92434\|\|full | Not-determined |
| S12_vs_k141_122801\|\|full | Not-determined |
| S12_vs_k141_154626\|\|full | Low-quality |
| S12_vs_k141_92443\|\|full | Low-quality |
| S12_vs_k141_20798\|\|full | Low-quality |
| S12_vs_k141_5735\|\|full | Low-quality |
| S12_vs_k141_177760\|\|full | Low-quality |
| S12_vs_k141_92453\|\|full | Not-determined |
| S12_vs_k141_186880\|\|full | Low-quality |
| S12_vs_k141_20806\|\|full | Low-quality |
| S12_vs_k141_75960\|\|full | Low-quality |
| S12_vs_k141_12310\|\|full | Low-quality |
| S12_vs_k141_186888\|\|full | Not-determined |
| S12_vs_k141_68190\|\|full | Not-determined |
| S12_vs_k141_177789\|\|full | Not-determined |
| S12_vs_k141_108065\|\|full | Low-quality |
| S12_vs_k141_100528\|\|full | Low-quality |
| S12_vs_k141_100533\|\|full | Low-quality |
| S12_vs_k141_76004\|\|full | Not-determined |
| S12_vs_k141_100538\|\|full | Not-determined |
| S12_vs_k141_108092\|\|full | Low-quality |
| S12_vs_k141_28939\|\|full | Low-quality |
| S12_vs_k141_5781\|\|full | Low-quality |
| S12_vs_k141_36252\|\|full | Not-determined |
| S12_vs_k141_76024\|\|full | Low-quality |
| S12_vs_k141_60439\|\|full | Low-quality |
| S12_vs_k141_139432\|\|full | Not-determined |
| S12_vs_k141_170482\|\|full | Low-quality |
| S12_vs_k141_92498\|\|full | Not-determined |
| S12_vs_k141_115153\|\|full | Low-quality |
| S12_vs_k141_147120\|\|full | Not-determined |
| S12_vs_k141_154674\|\|full | Low-quality |
| S12_vs_k141_100570\|\|full | Not-determined |
| S12_vs_k141_5790\|\|full | Low-quality |
| S12_vs_k141_122829\|\|full | Not-determined |
| S12_vs_k141_52318\|\|full | Not-determined |
| S12_vs_k141_20865\|\|full | Low-quality |
| S12_vs_k141_60442\|\|full | Low-quality |
| S12_vs_k141_139452\|\|full | Low-quality |
| S12_vs_k141_170493\|\|full | Low-quality |
| S12_vs_k141_60443\|\|full | Not-determined |
| S12_vs_k141_76045\|\|full | Not-determined |
| S12_vs_k141_5791\|\|full | Low-quality |
| S12_vs_k141_68255\|\|full | Not-determined |
| S12_vs_k141_147138\|\|full | Not-determined |
| S12_vs_k141_108125\|\|full | Not-determined |
| S12_vs_k141_122841\|\|full | Not-determined |
| S12_vs_k141_100587\|\|full | Low-quality |
| S12_vs_k141_68257\|\|full | Not-determined |
| S12_vs_k141_36268\|\|full | Not-determined |
| S12_vs_k141_154682\|\|full | Low-quality |
| S12_vs_k141_108128\|\|full | Low-quality |
| S12_vs_k141_139466\|\|full | Low-quality |
| S12_vs_k141_100599\|\|full | Low-quality |
| S12_vs_k141_139479\|\|full | Low-quality |
| S12_vs_k141_28971\|\|full | Low-quality |
| S12_vs_k141_177892\|\|full | Not-determined |
| S12_vs_k141_52350\|\|full | Low-quality |
| S12_vs_k141_52354\|\|full | Low-quality |
| S12_vs_k141_20920\|\|full | Low-quality |
| S12_vs_k141_68273\|\|full | Low-quality |
| S12_vs_k141_76060\|\|full | Low-quality |
| S12_vs_k141_52376\|\|full | Low-quality |
| S12_vs_k141_130977\|\|full | Not-determined |
| S12_vs_k141_115229\|\|full | Low-quality |
| S12_vs_k141_83763\|\|full | Low-quality |
| S12_vs_k141_12437\|\|full | Low-quality |
| S12_vs_k141_130988\|\|full | Not-determined |
| S12_vs_k141_130992\|\|full | Low-quality |
| S12_vs_k141_131000\|\|full | Not-determined |
| S12_vs_k141_177944\|\|full | Low-quality |
| S12_vs_k141_20963\|\|full | Not-determined |
| S12_vs_k141_187025\|\|full | Not-determined |
| S12_vs_k141_36312\|\|full | Low-quality |
| S12_vs_k141_52398\|\|full | Low-quality |
| S12_vs_k141_108166\|\|full | Low-quality |
| S12_vs_k141_36315\|\|full | Low-quality |
| S12_vs_k141_83771\|\|full | Low-quality |
| S12_vs_k141_131017\|\|full | Low-quality |
| S12_vs_k141_108177\|\|full | Low-quality |
| S12_vs_k141_29036\|\|full | Not-determined |
| S12_vs_k141_139540\|\|full | Low-quality |
| S12_vs_k141_60527\|\|full | Low-quality |
| S12_vs_k141_20969\|\|full | Low-quality |
| S12_vs_k141_177974\|\|full | Not-determined |
| S12_vs_k141_60537\|\|full | Low-quality |
| S12_vs_k141_20973\|\|full | Not-determined |
| S12_vs_k141_100677\|\|full | Not-determined |
| S12_vs_k141_83780\|\|full | Low-quality |
| S12_vs_k141_100679\|\|full | Low-quality |
| S12_vs_k141_45531\|\|full | Low-quality |
| S12_vs_k141_68305\|\|full | Not-determined |
| S12_vs_k141_131038\|\|full | Not-determined |
| S12_vs_k141_76072\|\|full | Low-quality |
| S10_vs_46328\|\|full | Medium-quality |
| S12_vs_k141_52432\|\|full | Low-quality |
| S12_vs_k141_76077\|\|full | Not-determined |
| S12_vs_k141_139552\|\|full | Low-quality |
| S12_vs_k141_52442\|\|full | Low-quality |
| S12_vs_k141_122922\|\|full | Low-quality |
| S14_vs_k141_100227\|\|full | Medium-quality |
| S12_vs_k141_52457\|\|full | Low-quality |
| S12_vs_k141_20993\|\|full | Low-quality |
| S12_vs_k141_29080\|\|full | Low-quality |
| S12_vs_k141_20998\|\|full | Not-determined |
| S12_vs_k141_21001\|\|full | Not-determined |
| S12_vs_k141_100709\|\|full | Not-determined |
| S12_vs_k141_52473\|\|full | Not-determined |
| S12_vs_k141_92553\|\|full | Not-determined |
| S12_vs_k141_76139\|\|full | Low-quality |
| S12_vs_k141_108194\|\|full | Low-quality |
| S12_vs_k141_139612\|\|full | Not-determined |
| S12_vs_k141_52479\|\|full | Not-determined |
| S12_vs_k141_187099\|\|full | Low-quality |
| S12_vs_k141_76145\|\|full | Not-determined |
| S12_vs_k141_29106\|\|full | Low-quality |
| S12_vs_k141_21018\|\|full | Low-quality |
| S12_vs_k141_100722\|\|full | Low-quality |
| S12_vs_k141_76148\|\|full | Not-determined |
| S12_vs_k141_139626\|\|full | Low-quality |
| S12_vs_k141_76151\|\|full | Not-determined |
| S12_vs_k141_147241\|\|full | Not-determined |
| S12_vs_k141_5895\|\|full | Not-determined |
| S12_vs_k141_162721\|\|full | Low-quality |
| S12_vs_k141_139634\|\|full | Not-determined |
| S12_vs_k141_92563\|\|full | Not-determined |
| S12_vs_k141_131083\|\|full | Not-determined |
| S12_vs_k141_122992\|\|full | Not-determined |
| S12_vs_k141_178076\|\|full | Low-quality |
| S12_vs_k141_162729\|\|full | Not-determined |
| S12_vs_k141_154828\|\|full | Low-quality |
| S12_vs_k141_5907\|\|full | Not-determined |
| S12_vs_k141_139653\|\|full | Low-quality |
| S12_vs_k141_92580\|\|full | Low-quality |
| S12_vs_k141_100762\|\|full | Low-quality |
| S12_vs_k141_60615\|\|full | Not-determined |
| S12_vs_k141_178087\|\|full | Low-quality |
| S12_vs_k141_108243\|\|full | Not-determined |
| S12_vs_k141_162744\|\|full | Not-determined |
| S12_vs_k141_178088\|\|full | Low-quality |
| S12_vs_k141_147256\|\|full | Not-determined |
| S12_vs_k141_45635\|\|full | Low-quality |
| S12_vs_k141_76209\|\|full | Not-determined |
| S12_vs_k141_83863\|\|full | Low-quality |
| S12_vs_k141_92594\|\|full | Low-quality |
| S12_vs_k141_12551\|\|full | Not-determined |
| S12_vs_k141_5954\|\|full | Not-determined |
| S12_vs_k141_45644\|\|full | Not-determined |
| S12_vs_k141_83867\|\|full | Low-quality |
| S12_vs_k141_162763\|\|full | Not-determined |
| S12_vs_k141_178102\|\|full | Low-quality |
| S12_vs_k141_36410\|\|full | Low-quality |
| S12_vs_k141_108264\|\|full | Not-determined |
| S12_vs_k141_21075\|\|full | Not-determined |
| S12_vs_k141_115369\|\|full | Low-quality |
| S12_vs_k141_139683\|\|full | Low-quality |
| S12_vs_k141_154885\|\|full | Not-determined |
| S12_vs_k141_147259\|\|full | Low-quality |
| S12_vs_k141_123041\|\|full | Not-determined |
| S12_vs_k141_83887\|\|full | Not-determined |
| S12_vs_k141_139696\|\|full | Low-quality |
| S12_vs_k141_178123\|\|full | Low-quality |
| S12_vs_k141_68396\|\|full | Not-determined |
| S12_vs_k141_83895\|\|full | Low-quality |
| S12_vs_k141_131110\|\|full | Low-quality |
| S12_vs_k141_178133\|\|full | Not-determined |
| S12_vs_k141_139705\|\|full | Low-quality |
| S12_vs_k141_147282\|\|full | Low-quality |
| S12_vs_k141_68410\|\|full | Low-quality |
| S12_vs_k141_147288\|\|full | Low-quality |
| S12_vs_k141_60676\|\|full | Low-quality |
| S12_vs_k141_115407\|\|full | Not-determined |
| S5_vs_k141_108086\|\|full | Medium-quality |
| S12_vs_k141_5983\|\|full | Low-quality |
| S12_vs_k141_29141\|\|full | Not-determined |
| S12_vs_k141_12611\|\|full | Low-quality |
| S12_vs_k141_187205\|\|full | Not-determined |
| S12_vs_k141_147323\|\|full | Not-determined |
| S12_vs_k141_115418\|\|full | Not-determined |
| S12_vs_k141_29157\|\|full | Not-determined |
| S12_vs_k141_108308\|\|full | Not-determined |
| S12_vs_k141_68448\|\|full | Low-quality |
| S12_vs_k141_29163\|\|full | Low-quality |
| S12_vs_k141_170641\|\|full | Low-quality |
| S12_vs_k141_162817\|\|full | Not-determined |
| S12_vs_k141_108315\|\|full | Low-quality |
| S12_vs_k141_12626\|\|full | Low-quality |
| S12_vs_k141_178185\|\|full | Not-determined |
| S12_vs_k141_60689\|\|full | Not-determined |
| S12_vs_k141_100876\|\|full | Not-determined |
| S12_vs_k141_131164\|\|full | Low-quality |
| S12_vs_k141_139754\|\|full | Not-determined |
| S12_vs_k141_170651\|\|full | Low-quality |
| S12_vs_k141_108336\|\|full | Low-quality |
| S12_vs_k141_6004\|\|full | Low-quality |
| S12_vs_k141_45724\|\|full | Low-quality |
| S12_vs_k141_115449\|\|full | Low-quality |
| S12_vs_k141_36445\|\|full | Low-quality |
| S12_vs_k141_170669\|\|full | Not-determined |
| S12_vs_k141_36450\|\|full | Low-quality |
| S12_vs_k141_108363\|\|full | Low-quality |
| S12_vs_k141_131185\|\|full | Not-determined |
| S12_vs_k141_21148\|\|full | Not-determined |
| S12_vs_k141_68486\|\|full | Low-quality |
| S12_vs_k141_139783\|\|full | Low-quality |
| S12_vs_k141_36457\|\|full | Not-determined |
| S12_vs_k141_170676\|\|full | Low-quality |
| S12_vs_k141_139787\|\|full | Low-quality |
| S4_extracted_2217 | Medium-quality |
| S12_vs_k141_147386\|\|full | Low-quality |
| S12_vs_k141_108382\|\|full | Not-determined |
| S12_vs_k141_83961\|\|full | Low-quality |
| S12_vs_k141_6038\|\|full | Low-quality |
| S12_vs_k141_187243\|\|full | Low-quality |
| S12_vs_k141_178236\|\|full | Low-quality |
| S12_vs_k141_92734\|\|full | Not-determined |
| S12_vs_k141_187254\|\|full | Not-determined |
| S12_vs_k141_162889\|\|full | Low-quality |
| S12_vs_k141_92743\|\|full | Not-determined |
| S12_vs_k141_68510\|\|full | Low-quality |
| S12_vs_k141_178258\|\|full | Not-determined |
| S12_vs_k141_162895\|\|full | Low-quality |
| S12_vs_k141_187265\|\|full | Low-quality |
| S12_vs_k141_68525\|\|full | Not-determined |
| S12_vs_k141_92755\|\|full | Not-determined |
| S12_vs_k141_76256\|\|full | Low-quality |
| S12_vs_k141_108433\|\|full | Low-quality |
| S12_vs_k141_131199\|\|full | Low-quality |
| S12_vs_k141_154981\|\|full | Not-determined |
| S12_vs_k141_92772\|\|full | Low-quality |
| S12_vs_k141_21220\|\|full | Not-determined |
| S12_vs_k141_60776\|\|full | Low-quality |
| S12_vs_k141_21225\|\|full | Low-quality |
| S12_vs_k141_131212\|\|full | Not-determined |
| S12_vs_k141_21226\|\|full | Low-quality |
| S12_vs_k141_6093\|\|full | Low-quality |
| S12_vs_k141_154991\|\|full | Low-quality |
| S12_vs_k141_6097\|\|full | Not-determined |
| S12_vs_k141_123192\|\|full | Low-quality |
| S12_vs_k141_84012\|\|full | Not-determined |
| S12_vs_k141_162905\|\|full | Not-determined |
| S12_vs_k141_68573\|\|full | Low-quality |
| S12_vs_k141_21233\|\|full | Low-quality |
| S12_vs_k141_100959\|\|full | Not-determined |
| S12_vs_k141_139842\|\|full | Not-determined |
| S12_vs_k141_139843\|\|full | Low-quality |
| S12_vs_k141_108460\|\|full | Not-determined |
| S12_vs_k141_12729\|\|full | Not-determined |
| S12_vs_k141_21250\|\|full | Low-quality |
| S12_vs_k141_131235\|\|full | Low-quality |
| S12_vs_k141_68590\|\|full | Low-quality |
| S12_vs_k141_131240\|\|full | Low-quality |
| S12_vs_k141_84045\|\|full | Not-determined |
| S12_vs_k141_123229\|\|full | Low-quality |
| S12_vs_k141_92826\|\|full | Low-quality |
| S12_vs_k141_6103\|\|full | Not-determined |
| S12_vs_k141_84063\|\|full | Low-quality |
| S12_vs_k141_139884\|\|full | Not-determined |
| S12_vs_k141_131278\|\|full | Low-quality |
| S12_vs_k141_139886\|\|full | Not-determined |
| S12_vs_k141_21292\|\|full | Not-determined |
| S12_vs_k141_147507\|\|full | Not-determined |
| S12_vs_k141_60860\|\|full | Not-determined |
| S12_vs_k141_123264\|\|full | Low-quality |
| S12_vs_k141_170848\|\|full | Low-quality |
| S12_vs_k141_45781\|\|full | Low-quality |
| S12_vs_k141_178351\|\|full | Low-quality |
| S12_vs_k141_101040\|\|full | Low-quality |
| S12_vs_k141_123269\|\|full | Low-quality |
| S12_vs_k141_147514\|\|full | Not-determined |
| S12_vs_k141_162976\|\|full | Low-quality |
| S12_vs_k141_60869\|\|full | Low-quality |
| S12_vs_k141_187304\|\|full | Not-determined |
| S12_vs_k141_178366\|\|full | Low-quality |
| S12_vs_k141_36532\|\|full | Low-quality |
| S12_vs_k141_12764\|\|full | Low-quality |
| S12_vs_k141_155056\|\|full | Low-quality |
| S12_vs_k141_123288\|\|full | Low-quality |
| S12_vs_k141_178379\|\|full | Low-quality |
| S12_vs_k141_101059\|\|full | Low-quality |
| S12_vs_k141_29223\|\|full | Low-quality |
| S12_vs_k141_187315\|\|full | Not-determined |
| S12_vs_k141_45821\|\|full | Not-determined |
| S12_vs_k141_36536\|\|full | Not-determined |
| S12_vs_k141_68619\|\|full | Low-quality |
| S12_vs_k141_170850\|\|full | Not-determined |
| S12_vs_k141_147547\|\|full | Not-determined |
| S12_vs_k141_45830\|\|full | Not-determined |
| S12_vs_k141_101075\|\|full | Low-quality |
| S12_vs_k141_163011\|\|full | Low-quality |
| S12_vs_k141_12788\|\|full | Not-determined |
| S12_vs_k141_147549\|\|full | Not-determined |
| S12_vs_k141_68641\|\|full | Not-determined |
| S12_vs_k141_60905\|\|full | Low-quality |
| S12_vs_k141_131296\|\|full | Not-determined |
| S12_vs_k141_84113\|\|full | Low-quality |
| S12_vs_k141_45845\|\|full | Low-quality |
| S12_vs_k141_101090\|\|full | Not-determined |
| S12_vs_k141_187336\|\|full | Low-quality |
| S12_vs_k141_12801\|\|full | Low-quality |
| S12_vs_k141_60925\|\|full | Not-determined |
| S12_vs_k141_108511\|\|full | Not-determined |
| S12_vs_k141_147567\|\|full | Not-determined |
| S12_vs_k141_139974\|\|full | Not-determined |
| S12_vs_k141_131326\|\|full | Not-determined |
| S12_vs_k141_108514\|\|full | Not-determined |
| S12_vs_k141_29277\|\|full | Not-determined |
| S12_vs_k141_131334\|\|full | Not-determined |
| S12_vs_k141_52648\|\|full | Not-determined |
| S12_vs_k141_123365\|\|full | Not-determined |
| S12_vs_k141_84139\|\|full | Low-quality |
| S12_vs_k141_36606\|\|full | Not-determined |
| S12_vs_k141_155074\|\|full | Not-determined |
| S12_vs_k141_101134\|\|full | Low-quality |
| S12_vs_k141_60954\|\|full | Low-quality |
| S12_vs_k141_45875\|\|full | Low-quality |
| S12_vs_k141_187386\|\|full | Low-quality |
| S12_vs_k141_29295\|\|full | Not-determined |
| S12_vs_k141_163075\|\|full | Low-quality |
| S12_vs_k141_178445\|\|full | Not-determined |
| S12_vs_k141_155111\|\|full | Low-quality |
| S12_vs_k141_84174\|\|full | Not-determined |
| S12_vs_k141_101156\|\|full | Low-quality |
| S12_vs_k141_123408\|\|full | Not-determined |
| S12_vs_k141_108568\|\|full | Not-determined |
| S12_vs_k141_76437\|\|full | Not-determined |
| S12_vs_k141_84188\|\|full | Low-quality |
| S4_vs_VIRSorter_2217-cat_2 | Medium-quality |
| S12_vs_k141_115598\|\|full | Not-determined |
| S12_vs_k141_187397\|\|full | Low-quality |
| S12_vs_k141_140048\|\|full | Low-quality |
| S12_vs_k141_21393\|\|full | Low-quality |
| S12_vs_k141_6264\|\|full | Low-quality |
| S12_vs_k141_163110\|\|full | Low-quality |
| S12_vs_k141_45938\|\|full | Low-quality |
| S12_vs_k141_21408\|\|full | Not-determined |
| S12_vs_k141_123437\|\|full | Low-quality |
| S12_vs_k141_131382\|\|full | Low-quality |
| S12_vs_k141_52723\|\|full | Not-determined |
| S12_vs_k141_187427\|\|full | Not-determined |
| S12_vs_k141_36652\|\|full | Not-determined |
| S12_vs_k141_21414\|\|full | Not-determined |
| S12_vs_k141_36654\|\|full | Low-quality |
| S12_vs_k141_147635\|\|full | Not-determined |
| S12_vs_k141_131389\|\|full | Not-determined |
| S12_vs_k141_147636\|\|full | Not-determined |
| S12_vs_k141_29353\|\|full | Low-quality |
| S12_vs_k141_131392\|\|full | Not-determined |
| S12_vs_k141_84233\|\|full | Not-determined |
| S12_vs_k141_187445\|\|full | Not-determined |
| S12_vs_k141_163134\|\|full | Low-quality |
| S12_vs_k141_187446\|\|full | Low-quality |
| S12_vs_k141_21425\|\|full | Not-determined |
| S12_vs_k141_12877\|\|full | Low-quality |
| S12_vs_k141_108638\|\|full | Not-determined |
| S12_vs_k141_21427\|\|full | Low-quality |
| S12_vs_k141_178519\|\|full | Low-quality |
| S12_vs_k141_45981\|\|full | Low-quality |
| S12_vs_k141_68758\|\|full | Not-determined |
| S12_vs_k141_140093\|\|full | Low-quality |
| S12_vs_k141_108646\|\|full | Low-quality |
| S12_vs_k141_76514\|\|full | Not-determined |
| S12_vs_k141_178527\|\|full | Not-determined |
| S12_vs_k141_52758\|\|full | Low-quality |
| S12_vs_k141_6307\|\|full | Not-determined |
| S12_vs_k141_45987\|\|full | Low-quality |
| S12_vs_k141_163158\|\|full | Low-quality |
| S12_vs_k141_61033\|\|full | Low-quality |
| S12_vs_k141_84248\|\|full | Not-determined |
| S12_vs_k141_52766\|\|full | Low-quality |
| S12_vs_k141_68768\|\|full | Low-quality |
| S12_vs_k141_123490\|\|full | Low-quality |
| S12_vs_k141_163168\|\|full | Low-quality |
| S12_vs_k141_84258\|\|full | Low-quality |
| S12_vs_k141_76534\|\|full | Low-quality |
| S12_vs_k141_46009\|\|full | Low-quality |
| S12_vs_k141_36694\|\|full | Not-determined |
| S12_vs_k141_123499\|\|full | Not-determined |
| S12_vs_k141_84276\|\|full | Not-determined |
| S12_vs_k141_108667\|\|full | Not-determined |
| S12_vs_k141_140147\|\|full | Not-determined |
| S12_vs_k141_140150\|\|full | Low-quality |
| S12_vs_k141_46037\|\|full | Not-determined |
| S12_vs_k141_21475\|\|full | Low-quality |
| S12_vs_k141_36721\|\|full | Low-quality |
| S12_vs_k141_163185\|\|full | Not-determined |
| S12_vs_k141_187464\|\|full | Low-quality |
| S12_vs_k141_178570\|\|full | Not-determined |
| S12_vs_k141_84301\|\|full | Low-quality |
| S12_vs_k141_163190\|\|full | Low-quality |
| S12_vs_k141_115669\|\|full | Low-quality |
| S12_vs_k141_12925\|\|full | Low-quality |
| S12_vs_k141_178580\|\|full | Low-quality |
| S12_vs_k141_46058\|\|full | Not-determined |
| S12_vs_k141_29446\|\|full | Not-determined |
| S12_vs_k141_108696\|\|full | Low-quality |
| S12_vs_k141_131502\|\|full | Low-quality |
| S12_vs_k141_84323\|\|full | Not-determined |
| S12_vs_k141_155199\|\|full | Low-quality |
| S12_vs_k141_115680\|\|full | Not-determined |
| S12_vs_k141_163199\|\|full | Low-quality |
| S12_vs_k141_29457\|\|full | Low-quality |
| S12_vs_k141_84329\|\|full | Not-determined |
| S12_vs_k141_36755\|\|full | Not-determined |
| S12_vs_k141_171012\|\|full | Low-quality |
| S12_vs_k141_123544\|\|full | Not-determined |
| S12_vs_k141_29471\|\|full | Low-quality |
| S12_vs_k141_12957\|\|full | Not-determined |
| S12_vs_k141_123546\|\|full | Not-determined |
| S12_vs_k141_76582\|\|full | Low-quality |
| S12_vs_k141_171019\|\|full | Low-quality |
| S12_vs_k141_147725\|\|full | Low-quality |
| S12_vs_k141_171020\|\|full | Low-quality |
| S12_vs_k141_131522\|\|full | Low-quality |
| S12_vs_k141_12974\|\|full | Not-determined |
| S12_vs_k141_61088\|\|full | Not-determined |
| S12_vs_k141_131530\|\|full | Low-quality |
| S12_vs_k141_101353\|\|full | Not-determined |
| S12_vs_k141_46092\|\|full | Low-quality |
| S12_vs_k141_61098\|\|full | Not-determined |
| S12_vs_k141_61099\|\|full | Low-quality |
| S12_vs_k141_115682\|\|full | Low-quality |
| S12_vs_k141_21534\|\|full | Not-determined |
| S12_vs_k141_131550\|\|full | Low-quality |
| S12_vs_k141_92897\|\|full | Low-quality |
| S12_vs_k141_92898\|\|full | Low-quality |
| S12_vs_k141_187531\|\|full | Low-quality |
| S12_vs_k141_178625\|\|full | Low-quality |
| S12_vs_k141_155239\|\|full | Low-quality |
| S12_vs_k141_6353\|\|full | Not-determined |
| S12_vs_k141_108751\|\|full | Low-quality |
| S12_vs_k141_187540\|\|full | Low-quality |
| S12_vs_k141_147776\|\|full | Not-determined |
| S12_vs_k141_6357\|\|full | Not-determined |
| S12_vs_k141_52882\|\|full | Not-determined |
| S12_vs_k141_29523\|\|full | Low-quality |
| S12_vs_k141_108777\|\|full | Low-quality |
| S12_vs_k141_155264\|\|full | Low-quality |
| S12_vs_k141_52892\|\|full | Low-quality |
| S12_vs_k141_76639\|\|full | Not-determined |
| S12_vs_k141_68859\|\|full | Not-determined |
| S12_vs_k141_171060\|\|full | Low-quality |
| S12_vs_k141_61142\|\|full | Not-determined |
| S12_vs_k141_68861\|\|full | Not-determined |
| S12_vs_k141_52893\|\|full | Low-quality |

S12_vs_k141_6379||full Low-quality
S12_vs_k141_84398||full Low-quality
S12_vs_k141_123588||full Not-determined
S12_vs_k141_36845||full Not-determined
S12_vs_k141_101417||full Low-quality
S12_vs_k141_76648||full Not-determined
S12_vs_k141_92944||full Low-quality
S12_vs_k141_140295||full Low-quality
S12_vs_k141_52910||full Not-determined
S12_vs_k141_163295||full Not-determined
S12_vs_k141_147809||full Low-quality
S12_vs_k141_52911||full Low-quality
S12_vs_k141_36871||full Low-quality
S12_vs_k141_155299||full Not-determined
S12_vs_k141_115750||full Not-determined
S12_vs_k141_92969||full Not-determined
S12_vs_k141_163319||full Not-determined
S12_vs_k141_84405||full Not-determined
S12_vs_k141_13075||full Not-determined
S12_vs_k141_52931||full Low-quality
S12_vs_k141_155319||full Low-quality
S12_vs_k141_140341||full Not-determined
S12_vs_k141_140342||full Low-quality
S12_vs_k141_187611||full Not-determined
S12_vs_k141_115780||full Not-determined
S12_vs_k141_178696||full Not-determined
S12_vs_k141_76682||full Low-quality
S12_vs_k141_36907||full Not-determined
S12_vs_k141_13098||full Not-determined
S12_vs_k141_84436||full Not-determined
S12_vs_k141_13100||full Not-determined
S12_vs_k141_178716||full Low-quality
S12_vs_k141_187645||full Not-determined
S12_vs_k141_13121||full Low-quality
S12_vs_k141_93008||full Low-quality
S12_vs_k141_171152||full Low-quality
S12_vs_k141_101490||full Not-determined
S12_vs_k141_101494||full Low-quality
S12_vs_k141_68963||full Not-determined
S12_vs_k141_36942||full Low-quality
S12_vs_k141_115832||full Low-quality
S12_vs_k141_163346||full Low-quality
S12_vs_k141_115834||full Low-quality
S12_vs_k141_21575||full Not-determined
S12_vs_k141_21577||full Low-quality
S12_vs_k141_147877||full Low-quality
S12_vs_k141_76740||full Not-determined
S12_vs_k141_21582||full Not-determined
S12_vs_k141_13151||full Not-determined
S12_vs_k141_76741||full Not-determined
S12_vs_k141_93047||full Low-quality
S12_vs_k141_93050||full Not-determined
S12_vs_k141_36962||full Low-quality
S12_vs_k141_171179||full Not-determined
S12_vs_k141_13170||full Not-determined
S12_vs_k141_93061||full Low-quality
S12_vs_k141_52973||full Not-determined
S12_vs_k141_84472||full Not-determined
S12_vs_k141_13183||full Low-quality
S12_vs_k141_46243||full Low-quality
S12_vs_k141_187703||full Not-determined
S12_vs_k141_29646||full Low-quality
S12_vs_k141_36981||full Not-determined
S12_vs_k141_36993||full Low-quality
S12_vs_k141_101547||full Not-determined
S12_vs_k141_69021||full Not-determined
S12_vs_k141_147918||full Low-quality
S12_vs_k141_131720||full Low-quality
S12_vs_k141_101552||full Low-quality
S12_vs_k141_61278||full Not-determined
S12_vs_k141_13236||full Low-quality
S12_vs_k141_46261||full Low-quality
S12_vs_k141_76773||full Low-quality
S12_vs_k141_13240||full Not-determined
S12_vs_k141_131729||full Low-quality
S5_vs_k141_80498||full Medium-quality
S12_vs_k141_147927||full Low-quality
S12_vs_k141_147931||full Low-quality
S12_vs_k141_76784||full Low-quality
S12_vs_k141_6511||full Low-quality
S12_vs_k141_13258||full Low-quality
S12_vs_k141_29664||full Not-determined
S12_vs_k141_163429||full Low-quality
S12_vs_k141_155373||full Low-quality
S12_vs_k141_29667||full Not-determined
S12_vs_k141_37046||full Not-determined
S12_vs_k141_13262||full Low-quality
S9_vs_k141_10424||full Medium-quality
S12_vs_k141_147958||full Low-quality
S12_vs_k141_163441||full Not-determined
S12_vs_k141_84494||full Low-quality
S12_vs_k141_69040||full Not-determined
S12_vs_k141_108921||full Not-determined
S12_vs_k141_131779||full Low-quality
S12_vs_k141_46280||full Not-determined
S12_vs_k141_171260||full Low-quality
S12_vs_k141_140503||full Low-quality
S12_vs_k141_101606||full Not-determined
S12_vs_k141_93159||full Not-determined
S12_vs_k141_6532||full Low-quality
S12_vs_k141_61327||full Low-quality
S12_vs_k141_46299||full Low-quality
S12_vs_k141_147964||full Low-quality
S12_vs_k141_93173||full Low-quality
S12_vs_k141_101616||full Not-determined
S12_vs_k141_171282||full Not-determined
S12_vs_k141_46314||full Not-determined
S12_vs_k141_21664||full Low-quality
S12_vs_k141_37086||full Low-quality
S12_vs_k141_108950||full Low-quality
S12_vs_k141_29739||full Low-quality
S12_vs_k141_101623||full Low-quality
S12_vs_k141_76839||full Not-determined
S12_vs_k141_53081||full Low-quality
S12_vs_k141_46321||full Not-determined
S12_vs_k141_115965||full Low-quality
S12_vs_k141_155443||full Low-quality
S12_vs_k141_76849||full Low-quality
S12_vs_k141_76853||full Not-determined
S12_vs_k141_115967||full Not-determined
S12_vs_k141_187796||full Not-determined
S12_vs_k141_84544||full Not-determined
S12_vs_k141_131808||full Low-quality
S12_vs_k141_29766||full Low-quality
S12_vs_k141_101649||full Not-determined
S12_vs_k141_131814||full Low-quality
S12_vs_k141_76871||full Not-determined
S12_vs_k141_69100||full Not-determined
S12_vs_k141_163478||full Low-quality
S12_vs_k141_84546||full Not-determined
S12_vs_k141_155463||full Not-determined
S12_vs_k141_163479||full Not-determined
S12_vs_k141_84550||full Low-quality
S12_vs_k141_46327||full Not-determined
S12_vs_k141_187812||full Not-determined
S12_vs_k141_178800||full Not-determined
S12_vs_k141_76879||full Not-determined
S12_vs_k141_29794||full Not-determined
S12_vs_k141_101670||full Not-determined
S12_vs_k141_84565||full Not-determined
S12_vs_k141_76892||full Low-quality
S12_vs_k141_148030||full Not-determined
S12_vs_k141_69120||full Low-quality
S12_vs_k141_6583||full Not-determined
S12_vs_k141_155493||full Not-determined
S12_vs_k141_37129||full Not-determined
S12_vs_k141_69133||full Low-quality
S12_vs_k141_109034||full Low-quality
S12_vs_k141_123769||full Low-quality
S12_vs_k141_178852||full Low-quality
S12_vs_k141_6615||full Low-quality
S12_vs_k141_131880||full Low-quality
S12_vs_k141_171343||full Low-quality
S12_vs_k141_76934||full Not-determined
S12_vs_k141_84619||full Not-determined
S12_vs_k141_93276||full Low-quality
S12_vs_k141_29853||full Not-determined
S12_vs_k141_109050||full Low-quality
S3_extracted_k141_34141 Medium-quality
S12_vs_k141_6625||full Not-determined
S12_vs_k141_187883||full Low-quality
S12_vs_k141_13337||full Not-determined
S12_vs_k141_116020||full Low-quality
S12_vs_k141_37166||full Low-quality
S12_vs_k141_6632||full Not-determined
S12_vs_k141_109063||full Low-quality
S3_vs_k141_34141||full Medium-quality
S12_vs_k141_187889||full Not-determined
S12_vs_k141_140580||full Low-quality
S12_vs_k141_140583||full Not-determined
S8_vs_k141_147670||full Medium-quality
S12_vs_k141_131912||full Not-determined
S12_vs_k141_171356||full Low-quality
S12_vs_k141_163587||full Not-determined
S12_vs_k141_53192||full Low-quality
S12_vs_k141_187911||full Low-quality
S12_vs_k141_93320||full Not-determined
S12_vs_k141_29862||full Low-quality
S12_vs_k141_61494||full Low-quality
S12_vs_k141_148065||full Low-quality
S12_vs_k141_69185||full Not-determined
S12_vs_k141_29867||full Low-quality
S12_vs_k141_37196||full Low-quality
S12_vs_k141_131937||full Not-determined
S12_vs_k141_61511||full Not-determined
S12_vs_k141_77030||full Low-quality
S12_vs_k141_116082||full Not-determined
S12_vs_k141_69199||full Not-determined
S12_vs_k141_29879||full Low-quality
S12_vs_k141_46406||full Not-determined
S12_vs_k141_109128||full Not-determined
S12_vs_k141_93353||full Not-determined
S12_vs_k141_131978||full Not-determined
S12_vs_k141_53255||full Not-determined
S12_vs_k141_6706||full Low-quality
S12_vs_k141_53260||full Low-quality
S12_vs_k141_148103||full Low-quality
S12_vs_k141_37254||full Low-quality
S12_vs_k141_132000||full Not-determined
S12_vs_k141_61570||full Not-determined
S12_vs_k141_148112||full Not-determined
S12_vs_k141_6731||full Low-quality
S12_vs_k141_109169||full Not-determined
S12_vs_k141_116123||full Low-quality
S12_vs_k141_29922||full Not-determined
S12_vs_k141_21721||full Low-quality
S12_vs_k141_178919||full Low-quality
S12_vs_k141_29929||full Low-quality
S12_vs_k141_123892||full Not-determined
S12_vs_k141_46446||full Low-quality
S12_vs_k141_123898||full Low-quality
S12_vs_k141_21723||full Not-determined
S12_vs_k141_13467||full Low-quality
S12_vs_k141_46452||full Not-determined
S8_extracted_k141_66528 Medium-quality
S12_vs_k141_140673||full Not-determined
S12_vs_k141_101813||full Low-quality
S12_vs_k141_21730||full Not-determined
S7_vs_87439||full Medium-quality
S12_vs_k141_37278||full Not-determined
S12_vs_k141_77044||full Low-quality
S12_vs_k141_46465||full Low-quality
S12_vs_k141_61602||full Not-determined
S12_vs_k141_101824||full Low-quality
S12_vs_k141_148141||full Not-determined
S12_vs_k141_84741||full Low-quality
S12_vs_k141_29960||full Not-determined
S12_vs_k141_53327||full Not-determined
S12_vs_k141_178944||full Low-quality
S12_vs_k141_84746||full Low-quality
S12_vs_k141_61616||full Not-determined
S12_vs_k141_140697||full Low-quality
S12_vs_k141_77072||full Low-quality
S12_vs_k141_148156||full Not-determined
S12_vs_k141_13503||full Not-determined
S12_vs_k141_123947||full Low-quality
S12_vs_k141_61643||full Not-determined
S12_vs_k141_171431||full Not-determined
S12_vs_k141_77088||full Not-determined
S12_vs_k141_69302||full Not-determined
S12_vs_k141_187991||full Low-quality
S12_vs_k141_163739||full Not-determined
S12_vs_k141_123950||full Not-determined
S12_vs_k141_61653||full Low-quality
S12_vs_k141_163742||full Low-quality
S12_vs_k141_53375||full Low-quality
S12_vs_k141_188000||full Low-quality
S12_vs_k141_171441||full Low-quality
S12_vs_k141_155639||full Low-quality
S12_vs_k141_116205||full Low-quality
S12_vs_k141_93408||full Low-quality
S12_vs_k141_178976||full Low-quality
S12_vs_k141_155650||full Low-quality
S12_vs_k141_109238||full Low-quality
S12_vs_k141_140766||full Low-quality
S12_vs_k141_109242||full Not-determined
S12_vs_k141_30055||full Low-quality
S12_vs_k141_140775||full Low-quality
S12_vs_k141_155670||full Low-quality
S12_vs_k141_179003||full Low-quality
S12_vs_k141_171481||full Low-quality
S12_vs_k141_109250||full Low-quality
S12_vs_k141_116216||full Not-determined
S12_vs_k141_30058||full Not-determined
S12_vs_k141_116219||full Not-determined
S12_vs_k141_140785||full Low-quality
S12_vs_k141_171485||full Low-quality
S12_vs_k141_188046||full Not-determined
S12_vs_k141_6829||full Not-determined
S12_vs_k141_163782||full Low-quality
S12_vs_k141_179012||full Low-quality
S12_vs_k141_101872||full Low-quality
S12_vs_k141_77167||full Not-determined
S12_vs_k141_132078||full Not-determined
S12_vs_k141_124018||full Not-determined
S12_vs_k141_93456||full Not-determined
S12_vs_k141_84826||full Not-determined
S12_vs_k141_155672||full Low-quality
S12_vs_k141_163795||full Not-determined
S12_vs_k141_171493||full Low-quality
S12_vs_k141_155676||full Low-quality
S12_vs_k141_69359||full Not-determined
S12_vs_k141_13577||full Low-quality
S12_vs_k141_21828||full Not-determined
S12_vs_k141_77191||full Low-quality
S12_vs_k141_179025||full Not-determined
S12_vs_k141_53451||full Low-quality
S12_vs_k141_163811||full Low-quality
S12_vs_k141_69372||full Low-quality
S12_vs_k141_124041||full Not-determined
S12_vs_k141_179042||full Not-determined
S12_vs_k141_163818||full Not-determined
S12_vs_k141_13609||full Low-quality
S14_vs_k141_102119||full Medium-quality
S12_vs_k141_93506||full Not-determined
S12_vs_k141_21852||full Low-quality
S12_vs_k141_116247||full Not-determined
S12_vs_k141_148276||full Low-quality
S12_vs_k141_46613||full Not-determined
S12_vs_k141_148277||full Not-determined
S12_vs_k141_13627||full Not-determined
S12_vs_k141_132111||full Low-quality
S12_vs_k141_13628||full Low-quality
S12_vs_k141_93516||full Low-quality
S12_vs_k141_116257||full Low-quality
S12_vs_k141_69400||full Not-determined
S12_vs_k141_132115||full Not-determined
S12_vs_k141_46626||full Low-quality
S12_vs_k141_69410||full Low-quality
S12_vs_k141_30142||full Low-quality
S12_vs_k141_109286||full Low-quality
S12_vs_k141_53502||full Low-quality
S12_vs_k141_101954||full Not-determined
S12_vs_k141_13640||full Low-quality
S12_vs_k141_46645||full Not-determined
S12_vs_k141_155696||full Low-quality
S12_vs_k141_132137||full Not-determined
S12_vs_k141_84885||full Low-quality
S12_vs_k141_124042||full Low-quality
S12_vs_k141_148318||full Low-quality
S12_vs_k141_46654||full Not-determined
S12_vs_k141_132153||full Not-determined
S12_vs_k141_155715||full Low-quality
S12_vs_k141_171530||full Not-determined
S12_vs_k141_109316||full Low-quality
S12_vs_k141_53520||full Not-determined
S12_vs_k141_155723||full Low-quality
S12_vs_k141_163889||full Low-quality
S12_vs_k141_132164||full Low-quality
S12_vs_k141_37433||full Low-quality
S12_vs_k141_163893||full Low-quality
S12_vs_k141_84920||full Low-quality
S12_vs_k141_140833||full Low-quality
S12_vs_k141_30193||full Not-determined
S12_vs_k141_188073||full Not-determined
S12_vs_k141_101988||full Low-quality
S12_vs_k141_30199||full Low-quality
S12_vs_k141_155743||full Not-determined
S12_vs_k141_93547||full Not-determined
S12_vs_k141_61712||full Low-quality
S12_vs_k141_163908||full Not-determined
S12_vs_k141_84941||full Low-quality
S12_vs_k141_140854||full Not-determined
S12_vs_k141_84943||full Not-determined
S12_vs_k141_102003||full Low-quality
S12_vs_k141_61723||full Not-determined
S12_vs_k141_155768||full Low-quality
S12_vs_k141_171545||full Not-determined
S12_vs_k141_124088||full Not-determined
S12_vs_k141_84956||full Not-determined
S12_vs_k141_6914||full Not-determined
S12_vs_k141_30225||full Low-quality
S12_vs_k141_132204||full Not-determined
S12_vs_k141_163939||full Low-quality
S12_vs_k141_148377||full Low-quality
S12_vs_k141_155784||full Not-determined
S12_vs_k141_37479||full Not-determined
S12_vs_k141_46695||full Not-determined
S12_vs_k141_69526||full Low-quality
S12_vs_k141_46702||full Not-determined
S12_vs_k141_13664||full Not-determined
S12_vs_k141_69531||full Low-quality
S12_vs_k141_13669||full Low-quality
S12_vs_k141_116337||full Low-quality
S12_vs_k141_6956||full Low-quality
S12_vs_k141_116346||full Not-determined
S12_vs_k141_163970||full Low-quality
S12_vs_k141_93614||full Not-determined
S12_vs_k141_69562||full Low-quality
S12_vs_k141_69563||full Low-quality
S12_vs_k141_148408||full Low-quality
S12_vs_k141_148410||full Low-quality
S12_vs_k141_171571||full Low-quality
S12_vs_k141_109432||full Low-quality
S12_vs_k141_85019||full Not-determined
S12_vs_k141_85026||full Low-quality
S12_vs_k141_13732||full Low-quality
S12_vs_k141_53638||full Low-quality
S12_vs_k141_163989||full Low-quality
S12_vs_k141_102047||full Low-quality
S12_vs_k141_132286||full Not-determined
S12_vs_k141_53651||full Not-determined
S12_vs_k141_109463||full Not-determined
S12_vs_k141_61784||full Low-quality
S12_vs_k141_102056||full Low-quality
S12_vs_k141_132289||full Low-quality
S12_vs_k141_116391||full Not-determined
S12_vs_k141_179149||full Not-determined
S12_vs_k141_37566||full Low-quality
S12_vs_k141_116395||full Not-determined
S12_vs_k141_171622||full Not-determined
S12_vs_k141_53681||full Not-determined
S12_vs_k141_179155||full Not-determined
S12_vs_k141_77377||full Low-quality
S12_vs_k141_85077||full Not-determined
S12_vs_k141_109482||full Low-quality
S12_vs_k141_188132||full Not-determined
S12_vs_k141_22016||full Not-determined
S12_vs_k141_148450||full Not-determined
S12_vs_k141_37579||full Low-quality
S12_vs_k141_13793||full Low-quality
S12_vs_k141_37585||full Low-quality
S12_vs_k141_155857||full Not-determined
S12_vs_k141_109500||full Low-quality
S12_vs_k141_148462||full Low-quality
S12_vs_k141_77407||full Low-quality
S12_vs_k141_69652||full Low-quality
S12_vs_k141_148464||full Low-quality
S12_vs_k141_30314||full Not-determined
S12_vs_k141_102079||full Low-quality
S12_vs_k141_22053||full Low-quality
S12_vs_k141_30322||full Not-determined
S12_vs_k141_109526||full Low-quality
S12_vs_k141_85119||full Low-quality
S12_vs_k141_148474||full Low-quality
S12_vs_k141_116416||full Low-quality
S12_vs_k141_140987||full Not-determined
S12_vs_k141_102090||full Not-determined
S12_vs_k141_109540||full Low-quality
S12_vs_k141_61885||full Low-quality
S12_vs_k141_155899||full Low-quality
S8_vs_k141_66528||full Medium-quality
S12_vs_k141_124205||full Low-quality
S12_vs_k141_13819||full Low-quality
S12_vs_k141_77438||full Not-determined
S12_vs_k141_179185||full Low-quality
S12_vs_k141_179186||full Low-quality
S12_vs_k141_37619||full Not-determined
S12_vs_k141_155903||full Low-quality
S12_vs_k141_37622||full Low-quality
S12_vs_k141_179191||full Not-determined
S12_vs_k141_148493||full Not-determined
S12_vs_k141_77450||full Low-quality
S12_vs_k141_46813||full Low-quality
S12_vs_k141_102112||full Not-determined
S12_vs_k141_116450||full Low-quality
S12_vs_k141_109556||full Low-quality
S12_vs_k141_93756||full Low-quality
S12_vs_k141_46816||full Low-quality
S12_vs_k141_109557||full Not-determined
S12_vs_k141_148498||full Not-determined
S12_vs_k141_13834||full Not-determined
S12_vs_k141_61913||full Not-determined
S12_vs_k141_61919||full Not-determined
S12_vs_k141_46821||full Not-determined
S12_vs_k141_179206||full Not-determined
S12_vs_k141_37641||full Low-quality
S12_vs_k141_13845||full Low-quality
S12_vs_k141_179209||full Low-quality
S12_vs_k141_188179||full Not-determined
S12_vs_k141_109567||full Not-determined
S12_vs_k141_53763||full Not-determined
S12_vs_k141_22107||full Not-determined
S12_vs_k141_13846||full Low-quality
S12_vs_k141_171696||full Low-quality
S12_vs_k141_85162||full Not-determined
S12_vs_k141_13851||full Low-quality
S12_vs_k141_124236||full Low-quality
S12_vs_k141_93798||full Not-determined
S12_vs_k141_37652||full Low-quality
S12_vs_k141_116484||full Low-quality
S12_vs_k141_164030||full Low-quality
S12_vs_k141_109591||full Low-quality
S12_vs_k141_188195||full Not-determined
S12_vs_k141_171713||full Not-determined
S12_vs_k141_171719||full Not-determined
S12_vs_k141_109600||full Low-quality
S12_vs_k141_77520||full Not-determined
S12_vs_k141_30376||full Not-determined
S12_vs_k141_69753||full Low-quality
S12_vs_k141_148550||full Not-determined
S12_vs_k141_171734||full Low-quality
S12_vs_k141_116498||full Not-determined
S12_vs_k141_132413||full Low-quality
S12_vs_k141_141064||full Not-determined
S12_vs_k141_53813||full Low-quality
S12_vs_k141_93836||full Not-determined
S12_vs_k141_141067||full Low-quality
S7_vs_129858||full Medium-quality
S12_vs_k141_46870||full Low-quality
S12_vs_k141_148575||full Low-quality
S12_vs_k141_171746||full Low-quality
S12_vs_k141_22179||full Not-determined
S12_vs_k141_77528||full Not-determined
S12_vs_k141_53827||full Low-quality
S12_vs_k141_7128||full Not-determined
S12_vs_k141_164076||full Not-determined
S12_vs_k141_37671||full Low-quality
S12_vs_k141_7130||full Low-quality
S12_vs_k141_171770||full Not-determined
S12_vs_k141_22201||full Low-quality
S12_vs_k141_53836||full Not-determined
S12_vs_k141_179269||full Not-determined
S12_vs_k141_171775||full Low-quality
S12_vs_k141_37691||full Low-quality
S12_vs_k141_179272||full Not-determined
S12_vs_k141_148620||full Low-quality
S12_vs_k141_93871||full Low-quality
S12_vs_k141_164102||full Low-quality
S12_vs_k141_77578||full Low-quality
S12_vs_k141_148630||full Low-quality
S12_vs_k141_37706||full Low-quality
S12_vs_k141_53847||full Low-quality
S12_vs_k141_124311||full Not-determined
S12_vs_k141_116530||full Not-determined
S12_vs_k141_93884||full Not-determined
S12_vs_k141_85300||full Low-quality
S12_vs_k141_155966||full Low-quality
S12_vs_k141_22241||full Low-quality
S12_vs_k141_132503||full Low-quality
S12_vs_k141_188268||full Low-quality
S12_vs_k141_7188||full Low-quality
S12_vs_k141_148654||full Low-quality
S12_vs_k141_188269||full Not-determined
S12_vs_k141_62023||full Low-quality
S12_vs_k141_93896||full Low-quality
S12_vs_k141_164152||full Not-determined
S12_vs_k141_13937||full Low-quality
S12_vs_k141_13941||full Low-quality
S12_vs_k141_37728||full Low-quality
S12_vs_k141_109633||full Low-quality
S12_vs_k141_124342||full Low-quality
S12_vs_k141_62044||full Low-quality
S12_vs_k141_188292||full Not-determined
S12_vs_k141_7207||full Not-determined
S12_vs_k141_22274||full Low-quality
S12_vs_k141_37741||full Low-quality
S12_vs_k141_7213||full Not-determined
S12_vs_k141_109641||full Low-quality
S12_vs_k141_124347||full Low-quality
S12_vs_k141_116559||full Low-quality
S12_vs_k141_69874||full Not-determined
S12_vs_k141_188310||full Not-determined
S12_vs_k141_109655||full Not-determined
S12_vs_k141_102230||full Not-determined
S12_vs_k141_148717||full Not-determined
S12_vs_k141_46968||full Low-quality
S12_vs_k141_46970||full Low-quality
S12_vs_k141_62087||full Not-determined
S12_vs_k141_109665||full Low-quality
S12_vs_k141_179354||full Low-quality
S12_vs_k141_22308||full Low-quality
S12_vs_k141_62099||full Not-determined
S12_vs_k141_148742||full Not-determined
S12_vs_k141_53884||full Not-determined
S12_vs_k141_7248||full Not-determined
S12_vs_k141_22317||full Low-quality
S12_vs_k141_148746||full Low-quality
S12_vs_k141_124397||full Not-determined
S12_vs_k141_85330||full Low-quality
S12_vs_k141_47014||full Low-quality
S12_vs_k141_7260||full Low-quality
S12_vs_k141_179375||full Not-determined
S12_vs_k141_85341||full Not-determined
S12_vs_k141_7266||full Low-quality
S12_vs_k141_22347||full Low-quality
S12_vs_k141_148769||full Low-quality
S12_vs_k141_62134||full Low-quality
S12_vs_k141_13980||full Not-determined
S12_vs_k141_179377||full Not-determined
S12_vs_k141_132600||full Low-quality
S12_vs_k141_13985||full Not-determined
S12_vs_k141_188387||full Not-determined
S12_vs_k141_7278||full Low-quality
S12_vs_k141_85380||full Not-determined
S12_vs_k141_109718||full Not-determined
S12_vs_k141_37773||full Low-quality
S12_vs_k141_77629||full Low-quality
S12_vs_k141_62169||full Low-quality
S12_vs_k141_22377||full Low-quality
S12_vs_k141_77639||full Not-determined
S12_vs_k141_37794||full Low-quality
S12_vs_k141_53920||full Not-determined
S12_vs_k141_179402||full Low-quality
S12_vs_k141_22395||full Low-quality
S12_vs_k141_132654||full Low-quality
S12_vs_k141_179406||full Low-quality
S12_vs_k141_164267||full Not-determined
S12_vs_k141_132657||full Low-quality
S12_vs_k141_77661||full Low-quality
S12_vs_k141_179420||full Low-quality
S12_vs_k141_85420||full Low-quality
S12_vs_k141_148819||full Low-quality
S12_vs_k141_14038||full Not-determined
S12_vs_k141_124441||full Low-quality
S12_vs_k141_116660||full Low-quality
S12_vs_k141_7346||full Low-quality
S12_vs_k141_141290||full Not-determined
S12_vs_k141_109766||full Low-quality

| | |
|---|---|
| S12_vs_k141_85437\|\|full | Low-quality |
| S12_vs_k141_7364\|\|full | Not-determined |
| S12_vs_k141_14062\|\|full | Low-quality |
| S12_vs_k141_37844\|\|full | Low-quality |
| S12_vs_k141_179457\|\|full | Not-determined |
| S12_vs_k141_141317\|\|full | Not-determined |
| S12_vs_k141_85444\|\|full | Not-determined |
| S12_vs_k141_93988\|\|full | Low-quality |
| S12_vs_k141_37857\|\|full | Not-determined |
| S12_vs_k141_124474\|\|full | Low-quality |
| S12_vs_k141_116679\|\|full | Not-determined |
| S12_vs_k141_37859\|\|full | Low-quality |
| S12_vs_k141_37863\|\|full | Low-quality |
| S12_vs_k141_164278\|\|full | Not-determined |
| S12_vs_k141_62222\|\|full | Low-quality |
| S12_vs_k141_85467\|\|full | Low-quality |
| S12_vs_k141_62228\|\|full | Low-quality |
| S12_vs_k141_109781\|\|full | Low-quality |
| S12_vs_k141_102306\|\|full | Not-determined |
| S12_vs_k141_124497\|\|full | Not-determined |
| S12_vs_k141_85472\|\|full | Low-quality |
| S12_vs_k141_30642\|\|full | Not-determined |
| S12_vs_k141_164289\|\|full | Not-determined |
| S12_vs_k141_62236\|\|full | Low-quality |
| S12_vs_k141_188496\|\|full | Low-quality |
| S4_vs_VIRSorter_21449-cat_2 | Medium-quality |
| S12_vs_k141_179505\|\|full | Low-quality |
| S12_vs_k141_7409\|\|full | Low-quality |
| S12_vs_k141_47141\|\|full | Not-determined |
| S12_vs_k141_22488\|\|full | Not-determined |
| S12_vs_k141_116722\|\|full | Low-quality |
| S12_vs_k141_179532\|\|full | Not-determined |
| S12_vs_k141_14132\|\|full | Not-determined |
| S12_vs_k141_14133\|\|full | Low-quality |
| S12_vs_k141_164309\|\|full | Not-determined |
| S12_vs_k141_109817\|\|full | Low-quality |
| S12_vs_k141_164316\|\|full | Not-determined |
| S12_vs_k141_54045\|\|full | Not-determined |
| S12_vs_k141_62295\|\|full | Not-determined |
| S12_vs_k141_132710\|\|full | Not-determined |
| S12_vs_k141_47162\|\|full | Not-determined |
| S12_vs_k141_109826\|\|full | Not-determined |
| S12_vs_k141_148915\|\|full | Low-quality |
| S12_vs_k141_58148\|\|full | Medium-quality |
| S12_vs_k141_22502\|\|full | Low-quality |
| S12_vs_k141_14156\|\|full | Not-determined |
| S12_vs_k141_132723\|\|full | Not-determined |
| S12_vs_k141_47172\|\|full | Low-quality |
| S12_vs_k141_54060\|\|full | Not-determined |
| S12_vs_k141_70087\|\|full | Low-quality |
| S12_vs_k141_54063\|\|full | Low-quality |
| S12_vs_k141_148925\|\|full | Low-quality |
| S12_vs_k141_132728\|\|full | Low-quality |
| S12_vs_k141_37906\|\|full | Not-determined |
| S12_vs_k141_7458\|\|full | Not-determined |
| S12_vs_k141_148930\|\|full | Low-quality |
| S12_vs_k141_85512\|\|full | Low-quality |
| S12_vs_k141_141361\|\|full | Not-determined |
| S12_vs_k141_85514\|\|full | Low-quality |
| S12_vs_k141_164340\|\|full | Low-quality |
| S12_vs_k141_179592\|\|full | Low-quality |
| S12_vs_k141_116782\|\|full | Low-quality |
| S12_vs_k141_85526\|\|full | Not-determined |
| S12_vs_k141_54092\|\|full | Low-quality |
| S12_vs_k141_14186\|\|full | Low-quality |
| S12_vs_k141_62319\|\|full | Low-quality |
| S12_vs_k141_109862\|\|full | Low-quality |
| S12_vs_k141_54107\|\|full | Not-determined |
| S12_vs_k141_22542\|\|full | Low-quality |
| S12_vs_k141_132777\|\|full | Not-determined |
| S12_vs_k141_70134\|\|full | Low-quality |
| S12_vs_k141_164347\|\|full | Low-quality |
| S12_vs_k141_85562\|\|full | Low-quality |
| S12_vs_k141_7508\|\|full | Not-determined |
| S12_vs_k141_164355\|\|full | Low-quality |
| S12_vs_k141_156013\|\|full | Low-quality |
| S12_vs_k141_7519\|\|full | Not-determined |
| S12_vs_k141_109893\|\|full | Not-determined |
| S12_vs_k141_62362\|\|full | Low-quality |
| S12_vs_k141_70145\|\|full | Low-quality |
| S12_vs_k141_14210\|\|full | Not-determined |
| S12_vs_k141_54138\|\|full | Low-quality |
| S12_vs_k141_116815\|\|full | Not-determined |
| S12_vs_k141_62367\|\|full | Not-determined |
| S12_vs_k141_124648\|\|full | Low-quality |
| S12_vs_k141_77863\|\|full | Low-quality |
| S12_vs_k141_188580\|\|full | Not-determined |
| S12_vs_k141_156032\|\|full | Not-determined |
| S12_vs_k141_62374\|\|full | Low-quality |
| S12_vs_k141_116825\|\|full | Not-determined |
| S12_vs_k141_124655\|\|full | Low-quality |
| S12_vs_k141_124659\|\|full | Low-quality |
| S12_vs_k141_132821\|\|full | Not-determined |
| S12_vs_k141_37977\|\|full | Not-determined |
| S12_vs_k141_7527\|\|full | Low-quality |
| S12_vs_k141_62395\|\|full | Not-determined |
| S12_vs_k141_37980\|\|full | Low-quality |
| S12_vs_k141_179654\|\|full | Low-quality |
| S12_vs_k141_109944\|\|full | Not-determined |
| S12_vs_k141_54184\|\|full | Low-quality |
| S12_vs_k141_156078\|\|full | Not-determined |
| S12_vs_k141_62418\|\|full | Not-determined |
| S12_vs_k141_14269\|\|full | Not-determined |
| S12_vs_k141_132855\|\|full | Not-determined |
| S12_vs_k141_132858\|\|full | Low-quality |
| S12_vs_k141_62421\|\|full | Low-quality |
| S12_vs_k141_62422\|\|full | Low-quality |
| S12_vs_k141_54209\|\|full | Low-quality |
| S12_vs_k141_38004\|\|full | Low-quality |
| S12_vs_k141_30761\|\|full | Low-quality |
| S12_vs_k141_188648\|\|full | Low-quality |
| S12_vs_k141_164471\|\|full | Low-quality |
| S12_vs_k141_85647\|\|full | Low-quality |
| S12_vs_k141_156087\|\|full | Low-quality |
| S12_vs_k141_188651\|\|full | Not-determined |
| S12_vs_k141_188653\|\|full | Not-determined |
| S12_vs_k141_132899\|\|full | Low-quality |
| S12_vs_k141_116875\|\|full | Low-quality |
| S12_vs_k141_156088\|\|full | Low-quality |
| S12_vs_k141_54231\|\|full | Not-determined |
| S12_vs_k141_77962\|\|full | Low-quality |
| S12_vs_k141_30768\|\|full | Not-determined |
| S12_vs_k141_62469\|\|full | Low-quality |
| S12_vs_k141_109994\|\|full | Low-quality |
| S5_extracted_k141_6506 | Medium-quality |
| S12_vs_k141_7562\|\|full | Not-determined |
| S12_vs_k141_179729\|\|full | Low-quality |
| S12_vs_k141_149119\|\|full | Not-determined |
| S12_vs_k141_110007\|\|full | Not-determined |
| S12_vs_k141_179732\|\|full | Low-quality |
| S12_vs_k141_141447\|\|full | Low-quality |
| S12_vs_k141_110010\|\|full | Low-quality |
| S12_vs_k141_110014\|\|full | Low-quality |
| S12_vs_k141_38034\|\|full | Not-determined |
| S12_vs_k141_124682\|\|full | Not-determined |
| S12_vs_k141_149136\|\|full | Low-quality |
| S12_vs_k141_179742\|\|full | Low-quality |
| S12_vs_k141_30811\|\|full | Low-quality |
| S12_vs_k141_110025\|\|full | Not-determined |
| S12_vs_k141_62484\|\|full | Not-determined |
| S12_vs_k141_179747\|\|full | Not-determined |
| S12_vs_k141_164490\|\|full | Not-determined |
| S12_vs_k141_164499\|\|full | Low-quality |
| S12_vs_k141_14370\|\|full | Not-determined |
| S12_vs_k141_179759\|\|full | Low-quality |
| S12_vs_k141_30812\|\|full | Low-quality |
| S12_vs_k141_77993\|\|full | Low-quality |
| S12_vs_k141_62509\|\|full | Low-quality |
| S12_vs_k141_14373\|\|full | Low-quality |
| S12_vs_k141_124699\|\|full | Low-quality |
| S1_vs_k141_145792\|\|full | Medium-quality |
| S12_vs_k141_149149\|\|full | Not-determined |
| S12_vs_k141_7621\|\|full | Low-quality |
| S12_vs_k141_124713\|\|full | Low-quality |
| S12_vs_k141_14378\|\|full | Not-determined |
| S12_vs_k141_78010\|\|full | Low-quality |
| S12_vs_k141_124718\|\|full | Not-determined |
| S12_vs_k141_179783\|\|full | Low-quality |
| S12_vs_k141_78012\|\|full | Low-quality |
| S12_vs_k141_14382\|\|full | Low-quality |
| S12_vs_k141_7632\|\|full | Not-determined |
| S12_vs_k141_124722\|\|full | Not-determined |
| S12_vs_k141_85728\|\|full | Not-determined |
| S12_vs_k141_7634\|\|full | Low-quality |
| S12_vs_k141_70305\|\|full | Low-quality |
| S12_vs_k141_179792\|\|full | Low-quality |
| S12_vs_k141_124732\|\|full | Low-quality |
| S12_vs_k141_30845\|\|full | Not-determined |
| S12_vs_k141_78016\|\|full | Not-determined |
| S12_vs_k141_164543\|\|full | Low-quality |
| S12_vs_k141_78024\|\|full | Not-determined |
| S12_vs_k141_141514\|\|full | Not-determined |
| S12_vs_k141_179806\|\|full | Low-quality |
| S12_vs_k141_94088\|\|full | Low-quality |
| S12_vs_k141_22640\|\|full | Not-determined |
| S12_vs_k141_149183\|\|full | Not-determined |
| S12_vs_k141_85753\|\|full | Low-quality |
| S12_vs_k141_110073\|\|full | Not-determined |
| S12_vs_k141_179814\|\|full | Low-quality |
| S12_vs_k141_94092\|\|full | Not-determined |
| S12_vs_k141_94097\|\|full | Not-determined |
| S12_vs_k141_7658\|\|full | Not-determined |
| S12_vs_k141_132967\|\|full | Not-determined |
| S12_vs_k141_116933\|\|full | Not-determined |
| S12_vs_k141_22664\|\|full | Low-quality |
| S12_vs_k141_78055\|\|full | Not-determined |
| S12_vs_k141_179844\|\|full | Low-quality |
| S12_vs_k141_116945\|\|full | Low-quality |
| S12_vs_k141_141550\|\|full | Not-determined |
| S12_vs_k141_179851\|\|full | Not-determined |
| S12_vs_k141_110109\|\|full | Not-determined |
| S12_vs_k141_70364\|\|full | Low-quality |
| S12_vs_k141_85794\|\|full | Low-quality |
| S12_vs_k141_70380\|\|full | Low-quality |
| S12_vs_k141_116967\|\|full | Low-quality |
| S12_vs_k141_38081\|\|full | Not-determined |
| S12_vs_k141_133022\|\|full | Not-determined |
| S12_vs_k141_179862\|\|full | Low-quality |
| S12_vs_k141_179864\|\|full | Not-determined |
| S12_vs_k141_22714\|\|full | Not-determined |
| S12_vs_k141_78082\|\|full | Not-determined |
| S12_vs_k141_141597\|\|full | Low-quality |
| S12_vs_k141_70404\|\|full | Low-quality |
| S12_vs_k141_110143\|\|full | Not-determined |
| S16_vs_54250\|\|full | Medium-quality |
| S12_vs_k141_94127\|\|full | Not-determined |
| S12_vs_k141_141604\|\|full | Low-quality |
| S12_vs_k141_179886\|\|full | Low-quality |
| S12_vs_k141_116997\|\|full | Low-quality |
| S12_vs_k141_164580\|\|full | Low-quality |
| S12_vs_k141_179890\|\|full | Low-quality |
| S12_vs_k141_124772\|\|full | Not-determined |
| S12_vs_k141_54391\|\|full | Low-quality |
| S12_vs_k141_179900\|\|full | Low-quality |
| S12_vs_k141_14489\|\|full | Low-quality |
| S12_vs_k141_171961\|\|full | Low-quality |
| S12_vs_k141_133065\|\|full | Not-determined |
| S12_vs_k141_22775\|\|full | Low-quality |
| S12_vs_k141_14504\|\|full | Not-determined |
| S12_vs_k141_156135\|\|full | Not-determined |
| S12_vs_k141_54397\|\|full | Low-quality |
| S12_vs_k141_85806\|\|full | Not-determined |
| S12_vs_k141_117018\|\|full | Low-quality |
| S12_vs_k141_171975\|\|full | Not-determined |
| S12_vs_k141_117019\|\|full | Low-quality |
| S12_vs_k141_62616\|\|full | Not-determined |
| S12_vs_k141_22789\|\|full | Low-quality |
| S12_vs_k141_30963\|\|full | Not-determined |
| S12_vs_k141_38145\|\|full | Not-determined |
| S12_vs_k141_164626\|\|full | Not-determined |
| S12_vs_k141_171981\|\|full | Low-quality |
| S12_vs_k141_141671\|\|full | Low-quality |
| S12_vs_k141_149287\|\|full | Low-quality |
| S12_vs_k141_7731\|\|full | Low-quality |
| S12_vs_k141_14525\|\|full | Low-quality |
| S12_vs_k141_30989\|\|full | Low-quality |
| S12_vs_k141_78140\|\|full | Low-quality |
| S12_vs_k141_172004\|\|full | Not-determined |
| S12_vs_k141_141695\|\|full | Not-determined |
| S12_vs_k141_124798\|\|full | Low-quality |
| S12_vs_k141_179951\|\|full | Not-determined |
| S12_vs_k141_22825\|\|full | Low-quality |
| S12_vs_k141_172020\|\|full | Low-quality |
| S12_vs_k141_179969\|\|full | Not-determined |
| S12_vs_k141_14548\|\|full | Not-determined |
| S12_vs_k141_94181\|\|full | Not-determined |
| S12_vs_k141_14557\|\|full | Not-determined |
| S12_vs_k141_22846\|\|full | Not-determined |
| S12_vs_k141_54433\|\|full | Low-quality |
| S12_vs_k141_156179\|\|full | Not-determined |
| S12_vs_k141_70484\|\|full | Low-quality |
| S12_vs_k141_85855\|\|full | Low-quality |
| S12_vs_k141_70486\|\|full | Not-determined |
| S12_vs_k141_85860\|\|full | Not-determined |
| S12_vs_k141_149353\|\|full | Low-quality |
| S12_vs_k141_85869\|\|full | Not-determined |
| S12_vs_k141_70504\|\|full | Not-determined |
| S12_vs_k141_156201\|\|full | Low-quality |
| S12_vs_k141_85877\|\|full | Low-quality |
| S12_vs_k141_141721\|\|full | Not-determined |
| S12_vs_k141_85881\|\|full | Low-quality |
| S12_vs_k141_54476\|\|full | Not-determined |
| S12_vs_k141_164679\|\|full | Low-quality |
| S12_vs_k141_94220\|\|full | Low-quality |
| S12_vs_k141_14602\|\|full | Low-quality |
| S12_vs_k141_124867\|\|full | Not-determined |
| S12_vs_k141_94223\|\|full | Low-quality |
| S12_vs_k141_94228\|\|full | Low-quality |
| S12_vs_k141_149370\|\|full | Low-quality |
| S10_extracted_62230 | Medium-quality |
| S12_vs_k141_22854\|\|full | Low-quality |
| S12_vs_k141_54496\|\|full | Not-determined |
| S12_vs_k141_78208\|\|full | Low-quality |
| S12_vs_k141_22857\|\|full | Low-quality |
| S12_vs_k141_22861\|\|full | Not-determined |
| S12_vs_k141_62752\|\|full | Low-quality |
| S12_vs_k141_164692\|\|full | Low-quality |
| S12_vs_k141_149383\|\|full | Not-determined |
| S12_vs_k141_78233\|\|full | Not-determined |
| S12_vs_k141_94245\|\|full | Low-quality |
| S12_vs_k141_85901\|\|full | Low-quality |
| S12_vs_k141_124902\|\|full | Not-determined |
| S12_vs_k141_149389\|\|full | Not-determined |
| S12_vs_k141_62762\|\|full | Low-quality |
| S12_vs_k141_117096\|\|full | Low-quality |
| S12_vs_k141_22875\|\|full | Low-quality |
| S12_vs_k141_94249\|\|full | Low-quality |
| S12_vs_k141_7845\|\|full | Low-quality |
| S12_vs_k141_164716\|\|full | Not-determined |
| S12_vs_k141_149394\|\|full | Low-quality |
| S12_vs_k141_188718\|\|full | Not-determined |
| S12_vs_k141_172135\|\|full | Low-quality |
| S12_vs_k141_7861\|\|full | Low-quality |
| S12_vs_k141_124923\|\|full | Not-determined |
| S12_vs_k141_188721\|\|full | Not-determined |
| S12_vs_k141_156235\|\|full | Low-quality |
| S12_vs_k141_156236\|\|full | Not-determined |
| S12_vs_k141_94266\|\|full | Not-determined |
| S12_vs_k141_149405\|\|full | Not-determined |
| S11_vs_k141_58930\|\|full | Medium-quality |
| S12_vs_k141_78262\|\|full | Low-quality |
| S12_vs_k141_94275\|\|full | Low-quality |
| S12_vs_k141_156264\|\|full | Not-determined |
| S12_vs_k141_164756\|\|full | Not-determined |
| S12_vs_k141_188747\|\|full | Low-quality |
| S12_vs_k141_14678\|\|full | Low-quality |
| S12_vs_k141_164760\|\|full | Not-determined |
| S12_vs_k141_14689\|\|full | Low-quality |
| S12_vs_k141_62831\|\|full | Not-determined |
| S12_vs_k141_85971\|\|full | Not-determined |
| S12_vs_k141_94302\|\|full | Low-quality |
| S12_vs_k141_31084\|\|full | Not-determined |
| S12_vs_k141_70589\|\|full | Not-determined |
| S12_vs_k141_149430\|\|full | Low-quality |
| S12_vs_k141_188774\|\|full | Low-quality |
| S12_vs_k141_54557\|\|full | Low-quality |
| S12_vs_k141_31093\|\|full | Low-quality |
| S12_vs_k141_149444\|\|full | Low-quality |
| S12_vs_k141_38294\|\|full | Low-quality |
| S12_vs_k141_133130\|\|full | Not-determined |
| S12_vs_k141_117193\|\|full | Not-determined |
| S12_vs_k141_22987\|\|full | Low-quality |
| S12_vs_k141_188795\|\|full | Low-quality |
| S12_vs_k141_38295\|\|full | Low-quality |
| S12_vs_k141_62855\|\|full | Not-determined |
| S12_vs_k141_164802\|\|full | Not-determined |
| S12_vs_k141_125006\|\|full | Not-determined |
| S12_vs_k141_31117\|\|full | Low-quality |
| S12_vs_k141_54583\|\|full | Not-determined |
| S12_vs_k141_117202\|\|full | Not-determined |
| S12_vs_k141_133144\|\|full | Not-determined |
| S12_vs_k141_62868\|\|full | Low-quality |
| S12_vs_k141_38313\|\|full | Low-quality |
| S12_vs_k141_54590\|\|full | Low-quality |
| S12_vs_k141_164809\|\|full | Low-quality |
| S12_vs_k141_78331\|\|full | Low-quality |
| S12_vs_k141_78338\|\|full | Not-determined |
| S12_vs_k141_14744\|\|full | Not-determined |
| S12_vs_k141_156349\|\|full | Low-quality |
| S12_vs_k141_14755\|\|full | Low-quality |
| S12_vs_k141_54605\|\|full | Not-determined |
| S12_vs_k141_164821\|\|full | Low-quality |
| S12_vs_k141_156359\|\|full | Not-determined |
| S12_vs_k141_172224\|\|full | Low-quality |
| S12_vs_k141_38336\|\|full | Not-determined |
| S12_vs_k141_62900\|\|full | Low-quality |
| S12_vs_k141_117231\|\|full | Not-determined |
| S12_vs_k141_94353\|\|full | Low-quality |
| S12_vs_k141_149507\|\|full | Low-quality |
| S12_vs_k141_117245\|\|full | Low-quality |
| S12_vs_k141_38351\|\|full | Not-determined |
| S12_vs_k141_180092\|\|full | Not-determined |
| S12_vs_k141_62922\|\|full | Not-determined |
| S12_vs_k141_62923\|\|full | Low-quality |
| S12_vs_k141_14777\|\|full | Not-determined |
| S12_vs_k141_156405\|\|full | Low-quality |
| S12_vs_k141_14778\|\|full | Low-quality |
| S12_vs_k141_149527\|\|full | Not-determined |
| S12_vs_k141_180112\|\|full | Not-determined |
| S12_vs_k141_133222\|\|full | Low-quality |
| S12_vs_k141_23060\|\|full | Low-quality |
| S12_vs_k141_62933\|\|full | Not-determined |
| S12_vs_k141_149539\|\|full | Not-determined |
| S12_vs_k141_70690\|\|full | Low-quality |
| S12_vs_k141_78422\|\|full | Low-quality |
| S12_vs_k141_180126\|\|full | Low-quality |
| S12_vs_k141_54649\|\|full | Not-determined |
| S12_vs_k141_86109\|\|full | Low-quality |
| S12_vs_k141_23078\|\|full | Low-quality |
| S12_vs_k141_149555\|\|full | Low-quality |
| S12_vs_k141_38383\|\|full | Not-determined |
| S12_vs_k141_180136\|\|full | Not-determined |
| S12_vs_k141_180137\|\|full | Not-determined |
| S12_vs_k141_78437\|\|full | Low-quality |
| S12_vs_k141_78448\|\|full | Low-quality |
| S12_vs_k141_38394\|\|full | Low-quality |
| S12_vs_k141_54670\|\|full | Not-determined |
| S12_vs_k141_23100\|\|full | Low-quality |
| S12_vs_k141_38402\|\|full | Low-quality |
| S12_vs_k141_38407\|\|full | Low-quality |
| S12_vs_k141_31179\|\|full | Not-determined |
| S12_vs_k141_149582\|\|full | Not-determined |
| S12_vs_k141_70709\|\|full | Not-determined |
| S12_vs_k141_31182\|\|full | Not-determined |
| S12_vs_k141_78471\|\|full | Not-determined |
| S12_vs_k141_149585\|\|full | Low-quality |
| S12_vs_k141_117310\|\|full | Not-determined |
| S12_vs_k141_172315\|\|full | Low-quality |
| S12_vs_k141_70725\|\|full | Low-quality |
| S12_vs_k141_62940\|\|full | Low-quality |
| S12_vs_k141_133246\|\|full | Not-determined |
| S12_vs_k141_31208\|\|full | Low-quality |
| S12_vs_k141_133251\|\|full | Low-quality |
| S12_vs_k141_70746\|\|full | Low-quality |
| S12_vs_k141_54717\|\|full | Low-quality |
| S12_vs_k141_62957\|\|full | Low-quality |
| S12_vs_k141_117343\|\|full | Low-quality |
| S12_vs_k141_31234\|\|full | Low-quality |
| S12_vs_k141_31240\|\|full | Low-quality |
| S12_vs_k141_86205\|\|full | Low-quality |
| S12_vs_k141_133271\|\|full | Not-determined |
| S12_vs_k141_38465\|\|full | Low-quality |
| S12_vs_k141_86209\|\|full | Not-determined |
| S12_vs_k141_31246\|\|full | Low-quality |
| S12_vs_k141_133274\|\|full | Low-quality |
| S12_vs_k141_164910\|\|full | Not-determined |
| S12_vs_k141_38481\|\|full | Not-determined |
| S12_vs_k141_78533\|\|full | Not-determined |
| S12_vs_k141_133294\|\|full | Not-determined |
| S12_vs_k141_70775\|\|full | Not-determined |
| S12_vs_k141_78539\|\|full | Not-determined |
| S12_vs_k141_54733\|\|full | Low-quality |
| S12_vs_k141_78550\|\|full | Not-determined |
| S12_vs_k141_180226\|\|full | Low-quality |
| S12_vs_k141_94487\|\|full | Not-determined |
| S12_vs_k141_156464\|\|full | Low-quality |
| S12_vs_k141_86259\|\|full | Not-determined |
| S12_vs_k141_125133\|\|full | Not-determined |
| S12_vs_k141_78571\|\|full | Low-quality |
| S12_vs_k141_23191\|\|full | Low-quality |
| S12_vs_k141_31327\|\|full | Low-quality |
| S12_vs_k141_117408\|\|full | Not-determined |
| S12_vs_k141_54789\|\|full | Not-determined |
| S12_vs_k141_156490\|\|full | Low-quality |
| S12_vs_k141_156494\|\|full | Low-quality |
| S12_vs_k141_38545\|\|full | Low-quality |
| S12_vs_k141_125183\|\|full | Low-quality |
| S12_vs_k141_38565\|\|full | Low-quality |
| S12_vs_k141_133346\|\|full | Not-determined |
| S12_vs_k141_78627\|\|full | Low-quality |
| S12_vs_k141_86325\|\|full | Low-quality |
| S12_vs_k141_156522\|\|full | Low-quality |
| S12_vs_k141_133349\|\|full | Low-quality |
| S12_vs_k141_133351\|\|full | Low-quality |
| S12_vs_k141_23238\|\|full | Low-quality |
| S12_vs_k141_31380\|\|full | Not-determined |
| S12_vs_k141_180271\|\|full | Not-determined |
| S12_vs_k141_14922\|\|full | Low-quality |
| S12_vs_k141_23244\|\|full | Not-determined |
| S12_vs_k141_165004\|\|full | Low-quality |
| S12_vs_k141_86345\|\|full | Low-quality |
| S12_vs_k141_165005\|\|full | Low-quality |
| S12_vs_k141_86349\|\|full | Not-determined |
| S12_vs_k141_165011\|\|full | Low-quality |
| S12_vs_k141_78664\|\|full | Low-quality |
| S12_vs_k141_180296\|\|full | Not-determined |
| S12_vs_k141_156552\|\|full | Low-quality |
| S12_vs_k141_78667\|\|full | Not-determined |
| S12_vs_k141_31408\|\|full | Not-determined |
| S12_vs_k141_31410\|\|full | Not-determined |
| S12_vs_k141_133385\|\|full | Not-determined |
| S12_vs_k141_86358\|\|full | Low-quality |
| S12_vs_k141_133401\|\|full | Not-determined |
| S12_vs_k141_38635\|\|full | Low-quality |
| S12_vs_k141_78692\|\|full | Low-quality |
| S12_vs_k141_86373\|\|full | Low-quality |
| S12_vs_k141_156579\|\|full | Not-determined |
| S12_vs_k141_86417\|\|full | Low-quality |
| S12_vs_k141_156612\|\|full | Low-quality |
| S12_vs_k141_180359\|\|full | Not-determined |
| S12_vs_k141_125306\|\|full | Low-quality |
| S8_extracted_k141_71214 | Medium-quality |
| S12_vs_k141_156630\|\|full | Not-determined |
| S12_vs_k141_172383\|\|full | Not-determined |
| S12_vs_k141_172418\|\|full | Not-determined |
| S12_vs_k141_86493\|\|full | Low-quality |
| S12_vs_k141_23381\|\|full | Low-quality |
| S12_vs_k141_86497\|\|full | Not-determined |
| S12_vs_k141_23383\|\|full | Low-quality |
| S12_vs_k141_133511\|\|full | Not-determined |
| S12_vs_k141_31457\|\|full | Not-determined |
| S11_vs_k141_93101\|\|full | High-quality |
| S12_vs_k141_180421\|\|full | Low-quality |
| S12_vs_k141_14928\|\|full | Not-determined |
| S16_vs_48714\|\|full | Medium-quality |
| S12_vs_k141_133524\|\|full | Low-quality |
| S12_vs_k141_14935\|\|full | Low-quality |
| S12_vs_k141_165124\|\|full | Low-quality |
| S12_vs_k141_165128\|\|full | Not-determined |
| S12_vs_k141_165134\|\|full | Low-quality |
| S12_vs_k141_165136\|\|full | Not-determined |
| S12_vs_k141_23392\|\|full | Low-quality |
| S12_vs_k141_165147\|\|full | Low-quality |
| S12_vs_k141_133550\|\|full | Low-quality |
| S12_vs_k141_125384\|\|full | Not-determined |
| S12_vs_k141_172523\|\|full | Low-quality |
| S12_vs_k141_55040\|\|full | Low-quality |
| S12_vs_k141_117518\|\|full | Not-determined |
| S12_vs_k141_14969\|\|full | Low-quality |
| S12_vs_k141_180475\|\|full | Not-determined |
| S12_vs_k141_165174\|\|full | Low-quality |
| S12_vs_k141_14974\|\|full | Low-quality |
| S12_vs_k141_117520\|\|full | Low-quality |
| S12_vs_k141_14981\|\|full | Not-determined |
| S12_vs_k141_125394\|\|full | Low-quality |
| S12_vs_k141_133610\|\|full | Not-determined |
| S12_vs_k141_172549\|\|full | Low-quality |
| S12_vs_k141_125410\|\|full | Low-quality |
| S12_vs_k141_38834\|\|full | Not-determined |
| S12_vs_k141_172584\|\|full | Low-quality |
| S12_vs_k141_86569\|\|full | Not-determined |
| S12_vs_k141_165216\|\|full | Not-determined |
| S12_vs_k141_180556\|\|full | Low-quality |
| S12_vs_k141_86580\|\|full | Not-determined |
| S12_vs_k141_23494\|\|full | Not-determined |
| S12_vs_k141_55075\|\|full | Low-quality |
| S12_vs_k141_117606\|\|full | Not-determined |
| S12_vs_k141_86604\|\|full | Not-determined |
| S12_vs_k141_180573\|\|full | Low-quality |
| S12_vs_k141_172646\|\|full | Low-quality |
| S12_vs_k141_172650\|\|full | Low-quality |
| S12_vs_k141_156797\|\|full | Low-quality |
| S12_vs_k141_15086\|\|full | Low-quality |
| S12_vs_k141_15092\|\|full | Low-quality |
| S12_vs_k141_180598\|\|full | Low-quality |
| S12_vs_k141_38912\|\|full | Low-quality |
| S12_vs_k141_172698\|\|full | Low-quality |
| S12_vs_k141_117690\|\|full | Not-determined |
| S12_vs_k141_23567\|\|full | Low-quality |
| S12_vs_k141_38939\|\|full | Not-determined |
| S12_vs_k141_156872\|\|full | Low-quality |
| S12_vs_k141_117719\|\|full | Low-quality |
| S12_vs_k141_172745\|\|full | Low-quality |
| S12_vs_k141_156896\|\|full | Not-determined |
| S12_vs_k141_133765\|\|full | Low-quality |
| S12_vs_k141_38990\|\|full | Low-quality |
| S12_vs_k141_23574\|\|full | Not-determined |
| S12_vs_k141_172790\|\|full | Not-determined |
| S12_vs_k141_15170\|\|full | Low-quality |
| S12_vs_k141_117759\|\|full | Not-determined |
| S12_vs_k141_133806\|\|full | Not-determined |
| S12_vs_k141_125482\|\|full | Low-quality |
| S12_vs_k141_15201\|\|full | Not-determined |
| S12_vs_k141_172835\|\|full | Low-quality |
| S12_vs_k141_125488\|\|full | Low-quality |
| S12_vs_k141_133821\|\|full | Low-quality |
| S12_vs_k141_117806\|\|full | Low-quality |
| S12_vs_k141_133839\|\|full | Not-determined |
| S12_vs_k141_172870\|\|full | Not-determined |
| S12_vs_k141_15250\|\|full | Low-quality |
| S12_vs_k141_172875\|\|full | Low-quality |
| S12_vs_k141_133843\|\|full | Not-determined |
| S12_vs_k141_117835\|\|full | Low-quality |
| S12_vs_k141_125520\|\|full | Not-determined |
| S12_vs_k141_172888\|\|full | Low-quality |
| S12_vs_k141_125527\|\|full | Not-determined |
| S12_vs_k141_39054\|\|full | Not-determined |
| S12_vs_k141_39079\|\|full | Not-determined |
| S12_vs_k141_39092\|\|full | Low-quality |
| S12_vs_k141_157003\|\|full | Not-determined |
| S12_vs_k141_39095\|\|full | Low-quality |
| S12_vs_k141_157016\|\|full | Not-determined |
| S12_vs_k141_15339\|\|full | Not-determined |
| S12_vs_k141_172942\|\|full | Low-quality |
| S12_vs_k141_125621\|\|full | Low-quality |
| S12_vs_k141_39116\|\|full | Not-determined |
| S12_vs_k141_180653\|\|full | Not-determined |
| S12_vs_k141_117938\|\|full | Low-quality |
| S12_vs_k141_117948\|\|full | Not-determined |
| S12_vs_k141_117951\|\|full | Not-determined |
| S12_vs_k141_39152\|\|full | Low-quality |
| S12_vs_k141_157075\|\|full | Low-quality |
| S12_vs_k141_125643\|\|full | Not-determined |
| S12_vs_k141_173002\|\|full | Not-determined |
| S12_vs_k141_125655\|\|full | Low-quality |
| S12_vs_k141_173007\|\|full | Not-determined |
| S12_vs_k141_125659\|\|full | Not-determined |
| S12_vs_k141_117972\|\|full | Low-quality |
| S12_vs_k141_125663\|\|full | Low-quality |
| S12_vs_k141_173020\|\|full | Not-determined |
| S12_vs_k141_117984\|\|full | Not-determined |
| S12_vs_k141_173025\|\|full | Not-determined |
| S12_vs_k141_39181\|\|full | Low-quality |
| S12_vs_k141_157107\|\|full | Low-quality |
| S12_vs_k141_173036\|\|full | Not-determined |
| S12_vs_k141_125692\|\|full | Low-quality |
| S12_vs_k141_180691\|\|full | Low-quality |
| S12_vs_k141_118003\|\|full | Low-quality |
| S12_vs_k141_180692\|\|full | Not-determined |
| S12_vs_k141_125694\|\|full | Low-quality |
| S12_vs_k141_15466\|\|full | Low-quality |
| S12_vs_k141_180700\|\|full | Low-quality |
| S12_vs_k141_125702\|\|full | Not-determined |
| S12_vs_k141_180712\|\|full | Low-quality |
| S12_vs_k141_39249\|\|full | Low-quality |
| S12_vs_k141_15494\|\|full | Low-quality |
| S12_vs_k141_157172\|\|full | Low-quality |
| S12_vs_k141_39260\|\|full | Not-determined |
| S12_vs_k141_15510\|\|full | Low-quality |

| | |
|---|---|
| S12_vs_k141_157183||full | Low-quality |
| S12_vs_k141_173132||full | Not-determined |
| S12_vs_k141_125714||full | Not-determined |
| S12_vs_k141_173141||full | Low-quality |
| S12_vs_k141_157200||full | Low-quality |
| S12_vs_k141_125739||full | Low-quality |
| S12_vs_k141_157212||full | Not-determined |
| S12_vs_k141_15561||full | Low-quality |
| S12_vs_k141_15562||full | Low-quality |
| S12_vs_k141_118045||full | Not-determined |
| S12_vs_k141_15596||full | Not-determined |
| S12_vs_k141_157221||full | Low-quality |
| S12_vs_k141_15621||full | Not-determined |
| S12_vs_k141_180844||full | Low-quality |
| S12_vs_k141_15638||full | Low-quality |
| S12_vs_k141_157267||full | Low-quality |
| S12_vs_k141_125798||full | Not-determined |
| S12_vs_k141_157278||full | Not-determined |
| S12_vs_k141_15667||full | Low-quality |
| S12_vs_k141_180863||full | Low-quality |
| S12_vs_k141_157288||full | Low-quality |
| S12_vs_k141_180878||full | Not-determined |
| S12_vs_k141_125820||full | Low-quality |
| S12_vs_k141_157344||full | Low-quality |
| S12_vs_k141_157350||full | Not-determined |
| S12_vs_k141_157359||full | Low-quality |
| S12_vs_k141_180915||full | Not-determined |
| S12_vs_k141_15708||full | Low-quality |
| S12_vs_k141_180930||full | Not-determined |
| S12_vs_k141_157387||full | Not-determined |
| S12_vs_k141_125897||full | Low-quality |
| S6_vs_k141_38250||full | Medium-quality |
| S12_vs_k141_180994||full | Low-quality |
| S8_vs_k141_71214||full | Medium-quality |
| S12_vs_k141_181039||full | Low-quality |
| S7_vs_23770||full | Medium-quality |
| S12_vs_k141_165729||0_partial | Low-quality |
| S12_vs_k141_31878||0_partial | Low-quality |
| S12_vs_k141_165858||0_partial | Not-determined |
| S12_vs_k141_158058||0_partial | Not-determined |
| S12_vs_k141_165925||0_partial | Low-quality |
| S12_vs_k141_87395||0_partial | Not-determined |
| S12_vs_k141_24899||0_partial | Low-quality |
| S12_vs_k141_103765||0_partial | Not-determined |
| S12_vs_k141_40748||0_partial | Low-quality |
| S12_vs_k141_48289||0_partial | Low-quality |
| S12_vs_k141_88221||0_partial | Low-quality |
| S17_extracted_k141_46388 | Medium-quality |
| S12_vs_k141_104347||0_partial | Low-quality |
| S12_vs_k141_25672||0_partial | Not-determined |
| S12_vs_k141_72635||0_partial | Low-quality |
| S12_vs_k141_183908||0_partial | Not-determined |
| S12_vs_k141_66035||0_partial | Not-determined |
| S12_vs_k141_175723||0_partial | Low-quality |
| S12_vs_k141_184313||0_partial | Low-quality |
| S12_vs_k141_90309||0_partial | Low-quality |
| S12_vs_k141_105885||0_partial | Low-quality |
| S12_vs_k141_58711||0_partial | Not-determined |
| S12_vs_k141_19581||0_partial | Not-determined |
| S12_vs_k141_106720||0_partial | Not-determined |
| S12_vs_k141_51104||0_partial | Low-quality |
| S12_vs_k141_35213||0_partial | Low-quality |
| S12_vs_k141_59958||0_partial | Low-quality |
| S12_vs_k141_60227||0_partial | Low-quality |
| S12_vs_k141_130864||0_partial | Not-determined |
| S12_vs_k141_92520||0_partial | Low-quality |
| S12_vs_k141_36441||0_partial | Not-determined |
| S12_vs_k141_6860||0_partial | Not-determined |
| S12_vs_k141_164274||0_partial | Not-determined |
| S12_vs_k141_171917||0_partial | Not-determined |
| S12_vs_k141_125063||0_partial | Not-determined |
| S12_vs_k141_23569||0_partial | Low-quality |
| S12_vs_k141_165294||0_partial | Not-determined |
| S12_vs_k141_110245||lt2gene | Low-quality |
| S12_vs_k141_63001||lt2gene | Low-quality |
| S12_vs_k141_133868||lt2gene | Low-quality |
| S12_vs_k141_15760||lt2gene | Low-quality |
| S12_vs_k141_125999||lt2gene | Low-quality |
| S12_vs_k141_55142||lt2gene | Not-determined |
| S12_vs_k141_31518||lt2gene | Low-quality |
| S12_vs_k141_86688||lt2gene | Low-quality |
| S12_vs_k141_181134||lt2gene | Not-determined |
| S12_vs_k141_102431||lt2gene | Low-quality |
| S12_vs_k141_100||lt2gene | Low-quality |
| S12_vs_k141_70951||lt2gene | Not-determined |
| S12_vs_k141_7954||lt2gene | Not-determined |
| S12_vs_k141_173312||lt2gene | Low-quality |
| S12_vs_k141_149691||lt2gene | Low-quality |
| S12_vs_k141_70976||lt2gene | Low-quality |
| S12_vs_k141_165477||lt2gene | Low-quality |
| S12_vs_k141_15813||lt2gene | Low-quality |
| S12_vs_k141_126130||lt2gene | Low-quality |
| S12_vs_k141_7987||lt2gene | Low-quality |
| S12_vs_k141_118202||lt2gene | Low-quality |
| S12_vs_k141_118203||lt2gene | Not-determined |
| S12_vs_k141_173347||lt2gene | Not-determined |
| S12_vs_k141_118214||lt2gene | Low-quality |
| S12_vs_k141_181222||lt2gene | Low-quality |
| S12_vs_k141_141815||lt2gene | Low-quality |
| S12_vs_k141_141820||lt2gene | Not-determined |
| S12_vs_k141_133980||lt2gene | Low-quality |
| S12_vs_k141_86802||lt2gene | Not-determined |
| S12_vs_k141_47334||lt2gene | Not-determined |
| S12_vs_k141_86814||lt2gene | Not-determined |
| S12_vs_k141_86815||lt2gene | Low-quality |
| S12_vs_k141_55210||lt2gene | Low-quality |
| S12_vs_k141_71048||lt2gene | Low-quality |
| S12_vs_k141_23787||lt2gene | Not-determined |
| S12_vs_k141_31617||lt2gene | Low-quality |
| S12_vs_k141_157617||lt2gene | Low-quality |
| S12_vs_k141_165571||lt2gene | Low-quality |
| S12_vs_k141_173424||lt2gene | Not-determined |
| S12_vs_k141_8065||lt2gene | Low-quality |
| S12_vs_k141_71076||lt2gene | Not-determined |
| S12_vs_k141_94718||lt2gene | Not-determined |
| S12_vs_k141_181308||lt2gene | Low-quality |
| S12_vs_k141_39508||lt2gene | Not-determined |
| S12_vs_k141_102604||lt2gene | Low-quality |
| S12_vs_k141_157684||lt2gene | Not-determined |
| S12_vs_k141_94750||lt2gene | Low-quality |
| S12_vs_k141_55255||lt2gene | Low-quality |
| S12_vs_k141_63222||lt2gene | Low-quality |
| S12_vs_k141_316||lt2gene | Low-quality |
| S12_vs_k141_325||lt2gene | Low-quality |
| S12_vs_k141_39546||lt2gene | Low-quality |
| S12_vs_k141_8113||lt2gene | Low-quality |
| S12_vs_k141_94784||lt2gene | Low-quality |
| S12_vs_k141_181372||lt2gene | Not-determined |
| S12_vs_k141_71151||lt2gene | Low-quality |
| S12_vs_k141_71156||lt2gene | Low-quality |
| S12_vs_k141_15897||lt2gene | Not-determined |
| S12_vs_k141_47422||lt2gene | Low-quality |
| S12_vs_k141_141956||lt2gene | Not-determined |
| S12_vs_k141_141978||lt2gene | Low-quality |
| S12_vs_k141_15933||lt2gene | Not-determined |
| S12_vs_k141_55354||lt2gene | Not-determined |
| S12_vs_k141_63376||lt2gene | Low-quality |
| S12_vs_k141_55413||lt2gene | Low-quality |
| S12_vs_k141_79091||lt2gene | Low-quality |
| S12_vs_k141_126411||lt2gene | Not-determined |
| S12_vs_k141_157796||lt2gene | Low-quality |
| S12_vs_k141_149890||lt2gene | Low-quality |
| S12_vs_k141_157801||lt2gene | Low-quality |
| S12_vs_k141_55436||lt2gene | Low-quality |
| S12_vs_k141_165725||lt2gene | Low-quality |
| S12_vs_k141_173554||lt2gene | Low-quality |
| S12_vs_k141_126452||lt2gene | Not-determined |
| S12_vs_k141_157834||lt2gene | Low-quality |
| S12_vs_k141_94917||lt2gene | Low-quality |
| S12_vs_k141_142159||lt2gene | Not-determined |
| S12_vs_k141_23801||lt2gene | Low-quality |
| S12_vs_k141_79165||lt2gene | Low-quality |
| S12_vs_k141_550||lt2gene | Not-determined |
| S12_vs_k141_23822||lt2gene | Low-quality |
| S12_vs_k141_23825||lt2gene | Low-quality |
| S12_vs_k141_71260||lt2gene | Low-quality |
| S12_vs_k141_79195||lt2gene | Low-quality |
| S12_vs_k141_79202||lt2gene | Low-quality |
| S12_vs_k141_134164||lt2gene | Low-quality |
| S12_vs_k141_16064||lt2gene | Not-determined |
| S12_vs_k141_157913||lt2gene | Low-quality |
| S12_vs_k141_134186||lt2gene | Low-quality |
| S12_vs_k141_47499||lt2gene | Low-quality |
| S12_vs_k141_142253||lt2gene | Low-quality |
| S12_vs_k141_23928||lt2gene | Not-determined |
| S12_vs_k141_647||lt2gene | Low-quality |
| S12_vs_k141_181534||lt2gene | Low-quality |
| S12_vs_k141_63538||lt2gene | Not-determined |
| S12_vs_k141_8288||lt2gene | Low-quality |
| S12_vs_k141_94971||lt2gene | Low-quality |
| S12_vs_k141_39853||lt2gene | Low-quality |
| S12_vs_k141_150065||lt2gene | Low-quality |
| S12_vs_k141_134204||lt2gene | Not-determined |
| S12_vs_k141_134226||lt2gene | Low-quality |
| S12_vs_k141_126593||lt2gene | Low-quality |
| S12_vs_k141_181599||lt2gene | Low-quality |
| S12_vs_k141_134238||lt2gene | Low-quality |
| S12_vs_k141_79323||lt2gene | Not-determined |
| S12_vs_k141_95007||lt2gene | Low-quality |
| S12_vs_k141_142353||lt2gene | Low-quality |
| S12_vs_k141_8308||lt2gene | Low-quality |
| S12_vs_k141_118693||lt2gene | Low-quality |
| S12_vs_k141_47559||lt2gene | Low-quality |
| S12_vs_k141_110723||lt2gene | Low-quality |
| S12_vs_k141_158045||lt2gene | Not-determined |
| S12_vs_k141_39896||lt2gene | Low-quality |
| S12_vs_k141_150129||lt2gene | Low-quality |
| S12_vs_k141_63677||lt2gene | Not-determined |
| S12_vs_k141_24099||lt2gene | Not-determined |
| S12_vs_k141_71446||lt2gene | Low-quality |
| S12_vs_k141_173642||lt2gene | Low-quality |
| S12_vs_k141_71454||lt2gene | Not-determined |
| S12_vs_k141_173654||lt2gene | Low-quality |
| S12_vs_k141_71464||lt2gene | Low-quality |
| S12_vs_k141_16122||lt2gene | Low-quality |
| S12_vs_k141_71492||lt2gene | Low-quality |
| S12_vs_k141_79411||lt2gene | Not-determined |
| S12_vs_k141_165953||lt2gene | Low-quality |
| S12_vs_k141_8444||lt2gene | Low-quality |
| S12_vs_k141_102988||lt2gene | Low-quality |
| S12_vs_k141_79440||lt2gene | Low-quality |
| S12_vs_k141_79445||lt2gene | Not-determined |
| S12_vs_k141_24156||lt2gene | Low-quality |
| S12_vs_k141_16182||lt2gene | Low-quality |
| S12_vs_k141_39995||lt2gene | Not-determined |
| S12_vs_k141_40004||lt2gene | Low-quality |
| S12_vs_k141_150239||lt2gene | Low-quality |
| S12_vs_k141_24225||lt2gene | Low-quality |
| S12_vs_k141_173709||lt2gene | Not-determined |
| S12_vs_k141_897||lt2gene | Low-quality |
| S12_vs_k141_150271||lt2gene | Low-quality |
| S12_vs_k141_8516||lt2gene | Low-quality |
| S12_vs_k141_24244||lt2gene | Not-determined |
| S12_vs_k141_47649||lt2gene | Low-quality |
| S12_vs_k141_173728||lt2gene | Not-determined |
| S12_vs_k141_173745||lt2gene | Not-determined |
| S12_vs_k141_173747||lt2gene | Low-quality |
| S12_vs_k141_103116||lt2gene | Low-quality |
| S12_vs_k141_158195||lt2gene | Not-determined |
| S12_vs_k141_55815||lt2gene | Low-quality |
| S12_vs_k141_8586||lt2gene | Not-determined |
| S12_vs_k141_32178||lt2gene | Not-determined |
| S12_vs_k141_181865||lt2gene | Low-quality |
| S12_vs_k141_966||lt2gene | Low-quality |
| S12_vs_k141_24334||lt2gene | Low-quality |
| S12_vs_k141_126736||lt2gene | Low-quality |
| S12_vs_k141_40149||lt2gene | Not-determined |
| S12_vs_k141_158246||lt2gene | Low-quality |
| S12_vs_k141_126746||lt2gene | Low-quality |
| S12_vs_k141_103203||lt2gene | Not-determined |
| S12_vs_k141_24362||lt2gene | Low-quality |
| S12_vs_k141_118933||lt2gene | Low-quality |
| S12_vs_k141_166154||lt2gene | Low-quality |
| S12_vs_k141_32262||lt2gene | Low-quality |
| S12_vs_k141_134482||lt2gene | Not-determined |
| S12_vs_k141_166160||lt2gene | Not-determined |
| S12_vs_k141_87419||lt2gene | Not-determined |
| S12_vs_k141_103292||lt2gene | Low-quality |
| S12_vs_k141_55939||lt2gene | Low-quality |
| S12_vs_k141_40229||lt2gene | Low-quality |
| S12_vs_k141_1021||lt2gene | Low-quality |
| S12_vs_k141_118973||lt2gene | Not-determined |
| S12_vs_k141_16400||lt2gene | Low-quality |
| S12_vs_k141_166255||lt2gene | Low-quality |
| S12_vs_k141_134576||lt2gene | Low-quality |
| S12_vs_k141_87501||lt2gene | Not-determined |
| S12_vs_k141_47872||lt2gene | Low-quality |
| S12_vs_k141_134591||lt2gene | Not-determined |
| S12_vs_k141_142693||lt2gene | Low-quality |
| S12_vs_k141_142694||lt2gene | Low-quality |
| S12_vs_k141_63974||lt2gene | Low-quality |
| S12_vs_k141_126840||lt2gene | Low-quality |
| S12_vs_k141_47889||lt2gene | Low-quality |
| S12_vs_k141_87524||lt2gene | Low-quality |
| S12_vs_k141_119029||lt2gene | Low-quality |
| S12_vs_k141_47896||lt2gene | Low-quality |
| S12_vs_k141_16462||lt2gene | Low-quality |
| S12_vs_k141_119048||lt2gene | Low-quality |
| S12_vs_k141_16465||lt2gene | Low-quality |
| S12_vs_k141_16468||lt2gene | Low-quality |
| S12_vs_k141_182056||lt2gene | Low-quality |
| S12_vs_k141_150490||lt2gene | Not-determined |
| S12_vs_k141_103423||lt2gene | Low-quality |
| S12_vs_k141_32363||lt2gene | Low-quality |
| S12_vs_k141_150497||lt2gene | Not-determined |
| S12_vs_k141_16496||lt2gene | Low-quality |
| S12_vs_k141_166328||lt2gene | Low-quality |
| S12_vs_k141_182083||lt2gene | Not-determined |
| S12_vs_k141_182087||lt2gene | Low-quality |
| S12_vs_k141_24519||lt2gene | Not-determined |
| S12_vs_k141_47977||lt2gene | Low-quality |
| S12_vs_k141_47978||lt2gene | Low-quality |
| S12_vs_k141_95462||lt2gene | Low-quality |
| S12_vs_k141_24580||lt2gene | Not-determined |
| S12_vs_k141_8721||lt2gene | Low-quality |
| S12_vs_k141_56128||lt2gene | Low-quality |
| S12_vs_k141_32434||lt2gene | Low-quality |
| S12_vs_k141_64107||lt2gene | Not-determined |
| S12_vs_k141_182183||lt2gene | Low-quality |
| S12_vs_k141_166419||lt2gene | Low-quality |
| S12_vs_k141_24604||lt2gene | Low-quality |
| S12_vs_k141_24645||lt2gene | Low-quality |
| S12_vs_k141_174083||lt2gene | Low-quality |
| S12_vs_k141_95591||lt2gene | Low-quality |
| S12_vs_k141_95594||lt2gene | Low-quality |
| S12_vs_k141_174105||lt2gene | Low-quality |
| S12_vs_k141_119186||lt2gene | Not-determined |
| S12_vs_k141_48093||lt2gene | Not-determined |
| S12_vs_k141_134690||lt2gene | Not-determined |
| S12_vs_k141_174162||lt2gene | Low-quality |
| S12_vs_k141_79686||lt2gene | Low-quality |
| S12_vs_k141_119230||lt2gene | Low-quality |
| S12_vs_k141_182321||lt2gene | Low-quality |
| S12_vs_k141_8819||lt2gene | Low-quality |
| S12_vs_k141_24793||lt2gene | Low-quality |
| S12_vs_k141_16699||lt2gene | Not-determined |
| S12_vs_k141_64256||lt2gene | Low-quality |
| S12_vs_k141_127045||lt2gene | Not-determined |
| S12_vs_k141_119271||lt2gene | Low-quality |
| S12_vs_k141_48169||lt2gene | Not-determined |
| S12_vs_k141_8861||lt2gene | Not-determined |
| S12_vs_k141_79765||lt2gene | Not-determined |
| S12_vs_k141_32532||lt2gene | Low-quality |
| S12_vs_k141_1293||lt2gene | Not-determined |
| S12_vs_k141_134759||lt2gene | Low-quality |
| S12_vs_k141_16744||lt2gene | Not-determined |
| S12_vs_k141_16750||lt2gene | Not-determined |
| S12_vs_k141_111165||lt2gene | Low-quality |
| S12_vs_k141_56380||lt2gene | Low-quality |
| S12_vs_k141_24852||lt2gene | Low-quality |
| S12_vs_k141_32581||lt2gene | Low-quality |
| S12_vs_k141_142992||lt2gene | Not-determined |
| S12_vs_k141_174257||lt2gene | Not-determined |
| S12_vs_k141_119342||lt2gene | Low-quality |
| S12_vs_k141_87865||lt2gene | Low-quality |
| S12_vs_k141_79789||lt2gene | Low-quality |
| S12_vs_k141_79792||lt2gene | Low-quality |
| S12_vs_k141_87872||lt2gene | Low-quality |
| S12_vs_k141_72079||lt2gene | Low-quality |
| S12_vs_k141_158517||lt2gene | Not-determined |
| S12_vs_k141_79822||lt2gene | Low-quality |
| S12_vs_k141_16829||lt2gene | Low-quality |
| S12_vs_k141_158520||lt2gene | Low-quality |
| S12_vs_k141_16837||lt2gene | Not-determined |
| S12_vs_k141_16849||lt2gene | Low-quality |
| S12_vs_k141_1382||lt2gene | Low-quality |
| S12_vs_k141_24903||lt2gene | Low-quality |
| S12_vs_k141_16860||lt2gene | Not-determined |
| S12_vs_k141_1447||lt2gene | Low-quality |
| S12_vs_k141_64401||lt2gene | Low-quality |
| S12_vs_k141_95888||lt2gene | Low-quality |
| S12_vs_k141_158611||lt2gene | Low-quality |
| S12_vs_k141_127297||lt2gene | Low-quality |
| S12_vs_k141_134948||lt2gene | Not-determined |
| S12_vs_k141_158640||lt2gene | Not-determined |
| S12_vs_k141_182519||lt2gene | Not-determined |
| S12_vs_k141_9072||lt2gene | Not-determined |
| S12_vs_k141_143087||lt2gene | Low-quality |
| S12_vs_k141_25004||lt2gene | Low-quality |
| S12_vs_k141_25010||lt2gene | Low-quality |
| S12_vs_k141_88057||lt2gene | Low-quality |
| S12_vs_k141_158678||lt2gene | Low-quality |
| S12_vs_k141_119480||lt2gene | Not-determined |
| S12_vs_k141_182577||lt2gene | Low-quality |
| S12_vs_k141_17016||lt2gene | Low-quality |
| S12_vs_k141_25070||lt2gene | Not-determined |
| S12_vs_k141_72275||lt2gene | Not-determined |
| S12_vs_k141_119538||lt2gene | Low-quality |
| S12_vs_k141_135090||lt2gene | Not-determined |
| S12_vs_k141_166835||lt2gene | Low-quality |
| S12_vs_k141_9141||lt2gene | Low-quality |
| S12_vs_k141_150811||lt2gene | Low-quality |
| S12_vs_k141_166851||lt2gene | Not-determined |
| S12_vs_k141_79971||lt2gene | Low-quality |
| S12_vs_k141_79972||lt2gene | Low-quality |
| S12_vs_k141_25134||lt2gene | Low-quality |
| S12_vs_k141_72355||lt2gene | Low-quality |
| S12_vs_k141_135131||lt2gene | Low-quality |
| S12_vs_k141_80002||lt2gene | Low-quality |
| S12_vs_k141_174378||lt2gene | Not-determined |
| S12_vs_k141_17061||lt2gene | Low-quality |
| S12_vs_k141_1598||lt2gene | Low-quality |
| S12_vs_k141_64619||lt2gene | Low-quality |
| S12_vs_k141_9232||lt2gene | Not-determined |
| S12_vs_k141_166956||lt2gene | Not-determined |
| S12_vs_k141_56751||lt2gene | Low-quality |
| S12_vs_k141_127395||lt2gene | Low-quality |
| S12_vs_k141_143228||lt2gene | Low-quality |
| S12_vs_k141_72437||lt2gene | Low-quality |
| S12_vs_k141_88174||lt2gene | Low-quality |
| S12_vs_k141_182682||lt2gene | Low-quality |
| S12_vs_k141_96144||lt2gene | Low-quality |
| S12_vs_k141_96157||lt2gene | Low-quality |
| S12_vs_k141_166983||lt2gene | Low-quality |
| S12_vs_k141_119779||lt2gene | Low-quality |
| S12_vs_k141_150943||lt2gene | Not-determined |
| S12_vs_k141_103918||lt2gene | Not-determined |
| S12_vs_k141_56809||lt2gene | Low-quality |
| S12_vs_k141_166996||lt2gene | Low-quality |
| S12_vs_k141_56819||lt2gene | Low-quality |
| S12_vs_k141_103931||lt2gene | Low-quality |
| S12_vs_k141_56831||lt2gene | Not-determined |
| S12_vs_k141_111462||lt2gene | Not-determined |
| S12_vs_k141_182741||lt2gene | Low-quality |
| S12_vs_k141_56846||lt2gene | Low-quality |
| S12_vs_k141_174482||lt2gene | Not-determined |
| S12_vs_k141_56852||lt2gene | Not-determined |
| S12_vs_k141_96250||lt2gene | Low-quality |
| S12_vs_k141_40833||lt2gene | Low-quality |
| S12_vs_k141_127571||lt2gene | Not-determined |
| S12_vs_k141_64735||lt2gene | Not-determined |
| S12_vs_k141_56879||lt2gene | Not-determined |
| S12_vs_k141_103977||lt2gene | Not-determined |
| S12_vs_k141_1743||lt2gene | Not-determined |
| S12_vs_k141_103989||lt2gene | Low-quality |
| S12_vs_k141_1769||lt2gene | Not-determined |
| S12_vs_k141_96308||lt2gene | Low-quality |
| S12_vs_k141_56929||lt2gene | Low-quality |
| S12_vs_k141_119882||lt2gene | Not-determined |
| S12_vs_k141_64771||lt2gene | Low-quality |
| S12_vs_k141_127630||lt2gene | Low-quality |
| S12_vs_k141_96357||lt2gene | Low-quality |
| S12_vs_k141_182779||lt2gene | Not-determined |
| S12_vs_k141_104056||lt2gene | Not-determined |
| S12_vs_k141_25352||lt2gene | Low-quality |
| S12_vs_k141_143400||lt2gene | Low-quality |
| S12_vs_k141_96398||lt2gene | Low-quality |
| S12_vs_k141_48450||lt2gene | Low-quality |
| S12_vs_k141_9348||lt2gene | Not-determined |
| S12_vs_k141_32698||lt2gene | Low-quality |
| S12_vs_k141_104105||lt2gene | Low-quality |
| S12_vs_k141_72617||lt2gene | Low-quality |
| S12_vs_k141_1866||lt2gene | Low-quality |
| S12_vs_k141_182805||lt2gene | Not-determined |
| S12_vs_k141_120048||lt2gene | Low-quality |
| S12_vs_k141_96483||lt2gene | Not-determined |
| S12_vs_k141_80106||lt2gene | Low-quality |
| S12_vs_k141_96498||lt2gene | Low-quality |
| S12_vs_k141_57094||lt2gene | Low-quality |
| S12_vs_k141_151044||lt2gene | Low-quality |
| S12_vs_k141_104188||lt2gene | Low-quality |
| S12_vs_k141_17310||lt2gene | Low-quality |
| S12_vs_k141_48506||lt2gene | Low-quality |
| S12_vs_k141_104209||lt2gene | Not-determined |
| S12_vs_k141_64828||lt2gene | Low-quality |
| S12_vs_k141_88391||lt2gene | Not-determined |
| S12_vs_k141_88421||lt2gene | Not-determined |
| S12_vs_k141_64858||lt2gene | Low-quality |
| S12_vs_k141_96573||lt2gene | Low-quality |
| S12_vs_k141_88424||lt2gene | Not-determined |
| S12_vs_k141_48523||lt2gene | Low-quality |
| S12_vs_k141_57134||lt2gene | Not-determined |
| S12_vs_k141_57140||lt2gene | Low-quality |
| S12_vs_k141_158819||lt2gene | Not-determined |
| S12_vs_k141_143568||lt2gene | Not-determined |
| S12_vs_k141_167185||lt2gene | Not-determined |
| S12_vs_k141_17400||lt2gene | Low-quality |
| S12_vs_k141_120118||lt2gene | Not-determined |
| S12_vs_k141_104312||lt2gene | Not-determined |
| S12_vs_k141_64900||lt2gene | Not-determined |
| S12_vs_k141_88492||lt2gene | Low-quality |
| S12_vs_k141_25532||lt2gene | Not-determined |
| S12_vs_k141_167233||lt2gene | Low-quality |
| S12_vs_k141_48581||lt2gene | Low-quality |
| S12_vs_k141_88530||lt2gene | Low-quality |
| S12_vs_k141_143659||lt2gene | Low-quality |
| S12_vs_k141_88550||lt2gene | Low-quality |
| S12_vs_k141_167295||lt2gene | Low-quality |
| S12_vs_k141_25608||lt2gene | Not-determined |
| S12_vs_k141_9520||lt2gene | Not-determined |
| S12_vs_k141_32810||lt2gene | Not-determined |
| S12_vs_k141_64976||lt2gene | Not-determined |
| S12_vs_k141_80226||lt2gene | Low-quality |
| S12_vs_k141_104374||lt2gene | Low-quality |
| S12_vs_k141_64995||lt2gene | Not-determined |
| S12_vs_k141_41276||lt2gene | Not-determined |
| S12_vs_k141_88602||lt2gene | Low-quality |
| S12_vs_k141_17550||lt2gene | Not-determined |
| S12_vs_k141_41292||lt2gene | Low-quality |
| S12_vs_k141_167323||lt2gene | Not-determined |
| S12_vs_k141_41318||lt2gene | Low-quality |
| S12_vs_k141_135540||lt2gene | Low-quality |
| S12_vs_k141_9613||lt2gene | Low-quality |
| S12_vs_k141_127833||lt2gene | Low-quality |
| S12_vs_k141_183076||lt2gene | Low-quality |
| S12_vs_k141_17580||lt2gene | Not-determined |
| S12_vs_k141_120269||lt2gene | Low-quality |
| S12_vs_k141_80313||lt2gene | Low-quality |
| S12_vs_k141_143767||lt2gene | Low-quality |
| S12_vs_k141_120310||lt2gene | Not-determined |
| S12_vs_k141_104430||lt2gene | Low-quality |
| S12_vs_k141_183114||lt2gene | Low-quality |
| S12_vs_k141_25710||lt2gene | Not-determined |
| S12_vs_k141_2126||lt2gene | Low-quality |
| S12_vs_k141_120316||lt2gene | Low-quality |
| S12_vs_k141_174687||lt2gene | Low-quality |
| S12_vs_k141_120324||lt2gene | Low-quality |
| S12_vs_k141_167390||lt2gene | Not-determined |
| S12_vs_k141_167406||lt2gene | Low-quality |
| S12_vs_k141_96846||lt2gene | Low-quality |
| S12_vs_k141_2159||lt2gene | Low-quality |
| S12_vs_k141_167416||lt2gene | Low-quality |
| S12_vs_k141_48803||lt2gene | Low-quality |
| S12_vs_k141_80456||lt2gene | Low-quality |
| S12_vs_k141_33071||lt2gene | Low-quality |
| S12_vs_k141_159155||lt2gene | Low-quality |
| S12_vs_k141_57410||lt2gene | Low-quality |
| S12_vs_k141_167453||lt2gene | Low-quality |
| S12_vs_k141_57420||lt2gene | Low-quality |
| S12_vs_k141_25835||lt2gene | Low-quality |
| S12_vs_k141_33164||lt2gene | Low-quality |
| S12_vs_k141_120458||lt2gene | Not-determined |
| S12_vs_k141_111828||lt2gene | Low-quality |
| S12_vs_k141_120486||lt2gene | Not-determined |
| S12_vs_k141_104571||lt2gene | Not-determined |
| S12_vs_k141_104585||lt2gene | Not-determined |
| S12_vs_k141_57529||lt2gene | Not-determined |
| S12_vs_k141_97033||lt2gene | Low-quality |
| S12_vs_k141_17834||lt2gene | Not-determined |
| S12_vs_k141_57541||lt2gene | Not-determined |
| S12_vs_k141_135775||lt2gene | Low-quality |
| S12_vs_k141_111867||lt2gene | Low-quality |
| S12_vs_k141_57545||lt2gene | Low-quality |
| S12_vs_k141_57547||lt2gene | Low-quality |
| S12_vs_k141_151360||lt2gene | Low-quality |
| S12_vs_k141_2262||lt2gene | Not-determined |
| S12_vs_k141_80668||lt2gene | Low-quality |
| S12_vs_k141_120535||lt2gene | Not-determined |
| S12_vs_k141_128068||lt2gene | Low-quality |
| S12_vs_k141_57581||lt2gene | Low-quality |
| S12_vs_k141_135852||lt2gene | Not-determined |
| S12_vs_k141_2328||lt2gene | Low-quality |
| S12_vs_k141_175023||lt2gene | Low-quality |
| S12_vs_k141_135881||lt2gene | Not-determined |
| S12_vs_k141_135895||lt2gene | Low-quality |
| S12_vs_k141_135906||lt2gene | Low-quality |
| S12_vs_k141_97139||lt2gene | Low-quality |
| S12_vs_k141_89015||lt2gene | Low-quality |
| S12_vs_k141_128161||lt2gene | Low-quality |
| S12_vs_k141_183380||lt2gene | Not-determined |
| S12_vs_k141_111997||lt2gene | Low-quality |
| S12_vs_k141_17950||lt2gene | Low-quality |
| S12_vs_k141_41801||lt2gene | Low-quality |
| S12_vs_k141_135940||lt2gene | Not-determined |
| S12_vs_k141_80749||lt2gene | Low-quality |
| S12_vs_k141_80750||lt2gene | Low-quality |
| S12_vs_k141_159309||lt2gene | Low-quality |
| S12_vs_k141_151487||lt2gene | Low-quality |
| S12_vs_k141_159331||lt2gene | Low-quality |
| S12_vs_k141_72660||lt2gene | Not-determined |
| S12_vs_k141_183399||lt2gene | Low-quality |
| S12_vs_k141_97190||lt2gene | Low-quality |
| S12_vs_k141_128223||lt2gene | Low-quality |
| S12_vs_k141_112073||lt2gene | Not-determined |
| S12_vs_k141_175071||lt2gene | Not-determined |
| S12_vs_k141_120617||lt2gene | Low-quality |
| S12_vs_k141_41850||lt2gene | Low-quality |
| S12_vs_k141_89111||lt2gene | Low-quality |
| S12_vs_k141_120638||lt2gene | Low-quality |
| S12_vs_k141_151555||lt2gene | Low-quality |
| S12_vs_k141_97239||lt2gene | Not-determined |
| S12_vs_k141_120656||lt2gene | Low-quality |
| S12_vs_k141_2431||lt2gene | Not-determined |
| S12_vs_k141_112101||lt2gene | Low-quality |
| S12_vs_k141_49128||lt2gene | Low-quality |
| S12_vs_k141_49134||lt2gene | Low-quality |
| S12_vs_k141_183487||lt2gene | Low-quality |
| S12_vs_k141_65542||lt2gene | Not-determined |
| S12_vs_k141_183492||lt2gene | Low-quality |
| S12_vs_k141_49166||lt2gene | Low-quality |
| S12_vs_k141_41912||lt2gene | Low-quality |
| S12_vs_k141_167481||lt2gene | Low-quality |
| S12_vs_k141_65565||lt2gene | Low-quality |
| S12_vs_k141_183531||lt2gene | Low-quality |
| S12_vs_k141_97366||lt2gene | Not-determined |
| S12_vs_k141_159417||lt2gene | Not-determined |
| S12_vs_k141_18075||lt2gene | Not-determined |
| S12_vs_k141_136097||lt2gene | Not-determined |
| S12_vs_k141_33446||lt2gene | Low-quality |
| S12_vs_k141_183559||lt2gene | Low-quality |
| S12_vs_k141_18108||lt2gene | Low-quality |
| S12_vs_k141_136137||lt2gene | Not-determined |
| S12_vs_k141_42018||lt2gene | Low-quality |
| S12_vs_k141_167549||lt2gene | Low-quality |
| S12_vs_k141_112186||lt2gene | Low-quality |
| S12_vs_k141_9821||lt2gene | Low-quality |
| S12_vs_k141_33492||lt2gene | Not-determined |
| S12_vs_k141_128324||lt2gene | Not-determined |
| S12_vs_k141_72834||lt2gene | Low-quality |
| S12_vs_k141_9835||lt2gene | Low-quality |
| S12_vs_k141_9840||lt2gene | Low-quality |
| S12_vs_k141_167598||lt2gene | Low-quality |
| S12_vs_k141_65648||lt2gene | Low-quality |
| S12_vs_k141_81017||lt2gene | Low-quality |
| S12_vs_k141_2593||lt2gene | Low-quality |
| S12_vs_k141_2611||lt2gene | Low-quality |
| S12_vs_k141_159521||lt2gene | Not-determined |
| S12_vs_k141_9868||lt2gene | Not-determined |
| S12_vs_k141_136212||lt2gene | Low-quality |
| S12_vs_k141_72867||lt2gene | Low-quality |
| S12_vs_k141_2666||lt2gene | Not-determined |
| S12_vs_k141_144303||lt2gene | Not-determined |
| S12_vs_k141_104973||lt2gene | Low-quality |
| S12_vs_k141_42129||lt2gene | Not-determined |
| S12_vs_k141_97486||lt2gene | Not-determined |
| S12_vs_k141_167684||lt2gene | Low-quality |
| S12_vs_k141_97490||lt2gene | Low-quality |
| S12_vs_k141_136261||lt2gene | Low-quality |
| S12_vs_k141_57954||lt2gene | Not-determined |

| | |
|---|---|
| S12_vs_k141_112261\|\|lt2gene | Low-quality |
| S12_vs_k141_112286\|\|lt2gene | Low-quality |
| S12_vs_k141_167723\|\|lt2gene | Low-quality |
| S12_vs_k141_9968\|\|lt2gene | Not-determined |
| S12_vs_k141_42197\|\|lt2gene | Low-quality |
| S12_vs_k141_105057\|\|lt2gene | Not-determined |
| S12_vs_k141_58016\|\|lt2gene | Not-determined |
| S12_vs_k141_49297\|\|lt2gene | Low-quality |
| S12_vs_k141_49298\|\|lt2gene | Not-determined |
| S12_vs_k141_183760\|\|lt2gene | Low-quality |
| S12_vs_k141_167761\|\|lt2gene | Low-quality |
| S12_vs_k141_2768\|\|lt2gene | Low-quality |
| S12_vs_k141_10035\|\|lt2gene | Low-quality |
| S12_vs_k141_72967\|\|lt2gene | Low-quality |
| S12_vs_k141_10045\|\|lt2gene | Low-quality |
| S12_vs_k141_58072\|\|lt2gene | Not-determined |
| S12_vs_k141_18362\|\|lt2gene | Low-quality |
| S12_vs_k141_105110\|\|lt2gene | Low-quality |
| S12_vs_k141_65830\|\|lt2gene | Not-determined |
| S12_vs_k141_58098\|\|lt2gene | Low-quality |
| S12_vs_k141_49375\|\|lt2gene | Not-determined |
| S12_vs_k141_18398\|\|lt2gene | Not-determined |
| S12_vs_k141_89574\|\|lt2gene | Low-quality |
| S12_vs_k141_18413\|\|lt2gene | Low-quality |
| S12_vs_k141_42352\|\|lt2gene | Low-quality |
| S12_vs_k141_65880\|\|lt2gene | Low-quality |
| S12_vs_k141_10122\|\|lt2gene | Low-quality |
| S12_vs_k141_183864\|\|lt2gene | Low-quality |
| S12_vs_k141_167892\|\|lt2gene | Low-quality |
| S12_vs_k141_105203\|\|lt2gene | Low-quality |
| S12_vs_k141_112458\|\|lt2gene | Low-quality |
| S12_vs_k141_18474\|\|lt2gene | Low-quality |
| S12_vs_k141_159761\|\|lt2gene | Low-quality |
| S12_vs_k141_128458\|\|lt2gene | Low-quality |
| S12_vs_k141_18484\|\|lt2gene | Low-quality |
| S12_vs_k141_18505\|\|lt2gene | Low-quality |
| S12_vs_k141_18508\|\|lt2gene | Low-quality |
| S12_vs_k141_65921\|\|lt2gene | Low-quality |
| S12_vs_k141_33766\|\|lt2gene | Not-determined |
| S12_vs_k141_18530\|\|lt2gene | Low-quality |
| S12_vs_k141_58219\|\|lt2gene | Low-quality |
| S12_vs_k141_151904\|\|lt2gene | Low-quality |
| S12_vs_k141_33802\|\|lt2gene | Low-quality |
| S12_vs_k141_58223\|\|lt2gene | Low-quality |
| S12_vs_k141_105302\|\|lt2gene | Not-determined |
| S12_vs_k141_167979\|\|lt2gene | Not-determined |
| S12_vs_k141_136513\|\|lt2gene | Low-quality |
| S12_vs_k141_136519\|\|lt2gene | Not-determined |
| S12_vs_k141_65994\|\|lt2gene | Low-quality |
| S12_vs_k141_112576\|\|lt2gene | Not-determined |
| S12_vs_k141_168014\|\|lt2gene | Not-determined |
| S12_vs_k141_105313\|\|lt2gene | Low-quality |
| S12_vs_k141_151960\|\|lt2gene | Low-quality |
| S12_vs_k141_159863\|\|lt2gene | Not-determined |
| S12_vs_k141_49563\|\|lt2gene | Low-quality |
| S12_vs_k141_136552\|\|lt2gene | Low-quality |
| S12_vs_k141_42517\|\|lt2gene | Not-determined |
| S12_vs_k141_66005\|\|lt2gene | Low-quality |
| S12_vs_k141_168035\|\|lt2gene | Not-determined |
| S12_vs_k141_136568\|\|lt2gene | Low-quality |
| S12_vs_k141_58293\|\|lt2gene | Not-determined |
| S12_vs_k141_58297\|\|lt2gene | Not-determined |
| S12_vs_k141_66015\|\|lt2gene | Low-quality |
| S12_vs_k141_3019\|\|lt2gene | Low-quality |
| S12_vs_k141_175533\|\|lt2gene | Low-quality |
| S12_vs_k141_26446\|\|lt2gene | Low-quality |
| S12_vs_k141_152021\|\|lt2gene | Not-determined |
| S12_vs_k141_159932\|\|lt2gene | Not-determined |
| S12_vs_k141_26467\|\|lt2gene | Low-quality |
| S12_vs_k141_105407\|\|lt2gene | Low-quality |
| S12_vs_k141_144546\|\|lt2gene | Not-determined |
| S12_vs_k141_144553\|\|lt2gene | Low-quality |
| S12_vs_k141_136595\|\|lt2gene | Low-quality |
| S12_vs_k141_49623\|\|lt2gene | Low-quality |
| S12_vs_k141_3064\|\|lt2gene | Low-quality |
| S12_vs_k141_81438\|\|lt2gene | Low-quality |
| S12_vs_k141_42591\|\|lt2gene | Low-quality |
| S12_vs_k141_120827\|\|lt2gene | Low-quality |
| S12_vs_k141_73244\|\|lt2gene | Not-determined |
| S12_vs_k141_3104\|\|lt2gene | Low-quality |
| S12_vs_k141_73269\|\|lt2gene | Low-quality |
| S12_vs_k141_49693\|\|lt2gene | Low-quality |
| S12_vs_k141_66063\|\|lt2gene | Low-quality |
| S12_vs_k141_49709\|\|lt2gene | Not-determined |
| S12_vs_k141_175600\|\|lt2gene | Not-determined |
| S12_vs_k141_175606\|\|lt2gene | Not-determined |
| S12_vs_k141_184053\|\|lt2gene | Not-determined |
| S12_vs_k141_168111\|\|lt2gene | Low-quality |
| S12_vs_k141_175615\|\|lt2gene | Low-quality |
| S12_vs_k141_18776\|\|lt2gene | Low-quality |
| S12_vs_k141_120899\|\|lt2gene | Not-determined |
| S12_vs_k141_175620\|\|lt2gene | Low-quality |
| S12_vs_k141_3113\|\|lt2gene | Low-quality |
| S12_vs_k141_89845\|\|lt2gene | Not-determined |
| S12_vs_k141_58395\|\|lt2gene | Low-quality |
| S12_vs_k141_136717\|\|lt2gene | Low-quality |
| S12_vs_k141_42676\|\|lt2gene | Low-quality |
| S12_vs_k141_66077\|\|lt2gene | Low-quality |
| S12_vs_k141_184107\|\|lt2gene | Not-determined |
| S12_vs_k141_128588\|\|lt2gene | Low-quality |
| S12_vs_k141_26566\|\|lt2gene | Low-quality |
| S12_vs_k141_98011\|\|lt2gene | Low-quality |
| S12_vs_k141_73332\|\|lt2gene | Not-determined |
| S12_vs_k141_136753\|\|lt2gene | Not-determined |
| S12_vs_k141_175705\|\|lt2gene | Low-quality |
| S12_vs_k141_49789\|\|lt2gene | Low-quality |
| S12_vs_k141_33984\|\|lt2gene | Not-determined |
| S12_vs_k141_10177\|\|lt2gene | Low-quality |
| S12_vs_k141_152159\|\|lt2gene | Low-quality |
| S12_vs_k141_66115\|\|lt2gene | Not-determined |
| S12_vs_k141_34017\|\|lt2gene | Low-quality |
| S12_vs_k141_73378\|\|lt2gene | Low-quality |
| S12_vs_k141_26641\|\|lt2gene | Not-determined |
| S12_vs_k141_98090\|\|lt2gene | Low-quality |
| S12_vs_k141_73399\|\|lt2gene | Low-quality |
| S12_vs_k141_168247\|\|lt2gene | Not-determined |
| S12_vs_k141_42774\|\|lt2gene | Low-quality |
| S12_vs_k141_136841\|\|lt2gene | Not-determined |
| S12_vs_k141_73409\|\|lt2gene | Low-quality |
| S12_vs_k141_184195\|\|lt2gene | Low-quality |
| S12_vs_k141_105552\|\|lt2gene | Low-quality |
| S12_vs_k141_90013\|\|lt2gene | Low-quality |
| S12_vs_k141_128664\|\|lt2gene | Low-quality |
| S12_vs_k141_81657\|\|lt2gene | Low-quality |
| S12_vs_k141_81660\|\|lt2gene | Low-quality |
| S12_vs_k141_81665\|\|lt2gene | Low-quality |
| S12_vs_k141_152230\|\|lt2gene | Low-quality |
| S12_vs_k141_152241\|\|lt2gene | Low-quality |
| S12_vs_k141_66174\|\|lt2gene | Low-quality |
| S12_vs_k141_49813\|\|lt2gene | Not-determined |
| S12_vs_k141_10266\|\|lt2gene | Low-quality |
| S12_vs_k141_128677\|\|lt2gene | Low-quality |
| S12_vs_k141_105606\|\|lt2gene | Low-quality |
| S12_vs_k141_3294\|\|lt2gene | Low-quality |
| S12_vs_k141_3307\|\|lt2gene | Low-quality |
| S12_vs_k141_3317\|\|lt2gene | Low-quality |
| S12_vs_k141_160135\|\|lt2gene | Low-quality |
| S12_vs_k141_160139\|\|lt2gene | Low-quality |
| S12_vs_k141_58536\|\|lt2gene | Low-quality |
| S12_vs_k141_168360\|\|lt2gene | Not-determined |
| S12_vs_k141_128759\|\|lt2gene | Not-determined |
| S12_vs_k141_105687\|\|lt2gene | Low-quality |
| S12_vs_k141_3369\|\|lt2gene | Low-quality |
| S12_vs_k141_160199\|\|lt2gene | Not-determined |
| S12_vs_k141_58563\|\|lt2gene | Low-quality |
| S12_vs_k141_66284\|\|lt2gene | Low-quality |
| S12_vs_k141_58581\|\|lt2gene | Low-quality |
| S12_vs_k141_136966\|\|lt2gene | Not-determined |
| S12_vs_k141_49911\|\|lt2gene | Low-quality |
| S12_vs_k141_18904\|\|lt2gene | Low-quality |
| S12_vs_k141_113039\|\|lt2gene | Not-determined |
| S12_vs_k141_152331\|\|lt2gene | Not-determined |
| S12_vs_k141_144956\|\|lt2gene | Low-quality |
| S12_vs_k141_136988\|\|lt2gene | Not-determined |
| S12_vs_k141_136995\|\|lt2gene | Low-quality |
| S12_vs_k141_113048\|\|lt2gene | Low-quality |
| S12_vs_k141_98233\|\|lt2gene | Not-determined |
| S12_vs_k141_160292\|\|lt2gene | Low-quality |
| S12_vs_k141_168495\|\|lt2gene | Low-quality |
| S12_vs_k141_98250\|\|lt2gene | Not-determined |
| S12_vs_k141_98252\|\|lt2gene | Low-quality |
| S12_vs_k141_58634\|\|lt2gene | Not-determined |
| S12_vs_k141_168516\|\|lt2gene | Not-determined |
| S12_vs_k141_81862\|\|lt2gene | Low-quality |
| S12_vs_k141_66310\|\|lt2gene | Low-quality |
| S12_vs_k141_43026\|\|lt2gene | Low-quality |
| S12_vs_k141_43028\|\|lt2gene | Not-determined |
| S12_vs_k141_137059\|\|lt2gene | Low-quality |
| S12_vs_k141_113100\|\|lt2gene | Low-quality |
| S12_vs_k141_98313\|\|lt2gene | Not-determined |
| S12_vs_k141_43061\|\|lt2gene | Low-quality |
| S12_vs_k141_152424\|\|lt2gene | Low-quality |
| S12_vs_k141_58700\|\|lt2gene | Low-quality |
| S12_vs_k141_105869\|\|lt2gene | Low-quality |
| S12_vs_k141_152434\|\|lt2gene | Low-quality |
| S12_vs_k141_137085\|\|lt2gene | Low-quality |
| S12_vs_k141_73639\|\|lt2gene | Low-quality |
| S12_vs_k141_184439\|\|lt2gene | Low-quality |
| S12_vs_k141_49974\|\|lt2gene | Low-quality |
| S12_vs_k141_10421\|\|lt2gene | Not-determined |
| S12_vs_k141_152470\|\|lt2gene | Low-quality |
| S12_vs_k141_3567\|\|lt2gene | Low-quality |
| S12_vs_k141_113160\|\|lt2gene | Low-quality |
| S12_vs_k141_81961\|\|lt2gene | Low-quality |
| S12_vs_k141_3591\|\|lt2gene | Not-determined |
| S12_vs_k141_98405\|\|lt2gene | Low-quality |
| S12_vs_k141_3599\|\|lt2gene | Low-quality |
| S12_vs_k141_90379\|\|lt2gene | Low-quality |
| S12_vs_k141_98441\|\|lt2gene | Low-quality |
| S12_vs_k141_3634\|\|lt2gene | Not-determined |
| S12_vs_k141_176009\|\|lt2gene | Low-quality |
| S12_vs_k141_90388\|\|lt2gene | Low-quality |
| S12_vs_k141_50086\|\|lt2gene | Low-quality |
| S12_vs_k141_10501\|\|lt2gene | Low-quality |
| S12_vs_k141_90415\|\|lt2gene | Low-quality |
| S12_vs_k141_184547\|\|lt2gene | Low-quality |
| S12_vs_k141_50104\|\|lt2gene | Not-determined |
| S12_vs_k141_90419\|\|lt2gene | Not-determined |
| S12_vs_k141_105981\|\|lt2gene | Low-quality |
| S12_vs_k141_82053\|\|lt2gene | Low-quality |
| S12_vs_k141_3689\|\|lt2gene | Low-quality |
| S12_vs_k141_3699\|\|lt2gene | Low-quality |
| S12_vs_k141_106009\|\|lt2gene | Not-determined |
| S12_vs_k141_50136\|\|lt2gene | Not-determined |
| S12_vs_k141_137169\|\|lt2gene | Low-quality |
| S12_vs_k141_50150\|\|lt2gene | Not-determined |
| S12_vs_k141_176093\|\|lt2gene | Low-quality |
| S12_vs_k141_98530\|\|lt2gene | Low-quality |
| S12_vs_k141_128954\|\|lt2gene | Low-quality |
| S12_vs_k141_82064\|\|lt2gene | Low-quality |
| S12_vs_k141_160509\|\|lt2gene | Low-quality |
| S12_vs_k141_184624\|\|lt2gene | Low-quality |
| S12_vs_k141_160539\|\|lt2gene | Not-determined |
| S12_vs_k141_82098\|\|lt2gene | Not-determined |
| S12_vs_k141_90508\|\|lt2gene | Not-determined |
| S12_vs_k141_160554\|\|lt2gene | Low-quality |
| S12_vs_k141_137215\|\|lt2gene | Not-determined |
| S12_vs_k141_10602\|\|lt2gene | Low-quality |
| S12_vs_k141_145084\|\|lt2gene | Low-quality |
| S12_vs_k141_106113\|\|lt2gene | Not-determined |
| S12_vs_k141_90542\|\|lt2gene | Low-quality |
| S12_vs_k141_90547\|\|lt2gene | Not-determined |
| S12_vs_k141_50243\|\|lt2gene | Not-determined |
| S12_vs_k141_50267\|\|lt2gene | Not-determined |
| S12_vs_k141_50272\|\|lt2gene | Low-quality |
| S12_vs_k141_121204\|\|lt2gene | Low-quality |
| S12_vs_k141_82151\|\|lt2gene | Low-quality |
| S12_vs_k141_106152\|\|lt2gene | Low-quality |
| S12_vs_k141_145137\|\|lt2gene | Low-quality |
| S12_vs_k141_19154\|\|lt2gene | Low-quality |
| S12_vs_k141_90593\|\|lt2gene | Low-quality |
| S12_vs_k141_90601\|\|lt2gene | Low-quality |
| S12_vs_k141_145151\|\|lt2gene | Not-determined |
| S12_vs_k141_129031\|\|lt2gene | Low-quality |
| S12_vs_k141_34399\|\|lt2gene | Low-quality |
| S12_vs_k141_50293\|\|lt2gene | Low-quality |
| S12_vs_k141_176241\|\|lt2gene | Not-determined |
| S12_vs_k141_152665\|\|lt2gene | Not-determined |
| S12_vs_k141_129066\|\|lt2gene | Low-quality |
| S12_vs_k141_10735\|\|lt2gene | Low-quality |
| S12_vs_k141_121247\|\|lt2gene | Low-quality |
| S12_vs_k141_43385\|\|lt2gene | Low-quality |
| S12_vs_k141_121251\|\|lt2gene | Low-quality |
| S12_vs_k141_27065\|\|lt2gene | Low-quality |
| S12_vs_k141_43405\|\|lt2gene | Not-determined |
| S12_vs_k141_43407\|\|lt2gene | Not-determined |
| S12_vs_k141_137375\|\|lt2gene | Low-quality |
| S12_vs_k141_43420\|\|lt2gene | Not-determined |
| S12_vs_k141_176295\|\|lt2gene | Low-quality |
| S12_vs_k141_152718\|\|lt2gene | Low-quality |
| S12_vs_k141_58833\|\|lt2gene | Low-quality |
| S12_vs_k141_10814\|\|lt2gene | Not-determined |
| S12_vs_k141_10818\|\|lt2gene | Low-quality |
| S12_vs_k141_184821\|\|lt2gene | Not-determined |
| S12_vs_k141_152776\|\|lt2gene | Low-quality |
| S12_vs_k141_176358\|\|lt2gene | Low-quality |
| S12_vs_k141_113405\|\|lt2gene | Low-quality |
| S12_vs_k141_176366\|\|lt2gene | Low-quality |
| S12_vs_k141_106219\|\|lt2gene | Low-quality |
| S12_vs_k141_176373\|\|lt2gene | Low-quality |
| S12_vs_k141_74013\|\|lt2gene | Low-quality |
| S12_vs_k141_152793\|\|lt2gene | Not-determined |
| S12_vs_k141_106232\|\|lt2gene | Low-quality |
| S12_vs_k141_121348\|\|lt2gene | Low-quality |
| S12_vs_k141_129199\|\|lt2gene | Low-quality |
| S12_vs_k141_10822\|\|lt2gene | Not-determined |
| S12_vs_k141_82246\|\|lt2gene | Low-quality |
| S12_vs_k141_106269\|\|lt2gene | Not-determined |
| S12_vs_k141_129247\|\|lt2gene | Low-quality |
| S12_vs_k141_152824\|\|lt2gene | Not-determined |
| S12_vs_k141_98838\|\|lt2gene | Low-quality |
| S12_vs_k141_58952\|\|lt2gene | Low-quality |
| S12_vs_k141_98847\|\|lt2gene | Low-quality |
| S12_vs_k141_27242\|\|lt2gene | Low-quality |
| S12_vs_k141_82279\|\|lt2gene | Low-quality |
| S12_vs_k141_145346\|\|lt2gene | Low-quality |
| S12_vs_k141_168808\|\|lt2gene | Low-quality |
| S12_vs_k141_19308\|\|lt2gene | Not-determined |
| S12_vs_k141_137553\|\|lt2gene | Low-quality |
| S12_vs_k141_176402\|\|lt2gene | Low-quality |
| S12_vs_k141_137571\|\|lt2gene | Low-quality |
| S12_vs_k141_137572\|\|lt2gene | Not-determined |
| S12_vs_k141_34556\|\|lt2gene | Low-quality |
| S12_vs_k141_168828\|\|lt2gene | Low-quality |
| S12_vs_k141_82329\|\|lt2gene | Not-determined |
| S12_vs_k141_50545\|\|lt2gene | Low-quality |
| S12_vs_k141_59004\|\|lt2gene | Low-quality |
| S12_vs_k141_184922\|\|lt2gene | Low-quality |
| S12_vs_k141_43615\|\|lt2gene | Low-quality |
| S12_vs_k141_10889\|\|lt2gene | Low-quality |
| S12_vs_k141_184926\|\|lt2gene | Low-quality |
| S12_vs_k141_74164\|\|lt2gene | Not-determined |
| S12_vs_k141_82368\|\|lt2gene | Low-quality |
| S12_vs_k141_113564\|\|lt2gene | Not-determined |
| S12_vs_k141_74184\|\|lt2gene | Not-determined |
| S12_vs_k141_50594\|\|lt2gene | Low-quality |
| S12_vs_k141_160749\|\|lt2gene | Low-quality |
| S12_vs_k141_10930\|\|lt2gene | Low-quality |
| S12_vs_k141_152940\|\|lt2gene | Low-quality |
| S12_vs_k141_66736\|\|lt2gene | Low-quality |
| S12_vs_k141_168927\|\|lt2gene | Low-quality |
| S12_vs_k141_34686\|\|lt2gene | Low-quality |
| S12_vs_k141_152994\|\|lt2gene | Low-quality |
| S12_vs_k141_74263\|\|lt2gene | Low-quality |
| S12_vs_k141_4061\|\|lt2gene | Low-quality |
| S12_vs_k141_121530\|\|lt2gene | Low-quality |
| S12_vs_k141_160818\|\|lt2gene | Low-quality |
| S12_vs_k141_121552\|\|lt2gene | Low-quality |
| S12_vs_k141_43738\|\|lt2gene | Low-quality |
| S12_vs_k141_74281\|\|lt2gene | Low-quality |
| S12_vs_k141_43757\|\|lt2gene | Low-quality |
| S12_vs_k141_74297\|\|lt2gene | Low-quality |
| S12_vs_k141_106469\|\|lt2gene | Not-determined |
| S12_vs_k141_106471\|\|lt2gene | Low-quality |
| S12_vs_k141_43764\|\|lt2gene | Low-quality |
| S12_vs_k141_4114\|\|lt2gene | Low-quality |
| S12_vs_k141_43773\|\|lt2gene | Low-quality |
| S12_vs_k141_90832\|\|lt2gene | Low-quality |
| S12_vs_k141_185101\|\|lt2gene | Low-quality |
| S12_vs_k141_99053\|\|lt2gene | Not-determined |
| S12_vs_k141_185129\|\|lt2gene | Low-quality |
| S12_vs_k141_113660\|\|lt2gene | Not-determined |
| S12_vs_k141_185136\|\|lt2gene | Low-quality |
| S12_vs_k141_99087\|\|lt2gene | Not-determined |
| S12_vs_k141_169051\|\|lt2gene | Low-quality |
| S12_vs_k141_153099\|\|lt2gene | Low-quality |
| S12_vs_k141_153143\|\|lt2gene | Low-quality |
| S12_vs_k141_34855\|\|lt2gene | Low-quality |
| S12_vs_k141_59249\|\|lt2gene | Low-quality |
| S12_vs_k141_74334\|\|lt2gene | Not-determined |
| S12_vs_k141_106600\|\|lt2gene | Low-quality |
| S12_vs_k141_99130\|\|lt2gene | Low-quality |
| S12_vs_k141_99137\|\|lt2gene | Low-quality |
| S12_vs_k141_129547\|\|lt2gene | Low-quality |
| S12_vs_k141_185311\|\|lt2gene | Not-determined |
| S12_vs_k141_137894\|\|lt2gene | Low-quality |
| S12_vs_k141_161059\|\|lt2gene | Low-quality |
| S12_vs_k141_161061\|\|lt2gene | Low-quality |
| S12_vs_k141_27693\|\|lt2gene | Low-quality |
| S12_vs_k141_11154\|\|lt2gene | Low-quality |
| S12_vs_k141_27695\|\|lt2gene | Low-quality |
| S12_vs_k141_66930\|\|lt2gene | Not-determined |
| S12_vs_k141_66941\|\|lt2gene | Not-determined |
| S12_vs_k141_169202\|\|lt2gene | Low-quality |
| S12_vs_k141_153269\|\|lt2gene | Low-quality |
| S12_vs_k141_50899\|\|lt2gene | Low-quality |
| S12_vs_k141_153272\|\|lt2gene | Not-determined |
| S12_vs_k141_4308\|\|lt2gene | Not-determined |
| S12_vs_k141_161135\|\|lt2gene | Low-quality |
| S12_vs_k141_43962\|\|lt2gene | Not-determined |
| S12_vs_k141_91013\|\|lt2gene | Low-quality |
| S12_vs_k141_27768\|\|lt2gene | Low-quality |
| S12_vs_k141_161198\|\|lt2gene | Low-quality |
| S12_vs_k141_11240\|\|lt2gene | Low-quality |
| S12_vs_k141_145812\|\|lt2gene | Not-determined |
| S12_vs_k141_138019\|\|lt2gene | Low-quality |
| S12_vs_k141_161230\|\|lt2gene | Low-quality |
| S12_vs_k141_153333\|\|lt2gene | Not-determined |
| S12_vs_k141_67047\|\|lt2gene | Low-quality |
| S12_vs_k141_35077\|\|lt2gene | Low-quality |
| S12_vs_k141_91128\|\|lt2gene | Low-quality |
| S12_vs_k141_145894\|\|lt2gene | Not-determined |
| S12_vs_k141_19595\|\|lt2gene | Low-quality |
| S12_vs_k141_82786\|\|lt2gene | Low-quality |
| S12_vs_k141_82794\|\|lt2gene | Not-determined |
| S12_vs_k141_176485\|\|lt2gene | Low-quality |
| S12_vs_k141_145928\|\|lt2gene | Not-determined |
| S12_vs_k141_51030\|\|lt2gene | Low-quality |
| S12_vs_k141_106743\|\|lt2gene | Low-quality |
| S12_vs_k141_91174\|\|lt2gene | Low-quality |
| S12_vs_k141_106754\|\|lt2gene | Not-determined |
| S12_vs_k141_99242\|\|lt2gene | Low-quality |
| S12_vs_k141_91186\|\|lt2gene | Low-quality |
| S12_vs_k141_99264\|\|lt2gene | Not-determined |
| S12_vs_k141_176518\|\|lt2gene | Low-quality |
| S12_vs_k141_91206\|\|lt2gene | Not-determined |
| S12_vs_k141_99277\|\|lt2gene | Not-determined |
| S12_vs_k141_153379\|\|lt2gene | Low-quality |
| S12_vs_k141_51093\|\|lt2gene | Low-quality |
| S12_vs_k141_59498\|\|lt2gene | Low-quality |
| S12_vs_k141_113906\|\|lt2gene | Low-quality |
| S12_vs_k141_44116\|\|lt2gene | Low-quality |
| S12_vs_k141_106799\|\|lt2gene | Low-quality |
| S12_vs_k141_44144\|\|lt2gene | Low-quality |
| S12_vs_k141_129649\|\|lt2gene | Low-quality |
| S12_vs_k141_129652\|\|lt2gene | Not-determined |
| S12_vs_k141_44152\|\|lt2gene | Low-quality |
| S12_vs_k141_35208\|\|lt2gene | Not-determined |
| S12_vs_k141_35209\|\|lt2gene | Low-quality |
| S12_vs_k141_113961\|\|lt2gene | Low-quality |
| S12_vs_k141_59586\|\|lt2gene | Low-quality |
| S12_vs_k141_91296\|\|lt2gene | Not-determined |
| S12_vs_k141_99322\|\|lt2gene | Low-quality |
| S12_vs_k141_11322\|\|lt2gene | Low-quality |
| S12_vs_k141_59636\|\|lt2gene | Not-determined |
| S12_vs_k141_82882\|\|lt2gene | Not-determined |
| S12_vs_k141_82896\|\|lt2gene | Not-determined |
| S12_vs_k141_74780\|\|lt2gene | Not-determined |
| S12_vs_k141_82904\|\|lt2gene | Low-quality |
| S12_vs_k141_44224\|\|lt2gene | Low-quality |
| S12_vs_k141_176645\|\|lt2gene | Low-quality |
| S12_vs_k141_74791\|\|lt2gene | Low-quality |
| S12_vs_k141_59640\|\|lt2gene | Low-quality |
| S12_vs_k141_19693\|\|lt2gene | Low-quality |
| S12_vs_k141_91336\|\|lt2gene | Not-determined |
| S12_vs_k141_153534\|\|lt2gene | Not-determined |
| S12_vs_k141_138258\|\|lt2gene | Low-quality |
| S12_vs_k141_106962\|\|lt2gene | Not-determined |
| S12_vs_k141_99380\|\|lt2gene | Low-quality |
| S12_vs_k141_106976\|\|lt2gene | Low-quality |
| S12_vs_k141_27974\|\|lt2gene | Low-quality |
| S12_vs_k141_67300\|\|lt2gene | Low-quality |
| S12_vs_k141_185694\|\|lt2gene | Not-determined |
| S12_vs_k141_19743\|\|lt2gene | Low-quality |
| S12_vs_k141_35328\|\|lt2gene | Low-quality |
| S12_vs_k141_59703\|\|lt2gene | Low-quality |
| S12_vs_k141_176726\|\|lt2gene | Low-quality |
| S12_vs_k141_169478\|\|lt2gene | Not-determined |
| S12_vs_k141_19809\|\|lt2gene | Low-quality |
| S12_vs_k141_99465\|\|lt2gene | Low-quality |
| S12_vs_k141_44364\|\|lt2gene | Low-quality |
| S12_vs_k141_146209\|\|lt2gene | Low-quality |
| S12_vs_k141_176760\|\|lt2gene | Low-quality |
| S12_vs_k141_129867\|\|lt2gene | Low-quality |
| S12_vs_k141_59760\|\|lt2gene | Low-quality |
| S12_vs_k141_185778\|\|lt2gene | Low-quality |
| S12_vs_k141_114211\|\|lt2gene | Not-determined |
| S12_vs_k141_44427\|\|lt2gene | Low-quality |
| S12_vs_k141_185817\|\|lt2gene | Low-quality |
| S12_vs_k141_99550\|\|lt2gene | Low-quality |
| S12_vs_k141_185830\|\|lt2gene | Low-quality |
| S12_vs_k141_67437\|\|lt2gene | Low-quality |
| S12_vs_k141_169583\|\|lt2gene | Low-quality |
| S12_vs_k141_67457\|\|lt2gene | Low-quality |
| S12_vs_k141_67460\|\|lt2gene | Low-quality |
| S12_vs_k141_11516\|\|lt2gene | Not-determined |
| S12_vs_k141_138370\|\|lt2gene | Low-quality |
| S12_vs_k141_169593\|\|lt2gene | Low-quality |
| S12_vs_k141_51292\|\|lt2gene | Not-determined |
| S12_vs_k141_176833\|\|lt2gene | Low-quality |
| S12_vs_k141_185864\|\|lt2gene | Low-quality |
| S12_vs_k141_59877\|\|lt2gene | Low-quality |
| S12_vs_k141_146281\|\|lt2gene | Low-quality |
| S12_vs_k141_44512\|\|lt2gene | Low-quality |
| S12_vs_k141_67506\|\|lt2gene | Low-quality |
| S12_vs_k141_67507\|\|lt2gene | Low-quality |
| S12_vs_k141_67514\|\|lt2gene | Not-determined |
| S12_vs_k141_169629\|\|lt2gene | Low-quality |
| S12_vs_k141_4807\|\|lt2gene | Not-determined |
| S12_vs_k141_185878\|\|lt2gene | Low-quality |
| S12_vs_k141_146314\|\|lt2gene | Not-determined |
| S12_vs_k141_176912\|\|lt2gene | Low-quality |
| S12_vs_k141_59921\|\|lt2gene | Low-quality |
| S12_vs_k141_35484\|\|lt2gene | Low-quality |
| S12_vs_k141_59957\|\|lt2gene | Low-quality |
| S12_vs_k141_130023\|\|lt2gene | Not-determined |
| S12_vs_k141_130027\|\|lt2gene | Low-quality |
| S12_vs_k141_107315\|\|lt2gene | Not-determined |
| S12_vs_k141_185954\|\|lt2gene | Low-quality |
| S12_vs_k141_28202\|\|lt2gene | Low-quality |
| S12_vs_k141_28215\|\|lt2gene | Not-determined |
| S12_vs_k141_138502\|\|lt2gene | Low-quality |
| S12_vs_k141_114417\|\|lt2gene | Low-quality |
| S12_vs_k141_75040\|\|lt2gene | Low-quality |
| S12_vs_k141_91664\|\|lt2gene | Low-quality |
| S12_vs_k141_169750\|\|lt2gene | Low-quality |
| S12_vs_k141_138533\|\|lt2gene | Low-quality |
| S12_vs_k141_153824\|\|lt2gene | Low-quality |
| S12_vs_k141_44650\|\|lt2gene | Low-quality |
| S12_vs_k141_44667\|\|lt2gene | Low-quality |
| S12_vs_k141_130107\|\|lt2gene | Low-quality |
| S12_vs_k141_107387\|\|lt2gene | Low-quality |
| S12_vs_k141_186044\|\|lt2gene | Not-determined |
| S12_vs_k141_35567\|\|lt2gene | Not-determined |
| S12_vs_k141_169790\|\|lt2gene | Low-quality |
| S12_vs_k141_99738\|\|lt2gene | Low-quality |
| S12_vs_k141_161925\|\|lt2gene | Not-determined |
| S12_vs_k141_186079\|\|lt2gene | Not-determined |
| S12_vs_k141_138605\|\|lt2gene | Low-quality |
| S12_vs_k141_153899\|\|lt2gene | Low-quality |
| S12_vs_k141_99760\|\|lt2gene | Low-quality |
| S12_vs_k141_186116\|\|lt2gene | Not-determined |
| S12_vs_k141_107460\|\|lt2gene | Low-quality |
| S12_vs_k141_91737\|\|lt2gene | Low-quality |
| S12_vs_k141_186120\|\|lt2gene | Low-quality |
| S12_vs_k141_67633\|\|lt2gene | Not-determined |
| S12_vs_k141_161990\|\|lt2gene | Not-determined |
| S12_vs_k141_114520\|\|lt2gene | Low-quality |
| S12_vs_k141_4949\|\|lt2gene | Not-determined |
| S12_vs_k141_20164\|\|lt2gene | Not-determined |
| S12_vs_k141_107482\|\|lt2gene | Low-quality |
| S12_vs_k141_122092\|\|lt2gene | Not-determined |
| S12_vs_k141_35658\|\|lt2gene | Low-quality |
| S12_vs_k141_51507\|\|lt2gene | Low-quality |
| S12_vs_k141_186159\|\|lt2gene | Not-determined |
| S12_vs_k141_51510\|\|lt2gene | Low-quality |
| S12_vs_k141_107509\|\|lt2gene | Not-determined |
| S12_vs_k141_177109\|\|lt2gene | Low-quality |
| S12_vs_k141_138688\|\|lt2gene | Low-quality |
| S12_vs_k141_75145\|\|lt2gene | Low-quality |
| S12_vs_k141_114556\|\|lt2gene | Low-quality |
| S12_vs_k141_5006\|\|lt2gene | Low-quality |
| S12_vs_k141_5019\|\|lt2gene | Low-quality |
| S12_vs_k141_177150\|\|lt2gene | Low-quality |
| S12_vs_k141_83312\|\|lt2gene | Low-quality |
| S12_vs_k141_5026\|\|lt2gene | Not-determined |
| S12_vs_k141_91842\|\|lt2gene | Low-quality |
| S12_vs_k141_177160\|\|lt2gene | Low-quality |
| S12_vs_k141_138723\|\|lt2gene | Not-determined |
| S12_vs_k141_162080\|\|lt2gene | Low-quality |
| S12_vs_k141_44845\|\|lt2gene | Low-quality |
| S12_vs_k141_75213\|\|lt2gene | Low-quality |
| S12_vs_k141_162100\|\|lt2gene | Low-quality |
| S12_vs_k141_91863\|\|lt2gene | Not-determined |
| S12_vs_k141_11703\|\|lt2gene | Low-quality |
| S12_vs_k141_20240\|\|lt2gene | Not-determined |
| S12_vs_k141_67720\|\|lt2gene | Low-quality |
| S12_vs_k141_162117\|\|lt2gene | Low-quality |
| S12_vs_k141_28441\|\|lt2gene | Not-determined |
| S12_vs_k141_138763\|\|lt2gene | Not-determined |
| S12_vs_k141_44880\|\|lt2gene | Low-quality |
| S12_vs_k141_130309\|\|lt2gene | Low-quality |
| S12_vs_k141_51649\|\|lt2gene | Low-quality |
| S12_vs_k141_59991\|\|lt2gene | Not-determined |
| S12_vs_k141_114707\|\|lt2gene | Not-determined |
| S12_vs_k141_35827\|\|lt2gene | Low-quality |
| S12_vs_k141_162157\|\|lt2gene | Not-determined |
| S12_vs_k141_186340\|\|lt2gene | Not-determined |
| S12_vs_k141_162161\|\|lt2gene | Low-quality |
| S12_vs_k141_83424\|\|lt2gene | Low-quality |
| S12_vs_k141_114731\|\|lt2gene | Low-quality |
| S12_vs_k141_122268\|\|lt2gene | Low-quality |
| S12_vs_k141_169924\|\|lt2gene | Low-quality |
| S12_vs_k141_146562\|\|lt2gene | Not-determined |
| S12_vs_k141_169929\|\|lt2gene | Not-determined |
| S12_vs_k141_11835\|\|lt2gene | Low-quality |
| S12_vs_k141_83475\|\|lt2gene | Not-determined |
| S12_vs_k141_11843\|\|lt2gene | Low-quality |
| S12_vs_k141_91965\|\|lt2gene | Low-quality |
| S12_vs_k141_114820\|\|lt2gene | Low-quality |
| S12_vs_k141_5199\|\|lt2gene | Low-quality |
| S12_vs_k141_5200\|\|lt2gene | Low-quality |
| S12_vs_k141_28638\|\|lt2gene | Low-quality |
| S12_vs_k141_130435\|\|lt2gene | Not-determined |
| S12_vs_k141_20478\|\|lt2gene | Low-quality |
| S12_vs_k141_162288\|\|lt2gene | Not-determined |
| S12_vs_k141_28658\|\|lt2gene | Low-quality |
| S12_vs_k141_5231\|\|lt2gene | Not-determined |
| S12_vs_k141_146677\|\|lt2gene | Low-quality |
| S12_vs_k141_60042\|\|lt2gene | Low-quality |
| S12_vs_k141_51741\|\|lt2gene | Low-quality |
| S12_vs_k141_92034\|\|lt2gene | Low-quality |
| S12_vs_k141_154202\|\|lt2gene | Low-quality |
| S12_vs_k141_92048\|\|lt2gene | Not-determined |
| S12_vs_k141_20516\|\|lt2gene | Low-quality |
| S12_vs_k141_60059\|\|lt2gene | Low-quality |
| S12_vs_k141_60081\|\|lt2gene | Low-quality |
| S12_vs_k141_130503\|\|lt2gene | Low-quality |
| S12_vs_k141_170053\|\|lt2gene | Low-quality |
| S12_vs_k141_20552\|\|lt2gene | Low-quality |
| S12_vs_k141_186506\|\|lt2gene | Low-quality |
| S12_vs_k141_83547\|\|lt2gene | Low-quality |
| S12_vs_k141_177362\|\|lt2gene | Low-quality |
| S12_vs_k141_177365\|\|lt2gene | Not-determined |
| S12_vs_k141_130561\|\|lt2gene | Low-quality |
| S12_vs_k141_162409\|\|lt2gene | Low-quality |
| S12_vs_k141_35946\|\|lt2gene | Not-determined |
| S12_vs_k141_146748\|\|lt2gene | Low-quality |
| S12_vs_k141_100202\|\|lt2gene | Not-determined |
| S12_vs_k141_100214\|\|lt2gene | Low-quality |
| S12_vs_k141_60172\|\|lt2gene | Low-quality |
| S12_vs_k141_100220\|\|lt2gene | Low-quality |
| S12_vs_k141_146782\|\|lt2gene | Low-quality |
| S12_vs_k141_51862\|\|lt2gene | Low-quality |
| S12_vs_k141_162493\|\|lt2gene | Low-quality |
| S12_vs_k141_162496\|\|lt2gene | Low-quality |
| S12_vs_k141_162513\|\|lt2gene | Low-quality |
| S12_vs_k141_177481\|\|lt2gene | Low-quality |
| S12_vs_k141_170122\|\|lt2gene | Low-quality |
| S12_vs_k141_12031\|\|lt2gene | Low-quality |
| S12_vs_k141_122581\|\|lt2gene | Low-quality |
| S12_vs_k141_51918\|\|lt2gene | Not-determined |
| S12_vs_k141_100322\|\|lt2gene | Low-quality |
| S12_vs_k141_114951\|\|lt2gene | Low-quality |
| S12_vs_k141_12052\|\|lt2gene | Low-quality |
| S12_vs_k141_154342\|\|lt2gene | Low-quality |
| S12_vs_k141_83602\|\|lt2gene | Low-quality |
| S12_vs_k141_146879\|\|lt2gene | Low-quality |
| S12_vs_k141_83620\|\|lt2gene | Not-determined |

| | |
|---|---|
| S12_vs_k141_67932||lt2gene | Low-quality |
| S12_vs_k141_36052||lt2gene | Low-quality |
| S12_vs_k141_100357||lt2gene | Low-quality |
| S12_vs_k141_36058||lt2gene | Low-quality |
| S12_vs_k141_45121||lt2gene | Low-quality |
| S12_vs_k141_51960||lt2gene | Low-quality |
| S12_vs_k141_170246||lt2gene | Low-quality |
| S12_vs_k141_75695||lt2gene | Not-determined |
| S12_vs_k141_139229||lt2gene | Low-quality |
| S12_vs_k141_67971||lt2gene | Low-quality |
| S12_vs_k141_170267||lt2gene | Low-quality |
| S12_vs_k141_170270||lt2gene | Low-quality |
| S12_vs_k141_75701||lt2gene | Low-quality |
| S12_vs_k141_28766||lt2gene | Low-quality |
| S12_vs_k141_146915||lt2gene | Not-determined |
| S12_vs_k141_5569||lt2gene | Not-determined |
| S12_vs_k141_154431||lt2gene | Low-quality |
| S12_vs_k141_75764||lt2gene | Low-quality |
| S12_vs_k141_130724||lt2gene | Low-quality |
| S12_vs_k141_12148||lt2gene | Low-quality |
| S12_vs_k141_45218||lt2gene | Low-quality |
| S12_vs_k141_12159||lt2gene | Low-quality |
| S12_vs_k141_107897||lt2gene | Low-quality |
| S12_vs_k141_20737||lt2gene | Low-quality |
| S12_vs_k141_100464||lt2gene | Low-quality |
| S12_vs_k141_162574||lt2gene | Low-quality |
| S12_vs_k141_92305||lt2gene | Not-determined |
| S12_vs_k141_139329||lt2gene | Low-quality |
| S12_vs_k141_68079||lt2gene | Low-quality |
| S12_vs_k141_68080||lt2gene | Not-determined |
| S12_vs_k141_28841||lt2gene | Not-determined |
| S12_vs_k141_177698||lt2gene | Low-quality |
| S12_vs_k141_170382||lt2gene | Low-quality |
| S12_vs_k141_36119||lt2gene | Low-quality |
| S12_vs_k141_92324||lt2gene | Low-quality |
| S12_vs_k141_75840||lt2gene | Not-determined |
| S12_vs_k141_147018||lt2gene | Low-quality |
| S12_vs_k141_147022||lt2gene | Low-quality |
| S12_vs_k141_107965||lt2gene | Not-determined |
| S12_vs_k141_130802||lt2gene | Not-determined |
| S12_vs_k141_186790||lt2gene | Low-quality |
| S12_vs_k141_115080||lt2gene | Not-determined |
| S12_vs_k141_107981||lt2gene | Low-quality |
| S12_vs_k141_154560||lt2gene | Low-quality |
| S12_vs_k141_60346||lt2gene | Low-quality |
| S12_vs_k141_45310||lt2gene | Low-quality |
| S12_vs_k141_92389||lt2gene | Not-determined |
| S12_vs_k141_108012||lt2gene | Low-quality |
| S12_vs_k141_139386||lt2gene | Low-quality |
| S12_vs_k141_139392||lt2gene | Not-determined |
| S12_vs_k141_122786||lt2gene | Not-determined |
| S12_vs_k141_154621||lt2gene | Not-determined |
| S12_vs_k141_92438||lt2gene | Low-quality |
| S12_vs_k141_92446||lt2gene | Low-quality |
| S12_vs_k141_5758||lt2gene | Low-quality |
| S12_vs_k141_68194||lt2gene | Not-determined |
| S12_vs_k141_75992||lt2gene | Low-quality |
| S12_vs_k141_170439||lt2gene | Low-quality |
| S12_vs_k141_76003||lt2gene | Low-quality |
| S12_vs_k141_154650||lt2gene | Low-quality |
| S12_vs_k141_100563||lt2gene | Not-determined |
| S12_vs_k141_76032||lt2gene | Low-quality |
| S12_vs_k141_177851||lt2gene | Low-quality |
| S12_vs_k141_147139||lt2gene | Low-quality |
| S12_vs_k141_60447||lt2gene | Low-quality |
| S12_vs_k141_177868||lt2gene | Low-quality |
| S12_vs_k141_12389||lt2gene | Low-quality |
| S12_vs_k141_100623||lt2gene | Low-quality |
| S12_vs_k141_5829||lt2gene | Low-quality |
| S12_vs_k141_28995||lt2gene | Low-quality |
| S12_vs_k141_60501||lt2gene | Low-quality |
| S12_vs_k141_130972||lt2gene | Low-quality |
| S12_vs_k141_12420||lt2gene | Not-determined |
| S12_vs_k141_12435||lt2gene | Low-quality |
| S12_vs_k141_45495||lt2gene | Low-quality |
| S12_vs_k141_115242||lt2gene | Not-determined |
| S12_vs_k141_68287||lt2gene | Low-quality |
| S12_vs_k141_115250||lt2gene | Not-determined |
| S12_vs_k141_115259||lt2gene | Low-quality |
| S12_vs_k141_154769||lt2gene | Not-determined |
| S12_vs_k141_177985||lt2gene | Low-quality |
| S12_vs_k141_29051||lt2gene | Low-quality |
| S12_vs_k141_139553||lt2gene | Low-quality |
| S12_vs_k141_83799||lt2gene | Low-quality |
| S12_vs_k141_178013||lt2gene | Low-quality |
| S12_vs_k141_139576||lt2gene | Not-determined |
| S12_vs_k141_29068||lt2gene | Not-determined |
| S12_vs_k141_139586||lt2gene | Low-quality |
| S12_vs_k141_12501||lt2gene | Low-quality |
| S12_vs_k141_131074||lt2gene | Low-quality |
| S12_vs_k141_45622||lt2gene | Low-quality |
| S12_vs_k141_100764||lt2gene | Low-quality |
| S12_vs_k141_68345||lt2gene | Low-quality |
| S12_vs_k141_100774||lt2gene | Low-quality |
| S12_vs_k141_5942||lt2gene | Not-determined |
| S12_vs_k141_83854||lt2gene | Low-quality |
| S12_vs_k141_52512||lt2gene | Low-quality |
| S12_vs_k141_115364||lt2gene | Not-determined |
| S12_vs_k141_92605||lt2gene | Not-determined |
| S12_vs_k141_123039||lt2gene | Low-quality |
| S12_vs_k141_147268||lt2gene | Low-quality |
| S12_vs_k141_83894||lt2gene | Not-determined |
| S12_vs_k141_60660||lt2gene | Low-quality |
| S12_vs_k141_68406||lt2gene | Low-quality |
| S12_vs_k141_139710||lt2gene | Low-quality |
| S12_vs_k141_178148||lt2gene | Low-quality |
| S12_vs_k141_123069||lt2gene | Low-quality |
| S12_vs_k141_29145||lt2gene | Low-quality |
| S12_vs_k141_29146||lt2gene | Low-quality |
| S12_vs_k141_131133||lt2gene | Low-quality |
| S12_vs_k141_115422||lt2gene | Low-quality |
| S12_vs_k141_29162||lt2gene | Not-determined |
| S12_vs_k141_29165||lt2gene | Not-determined |
| S12_vs_k141_83922||lt2gene | Low-quality |
| S12_vs_k141_21127||lt2gene | Low-quality |
| S12_vs_k141_147348||lt2gene | Low-quality |
| S12_vs_k141_45720||lt2gene | Not-determined |
| S12_vs_k141_45729||lt2gene | Low-quality |
| S12_vs_k141_170660||lt2gene | Low-quality |
| S12_vs_k141_12658||lt2gene | Not-determined |
| S12_vs_k141_170662||lt2gene | Low-quality |
| S12_vs_k141_108361||lt2gene | Low-quality |
| S12_vs_k141_92728||lt2gene | Low-quality |
| S12_vs_k141_68503||lt2gene | Not-determined |
| S12_vs_k141_147412||lt2gene | Not-determined |
| S12_vs_k141_108401||lt2gene | Not-determined |
| S12_vs_k141_92744||lt2gene | Low-quality |
| S12_vs_k141_60735||lt2gene | Low-quality |
| S12_vs_k141_187277||lt2gene | Not-determined |
| S12_vs_k141_187282||lt2gene | Low-quality |
| S12_vs_k141_108430||lt2gene | Low-quality |
| S12_vs_k141_178297||lt2gene | Low-quality |
| S12_vs_k141_68554||lt2gene | Low-quality |
| S12_vs_k141_170753||lt2gene | Not-determined |
| S12_vs_k141_154992||lt2gene | Low-quality |
| S12_vs_k141_92794||lt2gene | Low-quality |
| S12_vs_k141_108459||lt2gene | Low-quality |
| S12_vs_k141_84044||lt2gene | Low-quality |
| S12_vs_k141_52542||lt2gene | Low-quality |
| S12_vs_k141_108483||lt2gene | Low-quality |
| S12_vs_k141_147473||lt2gene | Low-quality |
| S12_vs_k141_92827||lt2gene | Low-quality |
| S12_vs_k141_60841||lt2gene | Low-quality |
| S12_vs_k141_123245||lt2gene | Low-quality |
| S12_vs_k141_162955||lt2gene | Low-quality |
| S12_vs_k141_123253||lt2gene | Low-quality |
| S12_vs_k141_101035||lt2gene | Not-determined |
| S12_vs_k141_139918||lt2gene | Low-quality |
| S12_vs_k141_123285||lt2gene | Low-quality |
| S12_vs_k141_36546||lt2gene | Low-quality |
| S12_vs_k141_163005||lt2gene | Low-quality |
| S12_vs_k141_101070||lt2gene | Low-quality |
| S12_vs_k141_36548||lt2gene | Not-determined |
| S12_vs_k141_76351||lt2gene | Not-determined |
| S12_vs_k141_139958||lt2gene | Low-quality |
| S12_vs_k141_21351||lt2gene | Low-quality |
| S12_vs_k141_123350||lt2gene | Low-quality |
| S12_vs_k141_12813||lt2gene | Not-determined |
| S12_vs_k141_52642||lt2gene | Low-quality |
| S12_vs_k141_36588||lt2gene | Low-quality |
| S12_vs_k141_163042||lt2gene | Low-quality |
| S12_vs_k141_187372||lt2gene | Not-determined |
| S12_vs_k141_21369||lt2gene | Low-quality |
| S12_vs_k141_52713||lt2gene | Low-quality |
| S12_vs_k141_108603||lt2gene | Low-quality |
| S12_vs_k141_140090||lt2gene | Low-quality |
| S12_vs_k141_170914||lt2gene | Not-determined |
| S12_vs_k141_131393||lt2gene | Low-quality |
| S12_vs_k141_108633||lt2gene | Not-determined |
| S12_vs_k141_6293||lt2gene | Low-quality |
| S12_vs_k141_163133||lt2gene | Not-determined |
| S12_vs_k141_45965||lt2gene | Low-quality |
| S12_vs_k141_115627||lt2gene | Not-determined |
| S12_vs_k141_6302||lt2gene | Low-quality |
| S12_vs_k141_61040||lt2gene | Not-determined |
| S12_vs_k141_68769||lt2gene | Low-quality |
| S12_vs_k141_170967||lt2gene | Low-quality |
| S12_vs_k141_147686||lt2gene | Low-quality |
| S12_vs_k141_46045||lt2gene | Low-quality |
| S12_vs_k141_163186||lt2gene | Low-quality |
| S12_vs_k141_29442||lt2gene | Low-quality |
| S12_vs_k141_147701||lt2gene | Low-quality |
| S12_vs_k141_131500||lt2gene | Not-determined |
| S12_vs_k141_163203||lt2gene | Low-quality |
| S12_vs_k141_36757||lt2gene | Low-quality |
| S12_vs_k141_101317||lt2gene | Low-quality |
| S12_vs_k141_12965||lt2gene | Low-quality |
| S12_vs_k141_84346||lt2gene | Low-quality |
| S12_vs_k141_131521||lt2gene | Low-quality |
| S12_vs_k141_171027||lt2gene | Low-quality |
| S12_vs_k141_61086||lt2gene | Low-quality |
| S12_vs_k141_6336||lt2gene | Low-quality |
| S12_vs_k141_171045||lt2gene | Low-quality |
| S12_vs_k141_131554||lt2gene | Not-determined |
| S12_vs_k141_108744||lt2gene | Low-quality |
| S12_vs_k141_115692||lt2gene | Not-determined |
| S12_vs_k141_101379||lt2gene | Low-quality |
| S12_vs_k141_140264||lt2gene | Low-quality |
| S12_vs_k141_6375||lt2gene | Low-quality |
| S12_vs_k141_92940||lt2gene | Low-quality |
| S12_vs_k141_61155||lt2gene | Low-quality |
| S12_vs_k141_108805||lt2gene | Low-quality |
| S12_vs_k141_187565||lt2gene | Low-quality |
| S12_vs_k141_155292||lt2gene | Low-quality |
| S12_vs_k141_13046||lt2gene | Low-quality |
| S12_vs_k141_178674||lt2gene | Not-determined |
| S12_vs_k141_101453||lt2gene | Not-determined |
| S12_vs_k141_178685||lt2gene | Not-determined |
| S12_vs_k141_29557||lt2gene | Low-quality |
| S12_vs_k141_68923||lt2gene | Low-quality |
| S12_vs_k141_36911||lt2gene | Low-quality |
| S12_vs_k141_6443||lt2gene | Not-determined |
| S12_vs_k141_13117||lt2gene | Low-quality |
| S12_vs_k141_68946||lt2gene | Low-quality |
| S12_vs_k141_29614||lt2gene | Low-quality |
| S12_vs_k141_187655||lt2gene | Low-quality |
| S12_vs_k141_140409||lt2gene | Not-determined |
| S12_vs_k141_131666||lt2gene | Low-quality |
| S12_vs_k141_61261||lt2gene | Low-quality |
| S12_vs_k141_52999||lt2gene | Low-quality |
| S12_vs_k141_131711||lt2gene | Not-determined |
| S12_vs_k141_93125||lt2gene | Not-determined |
| S12_vs_k141_123643||lt2gene | Not-determined |
| S12_vs_k141_21609||lt2gene | Low-quality |
| S12_vs_k141_131784||lt2gene | Low-quality |
| S12_vs_k141_155391||lt2gene | Not-determined |
| S12_vs_k141_171270||lt2gene | Low-quality |
| S12_vs_k141_37067||lt2gene | Not-determined |
| S12_vs_k141_21645||lt2gene | Not-determined |
| S12_vs_k141_76815||lt2gene | Low-quality |
| S12_vs_k141_163464||lt2gene | Low-quality |
| S12_vs_k141_131801||lt2gene | Not-determined |
| S12_vs_k141_21663||lt2gene | Low-quality |
| S12_vs_k141_108949||lt2gene | Low-quality |
| S12_vs_k141_155431||lt2gene | Low-quality |
| S12_vs_k141_123721||lt2gene | Not-determined |
| S12_vs_k141_84555||lt2gene | Low-quality |
| S12_vs_k141_93248||lt2gene | Low-quality |
| S12_vs_k141_187843||lt2gene | Low-quality |
| S12_vs_k141_76900||lt2gene | Low-quality |
| S12_vs_k141_131854||lt2gene | Low-quality |
| S12_vs_k141_148042||lt2gene | Low-quality |
| S12_vs_k141_131858||lt2gene | Low-quality |
| S12_vs_k141_101696||lt2gene | Low-quality |
| S12_vs_k141_178843||lt2gene | Low-quality |
| S12_vs_k141_37142||lt2gene | Not-determined |
| S12_vs_k141_123791||lt2gene | Low-quality |
| S12_vs_k141_53171||lt2gene | Not-determined |
| S12_vs_k141_171360||lt2gene | Not-determined |
| S12_vs_k141_93318||lt2gene | Low-quality |
| S12_vs_k141_171365||lt2gene | Low-quality |
| S12_vs_k141_46365||lt2gene | Not-determined |
| S12_vs_k141_187913||lt2gene | Not-determined |
| S12_vs_k141_109087||lt2gene | Low-quality |
| S12_vs_k141_53204||lt2gene | Low-quality |
| S12_vs_k141_171372||lt2gene | Low-quality |
| S12_vs_k141_37197||lt2gene | Low-quality |
| S12_vs_k141_163628||lt2gene | Low-quality |
| S12_vs_k141_123845||lt2gene | Low-quality |
| S12_vs_k141_77022||lt2gene | Not-determined |
| S12_vs_k141_69204||lt2gene | Low-quality |
| S12_vs_k141_84688||lt2gene | Not-determined |
| S12_vs_k141_37231||lt2gene | Low-quality |
| S12_vs_k141_61539||lt2gene | Low-quality |
| S12_vs_k141_61551||lt2gene | Low-quality |
| S12_vs_k141_131986||lt2gene | Not-determined |
| S12_vs_k141_6713||lt2gene | Not-determined |
| S12_vs_k141_155538||lt2gene | Not-determined |
| S12_vs_k141_37259||lt2gene | Not-determined |
| S12_vs_k141_29916||lt2gene | Low-quality |
| S12_vs_k141_21708||lt2gene | Not-determined |
| S12_vs_k141_178908||lt2gene | Low-quality |
| S12_vs_k141_53289||lt2gene | Low-quality |
| S12_vs_k141_116137||lt2gene | Low-quality |
| S12_vs_k141_29936||lt2gene | Low-quality |
| S12_vs_k141_29939||lt2gene | Not-determined |
| S12_vs_k141_77039||lt2gene | Low-quality |
| S12_vs_k141_77042||lt2gene | Low-quality |
| S12_vs_k141_53318||lt2gene | Low-quality |
| S12_vs_k141_37284||lt2gene | Not-determined |
| S12_vs_k141_178943||lt2gene | Low-quality |
| S12_vs_k141_46481||lt2gene | Low-quality |
| S12_vs_k141_140693||lt2gene | Not-determined |
| S12_vs_k141_61621||lt2gene | Low-quality |
| S12_vs_k141_171430||lt2gene | Low-quality |
| S12_vs_k141_163746||lt2gene | Not-determined |
| S12_vs_k141_21771||lt2gene | Low-quality |
| S12_vs_k141_148219||lt2gene | Low-quality |
| S12_vs_k141_101865||lt2gene | Low-quality |
| S12_vs_k141_93446||lt2gene | Low-quality |
| S12_vs_k141_13567||lt2gene | Low-quality |
| S12_vs_k141_53454||lt2gene | Low-quality |
| S12_vs_k141_132102||lt2gene | Low-quality |
| S12_vs_k141_155693||lt2gene | Not-determined |
| S12_vs_k141_101935||lt2gene | Low-quality |
| S12_vs_k141_77220||lt2gene | Low-quality |
| S12_vs_k141_21853||lt2gene | Low-quality |
| S12_vs_k141_21860||lt2gene | Low-quality |
| S12_vs_k141_61680||lt2gene | Not-determined |
| S12_vs_k141_101961||lt2gene | Not-determined |
| S12_vs_k141_109295||lt2gene | Not-determined |
| S12_vs_k141_132162||lt2gene | Not-determined |
| S12_vs_k141_6886||lt2gene | Low-quality |
| S12_vs_k141_148338||lt2gene | Low-quality |
| S12_vs_k141_140846||lt2gene | Low-quality |
| S12_vs_k141_109350||lt2gene | Low-quality |
| S12_vs_k141_6903||lt2gene | Low-quality |
| S12_vs_k141_155760||lt2gene | Low-quality |
| S12_vs_k141_163938||lt2gene | Not-determined |
| S12_vs_k141_155780||lt2gene | Low-quality |
| S12_vs_k141_46692||lt2gene | Not-determined |
| S12_vs_k141_69525||lt2gene | Not-determined |
| S12_vs_k141_13658||lt2gene | Not-determined |
| S12_vs_k141_53573||lt2gene | Low-quality |
| S12_vs_k141_46708||lt2gene | Low-quality |
| S12_vs_k141_77317||lt2gene | Low-quality |
| S12_vs_k141_13677||lt2gene | Low-quality |
| S12_vs_k141_6961||lt2gene | Not-determined |
| S12_vs_k141_116347||lt2gene | Low-quality |
| S12_vs_k141_93625||lt2gene | Low-quality |
| S12_vs_k141_6978||lt2gene | Not-determined |
| S12_vs_k141_93631||lt2gene | Not-determined |
| S12_vs_k141_102032||lt2gene | Low-quality |
| S12_vs_k141_171563||lt2gene | Low-quality |
| S12_vs_k141_132259||lt2gene | Not-determined |
| S12_vs_k141_155844||lt2gene | Low-quality |
| S12_vs_k141_132263||lt2gene | Low-quality |
| S12_vs_k141_93644||lt2gene | Not-determined |
| S12_vs_k141_109439||lt2gene | Not-determined |
| S12_vs_k141_148428||lt2gene | Low-quality |
| S12_vs_k141_116365||lt2gene | Not-determined |
| S12_vs_k141_69603||lt2gene | Low-quality |
| S12_vs_k141_77352||lt2gene | Low-quality |
| S12_vs_k141_61804||lt2gene | Not-determined |
| S12_vs_k141_77375||lt2gene | Not-determined |
| S12_vs_k141_69636||lt2gene | Low-quality |
| S12_vs_k141_132309||lt2gene | Low-quality |
| S12_vs_k141_85104||lt2gene | Low-quality |
| S12_vs_k141_7025||lt2gene | Not-determined |
| S12_vs_k141_102077||lt2gene | Not-determined |
| S12_vs_k141_69661||lt2gene | Low-quality |
| S12_vs_k141_37605||lt2gene | Low-quality |
| S12_vs_k141_109532||lt2gene | Low-quality |
| S12_vs_k141_22066||lt2gene | Low-quality |
| S12_vs_k141_61893||lt2gene | Low-quality |
| S12_vs_k141_46804||lt2gene | Low-quality |
| S12_vs_k141_171672||lt2gene | Low-quality |
| S12_vs_k141_141016||lt2gene | Low-quality |
| S12_vs_k141_93755||lt2gene | Low-quality |
| S12_vs_k141_116460||lt2gene | Not-determined |
| S12_vs_k141_77465||lt2gene | Low-quality |
| S12_vs_k141_61925||lt2gene | Not-determined |
| S12_vs_k141_116471||lt2gene | Low-quality |
| S12_vs_k141_7078||lt2gene | Low-quality |
| S12_vs_k141_102145||lt2gene | Not-determined |
| S12_vs_k141_164032||lt2gene | Not-determined |
| S12_vs_k141_141045||lt2gene | Low-quality |
| S12_vs_k141_30378||lt2gene | Low-quality |
| S12_vs_k141_53807||lt2gene | Not-determined |
| S12_vs_k141_116501||lt2gene | Low-quality |
| S12_vs_k141_53819||lt2gene | Low-quality |
| S12_vs_k141_102193||lt2gene | Low-quality |
| S12_vs_k141_148580||lt2gene | Low-quality |
| S12_vs_k141_141083||lt2gene | Low-quality |
| S12_vs_k141_53833||lt2gene | Low-quality |
| S12_vs_k141_22209||lt2gene | Low-quality |
| S12_vs_k141_22210||lt2gene | Not-determined |
| S12_vs_k141_7160||lt2gene | Low-quality |
| S12_vs_k141_85276||lt2gene | Not-determined |
| S12_vs_k141_46902||lt2gene | Low-quality |
| S12_vs_k141_46906||lt2gene | Not-determined |
| S12_vs_k141_148637||lt2gene | Low-quality |
| S12_vs_k141_46922||lt2gene | Low-quality |
| S12_vs_k141_69836||lt2gene | Not-determined |
| S12_vs_k141_22264||lt2gene | Low-quality |
| S12_vs_k141_179317||lt2gene | Low-quality |
| S12_vs_k141_109635||lt2gene | Low-quality |
| S12_vs_k141_62047||lt2gene | Not-determined |
| S12_vs_k141_46946||lt2gene | Not-determined |
| S12_vs_k141_46948||lt2gene | Low-quality |
| S12_vs_k141_46950||lt2gene | Low-quality |
| S12_vs_k141_171825||lt2gene | Low-quality |
| S12_vs_k141_93918||lt2gene | Low-quality |
| S12_vs_k141_164196||lt2gene | Not-determined |
| S12_vs_k141_132545||lt2gene | Not-determined |
| S12_vs_k141_132550||lt2gene | Low-quality |
| S12_vs_k141_132556||lt2gene | Not-determined |
| S12_vs_k141_69898||lt2gene | Low-quality |
| S12_vs_k141_148740||lt2gene | Low-quality |
| S12_vs_k141_85338||lt2gene | Low-quality |
| S12_vs_k141_69929||lt2gene | Not-determined |
| S12_vs_k141_69940||lt2gene | Low-quality |
| S12_vs_k141_141236||lt2gene | Low-quality |
| S12_vs_k141_109723||lt2gene | Low-quality |
| S12_vs_k141_62171||lt2gene | Not-determined |
| S12_vs_k141_37781||lt2gene | Low-quality |
| S12_vs_k141_85415||lt2gene | Low-quality |
| S12_vs_k141_148824||lt2gene | Low-quality |
| S12_vs_k141_37836||lt2gene | Low-quality |
| S12_vs_k141_37847||lt2gene | Low-quality |
| S12_vs_k141_14086||lt2gene | Low-quality |
| S12_vs_k141_188485||lt2gene | Low-quality |
| S12_vs_k141_148868||lt2gene | Low-quality |
| S12_vs_k141_164291||lt2gene | Low-quality |
| S12_vs_k141_62242||lt2gene | Low-quality |
| S12_vs_k141_47133||lt2gene | Low-quality |
| S12_vs_k141_179507||lt2gene | Not-determined |
| S12_vs_k141_148880||lt2gene | Low-quality |
| S12_vs_k141_179519||lt2gene | Not-determined |
| S12_vs_k141_124529||lt2gene | Low-quality |
| S12_vs_k141_179530||lt2gene | Low-quality |
| S12_vs_k141_54039||lt2gene | Low-quality |
| S12_vs_k141_132718||lt2gene | Low-quality |
| S12_vs_k141_141341||lt2gene | Not-determined |
| S12_vs_k141_116777||lt2gene | Not-determined |
| S12_vs_k141_132741||lt2gene | Low-quality |
| S12_vs_k141_132751||lt2gene | Low-quality |
| S12_vs_k141_62326||lt2gene | Low-quality |
| S12_vs_k141_30691||lt2gene | Low-quality |
| S12_vs_k141_109883||lt2gene | Low-quality |
| S12_vs_k141_124625||lt2gene | Low-quality |
| S12_vs_k141_77848||lt2gene | Not-determined |
| S12_vs_k141_132795||lt2gene | Low-quality |
| S12_vs_k141_124640||lt2gene | Low-quality |
| S12_vs_k141_70154||lt2gene | Low-quality |
| S12_vs_k141_132811||lt2gene | Not-determined |
| S12_vs_k141_85603||lt2gene | Not-determined |
| S12_vs_k141_54162||lt2gene | Low-quality |
| S12_vs_k141_30729||lt2gene | Low-quality |
| S12_vs_k141_85617||lt2gene | Low-quality |
| S12_vs_k141_132845||lt2gene | Low-quality |
| S12_vs_k141_77913||lt2gene | Not-determined |
| S12_vs_k141_14273||lt2gene | Low-quality |
| S12_vs_k141_179677||lt2gene | Low-quality |
| S12_vs_k141_77947||lt2gene | Low-quality |
| S12_vs_k141_62447||lt2gene | Low-quality |
| S12_vs_k141_14324||lt2gene | Low-quality |
| S12_vs_k141_141421||lt2gene | Low-quality |
| S12_vs_k141_30770||lt2gene | Low-quality |
| S12_vs_k141_179727||lt2gene | Low-quality |
| S12_vs_k141_116919||lt2gene | Low-quality |
| S12_vs_k141_62493||lt2gene | Low-quality |
| S12_vs_k141_179752||lt2gene | Not-determined |
| S12_vs_k141_141473||lt2gene | Low-quality |
| S12_vs_k141_14381||lt2gene | Low-quality |
| S12_vs_k141_78015||lt2gene | Low-quality |
| S12_vs_k141_141511||lt2gene | Low-quality |
| S12_vs_k141_54310||lt2gene | Low-quality |
| S12_vs_k141_132974||lt2gene | Not-determined |
| S12_vs_k141_156101||lt2gene | Low-quality |
| S12_vs_k141_30896||lt2gene | Low-quality |
| S12_vs_k141_54346||lt2gene | Low-quality |
| S12_vs_k141_14459||lt2gene | Low-quality |
| S12_vs_k141_54362||lt2gene | Low-quality |
| S12_vs_k141_124758||lt2gene | Low-quality |
| S12_vs_k141_164578||lt2gene | Low-quality |
| S12_vs_k141_141607||lt2gene | Low-quality |
| S12_vs_k141_124766||lt2gene | Not-determined |
| S12_vs_k141_70415||lt2gene | Low-quality |
| S12_vs_k141_22753||lt2gene | Not-determined |
| S12_vs_k141_38118||lt2gene | Low-quality |
| S12_vs_k141_7721||lt2gene | Low-quality |
| S12_vs_k141_156125||lt2gene | Low-quality |
| S12_vs_k141_179905||lt2gene | Low-quality |
| S12_vs_k141_70443||lt2gene | Not-determined |
| S12_vs_k141_164611||lt2gene | Low-quality |
| S12_vs_k141_179918||lt2gene | Not-determined |
| S12_vs_k141_85828||lt2gene | Low-quality |
| S12_vs_k141_124778||lt2gene | Not-determined |
| S12_vs_k141_110208||lt2gene | Low-quality |
| S12_vs_k141_110221||lt2gene | Low-quality |
| S12_vs_k141_14558||lt2gene | Not-determined |
| S12_vs_k141_54429||lt2gene | Low-quality |
| S12_vs_k141_62696||lt2gene | Low-quality |
| S12_vs_k141_31020||lt2gene | Low-quality |
| S12_vs_k141_14575||lt2gene | Not-determined |
| S12_vs_k141_156189||lt2gene | Low-quality |
| S12_vs_k141_164672||lt2gene | Not-determined |
| S12_vs_k141_164676||lt2gene | Low-quality |
| S12_vs_k141_180000||lt2gene | Low-quality |
| S12_vs_k141_14603||lt2gene | Low-quality |
| S12_vs_k141_7817||lt2gene | Low-quality |
| S12_vs_k141_22863||lt2gene | Low-quality |
| S12_vs_k141_164693||lt2gene | Low-quality |
| S12_vs_k141_22885||lt2gene | Low-quality |
| S12_vs_k141_7857||lt2gene | Low-quality |
| S12_vs_k141_62793||lt2gene | Not-determined |
| S12_vs_k141_70546||lt2gene | Not-determined |
| S12_vs_k141_180035||lt2gene | Low-quality |
| S12_vs_k141_117136||lt2gene | Low-quality |
| S12_vs_k141_164779||lt2gene | Low-quality |
| S12_vs_k141_38280||lt2gene | Low-quality |
| S12_vs_k141_188775||lt2gene | Not-determined |
| S12_vs_k141_188782||lt2gene | Low-quality |
| S12_vs_k141_149443||lt2gene | Low-quality |
| S12_vs_k141_62851||lt2gene | Not-determined |
| S12_vs_k141_54581||lt2gene | Not-determined |
| S12_vs_k141_22999||lt2gene | Low-quality |
| S12_vs_k141_125042||lt2gene | Low-quality |
| S12_vs_k141_133175||lt2gene | Not-determined |
| S12_vs_k141_156390||lt2gene | Not-determined |
| S12_vs_k141_149523||lt2gene | Not-determined |
| S12_vs_k141_23058||lt2gene | Low-quality |
| S12_vs_k141_94397||lt2gene | Low-quality |
| S12_vs_k141_180128||lt2gene | Low-quality |
| S12_vs_k141_54657||lt2gene | Low-quality |
| S12_vs_k141_14814||lt2gene | Not-determined |
| S12_vs_k141_94429||lt2gene | Low-quality |
| S12_vs_k141_117320||lt2gene | Low-quality |
| S12_vs_k141_62944||lt2gene | Low-quality |
| S12_vs_k141_70758||lt2gene | Low-quality |
| S12_vs_k141_78507||lt2gene | Not-determined |
| S12_vs_k141_86199||lt2gene | Not-determined |
| S12_vs_k141_172363||lt2gene | Low-quality |
| S12_vs_k141_31244||lt2gene | Low-quality |
| S12_vs_k141_164896||lt2gene | Not-determined |
| S12_vs_k141_180184||lt2gene | Not-determined |
| S12_vs_k141_31285||lt2gene | Low-quality |
| S12_vs_k141_31337||lt2gene | Low-quality |
| S12_vs_k141_70831||lt2gene | Not-determined |
| S12_vs_k141_125181||lt2gene | Low-quality |
| S12_vs_k141_78601||lt2gene | Low-quality |
| S12_vs_k141_14896||lt2gene | Low-quality |
| S12_vs_k141_156524||lt2gene | Not-determined |
| S12_vs_k141_180260||lt2gene | Low-quality |
| S12_vs_k141_180276||lt2gene | Low-quality |
| S12_vs_k141_117448||lt2gene | Low-quality |
| S12_vs_k141_165028||lt2gene | Low-quality |
| S12_vs_k141_133412||lt2gene | Not-determined |
| S12_vs_k141_133424||lt2gene | Low-quality |
| S12_vs_k141_54908||lt2gene | Low-quality |
| S12_vs_k141_165068||lt2gene | Low-quality |
| S12_vs_k141_86440||lt2gene | Low-quality |
| S12_vs_k141_180362||lt2gene | Low-quality |
| S12_vs_k141_165104||lt2gene | Low-quality |
| S12_vs_k141_54934||lt2gene | Low-quality |
| S12_vs_k141_165107||lt2gene | Low-quality |
| S12_vs_k141_172421||lt2gene | Not-determined |
| S12_vs_k141_172427||lt2gene | Low-quality |
| S12_vs_k141_133510||lt2gene | Not-determined |
| S12_vs_k141_14936||lt2gene | Low-quality |
| S12_vs_k141_78731||lt2gene | Low-quality |
| S12_vs_k141_125345||lt2gene | Low-quality |
| S12_vs_k141_55016||lt2gene | Not-determined |
| S12_vs_k141_180462||lt2gene | Low-quality |
| S12_vs_k141_86529||lt2gene | Not-determined |
| S12_vs_k141_172590||lt2gene | Low-quality |
| S12_vs_k141_180542||lt2gene | Low-quality |
| S12_vs_k141_172626||lt2gene | Low-quality |
| S12_vs_k141_172674||lt2gene | Low-quality |
| S12_vs_k141_55082||lt2gene | Low-quality |
| S12_vs_k141_133732||lt2gene | Low-quality |
| S12_vs_k141_38958||lt2gene | Low-quality |
| S12_vs_k141_172744||lt2gene | Not-determined |
| S12_vs_k141_172808||lt2gene | Low-quality |
| S12_vs_k141_15176||lt2gene | Low-quality |
| S12_vs_k141_15206||lt2gene | Not-determined |
| S12_vs_k141_15229||lt2gene | Not-determined |
| S12_vs_k141_125515||lt2gene | Low-quality |
| S12_vs_k141_117842||lt2gene | Not-determined |
| S12_vs_k141_156967||lt2gene | Low-quality |
| S12_vs_k141_157020||lt2gene | Low-quality |
| S12_vs_k141_15363||lt2gene | Low-quality |
| S12_vs_k141_172994||lt2gene | Low-quality |
| S12_vs_k141_173024||lt2gene | Not-determined |
| S12_vs_k141_157137||lt2gene | Low-quality |
| S12_vs_k141_125697||lt2gene | Not-determined |
| S12_vs_k141_173093||lt2gene | Low-quality |
| S12_vs_k141_180715||lt2gene | Low-quality |
| S12_vs_k141_173103||lt2gene | Low-quality |
| S12_vs_k141_180737||lt2gene | Low-quality |
| S12_vs_k141_118089||lt2gene | Low-quality |
| S12_vs_k141_118101||lt2gene | Low-quality |
| S12_vs_k141_157242||lt2gene | Not-determined |
| S12_vs_k141_157254||lt2gene | Low-quality |
| S12_vs_k141_15651||lt2gene | Low-quality |
| S12_vs_k141_125797||lt2gene | Low-quality |
| S12_vs_k141_180864||lt2gene | Low-quality |
| S12_vs_k141_125891||lt2gene | Low-quality |
| S12_vs_k141_173211||lt2gene | Low-quality |
| S12_vs_k141_125918||lt2gene | Low-quality |
| S12_vs_k141_125937||lt2gene | Low-quality |
| S15_extracted_6 | Not-determined |
| S15_extracted_11 | Low-quality |
| S15_extracted_14 | Not-determined |
| S15_extracted_23 | Not-determined |
| S15_extracted_27 | Low-quality |
| S15_extracted_31 | Low-quality |
| S15_extracted_47 | Not-determined |

| | |
|---|---|
| S1S_extracted_49 | Not-determined |
| S1S_extracted_71 | Not-determined |
| S1S_extracted_84 | Not-determined |
| S1S_extracted_91 | Not-determined |
| S1S_extracted_96 | Not-determined |
| S1S_extracted_97 | Not-determined |
| S1S_extracted_99 | Low-quality |
| S1S_extracted_100 | Not-determined |
| S1S_extracted_122 | Not-determined |
| S1S_extracted_124 | Not-determined |
| S1S_extracted_137 | Not-determined |
| S1S_extracted_144 | Not-determined |
| S1S_extracted_153 | Not-determined |
| S1S_extracted_160 | Low-quality |
| S1S_extracted_170 | Not-determined |
| S1S_extracted_171 | Not-determined |
| S1S_extracted_188 | Not-determined |
| S1S_extracted_193 | Not-determined |
| S1S_extracted_202 | Not-determined |
| S1S_extracted_230 | Not-determined |
| S1S_extracted_232 | Not-determined |
| S1S_extracted_236 | Not-determined |
| S1S_extracted_243 | Low-quality |
| S1S_extracted_257 | Low-quality |
| S1S_extracted_258 | Not-determined |
| S1S_extracted_265 | Not-determined |
| S1S_extracted_271 | Not-determined |
| S1S_extracted_291 | Not-determined |
| S1S_extracted_296 | Not-determined |
| S1S_extracted_330 | Not-determined |
| S1S_extracted_342 | Not-determined |
| S1S_extracted_344 | Low-quality |
| S1S_extracted_349 | Not-determined |
| S1S_extracted_354 | Not-determined |
| S1S_extracted_359 | Not-determined |
| S1S_extracted_369 | Not-determined |
| S1S_extracted_371 | Not-determined |
| S1S_extracted_383 | Not-determined |
| S1S_extracted_386 | Not-determined |
| S1S_extracted_391 | Not-determined |
| S1S_extracted_394 | Not-determined |
| S1S_extracted_411 | Not-determined |
| S1S_extracted_416 | Not-determined |
| S1S_extracted_420 | Low-quality |
| S1S_extracted_431 | Not-determined |
| S1S_extracted_441 | Not-determined |
| S1S_extracted_442 | Low-quality |
| S1S_extracted_443 | Not-determined |
| S1S_extracted_459 | Not-determined |
| S1S_extracted_477 | Low-quality |
| S1S_extracted_482 | Not-determined |
| S1S_extracted_486 | Not-determined |
| S1S_extracted_488 | Not-determined |
| S1S_extracted_497 | Not-determined |
| S1S_extracted_502 | Low-quality |
| S1S_extracted_511 | Not-determined |
| S1S_extracted_516 | Not-determined |
| S1S_extracted_539 | Not-determined |
| S1S_extracted_548 | Not-determined |
| S1S_extracted_554 | Not-determined |
| S1S_extracted_560 | Low-quality |
| S1S_extracted_571 | Not-determined |
| S1S_extracted_574 | Not-determined |
| S1S_extracted_584 | Not-determined |
| S1S_extracted_595 | Not-determined |
| S1S_extracted_610 | Low-quality |
| S1S_extracted_640 | Not-determined |
| S1S_extracted_647 | Not-determined |
| S1S_extracted_658 | Not-determined |
| S1S_extracted_660 | Not-determined |
| S1S_extracted_669 | Not-determined |
| S1S_extracted_671 | Not-determined |
| S1S_extracted_680 | Not-determined |
| S1S_extracted_683 | Not-determined |
| S1S_extracted_697 | Not-determined |
| S1S_extracted_712 | Not-determined |
| S1S_extracted_713 | Low-quality |
| S1S_extracted_717 | Not-determined |
| S1S_extracted_734 | Not-determined |
| S1S_extracted_737 | Not-determined |
| S1S_extracted_749 | Not-determined |
| S1S_extracted_752 | Not-determined |
| S1S_extracted_772 | Not-determined |
| S1S_extracted_776 | Low-quality |
| S1S_extracted_777 | Low-quality |
| S1S_extracted_781 | Low-quality |
| S1S_extracted_785 | Not-determined |
| S1S_extracted_790 | Not-determined |
| S1S_extracted_792 | Not-determined |
| S1S_extracted_799 | Not-determined |
| S1S_extracted_809 | Not-determined |
| S1S_extracted_829 | Not-determined |
| S1S_extracted_831 | Low-quality |
| S1S_extracted_847 | Not-determined |
| S1S_extracted_851 | Not-determined |
| S1S_extracted_857 | Not-determined |
| S1S_extracted_883 | Not-determined |
| S1S_extracted_884 | Not-determined |
| S1S_extracted_890 | Not-determined |
| S1S_extracted_907 | Not-determined |
| S1S_extracted_912 | Not-determined |
| S1S_extracted_917 | Not-determined |
| S1S_extracted_920 | Not-determined |
| S1S_extracted_928 | Low-quality |
| S1S_extracted_931 | Not-determined |
| S1S_extracted_933 | Not-determined |
| S1S_extracted_938 | Not-determined |
| S1S_extracted_944 | Not-determined |
| S1S_extracted_947 | Not-determined |
| S1S_extracted_952 | Low-quality |
| S1S_extracted_959 | Not-determined |
| S1S_extracted_974 | Not-determined |
| S1S_extracted_979 | Not-determined |
| S1S_extracted_1002 | Low-quality |
| S1S_extracted_1016 | Not-determined |
| S1S_extracted_1034 | Low-quality |
| S1S_extracted_1083 | Not-determined |
| S1S_extracted_1092 | Not-determined |
| S1S_extracted_1095 | Not-determined |
| S1S_extracted_1105 | Low-quality |
| S1S_extracted_1109 | Not-determined |
| S1S_extracted_1114 | Not-determined |
| S1S_extracted_1122 | Not-determined |
| S1S_extracted_1135 | Low-quality |
| S1S_extracted_1153 | Not-determined |
| S1S_extracted_1156 | Not-determined |
| S1S_extracted_1162 | Not-determined |
| S1S_extracted_1164 | Not-determined |
| S1S_extracted_1178 | Not-determined |
| S1S_extracted_1179 | Not-determined |
| S1S_extracted_1181 | Not-determined |
| S1S_extracted_1182 | Not-determined |
| S1S_extracted_1190 | Not-determined |
| S1S_extracted_1193 | Not-determined |
| S1S_extracted_1203 | Low-quality |
| S1S_extracted_1204 | Not-determined |
| S1S_extracted_1220 | Not-determined |
| S1S_extracted_1224 | Low-quality |
| S1S_extracted_1229 | Not-determined |
| S1S_extracted_1231 | Not-determined |
| S1S_extracted_1232 | Not-determined |
| S1S_extracted_1233 | Low-quality |
| S1S_extracted_1239 | Not-determined |
| S1S_extracted_1247 | Not-determined |
| S1S_extracted_1256 | Not-determined |
| S1S_extracted_1267 | Not-determined |
| S1S_extracted_1286 | Not-determined |
| S1S_extracted_1293 | Not-determined |
| S1S_extracted_1297 | Low-quality |
| S1S_extracted_1301 | Not-determined |
| S1S_extracted_1307 | Not-determined |
| S1S_extracted_1310 | Not-determined |
| S1S_extracted_1315 | Not-determined |
| S1S_extracted_1326 | Not-determined |
| S1S_extracted_1328 | Not-determined |
| S1S_extracted_1338 | Not-determined |
| S1S_extracted_1350 | Not-determined |
| S1S_extracted_1359 | Not-determined |
| S1S_extracted_1362 | Not-determined |
| S1S_extracted_1364 | Low-quality |
| S1S_extracted_1367 | Not-determined |
| S1S_extracted_1369 | Low-quality |
| S1S_extracted_1373 | Not-determined |
| S1S_extracted_1377 | Not-determined |
| S1S_extracted_1381 | Low-quality |
| S1S_extracted_1383 | Not-determined |
| S1S_extracted_1396 | Not-determined |
| S1S_extracted_1398 | Not-determined |
| S1S_extracted_1403 | Not-determined |
| S1S_extracted_1410 | Low-quality |
| S1S_extracted_1418 | Not-determined |
| S1S_extracted_1419 | Not-determined |
| S1S_extracted_1434 | Not-determined |
| S1S_extracted_1441 | Not-determined |
| S1S_extracted_1453 | Not-determined |
| S1S_extracted_1466 | Not-determined |
| S1S_extracted_1473 | Not-determined |
| S1S_extracted_1477 | Not-determined |
| S1S_extracted_1480 | Not-determined |
| S1S_extracted_1481 | Not-determined |
| S1S_extracted_1483 | Not-determined |
| S1S_extracted_1488 | Low-quality |
| S1S_extracted_1491 | Not-determined |
| S1S_extracted_1505 | Low-quality |
| S1S_extracted_1510 | Not-determined |
| S1S_extracted_1521 | Not-determined |
| S1S_extracted_1526 | Not-determined |
| S1S_extracted_1547 | Not-determined |
| S1S_extracted_1548 | Not-determined |
| S1S_extracted_1550 | Not-determined |
| S1S_extracted_1555 | Not-determined |
| S1S_extracted_1561 | Not-determined |
| S1S_extracted_1581 | Not-determined |
| S1S_extracted_1582 | Not-determined |
| S1S_extracted_1597 | Low-quality |
| S1S_extracted_1603 | Not-determined |
| S1S_extracted_1612 | Not-determined |
| S1S_extracted_1613 | Not-determined |
| S1S_extracted_1630 | Not-determined |
| S1S_extracted_1631 | Not-determined |
| S1S_extracted_1642 | Not-determined |
| S1S_extracted_1656 | Not-determined |
| S1S_extracted_1660 | Low-quality |
| S1S_extracted_1662 | Not-determined |
| S1S_extracted_1663 | Not-determined |
| S1S_extracted_1665 | Not-determined |
| S1S_extracted_1679 | Not-determined |
| S1S_extracted_1680 | Low-quality |
| S1S_extracted_1694 | Not-determined |
| S1S_extracted_1695 | Not-determined |
| S1S_extracted_1703 | Not-determined |
| S1S_extracted_1705 | Not-determined |
| S1S_extracted_1710 | Not-determined |
| S1S_extracted_1714 | Not-determined |
| S1S_extracted_1728 | Not-determined |
| S1S_extracted_1746 | Not-determined |
| S1S_extracted_1747 | Low-quality |
| S1S_extracted_1762 | Not-determined |
| S1S_extracted_1764 | Not-determined |
| S1S_extracted_1773 | Low-quality |
| S1S_extracted_1786 | Not-determined |
| S1S_extracted_1800 | Not-determined |
| S1S_extracted_1818 | Not-determined |
| S1S_extracted_1821 | Not-determined |
| S1S_extracted_1842 | Low-quality |
| S1S_extracted_1845 | Not-determined |
| S1S_extracted_1856 | Not-determined |
| S1S_extracted_1866 | Not-determined |
| S1S_extracted_1874 | Not-determined |
| S1S_extracted_1878 | Not-determined |
| S1S_extracted_1885 | Not-determined |
| S1S_extracted_1897 | Low-quality |
| S1S_extracted_1935 | Not-determined |
| S1S_extracted_1953 | Not-determined |
| S1S_extracted_1956 | Not-determined |
| S1S_extracted_1959 | Not-determined |
| S1S_extracted_1963 | Not-determined |
| S1S_extracted_1979 | Not-determined |
| S1S_extracted_1995 | Not-determined |
| S1S_extracted_2022 | Not-determined |
| S1S_extracted_2023 | Not-determined |
| S1S_extracted_2027 | Not-determined |
| S1S_extracted_2030 | Not-determined |
| S16_extracted_103938 | Medium-quality |
| S1S_extracted_2040 | Not-determined |
| S1S_extracted_2056 | Not-determined |
| S1S_extracted_2059 | Not-determined |
| S1S_extracted_2062 | Not-determined |
| S1S_extracted_2087 | Not-determined |
| S1S_extracted_2094 | Not-determined |
| S1S_extracted_2096 | Not-determined |
| S1S_extracted_2109 | Not-determined |
| S1S_extracted_2114 | Low-quality |
| S1S_extracted_2115 | Not-determined |
| S1S_extracted_2121 | Not-determined |
| S1S_extracted_2140 | Not-determined |
| S1S_extracted_2141 | Not-determined |
| S1S_extracted_2149 | Not-determined |
| S1S_extracted_2150 | Not-determined |
| S1S_extracted_2172 | Not-determined |
| S1S_extracted_2175 | Not-determined |
| S1S_extracted_2180 | Not-determined |
| S1S_extracted_2181 | Not-determined |
| S1S_extracted_2183 | Not-determined |
| S1S_extracted_2203 | Not-determined |
| S1S_extracted_2205 | Not-determined |
| S1S_extracted_2228 | Not-determined |
| S1S_extracted_2254 | Not-determined |
| S1S_extracted_2261 | Not-determined |
| S1S_extracted_2265 | Not-determined |
| S1S_extracted_2277 | Not-determined |
| S1S_extracted_2294 | Low-quality |
| S1S_extracted_2303 | Not-determined |
| S1S_extracted_2305 | Not-determined |
| S1S_extracted_2338 | Low-quality |
| S1S_extracted_2351 | Not-determined |
| S1S_extracted_2372 | Not-determined |
| S1S_extracted_2376 | Not-determined |
| S1S_extracted_2380 | Not-determined |
| S1S_extracted_2382 | Not-determined |
| S1S_extracted_2401 | Not-determined |
| S1S_extracted_2405 | Not-determined |
| S1S_extracted_2410 | Not-determined |
| S1S_extracted_2415 | Not-determined |
| S1S_extracted_2421 | Not-determined |
| S1S_extracted_2433 | Not-determined |
| S1S_extracted_2439 | Not-determined |
| S1S_extracted_2461 | Not-determined |
| S1S_extracted_2463 | Not-determined |
| S1S_extracted_2465 | Not-determined |
| S1S_extracted_2468 | Not-determined |
| S1S_extracted_2477 | Not-determined |
| S1S_extracted_2483 | Not-determined |
| S1S_extracted_2485 | Not-determined |
| S1S_extracted_2487 | Low-quality |
| S1S_extracted_2493 | Not-determined |
| S1S_extracted_2503 | Not-determined |
| SS_vs_k141_8042\|\|full | Medium-quality |
| S1S_extracted_2525 | Not-determined |
| S1S_extracted_2531 | Not-determined |
| S1S_extracted_2542 | Not-determined |
| S1S_extracted_2543 | Low-quality |
| S1S_extracted_2547 | Not-determined |
| S1S_extracted_2568 | Not-determined |
| S1S_extracted_2569 | Not-determined |
| S1S_extracted_2580 | Low-quality |
| S1S_extracted_2595 | Low-quality |
| S1S_extracted_2598 | Low-quality |
| S1S_extracted_2599 | Not-determined |
| S1S_extracted_2606 | Not-determined |
| S1S_extracted_2608 | Not-determined |
| S1S_extracted_2610 | Not-determined |
| S1S_extracted_2611 | Low-quality |
| S1S_extracted_2615 | Not-determined |
| S1S_extracted_2616 | Not-determined |
| S1S_extracted_2622 | Not-determined |
| S1S_extracted_2623 | Not-determined |
| S1S_extracted_2629 | Not-determined |
| S1S_extracted_2633 | Not-determined |
| S1S_extracted_2641 | Not-determined |
| S1S_extracted_2650 | Not-determined |
| S1S_extracted_2673 | Not-determined |
| S1S_extracted_2677 | Not-determined |
| S1S_extracted_2685 | Not-determined |
| S1S_extracted_2703 | Not-determined |
| S1S_extracted_2716 | Not-determined |
| S1S_extracted_2717 | Not-determined |
| S1S_extracted_2725 | Not-determined |
| S1S_extracted_2731 | Not-determined |
| S1S_extracted_2735 | Low-quality |
| S1S_extracted_2748 | Not-determined |
| S1S_extracted_2752 | Not-determined |
| S1S_extracted_2755 | Not-determined |
| S1S_extracted_2757 | Not-determined |
| S1S_extracted_2770 | Not-determined |
| S1S_extracted_2777 | Not-determined |
| S1S_extracted_2787 | Not-determined |
| S1S_extracted_2791 | Not-determined |
| S1S_extracted_2797 | Not-determined |
| S1S_extracted_2798 | Not-determined |
| S1S_extracted_2801 | Low-quality |
| S1S_extracted_2819 | Not-determined |
| S1S_extracted_2820 | Not-determined |
| S1S_extracted_2824 | Not-determined |
| S1S_extracted_2826 | Not-determined |
| S1S_extracted_2828 | Not-determined |
| S1S_extracted_2848 | Not-determined |
| S1S_extracted_2855 | Low-quality |
| S1S_extracted_2859 | Not-determined |
| S1S_extracted_2872 | Not-determined |
| S1S_extracted_2873 | Not-determined |
| S1S_extracted_2877 | Not-determined |
| S1S_extracted_2883 | Not-determined |
| S1S_extracted_2886 | Not-determined |
| S1S_extracted_2892 | Not-determined |
| S1S_extracted_2897 | Not-determined |
| S1S_extracted_2905 | Not-determined |
| S1S_extracted_2906 | Not-determined |
| S1S_extracted_2910 | Low-quality |
| S1S_extracted_2923 | Not-determined |
| S1S_extracted_2926 | Not-determined |
| S1S_extracted_2928 | Not-determined |
| S1S_extracted_2931 | Not-determined |
| S1S_extracted_2933 | Low-quality |
| S1S_extracted_2934 | Not-determined |
| S1S_extracted_2944 | Not-determined |
| S1S_extracted_2953 | Not-determined |
| S1S_extracted_2960 | Not-determined |
| S1S_extracted_2964 | Not-determined |
| S1S_extracted_2973 | Not-determined |
| S1S_extracted_2980 | Not-determined |
| S1S_extracted_2987 | Not-determined |
| S1S_extracted_2988 | Not-determined |
| S1S_extracted_2992 | Not-determined |
| S1S_extracted_2995 | Not-determined |
| S1S_extracted_2996 | Not-determined |
| S1S_extracted_2997 | Not-determined |
| S1S_extracted_2999 | Not-determined |
| S1S_extracted_3006 | Not-determined |
| S1S_extracted_3009 | Not-determined |
| S1S_extracted_3010 | Not-determined |
| S1S_extracted_3027 | Not-determined |
| S1S_extracted_3032 | Not-determined |
| S1S_extracted_3040 | Not-determined |
| S1S_extracted_3055 | Not-determined |
| S1S_extracted_3097 | Low-quality |
| S1S_extracted_3113 | Not-determined |
| S1S_extracted_3117 | Not-determined |
| S1S_extracted_3127 | Not-determined |
| S1S_extracted_3135 | Not-determined |
| S1S_extracted_3145 | Not-determined |
| S1S_extracted_3146 | Not-determined |
| S1S_extracted_3151 | Not-determined |
| S1S_extracted_3153 | Low-quality |
| S1S_extracted_3160 | Not-determined |
| S1S_extracted_3162 | Not-determined |
| S1S_extracted_3185 | Not-determined |
| S1S_extracted_3187 | Not-determined |
| S1S_extracted_3204 | Not-determined |
| S1S_extracted_3206 | Not-determined |
| S1S_extracted_3208 | Not-determined |
| S1S_extracted_3211 | Not-determined |
| S1S_extracted_3213 | Not-determined |
| S1S_extracted_3218 | Low-quality |
| S1S_extracted_3221 | Not-determined |
| S1S_extracted_3251 | Not-determined |
| S1S_extracted_3267 | Not-determined |
| S1S_extracted_3275 | Not-determined |
| S1S_extracted_3276 | Not-determined |
| S1S_extracted_3288 | Not-determined |
| S1S_extracted_3294 | Not-determined |
| S1S_extracted_3319 | Not-determined |
| S1S_extracted_3322 | Not-determined |
| S1S_extracted_3327 | Not-determined |
| S1S_extracted_3340 | Not-determined |
| S1S_extracted_3344 | Not-determined |
| S1S_extracted_3357 | Low-quality |
| S1S_extracted_3361 | Not-determined |
| S1S_extracted_3375 | Not-determined |
| S1S_extracted_3377 | Low-quality |
| S1S_extracted_3395 | Not-determined |
| S1S_extracted_3416 | Low-quality |
| S1S_extracted_3428 | Not-determined |
| S1S_extracted_3431 | Not-determined |
| S1S_extracted_3440 | Not-determined |
| S1S_extracted_3447 | Low-quality |
| S1S_extracted_3449 | Not-determined |
| S1S_extracted_3451 | Not-determined |
| S1S_extracted_3462 | Not-determined |
| S1S_extracted_3465 | Low-quality |
| S1S_extracted_3472 | Not-determined |
| S1S_extracted_3482 | Not-determined |
| S1S_extracted_3487 | Not-determined |
| S1S_extracted_3506 | Not-determined |
| S1S_extracted_3507 | Low-quality |
| S1S_extracted_3510 | Low-quality |
| S1S_extracted_3528 | Not-determined |
| S1S_extracted_3544 | Low-quality |
| S1S_extracted_3548 | Not-determined |
| S1S_extracted_3556 | Not-determined |
| S1S_extracted_3559 | Not-determined |
| S1S_extracted_3560 | Not-determined |
| S1S_extracted_3564 | Not-determined |
| S1S_extracted_3570 | Not-determined |
| S1S_extracted_3580 | Not-determined |
| S1S_extracted_3592 | Not-determined |
| S1S_extracted_3595 | Not-determined |
| S1S_extracted_3597 | Not-determined |
| S1S_extracted_3599 | Low-quality |
| S1S_extracted_3609 | Low-quality |
| S1S_extracted_3612 | Not-determined |
| S1S_extracted_3615 | Not-determined |
| S1S_extracted_3632 | Not-determined |
| S1S_extracted_3634 | Not-determined |
| S1S_extracted_3644 | Not-determined |
| S1S_extracted_3655 | Not-determined |
| S1S_extracted_3657 | Low-quality |
| S1S_extracted_3661 | Not-determined |
| S1S_extracted_3685 | Not-determined |
| S1S_extracted_3693 | Not-determined |
| S1S_extracted_3703 | Not-determined |
| S1S_extracted_3705 | Not-determined |
| S1S_extracted_3721 | Not-determined |
| S1S_extracted_3722 | Not-determined |
| S1S_extracted_3732 | Not-determined |
| S1S_extracted_3735 | Not-determined |
| S1S_extracted_3737 | Not-determined |
| S1S_extracted_3752 | Not-determined |
| S1S_extracted_3755 | Not-determined |
| S1S_extracted_3757 | Not-determined |
| S1S_extracted_3761 | Not-determined |
| S1S_extracted_3765 | Not-determined |
| S1S_extracted_3767 | Not-determined |
| S1S_extracted_3786 | Not-determined |
| S1S_extracted_3792 | Not-determined |
| S1S_extracted_3821 | Not-determined |
| S1S_extracted_3830 | Not-determined |
| S1S_extracted_3831 | Not-determined |
| S1S_extracted_3836 | Not-determined |
| S1S_extracted_3845 | Not-determined |
| S1S_extracted_3847 | Not-determined |
| S1S_extracted_3854 | Not-determined |
| S1S_extracted_3871 | Not-determined |
| S1S_extracted_3873 | Not-determined |
| S1S_extracted_3875 | Not-determined |
| S1S_extracted_3886 | Not-determined |
| S1S_extracted_3887 | Not-determined |
| S1S_extracted_3892 | Low-quality |
| S1S_extracted_3894 | Not-determined |
| S1S_extracted_3913 | Not-determined |
| S1S_extracted_3919 | Low-quality |
| S1S_extracted_3926 | Low-quality |
| S1S_extracted_3927 | Not-determined |
| S1S_extracted_3930 | Not-determined |
| S1S_extracted_3934 | Not-determined |
| S1S_extracted_3935 | Not-determined |
| S1S_extracted_3937 | Not-determined |
| S1S_extracted_3952 | Not-determined |
| S1S_extracted_3953 | Not-determined |
| S1S_extracted_3962 | Low-quality |
| S1S_extracted_3969 | Not-determined |
| S1S_extracted_3980 | Not-determined |
| S1S_extracted_4015 | Low-quality |
| S1S_extracted_4020 | Not-determined |
| S1S_extracted_4039 | Not-determined |
| S1S_extracted_4044 | Low-quality |
| S1S_extracted_4053 | Not-determined |
| S1S_extracted_4077 | Not-determined |
| S1S_extracted_4084 | Low-quality |
| S1S_extracted_4096 | Not-determined |
| S1S_extracted_4111 | Not-determined |
| S1S_extracted_4117 | Not-determined |
| S1S_extracted_4133 | Not-determined |
| S1S_extracted_4145 | Not-determined |
| S1S_extracted_4171 | Not-determined |
| S1S_extracted_4183 | Low-quality |
| S1S_extracted_4193 | Low-quality |
| S1S_extracted_4196 | Not-determined |
| S1S_extracted_4203 | Not-determined |
| S1S_extracted_4207 | Not-determined |
| S1S_extracted_4209 | Not-determined |
| S1S_extracted_4210 | Not-determined |
| S1S_extracted_4217 | Not-determined |
| S1S_extracted_4232 | Not-determined |
| S1S_extracted_4248 | Not-determined |
| S1S_extracted_4253 | Not-determined |
| S1S_extracted_4257 | Not-determined |
| S1S_extracted_4263 | Not-determined |
| S1S_extracted_4282 | Not-determined |
| S1S_extracted_4293 | Not-determined |
| S1S_extracted_4301 | Not-determined |
| S1S_extracted_4309 | Not-determined |
| S1S_extracted_4313 | Low-quality |
| S1S_extracted_4329 | Not-determined |
| S1S_extracted_4335 | Not-determined |
| S1S_extracted_4350 | Low-quality |
| S1S_extracted_4354 | Not-determined |
| S1S_extracted_4362 | Not-determined |
| S1S_extracted_4375 | Not-determined |
| S1S_extracted_4378 | Not-determined |
| S1S_extracted_4385 | Not-determined |
| S1S_extracted_4390 | Not-determined |
| S1S_extracted_4396 | Not-determined |
| S1S_extracted_4397 | Not-determined |
| S1S_extracted_4410 | Not-determined |
| S1S_extracted_4417 | Not-determined |
| S1S_extracted_4424 | Not-determined |
| S1S_extracted_4428 | Not-determined |
| S1S_extracted_4430 | Not-determined |
| S1S_extracted_4460 | Not-determined |
| S1S_extracted_4470 | Not-determined |
| S1S_extracted_4475 | Not-determined |
| S1S_extracted_4488 | Not-determined |
| S1S_extracted_4489 | Not-determined |
| S1S_extracted_4490 | Not-determined |
| S1S_extracted_4495 | Not-determined |
| S1S_extracted_4506 | Not-determined |
| S1S_extracted_4515 | Not-determined |
| S1S_extracted_4526 | Not-determined |
| S1S_extracted_4536 | Not-determined |
| S1S_extracted_4548 | Not-determined |
| S1S_extracted_4549 | Not-determined |
| S1S_extracted_4558 | Not-determined |
| S1S_extracted_4597 | Not-determined |
| S1S_extracted_4601 | Not-determined |
| S1S_extracted_4654 | Not-determined |
| S1S_extracted_4657 | Not-determined |
| S1S_extracted_4659 | Not-determined |
| S1S_extracted_4660 | Not-determined |
| S1S_extracted_4680 | Not-determined |
| S1S_extracted_4683 | Not-determined |
| S1S_extracted_4688 | Not-determined |
| S1S_extracted_4721 | Low-quality |
| S1S_extracted_4729 | Not-determined |
| S1S_extracted_4730 | Not-determined |
| S1S_extracted_4747 | Not-determined |
| S1S_extracted_4755 | Not-determined |
| S1S_extracted_4756 | Not-determined |
| S1S_extracted_4762 | Not-determined |
| S1S_extracted_4777 | Not-determined |
| S1S_extracted_4779 | Not-determined |
| S1S_extracted_4787 | Not-determined |
| S1S_extracted_4788 | Not-determined |
| S1S_extracted_4791 | Not-determined |
| S1S_extracted_4800 | Low-quality |
| S1S_extracted_4801 | Not-determined |
| S1S_extracted_4824 | Not-determined |
| S1S_extracted_4841 | Not-determined |
| S1S_extracted_4859 | Not-determined |

| | |
|---|---|
| S1S_extracted_4863 | Not-determined |
| S1S_extracted_4883 | Not-determined |
| S1S_extracted_4886 | Not-determined |
| S1S_extracted_4899 | Not-determined |
| S1S_extracted_4921 | Not-determined |
| S1S_extracted_4928 | Not-determined |
| S1S_extracted_4934 | Not-determined |
| S1S_extracted_4941 | Not-determined |
| S1S_extracted_4972 | Low-quality |
| S1S_extracted_4992 | Low-quality |
| S1S_extracted_5002 | Not-determined |
| S1S_extracted_5005 | Not-determined |
| S1S_extracted_5006 | Not-determined |
| S1S_extracted_5009 | Not-determined |
| S1S_extracted_5013 | Low-quality |
| S1S_extracted_5026 | Not-determined |
| S1S_extracted_5029 | Not-determined |
| S1S_extracted_5032 | Not-determined |
| S1S_extracted_5044 | Low-quality |
| S1S_extracted_5048 | Low-quality |
| S1S_extracted_5060 | Not-determined |
| S1S_extracted_5061 | Not-determined |
| S1S_extracted_5068 | Not-determined |
| S1S_extracted_5076 | Low-quality |
| S1S_extracted_5083 | Not-determined |
| S1S_extracted_5084 | Not-determined |
| S1S_extracted_5086 | Not-determined |
| S1S_extracted_5088 | Not-determined |
| S1S_extracted_5091 | Not-determined |
| S1S_extracted_5096 | Not-determined |
| S1S_extracted_5099 | Not-determined |
| S1S_extracted_5108 | Not-determined |
| S1S_extracted_5141 | Not-determined |
| S1S_extracted_5142 | Not-determined |
| S1S_extracted_5144 | Not-determined |
| S1S_extracted_5157 | Not-determined |
| S1S_extracted_5162 | Not-determined |
| S1S_extracted_5170 | Low-quality |
| S1S_extracted_5178 | Not-determined |
| S1S_extracted_5184 | Not-determined |
| S1S_extracted_5188 | Low-quality |
| S1S_extracted_5192 | Not-determined |
| S1S_extracted_5193 | Not-determined |
| S1S_extracted_5194 | Not-determined |
| S1S_extracted_5199 | Not-determined |
| S1S_extracted_5216 | Not-determined |
| S1S_extracted_5220 | Not-determined |
| S1S_extracted_5221 | Not-determined |
| S1S_extracted_5228 | Not-determined |
| S1S_extracted_5233 | Not-determined |
| S1S_extracted_5235 | Not-determined |
| S1S_extracted_5257 | Not-determined |
| S1S_extracted_5260 | Not-determined |
| S1S_extracted_5270 | Low-quality |
| S1S_extracted_5291 | Not-determined |
| S1S_extracted_5295 | Not-determined |
| S1S_extracted_5304 | Not-determined |
| S1S_extracted_5312 | Not-determined |
| S1S_extracted_5313 | Not-determined |
| S1S_extracted_5316 | Not-determined |
| S1S_extracted_5328 | Not-determined |
| S1S_extracted_5336 | Not-determined |
| S1S_extracted_5366 | Not-determined |
| S1S_extracted_5383 | Low-quality |
| S1S_extracted_5391 | Not-determined |
| S1S_extracted_5404 | Not-determined |
| S1S_extracted_5408 | Not-determined |
| S1S_extracted_5413 | Not-determined |
| S1S_extracted_5421 | Not-determined |
| S1S_extracted_5434 | Not-determined |
| S1S_extracted_5436 | Not-determined |
| S1S_extracted_5473 | Not-determined |
| S1S_extracted_5475 | Not-determined |
| S1S_extracted_5490 | Not-determined |
| S1S_extracted_5497 | Not-determined |
| S1S_extracted_5498 | Not-determined |
| S1S_extracted_5499 | Not-determined |
| S1S_extracted_5501 | Low-quality |
| S1S_extracted_5507 | Not-determined |
| S1S_extracted_5518 | Not-determined |
| S1S_extracted_5523 | Not-determined |
| S1S_extracted_5525 | Not-determined |
| S1S_extracted_5528 | Not-determined |
| S1S_extracted_5532 | Not-determined |
| S1S_extracted_5535 | Not-determined |
| S1S_extracted_5553 | Not-determined |
| S1S_extracted_5570 | Not-determined |
| S1S_extracted_5571 | Not-determined |
| S1S_extracted_5572 | Not-determined |
| S1S_extracted_5581 | Not-determined |
| S1S_extracted_5588 | Not-determined |
| S1S_extracted_5595 | Not-determined |
| S1S_extracted_5596 | Not-determined |
| S1S_extracted_5598 | Not-determined |
| S1S_extracted_5601 | Not-determined |
| S1S_extracted_5645 | Not-determined |
| S1S_extracted_5653 | Not-determined |
| S1S_extracted_5663 | Not-determined |
| S1S_extracted_5701 | Not-determined |
| S1S_extracted_5703 | Not-determined |
| S1S_extracted_5708 | Low-quality |
| S1S_extracted_5722 | Not-determined |
| S1S_extracted_5730 | Not-determined |
| S1S_extracted_5734 | Not-determined |
| S1S_extracted_5736 | Not-determined |
| S1S_extracted_5737 | Not-determined |
| S1S_extracted_5738 | Not-determined |
| S1S_extracted_5746 | Low-quality |
| S1S_extracted_5747 | Not-determined |
| S1S_extracted_5756 | Not-determined |
| S1S_extracted_5759 | Not-determined |
| S1S_extracted_5763 | Low-quality |
| S1S_extracted_5794 | Not-determined |
| S1S_extracted_5804 | Not-determined |
| S1S_extracted_5808 | Not-determined |
| S1S_extracted_5809 | Low-quality |
| S1S_extracted_5820 | Not-determined |
| S1S_extracted_5838 | Not-determined |
| S1S_extracted_5840 | Not-determined |
| S1S_extracted_5842 | Not-determined |
| S1S_extracted_5857 | Not-determined |
| S1S_extracted_5860 | Not-determined |
| S1S_extracted_5872 | Not-determined |
| S1S_extracted_5895 | Not-determined |
| S1S_extracted_5900 | Not-determined |
| S1S_extracted_5917 | Not-determined |
| S1S_extracted_5940 | Not-determined |
| S1S_extracted_5965 | Not-determined |
| S1S_extracted_5985 | Low-quality |
| S1S_extracted_6003 | Not-determined |
| S1S_extracted_6013 | Not-determined |
| S1S_extracted_6022 | Not-determined |
| S1S_extracted_6024 | Not-determined |
| S1S_extracted_6026 | Not-determined |
| S1S_extracted_6029 | Not-determined |
| S1S_extracted_6045 | Not-determined |
| S1S_extracted_6048 | Not-determined |
| S1S_extracted_6049 | Not-determined |
| S1S_extracted_6080 | Not-determined |
| S1S_extracted_6086 | Not-determined |
| S1S_extracted_6109 | Not-determined |
| S1S_extracted_6115 | Not-determined |
| S1S_extracted_6120 | Not-determined |
| S1S_extracted_6138 | Not-determined |
| S1S_extracted_6140 | Not-determined |
| S1S_extracted_6148 | Not-determined |
| S1S_extracted_6158 | Not-determined |
| S1S_extracted_6161 | Not-determined |
| S1S_extracted_6162 | Not-determined |
| S1S_extracted_6164 | Not-determined |
| S1S_extracted_6172 | Not-determined |
| S1S_extracted_6173 | Not-determined |
| S1S_extracted_6174 | Not-determined |
| S1S_extracted_6192 | Not-determined |
| S1S_extracted_6237 | Not-determined |
| S1S_extracted_6254 | Not-determined |
| S1S_extracted_6260 | Not-determined |
| S1S_extracted_6261 | Low-quality |
| S1S_extracted_6265 | Not-determined |
| S1S_extracted_6282 | Not-determined |
| S1S_extracted_6286 | Not-determined |
| S1S_extracted_6292 | Low-quality |
| S1S_extracted_6311 | Not-determined |
| S1S_extracted_6312 | Not-determined |
| S1S_extracted_6318 | Not-determined |
| S1S_extracted_6332 | Not-determined |
| S1S_extracted_6340 | Not-determined |
| S1S_extracted_6351 | Low-quality |
| S1S_extracted_6357 | Not-determined |
| S1S_extracted_6362 | Not-determined |
| S1S_extracted_6364 | Not-determined |
| S1S_extracted_6369 | Not-determined |
| S1S_extracted_6397 | Not-determined |
| S1S_extracted_6402 | Not-determined |
| S1S_extracted_6420 | Not-determined |
| S1S_extracted_6432 | Low-quality |
| S1S_extracted_6449 | Not-determined |
| S1S_extracted_6469 | Not-determined |
| S1S_extracted_6473 | Not-determined |
| S1S_extracted_6476 | Not-determined |
| S1S_extracted_6479 | Not-determined |
| S1S_extracted_6485 | Not-determined |
| S1S_extracted_6487 | Not-determined |
| S1S_extracted_6515 | Not-determined |
| S1S_extracted_6519 | Not-determined |
| S1S_extracted_6521 | Not-determined |
| S1S_extracted_6523 | Not-determined |
| S1S_extracted_6524 | Not-determined |
| S1S_extracted_6540 | Not-determined |
| S1S_extracted_6547 | Not-determined |
| S1S_extracted_6556 | Not-determined |
| S1S_extracted_6558 | Not-determined |
| S1S_extracted_6564 | Low-quality |
| S1S_extracted_6577 | Not-determined |
| S1S_extracted_6593 | Not-determined |
| S1S_extracted_6596 | Low-quality |
| S1S_extracted_6616 | Not-determined |
| S1S_extracted_6617 | Not-determined |
| S1S_extracted_6623 | Not-determined |
| S1S_extracted_6630 | Not-determined |
| S1S_extracted_6631 | Low-quality |
| S1S_extracted_6651 | Not-determined |
| S1S_extracted_6660 | Not-determined |
| S1S_extracted_6664 | Not-determined |
| S1S_extracted_6675 | Not-determined |
| S1S_extracted_6682 | Not-determined |
| S1S_extracted_6686 | Low-quality |
| S1S_extracted_6687 | Low-quality |
| S1S_extracted_6693 | Not-determined |
| S1S_extracted_6697 | Not-determined |
| S1S_extracted_6702 | Not-determined |
| S1S_extracted_6705 | Not-determined |
| S1S_extracted_6711 | Not-determined |
| S1S_extracted_6712 | Not-determined |
| S1S_extracted_6725 | Not-determined |
| S1S_extracted_6731 | Not-determined |
| S1S_extracted_6733 | Not-determined |
| S1S_extracted_6752 | Not-determined |
| S1S_extracted_6759 | Not-determined |
| S1S_extracted_6773 | Not-determined |
| S1S_extracted_6774 | Not-determined |
| S1S_extracted_6777 | Low-quality |
| S1S_extracted_6780 | Not-determined |
| S1S_extracted_6782 | Not-determined |
| S1S_extracted_6785 | Not-determined |
| S1S_extracted_6797 | Not-determined |
| S1S_extracted_6807 | Not-determined |
| S17_vs_k141_46388\|\|full | Medium-quality |
| S1S_extracted_6827 | Not-determined |
| S1S_extracted_6830 | Low-quality |
| S1S_extracted_6831 | Not-determined |
| S1S_extracted_6836 | Not-determined |
| S1S_extracted_6842 | Not-determined |
| S1S_extracted_6844 | Low-quality |
| S1S_extracted_6850 | Not-determined |
| S1S_extracted_6854 | Not-determined |
| S1S_extracted_6865 | Not-determined |
| S1S_extracted_6881 | Low-quality |
| S1S_extracted_6909 | Not-determined |
| S1S_extracted_6912 | Not-determined |
| S1S_extracted_6920 | Not-determined |
| S1S_extracted_6956 | Not-determined |
| S1S_extracted_6958 | Not-determined |
| S1S_extracted_6976 | Not-determined |
| S1S_extracted_6978 | Low-quality |
| S1S_extracted_6991 | Not-determined |
| S1S_extracted_6996 | Not-determined |
| S1S_extracted_6999 | Not-determined |
| S1S_extracted_7017 | Low-quality |
| S1S_extracted_7025 | Not-determined |
| S1S_extracted_7035 | Not-determined |
| S1S_extracted_7038 | Not-determined |
| S1S_extracted_7049 | Not-determined |
| S1S_extracted_7050 | Not-determined |
| S1S_extracted_7056 | Not-determined |
| S1S_extracted_7066 | Not-determined |
| S1S_extracted_7068 | Not-determined |
| S1S_extracted_7072 | Not-determined |
| S1S_extracted_7081 | Not-determined |
| S1S_extracted_7089 | Not-determined |
| S1S_extracted_7118 | Low-quality |
| S1S_extracted_7128 | Not-determined |
| S1S_extracted_7130 | Low-quality |
| S1S_extracted_7146 | Not-determined |
| S1S_extracted_7147 | Not-determined |
| S1S_extracted_7150 | Low-quality |
| S1S_extracted_7151 | Not-determined |
| S1S_extracted_7165 | Not-determined |
| S1S_extracted_7171 | Not-determined |
| S1S_extracted_7200 | Not-determined |
| S1S_extracted_7202 | Not-determined |
| S1S_extracted_7204 | Not-determined |
| S1S_extracted_7216 | Not-determined |
| S1S_extracted_7218 | Not-determined |
| S1S_extracted_7222 | Low-quality |
| S1S_extracted_7259 | Not-determined |
| S1S_extracted_7262 | Not-determined |
| S1S_extracted_7265 | Low-quality |
| S1S_extracted_7280 | Not-determined |
| S1S_extracted_7288 | Not-determined |
| S1S_extracted_7289 | Not-determined |
| S1S_extracted_7290 | Low-quality |
| S1S_extracted_7300 | Not-determined |
| S1S_extracted_7302 | Not-determined |
| S1S_extracted_7303 | Not-determined |
| S1S_extracted_7304 | Not-determined |
| S1S_extracted_7306 | Not-determined |
| S1S_extracted_7314 | Not-determined |
| S1S_extracted_7321 | Not-determined |
| S1S_extracted_7325 | Not-determined |
| S1S_extracted_7328 | Not-determined |
| S1S_extracted_7329 | Not-determined |
| S1S_extracted_7331 | Not-determined |
| S1S_extracted_7346 | Not-determined |
| S1S_extracted_7352 | Not-determined |
| S1S_extracted_7353 | Low-quality |
| S1S_extracted_7354 | Low-quality |
| S1S_extracted_7370 | Not-determined |
| S1S_extracted_7372 | Not-determined |
| S1S_extracted_7387 | Not-determined |
| S1S_extracted_7390 | Not-determined |
| S1S_extracted_7397 | Not-determined |
| S1S_extracted_7408 | Not-determined |
| S1S_extracted_7417 | Not-determined |
| S1S_extracted_7427 | Low-quality |
| S1S_extracted_7464 | Not-determined |
| S1S_extracted_7468 | Not-determined |
| S1S_extracted_7469 | Not-determined |
| S1S_extracted_7471 | Not-determined |
| S1S_extracted_7472 | Not-determined |
| S1S_extracted_7481 | Not-determined |
| S1S_extracted_7484 | Not-determined |
| S1S_extracted_7485 | Not-determined |
| S1S_extracted_7498 | Not-determined |
| S1S_extracted_7511 | Not-determined |
| S1S_extracted_7512 | Not-determined |
| S1S_extracted_7538 | Not-determined |
| S1S_extracted_7539 | Not-determined |
| S1S_extracted_7541 | Not-determined |
| S1S_extracted_7574 | Not-determined |
| S1S_extracted_7579 | Not-determined |
| S1S_extracted_7589 | Not-determined |
| S1S_extracted_7594 | Not-determined |
| S1S_extracted_7600 | Not-determined |
| S1S_extracted_7603 | Not-determined |
| S1S_extracted_7619 | Not-determined |
| S1S_extracted_7622 | Not-determined |
| S1S_extracted_7625 | Not-determined |
| S1S_extracted_7639 | Not-determined |
| S1S_extracted_7668 | Not-determined |
| S1S_extracted_7674 | Not-determined |
| S1S_extracted_7675 | Not-determined |
| S1S_extracted_7685 | Not-determined |
| S1S_extracted_7705 | Not-determined |
| S1S_extracted_7717 | Not-determined |
| S1S_extracted_7720 | Not-determined |
| S1S_extracted_7723 | Not-determined |
| S1S_extracted_7730 | Low-quality |
| S1S_extracted_7731 | Not-determined |
| S1S_extracted_7738 | Not-determined |
| S1S_extracted_7750 | Not-determined |
| S1S_extracted_7757 | Not-determined |
| S1S_extracted_7766 | Not-determined |
| S1S_extracted_7783 | Not-determined |
| S1S_extracted_7784 | Not-determined |
| S1S_extracted_7794 | Low-quality |
| S1S_extracted_7800 | Low-quality |
| S1S_extracted_7804 | Not-determined |
| S1S_extracted_7816 | Not-determined |
| S1S_extracted_7817 | Not-determined |
| S1S_extracted_7821 | Not-determined |
| S1S_extracted_7844 | Not-determined |
| S1S_extracted_7858 | Low-quality |
| S1S_extracted_7864 | Not-determined |
| S1S_extracted_7874 | Not-determined |
| S1S_extracted_7880 | Not-determined |
| S1S_extracted_7899 | Not-determined |
| S1S_extracted_7910 | Not-determined |
| S1S_extracted_7918 | Not-determined |
| S1S_extracted_7922 | Not-determined |
| S1S_extracted_7924 | Not-determined |
| S1S_extracted_7927 | Not-determined |
| S1S_extracted_7930 | Low-quality |
| S1S_extracted_7932 | Not-determined |
| S1S_extracted_7939 | Not-determined |
| S1S_extracted_7940 | Not-determined |
| S1S_extracted_7958 | Not-determined |
| S1S_extracted_7964 | Not-determined |
| S1S_extracted_7968 | Not-determined |
| S1S_extracted_7980 | Not-determined |
| S1S_extracted_7983 | Not-determined |
| S1S_extracted_7984 | Not-determined |
| S1S_extracted_7987 | Not-determined |
| S1S_extracted_8000 | Not-determined |
| S1S_extracted_8005 | Not-determined |
| S1S_extracted_8009 | Not-determined |
| S1S_extracted_8039 | Not-determined |
| S1S_extracted_8042 | Low-quality |
| S1S_extracted_8048 | Not-determined |
| S1S_extracted_8060 | Low-quality |
| S1S_extracted_8072 | Not-determined |
| S1S_extracted_8082 | Not-determined |
| S1S_extracted_8084 | Not-determined |
| S1S_extracted_8088 | Low-quality |
| S1S_extracted_8089 | Low-quality |
| S1S_extracted_8095 | Not-determined |
| S1S_extracted_8112 | Not-determined |
| S1S_extracted_8116 | Not-determined |
| S1S_extracted_8134 | Not-determined |
| S1S_extracted_8136 | Not-determined |
| S1S_extracted_8138 | Not-determined |
| S1S_extracted_8141 | Not-determined |
| S1S_extracted_8156 | Not-determined |
| S1S_extracted_8165 | Not-determined |
| S1S_extracted_8166 | Not-determined |
| S1S_extracted_8167 | Not-determined |
| S1S_extracted_8172 | Low-quality |
| S1S_extracted_8176 | Not-determined |
| S1S_extracted_8185 | Not-determined |
| S1S_extracted_8194 | Not-determined |
| S1S_extracted_8199 | Not-determined |
| S1S_extracted_8203 | Not-determined |
| S1S_extracted_8206 | Not-determined |
| S1S_extracted_8214 | Not-determined |
| S1S_extracted_8224 | Not-determined |
| S1S_extracted_8226 | Not-determined |
| S1S_extracted_8228 | Not-determined |
| S1S_extracted_8246 | Not-determined |
| S1S_extracted_8252 | Not-determined |
| S1S_extracted_8255 | Not-determined |
| S1S_extracted_8259 | Not-determined |
| S1S_extracted_8266 | Not-determined |
| S1S_extracted_8301 | Not-determined |
| S1S_extracted_8303 | Not-determined |
| S1S_extracted_8311 | Not-determined |
| S1S_extracted_8330 | Not-determined |
| S1S_extracted_8340 | Not-determined |
| S1S_extracted_8362 | Not-determined |
| S1S_extracted_8363 | Not-determined |
| S1S_extracted_8393 | Not-determined |
| S1S_extracted_8398 | Not-determined |
| S1S_extracted_8418 | Not-determined |
| S1S_extracted_8419 | Not-determined |
| S1S_extracted_8430 | Not-determined |
| S1S_extracted_8433 | Not-determined |
| S1S_extracted_8434 | Not-determined |
| S1S_extracted_8435 | Not-determined |
| S1S_extracted_8438 | Not-determined |
| S1S_extracted_8442 | Not-determined |
| S1S_extracted_8443 | Not-determined |
| S1S_extracted_8451 | Not-determined |
| S1S_extracted_8476 | Not-determined |
| S1S_extracted_8480 | Low-quality |
| S1S_extracted_8485 | Low-quality |
| S1S_extracted_8486 | Not-determined |
| S1S_extracted_8509 | Not-determined |
| S1S_extracted_8539 | Not-determined |
| S1S_extracted_8548 | Not-determined |
| S1S_extracted_8560 | Not-determined |
| S1S_extracted_8568 | Not-determined |
| S1S_extracted_8578 | Not-determined |
| S1S_extracted_8580 | Not-determined |
| S1S_extracted_8582 | Not-determined |
| S1S_extracted_8585 | Not-determined |
| S1S_extracted_8591 | Not-determined |
| S1S_extracted_8598 | Not-determined |
| S1S_extracted_8600 | Not-determined |
| S1S_extracted_8609 | Not-determined |
| S1S_extracted_8615 | Not-determined |
| S1S_extracted_8619 | Not-determined |
| S1S_extracted_8629 | Not-determined |
| S1S_extracted_8637 | Not-determined |
| S1S_extracted_8665 | Low-quality |
| S1S_extracted_8674 | Not-determined |
| S1S_extracted_8682 | Low-quality |
| S1S_extracted_8685 | Not-determined |
| S1S_extracted_8707 | Not-determined |
| S1S_extracted_8711 | Not-determined |
| S1S_extracted_8722 | Not-determined |
| S1S_extracted_8731 | Low-quality |
| S1S_extracted_8736 | Not-determined |
| S1S_extracted_8742 | Not-determined |
| S1S_extracted_8751 | Not-determined |
| S1S_extracted_8757 | Not-determined |
| S1S_extracted_8774 | Not-determined |
| S1S_extracted_8778 | Not-determined |
| S1S_extracted_8799 | Not-determined |
| S1S_extracted_8833 | Low-quality |
| S1S_extracted_8836 | Not-determined |
| S1S_extracted_8846 | Not-determined |
| S1S_extracted_8862 | Not-determined |
| S1S_extracted_8869 | Not-determined |
| S1S_extracted_8875 | Not-determined |
| S1S_extracted_8899 | Not-determined |
| S1S_extracted_8907 | Not-determined |
| S1S_extracted_8911 | Not-determined |
| S1S_extracted_8912 | Not-determined |
| S1S_extracted_8913 | Not-determined |
| S1S_extracted_8915 | Not-determined |
| S1S_extracted_8930 | Not-determined |
| S1S_extracted_8939 | Not-determined |
| S1S_extracted_8946 | Not-determined |
| S1S_extracted_8959 | Not-determined |
| S1S_extracted_8960 | Not-determined |
| S1S_extracted_8966 | Not-determined |
| S1S_extracted_8968 | Not-determined |
| S1S_extracted_8976 | Low-quality |
| S1S_extracted_8980 | Not-determined |
| S1S_extracted_8986 | Not-determined |
| S1S_extracted_8987 | Not-determined |
| S1S_extracted_8990 | Not-determined |
| S1S_extracted_9002 | Low-quality |
| S1S_extracted_9023 | Not-determined |
| S1S_extracted_9033 | Not-determined |
| S1S_extracted_9064 | Not-determined |
| S1S_extracted_9065 | Not-determined |
| S1S_extracted_9068 | Not-determined |
| S1S_extracted_9075 | Not-determined |
| S1S_extracted_9079 | Not-determined |
| S1S_extracted_9083 | Not-determined |
| S1S_extracted_9091 | Low-quality |
| S1S_extracted_9108 | Low-quality |
| S1S_extracted_9113 | Low-quality |
| S1S_extracted_9115 | Not-determined |
| S1S_extracted_9126 | Not-determined |
| S1S_extracted_9134 | Not-determined |
| S1S_extracted_9141 | Not-determined |
| S1S_extracted_9153 | Not-determined |
| S1S_extracted_9154 | Not-determined |
| S1S_extracted_9174 | Not-determined |
| S1S_extracted_9190 | Not-determined |
| S1S_extracted_9192 | Not-determined |
| S1S_extracted_9194 | Not-determined |
| S1S_extracted_9211 | Low-quality |
| S1S_extracted_9212 | Not-determined |
| S1S_extracted_9216 | Not-determined |
| S1S_extracted_9264 | Not-determined |
| S1S_extracted_9281 | Not-determined |
| S1S_extracted_9282 | Not-determined |
| S1S_extracted_9292 | Low-quality |
| S1S_extracted_9296 | Low-quality |
| S1S_extracted_9300 | Not-determined |
| S1S_extracted_9305 | Not-determined |
| S1S_extracted_9318 | Not-determined |
| S1S_extracted_9321 | Not-determined |
| S1S_extracted_9326 | Not-determined |
| S1S_extracted_9329 | Not-determined |
| S1S_extracted_9336 | Not-determined |
| S1S_extracted_9337 | Not-determined |
| S1S_extracted_9382 | Not-determined |
| S1S_extracted_9399 | Not-determined |
| S1S_extracted_9409 | Not-determined |
| S1S_extracted_9416 | Not-determined |
| S1S_extracted_9428 | Low-quality |
| S1S_extracted_9436 | Not-determined |
| S1S_extracted_9439 | Not-determined |
| S1S_extracted_9459 | Not-determined |
| S1S_extracted_9472 | Low-quality |
| S1S_extracted_9482 | Not-determined |
| S1S_extracted_9487 | Low-quality |
| S1S_extracted_9496 | Not-determined |
| S1S_extracted_9517 | Not-determined |
| S1S_extracted_9518 | Not-determined |
| S1S_extracted_9521 | Not-determined |
| S1S_extracted_9533 | Not-determined |
| S1S_extracted_9546 | Low-quality |
| S1S_extracted_9551 | Not-determined |
| S1S_extracted_9555 | Not-determined |
| S1S_extracted_9565 | Not-determined |
| S1S_extracted_9585 | Low-quality |
| S1S_extracted_9586 | Not-determined |
| S1S_extracted_9600 | Not-determined |
| S1S_extracted_9610 | Not-determined |
| S1S_extracted_9613 | Not-determined |
| S1S_extracted_9627 | Not-determined |
| S1S_extracted_9628 | Not-determined |
| S1S_extracted_9632 | Low-quality |
| S1S_extracted_9639 | Not-determined |
| S1S_extracted_9643 | Not-determined |
| S1S_extracted_9647 | Not-determined |
| S1S_extracted_9652 | Not-determined |
| S1S_extracted_9656 | Not-determined |
| S1S_extracted_9664 | Low-quality |
| S1S_extracted_9672 | Not-determined |
| S1S_extracted_9690 | Not-determined |
| S1S_extracted_9704 | Not-determined |
| S1S_extracted_9708 | Low-quality |
| S1S_extracted_9720 | Low-quality |
| S1S_extracted_9722 | Not-determined |
| S1S_extracted_9724 | Not-determined |
| S1S_extracted_9726 | Not-determined |
| S1S_extracted_9745 | Not-determined |
| S1S_extracted_9754 | Not-determined |
| S1S_extracted_9755 | Not-determined |
| S1S_extracted_9757 | Not-determined |
| S1S_extracted_9761 | Low-quality |
| S1S_extracted_9772 | Low-quality |
| S1S_extracted_9775 | Low-quality |
| S1S_extracted_9788 | Not-determined |
| S1S_extracted_9793 | Not-determined |
| S1S_extracted_9818 | Not-determined |
| S1S_extracted_9820 | Not-determined |
| S1S_extracted_9845 | Not-determined |
| S1S_extracted_9850 | Not-determined |
| S1S_extracted_9853 | Not-determined |
| S1S_extracted_9858 | Not-determined |
| S1S_extracted_9866 | Not-determined |
| S1S_extracted_9876 | Not-determined |
| S1S_extracted_9894 | Not-determined |

| | |
|---|---|
| S1S_extracted_9901 | Low-quality |
| S1S_extracted_9916 | Not-determined |
| S1S_extracted_9919 | Not-determined |
| S1S_extracted_9963 | Not-determined |
| S1S_extracted_9978 | Not-determined |
| S1S_extracted_9981 | Not-determined |
| S1S_extracted_9995 | Not-determined |
| S1S_extracted_9996 | Not-determined |
| S1S_extracted_9999 | Not-determined |
| S1S_extracted_10049 | Not-determined |
| S1S_extracted_10057 | Not-determined |
| S1S_extracted_10061 | Not-determined |
| S1S_extracted_10066 | Not-determined |
| S1S_extracted_10069 | Not-determined |
| S1S_extracted_10077 | Not-determined |
| S1S_extracted_10087 | Not-determined |
| S1S_extracted_10102 | Not-determined |
| S1S_extracted_10115 | Not-determined |
| S1S_extracted_10120 | Not-determined |
| S1S_extracted_10123 | Not-determined |
| S1S_extracted_10130 | Not-determined |
| S1S_extracted_10140 | Not-determined |
| S1S_extracted_10144 | Not-determined |
| S1S_extracted_10146 | Low-quality |
| S1S_extracted_10150 | Not-determined |
| S1S_extracted_10152 | Not-determined |
| S1S_extracted_10189 | Not-determined |
| S1S_extracted_10197 | Not-determined |
| S1S_extracted_10206 | Not-determined |
| S1S_extracted_10227 | Low-quality |
| S1S_extracted_10229 | Not-determined |
| S1S_extracted_10245 | Not-determined |
| S1S_extracted_10257 | Low-quality |
| S1S_extracted_10267 | Not-determined |
| S1S_extracted_10280 | Not-determined |
| S1S_extracted_10295 | Not-determined |
| S1S_extracted_10298 | Not-determined |
| S1S_extracted_10302 | Not-determined |
| S1S_extracted_10314 | Not-determined |
| S1S_extracted_10318 | Not-determined |
| S1S_extracted_10320 | Not-determined |
| S1S_extracted_10321 | Not-determined |
| S1S_extracted_10325 | Not-determined |
| S1S_extracted_10337 | Not-determined |
| S1S_extracted_10348 | Not-determined |
| S1S_extracted_10351 | Not-determined |
| S1S_extracted_10359 | Not-determined |
| S1S_extracted_10362 | Not-determined |
| S1S_extracted_10363 | Not-determined |
| S1S_extracted_10371 | Not-determined |
| S1S_extracted_10401 | Not-determined |
| S1S_extracted_10402 | Not-determined |
| S1S_extracted_10407 | Not-determined |
| S1S_extracted_10426 | Not-determined |
| S1S_extracted_10434 | Not-determined |
| S1S_extracted_10446 | Not-determined |
| S1S_extracted_10455 | Not-determined |
| S1S_extracted_10459 | Not-determined |
| S1S_extracted_10467 | Not-determined |
| S1S_extracted_10478 | Not-determined |
| S1S_extracted_10481 | Not-determined |
| S1S_extracted_10489 | Not-determined |
| S1S_extracted_10492 | Not-determined |
| S1S_extracted_10498 | Not-determined |
| S1S_extracted_10507 | Not-determined |
| S1S_extracted_10515 | Not-determined |
| S1S_extracted_10520 | Not-determined |
| S1S_extracted_10525 | Not-determined |
| S1S_extracted_10535 | Not-determined |
| S1S_extracted_10553 | Not-determined |
| S1S_extracted_10555 | Not-determined |
| S1S_extracted_10564 | Not-determined |
| S1S_extracted_10565 | Not-determined |
| S1S_extracted_10585 | Not-determined |
| S1S_extracted_10595 | Not-determined |
| S1S_extracted_10600 | Not-determined |
| S1S_extracted_10606 | Not-determined |
| S1S_extracted_10615 | Not-determined |
| S1S_extracted_10634 | Not-determined |
| S1S_extracted_10642 | Not-determined |
| S1S_extracted_10668 | Not-determined |
| S1S_extracted_10690 | Not-determined |
| S1S_extracted_10692 | Not-determined |
| S1S_extracted_10695 | Not-determined |
| S1S_extracted_10697 | Not-determined |
| S1S_extracted_10703 | Not-determined |
| S1S_extracted_10739 | Not-determined |
| S1S_extracted_10745 | Low-quality |
| S1S_extracted_10763 | Not-determined |
| S1S_extracted_10767 | Not-determined |
| S1S_extracted_10773 | Not-determined |
| S1S_extracted_10791 | Not-determined |
| S1S_extracted_10792 | Not-determined |
| S1S_extracted_10806 | Not-determined |
| S1S_extracted_10845 | Not-determined |
| S1S_extracted_10848 | Not-determined |
| S1S_extracted_10850 | Not-determined |
| S1S_extracted_10852 | Not-determined |
| S1S_extracted_10853 | Not-determined |
| S1S_extracted_10857 | Not-determined |
| S1S_extracted_10864 | Low-quality |
| S1S_extracted_10872 | Not-determined |
| S1S_extracted_10878 | Not-determined |
| S1S_extracted_10901 | Not-determined |
| S1S_extracted_10906 | Not-determined |
| S1S_extracted_10908 | Not-determined |
| S1S_extracted_10911 | Not-determined |
| S1S_extracted_10913 | Not-determined |
| S1S_extracted_10927 | Not-determined |
| S1S_extracted_10938 | Not-determined |
| S1S_extracted_10947 | Low-quality |
| S1S_extracted_10954 | Not-determined |
| S1S_extracted_10955 | Not-determined |
| S1S_extracted_10966 | Not-determined |
| S1S_extracted_10981 | Not-determined |
| S1S_extracted_10983 | Not-determined |
| S1S_extracted_10987 | Low-quality |
| S1S_extracted_10990 | Not-determined |
| S1S_extracted_10992 | Not-determined |
| S1S_extracted_11011 | Not-determined |
| S1S_extracted_11016 | Not-determined |
| S1S_extracted_11021 | Not-determined |
| S1S_extracted_11028 | Not-determined |
| S1S_extracted_11038 | Not-determined |
| S1S_extracted_11040 | Not-determined |
| S1S_extracted_11042 | Not-determined |
| S1S_extracted_11046 | Not-determined |
| S1S_extracted_11047 | Low-quality |
| S1S_extracted_11056 | Not-determined |
| S1S_extracted_11059 | Not-determined |
| S1S_extracted_11063 | Low-quality |
| S1S_extracted_11064 | Not-determined |
| S1S_extracted_11072 | Not-determined |
| S1S_extracted_11073 | Low-quality |
| S1S_extracted_11078 | Not-determined |
| S1S_extracted_11081 | Not-determined |
| S1S_extracted_11085 | Not-determined |
| S1S_extracted_11109 | Not-determined |
| S1S_extracted_11112 | Not-determined |
| S1S_extracted_11114 | Not-determined |
| S1S_extracted_11125 | Not-determined |
| S1S_extracted_11138 | Not-determined |
| S1S_extracted_11144 | Not-determined |
| S1S_extracted_11146 | Not-determined |
| S1S_extracted_11183 | Not-determined |
| S1S_extracted_11191 | Not-determined |
| S1S_extracted_11193 | Not-determined |
| S1S_extracted_11197 | Not-determined |
| S1S_extracted_11206 | Not-determined |
| S1S_extracted_11219 | Not-determined |
| S1S_extracted_11221 | Not-determined |
| S1S_extracted_11237 | Not-determined |
| S1S_extracted_11246 | Not-determined |
| S1S_extracted_11252 | Not-determined |
| S1S_extracted_11257 | Not-determined |
| S1S_extracted_11269 | Not-determined |
| S1S_extracted_11285 | Not-determined |
| S1S_extracted_11293 | Not-determined |
| S1S_extracted_11298 | Not-determined |
| S1S_extracted_11303 | Not-determined |
| S1S_extracted_11305 | Not-determined |
| S1S_extracted_11306 | Not-determined |
| S1S_extracted_11308 | Not-determined |
| S1S_extracted_11311 | Not-determined |
| S1S_extracted_11323 | Not-determined |
| S1S_extracted_11340 | Not-determined |
| S1S_extracted_11350 | Not-determined |
| S1S_extracted_11381 | Low-quality |
| S1S_extracted_11390 | Not-determined |
| S1S_extracted_11399 | Not-determined |
| S1S_extracted_11409 | Not-determined |
| S1S_extracted_11423 | Not-determined |
| S1S_extracted_11429 | Not-determined |
| S1S_extracted_11450 | Low-quality |
| S1S_extracted_11455 | Not-determined |
| S1S_extracted_11459 | Not-determined |
| S1S_extracted_11478 | Not-determined |
| S1S_extracted_11479 | Not-determined |
| S1S_extracted_11480 | Not-determined |
| S1S_extracted_11485 | Not-determined |
| S1S_extracted_11493 | Not-determined |
| S1S_extracted_11499 | Not-determined |
| S1S_extracted_11510 | Not-determined |
| S1S_extracted_11528 | Not-determined |
| S1S_extracted_11537 | Not-determined |
| S1S_extracted_11538 | Low-quality |
| S1S_extracted_11540 | Not-determined |
| S1S_extracted_11547 | Not-determined |
| S1S_extracted_11555 | Low-quality |
| S1S_extracted_11569 | Not-determined |
| S1S_extracted_11577 | Not-determined |
| S1S_extracted_11587 | Not-determined |
| S1S_extracted_11594 | Low-quality |
| S1S_extracted_11609 | Low-quality |
| S1S_extracted_11617 | Not-determined |
| S1S_extracted_11623 | Not-determined |
| S1S_extracted_11635 | Not-determined |
| S1S_extracted_11636 | Not-determined |
| S1S_extracted_11643 | Not-determined |
| S1S_extracted_11648 | Not-determined |
| S1S_extracted_11650 | Low-quality |
| S1S_extracted_11652 | Not-determined |
| S1S_extracted_11657 | Not-determined |
| S1S_extracted_11662 | Not-determined |
| S1S_extracted_11673 | Not-determined |
| S1S_extracted_11675 | Not-determined |
| S1S_extracted_11689 | Not-determined |
| S1S_extracted_11705 | Not-determined |
| S1S_extracted_11729 | Not-determined |
| S1S_extracted_11733 | Not-determined |
| S1S_extracted_11735 | Not-determined |
| S1S_extracted_11754 | Not-determined |
| S1S_extracted_11764 | Not-determined |
| S1S_extracted_11768 | Not-determined |
| S1S_extracted_11769 | Not-determined |
| S1S_extracted_11770 | Not-determined |
| S1S_extracted_11787 | Not-determined |
| S1S_extracted_11814 | Not-determined |
| SS_vs_k141_112452\|\|full | Medium-quality |
| S1S_extracted_11824 | Not-determined |
| S1S_extracted_11830 | Not-determined |
| S1S_extracted_11840 | Not-determined |
| S1S_extracted_11843 | Not-determined |
| S1S_extracted_11853 | Not-determined |
| S1S_extracted_11884 | Low-quality |
| S1S_extracted_11889 | Not-determined |
| S1S_extracted_11901 | Low-quality |
| S1S_extracted_11903 | Not-determined |
| S1S_extracted_11904 | Not-determined |
| S1S_extracted_11905 | Not-determined |
| S1S_extracted_11907 | Not-determined |
| S1S_extracted_11910 | Not-determined |
| S1S_extracted_11914 | Not-determined |
| S1S_extracted_11920 | Not-determined |
| S1S_extracted_11943 | Not-determined |
| S1S_extracted_11955 | Not-determined |
| S1S_extracted_11958 | Not-determined |
| S1S_extracted_11964 | Not-determined |
| S1S_extracted_11973 | Not-determined |
| S1S_extracted_11990 | Not-determined |
| S1S_extracted_11994 | Not-determined |
| S1S_extracted_12005 | Not-determined |
| S1S_extracted_12011 | Low-quality |
| S1S_extracted_12030 | Low-quality |
| S1S_extracted_12042 | Not-determined |
| S1S_extracted_12044 | Not-determined |
| S1S_extracted_12065 | Not-determined |
| S1S_extracted_12082 | Low-quality |
| S1S_extracted_12084 | Not-determined |
| S1S_extracted_12107 | Not-determined |
| S1S_extracted_12118 | Not-determined |
| S1S_extracted_12120 | Not-determined |
| S1S_extracted_12128 | Low-quality |
| S1S_extracted_12145 | Not-determined |
| S1S_extracted_12149 | Not-determined |
| S1S_extracted_12153 | Not-determined |
| S1S_extracted_12169 | Not-determined |
| S1S_extracted_12170 | Not-determined |
| S1S_extracted_12171 | Not-determined |
| S1S_extracted_12179 | Not-determined |
| S1S_extracted_12185 | Not-determined |
| S1S_extracted_12187 | Not-determined |
| S1S_extracted_12193 | Not-determined |
| S1S_extracted_12196 | Not-determined |
| S1S_extracted_12201 | Not-determined |
| S1S_extracted_12229 | Low-quality |
| S1S_extracted_12230 | Not-determined |
| S1S_extracted_12233 | Not-determined |
| S1S_extracted_12235 | Not-determined |
| S1S_extracted_12237 | Not-determined |
| S1S_extracted_12239 | Not-determined |
| S1S_extracted_12241 | Not-determined |
| S1S_extracted_12242 | Not-determined |
| S1S_extracted_12250 | Not-determined |
| S1S_extracted_12294 | Low-quality |
| S1S_extracted_12302 | Not-determined |
| S1S_extracted_12306 | Not-determined |
| S1S_extracted_12321 | Not-determined |
| S1S_extracted_12326 | Not-determined |
| S1S_extracted_12330 | Low-quality |
| S1S_extracted_12335 | Low-quality |
| S1S_extracted_12338 | Not-determined |
| S1S_extracted_12341 | Not-determined |
| S1S_extracted_12346 | Not-determined |
| S1S_extracted_12353 | Not-determined |
| S1S_extracted_12355 | Not-determined |
| S1S_extracted_12375 | Not-determined |
| S1S_extracted_12381 | Not-determined |
| S1S_extracted_12386 | Not-determined |
| S1S_extracted_12387 | Low-quality |
| S1S_extracted_12393 | Not-determined |
| S1S_extracted_12399 | Not-determined |
| S1S_extracted_12431 | Not-determined |
| S1S_extracted_12432 | Not-determined |
| S1S_extracted_12441 | Not-determined |
| S1S_extracted_12442 | Not-determined |
| S1S_extracted_12451 | Not-determined |
| S1S_extracted_12452 | Not-determined |
| S1S_extracted_12456 | Not-determined |
| S1S_extracted_12472 | Low-quality |
| S1S_extracted_12489 | Not-determined |
| S1S_extracted_12494 | Not-determined |
| S1S_extracted_12498 | Not-determined |
| S1S_extracted_12499 | Not-determined |
| S1S_extracted_12533 | Not-determined |
| S1S_extracted_12539 | Not-determined |
| S1S_extracted_12563 | Not-determined |
| S1S_extracted_12565 | Not-determined |
| S1S_extracted_12566 | Not-determined |
| S1S_extracted_12580 | Not-determined |
| S1S_extracted_12583 | Not-determined |
| S1S_extracted_12600 | Low-quality |
| S1S_extracted_12604 | Not-determined |
| S1S_extracted_12606 | Not-determined |
| S1S_extracted_12607 | Not-determined |
| S1S_extracted_12614 | Not-determined |
| S1S_extracted_12624 | Not-determined |
| S1S_extracted_12625 | Not-determined |
| S1S_extracted_12626 | Not-determined |
| S1S_extracted_12628 | Not-determined |
| S1S_extracted_12644 | Not-determined |
| S1S_extracted_12647 | Low-quality |
| S1S_extracted_12650 | Not-determined |
| S1S_extracted_12652 | Not-determined |
| S1S_extracted_12656 | Not-determined |
| S1S_extracted_12663 | Not-determined |
| S1S_extracted_12669 | Not-determined |
| S1S_extracted_12681 | Not-determined |
| S1S_extracted_12689 | Not-determined |
| S1S_extracted_12697 | Not-determined |
| S1S_extracted_12701 | Not-determined |
| S1S_extracted_12711 | Not-determined |
| S1S_extracted_12723 | Not-determined |
| S1S_extracted_12782 | Not-determined |
| S1S_extracted_12784 | Not-determined |
| S1S_extracted_12786 | Low-quality |
| S1S_extracted_12841 | Not-determined |
| S1S_extracted_12843 | Low-quality |
| S1S_extracted_12848 | Not-determined |
| S1S_extracted_12876 | Not-determined |
| S1S_extracted_12884 | Not-determined |
| S1S_extracted_12904 | Not-determined |
| S1S_extracted_12913 | Not-determined |
| S1S_extracted_12915 | Not-determined |
| S1S_extracted_12923 | Not-determined |
| S1S_extracted_12925 | Not-determined |
| S1S_extracted_12942 | Not-determined |
| S1S_extracted_12946 | Not-determined |
| S1S_extracted_12965 | Low-quality |
| S1S_extracted_12984 | Not-determined |
| S1S_extracted_12994 | Not-determined |
| S1S_extracted_12997 | Not-determined |
| S1S_extracted_13002 | Not-determined |
| S1S_extracted_13016 | Not-determined |
| S1S_extracted_13023 | Not-determined |
| S1S_extracted_13033 | Low-quality |
| S1S_extracted_13043 | Not-determined |
| S1S_extracted_13060 | Not-determined |
| S1S_extracted_13089 | Not-determined |
| S1S_extracted_13092 | Low-quality |
| S1S_extracted_13110 | Not-determined |
| S1S_extracted_13130 | Not-determined |
| S1S_extracted_13139 | Not-determined |
| S1S_extracted_13144 | Not-determined |
| S1S_extracted_13149 | Low-quality |
| S1S_extracted_13154 | Not-determined |
| S1S_extracted_13158 | Not-determined |
| S1S_extracted_13192 | Not-determined |
| S1S_extracted_13194 | Not-determined |
| S1S_extracted_13212 | Low-quality |
| S1S_extracted_13215 | Not-determined |
| S1S_extracted_13222 | Not-determined |
| S1S_extracted_13236 | Not-determined |
| S1S_extracted_13238 | Not-determined |
| S1S_extracted_13242 | Not-determined |
| S1S_extracted_13255 | Not-determined |
| S1S_extracted_13256 | Low-quality |
| S1S_extracted_13268 | Not-determined |
| S1S_extracted_13271 | Not-determined |
| S1S_extracted_13272 | Low-quality |
| S1S_extracted_13281 | Not-determined |
| S1S_extracted_13284 | Not-determined |
| S1S_extracted_13305 | Not-determined |
| S1S_extracted_13309 | Not-determined |
| S1S_extracted_13313 | Not-determined |
| S1S_extracted_13314 | Not-determined |
| S1S_extracted_13327 | Not-determined |
| S1S_extracted_13328 | Low-quality |
| S1S_extracted_13348 | Not-determined |
| S1S_extracted_13350 | Not-determined |
| S1S_extracted_13356 | Not-determined |
| S1S_extracted_13359 | Not-determined |
| S1S_extracted_13362 | Not-determined |
| S1S_extracted_13366 | Not-determined |
| S1S_extracted_13376 | Not-determined |
| S1S_extracted_13394 | Not-determined |
| S1S_extracted_13397 | Not-determined |
| S1S_extracted_13398 | Not-determined |
| S1S_extracted_13400 | Not-determined |
| S1S_extracted_13406 | Not-determined |
| S1S_extracted_13412 | Not-determined |
| S1S_extracted_13416 | Low-quality |
| S1S_extracted_13428 | Not-determined |
| S1S_extracted_13436 | Not-determined |
| S1S_extracted_13448 | Not-determined |
| S1S_extracted_13456 | Not-determined |
| S1S_extracted_13457 | Not-determined |
| S1S_extracted_13470 | Not-determined |
| S1S_extracted_13475 | Low-quality |
| S1S_extracted_13476 | Not-determined |
| S1S_extracted_13480 | Not-determined |
| S1S_extracted_13489 | Not-determined |
| S1S_extracted_13496 | Not-determined |
| S1S_extracted_13499 | Not-determined |
| S1S_extracted_13502 | Not-determined |
| S1S_extracted_13513 | Not-determined |
| S1S_extracted_13522 | Not-determined |
| S1S_extracted_13530 | Not-determined |
| S1S_extracted_13536 | Not-determined |
| S1S_extracted_13551 | Not-determined |
| S1S_extracted_13557 | Low-quality |
| S1S_extracted_13577 | Not-determined |
| S1S_extracted_13594 | Not-determined |
| S1S_extracted_13599 | Not-determined |
| S1S_extracted_13603 | Not-determined |
| S1S_extracted_13618 | Not-determined |
| S1S_extracted_13622 | Not-determined |
| S1S_extracted_13628 | Not-determined |
| S1S_extracted_13636 | Not-determined |
| S1S_extracted_13647 | Not-determined |
| S1S_extracted_13648 | Not-determined |
| S1S_extracted_13652 | Not-determined |
| S1S_extracted_13663 | Not-determined |
| S1S_extracted_13675 | Not-determined |
| S1S_extracted_13676 | Not-determined |
| S1S_extracted_13707 | Not-determined |
| S1S_extracted_13715 | Not-determined |
| S1S_extracted_13716 | Not-determined |
| S1S_extracted_13733 | Not-determined |
| S1S_extracted_13740 | Low-quality |
| S1S_extracted_13743 | Not-determined |
| S1S_extracted_13748 | Not-determined |
| S1S_extracted_13757 | Not-determined |
| S1S_extracted_13758 | Not-determined |
| S1S_extracted_13761 | Not-determined |
| S1S_extracted_13767 | Not-determined |
| S1S_extracted_13778 | Not-determined |
| S1S_extracted_13786 | Not-determined |
| S1S_extracted_13793 | Low-quality |
| S1S_extracted_13798 | Not-determined |
| S1S_extracted_13800 | Not-determined |
| S1S_extracted_13808 | Low-quality |
| S1S_extracted_13818 | Not-determined |
| S1S_extracted_13821 | Not-determined |
| S1S_extracted_13824 | Not-determined |
| S1S_extracted_13825 | Low-quality |
| S1S_extracted_13832 | Not-determined |
| S1S_extracted_13835 | Not-determined |
| S1S_extracted_13857 | Not-determined |
| S1S_extracted_13861 | Low-quality |
| S1S_extracted_13873 | Not-determined |
| S1S_extracted_13875 | Not-determined |
| S1S_extracted_13877 | Not-determined |
| S1S_extracted_13882 | Not-determined |
| S1S_extracted_13885 | Not-determined |
| S1S_extracted_13886 | Not-determined |
| S1S_extracted_13887 | Not-determined |
| S1S_extracted_13895 | Not-determined |
| S1S_extracted_13896 | Not-determined |
| S1S_extracted_13902 | Not-determined |
| S1S_extracted_13916 | Not-determined |
| S1S_extracted_13921 | Not-determined |
| S1S_extracted_13931 | Not-determined |
| S1S_extracted_13933 | Not-determined |
| S1S_extracted_13941 | Not-determined |
| S1S_extracted_13958 | Not-determined |
| S1S_extracted_13968 | Not-determined |
| S1S_extracted_13971 | Not-determined |
| S1S_extracted_13989 | Not-determined |
| S1S_extracted_13994 | Not-determined |
| S1S_extracted_14005 | Not-determined |
| S1S_extracted_14011 | Not-determined |
| S1S_extracted_14017 | Not-determined |
| S1S_extracted_14018 | Not-determined |
| S1S_extracted_14027 | Not-determined |
| S1S_extracted_14031 | Not-determined |
| S1S_extracted_14034 | Not-determined |
| S1S_extracted_14038 | Low-quality |
| S1S_extracted_14042 | Not-determined |
| S1S_extracted_14044 | Not-determined |
| S1S_extracted_14064 | Not-determined |
| S1S_extracted_14066 | Not-determined |
| S1S_extracted_14080 | Not-determined |
| S1S_extracted_14085 | Low-quality |
| S1S_extracted_14086 | Not-determined |
| S1S_extracted_14089 | Not-determined |
| S1S_extracted_14090 | Not-determined |
| S1S_extracted_14103 | Low-quality |
| S1S_extracted_14104 | Not-determined |
| S1S_extracted_14124 | Not-determined |
| S1S_extracted_14129 | Not-determined |
| S1S_extracted_14131 | Not-determined |
| S1S_extracted_14140 | Not-determined |
| S1S_extracted_14143 | Not-determined |
| S1S_extracted_14148 | Not-determined |
| S1S_extracted_14150 | Low-quality |
| S1S_extracted_14154 | Not-determined |
| S1S_extracted_14185 | Not-determined |
| S1S_extracted_14192 | Not-determined |
| S1S_extracted_14195 | Low-quality |
| S1S_extracted_14203 | Not-determined |
| S1S_extracted_14209 | Not-determined |
| S1S_extracted_14210 | Not-determined |
| S1S_extracted_14212 | Not-determined |
| S1S_extracted_14213 | Low-quality |
| S1S_extracted_14216 | Not-determined |
| S1S_extracted_14227 | Not-determined |
| S1S_extracted_14230 | Not-determined |
| S1S_extracted_14239 | Not-determined |
| S1S_extracted_14242 | Not-determined |
| S1S_extracted_14245 | Not-determined |
| S1S_extracted_14262 | Not-determined |
| S1S_extracted_14265 | Low-quality |
| S1S_extracted_14274 | Not-determined |
| S1S_extracted_14288 | Not-determined |
| S1S_extracted_14308 | Not-determined |
| S1S_extracted_14317 | Not-determined |
| S1S_extracted_14318 | Low-quality |
| S1S_extracted_14326 | Low-quality |
| S1S_extracted_14331 | Not-determined |
| S1S_extracted_14350 | Not-determined |
| S1S_extracted_14370 | Low-quality |
| S1S_extracted_14376 | Not-determined |
| S1S_extracted_14380 | Not-determined |
| S1S_extracted_14392 | Not-determined |
| S1S_extracted_14398 | Not-determined |
| S1S_extracted_14407 | Not-determined |
| S1S_extracted_14419 | Not-determined |
| S1S_extracted_14420 | Not-determined |
| S1S_extracted_14454 | Not-determined |
| S1S_extracted_14455 | Not-determined |
| S1S_extracted_14465 | Not-determined |
| S1S_extracted_14467 | Not-determined |
| S1S_extracted_14468 | Not-determined |
| S1S_extracted_14504 | Not-determined |
| S1S_extracted_14510 | Not-determined |
| S1S_extracted_14533 | Not-determined |
| S1S_extracted_14538 | Not-determined |
| S1S_extracted_14556 | Not-determined |
| S1S_extracted_14559 | Not-determined |
| S1S_extracted_14565 | Not-determined |
| S1S_extracted_14567 | Not-determined |
| S1S_extracted_14570 | Not-determined |
| S1S_extracted_14591 | Not-determined |
| S1S_extracted_14601 | Not-determined |
| S1S_extracted_14615 | Not-determined |
| S1S_extracted_14616 | Not-determined |
| S1S_extracted_14624 | Low-quality |
| S1S_extracted_14626 | Not-determined |
| S1S_extracted_14628 | Not-determined |
| S1S_extracted_14631 | Not-determined |
| S1S_extracted_14633 | Low-quality |
| S1S_extracted_14647 | Not-determined |
| S1S_extracted_14655 | Not-determined |
| S1S_extracted_14657 | Not-determined |
| S1S_extracted_14675 | Low-quality |
| S1S_extracted_14677 | Not-determined |
| S1S_extracted_14694 | Not-determined |
| S1S_extracted_14705 | Not-determined |
| S1S_extracted_14706 | Not-determined |
| S1S_extracted_14711 | Not-determined |
| S1S_extracted_14712 | Not-determined |
| S1S_extracted_14718 | Low-quality |
| S1S_extracted_14726 | Not-determined |
| S1S_extracted_14735 | Not-determined |
| S1S_extracted_14743 | Not-determined |
| S1S_extracted_14746 | Not-determined |
| S1S_extracted_14747 | Not-determined |
| S1S_extracted_14749 | Not-determined |
| S1S_extracted_14755 | Not-determined |
| S1S_extracted_14766 | Not-determined |
| S1S_extracted_14788 | Not-determined |
| S1S_extracted_14802 | Not-determined |
| S1S_extracted_14803 | Not-determined |
| S1S_extracted_14805 | Not-determined |
| S1S_extracted_14811 | Not-determined |
| S1S_extracted_14820 | Low-quality |
| S1S_extracted_14822 | Not-determined |
| S1S_extracted_14832 | Not-determined |

| | |
|---|---|
| S1S_extracted_14850 | Low-quality |
| S1S_extracted_14855 | Not-determined |
| S1S_extracted_14872 | Not-determined |
| S1S_extracted_14875 | Not-determined |
| S1S_extracted_14879 | Low-quality |
| S1S_extracted_14880 | Not-determined |
| S1S_extracted_14884 | Not-determined |
| S1S_extracted_14887 | Not-determined |
| S1S_extracted_14889 | Not-determined |
| S1S_extracted_14897 | Not-determined |
| S1S_extracted_14908 | Not-determined |
| S1S_extracted_14916 | Not-determined |
| S1S_extracted_14969 | Not-determined |
| S1S_extracted_14973 | Not-determined |
| S1S_extracted_14992 | Not-determined |
| S1S_extracted_14997 | Not-determined |
| S1S_extracted_14998 | Not-determined |
| S1S_extracted_15003 | Not-determined |
| S1S_extracted_15010 | Low-quality |
| S1S_extracted_15015 | Not-determined |
| S1S_extracted_15018 | Not-determined |
| S1S_extracted_15026 | Not-determined |
| S1S_extracted_15033 | Not-determined |
| S1S_extracted_15056 | Not-determined |
| S1S_extracted_15076 | Not-determined |
| S1S_extracted_15086 | Not-determined |
| S1S_extracted_15124 | Not-determined |
| S1S_extracted_15168 | Not-determined |
| S1S_extracted_15171 | Not-determined |
| S1S_extracted_15180 | Not-determined |
| S1S_extracted_15191 | Not-determined |
| S1S_extracted_15192 | Not-determined |
| S1S_extracted_15201 | Low-quality |
| S1S_extracted_15204 | Not-determined |
| S1S_extracted_15217 | Not-determined |
| S1S_extracted_15221 | Not-determined |
| S1S_extracted_15231 | Not-determined |
| S1S_extracted_15232 | Not-determined |
| S1S_extracted_15242 | Not-determined |
| S1S_extracted_15251 | Not-determined |
| S1S_extracted_15252 | Not-determined |
| S1S_extracted_15270 | Not-determined |
| S1S_extracted_15276 | Low-quality |
| S1S_extracted_15282 | Not-determined |
| S1S_extracted_15294 | Not-determined |
| S1S_extracted_15307 | Not-determined |
| S1S_extracted_15311 | Not-determined |
| S1S_extracted_15320 | Not-determined |
| S1S_extracted_15325 | Not-determined |
| S1S_extracted_15333 | Not-determined |
| S1S_extracted_15342 | Not-determined |
| S1S_extracted_15354 | Not-determined |
| S1S_extracted_15365 | Low-quality |
| S1S_extracted_15374 | Not-determined |
| S1S_extracted_15376 | Not-determined |
| S1S_extracted_15415 | Not-determined |
| S1S_extracted_15421 | Not-determined |
| S1S_extracted_15425 | Not-determined |
| S1S_extracted_15430 | Not-determined |
| S1S_extracted_15436 | Not-determined |
| S1S_extracted_15449 | Not-determined |
| S1S_extracted_15471 | Not-determined |
| S1S_extracted_15487 | Not-determined |
| S1S_extracted_15498 | Not-determined |
| S1S_extracted_15506 | Not-determined |
| S1S_extracted_15523 | Low-quality |
| S1S_extracted_15525 | Not-determined |
| S1S_extracted_15535 | Not-determined |
| S1S_extracted_15538 | Not-determined |
| S1S_extracted_15553 | Not-determined |
| S1S_extracted_15559 | Not-determined |
| S1S_extracted_15578 | Not-determined |
| S1S_extracted_15581 | Not-determined |
| S1S_extracted_15590 | Not-determined |
| S1S_extracted_15592 | Low-quality |
| S1S_extracted_15625 | Not-determined |
| S1S_extracted_15634 | Not-determined |
| S1S_extracted_15640 | Not-determined |
| S1S_extracted_15644 | Not-determined |
| S1S_extracted_15650 | Not-determined |
| S1S_extracted_15657 | Not-determined |
| S1S_extracted_15665 | Not-determined |
| S1S_extracted_15673 | Not-determined |
| S1S_extracted_15681 | Not-determined |
| S1S_extracted_15682 | Not-determined |
| S1S_extracted_15688 | Low-quality |
| S1S_extracted_15689 | Not-determined |
| S1S_extracted_15697 | Not-determined |
| S1S_extracted_15698 | Low-quality |
| S1S_extracted_15713 | Not-determined |
| S1S_extracted_15743 | Not-determined |
| S1S_extracted_15748 | Not-determined |
| S1S_extracted_15751 | Not-determined |
| S1S_extracted_15767 | Not-determined |
| S1S_extracted_15791 | Not-determined |
| S1S_extracted_15815 | Not-determined |
| S1S_extracted_15831 | Not-determined |
| S1S_extracted_15847 | Not-determined |
| S1S_extracted_15851 | Low-quality |
| S1S_extracted_15853 | Not-determined |
| S1S_extracted_15854 | Not-determined |
| S1S_extracted_15856 | Low-quality |
| S1S_extracted_15865 | Not-determined |
| S1S_extracted_15879 | Not-determined |
| S1S_extracted_15881 | Not-determined |
| S1S_extracted_15891 | Low-quality |
| S1S_extracted_15892 | Not-determined |
| S1S_extracted_15893 | Not-determined |
| S1S_extracted_15901 | Low-quality |
| S1S_extracted_15908 | Not-determined |
| S1S_extracted_15910 | Not-determined |
| S1S_extracted_15915 | Not-determined |
| S1S_extracted_15931 | Low-quality |
| S1S_extracted_15934 | Not-determined |
| S1S_extracted_15935 | Not-determined |
| S1S_extracted_15936 | Not-determined |
| S1S_extracted_15937 | Not-determined |
| S1S_extracted_15942 | Not-determined |
| S1S_extracted_15952 | Not-determined |
| S1S_extracted_15958 | Low-quality |
| S1S_extracted_15963 | Not-determined |
| S1S_extracted_15981 | Not-determined |
| S1S_extracted_15983 | Not-determined |
| S1S_extracted_15998 | Not-determined |
| S1S_extracted_16013 | Not-determined |
| S1S_extracted_16017 | Not-determined |
| S1S_extracted_16021 | Not-determined |
| S1S_extracted_16029 | Not-determined |
| S1S_extracted_16045 | Not-determined |
| S1S_extracted_16052 | Not-determined |
| S1S_extracted_16055 | Not-determined |
| S1S_extracted_16058 | Not-determined |
| S1S_extracted_16063 | Not-determined |
| S1S_extracted_16064 | Not-determined |
| S1S_extracted_16078 | Not-determined |
| S1S_extracted_16084 | Not-determined |
| S1S_extracted_16086 | Not-determined |
| S1S_extracted_16087 | Not-determined |
| S1S_extracted_16107 | Low-quality |
| S1S_extracted_16122 | Not-determined |
| S1S_extracted_16127 | Not-determined |
| S1S_extracted_16141 | Low-quality |
| S1S_extracted_16145 | Not-determined |
| S1S_extracted_16146 | Not-determined |
| S1S_extracted_16155 | Not-determined |
| S1S_extracted_16158 | Not-determined |
| S1S_extracted_16160 | Not-determined |
| S1S_extracted_16167 | Not-determined |
| S1S_extracted_16172 | Not-determined |
| S1S_extracted_16182 | Low-quality |
| S1S_extracted_16191 | Low-quality |
| S1S_extracted_16205 | Not-determined |
| S1S_extracted_16208 | Not-determined |
| S1S_extracted_16212 | Not-determined |
| S1S_extracted_16223 | Not-determined |
| S1S_extracted_16225 | Not-determined |
| S1S_extracted_16226 | Not-determined |
| S1S_extracted_16229 | Not-determined |
| S1S_extracted_16232 | Not-determined |
| S1S_extracted_16245 | Low-quality |
| S1S_extracted_16254 | Not-determined |
| S1S_extracted_16273 | Not-determined |
| S1S_extracted_16274 | Not-determined |
| S1S_extracted_16279 | Not-determined |
| S1S_extracted_16282 | Not-determined |
| S1S_extracted_16292 | Not-determined |
| S1S_extracted_16307 | Not-determined |
| S1S_extracted_16309 | Low-quality |
| S1S_extracted_16310 | Low-quality |
| S1S_extracted_16311 | Not-determined |
| S1S_extracted_16312 | Low-quality |
| S1S_extracted_16314 | Not-determined |
| S1S_extracted_16320 | Not-determined |
| S1S_extracted_16331 | Not-determined |
| S1S_extracted_16345 | Not-determined |
| S1S_extracted_16356 | Not-determined |
| S1S_extracted_16369 | Not-determined |
| S1S_extracted_16376 | Not-determined |
| S1S_extracted_16379 | Low-quality |
| S1S_extracted_16399 | Not-determined |
| S1S_extracted_16400 | Not-determined |
| S1S_extracted_16422 | Not-determined |
| S1S_extracted_16433 | Not-determined |
| S1S_extracted_16437 | Not-determined |
| S1S_extracted_16441 | Not-determined |
| S1S_extracted_16448 | Not-determined |
| S1S_extracted_16457 | Not-determined |
| S1S_extracted_16463 | Not-determined |
| S1S_extracted_16494 | Not-determined |
| S1S_extracted_16496 | Not-determined |
| S1S_extracted_16517 | Not-determined |
| S1S_extracted_16530 | Not-determined |
| S1S_extracted_16532 | Not-determined |
| S1S_extracted_16534 | Not-determined |
| S1S_extracted_16541 | Not-determined |
| S1S_extracted_16542 | Not-determined |
| S1S_extracted_16559 | Not-determined |
| S1S_extracted_16563 | Not-determined |
| S1S_extracted_16591 | Low-quality |
| S1S_extracted_16599 | Not-determined |
| S1S_extracted_16600 | Not-determined |
| S1S_extracted_16608 | Low-quality |
| S1S_extracted_16611 | Not-determined |
| S1S_extracted_16613 | Not-determined |
| S1S_extracted_16625 | Not-determined |
| S1S_extracted_16627 | Not-determined |
| S1S_extracted_16653 | Not-determined |
| S1S_extracted_16654 | Low-quality |
| S1S_extracted_16684 | Not-determined |
| S1S_extracted_16691 | Not-determined |
| S1S_extracted_16695 | Not-determined |
| S1S_extracted_16699 | Not-determined |
| S1S_extracted_16702 | Not-determined |
| S1S_extracted_16718 | Not-determined |
| S1S_extracted_16725 | Not-determined |
| S1S_extracted_16731 | Not-determined |
| S1S_extracted_16733 | Not-determined |
| S1S_extracted_16740 | Not-determined |
| S1S_extracted_16749 | Not-determined |
| S1S_extracted_16758 | Not-determined |
| S1S_extracted_16767 | Not-determined |
| S1S_extracted_16779 | Not-determined |
| S1S_extracted_16784 | Low-quality |
| S1S_extracted_16791 | Low-quality |
| S1S_extracted_16799 | Low-quality |
| S1S_extracted_16803 | Not-determined |
| S1S_extracted_16808 | Not-determined |
| S1S_extracted_16812 | Not-determined |
| S1S_extracted_16814 | Low-quality |
| S1S_extracted_16822 | Not-determined |
| S1S_extracted_16847 | Low-quality |
| S1S_extracted_16848 | Not-determined |
| S1S_extracted_16861 | Not-determined |
| S1S_extracted_16863 | Not-determined |
| S1S_extracted_16871 | Not-determined |
| S1S_extracted_16878 | Not-determined |
| S1S_extracted_16896 | Not-determined |
| S1S_extracted_16901 | Not-determined |
| S1S_extracted_16924 | Not-determined |
| S1S_extracted_16926 | Not-determined |
| S1S_extracted_16934 | Not-determined |
| S1S_extracted_16935 | Not-determined |
| S1S_extracted_16947 | Low-quality |
| S1S_extracted_16951 | Not-determined |
| S1S_extracted_16953 | Not-determined |
| S1S_extracted_16957 | Not-determined |
| S1S_extracted_16958 | Not-determined |
| S1S_extracted_16959 | Not-determined |
| S1S_extracted_16962 | Low-quality |
| S1S_extracted_16989 | Low-quality |
| S1S_extracted_17017 | Not-determined |
| S1S_extracted_17029 | Not-determined |
| S1S_extracted_17030 | Not-determined |
| S1S_extracted_17034 | Not-determined |
| S1S_extracted_17035 | Not-determined |
| S1S_extracted_17036 | Not-determined |
| S1S_extracted_17044 | Not-determined |
| S1S_extracted_17045 | Not-determined |
| S1S_extracted_17047 | Not-determined |
| S1S_extracted_17052 | Not-determined |
| S1S_extracted_17054 | Not-determined |
| S1S_extracted_17060 | Not-determined |
| S1S_extracted_17064 | Not-determined |
| S1S_extracted_17070 | Not-determined |
| S1S_extracted_17098 | Not-determined |
| S1S_extracted_17124 | Not-determined |
| S1S_extracted_17137 | Not-determined |
| S1S_extracted_17144 | Not-determined |
| S1S_extracted_17145 | Not-determined |
| S1S_extracted_17149 | Not-determined |
| S1S_extracted_17151 | Not-determined |
| S1S_extracted_17159 | Low-quality |
| S1S_extracted_17169 | Low-quality |
| S1S_extracted_17182 | Not-determined |
| S1S_extracted_17183 | Not-determined |
| S1S_extracted_17184 | Not-determined |
| S1S_extracted_17189 | Not-determined |
| S1S_extracted_17195 | Not-determined |
| S1S_extracted_17208 | Not-determined |
| S1S_extracted_17221 | Not-determined |
| S1S_extracted_17225 | Not-determined |
| S1S_extracted_17226 | Not-determined |
| S1S_extracted_17232 | Not-determined |
| S1S_extracted_17239 | Not-determined |
| S1S_extracted_17252 | Not-determined |
| S1S_extracted_17258 | Not-determined |
| S1S_extracted_17264 | Not-determined |
| S1S_extracted_17273 | Not-determined |
| S1S_extracted_17286 | Not-determined |
| S1S_extracted_17288 | Low-quality |
| S1S_extracted_17303 | Not-determined |
| S1S_extracted_17310 | Not-determined |
| S1S_extracted_17311 | Not-determined |
| S1S_extracted_17341 | Not-determined |
| S1S_extracted_17350 | Not-determined |
| S1S_extracted_17363 | Not-determined |
| S1S_extracted_17371 | Not-determined |
| S1S_extracted_17381 | Not-determined |
| S1S_extracted_17385 | Not-determined |
| S1S_extracted_17391 | Not-determined |
| S1S_extracted_17423 | Not-determined |
| S1S_extracted_17434 | Not-determined |
| S1S_extracted_17437 | Not-determined |
| S1S_extracted_17440 | Not-determined |
| S1S_extracted_17444 | Not-determined |
| S1S_extracted_17449 | Not-determined |
| S1S_extracted_17463 | Not-determined |
| S1S_extracted_17469 | Not-determined |
| S1S_extracted_17491 | Not-determined |
| S1S_extracted_17494 | Not-determined |
| S1S_extracted_17497 | Not-determined |
| S1S_extracted_17501 | Not-determined |
| S1S_extracted_17515 | Not-determined |
| S1S_extracted_17518 | Not-determined |
| S1S_extracted_17526 | Not-determined |
| S1S_extracted_17537 | Not-determined |
| S1S_extracted_17546 | Not-determined |
| S1S_extracted_17549 | Not-determined |
| S1S_extracted_17551 | Low-quality |
| S1S_extracted_17554 | Not-determined |
| S1S_extracted_17555 | Not-determined |
| S1S_extracted_17570 | Not-determined |
| S1S_extracted_17571 | Not-determined |
| S1S_extracted_17583 | Not-determined |
| S1S_extracted_17596 | Not-determined |
| S1S_extracted_17606 | Low-quality |
| S1S_extracted_17621 | Not-determined |
| S1S_extracted_17626 | Not-determined |
| S1S_extracted_17635 | Not-determined |
| S1S_extracted_17657 | Not-determined |
| S1S_extracted_17659 | Not-determined |
| S1S_extracted_17671 | Not-determined |
| S1S_extracted_17672 | Low-quality |
| S1S_extracted_17679 | Not-determined |
| S1S_extracted_17681 | Not-determined |
| S1S_extracted_17685 | Not-determined |
| S1S_extracted_17714 | Not-determined |
| S1S_extracted_17715 | Not-determined |
| S1S_extracted_17718 | Low-quality |
| S1S_extracted_17722 | Not-determined |
| S1S_extracted_17756 | Not-determined |
| S1S_extracted_17796 | Not-determined |
| S1S_extracted_17810 | Not-determined |
| S1S_extracted_17820 | Not-determined |
| S1S_extracted_17831 | Not-determined |
| S1S_extracted_17846 | Not-determined |
| S1S_extracted_17848 | Not-determined |
| S16_vs_103938\|\|full | Medium-quality |
| S1S_extracted_17861 | Not-determined |
| S1S_extracted_17863 | Not-determined |
| S1S_extracted_17881 | Not-determined |
| S1S_extracted_17882 | Not-determined |
| S1S_extracted_17885 | Not-determined |
| S1S_extracted_17889 | Not-determined |
| S1S_extracted_17897 | Not-determined |
| S1S_extracted_17903 | Not-determined |
| S1S_extracted_17909 | Not-determined |
| S1S_extracted_17917 | Not-determined |
| S1S_extracted_17926 | Not-determined |
| S1S_extracted_17931 | Not-determined |
| S1S_extracted_17937 | Not-determined |
| S1S_extracted_17950 | Not-determined |
| S1S_extracted_17952 | Not-determined |
| S1S_extracted_17955 | Not-determined |
| S1S_extracted_17963 | Not-determined |
| S1S_extracted_17969 | Low-quality |
| S1S_extracted_17978 | Low-quality |
| S1S_extracted_17987 | Not-determined |
| S1S_extracted_17992 | Not-determined |
| S1S_extracted_18025 | Not-determined |
| S1S_extracted_18029 | Low-quality |
| S1S_extracted_18038 | Not-determined |
| S1S_extracted_18051 | Not-determined |
| S1S_extracted_18076 | Low-quality |
| S1S_extracted_18084 | Not-determined |
| S1S_extracted_18092 | Not-determined |
| S1S_extracted_18110 | Not-determined |
| S1S_extracted_18124 | Not-determined |
| S1S_extracted_18129 | Not-determined |
| S1S_extracted_18139 | Not-determined |
| S1S_extracted_18146 | Low-quality |
| S1S_extracted_18148 | Not-determined |
| S1S_extracted_18171 | Low-quality |
| S1S_extracted_18188 | Not-determined |
| S1S_extracted_18195 | Not-determined |
| S1S_extracted_18199 | Not-determined |
| S1S_extracted_18200 | Low-quality |
| S1S_extracted_18201 | Not-determined |
| S1S_extracted_18210 | Not-determined |
| S1S_extracted_18217 | Not-determined |
| S1S_extracted_18219 | Not-determined |
| S1S_extracted_18220 | Not-determined |
| S1S_extracted_18277 | Not-determined |
| S1S_extracted_18282 | Low-quality |
| S1S_extracted_18289 | Not-determined |
| S1S_extracted_18303 | Not-determined |
| S1S_extracted_18317 | Not-determined |
| S1S_extracted_18322 | Not-determined |
| S1S_extracted_18330 | Not-determined |
| S1S_extracted_18331 | Not-determined |
| S1S_extracted_18336 | Not-determined |
| S1S_extracted_18345 | Not-determined |
| S1S_extracted_18347 | Not-determined |
| S1S_extracted_18361 | Not-determined |
| S1S_extracted_18362 | Not-determined |
| S1S_extracted_18370 | Not-determined |
| S1S_extracted_18372 | Not-determined |
| S1S_extracted_18383 | Not-determined |
| S1S_extracted_18390 | Not-determined |
| S1S_extracted_18391 | Not-determined |
| S1S_extracted_18395 | Not-determined |
| S1S_extracted_18401 | Not-determined |
| S1S_extracted_18403 | Not-determined |
| S1S_extracted_18428 | Not-determined |
| S1S_extracted_18432 | Not-determined |
| S1S_extracted_18433 | Low-quality |
| S1S_extracted_18435 | Not-determined |
| S1S_extracted_18442 | Not-determined |
| S1S_extracted_18456 | Not-determined |
| S1S_extracted_18459 | Not-determined |
| S1S_extracted_18465 | Not-determined |
| S1S_extracted_18476 | Low-quality |
| S1S_extracted_18481 | Not-determined |
| S1S_extracted_18487 | Low-quality |
| S1S_extracted_18489 | Not-determined |
| S1S_extracted_18494 | Not-determined |
| S1S_extracted_18500 | Not-determined |
| S1S_extracted_18506 | Low-quality |
| S1S_extracted_18515 | Not-determined |
| S1S_extracted_18517 | Not-determined |
| S1S_extracted_18548 | Not-determined |
| S1S_extracted_18549 | Not-determined |
| S1S_extracted_18557 | Not-determined |
| S1S_extracted_18563 | Low-quality |
| S1S_extracted_18573 | Not-determined |
| S1S_extracted_18574 | Not-determined |
| S1S_extracted_18576 | Not-determined |
| S1S_extracted_18581 | Not-determined |
| S1S_extracted_18598 | Not-determined |
| S1S_extracted_18618 | Not-determined |
| S1S_extracted_18619 | Not-determined |
| S1S_extracted_18635 | Not-determined |
| S1S_extracted_18643 | Not-determined |
| S1S_extracted_18644 | Not-determined |
| S1S_extracted_18670 | Low-quality |
| S1S_extracted_18685 | Not-determined |
| S1S_extracted_18695 | Low-quality |
| S1S_extracted_18708 | Not-determined |
| S1S_extracted_18718 | Not-determined |
| S1S_extracted_18719 | Not-determined |
| S1S_extracted_18724 | Not-determined |
| S1S_extracted_18739 | Not-determined |
| S1S_extracted_18741 | Not-determined |
| S1S_extracted_18747 | Not-determined |
| S1S_extracted_18764 | Not-determined |
| S1S_extracted_18765 | Not-determined |
| S1S_extracted_18790 | Not-determined |
| S1S_extracted_18805 | Not-determined |
| S1S_extracted_18808 | Not-determined |
| S1S_extracted_18813 | Not-determined |
| S1S_extracted_18817 | Not-determined |
| S1S_extracted_18823 | Low-quality |
| S1S_extracted_18824 | Not-determined |
| S1S_extracted_18826 | Not-determined |
| S1S_extracted_18827 | Not-determined |
| S1S_extracted_18851 | Not-determined |
| S1S_extracted_18855 | Not-determined |
| S1S_extracted_18870 | Not-determined |
| S1S_extracted_18881 | Not-determined |
| S1S_extracted_18910 | Not-determined |
| S1S_extracted_18948 | Not-determined |
| S1S_extracted_18982 | Not-determined |
| S1S_extracted_18989 | Low-quality |
| S1S_extracted_18991 | Not-determined |
| S1S_extracted_18992 | Not-determined |
| S1S_extracted_19003 | Not-determined |
| S1S_extracted_19015 | Not-determined |
| S1S_extracted_19031 | Not-determined |
| S1S_extracted_19032 | Not-determined |
| S1S_extracted_19037 | Not-determined |
| S1S_extracted_19042 | Not-determined |
| S1S_extracted_19045 | Low-quality |
| S1S_extracted_19059 | Not-determined |
| S1S_extracted_19066 | Not-determined |
| S1S_extracted_19070 | Not-determined |
| S1S_extracted_19071 | Not-determined |
| S1S_extracted_19097 | Not-determined |
| S1S_extracted_19104 | Not-determined |
| S1S_extracted_19108 | Not-determined |
| S1S_extracted_19122 | Not-determined |
| S1S_extracted_19124 | Not-determined |
| S1S_extracted_19136 | Not-determined |
| S1S_extracted_19145 | Not-determined |
| S1S_extracted_19167 | Not-determined |
| S1S_extracted_19186 | Not-determined |
| S1S_extracted_19188 | Not-determined |
| S1S_extracted_19189 | Low-quality |
| S1S_extracted_19190 | Not-determined |
| S1S_extracted_19192 | Not-determined |
| S1S_extracted_19207 | Low-quality |
| S1S_extracted_19245 | Low-quality |
| S1S_extracted_19263 | Not-determined |
| S1S_extracted_19275 | Not-determined |
| S1S_extracted_19283 | Not-determined |
| S1S_extracted_19287 | Not-determined |
| S1S_extracted_19312 | Not-determined |
| S1S_extracted_19318 | Not-determined |
| S1S_extracted_19320 | Not-determined |
| S1S_extracted_19323 | Not-determined |
| S1S_extracted_19331 | Not-determined |
| S1S_extracted_19352 | Not-determined |
| S1S_extracted_19356 | Not-determined |
| S1S_extracted_19357 | Not-determined |
| S1S_extracted_19359 | Not-determined |
| S1S_extracted_19367 | Not-determined |
| S1S_extracted_19378 | Not-determined |
| S1S_extracted_19381 | Low-quality |
| S1S_extracted_19382 | Not-determined |
| S1S_extracted_19385 | Not-determined |
| S1S_extracted_19403 | Low-quality |
| S1S_extracted_19407 | Low-quality |
| S1S_extracted_19412 | Not-determined |
| S1S_extracted_19417 | Not-determined |
| S1S_extracted_19421 | Not-determined |
| S1S_extracted_19430 | Not-determined |
| S1S_extracted_19434 | Not-determined |
| S1S_extracted_19439 | Not-determined |
| S1S_extracted_19458 | Not-determined |
| S1S_extracted_19475 | Not-determined |
| S1S_extracted_19477 | Low-quality |
| S1S_extracted_19488 | Not-determined |
| S1S_extracted_19491 | Low-quality |
| S1S_extracted_19498 | Not-determined |
| S1S_extracted_19501 | Not-determined |
| S1S_extracted_19521 | Low-quality |
| S1S_extracted_19523 | Not-determined |
| S1S_extracted_19532 | Low-quality |
| S1S_extracted_19533 | Low-quality |
| S1S_extracted_19536 | Not-determined |
| S1S_extracted_19541 | Low-quality |
| S1S_extracted_19551 | Low-quality |
| S1S_extracted_19573 | Not-determined |
| S1S_extracted_19587 | Not-determined |
| S1S_extracted_19593 | Not-determined |
| S1S_extracted_19594 | Low-quality |
| S1S_extracted_19605 | Low-quality |
| S1S_extracted_19607 | Not-determined |
| S1S_extracted_19609 | Not-determined |
| S1S_extracted_19645 | Not-determined |
| S1S_extracted_19650 | Low-quality |
| S1S_extracted_19658 | Not-determined |
| S1S_extracted_19662 | Not-determined |
| S1S_extracted_19669 | Not-determined |
| S1S_extracted_19671 | Not-determined |
| S1S_extracted_19674 | Not-determined |
| S1S_extracted_19676 | Not-determined |
| S1S_extracted_19684 | Not-determined |
| S1S_extracted_19690 | Low-quality |
| S1S_extracted_19698 | Not-determined |
| S1S_extracted_19712 | Not-determined |
| S1S_extracted_19718 | Not-determined |
| S1S_extracted_19748 | Not-determined |
| S1S_extracted_19757 | Low-quality |
| S1S_extracted_19765 | Not-determined |
| S1S_extracted_19767 | Not-determined |
| S1S_extracted_19768 | Not-determined |
| S1S_extracted_19782 | Not-determined |
| S1S_extracted_19784 | Not-determined |
| S1S_extracted_19790 | Not-determined |
| S1S_extracted_19796 | Low-quality |
| S1S_extracted_19797 | Low-quality |
| S1S_extracted_19799 | Not-determined |
| S1S_extracted_19802 | Not-determined |
| S1S_extracted_19805 | Not-determined |
| S1S_extracted_19806 | Not-determined |
| S1S_extracted_19811 | Not-determined |
| S1S_extracted_19814 | Not-determined |
| S1S_extracted_19821 | Not-determined |
| S1S_extracted_19829 | Not-determined |
| S1S_extracted_19843 | Not-determined |
| S1S_extracted_19849 | Not-determined |
| S1S_extracted_19850 | Not-determined |
| S1S_extracted_19854 | Not-determined |
| S1S_extracted_19857 | Not-determined |
| S1S_extracted_19859 | Not-determined |
| S1S_extracted_19860 | Not-determined |
| S1S_extracted_19864 | Not-determined |
| S1S_extracted_19866 | Low-quality |
| S1S_extracted_19873 | Not-determined |

| | |
|---|---|
| S15_extracted_19882 | Not-determined |
| S15_extracted_19905 | Not-determined |
| S15_extracted_19927 | Not-determined |
| S15_extracted_19928 | Not-determined |
| S15_extracted_19949 | Not-determined |
| S15_extracted_19956 | Not-determined |
| S15_extracted_19965 | Not-determined |
| S15_extracted_19969 | Not-determined |
| S15_extracted_19979 | Not-determined |
| S15_extracted_19983 | Not-determined |
| S15_extracted_19991 | Not-determined |
| S15_extracted_20005 | Not-determined |
| S15_extracted_20010 | Not-determined |
| S15_extracted_20022 | Not-determined |
| S15_extracted_20023 | Not-determined |
| S15_extracted_20034 | Not-determined |
| S15_extracted_20050 | Not-determined |
| S15_extracted_20055 | Not-determined |
| S15_extracted_20077 | Not-determined |
| S15_extracted_20079 | Not-determined |
| S15_extracted_20105 | Not-determined |
| S15_extracted_20118 | Not-determined |
| S15_extracted_20131 | Not-determined |
| S15_extracted_20146 | Not-determined |
| S15_extracted_20147 | Low-quality |
| S15_extracted_20150 | Not-determined |
| S15_extracted_20166 | Not-determined |
| S15_extracted_20174 | Not-determined |
| S15_extracted_20196 | Not-determined |
| S15_extracted_20198 | Low-quality |
| S15_extracted_20202 | Not-determined |
| S15_extracted_20208 | Not-determined |
| S15_extracted_20223 | Low-quality |
| S15_extracted_20231 | Not-determined |
| S15_extracted_20235 | Not-determined |
| S15_extracted_20237 | Not-determined |
| S15_extracted_20242 | Not-determined |
| S15_extracted_20244 | Not-determined |
| S15_extracted_20260 | Not-determined |
| S15_extracted_20301 | Not-determined |
| S15_extracted_20307 | Not-determined |
| S15_extracted_20313 | Not-determined |
| S15_extracted_20316 | Low-quality |
| S15_extracted_20350 | Not-determined |
| S15_extracted_20351 | Not-determined |
| S15_extracted_20352 | Not-determined |
| S15_extracted_20362 | Not-determined |
| S15_extracted_20367 | Not-determined |
| S15_extracted_20382 | Not-determined |
| S15_extracted_20384 | Not-determined |
| S15_extracted_20390 | Not-determined |
| S15_extracted_20406 | Not-determined |
| S15_extracted_20414 | Not-determined |
| S15_extracted_20431 | Not-determined |
| S15_extracted_20439 | Not-determined |
| S15_extracted_20450 | Not-determined |
| S15_extracted_20452 | Not-determined |
| S15_extracted_20482 | Not-determined |
| S15_extracted_20484 | Not-determined |
| S15_extracted_20488 | Not-determined |
| S15_extracted_20495 | Low-quality |
| S15_extracted_20508 | Not-determined |
| S15_extracted_20509 | Not-determined |
| S15_extracted_20520 | Not-determined |
| S15_extracted_20521 | Not-determined |
| S15_extracted_20529 | Not-determined |
| S15_extracted_20548 | Not-determined |
| S15_extracted_20563 | Not-determined |
| S15_extracted_20566 | Not-determined |
| S15_extracted_20567 | Not-determined |
| S15_extracted_20573 | Not-determined |
| S15_extracted_20574 | Not-determined |
| S15_extracted_20577 | Not-determined |
| S15_extracted_20588 | Not-determined |
| S15_extracted_20596 | Not-determined |
| S15_extracted_20616 | Not-determined |
| S15_extracted_20631 | Low-quality |
| S15_extracted_20642 | Not-determined |
| S9_vs_k141_26179\|\|full | Medium-quality |
| S15_extracted_20657 | Not-determined |
| S15_extracted_20672 | Not-determined |
| S15_extracted_20676 | Not-determined |
| S15_extracted_20680 | Not-determined |
| S15_extracted_20687 | Not-determined |
| S15_extracted_20694 | Not-determined |
| S15_extracted_20705 | Low-quality |
| S15_extracted_20711 | Not-determined |
| S15_extracted_20717 | Not-determined |
| S15_extracted_20731 | Not-determined |
| S15_extracted_20735 | Not-determined |
| S15_extracted_20743 | Not-determined |
| S15_extracted_20751 | Low-quality |
| S15_extracted_20768 | Not-determined |
| S15_extracted_20774 | Not-determined |
| S15_extracted_20778 | Not-determined |
| S15_extracted_20786 | Not-determined |
| S15_extracted_20787 | Not-determined |
| S15_extracted_20788 | Not-determined |
| S15_extracted_20790 | Not-determined |
| S15_extracted_20802 | Not-determined |
| S15_extracted_20804 | Not-determined |
| S15_extracted_20809 | Not-determined |
| S15_extracted_20820 | Not-determined |
| S15_extracted_20821 | Not-determined |
| S15_extracted_20822 | Not-determined |
| S15_extracted_20840 | Not-determined |
| S15_extracted_20851 | Not-determined |
| S15_extracted_20865 | Not-determined |
| S15_extracted_20867 | Not-determined |
| S15_extracted_20869 | Not-determined |
| S15_extracted_20885 | Not-determined |
| S15_extracted_20886 | Not-determined |
| S15_extracted_20895 | Not-determined |
| S15_extracted_20902 | Not-determined |
| S15_extracted_20924 | Not-determined |
| S15_extracted_20927 | Not-determined |
| S15_extracted_20931 | Not-determined |
| S15_extracted_20932 | Not-determined |
| S15_extracted_20934 | Not-determined |
| S15_extracted_20943 | Not-determined |
| S15_extracted_20979 | Low-quality |
| S15_extracted_21036 | Not-determined |
| S15_extracted_21046 | Not-determined |
| S15_extracted_21055 | Not-determined |
| S15_extracted_21059 | Not-determined |
| S15_extracted_21070 | Not-determined |
| S15_extracted_21081 | Not-determined |
| S15_extracted_21099 | Not-determined |
| S15_extracted_21102 | Not-determined |
| S15_extracted_21113 | Not-determined |
| S15_extracted_21125 | Not-determined |
| S15_extracted_21127 | Not-determined |
| S15_extracted_21129 | Not-determined |
| S15_extracted_21132 | Not-determined |
| S15_extracted_21167 | Not-determined |
| S15_extracted_21172 | Not-determined |
| S15_extracted_21190 | Not-determined |
| S15_extracted_21199 | Not-determined |
| S15_extracted_21200 | Low-quality |
| S15_extracted_21201 | Not-determined |
| S15_extracted_21203 | Not-determined |
| S15_extracted_21204 | Not-determined |
| S15_extracted_21223 | Not-determined |
| S15_extracted_21238 | Not-determined |
| S15_extracted_21253 | Not-determined |
| S15_extracted_21261 | Not-determined |
| S15_extracted_21263 | Not-determined |
| S15_extracted_21275 | Not-determined |
| S15_extracted_21284 | Not-determined |
| S15_extracted_21285 | Not-determined |
| S15_extracted_21289 | Not-determined |
| S15_extracted_21317 | Not-determined |
| S15_extracted_21343 | Not-determined |
| S15_extracted_21345 | Not-determined |
| S15_extracted_21347 | Not-determined |
| S15_extracted_21392 | Not-determined |
| S15_extracted_21394 | Not-determined |
| S15_extracted_21396 | Not-determined |
| S15_extracted_21411 | Not-determined |
| S15_extracted_21460 | Low-quality |
| S15_extracted_21463 | Not-determined |
| S15_extracted_21470 | Not-determined |
| S15_extracted_21477 | Not-determined |
| S15_extracted_21484 | Not-determined |
| S15_extracted_21504 | Not-determined |
| S15_extracted_21508 | Not-determined |
| S15_extracted_21538 | Not-determined |
| S15_extracted_21540 | Not-determined |
| S15_extracted_21547 | Not-determined |
| S15_extracted_21552 | Not-determined |
| S15_extracted_21560 | Not-determined |
| S15_extracted_21563 | Not-determined |
| S15_extracted_21570 | Not-determined |
| S15_extracted_21594 | Low-quality |
| S15_extracted_21598 | Not-determined |
| S15_extracted_21611 | Not-determined |
| S15_extracted_21619 | Not-determined |
| S15_extracted_21627 | Not-determined |
| S15_extracted_21628 | Not-determined |
| S15_extracted_21629 | Not-determined |
| S15_extracted_21642 | Not-determined |
| S15_extracted_21644 | Low-quality |
| S15_extracted_21648 | Not-determined |
| S15_extracted_21657 | Not-determined |
| S15_extracted_21680 | Low-quality |
| S15_extracted_21685 | Low-quality |
| S15_extracted_21705 | Not-determined |
| S15_extracted_21708 | Not-determined |
| S15_extracted_21730 | Not-determined |
| S15_extracted_21754 | Not-determined |
| S15_extracted_21757 | Not-determined |
| S15_extracted_21763 | Not-determined |
| S15_extracted_21766 | Not-determined |
| S15_extracted_21819 | Not-determined |
| S15_extracted_21839 | Not-determined |
| S15_extracted_21842 | Not-determined |
| S15_extracted_21843 | Not-determined |
| S15_extracted_21857 | Not-determined |
| S15_extracted_21866 | Not-determined |
| S15_extracted_21868 | Not-determined |
| S15_extracted_21873 | Not-determined |
| S15_extracted_21887 | Not-determined |
| S15_extracted_21889 | Not-determined |
| S15_extracted_21892 | Low-quality |
| S15_extracted_21904 | Not-determined |
| S15_extracted_21906 | Low-quality |
| S15_extracted_21921 | Not-determined |
| S15_extracted_21932 | Not-determined |
| S15_extracted_21945 | Not-determined |
| S15_extracted_21948 | Not-determined |
| S15_extracted_21957 | Not-determined |
| S15_extracted_21963 | Not-determined |
| S15_extracted_21965 | Not-determined |
| S15_extracted_21972 | Not-determined |
| S15_extracted_21976 | Not-determined |
| S15_extracted_21986 | Not-determined |
| S15_extracted_21987 | Not-determined |
| S15_extracted_21995 | Not-determined |
| S15_extracted_21999 | Not-determined |
| S15_extracted_22001 | Not-determined |
| S15_extracted_22014 | Not-determined |
| S15_extracted_22015 | Not-determined |
| S15_extracted_22027 | Not-determined |
| S15_extracted_22066 | Not-determined |
| S15_extracted_22073 | Low-quality |
| S15_extracted_22084 | Low-quality |
| S15_extracted_22089 | Not-determined |
| S15_extracted_22091 | Not-determined |
| S15_extracted_22101 | Not-determined |
| S15_extracted_22117 | Not-determined |
| S15_extracted_22119 | Not-determined |
| S15_extracted_22128 | Low-quality |
| S15_extracted_22142 | Not-determined |
| S15_extracted_22153 | Not-determined |
| S15_extracted_22174 | Not-determined |
| S15_extracted_22184 | Not-determined |
| S15_extracted_22187 | Not-determined |
| S15_extracted_22189 | Not-determined |
| S15_extracted_22218 | Not-determined |
| S15_extracted_22227 | Not-determined |
| S15_extracted_22228 | Not-determined |
| S15_extracted_22243 | Not-determined |
| S15_extracted_22263 | Not-determined |
| S15_extracted_22297 | Not-determined |
| S15_extracted_22298 | Not-determined |
| S15_extracted_22311 | Not-determined |
| S15_extracted_22318 | Not-determined |
| S15_extracted_22327 | Not-determined |
| S15_extracted_22346 | Low-quality |
| S15_extracted_22349 | Not-determined |
| S15_extracted_22356 | Not-determined |
| S15_extracted_22365 | Not-determined |
| S15_extracted_22381 | Not-determined |
| S15_extracted_22383 | Not-determined |
| S15_extracted_22393 | Low-quality |
| S15_extracted_22396 | Not-determined |
| S15_extracted_22414 | Low-quality |
| S15_extracted_22433 | Low-quality |
| S15_extracted_22438 | Not-determined |
| S15_extracted_22444 | Not-determined |
| S15_extracted_22453 | Not-determined |
| S15_extracted_22456 | Not-determined |
| S15_extracted_22458 | Not-determined |
| S15_extracted_22459 | Not-determined |
| S15_extracted_22462 | Not-determined |
| S15_extracted_22476 | Not-determined |
| S15_extracted_22479 | Not-determined |
| S15_extracted_22497 | Not-determined |
| S15_extracted_22502 | Low-quality |
| S15_extracted_22508 | Low-quality |
| S15_extracted_22510 | Not-determined |
| S15_extracted_22517 | Not-determined |
| S15_extracted_22523 | Not-determined |
| S15_extracted_22528 | Not-determined |
| S15_extracted_22536 | Not-determined |
| S15_extracted_22546 | Not-determined |
| S15_extracted_22548 | Not-determined |
| S15_extracted_22551 | Not-determined |
| S15_extracted_22581 | Not-determined |
| S15_extracted_22592 | Not-determined |
| S15_extracted_22595 | Not-determined |
| S15_extracted_22597 | Not-determined |
| S15_extracted_22609 | Not-determined |
| S15_extracted_22611 | Not-determined |
| S15_extracted_22618 | Not-determined |
| S15_extracted_22621 | Not-determined |
| S15_extracted_22641 | Not-determined |
| S15_extracted_22647 | Not-determined |
| S15_extracted_22651 | Not-determined |
| S15_extracted_22654 | Not-determined |
| S15_extracted_22659 | Not-determined |
| S15_extracted_22660 | Not-determined |
| S15_extracted_22684 | Low-quality |
| S15_extracted_22687 | Low-quality |
| S15_extracted_22699 | Low-quality |
| S15_extracted_22733 | Not-determined |
| S15_extracted_22742 | Not-determined |
| S15_extracted_22757 | Not-determined |
| S15_extracted_22759 | Not-determined |
| S15_extracted_22761 | Not-determined |
| S15_extracted_22779 | Not-determined |
| S15_extracted_22780 | Not-determined |
| S15_extracted_22783 | Low-quality |
| S15_extracted_22786 | Not-determined |
| S15_extracted_22787 | Not-determined |
| S15_extracted_22797 | Low-quality |
| S15_extracted_22806 | Not-determined |
| S15_extracted_22811 | Not-determined |
| S15_extracted_22816 | Not-determined |
| S15_extracted_22824 | Not-determined |
| S15_extracted_22831 | Not-determined |
| S15_extracted_22842 | Low-quality |
| S15_extracted_22869 | Not-determined |
| S15_extracted_22883 | Low-quality |
| S15_extracted_22889 | Not-determined |
| S15_extracted_22930 | Not-determined |
| S15_extracted_22939 | Not-determined |
| S15_extracted_22951 | Not-determined |
| S15_extracted_22954 | Not-determined |
| S15_extracted_22955 | Not-determined |
| S15_extracted_22959 | Not-determined |
| S15_extracted_22981 | Not-determined |
| S15_extracted_22984 | Not-determined |
| S15_extracted_23010 | Not-determined |
| S15_extracted_23049 | Not-determined |
| S15_extracted_23059 | Not-determined |
| S15_extracted_23076 | Low-quality |
| S15_extracted_23078 | Low-quality |
| S15_extracted_23080 | Not-determined |
| S15_extracted_23087 | Not-determined |
| S15_extracted_23089 | Not-determined |
| S15_extracted_23091 | Low-quality |
| S15_extracted_23094 | Not-determined |
| S15_extracted_23100 | Not-determined |
| S15_extracted_23107 | Not-determined |
| S15_extracted_23125 | Not-determined |
| S15_extracted_23127 | Not-determined |
| S15_extracted_23135 | Not-determined |
| S15_extracted_23149 | Not-determined |
| S15_extracted_23153 | Not-determined |
| S15_extracted_23161 | Not-determined |
| S15_extracted_23167 | Not-determined |
| S15_extracted_23171 | Not-determined |
| S15_extracted_23172 | Low-quality |
| S15_extracted_23173 | Not-determined |
| S15_extracted_23181 | Not-determined |
| S15_extracted_23185 | Not-determined |
| S15_extracted_23194 | Not-determined |
| S15_extracted_23196 | Not-determined |
| S15_extracted_23200 | Not-determined |
| S15_extracted_23209 | Not-determined |
| S15_extracted_23214 | Not-determined |
| S15_extracted_23237 | Not-determined |
| S15_extracted_23239 | Not-determined |
| S15_extracted_23240 | Not-determined |
| S15_extracted_23246 | Not-determined |
| S15_extracted_23254 | Not-determined |
| S15_extracted_23267 | Low-quality |
| S15_extracted_23277 | Not-determined |
| S15_extracted_23278 | Low-quality |
| S15_extracted_23293 | Not-determined |
| S15_extracted_23297 | Not-determined |
| S15_extracted_23308 | Not-determined |
| S15_extracted_23313 | Not-determined |
| S15_extracted_23321 | Not-determined |
| S15_extracted_23327 | Not-determined |
| S15_extracted_23331 | Not-determined |
| S15_extracted_23344 | Not-determined |
| S15_extracted_23346 | Not-determined |
| S15_extracted_23351 | Not-determined |
| S15_extracted_23356 | Not-determined |
| S15_extracted_23364 | Not-determined |
| S15_extracted_23370 | Not-determined |
| S15_extracted_23372 | Not-determined |
| S15_extracted_23382 | Not-determined |
| S15_extracted_23414 | Not-determined |
| S15_extracted_23415 | Not-determined |
| S15_extracted_23423 | Not-determined |
| S15_extracted_23426 | Not-determined |
| S15_extracted_23427 | Not-determined |
| S15_extracted_23428 | Not-determined |
| S15_extracted_23431 | Low-quality |
| S15_extracted_23463 | Not-determined |
| S15_extracted_23470 | Not-determined |
| S15_extracted_23475 | Not-determined |
| S15_extracted_23479 | Not-determined |
| S15_extracted_23480 | Not-determined |
| S15_extracted_23487 | Not-determined |
| S15_extracted_23499 | Not-determined |
| S15_extracted_23505 | Not-determined |
| S15_extracted_23508 | Not-determined |
| S15_extracted_23527 | Not-determined |
| S15_extracted_23529 | Low-quality |
| S15_extracted_23533 | Low-quality |
| S15_extracted_23538 | Not-determined |
| S15_extracted_23546 | Not-determined |
| S15_extracted_23547 | Not-determined |
| S15_extracted_23563 | Not-determined |
| S15_extracted_23566 | Not-determined |
| S15_extracted_23570 | Not-determined |
| S15_extracted_23580 | Not-determined |
| S15_extracted_23581 | Not-determined |
| S15_extracted_23588 | Low-quality |
| S15_extracted_23600 | Low-quality |
| S15_extracted_23603 | Not-determined |
| S15_extracted_23609 | Not-determined |
| S15_extracted_23612 | Not-determined |
| S15_extracted_23624 | Not-determined |
| S15_extracted_23632 | Low-quality |
| S15_extracted_23636 | Not-determined |
| S15_extracted_23655 | Not-determined |
| S15_extracted_23657 | Low-quality |
| S15_extracted_23659 | Not-determined |
| S15_extracted_23662 | Not-determined |
| S15_extracted_23665 | Low-quality |
| S15_extracted_23668 | Not-determined |
| S15_extracted_23673 | Not-determined |
| S15_extracted_23684 | Not-determined |
| S15_extracted_23693 | Low-quality |
| S15_extracted_23700 | Low-quality |
| S15_extracted_23702 | Low-quality |
| S15_extracted_23707 | Not-determined |
| S15_extracted_23715 | Low-quality |
| S15_extracted_23720 | Not-determined |
| S15_extracted_23730 | Not-determined |
| S15_extracted_23731 | Low-quality |
| S15_extracted_23738 | Not-determined |
| S15_extracted_23773 | Not-determined |
| S15_extracted_23809 | Not-determined |
| S15_extracted_23811 | Low-quality |
| S15_extracted_23817 | Not-determined |
| S15_extracted_23819 | Not-determined |
| S15_extracted_23821 | Not-determined |
| S15_extracted_23822 | Not-determined |
| S15_extracted_23836 | Not-determined |
| S15_extracted_23837 | Not-determined |
| S15_extracted_23847 | Not-determined |
| S15_extracted_23854 | Not-determined |
| S15_extracted_23860 | Not-determined |
| S15_extracted_23873 | Not-determined |
| S15_extracted_23882 | Not-determined |
| S15_extracted_23885 | Not-determined |
| S15_extracted_23891 | Not-determined |
| S15_extracted_23900 | Not-determined |
| S15_extracted_23904 | Not-determined |
| S15_extracted_23917 | Not-determined |
| S15_extracted_23927 | Not-determined |
| S15_extracted_23928 | Low-quality |
| S15_extracted_23934 | Not-determined |
| S15_extracted_23937 | Not-determined |
| S15_extracted_23941 | Not-determined |
| S15_extracted_23949 | Not-determined |
| S15_extracted_23950 | Not-determined |
| S15_extracted_23955 | Not-determined |
| S15_extracted_23965 | Not-determined |
| S15_extracted_23977 | Not-determined |
| S15_extracted_23993 | Not-determined |
| S15_extracted_24002 | Not-determined |
| S15_extracted_24016 | Not-determined |
| S15_extracted_24017 | Not-determined |
| S15_extracted_24024 | Not-determined |
| S15_extracted_24035 | Not-determined |
| S15_extracted_24037 | Low-quality |
| S15_extracted_24047 | Low-quality |
| S15_extracted_24049 | Low-quality |
| S15_extracted_24055 | Low-quality |
| S15_extracted_24063 | Not-determined |
| S15_extracted_24070 | Not-determined |
| S15_extracted_24076 | Not-determined |
| S15_extracted_24082 | Not-determined |
| S15_extracted_24083 | Not-determined |
| S15_extracted_24126 | Not-determined |
| S15_extracted_24127 | Not-determined |
| S15_extracted_24135 | Not-determined |
| S15_extracted_24137 | Not-determined |
| S15_extracted_24139 | Not-determined |
| S15_extracted_24143 | Not-determined |
| S15_extracted_24144 | Not-determined |
| S15_extracted_24146 | Low-quality |
| S15_extracted_24152 | Not-determined |
| S15_extracted_24153 | Not-determined |
| S15_extracted_24158 | Not-determined |
| S15_extracted_24160 | Not-determined |
| S15_extracted_24183 | Not-determined |
| S15_extracted_24191 | Not-determined |
| S15_extracted_24194 | Not-determined |
| S15_extracted_24202 | Not-determined |
| S15_extracted_24205 | Not-determined |
| S15_extracted_24223 | Not-determined |
| S15_extracted_24227 | Low-quality |
| S15_extracted_24267 | Not-determined |
| S15_extracted_24269 | Not-determined |
| S15_extracted_24270 | Low-quality |
| S15_extracted_24278 | Not-determined |
| S15_extracted_24288 | Not-determined |
| S15_extracted_24298 | Not-determined |
| S15_extracted_24299 | Not-determined |
| S15_extracted_24310 | Not-determined |
| S15_extracted_24323 | Not-determined |
| S15_extracted_24326 | Not-determined |
| S15_extracted_24357 | Not-determined |
| S15_extracted_24367 | Not-determined |
| S15_extracted_24383 | Not-determined |
| S15_extracted_24385 | Not-determined |
| S15_extracted_24388 | Not-determined |
| S15_extracted_24395 | Not-determined |
| S15_extracted_24398 | Not-determined |
| S15_extracted_24400 | Not-determined |
| S15_extracted_24404 | Not-determined |
| S15_extracted_24410 | Not-determined |
| S15_extracted_24411 | Low-quality |
| S15_extracted_24419 | Not-determined |
| S15_extracted_24423 | Not-determined |
| S15_extracted_24426 | Not-determined |
| S15_extracted_24447 | Not-determined |
| S15_extracted_24452 | Not-determined |
| S15_extracted_24460 | Not-determined |
| S15_extracted_24464 | Not-determined |
| S15_extracted_24474 | Low-quality |
| S15_extracted_24476 | Not-determined |
| S15_extracted_24481 | Not-determined |
| S15_extracted_24492 | Not-determined |
| S15_extracted_24496 | Not-determined |
| S15_extracted_24507 | Not-determined |
| S15_extracted_24520 | Not-determined |
| S15_extracted_24540 | Not-determined |
| S15_extracted_24546 | Not-determined |
| S15_extracted_24547 | Not-determined |
| S15_extracted_24555 | Low-quality |
| S15_extracted_24559 | Not-determined |
| S15_extracted_24583 | Low-quality |
| S15_extracted_24589 | Not-determined |
| S15_extracted_24591 | Not-determined |
| S15_extracted_24597 | Not-determined |
| S15_extracted_24625 | Low-quality |
| S15_extracted_24635 | Low-quality |
| S15_extracted_24640 | Not-determined |
| S15_extracted_24693 | Not-determined |
| S15_extracted_24705 | Not-determined |
| S15_extracted_24713 | Not-determined |
| S15_extracted_24720 | Not-determined |
| S15_extracted_24726 | Not-determined |
| S15_extracted_24757 | Not-determined |
| S15_extracted_24760 | Not-determined |
| S15_extracted_24772 | Not-determined |
| S15_extracted_24829 | Not-determined |
| S15_extracted_24833 | Not-determined |
| S15_extracted_24839 | Not-determined |
| S15_extracted_24840 | Not-determined |
| S15_extracted_24849 | Not-determined |
| S15_extracted_24877 | Low-quality |
| S15_extracted_24880 | Not-determined |
| S15_extracted_24881 | Not-determined |
| S15_extracted_24920 | Not-determined |
| S15_extracted_24930 | Not-determined |
| S15_extracted_24942 | Not-determined |
| S15_extracted_24945 | Not-determined |
| S15_extracted_24952 | Not-determined |
| S15_extracted_24958 | Low-quality |
| S15_extracted_24961 | Not-determined |
| S15_extracted_24962 | Not-determined |
| S15_extracted_24966 | Not-determined |
| S15_extracted_24977 | Not-determined |
| S15_extracted_24980 | Not-determined |
| S15_extracted_24991 | Low-quality |
| S15_extracted_25003 | Not-determined |
| S15_extracted_25009 | Not-determined |
| S15_extracted_25053 | Low-quality |
| S15_extracted_25054 | Not-determined |
| S15_extracted_25077 | Not-determined |
| S15_extracted_25082 | Not-determined |
| S15_extracted_25094 | Not-determined |
| S15_extracted_25095 | Not-determined |
| S15_extracted_25099 | Not-determined |
| S15_extracted_25100 | Not-determined |
| S15_extracted_25102 | Not-determined |
| S15_extracted_25110 | Not-determined |
| S15_extracted_25115 | Not-determined |
| S15_extracted_25123 | Not-determined |
| S15_extracted_25151 | Not-determined |
| S15_extracted_25159 | Not-determined |
| S15_extracted_25161 | Low-quality |
| S15_extracted_25170 | Not-determined |
| S15_extracted_25174 | Not-determined |
| S15_extracted_25182 | Not-determined |
| S15_extracted_25188 | Not-determined |
| S15_extracted_25205 | Not-determined |
| S15_extracted_25206 | Not-determined |
| S15_extracted_25212 | Not-determined |

| | |
|---|---|
| S1S_extracted_25222 | Not-determined |
| S1S_extracted_25230 | Not-determined |
| S1S_extracted_25243 | Low-quality |
| S1S_extracted_25266 | Not-determined |
| S1S_extracted_25267 | Not-determined |
| S1S_extracted_25289 | Not-determined |
| S1S_extracted_25294 | Not-determined |
| S1S_extracted_25307 | Not-determined |
| S1S_extracted_25325 | Not-determined |
| S1S_extracted_25362 | Not-determined |
| S1S_extracted_25363 | Not-determined |
| S1S_extracted_25365 | Not-determined |
| S1S_extracted_25366 | Low-quality |
| S1S_extracted_25376 | Not-determined |
| S1S_extracted_25385 | Not-determined |
| S1S_extracted_25393 | Not-determined |
| S1S_extracted_25408 | Not-determined |
| S1S_extracted_25420 | Low-quality |
| S1S_extracted_25425 | Not-determined |
| S1S_extracted_25429 | Low-quality |
| S1S_extracted_25445 | Low-quality |
| S1S_extracted_25460 | Not-determined |
| S1S_extracted_25466 | Not-determined |
| S1S_extracted_25468 | Not-determined |
| S1S_extracted_25477 | Not-determined |
| S1S_extracted_25490 | Not-determined |
| S1S_extracted_25492 | Not-determined |
| S1S_extracted_25497 | Not-determined |
| S1S_extracted_25500 | Not-determined |
| S1S_extracted_25510 | Not-determined |
| S1S_extracted_25511 | Not-determined |
| S1S_extracted_25513 | Not-determined |
| S1S_extracted_25529 | Not-determined |
| S1S_extracted_25530 | Low-quality |
| S1S_extracted_25532 | Not-determined |
| S1S_extracted_25533 | Not-determined |
| S1S_extracted_25534 | Low-quality |
| S1S_extracted_25535 | Not-determined |
| S1S_extracted_25551 | Low-quality |
| S1S_extracted_25563 | Not-determined |
| S1S_extracted_25566 | Not-determined |
| S1S_extracted_25568 | Not-determined |
| S1S_extracted_25573 | Not-determined |
| S1S_extracted_25574 | Not-determined |
| S1S_extracted_25577 | Not-determined |
| S1S_extracted_25584 | Low-quality |
| S1S_extracted_25593 | Not-determined |
| S1S_extracted_25616 | Not-determined |
| S1S_extracted_25622 | Not-determined |
| S1S_extracted_25638 | Not-determined |
| S1S_extracted_25687 | Not-determined |
| S1S_extracted_25700 | Not-determined |
| S1S_extracted_25718 | Not-determined |
| S1S_extracted_25732 | Not-determined |
| S1S_extracted_25741 | Low-quality |
| S1S_extracted_25742 | Not-determined |
| S1S_extracted_25749 | Not-determined |
| S1S_extracted_25753 | Low-quality |
| S1S_extracted_25756 | Not-determined |
| S1S_extracted_25757 | Not-determined |
| S1S_extracted_25769 | Not-determined |
| S1S_extracted_25777 | Not-determined |
| S1S_extracted_25800 | Not-determined |
| S1S_extracted_25806 | Not-determined |
| S1S_extracted_25814 | Not-determined |
| S1S_extracted_25823 | Low-quality |
| S1S_extracted_25829 | Not-determined |
| S1S_extracted_25832 | Not-determined |
| S1S_extracted_25849 | Not-determined |
| S1S_extracted_25851 | Not-determined |
| S1S_extracted_25857 | Not-determined |
| S1S_extracted_25873 | Low-quality |
| S1S_extracted_25878 | Not-determined |
| S1S_extracted_25881 | Not-determined |
| S1S_extracted_25893 | Not-determined |
| S1S_extracted_25901 | Not-determined |
| S1S_extracted_25915 | Not-determined |
| S1S_extracted_25916 | Not-determined |
| S1S_extracted_25936 | Not-determined |
| S1S_extracted_25940 | Not-determined |
| S1S_extracted_25966 | Not-determined |
| S1S_extracted_25990 | Not-determined |
| S1S_extracted_26002 | Not-determined |
| S1S_extracted_26009 | Not-determined |
| S1S_extracted_26011 | Not-determined |
| S1S_extracted_26017 | Not-determined |
| S1S_extracted_26021 | Not-determined |
| S1S_extracted_26030 | Not-determined |
| S1S_extracted_26042 | Not-determined |
| S1S_extracted_26043 | Not-determined |
| S1S_extracted_26047 | Not-determined |
| S1S_extracted_26049 | Not-determined |
| S1S_extracted_26053 | Not-determined |
| S1S_extracted_26060 | Not-determined |
| S1S_extracted_26088 | Not-determined |
| S1S_extracted_26095 | Not-determined |
| S1S_extracted_26146 | Not-determined |
| S1S_extracted_26150 | Not-determined |
| S1S_extracted_26161 | Not-determined |
| S1S_extracted_26163 | Not-determined |
| S1S_extracted_26166 | Not-determined |
| S1S_extracted_26167 | Not-determined |
| S1S_extracted_26180 | Not-determined |
| S1S_extracted_26190 | Not-determined |
| S1S_extracted_26195 | Not-determined |
| S1S_extracted_26224 | Not-determined |
| S1S_extracted_26231 | Not-determined |
| S1S_extracted_26240 | Not-determined |
| S1S_extracted_26258 | Not-determined |
| S1S_extracted_26287 | Not-determined |
| S1S_extracted_26288 | Not-determined |
| S1S_extracted_26291 | Not-determined |
| S1S_extracted_26292 | Not-determined |
| S1S_extracted_26299 | Not-determined |
| S1S_extracted_26306 | Low-quality |
| S1S_extracted_26323 | Not-determined |
| S1S_extracted_26348 | Not-determined |
| S1S_extracted_26350 | Not-determined |
| S1S_extracted_26361 | Not-determined |
| S1S_extracted_26365 | Low-quality |
| S1S_extracted_26369 | Not-determined |
| S1S_extracted_26381 | Not-determined |
| S1S_extracted_26390 | Not-determined |
| S1S_extracted_26400 | Not-determined |
| S1S_extracted_26427 | Not-determined |
| S1S_extracted_26436 | Not-determined |
| S1S_extracted_26439 | Not-determined |
| S1S_extracted_26460 | Not-determined |
| S1S_extracted_26462 | Low-quality |
| S1S_extracted_26466 | Not-determined |
| S1S_extracted_26479 | Not-determined |
| S1S_extracted_26480 | Not-determined |
| S1S_extracted_26481 | Not-determined |
| S1S_extracted_26498 | Not-determined |
| S1S_extracted_26500 | Not-determined |
| S1S_extracted_26507 | Not-determined |
| S1S_extracted_26517 | Not-determined |
| S1S_extracted_26519 | Not-determined |
| S1S_extracted_26532 | Not-determined |
| S1S_extracted_26595 | Not-determined |
| S1S_extracted_26601 | Low-quality |
| S1S_extracted_26609 | Not-determined |
| S1S_extracted_26640 | Not-determined |
| S1S_extracted_26644 | Not-determined |
| S1S_extracted_26655 | Not-determined |
| S1S_extracted_26670 | Not-determined |
| S1S_extracted_26687 | Not-determined |
| S1S_extracted_26692 | Low-quality |
| S1S_extracted_26693 | Not-determined |
| S1S_extracted_26695 | Not-determined |
| S1S_extracted_26704 | Not-determined |
| S1S_extracted_26705 | Not-determined |
| S1S_extracted_26709 | Low-quality |
| S1S_extracted_26720 | Not-determined |
| S1S_extracted_26721 | Not-determined |
| S1S_extracted_26723 | Not-determined |
| S1S_extracted_26729 | Not-determined |
| S1S_extracted_26737 | Low-quality |
| S1S_extracted_26742 | Not-determined |
| S1S_extracted_26751 | Not-determined |
| S1S_extracted_26754 | Not-determined |
| S1S_extracted_26768 | Not-determined |
| S1S_extracted_26779 | Not-determined |
| S1S_extracted_26780 | Not-determined |
| S1S_extracted_26792 | Not-determined |
| S1S_extracted_26800 | Not-determined |
| S1S_extracted_26809 | Not-determined |
| S1S_extracted_26827 | Not-determined |
| S1S_extracted_26830 | Low-quality |
| S1S_extracted_26835 | Low-quality |
| S1S_extracted_26848 | Not-determined |
| S1S_extracted_26849 | Not-determined |
| S1S_extracted_26853 | Low-quality |
| S1S_extracted_26866 | Not-determined |
| S1S_extracted_26869 | Not-determined |
| S1S_extracted_26880 | Not-determined |
| S1S_extracted_26884 | Not-determined |
| S1S_extracted_26887 | Not-determined |
| S1S_extracted_26913 | Low-quality |
| S1S_extracted_26925 | Not-determined |
| S1S_extracted_26928 | Not-determined |
| S1S_extracted_26938 | Not-determined |
| S1S_extracted_26967 | Not-determined |
| S1S_extracted_26970 | Not-determined |
| S1S_extracted_26977 | Not-determined |
| S1S_extracted_26981 | Not-determined |
| S1S_extracted_26982 | Low-quality |
| S1S_extracted_26991 | Not-determined |
| S1S_extracted_26992 | Not-determined |
| S1S_extracted_26997 | Not-determined |
| S1S_extracted_27002 | Not-determined |
| S1S_extracted_27013 | Not-determined |
| S1S_extracted_27020 | Not-determined |
| S1S_extracted_27026 | Low-quality |
| S1S_extracted_27037 | Not-determined |
| S1S_extracted_27058 | Low-quality |
| S1S_extracted_27060 | Not-determined |
| S1S_extracted_27062 | Not-determined |
| S1S_extracted_27067 | Not-determined |
| S1S_extracted_27074 | Not-determined |
| S1S_extracted_27081 | Not-determined |
| S1S_extracted_27089 | Not-determined |
| S1S_extracted_27093 | Not-determined |
| S1S_extracted_27096 | Not-determined |
| S1S_extracted_27115 | Not-determined |
| S1S_extracted_27117 | Not-determined |
| S1S_extracted_27129 | Not-determined |
| S1S_extracted_27130 | Not-determined |
| S1S_extracted_27133 | Not-determined |
| S1S_extracted_27136 | Not-determined |
| S1S_extracted_27145 | Not-determined |
| S1S_extracted_27146 | Not-determined |
| S1S_extracted_27149 | Not-determined |
| S1S_extracted_27158 | Not-determined |
| S1S_extracted_27175 | Low-quality |
| S1S_extracted_27184 | Not-determined |
| S1S_extracted_27194 | Not-determined |
| S1S_extracted_27196 | Low-quality |
| S1S_extracted_27217 | Not-determined |
| S1S_extracted_27224 | Not-determined |
| S1S_extracted_27228 | Not-determined |
| S1S_extracted_27231 | Not-determined |
| S1S_extracted_27235 | Not-determined |
| S1S_extracted_27259 | Not-determined |
| S1S_extracted_27260 | Not-determined |
| S1S_extracted_27262 | Not-determined |
| S1S_extracted_27267 | Not-determined |
| S1S_extracted_27274 | Not-determined |
| S1S_extracted_27275 | Not-determined |
| S1S_extracted_27276 | Not-determined |
| S1S_extracted_27280 | Not-determined |
| S1S_extracted_27285 | Low-quality |
| S1S_extracted_27294 | Not-determined |
| S1S_extracted_27298 | Not-determined |
| S1S_extracted_27299 | Not-determined |
| S1S_extracted_27305 | Not-determined |
| S1S_extracted_27312 | Not-determined |
| S1S_extracted_27339 | Not-determined |
| S1S_extracted_27354 | Not-determined |
| S1S_extracted_27363 | Not-determined |
| S1S_extracted_27370 | Low-quality |
| S1S_extracted_27378 | Low-quality |
| S1S_extracted_27381 | Not-determined |
| S1S_extracted_27385 | Low-quality |
| S1S_extracted_27392 | Not-determined |
| S1S_extracted_27399 | Not-determined |
| S1S_extracted_27405 | Not-determined |
| S1S_extracted_27422 | Not-determined |
| S1S_extracted_27423 | Not-determined |
| S1S_extracted_27424 | Not-determined |
| S1S_extracted_27435 | Not-determined |
| S1S_extracted_27451 | Not-determined |
| S1S_extracted_27456 | Not-determined |
| S1S_extracted_27457 | Not-determined |
| S1S_extracted_27462 | Not-determined |
| S1S_extracted_27465 | Not-determined |
| S1S_extracted_27478 | Not-determined |
| S1S_extracted_27484 | Not-determined |
| S1S_extracted_27507 | Not-determined |
| S1S_extracted_27508 | Not-determined |
| S1S_extracted_27511 | Not-determined |
| S1S_extracted_27535 | Not-determined |
| S1S_extracted_27542 | Not-determined |
| S1S_extracted_27567 | Not-determined |
| S1S_extracted_27569 | Not-determined |
| S1S_extracted_27571 | Not-determined |
| S1S_extracted_27574 | Not-determined |
| S1S_extracted_27628 | Not-determined |
| S1S_extracted_27634 | Not-determined |
| S1S_extracted_27636 | Not-determined |
| S1S_extracted_27646 | Not-determined |
| S1S_extracted_27648 | Not-determined |
| S1S_extracted_27650 | Not-determined |
| S1S_extracted_27657 | Not-determined |
| S1S_extracted_27667 | Not-determined |
| S1S_extracted_27677 | Not-determined |
| S1S_extracted_27683 | Low-quality |
| S1S_extracted_27687 | Not-determined |
| S1S_extracted_27701 | Not-determined |
| S1S_extracted_27705 | Not-determined |
| S1S_extracted_27708 | Not-determined |
| S1S_extracted_27732 | Low-quality |
| S1S_extracted_27738 | Not-determined |
| S1S_extracted_27768 | Not-determined |
| S1S_extracted_27769 | Not-determined |
| S1S_extracted_27831 | Not-determined |
| S1S_extracted_27838 | Not-determined |
| S1S_extracted_27847 | Not-determined |
| S1S_extracted_27850 | Not-determined |
| S1S_extracted_27854 | Not-determined |
| S1S_extracted_27857 | Not-determined |
| S1S_extracted_27858 | Not-determined |
| S1S_extracted_27860 | Not-determined |
| S1S_extracted_27876 | Not-determined |
| S1S_extracted_27877 | Not-determined |
| S1S_extracted_27882 | Not-determined |
| S1S_extracted_27891 | Not-determined |
| S1S_extracted_27920 | Not-determined |
| S1S_extracted_27926 | Not-determined |
| S1S_extracted_27932 | Not-determined |
| S1S_extracted_27937 | Not-determined |
| S1S_extracted_27939 | Not-determined |
| S1S_extracted_27950 | Not-determined |
| S1S_extracted_27953 | Not-determined |
| S1S_extracted_27959 | Not-determined |
| S1S_extracted_27973 | Not-determined |
| S1S_extracted_27974 | Not-determined |
| S1S_extracted_27980 | Low-quality |
| S1S_extracted_28018 | Not-determined |
| S1S_extracted_28046 | Not-determined |
| S1S_extracted_28052 | Not-determined |
| S1S_extracted_28060 | Not-determined |
| S1S_extracted_28067 | Not-determined |
| S1S_extracted_28070 | Not-determined |
| S1S_extracted_28080 | Not-determined |
| S1S_extracted_28085 | Not-determined |
| S1S_extracted_28098 | Not-determined |
| S1S_extracted_28107 | Low-quality |
| S1S_extracted_28114 | Not-determined |
| S1S_extracted_28126 | Not-determined |
| S1S_extracted_28141 | Not-determined |
| S1S_extracted_28145 | Not-determined |
| S1S_extracted_28150 | Low-quality |
| S1S_extracted_28166 | Not-determined |
| S1S_extracted_28168 | Not-determined |
| S1S_extracted_28183 | Not-determined |
| S1S_extracted_28185 | Not-determined |
| S1S_extracted_28188 | Not-determined |
| S1S_extracted_28190 | Not-determined |
| S1S_extracted_28193 | Not-determined |
| S1S_extracted_28197 | Not-determined |
| S1S_extracted_28207 | Not-determined |
| S1S_extracted_28211 | Not-determined |
| S1S_extracted_28222 | Not-determined |
| S1S_extracted_28227 | Low-quality |
| S1S_extracted_28251 | Not-determined |
| S1S_extracted_28253 | Not-determined |
| S1S_extracted_28277 | Not-determined |
| S1S_extracted_28278 | Not-determined |
| S1S_extracted_28281 | Low-quality |
| S1S_extracted_28282 | Not-determined |
| S1S_extracted_28290 | Low-quality |
| S1S_extracted_28294 | Not-determined |
| S1S_extracted_28309 | Not-determined |
| S1S_extracted_28313 | Not-determined |
| S1S_extracted_28315 | Not-determined |
| S1S_extracted_28337 | Not-determined |
| S1S_extracted_28339 | Not-determined |
| S1S_extracted_28349 | Not-determined |
| S1S_extracted_28354 | Not-determined |
| S1S_extracted_28372 | Not-determined |
| S1S_extracted_28374 | Not-determined |
| S1S_extracted_28396 | Not-determined |
| S1S_extracted_28401 | Not-determined |
| S1S_extracted_28410 | Not-determined |
| S1S_extracted_28430 | Not-determined |
| S1S_extracted_28435 | Low-quality |
| S1S_extracted_28448 | Not-determined |
| S1S_extracted_28462 | Not-determined |
| S1S_extracted_28466 | Low-quality |
| S1S_extracted_28479 | Not-determined |
| S1S_extracted_28483 | Not-determined |
| S1S_extracted_28490 | Not-determined |
| S1S_extracted_28501 | Not-determined |
| S1S_extracted_28502 | Low-quality |
| S1S_extracted_28524 | Not-determined |
| S1S_extracted_28535 | Not-determined |
| S1S_extracted_28568 | Not-determined |
| S1S_extracted_28571 | Not-determined |
| S1S_extracted_28573 | Not-determined |
| S1S_extracted_28595 | Not-determined |
| S1S_extracted_28608 | Not-determined |
| S1S_extracted_28618 | Low-quality |
| S1S_extracted_28621 | Not-determined |
| S1S_extracted_28623 | Not-determined |
| S1S_extracted_28638 | Not-determined |
| S1S_extracted_28647 | Not-determined |
| S1S_extracted_28656 | Low-quality |
| S1S_extracted_28659 | Not-determined |
| S1S_extracted_28679 | Not-determined |
| S1S_extracted_28693 | Not-determined |
| S1S_extracted_28704 | Not-determined |
| S1S_extracted_28712 | Low-quality |
| S1S_extracted_28717 | Not-determined |
| S1S_extracted_28732 | Not-determined |
| S1S_extracted_28737 | Not-determined |
| S1S_extracted_28739 | Not-determined |
| S1S_extracted_28749 | Not-determined |
| S1S_extracted_28752 | Not-determined |
| S1S_extracted_28763 | Not-determined |
| S1S_extracted_28766 | Not-determined |
| S1S_extracted_28771 | Not-determined |
| S1S_extracted_28778 | Not-determined |
| S1S_extracted_28781 | Not-determined |
| S1S_extracted_28784 | Not-determined |
| S1S_extracted_28786 | Not-determined |
| S1S_extracted_28802 | Not-determined |
| S1S_extracted_28812 | Not-determined |
| S1S_extracted_28821 | Not-determined |
| S1S_extracted_28825 | Not-determined |
| S1S_extracted_28848 | Not-determined |
| S1S_extracted_28852 | Not-determined |
| S1S_extracted_28860 | Not-determined |
| S1S_extracted_28889 | Low-quality |
| S1S_extracted_28903 | Not-determined |
| S1S_extracted_28919 | Not-determined |
| S1S_extracted_28929 | Not-determined |
| S1S_extracted_28947 | Not-determined |
| S1S_extracted_28950 | Not-determined |
| S1S_extracted_28953 | Not-determined |
| S1S_extracted_28954 | Not-determined |
| S1S_extracted_28967 | Not-determined |
| S1S_extracted_28970 | Not-determined |
| S1S_extracted_28973 | Low-quality |
| S1S_extracted_28981 | Not-determined |
| S1S_extracted_28983 | Not-determined |
| S1S_extracted_28985 | Low-quality |
| S1S_extracted_29005 | Not-determined |
| S1S_extracted_29009 | Not-determined |
| S1S_extracted_29012 | Not-determined |
| S1S_extracted_29027 | Not-determined |
| S1S_extracted_29051 | Not-determined |
| S1S_extracted_29056 | Not-determined |
| S1S_extracted_29069 | Not-determined |
| S1S_extracted_29074 | Not-determined |
| S1S_extracted_29087 | Not-determined |
| S1S_extracted_29105 | Not-determined |
| S1S_extracted_29114 | Not-determined |
| S1S_extracted_29125 | Not-determined |
| S1S_extracted_29126 | Not-determined |
| S1S_extracted_29127 | Not-determined |
| S1S_extracted_29141 | Not-determined |
| S1S_extracted_29151 | Not-determined |
| S1S_extracted_29160 | Low-quality |
| S1S_extracted_29161 | Not-determined |
| S1S_extracted_29170 | Not-determined |
| S1S_extracted_29171 | Low-quality |
| S1S_extracted_29172 | Not-determined |
| S1S_extracted_29173 | Not-determined |
| S1S_extracted_29175 | Not-determined |
| S1S_extracted_29185 | Low-quality |
| S1S_extracted_29186 | Not-determined |
| S1S_extracted_29209 | Not-determined |
| S1S_extracted_29220 | Not-determined |
| S1S_extracted_29226 | Not-determined |
| S1S_extracted_29228 | Not-determined |
| S1S_extracted_29249 | Low-quality |
| S1S_extracted_29258 | Not-determined |
| S1S_extracted_29276 | Not-determined |
| S1S_extracted_29283 | Not-determined |
| S1S_extracted_29298 | Not-determined |
| S1S_extracted_29305 | Not-determined |
| S1S_extracted_29307 | Not-determined |
| S1S_extracted_29308 | Not-determined |
| S1S_extracted_29329 | Not-determined |
| S1S_extracted_29353 | Not-determined |
| S1S_extracted_29358 | Not-determined |
| S1S_extracted_29370 | Not-determined |
| S1S_extracted_29371 | Not-determined |
| S1S_extracted_29387 | Low-quality |
| S1S_extracted_29388 | Not-determined |
| S1S_extracted_29394 | Not-determined |
| S1S_extracted_29395 | Not-determined |
| S1S_extracted_29397 | Not-determined |
| S1S_extracted_29404 | Not-determined |
| S1S_extracted_29416 | Not-determined |
| S1S_extracted_29417 | Not-determined |
| S1S_extracted_29420 | Not-determined |
| S1S_extracted_29442 | Low-quality |
| S1S_extracted_29448 | Not-determined |
| S1S_extracted_29457 | Not-determined |
| S1S_extracted_29460 | Not-determined |
| S1S_extracted_29462 | Not-determined |
| S1S_extracted_29464 | Not-determined |
| S1S_extracted_29466 | Not-determined |
| S1S_extracted_29479 | Not-determined |
| S1S_extracted_29481 | Not-determined |
| S1S_extracted_29489 | Not-determined |
| S1S_extracted_29510 | Not-determined |
| S1S_extracted_29523 | Not-determined |
| S1S_extracted_29535 | Not-determined |
| S1S_extracted_29552 | Not-determined |
| S1S_extracted_29563 | Not-determined |
| S1S_extracted_29568 | Not-determined |
| S1S_extracted_29571 | Not-determined |
| S1S_extracted_29573 | Low-quality |
| S1S_extracted_29579 | Not-determined |
| S1S_extracted_29596 | Low-quality |
| S1S_extracted_29597 | Not-determined |
| S1S_extracted_29619 | Not-determined |
| S1S_extracted_29627 | Low-quality |
| S1S_extracted_29637 | Not-determined |
| S1S_extracted_29639 | Not-determined |
| S1S_extracted_29642 | Not-determined |
| S1S_extracted_29650 | Not-determined |
| S1S_extracted_29655 | Low-quality |
| S1S_extracted_29660 | Not-determined |
| S1S_extracted_29663 | Not-determined |
| S1S_extracted_29667 | Not-determined |
| S1S_extracted_29681 | Not-determined |
| S1S_extracted_29686 | Not-determined |
| S1S_extracted_29687 | Not-determined |
| S1S_extracted_29691 | Not-determined |
| S1S_extracted_29709 | Not-determined |
| S1S_extracted_29719 | Not-determined |
| S1S_extracted_29747 | Not-determined |
| S1S_extracted_29754 | Not-determined |
| S1S_extracted_29765 | Not-determined |
| S1S_extracted_29769 | Not-determined |
| S1S_extracted_29775 | Not-determined |
| S1S_extracted_29776 | Not-determined |
| S1S_extracted_29778 | Not-determined |
| S1S_extracted_29786 | Not-determined |
| S1S_extracted_29796 | Low-quality |
| S1S_extracted_29802 | Not-determined |
| S1S_extracted_29812 | Not-determined |
| S1S_extracted_29818 | Not-determined |
| S1S_extracted_29826 | Not-determined |
| S1S_extracted_29838 | Not-determined |
| S1S_extracted_29839 | Not-determined |
| S1S_extracted_29847 | Not-determined |
| S1S_extracted_29849 | Not-determined |
| S1S_extracted_29854 | Not-determined |
| S1S_extracted_29856 | Not-determined |
| S1S_extracted_29872 | Not-determined |
| S1S_extracted_29888 | Not-determined |
| S1S_extracted_29897 | Low-quality |
| S1S_extracted_29928 | Not-determined |
| S1S_extracted_29939 | Not-determined |
| S1S_extracted_29941 | Not-determined |
| S1S_extracted_29950 | Not-determined |
| S1S_extracted_29959 | Not-determined |
| S1S_extracted_29960 | Not-determined |
| S1S_extracted_29967 | Not-determined |
| S1S_extracted_29975 | Low-quality |
| S1S_extracted_29977 | Not-determined |
| S1S_extracted_29984 | Not-determined |
| S1S_extracted_29987 | Not-determined |
| S1S_extracted_30022 | Not-determined |
| S1S_extracted_30030 | Not-determined |
| S1S_extracted_30039 | Low-quality |
| S1S_extracted_30056 | Not-determined |
| S1S_extracted_30057 | Not-determined |
| S1S_extracted_30059 | Not-determined |
| S1S_extracted_30073 | Not-determined |
| S1S_extracted_30080 | Not-determined |
| S1S_extracted_30082 | Not-determined |
| S1S_extracted_30124 | Not-determined |
| S1S_extracted_30131 | Not-determined |
| S1S_extracted_30137 | Not-determined |
| S1S_extracted_30144 | Not-determined |
| S1S_extracted_30150 | Not-determined |
| S1S_extracted_30151 | Not-determined |
| S1S_extracted_30153 | Not-determined |
| S1S_extracted_30173 | Not-determined |
| S1S_extracted_30198 | Not-determined |
| S1S_extracted_30219 | Not-determined |
| S1S_extracted_30224 | Low-quality |
| S1S_extracted_30235 | Not-determined |
| S1S_extracted_30261 | Not-determined |
| S1S_extracted_30264 | Not-determined |
| S1S_extracted_30274 | Not-determined |
| S1S_extracted_30283 | Not-determined |
| S1S_extracted_30289 | Not-determined |
| S1S_extracted_30300 | Not-determined |
| S1S_extracted_30306 | Not-determined |
| S1S_extracted_30323 | Not-determined |
| S1S_extracted_30328 | Not-determined |
| S1S_extracted_30336 | Low-quality |
| S1S_extracted_30340 | Not-determined |
| S1S_extracted_30351 | Not-determined |
| S1S_extracted_30352 | Not-determined |
| S1S_extracted_30374 | Not-determined |
| S1S_extracted_30379 | Not-determined |
| S1S_extracted_30382 | Not-determined |
| S1S_extracted_30385 | Not-determined |
| S1S_extracted_30421 | Not-determined |
| S1S_extracted_30435 | Not-determined |
| S1S_extracted_30442 | Not-determined |
| S1S_extracted_30462 | Not-determined |
| S1S_extracted_30481 | Not-determined |
| S1S_extracted_30495 | Not-determined |
| S1S_extracted_30496 | Not-determined |
| S1S_extracted_30515 | Low-quality |
| S1S_extracted_30525 | Not-determined |
| S1S_extracted_30526 | Not-determined |
| S1S_extracted_30533 | Not-determined |
| S1S_extracted_30542 | Not-determined |
| S1S_extracted_30544 | Not-determined |

| | |
|---|---|
| S15_extracted_30553 | Not-determined |
| S15_extracted_30559 | Not-determined |
| S15_extracted_30567 | Not-determined |
| S15_extracted_30577 | Not-determined |
| S15_extracted_30578 | Not-determined |
| S15_extracted_30587 | Low-quality |
| S15_extracted_30596 | Not-determined |
| S15_extracted_30616 | Not-determined |
| S15_extracted_30627 | Not-determined |
| S15_extracted_30632 | Not-determined |
| S15_extracted_30637 | Not-determined |
| S15_extracted_30638 | Not-determined |
| S15_extracted_30643 | Not-determined |
| S15_extracted_30651 | Not-determined |
| S15_extracted_30653 | Not-determined |
| S15_extracted_30669 | Not-determined |
| S15_extracted_30670 | Not-determined |
| S15_extracted_30674 | Not-determined |
| S15_extracted_30675 | Not-determined |
| S15_extracted_30676 | Not-determined |
| S15_extracted_30680 | Not-determined |
| S15_extracted_30681 | Not-determined |
| S15_extracted_30686 | Not-determined |
| S15_extracted_30692 | Not-determined |
| S15_extracted_30693 | Not-determined |
| S15_extracted_30703 | Low-quality |
| S15_extracted_30710 | Not-determined |
| S15_extracted_30714 | Not-determined |
| S15_extracted_30715 | Not-determined |
| S15_extracted_30719 | Low-quality |
| S15_extracted_30733 | Not-determined |
| S15_extracted_30741 | Not-determined |
| S15_extracted_30763 | Not-determined |
| S15_extracted_30768 | Not-determined |
| S15_extracted_30781 | Not-determined |
| S15_extracted_30792 | Low-quality |
| S15_extracted_30813 | Not-determined |
| S15_extracted_30814 | Not-determined |
| S15_extracted_30822 | Not-determined |
| S15_extracted_30827 | Not-determined |
| S15_extracted_30828 | Not-determined |
| S15_extracted_30838 | Not-determined |
| S15_extracted_30853 | Not-determined |
| S15_extracted_30855 | Not-determined |
| S15_extracted_30857 | Not-determined |
| S15_extracted_30876 | Not-determined |
| S15_extracted_30884 | Not-determined |
| S15_extracted_30895 | Not-determined |
| S15_extracted_30902 | Not-determined |
| S15_extracted_30906 | Not-determined |
| S15_extracted_30920 | Not-determined |
| S15_extracted_30925 | Low-quality |
| S15_extracted_30941 | Not-determined |
| S15_extracted_30959 | Not-determined |
| S15_extracted_30964 | Not-determined |
| S15_extracted_30975 | Not-determined |
| S15_extracted_30977 | Not-determined |
| S15_extracted_30985 | Not-determined |
| S15_extracted_30988 | Not-determined |
| S15_extracted_30989 | Not-determined |
| S15_extracted_31000 | Not-determined |
| S15_extracted_31010 | Not-determined |
| S15_extracted_31032 | Not-determined |
| S15_extracted_31038 | Not-determined |
| S15_extracted_31055 | Not-determined |
| S15_extracted_31066 | Not-determined |
| S15_extracted_31070 | Not-determined |
| S15_extracted_31083 | Not-determined |
| S15_extracted_31092 | Not-determined |
| S15_extracted_31096 | Not-determined |
| S15_extracted_31104 | Not-determined |
| S15_extracted_31112 | Not-determined |
| S15_extracted_31116 | Not-determined |
| S15_extracted_31119 | Not-determined |
| S15_extracted_31120 | Low-quality |
| S15_extracted_31137 | Not-determined |
| S15_extracted_31140 | Not-determined |
| S15_extracted_31169 | Low-quality |
| S15_extracted_31188 | Not-determined |
| S15_extracted_31197 | Not-determined |
| S15_extracted_31198 | Low-quality |
| S15_extracted_31203 | Not-determined |
| S15_extracted_31213 | Not-determined |
| S15_extracted_31218 | Not-determined |
| S15_extracted_31236 | Not-determined |
| S15_extracted_31248 | Not-determined |
| S15_extracted_31260 | Not-determined |
| S15_extracted_31277 | Not-determined |
| S15_extracted_31285 | Not-determined |
| S15_extracted_31287 | Not-determined |
| S15_extracted_31293 | Not-determined |
| S15_extracted_31294 | Not-determined |
| S15_extracted_31344 | Not-determined |
| S15_extracted_31352 | Not-determined |
| S15_extracted_31403 | Not-determined |
| S15_extracted_31410 | Not-determined |
| S15_extracted_31413 | Not-determined |
| S15_extracted_31415 | Not-determined |
| S15_extracted_31424 | Not-determined |
| S15_extracted_31437 | Not-determined |
| S15_extracted_31438 | Low-quality |
| S15_extracted_31448 | Not-determined |
| S15_extracted_31464 | Low-quality |
| S15_extracted_31465 | Not-determined |
| S15_extracted_31478 | Not-determined |
| S15_extracted_31481 | Not-determined |
| S15_extracted_31482 | Not-determined |
| S15_extracted_31484 | Low-quality |
| S15_extracted_31485 | Not-determined |
| S15_extracted_31492 | Not-determined |
| S15_extracted_31495 | Not-determined |
| S15_extracted_31513 | Not-determined |
| S15_extracted_31528 | Not-determined |
| S15_extracted_31542 | Not-determined |
| S15_extracted_31558 | Not-determined |
| S15_extracted_31590 | Not-determined |
| S15_extracted_31597 | Not-determined |
| S15_extracted_31606 | Not-determined |
| S15_extracted_31617 | Low-quality |
| S15_extracted_31635 | Not-determined |
| S15_extracted_31650 | Not-determined |
| S15_extracted_31651 | Not-determined |
| S15_extracted_31663 | Not-determined |
| S15_extracted_31668 | Low-quality |
| S15_extracted_31671 | Not-determined |
| S15_extracted_31683 | Not-determined |
| S15_extracted_31689 | Not-determined |
| S15_extracted_31698 | Not-determined |
| S15_extracted_31699 | Not-determined |
| S15_extracted_31716 | Not-determined |
| S15_extracted_31722 | Not-determined |
| S15_extracted_31728 | Not-determined |
| S15_extracted_31730 | Not-determined |
| S15_extracted_31735 | Not-determined |
| S15_extracted_31755 | Not-determined |
| S15_extracted_31785 | Not-determined |
| S15_extracted_31797 | Not-determined |
| S15_extracted_31800 | Low-quality |
| S15_extracted_31804 | Not-determined |
| S15_extracted_31817 | Not-determined |
| S15_extracted_31831 | Low-quality |
| S15_extracted_31863 | Low-quality |
| S15_extracted_31866 | Not-determined |
| S15_extracted_31872 | Not-determined |
| S15_extracted_31874 | Not-determined |
| S15_extracted_31875 | Not-determined |
| S15_extracted_31883 | Not-determined |
| S15_extracted_31897 | Not-determined |
| S15_extracted_31924 | Not-determined |
| S15_extracted_31930 | Not-determined |
| S15_extracted_31939 | Not-determined |
| S15_extracted_31941 | Not-determined |
| S15_extracted_31945 | Low-quality |
| S15_extracted_31950 | Not-determined |
| S15_extracted_31951 | Not-determined |
| S15_extracted_31983 | Not-determined |
| S15_extracted_32022 | Not-determined |
| S15_extracted_32024 | Not-determined |
| S15_extracted_32035 | Not-determined |
| S15_extracted_32042 | Not-determined |
| S15_extracted_32047 | Not-determined |
| S15_extracted_32048 | Low-quality |
| S15_extracted_32052 | Not-determined |
| S15_extracted_32063 | Not-determined |
| S15_extracted_32075 | Not-determined |
| S15_extracted_32084 | Low-quality |
| S15_extracted_32099 | Not-determined |
| S15_extracted_32103 | Not-determined |
| S15_extracted_32105 | Not-determined |
| S15_extracted_32108 | Not-determined |
| S15_extracted_32121 | Not-determined |
| S15_extracted_32123 | Not-determined |
| S15_extracted_32133 | Not-determined |
| S15_extracted_32153 | Not-determined |
| S15_extracted_32164 | Not-determined |
| S15_extracted_32168 | Not-determined |
| S15_extracted_32173 | Low-quality |
| S15_extracted_32174 | Not-determined |
| S15_extracted_32176 | Not-determined |
| S15_extracted_32177 | Not-determined |
| S15_extracted_32178 | Not-determined |
| S15_extracted_32180 | Not-determined |
| S15_extracted_32192 | Not-determined |
| S15_extracted_32197 | Not-determined |
| S15_extracted_32212 | Not-determined |
| S15_extracted_32217 | Not-determined |
| S15_extracted_32233 | Not-determined |
| S15_extracted_32240 | Not-determined |
| S15_extracted_32242 | Not-determined |
| S15_extracted_32245 | Not-determined |
| S15_extracted_32255 | Low-quality |
| S15_extracted_32263 | Not-determined |
| S15_extracted_32284 | Not-determined |
| S15_extracted_32286 | Low-quality |
| S15_extracted_32296 | Not-determined |
| S15_extracted_32307 | Not-determined |
| S15_extracted_32319 | Low-quality |
| S15_extracted_32323 | Not-determined |
| S15_extracted_32330 | Not-determined |
| S15_extracted_32332 | Not-determined |
| S15_extracted_32350 | Not-determined |
| S15_extracted_32356 | Not-determined |
| S15_extracted_32365 | Not-determined |
| S15_extracted_32368 | Not-determined |
| S15_extracted_32370 | Not-determined |
| S15_extracted_32399 | Low-quality |
| S15_extracted_32402 | Not-determined |
| S15_extracted_32406 | Not-determined |
| S15_extracted_32414 | Not-determined |
| S15_extracted_32423 | Not-determined |
| S15_extracted_32424 | Not-determined |
| S15_extracted_32426 | Not-determined |
| S15_extracted_32428 | Not-determined |
| S15_extracted_32434 | Not-determined |
| S15_extracted_32435 | Not-determined |
| S15_extracted_32438 | Not-determined |
| S15_extracted_32442 | Not-determined |
| S15_extracted_32443 | Not-determined |
| S15_extracted_32458 | Not-determined |
| S15_extracted_32460 | Not-determined |
| S15_extracted_32476 | Not-determined |
| S15_extracted_32484 | Not-determined |
| S15_extracted_32495 | Not-determined |
| S15_extracted_32500 | Not-determined |
| S15_extracted_32508 | Not-determined |
| S15_extracted_32514 | Low-quality |
| S15_extracted_32517 | Not-determined |
| S15_extracted_32527 | Not-determined |
| S15_extracted_32529 | Not-determined |
| S15_extracted_32539 | Not-determined |
| S15_extracted_32554 | Not-determined |
| S15_extracted_32560 | Not-determined |
| S15_extracted_32561 | Low-quality |
| S15_extracted_32563 | Not-determined |
| S15_extracted_32568 | Not-determined |
| S15_extracted_32587 | Not-determined |
| S15_extracted_32597 | Not-determined |
| S15_extracted_32616 | Not-determined |
| S15_extracted_32632 | Not-determined |
| S15_extracted_32638 | Not-determined |
| S15_extracted_32646 | Not-determined |
| S15_extracted_32657 | Not-determined |
| S15_extracted_32675 | Not-determined |
| S15_extracted_32681 | Not-determined |
| S15_extracted_32683 | Not-determined |
| S15_extracted_32691 | Not-determined |
| S15_extracted_32725 | Not-determined |
| S15_extracted_32736 | Not-determined |
| S15_extracted_32737 | Low-quality |
| S15_extracted_32764 | Not-determined |
| S15_extracted_32766 | Not-determined |
| S15_extracted_32796 | Not-determined |
| S15_extracted_32801 | Not-determined |
| S15_extracted_32809 | Low-quality |
| S15_extracted_32811 | Not-determined |
| S15_extracted_32813 | Not-determined |
| S15_extracted_32816 | Not-determined |
| S15_extracted_32831 | Not-determined |
| S15_extracted_32845 | Low-quality |
| S15_extracted_32879 | Low-quality |
| S15_extracted_32884 | Not-determined |
| S15_extracted_32887 | Not-determined |
| S15_extracted_32896 | Not-determined |
| S15_extracted_32910 | Not-determined |
| S15_extracted_32913 | Not-determined |
| S15_extracted_32916 | Not-determined |
| S15_extracted_32918 | Not-determined |
| S15_extracted_32929 | Not-determined |
| S15_extracted_32932 | Not-determined |
| S15_extracted_32939 | Not-determined |
| S15_extracted_32940 | Not-determined |
| S15_extracted_32954 | Not-determined |
| S15_extracted_32969 | Not-determined |
| S15_extracted_32972 | Not-determined |
| S15_extracted_32979 | Low-quality |
| S15_extracted_32988 | Not-determined |
| S15_extracted_32994 | Not-determined |
| S15_extracted_32995 | Not-determined |
| S15_extracted_33002 | Not-determined |
| S15_extracted_33004 | Not-determined |
| S15_extracted_33023 | Not-determined |
| S15_extracted_33049 | Not-determined |
| S15_extracted_33072 | Not-determined |
| S15_extracted_33073 | Not-determined |
| S15_extracted_33107 | Not-determined |
| S15_extracted_33109 | Not-determined |
| S15_extracted_33110 | Low-quality |
| S15_extracted_33112 | Not-determined |
| S15_extracted_33125 | Low-quality |
| S15_extracted_33134 | Not-determined |
| S15_extracted_33138 | Not-determined |
| S15_extracted_33168 | Not-determined |
| S15_extracted_33172 | Not-determined |
| S15_extracted_33178 | Not-determined |
| S15_extracted_33179 | Not-determined |
| S15_extracted_33192 | Not-determined |
| S15_extracted_33196 | Not-determined |
| S15_extracted_33197 | Not-determined |
| S15_extracted_33201 | Not-determined |
| S15_extracted_33205 | Low-quality |
| S15_extracted_33214 | Not-determined |
| S15_extracted_33215 | Not-determined |
| S15_extracted_33229 | Not-determined |
| S15_extracted_33236 | Not-determined |
| S15_extracted_33241 | Low-quality |
| S15_extracted_33256 | Not-determined |
| S15_extracted_33257 | Not-determined |
| S15_extracted_33260 | Not-determined |
| S15_extracted_33286 | Not-determined |
| S15_extracted_33290 | Not-determined |
| S15_extracted_33307 | Not-determined |
| S15_extracted_33308 | Low-quality |
| S15_extracted_33327 | Not-determined |
| S15_extracted_33329 | Not-determined |
| S15_extracted_33338 | Not-determined |
| S15_extracted_33342 | Not-determined |
| S15_extracted_33349 | Not-determined |
| S15_extracted_33350 | Not-determined |
| S15_extracted_33363 | Not-determined |
| S15_extracted_33371 | Not-determined |
| S15_extracted_33375 | Not-determined |
| S15_extracted_33376 | Not-determined |
| S15_extracted_33395 | Low-quality |
| S15_extracted_33408 | Not-determined |
| S15_extracted_33412 | Not-determined |
| S15_extracted_33414 | Not-determined |
| S15_extracted_33433 | Not-determined |
| S15_extracted_33445 | Not-determined |
| S15_extracted_33448 | Not-determined |
| S15_extracted_33464 | Not-determined |
| S15_extracted_33475 | Not-determined |
| S15_extracted_33482 | Low-quality |
| S15_extracted_33484 | Low-quality |
| S15_extracted_33495 | Low-quality |
| S15_extracted_33501 | Not-determined |
| S15_extracted_33502 | Not-determined |
| S10_vs_62230\|\|full | Medium-quality |
| S15_extracted_33517 | Not-determined |
| S15_extracted_33540 | Not-determined |
| S15_extracted_33549 | Low-quality |
| S15_extracted_33554 | Not-determined |
| S15_extracted_33560 | Not-determined |
| S15_extracted_33563 | Low-quality |
| S15_extracted_33567 | Not-determined |
| S15_extracted_33577 | Not-determined |
| S15_extracted_33598 | Not-determined |
| S15_extracted_33603 | Low-quality |
| S15_extracted_33608 | Not-determined |
| S15_extracted_33612 | Not-determined |
| S15_extracted_33623 | Not-determined |
| S15_extracted_33628 | Not-determined |
| S15_extracted_33631 | Not-determined |
| S15_extracted_33637 | Not-determined |
| S15_extracted_33644 | Not-determined |
| S15_extracted_33649 | Not-determined |
| S15_extracted_33664 | Not-determined |
| S15_extracted_33666 | Not-determined |
| S15_extracted_33667 | Not-determined |
| S15_extracted_33674 | Not-determined |
| S15_extracted_33683 | Not-determined |
| S15_extracted_33690 | Not-determined |
| S15_extracted_33720 | Not-determined |
| S15_extracted_33732 | Low-quality |
| S15_extracted_33741 | Not-determined |
| S15_extracted_33760 | Not-determined |
| S15_extracted_33761 | Not-determined |
| S15_extracted_33771 | Not-determined |
| S15_extracted_33773 | Not-determined |
| S15_extracted_33775 | Not-determined |
| S15_extracted_33779 | Not-determined |
| S15_extracted_33785 | Not-determined |
| S15_extracted_33789 | Not-determined |
| S15_extracted_33826 | Not-determined |
| S15_extracted_33830 | Not-determined |
| S15_extracted_33859 | Not-determined |
| S15_extracted_33867 | Not-determined |
| S15_extracted_33903 | Not-determined |
| S15_extracted_33914 | Not-determined |
| S15_extracted_33924 | Not-determined |
| S15_extracted_33932 | Not-determined |
| S15_extracted_33941 | Not-determined |
| S15_extracted_33942 | Not-determined |
| S15_extracted_33953 | Not-determined |
| S15_extracted_33961 | Low-quality |
| S15_extracted_33962 | Not-determined |
| S15_extracted_33972 | Not-determined |
| S15_extracted_33973 | Not-determined |
| S15_extracted_33987 | Not-determined |
| S15_extracted_33991 | Not-determined |
| S15_extracted_34005 | Not-determined |
| S15_extracted_34007 | Not-determined |
| S15_extracted_34009 | Not-determined |
| S15_extracted_34014 | Not-determined |
| S15_extracted_34021 | Not-determined |
| S15_extracted_34027 | Not-determined |
| S15_extracted_34040 | Not-determined |
| S15_extracted_34045 | Not-determined |
| S15_extracted_34051 | Low-quality |
| S15_extracted_34054 | Not-determined |
| S15_extracted_34063 | Low-quality |
| S15_extracted_34080 | Not-determined |
| S15_extracted_34089 | Not-determined |
| S15_extracted_34090 | Not-determined |
| S15_extracted_34104 | Not-determined |
| S15_extracted_34107 | Not-determined |
| S15_extracted_34118 | Not-determined |
| S15_extracted_34124 | Low-quality |
| S15_extracted_34158 | Not-determined |
| S15_extracted_34165 | Not-determined |
| S15_extracted_34169 | Not-determined |
| S15_extracted_34174 | Not-determined |
| S15_extracted_34178 | Not-determined |
| S15_extracted_34188 | Not-determined |
| S15_extracted_34190 | Not-determined |
| S15_extracted_34199 | Not-determined |
| S15_extracted_34214 | Not-determined |
| S15_extracted_34216 | Not-determined |
| S15_extracted_34223 | Not-determined |
| S15_extracted_34228 | Not-determined |
| S15_extracted_34237 | Not-determined |
| S15_extracted_34256 | Not-determined |
| S15_extracted_34270 | Not-determined |
| S15_extracted_34273 | Not-determined |
| S15_extracted_34303 | Not-determined |
| S15_extracted_34313 | Low-quality |
| S15_extracted_34316 | Not-determined |
| S15_extracted_34325 | Not-determined |
| S15_extracted_34333 | Low-quality |
| S15_extracted_34363 | Not-determined |
| S15_extracted_34368 | Low-quality |
| S15_extracted_34369 | Not-determined |
| S15_extracted_34370 | Low-quality |
| S15_extracted_34402 | Not-determined |
| S15_extracted_34404 | Not-determined |
| S15_extracted_34412 | Not-determined |
| S15_extracted_34425 | Low-quality |
| S15_extracted_34428 | Not-determined |
| S15_extracted_34430 | Not-determined |
| S15_extracted_34432 | Not-determined |
| S15_extracted_34440 | Not-determined |
| S15_extracted_34476 | Not-determined |
| S15_extracted_34477 | Not-determined |
| S15_extracted_34488 | Not-determined |
| S15_extracted_34491 | Not-determined |
| S15_extracted_34492 | Not-determined |
| S15_extracted_34494 | Not-determined |
| S15_extracted_34505 | Low-quality |
| S15_extracted_34508 | Low-quality |
| S15_extracted_34510 | Low-quality |
| S15_extracted_34512 | Not-determined |
| S15_extracted_34529 | Not-determined |
| S15_extracted_34534 | Not-determined |
| S15_extracted_34537 | Not-determined |
| S15_extracted_34548 | Not-determined |
| S15_extracted_34565 | Not-determined |
| S15_extracted_34567 | Not-determined |
| S15_extracted_34576 | Not-determined |
| S15_extracted_34577 | Not-determined |
| S15_extracted_34578 | Not-determined |
| S15_extracted_34600 | Not-determined |
| S15_extracted_34606 | Not-determined |
| S15_extracted_34617 | Low-quality |
| S15_extracted_34629 | Not-determined |
| S15_extracted_34647 | Not-determined |
| S15_extracted_34669 | Not-determined |
| S15_extracted_34671 | Not-determined |
| S15_extracted_34691 | Not-determined |
| S15_extracted_34714 | Not-determined |
| S15_extracted_34729 | Not-determined |
| S15_extracted_34732 | Not-determined |
| S15_extracted_34744 | Not-determined |
| S15_extracted_34751 | Not-determined |
| S15_extracted_34786 | Not-determined |
| S15_extracted_34795 | Not-determined |
| S15_extracted_34796 | Not-determined |
| S15_extracted_34800 | Low-quality |
| S15_extracted_34810 | Not-determined |
| S15_extracted_34832 | Not-determined |
| S15_extracted_34833 | Not-determined |
| S15_extracted_34839 | Not-determined |
| S15_extracted_34854 | Low-quality |
| S15_extracted_34855 | Not-determined |
| S15_extracted_34860 | Not-determined |
| S15_extracted_34890 | Not-determined |
| S15_extracted_34897 | Not-determined |
| S15_extracted_34900 | Not-determined |
| S15_extracted_34909 | Not-determined |
| S15_extracted_34910 | Not-determined |
| S15_extracted_34920 | Not-determined |
| S15_extracted_34931 | Not-determined |
| S15_extracted_34936 | Not-determined |
| S15_extracted_34945 | Low-quality |
| S15_extracted_34951 | Not-determined |
| S15_extracted_34955 | Not-determined |
| S15_extracted_34972 | Not-determined |
| S15_extracted_34978 | Low-quality |
| S15_extracted_35000 | Not-determined |
| S15_extracted_35004 | Not-determined |
| S15_extracted_35014 | Not-determined |
| S15_extracted_35024 | Not-determined |
| S15_extracted_35027 | Not-determined |
| S15_extracted_35031 | Not-determined |
| S15_extracted_35038 | Not-determined |
| S15_extracted_35045 | Not-determined |
| S15_extracted_35061 | Not-determined |
| S15_extracted_35066 | Not-determined |
| S15_extracted_35076 | Not-determined |
| S15_extracted_35084 | Low-quality |
| S15_extracted_35093 | Not-determined |
| S15_extracted_35096 | Not-determined |
| S15_extracted_35098 | Not-determined |
| S15_extracted_35135 | Low-quality |
| S15_extracted_35146 | Not-determined |
| S15_extracted_35157 | Not-determined |
| S15_extracted_35161 | Not-determined |
| S15_extracted_35168 | Not-determined |
| S15_extracted_35192 | Not-determined |
| S15_extracted_35217 | Not-determined |
| S15_extracted_35218 | Not-determined |
| S15_extracted_35228 | Not-determined |
| S15_extracted_35233 | Not-determined |
| S15_extracted_35246 | Low-quality |
| S15_extracted_35257 | Not-determined |
| S15_extracted_35268 | Low-quality |
| S15_extracted_35309 | Low-quality |
| S15_extracted_35324 | Not-determined |
| S15_extracted_35329 | Not-determined |
| S15_extracted_35333 | Not-determined |
| S15_extracted_35339 | Low-quality |
| S15_extracted_35357 | Not-determined |
| S15_extracted_35369 | Not-determined |
| S15_extracted_35382 | Not-determined |
| S15_extracted_35385 | Not-determined |
| S15_extracted_35390 | Not-determined |
| S15_extracted_35398 | Not-determined |
| S15_extracted_35408 | Not-determined |
| S15_extracted_35411 | Not-determined |
| S15_extracted_35423 | Not-determined |
| S15_extracted_35486 | Not-determined |
| S15_extracted_35487 | Low-quality |
| S15_extracted_35492 | Not-determined |
| S15_extracted_35500 | Not-determined |
| S15_extracted_35534 | Not-determined |
| S15_extracted_35539 | Not-determined |
| S15_extracted_35546 | Not-determined |
| S15_extracted_35565 | Not-determined |
| S15_extracted_35569 | Not-determined |
| S15_extracted_35599 | Not-determined |
| S15_extracted_35612 | Not-determined |
| S15_extracted_35613 | Not-determined |
| S15_extracted_35628 | Not-determined |
| S15_extracted_35629 | Not-determined |
| S15_extracted_35632 | Not-determined |
| S15_extracted_35635 | Not-determined |
| S15_extracted_35672 | Not-determined |
| S15_extracted_35690 | Not-determined |
| S15_extracted_35697 | Not-determined |
| S15_extracted_35700 | Not-determined |
| S15_extracted_35714 | Not-determined |
| S15_extracted_35725 | Not-determined |
| S15_extracted_35726 | Not-determined |
| S15_extracted_35731 | Not-determined |
| S15_extracted_35736 | Not-determined |
| S15_extracted_35739 | Not-determined |
| S15_extracted_35740 | Not-determined |
| S15_extracted_35750 | Not-determined |
| S15_extracted_35754 | Not-determined |
| S15_extracted_35762 | Not-determined |
| S15_extracted_35763 | Not-determined |
| S15_extracted_35767 | Low-quality |
| S15_extracted_35784 | Not-determined |
| S15_extracted_35796 | Not-determined |
| S15_extracted_35802 | Not-determined |
| S15_extracted_35812 | Low-quality |
| S15_extracted_35816 | Not-determined |
| S15_extracted_35820 | Not-determined |
| S15_extracted_35827 | Not-determined |
| S15_extracted_35839 | Not-determined |
| S15_extracted_35843 | Not-determined |
| S15_extracted_35844 | Not-determined |
| S15_extracted_35847 | Not-determined |
| S15_extracted_35860 | Low-quality |
| S15_extracted_35864 | Not-determined |
| S15_extracted_35870 | Not-determined |
| S15_extracted_35875 | Not-determined |
| S15_extracted_35877 | Not-determined |
| S15_extracted_35907 | Not-determined |
| S15_extracted_35927 | Not-determined |
| S15_extracted_35951 | Not-determined |

| | |
|---|---|
| S15_extracted_35958 | Not-determined |
| S15_extracted_35966 | Low-quality |
| S15_extracted_35980 | Low-quality |
| S15_extracted_35987 | Not-determined |
| S15_extracted_36001 | Not-determined |
| S15_extracted_36023 | Not-determined |
| S15_extracted_36042 | Not-determined |
| S15_extracted_36047 | Not-determined |
| S15_extracted_36049 | Not-determined |
| S15_extracted_36056 | Not-determined |
| S15_extracted_36065 | Not-determined |
| S15_extracted_36074 | Not-determined |
| S15_extracted_36077 | Not-determined |
| S15_extracted_36099 | Not-determined |
| S15_extracted_36110 | Not-determined |
| S15_extracted_36124 | Not-determined |
| S15_extracted_36153 | Not-determined |
| S15_extracted_36155 | Not-determined |
| S15_extracted_36165 | Not-determined |
| S15_extracted_36174 | Not-determined |
| S15_extracted_36178 | Not-determined |
| S15_extracted_36186 | Low-quality |
| S15_extracted_36187 | Not-determined |
| S15_extracted_36196 | Low-quality |
| S15_extracted_36210 | Low-quality |
| S15_extracted_36218 | Not-determined |
| S15_extracted_36230 | Not-determined |
| S15_extracted_36236 | Not-determined |
| S15_extracted_36244 | Not-determined |
| S15_extracted_36245 | Not-determined |
| S15_extracted_36246 | Not-determined |
| S15_extracted_36255 | Not-determined |
| S15_extracted_36256 | Not-determined |
| S15_extracted_36261 | Not-determined |
| S15_extracted_36274 | Not-determined |
| S15_extracted_36289 | Not-determined |
| S15_extracted_36291 | Not-determined |
| S15_extracted_36292 | Not-determined |
| S15_extracted_36296 | Low-quality |
| S15_extracted_36324 | Not-determined |
| S15_extracted_36329 | Low-quality |
| S15_extracted_36343 | Low-quality |
| S15_extracted_36346 | Not-determined |
| S15_extracted_36350 | Not-determined |
| S15_extracted_36358 | Not-determined |
| S15_extracted_36361 | Not-determined |
| S15_extracted_36373 | Low-quality |
| S15_extracted_36384 | Low-quality |
| S15_extracted_36389 | Not-determined |
| S15_extracted_36395 | Not-determined |
| S15_extracted_36399 | Not-determined |
| S15_extracted_36405 | Not-determined |
| S15_extracted_36410 | Not-determined |
| S15_extracted_36411 | Not-determined |
| S15_extracted_36416 | Not-determined |
| S15_extracted_36428 | Low-quality |
| S15_extracted_36431 | Not-determined |
| S15_extracted_36444 | Low-quality |
| S15_extracted_36450 | Low-quality |
| S15_extracted_36470 | Low-quality |
| S15_extracted_36473 | Not-determined |
| S15_extracted_36474 | Not-determined |
| S15_extracted_36483 | Not-determined |
| S15_extracted_36489 | Not-determined |
| S15_extracted_36504 | Low-quality |
| S15_extracted_36533 | Not-determined |
| S15_extracted_36548 | Not-determined |
| S15_extracted_36556 | Not-determined |
| S15_extracted_36558 | Not-determined |
| S15_extracted_36566 | Not-determined |
| S15_extracted_36576 | Low-quality |
| S15_extracted_36588 | Not-determined |
| S15_extracted_36593 | Not-determined |
| S15_extracted_36595 | Not-determined |
| S15_extracted_36598 | Not-determined |
| S15_extracted_36609 | Not-determined |
| S15_extracted_36614 | Not-determined |
| S15_extracted_36619 | Not-determined |
| S15_extracted_36633 | Not-determined |
| S15_extracted_36638 | Not-determined |
| S15_extracted_36640 | Not-determined |
| S15_extracted_36641 | Low-quality |
| S15_extracted_36651 | Not-determined |
| S15_extracted_36654 | Not-determined |
| S15_extracted_36656 | Low-quality |
| S15_extracted_36685 | Not-determined |
| S15_extracted_36692 | Not-determined |
| S15_extracted_36713 | Not-determined |
| S15_extracted_36732 | Not-determined |
| S15_extracted_36734 | Not-determined |
| S15_extracted_36735 | Not-determined |
| S15_extracted_36750 | Not-determined |
| S15_extracted_36767 | Not-determined |
| S15_extracted_36770 | Low-quality |
| S15_extracted_36775 | Not-determined |
| S15_extracted_36787 | Low-quality |
| S15_extracted_36788 | Not-determined |
| S15_extracted_36792 | Not-determined |
| S15_extracted_36793 | Not-determined |
| S15_extracted_36794 | Low-quality |
| S15_extracted_36800 | Not-determined |
| S15_extracted_36821 | Not-determined |
| S15_extracted_36823 | Not-determined |
| S15_extracted_36833 | Low-quality |
| S15_extracted_36836 | Not-determined |
| S15_extracted_36844 | Not-determined |
| S15_extracted_36852 | Not-determined |
| S15_extracted_36853 | Not-determined |
| S15_extracted_36864 | Not-determined |
| S15_extracted_36874 | Low-quality |
| S15_extracted_36894 | Not-determined |
| S15_extracted_36895 | Low-quality |
| S15_extracted_36900 | Not-determined |
| S15_extracted_36905 | Not-determined |
| S15_extracted_36909 | Not-determined |
| S15_extracted_36912 | Not-determined |
| S15_extracted_36918 | Not-determined |
| S15_extracted_36943 | Not-determined |
| S15_extracted_36946 | Not-determined |
| S15_extracted_36947 | Not-determined |
| S15_extracted_36949 | Not-determined |
| S15_extracted_36965 | Not-determined |
| S15_extracted_36983 | Not-determined |
| S15_extracted_36996 | Not-determined |
| S15_extracted_37001 | Not-determined |
| S15_extracted_37005 | Not-determined |
| S15_extracted_37008 | Not-determined |
| S15_extracted_37015 | Not-determined |
| S15_extracted_37019 | Not-determined |
| S15_extracted_37021 | Not-determined |
| S15_extracted_37032 | Not-determined |
| S15_extracted_37038 | Not-determined |
| S15_extracted_37052 | Not-determined |
| S15_extracted_37053 | Not-determined |
| S15_extracted_37057 | Low-quality |
| S15_extracted_37062 | Low-quality |
| S15_extracted_37075 | Not-determined |
| S15_extracted_37088 | Low-quality |
| S15_extracted_37100 | Not-determined |
| S15_extracted_37102 | Not-determined |
| S15_extracted_37103 | Not-determined |
| S15_extracted_37109 | Not-determined |
| S15_extracted_37110 | Not-determined |
| S15_extracted_37116 | Low-quality |
| S15_extracted_37123 | Not-determined |
| S15_extracted_37130 | Not-determined |
| S15_extracted_37135 | Not-determined |
| S15_extracted_37141 | Low-quality |
| S15_extracted_37146 | Not-determined |
| S15_extracted_37151 | Not-determined |
| S15_extracted_37161 | Not-determined |
| S15_extracted_37168 | Not-determined |
| S15_extracted_37171 | Not-determined |
| S15_extracted_37176 | Not-determined |
| S15_extracted_37180 | Not-determined |
| S15_extracted_37181 | Not-determined |
| S15_extracted_37204 | Not-determined |
| S15_extracted_37213 | Not-determined |
| S15_extracted_37221 | Not-determined |
| S15_extracted_37222 | Not-determined |
| S15_extracted_37237 | Not-determined |
| S15_extracted_37255 | Not-determined |
| S15_extracted_37256 | Not-determined |
| S15_extracted_37263 | Low-quality |
| S15_extracted_37283 | Not-determined |
| S15_extracted_37288 | Not-determined |
| S15_extracted_37289 | Not-determined |
| S15_extracted_37292 | Not-determined |
| S15_extracted_37312 | Not-determined |
| S15_extracted_37332 | Not-determined |
| S15_extracted_37334 | Low-quality |
| S15_extracted_37354 | Not-determined |
| S15_extracted_37355 | Not-determined |
| S15_extracted_37367 | Not-determined |
| S15_extracted_37370 | Not-determined |
| S15_extracted_37376 | Not-determined |
| S15_extracted_37377 | Not-determined |
| S15_extracted_37389 | Low-quality |
| S15_extracted_37417 | Low-quality |
| S15_extracted_37418 | Not-determined |
| S15_extracted_37422 | Not-determined |
| S15_extracted_37441 | Not-determined |
| S15_extracted_37451 | Not-determined |
| S15_extracted_37453 | Not-determined |
| S15_extracted_37475 | Not-determined |
| S15_extracted_37483 | Not-determined |
| S15_extracted_37508 | Not-determined |
| S15_extracted_37516 | Not-determined |
| S15_extracted_37519 | Not-determined |
| S15_extracted_37526 | Not-determined |
| S15_extracted_37528 | Not-determined |
| S15_extracted_37532 | Not-determined |
| S15_extracted_37540 | Not-determined |
| S15_extracted_37542 | Not-determined |
| S15_extracted_37558 | Not-determined |
| S15_extracted_37562 | Low-quality |
| S15_extracted_37567 | Not-determined |
| S15_extracted_37572 | Not-determined |
| S15_extracted_37574 | Not-determined |
| S15_extracted_37581 | Not-determined |
| S15_extracted_37584 | Low-quality |
| S15_extracted_37588 | Not-determined |
| S15_extracted_37592 | Not-determined |
| S15_extracted_37593 | Not-determined |
| S15_extracted_37594 | Low-quality |
| S15_extracted_37600 | Not-determined |
| S15_extracted_37622 | Not-determined |
| S15_extracted_37627 | Low-quality |
| S15_extracted_37632 | Not-determined |
| S15_extracted_37643 | Not-determined |
| S15_extracted_37654 | Not-determined |
| S15_extracted_37658 | Not-determined |
| S15_extracted_37666 | Low-quality |
| S15_extracted_37688 | Not-determined |
| S15_extracted_37691 | Not-determined |
| S15_extracted_37711 | Not-determined |
| S15_extracted_37721 | Not-determined |
| S15_extracted_37730 | Not-determined |
| S15_extracted_37733 | Not-determined |
| S15_extracted_37748 | Not-determined |
| S15_extracted_37749 | Not-determined |
| S15_extracted_37751 | Not-determined |
| S15_extracted_37754 | Not-determined |
| S15_extracted_37765 | Not-determined |
| S15_extracted_37768 | Not-determined |
| S15_extracted_37781 | Not-determined |
| S15_extracted_37784 | Not-determined |
| S15_extracted_37807 | Not-determined |
| S15_extracted_37816 | Not-determined |
| S15_extracted_37823 | Not-determined |
| S15_extracted_37824 | Not-determined |
| S15_extracted_37825 | Not-determined |
| S15_extracted_37833 | Not-determined |
| S15_extracted_37835 | Not-determined |
| S15_extracted_37840 | Not-determined |
| S15_extracted_37846 | Not-determined |
| S15_extracted_37852 | Not-determined |
| S15_extracted_37853 | Not-determined |
| S15_extracted_37857 | Not-determined |
| S15_extracted_37882 | Not-determined |
| S15_extracted_37884 | Not-determined |
| S15_extracted_37891 | Low-quality |
| S15_extracted_37893 | Not-determined |
| S15_extracted_37894 | Not-determined |
| S15_extracted_37898 | Not-determined |
| S15_extracted_37901 | Not-determined |
| S15_extracted_37919 | Not-determined |
| S15_extracted_37938 | Not-determined |
| S15_extracted_37943 | Not-determined |
| S15_extracted_37946 | Not-determined |
| S15_extracted_37957 | Not-determined |
| S15_extracted_37959 | Not-determined |
| S15_extracted_37962 | Not-determined |
| S15_extracted_37964 | Not-determined |
| S15_extracted_37971 | Not-determined |
| S15_extracted_37973 | Not-determined |
| S15_extracted_37976 | Not-determined |
| S15_extracted_37983 | Not-determined |
| S15_extracted_38007 | Not-determined |
| S15_extracted_38010 | Not-determined |
| S15_extracted_38023 | Not-determined |
| S15_extracted_38035 | Not-determined |
| S15_extracted_38045 | Low-quality |
| S15_extracted_38058 | Not-determined |
| S15_extracted_38079 | Not-determined |
| S15_extracted_38084 | Not-determined |
| S15_extracted_38091 | Not-determined |
| S15_extracted_38094 | Not-determined |
| S15_extracted_38102 | Not-determined |
| S15_extracted_38103 | Not-determined |
| S15_extracted_38106 | Not-determined |
| S15_extracted_38124 | Not-determined |
| S15_extracted_38134 | Not-determined |
| S15_extracted_38142 | Not-determined |
| S15_extracted_38146 | Not-determined |
| S15_extracted_38147 | Not-determined |
| S15_extracted_38158 | Not-determined |
| S15_extracted_38159 | Not-determined |
| S15_extracted_38161 | Not-determined |
| S15_extracted_38172 | Not-determined |
| S15_extracted_38177 | Not-determined |
| S15_extracted_38178 | Not-determined |
| S15_extracted_38183 | Low-quality |
| S15_extracted_38192 | Not-determined |
| S15_extracted_38193 | Not-determined |
| S15_extracted_38229 | Not-determined |
| S15_extracted_38231 | Not-determined |
| S15_extracted_38251 | Low-quality |
| S15_extracted_38270 | Low-quality |
| S15_extracted_38277 | Not-determined |
| S15_extracted_38282 | Not-determined |
| S15_extracted_38296 | Not-determined |
| S15_extracted_38318 | Not-determined |
| S15_extracted_38334 | Not-determined |
| S15_extracted_38338 | Not-determined |
| S15_extracted_38342 | Not-determined |
| S15_extracted_38345 | Low-quality |
| S15_extracted_38349 | Not-determined |
| S15_extracted_38362 | Not-determined |
| S15_extracted_38386 | Not-determined |
| S15_extracted_38387 | Not-determined |
| S15_extracted_38392 | Not-determined |
| S15_extracted_38412 | Not-determined |
| S15_extracted_38432 | Not-determined |
| S15_extracted_38434 | Not-determined |
| S15_extracted_38450 | Not-determined |
| S15_extracted_38454 | Not-determined |
| S15_extracted_38464 | Not-determined |
| S15_extracted_38475 | Not-determined |
| S15_extracted_38476 | Not-determined |
| S15_extracted_38478 | Not-determined |
| S15_extracted_38487 | Not-determined |
| S15_extracted_38497 | Not-determined |
| S15_extracted_38521 | Not-determined |
| S15_extracted_38522 | Not-determined |
| S15_extracted_38536 | Not-determined |
| S15_extracted_38539 | Not-determined |
| S15_extracted_38557 | Low-quality |
| S15_extracted_38560 | Not-determined |
| S15_extracted_38566 | Low-quality |
| S15_extracted_38568 | Not-determined |
| S15_extracted_38585 | Not-determined |
| S15_extracted_38587 | Not-determined |
| S15_extracted_38601 | Not-determined |
| S15_extracted_38606 | Not-determined |
| S15_extracted_38607 | Not-determined |
| S15_extracted_38626 | Low-quality |
| S15_extracted_38646 | Not-determined |
| S15_extracted_38652 | Not-determined |
| S15_extracted_38653 | Not-determined |
| S15_extracted_38654 | Not-determined |
| S15_extracted_38656 | Low-quality |
| S15_extracted_38661 | Not-determined |
| S15_extracted_38679 | Not-determined |
| S15_extracted_38680 | Not-determined |
| S15_extracted_38688 | Not-determined |
| S15_extracted_38690 | Not-determined |
| S15_extracted_38696 | Not-determined |
| S15_extracted_38714 | Not-determined |
| S15_extracted_38718 | Not-determined |
| S15_extracted_38722 | Not-determined |
| S15_extracted_38751 | Not-determined |
| S15_extracted_38754 | Low-quality |
| S15_extracted_38756 | Not-determined |
| S15_extracted_38765 | Not-determined |
| S15_extracted_38771 | Not-determined |
| S15_extracted_38774 | Not-determined |
| S15_extracted_38775 | Not-determined |
| S15_extracted_38778 | Not-determined |
| S15_extracted_38779 | Not-determined |
| S15_extracted_38780 | Low-quality |
| S15_extracted_38782 | Low-quality |
| S15_extracted_38791 | Not-determined |
| S15_extracted_38802 | Not-determined |
| S15_extracted_38815 | Not-determined |
| S15_extracted_38830 | Not-determined |
| S15_extracted_38831 | Not-determined |
| S15_extracted_38842 | Not-determined |
| S15_extracted_38844 | Not-determined |
| S15_extracted_38851 | Not-determined |
| S15_extracted_38863 | Not-determined |
| S15_extracted_38867 | Not-determined |
| S15_extracted_38869 | Not-determined |
| S15_extracted_38878 | Not-determined |
| S15_extracted_38881 | Low-quality |
| S15_extracted_38894 | Not-determined |
| S15_extracted_38895 | Not-determined |
| S15_extracted_38900 | Low-quality |
| S15_extracted_38916 | Not-determined |
| S15_extracted_38920 | Not-determined |
| S15_extracted_38966 | Low-quality |
| S15_extracted_38977 | Not-determined |
| S15_extracted_38984 | Not-determined |
| S15_extracted_38985 | Not-determined |
| S15_extracted_38991 | Not-determined |
| S15_extracted_39006 | Not-determined |
| S15_extracted_39017 | Not-determined |
| S15_extracted_39018 | Low-quality |
| S15_extracted_39023 | Not-determined |
| S15_extracted_39036 | Low-quality |
| S15_extracted_39051 | Not-determined |
| S15_extracted_39058 | Not-determined |
| S15_extracted_39063 | Not-determined |
| S15_extracted_39086 | Low-quality |
| S15_extracted_39091 | Not-determined |
| S15_extracted_39096 | Not-determined |
| S15_extracted_39104 | Not-determined |
| S15_extracted_39123 | Not-determined |
| S15_extracted_39129 | Not-determined |
| S15_extracted_39156 | Not-determined |
| S15_extracted_39167 | Not-determined |
| S15_extracted_39171 | Not-determined |
| S15_extracted_39174 | Not-determined |
| S15_extracted_39186 | Low-quality |
| S15_extracted_39192 | Low-quality |
| S15_extracted_39193 | Not-determined |
| S15_extracted_39200 | Not-determined |
| S15_extracted_39202 | Not-determined |
| S15_extracted_39211 | Not-determined |
| S15_extracted_39223 | Not-determined |
| S15_extracted_39225 | Not-determined |
| S15_extracted_39252 | Low-quality |
| S15_extracted_39264 | Not-determined |
| S15_extracted_39265 | Low-quality |
| S15_extracted_39267 | Not-determined |
| S15_extracted_39270 | Not-determined |
| S15_extracted_39273 | Not-determined |
| S15_extracted_39276 | Not-determined |
| S15_extracted_39284 | Low-quality |
| S15_extracted_39288 | Not-determined |
| S15_extracted_39294 | Not-determined |
| S15_extracted_39299 | Low-quality |
| S15_extracted_39317 | Not-determined |
| S15_extracted_39335 | Not-determined |
| S15_extracted_39347 | Not-determined |
| S15_extracted_39374 | Not-determined |
| S15_extracted_39376 | Not-determined |
| S15_extracted_39380 | Not-determined |
| S15_extracted_39383 | Not-determined |
| S15_extracted_39387 | Not-determined |
| S15_extracted_39388 | Not-determined |
| S15_extracted_39395 | Not-determined |
| S15_extracted_39406 | Not-determined |
| S15_extracted_39420 | Not-determined |
| S15_extracted_39421 | Not-determined |
| S15_extracted_39441 | Not-determined |
| S15_extracted_39443 | Not-determined |
| S15_extracted_39462 | Not-determined |
| S15_extracted_39466 | Not-determined |
| S15_extracted_39471 | Not-determined |
| S15_extracted_39474 | Not-determined |
| S15_extracted_39477 | Not-determined |
| S15_extracted_39483 | Not-determined |
| S15_extracted_39493 | Not-determined |
| S15_extracted_39504 | Low-quality |
| S15_extracted_39527 | Not-determined |
| S15_extracted_39531 | Low-quality |
| S15_extracted_39533 | Not-determined |
| S15_extracted_39540 | Not-determined |
| S15_extracted_39543 | Not-determined |
| S15_extracted_39545 | Not-determined |
| S15_extracted_39550 | Not-determined |
| S15_extracted_39557 | Low-quality |
| S15_extracted_39566 | Not-determined |
| S15_extracted_39578 | Not-determined |
| S15_extracted_39585 | Not-determined |
| S15_extracted_39597 | Not-determined |
| S15_extracted_39600 | Not-determined |
| S15_extracted_39610 | Not-determined |
| S15_extracted_39619 | Not-determined |
| S15_extracted_39636 | Not-determined |
| S15_extracted_39639 | Not-determined |
| S15_extracted_39644 | Not-determined |
| S15_extracted_39647 | Not-determined |
| S15_extracted_39655 | Not-determined |
| S15_extracted_39662 | Not-determined |
| S15_extracted_39663 | Not-determined |
| S15_extracted_39697 | Not-determined |
| S15_extracted_39725 | Not-determined |
| S15_extracted_39727 | Not-determined |
| S15_extracted_39775 | Not-determined |
| S15_extracted_39791 | Not-determined |
| S15_extracted_39809 | Not-determined |
| S15_extracted_39825 | Not-determined |
| S15_extracted_39831 | Not-determined |
| S15_extracted_39834 | Not-determined |
| S15_extracted_39854 | Low-quality |
| S15_extracted_39879 | Not-determined |
| S15_extracted_39901 | Not-determined |
| S15_extracted_39936 | Not-determined |
| S15_extracted_39964 | Not-determined |
| S15_extracted_39988 | Not-determined |
| S15_extracted_39990 | Not-determined |
| S15_extracted_40008 | Not-determined |
| S15_extracted_40060 | Not-determined |
| S15_extracted_40067 | Not-determined |
| S15_extracted_40072 | Low-quality |
| S15_extracted_40075 | Not-determined |
| S15_extracted_40086 | Not-determined |
| S15_extracted_40094 | Not-determined |
| S15_extracted_40111 | Not-determined |
| S15_extracted_40121 | Not-determined |
| S15_extracted_40129 | Not-determined |
| S15_extracted_40147 | Not-determined |
| S15_extracted_40148 | Not-determined |
| S15_extracted_40155 | Not-determined |
| S15_extracted_40166 | Not-determined |
| S15_extracted_40174 | Not-determined |
| S15_extracted_40180 | Not-determined |
| S15_extracted_40184 | Not-determined |
| S15_extracted_40191 | Not-determined |
| S15_extracted_40199 | Not-determined |
| S15_extracted_40200 | Not-determined |
| S15_extracted_40210 | Not-determined |
| S15_extracted_40217 | Not-determined |
| S15_extracted_40218 | Low-quality |
| S15_extracted_40232 | Low-quality |
| S15_extracted_40235 | Low-quality |
| S15_extracted_40245 | Not-determined |
| S15_extracted_40270 | Not-determined |
| S15_extracted_40276 | Not-determined |
| S15_extracted_40287 | Not-determined |
| S15_extracted_40297 | Not-determined |
| S15_extracted_40305 | Not-determined |
| S15_extracted_40308 | Not-determined |
| S15_extracted_40322 | Not-determined |
| S15_extracted_40339 | Not-determined |
| S15_extracted_40344 | Not-determined |
| S15_extracted_40367 | Not-determined |
| S15_extracted_40368 | Not-determined |
| S15_extracted_40372 | Not-determined |
| S15_extracted_40375 | Low-quality |
| S15_extracted_40377 | Not-determined |
| S15_extracted_40379 | Not-determined |
| S15_extracted_40385 | Not-determined |
| S15_extracted_40406 | Not-determined |
| S15_extracted_40412 | Not-determined |
| S15_extracted_40414 | Not-determined |
| S15_extracted_40417 | Not-determined |
| S15_extracted_40438 | Not-determined |
| S15_extracted_40454 | Not-determined |
| S15_extracted_40456 | Not-determined |
| S15_extracted_40465 | Not-determined |
| S15_extracted_40468 | Not-determined |
| S15_extracted_40469 | Not-determined |
| S15_extracted_40470 | Not-determined |
| S15_extracted_40479 | Low-quality |
| S15_extracted_40482 | Not-determined |
| S15_extracted_40516 | Not-determined |
| S15_extracted_40526 | Not-determined |
| S15_extracted_40531 | Low-quality |
| S15_extracted_40553 | Low-quality |
| S15_extracted_40558 | Not-determined |
| S15_extracted_40563 | Not-determined |
| S15_extracted_40581 | Not-determined |
| S15_extracted_40584 | Not-determined |
| S15_extracted_40591 | Not-determined |
| S15_extracted_40592 | Not-determined |
| S15_extracted_40605 | Low-quality |
| S15_extracted_40622 | Not-determined |
| S15_extracted_40655 | Not-determined |
| S15_extracted_40657 | Not-determined |
| S15_extracted_40667 | Not-determined |
| S15_extracted_40673 | Not-determined |
| S15_extracted_40678 | Not-determined |
| S15_extracted_40679 | Not-determined |
| S15_extracted_40682 | Not-determined |
| S15_extracted_40698 | Not-determined |
| S15_extracted_40700 | Not-determined |
| S15_extracted_40704 | Not-determined |
| S15_extracted_40706 | Not-determined |
| S15_extracted_40712 | Low-quality |
| S15_extracted_40718 | Not-determined |
| S15_extracted_40721 | Low-quality |
| S15_extracted_40722 | Not-determined |
| S15_extracted_40723 | Not-determined |
| S15_extracted_40733 | Not-determined |
| S15_extracted_40735 | Not-determined |
| S15_extracted_40746 | Not-determined |
| S15_extracted_40748 | Not-determined |
| S15_extracted_40753 | Not-determined |
| S15_extracted_40774 | Not-determined |
| S15_extracted_40782 | Not-determined |
| S15_extracted_40801 | Not-determined |
| S15_extracted_40804 | Not-determined |
| S15_extracted_40807 | Not-determined |
| S15_extracted_40808 | Not-determined |
| S15_extracted_40819 | Not-determined |
| S15_extracted_40826 | Not-determined |
| S15_extracted_40907 | Not-determined |
| S15_extracted_40911 | Not-determined |
| S15_extracted_40922 | Not-determined |
| S15_extracted_40931 | Not-determined |
| S15_extracted_40951 | Not-determined |
| S15_extracted_40956 | Not-determined |
| S15_extracted_40963 | Not-determined |
| S15_extracted_40977 | Not-determined |
| S15_extracted_40982 | Not-determined |
| S15_extracted_40983 | Not-determined |
| S15_extracted_41002 | Not-determined |
| S15_extracted_41004 | Not-determined |
| S15_extracted_41014 | Not-determined |
| S15_extracted_41017 | Not-determined |
| S15_extracted_41022 | Low-quality |
| S15_extracted_41023 | Not-determined |
| S15_extracted_41033 | Not-determined |
| S15_extracted_41034 | Low-quality |
| S15_extracted_41043 | Not-determined |
| S15_extracted_41046 | Not-determined |

| | |
|---|---|
| S15_extracted_41054 | Not-determined |
| S15_extracted_41077 | Not-determined |
| S15_extracted_41092 | Low-quality |
| S15_extracted_41118 | Not-determined |
| S15_extracted_41121 | Low-quality |
| S15_extracted_41157 | Not-determined |
| S15_extracted_41163 | Not-determined |
| S15_extracted_41170 | Not-determined |
| S15_extracted_41173 | Not-determined |
| S15_extracted_41182 | Not-determined |
| S15_extracted_41190 | Not-determined |
| S15_extracted_41194 | Not-determined |
| S15_extracted_41201 | Not-determined |
| S15_extracted_41202 | Not-determined |
| S15_extracted_41204 | Not-determined |
| S15_extracted_41210 | Not-determined |
| S15_extracted_41214 | Not-determined |
| S15_extracted_41221 | Not-determined |
| S15_extracted_41223 | Not-determined |
| S15_extracted_41231 | Not-determined |
| S15_extracted_41263 | Not-determined |
| S15_extracted_41266 | Not-determined |
| S15_extracted_41268 | Not-determined |
| S15_extracted_41272 | Not-determined |
| S15_extracted_41274 | Not-determined |
| S15_extracted_41282 | Not-determined |
| S15_extracted_41287 | Not-determined |
| S15_extracted_41290 | Not-determined |
| S15_extracted_41293 | Not-determined |
| S15_extracted_41295 | Not-determined |
| S15_extracted_41296 | Not-determined |
| S15_extracted_41322 | Not-determined |
| S15_extracted_41329 | Not-determined |
| S15_extracted_41330 | Not-determined |
| S15_extracted_41332 | Not-determined |
| S15_extracted_41340 | Low-quality |
| S15_extracted_41353 | Low-quality |
| S15_extracted_41357 | Low-quality |
| S15_extracted_41373 | Not-determined |
| S15_extracted_41374 | Not-determined |
| S15_extracted_41384 | Not-determined |
| S15_extracted_41387 | Not-determined |
| S15_extracted_41392 | Not-determined |
| S15_extracted_41406 | Not-determined |
| S15_extracted_41408 | Not-determined |
| S15_extracted_41418 | Not-determined |
| S15_extracted_41421 | Not-determined |
| S15_extracted_41424 | Not-determined |
| S15_extracted_41433 | Not-determined |
| S15_extracted_41465 | Not-determined |
| S15_extracted_41471 | Not-determined |
| S15_extracted_41480 | Not-determined |
| S15_extracted_41484 | Not-determined |
| S15_extracted_41491 | Not-determined |
| S15_extracted_41502 | Not-determined |
| S15_extracted_41521 | Not-determined |
| S15_extracted_41522 | Not-determined |
| S15_extracted_41525 | Not-determined |
| S15_extracted_41527 | Not-determined |
| S15_extracted_41530 | Low-quality |
| S15_extracted_41533 | Not-determined |
| S15_extracted_41542 | Not-determined |
| S15_extracted_41546 | Not-determined |
| S15_extracted_41557 | Not-determined |
| S15_extracted_41559 | Not-determined |
| S15_extracted_41565 | Low-quality |
| S15_extracted_41574 | Not-determined |
| S15_extracted_41581 | Not-determined |
| S15_extracted_41582 | Low-quality |
| S15_extracted_41584 | Not-determined |
| S15_extracted_41588 | Low-quality |
| S15_extracted_41605 | Not-determined |
| S15_extracted_41630 | Not-determined |
| S15_extracted_41638 | Not-determined |
| S15_extracted_41642 | Not-determined |
| S15_extracted_41649 | Not-determined |
| S15_extracted_41658 | Not-determined |
| S15_extracted_41675 | Not-determined |
| S15_extracted_41676 | Not-determined |
| S15_extracted_41681 | Not-determined |
| S15_extracted_41697 | Low-quality |
| S15_extracted_41706 | Not-determined |
| S15_extracted_41707 | Not-determined |
| S15_extracted_41708 | Not-determined |
| S15_extracted_41724 | Low-quality |
| S15_extracted_41729 | Low-quality |
| S15_extracted_41739 | Not-determined |
| S15_extracted_41747 | Not-determined |
| S15_extracted_41755 | Low-quality |
| S15_extracted_41769 | Not-determined |
| S15_extracted_41771 | Not-determined |
| S15_extracted_41777 | Not-determined |
| S15_extracted_41795 | Not-determined |
| S15_extracted_41796 | Not-determined |
| S15_extracted_41801 | Not-determined |
| S15_extracted_41821 | Low-quality |
| S15_extracted_41830 | Not-determined |
| S15_extracted_41837 | Not-determined |
| S15_extracted_41839 | Not-determined |
| S15_extracted_41864 | Not-determined |
| S15_extracted_41870 | Low-quality |
| S15_extracted_41886 | Not-determined |
| S15_extracted_41887 | Not-determined |
| S15_extracted_41897 | Not-determined |
| S15_extracted_41903 | Not-determined |
| S15_extracted_41908 | Not-determined |
| S15_extracted_41918 | Not-determined |
| S15_extracted_41921 | Low-quality |
| S15_extracted_41922 | Not-determined |
| S15_extracted_41926 | Not-determined |
| S15_extracted_41937 | Not-determined |
| S15_extracted_41945 | Not-determined |
| S15_extracted_41953 | Not-determined |
| S7_vs_944\|\|full | Medium-quality |
| S15_extracted_41958 | Not-determined |
| S15_extracted_41963 | Not-determined |
| S15_extracted_41967 | Not-determined |
| S15_extracted_41976 | Not-determined |
| S15_extracted_41991 | Not-determined |
| S15_extracted_42002 | Not-determined |
| S15_extracted_42019 | Not-determined |
| S15_extracted_42026 | Low-quality |
| S15_extracted_42034 | Not-determined |
| S15_extracted_42038 | Not-determined |
| S15_extracted_42040 | Not-determined |
| S15_extracted_42046 | Not-determined |
| S15_extracted_42058 | Not-determined |
| S15_extracted_42059 | Not-determined |
| S15_extracted_42061 | Not-determined |
| S15_extracted_42078 | Not-determined |
| S15_extracted_42083 | Not-determined |
| S15_extracted_42086 | Not-determined |
| S15_extracted_42091 | Not-determined |
| S15_extracted_42117 | Not-determined |
| S15_extracted_42118 | Low-quality |
| S15_extracted_42123 | Not-determined |
| S15_extracted_42134 | Not-determined |
| S15_extracted_42150 | Not-determined |
| S15_extracted_42155 | Not-determined |
| S15_extracted_42168 | Not-determined |
| S15_extracted_42169 | Not-determined |
| S15_extracted_42193 | Not-determined |
| S15_extracted_42198 | Not-determined |
| S15_extracted_42199 | Not-determined |
| S15_extracted_42201 | Not-determined |
| S15_extracted_42213 | Not-determined |
| S15_extracted_42225 | Not-determined |
| S15_extracted_42239 | Not-determined |
| S15_extracted_42240 | Not-determined |
| S15_extracted_42244 | Not-determined |
| S15_extracted_42247 | Not-determined |
| S15_extracted_42248 | Not-determined |
| S15_extracted_42249 | Not-determined |
| S15_extracted_42268 | Low-quality |
| S15_extracted_42269 | Not-determined |
| S15_extracted_42282 | Not-determined |
| S15_extracted_42306 | Not-determined |
| S15_extracted_42313 | Low-quality |
| S15_extracted_42316 | Not-determined |
| S15_extracted_42323 | Not-determined |
| S15_extracted_42328 | Not-determined |
| S15_extracted_42334 | Not-determined |
| S15_extracted_42339 | Low-quality |
| S15_extracted_42342 | Not-determined |
| S15_extracted_42344 | Low-quality |
| S15_extracted_42392 | Low-quality |
| S15_extracted_42401 | Not-determined |
| S15_extracted_42403 | Not-determined |
| S15_extracted_42408 | Not-determined |
| S15_extracted_42417 | Not-determined |
| S15_extracted_42423 | Not-determined |
| S15_extracted_42433 | Not-determined |
| S15_extracted_42434 | Low-quality |
| S15_extracted_42445 | Not-determined |
| S15_extracted_42448 | Not-determined |
| S15_extracted_42453 | Not-determined |
| S15_extracted_42461 | Not-determined |
| S15_extracted_42474 | Not-determined |
| S15_extracted_42475 | Not-determined |
| S15_extracted_42476 | Not-determined |
| S15_extracted_42477 | Not-determined |
| S15_extracted_42478 | Not-determined |
| S15_extracted_42497 | Not-determined |
| S15_extracted_42502 | Not-determined |
| S15_extracted_42511 | Not-determined |
| S15_extracted_42520 | Not-determined |
| S15_extracted_42526 | Not-determined |
| S15_extracted_42532 | Not-determined |
| S15_extracted_42538 | Not-determined |
| S15_extracted_42545 | Not-determined |
| S15_extracted_42581 | Not-determined |
| S15_extracted_42601 | Not-determined |
| S15_extracted_42617 | Not-determined |
| S15_extracted_42622 | Not-determined |
| S15_extracted_42641 | Not-determined |
| S15_extracted_42648 | Not-determined |
| S15_extracted_42651 | Not-determined |
| S15_extracted_42659 | Not-determined |
| S15_extracted_42663 | Not-determined |
| S15_extracted_42665 | Not-determined |
| S15_extracted_42682 | Not-determined |
| S15_extracted_42690 | Not-determined |
| S15_extracted_42696 | Not-determined |
| S15_extracted_42697 | Not-determined |
| S15_extracted_42699 | Not-determined |
| S15_extracted_42701 | Not-determined |
| S15_extracted_42707 | Not-determined |
| S15_extracted_42717 | Not-determined |
| S15_extracted_42719 | Not-determined |
| S15_extracted_42736 | Not-determined |
| S15_extracted_42742 | Not-determined |
| S15_extracted_42755 | Not-determined |
| S15_extracted_42768 | Not-determined |
| S15_extracted_42794 | Low-quality |
| S15_extracted_42796 | Not-determined |
| S15_extracted_42811 | Not-determined |
| S15_extracted_42860 | Not-determined |
| S15_extracted_42875 | Not-determined |
| S15_extracted_42902 | Not-determined |
| S15_extracted_42906 | Not-determined |
| S15_extracted_42908 | Not-determined |
| S15_extracted_42936 | Not-determined |
| S15_extracted_42941 | Not-determined |
| S15_extracted_42942 | Not-determined |
| S15_extracted_42943 | Low-quality |
| S15_extracted_42954 | Not-determined |
| S15_extracted_42960 | Not-determined |
| S15_extracted_42962 | Low-quality |
| S15_extracted_42974 | Not-determined |
| S15_extracted_42988 | Not-determined |
| S15_extracted_43001 | Not-determined |
| S15_extracted_43005 | Not-determined |
| S15_extracted_43028 | Not-determined |
| S15_extracted_43029 | Not-determined |
| S15_extracted_43038 | Not-determined |
| S15_extracted_43048 | Not-determined |
| S15_extracted_43061 | Not-determined |
| S15_extracted_43066 | Not-determined |
| S15_extracted_43070 | Not-determined |
| S15_extracted_43075 | Not-determined |
| S15_extracted_43080 | Not-determined |
| S15_extracted_43086 | Not-determined |
| S15_extracted_43094 | Not-determined |
| S15_extracted_43101 | Not-determined |
| S15_extracted_43107 | Not-determined |
| S15_extracted_43112 | Not-determined |
| S15_extracted_43116 | Not-determined |
| S15_extracted_43123 | Not-determined |
| S15_extracted_43126 | Not-determined |
| S15_extracted_43147 | Not-determined |
| S15_extracted_43152 | Not-determined |
| S15_extracted_43167 | Not-determined |
| S15_extracted_43178 | Not-determined |
| S15_extracted_43182 | Not-determined |
| S15_extracted_43210 | Not-determined |
| S15_extracted_43211 | Not-determined |
| S15_extracted_43212 | Not-determined |
| S15_extracted_43220 | Not-determined |
| S15_extracted_43232 | Not-determined |
| S15_extracted_43248 | Not-determined |
| S15_extracted_43256 | Not-determined |
| S15_extracted_43262 | Not-determined |
| S15_extracted_43272 | Not-determined |
| S15_extracted_43304 | Not-determined |
| S15_extracted_43312 | Not-determined |
| S15_extracted_43314 | Low-quality |
| S15_extracted_43320 | Not-determined |
| S15_extracted_43321 | Not-determined |
| S15_extracted_43343 | Low-quality |
| S15_extracted_43344 | Not-determined |
| S15_extracted_43360 | Low-quality |
| S15_extracted_43368 | Not-determined |
| S15_extracted_43372 | Not-determined |
| S15_extracted_43397 | Not-determined |
| S15_extracted_43402 | Not-determined |
| S15_extracted_43405 | Not-determined |
| S15_extracted_43434 | Not-determined |
| S15_extracted_43435 | Low-quality |
| S15_extracted_43446 | Not-determined |
| S15_extracted_43451 | Not-determined |
| S15_extracted_43452 | Low-quality |
| S15_extracted_43457 | Low-quality |
| S15_extracted_43484 | Low-quality |
| S15_extracted_43502 | Not-determined |
| S15_extracted_43515 | Not-determined |
| S15_extracted_43528 | Not-determined |
| S15_extracted_43534 | Low-quality |
| S15_extracted_43538 | Not-determined |
| S15_extracted_43553 | Not-determined |
| S15_extracted_43567 | Not-determined |
| S15_extracted_43568 | Not-determined |
| S15_extracted_43574 | Not-determined |
| S15_extracted_43575 | Not-determined |
| S15_extracted_43577 | Not-determined |
| S15_extracted_43582 | Not-determined |
| S15_extracted_43583 | Not-determined |
| S15_extracted_43610 | Not-determined |
| S15_extracted_43648 | Not-determined |
| S15_extracted_43661 | Not-determined |
| S15_extracted_43665 | Not-determined |
| S15_extracted_43687 | Not-determined |
| S15_extracted_43690 | Not-determined |
| S15_extracted_43722 | Not-determined |
| S15_extracted_43728 | Not-determined |
| S15_extracted_43737 | Low-quality |
| S15_extracted_43763 | Not-determined |
| S15_extracted_43777 | Not-determined |
| S15_extracted_43778 | Not-determined |
| S15_extracted_43779 | Not-determined |
| S15_extracted_43780 | Not-determined |
| S15_extracted_43791 | Not-determined |
| S15_extracted_43793 | Not-determined |
| S15_extracted_43803 | Not-determined |
| S15_extracted_43810 | Low-quality |
| S15_extracted_43818 | Not-determined |
| S15_extracted_43829 | Not-determined |
| S15_extracted_43844 | Not-determined |
| S15_extracted_43848 | Not-determined |
| S15_extracted_43881 | Not-determined |
| S15_extracted_43886 | Low-quality |
| S15_extracted_43888 | Not-determined |
| S15_extracted_43903 | Not-determined |
| S15_extracted_43924 | Not-determined |
| S15_extracted_43926 | Not-determined |
| S15_extracted_43927 | Low-quality |
| S15_extracted_43934 | Not-determined |
| S15_extracted_43936 | Not-determined |
| S15_extracted_43939 | Not-determined |
| S15_extracted_43941 | Not-determined |
| S15_extracted_43942 | Not-determined |
| S15_extracted_43953 | Not-determined |
| S15_extracted_43958 | Not-determined |
| S15_extracted_43962 | Not-determined |
| S15_extracted_43972 | Not-determined |
| S15_extracted_43985 | Not-determined |
| S15_extracted_43990 | Not-determined |
| S15_extracted_43991 | Not-determined |
| S15_extracted_43997 | Not-determined |
| S15_extracted_44024 | Not-determined |
| S15_extracted_44029 | Not-determined |
| S15_extracted_44031 | Not-determined |
| S15_extracted_44032 | Not-determined |
| S15_extracted_44037 | Not-determined |
| S15_extracted_44044 | Not-determined |
| S15_extracted_44046 | Not-determined |
| S15_extracted_44052 | Not-determined |
| S15_extracted_44054 | Not-determined |
| S15_extracted_44057 | Not-determined |
| S15_extracted_44064 | Not-determined |
| S15_extracted_44072 | Not-determined |
| S15_extracted_44075 | Not-determined |
| S15_extracted_44085 | Not-determined |
| S15_extracted_44095 | Low-quality |
| S15_extracted_44123 | Not-determined |
| S15_extracted_44137 | Not-determined |
| S15_extracted_44141 | Not-determined |
| S15_extracted_44146 | Not-determined |
| S15_extracted_44151 | Not-determined |
| S15_extracted_44153 | Low-quality |
| S15_extracted_44168 | Not-determined |
| S15_extracted_44172 | Not-determined |
| S15_extracted_44177 | Not-determined |
| S15_extracted_44186 | Not-determined |
| S15_extracted_44195 | Not-determined |
| S15_extracted_44200 | Not-determined |
| S15_extracted_44204 | Not-determined |
| S15_extracted_44209 | Not-determined |
| S15_extracted_44221 | Not-determined |
| S15_extracted_44224 | Not-determined |
| S15_extracted_44228 | Not-determined |
| S15_extracted_44237 | Not-determined |
| S15_extracted_44264 | Not-determined |
| S15_extracted_44266 | Not-determined |
| S15_extracted_44289 | Low-quality |
| S15_extracted_44290 | Low-quality |
| S15_extracted_44293 | Not-determined |
| S15_extracted_44297 | Not-determined |
| S15_extracted_44304 | Not-determined |
| S15_extracted_44313 | Not-determined |
| S15_extracted_44314 | Not-determined |
| S15_extracted_44319 | Low-quality |
| S15_extracted_44337 | Low-quality |
| S15_extracted_44349 | Not-determined |
| S15_extracted_44354 | Not-determined |
| S15_extracted_44357 | Not-determined |
| S15_extracted_44362 | Low-quality |
| S15_extracted_44371 | Not-determined |
| S15_extracted_44379 | Not-determined |
| S15_extracted_44388 | Not-determined |
| S15_extracted_44392 | Not-determined |
| S15_extracted_44428 | Not-determined |
| S15_extracted_44437 | Low-quality |
| S15_extracted_44438 | Not-determined |
| S15_extracted_44466 | Not-determined |
| S15_extracted_44469 | Not-determined |
| S15_extracted_44495 | Not-determined |
| S15_extracted_44516 | Not-determined |
| S15_extracted_44532 | Not-determined |
| S15_extracted_44533 | Not-determined |
| S15_extracted_44541 | Not-determined |
| S15_extracted_44544 | Low-quality |
| S15_extracted_44545 | Not-determined |
| S15_extracted_44547 | Low-quality |
| S15_extracted_44550 | Not-determined |
| S15_extracted_44555 | Not-determined |
| S15_extracted_44558 | Not-determined |
| S15_extracted_44573 | Not-determined |
| S15_extracted_44575 | Not-determined |
| S15_extracted_44578 | Not-determined |
| S15_extracted_44588 | Not-determined |
| S15_extracted_44595 | Not-determined |
| S15_extracted_44618 | Low-quality |
| S15_extracted_44623 | Not-determined |
| S15_extracted_44625 | Not-determined |
| S15_extracted_44672 | Not-determined |
| S15_extracted_44698 | Not-determined |
| S15_extracted_44708 | Not-determined |
| S15_extracted_44714 | Not-determined |
| S15_extracted_44763 | Not-determined |
| S15_extracted_44766 | Not-determined |
| S15_extracted_44800 | Not-determined |
| S15_extracted_44801 | Not-determined |
| S15_extracted_44805 | Not-determined |
| S15_extracted_44806 | Not-determined |
| S15_extracted_44819 | Not-determined |
| S15_extracted_44821 | Low-quality |
| S15_extracted_44822 | Not-determined |
| S15_extracted_44833 | Not-determined |
| S15_extracted_44843 | Low-quality |
| S15_extracted_44850 | Not-determined |
| S15_extracted_44852 | Not-determined |
| S15_extracted_44868 | Not-determined |
| S15_extracted_44875 | Not-determined |
| S15_extracted_44898 | Not-determined |
| S15_extracted_44918 | Not-determined |
| S15_extracted_44928 | Not-determined |
| S15_extracted_44931 | Not-determined |
| S15_extracted_44949 | Not-determined |
| S15_extracted_44951 | Not-determined |
| S15_extracted_44954 | Not-determined |
| S15_extracted_44960 | Not-determined |
| S15_extracted_44980 | Not-determined |
| S15_extracted_44999 | Not-determined |
| S15_extracted_45001 | Not-determined |
| S15_extracted_45002 | Not-determined |
| S15_extracted_45003 | Low-quality |
| S15_extracted_45011 | Not-determined |
| S15_extracted_45019 | Not-determined |
| S15_extracted_45024 | Not-determined |
| S15_extracted_45064 | Not-determined |
| S15_extracted_45077 | Not-determined |
| S15_extracted_45081 | Not-determined |
| S15_extracted_45103 | Not-determined |
| S15_extracted_45118 | Not-determined |
| S15_extracted_45120 | Low-quality |
| S15_extracted_45122 | Low-quality |
| S15_extracted_45125 | Not-determined |
| S15_extracted_45134 | Low-quality |
| S15_extracted_45142 | Not-determined |
| S15_extracted_45152 | Not-determined |
| S15_extracted_45160 | Not-determined |
| S15_extracted_45163 | Not-determined |
| S15_extracted_45166 | Low-quality |
| S15_extracted_45167 | Not-determined |
| S15_extracted_45174 | Not-determined |
| S15_extracted_45178 | Not-determined |
| S15_extracted_45180 | Not-determined |
| S15_extracted_45192 | Low-quality |
| S15_extracted_45227 | Not-determined |
| S15_extracted_45257 | Not-determined |
| S15_extracted_45262 | Not-determined |
| S15_extracted_45273 | Not-determined |
| S15_extracted_45284 | Not-determined |
| S15_extracted_45286 | Not-determined |
| S15_extracted_45287 | Not-determined |
| S15_extracted_45303 | Not-determined |
| S15_extracted_45304 | Low-quality |
| S15_extracted_45306 | Not-determined |
| S15_extracted_45327 | Not-determined |
| S15_extracted_45353 | Low-quality |
| S15_extracted_45354 | Not-determined |
| S15_extracted_45374 | Not-determined |
| S15_extracted_45381 | Not-determined |
| S15_extracted_45414 | Not-determined |
| S15_extracted_45431 | Not-determined |
| S15_extracted_45433 | Not-determined |
| S15_extracted_45443 | Not-determined |
| S15_extracted_45474 | Not-determined |
| S15_extracted_45488 | Low-quality |
| S15_extracted_45497 | Not-determined |
| S15_extracted_45529 | Not-determined |
| S15_extracted_45542 | Not-determined |
| S15_extracted_45572 | Not-determined |
| S15_extracted_45578 | Not-determined |
| S15_extracted_45594 | Not-determined |
| S15_extracted_45601 | Not-determined |
| S15_extracted_45603 | Not-determined |
| S15_extracted_45604 | Not-determined |
| S15_extracted_45615 | Not-determined |
| S15_extracted_45622 | Not-determined |
| S15_extracted_45627 | Not-determined |
| S15_extracted_45630 | Not-determined |
| S15_extracted_45636 | Low-quality |
| S15_extracted_45647 | Not-determined |
| S15_extracted_45690 | Low-quality |
| S15_extracted_45691 | Not-determined |
| S15_extracted_45693 | Not-determined |
| S15_extracted_45697 | Not-determined |
| S15_extracted_45712 | Not-determined |
| S15_extracted_45728 | Not-determined |
| S15_extracted_45743 | Not-determined |
| S15_extracted_45753 | Not-determined |
| S15_extracted_45756 | Low-quality |
| S15_extracted_45774 | Not-determined |
| S15_extracted_45780 | Not-determined |
| S15_extracted_45816 | Not-determined |
| S15_extracted_45818 | Not-determined |
| S15_extracted_45825 | Not-determined |
| S15_extracted_45834 | Not-determined |
| S15_extracted_45838 | Not-determined |
| S15_extracted_45859 | Not-determined |
| S15_extracted_45860 | Not-determined |
| S15_extracted_45873 | Not-determined |
| S15_extracted_45878 | Not-determined |
| S15_extracted_45879 | Not-determined |
| S15_extracted_45883 | Not-determined |
| S15_extracted_45902 | Not-determined |
| S15_extracted_45918 | Not-determined |
| S15_extracted_45922 | Not-determined |
| S15_extracted_45932 | Low-quality |
| S15_extracted_45941 | Not-determined |
| S15_extracted_45944 | Not-determined |
| S15_extracted_45947 | Not-determined |
| S15_extracted_45957 | Not-determined |
| S15_extracted_45962 | Not-determined |
| S15_extracted_45965 | Not-determined |
| S15_extracted_45974 | Not-determined |
| S15_extracted_45977 | Not-determined |
| S15_extracted_45979 | Not-determined |
| S15_extracted_45990 | Not-determined |
| S15_extracted_46001 | Not-determined |
| S15_extracted_46037 | Not-determined |
| S15_extracted_46045 | Not-determined |
| S15_extracted_46048 | Not-determined |
| S15_extracted_46054 | Not-determined |
| S15_extracted_46058 | Not-determined |
| S15_extracted_46065 | Not-determined |
| S15_extracted_46067 | Not-determined |
| S15_extracted_46070 | Not-determined |
| S15_extracted_46071 | Low-quality |
| S15_extracted_46090 | Not-determined |
| S15_extracted_46101 | Not-determined |
| S15_extracted_46106 | Not-determined |
| S15_extracted_46124 | Not-determined |
| S15_extracted_46147 | Not-determined |
| S15_extracted_46150 | Not-determined |
| S15_extracted_46153 | Not-determined |
| S15_extracted_46156 | Not-determined |
| S15_extracted_46158 | Low-quality |
| S15_extracted_46161 | Not-determined |
| S15_extracted_46164 | Not-determined |
| S15_extracted_46169 | Not-determined |
| S15_extracted_46171 | Not-determined |
| S15_extracted_46178 | Not-determined |
| S15_extracted_46180 | Not-determined |
| S15_extracted_46182 | Not-determined |
| S15_extracted_46190 | Not-determined |
| S15_extracted_46192 | Not-determined |
| S15_extracted_46201 | Not-determined |
| S15_extracted_46220 | Not-determined |
| S15_extracted_46223 | Low-quality |
| S15_extracted_46224 | Not-determined |
| S15_extracted_46228 | Not-determined |
| S15_extracted_46252 | Not-determined |
| S15_extracted_46266 | Not-determined |
| S15_extracted_46268 | Not-determined |
| S15_extracted_46307 | Not-determined |
| S15_extracted_46309 | Not-determined |
| S15_extracted_46316 | Not-determined |
| S15_extracted_46320 | Not-determined |
| S15_extracted_46321 | Not-determined |
| S15_extracted_46353 | Not-determined |
| S15_extracted_46357 | Low-quality |
| S15_extracted_46381 | Not-determined |
| S15_extracted_46382 | Not-determined |
| S15_extracted_46386 | Not-determined |
| S15_extracted_46399 | Not-determined |
| S15_extracted_46401 | Not-determined |
| S15_extracted_46402 | Not-determined |

| | |
|---|---|
| S15_extracted_46406 | Not-determined |
| S15_extracted_46416 | Not-determined |
| S15_extracted_46436 | Low-quality |
| S15_extracted_46451 | Not-determined |
| S15_extracted_46459 | Low-quality |
| S15_extracted_46467 | Not-determined |
| S15_extracted_46481 | Not-determined |
| S15_extracted_46497 | Not-determined |
| S15_extracted_46499 | Not-determined |
| S15_extracted_46503 | Not-determined |
| S15_extracted_46511 | Not-determined |
| S15_extracted_46517 | Not-determined |
| S15_extracted_46520 | Not-determined |
| S15_extracted_46532 | Not-determined |
| S15_extracted_46552 | Low-quality |
| S15_extracted_46554 | Not-determined |
| S15_extracted_46559 | Not-determined |
| S15_extracted_46560 | Not-determined |
| S15_extracted_46566 | Not-determined |
| S15_extracted_46567 | Not-determined |
| S15_extracted_46571 | Not-determined |
| S15_extracted_46578 | Not-determined |
| S15_extracted_46585 | Not-determined |
| S15_extracted_46589 | Low-quality |
| S15_extracted_46595 | Not-determined |
| S15_extracted_46608 | Not-determined |
| S15_extracted_46610 | Not-determined |
| S15_extracted_46615 | Not-determined |
| S15_extracted_46627 | Not-determined |
| S15_extracted_46633 | Not-determined |
| S15_extracted_46635 | Not-determined |
| S15_extracted_46651 | Not-determined |
| S15_extracted_46669 | Low-quality |
| S15_extracted_46674 | Not-determined |
| S15_extracted_46680 | Not-determined |
| S15_extracted_46682 | Not-determined |
| S15_extracted_46711 | Not-determined |
| S15_extracted_46717 | Not-determined |
| S15_extracted_46720 | Not-determined |
| S15_extracted_46725 | Not-determined |
| S15_extracted_46726 | Not-determined |
| S15_extracted_46734 | Not-determined |
| S15_extracted_46744 | Not-determined |
| S15_extracted_46755 | Not-determined |
| S15_extracted_46774 | Not-determined |
| S15_extracted_46784 | Not-determined |
| S15_extracted_46786 | Not-determined |
| S15_extracted_46791 | Not-determined |
| S15_extracted_46793 | Not-determined |
| S15_extracted_46795 | Not-determined |
| S15_extracted_46799 | Not-determined |
| S15_extracted_46801 | Not-determined |
| S15_extracted_46804 | Not-determined |
| S15_extracted_46809 | Low-quality |
| S15_extracted_46810 | Not-determined |
| S15_extracted_46818 | Low-quality |
| S15_extracted_46834 | Low-quality |
| S15_extracted_46847 | Not-determined |
| S15_extracted_46855 | Low-quality |
| S15_extracted_46864 | Not-determined |
| S15_extracted_46868 | Low-quality |
| S15_extracted_46877 | Not-determined |
| S15_extracted_46884 | Not-determined |
| S15_extracted_46899 | Not-determined |
| S15_extracted_46900 | Not-determined |
| S15_extracted_46920 | Not-determined |
| S15_extracted_46924 | Low-quality |
| S15_extracted_46925 | Not-determined |
| S15_extracted_46928 | Not-determined |
| S15_extracted_46941 | Not-determined |
| S15_extracted_46957 | Not-determined |
| S15_extracted_46991 | Not-determined |
| S15_extracted_47003 | Not-determined |
| S15_extracted_47009 | Low-quality |
| S15_extracted_47014 | Not-determined |
| S15_extracted_47017 | Not-determined |
| S15_extracted_47036 | Not-determined |
| S15_extracted_47040 | Not-determined |
| S15_extracted_47060 | Not-determined |
| S15_extracted_47093 | Not-determined |
| S15_extracted_47095 | Not-determined |
| S15_extracted_47111 | Not-determined |
| S15_extracted_47113 | Low-quality |
| S15_extracted_47118 | Not-determined |
| S15_extracted_47121 | Not-determined |
| S15_extracted_47144 | Low-quality |
| S15_extracted_47158 | Not-determined |
| S15_extracted_47161 | Not-determined |
| S15_extracted_47165 | Not-determined |
| S15_extracted_47186 | Not-determined |
| S15_extracted_47189 | Not-determined |
| S15_extracted_47195 | Not-determined |
| S15_extracted_47197 | Low-quality |
| S15_extracted_47198 | Low-quality |
| S15_extracted_47201 | Not-determined |
| S15_extracted_47203 | Low-quality |
| S15_extracted_47210 | Not-determined |
| S15_extracted_47223 | Not-determined |
| S15_extracted_47234 | Not-determined |
| S15_extracted_47241 | Not-determined |
| S15_extracted_47242 | Not-determined |
| S15_extracted_47248 | Not-determined |
| S15_extracted_47255 | Not-determined |
| S15_extracted_47270 | Not-determined |
| S15_extracted_47285 | Low-quality |
| S15_extracted_47290 | Not-determined |
| S15_extracted_47298 | Not-determined |
| S15_extracted_47299 | Low-quality |
| S15_extracted_47307 | Not-determined |
| S15_extracted_47311 | Not-determined |
| S15_extracted_47313 | Not-determined |
| S15_extracted_47315 | Low-quality |
| S15_extracted_47326 | Not-determined |
| S15_extracted_47334 | Not-determined |
| S15_extracted_47351 | Not-determined |
| S15_extracted_47366 | Not-determined |
| S15_extracted_47368 | Not-determined |
| S15_extracted_47372 | Not-determined |
| S15_extracted_47374 | Not-determined |
| S15_extracted_47380 | Not-determined |
| S15_extracted_47391 | Low-quality |
| S15_extracted_47413 | Not-determined |
| S15_extracted_47414 | Low-quality |
| S15_extracted_47417 | Not-determined |
| S15_extracted_47421 | Not-determined |
| S15_extracted_47436 | Not-determined |
| S15_extracted_47442 | Not-determined |
| S15_extracted_47458 | Not-determined |
| S15_extracted_47471 | Not-determined |
| S15_extracted_47491 | Not-determined |
| S15_extracted_47520 | Low-quality |
| S15_extracted_47527 | Not-determined |
| S15_extracted_47557 | Not-determined |
| S15_extracted_47558 | Not-determined |
| S15_extracted_47564 | Not-determined |
| S15_extracted_47580 | Low-quality |
| S15_extracted_47581 | Not-determined |
| S15_extracted_47611 | Not-determined |
| S15_extracted_47620 | Not-determined |
| S15_extracted_47621 | Not-determined |
| S15_extracted_47631 | Not-determined |
| S15_extracted_47635 | Not-determined |
| S15_extracted_47636 | Not-determined |
| S15_extracted_47640 | Not-determined |
| S15_extracted_47643 | Not-determined |
| S15_extracted_47646 | Not-determined |
| S15_extracted_47647 | Not-determined |
| S15_extracted_47653 | Not-determined |
| S15_extracted_47660 | Not-determined |
| S15_extracted_47663 | Not-determined |
| S15_extracted_47668 | Not-determined |
| S15_extracted_47671 | Not-determined |
| S15_extracted_47673 | Not-determined |
| S15_extracted_47679 | Not-determined |
| S15_extracted_47686 | Low-quality |
| S15_extracted_47693 | Not-determined |
| S15_extracted_47697 | Low-quality |
| S15_extracted_47701 | Not-determined |
| S15_extracted_47710 | Not-determined |
| S15_extracted_47719 | Not-determined |
| S15_extracted_47720 | Not-determined |
| S15_extracted_47746 | Not-determined |
| S15_extracted_47750 | Not-determined |
| S15_extracted_47751 | Not-determined |
| S15_extracted_47761 | Not-determined |
| S15_extracted_47765 | Not-determined |
| S15_extracted_47767 | Not-determined |
| S15_extracted_47768 | Not-determined |
| S15_extracted_47779 | Not-determined |
| S15_extracted_47782 | Not-determined |
| S15_extracted_47793 | Not-determined |
| S15_extracted_47803 | Not-determined |
| S15_extracted_47826 | Not-determined |
| S15_extracted_47829 | Not-determined |
| S15_extracted_47831 | Low-quality |
| S15_extracted_47832 | Not-determined |
| S15_extracted_47838 | Not-determined |
| S15_extracted_47864 | Not-determined |
| S15_extracted_47867 | Not-determined |
| S15_extracted_47873 | Not-determined |
| S15_extracted_47891 | Not-determined |
| S15_extracted_47909 | Not-determined |
| S15_extracted_47927 | Not-determined |
| S15_extracted_47929 | Not-determined |
| S15_extracted_47930 | Not-determined |
| S15_extracted_47940 | Not-determined |
| S15_extracted_47950 | Not-determined |
| S15_extracted_47951 | Not-determined |
| S15_extracted_47955 | Not-determined |
| S15_extracted_47960 | Not-determined |
| S15_extracted_47963 | Not-determined |
| S15_extracted_47978 | Not-determined |
| S15_extracted_48008 | Not-determined |
| S15_extracted_48022 | Not-determined |
| S15_extracted_48024 | Not-determined |
| S15_extracted_48047 | Not-determined |
| S15_extracted_48048 | Not-determined |
| S15_extracted_48052 | Not-determined |
| S15_extracted_48072 | Not-determined |
| S15_extracted_48073 | Not-determined |
| S15_extracted_48084 | Not-determined |
| S15_extracted_48091 | Not-determined |
| S15_extracted_48092 | Not-determined |
| S15_extracted_48112 | Not-determined |
| S15_extracted_48126 | Not-determined |
| S15_extracted_48135 | Not-determined |
| S15_extracted_48144 | Not-determined |
| S15_extracted_48154 | Not-determined |
| S15_extracted_48168 | Not-determined |
| S15_extracted_48171 | Not-determined |
| S15_extracted_48180 | Not-determined |
| S15_extracted_48189 | Not-determined |
| S15_extracted_48209 | Not-determined |
| S15_extracted_48220 | Not-determined |
| S15_extracted_48221 | Not-determined |
| S15_extracted_48242 | Not-determined |
| S15_extracted_48247 | Not-determined |
| S15_extracted_48263 | Not-determined |
| S15_extracted_48273 | Not-determined |
| S15_extracted_48274 | Low-quality |
| S15_extracted_48282 | Low-quality |
| S15_extracted_48315 | Not-determined |
| S15_extracted_48322 | Not-determined |
| S15_extracted_48325 | Not-determined |
| S15_extracted_48328 | Not-determined |
| S15_extracted_48333 | Not-determined |
| S15_extracted_48334 | Not-determined |
| S15_extracted_48353 | Not-determined |
| S15_extracted_48368 | Not-determined |
| S15_extracted_48380 | Low-quality |
| S15_extracted_48381 | Not-determined |
| S15_extracted_48397 | Not-determined |
| S15_extracted_48401 | Not-determined |
| S15_extracted_48407 | Not-determined |
| S15_extracted_48412 | Not-determined |
| S15_extracted_48414 | Low-quality |
| S15_extracted_48448 | Not-determined |
| S15_extracted_48467 | Not-determined |
| S15_extracted_48471 | Not-determined |
| S15_extracted_48473 | Not-determined |
| S15_extracted_48494 | Not-determined |
| S15_extracted_48505 | Not-determined |
| S15_extracted_48517 | Not-determined |
| S15_extracted_48527 | Not-determined |
| S15_extracted_48536 | Not-determined |
| S15_extracted_48538 | Not-determined |
| S15_extracted_48539 | Not-determined |
| S15_extracted_48542 | Not-determined |
| S15_extracted_48544 | Not-determined |
| S15_extracted_48546 | Not-determined |
| S15_extracted_48562 | Not-determined |
| S15_extracted_48563 | Not-determined |
| S15_extracted_48580 | Not-determined |
| S15_extracted_48585 | Not-determined |
| S15_extracted_48586 | Not-determined |
| S15_extracted_48590 | Not-determined |
| S15_extracted_48594 | Not-determined |
| S15_extracted_48597 | Not-determined |
| S15_extracted_48603 | Not-determined |
| S15_extracted_48609 | Not-determined |
| S15_extracted_48625 | Not-determined |
| S15_extracted_48648 | Not-determined |
| S15_extracted_48658 | Not-determined |
| S15_extracted_48695 | Not-determined |
| S15_extracted_48701 | Not-determined |
| S15_extracted_48706 | Low-quality |
| S15_extracted_48709 | Not-determined |
| S15_extracted_48729 | Low-quality |
| S15_extracted_48734 | Not-determined |
| S15_extracted_48742 | Not-determined |
| S15_extracted_48756 | Not-determined |
| S15_extracted_48766 | Not-determined |
| S15_extracted_48771 | Not-determined |
| S15_extracted_48776 | Not-determined |
| S15_extracted_48788 | Low-quality |
| S15_extracted_48805 | Not-determined |
| S15_extracted_48836 | Not-determined |
| S15_extracted_48859 | Not-determined |
| S15_extracted_48866 | Not-determined |
| S15_extracted_48869 | Not-determined |
| S15_extracted_48876 | Not-determined |
| S15_extracted_48888 | Not-determined |
| S15_extracted_48891 | Not-determined |
| S15_extracted_48899 | Not-determined |
| S15_extracted_48920 | Not-determined |
| S15_extracted_48923 | Not-determined |
| S15_extracted_48936 | Not-determined |
| S15_extracted_48937 | Not-determined |
| S15_extracted_48947 | Not-determined |
| S15_extracted_48956 | Not-determined |
| S15_extracted_48959 | Not-determined |
| S15_extracted_48960 | Not-determined |
| S15_extracted_48963 | Not-determined |
| S15_extracted_48964 | Not-determined |
| S15_extracted_48980 | Not-determined |
| S15_extracted_49003 | Not-determined |
| S15_extracted_49009 | Not-determined |
| S15_extracted_49045 | Not-determined |
| S15_extracted_49047 | Not-determined |
| S15_extracted_49055 | Not-determined |
| S15_extracted_49081 | Not-determined |
| S15_extracted_49087 | Not-determined |
| S15_extracted_49099 | Not-determined |
| S15_extracted_49105 | Not-determined |
| S15_extracted_49106 | Not-determined |
| S15_extracted_49111 | Not-determined |
| S15_extracted_49118 | Low-quality |
| S15_extracted_49140 | Not-determined |
| S15_extracted_49151 | Not-determined |
| S15_extracted_49167 | Not-determined |
| S15_extracted_49180 | Not-determined |
| S15_extracted_49182 | Not-determined |
| S15_extracted_49188 | Not-determined |
| S15_extracted_49193 | Not-determined |
| S15_extracted_49201 | Not-determined |
| S15_extracted_49210 | Not-determined |
| S15_extracted_49214 | Not-determined |
| S15_extracted_49233 | Not-determined |
| S15_extracted_49246 | Not-determined |
| S15_extracted_49247 | Not-determined |
| S15_extracted_49266 | Not-determined |
| S15_extracted_49267 | Not-determined |
| S15_extracted_49275 | Not-determined |
| S15_extracted_49288 | Not-determined |
| S15_extracted_49291 | Not-determined |
| S15_extracted_49298 | Not-determined |
| S15_extracted_49313 | Not-determined |
| S15_extracted_49320 | Low-quality |
| S15_extracted_49339 | Low-quality |
| S15_extracted_49351 | Low-quality |
| S15_extracted_49360 | Not-determined |
| S15_extracted_49361 | Not-determined |
| S15_extracted_49385 | Not-determined |
| S15_extracted_49386 | Not-determined |
| S15_extracted_49391 | Not-determined |
| S15_extracted_49394 | Not-determined |
| S15_extracted_49395 | Not-determined |
| S15_extracted_49411 | Low-quality |
| S15_extracted_49412 | Not-determined |
| S15_extracted_49423 | Not-determined |
| S15_extracted_49438 | Not-determined |
| S15_extracted_49440 | Not-determined |
| S15_extracted_49443 | Not-determined |
| S15_extracted_49469 | Not-determined |
| S15_extracted_49481 | Not-determined |
| S15_extracted_49486 | Low-quality |
| S15_extracted_49494 | Not-determined |
| S15_extracted_49507 | Not-determined |
| S15_extracted_49512 | Not-determined |
| S15_extracted_49516 | Not-determined |
| S15_extracted_49534 | Low-quality |
| S15_extracted_49539 | Not-determined |
| S15_extracted_49540 | Low-quality |
| S15_extracted_49542 | Not-determined |
| S15_extracted_49549 | Not-determined |
| S15_extracted_49553 | Not-determined |
| S15_extracted_49554 | Low-quality |
| S15_extracted_49556 | Not-determined |
| S15_extracted_49559 | Not-determined |
| S15_extracted_49560 | Not-determined |
| S15_extracted_49562 | Not-determined |
| S15_extracted_49570 | Not-determined |
| S15_extracted_49574 | Not-determined |
| S15_extracted_49575 | Not-determined |
| S15_extracted_49583 | Not-determined |
| S15_extracted_49587 | Not-determined |
| S15_extracted_49590 | Low-quality |
| S15_extracted_49598 | Not-determined |
| S15_extracted_49615 | Not-determined |
| S15_extracted_49620 | Not-determined |
| S15_extracted_49621 | Not-determined |
| S15_extracted_49636 | Not-determined |
| S15_extracted_49639 | Not-determined |
| S15_extracted_49641 | Not-determined |
| S15_extracted_49657 | Not-determined |
| S15_extracted_49678 | Not-determined |
| S15_extracted_49689 | Not-determined |
| S15_extracted_49706 | Not-determined |
| S15_extracted_49707 | Not-determined |
| S15_extracted_49717 | Low-quality |
| S15_extracted_49722 | Not-determined |
| S15_extracted_49735 | Not-determined |
| S15_extracted_49737 | Not-determined |
| S15_extracted_49742 | Low-quality |
| S15_extracted_49743 | Not-determined |
| S15_extracted_49753 | Low-quality |
| S15_extracted_49764 | Not-determined |
| S15_extracted_49774 | Not-determined |
| S15_extracted_49782 | Not-determined |
| S15_extracted_49796 | Not-determined |
| S15_extracted_49797 | Not-determined |
| S15_extracted_49799 | Not-determined |
| S15_extracted_49812 | Not-determined |
| S15_extracted_49818 | Not-determined |
| S15_extracted_49838 | Not-determined |
| S15_extracted_49840 | Not-determined |
| S15_extracted_49842 | Not-determined |
| S15_extracted_49853 | Not-determined |
| S15_extracted_49854 | Not-determined |
| S15_extracted_49856 | Not-determined |
| S15_extracted_49858 | Not-determined |
| S15_extracted_49859 | Not-determined |
| S15_extracted_49880 | Not-determined |
| S15_extracted_49881 | Not-determined |
| S15_extracted_49901 | Not-determined |
| S15_extracted_49919 | Low-quality |
| S15_extracted_49933 | Not-determined |
| S15_extracted_49938 | Not-determined |
| S15_extracted_49948 | Not-determined |
| S15_extracted_49959 | Not-determined |
| S15_extracted_49961 | Not-determined |
| S15_extracted_49962 | Not-determined |
| S15_extracted_49968 | Not-determined |
| S15_extracted_49983 | Not-determined |
| S15_extracted_49987 | Not-determined |
| S15_extracted_49988 | Not-determined |
| S15_extracted_49992 | Not-determined |
| S15_extracted_50015 | Not-determined |
| S15_extracted_50064 | Not-determined |
| S15_extracted_50075 | Not-determined |
| S15_extracted_50076 | Not-determined |
| S15_extracted_50086 | Not-determined |
| S15_extracted_50092 | Not-determined |
| S15_extracted_50096 | Low-quality |
| S15_extracted_50098 | Not-determined |
| S15_extracted_50103 | Not-determined |
| S15_extracted_50107 | Not-determined |
| S15_extracted_50109 | Not-determined |
| S15_extracted_50110 | Low-quality |
| S15_extracted_50130 | Not-determined |
| S15_extracted_50132 | Low-quality |
| S15_extracted_50133 | Not-determined |
| S15_extracted_50135 | Not-determined |
| S15_extracted_50138 | Not-determined |
| S15_extracted_50162 | Not-determined |
| S15_extracted_50163 | Low-quality |
| S15_extracted_50171 | Not-determined |
| S15_extracted_50180 | Not-determined |
| S15_extracted_50182 | Not-determined |
| S15_extracted_50185 | Not-determined |
| S15_extracted_50190 | Not-determined |
| S15_extracted_50197 | Not-determined |
| S15_extracted_50215 | Not-determined |
| S15_extracted_50219 | Not-determined |
| S15_extracted_50221 | Not-determined |
| S15_extracted_50223 | Not-determined |
| S15_extracted_50233 | Not-determined |
| S15_extracted_50236 | Not-determined |
| S15_extracted_50245 | Low-quality |
| S15_extracted_50258 | Low-quality |
| S15_extracted_50266 | Not-determined |
| S15_extracted_50271 | Not-determined |
| S15_extracted_50291 | Not-determined |
| S15_extracted_50292 | Not-determined |
| S15_extracted_50303 | Low-quality |
| S15_extracted_50315 | Not-determined |
| S15_extracted_50327 | Not-determined |
| S15_extracted_50343 | Not-determined |
| S15_extracted_50345 | Not-determined |
| S15_extracted_50351 | Not-determined |
| S15_extracted_50352 | Not-determined |
| S15_extracted_50354 | Not-determined |
| S15_extracted_50357 | Not-determined |
| S15_extracted_50361 | Not-determined |
| S15_extracted_50385 | Not-determined |
| S15_extracted_50387 | Not-determined |
| S15_extracted_50393 | Not-determined |
| S15_extracted_50444 | Not-determined |
| S15_extracted_50456 | Not-determined |
| S15_extracted_50459 | Not-determined |
| S15_extracted_50460 | Not-determined |
| S15_extracted_50471 | Not-determined |
| S15_extracted_50491 | Not-determined |
| S15_extracted_50496 | Not-determined |
| S15_extracted_50498 | Not-determined |
| S15_extracted_50506 | Not-determined |
| S15_extracted_50507 | Not-determined |
| S15_extracted_50518 | Low-quality |
| S15_extracted_50526 | Not-determined |
| S15_extracted_50544 | Not-determined |
| S15_extracted_50548 | Not-determined |
| S15_extracted_50552 | Not-determined |
| S15_extracted_50558 | Not-determined |
| S15_extracted_50559 | Low-quality |
| S15_extracted_50561 | Low-quality |
| S15_extracted_50562 | Not-determined |
| S15_extracted_50567 | Not-determined |
| S15_extracted_50574 | Not-determined |
| S15_extracted_50575 | Not-determined |
| S15_extracted_50578 | Not-determined |
| S15_extracted_50579 | Low-quality |
| S15_extracted_50592 | Not-determined |
| S15_extracted_50604 | Low-quality |
| S15_extracted_50605 | Low-quality |
| S15_extracted_50613 | Not-determined |
| S15_extracted_50625 | Not-determined |
| S15_extracted_50631 | Not-determined |
| S15_extracted_50652 | Low-quality |
| S15_extracted_50657 | Not-determined |
| S15_extracted_50680 | Not-determined |
| S15_extracted_50705 | Not-determined |
| S15_extracted_50709 | Not-determined |
| S15_extracted_50754 | Not-determined |
| S15_extracted_50779 | Not-determined |
| S15_extracted_50780 | Not-determined |
| S15_extracted_50785 | Not-determined |
| S15_extracted_50798 | Not-determined |
| S15_extracted_50809 | Not-determined |
| S15_extracted_50810 | Not-determined |
| S15_extracted_50818 | Not-determined |
| S15_extracted_50820 | Not-determined |
| S15_extracted_50834 | Not-determined |
| S15_extracted_50835 | Not-determined |
| S15_extracted_50838 | Not-determined |
| S15_extracted_50847 | Not-determined |
| S15_extracted_50856 | Not-determined |
| S15_extracted_50863 | Not-determined |
| S15_extracted_50875 | Not-determined |
| S15_extracted_50876 | Low-quality |
| S15_extracted_50880 | Not-determined |
| S15_extracted_50912 | Not-determined |
| S15_extracted_50918 | Not-determined |
| S15_extracted_50921 | Not-determined |
| S15_extracted_50938 | Not-determined |
| S15_extracted_50950 | Not-determined |
| S15_extracted_50957 | Not-determined |
| S15_extracted_50993 | Not-determined |
| S15_extracted_50994 | Not-determined |
| S15_extracted_50998 | Not-determined |
| S15_extracted_51031 | Not-determined |
| S15_extracted_51034 | Not-determined |
| S15_extracted_51037 | Not-determined |
| S15_extracted_51038 | Not-determined |
| S15_extracted_51043 | Not-determined |
| S15_extracted_51072 | Not-determined |
| S15_extracted_51100 | Not-determined |
| S15_extracted_51101 | Not-determined |
| S15_extracted_51106 | Not-determined |
| S15_extracted_51107 | Not-determined |
| S15_extracted_51111 | Not-determined |
| S15_extracted_51115 | Not-determined |
| S15_extracted_51122 | Not-determined |
| S15_extracted_51126 | Low-quality |
| S15_extracted_51135 | Low-quality |
| S15_extracted_51157 | Not-determined |
| S15_extracted_51163 | Not-determined |
| S15_extracted_51173 | Not-determined |
| S15_extracted_51178 | Not-determined |
| S15_extracted_51193 | Low-quality |
| S15_extracted_51195 | Not-determined |
| S15_extracted_51208 | Not-determined |
| S15_extracted_51217 | Not-determined |
| S15_extracted_51225 | Not-determined |
| S15_extracted_51226 | Not-determined |
| S15_extracted_51230 | Not-determined |
| S15_extracted_51233 | Not-determined |
| S15_extracted_51235 | Not-determined |
| S15_extracted_51240 | Not-determined |
| S15_extracted_51243 | Not-determined |
| S15_extracted_51253 | Not-determined |
| S15_extracted_51255 | Not-determined |
| S15_extracted_51258 | Low-quality |
| S15_extracted_51259 | Not-determined |
| S15_extracted_51269 | Low-quality |
| S15_extracted_51275 | Not-determined |
| S15_extracted_51276 | Not-determined |
| S15_extracted_51278 | Low-quality |
| S15_extracted_51279 | Not-determined |
| S15_extracted_51282 | Not-determined |
| S15_extracted_51307 | Not-determined |
| S15_extracted_51329 | Not-determined |
| S15_extracted_51331 | Not-determined |
| S15_extracted_51332 | Not-determined |
| S15_extracted_51333 | Not-determined |
| S15_extracted_51339 | Not-determined |
| S15_extracted_51359 | Low-quality |
| S15_extracted_51363 | Low-quality |
| S15_extracted_51371 | Not-determined |
| S15_extracted_51372 | Low-quality |
| S15_extracted_51399 | Not-determined |
| S15_extracted_51401 | Not-determined |
| S15_extracted_51416 | Not-determined |
| S15_extracted_51425 | Not-determined |
| S15_extracted_51427 | Not-determined |
| S15_extracted_51433 | Not-determined |
| S15_extracted_51434 | Not-determined |
| S15_extracted_51446 | Not-determined |
| S15_extracted_51455 | Not-determined |
| S15_extracted_51465 | Low-quality |

| | |
|---|---|
| S15_extracted_S1476 | Not-determined |
| S15_extracted_S1483 | Low-quality |
| S15_extracted_S1500 | Not-determined |
| S15_extracted_S1503 | Not-determined |
| S15_extracted_S1532 | Low-quality |
| S15_extracted_S1533 | Not-determined |
| S15_extracted_S1548 | Low-quality |
| S15_extracted_S1556 | Not-determined |
| S15_extracted_S1574 | Low-quality |
| S15_extracted_S1602 | Not-determined |
| S15_extracted_S1603 | Not-determined |
| S15_extracted_S1607 | Not-determined |
| S15_extracted_S1621 | Not-determined |
| S15_extracted_S1623 | Low-quality |
| S15_extracted_S1631 | Not-determined |
| S15_extracted_S1634 | Not-determined |
| S15_extracted_S1637 | Not-determined |
| S15_extracted_S1644 | Not-determined |
| S15_extracted_S1646 | Not-determined |
| S15_extracted_S1649 | Not-determined |
| S15_extracted_S1662 | Not-determined |
| S15_extracted_S1664 | Not-determined |
| S15_extracted_S1683 | Not-determined |
| S15_extracted_S1705 | Low-quality |
| S15_extracted_S1707 | Not-determined |
| S15_extracted_S1715 | Low-quality |
| S15_extracted_S1726 | Not-determined |
| S15_extracted_S1733 | Not-determined |
| S15_extracted_S1739 | Not-determined |
| S15_extracted_S1742 | Not-determined |
| S15_extracted_S1745 | Low-quality |
| S15_extracted_S1750 | Not-determined |
| S15_extracted_S1754 | Not-determined |
| S15_extracted_S1766 | Not-determined |
| S15_extracted_S1768 | Not-determined |
| S15_extracted_S1796 | Not-determined |
| S15_extracted_S1797 | Not-determined |
| S15_extracted_S1803 | Low-quality |
| S15_extracted_S1805 | Not-determined |
| S15_extracted_S1808 | Not-determined |
| S15_extracted_S1825 | Not-determined |
| S15_extracted_S1830 | Not-determined |
| S15_extracted_S1831 | Not-determined |
| S15_extracted_S1848 | Not-determined |
| S15_extracted_S1855 | Not-determined |
| S15_extracted_S1860 | Not-determined |
| S15_extracted_S1862 | Not-determined |
| S15_extracted_S1863 | Not-determined |
| S15_extracted_S1868 | Not-determined |
| S15_extracted_S1869 | Not-determined |
| S15_extracted_S1872 | Not-determined |
| S15_extracted_S1879 | Not-determined |
| S15_extracted_S1884 | Not-determined |
| S15_extracted_S1887 | Not-determined |
| S15_extracted_S1907 | Not-determined |
| S15_extracted_S1912 | Not-determined |
| S15_extracted_S1915 | Not-determined |
| S15_extracted_S1920 | Not-determined |
| S15_extracted_S1925 | Not-determined |
| S15_extracted_S1933 | Not-determined |
| S15_extracted_S1935 | Not-determined |
| S15_extracted_S1940 | Not-determined |
| S15_extracted_S1952 | Low-quality |
| S15_extracted_S1958 | Not-determined |
| S15_extracted_S1964 | Not-determined |
| S15_extracted_S1976 | Not-determined |
| S15_extracted_S1980 | Not-determined |
| S15_extracted_S1981 | Not-determined |
| S15_extracted_S1983 | Low-quality |
| S15_extracted_S1994 | Not-determined |
| S15_extracted_S2023 | Not-determined |
| S15_extracted_S2025 | Not-determined |
| S15_extracted_S2034 | Not-determined |
| S15_extracted_S2037 | Not-determined |
| S15_extracted_S2041 | Not-determined |
| S15_extracted_S2064 | Not-determined |
| S15_extracted_S2076 | Not-determined |
| S15_extracted_S2091 | Not-determined |
| S15_extracted_S2105 | Not-determined |
| S15_extracted_S2114 | Low-quality |
| S15_extracted_S2118 | Not-determined |
| S15_extracted_S2120 | Not-determined |
| S15_extracted_S2124 | Low-quality |
| S15_extracted_S2131 | Not-determined |
| S15_extracted_S2132 | Not-determined |
| S15_extracted_S2157 | Not-determined |
| S15_extracted_S2160 | Not-determined |
| S15_extracted_S2175 | Not-determined |
| S15_extracted_S2178 | Not-determined |
| S15_extracted_S2183 | Not-determined |
| S15_extracted_S2201 | Low-quality |
| S15_extracted_S2203 | Not-determined |
| S15_extracted_S2231 | Not-determined |
| S15_extracted_S2235 | Not-determined |
| S15_extracted_S2240 | Not-determined |
| S15_extracted_S2245 | Not-determined |
| S15_extracted_S2257 | Not-determined |
| S15_extracted_S2260 | Not-determined |
| S15_extracted_S2269 | Low-quality |
| S15_extracted_S2280 | Not-determined |
| S15_extracted_S2289 | Low-quality |
| S15_extracted_S2302 | Not-determined |
| S15_extracted_S2307 | Not-determined |
| S15_extracted_S2313 | Low-quality |
| S15_extracted_S2314 | Low-quality |
| S15_extracted_S2318 | Not-determined |
| S15_extracted_S2328 | Low-quality |
| S15_extracted_S2333 | Not-determined |
| S15_extracted_S2366 | Not-determined |
| S15_extracted_S2369 | Not-determined |
| S15_extracted_S2374 | Not-determined |
| S15_extracted_S2376 | Low-quality |
| S15_extracted_S2383 | Not-determined |
| S15_extracted_S2412 | Not-determined |
| S15_extracted_S2424 | Not-determined |
| S15_extracted_S2425 | Not-determined |
| S15_extracted_S2433 | Not-determined |
| S15_extracted_S2452 | Not-determined |
| S15_extracted_S2456 | Not-determined |
| S15_extracted_S2496 | Not-determined |
| S15_extracted_S2500 | Not-determined |
| S15_extracted_S2504 | Not-determined |
| S15_extracted_S2515 | Not-determined |
| S15_extracted_S2519 | Not-determined |
| S15_extracted_S2525 | Not-determined |
| S15_extracted_S2526 | Not-determined |
| S15_extracted_S2530 | Not-determined |
| S15_extracted_S2536 | Not-determined |
| S15_extracted_S2542 | Not-determined |
| S15_extracted_S2560 | Not-determined |
| S15_extracted_S2581 | Low-quality |
| S15_extracted_S2582 | Not-determined |
| S15_extracted_S2585 | Not-determined |
| S15_extracted_S2590 | Not-determined |
| S15_extracted_S2594 | Not-determined |
| S15_extracted_S2616 | Not-determined |
| S15_extracted_S2621 | Not-determined |
| S15_extracted_S2625 | Not-determined |
| S15_extracted_S2629 | Not-determined |
| S15_extracted_S2635 | Not-determined |
| S15_extracted_S2658 | Not-determined |
| S15_extracted_S2662 | Not-determined |
| S15_extracted_S2666 | Not-determined |
| S15_extracted_S2693 | Not-determined |
| S15_extracted_S2695 | Not-determined |
| S15_extracted_S2703 | Not-determined |
| S15_extracted_S2729 | Not-determined |
| S15_extracted_S2734 | Not-determined |
| S15_extracted_S2740 | Not-determined |
| S15_extracted_S2758 | Not-determined |
| S15_extracted_S2759 | Not-determined |
| S15_extracted_S2764 | Not-determined |
| S15_extracted_S2779 | Not-determined |
| S15_extracted_S2786 | Not-determined |
| S15_extracted_S2788 | Not-determined |
| S15_extracted_S2797 | Not-determined |
| S15_extracted_S2806 | Not-determined |
| S15_extracted_S2854 | Not-determined |
| S15_extracted_S2866 | Not-determined |
| S15_extracted_S2869 | Not-determined |
| S15_extracted_S2871 | Not-determined |
| S15_extracted_S2884 | Not-determined |
| S15_extracted_S2908 | Low-quality |
| S15_extracted_S2917 | Not-determined |
| S15_extracted_S2927 | Not-determined |
| S15_extracted_S2935 | Not-determined |
| S15_extracted_S2941 | Not-determined |
| S15_extracted_S2946 | Not-determined |
| S15_extracted_S2950 | Not-determined |
| S15_extracted_S2951 | Low-quality |
| S15_extracted_S2953 | Not-determined |
| S15_extracted_S2955 | Not-determined |
| S15_extracted_S2964 | Not-determined |
| S15_extracted_S2971 | Not-determined |
| S15_extracted_S2972 | Not-determined |
| S15_extracted_S2979 | Low-quality |
| S15_extracted_S2985 | Not-determined |
| S15_extracted_S2995 | Not-determined |
| S15_extracted_S2997 | Not-determined |
| S15_extracted_S2999 | Low-quality |
| S15_extracted_S3001 | Not-determined |
| S15_extracted_S3004 | Not-determined |
| S15_extracted_S3011 | Not-determined |
| S15_extracted_S3012 | Low-quality |
| S15_extracted_S3016 | Low-quality |
| S15_extracted_S3022 | Not-determined |
| S15_extracted_S3024 | Not-determined |
| S15_extracted_S3031 | Not-determined |
| S15_extracted_S3043 | Not-determined |
| S15_extracted_S3050 | Not-determined |
| S15_extracted_S3052 | Not-determined |
| S15_extracted_S3078 | Not-determined |
| S15_extracted_S3082 | Not-determined |
| S15_extracted_S3086 | Not-determined |
| S15_extracted_S3093 | Not-determined |
| S15_extracted_S3095 | Not-determined |
| S15_extracted_S3104 | Not-determined |
| S15_extracted_S3122 | Not-determined |
| S15_extracted_S3124 | Not-determined |
| S15_extracted_S3125 | Not-determined |
| S15_extracted_S3128 | Not-determined |
| S15_extracted_S3136 | Low-quality |
| S15_extracted_S3141 | Not-determined |
| S15_extracted_S3148 | Not-determined |
| S15_extracted_S3167 | Not-determined |
| S15_extracted_S3175 | Not-determined |
| S15_extracted_S3195 | Not-determined |
| S15_extracted_S3215 | Not-determined |
| S15_extracted_S3233 | Not-determined |
| S15_extracted_S3234 | Not-determined |
| S15_extracted_S3247 | Not-determined |
| S15_extracted_S3259 | Not-determined |
| S15_extracted_S3272 | Not-determined |
| S15_extracted_S3284 | Not-determined |
| S15_extracted_S3297 | Not-determined |
| S15_extracted_S3309 | Not-determined |
| S15_extracted_S3318 | Not-determined |
| S15_extracted_S3321 | Not-determined |
| S15_extracted_S3322 | Not-determined |
| S15_extracted_S3323 | Low-quality |
| S15_extracted_S3330 | Not-determined |
| S15_extracted_S3343 | Not-determined |
| S15_extracted_S3362 | Not-determined |
| S15_extracted_S3369 | Not-determined |
| S15_extracted_S3392 | Low-quality |
| S15_extracted_S3401 | Not-determined |
| S15_extracted_S3409 | Not-determined |
| S15_extracted_S3415 | Not-determined |
| S15_extracted_S3420 | Not-determined |
| S15_extracted_S3426 | Not-determined |
| S15_extracted_S3428 | Not-determined |
| S15_extracted_S3430 | Not-determined |
| S15_extracted_S3435 | Not-determined |
| S15_extracted_S3440 | Not-determined |
| S15_extracted_S3445 | Not-determined |
| S15_extracted_S3447 | Not-determined |
| S15_extracted_S3456 | Not-determined |
| S15_extracted_S3458 | Not-determined |
| S15_extracted_S3466 | Not-determined |
| S15_extracted_S3468 | Low-quality |
| S15_extracted_S3471 | Not-determined |
| S15_extracted_S3483 | Low-quality |
| S15_extracted_S3485 | Not-determined |
| S15_extracted_S3491 | Not-determined |
| S15_extracted_S3497 | Not-determined |
| S15_extracted_S3498 | Low-quality |
| S15_extracted_S3499 | Not-determined |
| S15_extracted_S3500 | Not-determined |
| S15_extracted_S3519 | Not-determined |
| S15_extracted_S3548 | Not-determined |
| S15_extracted_S3549 | Not-determined |
| S15_extracted_S3551 | Not-determined |
| S15_extracted_S3554 | Not-determined |
| S15_extracted_S3556 | Low-quality |
| S15_extracted_S3575 | Not-determined |
| S15_extracted_S3578 | Not-determined |
| S15_extracted_S3582 | Not-determined |
| S15_extracted_S3586 | Low-quality |
| S15_extracted_S3587 | Not-determined |
| S15_extracted_S3589 | Not-determined |
| S15_extracted_S3600 | Not-determined |
| S15_extracted_S3603 | Not-determined |
| S15_extracted_S3611 | Not-determined |
| S15_extracted_S3614 | Not-determined |
| S15_extracted_S3618 | Low-quality |
| S15_extracted_S3619 | Not-determined |
| S15_extracted_S3627 | Not-determined |
| S15_extracted_S3631 | Not-determined |
| S15_extracted_S3635 | Low-quality |
| S15_extracted_S3643 | Not-determined |
| S15_extracted_S3649 | Not-determined |
| S15_extracted_S3654 | Not-determined |
| S15_extracted_S3663 | Not-determined |
| S15_extracted_S3672 | Not-determined |
| S15_extracted_S3676 | Not-determined |
| S15_extracted_S3687 | Low-quality |
| S15_extracted_S3693 | Not-determined |
| S15_extracted_S3707 | Not-determined |
| S15_extracted_S3728 | Not-determined |
| S15_extracted_S3732 | Not-determined |
| S15_extracted_S3742 | Not-determined |
| S15_extracted_S3743 | Not-determined |
| S15_extracted_S3746 | Not-determined |
| S15_extracted_S3756 | Not-determined |
| S15_extracted_S3757 | Not-determined |
| S15_extracted_S3758 | Not-determined |
| S15_extracted_S3771 | Low-quality |
| S15_extracted_S3783 | Not-determined |
| S15_extracted_S3813 | Low-quality |
| S15_extracted_S3828 | Not-determined |
| S15_extracted_S3834 | Low-quality |
| S15_extracted_S3838 | Not-determined |
| S15_extracted_S3841 | Not-determined |
| S15_extracted_S3843 | Low-quality |
| S15_extracted_S3850 | Not-determined |
| S15_extracted_S3854 | Low-quality |
| S15_extracted_S3876 | Not-determined |
| S15_extracted_S3880 | Not-determined |
| S15_extracted_S3885 | Not-determined |
| S15_extracted_S3890 | Not-determined |
| S15_extracted_S3901 | Not-determined |
| S15_extracted_S3903 | Not-determined |
| S15_extracted_S3910 | Not-determined |
| S15_extracted_S3916 | Not-determined |
| S15_extracted_S3934 | Not-determined |
| S15_extracted_S3943 | Not-determined |
| S15_extracted_S3950 | Not-determined |
| S15_extracted_S3958 | Not-determined |
| S15_extracted_S3984 | Not-determined |
| S15_extracted_S3985 | Not-determined |
| S15_extracted_S3989 | Low-quality |
| S15_extracted_S3990 | Not-determined |
| S15_extracted_S3995 | Not-determined |
| S15_extracted_S3998 | Not-determined |
| S15_extracted_S4001 | Not-determined |
| S15_extracted_S4015 | Not-determined |
| S15_extracted_S4016 | Not-determined |
| S15_extracted_S4017 | Not-determined |
| S15_extracted_S4020 | Not-determined |
| S15_extracted_S4036 | Not-determined |
| S15_extracted_S4056 | Not-determined |
| S15_extracted_S4079 | Not-determined |
| S15_extracted_S4094 | Not-determined |
| S15_extracted_S4100 | Low-quality |
| S15_extracted_S4102 | Not-determined |
| S15_extracted_S4107 | Not-determined |
| S15_extracted_S4122 | Not-determined |
| S15_extracted_S4133 | Not-determined |
| S15_extracted_S4147 | Not-determined |
| S15_extracted_S4164 | Not-determined |
| S15_extracted_S4181 | Not-determined |
| S15_extracted_S4185 | Not-determined |
| S15_extracted_S4187 | Not-determined |
| S15_extracted_S4198 | Not-determined |
| S15_extracted_S4205 | Not-determined |
| S15_extracted_S4213 | Not-determined |
| S15_extracted_S4230 | Low-quality |
| S15_extracted_S4231 | Not-determined |
| S15_extracted_S4237 | Not-determined |
| S15_extracted_S4263 | Not-determined |
| S15_extracted_S4271 | Not-determined |
| S15_extracted_S4281 | Not-determined |
| S15_extracted_S4310 | Not-determined |
| S15_extracted_S4314 | Not-determined |
| S15_extracted_S4331 | Not-determined |
| S15_extracted_S4337 | Not-determined |
| S15_extracted_S4338 | Not-determined |
| S15_extracted_S4349 | Low-quality |
| S15_extracted_S4352 | Not-determined |
| S15_extracted_S4361 | Not-determined |
| S15_extracted_S4369 | Not-determined |
| S15_extracted_S4380 | Not-determined |
| S15_extracted_S4390 | Not-determined |
| S15_extracted_S4393 | Not-determined |
| S15_extracted_S4408 | Not-determined |
| S15_extracted_S4416 | Not-determined |
| S15_extracted_S4428 | Not-determined |
| S15_extracted_S4430 | Not-determined |
| S15_extracted_S4432 | Low-quality |
| S15_extracted_S4440 | Not-determined |
| S15_extracted_S4446 | Low-quality |
| S15_extracted_S4449 | Not-determined |
| S15_extracted_S4463 | Not-determined |
| S15_extracted_S4483 | Low-quality |
| S15_extracted_S4495 | Not-determined |
| S15_extracted_S4504 | Not-determined |
| S15_extracted_S4511 | Not-determined |
| S15_extracted_S4512 | Not-determined |
| S15_extracted_S4517 | Not-determined |
| S15_extracted_S4521 | Not-determined |
| S15_extracted_S4524 | Low-quality |
| S15_extracted_S4554 | Not-determined |
| S15_extracted_S4567 | Not-determined |
| S15_extracted_S4568 | Not-determined |
| S15_extracted_S4579 | Not-determined |
| S15_extracted_S4580 | Not-determined |
| S15_extracted_S4585 | Not-determined |
| S15_extracted_S4590 | Not-determined |
| S15_extracted_S4593 | Not-determined |
| S15_extracted_S4600 | Not-determined |
| S15_extracted_S4603 | Not-determined |
| S15_extracted_S4605 | Not-determined |
| S15_extracted_S4607 | Not-determined |
| S15_extracted_S4620 | Not-determined |
| S15_extracted_S4631 | Not-determined |
| S15_extracted_S4634 | Not-determined |
| S15_extracted_S4636 | Not-determined |
| S15_extracted_S4674 | Not-determined |
| S15_extracted_S4679 | Not-determined |
| S15_extracted_S4701 | Not-determined |
| S15_extracted_S4703 | Not-determined |
| S15_extracted_S4712 | Not-determined |
| S15_extracted_S4728 | Not-determined |
| S15_extracted_S4733 | Not-determined |
| S15_extracted_S4738 | Not-determined |
| S15_extracted_S4751 | Not-determined |
| S15_extracted_S4753 | Low-quality |
| S15_extracted_S4757 | Not-determined |
| S15_extracted_S4812 | Not-determined |
| S15_extracted_S4823 | Low-quality |
| S15_extracted_S4848 | Not-determined |
| S15_extracted_S4857 | Low-quality |
| S15_extracted_S4860 | Not-determined |
| S15_extracted_S4868 | Not-determined |
| S15_extracted_S4871 | Not-determined |
| S15_extracted_S4874 | Not-determined |
| S15_extracted_S4875 | Low-quality |
| S15_extracted_S4876 | Not-determined |
| S15_extracted_S4887 | Not-determined |
| S15_extracted_S4888 | Not-determined |
| S15_extracted_S4889 | Not-determined |
| S15_extracted_S4894 | Not-determined |
| S15_extracted_S4895 | Not-determined |
| S15_extracted_S4896 | Not-determined |
| S15_extracted_S4903 | Low-quality |
| S15_extracted_S4915 | Low-quality |
| S15_extracted_S4917 | Not-determined |
| S15_extracted_S4924 | Not-determined |
| S15_extracted_S4934 | Not-determined |
| S15_extracted_S4937 | Not-determined |
| S15_extracted_S4945 | Not-determined |
| S15_extracted_S4951 | Not-determined |
| S15_extracted_S4953 | Not-determined |
| S15_extracted_S4963 | Not-determined |
| S15_extracted_S4967 | Not-determined |
| S15_extracted_S4973 | Not-determined |
| S15_extracted_S4984 | Not-determined |
| S15_extracted_S5016 | Not-determined |
| S15_extracted_S5018 | Not-determined |
| S15_extracted_S5041 | Not-determined |
| S15_extracted_S5045 | Not-determined |
| S15_extracted_S5051 | Not-determined |
| S15_extracted_S5052 | Not-determined |
| S15_extracted_S5053 | Not-determined |
| S15_extracted_S5074 | Not-determined |
| S15_extracted_S5088 | Low-quality |
| S15_extracted_S5092 | Not-determined |
| S15_extracted_S5100 | Not-determined |
| S15_extracted_S5109 | Not-determined |
| S15_extracted_S5112 | Not-determined |
| S15_extracted_S5117 | Not-determined |
| S15_extracted_S5126 | Low-quality |
| S15_extracted_S5148 | Not-determined |
| S15_extracted_S5149 | Not-determined |
| S15_extracted_S5152 | Not-determined |
| S15_extracted_S5164 | Not-determined |
| S15_extracted_S5175 | Low-quality |
| S15_extracted_S5185 | Not-determined |
| S15_extracted_S5197 | Not-determined |
| S15_extracted_S5243 | Low-quality |
| S15_extracted_S5251 | Not-determined |
| S15_extracted_S5254 | Not-determined |
| S15_extracted_S5265 | Not-determined |
| S15_extracted_S5273 | Not-determined |
| S15_extracted_S5286 | Not-determined |
| S15_extracted_S5287 | Not-determined |
| S15_extracted_S5312 | Not-determined |
| S15_extracted_S5318 | Not-determined |
| S15_extracted_S5320 | Not-determined |
| S15_extracted_S5326 | Not-determined |
| S15_extracted_S5333 | Not-determined |
| S15_extracted_S5344 | Not-determined |
| S15_extracted_S5367 | Low-quality |
| S15_extracted_S5390 | Not-determined |
| S15_extracted_S5400 | Not-determined |
| S15_extracted_S5404 | Not-determined |
| S15_extracted_S5410 | Not-determined |
| S15_extracted_S5421 | Not-determined |
| S15_extracted_S5433 | Low-quality |
| S15_extracted_S5436 | Not-determined |
| S15_extracted_S5443 | Not-determined |
| S15_extracted_S5446 | Not-determined |
| S15_extracted_S5455 | Not-determined |
| S15_extracted_S5466 | Low-quality |
| S15_extracted_S5469 | Not-determined |
| S15_extracted_S5473 | Not-determined |
| S15_extracted_S5475 | Not-determined |
| S15_extracted_S5476 | Not-determined |
| S15_extracted_S5488 | Not-determined |
| S15_extracted_S5492 | Not-determined |
| S15_extracted_S5507 | Not-determined |
| S15_extracted_S5511 | Not-determined |
| S15_extracted_S5515 | Not-determined |
| S15_extracted_S5520 | Not-determined |
| S15_extracted_S5524 | Not-determined |
| S15_extracted_S5530 | Low-quality |
| S15_extracted_S5536 | Not-determined |
| S15_extracted_S5537 | Not-determined |
| S15_extracted_S5539 | Not-determined |
| S15_extracted_S5543 | Low-quality |
| S15_extracted_S5554 | Not-determined |
| S15_extracted_S5562 | Not-determined |
| S15_extracted_S5563 | Not-determined |
| S15_extracted_S5573 | Not-determined |
| S15_extracted_S5581 | Not-determined |
| S15_extracted_S5583 | Not-determined |
| S15_extracted_S5609 | Not-determined |
| S15_extracted_S5621 | Not-determined |
| S15_extracted_S5630 | Not-determined |
| S15_extracted_S5638 | Not-determined |
| S8_vs_k141_176720\|\|full | Medium-quality |
| S15_extracted_S5644 | Not-determined |
| S15_extracted_S5646 | Not-determined |
| S15_extracted_S5665 | Not-determined |
| S15_extracted_S5676 | Not-determined |
| S15_extracted_S5681 | Not-determined |
| S15_extracted_S5687 | Not-determined |
| S15_extracted_S5688 | Not-determined |
| S15_extracted_S5696 | Not-determined |
| S15_extracted_S5700 | Not-determined |
| S15_extracted_S5703 | Not-determined |
| S15_extracted_S5720 | Not-determined |
| S15_extracted_S5721 | Not-determined |
| S15_extracted_S5737 | Not-determined |
| S15_extracted_S5741 | Not-determined |
| S15_extracted_S5744 | Not-determined |
| S15_extracted_S5747 | Not-determined |
| S15_extracted_S5751 | Not-determined |
| S15_extracted_S5754 | Not-determined |
| S15_extracted_S5764 | Low-quality |
| S15_extracted_S5765 | Not-determined |
| S15_extracted_S5774 | Not-determined |
| S15_extracted_S5785 | Not-determined |
| S15_extracted_S5797 | Not-determined |
| S15_extracted_S5807 | Not-determined |
| S15_extracted_S5811 | Low-quality |
| S15_extracted_S5814 | Not-determined |
| S15_extracted_S5823 | Not-determined |
| S15_extracted_S5834 | Not-determined |
| S15_extracted_S5869 | Not-determined |
| S15_extracted_S5877 | Not-determined |
| S15_extracted_S5885 | Not-determined |
| S15_extracted_S5893 | Not-determined |
| S15_extracted_S5895 | Not-determined |
| S15_extracted_S5898 | Not-determined |
| S15_extracted_S5910 | Not-determined |
| S15_extracted_S5925 | Not-determined |
| S15_extracted_S5928 | Not-determined |
| S15_extracted_S5940 | Not-determined |
| S15_extracted_S5941 | Not-determined |
| S15_extracted_S5950 | Not-determined |
| S15_extracted_S5977 | Not-determined |
| S15_extracted_S6007 | Not-determined |
| S15_extracted_S6030 | Not-determined |
| S15_extracted_S6033 | Low-quality |
| S15_extracted_S6038 | Low-quality |
| S15_extracted_S6044 | Not-determined |
| S15_extracted_S6048 | Not-determined |
| S15_extracted_S6053 | Not-determined |
| S15_extracted_S6054 | Low-quality |
| S15_extracted_S6089 | Not-determined |
| S15_extracted_S6097 | Not-determined |
| S15_extracted_S6099 | Not-determined |
| S15_extracted_S6102 | Not-determined |
| S15_extracted_S6104 | Not-determined |
| S15_extracted_S6106 | Not-determined |
| S15_extracted_S6111 | Not-determined |
| S15_extracted_S6117 | Not-determined |
| S15_extracted_S6125 | Not-determined |
| S15_extracted_S6133 | Not-determined |
| S15_extracted_S6135 | Not-determined |
| S15_extracted_S6144 | Not-determined |
| S15_extracted_S6147 | Not-determined |
| S15_extracted_S6155 | Low-quality |
| S15_extracted_S6184 | Not-determined |
| S15_extracted_S6197 | Not-determined |
| S15_extracted_S6199 | Low-quality |
| S15_extracted_S6213 | Low-quality |
| S15_extracted_S6216 | Low-quality |
| S15_extracted_S6218 | Not-determined |
| S15_extracted_S6221 | Not-determined |
| S15_extracted_S6224 | Not-determined |
| S15_extracted_S6231 | Low-quality |
| S15_extracted_S6239 | Not-determined |
| S15_extracted_S6241 | Not-determined |
| S15_extracted_S6244 | Not-determined |
| S15_extracted_S6245 | Not-determined |
| S15_extracted_S6246 | Not-determined |
| S15_extracted_S6274 | Low-quality |
| S15_extracted_S6285 | Not-determined |
| S15_extracted_S6291 | Not-determined |
| S15_extracted_S6299 | Not-determined |
| S15_extracted_S6300 | Not-determined |
| S15_extracted_S6303 | Not-determined |
| S15_extracted_S6310 | Not-determined |
| S15_extracted_S6315 | Not-determined |
| S15_extracted_S6323 | Not-determined |
| S15_extracted_S6324 | Not-determined |
| S15_extracted_S6326 | Not-determined |

| | |
|---|---|
| S1S_extracted_56337 | Not-determined |
| S1S_extracted_56338 | Low-quality |
| S1S_extracted_56352 | Not-determined |
| S1S_extracted_56359 | Low-quality |
| S1S_extracted_56362 | Low-quality |
| S1S_extracted_56367 | Not-determined |
| S1S_extracted_56371 | Low-quality |
| S1S_extracted_56374 | Not-determined |
| S1S_extracted_56400 | Low-quality |
| S1S_extracted_56406 | Not-determined |
| S1S_extracted_56412 | Not-determined |
| S1S_extracted_56420 | Not-determined |
| S1S_extracted_56422 | Not-determined |
| S1S_extracted_56428 | Not-determined |
| S1S_extracted_56433 | Not-determined |
| S1S_extracted_56440 | Not-determined |
| S1S_extracted_56468 | Not-determined |
| S1S_extracted_56471 | Not-determined |
| S1S_extracted_56491 | Not-determined |
| S1S_extracted_56503 | Not-determined |
| S1S_extracted_56513 | Not-determined |
| S1S_extracted_56524 | Low-quality |
| S1S_extracted_56540 | Not-determined |
| S1S_extracted_56589 | Not-determined |
| S1S_extracted_56600 | Not-determined |
| S1S_extracted_56602 | Not-determined |
| S1S_extracted_56606 | Not-determined |
| S1S_extracted_56610 | Not-determined |
| S1S_extracted_56623 | Not-determined |
| S1S_extracted_56630 | Not-determined |
| S1S_extracted_56632 | Low-quality |
| S1S_extracted_56646 | Not-determined |
| S1S_extracted_56647 | Not-determined |
| S1S_extracted_56651 | Not-determined |
| S1S_extracted_56652 | Not-determined |
| S1S_extracted_56674 | Not-determined |
| S1S_extracted_56681 | Not-determined |
| S1S_extracted_56685 | Not-determined |
| S1S_extracted_56697 | Not-determined |
| S1S_extracted_56704 | Not-determined |
| S1S_extracted_56713 | Not-determined |
| S1S_extracted_56726 | Not-determined |
| S1S_extracted_56734 | Not-determined |
| S1S_extracted_56737 | Low-quality |
| S1S_extracted_56745 | Not-determined |
| S1S_extracted_56752 | Not-determined |
| S1S_extracted_56753 | Not-determined |
| S1S_extracted_56761 | Not-determined |
| S1S_extracted_56769 | Not-determined |
| S1S_extracted_56799 | Not-determined |
| S1S_extracted_56805 | Not-determined |
| S1S_extracted_56814 | Not-determined |
| S1S_extracted_56819 | Low-quality |
| S1S_extracted_56820 | Not-determined |
| S1S_extracted_56849 | Not-determined |
| S1S_extracted_56876 | Not-determined |
| S1S_extracted_56884 | Not-determined |
| S1S_extracted_56885 | Not-determined |
| S1S_extracted_56888 | Not-determined |
| S1S_extracted_56920 | Not-determined |
| S1S_extracted_56954 | Not-determined |
| S1S_extracted_56967 | Not-determined |
| S1S_extracted_56969 | Low-quality |
| S1S_extracted_56974 | Not-determined |
| S1S_extracted_56983 | Not-determined |
| S1S_extracted_57004 | Not-determined |
| S1S_extracted_57005 | Not-determined |
| S1S_extracted_57015 | Low-quality |
| S1S_extracted_57020 | Not-determined |
| S1S_extracted_57028 | Not-determined |
| S1S_extracted_57042 | Not-determined |
| S1S_extracted_57055 | Not-determined |
| S1S_extracted_57058 | Not-determined |
| S1S_extracted_57061 | Not-determined |
| S1S_extracted_57062 | Not-determined |
| S1S_extracted_57066 | Not-determined |
| S1S_extracted_57069 | Low-quality |
| S1S_extracted_57076 | Not-determined |
| S1S_extracted_57089 | Not-determined |
| S1S_extracted_57097 | Not-determined |
| S1S_extracted_57102 | Not-determined |
| S1S_extracted_57111 | Not-determined |
| S1S_extracted_57115 | Not-determined |
| S1S_extracted_57118 | Not-determined |
| S1S_extracted_57135 | Not-determined |
| S1S_extracted_57144 | Not-determined |
| S1S_extracted_57147 | Not-determined |
| S1S_extracted_57151 | Not-determined |
| S1S_extracted_57153 | Not-determined |
| S1S_extracted_57160 | Not-determined |
| S1S_extracted_57172 | Low-quality |
| S1S_extracted_57177 | Not-determined |
| S1S_extracted_57184 | Low-quality |
| S1S_extracted_57188 | Not-determined |
| S1S_extracted_57194 | Low-quality |
| S1S_extracted_57202 | Not-determined |
| S1S_extracted_57209 | Not-determined |
| S1S_extracted_57212 | Not-determined |
| S1S_extracted_57215 | Not-determined |
| S1S_extracted_57218 | Not-determined |
| S1S_extracted_57219 | Not-determined |
| S1S_extracted_57225 | Not-determined |
| S1S_extracted_57228 | Not-determined |
| S1S_extracted_57231 | Not-determined |
| S1S_extracted_57233 | Low-quality |
| S1S_extracted_57245 | Not-determined |
| S1S_extracted_57250 | Not-determined |
| S1S_extracted_57265 | Not-determined |
| S1S_extracted_57273 | Not-determined |
| S1S_extracted_57286 | Not-determined |
| S1S_extracted_57288 | Not-determined |
| S1S_extracted_57289 | Not-determined |
| S1S_extracted_57293 | Not-determined |
| S1S_extracted_57309 | Not-determined |
| S1S_extracted_57319 | Low-quality |
| S1S_extracted_57320 | Not-determined |
| S1S_extracted_57322 | Not-determined |
| S1S_extracted_57340 | Not-determined |
| S1S_extracted_57351 | Not-determined |
| S1S_extracted_57362 | Not-determined |
| S1S_extracted_57363 | Low-quality |
| S1S_extracted_57377 | Not-determined |
| S1S_extracted_57378 | Not-determined |
| S1S_extracted_57387 | Not-determined |
| S1S_extracted_57394 | Not-determined |
| S1S_extracted_57396 | Low-quality |
| S1S_extracted_57401 | Not-determined |
| S1S_extracted_57402 | Not-determined |
| S1S_extracted_57411 | Not-determined |
| S1S_extracted_57412 | Not-determined |
| S1S_extracted_57415 | Not-determined |
| S1S_extracted_57425 | Not-determined |
| S1S_extracted_57452 | Not-determined |
| S1S_extracted_57471 | Not-determined |
| S1S_extracted_57480 | Not-determined |
| S1S_extracted_57513 | Not-determined |
| S1S_extracted_57515 | Not-determined |
| S1S_extracted_57522 | Not-determined |
| S1S_extracted_57529 | Not-determined |
| S1S_extracted_57554 | Not-determined |
| S1S_extracted_57556 | Not-determined |
| S1S_extracted_57557 | Not-determined |
| S1S_extracted_57564 | Not-determined |
| S1S_extracted_57603 | Not-determined |
| S1S_extracted_57605 | Not-determined |
| S1S_extracted_57606 | Not-determined |
| S1S_extracted_57619 | Not-determined |
| S1S_extracted_57622 | Not-determined |
| S1S_extracted_57626 | Not-determined |
| S1S_extracted_57647 | Not-determined |
| S1S_extracted_57653 | Not-determined |
| S1S_extracted_57654 | Not-determined |
| S1S_extracted_57659 | Not-determined |
| S1S_extracted_57660 | Not-determined |
| S1S_extracted_57672 | Not-determined |
| S1S_extracted_57686 | Not-determined |
| S1S_extracted_57698 | Not-determined |
| S1S_extracted_57701 | Not-determined |
| S1S_extracted_57705 | Not-determined |
| S1S_extracted_57706 | Low-quality |
| S1S_extracted_57710 | Low-quality |
| S1S_extracted_57715 | Not-determined |
| S1S_extracted_57729 | Not-determined |
| S1S_extracted_57738 | Not-determined |
| S1S_extracted_57747 | Not-determined |
| S1S_extracted_57755 | Not-determined |
| S1S_extracted_57765 | Not-determined |
| S1S_extracted_57773 | Not-determined |
| S1S_extracted_57786 | Not-determined |
| S1S_extracted_57810 | Not-determined |
| S1S_extracted_57815 | Not-determined |
| S1S_extracted_57822 | Not-determined |
| S1S_extracted_57826 | Not-determined |
| S1S_extracted_57838 | Not-determined |
| S1S_extracted_57839 | Not-determined |
| S1S_extracted_57841 | Not-determined |
| S1S_extracted_57853 | Not-determined |
| S1S_extracted_57856 | Not-determined |
| S1S_extracted_57860 | Not-determined |
| S1S_extracted_57864 | Low-quality |
| S1S_extracted_57871 | Not-determined |
| S1S_extracted_57878 | Not-determined |
| S1S_extracted_57881 | Not-determined |
| S1S_extracted_57896 | Not-determined |
| S1S_extracted_57903 | Not-determined |
| S1S_extracted_57904 | Not-determined |
| S1S_extracted_57911 | Not-determined |
| S1S_extracted_57915 | Not-determined |
| S1S_extracted_57926 | Not-determined |
| S1S_extracted_57932 | Not-determined |
| S1S_extracted_57934 | Low-quality |
| S1S_extracted_57938 | Not-determined |
| S1S_extracted_57944 | Low-quality |
| S1S_extracted_57950 | Not-determined |
| S1S_extracted_57962 | Not-determined |
| S1S_extracted_57966 | Not-determined |
| S1S_extracted_57976 | Not-determined |
| S1S_extracted_57978 | Not-determined |
| S1S_extracted_57993 | Not-determined |
| S1S_extracted_58006 | Not-determined |
| S1S_extracted_58025 | Not-determined |
| S1S_extracted_58030 | Not-determined |
| S1S_extracted_58042 | Not-determined |
| S1S_extracted_58043 | Not-determined |
| S1S_extracted_58073 | Not-determined |
| S1S_extracted_58078 | Low-quality |
| S1S_extracted_58085 | Not-determined |
| S1S_extracted_58090 | Not-determined |
| S1S_extracted_58097 | Not-determined |
| S1S_extracted_58102 | Not-determined |
| S1S_extracted_58107 | Not-determined |
| S1S_extracted_58110 | Low-quality |
| S1S_extracted_58117 | Not-determined |
| S1S_extracted_58126 | Not-determined |
| S1S_extracted_58135 | Not-determined |
| S1S_extracted_58137 | Not-determined |
| S1S_extracted_58138 | Not-determined |
| S1S_extracted_58149 | Not-determined |
| S1S_extracted_58158 | Not-determined |
| S1S_extracted_58165 | Not-determined |
| S1S_extracted_58166 | Not-determined |
| S1S_extracted_58170 | Not-determined |
| S1S_extracted_58176 | Low-quality |
| S1S_extracted_58179 | Low-quality |
| S1S_extracted_58180 | Not-determined |
| S1S_extracted_58187 | Low-quality |
| S1S_extracted_58196 | Not-determined |
| S1S_extracted_58201 | Not-determined |
| S1S_extracted_58238 | Not-determined |
| S1S_extracted_58254 | Low-quality |
| S1S_extracted_58259 | Not-determined |
| S1S_extracted_58263 | Not-determined |
| S1S_extracted_58289 | Not-determined |
| S1S_extracted_58297 | Low-quality |
| S1S_extracted_58304 | Not-determined |
| S1S_extracted_58312 | Not-determined |
| S1S_extracted_58314 | Not-determined |
| S1S_extracted_58329 | Not-determined |
| S1S_extracted_58348 | Not-determined |
| S1S_extracted_58350 | Not-determined |
| S1S_extracted_58362 | Not-determined |
| S1S_extracted_58371 | Not-determined |
| S1S_extracted_58373 | Not-determined |
| S1S_extracted_58380 | Not-determined |
| S1S_extracted_58383 | Not-determined |
| S1S_extracted_58392 | Not-determined |
| S1S_extracted_58404 | Not-determined |
| S1S_extracted_58421 | Not-determined |
| S1S_extracted_58449 | Not-determined |
| S11_vs_k141_13652\|\|full | Medium-quality |
| S1S_extracted_58480 | Not-determined |
| S1S_extracted_58498 | Not-determined |
| S1S_extracted_58513 | Not-determined |
| S1S_extracted_58534 | Not-determined |
| S1S_extracted_58549 | Not-determined |
| S1S_extracted_58551 | Not-determined |
| S1S_extracted_58565 | Not-determined |
| S1S_extracted_58583 | Not-determined |
| S1S_extracted_58601 | Not-determined |
| S1S_extracted_58610 | Not-determined |
| S1S_extracted_58615 | Not-determined |
| S1S_extracted_58616 | Not-determined |
| S1S_extracted_58621 | Not-determined |
| S1S_extracted_58626 | Not-determined |
| S1S_extracted_58633 | Low-quality |
| S1S_extracted_58635 | Not-determined |
| S1S_extracted_58646 | Not-determined |
| S1S_extracted_58647 | Not-determined |
| S1S_extracted_58652 | Not-determined |
| S1S_extracted_58665 | Not-determined |
| S1S_extracted_58687 | Not-determined |
| S1S_extracted_58692 | Not-determined |
| S1S_extracted_58697 | Not-determined |
| S1S_extracted_58711 | Not-determined |
| S1S_extracted_58712 | Low-quality |
| S1S_extracted_58726 | Low-quality |
| S1S_extracted_58736 | Not-determined |
| S1S_extracted_58750 | Not-determined |
| S1S_extracted_58752 | Not-determined |
| S1S_extracted_58754 | Not-determined |
| S1S_extracted_58756 | Not-determined |
| S1S_extracted_58760 | Not-determined |
| S1S_extracted_58781 | Not-determined |
| S1S_extracted_58789 | Low-quality |
| S1S_extracted_58805 | Not-determined |
| S1S_extracted_58818 | Not-determined |
| S1S_extracted_58852 | Not-determined |
| S1S_extracted_58858 | Not-determined |
| S1S_extracted_58870 | Not-determined |
| S1S_extracted_58871 | Not-determined |
| S1S_extracted_58919 | Not-determined |
| S1S_extracted_58929 | Low-quality |
| S1S_extracted_58932 | Not-determined |
| S1S_extracted_58935 | Not-determined |
| S1S_extracted_58936 | Not-determined |
| S1S_extracted_58937 | Not-determined |
| S1S_extracted_58941 | Low-quality |
| S1S_extracted_58943 | Not-determined |
| S1S_extracted_58944 | Not-determined |
| S1S_extracted_58948 | Not-determined |
| S1S_extracted_58971 | Not-determined |
| S1S_extracted_58972 | Not-determined |
| S1S_extracted_58978 | Not-determined |
| S1S_extracted_58983 | Not-determined |
| S1S_extracted_58988 | Not-determined |
| S1S_extracted_58999 | Not-determined |
| S1S_extracted_59005 | Not-determined |
| S1S_extracted_59009 | Not-determined |
| S1S_extracted_59041 | Not-determined |
| S1S_extracted_59043 | Not-determined |
| S1S_extracted_59044 | Not-determined |
| S1S_extracted_59045 | Not-determined |
| S1S_extracted_59049 | Not-determined |
| S1S_extracted_59061 | Not-determined |
| S1S_extracted_59079 | Not-determined |
| S1S_extracted_59084 | Not-determined |
| S1S_extracted_59093 | Not-determined |
| S1S_extracted_59096 | Not-determined |
| S1S_extracted_59098 | Not-determined |
| S1S_extracted_59106 | Not-determined |
| S1S_extracted_59108 | Not-determined |
| S1S_extracted_59118 | Not-determined |
| S1S_extracted_59123 | Not-determined |
| S1S_extracted_59128 | Not-determined |
| S1S_extracted_59129 | Not-determined |
| S1S_extracted_59134 | Not-determined |
| S1S_extracted_59155 | Not-determined |
| S1S_extracted_59160 | Not-determined |
| S1S_extracted_59161 | Not-determined |
| S1S_extracted_59165 | Low-quality |
| S1S_extracted_59186 | Low-quality |
| S1S_extracted_59191 | Not-determined |
| S1S_extracted_59206 | Low-quality |
| S1S_extracted_59209 | Not-determined |
| S1S_extracted_59212 | Not-determined |
| S1S_extracted_59231 | Not-determined |
| S1S_extracted_59235 | Not-determined |
| S1S_extracted_59248 | Not-determined |
| S1S_extracted_59253 | Low-quality |
| S1S_extracted_59257 | Not-determined |
| S1S_extracted_59259 | Not-determined |
| S1S_extracted_59262 | Low-quality |
| S1S_extracted_59266 | Not-determined |
| S1S_extracted_59270 | Not-determined |
| S1S_extracted_59286 | Not-determined |
| S1S_extracted_59289 | Not-determined |
| S1S_extracted_59311 | Not-determined |
| S1S_extracted_59313 | Not-determined |
| S1S_extracted_59323 | Not-determined |
| S1S_extracted_59334 | Not-determined |
| S1S_extracted_59335 | Not-determined |
| S1S_extracted_59341 | Not-determined |
| S1S_extracted_59342 | Not-determined |
| S1S_extracted_59344 | Low-quality |
| S1S_extracted_59350 | Not-determined |
| S1S_extracted_59352 | Not-determined |
| S1S_extracted_59372 | Not-determined |
| S1S_extracted_59379 | Not-determined |
| S1S_extracted_59383 | Low-quality |
| S1S_extracted_59391 | Not-determined |
| S1S_extracted_59400 | Not-determined |
| S1S_extracted_59418 | Not-determined |
| S1S_extracted_59424 | Not-determined |
| S1S_extracted_59427 | Not-determined |
| S1S_extracted_59437 | Low-quality |
| S1S_extracted_59438 | Not-determined |
| S1S_extracted_59440 | Low-quality |
| S1S_extracted_59443 | Not-determined |
| S1S_extracted_59458 | Not-determined |
| S1S_extracted_59468 | Not-determined |
| S1S_extracted_59472 | Not-determined |
| S1S_extracted_59484 | Not-determined |
| S1S_extracted_59497 | Not-determined |
| S1S_extracted_59509 | Not-determined |
| S1S_extracted_59513 | Not-determined |
| S1S_extracted_59531 | Low-quality |
| S1S_extracted_59532 | Not-determined |
| S1S_extracted_59535 | Not-determined |
| S1S_extracted_59546 | Not-determined |
| S1S_extracted_59549 | Not-determined |
| S1S_extracted_59553 | Not-determined |
| S1S_extracted_59566 | Low-quality |
| S1S_extracted_59578 | Not-determined |
| S1S_extracted_59593 | Not-determined |
| S1S_extracted_59596 | Not-determined |
| S1S_extracted_59599 | Not-determined |
| S1S_extracted_59605 | Not-determined |
| S1S_extracted_59609 | Not-determined |
| S1S_extracted_59649 | Not-determined |
| S1S_extracted_59653 | Not-determined |
| S1S_extracted_59658 | Not-determined |
| S1S_extracted_59670 | Not-determined |
| S1S_extracted_59675 | Not-determined |
| S1S_extracted_59676 | Not-determined |
| S1S_extracted_59681 | Not-determined |
| S1S_extracted_59688 | Not-determined |
| S1S_extracted_59693 | Not-determined |
| S1S_extracted_59699 | Not-determined |
| S1S_extracted_59700 | Not-determined |
| S1S_extracted_59708 | Not-determined |
| S1S_extracted_59711 | Not-determined |
| S1S_extracted_59728 | Not-determined |
| S1S_extracted_59737 | Not-determined |
| S1S_extracted_59740 | Not-determined |
| S1S_extracted_59741 | Low-quality |
| S1S_extracted_59770 | Low-quality |
| S1S_extracted_59777 | Not-determined |
| S1S_extracted_59787 | Low-quality |
| S1S_extracted_59801 | Not-determined |
| S1S_extracted_59802 | Not-determined |
| S1S_extracted_59803 | Not-determined |
| S1S_extracted_59814 | Not-determined |
| S1S_extracted_59816 | Not-determined |
| S1S_extracted_59818 | Low-quality |
| S1S_extracted_59839 | Not-determined |
| S1S_extracted_59840 | Not-determined |
| S1S_extracted_59841 | Not-determined |
| S1S_extracted_59843 | Not-determined |
| S1S_extracted_59848 | Not-determined |
| S1S_extracted_59851 | Not-determined |
| S1S_extracted_59862 | Not-determined |
| S1S_extracted_59864 | Not-determined |
| S1S_extracted_59865 | Not-determined |
| S1S_extracted_59870 | Not-determined |
| S1S_extracted_59874 | Low-quality |
| S1S_extracted_59880 | Not-determined |
| S1S_extracted_59885 | Not-determined |
| S1S_extracted_59892 | Not-determined |
| S1S_extracted_59903 | Not-determined |
| S1S_extracted_59910 | Not-determined |
| S1S_extracted_59911 | Not-determined |
| S1S_extracted_59924 | Not-determined |
| S1S_extracted_59937 | Low-quality |
| S1S_extracted_59941 | Not-determined |
| S1S_extracted_59943 | Not-determined |
| S1S_extracted_59944 | Not-determined |
| S1S_extracted_59946 | Low-quality |
| S1S_extracted_59947 | Not-determined |
| S1S_extracted_59957 | Low-quality |
| S1S_extracted_59958 | Not-determined |
| S1S_extracted_59966 | Not-determined |
| S1S_extracted_59968 | Not-determined |
| S1S_extracted_59981 | Not-determined |
| S1S_extracted_59985 | Not-determined |
| S1S_extracted_59988 | Not-determined |
| S1S_extracted_59998 | Not-determined |
| S1S_extracted_60004 | Not-determined |
| S1S_extracted_60005 | Not-determined |
| S1S_extracted_60021 | Not-determined |
| S1S_extracted_60028 | Not-determined |
| S1S_extracted_60034 | Not-determined |
| S1S_extracted_60040 | Not-determined |
| S1S_extracted_60060 | Not-determined |
| S1S_extracted_60071 | Not-determined |
| S1S_extracted_60078 | Not-determined |
| S1S_extracted_60082 | Not-determined |
| S1S_extracted_60086 | Not-determined |
| S1S_extracted_60088 | Not-determined |
| S1S_extracted_60103 | Not-determined |
| S1S_extracted_60105 | Not-determined |
| S1S_extracted_60115 | Not-determined |
| S1S_extracted_60119 | Not-determined |
| S1S_extracted_60120 | Not-determined |
| S1S_extracted_60124 | Not-determined |
| S1S_extracted_60125 | Not-determined |
| S1S_extracted_60130 | Low-quality |
| S1S_extracted_60139 | Not-determined |
| S1S_extracted_60153 | Low-quality |
| S1S_extracted_60159 | Not-determined |
| S1S_extracted_60169 | Not-determined |
| S1S_extracted_60171 | Not-determined |
| S1S_extracted_60180 | Not-determined |
| S1S_extracted_60184 | Not-determined |
| S1S_extracted_60187 | Not-determined |
| S1S_extracted_60198 | Not-determined |
| S1S_extracted_60207 | Not-determined |
| S1S_extracted_60212 | Not-determined |
| S1S_extracted_60238 | Not-determined |
| S1S_extracted_60248 | Not-determined |
| S1S_extracted_60253 | Not-determined |
| S1S_extracted_60257 | Not-determined |
| S1S_extracted_60260 | Low-quality |
| S1S_extracted_60261 | Not-determined |
| S1S_extracted_60263 | Not-determined |
| S1S_extracted_60275 | Low-quality |
| S1S_extracted_60278 | Not-determined |
| S1S_extracted_60282 | Not-determined |
| S1S_extracted_60291 | Not-determined |
| S1S_extracted_60323 | Not-determined |
| S1S_extracted_60334 | Not-determined |
| S1S_extracted_60353 | Not-determined |
| S1S_extracted_60359 | Not-determined |
| S1S_extracted_60362 | Not-determined |
| S1S_extracted_60372 | Not-determined |
| S1S_extracted_60374 | Not-determined |
| S1S_extracted_60384 | Not-determined |
| S1S_extracted_60387 | Not-determined |
| S1S_extracted_60398 | Not-determined |
| S1S_extracted_60403 | Low-quality |
| S1S_extracted_60414 | Not-determined |
| S1S_extracted_60417 | Not-determined |
| S1S_extracted_60423 | Not-determined |
| S1S_extracted_60424 | Not-determined |
| S1S_extracted_60428 | Not-determined |
| S1S_extracted_60437 | Not-determined |
| S1S_extracted_60450 | Not-determined |
| S1S_extracted_60453 | Not-determined |
| S1S_extracted_60470 | Not-determined |
| S1S_extracted_60482 | Low-quality |
| S1S_extracted_60504 | Not-determined |
| S1S_extracted_60510 | Not-determined |
| S1S_extracted_60513 | Not-determined |
| S1S_extracted_60530 | Not-determined |
| S1S_extracted_60534 | Not-determined |
| S1S_extracted_60538 | Not-determined |
| S1S_extracted_60543 | Not-determined |
| S1S_extracted_60545 | Not-determined |
| S1S_extracted_60569 | Not-determined |
| S1S_extracted_60572 | Not-determined |
| S1S_extracted_60591 | Not-determined |
| S1S_extracted_60593 | Not-determined |
| S1S_extracted_60611 | Not-determined |
| S1S_extracted_60616 | Not-determined |
| S1S_extracted_60625 | Not-determined |
| S1S_extracted_60638 | Not-determined |
| S1S_extracted_60647 | Low-quality |
| S1S_extracted_60648 | Not-determined |
| S1S_extracted_60655 | Not-determined |
| S1S_extracted_60664 | Not-determined |
| S1S_extracted_60666 | Not-determined |
| S1S_extracted_60668 | Not-determined |
| S1S_extracted_60670 | Low-quality |
| S1S_extracted_60675 | Not-determined |
| S1S_extracted_60677 | Not-determined |
| S1S_extracted_60680 | Not-determined |
| S1S_extracted_60689 | Low-quality |
| S1S_extracted_60696 | Low-quality |
| S1S_extracted_60699 | Not-determined |
| S1S_extracted_60705 | Not-determined |
| S1S_extracted_60727 | Low-quality |
| S1S_extracted_60731 | Not-determined |
| S1S_extracted_60755 | Low-quality |
| S1S_extracted_60759 | Not-determined |
| S1S_extracted_60762 | Not-determined |
| S1S_extracted_60764 | Not-determined |
| S1S_extracted_60778 | Not-determined |
| S1S_extracted_60786 | Not-determined |
| S1S_extracted_60798 | Not-determined |
| S1S_extracted_60799 | Not-determined |
| S1S_extracted_60803 | Not-determined |
| S1S_extracted_60813 | Not-determined |
| S1S_extracted_60820 | Not-determined |
| S1S_extracted_60825 | Not-determined |
| S1S_extracted_60831 | Not-determined |
| S1S_extracted_60833 | Not-determined |
| S1S_extracted_60845 | Low-quality |
| S1S_extracted_60869 | Not-determined |
| S1S_extracted_60897 | Not-determined |
| S1S_extracted_60918 | Not-determined |
| S1S_extracted_60930 | Not-determined |
| S1S_extracted_60951 | Not-determined |
| S1S_extracted_60953 | Not-determined |
| S1S_extracted_60959 | Low-quality |
| S1S_extracted_60974 | Not-determined |
| S1S_extracted_61003 | Not-determined |
| S1S_extracted_61013 | Not-determined |
| S1S_extracted_61021 | Not-determined |
| S1S_extracted_61033 | Not-determined |
| S1S_extracted_61040 | Not-determined |
| S1S_extracted_61046 | Not-determined |
| S1S_extracted_61049 | Not-determined |
| S1S_extracted_61081 | Not-determined |
| S1S_extracted_61082 | Not-determined |
| S1S_extracted_61093 | Low-quality |
| S1S_extracted_61099 | Not-determined |
| S1S_extracted_61104 | Not-determined |
| S1S_extracted_61111 | Not-determined |
| S1S_extracted_61116 | Not-determined |
| S1S_extracted_61122 | Not-determined |
| S1S_extracted_61134 | Not-determined |
| S1S_extracted_61138 | Not-determined |
| S1S_extracted_61142 | Not-determined |
| S1S_extracted_61143 | Not-determined |
| S1S_extracted_61152 | Not-determined |
| S1S_extracted_61154 | Not-determined |
| S1S_extracted_61157 | Not-determined |
| S1S_extracted_61170 | Not-determined |
| S1S_extracted_61172 | Not-determined |
| S1S_extracted_61174 | Low-quality |

| | |
|---|---|
| S15_extracted_61189 | Not-determined |
| S15_extracted_61198 | Not-determined |
| S15_extracted_61201 | Not-determined |
| S15_extracted_61214 | Not-determined |
| S15_extracted_61216 | Not-determined |
| S15_extracted_61218 | Not-determined |
| S15_extracted_61219 | Not-determined |
| S15_extracted_61223 | Not-determined |
| S15_extracted_61248 | Not-determined |
| S15_extracted_61255 | Not-determined |
| S15_extracted_61268 | Not-determined |
| S15_extracted_61277 | Not-determined |
| S15_extracted_61292 | Not-determined |
| S15_extracted_61296 | Not-determined |
| S15_extracted_61318 | Not-determined |
| S15_extracted_61320 | Low-quality |
| S15_extracted_61322 | Not-determined |
| S15_extracted_61326 | Not-determined |
| S15_extracted_61330 | Not-determined |
| S15_extracted_61333 | Not-determined |
| S15_extracted_61334 | Low-quality |
| S15_extracted_61339 | Not-determined |
| S15_extracted_61348 | Not-determined |
| S15_extracted_61356 | Not-determined |
| S15_extracted_61371 | Not-determined |
| S15_extracted_61376 | Not-determined |
| S15_extracted_61391 | Not-determined |
| S15_extracted_61394 | Not-determined |
| S15_extracted_61404 | Not-determined |
| S15_extracted_61408 | Not-determined |
| S15_extracted_61419 | Not-determined |
| S15_extracted_61430 | Not-determined |
| S15_extracted_61443 | Low-quality |
| S15_extracted_61447 | Not-determined |
| S15_extracted_61451 | Not-determined |
| S15_extracted_61454 | Not-determined |
| S15_extracted_61458 | Not-determined |
| S15_extracted_61466 | Not-determined |
| S15_extracted_61483 | Not-determined |
| S15_extracted_61490 | Not-determined |
| S15_extracted_61501 | Not-determined |
| S15_extracted_61513 | Not-determined |
| S15_extracted_61516 | Not-determined |
| S15_extracted_61527 | Not-determined |
| S15_extracted_61529 | Not-determined |
| S15_extracted_61532 | Not-determined |
| S15_extracted_61539 | Not-determined |
| S15_extracted_61560 | Not-determined |
| S15_extracted_61562 | Not-determined |
| S15_extracted_61564 | Not-determined |
| S15_extracted_61585 | Not-determined |
| S15_extracted_61610 | Not-determined |
| S15_extracted_61613 | Not-determined |
| S15_extracted_61618 | Not-determined |
| S15_extracted_61626 | Not-determined |
| S15_extracted_61627 | Not-determined |
| S15_extracted_61652 | Not-determined |
| S15_extracted_61655 | Low-quality |
| S15_extracted_61664 | Not-determined |
| S15_extracted_61667 | Not-determined |
| S15_extracted_61676 | Not-determined |
| S15_extracted_61698 | Not-determined |
| S15_extracted_61702 | Not-determined |
| S15_extracted_61712 | Not-determined |
| S15_extracted_61713 | Not-determined |
| S15_extracted_61731 | Not-determined |
| S15_extracted_61733 | Not-determined |
| S15_extracted_61737 | Not-determined |
| S15_extracted_61739 | Not-determined |
| S15_extracted_61753 | Not-determined |
| S15_extracted_61768 | Not-determined |
| S15_extracted_61772 | Not-determined |
| S15_extracted_61777 | Not-determined |
| S15_extracted_61791 | Low-quality |
| S15_extracted_61798 | Not-determined |
| S15_extracted_61807 | Not-determined |
| S15_extracted_61833 | Not-determined |
| S15_extracted_61840 | Not-determined |
| S15_extracted_61850 | Low-quality |
| S15_extracted_61858 | Not-determined |
| S15_extracted_61881 | Not-determined |
| S15_extracted_61908 | Not-determined |
| S15_extracted_61910 | Not-determined |
| S15_extracted_61913 | Not-determined |
| S15_extracted_61917 | Not-determined |
| S15_extracted_61945 | Not-determined |
| S15_extracted_61950 | Not-determined |
| S15_extracted_61952 | Not-determined |
| S15_extracted_61984 | Not-determined |
| S15_extracted_61989 | Not-determined |
| S15_extracted_61992 | Not-determined |
| S15_extracted_62011 | Low-quality |
| S15_extracted_62013 | Low-quality |
| S15_extracted_62016 | Low-quality |
| S15_extracted_62042 | Not-determined |
| S15_extracted_62060 | Not-determined |
| S15_extracted_62067 | Not-determined |
| S15_extracted_62076 | Not-determined |
| S15_extracted_62078 | Low-quality |
| S15_extracted_62083 | Not-determined |
| S15_extracted_62084 | Not-determined |
| S15_extracted_62088 | Not-determined |
| S15_extracted_62105 | Not-determined |
| S15_extracted_62114 | Not-determined |
| S15_extracted_62124 | Not-determined |
| S15_extracted_62126 | Not-determined |
| S15_extracted_62134 | Not-determined |
| S15_extracted_62148 | Not-determined |
| S15_extracted_62150 | Not-determined |
| S15_extracted_62151 | Not-determined |
| S15_extracted_62162 | Low-quality |
| S15_extracted_62166 | Not-determined |
| S15_extracted_62186 | Not-determined |
| S15_extracted_62190 | Not-determined |
| S15_extracted_62213 | Not-determined |
| S15_extracted_62231 | Not-determined |
| S15_extracted_62234 | Not-determined |
| S15_extracted_62238 | Not-determined |
| S15_extracted_62240 | Not-determined |
| S15_extracted_62250 | Not-determined |
| S15_extracted_62263 | Not-determined |
| S15_extracted_62278 | Not-determined |
| S15_extracted_62284 | Not-determined |
| S15_extracted_62285 | Not-determined |
| S15_extracted_62298 | Not-determined |
| S15_extracted_62327 | Not-determined |
| S15_extracted_62340 | Not-determined |
| S15_extracted_62366 | Not-determined |
| S15_extracted_62369 | Not-determined |
| S15_extracted_62395 | Low-quality |
| S15_extracted_62405 | Not-determined |
| S15_extracted_62410 | Not-determined |
| S15_extracted_62411 | Not-determined |
| S15_extracted_62427 | Not-determined |
| S15_extracted_62453 | Low-quality |
| S15_extracted_62462 | Not-determined |
| S15_extracted_62496 | Not-determined |
| S15_extracted_62505 | Not-determined |
| S15_extracted_62509 | Not-determined |
| S15_extracted_62528 | Not-determined |
| S15_extracted_62531 | Not-determined |
| S15_extracted_62564 | Not-determined |
| S15_extracted_62566 | Not-determined |
| S15_extracted_62568 | Not-determined |
| S15_extracted_62572 | Not-determined |
| S15_extracted_62595 | Not-determined |
| S15_extracted_62621 | Not-determined |
| S15_extracted_62625 | Not-determined |
| S15_extracted_62627 | Not-determined |
| S15_extracted_62628 | Not-determined |
| S15_extracted_62631 | Not-determined |
| S15_extracted_62637 | Not-determined |
| S15_extracted_62649 | Not-determined |
| S15_extracted_62657 | Not-determined |
| S15_extracted_62660 | Not-determined |
| S15_extracted_62662 | Not-determined |
| S15_extracted_62681 | Not-determined |
| S15_extracted_62696 | Not-determined |
| S15_extracted_62707 | Not-determined |
| S15_extracted_62714 | Not-determined |
| S15_extracted_62723 | Not-determined |
| S15_extracted_62724 | Not-determined |
| S15_extracted_62726 | Not-determined |
| S15_extracted_62727 | Low-quality |
| S15_extracted_62728 | Not-determined |
| S15_extracted_62733 | Not-determined |
| S15_extracted_62746 | Not-determined |
| S15_extracted_62752 | Not-determined |
| S15_extracted_62766 | Low-quality |
| S15_extracted_62768 | Not-determined |
| S15_extracted_62778 | Not-determined |
| S15_extracted_62781 | Low-quality |
| S15_extracted_62791 | Not-determined |
| S15_extracted_62802 | Not-determined |
| S15_extracted_62803 | Not-determined |
| S15_extracted_62808 | Not-determined |
| S15_extracted_62811 | Not-determined |
| S15_extracted_62829 | Not-determined |
| S15_extracted_62837 | Low-quality |
| S15_extracted_62854 | Not-determined |
| S15_extracted_62855 | Not-determined |
| S15_extracted_62883 | Not-determined |
| S15_extracted_62889 | Not-determined |
| S15_extracted_62896 | Not-determined |
| S15_extracted_62902 | Not-determined |
| S15_extracted_62922 | Not-determined |
| S15_extracted_62925 | Not-determined |
| S15_extracted_62927 | Not-determined |
| S15_extracted_62930 | Not-determined |
| S15_extracted_62936 | Not-determined |
| S15_extracted_62937 | Not-determined |
| S15_extracted_62943 | Not-determined |
| S15_extracted_62959 | Not-determined |
| S15_extracted_62995 | Not-determined |
| S15_extracted_62998 | Not-determined |
| S15_extracted_63013 | Not-determined |
| S15_extracted_63022 | Not-determined |
| S15_extracted_63037 | Not-determined |
| S15_extracted_63043 | Not-determined |
| S15_extracted_63050 | Not-determined |
| S15_extracted_63052 | Not-determined |
| S15_extracted_63056 | Not-determined |
| S15_extracted_63072 | Not-determined |
| S15_extracted_63073 | Not-determined |
| S15_extracted_63077 | Not-determined |
| S15_extracted_63078 | Not-determined |
| S15_extracted_63091 | Not-determined |
| S15_extracted_63097 | Low-quality |
| S15_extracted_63120 | Not-determined |
| S15_extracted_63125 | Not-determined |
| S15_extracted_63146 | Not-determined |
| S5_vs_k141_135750\|\|full | Medium-quality |
| S15_extracted_63178 | Not-determined |
| S15_extracted_63179 | Not-determined |
| S15_extracted_63193 | Not-determined |
| S15_extracted_63194 | Not-determined |
| S15_extracted_63196 | Not-determined |
| S15_extracted_63199 | Low-quality |
| S15_extracted_63209 | Not-determined |
| S15_extracted_63210 | Not-determined |
| S15_extracted_63214 | Not-determined |
| S15_extracted_63215 | Not-determined |
| S15_extracted_63218 | Not-determined |
| S15_extracted_63219 | Not-determined |
| S15_extracted_63227 | Not-determined |
| S15_extracted_63228 | Not-determined |
| S15_extracted_63247 | Not-determined |
| S15_extracted_63254 | Not-determined |
| S15_extracted_63267 | Not-determined |
| S15_extracted_63274 | Not-determined |
| S15_extracted_63280 | Not-determined |
| S15_extracted_63292 | Not-determined |
| S15_extracted_63295 | Not-determined |
| S15_extracted_63297 | Not-determined |
| S15_extracted_63306 | Not-determined |
| S15_extracted_63314 | Low-quality |
| S15_extracted_63325 | Low-quality |
| S15_extracted_63329 | Not-determined |
| S15_extracted_63340 | Not-determined |
| S15_extracted_63342 | Not-determined |
| S15_extracted_63349 | Not-determined |
| S15_extracted_63371 | Not-determined |
| S15_extracted_63373 | Not-determined |
| S15_extracted_63375 | Not-determined |
| S15_extracted_63376 | Not-determined |
| S15_extracted_63384 | Not-determined |
| S15_extracted_63389 | Not-determined |
| S15_extracted_63416 | Not-determined |
| S15_extracted_63423 | Not-determined |
| S15_extracted_63436 | Low-quality |
| S15_extracted_63439 | Low-quality |
| S15_extracted_63441 | Not-determined |
| S15_extracted_63444 | Not-determined |
| S15_extracted_63448 | Not-determined |
| S15_extracted_63469 | Not-determined |
| S15_extracted_63480 | Not-determined |
| S15_extracted_63493 | Low-quality |
| S15_extracted_63509 | Not-determined |
| S15_extracted_63520 | Not-determined |
| S15_extracted_63533 | Not-determined |
| S15_extracted_63538 | Not-determined |
| S15_extracted_63540 | Not-determined |
| S15_extracted_63549 | Not-determined |
| S15_extracted_63560 | Not-determined |
| S15_extracted_63587 | Not-determined |
| S15_extracted_63600 | Low-quality |
| S15_extracted_63603 | Not-determined |
| S15_extracted_63617 | Not-determined |
| S15_extracted_63620 | Not-determined |
| S15_extracted_63623 | Not-determined |
| S15_extracted_63631 | Not-determined |
| S15_extracted_63654 | Low-quality |
| S15_extracted_63665 | Not-determined |
| S15_extracted_63672 | Not-determined |
| S15_extracted_63674 | Not-determined |
| S15_extracted_63698 | Not-determined |
| S15_extracted_63742 | Not-determined |
| S15_extracted_63746 | Not-determined |
| S15_extracted_63749 | Low-quality |
| S15_extracted_63752 | Not-determined |
| S15_extracted_63755 | Not-determined |
| S15_extracted_63763 | Not-determined |
| S15_extracted_63768 | Not-determined |
| S15_extracted_63774 | Not-determined |
| S15_extracted_63784 | Not-determined |
| S15_extracted_63789 | Low-quality |
| S15_extracted_63796 | Low-quality |
| S15_extracted_63805 | Not-determined |
| S15_extracted_63820 | Low-quality |
| S15_extracted_63827 | Not-determined |
| S15_extracted_63838 | Not-determined |
| S15_extracted_63854 | Not-determined |
| S15_extracted_63880 | Not-determined |
| S15_extracted_63883 | Not-determined |
| S15_extracted_63884 | Not-determined |
| S15_extracted_63891 | Not-determined |
| S15_extracted_63896 | Not-determined |
| S15_extracted_63901 | Not-determined |
| S15_extracted_63905 | Not-determined |
| S15_extracted_63911 | Not-determined |
| S15_extracted_63916 | Not-determined |
| S15_extracted_63918 | Not-determined |
| S15_extracted_63920 | Not-determined |
| S15_extracted_63930 | Not-determined |
| S15_extracted_63931 | Not-determined |
| S15_extracted_63932 | Not-determined |
| S15_extracted_63933 | Not-determined |
| S15_extracted_63938 | Low-quality |
| S15_extracted_63943 | Not-determined |
| S15_extracted_63944 | Not-determined |
| S15_extracted_63954 | Not-determined |
| S15_extracted_63962 | Not-determined |
| S15_extracted_63964 | Not-determined |
| S15_extracted_63971 | Not-determined |
| S15_extracted_63976 | Not-determined |
| S15_extracted_63989 | Not-determined |
| S15_extracted_64001 | Not-determined |
| S15_extracted_64002 | Not-determined |
| S15_extracted_64008 | Low-quality |
| S15_extracted_64011 | Not-determined |
| S15_extracted_64014 | Not-determined |
| S15_extracted_64018 | Not-determined |
| S15_extracted_64024 | Low-quality |
| S15_extracted_64028 | Not-determined |
| S15_extracted_64041 | Not-determined |
| S15_extracted_64054 | Not-determined |
| S15_extracted_64070 | Not-determined |
| S15_extracted_64082 | Not-determined |
| S15_extracted_64084 | Not-determined |
| S15_extracted_64089 | Low-quality |
| S15_extracted_64097 | Not-determined |
| S15_extracted_64098 | Not-determined |
| S15_extracted_64105 | Not-determined |
| S15_extracted_64124 | Not-determined |
| S15_extracted_64139 | Not-determined |
| S15_extracted_64140 | Not-determined |
| S15_extracted_64148 | Not-determined |
| S15_extracted_64174 | Not-determined |
| S15_extracted_64175 | Not-determined |
| S15_extracted_64179 | Not-determined |
| S15_extracted_64182 | Not-determined |
| S15_extracted_64188 | Not-determined |
| S15_extracted_64189 | Not-determined |
| S15_extracted_64190 | Not-determined |
| S15_extracted_64193 | Not-determined |
| S15_extracted_64194 | Not-determined |
| S15_extracted_64196 | Not-determined |
| S15_extracted_64208 | Not-determined |
| S15_extracted_64210 | Not-determined |
| S15_extracted_64213 | Not-determined |
| S15_extracted_64223 | Not-determined |
| S15_extracted_64224 | Not-determined |
| S15_extracted_64245 | Not-determined |
| S15_extracted_64253 | Not-determined |
| S15_extracted_64267 | Not-determined |
| S15_extracted_64293 | Not-determined |
| S15_extracted_64295 | Not-determined |
| S15_extracted_64296 | Not-determined |
| S15_extracted_64311 | Not-determined |
| S15_extracted_64318 | Not-determined |
| S15_extracted_64330 | Not-determined |
| S15_extracted_64340 | Not-determined |
| S15_extracted_64342 | Not-determined |
| S15_extracted_64358 | Not-determined |
| S15_extracted_64370 | Not-determined |
| S15_extracted_64377 | Not-determined |
| S15_extracted_64381 | Not-determined |
| S15_extracted_64390 | Not-determined |
| S15_extracted_64398 | Not-determined |
| S15_extracted_64401 | Not-determined |
| S15_extracted_64422 | Not-determined |
| S15_extracted_64428 | Not-determined |
| S15_extracted_64430 | Not-determined |
| S15_extracted_64434 | Not-determined |
| S15_extracted_64437 | Not-determined |
| S15_extracted_64443 | Not-determined |
| S15_extracted_64464 | Not-determined |
| S15_extracted_64467 | Not-determined |
| S15_extracted_64475 | Not-determined |
| S15_extracted_64483 | Not-determined |
| S15_extracted_64489 | Not-determined |
| S15_extracted_64497 | Not-determined |
| S15_extracted_64500 | Not-determined |
| S15_extracted_64506 | Not-determined |
| S15_extracted_64514 | Not-determined |
| S15_extracted_64516 | Not-determined |
| S15_extracted_64525 | Not-determined |
| S15_extracted_64527 | Not-determined |
| S15_extracted_64533 | Not-determined |
| S15_extracted_64541 | Not-determined |
| S15_extracted_64544 | Not-determined |
| S15_extracted_64548 | Not-determined |
| S15_extracted_64554 | Not-determined |
| S15_extracted_64555 | Not-determined |
| S15_extracted_64560 | Not-determined |
| S15_extracted_64564 | Not-determined |
| S15_extracted_64576 | Low-quality |
| S15_extracted_64596 | Not-determined |
| S15_extracted_64598 | Not-determined |
| S15_extracted_64601 | Not-determined |
| S15_extracted_64609 | Low-quality |
| S15_extracted_64613 | Not-determined |
| S15_extracted_64617 | Not-determined |
| S15_extracted_64620 | Not-determined |
| S15_extracted_64625 | Not-determined |
| S15_extracted_64629 | Not-determined |
| S15_extracted_64636 | Not-determined |
| S15_extracted_64647 | Not-determined |
| S15_extracted_64652 | Not-determined |
| S15_extracted_64657 | Not-determined |
| S15_extracted_64660 | Not-determined |
| S15_extracted_64667 | Low-quality |
| S15_extracted_64669 | Low-quality |
| S15_extracted_64686 | Not-determined |
| S15_extracted_64690 | Not-determined |
| S15_extracted_64691 | Not-determined |
| S15_extracted_64701 | Not-determined |
| S15_extracted_64706 | Not-determined |
| S15_extracted_64711 | Not-determined |
| S15_extracted_64721 | Not-determined |
| S15_extracted_64722 | Not-determined |
| S15_extracted_64725 | Not-determined |
| S15_extracted_64734 | Not-determined |
| S15_extracted_64755 | Not-determined |
| S15_extracted_64756 | Not-determined |
| S15_extracted_64759 | Low-quality |
| S15_extracted_64762 | Not-determined |
| S15_extracted_64797 | Not-determined |
| S15_extracted_64798 | Not-determined |
| S15_extracted_64817 | Not-determined |
| S15_extracted_64822 | Not-determined |
| S15_extracted_64841 | Not-determined |
| S15_extracted_64857 | Not-determined |
| S15_extracted_64871 | Not-determined |
| S15_extracted_64881 | Low-quality |
| S15_extracted_64889 | Not-determined |
| S15_extracted_64902 | Not-determined |
| S15_extracted_64909 | Not-determined |
| S15_extracted_64911 | Not-determined |
| S15_extracted_64920 | Not-determined |
| S15_extracted_64926 | Not-determined |
| S15_extracted_64933 | Not-determined |
| S15_extracted_64934 | Not-determined |
| S15_extracted_64938 | Not-determined |
| S15_extracted_64940 | Not-determined |
| S15_extracted_64949 | Not-determined |
| S15_extracted_64954 | Not-determined |
| S15_extracted_64955 | Not-determined |
| S15_extracted_64961 | Not-determined |
| S15_extracted_64969 | Not-determined |
| S15_extracted_64972 | Not-determined |
| S15_extracted_64976 | Low-quality |
| S15_extracted_64983 | Not-determined |
| S15_extracted_65013 | Not-determined |
| S15_extracted_65052 | Not-determined |
| S15_extracted_65053 | Not-determined |
| S15_extracted_65067 | Not-determined |
| S15_extracted_65068 | Not-determined |
| S15_extracted_65081 | Not-determined |
| S15_extracted_65104 | Not-determined |
| S15_extracted_65105 | Not-determined |
| S15_extracted_65115 | Not-determined |
| S15_extracted_65120 | Not-determined |
| S15_extracted_65124 | Not-determined |
| S15_extracted_65127 | Not-determined |
| S15_extracted_65133 | Not-determined |
| S15_extracted_65136 | Not-determined |
| S15_extracted_65154 | Not-determined |
| S15_extracted_65171 | Not-determined |
| S15_extracted_65172 | Not-determined |
| S15_extracted_65183 | Low-quality |
| S15_extracted_65200 | Not-determined |
| S15_extracted_65201 | Not-determined |
| S15_extracted_65204 | Not-determined |
| S15_extracted_65205 | Low-quality |
| S15_extracted_65209 | Not-determined |
| S15_extracted_65227 | Not-determined |
| S15_extracted_65230 | Not-determined |
| S15_extracted_65235 | Low-quality |
| S15_extracted_65257 | Not-determined |
| S15_extracted_65273 | Not-determined |
| S15_extracted_65275 | Not-determined |
| S15_extracted_65276 | Not-determined |
| S15_extracted_65283 | Not-determined |
| S15_extracted_65295 | Not-determined |
| S15_extracted_65306 | Not-determined |
| S15_extracted_65307 | Not-determined |
| S15_extracted_65329 | Not-determined |
| S15_extracted_65331 | Not-determined |
| S15_extracted_65333 | Not-determined |
| S15_extracted_65340 | Not-determined |
| S15_extracted_65343 | Not-determined |
| S15_extracted_65355 | Not-determined |
| S15_extracted_65367 | Not-determined |
| S15_extracted_65376 | Not-determined |
| S15_extracted_65409 | Not-determined |
| S15_extracted_65410 | Not-determined |
| S15_extracted_65412 | Not-determined |
| S15_extracted_65419 | Not-determined |
| S15_extracted_65421 | Not-determined |
| S15_extracted_65430 | Not-determined |
| S15_extracted_65431 | Low-quality |
| S15_extracted_65437 | Not-determined |
| S15_extracted_65442 | Not-determined |
| S15_extracted_65445 | Not-determined |
| S15_extracted_65450 | Not-determined |
| S15_extracted_65462 | Not-determined |
| S15_extracted_65470 | Not-determined |
| S15_extracted_65481 | Not-determined |
| S15_extracted_65482 | Not-determined |
| S15_extracted_65493 | Not-determined |
| S15_extracted_65507 | Not-determined |
| S15_extracted_65538 | Not-determined |
| S15_extracted_65546 | Not-determined |
| S15_extracted_65547 | Not-determined |
| S15_extracted_65549 | Not-determined |
| S15_extracted_65566 | Not-determined |
| S15_extracted_65572 | Low-quality |
| S15_extracted_65581 | Not-determined |
| S15_extracted_65587 | Not-determined |
| S15_extracted_65593 | Not-determined |
| S15_extracted_65598 | Not-determined |
| S15_extracted_65616 | Not-determined |
| S15_extracted_65620 | Not-determined |
| S15_extracted_65626 | Low-quality |
| S15_extracted_65640 | Not-determined |
| S15_extracted_65676 | Not-determined |
| S15_extracted_65677 | Not-determined |
| S15_extracted_65682 | Not-determined |
| S15_extracted_65686 | Not-determined |
| S15_extracted_65696 | Not-determined |
| S15_extracted_65705 | Not-determined |
| S15_extracted_65710 | Not-determined |
| S15_extracted_65715 | Not-determined |
| S15_extracted_65720 | Not-determined |
| S15_extracted_65768 | Not-determined |
| S15_extracted_65770 | Not-determined |
| S15_extracted_65773 | Not-determined |
| S15_extracted_65824 | Not-determined |
| S15_extracted_65835 | Not-determined |
| S15_extracted_65843 | Not-determined |
| S15_extracted_65855 | Not-determined |
| S15_extracted_65858 | Not-determined |
| S15_extracted_65867 | Not-determined |
| S15_extracted_65869 | Not-determined |
| S15_extracted_65883 | Low-quality |
| S15_extracted_65886 | Not-determined |
| S15_extracted_65899 | Low-quality |
| S15_extracted_65906 | Not-determined |
| S15_extracted_65910 | Not-determined |
| S15_extracted_65915 | Not-determined |
| S15_extracted_65920 | Not-determined |
| S15_extracted_65923 | Not-determined |
| S15_extracted_65925 | Not-determined |
| S15_extracted_65929 | Not-determined |
| S15_extracted_65966 | Not-determined |
| S15_extracted_65967 | Low-quality |
| S15_extracted_65968 | Not-determined |
| S15_extracted_65970 | Not-determined |
| S15_extracted_65976 | Not-determined |
| S15_extracted_65987 | Not-determined |
| S15_extracted_65989 | Not-determined |
| S15_extracted_65990 | Not-determined |
| S15_extracted_65992 | Not-determined |
| S15_extracted_65997 | Not-determined |
| S15_extracted_66003 | Not-determined |
| S15_extracted_66008 | Not-determined |
| S15_extracted_66012 | Not-determined |
| S15_extracted_66016 | Not-determined |
| S15_extracted_66028 | Not-determined |
| S15_extracted_66046 | Not-determined |
| S15_extracted_66050 | Not-determined |
| S15_extracted_66054 | Not-determined |
| S15_extracted_66074 | Not-determined |
| S15_extracted_66084 | Not-determined |
| S15_extracted_66108 | Not-determined |
| S15_extracted_66124 | Low-quality |
| S15_extracted_66138 | Not-determined |
| S15_extracted_66150 | Not-determined |
| S15_extracted_66151 | Not-determined |
| S15_extracted_66159 | Not-determined |
| S15_extracted_66162 | Low-quality |
| S15_extracted_66163 | Not-determined |
| S15_extracted_66173 | Not-determined |
| S15_extracted_66185 | Not-determined |
| S15_extracted_66188 | Low-quality |
| S15_extracted_66199 | Not-determined |
| S15_extracted_66205 | Not-determined |
| S15_extracted_66212 | Not-determined |

| | |
|---|---|
| S1S_extracted_66218 | Not-determined |
| S1S_extracted_66224 | Not-determined |
| S1S_extracted_66227 | Not-determined |
| S1S_extracted_66231 | Not-determined |
| S1S_extracted_66238 | Not-determined |
| S1S_extracted_66252 | Not-determined |
| S1S_extracted_66256 | Low-quality |
| S1S_extracted_66257 | Not-determined |
| S1S_extracted_66289 | Low-quality |
| S1S_extracted_66295 | Not-determined |
| S1S_extracted_66314 | Low-quality |
| S1S_extracted_66332 | Not-determined |
| S1S_extracted_66338 | Low-quality |
| S1S_extracted_66347 | Not-determined |
| S1S_extracted_66348 | Low-quality |
| S1S_extracted_66357 | Not-determined |
| S1S_extracted_66358 | Not-determined |
| S1S_extracted_66363 | Not-determined |
| S1S_extracted_66367 | Low-quality |
| S1S_extracted_66374 | Not-determined |
| S1S_extracted_66377 | Low-quality |
| S1S_extracted_66383 | Not-determined |
| S1S_extracted_66386 | Not-determined |
| S1S_extracted_66387 | Not-determined |
| S1S_extracted_66388 | Not-determined |
| S1S_extracted_66395 | Not-determined |
| S1S_extracted_66401 | Not-determined |
| S1S_extracted_66419 | Not-determined |
| S1S_extracted_66423 | Not-determined |
| S1S_extracted_66476 | Not-determined |
| S1S_extracted_66483 | Low-quality |
| S1S_extracted_66493 | Not-determined |
| S1S_extracted_66494 | Not-determined |
| S1S_extracted_66501 | Not-determined |
| S1S_extracted_66503 | Low-quality |
| S1S_extracted_66508 | Not-determined |
| S1S_extracted_66515 | Not-determined |
| S1S_extracted_66519 | Not-determined |
| S1S_extracted_66522 | Low-quality |
| S1S_extracted_66527 | Not-determined |
| S1S_extracted_66531 | Not-determined |
| S1S_extracted_66541 | Not-determined |
| S1S_extracted_66550 | Low-quality |
| S1S_extracted_66572 | Not-determined |
| S1S_extracted_66578 | Not-determined |
| S1S_extracted_66594 | Low-quality |
| S1S_extracted_66597 | Not-determined |
| S1S_extracted_66605 | Low-quality |
| S1S_extracted_66610 | Low-quality |
| S1S_extracted_66616 | Not-determined |
| S1S_extracted_66619 | Low-quality |
| S1S_extracted_66620 | Not-determined |
| S1S_extracted_66627 | Not-determined |
| S1S_extracted_66629 | Not-determined |
| S1S_extracted_66635 | Not-determined |
| S1S_extracted_66641 | Not-determined |
| S1S_extracted_66655 | Not-determined |
| S1S_extracted_66656 | Not-determined |
| S1S_extracted_66659 | Not-determined |
| S1S_extracted_66661 | Not-determined |
| S1S_extracted_66667 | Not-determined |
| S1S_extracted_66682 | Not-determined |
| S1S_extracted_66684 | Not-determined |
| S1S_extracted_66686 | Not-determined |
| S1S_extracted_66694 | Not-determined |
| S1S_extracted_66719 | Not-determined |
| S1S_extracted_66727 | Low-quality |
| S1S_extracted_66735 | Not-determined |
| S1S_extracted_66758 | Not-determined |
| S1S_extracted_66770 | Not-determined |
| S1S_extracted_66783 | Not-determined |
| S1S_extracted_66785 | Not-determined |
| S1S_extracted_66789 | Low-quality |
| S1S_extracted_66793 | Low-quality |
| S1S_extracted_66814 | Not-determined |
| S1S_extracted_66815 | Not-determined |
| S1S_extracted_66818 | Not-determined |
| S1S_extracted_66829 | Low-quality |
| S1S_extracted_66839 | Low-quality |
| S1S_extracted_66847 | Low-quality |
| S1S_extracted_66867 | Not-determined |
| S1S_extracted_66870 | Not-determined |
| S1S_extracted_66881 | Not-determined |
| S1S_extracted_66890 | Not-determined |
| S1S_extracted_66909 | Not-determined |
| S1S_extracted_66915 | Not-determined |
| S1S_extracted_66916 | Not-determined |
| S1S_extracted_66921 | Not-determined |
| S1S_extracted_66937 | Not-determined |
| S1S_extracted_66945 | Not-determined |
| S1S_extracted_66954 | Low-quality |
| S1S_extracted_66957 | Low-quality |
| S1S_extracted_66970 | Not-determined |
| S1S_extracted_66973 | Not-determined |
| S1S_extracted_66979 | Not-determined |
| S1S_extracted_67018 | Not-determined |
| S1S_extracted_67023 | Not-determined |
| S1S_extracted_67035 | Not-determined |
| S1S_extracted_67044 | Not-determined |
| S1S_extracted_67062 | Not-determined |
| S1S_extracted_67082 | Not-determined |
| S1S_extracted_67088 | Not-determined |
| S1S_extracted_67107 | Not-determined |
| S1S_extracted_67117 | Not-determined |
| S1S_extracted_67118 | Not-determined |
| S1S_extracted_67131 | Not-determined |
| S1S_extracted_67135 | Not-determined |
| S1S_extracted_67143 | Not-determined |
| S1S_extracted_67160 | Not-determined |
| S1S_extracted_67167 | Not-determined |
| S1S_extracted_67170 | Not-determined |
| S1S_extracted_67194 | Not-determined |
| S1S_extracted_67197 | Not-determined |
| S1S_extracted_67201 | Not-determined |
| S1S_extracted_67215 | Not-determined |
| S1S_extracted_67219 | Not-determined |
| S1S_extracted_67221 | Not-determined |
| S1S_extracted_67223 | Not-determined |
| S1S_extracted_67224 | Not-determined |
| S1S_extracted_67226 | Not-determined |
| S1S_extracted_67236 | Not-determined |
| S1S_extracted_67242 | Not-determined |
| S1S_extracted_67243 | Not-determined |
| S1S_extracted_67246 | Not-determined |
| S1S_extracted_67247 | Not-determined |
| S1S_extracted_67248 | Not-determined |
| S1S_extracted_67249 | Not-determined |
| S1S_extracted_67259 | Not-determined |
| S1S_extracted_67270 | Not-determined |
| S1S_extracted_67274 | Not-determined |
| S1S_extracted_67287 | Not-determined |
| S1S_extracted_67291 | Not-determined |
| S1S_extracted_67293 | Not-determined |
| S1S_extracted_67297 | Not-determined |
| S1S_extracted_67316 | Low-quality |
| S1S_extracted_67319 | Not-determined |
| S1S_extracted_67328 | Not-determined |
| S1S_extracted_67330 | Not-determined |
| S1S_extracted_67334 | Not-determined |
| S1S_extracted_67338 | Not-determined |
| S1S_extracted_67360 | Not-determined |
| S1S_extracted_67376 | Not-determined |
| S1S_extracted_67382 | Not-determined |
| S1S_extracted_67400 | Not-determined |
| S1S_extracted_67402 | Not-determined |
| S1S_extracted_67405 | Not-determined |
| S1S_extracted_67416 | Not-determined |
| S1S_extracted_67417 | Low-quality |
| S1S_extracted_67427 | Not-determined |
| S1S_extracted_67437 | Not-determined |
| S1S_extracted_67444 | Not-determined |
| S1S_extracted_67450 | Not-determined |
| S1S_extracted_67455 | Not-determined |
| S1S_extracted_67457 | Not-determined |
| S1S_extracted_67473 | Not-determined |
| S1S_extracted_67478 | Low-quality |
| S1S_extracted_67493 | Not-determined |
| S1S_extracted_67502 | Not-determined |
| S1S_extracted_67503 | Not-determined |
| S1S_extracted_67507 | Low-quality |
| S1S_extracted_67508 | Low-quality |
| S1S_extracted_67511 | Not-determined |
| S1S_extracted_67514 | Low-quality |
| S1S_extracted_67529 | Not-determined |
| S1S_extracted_67534 | Not-determined |
| S1S_extracted_67546 | Low-quality |
| S1S_extracted_67555 | Not-determined |
| S1S_extracted_67556 | Not-determined |
| S1S_extracted_67563 | Not-determined |
| S1S_extracted_67566 | Not-determined |
| S1S_extracted_67569 | Not-determined |
| S1S_extracted_67574 | Not-determined |
| S1S_extracted_67580 | Not-determined |
| S1S_extracted_67585 | Not-determined |
| S1S_extracted_67608 | Not-determined |
| S1S_extracted_67626 | Not-determined |
| S1S_extracted_67634 | Not-determined |
| S1S_extracted_67647 | Not-determined |
| S1S_extracted_67652 | Not-determined |
| S1S_extracted_67654 | Not-determined |
| S1S_extracted_67670 | Not-determined |
| S1S_extracted_67675 | Low-quality |
| S1S_extracted_67677 | Low-quality |
| S1S_extracted_67678 | Not-determined |
| S1S_extracted_67690 | Not-determined |
| S1S_extracted_67711 | Not-determined |
| S1S_extracted_67717 | Not-determined |
| S1S_extracted_67721 | Not-determined |
| S1S_extracted_67733 | Not-determined |
| S1S_extracted_67736 | Not-determined |
| S1S_extracted_67741 | Low-quality |
| S1S_extracted_67744 | Not-determined |
| S1S_extracted_67757 | Not-determined |
| S1S_extracted_67759 | Not-determined |
| S1S_extracted_67761 | Not-determined |
| S1S_extracted_67763 | Not-determined |
| S1S_extracted_67787 | Not-determined |
| S1S_extracted_67790 | Not-determined |
| S1S_extracted_67792 | Not-determined |
| S1S_extracted_67797 | Not-determined |
| S1S_extracted_67825 | Not-determined |
| S1S_extracted_67828 | Not-determined |
| S1S_extracted_67842 | Not-determined |
| S1S_extracted_67849 | Not-determined |
| S1S_extracted_67852 | Not-determined |
| S1S_extracted_67854 | Low-quality |
| S1S_extracted_67884 | Not-determined |
| S1S_extracted_67893 | Not-determined |
| S1S_extracted_67895 | Not-determined |
| S1S_extracted_67909 | Low-quality |
| S1S_extracted_67918 | Low-quality |
| S1S_extracted_67920 | Not-determined |
| S1S_extracted_67922 | Not-determined |
| S1S_extracted_67934 | Not-determined |
| S1S_extracted_67938 | Low-quality |
| S1S_extracted_67940 | Not-determined |
| S1S_extracted_67941 | Low-quality |
| S1S_extracted_67942 | Not-determined |
| S1S_extracted_67947 | Not-determined |
| S1S_extracted_67949 | Not-determined |
| S1S_extracted_67951 | Not-determined |
| S1S_extracted_67953 | Not-determined |
| S1S_extracted_67957 | Not-determined |
| S1S_extracted_67959 | Not-determined |
| S1S_extracted_67962 | Not-determined |
| S1S_extracted_67964 | Not-determined |
| S1S_extracted_67982 | Not-determined |
| S1S_extracted_67989 | Not-determined |
| S1S_extracted_67990 | Not-determined |
| S1S_extracted_68003 | Not-determined |
| S1S_extracted_68028 | Not-determined |
| S1S_extracted_68030 | Not-determined |
| S1S_extracted_68037 | Low-quality |
| S1S_extracted_68042 | Not-determined |
| S1S_extracted_68043 | Not-determined |
| S1S_extracted_68044 | Not-determined |
| S1S_extracted_68049 | Not-determined |
| S1S_extracted_68050 | Not-determined |
| S1S_extracted_68051 | Not-determined |
| S1S_extracted_68057 | Not-determined |
| S1S_extracted_68073 | Not-determined |
| S1S_extracted_68079 | Not-determined |
| S1S_extracted_68087 | Not-determined |
| S1S_extracted_68103 | Not-determined |
| S1S_extracted_68107 | Not-determined |
| S1S_extracted_68114 | Low-quality |
| S1S_extracted_68119 | Not-determined |
| S1S_extracted_68130 | Not-determined |
| S1S_extracted_68136 | Not-determined |
| S1S_extracted_68151 | Not-determined |
| S1S_extracted_68171 | Not-determined |
| S1S_extracted_68179 | Not-determined |
| S1S_extracted_68190 | Low-quality |
| S1S_extracted_68197 | Not-determined |
| S1S_extracted_68198 | Not-determined |
| S1S_extracted_68206 | Not-determined |
| S1S_extracted_68224 | Low-quality |
| S1S_extracted_68241 | Low-quality |
| S1S_extracted_68243 | Not-determined |
| S1S_extracted_68244 | Not-determined |
| S1S_extracted_68246 | Low-quality |
| S1S_extracted_68256 | Not-determined |
| S1S_extracted_68257 | Not-determined |
| S1S_extracted_68265 | Low-quality |
| S1S_extracted_68278 | Not-determined |
| S1S_extracted_68280 | Not-determined |
| S1S_extracted_68288 | Not-determined |
| S1S_extracted_68309 | Not-determined |
| S1S_extracted_68310 | Not-determined |
| S1S_extracted_68326 | Not-determined |
| S1S_extracted_68329 | Not-determined |
| S1S_extracted_68339 | Not-determined |
| S1S_extracted_68343 | Not-determined |
| S1S_extracted_68349 | Not-determined |
| S1S_extracted_68356 | Not-determined |
| S1S_extracted_68373 | Not-determined |
| S1S_extracted_68376 | Not-determined |
| S1S_extracted_68378 | Not-determined |
| S1S_extracted_68390 | Not-determined |
| S1S_extracted_68396 | Not-determined |
| S1S_extracted_68403 | Low-quality |
| S1S_extracted_68407 | Not-determined |
| S1S_extracted_68414 | Not-determined |
| S1S_extracted_68431 | Not-determined |
| S1S_extracted_68434 | Not-determined |
| S1S_extracted_68441 | Not-determined |
| S1S_extracted_68449 | Not-determined |
| S1S_extracted_68466 | Not-determined |
| S1S_extracted_68485 | Not-determined |
| S1S_extracted_68486 | Not-determined |
| S1S_extracted_68492 | Not-determined |
| S1S_extracted_68494 | Low-quality |
| S1S_extracted_68497 | Not-determined |
| S1S_extracted_68507 | Not-determined |
| S1S_extracted_68508 | Not-determined |
| S1S_extracted_68513 | Low-quality |
| S1S_extracted_68520 | Not-determined |
| S1S_extracted_68543 | Not-determined |
| S1S_extracted_68544 | Not-determined |
| S1S_extracted_68549 | Not-determined |
| S1S_extracted_68557 | Not-determined |
| S1S_extracted_68563 | Not-determined |
| S1S_extracted_68577 | Not-determined |
| S1S_extracted_68584 | Not-determined |
| S1S_extracted_68588 | Low-quality |
| S1S_extracted_68596 | Not-determined |
| S1S_extracted_68598 | Not-determined |
| S1S_extracted_68601 | Not-determined |
| S1S_extracted_68629 | Not-determined |
| S1S_extracted_68640 | Not-determined |
| S1S_extracted_68641 | Not-determined |
| S1S_extracted_68650 | Not-determined |
| S1S_extracted_68659 | Not-determined |
| S1S_extracted_68661 | Not-determined |
| S1S_extracted_68676 | Not-determined |
| S1S_extracted_68681 | Not-determined |
| S1S_extracted_68686 | Low-quality |
| S1S_extracted_68689 | Not-determined |
| S1S_extracted_68691 | Not-determined |
| S1S_extracted_68704 | Not-determined |
| S1S_extracted_68707 | Low-quality |
| S1S_extracted_68718 | Not-determined |
| S1S_extracted_68719 | Not-determined |
| S1S_extracted_68721 | Low-quality |
| S1S_extracted_68725 | Low-quality |
| S1S_extracted_68728 | Not-determined |
| S1S_extracted_68730 | Not-determined |
| S1S_extracted_68754 | Not-determined |
| S1S_extracted_68770 | Not-determined |
| S1S_extracted_68781 | Not-determined |
| S1S_extracted_68798 | Not-determined |
| S1S_extracted_68801 | Not-determined |
| S1S_extracted_68808 | Not-determined |
| S1S_extracted_68825 | Not-determined |
| S1S_extracted_68832 | Not-determined |
| S1S_extracted_68890 | Not-determined |
| S1S_extracted_68896 | Not-determined |
| S1S_extracted_68908 | Low-quality |
| S1S_extracted_68922 | Not-determined |
| S1S_extracted_68926 | Not-determined |
| S1S_extracted_68927 | Not-determined |
| S1S_extracted_68944 | Not-determined |
| S1S_extracted_68950 | Not-determined |
| S1S_extracted_68953 | Not-determined |
| S1S_extracted_68961 | Not-determined |
| S1S_extracted_68963 | Not-determined |
| S1S_extracted_68968 | Not-determined |
| S1S_extracted_68972 | Not-determined |
| S1S_extracted_68976 | Not-determined |
| S1S_extracted_68980 | Not-determined |
| S1S_extracted_68987 | Not-determined |
| S1S_extracted_68996 | Not-determined |
| S1S_extracted_68999 | Not-determined |
| S1S_extracted_69000 | Not-determined |
| S1S_extracted_69009 | Not-determined |
| S1S_extracted_69014 | Not-determined |
| S1S_extracted_69024 | Not-determined |
| S1S_extracted_69043 | Not-determined |
| S1S_extracted_69070 | Not-determined |
| S1S_extracted_69095 | Not-determined |
| S1S_extracted_69098 | Not-determined |
| S1S_extracted_69100 | Not-determined |
| S1S_extracted_69105 | Not-determined |
| S1S_extracted_69113 | Not-determined |
| S1S_extracted_69122 | Not-determined |
| S1S_extracted_69128 | Low-quality |
| S1S_extracted_69151 | Not-determined |
| S1S_extracted_69165 | Not-determined |
| S1S_extracted_69170 | Not-determined |
| S1S_extracted_69177 | Low-quality |
| S1S_extracted_69185 | Not-determined |
| S1S_extracted_69186 | Not-determined |
| S1S_extracted_69205 | Not-determined |
| S1S_extracted_69213 | Not-determined |
| S1S_extracted_69243 | Not-determined |
| S1S_extracted_69246 | Low-quality |
| S1S_extracted_69250 | Not-determined |
| S1S_extracted_69259 | Not-determined |
| S1S_extracted_69262 | Not-determined |
| S1S_extracted_69282 | Not-determined |
| S1S_extracted_69289 | Not-determined |
| S1S_extracted_69300 | Not-determined |
| S1S_extracted_69312 | Not-determined |
| S1S_extracted_69315 | Low-quality |
| S1S_extracted_69342 | Not-determined |
| S1S_extracted_69346 | Not-determined |
| S1S_extracted_69350 | Not-determined |
| S1S_extracted_69353 | Not-determined |
| S1S_extracted_69354 | Not-determined |
| S1S_extracted_69355 | Not-determined |
| S1S_extracted_69359 | Not-determined |
| S1S_extracted_69360 | Not-determined |
| S1S_extracted_69364 | Not-determined |
| S1S_extracted_69375 | Not-determined |
| S1S_extracted_69383 | Not-determined |
| S1S_extracted_69389 | Not-determined |
| S1S_extracted_69399 | Not-determined |
| S1S_extracted_69400 | Not-determined |
| S1S_extracted_69402 | Not-determined |
| S1S_extracted_69405 | Not-determined |
| S1S_extracted_69417 | Not-determined |
| S1S_extracted_69440 | Not-determined |
| S1S_extracted_69442 | Not-determined |
| S1S_extracted_69446 | Not-determined |
| S1S_extracted_69454 | Not-determined |
| S1S_extracted_69468 | Not-determined |
| S1S_extracted_69477 | Not-determined |
| S1S_extracted_69480 | Not-determined |
| S1S_extracted_69488 | Not-determined |
| S1S_extracted_69494 | Not-determined |
| S1S_extracted_69505 | Not-determined |
| S1S_extracted_69510 | Not-determined |
| S1S_extracted_69518 | Not-determined |
| S1S_extracted_69531 | Low-quality |
| S1S_extracted_69545 | Not-determined |
| S1S_extracted_69551 | Not-determined |
| S1S_extracted_69558 | Not-determined |
| S1S_extracted_69564 | Low-quality |
| S1S_extracted_69566 | Not-determined |
| S1S_extracted_69575 | Not-determined |
| S1S_extracted_69583 | Not-determined |
| S1S_extracted_69584 | Not-determined |
| S1S_extracted_69595 | Low-quality |
| S1S_extracted_69598 | Not-determined |
| S1S_extracted_69644 | Not-determined |
| S1S_extracted_69663 | Not-determined |
| S1S_extracted_69680 | Not-determined |
| S1S_extracted_69686 | Not-determined |
| S1S_extracted_69703 | Not-determined |
| S1S_extracted_69706 | Not-determined |
| S1S_extracted_69713 | Not-determined |
| S1S_extracted_69715 | Not-determined |
| S1S_extracted_69716 | Low-quality |
| S1S_extracted_69717 | Low-quality |
| S1S_extracted_69718 | Not-determined |
| S1S_extracted_69719 | Not-determined |
| S1S_extracted_69727 | Not-determined |
| S1S_extracted_69737 | Low-quality |
| S1S_extracted_69744 | Not-determined |
| S1S_extracted_69745 | Not-determined |
| S1S_extracted_69747 | Not-determined |
| S1S_extracted_69763 | Low-quality |
| S1S_extracted_69774 | Not-determined |
| S1S_extracted_69775 | Not-determined |
| S1S_extracted_69777 | Low-quality |
| S1S_extracted_69786 | Not-determined |
| S1S_extracted_69803 | Not-determined |
| S1S_extracted_69813 | Not-determined |
| S1S_extracted_69817 | Not-determined |
| S1S_extracted_69818 | Not-determined |
| S1S_extracted_69819 | Not-determined |
| S1S_extracted_69821 | Not-determined |
| S1S_extracted_69840 | Not-determined |
| S1S_extracted_69849 | Not-determined |
| S1S_extracted_69850 | Not-determined |
| S1S_extracted_69852 | Not-determined |
| S1S_extracted_69878 | Low-quality |
| S1S_extracted_69887 | Not-determined |
| S1S_extracted_69894 | Not-determined |
| S1S_extracted_69905 | Not-determined |
| S1S_extracted_69909 | Not-determined |
| S1S_extracted_69926 | Not-determined |
| S1S_extracted_69930 | Not-determined |
| S1S_extracted_69934 | Not-determined |
| S1S_extracted_69936 | Low-quality |
| S1S_extracted_69939 | Not-determined |
| S1S_extracted_69952 | Not-determined |
| S1S_extracted_69994 | Not-determined |
| S1S_extracted_70001 | Not-determined |
| S1S_extracted_70006 | Not-determined |
| S1S_extracted_70015 | Not-determined |
| S1S_extracted_70023 | Not-determined |
| S1S_extracted_70030 | Not-determined |
| S1S_extracted_70046 | Not-determined |
| S1S_extracted_70048 | Not-determined |
| S1S_extracted_70055 | Low-quality |
| S1S_extracted_70068 | Not-determined |
| S1S_extracted_70071 | Low-quality |
| S1S_extracted_70074 | Not-determined |
| S1S_extracted_70080 | Not-determined |
| S1S_extracted_70099 | Not-determined |
| S1S_extracted_70105 | Low-quality |
| S1S_extracted_70130 | Not-determined |
| S1S_extracted_70151 | Low-quality |
| S1S_extracted_70165 | Not-determined |
| S1S_extracted_70189 | Not-determined |
| S1S_extracted_70199 | Not-determined |
| S1S_extracted_70209 | Not-determined |
| S1S_extracted_70210 | Low-quality |
| S1S_extracted_70214 | Not-determined |
| S1S_extracted_70215 | Not-determined |
| S1S_extracted_70230 | Not-determined |
| S1S_extracted_70234 | Not-determined |
| S1S_extracted_70243 | Not-determined |
| S1S_extracted_70252 | Low-quality |
| S1S_extracted_70253 | Not-determined |
| S1S_extracted_70260 | Not-determined |
| S1S_extracted_70271 | Not-determined |
| S1S_extracted_70280 | Not-determined |
| S1S_extracted_70288 | Not-determined |
| S1S_extracted_70293 | Not-determined |
| S1S_extracted_70312 | Not-determined |
| S1S_extracted_70346 | Not-determined |
| S1S_extracted_70349 | Not-determined |
| S1S_extracted_70350 | Not-determined |
| S1S_extracted_70363 | Not-determined |
| S1S_extracted_70374 | Low-quality |
| S1S_extracted_70407 | Not-determined |
| S1S_extracted_70412 | Not-determined |
| S1S_extracted_70421 | Low-quality |
| S1S_extracted_70422 | Not-determined |
| S1S_extracted_70431 | Not-determined |
| S1S_extracted_70507 | Not-determined |
| S1S_extracted_70509 | Not-determined |
| S1S_extracted_70514 | Not-determined |
| S1S_extracted_70520 | Not-determined |
| S1S_extracted_70525 | Not-determined |
| S1S_extracted_70563 | Not-determined |
| S1S_extracted_70568 | Not-determined |
| S1S_extracted_70572 | Not-determined |
| S1S_extracted_70577 | Low-quality |
| S1S_extracted_70581 | Low-quality |
| S1S_extracted_70585 | Not-determined |
| S1S_extracted_70609 | Not-determined |
| S1S_extracted_70614 | Not-determined |
| S1S_extracted_70625 | Not-determined |
| S1S_extracted_70630 | Not-determined |
| S1S_extracted_70632 | Not-determined |
| S1S_extracted_70660 | Not-determined |
| S1S_extracted_70662 | Not-determined |
| S1S_extracted_70663 | Not-determined |
| S1S_extracted_70669 | Low-quality |
| S1S_extracted_70670 | Not-determined |
| S1S_extracted_70679 | Not-determined |
| S1S_extracted_70688 | Not-determined |
| S1S_extracted_70695 | Not-determined |
| S1S_extracted_70710 | Not-determined |
| S1S_extracted_70713 | Not-determined |
| S1S_extracted_70714 | Not-determined |
| S1S_extracted_70730 | Low-quality |
| S1S_extracted_70733 | Not-determined |
| S1S_extracted_70739 | Not-determined |
| S1S_extracted_70740 | Not-determined |
| S1S_extracted_70743 | Not-determined |
| S1S_extracted_70745 | Not-determined |
| S1S_extracted_70760 | Not-determined |
| S1S_extracted_70767 | Not-determined |
| S1S_extracted_70769 | Not-determined |
| S1S_extracted_70773 | Not-determined |
| S1S_extracted_70786 | Not-determined |
| S1S_extracted_70795 | Not-determined |
| S1S_extracted_70797 | Low-quality |
| S1S_extracted_70802 | Not-determined |
| S1S_extracted_70810 | Low-quality |
| S1S_extracted_70815 | Not-determined |
| S1S_extracted_70818 | Not-determined |
| S1S_extracted_70822 | Not-determined |
| S1S_extracted_70828 | Not-determined |
| S1S_extracted_70829 | Not-determined |
| S1S_extracted_70855 | Not-determined |
| S1S_extracted_70866 | Not-determined |
| S1S_extracted_70872 | Not-determined |
| S1S_extracted_70874 | Not-determined |
| S1S_extracted_70896 | Not-determined |
| S1S_extracted_70900 | Not-determined |
| S1S_extracted_70916 | Not-determined |
| S1S_extracted_70919 | Not-determined |
| S1S_extracted_70924 | Not-determined |
| S1S_extracted_70933 | Not-determined |
| S1S_extracted_70935 | Not-determined |
| S1S_extracted_70936 | Not-determined |
| S1S_extracted_70939 | Not-determined |
| S1S_extracted_70948 | Not-determined |
| S1S_extracted_70953 | Not-determined |
| S1S_extracted_70960 | Not-determined |
| S1S_extracted_70970 | Not-determined |
| S1S_extracted_70973 | Not-determined |
| S1S_extracted_70977 | Not-determined |
| S1S_extracted_70999 | Not-determined |
| S1S_extracted_71017 | Not-determined |
| S1S_extracted_71024 | Not-determined |
| S1S_extracted_71037 | Not-determined |
| S1S_extracted_71052 | Not-determined |
| S1S_extracted_71055 | Not-determined |
| S1S_extracted_71060 | Not-determined |
| S1S_extracted_71062 | Not-determined |
| S1S_extracted_71090 | Not-determined |
| S1S_extracted_71096 | Not-determined |
| S1S_extracted_71100 | Not-determined |

| | |
|---|---|
| S15_extracted_71105 | Not-determined |
| S15_extracted_71112 | Not-determined |
| S15_extracted_71116 | Not-determined |
| S15_extracted_71121 | Not-determined |
| S15_extracted_71124 | Not-determined |
| S15_extracted_71125 | Not-determined |
| S15_extracted_71127 | Not-determined |
| S15_extracted_71134 | Not-determined |
| S15_extracted_71152 | Low-quality |
| S15_extracted_71165 | Not-determined |
| S15_extracted_71167 | Not-determined |
| S15_extracted_71169 | Not-determined |
| S15_extracted_71181 | Not-determined |
| S15_extracted_71188 | Not-determined |
| S15_extracted_71195 | Not-determined |
| S15_extracted_71200 | Not-determined |
| S15_extracted_71204 | Not-determined |
| S15_extracted_71214 | Not-determined |
| S15_extracted_71215 | Not-determined |
| S15_extracted_71230 | Not-determined |
| S15_extracted_71247 | Not-determined |
| S15_extracted_71256 | Not-determined |
| S15_extracted_71262 | Not-determined |
| S15_extracted_71268 | Low-quality |
| S15_extracted_71280 | Not-determined |
| S15_extracted_71293 | Low-quality |
| S15_extracted_71296 | Not-determined |
| S15_extracted_71299 | Not-determined |
| S15_extracted_71316 | Not-determined |
| S15_extracted_71317 | Not-determined |
| S15_extracted_71332 | Not-determined |
| S15_extracted_71336 | Not-determined |
| S15_extracted_71348 | Not-determined |
| S15_extracted_71354 | Not-determined |
| S15_extracted_71369 | Not-determined |
| S15_extracted_71388 | Not-determined |
| S15_extracted_71392 | Not-determined |
| S15_extracted_71394 | Low-quality |
| S15_extracted_71400 | Not-determined |
| S15_extracted_71428 | Not-determined |
| S15_extracted_71429 | Not-determined |
| S15_extracted_71439 | Not-determined |
| S15_extracted_71453 | Not-determined |
| S15_extracted_71454 | Not-determined |
| S15_extracted_71467 | Not-determined |
| S15_extracted_71470 | Not-determined |
| S15_extracted_71475 | Not-determined |
| S15_extracted_71476 | Low-quality |
| S15_extracted_71478 | Not-determined |
| S15_extracted_71481 | Not-determined |
| S15_extracted_71490 | Not-determined |
| S15_extracted_71495 | Not-determined |
| S15_extracted_71498 | Not-determined |
| S15_extracted_71530 | Not-determined |
| S15_extracted_71534 | Not-determined |
| S15_extracted_71553 | Not-determined |
| S15_extracted_71555 | Not-determined |
| S15_extracted_71557 | Low-quality |
| S15_extracted_71570 | Not-determined |
| S15_extracted_71583 | Not-determined |
| S15_extracted_71598 | Low-quality |
| S15_extracted_71608 | Not-determined |
| S7_extracted_72493 | Medium-quality |
| S15_extracted_71629 | Low-quality |
| S15_extracted_71634 | Not-determined |
| S15_extracted_71637 | Not-determined |
| S15_extracted_71647 | Not-determined |
| S15_extracted_71648 | Not-determined |
| S15_extracted_71651 | Not-determined |
| S15_extracted_71652 | Not-determined |
| S15_extracted_71656 | Not-determined |
| S15_extracted_71658 | Low-quality |
| S15_extracted_71659 | Not-determined |
| S15_extracted_71663 | Not-determined |
| S15_extracted_71668 | Not-determined |
| S15_extracted_71672 | Not-determined |
| S15_extracted_71676 | Not-determined |
| S15_extracted_71679 | Not-determined |
| S15_extracted_71689 | Not-determined |
| S15_extracted_71692 | Not-determined |
| S15_extracted_71710 | Not-determined |
| S15_extracted_71731 | Not-determined |
| S15_extracted_71741 | Not-determined |
| S15_extracted_71751 | Not-determined |
| S15_extracted_71757 | Not-determined |
| S15_extracted_71769 | Not-determined |
| S15_extracted_71773 | Not-determined |
| S15_extracted_71774 | Not-determined |
| S15_extracted_71777 | Not-determined |
| S15_extracted_71785 | Not-determined |
| S15_extracted_71788 | Not-determined |
| S15_extracted_71790 | Not-determined |
| S15_extracted_71793 | Not-determined |
| S15_extracted_71803 | Not-determined |
| S15_extracted_71816 | Low-quality |
| S15_extracted_71823 | Low-quality |
| S15_extracted_71834 | Not-determined |
| S15_extracted_71835 | Not-determined |
| S15_extracted_71841 | Not-determined |
| S15_extracted_71849 | Low-quality |
| S15_extracted_71850 | Not-determined |
| S15_extracted_71863 | Not-determined |
| S15_extracted_71871 | Not-determined |
| S15_extracted_71875 | Not-determined |
| S15_extracted_71904 | Not-determined |
| S15_extracted_71907 | Not-determined |
| S15_extracted_71939 | Not-determined |
| S15_extracted_71943 | Low-quality |
| S15_extracted_71949 | Not-determined |
| S15_extracted_71958 | Low-quality |
| S15_extracted_71964 | Not-determined |
| S15_extracted_71966 | Not-determined |
| S15_extracted_71976 | Not-determined |
| S15_extracted_71987 | Low-quality |
| S15_extracted_72003 | Not-determined |
| S15_extracted_72018 | Not-determined |
| S15_extracted_72064 | Not-determined |
| S15_extracted_72068 | Low-quality |
| S15_extracted_72069 | Not-determined |
| S15_extracted_72075 | Not-determined |
| S15_extracted_72081 | Not-determined |
| S15_extracted_72097 | Not-determined |
| S15_extracted_72098 | Not-determined |
| S15_extracted_72102 | Not-determined |
| S15_extracted_72118 | Not-determined |
| S15_extracted_72137 | Not-determined |
| S15_extracted_72142 | Not-determined |
| S15_extracted_72153 | Not-determined |
| S15_extracted_72156 | Not-determined |
| S15_extracted_72161 | Not-determined |
| S15_extracted_72165 | Not-determined |
| S15_extracted_72183 | Not-determined |
| S15_extracted_72187 | Low-quality |
| S15_extracted_72189 | Not-determined |
| S15_extracted_72190 | Not-determined |
| S15_extracted_72221 | Not-determined |
| S15_extracted_72243 | Not-determined |
| S15_extracted_72245 | Not-determined |
| S15_extracted_72246 | Low-quality |
| S15_extracted_72251 | Low-quality |
| S15_extracted_72268 | Not-determined |
| S15_extracted_72295 | Not-determined |
| S15_extracted_72309 | Not-determined |
| S15_extracted_72327 | Not-determined |
| S15_extracted_72337 | Not-determined |
| S15_extracted_72352 | Not-determined |
| S15_extracted_72411 | Not-determined |
| S15_extracted_72417 | Not-determined |
| S15_extracted_72421 | Not-determined |
| S15_extracted_72422 | Not-determined |
| S15_extracted_72424 | Not-determined |
| S15_extracted_72427 | Not-determined |
| S15_extracted_72429 | Not-determined |
| S15_extracted_72435 | Not-determined |
| S15_extracted_72437 | Not-determined |
| S15_extracted_72441 | Not-determined |
| S15_extracted_72443 | Not-determined |
| S15_extracted_72453 | Low-quality |
| S15_extracted_72454 | Not-determined |
| S15_extracted_72455 | Not-determined |
| S15_extracted_72459 | Not-determined |
| S15_extracted_72470 | Low-quality |
| S15_extracted_72479 | Not-determined |
| S15_extracted_72490 | Not-determined |
| S15_extracted_72493 | Not-determined |
| S15_extracted_72518 | Not-determined |
| S15_extracted_72527 | Not-determined |
| S15_extracted_72533 | Not-determined |
| S15_extracted_72536 | Not-determined |
| S15_extracted_72537 | Not-determined |
| S15_extracted_72546 | Not-determined |
| S15_extracted_72547 | Not-determined |
| S15_extracted_72558 | Not-determined |
| S15_extracted_72563 | Low-quality |
| S15_extracted_72575 | Not-determined |
| S15_extracted_72590 | Not-determined |
| S15_extracted_72599 | Not-determined |
| S15_extracted_72601 | Not-determined |
| S15_extracted_72604 | Not-determined |
| S15_extracted_72605 | Not-determined |
| S15_extracted_72608 | Not-determined |
| S15_extracted_72609 | Not-determined |
| S15_extracted_72616 | Not-determined |
| S15_extracted_72634 | Not-determined |
| S15_extracted_72636 | Not-determined |
| S15_extracted_72655 | Not-determined |
| S15_extracted_72657 | Not-determined |
| S15_extracted_72676 | Not-determined |
| S15_extracted_72686 | Not-determined |
| S15_extracted_72706 | Not-determined |
| S15_extracted_72707 | Not-determined |
| S15_extracted_72721 | Low-quality |
| S15_extracted_72724 | Not-determined |
| S15_extracted_72736 | Not-determined |
| S15_extracted_72737 | Not-determined |
| S15_extracted_72739 | Low-quality |
| S15_extracted_72744 | Not-determined |
| S15_extracted_72747 | Not-determined |
| S15_extracted_72748 | Not-determined |
| S15_extracted_72752 | Not-determined |
| S15_extracted_72758 | Low-quality |
| S15_extracted_72770 | Low-quality |
| S15_extracted_72779 | Not-determined |
| S15_extracted_72783 | Not-determined |
| S15_extracted_72784 | Not-determined |
| S15_extracted_72786 | Not-determined |
| S15_extracted_72802 | Not-determined |
| S15_extracted_72818 | Not-determined |
| S15_extracted_72821 | Low-quality |
| S15_extracted_72823 | Low-quality |
| S15_extracted_72829 | Not-determined |
| S15_extracted_72842 | Not-determined |
| S15_extracted_72843 | Not-determined |
| S15_extracted_72852 | Not-determined |
| S15_extracted_72876 | Low-quality |
| S15_extracted_72879 | Not-determined |
| S15_extracted_72883 | Not-determined |
| S15_extracted_72885 | Not-determined |
| S15_extracted_72890 | Not-determined |
| S15_extracted_72892 | Low-quality |
| S15_extracted_72896 | Not-determined |
| S15_extracted_72899 | Not-determined |
| S15_extracted_72901 | Not-determined |
| S15_extracted_72907 | Not-determined |
| S15_extracted_72913 | Low-quality |
| S15_extracted_72923 | Low-quality |
| S15_extracted_72927 | Not-determined |
| S15_extracted_72935 | Not-determined |
| S15_extracted_72938 | Low-quality |
| S15_extracted_72961 | Low-quality |
| S15_extracted_72978 | Not-determined |
| S15_extracted_72980 | Not-determined |
| S15_extracted_72987 | Not-determined |
| S15_extracted_72999 | Not-determined |
| S15_extracted_73005 | Not-determined |
| S15_extracted_73040 | Not-determined |
| S15_extracted_73048 | Low-quality |
| S15_extracted_73054 | Low-quality |
| S15_extracted_73060 | Not-determined |
| S15_extracted_73089 | Not-determined |
| S15_extracted_73094 | Not-determined |
| S15_extracted_73105 | Not-determined |
| S15_extracted_73110 | Not-determined |
| S15_extracted_73117 | Not-determined |
| S15_extracted_73118 | Not-determined |
| S15_extracted_73119 | Low-quality |
| S15_extracted_73126 | Not-determined |
| S15_extracted_73129 | Not-determined |
| S15_extracted_73145 | Not-determined |
| S15_extracted_73151 | Not-determined |
| S15_extracted_73164 | Not-determined |
| S15_extracted_73168 | Not-determined |
| S15_extracted_73196 | Not-determined |
| S15_extracted_73201 | Not-determined |
| S15_extracted_73205 | Not-determined |
| S15_extracted_73220 | Not-determined |
| S15_extracted_73229 | Not-determined |
| S15_extracted_73241 | Not-determined |
| S15_extracted_73245 | Not-determined |
| S15_extracted_73249 | Not-determined |
| S15_extracted_73250 | Not-determined |
| S15_extracted_73251 | Not-determined |
| S15_extracted_73267 | Not-determined |
| S15_extracted_73269 | Not-determined |
| S15_extracted_73295 | Low-quality |
| S15_extracted_73302 | Low-quality |
| S15_extracted_73304 | Not-determined |
| S15_extracted_73309 | Not-determined |
| S15_extracted_73313 | Not-determined |
| S15_extracted_73325 | Not-determined |
| S15_extracted_73331 | Not-determined |
| S15_extracted_73335 | Not-determined |
| S15_extracted_73349 | Not-determined |
| S15_extracted_73352 | Not-determined |
| S15_extracted_73362 | Not-determined |
| S15_extracted_73380 | Not-determined |
| S15_extracted_73388 | Not-determined |
| S15_extracted_73398 | Not-determined |
| S15_extracted_73404 | Not-determined |
| S15_extracted_73412 | Not-determined |
| S15_extracted_73413 | Low-quality |
| S15_extracted_73414 | Not-determined |
| S15_extracted_73416 | Low-quality |
| S15_extracted_73424 | Not-determined |
| S15_extracted_73427 | Not-determined |
| S15_extracted_73433 | Not-determined |
| S15_extracted_73440 | Not-determined |
| S15_extracted_73447 | Not-determined |
| S15_extracted_73449 | Not-determined |
| S15_extracted_73450 | Not-determined |
| S15_extracted_73468 | Low-quality |
| S15_extracted_73471 | Not-determined |
| S15_extracted_73478 | Not-determined |
| S15_extracted_73494 | Not-determined |
| S15_extracted_73502 | Not-determined |
| S15_extracted_73505 | Low-quality |
| S15_extracted_73511 | Not-determined |
| S15_extracted_73523 | Not-determined |
| S15_extracted_73528 | Not-determined |
| S15_extracted_73531 | Not-determined |
| S15_extracted_73549 | Not-determined |
| S15_extracted_73562 | Not-determined |
| S15_extracted_73565 | Not-determined |
| S15_extracted_73577 | Not-determined |
| S15_extracted_73586 | Not-determined |
| S15_extracted_73591 | Low-quality |
| S15_extracted_73603 | Not-determined |
| S15_extracted_73607 | Low-quality |
| S15_extracted_73608 | Not-determined |
| S15_extracted_73618 | Not-determined |
| S15_extracted_73621 | Not-determined |
| S15_extracted_73622 | Not-determined |
| S15_extracted_73641 | Low-quality |
| S15_extracted_73656 | Not-determined |
| S15_extracted_73666 | Not-determined |
| S15_extracted_73669 | Not-determined |
| S15_extracted_73672 | Not-determined |
| S15_extracted_73683 | Not-determined |
| S15_extracted_73684 | Low-quality |
| S15_extracted_73687 | Not-determined |
| S15_extracted_73701 | Not-determined |
| S15_extracted_73717 | Not-determined |
| S15_extracted_73738 | Not-determined |
| S15_extracted_73741 | Not-determined |
| S15_extracted_73754 | Not-determined |
| S15_extracted_73777 | Low-quality |
| S15_extracted_73784 | Not-determined |
| S15_extracted_73788 | Not-determined |
| S15_extracted_73793 | Not-determined |
| S15_extracted_73794 | Not-determined |
| S15_extracted_73799 | Not-determined |
| S15_extracted_73811 | Not-determined |
| S15_extracted_73835 | Not-determined |
| S15_extracted_73839 | Not-determined |
| S15_extracted_73841 | Not-determined |
| S15_extracted_73850 | Not-determined |
| S15_extracted_73856 | Not-determined |
| S15_extracted_73867 | Low-quality |
| S15_extracted_73889 | Low-quality |
| S15_extracted_73898 | Low-quality |
| S15_extracted_73909 | Not-determined |
| S15_extracted_73918 | Not-determined |
| S15_extracted_73924 | Low-quality |
| S15_extracted_73930 | Not-determined |
| S15_extracted_73941 | Not-determined |
| S15_extracted_73947 | Not-determined |
| S15_extracted_73955 | Not-determined |
| S15_extracted_73974 | Not-determined |
| S15_extracted_73986 | Low-quality |
| S15_extracted_73994 | Not-determined |
| S15_extracted_74004 | Not-determined |
| S15_extracted_74005 | Not-determined |
| S15_extracted_74013 | Not-determined |
| S15_extracted_74017 | Not-determined |
| S15_extracted_74024 | Not-determined |
| S15_extracted_74036 | Low-quality |
| S15_extracted_74043 | Not-determined |
| S15_extracted_74045 | Not-determined |
| S15_extracted_74051 | Not-determined |
| S15_extracted_74057 | Not-determined |
| S15_extracted_74059 | Low-quality |
| S15_extracted_74074 | Not-determined |
| S15_extracted_74088 | Not-determined |
| S15_extracted_74101 | Low-quality |
| S15_extracted_74111 | Low-quality |
| S15_extracted_74117 | Not-determined |
| S15_extracted_74125 | Not-determined |
| S15_extracted_74138 | Not-determined |
| S15_extracted_74141 | Not-determined |
| S15_extracted_74143 | Not-determined |
| S15_extracted_74148 | Not-determined |
| S15_extracted_74154 | Not-determined |
| S15_extracted_74167 | Low-quality |
| S15_extracted_74177 | Not-determined |
| S15_extracted_74187 | Not-determined |
| S15_extracted_74209 | Not-determined |
| S15_extracted_74219 | Not-determined |
| S15_extracted_74220 | Low-quality |
| S15_extracted_74223 | Not-determined |
| S15_extracted_74224 | Not-determined |
| S15_extracted_74227 | Low-quality |
| S15_extracted_74228 | Not-determined |
| S15_extracted_74230 | Not-determined |
| S15_extracted_74232 | Not-determined |
| S15_extracted_74241 | Not-determined |
| S15_extracted_74257 | Not-determined |
| S15_extracted_74260 | Low-quality |
| S15_extracted_74262 | Not-determined |
| S15_extracted_74286 | Not-determined |
| S15_extracted_74294 | Low-quality |
| S15_extracted_74304 | Not-determined |
| S15_extracted_74311 | Not-determined |
| S15_extracted_74328 | Not-determined |
| S15_extracted_74351 | Not-determined |
| S15_extracted_74355 | Not-determined |
| S15_extracted_74358 | Not-determined |
| S15_extracted_74362 | Not-determined |
| S15_extracted_74376 | Not-determined |
| S15_extracted_74382 | Low-quality |
| S15_extracted_74385 | Not-determined |
| S15_extracted_74386 | Not-determined |
| S15_extracted_74387 | Low-quality |
| S15_extracted_74399 | Not-determined |
| S15_extracted_74403 | Not-determined |
| S15_extracted_74405 | Not-determined |
| S15_extracted_74438 | Not-determined |
| S15_extracted_74449 | Not-determined |
| S15_extracted_74454 | Not-determined |
| S15_extracted_74469 | Not-determined |
| S15_extracted_74470 | Not-determined |
| S15_extracted_74488 | Not-determined |
| S15_extracted_74496 | Not-determined |
| S15_extracted_74515 | Not-determined |
| S15_extracted_74517 | Not-determined |
| S15_extracted_74518 | Low-quality |
| S15_extracted_74526 | Low-quality |
| S15_extracted_74549 | Not-determined |
| S15_extracted_74556 | Not-determined |
| S15_extracted_74557 | Low-quality |
| S15_extracted_74558 | Not-determined |
| S15_extracted_74570 | Not-determined |
| S15_extracted_74571 | Not-determined |
| S15_extracted_74573 | Low-quality |
| S15_extracted_74578 | Not-determined |
| S16_vs_88477\|\|full | Medium-quality |
| S15_extracted_74587 | Not-determined |
| S15_extracted_74605 | Not-determined |
| S15_extracted_74609 | Not-determined |
| S15_extracted_74611 | Low-quality |
| S15_extracted_74612 | Not-determined |
| S15_extracted_74617 | Not-determined |
| S15_extracted_74627 | Not-determined |
| S15_extracted_74639 | Not-determined |
| S15_extracted_74644 | Not-determined |
| S15_extracted_74651 | Not-determined |
| S15_extracted_74678 | Not-determined |
| S15_extracted_74688 | Not-determined |
| S15_extracted_74690 | Not-determined |
| S15_extracted_74691 | Not-determined |
| S15_extracted_74717 | Not-determined |
| S15_extracted_74719 | Not-determined |
| S15_extracted_74722 | Not-determined |
| S15_extracted_74730 | Not-determined |
| S15_extracted_74750 | Not-determined |
| S15_extracted_74753 | Low-quality |
| S15_extracted_74754 | Not-determined |
| S15_extracted_74764 | Not-determined |
| S15_extracted_74765 | Not-determined |
| S15_extracted_74792 | Not-determined |
| S15_extracted_74807 | Not-determined |
| S15_extracted_74810 | Not-determined |
| S15_extracted_74812 | Not-determined |
| S15_extracted_74816 | Not-determined |
| S15_extracted_74819 | Not-determined |
| S15_extracted_74825 | Not-determined |
| S15_extracted_74831 | Not-determined |
| S15_extracted_74843 | Not-determined |
| S15_extracted_74860 | Not-determined |
| S15_extracted_74871 | Low-quality |
| S15_extracted_74880 | Not-determined |
| S15_extracted_74886 | Not-determined |
| S15_extracted_74895 | Not-determined |
| S15_extracted_74903 | Not-determined |
| S15_extracted_74923 | Not-determined |
| S15_extracted_74930 | Not-determined |
| S15_extracted_74948 | Not-determined |
| S15_extracted_74953 | Not-determined |
| S15_extracted_74957 | Not-determined |
| S15_extracted_74963 | Low-quality |
| S15_extracted_74970 | Not-determined |
| S15_extracted_74979 | Not-determined |
| S15_extracted_74992 | Low-quality |
| S15_extracted_75006 | Not-determined |
| S15_extracted_75011 | Not-determined |
| S15_extracted_75041 | Not-determined |
| S15_extracted_75043 | Not-determined |
| S15_extracted_75054 | Low-quality |
| S15_extracted_75064 | Not-determined |
| S15_extracted_75072 | Not-determined |
| S15_extracted_75073 | Not-determined |
| S15_extracted_75077 | Not-determined |
| S15_extracted_75082 | Not-determined |
| S15_extracted_75084 | Low-quality |
| S15_extracted_75092 | Not-determined |
| S15_extracted_75100 | Not-determined |
| S15_extracted_75106 | Not-determined |
| S15_extracted_75114 | Not-determined |
| S15_extracted_75139 | Not-determined |
| S15_extracted_75140 | Not-determined |
| S15_extracted_75142 | Not-determined |
| S15_extracted_75144 | Not-determined |
| S15_extracted_75175 | Not-determined |
| S15_extracted_75195 | Not-determined |
| S15_extracted_75198 | Not-determined |
| S15_extracted_75201 | Not-determined |
| S15_extracted_75204 | Not-determined |
| S15_extracted_75211 | Low-quality |
| S15_extracted_75260 | Not-determined |
| S15_extracted_75267 | Not-determined |
| S15_extracted_75284 | Not-determined |
| S15_extracted_75287 | Not-determined |
| S15_extracted_75295 | Not-determined |
| S15_extracted_75297 | Low-quality |
| S15_extracted_75313 | Not-determined |
| S15_extracted_75320 | Not-determined |
| S15_extracted_75337 | Not-determined |
| S15_extracted_75347 | Not-determined |
| S15_extracted_75348 | Low-quality |
| S15_extracted_75349 | Not-determined |
| S15_extracted_75360 | Not-determined |
| S15_extracted_75361 | Not-determined |
| S15_extracted_75366 | Not-determined |
| S15_extracted_75378 | Not-determined |
| S15_extracted_75386 | Not-determined |
| S15_extracted_75405 | Not-determined |
| S15_extracted_75406 | Low-quality |
| S15_extracted_75409 | Not-determined |
| S15_extracted_75413 | Not-determined |
| S15_extracted_75416 | Not-determined |
| S15_extracted_75422 | Not-determined |
| S15_extracted_75425 | Not-determined |
| S15_extracted_75433 | Not-determined |
| S15_extracted_75437 | Not-determined |
| S15_extracted_75443 | Not-determined |
| S15_extracted_75461 | Not-determined |
| S15_extracted_75468 | Not-determined |
| S15_extracted_75477 | Not-determined |
| S15_extracted_75480 | Not-determined |
| S15_extracted_75482 | Not-determined |
| S15_extracted_75486 | Not-determined |
| S15_extracted_75497 | Not-determined |
| S15_extracted_75506 | Not-determined |
| S15_extracted_75509 | Not-determined |
| S15_extracted_75515 | Not-determined |
| S15_extracted_75521 | Not-determined |
| S15_extracted_75533 | Not-determined |
| S15_extracted_75538 | Not-determined |
| S15_extracted_75544 | Not-determined |
| S15_extracted_75550 | Not-determined |
| S15_extracted_75555 | Not-determined |
| S15_extracted_75556 | Not-determined |
| S15_extracted_75561 | Not-determined |
| S15_extracted_75576 | Not-determined |
| S15_extracted_75580 | Not-determined |
| S15_extracted_75589 | Not-determined |
| S15_extracted_75591 | Not-determined |
| S15_extracted_75594 | Not-determined |
| S15_extracted_75606 | Not-determined |
| S15_extracted_75622 | Not-determined |
| S15_extracted_75623 | Not-determined |
| S15_extracted_75642 | Not-determined |
| S15_extracted_75644 | Not-determined |
| S15_extracted_75673 | Not-determined |
| S15_extracted_75674 | Not-determined |
| S15_extracted_75682 | Low-quality |
| S15_extracted_75698 | Not-determined |
| S15_extracted_75739 | Not-determined |
| S15_extracted_75752 | Not-determined |
| S15_extracted_75768 | Not-determined |
| S15_extracted_75769 | Not-determined |
| S15_extracted_75772 | Not-determined |
| S15_extracted_75773 | Not-determined |
| S15_extracted_75779 | Not-determined |
| S15_extracted_75797 | Not-determined |
| S15_extracted_75806 | Low-quality |
| S15_extracted_75814 | Not-determined |
| S15_extracted_75827 | Not-determined |
| S15_extracted_75829 | Not-determined |
| S15_extracted_75831 | Not-determined |
| S15_extracted_75836 | Not-determined |
| S15_extracted_75848 | Not-determined |
| S15_extracted_75857 | Not-determined |
| S15_extracted_75861 | Not-determined |
| S15_extracted_75873 | Not-determined |
| S15_extracted_75874 | Not-determined |
| S15_extracted_75891 | Not-determined |
| S15_extracted_75892 | Not-determined |
| S15_extracted_75896 | Not-determined |
| S15_extracted_75902 | Low-quality |
| S15_extracted_75905 | Not-determined |
| S15_extracted_75914 | Not-determined |
| S15_extracted_75917 | Not-determined |
| S8_vs_k141_76852\|\|full | Medium-quality |
| S15_extracted_75928 | Not-determined |
| S15_extracted_75934 | Not-determined |
| S15_extracted_75939 | Not-determined |
| S15_extracted_75946 | Not-determined |
| S15_extracted_75950 | Not-determined |
| S15_extracted_75952 | Not-determined |

| | |
|---|---|
| S15_extracted_75971 | Low-quality |
| S15_extracted_75995 | Low-quality |
| S15_extracted_76020 | Not-determined |
| S15_extracted_76035 | Not-determined |
| S15_extracted_76037 | Low-quality |
| S15_extracted_76041 | Not-determined |
| S15_extracted_76048 | Not-determined |
| S15_extracted_76049 | Not-determined |
| S15_extracted_76063 | Not-determined |
| S15_extracted_76064 | Not-determined |
| S15_extracted_76071 | Low-quality |
| S15_extracted_76072 | Not-determined |
| S15_extracted_76074 | Low-quality |
| S15_extracted_76075 | Not-determined |
| S15_extracted_76080 | Not-determined |
| S15_extracted_76084 | Not-determined |
| S15_extracted_76089 | Not-determined |
| S15_extracted_76093 | Not-determined |
| S15_extracted_76097 | Not-determined |
| S15_extracted_76098 | Not-determined |
| S15_extracted_76102 | Not-determined |
| S15_extracted_76105 | Not-determined |
| S15_extracted_76108 | Not-determined |
| S15_extracted_76136 | Low-quality |
| S15_extracted_76144 | Not-determined |
| S15_extracted_76149 | Not-determined |
| S15_extracted_76156 | Not-determined |
| S15_extracted_76178 | Not-determined |
| S15_extracted_76182 | Not-determined |
| S15_extracted_76186 | Low-quality |
| S15_extracted_76193 | Not-determined |
| S15_extracted_76196 | Not-determined |
| S15_extracted_76214 | Not-determined |
| S15_extracted_76216 | Not-determined |
| S15_extracted_76224 | Not-determined |
| S15_extracted_76229 | Not-determined |
| S15_extracted_76243 | Not-determined |
| S15_extracted_76246 | Not-determined |
| S15_extracted_76253 | Low-quality |
| S15_extracted_76262 | Low-quality |
| S15_extracted_76264 | Not-determined |
| S15_extracted_76265 | Not-determined |
| S15_extracted_76271 | Low-quality |
| S15_extracted_76274 | Not-determined |
| S15_extracted_76275 | Not-determined |
| S15_extracted_76276 | Not-determined |
| S15_extracted_76282 | Not-determined |
| S15_extracted_76283 | Low-quality |
| S15_extracted_76289 | Not-determined |
| S15_extracted_76295 | Low-quality |
| S15_extracted_76300 | Not-determined |
| S15_extracted_76314 | Low-quality |
| S15_extracted_76329 | Not-determined |
| S15_extracted_76337 | Not-determined |
| S15_extracted_76352 | Not-determined |
| S15_extracted_76365 | Not-determined |
| S15_extracted_76374 | Not-determined |
| S15_extracted_76376 | Not-determined |
| S15_extracted_76387 | Low-quality |
| S15_extracted_76405 | Low-quality |
| S15_extracted_76410 | Not-determined |
| S15_extracted_76413 | Not-determined |
| S15_extracted_76421 | Not-determined |
| S15_extracted_76430 | Not-determined |
| S15_extracted_76431 | Not-determined |
| S15_extracted_76451 | Not-determined |
| S15_extracted_76459 | Not-determined |
| S15_extracted_76462 | Not-determined |
| S15_extracted_76463 | Not-determined |
| S15_extracted_76486 | Not-determined |
| S15_extracted_76496 | Low-quality |
| S15_extracted_76504 | Not-determined |
| S15_extracted_76507 | Not-determined |
| S15_extracted_76517 | Not-determined |
| S15_extracted_76526 | Not-determined |
| S15_extracted_76527 | Not-determined |
| S15_extracted_76529 | Low-quality |
| S15_extracted_76532 | Not-determined |
| S15_extracted_76538 | Not-determined |
| S15_extracted_76545 | Not-determined |
| S15_extracted_76558 | Not-determined |
| S15_extracted_76561 | Not-determined |
| S15_extracted_76574 | Not-determined |
| S15_extracted_76576 | Not-determined |
| S15_extracted_76580 | Not-determined |
| S15_extracted_76590 | Not-determined |
| S15_extracted_76591 | Not-determined |
| S15_extracted_76614 | Not-determined |
| S15_extracted_76617 | Not-determined |
| S15_extracted_76618 | Not-determined |
| S15_extracted_76638 | Not-determined |
| S15_extracted_76647 | Not-determined |
| S15_extracted_76648 | Not-determined |
| S15_extracted_76650 | Low-quality |
| S15_extracted_76661 | Not-determined |
| S15_extracted_76662 | Not-determined |
| S15_extracted_76668 | Low-quality |
| S15_extracted_76693 | Not-determined |
| S15_extracted_76700 | Not-determined |
| S15_extracted_76706 | Not-determined |
| S15_extracted_76717 | Not-determined |
| S15_extracted_76723 | Low-quality |
| S15_extracted_76726 | Not-determined |
| S15_extracted_76732 | Not-determined |
| S15_extracted_76735 | Not-determined |
| S15_extracted_76744 | Not-determined |
| S15_extracted_76746 | Not-determined |
| S15_extracted_76750 | Not-determined |
| S15_extracted_76783 | Not-determined |
| S15_extracted_76787 | Not-determined |
| S15_extracted_76837 | Not-determined |
| S15_extracted_76841 | Low-quality |
| S15_extracted_76857 | Not-determined |
| S15_extracted_76867 | Low-quality |
| S15_extracted_76869 | Low-quality |
| S15_extracted_76884 | Not-determined |
| S15_extracted_76896 | Not-determined |
| S15_extracted_76958 | Not-determined |
| S15_extracted_76960 | Not-determined |
| S15_extracted_76961 | Low-quality |
| S15_extracted_76970 | Not-determined |
| S15_extracted_76972 | Not-determined |
| S15_extracted_76976 | Low-quality |
| S15_extracted_76982 | Not-determined |
| S15_extracted_76992 | Not-determined |
| S15_extracted_76997 | Not-determined |
| S15_extracted_77012 | Not-determined |
| S15_extracted_77039 | Not-determined |
| S15_extracted_77047 | Not-determined |
| S15_extracted_77091 | Not-determined |
| S15_extracted_77096 | Not-determined |
| S15_extracted_77103 | Not-determined |
| S15_extracted_77110 | Not-determined |
| S15_extracted_77111 | Not-determined |
| S15_extracted_77113 | Not-determined |
| S15_extracted_77116 | Not-determined |
| S15_extracted_77130 | Not-determined |
| S15_extracted_77140 | Not-determined |
| S15_extracted_77160 | Not-determined |
| S15_extracted_77162 | Low-quality |
| S15_extracted_77189 | Not-determined |
| S15_extracted_77201 | Not-determined |
| S15_extracted_77224 | Low-quality |
| S15_extracted_77231 | Low-quality |
| S15_extracted_77232 | Not-determined |
| S15_extracted_77240 | Not-determined |
| S15_extracted_77244 | Not-determined |
| S15_extracted_77249 | Not-determined |
| S15_extracted_77258 | Not-determined |
| S15_extracted_77261 | Not-determined |
| S15_extracted_77275 | Not-determined |
| S15_extracted_77284 | Not-determined |
| S15_extracted_77307 | Not-determined |
| S15_extracted_77314 | Not-determined |
| S15_extracted_77315 | Not-determined |
| S15_extracted_77323 | Not-determined |
| S15_extracted_77333 | Not-determined |
| S15_extracted_77335 | Not-determined |
| S15_extracted_77345 | Low-quality |
| S15_extracted_77350 | Not-determined |
| S15_extracted_77356 | Not-determined |
| S15_extracted_77368 | Not-determined |
| S15_extracted_77378 | Not-determined |
| S15_extracted_77380 | Not-determined |
| S15_extracted_77382 | Not-determined |
| S15_extracted_77391 | Not-determined |
| S15_extracted_77394 | Not-determined |
| S15_extracted_77413 | Low-quality |
| S15_extracted_77423 | Not-determined |
| S15_extracted_77426 | Not-determined |
| S15_extracted_77434 | Not-determined |
| S15_extracted_77438 | Low-quality |
| S15_extracted_77440 | Low-quality |
| S15_extracted_77451 | Low-quality |
| S15_extracted_77454 | Not-determined |
| S15_extracted_77455 | Not-determined |
| S15_extracted_77463 | Not-determined |
| S15_extracted_77475 | Not-determined |
| S15_extracted_77509 | Not-determined |
| S15_extracted_77511 | Not-determined |
| S15_extracted_77515 | Not-determined |
| S5_extracted_k141_8965 | Medium-quality |
| S12_vs_k141_98175\|\|full | Medium-quality |
| S17_vs_k141_143748\|\|full | Medium-quality |
| S5_extracted_k141_8808 | Medium-quality |
| S14_vs_k141_34010\|\|full | Medium-quality |
| S14_vs_k141_77716\|\|full | Medium-quality |
| S15_vs_32599\|\|full | Low-quality |
| S8_vs_k141_208129\|\|full | High-quality |
| S11_vs_k141_58702\|\|full | Medium-quality |
| S14_vs_k141_112252\|\|full | Medium-quality |
| S5_vs_k141_8808\|\|full | Medium-quality |
| S3_vs_k141_95574\|\|full | Medium-quality |
| S15_vs_76400\|\|full | Low-quality |
| S15_vs_76435\|\|full | Not-determined |
| S15_vs_76464\|\|full | Not-determined |
| S15_vs_76482\|\|full | Not-determined |
| S15_vs_76554\|\|full | Not-determined |
| S15_vs_76598\|\|full | Not-determined |
| S15_vs_49\|\|full | Not-determined |
| S15_vs_151\|\|full | Not-determined |
| S15_vs_208\|\|full | Low-quality |
| S15_vs_210\|\|full | Low-quality |
| S15_vs_256\|\|full | Not-determined |
| S15_vs_281\|\|full | Not-determined |
| S15_vs_344\|\|full | Low-quality |
| S15_vs_360\|\|full | Not-determined |
| S15_vs_469\|\|full | Low-quality |
| S15_vs_477\|\|full | Low-quality |
| S15_vs_574\|\|full | Not-determined |
| S15_vs_605\|\|full | Low-quality |
| S15_vs_616\|\|full | Low-quality |
| S15_vs_859\|\|full | Not-determined |
| S15_vs_907\|\|full | Low-quality |
| S15_vs_913\|\|full | Low-quality |
| S15_vs_948\|\|full | Not-determined |
| S15_vs_1002\|\|full | Low-quality |
| S15_vs_1034\|\|full | Low-quality |
| S15_vs_1051\|\|full | Low-quality |
| S15_vs_1056\|\|full | Low-quality |
| S5_vs_k141_69980\|\|full | Medium-quality |
| S15_vs_1143\|\|full | Low-quality |
| S15_vs_1219\|\|full | Not-determined |
| S15_vs_1224\|\|full | Low-quality |
| S15_vs_1229\|\|full | Not-determined |
| S15_vs_1290\|\|full | Low-quality |
| S15_vs_1450\|\|full | Not-determined |
| S15_vs_1488\|\|full | Low-quality |
| S15_vs_1554\|\|full | Not-determined |
| S15_vs_1576\|\|full | Low-quality |
| S15_vs_1590\|\|full | Not-determined |
| S15_vs_1621\|\|full | Low-quality |
| S15_vs_1628\|\|full | Low-quality |
| S15_vs_1648\|\|full | Not-determined |
| S15_vs_1706\|\|full | Low-quality |
| S15_vs_1734\|\|full | Not-determined |
| S15_vs_1794\|\|full | Low-quality |
| S15_vs_1814\|\|full | Low-quality |
| S15_vs_1835\|\|full | Low-quality |
| S15_vs_1897\|\|full | Low-quality |
| S15_vs_1978\|\|full | Not-determined |
| S15_vs_1983\|\|full | Not-determined |
| S15_vs_2067\|\|full | Not-determined |
| S15_vs_2153\|\|full | Not-determined |
| S15_vs_2424\|\|full | Low-quality |
| S15_vs_2543\|\|full | Low-quality |
| S15_vs_2598\|\|full | Low-quality |
| S15_vs_2662\|\|full | Low-quality |
| S15_vs_2762\|\|full | Not-determined |
| S15_vs_2801\|\|full | Low-quality |
| S15_vs_2862\|\|full | Low-quality |
| S15_vs_2928\|\|full | Not-determined |
| S15_vs_2933\|\|full | Low-quality |
| S15_vs_2992\|\|full | Not-determined |
| S15_vs_3038\|\|full | Low-quality |
| S15_vs_3131\|\|full | Low-quality |
| S15_vs_3153\|\|full | Low-quality |
| S15_vs_3305\|\|full | Low-quality |
| S15_vs_3356\|\|full | Not-determined |
| S15_vs_3377\|\|full | Low-quality |
| S15_vs_3416\|\|full | Low-quality |
| S15_vs_3447\|\|full | Low-quality |
| S15_vs_3474\|\|full | Low-quality |
| S15_vs_3480\|\|full | Low-quality |
| S15_vs_3507\|\|full | Low-quality |
| S15_vs_3510\|\|full | Low-quality |
| S15_vs_3527\|\|full | Low-quality |
| S15_vs_3575\|\|full | Low-quality |
| S15_vs_3579\|\|full | Not-determined |
| S15_vs_3609\|\|full | Low-quality |
| S15_vs_3669\|\|full | Low-quality |
| S15_vs_3726\|\|full | Low-quality |
| S15_vs_3757\|\|full | Not-determined |
| S15_vs_3791\|\|full | Low-quality |
| S15_vs_3831\|\|full | Not-determined |
| S15_vs_3835\|\|full | Low-quality |
| S15_vs_3863\|\|full | Low-quality |
| S7_vs_72493\|\|full | Medium-quality |
| S15_vs_3951\|\|full | Low-quality |
| S15_vs_3962\|\|full | Low-quality |
| S15_vs_3975\|\|full | Not-determined |
| S15_vs_3997\|\|full | Not-determined |
| S15_vs_4001\|\|full | Low-quality |
| S15_vs_4061\|\|full | Low-quality |
| S15_vs_4084\|\|full | Low-quality |
| S15_vs_4095\|\|full | Low-quality |
| S15_vs_4097\|\|full | Low-quality |
| S15_vs_4132\|\|full | Low-quality |
| S15_vs_4193\|\|full | Low-quality |
| S15_vs_4199\|\|full | Not-determined |
| S15_vs_4299\|\|full | Not-determined |
| S15_vs_4455\|\|full | Not-determined |
| S15_vs_4482\|\|full | Low-quality |
| S15_vs_4555\|\|full | Low-quality |
| S15_vs_4562\|\|full | Low-quality |
| S15_vs_4606\|\|full | Not-determined |
| S15_vs_4621\|\|full | Low-quality |
| S15_vs_4664\|\|full | Low-quality |
| S15_vs_4751\|\|full | Low-quality |
| S15_vs_4774\|\|full | Low-quality |
| S15_vs_4846\|\|full | Not-determined |
| S15_vs_4901\|\|full | Low-quality |
| S15_vs_4913\|\|full | Low-quality |
| S15_vs_4966\|\|full | Low-quality |
| S15_vs_5052\|\|full | Not-determined |
| S15_vs_5070\|\|full | Low-quality |
| S15_vs_5090\|\|full | Not-determined |
| S15_vs_5102\|\|full | Low-quality |
| S15_vs_5155\|\|full | Low-quality |
| S15_vs_5173\|\|full | Low-quality |
| S15_vs_5188\|\|full | Low-quality |
| S15_vs_5236\|\|full | Low-quality |
| S15_vs_5322\|\|full | Low-quality |
| S15_vs_5369\|\|full | Not-determined |
| S15_vs_5383\|\|full | Low-quality |
| S15_vs_5397\|\|full | Not-determined |
| S15_vs_5425\|\|full | Low-quality |
| S15_vs_5510\|\|full | Not-determined |
| S15_vs_5582\|\|full | Low-quality |
| S15_vs_5634\|\|full | Not-determined |
| S15_vs_5636\|\|full | Not-determined |
| S15_vs_5652\|\|full | Low-quality |
| S15_vs_5708\|\|full | Low-quality |
| S15_vs_5718\|\|full | Low-quality |
| S15_vs_5720\|\|full | Not-determined |
| S15_vs_5768\|\|full | Low-quality |
| S15_vs_5851\|\|full | Low-quality |
| S15_vs_5879\|\|full | Low-quality |
| S15_vs_5944\|\|full | Low-quality |
| S15_vs_5976\|\|full | Not-determined |
| S15_vs_6006\|\|full | Low-quality |
| S15_vs_6079\|\|full | Not-determined |
| S15_vs_6104\|\|full | Low-quality |
| S15_vs_6129\|\|full | Low-quality |
| S15_vs_6132\|\|full | Not-determined |
| S15_vs_6395\|\|full | Low-quality |
| S15_vs_6432\|\|full | Low-quality |
| S15_vs_6441\|\|full | Low-quality |
| S15_vs_6454\|\|full | Not-determined |
| S15_vs_6458\|\|full | Not-determined |
| S15_vs_6516\|\|full | Not-determined |
| S15_vs_6562\|\|full | Low-quality |
| S15_vs_6564\|\|full | Low-quality |
| S15_vs_6644\|\|full | Not-determined |
| S15_vs_6649\|\|full | Low-quality |
| S15_vs_6743\|\|full | Not-determined |
| S15_vs_6747\|\|full | Low-quality |
| S15_vs_6766\|\|full | Not-determined |
| S15_vs_6777\|\|full | Low-quality |
| S15_vs_6787\|\|full | Not-determined |
| S15_vs_6866\|\|full | Not-determined |
| S15_vs_7083\|\|full | Low-quality |
| S15_vs_7130\|\|full | Low-quality |
| S15_vs_7138\|\|full | Low-quality |
| S15_vs_7183\|\|full | Not-determined |
| S15_vs_7222\|\|full | Low-quality |
| S15_vs_7265\|\|full | Low-quality |
| S15_vs_7280\|\|full | Not-determined |
| S15_vs_7427\|\|full | Low-quality |
| S15_vs_7715\|\|full | Low-quality |
| S15_vs_7741\|\|full | Low-quality |
| S15_vs_7771\|\|full | Low-quality |
| S15_vs_7794\|\|full | Low-quality |
| S15_vs_7800\|\|full | Low-quality |
| S15_vs_7858\|\|full | Low-quality |
| S15_vs_8038\|\|full | Low-quality |
| S15_vs_8089\|\|full | Low-quality |
| S15_vs_8092\|\|full | Not-determined |
| S15_vs_8159\|\|full | Low-quality |
| S15_vs_8255\|\|full | Not-determined |
| S15_vs_8446\|\|full | Not-determined |
| S15_vs_8485\|\|full | Low-quality |
| S15_vs_8490\|\|full | Low-quality |
| S15_vs_8517\|\|full | Low-quality |
| S15_vs_8533\|\|full | Low-quality |
| S15_vs_8549\|\|full | Low-quality |
| S15_vs_8571\|\|full | Low-quality |
| S15_vs_8613\|\|full | Not-determined |
| S15_vs_8682\|\|full | Low-quality |
| S15_vs_8699\|\|full | Not-determined |
| S15_vs_8883\|\|full | Low-quality |
| S15_vs_8929\|\|full | Not-determined |
| S15_vs_8938\|\|full | Low-quality |
| S15_vs_8977\|\|full | Low-quality |
| S15_vs_8987\|\|full | Not-determined |
| S15_vs_9109\|\|full | Not-determined |
| S15_vs_9111\|\|full | Low-quality |
| S15_vs_9198\|\|full | Not-determined |
| S15_vs_9211\|\|full | Low-quality |
| S15_vs_9270\|\|full | Not-determined |
| S15_vs_9296\|\|full | Low-quality |
| S15_vs_9328\|\|full | Low-quality |
| S15_vs_9344\|\|full | Low-quality |
| S15_vs_9386\|\|full | Low-quality |
| S15_vs_9427\|\|full | Low-quality |
| S15_vs_9428\|\|full | Low-quality |
| S15_vs_9449\|\|full | Not-determined |
| S15_vs_9465\|\|full | Not-determined |
| S15_vs_9532\|\|full | Low-quality |
| S15_vs_9554\|\|full | Low-quality |
| S15_vs_9660\|\|full | Not-determined |
| S15_vs_9708\|\|full | Low-quality |
| S15_vs_9776\|\|full | Not-determined |
| S15_vs_9780\|\|full | Low-quality |
| S15_vs_9848\|\|full | Not-determined |
| S15_vs_9852\|\|full | Not-determined |
| S15_vs_9888\|\|full | Not-determined |
| S15_vs_9901\|\|full | Low-quality |
| S15_vs_9934\|\|full | Not-determined |
| S15_vs_9992\|\|full | Low-quality |
| S15_vs_10014\|\|full | Low-quality |
| S15_vs_10023\|\|full | Low-quality |
| S15_vs_10147\|\|full | Low-quality |
| S15_vs_10181\|\|full | Low-quality |
| S15_vs_10227\|\|full | Low-quality |
| S15_vs_10238\|\|full | Not-determined |
| S15_vs_10239\|\|full | Not-determined |
| S15_vs_10245\|\|full | Not-determined |
| S15_vs_10380\|\|full | Not-determined |
| S15_vs_10411\|\|full | Not-determined |
| S14_vs_k141_84697\|\|full | Medium-quality |
| S15_vs_10521\|\|full | Not-determined |
| S15_vs_10532\|\|full | Not-determined |
| S15_vs_10560\|\|full | Low-quality |
| S15_vs_10612\|\|full | Low-quality |
| S15_vs_10652\|\|full | Low-quality |
| S15_vs_10674\|\|full | Not-determined |
| S15_vs_10735\|\|full | Low-quality |
| S15_vs_10818\|\|full | Not-determined |
| S15_vs_10825\|\|full | Low-quality |
| S15_vs_10864\|\|full | Low-quality |
| S15_vs_10868\|\|full | Low-quality |
| S15_vs_10896\|\|full | Low-quality |
| S15_vs_10947\|\|full | Low-quality |
| S15_vs_11047\|\|full | Low-quality |
| S15_vs_11101\|\|full | Low-quality |
| S15_vs_11206\|\|full | Not-determined |
| S15_vs_11207\|\|full | Not-determined |
| S15_vs_11255\|\|full | Low-quality |
| S15_vs_11297\|\|full | Not-determined |
| S15_vs_11321\|\|full | Low-quality |
| S15_vs_11326\|\|full | Low-quality |
| S15_vs_11381\|\|full | Low-quality |
| S15_vs_11435\|\|full | Not-determined |
| S15_vs_11450\|\|full | Low-quality |
| S15_vs_11493\|\|full | Not-determined |
| S15_vs_11546\|\|full | Not-determined |
| S15_vs_11650\|\|full | Low-quality |
| S15_vs_11682\|\|full | Not-determined |
| S15_vs_11738\|\|full | Low-quality |
| S15_vs_11745\|\|full | Not-determined |
| S15_vs_11767\|\|full | Low-quality |
| S15_vs_11819\|\|full | Low-quality |
| S15_vs_76390\|\|full | High-quality |
| S15_vs_11876\|\|full | Not-determined |
| S15_vs_11930\|\|full | Not-determined |
| S15_vs_11949\|\|full | Low-quality |
| S15_vs_11986\|\|full | Low-quality |
| S15_vs_12115\|\|full | Low-quality |
| S15_vs_12152\|\|full | Low-quality |
| S15_vs_12168\|\|full | Low-quality |
| S15_vs_12174\|\|full | Not-determined |
| S15_vs_12307\|\|full | Low-quality |
| S15_vs_12335\|\|full | Low-quality |
| S15_vs_12424\|\|full | Low-quality |
| S15_vs_12437\|\|full | Low-quality |
| S15_vs_12451\|\|full | Not-determined |
| S15_vs_12537\|\|full | Not-determined |
| S15_vs_12579\|\|full | Low-quality |
| S15_vs_12622\|\|full | Not-determined |
| S15_vs_12688\|\|full | Not-determined |
| S15_vs_12710\|\|full | Low-quality |
| S15_vs_12787\|\|full | Low-quality |
| S15_vs_12858\|\|full | Not-determined |
| S15_vs_13046\|\|full | Low-quality |
| S15_vs_13119\|\|full | Low-quality |
| S15_vs_13171\|\|full | Low-quality |
| S15_vs_13256\|\|full | Low-quality |
| S15_vs_13285\|\|full | Not-determined |
| S15_vs_13298\|\|full | Not-determined |
| S15_vs_13401\|\|full | Not-determined |
| S15_vs_13416\|\|full | Low-quality |
| S15_vs_13438\|\|full | Not-determined |
| S15_vs_13455\|\|full | Not-determined |
| S15_vs_13610\|\|full | Low-quality |
| S15_vs_13621\|\|full | Low-quality |
| S15_vs_13681\|\|full | Low-quality |
| S15_vs_13861\|\|full | Low-quality |
| S15_vs_13914\|\|full | Not-determined |
| S15_vs_13915\|\|full | Low-quality |
| S15_vs_13967\|\|full | Not-determined |
| S15_vs_13970\|\|full | Not-determined |
| S15_vs_14110\|\|full | Not-determined |
| S15_vs_14150\|\|full | Low-quality |
| S15_vs_14213\|\|full | Low-quality |
| S15_vs_14234\|\|full | Low-quality |
| S15_vs_14252\|\|full | Not-determined |
| S15_vs_14268\|\|full | Low-quality |
| S15_vs_14318\|\|full | Low-quality |
| S15_vs_14526\|\|full | Low-quality |
| S15_vs_14624\|\|full | Low-quality |
| S15_vs_14667\|\|full | Low-quality |
| S15_vs_14743\|\|full | Not-determined |
| S15_vs_14866\|\|full | Not-determined |
| S15_vs_14882\|\|full | Not-determined |
| S15_vs_14902\|\|full | Low-quality |
| S15_vs_15117\|\|full | Not-determined |
| S15_vs_15196\|\|full | Not-determined |
| S15_vs_15276\|\|full | Low-quality |
| S15_vs_15345\|\|full | Not-determined |
| S15_vs_15419\|\|full | Low-quality |
| S15_vs_15592\|\|full | Low-quality |
| S15_vs_15726\|\|full | Low-quality |
| S15_vs_15728\|\|full | Not-determined |
| S15_vs_15738\|\|full | Low-quality |
| S15_vs_15949\|\|full | Not-determined |
| S15_vs_16001\|\|full | Low-quality |
| S15_vs_16033\|\|full | Low-quality |
| S15_vs_16099\|\|full | Low-quality |
| S15_vs_16134\|\|full | Not-determined |
| S15_vs_16156\|\|full | Low-quality |
| S15_vs_16182\|\|full | Low-quality |
| S15_vs_16191\|\|full | Low-quality |
| S15_vs_16199\|\|full | Not-determined |
| S15_vs_16220\|\|full | Low-quality |
| S15_vs_16231\|\|full | Not-determined |
| S15_vs_16245\|\|full | Low-quality |
| S15_vs_16308\|\|full | Low-quality |
| S15_vs_16363\|\|full | Not-determined |
| S15_vs_16407\|\|full | Low-quality |
| S15_vs_16514\|\|full | Low-quality |
| S15_vs_16619\|\|full | Not-determined |
| S15_vs_16654\|\|full | Low-quality |
| S15_vs_16704\|\|full | Low-quality |
| S15_vs_16786\|\|full | Not-determined |
| S15_vs_16847\|\|full | Low-quality |
| S15_vs_16872\|\|full | Not-determined |
| S15_vs_16892\|\|full | Low-quality |
| S15_vs_17009\|\|full | Low-quality |
| S15_vs_17079\|\|full | Not-determined |
| S15_vs_17104\|\|full | Not-determined |
| S15_vs_17114\|\|full | Not-determined |
| S15_vs_17194\|\|full | Not-determined |
| S15_vs_17228\|\|full | Low-quality |
| S15_vs_17238\|\|full | Not-determined |
| S15_vs_17342\|\|full | Low-quality |
| S15_vs_17361\|\|full | Not-determined |
| S15_vs_17403\|\|full | Not-determined |
| S15_vs_17424\|\|full | Low-quality |
| S15_vs_17471\|\|full | Not-determined |
| S15_vs_17478\|\|full | Low-quality |
| S15_vs_17594\|\|full | Not-determined |
| S15_vs_17606\|\|full | Low-quality |
| S15_vs_17664\|\|full | Not-determined |
| S15_vs_17788\|\|full | Low-quality |
| S15_vs_17816\|\|full | Low-quality |
| S15_vs_17827\|\|full | Low-quality |
| S5_vs_k141_2876\|\|full | Medium-quality |
| S15_vs_17864\|\|full | Low-quality |
| S15_vs_17874\|\|full | Low-quality |
| S15_vs_17886\|\|full | Low-quality |
| S15_vs_17954\|\|full | Not-determined |
| S15_vs_17969\|\|full | Low-quality |
| S15_vs_17990\|\|full | Low-quality |
| S15_vs_18006\|\|full | Low-quality |
| S15_vs_18138\|\|full | Low-quality |
| S15_vs_18146\|\|full | Low-quality |
| S15_vs_18250\|\|full | Low-quality |
| S15_vs_18385\|\|full | Low-quality |
| S15_vs_18395\|\|full | Not-determined |
| S15_vs_18399\|\|full | Low-quality |
| S15_vs_18413\|\|full | Low-quality |
| S15_vs_18522\|\|full | Not-determined |
| S15_vs_18555\|\|full | Not-determined |
| S15_vs_18587\|\|full | Not-determined |
| S15_vs_18604\|\|full | Not-determined |
| S15_vs_18620\|\|full | Low-quality |
| S15_vs_18655\|\|full | Not-determined |
| S15_vs_18686\|\|full | Low-quality |
| S15_vs_18789\|\|full | Not-determined |
| S15_vs_18837\|\|full | Low-quality |
| S15_vs_18926\|\|full | Not-determined |
| S15_vs_18949\|\|full | Not-determined |
| S15_vs_19140\|\|full | Low-quality |
| S15_vs_19184\|\|full | Not-determined |
| S15_vs_19235\|\|full | Low-quality |
| S15_vs_19261\|\|full | Low-quality |
| S15_vs_19341\|\|full | Not-determined |
| S15_vs_19381\|\|full | Low-quality |

| | |
|---|---|
| S15_vs_19399\|\|full | Not-determined |
| S15_vs_19403\|\|full | Low-quality |
| S15_vs_19521\|\|full | Low-quality |
| S15_vs_19561\|\|full | Low-quality |
| S15_vs_19629\|\|full | Not-determined |
| S15_vs_19643\|\|full | Low-quality |
| S15_vs_19647\|\|full | Low-quality |
| S15_vs_19654\|\|full | Not-determined |
| S15_vs_19672\|\|full | Not-determined |
| S15_vs_19690\|\|full | Low-quality |
| S15_vs_19727\|\|full | Not-determined |
| S15_vs_19797\|\|full | Low-quality |
| S15_vs_19840\|\|full | Low-quality |
| S15_vs_19881\|\|full | Low-quality |
| S15_vs_19908\|\|full | Low-quality |
| S15_vs_19939\|\|full | Low-quality |
| S15_vs_19964\|\|full | Not-determined |
| S15_vs_20030\|\|full | Not-determined |
| S15_vs_20090\|\|full | Not-determined |
| S15_vs_20147\|\|full | Low-quality |
| S15_vs_20149\|\|full | Not-determined |
| S15_vs_20193\|\|full | Not-determined |
| S15_vs_20303\|\|full | Not-determined |
| S15_vs_20322\|\|full | Low-quality |
| S15_vs_20366\|\|full | Low-quality |
| S15_vs_20380\|\|full | Low-quality |
| S15_vs_20381\|\|full | Low-quality |
| S15_vs_20445\|\|full | Not-determined |
| S15_vs_20570\|\|full | Low-quality |
| S15_vs_20606\|\|full | Not-determined |
| S15_vs_20610\|\|full | Not-determined |
| S14_vs_k141_102698\|\|full | Medium-quality |
| S15_vs_20676\|\|full | Not-determined |
| S15_vs_20762\|\|full | Not-determined |
| S15_vs_20834\|\|full | Not-determined |
| S15_vs_20880\|\|full | Low-quality |
| S15_vs_20908\|\|full | Low-quality |
| S15_vs_20909\|\|full | Low-quality |
| S15_vs_20917\|\|full | Low-quality |
| S15_vs_20952\|\|full | Not-determined |
| S15_vs_20993\|\|full | Low-quality |
| S15_vs_21014\|\|full | Low-quality |
| S15_vs_21051\|\|full | Not-determined |
| S15_vs_21054\|\|full | Low-quality |
| S15_vs_21068\|\|full | Low-quality |
| S15_vs_21078\|\|full | Not-determined |
| S15_vs_21097\|\|full | Low-quality |
| S15_vs_21105\|\|full | Not-determined |
| S15_vs_21122\|\|full | Not-determined |
| S15_vs_21198\|\|full | Low-quality |
| S15_vs_21278\|\|full | Low-quality |
| S15_vs_21281\|\|full | Not-determined |
| S15_vs_21309\|\|full | Not-determined |
| S15_vs_21341\|\|full | Low-quality |
| S15_vs_21454\|\|full | Low-quality |
| S15_vs_21519\|\|full | Not-determined |
| S15_vs_21520\|\|full | Low-quality |
| S15_vs_21557\|\|full | Low-quality |
| S15_vs_21640\|\|full | Not-determined |
| S15_vs_21644\|\|full | Low-quality |
| S15_vs_21685\|\|full | Low-quality |
| S15_vs_21804\|\|full | Low-quality |
| S15_vs_21852\|\|full | Low-quality |
| S15_vs_21865\|\|full | Low-quality |
| S15_vs_21873\|\|full | Not-determined |
| S15_vs_21947\|\|full | Low-quality |
| S15_vs_21974\|\|full | Low-quality |
| S15_vs_22033\|\|full | Not-determined |
| S15_vs_22040\|\|full | Low-quality |
| S15_vs_22048\|\|full | Not-determined |
| S15_vs_22128\|\|full | Low-quality |
| S15_vs_22207\|\|full | Not-determined |
| S15_vs_22299\|\|full | Not-determined |
| S15_vs_22351\|\|full | Low-quality |
| S15_vs_22393\|\|full | Low-quality |
| S15_vs_22407\|\|full | Low-quality |
| S15_vs_22508\|\|full | Low-quality |
| S15_vs_22803\|\|full | Low-quality |
| S15_vs_22807\|\|full | Low-quality |
| S15_vs_23235\|\|full | Low-quality |
| S15_vs_23250\|\|full | Not-determined |
| S15_vs_23273\|\|full | Not-determined |
| S15_vs_23279\|\|full | Low-quality |
| S15_vs_23418\|\|full | Not-determined |
| S15_vs_23560\|\|full | Low-quality |
| S15_vs_23625\|\|full | Low-quality |
| S15_vs_23632\|\|full | Low-quality |
| S15_vs_23702\|\|full | Low-quality |
| S15_vs_23715\|\|full | Low-quality |
| S15_vs_23752\|\|full | Not-determined |
| S15_vs_23832\|\|full | Not-determined |
| S15_vs_23928\|\|full | Low-quality |
| S15_vs_24032\|\|full | Low-quality |
| S15_vs_24049\|\|full | Low-quality |
| S15_vs_24061\|\|full | Not-determined |
| S15_vs_24092\|\|full | Low-quality |
| S15_vs_24146\|\|full | Low-quality |
| S15_vs_24345\|\|full | Not-determined |
| S15_vs_24411\|\|full | Low-quality |
| S15_vs_24417\|\|full | Low-quality |
| S15_vs_24474\|\|full | Low-quality |
| S15_vs_24480\|\|full | Not-determined |
| S15_vs_24493\|\|full | Low-quality |
| S15_vs_24539\|\|full | Low-quality |
| S15_vs_24602\|\|full | Low-quality |
| S15_vs_24781\|\|full | Low-quality |
| S15_vs_24792\|\|full | Not-determined |
| S15_vs_24913\|\|full | Low-quality |
| S15_vs_24921\|\|full | Not-determined |
| S15_vs_25053\|\|full | Low-quality |
| S15_vs_25104\|\|full | Not-determined |
| S15_vs_25133\|\|full | Not-determined |
| S15_vs_25195\|\|full | Not-determined |
| S15_vs_25216\|\|full | Low-quality |
| S15_vs_25234\|\|full | Low-quality |
| S15_vs_25244\|\|full | Not-determined |
| S15_vs_25291\|\|full | Not-determined |
| S15_vs_25493\|\|full | Not-determined |
| S15_vs_25502\|\|full | Low-quality |
| S15_vs_25514\|\|full | Low-quality |
| S15_vs_25534\|\|full | Low-quality |
| S15_vs_25551\|\|full | Low-quality |
| S15_vs_25606\|\|full | Not-determined |
| S15_vs_25652\|\|full | Low-quality |
| S15_vs_25673\|\|full | Not-determined |
| S15_vs_25776\|\|full | Low-quality |
| S15_vs_25780\|\|full | Not-determined |
| S15_vs_25785\|\|full | Not-determined |
| S15_vs_25791\|\|full | Low-quality |
| S15_vs_25823\|\|full | Low-quality |
| S15_vs_25848\|\|full | Low-quality |
| S15_vs_25859\|\|full | Not-determined |
| S15_vs_26015\|\|full | Not-determined |
| S15_vs_26044\|\|full | Not-determined |
| S15_vs_26080\|\|full | Low-quality |
| S15_vs_26098\|\|full | Low-quality |
| S15_vs_26136\|\|full | Low-quality |
| S15_vs_26235\|\|full | Not-determined |
| S15_vs_26282\|\|full | Low-quality |
| S15_vs_26306\|\|full | Low-quality |
| S15_vs_26392\|\|full | Low-quality |
| S15_vs_26460\|\|full | Not-determined |
| S15_vs_26462\|\|full | Low-quality |
| S15_vs_26463\|\|full | Not-determined |
| S15_vs_26692\|\|full | Low-quality |
| S15_vs_26781\|\|full | Low-quality |
| S15_vs_26853\|\|full | Low-quality |
| S15_vs_26968\|\|full | Low-quality |
| S15_vs_27004\|\|full | Low-quality |
| S15_vs_27049\|\|full | Low-quality |
| S15_vs_27088\|\|full | Low-quality |
| S15_vs_27175\|\|full | Low-quality |
| S15_vs_27234\|\|full | Low-quality |
| S15_vs_27285\|\|full | Low-quality |
| S15_vs_27291\|\|full | Not-determined |
| S15_vs_27339\|\|full | Not-determined |
| S15_vs_27390\|\|full | Not-determined |
| S15_vs_27464\|\|full | Low-quality |
| S5_vs_k141_89540\|\|full | Medium-quality |
| S15_vs_27530\|\|full | Not-determined |
| S15_vs_27533\|\|full | Low-quality |
| S15_vs_27631\|\|full | Low-quality |
| S15_vs_27682\|\|full | Not-determined |
| S15_vs_27718\|\|full | Low-quality |
| S15_vs_27813\|\|full | Not-determined |
| S15_vs_27872\|\|full | Low-quality |
| S15_vs_27883\|\|full | Not-determined |
| S15_vs_27892\|\|full | Not-determined |
| S15_vs_27904\|\|full | Not-determined |
| S15_vs_27913\|\|full | Low-quality |
| S15_vs_27961\|\|full | Low-quality |
| S15_vs_27971\|\|full | Low-quality |
| S15_vs_27992\|\|full | Not-determined |
| S15_vs_28020\|\|full | Low-quality |
| S15_vs_28107\|\|full | Low-quality |
| S15_vs_28129\|\|full | Low-quality |
| S15_vs_28280\|\|full | Low-quality |
| S15_vs_28304\|\|full | Not-determined |
| S15_vs_28329\|\|full | Not-determined |
| S15_vs_28556\|\|full | Low-quality |
| S15_vs_28695\|\|full | Low-quality |
| S15_vs_28712\|\|full | Low-quality |
| S15_vs_28741\|\|full | Low-quality |
| S15_vs_28877\|\|full | Low-quality |
| S15_vs_28984\|\|full | Low-quality |
| S15_vs_29001\|\|full | Not-determined |
| S15_vs_29035\|\|full | Low-quality |
| S15_vs_29062\|\|full | Low-quality |
| S15_vs_29160\|\|full | Low-quality |
| S15_vs_29221\|\|full | Not-determined |
| S15_vs_29317\|\|full | Not-determined |
| S15_vs_29374\|\|full | Low-quality |
| S15_vs_29461\|\|full | Not-determined |
| S15_vs_29487\|\|full | Not-determined |
| S15_vs_29492\|\|full | Not-determined |
| S15_vs_29512\|\|full | Low-quality |
| S15_vs_29613\|\|full | Low-quality |
| S15_vs_29615\|\|full | Not-determined |
| S15_vs_29627\|\|full | Low-quality |
| S15_vs_29646\|\|full | Not-determined |
| S15_vs_29720\|\|full | Not-determined |
| S15_vs_29756\|\|full | Low-quality |
| S15_vs_29805\|\|full | Not-determined |
| S15_vs_29831\|\|full | Not-determined |
| S15_vs_29839\|\|full | Not-determined |
| S15_vs_29841\|\|full | Low-quality |
| S15_vs_30158\|\|full | Not-determined |
| S15_vs_30173\|\|full | Not-determined |
| S15_vs_30178\|\|full | Not-determined |
| S15_vs_30195\|\|full | Low-quality |
| S15_vs_30206\|\|full | Not-determined |
| S15_vs_30217\|\|full | Not-determined |
| S15_vs_30221\|\|full | Not-determined |
| S15_vs_30224\|\|full | Low-quality |
| S15_vs_30334\|\|full | Not-determined |
| S15_vs_30350\|\|full | Low-quality |
| S15_vs_30515\|\|full | Low-quality |
| S15_vs_30558\|\|full | Not-determined |
| S15_vs_30642\|\|full | Not-determined |
| S15_vs_30737\|\|full | Low-quality |
| S15_vs_30738\|\|full | Not-determined |
| S15_vs_30908\|\|full | Low-quality |
| S15_vs_30933\|\|full | Low-quality |
| S15_vs_30967\|\|full | Not-determined |
| S15_vs_30983\|\|full | Low-quality |
| S15_vs_31088\|\|full | Low-quality |
| S15_vs_31136\|\|full | Not-determined |
| S15_vs_31150\|\|full | Low-quality |
| S15_vs_31152\|\|full | Low-quality |
| S15_vs_31155\|\|full | Not-determined |
| S15_vs_31246\|\|full | Not-determined |
| S15_vs_31290\|\|full | Low-quality |
| S15_vs_31298\|\|full | Not-determined |
| S15_vs_31318\|\|full | Not-determined |
| S15_vs_31351\|\|full | Low-quality |
| S15_vs_31353\|\|full | Not-determined |
| S15_vs_31495\|\|full | Not-determined |
| S15_vs_31514\|\|full | Low-quality |
| S15_vs_31536\|\|full | Not-determined |
| S15_vs_31591\|\|full | Not-determined |
| S15_vs_31594\|\|full | Not-determined |
| S15_vs_31600\|\|full | Not-determined |
| S15_vs_31611\|\|full | Low-quality |
| S15_vs_31634\|\|full | Low-quality |
| S15_vs_31640\|\|full | Low-quality |
| S15_vs_31653\|\|full | Not-determined |
| S15_vs_31694\|\|full | Low-quality |
| S15_vs_31709\|\|full | Not-determined |
| S15_vs_31732\|\|full | Low-quality |
| S15_vs_31929\|\|full | Low-quality |
| S15_vs_31945\|\|full | Low-quality |
| S15_vs_31946\|\|full | Not-determined |
| S15_vs_31992\|\|full | Not-determined |
| S15_vs_32044\|\|full | Not-determined |
| S15_vs_32085\|\|full | Low-quality |
| S15_vs_32113\|\|full | Low-quality |
| S15_vs_32136\|\|full | Not-determined |
| S15_vs_32189\|\|full | Not-determined |
| S15_vs_32204\|\|full | Not-determined |
| S15_vs_32352\|\|full | Low-quality |
| S15_vs_32441\|\|full | Low-quality |
| S1_extracted_k141_138157 | Medium-quality |
| S15_vs_32561\|\|full | Low-quality |
| S15_vs_32589\|\|full | Low-quality |
| S15_vs_32690\|\|full | Low-quality |
| S15_vs_32726\|\|full | Not-determined |
| S15_vs_32737\|\|full | Low-quality |
| S15_vs_32803\|\|full | Not-determined |
| S15_vs_32809\|\|full | Low-quality |
| S15_vs_32834\|\|full | Not-determined |
| S15_vs_32857\|\|full | Low-quality |
| S15_vs_32933\|\|full | Low-quality |
| S15_vs_33087\|\|full | Not-determined |
| S15_vs_33110\|\|full | Low-quality |
| S15_vs_33158\|\|full | Not-determined |
| S15_vs_33273\|\|full | Low-quality |
| S15_vs_33358\|\|full | Not-determined |
| S15_vs_33495\|\|full | Low-quality |
| S15_vs_33549\|\|full | Low-quality |
| S15_vs_33563\|\|full | Low-quality |
| S15_vs_33642\|\|full | Low-quality |
| S15_vs_33648\|\|full | Not-determined |
| S15_vs_33663\|\|full | Low-quality |
| S15_vs_33665\|\|full | Low-quality |
| S15_vs_33686\|\|full | Not-determined |
| S15_vs_33756\|\|full | Low-quality |
| S15_vs_33763\|\|full | Low-quality |
| S15_vs_33969\|\|full | Not-determined |
| S15_vs_33970\|\|full | Low-quality |
| S15_vs_33979\|\|full | Low-quality |
| S15_vs_34012\|\|full | Low-quality |
| S15_vs_34108\|\|full | Not-determined |
| S15_vs_34193\|\|full | Not-determined |
| S15_vs_34274\|\|full | Low-quality |
| S15_vs_34305\|\|full | Low-quality |
| S15_vs_34368\|\|full | Low-quality |
| S15_vs_34378\|\|full | Not-determined |
| S15_vs_34425\|\|full | Low-quality |
| S15_vs_34508\|\|full | Low-quality |
| S15_vs_34581\|\|full | Not-determined |
| S15_vs_34584\|\|full | Low-quality |
| S15_vs_34704\|\|full | Low-quality |
| S15_vs_34749\|\|full | Not-determined |
| S15_vs_34800\|\|full | Low-quality |
| S15_vs_34879\|\|full | Low-quality |
| S15_vs_35113\|\|full | Low-quality |
| S15_vs_35135\|\|full | Low-quality |
| S15_vs_35139\|\|full | Low-quality |
| S15_vs_35246\|\|full | Low-quality |
| S11_vs_k141_89911\|\|full | Medium-quality |
| S15_vs_35452\|\|full | Low-quality |
| S15_vs_35473\|\|full | Not-determined |
| S15_vs_35544\|\|full | Not-determined |
| S15_vs_35563\|\|full | Low-quality |
| S15_vs_35639\|\|full | Low-quality |
| S15_vs_35681\|\|full | Low-quality |
| S15_vs_35713\|\|full | Not-determined |
| S15_vs_35733\|\|full | Low-quality |
| S15_vs_35812\|\|full | Low-quality |
| S15_vs_35860\|\|full | Low-quality |
| S15_vs_35861\|\|full | Not-determined |
| S15_vs_35966\|\|full | Low-quality |
| S15_vs_36010\|\|full | Not-determined |
| S15_vs_36014\|\|full | Low-quality |
| S15_vs_36070\|\|full | Not-determined |
| S15_vs_36094\|\|full | Low-quality |
| S15_vs_36121\|\|full | Low-quality |
| S15_vs_36125\|\|full | Low-quality |
| S15_vs_36194\|\|full | Not-determined |
| S15_vs_36302\|\|full | Low-quality |
| S15_vs_36375\|\|full | Low-quality |
| S15_vs_36428\|\|full | Low-quality |
| S15_vs_36432\|\|full | Low-quality |
| S15_vs_36471\|\|full | Not-determined |
| S15_vs_36504\|\|full | Low-quality |
| S15_vs_36526\|\|full | Low-quality |
| S15_vs_36576\|\|full | Low-quality |
| S15_vs_36583\|\|full | Not-determined |
| S15_vs_36656\|\|full | Low-quality |
| S15_vs_36687\|\|full | Low-quality |
| S15_vs_36770\|\|full | Low-quality |
| S15_vs_36779\|\|full | Not-determined |
| S15_vs_36865\|\|full | Not-determined |
| S15_vs_36895\|\|full | Low-quality |
| S15_vs_36952\|\|full | Not-determined |
| S15_vs_36967\|\|full | Not-determined |
| S15_vs_37062\|\|full | Low-quality |
| S15_vs_37086\|\|full | Low-quality |
| S15_vs_37124\|\|full | Not-determined |
| S15_vs_37193\|\|full | Low-quality |
| S15_vs_37214\|\|full | Not-determined |
| S15_vs_37284\|\|full | Low-quality |
| S15_vs_37317\|\|full | Low-quality |
| S15_vs_37523\|\|full | Not-determined |
| S15_vs_37530\|\|full | Not-determined |
| S15_vs_37560\|\|full | Low-quality |
| S15_vs_37562\|\|full | Low-quality |
| S15_vs_37573\|\|full | Low-quality |
| S15_vs_37627\|\|full | Low-quality |
| S15_vs_37645\|\|full | Low-quality |
| S15_vs_37666\|\|full | Low-quality |
| S15_vs_37866\|\|full | Not-determined |
| S15_vs_37871\|\|full | Low-quality |
| S15_vs_37891\|\|full | Low-quality |
| S15_vs_37950\|\|full | Low-quality |
| S15_vs_38059\|\|full | Low-quality |
| S15_vs_38075\|\|full | Not-determined |
| S15_vs_38081\|\|full | Low-quality |
| S15_vs_38087\|\|full | Low-quality |
| S15_vs_38208\|\|full | Low-quality |
| S15_vs_38251\|\|full | Low-quality |
| S15_vs_38280\|\|full | Not-determined |
| S15_vs_38296\|\|full | Not-determined |
| S15_vs_38347\|\|full | Low-quality |
| S15_vs_38382\|\|full | Not-determined |
| S15_vs_38408\|\|full | Low-quality |
| S15_vs_38445\|\|full | Low-quality |
| S15_vs_38465\|\|full | Not-determined |
| S15_vs_38471\|\|full | Not-determined |
| S15_vs_38484\|\|full | Low-quality |
| S15_vs_38503\|\|full | Low-quality |
| S15_vs_38537\|\|full | Low-quality |
| S15_vs_38602\|\|full | Not-determined |
| S15_vs_38614\|\|full | Not-determined |
| S15_vs_38709\|\|full | Low-quality |
| S15_vs_38710\|\|full | Not-determined |
| S15_vs_38874\|\|full | Not-determined |
| S15_vs_38925\|\|full | Low-quality |
| S15_vs_39009\|\|full | Not-determined |
| S15_vs_39111\|\|full | Low-quality |
| S15_vs_39250\|\|full | Low-quality |
| S15_vs_39284\|\|full | Low-quality |
| S15_vs_39557\|\|full | Low-quality |
| S5_vs_k141_8965\|\|full | Medium-quality |
| S15_vs_39806\|\|full | Low-quality |
| S15_vs_39854\|\|full | Low-quality |
| S15_vs_39897\|\|full | Low-quality |
| S15_vs_39943\|\|full | Low-quality |
| S15_vs_39960\|\|full | Low-quality |
| S15_vs_40020\|\|full | Not-determined |
| S15_vs_40175\|\|full | Low-quality |
| S15_vs_40218\|\|full | Low-quality |
| S15_vs_40221\|\|full | Low-quality |
| S15_vs_40232\|\|full | Low-quality |
| S15_vs_40239\|\|full | Low-quality |
| S15_vs_40277\|\|full | Not-determined |
| S15_vs_40284\|\|full | Not-determined |
| S15_vs_40291\|\|full | Low-quality |
| S15_vs_40326\|\|full | Low-quality |
| S15_vs_40405\|\|full | Low-quality |
| S15_vs_40498\|\|full | Not-determined |
| S15_vs_40539\|\|full | Not-determined |
| S15_vs_40662\|\|full | Low-quality |
| S15_vs_40758\|\|full | Low-quality |
| S15_vs_40773\|\|full | Low-quality |
| S15_vs_40775\|\|full | Not-determined |
| S15_vs_40809\|\|full | Not-determined |
| S15_vs_40975\|\|full | Low-quality |
| S15_vs_41013\|\|full | Not-determined |
| S15_vs_41022\|\|full | Low-quality |
| S15_vs_41092\|\|full | Low-quality |
| S15_vs_41291\|\|full | Not-determined |
| S15_vs_41340\|\|full | Low-quality |
| S15_vs_41353\|\|full | Low-quality |
| S15_vs_41411\|\|full | Not-determined |
| S15_vs_41432\|\|full | Low-quality |
| S15_vs_41441\|\|full | Low-quality |
| S15_vs_41475\|\|full | Not-determined |
| S15_vs_41539\|\|full | Low-quality |
| S15_vs_41582\|\|full | Low-quality |
| S15_vs_41697\|\|full | Low-quality |
| S15_vs_41725\|\|full | Low-quality |
| S15_vs_41862\|\|full | Low-quality |
| S15_vs_41863\|\|full | Low-quality |
| S15_vs_41921\|\|full | Low-quality |
| S15_vs_41929\|\|full | Low-quality |
| S15_vs_41930\|\|full | Low-quality |
| S15_vs_41946\|\|full | Low-quality |
| S1_vs_k141_138157\|\|full | Medium-quality |
| S15_vs_41980\|\|full | Not-determined |
| S15_vs_42026\|\|full | Low-quality |
| S7_vs_82140\|\|full | Medium-quality |
| S15_vs_42109\|\|full | Not-determined |
| S15_vs_42255\|\|full | Low-quality |
| S15_vs_42295\|\|full | Low-quality |
| S15_vs_42528\|\|full | Low-quality |
| S15_vs_42644\|\|full | Not-determined |
| S15_vs_42676\|\|full | Low-quality |
| S15_vs_42680\|\|full | Not-determined |
| S15_vs_42681\|\|full | Not-determined |
| S15_vs_42689\|\|full | Low-quality |
| S15_vs_42725\|\|full | Not-determined |
| S15_vs_42726\|\|full | Not-determined |
| S15_vs_42783\|\|full | Low-quality |
| S15_vs_42805\|\|full | Not-determined |
| S15_vs_42872\|\|full | Not-determined |
| S15_vs_42880\|\|full | Low-quality |
| S15_vs_42968\|\|full | Low-quality |
| S15_vs_42974\|\|full | Not-determined |
| S15_vs_42999\|\|full | Low-quality |
| S15_vs_43005\|\|full | Not-determined |
| S15_vs_43006\|\|full | Not-determined |
| S15_vs_43014\|\|full | Not-determined |
| S15_vs_43017\|\|full | Not-determined |
| S15_vs_43043\|\|full | Low-quality |
| S15_vs_43068\|\|full | Low-quality |
| S15_vs_43204\|\|full | Low-quality |
| S15_vs_43215\|\|full | Low-quality |
| S15_vs_43454\|\|full | Not-determined |
| S15_vs_43484\|\|full | Low-quality |
| S15_vs_43515\|\|full | Not-determined |
| S15_vs_43524\|\|full | Low-quality |
| S15_vs_43690\|\|full | Not-determined |
| S15_vs_43709\|\|full | Not-determined |
| S15_vs_43721\|\|full | Low-quality |
| S15_vs_43804\|\|full | Not-determined |
| S15_vs_43904\|\|full | Low-quality |
| S1_vs_k141_7418\|\|full | Medium-quality |
| S15_vs_43948\|\|full | Not-determined |
| S15_vs_44265\|\|full | Low-quality |
| S15_vs_44290\|\|full | Low-quality |
| S15_vs_44308\|\|full | Not-determined |
| S15_vs_44347\|\|full | Low-quality |
| S15_vs_44386\|\|full | Low-quality |
| S15_vs_44402\|\|full | Low-quality |
| S15_vs_44437\|\|full | Low-quality |
| S15_vs_44526\|\|full | Not-determined |
| S15_vs_44600\|\|full | Low-quality |
| S15_vs_44618\|\|full | Low-quality |
| S15_vs_44721\|\|full | Low-quality |
| S15_vs_44742\|\|full | Low-quality |
| S15_vs_44837\|\|full | Low-quality |
| S15_vs_44914\|\|full | Low-quality |
| S15_vs_44947\|\|full | Low-quality |
| S15_vs_44957\|\|full | Not-determined |
| S15_vs_45085\|\|full | Low-quality |
| S15_vs_45091\|\|full | Not-determined |
| S15_vs_45154\|\|full | Low-quality |
| S15_vs_45166\|\|full | Low-quality |
| S15_vs_45181\|\|full | Not-determined |
| S15_vs_45192\|\|full | Low-quality |
| S15_vs_45266\|\|full | Low-quality |
| S15_vs_45380\|\|full | Not-determined |
| S15_vs_45391\|\|full | Low-quality |
| S15_vs_45417\|\|full | Low-quality |
| S15_vs_45428\|\|full | Low-quality |
| S15_vs_45437\|\|full | Low-quality |
| S15_vs_45631\|\|full | Low-quality |
| S15_vs_45653\|\|full | Low-quality |
| S15_vs_45658\|\|full | Not-determined |
| S15_vs_45694\|\|full | Low-quality |
| S15_vs_45734\|\|full | Not-determined |
| S15_vs_45767\|\|full | Low-quality |
| S15_vs_45781\|\|full | Low-quality |
| S15_vs_45836\|\|full | Not-determined |
| S15_vs_45881\|\|full | Low-quality |
| S15_vs_46128\|\|full | Not-determined |
| S15_vs_46158\|\|full | Low-quality |
| S15_vs_46223\|\|full | Low-quality |
| S15_vs_46230\|\|full | Low-quality |
| S15_vs_46242\|\|full | Low-quality |
| S15_vs_46257\|\|full | Low-quality |
| S15_vs_46265\|\|full | Low-quality |
| S15_vs_46275\|\|full | Not-determined |
| S15_vs_46331\|\|full | Not-determined |
| S15_vs_46332\|\|full | Not-determined |
| S15_vs_46336\|\|full | Not-determined |
| S15_vs_46355\|\|full | Not-determined |
| S15_vs_46459\|\|full | Low-quality |
| S15_vs_46478\|\|full | Not-determined |
| S15_vs_46579\|\|full | Low-quality |
| S15_vs_46593\|\|full | Not-determined |
| S15_vs_46617\|\|full | Not-determined |
| S15_vs_46669\|\|full | Low-quality |
| S15_vs_46749\|\|full | Not-determined |
| S15_vs_46828\|\|full | Low-quality |
| S15_vs_46838\|\|full | Not-determined |
| S15_vs_46854\|\|full | Not-determined |
| S15_vs_46867\|\|full | Not-determined |
| S15_vs_46889\|\|full | Low-quality |
| S15_vs_46924\|\|full | Low-quality |
| S15_vs_46978\|\|full | Low-quality |
| S15_vs_47220\|\|full | Low-quality |
| S15_vs_47232\|\|full | Not-determined |
| S15_vs_47237\|\|full | Not-determined |
| S15_vs_47285\|\|full | Low-quality |
| S15_vs_47315\|\|full | Low-quality |
| S15_vs_47316\|\|full | Not-determined |
| S15_vs_47391\|\|full | Low-quality |
| S15_vs_47400\|\|full | Low-quality |
| S15_vs_47402\|\|full | Not-determined |
| S15_vs_47403\|\|full | Not-determined |
| S15_vs_47505\|\|full | Low-quality |
| S15_vs_47525\|\|full | Low-quality |
| S15_vs_47531\|\|full | Low-quality |
| S15_vs_47596\|\|full | Low-quality |
| S15_vs_47603\|\|full | Not-determined |
| S15_vs_47686\|\|full | Low-quality |
| S15_vs_47697\|\|full | Low-quality |
| S15_vs_47726\|\|full | Low-quality |
| S15_vs_47744\|\|full | Not-determined |
| S15_vs_47781\|\|full | Low-quality |
| S15_vs_47869\|\|full | Not-determined |
| S15_vs_47924\|\|full | Low-quality |
| S15_vs_48003\|\|full | Not-determined |
| S15_vs_48051\|\|full | Low-quality |
| S15_vs_48132\|\|full | Not-determined |
| S15_vs_48146\|\|full | Low-quality |
| S15_vs_48172\|\|full | Low-quality |
| S15_vs_48292\|\|full | Low-quality |
| S15_vs_48332\|\|full | Not-determined |
| S15_vs_48358\|\|full | Low-quality |
| S15_vs_48380\|\|full | Low-quality |
| S15_vs_48389\|\|full | Low-quality |
| S15_vs_48414\|\|full | Low-quality |
| S15_vs_48426\|\|full | Low-quality |
| S15_vs_48456\|\|full | Not-determined |
| S15_vs_48613\|\|full | Not-determined |
| S15_vs_48661\|\|full | Low-quality |
| S15_vs_48667\|\|full | Not-determined |
| S15_vs_48679\|\|full | Low-quality |
| S15_vs_48749\|\|full | Not-determined |
| S15_vs_48788\|\|full | Low-quality |
| S15_vs_48884\|\|full | Low-quality |
| S15_vs_48892\|\|full | Not-determined |
| S15_vs_48930\|\|full | Not-determined |
| S15_vs_48935\|\|full | Not-determined |
| S15_vs_48951\|\|full | Not-determined |
| S15_vs_48953\|\|full | Not-determined |

| | |
|---|---|
| S15_vs_48969\|\|full | Low-quality |
| S15_vs_49004\|\|full | Not-determined |
| S15_vs_49040\|\|full | Low-quality |
| S15_vs_49110\|\|full | Not-determined |
| S15_vs_49264\|\|full | Not-determined |
| S15_vs_49266\|\|full | Not-determined |
| S15_vs_49433\|\|full | Low-quality |
| S15_vs_49452\|\|full | Low-quality |
| S15_vs_49455\|\|full | Not-determined |
| S15_vs_49515\|\|full | Low-quality |
| S15_vs_49540\|\|full | Low-quality |
| S15_vs_49552\|\|full | Low-quality |
| S15_vs_49563\|\|full | Low-quality |
| S15_vs_49573\|\|full | Not-determined |
| S15_vs_49686\|\|full | Not-determined |
| S15_vs_49694\|\|full | Low-quality |
| S15_vs_49712\|\|full | Low-quality |
| S15_vs_49753\|\|full | Low-quality |
| S15_vs_49820\|\|full | Low-quality |
| S15_vs_49919\|\|full | Low-quality |
| S15_vs_49935\|\|full | Low-quality |
| S15_vs_49993\|\|full | Not-determined |
| S15_vs_50081\|\|full | Low-quality |
| S15_vs_50112\|\|full | Low-quality |
| S15_vs_50132\|\|full | Low-quality |
| S15_vs_50157\|\|full | Low-quality |
| S15_vs_50258\|\|full | Low-quality |
| S15_vs_50440\|\|full | Low-quality |
| S15_vs_50516\|\|full | Low-quality |
| S15_vs_50559\|\|full | Low-quality |
| S15_vs_50561\|\|full | Low-quality |
| S15_vs_50579\|\|full | Low-quality |
| S15_vs_50604\|\|full | Low-quality |
| S15_vs_50605\|\|full | Low-quality |
| S15_vs_50659\|\|full | Low-quality |
| S15_vs_50708\|\|full | Not-determined |
| S15_vs_50733\|\|full | Low-quality |
| S15_vs_50876\|\|full | Low-quality |
| S15_vs_50904\|\|full | Not-determined |
| S15_vs_50916\|\|full | Low-quality |
| S15_vs_50927\|\|full | Not-determined |
| S15_vs_50947\|\|full | Not-determined |
| S15_vs_50970\|\|full | Low-quality |
| S15_vs_50981\|\|full | Low-quality |
| S15_vs_50996\|\|full | Low-quality |
| S15_vs_51013\|\|full | Not-determined |
| S15_vs_51052\|\|full | Not-determined |
| S15_vs_51067\|\|full | Low-quality |
| S15_vs_51090\|\|full | Not-determined |
| S15_vs_51250\|\|full | Not-determined |
| S15_vs_51281\|\|full | Low-quality |
| S15_vs_51303\|\|full | Not-determined |
| S15_vs_51324\|\|full | Low-quality |
| S15_vs_51337\|\|full | Low-quality |
| S15_vs_51378\|\|full | Not-determined |
| S15_vs_51387\|\|full | Low-quality |
| S15_vs_51420\|\|full | Low-quality |
| S15_vs_51438\|\|full | Not-determined |
| S15_vs_51451\|\|full | Not-determined |
| S15_vs_51465\|\|full | Low-quality |
| S15_vs_51483\|\|full | Low-quality |
| S15_vs_51515\|\|full | Not-determined |
| S15_vs_51548\|\|full | Low-quality |
| S15_vs_51553\|\|full | Low-quality |
| S15_vs_51604\|\|full | Low-quality |
| S15_vs_51606\|\|full | Low-quality |
| S15_vs_51623\|\|full | Low-quality |
| S15_vs_51642\|\|full | Low-quality |
| S15_vs_51691\|\|full | Not-determined |
| S15_vs_51780\|\|full | Not-determined |
| S15_vs_51798\|\|full | Not-determined |
| S15_vs_51849\|\|full | Low-quality |
| S15_vs_51952\|\|full | Low-quality |
| S15_vs_51983\|\|full | Low-quality |
| S15_vs_51984\|\|full | Low-quality |
| S15_vs_52011\|\|full | Not-determined |
| S15_vs_52114\|\|full | Low-quality |
| S15_vs_52124\|\|full | Low-quality |
| S15_vs_52145\|\|full | Not-determined |
| S15_vs_52153\|\|full | Low-quality |
| S15_vs_52164\|\|full | Low-quality |
| S15_vs_52311\|\|full | Not-determined |
| S15_vs_52314\|\|full | Low-quality |
| S15_vs_52328\|\|full | Low-quality |
| S15_vs_52405\|\|full | Not-determined |
| S15_vs_52422\|\|full | Low-quality |
| S15_vs_52428\|\|full | Low-quality |
| S15_vs_52653\|\|full | Not-determined |
| S15_vs_52680\|\|full | Low-quality |
| S15_vs_52882\|\|full | Not-determined |
| S15_vs_52948\|\|full | Low-quality |
| S15_vs_52951\|\|full | Low-quality |
| S15_vs_53016\|\|full | Low-quality |
| S15_vs_53132\|\|full | Not-determined |
| S15_vs_53136\|\|full | Low-quality |
| S15_vs_53139\|\|full | Low-quality |
| S15_vs_53150\|\|full | Low-quality |
| S15_vs_53211\|\|full | Not-determined |
| S15_vs_53281\|\|full | Not-determined |
| S15_vs_53284\|\|full | Not-determined |
| S15_vs_53388\|\|full | Low-quality |
| S15_vs_53464\|\|full | Low-quality |
| S15_vs_53563\|\|full | Low-quality |
| S15_vs_53573\|\|full | Low-quality |
| S15_vs_53590\|\|full | Low-quality |
| S15_vs_53624\|\|full | Low-quality |
| S15_vs_53700\|\|full | Low-quality |
| S15_vs_53813\|\|full | Low-quality |
| S15_vs_53834\|\|full | Low-quality |
| S15_vs_53854\|\|full | Low-quality |
| S15_vs_53862\|\|full | Low-quality |
| S15_vs_53867\|\|full | Low-quality |
| S15_vs_53881\|\|full | Not-determined |
| S15_vs_53911\|\|full | Not-determined |
| S15_vs_53914\|\|full | Low-quality |
| S15_vs_54049\|\|full | Not-determined |
| S15_vs_54176\|\|full | Low-quality |
| S15_vs_54190\|\|full | Low-quality |
| S15_vs_54214\|\|full | Low-quality |
| S15_vs_54230\|\|full | Low-quality |
| S15_vs_54236\|\|full | Low-quality |
| S15_vs_54399\|\|full | Not-determined |
| S15_vs_54432\|\|full | Low-quality |
| S15_vs_54489\|\|full | Not-determined |
| S15_vs_54501\|\|full | Not-determined |
| S15_vs_54578\|\|full | Low-quality |
| S15_vs_54675\|\|full | Not-determined |
| S15_vs_54756\|\|full | Not-determined |
| S15_vs_54767\|\|full | Not-determined |
| S15_vs_54823\|\|full | Low-quality |
| S15_vs_54855\|\|full | Not-determined |
| S15_vs_54942\|\|full | Low-quality |
| S15_vs_55034\|\|full | Not-determined |
| S15_vs_55062\|\|full | Not-determined |
| S15_vs_55071\|\|full | Not-determined |
| S15_vs_55079\|\|full | Not-determined |
| S15_vs_55088\|\|full | Low-quality |
| S15_vs_55175\|\|full | Low-quality |
| S15_vs_55196\|\|full | Not-determined |
| S15_vs_55212\|\|full | Not-determined |
| S15_vs_55584\|\|full | Low-quality |
| S9_vs_k141_58402\|\|full | Medium-quality |
| S15_vs_55666\|\|full | Low-quality |
| S15_vs_55670\|\|full | Low-quality |
| S15_vs_55764\|\|full | Low-quality |
| S15_vs_55853\|\|full | Low-quality |
| S15_vs_55880\|\|full | Low-quality |
| S15_vs_55976\|\|full | Low-quality |
| S15_vs_56033\|\|full | Low-quality |
| S15_vs_56054\|\|full | Low-quality |
| S15_vs_56112\|\|full | Not-determined |
| S15_vs_56155\|\|full | Low-quality |
| S15_vs_56213\|\|full | Low-quality |
| S15_vs_56231\|\|full | Low-quality |
| S15_vs_56252\|\|full | Low-quality |
| S15_vs_56272\|\|full | Not-determined |
| S15_vs_56285\|\|full | Not-determined |
| S15_vs_56340\|\|full | Not-determined |
| S15_vs_56359\|\|full | Low-quality |
| S15_vs_56362\|\|full | Low-quality |
| S15_vs_56371\|\|full | Low-quality |
| S15_vs_56378\|\|full | Low-quality |
| S15_vs_56470\|\|full | Low-quality |
| S15_vs_56516\|\|full | Low-quality |
| S15_vs_56551\|\|full | Low-quality |
| S15_vs_56620\|\|full | Low-quality |
| S15_vs_56735\|\|full | Not-determined |
| S15_vs_56740\|\|full | Low-quality |
| S15_vs_56819\|\|full | Low-quality |
| S15_vs_56942\|\|full | Low-quality |
| S15_vs_56961\|\|full | Not-determined |
| S15_vs_56979\|\|full | Low-quality |
| S15_vs_57018\|\|full | Not-determined |
| S15_vs_57069\|\|full | Low-quality |
| S15_vs_57083\|\|full | Low-quality |
| S15_vs_57111\|\|full | Not-determined |
| S15_vs_57182\|\|full | Not-determined |
| S15_vs_57233\|\|full | Low-quality |
| S15_vs_57266\|\|full | Not-determined |
| S15_vs_57353\|\|full | Not-determined |
| S15_vs_57434\|\|full | Low-quality |
| S15_vs_57503\|\|full | Low-quality |
| S15_vs_57572\|\|full | Not-determined |
| S15_vs_57710\|\|full | Low-quality |
| S15_vs_57887\|\|full | Low-quality |
| S15_vs_57893\|\|full | Not-determined |
| S15_vs_57898\|\|full | Low-quality |
| S15_vs_57944\|\|full | Low-quality |
| S15_vs_58117\|\|full | Not-determined |
| S15_vs_58179\|\|full | Low-quality |
| S15_vs_58187\|\|full | Low-quality |
| S15_vs_58209\|\|full | Low-quality |
| S15_vs_58227\|\|full | Not-determined |
| S15_vs_58228\|\|full | Low-quality |
| S15_vs_58254\|\|full | Low-quality |
| S15_vs_58264\|\|full | Not-determined |
| S15_vs_58311\|\|full | Not-determined |
| S15_vs_58347\|\|full | Low-quality |
| S15_vs_58351\|\|full | Not-determined |
| S8_vs_k141_182331\|\|full | Medium-quality |
| S15_vs_58529\|\|full | Not-determined |
| S15_vs_58570\|\|full | Low-quality |
| S15_vs_58586\|\|full | Not-determined |
| S15_vs_58638\|\|full | Not-determined |
| S15_vs_58683\|\|full | Low-quality |
| S15_vs_58726\|\|full | Low-quality |
| S15_vs_58759\|\|full | Not-determined |
| S15_vs_58778\|\|full | Not-determined |
| S15_vs_58790\|\|full | Low-quality |
| S15_vs_58836\|\|full | Not-determined |
| S15_vs_58917\|\|full | Not-determined |
| S15_vs_58963\|\|full | Not-determined |
| S15_vs_59012\|\|full | Low-quality |
| S15_vs_59130\|\|full | Low-quality |
| S15_vs_59162\|\|full | Low-quality |
| S15_vs_59184\|\|full | Not-determined |
| S15_vs_59214\|\|full | Low-quality |
| S15_vs_59232\|\|full | Low-quality |
| S15_vs_59253\|\|full | Low-quality |
| S15_vs_59262\|\|full | Low-quality |
| S15_vs_59273\|\|full | Not-determined |
| S15_vs_59350\|\|full | Not-determined |
| S15_vs_59364\|\|full | Low-quality |
| S15_vs_59381\|\|full | Low-quality |
| S15_vs_59383\|\|full | Low-quality |
| S15_vs_59401\|\|full | Low-quality |
| S15_vs_59451\|\|full | Not-determined |
| S15_vs_59488\|\|full | Not-determined |
| S15_vs_59636\|\|full | Low-quality |
| S15_vs_59704\|\|full | Not-determined |
| S15_vs_59856\|\|full | Low-quality |
| S15_vs_59874\|\|full | Low-quality |
| S15_vs_59957\|\|full | Low-quality |
| S15_vs_60001\|\|full | Not-determined |
| S15_vs_60275\|\|full | Low-quality |
| S15_vs_60277\|\|full | Low-quality |
| S15_vs_60316\|\|full | Low-quality |
| S15_vs_60322\|\|full | Low-quality |
| S15_vs_60331\|\|full | Not-determined |
| S15_vs_60336\|\|full | Not-determined |
| S15_vs_60370\|\|full | Not-determined |
| S15_vs_60393\|\|full | Low-quality |
| S15_vs_60479\|\|full | Low-quality |
| S15_vs_60482\|\|full | Low-quality |
| S15_vs_60542\|\|full | Not-determined |
| S15_vs_60550\|\|full | Not-determined |
| S15_vs_60766\|\|full | Low-quality |
| S15_vs_60832\|\|full | Not-determined |
| S15_vs_60845\|\|full | Low-quality |
| S15_vs_60944\|\|full | Low-quality |
| S15_vs_60948\|\|full | Not-determined |
| S15_vs_60963\|\|full | Low-quality |
| S15_vs_60970\|\|full | Low-quality |
| S15_vs_61050\|\|full | Low-quality |
| S15_vs_61073\|\|full | Low-quality |
| S15_vs_61207\|\|full | Not-determined |
| S15_vs_61245\|\|full | Not-determined |
| S15_vs_61322\|\|full | Low-quality |
| S15_vs_61334\|\|full | Low-quality |
| S15_vs_61337\|\|full | Not-determined |
| S15_vs_61379\|\|full | Not-determined |
| S15_vs_61456\|\|full | Low-quality |
| S15_vs_61472\|\|full | Low-quality |
| S15_vs_61479\|\|full | Low-quality |
| S15_vs_61521\|\|full | Low-quality |
| S15_vs_61638\|\|full | Low-quality |
| S15_vs_61653\|\|full | Not-determined |
| S15_vs_61655\|\|full | Low-quality |
| S15_vs_61677\|\|full | Low-quality |
| S15_vs_61757\|\|full | Low-quality |
| S15_vs_61791\|\|full | Low-quality |
| S15_vs_61844\|\|full | Low-quality |
| S15_vs_61965\|\|full | Low-quality |
| S15_vs_61973\|\|full | Not-determined |
| S15_vs_62008\|\|full | Low-quality |
| S15_vs_62013\|\|full | Low-quality |
| S15_vs_62039\|\|full | Not-determined |
| S15_vs_62041\|\|full | Low-quality |
| S15_vs_62111\|\|full | Not-determined |
| S15_vs_62131\|\|full | Low-quality |
| S15_vs_62178\|\|full | Not-determined |
| S15_vs_62302\|\|full | Not-determined |
| S15_vs_62354\|\|full | Not-determined |
| S15_vs_62431\|\|full | Not-determined |
| S15_vs_62638\|\|full | Not-determined |
| S15_vs_62686\|\|full | Low-quality |
| S15_vs_62764\|\|full | Low-quality |
| S15_vs_63053\|\|full | Not-determined |
| S15_vs_63097\|\|full | Low-quality |
| S15_vs_63143\|\|full | Low-quality |
| S3_extracted_k141_96222 | Medium-quality |
| S15_vs_63264\|\|full | Low-quality |
| S15_vs_63325\|\|full | Low-quality |
| S15_vs_63489\|\|full | Not-determined |
| S15_vs_63568\|\|full | Not-determined |
| S15_vs_63570\|\|full | Low-quality |
| S15_vs_63580\|\|full | Low-quality |
| S15_vs_63654\|\|full | Low-quality |
| S15_vs_63661\|\|full | Low-quality |
| S15_vs_63677\|\|full | Low-quality |
| S15_vs_63718\|\|full | Not-determined |
| S15_vs_63789\|\|full | Low-quality |
| S15_vs_63917\|\|full | Not-determined |
| S15_vs_63955\|\|full | Low-quality |
| S15_vs_63999\|\|full | Not-determined |
| S15_vs_64024\|\|full | Low-quality |
| S15_vs_64025\|\|full | Low-quality |
| S15_vs_64027\|\|full | Low-quality |
| S15_vs_64043\|\|full | Not-determined |
| S15_vs_64066\|\|full | Not-determined |
| S15_vs_64071\|\|full | Low-quality |
| S15_vs_64232\|\|full | Low-quality |
| S15_vs_64236\|\|full | Not-determined |
| S15_vs_64361\|\|full | Not-determined |
| S15_vs_64417\|\|full | Not-determined |
| S15_vs_64473\|\|full | Low-quality |
| S15_vs_64520\|\|full | Not-determined |
| S15_vs_64695\|\|full | Low-quality |
| S15_vs_64714\|\|full | Not-determined |
| S15_vs_64759\|\|full | Low-quality |
| S15_vs_64888\|\|full | Not-determined |
| S15_vs_64960\|\|full | Low-quality |
| S15_vs_65116\|\|full | Low-quality |
| S15_vs_65128\|\|full | Low-quality |
| S15_vs_65235\|\|full | Low-quality |
| S15_vs_65248\|\|full | Not-determined |
| S15_vs_65294\|\|full | Low-quality |
| S15_vs_65386\|\|full | Low-quality |
| S15_vs_65428\|\|full | Not-determined |
| S15_vs_65456\|\|full | Low-quality |
| S15_vs_65671\|\|full | Low-quality |
| S15_vs_65883\|\|full | Low-quality |
| S15_vs_65903\|\|full | Low-quality |
| S15_vs_65948\|\|full | Not-determined |
| S15_vs_65949\|\|full | Not-determined |
| S15_vs_65970\|\|full | Not-determined |
| S15_vs_65971\|\|full | Low-quality |
| S15_vs_66020\|\|full | Low-quality |
| S15_vs_66077\|\|full | Not-determined |
| S15_vs_66188\|\|full | Low-quality |
| S15_vs_66233\|\|full | Low-quality |
| S15_vs_66269\|\|full | Not-determined |
| S15_vs_66284\|\|full | Low-quality |
| S15_vs_66314\|\|full | Low-quality |
| S15_vs_66356\|\|full | Not-determined |
| S15_vs_66377\|\|full | Low-quality |
| S15_vs_66652\|\|full | Low-quality |
| S15_vs_66657\|\|full | Low-quality |
| S15_vs_66750\|\|full | Low-quality |
| S15_vs_66778\|\|full | Not-determined |
| S15_vs_66794\|\|full | Not-determined |
| S15_vs_66802\|\|full | Not-determined |
| S15_vs_66979\|\|full | Not-determined |
| S15_vs_67178\|\|full | Not-determined |
| S15_vs_67323\|\|full | Not-determined |
| S15_vs_67508\|\|full | Low-quality |
| S15_vs_67510\|\|full | Low-quality |
| S15_vs_67545\|\|full | Low-quality |
| S15_vs_67665\|\|full | Not-determined |
| S15_vs_67737\|\|full | Low-quality |
| S15_vs_67754\|\|full | Not-determined |
| S15_vs_67843\|\|full | Low-quality |
| S15_vs_67911\|\|full | Low-quality |
| S15_vs_68169\|\|full | Low-quality |
| S15_vs_68418\|\|full | Not-determined |
| S15_vs_68420\|\|full | Not-determined |
| S15_vs_68444\|\|full | Not-determined |
| S15_vs_68466\|\|full | Not-determined |
| S15_vs_68490\|\|full | Not-determined |
| S15_vs_68494\|\|full | Low-quality |
| S15_vs_68531\|\|full | Not-determined |
| S15_vs_68707\|\|full | Low-quality |
| S15_vs_68721\|\|full | Low-quality |
| S15_vs_68725\|\|full | Low-quality |
| S15_vs_68783\|\|full | Not-determined |
| S15_vs_68802\|\|full | Low-quality |
| S15_vs_68814\|\|full | Not-determined |
| S15_vs_68827\|\|full | Not-determined |
| S15_vs_68831\|\|full | Not-determined |
| S15_vs_68898\|\|full | Not-determined |
| S15_vs_68963\|\|full | Not-determined |
| S15_vs_68973\|\|full | Low-quality |
| S15_vs_69194\|\|full | Low-quality |
| S15_vs_69206\|\|full | Low-quality |
| S15_vs_69229\|\|full | Low-quality |
| S15_vs_69244\|\|full | Not-determined |
| S15_vs_69323\|\|full | Low-quality |
| S15_vs_69544\|\|full | Not-determined |
| S15_vs_69553\|\|full | Not-determined |
| S15_vs_69644\|\|full | Not-determined |
| S15_vs_69665\|\|full | Not-determined |
| S15_vs_69708\|\|full | Not-determined |
| S15_vs_69876\|\|full | Low-quality |
| S15_vs_69902\|\|full | Not-determined |
| S15_vs_69904\|\|full | Not-determined |
| S15_vs_69936\|\|full | Low-quality |
| S15_vs_69950\|\|full | Not-determined |
| S15_vs_70062\|\|full | Not-determined |
| S15_vs_70096\|\|full | Low-quality |
| S15_vs_70100\|\|full | Low-quality |
| S15_vs_70116\|\|full | Low-quality |
| S15_vs_70301\|\|full | Low-quality |
| S15_vs_70325\|\|full | Low-quality |
| S15_vs_70340\|\|full | Not-determined |
| S15_vs_70374\|\|full | Low-quality |
| S15_vs_70415\|\|full | Low-quality |
| S15_vs_70421\|\|full | Low-quality |
| S15_vs_70458\|\|full | Not-determined |
| S15_vs_70494\|\|full | Low-quality |
| S15_vs_70524\|\|full | Low-quality |
| S15_vs_70553\|\|full | Low-quality |
| S15_vs_70569\|\|full | Low-quality |
| S15_vs_70619\|\|full | Low-quality |
| S15_vs_70737\|\|full | Low-quality |
| S15_vs_70744\|\|full | Not-determined |
| S15_vs_70783\|\|full | Not-determined |
| S15_vs_70800\|\|full | Low-quality |
| S15_vs_70985\|\|full | Not-determined |
| S15_vs_71163\|\|full | Low-quality |
| S15_vs_71184\|\|full | Not-determined |
| S15_vs_71224\|\|full | Not-determined |
| S15_vs_71291\|\|full | Low-quality |
| S15_vs_71297\|\|full | Low-quality |
| S15_vs_71368\|\|full | Not-determined |
| S15_vs_71538\|\|full | Low-quality |
| S15_vs_71557\|\|full | Low-quality |
| S15_vs_71598\|\|full | Low-quality |
| S1_vs_k141_56489\|\|full | Medium-quality |
| S15_vs_71658\|\|full | Low-quality |
| S15_vs_71673\|\|full | Low-quality |
| S15_vs_71697\|\|full | Low-quality |
| S15_vs_71717\|\|full | Low-quality |
| S15_vs_71725\|\|full | Not-determined |
| S15_vs_71734\|\|full | Not-determined |
| S15_vs_71787\|\|full | Not-determined |
| S15_vs_71823\|\|full | Low-quality |
| S15_vs_71825\|\|full | Low-quality |
| S15_vs_71849\|\|full | Low-quality |
| S15_vs_71892\|\|full | Low-quality |
| S15_vs_71927\|\|full | Low-quality |
| S15_vs_71951\|\|full | Low-quality |
| S15_vs_71953\|\|full | Not-determined |
| S3_vs_k141_96222\|\|full | Medium-quality |
| S15_vs_71987\|\|full | Low-quality |
| S15_vs_72087\|\|full | Low-quality |
| S15_vs_72143\|\|full | Not-determined |
| S15_vs_72187\|\|full | Low-quality |
| S15_vs_72230\|\|full | Not-determined |
| S15_vs_72251\|\|full | Low-quality |
| S15_vs_72262\|\|full | Low-quality |
| S15_vs_72384\|\|full | Not-determined |
| S15_vs_72459\|\|full | Not-determined |
| S15_vs_72470\|\|full | Low-quality |
| S15_vs_72544\|\|full | Low-quality |
| S15_vs_72648\|\|full | Not-determined |
| S15_vs_72688\|\|full | Low-quality |
| S15_vs_72739\|\|full | Low-quality |
| S15_vs_72793\|\|full | Not-determined |
| S15_vs_72796\|\|full | Not-determined |
| S15_vs_72821\|\|full | Low-quality |
| S15_vs_72876\|\|full | Low-quality |
| S15_vs_72892\|\|full | Low-quality |
| S15_vs_72923\|\|full | Low-quality |
| S15_vs_73048\|\|full | Low-quality |
| S15_vs_73167\|\|full | Low-quality |
| S15_vs_73233\|\|full | Low-quality |
| S15_vs_73264\|\|full | Low-quality |
| S15_vs_73281\|\|full | Low-quality |
| S15_vs_73301\|\|full | Not-determined |
| S15_vs_73406\|\|full | Low-quality |
| S8_extracted_k141_137836 | Medium-quality |
| S15_vs_73475\|\|full | Not-determined |
| S15_vs_73556\|\|full | Not-determined |
| S15_vs_73595\|\|full | Low-quality |
| S15_vs_73644\|\|full | Low-quality |
| S15_vs_73789\|\|full | Not-determined |
| S15_vs_73891\|\|full | Low-quality |
| S15_vs_73924\|\|full | Low-quality |
| S15_vs_73947\|\|full | Not-determined |
| S15_vs_73982\|\|full | Low-quality |
| S15_vs_74148\|\|full | Not-determined |
| S15_vs_74327\|\|full | Not-determined |
| S15_vs_74349\|\|full | Low-quality |
| S15_vs_74371\|\|full | Low-quality |
| S15_vs_74374\|\|full | Low-quality |
| S15_vs_74410\|\|full | Low-quality |
| S15_vs_74461\|\|full | Low-quality |
| S15_vs_74463\|\|full | Not-determined |
| S15_vs_74477\|\|full | Not-determined |
| S15_vs_74496\|\|full | Not-determined |
| S15_vs_74565\|\|full | Low-quality |
| S15_vs_74573\|\|full | Low-quality |
| S15_vs_74590\|\|full | Not-determined |
| S15_vs_74611\|\|full | Low-quality |
| S15_vs_74710\|\|full | Low-quality |
| S15_vs_74723\|\|full | Low-quality |
| S15_vs_74816\|\|full | Not-determined |
| S15_vs_74871\|\|full | Low-quality |
| S15_vs_74936\|\|full | Low-quality |
| S15_vs_74964\|\|full | Low-quality |
| S15_vs_75016\|\|full | Not-determined |
| S15_vs_75030\|\|full | Not-determined |
| S15_vs_75206\|\|full | Low-quality |
| S15_vs_75407\|\|full | Not-determined |
| S15_vs_75420\|\|full | Not-determined |
| S15_vs_75463\|\|full | Low-quality |
| S15_vs_75474\|\|full | Not-determined |
| S15_vs_75487\|\|full | Low-quality |
| S15_vs_75507\|\|full | Not-determined |
| S15_vs_75566\|\|full | Not-determined |
| S15_vs_75571\|\|full | Not-determined |
| S15_vs_75590\|\|full | Low-quality |
| S15_vs_75640\|\|full | Low-quality |
| S15_vs_75680\|\|full | Low-quality |
| S15_vs_75734\|\|full | Low-quality |
| S15_vs_75941\|\|full | Low-quality |
| S15_vs_75949\|\|full | Low-quality |
| S15_vs_75970\|\|full | Not-determined |
| S15_vs_75971\|\|full | Low-quality |
| S15_vs_76010\|\|full | Low-quality |
| S15_vs_76037\|\|full | Low-quality |
| S15_vs_76153\|\|full | Low-quality |
| S15_vs_76232\|\|full | Low-quality |
| S15_vs_76235\|\|full | Low-quality |
| S15_vs_76253\|\|full | Low-quality |
| S15_vs_76259\|\|full | Low-quality |
| S15_vs_76404\|\|full | Low-quality |
| S15_vs_76427\|\|full | Low-quality |
| S15_vs_76553\|\|full | Low-quality |
| S15_vs_76633\|\|full | Low-quality |
| S15_vs_76787\|\|full | Not-determined |
| S15_vs_76804\|\|full | Low-quality |
| S15_vs_76845\|\|full | Low-quality |
| S15_vs_76905\|\|full | Low-quality |
| S15_vs_76924\|\|full | Low-quality |
| S15_vs_76957\|\|full | Low-quality |
| S15_vs_76961\|\|full | Low-quality |
| S15_vs_76990\|\|full | Low-quality |
| S15_vs_77010\|\|full | Low-quality |
| S15_vs_77065\|\|full | Not-determined |
| S15_vs_77074\|\|full | Not-determined |
| S15_vs_77093\|\|full | Low-quality |
| S15_vs_77154\|\|full | Not-determined |
| S15_vs_77171\|\|full | Not-determined |
| S15_vs_77255\|\|full | Not-determined |
| S15_vs_77345\|\|full | Low-quality |
| S15_vs_77413\|\|full | Low-quality |
| S15_vs_77415\|\|full | Not-determined |
| S15_vs_12545\|\|0_partial | Not-determined |
| S15_vs_14765\|\|0_partial | Not-determined |
| S15_vs_20682\|\|0_partial | Not-determined |
| S15_vs_28139\|\|0_partial | Low-quality |
| S15_vs_33541\|\|0_partial | Not-determined |
| S15_vs_42979\|\|0_partial | Not-determined |
| S15_vs_46546\|\|0_partial | Not-determined |
| S15_vs_58840\|\|0_partial | Not-determined |
| S15_vs_316\|\|lt2gene | Low-quality |
| S15_vs_442\|\|lt2gene | Low-quality |
| S15_vs_502\|\|lt2gene | Low-quality |
| S15_vs_583\|\|lt2gene | Low-quality |
| S15_vs_664\|\|lt2gene | Low-quality |
| S15_vs_688\|\|lt2gene | Low-quality |
| S15_vs_695\|\|lt2gene | Low-quality |
| S15_vs_761\|\|lt2gene | Low-quality |
| S15_vs_780\|\|lt2gene | Low-quality |
| S15_vs_928\|\|lt2gene | Low-quality |
| S15_vs_960\|\|lt2gene | Not-determined |
| S15_vs_1110\|\|lt2gene | Not-determined |
| S15_vs_1297\|\|lt2gene | Low-quality |
| S15_vs_1364\|\|lt2gene | Low-quality |
| S15_vs_1497\|\|lt2gene | Not-determined |
| S15_vs_1505\|\|lt2gene | Low-quality |
| S15_vs_1660\|\|lt2gene | Low-quality |
| S15_vs_1670\|\|lt2gene | Low-quality |
| S15_vs_1841\|\|lt2gene | Not-determined |
| S15_vs_1975\|\|lt2gene | Low-quality |
| S15_vs_2008\|\|lt2gene | Not-determined |
| S15_vs_2057\|\|lt2gene | Not-determined |
| S15_vs_2114\|\|lt2gene | Low-quality |
| S15_vs_2146\|\|lt2gene | Not-determined |
| S15_vs_2209\|\|lt2gene | Low-quality |
| S15_vs_2427\|\|lt2gene | Low-quality |
| S15_vs_2459\|\|lt2gene | Low-quality |
| S15_vs_2580\|\|lt2gene | Low-quality |
| S15_vs_2595\|\|lt2gene | Low-quality |
| S15_vs_2611\|\|lt2gene | Low-quality |
| S15_vs_2612\|\|lt2gene | Low-quality |
| S15_vs_2691\|\|lt2gene | Not-determined |
| S15_vs_2701\|\|lt2gene | Low-quality |

S1S_vs_2805||lt2gene Not-determined
S1S_vs_2807||lt2gene Not-determined
S1S_vs_3058||lt2gene Low-quality
S1S_vs_3161||lt2gene Low-quality
S1S_vs_3349||lt2gene Low-quality
S1S_vs_3437||lt2gene Low-quality
S1S_vs_3534||lt2gene Not-determined
S1S_vs_3548||lt2gene Not-determined
S1S_vs_3560||lt2gene Not-determined
S1S_vs_3611||lt2gene Low-quality
S1S_vs_3815||lt2gene Low-quality
S1S_vs_3892||lt2gene Low-quality
S1S_vs_3926||lt2gene Low-quality
S1S_vs_4006||lt2gene Low-quality
S1S_vs_4170||lt2gene Not-determined
S1S_vs_4313||lt2gene Low-quality
S1S_vs_4382||lt2gene Low-quality
S1S_vs_4481||lt2gene Low-quality
S1S_vs_4483||lt2gene Not-determined
S1S_vs_4804||lt2gene Low-quality
S1S_vs_4969||lt2gene Not-determined
S1S_vs_4970||lt2gene Not-determined
S1S_vs_5036||lt2gene Low-quality
S1S_vs_5161||lt2gene Low-quality
S1S_vs_5302||lt2gene Low-quality
S1S_vs_5454||lt2gene Not-determined
S1S_vs_5551||lt2gene Not-determined
S1S_vs_5558||lt2gene Not-determined
S1S_vs_5763||lt2gene Low-quality
S1S_vs_5775||lt2gene Low-quality
S1S_vs_5793||lt2gene Low-quality
S1S_vs_5847||lt2gene Low-quality
S1S_vs_6149||lt2gene Not-determined
S1S_vs_6156||lt2gene Low-quality
S1S_vs_6308||lt2gene Low-quality
S1S_vs_6384||lt2gene Low-quality
S1S_vs_6470||lt2gene Low-quality
S1S_vs_6510||lt2gene Low-quality
S1S_vs_6631||lt2gene Low-quality
S1S_vs_6687||lt2gene Low-quality
S1S_vs_6830||lt2gene Low-quality
S1S_vs_6844||lt2gene Low-quality
S1S_vs_6942||lt2gene Low-quality
S1S_vs_7036||lt2gene Low-quality
S1S_vs_7045||lt2gene Not-determined
S1S_vs_7112||lt2gene Not-determined
S1S_vs_7118||lt2gene Low-quality
S1S_vs_7134||lt2gene Low-quality
S1S_vs_7410||lt2gene Not-determined
S1S_vs_7421||lt2gene Not-determined
S1S_vs_7509||lt2gene Low-quality
S1S_vs_7602||lt2gene Not-determined
S1S_vs_7663||lt2gene Low-quality
S1S_vs_7852||lt2gene Not-determined
S1S_vs_7903||lt2gene Low-quality
S1S_vs_7928||lt2gene Low-quality
S1S_vs_7984||lt2gene Not-determined
S1S_vs_8275||lt2gene Low-quality
S1S_vs_8431||lt2gene Not-determined
S1S_vs_8487||lt2gene Low-quality
S1S_vs_8502||lt2gene Low-quality
S1S_vs_8621||lt2gene Low-quality
S1S_vs_8665||lt2gene Low-quality
S1S_vs_8719||lt2gene Low-quality
S1S_vs_8787||lt2gene Low-quality
S1S_vs_8880||lt2gene Not-determined
S1S_vs_8915||lt2gene Not-determined
S1S_vs_9002||lt2gene Low-quality
S1S_vs_9046||lt2gene Not-determined
S1S_vs_9057||lt2gene Low-quality
S1S_vs_9080||lt2gene Low-quality
S1S_vs_9103||lt2gene Low-quality
S1S_vs_9232||lt2gene Low-quality
S1S_vs_9374||lt2gene Low-quality
S1S_vs_9375||lt2gene Not-determined
S1S_vs_9379||lt2gene Low-quality
S1S_vs_9576||lt2gene Low-quality
S1S_vs_9761||lt2gene Low-quality
S1S_vs_9772||lt2gene Low-quality
S1S_vs_9775||lt2gene Low-quality
S1S_vs_9957||lt2gene Not-determined
S1S_vs_10290||lt2gene Low-quality
S1S_vs_10420||lt2gene Low-quality
S1S_vs_10659||lt2gene Low-quality
S1S_vs_10667||lt2gene Low-quality
S1S_vs_10686||lt2gene Low-quality
S1S_vs_10714||lt2gene Not-determined
S1S_vs_10870||lt2gene Low-quality
S1S_vs_10876||lt2gene Low-quality
S1S_vs_11153||lt2gene Low-quality
S1S_vs_11414||lt2gene Low-quality
S1S_vs_11527||lt2gene Low-quality
S1S_vs_11594||lt2gene Low-quality
S1S_vs_11609||lt2gene Low-quality
S1S_vs_11664||lt2gene Low-quality
S1S_vs_11901||lt2gene Low-quality
S1S_vs_12011||lt2gene Low-quality
S1S_vs_12128||lt2gene Low-quality
S1S_vs_12647||lt2gene Low-quality
S1S_vs_12772||lt2gene Low-quality
S1S_vs_12954||lt2gene Low-quality
S1S_vs_12970||lt2gene Low-quality
S1S_vs_13387||lt2gene Low-quality
S1S_vs_13432||lt2gene Low-quality
S1S_vs_13468||lt2gene Low-quality
S1S_vs_13740||lt2gene Low-quality
S1S_vs_14038||lt2gene Low-quality
S1S_vs_14088||lt2gene Low-quality
S1S_vs_14108||lt2gene Low-quality
S1S_vs_14370||lt2gene Low-quality
S1S_vs_14536||lt2gene Not-determined
S1S_vs_14566||lt2gene Low-quality
S1S_vs_14718||lt2gene Low-quality
S1S_vs_14722||lt2gene Low-quality
S1S_vs_14752||lt2gene Not-determined
S1S_vs_14815||lt2gene Not-determined
S1S_vs_14821||lt2gene Low-quality
S1S_vs_14994||lt2gene Low-quality
S1S_vs_15007||lt2gene Not-determined
S1S_vs_15017||lt2gene Low-quality
S1S_vs_15294||lt2gene Not-determined
S1S_vs_15353||lt2gene Low-quality
S1S_vs_15387||lt2gene Not-determined
S1S_vs_15523||lt2gene Low-quality
S1S_vs_15534||lt2gene Not-determined
S1S_vs_15625||lt2gene Not-determined
S1S_vs_15640||lt2gene Not-determined
S1S_vs_15698||lt2gene Low-quality
S1S_vs_15826||lt2gene Low-quality
S1S_vs_15919||lt2gene Low-quality
S1S_vs_16066||lt2gene Low-quality
S1S_vs_16090||lt2gene Low-quality
S1S_vs_16093||lt2gene Not-determined
S1S_vs_16312||lt2gene Low-quality
S1S_vs_16379||lt2gene Low-quality
S1S_vs_16550||lt2gene Not-determined
S1S_vs_16627||lt2gene Not-determined
S1S_vs_16676||lt2gene Not-determined
S1S_vs_16746||lt2gene Low-quality
S1S_vs_16963||lt2gene Low-quality
S1S_vs_17137||lt2gene Not-determined
S1S_vs_17144||lt2gene Not-determined
S1S_vs_17242||lt2gene Not-determined
S1S_vs_17288||lt2gene Low-quality
S1S_vs_17472||lt2gene Low-quality
S1S_vs_17557||lt2gene Low-quality
S1S_vs_17655||lt2gene Low-quality
S1S_vs_17818||lt2gene Not-determined
S1S_vs_17942||lt2gene Low-quality
S1S_vs_18029||lt2gene Low-quality
S1S_vs_18031||lt2gene Low-quality
S1S_vs_18034||lt2gene Low-quality
S1S_vs_18227||lt2gene Low-quality
S1S_vs_18318||lt2gene Low-quality
S1S_vs_18433||lt2gene Low-quality
S1S_vs_18593||lt2gene Not-determined
S1S_vs_18788||lt2gene Low-quality
S1S_vs_18863||lt2gene Not-determined
S1S_vs_18970||lt2gene Low-quality
S1S_vs_19302||lt2gene Not-determined
S1S_vs_19317||lt2gene Not-determined
S1S_vs_19345||lt2gene Low-quality
S1S_vs_19498||lt2gene Not-determined
S1S_vs_19575||lt2gene Low-quality
S1S_vs_19605||lt2gene Low-quality
S1S_vs_20078||lt2gene Not-determined
S1S_vs_20300||lt2gene Low-quality
S1S_vs_20312||lt2gene Not-determined
S1S_vs_20705||lt2gene Low-quality
S1S_vs_20776||lt2gene Low-quality
S1S_vs_20828||lt2gene Low-quality
S1S_vs_20946||lt2gene Low-quality
S1S_vs_21013||lt2gene Not-determined
S1S_vs_21115||lt2gene Not-determined
S1S_vs_21226||lt2gene Low-quality
S1S_vs_21295||lt2gene Low-quality
S1S_vs_21460||lt2gene Low-quality
S1S_vs_21555||lt2gene Low-quality
S1S_vs_21592||lt2gene Low-quality
S1S_vs_21761||lt2gene Not-determined
S1S_vs_21784||lt2gene Low-quality
S1S_vs_21824||lt2gene Low-quality
S1S_vs_21892||lt2gene Low-quality
S1S_vs_21913||lt2gene Low-quality
S1S_vs_22079||lt2gene Low-quality
S1S_vs_22084||lt2gene Low-quality
S1S_vs_22346||lt2gene Low-quality
S1S_vs_22433||lt2gene Low-quality
S1S_vs_22436||lt2gene Low-quality
S1S_vs_22544||lt2gene Low-quality
S1S_vs_22792||lt2gene Not-determined
S1S_vs_22883||lt2gene Low-quality
S1S_vs_23066||lt2gene Not-determined
S1S_vs_23205||lt2gene Not-determined
S1S_vs_23370||lt2gene Not-determined
S1S_vs_23688||lt2gene Not-determined
S1S_vs_23740||lt2gene Not-determined
S1S_vs_24083||lt2gene Not-determined
S1S_vs_24145||lt2gene Low-quality
S1S_vs_24342||lt2gene Low-quality
S1S_vs_24555||lt2gene Low-quality
S1S_vs_24742||lt2gene Not-determined
S1S_vs_24743||lt2gene Low-quality
S1S_vs_24886||lt2gene Low-quality
S1S_vs_24939||lt2gene Low-quality
S1S_vs_24991||lt2gene Low-quality
S1S_vs_25101||lt2gene Low-quality
S1S_vs_25505||lt2gene Low-quality
S1S_vs_25506||lt2gene Low-quality
S1S_vs_25511||lt2gene Not-determined
S1S_vs_25594||lt2gene Not-determined
S1S_vs_25639||lt2gene Low-quality
S1S_vs_25681||lt2gene Not-determined
S1S_vs_25857||lt2gene Not-determined
S1S_vs_25948||lt2gene Low-quality
S1S_vs_26016||lt2gene Low-quality
S1S_vs_26150||lt2gene Not-determined
S1S_vs_26190||lt2gene Not-determined
S1S_vs_26264||lt2gene Low-quality
S1S_vs_26267||lt2gene Not-determined
S1S_vs_26294||lt2gene Not-determined
S1S_vs_26601||lt2gene Low-quality
S1S_vs_26604||lt2gene Low-quality
S1S_vs_26702||lt2gene Low-quality
S1S_vs_26737||lt2gene Low-quality
S1S_vs_26982||lt2gene Low-quality
S1S_vs_27022||lt2gene Low-quality
S1S_vs_27058||lt2gene Low-quality
S1S_vs_27135||lt2gene Low-quality
S1S_vs_27370||lt2gene Low-quality
S1S_vs_27436||lt2gene Low-quality
S1S_vs_27469||lt2gene Low-quality
S1S_vs_27490||lt2gene Low-quality
S1S_vs_27696||lt2gene Not-determined
S1S_vs_27721||lt2gene Not-determined
S1S_vs_28050||lt2gene Low-quality
S1S_vs_28339||lt2gene Not-determined
S1S_vs_28469||lt2gene Low-quality
S1S_vs_28602||lt2gene Not-determined
S1S_vs_28618||lt2gene Low-quality
S1S_vs_28772||lt2gene Low-quality
S1S_vs_28817||lt2gene Low-quality
S1S_vs_28879||lt2gene Low-quality
S1S_vs_28906||lt2gene Low-quality
S1S_vs_28972||lt2gene Not-determined
S1S_vs_29257||lt2gene Low-quality
S1S_vs_29258||lt2gene Not-determined
S1S_vs_29297||lt2gene Low-quality
S1S_vs_29383||lt2gene Low-quality
S1S_vs_29403||lt2gene Not-determined
S1S_vs_29426||lt2gene Low-quality
S1S_vs_29633||lt2gene Low-quality
S1S_vs_29655||lt2gene Low-quality
S1S_vs_29696||lt2gene Not-determined
S1S_vs_29714||lt2gene Low-quality
S1S_vs_29880||lt2gene Low-quality
S1S_vs_29901||lt2gene Low-quality
S1S_vs_30314||lt2gene Not-determined
S1S_vs_30635||lt2gene Low-quality
S1S_vs_30877||lt2gene Low-quality
S1S_vs_31122||lt2gene Low-quality
S1S_vs_31169||lt2gene Low-quality
S1S_vs_31287||lt2gene Not-determined
S1S_vs_31332||lt2gene Low-quality
S1S_vs_31404||lt2gene Not-determined
S1S_vs_31484||lt2gene Low-quality
S1S_vs_31537||lt2gene Low-quality
S1S_vs_31617||lt2gene Low-quality
S1S_vs_31631||lt2gene Low-quality
S1S_vs_31768||lt2gene Not-determined
S1S_vs_31772||lt2gene Low-quality
S1S_vs_31800||lt2gene Low-quality
S1S_vs_31816||lt2gene Low-quality
S1S_vs_31853||lt2gene Not-determined
S1S_vs_31906||lt2gene Not-determined
S1S_vs_32020||lt2gene Not-determined
S1S_vs_32084||lt2gene Low-quality
S1S_vs_32159||lt2gene Not-determined
S1S_vs_32201||lt2gene Low-quality
S1S_vs_32443||lt2gene Not-determined
S1S_vs_32505||lt2gene Not-determined
S1S_vs_32662||lt2gene Low-quality
S1S_vs_32768||lt2gene Low-quality
S1S_vs_32822||lt2gene Low-quality
S1S_vs_32973||lt2gene Low-quality
S1S_vs_33005||lt2gene Low-quality
S1S_vs_33241||lt2gene Low-quality
S1S_vs_33489||lt2gene Low-quality
S1S_vs_33603||lt2gene Low-quality
S1S_vs_33688||lt2gene Low-quality
S1S_vs_33732||lt2gene Low-quality
S1S_vs_33858||lt2gene Low-quality
S1S_vs_33961||lt2gene Low-quality
S1S_vs_34071||lt2gene Low-quality
S1S_vs_34080||lt2gene Not-determined
S1S_vs_34152||lt2gene Not-determined
S1S_vs_34179||lt2gene Low-quality
S1S_vs_34181||lt2gene Not-determined
S1S_vs_34192||lt2gene Low-quality
S1S_vs_34226||lt2gene Not-determined
S1S_vs_34270||lt2gene Not-determined
S1S_vs_34291||lt2gene Low-quality
S1S_vs_34306||lt2gene Low-quality
S1S_vs_34426||lt2gene Low-quality
S1S_vs_34482||lt2gene Low-quality
S1S_vs_34677||lt2gene Low-quality
S1S_vs_34781||lt2gene Low-quality
S1S_vs_34963||lt2gene Not-determined
S1S_vs_34990||lt2gene Not-determined
S1S_vs_35309||lt2gene Low-quality
S1S_vs_35339||lt2gene Low-quality
S1S_vs_35433||lt2gene Low-quality
S1S_vs_35624||lt2gene Not-determined
S1S_vs_35718||lt2gene Low-quality
S1S_vs_35816||lt2gene Not-determined
S1S_vs_36027||lt2gene Low-quality
S1S_vs_36139||lt2gene Low-quality
S1S_vs_36195||lt2gene Low-quality
S1S_vs_36207||lt2gene Low-quality
S1S_vs_36208||lt2gene Not-determined
S1S_vs_36450||lt2gene Low-quality
S1S_vs_36498||lt2gene Not-determined
S1S_vs_36610||lt2gene Not-determined
S1S_vs_36708||lt2gene Low-quality
S1S_vs_36736||lt2gene Low-quality
S1S_vs_36833||lt2gene Low-quality
S1S_vs_36851||lt2gene Not-determined
S1S_vs_36874||lt2gene Low-quality
S1S_vs_36982||lt2gene Low-quality
S1S_vs_37007||lt2gene Low-quality
S1S_vs_37116||lt2gene Low-quality
S1S_vs_37141||lt2gene Low-quality
S1S_vs_37181||lt2gene Not-determined
S1S_vs_37255||lt2gene Not-determined
S1S_vs_37398||lt2gene Low-quality
S1S_vs_37789||lt2gene Low-quality
S1S_vs_37832||lt2gene Low-quality
S1S_vs_37943||lt2gene Not-determined
S1S_vs_38011||lt2gene Low-quality
S1S_vs_38045||lt2gene Low-quality
S1S_vs_38066||lt2gene Not-determined
S1S_vs_38327||lt2gene Low-quality
S1S_vs_38606||lt2gene Not-determined
S1S_vs_38626||lt2gene Low-quality
S1S_vs_38754||lt2gene Low-quality
S1S_vs_38790||lt2gene Low-quality
S1S_vs_38811||lt2gene Low-quality
S1S_vs_38855||lt2gene Not-determined
S1S_vs_38927||lt2gene Low-quality
S1S_vs_39113||lt2gene Low-quality
S1S_vs_39189||lt2gene Not-determined
S1S_vs_39249||lt2gene Low-quality
S1S_vs_39252||lt2gene Low-quality
S1S_vs_39415||lt2gene Low-quality
S1S_vs_39978||lt2gene Not-determined
S1S_vs_39983||lt2gene Low-quality
S1S_vs_40195||lt2gene Low-quality
S1S_vs_40212||lt2gene Low-quality
S1S_vs_40235||lt2gene Low-quality
S1S_vs_40294||lt2gene Low-quality
S1S_vs_40331||lt2gene Low-quality
S1S_vs_40361||lt2gene Not-determined
S1S_vs_40377||lt2gene Not-determined
S1S_vs_40400||lt2gene Low-quality
S1S_vs_40513||lt2gene Low-quality
S1S_vs_40553||lt2gene Low-quality
S1S_vs_40721||lt2gene Low-quality
S1S_vs_40742||lt2gene Low-quality
S1S_vs_40951||lt2gene Not-determined
S1S_vs_40989||lt2gene Not-determined
S1S_vs_41287||lt2gene Not-determined
S1S_vs_41398||lt2gene Not-determined
S1S_vs_41744||lt2gene Not-determined
S1S_vs_41870||lt2gene Low-quality
S1S_vs_41909||lt2gene Low-quality
S1S_vs_41955||lt2gene Not-determined
S1S_vs_42057||lt2gene Low-quality
S1S_vs_42177||lt2gene Low-quality
S1S_vs_42268||lt2gene Low-quality
S1S_vs_42339||lt2gene Low-quality
S1S_vs_42434||lt2gene Low-quality
S1S_vs_42466||lt2gene Not-determined
S1S_vs_42571||lt2gene Not-determined
S1S_vs_42612||lt2gene Low-quality
S1S_vs_42640||lt2gene Low-quality
S1S_vs_42650||lt2gene Low-quality
S1S_vs_42734||lt2gene Not-determined
S1S_vs_42837||lt2gene Not-determined
S1S_vs_42853||lt2gene Not-determined
S1S_vs_42939||lt2gene Low-quality
S1S_vs_42957||lt2gene Low-quality
S1S_vs_43108||lt2gene Not-determined
S1S_vs_43233||lt2gene Not-determined
S1S_vs_43249||lt2gene Not-determined
S1S_vs_43307||lt2gene Low-quality
S1S_vs_43314||lt2gene Low-quality
S1S_vs_43343||lt2gene Low-quality
S1S_vs_43435||lt2gene Low-quality
S1S_vs_43717||lt2gene Not-determined
S1S_vs_43775||lt2gene Low-quality
S1S_vs_43873||lt2gene Low-quality
S1S_vs_43906||lt2gene Low-quality
S1S_vs_44516||lt2gene Not-determined
S1S_vs_44563||lt2gene Low-quality
S1S_vs_44645||lt2gene Low-quality
S1S_vs_44698||lt2gene Not-determined
S1S_vs_44789||lt2gene Low-quality
S1S_vs_44821||lt2gene Low-quality
S1S_vs_44866||lt2gene Not-determined
S1S_vs_44927||lt2gene Low-quality
S1S_vs_45237||lt2gene Low-quality
S1S_vs_45274||lt2gene Not-determined
S1S_vs_45419||lt2gene Not-determined
S1S_vs_45425||lt2gene Low-quality
S1S_vs_45465||lt2gene Low-quality
S1S_vs_45483||lt2gene Low-quality
S1S_vs_45667||lt2gene Low-quality
S1S_vs_45691||lt2gene Not-determined
S1S_vs_45853||lt2gene Low-quality
S1S_vs_45882||lt2gene Not-determined
S1S_vs_45952||lt2gene Not-determined
S1S_vs_46295||lt2gene Low-quality
S1S_vs_46357||lt2gene Low-quality
S1S_vs_46359||lt2gene Low-quality
S1S_vs_46765||lt2gene Low-quality
S1S_vs_46982||lt2gene Not-determined
S1S_vs_46983||lt2gene Not-determined
S1S_vs_47003||lt2gene Not-determined
S1S_vs_47065||lt2gene Low-quality
S1S_vs_47120||lt2gene Low-quality
S1S_vs_47197||lt2gene Low-quality
S1S_vs_47299||lt2gene Low-quality
S1S_vs_47362||lt2gene Low-quality
S1S_vs_47419||lt2gene Low-quality
S1S_vs_47827||lt2gene Low-quality
S1S_vs_48379||lt2gene Not-determined
S1S_vs_48472||lt2gene Low-quality
S1S_vs_48608||lt2gene Not-determined
S1S_vs_48891||lt2gene Not-determined
S1S_vs_48895||lt2gene Not-determined
S1S_vs_48936||lt2gene Not-determined
S1S_vs_49298||lt2gene Not-determined
S1S_vs_49354||lt2gene Low-quality
S1S_vs_49411||lt2gene Low-quality
S1S_vs_49572||lt2gene Low-quality
S1S_vs_49635||lt2gene Low-quality
S1S_vs_49785||lt2gene Not-determined
S1S_vs_49819||lt2gene Low-quality
S1S_vs_49843||lt2gene Low-quality
S1S_vs_49875||lt2gene Low-quality
S1S_vs_49950||lt2gene Low-quality
S1S_vs_50276||lt2gene Not-determined
S1S_vs_50469||lt2gene Not-determined
S1S_vs_50630||lt2gene Low-quality
S1S_vs_50870||lt2gene Low-quality
S1S_vs_50919||lt2gene Low-quality
S1S_vs_51158||lt2gene Not-determined
S1S_vs_51277||lt2gene Not-determined
S1S_vs_51319||lt2gene Low-quality
S1S_vs_51359||lt2gene Low-quality
S1S_vs_51373||lt2gene Low-quality
S1S_vs_51575||lt2gene Not-determined
S1S_vs_51667||lt2gene Low-quality
S1S_vs_51705||lt2gene Low-quality
S1S_vs_52125||lt2gene Low-quality
S1S_vs_52140||lt2gene Low-quality
S1S_vs_52190||lt2gene Not-determined
S1S_vs_52235||lt2gene Not-determined
S1S_vs_52285||lt2gene Not-determined
S1S_vs_52289||lt2gene Low-quality
S1S_vs_52313||lt2gene Low-quality
S1S_vs_52481||lt2gene Low-quality
S1S_vs_52498||lt2gene Low-quality
S1S_vs_52581||lt2gene Low-quality
S1S_vs_52612||lt2gene Not-determined
S1S_vs_52718||lt2gene Low-quality
S1S_vs_53108||lt2gene Not-determined
S1S_vs_53340||lt2gene Not-determined
S1S_vs_53430||lt2gene Not-determined
S1S_vs_53468||lt2gene Low-quality
S1S_vs_53523||lt2gene Not-determined
S1S_vs_53528||lt2gene Not-determined
S1S_vs_53556||lt2gene Low-quality
S1S_vs_53577||lt2gene Low-quality
S1S_vs_53586||lt2gene Low-quality
S1S_vs_53635||lt2gene Low-quality
S1S_vs_53687||lt2gene Low-quality
S1S_vs_53771||lt2gene Low-quality
S1S_vs_53847||lt2gene Low-quality
S1S_vs_54073||lt2gene Low-quality
S1S_vs_54101||lt2gene Low-quality
S1S_vs_54105||lt2gene Not-determined
S1S_vs_54143||lt2gene Low-quality
S1S_vs_54266||lt2gene Low-quality
S1S_vs_54374||lt2gene Low-quality
S1S_vs_54414||lt2gene Low-quality
S1S_vs_54483||lt2gene Low-quality
S1S_vs_54488||lt2gene Not-determined
S1S_vs_54740||lt2gene Not-determined
S1S_vs_54781||lt2gene Not-determined
S1S_vs_54875||lt2gene Low-quality
S1S_vs_55123||lt2gene Low-quality
S1S_vs_55243||lt2gene Low-quality
S1S_vs_55270||lt2gene Low-quality
S1S_vs_55331||lt2gene Low-quality
S1S_vs_55414||lt2gene Not-determined
S1S_vs_55426||lt2gene Low-quality
S1S_vs_55428||lt2gene Low-quality
S1S_vs_55456||lt2gene Low-quality
S1S_vs_55540||lt2gene Low-quality
S1S_vs_55890||lt2gene Low-quality
S1S_vs_55968||lt2gene Low-quality
S1S_vs_56300||lt2gene Not-determined
S1S_vs_56524||lt2gene Low-quality
S1S_vs_56583||lt2gene Low-quality
S1S_vs_56836||lt2gene Low-quality
S1S_vs_56928||lt2gene Low-quality
S1S_vs_57179||lt2gene Low-quality
S1S_vs_57216||lt2gene Low-quality
S1S_vs_57319||lt2gene Low-quality
S1S_vs_57467||lt2gene Low-quality
S1S_vs_57700||lt2gene Not-determined
S1S_vs_57811||lt2gene Low-quality
S1S_vs_57841||lt2gene Not-determined
S1S_vs_57846||lt2gene Low-quality
S1S_vs_57934||lt2gene Low-quality
S1S_vs_58021||lt2gene Low-quality
S1S_vs_58032||lt2gene Not-determined
S1S_vs_58110||lt2gene Low-quality
S1S_vs_58176||lt2gene Low-quality
S1S_vs_58214||lt2gene Low-quality
S1S_vs_58380||lt2gene Not-determined
S1S_vs_58419||lt2gene Low-quality
S1S_vs_58536||lt2gene Low-quality
S1S_vs_58627||lt2gene Low-quality
S1S_vs_58735||lt2gene Low-quality
S1S_vs_58789||lt2gene Low-quality
S1S_vs_58818||lt2gene Not-determined
S1S_vs_58847||lt2gene Low-quality
S1S_vs_58929||lt2gene Low-quality
S1S_vs_58961||lt2gene Low-quality
S1S_vs_59029||lt2gene Low-quality
S1S_vs_59212||lt2gene Not-determined
S1S_vs_59272||lt2gene Not-determined
S1S_vs_59316||lt2gene Low-quality
S1S_vs_59347||lt2gene Low-quality
S1S_vs_59440||lt2gene Low-quality
S1S_vs_59468||lt2gene Not-determined
S1S_vs_59770||lt2gene Low-quality
S1S_vs_59859||lt2gene Low-quality
S1S_vs_59937||lt2gene Low-quality
S1S_vs_60020||lt2gene Low-quality
S1S_vs_60153||lt2gene Low-quality
S1S_vs_60219||lt2gene Not-determined
S1S_vs_60307||lt2gene Low-quality
S1S_vs_60335||lt2gene Low-quality
S1S_vs_60412||lt2gene Low-quality
S1S_vs_60483||lt2gene Low-quality
S1S_vs_60675||lt2gene Not-determined
S1S_vs_60727||lt2gene Low-quality
S1S_vs_60757||lt2gene Low-quality
S1S_vs_60993||lt2gene Not-determined
S1S_vs_61210||lt2gene Low-quality
S1S_vs_61253||lt2gene Low-quality
S1S_vs_61269||lt2gene Low-quality
S1S_vs_61304||lt2gene Low-quality
S1S_vs_61852||lt2gene Low-quality
S1S_vs_61857||lt2gene Low-quality
S1S_vs_61913||lt2gene Not-determined
S1S_vs_61968||lt2gene Low-quality
S1S_vs_61972||lt2gene Not-determined
S1S_vs_61998||lt2gene Not-determined
S1S_vs_62134||lt2gene Not-determined
S1S_vs_62296||lt2gene Not-determined
S1S_vs_62362||lt2gene Low-quality
S1S_vs_62405||lt2gene Not-determined
S1S_vs_62540||lt2gene Low-quality
S1S_vs_62741||lt2gene Not-determined
S1S_vs_62781||lt2gene Low-quality
S1S_vs_62994||lt2gene Low-quality
S1S_vs_63199||lt2gene Low-quality
S1S_vs_63200||lt2gene Low-quality
S1S_vs_63334||lt2gene Low-quality
S1S_vs_63452||lt2gene Low-quality
S1S_vs_63708||lt2gene Low-quality
S1S_vs_63749||lt2gene Low-quality
S1S_vs_63897||lt2gene Not-determined
S1S_vs_63992||lt2gene Not-determined
S1S_vs_64072||lt2gene Low-quality

| | |
|---|---|
| S1S_vs_64089\|\|lt2gene | Low-quality |
| S1S_vs_64284\|\|lt2gene | Low-quality |
| S1S_vs_64333\|\|lt2gene | Low-quality |
| S1S_vs_64447\|\|lt2gene | Not-determined |
| S1S_vs_64579\|\|lt2gene | Not-determined |
| S1S_vs_64669\|\|lt2gene | Low-quality |
| S1S_vs_64902\|\|lt2gene | Not-determined |
| S1S_vs_64911\|\|lt2gene | Not-determined |
| S1S_vs_65005\|\|lt2gene | Low-quality |
| S1S_vs_65090\|\|lt2gene | Low-quality |
| S1S_vs_65332\|\|lt2gene | Low-quality |
| S1S_vs_65347\|\|lt2gene | Not-determined |
| S1S_vs_65492\|\|lt2gene | Low-quality |
| S1S_vs_65623\|\|lt2gene | Low-quality |
| S1S_vs_65626\|\|lt2gene | Low-quality |
| S1S_vs_65764\|\|lt2gene | Low-quality |
| S1S_vs_65892\|\|lt2gene | Not-determined |
| S1S_vs_66189\|\|lt2gene | Low-quality |
| S1S_vs_66435\|\|lt2gene | Low-quality |
| S1S_vs_66503\|\|lt2gene | Low-quality |
| S1S_vs_66522\|\|lt2gene | Low-quality |
| S1S_vs_66524\|\|lt2gene | Low-quality |
| S1S_vs_66568\|\|lt2gene | Low-quality |
| S1S_vs_66572\|\|lt2gene | Not-determined |
| S1S_vs_66605\|\|lt2gene | Low-quality |
| S1S_vs_66630\|\|lt2gene | Low-quality |
| S1S_vs_66793\|\|lt2gene | Low-quality |
| S1S_vs_66825\|\|lt2gene | Not-determined |
| S1S_vs_66839\|\|lt2gene | Low-quality |
| S1S_vs_66847\|\|lt2gene | Low-quality |
| S1S_vs_66915\|\|lt2gene | Not-determined |
| S1S_vs_66941\|\|lt2gene | Low-quality |
| S1S_vs_66951\|\|lt2gene | Low-quality |
| S1S_vs_66954\|\|lt2gene | Low-quality |
| S1S_vs_66957\|\|lt2gene | Low-quality |
| S1S_vs_67276\|\|lt2gene | Low-quality |
| S1S_vs_67478\|\|lt2gene | Low-quality |
| S1S_vs_67512\|\|lt2gene | Low-quality |
| S1S_vs_67526\|\|lt2gene | Low-quality |
| S1S_vs_67541\|\|lt2gene | Not-determined |
| S1S_vs_67654\|\|lt2gene | Not-determined |
| S1S_vs_67721\|\|lt2gene | Not-determined |
| S1S_vs_67822\|\|lt2gene | Low-quality |
| S1S_vs_67909\|\|lt2gene | Low-quality |
| S1S_vs_68187\|\|lt2gene | Low-quality |
| S1S_vs_68265\|\|lt2gene | Low-quality |
| S1S_vs_68286\|\|lt2gene | Low-quality |
| S1S_vs_68588\|\|lt2gene | Low-quality |
| S1S_vs_68632\|\|lt2gene | Low-quality |
| S1S_vs_68676\|\|lt2gene | Not-determined |
| S1S_vs_68723\|\|lt2gene | Low-quality |
| S1S_vs_68824\|\|lt2gene | Not-determined |
| S1S_vs_69026\|\|lt2gene | Low-quality |
| S1S_vs_69121\|\|lt2gene | Low-quality |
| S1S_vs_69284\|\|lt2gene | Low-quality |
| S1S_vs_69500\|\|lt2gene | Low-quality |
| S1S_vs_69547\|\|lt2gene | Not-determined |
| S1S_vs_69564\|\|lt2gene | Low-quality |
| S1S_vs_69573\|\|lt2gene | Not-determined |
| S1S_vs_69578\|\|lt2gene | Not-determined |
| S1S_vs_69589\|\|lt2gene | Not-determined |
| S1S_vs_69771\|\|lt2gene | Not-determined |
| S1S_vs_70105\|\|lt2gene | Low-quality |
| S1S_vs_70284\|\|lt2gene | Not-determined |
| S1S_vs_70397\|\|lt2gene | Low-quality |
| S1S_vs_70489\|\|lt2gene | Not-determined |
| S1S_vs_70770\|\|lt2gene | Low-quality |
| S1S_vs_70844\|\|lt2gene | Low-quality |
| S1S_vs_71093\|\|lt2gene | Low-quality |
| S1S_vs_71161\|\|lt2gene | Not-determined |
| S1S_vs_71260\|\|lt2gene | Low-quality |
| S1S_vs_71352\|\|lt2gene | Not-determined |
| S1S_vs_71476\|\|lt2gene | Low-quality |
| S1S_vs_71527\|\|lt2gene | Not-determined |
| S1S_vs_71537\|\|lt2gene | Not-determined |
| S1S_vs_71773\|\|lt2gene | Not-determined |
| S1S_vs_71839\|\|lt2gene | Low-quality |
| S1S_vs_72068\|\|lt2gene | Low-quality |
| S1S_vs_72142\|\|lt2gene | Not-determined |
| S1S_vs_72246\|\|lt2gene | Low-quality |
| S1S_vs_72272\|\|lt2gene | Low-quality |
| S1S_vs_72365\|\|lt2gene | Not-determined |
| S1S_vs_72523\|\|lt2gene | Low-quality |
| S1S_vs_72670\|\|lt2gene | Low-quality |
| S1S_vs_72770\|\|lt2gene | Low-quality |
| S1S_vs_72880\|\|lt2gene | Low-quality |
| S1S_vs_73021\|\|lt2gene | Not-determined |
| S1S_vs_73105\|\|lt2gene | Not-determined |
| S1S_vs_73215\|\|lt2gene | Not-determined |
| S1S_vs_73385\|\|lt2gene | Not-determined |
| S1S_vs_73610\|\|lt2gene | Low-quality |
| S1S_vs_73873\|\|lt2gene | Not-determined |
| S1S_vs_73875\|\|lt2gene | Low-quality |
| S1S_vs_73883\|\|lt2gene | Low-quality |
| S1S_vs_73898\|\|lt2gene | Low-quality |
| S1S_vs_73918\|\|lt2gene | Not-determined |
| S1S_vs_73999\|\|lt2gene | Low-quality |
| S1S_vs_74145\|\|lt2gene | Low-quality |
| S1S_vs_74236\|\|lt2gene | Low-quality |
| S1S_vs_74439\|\|lt2gene | Not-determined |
| S1S_vs_74549\|\|lt2gene | Not-determined |
| S1S_vs_74557\|\|lt2gene | Low-quality |
| S1S_vs_74622\|\|lt2gene | Low-quality |
| S1S_vs_74939\|\|lt2gene | Low-quality |
| S1S_vs_74975\|\|lt2gene | Low-quality |
| S1S_vs_74992\|\|lt2gene | Low-quality |
| S1S_vs_75085\|\|lt2gene | Low-quality |
| S1S_vs_75131\|\|lt2gene | Low-quality |
| S1S_vs_75215\|\|lt2gene | Low-quality |
| S1S_vs_75370\|\|lt2gene | Low-quality |
| S1S_vs_75492\|\|lt2gene | Not-determined |
| S1S_vs_75619\|\|lt2gene | Not-determined |
| S1S_vs_75699\|\|lt2gene | Low-quality |
| S1S_vs_76017\|\|lt2gene | Low-quality |
| S1S_vs_76295\|\|lt2gene | Low-quality |
| S1S_vs_76491\|\|lt2gene | Low-quality |
| S1S_vs_76496\|\|lt2gene | Low-quality |
| S1S_vs_76521\|\|lt2gene | Low-quality |
| S1S_vs_76529\|\|lt2gene | Low-quality |
| S1S_vs_76618\|\|lt2gene | Not-determined |
| S1S_vs_76685\|\|lt2gene | Low-quality |
| S1S_vs_76879\|\|lt2gene | Not-determined |
| S1S_vs_76903\|\|lt2gene | Low-quality |
| S1S_vs_77027\|\|lt2gene | Low-quality |
| S1S_vs_77043\|\|lt2gene | Low-quality |
| S1S_vs_77128\|\|lt2gene | Low-quality |
| S1S_vs_77139\|\|lt2gene | Low-quality |
| S1S_vs_77231\|\|lt2gene | Low-quality |
| S1S_vs_77301\|\|lt2gene | Low-quality |
| S1S_vs_77456\|\|lt2gene | Not-determined |
| S1S_vs_77462\|\|lt2gene | Not-determined |
| S2_extracted_k141_4059 | Not-determined |
| S2_extracted_k141_1353 | Low-quality |
| S2_extracted_k141_2706 | Not-determined |
| S2_extracted_k141_18938 | Low-quality |
| S2_extracted_k141_9472 | Not-determined |
| S2_extracted_k141_18942 | Not-determined |
| S2_extracted_k141_2711 | Not-determined |
| S2_extracted_k141_8122 | Not-determined |
| S2_extracted_k141_12180 | Not-determined |
| S2_extracted_k141_2718 | Not-determined |
| S2_extracted_k141_14886 | Not-determined |
| S2_extracted_k141_9477 | Not-determined |
| S2_extracted_k141_16242 | Not-determined |
| S2_extracted_k141_16243 | Not-determined |
| S2_extracted_k141_20296 | Not-determined |
| S2_extracted_k141_16245 | Low-quality |
| S2_extracted_k141_16246 | Low-quality |
| S2_extracted_k141_2726 | Low-quality |
| S2_extracted_k141_16249 | Not-determined |
| S2_extracted_k141_19 | Not-determined |
| S2_extracted_k141_16256 | Not-determined |
| S2_extracted_k141_16258 | Not-determined |
| S2_extracted_k141_8129 | Low-quality |
| S2_extracted_k141_5418 | Low-quality |
| S2_extracted_k141_8131 | Not-determined |
| S2_extracted_k141_17595 | Not-determined |
| S2_extracted_k141_9482 | Not-determined |
| S2_extracted_k141_20303 | Low-quality |
| S2_extracted_k141_18949 | Low-quality |
| S2_extracted_k141_16261 | Not-determined |
| S2_extracted_k141_10825 | Low-quality |
| S2_extracted_k141_18950 | Not-determined |
| S2_extracted_k141_9489 | Not-determined |
| S2_extracted_k141_18951 | Low-quality |
| S2_extracted_k141_13533 | Not-determined |
| S2_extracted_k141_8138 | Not-determined |
| S2_extracted_k141_9491 | Low-quality |
| S2_extracted_k141_17598 | Not-determined |
| S2_extracted_k141_10828 | Not-determined |
| S2_extracted_k141_18959 | Not-determined |
| S2_extracted_k141_9493 | Not-determined |
| S2_extracted_k141_9495 | Not-determined |
| S2_extracted_k141_4073 | Low-quality |
| S2_extracted_k141_1356 | Not-determined |
| S2_extracted_k141_1358 | Not-determined |
| S2_extracted_k141_1359 | Not-determined |
| S2_extracted_k141_8141 | Low-quality |
| S2_extracted_k141_21 | Not-determined |
| S2_extracted_k141_8143 | Not-determined |
| S2_extracted_k141_2737 | Not-determined |
| S2_extracted_k141_6773 | Not-determined |
| S2_extracted_k141_8144 | Low-quality |
| S2_extracted_k141_1364 | Not-determined |
| S2_extracted_k141_18970 | Not-determined |
| S2_extracted_k141_6774 | Low-quality |
| S2_extracted_k141_12194 | Low-quality |
| S2_extracted_k141_2742 | Not-determined |
| S2_extracted_k141_12195 | Not-determined |
| S2_extracted_k141_23 | Not-determined |
| S2_extracted_k141_4085 | Low-quality |
| S2_extracted_k141_13548 | Not-determined |
| S2_extracted_k141_2745 | Low-quality |
| S2_extracted_k141_4088 | Not-determined |
| S2_extracted_k141_9500 | Low-quality |
| S2_extracted_k141_8149 | Not-determined |
| S2_extracted_k141_9501 | Not-determined |
| S2_extracted_k141_9503 | Low-quality |
| S2_extracted_k141_26 | Not-determined |
| S2_extracted_k141_12202 | Not-determined |
| S2_extracted_k141_16266 | Low-quality |
| S2_extracted_k141_17612 | Not-determined |
| S2_extracted_k141_13557 | Not-determined |
| S2_extracted_k141_1373 | Not-determined |
| S2_extracted_k141_12205 | Not-determined |
| S2_extracted_k141_1376 | Not-determined |
| S2_extracted_k141_33 | Not-determined |
| S2_extracted_k141_8151 | Not-determined |
| S2_extracted_k141_2754 | Not-determined |
| S2_extracted_k141_4098 | Not-determined |
| S2_extracted_k141_13562 | Not-determined |
| S2_extracted_k141_9509 | Low-quality |
| S2_extracted_k141_8155 | Not-determined |
| S2_extracted_k141_16271 | Not-determined |
| S2_extracted_k141_4101 | Not-determined |
| S2_extracted_k141_20309 | Not-determined |
| S2_extracted_k141_9512 | Not-determined |
| S2_extracted_k141_4104 | Not-determined |
| S2_extracted_k141_20313 | Not-determined |
| S2_extracted_k141_12212 | Not-determined |
| S2_extracted_k141_20316 | Not-determined |
| S2_extracted_k141_20317 | Not-determined |
| S2_extracted_k141_38 | Not-determined |
| S2_extracted_k141_4111 | Not-determined |
| S2_extracted_k141_1380 | Not-determined |
| S2_extracted_k141_6793 | Not-determined |
| S2_extracted_k141_43 | Not-determined |
| S2_extracted_k141_44 | Not-determined |
| S2_extracted_k141_46 | Low-quality |
| S2_extracted_k141_16277 | Low-quality |
| S2_extracted_k141_14905 | Not-determined |
| S2_extracted_k141_14906 | Not-determined |
| S2_extracted_k141_17628 | Not-determined |
| S2_extracted_k141_6795 | Low-quality |
| S2_extracted_k141_6798 | Not-determined |
| S2_extracted_k141_12218 | Low-quality |
| S2_extracted_k141_9527 | Not-determined |
| S2_extracted_k141_5433 | Not-determined |
| S2_extracted_k141_20325 | Low-quality |
| S2_extracted_k141_9528 | Not-determined |
| S2_extracted_k141_6802 | Not-determined |
| S2_extracted_k141_2768 | Low-quality |
| S2_extracted_k141_8162 | Low-quality |
| S2_extracted_k141_8165 | Not-determined |
| S2_extracted_k141_8166 | Not-determined |
| S2_extracted_k141_20329 | Not-determined |
| S2_extracted_k141_6805 | Low-quality |
| S2_extracted_k141_18990 | Not-determined |
| S2_extracted_k141_17635 | Not-determined |
| S2_extracted_k141_9529 | Low-quality |
| S2_extracted_k141_13585 | Not-determined |
| S2_extracted_k141_1382 | Not-determined |
| S2_extracted_k141_1383 | Low-quality |
| S2_extracted_k141_6810 | Not-determined |
| S2_extracted_k141_8168 | Low-quality |
| S2_extracted_k141_17639 | Not-determined |
| S2_extracted_k141_20338 | Not-determined |
| S2_extracted_k141_14916 | Not-determined |
| S2_extracted_k141_14917 | Low-quality |
| S2_extracted_k141_6812 | Not-determined |
| S2_extracted_k141_5442 | Not-determined |
| S2_extracted_k141_20345 | Not-determined |
| S2_extracted_k141_20348 | Not-determined |
| S2_extracted_k141_8176 | Not-determined |
| S2_extracted_k141_20349 | Not-determined |
| S2_extracted_k141_20351 | Not-determined |
| S2_extracted_k141_20352 | Not-determined |
| S2_extracted_k141_10852 | Low-quality |
| S2_extracted_k141_5446 | Low-quality |
| S2_extracted_k141_10853 | Not-determined |
| S2_extracted_k141_60 | Not-determined |
| S2_extracted_k141_20359 | Not-determined |
| S2_extracted_k141_20364 | Not-determined |
| S2_extracted_k141_5455 | Not-determined |
| S2_extracted_k141_16302 | Not-determined |
| S2_extracted_k141_65 | Not-determined |
| S2_extracted_k141_17650 | Not-determined |
| S2_extracted_k141_6813 | Not-determined |
| S2_extracted_k141_10854 | Low-quality |
| S2_extracted_k141_20376 | Not-determined |
| S2_extracted_k141_10855 | Not-determined |
| S2_extracted_k141_20377 | Not-determined |
| S2_extracted_k141_6820 | Low-quality |
| S2_extracted_k141_17659 | Not-determined |
| S2_extracted_k141_17663 | Not-determined |
| S2_extracted_k141_14948 | Not-determined |
| S2_extracted_k141_19004 | Not-determined |
| S2_extracted_k141_17664 | Not-determined |
| S2_extracted_k141_13592 | Low-quality |
| S2_extracted_k141_14953 | Not-determined |
| S2_extracted_k141_8198 | Low-quality |
| S2_extracted_k141_17667 | Not-determined |
| S2_extracted_k141_10861 | Low-quality |
| S2_extracted_k141_19008 | Low-quality |
| S2_extracted_k141_8201 | Not-determined |
| S2_extracted_k141_13596 | Not-determined |
| S2_extracted_k141_9533 | Not-determined |
| S2_extracted_k141_9535 | Not-determined |
| S2_extracted_k141_19014 | Not-determined |
| S2_extracted_k141_13597 | Low-quality |
| S2_extracted_k141_14961 | Not-determined |
| S2_extracted_k141_14964 | Low-quality |
| S2_extracted_k141_10864 | Low-quality |
| S2_extracted_k141_20391 | Not-determined |
| S2_extracted_k141_14967 | Low-quality |
| S2_extracted_k141_19017 | Not-determined |
| S2_extracted_k141_19018 | Not-determined |
| S2_extracted_k141_10867 | Not-determined |
| S2_extracted_k141_14971 | Not-determined |
| S2_extracted_k141_13601 | Not-determined |
| S2_extracted_k141_20395 | Not-determined |
| S2_extracted_k141_20396 | Not-determined |
| S2_extracted_k141_20398 | Not-determined |
| S2_extracted_k141_17681 | Low-quality |
| S2_extracted_k141_2791 | Not-determined |
| S2_extracted_k141_20402 | Not-determined |
| S2_extracted_k141_17686 | Not-determined |
| S2_extracted_k141_13606 | Not-determined |
| S2_extracted_k141_17688 | Not-determined |
| S2_extracted_k141_81 | Not-determined |
| S2_extracted_k141_10874 | Not-determined |
| S2_extracted_k141_19025 | Not-determined |
| S2_extracted_k141_88 | Not-determined |
| S2_extracted_k141_13613 | Not-determined |
| S2_extracted_k141_14975 | Low-quality |
| S2_extracted_k141_17693 | Not-determined |
| S2_extracted_k141_13614 | Not-determined |
| S2_extracted_k141_5470 | Not-determined |
| S2_extracted_k141_20417 | Low-quality |
| S2_extracted_k141_2795 | Not-determined |
| S2_extracted_k141_2796 | Not-determined |
| S2_extracted_k141_9551 | Not-determined |
| S2_extracted_k141_17696 | Not-determined |
| S2_extracted_k141_2798 | Not-determined |
| S2_extracted_k141_2799 | Not-determined |
| S2_extracted_k141_17699 | Not-determined |
| S2_extracted_k141_1396 | Low-quality |
| S2_extracted_k141_14978 | Low-quality |
| S2_extracted_k141_14980 | Not-determined |
| S2_extracted_k141_1398 | Low-quality |
| S2_extracted_k141_19038 | Not-determined |
| S2_extracted_k141_19040 | Not-determined |
| S2_extracted_k141_17707 | Not-determined |
| S2_extracted_k141_2803 | Not-determined |
| S2_extracted_k141_9559 | Not-determined |
| S2_extracted_k141_9560 | Not-determined |
| S2_extracted_k141_9561 | Not-determined |
| S2_extracted_k141_17715 | Not-determined |
| S2_extracted_k141_9564 | Not-determined |
| S2_extracted_k141_17717 | Low-quality |
| S2_extracted_k141_6842 | Not-determined |
| S2_extracted_k141_6843 | Not-determined |
| S2_extracted_k141_9567 | Not-determined |
| S2_extracted_k141_6845 | Not-determined |
| S2_extracted_k141_5478 | Not-determined |
| S2_extracted_k141_2820 | Not-determined |
| S2_extracted_k141_9572 | Not-determined |
| S2_extracted_k141_10889 | Not-determined |
| S2_extracted_k141_14987 | Not-determined |
| S2_extracted_k141_9577 | Not-determined |
| S2_extracted_k141_2825 | Not-determined |
| S2_extracted_k141_20428 | Not-determined |
| S2_extracted_k141_14988 | Low-quality |
| S2_extracted_k141_10891 | Not-determined |
| S2_extracted_k141_14991 | Low-quality |
| S2_extracted_k141_9579 | Not-determined |
| S2_extracted_k141_2826 | Not-determined |
| S2_extracted_k141_2829 | Not-determined |
| S2_extracted_k141_6866 | Not-determined |
| S2_extracted_k141_6867 | Not-determined |
| S2_extracted_k141_2836 | Not-determined |
| S2_extracted_k141_10901 | Not-determined |
| S2_extracted_k141_12223 | Not-determined |
| S2_extracted_k141_13623 | Low-quality |
| S2_extracted_k141_13624 | Not-determined |
| S2_extracted_k141_10907 | Not-determined |
| S2_extracted_k141_20444 | Not-determined |
| S2_extracted_k141_9591 | Low-quality |
| S2_extracted_k141_14998 | Low-quality |
| S2_extracted_k141_16313 | Not-determined |
| S2_extracted_k141_15003 | Not-determined |
| S2_extracted_k141_9596 | Low-quality |
| S2_extracted_k141_2855 | Low-quality |
| S2_extracted_k141_2856 | Not-determined |
| S2_extracted_k141_13632 | Not-determined |
| S2_extracted_k141_9599 | Not-determined |
| S2_extracted_k141_12239 | Not-determined |
| S2_extracted_k141_13634 | Not-determined |
| S2_extracted_k141_112 | Not-determined |
| S2_extracted_k141_2862 | Not-determined |
| S2_extracted_k141_12243 | Not-determined |
| S2_extracted_k141_8238 | Low-quality |
| S2_extracted_k141_13638 | Not-determined |
| S2_extracted_k141_16315 | Not-determined |
| S2_extracted_k141_19048 | Not-determined |
| S2_extracted_k141_8241 | Not-determined |
| S2_extracted_k141_6876 | Not-determined |
| S2_extracted_k141_2870 | Not-determined |
| S2_extracted_k141_19051 | Low-quality |
| S2_extracted_k141_8244 | Low-quality |
| S2_extracted_k141_2871 | Not-determined |
| S2_extracted_k141_16322 | Low-quality |
| S2_extracted_k141_8248 | Low-quality |
| S2_extracted_k141_10917 | Not-determined |
| S2_extracted_k141_19056 | Not-determined |
| S2_extracted_k141_19059 | Not-determined |
| S2_extracted_k141_12252 | Not-determined |
| S2_extracted_k141_13646 | Not-determined |
| S2_extracted_k141_20467 | Not-determined |
| S2_extracted_k141_6878 | Not-determined |
| S2_extracted_k141_9613 | Low-quality |
| S8_vs_k141_137836\|\|full | Medium-quality |
| S2_extracted_k141_15008 | Low-quality |
| S2_extracted_k141_13654 | Not-determined |
| S2_extracted_k141_6884 | Not-determined |
| S2_extracted_k141_10931 | Not-determined |
| S2_extracted_k141_6886 | Not-determined |
| S2_extracted_k141_15011 | Low-quality |
| S2_extracted_k141_6888 | Not-determined |
| S2_extracted_k141_17727 | Low-quality |
| S2_extracted_k141_6889 | Not-determined |
| S2_extracted_k141_4137 | Low-quality |
| S2_extracted_k141_15016 | Not-determined |
| S2_extracted_k141_4140 | Not-determined |
| S2_extracted_k141_4141 | Not-determined |
| S2_extracted_k141_19066 | Not-determined |
| S2_extracted_k141_6895 | Not-determined |
| S2_extracted_k141_19068 | Low-quality |
| S2_extracted_k141_15021 | Not-determined |
| S2_extracted_k141_15024 | Low-quality |
| S2_extracted_k141_15025 | Not-determined |
| S2_extracted_k141_4147 | Not-determined |
| S2_extracted_k141_19073 | Not-determined |
| S2_extracted_k141_5505 | Not-determined |
| S2_extracted_k141_4149 | Not-determined |
| S2_extracted_k141_4151 | Not-determined |
| S2_extracted_k141_15028 | Not-determined |
| S2_extracted_k141_15030 | Not-determined |
| S2_extracted_k141_16342 | Not-determined |
| S2_extracted_k141_6907 | Not-determined |
| S2_extracted_k141_13666 | Not-determined |
| S2_extracted_k141_16346 | Not-determined |
| S2_extracted_k141_4160 | Not-determined |
| S2_extracted_k141_137 | Not-determined |
| S2_extracted_k141_15045 | Low-quality |
| S2_extracted_k141_15046 | Not-determined |
| S2_extracted_k141_13677 | Not-determined |
| S2_extracted_k141_6914 | Low-quality |
| S2_extracted_k141_15051 | Not-determined |
| S2_extracted_k141_13680 | Not-determined |
| S2_extracted_k141_16359 | Not-determined |
| S2_extracted_k141_15052 | Not-determined |
| S2_extracted_k141_10948 | Not-determined |
| S2_extracted_k141_17728 | Not-determined |
| S2_extracted_k141_141 | Not-determined |
| S2_extracted_k141_16360 | Not-determined |
| S2_extracted_k141_6920 | Low-quality |
| S2_extracted_k141_17731 | Not-determined |
| S2_extracted_k141_4171 | Not-determined |
| S2_extracted_k141_13688 | Not-determined |
| S2_extracted_k141_10954 | Not-determined |
| S2_extracted_k141_17733 | Not-determined |
| S2_extracted_k141_17735 | Not-determined |
| S2_extracted_k141_10959 | Not-determined |
| S2_extracted_k141_4175 | Not-determined |
| S2_extracted_k141_17738 | Not-determined |
| S2_extracted_k141_10963 | Not-determined |
| S2_extracted_k141_10967 | Not-determined |
| S2_extracted_k141_144 | Not-determined |
| S2_extracted_k141_10968 | Low-quality |
| S2_extracted_k141_10969 | Not-determined |
| S2_extracted_k141_17739 | Low-quality |
| S2_extracted_k141_12269 | Not-determined |
| S2_extracted_k141_9618 | Not-determined |
| S2_extracted_k141_15069 | Low-quality |
| S2_extracted_k141_146 | Low-quality |
| S2_extracted_k141_9621 | Not-determined |
| S2_extracted_k141_20472 | Not-determined |
| S2_extracted_k141_15071 | Low-quality |
| S2_extracted_k141_20473 | Not-determined |
| S2_extracted_k141_4186 | Not-determined |
| S2_extracted_k141_4187 | Not-determined |
| S2_extracted_k141_15077 | Low-quality |
| S2_extracted_k141_12274 | Low-quality |
| S2_extracted_k141_4190 | Low-quality |
| S2_extracted_k141_17741 | Not-determined |
| S2_extracted_k141_161 | Not-determined |
| S2_extracted_k141_16386 | Not-determined |
| S2_extracted_k141_163 | Not-determined |
| S2_extracted_k141_17745 | Not-determined |
| S3_vs_k141_122706\|\|full | Medium-quality |
| S2_extracted_k141_9630 | Not-determined |
| S2_extracted_k141_12285 | Low-quality |
| S2_extracted_k141_4195 | Not-determined |
| S2_extracted_k141_19093 | Not-determined |
| S2_extracted_k141_4198 | Not-determined |
| S2_extracted_k141_5518 | Not-determined |
| S2_extracted_k141_1403 | Not-determined |
| S2_extracted_k141_12291 | Not-determined |
| S2_extracted_k141_10978 | Not-determined |
| S2_extracted_k141_19096 | Not-determined |
| S2_extracted_k141_20488 | Not-determined |
| S2_extracted_k141_19099 | Not-determined |
| S2_extracted_k141_2875 | Not-determined |
| S2_extracted_k141_10983 | Not-determined |
| S2_extracted_k141_2881 | Not-determined |
| S2_extracted_k141_13715 | Not-determined |
| S2_extracted_k141_2883 | Not-determined |
| S2_extracted_k141_19102 | Not-determined |
| S2_extracted_k141_13719 | Low-quality |
| S2_extracted_k141_20497 | Not-determined |
| S2_extracted_k141_9631 | Not-determined |
| S2_extracted_k141_20498 | Not-determined |
| S2_extracted_k141_6941 | Not-determined |
| S2_extracted_k141_13720 | Not-determined |
| S2_extracted_k141_9634 | Not-determined |
| S2_extracted_k141_20504 | Low-quality |
| S2_extracted_k141_13722 | Not-determined |
| S2_extracted_k141_5524 | Low-quality |
| S2_extracted_k141_10985 | Not-determined |
| S2_extracted_k141_2891 | Low-quality |
| S2_extracted_k141_2892 | Not-determined |
| S2_extracted_k141_10987 | Low-quality |
| S2_extracted_k141_2893 | Low-quality |
| S2_extracted_k141_13727 | Not-determined |
| S2_extracted_k141_20511 | Not-determined |
| S2_extracted_k141_1413 | Not-determined |
| S2_extracted_k141_13729 | Low-quality |
| S2_extracted_k141_16413 | Not-determined |
| S2_extracted_k141_5536 | Not-determined |
| S2_extracted_k141_5537 | Not-determined |
| S2_extracted_k141_9643 | Not-determined |
| S2_extracted_k141_13731 | Low-quality |
| S2_extracted_k141_1422 | Not-determined |
| S2_extracted_k141_16418 | Not-determined |
| S2_extracted_k141_2903 | Not-determined |
| S2_extracted_k141_17756 | Not-determined |
| S2_extracted_k141_10993 | Not-determined |
| S2_extracted_k141_17759 | Not-determined |
| S2_extracted_k141_16420 | Not-determined |
| S2_extracted_k141_6949 | Not-determined |
| S2_extracted_k141_181 | Low-quality |
| S2_extracted_k141_182 | Not-determined |
| S2_extracted_k141_6950 | Not-determined |
| S2_extracted_k141_9654 | Not-determined |
| S2_extracted_k141_16424 | Not-determined |
| S2_extracted_k141_13741 | Not-determined |
| S2_extracted_k141_16425 | Low-quality |
| S2_extracted_k141_1432 | Low-quality |
| S2_extracted_k141_9658 | Not-determined |
| S2_extracted_k141_13744 | Not-determined |
| S2_extracted_k141_6955 | Not-determined |
| S2_extracted_k141_10995 | Low-quality |
| S2_extracted_k141_9662 | Not-determined |
| S2_extracted_k141_6959 | Not-determined |
| S2_extracted_k141_1433 | Low-quality |
| S2_extracted_k141_13747 | Not-determined |
| S2_extracted_k141_1437 | Not-determined |
| S2_extracted_k141_6963 | Low-quality |
| S2_extracted_k141_11001 | Low-quality |
| S2_extracted_k141_16435 | Not-determined |
| S2_extracted_k141_9669 | Not-determined |
| S2_extracted_k141_13750 | Not-determined |
| S2_extracted_k141_6967 | Not-determined |
| S2_extracted_k141_6968 | Not-determined |
| S2_extracted_k141_4210 | Not-determined |
| S2_extracted_k141_19123 | Low-quality |
| S2_extracted_k141_2927 | Not-determined |
| S2_extracted_k141_11004 | Not-determined |
| S2_extracted_k141_16437 | Not-determined |
| S2_extracted_k141_17774 | Low-quality |
| S2_extracted_k141_16439 | Not-determined |
| S2_extracted_k141_12300 | Low-quality |
| S2_extracted_k141_11006 | Not-determined |
| S2_extracted_k141_4216 | Low-quality |
| S2_extracted_k141_13755 | Not-determined |
| S2_extracted_k141_11007 | Not-determined |
| S2_extracted_k141_4218 | Not-determined |
| S2_extracted_k141_4219 | Not-determined |
| S2_extracted_k141_2933 | Not-determined |
| S2_extracted_k141_13758 | Not-determined |
| S2_extracted_k141_5544 | Not-determined |
| S2_extracted_k141_16442 | Not-determined |
| S2_extracted_k141_13761 | Low-quality |
| S2_extracted_k141_4224 | Not-determined |
| S2_extracted_k141_2937 | Not-determined |
| S2_extracted_k141_6973 | Not-determined |
| S2_extracted_k141_19127 | Not-determined |
| S2_extracted_k141_12311 | Not-determined |

| | |
|---|---|
| S2_extracted_k141_20522 | Low-quality |
| S2_extracted_k141_1442 | Not-determined |
| S2_extracted_k141_2945 | Not-determined |
| S12_vs_k141_110623\|\|full | Medium-quality |
| S2_extracted_k141_5551 | Low-quality |
| S2_extracted_k141_186 | Not-determined |
| S2_extracted_k141_19139 | Not-determined |
| S2_extracted_k141_4230 | Not-determined |
| S2_extracted_k141_5554 | Not-determined |
| S2_extracted_k141_16453 | Not-determined |
| S2_extracted_k141_2948 | Low-quality |
| S2_extracted_k141_19140 | Not-determined |
| S2_extracted_k141_4234 | Not-determined |
| S2_extracted_k141_5557 | Low-quality |
| S2_extracted_k141_19144 | Not-determined |
| S2_extracted_k141_5559 | Low-quality |
| S2_extracted_k141_12328 | Not-determined |
| S2_extracted_k141_11028 | Low-quality |
| S2_extracted_k141_1446 | Not-determined |
| S2_extracted_k141_6981 | Low-quality |
| S2_extracted_k141_1449 | Not-determined |
| S2_extracted_k141_1452 | Low-quality |
| S2_extracted_k141_1453 | Not-determined |
| S2_extracted_k141_11033 | Not-determined |
| S2_extracted_k141_4244 | Not-determined |
| S2_extracted_k141_5573 | Not-determined |
| S2_extracted_k141_11034 | Low-quality |
| S2_extracted_k141_6990 | Not-determined |
| S2_extracted_k141_4246 | Low-quality |
| S2_extracted_k141_4247 | Not-determined |
| S2_extracted_k141_12332 | Low-quality |
| S2_extracted_k141_20527 | Not-determined |
| S2_extracted_k141_1460 | Not-determined |
| S2_extracted_k141_12335 | Low-quality |
| S2_extracted_k141_4252 | Low-quality |
| S2_extracted_k141_6993 | Low-quality |
| S2_extracted_k141_12337 | Not-determined |
| S2_extracted_k141_13782 | Not-determined |
| S2_extracted_k141_15094 | Not-determined |
| S2_extracted_k141_4253 | Not-determined |
| S2_extracted_k141_1466 | Not-determined |
| S2_extracted_k141_17781 | Not-determined |
| S2_extracted_k141_2956 | Not-determined |
| S2_extracted_k141_17782 | Not-determined |
| S2_extracted_k141_16463 | Not-determined |
| S2_extracted_k141_17784 | Not-determined |
| S2_extracted_k141_16464 | Not-determined |
| S2_extracted_k141_1471 | Low-quality |
| S2_extracted_k141_15098 | Not-determined |
| S2_extracted_k141_2959 | Not-determined |
| S2_extracted_k141_1472 | Not-determined |
| S2_extracted_k141_12345 | Low-quality |
| S2_extracted_k141_4257 | Low-quality |
| S2_extracted_k141_17786 | Low-quality |
| S2_extracted_k141_1473 | Not-determined |
| S2_extracted_k141_9673 | Not-determined |
| S2_extracted_k141_16466 | Low-quality |
| S2_extracted_k141_5585 | Low-quality |
| S2_extracted_k141_19156 | Low-quality |
| S2_extracted_k141_1476 | Not-determined |
| S2_extracted_k141_19157 | Not-determined |
| S2_extracted_k141_9676 | Not-determined |
| S2_extracted_k141_9677 | Not-determined |
| S2_extracted_k141_8257 | Not-determined |
| S2_extracted_k141_15105 | Low-quality |
| S2_extracted_k141_16470 | Not-determined |
| S2_extracted_k141_15106 | Not-determined |
| S2_extracted_k141_13790 | Not-determined |
| S2_extracted_k141_20532 | Low-quality |
| S2_extracted_k141_13791 | Not-determined |
| S2_extracted_k141_15110 | Not-determined |
| S2_extracted_k141_20533 | Low-quality |
| S2_extracted_k141_16472 | Low-quality |
| S2_extracted_k141_9682 | Not-determined |
| S2_extracted_k141_19163 | Not-determined |
| S2_extracted_k141_7003 | Not-determined |
| S2_extracted_k141_19164 | Not-determined |
| S2_extracted_k141_20535 | Not-determined |
| S2_extracted_k141_16474 | Not-determined |
| S2_extracted_k141_11040 | Not-determined |
| S2_extracted_k141_9683 | Not-determined |
| S2_extracted_k141_7005 | Not-determined |
| S2_extracted_k141_15114 | Not-determined |
| S2_extracted_k141_15115 | Not-determined |
| S2_extracted_k141_8264 | Not-determined |
| S2_extracted_k141_12350 | Not-determined |
| S2_extracted_k141_12351 | Not-determined |
| S2_extracted_k141_12352 | Not-determined |
| S2_extracted_k141_15121 | Not-determined |
| S2_extracted_k141_12355 | Not-determined |
| S2_extracted_k141_4262 | Not-determined |
| S2_extracted_k141_13793 | Not-determined |
| S2_extracted_k141_15128 | Not-determined |
| S2_extracted_k141_2966 | Low-quality |
| S2_extracted_k141_8268 | Not-determined |
| S2_extracted_k141_11048 | Low-quality |
| S2_extracted_k141_8269 | Not-determined |
| S2_extracted_k141_13797 | Not-determined |
| S2_extracted_k141_11049 | Low-quality |
| S2_extracted_k141_7012 | Low-quality |
| S2_extracted_k141_4272 | Not-determined |
| S2_extracted_k141_5592 | Not-determined |
| S2_extracted_k141_16486 | Not-determined |
| S2_extracted_k141_5594 | Not-determined |
| S2_extracted_k141_5598 | Not-determined |
| S2_extracted_k141_15138 | Not-determined |
| S2_extracted_k141_15139 | Not-determined |
| S2_extracted_k141_9687 | Low-quality |
| S2_extracted_k141_16492 | Not-determined |
| S2_extracted_k141_19170 | Not-determined |
| S2_extracted_k141_5605 | Not-determined |
| S2_extracted_k141_19174 | Not-determined |
| S2_extracted_k141_5610 | Not-determined |
| S2_extracted_k141_19178 | Not-determined |
| S2_extracted_k141_15147 | Not-determined |
| S2_extracted_k141_5615 | Not-determined |
| S2_extracted_k141_16502 | Low-quality |
| S2_extracted_k141_20545 | Not-determined |
| S2_extracted_k141_13820 | Low-quality |
| S2_extracted_k141_8293 | Low-quality |
| S2_extracted_k141_19179 | Low-quality |
| S2_extracted_k141_19182 | Not-determined |
| S2_extracted_k141_15152 | Low-quality |
| S2_extracted_k141_20549 | Not-determined |
| S2_extracted_k141_5618 | Low-quality |
| S2_extracted_k141_15153 | Not-determined |
| S2_extracted_k141_9693 | Low-quality |
| S2_extracted_k141_11075 | Not-determined |
| S2_extracted_k141_11076 | Not-determined |
| S2_extracted_k141_5621 | Not-determined |
| S2_extracted_k141_5623 | Not-determined |
| S2_extracted_k141_5624 | Not-determined |
| S2_extracted_k141_20558 | Not-determined |
| S2_extracted_k141_12367 | Not-determined |
| S2_extracted_k141_5626 | Low-quality |
| S2_extracted_k141_5627 | Low-quality |
| S2_extracted_k141_12370 | Low-quality |
| S2_extracted_k141_9704 | Not-determined |
| S2_extracted_k141_13835 | Not-determined |
| S2_extracted_k141_12371 | Not-determined |
| S2_extracted_k141_4283 | Not-determined |
| S2_extracted_k141_20563 | Not-determined |
| S2_extracted_k141_4287 | Low-quality |
| S2_extracted_k141_2994 | Not-determined |
| S2_extracted_k141_2995 | Not-determined |
| S2_extracted_k141_16522 | Not-determined |
| S2_extracted_k141_12373 | Not-determined |
| S2_extracted_k141_5637 | Not-determined |
| S2_extracted_k141_2998 | Not-determined |
| S2_extracted_k141_5638 | Low-quality |
| S2_extracted_k141_2999 | Not-determined |
| S2_extracted_k141_9707 | Not-determined |
| S2_extracted_k141_3000 | Not-determined |
| S2_extracted_k141_16528 | Not-determined |
| S2_extracted_k141_3002 | Not-determined |
| S2_extracted_k141_13844 | Not-determined |
| S2_extracted_k141_8312 | Not-determined |
| S2_extracted_k141_15155 | Low-quality |
| S2_extracted_k141_5644 | Not-determined |
| S2_extracted_k141_208 | Not-determined |
| S2_extracted_k141_15159 | Not-determined |
| S2_extracted_k141_17805 | Not-determined |
| S2_extracted_k141_3006 | Not-determined |
| S2_extracted_k141_3008 | Not-determined |
| S2_extracted_k141_16536 | Low-quality |
| S2_extracted_k141_11085 | Not-determined |
| S2_extracted_k141_15165 | Not-determined |
| S2_extracted_k141_16538 | Not-determined |
| S2_extracted_k141_9715 | Not-determined |
| S2_extracted_k141_17810 | Not-determined |
| S2_extracted_k141_16539 | Low-quality |
| S2_extracted_k141_19196 | Not-determined |
| S2_extracted_k141_9716 | Not-determined |
| S2_extracted_k141_5650 | Low-quality |
| S2_extracted_k141_15168 | Not-determined |
| S2_extracted_k141_16541 | Not-determined |
| S2_extracted_k141_19199 | Not-determined |
| S2_extracted_k141_17813 | Not-determined |
| S2_extracted_k141_9725 | Not-determined |
| S2_extracted_k141_5657 | Not-determined |
| S2_extracted_k141_13852 | Not-determined |
| S2_extracted_k141_15174 | Not-determined |
| S2_extracted_k141_15175 | Not-determined |
| S2_extracted_k141_12385 | Low-quality |
| S2_extracted_k141_9726 | Not-determined |
| S2_extracted_k141_12387 | Not-determined |
| S2_extracted_k141_8324 | Not-determined |
| S2_extracted_k141_5661 | Not-determined |
| S2_extracted_k141_8325 | Not-determined |
| S2_extracted_k141_17828 | Low-quality |
| S2_extracted_k141_7036 | Low-quality |
| S2_extracted_k141_19203 | Not-determined |
| S2_extracted_k141_13857 | Not-determined |
| S2_extracted_k141_13858 | Not-determined |
| S2_extracted_k141_16549 | Low-quality |
| S2_extracted_k141_12394 | Not-determined |
| S2_extracted_k141_9730 | Not-determined |
| S2_extracted_k141_15185 | Not-determined |
| S2_extracted_k141_3024 | Not-determined |
| S2_extracted_k141_16550 | Low-quality |
| S2_extracted_k141_8332 | Low-quality |
| S2_extracted_k141_19205 | Not-determined |
| S2_extracted_k141_8333 | Not-determined |
| S2_extracted_k141_20571 | Not-determined |
| S2_extracted_k141_12399 | Not-determined |
| S2_extracted_k141_19206 | Not-determined |
| S2_extracted_k141_15191 | Not-determined |
| S2_extracted_k141_19207 | Not-determined |
| S2_extracted_k141_20572 | Not-determined |
| S2_extracted_k141_229 | Not-determined |
| S2_extracted_k141_11095 | Low-quality |
| S2_extracted_k141_13864 | Not-determined |
| S2_extracted_k141_8339 | Not-determined |
| S2_extracted_k141_13865 | Not-determined |
| S2_extracted_k141_8341 | Low-quality |
| S2_extracted_k141_8342 | Low-quality |
| S2_extracted_k141_19215 | Not-determined |
| S2_extracted_k141_15199 | Low-quality |
| S2_extracted_k141_12409 | Low-quality |
| S2_extracted_k141_3027 | Low-quality |
| S2_extracted_k141_11104 | Low-quality |
| S2_extracted_k141_12410 | Low-quality |
| S2_extracted_k141_13873 | Not-determined |
| S2_extracted_k141_12412 | Not-determined |
| S2_extracted_k141_12413 | Not-determined |
| S2_extracted_k141_13875 | Low-quality |
| S2_extracted_k141_7040 | Low-quality |
| S2_extracted_k141_17844 | Not-determined |
| S2_extracted_k141_15207 | Not-determined |
| S2_extracted_k141_4307 | Low-quality |
| S2_extracted_k141_12417 | Low-quality |
| S2_extracted_k141_20578 | Not-determined |
| S2_extracted_k141_12418 | Low-quality |
| S2_extracted_k141_3034 | Not-determined |
| S2_extracted_k141_7048 | Not-determined |
| S2_extracted_k141_7049 | Not-determined |
| S2_extracted_k141_16567 | Not-determined |
| S2_extracted_k141_13879 | Not-determined |
| S2_extracted_k141_4308 | Not-determined |
| S2_extracted_k141_16568 | Not-determined |
| S2_extracted_k141_9742 | Not-determined |
| S2_extracted_k141_9743 | Not-determined |
| S2_extracted_k141_13882 | Not-determined |
| S2_extracted_k141_4313 | Not-determined |
| S2_extracted_k141_12426 | Not-determined |
| S2_extracted_k141_16572 | Low-quality |
| S2_extracted_k141_9748 | Not-determined |
| S2_extracted_k141_12428 | Not-determined |
| S2_extracted_k141_12429 | Not-determined |
| S2_extracted_k141_4316 | Not-determined |
| S2_extracted_k141_12431 | Not-determined |
| S2_extracted_k141_9753 | Not-determined |
| S2_extracted_k141_15223 | Not-determined |
| S2_extracted_k141_12434 | Not-determined |
| S2_extracted_k141_12435 | Not-determined |
| S2_extracted_k141_12437 | Not-determined |
| S2_extracted_k141_7064 | Low-quality |
| S2_extracted_k141_5679 | Not-determined |
| S2_extracted_k141_8353 | Not-determined |
| S2_extracted_k141_19221 | Not-determined |
| S2_extracted_k141_19223 | Low-quality |
| S2_extracted_k141_13894 | Low-quality |
| S2_extracted_k141_19224 | Not-determined |
| S2_extracted_k141_3042 | Low-quality |
| S2_extracted_k141_3044 | Not-determined |
| S2_extracted_k141_12445 | Not-determined |
| S2_extracted_k141_15227 | Low-quality |
| S2_extracted_k141_9754 | Not-determined |
| S2_extracted_k141_15228 | Low-quality |
| S2_extracted_k141_9756 | Not-determined |
| S2_extracted_k141_13898 | Low-quality |
| S2_extracted_k141_12446 | Low-quality |
| S2_extracted_k141_15231 | Low-quality |
| S2_extracted_k141_19227 | Low-quality |
| S2_extracted_k141_9759 | Not-determined |
| S2_extracted_k141_13904 | Not-determined |
| S2_extracted_k141_19230 | Low-quality |
| S2_extracted_k141_7074 | Not-determined |
| S2_extracted_k141_12452 | Low-quality |
| S2_extracted_k141_8359 | Not-determined |
| S2_extracted_k141_12453 | Low-quality |
| S2_extracted_k141_15238 | Not-determined |
| S5_vs_k141_58859\|\|full | Medium-quality |
| S2_extracted_k141_7077 | Not-determined |
| S2_extracted_k141_12454 | Not-determined |
| S2_extracted_k141_8361 | Not-determined |
| S2_extracted_k141_7078 | Not-determined |
| S2_extracted_k141_9763 | Not-determined |
| S2_extracted_k141_19234 | Not-determined |
| S2_extracted_k141_230 | Not-determined |
| S2_extracted_k141_7083 | Not-determined |
| S2_extracted_k141_7084 | Not-determined |
| S2_extracted_k141_11108 | Not-determined |
| S2_extracted_k141_19238 | Not-determined |
| S2_extracted_k141_11110 | Not-determined |
| S2_extracted_k141_7086 | Not-determined |
| S2_extracted_k141_20589 | Not-determined |
| S2_extracted_k141_20590 | Low-quality |
| S2_extracted_k141_11115 | Not-determined |
| S2_extracted_k141_15244 | Low-quality |
| S2_extracted_k141_11116 | Not-determined |
| S2_extracted_k141_236 | Not-determined |
| S2_extracted_k141_19251 | Not-determined |
| S2_extracted_k141_15247 | Not-determined |
| S2_extracted_k141_7097 | Not-determined |
| S2_extracted_k141_15248 | Not-determined |
| S2_extracted_k141_19252 | Not-determined |
| S2_extracted_k141_19254 | Low-quality |
| S2_extracted_k141_13918 | Not-determined |
| S2_extracted_k141_7102 | Not-determined |
| S2_extracted_k141_17854 | Low-quality |
| S2_extracted_k141_13920 | Not-determined |
| S2_extracted_k141_7105 | Not-determined |
| S2_extracted_k141_5722 | Low-quality |
| S2_extracted_k141_17861 | Not-determined |
| S2_extracted_k141_5724 | Not-determined |
| S2_extracted_k141_8367 | Not-determined |
| S2_extracted_k141_7110 | Not-determined |
| S2_extracted_k141_7111 | Not-determined |
| S2_extracted_k141_12457 | Not-determined |
| S2_extracted_k141_9777 | Not-determined |
| S2_extracted_k141_12458 | Low-quality |
| S2_extracted_k141_19273 | Not-determined |
| S2_extracted_k141_9778 | Low-quality |
| S2_extracted_k141_20603 | Not-determined |
| S2_extracted_k141_9779 | Low-quality |
| S2_extracted_k141_12460 | Low-quality |
| S2_extracted_k141_20606 | Low-quality |
| S2_extracted_k141_16583 | Not-determined |
| S2_extracted_k141_9785 | Low-quality |
| S2_extracted_k141_8374 | Not-determined |
| S2_extracted_k141_12466 | Not-determined |
| S2_extracted_k141_8376 | Not-determined |
| S2_extracted_k141_12467 | Not-determined |
| S2_extracted_k141_8377 | Low-quality |
| S2_extracted_k141_13937 | Low-quality |
| S2_extracted_k141_20614 | Low-quality |
| S2_extracted_k141_12470 | Not-determined |
| S2_extracted_k141_4340 | Not-determined |
| S2_extracted_k141_12471 | Not-determined |
| S2_extracted_k141_12472 | Not-determined |
| S2_extracted_k141_246 | Not-determined |
| S2_extracted_k141_20619 | Not-determined |
| S2_extracted_k141_12476 | Not-determined |
| S2_extracted_k141_16591 | Low-quality |
| S2_extracted_k141_9793 | Not-determined |
| S2_extracted_k141_13941 | Not-determined |
| S2_extracted_k141_16592 | Low-quality |
| S2_extracted_k141_16593 | Not-determined |
| S2_extracted_k141_11149 | Not-determined |
| S2_extracted_k141_19276 | Low-quality |
| S2_extracted_k141_13944 | Not-determined |
| S2_extracted_k141_16596 | Not-determined |
| S2_extracted_k141_9797 | Not-determined |
| S2_extracted_k141_17871 | Not-determined |
| S2_extracted_k141_13946 | Low-quality |
| S2_extracted_k141_19280 | Low-quality |
| S2_extracted_k141_19281 | Not-determined |
| S2_extracted_k141_17876 | Not-determined |
| S2_extracted_k141_19282 | Not-determined |
| S2_extracted_k141_19286 | Not-determined |
| S2_extracted_k141_19289 | Not-determined |
| S2_extracted_k141_17884 | Not-determined |
| S2_extracted_k141_19290 | Low-quality |
| S2_extracted_k141_7117 | Not-determined |
| S2_extracted_k141_9806 | Low-quality |
| S2_extracted_k141_19292 | Not-determined |
| S2_extracted_k141_9807 | Not-determined |
| S2_extracted_k141_19294 | Not-determined |
| S2_extracted_k141_19295 | Not-determined |
| S2_extracted_k141_15263 | Not-determined |
| S2_extracted_k141_19296 | Low-quality |
| S2_extracted_k141_20623 | Low-quality |
| S2_extracted_k141_15264 | Not-determined |
| S2_extracted_k141_20624 | Not-determined |
| S2_extracted_k141_11163 | Not-determined |
| S2_extracted_k141_13953 | Low-quality |
| S2_extracted_k141_20625 | Not-determined |
| S2_extracted_k141_13954 | Not-determined |
| S2_extracted_k141_4363 | Not-determined |
| S2_extracted_k141_19301 | Not-determined |
| S2_extracted_k141_16615 | Low-quality |
| S2_extracted_k141_7129 | Not-determined |
| S2_extracted_k141_20631 | Not-determined |
| S2_extracted_k141_13964 | Not-determined |
| S2_extracted_k141_12494 | Not-determined |
| S2_extracted_k141_9817 | Low-quality |
| S2_extracted_k141_16616 | Not-determined |
| S2_extracted_k141_16618 | Not-determined |
| S2_extracted_k141_12500 | Not-determined |
| S2_extracted_k141_12501 | Not-determined |
| S2_extracted_k141_7132 | Not-determined |
| S2_extracted_k141_16621 | Not-determined |
| S2_extracted_k141_12502 | Low-quality |
| S2_extracted_k141_16623 | Not-determined |
| S2_extracted_k141_12503 | Not-determined |
| S2_extracted_k141_16624 | Not-determined |
| S2_extracted_k141_12506 | Not-determined |
| S2_extracted_k141_13971 | Low-quality |
| S2_extracted_k141_15277 | Not-determined |
| S2_extracted_k141_12509 | Not-determined |
| S2_extracted_k141_11174 | Not-determined |
| S2_extracted_k141_12513 | Low-quality |
| S2_extracted_k141_12515 | Not-determined |
| S2_extracted_k141_8388 | Not-determined |
| S2_extracted_k141_8389 | Not-determined |
| S2_extracted_k141_7152 | Not-determined |
| S2_extracted_k141_1512 | Not-determined |
| S2_extracted_k141_9830 | Not-determined |
| S2_extracted_k141_1513 | Not-determined |
| S2_extracted_k141_5734 | Low-quality |
| S2_extracted_k141_15290 | Not-determined |
| S2_extracted_k141_9835 | Not-determined |
| S2_extracted_k141_5735 | Low-quality |
| S2_extracted_k141_15294 | Not-determined |
| S2_extracted_k141_5736 | Not-determined |
| S2_extracted_k141_5737 | Not-determined |
| S2_extracted_k141_16634 | Not-determined |
| S2_extracted_k141_5738 | Low-quality |
| S2_extracted_k141_253 | Not-determined |
| S2_extracted_k141_254 | Not-determined |
| S2_extracted_k141_15299 | Not-determined |
| S2_extracted_k141_12529 | Low-quality |
| S2_extracted_k141_16639 | Not-determined |
| S2_extracted_k141_5743 | Not-determined |
| S2_extracted_k141_16641 | Not-determined |
| S2_extracted_k141_5745 | Not-determined |
| S2_extracted_k141_7167 | Not-determined |
| S2_extracted_k141_7168 | Not-determined |
| S2_extracted_k141_7169 | Not-determined |
| S2_extracted_k141_17891 | Not-determined |
| S2_extracted_k141_13975 | Low-quality |
| S2_extracted_k141_16645 | Low-quality |
| S2_extracted_k141_11184 | Not-determined |
| S2_extracted_k141_13978 | Not-determined |
| S2_extracted_k141_4375 | Not-determined |
| S2_extracted_k141_7174 | Not-determined |
| S2_extracted_k141_13980 | Not-determined |
| S2_extracted_k141_20639 | Low-quality |
| S2_extracted_k141_13983 | Low-quality |
| S2_extracted_k141_20642 | Not-determined |
| S2_extracted_k141_7180 | Not-determined |
| S2_extracted_k141_15309 | Not-determined |
| S2_extracted_k141_1523 | Not-determined |
| S2_extracted_k141_19325 | Low-quality |
| S2_extracted_k141_19326 | Not-determined |
| S2_extracted_k141_1525 | Not-determined |
| S2_extracted_k141_13988 | Not-determined |
| S2_extracted_k141_7188 | Not-determined |
| S2_extracted_k141_13989 | Low-quality |
| S2_extracted_k141_7190 | Not-determined |
| S2_extracted_k141_7191 | Not-determined |
| S2_extracted_k141_13990 | Low-quality |
| S2_extracted_k141_11195 | Not-determined |
| S2_extracted_k141_13991 | Not-determined |
| S2_extracted_k141_16653 | Not-determined |
| S2_extracted_k141_1530 | Low-quality |
| S2_extracted_k141_12538 | Low-quality |
| S2_extracted_k141_20655 | Low-quality |
| S2_extracted_k141_12542 | Not-determined |
| S2_extracted_k141_20656 | Not-determined |
| S2_extracted_k141_1532 | Low-quality |
| S2_extracted_k141_9840 | Not-determined |
| S2_extracted_k141_19343 | Not-determined |
| S2_extracted_k141_1536 | Not-determined |
| S2_extracted_k141_1537 | Not-determined |
| S2_extracted_k141_9841 | Not-determined |
| S2_extracted_k141_5751 | Not-determined |
| S2_extracted_k141_1539 | Not-determined |
| S2_extracted_k141_13995 | Low-quality |
| S2_extracted_k141_7196 | Not-determined |
| S2_extracted_k141_5756 | Not-determined |
| S2_extracted_k141_7198 | Not-determined |
| S2_extracted_k141_9847 | Not-determined |
| S2_extracted_k141_7199 | Low-quality |
| S2_extracted_k141_16668 | Not-determined |
| S2_extracted_k141_19350 | Not-determined |
| S2_extracted_k141_9850 | Not-determined |
| S2_extracted_k141_4383 | Low-quality |
| S2_extracted_k141_14000 | Not-determined |
| S2_extracted_k141_5760 | Not-determined |
| S2_extracted_k141_7201 | Not-determined |
| S2_extracted_k141_5763 | Not-determined |
| S2_extracted_k141_9854 | Low-quality |
| S2_extracted_k141_1547 | Low-quality |
| S2_extracted_k141_16674 | Not-determined |
| S2_extracted_k141_5766 | Low-quality |
| S2_extracted_k141_12560 | Not-determined |
| S2_extracted_k141_1551 | Not-determined |
| S2_extracted_k141_8396 | Not-determined |
| S2_extracted_k141_15325 | Not-determined |
| S2_extracted_k141_267 | Not-determined |
| S2_extracted_k141_16677 | Not-determined |
| S2_extracted_k141_4392 | Not-determined |
| S2_extracted_k141_269 | Low-quality |
| S2_extracted_k141_16678 | Low-quality |
| S2_extracted_k141_12563 | Not-determined |
| S2_extracted_k141_19360 | Not-determined |
| S2_extracted_k141_15329 | Not-determined |
| S2_extracted_k141_15333 | Not-determined |
| S2_extracted_k141_12570 | Not-determined |
| S2_extracted_k141_19365 | Not-determined |
| S2_extracted_k141_12572 | Not-determined |
| S2_extracted_k141_15336 | Not-determined |
| S2_extracted_k141_15337 | Not-determined |
| S2_extracted_k141_12574 | Not-determined |
| S2_extracted_k141_16682 | Not-determined |
| S2_extracted_k141_12578 | Not-determined |
| S2_extracted_k141_11221 | Low-quality |
| S2_extracted_k141_16684 | Not-determined |
| S2_extracted_k141_4402 | Not-determined |
| S2_extracted_k141_9862 | Not-determined |
| S2_extracted_k141_275 | Not-determined |
| S2_extracted_k141_11227 | Not-determined |
| S2_extracted_k141_16687 | Not-determined |
| S2_extracted_k141_19376 | Not-determined |
| S2_extracted_k141_277 | Not-determined |
| S2_extracted_k141_16689 | Not-determined |
| S2_extracted_k141_19381 | Not-determined |
| S2_extracted_k141_1553 | Not-determined |
| S2_extracted_k141_19382 | Not-determined |
| S2_extracted_k141_20668 | Not-determined |
| S2_extracted_k141_7207 | Not-determined |
| S2_extracted_k141_15352 | Not-determined |
| S2_extracted_k141_20670 | Not-determined |
| S2_extracted_k141_9875 | Not-determined |
| S2_extracted_k141_3067 | Not-determined |
| S2_extracted_k141_4422 | Not-determined |
| S2_extracted_k141_16696 | Not-determined |
| S2_extracted_k141_20675 | Not-determined |
| S2_extracted_k141_16697 | Not-determined |
| S2_extracted_k141_11239 | Not-determined |
| S2_extracted_k141_12598 | Not-determined |
| S2_extracted_k141_15361 | Not-determined |
| S2_extracted_k141_1559 | Not-determined |
| S2_extracted_k141_4426 | Low-quality |
| S2_extracted_k141_4428 | Not-determined |
| S2_extracted_k141_3077 | Low-quality |
| S2_extracted_k141_19391 | Low-quality |
| S2_extracted_k141_12606 | Not-determined |
| S2_extracted_k141_7221 | Low-quality |
| S2_extracted_k141_3080 | Low-quality |
| S2_extracted_k141_8413 | Low-quality |
| S2_extracted_k141_16707 | Low-quality |
| S2_extracted_k141_14007 | Not-determined |
| S2_extracted_k141_19394 | Low-quality |
| S2_extracted_k141_20685 | Not-determined |
| S2_extracted_k141_7224 | Not-determined |
| S2_extracted_k141_20686 | Not-determined |
| S2_extracted_k141_11243 | Low-quality |
| S2_extracted_k141_1568 | Not-determined |
| S2_extracted_k141_9884 | Not-determined |
| S2_extracted_k141_20689 | Not-determined |
| S2_extracted_k141_1571 | Low-quality |
| S2_extracted_k141_9887 | Not-determined |
| S2_extracted_k141_12614 | Not-determined |
| S2_extracted_k141_12615 | Not-determined |
| S2_extracted_k141_286 | Low-quality |
| S2_extracted_k141_12616 | Low-quality |
| S2_extracted_k141_15376 | Not-determined |
| S2_extracted_k141_4434 | Low-quality |
| S2_extracted_k141_17919 | Not-determined |
| S2_extracted_k141_19406 | Low-quality |
| S2_extracted_k141_16713 | Low-quality |
| S2_extracted_k141_12620 | Not-determined |
| S2_extracted_k141_4437 | Low-quality |
| S2_extracted_k141_4438 | Not-determined |
| S2_extracted_k141_16717 | Low-quality |
| S2_extracted_k141_19408 | Not-determined |
| S2_extracted_k141_291 | Not-determined |
| S2_extracted_k141_15382 | Not-determined |
| S2_extracted_k141_11250 | Not-determined |
| S2_extracted_k141_7228 | Not-determined |
| S2_extracted_k141_12622 | Not-determined |
| S2_extracted_k141_15383 | Not-determined |
| S2_extracted_k141_9894 | Not-determined |
| S2_extracted_k141_9895 | Not-determined |
| S2_extracted_k141_4447 | Low-quality |
| S2_extracted_k141_4448 | Not-determined |
| S2_extracted_k141_7231 | Not-determined |
| S2_extracted_k141_8433 | Not-determined |
| S2_extracted_k141_15386 | Low-quality |
| S2_extracted_k141_17926 | Not-determined |
| S2_extracted_k141_4450 | Low-quality |
| S2_extracted_k141_4451 | Not-determined |
| S2_extracted_k141_9896 | Low-quality |
| S2_extracted_k141_9897 | Not-determined |
| S2_extracted_k141_11255 | Not-determined |
| S2_extracted_k141_16722 | Low-quality |

| | |
|---|---|
| S2_extracted_k141_5775 | Not-determined |
| S2_extracted_k141_12631 | Not-determined |
| S2_extracted_k141_4452 | Low-quality |
| S2_extracted_k141_19419 | Low-quality |
| S2_extracted_k141_19420 | Not-determined |
| S2_extracted_k141_9902 | Not-determined |
| S2_extracted_k141_15394 | Not-determined |
| S2_extracted_k141_12636 | Not-determined |
| S2_extracted_k141_20692 | Not-determined |
| S2_extracted_k141_12637 | Not-determined |
| S2_extracted_k141_9905 | Not-determined |
| S2_extracted_k141_9906 | Not-determined |
| S2_extracted_k141_11265 | Not-determined |
| S2_extracted_k141_14019 | Low-quality |
| S2_extracted_k141_14020 | Not-determined |
| S2_extracted_k141_4458 | Not-determined |
| S2_extracted_k141_19427 | Low-quality |
| S2_extracted_k141_12648 | Not-determined |
| S2_extracted_k141_4460 | Not-determined |
| S2_extracted_k141_20702 | Not-determined |
| S2_extracted_k141_3089 | Not-determined |
| S2_extracted_k141_20705 | Not-determined |
| S2_extracted_k141_17946 | Not-determined |
| S2_extracted_k141_19433 | Not-determined |
| S2_extracted_k141_15408 | Not-determined |
| S2_extracted_k141_20708 | Not-determined |
| S2_extracted_k141_9909 | Not-determined |
| S2_extracted_k141_19435 | Not-determined |
| S2_extracted_k141_11273 | Not-determined |
| S2_extracted_k141_15412 | Low-quality |
| S2_extracted_k141_7234 | Not-determined |
| S2_extracted_k141_302 | Not-determined |
| S2_extracted_k141_19440 | Not-determined |
| S2_extracted_k141_17954 | Low-quality |
| S2_extracted_k141_11281 | Low-quality |
| S2_extracted_k141_11283 | Not-determined |
| S2_extracted_k141_17957 | Not-determined |
| S2_extracted_k141_15416 | Not-determined |
| S2_extracted_k141_4474 | Not-determined |
| S2_extracted_k141_15418 | Not-determined |
| S2_extracted_k141_14036 | Not-determined |
| S2_extracted_k141_16739 | Not-determined |
| S2_extracted_k141_16740 | Low-quality |
| S2_extracted_k141_4480 | Not-determined |
| S2_extracted_k141_4484 | Low-quality |
| S2_extracted_k141_9911 | Low-quality |
| S2_extracted_k141_14039 | Not-determined |
| S2_extracted_k141_12677 | Not-determined |
| S2_extracted_k141_15426 | Not-determined |
| S2_extracted_k141_4487 | Low-quality |
| S2_extracted_k141_12679 | Not-determined |
| S2_extracted_k141_9914 | Not-determined |
| S2_extracted_k141_12680 | Not-determined |
| S2_extracted_k141_17965 | Not-determined |
| S2_extracted_k141_9915 | Not-determined |
| S2_extracted_k141_17966 | Not-determined |
| S2_extracted_k141_4493 | Not-determined |
| S2_extracted_k141_12685 | Not-determined |
| S2_extracted_k141_19456 | Not-determined |
| S2_extracted_k141_17970 | Not-determined |
| S2_extracted_k141_19457 | Not-determined |
| S2_extracted_k141_16747 | Not-determined |
| S2_extracted_k141_15430 | Not-determined |
| S2_extracted_k141_9922 | Not-determined |
| S2_extracted_k141_4500 | Not-determined |
| S2_extracted_k141_9923 | Not-determined |
| S2_extracted_k141_4501 | Not-determined |
| S2_extracted_k141_17977 | Not-determined |
| S2_extracted_k141_9925 | Low-quality |
| S2_extracted_k141_17979 | Not-determined |
| S2_extracted_k141_1593 | Low-quality |
| S2_extracted_k141_19467 | Not-determined |
| S2_extracted_k141_16754 | Not-determined |
| S2_extracted_k141_9927 | Not-determined |
| S2_extracted_k141_17982 | Not-determined |
| S2_extracted_k141_17985 | Not-determined |
| S2_extracted_k141_4513 | Not-determined |
| S2_extracted_k141_17986 | Low-quality |
| S2_extracted_k141_20711 | Not-determined |
| S2_extracted_k141_15440 | Not-determined |
| S2_extracted_k141_16759 | Not-determined |
| S2_extracted_k141_20714 | Low-quality |
| S2_extracted_k141_3107 | Not-determined |
| S2_extracted_k141_7242 | Not-determined |
| S2_extracted_k141_7244 | Not-determined |
| S2_extracted_k141_8446 | Not-determined |
| S2_extracted_k141_20720 | Not-determined |
| S2_extracted_k141_12705 | Low-quality |
| S2_extracted_k141_8448 | Not-determined |
| S2_extracted_k141_16762 | Not-determined |
| S2_extracted_k141_20722 | Not-determined |
| S2_extracted_k141_4525 | Not-determined |
| S2_extracted_k141_11312 | Not-determined |
| S2_extracted_k141_7249 | Low-quality |
| S2_extracted_k141_8453 | Low-quality |
| S2_extracted_k141_11315 | Low-quality |
| S2_extracted_k141_8457 | Not-determined |
| S2_extracted_k141_3117 | Not-determined |
| S2_extracted_k141_7253 | Not-determined |
| S2_extracted_k141_19480 | Low-quality |
| S2_extracted_k141_3119 | Not-determined |
| S2_extracted_k141_19483 | Not-determined |
| S2_extracted_k141_19484 | Low-quality |
| S2_extracted_k141_7257 | Not-determined |
| S2_extracted_k141_14057 | Not-determined |
| S2_extracted_k141_18004 | Not-determined |
| S2_extracted_k141_12716 | Not-determined |
| S2_extracted_k141_8464 | Not-determined |
| S2_extracted_k141_14059 | Not-determined |
| S2_extracted_k141_18005 | Not-determined |
| S2_extracted_k141_14060 | Not-determined |
| S2_extracted_k141_20738 | Not-determined |
| S2_extracted_k141_19490 | Not-determined |
| S2_extracted_k141_16773 | Not-determined |
| S2_extracted_k141_18009 | Not-determined |
| S2_extracted_k141_19492 | Not-determined |
| S2_extracted_k141_18010 | Low-quality |
| S2_extracted_k141_1596 | Not-determined |
| S2_extracted_k141_12728 | Not-determined |
| S2_extracted_k141_16775 | Not-determined |
| S2_extracted_k141_9930 | Not-determined |
| S2_extracted_k141_4532 | Not-determined |
| S2_extracted_k141_16778 | Low-quality |
| S2_extracted_k141_14064 | Not-determined |
| S2_extracted_k141_4534 | Not-determined |
| S2_extracted_k141_4536 | Not-determined |
| S2_extracted_k141_16784 | Not-determined |
| S2_extracted_k141_1606 | Not-determined |
| S2_extracted_k141_1607 | Not-determined |
| S2_extracted_k141_1611 | Not-determined |
| S2_extracted_k141_18023 | Not-determined |
| S2_extracted_k141_12741 | Not-determined |
| S2_extracted_k141_19509 | Low-quality |
| S2_extracted_k141_16789 | Low-quality |
| S2_extracted_k141_1615 | Not-determined |
| S2_extracted_k141_4543 | Not-determined |
| S2_extracted_k141_16791 | Not-determined |
| S2_extracted_k141_3135 | Not-determined |
| S2_extracted_k141_12745 | Not-determined |
| S2_extracted_k141_12747 | Low-quality |
| S2_extracted_k141_18029 | Low-quality |
| S2_extracted_k141_16796 | Not-determined |
| S2_extracted_k141_16798 | Not-determined |
| S2_extracted_k141_1622 | Not-determined |
| S2_extracted_k141_317 | Not-determined |
| S2_extracted_k141_11325 | Not-determined |
| S2_extracted_k141_18036 | Not-determined |
| S2_extracted_k141_18037 | Not-determined |
| S2_extracted_k141_18038 | Not-determined |
| S2_extracted_k141_1627 | Not-determined |
| S2_extracted_k141_16802 | Not-determined |
| S2_extracted_k141_1630 | Not-determined |
| S2_extracted_k141_11329 | Not-determined |
| S2_extracted_k141_11333 | Not-determined |
| S2_extracted_k141_1635 | Not-determined |
| S2_extracted_k141_16809 | Low-quality |
| S2_extracted_k141_18046 | Low-quality |
| S2_extracted_k141_12752 | Low-quality |
| S2_extracted_k141_330 | Not-determined |
| S2_extracted_k141_18048 | Not-determined |
| S2_extracted_k141_9947 | Not-determined |
| S2_extracted_k141_8470 | Not-determined |
| S2_extracted_k141_12754 | Not-determined |
| S2_extracted_k141_18051 | Low-quality |
| S2_extracted_k141_18052 | Not-determined |
| S2_extracted_k141_14067 | Low-quality |
| S2_extracted_k141_8473 | Low-quality |
| S2_extracted_k141_8476 | Not-determined |
| S2_extracted_k141_337 | Low-quality |
| S2_extracted_k141_8481 | Not-determined |
| S2_extracted_k141_18061 | Not-determined |
| S2_extracted_k141_16815 | Not-determined |
| S2_extracted_k141_8487 | Not-determined |
| S2_extracted_k141_343 | Not-determined |
| S2_extracted_k141_18064 | Not-determined |
| S2_extracted_k141_18065 | Not-determined |
| S2_extracted_k141_1646 | Not-determined |
| S2_extracted_k141_9954 | Not-determined |
| S2_extracted_k141_18067 | Low-quality |
| S2_extracted_k141_16822 | Not-determined |
| S2_extracted_k141_16824 | Not-determined |
| S2_extracted_k141_8491 | Not-determined |
| S2_extracted_k141_16825 | Not-determined |
| S2_extracted_k141_4545 | Low-quality |
| S2_extracted_k141_1653 | Not-determined |
| S2_extracted_k141_12767 | Low-quality |
| S2_extracted_k141_1654 | Not-determined |
| S2_extracted_k141_19528 | Not-determined |
| S2_extracted_k141_18073 | Not-determined |
| S2_extracted_k141_20743 | Low-quality |
| S2_extracted_k141_19531 | Not-determined |
| S2_extracted_k141_18075 | Not-determined |
| S2_extracted_k141_18077 | Not-determined |
| S2_extracted_k141_12769 | Not-determined |
| S2_extracted_k141_8498 | Not-determined |
| S2_extracted_k141_18079 | Low-quality |
| S2_extracted_k141_19534 | Not-determined |
| S2_extracted_k141_18080 | Not-determined |
| S2_extracted_k141_1662 | Not-determined |
| S2_extracted_k141_12772 | Not-determined |
| S2_extracted_k141_4555 | Not-determined |
| S2_extracted_k141_18083 | Low-quality |
| S2_extracted_k141_4557 | Not-determined |
| S2_extracted_k141_8506 | Not-determined |
| S2_extracted_k141_18084 | Not-determined |
| S2_extracted_k141_12776 | Not-determined |
| S2_extracted_k141_4566 | Not-determined |
| S2_extracted_k141_11341 | Not-determined |
| S2_extracted_k141_18095 | Not-determined |
| S2_extracted_k141_8513 | Not-determined |
| S2_extracted_k141_8516 | Not-determined |
| S2_extracted_k141_9957 | Low-quality |
| S2_extracted_k141_7292 | Not-determined |
| S2_extracted_k141_357 | Not-determined |
| S2_extracted_k141_18103 | Not-determined |
| S2_extracted_k141_14072 | Not-determined |
| S2_extracted_k141_4578 | Not-determined |
| S2_extracted_k141_3146 | Not-determined |
| S2_extracted_k141_1672 | Not-determined |
| S9_vs_k141_14235\|\|full | Medium-quality |
| S2_extracted_k141_3148 | Not-determined |
| S2_extracted_k141_11357 | Not-determined |
| S2_extracted_k141_12786 | Not-determined |
| S2_extracted_k141_15453 | Low-quality |
| S2_extracted_k141_15454 | Not-determined |
| S2_extracted_k141_8521 | Low-quality |
| S2_extracted_k141_12790 | Not-determined |
| S2_extracted_k141_9972 | Not-determined |
| S2_extracted_k141_12791 | Not-determined |
| S2_extracted_k141_15458 | Low-quality |
| S2_extracted_k141_12797 | Not-determined |
| S2_extracted_k141_12798 | Not-determined |
| S2_extracted_k141_18118 | Not-determined |
| S2_extracted_k141_14084 | Not-determined |
| S2_extracted_k141_9977 | Not-determined |
| S2_extracted_k141_11364 | Low-quality |
| S2_extracted_k141_5788 | Not-determined |
| S2_extracted_k141_9980 | Low-quality |
| S2_extracted_k141_11365 | Low-quality |
| S2_extracted_k141_11366 | Not-determined |
| S2_extracted_k141_11367 | Not-determined |
| S2_extracted_k141_18125 | Low-quality |
| S2_extracted_k141_5792 | Not-determined |
| S2_extracted_k141_18126 | Not-determined |
| S2_extracted_k141_18129 | Not-determined |
| S2_extracted_k141_9996 | Not-determined |
| S2_extracted_k141_9997 | Not-determined |
| S2_extracted_k141_14096 | Not-determined |
| S2_extracted_k141_12822 | Not-determined |
| S2_extracted_k141_14098 | Not-determined |
| S2_extracted_k141_18137 | Low-quality |
| S2_extracted_k141_3187 | Not-determined |
| S2_extracted_k141_3189 | Not-determined |
| S2_extracted_k141_15469 | Not-determined |
| S2_extracted_k141_12826 | Not-determined |
| S2_extracted_k141_12828 | Not-determined |
| S2_extracted_k141_20770 | Not-determined |
| S2_extracted_k141_10005 | Not-determined |
| S2_extracted_k141_358 | Not-determined |
| S2_extracted_k141_360 | Not-determined |
| S2_extracted_k141_4593 | Not-determined |
| S2_extracted_k141_15477 | Not-determined |
| S2_extracted_k141_4595 | Not-determined |
| S2_extracted_k141_365 | Not-determined |
| S2_extracted_k141_372 | Not-determined |
| S2_extracted_k141_4604 | Not-determined |
| S2_extracted_k141_15486 | Low-quality |
| S2_extracted_k141_10019 | Not-determined |
| S2_extracted_k141_10020 | Not-determined |
| S2_extracted_k141_15492 | Not-determined |
| S2_extracted_k141_11390 | Not-determined |
| S2_extracted_k141_11391 | Not-determined |
| S2_extracted_k141_12849 | Not-determined |
| S2_extracted_k141_4611 | Low-quality |
| S2_extracted_k141_15497 | Low-quality |
| S2_extracted_k141_15499 | Not-determined |
| S2_extracted_k141_1682 | Not-determined |
| S2_extracted_k141_3192 | Low-quality |
| S2_extracted_k141_18168 | Not-determined |
| S2_extracted_k141_18174 | Not-determined |
| S2_extracted_k141_1689 | Not-determined |
| S2_extracted_k141_1691 | Not-determined |
| S2_extracted_k141_15507 | Not-determined |
| S2_extracted_k141_1692 | Low-quality |
| S2_extracted_k141_15508 | Not-determined |
| S2_extracted_k141_15511 | Low-quality |
| S2_extracted_k141_15512 | Not-determined |
| S2_extracted_k141_15517 | Not-determined |
| S2_extracted_k141_8542 | Not-determined |
| S2_extracted_k141_5796 | Not-determined |
| S2_extracted_k141_15522 | Not-determined |
| S2_extracted_k141_10026 | Low-quality |
| S2_extracted_k141_14117 | Not-determined |
| S2_extracted_k141_14120 | Not-determined |
| S2_extracted_k141_5806 | Not-determined |
| S2_extracted_k141_16846 | Not-determined |
| S2_extracted_k141_15529 | Not-determined |
| S2_extracted_k141_5809 | Low-quality |
| S2_extracted_k141_15533 | Not-determined |
| S2_extracted_k141_14126 | Low-quality |
| S2_extracted_k141_14127 | Not-determined |
| S2_extracted_k141_14128 | Low-quality |
| S11_vs_k141_30739\|\|full | Medium-quality |
| S2_extracted_k141_16853 | Not-determined |
| S2_extracted_k141_5819 | Not-determined |
| S2_extracted_k141_16854 | Not-determined |
| S2_extracted_k141_19552 | Not-determined |
| S2_extracted_k141_14135 | Not-determined |
| S2_extracted_k141_19554 | Not-determined |
| S2_extracted_k141_15537 | Not-determined |
| S2_extracted_k141_14136 | Not-determined |
| S2_extracted_k141_14137 | Not-determined |
| S2_extracted_k141_4631 | Not-determined |
| S2_extracted_k141_19557 | Low-quality |
| S2_extracted_k141_5826 | Low-quality |
| S2_extracted_k141_14139 | Not-determined |
| S2_extracted_k141_10031 | Not-determined |
| S2_extracted_k141_12863 | Not-determined |
| S2_extracted_k141_16858 | Not-determined |
| S2_extracted_k141_5827 | Not-determined |
| S2_extracted_k141_12864 | Low-quality |
| S2_extracted_k141_8557 | Not-determined |
| S2_extracted_k141_14145 | Not-determined |
| S2_extracted_k141_8562 | Not-determined |
| S2_extracted_k141_14149 | Not-determined |
| S2_extracted_k141_10046 | Not-determined |
| S2_extracted_k141_10047 | Not-determined |
| S2_extracted_k141_12876 | Low-quality |
| S2_extracted_k141_14150 | Low-quality |
| S2_extracted_k141_12878 | Low-quality |
| S2_extracted_k141_4640 | Not-determined |
| S2_extracted_k141_4642 | Not-determined |
| S2_extracted_k141_7312 | Not-determined |
| S2_extracted_k141_12881 | Not-determined |
| S2_extracted_k141_381 | Not-determined |
| S2_extracted_k141_10054 | Low-quality |
| S2_extracted_k141_3215 | Not-determined |
| S2_extracted_k141_8564 | Not-determined |
| S2_extracted_k141_8565 | Not-determined |
| S2_extracted_k141_5840 | Not-determined |
| S2_extracted_k141_12889 | Low-quality |
| S2_extracted_k141_14158 | Not-determined |
| S2_extracted_k141_8568 | Not-determined |
| S2_extracted_k141_8569 | Not-determined |
| S2_extracted_k141_14161 | Not-determined |
| S2_extracted_k141_10061 | Low-quality |
| S2_extracted_k141_1699 | Not-determined |
| S2_extracted_k141_5845 | Not-determined |
| S2_extracted_k141_11395 | Not-determined |
| S2_extracted_k141_10063 | Not-determined |
| S2_extracted_k141_14164 | Not-determined |
| S2_extracted_k141_12892 | Low-quality |
| S2_extracted_k141_14167 | Low-quality |
| S2_extracted_k141_1706 | Not-determined |
| S2_extracted_k141_16879 | Not-determined |
| S2_extracted_k141_8577 | Not-determined |
| S2_extracted_k141_16880 | Not-determined |
| S2_extracted_k141_15556 | Low-quality |
| S2_extracted_k141_14169 | Not-determined |
| S2_extracted_k141_19570 | Not-determined |
| S2_extracted_k141_3230 | Low-quality |
| S2_extracted_k141_12899 | Not-determined |
| S2_extracted_k141_19571 | Not-determined |
| S2_extracted_k141_15562 | Not-determined |
| S2_extracted_k141_12903 | Low-quality |
| S2_extracted_k141_401 | Low-quality |
| S2_extracted_k141_1712 | Not-determined |
| S2_extracted_k141_15563 | Not-determined |
| S2_extracted_k141_11406 | Not-determined |
| S2_extracted_k141_5858 | Low-quality |
| S2_extracted_k141_16884 | Not-determined |
| S2_extracted_k141_19578 | Not-determined |
| S2_extracted_k141_19579 | Not-determined |
| S2_extracted_k141_12907 | Not-determined |
| S2_extracted_k141_7325 | Not-determined |
| S2_extracted_k141_11412 | Not-determined |
| S2_extracted_k141_16888 | Not-determined |
| S2_extracted_k141_16889 | Not-determined |
| S2_extracted_k141_8590 | Not-determined |
| S2_extracted_k141_19590 | Not-determined |
| S2_extracted_k141_409 | Not-determined |
| S2_extracted_k141_12914 | Not-determined |
| S2_extracted_k141_19593 | Not-determined |
| S2_extracted_k141_16895 | Low-quality |
| S2_extracted_k141_4648 | Not-determined |
| S2_extracted_k141_413 | Not-determined |
| S2_extracted_k141_3236 | Not-determined |
| S2_extracted_k141_14177 | Not-determined |
| S2_extracted_k141_19600 | Not-determined |
| S2_extracted_k141_18214 | Not-determined |
| S2_extracted_k141_4651 | Not-determined |
| S2_extracted_k141_8599 | Not-determined |
| S2_extracted_k141_7336 | Not-determined |
| S2_extracted_k141_4653 | Not-determined |
| S2_extracted_k141_10069 | Low-quality |
| S2_extracted_k141_18219 | Not-determined |
| S2_extracted_k141_7339 | Not-determined |
| S2_extracted_k141_14184 | Low-quality |
| S2_extracted_k141_18222 | Not-determined |
| S2_extracted_k141_16899 | Not-determined |
| S2_extracted_k141_5864 | Not-determined |
| S2_extracted_k141_15583 | Not-determined |
| S2_extracted_k141_12929 | Not-determined |
| S2_extracted_k141_5869 | Low-quality |
| S2_extracted_k141_15584 | Low-quality |
| S2_extracted_k141_10079 | Not-determined |
| S2_extracted_k141_12932 | Not-determined |
| S2_extracted_k141_11419 | Not-determined |
| S2_extracted_k141_5873 | Not-determined |
| S2_extracted_k141_12934 | Not-determined |
| S2_extracted_k141_12935 | Low-quality |
| S2_extracted_k141_3243 | Not-determined |
| S2_extracted_k141_5879 | Low-quality |
| S2_extracted_k141_18240 | Not-determined |
| S2_extracted_k141_5882 | Not-determined |
| S2_extracted_k141_3247 | Low-quality |
| S2_extracted_k141_15590 | Not-determined |
| S2_extracted_k141_20775 | Low-quality |
| S2_extracted_k141_12937 | Not-determined |
| S2_extracted_k141_15594 | Not-determined |
| S2_extracted_k141_3253 | Not-determined |
| S2_extracted_k141_7355 | Low-quality |
| S2_extracted_k141_3254 | Not-determined |
| S2_extracted_k141_15599 | Low-quality |
| S2_extracted_k141_16900 | Low-quality |
| S2_extracted_k141_15602 | Not-determined |
| S2_extracted_k141_15603 | Not-determined |
| S2_extracted_k141_19621 | Low-quality |
| S2_extracted_k141_426 | Low-quality |
| S2_extracted_k141_7361 | Low-quality |
| S2_extracted_k141_427 | Low-quality |
| S2_extracted_k141_1735 | Low-quality |
| S2_extracted_k141_16902 | Low-quality |
| S2_extracted_k141_10089 | Not-determined |
| S2_extracted_k141_4674 | Not-determined |
| S2_extracted_k141_5897 | Low-quality |
| S2_extracted_k141_10096 | Low-quality |
| S2_extracted_k141_4677 | Not-determined |
| S2_extracted_k141_4679 | Not-determined |
| S2_extracted_k141_15614 | Not-determined |
| S2_extracted_k141_15616 | Not-determined |
| S2_extracted_k141_18246 | Not-determined |
| S2_extracted_k141_19626 | Low-quality |
| S2_extracted_k141_4684 | Low-quality |
| S2_extracted_k141_5909 | Low-quality |
| S2_extracted_k141_4685 | Low-quality |
| S2_extracted_k141_12948 | Not-determined |
| S2_extracted_k141_7367 | Not-determined |
| S2_extracted_k141_3276 | Low-quality |
| S2_extracted_k141_20776 | Not-determined |
| S2_extracted_k141_3277 | Not-determined |
| S2_extracted_k141_20779 | Not-determined |
| S2_extracted_k141_3282 | Low-quality |
| S2_extracted_k141_11422 | Low-quality |
| S2_extracted_k141_15629 | Not-determined |
| S2_extracted_k141_7369 | Not-determined |
| S2_extracted_k141_437 | Low-quality |
| S2_extracted_k141_12953 | Not-determined |
| S2_extracted_k141_5918 | Not-determined |
| S2_extracted_k141_439 | Not-determined |
| S2_extracted_k141_5919 | Not-determined |
| S2_extracted_k141_11431 | Not-determined |
| S2_extracted_k141_447 | Low-quality |
| S2_extracted_k141_448 | Not-determined |
| S2_extracted_k141_449 | Not-determined |
| S2_extracted_k141_10111 | Not-determined |
| S2_extracted_k141_4696 | Not-determined |
| S2_extracted_k141_19636 | Low-quality |
| S2_extracted_k141_5927 | Not-determined |
| S2_extracted_k141_14213 | Not-determined |
| S2_extracted_k141_15643 | Low-quality |
| S2_extracted_k141_5932 | Not-determined |
| S2_extracted_k141_11437 | Not-determined |
| S2_extracted_k141_15647 | Not-determined |
| S2_extracted_k141_4699 | Not-determined |
| S2_extracted_k141_14217 | Low-quality |
| S2_extracted_k141_19645 | Low-quality |
| S2_extracted_k141_4700 | Not-determined |
| S2_extracted_k141_19647 | Not-determined |
| S2_extracted_k141_16913 | Not-determined |
| S2_extracted_k141_15652 | Not-determined |
| S2_extracted_k141_8624 | Not-determined |
| S2_extracted_k141_19648 | Low-quality |
| S2_extracted_k141_11441 | Not-determined |
| S2_extracted_k141_4704 | Not-determined |
| S2_extracted_k141_14224 | Not-determined |
| S2_extracted_k141_14226 | Not-determined |
| S2_extracted_k141_19652 | Low-quality |
| S2_extracted_k141_15660 | Not-determined |
| S2_extracted_k141_12965 | Not-determined |
| S2_extracted_k141_14227 | Low-quality |
| S2_extracted_k141_1737 | Not-determined |
| S2_extracted_k141_8634 | Not-determined |
| S2_extracted_k141_8635 | Low-quality |
| S2_extracted_k141_14228 | Not-determined |
| S2_extracted_k141_18274 | Low-quality |
| S2_extracted_k141_19661 | Not-determined |
| S2_extracted_k141_4710 | Low-quality |
| S2_extracted_k141_12971 | Not-determined |
| S2_extracted_k141_7378 | Not-determined |
| S2_extracted_k141_14229 | Not-determined |
| S2_extracted_k141_4712 | Not-determined |
| S2_extracted_k141_12974 | Not-determined |
| S2_extracted_k141_5949 | Not-determined |
| S2_extracted_k141_12977 | Not-determined |
| S2_extracted_k141_15666 | Not-determined |
| S2_extracted_k141_471 | Not-determined |
| S2_extracted_k141_1745 | Not-determined |
| S2_extracted_k141_5952 | Not-determined |
| S2_extracted_k141_12981 | Not-determined |
| S2_extracted_k141_15667 | Not-determined |
| S2_extracted_k141_18287 | Not-determined |
| S2_extracted_k141_18288 | Not-determined |
| S2_extracted_k141_8647 | Not-determined |
| S2_extracted_k141_11461 | Not-determined |
| S2_extracted_k141_7385 | Not-determined |
| S2_extracted_k141_15675 | Not-determined |
| S2_extracted_k141_18292 | Not-determined |
| S2_extracted_k141_8650 | Not-determined |
| S2_extracted_k141_8651 | Not-determined |
| S2_extracted_k141_15676 | Not-determined |
| S2_extracted_k141_5965 | Not-determined |
| S2_extracted_k141_19681 | Not-determined |
| S2_extracted_k141_4720 | Not-determined |
| S2_extracted_k141_19683 | Not-determined |
| S2_extracted_k141_18295 | Not-determined |
| S2_extracted_k141_5967 | Not-determined |
| S2_extracted_k141_16915 | Low-quality |
| S2_extracted_k141_7389 | Not-determined |
| S2_extracted_k141_5969 | Low-quality |
| S2_extracted_k141_11464 | Not-determined |
| S2_extracted_k141_3286 | Not-determined |
| S2_extracted_k141_11466 | Not-determined |
| S2_extracted_k141_11467 | Not-determined |
| S2_extracted_k141_16920 | Not-determined |
| S2_extracted_k141_11468 | Not-determined |
| S2_extracted_k141_5970 | Not-determined |
| S2_extracted_k141_8660 | Not-determined |
| S2_extracted_k141_3294 | Not-determined |
| S2_extracted_k141_3295 | Not-determined |
| S2_extracted_k141_5975 | Not-determined |
| S2_extracted_k141_3297 | Not-determined |
| S2_extracted_k141_18306 | Low-quality |
| S2_extracted_k141_19690 | Not-determined |
| S2_extracted_k141_5976 | Low-quality |
| S2_extracted_k141_15688 | Not-determined |
| S2_extracted_k141_7399 | Not-determined |
| S2_extracted_k141_3298 | Not-determined |
| S2_extracted_k141_19693 | Not-determined |
| S2_extracted_k141_3299 | Not-determined |
| S2_extracted_k141_4721 | Low-quality |
| S2_extracted_k141_5978 | Not-determined |
| S2_extracted_k141_4722 | Low-quality |
| S2_extracted_k141_19696 | Not-determined |
| S2_extracted_k141_15697 | Not-determined |
| S2_extracted_k141_14241 | Low-quality |
| S2_extracted_k141_16930 | Not-determined |
| S2_extracted_k141_5982 | Not-determined |
| S2_extracted_k141_3305 | Not-determined |
| S2_extracted_k141_19701 | Not-determined |
| S2_extracted_k141_5983 | Low-quality |
| S2_extracted_k141_15702 | Not-determined |
| S2_extracted_k141_1756 | Not-determined |
| S2_extracted_k141_5987 | Not-determined |
| S2_extracted_k141_19706 | Low-quality |
| S2_extracted_k141_4728 | Low-quality |
| S2_extracted_k141_20786 | Not-determined |
| S2_extracted_k141_18310 | Not-determined |
| S2_extracted_k141_18311 | Not-determined |
| S2_extracted_k141_18312 | Not-determined |
| S2_extracted_k141_11484 | Not-determined |
| S2_extracted_k141_16938 | Low-quality |
| S2_extracted_k141_3311 | Not-determined |
| S2_extracted_k141_4729 | Not-determined |
| S2_extracted_k141_20789 | Low-quality |
| S2_extracted_k141_4730 | Low-quality |
| S2_extracted_k141_16940 | Not-determined |
| S2_extracted_k141_11486 | Not-determined |
| S2_extracted_k141_14248 | Low-quality |
| S2_extracted_k141_16941 | Not-determined |
| S2_extracted_k141_3316 | Not-determined |
| S2_extracted_k141_8690 | Not-determined |
| S2_extracted_k141_8691 | Not-determined |
| S2_extracted_k141_15713 | Not-determined |
| S2_extracted_k141_12988 | Not-determined |
| S2_extracted_k141_18321 | Low-quality |
| S2_extracted_k141_15714 | Low-quality |
| S2_extracted_k141_3320 | Not-determined |
| S2_extracted_k141_18324 | Not-determined |

| | |
|---|---|
| S2_extracted_k141_20796 | Low-quality |
| S2_extracted_k141_14252 | Not-determined |
| S2_extracted_k141_15718 | Not-determined |
| S2_extracted_k141_20799 | Not-determined |
| S2_extracted_k141_11491 | Not-determined |
| S2_extracted_k141_19717 | Not-determined |
| S2_extracted_k141_3323 | Not-determined |
| S2_extracted_k141_14255 | Not-determined |
| S2_extracted_k141_4739 | Not-determined |
| S2_extracted_k141_7417 | Not-determined |
| S2_extracted_k141_4741 | Not-determined |
| S2_extracted_k141_14258 | Not-determined |
| S2_extracted_k141_19722 | Not-determined |
| S2_extracted_k141_19723 | Not-determined |
| S2_extracted_k141_15723 | Not-determined |
| S2_extracted_k141_14259 | Not-determined |
| S2_extracted_k141_18335 | Low-quality |
| S2_extracted_k141_6007 | Not-determined |
| S2_extracted_k141_6010 | Not-determined |
| S2_extracted_k141_11498 | Not-determined |
| S2_extracted_k141_1776 | Low-quality |
| S2_extracted_k141_12999 | Low-quality |
| S2_extracted_k141_14262 | Not-determined |
| S2_extracted_k141_6015 | Not-determined |
| S2_extracted_k141_8703 | Not-determined |
| S2_extracted_k141_7420 | Not-determined |
| S2_extracted_k141_11502 | Not-determined |
| S2_extracted_k141_4743 | Not-determined |
| S2_extracted_k141_18344 | Not-determined |
| S2_extracted_k141_18346 | Not-determined |
| S2_extracted_k141_19740 | Not-determined |
| S2_extracted_k141_20809 | Low-quality |
| S2_extracted_k141_480 | Not-determined |
| S2_extracted_k141_14266 | Low-quality |
| S2_extracted_k141_20811 | Not-determined |
| S2_extracted_k141_14267 | Not-determined |
| S2_extracted_k141_16947 | Low-quality |
| S2_extracted_k141_19745 | Not-determined |
| S2_extracted_k141_4749 | Low-quality |
| S2_extracted_k141_16948 | Low-quality |
| S2_extracted_k141_20815 | Not-determined |
| S2_extracted_k141_8714 | Not-determined |
| S2_extracted_k141_14271 | Not-determined |
| S2_extracted_k141_14272 | Not-determined |
| S2_extracted_k141_6025 | Not-determined |
| S2_extracted_k141_1794 | Not-determined |
| S2_extracted_k141_1795 | Not-determined |
| S2_extracted_k141_11512 | Not-determined |
| S2_extracted_k141_11513 | Not-determined |
| S2_extracted_k141_4755 | Not-determined |
| S2_extracted_k141_15730 | Not-determined |
| S2_extracted_k141_8721 | Not-determined |
| S2_extracted_k141_1799 | Low-quality |
| S2_extracted_k141_4757 | Not-determined |
| S2_extracted_k141_20824 | Low-quality |
| S2_extracted_k141_16951 | Not-determined |
| S2_extracted_k141_6035 | Low-quality |
| S2_extracted_k141_8726 | Not-determined |
| S2_extracted_k141_13014 | Not-determined |
| S2_extracted_k141_8727 | Low-quality |
| S2_extracted_k141_11518 | Low-quality |
| S2_extracted_k141_16954 | Not-determined |
| S2_extracted_k141_18356 | Low-quality |
| S2_extracted_k141_20829 | Not-determined |
| S2_extracted_k141_3342 | Not-determined |
| S2_extracted_k141_497 | Low-quality |
| S2_extracted_k141_16959 | Not-determined |
| S2_extracted_k141_20832 | Not-determined |
| S2_extracted_k141_6045 | Not-determined |
| S2_extracted_k141_18361 | Not-determined |
| S2_extracted_k141_18363 | Not-determined |
| S2_extracted_k141_6048 | Not-determined |
| S2_extracted_k141_18364 | Not-determined |
| S2_extracted_k141_11526 | Not-determined |
| S2_extracted_k141_506 | Not-determined |
| S2_extracted_k141_13024 | Not-determined |
| S2_extracted_k141_11527 | Low-quality |
| S2_extracted_k141_14284 | Not-determined |
| S2_extracted_k141_6055 | Not-determined |
| S2_extracted_k141_511 | Not-determined |
| S2_extracted_k141_18370 | Not-determined |
| S2_extracted_k141_11532 | Low-quality |
| S2_extracted_k141_18371 | Not-determined |
| S2_extracted_k141_18372 | Low-quality |
| S2_extracted_k141_13025 | Low-quality |
| S2_extracted_k141_8746 | Not-determined |
| S2_extracted_k141_13029 | Not-determined |
| S2_extracted_k141_8751 | Not-determined |
| S2_extracted_k141_18377 | Low-quality |
| S2_extracted_k141_15747 | Not-determined |
| S2_extracted_k141_14289 | Not-determined |
| S2_extracted_k141_15750 | Low-quality |
| S2_extracted_k141_18380 | Not-determined |
| S2_extracted_k141_18381 | Not-determined |
| S2_extracted_k141_7442 | Not-determined |
| S2_extracted_k141_8755 | Low-quality |
| S2_extracted_k141_13035 | Not-determined |
| S2_extracted_k141_19752 | Not-determined |
| S2_extracted_k141_7443 | Not-determined |
| S2_extracted_k141_19754 | Not-determined |
| S2_extracted_k141_14293 | Not-determined |
| S2_extracted_k141_13039 | Low-quality |
| S2_extracted_k141_19757 | Not-determined |
| S2_extracted_k141_7446 | Not-determined |
| S2_extracted_k141_18386 | Not-determined |
| S2_extracted_k141_8758 | Not-determined |
| S2_extracted_k141_8760 | Not-determined |
| S2_extracted_k141_6065 | Not-determined |
| S2_extracted_k141_515 | Low-quality |
| S2_extracted_k141_13042 | Not-determined |
| S2_extracted_k141_13043 | Low-quality |
| S2_extracted_k141_13044 | Not-determined |
| S2_extracted_k141_13045 | Not-determined |
| S2_extracted_k141_14299 | Not-determined |
| S2_extracted_k141_13046 | Not-determined |
| S2_extracted_k141_13047 | Not-determined |
| S2_extracted_k141_8769 | Not-determined |
| S2_extracted_k141_19764 | Not-determined |
| S2_extracted_k141_13049 | Not-determined |
| S2_extracted_k141_520 | Not-determined |
| S2_extracted_k141_4769 | Not-determined |
| S2_extracted_k141_13051 | Low-quality |
| S2_extracted_k141_13053 | Not-determined |
| S2_extracted_k141_525 | Not-determined |
| S2_extracted_k141_4772 | Not-determined |
| S16_vs_165662\|\|full | Medium-quality |
| S2_extracted_k141_3357 | Not-determined |
| S2_extracted_k141_19771 | Not-determined |
| S2_extracted_k141_6078 | Not-determined |
| S2_extracted_k141_4774 | Low-quality |
| S2_extracted_k141_15754 | Not-determined |
| S2_extracted_k141_7455 | Not-determined |
| S2_extracted_k141_6084 | Not-determined |
| S2_extracted_k141_4778 | Not-determined |
| S2_extracted_k141_3362 | Low-quality |
| S2_extracted_k141_8784 | Not-determined |
| S2_extracted_k141_8785 | Not-determined |
| S2_extracted_k141_13064 | Not-determined |
| S2_extracted_k141_3365 | Not-determined |
| S2_extracted_k141_11546 | Not-determined |
| S2_extracted_k141_18390 | Not-determined |
| S2_extracted_k141_20834 | Not-determined |
| S2_extracted_k141_20835 | Not-determined |
| S2_extracted_k141_7462 | Not-determined |
| S2_extracted_k141_3366 | Low-quality |
| S2_extracted_k141_3367 | Not-determined |
| S2_extracted_k141_6096 | Low-quality |
| S2_extracted_k141_10125 | Low-quality |
| S2_extracted_k141_6097 | Not-determined |
| S2_extracted_k141_18396 | Not-determined |
| S2_extracted_k141_6098 | Not-determined |
| S2_extracted_k141_18397 | Not-determined |
| S2_extracted_k141_8799 | Not-determined |
| S2_extracted_k141_15764 | Low-quality |
| S2_extracted_k141_7469 | Not-determined |
| S2_extracted_k141_7470 | Low-quality |
| S2_extracted_k141_13069 | Not-determined |
| S2_extracted_k141_20847 | Not-determined |
| S2_extracted_k141_20848 | Not-determined |
| S2_extracted_k141_8803 | Low-quality |
| S2_extracted_k141_14305 | Not-determined |
| S2_extracted_k141_18403 | Not-determined |
| S2_extracted_k141_14306 | Not-determined |
| S2_extracted_k141_11565 | Low-quality |
| S2_extracted_k141_16962 | Low-quality |
| S2_extracted_k141_559 | Not-determined |
| S2_extracted_k141_20853 | Not-determined |
| S2_extracted_k141_13075 | Not-determined |
| S2_extracted_k141_562 | Not-determined |
| S2_extracted_k141_20855 | Not-determined |
| S2_extracted_k141_18409 | Not-determined |
| S2_extracted_k141_4798 | Low-quality |
| S2_extracted_k141_13079 | Not-determined |
| S2_extracted_k141_13080 | Low-quality |
| S2_extracted_k141_8814 | Not-determined |
| S2_extracted_k141_14311 | Not-determined |
| S2_extracted_k141_14312 | Not-determined |
| S2_extracted_k141_16968 | Not-determined |
| S2_extracted_k141_14313 | Not-determined |
| S2_extracted_k141_14314 | Not-determined |
| S2_extracted_k141_13084 | Not-determined |
| S2_extracted_k141_6102 | Low-quality |
| S2_extracted_k141_20863 | Not-determined |
| S2_extracted_k141_1808 | Low-quality |
| S2_extracted_k141_15768 | Low-quality |
| S2_extracted_k141_8823 | Not-determined |
| S2_extracted_k141_15769 | Not-determined |
| S2_extracted_k141_16979 | Not-determined |
| S2_extracted_k141_16980 | Not-determined |
| S2_extracted_k141_577 | Low-quality |
| S2_extracted_k141_16981 | Not-determined |
| S2_extracted_k141_16983 | Not-determined |
| S2_extracted_k141_15773 | Low-quality |
| S2_extracted_k141_6111 | Low-quality |
| S2_extracted_k141_15776 | Not-determined |
| S2_extracted_k141_16985 | Low-quality |
| S2_extracted_k141_14323 | Not-determined |
| S2_extracted_k141_587 | Not-determined |
| S2_extracted_k141_588 | Low-quality |
| S2_extracted_k141_8827 | Not-determined |
| S2_extracted_k141_4814 | Not-determined |
| S2_extracted_k141_8829 | Not-determined |
| S2_extracted_k141_6119 | Not-determined |
| S2_extracted_k141_8831 | Not-determined |
| S2_extracted_k141_6122 | Not-determined |
| S2_extracted_k141_11591 | Not-determined |
| S2_extracted_k141_15788 | Low-quality |
| S2_extracted_k141_8837 | Not-determined |
| S2_extracted_k141_8839 | Not-determined |
| S2_extracted_k141_11600 | Not-determined |
| S2_extracted_k141_4819 | Low-quality |
| S2_extracted_k141_6135 | Not-determined |
| S2_extracted_k141_591 | Low-quality |
| S2_extracted_k141_15796 | Not-determined |
| S2_extracted_k141_15797 | Not-determined |
| S2_extracted_k141_10129 | Not-determined |
| S2_extracted_k141_15800 | Not-determined |
| S2_extracted_k141_11607 | Low-quality |
| S2_extracted_k141_595 | Not-determined |
| S2_extracted_k141_16995 | Not-determined |
| S2_extracted_k141_8847 | Not-determined |
| S2_extracted_k141_19795 | Not-determined |
| S2_extracted_k141_16998 | Not-determined |
| S2_extracted_k141_16999 | Not-determined |
| S2_extracted_k141_596 | Low-quality |
| S2_extracted_k141_3380 | Not-determined |
| S2_extracted_k141_6145 | Low-quality |
| S2_extracted_k141_11610 | Low-quality |
| S2_extracted_k141_597 | Low-quality |
| S2_extracted_k141_600 | Not-determined |
| S2_extracted_k141_19805 | Not-determined |
| S2_extracted_k141_601 | Not-determined |
| S2_extracted_k141_8852 | Low-quality |
| S2_extracted_k141_8853 | Not-determined |
| S2_extracted_k141_17002 | Not-determined |
| S2_extracted_k141_6152 | Not-determined |
| S2_extracted_k141_20878 | Low-quality |
| S2_extracted_k141_3388 | Not-determined |
| S2_extracted_k141_11618 | Not-determined |
| S2_extracted_k141_3389 | Not-determined |
| S2_extracted_k141_8859 | Low-quality |
| S2_extracted_k141_18435 | Not-determined |
| S2_extracted_k141_11619 | Low-quality |
| S2_extracted_k141_6159 | Not-determined |
| S2_extracted_k141_3392 | Low-quality |
| S2_extracted_k141_11621 | Not-determined |
| S2_extracted_k141_8862 | Not-determined |
| S2_extracted_k141_6160 | Low-quality |
| S2_extracted_k141_612 | Not-determined |
| S2_extracted_k141_20883 | Not-determined |
| S2_extracted_k141_18439 | Low-quality |
| S2_extracted_k141_8867 | Not-determined |
| S2_extracted_k141_17011 | Not-determined |
| S2_extracted_k141_14339 | Not-determined |
| S2_extracted_k141_8868 | Low-quality |
| S2_extracted_k141_1840 | Not-determined |
| S2_extracted_k141_17012 | Not-determined |
| S2_extracted_k141_20887 | Not-determined |
| S2_extracted_k141_4833 | Low-quality |
| S2_extracted_k141_20888 | Not-determined |
| S2_extracted_k141_6165 | Not-determined |
| S2_extracted_k141_8873 | Low-quality |
| S2_extracted_k141_6169 | Low-quality |
| S2_extracted_k141_3399 | Not-determined |
| S2_extracted_k141_6171 | Low-quality |
| S2_extracted_k141_17017 | Not-determined |
| S2_extracted_k141_14342 | Low-quality |
| S2_extracted_k141_20894 | Not-determined |
| S2_extracted_k141_6175 | Not-determined |
| S2_extracted_k141_8877 | Not-determined |
| S2_extracted_k141_13094 | Low-quality |
| S2_extracted_k141_3404 | Not-determined |
| S2_extracted_k141_14344 | Not-determined |
| S2_extracted_k141_6179 | Not-determined |
| S2_extracted_k141_8884 | Low-quality |
| S2_extracted_k141_18454 | Not-determined |
| S2_extracted_k141_11643 | Low-quality |
| S2_extracted_k141_20900 | Not-determined |
| S2_extracted_k141_19817 | Not-determined |
| S2_extracted_k141_11645 | Not-determined |
| S2_extracted_k141_14352 | Not-determined |
| S2_extracted_k141_18458 | Not-determined |
| S2_extracted_k141_15812 | Not-determined |
| S2_extracted_k141_15813 | Not-determined |
| S2_extracted_k141_17023 | Not-determined |
| S2_extracted_k141_6186 | Not-determined |
| S2_extracted_k141_19821 | Not-determined |
| S2_extracted_k141_8890 | Not-determined |
| S2_extracted_k141_19824 | Not-determined |
| S2_extracted_k141_19825 | Low-quality |
| S2_extracted_k141_10143 | Not-determined |
| S2_extracted_k141_15819 | Not-determined |
| S2_extracted_k141_10144 | Not-determined |
| S2_extracted_k141_11650 | Low-quality |
| S2_extracted_k141_10146 | Low-quality |
| S2_extracted_k141_11651 | Not-determined |
| S2_extracted_k141_10147 | Not-determined |
| S2_extracted_k141_6194 | Not-determined |
| S2_extracted_k141_639 | Not-determined |
| S2_extracted_k141_8897 | Not-determined |
| S2_extracted_k141_10152 | Low-quality |
| S2_extracted_k141_17032 | Not-determined |
| S2_extracted_k141_17033 | Not-determined |
| S2_extracted_k141_6197 | Low-quality |
| S2_extracted_k141_643 | Not-determined |
| S2_extracted_k141_1863 | Low-quality |
| S2_extracted_k141_17036 | Not-determined |
| S2_extracted_k141_14363 | Low-quality |
| S2_extracted_k141_4849 | Not-determined |
| S2_extracted_k141_650 | Low-quality |
| S2_extracted_k141_651 | Not-determined |
| S2_extracted_k141_18462 | Low-quality |
| S2_extracted_k141_1868 | Not-determined |
| S2_extracted_k141_19832 | Not-determined |
| S2_extracted_k141_6200 | Not-determined |
| S2_extracted_k141_17040 | Low-quality |
| S2_extracted_k141_11662 | Low-quality |
| S2_extracted_k141_1871 | Low-quality |
| S2_extracted_k141_13110 | Low-quality |
| S2_extracted_k141_18468 | Not-determined |
| S2_extracted_k141_653 | Not-determined |
| S2_extracted_k141_19839 | Not-determined |
| S2_extracted_k141_7486 | Not-determined |
| S2_extracted_k141_11667 | Low-quality |
| S2_extracted_k141_7488 | Not-determined |
| S2_extracted_k141_18475 | Not-determined |
| S2_extracted_k141_659 | Not-determined |
| S2_extracted_k141_19845 | Not-determined |
| S2_extracted_k141_17048 | Low-quality |
| S2_extracted_k141_18477 | Not-determined |
| S2_extracted_k141_18478 | Not-determined |
| S2_extracted_k141_1883 | Not-determined |
| S2_extracted_k141_17049 | Low-quality |
| S2_extracted_k141_3416 | Not-determined |
| S2_extracted_k141_7492 | Not-determined |
| S2_extracted_k141_11675 | Not-determined |
| S2_extracted_k141_4854 | Not-determined |
| S2_extracted_k141_17053 | Not-determined |
| S2_extracted_k141_10160 | Low-quality |
| S2_extracted_k141_11679 | Not-determined |
| S2_extracted_k141_18487 | Not-determined |
| S2_extracted_k141_1887 | Not-determined |
| S4_vs_VIRSorter_29332-cat_2 | Medium-quality |
| S2_extracted_k141_8899 | Low-quality |
| S2_extracted_k141_1890 | Not-determined |
| S2_extracted_k141_4862 | Not-determined |
| S2_extracted_k141_10164 | Not-determined |
| S2_extracted_k141_14372 | Not-determined |
| S2_extracted_k141_14374 | Not-determined |
| S2_extracted_k141_15837 | Not-determined |
| S2_extracted_k141_18493 | Low-quality |
| S2_extracted_k141_11685 | Not-determined |
| S2_extracted_k141_19850 | Not-determined |
| S2_extracted_k141_18495 | Not-determined |
| S2_extracted_k141_20917 | Not-determined |
| S2_extracted_k141_13117 | Not-determined |
| S2_extracted_k141_19851 | Not-determined |
| S2_extracted_k141_18496 | Not-determined |
| S2_extracted_k141_13119 | Low-quality |
| S2_extracted_k141_19856 | Low-quality |
| S2_extracted_k141_20919 | Not-determined |
| S2_extracted_k141_13123 | Not-determined |
| S2_extracted_k141_20920 | Not-determined |
| S2_extracted_k141_1904 | Not-determined |
| S2_extracted_k141_11687 | Low-quality |
| S2_extracted_k141_1907 | Not-determined |
| S2_extracted_k141_4875 | Not-determined |
| S2_extracted_k141_15838 | Low-quality |
| S2_extracted_k141_20925 | Not-determined |
| S16_vs_26078\|\|full | Medium-quality |
| S2_extracted_k141_19864 | Not-determined |
| S2_extracted_k141_19867 | Not-determined |
| S2_extracted_k141_13136 | Not-determined |
| S2_extracted_k141_11694 | Not-determined |
| S2_extracted_k141_18513 | Not-determined |
| S2_extracted_k141_6215 | Not-determined |
| S2_extracted_k141_13138 | Not-determined |
| S2_extracted_k141_6216 | Low-quality |
| S2_extracted_k141_6217 | Not-determined |
| S2_extracted_k141_17072 | Not-determined |
| S2_extracted_k141_11696 | Not-determined |
| S2_extracted_k141_7508 | Low-quality |
| S2_extracted_k141_10170 | Not-determined |
| S2_extracted_k141_13142 | Not-determined |
| S2_extracted_k141_7509 | Not-determined |
| S2_extracted_k141_6221 | Low-quality |
| S2_extracted_k141_13144 | Not-determined |
| S2_extracted_k141_19876 | Not-determined |
| S2_extracted_k141_4881 | Not-determined |
| S2_extracted_k141_17074 | Not-determined |
| S2_extracted_k141_8925 | Not-determined |
| S2_extracted_k141_7512 | Low-quality |
| S2_extracted_k141_13146 | Low-quality |
| S2_extracted_k141_6223 | Low-quality |
| S2_extracted_k141_672 | Not-determined |
| S2_extracted_k141_18521 | Low-quality |
| S2_extracted_k141_11700 | Low-quality |
| S2_extracted_k141_10176 | Not-determined |
| S2_extracted_k141_7514 | Not-determined |
| S2_extracted_k141_15844 | Not-determined |
| S2_extracted_k141_11703 | Not-determined |
| S2_extracted_k141_17081 | Not-determined |
| S2_extracted_k141_3436 | Low-quality |
| S2_extracted_k141_6227 | Not-determined |
| S2_extracted_k141_20929 | Not-determined |
| S2_extracted_k141_4887 | Not-determined |
| S2_extracted_k141_3441 | Not-determined |
| S2_extracted_k141_18531 | Not-determined |
| S2_extracted_k141_4888 | Not-determined |
| S2_extracted_k141_17083 | Not-determined |
| S2_extracted_k141_18534 | Not-determined |
| S2_extracted_k141_4890 | Not-determined |
| S2_extracted_k141_18535 | Not-determined |
| S2_extracted_k141_11707 | Not-determined |
| S2_extracted_k141_14377 | Not-determined |
| S2_extracted_k141_8938 | Not-determined |
| S2_extracted_k141_15848 | Not-determined |
| S2_extracted_k141_1942 | Not-determined |
| S2_extracted_k141_6233 | Not-determined |
| S2_extracted_k141_13163 | Not-determined |
| S2_extracted_k141_13164 | Not-determined |
| S2_extracted_k141_10181 | Not-determined |
| S2_extracted_k141_13165 | Not-determined |
| S2_extracted_k141_17089 | Not-determined |
| S2_extracted_k141_17091 | Not-determined |
| S2_extracted_k141_10183 | Not-determined |
| S2_extracted_k141_14381 | Not-determined |
| S2_extracted_k141_17095 | Not-determined |
| S2_extracted_k141_1947 | Not-determined |
| S2_extracted_k141_14382 | Low-quality |
| S2_extracted_k141_18540 | Low-quality |
| S2_extracted_k141_14384 | Low-quality |
| S2_extracted_k141_13178 | Not-determined |
| S2_extracted_k141_8953 | Not-determined |
| S2_extracted_k141_10192 | Not-determined |
| S2_extracted_k141_1951 | Not-determined |
| S2_extracted_k141_8955 | Not-determined |
| S2_extracted_k141_15860 | Not-determined |
| S2_extracted_k141_13181 | Not-determined |
| S2_extracted_k141_15862 | Not-determined |
| S2_extracted_k141_15864 | Low-quality |
| S2_extracted_k141_13187 | Low-quality |
| S2_extracted_k141_15866 | Low-quality |
| S2_extracted_k141_4902 | Not-determined |
| S2_extracted_k141_15868 | Not-determined |
| S2_extracted_k141_13191 | Low-quality |
| S2_extracted_k141_18557 | Not-determined |
| S2_extracted_k141_20938 | Not-determined |
| S2_extracted_k141_11726 | Not-determined |
| S2_extracted_k141_14398 | Low-quality |
| S2_extracted_k141_13195 | Not-determined |
| S2_extracted_k141_15873 | Not-determined |
| S2_extracted_k141_4907 | Low-quality |
| S2_extracted_k141_20945 | Not-determined |
| S2_extracted_k141_4908 | Not-determined |
| S2_extracted_k141_19889 | Not-determined |
| S2_extracted_k141_13197 | Low-quality |
| S2_extracted_k141_14402 | Not-determined |
| S2_extracted_k141_20950 | Not-determined |
| S2_extracted_k141_19895 | Not-determined |
| S2_extracted_k141_17109 | Not-determined |
| S2_extracted_k141_680 | Not-determined |
| S2_extracted_k141_18577 | Not-determined |
| S2_extracted_k141_20957 | Not-determined |
| S2_extracted_k141_13206 | Not-determined |
| S2_extracted_k141_13207 | Not-determined |
| S2_extracted_k141_20959 | Low-quality |
| S2_extracted_k141_13211 | Not-determined |
| S2_extracted_k141_14411 | Not-determined |
| S2_extracted_k141_20962 | Not-determined |
| S2_extracted_k141_17117 | Not-determined |
| S2_extracted_k141_20964 | Not-determined |
| S2_extracted_k141_17122 | Not-determined |
| S2_extracted_k141_20967 | Low-quality |
| S2_extracted_k141_17125 | Not-determined |
| S2_extracted_k141_3457 | Not-determined |
| S2_extracted_k141_17129 | Low-quality |
| S2_extracted_k141_20974 | Not-determined |
| S2_extracted_k141_17130 | Not-determined |
| S2_extracted_k141_14422 | Not-determined |
| S2_extracted_k141_19917 | Not-determined |
| S2_extracted_k141_20979 | Not-determined |
| S2_extracted_k141_8969 | Not-determined |
| S2_extracted_k141_20984 | Not-determined |
| S2_extracted_k141_15888 | Not-determined |
| S2_extracted_k141_17137 | Not-determined |
| S2_extracted_k141_3468 | Not-determined |
| S2_extracted_k141_15890 | Not-determined |
| S2_extracted_k141_15891 | Not-determined |
| S2_extracted_k141_15893 | Not-determined |
| S2_extracted_k141_10209 | Not-determined |
| S2_extracted_k141_8976 | Not-determined |
| S2_extracted_k141_18588 | Not-determined |
| S2_extracted_k141_20991 | Not-determined |
| S2_extracted_k141_20994 | Not-determined |
| S13_extracted_107195 | Medium-quality |
| S2_extracted_k141_8978 | Not-determined |
| S2_extracted_k141_11738 | Low-quality |
| S2_extracted_k141_3476 | Not-determined |
| S2_extracted_k141_11739 | Low-quality |
| S2_extracted_k141_19927 | Not-determined |
| S2_extracted_k141_17147 | Low-quality |
| S2_extracted_k141_21001 | Low-quality |
| S2_extracted_k141_699 | Not-determined |
| S2_extracted_k141_701 | Low-quality |
| S2_extracted_k141_21004 | Low-quality |
| S2_extracted_k141_702 | Not-determined |
| S2_extracted_k141_17149 | Low-quality |
| S2_extracted_k141_19933 | Not-determined |
| S2_extracted_k141_7526 | Not-determined |
| S2_extracted_k141_704 | Not-determined |
| S2_extracted_k141_8985 | Not-determined |
| S2_extracted_k141_8986 | Low-quality |
| S2_extracted_k141_10220 | Not-determined |
| S2_extracted_k141_4922 | Low-quality |
| S2_extracted_k141_14433 | Not-determined |
| S2_extracted_k141_14434 | Not-determined |
| S2_extracted_k141_709 | Low-quality |
| S2_extracted_k141_21011 | Not-determined |
| S2_extracted_k141_19940 | Not-determined |
| S2_extracted_k141_21012 | Not-determined |
| S2_extracted_k141_711 | Low-quality |
| S2_extracted_k141_21014 | Not-determined |
| S2_extracted_k141_21015 | Not-determined |
| S2_extracted_k141_11753 | Low-quality |
| S2_extracted_k141_14440 | Not-determined |
| S2_extracted_k141_717 | Not-determined |
| S2_extracted_k141_19946 | Not-determined |
| S2_extracted_k141_21018 | Not-determined |
| S2_extracted_k141_18597 | Not-determined |
| S2_extracted_k141_18598 | Not-determined |
| S2_extracted_k141_21020 | Not-determined |
| S2_extracted_k141_4932 | Not-determined |
| S2_extracted_k141_19949 | Not-determined |
| S2_extracted_k141_11761 | Not-determined |
| S2_extracted_k141_720 | Low-quality |
| S2_extracted_k141_3491 | Not-determined |
| S2_extracted_k141_4933 | Not-determined |
| S2_extracted_k141_11763 | Not-determined |
| S2_extracted_k141_18600 | Low-quality |
| S2_extracted_k141_19951 | Not-determined |
| S2_extracted_k141_11764 | Low-quality |
| S2_extracted_k141_3494 | Not-determined |
| S2_extracted_k141_3495 | Not-determined |
| S2_extracted_k141_1974 | Not-determined |
| S2_extracted_k141_14446 | Not-determined |
| S2_extracted_k141_21027 | Low-quality |
| S2_extracted_k141_4937 | Not-determined |
| S2_extracted_k141_21028 | Not-determined |
| S2_extracted_k141_13228 | Not-determined |
| S2_extracted_k141_1977 | Low-quality |
| S2_extracted_k141_1978 | Low-quality |
| S2_extracted_k141_3498 | Not-determined |
| S2_extracted_k141_1980 | Not-determined |
| S2_extracted_k141_1982 | Not-determined |
| S2_extracted_k141_3503 | Low-quality |
| S2_extracted_k141_1990 | Not-determined |
| S2_extracted_k141_21033 | Not-determined |
| S2_extracted_k141_21034 | Not-determined |
| S2_extracted_k141_11775 | Not-determined |
| S2_extracted_k141_1991 | Low-quality |
| S2_extracted_k141_7548 | Low-quality |
| S2_extracted_k141_13242 | Not-determined |
| S2_extracted_k141_7552 | Not-determined |
| S2_extracted_k141_10243 | Not-determined |
| S2_extracted_k141_732 | Not-determined |
| S2_extracted_k141_13247 | Not-determined |
| S2_extracted_k141_13249 | Not-determined |
| S2_extracted_k141_3510 | Low-quality |
| S2_extracted_k141_735 | Not-determined |
| S2_extracted_k141_736 | Not-determined |
| S2_extracted_k141_19959 | Not-determined |
| S2_extracted_k141_7558 | Not-determined |
| S2_extracted_k141_18611 | Not-determined |
| S2_extracted_k141_14463 | Low-quality |
| S2_extracted_k141_19962 | Not-determined |
| S2_extracted_k141_741 | Not-determined |
| S2_extracted_k141_18615 | Not-determined |
| S2_extracted_k141_18616 | Low-quality |
| S2_extracted_k141_15946 | Not-determined |
| S2_extracted_k141_2007 | Not-determined |
| S2_extracted_k141_750 | Not-determined |
| S2_extracted_k141_11781 | Low-quality |
| S2_extracted_k141_9009 | Low-quality |
| S2_extracted_k141_13268 | Not-determined |
| S2_extracted_k141_9011 | Not-determined |
| S2_extracted_k141_3526 | Not-determined |
| S2_extracted_k141_756 | Not-determined |
| S2_extracted_k141_757 | Not-determined |

| | |
|---|---|
| S2_extracted_k141_3527 | Low-quality |
| S2_extracted_k141_13272 | Not-determined |
| S2_extracted_k141_11785 | Low-quality |
| S2_extracted_k141_7570 | Not-determined |
| S2_extracted_k141_3529 | Not-determined |
| S2_extracted_k141_763 | Not-determined |
| S2_extracted_k141_13280 | Not-determined |
| S2_extracted_k141_9021 | Low-quality |
| S2_extracted_k141_18628 | Low-quality |
| S2_extracted_k141_18630 | Not-determined |
| S2_extracted_k141_10257 | Not-determined |
| S2_extracted_k141_21054 | Not-determined |
| S2_extracted_k141_10259 | Not-determined |
| S2_extracted_k141_21056 | Not-determined |
| S2_extracted_k141_13284 | Low-quality |
| S2_extracted_k141_3536 | Low-quality |
| S2_extracted_k141_10262 | Not-determined |
| S2_extracted_k141_3537 | Not-determined |
| S2_extracted_k141_10265 | Not-determined |
| S2_extracted_k141_3540 | Not-determined |
| S2_extracted_k141_21059 | Low-quality |
| S2_extracted_k141_11798 | Not-determined |
| S2_extracted_k141_14488 | Low-quality |
| S2_extracted_k141_775 | Not-determined |
| S2_extracted_k141_10267 | Not-determined |
| S2_extracted_k141_10268 | Not-determined |
| S2_extracted_k141_782 | Low-quality |
| S2_extracted_k141_10273 | Not-determined |
| S2_extracted_k141_17155 | Not-determined |
| S2_extracted_k141_21078 | Not-determined |
| S2_extracted_k141_14498 | Not-determined |
| S2_extracted_k141_14499 | Low-quality |
| S2_extracted_k141_3549 | Not-determined |
| S2_extracted_k141_21081 | Not-determined |
| S2_extracted_k141_14500 | Low-quality |
| S2_extracted_k141_14501 | Not-determined |
| S2_extracted_k141_19975 | Low-quality |
| S2_extracted_k141_21085 | Not-determined |
| S2_extracted_k141_14507 | Low-quality |
| S2_extracted_k141_2043 | Low-quality |
| S2_extracted_k141_21087 | Not-determined |
| S2_extracted_k141_790 | Not-determined |
| S2_extracted_k141_10277 | Not-determined |
| S2_extracted_k141_2046 | Not-determined |
| S2_extracted_k141_791 | Not-determined |
| S2_extracted_k141_3558 | Not-determined |
| S2_extracted_k141_11808 | Not-determined |
| S2_extracted_k141_7586 | Not-determined |
| S2_extracted_k141_11810 | Not-determined |
| S2_extracted_k141_4951 | Low-quality |
| S2_extracted_k141_2052 | Not-determined |
| S2_extracted_k141_795 | Not-determined |
| S2_extracted_k141_21090 | Low-quality |
| S2_extracted_k141_9043 | Not-determined |
| S2_extracted_k141_2055 | Low-quality |
| S2_extracted_k141_19988 | Not-determined |
| S2_extracted_k141_800 | Not-determined |
| S2_extracted_k141_802 | Not-determined |
| S2_extracted_k141_19991 | Not-determined |
| S2_extracted_k141_17170 | Not-determined |
| S2_extracted_k141_4958 | Not-determined |
| S2_extracted_k141_14509 | Not-determined |
| S2_extracted_k141_2059 | Not-determined |
| S2_extracted_k141_806 | Not-determined |
| S2_extracted_k141_14510 | Not-determined |
| S2_extracted_k141_10286 | Low-quality |
| S2_extracted_k141_19993 | Not-determined |
| S2_extracted_k141_10287 | Not-determined |
| S2_extracted_k141_17171 | Low-quality |
| S2_extracted_k141_3570 | Not-determined |
| S2_extracted_k141_3571 | Not-determined |
| S2_extracted_k141_810 | Not-determined |
| S2_extracted_k141_14511 | Not-determined |
| S2_extracted_k141_15951 | Low-quality |
| S2_extracted_k141_3577 | Not-determined |
| S2_extracted_k141_817 | Not-determined |
| S2_extracted_k141_818 | Not-determined |
| S2_extracted_k141_19997 | Not-determined |
| S2_extracted_k141_14516 | Not-determined |
| S2_extracted_k141_13288 | Not-determined |
| S2_extracted_k141_19998 | Not-determined |
| S2_extracted_k141_2069 | Not-determined |
| S2_extracted_k141_15958 | Not-determined |
| S2_extracted_k141_20000 | Not-determined |
| S2_extracted_k141_11822 | Low-quality |
| S2_extracted_k141_13290 | Not-determined |
| S2_extracted_k141_4970 | Not-determined |
| S2_extracted_k141_9048 | Not-determined |
| S2_extracted_k141_3585 | Low-quality |
| S2_extracted_k141_17177 | Not-determined |
| S2_extracted_k141_9049 | Low-quality |
| S2_extracted_k141_15964 | Not-determined |
| S2_extracted_k141_11824 | Not-determined |
| S2_extracted_k141_20004 | Low-quality |
| S2_extracted_k141_15966 | Low-quality |
| S2_extracted_k141_9054 | Not-determined |
| S2_extracted_k141_824 | Low-quality |
| S2_extracted_k141_11834 | Not-determined |
| S2_extracted_k141_828 | Not-determined |
| S2_extracted_k141_7597 | Low-quality |
| S2_extracted_k141_4981 | Not-determined |
| S2_extracted_k141_9059 | Low-quality |
| S2_extracted_k141_9060 | Not-determined |
| S2_extracted_k141_833 | Not-determined |
| S2_extracted_k141_9064 | Not-determined |
| S2_extracted_k141_834 | Low-quality |
| S2_extracted_k141_3598 | Not-determined |
| S2_extracted_k141_9068 | Not-determined |
| S2_extracted_k141_15976 | Low-quality |
| S2_extracted_k141_3602 | Not-determined |
| S2_extracted_k141_18636 | Low-quality |
| S2_extracted_k141_9069 | Not-determined |
| S2_extracted_k141_15977 | Not-determined |
| S2_extracted_k141_14548 | Not-determined |
| S2_extracted_k141_11837 | Low-quality |
| S2_extracted_k141_9072 | Not-determined |
| S2_extracted_k141_14550 | Not-determined |
| S2_extracted_k141_3607 | Not-determined |
| S2_extracted_k141_840 | Not-determined |
| S2_extracted_k141_9074 | Low-quality |
| S2_extracted_k141_15983 | Low-quality |
| S2_extracted_k141_17201 | Low-quality |
| S2_extracted_k141_845 | Not-determined |
| S2_extracted_k141_15988 | Not-determined |
| S2_extracted_k141_848 | Not-determined |
| S2_extracted_k141_17204 | Not-determined |
| S2_extracted_k141_2071 | Not-determined |
| S2_extracted_k141_11845 | Not-determined |
| S2_extracted_k141_18647 | Low-quality |
| S2_extracted_k141_3621 | Not-determined |
| S2_extracted_k141_15989 | Not-determined |
| S2_extracted_k141_7616 | Not-determined |
| S2_extracted_k141_855 | Not-determined |
| S2_extracted_k141_15991 | Not-determined |
| S2_extracted_k141_13299 | Not-determined |
| S2_extracted_k141_15993 | Not-determined |
| S2_extracted_k141_9084 | Not-determined |
| S2_extracted_k141_17216 | Not-determined |
| S2_extracted_k141_3627 | Low-quality |
| S2_extracted_k141_9089 | Low-quality |
| S2_extracted_k141_9094 | Not-determined |
| S2_extracted_k141_17221 | Low-quality |
| S2_extracted_k141_17222 | Not-determined |
| S2_extracted_k141_9097 | Low-quality |
| S2_extracted_k141_9098 | Not-determined |
| S2_extracted_k141_3633 | Not-determined |
| S2_extracted_k141_14557 | Not-determined |
| S2_extracted_k141_9100 | Not-determined |
| S2_extracted_k141_9101 | Not-determined |
| S2_extracted_k141_9102 | Not-determined |
| S2_extracted_k141_3637 | Low-quality |
| S2_extracted_k141_9103 | Not-determined |
| S2_extracted_k141_9104 | Not-determined |
| S2_extracted_k141_17224 | Low-quality |
| S2_extracted_k141_14562 | Not-determined |
| S2_extracted_k141_2089 | Not-determined |
| S2_extracted_k141_869 | Not-determined |
| S2_extracted_k141_2090 | Not-determined |
| S2_extracted_k141_4994 | Low-quality |
| S2_extracted_k141_2093 | Low-quality |
| S2_extracted_k141_9105 | Low-quality |
| S2_extracted_k141_17231 | Not-determined |
| S2_extracted_k141_14568 | Not-determined |
| S2_extracted_k141_7636 | Not-determined |
| S2_extracted_k141_3650 | Not-determined |
| S2_extracted_k141_14571 | Not-determined |
| S2_extracted_k141_2105 | Not-determined |
| S2_extracted_k141_17236 | Not-determined |
| S2_extracted_k141_3657 | Not-determined |
| S2_extracted_k141_2108 | Not-determined |
| S2_extracted_k141_17238 | Low-quality |
| S2_extracted_k141_14575 | Low-quality |
| S2_extracted_k141_20040 | Not-determined |
| S2_extracted_k141_9118 | Low-quality |
| S2_extracted_k141_17240 | Not-determined |
| S2_extracted_k141_20044 | Not-determined |
| S2_extracted_k141_2114 | Low-quality |
| S2_extracted_k141_13341 | Low-quality |
| S2_extracted_k141_11859 | Low-quality |
| S2_extracted_k141_20047 | Not-determined |
| S2_extracted_k141_20048 | Not-determined |
| S2_extracted_k141_20049 | Not-determined |
| S2_extracted_k141_9126 | Not-determined |
| S2_extracted_k141_14583 | Not-determined |
| S2_extracted_k141_21105 | Not-determined |
| S2_extracted_k141_20053 | Low-quality |
| S2_extracted_k141_897 | Not-determined |
| S2_extracted_k141_11863 | Not-determined |
| S2_extracted_k141_2123 | Not-determined |
| S2_extracted_k141_15997 | Not-determined |
| S2_extracted_k141_20054 | Low-quality |
| S2_extracted_k141_20059 | Not-determined |
| S2_extracted_k141_7645 | Not-determined |
| S2_extracted_k141_13358 | Low-quality |
| S2_extracted_k141_21109 | Low-quality |
| S2_extracted_k141_14593 | Not-determined |
| S2_extracted_k141_21116 | Low-quality |
| S2_extracted_k141_10290 | Not-determined |
| S2_extracted_k141_10292 | Not-determined |
| S2_extracted_k141_14600 | Not-determined |
| S2_extracted_k141_18653 | Low-quality |
| S2_extracted_k141_18654 | Not-determined |
| S2_extracted_k141_14602 | Not-determined |
| S2_extracted_k141_3661 | Low-quality |
| S2_extracted_k141_20068 | Not-determined |
| S2_extracted_k141_20069 | Not-determined |
| S2_extracted_k141_13372 | Not-determined |
| S2_extracted_k141_18663 | Not-determined |
| S2_extracted_k141_7657 | Not-determined |
| S2_extracted_k141_3668 | Low-quality |
| S2_extracted_k141_18666 | Not-determined |
| S2_extracted_k141_7659 | Not-determined |
| S2_extracted_k141_11883 | Not-determined |
| S2_extracted_k141_20076 | Not-determined |
| S2_extracted_k141_21122 | Not-determined |
| S2_extracted_k141_910 | Not-determined |
| S2_extracted_k141_20081 | Not-determined |
| S2_extracted_k141_13376 | Low-quality |
| S2_extracted_k141_9151 | Not-determined |
| S2_extracted_k141_5014 | Not-determined |
| S2_extracted_k141_18674 | Not-determined |
| S2_extracted_k141_11889 | Not-determined |
| S2_extracted_k141_17251 | Low-quality |
| S2_extracted_k141_18675 | Not-determined |
| S2_extracted_k141_5016 | Not-determined |
| S2_extracted_k141_9157 | Not-determined |
| S2_extracted_k141_18678 | Low-quality |
| S2_extracted_k141_2125 | Low-quality |
| S2_extracted_k141_18680 | Low-quality |
| S2_extracted_k141_2127 | Low-quality |
| S2_extracted_k141_9163 | Low-quality |
| S2_extracted_k141_3681 | Low-quality |
| S2_extracted_k141_2128 | Not-determined |
| S2_extracted_k141_9167 | Low-quality |
| S2_extracted_k141_7674 | Low-quality |
| S2_extracted_k141_18690 | Low-quality |
| S2_extracted_k141_9172 | Low-quality |
| S2_extracted_k141_18693 | Low-quality |
| S2_extracted_k141_20096 | Low-quality |
| S2_extracted_k141_11901 | Not-determined |
| S2_extracted_k141_11902 | Not-determined |
| S2_extracted_k141_3694 | Not-determined |
| S2_extracted_k141_5035 | Low-quality |
| S2_extracted_k141_11905 | Not-determined |
| S2_extracted_k141_2137 | Not-determined |
| S2_extracted_k141_18702 | Not-determined |
| S2_extracted_k141_9183 | Not-determined |
| S2_extracted_k141_14608 | Low-quality |
| S2_extracted_k141_16002 | Not-determined |
| S2_extracted_k141_9184 | Not-determined |
| S2_extracted_k141_3696 | Low-quality |
| S2_extracted_k141_3700 | Not-determined |
| S2_extracted_k141_14613 | Low-quality |
| S2_extracted_k141_3701 | Not-determined |
| S2_extracted_k141_5044 | Low-quality |
| S2_extracted_k141_10297 | Not-determined |
| S2_extracted_k141_16013 | Low-quality |
| S2_extracted_k141_11914 | Not-determined |
| S2_extracted_k141_18712 | Not-determined |
| S2_extracted_k141_16019 | Not-determined |
| S2_extracted_k141_5050 | Not-determined |
| S2_extracted_k141_18719 | Not-determined |
| S2_extracted_k141_9205 | Not-determined |
| S2_extracted_k141_5053 | Not-determined |
| S2_extracted_k141_21160 | Not-determined |
| S2_extracted_k141_21161 | Not-determined |
| S2_extracted_k141_7693 | Not-determined |
| S2_extracted_k141_21162 | Not-determined |
| S2_extracted_k141_3706 | Not-determined |
| S2_extracted_k141_11930 | Not-determined |
| S2_extracted_k141_21163 | Low-quality |
| S2_extracted_k141_11933 | Not-determined |
| S2_extracted_k141_920 | Not-determined |
| S2_extracted_k141_3712 | Not-determined |
| S2_extracted_k141_11935 | Not-determined |
| S2_extracted_k141_3713 | Not-determined |
| S2_extracted_k141_5065 | Not-determined |
| S2_extracted_k141_2144 | Not-determined |
| S2_extracted_k141_7702 | Not-determined |
| S2_extracted_k141_14620 | Not-determined |
| S2_extracted_k141_3723 | Not-determined |
| S2_extracted_k141_9213 | Not-determined |
| S2_extracted_k141_11941 | Not-determined |
| S2_extracted_k141_928 | Not-determined |
| S2_extracted_k141_14623 | Not-determined |
| S2_extracted_k141_3731 | Not-determined |
| S2_extracted_k141_17258 | Not-determined |
| S2_extracted_k141_14625 | Not-determined |
| S2_extracted_k141_9219 | Not-determined |
| S2_extracted_k141_934 | Not-determined |
| S2_extracted_k141_21179 | Not-determined |
| S2_extracted_k141_11945 | Not-determined |
| S2_extracted_k141_14629 | Not-determined |
| S2_extracted_k141_5076 | Low-quality |
| S2_extracted_k141_11948 | Low-quality |
| S2_extracted_k141_21184 | Not-determined |
| S2_extracted_k141_9227 | Not-determined |
| S2_extracted_k141_17262 | Not-determined |
| S2_extracted_k141_11954 | Not-determined |
| S2_extracted_k141_17265 | Not-determined |
| S2_extracted_k141_16032 | Not-determined |
| S2_extracted_k141_9228 | Low-quality |
| S2_extracted_k141_7722 | Not-determined |
| S2_extracted_k141_2166 | Not-determined |
| S2_extracted_k141_9229 | Not-determined |
| S2_extracted_k141_7723 | Low-quality |
| S2_extracted_k141_11959 | Low-quality |
| S2_extracted_k141_9236 | Low-quality |
| S2_extracted_k141_946 | Not-determined |
| S2_extracted_k141_5094 | Low-quality |
| S2_extracted_k141_947 | Not-determined |
| S2_extracted_k141_9238 | Not-determined |
| S2_extracted_k141_3762 | Not-determined |
| S2_extracted_k141_16036 | Not-determined |
| S2_extracted_k141_5097 | Low-quality |
| S2_extracted_k141_9244 | Not-determined |
| S2_extracted_k141_11965 | Low-quality |
| S2_extracted_k141_951 | Not-determined |
| S2_extracted_k141_21189 | Not-determined |
| S2_extracted_k141_17279 | Not-determined |
| S2_extracted_k141_17282 | Not-determined |
| S2_extracted_k141_954 | Not-determined |
| S2_extracted_k141_20149 | Low-quality |
| S2_extracted_k141_955 | Not-determined |
| S2_extracted_k141_9249 | Not-determined |
| S2_extracted_k141_957 | Low-quality |
| S2_extracted_k141_20156 | Not-determined |
| S2_extracted_k141_9253 | Not-determined |
| S2_extracted_k141_3779 | Not-determined |
| S2_extracted_k141_20158 | Not-determined |
| S2_extracted_k141_17288 | Not-determined |
| S2_extracted_k141_20159 | Not-determined |
| S2_extracted_k141_3780 | Not-determined |
| S2_extracted_k141_5109 | Low-quality |
| S2_extracted_k141_5110 | Low-quality |
| S2_extracted_k141_3782 | Low-quality |
| S2_extracted_k141_9257 | Not-determined |
| S2_extracted_k141_20162 | Not-determined |
| S2_extracted_k141_17291 | Not-determined |
| S2_extracted_k141_13383 | Not-determined |
| S2_extracted_k141_13384 | Low-quality |
| S2_extracted_k141_13386 | Not-determined |
| S2_extracted_k141_9262 | Not-determined |
| S2_extracted_k141_7747 | Not-determined |
| S2_extracted_k141_967 | Low-quality |
| S2_extracted_k141_18742 | Not-determined |
| S2_extracted_k141_9265 | Not-determined |
| S2_extracted_k141_13393 | Not-determined |
| S2_extracted_k141_13394 | Not-determined |
| S2_extracted_k141_7751 | Not-determined |
| S2_extracted_k141_9272 | Low-quality |
| S2_extracted_k141_11985 | Low-quality |
| S2_extracted_k141_16052 | Low-quality |
| S2_extracted_k141_2210 | Not-determined |
| S2_extracted_k141_982 | Not-determined |
| S2_extracted_k141_16054 | Not-determined |
| S2_extracted_k141_13404 | Not-determined |
| S2_extracted_k141_11993 | Low-quality |
| S2_extracted_k141_985 | Not-determined |
| S2_extracted_k141_16056 | Not-determined |
| S2_extracted_k141_13406 | Not-determined |
| S2_extracted_k141_18759 | Low-quality |
| S2_extracted_k141_16057 | Low-quality |
| S2_extracted_k141_11996 | Not-determined |
| S2_extracted_k141_18761 | Low-quality |
| S2_extracted_k141_988 | Not-determined |
| S2_extracted_k141_16061 | Not-determined |
| S2_extracted_k141_2218 | Not-determined |
| S2_extracted_k141_13411 | Low-quality |
| S2_extracted_k141_12000 | Not-determined |
| S2_extracted_k141_7761 | Not-determined |
| S2_extracted_k141_9282 | Low-quality |
| S2_extracted_k141_12004 | Not-determined |
| S2_extracted_k141_2225 | Not-determined |
| S2_extracted_k141_21207 | Low-quality |
| S2_extracted_k141_14644 | Not-determined |
| S2_extracted_k141_6237 | Not-determined |
| S2_extracted_k141_16069 | Low-quality |
| S2_extracted_k141_13420 | Low-quality |
| S2_extracted_k141_5119 | Not-determined |
| S2_extracted_k141_16071 | Low-quality |
| S2_extracted_k141_20180 | Not-determined |
| S2_extracted_k141_20181 | Not-determined |
| S2_extracted_k141_13424 | Not-determined |
| S2_extracted_k141_7769 | Not-determined |
| S2_extracted_k141_14647 | Not-determined |
| S2_extracted_k141_18768 | Not-determined |
| S2_extracted_k141_14648 | Not-determined |
| S2_extracted_k141_7770 | Not-determined |
| S2_extracted_k141_9292 | Low-quality |
| S2_extracted_k141_10310 | Not-determined |
| S2_extracted_k141_5124 | Not-determined |
| S2_extracted_k141_14649 | Not-determined |
| S2_extracted_k141_2233 | Not-determined |
| S2_extracted_k141_5125 | Not-determined |
| S2_extracted_k141_21211 | Not-determined |
| S2_extracted_k141_10312 | Not-determined |
| S2_extracted_k141_13426 | Not-determined |
| S2_extracted_k141_14650 | Not-determined |
| S2_extracted_k141_14651 | Not-determined |
| S2_extracted_k141_13430 | Not-determined |
| S2_extracted_k141_14652 | Low-quality |
| S2_extracted_k141_2241 | Not-determined |
| S2_extracted_k141_3804 | Low-quality |
| S2_extracted_k141_2243 | Not-determined |
| S2_extracted_k141_14653 | Not-determined |
| S2_extracted_k141_16081 | Low-quality |
| S2_extracted_k141_16082 | Not-determined |
| S2_extracted_k141_13435 | Not-determined |
| S2_extracted_k141_18781 | Not-determined |
| S2_extracted_k141_16085 | Not-determined |
| S2_extracted_k141_2248 | Not-determined |
| S2_extracted_k141_20194 | Low-quality |
| S2_extracted_k141_16090 | Not-determined |
| S2_extracted_k141_16091 | Not-determined |
| S2_extracted_k141_7785 | Not-determined |
| S2_extracted_k141_17317 | Not-determined |
| S2_extracted_k141_20198 | Not-determined |
| S2_extracted_k141_18789 | Low-quality |
| S2_extracted_k141_16095 | Not-determined |
| S2_extracted_k141_12008 | Not-determined |
| S2_extracted_k141_2252 | Not-determined |
| S2_extracted_k141_9309 | Low-quality |
| S2_extracted_k141_14658 | Not-determined |
| S2_extracted_k141_20202 | Not-determined |
| S2_extracted_k141_9311 | Not-determined |
| S2_extracted_k141_21221 | Low-quality |
| S2_extracted_k141_9313 | Not-determined |
| S2_extracted_k141_9314 | Not-determined |
| S2_extracted_k141_21222 | Low-quality |
| S2_extracted_k141_20205 | Not-determined |
| S2_extracted_k141_13447 | Not-determined |
| S2_extracted_k141_6249 | Not-determined |
| S2_extracted_k141_2254 | Not-determined |
| S2_extracted_k141_13450 | Not-determined |
| S2_extracted_k141_14662 | Low-quality |
| S2_extracted_k141_9319 | Low-quality |
| S2_extracted_k141_21226 | Low-quality |
| S2_extracted_k141_2259 | Low-quality |
| S2_extracted_k141_9323 | Not-determined |
| S2_extracted_k141_21228 | Low-quality |
| S2_extracted_k141_9326 | Low-quality |
| S2_extracted_k141_2263 | Not-determined |
| S2_extracted_k141_6263 | Low-quality |
| S2_extracted_k141_12016 | Not-determined |
| S2_extracted_k141_6265 | Not-determined |
| S2_extracted_k141_6266 | Low-quality |
| S2_extracted_k141_6267 | Not-determined |
| S2_extracted_k141_14673 | Not-determined |
| S2_extracted_k141_7800 | Low-quality |
| S2_extracted_k141_7801 | Not-determined |
| S2_extracted_k141_6273 | Not-determined |
| S2_extracted_k141_13460 | Not-determined |
| S2_extracted_k141_2273 | Low-quality |
| S2_extracted_k141_12024 | Not-determined |
| S2_extracted_k141_2275 | Not-determined |
| S2_extracted_k141_5137 | Not-determined |
| S2_extracted_k141_7807 | Not-determined |
| S2_extracted_k141_20227 | Low-quality |
| S2_extracted_k141_14678 | Not-determined |
| S2_extracted_k141_12030 | Low-quality |
| S2_extracted_k141_1001 | Low-quality |
| S2_extracted_k141_6282 | Not-determined |
| S2_extracted_k141_20229 | Low-quality |
| S2_extracted_k141_20231 | Low-quality |
| S2_extracted_k141_20234 | Not-determined |
| S2_extracted_k141_5142 | Not-determined |
| S2_extracted_k141_7814 | Not-determined |
| S2_extracted_k141_21235 | Not-determined |
| S2_extracted_k141_6292 | Not-determined |
| S2_extracted_k141_17334 | Not-determined |
| S2_extracted_k141_21237 | Not-determined |
| S2_extracted_k141_17338 | Low-quality |
| S2_extracted_k141_6298 | Not-determined |
| S2_extracted_k141_17340 | Not-determined |
| S2_extracted_k141_20239 | Not-determined |
| S2_extracted_k141_7824 | Not-determined |
| S2_extracted_k141_5155 | Not-determined |
| S2_extracted_k141_3851 | Not-determined |
| S2_extracted_k141_5158 | Low-quality |
| S2_extracted_k141_17342 | Not-determined |
| S2_extracted_k141_13463 | Not-determined |
| S2_extracted_k141_3854 | Low-quality |
| S2_extracted_k141_13464 | Not-determined |
| S2_extracted_k141_5161 | Not-determined |
| S2_extracted_k141_16106 | Low-quality |
| S2_extracted_k141_18818 | Low-quality |
| S2_extracted_k141_10325 | Low-quality |
| S2_extracted_k141_5164 | Not-determined |
| S2_extracted_k141_20250 | Not-determined |
| S2_extracted_k141_12047 | Not-determined |
| S2_extracted_k141_12048 | Not-determined |
| S2_extracted_k141_17346 | Not-determined |
| S2_extracted_k141_17347 | Not-determined |
| S2_extracted_k141_16112 | Not-determined |
| S2_extracted_k141_10330 | Not-determined |
| S2_extracted_k141_16115 | Not-determined |
| S2_extracted_k141_16116 | Not-determined |
| S2_extracted_k141_20256 | Not-determined |
| S2_extracted_k141_7831 | Not-determined |
| S2_extracted_k141_20259 | Not-determined |
| S2_extracted_k141_5168 | Not-determined |
| S2_extracted_k141_14693 | Not-determined |
| S2_extracted_k141_3859 | Low-quality |
| S2_extracted_k141_16117 | Not-determined |
| S2_extracted_k141_3860 | Not-determined |
| S2_extracted_k141_5169 | Low-quality |
| S2_extracted_k141_7833 | Not-determined |
| S2_extracted_k141_5170 | Not-determined |
| S2_extracted_k141_10333 | Not-determined |
| S2_extracted_k141_14694 | Not-determined |
| S2_extracted_k141_16118 | Not-determined |
| S2_extracted_k141_6309 | Low-quality |
| S2_extracted_k141_20263 | Not-determined |
| S2_extracted_k141_3862 | Low-quality |
| S2_extracted_k141_16119 | Low-quality |
| S2_extracted_k141_20264 | Not-determined |
| S2_extracted_k141_21247 | Low-quality |
| S2_extracted_k141_7836 | Low-quality |
| S2_extracted_k141_12054 | Not-determined |
| S2_extracted_k141_21248 | Not-determined |
| S2_extracted_k141_21249 | Not-determined |
| S2_extracted_k141_21250 | Not-determined |
| S2_extracted_k141_16123 | Not-determined |
| S2_extracted_k141_20267 | Not-determined |
| S2_extracted_k141_20268 | Low-quality |
| S2_extracted_k141_14700 | Not-determined |
| S2_extracted_k141_21253 | Not-determined |
| S2_extracted_k141_7839 | Not-determined |
| S2_extracted_k141_20270 | Not-determined |
| S2_extracted_k141_17357 | Low-quality |
| S2_extracted_k141_16125 | Low-quality |
| S2_extracted_k141_10340 | Not-determined |
| S2_extracted_k141_21258 | Low-quality |
| S2_extracted_k141_14704 | Not-determined |
| S2_extracted_k141_2285 | Low-quality |
| S2_extracted_k141_3864 | Not-determined |
| S2_extracted_k141_13475 | Not-determined |
| S2_extracted_k141_6322 | Low-quality |
| S2_extracted_k141_5175 | Not-determined |
| S2_extracted_k141_2287 | Not-determined |
| S2_extracted_k141_7845 | Not-determined |
| S2_extracted_k141_3868 | Not-determined |
| S2_extracted_k141_5176 | Low-quality |
| S2_extracted_k141_13477 | Not-determined |
| S2_extracted_k141_20281 | Not-determined |
| S2_extracted_k141_13478 | Not-determined |
| S2_extracted_k141_17363 | Not-determined |
| S2_extracted_k141_21266 | Low-quality |
| S2_extracted_k141_13480 | Not-determined |
| S2_extracted_k141_6326 | Not-determined |
| S2_extracted_k141_21269 | Not-determined |
| S2_extracted_k141_17366 | Not-determined |
| S2_extracted_k141_20287 | Not-determined |
| S2_extracted_k141_17367 | Not-determined |
| S2_extracted_k141_20288 | Not-determined |
| S2_extracted_k141_7850 | Not-determined |
| S2_extracted_k141_17368 | Not-determined |
| S2_extracted_k141_18832 | Low-quality |
| S2_extracted_k141_9348 | Low-quality |
| S2_extracted_k141_13483 | Low-quality |
| S2_extracted_k141_6333 | Not-determined |
| S2_extracted_k141_17371 | Not-determined |
| S2_extracted_k141_13486 | Not-determined |
| S2_extracted_k141_12072 | Low-quality |
| S2_extracted_k141_12073 | Not-determined |
| S2_extracted_k141_5185 | Not-determined |
| S2_extracted_k141_21277 | Not-determined |
| S2_extracted_k141_18839 | Not-determined |
| S2_extracted_k141_18840 | Low-quality |
| S2_extracted_k141_6337 | Low-quality |
| S2_extracted_k141_18841 | Not-determined |
| S2_extracted_k141_13492 | Not-determined |
| S2_extracted_k141_6340 | Not-determined |
| S2_extracted_k141_5193 | Not-determined |
| S2_extracted_k141_16144 | Low-quality |
| S2_extracted_k141_13496 | Not-determined |
| S2_extracted_k141_3883 | Not-determined |
| S2_extracted_k141_14708 | Not-determined |
| S2_extracted_k141_21280 | Low-quality |
| S2_extracted_k141_21282 | Low-quality |
| S2_extracted_k141_18851 | Not-determined |
| S2_extracted_k141_18852 | Not-determined |
| S2_extracted_k141_6350 | Not-determined |
| S2_extracted_k141_1017 | Not-determined |
| S2_extracted_k141_21285 | Low-quality |
| S2_extracted_k141_5201 | Not-determined |
| S2_extracted_k141_7856 | Low-quality |
| S2_extracted_k141_6353 | Not-determined |

| | |
|---|---|
| S2_extracted_k141_9370 | Low-quality |
| S2_extracted_k141_7858 | Not-determined |
| S2_extracted_k141_18857 | Not-determined |
| S2_extracted_k141_14719 | Not-determined |
| S2_extracted_k141_18859 | Not-determined |
| S2_extracted_k141_1020 | Low-quality |
| S2_extracted_k141_12089 | Not-determined |
| S2_extracted_k141_6358 | Not-determined |
| S2_extracted_k141_16150 | Not-determined |
| S2_extracted_k141_6360 | Low-quality |
| S2_extracted_k141_9375 | Not-determined |
| S2_extracted_k141_13508 | Not-determined |
| S2_extracted_k141_21300 | Not-determined |
| S2_extracted_k141_16153 | Not-determined |
| S2_extracted_k141_9380 | Not-determined |
| S2_extracted_k141_13513 | Not-determined |
| S2_extracted_k141_17385 | Not-determined |
| S2_extracted_k141_21309 | Not-determined |
| S2_extracted_k141_17390 | Not-determined |
| S2_extracted_k141_21314 | Not-determined |
| S2_extracted_k141_21315 | Not-determined |
| S2_extracted_k141_14732 | Not-determined |
| S2_extracted_k141_3893 | Low-quality |
| S2_extracted_k141_14733 | Low-quality |
| S2_extracted_k141_21316 | Not-determined |
| S2_extracted_k141_17394 | Not-determined |
| S2_extracted_k141_13521 | Not-determined |
| S2_extracted_k141_14735 | Not-determined |
| S2_extracted_k141_13522 | Not-determined |
| S2_extracted_k141_16169 | Not-determined |
| S2_extracted_k141_9392 | Not-determined |
| S2_extracted_k141_16170 | Low-quality |
| S2_extracted_k141_5217 | Low-quality |
| S2_extracted_k141_21325 | Low-quality |
| S2_extracted_k141_16172 | Not-determined |
| S2_extracted_k141_1023 | Low-quality |
| S2_extracted_k141_13526 | Low-quality |
| S2_extracted_k141_2298 | Not-determined |
| S2_extracted_k141_9401 | Not-determined |
| S2_extracted_k141_2300 | Not-determined |
| S2_extracted_k141_6375 | Not-determined |
| S2_extracted_k141_17403 | Low-quality |
| S2_extracted_k141_6378 | Not-determined |
| S2_extracted_k141_9405 | Not-determined |
| S2_extracted_k141_2305 | Not-determined |
| S2_extracted_k141_16185 | Not-determined |
| S2_extracted_k141_21331 | Low-quality |
| S2_extracted_k141_16186 | Not-determined |
| S2_extracted_k141_7883 | Low-quality |
| S2_extracted_k141_14752 | Not-determined |
| S2_extracted_k141_16188 | Not-determined |
| S2_extracted_k141_2311 | Low-quality |
| S2_extracted_k141_14757 | Not-determined |
| S2_extracted_k141_21336 | Not-determined |
| S2_extracted_k141_14758 | Low-quality |
| S2_extracted_k141_16202 | Not-determined |
| S2_extracted_k141_9416 | Not-determined |
| S2_extracted_k141_9417 | Not-determined |
| S2_extracted_k141_14764 | Not-determined |
| S2_extracted_k141_5222 | Low-quality |
| S2_extracted_k141_7895 | Not-determined |
| S2_extracted_k141_16206 | Low-quality |
| S2_extracted_k141_16207 | Not-determined |
| S2_extracted_k141_2319 | Not-determined |
| S2_extracted_k141_18862 | Low-quality |
| S2_extracted_k141_6400 | Low-quality |
| S2_extracted_k141_9424 | Low-quality |
| S2_extracted_k141_14771 | Not-determined |
| S2_extracted_k141_9426 | Not-determined |
| S2_extracted_k141_5227 | Not-determined |
| S2_extracted_k141_14774 | Low-quality |
| S2_extracted_k141_12095 | Not-determined |
| S2_extracted_k141_16214 | Not-determined |
| S2_extracted_k141_21358 | Not-determined |
| S2_extracted_k141_1047 | Not-determined |
| S2_extracted_k141_21359 | Low-quality |
| S2_extracted_k141_1051 | Not-determined |
| S2_extracted_k141_12102 | Not-determined |
| S2_extracted_k141_1054 | Low-quality |
| S2_extracted_k141_12103 | Not-determined |
| S2_extracted_k141_21366 | Not-determined |
| S2_extracted_k141_16223 | Not-determined |
| S2_extracted_k141_1056 | Not-determined |
| S2_extracted_k141_21369 | Not-determined |
| S2_extracted_k141_14779 | Low-quality |
| S2_extracted_k141_1057 | Not-determined |
| S2_extracted_k141_1059 | Low-quality |
| S2_extracted_k141_16227 | Not-determined |
| S2_extracted_k141_18884 | Not-determined |
| S2_extracted_k141_1062 | Not-determined |
| S2_extracted_k141_21374 | Not-determined |
| S2_extracted_k141_16228 | Not-determined |
| S2_extracted_k141_12106 | Not-determined |
| S2_extracted_k141_10352 | Not-determined |
| S2_extracted_k141_9442 | Low-quality |
| S2_extracted_k141_10353 | Low-quality |
| S2_extracted_k141_12111 | Not-determined |
| S2_extracted_k141_10356 | Low-quality |
| S2_extracted_k141_5235 | Not-determined |
| S2_extracted_k141_6406 | Not-determined |
| S2_extracted_k141_1073 | Low-quality |
| S2_extracted_k141_9446 | Not-determined |
| S2_extracted_k141_17414 | Not-determined |
| S2_extracted_k141_14793 | Not-determined |
| S2_extracted_k141_2348 | Not-determined |
| S2_extracted_k141_6413 | Not-determined |
| S2_extracted_k141_3904 | Not-determined |
| S2_extracted_k141_2350 | Not-determined |
| S2_extracted_k141_21383 | Not-determined |
| S2_extracted_k141_21384 | Low-quality |
| S2_extracted_k141_2353 | Not-determined |
| S2_extracted_k141_18890 | Not-determined |
| S2_extracted_k141_3906 | Not-determined |
| S2_extracted_k141_6418 | Not-determined |
| S2_extracted_k141_21386 | Not-determined |
| S2_extracted_k141_5245 | Not-determined |
| S2_extracted_k141_12112 | Low-quality |
| S2_extracted_k141_18896 | Not-determined |
| S2_extracted_k141_17420 | Not-determined |
| S2_extracted_k141_18898 | Low-quality |
| S2_extracted_k141_5248 | Low-quality |
| S2_extracted_k141_5249 | Not-determined |
| S2_extracted_k141_17423 | Not-determined |
| S2_extracted_k141_10363 | Low-quality |
| S2_extracted_k141_9450 | Not-determined |
| S2_extracted_k141_10365 | Not-determined |
| S2_extracted_k141_18900 | Not-determined |
| S2_extracted_k141_14808 | Not-determined |
| S2_extracted_k141_1084 | Not-determined |
| S2_extracted_k141_9457 | Low-quality |
| S2_extracted_k141_12123 | Not-determined |
| S2_extracted_k141_2376 | Not-determined |
| S2_extracted_k141_5257 | Low-quality |
| S2_extracted_k141_2379 | Not-determined |
| S2_extracted_k141_18905 | Not-determined |
| S2_extracted_k141_14813 | Not-determined |
| S2_extracted_k141_1089 | Low-quality |
| S2_extracted_k141_2381 | Not-determined |
| S2_extracted_k141_6427 | Low-quality |
| S2_extracted_k141_2384 | Not-determined |
| S2_extracted_k141_17430 | Low-quality |
| S2_extracted_k141_1093 | Not-determined |
| S2_extracted_k141_17434 | Low-quality |
| S2_extracted_k141_6432 | Not-determined |
| S2_extracted_k141_1097 | Not-determined |
| S2_extracted_k141_21398 | Not-determined |
| S2_extracted_k141_14819 | Not-determined |
| S2_extracted_k141_2389 | Not-determined |
| S2_extracted_k141_5265 | Not-determined |
| S2_extracted_k141_21400 | Not-determined |
| S2_extracted_k141_3912 | Low-quality |
| S2_extracted_k141_3913 | Not-determined |
| S2_extracted_k141_12135 | Low-quality |
| S2_extracted_k141_21401 | Not-determined |
| S2_extracted_k141_5266 | Low-quality |
| S2_extracted_k141_9467 | Not-determined |
| S2_extracted_k141_1107 | Not-determined |
| S2_extracted_k141_6443 | Not-determined |
| S2_extracted_k141_5269 | Not-determined |
| S2_extracted_k141_1112 | Not-determined |
| S2_extracted_k141_2398 | Low-quality |
| S2_extracted_k141_7913 | Not-determined |
| S2_extracted_k141_18931 | Not-determined |
| S2_extracted_k141_17438 | Not-determined |
| S2_extracted_k141_6448 | Low-quality |
| S2_extracted_k141_14831 | Not-determined |
| S2_extracted_k141_6451 | Not-determined |
| S2_extracted_k141_7918 | Not-determined |
| S2_extracted_k141_7920 | Not-determined |
| S2_extracted_k141_14834 | Low-quality |
| S2_extracted_k141_14836 | Not-determined |
| S2_extracted_k141_2407 | Not-determined |
| S2_extracted_k141_7922 | Not-determined |
| S2_extracted_k141_7924 | Not-determined |
| S2_extracted_k141_14840 | Not-determined |
| S2_extracted_k141_6463 | Low-quality |
| S2_extracted_k141_6466 | Not-determined |
| S2_extracted_k141_6467 | Low-quality |
| S2_extracted_k141_7930 | Not-determined |
| S2_extracted_k141_6468 | Not-determined |
| S2_extracted_k141_21407 | Low-quality |
| S2_extracted_k141_14842 | Low-quality |
| S2_extracted_k141_6470 | Not-determined |
| S2_extracted_k141_1126 | Not-determined |
| S2_extracted_k141_1129 | Low-quality |
| S2_extracted_k141_7934 | Not-determined |
| S2_extracted_k141_2418 | Low-quality |
| S2_extracted_k141_7936 | Not-determined |
| S2_extracted_k141_7937 | Not-determined |
| S2_extracted_k141_14849 | Not-determined |
| S2_extracted_k141_17456 | Not-determined |
| S2_extracted_k141_1135 | Low-quality |
| S2_extracted_k141_21415 | Not-determined |
| S2_extracted_k141_21419 | Not-determined |
| S2_extracted_k141_6483 | Not-determined |
| S2_extracted_k141_6484 | Not-determined |
| S2_extracted_k141_10372 | Low-quality |
| S2_extracted_k141_14853 | Not-determined |
| S2_extracted_k141_10373 | Not-determined |
| S2_extracted_k141_2428 | Not-determined |
| S2_extracted_k141_1140 | Not-determined |
| S2_extracted_k141_21422 | Not-determined |
| S2_extracted_k141_2434 | Not-determined |
| S2_extracted_k141_6490 | Low-quality |
| S2_extracted_k141_6492 | Not-determined |
| S2_extracted_k141_2438 | Not-determined |
| S2_extracted_k141_21425 | Not-determined |
| S2_extracted_k141_21426 | Not-determined |
| S2_extracted_k141_1151 | Not-determined |
| S2_extracted_k141_1154 | Not-determined |
| S2_extracted_k141_10383 | Not-determined |
| S2_extracted_k141_17460 | Low-quality |
| S2_extracted_k141_10384 | Low-quality |
| S2_extracted_k141_10385 | Not-determined |
| S2_extracted_k141_17464 | Not-determined |
| S2_extracted_k141_1158 | Not-determined |
| S2_extracted_k141_21431 | Not-determined |
| S2_extracted_k141_1160 | Not-determined |
| S2_extracted_k141_10389 | Not-determined |
| S2_extracted_k141_5274 | Not-determined |
| S2_extracted_k141_12174 | Not-determined |
| S2_extracted_k141_10397 | Low-quality |
| S2_extracted_k141_14869 | Not-determined |
| S2_extracted_k141_5278 | Low-quality |
| S2_extracted_k141_10399 | Not-determined |
| S2_extracted_k141_12176 | Low-quality |
| S2_extracted_k141_3915 | Not-determined |
| S2_extracted_k141_17473 | Not-determined |
| S2_extracted_k141_21444 | Not-determined |
| S2_extracted_k141_14871 | Not-determined |
| S2_extracted_k141_1173 | Low-quality |
| S2_extracted_k141_6506 | Not-determined |
| S2_extracted_k141_21447 | Not-determined |
| S2_extracted_k141_6508 | Not-determined |
| S2_extracted_k141_17480 | Not-determined |
| S2_extracted_k141_1181 | Not-determined |
| S2_extracted_k141_1184 | Not-determined |
| S2_extracted_k141_6515 | Low-quality |
| S2_extracted_k141_6518 | Not-determined |
| S2_extracted_k141_7964 | Not-determined |
| S2_extracted_k141_21461 | Not-determined |
| S2_extracted_k141_6520 | Low-quality |
| S2_extracted_k141_2467 | Not-determined |
| S2_extracted_k141_2469 | Not-determined |
| S2_extracted_k141_2473 | Not-determined |
| S2_extracted_k141_1212 | Not-determined |
| S2_extracted_k141_1213 | Not-determined |
| S2_extracted_k141_17504 | Not-determined |
| S2_extracted_k141_3944 | Not-determined |
| S2_extracted_k141_17505 | Not-determined |
| S2_extracted_k141_1215 | Not-determined |
| S2_extracted_k141_5281 | Low-quality |
| S2_extracted_k141_1217 | Not-determined |
| S2_extracted_k141_17511 | Not-determined |
| S2_extracted_k141_17512 | Not-determined |
| S2_extracted_k141_6541 | Low-quality |
| S2_extracted_k141_6542 | Not-determined |
| S2_extracted_k141_1226 | Not-determined |
| S2_extracted_k141_7975 | Low-quality |
| S2_extracted_k141_17519 | Not-determined |
| S2_extracted_k141_3962 | Not-determined |
| S2_extracted_k141_5288 | Low-quality |
| S2_extracted_k141_7977 | Not-determined |
| S2_extracted_k141_3966 | Not-determined |
| S2_extracted_k141_3968 | Low-quality |
| S2_extracted_k141_5290 | Not-determined |
| S2_extracted_k141_3970 | Not-determined |
| S2_extracted_k141_3971 | Not-determined |
| S2_extracted_k141_21488 | Not-determined |
| S2_extracted_k141_21492 | Not-determined |
| S2_extracted_k141_1233 | Low-quality |
| S2_extracted_k141_21495 | Not-determined |
| S2_extracted_k141_17523 | Not-determined |
| S2_extracted_k141_5296 | Not-determined |
| S2_extracted_k141_5298 | Low-quality |
| S2_extracted_k141_3981 | Low-quality |
| S2_extracted_k141_3983 | Not-determined |
| S2_extracted_k141_3985 | Not-determined |
| S2_extracted_k141_21503 | Not-determined |
| S2_extracted_k141_21505 | Not-determined |
| S2_extracted_k141_1249 | Low-quality |
| S2_extracted_k141_1250 | Not-determined |
| S2_extracted_k141_17531 | Not-determined |
| S2_extracted_k141_1253 | Not-determined |
| S2_extracted_k141_21510 | Low-quality |
| S2_extracted_k141_3989 | Not-determined |
| S2_extracted_k141_2482 | Not-determined |
| S2_extracted_k141_17535 | Not-determined |
| S2_extracted_k141_2483 | Not-determined |
| S2_extracted_k141_17536 | Not-determined |
| S2_extracted_k141_3992 | Not-determined |
| S2_extracted_k141_1258 | Not-determined |
| S2_extracted_k141_1261 | Low-quality |
| S2_extracted_k141_1263 | Not-determined |
| S2_extracted_k141_1266 | Not-determined |
| S2_extracted_k141_1267 | Low-quality |
| S2_extracted_k141_1268 | Not-determined |
| S2_extracted_k141_10425 | Not-determined |
| S2_extracted_k141_5303 | Not-determined |
| S2_extracted_k141_2495 | Not-determined |
| S2_extracted_k141_17541 | Not-determined |
| S2_extracted_k141_1273 | Not-determined |
| S2_extracted_k141_10431 | Not-determined |
| S2_extracted_k141_17546 | Not-determined |
| S2_extracted_k141_2503 | Not-determined |
| S2_extracted_k141_1284 | Low-quality |
| S2_extracted_k141_10438 | Not-determined |
| S2_extracted_k141_1288 | Not-determined |
| S2_extracted_k141_17551 | Not-determined |
| S2_extracted_k141_7985 | Not-determined |
| S2_extracted_k141_4008 | Not-determined |
| S2_extracted_k141_17552 | Not-determined |
| S2_extracted_k141_1292 | Not-determined |
| S2_extracted_k141_10443 | Not-determined |
| S2_extracted_k141_1300 | Not-determined |
| S2_extracted_k141_4017 | Not-determined |
| S2_extracted_k141_5310 | Not-determined |
| S2_extracted_k141_2518 | Low-quality |
| S2_extracted_k141_4021 | Not-determined |
| S2_extracted_k141_10451 | Low-quality |
| S2_extracted_k141_5311 | Low-quality |
| S2_extracted_k141_2520 | Low-quality |
| S2_extracted_k141_2521 | Not-determined |
| S2_extracted_k141_17566 | Not-determined |
| S2_extracted_k141_1309 | Not-determined |
| S2_extracted_k141_2522 | Low-quality |
| S2_extracted_k141_1310 | Not-determined |
| S2_extracted_k141_10455 | Not-determined |
| S2_extracted_k141_4030 | Low-quality |
| S2_extracted_k141_1311 | Not-determined |
| S2_extracted_k141_1313 | Not-determined |
| S2_extracted_k141_17569 | Not-determined |
| S2_extracted_k141_1316 | Not-determined |
| S2_extracted_k141_1318 | Not-determined |
| S2_extracted_k141_10460 | Not-determined |
| S2_extracted_k141_4043 | Not-determined |
| S2_extracted_k141_1326 | Not-determined |
| S2_extracted_k141_10468 | Low-quality |
| S2_extracted_k141_1330 | Not-determined |
| S2_extracted_k141_2535 | Low-quality |
| S2_extracted_k141_2536 | Not-determined |
| S2_extracted_k141_6555 | Not-determined |
| S2_extracted_k141_1332 | Not-determined |
| S2_extracted_k141_4048 | Not-determined |
| S2_extracted_k141_1333 | Not-determined |
| S2_extracted_k141_8005 | Not-determined |
| S2_extracted_k141_17583 | Not-determined |
| S2_extracted_k141_8008 | Low-quality |
| S2_extracted_k141_2542 | Not-determined |
| S2_extracted_k141_8009 | Low-quality |
| S2_extracted_k141_2545 | Not-determined |
| S2_extracted_k141_10479 | Not-determined |
| S2_extracted_k141_8013 | Not-determined |
| S2_extracted_k141_6568 | Low-quality |
| S2_extracted_k141_2551 | Not-determined |
| S2_extracted_k141_2552 | Not-determined |
| S2_extracted_k141_6572 | Low-quality |
| S2_extracted_k141_6574 | Not-determined |
| S2_extracted_k141_2557 | Low-quality |
| S2_extracted_k141_8021 | Not-determined |
| S2_extracted_k141_6579 | Not-determined |
| S2_extracted_k141_6580 | Not-determined |
| S2_extracted_k141_6582 | Not-determined |
| S2_extracted_k141_10490 | Not-determined |
| S2_extracted_k141_1338 | Not-determined |
| S2_extracted_k141_6592 | Not-determined |
| S2_extracted_k141_1341 | Not-determined |
| S2_extracted_k141_6595 | Low-quality |
| S2_extracted_k141_4058 | Low-quality |
| S2_extracted_k141_10500 | Not-determined |
| S2_extracted_k141_6598 | Not-determined |
| S2_extracted_k141_10501 | Not-determined |
| S2_extracted_k141_10502 | Not-determined |
| S2_extracted_k141_6601 | Not-determined |
| S2_extracted_k141_10504 | Low-quality |
| S2_extracted_k141_5330 | Not-determined |
| S2_extracted_k141_2578 | Not-determined |
| S2_extracted_k141_2581 | Not-determined |
| S2_extracted_k141_5338 | Not-determined |
| S2_extracted_k141_8041 | Not-determined |
| S2_extracted_k141_10513 | Not-determined |
| S2_extracted_k141_2586 | Not-determined |
| S2_extracted_k141_10516 | Low-quality |
| S2_extracted_k141_2590 | Low-quality |
| S2_extracted_k141_1351 | Not-determined |
| S2_extracted_k141_6613 | Not-determined |
| S2_extracted_k141_1352 | Not-determined |
| S2_extracted_k141_8045 | Not-determined |
| S2_extracted_k141_6616 | Low-quality |
| S4_extracted_13759 | Medium-quality |
| S2_extracted_k141_5351 | Not-determined |
| S2_extracted_k141_5352 | Not-determined |
| S2_extracted_k141_8053 | Not-determined |
| S2_extracted_k141_5354 | Low-quality |
| S2_extracted_k141_8055 | Not-determined |
| S2_extracted_k141_2609 | Not-determined |
| S2_extracted_k141_8056 | Not-determined |
| S2_extracted_k141_5357 | Not-determined |
| S2_extracted_k141_8057 | Low-quality |
| S2_extracted_k141_2611 | Not-determined |
| S2_extracted_k141_10529 | Not-determined |
| S2_extracted_k141_2613 | Not-determined |
| S2_extracted_k141_2614 | Not-determined |
| S2_extracted_k141_10539 | Not-determined |
| S2_extracted_k141_8069 | Not-determined |
| S2_extracted_k141_8070 | Not-determined |
| S2_extracted_k141_5360 | Low-quality |
| S2_extracted_k141_6631 | Not-determined |
| S2_extracted_k141_10546 | Not-determined |
| S2_extracted_k141_5363 | Not-determined |
| S2_extracted_k141_8072 | Not-determined |
| S2_extracted_k141_10548 | Not-determined |
| S2_extracted_k141_8074 | Not-determined |
| S2_extracted_k141_8077 | Low-quality |
| S2_extracted_k141_5367 | Low-quality |
| S2_extracted_k141_8078 | Not-determined |
| S2_extracted_k141_10556 | Not-determined |
| S2_extracted_k141_5375 | Not-determined |
| S2_extracted_k141_6634 | Not-determined |
| S2_extracted_k141_21519 | Not-determined |
| S2_extracted_k141_8091 | Not-determined |
| S2_extracted_k141_2628 | Low-quality |
| S2_extracted_k141_21523 | Not-determined |
| S2_extracted_k141_21524 | Not-determined |
| S2_extracted_k141_6638 | Low-quality |
| S2_extracted_k141_2631 | Not-determined |
| S2_extracted_k141_10568 | Low-quality |
| S2_extracted_k141_10569 | Not-determined |
| S2_extracted_k141_2633 | Low-quality |
| S2_extracted_k141_8098 | Not-determined |
| S2_extracted_k141_8101 | Not-determined |
| S2_extracted_k141_8103 | Not-determined |
| S2_extracted_k141_6645 | Not-determined |
| S2_extracted_k141_5390 | Low-quality |
| S2_extracted_k141_8108 | Not-determined |
| S2_extracted_k141_6653 | Not-determined |
| S2_extracted_k141_10579 | Not-determined |
| S2_extracted_k141_5394 | Not-determined |
| S2_extracted_k141_6662 | Low-quality |
| S2_extracted_k141_10584 | Not-determined |
| S2_extracted_k141_10588 | Not-determined |
| S2_extracted_k141_10592 | Not-determined |
| S2_extracted_k141_8117 | Low-quality |
| S2_extracted_k141_5402 | Not-determined |
| S2_extracted_k141_10597 | Low-quality |
| S2_extracted_k141_5404 | Low-quality |
| S2_extracted_k141_21533 | Not-determined |
| S2_extracted_k141_10609 | Not-determined |
| S2_extracted_k141_10610 | Not-determined |
| S2_extracted_k141_6671 | Not-determined |
| S2_extracted_k141_5410 | Not-determined |
| S2_extracted_k141_6673 | Not-determined |
| S2_extracted_k141_21543 | Not-determined |
| S2_extracted_k141_21545 | Not-determined |
| S2_extracted_k141_6676 | Not-determined |
| S2_extracted_k141_6677 | Not-determined |
| S2_extracted_k141_6679 | Not-determined |
| S2_extracted_k141_6680 | Not-determined |
| S2_extracted_k141_10624 | Not-determined |
| S2_extracted_k141_10625 | Low-quality |
| S2_extracted_k141_6700 | Not-determined |
| S2_extracted_k141_6701 | Not-determined |
| S2_extracted_k141_10645 | Not-determined |
| S2_extracted_k141_10647 | Not-determined |
| S2_extracted_k141_21551 | Not-determined |
| S2_extracted_k141_21554 | Low-quality |
| S2_extracted_k141_6717 | Low-quality |
| S2_extracted_k141_21561 | Not-determined |
| S2_extracted_k141_10656 | Not-determined |
| S2_extracted_k141_6723 | Low-quality |
| S2_extracted_k141_10664 | Not-determined |
| S2_extracted_k141_10665 | Not-determined |
| S2_extracted_k141_6725 | Not-determined |
| S2_extracted_k141_10668 | Not-determined |
| S2_extracted_k141_6726 | Not-determined |
| S2_extracted_k141_10671 | Not-determined |
| S2_extracted_k141_21571 | Not-determined |
| S2_extracted_k141_21573 | Not-determined |
| S2_extracted_k141_21574 | Low-quality |
| S2_extracted_k141_6734 | Low-quality |
| S2_extracted_k141_10678 | Not-determined |
| S2_extracted_k141_21580 | Not-determined |
| S2_extracted_k141_21581 | Not-determined |
| S2_extracted_k141_6741 | Low-quality |
| S2_extracted_k141_6743 | Low-quality |
| S2_extracted_k141_21587 | Low-quality |
| S2_extracted_k141_6748 | Not-determined |
| S2_extracted_k141_21588 | Not-determined |
| S2_extracted_k141_21589 | Not-determined |
| S2_extracted_k141_6751 | Not-determined |
| S2_extracted_k141_21596 | Not-determined |
| S2_extracted_k141_6756 | Not-determined |
| S2_extracted_k141_10706 | Not-determined |
| S2_extracted_k141_10707 | Not-determined |
| S2_extracted_k141_21608 | Not-determined |
| S2_extracted_k141_10723 | Low-quality |
| S2_extracted_k141_21612 | Not-determined |
| S2_extracted_k141_21614 | Not-determined |
| S2_extracted_k141_10732 | Low-quality |
| S2_extracted_k141_10735 | Not-determined |
| S2_extracted_k141_21623 | Not-determined |
| S2_extracted_k141_10742 | Low-quality |
| S2_extracted_k141_10750 | Not-determined |
| S2_extracted_k141_10761 | Not-determined |
| S2_extracted_k141_10762 | Not-determined |
| S2_extracted_k141_10765 | Not-determined |
| S2_extracted_k141_10768 | Not-determined |
| S2_extracted_k141_10769 | Not-determined |
| S2_extracted_k141_10784 | Not-determined |
| S2_extracted_k141_10790 | Not-determined |
| S2_extracted_k141_10793 | Not-determined |
| S2_extracted_k141_10808 | Not-determined |
| S4_vs_VIRSorter_13759-cat_2 | Medium-quality |
| S7_vs_83474\|\|full | Medium-quality |
| S1_extracted_k141_50591 | Medium-quality |
| S2_extracted_k141_21634 | Low-quality |
| S2_extracted_k141_21635 | Not-determined |
| S2_extracted_k141_21639 | Not-determined |
| S2_vs_k141_2774\|\|full | Low-quality |
| S5_vs_k141_130476\|\|full | Medium-quality |
| S2_vs_k141_16812\|\|full | Not-determined |
| S2_vs_k141_4368\|\|full | Medium-quality |
| S2_vs_k141_10280\|\|full | Not-determined |
| S5_vs_k141_58766\|\|full | Medium-quality |
| S2_vs_k141_21629\|\|full | Not-determined |
| S1_vs_k141_50591\|\|full | Medium-quality |
| S9_vs_k141_46498\|\|full | Medium-quality |
| S2_vs_k141_21632\|\|full | Not-determined |
| S2_vs_k141_21633\|\|full | Low-quality |
| S2_vs_k141_18943\|\|full | Low-quality |
| S2_vs_k141_9477\|\|full | Not-determined |
| S2_vs_k141_8129\|\|full | Low-quality |
| S2_vs_k141_5418\|\|full | Low-quality |
| S2_vs_k141_13530\|\|full | Not-determined |
| S2_vs_k141_5419\|\|full | Not-determined |
| S2_vs_k141_5420\|\|full | Not-determined |
| S2_vs_k141_16263\|\|full | Low-quality |
| S2_vs_k141_17600\|\|full | Low-quality |
| S2_vs_k141_8144\|\|full | Low-quality |
| S2_vs_k141_4085\|\|full | Low-quality |
| S2_vs_k141_20306\|\|full | Low-quality |
| S2_vs_k141_16265\|\|full | Not-determined |
| S2_vs_k141_9503\|\|full | Low-quality |
| S2_vs_k141_13558\|\|full | Not-determined |
| S2_vs_k141_13562\|\|full | Not-determined |
| S2_vs_k141_2755\|\|full | Low-quality |
| S2_vs_k141_9509\|\|full | Low-quality |
| S2_vs_k141_10837\|\|full | Not-determined |
| S2_vs_k141_13566\|\|full | Low-quality |
| S2_vs_k141_5429\|\|full | Low-quality |
| S2_vs_k141_16277\|\|full | Low-quality |
| S2_vs_k141_6795\|\|full | Low-quality |
| S2_vs_k141_14907\|\|full | Not-determined |
| S2_vs_k141_20327\|\|full | Low-quality |
| S2_vs_k141_49\|\|full | Not-determined |
| S2_vs_k141_9529\|\|full | Low-quality |
| S2_vs_k141_8168\|\|full | Low-quality |
| S2_vs_k141_50\|\|full | Not-determined |
| S2_vs_k141_8170\|\|full | Low-quality |
| S2_vs_k141_8171\|\|full | Not-determined |
| S2_vs_k141_5446\|\|full | Low-quality |
| S2_vs_k141_8185\|\|full | Not-determined |
| S2_vs_k141_10854\|\|full | Low-quality |
| S2_vs_k141_8196\|\|full | Low-quality |
| S2_vs_k141_20383\|\|full | Low-quality |
| S2_vs_k141_19008\|\|full | Low-quality |
| S2_vs_k141_13597\|\|full | Low-quality |
| S2_vs_k141_9536\|\|full | Not-determined |
| S2_vs_k141_19016\|\|full | Low-quality |
| S2_vs_k141_10864\|\|full | Low-quality |
| S2_vs_k141_14970\|\|full | Low-quality |
| S2_vs_k141_2785\|\|full | Low-quality |
| S2_vs_k141_8218\|\|full | Low-quality |
| S2_vs_k141_17681\|\|full | Low-quality |
| S2_vs_k141_20401\|\|full | Low-quality |
| S2_vs_k141_6824\|\|full | Low-quality |
| S2_vs_k141_17690\|\|full | Low-quality |
| S2_vs_k141_9550\|\|full | Low-quality |
| S2_vs_k141_17698\|\|full | Low-quality |
| S2_vs_k141_8230\|\|full | Low-quality |
| S2_vs_k141_97\|\|full | Low-quality |
| S2_vs_k141_6839\|\|full | Not-determined |
| S2_vs_k141_2808\|\|full | Low-quality |
| S2_vs_k141_14988\|\|full | Low-quality |
| S2_vs_k141_9583\|\|full | Low-quality |
| S2_vs_k141_13623\|\|full | Low-quality |
| S2_vs_k141_20443\|\|full | Low-quality |
| S2_vs_k141_2849\|\|full | Not-determined |
| S2_vs_k141_10914\|\|full | Not-determined |

S2_vs_k141_12248||full Low-quality
S2_vs_k141_118||full Not-determined
S8_extracted_k141_195032 Medium-quality
S2_vs_k141_15008||full Low-quality
S2_vs_k141_17727||full Low-quality
S2_vs_k141_4127||full Low-quality
S2_vs_k141_19068||full Low-quality
S2_vs_k141_133||full Not-determined
S2_vs_k141_16349||full Low-quality
S2_vs_k141_13673||full Low-quality
S2_vs_k141_6914||full Low-quality
S2_vs_k141_6916||full Low-quality
S2_vs_k141_139||full Low-quality
S2_vs_k141_4167||full Not-determined
S2_vs_k141_6922||full Not-determined
S2_vs_k141_10956||full Low-quality
S2_vs_k141_13694||full Low-quality
S2_vs_k141_146||full Low-quality
S2_vs_k141_4182||full Low-quality
S2_vs_k141_15070||full Low-quality
S2_vs_k141_15074||full Low-quality
S2_vs_k141_12272||full Low-quality
S2_vs_k141_15081||full Low-quality
S2_vs_k141_4190||full Low-quality
S2_vs_k141_16389||full Not-determined
S2_vs_k141_20480||full Low-quality
S2_vs_k141_4194||full Low-quality
S2_vs_k141_1401||full Low-quality
S8_vs_k141_195032||full Medium-quality
S2_vs_k141_13711||full Low-quality
S2_vs_k141_4205||full Low-quality
S2_vs_k141_13717||full Low-quality
S2_vs_k141_9634||full Not-determined
S2_vs_k141_9641||full Not-determined
S2_vs_k141_2897||full Low-quality
S2_vs_k141_13739||full Low-quality
S2_vs_k141_1433||full Low-quality
S2_vs_k141_11001||full Low-quality
S2_vs_k141_9668||full Low-quality
S2_vs_k141_13749||full Low-quality
S2_vs_k141_11005||full Low-quality
S2_vs_k141_12300||full Low-quality
S2_vs_k141_2930||full Low-quality
S2_vs_k141_11013||full Not-determined
S8_extracted_k141_13758 Medium-quality
S2_vs_k141_5551||full Low-quality
S2_vs_k141_4229||full Low-quality
S2_vs_k141_2948||full Low-quality
S2_vs_k141_13777||full Low-quality
S2_vs_k141_6987||full Low-quality
S2_vs_k141_15085||full Not-determined
S2_vs_k141_12332||full Low-quality
S2_vs_k141_5585||full Low-quality
S2_vs_k141_19156||full Low-quality
S2_vs_k141_15108||full Low-quality
S2_vs_k141_20533||full Low-quality
S2_vs_k141_16472||full Low-quality
S2_vs_k141_11046||full Not-determined
S2_vs_k141_11049||full Low-quality
S2_vs_k141_15132||full Low-quality
S2_vs_k141_7012||full Low-quality
S2_vs_k141_16491||full Not-determined
S2_vs_k141_15140||full Low-quality
S2_vs_k141_2979||full Low-quality
S2_vs_k141_8297||full Low-quality
S2_vs_k141_12362||full Low-quality
S2_vs_k141_8301||full Not-determined
S2_vs_k141_5626||full Low-quality
S2_vs_k141_12373||full Not-determined
S2_vs_k141_19187||full Low-quality
S2_vs_k141_7028||full Low-quality
S2_vs_k141_15155||full Low-quality
S2_vs_k141_3010||full Low-quality
S2_vs_k141_5651||full Low-quality
S2_vs_k141_11088||full Not-determined
S2_vs_k141_12385||full Low-quality
S2_vs_k141_11089||full Low-quality
S2_vs_k141_15179||full Low-quality
S2_vs_k141_7037||full Low-quality
S2_vs_k141_16549||full Low-quality
S2_vs_k141_16550||full Low-quality
S2_vs_k141_11095||full Low-quality
S2_vs_k141_3027||full Low-quality
S2_vs_k141_7040||full Low-quality
S2_vs_k141_12417||full Low-quality
S2_vs_k141_12419||full Low-quality
S2_vs_k141_3039||full Low-quality
S2_vs_k141_15228||full Low-quality
S2_vs_k141_3047||full Not-determined
S2_vs_k141_19227||full Low-quality
S2_vs_k141_5688||full Low-quality
S2_vs_k141_3049||full Low-quality
S2_vs_k141_13906||full Low-quality
S2_vs_k141_15235||full Not-determined
S8_vs_k141_13758||full Medium-quality
S2_vs_k141_11105||full Low-quality
S2_vs_k141_5709||full Low-quality
S2_vs_k141_15253||full Low-quality
S2_vs_k141_7099||full Low-quality
S2_vs_k141_20597||full Not-determined
S2_vs_k141_13916||full Low-quality
S2_vs_k141_8371||full Low-quality
S2_vs_k141_20606||full Low-quality
S2_vs_k141_4332||full Low-quality
S2_vs_k141_20614||full Low-quality
S2_vs_k141_11143||full Low-quality
S2_vs_k141_16591||full Low-quality
S2_vs_k141_4343||full Not-determined
S2_vs_k141_19276||full Low-quality
S2_vs_k141_9803||full Not-determined
S2_vs_k141_19290||full Low-quality
S2_vs_k141_7119||full Not-determined
S2_vs_k141_20623||full Low-quality
S2_vs_k141_4366||full Not-determined
S2_vs_k141_9815||full Not-determined
S2_vs_k141_13963||full Low-quality
S2_vs_k141_1496||full Low-quality
S2_vs_k141_247||full Low-quality
S2_vs_k141_12502||full Low-quality
S2_vs_k141_7138||full Not-determined
S2_vs_k141_19319||full Low-quality
S2_vs_k141_11176||full Low-quality
S2_vs_k141_16627||full Low-quality
S2_vs_k141_5734||full Low-quality
S2_vs_k141_5735||full Low-quality
S2_vs_k141_9836||full Low-quality
S2_vs_k141_255||full Low-quality
S2_vs_k141_16638||full Low-quality
S2_vs_k141_257||full Not-determined
S17_vs_k141_116640||full Medium-quality
S2_vs_k141_7165||full Low-quality
S2_vs_k141_13975||full Low-quality
S2_vs_k141_7170||full Low-quality
S2_vs_k141_4377||full Low-quality
S2_vs_k141_15307||full Low-quality
S2_vs_k141_13986||full Low-quality
S2_vs_k141_13989||full Low-quality
S2_vs_k141_16649||full Not-determined
S2_vs_k141_12538||full Low-quality
S2_vs_k141_20655||full Low-quality
S2_vs_k141_1532||full Low-quality
S2_vs_k141_13995||full Low-quality
S2_vs_k141_12550||full Low-quality
S2_vs_k141_4383||full Low-quality
S2_vs_k141_11211||full Low-quality
S2_vs_k141_5766||full Low-quality
S2_vs_k141_15324||full Not-determined
S2_vs_k141_19361||full Low-quality
S2_vs_k141_16679||full Low-quality
S2_vs_k141_20667||full Low-quality
S2_vs_k141_9871||full Low-quality
S2_vs_k141_20675||full Not-determined
S2_vs_k141_7221||full Low-quality
S2_vs_k141_3080||full Low-quality
S2_vs_k141_20681||full Low-quality
S2_vs_k141_19393||full Not-determined
S2_vs_k141_15364||full Low-quality
S2_vs_k141_19397||full Low-quality
S2_vs_k141_4434||full Low-quality
S2_vs_k141_1572||full Low-quality
S2_vs_k141_4437||full Low-quality
S2_vs_k141_7229||full Not-determined
S2_vs_k141_293||full Low-quality
S2_vs_k141_15387||full Low-quality
S2_vs_k141_17935||full Low-quality
S2_vs_k141_20694||full Low-quality
S2_vs_k141_11281||full Low-quality
S2_vs_k141_15413||full Not-determined
S2_vs_k141_3096||full Low-quality
S2_vs_k141_4482||full Not-determined
S2_vs_k141_11296||full Not-determined
S2_vs_k141_14049||full Not-determined
S2_vs_k141_310||full Low-quality
S2_vs_k141_8440||full Low-quality
S2_vs_k141_4521||full Low-quality
S2_vs_k141_12708||full Low-quality
S2_vs_k141_19480||full Low-quality
S2_vs_k141_14056||full Low-quality
S2_vs_k141_4528||full Not-determined
S2_vs_k141_12726||full Low-quality
S2_vs_k141_16778||full Low-quality
S2_vs_k141_12740||full Not-determined
S2_vs_k141_16789||full Low-quality
S2_vs_k141_18029||full Low-quality
S2_vs_k141_16802||full Not-determined
S2_vs_k141_12751||full Low-quality
S2_vs_k141_18046||full Low-quality
S2_vs_k141_12757||full Low-quality
S2_vs_k141_337||full Low-quality
S2_vs_k141_4545||full Low-quality
S2_vs_k141_20743||full Low-quality
S2_vs_k141_18076||full Low-quality
S2_vs_k141_11335||full Low-quality
S2_vs_k141_9957||full Low-quality
S2_vs_k141_14070||full Low-quality
S17_vs_k141_138036||full High-quality
S2_vs_k141_12785||full Not-determined
S2_vs_k141_18111||full Not-determined
S2_vs_k141_8521||full Low-quality
S2_vs_k141_7298||full Low-quality
S2_vs_k141_15458||full Low-quality
S2_vs_k141_11364||full Low-quality
S2_vs_k141_11365||full Low-quality
S2_vs_k141_18131||full Not-determined
S2_vs_k141_12822||full Not-determined
S2_vs_k141_15486||full Low-quality
S2_vs_k141_12843||full Low-quality
S2_vs_k141_1681||full Low-quality
S2_vs_k141_15511||full Low-quality
S2_vs_k141_8541||full Low-quality
S2_vs_k141_18185||full Low-quality
S2_vs_k141_15521||full Low-quality
S2_vs_k141_8546||full Low-quality
S2_vs_k141_5809||full Low-quality
S10_vs_20400||full Medium-quality
S2_vs_k141_14135||full Low-quality
S2_vs_k141_12861||full Low-quality
S2_vs_k141_8551||full Low-quality
S2_vs_k141_10032||full Low-quality
S2_vs_k141_16861||full Low-quality
S2_vs_k141_7309||full Low-quality
S2_vs_k141_14150||full Low-quality
S2_vs_k141_374||full Low-quality
S2_vs_k141_12878||full Low-quality
S2_vs_k141_16869||full Not-determined
S2_vs_k141_10061||full Low-quality
S2_vs_k141_16876||full Low-quality
S2_vs_k141_3222||full Not-determined
S2_vs_k141_396||full Low-quality
S2_vs_k141_11398||full Low-quality
S2_vs_k141_15556||full Low-quality
S2_vs_k141_12901||full Low-quality
S2_vs_k141_5858||full Low-quality
S2_vs_k141_18201||full Not-determined
S2_vs_k141_412||full Low-quality
S2_vs_k141_8593||full Low-quality
S2_vs_k141_10066||full Low-quality
S2_vs_k141_16899||full Not-determined
S2_vs_k141_15593||full Low-quality
S2_vs_k141_7355||full Low-quality
S2_vs_k141_14187||full Low-quality
S2_vs_k141_4669||full Low-quality
S2_vs_k141_426||full Low-quality
S2_vs_k141_19622||full Low-quality
S2_vs_k141_5897||full Low-quality
S2_vs_k141_4675||full Low-quality
S2_vs_k141_10096||full Low-quality
S2_vs_k141_14196||full Low-quality
S2_vs_k141_19626||full Low-quality
S2_vs_k141_4684||full Low-quality
S2_vs_k141_4685||full Low-quality
S2_vs_k141_437||full Low-quality
S2_vs_k141_18252||full Not-determined
S2_vs_k141_4692||full Low-quality
S2_vs_k141_7375||full Not-determined
S2_vs_k141_18258||full Low-quality
S2_vs_k141_15643||full Low-quality
S2_vs_k141_19648||full Low-quality
S2_vs_k141_11443||full Low-quality
S2_vs_k141_14227||full Low-quality
S2_vs_k141_4710||full Low-quality
S2_vs_k141_4714||full Low-quality
S2_vs_k141_15667||full Not-determined
S2_vs_k141_472||full Low-quality
S2_vs_k141_7384||full Low-quality
S2_vs_k141_11462||full Not-determined
S2_vs_k141_16915||full Low-quality
S2_vs_k141_18296||full Low-quality
S2_vs_k141_18306||full Low-quality
S2_vs_k141_3298||full Low-quality
S2_vs_k141_4721||full Low-quality
S2_vs_k141_5981||full Low-quality
S2_vs_k141_15704||full Low-quality
S2_vs_k141_18310||full Not-determined
S2_vs_k141_20789||full Low-quality
S2_vs_k141_14247||full Low-quality
S2_vs_k141_1771||full Low-quality
S2_vs_k141_12993||full Low-quality
S2_vs_k141_18327||full Low-quality
S2_vs_k141_12999||full Low-quality
S2_vs_k141_479||full Low-quality
S2_vs_k141_20809||full Low-quality
S2_vs_k141_7421||full Low-quality
S2_vs_k141_14266||full Low-quality
S2_vs_k141_16947||full Low-quality
S2_vs_k141_1793||full Low-quality
S2_vs_k141_13015||full Not-determined
S2_vs_k141_18356||full Low-quality
S2_vs_k141_15731||full Low-quality
S2_vs_k141_10117||full Low-quality
S2_vs_k141_14285||full Not-determined
S2_vs_k141_18378||full Not-determined
S2_vs_k141_7449||full Low-quality
S2_vs_k141_13055||full Low-quality
S2_vs_k141_6074||full Low-quality
S16_vs_120149||full Medium-quality
S2_vs_k141_8783||full Not-determined
S2_vs_k141_3362||full Low-quality
S2_vs_k141_6085||full Low-quality
S2_vs_k141_3364||full Low-quality
S2_vs_k141_7460||full Low-quality
S2_vs_k141_543||full Not-determined
S2_vs_k141_3366||full Low-quality
S2_vs_k141_6096||full Low-quality
S2_vs_k141_18395||full Low-quality
S2_vs_k141_13065||full Not-determined
S2_vs_k141_4784||full Low-quality
S2_vs_k141_11559||full Low-quality
S2_vs_k141_552||full Low-quality
S2_vs_k141_14304||full Low-quality
S2_vs_k141_20850||full Low-quality
S2_vs_k141_8803||full Low-quality
S2_vs_k141_18405||full Not-determined
S2_vs_k141_16962||full Low-quality
S2_vs_k141_570||full Not-determined
S2_vs_k141_14316||full Low-quality
S2_vs_k141_20865||full Low-quality
S2_vs_k141_1814||full Low-quality
S2_vs_k141_8825||full Not-determined
S2_vs_k141_588||full Low-quality
S2_vs_k141_15781||full Low-quality
S2_vs_k141_6123||full Low-quality
S2_vs_k141_4816||full Low-quality
S2_vs_k141_591||full Low-quality
S2_vs_k141_597||full Low-quality
S2_vs_k141_4826||full Low-quality
S2_vs_k141_14331||full Not-determined
S2_vs_k141_18431||full Low-quality
S2_vs_k141_20878||full Low-quality
S2_vs_k141_17007||full Low-quality
S2_vs_k141_1834||full Low-quality
S2_vs_k141_18439||full Low-quality
S2_vs_k141_8868||full Low-quality
S2_vs_k141_13092||full Low-quality
S2_vs_k141_18451||full Low-quality
S2_vs_k141_17019||full Low-quality
S2_vs_k141_8886||full Not-determined
S2_vs_k141_6184||full Not-determined
S2_vs_k141_15821||full Low-quality
S2_vs_k141_10152||full Low-quality
S2_vs_k141_4847||full Not-determined
S2_vs_k141_11654||full Low-quality
S2_vs_k141_7485||full Low-quality
S2_vs_k141_653||full Not-determined
S2_vs_k141_7487||full Low-quality
S2_vs_k141_4850||full Low-quality
S2_vs_k141_8898||full Not-determined
S2_vs_k141_10160||full Low-quality
S11_vs_k141_53675||full Medium-quality
S2_vs_k141_8899||full Low-quality
S2_vs_k141_18493||full Low-quality
S2_vs_k141_1894||full Not-determined
S2_vs_k141_3423||full Low-quality
S2_vs_k141_11687||full Low-quality
S2_vs_k141_3426||full Low-quality
S2_vs_k141_13128||full Not-determined
S7_vs_38834||full Medium-quality
S2_vs_k141_11697||full Low-quality
S2_vs_k141_669||full Not-determined
S2_vs_k141_13146||full Low-quality
S2_vs_k141_18521||full Low-quality
S2_vs_k141_7516||full Low-quality
S2_vs_k141_3436||full Low-quality
S2_vs_k141_6230||full Low-quality
S2_vs_k141_4891||full Low-quality
S2_vs_k141_14379||full Low-quality
S2_vs_k141_4893||full Low-quality
S2_vs_k141_14382||full Low-quality
S2_vs_k141_4897||full Not-determined
S2_vs_k141_1955||full Low-quality
S2_vs_k141_17100||full Low-quality
S2_vs_k141_14398||full Low-quality
S2_vs_k141_4907||full Low-quality
S2_vs_k141_19888||full Not-determined
S2_vs_k141_14401||full Low-quality
S2_vs_k141_13197||full Low-quality
S2_vs_k141_11732||full Low-quality
S2_vs_k141_15878||full Low-quality
S2_vs_k141_1964||full Low-quality
S2_vs_k141_18578||full Low-quality
S2_vs_k141_13212||full Low-quality
S3_vs_k141_70093||full Medium-quality
S2_vs_k141_11738||full Low-quality
S2_vs_k141_17146||full Not-determined
S2_vs_k141_17149||full Low-quality
S2_vs_k141_11747||full Not-determined
S2_vs_k141_21017||full Low-quality
S2_vs_k141_11753||full Low-quality
S2_vs_k141_11759||full Not-determined
S2_vs_k141_13230||full Low-quality
S2_vs_k141_4940||full Low-quality
S2_vs_k141_3501||full Not-determined
S2_vs_k141_14448||full Low-quality
S2_vs_k141_1991||full Low-quality
S2_vs_k141_7548||full Low-quality
S2_vs_k141_21039||full Low-quality
S2_vs_k141_3508||full Low-quality
S2_vs_k141_11781||full Low-quality
S2_vs_k141_755||full Not-determined
S2_vs_k141_9011||full Not-determined
S2_vs_k141_15947||full Low-quality
S2_vs_k141_13275||full Not-determined
S2_vs_k141_10256||full Not-determined
S2_vs_k141_21047||full Not-determined
S2_vs_k141_3534||full Low-quality
S2_vs_k141_21053||full Low-quality
S2_vs_k141_3535||full Low-quality
S2_vs_k141_13284||full Low-quality
S2_vs_k141_21059||full Low-quality
S2_vs_k141_10266||full Low-quality
S2_vs_k141_21078||full Not-determined
S2_vs_k141_18631||full Low-quality
S2_vs_k141_9031||full Low-quality
S2_vs_k141_11805||full Low-quality
S2_vs_k141_21090||full Low-quality
S2_vs_k141_2055||full Low-quality
S2_vs_k141_21093||full Low-quality
S2_vs_k141_14509||full Not-determined
S2_vs_k141_19992||full Low-quality
S2_vs_k141_19995||full Low-quality
S2_vs_k141_15951||full Low-quality
S2_vs_k141_11817||full Low-quality
S2_vs_k141_17173||full Not-determined
S2_vs_k141_3585||full Low-quality
S2_vs_k141_9049||full Low-quality
S2_vs_k141_3588||full Not-determined
S2_vs_k141_824||full Low-quality
S2_vs_k141_11836||full Low-quality
S2_vs_k141_834||full Low-quality
S2_vs_k141_835||full Low-quality
S2_vs_k141_18637||full Low-quality
S2_vs_k141_11837||full Low-quality
S2_vs_k141_7610||full Low-quality
S2_vs_k141_15983||full Low-quality
S2_vs_k141_11843||full Low-quality
S2_vs_k141_17206||full Low-quality
S2_vs_k141_18647||full Low-quality
S2_vs_k141_3627||full Low-quality
S2_vs_k141_20017||full Low-quality
S2_vs_k141_9094||full Not-determined
S2_vs_k141_14555||full Not-determined
S2_vs_k141_17221||full Low-quality
S2_vs_k141_3637||full Low-quality
S2_vs_k141_17224||full Low-quality
S2_vs_k141_4994||full Low-quality
S2_vs_k141_3645||full Low-quality
S2_vs_k141_9105||full Low-quality
S2_vs_k141_2100||full Low-quality
S2_vs_k141_883||full Low-quality
S2_vs_k141_5003||full Not-determined
S2_vs_k141_20040||full Not-determined
S2_vs_k141_11859||full Low-quality
S2_vs_k141_20051||full Not-determined
S2_vs_k141_20053||full Low-quality
S2_vs_k141_11865||full Low-quality
S2_vs_k141_21108||full Low-quality
S2_vs_k141_11866||full Low-quality
S2_vs_k141_21109||full Low-quality
S2_vs_k141_20062||full Low-quality
S4_vs_VIRSorter_2744-cat_3 Medium-quality
S2_vs_k141_21121||full Not-determined
S2_vs_k141_3661||full Low-quality
S2_vs_k141_11878||full Low-quality
S2_vs_k141_7660||full Not-determined
S2_vs_k141_21122||full Not-determined
S2_vs_k141_11886||full Low-quality
S2_vs_k141_17251||full Low-quality
S2_vs_k141_2125||full Low-quality
S2_vs_k141_14603||full Low-quality
S2_vs_k141_3681||full Low-quality
S2_vs_k141_21136||full Low-quality
S2_vs_k141_10293||full Low-quality
S2_vs_k141_20096||full Low-quality
S2_vs_k141_18700||full Low-quality
S2_vs_k141_14608||full Low-quality
S2_vs_k141_14613||full Low-quality
S2_vs_k141_5044||full Low-quality
S2_vs_k141_21163||full Low-quality
S2_vs_k141_16020||full Not-determined
S2_vs_k141_7696||full Low-quality
S2_vs_k141_9210||full Not-determined
S2_vs_k141_7701||full Low-quality
S2_vs_k141_11939||full Low-quality
S2_vs_k141_9213||full Not-determined
S2_vs_k141_21178||full Low-quality
S2_vs_k141_10306||full Not-determined
S2_vs_k141_11952||full Low-quality
S2_vs_k141_2165||full Not-determined
S2_vs_k141_9228||full Low-quality
S2_vs_k141_9229||full Not-determined
S2_vs_k141_9236||full Low-quality
S2_vs_k141_20149||full Low-quality
S2_vs_k141_956||full Low-quality
S2_vs_k141_20157||full Low-quality
S2_vs_k141_2187||full Low-quality
S2_vs_k141_5105||full Not-determined
S2_vs_k141_11977||full Not-determined
S2_vs_k141_16048||full Not-determined
S2_vs_k141_7754||full Low-quality
S2_vs_k141_13401||full Low-quality
S2_vs_k141_9277||full Not-determined
S2_vs_k141_9281||full Low-quality
S2_vs_k141_18767||full Not-determined
S2_vs_k141_9287||full Low-quality
S2_vs_k141_14647||full Not-determined
S2_vs_k141_13425||full Low-quality
S2_vs_k141_3802||full Not-determined
S2_vs_k141_990||full Low-quality
S2_vs_k141_16078||full Not-determined
S2_vs_k141_16079||full Low-quality
S2_vs_k141_14652||full Low-quality
S2_vs_k141_991||full Not-determined
S2_vs_k141_13441||full Low-quality
S2_vs_k141_16092||full Low-quality
S2_vs_k141_3808||full Low-quality
S2_vs_k141_16100||full Low-quality
S2_vs_k141_3816||full Low-quality
S2_vs_k141_7800||full Low-quality
S2_vs_k141_20220||full Low-quality
S2_vs_k141_17325||full Low-quality
S2_vs_k141_2273||full Low-quality
S2_vs_k141_12030||full Low-quality
S2_vs_k141_1001||full Low-quality
S2_vs_k141_20229||full Low-quality
S2_vs_k141_1002||full Low-quality
S2_vs_k141_17330||full Not-determined
S2_vs_k141_3843||full Not-determined
S2_vs_k141_7821||full Not-determined
S2_vs_k141_13462||full Low-quality
S2_vs_k141_10325||full Low-quality
S2_vs_k141_21238||full Low-quality
S2_vs_k141_6303||full Not-determined
S2_vs_k141_20255||full Low-quality
S2_vs_k141_5166||full Not-determined
S2_vs_k141_3862||full Low-quality
S2_vs_k141_16119||full Low-quality
S2_vs_k141_14696||full Low-quality
S2_vs_k141_6311||full Not-determined
S2_vs_k141_6313||full Low-quality
S2_vs_k141_5173||full Low-quality
S2_vs_k141_13473||full Low-quality
S2_vs_k141_21257||full Not-determined
S2_vs_k141_2285||full Low-quality
S2_vs_k141_5176||full Low-quality
S2_vs_k141_3870||full Low-quality
S2_vs_k141_21270||full Low-quality
S2_vs_k141_9348||full Low-quality
S2_vs_k141_13483||full Low-quality
S2_vs_k141_18840||full Low-quality
S2_vs_k141_17376||full Not-determined
S2_vs_k141_21280||full Low-quality
S2_vs_k141_16146||full Low-quality
S2_vs_k141_17377||full Not-determined
S2_vs_k141_14725||full Low-quality
S2_vs_k141_3890||full Not-determined
S2_vs_k141_16159||full Low-quality
S2_vs_k141_3893||full Low-quality
S2_vs_k141_14733||full Low-quality
S2_vs_k141_13523||full Low-quality
S2_vs_k141_16170||full Low-quality
S2_vs_k141_5217||full Low-quality
S2_vs_k141_2295||full Low-quality
S2_vs_k141_9399||full Low-quality
S2_vs_k141_6374||full Not-determined
S2_vs_k141_21330||full Low-quality
S2_vs_k141_2311||full Low-quality
S2_vs_k141_14758||full Low-quality
S2_vs_k141_9415||full Low-quality
S2_vs_k141_5222||full Low-quality
S2_vs_k141_18862||full Low-quality
S2_vs_k141_21353||full Low-quality
S2_vs_k141_14774||full Low-quality
S2_vs_k141_5233||full Low-quality
S2_vs_k141_17411||full Low-quality
S2_vs_k141_3906||full Not-determined
S2_vs_k141_14799||full Low-quality
S2_vs_k141_12112||full Low-quality
S2_vs_k141_18898||full Low-quality
S2_vs_k141_10363||full Low-quality
S2_vs_k141_1084||full Not-determined
S2_vs_k141_1088||full Low-quality
S2_vs_k141_6427||full Low-quality
S2_vs_k141_5259||full Low-quality
S2_vs_k141_1098||full Low-quality
S2_vs_k141_12129||full Low-quality
S2_vs_k141_2394||full Not-determined
S2_vs_k141_5272||full Low-quality
S2_vs_k141_17438||full Not-determined
S2_vs_k141_2400||full Low-quality

| | |
|---|---|
| S2_vs_k141_6463||full | Low-quality |
| S2_vs_k141_6467||full | Low-quality |
| S2_vs_k141_14842||full | Low-quality |
| S2_vs_k141_21408||full | Low-quality |
| S2_vs_k141_21411||full | Low-quality |
| S2_vs_k141_21412||full | Low-quality |
| S2_vs_k141_1135||full | Low-quality |
| S2_vs_k141_6487||full | Not-determined |
| S2_vs_k141_10380||full | Low-quality |
| S2_vs_k141_12161||full | Low-quality |
| S2_vs_k141_17460||full | Low-quality |
| S2_vs_k141_1163||full | Low-quality |
| S2_vs_k141_3914||full | Low-quality |
| S2_vs_k141_7958||full | Low-quality |
| S2_vs_k141_6514||full | Not-determined |
| S2_vs_k141_1190||full | Low-quality |
| S2_vs_k141_6523||full | Not-determined |
| S2_vs_k141_3940||full | Low-quality |
| S2_vs_k141_1207||full | Not-determined |
| S2_vs_k141_6541||full | Low-quality |
| S2_vs_k141_7975||full | Low-quality |
| S2_vs_k141_21471||full | Low-quality |
| S2_vs_k141_5288||full | Low-quality |
| S2_vs_k141_3968||full | Low-quality |
| S2_vs_k141_17520||full | Low-quality |
| S2_vs_k141_3981||full | Low-quality |
| S2_vs_k141_21502||full | Low-quality |
| S2_vs_k141_1248||full | Low-quality |
| S2_vs_k141_21510||full | Low-quality |
| S2_vs_k141_7978||full | Low-quality |
| S2_vs_k141_4003||full | Low-quality |
| S2_vs_k141_1287||full | Low-quality |
| S2_vs_k141_10439||full | Low-quality |
| S2_vs_k141_10451||full | Low-quality |
| S2_vs_k141_5311||full | Low-quality |
| S2_vs_k141_10452||full | Not-determined |
| S2_vs_k141_1312||full | Low-quality |
| S2_vs_k141_10456||full | Low-quality |
| S2_vs_k141_4045||full | Low-quality |
| S2_vs_k141_10468||full | Low-quality |
| S2_vs_k141_2535||full | Low-quality |
| S2_vs_k141_5325||full | Low-quality |
| S2_vs_k141_6563||full | Not-determined |
| S2_vs_k141_6564||full | Low-quality |
| S2_vs_k141_8012||full | Not-determined |
| S2_vs_k141_6568||full | Low-quality |
| S2_vs_k141_1334||full | Low-quality |
| S2_vs_k141_4058||full | Low-quality |
| S2_vs_k141_6602||full | Not-determined |
| S2_vs_k141_10519||full | Not-determined |
| S13_vs_107195||full | Medium-quality |
| S2_vs_k141_8061||full | Low-quality |
| S2_vs_k141_6628||full | Not-determined |
| S2_vs_k141_5360||full | Low-quality |
| S2_vs_k141_10552||full | Not-determined |
| S2_vs_k141_8077||full | Low-quality |
| S2_vs_k141_2626||full | Low-quality |
| S2_vs_k141_2628||full | Low-quality |
| S2_vs_k141_6640||full | Low-quality |
| S2_vs_k141_6642||full | Low-quality |
| S2_vs_k141_2642||full | Not-determined |
| S2_vs_k141_8109||full | Low-quality |
| S2_vs_k141_8117||full | Low-quality |
| S2_vs_k141_21540||full | Low-quality |
| S2_vs_k141_6717||full | Low-quality |
| S2_vs_k141_6720||full | Low-quality |
| S2_vs_k141_21574||full | Low-quality |
| S2_vs_k141_10694||full | Low-quality |
| S2_vs_k141_10723||full | Low-quality |
| S2_vs_k141_10732||full | Low-quality |
| S2_vs_k141_21615||full | Low-quality |
| S2_vs_k141_10742||full | Low-quality |
| S2_vs_k141_10791||full | Low-quality |
| S2_vs_k141_10798||full | Not-determined |
| S2_vs_k141_3051||0_partial | Not-determined |
| S2_vs_k141_18938||lt2gene | Low-quality |
| S2_vs_k141_8119||lt2gene | Not-determined |
| S2_vs_k141_16237||lt2gene | Low-quality |
| S2_vs_k141_9474||lt2gene | Low-quality |
| S2_vs_k141_16239||lt2gene | Low-quality |
| S2_vs_k141_16246||lt2gene | Low-quality |
| S2_vs_k141_18951||lt2gene | Low-quality |
| S2_vs_k141_8136||lt2gene | Low-quality |
| S2_vs_k141_4067||lt2gene | Not-determined |
| S2_vs_k141_8141||lt2gene | Low-quality |
| S2_vs_k141_14891||lt2gene | Not-determined |
| S2_vs_k141_12194||lt2gene | Low-quality |
| S2_vs_k141_4087||lt2gene | Low-quality |
| S2_vs_k141_2745||lt2gene | Low-quality |
| S2_vs_k141_13551||lt2gene | Not-determined |
| S2_vs_k141_10834||lt2gene | Low-quality |
| S2_vs_k141_9504||lt2gene | Low-quality |
| S2_vs_k141_1374||lt2gene | Low-quality |
| S2_vs_k141_34||lt2gene | Low-quality |
| S2_vs_k141_46||lt2gene | Low-quality |
| S2_vs_k141_2764||lt2gene | Not-determined |
| S2_vs_k141_20325||lt2gene | Low-quality |
| S2_vs_k141_16282||lt2gene | Low-quality |
| S2_vs_k141_10843||lt2gene | Low-quality |
| S2_vs_k141_8157||lt2gene | Low-quality |
| S2_vs_k141_4118||lt2gene | Not-determined |
| S2_vs_k141_8162||lt2gene | Low-quality |
| S2_vs_k141_16301||lt2gene | Low-quality |
| S2_vs_k141_16302||lt2gene | Not-determined |
| S2_vs_k141_20368||lt2gene | Low-quality |
| S2_vs_k141_17648||lt2gene | Not-determined |
| S2_vs_k141_17651||lt2gene | Low-quality |
| S2_vs_k141_5462||lt2gene | Low-quality |
| S2_vs_k141_9534||lt2gene | Not-determined |
| S2_vs_k141_8210||lt2gene | Low-quality |
| S2_vs_k141_14964||lt2gene | Low-quality |
| S2_vs_k141_14967||lt2gene | Low-quality |
| S2_vs_k141_17675||lt2gene | Not-determined |
| S2_vs_k141_2789||lt2gene | Low-quality |
| S2_vs_k141_8222||lt2gene | Not-determined |
| S2_vs_k141_8223||lt2gene | Not-determined |
| S2_vs_k141_5474||lt2gene | Low-quality |
| S2_vs_k141_14991||lt2gene | Low-quality |
| S2_vs_k141_2871||lt2gene | Not-determined |
| S2_vs_k141_9613||lt2gene | Low-quality |
| S2_vs_k141_8252||lt2gene | Low-quality |
| S2_vs_k141_10936||lt2gene | Low-quality |
| S2_vs_k141_10938||lt2gene | Low-quality |
| S2_vs_k141_15024||lt2gene | Low-quality |
| S2_vs_k141_10941||lt2gene | Not-determined |
| S2_vs_k141_4154||lt2gene | Not-determined |
| S2_vs_k141_15038||lt2gene | Not-determined |
| S2_vs_k141_6920||lt2gene | Low-quality |
| S2_vs_k141_13686||lt2gene | Low-quality |
| S2_vs_k141_145||lt2gene | Not-determined |
| S2_vs_k141_10968||lt2gene | Low-quality |
| S2_vs_k141_15067||lt2gene | Low-quality |
| S2_vs_k141_10970||lt2gene | Low-quality |
| S2_vs_k141_15069||lt2gene | Low-quality |
| S2_vs_k141_9621||lt2gene | Not-determined |
| S2_vs_k141_150||lt2gene | Low-quality |
| S2_vs_k141_15073||lt2gene | Not-determined |
| S2_vs_k141_12274||lt2gene | Low-quality |
| S2_vs_k141_163||lt2gene | Not-determined |
| S2_vs_k141_2873||lt2gene | Low-quality |
| S2_vs_k141_20489||lt2gene | Low-quality |
| S2_vs_k141_20492||lt2gene | Low-quality |
| S2_vs_k141_4208||lt2gene | Not-determined |
| S2_vs_k141_13719||lt2gene | Low-quality |
| S2_vs_k141_2890||lt2gene | Not-determined |
| S2_vs_k141_2891||lt2gene | Low-quality |
| S2_vs_k141_168||lt2gene | Low-quality |
| S2_vs_k141_2893||lt2gene | Low-quality |
| S2_vs_k141_13729||lt2gene | Low-quality |
| S2_vs_k141_180||lt2gene | Not-determined |
| S2_vs_k141_181||lt2gene | Low-quality |
| S2_vs_k141_17766||lt2gene | Low-quality |
| S2_vs_k141_13747||lt2gene | Not-determined |
| S2_vs_k141_4217||lt2gene | Not-determined |
| S2_vs_k141_13761||lt2gene | Low-quality |
| S2_vs_k141_19139||lt2gene | Not-determined |
| S2_vs_k141_4230||lt2gene | Not-determined |
| S2_vs_k141_12319||lt2gene | Not-determined |
| S2_vs_k141_11020||lt2gene | Not-determined |
| S2_vs_k141_19144||lt2gene | Not-determined |
| S2_vs_k141_5559||lt2gene | Low-quality |
| S2_vs_k141_11028||lt2gene | Low-quality |
| S2_vs_k141_15091||lt2gene | Not-determined |
| S2_vs_k141_4252||lt2gene | Low-quality |
| S2_vs_k141_17785||lt2gene | Not-determined |
| S2_vs_k141_16470||lt2gene | Not-determined |
| S2_vs_k141_7008||lt2gene | Low-quality |
| S2_vs_k141_20543||lt2gene | Not-determined |
| S2_vs_k141_13817||lt2gene | Not-determined |
| S2_vs_k141_13820||lt2gene | Low-quality |
| S2_vs_k141_8293||lt2gene | Low-quality |
| S2_vs_k141_19179||lt2gene | Low-quality |
| S2_vs_k141_15152||lt2gene | Low-quality |
| S2_vs_k141_16512||lt2gene | Low-quality |
| S2_vs_k141_12370||lt2gene | Low-quality |
| S2_vs_k141_2996||lt2gene | Low-quality |
| S2_vs_k141_207||lt2gene | Low-quality |
| S2_vs_k141_16536||lt2gene | Low-quality |
| S2_vs_k141_16538||lt2gene | Not-determined |
| S2_vs_k141_5650||lt2gene | Low-quality |
| S2_vs_k141_17820||lt2gene | Low-quality |
| S2_vs_k141_17828||lt2gene | Low-quality |
| S2_vs_k141_7036||lt2gene | Low-quality |
| S2_vs_k141_8341||lt2gene | Low-quality |
| S2_vs_k141_15199||lt2gene | Low-quality |
| S2_vs_k141_16560||lt2gene | Low-quality |
| S2_vs_k141_13875||lt2gene | Low-quality |
| S2_vs_k141_12418||lt2gene | Low-quality |
| S2_vs_k141_7065||lt2gene | Not-determined |
| S2_vs_k141_12440||lt2gene | Low-quality |
| S2_vs_k141_12446||lt2gene | Low-quality |
| S2_vs_k141_12453||lt2gene | Low-quality |
| S2_vs_k141_20583||lt2gene | Not-determined |
| S2_vs_k141_9773||lt2gene | Not-determined |
| S2_vs_k141_11119||lt2gene | Low-quality |
| S2_vs_k141_11120||lt2gene | Low-quality |
| S2_vs_k141_241||lt2gene | Not-determined |
| S2_vs_k141_11123||lt2gene | Not-determined |
| S2_vs_k141_17853||lt2gene | Not-determined |
| S2_vs_k141_17854||lt2gene | Low-quality |
| S2_vs_k141_11134||lt2gene | Low-quality |
| S2_vs_k141_9778||lt2gene | Low-quality |
| S2_vs_k141_12460||lt2gene | Low-quality |
| S2_vs_k141_20608||lt2gene | Low-quality |
| S2_vs_k141_4333||lt2gene | Low-quality |
| S2_vs_k141_4335||lt2gene | Low-quality |
| S2_vs_k141_20615||lt2gene | Not-determined |
| S2_vs_k141_11144||lt2gene | Not-determined |
| S2_vs_k141_9793||lt2gene | Not-determined |
| S2_vs_k141_4352||lt2gene | Not-determined |
| S2_vs_k141_11157||lt2gene | Low-quality |
| S2_vs_k141_16609||lt2gene | Low-quality |
| S2_vs_k141_16611||lt2gene | Not-determined |
| S2_vs_k141_7122||lt2gene | Not-determined |
| S2_vs_k141_7126||lt2gene | Not-determined |
| S2_vs_k141_20629||lt2gene | Not-determined |
| S2_vs_k141_12513||lt2gene | Low-quality |
| S2_vs_k141_5737||lt2gene | Not-determined |
| S2_vs_k141_12529||lt2gene | Low-quality |
| S2_vs_k141_11188||lt2gene | Not-determined |
| S2_vs_k141_13983||lt2gene | Low-quality |
| S2_vs_k141_20642||lt2gene | Not-determined |
| S2_vs_k141_11194||lt2gene | Low-quality |
| S2_vs_k141_3056||lt2gene | Low-quality |
| S2_vs_k141_7201||lt2gene | Not-determined |
| S2_vs_k141_1545||lt2gene | Not-determined |
| S2_vs_k141_3062||lt2gene | Low-quality |
| S2_vs_k141_9864||lt2gene | Low-quality |
| S2_vs_k141_4412||lt2gene | Not-determined |
| S2_vs_k141_15356||lt2gene | Not-determined |
| S2_vs_k141_3073||lt2gene | Not-determined |
| S2_vs_k141_19388||lt2gene | Low-quality |
| S2_vs_k141_8413||lt2gene | Low-quality |
| S2_vs_k141_16707||lt2gene | Low-quality |
| S2_vs_k141_4433||lt2gene | Not-determined |
| S2_vs_k141_19406||lt2gene | Low-quality |
| S2_vs_k141_16713||lt2gene | Low-quality |
| S2_vs_k141_288||lt2gene | Low-quality |
| S2_vs_k141_16717||lt2gene | Low-quality |
| S2_vs_k141_19415||lt2gene | Low-quality |
| S2_vs_k141_16722||lt2gene | Low-quality |
| S2_vs_k141_20696||lt2gene | Low-quality |
| S2_vs_k141_19427||lt2gene | Low-quality |
| S2_vs_k141_9911||lt2gene | Low-quality |
| S2_vs_k141_12677||lt2gene | Not-determined |
| S2_vs_k141_4498||lt2gene | Low-quality |
| S2_vs_k141_9925||lt2gene | Low-quality |
| S2_vs_k141_9926||lt2gene | Low-quality |
| S2_vs_k141_15435||lt2gene | Low-quality |
| S2_vs_k141_8451||lt2gene | Low-quality |
| S2_vs_k141_7251||lt2gene | Low-quality |
| S2_vs_k141_12732||lt2gene | Not-determined |
| S2_vs_k141_3124||lt2gene | Low-quality |
| S2_vs_k141_3133||lt2gene | Low-quality |
| S2_vs_k141_3136||lt2gene | Low-quality |
| S2_vs_k141_12747||lt2gene | Low-quality |
| S2_vs_k141_3142||lt2gene | Low-quality |
| S2_vs_k141_18051||lt2gene | Low-quality |
| S2_vs_k141_335||lt2gene | Not-determined |
| S2_vs_k141_341||lt2gene | Low-quality |
| S2_vs_k141_343||lt2gene | Not-determined |
| S2_vs_k141_18064||lt2gene | Not-determined |
| S2_vs_k141_19518||lt2gene | Low-quality |
| S2_vs_k141_19519||lt2gene | Not-determined |
| S2_vs_k141_12769||lt2gene | Not-determined |
| S2_vs_k141_18083||lt2gene | Low-quality |
| S2_vs_k141_8507||lt2gene | Low-quality |
| S2_vs_k141_18107||lt2gene | Not-determined |
| S2_vs_k141_1674||lt2gene | Low-quality |
| S2_vs_k141_15453||lt2gene | Low-quality |
| S2_vs_k141_18127||lt2gene | Low-quality |
| S2_vs_k141_12818||lt2gene | Low-quality |
| S2_vs_k141_11371||lt2gene | Low-quality |
| S2_vs_k141_18137||lt2gene | Low-quality |
| S2_vs_k141_15471||lt2gene | Low-quality |
| S2_vs_k141_4610||lt2gene | Not-determined |
| S2_vs_k141_4611||lt2gene | Low-quality |
| S2_vs_k141_15497||lt2gene | Low-quality |
| S2_vs_k141_3192||lt2gene | Low-quality |
| S2_vs_k141_1692||lt2gene | Low-quality |
| S2_vs_k141_8545||lt2gene | Low-quality |
| S2_vs_k141_15531||lt2gene | Not-determined |
| S2_vs_k141_14130||lt2gene | Low-quality |
| S2_vs_k141_10030||lt2gene | Not-determined |
| S2_vs_k141_12864||lt2gene | Low-quality |
| S2_vs_k141_3202||lt2gene | Low-quality |
| S2_vs_k141_4638||lt2gene | Not-determined |
| S2_vs_k141_15548||lt2gene | Not-determined |
| S2_vs_k141_7313||lt2gene | Not-determined |
| S2_vs_k141_16871||lt2gene | Not-determined |
| S2_vs_k141_390||lt2gene | Not-determined |
| S2_vs_k141_1700||lt2gene | Low-quality |
| S2_vs_k141_10063||lt2gene | Not-determined |
| S2_vs_k141_12892||lt2gene | Low-quality |
| S2_vs_k141_19573||lt2gene | Low-quality |
| S2_vs_k141_12903||lt2gene | Low-quality |
| S2_vs_k141_401||lt2gene | Low-quality |
| S2_vs_k141_7323||lt2gene | Not-determined |
| S2_vs_k141_18196||lt2gene | Not-determined |
| S2_vs_k141_19582||lt2gene | Low-quality |
| S2_vs_k141_7328||lt2gene | Not-determined |
| S2_vs_k141_19599||lt2gene | Low-quality |
| S2_vs_k141_10075||lt2gene | Low-quality |
| S2_vs_k141_15583||lt2gene | Not-determined |
| S2_vs_k141_10081||lt2gene | Low-quality |
| S2_vs_k141_4665||lt2gene | Low-quality |
| S2_vs_k141_15599||lt2gene | Low-quality |
| S2_vs_k141_19621||lt2gene | Low-quality |
| S2_vs_k141_18245||lt2gene | Low-quality |
| S2_vs_k141_3265||lt2gene | Low-quality |
| S2_vs_k141_5912||lt2gene | Not-determined |
| S2_vs_k141_4688||lt2gene | Not-determined |
| S2_vs_k141_3282||lt2gene | Low-quality |
| S2_vs_k141_10106||lt2gene | Low-quality |
| S2_vs_k141_447||lt2gene | Low-quality |
| S2_vs_k141_18259||lt2gene | Not-determined |
| S2_vs_k141_458||lt2gene | Low-quality |
| S2_vs_k141_4701||lt2gene | Low-quality |
| S2_vs_k141_15653||lt2gene | Low-quality |
| S2_vs_k141_15657||lt2gene | Not-determined |
| S2_vs_k141_18267||lt2gene | Low-quality |
| S2_vs_k141_8635||lt2gene | Low-quality |
| S2_vs_k141_471||lt2gene | Not-determined |
| S2_vs_k141_4715||lt2gene | Low-quality |
| S2_vs_k141_5973||lt2gene | Low-quality |
| S2_vs_k141_4723||lt2gene | Not-determined |
| S2_vs_k141_16927||lt2gene | Low-quality |
| S2_vs_k141_16938||lt2gene | Low-quality |
| S2_vs_k141_3313||lt2gene | Low-quality |
| S2_vs_k141_18319||lt2gene | Low-quality |
| S2_vs_k141_1769||lt2gene | Low-quality |
| S2_vs_k141_3323||lt2gene | Not-determined |
| S2_vs_k141_3327||lt2gene | Low-quality |
| S2_vs_k141_6010||lt2gene | Not-determined |
| S2_vs_k141_1781||lt2gene | Not-determined |
| S2_vs_k141_6021||lt2gene | Low-quality |
| S2_vs_k141_13008||lt2gene | Not-determined |
| S2_vs_k141_16948||lt2gene | Low-quality |
| S2_vs_k141_11510||lt2gene | Not-determined |
| S2_vs_k141_20824||lt2gene | Low-quality |
| S2_vs_k141_11518||lt2gene | Low-quality |
| S2_vs_k141_494||lt2gene | Low-quality |
| S2_vs_k141_497||lt2gene | Low-quality |
| S2_vs_k141_11527||lt2gene | Low-quality |
| S2_vs_k141_11532||lt2gene | Low-quality |
| S2_vs_k141_13026||lt2gene | Low-quality |
| S2_vs_k141_18375||lt2gene | Low-quality |
| S2_vs_k141_18377||lt2gene | Low-quality |
| S2_vs_k141_8757||lt2gene | Low-quality |
| S2_vs_k141_19755||lt2gene | Low-quality |
| S2_vs_k141_13038||lt2gene | Low-quality |
| S2_vs_k141_13044||lt2gene | Not-determined |
| S2_vs_k141_11541||lt2gene | Not-determined |
| S2_vs_k141_4774||lt2gene | Low-quality |
| S2_vs_k141_7465||lt2gene | Low-quality |
| S2_vs_k141_10125||lt2gene | Low-quality |
| S2_vs_k141_8798||lt2gene | Low-quality |
| S2_vs_k141_7472||lt2gene | Not-determined |
| S2_vs_k141_11565||lt2gene | Low-quality |
| S2_vs_k141_8809||lt2gene | Low-quality |
| S2_vs_k141_18409||lt2gene | Not-determined |
| S2_vs_k141_4798||lt2gene | Low-quality |
| S2_vs_k141_4804||lt2gene | Low-quality |
| S2_vs_k141_15768||lt2gene | Low-quality |
| S2_vs_k141_15777||lt2gene | Low-quality |
| S2_vs_k141_15784||lt2gene | Low-quality |
| S2_vs_k141_11607||lt2gene | Low-quality |
| S2_vs_k141_11610||lt2gene | Low-quality |
| S2_vs_k141_599||lt2gene | Low-quality |
| S2_vs_k141_8859||lt2gene | Low-quality |
| S2_vs_k141_1835||lt2gene | Not-determined |
| S2_vs_k141_14339||lt2gene | Not-determined |
| S2_vs_k141_4834||lt2gene | Not-determined |
| S2_vs_k141_11627||lt2gene | Not-determined |
| S2_vs_k141_8873||lt2gene | Low-quality |
| S2_vs_k141_6169||lt2gene | Low-quality |
| S2_vs_k141_6171||lt2gene | Low-quality |
| S2_vs_k141_619||lt2gene | Low-quality |
| S2_vs_k141_20895||lt2gene | Not-determined |
| S2_vs_k141_20896||lt2gene | Not-determined |
| S2_vs_k141_6177||lt2gene | Low-quality |
| S2_vs_k141_13094||lt2gene | Low-quality |
| S2_vs_k141_13096||lt2gene | Low-quality |
| S2_vs_k141_10131||lt2gene | Low-quality |
| S2_vs_k141_628||lt2gene | Low-quality |
| S2_vs_k141_17026||lt2gene | Not-determined |
| S2_vs_k141_6200||lt2gene | Not-determined |
| S2_vs_k141_14370||lt2gene | Not-determined |
| S2_vs_k141_18479||lt2gene | Not-determined |
| S2_vs_k141_11676||lt2gene | Low-quality |
| S2_vs_k141_11686||lt2gene | Not-determined |
| S2_vs_k141_1900||lt2gene | Not-determined |
| S2_vs_k141_4873||lt2gene | Low-quality |
| S2_vs_k141_19868||lt2gene | Low-quality |
| S2_vs_k141_19869||lt2gene | Not-determined |
| S2_vs_k141_19873||lt2gene | Low-quality |
| S2_vs_k141_10174||lt2gene | Low-quality |
| S2_vs_k141_18519||lt2gene | Low-quality |
| S2_vs_k141_1926||lt2gene | Not-determined |
| S2_vs_k141_6223||lt2gene | Low-quality |
| S2_vs_k141_6225||lt2gene | Low-quality |
| S2_vs_k141_8933||lt2gene | Low-quality |
| S2_vs_k141_17083||lt2gene | Not-determined |
| S2_vs_k141_18540||lt2gene | Low-quality |
| S2_vs_k141_17098||lt2gene | Low-quality |
| S2_vs_k141_8966||lt2gene | Low-quality |
| S2_vs_k141_14389||lt2gene | Low-quality |
| S2_vs_k141_13187||lt2gene | Low-quality |
| S2_vs_k141_17101||lt2gene | Not-determined |
| S2_vs_k141_15866||lt2gene | Low-quality |
| S2_vs_k141_683||lt2gene | Low-quality |
| S2_vs_k141_17116||lt2gene | Low-quality |
| S2_vs_k141_15884||lt2gene | Low-quality |
| S2_vs_k141_10206||lt2gene | Low-quality |
| S2_vs_k141_14426||lt2gene | Low-quality |
| S2_vs_k141_11739||lt2gene | Low-quality |
| S2_vs_k141_8998||lt2gene | Low-quality |
| S2_vs_k141_720||lt2gene | Low-quality |
| S2_vs_k141_18600||lt2gene | Low-quality |
| S2_vs_k141_11765||lt2gene | Low-quality |
| S2_vs_k141_21028||lt2gene | Not-determined |
| S2_vs_k141_3503||lt2gene | Low-quality |
| S2_vs_k141_7543||lt2gene | Low-quality |
| S2_vs_k141_7546||lt2gene | Low-quality |
| S2_vs_k141_746||lt2gene | Low-quality |
| S2_vs_k141_15946||lt2gene | Not-determined |
| S2_vs_k141_2016||lt2gene | Low-quality |
| S2_vs_k141_11785||lt2gene | Low-quality |
| S2_vs_k141_11788||lt2gene | Not-determined |
| S2_vs_k141_14488||lt2gene | Low-quality |
| S2_vs_k141_21065||lt2gene | Low-quality |
| S2_vs_k141_10272||lt2gene | Low-quality |
| S2_vs_k141_19975||lt2gene | Low-quality |
| S2_vs_k141_19987||lt2gene | Low-quality |
| S2_vs_k141_802||lt2gene | Not-determined |
| S2_vs_k141_17171||lt2gene | Low-quality |
| S2_vs_k141_21096||lt2gene | Low-quality |
| S2_vs_k141_14516||lt2gene | Not-determined |
| S2_vs_k141_3584||lt2gene | Low-quality |
| S2_vs_k141_819||lt2gene | Low-quality |
| S2_vs_k141_4970||lt2gene | Not-determined |
| S2_vs_k141_14531||lt2gene | Low-quality |
| S2_vs_k141_9059||lt2gene | Low-quality |
| S2_vs_k141_17189||lt2gene | Low-quality |
| S2_vs_k141_4989||lt2gene | Low-quality |
| S2_vs_k141_17201||lt2gene | Low-quality |
| S2_vs_k141_3626||lt2gene | Low-quality |
| S2_vs_k141_9098||lt2gene | Not-determined |
| S2_vs_k141_20024||lt2gene | Low-quality |
| S2_vs_k141_2093||lt2gene | Low-quality |
| S2_vs_k141_875||lt2gene | Low-quality |
| S2_vs_k141_7636||lt2gene | Not-determined |
| S2_vs_k141_879||lt2gene | Low-quality |
| S2_vs_k141_14570||lt2gene | Not-determined |
| S2_vs_k141_20039||lt2gene | Low-quality |
| S2_vs_k141_9126||lt2gene | Not-determined |
| S2_vs_k141_20054||lt2gene | Low-quality |
| S2_vs_k141_7652||lt2gene | Low-quality |
| S2_vs_k141_18665||lt2gene | Low-quality |
| S2_vs_k141_18676||lt2gene | Low-quality |
| S2_vs_k141_20087||lt2gene | Low-quality |
| S2_vs_k141_2127||lt2gene | Low-quality |
| S2_vs_k141_7674||lt2gene | Low-quality |
| S2_vs_k141_16001||lt2gene | Low-quality |
| S2_vs_k141_5055||lt2gene | Low-quality |
| S2_vs_k141_10305||lt2gene | Low-quality |
| S2_vs_k141_3716||lt2gene | Low-quality |
| S2_vs_k141_11944||lt2gene | Low-quality |
| S2_vs_k141_11946||lt2gene | Low-quality |
| S2_vs_k141_5079||lt2gene | Low-quality |
| S2_vs_k141_11953||lt2gene | Low-quality |
| S2_vs_k141_11958||lt2gene | Low-quality |
| S2_vs_k141_9231||lt2gene | Not-determined |
| S2_vs_k141_11960||lt2gene | Low-quality |
| S2_vs_k141_7743||lt2gene | Low-quality |
| S2_vs_k141_18736||lt2gene | Low-quality |
| S2_vs_k141_13384||lt2gene | Low-quality |
| S2_vs_k141_18738||lt2gene | Low-quality |
| S2_vs_k141_2201||lt2gene | Low-quality |
| S2_vs_k141_9272||lt2gene | Low-quality |
| S2_vs_k141_14636||lt2gene | Low-quality |
| S2_vs_k141_9280||lt2gene | Not-determined |
| S2_vs_k141_11995||lt2gene | Low-quality |
| S2_vs_k141_13411||lt2gene | Low-quality |
| S2_vs_k141_13421||lt2gene | Not-determined |
| S2_vs_k141_2241||lt2gene | Not-determined |
| S2_vs_k141_3804||lt2gene | Low-quality |
| S2_vs_k141_5131||lt2gene | Low-quality |
| S2_vs_k141_16081||lt2gene | Low-quality |
| S2_vs_k141_18783||lt2gene | Not-determined |
| S2_vs_k141_16089||lt2gene | Low-quality |
| S2_vs_k141_9309||lt2gene | Low-quality |
| S2_vs_k141_21222||lt2gene | Low-quality |
| S2_vs_k141_7793||lt2gene | Low-quality |
| S2_vs_k141_2259||lt2gene | Low-quality |
| S2_vs_k141_21231||lt2gene | Not-determined |
| S2_vs_k141_9326||lt2gene | Low-quality |
| S2_vs_k141_20217||lt2gene | Low-quality |
| S2_vs_k141_1000||lt2gene | Low-quality |
| S2_vs_k141_6266||lt2gene | Low-quality |
| S2_vs_k141_20230||lt2gene | Not-determined |
| S2_vs_k141_6288||lt2gene | Low-quality |
| S2_vs_k141_12042||lt2gene | Low-quality |
| S2_vs_k141_5158||lt2gene | Low-quality |
| S2_vs_k141_10329||lt2gene | Low-quality |
| S2_vs_k141_21247||lt2gene | Low-quality |
| S2_vs_k141_16121||lt2gene | Not-determined |
| S2_vs_k141_9338||lt2gene | Not-determined |
| S2_vs_k141_7841||lt2gene | Not-determined |
| S2_vs_k141_13474||lt2gene | Low-quality |
| S2_vs_k141_13475||lt2gene | Not-determined |
| S2_vs_k141_6322||lt2gene | Low-quality |
| S2_vs_k141_7849||lt2gene | Low-quality |
| S2_vs_k141_17370||lt2gene | Low-quality |
| S2_vs_k141_13501||lt2gene | Low-quality |
| S2_vs_k141_21285||lt2gene | Low-quality |
| S2_vs_k141_1020||lt2gene | Low-quality |
| S2_vs_k141_14727||lt2gene | Not-determined |
| S2_vs_k141_5214||lt2gene | Not-determined |
| S2_vs_k141_21325||lt2gene | Low-quality |
| S2_vs_k141_9408||lt2gene | Low-quality |
| S2_vs_k141_7883||lt2gene | Low-quality |
| S2_vs_k141_16188||lt2gene | Not-determined |
| S2_vs_k141_16196||lt2gene | Not-determined |
| S2_vs_k141_21342||lt2gene | Low-quality |
| S2_vs_k141_18863||lt2gene | Low-quality |
| S2_vs_k141_6401||lt2gene | Not-determined |
| S2_vs_k141_2330||lt2gene | Low-quality |
| S2_vs_k141_1054||lt2gene | Low-quality |
| S2_vs_k141_18877||lt2gene | Low-quality |
| S2_vs_k141_18885||lt2gene | Low-quality |
| S2_vs_k141_1064||lt2gene | Low-quality |
| S2_vs_k141_10352||lt2gene | Not-determined |
| S2_vs_k141_10356||lt2gene | Low-quality |
| S2_vs_k141_14787||lt2gene | Not-determined |
| S2_vs_k141_1073||lt2gene | Low-quality |
| S2_vs_k141_14788||lt2gene | Low-quality |
| S2_vs_k141_10365||lt2gene | Not-determined |
| S2_vs_k141_5255||lt2gene | Not-determined |
| S2_vs_k141_5257||lt2gene | Low-quality |
| S2_vs_k141_17430||lt2gene | Low-quality |
| S2_vs_k141_17432||lt2gene | Low-quality |
| S2_vs_k141_2398||lt2gene | Low-quality |
| S2_vs_k141_18933||lt2gene | Low-quality |
| S2_vs_k141_6448||lt2gene | Low-quality |
| S2_vs_k141_12145||lt2gene | Low-quality |
| S2_vs_k141_17443||lt2gene | Low-quality |
| S2_vs_k141_10372||lt2gene | Low-quality |
| S2_vs_k141_10399||lt2gene | Not-determined |
| S2_vs_k141_6508||lt2gene | Not-determined |
| S2_vs_k141_14878||lt2gene | Low-quality |
| S2_vs_k141_1198||lt2gene | Low-quality |
| S2_vs_k141_10417||lt2gene | Low-quality |
| S2_vs_k141_3939||lt2gene | Not-determined |
| S2_vs_k141_17509||lt2gene | Low-quality |
| S2_vs_k141_1257||lt2gene | Not-determined |
| S2_vs_k141_1261||lt2gene | Low-quality |
| S2_vs_k141_1275||lt2gene | Not-determined |
| S2_vs_k141_10438||lt2gene | Not-determined |
| S2_vs_k141_7988||lt2gene | Low-quality |
| S2_vs_k141_4009||lt2gene | Low-quality |
| S2_vs_k141_1300||lt2gene | Not-determined |
| S2_vs_k141_1304||lt2gene | Low-quality |
| S2_vs_k141_1321||lt2gene | Low-quality |
| S2_vs_k141_2533||lt2gene | Not-determined |
| S2_vs_k141_6572||lt2gene | Low-quality |
| S2_vs_k141_10488||lt2gene | Not-determined |
| S2_vs_k141_1338||lt2gene | Not-determined |
| S2_vs_k141_10516||lt2gene | Low-quality |
| S2_vs_k141_5346||lt2gene | Low-quality |
| S2_vs_k141_5347||lt2gene | Low-quality |
| S2_vs_k141_6616||lt2gene | Low-quality |
| S2_vs_k141_8058||lt2gene | Not-determined |
| S2_vs_k141_8060||lt2gene | Not-determined |
| S2_vs_k141_10563||lt2gene | Low-quality |
| S2_vs_k141_10568||lt2gene | Low-quality |
| S2_vs_k141_8098||lt2gene | Not-determined |
| S2_vs_k141_5387||lt2gene | Low-quality |
| S2_vs_k141_10573||lt2gene | Low-quality |
| S2_vs_k141_6657||lt2gene | Low-quality |
| S2_vs_k141_6662||lt2gene | Low-quality |
| S2_vs_k141_21529||lt2gene | Low-quality |
| S2_vs_k141_10597||lt2gene | Low-quality |
| S2_vs_k141_5404||lt2gene | Low-quality |
| S2_vs_k141_10603||lt2gene | Not-determined |
| S2_vs_k141_10621||lt2gene | Low-quality |
| S2_vs_k141_10625||lt2gene | Low-quality |
| S2_vs_k141_2675||lt2gene | Not-determined |
| S2_vs_k141_2681||lt2gene | Not-determined |

| | |
|---|---|
| S2_vs_k141_6741\|\|lt2gene | Low-quality |
| S2_vs_k141_6758\|\|lt2gene | Not-determined |
| S2_vs_k141_10729\|\|lt2gene | Low-quality |
| S2_vs_k141_10735\|\|lt2gene | Not-determined |
| S2_vs_k141_10744\|\|lt2gene | Low-quality |
| S2_vs_k141_10777\|\|lt2gene | Not-determined |
| S2_vs_k141_10785\|\|lt2gene | Not-determined |
| S4_extracted_2 | Not-determined |
| S4_extracted_11 | Not-determined |
| S4_extracted_28 | Not-determined |
| S4_extracted_49 | Not-determined |
| S4_extracted_55 | Not-determined |
| S4_extracted_65 | Not-determined |
| S4_extracted_66 | Low-quality |
| S4_extracted_87 | Not-determined |
| S4_extracted_91 | Low-quality |
| S4_extracted_93 | Not-determined |
| S4_extracted_100 | Low-quality |
| S4_extracted_101 | Not-determined |
| S4_extracted_105 | Not-determined |
| S4_extracted_116 | Not-determined |
| S4_extracted_120 | Low-quality |
| S4_extracted_134 | Low-quality |
| S4_extracted_149 | Not-determined |
| S4_extracted_152 | Not-determined |
| S4_extracted_159 | Not-determined |
| S4_extracted_162 | Not-determined |
| S4_extracted_208 | Not-determined |
| S4_extracted_211 | Not-determined |
| S4_extracted_212 | Not-determined |
| S4_extracted_222 | Low-quality |
| S4_extracted_230 | Low-quality |
| S4_extracted_231 | Not-determined |
| S4_extracted_233 | Not-determined |
| S4_extracted_239 | Not-determined |
| S4_extracted_245 | Not-determined |
| S4_extracted_267 | Not-determined |
| S4_extracted_269 | Not-determined |
| S4_extracted_271 | Not-determined |
| S4_extracted_274 | Not-determined |
| S4_extracted_305 | Not-determined |
| S4_extracted_307 | Not-determined |
| S4_extracted_311 | Not-determined |
| S4_extracted_313 | Not-determined |
| S4_extracted_317 | Not-determined |
| S4_extracted_320 | Not-determined |
| S4_extracted_392 | Not-determined |
| S4_extracted_397 | Not-determined |
| S4_extracted_408 | Not-determined |
| S4_extracted_410 | Not-determined |
| S4_extracted_416 | Not-determined |
| S4_extracted_423 | Not-determined |
| S4_extracted_427 | Not-determined |
| S4_extracted_447 | Not-determined |
| S4_extracted_450 | Not-determined |
| S4_extracted_451 | Not-determined |
| S4_extracted_456 | Not-determined |
| S4_extracted_459 | Low-quality |
| S4_extracted_465 | Not-determined |
| S4_extracted_482 | Not-determined |
| S4_extracted_486 | Not-determined |
| S4_extracted_493 | Not-determined |
| S4_extracted_497 | Not-determined |
| S4_extracted_506 | Not-determined |
| S4_extracted_508 | Not-determined |
| S4_extracted_509 | Not-determined |
| S4_extracted_517 | Not-determined |
| S4_extracted_525 | Low-quality |
| S4_extracted_552 | Not-determined |
| S4_extracted_558 | Low-quality |
| S4_extracted_559 | Not-determined |
| S4_extracted_566 | Not-determined |
| S4_extracted_568 | Not-determined |
| S4_extracted_569 | Not-determined |
| S4_extracted_575 | Not-determined |
| S4_extracted_576 | Not-determined |
| S4_extracted_592 | Low-quality |
| S4_extracted_595 | Not-determined |
| S4_extracted_608 | Not-determined |
| S4_extracted_622 | Not-determined |
| S4_extracted_624 | Low-quality |
| S4_extracted_641 | Low-quality |
| S4_extracted_642 | Low-quality |
| S4_extracted_648 | Not-determined |
| S4_extracted_651 | Not-determined |
| S4_extracted_684 | Low-quality |
| S4_extracted_685 | Low-quality |
| S4_extracted_692 | Low-quality |
| S4_extracted_696 | Not-determined |
| S4_extracted_702 | Not-determined |
| S4_extracted_708 | Not-determined |
| S4_extracted_714 | Not-determined |
| S4_extracted_716 | Not-determined |
| S4_extracted_719 | Not-determined |
| S4_extracted_739 | Not-determined |
| S4_extracted_751 | Not-determined |
| S4_extracted_757 | Low-quality |
| S4_extracted_759 | Low-quality |
| S4_extracted_770 | Low-quality |
| S4_extracted_771 | Low-quality |
| S4_extracted_784 | Low-quality |
| S4_extracted_793 | Not-determined |
| S4_extracted_799 | Not-determined |
| S4_extracted_802 | Not-determined |
| S4_extracted_813 | Not-determined |
| S4_extracted_816 | Not-determined |
| S4_extracted_819 | Not-determined |
| S4_extracted_827 | Low-quality |
| S4_extracted_859 | Low-quality |
| S4_extracted_864 | Not-determined |
| S4_extracted_867 | Not-determined |
| S4_extracted_874 | Low-quality |
| S4_extracted_876 | Not-determined |
| S4_extracted_879 | Not-determined |
| S4_extracted_891 | Not-determined |
| S4_extracted_898 | Not-determined |
| S4_extracted_900 | Not-determined |
| S4_extracted_905 | Not-determined |
| S4_extracted_931 | Not-determined |
| S4_extracted_934 | Not-determined |
| S4_extracted_938 | Not-determined |
| S4_extracted_952 | Low-quality |
| S4_extracted_955 | Not-determined |
| S4_extracted_958 | Low-quality |
| S4_extracted_966 | Not-determined |
| S4_extracted_968 | Not-determined |
| S4_extracted_971 | Not-determined |
| S4_extracted_977 | Not-determined |
| S4_extracted_981 | Not-determined |
| S4_extracted_983 | Low-quality |
| S4_extracted_993 | Not-determined |
| S4_extracted_994 | Low-quality |
| S4_extracted_996 | Not-determined |
| S4_extracted_1000 | Not-determined |
| S4_extracted_1001 | Not-determined |
| S4_extracted_1016 | Not-determined |
| S4_extracted_1020 | Low-quality |
| S4_extracted_1036 | Low-quality |
| S4_extracted_1039 | Not-determined |
| S4_extracted_1040 | Not-determined |
| S4_extracted_1045 | Not-determined |
| S4_extracted_1047 | Not-determined |
| S4_extracted_1067 | Not-determined |
| S4_extracted_1086 | Not-determined |
| S4_extracted_1093 | Not-determined |
| S4_extracted_1098 | Low-quality |
| S4_extracted_1100 | Low-quality |
| S4_extracted_1106 | Not-determined |
| S4_extracted_1108 | Low-quality |
| S4_extracted_1124 | Low-quality |
| S4_extracted_1134 | Low-quality |
| S4_extracted_1137 | Not-determined |
| S4_extracted_1143 | Not-determined |
| S4_extracted_1144 | Not-determined |
| S4_extracted_1155 | Low-quality |
| S4_extracted_1162 | Low-quality |
| S4_extracted_1164 | Not-determined |
| S4_extracted_1188 | Not-determined |
| S4_extracted_1191 | Not-determined |
| S4_extracted_1199 | Low-quality |
| S4_extracted_1207 | Not-determined |
| S4_extracted_1211 | Low-quality |
| S4_extracted_1213 | Not-determined |
| S4_extracted_1219 | Not-determined |
| S4_extracted_1223 | Not-determined |
| S4_extracted_1237 | Low-quality |
| S4_extracted_1244 | Not-determined |
| S4_extracted_1255 | Not-determined |
| S4_extracted_1268 | Low-quality |
| S4_extracted_1269 | Not-determined |
| S4_extracted_1271 | Low-quality |
| S4_extracted_1279 | Low-quality |
| S4_extracted_1280 | Not-determined |
| S4_extracted_1283 | Not-determined |
| S4_extracted_1284 | Not-determined |
| S4_extracted_1287 | Low-quality |
| S4_extracted_1290 | Not-determined |
| S4_extracted_1293 | Not-determined |
| S4_extracted_1313 | Not-determined |
| S4_extracted_1360 | Not-determined |
| S4_extracted_1395 | Not-determined |
| S4_extracted_1405 | Not-determined |
| S4_extracted_1426 | Not-determined |
| S4_extracted_1431 | Not-determined |
| S4_extracted_1432 | Not-determined |
| S4_extracted_1445 | Not-determined |
| S4_extracted_1447 | Not-determined |
| S4_extracted_1458 | Not-determined |
| S4_extracted_1469 | Not-determined |
| S4_extracted_1473 | Low-quality |
| S4_extracted_1483 | Not-determined |
| S4_extracted_1489 | Not-determined |
| S4_extracted_1510 | Not-determined |
| S4_extracted_1519 | Not-determined |
| S4_extracted_1541 | Not-determined |
| S4_extracted_1544 | Not-determined |
| S4_extracted_1569 | Not-determined |
| S4_extracted_1575 | Not-determined |
| S4_extracted_1576 | Not-determined |
| S4_extracted_1583 | Not-determined |
| S4_extracted_1586 | Low-quality |
| S4_extracted_1587 | Not-determined |
| S4_extracted_1591 | Not-determined |
| S4_extracted_1592 | Not-determined |
| S4_extracted_1595 | Not-determined |
| S4_extracted_1601 | Not-determined |
| S4_extracted_1615 | Not-determined |
| S4_extracted_1625 | Low-quality |
| S4_extracted_1627 | Low-quality |
| S4_extracted_1632 | Not-determined |
| S4_extracted_1635 | Not-determined |
| S4_extracted_1644 | Not-determined |
| S4_extracted_1655 | Not-determined |
| S4_extracted_1659 | Not-determined |
| S4_extracted_1673 | Not-determined |
| S4_extracted_1675 | Not-determined |
| S4_extracted_1678 | Not-determined |
| S4_extracted_1708 | Not-determined |
| S4_extracted_1713 | Not-determined |
| S4_extracted_1719 | Low-quality |
| S4_extracted_1736 | Not-determined |
| S4_extracted_1742 | Not-determined |
| S4_extracted_1748 | Not-determined |
| S4_extracted_1753 | Low-quality |
| S4_extracted_1754 | Not-determined |
| S4_extracted_1757 | Not-determined |
| S4_extracted_1759 | Not-determined |
| S4_extracted_1763 | Not-determined |
| S4_extracted_1768 | Low-quality |
| S4_extracted_1777 | Low-quality |
| S4_extracted_1784 | Not-determined |
| S4_extracted_1791 | Not-determined |
| S4_extracted_1803 | Not-determined |
| S4_extracted_1816 | Low-quality |
| S4_extracted_1865 | Not-determined |
| S4_extracted_1882 | Not-determined |
| S4_extracted_1886 | Not-determined |
| S4_extracted_1895 | Not-determined |
| S4_extracted_1896 | Low-quality |
| S4_extracted_1912 | Not-determined |
| S4_extracted_1918 | Not-determined |
| S4_extracted_1936 | Not-determined |
| S4_extracted_1945 | Not-determined |
| S4_extracted_1949 | Not-determined |
| S4_extracted_1961 | Low-quality |
| S4_extracted_1970 | Low-quality |
| S4_extracted_1979 | Not-determined |
| S4_extracted_1983 | Not-determined |
| S4_extracted_1989 | Not-determined |
| S4_extracted_2012 | Low-quality |
| S4_extracted_2015 | Not-determined |
| S4_extracted_2019 | Not-determined |
| S4_extracted_2024 | Not-determined |
| S4_extracted_2027 | Not-determined |
| S4_extracted_2032 | Not-determined |
| S4_extracted_2035 | Not-determined |
| S4_extracted_2038 | Low-quality |
| S4_extracted_2050 | Not-determined |
| S4_extracted_2051 | Low-quality |
| S4_extracted_2057 | Not-determined |
| S4_extracted_2064 | Low-quality |
| S4_extracted_2081 | Not-determined |
| S4_extracted_2088 | Low-quality |
| S4_extracted_2090 | Not-determined |
| S4_extracted_2091 | Not-determined |
| S4_extracted_2102 | Not-determined |
| S4_extracted_2104 | Low-quality |
| S4_extracted_2117 | Low-quality |
| S4_extracted_2119 | Low-quality |
| S4_extracted_2124 | Not-determined |
| S4_extracted_2130 | Not-determined |
| S4_extracted_2131 | Not-determined |
| S4_extracted_2139 | Low-quality |
| S4_extracted_2143 | Not-determined |
| S4_extracted_2146 | Not-determined |
| S4_extracted_2171 | Not-determined |
| S4_extracted_2179 | Not-determined |
| S4_extracted_2201 | Not-determined |
| S4_extracted_2204 | Not-determined |
| S8_vs_k141_132472\|\|full | Medium-quality |
| S4_extracted_2218 | Not-determined |
| S4_extracted_2219 | Not-determined |
| S4_extracted_2224 | Not-determined |
| S4_extracted_2256 | Not-determined |
| S4_extracted_2261 | Not-determined |
| S4_extracted_2264 | Not-determined |
| S4_extracted_2287 | Not-determined |
| S4_extracted_2296 | Not-determined |
| S4_extracted_2319 | Not-determined |
| S4_extracted_2326 | Not-determined |
| S4_extracted_2327 | Not-determined |
| S4_extracted_2331 | Not-determined |
| S4_extracted_2334 | Not-determined |
| S4_extracted_2348 | Low-quality |
| S4_extracted_2351 | Not-determined |
| S4_extracted_2357 | Not-determined |
| S4_extracted_2368 | Not-determined |
| S4_extracted_2371 | Low-quality |
| S4_extracted_2379 | Not-determined |
| S4_extracted_2386 | Low-quality |
| S4_extracted_2388 | Not-determined |
| S4_extracted_2391 | Not-determined |
| S4_extracted_2399 | Not-determined |
| S4_extracted_2400 | Not-determined |
| S4_extracted_2408 | Not-determined |
| S4_extracted_2413 | Not-determined |
| S4_extracted_2415 | Not-determined |
| S4_extracted_2421 | Not-determined |
| S4_extracted_2435 | Low-quality |
| S4_extracted_2436 | Low-quality |
| S16_vs_140468\|\|full | Medium-quality |
| S4_extracted_2442 | Low-quality |
| S4_extracted_2447 | Not-determined |
| S4_extracted_2464 | Not-determined |
| S4_extracted_2465 | Not-determined |
| S4_extracted_2468 | Not-determined |
| S4_extracted_2469 | Low-quality |
| S4_extracted_2470 | Not-determined |
| S4_extracted_2475 | Not-determined |
| S4_extracted_2493 | Not-determined |
| S4_extracted_2497 | Not-determined |
| S4_extracted_2500 | Not-determined |
| S4_extracted_2505 | Not-determined |
| S4_extracted_2519 | Not-determined |
| S4_extracted_2520 | Not-determined |
| S4_extracted_2521 | Not-determined |
| S4_extracted_2523 | Low-quality |
| S13_vs_121307\|\|full | Medium-quality |
| S4_extracted_2528 | Low-quality |
| S4_extracted_2533 | Not-determined |
| S4_extracted_2544 | Low-quality |
| S4_extracted_2552 | Not-determined |
| S4_extracted_2554 | Not-determined |
| S4_extracted_2560 | Not-determined |
| S4_extracted_2577 | Not-determined |
| S4_extracted_2592 | Not-determined |
| S4_extracted_2594 | Not-determined |
| S4_extracted_2602 | Not-determined |
| S4_extracted_2604 | Not-determined |
| S4_extracted_2612 | Low-quality |
| S4_extracted_2632 | Not-determined |
| S4_extracted_2635 | Not-determined |
| S4_extracted_2638 | Low-quality |
| S3_vs_k141_94577\|\|full | Medium-quality |
| S4_extracted_2663 | Low-quality |
| S4_extracted_2672 | Low-quality |
| S4_extracted_2683 | Not-determined |
| S4_extracted_2685 | Low-quality |
| S4_extracted_2686 | Not-determined |
| S4_extracted_2688 | Not-determined |
| S4_extracted_2692 | Not-determined |
| S4_extracted_2706 | Not-determined |
| S4_extracted_2712 | Not-determined |
| S4_extracted_2714 | Not-determined |
| S4_extracted_2724 | Not-determined |
| S4_extracted_2735 | Not-determined |
| S4_extracted_2742 | Not-determined |
| S4_extracted_2743 | Not-determined |
| S4_extracted_2750 | Low-quality |
| S4_extracted_2754 | Not-determined |
| S4_extracted_2759 | Not-determined |
| S4_extracted_2766 | Not-determined |
| S4_extracted_2778 | Not-determined |
| S4_extracted_2779 | Not-determined |
| S4_extracted_2783 | Low-quality |
| S4_extracted_2789 | Not-determined |
| S4_extracted_2793 | Not-determined |
| S4_extracted_2795 | Not-determined |
| S4_extracted_2797 | Not-determined |
| S4_extracted_2802 | Not-determined |
| S4_extracted_2813 | Not-determined |
| S4_extracted_2822 | Not-determined |
| S4_extracted_2876 | Not-determined |
| S4_extracted_2885 | Not-determined |
| S4_extracted_2889 | Not-determined |
| S4_extracted_2900 | Not-determined |
| S4_extracted_2910 | Not-determined |
| S4_extracted_2926 | Not-determined |
| S4_extracted_2930 | Low-quality |
| S4_extracted_2931 | Not-determined |
| S4_extracted_2940 | Not-determined |
| S4_extracted_2946 | Not-determined |
| S4_extracted_2947 | Not-determined |
| S4_extracted_2954 | Low-quality |
| S4_extracted_2956 | Not-determined |
| S4_extracted_2962 | Not-determined |
| S4_extracted_2963 | Not-determined |
| S4_extracted_2966 | Low-quality |
| S4_extracted_2987 | Not-determined |
| S4_extracted_3001 | Not-determined |
| S4_extracted_3002 | Low-quality |
| S4_extracted_3005 | Not-determined |
| S4_extracted_3013 | Not-determined |
| S4_extracted_3020 | Not-determined |
| S4_extracted_3032 | Low-quality |
| S4_extracted_3035 | Not-determined |
| S4_extracted_3037 | Not-determined |
| S4_extracted_3057 | Not-determined |
| S4_extracted_3062 | Not-determined |
| S4_extracted_3072 | Not-determined |
| S4_extracted_3075 | Not-determined |
| S4_extracted_3078 | Low-quality |
| S4_extracted_3088 | Low-quality |
| S4_extracted_3091 | Not-determined |
| S4_extracted_3093 | Not-determined |
| S4_extracted_3096 | Not-determined |
| S4_extracted_3105 | Not-determined |
| S4_extracted_3107 | Not-determined |
| S4_extracted_3109 | Not-determined |
| S4_extracted_3118 | Low-quality |
| S4_extracted_3125 | Not-determined |
| S4_extracted_3126 | Not-determined |
| S4_extracted_3161 | Not-determined |
| S4_extracted_3168 | Not-determined |
| S4_extracted_3170 | Not-determined |
| S4_extracted_3182 | Not-determined |
| S4_extracted_3195 | Not-determined |
| S4_extracted_3203 | Not-determined |
| S4_extracted_3216 | Not-determined |
| S4_extracted_3217 | Not-determined |
| S4_extracted_3221 | Low-quality |
| S4_extracted_3223 | Not-determined |
| S4_extracted_3236 | Not-determined |
| S4_extracted_3237 | Low-quality |
| S4_extracted_3245 | Not-determined |
| S4_extracted_3246 | Not-determined |
| S4_extracted_3252 | Low-quality |
| S4_extracted_3262 | Not-determined |
| S4_extracted_3270 | Not-determined |
| S4_extracted_3272 | Not-determined |
| S4_extracted_3276 | Not-determined |
| S4_extracted_3284 | Low-quality |
| S4_extracted_3285 | Not-determined |
| S4_extracted_3289 | Not-determined |
| S4_extracted_3295 | Not-determined |
| S4_extracted_3302 | Low-quality |
| S4_extracted_3324 | Not-determined |
| S4_extracted_3346 | Low-quality |
| S4_extracted_3348 | Low-quality |
| S4_extracted_3349 | Not-determined |
| S4_extracted_3353 | Not-determined |
| S4_extracted_3357 | Not-determined |
| S4_extracted_3379 | Not-determined |
| S4_extracted_3394 | Not-determined |
| S4_extracted_3410 | Not-determined |
| S4_extracted_3415 | Not-determined |
| S4_extracted_3417 | Not-determined |
| S4_extracted_3420 | Low-quality |
| S4_extracted_3421 | Not-determined |
| S4_extracted_3425 | Not-determined |
| S4_extracted_3427 | Low-quality |
| S4_extracted_3430 | Not-determined |
| S4_extracted_3436 | Low-quality |
| S4_extracted_3441 | Low-quality |
| S4_extracted_3443 | Not-determined |
| S4_extracted_3452 | Not-determined |
| S4_extracted_3455 | Not-determined |
| S4_extracted_3459 | Not-determined |
| S4_extracted_3473 | Not-determined |
| S4_extracted_3474 | Not-determined |
| S4_extracted_3475 | Not-determined |
| S4_extracted_3483 | Not-determined |
| S4_extracted_3490 | Low-quality |
| S4_extracted_3505 | Not-determined |
| S4_extracted_3509 | Not-determined |
| S4_extracted_3522 | Not-determined |
| S4_extracted_3535 | Not-determined |
| S4_extracted_3539 | Low-quality |
| S4_extracted_3541 | Not-determined |
| S4_extracted_3542 | Not-determined |
| S4_extracted_3543 | Low-quality |
| S4_extracted_3550 | Low-quality |
| S4_extracted_3551 | Not-determined |
| S4_extracted_3560 | Not-determined |
| S4_extracted_3590 | Not-determined |
| S4_extracted_3600 | Not-determined |
| S4_extracted_3604 | Not-determined |
| S4_extracted_3612 | Low-quality |
| S4_extracted_3613 | Not-determined |
| S4_extracted_3617 | Not-determined |
| S4_extracted_3628 | Not-determined |
| S4_extracted_3630 | Not-determined |
| S4_extracted_3640 | Not-determined |
| S4_extracted_3647 | Not-determined |
| S4_extracted_3659 | Low-quality |
| S4_extracted_3678 | Not-determined |
| S4_extracted_3686 | Not-determined |
| S4_extracted_3706 | Not-determined |
| S4_extracted_3709 | Not-determined |
| S4_extracted_3715 | Not-determined |
| S4_extracted_3727 | Not-determined |
| S4_extracted_3732 | Low-quality |
| S4_extracted_3735 | Low-quality |
| S4_extracted_3743 | Not-determined |
| S4_extracted_3750 | Low-quality |
| S4_extracted_3757 | Low-quality |
| S4_extracted_3763 | Not-determined |
| S4_extracted_3765 | Not-determined |
| S4_extracted_3768 | Not-determined |
| S4_extracted_3773 | Not-determined |
| S4_extracted_3774 | Not-determined |
| S4_extracted_3775 | Not-determined |
| S4_extracted_3780 | Low-quality |
| S4_extracted_3795 | Not-determined |
| S4_extracted_3802 | Not-determined |
| S4_extracted_3805 | Not-determined |
| S4_extracted_3810 | Not-determined |
| S4_extracted_3811 | Not-determined |
| S4_extracted_3817 | Not-determined |
| S4_extracted_3821 | Not-determined |
| S4_extracted_3833 | Not-determined |
| S4_extracted_3850 | Not-determined |
| S4_extracted_3855 | Not-determined |
| S4_extracted_3857 | Not-determined |
| S4_extracted_3858 | Not-determined |
| S4_extracted_3862 | Not-determined |
| S4_extracted_3876 | Not-determined |
| S4_extracted_3881 | Low-quality |
| S4_extracted_3887 | Not-determined |
| S4_extracted_3897 | Not-determined |
| S4_extracted_3904 | Not-determined |
| S4_extracted_3906 | Not-determined |
| S4_extracted_3912 | Not-determined |
| S4_extracted_3917 | Not-determined |
| S4_extracted_3919 | Not-determined |
| S4_extracted_3950 | Not-determined |
| S4_extracted_3954 | Not-determined |
| S4_extracted_3955 | Not-determined |
| S4_extracted_3960 | Not-determined |
| S4_extracted_3965 | Not-determined |
| S4_extracted_3973 | Low-quality |
| S4_extracted_3998 | Low-quality |
| S4_extracted_4004 | Not-determined |
| S4_extracted_4013 | Not-determined |
| S4_extracted_4023 | Low-quality |
| S4_extracted_4032 | Not-determined |
| S4_extracted_4037 | Not-determined |
| S4_extracted_4042 | Low-quality |
| S4_extracted_4046 | Not-determined |
| S4_extracted_4051 | Not-determined |
| S4_extracted_4052 | Not-determined |
| S4_extracted_4055 | Not-determined |
| S4_extracted_4074 | Not-determined |
| S4_extracted_4097 | Not-determined |
| S4_extracted_4102 | Low-quality |
| S4_extracted_4120 | Low-quality |
| S4_extracted_4122 | Not-determined |
| S4_extracted_4132 | Not-determined |
| S4_extracted_4142 | Low-quality |
| S4_extracted_4148 | Not-determined |
| S4_extracted_4150 | Not-determined |
| S4_extracted_4156 | Not-determined |
| S4_extracted_4162 | Not-determined |
| S4_extracted_4172 | Not-determined |
| S4_extracted_4176 | Not-determined |
| S4_extracted_4189 | Not-determined |
| S4_extracted_4190 | Not-determined |
| S4_extracted_4192 | Low-quality |
| S4_extracted_4201 | Low-quality |
| S4_extracted_4208 | Not-determined |
| S4_extracted_4209 | Not-determined |
| S4_extracted_4213 | Not-determined |
| S4_extracted_4215 | Low-quality |
| S4_extracted_4230 | Not-determined |
| S4_extracted_4234 | Low-quality |
| S4_extracted_4244 | Not-determined |
| S4_extracted_4249 | Not-determined |
| S4_extracted_4257 | Not-determined |
| S4_extracted_4266 | Not-determined |
| S4_extracted_4268 | Not-determined |
| S4_extracted_4275 | Not-determined |
| S4_extracted_4283 | Low-quality |
| S4_extracted_4284 | Not-determined |
| S4_extracted_4323 | Low-quality |
| S4_extracted_4329 | Low-quality |
| S4_extracted_4333 | Not-determined |
| S4_extracted_4339 | Not-determined |
| S4_extracted_4348 | Not-determined |
| S4_extracted_4360 | Not-determined |
| S4_extracted_4370 | Low-quality |
| S4_extracted_4396 | Low-quality |
| S4_extracted_4398 | Not-determined |
| S4_extracted_4426 | Not-determined |
| S4_extracted_4427 | Not-determined |
| S4_extracted_4444 | Not-determined |
| S4_extracted_4451 | Not-determined |
| S4_extracted_4456 | Not-determined |
| S4_extracted_4458 | Low-quality |
| S4_extracted_4459 | Not-determined |

| | |
|---|---|
| S4_extracted_4462 | Not-determined |
| S4_extracted_4468 | Not-determined |
| S4_extracted_4484 | Not-determined |
| S4_extracted_4489 | Not-determined |
| S4_extracted_4492 | Not-determined |
| S4_extracted_4499 | Low-quality |
| S4_extracted_4504 | Not-determined |
| S4_extracted_4527 | Not-determined |
| S4_extracted_4533 | Not-determined |
| S4_extracted_4551 | Low-quality |
| S4_extracted_4557 | Low-quality |
| S4_extracted_4562 | Low-quality |
| S4_extracted_4571 | Not-determined |
| S4_extracted_4593 | Not-determined |
| S4_extracted_4612 | Not-determined |
| S4_extracted_4649 | Not-determined |
| S4_extracted_4671 | Not-determined |
| S4_extracted_4674 | Not-determined |
| S4_extracted_4675 | Not-determined |
| S4_extracted_4676 | Not-determined |
| S4_extracted_4677 | Low-quality |
| S4_extracted_4692 | Not-determined |
| S4_extracted_4700 | Not-determined |
| S4_extracted_4703 | Not-determined |
| S4_extracted_4704 | Not-determined |
| S4_extracted_4708 | Not-determined |
| S4_extracted_4719 | Low-quality |
| S4_extracted_4734 | Not-determined |
| S4_extracted_4766 | Not-determined |
| S4_extracted_4769 | Not-determined |
| S4_extracted_4789 | Low-quality |
| S4_extracted_4801 | Low-quality |
| S4_extracted_4804 | Not-determined |
| S4_extracted_4807 | Not-determined |
| S4_extracted_4826 | Not-determined |
| S14_vs_k141_101482\|\|full | Medium-quality |
| S4_extracted_4846 | Not-determined |
| S4_extracted_4858 | Not-determined |
| S4_extracted_4879 | Not-determined |
| S4_extracted_4887 | Not-determined |
| S8_vs_k141_190902\|\|full | Medium-quality |
| S4_extracted_4904 | Not-determined |
| S4_extracted_4921 | Not-determined |
| S4_extracted_4932 | Not-determined |
| S4_extracted_4943 | Not-determined |
| S4_extracted_4947 | Not-determined |
| S4_extracted_4948 | Not-determined |
| S4_extracted_4955 | Low-quality |
| S4_extracted_4978 | Low-quality |
| S4_extracted_4979 | Not-determined |
| S4_extracted_4987 | Not-determined |
| S4_extracted_4997 | Low-quality |
| S4_extracted_4998 | Not-determined |
| S4_extracted_5004 | Not-determined |
| S5_vs_k141_113386\|\|full | Medium-quality |
| S4_extracted_5025 | Not-determined |
| S4_extracted_5027 | Not-determined |
| S4_extracted_5053 | Not-determined |
| S4_extracted_5060 | Not-determined |
| S4_extracted_5070 | Not-determined |
| S4_extracted_5081 | Not-determined |
| S4_extracted_5095 | Not-determined |
| S4_extracted_5099 | Not-determined |
| S4_extracted_5105 | Not-determined |
| S4_extracted_5106 | Not-determined |
| S4_extracted_5110 | Not-determined |
| S4_extracted_5121 | Not-determined |
| S4_extracted_5130 | Not-determined |
| S4_extracted_5131 | Not-determined |
| S4_extracted_5132 | Not-determined |
| S4_extracted_5135 | Not-determined |
| S4_extracted_5141 | Not-determined |
| S4_extracted_5151 | Not-determined |
| S4_extracted_5153 | Not-determined |
| S4_extracted_5165 | Not-determined |
| S4_extracted_5168 | Not-determined |
| S4_extracted_5173 | Not-determined |
| S4_extracted_5175 | Not-determined |
| S4_extracted_5183 | Low-quality |
| S4_extracted_5192 | Low-quality |
| S4_extracted_5195 | Low-quality |
| S4_extracted_5223 | Not-determined |
| S4_extracted_5235 | Not-determined |
| S4_extracted_5238 | Not-determined |
| S4_extracted_5245 | Not-determined |
| S4_extracted_5248 | Not-determined |
| S4_extracted_5284 | Not-determined |
| S4_extracted_5296 | Not-determined |
| S4_extracted_5314 | Not-determined |
| S4_extracted_5315 | Not-determined |
| S4_extracted_5336 | Not-determined |
| S4_extracted_5342 | Low-quality |
| S4_extracted_5349 | Low-quality |
| S4_extracted_5353 | Not-determined |
| S4_extracted_5359 | Not-determined |
| S4_extracted_5360 | Low-quality |
| S4_extracted_5362 | Not-determined |
| S4_extracted_5365 | Not-determined |
| S4_extracted_5368 | Low-quality |
| S4_extracted_5378 | Not-determined |
| S4_extracted_5387 | Not-determined |
| S4_extracted_5392 | Not-determined |
| S4_extracted_5394 | Not-determined |
| S4_extracted_5409 | Not-determined |
| S4_extracted_5412 | Low-quality |
| S4_extracted_5416 | Not-determined |
| S4_extracted_5426 | Not-determined |
| S4_extracted_5433 | Low-quality |
| S4_extracted_5438 | Low-quality |
| S4_extracted_5440 | Not-determined |
| S4_extracted_5441 | Not-determined |
| S4_extracted_5445 | Not-determined |
| S4_extracted_5449 | Not-determined |
| S4_extracted_5466 | Not-determined |
| S4_extracted_5490 | Low-quality |
| S4_extracted_5491 | Not-determined |
| S4_extracted_5497 | Not-determined |
| S4_extracted_5502 | Not-determined |
| S4_extracted_5515 | Low-quality |
| S4_extracted_5519 | Low-quality |
| S4_extracted_5520 | Not-determined |
| S4_extracted_5533 | Not-determined |
| S4_extracted_5537 | Not-determined |
| S4_extracted_5546 | Not-determined |
| S4_extracted_5549 | Not-determined |
| S4_extracted_5557 | Not-determined |
| S4_extracted_5566 | Not-determined |
| S4_extracted_5569 | Not-determined |
| S4_extracted_5572 | Low-quality |
| S4_extracted_5586 | Low-quality |
| S4_extracted_5597 | Not-determined |
| S4_extracted_5598 | Not-determined |
| S4_extracted_5608 | Not-determined |
| S4_extracted_5626 | Not-determined |
| S4_extracted_5629 | Not-determined |
| S4_extracted_5633 | Not-determined |
| S4_extracted_5639 | Not-determined |
| S4_extracted_5643 | Low-quality |
| S4_extracted_5653 | Not-determined |
| S4_extracted_5654 | Not-determined |
| S4_extracted_5655 | Low-quality |
| S4_extracted_5663 | Low-quality |
| S4_extracted_5664 | Not-determined |
| S4_extracted_5673 | Low-quality |
| S4_extracted_5676 | Not-determined |
| S4_extracted_5677 | Low-quality |
| S4_extracted_5684 | Not-determined |
| S4_extracted_5695 | Not-determined |
| S4_extracted_5704 | Not-determined |
| S4_extracted_5711 | Low-quality |
| S4_extracted_5768 | Not-determined |
| S4_extracted_5770 | Not-determined |
| S4_extracted_5775 | Low-quality |
| S4_extracted_5778 | Not-determined |
| S4_extracted_5780 | Not-determined |
| S4_extracted_5792 | Not-determined |
| S4_extracted_5793 | Not-determined |
| S4_extracted_5797 | Not-determined |
| S4_extracted_5810 | Low-quality |
| S4_extracted_5814 | Not-determined |
| S4_extracted_5817 | Not-determined |
| S4_extracted_5818 | Low-quality |
| S4_extracted_5822 | Low-quality |
| S4_extracted_5823 | Not-determined |
| S4_extracted_5827 | Not-determined |
| S4_extracted_5828 | Not-determined |
| S4_extracted_5829 | Not-determined |
| S4_extracted_5845 | Not-determined |
| S4_extracted_5853 | Not-determined |
| S4_extracted_5859 | Not-determined |
| S4_extracted_5861 | Not-determined |
| S4_extracted_5869 | Not-determined |
| S4_extracted_5872 | Not-determined |
| S4_extracted_5877 | Not-determined |
| S4_extracted_5881 | Low-quality |
| S4_extracted_5892 | Low-quality |
| S4_extracted_5913 | Low-quality |
| S4_extracted_5923 | Not-determined |
| S4_extracted_5938 | Not-determined |
| S4_extracted_5949 | Not-determined |
| S4_extracted_5951 | Not-determined |
| S4_extracted_5966 | Not-determined |
| S4_extracted_5985 | Not-determined |
| S4_extracted_6000 | Not-determined |
| S4_extracted_6004 | Not-determined |
| S4_extracted_6006 | Not-determined |
| S4_extracted_6021 | Low-quality |
| S4_extracted_6023 | Low-quality |
| S4_extracted_6032 | Not-determined |
| S4_extracted_6033 | Not-determined |
| S4_extracted_6037 | Not-determined |
| S4_extracted_6046 | Not-determined |
| S4_extracted_6047 | Not-determined |
| S4_extracted_6048 | Not-determined |
| S4_extracted_6052 | Low-quality |
| S4_extracted_6060 | Low-quality |
| S4_extracted_6070 | Not-determined |
| S4_extracted_6079 | Not-determined |
| S4_extracted_6091 | Not-determined |
| S4_extracted_6106 | Not-determined |
| S4_extracted_6112 | Low-quality |
| S4_extracted_6118 | Not-determined |
| S4_extracted_6128 | Not-determined |
| S4_extracted_6129 | Not-determined |
| S4_extracted_6134 | Not-determined |
| S4_extracted_6151 | Not-determined |
| S4_extracted_6154 | Not-determined |
| S4_extracted_6163 | Not-determined |
| S5_extracted_k141_116300 | Medium-quality |
| S4_extracted_6170 | Low-quality |
| S4_extracted_6175 | Not-determined |
| S4_extracted_6185 | Not-determined |
| S4_extracted_6192 | Low-quality |
| S4_extracted_6197 | Not-determined |
| S4_extracted_6211 | Not-determined |
| S4_extracted_6227 | Not-determined |
| S4_extracted_6237 | Not-determined |
| S4_extracted_6240 | Not-determined |
| S4_extracted_6248 | Not-determined |
| S4_extracted_6256 | Not-determined |
| S4_extracted_6258 | Not-determined |
| S4_extracted_6266 | Low-quality |
| S4_extracted_6269 | Not-determined |
| S4_extracted_6272 | Not-determined |
| S4_extracted_6273 | Not-determined |
| S4_extracted_6296 | Not-determined |
| S4_extracted_6298 | Not-determined |
| S4_extracted_6305 | Not-determined |
| S4_extracted_6307 | Not-determined |
| S4_extracted_6309 | Not-determined |
| S4_extracted_6320 | Low-quality |
| S4_extracted_6330 | Low-quality |
| S4_extracted_6348 | Not-determined |
| S4_extracted_6364 | Low-quality |
| S4_extracted_6368 | Low-quality |
| S4_extracted_6371 | Not-determined |
| S4_extracted_6383 | Low-quality |
| S4_extracted_6396 | Not-determined |
| S4_extracted_6411 | Not-determined |
| S4_extracted_6412 | Not-determined |
| S4_extracted_6421 | Not-determined |
| S4_extracted_6437 | Not-determined |
| S4_extracted_6441 | Not-determined |
| S4_extracted_6446 | Not-determined |
| S4_extracted_6462 | Low-quality |
| S4_extracted_6477 | Low-quality |
| S4_extracted_6483 | Low-quality |
| S4_extracted_6488 | Not-determined |
| S4_extracted_6511 | Not-determined |
| S4_extracted_6516 | Not-determined |
| S4_extracted_6540 | Not-determined |
| S4_extracted_6560 | Not-determined |
| S4_extracted_6573 | Low-quality |
| S4_extracted_6576 | Not-determined |
| S4_extracted_6582 | Not-determined |
| S4_extracted_6584 | Not-determined |
| S4_extracted_6585 | Not-determined |
| S4_extracted_6597 | Not-determined |
| S4_extracted_6598 | Low-quality |
| S4_extracted_6601 | Not-determined |
| S4_extracted_6616 | Not-determined |
| S4_extracted_6639 | Not-determined |
| S4_extracted_6660 | Not-determined |
| S4_extracted_6662 | Low-quality |
| S4_extracted_6674 | Low-quality |
| S4_extracted_6675 | Not-determined |
| S4_extracted_6679 | Not-determined |
| S4_extracted_6691 | Low-quality |
| S4_extracted_6692 | Not-determined |
| S4_extracted_6710 | Low-quality |
| S4_extracted_6713 | Not-determined |
| S4_extracted_6718 | Not-determined |
| S4_extracted_6727 | Low-quality |
| S4_extracted_6742 | Low-quality |
| S4_extracted_6744 | Not-determined |
| S4_extracted_6759 | Not-determined |
| S4_extracted_6763 | Low-quality |
| S4_extracted_6781 | Not-determined |
| S4_extracted_6788 | Not-determined |
| S4_extracted_6791 | Not-determined |
| S4_extracted_6818 | Not-determined |
| S4_extracted_6819 | Low-quality |
| S4_extracted_6833 | Low-quality |
| S4_extracted_6841 | Low-quality |
| S4_extracted_6851 | Low-quality |
| S4_extracted_6854 | Not-determined |
| S4_extracted_6855 | Not-determined |
| S4_extracted_6858 | Not-determined |
| S4_extracted_6860 | Not-determined |
| S4_extracted_6869 | Low-quality |
| S4_extracted_6879 | Not-determined |
| S4_extracted_6899 | Not-determined |
| S4_extracted_6900 | Not-determined |
| S4_extracted_6904 | Not-determined |
| S4_extracted_6912 | Not-determined |
| S4_extracted_6921 | Not-determined |
| S4_extracted_6957 | Not-determined |
| S4_extracted_6964 | Not-determined |
| S4_extracted_6965 | Low-quality |
| S4_extracted_6990 | Not-determined |
| S4_extracted_6996 | Low-quality |
| S4_extracted_7010 | Not-determined |
| S4_extracted_7014 | Not-determined |
| S4_extracted_7022 | Not-determined |
| S4_extracted_7023 | Not-determined |
| S4_extracted_7024 | Not-determined |
| S4_extracted_7034 | Not-determined |
| S4_extracted_7040 | Not-determined |
| S4_extracted_7048 | Not-determined |
| S4_extracted_7056 | Low-quality |
| S4_extracted_7057 | Low-quality |
| S4_extracted_7061 | Not-determined |
| S4_extracted_7069 | Not-determined |
| S4_extracted_7070 | Not-determined |
| S4_extracted_7074 | Not-determined |
| S4_extracted_7085 | Low-quality |
| S4_extracted_7088 | Not-determined |
| S4_extracted_7089 | Low-quality |
| S4_extracted_7112 | Not-determined |
| S4_extracted_7129 | Not-determined |
| S4_extracted_7136 | Not-determined |
| S4_extracted_7147 | Not-determined |
| S4_extracted_7153 | Low-quality |
| S4_extracted_7172 | Not-determined |
| S4_extracted_7180 | Low-quality |
| S4_extracted_7199 | Not-determined |
| S4_extracted_7205 | Low-quality |
| S4_extracted_7211 | Not-determined |
| S4_extracted_7214 | Not-determined |
| S4_extracted_7227 | Not-determined |
| S4_extracted_7228 | Not-determined |
| S4_extracted_7231 | Low-quality |
| S4_extracted_7233 | Low-quality |
| S4_extracted_7244 | Low-quality |
| S4_extracted_7253 | Not-determined |
| S4_extracted_7263 | Not-determined |
| S4_extracted_7274 | Not-determined |
| S4_extracted_7286 | Not-determined |
| S4_extracted_7289 | Low-quality |
| S4_extracted_7294 | Not-determined |
| S4_extracted_7315 | Not-determined |
| S3_vs_k141_39940\|\|full | Medium-quality |
| S4_extracted_7320 | Not-determined |
| S4_extracted_7323 | Not-determined |
| S4_extracted_7325 | Low-quality |
| S4_extracted_7327 | Not-determined |
| S4_extracted_7355 | Low-quality |
| S4_extracted_7376 | Not-determined |
| S4_extracted_7381 | Not-determined |
| S4_extracted_7389 | Not-determined |
| S4_extracted_7404 | Not-determined |
| S4_extracted_7410 | Not-determined |
| S4_extracted_7415 | Low-quality |
| S4_extracted_7432 | Not-determined |
| S4_extracted_7446 | Not-determined |
| S4_extracted_7451 | Low-quality |
| S4_extracted_7465 | Not-determined |
| S4_extracted_7476 | Not-determined |
| S4_extracted_7485 | Low-quality |
| S4_extracted_7493 | Not-determined |
| S4_extracted_7510 | Low-quality |
| S4_extracted_7516 | Not-determined |
| S4_extracted_7538 | Not-determined |
| S4_extracted_7541 | Not-determined |
| S4_extracted_7545 | Not-determined |
| S4_extracted_7547 | Low-quality |
| S4_extracted_7565 | Low-quality |
| S4_extracted_7571 | Not-determined |
| S4_extracted_7577 | Not-determined |
| S4_extracted_7579 | Not-determined |
| S4_extracted_7581 | Not-determined |
| S4_extracted_7594 | Not-determined |
| S4_extracted_7607 | Low-quality |
| S4_extracted_7610 | Not-determined |
| S4_extracted_7629 | Not-determined |
| S4_extracted_7632 | Not-determined |
| S4_extracted_7634 | Not-determined |
| S4_extracted_7637 | Not-determined |
| S4_extracted_7638 | Not-determined |
| S4_extracted_7639 | Low-quality |
| S4_extracted_7643 | Not-determined |
| S4_extracted_7681 | Not-determined |
| S4_extracted_7682 | Not-determined |
| S4_extracted_7692 | Not-determined |
| S4_extracted_7709 | Not-determined |
| S4_extracted_7732 | Low-quality |
| S4_extracted_7734 | Not-determined |
| S4_extracted_7758 | Not-determined |
| S4_extracted_7761 | Not-determined |
| S4_extracted_7763 | Not-determined |
| S4_extracted_7764 | Not-determined |
| S4_extracted_7769 | Not-determined |
| S4_extracted_7772 | Not-determined |
| S4_extracted_7793 | Not-determined |
| S4_extracted_7798 | Not-determined |
| S4_extracted_7803 | Low-quality |
| S4_extracted_7804 | Not-determined |
| S4_extracted_7828 | Not-determined |
| S4_extracted_7837 | Low-quality |
| S4_extracted_7845 | Not-determined |
| S4_extracted_7849 | Not-determined |
| S4_extracted_7855 | Not-determined |
| S4_extracted_7858 | Low-quality |
| S4_extracted_7869 | Not-determined |
| S4_extracted_7875 | Low-quality |
| S4_extracted_7889 | Low-quality |
| S4_extracted_7896 | Low-quality |
| S4_extracted_7901 | Low-quality |
| S4_extracted_7905 | Low-quality |
| S4_extracted_7916 | Low-quality |
| S4_extracted_7922 | Not-determined |
| S4_extracted_7927 | Not-determined |
| S4_extracted_7936 | Low-quality |
| S4_extracted_7946 | Not-determined |
| S4_extracted_7947 | Not-determined |
| S4_extracted_7965 | Not-determined |
| S4_extracted_7970 | Not-determined |
| S4_extracted_7996 | Not-determined |
| S4_extracted_8002 | Low-quality |
| S4_extracted_8008 | Not-determined |
| S4_extracted_8018 | Not-determined |
| S4_extracted_8023 | Not-determined |
| S4_extracted_8024 | Not-determined |
| S4_extracted_8027 | Not-determined |
| S4_extracted_8040 | Low-quality |
| S4_extracted_8044 | Not-determined |
| S4_extracted_8048 | Low-quality |
| S4_extracted_8060 | Low-quality |
| S4_extracted_8062 | Low-quality |
| S4_extracted_8064 | Not-determined |
| S4_extracted_8076 | Not-determined |
| S4_extracted_8077 | Low-quality |
| S4_extracted_8079 | Not-determined |
| S4_extracted_8080 | Not-determined |
| S4_extracted_8084 | Low-quality |
| S4_extracted_8099 | Not-determined |
| S4_extracted_8100 | Not-determined |
| S4_extracted_8104 | Not-determined |
| S4_extracted_8115 | Not-determined |
| S4_extracted_8116 | Low-quality |
| S4_extracted_8144 | Low-quality |
| S4_extracted_8145 | Not-determined |
| S4_extracted_8156 | Not-determined |
| S4_extracted_8157 | Not-determined |
| S4_extracted_8179 | Not-determined |
| S4_extracted_8195 | Not-determined |
| S4_extracted_8204 | Not-determined |
| S4_extracted_8210 | Not-determined |
| S4_extracted_8213 | Not-determined |
| S4_extracted_8215 | Not-determined |
| S4_extracted_8226 | Not-determined |
| S4_extracted_8231 | Not-determined |
| S4_extracted_8239 | Not-determined |
| S4_extracted_8240 | Not-determined |
| S4_extracted_8244 | Not-determined |
| S4_extracted_8256 | Not-determined |
| S4_extracted_8263 | Low-quality |
| S4_extracted_8265 | Not-determined |
| S4_extracted_8274 | Not-determined |
| S4_extracted_8279 | Low-quality |
| S4_extracted_8281 | Not-determined |
| S4_extracted_8284 | Low-quality |
| S4_extracted_8291 | Low-quality |
| S4_extracted_8300 | Not-determined |
| S4_extracted_8301 | Not-determined |
| S4_extracted_8310 | Low-quality |
| S4_extracted_8315 | Not-determined |
| S4_extracted_8332 | Not-determined |
| S4_extracted_8334 | Not-determined |
| S4_extracted_8336 | Low-quality |
| S4_extracted_8346 | Low-quality |
| S4_extracted_8355 | Low-quality |
| S4_extracted_8356 | Not-determined |
| S4_extracted_8387 | Low-quality |
| S4_extracted_8392 | Low-quality |
| S4_extracted_8395 | Not-determined |
| S4_extracted_8400 | Low-quality |
| S4_extracted_8402 | Not-determined |
| S4_extracted_8407 | Not-determined |
| S4_extracted_8418 | Not-determined |
| S4_extracted_8439 | Low-quality |
| S4_extracted_8448 | Not-determined |
| S4_extracted_8468 | Low-quality |
| S4_extracted_8474 | Not-determined |
| S4_extracted_8475 | Low-quality |
| S4_extracted_8476 | Low-quality |
| S4_extracted_8483 | Not-determined |
| S4_extracted_8487 | Low-quality |
| S4_extracted_8497 | Not-determined |
| S4_extracted_8501 | Not-determined |
| S4_extracted_8518 | Not-determined |
| S4_extracted_8523 | Not-determined |
| S4_extracted_8531 | Not-determined |
| S4_extracted_8541 | Not-determined |
| S4_extracted_8544 | Low-quality |
| S4_extracted_8555 | Not-determined |
| S4_extracted_8570 | Not-determined |
| S4_extracted_8579 | Not-determined |
| S4_extracted_8588 | Not-determined |
| S4_extracted_8596 | Not-determined |
| S4_extracted_8598 | Not-determined |
| S4_extracted_8599 | Not-determined |
| S4_extracted_8614 | Not-determined |
| S4_extracted_8616 | Not-determined |
| S4_extracted_8617 | Not-determined |
| S4_extracted_8626 | Low-quality |
| S4_extracted_8634 | Not-determined |
| S4_extracted_8646 | Not-determined |
| S4_extracted_8655 | Low-quality |
| S4_extracted_8658 | Low-quality |
| S4_extracted_8680 | Low-quality |
| S4_extracted_8705 | Low-quality |
| S4_extracted_8724 | Not-determined |
| S4_extracted_8729 | Not-determined |
| S4_extracted_8738 | Not-determined |
| S4_extracted_8746 | Low-quality |
| S4_extracted_8752 | Low-quality |
| S4_extracted_8753 | Not-determined |
| S4_extracted_8767 | Not-determined |
| S4_extracted_8795 | Not-determined |
| S4_extracted_8802 | Not-determined |
| S4_extracted_8809 | Not-determined |
| S4_extracted_8821 | Not-determined |
| S4_extracted_8830 | Not-determined |
| S4_extracted_8832 | Not-determined |
| S4_extracted_8841 | Not-determined |
| S4_extracted_8854 | Low-quality |
| S4_extracted_8864 | Not-determined |
| S4_extracted_8874 | Low-quality |
| S4_extracted_8876 | Not-determined |
| S4_extracted_8877 | Not-determined |
| S4_extracted_8880 | Not-determined |
| S4_extracted_8886 | Not-determined |
| S4_extracted_8894 | Low-quality |
| S4_extracted_8899 | Not-determined |
| S4_extracted_8906 | Not-determined |
| S4_extracted_8907 | Not-determined |
| S4_extracted_8921 | Not-determined |
| S3_vs_k141_12973\|\|full | Medium-quality |
| S4_extracted_8939 | Low-quality |
| S4_extracted_8940 | Low-quality |
| S4_extracted_8942 | Not-determined |
| S4_extracted_8947 | Not-determined |
| S4_extracted_8959 | Not-determined |
| S4_extracted_8960 | Not-determined |
| S4_extracted_8964 | Not-determined |
| S4_extracted_8990 | Not-determined |
| S4_extracted_8991 | Low-quality |
| S4_extracted_8998 | Not-determined |
| S4_extracted_9010 | Not-determined |
| S4_extracted_9013 | Not-determined |
| S4_extracted_9020 | Not-determined |
| S4_extracted_9022 | Not-determined |
| S4_extracted_9033 | Not-determined |
| S4_extracted_9037 | Not-determined |
| S4_extracted_9039 | Not-determined |
| S4_extracted_9040 | Low-quality |
| S4_extracted_9048 | Low-quality |
| S4_extracted_9053 | Not-determined |
| S4_extracted_9057 | Low-quality |
| S4_extracted_9098 | Not-determined |
| S4_extracted_9101 | Not-determined |
| S4_extracted_9103 | Low-quality |
| S4_extracted_9117 | Not-determined |
| S4_extracted_9126 | Not-determined |
| S4_extracted_9128 | Low-quality |
| S4_extracted_9130 | Low-quality |
| S4_extracted_9143 | Not-determined |
| S4_extracted_9149 | Not-determined |
| S4_extracted_9154 | Low-quality |
| S4_extracted_9157 | Low-quality |
| S4_extracted_9162 | Not-determined |
| S4_extracted_9164 | Not-determined |
| S4_extracted_9167 | Not-determined |
| S4_extracted_9169 | Not-determined |
| S4_extracted_9174 | Not-determined |
| S4_extracted_9184 | Low-quality |
| S4_extracted_9185 | Not-determined |
| S4_extracted_9186 | Low-quality |
| S4_extracted_9190 | Low-quality |
| S4_extracted_9193 | Low-quality |
| S4_extracted_9198 | Not-determined |
| S4_extracted_9205 | Not-determined |
| S4_extracted_9208 | Not-determined |
| S4_extracted_9211 | Not-determined |
| S4_extracted_9220 | Not-determined |
| S4_extracted_9226 | Low-quality |
| S4_extracted_9229 | Not-determined |
| S4_extracted_9243 | Not-determined |

| | |
|---|---|
| S4_extracted_9255 | Not-determined |
| S4_extracted_9272 | Not-determined |
| S4_extracted_9289 | Low-quality |
| S4_extracted_9301 | Not-determined |
| S4_extracted_9306 | Not-determined |
| S4_extracted_9308 | Not-determined |
| S4_extracted_9311 | Not-determined |
| S4_extracted_9317 | Not-determined |
| S4_extracted_9319 | Not-determined |
| S4_extracted_9328 | Not-determined |
| S4_extracted_9353 | Not-determined |
| S4_extracted_9366 | Not-determined |
| S4_extracted_9371 | Not-determined |
| S4_extracted_9384 | Low-quality |
| S4_extracted_9385 | Low-quality |
| S4_extracted_9401 | Not-determined |
| S4_extracted_9403 | Not-determined |
| S4_extracted_9411 | Low-quality |
| S4_extracted_9416 | Not-determined |
| S4_extracted_9428 | Low-quality |
| S4_extracted_9429 | Not-determined |
| S4_extracted_9441 | Not-determined |
| S4_extracted_9442 | Not-determined |
| S14_vs_k141_56410\|\|full | Medium-quality |
| S4_extracted_9445 | Not-determined |
| S4_extracted_9447 | Not-determined |
| S4_extracted_9455 | Not-determined |
| S4_extracted_9461 | Not-determined |
| S4_extracted_9475 | Low-quality |
| S4_extracted_9485 | Low-quality |
| S4_extracted_9486 | Not-determined |
| S4_extracted_9492 | Low-quality |
| S4_extracted_9496 | Not-determined |
| S4_extracted_9504 | Not-determined |
| S4_extracted_9515 | Not-determined |
| S4_extracted_9524 | Not-determined |
| S4_extracted_9530 | Not-determined |
| S4_extracted_9532 | Not-determined |
| S4_extracted_9557 | Not-determined |
| S4_extracted_9564 | Low-quality |
| S4_extracted_9583 | Low-quality |
| S4_extracted_9595 | Not-determined |
| S4_extracted_9599 | Not-determined |
| S4_extracted_9601 | Low-quality |
| S4_extracted_9619 | Not-determined |
| S4_extracted_9623 | Low-quality |
| S4_extracted_9630 | Not-determined |
| S4_extracted_9631 | Not-determined |
| S4_extracted_9634 | Not-determined |
| S4_extracted_9637 | Not-determined |
| S4_extracted_9651 | Not-determined |
| S4_extracted_9669 | Not-determined |
| S4_extracted_9681 | Not-determined |
| S4_extracted_9698 | Not-determined |
| S4_extracted_9705 | Not-determined |
| S4_extracted_9706 | Not-determined |
| S4_extracted_9719 | Not-determined |
| S4_extracted_9722 | Not-determined |
| S4_extracted_9724 | Not-determined |
| S4_extracted_9725 | Not-determined |
| S4_extracted_9731 | Not-determined |
| S4_extracted_9733 | Low-quality |
| S4_extracted_9736 | Not-determined |
| S4_extracted_9749 | Low-quality |
| S4_extracted_9758 | Not-determined |
| S4_extracted_9769 | Low-quality |
| S4_extracted_9793 | Not-determined |
| S4_extracted_9808 | Not-determined |
| S4_extracted_9838 | Not-determined |
| S4_extracted_9844 | Not-determined |
| S4_extracted_9861 | Not-determined |
| S4_extracted_9870 | Not-determined |
| S4_extracted_9877 | Not-determined |
| S4_extracted_9879 | Not-determined |
| S4_extracted_9899 | Not-determined |
| S4_extracted_9902 | Low-quality |
| S4_extracted_9912 | Not-determined |
| S4_extracted_9919 | Low-quality |
| S4_extracted_9924 | Not-determined |
| S4_extracted_9928 | Not-determined |
| S4_extracted_9948 | Not-determined |
| S4_extracted_9950 | Not-determined |
| S4_extracted_9951 | Not-determined |
| S4_extracted_9960 | Not-determined |
| S4_extracted_9963 | Not-determined |
| S4_extracted_9974 | Low-quality |
| S4_extracted_9984 | Not-determined |
| S4_extracted_9994 | Not-determined |
| S4_extracted_9996 | Not-determined |
| S4_extracted_10011 | Not-determined |
| S4_extracted_10031 | Not-determined |
| S4_extracted_10046 | Low-quality |
| S4_extracted_10058 | Not-determined |
| S4_extracted_10059 | Low-quality |
| S4_extracted_10066 | Not-determined |
| S4_extracted_10071 | Not-determined |
| S4_extracted_10072 | Low-quality |
| S4_extracted_10081 | Not-determined |
| S4_extracted_10093 | Low-quality |
| S4_extracted_10098 | Not-determined |
| S4_extracted_10099 | Not-determined |
| S4_extracted_10124 | Not-determined |
| S4_extracted_10127 | Not-determined |
| S4_extracted_10130 | Low-quality |
| S4_extracted_10141 | Not-determined |
| S4_extracted_10152 | Not-determined |
| S4_extracted_10172 | Not-determined |
| S4_extracted_10173 | Not-determined |
| S4_extracted_10181 | Low-quality |
| S4_extracted_10200 | Not-determined |
| S4_extracted_10202 | Low-quality |
| S4_extracted_10203 | Not-determined |
| S4_extracted_10234 | Not-determined |
| S4_extracted_10237 | Not-determined |
| S4_extracted_10243 | Not-determined |
| S4_extracted_10250 | Not-determined |
| S4_extracted_10252 | Not-determined |
| S4_extracted_10254 | Not-determined |
| S4_extracted_10262 | Low-quality |
| S4_extracted_10266 | Not-determined |
| S4_extracted_10270 | Not-determined |
| S4_extracted_10272 | Not-determined |
| S4_extracted_10277 | Not-determined |
| S4_extracted_10278 | Not-determined |
| S4_extracted_10291 | Not-determined |
| S4_extracted_10321 | Low-quality |
| S4_extracted_10330 | Low-quality |
| S4_extracted_10344 | Low-quality |
| S4_extracted_10359 | Not-determined |
| S4_extracted_10361 | Not-determined |
| S4_extracted_10362 | Low-quality |
| S4_extracted_10367 | Not-determined |
| S4_extracted_10378 | Not-determined |
| S4_extracted_10388 | Low-quality |
| S4_extracted_10391 | Not-determined |
| S4_extracted_10395 | Not-determined |
| S4_extracted_10405 | Not-determined |
| S4_extracted_10417 | Low-quality |
| S4_extracted_10420 | Low-quality |
| S4_extracted_10422 | Not-determined |
| S4_extracted_10423 | Not-determined |
| S4_extracted_10425 | Not-determined |
| S4_extracted_10441 | Not-determined |
| S4_extracted_10445 | Not-determined |
| S4_extracted_10458 | Not-determined |
| S16_vs_34751\|\|full | Medium-quality |
| S4_extracted_10508 | Low-quality |
| S4_extracted_10510 | Low-quality |
| S4_extracted_10511 | Not-determined |
| S4_extracted_10523 | Low-quality |
| S4_extracted_10548 | Not-determined |
| S4_extracted_10558 | Low-quality |
| S4_extracted_10571 | Not-determined |
| S4_extracted_10592 | Not-determined |
| S4_extracted_10609 | Not-determined |
| S4_extracted_10611 | Low-quality |
| S4_extracted_10614 | Not-determined |
| S4_extracted_10622 | Low-quality |
| S4_extracted_10627 | Low-quality |
| S4_extracted_10642 | Not-determined |
| S4_extracted_10643 | Not-determined |
| S4_extracted_10644 | Not-determined |
| S4_extracted_10646 | Not-determined |
| S4_extracted_10653 | Not-determined |
| S4_extracted_10667 | Not-determined |
| S4_extracted_10692 | Low-quality |
| S4_extracted_10694 | Not-determined |
| S4_extracted_10697 | Not-determined |
| S4_extracted_10699 | Not-determined |
| S4_extracted_10708 | Not-determined |
| S4_extracted_10720 | Not-determined |
| S4_extracted_10730 | Low-quality |
| S4_extracted_10745 | Not-determined |
| S4_extracted_10748 | Not-determined |
| S4_extracted_10751 | Not-determined |
| S4_extracted_10754 | Not-determined |
| S4_extracted_10759 | Not-determined |
| S4_extracted_10768 | Not-determined |
| S4_extracted_10780 | Not-determined |
| S4_extracted_10789 | Low-quality |
| S4_extracted_10795 | Not-determined |
| S4_extracted_10818 | Not-determined |
| S4_extracted_10819 | Not-determined |
| S4_extracted_10820 | Not-determined |
| S4_extracted_10822 | Not-determined |
| S4_extracted_10823 | Low-quality |
| S4_extracted_10825 | Not-determined |
| S4_extracted_10831 | Low-quality |
| S4_extracted_10834 | Low-quality |
| S4_extracted_10835 | Low-quality |
| S4_extracted_10843 | Not-determined |
| S4_extracted_10849 | Low-quality |
| S4_extracted_10854 | Not-determined |
| S4_extracted_10861 | Not-determined |
| S4_extracted_10864 | Low-quality |
| S4_extracted_10866 | Low-quality |
| S4_extracted_10868 | Not-determined |
| S4_extracted_10878 | Low-quality |
| S4_extracted_10888 | Not-determined |
| S4_extracted_10892 | Not-determined |
| S4_extracted_10895 | Low-quality |
| S4_extracted_10896 | Not-determined |
| S4_extracted_10917 | Not-determined |
| S4_extracted_10920 | Low-quality |
| S4_extracted_10923 | Not-determined |
| S4_extracted_10946 | Not-determined |
| S4_extracted_10950 | Not-determined |
| S4_extracted_10967 | Not-determined |
| S4_extracted_10979 | Low-quality |
| S4_extracted_10988 | Not-determined |
| S4_extracted_10991 | Low-quality |
| S4_extracted_10997 | Not-determined |
| S4_extracted_11000 | Not-determined |
| S4_extracted_11001 | Not-determined |
| S4_extracted_11008 | Low-quality |
| S4_extracted_11014 | Low-quality |
| S4_extracted_11015 | Not-determined |
| S4_extracted_11016 | Low-quality |
| S4_extracted_11017 | Not-determined |
| S4_extracted_11023 | Not-determined |
| S4_extracted_11036 | Low-quality |
| S4_extracted_11042 | Low-quality |
| S4_extracted_11045 | Not-determined |
| S4_extracted_11051 | Not-determined |
| S4_extracted_11071 | Not-determined |
| S4_extracted_11084 | Not-determined |
| S4_extracted_11087 | Not-determined |
| S4_extracted_11088 | Not-determined |
| S4_extracted_11092 | Not-determined |
| S4_extracted_11093 | Not-determined |
| S4_extracted_11097 | Not-determined |
| S4_extracted_11101 | Low-quality |
| S4_extracted_11117 | Not-determined |
| S4_extracted_11122 | Not-determined |
| S4_extracted_11168 | Not-determined |
| S4_extracted_11169 | Not-determined |
| S4_extracted_11179 | Not-determined |
| S4_extracted_11192 | Not-determined |
| S4_extracted_11193 | Not-determined |
| S4_extracted_11199 | Low-quality |
| S4_extracted_11203 | Not-determined |
| S4_extracted_11210 | Not-determined |
| S4_extracted_11215 | Not-determined |
| S4_extracted_11222 | Low-quality |
| S4_extracted_11230 | Low-quality |
| S4_extracted_11231 | Not-determined |
| S4_extracted_11237 | Not-determined |
| S4_extracted_11238 | Not-determined |
| S4_extracted_11263 | Not-determined |
| S4_extracted_11264 | Low-quality |
| S4_extracted_11289 | Not-determined |
| S4_extracted_11300 | Not-determined |
| S4_extracted_11303 | Low-quality |
| S4_extracted_11304 | Low-quality |
| S14_vs_k141_108376\|\|full | Medium-quality |
| S4_extracted_11310 | Low-quality |
| S4_extracted_11311 | Not-determined |
| S4_extracted_11320 | Low-quality |
| S4_extracted_11335 | Not-determined |
| S4_extracted_11340 | Not-determined |
| S4_extracted_11356 | Not-determined |
| S4_extracted_11366 | Not-determined |
| S4_extracted_11371 | Not-determined |
| S4_extracted_11372 | Not-determined |
| S4_extracted_11381 | Not-determined |
| S4_extracted_11391 | Low-quality |
| S4_extracted_11392 | Low-quality |
| S4_extracted_11394 | Not-determined |
| S4_extracted_11411 | Not-determined |
| S4_extracted_11424 | Not-determined |
| S4_extracted_11430 | Low-quality |
| S4_extracted_11438 | Not-determined |
| S4_extracted_11443 | Not-determined |
| S4_extracted_11447 | Not-determined |
| S4_extracted_11448 | Not-determined |
| S4_extracted_11458 | Not-determined |
| S4_extracted_11476 | Not-determined |
| S4_extracted_11478 | Not-determined |
| S4_extracted_11494 | Not-determined |
| S4_extracted_11500 | Low-quality |
| S4_extracted_11511 | Not-determined |
| S4_extracted_11512 | Low-quality |
| S4_extracted_11513 | Not-determined |
| S4_extracted_11523 | Not-determined |
| S4_extracted_11542 | Not-determined |
| S4_extracted_11547 | Not-determined |
| S4_extracted_11549 | Low-quality |
| S4_extracted_11552 | Low-quality |
| S4_extracted_11558 | Low-quality |
| S4_extracted_11562 | Not-determined |
| S4_extracted_11584 | Low-quality |
| S4_extracted_11610 | Low-quality |
| S4_extracted_11611 | Not-determined |
| S4_extracted_11616 | Not-determined |
| S4_extracted_11639 | Not-determined |
| S4_extracted_11640 | Not-determined |
| S4_extracted_11642 | Not-determined |
| S4_extracted_11647 | Low-quality |
| S4_extracted_11649 | Low-quality |
| S4_extracted_11651 | Not-determined |
| S4_extracted_11667 | Low-quality |
| S4_extracted_11682 | Not-determined |
| S4_extracted_11698 | Not-determined |
| S4_extracted_11700 | Not-determined |
| S4_extracted_11701 | Not-determined |
| S4_extracted_11721 | Not-determined |
| S4_extracted_11724 | Low-quality |
| S4_extracted_11727 | Not-determined |
| S4_extracted_11736 | Not-determined |
| S4_extracted_11749 | Not-determined |
| S4_extracted_11755 | Not-determined |
| S4_extracted_11757 | Not-determined |
| S4_extracted_11758 | Not-determined |
| S4_extracted_11772 | Not-determined |
| S4_extracted_11777 | Not-determined |
| S4_extracted_11784 | Not-determined |
| S4_extracted_11799 | Not-determined |
| S4_extracted_11805 | Not-determined |
| S4_extracted_11806 | Not-determined |
| S4_extracted_11810 | Not-determined |
| S4_extracted_11813 | Not-determined |
| S4_extracted_11822 | Not-determined |
| S4_extracted_11828 | Not-determined |
| S4_extracted_11847 | Not-determined |
| S4_extracted_11851 | Not-determined |
| S4_extracted_11853 | Not-determined |
| S4_extracted_11854 | Not-determined |
| S4_extracted_11864 | Not-determined |
| S4_extracted_11899 | Not-determined |
| S4_extracted_11900 | Not-determined |
| S4_extracted_11915 | Low-quality |
| S4_extracted_11919 | Low-quality |
| S4_extracted_11929 | Not-determined |
| S4_extracted_11933 | Low-quality |
| S4_extracted_11936 | Not-determined |
| S4_extracted_11947 | Not-determined |
| S4_extracted_11964 | Low-quality |
| S4_extracted_11977 | Not-determined |
| S4_extracted_11978 | Low-quality |
| S4_extracted_11989 | Not-determined |
| S4_extracted_11993 | Not-determined |
| S4_extracted_12014 | Not-determined |
| S4_extracted_12017 | Not-determined |
| S4_extracted_12023 | Not-determined |
| S4_extracted_12040 | Not-determined |
| S4_extracted_12041 | Not-determined |
| S4_extracted_12060 | Not-determined |
| S4_extracted_12072 | Not-determined |
| S4_extracted_12105 | Not-determined |
| S4_extracted_12125 | Not-determined |
| S4_extracted_12126 | Not-determined |
| S4_extracted_12133 | Not-determined |
| S4_extracted_12142 | Not-determined |
| S4_extracted_12180 | Low-quality |
| S4_extracted_12190 | Not-determined |
| S4_extracted_12198 | Not-determined |
| S4_extracted_12209 | Not-determined |
| S4_extracted_12210 | Not-determined |
| S4_extracted_12213 | Not-determined |
| S4_extracted_12214 | Not-determined |
| S4_extracted_12219 | Not-determined |
| S4_extracted_12243 | Not-determined |
| S4_extracted_12246 | Not-determined |
| S4_extracted_12250 | Not-determined |
| S4_extracted_12252 | Not-determined |
| S4_extracted_12254 | Low-quality |
| S4_extracted_12305 | Not-determined |
| S4_extracted_12328 | Not-determined |
| S4_extracted_12333 | Not-determined |
| S4_extracted_12355 | Not-determined |
| S4_extracted_12356 | Not-determined |
| S4_extracted_12370 | Not-determined |
| S4_extracted_12371 | Not-determined |
| S4_extracted_12376 | Not-determined |
| S4_extracted_12378 | Not-determined |
| S4_extracted_12381 | Not-determined |
| S4_extracted_12386 | Not-determined |
| S4_extracted_12405 | Not-determined |
| S4_extracted_12407 | Not-determined |
| S4_extracted_12418 | Not-determined |
| S4_extracted_12422 | Low-quality |
| S4_extracted_12426 | Not-determined |
| S4_extracted_12433 | Not-determined |
| S4_extracted_12451 | Not-determined |
| S4_extracted_12460 | Not-determined |
| S4_extracted_12472 | Not-determined |
| S4_extracted_12480 | Not-determined |
| S4_extracted_12496 | Not-determined |
| S4_extracted_12513 | Not-determined |
| S4_extracted_12515 | Not-determined |
| S4_extracted_12527 | Not-determined |
| S4_extracted_12535 | Not-determined |
| S4_extracted_12542 | Low-quality |
| S4_extracted_12547 | Not-determined |
| S4_extracted_12560 | Low-quality |
| S4_extracted_12563 | Not-determined |
| S4_extracted_12569 | Low-quality |
| S4_extracted_12572 | Low-quality |
| S4_extracted_12574 | Low-quality |
| S4_extracted_12575 | Low-quality |
| S4_extracted_12576 | Not-determined |
| S4_extracted_12588 | Not-determined |
| S4_extracted_12600 | Not-determined |
| S17_extracted_k141_97671 | Medium-quality |
| S15_vs_10460\|\|full | Medium-quality |
| S4_extracted_12628 | Low-quality |
| S4_extracted_12657 | Not-determined |
| S4_extracted_12658 | Not-determined |
| S4_extracted_12665 | Not-determined |
| S4_extracted_12677 | Not-determined |
| S4_extracted_12686 | Not-determined |
| S4_extracted_12699 | Low-quality |
| S4_extracted_12714 | Not-determined |
| S4_extracted_12724 | Not-determined |
| S4_extracted_12728 | Not-determined |
| S4_extracted_12736 | Not-determined |
| S4_extracted_12741 | Not-determined |
| S4_extracted_12747 | Not-determined |
| S4_extracted_12772 | Not-determined |
| S4_extracted_12798 | Not-determined |
| S4_extracted_12801 | Low-quality |
| S4_extracted_12803 | Not-determined |
| S4_extracted_12813 | Not-determined |
| S4_extracted_12818 | Not-determined |
| S4_extracted_12821 | Low-quality |
| S4_extracted_12833 | Not-determined |
| S4_extracted_12852 | Low-quality |
| S4_extracted_12861 | Not-determined |
| S4_extracted_12863 | Low-quality |
| S4_extracted_12874 | Not-determined |
| S4_extracted_12891 | Not-determined |
| S4_extracted_12904 | Low-quality |
| S4_extracted_12907 | Not-determined |
| S4_extracted_12911 | Not-determined |
| S4_extracted_12917 | Not-determined |
| S4_extracted_12927 | Not-determined |
| S4_extracted_12930 | Not-determined |
| S4_extracted_12936 | Not-determined |
| S4_extracted_12937 | Not-determined |
| S4_extracted_12940 | Not-determined |
| S4_extracted_12943 | Not-determined |
| S4_extracted_12946 | Not-determined |
| S4_extracted_12957 | Not-determined |
| S4_extracted_12962 | Not-determined |
| S4_extracted_12970 | Not-determined |
| S4_extracted_12977 | Not-determined |
| S4_extracted_13003 | Not-determined |
| S4_extracted_13007 | Not-determined |
| S4_extracted_13014 | Not-determined |
| S4_extracted_13033 | Not-determined |
| S4_extracted_13039 | Not-determined |
| S4_extracted_13048 | Not-determined |
| S4_extracted_13057 | Low-quality |
| S4_extracted_13062 | Not-determined |
| S4_extracted_13086 | Not-determined |
| S4_extracted_13099 | Not-determined |
| S4_extracted_13116 | Not-determined |
| S4_extracted_13121 | Not-determined |
| S4_extracted_13141 | Not-determined |
| S4_extracted_13150 | Not-determined |
| S4_extracted_13151 | Not-determined |
| S4_extracted_13156 | Not-determined |
| S4_extracted_13159 | Not-determined |
| S4_extracted_13160 | Not-determined |
| S4_extracted_13167 | Not-determined |
| S4_extracted_13173 | Not-determined |
| S4_extracted_13194 | Low-quality |
| S4_extracted_13195 | Not-determined |
| S4_extracted_13201 | Not-determined |
| S4_extracted_13205 | Not-determined |
| S4_extracted_13212 | Not-determined |
| S4_extracted_13215 | Not-determined |
| S4_extracted_13221 | Not-determined |
| S4_extracted_13223 | Low-quality |
| S4_extracted_13226 | Not-determined |
| S4_extracted_13233 | Not-determined |
| S4_extracted_13238 | Not-determined |
| S4_extracted_13246 | Not-determined |
| S4_extracted_13252 | Not-determined |
| S4_extracted_13263 | Not-determined |
| S4_extracted_13278 | Not-determined |
| S4_extracted_13296 | Not-determined |
| S4_extracted_13300 | Not-determined |
| S4_extracted_13302 | Not-determined |
| S4_extracted_13317 | Not-determined |
| S4_extracted_13324 | Not-determined |
| S4_extracted_13325 | Not-determined |
| S4_extracted_13327 | Not-determined |
| S4_extracted_13336 | Low-quality |
| S4_extracted_13337 | Not-determined |
| S4_extracted_13344 | Not-determined |
| S7_extracted_11744 | Medium-quality |
| S4_extracted_13357 | Low-quality |
| S4_extracted_13374 | Not-determined |
| S4_extracted_13375 | Not-determined |
| S4_extracted_13377 | Not-determined |
| S4_extracted_13380 | Low-quality |
| S4_extracted_13386 | Not-determined |
| S4_extracted_13394 | Not-determined |
| S4_extracted_13395 | Not-determined |
| S4_extracted_13404 | Not-determined |
| S4_extracted_13407 | Not-determined |
| S4_extracted_13421 | Not-determined |
| S4_extracted_13430 | Not-determined |
| S4_extracted_13436 | Not-determined |
| S4_extracted_13440 | Not-determined |
| S4_extracted_13441 | Not-determined |
| S4_extracted_13444 | Not-determined |
| S4_extracted_13449 | Not-determined |
| S4_extracted_13455 | Not-determined |
| S4_extracted_13457 | Not-determined |
| S4_extracted_13476 | Low-quality |
| S4_extracted_13505 | Low-quality |
| S4_extracted_13507 | Not-determined |
| S4_extracted_13510 | Not-determined |
| S4_extracted_13519 | Not-determined |
| S4_extracted_13531 | Not-determined |
| S4_extracted_13539 | Not-determined |
| S4_extracted_13541 | Low-quality |
| S4_extracted_13549 | Not-determined |
| S4_extracted_13552 | Low-quality |
| S4_extracted_13554 | Not-determined |
| S4_extracted_13568 | Low-quality |
| S4_extracted_13570 | Not-determined |
| S4_extracted_13572 | Not-determined |
| S4_extracted_13576 | Not-determined |
| S4_extracted_13582 | Not-determined |
| S4_extracted_13590 | Not-determined |
| S4_extracted_13591 | Not-determined |
| S4_extracted_13604 | Not-determined |
| S4_extracted_13610 | Not-determined |
| S4_extracted_13612 | Not-determined |
| S4_extracted_13616 | Not-determined |
| S4_extracted_13622 | Not-determined |
| S7_vs_11744\|\|full | Medium-quality |
| S4_extracted_13631 | Not-determined |
| S4_extracted_13639 | Not-determined |
| S4_extracted_13657 | Not-determined |
| S4_extracted_13661 | Not-determined |
| S4_extracted_13665 | Not-determined |
| S4_extracted_13682 | Not-determined |
| S4_extracted_13686 | Low-quality |
| S4_extracted_13687 | Not-determined |
| S4_extracted_13691 | Not-determined |
| S4_extracted_13724 | Not-determined |
| S4_extracted_13726 | Not-determined |
| S4_extracted_13731 | Low-quality |
| S4_extracted_13738 | Not-determined |
| S4_extracted_13740 | Not-determined |
| S4_extracted_13742 | Low-quality |
| S4_extracted_13743 | Low-quality |
| S4_extracted_13747 | Not-determined |
| S4_extracted_13749 | Low-quality |
| S17_vs_k141_97671\|\|full | Medium-quality |
| S4_extracted_13763 | Not-determined |
| S4_extracted_13770 | Not-determined |
| S4_extracted_13771 | Low-quality |
| S4_extracted_13773 | Not-determined |
| S4_extracted_13783 | Not-determined |
| S4_extracted_13803 | Not-determined |
| S4_extracted_13810 | Low-quality |
| S8_vs_k141_110692\|\|full | Medium-quality |
| S4_extracted_13834 | Low-quality |
| S4_extracted_13839 | Low-quality |
| S4_extracted_13857 | Not-determined |
| S4_extracted_13863 | Low-quality |
| S4_extracted_13879 | Not-determined |
| S4_extracted_13886 | Not-determined |
| S4_extracted_13888 | Not-determined |
| S4_extracted_13906 | Not-determined |
| S4_extracted_13911 | Not-determined |
| S4_extracted_13916 | Not-determined |
| S4_extracted_13917 | Low-quality |
| S4_extracted_13933 | Not-determined |
| S4_extracted_13939 | Not-determined |
| S4_extracted_13941 | Not-determined |
| S4_extracted_13942 | Not-determined |
| S4_extracted_13944 | Not-determined |
| S4_extracted_13953 | Low-quality |
| S4_extracted_13954 | Low-quality |
| S4_extracted_13958 | Not-determined |
| S4_extracted_13976 | Not-determined |
| S4_extracted_13978 | Not-determined |
| S4_extracted_13981 | Not-determined |
| S4_extracted_13994 | Low-quality |
| S4_extracted_13996 | Not-determined |
| S4_extracted_14000 | Not-determined |
| S4_extracted_14006 | Not-determined |
| S4_extracted_14011 | Not-determined |

| | |
|---|---|
| S4_extracted_14012 | Not-determined |
| S4_extracted_14013 | Not-determined |
| S4_extracted_14033 | Not-determined |
| S4_extracted_14043 | Not-determined |
| S4_extracted_14053 | Not-determined |
| S4_extracted_14061 | Low-quality |
| S4_extracted_14063 | Low-quality |
| S4_extracted_14078 | Not-determined |
| S4_extracted_14086 | Not-determined |
| S4_extracted_14089 | Not-determined |
| S4_extracted_14093 | Not-determined |
| S4_extracted_14095 | Not-determined |
| S4_extracted_14102 | Not-determined |
| S4_extracted_14103 | Not-determined |
| S4_extracted_14118 | Low-quality |
| S4_extracted_14129 | Not-determined |
| S4_extracted_14136 | Not-determined |
| S4_extracted_14144 | Low-quality |
| S4_extracted_14146 | Not-determined |
| S4_extracted_14149 | Not-determined |
| S4_extracted_14167 | Not-determined |
| S4_extracted_14169 | Not-determined |
| S4_extracted_14184 | Low-quality |
| S4_extracted_14191 | Not-determined |
| S4_extracted_14195 | Not-determined |
| S4_extracted_14197 | Low-quality |
| S4_extracted_14198 | Not-determined |
| S4_extracted_14201 | Not-determined |
| S4_extracted_14204 | Not-determined |
| S4_extracted_14211 | Not-determined |
| S4_extracted_14212 | Low-quality |
| S4_extracted_14222 | Low-quality |
| S4_extracted_14228 | Not-determined |
| S4_extracted_14229 | Not-determined |
| S4_extracted_14238 | Not-determined |
| S4_extracted_14242 | Not-determined |
| S4_extracted_14255 | Not-determined |
| S4_extracted_14259 | Not-determined |
| S4_extracted_14269 | Low-quality |
| S4_extracted_14270 | Not-determined |
| S4_extracted_14271 | Not-determined |
| S4_extracted_14276 | Low-quality |
| S4_extracted_14283 | Not-determined |
| S4_extracted_14291 | Not-determined |
| S4_extracted_14294 | Not-determined |
| S4_extracted_14296 | Low-quality |
| S4_extracted_14312 | Not-determined |
| S4_extracted_14322 | Not-determined |
| S4_extracted_14327 | Not-determined |
| S4_extracted_14331 | Not-determined |
| S4_extracted_14337 | Not-determined |
| S4_extracted_14340 | Not-determined |
| S4_extracted_14341 | Not-determined |
| S4_extracted_14346 | Not-determined |
| S4_extracted_14349 | Not-determined |
| S4_extracted_14353 | Not-determined |
| S4_extracted_14369 | Low-quality |
| S4_extracted_14373 | Not-determined |
| S4_extracted_14378 | Not-determined |
| S4_extracted_14389 | Not-determined |
| S4_extracted_14393 | Low-quality |
| S4_extracted_14419 | Low-quality |
| S4_extracted_14421 | Not-determined |
| S4_extracted_14439 | Low-quality |
| S4_extracted_14446 | Not-determined |
| S4_extracted_14447 | Not-determined |
| S4_extracted_14454 | Low-quality |
| S4_extracted_14477 | Low-quality |
| S4_extracted_14485 | Not-determined |
| S4_extracted_14498 | Not-determined |
| S4_extracted_14499 | Not-determined |
| S4_extracted_14504 | Low-quality |
| S4_extracted_14515 | Low-quality |
| S4_extracted_14516 | Not-determined |
| S4_extracted_14528 | Not-determined |
| S4_extracted_14530 | Not-determined |
| S4_extracted_14532 | Not-determined |
| S4_extracted_14534 | Not-determined |
| S4_extracted_14563 | Not-determined |
| S4_extracted_14573 | Not-determined |
| S4_extracted_14579 | Not-determined |
| S4_extracted_14587 | Not-determined |
| S4_extracted_14594 | Not-determined |
| S4_extracted_14623 | Not-determined |
| S4_extracted_14648 | Not-determined |
| S4_extracted_14657 | Low-quality |
| S4_extracted_14660 | Not-determined |
| S4_extracted_14679 | Low-quality |
| S4_extracted_14683 | Not-determined |
| S4_extracted_14706 | Not-determined |
| S4_extracted_14707 | Not-determined |
| S4_extracted_14716 | Not-determined |
| S4_extracted_14724 | Not-determined |
| S4_extracted_14736 | Low-quality |
| S4_extracted_14739 | Low-quality |
| S4_extracted_14750 | Low-quality |
| S4_extracted_14763 | Low-quality |
| S4_extracted_14767 | Not-determined |
| S4_extracted_14772 | Not-determined |
| S4_extracted_14778 | Not-determined |
| S4_extracted_14796 | Not-determined |
| S4_extracted_14797 | Not-determined |
| S4_extracted_14799 | Not-determined |
| S4_extracted_14806 | Not-determined |
| S4_extracted_14855 | Not-determined |
| S4_extracted_14860 | Not-determined |
| S4_extracted_14875 | Not-determined |
| S4_extracted_14876 | Not-determined |
| S4_extracted_14878 | Not-determined |
| S4_extracted_14906 | Not-determined |
| S4_extracted_14909 | Not-determined |
| S4_extracted_14914 | Not-determined |
| S4_extracted_14915 | Not-determined |
| S4_extracted_14917 | Low-quality |
| S4_extracted_14924 | Not-determined |
| S4_extracted_14925 | Not-determined |
| S4_extracted_14933 | Not-determined |
| S4_extracted_14934 | Not-determined |
| S4_extracted_14935 | Not-determined |
| S4_extracted_14937 | Low-quality |
| S4_extracted_14947 | Not-determined |
| S4_extracted_14979 | Not-determined |
| S4_extracted_14990 | Not-determined |
| S4_extracted_14998 | Not-determined |
| S4_extracted_15002 | Low-quality |
| S4_extracted_15011 | Low-quality |
| S4_extracted_15016 | Not-determined |
| S4_extracted_15029 | Not-determined |
| S4_extracted_15031 | Not-determined |
| S4_extracted_15042 | Low-quality |
| S4_extracted_15043 | Not-determined |
| S4_extracted_15048 | Low-quality |
| S4_extracted_15054 | Not-determined |
| S4_extracted_15064 | Not-determined |
| S4_extracted_15067 | Low-quality |
| S4_extracted_15099 | Not-determined |
| S4_extracted_15107 | Low-quality |
| S4_extracted_15108 | Low-quality |
| S4_extracted_15111 | Not-determined |
| S4_extracted_15114 | Not-determined |
| S4_extracted_15117 | Low-quality |
| S4_extracted_15125 | Low-quality |
| S4_extracted_15126 | Low-quality |
| S4_extracted_15132 | Not-determined |
| S4_extracted_15133 | Low-quality |
| S4_extracted_15136 | Not-determined |
| S4_extracted_15137 | Low-quality |
| S4_extracted_15141 | Not-determined |
| S4_extracted_15170 | Not-determined |
| S4_extracted_15179 | Not-determined |
| S4_extracted_15186 | Not-determined |
| S4_extracted_15200 | Not-determined |
| S4_extracted_15201 | Not-determined |
| S4_extracted_15202 | Not-determined |
| S4_extracted_15204 | Low-quality |
| S4_extracted_15211 | Low-quality |
| S4_extracted_15245 | Low-quality |
| S4_extracted_15249 | Not-determined |
| S4_extracted_15250 | Not-determined |
| S4_extracted_15261 | Low-quality |
| S4_extracted_15263 | Not-determined |
| S4_extracted_15271 | Not-determined |
| S4_extracted_15274 | Low-quality |
| S4_extracted_15301 | Not-determined |
| S4_extracted_15315 | Not-determined |
| S4_extracted_15322 | Not-determined |
| S4_extracted_15344 | Not-determined |
| S4_extracted_15355 | Low-quality |
| S4_extracted_15358 | Not-determined |
| S4_extracted_15360 | Not-determined |
| S4_extracted_15363 | Not-determined |
| S5_vs_k141_117417\|\|full | Medium-quality |
| S4_extracted_15384 | Not-determined |
| S4_extracted_15397 | Not-determined |
| S4_extracted_15400 | Not-determined |
| S4_extracted_15404 | Not-determined |
| S4_extracted_15410 | Low-quality |
| S4_extracted_15414 | Low-quality |
| S4_extracted_15421 | Low-quality |
| S4_extracted_15441 | Not-determined |
| S4_extracted_15453 | Not-determined |
| S4_extracted_15456 | Low-quality |
| S4_extracted_15465 | Not-determined |
| S4_extracted_15467 | Not-determined |
| S4_extracted_15478 | Not-determined |
| S4_extracted_15488 | Not-determined |
| S4_extracted_15520 | Not-determined |
| S4_extracted_15527 | Not-determined |
| S4_extracted_15533 | Not-determined |
| S4_extracted_15537 | Low-quality |
| S4_extracted_15544 | Low-quality |
| S4_extracted_15567 | Not-determined |
| S4_extracted_15578 | Low-quality |
| S4_extracted_15583 | Low-quality |
| S4_extracted_15593 | Not-determined |
| S4_extracted_15594 | Not-determined |
| S4_extracted_15619 | Not-determined |
| S4_extracted_15625 | Low-quality |
| S4_extracted_15657 | Not-determined |
| S4_extracted_15663 | Not-determined |
| S4_extracted_15667 | Not-determined |
| S4_extracted_15681 | Low-quality |
| S4_extracted_15689 | Not-determined |
| S4_extracted_15697 | Not-determined |
| S4_extracted_15710 | Not-determined |
| S4_extracted_15715 | Not-determined |
| S4_extracted_15723 | Not-determined |
| S4_extracted_15731 | Not-determined |
| S4_extracted_15738 | Not-determined |
| S4_extracted_15740 | Low-quality |
| S4_extracted_15746 | Not-determined |
| S4_extracted_15758 | Low-quality |
| S4_extracted_15774 | Not-determined |
| S4_extracted_15780 | Not-determined |
| S4_extracted_15783 | Not-determined |
| S4_extracted_15786 | Not-determined |
| S4_extracted_15787 | Not-determined |
| S4_extracted_15798 | Not-determined |
| S4_extracted_15808 | Not-determined |
| S4_extracted_15839 | Not-determined |
| S4_extracted_15842 | Low-quality |
| S4_extracted_15853 | Not-determined |
| S4_extracted_15855 | Low-quality |
| S4_extracted_15856 | Not-determined |
| S4_extracted_15858 | Not-determined |
| S4_extracted_15865 | Low-quality |
| S4_extracted_15869 | Not-determined |
| S4_extracted_15877 | Not-determined |
| S4_extracted_15886 | Low-quality |
| S4_extracted_15891 | Low-quality |
| S4_extracted_15892 | Low-quality |
| S4_extracted_15895 | Not-determined |
| S4_extracted_15901 | Not-determined |
| S4_extracted_15922 | Low-quality |
| S4_extracted_15929 | Not-determined |
| S4_extracted_15930 | Low-quality |
| S4_extracted_15934 | Low-quality |
| S4_extracted_15937 | Not-determined |
| S4_extracted_15963 | Low-quality |
| S4_extracted_15970 | Not-determined |
| S4_extracted_15976 | Low-quality |
| S4_extracted_16015 | Not-determined |
| S4_extracted_16042 | Not-determined |
| S4_extracted_16054 | Not-determined |
| S4_extracted_16059 | Not-determined |
| S4_extracted_16081 | Not-determined |
| S4_extracted_16088 | Not-determined |
| S4_extracted_16093 | Not-determined |
| S4_extracted_16098 | Not-determined |
| S4_extracted_16102 | Low-quality |
| S4_extracted_16107 | Not-determined |
| S4_extracted_16108 | Low-quality |
| S4_extracted_16122 | Not-determined |
| S4_extracted_16127 | Not-determined |
| S4_extracted_16131 | Low-quality |
| S4_extracted_16136 | Not-determined |
| S4_extracted_16137 | Not-determined |
| S4_extracted_16142 | Not-determined |
| S4_extracted_16150 | Not-determined |
| S4_extracted_16154 | Low-quality |
| S4_extracted_16157 | Not-determined |
| S4_extracted_16162 | Low-quality |
| S4_extracted_16167 | Not-determined |
| S4_extracted_16188 | Not-determined |
| S4_extracted_16190 | Not-determined |
| S4_extracted_16194 | Low-quality |
| S4_extracted_16195 | Low-quality |
| S4_extracted_16200 | Low-quality |
| S4_extracted_16202 | Low-quality |
| S4_extracted_16203 | Not-determined |
| S4_extracted_16212 | Not-determined |
| S4_extracted_16214 | Not-determined |
| S4_extracted_16218 | Not-determined |
| S4_extracted_16226 | Not-determined |
| S4_extracted_16235 | Not-determined |
| S4_extracted_16239 | Not-determined |
| S4_extracted_16241 | Not-determined |
| S4_extracted_16254 | Not-determined |
| S4_extracted_16267 | Not-determined |
| S4_extracted_16270 | Low-quality |
| S4_extracted_16272 | Low-quality |
| S4_extracted_16276 | Low-quality |
| S4_extracted_16288 | Low-quality |
| S4_extracted_16289 | Not-determined |
| S4_extracted_16298 | Not-determined |
| S4_extracted_16306 | Not-determined |
| S4_extracted_16311 | Not-determined |
| S4_extracted_16337 | Not-determined |
| S4_extracted_16347 | Low-quality |
| S4_extracted_16363 | Low-quality |
| S4_extracted_16367 | Not-determined |
| S4_extracted_16369 | Not-determined |
| S4_extracted_16379 | Not-determined |
| S4_extracted_16382 | Not-determined |
| S4_extracted_16388 | Low-quality |
| S4_extracted_16400 | Low-quality |
| S4_extracted_16401 | Not-determined |
| S4_extracted_16405 | Not-determined |
| S4_extracted_16410 | Not-determined |
| S4_extracted_16411 | Not-determined |
| S4_extracted_16415 | Not-determined |
| S4_extracted_16427 | Not-determined |
| S4_extracted_16438 | Low-quality |
| S4_extracted_16439 | Not-determined |
| S4_extracted_16443 | Low-quality |
| S4_extracted_16452 | Low-quality |
| S4_extracted_16463 | Not-determined |
| S4_extracted_16466 | Low-quality |
| S4_extracted_16483 | Not-determined |
| S4_extracted_16484 | Not-determined |
| S13_vs_149860\|\|full | High-quality |
| S4_extracted_16490 | Not-determined |
| S4_extracted_16491 | Not-determined |
| S4_extracted_16510 | Not-determined |
| S4_extracted_16515 | Not-determined |
| S4_extracted_16518 | Not-determined |
| S4_extracted_16522 | Not-determined |
| S4_extracted_16545 | Low-quality |
| S4_extracted_16550 | Not-determined |
| S4_extracted_16569 | Not-determined |
| S4_extracted_16586 | Not-determined |
| S4_extracted_16588 | Not-determined |
| S4_extracted_16593 | Not-determined |
| S4_extracted_16594 | Not-determined |
| S4_extracted_16595 | Not-determined |
| S4_extracted_16602 | Not-determined |
| S4_extracted_16603 | Not-determined |
| S4_extracted_16607 | Not-determined |
| S4_extracted_16615 | Not-determined |
| S4_extracted_16645 | Not-determined |
| S4_extracted_16662 | Not-determined |
| S4_extracted_16674 | Low-quality |
| S4_extracted_16677 | Not-determined |
| S4_extracted_16682 | Not-determined |
| S4_extracted_16707 | Not-determined |
| S4_extracted_16712 | Not-determined |
| S4_extracted_16719 | Not-determined |
| S4_extracted_16723 | Not-determined |
| S4_extracted_16740 | Not-determined |
| S4_extracted_16742 | Not-determined |
| S4_extracted_16751 | Not-determined |
| S4_extracted_16760 | Not-determined |
| S4_extracted_16761 | Not-determined |
| S4_extracted_16765 | Low-quality |
| S4_extracted_16770 | Not-determined |
| S4_extracted_16773 | Not-determined |
| S4_extracted_16774 | Not-determined |
| S4_extracted_16783 | Low-quality |
| S4_extracted_16793 | Not-determined |
| S4_extracted_16830 | Low-quality |
| S4_extracted_16847 | Not-determined |
| S4_extracted_16855 | Not-determined |
| S4_extracted_16872 | Not-determined |
| S4_extracted_16926 | Not-determined |
| S4_extracted_16934 | Not-determined |
| S4_extracted_16935 | Not-determined |
| S4_extracted_16942 | Not-determined |
| S4_extracted_16943 | Not-determined |
| S4_extracted_16954 | Not-determined |
| S4_extracted_16958 | Not-determined |
| S4_extracted_16973 | Not-determined |
| S4_extracted_16984 | Not-determined |
| S4_extracted_16991 | Not-determined |
| S4_extracted_17010 | Not-determined |
| S4_extracted_17012 | Low-quality |
| S4_extracted_17014 | Not-determined |
| S4_extracted_17017 | Not-determined |
| S4_extracted_17023 | Not-determined |
| S4_extracted_17032 | Not-determined |
| S4_extracted_17033 | Low-quality |
| S4_extracted_17046 | Low-quality |
| S4_extracted_17052 | Not-determined |
| S4_extracted_17063 | Not-determined |
| S4_extracted_17072 | Not-determined |
| S4_extracted_17078 | Not-determined |
| S4_extracted_17085 | Not-determined |
| S4_extracted_17106 | Not-determined |
| S4_extracted_17109 | Not-determined |
| S4_extracted_17111 | Not-determined |
| S4_extracted_17117 | Not-determined |
| S4_extracted_17143 | Not-determined |
| S4_extracted_17150 | Not-determined |
| S4_extracted_17155 | Not-determined |
| S4_extracted_17157 | Not-determined |
| S4_extracted_17162 | Not-determined |
| S4_extracted_17164 | Low-quality |
| S4_extracted_17173 | Not-determined |
| S4_extracted_17180 | Not-determined |
| S4_extracted_17192 | Not-determined |
| S4_extracted_17208 | Low-quality |
| S4_extracted_17218 | Not-determined |
| S4_extracted_17236 | Not-determined |
| S4_extracted_17244 | Not-determined |
| S4_extracted_17248 | Not-determined |
| S4_extracted_17250 | Not-determined |
| S4_extracted_17264 | Not-determined |
| S4_extracted_17270 | Not-determined |
| S4_extracted_17276 | Not-determined |
| S4_extracted_17278 | Low-quality |
| S4_extracted_17300 | Low-quality |
| S4_extracted_17302 | Not-determined |
| S4_extracted_17315 | Not-determined |
| S4_extracted_17324 | Not-determined |
| S4_extracted_17325 | Low-quality |
| S4_extracted_17327 | Not-determined |
| S4_extracted_17343 | Low-quality |
| S4_extracted_17347 | Low-quality |
| S4_extracted_17367 | Not-determined |
| S4_extracted_17372 | Not-determined |
| S4_extracted_17385 | Not-determined |
| S4_extracted_17404 | Not-determined |
| S4_extracted_17408 | Not-determined |
| S4_extracted_17419 | Not-determined |
| S4_extracted_17432 | Low-quality |
| S4_extracted_17457 | Low-quality |
| S4_extracted_17473 | Low-quality |
| S4_extracted_17475 | Not-determined |
| S4_extracted_17499 | Not-determined |
| S4_extracted_17503 | Not-determined |
| S4_extracted_17507 | Low-quality |
| S4_extracted_17508 | Not-determined |
| S4_extracted_17514 | Low-quality |
| S4_extracted_17517 | Not-determined |
| S4_extracted_17521 | Not-determined |
| S4_extracted_17533 | Not-determined |
| S4_extracted_17542 | Low-quality |
| S4_extracted_17559 | Not-determined |
| S4_extracted_17563 | Not-determined |
| S4_extracted_17583 | Not-determined |
| S4_extracted_17597 | Not-determined |
| S4_extracted_17599 | Not-determined |
| S4_extracted_17602 | Not-determined |
| S4_extracted_17604 | Not-determined |
| S4_extracted_17605 | Not-determined |
| S4_extracted_17610 | Not-determined |
| S4_extracted_17629 | Not-determined |
| S4_extracted_17635 | Not-determined |
| S4_extracted_17645 | Not-determined |
| S4_extracted_17666 | Not-determined |
| S4_extracted_17679 | Not-determined |
| S4_extracted_17692 | Not-determined |
| S4_extracted_17705 | Not-determined |
| S4_extracted_17709 | Not-determined |
| S4_extracted_17711 | Not-determined |
| S4_extracted_17717 | Not-determined |
| S4_extracted_17720 | Low-quality |
| S4_extracted_17731 | Low-quality |
| S4_extracted_17735 | Not-determined |
| S4_extracted_17760 | Not-determined |
| S4_extracted_17761 | Not-determined |
| S4_extracted_17768 | Low-quality |
| S4_extracted_17771 | Not-determined |
| S4_extracted_17774 | Low-quality |
| S4_extracted_17778 | Not-determined |
| S4_extracted_17786 | Not-determined |
| S4_extracted_17788 | Not-determined |
| S4_extracted_17795 | Not-determined |
| S4_extracted_17822 | Low-quality |
| S4_extracted_17852 | Not-determined |
| S4_extracted_17855 | Not-determined |
| S4_extracted_17871 | Low-quality |
| S4_extracted_17884 | Not-determined |
| S4_extracted_17887 | Not-determined |
| S4_extracted_17892 | Not-determined |
| S4_extracted_17893 | Not-determined |
| S4_extracted_17897 | Low-quality |
| S4_extracted_17908 | Not-determined |
| S14_vs_k141_70999\|\|full | Medium-quality |
| S4_extracted_17957 | Not-determined |
| S4_extracted_17964 | Low-quality |
| S4_extracted_17965 | Not-determined |
| S4_extracted_17966 | Not-determined |
| S4_extracted_17967 | Not-determined |
| S4_extracted_17969 | Not-determined |
| S4_extracted_17971 | Not-determined |
| S4_extracted_17973 | Not-determined |
| S4_extracted_17976 | Low-quality |
| S4_extracted_17989 | Not-determined |
| S4_extracted_17995 | Low-quality |
| S4_extracted_18013 | Not-determined |
| S4_extracted_18015 | Not-determined |
| S4_extracted_18016 | Not-determined |
| S4_extracted_18022 | Not-determined |
| S4_extracted_18025 | Not-determined |
| S4_extracted_18026 | Low-quality |
| S4_extracted_18052 | Low-quality |
| S4_extracted_18059 | Not-determined |
| S4_extracted_18061 | Not-determined |
| S4_extracted_18066 | Not-determined |
| S4_extracted_18067 | Not-determined |
| S4_extracted_18073 | Not-determined |
| S4_extracted_18077 | Not-determined |
| S4_extracted_18085 | Not-determined |
| S4_extracted_18089 | Not-determined |
| S4_extracted_18094 | Not-determined |
| S4_extracted_18099 | Low-quality |
| S4_extracted_18100 | Not-determined |
| S4_extracted_18101 | Not-determined |
| S4_extracted_18103 | Low-quality |
| S4_extracted_18104 | Not-determined |
| S4_extracted_18110 | Not-determined |
| S4_extracted_18120 | Not-determined |
| S4_extracted_18121 | Not-determined |
| S4_extracted_18130 | Not-determined |
| S4_extracted_18145 | Low-quality |
| S4_extracted_18170 | Low-quality |
| S4_extracted_18174 | Low-quality |
| S4_extracted_18181 | Not-determined |
| S4_extracted_18184 | Not-determined |
| S4_extracted_18190 | Not-determined |
| S4_extracted_18196 | Not-determined |
| S4_extracted_18204 | Not-determined |
| S4_extracted_18205 | Not-determined |
| S4_extracted_18207 | Low-quality |
| S4_extracted_18213 | Low-quality |
| S4_extracted_18220 | Not-determined |
| S4_extracted_18233 | Not-determined |
| S4_extracted_18239 | Not-determined |
| S4_extracted_18249 | Low-quality |
| S4_extracted_18262 | Low-quality |
| S4_extracted_18266 | Not-determined |
| S4_extracted_18268 | Not-determined |
| S4_extracted_18279 | Low-quality |
| S4_extracted_18280 | Not-determined |
| S4_extracted_18294 | Not-determined |
| S4_extracted_18296 | Not-determined |
| S4_extracted_18299 | Not-determined |
| S4_extracted_18303 | Not-determined |
| S4_extracted_18304 | Not-determined |
| S4_extracted_18326 | Low-quality |
| S4_extracted_18328 | Not-determined |
| S4_extracted_18330 | Low-quality |
| S4_extracted_18346 | Not-determined |
| S4_extracted_18355 | Not-determined |
| S4_extracted_18363 | Not-determined |
| S4_extracted_18365 | Not-determined |
| S4_extracted_18370 | Low-quality |
| S4_extracted_18376 | Low-quality |
| S4_extracted_18386 | Not-determined |
| S4_extracted_18390 | Not-determined |
| S4_extracted_18392 | Not-determined |
| S4_extracted_18394 | Not-determined |
| S4_extracted_18402 | Not-determined |
| S4_extracted_18403 | Not-determined |
| S4_extracted_18414 | Not-determined |
| S4_extracted_18420 | Not-determined |
| S4_extracted_18437 | Not-determined |
| S4_extracted_18450 | Low-quality |
| S4_extracted_18451 | Not-determined |
| S4_extracted_18453 | Not-determined |
| S4_extracted_18455 | Not-determined |
| S4_extracted_18463 | Low-quality |
| S4_extracted_18480 | Not-determined |
| S4_extracted_18489 | Not-determined |
| S4_extracted_18498 | Not-determined |
| S4_extracted_18506 | Not-determined |
| S4_extracted_18512 | Not-determined |
| S4_extracted_18531 | Not-determined |
| S4_extracted_18534 | Not-determined |
| S4_extracted_18543 | Not-determined |
| S4_extracted_18544 | Not-determined |
| S4_extracted_18547 | Not-determined |
| S4_extracted_18555 | Low-quality |
| S4_extracted_18566 | Not-determined |
| S4_extracted_18567 | Low-quality |
| S4_extracted_18569 | Not-determined |
| S4_extracted_18586 | Not-determined |
| S4_extracted_18591 | Low-quality |
| S4_extracted_18601 | Not-determined |
| S4_extracted_18621 | Not-determined |
| S4_extracted_18626 | Not-determined |
| S4_extracted_18628 | Not-determined |
| S4_extracted_18630 | Not-determined |
| S4_extracted_18632 | Not-determined |
| S4_extracted_18651 | Not-determined |
| S4_extracted_18658 | Not-determined |
| S4_extracted_18678 | Not-determined |
| S4_extracted_18688 | Not-determined |
| S4_extracted_18690 | Not-determined |
| S4_extracted_18699 | Not-determined |
| S4_extracted_18718 | Low-quality |
| S4_extracted_18725 | Not-determined |
| S4_extracted_18734 | Not-determined |
| S4_extracted_18756 | Not-determined |
| S4_extracted_18772 | Not-determined |
| S4_extracted_18776 | Not-determined |
| S4_extracted_18783 | Low-quality |
| S4_extracted_18784 | Low-quality |
| S4_extracted_18789 | Not-determined |

| | |
|---|---|
| S4_extracted_18793 | Not-determined |
| S4_extracted_18796 | Low-quality |
| S1_vs_k141_112116\|\|full | High-quality |
| S4_extracted_18804 | Low-quality |
| S4_extracted_18807 | Low-quality |
| S4_extracted_18817 | Not-determined |
| S4_extracted_18844 | Not-determined |
| S4_extracted_18854 | Not-determined |
| S4_extracted_18860 | Not-determined |
| S4_extracted_18874 | Low-quality |
| S4_extracted_18881 | Not-determined |
| S4_extracted_18894 | Not-determined |
| S4_extracted_18896 | Not-determined |
| S4_extracted_18899 | Low-quality |
| S4_extracted_18905 | Not-determined |
| S4_extracted_18914 | Not-determined |
| S4_extracted_18916 | Not-determined |
| S4_extracted_18926 | Not-determined |
| S4_extracted_18940 | Not-determined |
| S4_extracted_18945 | Low-quality |
| S4_extracted_18962 | Not-determined |
| S4_extracted_18965 | Not-determined |
| S4_extracted_18974 | Not-determined |
| S4_extracted_18976 | Not-determined |
| S4_extracted_18983 | Not-determined |
| S4_extracted_18987 | Not-determined |
| S4_extracted_18990 | Low-quality |
| S4_extracted_18995 | Not-determined |
| S4_extracted_18997 | Low-quality |
| S4_extracted_18999 | Not-determined |
| S4_extracted_19000 | Not-determined |
| S4_extracted_19001 | Not-determined |
| S4_extracted_19013 | Not-determined |
| S4_extracted_19014 | Not-determined |
| S4_extracted_19018 | Not-determined |
| S4_extracted_19020 | Not-determined |
| S4_extracted_19021 | Low-quality |
| S4_extracted_19027 | Not-determined |
| S4_extracted_19028 | Not-determined |
| S4_extracted_19035 | Not-determined |
| S4_extracted_19038 | Not-determined |
| S4_extracted_19073 | Not-determined |
| S4_extracted_19081 | Not-determined |
| S4_extracted_19084 | Low-quality |
| S4_extracted_19085 | Low-quality |
| S4_extracted_19089 | Low-quality |
| S4_extracted_19106 | Low-quality |
| S4_extracted_19114 | Low-quality |
| S4_extracted_19122 | Not-determined |
| S4_extracted_19124 | Not-determined |
| S4_extracted_19132 | Low-quality |
| S4_extracted_19148 | Low-quality |
| S4_extracted_19155 | Not-determined |
| S4_extracted_19174 | Not-determined |
| S4_extracted_19179 | Not-determined |
| S4_extracted_19192 | Not-determined |
| S4_extracted_19203 | Not-determined |
| S4_extracted_19216 | Not-determined |
| S4_extracted_19232 | Not-determined |
| S4_extracted_19248 | Not-determined |
| S4_extracted_19252 | Not-determined |
| S4_extracted_19253 | Not-determined |
| S4_extracted_19254 | Not-determined |
| S4_extracted_19261 | Not-determined |
| S4_extracted_19265 | Low-quality |
| S4_extracted_19282 | Not-determined |
| S4_extracted_19299 | Not-determined |
| S4_extracted_19305 | Low-quality |
| S4_extracted_19313 | Low-quality |
| S4_extracted_19314 | Low-quality |
| S4_extracted_19325 | Not-determined |
| S4_extracted_19328 | Not-determined |
| S4_extracted_19339 | Not-determined |
| S4_extracted_19349 | Not-determined |
| S4_extracted_19373 | Low-quality |
| S4_extracted_19390 | Not-determined |
| S4_extracted_19397 | Not-determined |
| S4_extracted_19403 | Not-determined |
| S4_extracted_19408 | Not-determined |
| S4_extracted_19410 | Not-determined |
| S4_extracted_19439 | Not-determined |
| S4_extracted_19441 | Not-determined |
| S4_extracted_19443 | Not-determined |
| S4_extracted_19444 | Not-determined |
| S4_extracted_19449 | Not-determined |
| S4_extracted_19450 | Not-determined |
| S4_extracted_19451 | Not-determined |
| S4_extracted_19457 | Not-determined |
| S4_extracted_19463 | Low-quality |
| S4_extracted_19486 | Not-determined |
| S4_extracted_19496 | Not-determined |
| S4_extracted_19513 | Not-determined |
| S4_extracted_19526 | Low-quality |
| S4_extracted_19539 | Not-determined |
| S4_extracted_19551 | Not-determined |
| S4_extracted_19554 | Low-quality |
| S4_extracted_19565 | Low-quality |
| S4_extracted_19580 | Not-determined |
| S4_extracted_19583 | Not-determined |
| S4_extracted_19596 | Low-quality |
| S4_extracted_19599 | Low-quality |
| S4_extracted_19602 | Not-determined |
| S4_extracted_19629 | Not-determined |
| S4_extracted_19632 | Not-determined |
| S4_extracted_19634 | Low-quality |
| S4_extracted_19638 | Not-determined |
| S4_extracted_19641 | Not-determined |
| S4_extracted_19646 | Not-determined |
| S4_extracted_19649 | Not-determined |
| S4_extracted_19656 | Not-determined |
| S4_extracted_19666 | Not-determined |
| S4_extracted_19687 | Not-determined |
| S4_extracted_19690 | Not-determined |
| S4_extracted_19691 | Not-determined |
| S4_extracted_19701 | Not-determined |
| S4_extracted_19719 | Not-determined |
| S4_extracted_19736 | Not-determined |
| S4_extracted_19747 | Not-determined |
| S4_extracted_19756 | Not-determined |
| S4_extracted_19758 | Low-quality |
| S4_extracted_19765 | Low-quality |
| S4_extracted_19768 | Low-quality |
| S4_extracted_19773 | Not-determined |
| S4_extracted_19778 | Not-determined |
| S4_extracted_19785 | Not-determined |
| S4_extracted_19807 | Low-quality |
| S4_extracted_19815 | Not-determined |
| S4_extracted_19817 | Not-determined |
| S4_extracted_19844 | Not-determined |
| S4_extracted_19848 | Not-determined |
| S4_extracted_19863 | Not-determined |
| S4_extracted_19864 | Not-determined |
| S4_extracted_19865 | Not-determined |
| S4_extracted_19869 | Not-determined |
| S4_extracted_19870 | Not-determined |
| S4_extracted_19872 | Not-determined |
| S4_extracted_19874 | Not-determined |
| S4_extracted_19885 | Not-determined |
| S4_extracted_19890 | Not-determined |
| S4_extracted_19898 | Not-determined |
| S4_extracted_19901 | Not-determined |
| S4_extracted_19907 | Not-determined |
| S4_extracted_19924 | Not-determined |
| S4_extracted_19928 | Not-determined |
| S4_extracted_19934 | Low-quality |
| S4_extracted_19944 | Low-quality |
| S4_extracted_19951 | Low-quality |
| S4_extracted_19954 | Low-quality |
| S4_extracted_19972 | Not-determined |
| S4_extracted_19978 | Not-determined |
| S4_extracted_19981 | Low-quality |
| S4_extracted_19984 | Not-determined |
| S4_extracted_19989 | Low-quality |
| S4_extracted_19999 | Not-determined |
| S4_extracted_20010 | Not-determined |
| S4_extracted_20020 | Low-quality |
| S4_extracted_20030 | Not-determined |
| S4_extracted_20038 | Not-determined |
| S4_extracted_20049 | Not-determined |
| S4_extracted_20052 | Low-quality |
| S4_extracted_20057 | Not-determined |
| S4_extracted_20059 | Not-determined |
| S4_extracted_20072 | Not-determined |
| S4_extracted_20077 | Not-determined |
| S4_extracted_20100 | Low-quality |
| S4_extracted_20105 | Not-determined |
| S4_extracted_20106 | Not-determined |
| S4_extracted_20109 | Not-determined |
| S4_extracted_20114 | Low-quality |
| S4_extracted_20125 | Not-determined |
| S4_extracted_20131 | Low-quality |
| S4_extracted_20132 | Not-determined |
| S4_extracted_20158 | Low-quality |
| S4_extracted_20163 | Not-determined |
| S4_extracted_20188 | Not-determined |
| S4_extracted_20196 | Not-determined |
| S4_extracted_20217 | Not-determined |
| S4_extracted_20227 | Low-quality |
| S4_extracted_20256 | Not-determined |
| S4_extracted_20267 | Low-quality |
| S4_extracted_20268 | Not-determined |
| S4_extracted_20269 | Not-determined |
| S4_extracted_20273 | Low-quality |
| S4_extracted_20276 | Not-determined |
| S4_extracted_20291 | Not-determined |
| S4_extracted_20305 | Not-determined |
| S4_extracted_20309 | Not-determined |
| S4_extracted_20313 | Not-determined |
| S4_extracted_20323 | Not-determined |
| S4_extracted_20324 | Low-quality |
| S4_extracted_20339 | Not-determined |
| S4_extracted_20386 | Not-determined |
| S4_extracted_20396 | Not-determined |
| S4_extracted_20398 | Not-determined |
| S4_extracted_20406 | Low-quality |
| S4_extracted_20408 | Not-determined |
| S4_extracted_20412 | Low-quality |
| S4_extracted_20435 | Not-determined |
| S12_vs_k141_14467\|\|full | Medium-quality |
| S4_extracted_20445 | Not-determined |
| S4_extracted_20458 | Not-determined |
| S4_extracted_20471 | Not-determined |
| S4_extracted_20483 | Low-quality |
| S4_extracted_20498 | Not-determined |
| S4_extracted_20544 | Low-quality |
| S4_extracted_20558 | Not-determined |
| S4_extracted_20577 | Not-determined |
| S4_extracted_20582 | Low-quality |
| S4_extracted_20585 | Not-determined |
| S4_extracted_20603 | Not-determined |
| S4_extracted_20624 | Low-quality |
| S4_extracted_20632 | Not-determined |
| S4_extracted_20643 | Not-determined |
| S4_extracted_20645 | Not-determined |
| S4_extracted_20648 | Low-quality |
| S4_extracted_20658 | Not-determined |
| S4_extracted_20659 | Not-determined |
| S4_extracted_20660 | Not-determined |
| S4_extracted_20673 | Not-determined |
| S4_extracted_20691 | Not-determined |
| S4_extracted_20698 | Not-determined |
| S4_extracted_20708 | Not-determined |
| S4_extracted_20722 | Not-determined |
| S4_extracted_20736 | Not-determined |
| S4_extracted_20737 | Not-determined |
| S4_extracted_20743 | Not-determined |
| S4_extracted_20758 | Not-determined |
| S4_extracted_20767 | Low-quality |
| S4_extracted_20771 | Not-determined |
| S4_extracted_20778 | Low-quality |
| S4_extracted_20781 | Low-quality |
| S4_extracted_20783 | Low-quality |
| S4_extracted_20786 | Not-determined |
| S4_extracted_20787 | Not-determined |
| S4_extracted_20791 | Not-determined |
| S4_extracted_20799 | Not-determined |
| S4_extracted_20801 | Not-determined |
| S4_extracted_20803 | Not-determined |
| S4_extracted_20806 | Not-determined |
| S4_extracted_20814 | Not-determined |
| S4_extracted_20818 | Low-quality |
| S4_extracted_20820 | Not-determined |
| S4_extracted_20823 | Not-determined |
| S4_extracted_20834 | Not-determined |
| S4_extracted_20835 | Low-quality |
| S4_extracted_20848 | Not-determined |
| S4_extracted_20852 | Not-determined |
| S4_extracted_20862 | Not-determined |
| S4_extracted_20865 | Not-determined |
| S4_extracted_20867 | Not-determined |
| S4_extracted_20881 | Not-determined |
| S4_extracted_20891 | Not-determined |
| S4_extracted_20897 | Not-determined |
| S4_extracted_20905 | Not-determined |
| S4_extracted_20907 | Not-determined |
| S4_extracted_20931 | Not-determined |
| S4_extracted_20932 | Not-determined |
| S4_extracted_20937 | Not-determined |
| S4_extracted_20946 | Not-determined |
| S4_extracted_20950 | Low-quality |
| S4_extracted_20956 | Not-determined |
| S4_extracted_20957 | Not-determined |
| S4_extracted_20964 | Not-determined |
| S4_extracted_20965 | Not-determined |
| S4_extracted_20972 | Not-determined |
| S4_extracted_20976 | Not-determined |
| S4_extracted_20981 | Not-determined |
| S4_extracted_20987 | Low-quality |
| S4_extracted_20988 | Not-determined |
| S4_extracted_20999 | Not-determined |
| S4_extracted_21006 | Low-quality |
| S4_extracted_21022 | Not-determined |
| S4_extracted_21034 | Not-determined |
| S4_extracted_21036 | Not-determined |
| S4_extracted_21041 | Not-determined |
| S4_extracted_21042 | Not-determined |
| S4_extracted_21056 | Not-determined |
| S4_extracted_21065 | Low-quality |
| S4_extracted_21074 | Not-determined |
| S4_extracted_21083 | Low-quality |
| S4_extracted_21101 | Not-determined |
| S4_extracted_21106 | Low-quality |
| S4_extracted_21111 | Not-determined |
| S4_extracted_21117 | Not-determined |
| S4_extracted_21119 | Not-determined |
| S4_extracted_21122 | Not-determined |
| S4_extracted_21133 | Not-determined |
| S4_extracted_21135 | Not-determined |
| S4_extracted_21152 | Low-quality |
| S4_extracted_21162 | Not-determined |
| S4_extracted_21183 | Not-determined |
| S4_extracted_21185 | Low-quality |
| S4_extracted_21193 | Low-quality |
| S4_extracted_21208 | Not-determined |
| S4_extracted_21212 | Not-determined |
| S4_extracted_21214 | Not-determined |
| S4_extracted_21217 | Not-determined |
| S4_extracted_21220 | Not-determined |
| S4_extracted_21223 | Not-determined |
| S4_extracted_21226 | Not-determined |
| S4_extracted_21231 | Not-determined |
| S4_extracted_21232 | Low-quality |
| S4_extracted_21274 | Not-determined |
| S4_extracted_21280 | Low-quality |
| S4_extracted_21309 | Low-quality |
| S4_extracted_21313 | Not-determined |
| S4_extracted_21317 | Not-determined |
| S4_extracted_21319 | Not-determined |
| S4_extracted_21323 | Not-determined |
| S4_extracted_21331 | Low-quality |
| S4_extracted_21345 | Not-determined |
| S4_extracted_21347 | Not-determined |
| S4_extracted_21373 | Not-determined |
| S4_extracted_21375 | Not-determined |
| S4_extracted_21379 | Not-determined |
| S4_extracted_21383 | Not-determined |
| S4_extracted_21394 | Not-determined |
| S4_extracted_21395 | Not-determined |
| S4_extracted_21404 | Low-quality |
| S4_extracted_21415 | Low-quality |
| S4_extracted_21421 | Low-quality |
| S4_extracted_21436 | Not-determined |
| S4_extracted_21438 | Low-quality |
| S4_extracted_21445 | Not-determined |
| S4_extracted_21446 | Not-determined |
| S4_extracted_21457 | Low-quality |
| S4_extracted_21470 | Not-determined |
| S4_extracted_21489 | Not-determined |
| S4_extracted_21490 | Not-determined |
| S4_extracted_21493 | Not-determined |
| S4_extracted_21498 | Not-determined |
| S4_extracted_21501 | Not-determined |
| S4_extracted_21507 | Low-quality |
| S4_extracted_21508 | Not-determined |
| S4_extracted_21512 | Low-quality |
| S4_extracted_21520 | Not-determined |
| S4_extracted_21522 | Not-determined |
| S4_extracted_21530 | Not-determined |
| S4_extracted_21533 | Not-determined |
| S4_extracted_21536 | Not-determined |
| S4_extracted_21540 | Not-determined |
| S4_extracted_21542 | Not-determined |
| S4_extracted_21545 | Not-determined |
| S4_extracted_21549 | Not-determined |
| S4_extracted_21553 | Low-quality |
| S4_extracted_21554 | Not-determined |
| S4_extracted_21559 | Low-quality |
| S4_extracted_21569 | Not-determined |
| S4_extracted_21577 | Not-determined |
| S4_extracted_21586 | Not-determined |
| S4_extracted_21590 | Not-determined |
| S4_extracted_21593 | Low-quality |
| S4_extracted_21601 | Not-determined |
| S4_extracted_21602 | Not-determined |
| S4_extracted_21614 | Not-determined |
| S4_extracted_21616 | Not-determined |
| S4_extracted_21617 | Not-determined |
| S4_extracted_21619 | Not-determined |
| S4_extracted_21625 | Low-quality |
| S4_extracted_21630 | Not-determined |
| S4_extracted_21633 | Not-determined |
| S4_extracted_21644 | Not-determined |
| S4_extracted_21647 | Not-determined |
| S4_extracted_21692 | Not-determined |
| S4_extracted_21695 | Not-determined |
| S4_extracted_21700 | Low-quality |
| S4_extracted_21705 | Not-determined |
| S4_extracted_21706 | Not-determined |
| S4_extracted_21707 | Not-determined |
| S4_extracted_21710 | Not-determined |
| S4_extracted_21713 | Not-determined |
| S4_extracted_21716 | Not-determined |
| S4_extracted_21732 | Not-determined |
| S4_extracted_21744 | Not-determined |
| S4_extracted_21746 | Not-determined |
| S4_extracted_21760 | Not-determined |
| S4_extracted_21762 | Not-determined |
| S4_extracted_21768 | Not-determined |
| S4_extracted_21774 | Not-determined |
| S4_extracted_21779 | Not-determined |
| S4_extracted_21789 | Not-determined |
| S4_extracted_21792 | Not-determined |
| S4_extracted_21800 | Low-quality |
| S4_extracted_21808 | Low-quality |
| S4_extracted_21809 | Low-quality |
| S4_extracted_21824 | Not-determined |
| S4_extracted_21830 | Not-determined |
| S4_extracted_21833 | Not-determined |
| S4_extracted_21846 | Low-quality |
| S4_extracted_21847 | Not-determined |
| S4_extracted_21853 | Low-quality |
| S4_extracted_21862 | Not-determined |
| S4_extracted_21866 | Low-quality |
| S4_extracted_21869 | Low-quality |
| S4_extracted_21870 | Not-determined |
| S4_extracted_21899 | Not-determined |
| S4_extracted_21915 | Not-determined |
| S4_extracted_21925 | Not-determined |
| S4_extracted_21934 | Low-quality |
| S4_extracted_21939 | Not-determined |
| S4_extracted_21945 | Not-determined |
| S4_extracted_21946 | Not-determined |
| S4_extracted_21966 | Not-determined |
| S4_extracted_21973 | Low-quality |
| S4_extracted_21990 | Low-quality |
| S4_extracted_22007 | Not-determined |
| S4_extracted_22016 | Low-quality |
| S4_extracted_22020 | Not-determined |
| S4_extracted_22022 | Not-determined |
| S4_extracted_22025 | Not-determined |
| S4_extracted_22035 | Not-determined |
| S4_extracted_22037 | Not-determined |
| S4_extracted_22040 | Low-quality |
| S4_extracted_22047 | Not-determined |
| S4_extracted_22049 | Low-quality |
| S4_extracted_22050 | Low-quality |
| S4_extracted_22074 | Not-determined |
| S4_extracted_22083 | Low-quality |
| S4_extracted_22092 | Not-determined |
| S4_extracted_22096 | Not-determined |
| S4_extracted_22098 | Not-determined |
| S4_extracted_22103 | Not-determined |
| S4_extracted_22105 | Not-determined |
| S4_extracted_22110 | Low-quality |
| S4_extracted_22111 | Not-determined |
| S4_extracted_22116 | Low-quality |
| S4_extracted_22141 | Not-determined |
| S4_extracted_22153 | Not-determined |
| S4_extracted_22159 | Low-quality |
| S4_extracted_22169 | Not-determined |
| S4_extracted_22197 | Not-determined |
| S4_extracted_22202 | Low-quality |
| S4_extracted_22206 | Not-determined |
| S4_extracted_22208 | Low-quality |
| S4_extracted_22209 | Low-quality |
| S4_extracted_22210 | Low-quality |
| S14_vs_k141_66841\|\|full | Medium-quality |
| S4_extracted_22222 | Not-determined |
| S4_extracted_22230 | Not-determined |
| S4_extracted_22231 | Low-quality |
| S4_extracted_22234 | Not-determined |
| S4_extracted_22235 | Not-determined |
| S4_extracted_22236 | Not-determined |
| S4_extracted_22239 | Not-determined |
| S4_extracted_22244 | Low-quality |
| S4_extracted_22283 | Not-determined |
| S4_extracted_22284 | Not-determined |
| S4_extracted_22286 | Not-determined |
| S4_extracted_22291 | Low-quality |
| S4_extracted_22303 | Not-determined |
| S4_extracted_22314 | Not-determined |
| S4_extracted_22330 | Not-determined |
| S4_extracted_22331 | Not-determined |
| S4_extracted_22339 | Not-determined |
| S4_extracted_22340 | Not-determined |
| S4_extracted_22342 | Not-determined |
| S4_extracted_22369 | Not-determined |
| S4_extracted_22392 | Low-quality |
| S4_extracted_22393 | Not-determined |
| S4_extracted_22399 | Not-determined |
| S4_extracted_22403 | Not-determined |
| S4_extracted_22406 | Not-determined |
| S4_extracted_22418 | Not-determined |
| S4_extracted_22432 | Not-determined |
| S4_extracted_22435 | Not-determined |
| S4_extracted_22440 | Not-determined |
| S4_extracted_22443 | Not-determined |
| S4_extracted_22444 | Low-quality |
| S4_extracted_22447 | Not-determined |
| S4_extracted_22456 | Not-determined |
| S4_extracted_22457 | Not-determined |
| S4_extracted_22462 | Low-quality |
| S4_extracted_22473 | Not-determined |
| S4_extracted_22474 | Not-determined |
| S4_extracted_22480 | Not-determined |
| S4_extracted_22482 | Not-determined |
| S4_extracted_22488 | Not-determined |
| S4_extracted_22489 | Not-determined |
| S4_extracted_22502 | Not-determined |
| S4_extracted_22506 | Not-determined |
| S4_extracted_22510 | Not-determined |
| S4_extracted_22514 | Not-determined |
| S4_extracted_22515 | Low-quality |
| S4_extracted_22531 | Not-determined |
| S4_extracted_22533 | Not-determined |
| S4_extracted_22536 | Not-determined |
| S4_extracted_22537 | Not-determined |
| S4_extracted_22544 | Not-determined |
| S4_extracted_22547 | Not-determined |
| S4_extracted_22548 | Low-quality |
| S4_extracted_22549 | Low-quality |
| S4_extracted_22551 | Not-determined |
| S4_extracted_22558 | Not-determined |
| S4_extracted_22560 | Not-determined |
| S4_extracted_22561 | Not-determined |
| S4_extracted_22565 | Not-determined |
| S4_extracted_22573 | Not-determined |
| S4_extracted_22575 | Not-determined |
| S4_extracted_22580 | Not-determined |
| S4_extracted_22587 | Not-determined |
| S4_extracted_22588 | Not-determined |
| S4_extracted_22597 | Not-determined |
| S4_extracted_22612 | Not-determined |
| S4_extracted_22617 | Not-determined |
| S4_extracted_22625 | Not-determined |
| S4_extracted_22630 | Not-determined |
| S4_extracted_22633 | Not-determined |
| S4_extracted_22644 | Low-quality |
| S4_extracted_22652 | Not-determined |
| S4_extracted_22666 | Low-quality |
| S4_extracted_22668 | Low-quality |
| S4_extracted_22669 | Not-determined |
| S4_extracted_22670 | Not-determined |
| S4_extracted_22672 | Not-determined |
| S4_extracted_22682 | Low-quality |
| S4_extracted_22696 | Not-determined |
| S4_extracted_22698 | Not-determined |
| S4_extracted_22700 | Not-determined |
| S4_extracted_22706 | Not-determined |
| S4_extracted_22707 | Not-determined |
| S4_extracted_22722 | Not-determined |
| S4_extracted_22732 | Not-determined |
| S4_extracted_22735 | Not-determined |
| S4_extracted_22752 | Low-quality |
| S4_extracted_22755 | Low-quality |
| S4_extracted_22760 | Not-determined |
| S4_extracted_22765 | Not-determined |
| S4_extracted_22776 | Not-determined |
| S4_extracted_22778 | Not-determined |
| S4_extracted_22783 | Not-determined |
| S4_extracted_22784 | Low-quality |
| S4_extracted_22800 | Not-determined |
| S4_extracted_22813 | Not-determined |
| S4_extracted_22817 | Low-quality |
| S4_extracted_22831 | Not-determined |
| S4_extracted_22834 | Not-determined |
| S4_extracted_22842 | Not-determined |
| S4_extracted_22847 | Not-determined |
| S4_extracted_22855 | Not-determined |
| S4_extracted_22860 | Not-determined |
| S4_extracted_22886 | Not-determined |
| S4_extracted_22899 | Low-quality |
| S4_extracted_22902 | Not-determined |
| S4_extracted_22907 | Not-determined |
| S4_extracted_22925 | Not-determined |
| S4_extracted_22952 | Not-determined |
| S4_extracted_22953 | Not-determined |
| S4_extracted_22955 | Not-determined |
| S4_extracted_22962 | Not-determined |
| S4_extracted_22966 | Not-determined |
| S4_extracted_22974 | Low-quality |
| S4_extracted_22978 | Not-determined |
| S4_extracted_22981 | Low-quality |
| S4_extracted_22988 | Not-determined |
| S4_extracted_23000 | Not-determined |
| S4_extracted_23008 | Not-determined |
| S4_extracted_23012 | Low-quality |
| S4_extracted_23013 | Low-quality |
| S4_extracted_23016 | Low-quality |
| S4_extracted_23020 | Not-determined |
| S4_extracted_23021 | Low-quality |
| S4_extracted_23022 | Not-determined |
| S4_extracted_23031 | Not-determined |
| S4_extracted_23057 | Not-determined |
| S4_extracted_23058 | Not-determined |
| S4_extracted_23060 | Not-determined |
| S4_extracted_23067 | Not-determined |
| S4_extracted_23078 | Not-determined |
| S4_extracted_23087 | Not-determined |
| S4_extracted_23088 | Not-determined |
| S4_extracted_23090 | Not-determined |
| S4_extracted_23098 | Low-quality |
| S4_extracted_23100 | Not-determined |
| S4_extracted_23104 | Not-determined |
| S4_extracted_23105 | Low-quality |
| S4_extracted_23118 | Not-determined |
| S4_extracted_23122 | Not-determined |
| S4_extracted_23131 | Not-determined |
| S4_extracted_23144 | Not-determined |
| S4_extracted_23161 | Not-determined |
| S4_extracted_23168 | Not-determined |
| S4_extracted_23174 | Low-quality |
| S4_extracted_23185 | Not-determined |
| S4_extracted_23188 | Low-quality |
| S4_extracted_23191 | Low-quality |
| S4_extracted_23199 | Low-quality |
| S4_extracted_23213 | Low-quality |
| S4_extracted_23215 | Not-determined |
| S4_extracted_23222 | Not-determined |

| | |
|---|---|
| S4_extracted_23233 | Not-determined |
| S4_extracted_23237 | Not-determined |
| S4_extracted_23238 | Low-quality |
| S4_extracted_23243 | Not-determined |
| S4_extracted_23250 | Not-determined |
| S4_extracted_23255 | Not-determined |
| S4_extracted_23256 | Not-determined |
| S4_extracted_23276 | Low-quality |
| S4_extracted_23277 | Not-determined |
| S4_extracted_23284 | Not-determined |
| S4_extracted_23290 | Not-determined |
| S4_extracted_23292 | Not-determined |
| S4_extracted_23299 | Not-determined |
| S4_extracted_23307 | Not-determined |
| S4_extracted_23312 | Not-determined |
| S4_extracted_23313 | Not-determined |
| S4_extracted_23316 | Not-determined |
| S4_extracted_23318 | Not-determined |
| S4_extracted_23322 | Not-determined |
| S4_extracted_23324 | Low-quality |
| S4_extracted_23325 | Low-quality |
| S4_extracted_23326 | Not-determined |
| S4_extracted_23333 | Not-determined |
| S4_extracted_23337 | Not-determined |
| S4_extracted_23341 | Not-determined |
| S4_extracted_23351 | Not-determined |
| S4_extracted_23355 | Not-determined |
| S4_extracted_23356 | Not-determined |
| S4_extracted_23361 | Low-quality |
| S4_extracted_23362 | Not-determined |
| S4_extracted_23369 | Not-determined |
| S4_extracted_23371 | Not-determined |
| S4_extracted_23382 | Low-quality |
| S4_extracted_23383 | Not-determined |
| S4_extracted_23389 | Not-determined |
| S4_extracted_23392 | Not-determined |
| S4_extracted_23417 | Not-determined |
| S4_extracted_23422 | Not-determined |
| S4_extracted_23423 | Low-quality |
| S4_extracted_23449 | Not-determined |
| S4_extracted_23450 | Not-determined |
| S4_extracted_23465 | Not-determined |
| S4_extracted_23475 | Not-determined |
| S4_extracted_23476 | Not-determined |
| S4_extracted_23478 | Not-determined |
| S4_extracted_23482 | Not-determined |
| S4_extracted_23489 | Not-determined |
| S4_extracted_23506 | Low-quality |
| S4_extracted_23507 | Not-determined |
| S4_extracted_23514 | Low-quality |
| S4_extracted_23519 | Not-determined |
| S4_extracted_23527 | Not-determined |
| S4_extracted_23528 | Not-determined |
| S4_extracted_23530 | Not-determined |
| S4_extracted_23534 | Not-determined |
| S4_extracted_23548 | Not-determined |
| S4_extracted_23551 | Not-determined |
| S4_extracted_23555 | Not-determined |
| S4_extracted_23576 | Not-determined |
| S4_extracted_23581 | Low-quality |
| S4_extracted_23599 | Not-determined |
| S4_extracted_23607 | Not-determined |
| S4_extracted_23608 | Not-determined |
| S4_extracted_23609 | Not-determined |
| S4_extracted_23615 | Not-determined |
| S4_extracted_23627 | Not-determined |
| S4_extracted_23631 | Not-determined |
| S4_extracted_23655 | Not-determined |
| S4_extracted_23657 | Not-determined |
| S4_extracted_23659 | Not-determined |
| S4_extracted_23661 | Not-determined |
| S4_extracted_23662 | Not-determined |
| S4_extracted_23674 | Low-quality |
| S4_extracted_23677 | Not-determined |
| S4_extracted_23691 | Not-determined |
| S4_extracted_23704 | Not-determined |
| S4_extracted_23705 | Low-quality |
| S4_extracted_23711 | Not-determined |
| S4_extracted_23718 | Low-quality |
| S4_extracted_23723 | Not-determined |
| S4_extracted_23726 | Not-determined |
| S4_extracted_23727 | Not-determined |
| S4_extracted_23730 | Not-determined |
| S4_extracted_23739 | Not-determined |
| S4_extracted_23741 | Low-quality |
| S4_extracted_23744 | Low-quality |
| S4_extracted_23755 | Not-determined |
| S4_extracted_23767 | Not-determined |
| S4_extracted_23774 | Not-determined |
| S4_extracted_23777 | Not-determined |
| S4_extracted_23798 | Not-determined |
| S4_extracted_23803 | Low-quality |
| S4_extracted_23810 | Not-determined |
| S4_extracted_23817 | Not-determined |
| S4_extracted_23819 | Not-determined |
| S4_extracted_23840 | Not-determined |
| S4_extracted_23842 | Not-determined |
| S4_extracted_23846 | Not-determined |
| S4_extracted_23847 | Not-determined |
| S4_extracted_23853 | Low-quality |
| S4_extracted_23869 | Not-determined |
| S4_extracted_23881 | Not-determined |
| S4_extracted_23882 | Low-quality |
| S4_extracted_23910 | Low-quality |
| S4_extracted_23912 | Not-determined |
| S4_extracted_23943 | Low-quality |
| S4_extracted_23946 | Low-quality |
| S4_extracted_23954 | Not-determined |
| S4_extracted_23989 | Low-quality |
| S4_extracted_23999 | Not-determined |
| S4_extracted_24018 | Not-determined |
| S4_extracted_24023 | Not-determined |
| S4_extracted_24030 | Not-determined |
| S4_extracted_24032 | Not-determined |
| S4_extracted_24046 | Not-determined |
| S4_extracted_24047 | Not-determined |
| S4_extracted_24050 | Not-determined |
| S4_extracted_24056 | Not-determined |
| S4_extracted_24058 | Not-determined |
| S4_extracted_24074 | Not-determined |
| S4_extracted_24082 | Not-determined |
| S4_extracted_24087 | Low-quality |
| S4_extracted_24095 | Not-determined |
| S4_extracted_24099 | Not-determined |
| S4_extracted_24109 | Not-determined |
| S4_extracted_24118 | Low-quality |
| S4_extracted_24135 | Not-determined |
| S4_extracted_24138 | Low-quality |
| S4_extracted_24143 | Not-determined |
| S4_extracted_24145 | Not-determined |
| S4_extracted_24147 | Not-determined |
| S4_extracted_24151 | Not-determined |
| S4_extracted_24155 | Not-determined |
| S4_extracted_24170 | Not-determined |
| S4_extracted_24176 | Not-determined |
| S4_extracted_24181 | Not-determined |
| S4_extracted_24195 | Low-quality |
| S4_extracted_24203 | Low-quality |
| S4_extracted_24213 | Not-determined |
| S4_extracted_24223 | Not-determined |
| S4_extracted_24233 | Not-determined |
| S4_extracted_24234 | Not-determined |
| S4_extracted_24258 | Low-quality |
| S4_extracted_24266 | Not-determined |
| S4_extracted_24274 | Not-determined |
| S4_extracted_24279 | Not-determined |
| S4_extracted_24283 | Not-determined |
| S4_extracted_24284 | Not-determined |
| S4_extracted_24286 | Not-determined |
| S4_extracted_24292 | Not-determined |
| S4_extracted_24301 | Not-determined |
| S4_extracted_24302 | Not-determined |
| S4_extracted_24305 | Not-determined |
| S4_extracted_24322 | Not-determined |
| S4_extracted_24329 | Not-determined |
| S4_extracted_24330 | Not-determined |
| S4_extracted_24335 | Not-determined |
| S4_extracted_24337 | Not-determined |
| S4_extracted_24359 | Low-quality |
| S4_extracted_24380 | Not-determined |
| S4_extracted_24391 | Not-determined |
| S4_extracted_24394 | Low-quality |
| S4_extracted_24408 | Not-determined |
| S4_extracted_24412 | Not-determined |
| S4_extracted_24436 | Not-determined |
| S4_extracted_24443 | Not-determined |
| S4_extracted_24447 | Not-determined |
| S4_extracted_24465 | Not-determined |
| S4_extracted_24469 | Not-determined |
| S4_extracted_24527 | Not-determined |
| S4_extracted_24531 | Not-determined |
| S4_extracted_24532 | Low-quality |
| S4_extracted_24546 | Low-quality |
| S4_extracted_24557 | Not-determined |
| S4_extracted_24559 | Low-quality |
| S4_extracted_24573 | Not-determined |
| S4_extracted_24574 | Not-determined |
| S4_extracted_24576 | Not-determined |
| S4_extracted_24580 | Not-determined |
| S4_extracted_24611 | Not-determined |
| S4_extracted_24614 | Not-determined |
| S4_extracted_24617 | Low-quality |
| S4_extracted_24641 | Not-determined |
| S4_extracted_24644 | Not-determined |
| S4_extracted_24648 | Not-determined |
| S4_extracted_24656 | Not-determined |
| S4_extracted_24659 | Not-determined |
| S4_extracted_24660 | Not-determined |
| S4_extracted_24665 | Not-determined |
| S4_extracted_24670 | Not-determined |
| S4_extracted_24680 | Low-quality |
| S4_extracted_24692 | Not-determined |
| S4_extracted_24698 | Not-determined |
| S4_extracted_24720 | Not-determined |
| S4_extracted_24766 | Not-determined |
| S4_extracted_24782 | Not-determined |
| S4_extracted_24820 | Low-quality |
| S4_extracted_24835 | Low-quality |
| S4_extracted_24839 | Not-determined |
| S4_extracted_24845 | Not-determined |
| S4_extracted_24863 | Not-determined |
| S4_extracted_24867 | Not-determined |
| S4_extracted_24870 | Not-determined |
| S4_extracted_24878 | Not-determined |
| S4_extracted_24879 | Not-determined |
| S4_extracted_24889 | Not-determined |
| S4_extracted_24903 | Not-determined |
| S4_extracted_24904 | Not-determined |
| S4_extracted_24905 | Low-quality |
| S4_extracted_24909 | Low-quality |
| S4_extracted_24916 | Not-determined |
| S4_extracted_24924 | Not-determined |
| S4_extracted_24926 | Low-quality |
| S4_extracted_24935 | Not-determined |
| S4_extracted_24945 | Not-determined |
| S4_extracted_24948 | Not-determined |
| S4_extracted_24970 | Low-quality |
| S4_extracted_24971 | Not-determined |
| S4_extracted_24972 | Not-determined |
| S4_extracted_24978 | Not-determined |
| S4_extracted_24979 | Not-determined |
| S4_extracted_24980 | Not-determined |
| S4_extracted_24981 | Not-determined |
| S4_extracted_24986 | Low-quality |
| S4_extracted_24991 | Not-determined |
| S4_extracted_25004 | Not-determined |
| S4_extracted_25009 | Not-determined |
| S4_extracted_25010 | Not-determined |
| S4_extracted_25011 | Not-determined |
| S4_extracted_25034 | Not-determined |
| S4_extracted_25041 | Not-determined |
| S4_extracted_25044 | Not-determined |
| S4_extracted_25049 | Not-determined |
| S4_extracted_25051 | Low-quality |
| S4_extracted_25054 | Not-determined |
| S4_extracted_25058 | Not-determined |
| S4_extracted_25071 | Not-determined |
| S4_extracted_25081 | Low-quality |
| S4_extracted_25092 | Not-determined |
| S4_extracted_25103 | Not-determined |
| S4_extracted_25104 | Not-determined |
| S4_extracted_25123 | Not-determined |
| S4_extracted_25126 | Low-quality |
| S4_extracted_25135 | Not-determined |
| S4_extracted_25148 | Not-determined |
| S4_extracted_25161 | Not-determined |
| S4_extracted_25162 | Low-quality |
| S4_extracted_25165 | Not-determined |
| S4_extracted_25169 | Not-determined |
| S4_extracted_25174 | Low-quality |
| S4_extracted_25184 | Not-determined |
| S4_extracted_25198 | Not-determined |
| S4_extracted_25200 | Not-determined |
| S4_extracted_25206 | Not-determined |
| S4_extracted_25211 | Low-quality |
| S4_extracted_25217 | Not-determined |
| S4_extracted_25221 | Not-determined |
| S4_extracted_25233 | Not-determined |
| S4_extracted_25238 | Not-determined |
| S4_extracted_25239 | Not-determined |
| S4_extracted_25281 | Low-quality |
| S4_extracted_25284 | Not-determined |
| S4_extracted_25290 | Not-determined |
| S4_extracted_25293 | Low-quality |
| S4_extracted_25301 | Low-quality |
| S4_extracted_25304 | Not-determined |
| S4_extracted_25307 | Not-determined |
| S4_extracted_25324 | Not-determined |
| S4_extracted_25328 | Not-determined |
| S4_extracted_25336 | Not-determined |
| S4_extracted_25351 | Not-determined |
| S4_extracted_25375 | Not-determined |
| S4_extracted_25377 | Not-determined |
| S4_extracted_25391 | Low-quality |
| S4_extracted_25392 | Low-quality |
| S4_extracted_25407 | Not-determined |
| S4_extracted_25436 | Low-quality |
| S4_extracted_25446 | Not-determined |
| S4_extracted_25478 | Not-determined |
| S4_extracted_25487 | Not-determined |
| S4_extracted_25491 | Not-determined |
| S4_extracted_25497 | Not-determined |
| S4_extracted_25499 | Not-determined |
| S4_extracted_25504 | Low-quality |
| S4_extracted_25517 | Low-quality |
| S4_extracted_25520 | Not-determined |
| S4_extracted_25533 | Low-quality |
| S4_extracted_25549 | Low-quality |
| S4_extracted_25567 | Low-quality |
| S4_extracted_25575 | Not-determined |
| S4_extracted_25580 | Not-determined |
| S4_extracted_25585 | Low-quality |
| S4_extracted_25589 | Not-determined |
| S4_extracted_25590 | Not-determined |
| S4_extracted_25597 | Not-determined |
| S4_extracted_25609 | Not-determined |
| S4_extracted_25612 | Not-determined |
| S4_extracted_25613 | Not-determined |
| S4_extracted_25626 | Low-quality |
| S4_extracted_25642 | Not-determined |
| S4_extracted_25655 | Not-determined |
| S4_extracted_25680 | Not-determined |
| S4_extracted_25681 | Not-determined |
| S4_extracted_25682 | Low-quality |
| S4_extracted_25701 | Low-quality |
| S4_extracted_25703 | Not-determined |
| S4_extracted_25704 | Not-determined |
| S4_extracted_25707 | Low-quality |
| S4_extracted_25712 | Not-determined |
| S4_extracted_25728 | Low-quality |
| S4_extracted_25733 | Not-determined |
| S4_extracted_25781 | Not-determined |
| S4_extracted_25783 | Low-quality |
| S4_extracted_25791 | Not-determined |
| S4_extracted_25797 | Not-determined |
| S4_extracted_25816 | Low-quality |
| S4_extracted_25818 | Not-determined |
| S4_extracted_25828 | Low-quality |
| S4_extracted_25838 | Not-determined |
| S4_extracted_25846 | Low-quality |
| S4_extracted_25859 | Not-determined |
| S4_extracted_25864 | Not-determined |
| S4_extracted_25876 | Not-determined |
| S4_extracted_25927 | Not-determined |
| S4_extracted_25931 | Not-determined |
| S4_extracted_25942 | Not-determined |
| S4_extracted_25954 | Not-determined |
| S4_extracted_25959 | Not-determined |
| S4_extracted_25965 | Low-quality |
| S4_extracted_25975 | Not-determined |
| S4_extracted_25989 | Not-determined |
| S4_extracted_25997 | Not-determined |
| S4_extracted_26002 | Not-determined |
| S4_extracted_26004 | Low-quality |
| S4_extracted_26012 | Not-determined |
| S4_extracted_26019 | Not-determined |
| S4_extracted_26026 | Not-determined |
| S4_extracted_26027 | Not-determined |
| S4_extracted_26044 | Not-determined |
| S4_extracted_26065 | Not-determined |
| S4_extracted_26092 | Not-determined |
| S4_extracted_26095 | Low-quality |
| S4_extracted_26097 | Not-determined |
| S4_extracted_26102 | Not-determined |
| S4_extracted_26105 | Not-determined |
| S4_extracted_26122 | Low-quality |
| S4_extracted_26124 | Low-quality |
| S4_extracted_26125 | Not-determined |
| S4_extracted_26130 | Not-determined |
| S4_extracted_26148 | Not-determined |
| S4_extracted_26150 | Low-quality |
| S4_extracted_26167 | Low-quality |
| S4_extracted_26171 | Not-determined |
| S4_extracted_26176 | Not-determined |
| S4_extracted_26178 | Low-quality |
| S4_extracted_26180 | Not-determined |
| S4_extracted_26201 | Not-determined |
| S4_extracted_26202 | Not-determined |
| S4_extracted_26208 | Not-determined |
| S4_extracted_26214 | Not-determined |
| S4_extracted_26235 | Not-determined |
| S4_extracted_26243 | Not-determined |
| S4_extracted_26250 | Low-quality |
| S4_extracted_26255 | Low-quality |
| S4_extracted_26259 | Not-determined |
| S4_extracted_26277 | Not-determined |
| S4_extracted_26281 | Not-determined |
| S4_extracted_26286 | Not-determined |
| S4_extracted_26315 | Not-determined |
| S4_extracted_26323 | Not-determined |
| S4_extracted_26350 | Not-determined |
| S4_extracted_26353 | Not-determined |
| S4_extracted_26359 | Low-quality |
| S4_extracted_26363 | Low-quality |
| S3_extracted_k141_77792 | Medium-quality |
| S4_extracted_26420 | Not-determined |
| S4_extracted_26421 | Not-determined |
| S4_extracted_26425 | Not-determined |
| S4_extracted_26438 | Low-quality |
| S4_extracted_26452 | Not-determined |
| S4_extracted_26464 | Not-determined |
| S4_extracted_26466 | Not-determined |
| S4_extracted_26470 | Not-determined |
| S4_extracted_26472 | Not-determined |
| S4_extracted_26481 | Not-determined |
| S4_extracted_26489 | Not-determined |
| S4_extracted_26505 | Low-quality |
| S4_extracted_26513 | Not-determined |
| S4_extracted_26528 | Low-quality |
| S4_extracted_26532 | Not-determined |
| S4_extracted_26533 | Not-determined |
| S4_extracted_26536 | Low-quality |
| S4_extracted_26540 | Low-quality |
| S4_extracted_26566 | Not-determined |
| S4_extracted_26573 | Not-determined |
| S4_extracted_26576 | Not-determined |
| S4_extracted_26583 | Not-determined |
| S4_extracted_26586 | Low-quality |
| S4_extracted_26618 | Not-determined |
| S4_extracted_26626 | Low-quality |
| S4_extracted_26627 | Not-determined |
| S4_extracted_26647 | Low-quality |
| S4_extracted_26656 | Not-determined |
| S4_extracted_26660 | Not-determined |
| S4_extracted_26664 | Not-determined |
| S4_extracted_26670 | Low-quality |
| S4_extracted_26672 | Low-quality |
| S4_extracted_26681 | Not-determined |
| S4_extracted_26686 | Not-determined |
| S4_extracted_26694 | Not-determined |
| S4_extracted_26701 | Not-determined |
| S4_extracted_26703 | Not-determined |
| S4_extracted_26711 | Low-quality |
| S4_extracted_26716 | Not-determined |
| S4_extracted_26717 | Not-determined |
| S4_extracted_26718 | Not-determined |
| S4_extracted_26731 | Not-determined |
| S4_extracted_26732 | Not-determined |
| S4_extracted_26739 | Not-determined |
| S4_extracted_26752 | Low-quality |
| S4_extracted_26777 | Low-quality |
| S4_extracted_26778 | Not-determined |
| S4_extracted_26782 | Not-determined |
| S4_extracted_26791 | Not-determined |
| S4_extracted_26806 | Not-determined |
| S4_extracted_26823 | Not-determined |
| S4_extracted_26834 | Not-determined |
| S4_extracted_26836 | Not-determined |
| S4_extracted_26841 | Not-determined |
| S4_extracted_26843 | Low-quality |
| S4_extracted_26853 | Not-determined |
| S4_extracted_26854 | Not-determined |
| S4_extracted_26861 | Low-quality |
| S4_extracted_26868 | Low-quality |
| S4_extracted_26890 | Not-determined |
| S4_extracted_26904 | Not-determined |
| S4_extracted_26907 | Not-determined |
| S4_extracted_26909 | Low-quality |
| S4_extracted_26918 | Not-determined |
| S4_extracted_26933 | Not-determined |
| S4_extracted_26938 | Low-quality |
| S4_extracted_26941 | Not-determined |
| S4_extracted_26954 | Low-quality |
| S4_extracted_26969 | Not-determined |
| S4_extracted_26971 | Low-quality |
| S4_extracted_26977 | Not-determined |
| S4_extracted_26978 | Not-determined |
| S4_extracted_26981 | Not-determined |
| S4_extracted_26984 | Not-determined |
| S4_extracted_26995 | Not-determined |
| S4_extracted_27009 | Not-determined |
| S4_extracted_27016 | Not-determined |
| S4_extracted_27023 | Low-quality |
| S4_extracted_27025 | Not-determined |
| S4_extracted_27071 | Not-determined |
| S4_extracted_27078 | Not-determined |
| S4_extracted_27079 | Not-determined |
| S4_extracted_27091 | Not-determined |
| S4_extracted_27097 | Not-determined |
| S4_extracted_27105 | Low-quality |
| S4_extracted_27111 | Not-determined |
| S4_extracted_27144 | Low-quality |
| S4_extracted_27152 | Low-quality |
| S4_extracted_27166 | Not-determined |
| S4_extracted_27168 | Not-determined |
| S4_extracted_27183 | Low-quality |
| S4_extracted_27186 | Not-determined |
| S4_extracted_27193 | Not-determined |
| S4_extracted_27194 | Not-determined |
| S4_extracted_27209 | Not-determined |
| S4_extracted_27226 | Not-determined |
| S4_extracted_27230 | Not-determined |
| S4_extracted_27238 | Not-determined |
| S4_extracted_27248 | Low-quality |
| S4_extracted_27251 | Not-determined |
| S4_extracted_27259 | Not-determined |
| S4_extracted_27263 | Not-determined |
| S4_extracted_27265 | Not-determined |
| S4_extracted_27275 | Not-determined |
| S4_extracted_27282 | Not-determined |
| S4_extracted_27283 | Low-quality |
| S4_extracted_27284 | Low-quality |
| S4_extracted_27298 | Not-determined |
| S4_extracted_27308 | Not-determined |
| S4_extracted_27314 | Low-quality |
| S4_extracted_27341 | Not-determined |
| S4_extracted_27354 | Not-determined |
| S4_extracted_27355 | Not-determined |
| S4_extracted_27364 | Not-determined |
| S4_extracted_27367 | Not-determined |
| S4_extracted_27381 | Not-determined |
| S4_extracted_27390 | Not-determined |
| S4_extracted_27394 | Not-determined |
| S4_extracted_27396 | Not-determined |
| S4_extracted_27413 | Not-determined |
| S4_extracted_27421 | Not-determined |
| S4_extracted_27427 | Not-determined |
| S4_extracted_27444 | Low-quality |
| S4_extracted_27448 | Not-determined |
| S4_extracted_27454 | Not-determined |
| S4_extracted_27459 | Not-determined |
| S4_extracted_27460 | Not-determined |
| S4_extracted_27473 | Not-determined |
| S4_extracted_27476 | Not-determined |
| S4_extracted_27490 | Not-determined |
| S4_extracted_27496 | Not-determined |
| S4_extracted_27504 | Low-quality |
| S4_extracted_27510 | Not-determined |
| S4_extracted_27522 | Not-determined |
| S4_extracted_27529 | Not-determined |
| S4_extracted_27532 | Not-determined |
| S4_extracted_27538 | Not-determined |
| S4_extracted_27540 | Low-quality |
| S4_extracted_27550 | Not-determined |
| S4_extracted_27579 | Not-determined |
| S4_extracted_27595 | Not-determined |
| S4_extracted_27624 | Low-quality |
| S4_extracted_27625 | Low-quality |
| S4_extracted_27628 | Not-determined |
| S4_extracted_27630 | Not-determined |
| S4_extracted_27635 | Not-determined |
| S4_extracted_27656 | Not-determined |
| S4_extracted_27672 | Not-determined |
| S4_extracted_27686 | Not-determined |
| S4_extracted_27692 | Not-determined |
| S4_extracted_27701 | Not-determined |
| S4_extracted_27702 | Not-determined |
| S4_extracted_27711 | Not-determined |
| S4_extracted_27723 | Not-determined |
| S4_extracted_27727 | Not-determined |
| S4_extracted_27729 | Not-determined |
| S4_extracted_27740 | Not-determined |
| S4_extracted_27748 | Not-determined |
| S4_extracted_27753 | Not-determined |
| S4_extracted_27766 | Not-determined |
| S4_extracted_27799 | Not-determined |
| S4_extracted_27806 | Low-quality |
| S4_extracted_27813 | Not-determined |
| S4_extracted_27821 | Not-determined |
| S4_extracted_27832 | Not-determined |
| S7_vs_100640\|\|full | Medium-quality |
| S4_extracted_27892 | Low-quality |
| S4_extracted_27909 | Not-determined |
| S4_extracted_27919 | Not-determined |
| S4_extracted_27926 | Not-determined |
| S4_extracted_27947 | Not-determined |
| S4_extracted_27949 | Not-determined |
| S4_extracted_27956 | Not-determined |
| S4_extracted_27966 | Low-quality |
| S4_extracted_27978 | Not-determined |
| S4_extracted_27979 | Not-determined |
| S4_extracted_27980 | Not-determined |
| S4_extracted_27984 | Low-quality |
| S4_extracted_27990 | Not-determined |
| S4_extracted_28007 | Low-quality |
| S4_extracted_28008 | Not-determined |
| S4_extracted_28023 | Not-determined |
| S4_extracted_28030 | Not-determined |
| S4_extracted_28033 | Not-determined |
| S4_extracted_28040 | Not-determined |
| S4_extracted_28050 | Not-determined |
| S4_extracted_28055 | Not-determined |
| S4_extracted_28056 | Not-determined |
| S4_extracted_28061 | Not-determined |
| S4_extracted_28063 | Not-determined |
| S4_extracted_28072 | Not-determined |
| S4_extracted_28083 | Not-determined |
| S4_extracted_28087 | Not-determined |
| S4_extracted_28091 | Not-determined |
| S4_extracted_28108 | Low-quality |
| S4_extracted_28111 | Not-determined |
| S4_extracted_28112 | Not-determined |
| S4_extracted_28121 | Not-determined |
| S4_extracted_28126 | Not-determined |
| S4_extracted_28148 | Not-determined |
| S4_extracted_28154 | Not-determined |
| S4_extracted_28160 | Not-determined |
| S4_extracted_28162 | Not-determined |
| S4_extracted_28168 | Low-quality |
| S4_extracted_28181 | Not-determined |
| S4_extracted_28182 | Not-determined |
| S4_extracted_28187 | Not-determined |
| S4_extracted_28193 | Not-determined |
| S4_extracted_28195 | Not-determined |
| S4_extracted_28199 | Not-determined |
| S4_extracted_28227 | Not-determined |
| S4_extracted_28240 | Not-determined |
| S4_extracted_28255 | Low-quality |
| S4_extracted_28268 | Not-determined |

| | |
|---|---|
| S4_extracted_28276 | Not-determined |
| S4_extracted_28282 | Not-determined |
| S4_extracted_28287 | Not-determined |
| S4_extracted_28301 | Not-determined |
| S4_extracted_28313 | Not-determined |
| S4_extracted_28323 | Low-quality |
| S4_extracted_28346 | Not-determined |
| S4_extracted_28347 | Not-determined |
| S4_extracted_28349 | Not-determined |
| S4_extracted_28353 | Not-determined |
| S4_extracted_28357 | Not-determined |
| S4_extracted_28359 | Not-determined |
| S4_extracted_28373 | Not-determined |
| S4_extracted_28403 | Not-determined |
| S4_extracted_28418 | Not-determined |
| S4_extracted_28429 | Not-determined |
| S4_extracted_28434 | Not-determined |
| S4_extracted_28442 | Not-determined |
| S4_extracted_28444 | Not-determined |
| S4_extracted_28461 | Not-determined |
| S4_extracted_28464 | Not-determined |
| S4_extracted_28478 | Not-determined |
| S4_extracted_28487 | Not-determined |
| S4_extracted_28504 | Not-determined |
| S4_extracted_28507 | Not-determined |
| S4_extracted_28511 | Low-quality |
| S4_extracted_28514 | Not-determined |
| S4_extracted_28530 | Not-determined |
| S4_extracted_28542 | Not-determined |
| S4_extracted_28558 | Not-determined |
| S4_extracted_28574 | Not-determined |
| S4_extracted_28576 | Low-quality |
| S4_extracted_28584 | Not-determined |
| S4_extracted_28590 | Not-determined |
| S4_extracted_28595 | Not-determined |
| S7_extracted_29941 | Medium-quality |
| S4_extracted_28611 | Not-determined |
| S4_extracted_28614 | Not-determined |
| S4_extracted_28633 | Not-determined |
| S4_extracted_28643 | Not-determined |
| S4_extracted_28647 | Not-determined |
| S4_extracted_28664 | Not-determined |
| S4_extracted_28670 | Not-determined |
| S4_extracted_28673 | Low-quality |
| S4_extracted_28678 | Low-quality |
| S4_extracted_28679 | Not-determined |
| S4_extracted_28688 | Not-determined |
| S4_extracted_28690 | Not-determined |
| S4_extracted_28699 | Not-determined |
| S4_extracted_28703 | Low-quality |
| S4_extracted_28709 | Not-determined |
| S4_extracted_28710 | Not-determined |
| S4_extracted_28713 | Low-quality |
| S4_extracted_28722 | Not-determined |
| S4_extracted_28724 | Not-determined |
| S4_extracted_28728 | Not-determined |
| S4_extracted_28737 | Not-determined |
| S4_extracted_28743 | Low-quality |
| S4_extracted_28750 | Low-quality |
| S4_extracted_28751 | Not-determined |
| S4_extracted_28756 | Not-determined |
| S4_extracted_28762 | Low-quality |
| S4_extracted_28770 | Low-quality |
| S4_extracted_28779 | Low-quality |
| S4_extracted_28780 | Not-determined |
| S4_extracted_28782 | Not-determined |
| S4_extracted_28783 | Not-determined |
| S4_extracted_28784 | Not-determined |
| S4_extracted_28822 | Not-determined |
| S4_extracted_28825 | Not-determined |
| S4_extracted_28835 | Not-determined |
| S4_extracted_28836 | Low-quality |
| S4_extracted_28839 | Not-determined |
| S4_extracted_28846 | Not-determined |
| S4_extracted_28857 | Not-determined |
| S4_extracted_28859 | Low-quality |
| S4_extracted_28862 | Low-quality |
| S4_extracted_28864 | Low-quality |
| S4_extracted_28877 | Not-determined |
| S4_extracted_28882 | Not-determined |
| S4_extracted_28886 | Low-quality |
| S4_extracted_28900 | Not-determined |
| S4_extracted_28903 | Not-determined |
| S4_extracted_28930 | Not-determined |
| S4_extracted_28938 | Not-determined |
| S4_extracted_28940 | Low-quality |
| S4_extracted_28965 | Not-determined |
| S4_extracted_28975 | Not-determined |
| S4_extracted_28977 | Not-determined |
| S4_extracted_28983 | Not-determined |
| S4_extracted_28984 | Low-quality |
| S4_extracted_29004 | Not-determined |
| S4_extracted_29008 | Not-determined |
| S4_extracted_29010 | Low-quality |
| S4_extracted_29021 | Not-determined |
| S4_extracted_29022 | Not-determined |
| S4_extracted_29030 | Not-determined |
| S4_extracted_29035 | Not-determined |
| S4_extracted_29038 | Low-quality |
| S4_extracted_29041 | Not-determined |
| S4_extracted_29049 | Low-quality |
| S4_extracted_29061 | Not-determined |
| S4_extracted_29066 | Not-determined |
| S4_extracted_29071 | Not-determined |
| S4_extracted_29075 | Low-quality |
| S4_extracted_29089 | Not-determined |
| S4_extracted_29093 | Not-determined |
| S4_extracted_29098 | Low-quality |
| S4_extracted_29115 | Not-determined |
| S4_extracted_29128 | Low-quality |
| S4_extracted_29129 | Low-quality |
| S4_extracted_29133 | Not-determined |
| S4_extracted_29135 | Not-determined |
| S4_extracted_29136 | Not-determined |
| S4_extracted_29156 | Not-determined |
| S4_extracted_29171 | Not-determined |
| S4_extracted_29175 | Not-determined |
| S4_extracted_29183 | Low-quality |
| S4_extracted_29209 | Low-quality |
| S4_extracted_29220 | Not-determined |
| S4_extracted_29221 | Not-determined |
| S4_extracted_29231 | Low-quality |
| S4_extracted_29232 | Not-determined |
| S4_extracted_29251 | Not-determined |
| S4_extracted_29256 | Not-determined |
| S4_extracted_29257 | Low-quality |
| S4_extracted_29261 | Not-determined |
| S4_extracted_29264 | Not-determined |
| S4_extracted_29269 | Not-determined |
| S4_extracted_29275 | Not-determined |
| S4_extracted_29284 | Not-determined |
| S4_extracted_29286 | Low-quality |
| S4_extracted_29291 | Not-determined |
| S4_extracted_29295 | Not-determined |
| S4_extracted_29298 | Not-determined |
| S4_extracted_29300 | Not-determined |
| S4_extracted_29314 | Low-quality |
| S4_extracted_29326 | Not-determined |
| S4_extracted_29331 | Not-determined |
| S4_extracted_29367 | Low-quality |
| S4_extracted_29380 | Not-determined |
| S4_extracted_29385 | Not-determined |
| S4_extracted_29392 | Not-determined |
| S4_extracted_29393 | Not-determined |
| S4_extracted_29404 | Not-determined |
| S4_extracted_29412 | Not-determined |
| S4_extracted_29415 | Not-determined |
| S4_extracted_29424 | Not-determined |
| S4_extracted_29425 | Not-determined |
| S4_extracted_29443 | Low-quality |
| S4_extracted_29452 | Not-determined |
| S4_extracted_29466 | Low-quality |
| S4_extracted_29471 | Not-determined |
| S4_extracted_29475 | Not-determined |
| S4_extracted_29480 | Not-determined |
| S4_extracted_29491 | Low-quality |
| S4_extracted_29498 | Low-quality |
| S4_extracted_29499 | Not-determined |
| S4_extracted_29501 | Not-determined |
| S4_extracted_29507 | Not-determined |
| S4_extracted_29518 | Not-determined |
| S4_extracted_29523 | Low-quality |
| S4_extracted_29533 | Not-determined |
| S4_extracted_29536 | Not-determined |
| S4_extracted_29537 | Not-determined |
| S4_extracted_29538 | Not-determined |
| S4_extracted_29576 | Not-determined |
| S4_extracted_29581 | Not-determined |
| S4_extracted_29592 | Not-determined |
| S4_extracted_29599 | Not-determined |
| S4_extracted_29605 | Not-determined |
| S4_extracted_29645 | Not-determined |
| S4_extracted_29650 | Not-determined |
| S4_extracted_29661 | Low-quality |
| S4_extracted_29662 | Not-determined |
| S4_extracted_29667 | Low-quality |
| S4_extracted_29673 | Low-quality |
| S4_extracted_29677 | Not-determined |
| S4_extracted_29678 | Low-quality |
| S4_extracted_29687 | Not-determined |
| S4_extracted_29695 | Low-quality |
| S4_extracted_29702 | Not-determined |
| S4_extracted_29703 | Not-determined |
| S4_extracted_29704 | Low-quality |
| S4_extracted_29718 | Not-determined |
| S4_extracted_29719 | Not-determined |
| S4_extracted_29721 | Not-determined |
| S4_extracted_29751 | Not-determined |
| S4_extracted_29755 | Not-determined |
| S4_extracted_29767 | Not-determined |
| S4_extracted_29770 | Not-determined |
| S4_extracted_29776 | Not-determined |
| S4_extracted_29781 | Not-determined |
| S4_extracted_29789 | Not-determined |
| S4_extracted_29799 | Not-determined |
| S4_extracted_29810 | Low-quality |
| S4_extracted_29829 | Not-determined |
| S4_extracted_29834 | Not-determined |
| S4_extracted_29836 | Not-determined |
| S4_extracted_29848 | Low-quality |
| S4_extracted_29852 | Not-determined |
| S4_extracted_29855 | Not-determined |
| S4_extracted_29862 | Not-determined |
| S4_extracted_29870 | Not-determined |
| S4_extracted_29880 | Not-determined |
| S4_extracted_29894 | Not-determined |
| S4_extracted_29900 | Low-quality |
| S4_extracted_29911 | Low-quality |
| S4_extracted_29912 | Not-determined |
| S4_extracted_29914 | Not-determined |
| S4_extracted_29930 | Low-quality |
| S4_extracted_29931 | Not-determined |
| S4_extracted_29932 | Not-determined |
| S4_extracted_29936 | Not-determined |
| S4_extracted_29945 | Not-determined |
| S4_extracted_29947 | Not-determined |
| S4_extracted_29951 | Not-determined |
| S4_extracted_29960 | Not-determined |
| S4_extracted_29964 | Not-determined |
| S4_extracted_29965 | Not-determined |
| S4_extracted_29971 | Not-determined |
| S4_extracted_29972 | Not-determined |
| S4_extracted_29974 | Not-determined |
| S4_extracted_29985 | Not-determined |
| S4_extracted_29987 | Not-determined |
| S4_extracted_30001 | Not-determined |
| S4_extracted_30022 | Not-determined |
| S4_extracted_30041 | Not-determined |
| S4_extracted_30047 | Not-determined |
| S4_extracted_30070 | Not-determined |
| S4_extracted_30074 | Not-determined |
| S7_vs_29941\|\|full | Medium-quality |
| S4_extracted_30105 | Not-determined |
| S4_extracted_30107 | Not-determined |
| S4_extracted_30110 | Not-determined |
| S4_extracted_30116 | Not-determined |
| S4_extracted_30126 | Low-quality |
| S4_extracted_30139 | Not-determined |
| S4_extracted_30153 | Not-determined |
| S4_extracted_30169 | Not-determined |
| S4_extracted_30177 | Not-determined |
| S4_extracted_30197 | Not-determined |
| S4_extracted_30202 | Not-determined |
| S4_extracted_30215 | Not-determined |
| S4_extracted_30228 | Low-quality |
| S4_extracted_30231 | Not-determined |
| S4_extracted_30236 | Low-quality |
| S4_extracted_30238 | Not-determined |
| S4_extracted_30241 | Low-quality |
| S4_extracted_30248 | Not-determined |
| S4_extracted_30259 | Not-determined |
| S4_extracted_30280 | Not-determined |
| S4_extracted_30283 | Low-quality |
| S4_extracted_30286 | Not-determined |
| S4_extracted_30288 | Not-determined |
| S4_extracted_30295 | Not-determined |
| S4_extracted_30300 | Not-determined |
| S4_extracted_30303 | Not-determined |
| S4_extracted_30312 | Not-determined |
| S4_extracted_30315 | Not-determined |
| S4_extracted_30326 | Not-determined |
| S4_extracted_30336 | Not-determined |
| S4_extracted_30354 | Low-quality |
| S4_extracted_30381 | Not-determined |
| S4_extracted_30382 | Not-determined |
| S4_extracted_30385 | Not-determined |
| S4_extracted_30401 | Low-quality |
| S4_extracted_30418 | Not-determined |
| S4_extracted_30426 | Not-determined |
| S4_extracted_30428 | Not-determined |
| S4_extracted_30434 | Low-quality |
| S4_extracted_30449 | Not-determined |
| S4_extracted_30455 | Low-quality |
| S4_extracted_30489 | Not-determined |
| S4_extracted_30507 | Not-determined |
| S4_extracted_30512 | Low-quality |
| S4_extracted_30517 | Not-determined |
| S4_extracted_30523 | Not-determined |
| S4_extracted_30524 | Not-determined |
| S4_extracted_30528 | Not-determined |
| S4_extracted_30551 | Not-determined |
| S4_extracted_30556 | Not-determined |
| S4_extracted_30558 | Not-determined |
| S4_extracted_30560 | Low-quality |
| S4_extracted_30563 | Low-quality |
| S4_extracted_30564 | Not-determined |
| S4_extracted_30570 | Not-determined |
| S4_extracted_30572 | Not-determined |
| S4_extracted_30579 | Not-determined |
| S4_extracted_30583 | Not-determined |
| S4_extracted_30593 | Not-determined |
| S4_extracted_30618 | Not-determined |
| S4_extracted_30619 | Not-determined |
| S4_extracted_30620 | Low-quality |
| S4_extracted_30622 | Not-determined |
| S4_extracted_30624 | Not-determined |
| S4_extracted_30640 | Not-determined |
| S4_extracted_30654 | Low-quality |
| S4_extracted_30658 | Not-determined |
| S4_extracted_30700 | Not-determined |
| S4_extracted_30710 | Not-determined |
| S4_extracted_30734 | Not-determined |
| S4_extracted_30738 | Not-determined |
| S4_extracted_30748 | Not-determined |
| S4_extracted_30750 | Not-determined |
| S4_extracted_30751 | Not-determined |
| S4_extracted_30757 | Low-quality |
| S4_extracted_30765 | Not-determined |
| S4_extracted_30768 | Not-determined |
| S4_extracted_30773 | Not-determined |
| S4_extracted_30776 | Not-determined |
| S4_extracted_30791 | Low-quality |
| S4_extracted_30808 | Not-determined |
| S4_extracted_30815 | Not-determined |
| S4_extracted_30824 | Not-determined |
| S4_extracted_30826 | Low-quality |
| S4_extracted_30829 | Not-determined |
| S4_extracted_30832 | Not-determined |
| S4_extracted_30834 | Not-determined |
| S4_extracted_30838 | Not-determined |
| S4_extracted_30840 | Not-determined |
| S4_extracted_30847 | Low-quality |
| S4_extracted_30851 | Not-determined |
| S4_extracted_30862 | Low-quality |
| S4_extracted_30878 | Low-quality |
| S4_extracted_30890 | Low-quality |
| S4_extracted_30892 | Not-determined |
| S4_extracted_30894 | Not-determined |
| S4_extracted_30898 | Not-determined |
| S4_extracted_30904 | Low-quality |
| S4_extracted_30931 | Not-determined |
| S4_extracted_30939 | Not-determined |
| S4_extracted_30942 | Low-quality |
| S4_extracted_30947 | Not-determined |
| S4_extracted_30960 | Not-determined |
| S4_extracted_30967 | Not-determined |
| S4_extracted_30974 | Not-determined |
| S4_extracted_30976 | Not-determined |
| S4_extracted_31000 | Not-determined |
| S4_extracted_31011 | Not-determined |
| S4_extracted_31012 | Not-determined |
| S4_extracted_31025 | Not-determined |
| S4_extracted_31043 | Not-determined |
| S4_extracted_31059 | Not-determined |
| S4_extracted_31063 | Low-quality |
| S4_extracted_31069 | Not-determined |
| S4_extracted_31076 | Not-determined |
| S4_extracted_31080 | Not-determined |
| S4_extracted_31081 | Not-determined |
| S4_extracted_31085 | Low-quality |
| S4_extracted_31090 | Not-determined |
| S4_extracted_31092 | Low-quality |
| S4_extracted_31093 | Not-determined |
| S4_extracted_31113 | Not-determined |
| S4_extracted_31135 | Not-determined |
| S4_extracted_31140 | Low-quality |
| S4_extracted_31149 | Not-determined |
| S4_extracted_31155 | Low-quality |
| S4_extracted_31212 | Not-determined |
| S4_extracted_31214 | Not-determined |
| S4_extracted_31217 | Not-determined |
| S4_extracted_31222 | Not-determined |
| S4_extracted_31239 | Not-determined |
| S4_extracted_31244 | Not-determined |
| S4_extracted_31268 | Not-determined |
| S4_extracted_31274 | Not-determined |
| S4_extracted_31276 | Not-determined |
| S4_extracted_31286 | Not-determined |
| S4_extracted_31309 | Not-determined |
| S4_extracted_31328 | Not-determined |
| S4_extracted_31348 | Not-determined |
| S4_extracted_31349 | Not-determined |
| S4_extracted_31354 | Not-determined |
| S4_extracted_31359 | Not-determined |
| S4_extracted_31363 | Low-quality |
| S4_extracted_31376 | Not-determined |
| S4_extracted_31377 | Not-determined |
| S4_extracted_31378 | Not-determined |
| S4_extracted_31388 | Not-determined |
| S4_extracted_31403 | Not-determined |
| S4_extracted_31405 | Not-determined |
| S4_extracted_31420 | Low-quality |
| S4_extracted_31423 | Low-quality |
| S4_extracted_31434 | Low-quality |
| S4_extracted_31436 | Not-determined |
| S4_extracted_31446 | Not-determined |
| S4_extracted_31450 | Not-determined |
| S4_extracted_31457 | Not-determined |
| S4_extracted_31477 | Not-determined |
| S4_extracted_31498 | Low-quality |
| S4_extracted_31503 | Not-determined |
| S4_extracted_31518 | Low-quality |
| S4_extracted_31525 | Low-quality |
| S4_extracted_31535 | Not-determined |
| S4_extracted_31544 | Not-determined |
| S4_extracted_31546 | Not-determined |
| S4_extracted_31566 | Not-determined |
| S4_extracted_31568 | Not-determined |
| S4_extracted_31598 | Not-determined |
| S4_extracted_31601 | Not-determined |
| S4_extracted_31611 | Not-determined |
| S4_extracted_31615 | Not-determined |
| S4_extracted_31616 | Not-determined |
| S4_extracted_31625 | Not-determined |
| S4_extracted_31636 | Not-determined |
| S4_extracted_31652 | Not-determined |
| S4_extracted_31653 | Not-determined |
| S4_extracted_31669 | Not-determined |
| S5_extracted_k141_20071 | Medium-quality |
| S4_extracted_31689 | Not-determined |
| S4_extracted_31694 | Not-determined |
| S4_extracted_31699 | Not-determined |
| S4_extracted_31714 | Not-determined |
| S4_extracted_31717 | Not-determined |
| S4_extracted_31722 | Low-quality |
| S4_extracted_31725 | Not-determined |
| S4_extracted_31738 | Low-quality |
| S4_extracted_31742 | Not-determined |
| S4_extracted_31745 | Not-determined |
| S4_extracted_31760 | Low-quality |
| S4_extracted_31769 | Low-quality |
| S4_extracted_31780 | Not-determined |
| S4_extracted_31785 | Low-quality |
| S4_extracted_31800 | Not-determined |
| S4_extracted_31819 | Not-determined |
| S4_extracted_31824 | Not-determined |
| S5_vs_k141_20071\|\|full | Medium-quality |
| S4_extracted_31829 | Not-determined |
| S4_extracted_31838 | Not-determined |
| S4_extracted_31841 | Not-determined |
| S4_extracted_31849 | Low-quality |
| S4_extracted_31855 | Not-determined |
| S4_extracted_31858 | Not-determined |
| S4_extracted_31885 | Not-determined |
| S4_extracted_31892 | Not-determined |
| S4_extracted_31896 | Not-determined |
| S4_extracted_31900 | Not-determined |
| S4_extracted_31903 | Low-quality |
| S4_extracted_31905 | Not-determined |
| S4_extracted_31908 | Not-determined |
| S4_extracted_31917 | Not-determined |
| S4_extracted_31929 | Not-determined |
| S4_extracted_31933 | Not-determined |
| S4_extracted_31941 | Not-determined |
| S4_extracted_31946 | Not-determined |
| S4_extracted_31948 | Not-determined |
| S4_extracted_31964 | Low-quality |
| S4_extracted_31965 | Not-determined |
| S4_extracted_31967 | Not-determined |
| S4_extracted_31971 | Not-determined |
| S3_extracted_k141_23042 | Medium-quality |
| S3_vs_k141_77792\|\|full | Medium-quality |
| S3_vs_k141_12985\|\|full | Medium-quality |
| S14_vs_k141_69422\|\|full | Medium-quality |
| S5_vs_k141_3183\|\|full | Medium-quality |
| S4_extracted_31984 | Not-determined |
| S4_extracted_31985 | Low-quality |
| S4_extracted_32008 | Not-determined |
| S4_extracted_32012 | Not-determined |
| S4_extracted_32014 | Not-determined |
| S4_extracted_32016 | Not-determined |
| S4_extracted_32023 | Not-determined |
| S4_extracted_32030 | Not-determined |
| S4_extracted_32033 | Not-determined |
| S4_extracted_32034 | Not-determined |
| S4_extracted_32043 | Not-determined |
| S4_extracted_32048 | Not-determined |
| S4_extracted_32057 | Not-determined |
| S5_vs_k141_141621\|\|full | Medium-quality |
| S3_vs_k141_23042\|\|full | Medium-quality |
| S11_vs_k141_11267\|\|full | Medium-quality |
| S6_vs_k141_44070\|\|full | Medium-quality |
| S8_vs_k141_143608\|\|full | Medium-quality |
| S5_vs_k141_122633\|\|full | Medium-quality |
| S14_vs_k141_50020\|\|full | Medium-quality |
| S4_vs_VIRSorter_23242-cat_1 | Low-quality |
| S4_vs_VIRSorter_23805-cat_1 | Low-quality |
| S4_vs_VIRSorter_16830-cat_1 | Low-quality |
| S4_vs_VIRSorter_27624-cat_1 | Low-quality |
| S4_vs_VIRSorter_18784-cat_1 | Low-quality |
| S4_vs_VIRSorter_20324-cat_1 | Low-quality |
| S4_vs_VIRSorter_2932-cat_1 | Low-quality |
| S4_vs_VIRSorter_3461-cat_1 | Low-quality |
| S4_vs_VIRSorter_23346-cat_1 | Low-quality |
| S4_vs_VIRSorter_31146-cat_1 | Low-quality |
| S4_vs_VIRSorter_4102-cat_1 | Low-quality |
| S4_vs_VIRSorter_10786-cat_1 | Low-quality |
| S4_vs_VIRSorter_16102-cat_1 | Low-quality |
| S4_vs_VIRSorter_5881-cat_1 | Low-quality |
| S4_vs_VIRSorter_17164-cat_1 | Low-quality |
| S4_vs_VIRSorter_3002-cat_1 | Low-quality |
| S4_vs_VIRSorter_10386-cat_1 | Low-quality |
| S4_vs_VIRSorter_31970-cat_1 | Low-quality |
| S4_vs_VIRSorter_21808-cat_1 | Low-quality |
| S4_vs_VIRSorter_21559-cat_1 | Low-quality |
| S5_extracted_k141_103206 | High-quality |
| S3_vs_k141_52937\|\|full | Medium-quality |
| S7_vs_131314\|\|full | Medium-quality |
| S8_vs_k141_63277\|\|full | Medium-quality |
| S1_vs_k141_91270\|\|full | Medium-quality |
| S5_vs_k141_137154\|\|full | Medium-quality |
| S12_extracted_k141_173208 | Medium-quality |
| S12_vs_k141_173208\|\|full | Medium-quality |
| S16_extracted_100235 | Medium-quality |
| S12_extracted_k141_84625 | Medium-quality |
| S12_vs_k141_84625\|\|full | Medium-quality |
| S7_vs_70265\|\|full | Medium-quality |
| S16_vs_146813\|\|full | Medium-quality |
| S16_vs_54616\|\|full | Medium-quality |
| S3_extracted_k141_33389 | High-quality |
| S17_vs_k141_771\|\|full | Medium-quality |
| S11_extracted_k141_3745 | Medium-quality |
| S5_vs_k141_103206\|\|full | High-quality |
| S3_vs_k141_33389\|\|full | High-quality |
| S16_vs_100235\|\|full | Medium-quality |
| S11_vs_k141_3745\|\|full | Medium-quality |
| S8_extracted_k141_203044 | Medium-quality |
| S16_extracted_169895 | Medium-quality |
| S17_extracted_k141_28210 | Medium-quality |
| S17_vs_k141_8690\|\|0_partial | High-quality |
| S16_extracted_33218 | Medium-quality |
| S16_vs_33218\|\|full | Medium-quality |
| S2_extracted_k141_16829 | Medium-quality |
| S5_extracted_k141_123990 | Medium-quality |
| S6_extracted_k141_2128 | Medium-quality |
| S5_vs_k141_123990\|\|full | Medium-quality |
| S6_vs_k141_2128\|\|full | Medium-quality |
| S2_extracted_k141_4910 | Medium-quality |
| S17_vs_k141_28210\|\|full | Medium-quality |
| S2_vs_k141_16829\|\|full | Medium-quality |
| S8_extracted_k141_52050 | Medium-quality |
| S13_extracted_107349 | Medium-quality |
| S4_vs_VIRSorter_23724-cat_2 | Low-quality |
| S7_extracted_76382 | Medium-quality |
| S7_vs_38113\|\|full | Medium-quality |
| S16_extracted_49628 | Medium-quality |
| S17_extracted_k141_67812 | Medium-quality |
| S16_extracted_164913 | High-quality |
| S17_extracted_k141_111931 | Medium-quality |
| S16_vs_169895\|\|full | Medium-quality |
| S7_extracted_89197 | Medium-quality |
| S7_vs_89197\|\|full | Medium-quality |
| S6_extracted_k141_24553 | Medium-quality |
| S16_vs_49628\|\|full | Medium-quality |
| S4_vs_VIRSorter_9564-cat_2 | Low-quality |
| S4_vs_VIRSorter_24856-cat_2 | Low-quality |
| S4_vs_VIRSorter_10448-cat_2 | Low-quality |
| S4_vs_VIRSorter_4828-cat_2 | Low-quality |
| S4_vs_VIRSorter_8597-cat_2 | Low-quality |
| S4_vs_VIRSorter_26777-cat_2 | Low-quality |
| S4_vs_VIRSorter_12852-cat_2 | Low-quality |
| S17_vs_k141_67812\|\|full | Medium-quality |
| S7_extracted_89668 | Medium-quality |
| S14_extracted_k141_62888 | Medium-quality |
| S1_vs_k141_62409\|\|full | Medium-quality |
| S4_vs_VIRSorter_1863-cat_2 | Low-quality |
| S4_vs_VIRSorter_4677-cat_2 | Low-quality |
| S8_vs_k141_203044\|\|full | Medium-quality |
| S16_extracted_94866 | Medium-quality |
| S4_vs_VIRSorter_1062-cat_2 | Low-quality |
| S4_vs_VIRSorter_7479-cat_2 | Low-quality |
| S4_vs_VIRSorter_29443-cat_2 | Low-quality |
| S4_vs_VIRSorter_27216-cat_2 | Low-quality |
| S4_vs_VIRSorter_29007-cat_2 | Low-quality |
| S4_vs_VIRSorter_1816-cat_2 | Low-quality |
| S4_vs_VIRSorter_18555-cat_2 | Low-quality |
| S4_vs_VIRSorter_20401-cat_2 | Low-quality |
| S6_vs_k141_24553\|\|full | Medium-quality |
| S4_vs_VIRSorter_14679-cat_2 | Low-quality |
| S4_vs_VIRSorter_30848-cat_2 | Low-quality |
| S4_vs_VIRSorter_24513-cat_2 | Low-quality |
| S4_vs_VIRSorter_8400-cat_2 | Low-quality |
| S4_vs_VIRSorter_15245-cat_2 | Low-quality |
| S4_vs_VIRSorter_3742-cat_2 | Low-quality |
| S4_vs_VIRSorter_25549-cat_2 | Low-quality |
| S4_vs_VIRSorter_4323-cat_2 | Low-quality |
| S4_vs_VIRSorter_26037-cat_2 | Low-quality |
| S4_vs_VIRSorter_9618-cat_2 | Low-quality |
| S4_vs_VIRSorter_20624-cat_2 | Low-quality |
| S4_vs_VIRSorter_7896-cat_2 | Low-quality |
| S4_vs_VIRSorter_7325-cat_2 | Low-quality |
| S4_vs_VIRSorter_8544-cat_2 | Low-quality |
| S4_vs_VIRSorter_2064-cat_2 | Low-quality |
| S4_vs_VIRSorter_9974-cat_2 | Low-quality |
| S4_vs_VIRSorter_17822-cat_2 | Low-quality |
| S4_vs_VIRSorter_12183-cat_2 | Low-quality |
| S4_vs_VIRSorter_4142-cat_2 | Low-quality |
| S4_vs_VIRSorter_13524-cat_2 | Low-quality |
| S4_vs_VIRSorter_24249-cat_2 | Low-quality |

| | |
|---|---|
| S4_vs_VIRSorter_759-cat_2 | Low-quality |
| S4_vs_VIRSorter_29122-cat_2 | Low-quality |
| S4_vs_VIRSorter_10093-cat_2 | Low-quality |
| S4_vs_VIRSorter_2992-cat_2 | Low-quality |
| S4_vs_VIRSorter_9186-cat_2 | Low-quality |
| S4_vs_VIRSorter_8060-cat_2 | Low-quality |
| S4_vs_VIRSorter_1584-cat_2 | Low-quality |
| S4_vs_VIRSorter_2435-cat_2 | Low-quality |
| S4_vs_VIRSorter_2798-cat_2 | Low-quality |
| S4_vs_VIRSorter_1961-cat_2 | Low-quality |
| S4_vs_VIRSorter_20666-cat_2 | Low-quality |
| S4_vs_VIRSorter_28108-cat_2 | Low-quality |
| S4_vs_VIRSorter_10523-cat_2 | Low-quality |
| S4_vs_VIRSorter_17208-cat_2 | Low-quality |
| S4_vs_VIRSorter_22974-cat_2 | Low-quality |
| S4_vs_VIRSorter_8310-cat_2 | Low-quality |
| S4_vs_VIRSorter_3973-cat_2 | Low-quality |
| S4_vs_VIRSorter_15625-cat_2 | Low-quality |
| S4_vs_VIRSorter_25783-cat_2 | Low-quality |
| S4_vs_VIRSorter_26379-cat_2 | Low-quality |
| S4_vs_VIRSorter_22444-cat_2 | Low-quality |
| S4_vs_VIRSorter_3284-cat_2 | Low-quality |
| S4_vs_VIRSorter_30053-cat_2 | Low-quality |
| S4_vs_VIRSorter_20835-cat_2 | Low-quality |
| S4_vs_VIRSorter_8752-cat_2 | Low-quality |
| S4_vs_VIRSorter_4640-cat_2 | Low-quality |
| S4_vs_VIRSorter_6763-cat_2 | Low-quality |
| S4_vs_VIRSorter_14692-cat_2 | Low-quality |
| S4_vs_VIRSorter_24926-cat_2 | Low-quality |
| S4_vs_VIRSorter_28837-cat_2 | Low-quality |
| S4_vs_VIRSorter_11382-cat_2 | Low-quality |
| S4_vs_VIRSorter_525-cat_2 | Low-quality |
| S4_vs_VIRSorter_1970-cat_2 | Low-quality |
| S4_vs_VIRSorter_9937-cat_2 | Low-quality |
| S4_vs_VIRSorter_16194-cat_2 | Low-quality |
| S4_vs_VIRSorter_22024-cat_2 | Low-quality |
| S4_vs_VIRSorter_3735-cat_2 | Low-quality |
| S4_vs_VIRSorter_18567-cat_2 | Low-quality |
| S4_vs_VIRSorter_18816-cat_2 | Low-quality |
| S4_vs_VIRSorter_6023-cat_2 | Low-quality |
| S4_vs_VIRSorter_9973-cat_2 | Low-quality |
| S4_vs_VIRSorter_13276-cat_2 | Low-quality |
| S4_vs_VIRSorter_15892-cat_2 | Low-quality |
| S4_vs_VIRSorter_22392-cat_2 | Low-quality |
| S4_vs_VIRSorter_26505-cat_2 | Low-quality |
| S4_vs_VIRSorter_1279-cat_2 | Low-quality |
| S4_vs_VIRSorter_3612-cat_2 | Low-quality |
| S4_vs_VIRSorter_21793-cat_2 | Low-quality |
| S4_vs_VIRSorter_7485-cat_2 | Low-quality |
| S4_vs_VIRSorter_16200-cat_2 | Low-quality |
| S4_vs_VIRSorter_10627-cat_2 | Low-quality |
| S4_extracted_13627 | Medium-quality |
| S4_vs_VIRSorter_13627-cat_2 | Medium-quality |
| S9_extracted_k141_85495 | Medium-quality |
| S7_vs_76382\|\|full | Medium-quality |
| S4_extracted_13831 | Medium-quality |
| S4_vs_VIRSorter_13831-cat_2 | Medium-quality |
| S4_vs_VIRSorter_9769-cat_3 | Low-quality |
| S2_vs_k141_4910\|\|full | Medium-quality |
| S8_extracted_k141_3410 | High-quality |
| S4_vs_VIRSorter_16270-cat_3 | Low-quality |
| S4_vs_VIRSorter_30283-cat_3 | Low-quality |
| S4_vs_VIRSorter_17514-cat_3 | Low-quality |
| S4_vs_VIRSorter_31698-cat_3 | Low-quality |
| S14_vs_k141_93565\|\|full | Medium-quality |
| S8_vs_k141_52050\|\|full | Medium-quality |
| S13_vs_107349\|\|full | Medium-quality |
| S6_extracted_k141_14142 | Not-determined |
| S6_extracted_k141_42416 | Not-determined |
| S6_extracted_k141_40062 | Low-quality |
| S6_extracted_k141_1 | Not-determined |
| S6_extracted_k141_37703 | Not-determined |
| S6_extracted_k141_4715 | Not-determined |
| S6_extracted_k141_37707 | Low-quality |
| S6_extracted_k141_37708 | Not-determined |
| S6_extracted_k141_14147 | Low-quality |
| S6_extracted_k141_7073 | Not-determined |
| S6_extracted_k141_16504 | Low-quality |
| S6_extracted_k141_14150 | Low-quality |
| S6_extracted_k141_4721 | Not-determined |
| S6_extracted_k141_18858 | Not-determined |
| S6_extracted_k141_23571 | Not-determined |
| S6_extracted_k141_21216 | Not-determined |
| S6_extracted_k141_9430 | Not-determined |
| S6_extracted_k141_23574 | Not-determined |
| S6_extracted_k141_11794 | Not-determined |
| S6_extracted_k141_28289 | Not-determined |
| S6_extracted_k141_47134 | Not-determined |
| S6_extracted_k141_42430 | Not-determined |
| S6_extracted_k141_47135 | Not-determined |
| S6_extracted_k141_4723 | Not-determined |
| S6_extracted_k141_23580 | Not-determined |
| S6_extracted_k141_11801 | Not-determined |
| S6_extracted_k141_33005 | Not-determined |
| S6_extracted_k141_42432 | Low-quality |
| S6_extracted_k141_4729 | Not-determined |
| S6_extracted_k141_16511 | Not-determined |
| S6_extracted_k141_47139 | Not-determined |
| S6_extracted_k141_47141 | Not-determined |
| S6_extracted_k141_49495 | Not-determined |
| S6_extracted_k141_54199 | Not-determined |
| S6_extracted_k141_16515 | Low-quality |
| S6_extracted_k141_51855 | Not-determined |
| S6_extracted_k141_16517 | Not-determined |
| S6_extracted_k141_4733 | Low-quality |
| S6_extracted_k141_37715 | Not-determined |
| S6_extracted_k141_11808 | Low-quality |
| S6_extracted_k141_49501 | Not-determined |
| S6_extracted_k141_18868 | Not-determined |
| S6_extracted_k141_47148 | Not-determined |
| S6_extracted_k141_18871 | Not-determined |
| S6_extracted_k141_16521 | Not-determined |
| S6_extracted_k141_16523 | Not-determined |
| S6_extracted_k141_18872 | Not-determined |
| S6_extracted_k141_44782 | Not-determined |
| S6_extracted_k141_16526 | Not-determined |
| S6_extracted_k141_18873 | Not-determined |
| S6_extracted_k141_16527 | Not-determined |
| S6_extracted_k141_16528 | Low-quality |
| S6_extracted_k141_44786 | Not-determined |
| S6_extracted_k141_25933 | Not-determined |
| S6_extracted_k141_16533 | Not-determined |
| S6_extracted_k141_23598 | Not-determined |
| S6_extracted_k141_33017 | Not-determined |
| S6_extracted_k141_11814 | Low-quality |
| S6_extracted_k141_33019 | Not-determined |
| S6_extracted_k141_54222 | Low-quality |
| S6_extracted_k141_5 | Not-determined |
| S6_extracted_k141_37718 | Not-determined |
| S6_extracted_k141_51857 | Low-quality |
| S6_extracted_k141_42451 | Low-quality |
| S6_extracted_k141_18877 | Not-determined |
| S6_extracted_k141_18878 | Not-determined |
| S6_extracted_k141_49506 | Not-determined |
| S6_extracted_k141_42458 | Not-determined |
| S6_extracted_k141_21241 | Not-determined |
| S6_extracted_k141_12 | Not-determined |
| S6_extracted_k141_16539 | Low-quality |
| S6_extracted_k141_40076 | Not-determined |
| S6_extracted_k141_42460 | Low-quality |
| S6_extracted_k141_25948 | Low-quality |
| S6_extracted_k141_40079 | Low-quality |
| S6_extracted_k141_42463 | Not-determined |
| S6_extracted_k141_40080 | Not-determined |
| S6_extracted_k141_40083 | Low-quality |
| S6_extracted_k141_14154 | Not-determined |
| S6_extracted_k141_49513 | Not-determined |
| S6_extracted_k141_51868 | Low-quality |
| S6_extracted_k141_47166 | Not-determined |
| S6_extracted_k141_40087 | Not-determined |
| S6_extracted_k141_16550 | Not-determined |
| S6_extracted_k141_24 | Not-determined |
| S6_extracted_k141_51871 | Low-quality |
| S6_extracted_k141_47170 | Not-determined |
| S6_extracted_k141_51872 | Not-determined |
| S6_extracted_k141_49518 | Not-determined |
| S6_extracted_k141_35370 | Not-determined |
| S6_extracted_k141_40095 | Not-determined |
| S6_extracted_k141_47172 | Not-determined |
| S6_extracted_k141_51876 | Not-determined |
| S6_extracted_k141_47175 | Not-determined |
| S6_extracted_k141_37727 | Not-determined |
| S6_extracted_k141_30648 | Low-quality |
| S6_extracted_k141_18891 | Not-determined |
| S6_extracted_k141_54230 | Low-quality |
| S6_extracted_k141_30656 | Not-determined |
| S6_extracted_k141_33 | Not-determined |
| S6_extracted_k141_49534 | Not-determined |
| S6_extracted_k141_49535 | Not-determined |
| S6_extracted_k141_51882 | Not-determined |
| S6_extracted_k141_51883 | Not-determined |
| S6_extracted_k141_49538 | Not-determined |
| S6_extracted_k141_35380 | Not-determined |
| S6_extracted_k141_30667 | Not-determined |
| S6_extracted_k141_33033 | Not-determined |
| S6_extracted_k141_40122 | Not-determined |
| S6_extracted_k141_30668 | Not-determined |
| S6_extracted_k141_18897 | Low-quality |
| S6_extracted_k141_30673 | Not-determined |
| S6_extracted_k141_30674 | Not-determined |
| S6_extracted_k141_49544 | Not-determined |
| S6_extracted_k141_42474 | Not-determined |
| S6_extracted_k141_40 | Not-determined |
| S6_extracted_k141_54238 | Not-determined |
| S6_extracted_k141_49558 | Not-determined |
| S6_extracted_k141_30688 | Not-determined |
| S6_extracted_k141_49561 | Not-determined |
| S6_extracted_k141_51892 | Not-determined |
| S6_extracted_k141_30693 | Low-quality |
| S6_extracted_k141_30695 | Not-determined |
| S6_extracted_k141_54243 | Low-quality |
| S6_extracted_k141_49 | Not-determined |
| S6_extracted_k141_18909 | Not-determined |
| S6_extracted_k141_30700 | Not-determined |
| S6_extracted_k141_30702 | Not-determined |
| S6_extracted_k141_54246 | Not-determined |
| S6_extracted_k141_30703 | Not-determined |
| S6_extracted_k141_42494 | Not-determined |
| S6_extracted_k141_30707 | Not-determined |
| S6_extracted_k141_51916 | Low-quality |
| S6_extracted_k141_14161 | Not-determined |
| S6_extracted_k141_47188 | Not-determined |
| S6_extracted_k141_42497 | Not-determined |
| S6_extracted_k141_30710 | Not-determined |
| S6_extracted_k141_51927 | Not-determined |
| S6_extracted_k141_54248 | Low-quality |
| S6_extracted_k141_42509 | Not-determined |
| S6_extracted_k141_42510 | Not-determined |
| S6_extracted_k141_16587 | Not-determined |
| S6_extracted_k141_42511 | Not-determined |
| S6_extracted_k141_30719 | Not-determined |
| S6_extracted_k141_16589 | Not-determined |
| S6_extracted_k141_30721 | Not-determined |
| S6_extracted_k141_42518 | Not-determined |
| S6_extracted_k141_16600 | Not-determined |
| S6_extracted_k141_54253 | Not-determined |
| S6_extracted_k141_16603 | Not-determined |
| S6_extracted_k141_18920 | Not-determined |
| S6_extracted_k141_28305 | Not-determined |
| S6_extracted_k141_14172 | Not-determined |
| S6_extracted_k141_63 | Not-determined |
| S6_extracted_k141_64 | Not-determined |
| S6_extracted_k141_42532 | Not-determined |
| S6_extracted_k141_28315 | Not-determined |
| S6_extracted_k141_28318 | Low-quality |
| S6_extracted_k141_42545 | Low-quality |
| S6_extracted_k141_28319 | Not-determined |
| S6_extracted_k141_47204 | Low-quality |
| S6_extracted_k141_47205 | Not-determined |
| S6_extracted_k141_69 | Low-quality |
| S6_extracted_k141_16607 | Not-determined |
| S6_extracted_k141_28326 | Low-quality |
| S6_extracted_k141_9442 | Not-determined |
| S6_extracted_k141_14183 | Not-determined |
| S6_extracted_k141_33048 | Not-determined |
| S6_extracted_k141_28331 | Low-quality |
| S6_extracted_k141_28332 | Not-determined |
| S6_extracted_k141_14190 | Low-quality |
| S6_extracted_k141_14195 | Not-determined |
| S6_extracted_k141_14197 | Not-determined |
| S6_extracted_k141_35391 | Low-quality |
| S6_extracted_k141_33058 | Not-determined |
| S6_extracted_k141_35393 | Not-determined |
| S6_extracted_k141_25956 | Not-determined |
| S6_extracted_k141_18927 | Not-determined |
| S6_extracted_k141_21251 | Low-quality |
| S6_extracted_k141_30744 | Not-determined |
| S6_extracted_k141_9452 | Not-determined |
| S6_extracted_k141_33065 | Not-determined |
| S6_extracted_k141_33067 | Not-determined |
| S6_extracted_k141_14205 | Not-determined |
| S6_extracted_k141_30750 | Not-determined |
| S6_extracted_k141_51946 | Not-determined |
| S6_extracted_k141_18939 | Not-determined |
| S6_extracted_k141_18940 | Not-determined |
| S6_extracted_k141_47218 | Not-determined |
| S6_extracted_k141_35403 | Not-determined |
| S6_extracted_k141_4737 | Not-determined |
| S6_extracted_k141_33075 | Not-determined |
| S6_extracted_k141_4739 | Not-determined |
| S6_extracted_k141_4740 | Not-determined |
| S6_extracted_k141_35407 | Not-determined |
| S6_extracted_k141_28341 | Not-determined |
| S6_extracted_k141_49574 | Not-determined |
| S6_extracted_k141_89 | Not-determined |
| S6_extracted_k141_21261 | Not-determined |
| S6_extracted_k141_37743 | Not-determined |
| S6_extracted_k141_37746 | Not-determined |
| S6_extracted_k141_33079 | Not-determined |
| S6_extracted_k141_4743 | Not-determined |
| S6_extracted_k141_28349 | Not-determined |
| S6_extracted_k141_93 | Low-quality |
| S6_extracted_k141_9457 | Not-determined |
| S6_extracted_k141_95 | Not-determined |
| S6_extracted_k141_49584 | Not-determined |
| S6_extracted_k141_96 | Not-determined |
| S6_extracted_k141_21263 | Not-determined |
| S6_extracted_k141_9462 | Not-determined |
| S6_extracted_k141_28351 | Not-determined |
| S6_extracted_k141_16619 | Low-quality |
| S6_extracted_k141_33090 | Not-determined |
| S6_extracted_k141_51962 | Not-determined |
| S6_extracted_k141_37765 | Not-determined |
| S6_extracted_k141_37768 | Not-determined |
| S6_extracted_k141_4759 | Not-determined |
| S6_extracted_k141_37771 | Not-determined |
| S6_extracted_k141_33099 | Not-determined |
| S6_extracted_k141_51969 | Not-determined |
| S6_extracted_k141_16624 | Not-determined |
| S6_extracted_k141_33100 | Not-determined |
| S6_extracted_k141_18952 | Not-determined |
| S6_extracted_k141_33101 | Not-determined |
| S6_extracted_k141_51973 | Not-determined |
| S6_extracted_k141_103 | Low-quality |
| S6_extracted_k141_14239 | Not-determined |
| S6_extracted_k141_42562 | Not-determined |
| S6_extracted_k141_18954 | Low-quality |
| S6_extracted_k141_16630 | Not-determined |
| S6_extracted_k141_33107 | Not-determined |
| S6_extracted_k141_42564 | Not-determined |
| S6_extracted_k141_16637 | Not-determined |
| S6_extracted_k141_18963 | Low-quality |
| S6_extracted_k141_16638 | Low-quality |
| S6_extracted_k141_33114 | Low-quality |
| S6_extracted_k141_16641 | Not-determined |
| S6_extracted_k141_14256 | Not-determined |
| S6_extracted_k141_51981 | Not-determined |
| S6_extracted_k141_44822 | Not-determined |
| S6_extracted_k141_44824 | Not-determined |
| S6_extracted_k141_40135 | Not-determined |
| S6_extracted_k141_21273 | Not-determined |
| S6_extracted_k141_44834 | Not-determined |
| S6_extracted_k141_28371 | Low-quality |
| S6_extracted_k141_136 | Not-determined |
| S6_extracted_k141_4791 | Not-determined |
| S6_extracted_k141_28373 | Not-determined |
| S6_extracted_k141_33124 | Not-determined |
| S6_extracted_k141_44842 | Not-determined |
| S6_extracted_k141_33126 | Not-determined |
| S6_extracted_k141_139 | Not-determined |
| S6_extracted_k141_28376 | Not-determined |
| S6_extracted_k141_44846 | Low-quality |
| S6_extracted_k141_33134 | Not-determined |
| S6_extracted_k141_148 | Not-determined |
| S6_extracted_k141_40157 | Not-determined |
| S6_extracted_k141_9501 | Not-determined |
| S6_extracted_k141_4800 | Low-quality |
| S6_extracted_k141_42580 | Not-determined |
| S6_extracted_k141_42584 | Not-determined |
| S6_extracted_k141_9505 | Not-determined |
| S6_extracted_k141_44861 | Low-quality |
| S6_extracted_k141_28380 | Low-quality |
| S4_extracted_12624 | High-quality |
| S6_extracted_k141_23632 | Low-quality |
| S6_extracted_k141_42593 | Not-determined |
| S6_extracted_k141_28383 | Not-determined |
| S6_extracted_k141_11823 | Not-determined |
| S6_extracted_k141_49619 | Not-determined |
| S6_extracted_k141_160 | Low-quality |
| S6_extracted_k141_18984 | Not-determined |
| S6_extracted_k141_44872 | Not-determined |
| S6_extracted_k141_2362 | Not-determined |
| S6_extracted_k141_42595 | Not-determined |
| S6_extracted_k141_28390 | Not-determined |
| S6_extracted_k141_44875 | Not-determined |
| S6_extracted_k141_2367 | Low-quality |
| S6_extracted_k141_35440 | Not-determined |
| S6_extracted_k141_28391 | Low-quality |
| S6_extracted_k141_166 | Not-determined |
| S6_extracted_k141_167 | Not-determined |
| S6_extracted_k141_11828 | Not-determined |
| S6_extracted_k141_49621 | Not-determined |
| S6_extracted_k141_37814 | Not-determined |
| S6_extracted_k141_52019 | Not-determined |
| S6_extracted_k141_4817 | Not-determined |
| S6_extracted_k141_37815 | Not-determined |
| S6_extracted_k141_37816 | Not-determined |
| S6_extracted_k141_11831 | Not-determined |
| S6_extracted_k141_11832 | Low-quality |
| S6_extracted_k141_37823 | Not-determined |
| S6_extracted_k141_35451 | Not-determined |
| S6_extracted_k141_11839 | Not-determined |
| S6_extracted_k141_35454 | Low-quality |
| S6_extracted_k141_37828 | Low-quality |
| S6_extracted_k141_9515 | Not-determined |
| S6_extracted_k141_44886 | Not-determined |
| S6_extracted_k141_33150 | Not-determined |
| S6_extracted_k141_28402 | Not-determined |
| S6_extracted_k141_44892 | Not-determined |
| S6_extracted_k141_11847 | Not-determined |
| S6_extracted_k141_23648 | Not-determined |
| S6_extracted_k141_7081 | Not-determined |
| S6_extracted_k141_44894 | Not-determined |
| S6_extracted_k141_7084 | Not-determined |
| S6_extracted_k141_14275 | Not-determined |
| S6_extracted_k141_28406 | Not-determined |
| S6_extracted_k141_37849 | Not-determined |
| S6_extracted_k141_35457 | Low-quality |
| S6_extracted_k141_2382 | Not-determined |
| S6_extracted_k141_23651 | Not-determined |
| S6_extracted_k141_28413 | Not-determined |
| S6_extracted_k141_49623 | Not-determined |
| S6_extracted_k141_28414 | Not-determined |
| S6_extracted_k141_52040 | Not-determined |
| S6_extracted_k141_42607 | Not-determined |
| S6_extracted_k141_49629 | Not-determined |
| S6_extracted_k141_40174 | Low-quality |
| S6_extracted_k141_40175 | Not-determined |
| S6_extracted_k141_21278 | Not-determined |
| S6_extracted_k141_14296 | Low-quality |
| S6_extracted_k141_21283 | Low-quality |
| S6_extracted_k141_42613 | Not-determined |
| S6_extracted_k141_21285 | Not-determined |
| S6_extracted_k141_21287 | Not-determined |
| S6_extracted_k141_42616 | Not-determined |
| S6_extracted_k141_21290 | Not-determined |
| S6_extracted_k141_52043 | Not-determined |
| S6_extracted_k141_9521 | Not-determined |
| S6_extracted_k141_49646 | Not-determined |
| S6_extracted_k141_49647 | Not-determined |
| S6_extracted_k141_40182 | Not-determined |
| S6_extracted_k141_52050 | Not-determined |
| S6_extracted_k141_52051 | Not-determined |
| S6_extracted_k141_52053 | Not-determined |
| S6_extracted_k141_42627 | Low-quality |
| S6_extracted_k141_42628 | Low-quality |
| S6_extracted_k141_37874 | Not-determined |
| S6_extracted_k141_49651 | Not-determined |
| S6_extracted_k141_14312 | Not-determined |
| S6_extracted_k141_52063 | Not-determined |
| S6_extracted_k141_37887 | Not-determined |
| S6_extracted_k141_52066 | Low-quality |
| S6_extracted_k141_44904 | Not-determined |
| S6_extracted_k141_42635 | Not-determined |
| S6_extracted_k141_2409 | Not-determined |
| S6_extracted_k141_37892 | Not-determined |
| S6_extracted_k141_14320 | Not-determined |
| S6_extracted_k141_42638 | Not-determined |
| S6_extracted_k141_14322 | Not-determined |
| S6_extracted_k141_40190 | Not-determined |
| S6_extracted_k141_16653 | Not-determined |
| S6_extracted_k141_216 | Not-determined |
| S6_extracted_k141_37901 | Not-determined |
| S6_extracted_k141_9536 | Not-determined |
| S6_extracted_k141_52086 | Not-determined |
| S6_extracted_k141_52087 | Not-determined |
| S6_extracted_k141_52090 | Not-determined |
| S6_extracted_k141_9542 | Not-determined |
| S6_extracted_k141_2418 | Not-determined |
| S6_extracted_k141_44917 | Not-determined |
| S6_extracted_k141_44919 | Not-determined |
| S6_extracted_k141_52095 | Not-determined |
| S6_extracted_k141_16660 | Not-determined |
| S6_extracted_k141_37906 | Low-quality |
| S6_extracted_k141_9545 | Not-determined |
| S6_extracted_k141_225 | Not-determined |
| S6_extracted_k141_226 | Not-determined |
| S6_extracted_k141_52098 | Not-determined |
| S6_extracted_k141_28451 | Not-determined |
| S6_extracted_k141_16662 | Low-quality |
| S6_extracted_k141_227 | Not-determined |
| S6_extracted_k141_44926 | Not-determined |
| S6_extracted_k141_28454 | Not-determined |
| S6_extracted_k141_28455 | Low-quality |
| S6_extracted_k141_49661 | Not-determined |
| S6_extracted_k141_49662 | Not-determined |
| S6_extracted_k141_37916 | Low-quality |
| S6_extracted_k141_28457 | Not-determined |
| S6_extracted_k141_49665 | Not-determined |
| S6_extracted_k141_49667 | Low-quality |
| S6_extracted_k141_11860 | Not-determined |
| S6_extracted_k141_52106 | Low-quality |
| S6_extracted_k141_11862 | Not-determined |
| S6_extracted_k141_30783 | Not-determined |
| S6_extracted_k141_26000 | Not-determined |
| S6_extracted_k141_26002 | Not-determined |
| S6_extracted_k141_49672 | Not-determined |
| S6_extracted_k141_52108 | Low-quality |
| S6_extracted_k141_26003 | Low-quality |
| S6_extracted_k141_230 | Not-determined |
| S6_extracted_k141_231 | Not-determined |
| S6_extracted_k141_2424 | Not-determined |
| S6_extracted_k141_2426 | Not-determined |
| S6_extracted_k141_44944 | Not-determined |
| S6_extracted_k141_26005 | Not-determined |
| S6_extracted_k141_16670 | Not-determined |
| S6_extracted_k141_21303 | Not-determined |
| S6_extracted_k141_21305 | Not-determined |
| S6_extracted_k141_52122 | Low-quality |
| S6_extracted_k141_37934 | Not-determined |
| S6_extracted_k141_235 | Not-determined |
| S6_extracted_k141_49687 | Not-determined |
| S6_extracted_k141_9548 | Low-quality |
| S6_extracted_k141_2432 | Not-determined |
| S6_extracted_k141_2434 | Not-determined |
| S6_extracted_k141_242 | Not-determined |
| S6_extracted_k141_44953 | Not-determined |
| S6_extracted_k141_30799 | Not-determined |
| S6_extracted_k141_9554 | Not-determined |
| S6_extracted_k141_2439 | Not-determined |
| S6_extracted_k141_9555 | Not-determined |
| S6_extracted_k141_16678 | Not-determined |
| S6_extracted_k141_30800 | Not-determined |
| S6_extracted_k141_16679 | Not-determined |
| S6_extracted_k141_44954 | Not-determined |
| S6_extracted_k141_40206 | Not-determined |
| S6_extracted_k141_30805 | Not-determined |
| S6_extracted_k141_11879 | Not-determined |
| S6_extracted_k141_42648 | Not-determined |
| S6_extracted_k141_44957 | Low-quality |
| S6_extracted_k141_37945 | Not-determined |
| S6_extracted_k141_11883 | Low-quality |
| S6_extracted_k141_37948 | Not-determined |
| S6_extracted_k141_33172 | Not-determined |
| S6_extracted_k141_23666 | Not-determined |
| S6_extracted_k141_11890 | Not-determined |
| S6_extracted_k141_2453 | Not-determined |
| S6_extracted_k141_2454 | Not-determined |
| S6_extracted_k141_44962 | Low-quality |
| S6_extracted_k141_252 | Not-determined |
| S6_extracted_k141_255 | Not-determined |
| S6_extracted_k141_14335 | Not-determined |
| S6_extracted_k141_37963 | Not-determined |
| S6_extracted_k141_23674 | Not-determined |
| S6_extracted_k141_14337 | Not-determined |
| S6_extracted_k141_49697 | Not-determined |
| S6_extracted_k141_14338 | Not-determined |
| S6_extracted_k141_23676 | Low-quality |
| S6_extracted_k141_49698 | Low-quality |
| S6_extracted_k141_44966 | Not-determined |
| S6_extracted_k141_42661 | Low-quality |
| S6_extracted_k141_30810 | Low-quality |
| S6_extracted_k141_49703 | Not-determined |
| S6_extracted_k141_14342 | Not-determined |
| S6_extracted_k141_258 | Not-determined |
| S6_extracted_k141_49707 | Not-determined |
| S6_extracted_k141_14346 | Not-determined |
| S6_extracted_k141_35487 | Not-determined |
| S6_extracted_k141_16689 | Not-determined |
| S6_extracted_k141_26028 | Low-quality |
| S6_extracted_k141_16691 | Not-determined |
| S6_extracted_k141_44980 | Not-determined |
| S6_extracted_k141_37974 | Not-determined |
| S6_extracted_k141_16692 | Not-determined |
| S6_extracted_k141_35493 | Not-determined |
| S6_extracted_k141_35496 | Not-determined |
| S6_extracted_k141_2475 | Not-determined |
| S6_extracted_k141_44984 | Not-determined |
| S6_extracted_k141_265 | Not-determined |
| S6_extracted_k141_35497 | Not-determined |
| S6_extracted_k141_44988 | Not-determined |
| S6_extracted_k141_40217 | Not-determined |
| S6_extracted_k141_35500 | Not-determined |
| S6_extracted_k141_23687 | Not-determined |
| S6_extracted_k141_35506 | Not-determined |
| S6_extracted_k141_11894 | Low-quality |
| S6_extracted_k141_44992 | Not-determined |
| S6_extracted_k141_33200 | Not-determined |
| S6_extracted_k141_40226 | Not-determined |
| S6_extracted_k141_11898 | Not-determined |
| S6_extracted_k141_2483 | Not-determined |
| S6_extracted_k141_2485 | Not-determined |
| S6_extracted_k141_16698 | Not-determined |
| S6_extracted_k141_16699 | Not-determined |
| S6_extracted_k141_28460 | Low-quality |
| S6_extracted_k141_16701 | Not-determined |
| S6_extracted_k141_274 | Low-quality |
| S6_extracted_k141_44997 | Low-quality |
| S6_extracted_k141_28463 | Not-determined |
| S6_extracted_k141_16704 | Not-determined |
| S6_extracted_k141_33204 | Not-determined |
| S6_extracted_k141_42671 | Not-determined |
| S6_extracted_k141_26041 | Not-determined |
| S6_extracted_k141_4845 | Low-quality |
| S6_extracted_k141_16709 | Not-determined |
| S6_extracted_k141_26045 | Not-determined |
| S6_extracted_k141_4847 | Not-determined |
| S6_extracted_k141_28468 | Not-determined |
| S6_extracted_k141_287 | Low-quality |
| S6_extracted_k141_42682 | Low-quality |
| S6_extracted_k141_28470 | Not-determined |
| S6_extracted_k141_7132 | Not-determined |
| S6_extracted_k141_35526 | Not-determined |
| S6_extracted_k141_26050 | Not-determined |
| S6_extracted_k141_4848 | Not-determined |
| S6_extracted_k141_9563 | Low-quality |
| S6_extracted_k141_11914 | Low-quality |
| S6_extracted_k141_26054 | Not-determined |
| S6_extracted_k141_7135 | Not-determined |
| S6_extracted_k141_2495 | Not-determined |
| S6_extracted_k141_4852 | Not-determined |
| S6_extracted_k141_7141 | Not-determined |
| S6_extracted_k141_299 | Not-determined |
| S6_extracted_k141_26063 | Not-determined |
| S6_extracted_k141_11924 | Not-determined |
| S6_extracted_k141_14363 | Not-determined |
| S6_extracted_k141_14365 | Not-determined |
| S6_extracted_k141_7145 | Low-quality |
| S6_extracted_k141_2503 | Not-determined |
| S6_extracted_k141_4854 | Low-quality |
| S6_extracted_k141_304 | Not-determined |
| S6_extracted_k141_35530 | Not-determined |
| S6_extracted_k141_306 | Not-determined |
| S6_extracted_k141_9580 | Not-determined |
| S6_extracted_k141_11938 | Not-determined |
| S6_extracted_k141_7154 | Not-determined |
| S6_extracted_k141_37999 | Not-determined |
| S6_extracted_k141_40241 | Not-determined |
| S6_extracted_k141_4870 | Low-quality |

| | |
|---|---|
| SG_extracted_k141_2513 | Not-determined |
| SG_extracted_k141_38001 | Not-determined |
| SG_extracted_k141_11942 | Not-determined |
| SG_extracted_k141_2514 | Not-determined |
| SG_extracted_k141_38004 | Not-determined |
| SG_extracted_k141_7160 | Not-determined |
| SG_extracted_k141_7161 | Not-determined |
| SG_extracted_k141_40247 | Not-determined |
| SG_extracted_k141_2522 | Not-determined |
| SG_extracted_k141_40249 | Not-determined |
| SG_extracted_k141_7172 | Not-determined |
| SG_extracted_k141_7175 | Not-determined |
| SG_extracted_k141_14371 | Low-quality |
| SG_extracted_k141_40250 | Not-determined |
| SG_extracted_k141_35549 | Not-determined |
| SG_extracted_k141_28491 | Not-determined |
| SG_extracted_k141_7180 | Not-determined |
| SG_extracted_k141_26098 | Not-determined |
| SG_extracted_k141_26099 | Not-determined |
| SG_extracted_k141_2532 | Not-determined |
| SG_extracted_k141_38009 | Not-determined |
| SG_extracted_k141_26102 | Low-quality |
| SG_extracted_k141_9589 | Not-determined |
| SG_extracted_k141_38011 | Low-quality |
| SG_extracted_k141_35555 | Not-determined |
| SG_extracted_k141_331 | Low-quality |
| SG_extracted_k141_14375 | Low-quality |
| SG_extracted_k141_35559 | Not-determined |
| SG_extracted_k141_16725 | Low-quality |
| SG_extracted_k141_38015 | Not-determined |
| SG_extracted_k141_38017 | Low-quality |
| SG_extracted_k141_45005 | Not-determined |
| SG_extracted_k141_7186 | Not-determined |
| SG_extracted_k141_11958 | Not-determined |
| SG_extracted_k141_9592 | Not-determined |
| SG_extracted_k141_7188 | Not-determined |
| SG_extracted_k141_23702 | Not-determined |
| SG_extracted_k141_26116 | Not-determined |
| SG_extracted_k141_4884 | Not-determined |
| SG_extracted_k141_7191 | Not-determined |
| SG_extracted_k141_11966 | Not-determined |
| SG_extracted_k141_2542 | Not-determined |
| SG_extracted_k141_38025 | Not-determined |
| SG_extracted_k141_28510 | Not-determined |
| SG_extracted_k141_28511 | Low-quality |
| SG_extracted_k141_38027 | Not-determined |
| SG_extracted_k141_28512 | Not-determined |
| SG_extracted_k141_38032 | Not-determined |
| SG_extracted_k141_2547 | Not-determined |
| SG_extracted_k141_38036 | Not-determined |
| SG_extracted_k141_9599 | Not-determined |
| SG_extracted_k141_38040 | Not-determined |
| SG_extracted_k141_33243 | Not-determined |
| SG_extracted_k141_2555 | Not-determined |
| SG_extracted_k141_21319 | Not-determined |
| SG_extracted_k141_45017 | Not-determined |
| SG_extracted_k141_21320 | Not-determined |
| SG_extracted_k141_38044 | Not-determined |
| SG_extracted_k141_38045 | Not-determined |
| SG_extracted_k141_54272 | Not-determined |
| SG_extracted_k141_9606 | Not-determined |
| SG_extracted_k141_54275 | Not-determined |
| SG_extracted_k141_11980 | Low-quality |
| SG_extracted_k141_38049 | Not-determined |
| SG_extracted_k141_33251 | Low-quality |
| SG_extracted_k141_2569 | Not-determined |
| SG_extracted_k141_45020 | Not-determined |
| SG_extracted_k141_2573 | Not-determined |
| SG_extracted_k141_45025 | Not-determined |
| SG_extracted_k141_40271 | Not-determined |
| SG_extracted_k141_2578 | Not-determined |
| SG_extracted_k141_45027 | Not-determined |
| SG_extracted_k141_40276 | Not-determined |
| SG_extracted_k141_42706 | Low-quality |
| SG_extracted_k141_40279 | Not-determined |
| SG_extracted_k141_42708 | Low-quality |
| SG_extracted_k141_42711 | Not-determined |
| SG_extracted_k141_40284 | Not-determined |
| SG_extracted_k141_38062 | Not-determined |
| SG_extracted_k141_367 | Not-determined |
| SG_extracted_k141_38063 | Not-determined |
| SG_extracted_k141_40288 | Not-determined |
| SG_extracted_k141_52129 | Not-determined |
| SG_extracted_k141_12002 | Not-determined |
| SG_extracted_k141_52130 | Not-determined |
| SG_extracted_k141_14404 | Not-determined |
| SG_extracted_k141_38072 | Not-determined |
| SG_extracted_k141_38073 | Not-determined |
| SG_extracted_k141_376 | Not-determined |
| SG_extracted_k141_33279 | Not-determined |
| SG_extracted_k141_2607 | Low-quality |
| SG_extracted_k141_2609 | Low-quality |
| SG_extracted_k141_12008 | Not-determined |
| SG_extracted_k141_14412 | Not-determined |
| SG_extracted_k141_45032 | Not-determined |
| SG_extracted_k141_14413 | Not-determined |
| SG_extracted_k141_47230 | Not-determined |
| SG_extracted_k141_12012 | Not-determined |
| SG_extracted_k141_52138 | Not-determined |
| SG_extracted_k141_38085 | Low-quality |
| SG_extracted_k141_38086 | Not-determined |
| SG_extracted_k141_14420 | Not-determined |
| SG_extracted_k141_16730 | Not-determined |
| SG_extracted_k141_33283 | Not-determined |
| SG_extracted_k141_9613 | Not-determined |
| SG_extracted_k141_52145 | Not-determined |
| SG_extracted_k141_23731 | Not-determined |
| SG_extracted_k141_45042 | Not-determined |
| SG_extracted_k141_391 | Low-quality |
| SG_extracted_k141_12027 | Low-quality |
| SG_extracted_k141_45051 | Low-quality |
| SG_extracted_k141_12028 | Low-quality |
| SG_extracted_k141_33296 | Not-determined |
| SG_extracted_k141_16743 | Not-determined |
| SG_extracted_k141_38103 | Not-determined |
| SG_extracted_k141_394 | Low-quality |
| SG_extracted_k141_33298 | Not-determined |
| SG_extracted_k141_16744 | Not-determined |
| SG_extracted_k141_33299 | Not-determined |
| SG_extracted_k141_398 | Not-determined |
| SG_extracted_k141_9624 | Not-determined |
| SG_extracted_k141_33303 | Not-determined |
| SG_extracted_k141_9626 | Low-quality |
| SG_extracted_k141_16751 | Not-determined |
| SG_extracted_k141_2640 | Not-determined |
| SG_extracted_k141_404 | Not-determined |
| SG_extracted_k141_40296 | Low-quality |
| SG_extracted_k141_12041 | Not-determined |
| SG_extracted_k141_40297 | Not-determined |
| SG_extracted_k141_16758 | Not-determined |
| SG_extracted_k141_45069 | Not-determined |
| SG_extracted_k141_2647 | Not-determined |
| SG_extracted_k141_4896 | Not-determined |
| SG_extracted_k141_45072 | Low-quality |
| SG_extracted_k141_2653 | Not-determined |
| SG_extracted_k141_45073 | Not-determined |
| SG_extracted_k141_45074 | Not-determined |
| SG_extracted_k141_33317 | Low-quality |
| SG_extracted_k141_4899 | Not-determined |
| SG_extracted_k141_2656 | Low-quality |
| SG_extracted_k141_33320 | Low-quality |
| SG_extracted_k141_40299 | Not-determined |
| SG_extracted_k141_33322 | Not-determined |
| SG_extracted_k141_45077 | Not-determined |
| SG_extracted_k141_45078 | Not-determined |
| SG_extracted_k141_40306 | Not-determined |
| SG_extracted_k141_416 | Low-quality |
| SG_extracted_k141_45083 | Low-quality |
| SG_extracted_k141_2669 | Not-determined |
| SG_extracted_k141_2670 | Not-determined |
| SG_extracted_k141_2671 | Not-determined |
| SG_extracted_k141_40316 | Not-determined |
| SG_extracted_k141_38138 | Not-determined |
| SG_extracted_k141_18991 | Not-determined |
| SG_extracted_k141_2674 | Low-quality |
| SG_extracted_k141_7212 | Not-determined |
| SG_extracted_k141_45090 | Not-determined |
| S4_vs_VIRSorter_12624-cat_2 | High-quality |
| SG_extracted_k141_35563 | Not-determined |
| SG_extracted_k141_18994 | Not-determined |
| SG_extracted_k141_12055 | Low-quality |
| SG_extracted_k141_38142 | Low-quality |
| SG_extracted_k141_38144 | Not-determined |
| SG_extracted_k141_35569 | Not-determined |
| SG_extracted_k141_40325 | Not-determined |
| SG_extracted_k141_4910 | Not-determined |
| SG_extracted_k141_38155 | Not-determined |
| SG_extracted_k141_2685 | Not-determined |
| SG_extracted_k141_9639 | Not-determined |
| SG_extracted_k141_45095 | Low-quality |
| SG_extracted_k141_12063 | Not-determined |
| SG_extracted_k141_52159 | Not-determined |
| SG_extracted_k141_40339 | Low-quality |
| SG_extracted_k141_432 | Not-determined |
| SG_extracted_k141_433 | Not-determined |
| SG_extracted_k141_33334 | Not-determined |
| SG_extracted_k141_16771 | Not-determined |
| SG_extracted_k141_14474 | Low-quality |
| SG_extracted_k141_38159 | Not-determined |
| SG_extracted_k141_23790 | Not-determined |
| SG_extracted_k141_23791 | Not-determined |
| SG_extracted_k141_16774 | Not-determined |
| SG_extracted_k141_14478 | Not-determined |
| SG_extracted_k141_33340 | Not-determined |
| SG_extracted_k141_4921 | Low-quality |
| SG_extracted_k141_4923 | Not-determined |
| SG_extracted_k141_2700 | Not-determined |
| SG_extracted_k141_4926 | Not-determined |
| SG_extracted_k141_9648 | Not-determined |
| SG_extracted_k141_2705 | Not-determined |
| SG_extracted_k141_23800 | Low-quality |
| SG_extracted_k141_35570 | Not-determined |
| SG_extracted_k141_16787 | Not-determined |
| SG_extracted_k141_47248 | Not-determined |
| SG_extracted_k141_23806 | Not-determined |
| SG_extracted_k141_16788 | Not-determined |
| SG_extracted_k141_28535 | Low-quality |
| SG_extracted_k141_47255 | Low-quality |
| SG_extracted_k141_28540 | Not-determined |
| SG_extracted_k141_23809 | Not-determined |
| SG_extracted_k141_9659 | Low-quality |
| SG_extracted_k141_9660 | Not-determined |
| SG_extracted_k141_40366 | Not-determined |
| SG_extracted_k141_12081 | Not-determined |
| SG_extracted_k141_23813 | Not-determined |
| SG_extracted_k141_16802 | Not-determined |
| SG_extracted_k141_47259 | Not-determined |
| SG_extracted_k141_454 | Not-determined |
| SG_extracted_k141_47261 | Not-determined |
| SG_extracted_k141_40369 | Low-quality |
| SG_extracted_k141_457 | Not-determined |
| SG_extracted_k141_40372 | Not-determined |
| SG_extracted_k141_23820 | Low-quality |
| SG_extracted_k141_16806 | Not-determined |
| SG_extracted_k141_461 | Low-quality |
| SG_extracted_k141_12085 | Low-quality |
| SG_extracted_k141_40378 | Not-determined |
| SG_extracted_k141_464 | Not-determined |
| SG_extracted_k141_465 | Not-determined |
| SG_extracted_k141_28551 | Not-determined |
| SG_extracted_k141_7221 | Not-determined |
| SG_extracted_k141_12091 | Not-determined |
| SG_extracted_k141_28557 | Not-determined |
| SG_extracted_k141_33373 | Not-determined |
| SG_extracted_k141_33374 | Low-quality |
| SG_extracted_k141_26122 | Not-determined |
| SG_extracted_k141_7225 | Not-determined |
| SG_extracted_k141_472 | Low-quality |
| SG_extracted_k141_23825 | Not-determined |
| SG_extracted_k141_7227 | Not-determined |
| SG_extracted_k141_33379 | Not-determined |
| SG_extracted_k141_9679 | Low-quality |
| SG_extracted_k141_23828 | Not-determined |
| SG_extracted_k141_26128 | Not-determined |
| SG_extracted_k141_9681 | Not-determined |
| SG_extracted_k141_52177 | Not-determined |
| SG_extracted_k141_16820 | Not-determined |
| SG_extracted_k141_38169 | Not-determined |
| SG_extracted_k141_23833 | Low-quality |
| SG_extracted_k141_23834 | Not-determined |
| SG_extracted_k141_38172 | Low-quality |
| SG_extracted_k141_33388 | Not-determined |
| SG_extracted_k141_38173 | Not-determined |
| SG_extracted_k141_7234 | Low-quality |
| SG_extracted_k141_35586 | Not-determined |
| SG_extracted_k141_7236 | Not-determined |
| SG_extracted_k141_7237 | Not-determined |
| SG_extracted_k141_26131 | Not-determined |
| SG_extracted_k141_23841 | Not-determined |
| SG_extracted_k141_35588 | Not-determined |
| SG_extracted_k141_33391 | Not-determined |
| SG_extracted_k141_7241 | Not-determined |
| SG_extracted_k141_23842 | Not-determined |
| SG_extracted_k141_38174 | Low-quality |
| SG_extracted_k141_9692 | Not-determined |
| SG_extracted_k141_28562 | Not-determined |
| SG_extracted_k141_16831 | Low-quality |
| SG_extracted_k141_26136 | Not-determined |
| SG_extracted_k141_26140 | Not-determined |
| SG_extracted_k141_28568 | Not-determined |
| SG_extracted_k141_33394 | Not-determined |
| SG_extracted_k141_33396 | Not-determined |
| SG_extracted_k141_16835 | Low-quality |
| SG_extracted_k141_7249 | Not-determined |
| SG_extracted_k141_38192 | Not-determined |
| SG_extracted_k141_38195 | Not-determined |
| SG_extracted_k141_21326 | Not-determined |
| SG_extracted_k141_21327 | Not-determined |
| SG_extracted_k141_33403 | Low-quality |
| SG_extracted_k141_486 | Not-determined |
| SG_extracted_k141_7253 | Not-determined |
| SG_extracted_k141_12112 | Not-determined |
| SG_extracted_k141_52207 | Not-determined |
| SG_extracted_k141_52209 | Not-determined |
| SG_extracted_k141_40397 | Low-quality |
| SG_extracted_k141_487 | Not-determined |
| SG_extracted_k141_35595 | Not-determined |
| SG_extracted_k141_52213 | Not-determined |
| SG_extracted_k141_489 | Not-determined |
| SG_extracted_k141_7260 | Not-determined |
| SG_extracted_k141_12118 | Not-determined |
| SG_extracted_k141_16843 | Not-determined |
| SG_extracted_k141_40404 | Not-determined |
| SG_extracted_k141_23860 | Not-determined |
| SG_extracted_k141_40405 | Low-quality |
| SG_extracted_k141_40406 | Not-determined |
| SG_extracted_k141_30816 | Not-determined |
| SG_extracted_k141_30817 | Low-quality |
| SG_extracted_k141_52226 | Not-determined |
| SG_extracted_k141_495 | Low-quality |
| SG_extracted_k141_35604 | Not-determined |
| SG_extracted_k141_42719 | Not-determined |
| SG_extracted_k141_47289 | Not-determined |
| SG_extracted_k141_52229 | Not-determined |
| SG_extracted_k141_42730 | Low-quality |
| SG_extracted_k141_14485 | Not-determined |
| SG_extracted_k141_2720 | Not-determined |
| SG_extracted_k141_47296 | Not-determined |
| SG_extracted_k141_23869 | Low-quality |
| SG_extracted_k141_18999 | Not-determined |
| SG_extracted_k141_514 | Low-quality |
| SG_extracted_k141_42736 | Not-determined |
| SG_extracted_k141_19007 | Not-determined |
| SG_extracted_k141_19008 | Not-determined |
| SG_extracted_k141_26146 | Not-determined |
| SG_extracted_k141_19009 | Not-determined |
| SG_extracted_k141_26147 | Low-quality |
| SG_extracted_k141_52238 | Not-determined |
| SG_extracted_k141_26149 | Low-quality |
| SG_extracted_k141_2732 | Not-determined |
| SG_extracted_k141_35615 | Not-determined |
| SG_extracted_k141_23873 | Not-determined |
| SG_extracted_k141_9703 | Low-quality |
| SG_extracted_k141_52243 | Low-quality |
| SG_extracted_k141_35622 | Not-determined |
| SG_extracted_k141_23878 | Not-determined |
| SG_extracted_k141_9707 | Not-determined |
| SG_extracted_k141_52253 | Not-determined |
| SG_extracted_k141_30833 | Not-determined |
| SG_extracted_k141_26159 | Not-determined |
| SG_extracted_k141_23882 | Not-determined |
| SG_extracted_k141_30836 | Not-determined |
| SG_extracted_k141_35625 | Not-determined |
| SG_extracted_k141_30838 | Low-quality |
| SG_extracted_k141_26164 | Not-determined |
| SG_extracted_k141_2739 | Not-determined |
| SG_extracted_k141_14497 | Low-quality |
| SG_extracted_k141_42757 | Not-determined |
| SG_extracted_k141_52258 | Low-quality |
| SG_extracted_k141_40424 | Not-determined |
| SG_extracted_k141_14504 | Not-determined |
| SG_extracted_k141_21347 | Not-determined |
| SG_extracted_k141_52269 | Low-quality |
| SG_extracted_k141_21353 | Not-determined |
| SG_extracted_k141_21354 | Not-determined |
| SG_extracted_k141_21355 | Not-determined |
| SG_extracted_k141_52272 | Low-quality |
| SG_extracted_k141_52273 | Not-determined |
| SG_extracted_k141_26172 | Low-quality |
| SG_extracted_k141_52274 | Not-determined |
| SG_extracted_k141_14510 | Not-determined |
| SG_extracted_k141_19021 | Not-determined |
| SG_extracted_k141_40438 | Not-determined |
| SG_extracted_k141_33419 | Not-determined |
| SG_extracted_k141_26179 | Not-determined |
| SG_extracted_k141_33424 | Low-quality |
| SG_extracted_k141_2758 | Not-determined |
| SG_extracted_k141_19027 | Not-determined |
| SG_extracted_k141_33427 | Not-determined |
| SG_extracted_k141_33428 | Not-determined |
| SG_extracted_k141_33433 | Not-determined |
| S5_extracted_k141_74948 | Medium-quality |
| SG_extracted_k141_33434 | Not-determined |
| SG_extracted_k141_7269 | Not-determined |
| SG_extracted_k141_527 | Not-determined |
| SG_extracted_k141_529 | Not-determined |
| SG_extracted_k141_30854 | Not-determined |
| SG_extracted_k141_30855 | Not-determined |
| SG_extracted_k141_33437 | Not-determined |
| SG_extracted_k141_42777 | Not-determined |
| SG_extracted_k141_2769 | Not-determined |
| SG_extracted_k141_35633 | Not-determined |
| SG_extracted_k141_2776 | Not-determined |
| SG_extracted_k141_33439 | Not-determined |
| SG_extracted_k141_2778 | Not-determined |
| SG_extracted_k141_33440 | Not-determined |
| SG_extracted_k141_35638 | Not-determined |
| SG_extracted_k141_12134 | Not-determined |
| SG_extracted_k141_33443 | Not-determined |
| SG_extracted_k141_12136 | Not-determined |
| SG_extracted_k141_42782 | Not-determined |
| SG_extracted_k141_12139 | Not-determined |
| SG_extracted_k141_19031 | Not-determined |
| SG_extracted_k141_19033 | Not-determined |
| SG_extracted_k141_16849 | Not-determined |
| SG_extracted_k141_12144 | Low-quality |
| SG_extracted_k141_19036 | Not-determined |
| SG_extracted_k141_19037 | Not-determined |
| SG_extracted_k141_26203 | Not-determined |
| SG_extracted_k141_33452 | Low-quality |
| SG_extracted_k141_16852 | Not-determined |
| SG_extracted_k141_9716 | Low-quality |
| SG_extracted_k141_2792 | Low-quality |
| SG_extracted_k141_16853 | Not-determined |
| SG_extracted_k141_40444 | Not-determined |
| SG_extracted_k141_26205 | Not-determined |
| SG_extracted_k141_23884 | Not-determined |
| SG_extracted_k141_12146 | Not-determined |
| SG_extracted_k141_16857 | Not-determined |
| SG_extracted_k141_33459 | Not-determined |
| SG_extracted_k141_45112 | Not-determined |
| SG_extracted_k141_2801 | Not-determined |
| SG_extracted_k141_40450 | Not-determined |
| SG_extracted_k141_19049 | Not-determined |
| SG_extracted_k141_23894 | Not-determined |
| SG_extracted_k141_42797 | Not-determined |
| SG_extracted_k141_19053 | Not-determined |
| SG_extracted_k141_40453 | Low-quality |
| SG_extracted_k141_16858 | Not-determined |
| SG_extracted_k141_52295 | Not-determined |
| SG_extracted_k141_42802 | Low-quality |
| SG_extracted_k141_26218 | Not-determined |
| SG_extracted_k141_2816 | Not-determined |
| SG_extracted_k141_2819 | Not-determined |
| SG_extracted_k141_23909 | Not-determined |
| SG_extracted_k141_40457 | Low-quality |
| SG_extracted_k141_52304 | Not-determined |
| SG_extracted_k141_2825 | Not-determined |
| SG_extracted_k141_9737 | Low-quality |
| SG_extracted_k141_14535 | Not-determined |
| SG_extracted_k141_9738 | Not-determined |
| SG_extracted_k141_16870 | Low-quality |
| SG_extracted_k141_52313 | Low-quality |
| SG_extracted_k141_38212 | Not-determined |
| SG_extracted_k141_52316 | Not-determined |
| SG_extracted_k141_12174 | Not-determined |
| SG_extracted_k141_26226 | Not-determined |
| SG_extracted_k141_2834 | Low-quality |
| SG_extracted_k141_45120 | Not-determined |
| SG_extracted_k141_2837 | Low-quality |
| SG_extracted_k141_2838 | Not-determined |
| SG_extracted_k141_2841 | Not-determined |
| SG_extracted_k141_40472 | Not-determined |
| SG_extracted_k141_42832 | Not-determined |
| SG_extracted_k141_35650 | Not-determined |
| SG_extracted_k141_2847 | Not-determined |
| SG_extracted_k141_42836 | Not-determined |
| SG_extracted_k141_42838 | Not-determined |
| SG_extracted_k141_2854 | Not-determined |
| SG_extracted_k141_40487 | Not-determined |
| SG_extracted_k141_2859 | Not-determined |
| SG_extracted_k141_9740 | Not-determined |
| SG_extracted_k141_9741 | Not-determined |
| SG_extracted_k141_9743 | Not-determined |
| SG_extracted_k141_52332 | Low-quality |
| SG_extracted_k141_40494 | Not-determined |
| SG_extracted_k141_42844 | Low-quality |
| SG_extracted_k141_52338 | Not-determined |
| SG_extracted_k141_52339 | Not-determined |
| SG_extracted_k141_2868 | Not-determined |
| SG_extracted_k141_52342 | Not-determined |
| SG_extracted_k141_40502 | Not-determined |
| SG_extracted_k141_40504 | Not-determined |
| SG_extracted_k141_14556 | Not-determined |
| SG_extracted_k141_42848 | Not-determined |
| SG_extracted_k141_2875 | Not-determined |
| SG_extracted_k141_2876 | Not-determined |
| SG_extracted_k141_52349 | Low-quality |
| SG_extracted_k141_52350 | Not-determined |
| SG_extracted_k141_40515 | Not-determined |
| SG_extracted_k141_7275 | Not-determined |
| SG_extracted_k141_52356 | Not-determined |
| SG_extracted_k141_40522 | Not-determined |
| SG_extracted_k141_40524 | Not-determined |
| SG_extracted_k141_7280 | Not-determined |
| SG_extracted_k141_2885 | Not-determined |
| SG_extracted_k141_23921 | Not-determined |
| SG_extracted_k141_52366 | Not-determined |
| SG_extracted_k141_7284 | Not-determined |
| SG_extracted_k141_52370 | Not-determined |
| SG_extracted_k141_45152 | Low-quality |
| SG_extracted_k141_52371 | Not-determined |
| SG_extracted_k141_23927 | Not-determined |
| SG_extracted_k141_7287 | Not-determined |
| SG_extracted_k141_45162 | Not-determined |
| SG_extracted_k141_40535 | Not-determined |
| SG_extracted_k141_2897 | Not-determined |
| SG_extracted_k141_23938 | Not-determined |
| SG_extracted_k141_40543 | Not-determined |
| SG_extracted_k141_52380 | Not-determined |
| SG_extracted_k141_28618 | Not-determined |
| SG_extracted_k141_12187 | Not-determined |
| SG_extracted_k141_16882 | Not-determined |
| SG_extracted_k141_16883 | Not-determined |
| SG_extracted_k141_40552 | Not-determined |
| SG_extracted_k141_4960 | Not-determined |
| SG_extracted_k141_23944 | Not-determined |
| SG_extracted_k141_16891 | Not-determined |
| SG_extracted_k141_12196 | Not-determined |
| SG_extracted_k141_47314 | Not-determined |
| SG_extracted_k141_23949 | Not-determined |
| SG_extracted_k141_23952 | Not-determined |
| SG_extracted_k141_35667 | Low-quality |
| SG_extracted_k141_23955 | Not-determined |
| SG_extracted_k141_16896 | Not-determined |
| SG_extracted_k141_9774 | Not-determined |
| SG_extracted_k141_45165 | Not-determined |
| SG_extracted_k141_14579 | Not-determined |
| SG_extracted_k141_9780 | Not-determined |
| SG_extracted_k141_12208 | Not-determined |
| SG_extracted_k141_52400 | Not-determined |
| SG_extracted_k141_9783 | Not-determined |
| SG_extracted_k141_19066 | Not-determined |
| SG_extracted_k141_52404 | Not-determined |
| SG_extracted_k141_52410 | Not-determined |
| SG_extracted_k141_12217 | Not-determined |
| SG_extracted_k141_45179 | Not-determined |
| SG_extracted_k141_45180 | Not-determined |
| SG_extracted_k141_35678 | Not-determined |
| SG_extracted_k141_45184 | Not-determined |
| SG_extracted_k141_45189 | Not-determined |
| SG_extracted_k141_45193 | Not-determined |
| SG_extracted_k141_52412 | Low-quality |
| SG_extracted_k141_45194 | Not-determined |
| SG_extracted_k141_38239 | Not-determined |
| SG_extracted_k141_35681 | Low-quality |
| SG_extracted_k141_35682 | Not-determined |
| SG_extracted_k141_45202 | Not-determined |
| SG_extracted_k141_33466 | Not-determined |
| SG_extracted_k141_544 | Not-determined |
| SG_extracted_k141_546 | Not-determined |
| SG_extracted_k141_23976 | Low-quality |
| SG_extracted_k141_23977 | Low-quality |
| SG_extracted_k141_42882 | Not-determined |
| SG_extracted_k141_42883 | Not-determined |
| SG_extracted_k141_7322 | Not-determined |
| SG_extracted_k141_19069 | Not-determined |
| SG_extracted_k141_7326 | Not-determined |
| SG_extracted_k141_23985 | Not-determined |
| SG_extracted_k141_33469 | Not-determined |
| SG_extracted_k141_33471 | Not-determined |
| SG_extracted_k141_12232 | Not-determined |
| SG_extracted_k141_14617 | Not-determined |
| SG_extracted_k141_47331 | Not-determined |
| SG_extracted_k141_47332 | Not-determined |
| SG_extracted_k141_19076 | Not-determined |
| SG_extracted_k141_14620 | Not-determined |
| SG_extracted_k141_558 | Low-quality |
| SG_extracted_k141_12240 | Not-determined |
| SG_extracted_k141_14625 | Not-determined |
| SG_extracted_k141_7330 | Not-determined |
| SG_extracted_k141_42896 | Not-determined |
| SG_extracted_k141_16923 | Not-determined |
| SG_extracted_k141_16924 | Not-determined |
| SG_extracted_k141_7331 | Low-quality |
| SG_extracted_k141_52430 | Not-determined |
| SG_extracted_k141_24006 | Not-determined |
| SG_extracted_k141_24007 | Not-determined |
| SG_extracted_k141_4985 | Not-determined |
| SG_extracted_k141_52434 | Low-quality |
| SG_extracted_k141_569 | Not-determined |
| SG_extracted_k141_42902 | Not-determined |
| SG_extracted_k141_570 | Not-determined |
| SG_extracted_k141_28644 | Not-determined |
| SG_extracted_k141_52441 | Not-determined |
| SG_extracted_k141_571 | Not-determined |
| SG_extracted_k141_2925 | Not-determined |
| SG_extracted_k141_12249 | Low-quality |
| SG_extracted_k141_573 | Not-determined |
| SG_extracted_k141_12250 | Not-determined |
| SG_extracted_k141_35688 | Not-determined |
| SG_extracted_k141_52446 | Not-determined |
| SG_extracted_k141_47345 | Low-quality |
| SG_extracted_k141_4992 | Not-determined |
| SG_extracted_k141_28652 | Not-determined |
| SG_extracted_k141_14632 | Not-determined |
| SG_extracted_k141_52448 | Not-determined |
| SG_extracted_k141_12255 | Low-quality |
| SG_extracted_k141_28653 | Low-quality |
| SG_extracted_k141_4994 | Not-determined |
| SG_extracted_k141_33481 | Low-quality |
| SG_extracted_k141_49716 | Not-determined |
| SG_extracted_k141_2934 | Low-quality |
| SG_extracted_k141_49720 | Not-determined |
| SG_extracted_k141_14636 | Low-quality |
| SG_extracted_k141_49722 | Not-determined |
| SG_extracted_k141_16939 | Not-determined |
| SG_extracted_k141_33488 | Not-determined |
| SG_extracted_k141_16940 | Not-determined |
| SG_extracted_k141_33489 | Not-determined |
| SG_extracted_k141_52452 | Not-determined |
| SG_extracted_k141_14642 | Low-quality |
| SG_extracted_k141_2942 | Low-quality |
| SG_extracted_k141_28660 | Low-quality |
| SG_extracted_k141_28664 | Not-determined |
| SG_extracted_k141_16944 | Not-determined |
| SG_extracted_k141_2950 | Not-determined |
| SG_extracted_k141_16947 | Not-determined |
| SG_extracted_k141_2953 | Low-quality |
| SG_extracted_k141_47356 | Not-determined |
| SG_extracted_k141_52461 | Not-determined |
| SG_extracted_k141_38253 | Not-determined |
| SG_extracted_k141_49733 | Not-determined |
| SG_extracted_k141_16953 | Not-determined |
| SG_extracted_k141_28677 | Not-determined |
| SG_extracted_k141_16954 | Not-determined |
| SG_extracted_k141_47359 | Not-determined |
| SG_extracted_k141_14660 | Not-determined |
| SG_extracted_k141_2966 | Not-determined |

| | |
|---|---|
| S6_extracted_k141_28681 | Not-determined |
| S6_extracted_k141_16960 | Not-determined |
| S6_extracted_k141_26230 | Not-determined |
| S6_extracted_k141_33492 | Not-determined |
| S6_extracted_k141_12270 | Not-determined |
| S6_extracted_k141_7333 | Low-quality |
| S6_extracted_k141_33499 | Not-determined |
| S6_extracted_k141_16963 | Not-determined |
| S6_extracted_k141_582 | Not-determined |
| S6_extracted_k141_16964 | Not-determined |
| S6_extracted_k141_7339 | Not-determined |
| S6_extracted_k141_12278 | Not-determined |
| S6_extracted_k141_38259 | Low-quality |
| S6_extracted_k141_7346 | Not-determined |
| S6_extracted_k141_47365 | Not-determined |
| S6_extracted_k141_52482 | Not-determined |
| S6_extracted_k141_12287 | Not-determined |
| S6_extracted_k141_12288 | Not-determined |
| S6_extracted_k141_52488 | Not-determined |
| S6_extracted_k141_2980 | Not-determined |
| S6_extracted_k141_45213 | Not-determined |
| S6_extracted_k141_26244 | Not-determined |
| S6_extracted_k141_596 | Not-determined |
| S6_extracted_k141_7353 | Not-determined |
| S6_extracted_k141_2986 | Not-determined |
| S6_extracted_k141_2988 | Not-determined |
| S6_extracted_k141_2990 | Not-determined |
| S6_extracted_k141_2992 | Low-quality |
| S6_extracted_k141_12296 | Not-determined |
| S6_extracted_k141_603 | Not-determined |
| S6_extracted_k141_605 | Not-determined |
| S6_extracted_k141_45219 | Low-quality |
| S6_extracted_k141_28707 | Not-determined |
| S6_extracted_k141_9801 | Not-determined |
| S6_extracted_k141_47376 | Not-determined |
| S6_extracted_k141_7369 | Low-quality |
| S6_extracted_k141_47379 | Not-determined |
| S6_extracted_k141_612 | Not-determined |
| S6_extracted_k141_9810 | Not-determined |
| S6_extracted_k141_45227 | Not-determined |
| S6_extracted_k141_9813 | Not-determined |
| S6_extracted_k141_47387 | Not-determined |
| S6_extracted_k141_16992 | Not-determined |
| S6_extracted_k141_24015 | Not-determined |
| S6_extracted_k141_24017 | Low-quality |
| S6_extracted_k141_24019 | Not-determined |
| S6_extracted_k141_5023 | Low-quality |
| S6_extracted_k141_5024 | Not-determined |
| S6_extracted_k141_47395 | Not-determined |
| S6_extracted_k141_47398 | Not-determined |
| S6_extracted_k141_9825 | Not-determined |
| S6_extracted_k141_5030 | Not-determined |
| S6_extracted_k141_7378 | Not-determined |
| S6_extracted_k141_28708 | Not-determined |
| S6_extracted_k141_26272 | Not-determined |
| S6_extracted_k141_9833 | Low-quality |
| S6_extracted_k141_9835 | Low-quality |
| S6_extracted_k141_7386 | Not-determined |
| S6_extracted_k141_7387 | Not-determined |
| S6_extracted_k141_5037 | Not-determined |
| S6_extracted_k141_26275 | Not-determined |
| S7_extracted_19866 | Medium-quality |
| S6_extracted_k141_5039 | Not-determined |
| S6_extracted_k141_12336 | Low-quality |
| S6_extracted_k141_47410 | Not-determined |
| S6_extracted_k141_26278 | Low-quality |
| S6_extracted_k141_26279 | Low-quality |
| S6_extracted_k141_52547 | Low-quality |
| S6_extracted_k141_45245 | Not-determined |
| S6_extracted_k141_52548 | Not-determined |
| S6_extracted_k141_52549 | Low-quality |
| S6_extracted_k141_26280 | Not-determined |
| S6_extracted_k141_24034 | Not-determined |
| S6_extracted_k141_17017 | Not-determined |
| S6_extracted_k141_26282 | Not-determined |
| S6_extracted_k141_30873 | Not-determined |
| S6_extracted_k141_33564 | Not-determined |
| S6_extracted_k141_52556 | Not-determined |
| S6_extracted_k141_12346 | Not-determined |
| S6_extracted_k141_52558 | Low-quality |
| S6_extracted_k141_28721 | Not-determined |
| S6_extracted_k141_628 | Not-determined |
| S6_extracted_k141_24037 | Low-quality |
| S6_extracted_k141_30879 | Low-quality |
| S6_extracted_k141_17021 | Not-determined |
| S6_extracted_k141_28722 | Not-determined |
| S6_extracted_k141_52560 | Not-determined |
| S6_extracted_k141_52561 | Not-determined |
| S6_extracted_k141_33568 | Not-determined |
| S6_extracted_k141_52562 | Low-quality |
| S6_extracted_k141_47423 | Not-determined |
| S6_extracted_k141_47424 | Not-determined |
| S6_extracted_k141_12352 | Low-quality |
| S6_extracted_k141_28724 | Not-determined |
| S6_extracted_k141_47425 | Low-quality |
| S6_extracted_k141_12355 | Not-determined |
| S6_extracted_k141_3011 | Not-determined |
| S6_extracted_k141_33571 | Not-determined |
| S6_extracted_k141_30881 | Not-determined |
| S6_extracted_k141_47426 | Low-quality |
| S6_extracted_k141_12357 | Not-determined |
| S6_extracted_k141_5049 | Not-determined |
| S6_extracted_k141_5050 | Not-determined |
| S6_extracted_k141_7414 | Not-determined |
| S6_extracted_k141_54291 | Low-quality |
| S6_extracted_k141_12361 | Not-determined |
| S6_extracted_k141_52575 | Not-determined |
| S6_extracted_k141_52576 | Low-quality |
| S6_extracted_k141_3017 | Low-quality |
| S6_extracted_k141_12364 | Not-determined |
| S6_extracted_k141_52578 | Not-determined |
| S6_extracted_k141_54296 | Not-determined |
| S6_extracted_k141_30888 | Not-determined |
| S6_extracted_k141_28728 | Not-determined |
| S6_extracted_k141_28729 | Not-determined |
| S6_extracted_k141_12366 | Not-determined |
| S6_extracted_k141_644 | Low-quality |
| S6_extracted_k141_52589 | Not-determined |
| S6_extracted_k141_12373 | Not-determined |
| S6_extracted_k141_52590 | Not-determined |
| S6_extracted_k141_3030 | Not-determined |
| S6_extracted_k141_40602 | Not-determined |
| S6_extracted_k141_30896 | Low-quality |
| S6_extracted_k141_26292 | Not-determined |
| S6_extracted_k141_30897 | Low-quality |
| S6_extracted_k141_21393 | Not-determined |
| S6_extracted_k141_47450 | Not-determined |
| S6_extracted_k141_17038 | Not-determined |
| S6_extracted_k141_28741 | Not-determined |
| S6_extracted_k141_45275 | Not-determined |
| S6_extracted_k141_28743 | Low-quality |
| S6_extracted_k141_30907 | Not-determined |
| S6_extracted_k141_38301 | Not-determined |
| S6_extracted_k141_54312 | Not-determined |
| S6_extracted_k141_33589 | Not-determined |
| S6_extracted_k141_33591 | Not-determined |
| S6_extracted_k141_54315 | Not-determined |
| S6_extracted_k141_21399 | Not-determined |
| S6_extracted_k141_30915 | Not-determined |
| S6_extracted_k141_54319 | Not-determined |
| S6_extracted_k141_45287 | Low-quality |
| S6_extracted_k141_54327 | Not-determined |
| S6_extracted_k141_3047 | Not-determined |
| S6_extracted_k141_33595 | Not-determined |
| S6_extracted_k141_33596 | Not-determined |
| S6_extracted_k141_3048 | Not-determined |
| S6_extracted_k141_45291 | Not-determined |
| S6_extracted_k141_30923 | Not-determined |
| S6_extracted_k141_30924 | Not-determined |
| S6_extracted_k141_38305 | Low-quality |
| S6_extracted_k141_47472 | Not-determined |
| S6_extracted_k141_47475 | Not-determined |
| S6_extracted_k141_38309 | Not-determined |
| S6_extracted_k141_28745 | Not-determined |
| S6_extracted_k141_30928 | Not-determined |
| S6_extracted_k141_28748 | Not-determined |
| S6_extracted_k141_3050 | Low-quality |
| S6_extracted_k141_30931 | Not-determined |
| S6_extracted_k141_38314 | Not-determined |
| S6_extracted_k141_33606 | Not-determined |
| S6_extracted_k141_49744 | Low-quality |
| S6_extracted_k141_12397 | Not-determined |
| S6_extracted_k141_3054 | Not-determined |
| S6_extracted_k141_9853 | Not-determined |
| S6_extracted_k141_9854 | Not-determined |
| S6_extracted_k141_54342 | Not-determined |
| S6_extracted_k141_3057 | Not-determined |
| S6_extracted_k141_28756 | Not-determined |
| S6_extracted_k141_26321 | Low-quality |
| S6_extracted_k141_49752 | Not-determined |
| S6_extracted_k141_38323 | Low-quality |
| S6_extracted_k141_42925 | Not-determined |
| S6_extracted_k141_38326 | Not-determined |
| S6_extracted_k141_28766 | Not-determined |
| S6_extracted_k141_54349 | Not-determined |
| S6_extracted_k141_38328 | Not-determined |
| S6_extracted_k141_54350 | Low-quality |
| S6_extracted_k141_24091 | Low-quality |
| S6_extracted_k141_47503 | Not-determined |
| S6_extracted_k141_677 | Not-determined |
| S6_extracted_k141_678 | Not-determined |
| S6_extracted_k141_9869 | Not-determined |
| S6_extracted_k141_680 | Not-determined |
| S6_extracted_k141_47510 | Not-determined |
| S6_extracted_k141_47513 | Not-determined |
| S6_extracted_k141_681 | Low-quality |
| S6_extracted_k141_54357 | Not-determined |
| S6_extracted_k141_38333 | Low-quality |
| S6_extracted_k141_683 | Not-determined |
| S6_extracted_k141_685 | Not-determined |
| S6_extracted_k141_30944 | Not-determined |
| S6_extracted_k141_24103 | Not-determined |
| S12_extracted_k141_187882 | Medium-quality |
| S6_extracted_k141_49757 | Low-quality |
| S6_extracted_k141_38337 | Not-determined |
| S6_extracted_k141_52606 | Not-determined |
| S6_extracted_k141_12423 | Not-determined |
| S6_extracted_k141_30948 | Not-determined |
| S6_extracted_k141_52611 | Not-determined |
| S6_extracted_k141_12427 | Not-determined |
| S6_extracted_k141_24110 | Not-determined |
| S6_extracted_k141_49770 | Not-determined |
| S6_extracted_k141_17058 | Not-determined |
| S6_extracted_k141_12433 | Not-determined |
| S6_extracted_k141_30954 | Not-determined |
| S6_extracted_k141_49772 | Low-quality |
| S6_extracted_k141_38342 | Not-determined |
| S6_extracted_k141_49773 | Low-quality |
| S6_extracted_k141_9889 | Not-determined |
| S6_extracted_k141_45335 | Not-determined |
| S6_extracted_k141_45337 | Not-determined |
| S6_extracted_k141_52621 | Not-determined |
| S6_extracted_k141_9893 | Not-determined |
| S6_extracted_k141_9895 | Not-determined |
| S6_extracted_k141_52623 | Not-determined |
| S6_extracted_k141_9896 | Not-determined |
| S6_extracted_k141_9897 | Not-determined |
| S6_extracted_k141_9899 | Not-determined |
| S6_extracted_k141_30969 | Not-determined |
| S6_extracted_k141_12448 | Not-determined |
| S6_extracted_k141_38348 | Not-determined |
| S6_extracted_k141_24121 | Not-determined |
| S6_extracted_k141_26328 | Not-determined |
| S6_extracted_k141_12452 | Not-determined |
| S6_extracted_k141_30977 | Not-determined |
| S6_extracted_k141_17071 | Not-determined |
| S6_extracted_k141_12456 | Not-determined |
| S6_extracted_k141_47522 | Not-determined |
| S6_extracted_k141_26336 | Not-determined |
| S6_extracted_k141_54367 | Low-quality |
| S6_extracted_k141_49797 | Not-determined |
| S6_extracted_k141_54371 | Not-determined |
| S6_extracted_k141_12470 | Not-determined |
| S6_extracted_k141_47531 | Not-determined |
| S6_extracted_k141_5080 | Not-determined |
| S6_extracted_k141_21406 | Not-determined |
| S6_extracted_k141_42944 | Not-determined |
| S6_extracted_k141_49804 | Not-determined |
| S6_extracted_k141_24139 | Low-quality |
| S6_extracted_k141_45343 | Not-determined |
| S6_extracted_k141_14693 | Low-quality |
| S6_extracted_k141_26345 | Low-quality |
| S6_extracted_k141_47540 | Low-quality |
| S6_extracted_k141_21411 | Not-determined |
| S6_extracted_k141_54378 | Not-determined |
| S6_extracted_k141_14695 | Not-determined |
| S6_extracted_k141_30992 | Not-determined |
| S6_extracted_k141_718 | Not-determined |
| S6_extracted_k141_7423 | Not-determined |
| S6_extracted_k141_719 | Not-determined |
| S6_extracted_k141_42952 | Not-determined |
| S6_extracted_k141_52644 | Not-determined |
| S6_extracted_k141_721 | Low-quality |
| S6_extracted_k141_54388 | Not-determined |
| S6_extracted_k141_21422 | Low-quality |
| S6_extracted_k141_24154 | Not-determined |
| S6_extracted_k141_54391 | Not-determined |
| S6_extracted_k141_54392 | Not-determined |
| S6_extracted_k141_28778 | Not-determined |
| S6_extracted_k141_725 | Not-determined |
| S6_extracted_k141_24157 | Not-determined |
| S6_extracted_k141_726 | Not-determined |
| S6_extracted_k141_17087 | Not-determined |
| S6_extracted_k141_49824 | Not-determined |
| S6_extracted_k141_52656 | Not-determined |
| S6_extracted_k141_14707 | Not-determined |
| S6_extracted_k141_42957 | Not-determined |
| S6_extracted_k141_49832 | Low-quality |
| S6_extracted_k141_26350 | Not-determined |
| S6_extracted_k141_736 | Not-determined |
| S6_extracted_k141_54413 | Not-determined |
| S6_extracted_k141_49843 | Not-determined |
| S6_extracted_k141_35768 | Not-determined |
| S6_extracted_k141_26354 | Not-determined |
| S6_extracted_k141_54416 | Not-determined |
| S6_extracted_k141_12496 | Not-determined |
| S6_extracted_k141_24167 | Low-quality |
| S6_extracted_k141_12497 | Not-determined |
| S6_extracted_k141_26356 | Low-quality |
| S6_extracted_k141_14721 | Not-determined |
| S6_extracted_k141_14722 | Not-determined |
| S6_extracted_k141_12498 | Not-determined |
| S6_extracted_k141_54421 | Not-determined |
| S6_extracted_k141_14723 | Low-quality |
| S6_extracted_k141_5084 | Not-determined |
| S6_extracted_k141_28787 | Low-quality |
| S6_extracted_k141_5088 | Not-determined |
| S6_extracted_k141_741 | Not-determined |
| S6_extracted_k141_42978 | Not-determined |
| S6_extracted_k141_42985 | Low-quality |
| S6_extracted_k141_35784 | Low-quality |
| S6_extracted_k141_28795 | Not-determined |
| S6_extracted_k141_14731 | Not-determined |
| S6_extracted_k141_42989 | Not-determined |
| S6_extracted_k141_14733 | Low-quality |
| S6_extracted_k141_26366 | Not-determined |
| S6_extracted_k141_5105 | Not-determined |
| S6_extracted_k141_5106 | Low-quality |
| S6_extracted_k141_42993 | Low-quality |
| S6_extracted_k141_5109 | Not-determined |
| S6_extracted_k141_14738 | Not-determined |
| S6_extracted_k141_33619 | Not-determined |
| S6_extracted_k141_12524 | Not-determined |
| S6_extracted_k141_12526 | Not-determined |
| S6_extracted_k141_14740 | Not-determined |
| S6_extracted_k141_35790 | Low-quality |
| S6_extracted_k141_7442 | Not-determined |
| S6_extracted_k141_21441 | Not-determined |
| S6_extracted_k141_35791 | Not-determined |
| S6_extracted_k141_35793 | Not-determined |
| S6_extracted_k141_26368 | Low-quality |
| S6_extracted_k141_14744 | Not-determined |
| S6_extracted_k141_17103 | Low-quality |
| S6_extracted_k141_7445 | Not-determined |
| S6_extracted_k141_35799 | Not-determined |
| S6_extracted_k141_35802 | Not-determined |
| S6_extracted_k141_43005 | Not-determined |
| S6_extracted_k141_14751 | Low-quality |
| S6_extracted_k141_43006 | Not-determined |
| S6_extracted_k141_12532 | Not-determined |
| S6_extracted_k141_7455 | Not-determined |
| S6_extracted_k141_21448 | Low-quality |
| S6_extracted_k141_14760 | Low-quality |
| S6_extracted_k141_49875 | Not-determined |
| S6_extracted_k141_7461 | Not-determined |
| S6_extracted_k141_26387 | Not-determined |
| S6_extracted_k141_49876 | Not-determined |
| S6_extracted_k141_43016 | Not-determined |
| S6_extracted_k141_49879 | Not-determined |
| S6_extracted_k141_21452 | Low-quality |
| S6_extracted_k141_26389 | Not-determined |
| S6_extracted_k141_43019 | Not-determined |
| S6_extracted_k141_5121 | Low-quality |
| S6_extracted_k141_5122 | Not-determined |
| S6_extracted_k141_49883 | Not-determined |
| S6_extracted_k141_14763 | Not-determined |
| S6_extracted_k141_49884 | Not-determined |
| S6_extracted_k141_5127 | Not-determined |
| S6_extracted_k141_7468 | Not-determined |
| S6_extracted_k141_5133 | Low-quality |
| S6_extracted_k141_12545 | Low-quality |
| S6_extracted_k141_26395 | Not-determined |
| S6_extracted_k141_7471 | Not-determined |
| S6_extracted_k141_52696 | Not-determined |
| S6_extracted_k141_19094 | Low-quality |
| S6_extracted_k141_52697 | Not-determined |
| S6_extracted_k141_12553 | Not-determined |
| S6_extracted_k141_21468 | Not-determined |
| S6_extracted_k141_26398 | Not-determined |
| S6_extracted_k141_7480 | Not-determined |
| S6_extracted_k141_21471 | Not-determined |
| S6_extracted_k141_19101 | Not-determined |
| S6_extracted_k141_753 | Not-determined |
| S6_extracted_k141_35827 | Not-determined |
| S6_extracted_k141_54435 | Not-determined |
| S6_extracted_k141_7486 | Not-determined |
| S6_extracted_k141_35830 | Low-quality |
| S6_extracted_k141_7488 | Not-determined |
| S6_extracted_k141_7489 | Not-determined |
| S6_extracted_k141_35831 | Not-determined |
| S6_extracted_k141_45373 | Not-determined |
| S6_extracted_k141_54440 | Not-determined |
| S6_extracted_k141_45378 | Not-determined |
| S6_extracted_k141_35835 | Not-determined |
| S6_extracted_k141_33630 | Not-determined |
| S6_extracted_k141_54446 | Not-determined |
| S6_extracted_k141_26404 | Not-determined |
| S6_extracted_k141_49902 | Not-determined |
| S6_extracted_k141_33632 | Not-determined |
| S6_extracted_k141_54449 | Low-quality |
| S6_extracted_k141_24177 | Not-determined |
| S6_extracted_k141_21487 | Low-quality |
| S6_extracted_k141_47554 | Low-quality |
| S6_extracted_k141_54450 | Not-determined |
| S6_extracted_k141_24178 | Not-determined |
| S6_extracted_k141_47560 | Low-quality |
| S6_extracted_k141_24179 | Not-determined |
| S6_extracted_k141_24180 | Not-determined |
| S6_extracted_k141_47563 | Not-determined |
| S6_extracted_k141_47564 | Not-determined |
| S6_extracted_k141_24182 | Not-determined |
| S6_extracted_k141_43024 | Not-determined |
| S6_extracted_k141_49914 | Not-determined |
| S6_extracted_k141_24186 | Not-determined |
| S6_extracted_k141_47569 | Not-determined |
| S6_extracted_k141_54456 | Not-determined |
| S6_extracted_k141_54458 | Not-determined |
| S6_extracted_k141_24189 | Not-determined |
| S6_extracted_k141_54462 | Low-quality |
| S6_extracted_k141_21497 | Low-quality |
| S6_extracted_k141_54465 | Not-determined |
| S6_extracted_k141_21498 | Not-determined |
| S6_extracted_k141_21509 | Not-determined |
| S6_extracted_k141_31017 | Not-determined |
| S6_extracted_k141_45388 | Not-determined |
| S6_extracted_k141_24201 | Not-determined |
| S6_extracted_k141_31018 | Not-determined |
| S6_extracted_k141_24204 | Not-determined |
| S6_extracted_k141_26417 | Low-quality |
| S6_extracted_k141_26418 | Not-determined |
| S6_extracted_k141_24211 | Not-determined |
| S6_extracted_k141_17109 | Low-quality |
| S6_extracted_k141_33661 | Not-determined |
| S6_extracted_k141_31026 | Not-determined |
| S6_extracted_k141_24218 | Not-determined |
| S6_extracted_k141_33668 | Not-determined |
| S6_extracted_k141_19105 | Not-determined |
| S6_extracted_k141_24226 | Low-quality |
| S6_extracted_k141_24227 | Not-determined |
| S6_extracted_k141_24229 | Not-determined |
| S6_extracted_k141_26430 | Not-determined |
| S6_extracted_k141_26431 | Not-determined |
| S6_extracted_k141_45405 | Low-quality |
| S6_extracted_k141_45408 | Not-determined |
| S6_extracted_k141_24234 | Not-determined |
| S6_extracted_k141_12567 | Low-quality |
| S6_extracted_k141_43029 | Not-determined |
| S6_extracted_k141_49922 | Not-determined |
| S6_extracted_k141_31039 | Not-determined |
| S6_extracted_k141_19111 | Not-determined |
| S6_extracted_k141_26436 | Not-determined |
| S6_extracted_k141_12570 | Not-determined |
| S6_extracted_k141_24236 | Not-determined |
| S6_extracted_k141_26438 | Not-determined |
| S6_extracted_k141_24238 | Low-quality |
| S6_extracted_k141_45417 | Not-determined |
| S6_extracted_k141_33678 | Not-determined |
| S6_extracted_k141_12576 | Low-quality |
| S6_extracted_k141_38366 | Not-determined |
| S6_extracted_k141_26446 | Not-determined |
| S6_extracted_k141_35846 | Not-determined |
| S6_extracted_k141_47598 | Not-determined |
| S6_extracted_k141_767 | Not-determined |
| S6_extracted_k141_40628 | Not-determined |
| S6_extracted_k141_19118 | Low-quality |
| S6_extracted_k141_54482 | Not-determined |
| S6_extracted_k141_33691 | Not-determined |
| S6_extracted_k141_43042 | Low-quality |
| S6_extracted_k141_31052 | Low-quality |
| S6_extracted_k141_21516 | Not-determined |
| S6_extracted_k141_24246 | Not-determined |
| S6_extracted_k141_49935 | Low-quality |
| S6_extracted_k141_21522 | Low-quality |
| S6_extracted_k141_12579 | Not-determined |
| S6_extracted_k141_49937 | Not-determined |
| S6_extracted_k141_35857 | Low-quality |
| S6_extracted_k141_21526 | Not-determined |
| S6_extracted_k141_21528 | Not-determined |
| S6_extracted_k141_47605 | Not-determined |
| S6_extracted_k141_24251 | Not-determined |
| S6_extracted_k141_5145 | Low-quality |
| S6_extracted_k141_49943 | Not-determined |
| S6_extracted_k141_38371 | Not-determined |
| S6_extracted_k141_47609 | Not-determined |
| S6_extracted_k141_19129 | Not-determined |
| S6_extracted_k141_47611 | Not-determined |
| S6_extracted_k141_43049 | Not-determined |
| S6_extracted_k141_24260 | Not-determined |
| S6_extracted_k141_43051 | Not-determined |
| S6_extracted_k141_49948 | Not-determined |
| S6_extracted_k141_43053 | Not-determined |
| S6_extracted_k141_5149 | Low-quality |
| S6_extracted_k141_49951 | Not-determined |
| S6_extracted_k141_7508 | Not-determined |
| S6_extracted_k141_28805 | Not-determined |
| S6_extracted_k141_19137 | Not-determined |
| S6_extracted_k141_19139 | Not-determined |
| S6_extracted_k141_31075 | Low-quality |
| S6_extracted_k141_7514 | Not-determined |
| S6_extracted_k141_31078 | Low-quality |
| S6_extracted_k141_7517 | Not-determined |
| S6_extracted_k141_12593 | Not-determined |
| S6_extracted_k141_17125 | Not-determined |
| S6_extracted_k141_17126 | Not-determined |
| S6_extracted_k141_790 | Not-determined |
| S6_extracted_k141_24283 | Not-determined |
| S6_extracted_k141_12597 | Low-quality |
| S6_extracted_k141_7528 | Not-determined |
| S6_extracted_k141_7529 | Not-determined |
| S6_extracted_k141_5157 | Not-determined |
| S6_extracted_k141_52752 | Not-determined |
| S6_extracted_k141_52754 | Not-determined |
| S6_extracted_k141_49960 | Not-determined |
| S6_extracted_k141_21559 | Not-determined |
| S6_extracted_k141_7538 | Not-determined |
| S6_extracted_k141_794 | Not-determined |
| S6_extracted_k141_17142 | Not-determined |
| S6_extracted_k141_52766 | Not-determined |
| S6_extracted_k141_796 | Not-determined |
| S6_extracted_k141_800 | Low-quality |
| S6_extracted_k141_52772 | Not-determined |
| S6_extracted_k141_3080 | Not-determined |
| S6_extracted_k141_7542 | Not-determined |
| S6_extracted_k141_24299 | Not-determined |
| S6_extracted_k141_7545 | Not-determined |
| S6_extracted_k141_54497 | Not-determined |
| S6_extracted_k141_49981 | Not-determined |
| S6_extracted_k141_5182 | Not-determined |
| S6_extracted_k141_49982 | Low-quality |
| S6_extracted_k141_807 | Not-determined |
| S6_extracted_k141_9954 | Not-determined |
| S6_extracted_k141_17150 | Low-quality |
| S6_extracted_k141_24303 | Not-determined |
| S6_extracted_k141_49984 | Not-determined |
| S6_extracted_k141_3082 | Not-determined |
| S6_extracted_k141_52780 | Not-determined |
| S6_extracted_k141_7549 | Not-determined |
| S6_extracted_k141_52781 | Not-determined |
| S6_extracted_k141_3085 | Not-determined |
| S6_extracted_k141_49994 | Low-quality |
| S6_extracted_k141_7551 | Not-determined |
| S6_extracted_k141_49995 | Not-determined |
| S6_extracted_k141_7552 | Not-determined |
| S6_extracted_k141_17157 | Not-determined |
| S6_extracted_k141_45423 | Not-determined |
| S6_extracted_k141_9971 | Not-determined |
| S6_extracted_k141_38396 | Low-quality |
| S6_extracted_k141_52789 | Not-determined |
| S6_extracted_k141_38404 | Not-determined |
| S6_extracted_k141_35884 | Not-determined |
| S6_extracted_k141_38407 | Not-determined |
| S6_extracted_k141_52796 | Low-quality |
| S6_extracted_k141_52797 | Low-quality |
| S6_extracted_k141_50009 | Low-quality |
| S6_extracted_k141_821 | Not-determined |
| S6_extracted_k141_52800 | Not-determined |
| S6_extracted_k141_31083 | Not-determined |
| S6_extracted_k141_35892 | Not-determined |
| S6_extracted_k141_52807 | Not-determined |
| S6_extracted_k141_38411 | Not-determined |
| S6_extracted_k141_45434 | Not-determined |
| S6_extracted_k141_52808 | Not-determined |
| S6_extracted_k141_45436 | Not-determined |
| S6_extracted_k141_35894 | Not-determined |
| S6_extracted_k141_50027 | Not-determined |
| S6_extracted_k141_35903 | Not-determined |
| S6_extracted_k141_45443 | Not-determined |
| S6_extracted_k141_45445 | Not-determined |
| S6_extracted_k141_50028 | Not-determined |
| S6_extracted_k141_45446 | Not-determined |
| S6_extracted_k141_824 | Low-quality |
| S6_extracted_k141_50031 | Not-determined |
| S6_extracted_k141_24310 | Not-determined |
| S6_extracted_k141_826 | Not-determined |
| S6_extracted_k141_45453 | Not-determined |
| S6_extracted_k141_45455 | Not-determined |
| S6_extracted_k141_28822 | Not-determined |
| S6_extracted_k141_35909 | Not-determined |
| S6_extracted_k141_3089 | Not-determined |
| S6_extracted_k141_24316 | Not-determined |
| S6_extracted_k141_45460 | Not-determined |
| S6_extracted_k141_5189 | Not-determined |
| S6_extracted_k141_835 | Not-determined |
| S6_extracted_k141_45462 | Not-determined |
| S6_extracted_k141_840 | Not-determined |
| S6_extracted_k141_24320 | Not-determined |
| S6_extracted_k141_35916 | Not-determined |
| S6_extracted_k141_5206 | Low-quality |
| S6_extracted_k141_45469 | Low-quality |
| S6_extracted_k141_28832 | Not-determined |
| S6_extracted_k141_24334 | Low-quality |
| S6_extracted_k141_5212 | Not-determined |
| S6_extracted_k141_24335 | Not-determined |
| S6_extracted_k141_5215 | Not-determined |
| S6_extracted_k141_17204 | Not-determined |
| S6_extracted_k141_47615 | Not-determined |
| S6_extracted_k141_40703 | Not-determined |
| S6_extracted_k141_28841 | Not-determined |
| S6_extracted_k141_843 | Not-determined |
| S6_extracted_k141_17205 | Low-quality |
| S6_extracted_k141_17206 | Not-determined |
| S6_extracted_k141_9981 | Not-determined |
| S6_extracted_k141_50054 | Not-determined |
| S6_extracted_k141_40708 | Not-determined |
| S6_extracted_k141_40710 | Not-determined |
| S6_extracted_k141_40711 | Not-determined |
| S6_extracted_k141_9985 | Not-determined |
| S6_extracted_k141_40713 | Not-determined |
| S6_extracted_k141_40717 | Not-determined |
| S6_extracted_k141_9992 | Not-determined |
| S6_extracted_k141_24355 | Not-determined |
| S6_extracted_k141_9993 | Not-determined |
| S6_extracted_k141_35943 | Not-determined |
| S6_extracted_k141_50064 | Not-determined |
| S6_extracted_k141_31096 | Not-determined |
| S6_extracted_k141_50065 | Not-determined |
| S6_extracted_k141_31097 | Not-determined |
| S6_extracted_k141_24361 | Not-determined |
| S6_extracted_k141_10003 | Not-determined |

| | |
|---|---|
| SG_extracted_k141_31100 | Not-determined |
| SG_extracted_k141_28858 | Not-determined |
| SG_extracted_k141_10008 | Low-quality |
| SG_extracted_k141_28862 | Low-quality |
| SG_extracted_k141_7578 | Low-quality |
| SG_extracted_k141_855 | Not-determined |
| SG_extracted_k141_28866 | Not-determined |
| SG_extracted_k141_10014 | Not-determined |
| SG_extracted_k141_28867 | Not-determined |
| SG_extracted_k141_28868 | Not-determined |
| SG_extracted_k141_31107 | Not-determined |
| SG_extracted_k141_24370 | Low-quality |
| SG_extracted_k141_3120 | Not-determined |
| SG_extracted_k141_3121 | Not-determined |
| SG_extracted_k141_28875 | Low-quality |
| SG_extracted_k141_7585 | Not-determined |
| SG_extracted_k141_24377 | Low-quality |
| SG_extracted_k141_40729 | Low-quality |
| SG_extracted_k141_47625 | Not-determined |
| SG_extracted_k141_864 | Not-determined |
| SG_extracted_k141_47626 | Not-determined |
| SG_extracted_k141_866 | Not-determined |
| SG_extracted_k141_868 | Not-determined |
| SG_extracted_k141_47629 | Not-determined |
| SG_extracted_k141_5259 | Not-determined |
| SG_extracted_k141_10033 | Not-determined |
| SG_extracted_k141_40735 | Not-determined |
| SG_extracted_k141_3132 | Not-determined |
| SG_extracted_k141_870 | Not-determined |
| SG_extracted_k141_3133 | Low-quality |
| SG_extracted_k141_21632 | Not-determined |
| SG_extracted_k141_50082 | Not-determined |
| SG_extracted_k141_875 | Low-quality |
| SG_extracted_k141_7601 | Not-determined |
| SG_extracted_k141_50089 | Not-determined |
| SG_extracted_k141_31131 | Not-determined |
| SG_extracted_k141_31132 | Not-determined |
| SG_extracted_k141_31133 | Not-determined |
| SG_extracted_k141_3136 | Not-determined |
| SG_extracted_k141_47645 | Not-determined |
| SG_extracted_k141_47646 | Not-determined |
| SG_extracted_k141_7609 | Not-determined |
| SG_extracted_k141_33712 | Not-determined |
| SG_extracted_k141_43064 | Not-determined |
| SG_extracted_k141_43065 | Not-determined |
| SG_extracted_k141_7614 | Not-determined |
| SG_extracted_k141_5273 | Not-determined |
| SG_extracted_k141_3139 | Not-determined |
| SG_extracted_k141_7616 | Not-determined |
| SG_extracted_k141_5275 | Not-determined |
| SG_extracted_k141_40748 | Low-quality |
| SG_extracted_k141_14819 | Not-determined |
| SG_extracted_k141_14821 | Not-determined |
| SG_extracted_k141_14823 | Not-determined |
| SG_extracted_k141_14824 | Not-determined |
| SG_extracted_k141_24382 | Not-determined |
| SG_extracted_k141_884 | Not-determined |
| SG_extracted_k141_35967 | Not-determined |
| SG_extracted_k141_3145 | Not-determined |
| SG_extracted_k141_31145 | Low-quality |
| SG_extracted_k141_3146 | Low-quality |
| SG_extracted_k141_35970 | Not-determined |
| SG_extracted_k141_40754 | Not-determined |
| SG_extracted_k141_5286 | Not-determined |
| SG_extracted_k141_43076 | Not-determined |
| SG_extracted_k141_31153 | Not-determined |
| SG_extracted_k141_38433 | Not-determined |
| SG_extracted_k141_14827 | Low-quality |
| SG_extracted_k141_21638 | Not-determined |
| SG_extracted_k141_43077 | Not-determined |
| SG_extracted_k141_43078 | Not-determined |
| SG_extracted_k141_3149 | Not-determined |
| SG_extracted_k141_14830 | Low-quality |
| SG_extracted_k141_7628 | Not-determined |
| SG_extracted_k141_31156 | Not-determined |
| SG_extracted_k141_43081 | Not-determined |
| SG_extracted_k141_7629 | Not-determined |
| SG_extracted_k141_38439 | Not-determined |
| SG_extracted_k141_35976 | Not-determined |
| SG_extracted_k141_31161 | Not-determined |
| SG_extracted_k141_21652 | Not-determined |
| SG_extracted_k141_898 | Not-determined |
| SG_extracted_k141_21653 | Not-determined |
| SG_extracted_k141_43086 | Not-determined |
| SG_extracted_k141_43088 | Low-quality |
| SG_extracted_k141_5297 | Not-determined |
| SG_extracted_k141_31168 | Not-determined |
| SG_extracted_k141_47661 | Not-determined |
| SG_extracted_k141_5299 | Not-determined |
| SG_extracted_k141_14840 | Not-determined |
| SG_extracted_k141_21656 | Not-determined |
| SG_extracted_k141_7643 | Not-determined |
| SG_extracted_k141_906 | Not-determined |
| SG_extracted_k141_35990 | Not-determined |
| SG_extracted_k141_33729 | Not-determined |
| SG_extracted_k141_31176 | Not-determined |
| SG_extracted_k141_31178 | Not-determined |
| SG_extracted_k141_21660 | Low-quality |
| SG_extracted_k141_33733 | Not-determined |
| SG_extracted_k141_7646 | Not-determined |
| SG_extracted_k141_47665 | Not-determined |
| SG_extracted_k141_915 | Low-quality |
| SG_extracted_k141_17267 | Not-determined |
| SG_extracted_k141_38449 | Low-quality |
| SG_extracted_k141_3162 | Low-quality |
| SG_extracted_k141_47672 | Not-determined |
| SG_extracted_k141_922 | Not-determined |
| SG_extracted_k141_923 | Not-determined |
| SG_extracted_k141_7659 | Low-quality |
| SG_extracted_k141_3168 | Not-determined |
| SG_extracted_k141_5306 | Not-determined |
| SG_extracted_k141_33744 | Not-determined |
| SG_extracted_k141_45525 | Not-determined |
| SG_extracted_k141_33747 | Not-determined |
| SG_extracted_k141_45527 | Not-determined |
| SG_extracted_k141_47679 | Not-determined |
| SG_extracted_k141_47681 | Not-determined |
| SG_extracted_k141_12603 | Not-determined |
| SG_extracted_k141_31202 | Not-determined |
| SG_extracted_k141_45534 | Not-determined |
| SG_extracted_k141_5317 | Not-determined |
| SG_extracted_k141_45535 | Not-determined |
| SG_extracted_k141_45538 | Not-determined |
| SG_extracted_k141_7672 | Not-determined |
| SG_extracted_k141_45547 | Not-determined |
| SG_extracted_k141_12608 | Not-determined |
| SG_extracted_k141_45552 | Low-quality |
| SG_extracted_k141_40760 | Not-determined |
| SG_extracted_k141_45556 | Not-determined |
| SG_extracted_k141_3192 | Not-determined |
| SG_extracted_k141_24395 | Not-determined |
| SG_extracted_k141_21672 | Low-quality |
| SG_extracted_k141_47690 | Low-quality |
| SG_extracted_k141_45561 | Low-quality |
| SG_extracted_k141_45562 | Not-determined |
| SG_extracted_k141_33766 | Low-quality |
| SG_extracted_k141_38458 | Not-determined |
| SG_extracted_k141_12618 | Low-quality |
| SG_extracted_k141_47693 | Not-determined |
| SG_extracted_k141_19152 | Not-determined |
| SG_extracted_k141_28901 | Low-quality |
| SG_extracted_k141_21684 | Not-determined |
| SG_extracted_k141_24404 | Not-determined |
| SG_extracted_k141_21685 | Not-determined |
| SG_extracted_k141_33774 | Not-determined |
| SG_extracted_k141_21686 | Not-determined |
| SG_extracted_k141_21687 | Not-determined |
| SG_extracted_k141_33776 | Not-determined |
| SG_extracted_k141_38465 | Not-determined |
| SG_extracted_k141_17300 | Not-determined |
| SG_extracted_k141_935 | Low-quality |
| SG_extracted_k141_24414 | Not-determined |
| SG_extracted_k141_12625 | Low-quality |
| SG_extracted_k141_19155 | Low-quality |
| SG_extracted_k141_17304 | Not-determined |
| SG_extracted_k141_17305 | Not-determined |
| SG_extracted_k141_12631 | Not-determined |
| SG_extracted_k141_50137 | Not-determined |
| SG_extracted_k141_45567 | Not-determined |
| SG_extracted_k141_943 | Not-determined |
| SG_extracted_k141_14863 | Not-determined |
| SG_extracted_k141_33783 | Not-determined |
| SG_extracted_k141_19162 | Not-determined |
| SG_extracted_k141_946 | Not-determined |
| SG_extracted_k141_47716 | Not-determined |
| SG_extracted_k141_45572 | Not-determined |
| SG_extracted_k141_14868 | Not-determined |
| SG_extracted_k141_19165 | Low-quality |
| SG_extracted_k141_3212 | Not-determined |
| SG_extracted_k141_38467 | Not-determined |
| SG_extracted_k141_17310 | Low-quality |
| SG_extracted_k141_3214 | Not-determined |
| SG_extracted_k141_19169 | Not-determined |
| SG_extracted_k141_31208 | Not-determined |
| SG_extracted_k141_3219 | Not-determined |
| SG_extracted_k141_47725 | Not-determined |
| SG_extracted_k141_3222 | Not-determined |
| SG_extracted_k141_31218 | Not-determined |
| SG_extracted_k141_961 | Low-quality |
| SG_extracted_k141_24425 | Low-quality |
| SG_extracted_k141_50147 | Not-determined |
| SG_extracted_k141_47735 | Not-determined |
| SG_extracted_k141_28936 | Not-determined |
| SG_extracted_k141_967 | Not-determined |
| SG_extracted_k141_968 | Not-determined |
| SG_extracted_k141_45612 | Not-determined |
| SG_extracted_k141_3243 | Not-determined |
| SG_extracted_k141_7704 | Low-quality |
| SG_extracted_k141_970 | Not-determined |
| SG_extracted_k141_47744 | Not-determined |
| SG_extracted_k141_33812 | Not-determined |
| SG_extracted_k141_7708 | Not-determined |
| SG_extracted_k141_31229 | Not-determined |
| SG_extracted_k141_19184 | Not-determined |
| SG_extracted_k141_7710 | Not-determined |
| SG_extracted_k141_31233 | Not-determined |
| SG_extracted_k141_33819 | Not-determined |
| SG_extracted_k141_31235 | Low-quality |
| SG_extracted_k141_19187 | Low-quality |
| SG_extracted_k141_33823 | Low-quality |
| SG_extracted_k141_19189 | Not-determined |
| SG_extracted_k141_19190 | Not-determined |
| SG_extracted_k141_24465 | Not-determined |
| SG_extracted_k141_24466 | Not-determined |
| SG_extracted_k141_33825 | Not-determined |
| SG_extracted_k141_31243 | Not-determined |
| SG_extracted_k141_28939 | Low-quality |
| SG_extracted_k141_17321 | Not-determined |
| SG_extracted_k141_19201 | Not-determined |
| SG_extracted_k141_7713 | Low-quality |
| SG_extracted_k141_3249 | Not-determined |
| SG_extracted_k141_33836 | Not-determined |
| SG_extracted_k141_3251 | Not-determined |
| SG_extracted_k141_5356 | Low-quality |
| SG_extracted_k141_3252 | Not-determined |
| SG_extracted_k141_5357 | Not-determined |
| SG_extracted_k141_5361 | Low-quality |
| SG_extracted_k141_33845 | Not-determined |
| SG_extracted_k141_40785 | Not-determined |
| SG_extracted_k141_40787 | Low-quality |
| SG_extracted_k141_17332 | Not-determined |
| SG_extracted_k141_33854 | Low-quality |
| SG_extracted_k141_28956 | Not-determined |
| SG_extracted_k141_24480 | Low-quality |
| SG_extracted_k141_33856 | Not-determined |
| SG_extracted_k141_17334 | Not-determined |
| SG_extracted_k141_28958 | Not-determined |
| SG_extracted_k141_17337 | Not-determined |
| SG_extracted_k141_28960 | Low-quality |
| SG_extracted_k141_3262 | Not-determined |
| SG_extracted_k141_17339 | Not-determined |
| S7_extracted_32396 | Medium-quality |
| SG_extracted_k141_28965 | Not-determined |
| SG_extracted_k141_50164 | Not-determined |
| SG_extracted_k141_31245 | Low-quality |
| SG_extracted_k141_50169 | Low-quality |
| SG_extracted_k141_50171 | Not-determined |
| SG_extracted_k141_28968 | Not-determined |
| S8_extracted_k141_153696 | Medium-quality |
| SG_extracted_k141_3279 | Not-determined |
| SG_extracted_k141_24489 | Low-quality |
| SG_extracted_k141_24495 | Not-determined |
| SG_extracted_k141_36017 | Low-quality |
| SG_extracted_k141_26449 | Low-quality |
| SG_extracted_k141_40795 | Not-determined |
| SG_extracted_k141_40796 | Not-determined |
| SG_extracted_k141_33873 | Not-determined |
| SG_extracted_k141_54514 | Not-determined |
| SG_extracted_k141_54515 | Not-determined |
| SG_extracted_k141_5378 | Not-determined |
| SG_extracted_k141_10069 | Not-determined |
| SG_extracted_k141_10070 | Not-determined |
| SG_extracted_k141_26458 | Not-determined |
| SG_extracted_k141_5382 | Not-determined |
| SG_extracted_k141_40804 | Not-determined |
| SG_extracted_k141_45624 | Not-determined |
| SG_extracted_k141_45626 | Low-quality |
| SG_extracted_k141_33881 | Not-determined |
| SG_extracted_k141_45629 | Not-determined |
| SG_extracted_k141_38481 | Not-determined |
| SG_extracted_k141_38484 | Not-determined |
| SG_extracted_k141_50180 | Low-quality |
| SG_extracted_k141_38485 | Not-determined |
| SG_extracted_k141_33884 | Not-determined |
| SG_extracted_k141_38486 | Not-determined |
| SG_extracted_k141_38488 | Not-determined |
| SG_extracted_k141_38490 | Not-determined |
| SG_extracted_k141_54519 | Not-determined |
| SG_extracted_k141_45636 | Not-determined |
| SG_extracted_k141_54522 | Not-determined |
| SG_extracted_k141_54523 | Not-determined |
| SG_extracted_k141_3289 | Not-determined |
| SG_extracted_k141_54534 | Not-determined |
| SG_extracted_k141_17354 | Not-determined |
| SG_extracted_k141_24514 | Not-determined |
| SG_extracted_k141_17357 | Low-quality |
| SG_extracted_k141_43091 | Not-determined |
| SG_extracted_k141_38493 | Not-determined |
| SG_extracted_k141_33899 | Low-quality |
| SG_extracted_k141_33900 | Low-quality |
| SG_extracted_k141_43093 | Not-determined |
| SG_extracted_k141_10091 | Not-determined |
| SG_extracted_k141_19210 | Not-determined |
| SG_extracted_k141_40826 | Low-quality |
| SG_extracted_k141_17364 | Low-quality |
| SG_extracted_k141_40831 | Not-determined |
| SG_extracted_k141_12663 | Not-determined |
| SG_extracted_k141_54540 | Not-determined |
| SG_extracted_k141_26474 | Low-quality |
| SG_extracted_k141_12665 | Low-quality |
| SG_extracted_k141_12670 | Low-quality |
| SG_extracted_k141_28982 | Low-quality |
| SG_extracted_k141_33911 | Not-determined |
| SG_extracted_k141_28988 | Not-determined |
| SG_extracted_k141_28989 | Not-determined |
| SG_extracted_k141_26482 | Not-determined |
| SG_extracted_k141_10110 | Not-determined |
| SG_extracted_k141_28992 | Not-determined |
| SG_extracted_k141_40840 | Low-quality |
| SG_extracted_k141_43110 | Low-quality |
| SG_extracted_k141_10112 | Not-determined |
| SG_extracted_k141_3319 | Not-determined |
| SG_extracted_k141_31264 | Not-determined |
| SG_extracted_k141_29000 | Not-determined |
| SG_extracted_k141_24524 | Not-determined |
| SG_extracted_k141_3324 | Low-quality |
| SG_extracted_k141_29002 | Not-determined |
| SG_extracted_k141_10118 | Not-determined |
| SG_extracted_k141_10119 | Not-determined |
| SG_extracted_k141_17372 | Not-determined |
| SG_extracted_k141_43116 | Not-determined |
| SG_extracted_k141_10121 | Not-determined |
| SG_extracted_k141_17375 | Not-determined |
| SG_extracted_k141_24530 | Low-quality |
| SG_extracted_k141_31279 | Not-determined |
| SG_extracted_k141_31280 | Not-determined |
| SG_extracted_k141_36062 | Not-determined |
| SG_extracted_k141_31282 | Low-quality |
| SG_extracted_k141_24535 | Not-determined |
| SG_extracted_k141_10129 | Not-determined |
| SG_extracted_k141_10132 | Not-determined |
| SG_extracted_k141_26490 | Not-determined |
| SG_extracted_k141_14882 | Not-determined |
| SG_extracted_k141_26494 | Not-determined |
| SG_extracted_k141_26495 | Not-determined |
| SG_extracted_k141_14892 | Not-determined |
| SG_extracted_k141_14894 | Not-determined |
| SG_extracted_k141_17397 | Not-determined |
| SG_extracted_k141_24549 | Not-determined |
| SG_extracted_k141_26505 | Not-determined |
| SG_extracted_k141_26506 | Low-quality |
| SG_extracted_k141_12720 | Not-determined |
| SG_extracted_k141_26507 | Low-quality |
| SG_extracted_k141_7738 | Not-determined |
| SG_extracted_k141_26508 | Not-determined |
| SG_extracted_k141_36068 | Not-determined |
| SG_extracted_k141_36069 | Not-determined |
| SG_extracted_k141_10146 | Not-determined |
| SG_extracted_k141_12725 | Not-determined |
| SG_extracted_k141_7742 | Low-quality |
| SG_extracted_k141_29018 | Not-determined |
| SG_extracted_k141_7745 | Not-determined |
| SG_extracted_k141_40841 | Not-determined |
| SG_extracted_k141_29023 | Not-determined |
| SG_extracted_k141_17416 | Not-determined |
| SG_extracted_k141_12737 | Not-determined |
| SG_extracted_k141_45663 | Low-quality |
| SG_extracted_k141_12738 | Not-determined |
| SG_extracted_k141_17420 | Not-determined |
| SG_extracted_k141_26520 | Not-determined |
| SG_extracted_k141_17423 | Not-determined |
| SG_extracted_k141_12742 | Not-determined |
| SG_extracted_k141_12747 | Not-determined |
| S11_extracted_k141_39861 | Medium-quality |
| SG_extracted_k141_24556 | Not-determined |
| SG_extracted_k141_40856 | Low-quality |
| SG_extracted_k141_45673 | Not-determined |
| SG_extracted_k141_24557 | Low-quality |
| SG_extracted_k141_24559 | Not-determined |
| SG_extracted_k141_24560 | Not-determined |
| SG_extracted_k141_45675 | Not-determined |
| SG_extracted_k141_45677 | Not-determined |
| SG_extracted_k141_24564 | Not-determined |
| SG_extracted_k141_14905 | Not-determined |
| SG_extracted_k141_14906 | Not-determined |
| SG_extracted_k141_45685 | Not-determined |
| SG_extracted_k141_45686 | Low-quality |
| SG_extracted_k141_17436 | Low-quality |
| SG_extracted_k141_24569 | Not-determined |
| SG_extracted_k141_7767 | Not-determined |
| SG_extracted_k141_17446 | Not-determined |
| SG_extracted_k141_10180 | Not-determined |
| SG_extracted_k141_38512 | Low-quality |
| SG_extracted_k141_7770 | Not-determined |
| SG_extracted_k141_45695 | Not-determined |
| SG_extracted_k141_45696 | Not-determined |
| SG_extracted_k141_43128 | Not-determined |
| SG_extracted_k141_24584 | Not-determined |
| SG_extracted_k141_45707 | Not-determined |
| SG_extracted_k141_52834 | Not-determined |
| SG_extracted_k141_45710 | Not-determined |
| SG_extracted_k141_36075 | Not-determined |
| SG_extracted_k141_52837 | Not-determined |
| SG_extracted_k141_3335 | Low-quality |
| SG_extracted_k141_7780 | Not-determined |
| SG_extracted_k141_7784 | Not-determined |
| SG_extracted_k141_14913 | Not-determined |
| SG_extracted_k141_52844 | Not-determined |
| SG_extracted_k141_45714 | Not-determined |
| SG_extracted_k141_33955 | Not-determined |
| SG_extracted_k141_29033 | Not-determined |
| SG_extracted_k141_17464 | Not-determined |
| SG_extracted_k141_24591 | Not-determined |
| SG_extracted_k141_12754 | Not-determined |
| SG_extracted_k141_43145 | Not-determined |
| SG_extracted_k141_38522 | Low-quality |
| SG_extracted_k141_24598 | Not-determined |
| SG_extracted_k141_29041 | Not-determined |
| SG_extracted_k141_43150 | Not-determined |
| SG_extracted_k141_7794 | Low-quality |
| SG_extracted_k141_14926 | Not-determined |
| SG_extracted_k141_38528 | Not-determined |
| SG_extracted_k141_3345 | Not-determined |
| SG_extracted_k141_40865 | Not-determined |
| SG_extracted_k141_29048 | Not-determined |
| SG_extracted_k141_40866 | Not-determined |
| SG_extracted_k141_3346 | Low-quality |
| SG_extracted_k141_52863 | Not-determined |
| SG_extracted_k141_3347 | Not-determined |
| SG_extracted_k141_29049 | Not-determined |
| SG_extracted_k141_40868 | Not-determined |
| SG_extracted_k141_19231 | Low-quality |
| SG_extracted_k141_33963 | Not-determined |
| SG_extracted_k141_29052 | Not-determined |
| SG_extracted_k141_17478 | Not-determined |
| SG_extracted_k141_29053 | Low-quality |
| SG_extracted_k141_45732 | Not-determined |
| SG_extracted_k141_29055 | Not-determined |
| SG_extracted_k141_52866 | Not-determined |
| SG_extracted_k141_7802 | Not-determined |
| SG_extracted_k141_40869 | Low-quality |
| SG_extracted_k141_38538 | Not-determined |
| SG_extracted_k141_52870 | Not-determined |
| SG_extracted_k141_24602 | Not-determined |
| SG_extracted_k141_10220 | Low-quality |
| SG_extracted_k141_33968 | Low-quality |
| SG_extracted_k141_10221 | Low-quality |
| SG_extracted_k141_29058 | Not-determined |
| SG_extracted_k141_43162 | Low-quality |
| SG_extracted_k141_10225 | Not-determined |
| SG_extracted_k141_10226 | Not-determined |
| SG_extracted_k141_45749 | Not-determined |
| SG_extracted_k141_17492 | Not-determined |
| SG_extracted_k141_43175 | Not-determined |
| SG_extracted_k141_24614 | Low-quality |
| SG_extracted_k141_45758 | Low-quality |
| SG_extracted_k141_24619 | Not-determined |
| SG_extracted_k141_43178 | Not-determined |
| SG_extracted_k141_24623 | Not-determined |
| SG_extracted_k141_14949 | Low-quality |
| SG_extracted_k141_14950 | Not-determined |
| SG_extracted_k141_29070 | Not-determined |
| SG_extracted_k141_43182 | Not-determined |
| SG_extracted_k141_7827 | Low-quality |
| SG_extracted_k141_52886 | Low-quality |
| SG_extracted_k141_7831 | Not-determined |
| SG_extracted_k141_45766 | Not-determined |
| SG_extracted_k141_7835 | Low-quality |
| SG_extracted_k141_29078 | Not-determined |
| SG_extracted_k141_24635 | Not-determined |
| SG_extracted_k141_10239 | Not-determined |
| SG_extracted_k141_47755 | Low-quality |
| SG_extracted_k141_14961 | Not-determined |
| SG_extracted_k141_47758 | Not-determined |
| SG_extracted_k141_10240 | Low-quality |
| SG_extracted_k141_31298 | Not-determined |
| SG_extracted_k141_43191 | Low-quality |
| SG_extracted_k141_36078 | Not-determined |
| SG_extracted_k141_31301 | Not-determined |
| SG_extracted_k141_43192 | Not-determined |
| SG_extracted_k141_3366 | Not-determined |
| SG_extracted_k141_47763 | Not-determined |
| SG_extracted_k141_7845 | Not-determined |
| SG_extracted_k141_36080 | Not-determined |
| SG_extracted_k141_31303 | Low-quality |
| SG_extracted_k141_24640 | Not-determined |
| SG_extracted_k141_31305 | Not-determined |
| SG_extracted_k141_24641 | Not-determined |
| SG_extracted_k141_10248 | Not-determined |
| SG_extracted_k141_38554 | Not-determined |
| SG_extracted_k141_3374 | Not-determined |
| SG_extracted_k141_54557 | Not-determined |
| SG_extracted_k141_7852 | Not-determined |
| SG_extracted_k141_45780 | Not-determined |
| SG_extracted_k141_19264 | Not-determined |
| SG_extracted_k141_14968 | Not-determined |
| SG_extracted_k141_10255 | Not-determined |
| SG_extracted_k141_38564 | Not-determined |
| SG_extracted_k141_19267 | Not-determined |
| SG_extracted_k141_31313 | Not-determined |
| SG_extracted_k141_45786 | Low-quality |
| SG_extracted_k141_54569 | Not-determined |
| SG_extracted_k141_24655 | Not-determined |
| SG_extracted_k141_45792 | Low-quality |
| SG_extracted_k141_5407 | Low-quality |
| SG_extracted_k141_24659 | Not-determined |
| SG_extracted_k141_14981 | Not-determined |
| SG_extracted_k141_31324 | Not-determined |
| SG_extracted_k141_3384 | Not-determined |
| SG_extracted_k141_10267 | Not-determined |
| SG_extracted_k141_24665 | Not-determined |
| SG_extracted_k141_5419 | Not-determined |
| SG_extracted_k141_10269 | Not-determined |
| SG_extracted_k141_17539 | Not-determined |
| SG_extracted_k141_31331 | Not-determined |
| SG_extracted_k141_17541 | Not-determined |
| SG_extracted_k141_47777 | Not-determined |
| SG_extracted_k141_7873 | Not-determined |
| SG_extracted_k141_33971 | Low-quality |
| SG_extracted_k141_5425 | Not-determined |
| SG_extracted_k141_38581 | Not-determined |
| SG_extracted_k141_7877 | Not-determined |
| SG_extracted_k141_24676 | Not-determined |
| SG_extracted_k141_7880 | Not-determined |
| SG_extracted_k141_45803 | Not-determined |
| SG_extracted_k141_31354 | Not-determined |
| SG_extracted_k141_36106 | Not-determined |
| SG_extracted_k141_24682 | Not-determined |
| SG_extracted_k141_7889 | Not-determined |
| SG_extracted_k141_7890 | Not-determined |
| SG_extracted_k141_7892 | Low-quality |
| SG_extracted_k141_33980 | Not-determined |
| SG_extracted_k141_45809 | Not-determined |
| SG_extracted_k141_7893 | Not-determined |
| SG_extracted_k141_54585 | Not-determined |
| SG_extracted_k141_54586 | Not-determined |
| SG_extracted_k141_24688 | Not-determined |
| SG_extracted_k141_33982 | Not-determined |
| SG_extracted_k141_981 | Not-determined |
| SG_extracted_k141_54594 | Not-determined |
| SG_extracted_k141_45825 | Not-determined |
| SG_extracted_k141_10270 | Not-determined |
| SG_extracted_k141_47786 | Not-determined |
| SG_extracted_k141_990 | Not-determined |
| SG_extracted_k141_3405 | Low-quality |
| SG_extracted_k141_991 | Low-quality |
| SG_extracted_k141_47788 | Not-determined |
| SG_extracted_k141_45829 | Not-determined |
| SG_extracted_k141_33990 | Not-determined |
| SG_extracted_k141_998 | Not-determined |
| SG_extracted_k141_45837 | Not-determined |
| SG_extracted_k141_36127 | Not-determined |
| SG_extracted_k141_5461 | Not-determined |
| SG_extracted_k141_1002 | Not-determined |
| SG_extracted_k141_33999 | Low-quality |
| SG_extracted_k141_38584 | Low-quality |
| SG_extracted_k141_34002 | Not-determined |
| SG_extracted_k141_1006 | Not-determined |
| SG_extracted_k141_47793 | Not-determined |
| SG_extracted_k141_10277 | Not-determined |
| SG_extracted_k141_10278 | Not-determined |
| SG_extracted_k141_47795 | Not-determined |
| SG_extracted_k141_43204 | Not-determined |
| SG_extracted_k141_1009 | Not-determined |
| SG_extracted_k141_1011 | Not-determined |
| SG_extracted_k141_1014 | Not-determined |
| SG_extracted_k141_43209 | Not-determined |
| SG_extracted_k141_7918 | Low-quality |
| SG_extracted_k141_34014 | Not-determined |
| SG_extracted_k141_45846 | Not-determined |
| SG_extracted_k141_47802 | Not-determined |
| SG_extracted_k141_47804 | Not-determined |
| SG_extracted_k141_47806 | Not-determined |
| SG_extracted_k141_38588 | Not-determined |
| SG_extracted_k141_38590 | Not-determined |
| SG_extracted_k141_34018 | Low-quality |
| SG_extracted_k141_10281 | Low-quality |
| SG_extracted_k141_31372 | Not-determined |
| SG_extracted_k141_36143 | Not-determined |
| SG_extracted_k141_36146 | Not-determined |
| SG_extracted_k141_34027 | Not-determined |
| SG_extracted_k141_7925 | Not-determined |
| SG_extracted_k141_36149 | Not-determined |
| SG_extracted_k141_34030 | Not-determined |
| SG_extracted_k141_31376 | Not-determined |
| SG_extracted_k141_7934 | Not-determined |
| SG_extracted_k141_34033 | Not-determined |
| SG_extracted_k141_38602 | Not-determined |
| SG_extracted_k141_36155 | Low-quality |
| SG_extracted_k141_36156 | Not-determined |
| SG_extracted_k141_43219 | Not-determined |
| SG_extracted_k141_7944 | Low-quality |
| SG_extracted_k141_43221 | Not-determined |
| SG_extracted_k141_34040 | Not-determined |
| SG_extracted_k141_47830 | Not-determined |
| SG_extracted_k141_34042 | Not-determined |
| SG_extracted_k141_14993 | Not-determined |
| SG_extracted_k141_34043 | Not-determined |
| S16_vs_164913\|\|full | High-quality |
| SG_extracted_k141_7947 | Low-quality |
| SG_extracted_k141_29112 | Not-determined |
| SG_extracted_k141_29113 | Not-determined |
| SG_extracted_k141_15003 | Not-determined |
| SG_extracted_k141_5466 | Not-determined |
| SG_extracted_k141_15006 | Low-quality |
| SG_extracted_k141_40872 | Not-determined |
| SG_extracted_k141_34051 | Not-determined |
| SG_extracted_k141_34054 | Not-determined |
| SG_extracted_k141_5474 | Not-determined |

| | |
|---|---|
| SG_extracted_k141_7965 | Not-determined |
| SG_extracted_k141_5476 | Not-determined |
| SG_extracted_k141_5478 | Not-determined |
| SG_extracted_k141_5479 | Low-quality |
| SG_extracted_k141_45869 | Not-determined |
| SG_extracted_k141_31388 | Not-determined |
| SG_extracted_k141_7977 | Not-determined |
| SG_extracted_k141_15023 | Not-determined |
| SG_extracted_k141_34069 | Not-determined |
| SG_extracted_k141_45881 | Not-determined |
| SG_extracted_k141_47838 | Not-determined |
| SG_extracted_k141_52901 | Not-determined |
| SG_extracted_k141_52904 | Not-determined |
| SG_extracted_k141_45885 | Low-quality |
| SG_extracted_k141_7986 | Low-quality |
| SG_extracted_k141_43230 | Not-determined |
| SG_extracted_k141_3443 | Not-determined |
| SG_extracted_k141_36164 | Low-quality |
| SG_extracted_k141_29123 | Not-determined |
| SG_extracted_k141_3446 | Not-determined |
| SG_extracted_k141_29124 | Low-quality |
| SG_extracted_k141_38631 | Not-determined |
| SG_extracted_k141_45891 | Not-determined |
| SG_extracted_k141_38634 | Not-determined |
| SG_extracted_k141_24694 | Not-determined |
| SG_extracted_k141_45893 | Not-determined |
| SG_extracted_k141_52909 | Low-quality |
| SG_extracted_k141_52910 | Not-determined |
| SG_extracted_k141_15039 | Not-determined |
| SG_extracted_k141_36169 | Low-quality |
| SG_extracted_k141_40885 | Not-determined |
| SG_extracted_k141_45901 | Not-determined |
| SG_extracted_k141_43242 | Not-determined |
| SG_extracted_k141_29134 | Not-determined |
| SG_extracted_k141_45904 | Low-quality |
| SG_extracted_k141_17561 | Not-determined |
| SG_extracted_k141_10290 | Not-determined |
| SG_extracted_k141_47848 | Not-determined |
| SG_extracted_k141_17562 | Low-quality |
| SG_extracted_k141_15044 | Not-determined |
| SG_extracted_k141_17563 | Low-quality |
| SG_extracted_k141_17566 | Not-determined |
| SG_extracted_k141_17569 | Not-determined |
| SG_extracted_k141_47855 | Not-determined |
| SG_extracted_k141_38642 | Not-determined |
| SG_extracted_k141_15046 | Low-quality |
| SG_extracted_k141_38643 | Not-determined |
| SG_extracted_k141_38645 | Not-determined |
| SG_extracted_k141_15047 | Low-quality |
| SG_extracted_k141_52923 | Not-determined |
| SG_extracted_k141_26537 | Not-determined |
| SG_extracted_k141_45924 | Not-determined |
| SG_extracted_k141_47868 | Not-determined |
| SG_extracted_k141_17589 | Not-determined |
| SG_extracted_k141_40901 | Not-determined |
| SG_extracted_k141_45932 | Not-determined |
| SG_extracted_k141_10295 | Not-determined |
| SG_extracted_k141_7997 | Not-determined |
| SG_extracted_k141_15053 | Not-determined |
| SG_extracted_k141_1059 | Low-quality |
| SG_extracted_k141_15056 | Not-determined |
| SG_extracted_k141_38670 | Low-quality |
| SG_extracted_k141_38671 | Not-determined |
| SG_extracted_k141_40913 | Not-determined |
| SG_extracted_k141_38672 | Low-quality |
| SG_extracted_k141_21701 | Not-determined |
| SG_extracted_k141_10297 | Not-determined |
| SG_extracted_k141_38675 | Not-determined |
| SG_extracted_k141_40919 | Not-determined |
| SG_extracted_k141_38676 | Not-determined |
| SG_extracted_k141_21705 | Not-determined |
| SG_extracted_k141_40922 | Low-quality |
| SG_extracted_k141_38680 | Not-determined |
| SG_extracted_k141_1062 | Not-determined |
| SG_extracted_k141_5496 | Not-determined |
| SG_extracted_k141_15075 | Not-determined |
| SG_extracted_k141_5497 | Low-quality |
| SG_extracted_k141_40924 | Not-determined |
| SG_extracted_k141_54614 | Not-determined |
| SG_extracted_k141_38687 | Low-quality |
| SG_extracted_k141_38690 | Low-quality |
| SG_extracted_k141_36178 | Low-quality |
| SG_extracted_k141_5504 | Not-determined |
| SG_extracted_k141_26547 | Low-quality |
| SG_extracted_k141_1075 | Not-determined |
| SG_extracted_k141_47876 | Not-determined |
| SG_extracted_k141_5510 | Not-determined |
| SG_extracted_k141_1079 | Not-determined |
| SG_extracted_k141_36185 | Not-determined |
| SG_extracted_k141_1081 | Not-determined |
| SG_extracted_k141_34099 | Not-determined |
| SG_extracted_k141_21715 | Not-determined |
| SG_extracted_k141_12784 | Not-determined |
| SG_extracted_k141_21716 | Low-quality |
| SG_extracted_k141_12786 | Not-determined |
| SG_extracted_k141_52943 | Low-quality |
| SG_extracted_k141_54628 | Not-determined |
| SG_extracted_k141_54631 | Not-determined |
| SG_extracted_k141_26558 | Not-determined |
| SG_extracted_k141_26559 | Not-determined |
| SG_extracted_k141_8024 | Not-determined |
| SG_extracted_k141_34109 | Not-determined |
| SG_extracted_k141_26562 | Low-quality |
| SG_extracted_k141_26564 | Not-determined |
| SG_extracted_k141_34113 | Not-determined |
| SG_extracted_k141_34115 | Not-determined |
| SG_extracted_k141_34117 | Not-determined |
| SG_extracted_k141_10312 | Not-determined |
| SG_extracted_k141_52951 | Not-determined |
| SG_extracted_k141_8028 | Low-quality |
| SG_extracted_k141_52953 | Not-determined |
| SG_extracted_k141_52958 | Low-quality |
| SG_extracted_k141_43273 | Not-determined |
| SG_extracted_k141_52966 | Not-determined |
| SG_extracted_k141_3460 | Not-determined |
| SG_extracted_k141_3461 | Not-determined |
| SG_extracted_k141_3462 | Not-determined |
| SG_extracted_k141_38706 | Not-determined |
| SG_extracted_k141_40944 | Not-determined |
| SG_extracted_k141_12801 | Not-determined |
| SG_extracted_k141_21735 | Not-determined |
| SG_extracted_k141_50196 | Low-quality |
| SG_extracted_k141_21738 | Low-quality |
| SG_extracted_k141_34138 | Not-determined |
| SG_extracted_k141_38720 | Not-determined |
| SG_extracted_k141_38724 | Not-determined |
| SG_extracted_k141_5528 | Not-determined |
| SG_extracted_k141_21749 | Not-determined |
| SG_extracted_k141_47895 | Not-determined |
| SG_extracted_k141_21753 | Not-determined |
| SG_extracted_k141_47896 | Not-determined |
| SG_extracted_k141_24706 | Low-quality |
| SG_extracted_k141_36191 | Not-determined |
| SG_extracted_k141_12818 | Not-determined |
| SG_extracted_k141_50205 | Not-determined |
| SG_extracted_k141_47907 | Not-determined |
| SG_extracted_k141_50207 | Not-determined |
| SG_extracted_k141_43283 | Not-determined |
| SG_extracted_k141_24716 | Not-determined |
| SG_extracted_k141_45941 | Not-determined |
| SG_extracted_k141_52975 | Not-determined |
| SG_extracted_k141_50210 | Not-determined |
| SG_extracted_k141_54667 | Not-determined |
| SG_extracted_k141_24717 | Not-determined |
| SG_extracted_k141_52976 | Not-determined |
| SG_extracted_k141_36213 | Not-determined |
| SG_extracted_k141_50220 | Not-determined |
| SG_extracted_k141_43294 | Not-determined |
| SG_extracted_k141_54680 | Not-determined |
| SG_extracted_k141_19291 | Not-determined |
| SG_extracted_k141_43297 | Not-determined |
| SG_extracted_k141_43300 | Not-determined |
| SG_extracted_k141_8036 | Not-determined |
| SG_extracted_k141_36226 | Not-determined |
| SG_extracted_k141_54684 | Low-quality |
| SG_extracted_k141_12833 | Not-determined |
| SG_extracted_k141_15104 | Not-determined |
| SG_extracted_k141_19298 | Low-quality |
| SG_extracted_k141_50228 | Not-determined |
| SG_extracted_k141_21780 | Not-determined |
| SG_extracted_k141_21781 | Not-determined |
| SG_extracted_k141_43305 | Not-determined |
| SG_extracted_k141_36232 | Low-quality |
| SG_extracted_k141_19300 | Not-determined |
| SG_extracted_k141_29163 | Not-determined |
| SG_extracted_k141_19301 | Not-determined |
| SG_extracted_k141_47919 | Not-determined |
| SG_extracted_k141_21793 | Not-determined |
| SG_extracted_k141_5546 | Not-determined |
| SG_extracted_k141_19306 | Not-determined |
| SG_extracted_k141_21797 | Not-determined |
| SG_extracted_k141_29172 | Not-determined |
| SG_extracted_k141_50231 | Not-determined |
| SG_extracted_k141_50232 | Not-determined |
| SG_extracted_k141_50233 | Not-determined |
| SG_extracted_k141_47928 | Not-determined |
| SG_extracted_k141_19316 | Not-determined |
| S11_extracted_k141_58587 | Medium-quality |
| SG_extracted_k141_36237 | Not-determined |
| SG_extracted_k141_24744 | Not-determined |
| SG_extracted_k141_5553 | Not-determined |
| SG_extracted_k141_40955 | Not-determined |
| SG_extracted_k141_52983 | Not-determined |
| SG_extracted_k141_21809 | Not-determined |
| SG_extracted_k141_19321 | Not-determined |
| SG_extracted_k141_52987 | Low-quality |
| SG_extracted_k141_19322 | Low-quality |
| SG_extracted_k141_19324 | Not-determined |
| SG_extracted_k141_52990 | Not-determined |
| SG_extracted_k141_43310 | Not-determined |
| SG_extracted_k141_36239 | Low-quality |
| SG_extracted_k141_24755 | Low-quality |
| SG_extracted_k141_52996 | Not-determined |
| SG_extracted_k141_50247 | Not-determined |
| SG_extracted_k141_45962 | Low-quality |
| SG_extracted_k141_50249 | Not-determined |
| SG_extracted_k141_24758 | Not-determined |
| SG_extracted_k141_3502 | Not-determined |
| SG_extracted_k141_5559 | Not-determined |
| SG_extracted_k141_8046 | Not-determined |
| SG_extracted_k141_36245 | Not-determined |
| SG_extracted_k141_54689 | Not-determined |
| SG_extracted_k141_54690 | Not-determined |
| SG_extracted_k141_50255 | Not-determined |
| SG_extracted_k141_24768 | Not-determined |
| SG_extracted_k141_3510 | Not-determined |
| SG_extracted_k141_43317 | Not-determined |
| SG_extracted_k141_24770 | Not-determined |
| SG_extracted_k141_31396 | Not-determined |
| SG_extracted_k141_21822 | Low-quality |
| SG_extracted_k141_38751 | Not-determined |
| SG_extracted_k141_31398 | Not-determined |
| SG_extracted_k141_38752 | Not-determined |
| SG_extracted_k141_36259 | Not-determined |
| SG_extracted_k141_29205 | Low-quality |
| SG_extracted_k141_29212 | Not-determined |
| SG_extracted_k141_15122 | Not-determined |
| SG_extracted_k141_12855 | Low-quality |
| SG_extracted_k141_45970 | Not-determined |
| SG_extracted_k141_34169 | Not-determined |
| SG_extracted_k141_21834 | Low-quality |
| SG_extracted_k141_29220 | Not-determined |
| SG_extracted_k141_15127 | Not-determined |
| SG_extracted_k141_21836 | Low-quality |
| SG_extracted_k141_5575 | Not-determined |
| SG_extracted_k141_12860 | Low-quality |
| SG_extracted_k141_15130 | Not-determined |
| SG_extracted_k141_12862 | Not-determined |
| SG_extracted_k141_54719 | Not-determined |
| SG_extracted_k141_47941 | Not-determined |
| SG_extracted_k141_50263 | Not-determined |
| SG_extracted_k141_19344 | Not-determined |
| SG_extracted_k141_29229 | Not-determined |
| SG_extracted_k141_47951 | Not-determined |
| SG_extracted_k141_47953 | Not-determined |
| SG_extracted_k141_29233 | Not-determined |
| SG_extracted_k141_21850 | Not-determined |
| SG_extracted_k141_54731 | Not-determined |
| SG_extracted_k141_47955 | Not-determined |
| SG_extracted_k141_21853 | Not-determined |
| SG_extracted_k141_36272 | Low-quality |
| SG_extracted_k141_15139 | Not-determined |
| SG_extracted_k141_31410 | Not-determined |
| SG_extracted_k141_15141 | Low-quality |
| SG_extracted_k141_5584 | Low-quality |
| SG_extracted_k141_29241 | Not-determined |
| SG_extracted_k141_15143 | Not-determined |
| SG_extracted_k141_43336 | Not-determined |
| SG_extracted_k141_15146 | Not-determined |
| SG_extracted_k141_34171 | Not-determined |
| SG_extracted_k141_29245 | Not-determined |
| SG_extracted_k141_47970 | Not-determined |
| SG_extracted_k141_31419 | Not-determined |
| SG_extracted_k141_47971 | Not-determined |
| SG_extracted_k141_29250 | Not-determined |
| SG_extracted_k141_50277 | Not-determined |
| SG_extracted_k141_36274 | Not-determined |
| SG_extracted_k141_31420 | Low-quality |
| SG_extracted_k141_3524 | Not-determined |
| SG_extracted_k141_29254 | Not-determined |
| SG_extracted_k141_31422 | Not-determined |
| SG_extracted_k141_29256 | Not-determined |
| SG_extracted_k141_24812 | Not-determined |
| SG_extracted_k141_3528 | Not-determined |
| SG_extracted_k141_15158 | Low-quality |
| SG_extracted_k141_34184 | Not-determined |
| SG_extracted_k141_43347 | Low-quality |
| SG_extracted_k141_34187 | Low-quality |
| SG_extracted_k141_43348 | Not-determined |
| SG_extracted_k141_24816 | Not-determined |
| SG_extracted_k141_17596 | Not-determined |
| SG_extracted_k141_46001 | Not-determined |
| SG_extracted_k141_53044 | Low-quality |
| SG_extracted_k141_3536 | Low-quality |
| SG_extracted_k141_31431 | Not-determined |
| SG_extracted_k141_21860 | Low-quality |
| SG_extracted_k141_34191 | Not-determined |
| SG_extracted_k141_31432 | Not-determined |
| SG_extracted_k141_21864 | Not-determined |
| SG_extracted_k141_36282 | Not-determined |
| SG_extracted_k141_17601 | Not-determined |
| SG_extracted_k141_47975 | Not-determined |
| SG_extracted_k141_43354 | Not-determined |
| SG_extracted_k141_47976 | Not-determined |
| SG_extracted_k141_3542 | Not-determined |
| SG_extracted_k141_24820 | Low-quality |
| SG_extracted_k141_47983 | Not-determined |
| SG_extracted_k141_1110 | Not-determined |
| SG_extracted_k141_31447 | Not-determined |
| SG_extracted_k141_34206 | Not-determined |
| SG_extracted_k141_34207 | Not-determined |
| SG_extracted_k141_31449 | Not-determined |
| SG_extracted_k141_31450 | Not-determined |
| SG_extracted_k141_3544 | Low-quality |
| SG_extracted_k141_43365 | Not-determined |
| SG_extracted_k141_24830 | Not-determined |
| SG_extracted_k141_43372 | Low-quality |
| SG_extracted_k141_46005 | Not-determined |
| SG_extracted_k141_3546 | Low-quality |
| SG_extracted_k141_34208 | Not-determined |
| SG_extracted_k141_17615 | Not-determined |
| SG_extracted_k141_38756 | Low-quality |
| SG_extracted_k141_50290 | Not-determined |
| SG_extracted_k141_17620 | Not-determined |
| SG_extracted_k141_15167 | Not-determined |
| SG_extracted_k141_50291 | Not-determined |
| SG_extracted_k141_38763 | Not-determined |
| SG_extracted_k141_43382 | Not-determined |
| SG_extracted_k141_34220 | Not-determined |
| SG_extracted_k141_38769 | Not-determined |
| SG_extracted_k141_31462 | Low-quality |
| SG_extracted_k141_36302 | Not-determined |
| SG_extracted_k141_54747 | Not-determined |
| SG_extracted_k141_3562 | Not-determined |
| SG_extracted_k141_31470 | Not-determined |
| SG_extracted_k141_1122 | Not-determined |
| SG_extracted_k141_1123 | Not-determined |
| SG_extracted_k141_47996 | Not-determined |
| SG_extracted_k141_38772 | Not-determined |
| SG_extracted_k141_17626 | Not-determined |
| SG_extracted_k141_54754 | Not-determined |
| SG_extracted_k141_1125 | Not-determined |
| SG_extracted_k141_10318 | Not-determined |
| SG_extracted_k141_3567 | Not-determined |
| SG_extracted_k141_46034 | Not-determined |
| SG_extracted_k141_31482 | Not-determined |
| SG_extracted_k141_54759 | Not-determined |
| SG_extracted_k141_3569 | Not-determined |
| SG_extracted_k141_26597 | Not-determined |
| SG_extracted_k141_10327 | Not-determined |
| SG_extracted_k141_46038 | Not-determined |
| SG_extracted_k141_26602 | Not-determined |
| SG_extracted_k141_34228 | Not-determined |
| SG_extracted_k141_31491 | Not-determined |
| SG_extracted_k141_54766 | Not-determined |
| SG_extracted_k141_26605 | Not-determined |
| SG_extracted_k141_50302 | Not-determined |
| SG_extracted_k141_21883 | Low-quality |
| SG_extracted_k141_50303 | Not-determined |
| SG_extracted_k141_3574 | Not-determined |
| SG_extracted_k141_50304 | Not-determined |
| SG_extracted_k141_1132 | Not-determined |
| SG_extracted_k141_31495 | Not-determined |
| SG_extracted_k141_21888 | Not-determined |
| SG_extracted_k141_50308 | Not-determined |
| SG_extracted_k141_1135 | Not-determined |
| SG_extracted_k141_50309 | Not-determined |
| SG_extracted_k141_50314 | Not-determined |
| SG_extracted_k141_53061 | Not-determined |
| SG_extracted_k141_40979 | Not-determined |
| SG_extracted_k141_53066 | Not-determined |
| SG_extracted_k141_31497 | Not-determined |
| SG_extracted_k141_26613 | Not-determined |
| SG_extracted_k141_26614 | Not-determined |
| SG_extracted_k141_34243 | Not-determined |
| SG_extracted_k141_26615 | Not-determined |
| SG_extracted_k141_3585 | Not-determined |
| SG_extracted_k141_3587 | Not-determined |
| SG_extracted_k141_54781 | Not-determined |
| SG_extracted_k141_54782 | Not-determined |
| SG_extracted_k141_54784 | Not-determined |
| SG_extracted_k141_3589 | Low-quality |
| SG_extracted_k141_31501 | Not-determined |
| SG_extracted_k141_34250 | Not-determined |
| SG_extracted_k141_40981 | Not-determined |
| SG_extracted_k141_31502 | Not-determined |
| SG_extracted_k141_53072 | Not-determined |
| SG_extracted_k141_50326 | Not-determined |
| SG_extracted_k141_50327 | Not-determined |
| SG_extracted_k141_54790 | Low-quality |
| SG_extracted_k141_54792 | Not-determined |
| SG_extracted_k141_40989 | Low-quality |
| SG_extracted_k141_10350 | Low-quality |
| SG_extracted_k141_53082 | Not-determined |
| SG_extracted_k141_34257 | Not-determined |
| SG_extracted_k141_53085 | Not-determined |
| SG_extracted_k141_53086 | Not-determined |
| SG_extracted_k141_53087 | Not-determined |
| SG_extracted_k141_5638 | Not-determined |
| SG_extracted_k141_34258 | Not-determined |
| SG_extracted_k141_36314 | Not-determined |
| SG_extracted_k141_34260 | Not-determined |
| SG_extracted_k141_43384 | Not-determined |
| SG_extracted_k141_54803 | Not-determined |
| SG_extracted_k141_38778 | Not-determined |
| SG_extracted_k141_38780 | Low-quality |
| SG_extracted_k141_34268 | Not-determined |
| SG_extracted_k141_3597 | Not-determined |
| SG_extracted_k141_38785 | Low-quality |
| SG_extracted_k141_38789 | Not-determined |
| SG_extracted_k141_43391 | Not-determined |
| SG_extracted_k141_10361 | Not-determined |
| SG_extracted_k141_31516 | Not-determined |
| SG_extracted_k141_12876 | Not-determined |
| SG_extracted_k141_3603 | Low-quality |
| SG_extracted_k141_12878 | Not-determined |
| SG_extracted_k141_12879 | Not-determined |
| SG_extracted_k141_5649 | Not-determined |
| SG_extracted_k141_12881 | Not-determined |
| SG_extracted_k141_10363 | Low-quality |
| SG_extracted_k141_12882 | Low-quality |
| SG_extracted_k141_12883 | Low-quality |
| SG_extracted_k141_5655 | Not-determined |
| S7_vs_89668\|\|full | Medium-quality |
| SG_extracted_k141_10365 | Not-determined |
| SG_extracted_k141_10366 | Not-determined |
| SG_extracted_k141_5658 | Not-determined |
| SG_extracted_k141_3609 | Low-quality |
| S5_extracted_k141_38011 | Medium-quality |
| SG_extracted_k141_12887 | Low-quality |
| SG_extracted_k141_38812 | Not-determined |
| SG_extracted_k141_17628 | Not-determined |
| SG_extracted_k141_17629 | Not-determined |
| SG_extracted_k141_5665 | Not-determined |
| SG_extracted_k141_5668 | Not-determined |
| SG_extracted_k141_29294 | Not-determined |
| SG_extracted_k141_10384 | Not-determined |
| SG_extracted_k141_41009 | Not-determined |
| SG_extracted_k141_5673 | Not-determined |
| SG_extracted_k141_17634 | Not-determined |
| SG_extracted_k141_41015 | Not-determined |
| SG_extracted_k141_31533 | Low-quality |
| SG_extracted_k141_17637 | Not-determined |
| SG_extracted_k141_31538 | Not-determined |
| SG_extracted_k141_17641 | Not-determined |
| SG_extracted_k141_17643 | Not-determined |
| SG_extracted_k141_10393 | Not-determined |
| SG_extracted_k141_31543 | Not-determined |
| SG_extracted_k141_17645 | Not-determined |
| SG_extracted_k141_3630 | Not-determined |
| SG_extracted_k141_38818 | Low-quality |
| SG_extracted_k141_21904 | Not-determined |
| SG_extracted_k141_10396 | Not-determined |
| SG_extracted_k141_10398 | Not-determined |
| SG_extracted_k141_43410 | Low-quality |
| SG_extracted_k141_17648 | Not-determined |
| SG_extracted_k141_43411 | Low-quality |
| SG_extracted_k141_43412 | Not-determined |
| SG_extracted_k141_12908 | Not-determined |
| SG_extracted_k141_31546 | Not-determined |
| SG_extracted_k141_31547 | Not-determined |
| SG_extracted_k141_43415 | Not-determined |
| SG_extracted_k141_54842 | Low-quality |
| SG_extracted_k141_17652 | Not-determined |
| SG_extracted_k141_43419 | Not-determined |
| SG_extracted_k141_10408 | Not-determined |
| SG_extracted_k141_5684 | Low-quality |
| SG_extracted_k141_10410 | Not-determined |
| SG_extracted_k141_50339 | Low-quality |
| SG_extracted_k141_50340 | Not-determined |
| SG_extracted_k141_3634 | Not-determined |
| SG_extracted_k141_19400 | Not-determined |
| SG_extracted_k141_50342 | Not-determined |
| SG_extracted_k141_3636 | Not-determined |
| SG_extracted_k141_19401 | Not-determined |
| SG_extracted_k141_5690 | Not-determined |
| SG_extracted_k141_50345 | Not-determined |
| SG_extracted_k141_21908 | Not-determined |
| SG_extracted_k141_17661 | Not-determined |
| SG_extracted_k141_21910 | Not-determined |
| SG_extracted_k141_34278 | Not-determined |
| SG_extracted_k141_5691 | Not-determined |
| SG_extracted_k141_19403 | Not-determined |
| SG_extracted_k141_41030 | Not-determined |
| SG_extracted_k141_21916 | Not-determined |
| SG_extracted_k141_12925 | Low-quality |
| SG_extracted_k141_21919 | Not-determined |
| SG_extracted_k141_1147 | Not-determined |
| SG_extracted_k141_10426 | Not-determined |
| SG_extracted_k141_31565 | Not-determined |
| SG_extracted_k141_12933 | Not-determined |
| SG_extracted_k141_38834 | Not-determined |
| SG_extracted_k141_38835 | Not-determined |
| SG_extracted_k141_12936 | Not-determined |
| SG_extracted_k141_12937 | Not-determined |
| SG_extracted_k141_12939 | Low-quality |
| SG_extracted_k141_43430 | Not-determined |
| SG_extracted_k141_26646 | Low-quality |
| SG_extracted_k141_3645 | Not-determined |
| SG_extracted_k141_21930 | Not-determined |
| SG_extracted_k141_21931 | Not-determined |
| SG_extracted_k141_21932 | Not-determined |
| SG_extracted_k141_21935 | Not-determined |
| SG_extracted_k141_50350 | Not-determined |
| SG_extracted_k141_17672 | Not-determined |
| SG_extracted_k141_54858 | Low-quality |
| SG_extracted_k141_31575 | Low-quality |
| SG_extracted_k141_50355 | Low-quality |
| SG_extracted_k141_29351 | Not-determined |
| SG_extracted_k141_50356 | Not-determined |
| SG_extracted_k141_38839 | Not-determined |
| SG_extracted_k141_53113 | Not-determined |
| SG_extracted_k141_50360 | Not-determined |
| SG_extracted_k141_54863 | Not-determined |
| SG_extracted_k141_3649 | Not-determined |
| SG_extracted_k141_50367 | Not-determined |
| SG_extracted_k141_54870 | Not-determined |
| SG_extracted_k141_17685 | Not-determined |
| SG_extracted_k141_54871 | Not-determined |
| SG_extracted_k141_5696 | Low-quality |
| SG_extracted_k141_53122 | Not-determined |
| SG_extracted_k141_8060 | Not-determined |
| SG_extracted_k141_21950 | Not-determined |
| SG_extracted_k141_29366 | Not-determined |
| SG_extracted_k141_36319 | Not-determined |
| SG_extracted_k141_38861 | Not-determined |
| SG_extracted_k141_15201 | Not-determined |
| SG_extracted_k141_29368 | Not-determined |
| SG_extracted_k141_8067 | Low-quality |
| SG_extracted_k141_38866 | Not-determined |
| SG_extracted_k141_36321 | Not-determined |
| SG_extracted_k141_3660 | Not-determined |
| SG_extracted_k141_43454 | Not-determined |
| SG_extracted_k141_34284 | Not-determined |
| SG_extracted_k141_10446 | Low-quality |
| SG_extracted_k141_34285 | Low-quality |
| SG_extracted_k141_34289 | Low-quality |
| SG_extracted_k141_3667 | Not-determined |
| SG_extracted_k141_8087 | Not-determined |
| SG_extracted_k141_3670 | Low-quality |
| SG_extracted_k141_10453 | Not-determined |
| SG_extracted_k141_3681 | Not-determined |
| SG_extracted_k141_3684 | Not-determined |
| SG_extracted_k141_50379 | Not-determined |
| SG_extracted_k141_3686 | Low-quality |
| SG_extracted_k141_17691 | Low-quality |
| SG_extracted_k141_53138 | Not-determined |
| SG_extracted_k141_10465 | Not-determined |
| SG_extracted_k141_10467 | Not-determined |
| SG_extracted_k141_53145 | Low-quality |
| SG_extracted_k141_43459 | Not-determined |
| SG_extracted_k141_43461 | Not-determined |
| SG_extracted_k141_3694 | Low-quality |
| SG_extracted_k141_50389 | Not-determined |
| SG_extracted_k141_10468 | Low-quality |
| SG_extracted_k141_15208 | Low-quality |
| SG_extracted_k141_10469 | Not-determined |
| SG_extracted_k141_12947 | Not-determined |
| SG_extracted_k141_34312 | Not-determined |
| SG_extracted_k141_1155 | Not-determined |
| SG_extracted_k141_3698 | Not-determined |
| SG_extracted_k141_10474 | Not-determined |
| SG_extracted_k141_50396 | Not-determined |
| SG_extracted_k141_15213 | Not-determined |
| SG_extracted_k141_15215 | Not-determined |
| SG_extracted_k141_50400 | Not-determined |
| SG_extracted_k141_15219 | Not-determined |
| SG_extracted_k141_43465 | Not-determined |
| SG_extracted_k141_43467 | Not-determined |
| SG_extracted_k141_46053 | Low-quality |
| SG_extracted_k141_50407 | Not-determined |
| SG_extracted_k141_19412 | Not-determined |
| SG_extracted_k141_38884 | Not-determined |
| SG_extracted_k141_38885 | Not-determined |
| SG_extracted_k141_38889 | Not-determined |
| SG_extracted_k141_15226 | Not-determined |
| SG_extracted_k141_19413 | Not-determined |
| SG_extracted_k141_19414 | Not-determined |
| SG_extracted_k141_12957 | Not-determined |
| SG_extracted_k141_3710 | Not-determined |
| SG_extracted_k141_5711 | Low-quality |
| SG_extracted_k141_10489 | Not-determined |
| SG_extracted_k141_26651 | Low-quality |
| SG_extracted_k141_5713 | Not-determined |
| SG_extracted_k141_10492 | Not-determined |
| SG_extracted_k141_10493 | Not-determined |
| SG_extracted_k141_50420 | Not-determined |
| SG_extracted_k141_43470 | Not-determined |
| SG_extracted_k141_15237 | Not-determined |
| SG_extracted_k141_53147 | Low-quality |
| SG_extracted_k141_12967 | Not-determined |
| SG_extracted_k141_43476 | Not-determined |
| SG_extracted_k141_19416 | Not-determined |
| SG_extracted_k141_5720 | Low-quality |
| SG_extracted_k141_19419 | Not-determined |
| SG_extracted_k141_31579 | Not-determined |
| SG_extracted_k141_26657 | Not-determined |
| SG_extracted_k141_12978 | Not-determined |
| SG_extracted_k141_17716 | Not-determined |
| SG_extracted_k141_10499 | Not-determined |
| SG_extracted_k141_21972 | Not-determined |
| SG_extracted_k141_15248 | Not-determined |
| SG_extracted_k141_17719 | Not-determined |
| SG_extracted_k141_15249 | Not-determined |
| SG_extracted_k141_15250 | Low-quality |
| SG_extracted_k141_5724 | Not-determined |
| SG_extracted_k141_43488 | Not-determined |
| SG_extracted_k141_5725 | Not-determined |
| SG_extracted_k141_12986 | Low-quality |

| | |
|---|---|
| SG_extracted_k141_12987 | Not-determined |
| SG_extracted_k141_17722 | Not-determined |
| SG_extracted_k141_34331 | Not-determined |
| SG_extracted_k141_26661 | Low-quality |
| SG_extracted_k141_15258 | Not-determined |
| SG_extracted_k141_34339 | Not-determined |
| SG_extracted_k141_19425 | Low-quality |
| SG_extracted_k141_5737 | Not-determined |
| SG_extracted_k141_19427 | Not-determined |
| SG_extracted_k141_15262 | Not-determined |
| SG_extracted_k141_3739 | Not-determined |
| SG_extracted_k141_31587 | Not-determined |
| SG_extracted_k141_3742 | Not-determined |
| SG_extracted_k141_17735 | Low-quality |
| SG_extracted_k141_8115 | Not-determined |
| SG_extracted_k141_34350 | Not-determined |
| SG_extracted_k141_54873 | Not-determined |
| SG_extracted_k141_3747 | Not-determined |
| SG_extracted_k141_15268 | Not-determined |
| SG_extracted_k141_13013 | Not-determined |
| SG_extracted_k141_31593 | Not-determined |
| SG_extracted_k141_17739 | Not-determined |
| SG_extracted_k141_17741 | Not-determined |
| SG_extracted_k141_34356 | Not-determined |
| SG_extracted_k141_34357 | Not-determined |
| SG_extracted_k141_31597 | Low-quality |
| SG_extracted_k141_34360 | Not-determined |
| SG_extracted_k141_34361 | Not-determined |
| SG_extracted_k141_29407 | Not-determined |
| SG_extracted_k141_29410 | Not-determined |
| SG_extracted_k141_31604 | Not-determined |
| SG_extracted_k141_43500 | Not-determined |
| SG_extracted_k141_53169 | Not-determined |
| SG_extracted_k141_15280 | Not-determined |
| SG_extracted_k141_24849 | Not-determined |
| SG_extracted_k141_24850 | Not-determined |
| SG_extracted_k141_24851 | Not-determined |
| SG_extracted_k141_15285 | Not-determined |
| SG_extracted_k141_43507 | Not-determined |
| SG_extracted_k141_26673 | Not-determined |
| SG_extracted_k141_31618 | Not-determined |
| SG_extracted_k141_36336 | Not-determined |
| SG_extracted_k141_15291 | Not-determined |
| SG_extracted_k141_34377 | Not-determined |
| SG_extracted_k141_43517 | Not-determined |
| SG_extracted_k141_34381 | Low-quality |
| SG_extracted_k141_8128 | Low-quality |
| SG_extracted_k141_15296 | Not-determined |
| SG_extracted_k141_43520 | Not-determined |
| SG_extracted_k141_54887 | Not-determined |
| SG_extracted_k141_26686 | Not-determined |
| SG_extracted_k141_43524 | Not-determined |
| SG_extracted_k141_34390 | Not-determined |
| SG_extracted_k141_54895 | Low-quality |
| SG_extracted_k141_36344 | Not-determined |
| [illegible] | [illegible] |

| | |
|---|---|
| SG_extracted_k141_36562 | Not-determined |
| SG_extracted_k141_34571 | Low-quality |
| SG_extracted_k141_8319 | Not-determined |
| SG_extracted_k141_53314 | Low-quality |
| SG_extracted_k141_50610 | Not-determined |
| SG_extracted_k141_8320 | Low-quality |
| SG_extracted_k141_26921 | Not-determined |
| SG_extracted_k141_26923 | Low-quality |
| SG_extracted_k141_41264 | Not-determined |
| SG_extracted_k141_34579 | Not-determined |
| SG_extracted_k141_8323 | Low-quality |
| SG_extracted_k141_50615 | Low-quality |
| SG_extracted_k141_36565 | Not-determined |
| SG_extracted_k141_34585 | Not-determined |
| SG_extracted_k141_34588 | Low-quality |
| SG_extracted_k141_53321 | Not-determined |
| SG_extracted_k141_53322 | Not-determined |
| SG_extracted_k141_10796 | Not-determined |
| SG_extracted_k141_46126 | Not-determined |
| SG_extracted_k141_41271 | Not-determined |
| SG_extracted_k141_50620 | Not-determined |
| SG_extracted_k141_8334 | Not-determined |
| SG_extracted_k141_50624 | Not-determined |
| SG_extracted_k141_48242 | Not-determined |
| SG_extracted_k141_25118 | Low-quality |
| SG_extracted_k141_41279 | Not-determined |
| SG_extracted_k141_10806 | Not-determined |
| SG_extracted_k141_41280 | Not-determined |
| SG_extracted_k141_26927 | Not-determined |
| SG_extracted_k141_1331 | Not-determined |
| SG_extracted_k141_26930 | Not-determined |
| SG_extracted_k141_41287 | Not-determined |
| SG_extracted_k141_25119 | Low-quality |
| SG_extracted_k141_1337 | Not-determined |
| SG_extracted_k141_41289 | Not-determined |
| SG_extracted_k141_41290 | Not-determined |
| SG_extracted_k141_1341 | Not-determined |
| SG_extracted_k141_34616 | Not-determined |
| SG_extracted_k141_41298 | Not-determined |
| SG_extracted_k141_34622 | Not-determined |
| SG_extracted_k141_3986 | Not-determined |
| SG_extracted_k141_3988 | Not-determined |
| SG_extracted_k141_34631 | Not-determined |
| SG_extracted_k141_5944 | Not-determined |
| SG_extracted_k141_3993 | Not-determined |
| SG_extracted_k141_46156 | Low-quality |
| SG_extracted_k141_10815 | Low-quality |
| SG_extracted_k141_48249 | Not-determined |
| SG_extracted_k141_46157 | Low-quality |
| SG_extracted_k141_41310 | Not-determined |
| SG_extracted_k141_5950 | Not-determined |
| SG_extracted_k141_3996 | Not-determined |
| SG_extracted_k141_1355 | Not-determined |
| SG_extracted_k141_53349 | Low-quality |
| SG_extracted_k141_43572 | Low-quality |
| SG_extracted_k141_53352 | Not-determined |
| SG_extracted_k141_1356 | Not-determined |
| SG_extracted_k141_50630 | Not-determined |
| SG_extracted_k141_26939 | Not-determined |
| SG_extracted_k141_46169 | Not-determined |
| SG_extracted_k141_3997 | Not-determined |
| SG_extracted_k141_26940 | Low-quality |
| SG_extracted_k141_10827 | Not-determined |
| SG_extracted_k141_31880 | Low-quality |
| SG_extracted_k141_54965 | Low-quality |
| SG_extracted_k141_4001 | Not-determined |
| SG_extracted_k141_31884 | Not-determined |
| SG_extracted_k141_10829 | Low-quality |
| SG_extracted_k141_34657 | Not-determined |
| SG_extracted_k141_54969 | Not-determined |
| SG_extracted_k141_54972 | Not-determined |
| SG_extracted_k141_26947 | Not-determined |
| SG_extracted_k141_43592 | Not-determined |
| SG_extracted_k141_34661 | Low-quality |
| SG_extracted_k141_26950 | Not-determined |
| SG_extracted_k141_36588 | Not-determined |
| SG_extracted_k141_31888 | Low-quality |
| SG_extracted_k141_22224 | Not-determined |
| SG_extracted_k141_22230 | Not-determined |
| SG_extracted_k141_41318 | Not-determined |
| SG_extracted_k141_22232 | Low-quality |
| SG_extracted_k141_43605 | Low-quality |
| SG_extracted_k141_34672 | Not-determined |
| SG_extracted_k141_22233 | Not-determined |
| SG_extracted_k141_25125 | Not-determined |
| SG_extracted_k141_25126 | Low-quality |
| SG_extracted_k141_15489 | Not-determined |
| SG_extracted_k141_22239 | Not-determined |
| SG_extracted_k141_43614 | Not-determined |
| SG_extracted_k141_8338 | Not-determined |
| SG_extracted_k141_15497 | Not-determined |
| SG_extracted_k141_13317 | Low-quality |
| SG_extracted_k141_41328 | Low-quality |
| SG_extracted_k141_8342 | Not-determined |
| SG_extracted_k141_50644 | Not-determined |
| SG_extracted_k141_15502 | Low-quality |
| SG_extracted_k141_34687 | Low-quality |
| SG_extracted_k141_5970 | Not-determined |
| SG_extracted_k141_15506 | Not-determined |
| SG_extracted_k141_5972 | Not-determined |
| SG_extracted_k141_48253 | Not-determined |
| SG_extracted_k141_13327 | Not-determined |
| SG_extracted_k141_31911 | Not-determined |
| SG_extracted_k141_8352 | Not-determined |
| SG_extracted_k141_36604 | Low-quality |
| SG_extracted_k141_48257 | Not-determined |
| SG_extracted_k141_50649 | Not-determined |
| SG_extracted_k141_5984 | Not-determined |
| SG_extracted_k141_43627 | Not-determined |
| SG_extracted_k141_25148 | Low-quality |
| SG_extracted_k141_4023 | Not-determined |
| SG_extracted_k141_13332 | Not-determined |
| SG_extracted_k141_41346 | Low-quality |
| SG_extracted_k141_41348 | Low-quality |
| SG_extracted_k141_4026 | Not-determined |
| SG_extracted_k141_55025 | Not-determined |
| SG_extracted_k141_55026 | Not-determined |
| SG_extracted_k141_50656 | Not-determined |
| SG_extracted_k141_50657 | Not-determined |
| SG_extracted_k141_4029 | Not-determined |
| SG_extracted_k141_36615 | Not-determined |
| SG_extracted_k141_5995 | Not-determined |
| SG_extracted_k141_5996 | Low-quality |
| SG_extracted_k141_15518 | Not-determined |
| SG_extracted_k141_15519 | Not-determined |
| SG_extracted_k141_50658 | Not-determined |
| SG_extracted_k141_13345 | Not-determined |
| SG_extracted_k141_4038 | Not-determined |
| SG_extracted_k141_43649 | Not-determined |
| SG_extracted_k141_55033 | Not-determined |
| SG_extracted_k141_8367 | Not-determined |
| SG_extracted_k141_18107 | Not-determined |
| SG_extracted_k141_13350 | Low-quality |
| SG_extracted_k141_10888 | Not-determined |
| SG_extracted_k141_53381 | Not-determined |
| SG_extracted_k141_10889 | Not-determined |
| SG_extracted_k141_50667 | Not-determined |
| SG_extracted_k141_53383 | Not-determined |
| SG_extracted_k141_6010 | Not-determined |
| SG_extracted_k141_8373 | Not-determined |
| SG_extracted_k141_53386 | Not-determined |
| SG_extracted_k141_36624 | Not-determined |
| SG_extracted_k141_55041 | Low-quality |
| SG_extracted_k141_10899 | Low-quality |
| SG_extracted_k141_36628 | Not-determined |
| SG_extracted_k141_48265 | Low-quality |
| SG_extracted_k141_55046 | Not-determined |
| SG_extracted_k141_4054 | Not-determined |
| SG_extracted_k141_25183 | Not-determined |
| SG_extracted_k141_10904 | Not-determined |
| SG_extracted_k141_4055 | Not-determined |
| SG_extracted_k141_4058 | Not-determined |
| SG_extracted_k141_6020 | Not-determined |
| SG_extracted_k141_25186 | Low-quality |
| SG_extracted_k141_4060 | Not-determined |
| SG_extracted_k141_10905 | Low-quality |
| SG_extracted_k141_36633 | Not-determined |
| SG_extracted_k141_19555 | Not-determined |
| SG_extracted_k141_18125 | Low-quality |
| SG_extracted_k141_8387 | Not-determined |
| SG_extracted_k141_46182 | Not-determined |
| SG_extracted_k141_46183 | Not-determined |
| SG_extracted_k141_19563 | Not-determined |
| SG_extracted_k141_8390 | Not-determined |
| SG_extracted_k141_6030 | Not-determined |
| SG_extracted_k141_18133 | Not-determined |
| SG_extracted_k141_8393 | Not-determined |
| SG_extracted_k141_6034 | Not-determined |
| SG_extracted_k141_8396 | Not-determined |
| SG_extracted_k141_41366 | Not-determined |
| SG_extracted_k141_46192 | Not-determined |
| SG_extracted_k141_31930 | Not-determined |
| SG_extracted_k141_46193 | Not-determined |
| SG_extracted_k141_18137 | Not-determined |
| SG_extracted_k141_31935 | Low-quality |
| SG_extracted_k141_13368 | Low-quality |
| SG_extracted_k141_19580 | Not-determined |
| SG_extracted_k141_18143 | Not-determined |
| SG_extracted_k141_50668 | Not-determined |
| SG_extracted_k141_25208 | Not-determined |
| SG_extracted_k141_46207 | Not-determined |
| SG_extracted_k141_41384 | Low-quality |
| SG_extracted_k141_50676 | Not-determined |
| SG_extracted_k141_19593 | Not-determined |
| SG_extracted_k141_19594 | Not-determined |
| SG_extracted_k141_50679 | Low-quality |
| SG_extracted_k141_25213 | Not-determined |
| SG_extracted_k141_36640 | Low-quality |
| SG_extracted_k141_36641 | Low-quality |
| SG_extracted_k141_26970 | Not-determined |
| SG_extracted_k141_8411 | Not-determined |
| SG_extracted_k141_15535 | Not-determined |
| SG_extracted_k141_46216 | Not-determined |
| SG_extracted_k141_50686 | Not-determined |
| SG_extracted_k141_46221 | Not-determined |
| SG_extracted_k141_4092 | Not-determined |
| SG_extracted_k141_19601 | Not-determined |
| SG_extracted_k141_8420 | Low-quality |
| SG_extracted_k141_6044 | Not-determined |
| SG_extracted_k141_50692 | Not-determined |
| SG_extracted_k141_50695 | Not-determined |
| SG_extracted_k141_26981 | Low-quality |
| SG_extracted_k141_13385 | Low-quality |
| SG_extracted_k141_8428 | Not-determined |
| SG_extracted_k141_15546 | Not-determined |
| SG_extracted_k141_18154 | Not-determined |
| SG_extracted_k141_55053 | Not-determined |
| SG_extracted_k141_4098 | Not-determined |
| SG_extracted_k141_15549 | Low-quality |
| SG_extracted_k141_19611 | Not-determined |
| SG_extracted_k141_19613 | Not-determined |
| SG_extracted_k141_6053 | Not-determined |
| SG_extracted_k141_48283 | Not-determined |
| SG_extracted_k141_29754 | Not-determined |
| SG_extracted_k141_19621 | Not-determined |
| SG_extracted_k141_43657 | Not-determined |
| SG_extracted_k141_29758 | Not-determined |
| SG_extracted_k141_19625 | Not-determined |
| SG_extracted_k141_15559 | Low-quality |
| SG_extracted_k141_26998 | Not-determined |
| SG_extracted_k141_55062 | Not-determined |
| SG_extracted_k141_34713 | Not-determined |
| SG_extracted_k141_34714 | Not-determined |
| SG_extracted_k141_13394 | Not-determined |
| SG_extracted_k141_19633 | Not-determined |
| SG_extracted_k141_41420 | Not-determined |
| SG_extracted_k141_18175 | Not-determined |
| SG_extracted_k141_13400 | Not-determined |
| SG_extracted_k141_13401 | Not-determined |
| SG_extracted_k141_19634 | Not-determined |
| SG_extracted_k141_29766 | Not-determined |
| SG_extracted_k141_19635 | Not-determined |
| SG_extracted_k141_8456 | Not-determined |
| SG_extracted_k141_8458 | Low-quality |
| SG_extracted_k141_19638 | Not-determined |
| SG_extracted_k141_50699 | Not-determined |
| SG_extracted_k141_6073 | Not-determined |
| SG_extracted_k141_55073 | Not-determined |
| SG_extracted_k141_6074 | Not-determined |
| SG_extracted_k141_15569 | Low-quality |
| SG_extracted_k141_34728 | Not-determined |
| SG_extracted_k141_4105 | Not-determined |
| SG_extracted_k141_34730 | Not-determined |
| SG_extracted_k141_31963 | Low-quality |
| SG_extracted_k141_34731 | Not-determined |
| SG_extracted_k141_31964 | Not-determined |
| SG_extracted_k141_19656 | Low-quality |
| SG_extracted_k141_15574 | Not-determined |
| SG_extracted_k141_18195 | Not-determined |
| SG_extracted_k141_41431 | Low-quality |
| SG_extracted_k141_27012 | Not-determined |
| SG_extracted_k141_25222 | Not-determined |
| SG_extracted_k141_27015 | Low-quality |
| SG_extracted_k141_25226 | Not-determined |
| SG_extracted_k141_25227 | Not-determined |
| SG_extracted_k141_19667 | Not-determined |
| SG_extracted_k141_22286 | Not-determined |
| SG_extracted_k141_18198 | Not-determined |
| SG_extracted_k141_48299 | Low-quality |
| SG_extracted_k141_50702 | Not-determined |
| SG_extracted_k141_34737 | Low-quality |
| SG_extracted_k141_4111 | Not-determined |
| SG_extracted_k141_34738 | Low-quality |
| SG_extracted_k141_15581 | Not-determined |
| SG_extracted_k141_25233 | Low-quality |
| SG_extracted_k141_8481 | Not-determined |
| SG_extracted_k141_8482 | Not-determined |
| SG_extracted_k141_34744 | Not-determined |
| SG_extracted_k141_41448 | Not-determined |
| SG_extracted_k141_48303 | Not-determined |
| SG_extracted_k141_34754 | Not-determined |
| SG_extracted_k141_22298 | Low-quality |
| SG_extracted_k141_15586 | Low-quality |
| SG_extracted_k141_41460 | Not-determined |
| SG_extracted_k141_34762 | Not-determined |
| SG_extracted_k141_22308 | Not-determined |
| SG_extracted_k141_46226 | Not-determined |
| SG_extracted_k141_22313 | Not-determined |
| SG_extracted_k141_8490 | Low-quality |
| SG_extracted_k141_25243 | Not-determined |
| SG_extracted_k141_13418 | Not-determined |
| SG_extracted_k141_46231 | Not-determined |
| SG_extracted_k141_46232 | Not-determined |
| SG_extracted_k141_25249 | Low-quality |
| SG_extracted_k141_15600 | Not-determined |
| SG_extracted_k141_36672 | Not-determined |
| SG_extracted_k141_13422 | Not-determined |
| SG_extracted_k141_15601 | Low-quality |
| SG_extracted_k141_34772 | Not-determined |
| SG_extracted_k141_15604 | Not-determined |
| SG_extracted_k141_13428 | Low-quality |
| SG_extracted_k141_36680 | Low-quality |
| SG_extracted_k141_8508 | Not-determined |
| SG_extracted_k141_19671 | Low-quality |
| SG_extracted_k141_36681 | Low-quality |
| SG_extracted_k141_25251 | Not-determined |
| SG_extracted_k141_36683 | Not-determined |
| SG_extracted_k141_13434 | Not-determined |
| SG_extracted_k141_13435 | Not-determined |
| SG_extracted_k141_19678 | Not-determined |
| SG_extracted_k141_4147 | Not-determined |
| SG_extracted_k141_8517 | Low-quality |
| SG_extracted_k141_4153 | Not-determined |
| SG_extracted_k141_34779 | Not-determined |
| SG_extracted_k141_15622 | Low-quality |
| SG_extracted_k141_41477 | Not-determined |
| SG_extracted_k141_36696 | Low-quality |
| SG_extracted_k141_15628 | Not-determined |
| SG_extracted_k141_19695 | Not-determined |
| SG_extracted_k141_48333 | Not-determined |
| SG_extracted_k141_48336 | Not-determined |
| SG_extracted_k141_8525 | Not-determined |
| SG_extracted_k141_41481 | Low-quality |
| SG_extracted_k141_4162 | Not-determined |
| SG_extracted_k141_8530 | Not-determined |
| SG_extracted_k141_13447 | Low-quality |
| SG_extracted_k141_4165 | Low-quality |
| SG_extracted_k141_19708 | Not-determined |
| SG_extracted_k141_34802 | Not-determined |
| SG_extracted_k141_19711 | Not-determined |
| SG_extracted_k141_25268 | Not-determined |
| SG_extracted_k141_13448 | Not-determined |
| SG_extracted_k141_43669 | Not-determined |
| SG_extracted_k141_27048 | Not-determined |
| SG_extracted_k141_25270 | Not-determined |
| SG_extracted_k141_13451 | Low-quality |
| SG_extracted_k141_8535 | Not-determined |
| SG_extracted_k141_41487 | Not-determined |
| SG_extracted_k141_15646 | Low-quality |
| SG_extracted_k141_8537 | Not-determined |
| SG_extracted_k141_19722 | Not-determined |
| SG_extracted_k141_19725 | Not-determined |
| SG_extracted_k141_27058 | Not-determined |
| SG_extracted_k141_41492 | Not-determined |
| SG_extracted_k141_27059 | Not-determined |
| SG_extracted_k141_4172 | Not-determined |
| SG_extracted_k141_19727 | Not-determined |
| SG_extracted_k141_15654 | Not-determined |
| SG_extracted_k141_8539 | Not-determined |
| SG_extracted_k141_13460 | Low-quality |
| SG_extracted_k141_19731 | Not-determined |
| SG_extracted_k141_15656 | Not-determined |
| SG_extracted_k141_46265 | Low-quality |
| SG_extracted_k141_8542 | Not-determined |
| SG_extracted_k141_13467 | Not-determined |
| SG_extracted_k141_27063 | Low-quality |
| SG_extracted_k141_8546 | Not-determined |
| SG_extracted_k141_34823 | Not-determined |
| SG_extracted_k141_46272 | Not-determined |
| SG_extracted_k141_8548 | Not-determined |
| SG_extracted_k141_13474 | Not-determined |
| SG_extracted_k141_4181 | Not-determined |
| SG_extracted_k141_13477 | Not-determined |
| SG_extracted_k141_25293 | Not-determined |
| SG_extracted_k141_27067 | Not-determined |
| SG_extracted_k141_34832 | Not-determined |
| SG_extracted_k141_13480 | Not-determined |
| SG_extracted_k141_13481 | Not-determined |
| SG_extracted_k141_41510 | Not-determined |
| SG_extracted_k141_13482 | Low-quality |
| SG_extracted_k141_13483 | Not-determined |
| SG_extracted_k141_34837 | Not-determined |
| SG_extracted_k141_15670 | Not-determined |
| SG_extracted_k141_6085 | Not-determined |
| SG_extracted_k141_15676 | Not-determined |
| SG_extracted_k141_41519 | Low-quality |
| SG_extracted_k141_25305 | Low-quality |
| SG_extracted_k141_15679 | Not-determined |
| SG_extracted_k141_1389 | Not-determined |
| SG_extracted_k141_46292 | Not-determined |
| SG_extracted_k141_43679 | Not-determined |
| SG_extracted_k141_29786 | Not-determined |
| SG_extracted_k141_48360 | Not-determined |
| SG_extracted_k141_15680 | Not-determined |
| SG_extracted_k141_19738 | Not-determined |
| SG_extracted_k141_41524 | Not-determined |
| SG_extracted_k141_27084 | Not-determined |
| SG_extracted_k141_41525 | Not-determined |
| SG_extracted_k141_27087 | Not-determined |
| SG_extracted_k141_1392 | Not-determined |
| SG_extracted_k141_1393 | Not-determined |
| SG_extracted_k141_6090 | Low-quality |
| SG_extracted_k141_4193 | Not-determined |
| SG_extracted_k141_48375 | Not-determined |
| SG_extracted_k141_8571 | Not-determined |
| SG_extracted_k141_19749 | Not-determined |
| SG_extracted_k141_4196 | Not-determined |
| SG_extracted_k141_25311 | Not-determined |
| SG_extracted_k141_34846 | Not-determined |
| SG_extracted_k141_34847 | Not-determined |
| SG_extracted_k141_15689 | Not-determined |
| SG_extracted_k141_41532 | Not-determined |
| SG_extracted_k141_1400 | Not-determined |
| SG_extracted_k141_41535 | Low-quality |
| SG_extracted_k141_25312 | Low-quality |
| SG_extracted_k141_19756 | Not-determined |
| SG_extracted_k141_15692 | Not-determined |
| SG_extracted_k141_27089 | Not-determined |
| SG_extracted_k141_4210 | Not-determined |
| SG_extracted_k141_41543 | Not-determined |
| SG_extracted_k141_41544 | Not-determined |
| SG_extracted_k141_1404 | Not-determined |
| SG_extracted_k141_22324 | Not-determined |
| SG_extracted_k141_4214 | Not-determined |
| SG_extracted_k141_19765 | Not-determined |
| SG_extracted_k141_1405 | Not-determined |
| SG_extracted_k141_8590 | Not-determined |
| SG_extracted_k141_4215 | Low-quality |
| SG_extracted_k141_1407 | Not-determined |
| SG_extracted_k141_1408 | Not-determined |
| SG_extracted_k141_46313 | Not-determined |
| SG_extracted_k141_46316 | Not-determined |
| SG_extracted_k141_4217 | Not-determined |
| SG_extracted_k141_22328 | Not-determined |
| SG_extracted_k141_53394 | Not-determined |
| SG_extracted_k141_46317 | Not-determined |
| SG_extracted_k141_27098 | Not-determined |
| SG_extracted_k141_1414 | Not-determined |
| SG_extracted_k141_22331 | Not-determined |
| SG_extracted_k141_27100 | Not-determined |
| SG_extracted_k141_8599 | Not-determined |
| SG_extracted_k141_46320 | Not-determined |
| SG_extracted_k141_41557 | Not-determined |
| SG_extracted_k141_22336 | Not-determined |
| SG_extracted_k141_46323 | Not-determined |
| SG_extracted_k141_41560 | Not-determined |
| SG_extracted_k141_25322 | Low-quality |
| SG_extracted_k141_4228 | Not-determined |
| SG_extracted_k141_27107 | Not-determined |
| SG_extracted_k141_4229 | Not-determined |
| SG_extracted_k141_8612 | Not-determined |
| SG_extracted_k141_46331 | Not-determined |
| SG_extracted_k141_1421 | Not-determined |
| SG_extracted_k141_15720 | Not-determined |
| SG_extracted_k141_46334 | Low-quality |
| SG_extracted_k141_10912 | Not-determined |
| SG_extracted_k141_10915 | Not-determined |
| SG_extracted_k141_25329 | Not-determined |
| SG_extracted_k141_15728 | Not-determined |
| SG_extracted_k141_13499 | Not-determined |
| SG_extracted_k141_46339 | Low-quality |
| SG_extracted_k141_13503 | Not-determined |
| SG_extracted_k141_13506 | Not-determined |
| SG_extracted_k141_13509 | Low-quality |
| SG_extracted_k141_29833 | Not-determined |
| SG_extracted_k141_10929 | Not-determined |
| SG_extracted_k141_46351 | Not-determined |
| SG_extracted_k141_25345 | Not-determined |
| SG_extracted_k141_6127 | Not-determined |
| SG_extracted_k141_13521 | Not-determined |
| SG_extracted_k141_46358 | Not-determined |
| SG_extracted_k141_4254 | Not-determined |
| SG_extracted_k141_6130 | Not-determined |
| SG_extracted_k141_6132 | Low-quality |
| SG_extracted_k141_13525 | Not-determined |
| SG_extracted_k141_4258 | Low-quality |
| SG_extracted_k141_27124 | Low-quality |
| SG_extracted_k141_25353 | Low-quality |
| SG_extracted_k141_27127 | Not-determined |
| SG_extracted_k141_25354 | Not-determined |
| SG_extracted_k141_43699 | Not-determined |
| SG_extracted_k141_25357 | Not-determined |
| SG_extracted_k141_10949 | Low-quality |
| SG_extracted_k141_19801 | Not-determined |
| SG_extracted_k141_27135 | Not-determined |
| SG_extracted_k141_15741 | Not-determined |
| SG_extracted_k141_13536 | Not-determined |
| SG_extracted_k141_46369 | Low-quality |
| SG_extracted_k141_19804 | Low-quality |
| SG_extracted_k141_13538 | Not-determined |
| SG_extracted_k141_19806 | Not-determined |
| SG_extracted_k141_13540 | Not-determined |
| SG_extracted_k141_13542 | Not-determined |
| SG_extracted_k141_46370 | Not-determined |
| SG_extracted_k141_10958 | Low-quality |
| SG_extracted_k141_1427 | Not-determined |
| SG_extracted_k141_13546 | Low-quality |
| SG_extracted_k141_15748 | Not-determined |
| SG_extracted_k141_10963 | Not-determined |
| SG_extracted_k141_46376 | Not-determined |
| SG_extracted_k141_29861 | Not-determined |
| SG_extracted_k141_19809 | Not-determined |
| SG_extracted_k141_13548 | Not-determined |
| SG_extracted_k141_22341 | Not-determined |
| SG_extracted_k141_32035 | Not-determined |
| SG_extracted_k141_41608 | Not-determined |
| SG_extracted_k141_4279 | Low-quality |
| SG_extracted_k141_27142 | Low-quality |
| SG_extracted_k141_13558 | Not-determined |
| SG_extracted_k141_29869 | Not-determined |
| SG_extracted_k141_6146 | Not-determined |
| SG_extracted_k141_29870 | Not-determined |
| SG_extracted_k141_43707 | Not-determined |
| SG_extracted_k141_41613 | Not-determined |
| SG_extracted_k141_41615 | Low-quality |
| SG_extracted_k141_29872 | Not-determined |
| SG_extracted_k141_41619 | Not-determined |
| SG_extracted_k141_15756 | Not-determined |
| SG_extracted_k141_29878 | Not-determined |
| SG_extracted_k141_1436 | Not-determined |
| SG_extracted_k141_1437 | Not-determined |
| SG_extracted_k141_15758 | Not-determined |
| SG_extracted_k141_22364 | Low-quality |
| SG_extracted_k141_43716 | Low-quality |
| SG_extracted_k141_32040 | Low-quality |
| SG_extracted_k141_43717 | Low-quality |
| SG_extracted_k141_13561 | Not-determined |
| SG_extracted_k141_32041 | Not-determined |
| SG_extracted_k141_43718 | Not-determined |
| SG_extracted_k141_1448 | Not-determined |
| SG_extracted_k141_43722 | Not-determined |
| SG_extracted_k141_29887 | Not-determined |
| SG_extracted_k141_8649 | Not-determined |
| SG_extracted_k141_13569 | Not-determined |
| SG_extracted_k141_27154 | Not-determined |
| SG_extracted_k141_43726 | Not-determined |
| SG_extracted_k141_8654 | Not-determined |
| SG_extracted_k141_41630 | Low-quality |
| SG_extracted_k141_1461 | Not-determined |
| SG_extracted_k141_13574 | Not-determined |
| SG_extracted_k141_13575 | Not-determined |
| SG_extracted_k141_43730 | Not-determined |
| SG_extracted_k141_27161 | Not-determined |
| SG_extracted_k141_13577 | Not-determined |
| SG_extracted_k141_15766 | Not-determined |
| SG_extracted_k141_22379 | Not-determined |
| SG_extracted_k141_4312 | Low-quality |
| SG_extracted_k141_13582 | Not-determined |
| SG_extracted_k141_13583 | Low-quality |
| SG_extracted_k141_13584 | Not-determined |
| SG_extracted_k141_43741 | Low-quality |
| SG_extracted_k141_6158 | Not-determined |
| SG_extracted_k141_1474 | Not-determined |
| SG_extracted_k141_6162 | Not-determined |
| SG_extracted_k141_34878 | Low-quality |
| SG_extracted_k141_4313 | Low-quality |
| SG_extracted_k141_53413 | Not-determined |
| SG_extracted_k141_6164 | Not-determined |
| SG_extracted_k141_4315 | Not-determined |
| SG_extracted_k141_8670 | Low-quality |
| SG_extracted_k141_27163 | Not-determined |
| SG_extracted_k141_34881 | Not-determined |
| SG_extracted_k141_27166 | Not-determined |
| SG_extracted_k141_22397 | Not-determined |
| SG_extracted_k141_6170 | Not-determined |
| SG_extracted_k141_22402 | Not-determined |
| SG_extracted_k141_6175 | Low-quality |
| SG_extracted_k141_19816 | Not-determined |
| SG_extracted_k141_19817 | Low-quality |
| SG_extracted_k141_15783 | Not-determined |
| SG_extracted_k141_22407 | Not-determined |
| SG_extracted_k141_8684 | Not-determined |
| SG_extracted_k141_22411 | Not-determined |
| SG_extracted_k141_50729 | Not-determined |
| SG_extracted_k141_8687 | Not-determined |
| SG_extracted_k141_50732 | Not-determined |
| SG_extracted_k141_6179 | Low-quality |
| SG_extracted_k141_32053 | Not-determined |
| SG_extracted_k141_53424 | Not-determined |
| SG_extracted_k141_13594 | Not-determined |
| SG_extracted_k141_55098 | Low-quality |
| SG_extracted_k141_6188 | Not-determined |
| SG_extracted_k141_15794 | Not-determined |
| SG_extracted_k141_55101 | Not-determined |
| SG_extracted_k141_1501 | Not-determined |
| SG_extracted_k141_15797 | Not-determined |
| SG_extracted_k141_55104 | Not-determined |
| SG_extracted_k141_18225 | Not-determined |
| SG_extracted_k141_18228 | Not-determined |
| SG_extracted_k141_13605 | Not-determined |
| SG_extracted_k141_15801 | Not-determined |
| SG_extracted_k141_32070 | Not-determined |
| SG_extracted_k141_55108 | Not-determined |
| SG_extracted_k141_27190 | Not-determined |
| SG_extracted_k141_13611 | Not-determined |
| SG_extracted_k141_6194 | Low-quality |
| SG_extracted_k141_27193 | Not-determined |
| SG_extracted_k141_6197 | Not-determined |
| SG_extracted_k141_19822 | Not-determined |
| SG_extracted_k141_29901 | Not-determined |
| SG_extracted_k141_4337 | Low-quality |
| SG_extracted_k141_53442 | Not-determined |
| SG_extracted_k141_4339 | Low-quality |
| SG_extracted_k141_18244 | Not-determined |
| SG_extracted_k141_4341 | Not-determined |
| SG_extracted_k141_18247 | Not-determined |
| SG_extracted_k141_19832 | Not-determined |
| SG_extracted_k141_29905 | Not-determined |
| SG_extracted_k141_32084 | Not-determined |

| | |
|---|---|
| SG_extracted_k141_29911 | Not-determined |
| SG_extracted_k141_6206 | Low-quality |
| SG_extracted_k141_19834 | Not-determined |
| SG_extracted_k141_6207 | Not-determined |
| SG_extracted_k141_43760 | Not-determined |
| SG_extracted_k141_18257 | Not-determined |
| SG_extracted_k141_8711 | Low-quality |
| SG_extracted_k141_13631 | Not-determined |
| SG_extracted_k141_8717 | Not-determined |
| SG_extracted_k141_41651 | Not-determined |
| SG_extracted_k141_8718 | Not-determined |
| SG_extracted_k141_50750 | Not-determined |
| SG_extracted_k141_41654 | Not-determined |
| SG_extracted_k141_13639 | Low-quality |
| SG_extracted_k141_8726 | Low-quality |
| SG_extracted_k141_50758 | Low-quality |
| SG_extracted_k141_13642 | Not-determined |
| SG_extracted_k141_18265 | Not-determined |
| SG_extracted_k141_55126 | Not-determined |
| SG_extracted_k141_27200 | Low-quality |
| SG_extracted_k141_8731 | Low-quality |
| SG_extracted_k141_29925 | Low-quality |
| SG_extracted_k141_41671 | Not-determined |
| SG_extracted_k141_8735 | Not-determined |
| SG_extracted_k141_29928 | Low-quality |
| SG_extracted_k141_6231 | Not-determined |
| SG_extracted_k141_8737 | Not-determined |
| SG_extracted_k141_41675 | Low-quality |
| SG_extracted_k141_29932 | Not-determined |
| SG_extracted_k141_32110 | Not-determined |
| SG_extracted_k141_32111 | Not-determined |
| SG_extracted_k141_32112 | Low-quality |
| SG_extracted_k141_13656 | Low-quality |
| SG_extracted_k141_41681 | Not-determined |
| SG_extracted_k141_55136 | Not-determined |
| SG_extracted_k141_27206 | Not-determined |
| SG_extracted_k141_36710 | Not-determined |
| SG_extracted_k141_29942 | Not-determined |
| SG_extracted_k141_27210 | Not-determined |
| SG_extracted_k141_36715 | Not-determined |
| SG_extracted_k141_13679 | Not-determined |
| SG_extracted_k141_18277 | Not-determined |
| SG_extracted_k141_27214 | Not-determined |
| SG_extracted_k141_13680 | Not-determined |
| SG_extracted_k141_6248 | Not-determined |
| SG_extracted_k141_55144 | Not-determined |
| SG_extracted_k141_46389 | Not-determined |
| SG_extracted_k141_6250 | Not-determined |
| SG_extracted_k141_6252 | Not-determined |
| SG_extracted_k141_50763 | Not-determined |
| SG_extracted_k141_19847 | Not-determined |
| SG_extracted_k141_18288 | Not-determined |
| SG_extracted_k141_36724 | Not-determined |
| SG_extracted_k141_27227 | Low-quality |
| SG_extracted_k141_55151 | Low-quality |
| SG_extracted_k141_25373 | Not-determined |
| SG_extracted_k141_18294 | Not-determined |
| SG_extracted_k141_41689 | Not-determined |
| SG_extracted_k141_41692 | Not-determined |
| SG_extracted_k141_32119 | Not-determined |
| SG_extracted_k141_41695 | Not-determined |
| SG_extracted_k141_46395 | Not-determined |
| SG_extracted_k141_32123 | Low-quality |
| SG_extracted_k141_22429 | Not-determined |
| SG_extracted_k141_29968 | Not-determined |
| SG_extracted_k141_46399 | Not-determined |
| SG_extracted_k141_25394 | Not-determined |
| SG_extracted_k141_19877 | Low-quality |
| SG_extracted_k141_19878 | Not-determined |
| SG_extracted_k141_41713 | Low-quality |
| SG_extracted_k141_46408 | Not-determined |
| SG_extracted_k141_41715 | Low-quality |
| SG_extracted_k141_25404 | Low-quality |
| SG_extracted_k141_19885 | Low-quality |
| SG_extracted_k141_50774 | Not-determined |
| SG_extracted_k141_25406 | Not-determined |
| SG_extracted_k141_19886 | Not-determined |
| SG_extracted_k141_18318 | Not-determined |
| SG_extracted_k141_41719 | Not-determined |
| SG_extracted_k141_25408 | Low-quality |
| SG_extracted_k141_15819 | Not-determined |
| SG_extracted_k141_53482 | Low-quality |
| SG_extracted_k141_29982 | Not-determined |
| SG_extracted_k141_55171 | Not-determined |
| SG_extracted_k141_4362 | Not-determined |
| SG_extracted_k141_15825 | Not-determined |
| SG_extracted_k141_53484 | Not-determined |
| SG_extracted_k141_22438 | Low-quality |
| SG_extracted_k141_15832 | Not-determined |
| SG_extracted_k141_41728 | Low-quality |
| SG_extracted_k141_46417 | Not-determined |
| SG_extracted_k141_41729 | Not-determined |
| SG_extracted_k141_46419 | Not-determined |
| SG_extracted_k141_15836 | Not-determined |
| SG_extracted_k141_53497 | Not-determined |
| SG_extracted_k141_29998 | Not-determined |
| SG_extracted_k141_53501 | Not-determined |
| SG_extracted_k141_55186 | Not-determined |
| SG_extracted_k141_4370 | Not-determined |
| SG_extracted_k141_22451 | Not-determined |
| SG_extracted_k141_22453 | Not-determined |
| SG_extracted_k141_55189 | Not-determined |
| SG_extracted_k141_55191 | Not-determined |
| SG_extracted_k141_18333 | Not-determined |
| SG_extracted_k141_55192 | Not-determined |
| SG_extracted_k141_55193 | Not-determined |
| SG_extracted_k141_55194 | Not-determined |
| SG_extracted_k141_55195 | Not-determined |
| SG_extracted_k141_22461 | Not-determined |
| SG_extracted_k141_55198 | Not-determined |
| SG_extracted_k141_18334 | Not-determined |
| SG_extracted_k141_18335 | Not-determined |
| SG_extracted_k141_55199 | Low-quality |
| SG_extracted_k141_46430 | Not-determined |
| SG_extracted_k141_30010 | Not-determined |
| SG_extracted_k141_53515 | Not-determined |
| SG_extracted_k141_11010 | Not-determined |
| SG_extracted_k141_11012 | Not-determined |
| SG_extracted_k141_53525 | Not-determined |
| SG_extracted_k141_41749 | Not-determined |
| SG_extracted_k141_53526 | Not-determined |
| SG_extracted_k141_36732 | Not-determined |
| SG_extracted_k141_8782 | Not-determined |
| SG_extracted_k141_41752 | Not-determined |
| SG_extracted_k141_36733 | Not-determined |
| SG_extracted_k141_36734 | Low-quality |
| SG_extracted_k141_53531 | Not-determined |
| SG_extracted_k141_8788 | Not-determined |
| SG_extracted_k141_36741 | Not-determined |
| SG_extracted_k141_18350 | Not-determined |
| SG_extracted_k141_18352 | Low-quality |
| SG_extracted_k141_53539 | Not-determined |
| SG_extracted_k141_4382 | Not-determined |
| SG_extracted_k141_15871 | Low-quality |
| SG_extracted_k141_22483 | Not-determined |
| SG_extracted_k141_46439 | Not-determined |
| SG_extracted_k141_55220 | Not-determined |
| SG_extracted_k141_11027 | Low-quality |
| SG_extracted_k141_15878 | Not-determined |
| SG_extracted_k141_46442 | Not-determined |
| SG_extracted_k141_50789 | Not-determined |
| SG_extracted_k141_15879 | Low-quality |
| SG_extracted_k141_46447 | Not-determined |
| SG_extracted_k141_39081 | Not-determined |
| SG_extracted_k141_41774 | Not-determined |
| SG_extracted_k141_46453 | Not-determined |
| SG_extracted_k141_27250 | Not-determined |
| SG_extracted_k141_55237 | Not-determined |
| SG_extracted_k141_4397 | Low-quality |
| SG_extracted_k141_41779 | Not-determined |
| SG_extracted_k141_27254 | Not-determined |
| SG_extracted_k141_41784 | Not-determined |
| SG_extracted_k141_27256 | Low-quality |
| SG_extracted_k141_32126 | Low-quality |
| SG_extracted_k141_30019 | Not-determined |
| SG_extracted_k141_46465 | Not-determined |
| SG_extracted_k141_55242 | Not-determined |
| SG_extracted_k141_32130 | Not-determined |
| SG_extracted_k141_4401 | Not-determined |
| SG_extracted_k141_34908 | Not-determined |
| SG_extracted_k141_27261 | Low-quality |
| SG_extracted_k141_6299 | Not-determined |
| SG_extracted_k141_46469 | Not-determined |
| SG_extracted_k141_55247 | Low-quality |
| SG_extracted_k141_55249 | Not-determined |
| SG_extracted_k141_46474 | Not-determined |
| SG_extracted_k141_30032 | Not-determined |
| SG_extracted_k141_4407 | Not-determined |
| SG_extracted_k141_22497 | Not-determined |
| SG_extracted_k141_50797 | Low-quality |
| SG_extracted_k141_43774 | Not-determined |
| SG_extracted_k141_22500 | Not-determined |
| SG_extracted_k141_32145 | Not-determined |
| SG_extracted_k141_4414 | Not-determined |
| SG_extracted_k141_46477 | Not-determined |
| SG_extracted_k141_30042 | Not-determined |
| SG_extracted_k141_22502 | Not-determined |
| SG_extracted_k141_43781 | Not-determined |
| SG_extracted_k141_32157 | Not-determined |
| SG_extracted_k141_4421 | Not-determined |
| SG_extracted_k141_4422 | Low-quality |
| SG_extracted_k141_22510 | Not-determined |
| SG_extracted_k141_43785 | Not-determined |
| SG_extracted_k141_55261 | Not-determined |
| SG_extracted_k141_4426 | Low-quality |
| SG_extracted_k141_46485 | Low-quality |
| SG_extracted_k141_22516 | Not-determined |
| SG_extracted_k141_4429 | Not-determined |
| SG_extracted_k141_46489 | Not-determined |
| SG_extracted_k141_34918 | Low-quality |
| SG_extracted_k141_30051 | Not-determined |
| SG_extracted_k141_32169 | Low-quality |
| SG_extracted_k141_15901 | Not-determined |
| SG_extracted_k141_30052 | Not-determined |
| SG_extracted_k141_36784 | Not-determined |
| SG_extracted_k141_27276 | Not-determined |
| SG_extracted_k141_32174 | Low-quality |
| SG_extracted_k141_30057 | Not-determined |
| SG_extracted_k141_41798 | Not-determined |
| SG_extracted_k141_30058 | Low-quality |
| SG_extracted_k141_53568 | Not-determined |
| SG_extracted_k141_30063 | Not-determined |
| SG_extracted_k141_4437 | Not-determined |
| SG_extracted_k141_46501 | Not-determined |
| SG_extracted_k141_32180 | Not-determined |
| S9_vs_k141_85495\|\|full | Medium-quality |
| SG_extracted_k141_55273 | Not-determined |
| SG_extracted_k141_55274 | Low-quality |
| SG_extracted_k141_53575 | Not-determined |
| SG_extracted_k141_43789 | Not-determined |
| SG_extracted_k141_32184 | Low-quality |
| SG_extracted_k141_43791 | Not-determined |
| SG_extracted_k141_43792 | Not-determined |
| SG_extracted_k141_55278 | Not-determined |
| SG_extracted_k141_34935 | Not-determined |
| SG_extracted_k141_36789 | Low-quality |
| SG_extracted_k141_11070 | Not-determined |
| SG_extracted_k141_30070 | Not-determined |
| SG_extracted_k141_22536 | Not-determined |
| SG_extracted_k141_43802 | Not-determined |
| SG_extracted_k141_36797 | Not-determined |
| SG_extracted_k141_43805 | Low-quality |
| SG_extracted_k141_39107 | Not-determined |
| SG_extracted_k141_13701 | Not-determined |
| SG_extracted_k141_32194 | Not-determined |
| SG_extracted_k141_22545 | Not-determined |
| SG_extracted_k141_32197 | Not-determined |
| SG_extracted_k141_55287 | Not-determined |
| SG_extracted_k141_39118 | Not-determined |
| SG_extracted_k141_13708 | Not-determined |
| SG_extracted_k141_55292 | Not-determined |
| SG_extracted_k141_34945 | Not-determined |
| SG_extracted_k141_27299 | Not-determined |
| SG_extracted_k141_36807 | Low-quality |
| SG_extracted_k141_36808 | Not-determined |
| SG_extracted_k141_41815 | Not-determined |
| SG_extracted_k141_22553 | Not-determined |
| SG_extracted_k141_39121 | Not-determined |
| SG_extracted_k141_39123 | Low-quality |
| SG_extracted_k141_15911 | Not-determined |
| SG_extracted_k141_30076 | Not-determined |
| SG_extracted_k141_13717 | Not-determined |
| SG_extracted_k141_32210 | Not-determined |
| SG_extracted_k141_15912 | Low-quality |
| SG_extracted_k141_13720 | Low-quality |
| SG_extracted_k141_30078 | Not-determined |
| SG_extracted_k141_41822 | Not-determined |
| SG_extracted_k141_32213 | Not-determined |
| SG_extracted_k141_22561 | Not-determined |
| SG_extracted_k141_41824 | Not-determined |
| SG_extracted_k141_41825 | Low-quality |
| SG_extracted_k141_18384 | Not-determined |
| SG_extracted_k141_36818 | Not-determined |
| SG_extracted_k141_55303 | Not-determined |
| SG_extracted_k141_22567 | Low-quality |
| SG_extracted_k141_55306 | Not-determined |
| SG_extracted_k141_55309 | Not-determined |
| SG_extracted_k141_18394 | Not-determined |
| SG_extracted_k141_46539 | Low-quality |
| SG_extracted_k141_30082 | Not-determined |
| SG_extracted_k141_46540 | Not-determined |
| SG_extracted_k141_15922 | Not-determined |
| SG_extracted_k141_46541 | Not-determined |
| SG_extracted_k141_55313 | Not-determined |
| SG_extracted_k141_41845 | Not-determined |
| SG_extracted_k141_30087 | Not-determined |
| SG_extracted_k141_22579 | Not-determined |
| SG_extracted_k141_22580 | Not-determined |
| SG_extracted_k141_50823 | Low-quality |
| SG_extracted_k141_55323 | Not-determined |
| SG_extracted_k141_22588 | Not-determined |
| SG_extracted_k141_55336 | Not-determined |
| SG_extracted_k141_39127 | Not-determined |
| SG_extracted_k141_55338 | Low-quality |
| SG_extracted_k141_11098 | Low-quality |
| SG_extracted_k141_11099 | Not-determined |
| SG_extracted_k141_39132 | Not-determined |
| SG_extracted_k141_30097 | Not-determined |
| SG_extracted_k141_39138 | Low-quality |
| SG_extracted_k141_22591 | Low-quality |
| SG_extracted_k141_22592 | Not-determined |
| SG_extracted_k141_30107 | Not-determined |
| SG_extracted_k141_30108 | Not-determined |
| SG_extracted_k141_30110 | Not-determined |
| SG_extracted_k141_50848 | Not-determined |
| SG_extracted_k141_36824 | Not-determined |
| SG_extracted_k141_50854 | Not-determined |
| SG_extracted_k141_18396 | Not-determined |
| SG_extracted_k141_22596 | Not-determined |
| SG_extracted_k141_30118 | Not-determined |
| SG_extracted_k141_41864 | Not-determined |
| SG_extracted_k141_41865 | Not-determined |
| SG_extracted_k141_41866 | Not-determined |
| SG_extracted_k141_15929 | Not-determined |
| SG_extracted_k141_15930 | Not-determined |
| SG_extracted_k141_6368 | Not-determined |
| SG_extracted_k141_18402 | Not-determined |
| SG_extracted_k141_6378 | Low-quality |
| SG_extracted_k141_55361 | Not-determined |
| SG_extracted_k141_41881 | Not-determined |
| SG_extracted_k141_15941 | Low-quality |
| SG_extracted_k141_6383 | Not-determined |
| SG_extracted_k141_18408 | Not-determined |
| SG_extracted_k141_41883 | Not-determined |
| SG_extracted_k141_41884 | Not-determined |
| SG_extracted_k141_41887 | Not-determined |
| SG_extracted_k141_34962 | Not-determined |
| SG_extracted_k141_30141 | Not-determined |
| SG_extracted_k141_53619 | Not-determined |
| SG_extracted_k141_6390 | Not-determined |
| SG_extracted_k141_22605 | Not-determined |
| SG_extracted_k141_53620 | Not-determined |
| SG_extracted_k141_1549 | Not-determined |
| SG_extracted_k141_30150 | Not-determined |
| SG_extracted_k141_50866 | Not-determined |
| SG_extracted_k141_50868 | Not-determined |
| SG_extracted_k141_22610 | Low-quality |
| SG_extracted_k141_15949 | Low-quality |
| SG_extracted_k141_50873 | Not-determined |
| SG_extracted_k141_32254 | Not-determined |
| SG_extracted_k141_15955 | Not-determined |
| SG_extracted_k141_32256 | Low-quality |
| SG_extracted_k141_55379 | Not-determined |
| SG_extracted_k141_50879 | Not-determined |
| SG_extracted_k141_32262 | Not-determined |
| SG_extracted_k141_22622 | Not-determined |
| SG_extracted_k141_30153 | Low-quality |
| SG_extracted_k141_15959 | Not-determined |
| SG_extracted_k141_36830 | Not-determined |
| SG_extracted_k141_53624 | Not-determined |
| SG_extracted_k141_30156 | Not-determined |
| SG_extracted_k141_15961 | Not-determined |
| SG_extracted_k141_15962 | Not-determined |
| SG_extracted_k141_6404 | Not-determined |
| SG_extracted_k141_53630 | Not-determined |
| SG_extracted_k141_4466 | Not-determined |
| SG_extracted_k141_6406 | Not-determined |
| SG_extracted_k141_18418 | Not-determined |
| SG_extracted_k141_53632 | Not-determined |
| SG_extracted_k141_30169 | Not-determined |
| SG_extracted_k141_22630 | Not-determined |
| SG_extracted_k141_53634 | Not-determined |
| SG_extracted_k141_22631 | Low-quality |
| SG_extracted_k141_55398 | Not-determined |
| SG_extracted_k141_55404 | Low-quality |
| SG_extracted_k141_15968 | Not-determined |
| SG_extracted_k141_6412 | Not-determined |
| SG_extracted_k141_55407 | Not-determined |
| SG_extracted_k141_6416 | Not-determined |
| SG_extracted_k141_53641 | Not-determined |
| SG_extracted_k141_36832 | Low-quality |
| SG_extracted_k141_53643 | Not-determined |
| SG_extracted_k141_6419 | Not-determined |
| SG_extracted_k141_19899 | Not-determined |
| SG_extracted_k141_13730 | Not-determined |
| SG_extracted_k141_15971 | Not-determined |
| SG_extracted_k141_19901 | Not-determined |
| SG_extracted_k141_13731 | Low-quality |
| SG_extracted_k141_35001 | Not-determined |
| SG_extracted_k141_39179 | Not-determined |
| SG_extracted_k141_55416 | Low-quality |
| SG_extracted_k141_53650 | Not-determined |
| SG_extracted_k141_35003 | Not-determined |
| SG_extracted_k141_6425 | Not-determined |
| SG_extracted_k141_53652 | Not-determined |
| SG_extracted_k141_35006 | Low-quality |
| SG_extracted_k141_15976 | Not-determined |
| SG_extracted_k141_55422 | Not-determined |
| SG_extracted_k141_35009 | Not-determined |
| SG_extracted_k141_50892 | Not-determined |
| SG_extracted_k141_50893 | Not-determined |
| SG_extracted_k141_19908 | Not-determined |
| SG_extracted_k141_55429 | Not-determined |
| SG_extracted_k141_13746 | Not-determined |
| SG_extracted_k141_46561 | Not-determined |
| SG_extracted_k141_46564 | Not-determined |
| SG_extracted_k141_55436 | Low-quality |
| SG_extracted_k141_6433 | Not-determined |
| SG_extracted_k141_1558 | Not-determined |
| SG_extracted_k141_32265 | Not-determined |
| SG_extracted_k141_13755 | Not-determined |
| SG_extracted_k141_32266 | Low-quality |
| SG_extracted_k141_32268 | Not-determined |
| SG_extracted_k141_32269 | Not-determined |
| SG_extracted_k141_36840 | Not-determined |
| SG_extracted_k141_46571 | Not-determined |
| SG_extracted_k141_18424 | Not-determined |
| SG_extracted_k141_13760 | Not-determined |
| SG_extracted_k141_22639 | Not-determined |
| SG_extracted_k141_35022 | Not-determined |
| SG_extracted_k141_15991 | Not-determined |
| SG_extracted_k141_35023 | Low-quality |
| SG_extracted_k141_53661 | Not-determined |
| SG_extracted_k141_53662 | Not-determined |
| SG_extracted_k141_55451 | Not-determined |
| SG_extracted_k141_6453 | Not-determined |
| SG_extracted_k141_53666 | Not-determined |
| SG_extracted_k141_46582 | Not-determined |
| SG_extracted_k141_53669 | Low-quality |
| SG_extracted_k141_46583 | Not-determined |
| SG_extracted_k141_46585 | Not-determined |
| SG_extracted_k141_22658 | Not-determined |
| SG_extracted_k141_6459 | Not-determined |
| SG_extracted_k141_13773 | Not-determined |
| SG_extracted_k141_53677 | Low-quality |
| SG_extracted_k141_32302 | Not-determined |
| SG_extracted_k141_35044 | Not-determined |
| SG_extracted_k141_46597 | Low-quality |
| SG_extracted_k141_41907 | Not-determined |
| SG_extracted_k141_36844 | Not-determined |
| SG_extracted_k141_25414 | Not-determined |
| SG_extracted_k141_19924 | Not-determined |
| SG_extracted_k141_18449 | Not-determined |
| SG_extracted_k141_32305 | Low-quality |
| SG_extracted_k141_22666 | Low-quality |
| SG_extracted_k141_25419 | Not-determined |
| SG_extracted_k141_25420 | Not-determined |
| SG_extracted_k141_46605 | Not-determined |
| SG_extracted_k141_22669 | Not-determined |
| SG_extracted_k141_25424 | Low-quality |
| SG_extracted_k141_53692 | Low-quality |
| SG_extracted_k141_27317 | Not-determined |
| SG_extracted_k141_27318 | Not-determined |
| SG_extracted_k141_36852 | Low-quality |
| SG_extracted_k141_18459 | Not-determined |
| SG_extracted_k141_27320 | Low-quality |
| SG_extracted_k141_22671 | Low-quality |
| SG_extracted_k141_4503 | Not-determined |
| SG_extracted_k141_22674 | Not-determined |
| SG_extracted_k141_4504 | Not-determined |
| SG_extracted_k141_36855 | Low-quality |
| SG_extracted_k141_46613 | Low-quality |
| SG_extracted_k141_32322 | Not-determined |
| SG_extracted_k141_41917 | Low-quality |
| SG_extracted_k141_27333 | Low-quality |
| SG_extracted_k141_53702 | Not-determined |
| SG_extracted_k141_41923 | Not-determined |
| SG_extracted_k141_46621 | Not-determined |
| SG_extracted_k141_46622 | Not-determined |
| SG_extracted_k141_41929 | Not-determined |
| SG_extracted_k141_32324 | Not-determined |
| SG_extracted_k141_6466 | Not-determined |
| SG_extracted_k141_6467 | Not-determined |
| SG_extracted_k141_4522 | Not-determined |
| SG_extracted_k141_4524 | Not-determined |
| SG_extracted_k141_41943 | Not-determined |
| SG_extracted_k141_4527 | Not-determined |
| SG_extracted_k141_4530 | Not-determined |
| SG_extracted_k141_25440 | Not-determined |
| SG_extracted_k141_41951 | Not-determined |
| SG_extracted_k141_32336 | Not-determined |
| SG_extracted_k141_41952 | Low-quality |
| SG_extracted_k141_19938 | Not-determined |
| SG_extracted_k141_16005 | Not-determined |
| SG_extracted_k141_50922 | Not-determined |
| SG_extracted_k141_32340 | Not-determined |
| SG_extracted_k141_48383 | Not-determined |
| SG_extracted_k141_25446 | Not-determined |
| SG_extracted_k141_36873 | Not-determined |
| SG_extracted_k141_27348 | Not-determined |
| SG_extracted_k141_27349 | Low-quality |
| SG_extracted_k141_16011 | Not-determined |
| SG_extracted_k141_48393 | Low-quality |
| SG_extracted_k141_48394 | Not-determined |
| SG_extracted_k141_36875 | Not-determined |
| SG_extracted_k141_48396 | Low-quality |
| SG_extracted_k141_25454 | Low-quality |
| SG_extracted_k141_46636 | Not-determined |
| SG_extracted_k141_46637 | Not-determined |
| SG_extracted_k141_18484 | Not-determined |
| SG_extracted_k141_41962 | Not-determined |
| SG_extracted_k141_18486 | Not-determined |
| SG_extracted_k141_19943 | Not-determined |
| SG_extracted_k141_27357 | Not-determined |
| SG_extracted_k141_35070 | Not-determined |
| SG_extracted_k141_19944 | Not-determined |
| SG_extracted_k141_41968 | Not-determined |
| SG_extracted_k141_18492 | Not-determined |
| SG_extracted_k141_35072 | Not-determined |
| SG_extracted_k141_35073 | Not-determined |
| SG_extracted_k141_30184 | Not-determined |
| SG_extracted_k141_48405 | Not-determined |
| SG_extracted_k141_41974 | Not-determined |
| SG_extracted_k141_48411 | Not-determined |
| SG_extracted_k141_35082 | Not-determined |
| SG_extracted_k141_55488 | Not-determined |
| SG_extracted_k141_11118 | Low-quality |
| SG_extracted_k141_22681 | Low-quality |
| SG_extracted_k141_18497 | Not-determined |
| SG_extracted_k141_13793 | Not-determined |
| SG_extracted_k141_18498 | Not-determined |
| SG_extracted_k141_41977 | Not-determined |
| SG_extracted_k141_53724 | Not-determined |
| SG_extracted_k141_13798 | Not-determined |
| SG_extracted_k141_48426 | Low-quality |
| SG_extracted_k141_46642 | Not-determined |
| SG_extracted_k141_35093 | Not-determined |
| SG_extracted_k141_18504 | Not-determined |
| SG_extracted_k141_27376 | Not-determined |
| SG_extracted_k141_35097 | Not-determined |
| SG_extracted_k141_46646 | Not-determined |
| SG_extracted_k141_53730 | Not-determined |
| SG_extracted_k141_48435 | Not-determined |
| SG_extracted_k141_30192 | Low-quality |
| SG_extracted_k141_53735 | Low-quality |
| SG_extracted_k141_53741 | Not-determined |
| SG_extracted_k141_53742 | Not-determined |
| SG_extracted_k141_35108 | Not-determined |
| SG_extracted_k141_35109 | Not-determined |
| SG_extracted_k141_53744 | Not-determined |
| SG_extracted_k141_53746 | Not-determined |
| SG_extracted_k141_22692 | Not-determined |
| SG_extracted_k141_50943 | Not-determined |
| SG_extracted_k141_27389 | Not-determined |
| SG_extracted_k141_55500 | Low-quality |
| SG_extracted_k141_13806 | Not-determined |
| SG_extracted_k141_50946 | Not-determined |
| SG_extracted_k141_55502 | Low-quality |
| SG_extracted_k141_22700 | Not-determined |
| SG_extracted_k141_13808 | Low-quality |
| SG_extracted_k141_13809 | Not-determined |
| SG_extracted_k141_13810 | Not-determined |
| SG_extracted_k141_13811 | Not-determined |
| SG_extracted_k141_19951 | Low-quality |
| SG_extracted_k141_39187 | Not-determined |
| SG_extracted_k141_35129 | Not-determined |
| SG_extracted_k141_39189 | Not-determined |
| SG_extracted_k141_39190 | Not-determined |
| SG_extracted_k141_27399 | Not-determined |
| SG_extracted_k141_39191 | Low-quality |
| SG_extracted_k141_39192 | Not-determined |
| SG_extracted_k141_39193 | Not-determined |
| SG_extracted_k141_13818 | Not-determined |
| SG_extracted_k141_18511 | Not-determined |
| SG_extracted_k141_27400 | Not-determined |
| SG_extracted_k141_35131 | Not-determined |
| SG_extracted_k141_22704 | Not-determined |
| SG_extracted_k141_35135 | Not-determined |
| SG_extracted_k141_46654 | Not-determined |
| SG_extracted_k141_42003 | Low-quality |
| SG_extracted_k141_19964 | Not-determined |
| SG_extracted_k141_42004 | Not-determined |
| SG_extracted_k141_13826 | Not-determined |
| SG_extracted_k141_19970 | Not-determined |
| SG_extracted_k141_27408 | Not-determined |
| SG_extracted_k141_18525 | Not-determined |
| SG_extracted_k141_27411 | Not-determined |
| SG_extracted_k141_13829 | Not-determined |
| SG_extracted_k141_18529 | Low-quality |
| SG_extracted_k141_22708 | Not-determined |
| SG_extracted_k141_22709 | Not-determined |
| SG_extracted_k141_11134 | Low-quality |
| SG_extracted_k141_18533 | Not-determined |
| SG_extracted_k141_50951 | Not-determined |
| SG_extracted_k141_18534 | Low-quality |
| SG_extracted_k141_18535 | Not-determined |
| SG_extracted_k141_42013 | Low-quality |
| SG_extracted_k141_11142 | Not-determined |
| SG_extracted_k141_50954 | Not-determined |
| SG_extracted_k141_50955 | Not-determined |
| SG_extracted_k141_22720 | Not-determined |
| SG_extracted_k141_13848 | Not-determined |
| SG_extracted_k141_42014 | Not-determined |
| SG_extracted_k141_11150 | Not-determined |
| SG_extracted_k141_6505 | Not-determined |
| SG_extracted_k141_6507 | Not-determined |
| SG_extracted_k141_22729 | Not-determined |
| SG_extracted_k141_19985 | Not-determined |
| SG_extracted_k141_19990 | Not-determined |
| SG_extracted_k141_13855 | Not-determined |
| SG_extracted_k141_11160 | Not-determined |
| SG_extracted_k141_13860 | Not-determined |
| SG_extracted_k141_39223 | Low-quality |
| SG_extracted_k141_4540 | Not-determined |
| SG_extracted_k141_39226 | Low-quality |
| SG_extracted_k141_13864 | Not-determined |
| SG_extracted_k141_4544 | Not-determined |
| SG_extracted_k141_32359 | Not-determined |
| SG_extracted_k141_27423 | Low-quality |
| SG_extracted_k141_13868 | Not-determined |
| SG_extracted_k141_32363 | Not-determined |
| SG_extracted_k141_4550 | Not-determined |
| SG_extracted_k141_50966 | Not-determined |
| SG_extracted_k141_32365 | Not-determined |
| SG_extracted_k141_32369 | Not-determined |
| SG_extracted_k141_27429 | Not-determined |
| SG_extracted_k141_13878 | Low-quality |
| SG_extracted_k141_27438 | Not-determined |
| SG_extracted_k141_18541 | Not-determined |

| | |
|---|---|
| SG_extracted_k141_22746 | Not-determined |
| SG_extracted_k141_20001 | Low-quality |
| SG_extracted_k141_27444 | Not-determined |
| SG_extracted_k141_11174 | Not-determined |
| SG_extracted_k141_35169 | Not-determined |
| SG_extracted_k141_32394 | Not-determined |
| SG_extracted_k141_27447 | Not-determined |
| SG_extracted_k141_20007 | Not-determined |
| SG_extracted_k141_35171 | Not-determined |
| SG_extracted_k141_42032 | Not-determined |
| SG_extracted_k141_42038 | Not-determined |
| SG_extracted_k141_11184 | Low-quality |
| SG_extracted_k141_42042 | Not-determined |
| SG_extracted_k141_30236 | Not-determined |
| SG_extracted_k141_48443 | Not-determined |
| SG_extracted_k141_18551 | Low-quality |
| SG_extracted_k141_46665 | Not-determined |
| SG_extracted_k141_4585 | Not-determined |
| SG_extracted_k141_27460 | Not-determined |
| SG_extracted_k141_11193 | Not-determined |
| SG_extracted_k141_13900 | Not-determined |
| SG_extracted_k141_22758 | Low-quality |
| SG_extracted_k141_13903 | Not-determined |
| SG_extracted_k141_42046 | Low-quality |
| SG_extracted_k141_8803 | Low-quality |
| SG_extracted_k141_32399 | Not-determined |
| SG_extracted_k141_11196 | Not-determined |
| SG_extracted_k141_27468 | Low-quality |
| SG_extracted_k141_13911 | Not-determined |
| SG_extracted_k141_32402 | Not-determined |
| SG_extracted_k141_48448 | Not-determined |
| SG_extracted_k141_35176 | Not-determined |
| SG_extracted_k141_39262 | Not-determined |
| SG_extracted_k141_46668 | Low-quality |
| S16_vs_94866\|\|full | Medium-quality |
| SG_extracted_k141_48456 | Not-determined |
| SG_extracted_k141_18572 | Not-determined |
| SG_extracted_k141_32413 | Not-determined |
| SG_extracted_k141_43816 | Not-determined |
| SG_extracted_k141_32417 | Not-determined |
| SG_extracted_k141_46674 | Not-determined |
| SG_extracted_k141_18584 | Not-determined |
| SG_extracted_k141_35182 | Low-quality |
| SG_extracted_k141_22770 | Not-determined |
| SG_extracted_k141_13923 | Not-determined |
| SG_extracted_k141_32423 | Not-determined |
| SG_extracted_k141_48466 | Low-quality |
| SG_extracted_k141_48467 | Low-quality |
| SG_extracted_k141_6512 | Not-determined |
| SG_extracted_k141_13927 | Low-quality |
| SG_extracted_k141_32426 | Not-determined |
| SG_extracted_k141_32427 | Low-quality |
| SG_extracted_k141_50986 | Not-determined |
| SG_extracted_k141_13930 | Not-determined |
| SG_extracted_k141_50988 | Low-quality |
| SG_extracted_k141_46683 | Not-determined |
| SG_extracted_k141_13931 | Low-quality |
| SG_extracted_k141_13932 | Not-determined |
| SG_extracted_k141_48484 | Not-determined |
| SG_extracted_k141_32446 | Not-determined |
| SG_extracted_k141_42059 | Low-quality |
| SG_extracted_k141_32449 | Low-quality |
| SG_extracted_k141_32450 | Not-determined |
| SG_extracted_k141_13936 | Low-quality |
| SG_extracted_k141_48488 | Low-quality |
| SG_extracted_k141_48490 | Low-quality |
| SG_extracted_k141_42066 | Not-determined |
| SG_extracted_k141_27478 | Low-quality |
| SG_extracted_k141_32457 | Not-determined |
| SG_extracted_k141_6529 | Not-determined |
| SG_extracted_k141_8814 | Not-determined |
| SG_extracted_k141_6534 | Low-quality |
| SG_extracted_k141_6535 | Not-determined |
| SG_extracted_k141_43840 | Not-determined |
| SG_extracted_k141_6541 | Low-quality |
| SG_extracted_k141_43841 | Not-determined |
| SG_extracted_k141_27485 | Low-quality |
| SG_extracted_k141_27486 | Not-determined |
| SG_extracted_k141_32471 | Not-determined |
| SG_extracted_k141_13951 | Not-determined |
| SG_extracted_k141_22806 | Not-determined |
| SG_extracted_k141_16052 | Low-quality |
| SG_extracted_k141_43854 | Low-quality |
| SG_extracted_k141_46700 | Not-determined |
| SG_extracted_k141_16055 | Not-determined |
| SG_extracted_k141_43860 | Not-determined |
| SG_extracted_k141_27491 | Low-quality |
| SG_extracted_k141_4612 | Not-determined |
| SG_extracted_k141_42096 | Not-determined |
| SG_extracted_k141_48506 | Not-determined |
| SG_extracted_k141_42101 | Not-determined |
| SG_extracted_k141_42102 | Not-determined |
| SG_extracted_k141_48509 | Not-determined |
| SG_extracted_k141_48513 | Not-determined |
| SG_extracted_k141_30251 | Not-determined |
| SG_extracted_k141_16063 | Not-determined |
| SG_extracted_k141_32478 | Not-determined |
| SG_extracted_k141_27503 | Not-determined |
| SG_extracted_k141_42106 | Not-determined |
| SG_extracted_k141_46715 | Low-quality |
| SG_extracted_k141_42110 | Not-determined |
| SG_extracted_k141_42111 | Not-determined |
| SG_extracted_k141_32489 | Low-quality |
| SG_extracted_k141_39264 | Not-determined |
| SG_extracted_k141_32492 | Not-determined |
| SG_extracted_k141_43877 | Low-quality |
| SG_extracted_k141_43882 | Not-determined |
| SG_extracted_k141_30272 | Not-determined |
| SG_extracted_k141_43884 | Not-determined |
| SG_extracted_k141_39268 | Not-determined |
| SG_extracted_k141_22824 | Not-determined |
| SG_extracted_k141_39270 | Not-determined |
| SG_extracted_k141_32499 | Low-quality |
| SG_extracted_k141_32501 | Not-determined |
| SG_extracted_k141_46722 | Not-determined |
| SG_extracted_k141_8819 | Not-determined |
| SG_extracted_k141_27510 | Not-determined |
| SG_extracted_k141_39276 | Low-quality |
| SG_extracted_k141_27513 | Not-determined |
| SG_extracted_k141_22839 | Not-determined |
| SG_extracted_k141_8821 | Low-quality |
| SG_extracted_k141_53756 | Not-determined |
| SG_extracted_k141_32510 | Low-quality |
| SG_extracted_k141_8823 | Not-determined |
| SG_extracted_k141_32512 | Not-determined |
| SG_extracted_k141_27517 | Not-determined |
| SG_extracted_k141_27518 | Not-determined |
| SG_extracted_k141_27520 | Not-determined |
| SG_extracted_k141_22843 | Low-quality |
| SG_extracted_k141_53761 | Low-quality |
| SG_extracted_k141_42136 | Not-determined |
| SG_extracted_k141_4643 | Low-quality |
| SG_extracted_k141_13972 | Low-quality |
| SG_extracted_k141_8829 | Not-determined |
| SG_extracted_k141_27523 | Not-determined |
| SG_extracted_k141_42141 | Not-determined |
| SG_extracted_k141_50995 | Not-determined |
| SG_extracted_k141_48531 | Not-determined |
| SG_extracted_k141_13976 | Low-quality |
| SG_extracted_k141_13978 | Not-determined |
| SG_extracted_k141_13980 | Not-determined |
| SG_extracted_k141_48533 | Low-quality |
| SG_extracted_k141_46740 | Not-determined |
| SG_extracted_k141_53766 | Not-determined |
| SG_extracted_k141_8836 | Low-quality |
| SG_extracted_k141_32520 | Not-determined |
| SG_extracted_k141_13983 | Not-determined |
| SG_extracted_k141_48540 | Not-determined |
| SG_extracted_k141_13985 | Not-determined |
| SG_extracted_k141_8842 | Not-determined |
| SG_extracted_k141_4646 | Not-determined |
| SG_extracted_k141_8844 | Not-determined |
| SG_extracted_k141_13990 | Not-determined |
| SG_extracted_k141_48544 | Not-determined |
| SG_extracted_k141_48546 | Not-determined |
| SG_extracted_k141_6558 | Low-quality |
| SG_extracted_k141_8845 | Not-determined |
| SG_extracted_k141_8846 | Not-determined |
| SG_extracted_k141_6560 | Not-determined |
| SG_extracted_k141_53773 | Low-quality |
| SG_extracted_k141_30275 | Not-determined |
| SG_extracted_k141_39287 | Not-determined |
| SG_extracted_k141_39288 | Not-determined |
| SG_extracted_k141_35209 | Not-determined |
| SG_extracted_k141_53780 | Low-quality |
| SG_extracted_k141_20018 | Low-quality |
| SG_extracted_k141_4658 | Not-determined |
| SG_extracted_k141_48550 | Low-quality |
| SG_extracted_k141_6569 | Not-determined |
| SG_extracted_k141_51005 | Not-determined |
| SG_extracted_k141_39297 | Low-quality |
| SG_extracted_k141_53787 | Not-determined |
| SG_extracted_k141_42169 | Not-determined |
| SG_extracted_k141_53788 | Not-determined |
| SG_extracted_k141_42174 | Not-determined |
| SG_extracted_k141_53792 | Not-determined |
| SG_extracted_k141_6575 | Not-determined |
| SG_extracted_k141_53795 | Not-determined |
| SG_extracted_k141_27539 | Not-determined |
| SG_extracted_k141_14002 | Not-determined |
| SG_extracted_k141_32557 | Low-quality |
| SG_extracted_k141_20041 | Not-determined |
| SG_extracted_k141_32560 | Not-determined |
| SG_extracted_k141_6590 | Not-determined |
| SG_extracted_k141_48564 | Not-determined |
| SG_extracted_k141_20043 | Not-determined |
| SG_extracted_k141_20047 | Not-determined |
| SG_extracted_k141_20049 | Low-quality |
| SG_extracted_k141_20052 | Not-determined |
| SG_extracted_k141_6593 | Not-determined |
| SG_extracted_k141_6597 | Not-determined |
| SG_extracted_k141_42204 | Low-quality |
| SG_extracted_k141_22855 | Not-determined |
| SG_extracted_k141_22858 | Not-determined |
| SG_extracted_k141_22859 | Not-determined |
| SG_extracted_k141_42210 | Not-determined |
| SG_extracted_k141_42216 | Not-determined |
| SG_extracted_k141_42219 | Not-determined |
| SG_extracted_k141_20061 | Not-determined |
| SG_extracted_k141_22876 | Low-quality |
| SG_extracted_k141_22880 | Not-determined |
| SG_extracted_k141_22881 | Not-determined |
| SG_extracted_k141_42228 | Not-determined |
| SG_extracted_k141_39322 | Not-determined |
| SG_extracted_k141_22886 | Not-determined |
| SG_extracted_k141_22888 | Low-quality |
| SG_extracted_k141_42234 | Not-determined |
| SG_extracted_k141_53811 | Low-quality |
| SG_extracted_k141_53815 | Not-determined |
| SG_extracted_k141_39325 | Not-determined |
| SG_extracted_k141_6607 | Low-quality |
| SG_extracted_k141_25479 | Low-quality |
| SG_extracted_k141_16094 | Not-determined |
| SG_extracted_k141_42244 | Not-determined |
| SG_extracted_k141_16099 | Not-determined |
| SG_extracted_k141_16100 | Not-determined |
| SG_extracted_k141_1633 | Low-quality |
| SG_extracted_k141_16101 | Not-determined |
| SG_extracted_k141_51014 | Not-determined |
| SG_extracted_k141_22896 | Not-determined |
| SG_extracted_k141_22898 | Not-determined |
| SG_extracted_k141_39333 | Not-determined |
| SG_extracted_k141_53823 | Not-determined |
| SG_extracted_k141_20069 | Not-determined |
| SG_extracted_k141_42259 | Low-quality |
| SG_extracted_k141_1642 | Not-determined |
| SG_extracted_k141_16108 | Not-determined |
| SG_extracted_k141_32577 | Low-quality |
| SG_extracted_k141_43911 | Not-determined |
| SG_extracted_k141_1646 | Not-determined |
| SG_extracted_k141_39344 | Not-determined |
| SG_extracted_k141_14008 | Not-determined |
| SG_extracted_k141_16115 | Low-quality |
| SG_extracted_k141_16117 | Not-determined |
| SG_extracted_k141_39349 | Not-determined |
| SG_extracted_k141_32586 | Not-determined |
| SG_extracted_k141_39353 | Not-determined |
| SG_extracted_k141_51027 | Not-determined |
| SG_extracted_k141_32589 | Not-determined |
| SG_extracted_k141_25492 | Not-determined |
| SG_extracted_k141_39357 | Not-determined |
| SG_extracted_k141_39358 | Not-determined |
| SG_extracted_k141_16131 | Not-determined |
| SG_extracted_k141_39360 | Not-determined |
| SG_extracted_k141_6640 | Not-determined |
| SG_extracted_k141_32592 | Not-determined |
| SG_extracted_k141_22913 | Not-determined |
| SG_extracted_k141_25502 | Not-determined |
| SG_extracted_k141_32593 | Low-quality |
| SG_extracted_k141_32594 | Not-determined |
| SG_extracted_k141_32598 | Not-determined |
| SG_extracted_k141_42273 | Low-quality |
| SG_extracted_k141_22923 | Not-determined |
| SG_extracted_k141_22928 | Not-determined |
| SG_extracted_k141_22929 | Low-quality |
| SG_extracted_k141_22934 | Not-determined |
| SG_extracted_k141_20076 | Not-determined |
| SG_extracted_k141_6646 | Not-determined |
| SG_extracted_k141_53828 | Low-quality |
| SG_extracted_k141_25507 | Not-determined |
| SG_extracted_k141_22939 | Not-determined |
| SG_extracted_k141_22940 | Not-determined |
| SG_extracted_k141_4691 | Low-quality |
| SG_extracted_k141_1667 | Not-determined |
| SG_extracted_k141_25513 | Not-determined |
| SG_extracted_k141_32604 | Not-determined |
| SG_extracted_k141_25515 | Not-determined |
| SG_extracted_k141_53832 | Not-determined |
| S13_extracted_97702 | Complete |
| SG_extracted_k141_42303 | Not-determined |
| SG_extracted_k141_4696 | Not-determined |
| SG_extracted_k141_20083 | Low-quality |
| SG_extracted_k141_20084 | Not-determined |
| SG_extracted_k141_18635 | Low-quality |
| SG_extracted_k141_14026 | Low-quality |
| SG_extracted_k141_18637 | Low-quality |
| SG_extracted_k141_14032 | Low-quality |
| SG_extracted_k141_1673 | Not-determined |
| SG_extracted_k141_14033 | Not-determined |
| SG_extracted_k141_25538 | Not-determined |
| SG_extracted_k141_20092 | Not-determined |
| SG_extracted_k141_25540 | Not-determined |
| SG_extracted_k141_20099 | Not-determined |
| SG_extracted_k141_25548 | Not-determined |
| SG_extracted_k141_32622 | Low-quality |
| SG_extracted_k141_18644 | Not-determined |
| SG_extracted_k141_25551 | Not-determined |
| SG_extracted_k141_1689 | Not-determined |
| SG_extracted_k141_14045 | Not-determined |
| SG_extracted_k141_6689 | Not-determined |
| SG_extracted_k141_30308 | Not-determined |
| SG_extracted_k141_4706 | Not-determined |
| SG_extracted_k141_18649 | Low-quality |
| SG_extracted_k141_4707 | Not-determined |
| SG_extracted_k141_4710 | Not-determined |
| SG_extracted_k141_39401 | Not-determined |
| SG_extracted_k141_25570 | Low-quality |
| SG_extracted_k141_32628 | Not-determined |
| SG_extracted_k141_11254 | Low-quality |
| SG_extracted_k141_25573 | Not-determined |
| SG_extracted_k141_25579 | Not-determined |
| SG_extracted_k141_16139 | Not-determined |
| SG_extracted_k141_20115 | Not-determined |
| SG_extracted_k141_6693 | Low-quality |
| SG_extracted_k141_30312 | Not-determined |
| SG_extracted_k141_1705 | Not-determined |
| SG_extracted_k141_6696 | Low-quality |
| SG_extracted_k141_20119 | Not-determined |
| SG_extracted_k141_22971 | Not-determined |
| SG_extracted_k141_32631 | Not-determined |
| SG_extracted_k141_1710 | Not-determined |
| SG_extracted_k141_11264 | Not-determined |
| SG_extracted_k141_18663 | Not-determined |
| SG_extracted_k141_18664 | Not-determined |
| SG_extracted_k141_32634 | Not-determined |
| SG_extracted_k141_20124 | Not-determined |
| SG_extracted_k141_11267 | Not-determined |
| SG_extracted_k141_1713 | Low-quality |
| SG_extracted_k141_16146 | Not-determined |
| SG_extracted_k141_18668 | Not-determined |
| SG_extracted_k141_32639 | Low-quality |
| SG_extracted_k141_20128 | Not-determined |
| SG_extracted_k141_20129 | Not-determined |
| SG_extracted_k141_30321 | Not-determined |
| SG_extracted_k141_22979 | Not-determined |
| SG_extracted_k141_32642 | Not-determined |
| SG_extracted_k141_22980 | Not-determined |
| SG_extracted_k141_32643 | Not-determined |
| SG_extracted_k141_20135 | Not-determined |
| SG_extracted_k141_1715 | Not-determined |
| SG_extracted_k141_32646 | Not-determined |
| SG_extracted_k141_39412 | Not-determined |
| SG_extracted_k141_39413 | Not-determined |
| SG_extracted_k141_32649 | Not-determined |
| SG_extracted_k141_1722 | Not-determined |
| SG_extracted_k141_55513 | Not-determined |
| SG_extracted_k141_1723 | Not-determined |
| SG_extracted_k141_39419 | Not-determined |
| SG_extracted_k141_39420 | Not-determined |
| SG_extracted_k141_11282 | Not-determined |
| SG_extracted_k141_32655 | Low-quality |
| SG_extracted_k141_25607 | Not-determined |
| SG_extracted_k141_32657 | Not-determined |
| SG_extracted_k141_32658 | Not-determined |
| SG_extracted_k141_39424 | Not-determined |
| SG_extracted_k141_1725 | Not-determined |
| SG_extracted_k141_20148 | Not-determined |
| SG_extracted_k141_39426 | Not-determined |
| SG_extracted_k141_25612 | Not-determined |
| SG_extracted_k141_35228 | Low-quality |
| SG_extracted_k141_11284 | Low-quality |
| SG_extracted_k141_35230 | Not-determined |
| SG_extracted_k141_39427 | Not-determined |
| SG_extracted_k141_32665 | Not-determined |
| SG_extracted_k141_25618 | Not-determined |
| SG_extracted_k141_39430 | Not-determined |
| SG_extracted_k141_1734 | Not-determined |
| SG_extracted_k141_16155 | Not-determined |
| SG_extracted_k141_14061 | Not-determined |
| SG_extracted_k141_16156 | Not-determined |
| SG_extracted_k141_14062 | Not-determined |
| SG_extracted_k141_25622 | Not-determined |
| SG_extracted_k141_39434 | Not-determined |
| SG_extracted_k141_25624 | Not-determined |
| SG_extracted_k141_11288 | Low-quality |
| SG_extracted_k141_1742 | Low-quality |
| SG_extracted_k141_39439 | Not-determined |
| SG_extracted_k141_53871 | Not-determined |
| SG_extracted_k141_14067 | Not-determined |
| SG_extracted_k141_32678 | Low-quality |
| SG_extracted_k141_11290 | Not-determined |
| SG_extracted_k141_25627 | Not-determined |
| SG_extracted_k141_16163 | Not-determined |
| SG_extracted_k141_14076 | Not-determined |
| SG_extracted_k141_25633 | Not-determined |
| SG_extracted_k141_39446 | Not-determined |
| SG_extracted_k141_1749 | Not-determined |
| SG_extracted_k141_25635 | Not-determined |
| SG_extracted_k141_1754 | Not-determined |
| SG_extracted_k141_14088 | Not-determined |
| SG_extracted_k141_20162 | Low-quality |
| SG_extracted_k141_18679 | Not-determined |
| SG_extracted_k141_14092 | Not-determined |
| SG_extracted_k141_11309 | Not-determined |
| SG_extracted_k141_18682 | Not-determined |
| SG_extracted_k141_11312 | Not-determined |
| SG_extracted_k141_39453 | Not-determined |
| SG_extracted_k141_11316 | Not-determined |
| SG_extracted_k141_43928 | Not-determined |
| SG_extracted_k141_1765 | Not-determined |
| SG_extracted_k141_16187 | Not-determined |
| SG_extracted_k141_53876 | Not-determined |
| SG_extracted_k141_32698 | Not-determined |
| SG_extracted_k141_30342 | Not-determined |
| SG_extracted_k141_23002 | Not-determined |
| SG_extracted_k141_48584 | Not-determined |
| SG_extracted_k141_1774 | Low-quality |
| SG_extracted_k141_48588 | Low-quality |
| SG_extracted_k141_23005 | Not-determined |
| SG_extracted_k141_18704 | Low-quality |
| SG_extracted_k141_43961 | Not-determined |
| SG_extracted_k141_18706 | Not-determined |
| SG_extracted_k141_53880 | Low-quality |
| SG_extracted_k141_23007 | Not-determined |
| SG_extracted_k141_39465 | Low-quality |
| SG_extracted_k141_25648 | Not-determined |
| SG_extracted_k141_39468 | Not-determined |
| SG_extracted_k141_43968 | Not-determined |
| SG_extracted_k141_16199 | Low-quality |
| SG_extracted_k141_43970 | Low-quality |
| SG_extracted_k141_48593 | Low-quality |
| SG_extracted_k141_16202 | Not-determined |
| SG_extracted_k141_53886 | Low-quality |
| SG_extracted_k141_16206 | Not-determined |
| SG_extracted_k141_32717 | Not-determined |
| SG_extracted_k141_23021 | Not-determined |
| SG_extracted_k141_16207 | Not-determined |
| SG_extracted_k141_43978 | Not-determined |
| SG_extracted_k141_39472 | Not-determined |
| SG_extracted_k141_16208 | Not-determined |
| SG_extracted_k141_23024 | Not-determined |
| SG_extracted_k141_53893 | Not-determined |
| SG_extracted_k141_43983 | Not-determined |
| SG_extracted_k141_39477 | Not-determined |
| SG_extracted_k141_23026 | Low-quality |
| SG_extracted_k141_43987 | Not-determined |
| SG_extracted_k141_43989 | Not-determined |
| SG_extracted_k141_48610 | Not-determined |
| SG_extracted_k141_20181 | Not-determined |
| SG_extracted_k141_23040 | Not-determined |
| SG_extracted_k141_20184 | Not-determined |
| SG_extracted_k141_16217 | Not-determined |
| SG_extracted_k141_20185 | Not-determined |
| SG_extracted_k141_44003 | Not-determined |
| SG_extracted_k141_20187 | Not-determined |
| SG_extracted_k141_23049 | Not-determined |
| SG_extracted_k141_44006 | Not-determined |
| SG_extracted_k141_44007 | Not-determined |
| SG_extracted_k141_30360 | Not-determined |
| SG_extracted_k141_23051 | Not-determined |
| SG_extracted_k141_32743 | Low-quality |
| SG_extracted_k141_16220 | Not-determined |
| SG_extracted_k141_35257 | Low-quality |
| SG_extracted_k141_48624 | Not-determined |
| SG_extracted_k141_30368 | Not-determined |
| SG_extracted_k141_44016 | Not-determined |
| SG_extracted_k141_6709 | Not-determined |
| SG_extracted_k141_32762 | Not-determined |
| SG_extracted_k141_44022 | Not-determined |
| SG_extracted_k141_32763 | Not-determined |
| SG_extracted_k141_32765 | Not-determined |
| SG_extracted_k141_23072 | Not-determined |
| SG_extracted_k141_48646 | Not-determined |
| SG_extracted_k141_16238 | Not-determined |
| SG_extracted_k141_44029 | Low-quality |
| SG_extracted_k141_32771 | Not-determined |
| SG_extracted_k141_55531 | Not-determined |
| SG_extracted_k141_42340 | Not-determined |
| SG_extracted_k141_23078 | Not-determined |
| SG_extracted_k141_18726 | Not-determined |
| SG_extracted_k141_16243 | Low-quality |
| SG_extracted_k141_30382 | Not-determined |
| SG_extracted_k141_18729 | Not-determined |
| SG_extracted_k141_23082 | Not-determined |
| SG_extracted_k141_53908 | Not-determined |
| SG_extracted_k141_48654 | Not-determined |
| SG_extracted_k141_53912 | Not-determined |
| SG_extracted_k141_48656 | Not-determined |
| SG_extracted_k141_42345 | Not-determined |
| SG_extracted_k141_44035 | Low-quality |
| SG_extracted_k141_20201 | Not-determined |
| SG_extracted_k141_53916 | Not-determined |
| SG_extracted_k141_46742 | Not-determined |
| SG_extracted_k141_55536 | Not-determined |
| SG_extracted_k141_46743 | Not-determined |
| SG_extracted_k141_46744 | Not-determined |
| SG_extracted_k141_46745 | Low-quality |
| SG_extracted_k141_23089 | Not-determined |
| SG_extracted_k141_32779 | Low-quality |
| SG_extracted_k141_23091 | Not-determined |
| SG_extracted_k141_35263 | Not-determined |
| SG_extracted_k141_53918 | Not-determined |
| SG_extracted_k141_23096 | Not-determined |
| SG_extracted_k141_48657 | Low-quality |
| SG_extracted_k141_55545 | Not-determined |
| SG_extracted_k141_30400 | Not-determined |
| SG_extracted_k141_55548 | Not-determined |
| SG_extracted_k141_42364 | Not-determined |
| SG_extracted_k141_16259 | Not-determined |
| SG_extracted_k141_46757 | Not-determined |
| SG_extracted_k141_11341 | Not-determined |
| SG_extracted_k141_6716 | Not-determined |
| SG_extracted_k141_11343 | Low-quality |
| SG_extracted_k141_20228 | Not-determined |
| SG_extracted_k141_16263 | Not-determined |
| SG_extracted_k141_53931 | Not-determined |
| SG_extracted_k141_42381 | Not-determined |
| SG_extracted_k141_20240 | Low-quality |
| SG_extracted_k141_6726 | Not-determined |
| SG_extracted_k141_20241 | Not-determined |
| SG_extracted_k141_46766 | Low-quality |
| SG_extracted_k141_48662 | Not-determined |
| SG_extracted_k141_32788 | Low-quality |
| SG_extracted_k141_20246 | Not-determined |
| SG_extracted_k141_16274 | Not-determined |
| SG_extracted_k141_16275 | Not-determined |
| SG_extracted_k141_48665 | Not-determined |
| SG_extracted_k141_23107 | Low-quality |
| SG_extracted_k141_6733 | Not-determined |
| SG_extracted_k141_27558 | Not-determined |
| SG_extracted_k141_23112 | Low-quality |
| SG_extracted_k141_18738 | Not-determined |
| SG_extracted_k141_23115 | Low-quality |
| SG_extracted_k141_23116 | Low-quality |
| SG_extracted_k141_23118 | Not-determined |
| SG_extracted_k141_53940 | Not-determined |
| SG_extracted_k141_53943 | Not-determined |
| SG_extracted_k141_44054 | Not-determined |
| SG_extracted_k141_42392 | Not-determined |
| SG_extracted_k141_32798 | Not-determined |
| SG_extracted_k141_53946 | Not-determined |
| SG_extracted_k141_48679 | Low-quality |
| SG_extracted_k141_44061 | Not-determined |
| SG_extracted_k141_53949 | Low-quality |
| SG_extracted_k141_27562 | Not-determined |
| SG_extracted_k141_8874 | Low-quality |
| SG_extracted_k141_8876 | Not-determined |
| SG_extracted_k141_48687 | Not-determined |
| SG_extracted_k141_8878 | Not-determined |
| SG_extracted_k141_42402 | Not-determined |
| SG_extracted_k141_8880 | Not-determined |
| SG_extracted_k141_27567 | Not-determined |
| SG_extracted_k141_16289 | Low-quality |
| SG_extracted_k141_6743 | Not-determined |
| SG_extracted_k141_32814 | Not-determined |
| SG_extracted_k141_18745 | Not-determined |
| SG_extracted_k141_16301 | Not-determined |
| SG_extracted_k141_16302 | Not-determined |
| SG_extracted_k141_18748 | Low-quality |
| SG_extracted_k141_32821 | Not-determined |
| SG_extracted_k141_48699 | Not-determined |
| SG_extracted_k141_32825 | Low-quality |
| SG_extracted_k141_53959 | Not-determined |
| SG_extracted_k141_32827 | Not-determined |
| SG_extracted_k141_36878 | Not-determined |
| SG_extracted_k141_36881 | Not-determined |
| SG_extracted_k141_44071 | Low-quality |
| SG_extracted_k141_16311 | Not-determined |
| SG_extracted_k141_36882 | Not-determined |
| SG_extracted_k141_36885 | Not-determined |
| SG_extracted_k141_32842 | Low-quality |
| SG_extracted_k141_11352 | Not-determined |
| SG_extracted_k141_36888 | Not-determined |
| SG_extracted_k141_35266 | Low-quality |
| SG_extracted_k141_23145 | Not-determined |
| SG_extracted_k141_35268 | Not-determined |
| SG_extracted_k141_8888 | Not-determined |
| SG_extracted_k141_44089 | Not-determined |
| SG_extracted_k141_32848 | Not-determined |
| SG_extracted_k141_11362 | Not-determined |
| SG_extracted_k141_36898 | Not-determined |
| SG_extracted_k141_39503 | Not-determined |
| SG_extracted_k141_39506 | Not-determined |
| SG_extracted_k141_11367 | Not-determined |
| SG_extracted_k141_36905 | Low-quality |
| SG_extracted_k141_32849 | Low-quality |
| SG_extracted_k141_18752 | Not-determined |
| SG_extracted_k141_32850 | Not-determined |
| SG_extracted_k141_27590 | Not-determined |
| SG_extracted_k141_18755 | Not-determined |
| SG_extracted_k141_48707 | Not-determined |
| SG_extracted_k141_44109 | Not-determined |
| SG_extracted_k141_30408 | Low-quality |
| SG_extracted_k141_30410 | Not-determined |
| SG_extracted_k141_27597 | Not-determined |
| SG_extracted_k141_27599 | Not-determined |
| SG_extracted_k141_23149 | Not-determined |
| SG_extracted_k141_18764 | Not-determined |
| SG_extracted_k141_27604 | Not-determined |
| SG_extracted_k141_8899 | Not-determined |
| SG_extracted_k141_39520 | Not-determined |
| SG_extracted_k141_6756 | Not-determined |
| SG_extracted_k141_11382 | Not-determined |
| SG_extracted_k141_53976 | Not-determined |
| SG_extracted_k141_35274 | Not-determined |
| SG_extracted_k141_44131 | Not-determined |
| SG_extracted_k141_23160 | Not-determined |
| SG_extracted_k141_53984 | Low-quality |

| | |
|---|---|
| S6_extracted_k141_11385 | Not-determined |
| S6_extracted_k141_8906 | Not-determined |
| S6_extracted_k141_53987 | Not-determined |
| S6_extracted_k141_20275 | Not-determined |
| S6_extracted_k141_32860 | Not-determined |
| S6_extracted_k141_44143 | Not-determined |
| S6_extracted_k141_11393 | Not-determined |
| S6_extracted_k141_18768 | Not-determined |
| S6_extracted_k141_11394 | Not-determined |
| S6_extracted_k141_53993 | Not-determined |
| S6_extracted_k141_18771 | Not-determined |
| S6_extracted_k141_53996 | Not-determined |
| S6_extracted_k141_8916 | Not-determined |
| S6_extracted_k141_18775 | Low-quality |
| S6_extracted_k141_8922 | Not-determined |
| S6_extracted_k141_54012 | Not-determined |
| S6_extracted_k141_8925 | Low-quality |
| S6_extracted_k141_25663 | Not-determined |
| S6_extracted_k141_6776 | Low-quality |
| S6_extracted_k141_54014 | Not-determined |
| S6_extracted_k141_25667 | Not-determined |
| S6_extracted_k141_32873 | Low-quality |
| S6_extracted_k141_8927 | Not-determined |
| S6_extracted_k141_6785 | Not-determined |
| S6_extracted_k141_32876 | Not-determined |
| S6_extracted_k141_39550 | Not-determined |
| S6_extracted_k141_30437 | Low-quality |
| S6_extracted_k141_25670 | Not-determined |
| S6_extracted_k141_8932 | Not-determined |
| S6_extracted_k141_18781 | Not-determined |
| S6_extracted_k141_20295 | Not-determined |
| S6_extracted_k141_20300 | Not-determined |
| S6_extracted_k141_8939 | Not-determined |
| S6_extracted_k141_20302 | Not-determined |
| S6_extracted_k141_39564 | Not-determined |
| S6_extracted_k141_23183 | Not-determined |
| S6_extracted_k141_39565 | Not-determined |
| S6_extracted_k141_6798 | Low-quality |
| S6_extracted_k141_36926 | Not-determined |
| S6_extracted_k141_6799 | Not-determined |
| S6_extracted_k141_30445 | Low-quality |
| S6_extracted_k141_14111 | Not-determined |
| S6_extracted_k141_20310 | Not-determined |
| S6_extracted_k141_35292 | Not-determined |
| S6_extracted_k141_20311 | Not-determined |
| S6_extracted_k141_18789 | Low-quality |
| S6_extracted_k141_35294 | Not-determined |
| S6_extracted_k141_14114 | Not-determined |
| S6_extracted_k141_14115 | Not-determined |
| S6_extracted_k141_6816 | Low-quality |
| S6_extracted_k141_48720 | Not-determined |
| S6_extracted_k141_35300 | Not-determined |
| S6_extracted_k141_23197 | Low-quality |
| S6_extracted_k141_36945 | Not-determined |
| S6_extracted_k141_20324 | Not-determined |
| S6_extracted_k141_48726 | Not-determined |
| S6_extracted_k141_6831 | Not-determined |
| S6_extracted_k141_35305 | Not-determined |
| S6_extracted_k141_36954 | Not-determined |
| S6_extracted_k141_27615 | Low-quality |
| S6_extracted_k141_44199 | Not-determined |
| S6_extracted_k141_48734 | Not-determined |
| S6_extracted_k141_54046 | Not-determined |
| S6_extracted_k141_54048 | Low-quality |
| S6_extracted_k141_20335 | Not-determined |
| S6_extracted_k141_39574 | Low-quality |
| S6_extracted_k141_20337 | Not-determined |
| S6_extracted_k141_14128 | Low-quality |
| S6_extracted_k141_39576 | Not-determined |
| S6_extracted_k141_54051 | Not-determined |
| S6_extracted_k141_36960 | Not-determined |
| S6_extracted_k141_36961 | Not-determined |
| S6_extracted_k141_48737 | Not-determined |
| S6_extracted_k141_20342 | Not-determined |
| S6_extracted_k141_46795 | Not-determined |
| S6_extracted_k141_48741 | Not-determined |
| S6_extracted_k141_27625 | Not-determined |
| S6_extracted_k141_14136 | Not-determined |
| S6_extracted_k141_11422 | Not-determined |
| S6_extracted_k141_27626 | Not-determined |
| S6_extracted_k141_23215 | Low-quality |
| S6_extracted_k141_46802 | Not-determined |
| S6_extracted_k141_46803 | Not-determined |
| S6_extracted_k141_23218 | Not-determined |
| S6_extracted_k141_39590 | Not-determined |
| S6_extracted_k141_39591 | Not-determined |
| S6_extracted_k141_46810 | Not-determined |
| S6_extracted_k141_23224 | Low-quality |
| S6_extracted_k141_39598 | Not-determined |
| S6_extracted_k141_23229 | Not-determined |
| S6_extracted_k141_1805 | Low-quality |
| S6_extracted_k141_46815 | Low-quality |
| S6_extracted_k141_46817 | Not-determined |
| S6_extracted_k141_1808 | Not-determined |
| S6_extracted_k141_55560 | Not-determined |
| S6_extracted_k141_18800 | Not-determined |
| S6_extracted_k141_55563 | Not-determined |
| S6_extracted_k141_55564 | Not-determined |
| S6_extracted_k141_46820 | Low-quality |
| S6_extracted_k141_39605 | Not-determined |
| S6_extracted_k141_16328 | Low-quality |
| S6_extracted_k141_46822 | Not-determined |
| S6_extracted_k141_6838 | Not-determined |
| S6_extracted_k141_36976 | Not-determined |
| S6_extracted_k141_46827 | Not-determined |
| S6_extracted_k141_46829 | Not-determined |
| S6_extracted_k141_30462 | Not-determined |
| S6_extracted_k141_39614 | Not-determined |
| S6_extracted_k141_54063 | Not-determined |
| S6_extracted_k141_36981 | Not-determined |
| S6_extracted_k141_39617 | Low-quality |
| S6_extracted_k141_23240 | Not-determined |
| S6_extracted_k141_46835 | Not-determined |
| S6_extracted_k141_32906 | Not-determined |
| S6_extracted_k141_48748 | Not-determined |
| S6_extracted_k141_39619 | Not-determined |
| S6_extracted_k141_44215 | Low-quality |
| S6_extracted_k141_48750 | Not-determined |
| S6_extracted_k141_54067 | Not-determined |
| S6_extracted_k141_48751 | Not-determined |
| S6_extracted_k141_39621 | Low-quality |
| S6_extracted_k141_36987 | Low-quality |
| S6_extracted_k141_46840 | Not-determined |
| S6_extracted_k141_48760 | Not-determined |
| S6_extracted_k141_23247 | Not-determined |
| S6_extracted_k141_11448 | Not-determined |
| S6_extracted_k141_46845 | Not-determined |
| S6_extracted_k141_30472 | Not-determined |
| S6_extracted_k141_48763 | Not-determined |
| S6_extracted_k141_11459 | Low-quality |
| S6_extracted_k141_36992 | Not-determined |
| S6_extracted_k141_27647 | Not-determined |
| S6_extracted_k141_23255 | Not-determined |
| S6_extracted_k141_27649 | Not-determined |
| S6_extracted_k141_54072 | Low-quality |
| S6_extracted_k141_11467 | Not-determined |
| S6_extracted_k141_11471 | Low-quality |
| S6_extracted_k141_54078 | Not-determined |
| S6_extracted_k141_54080 | Low-quality |
| S6_extracted_k141_54083 | Low-quality |
| S6_extracted_k141_54085 | Not-determined |
| S6_extracted_k141_36999 | Low-quality |
| S6_extracted_k141_48779 | Not-determined |
| S6_extracted_k141_48781 | Low-quality |
| S6_extracted_k141_25680 | Not-determined |
| S6_extracted_k141_46847 | Not-determined |
| S6_extracted_k141_46848 | Not-determined |
| S6_extracted_k141_32942 | Low-quality |
| S6_extracted_k141_25683 | Not-determined |
| S6_extracted_k141_37007 | Not-determined |
| S6_extracted_k141_18834 | Not-determined |
| S6_extracted_k141_11487 | Not-determined |
| S6_extracted_k141_20379 | Not-determined |
| S6_extracted_k141_54100 | Not-determined |
| S6_extracted_k141_27662 | Not-determined |
| S6_extracted_k141_25693 | Not-determined |
| S6_extracted_k141_20382 | Low-quality |
| S6_extracted_k141_20386 | Not-determined |
| S6_extracted_k141_46849 | Not-determined |
| S6_extracted_k141_35344 | Not-determined |
| S6_extracted_k141_23287 | Low-quality |
| S6_extracted_k141_25696 | Not-determined |
| S6_extracted_k141_54117 | Not-determined |
| S6_extracted_k141_23290 | Not-determined |
| S6_extracted_k141_23295 | Not-determined |
| S6_extracted_k141_23296 | Not-determined |
| S6_extracted_k141_23302 | Low-quality |
| S6_extracted_k141_37018 | Not-determined |
| S6_extracted_k141_18849 | Not-determined |
| S6_extracted_k141_37020 | Low-quality |
| S6_extracted_k141_23310 | Not-determined |
| S6_extracted_k141_23311 | Not-determined |
| S6_extracted_k141_46856 | Not-determined |
| S6_extracted_k141_11515 | Not-determined |
| S6_extracted_k141_20397 | Not-determined |
| S6_extracted_k141_46860 | Not-determined |
| S6_extracted_k141_54139 | Not-determined |
| S6_extracted_k141_46863 | Not-determined |
| S6_extracted_k141_27678 | Low-quality |
| S6_extracted_k141_23316 | Not-determined |
| S6_extracted_k141_23318 | Not-determined |
| S6_extracted_k141_27681 | Not-determined |
| S6_extracted_k141_54149 | Not-determined |
| S6_extracted_k141_6869 | Low-quality |
| S6_extracted_k141_54151 | Not-determined |
| S6_extracted_k141_20412 | Not-determined |
| S6_extracted_k141_54152 | Not-determined |
| S6_extracted_k141_54153 | Not-determined |
| S6_extracted_k141_27687 | Not-determined |
| S6_extracted_k141_6877 | Low-quality |
| S6_extracted_k141_11523 | Not-determined |
| S6_extracted_k141_27692 | Not-determined |
| S6_extracted_k141_27694 | Not-determined |
| S6_extracted_k141_27697 | Not-determined |
| S6_extracted_k141_54159 | Not-determined |
| S6_extracted_k141_23332 | Not-determined |
| S6_extracted_k141_11532 | Low-quality |
| S6_extracted_k141_23334 | Not-determined |
| S6_extracted_k141_54174 | Not-determined |
| S6_extracted_k141_46889 | Not-determined |
| S6_extracted_k141_48793 | Not-determined |
| S6_extracted_k141_48795 | Not-determined |
| S6_extracted_k141_55575 | Not-determined |
| S6_extracted_k141_20420 | Not-determined |
| S6_extracted_k141_11536 | Low-quality |
| S6_extracted_k141_11537 | Not-determined |
| S6_extracted_k141_11538 | Not-determined |
| S6_extracted_k141_32978 | Not-determined |
| S6_extracted_k141_6889 | Not-determined |
| S6_extracted_k141_32979 | Not-determined |
| S14_vs_k141_62888\|\|full | Medium-quality |
| S6_extracted_k141_32980 | Not-determined |
| S6_extracted_k141_25711 | Not-determined |
| S6_extracted_k141_54179 | Not-determined |
| S6_extracted_k141_6891 | Not-determined |
| S6_extracted_k141_11545 | Not-determined |
| S6_extracted_k141_32984 | Not-determined |
| S6_extracted_k141_11547 | Low-quality |
| S6_extracted_k141_23363 | Not-determined |
| S6_extracted_k141_23364 | Not-determined |
| S6_extracted_k141_11549 | Not-determined |
| S6_extracted_k141_25716 | Not-determined |
| S6_extracted_k141_6900 | Not-determined |
| S6_extracted_k141_48813 | Not-determined |
| S6_extracted_k141_11554 | Not-determined |
| S6_extracted_k141_32989 | Not-determined |
| S6_extracted_k141_23369 | Not-determined |
| S6_extracted_k141_48815 | Not-determined |
| S6_extracted_k141_30484 | Not-determined |
| S6_extracted_k141_30485 | Not-determined |
| S6_extracted_k141_48819 | Not-determined |
| S6_extracted_k141_6909 | Low-quality |
| S6_extracted_k141_25726 | Low-quality |
| S6_extracted_k141_23378 | Not-determined |
| S6_extracted_k141_23380 | Not-determined |
| S17_extracted_k141_159527 | Complete |
| S6_extracted_k141_25730 | Not-determined |
| S6_extracted_k141_48827 | Low-quality |
| S6_extracted_k141_39626 | Low-quality |
| S6_extracted_k141_23387 | Not-determined |
| S6_extracted_k141_16341 | Low-quality |
| S6_extracted_k141_48831 | Low-quality |
| S6_extracted_k141_11574 | Not-determined |
| S6_extracted_k141_30502 | Not-determined |
| S6_extracted_k141_46930 | Low-quality |
| S6_extracted_k141_39633 | Not-determined |
| S6_extracted_k141_48846 | Not-determined |
| S6_extracted_k141_39635 | Not-determined |
| S6_extracted_k141_48847 | Low-quality |
| S6_extracted_k141_11581 | Low-quality |
| S6_extracted_k141_39638 | Low-quality |
| S6_extracted_k141_39639 | Not-determined |
| S6_extracted_k141_16352 | Not-determined |
| S6_extracted_k141_16353 | Not-determined |
| S6_extracted_k141_11587 | Not-determined |
| S6_extracted_k141_48857 | Low-quality |
| S6_extracted_k141_48858 | Not-determined |
| S6_extracted_k141_48865 | Low-quality |
| S6_extracted_k141_46945 | Low-quality |
| S6_extracted_k141_46948 | Not-determined |
| S6_extracted_k141_11599 | Not-determined |
| S6_extracted_k141_30507 | Low-quality |
| S6_extracted_k141_23399 | Not-determined |
| S6_extracted_k141_46958 | Not-determined |
| S6_extracted_k141_6916 | Not-determined |
| S6_extracted_k141_55592 | Low-quality |
| S6_extracted_k141_39655 | Not-determined |
| S6_extracted_k141_6925 | Not-determined |
| S6_extracted_k141_6927 | Not-determined |
| S6_extracted_k141_11611 | Not-determined |
| S6_extracted_k141_48879 | Not-determined |
| S6_extracted_k141_48881 | Not-determined |
| S6_extracted_k141_1825 | Not-determined |
| S6_extracted_k141_23401 | Not-determined |
| S6_extracted_k141_11617 | Low-quality |
| S6_extracted_k141_37024 | Not-determined |
| S6_extracted_k141_11624 | Not-determined |
| S6_extracted_k141_48901 | Not-determined |
| S6_extracted_k141_23407 | Low-quality |
| S6_extracted_k141_48903 | Not-determined |
| S6_extracted_k141_30512 | Low-quality |
| S6_extracted_k141_55599 | Low-quality |
| S6_extracted_k141_11633 | Not-determined |
| S6_extracted_k141_23416 | Not-determined |
| S6_extracted_k141_23417 | Not-determined |
| S6_extracted_k141_44230 | Not-determined |
| S6_extracted_k141_25777 | Not-determined |
| S6_extracted_k141_11637 | Not-determined |
| S6_extracted_k141_30523 | Not-determined |
| S6_extracted_k141_23425 | Low-quality |
| S6_extracted_k141_44239 | Not-determined |
| S6_extracted_k141_55608 | Low-quality |
| S6_extracted_k141_23430 | Not-determined |
| S6_extracted_k141_11641 | Not-determined |
| S6_extracted_k141_11642 | Not-determined |
| S6_extracted_k141_23433 | Not-determined |
| S6_extracted_k141_30534 | Not-determined |
| S6_extracted_k141_30536 | Not-determined |
| S6_extracted_k141_55615 | Not-determined |
| S6_extracted_k141_25790 | Not-determined |
| S6_extracted_k141_23434 | Not-determined |
| S6_extracted_k141_11651 | Not-determined |
| S6_extracted_k141_30538 | Not-determined |
| S6_extracted_k141_30539 | Not-determined |
| S6_extracted_k141_25793 | Not-determined |
| S6_extracted_k141_11656 | Not-determined |
| S6_extracted_k141_55617 | Not-determined |
| S6_extracted_k141_11660 | Not-determined |
| S6_extracted_k141_55620 | Not-determined |
| S6_extracted_k141_11661 | Low-quality |
| S6_extracted_k141_55625 | Not-determined |
| S6_extracted_k141_39693 | Not-determined |
| S6_extracted_k141_55630 | Not-determined |
| S6_extracted_k141_6955 | Not-determined |
| S6_extracted_k141_39699 | Not-determined |
| S6_extracted_k141_44246 | Not-determined |
| S6_extracted_k141_39701 | Not-determined |
| S6_extracted_k141_48930 | Not-determined |
| S6_extracted_k141_39710 | Not-determined |
| S6_extracted_k141_30553 | Not-determined |
| S6_extracted_k141_11663 | Not-determined |
| S6_extracted_k141_30557 | Not-determined |
| S6_extracted_k141_16375 | Not-determined |
| S6_extracted_k141_48936 | Not-determined |
| S6_extracted_k141_48938 | Not-determined |
| S6_extracted_k141_48940 | Low-quality |
| S6_extracted_k141_39724 | Not-determined |
| S6_extracted_k141_16385 | Not-determined |
| S6_extracted_k141_6981 | Not-determined |
| S6_extracted_k141_16386 | Not-determined |
| S6_extracted_k141_6982 | Low-quality |
| S6_extracted_k141_39727 | Not-determined |
| S6_extracted_k141_23464 | Not-determined |
| S6_extracted_k141_6985 | Not-determined |
| S6_extracted_k141_23465 | Not-determined |
| S6_extracted_k141_23467 | Not-determined |
| S6_extracted_k141_30570 | Not-determined |
| S6_extracted_k141_55657 | Not-determined |
| S6_extracted_k141_46977 | Not-determined |
| S6_extracted_k141_55663 | Not-determined |
| S6_extracted_k141_25800 | Not-determined |
| S6_extracted_k141_37065 | Not-determined |
| S6_extracted_k141_37068 | Not-determined |
| S6_extracted_k141_37069 | Low-quality |
| S6_extracted_k141_48946 | Not-determined |
| S6_extracted_k141_7003 | Not-determined |
| S6_extracted_k141_55668 | Not-determined |
| S6_extracted_k141_30576 | Not-determined |
| S6_extracted_k141_48950 | Not-determined |
| S6_extracted_k141_48952 | Not-determined |
| S6_extracted_k141_44262 | Not-determined |
| S6_extracted_k141_25809 | Not-determined |
| S6_extracted_k141_55677 | Not-determined |
| S6_extracted_k141_7012 | Not-determined |
| S6_extracted_k141_44267 | Low-quality |
| S6_extracted_k141_7014 | Not-determined |
| S6_extracted_k141_44270 | Low-quality |
| S6_extracted_k141_46982 | Not-determined |
| S6_extracted_k141_46983 | Not-determined |
| S6_extracted_k141_30589 | Not-determined |
| S6_extracted_k141_7033 | Not-determined |
| S6_extracted_k141_46987 | Not-determined |
| S6_extracted_k141_30595 | Not-determined |
| S6_extracted_k141_48962 | Not-determined |
| S6_extracted_k141_7043 | Not-determined |
| S6_extracted_k141_37092 | Not-determined |
| S6_extracted_k141_55690 | Not-determined |
| S6_extracted_k141_7049 | Low-quality |
| S6_extracted_k141_7051 | Low-quality |
| S6_extracted_k141_48969 | Low-quality |
| S6_extracted_k141_48972 | Not-determined |
| S6_extracted_k141_55697 | Low-quality |
| S6_extracted_k141_11681 | Not-determined |
| S6_extracted_k141_39779 | Not-determined |
| S6_extracted_k141_7058 | Not-determined |
| S6_extracted_k141_39781 | Not-determined |
| S6_extracted_k141_55702 | Not-determined |
| S6_extracted_k141_11683 | Not-determined |
| S6_extracted_k141_7060 | Not-determined |
| S6_extracted_k141_48973 | Not-determined |
| S6_extracted_k141_48976 | Not-determined |
| S6_extracted_k141_55705 | Not-determined |
| S6_extracted_k141_48978 | Not-determined |
| S6_extracted_k141_23471 | Not-determined |
| S6_extracted_k141_7063 | Not-determined |
| S6_extracted_k141_25828 | Not-determined |
| S6_extracted_k141_39785 | Not-determined |
| S6_extracted_k141_7068 | Not-determined |
| S6_extracted_k141_16392 | Not-determined |
| S6_extracted_k141_25830 | Not-determined |
| S6_extracted_k141_55712 | Low-quality |
| S6_extracted_k141_37114 | Not-determined |
| S6_extracted_k141_16401 | Not-determined |
| S6_extracted_k141_23483 | Low-quality |
| S6_extracted_k141_25838 | Not-determined |
| S6_extracted_k141_23484 | Not-determined |
| S6_extracted_k141_25839 | Not-determined |
| S6_extracted_k141_25840 | Low-quality |
| S6_extracted_k141_48991 | Not-determined |
| S6_extracted_k141_48994 | Not-determined |
| S6_extracted_k141_48997 | Low-quality |
| S16_extracted_8493 | Complete |
| S6_extracted_k141_49007 | Not-determined |
| S6_extracted_k141_49008 | Not-determined |
| S6_extracted_k141_25844 | Not-determined |
| S6_extracted_k141_25846 | Low-quality |
| S6_extracted_k141_49013 | Not-determined |
| S6_extracted_k141_49014 | Not-determined |
| S6_extracted_k141_23502 | Not-determined |
| S6_extracted_k141_11706 | Not-determined |
| S6_extracted_k141_23507 | Not-determined |
| S6_extracted_k141_25855 | Not-determined |
| S6_extracted_k141_49018 | Low-quality |
| S6_extracted_k141_30608 | Not-determined |
| S6_extracted_k141_25858 | Not-determined |
| S6_extracted_k141_23512 | Low-quality |
| S6_extracted_k141_44317 | Not-determined |
| S6_extracted_k141_39793 | Low-quality |
| S6_extracted_k141_44318 | Not-determined |
| S6_extracted_k141_44320 | Not-determined |
| S6_extracted_k141_49026 | Low-quality |
| S6_extracted_k141_1834 | Not-determined |
| S6_extracted_k141_23519 | Not-determined |
| S6_extracted_k141_1837 | Not-determined |
| S6_extracted_k141_23520 | Not-determined |
| S6_extracted_k141_39802 | Low-quality |
| S6_extracted_k141_55739 | Not-determined |
| S6_extracted_k141_39807 | Not-determined |
| S6_extracted_k141_25877 | Not-determined |
| S6_extracted_k141_39810 | Not-determined |
| S6_extracted_k141_11724 | Low-quality |
| S6_extracted_k141_1842 | Not-determined |
| S6_extracted_k141_25882 | Low-quality |
| S6_extracted_k141_25886 | Not-determined |
| S6_extracted_k141_55752 | Low-quality |
| S6_extracted_k141_1848 | Not-determined |
| S6_extracted_k141_39819 | Low-quality |
| S6_extracted_k141_55753 | Not-determined |
| S6_extracted_k141_46994 | Low-quality |
| S6_extracted_k141_30626 | Not-determined |
| S6_extracted_k141_23528 | Not-determined |
| S6_extracted_k141_39827 | Not-determined |
| S6_extracted_k141_1863 | Not-determined |
| S6_extracted_k141_25897 | Not-determined |
| S6_extracted_k141_20425 | Not-determined |
| S6_extracted_k141_16408 | Low-quality |
| S6_extracted_k141_20428 | Not-determined |
| S6_extracted_k141_1877 | Not-determined |
| S6_extracted_k141_16413 | Not-determined |
| S6_extracted_k141_1878 | Not-determined |
| S6_extracted_k141_47000 | Low-quality |
| S6_extracted_k141_37150 | Not-determined |
| S6_extracted_k141_1880 | Not-determined |
| S6_extracted_k141_16417 | Not-determined |
| S6_extracted_k141_47008 | Low-quality |
| S6_extracted_k141_25902 | Not-determined |
| S6_extracted_k141_16426 | Not-determined |
| S6_extracted_k141_1895 | Not-determined |
| S6_extracted_k141_20447 | Not-determined |
| S6_extracted_k141_1896 | Not-determined |
| S6_extracted_k141_20448 | Not-determined |
| S6_extracted_k141_37159 | Not-determined |
| S6_extracted_k141_39831 | Not-determined |
| S6_extracted_k141_16436 | Not-determined |
| S6_extracted_k141_39833 | Low-quality |
| S6_extracted_k141_23548 | Not-determined |
| S6_extracted_k141_44346 | Not-determined |
| S6_extracted_k141_1907 | Not-determined |
| S6_extracted_k141_8982 | Not-determined |
| S6_extracted_k141_23552 | Low-quality |
| S6_extracted_k141_1912 | Low-quality |
| S6_extracted_k141_8985 | Not-determined |
| S6_extracted_k141_20456 | Low-quality |
| S6_extracted_k141_8987 | Not-determined |
| S6_extracted_k141_20460 | Not-determined |
| S6_extracted_k141_8989 | Not-determined |
| S6_extracted_k141_44356 | Not-determined |
| S6_extracted_k141_39842 | Low-quality |
| S6_extracted_k141_44357 | Not-determined |
| S6_extracted_k141_1916 | Not-determined |
| S6_extracted_k141_20464 | Not-determined |
| S6_extracted_k141_11746 | Not-determined |
| S6_extracted_k141_44362 | Not-determined |
| S6_extracted_k141_1921 | Not-determined |
| S6_extracted_k141_55779 | Not-determined |
| S6_extracted_k141_16450 | Not-determined |
| S6_extracted_k141_23558 | Not-determined |
| S6_extracted_k141_1927 | Not-determined |
| S6_extracted_k141_55785 | Not-determined |
| S6_extracted_k141_16457 | Not-determined |
| S6_extracted_k141_23561 | Not-determined |
| S6_extracted_k141_39857 | Not-determined |
| S6_extracted_k141_20476 | Not-determined |
| S6_extracted_k141_55800 | Not-determined |
| S6_extracted_k141_44374 | Not-determined |
| S6_extracted_k141_20479 | Not-determined |
| S6_extracted_k141_9005 | Low-quality |
| S6_extracted_k141_9006 | Low-quality |
| S6_extracted_k141_20481 | Not-determined |
| S6_extracted_k141_37170 | Not-determined |
| S6_extracted_k141_37171 | Not-determined |
| S6_extracted_k141_37172 | Not-determined |
| S6_extracted_k141_20483 | Not-determined |
| S6_extracted_k141_23566 | Not-determined |
| S6_extracted_k141_39859 | Not-determined |
| S6_extracted_k141_37175 | Not-determined |
| S6_extracted_k141_37177 | Not-determined |
| S6_extracted_k141_55813 | Not-determined |
| S6_extracted_k141_9015 | Not-determined |
| S6_extracted_k141_55814 | Not-determined |
| S6_extracted_k141_9016 | Not-determined |
| S6_extracted_k141_39865 | Not-determined |
| S6_extracted_k141_49049 | Not-determined |
| S6_extracted_k141_1936 | Not-determined |
| S6_extracted_k141_39871 | Low-quality |
| S6_extracted_k141_55822 | Low-quality |
| S6_extracted_k141_39874 | Not-determined |
| S6_extracted_k141_1942 | Low-quality |
| S6_extracted_k141_37181 | Not-determined |
| S6_extracted_k141_39876 | Not-determined |
| S6_extracted_k141_37182 | Not-determined |
| S6_extracted_k141_16460 | Not-determined |
| S6_extracted_k141_9030 | Not-determined |
| S6_extracted_k141_9031 | Not-determined |
| S6_extracted_k141_49070 | Low-quality |
| S6_extracted_k141_1947 | Low-quality |
| S6_extracted_k141_39886 | Not-determined |
| S6_extracted_k141_20499 | Not-determined |
| S6_extracted_k141_39887 | Not-determined |
| S6_extracted_k141_39889 | Not-determined |
| S6_extracted_k141_37191 | Not-determined |
| S6_extracted_k141_39891 | Not-determined |
| S6_extracted_k141_39892 | Not-determined |
| S6_extracted_k141_9043 | Not-determined |
| S6_extracted_k141_39894 | Not-determined |
| S6_extracted_k141_37198 | Not-determined |
| S6_extracted_k141_49080 | Low-quality |
| S6_extracted_k141_39899 | Low-quality |
| S6_extracted_k141_9048 | Not-determined |
| S6_extracted_k141_9055 | Not-determined |
| S6_extracted_k141_1952 | Low-quality |
| S6_extracted_k141_9057 | Not-determined |
| S6_extracted_k141_49086 | Not-determined |
| S6_extracted_k141_9062 | Not-determined |
| S6_extracted_k141_9067 | Not-determined |
| S6_extracted_k141_49096 | Not-determined |
| S6_extracted_k141_20517 | Not-determined |
| S6_extracted_k141_37216 | Not-determined |
| S6_extracted_k141_20521 | Not-determined |
| S6_extracted_k141_37218 | Not-determined |
| S6_extracted_k141_44401 | Not-determined |
| S6_extracted_k141_20523 | Not-determined |
| S6_extracted_k141_39912 | Not-determined |
| S6_extracted_k141_37223 | Low-quality |
| S6_extracted_k141_20529 | Not-determined |
| S6_extracted_k141_49104 | Not-determined |
| S6_extracted_k141_49107 | Not-determined |
| S6_extracted_k141_39930 | Not-determined |
| S6_extracted_k141_39934 | Not-determined |
| S6_extracted_k141_1992 | Not-determined |
| S6_extracted_k141_9069 | Not-determined |
| S6_extracted_k141_51040 | Low-quality |
| S6_extracted_k141_9070 | Not-determined |
| S6_extracted_k141_37226 | Not-determined |
| S6_extracted_k141_20537 | Not-determined |
| S6_extracted_k141_51046 | Low-quality |
| S6_extracted_k141_2000 | Not-determined |
| S6_extracted_k141_37230 | Not-determined |
| S6_extracted_k141_51050 | Not-determined |
| S6_extracted_k141_9076 | Not-determined |
| S6_extracted_k141_39951 | Not-determined |
| S6_extracted_k141_44434 | Not-determined |
| S6_extracted_k141_39953 | Not-determined |
| S6_extracted_k141_37232 | Low-quality |
| S6_extracted_k141_9081 | Not-determined |
| S6_extracted_k141_39954 | Not-determined |
| S6_extracted_k141_20542 | Low-quality |
| S6_extracted_k141_51056 | Not-determined |
| S6_extracted_k141_39959 | Low-quality |
| S6_extracted_k141_44442 | Not-determined |
| S6_extracted_k141_51060 | Not-determined |
| S6_extracted_k141_20554 | Not-determined |
| S6_extracted_k141_9090 | Not-determined |
| S6_extracted_k141_9093 | Not-determined |
| S6_extracted_k141_9095 | Not-determined |
| S6_extracted_k141_51067 | Not-determined |
| S6_extracted_k141_39977 | Low-quality |
| S6_extracted_k141_49136 | Not-determined |
| S6_extracted_k141_44450 | Not-determined |

| | |
|---|---|
| S6_extracted_k141_44453 | Not-determined |
| S6_extracted_k141_37247 | Not-determined |
| S6_extracted_k141_20567 | Not-determined |
| S6_extracted_k141_37255 | Not-determined |
| S6_extracted_k141_44457 | Not-determined |
| S6_extracted_k141_37260 | Not-determined |
| S6_extracted_k141_37264 | Low-quality |
| S6_extracted_k141_51079 | Not-determined |
| S6_extracted_k141_51081 | Not-determined |
| S6_extracted_k141_9119 | Not-determined |
| S6_extracted_k141_51083 | Not-determined |
| S6_extracted_k141_40004 | Not-determined |
| S6_extracted_k141_40012 | Not-determined |
| S6_extracted_k141_49157 | Low-quality |
| S6_extracted_k141_9138 | Not-determined |
| S6_extracted_k141_9143 | Not-determined |
| S6_extracted_k141_9144 | Not-determined |
| S6_extracted_k141_16471 | Not-determined |
| S6_extracted_k141_9146 | Low-quality |
| S6_extracted_k141_37287 | Not-determined |
| S6_extracted_k141_16477 | Not-determined |
| S6_extracted_k141_44474 | Not-determined |
| S6_extracted_k141_16478 | Not-determined |
| S6_extracted_k141_9163 | Not-determined |
| S6_extracted_k141_9164 | Not-determined |
| S6_extracted_k141_16479 | Not-determined |
| S6_extracted_k141_9166 | Not-determined |
| S6_extracted_k141_51098 | Low-quality |
| S6_extracted_k141_44482 | Not-determined |
| S6_extracted_k141_44483 | Not-determined |
| S6_extracted_k141_2012 | Not-determined |
| S6_extracted_k141_16485 | Not-determined |
| S6_extracted_k141_37306 | Not-determined |
| S6_extracted_k141_2019 | Not-determined |
| S6_extracted_k141_55825 | Not-determined |
| S6_extracted_k141_37312 | Not-determined |
| S6_extracted_k141_2020 | Low-quality |
| S6_extracted_k141_2023 | Not-determined |
| S6_extracted_k141_2024 | Not-determined |
| S6_extracted_k141_55834 | Not-determined |
| S6_extracted_k141_9178 | Not-determined |
| S6_extracted_k141_2030 | Not-determined |
| S6_extracted_k141_9185 | Not-determined |
| S6_extracted_k141_55842 | Not-determined |
| S6_extracted_k141_37325 | Not-determined |
| S6_extracted_k141_20600 | Not-determined |
| S6_extracted_k141_55846 | Not-determined |
| S6_extracted_k141_2039 | Not-determined |
| S6_extracted_k141_2042 | Not-determined |
| S6_extracted_k141_20605 | Not-determined |
| S6_extracted_k141_37332 | Low-quality |
| S6_extracted_k141_49182 | Not-determined |
| S6_extracted_k141_9202 | Not-determined |
| S6_extracted_k141_49184 | Not-determined |
| S6_extracted_k141_2055 | Not-determined |
| S6_extracted_k141_37339 | Not-determined |
| S6_extracted_k141_55879 | Not-determined |
| S6_extracted_k141_2059 | Not-determined |
| S6_extracted_k141_55881 | Not-determined |
| S6_extracted_k141_9212 | Not-determined |
| S6_extracted_k141_9214 | Not-determined |
| S6_extracted_k141_2069 | Not-determined |
| S6_extracted_k141_44508 | Not-determined |
| S6_extracted_k141_20615 | Low-quality |
| S6_extracted_k141_44516 | Not-determined |
| S6_extracted_k141_49197 | Not-determined |
| S6_extracted_k141_44521 | Not-determined |
| S6_extracted_k141_9229 | Not-determined |
| S6_extracted_k141_44524 | Not-determined |
| S6_extracted_k141_49204 | Not-determined |
| S6_extracted_k141_49208 | Not-determined |
| S6_extracted_k141_27700 | Not-determined |
| S6_extracted_k141_27702 | Not-determined |
| S6_extracted_k141_55916 | Not-determined |
| S6_extracted_k141_55918 | Not-determined |
| S6_extracted_k141_27707 | Not-determined |
| S6_extracted_k141_37342 | Low-quality |
| S6_extracted_k141_37343 | Low-quality |
| S6_extracted_k141_2080 | Not-determined |
| S6_extracted_k141_27708 | Not-determined |
| S6_extracted_k141_55924 | Not-determined |
| S6_extracted_k141_55925 | Not-determined |
| S6_extracted_k141_27709 | Low-quality |
| S6_extracted_k141_49229 | Low-quality |
| S6_extracted_k141_27711 | Not-determined |
| S6_extracted_k141_51134 | Not-determined |
| S6_extracted_k141_27715 | Not-determined |
| S6_extracted_k141_51140 | Not-determined |
| S6_extracted_k141_49238 | Not-determined |
| S6_extracted_k141_49241 | Not-determined |
| S6_extracted_k141_2098 | Not-determined |
| S6_extracted_k141_2099 | Not-determined |
| S6_extracted_k141_55942 | Not-determined |
| S6_extracted_k141_27722 | Low-quality |
| S6_extracted_k141_51149 | Low-quality |
| S6_extracted_k141_27726 | Not-determined |
| S6_extracted_k141_55946 | Not-determined |
| S6_extracted_k141_27729 | Not-determined |
| S6_extracted_k141_2102 | Not-determined |
| S6_extracted_k141_2104 | Not-determined |
| S6_extracted_k141_2106 | Not-determined |
| S6_extracted_k141_37353 | Not-determined |
| S6_extracted_k141_51158 | Not-determined |
| S6_extracted_k141_37355 | Not-determined |
| S6_extracted_k141_55954 | Not-determined |
| S6_extracted_k141_37357 | Not-determined |
| S6_extracted_k141_37358 | Not-determined |
| S6_extracted_k141_51164 | Not-determined |
| S6_extracted_k141_55964 | Low-quality |
| S6_extracted_k141_55966 | Low-quality |
| S6_extracted_k141_55967 | Not-determined |
| S6_extracted_k141_51170 | Not-determined |
| S6_extracted_k141_51171 | Low-quality |
| S6_extracted_k141_51176 | Not-determined |
| S6_extracted_k141_51177 | Not-determined |
| S6_extracted_k141_49260 | Not-determined |
| S6_extracted_k141_51178 | Not-determined |
| S6_extracted_k141_37363 | Not-determined |
| S6_extracted_k141_51179 | Not-determined |
| S6_extracted_k141_51187 | Low-quality |
| S6_extracted_k141_49266 | Not-determined |
| S6_extracted_k141_51191 | Not-determined |
| S6_extracted_k141_51194 | Not-determined |
| S6_extracted_k141_49280 | Low-quality |
| S6_extracted_k141_49283 | Not-determined |
| S6_extracted_k141_55982 | Not-determined |
| S6_extracted_k141_51209 | Not-determined |
| S6_extracted_k141_9270 | Not-determined |
| S6_extracted_k141_51210 | Not-determined |
| S6_extracted_k141_37377 | Low-quality |
| S7_vs_107876\|\|full | High-quality |
| S6_extracted_k141_9276 | Not-determined |
| S6_extracted_k141_9278 | Not-determined |
| S6_extracted_k141_2132 | Not-determined |
| S6_extracted_k141_49301 | Not-determined |
| S6_extracted_k141_37381 | Not-determined |
| S6_extracted_k141_49303 | Not-determined |
| S6_extracted_k141_9283 | Not-determined |
| S6_extracted_k141_49304 | Not-determined |
| S6_extracted_k141_9288 | Not-determined |
| S6_extracted_k141_9295 | Not-determined |
| S6_extracted_k141_55988 | Low-quality |
| S6_extracted_k141_44530 | Not-determined |
| S6_extracted_k141_37383 | Low-quality |
| S6_extracted_k141_44532 | Not-determined |
| S6_extracted_k141_47029 | Not-determined |
| S6_extracted_k141_47034 | Not-determined |
| S6_extracted_k141_44535 | Not-determined |
| S6_extracted_k141_9311 | Not-determined |
| S6_extracted_k141_37389 | Not-determined |
| S6_extracted_k141_9315 | Not-determined |
| S6_extracted_k141_51248 | Not-determined |
| S6_extracted_k141_51252 | Not-determined |
| S6_extracted_k141_2149 | Not-determined |
| S6_extracted_k141_9322 | Not-determined |
| S6_extracted_k141_2150 | Not-determined |
| S6_extracted_k141_9328 | Low-quality |
| S6_extracted_k141_9329 | Low-quality |
| S6_extracted_k141_37391 | Not-determined |
| S6_extracted_k141_47039 | Not-determined |
| S6_extracted_k141_47042 | Not-determined |
| S6_extracted_k141_44550 | Not-determined |
| S6_extracted_k141_55997 | Not-determined |
| S6_extracted_k141_51260 | Not-determined |
| S6_extracted_k141_47047 | Not-determined |
| S6_extracted_k141_51265 | Low-quality |
| S6_extracted_k141_2161 | Not-determined |
| S6_extracted_k141_51268 | Not-determined |
| S6_extracted_k141_56010 | Low-quality |
| S6_extracted_k141_9350 | Not-determined |
| S6_extracted_k141_49324 | Not-determined |
| S6_extracted_k141_56014 | Not-determined |
| S6_extracted_k141_49328 | Low-quality |
| S6_extracted_k141_20624 | Not-determined |
| S6_extracted_k141_47063 | Not-determined |
| S6_extracted_k141_20625 | Not-determined |
| S6_extracted_k141_37398 | Low-quality |
| S6_extracted_k141_44583 | Not-determined |
| S6_extracted_k141_37404 | Not-determined |
| S6_extracted_k141_51278 | Not-determined |
| S6_extracted_k141_47073 | Not-determined |
| S6_extracted_k141_2169 | Not-determined |
| S6_extracted_k141_47076 | Not-determined |
| S6_extracted_k141_20651 | Not-determined |
| S6_extracted_k141_20654 | Not-determined |
| S6_extracted_k141_20657 | Not-determined |
| S6_extracted_k141_20659 | Not-determined |
| S6_extracted_k141_49357 | Not-determined |
| S6_extracted_k141_37426 | Not-determined |
| S6_extracted_k141_47085 | Not-determined |
| S6_extracted_k141_56022 | Low-quality |
| S6_extracted_k141_47087 | Not-determined |
| S6_extracted_k141_47091 | Not-determined |
| S6_extracted_k141_47092 | Not-determined |
| S6_extracted_k141_56026 | Not-determined |
| S6_extracted_k141_20667 | Not-determined |
| S6_extracted_k141_44605 | Not-determined |
| S6_extracted_k141_44608 | Low-quality |
| S6_extracted_k141_49377 | Not-determined |
| S6_extracted_k141_49378 | Not-determined |
| S6_extracted_k141_47099 | Not-determined |
| S6_extracted_k141_49384 | Not-determined |
| S6_extracted_k141_49385 | Not-determined |
| S6_extracted_k141_51287 | Low-quality |
| S6_extracted_k141_51289 | Low-quality |
| S6_extracted_k141_51291 | Not-determined |
| S6_extracted_k141_37439 | Not-determined |
| S6_extracted_k141_49391 | Not-determined |
| S6_extracted_k141_49393 | Not-determined |
| S6_extracted_k141_37446 | Not-determined |
| S6_extracted_k141_49401 | Not-determined |
| S6_extracted_k141_37453 | Not-determined |
| S6_extracted_k141_37454 | Low-quality |
| S6_extracted_k141_40036 | Not-determined |
| S6_extracted_k141_37462 | Not-determined |
| S6_extracted_k141_51297 | Low-quality |
| S6_extracted_k141_37464 | Not-determined |
| S6_extracted_k141_40040 | Not-determined |
| S6_extracted_k141_40043 | Not-determined |
| S6_extracted_k141_51299 | Not-determined |
| S6_extracted_k141_40049 | Not-determined |
| S6_extracted_k141_51305 | Not-determined |
| S6_extracted_k141_37472 | Low-quality |
| S6_extracted_k141_51308 | Low-quality |
| S6_extracted_k141_40055 | Not-determined |
| S6_extracted_k141_9372 | Not-determined |
| S6_extracted_k141_20676 | Not-determined |
| S6_extracted_k141_20679 | Not-determined |
| S6_extracted_k141_9378 | Not-determined |
| S6_extracted_k141_51319 | Not-determined |
| S6_extracted_k141_20683 | Not-determined |
| S6_extracted_k141_51321 | Not-determined |
| S6_extracted_k141_51323 | Not-determined |
| S6_extracted_k141_20688 | Not-determined |
| S6_extracted_k141_51332 | Not-determined |
| S6_extracted_k141_51334 | Not-determined |
| S6_extracted_k141_20692 | Not-determined |
| S6_extracted_k141_51336 | Not-determined |
| S6_extracted_k141_51337 | Not-determined |
| S6_extracted_k141_51338 | Not-determined |
| S6_extracted_k141_2204 | Not-determined |
| S6_extracted_k141_51345 | Not-determined |
| S6_extracted_k141_49413 | Not-determined |
| S6_extracted_k141_49419 | Not-determined |
| S6_extracted_k141_44640 | Not-determined |
| S6_extracted_k141_44642 | Not-determined |
| S6_extracted_k141_20703 | Not-determined |
| S6_extracted_k141_20704 | Not-determined |
| S6_extracted_k141_9388 | Not-determined |
| S6_extracted_k141_44644 | Not-determined |
| S6_extracted_k141_56031 | Not-determined |
| S6_extracted_k141_20714 | Low-quality |
| S6_extracted_k141_20715 | Low-quality |
| S6_extracted_k141_51370 | Low-quality |
| S6_extracted_k141_44653 | Not-determined |
| S6_extracted_k141_9412 | Not-determined |
| S6_extracted_k141_20718 | Not-determined |
| S6_extracted_k141_37484 | Not-determined |
| S6_extracted_k141_9418 | Low-quality |
| S6_extracted_k141_20723 | Not-determined |
| S6_extracted_k141_56049 | Not-determined |
| S6_extracted_k141_56050 | Low-quality |
| S6_extracted_k141_47121 | Not-determined |
| S6_extracted_k141_9427 | Not-determined |
| S6_extracted_k141_37487 | Not-determined |
| S6_extracted_k141_20729 | Not-determined |
| S6_extracted_k141_44659 | Not-determined |
| S6_extracted_k141_56060 | Not-determined |
| S6_extracted_k141_56064 | Not-determined |
| S6_extracted_k141_49434 | Not-determined |
| S6_extracted_k141_44666 | Not-determined |
| S6_extracted_k141_56072 | Not-determined |
| S6_extracted_k141_2242 | Not-determined |
| S6_extracted_k141_56084 | Not-determined |
| S6_extracted_k141_37521 | Not-determined |
| S6_extracted_k141_37522 | Not-determined |
| S6_extracted_k141_56091 | Not-determined |
| S6_extracted_k141_56095 | Not-determined |
| S6_extracted_k141_37527 | Not-determined |
| S6_extracted_k141_49452 | Not-determined |
| S6_extracted_k141_56109 | Low-quality |
| S6_extracted_k141_44682 | Not-determined |
| S6_extracted_k141_2257 | Low-quality |
| S6_extracted_k141_51375 | Not-determined |
| S6_extracted_k141_37543 | Not-determined |
| S6_extracted_k141_44689 | Not-determined |
| S6_extracted_k141_51389 | Not-determined |
| S6_extracted_k141_56115 | Not-determined |
| S6_extracted_k141_49468 | Not-determined |
| S6_extracted_k141_51408 | Low-quality |
| S6_extracted_k141_56126 | Not-determined |
| S6_extracted_k141_44712 | Not-determined |
| S6_extracted_k141_44713 | Low-quality |
| S6_extracted_k141_56128 | Low-quality |
| S6_extracted_k141_56129 | Not-determined |
| S6_extracted_k141_37556 | Low-quality |
| S6_extracted_k141_51418 | Not-determined |
| S6_extracted_k141_44714 | Not-determined |
| S6_extracted_k141_51420 | Not-determined |
| S6_extracted_k141_37562 | Low-quality |
| S6_extracted_k141_56137 | Not-determined |
| S6_extracted_k141_51429 | Not-determined |
| S6_extracted_k141_37567 | Not-determined |
| S6_extracted_k141_44719 | Not-determined |
| S6_extracted_k141_2262 | Not-determined |
| S6_extracted_k141_2264 | Low-quality |
| S6_extracted_k141_51440 | Not-determined |
| S6_extracted_k141_44731 | Not-determined |
| S6_extracted_k141_56149 | Not-determined |
| S6_extracted_k141_2270 | Not-determined |
| S6_extracted_k141_56159 | Not-determined |
| S6_extracted_k141_44741 | Not-determined |
| S6_extracted_k141_44744 | Not-determined |
| S6_extracted_k141_37583 | Not-determined |
| S6_extracted_k141_2272 | Not-determined |
| S6_extracted_k141_56173 | Not-determined |
| S6_extracted_k141_44748 | Not-determined |
| S6_extracted_k141_56174 | Not-determined |
| S6_extracted_k141_51460 | Not-determined |
| S6_extracted_k141_44749 | Not-determined |
| S6_extracted_k141_56175 | Not-determined |
| S6_extracted_k141_44752 | Not-determined |
| S6_extracted_k141_51467 | Not-determined |
| S6_extracted_k141_51470 | Not-determined |
| S6_extracted_k141_37591 | Not-determined |
| S6_extracted_k141_44756 | Low-quality |
| S6_extracted_k141_37592 | Not-determined |
| S6_extracted_k141_51476 | Not-determined |
| S6_extracted_k141_2284 | Not-determined |
| S6_extracted_k141_44758 | Not-determined |
| S6_extracted_k141_2287 | Not-determined |
| S6_extracted_k141_56185 | Not-determined |
| S6_extracted_k141_2288 | Not-determined |
| S6_extracted_k141_51480 | Not-determined |
| S6_extracted_k141_56195 | Not-determined |
| S6_extracted_k141_37595 | Not-determined |
| S6_extracted_k141_51484 | Not-determined |
| S6_extracted_k141_51485 | Not-determined |
| S6_extracted_k141_51489 | Not-determined |
| S6_extracted_k141_51494 | Low-quality |
| S6_extracted_k141_51496 | Not-determined |
| S6_extracted_k141_56211 | Low-quality |
| S6_extracted_k141_56212 | Not-determined |
| S6_extracted_k141_44770 | Not-determined |
| S6_extracted_k141_51505 | Not-determined |
| S6_extracted_k141_20761 | Not-determined |
| S6_extracted_k141_20763 | Not-determined |
| S6_extracted_k141_37630 | Not-determined |
| S6_extracted_k141_20776 | Not-determined |
| S6_extracted_k141_20777 | Not-determined |
| S6_extracted_k141_20781 | Low-quality |
| S6_extracted_k141_56226 | Low-quality |
| S6_extracted_k141_51518 | Not-determined |
| S6_extracted_k141_56237 | Not-determined |
| S6_extracted_k141_20801 | Not-determined |
| S6_extracted_k141_56240 | Not-determined |
| S6_extracted_k141_20808 | Not-determined |
| S6_extracted_k141_20809 | Not-determined |
| S6_extracted_k141_56246 | Not-determined |
| S6_extracted_k141_56253 | Not-determined |
| S6_extracted_k141_37654 | Not-determined |
| S6_extracted_k141_51523 | Not-determined |
| S6_extracted_k141_51525 | Not-determined |
| S6_extracted_k141_37662 | Not-determined |
| S6_extracted_k141_37664 | Not-determined |
| S6_extracted_k141_51529 | Low-quality |
| S6_extracted_k141_37665 | Not-determined |
| S6_extracted_k141_20818 | Not-determined |
| S6_extracted_k141_27747 | Not-determined |
| S6_extracted_k141_27756 | Not-determined |
| S6_extracted_k141_2296 | Not-determined |
| S6_extracted_k141_2298 | Not-determined |
| S6_extracted_k141_2300 | Not-determined |
| S6_extracted_k141_20825 | Low-quality |
| S6_extracted_k141_20828 | Not-determined |
| S6_extracted_k141_27767 | Not-determined |
| S6_extracted_k141_37680 | Not-determined |
| S6_extracted_k141_27768 | Low-quality |
| S6_extracted_k141_37681 | Low-quality |
| S6_extracted_k141_20834 | Not-determined |
| S6_extracted_k141_37688 | Not-determined |
| S6_extracted_k141_27785 | Low-quality |
| S6_extracted_k141_27786 | Not-determined |
| S6_extracted_k141_20854 | Not-determined |
| S6_extracted_k141_20856 | Low-quality |
| S6_extracted_k141_20858 | Not-determined |
| S6_extracted_k141_37699 | Not-determined |
| S6_extracted_k141_37700 | Not-determined |
| S6_extracted_k141_20867 | Not-determined |
| S6_extracted_k141_20870 | Not-determined |
| S6_extracted_k141_51553 | Low-quality |
| S6_extracted_k141_20873 | Not-determined |
| S6_extracted_k141_20879 | Not-determined |
| S6_extracted_k141_20880 | Not-determined |
| S6_extracted_k141_20881 | Low-quality |
| S6_extracted_k141_20882 | Not-determined |
| S6_extracted_k141_20887 | Low-quality |
| S6_extracted_k141_20901 | Low-quality |
| S6_extracted_k141_20902 | Not-determined |
| S6_extracted_k141_20906 | Low-quality |
| S6_extracted_k141_51585 | Low-quality |
| S6_extracted_k141_2343 | Low-quality |
| S6_extracted_k141_2345 | Not-determined |
| S6_extracted_k141_20914 | Not-determined |
| S6_extracted_k141_2349 | Not-determined |
| S6_extracted_k141_2351 | Not-determined |
| S6_extracted_k141_20921 | Not-determined |
| S6_extracted_k141_51600 | Not-determined |
| S6_extracted_k141_20929 | Not-determined |
| S6_extracted_k141_51606 | Not-determined |
| S6_extracted_k141_27805 | Low-quality |
| S6_extracted_k141_27808 | Not-determined |
| S6_extracted_k141_20941 | Low-quality |
| S6_extracted_k141_51615 | Not-determined |
| S6_extracted_k141_27816 | Not-determined |
| S6_extracted_k141_56293 | Not-determined |
| S6_extracted_k141_56297 | Not-determined |
| S6_extracted_k141_27837 | Not-determined |
| S6_extracted_k141_27838 | Low-quality |
| S6_extracted_k141_27839 | Not-determined |
| S6_extracted_k141_27845 | Not-determined |
| S6_extracted_k141_56308 | Not-determined |
| S6_extracted_k141_56309 | Not-determined |
| S6_extracted_k141_51639 | Not-determined |
| S6_extracted_k141_20968 | Not-determined |
| S6_extracted_k141_27871 | Not-determined |
| S6_extracted_k141_56312 | Not-determined |
| S6_extracted_k141_27930 | Not-determined |
| S6_extracted_k141_27934 | Low-quality |
| S6_extracted_k141_20984 | Not-determined |
| S6_extracted_k141_27937 | Low-quality |
| S6_extracted_k141_56316 | Not-determined |
| S6_extracted_k141_56320 | Low-quality |
| S6_extracted_k141_27942 | Not-determined |
| S6_extracted_k141_51665 | Not-determined |
| S6_extracted_k141_21008 | Not-determined |
| S6_extracted_k141_51669 | Not-determined |
| S6_extracted_k141_51673 | Low-quality |
| S6_extracted_k141_51674 | Not-determined |
| S6_extracted_k141_51683 | Low-quality |
| S6_extracted_k141_51684 | Low-quality |
| S6_extracted_k141_51690 | Not-determined |
| S6_extracted_k141_51694 | Low-quality |
| S6_extracted_k141_51697 | Not-determined |
| S6_extracted_k141_51712 | Not-determined |
| S6_extracted_k141_56333 | Not-determined |
| S6_extracted_k141_56343 | Not-determined |
| S6_extracted_k141_56344 | Not-determined |
| S6_extracted_k141_56345 | Not-determined |
| S6_extracted_k141_51731 | Not-determined |
| S6_extracted_k141_56353 | Not-determined |
| S6_extracted_k141_56354 | Not-determined |
| S6_extracted_k141_56356 | Not-determined |
| S6_extracted_k141_56360 | Not-determined |
| S6_extracted_k141_51739 | Not-determined |
| S6_extracted_k141_27997 | Not-determined |
| S6_extracted_k141_51742 | Not-determined |
| S6_extracted_k141_51743 | Not-determined |
| S6_extracted_k141_51746 | Not-determined |
| S17_vs_k141_111931\|\|full | Medium-quality |
| S6_extracted_k141_28016 | Not-determined |
| S6_extracted_k141_56370 | Low-quality |
| S6_extracted_k141_51759 | Not-determined |
| S6_extracted_k141_56390 | Not-determined |
| S6_extracted_k141_51774 | Not-determined |
| S6_extracted_k141_21017 | Not-determined |
| S6_extracted_k141_51780 | Low-quality |
| S6_extracted_k141_56400 | Not-determined |
| S6_extracted_k141_56401 | Not-determined |
| S6_extracted_k141_21025 | Not-determined |
| S6_extracted_k141_21028 | Low-quality |
| S6_extracted_k141_21029 | Low-quality |
| S6_extracted_k141_28051 | Not-determined |
| S6_extracted_k141_28054 | Low-quality |
| S6_extracted_k141_21039 | Not-determined |
| S6_extracted_k141_28065 | Not-determined |
| S6_extracted_k141_56412 | Low-quality |
| S6_extracted_k141_56413 | Not-determined |
| S6_extracted_k141_56428 | Not-determined |
| S6_extracted_k141_28085 | Not-determined |
| S6_extracted_k141_51803 | Not-determined |
| S6_extracted_k141_51808 | Low-quality |
| S6_extracted_k141_51815 | Not-determined |
| S6_extracted_k141_56447 | Low-quality |
| S6_extracted_k141_21050 | Not-determined |
| S6_extracted_k141_21055 | Not-determined |
| S6_extracted_k141_51826 | Not-determined |
| S6_extracted_k141_51827 | Not-determined |
| S6_extracted_k141_56450 | Not-determined |
| S6_extracted_k141_51834 | Low-quality |
| S6_extracted_k141_28113 | Not-determined |
| S6_extracted_k141_56470 | Low-quality |
| S6_extracted_k141_56475 | Not-determined |
| S6_extracted_k141_28120 | Not-determined |
| S6_extracted_k141_21061 | Not-determined |
| S6_extracted_k141_21065 | Not-determined |
| S6_extracted_k141_21068 | Not-determined |
| S6_extracted_k141_28137 | Not-determined |
| S6_extracted_k141_21075 | Not-determined |
| S6_extracted_k141_28140 | Not-determined |
| S6_extracted_k141_21077 | Not-determined |
| S6_extracted_k141_21081 | Not-determined |
| S6_extracted_k141_21083 | Not-determined |
| S6_extracted_k141_28144 | Low-quality |
| S6_extracted_k141_21088 | Not-determined |
| S6_extracted_k141_21092 | Not-determined |
| S6_extracted_k141_21099 | Not-determined |
| S12_extracted_k141_61299 | Medium-quality |
| S6_extracted_k141_28165 | Not-determined |
| S6_extracted_k141_28176 | Not-determined |
| S6_extracted_k141_28187 | Not-determined |
| S6_extracted_k141_28198 | Low-quality |
| S6_extracted_k141_21110 | Not-determined |
| S6_extracted_k141_28209 | Not-determined |
| S6_extracted_k141_28215 | Not-determined |
| S11_vs_k141_75554\|\|full | Medium-quality |
| S6_extracted_k141_21138 | Not-determined |
| S6_extracted_k141_21156 | Not-determined |
| S6_extracted_k141_28235 | Low-quality |
| S6_extracted_k141_28238 | Low-quality |
| S6_extracted_k141_21173 | Not-determined |
| S6_extracted_k141_21180 | Low-quality |
| S6_extracted_k141_28243 | Not-determined |
| S6_extracted_k141_28247 | Low-quality |
| S8_extracted_k141_91033 | Medium-quality |
| S6_extracted_k141_28273 | Not-determined |
| S6_extracted_k141_21186 | Not-determined |
| S6_extracted_k141_28278 | Low-quality |
| S6_extracted_k141_21190 | Not-determined |
| S6_extracted_k141_21194 | Not-determined |
| S6_extracted_k141_21203 | Not-determined |
| S6_extracted_k141_21205 | Not-determined |
| S5_vs_k141_74948\|\|full | Medium-quality |
| S7_extracted_20129 | Medium-quality |
| S6_extracted_k141_56499 | Low-quality |
| S6_extracted_k141_56501 | Not-determined |
| S6_extracted_k141_56508 | Low-quality |
| S6_extracted_k141_56509 | Low-quality |
| S6_extracted_k141_56510 | Not-determined |
| S6_extracted_k141_56513 | Not-determined |
| S6_extracted_k141_56516 | Not-determined |
| S6_extracted_k141_56521 | Not-determined |
| S6_extracted_k141_56522 | Not-determined |
| S6_extracted_k141_56532 | Not-determined |
| S6_extracted_k141_56534 | Not-determined |
| S6_extracted_k141_56539 | Not-determined |
| S6_extracted_k141_56544 | Not-determined |
| S3_extracted_k141_111776 | Medium-quality |
| S11_extracted_k141_26593 | Medium-quality |
| S7_extracted_83624 | Medium-quality |
| S8_extracted_k141_142700 | Medium-quality |
| S5_vs_k141_38011\|\|full | Medium-quality |
| S6_vs_k141_36178\|\|full | Low-quality |
| S6_vs_k141_50298\|\|full | Not-determined |
| S6_vs_k141_22127\|\|full | Medium-quality |
| S8_extracted_k141_174559 | Medium-quality |
| S7_vs_19866\|\|full | Medium-quality |
| S12_vs_k141_187882\|\|full | Medium-quality |
| S11_vs_k141_39861\|\|full | Medium-quality |
| S7_vs_32396\|\|full | Medium-quality |
| S8_extracted_k141_159173 | Medium-quality |
| S6_vs_k141_6916\|\|full | Not-determined |
| S17_extracted_k141_36452 | Medium-quality |
| S6_vs_k141_49122\|\|full | Not-determined |
| S12_extracted_k141_170614 | Medium-quality |
| S8_vs_k141_153696\|\|full | Medium-quality |
| S16_vs_96874\|\|full | High-quality |
| S17_extracted_k141_79151 | Medium-quality |
| S5_vs_k141_110072\|\|full | High-quality |
| S6_vs_k141_36820\|\|full | High-quality |
| S5_extracted_k141_137260 | Complete |
| S6_extracted_k141_56361 | Complete |
| S7_extracted_21981 | Medium-quality |
| S16_extracted_100473 | Medium-quality |
| S12_vs_k141_61299\|\|full | Medium-quality |
| S11_vs_k141_58587\|\|full | Medium-quality |
| S11_vs_k141_85081\|\|full | High-quality |
| S3_extracted_k141_57422 | Medium-quality |
| S6_vs_k141_56500\|\|full | Low-quality |
| S6_vs_k141_56504\|\|full | Not-determined |
| S6_vs_k141_56511\|\|full | Not-determined |
| S6_vs_k141_7071\|\|full | Low-quality |
| S6_vs_k141_42420\|\|full | Not-determined |
| S6_vs_k141_14147\|\|full | Low-quality |
| S6_vs_k141_16504\|\|full | Low-quality |
| S6_vs_k141_51844\|\|full | Not-determined |
| S6_vs_k141_28286\|\|full | Low-quality |
| S6_vs_k141_51846\|\|full | Not-determined |
| S6_vs_k141_42432\|\|full | Low-quality |

| | |
|---|---|
| SG_vs_k141_21228\|\|full | Low-quality |
| SG_vs_k141_44777\|\|full | Low-quality |
| SG_vs_k141_4733\|\|full | Low-quality |
| SG_vs_k141_44781\|\|full | Not-determined |
| SG_vs_k141_11813\|\|full | Low-quality |
| SG_vs_k141_25938\|\|full | Low-quality |
| SB_vs_k141_142097\|\|full | Medium-quality |
| SG_vs_k141_25941\|\|full | Low-quality |
| SG_vs_k141_23612\|\|full | Not-determined |
| SG_vs_k141_49511\|\|full | Low-quality |
| SG_vs_k141_23613\|\|full | Low-quality |
| SG_vs_k141_14156\|\|full | Low-quality |
| SG_vs_k141_42464\|\|full | Not-determined |
| SG_vs_k141_51871\|\|full | Low-quality |
| SG_vs_k141_47177\|\|full | Low-quality |
| SG_vs_k141_30648\|\|full | Low-quality |
| SG_vs_k141_30651\|\|full | Not-determined |
| SG_vs_k141_54231\|\|full | Low-quality |
| SG_vs_k141_33032\|\|full | Low-quality |
| SG_vs_k141_38\|\|full | Not-determined |
| SG_vs_k141_16559\|\|full | Low-quality |
| SG_vs_k141_42475\|\|full | Low-quality |
| SG_vs_k141_54241\|\|full | Not-determined |
| SG_vs_k141_18898\|\|full | Low-quality |
| SG_vs_k141_49567\|\|full | Low-quality |
| SG_vs_k141_49568\|\|full | Low-quality |
| SG_vs_k141_42491\|\|full | Not-determined |
| SG_vs_k141_47189\|\|full | Not-determined |
| SG_vs_k141_42504\|\|full | Low-quality |
| SG_vs_k141_47190\|\|full | Low-quality |
| SG_vs_k141_42505\|\|full | Not-determined |
| SG_vs_k141_56\|\|full | Low-quality |
| SG_vs_k141_54248\|\|full | Low-quality |
| SG_vs_k141_18917\|\|full | Not-determined |
| SG_vs_k141_51939\|\|full | Not-determined |
| SG_vs_k141_30736\|\|full | Not-determined |
| SG_vs_k141_9436\|\|full | Low-quality |
| SG_vs_k141_18926\|\|full | Low-quality |
| SG_vs_k141_9439\|\|full | Low-quality |
| SG_vs_k141_75\|\|full | Low-quality |
| SG_vs_k141_78\|\|full | Low-quality |
| SG_vs_k141_25965\|\|full | Low-quality |
| SG_vs_k141_93\|\|full | Low-quality |
| SG_vs_k141_25973\|\|full | Not-determined |
| SG_vs_k141_21262\|\|full | Not-determined |
| SG_vs_k141_14233\|\|full | Low-quality |
| SG_vs_k141_18954\|\|full | Low-quality |
| SG_vs_k141_37781\|\|full | Low-quality |
| SG_vs_k141_110\|\|full | Not-determined |
| SG_vs_k141_4781\|\|full | Not-determined |
| SG_vs_k141_51975\|\|full | Low-quality |
| SG_vs_k141_14253\|\|full | Not-determined |
| SG_vs_k141_14261\|\|full | Low-quality |
| SG_vs_k141_49605\|\|full | Not-determined |
| SG_vs_k141_40130\|\|full | Not-determined |
| SG_vs_k141_33117\|\|full | Low-quality |
| SG_vs_k141_49607\|\|full | Low-quality |
| SG_vs_k141_44835\|\|full | Low-quality |
| SG_vs_k141_51996\|\|full | Not-determined |
| SG_vs_k141_40145\|\|full | Low-quality |
| SG_vs_k141_44846\|\|full | Low-quality |
| SG_vs_k141_40151\|\|full | Low-quality |
| SG_vs_k141_149\|\|full | Not-determined |
| SG_vs_k141_52002\|\|full | Not-determined |
| SG_vs_k141_42580\|\|full | Not-determined |
| SG_vs_k141_28377\|\|full | Low-quality |
| SG_vs_k141_44861\|\|full | Low-quality |
| SG_vs_k141_52005\|\|full | Low-quality |
| SG_vs_k141_49620\|\|full | Low-quality |
| SG_vs_k141_52014\|\|full | Low-quality |
| SB_vs_k141_142700\|\|full | Medium-quality |
| SG_vs_k141_44882\|\|full | Low-quality |
| SG_vs_k141_23645\|\|full | Low-quality |
| SG_vs_k141_44884\|\|full | Low-quality |
| SG_vs_k141_44885\|\|full | Not-determined |
| SG_vs_k141_11842\|\|full | Low-quality |
| SG_vs_k141_28403\|\|full | Low-quality |
| SG_vs_k141_14270\|\|full | Low-quality |
| SG_vs_k141_33151\|\|full | Not-determined |
| SG_vs_k141_35457\|\|full | Low-quality |
| SG_vs_k141_2382\|\|full | Not-determined |
| SG_vs_k141_14281\|\|full | Low-quality |
| SG_vs_k141_52038\|\|full | Low-quality |
| SG_vs_k141_14288\|\|full | Not-determined |
| SG_vs_k141_52042\|\|full | Not-determined |
| SG_vs_k141_42624\|\|full | Not-determined |
| SG_vs_k141_9524\|\|full | Not-determined |
| SG_vs_k141_2399\|\|full | Low-quality |
| SG_vs_k141_37875\|\|full | Low-quality |
| SG_vs_k141_37881\|\|full | Low-quality |
| SG_vs_k141_44900\|\|full | Not-determined |
| SG_vs_k141_52066\|\|full | Low-quality |
| SG_vs_k141_42635\|\|full | Not-determined |
| SG_vs_k141_9536\|\|full | Not-determined |
| SG_vs_k141_44915\|\|full | Not-determined |
| SG_vs_k141_25987\|\|full | Low-quality |
| SG_vs_k141_52097\|\|full | Not-determined |
| SG_vs_k141_40194\|\|full | Not-determined |
| SG_vs_k141_16662\|\|full | Low-quality |
| SG_vs_k141_30777\|\|full | Low-quality |
| SG_vs_k141_42642\|\|full | Low-quality |
| SG_vs_k141_37919\|\|full | Low-quality |
| SG_vs_k141_52106\|\|full | Low-quality |
| SG_vs_k141_14331\|\|full | Low-quality |
| SG_vs_k141_52108\|\|full | Low-quality |
| SG_vs_k141_232\|\|full | Low-quality |
| SG_vs_k141_52122\|\|full | Low-quality |
| SG_vs_k141_16671\|\|full | Not-determined |
| SG_vs_k141_235\|\|full | Not-determined |
| SG_vs_k141_23658\|\|full | Low-quality |
| SG_vs_k141_23659\|\|full | Not-determined |
| SG_vs_k141_4821\|\|full | Low-quality |
| SG_vs_k141_42653\|\|full | Low-quality |
| SG_vs_k141_244\|\|full | Not-determined |
| SG_vs_k141_37949\|\|full | Not-determined |
| SG_vs_k141_23667\|\|full | Not-determined |
| SG_vs_k141_26013\|\|full | Not-determined |
| SG_vs_k141_44965\|\|full | Low-quality |
| SG_vs_k141_44967\|\|full | Not-determined |
| SG_vs_k141_42663\|\|full | Low-quality |
| SG_vs_k141_30810\|\|full | Low-quality |
| SG_vs_k141_2466\|\|full | Low-quality |
| SG_vs_k141_37967\|\|full | Not-determined |
| SG_vs_k141_37972\|\|full | Low-quality |
| SG_vs_k141_16693\|\|full | Low-quality |
| SG_vs_k141_11894\|\|full | Low-quality |
| SG_vs_k141_23688\|\|full | Low-quality |
| SG_vs_k141_11897\|\|full | Not-determined |
| SG_vs_k141_28460\|\|full | Low-quality |
| SG_vs_k141_40228\|\|full | Not-determined |
| SG_vs_k141_11900\|\|full | Not-determined |
| SG_vs_k141_7116\|\|full | Low-quality |
| SG_vs_k141_2490\|\|full | Low-quality |
| SG_vs_k141_4843\|\|full | Low-quality |
| SG_vs_k141_42678\|\|full | Not-determined |
| SG_vs_k141_2492\|\|full | Not-determined |
| SG_vs_k141_11912\|\|full | Low-quality |
| SG_vs_k141_286\|\|full | Not-determined |
| SG_vs_k141_37986\|\|full | Not-determined |
| SG_vs_k141_40239\|\|full | Low-quality |
| SG_vs_k141_4848\|\|full | Not-determined |
| SG_vs_k141_9563\|\|full | Low-quality |
| SG_vs_k141_11914\|\|full | Low-quality |
| SG_vs_k141_37989\|\|full | Not-determined |
| SG_vs_k141_14364\|\|full | Low-quality |
| SG_vs_k141_26066\|\|full | Low-quality |
| SG_vs_k141_33214\|\|full | Low-quality |
| SG_vs_k141_28481\|\|full | Low-quality |
| SG_vs_k141_26067\|\|full | Low-quality |
| SG_vs_k141_7146\|\|full | Low-quality |
| SG_vs_k141_4860\|\|full | Not-determined |
| SG_vs_k141_28484\|\|full | Low-quality |
| SG_vs_k141_11942\|\|full | Not-determined |
| SG_vs_k141_4871\|\|full | Low-quality |
| SG_vs_k141_40244\|\|full | Not-determined |
| SG_vs_k141_14371\|\|full | Low-quality |
| SG_vs_k141_11949\|\|full | Low-quality |
| SG_vs_k141_14373\|\|full | Low-quality |
| SG_vs_k141_14375\|\|full | Low-quality |
| SG_vs_k141_16725\|\|full | Low-quality |
| SG_vs_k141_23696\|\|full | Low-quality |
| SG_vs_k141_16727\|\|full | Low-quality |
| SG_vs_k141_26110\|\|full | Low-quality |
| SG_vs_k141_21312\|\|full | Not-determined |
| SG_vs_k141_45011\|\|full | Not-determined |
| SG_vs_k141_38028\|\|full | Low-quality |
| SG_vs_k141_9601\|\|full | Low-quality |
| SG_vs_k141_352\|\|full | Low-quality |
| SG_vs_k141_2560\|\|full | Not-determined |
| SG_vs_k141_11979\|\|full | Low-quality |
| SG_vs_k141_40269\|\|full | Low-quality |
| SG_vs_k141_40273\|\|full | Low-quality |
| SG_vs_k141_33255\|\|full | Not-determined |
| SG_vs_k141_42708\|\|full | Low-quality |
| SG_vs_k141_28524\|\|full | Low-quality |
| SG_vs_k141_52130\|\|full | Not-determined |
| SG_vs_k141_14405\|\|full | Low-quality |
| SG_vs_k141_9609\|\|full | Not-determined |
| SG_vs_k141_33283\|\|full | Not-determined |
| SG_vs_k141_12025\|\|full | Low-quality |
| SG_vs_k141_40295\|\|full | Not-determined |
| SG_vs_k141_9619\|\|full | Low-quality |
| SG_vs_k141_14433\|\|full | Not-determined |
| SG_vs_k141_391\|\|full | Low-quality |
| SG_vs_k141_12027\|\|full | Low-quality |
| SG_vs_k141_16741\|\|full | Low-quality |
| SG_vs_k141_12028\|\|full | Low-quality |
| SG_vs_k141_33298\|\|full | Not-determined |
| SG_vs_k141_14439\|\|full | Low-quality |
| SG_vs_k141_9626\|\|full | Low-quality |
| SG_vs_k141_33307\|\|full | Low-quality |
| SG_vs_k141_402\|\|full | Not-determined |
| SG_vs_k141_45071\|\|full | Low-quality |
| SG_vs_k141_45072\|\|full | Low-quality |
| SG_vs_k141_23752\|\|full | Low-quality |
| SG_vs_k141_33315\|\|full | Not-determined |
| SG_vs_k141_23754\|\|full | Low-quality |
| SG_vs_k141_47236\|\|full | Not-determined |
| SG_vs_k141_7198\|\|full | Low-quality |
| SG_vs_k141_45083\|\|full | Low-quality |
| SG_vs_k141_23767\|\|full | Low-quality |
| S4_vs_VIRSorter_4746-cat_2 | Medium-quality |
| SG_vs_k141_12055\|\|full | Low-quality |
| SG_vs_k141_40320\|\|full | Low-quality |
| SG_vs_k141_38142\|\|full | Low-quality |
| SG_vs_k141_4912\|\|full | Low-quality |
| SG_vs_k141_12058\|\|full | Not-determined |
| SG_vs_k141_16769\|\|full | Low-quality |
| SG_vs_k141_431\|\|full | Low-quality |
| SG_vs_k141_12066\|\|full | Low-quality |
| SG_vs_k141_33333\|\|full | Not-determined |
| SG_vs_k141_23792\|\|full | Not-determined |
| SG_vs_k141_23793\|\|full | Not-determined |
| SG_vs_k141_12068\|\|full | Not-determined |
| SG_vs_k141_40353\|\|full | Low-quality |
| SG_vs_k141_23802\|\|full | Low-quality |
| SG_vs_k141_437\|\|full | Low-quality |
| SG_vs_k141_28535\|\|full | Low-quality |
| SG_vs_k141_47255\|\|full | Low-quality |
| SG_vs_k141_4930\|\|full | Not-determined |
| SG_vs_k141_12078\|\|full | Not-determined |
| SG_vs_k141_28543\|\|full | Not-determined |
| SG_vs_k141_23818\|\|full | Low-quality |
| SG_vs_k141_40369\|\|full | Low-quality |
| SG_vs_k141_47262\|\|full | Low-quality |
| SG_vs_k141_23820\|\|full | Low-quality |
| SG_vs_k141_16811\|\|full | Low-quality |
| SG_vs_k141_472\|\|full | Low-quality |
| SG_vs_k141_52172\|\|full | Low-quality |
| SG_vs_k141_23828\|\|full | Not-determined |
| SG_vs_k141_35580\|\|full | Not-determined |
| SG_vs_k141_23833\|\|full | Low-quality |
| SG_vs_k141_478\|\|full | Not-determined |
| SG_vs_k141_9694\|\|full | Not-determined |
| SG_vs_k141_7247\|\|full | Not-determined |
| SG_vs_k141_481\|\|full | Low-quality |
| SG_vs_k141_47282\|\|full | Not-determined |
| SG_vs_k141_16838\|\|full | Low-quality |
| SG_vs_k141_23856\|\|full | Low-quality |
| SG_vs_k141_40397\|\|full | Low-quality |
| SG_vs_k141_487\|\|full | Not-determined |
| SG_vs_k141_7257\|\|full | Not-determined |
| SG_vs_k141_38196\|\|full | Low-quality |
| SG_vs_k141_40405\|\|full | Low-quality |
| SG_vs_k141_12125\|\|full | Low-quality |
| SG_vs_k141_495\|\|full | Low-quality |
| SG_vs_k141_42726\|\|full | Low-quality |
| SG_vs_k141_35612\|\|full | Low-quality |
| SG_vs_k141_42730\|\|full | Low-quality |
| SG_vs_k141_42734\|\|full | Low-quality |
| SG_vs_k141_2730\|\|full | Not-determined |
| SG_vs_k141_26149\|\|full | Low-quality |
| SG_vs_k141_23872\|\|full | Low-quality |
| SG_vs_k141_26152\|\|full | Low-quality |
| SG_vs_k141_14494\|\|full | Low-quality |
| SG_vs_k141_2741\|\|full | Not-determined |
| SG_vs_k141_26165\|\|full | Low-quality |
| SG_vs_k141_14502\|\|full | Low-quality |
| SG_vs_k141_33410\|\|full | Not-determined |
| SG_vs_k141_26172\|\|full | Low-quality |
| SG_vs_k141_2755\|\|full | Low-quality |
| SG_vs_k141_2759\|\|full | Not-determined |
| SG_vs_k141_52275\|\|full | Not-determined |
| SG_vs_k141_7270\|\|full | Not-determined |
| SG_vs_k141_42776\|\|full | Low-quality |
| SG_vs_k141_2768\|\|full | Low-quality |
| SG_vs_k141_33438\|\|full | Not-determined |
| SG_vs_k141_537\|\|full | Low-quality |
| SG_vs_k141_14523\|\|full | Low-quality |
| SG_vs_k141_9716\|\|full | Low-quality |
| SG_vs_k141_19041\|\|full | Low-quality |
| SG_vs_k141_42802\|\|full | Low-quality |
| SG_vs_k141_23903\|\|full | Low-quality |
| SG_vs_k141_12162\|\|full | Low-quality |
| SG_vs_k141_19055\|\|full | Low-quality |
| SG_vs_k141_42820\|\|full | Low-quality |
| SG_vs_k141_2834\|\|full | Low-quality |
| SG_vs_k141_2837\|\|full | Low-quality |
| SG_vs_k141_38220\|\|full | Not-determined |
| SG_vs_k141_42839\|\|full | Low-quality |
| SG_vs_k141_52332\|\|full | Low-quality |
| SG_vs_k141_42842\|\|full | Low-quality |
| SG_vs_k141_14553\|\|full | Not-determined |
| SG_vs_k141_45134\|\|full | Not-determined |
| SG_vs_k141_28612\|\|full | Not-determined |
| SG_vs_k141_52349\|\|full | Low-quality |
| SG_vs_k141_40510\|\|full | Low-quality |
| SG_vs_k141_52354\|\|full | Low-quality |
| SG_vs_k141_40516\|\|full | Low-quality |
| SG_vs_k141_52358\|\|full | Low-quality |
| SG_vs_k141_9769\|\|full | Not-determined |
| S7_vs_13903\|\|full | High-quality |
| SG_vs_k141_52364\|\|full | Not-determined |
| SG_vs_k141_7282\|\|full | Not-determined |
| SG_vs_k141_2894\|\|full | Low-quality |
| SG_vs_k141_40550\|\|full | Low-quality |
| SG_vs_k141_4960\|\|full | Not-determined |
| SG_vs_k141_35667\|\|full | Low-quality |
| SG_vs_k141_16898\|\|full | Not-determined |
| SG_vs_k141_7300\|\|full | Low-quality |
| SG_vs_k141_14586\|\|full | Not-determined |
| SG_vs_k141_35676\|\|full | Not-determined |
| SG_vs_k141_14593\|\|full | Low-quality |
| SG_vs_k141_38230\|\|full | Low-quality |
| SG_vs_k141_35681\|\|full | Low-quality |
| SG_vs_k141_16914\|\|full | Not-determined |
| SG_vs_k141_42874\|\|full | Low-quality |
| SG_vs_k141_38247\|\|full | Low-quality |
| SG_vs_k141_23976\|\|full | Low-quality |
| SG_vs_k141_19068\|\|full | Low-quality |
| SG_vs_k141_12239\|\|full | Low-quality |
| SG_vs_k141_24001\|\|full | Low-quality |
| SG_vs_k141_47337\|\|full | Low-quality |
| SG_vs_k141_28642\|\|full | Not-determined |
| SG_vs_k141_24009\|\|full | Low-quality |
| SG_vs_k141_28653\|\|full | Low-quality |
| S3_extracted_k141_7137 | Medium-quality |
| SG_vs_k141_33481\|\|full | Low-quality |
| SG_vs_k141_47349\|\|full | Not-determined |
| SG_vs_k141_12265\|\|full | Not-determined |
| SG_vs_k141_579\|\|full | Low-quality |
| SG_vs_k141_33490\|\|full | Not-determined |
| S5_extracted_k141_86016 | Medium-quality |
| SG_vs_k141_52459\|\|full | Low-quality |
| SG_vs_k141_16952\|\|full | Low-quality |
| SG_vs_k141_47358\|\|full | Low-quality |
| SG_vs_k141_2965\|\|full | Low-quality |
| SG_vs_k141_2967\|\|full | Not-determined |
| SG_vs_k141_7333\|\|full | Low-quality |
| SG_vs_k141_7338\|\|full | Low-quality |
| SG_vs_k141_38259\|\|full | Low-quality |
| S7_extracted_52002 | Medium-quality |
| SG_vs_k141_45209\|\|full | Low-quality |
| SG_vs_k141_595\|\|full | Not-determined |
| SG_vs_k141_7352\|\|full | Low-quality |
| SG_vs_k141_52494\|\|full | Not-determined |
| SG_vs_k141_42909\|\|full | Not-determined |
| SG_vs_k141_42910\|\|full | Low-quality |
| SG_vs_k141_28699\|\|full | Low-quality |
| SG_vs_k141_7357\|\|full | Not-determined |
| SG_vs_k141_52505\|\|full | Low-quality |
| SG_vs_k141_2993\|\|full | Not-determined |
| SG_vs_k141_45219\|\|full | Low-quality |
| SG_vs_k141_38276\|\|full | Not-determined |
| SG_vs_k141_38284\|\|full | Not-determined |
| SG_vs_k141_9814\|\|full | Low-quality |
| SG_vs_k141_33534\|\|full | Not-determined |
| SG_vs_k141_5023\|\|full | Low-quality |
| SG_vs_k141_7385\|\|full | Low-quality |
| S12_extracted_k141_38725 | Medium-quality |
| SG_vs_k141_28711\|\|full | Low-quality |
| SG_vs_k141_7394\|\|full | Not-determined |
| SG_vs_k141_28712\|\|full | Low-quality |
| SG_vs_k141_30879\|\|full | Low-quality |
| SG_vs_k141_40592\|\|full | Not-determined |
| SG_vs_k141_47425\|\|full | Low-quality |
| SG_vs_k141_17023\|\|full | Low-quality |
| SG_vs_k141_24044\|\|full | Not-determined |
| SG_vs_k141_26287\|\|full | Not-determined |
| SG_vs_k141_47426\|\|full | Low-quality |
| SG_vs_k141_54291\|\|full | Low-quality |
| SG_vs_k141_47428\|\|full | Low-quality |
| SG_vs_k141_3017\|\|full | Low-quality |
| SG_vs_k141_5052\|\|full | Low-quality |
| SG_vs_k141_644\|\|full | Low-quality |
| SG_vs_k141_54297\|\|full | Not-determined |
| SG_vs_k141_45266\|\|full | Low-quality |
| SG_vs_k141_54305\|\|full | Not-determined |
| SG_vs_k141_38290\|\|full | Not-determined |
| SG_vs_k141_52596\|\|full | Not-determined |
| SG_vs_k141_26294\|\|full | Not-determined |
| SG_vs_k141_33588\|\|full | Not-determined |
| SG_vs_k141_12383\|\|full | Low-quality |
| SG_vs_k141_3042\|\|full | Low-quality |
| SG_vs_k141_26306\|\|full | Low-quality |
| SG_vs_k141_30917\|\|full | Low-quality |
| SG_vs_k141_45289\|\|full | Not-determined |
| SG_vs_k141_12391\|\|full | Low-quality |
| SG_vs_k141_12395\|\|full | Not-determined |
| SG_vs_k141_3050\|\|full | Low-quality |
| SG_vs_k141_14664\|\|full | Low-quality |
| SG_vs_k141_33604\|\|full | Not-determined |
| SG_vs_k141_35734\|\|full | Low-quality |
| SG_vs_k141_3053\|\|full | Low-quality |
| SG_vs_k141_26317\|\|full | Low-quality |
| SG_vs_k141_24078\|\|full | Not-determined |
| SG_vs_k141_38320\|\|full | Low-quality |
| SG_vs_k141_26321\|\|full | Low-quality |
| SG_vs_k141_42924\|\|full | Low-quality |
| SG_vs_k141_38329\|\|full | Low-quality |
| SG_vs_k141_54350\|\|full | Low-quality |
| SG_vs_k141_12409\|\|full | Low-quality |
| SG_vs_k141_681\|\|full | Low-quality |
| SG_vs_k141_35740\|\|full | Not-determined |
| SG_vs_k141_24098\|\|full | Not-determined |
| SG_vs_k141_54362\|\|full | Low-quality |
| SG_vs_k141_38334\|\|full | Not-determined |
| SG_vs_k141_49756\|\|full | Low-quality |
| SG_vs_k141_690\|\|full | Low-quality |
| SG_vs_k141_24107\|\|full | Low-quality |
| SG_vs_k141_38338\|\|full | Low-quality |
| SG_vs_k141_38340\|\|full | Low-quality |
| SG_vs_k141_17058\|\|full | Not-determined |
| SG_vs_k141_35758\|\|full | Low-quality |
| SG_vs_k141_9901\|\|full | Low-quality |
| SG_vs_k141_26325\|\|full | Low-quality |
| SG_vs_k141_9905\|\|full | Low-quality |
| SG_vs_k141_38353\|\|full | Low-quality |
| SG_vs_k141_24123\|\|full | Low-quality |
| SG_vs_k141_14687\|\|full | Low-quality |
| SG_vs_k141_17079\|\|full | Low-quality |
| SG_vs_k141_52634\|\|full | Low-quality |
| SG_vs_k141_5079\|\|full | Not-determined |
| SG_vs_k141_708\|\|full | Low-quality |
| SG_vs_k141_45342\|\|full | Low-quality |
| SG_vs_k141_711\|\|full | Low-quality |
| SG_vs_k141_26345\|\|full | Low-quality |
| SG_vs_k141_42950\|\|full | Low-quality |
| SG_vs_k141_17081\|\|full | Low-quality |
| SG_vs_k141_21422\|\|full | Low-quality |
| SG_vs_k141_14705\|\|full | Low-quality |
| SG_vs_k141_38360\|\|full | Not-determined |
| SG_vs_k141_7426\|\|full | Low-quality |
| SG_vs_k141_12491\|\|full | Low-quality |
| SG_vs_k141_54409\|\|full | Not-determined |
| SG_vs_k141_14713\|\|full | Low-quality |
| SG_vs_k141_26351\|\|full | Not-determined |
| SG_vs_k141_54414\|\|full | Low-quality |
| SG_vs_k141_24167\|\|full | Low-quality |
| SG_vs_k141_54418\|\|full | Low-quality |
| SG_vs_k141_7433\|\|full | Not-determined |
| SG_vs_k141_42979\|\|full | Low-quality |
| SG_vs_k141_17095\|\|full | Low-quality |
| SG_vs_k141_5100\|\|full | Low-quality |
| SG_vs_k141_17099\|\|full | Low-quality |
| SG_vs_k141_43000\|\|full | Not-determined |
| SG_vs_k141_26368\|\|full | Low-quality |
| SG_vs_k141_17103\|\|full | Low-quality |
| SG_vs_k141_43001\|\|full | Not-determined |
| SG_vs_k141_26370\|\|full | Not-determined |
| SG_vs_k141_35808\|\|full | Low-quality |
| SG_vs_k141_26381\|\|full | Not-determined |
| SG_vs_k141_5121\|\|full | Low-quality |
| SG_vs_k141_49887\|\|full | Not-determined |
| SG_vs_k141_21458\|\|full | Not-determined |
| SG_vs_k141_35825\|\|full | Low-quality |
| SG_vs_k141_45369\|\|full | Low-quality |
| SG_vs_k141_49895\|\|full | Not-determined |
| SG_vs_k141_47547\|\|full | Low-quality |
| SG_vs_k141_26403\|\|full | Low-quality |
| SG_vs_k141_52715\|\|full | Low-quality |
| SG_vs_k141_49906\|\|full | Low-quality |
| SG_vs_k141_49912\|\|full | Low-quality |
| SG_vs_k141_24187\|\|full | Low-quality |
| SG_vs_k141_21497\|\|full | Low-quality |
| SG_vs_k141_26406\|\|full | Low-quality |
| SG_vs_k141_24207\|\|full | Low-quality |
| SG_vs_k141_31025\|\|full | Not-determined |
| SG_vs_k141_24219\|\|full | Not-determined |
| S8_extracted_k141_81691 | Medium-quality |
| S2_extracted_k141_20910 | Medium-quality |
| SG_vs_k141_5142\|\|full | Low-quality |
| SG_vs_k141_35840\|\|full | Low-quality |
| SG_vs_k141_12576\|\|full | Low-quality |
| SG_vs_k141_43033\|\|full | Low-quality |
| SG_vs_k141_763\|\|full | Low-quality |
| SG_vs_k141_19112\|\|full | Not-determined |
| SG_vs_k141_47596\|\|full | Low-quality |
| SG_vs_k141_49924\|\|full | Not-determined |
| SG_vs_k141_43038\|\|full | Low-quality |
| SG_vs_k141_54481\|\|full | Low-quality |
| SG_vs_k141_19126\|\|full | Not-determined |
| SG_vs_k141_40643\|\|full | Low-quality |
| SG_vs_k141_24248\|\|full | Not-determined |
| SG_vs_k141_5145\|\|full | Low-quality |
| SG_vs_k141_35866\|\|full | Low-quality |
| SG_vs_k141_31071\|\|full | Not-determined |
| SG_vs_k141_7504\|\|full | Not-determined |
| SG_vs_k141_35869\|\|full | Not-determined |
| SG_vs_k141_5149\|\|full | Low-quality |
| SG_vs_k141_43055\|\|full | Low-quality |
| SG_vs_k141_24269\|\|full | Low-quality |
| SG_vs_k141_31078\|\|full | Low-quality |
| SG_vs_k141_35873\|\|full | Low-quality |
| SG_vs_k141_17124\|\|full | Low-quality |
| SG_vs_k141_24279\|\|full | Low-quality |
| SG_vs_k141_17125\|\|full | Not-determined |
| SG_vs_k141_43058\|\|full | Low-quality |
| SG_vs_k141_804\|\|full | Not-determined |
| SG_vs_k141_49979\|\|full | Low-quality |
| SG_vs_k141_808\|\|full | Low-quality |
| SG_vs_k141_9961\|\|full | Low-quality |
| SG_vs_k141_40682\|\|full | Low-quality |
| SG_vs_k141_49994\|\|full | Low-quality |
| SG_vs_k141_40685\|\|full | Low-quality |
| SG_vs_k141_52783\|\|full | Low-quality |
| SG_vs_k141_31079\|\|full | Not-determined |
| SG_vs_k141_49998\|\|full | Low-quality |
| SG_vs_k141_7555\|\|full | Low-quality |
| SG_vs_k141_17161\|\|full | Not-determined |
| SG_vs_k141_21574\|\|full | Not-determined |
| SG_vs_k141_40689\|\|full | Not-determined |
| SG_vs_k141_28818\|\|full | Not-determined |
| SG_vs_k141_45441\|\|full | Low-quality |
| SG_vs_k141_824\|\|full | Low-quality |
| SG_vs_k141_45454\|\|full | Low-quality |
| SG_vs_k141_40701\|\|full | Not-determined |
| SG_vs_k141_35907\|\|full | Not-determined |
| SG_vs_k141_5186\|\|full | Low-quality |
| SG_vs_k141_24325\|\|full | Low-quality |
| SG_vs_k141_45467\|\|full | Low-quality |
| SG_vs_k141_24334\|\|full | Low-quality |
| SG_vs_k141_45473\|\|full | Not-determined |
| SG_vs_k141_47616\|\|full | Low-quality |
| SG_vs_k141_17207\|\|full | Low-quality |
| SG_vs_k141_7563\|\|full | Low-quality |
| SG_vs_k141_24342\|\|full | Low-quality |
| SG_vs_k141_17214\|\|full | Low-quality |
| SG_vs_k141_45479\|\|full | Not-determined |
| SG_vs_k141_17220\|\|full | Not-determined |
| SG_vs_k141_10008\|\|full | Low-quality |
| SG_vs_k141_45491\|\|full | Low-quality |
| SG_vs_k141_45493\|\|full | Low-quality |
| SG_vs_k141_10009\|\|full | Not-determined |
| SG_vs_k141_5249\|\|full | Low-quality |
| SG_vs_k141_31107\|\|full | Not-determined |
| SG_vs_k141_28871\|\|full | Low-quality |
| SG_vs_k141_24377\|\|full | Low-quality |
| SG_vs_k141_862\|\|full | Low-quality |
| SG_vs_k141_7587\|\|full | Not-determined |
| SG_vs_k141_40737\|\|full | Low-quality |
| SG_vs_k141_3133\|\|full | Low-quality |
| SG_vs_k141_47634\|\|full | Low-quality |
| SG_vs_k141_7596\|\|full | Low-quality |
| SG_vs_k141_45494\|\|full | Not-determined |
| SG_vs_k141_28888\|\|full | Low-quality |
| SG_vs_k141_7598\|\|full | Low-quality |
| SG_vs_k141_875\|\|full | Low-quality |
| SG_vs_k141_21636\|\|full | Not-determined |
| SG_vs_k141_7602\|\|full | Not-determined |
| SG_vs_k141_47639\|\|full | Not-determined |
| SG_vs_k141_14811\|\|full | Low-quality |
| SG_vs_k141_7608\|\|full | Low-quality |
| S7_vs_83624\|\|full | Medium-quality |
| SG_vs_k141_31135\|\|full | Not-determined |
| SG_vs_k141_31136\|\|full | Not-determined |
| SG_vs_k141_14816\|\|full | Not-determined |
| SG_vs_k141_43074\|\|full | Low-quality |
| SG_vs_k141_35972\|\|full | Low-quality |
| SG_vs_k141_43077\|\|full | Not-determined |
| SG_vs_k141_889\|\|full | Low-quality |
| SG_vs_k141_14830\|\|full | Low-quality |
| SG_vs_k141_35975\|\|full | Low-quality |
| SG_vs_k141_43082\|\|full | Not-determined |
| SG_vs_k141_47659\|\|full | Low-quality |
| SG_vs_k141_47660\|\|full | Low-quality |
| SG_vs_k141_7642\|\|full | Not-determined |
| SG_vs_k141_47665\|\|full | Not-determined |
| SG_vs_k141_21665\|\|full | Not-determined |
| SG_vs_k141_17266\|\|full | Low-quality |
| SG_vs_k141_33736\|\|full | Low-quality |
| SG_vs_k141_35995\|\|full | Not-determined |
| SG_vs_k141_47675\|\|full | Low-quality |
| SG_vs_k141_31200\|\|full | Low-quality |
| SG_vs_k141_40758\|\|full | Low-quality |
| SG_vs_k141_45531\|\|full | Not-determined |
| SG_vs_k141_5317\|\|full | Not-determined |
| SG_vs_k141_12605\|\|full | Not-determined |
| SG_vs_k141_45552\|\|full | Low-quality |
| SG_vs_k141_7675\|\|full | Low-quality |
| SG_vs_k141_40761\|\|full | Low-quality |
| SG_vs_k141_50118\|\|full | Low-quality |
| SG_vs_k141_40762\|\|full | Not-determined |
| SG_vs_k141_45561\|\|full | Low-quality |
| SG_vs_k141_28901\|\|full | Low-quality |
| SG_vs_k141_47700\|\|full | Not-determined |
| SG_vs_k141_935\|\|full | Low-quality |
| SG_vs_k141_50126\|\|full | Low-quality |
| SG_vs_k141_33779\|\|full | Not-determined |
| SG_vs_k141_12625\|\|full | Low-quality |
| SG_vs_k141_7694\|\|full | Not-determined |
| SG_vs_k141_17302\|\|full | Not-determined |
| SG_vs_k141_19155\|\|full | Low-quality |
| SG_vs_k141_17305\|\|full | Not-determined |
| SG_vs_k141_3203\|\|full | Low-quality |
| SG_vs_k141_45573\|\|full | Low-quality |
| SG_vs_k141_19165\|\|full | Low-quality |
| SG_vs_k141_31212\|\|full | Low-quality |
| SG_vs_k141_50145\|\|full | Low-quality |

| | |
|---|---|
| S6_vs_k141_33801\|\|full | Low-quality |
| S6_vs_k141_50153\|\|full | Low-quality |
| S6_vs_k141_33807\|\|full | Not-determined |
| S6_vs_k141_45613\|\|full | Low-quality |
| S6_vs_k141_50154\|\|full | Not-determined |
| S6_vs_k141_971\|\|full | Low-quality |
| S6_vs_k141_33811\|\|full | Low-quality |
| S6_vs_k141_24449\|\|full | Low-quality |
| S6_vs_k141_5336\|\|full | Low-quality |
| S6_vs_k141_33822\|\|full | Low-quality |
| S6_vs_k141_19197\|\|full | Not-determined |
| S6_vs_k141_28939\|\|full | Low-quality |
| S6_vs_k141_7713\|\|full | Low-quality |
| S6_vs_k141_33835\|\|full | Low-quality |
| S6_vs_k141_5356\|\|full | Low-quality |
| S6_vs_k141_28960\|\|full | Low-quality |
| S6_vs_k141_38472\|\|full | Low-quality |
| S6_vs_k141_10047\|\|full | Low-quality |
| S6_vs_k141_31245\|\|full | Low-quality |
| S6_vs_k141_50169\|\|full | Low-quality |
| S6_vs_k141_17350\|\|full | Not-determined |
| S6_vs_k141_33866\|\|full | Low-quality |
| S6_vs_k141_36017\|\|full | Low-quality |
| S6_vs_k141_50176\|\|full | Low-quality |
| S6_vs_k141_50180\|\|full | Low-quality |
| S6_vs_k141_40805\|\|full | Not-determined |
| S6_vs_k141_24509\|\|full | Low-quality |
| S6_vs_k141_26466\|\|full | Low-quality |
| S6_vs_k141_24513\|\|full | Low-quality |
| S6_vs_k141_3293\|\|full | Not-determined |
| S6_vs_k141_38492\|\|full | Low-quality |
| S6_vs_k141_17360\|\|full | Not-determined |
| S6_vs_k141_31254\|\|full | Low-quality |
| S6_vs_k141_17361\|\|full | Low-quality |
| S6_vs_k141_43097\|\|full | Not-determined |
| S6_vs_k141_26474\|\|full | Low-quality |
| S6_vs_k141_3305\|\|full | Low-quality |
| S6_vs_k141_26475\|\|full | Low-quality |
| S6_vs_k141_28982\|\|full | Low-quality |
| S6_vs_k141_12674\|\|full | Not-determined |
| S6_vs_k141_40840\|\|full | Low-quality |
| S6_vs_k141_24525\|\|full | Low-quality |
| S6_vs_k141_24530\|\|full | Low-quality |
| S6_vs_k141_14877\|\|full | Not-determined |
| S6_vs_k141_10133\|\|full | Not-determined |
| S6_vs_k141_12707\|\|full | Low-quality |
| S6_vs_k141_12711\|\|full | Not-determined |
| S6_vs_k141_10143\|\|full | Low-quality |
| S6_vs_k141_26507\|\|full | Low-quality |
| S6_vs_k141_7739\|\|full | Low-quality |
| S6_vs_k141_17408\|\|full | Low-quality |
| S6_vs_k141_26518\|\|full | Low-quality |
| S17_extracted_k141_148303 | Medium-quality |
| S6_vs_k141_45672\|\|full | Low-quality |
| S6_vs_k141_40856\|\|full | Low-quality |
| S6_vs_k141_24557\|\|full | Low-quality |
| S6_vs_k141_40858\|\|full | Low-quality |
| S6_vs_k141_17441\|\|full | Low-quality |
| S6_vs_k141_7765\|\|full | Low-quality |
| S6_vs_k141_52819\|\|full | Not-determined |
| S6_vs_k141_7766\|\|full | Low-quality |
| S6_vs_k141_45692\|\|full | Not-determined |
| S6_vs_k141_24582\|\|full | Low-quality |
| S6_vs_k141_52849\|\|full | Low-quality |
| S6_vs_k141_24590\|\|full | Low-quality |
| S6_vs_k141_19225\|\|full | Low-quality |
| S6_vs_k141_10208\|\|full | Not-determined |
| S6_vs_k141_45722\|\|full | Low-quality |
| S6_vs_k141_45725\|\|full | Not-determined |
| S6_vs_k141_52862\|\|full | Low-quality |
| S6_vs_k141_19231\|\|full | Low-quality |
| S6_vs_k141_52864\|\|full | Low-quality |
| S6_vs_k141_24600\|\|full | Not-determined |
| S6_vs_k141_19233\|\|full | Not-determined |
| S6_vs_k141_24602\|\|full | Not-determined |
| S6_vs_k141_29057\|\|full | Not-determined |
| S6_vs_k141_52872\|\|full | Not-determined |
| S6_vs_k141_7816\|\|full | Low-quality |
| S6_vs_k141_24609\|\|full | Not-determined |
| S6_vs_k141_24614\|\|full | Low-quality |
| S6_vs_k141_45758\|\|full | Low-quality |
| S6_vs_k141_17497\|\|full | Low-quality |
| S6_vs_k141_14948\|\|full | Low-quality |
| S6_vs_k141_24632\|\|full | Low-quality |
| S6_vs_k141_10240\|\|full | Low-quality |
| S6_vs_k141_31298\|\|full | Low-quality |
| S6_vs_k141_36078\|\|full | Not-determined |
| S6_vs_k141_31303\|\|full | Low-quality |
| S6_vs_k141_31308\|\|full | Low-quality |
| S6_vs_k141_3378\|\|full | Low-quality |
| S6_vs_k141_19270\|\|full | Low-quality |
| S6_vs_k141_31317\|\|full | Not-determined |
| S6_vs_k141_5407\|\|full | Low-quality |
| S6_vs_k141_17533\|\|full | Low-quality |
| S6_vs_k141_38573\|\|full | Low-quality |
| S6_vs_k141_5420\|\|full | Not-determined |
| S6_vs_k141_33971\|\|full | Low-quality |
| S6_vs_k141_31341\|\|full | Low-quality |
| S6_vs_k141_54578\|\|full | Low-quality |
| S6_vs_k141_45806\|\|full | Not-determined |
| S6_vs_k141_7892\|\|full | Low-quality |
| S6_vs_k141_45809\|\|full | Not-determined |
| S6_vs_k141_980\|\|full | Low-quality |
| S6_vs_k141_3395\|\|full | Low-quality |
| S6_vs_k141_24689\|\|full | Low-quality |
| S6_vs_k141_3396\|\|full | Low-quality |
| S6_vs_k141_3405\|\|full | Low-quality |
| S6_vs_k141_5449\|\|full | Low-quality |
| S6_vs_k141_33995\|\|full | Low-quality |
| S6_vs_k141_1000\|\|full | Not-determined |
| S6_vs_k141_36136\|\|full | Low-quality |
| S6_vs_k141_31368\|\|full | Low-quality |
| S6_vs_k141_29086\|\|full | Low-quality |
| S6_vs_k141_7918\|\|full | Low-quality |
| S6_vs_k141_43211\|\|full | Low-quality |
| S6_vs_k141_1017\|\|full | Low-quality |
| S6_vs_k141_34018\|\|full | Low-quality |
| S6_vs_k141_36145\|\|full | Low-quality |
| S6_vs_k141_31373\|\|full | Not-determined |
| S6_vs_k141_29098\|\|full | Low-quality |
| S6_vs_k141_34032\|\|full | Low-quality |
| S6_vs_k141_38600\|\|full | Not-determined |
| S6_vs_k141_47824\|\|full | Low-quality |
| S6_vs_k141_14991\|\|full | Low-quality |
| S6_vs_k141_7945\|\|full | Not-determined |
| S17_vs_k141_148303\|\|full | Medium-quality |
| S6_vs_k141_15002\|\|full | Not-determined |
| S6_vs_k141_34050\|\|full | Not-determined |
| S6_vs_k141_45859\|\|full | Not-determined |
| S6_vs_k141_15030\|\|full | Low-quality |
| S6_vs_k141_47842\|\|full | Not-determined |
| S6_vs_k141_7986\|\|full | Not-determined |
| S6_vs_k141_36164\|\|full | Low-quality |
| S14_extracted_k141_114491 | Medium-quality |
| S6_vs_k141_52909\|\|full | Low-quality |
| S6_vs_k141_40882\|\|full | Not-determined |
| S6_vs_k141_36169\|\|full | Low-quality |
| S6_vs_k141_45905\|\|full | Not-determined |
| S6_vs_k141_17562\|\|full | Low-quality |
| S6_vs_k141_17572\|\|full | Not-determined |
| S6_vs_k141_15047\|\|full | Low-quality |
| S6_vs_k141_43247\|\|full | Not-determined |
| S6_vs_k141_45927\|\|full | Low-quality |
| S6_vs_k141_52928\|\|full | Low-quality |
| S6_vs_k141_1055\|\|full | Low-quality |
| S7_extracted_82801 | Medium-quality |
| S6_vs_k141_43261\|\|full | Not-determined |
| S6_vs_k141_34080\|\|full | Low-quality |
| S6_vs_k141_8009\|\|full | Low-quality |
| S6_vs_k141_38687\|\|full | Low-quality |
| S6_vs_k141_36177\|\|full | Low-quality |
| S6_vs_k141_5511\|\|full | Low-quality |
| S6_vs_k141_38694\|\|full | Low-quality |
| S6_vs_k141_12787\|\|full | Low-quality |
| S6_vs_k141_52949\|\|full | Not-determined |
| S6_vs_k141_10309\|\|full | Not-determined |
| S6_vs_k141_38699\|\|full | Low-quality |
| S6_vs_k141_52958\|\|full | Low-quality |
| S6_vs_k141_38702\|\|full | Low-quality |
| S6_vs_k141_54645\|\|full | Not-determined |
| S6_vs_k141_50192\|\|full | Low-quality |
| S6_vs_k141_54648\|\|full | Low-quality |
| S6_vs_k141_21738\|\|full | Low-quality |
| S6_vs_k141_21739\|\|full | Not-determined |
| S6_vs_k141_47885\|\|full | Low-quality |
| S6_vs_k141_34142\|\|full | Not-determined |
| S6_vs_k141_3469\|\|full | Low-quality |
| S6_vs_k141_12810\|\|full | Low-quality |
| S6_vs_k141_38734\|\|full | Not-determined |
| S6_vs_k141_24715\|\|full | Not-determined |
| S6_vs_k141_54670\|\|full | Not-determined |
| S6_vs_k141_24720\|\|full | Low-quality |
| S6_vs_k141_21768\|\|full | Not-determined |
| S6_vs_k141_19295\|\|full | Low-quality |
| S7_vs_82801\|\|full | Medium-quality |
| S6_vs_k141_19298\|\|full | Low-quality |
| S6_vs_k141_45950\|\|full | Low-quality |
| S6_vs_k141_21784\|\|full | Low-quality |
| S6_vs_k141_24728\|\|full | Low-quality |
| S6_vs_k141_12842\|\|full | Not-determined |
| S2_extracted_k141_661 | Medium-quality |
| S6_vs_k141_36239\|\|full | Low-quality |
| S6_vs_k141_50243\|\|full | Low-quality |
| S6_vs_k141_15113\|\|full | Low-quality |
| S6_vs_k141_24755\|\|full | Low-quality |
| S6_vs_k141_50247\|\|full | Not-determined |
| S6_vs_k141_5557\|\|full | Not-determined |
| S6_vs_k141_53000\|\|full | Low-quality |
| S6_vs_k141_53002\|\|full | Not-determined |
| S6_vs_k141_5560\|\|full | Not-determined |
| S6_vs_k141_3506\|\|full | Not-determined |
| S6_vs_k141_54688\|\|full | Not-determined |
| S6_vs_k141_53006\|\|full | Not-determined |
| S6_vs_k141_24772\|\|full | Low-quality |
| S6_vs_k141_29200\|\|full | Low-quality |
| S6_vs_k141_15119\|\|full | Not-determined |
| S6_vs_k141_29205\|\|full | Low-quality |
| S6_vs_k141_15121\|\|full | Not-determined |
| S6_vs_k141_34166\|\|full | Not-determined |
| S6_vs_k141_12860\|\|full | Low-quality |
| S6_vs_k141_1082\|\|full | Low-quality |
| S6_vs_k141_50271\|\|full | Low-quality |
| S6_vs_k141_34170\|\|full | Low-quality |
| S6_vs_k141_3522\|\|full | Low-quality |
| S6_vs_k141_24811\|\|full | Low-quality |
| S6_vs_k141_5597\|\|full | Low-quality |
| S6_vs_k141_15158\|\|full | Low-quality |
| S6_vs_k141_34193\|\|full | Low-quality |
| S6_vs_k141_24820\|\|full | Low-quality |
| S6_vs_k141_24821\|\|full | Not-determined |
| S6_vs_k141_34204\|\|full | Not-determined |
| S6_vs_k141_3544\|\|full | Low-quality |
| S6_vs_k141_17609\|\|full | Not-determined |
| S11_extracted_k141_92391 | Medium-quality |
| S6_vs_k141_17611\|\|full | Low-quality |
| S6_vs_k141_3545\|\|full | Low-quality |
| S6_vs_k141_3546\|\|full | Low-quality |
| S6_vs_k141_38756\|\|full | Low-quality |
| S6_vs_k141_29279\|\|full | Low-quality |
| S6_vs_k141_43376\|\|full | Not-determined |
| S6_vs_k141_3554\|\|full | Low-quality |
| S6_vs_k141_21868\|\|full | Low-quality |
| S6_vs_k141_53051\|\|full | Not-determined |
| S6_vs_k141_34227\|\|full | Not-determined |
| S6_vs_k141_50300\|\|full | Not-determined |
| S6_vs_k141_34229\|\|full | Low-quality |
| S6_vs_k141_31494\|\|full | Low-quality |
| S6_vs_k141_1140\|\|full | Low-quality |
| S6_vs_k141_1141\|\|full | Low-quality |
| S6_vs_k141_5624\|\|full | Low-quality |
| S6_vs_k141_53067\|\|full | Not-determined |
| S6_vs_k141_3589\|\|full | Low-quality |
| S6_vs_k141_10348\|\|full | Low-quality |
| S6_vs_k141_40982\|\|full | Low-quality |
| S6_vs_k141_40987\|\|full | Not-determined |
| S6_vs_k141_5632\|\|full | Not-determined |
| S6_vs_k141_40992\|\|full | Not-determined |
| S6_vs_k141_36309\|\|full | Low-quality |
| S6_vs_k141_38776\|\|full | Low-quality |
| S6_vs_k141_3592\|\|full | Not-determined |
| S6_vs_k141_34271\|\|full | Low-quality |
| S6_vs_k141_31515\|\|full | Not-determined |
| S6_vs_k141_3602\|\|full | Not-determined |
| S6_vs_k141_43398\|\|full | Low-quality |
| S6_vs_k141_31519\|\|full | Low-quality |
| S6_vs_k141_10362\|\|full | Not-determined |
| S6_vs_k141_5658\|\|full | Not-determined |
| S6_vs_k141_3609\|\|full | Low-quality |
| S6_vs_k141_29293\|\|full | Low-quality |
| S6_vs_k141_29299\|\|full | Low-quality |
| S6_vs_k141_38813\|\|full | Low-quality |
| S6_vs_k141_54831\|\|full | Not-determined |
| S6_vs_k141_5678\|\|full | Not-determined |
| S6_vs_k141_38816\|\|full | Low-quality |
| S6_vs_k141_38818\|\|full | Low-quality |
| S6_vs_k141_43410\|\|full | Low-quality |
| S6_vs_k141_31545\|\|full | Low-quality |
| S6_vs_k141_54842\|\|full | Low-quality |
| S6_vs_k141_10404\|\|full | Low-quality |
| S6_vs_k141_19391\|\|full | Low-quality |
| S6_vs_k141_26637\|\|full | Not-determined |
| S6_vs_k141_50339\|\|full | Low-quality |
| S6_vs_k141_5687\|\|full | Low-quality |
| S6_vs_k141_41024\|\|full | Not-determined |
| S6_vs_k141_41028\|\|full | Not-determined |
| S6_vs_k141_12922\|\|full | Low-quality |
| S8_extracted_k141_136235 | Medium-quality |
| S6_vs_k141_31569\|\|full | Not-determined |
| S6_vs_k141_26646\|\|full | Low-quality |
| S6_vs_k141_15188\|\|full | Low-quality |
| S6_vs_k141_38838\|\|full | Low-quality |
| S6_vs_k141_29353\|\|full | Low-quality |
| S6_vs_k141_21940\|\|full | Not-determined |
| S6_vs_k141_5696\|\|full | Low-quality |
| S6_vs_k141_15196\|\|full | Low-quality |
| S6_vs_k141_15197\|\|full | Low-quality |
| S6_vs_k141_38859\|\|full | Low-quality |
| S6_vs_k141_8067\|\|full | Low-quality |
| S6_vs_k141_3652\|\|full | Low-quality |
| S6_vs_k141_34279\|\|full | Low-quality |
| S6_vs_k141_10446\|\|full | Not-determined |
| S6_vs_k141_34285\|\|full | Low-quality |
| S6_vs_k141_34288\|\|full | Not-determined |
| S8_vs_k141_91033\|\|full | Medium-quality |
| S6_vs_k141_3670\|\|full | Low-quality |
| S6_vs_k141_8092\|\|full | Not-determined |
| S6_vs_k141_17691\|\|full | Low-quality |
| S6_vs_k141_53137\|\|full | Not-determined |
| S6_vs_k141_3687\|\|full | Low-quality |
| S6_vs_k141_34301\|\|full | Not-determined |
| S6_vs_k141_10460\|\|full | Not-determined |
| S6_vs_k141_53145\|\|full | Low-quality |
| S6_vs_k141_15208\|\|full | Low-quality |
| S3_vs_k141_87728\|\|0_partial | Medium-quality |
| S6_vs_k141_19406\|\|full | Low-quality |
| S6_vs_k141_34315\|\|full | Not-determined |
| S6_vs_k141_5703\|\|full | Low-quality |
| S6_vs_k141_19415\|\|full | Not-determined |
| S6_vs_k141_12973\|\|full | Not-determined |
| S6_vs_k141_5720\|\|full | Low-quality |
| S6_vs_k141_10498\|\|full | Low-quality |
| S6_vs_k141_31584\|\|full | Not-determined |
| S6_vs_k141_8104\|\|full | Low-quality |
| S6_vs_k141_12986\|\|full | Low-quality |
| S6_vs_k141_15254\|\|full | Not-determined |
| S6_vs_k141_29391\|\|full | Not-determined |
| S6_vs_k141_15257\|\|full | Low-quality |
| S6_vs_k141_17723\|\|full | Low-quality |
| S6_vs_k141_19425\|\|full | Low-quality |
| S6_vs_k141_15263\|\|full | Not-determined |
| S6_vs_k141_5743\|\|full | Not-determined |
| S6_vs_k141_29403\|\|full | Not-determined |
| S6_vs_k141_31597\|\|full | Low-quality |
| S6_vs_k141_8119\|\|full | Not-determined |
| S6_vs_k141_53162\|\|full | Low-quality |
| S6_vs_k141_3760\|\|full | Low-quality |
| S6_vs_k141_31610\|\|full | Not-determined |
| S6_vs_k141_31624\|\|full | Low-quality |
| S6_vs_k141_29424\|\|full | Not-determined |
| S6_vs_k141_8128\|\|full | Low-quality |
| S6_vs_k141_5762\|\|full | Not-determined |
| S6_vs_k141_54889\|\|full | Low-quality |
| S6_vs_k141_36343\|\|full | Not-determined |
| S6_vs_k141_26689\|\|full | Not-determined |
| S6_vs_k141_38918\|\|full | Low-quality |
| S6_vs_k141_1156\|\|full | Low-quality |
| S6_vs_k141_31634\|\|full | Low-quality |
| S6_vs_k141_10539\|\|full | Low-quality |
| S6_vs_k141_5785\|\|full | Not-determined |
| S6_vs_k141_38924\|\|full | Not-determined |
| S6_vs_k141_5790\|\|full | Low-quality |
| S6_vs_k141_36360\|\|full | Low-quality |
| S6_vs_k141_34424\|\|full | Not-determined |
| S6_vs_k141_29444\|\|full | Low-quality |
| S6_vs_k141_8166\|\|full | Not-determined |
| S6_vs_k141_10561\|\|full | Low-quality |
| S6_vs_k141_21981\|\|full | Not-determined |
| S6_vs_k141_48065\|\|full | Low-quality |
| S6_vs_k141_8174\|\|full | Low-quality |
| S6_vs_k141_48085\|\|full | Low-quality |
| S6_vs_k141_48086\|\|full | Low-quality |
| S6_vs_k141_43550\|\|full | Low-quality |
| S6_vs_k141_22003\|\|full | Low-quality |
| S6_vs_k141_5812\|\|full | Not-determined |
| S6_vs_k141_22005\|\|full | Not-determined |
| S6_vs_k141_5814\|\|full | Low-quality |
| S6_vs_k141_1178\|\|full | Low-quality |
| S6_vs_k141_43560\|\|full | Not-determined |
| S6_vs_k141_1179\|\|full | Low-quality |
| S6_vs_k141_17805\|\|full | Low-quality |
| S6_vs_k141_48116\|\|full | Low-quality |
| S6_vs_k141_3834\|\|full | Not-determined |
| S6_vs_k141_46062\|\|full | Low-quality |
| S6_vs_k141_8216\|\|full | Low-quality |
| S6_vs_k141_29493\|\|full | Not-determined |
| S6_vs_k141_10591\|\|full | Low-quality |
| S6_vs_k141_26725\|\|full | Not-determined |
| S6_vs_k141_10598\|\|full | Low-quality |
| S6_vs_k141_10604\|\|full | Not-determined |
| S6_vs_k141_19473\|\|full | Low-quality |
| S6_vs_k141_19475\|\|full | Low-quality |
| S6_vs_k141_17847\|\|full | Low-quality |
| S6_vs_k141_41050\|\|full | Low-quality |
| S6_vs_k141_41052\|\|full | Low-quality |
| S17_extracted_k141_174203 | Medium-quality |
| S6_vs_k141_13046\|\|full | Low-quality |
| S6_vs_k141_36418\|\|full | Low-quality |
| S6_vs_k141_19489\|\|full | Low-quality |
| S6_vs_k141_50459\|\|full | Low-quality |
| S7_extracted_66177 | Medium-quality |
| S6_vs_k141_29554\|\|full | Low-quality |
| S6_vs_k141_53228\|\|full | Not-determined |
| S6_vs_k141_19505\|\|full | Low-quality |
| S6_vs_k141_31756\|\|full | Low-quality |
| S6_vs_k141_10630\|\|full | Low-quality |
| S6_vs_k141_24919\|\|full | Low-quality |
| S6_vs_k141_50471\|\|full | Not-determined |
| S6_vs_k141_24921\|\|full | Low-quality |
| S6_vs_k141_10631\|\|full | Low-quality |
| S6_vs_k141_13072\|\|full | Low-quality |
| S6_vs_k141_22069\|\|full | Low-quality |
| S6_vs_k141_36443\|\|full | Not-determined |
| S6_vs_k141_17882\|\|full | Low-quality |
| S2_vs_k141_20910\|\|full | Medium-quality |
| S6_vs_k141_5868\|\|full | Not-determined |
| S6_vs_k141_5869\|\|full | Low-quality |
| S6_vs_k141_38997\|\|full | Low-quality |
| S6_vs_k141_1232\|\|full | Low-quality |
| S6_vs_k141_26769\|\|full | Low-quality |
| S6_vs_k141_24951\|\|full | Low-quality |
| S6_vs_k141_13116\|\|full | Low-quality |
| S6_vs_k141_22091\|\|full | Low-quality |
| S6_vs_k141_50480\|\|full | Not-determined |
| S6_vs_k141_36466\|\|full | Low-quality |
| S6_vs_k141_41115\|\|full | Not-determined |
| S6_vs_k141_39021\|\|full | Low-quality |
| S6_vs_k141_26792\|\|full | Low-quality |
| S6_vs_k141_15407\|\|full | Not-determined |
| S6_vs_k141_24983\|\|full | Low-quality |
| S6_vs_k141_17921\|\|full | Low-quality |
| S6_vs_k141_15418\|\|full | Not-determined |
| S6_vs_k141_22108\|\|full | Not-determined |
| S6_vs_k141_3942\|\|full | Not-determined |
| S6_vs_k141_34474\|\|full | Low-quality |
| S6_vs_k141_26812\|\|full | Not-determined |
| S6_vs_k141_41135\|\|full | Low-quality |
| S6_vs_k141_10674\|\|full | Low-quality |
| S6_vs_k141_17938\|\|full | Not-determined |
| S6_vs_k141_29628\|\|full | Low-quality |
| S6_vs_k141_34497\|\|full | Low-quality |
| S6_vs_k141_1245\|\|full | Low-quality |
| S6_vs_k141_24999\|\|full | Low-quality |
| S6_vs_k141_15452\|\|full | Low-quality |
| S6_vs_k141_1246\|\|full | Low-quality |
| S6_vs_k141_50509\|\|full | Not-determined |
| S6_vs_k141_1247\|\|full | Low-quality |
| S6_vs_k141_17960\|\|full | Low-quality |
| S6_vs_k141_36493\|\|full | Low-quality |
| S6_vs_k141_5910\|\|full | Low-quality |
| S6_vs_k141_48163\|\|full | Low-quality |
| S6_vs_k141_41193\|\|full | Not-determined |
| S6_vs_k141_17977\|\|full | Low-quality |
| S12_extracted_k141_42408 | Medium-quality |
| S6_vs_k141_10715\|\|full | Low-quality |
| S11_vs_k141_26593\|\|full | Medium-quality |
| S7_vs_20129\|\|full | Medium-quality |
| S6_vs_k141_29689\|\|full | Low-quality |
| S6_vs_k141_10726\|\|full | Low-quality |
| S6_vs_k141_39030\|\|full | Low-quality |
| S6_vs_k141_17983\|\|full | Low-quality |
| S6_vs_k141_17988\|\|full | Low-quality |
| S6_vs_k141_22157\|\|full | Low-quality |
| S6_vs_k141_36525\|\|full | Low-quality |
| S6_vs_k141_34519\|\|full | Not-determined |
| S6_vs_k141_17995\|\|full | Not-determined |
| S6_vs_k141_25059\|\|full | Low-quality |
| S6_vs_k141_26835\|\|full | Low-quality |
| S6_vs_k141_34534\|\|full | Not-determined |
| S6_vs_k141_31824\|\|full | Low-quality |
| S6_vs_k141_13244\|\|full | Low-quality |
| S6_vs_k141_29695\|\|full | Not-determined |
| S6_vs_k141_1265\|\|full | Low-quality |
| S6_vs_k141_13248\|\|full | Low-quality |
| S6_vs_k141_39048\|\|full | Low-quality |
| S6_vs_k141_10755\|\|full | Low-quality |
| S6_vs_k141_1272\|\|full | Low-quality |
| S6_vs_k141_31844\|\|full | Low-quality |
| S6_vs_k141_13250\|\|full | Low-quality |
| S6_vs_k141_41212\|\|full | Not-determined |
| S6_vs_k141_26858\|\|full | Not-determined |
| S6_vs_k141_18032\|\|full | Low-quality |
| S6_vs_k141_41217\|\|full | Low-quality |
| S6_vs_k141_48199\|\|full | Low-quality |
| S6_vs_k141_48205\|\|full | Low-quality |
| S8_vs_k141_3410\|\|full | High-quality |
| S6_vs_k141_13275\|\|full | Low-quality |
| S6_vs_k141_13281\|\|full | Low-quality |
| S6_vs_k141_48211\|\|full | Low-quality |
| S6_vs_k141_13287\|\|full | Low-quality |
| S6_vs_k141_46102\|\|full | Not-determined |
| S6_vs_k141_50580\|\|full | Not-determined |
| S6_vs_k141_18056\|\|full | Low-quality |
| S6_vs_k141_22193\|\|full | Low-quality |
| S6_vs_k141_26883\|\|full | Low-quality |
| S6_vs_k141_1302\|\|full | Not-determined |
| S6_vs_k141_18063\|\|full | Not-determined |
| S6_vs_k141_48232\|\|full | Low-quality |
| S6_vs_k141_34549\|\|full | Low-quality |
| S6_vs_k141_18067\|\|full | Not-determined |
| S6_vs_k141_53280\|\|full | Not-determined |
| S6_vs_k141_36550\|\|full | Low-quality |
| S6_vs_k141_53291\|\|full | Not-determined |
| S6_vs_k141_36552\|\|full | Not-determined |
| S6_vs_k141_26894\|\|full | Not-determined |
| S6_vs_k141_8293\|\|full | Low-quality |
| S6_vs_k141_25088\|\|full | Not-determined |
| S6_vs_k141_41234\|\|full | Not-determined |
| S6_vs_k141_26916\|\|full | Low-quality |
| S6_vs_k141_41255\|\|full | Low-quality |
| S6_vs_k141_53314\|\|full | Low-quality |
| S6_vs_k141_26923\|\|full | Low-quality |
| S6_vs_k141_25105\|\|full | Low-quality |
| S14_extracted_k141_99344 | Medium-quality |
| S6_vs_k141_22219\|\|full | Not-determined |
| S6_vs_k141_36573\|\|full | Low-quality |
| S6_vs_k141_41266\|\|full | Low-quality |
| S6_vs_k141_5924\|\|full | Not-determined |
| S6_vs_k141_41272\|\|full | Not-determined |
| S6_vs_k141_25119\|\|full | Low-quality |
| S6_vs_k141_5934\|\|full | Low-quality |
| S6_vs_k141_46149\|\|full | Low-quality |
| S6_vs_k141_46151\|\|full | Low-quality |
| S6_vs_k141_18087\|\|full | Not-determined |
| S6_vs_k141_5940\|\|full | Low-quality |
| S6_vs_k141_46156\|\|full | Low-quality |
| S6_vs_k141_53340\|\|full | Not-determined |
| S6_vs_k141_46160\|\|full | Low-quality |
| S6_vs_k141_10824\|\|full | Not-determined |
| S6_vs_k141_10826\|\|full | Not-determined |
| S6_vs_k141_31880\|\|full | Low-quality |
| S6_vs_k141_26942\|\|full | Not-determined |
| S6_vs_k141_34661\|\|full | Low-quality |
| S6_vs_k141_31888\|\|full | Low-quality |
| S6_vs_k141_54986\|\|full | Not-determined |
| S6_vs_k141_43605\|\|full | Low-quality |
| S6_vs_k141_25126\|\|full | Low-quality |
| S6_vs_k141_31896\|\|full | Not-determined |
| S6_vs_k141_22247\|\|full | Low-quality |
| S6_vs_k141_41333\|\|full | Not-determined |
| S6_vs_k141_41341\|\|full | Low-quality |
| S6_vs_k141_31915\|\|full | Low-quality |
| S6_vs_k141_25148\|\|full | Low-quality |
| S6_vs_k141_31916\|\|full | Not-determined |
| S6_vs_k141_15514\|\|full | Not-determined |
| S6_vs_k141_5996\|\|full | Low-quality |
| S6_vs_k141_18102\|\|full | Not-determined |
| S6_vs_k141_46179\|\|full | Low-quality |
| S6_vs_k141_13350\|\|full | Low-quality |
| S6_vs_k141_43652\|\|full | Low-quality |
| S6_vs_k141_53387\|\|full | Low-quality |
| S6_vs_k141_55044\|\|full | Not-determined |
| S6_vs_k141_18125\|\|full | Low-quality |
| S6_vs_k141_13362\|\|full | Not-determined |
| S6_vs_k141_48278\|\|full | Low-quality |
| S6_vs_k141_19568\|\|full | Low-quality |
| S6_vs_k141_34700\|\|full | Low-quality |
| S6_vs_k141_31928\|\|full | Not-determined |
| S6_vs_k141_31935\|\|full | Low-quality |
| S6_vs_k141_25201\|\|full | Low-quality |
| S6_vs_k141_41376\|\|full | Low-quality |
| S6_vs_k141_18142\|\|full | Low-quality |
| S6_vs_k141_48279\|\|full | Not-determined |
| S6_vs_k141_15532\|\|full | Not-determined |
| S6_vs_k141_8401\|\|full | Not-determined |
| S6_vs_k141_41384\|\|full | Low-quality |
| S6_vs_k141_8404\|\|full | Low-quality |
| S6_vs_k141_13378\|\|full | Low-quality |
| S6_vs_k141_4092\|\|full | Not-determined |
| S6_vs_k141_8420\|\|full | Low-quality |
| S6_vs_k141_41397\|\|full | Not-determined |
| S6_vs_k141_50696\|\|full | Not-determined |
| S6_vs_k141_13385\|\|full | Low-quality |
| S6_vs_k141_8430\|\|full | Low-quality |
| S6_vs_k141_41400\|\|full | Not-determined |
| S6_vs_k141_41420\|\|full | Not-determined |
| S6_vs_k141_48295\|\|full | Low-quality |
| S6_vs_k141_27002\|\|full | Low-quality |
| S6_vs_k141_13404\|\|full | Not-determined |
| S6_vs_k141_55067\|\|full | Not-determined |
| S6_vs_k141_15567\|\|full | Not-determined |
| S6_vs_k141_18191\|\|full | Not-determined |
| S6_vs_k141_34729\|\|full | Not-determined |
| S6_vs_k141_27009\|\|full | Low-quality |
| S6_vs_k141_31963\|\|full | Low-quality |
| S6_vs_k141_27010\|\|full | Not-determined |
| S6_vs_k141_25219\|\|full | Not-determined |
| S6_vs_k141_41431\|\|full | Low-quality |
| S6_vs_k141_22283\|\|full | Not-determined |
| S6_vs_k141_48299\|\|full | Low-quality |
| S6_vs_k141_34737\|\|full | Low-quality |
| S6_vs_k141_8476\|\|full | Not-determined |
| S6_vs_k141_27017\|\|full | Low-quality |
| S6_vs_k141_25233\|\|full | Low-quality |
| S6_vs_k141_8488\|\|full | Low-quality |
| S6_vs_k141_15586\|\|full | Low-quality |
| S6_vs_k141_50722\|\|full | Low-quality |
| S6_vs_k141_34765\|\|full | Not-determined |
| S6_vs_k141_8490\|\|full | Low-quality |
| S6_vs_k141_25242\|\|full | Not-determined |
| S6_vs_k141_25244\|\|full | Not-determined |
| S6_vs_k141_34768\|\|full | Low-quality |
| S6_vs_k141_13424\|\|full | Low-quality |
| S6_vs_k141_36677\|\|full | Low-quality |
| S6_vs_k141_13428\|\|full | Low-quality |
| S6_vs_k141_4135\|\|full | Not-determined |
| S6_vs_k141_36680\|\|full | Low-quality |
| S6_vs_k141_8517\|\|full | Low-quality |
| S6_vs_k141_15622\|\|full | Low-quality |
| S6_vs_k141_15635\|\|full | Low-quality |
| S6_vs_k141_19714\|\|full | Not-determined |
| S6_vs_k141_15649\|\|full | Not-determined |
| S6_vs_k141_34813\|\|full | Low-quality |
| S6_vs_k141_13460\|\|full | Low-quality |
| S6_vs_k141_8540\|\|full | Low-quality |
| S6_vs_k141_27063\|\|full | Low-quality |
| S6_vs_k141_8549\|\|full | Not-determined |
| S6_vs_k141_34831\|\|full | Not-determined |
| S6_vs_k141_25302\|\|full | Not-determined |
| S6_vs_k141_6090\|\|full | Low-quality |

| | |
|---|---|
| S6_vs_k141_29795\|\|full | Not-determined |
| S6_vs_k141_1395\|\|full | Not-determined |
| S6_vs_k141_29806\|\|full | Low-quality |
| S6_vs_k141_34848\|\|full | Low-quality |
| S6_vs_k141_22319\|\|full | Not-determined |
| S6_vs_k141_8587\|\|full | Not-determined |
| S6_vs_k141_29819\|\|full | Not-determined |
| S6_vs_k141_46310\|\|full | Low-quality |
| S6_vs_k141_41546\|\|full | Not-determined |
| S17_vs_k141_36452\|\|full | Medium-quality |
| S6_vs_k141_6106\|\|full | Not-determined |
| S6_vs_k141_53395\|\|full | Not-determined |
| S6_vs_k141_25317\|\|full | Low-quality |
| S6_vs_k141_15712\|\|full | Low-quality |
| S6_vs_k141_25322\|\|full | Low-quality |
| S6_vs_k141_8614\|\|full | Not-determined |
| S6_vs_k141_25324\|\|full | Low-quality |
| S6_vs_k141_29828\|\|full | Low-quality |
| S6_vs_k141_15729\|\|full | Not-determined |
| S6_vs_k141_41579\|\|full | Low-quality |
| S6_vs_k141_13505\|\|full | Not-determined |
| S6_vs_k141_25337\|\|full | Low-quality |
| S6_vs_k141_13513\|\|full | Low-quality |
| S6_vs_k141_46350\|\|full | Not-determined |
| S6_vs_k141_10932\|\|full | Low-quality |
| S6_vs_k141_13517\|\|full | Low-quality |
| S6_vs_k141_8625\|\|full | Not-determined |
| S6_vs_k141_8628\|\|full | Low-quality |
| S6_vs_k141_43694\|\|full | Low-quality |
| S6_vs_k141_25353\|\|full | Low-quality |
| S6_vs_k141_10944\|\|full | Not-determined |
| S6_vs_k141_4264\|\|full | Not-determined |
| S6_vs_k141_19801\|\|full | Not-determined |
| S6_vs_k141_46369\|\|full | Low-quality |
| S6_vs_k141_10958\|\|full | Low-quality |
| S6_vs_k141_13546\|\|full | Low-quality |
| S6_vs_k141_10962\|\|full | Low-quality |
| S6_vs_k141_13548\|\|full | Not-determined |
| S6_vs_k141_4278\|\|full | Low-quality |
| S6_vs_k141_13550\|\|full | Not-determined |
| S6_vs_k141_8634\|\|full | Low-quality |
| S6_vs_k141_29879\|\|full | Low-quality |
| S6_vs_k141_32040\|\|full | Low-quality |
| S6_vs_k141_4303\|\|full | Low-quality |
| S6_vs_k141_8650\|\|full | Not-determined |
| S6_vs_k141_4304\|\|full | Not-determined |
| S6_vs_k141_4310\|\|full | Not-determined |
| S6_vs_k141_55090\|\|full | Low-quality |
| S6_vs_k141_13583\|\|full | Low-quality |
| S6_vs_k141_43742\|\|full | Low-quality |
| S6_vs_k141_15776\|\|full | Low-quality |
| S6_vs_k141_8672\|\|full | Low-quality |
| S6_vs_k141_6167\|\|full | Low-quality |
| S6_vs_k141_6169\|\|full | Not-determined |
| S6_vs_k141_13587\|\|full | Low-quality |
| S6_vs_k141_4319\|\|full | Not-determined |
| S6_vs_k141_22403\|\|full | Not-determined |
| S6_vs_k141_18206\|\|full | Not-determined |
| S6_vs_k141_6179\|\|full | Low-quality |
| S6_vs_k141_4329\|\|full | Not-determined |
| S6_vs_k141_55096\|\|full | Not-determined |
| S6_vs_k141_8688\|\|full | Not-determined |
| S6_vs_k141_55105\|\|full | Not-determined |
| S6_vs_k141_13603\|\|full | Low-quality |
| S6_vs_k141_6194\|\|full | Low-quality |
| S6_vs_k141_27191\|\|full | Low-quality |
| S6_vs_k141_6195\|\|full | Low-quality |
| S6_vs_k141_8702\|\|full | Low-quality |
| S6_vs_k141_13618\|\|full | Low-quality |
| S6_vs_k141_41636\|\|full | Not-determined |
| S6_vs_k141_6203\|\|full | Low-quality |
| S6_vs_k141_8708\|\|full | Low-quality |
| S6_vs_k141_6206\|\|full | Low-quality |
| S6_vs_k141_6211\|\|full | Low-quality |
| S6_vs_k141_8726\|\|full | Low-quality |
| S6_vs_k141_13648\|\|full | Low-quality |
| S6_vs_k141_6224\|\|full | Low-quality |
| S6_vs_k141_18266\|\|full | Low-quality |
| S6_vs_k141_13650\|\|full | Low-quality |
| S6_vs_k141_29925\|\|full | Low-quality |
| S6_vs_k141_41669\|\|full | Low-quality |
| S6_vs_k141_41675\|\|full | Low-quality |
| S6_vs_k141_36704\|\|full | Not-determined |
| S6_vs_k141_29934\|\|full | Not-determined |
| S6_vs_k141_4345\|\|full | Low-quality |
| S6_vs_k141_55139\|\|full | Low-quality |
| S6_vs_k141_55146\|\|full | Low-quality |
| S6_vs_k141_18287\|\|full | Low-quality |
| S6_vs_k141_50769\|\|full | Low-quality |
| S6_vs_k141_8754\|\|full | Low-quality |
| S6_vs_k141_27229\|\|full | Low-quality |
| S6_vs_k141_29956\|\|full | Low-quality |
| S6_vs_k141_29972\|\|full | Low-quality |
| S6_vs_k141_19877\|\|full | Low-quality |
| S6_vs_k141_25395\|\|full | Low-quality |
| S13_vs_68102\|\|full | Medium-quality |
| S6_vs_k141_4356\|\|full | Low-quality |
| S6_vs_k141_18315\|\|full | Low-quality |
| S6_vs_k141_25409\|\|full | Low-quality |
| S7_extracted_112914 | Medium-quality |
| S6_vs_k141_41721\|\|full | Low-quality |
| S6_vs_k141_4362\|\|full | Not-determined |
| S6_vs_k141_4363\|\|full | Low-quality |
| S6_vs_k141_41726\|\|full | Low-quality |
| S6_vs_k141_22443\|\|full | Low-quality |
| S6_vs_k141_46420\|\|full | Low-quality |
| S6_vs_k141_22449\|\|full | Low-quality |
| S6_vs_k141_15838\|\|full | Low-quality |
| S6_vs_k141_55188\|\|full | Not-determined |
| S6_vs_k141_53507\|\|full | Low-quality |
| S6_vs_k141_55199\|\|full | Low-quality |
| S6_vs_k141_6271\|\|full | Low-quality |
| S6_vs_k141_41743\|\|full | Low-quality |
| S6_vs_k141_11016\|\|full | Not-determined |
| S6_vs_k141_18345\|\|full | Not-determined |
| S6_vs_k141_18350\|\|full | Not-determined |
| S6_vs_k141_41762\|\|full | Low-quality |
| S6_vs_k141_39067\|\|full | Low-quality |
| S6_vs_k141_53546\|\|full | Not-determined |
| S6_vs_k141_11031\|\|full | Not-determined |
| S6_vs_k141_55226\|\|full | Low-quality |
| S6_vs_k141_15879\|\|full | Low-quality |
| S6_vs_k141_11037\|\|full | Low-quality |
| S6_vs_k141_55233\|\|full | Not-determined |
| S6_vs_k141_30015\|\|full | Low-quality |
| S6_vs_k141_41780\|\|full | Low-quality |
| S6_vs_k141_53564\|\|full | Not-determined |
| S6_vs_k141_30018\|\|full | Low-quality |
| S6_vs_k141_32125\|\|full | Low-quality |
| S6_vs_k141_27256\|\|full | Low-quality |
| S6_vs_k141_30021\|\|full | Low-quality |
| S6_vs_k141_30022\|\|full | Not-determined |
| S6_vs_k141_11055\|\|full | Low-quality |
| S6_vs_k141_36764\|\|full | Not-determined |
| S6_vs_k141_4416\|\|full | Low-quality |
| S6_vs_k141_55255\|\|full | Low-quality |
| S6_vs_k141_30048\|\|full | Low-quality |
| S6_vs_k141_46480\|\|full | Low-quality |
| S6_vs_k141_32162\|\|full | Not-determined |
| S6_vs_k141_4426\|\|full | Low-quality |
| S6_vs_k141_41794\|\|full | Not-determined |
| S6_vs_k141_22524\|\|full | Low-quality |
| S6_vs_k141_32178\|\|full | Not-determined |
| S6_vs_k141_22525\|\|full | Low-quality |
| S16_extracted_119834 | Medium-quality |
| S6_vs_k141_53575\|\|full | Not-determined |
| S6_vs_k141_36789\|\|full | Low-quality |
| S6_vs_k141_43800\|\|full | Not-determined |
| S6_vs_k141_36798\|\|full | Low-quality |
| S6_vs_k141_6329\|\|full | Not-determined |
| S6_vs_k141_13705\|\|full | Not-determined |
| S6_vs_k141_34945\|\|full | Not-determined |
| S6_vs_k141_34951\|\|full | Not-determined |
| S6_vs_k141_30077\|\|full | Not-determined |
| S6_vs_k141_15912\|\|full | Low-quality |
| S6_vs_k141_13720\|\|full | Low-quality |
| S6_vs_k141_53593\|\|full | Not-determined |
| S6_vs_k141_41831\|\|full | Low-quality |
| S6_vs_k141_15916\|\|full | Low-quality |
| S6_vs_k141_6339\|\|full | Low-quality |
| S6_vs_k141_15919\|\|full | Not-determined |
| S6_vs_k141_30079\|\|full | Low-quality |
| S6_vs_k141_22572\|\|full | Low-quality |
| S6_vs_k141_27310\|\|full | Low-quality |
| S6_vs_k141_1511\|\|full | Low-quality |
| S6_vs_k141_41848\|\|full | Low-quality |
| S6_vs_k141_55338\|\|full | Low-quality |
| S6_vs_k141_46556\|\|full | Not-determined |
| S11_vs_k141_92391\|\|full | Medium-quality |
| S6_vs_k141_32221\|\|full | Not-determined |
| S6_vs_k141_22596\|\|full | Not-determined |
| S6_vs_k141_53605\|\|full | Not-determined |
| S6_vs_k141_50856\|\|full | Not-determined |
| S6_vs_k141_18399\|\|full | Low-quality |
| S6_vs_k141_30126\|\|full | Low-quality |
| S6_vs_k141_15941\|\|full | Low-quality |
| S6_vs_k141_1546\|\|full | Not-determined |
| S6_vs_k141_15949\|\|full | Low-quality |
| S6_vs_k141_55372\|\|full | Not-determined |
| S6_vs_k141_50874\|\|full | Not-determined |
| S6_vs_k141_30153\|\|full | Low-quality |
| S6_vs_k141_18415\|\|full | Low-quality |
| S6_vs_k141_53628\|\|full | Low-quality |
| S6_vs_k141_4465\|\|full | Low-quality |
| S6_vs_k141_6405\|\|full | Low-quality |
| S6_vs_k141_22631\|\|full | Low-quality |
| S6_vs_k141_4467\|\|full | Low-quality |
| S6_vs_k141_53640\|\|full | Low-quality |
| S6_vs_k141_36832\|\|full | Low-quality |
| S6_vs_k141_34997\|\|full | Low-quality |
| S6_vs_k141_13728\|\|full | Low-quality |
| S6_vs_k141_13731\|\|full | Low-quality |
| S6_vs_k141_53647\|\|full | Low-quality |
| S6_vs_k141_55416\|\|full | Low-quality |
| S6_vs_k141_39180\|\|full | Low-quality |
| S6_vs_k141_35008\|\|full | Low-quality |
| S6_vs_k141_19912\|\|full | Not-determined |
| S6_vs_k141_18422\|\|full | Not-determined |
| S6_vs_k141_15986\|\|full | Not-determined |
| S8_vs_k141_159173\|\|full | Medium-quality |
| S6_vs_k141_15990\|\|full | Low-quality |
| S6_vs_k141_15993\|\|full | Not-determined |
| S6_vs_k141_53677\|\|full | Low-quality |
| S6_vs_k141_19919\|\|full | Low-quality |
| S6_vs_k141_19921\|\|full | Low-quality |
| S6_vs_k141_4492\|\|full | Low-quality |
| S6_vs_k141_22666\|\|full | Low-quality |
| S6_vs_k141_53686\|\|full | Not-determined |
| S6_vs_k141_6462\|\|full | Low-quality |
| S6_vs_k141_53693\|\|full | Not-determined |
| S6_vs_k141_32313\|\|full | Low-quality |
| S6_vs_k141_46614\|\|full | Low-quality |
| S6_vs_k141_18467\|\|full | Low-quality |
| S6_vs_k141_27333\|\|full | Low-quality |
| S6_vs_k141_32329\|\|full | Low-quality |
| S6_vs_k141_32332\|\|full | Low-quality |
| S6_vs_k141_27339\|\|full | Low-quality |
| S6_vs_k141_19935\|\|full | Not-determined |
| S6_vs_k141_50923\|\|full | Not-determined |
| S6_vs_k141_18481\|\|full | Not-determined |
| S6_vs_k141_6483\|\|full | Not-determined |
| S6_vs_k141_27350\|\|full | Low-quality |
| S6_vs_k141_48396\|\|full | Low-quality |
| S6_vs_k141_25454\|\|full | Low-quality |
| S6_vs_k141_18483\|\|full | Low-quality |
| S6_vs_k141_1586\|\|full | Low-quality |
| S6_vs_k141_35069\|\|full | Low-quality |
| S6_vs_k141_41965\|\|full | Low-quality |
| S6_vs_k141_19945\|\|full | Not-determined |
| S6_vs_k141_11118\|\|full | Low-quality |
| S6_vs_k141_18496\|\|full | Not-determined |
| S6_vs_k141_27367\|\|full | Low-quality |
| S6_vs_k141_41979\|\|full | Not-determined |
| S6_vs_k141_30192\|\|full | Low-quality |
| S6_vs_k141_41992\|\|full | Low-quality |
| S6_vs_k141_46650\|\|full | Low-quality |
| S6_vs_k141_39191\|\|full | Low-quality |
| S6_vs_k141_22706\|\|full | Low-quality |
| S6_vs_k141_42003\|\|full | Low-quality |
| S6_vs_k141_13824\|\|full | Low-quality |
| S6_vs_k141_19971\|\|full | Low-quality |
| S6_vs_k141_35143\|\|full | Low-quality |
| S6_vs_k141_6492\|\|full | Low-quality |
| S6_vs_k141_18528\|\|full | Low-quality |
| S6_vs_k141_27415\|\|full | Low-quality |
| S6_vs_k141_35151\|\|full | Low-quality |
| S6_vs_k141_8798\|\|full | Low-quality |
| S6_vs_k141_27423\|\|full | Low-quality |
| S6_vs_k141_27424\|\|full | Low-quality |
| S6_vs_k141_32370\|\|full | Low-quality |
| S6_vs_k141_13878\|\|full | Low-quality |
| S6_vs_k141_27438\|\|full | Not-determined |
| S6_vs_k141_32386\|\|full | Low-quality |
| S6_vs_k141_27445\|\|full | Low-quality |
| S6_vs_k141_18545\|\|full | Low-quality |
| S6_vs_k141_18546\|\|full | Low-quality |
| S6_vs_k141_48439\|\|full | Not-determined |
| S6_vs_k141_11180\|\|full | Low-quality |
| S6_vs_k141_11182\|\|full | Low-quality |
| S6_vs_k141_32397\|\|full | Low-quality |
| S6_vs_k141_18547\|\|full | Not-determined |
| S6_vs_k141_39237\|\|full | Not-determined |
| S6_vs_k141_48443\|\|full | Not-determined |
| S6_vs_k141_46664\|\|full | Not-determined |
| S6_vs_k141_18551\|\|full | Low-quality |
| S6_vs_k141_48444\|\|full | Not-determined |
| S6_vs_k141_16045\|\|full | Low-quality |
| S6_vs_k141_4592\|\|full | Low-quality |
| S6_vs_k141_11203\|\|full | Low-quality |
| S6_vs_k141_46668\|\|full | Low-quality |
| S17_extracted_k141_77057 | Medium-quality |
| S6_vs_k141_13918\|\|full | Low-quality |
| S6_vs_k141_48459\|\|full | Not-determined |
| S6_vs_k141_43818\|\|full | Low-quality |
| S6_vs_k141_35182\|\|full | Low-quality |
| S6_vs_k141_4595\|\|full | Low-quality |
| S6_vs_k141_13931\|\|full | Low-quality |
| S6_vs_k141_18586\|\|full | Low-quality |
| S6_vs_k141_18590\|\|full | Not-determined |
| S6_vs_k141_43828\|\|full | Low-quality |
| S6_vs_k141_32449\|\|full | Low-quality |
| S6_vs_k141_13936\|\|full | Low-quality |
| S6_vs_k141_48485\|\|full | Low-quality |
| S6_vs_k141_48490\|\|full | Low-quality |
| S6_vs_k141_35183\|\|full | Low-quality |
| S6_vs_k141_13941\|\|full | Low-quality |
| S6_vs_k141_35184\|\|full | Not-determined |
| S6_vs_k141_42073\|\|full | Not-determined |
| S6_vs_k141_46687\|\|full | Not-determined |
| S6_vs_k141_46691\|\|full | Low-quality |
| S6_vs_k141_42083\|\|full | Low-quality |
| S6_vs_k141_4604\|\|full | Low-quality |
| S6_vs_k141_22807\|\|full | Low-quality |
| S6_vs_k141_13956\|\|full | Low-quality |
| S6_vs_k141_42094\|\|full | Not-determined |
| S6_vs_k141_27491\|\|full | Low-quality |
| S6_vs_k141_46711\|\|full | Not-determined |
| S6_vs_k141_4624\|\|full | Low-quality |
| S6_vs_k141_48522\|\|full | Not-determined |
| S6_vs_k141_43871\|\|full | Low-quality |
| S6_vs_k141_32489\|\|full | Low-quality |
| S6_vs_k141_16077\|\|full | Low-quality |
| S6_vs_k141_4628\|\|full | Not-determined |
| S6_vs_k141_32508\|\|full | Low-quality |
| S6_vs_k141_22840\|\|full | Low-quality |
| S6_vs_k141_53761\|\|full | Low-quality |
| S6_vs_k141_13972\|\|full | Low-quality |
| S6_vs_k141_8832\|\|full | Low-quality |
| S6_vs_k141_46739\|\|full | Not-determined |
| S6_vs_k141_8836\|\|full | Low-quality |
| S6_vs_k141_32519\|\|full | Not-determined |
| S6_vs_k141_27528\|\|full | Low-quality |
| S6_vs_k141_35194\|\|full | Low-quality |
| S6_vs_k141_8845\|\|full | Not-determined |
| S6_vs_k141_35200\|\|full | Low-quality |
| S6_vs_k141_30275\|\|full | Not-determined |
| S6_vs_k141_4647\|\|full | Low-quality |
| S6_vs_k141_53780\|\|full | Low-quality |
| S6_vs_k141_53785\|\|full | Low-quality |
| S6_vs_k141_39297\|\|full | Low-quality |
| S6_vs_k141_53789\|\|full | Not-determined |
| S6_vs_k141_42179\|\|full | Not-determined |
| S6_vs_k141_42182\|\|full | Low-quality |
| S6_vs_k141_48559\|\|full | Not-determined |
| S6_vs_k141_20043\|\|full | Not-determined |
| S6_vs_k141_4670\|\|full | Not-determined |
| S6_vs_k141_42198\|\|full | Not-determined |
| S6_vs_k141_39316\|\|full | Not-determined |
| S6_vs_k141_53811\|\|full | Low-quality |
| S6_vs_k141_25479\|\|full | Low-quality |
| S6_vs_k141_43900\|\|full | Not-determined |
| S6_vs_k141_1633\|\|full | Low-quality |
| S6_vs_k141_51013\|\|full | Not-determined |
| S6_vs_k141_42259\|\|full | Low-quality |
| S6_vs_k141_51024\|\|full | Low-quality |
| S6_vs_k141_42271\|\|full | Low-quality |
| S6_vs_k141_42274\|\|full | Low-quality |
| S6_vs_k141_42278\|\|full | Not-determined |
| S6_vs_k141_4681\|\|full | Not-determined |
| S6_vs_k141_53828\|\|full | Low-quality |
| S6_vs_k141_53832\|\|full | Not-determined |
| S6_vs_k141_18633\|\|full | Low-quality |
| S6_vs_k141_18637\|\|full | Low-quality |
| S6_vs_k141_6681\|\|full | Low-quality |
| S6_vs_k141_18645\|\|full | Low-quality |
| S6_vs_k141_30301\|\|full | Not-determined |
| S6_vs_k141_39394\|\|full | Not-determined |
| S6_vs_k141_18649\|\|full | Low-quality |
| S6_vs_k141_14055\|\|full | Low-quality |
| S6_vs_k141_11250\|\|full | Not-determined |
| S6_vs_k141_53852\|\|full | Not-determined |
| S6_vs_k141_22957\|\|full | Low-quality |
| S6_vs_k141_18654\|\|full | Low-quality |
| S6_vs_k141_1703\|\|full | Low-quality |
| S6_vs_k141_4713\|\|full | Low-quality |
| S6_vs_k141_11262\|\|full | Low-quality |
| S6_vs_k141_25592\|\|full | Low-quality |
| S6_vs_k141_25593\|\|full | Low-quality |
| S6_vs_k141_32639\|\|full | Low-quality |
| S6_vs_k141_20143\|\|full | Low-quality |
| S6_vs_k141_22986\|\|full | Low-quality |
| S6_vs_k141_22992\|\|full | Not-determined |
| S6_vs_k141_35228\|\|full | Low-quality |
| S6_vs_k141_11288\|\|full | Low-quality |
| S6_vs_k141_43922\|\|full | Low-quality |
| S6_vs_k141_43926\|\|full | Low-quality |
| S6_vs_k141_14080\|\|full | Low-quality |
| S6_vs_k141_25635\|\|full | Low-quality |
| S6_vs_k141_11300\|\|full | Not-determined |
| S6_vs_k141_20162\|\|full | Low-quality |
| S6_vs_k141_20165\|\|full | Low-quality |
| S6_vs_k141_16189\|\|full | Low-quality |
| S6_vs_k141_48588\|\|full | Low-quality |
| S6_vs_k141_32703\|\|full | Low-quality |
| S6_vs_k141_53880\|\|full | Low-quality |
| S6_vs_k141_30346\|\|full | Low-quality |
| S6_vs_k141_16196\|\|full | Not-determined |
| S6_vs_k141_43970\|\|full | Low-quality |
| S6_vs_k141_48593\|\|full | Low-quality |
| S6_vs_k141_32710\|\|full | Not-determined |
| S6_vs_k141_53886\|\|full | Low-quality |
| S6_vs_k141_16205\|\|full | Low-quality |
| S6_vs_k141_43977\|\|full | Not-determined |
| S6_vs_k141_39471\|\|full | Low-quality |
| S6_vs_k141_1784\|\|full | Not-determined |
| S6_vs_k141_30353\|\|full | Not-determined |
| S6_vs_k141_43996\|\|full | Not-determined |
| S6_vs_k141_32736\|\|full | Low-quality |
| S6_vs_k141_48617\|\|full | Not-determined |
| S6_vs_k141_23047\|\|full | Low-quality |
| S6_vs_k141_32745\|\|full | Low-quality |
| S6_vs_k141_48631\|\|full | Low-quality |
| S6_vs_k141_32762\|\|full | Not-determined |
| S6_vs_k141_44021\|\|full | Low-quality |
| S6_vs_k141_32772\|\|full | Low-quality |
| S6_vs_k141_18726\|\|full | Not-determined |
| S6_vs_k141_44032\|\|full | Not-determined |
| S6_vs_k141_44033\|\|full | Low-quality |
| S6_vs_k141_55535\|\|full | Not-determined |
| S6_vs_k141_23093\|\|full | Low-quality |
| S6_vs_k141_11337\|\|full | Low-quality |
| S6_vs_k141_27548\|\|full | Not-determined |
| S6_vs_k141_42379\|\|full | Low-quality |
| S6_vs_k141_32788\|\|full | Low-quality |
| S6_vs_k141_48666\|\|full | Low-quality |
| S6_vs_k141_46776\|\|full | Low-quality |
| S6_vs_k141_48679\|\|full | Low-quality |
| S6_vs_k141_18741\|\|full | Not-determined |
| S6_vs_k141_42395\|\|full | Low-quality |
| S6_vs_k141_53949\|\|full | Low-quality |
| S6_vs_k141_8872\|\|full | Low-quality |
| S6_vs_k141_48689\|\|full | Low-quality |
| S6_vs_k141_27567\|\|full | Not-determined |
| S6_vs_k141_6743\|\|full | Not-determined |
| S6_vs_k141_32813\|\|full | Not-determined |
| S6_vs_k141_16296\|\|full | Not-determined |
| S6_vs_k141_32817\|\|full | Low-quality |
| S6_vs_k141_42412\|\|full | Not-determined |
| S3_vs_k141_111776\|\|full | Medium-quality |
| S6_vs_k141_48702\|\|full | Not-determined |
| S6_vs_k141_36883\|\|full | Not-determined |
| S6_vs_k141_36886\|\|full | Not-determined |
| S6_vs_k141_27585\|\|full | Low-quality |
| S6_vs_k141_35269\|\|full | Not-determined |
| S6_vs_k141_44092\|\|full | Low-quality |
| S6_vs_k141_32849\|\|full | Low-quality |
| S6_vs_k141_48708\|\|full | Low-quality |
| S7_extracted_93097 | Medium-quality |
| S6_vs_k141_32856\|\|full | Not-determined |
| S6_vs_k141_6766\|\|full | Low-quality |
| S6_vs_k141_18774\|\|full | Low-quality |
| S6_vs_k141_30418\|\|full | Not-determined |
| S6_vs_k141_54010\|\|full | Low-quality |
| S6_vs_k141_44161\|\|full | Not-determined |
| S6_vs_k141_20285\|\|full | Low-quality |
| S6_vs_k141_8926\|\|full | Not-determined |
| S6_vs_k141_44165\|\|full | Low-quality |
| S6_vs_k141_39550\|\|full | Not-determined |
| S6_vs_k141_30437\|\|full | Low-quality |
| S6_vs_k141_54019\|\|full | Not-determined |
| S6_vs_k141_54023\|\|full | Not-determined |
| S6_vs_k141_36924\|\|full | Low-quality |
| S6_vs_k141_30445\|\|full | Low-quality |
| S6_vs_k141_6808\|\|full | Not-determined |
| S6_vs_k141_18789\|\|full | Low-quality |
| S6_vs_k141_36940\|\|full | Not-determined |
| S6_vs_k141_20317\|\|full | Low-quality |
| S6_vs_k141_39568\|\|full | Low-quality |
| S6_vs_k141_36951\|\|full | Not-determined |
| S6_vs_k141_48732\|\|full | Low-quality |
| S6_vs_k141_20329\|\|full | Low-quality |
| S6_vs_k141_35307\|\|full | Not-determined |
| S6_vs_k141_48734\|\|full | Not-determined |
| S6_vs_k141_54046\|\|full | Not-determined |
| S6_vs_k141_36957\|\|full | Low-quality |
| S6_vs_k141_14128\|\|full | Low-quality |
| S6_vs_k141_14134\|\|full | Low-quality |
| S6_vs_k141_11422\|\|full | Not-determined |
| S6_vs_k141_23211\|\|full | Not-determined |
| S6_vs_k141_35318\|\|full | Not-determined |
| S6_vs_k141_27633\|\|full | Not-determined |
| S6_vs_k141_1805\|\|full | Low-quality |
| S6_vs_k141_36973\|\|full | Not-determined |
| S6_vs_k141_39600\|\|full | Low-quality |
| S6_vs_k141_20357\|\|full | Not-determined |
| S6_vs_k141_16328\|\|full | Low-quality |
| S6_vs_k141_1813\|\|full | Low-quality |
| S6_vs_k141_30467\|\|full | Low-quality |
| S6_vs_k141_48757\|\|full | Not-determined |
| S6_vs_k141_48762\|\|full | Not-determined |
| S6_vs_k141_11455\|\|full | Low-quality |
| S6_vs_k141_11459\|\|full | Low-quality |
| S6_vs_k141_36992\|\|full | Not-determined |
| S6_vs_k141_30474\|\|full | Low-quality |
| S6_vs_k141_48769\|\|full | Low-quality |
| S6_vs_k141_18819\|\|full | Low-quality |
| S6_vs_k141_54072\|\|full | Low-quality |
| S6_vs_k141_11471\|\|full | Low-quality |
| S6_vs_k141_20365\|\|full | Low-quality |
| S6_vs_k141_35337\|\|full | Not-determined |
| S6_vs_k141_54094\|\|full | Low-quality |
| S6_vs_k141_11485\|\|full | Low-quality |
| S6_vs_k141_11486\|\|full | Low-quality |
| S6_vs_k141_32951\|\|full | Low-quality |
| S6_vs_k141_6842\|\|full | Low-quality |
| S6_vs_k141_37013\|\|full | Low-quality |
| S6_vs_k141_25704\|\|full | Low-quality |
| S6_vs_k141_18843\|\|full | Low-quality |
| S6_vs_k141_27670\|\|full | Not-determined |
| S6_vs_k141_6865\|\|full | Low-quality |
| S6_vs_k141_23315\|\|full | Not-determined |
| S6_vs_k141_27678\|\|full | Low-quality |
| S6_vs_k141_27679\|\|full | Not-determined |
| S6_vs_k141_20405\|\|full | Low-quality |
| S6_vs_k141_23319\|\|full | Not-determined |
| S6_vs_k141_20409\|\|full | Not-determined |
| S6_vs_k141_11521\|\|full | Low-quality |
| S6_vs_k141_6875\|\|full | Low-quality |
| S6_vs_k141_46880\|\|full | Low-quality |
| S6_vs_k141_11532\|\|full | Low-quality |
| S6_vs_k141_11536\|\|full | Low-quality |
| S6_vs_k141_48804\|\|full | Low-quality |
| S6_vs_k141_11539\|\|full | Not-determined |
| S6_vs_k141_32979\|\|full | Not-determined |
| S7_vs_41534\|\|full | Medium-quality |
| S6_vs_k141_48811\|\|full | Low-quality |
| S6_vs_k141_11564\|\|full | Low-quality |
| S6_vs_k141_25726\|\|full | Low-quality |
| S6_vs_k141_25727\|\|full | Not-determined |
| S5_vs_k141_137260\|\|full | Medium-quality |
| S6_vs_k141_11570\|\|full | Low-quality |
| S6_vs_k141_39628\|\|full | Low-quality |
| S6_vs_k141_48837\|\|full | Not-determined |
| S6_vs_k141_39637\|\|full | Not-determined |
| S6_vs_k141_11588\|\|full | Not-determined |
| S6_vs_k141_11589\|\|full | Low-quality |
| S6_vs_k141_39658\|\|full | Low-quality |
| S6_vs_k141_25769\|\|full | Low-quality |
| S6_vs_k141_55599\|\|full | Low-quality |
| S6_vs_k141_6931\|\|full | Not-determined |
| S6_vs_k141_11636\|\|full | Not-determined |
| S6_vs_k141_48906\|\|full | Low-quality |
| S6_vs_k141_23425\|\|full | Low-quality |
| S6_vs_k141_25781\|\|full | Not-determined |
| S6_vs_k141_55606\|\|full | Low-quality |
| S6_vs_k141_16365\|\|full | Low-quality |
| S6_vs_k141_37042\|\|full | Low-quality |
| S6_vs_k141_23448\|\|full | Not-determined |
| S6_vs_k141_30561\|\|full | Low-quality |
| S6_vs_k141_23454\|\|full | Not-determined |
| S6_vs_k141_39721\|\|full | Not-determined |
| S6_vs_k141_55652\|\|full | Not-determined |
| S6_vs_k141_37061\|\|full | Low-quality |
| S6_vs_k141_37069\|\|full | Low-quality |
| S6_vs_k141_48947\|\|full | Low-quality |
| S6_vs_k141_39749\|\|full | Not-determined |
| S6_vs_k141_48956\|\|full | Low-quality |
| S6_vs_k141_44267\|\|full | Low-quality |
| S6_vs_k141_37079\|\|full | Low-quality |
| S6_vs_k141_37085\|\|full | Not-determined |
| S6_vs_k141_48960\|\|full | Low-quality |
| S6_vs_k141_48961\|\|full | Low-quality |
| S6_vs_k141_30594\|\|full | Low-quality |
| S6_vs_k141_11671\|\|full | Low-quality |
| S6_vs_k141_48967\|\|full | Low-quality |
| S6_vs_k141_46990\|\|full | Low-quality |
| S6_vs_k141_11680\|\|full | Low-quality |
| S6_vs_k141_44288\|\|full | Low-quality |
| S6_vs_k141_25824\|\|full | Low-quality |
| S6_vs_k141_23481\|\|full | Not-determined |
| S6_vs_k141_37126\|\|full | Low-quality |
| S6_vs_k141_11704\|\|full | Low-quality |
| S6_vs_k141_37130\|\|full | Low-quality |
| S6_vs_k141_49018\|\|full | Low-quality |
| S6_vs_k141_23512\|\|full | Low-quality |
| S6_vs_k141_39793\|\|full | Low-quality |
| S6_vs_k141_1832\|\|full | Not-determined |
| S6_vs_k141_44325\|\|full | Low-quality |
| S6_vs_k141_25871\|\|full | Low-quality |
| S6_vs_k141_39802\|\|full | Low-quality |
| S6_vs_k141_46994\|\|full | Low-quality |
| S6_vs_k141_1868\|\|full | Low-quality |
| S6_vs_k141_16408\|\|full | Low-quality |
| S6_vs_k141_1874\|\|full | Low-quality |
| S6_vs_k141_47000\|\|full | Low-quality |
| S6_vs_k141_37151\|\|full | Low-quality |
| S6_vs_k141_25900\|\|full | Low-quality |
| S6_vs_k141_25903\|\|full | Low-quality |
| S6_vs_k141_8961\|\|full | Not-determined |
| S6_vs_k141_1893\|\|full | Low-quality |
| S6_vs_k141_8970\|\|full | Not-determined |
| S6_vs_k141_23540\|\|full | Not-determined |
| S6_vs_k141_37163\|\|full | Low-quality |
| S6_vs_k141_39838\|\|full | Low-quality |
| S6_vs_k141_16440\|\|full | Low-quality |
| S6_vs_k141_44348\|\|full | Low-quality |
| S6_vs_k141_8983\|\|full | Low-quality |
| S6_vs_k141_55759\|\|full | Not-determined |
| S6_vs_k141_11740\|\|full | Low-quality |
| S6_vs_k141_44359\|\|full | Low-quality |
| S6_vs_k141_8993\|\|full | Low-quality |
| S6_vs_k141_11758\|\|full | Not-determined |
| S6_vs_k141_55793\|\|full | Low-quality |
| S6_vs_k141_9006\|\|full | Low-quality |
| S6_vs_k141_1936\|\|full | Not-determined |
| S6_vs_k141_1942\|\|full | Low-quality |
| S6_vs_k141_49065\|\|full | Not-determined |
| S6_vs_k141_49078\|\|full | Low-quality |
| S6_vs_k141_39899\|\|full | Low-quality |

| | |
|---|---|
| S6_vs_k141_37205||full | Low-quality |
| S6_vs_k141_49090||full | Low-quality |
| S6_vs_k141_9065||full | Low-quality |
| S6_vs_k141_44388||full | Low-quality |
| S6_vs_k141_20516||full | Not-determined |
| S6_vs_k141_1983||full | Low-quality |
| S6_vs_k141_39924||full | Not-determined |
| S6_vs_k141_49108||full | Low-quality |
| S6_vs_k141_20536||full | Low-quality |
| S6_vs_k141_39938||full | Low-quality |
| S6_vs_k141_37232||full | Low-quality |
| S6_vs_k141_20542||full | Low-quality |
| S6_vs_k141_20553||full | Not-determined |
| S6_vs_k141_49137||full | Low-quality |
| S6_vs_k141_9102||full | Not-determined |
| S6_vs_k141_20569||full | Low-quality |
| S6_vs_k141_44467||full | Low-quality |
| S6_vs_k141_44469||full | Low-quality |
| S6_vs_k141_37264||full | Low-quality |
| S6_vs_k141_37276||full | Low-quality |
| S6_vs_k141_16470||full | Low-quality |
| S6_vs_k141_20579||full | Not-determined |
| S6_vs_k141_51102||full | Low-quality |
| S6_vs_k141_51106||full | Not-determined |
| S6_vs_k141_51107||full | Low-quality |
| S6_vs_k141_20589||full | Low-quality |
| S6_vs_k141_44495||full | Low-quality |
| S6_vs_k141_2032||full | Not-determined |
| S6_vs_k141_2041||full | Low-quality |
| S6_vs_k141_20604||full | Low-quality |
| S6_vs_k141_2046||full | Low-quality |
| S6_vs_k141_44502||full | Low-quality |
| S6_vs_k141_55874||full | Low-quality |
| S6_vs_k141_49185||full | Low-quality |
| S6_vs_k141_9217||full | Low-quality |
| S6_vs_k141_44505||full | Not-determined |
| S6_vs_k141_2076||full | Low-quality |
| S6_vs_k141_9221||full | Not-determined |
| S6_vs_k141_9231||full | Low-quality |
| S6_vs_k141_55910||full | Low-quality |
| S6_vs_k141_55911||full | Not-determined |
| S6_vs_k141_9233||full | Not-determined |
| S6_vs_k141_37342||full | Low-quality |
| S6_vs_k141_27710||full | Low-quality |
| S6_vs_k141_27722||full | Low-quality |
| S6_vs_k141_51166||full | Low-quality |
| S6_vs_k141_2120||full | Low-quality |
| S6_vs_k141_49257||full | Low-quality |
| S6_vs_k141_51187||full | Low-quality |
| S6_vs_k141_49278||full | Not-determined |
| S6_vs_k141_9256||full | Low-quality |
| S6_vs_k141_49280||full | Low-quality |
| S6_vs_k141_56361||full | Medium-quality |
| S6_vs_k141_49295||full | Low-quality |
| S6_vs_k141_2139||full | Not-determined |
| S6_vs_k141_9300||full | Not-determined |
| S6_vs_k141_55988||full | Low-quality |
| S6_vs_k141_37383||full | Low-quality |
| S6_vs_k141_2147||full | Low-quality |
| S6_vs_k141_9334||full | Low-quality |
| S6_vs_k141_49311||full | Low-quality |
| S6_vs_k141_9348||full | Not-determined |
| S6_vs_k141_9349||full | Not-determined |
| S6_vs_k141_47055||full | Not-determined |
| S6_vs_k141_47059||full | Low-quality |
| S6_vs_k141_20622||full | Not-determined |
| S6_vs_k141_47065||full | Low-quality |
| S6_vs_k141_37402||full | Not-determined |
| S6_vs_k141_37418||full | Low-quality |
| S6_vs_k141_2176||full | Low-quality |
| S6_vs_k141_56022||full | Low-quality |
| S6_vs_k141_20665||full | Low-quality |
| S6_vs_k141_44608||full | Low-quality |
| S6_vs_k141_47105||full | Low-quality |
| S6_vs_k141_51287||full | Low-quality |
| S6_vs_k141_51289||full | Low-quality |
| S6_vs_k141_49402||full | Low-quality |
| S6_vs_k141_40030||full | Low-quality |
| S6_vs_k141_40039||full | Not-determined |
| S17_vs_k141_79151||full | Medium-quality |
| S6_vs_k141_9372||full | Not-determined |
| S6_vs_k141_20680||full | Low-quality |
| S6_vs_k141_49423||full | Not-determined |
| S6_vs_k141_44648||full | Not-determined |
| S6_vs_k141_49428||full | Low-quality |
| S6_vs_k141_9409||full | Low-quality |
| S6_vs_k141_37485||full | Not-determined |
| S6_vs_k141_56051||full | Low-quality |
| S6_vs_k141_56057||full | Low-quality |
| S6_vs_k141_49433||full | Not-determined |
| S6_vs_k141_49444||full | Not-determined |
| S7_vs_21981||full | Medium-quality |
| S6_vs_k141_44678||full | Low-quality |
| S6_vs_k141_37537||full | Low-quality |
| S6_vs_k141_37541||full | Low-quality |
| S6_vs_k141_51390||full | Not-determined |
| S6_vs_k141_44694||full | Low-quality |
| S6_vs_k141_49458||full | Not-determined |
| S6_vs_k141_44700||full | Low-quality |
| S6_vs_k141_51393||full | Not-determined |
| S6_vs_k141_49462||full | Not-determined |
| S6_vs_k141_51408||full | Low-quality |
| S6_vs_k141_44711||full | Low-quality |
| S6_vs_k141_56128||full | Low-quality |
| S6_vs_k141_51433||full | Low-quality |
| S6_vs_k141_56155||full | Low-quality |
| S6_vs_k141_56160||full | Low-quality |
| S6_vs_k141_49475||full | Low-quality |
| S6_vs_k141_2281||full | Low-quality |
| S6_vs_k141_51492||full | Not-determined |
| S6_vs_k141_44765||full | Low-quality |
| S6_vs_k141_20781||full | Low-quality |
| S6_vs_k141_56248||full | Low-quality |
| S6_vs_k141_51522||full | Not-determined |
| S6_vs_k141_20825||full | Low-quality |
| S6_vs_k141_37686||full | Low-quality |
| S6_vs_k141_27783||full | Low-quality |
| S6_vs_k141_51541||full | Low-quality |
| S6_vs_k141_37696||full | Low-quality |
| S6_vs_k141_20863||full | Low-quality |
| S6_vs_k141_20881||full | Low-quality |
| S6_vs_k141_20887||full | Low-quality |
| S6_vs_k141_51578||full | Not-determined |
| S6_vs_k141_20901||full | Low-quality |
| S6_vs_k141_2349||full | Not-determined |
| S6_vs_k141_51598||full | Not-determined |
| S6_vs_k141_51601||full | Not-determined |
| S6_vs_k141_27803||full | Low-quality |
| S6_vs_k141_20930||full | Low-quality |
| S6_vs_k141_27805||full | Low-quality |
| S6_vs_k141_51614||full | Not-determined |
| S6_vs_k141_56278||full | Not-determined |
| S6_vs_k141_20952||full | Not-determined |
| S6_vs_k141_27838||full | Low-quality |
| S6_vs_k141_20959||full | Not-determined |
| S6_vs_k141_51645||full | Low-quality |
| S6_vs_k141_27915||full | Low-quality |
| S9_extracted_k141_70236 | Medium-quality |
| S6_vs_k141_27931||full | Low-quality |
| S6_vs_k141_20975||full | Low-quality |
| S6_vs_k141_27934||full | Low-quality |
| S6_vs_k141_56320||full | Low-quality |
| S6_vs_k141_21003||full | Low-quality |
| S6_vs_k141_27953||full | Low-quality |
| S6_vs_k141_27962||full | Not-determined |
| S6_vs_k141_27980||full | Not-determined |
| S6_vs_k141_56331||full | Not-determined |
| S6_vs_k141_28001||full | Low-quality |
| S12_extracted_k141_176437 | Medium-quality |
| S6_vs_k141_56376||full | Low-quality |
| S6_vs_k141_51780||full | Low-quality |
| S6_vs_k141_56410||full | Not-determined |
| S6_vs_k141_56412||full | Low-quality |
| S6_vs_k141_28077||full | Not-determined |
| S6_vs_k141_51807||full | Not-determined |
| S6_vs_k141_51812||full | Low-quality |
| S6_vs_k141_56444||full | Not-determined |
| S6_vs_k141_28099||full | Not-determined |
| S6_vs_k141_56461||full | Not-determined |
| S6_vs_k141_56463||full | Low-quality |
| S6_vs_k141_56469||full | Not-determined |
| S6_vs_k141_21068||full | Not-determined |
| S6_vs_k141_28135||full | Not-determined |
| S6_vs_k141_28144||full | Low-quality |
| S6_vs_k141_21106||full | Not-determined |
| S6_vs_k141_21128||full | Low-quality |
| S6_vs_k141_28238||full | Low-quality |
| S6_vs_k141_28253||full | Low-quality |
| S6_vs_k141_28260||full | Low-quality |
| S6_vs_k141_28278||full | Low-quality |
| S6_vs_k141_14480||0_partial | Not-determined |
| S6_vs_k141_25980||0_partial | Not-determined |
| S6_vs_k141_38470||0_partial | Not-determined |
| S6_vs_k141_43120||0_partial | Low-quality |
| S6_vs_k141_47754||0_partial | Not-determined |
| S6_vs_k141_52891||0_partial | Low-quality |
| S6_vs_k141_50185||0_partial | Not-determined |
| S6_vs_k141_50185||1_partial | Low-quality |
| S6_vs_k141_54925||0_partial | Not-determined |
| S6_vs_k141_10776||0_partial | Low-quality |
| S6_vs_k141_43653||0_partial | Low-quality |
| S6_vs_k141_10908||0_partial | Not-determined |
| S6_vs_k141_55084||0_partial | Not-determined |
| S17_vs_k141_174203||full | Medium-quality |
| S6_vs_k141_1504||0_partial | Not-determined |
| S6_vs_k141_4457||0_partial | Low-quality |
| S6_vs_k141_39232||0_partial | Low-quality |
| S6_vs_k141_39286||0_partial | Not-determined |
| S6_vs_k141_30297||0_partial | Not-determined |
| S6_vs_k141_55570||0_partial | Not-determined |
| S6_vs_k141_20421||0_partial | Low-quality |
| S6_vs_k141_40014||0_partial | Low-quality |
| S6_vs_k141_42422||lt2gene | Low-quality |
| S6_vs_k141_18859||lt2gene | Not-determined |
| S6_vs_k141_11803||lt2gene | Low-quality |
| S6_vs_k141_42439||lt2gene | Low-quality |
| S6_vs_k141_16516||lt2gene | Low-quality |
| S6_vs_k141_44786||lt2gene | Not-determined |
| S6_vs_k141_49507||lt2gene | Not-determined |
| S6_vs_k141_16539||lt2gene | Low-quality |
| S6_vs_k141_40090||lt2gene | Low-quality |
| S6_vs_k141_37728||lt2gene | Not-determined |
| S6_vs_k141_40099||lt2gene | Low-quality |
| S6_vs_k141_42469||lt2gene | Low-quality |
| S6_vs_k141_49535||lt2gene | Not-determined |
| S6_vs_k141_42474||lt2gene | Not-determined |
| S6_vs_k141_42481||lt2gene | Low-quality |
| S6_vs_k141_42488||lt2gene | Low-quality |
| S6_vs_k141_35384||lt2gene | Not-determined |
| S6_vs_k141_54255||lt2gene | Low-quality |
| S6_vs_k141_42520||lt2gene | Not-determined |
| S6_vs_k141_16604||lt2gene | Low-quality |
| S6_vs_k141_47193||lt2gene | Low-quality |
| S6_vs_k141_42534||lt2gene | Low-quality |
| S6_vs_k141_28318||lt2gene | Low-quality |
| S6_vs_k141_68||lt2gene | Low-quality |
| S6_vs_k141_69||lt2gene | Low-quality |
| S6_vs_k141_35395||lt2gene | Low-quality |
| S6_vs_k141_37744||lt2gene | Not-determined |
| S6_vs_k141_4747||lt2gene | Not-determined |
| S6_vs_k141_37762||lt2gene | Low-quality |
| S6_vs_k141_18949||lt2gene | Low-quality |
| S6_vs_k141_14240||lt2gene | Low-quality |
| S6_vs_k141_33116||lt2gene | Low-quality |
| S6_vs_k141_28371||lt2gene | Low-quality |
| S6_vs_k141_40155||lt2gene | Low-quality |
| S6_vs_k141_157||lt2gene | Low-quality |
| S6_vs_k141_2365||lt2gene | Low-quality |
| S6_vs_k141_14272||lt2gene | Not-determined |
| S6_vs_k141_52023||lt2gene | Low-quality |
| S6_vs_k141_23656||lt2gene | Not-determined |
| S6_vs_k141_42605||lt2gene | Low-quality |
| S6_vs_k141_14296||lt2gene | Low-quality |
| S6_vs_k141_14301||lt2gene | Low-quality |
| S6_vs_k141_52046||lt2gene | Not-determined |
| S6_vs_k141_42622||lt2gene | Low-quality |
| S6_vs_k141_42628||lt2gene | Low-quality |
| S6_vs_k141_44902||lt2gene | Low-quality |
| S6_vs_k141_30780||lt2gene | Not-determined |
| S6_vs_k141_49665||lt2gene | Not-determined |
| S6_vs_k141_49671||lt2gene | Low-quality |
| S6_vs_k141_44936||lt2gene | Not-determined |
| S6_vs_k141_44938||lt2gene | Not-determined |
| S6_vs_k141_37930||lt2gene | Low-quality |
| S6_vs_k141_49681||lt2gene | Low-quality |
| S6_vs_k141_9548||lt2gene | Low-quality |
| S6_vs_k141_2452||lt2gene | Not-determined |
| S6_vs_k141_2454||lt2gene | Not-determined |
| S6_vs_k141_11893||lt2gene | Low-quality |
| S6_vs_k141_49698||lt2gene | Low-quality |
| S6_vs_k141_40209||lt2gene | Low-quality |
| S6_vs_k141_2475||lt2gene | Not-determined |
| S6_vs_k141_2478||lt2gene | Low-quality |
| S6_vs_k141_54265||lt2gene | Low-quality |
| S6_vs_k141_26037||lt2gene | Low-quality |
| S6_vs_k141_42672||lt2gene | Not-determined |
| S6_vs_k141_35521||lt2gene | Low-quality |
| S6_vs_k141_14353||lt2gene | Not-determined |
| S6_vs_k141_26059||lt2gene | Low-quality |
| S6_vs_k141_14367||lt2gene | Low-quality |
| S6_vs_k141_26069||lt2gene | Low-quality |
| S6_vs_k141_9583||lt2gene | Not-determined |
| S6_vs_k141_4873||lt2gene | Low-quality |
| S6_vs_k141_4876||lt2gene | Low-quality |
| S6_vs_k141_321||lt2gene | Not-determined |
| S6_vs_k141_38008||lt2gene | Low-quality |
| S6_vs_k141_38017||lt2gene | Low-quality |
| S6_vs_k141_336||lt2gene | Low-quality |
| S6_vs_k141_33229||lt2gene | Not-determined |
| S6_vs_k141_26112||lt2gene | Not-determined |
| S6_vs_k141_38021||lt2gene | Low-quality |
| S6_vs_k141_33239||lt2gene | Low-quality |
| S6_vs_k141_33251||lt2gene | Low-quality |
| S6_vs_k141_33280||lt2gene | Low-quality |
| S6_vs_k141_38085||lt2gene | Low-quality |
| S6_vs_k141_45043||lt2gene | Not-determined |
| S6_vs_k141_394||lt2gene | Low-quality |
| S6_vs_k141_14442||lt2gene | Low-quality |
| S6_vs_k141_45075||lt2gene | Low-quality |
| S6_vs_k141_2656||lt2gene | Low-quality |
| S6_vs_k141_411||lt2gene | Not-determined |
| S6_vs_k141_416||lt2gene | Low-quality |
| S6_vs_k141_14474||lt2gene | Low-quality |
| S6_vs_k141_52165||lt2gene | Low-quality |
| S6_vs_k141_16777||lt2gene | Not-determined |
| S6_vs_k141_4921||lt2gene | Low-quality |
| S6_vs_k141_447||lt2gene | Low-quality |
| S6_vs_k141_9659||lt2gene | Low-quality |
| S6_vs_k141_23819||lt2gene | Not-determined |
| S6_vs_k141_461||lt2gene | Low-quality |
| S6_vs_k141_33374||lt2gene | Low-quality |
| S6_vs_k141_16822||lt2gene | Low-quality |
| S6_vs_k141_4933||lt2gene | Not-determined |
| S6_vs_k141_16831||lt2gene | Low-quality |
| S6_vs_k141_30817||lt2gene | Low-quality |
| S6_vs_k141_52228||lt2gene | Low-quality |
| S6_vs_k141_42736||lt2gene | Not-determined |
| S6_vs_k141_35617||lt2gene | Low-quality |
| S6_vs_k141_21351||lt2gene | Low-quality |
| S6_vs_k141_42767||lt2gene | Low-quality |
| S6_vs_k141_52272||lt2gene | Low-quality |
| S6_vs_k141_14507||lt2gene | Low-quality |
| S6_vs_k141_21361||lt2gene | Low-quality |
| S6_vs_k141_21362||lt2gene | Low-quality |
| S6_vs_k141_33424||lt2gene | Low-quality |
| S6_vs_k141_21364||lt2gene | Low-quality |
| S6_vs_k141_2774||lt2gene | Low-quality |
| S6_vs_k141_2782||lt2gene | Low-quality |
| S6_vs_k141_2789||lt2gene | Low-quality |
| S6_vs_k141_33452||lt2gene | Low-quality |
| S6_vs_k141_2799||lt2gene | Not-determined |
| S6_vs_k141_2814||lt2gene | Low-quality |
| S6_vs_k141_16862||lt2gene | Low-quality |
| S6_vs_k141_9733||lt2gene | Low-quality |
| S6_vs_k141_16870||lt2gene | Low-quality |
| S6_vs_k141_40471||lt2gene | Low-quality |
| S6_vs_k141_4950||lt2gene | Low-quality |
| S6_vs_k141_35650||lt2gene | Not-determined |
| S6_vs_k141_52326||lt2gene | Low-quality |
| S6_vs_k141_42844||lt2gene | Low-quality |
| S6_vs_k141_23931||lt2gene | Low-quality |
| S6_vs_k141_12190||lt2gene | Low-quality |
| S6_vs_k141_23956||lt2gene | Not-determined |
| S6_vs_k141_14584||lt2gene | Low-quality |
| S6_vs_k141_7314||lt2gene | Not-determined |
| S6_vs_k141_28634||lt2gene | Low-quality |
| S6_vs_k141_45192||lt2gene | Low-quality |
| S6_vs_k141_14608||lt2gene | Low-quality |
| S6_vs_k141_47340||lt2gene | Low-quality |
| S6_vs_k141_4983||lt2gene | Low-quality |
| S6_vs_k141_2924||lt2gene | Low-quality |
| S6_vs_k141_14629||lt2gene | Not-determined |
| S6_vs_k141_47345||lt2gene | Low-quality |
| S6_vs_k141_5000||lt2gene | Low-quality |
| S6_vs_k141_2942||lt2gene | Low-quality |
| S6_vs_k141_28660||lt2gene | Low-quality |
| S6_vs_k141_14648||lt2gene | Low-quality |
| S6_vs_k141_52474||lt2gene | Low-quality |
| S6_vs_k141_40575||lt2gene | Not-determined |
| S6_vs_k141_9799||lt2gene | Low-quality |
| S6_vs_k141_24012||lt2gene | Not-determined |
| S6_vs_k141_26265||lt2gene | Low-quality |
| S6_vs_k141_33537||lt2gene | Not-determined |
| S6_vs_k141_9835||lt2gene | Low-quality |
| S6_vs_k141_5038||lt2gene | Low-quality |
| S6_vs_k141_52528||lt2gene | Low-quality |
| S6_vs_k141_5041||lt2gene | Not-determined |
| S6_vs_k141_33556||lt2gene | Not-determined |
| S6_vs_k141_30883||lt2gene | Low-quality |
| S6_vs_k141_54294||lt2gene | Low-quality |
| S6_vs_k141_3019||lt2gene | Low-quality |
| S6_vs_k141_3028||lt2gene | Not-determined |
| S6_vs_k141_47456||lt2gene | Low-quality |
| S6_vs_k141_3040||lt2gene | Low-quality |
| S6_vs_k141_30918||lt2gene | Not-determined |
| S6_vs_k141_24072||lt2gene | Not-determined |
| S6_vs_k141_49744||lt2gene | Low-quality |
| S6_vs_k141_45303||lt2gene | Not-determined |
| S6_vs_k141_26316||lt2gene | Low-quality |
| S6_vs_k141_47495||lt2gene | Low-quality |
| S6_vs_k141_38322||lt2gene | Low-quality |
| S6_vs_k141_38323||lt2gene | Low-quality |
| S6_vs_k141_24085||lt2gene | Low-quality |
| S6_vs_k141_9867||lt2gene | Low-quality |
| S6_vs_k141_9895||lt2gene | Not-determined |
| S6_vs_k141_30979||lt2gene | Not-determined |
| S6_vs_k141_9918||lt2gene | Low-quality |
| S6_vs_k141_12463||lt2gene | Low-quality |
| S6_vs_k141_54367||lt2gene | Low-quality |
| S6_vs_k141_54373||lt2gene | Low-quality |
| S6_vs_k141_26340||lt2gene | Not-determined |
| S6_vs_k141_47540||lt2gene | Low-quality |
| S6_vs_k141_54381||lt2gene | Low-quality |
| S6_vs_k141_14718||lt2gene | Low-quality |
| S6_vs_k141_9945||lt2gene | Low-quality |
| S6_vs_k141_26366||lt2gene | Not-determined |
| S6_vs_k141_5106||lt2gene | Low-quality |
| S6_vs_k141_12521||lt2gene | Low-quality |
| S6_vs_k141_7441||lt2gene | Low-quality |
| S6_vs_k141_35790||lt2gene | Low-quality |
| S6_vs_k141_7442||lt2gene | Not-determined |
| S6_vs_k141_12545||lt2gene | Low-quality |
| S6_vs_k141_7472||lt2gene | Low-quality |
| S6_vs_k141_19094||lt2gene | Low-quality |
| S6_vs_k141_52708||lt2gene | Low-quality |
| S6_vs_k141_35829||lt2gene | Low-quality |
| S6_vs_k141_35830||lt2gene | Low-quality |
| S6_vs_k141_45379||lt2gene | Low-quality |
| S6_vs_k141_49904||lt2gene | Low-quality |
| S6_vs_k141_52719||lt2gene | Low-quality |
| S6_vs_k141_24190||lt2gene | Low-quality |
| S6_vs_k141_14783||lt2gene | Not-determined |
| S6_vs_k141_24226||lt2gene | Low-quality |
| S6_vs_k141_35841||lt2gene | Not-determined |
| S6_vs_k141_19117||lt2gene | Low-quality |
| S6_vs_k141_14802||lt2gene | Low-quality |
| S6_vs_k141_31057||lt2gene | Not-determined |
| S6_vs_k141_21521||lt2gene | Low-quality |
| S6_vs_k141_12580||lt2gene | Low-quality |
| S6_vs_k141_43044||lt2gene | Low-quality |
| S6_vs_k141_19135||lt2gene | Low-quality |
| S6_vs_k141_21534||lt2gene | Low-quality |
| S6_vs_k141_24281||lt2gene | Low-quality |
| S6_vs_k141_21543||lt2gene | Low-quality |
| S6_vs_k141_43059||lt2gene | Not-determined |
| S6_vs_k141_52764||lt2gene | Low-quality |
| S6_vs_k141_5171||lt2gene | Low-quality |
| S6_vs_k141_24300||lt2gene | Low-quality |
| S6_vs_k141_805||lt2gene | Not-determined |
| S6_vs_k141_9959||lt2gene | Low-quality |
| S6_vs_k141_7556||lt2gene | Low-quality |
| S6_vs_k141_50009||lt2gene | Low-quality |
| S6_vs_k141_52806||lt2gene | Low-quality |
| S6_vs_k141_21587||lt2gene | Low-quality |
| S6_vs_k141_45469||lt2gene | Low-quality |
| S6_vs_k141_50047||lt2gene | Low-quality |
| S6_vs_k141_24343||lt2gene | Low-quality |
| S6_vs_k141_24347||lt2gene | Low-quality |
| S6_vs_k141_5231||lt2gene | Not-determined |
| S6_vs_k141_3120||lt2gene | Not-determined |
| S6_vs_k141_21626||lt2gene | Not-determined |
| S6_vs_k141_17259||lt2gene | Not-determined |
| S6_vs_k141_50098||lt2gene | Not-determined |
| S6_vs_k141_35963||lt2gene | Low-quality |
| S6_vs_k141_47655||lt2gene | Not-determined |
| S6_vs_k141_14827||lt2gene | Low-quality |
| S6_vs_k141_43079||lt2gene | Low-quality |
| S6_vs_k141_31161||lt2gene | Not-determined |
| S6_vs_k141_21657||lt2gene | Low-quality |
| S6_vs_k141_45523||lt2gene | Low-quality |
| S6_vs_k141_17291||lt2gene | Low-quality |
| S6_vs_k141_45555||lt2gene | Not-determined |
| S6_vs_k141_24396||lt2gene | Low-quality |
| S6_vs_k141_21672||lt2gene | Low-quality |
| S6_vs_k141_5326||lt2gene | Not-determined |
| S6_vs_k141_17310||lt2gene | Low-quality |
| S6_vs_k141_45581||lt2gene | Low-quality |
| S6_vs_k141_45593||lt2gene | Low-quality |
| S6_vs_k141_50150||lt2gene | Low-quality |
| S6_vs_k141_45605||lt2gene | Low-quality |
| S6_vs_k141_31236||lt2gene | Low-quality |
| S6_vs_k141_5365||lt2gene | Not-determined |
| S6_vs_k141_28963||lt2gene | Low-quality |
| S6_vs_k141_3269||lt2gene | Low-quality |
| S6_vs_k141_38474||lt2gene | Low-quality |
| S6_vs_k141_17364||lt2gene | Low-quality |
| S6_vs_k141_54547||lt2gene | Low-quality |
| S6_vs_k141_10108||lt2gene | Low-quality |
| S6_vs_k141_10109||lt2gene | Not-determined |
| S6_vs_k141_12690||lt2gene | Low-quality |
| S6_vs_k141_31282||lt2gene | Low-quality |
| S6_vs_k141_12703||lt2gene | Not-determined |
| S6_vs_k141_31286||lt2gene | Low-quality |
| S6_vs_k141_12739||lt2gene | Not-determined |
| S6_vs_k141_38512||lt2gene | Low-quality |
| S6_vs_k141_12754||lt2gene | Not-determined |
| S6_vs_k141_10214||lt2gene | Low-quality |
| S6_vs_k141_38528||lt2gene | Not-determined |
| S6_vs_k141_45732||lt2gene | Not-determined |
| S6_vs_k141_7827||lt2gene | Low-quality |
| S6_vs_k141_29073||lt2gene | Low-quality |
| S6_vs_k141_52886||lt2gene | Low-quality |
| S6_vs_k141_47755||lt2gene | Low-quality |
| S6_vs_k141_43191||lt2gene | Low-quality |
| S6_vs_k141_17521||lt2gene | Low-quality |
| S6_vs_k141_36083||lt2gene | Low-quality |
| S6_vs_k141_7852||lt2gene | Not-determined |
| S6_vs_k141_54562||lt2gene | Not-determined |
| S6_vs_k141_45788||lt2gene | Not-determined |
| S6_vs_k141_14985||lt2gene | Not-determined |
| S6_vs_k141_7887||lt2gene | Not-determined |
| S6_vs_k141_24682||lt2gene | Not-determined |
| S6_vs_k141_54583||lt2gene | Low-quality |
| S6_vs_k141_7893||lt2gene | Not-determined |
| S6_vs_k141_36110||lt2gene | Not-determined |
| S6_vs_k141_36114||lt2gene | Not-determined |
| S6_vs_k141_54591||lt2gene | Low-quality |
| S6_vs_k141_45823||lt2gene | Low-quality |
| S6_vs_k141_987||lt2gene | Low-quality |
| S6_vs_k141_34015||lt2gene | Low-quality |
| S6_vs_k141_10281||lt2gene | Low-quality |
| S6_vs_k141_34043||lt2gene | Not-determined |
| S6_vs_k141_29114||lt2gene | Low-quality |
| S6_vs_k141_15006||lt2gene | Low-quality |
| S6_vs_k141_3437||lt2gene | Low-quality |
| S6_vs_k141_3441||lt2gene | Low-quality |
| S6_vs_k141_7993||lt2gene | Low-quality |
| S6_vs_k141_17563||lt2gene | Low-quality |
| S6_vs_k141_29144||lt2gene | Low-quality |
| S6_vs_k141_15046||lt2gene | Low-quality |
| S6_vs_k141_38645||lt2gene | Not-determined |
| S6_vs_k141_1059||lt2gene | Low-quality |
| S6_vs_k141_15057||lt2gene | Low-quality |
| S6_vs_k141_38672||lt2gene | Low-quality |
| S6_vs_k141_40922||lt2gene | Low-quality |
| S6_vs_k141_5497||lt2gene | Low-quality |
| S6_vs_k141_54613||lt2gene | Not-determined |
| S6_vs_k141_36180||lt2gene | Low-quality |
| S6_vs_k141_26547||lt2gene | Low-quality |
| S6_vs_k141_26554||lt2gene | Not-determined |
| S6_vs_k141_52943||lt2gene | Low-quality |
| S6_vs_k141_8031||lt2gene | Low-quality |
| S6_vs_k141_43274||lt2gene | Low-quality |
| S6_vs_k141_21724||lt2gene | Low-quality |
| S6_vs_k141_21728||lt2gene | Low-quality |
| S6_vs_k141_12817||lt2gene | Not-determined |
| S6_vs_k141_54663||lt2gene | Low-quality |
| S6_vs_k141_24714||lt2gene | Low-quality |
| S6_vs_k141_12835||lt2gene | Not-determined |
| S6_vs_k141_50244||lt2gene | Low-quality |
| S6_vs_k141_43313||lt2gene | Not-determined |
| S6_vs_k141_3510||lt2gene | Not-determined |
| S6_vs_k141_31398||lt2gene | Not-determined |
| S6_vs_k141_36259||lt2gene | Not-determined |
| S6_vs_k141_21836||lt2gene | Low-quality |
| S6_vs_k141_24790||lt2gene | Not-determined |
| S6_vs_k141_53038||lt2gene | Low-quality |
| S6_vs_k141_54720||lt2gene | Low-quality |
| S6_vs_k141_12867||lt2gene | Low-quality |
| S6_vs_k141_21850||lt2gene | Not-determined |
| S6_vs_k141_29237||lt2gene | Low-quality |
| S6_vs_k141_1098||lt2gene | Low-quality |
| S6_vs_k141_1103||lt2gene | Low-quality |
| S6_vs_k141_31429||lt2gene | Low-quality |
| S6_vs_k141_3536||lt2gene | Low-quality |
| S6_vs_k141_3538||lt2gene | Not-determined |
| S6_vs_k141_15160||lt2gene | Low-quality |
| S6_vs_k141_1110||lt2gene | Not-determined |
| S6_vs_k141_17607||lt2gene | Low-quality |
| S6_vs_k141_26579||lt2gene | Low-quality |
| S6_vs_k141_1116||lt2gene | Low-quality |
| S6_vs_k141_54745||lt2gene | Low-quality |
| S6_vs_k141_34220||lt2gene | Not-determined |
| S6_vs_k141_3558||lt2gene | Low-quality |
| S6_vs_k141_36305||lt2gene | Not-determined |
| S6_vs_k141_21875||lt2gene | Low-quality |
| S6_vs_k141_47998||lt2gene | Not-determined |
| S6_vs_k141_1128||lt2gene | Low-quality |
| S6_vs_k141_50325||lt2gene | Low-quality |
| S6_vs_k141_40985||lt2gene | Low-quality |
| S6_vs_k141_40990||lt2gene | Not-determined |
| S6_vs_k141_38785||lt2gene | Low-quality |
| S6_vs_k141_3600||lt2gene | Low-quality |
| S6_vs_k141_43389||lt2gene | Low-quality |
| S6_vs_k141_10363||lt2gene | Low-quality |
| S6_vs_k141_31533||lt2gene | Low-quality |
| S6_vs_k141_10391||lt2gene | Low-quality |
| S6_vs_k141_19386||lt2gene | Low-quality |
| S6_vs_k141_41025||lt2gene | Low-quality |
| S6_vs_k141_31558||lt2gene | Low-quality |
| S6_vs_k141_15183||lt2gene | Low-quality |
| S6_vs_k141_17677||lt2gene | Not-determined |
| S6_vs_k141_54869||lt2gene | Low-quality |
| S6_vs_k141_43455||lt2gene | Low-quality |
| S6_vs_k141_10444||lt2gene | Low-quality |
| S6_vs_k141_3694||lt2gene | Low-quality |
| S6_vs_k141_38872||lt2gene | Not-determined |
| S6_vs_k141_50389||lt2gene | Not-determined |
| S6_vs_k141_34312||lt2gene | Not-determined |
| S6_vs_k141_3706||lt2gene | Low-quality |
| S6_vs_k141_31580||lt2gene | Low-quality |
| S6_vs_k141_12996||lt2gene | Low-quality |
| S6_vs_k141_54878||lt2gene | Low-quality |
| S6_vs_k141_36335||lt2gene | Low-quality |
| S6_vs_k141_36339||lt2gene | Low-quality |
| S6_vs_k141_19444||lt2gene | Low-quality |
| S6_vs_k141_10529||lt2gene | Low-quality |
| S6_vs_k141_34400||lt2gene | Not-determined |
| S6_vs_k141_29435||lt2gene | Not-determined |
| S6_vs_k141_48036||lt2gene | Low-quality |
| S6_vs_k141_48045||lt2gene | Low-quality |
| S6_vs_k141_48049||lt2gene | Low-quality |
| S6_vs_k141_31653||lt2gene | Low-quality |
| S6_vs_k141_31656||lt2gene | Low-quality |
| S6_vs_k141_10568||lt2gene | Not-determined |
| S6_vs_k141_15330||lt2gene | Low-quality |
| S6_vs_k141_21995||lt2gene | Low-quality |
| S6_vs_k141_21999||lt2gene | Not-determined |
| S6_vs_k141_3807||lt2gene | Low-quality |
| S6_vs_k141_8179||lt2gene | Not-determined |
| S6_vs_k141_8182||lt2gene | Low-quality |
| S6_vs_k141_8187||lt2gene | Low-quality |
| S6_vs_k141_36384||lt2gene | Not-determined |
| S6_vs_k141_1171||lt2gene | Low-quality |
| S6_vs_k141_15345||lt2gene | Not-determined |
| S6_vs_k141_22020||lt2gene | Low-quality |
| S6_vs_k141_3835||lt2gene | Not-determined |
| S6_vs_k141_13048||lt2gene | Not-determined |
| S6_vs_k141_29541||lt2gene | Low-quality |

SG_vs_k141_38947||lt2gene Not-determined
SG_vs_k141_31750||lt2gene Low-quality
SG_vs_k141_3900||lt2gene Low-quality
SG_vs_k141_3903||lt2gene Low-quality
SG_vs_k141_29591||lt2gene Low-quality
SG_vs_k141_41114||lt2gene Low-quality
SG_vs_k141_22090||lt2gene Low-quality
SG_vs_k141_29605||lt2gene Low-quality
SG_vs_k141_39016||lt2gene Low-quality
SG_vs_k141_22103||lt2gene Not-determined
SG_vs_k141_15421||lt2gene Low-quality
SG_vs_k141_53268||lt2gene Low-quality
SG_vs_k141_13179||lt2gene Low-quality
SG_vs_k141_25040||lt2gene Low-quality
SG_vs_k141_13215||lt2gene Low-quality
SG_vs_k141_34506||lt2gene Low-quality
SG_vs_k141_22147||lt2gene Low-quality
SG_vs_k141_10737||lt2gene Low-quality
SG_vs_k141_25070||lt2gene Low-quality
SG_vs_k141_18013||lt2gene Not-determined
SG_vs_k141_31839||lt2gene Not-determined
SG_vs_k141_50586||lt2gene Low-quality
SG_vs_k141_36532||lt2gene Not-determined
SG_vs_k141_29731||lt2gene Low-quality
SG_vs_k141_48236||lt2gene Low-quality
SG_vs_k141_1315||lt2gene Low-quality
SG_vs_k141_34599||lt2gene Low-quality
SG_vs_k141_41271||lt2gene Not-determined
SG_vs_k141_34609||lt2gene Low-quality
SG_vs_k141_1338||lt2gene Not-determined
SG_vs_k141_41299||lt2gene Low-quality
SG_vs_k141_3987||lt2gene Not-determined
SG_vs_k141_34630||lt2gene Low-quality
SG_vs_k141_46157||lt2gene Low-quality
SG_vs_k141_10818||lt2gene Low-quality
SG_vs_k141_34648||lt2gene Low-quality
SG_vs_k141_22243||lt2gene Low-quality
SG_vs_k141_13317||lt2gene Low-quality
SG_vs_k141_15502||lt2gene Low-quality
SG_vs_k141_5972||lt2gene Not-determined
SG_vs_k141_41336||lt2gene Not-determined
SG_vs_k141_15512||lt2gene Low-quality
SG_vs_k141_13334||lt2gene Not-determined
SG_vs_k141_41347||lt2gene Not-determined
SG_vs_k141_15517||lt2gene Not-determined
SG_vs_k141_15519||lt2gene Not-determined
SG_vs_k141_25170||lt2gene Low-quality
SG_vs_k141_50666||lt2gene Low-quality
SG_vs_k141_18121||lt2gene Low-quality
SG_vs_k141_4062||lt2gene Low-quality
SG_vs_k141_46199||lt2gene Low-quality
SG_vs_k141_31937||lt2gene Not-determined
SG_vs_k141_41379||lt2gene Low-quality
SG_vs_k141_19599||lt2gene Not-determined
SG_vs_k141_19602||lt2gene Low-quality
SG_vs_k141_13383||lt2gene Low-quality
SG_vs_k141_50697||lt2gene Low-quality
SG_vs_k141_26986||lt2gene Low-quality
SG_vs_k141_15549||lt2gene Low-quality
SG_vs_k141_8436||lt2gene Low-quality
SG_vs_k141_19620||lt2gene Low-quality
SG_vs_k141_18178||lt2gene Low-quality
SG_vs_k141_31960||lt2gene Low-quality
SG_vs_k141_15571||lt2gene Low-quality
SG_vs_k141_19656||lt2gene Low-quality
SG_vs_k141_25222||lt2gene Not-determined
SG_vs_k141_27015||lt2gene Low-quality
SG_vs_k141_41449||lt2gene Low-quality
SG_vs_k141_22298||lt2gene Low-quality
SG_vs_k141_41459||lt2gene Low-quality
SG_vs_k141_41460||lt2gene Not-determined
SG_vs_k141_15600||lt2gene Not-determined
SG_vs_k141_27030||lt2gene Low-quality
SG_vs_k141_32013||lt2gene Low-quality
SG_vs_k141_4143||lt2gene Low-quality
SG_vs_k141_27040||lt2gene Low-quality
SG_vs_k141_8528||lt2gene Low-quality
SG_vs_k141_41492||lt2gene Not-determined
SG_vs_k141_46272||lt2gene Not-determined
SG_vs_k141_6082||lt2gene Low-quality
SG_vs_k141_6085||lt2gene Not-determined
SG_vs_k141_48359||lt2gene Low-quality
SG_vs_k141_43687||lt2gene Low-quality
SG_vs_k141_1399||lt2gene Low-quality
SG_vs_k141_4202||lt2gene Low-quality
SG_vs_k141_19790||lt2gene Low-quality
SG_vs_k141_19792||lt2gene Low-quality
SG_vs_k141_15727||lt2gene Not-determined
SG_vs_k141_4239||lt2gene Low-quality
SG_vs_k141_46347||lt2gene Low-quality
SG_vs_k141_13509||lt2gene Low-quality
SG_vs_k141_4246||lt2gene Low-quality
SG_vs_k141_6123||lt2gene Low-quality
SG_vs_k141_4253||lt2gene Low-quality
SG_vs_k141_19797||lt2gene Low-quality
SG_vs_k141_10949||lt2gene Low-quality
SG_vs_k141_8633||lt2gene Low-quality
SG_vs_k141_41603||lt2gene Not-determined
SG_vs_k141_41622||lt2gene Low-quality
SG_vs_k141_22364||lt2gene Low-quality
SG_vs_k141_8642||lt2gene Low-quality
SG_vs_k141_8655||lt2gene Not-determined
SG_vs_k141_15764||lt2gene Low-quality
SG_vs_k141_55091||lt2gene Low-quality
SG_vs_k141_34875||lt2gene Low-quality
SG_vs_k141_1479||lt2gene Low-quality
SG_vs_k141_19817||lt2gene Low-quality
SG_vs_k141_27172||lt2gene Not-determined
SG_vs_k141_32050||lt2gene Not-determined
SG_vs_k141_34893||lt2gene Not-determined
SG_vs_k141_50735||lt2gene Low-quality
SG_vs_k141_55098||lt2gene Low-quality
SG_vs_k141_15797||lt2gene Not-determined
SG_vs_k141_53428||lt2gene Low-quality
SG_vs_k141_27187||lt2gene Low-quality
SG_vs_k141_19826||lt2gene Low-quality
SG_vs_k141_8722||lt2gene Low-quality
SG_vs_k141_13641||lt2gene Not-determined
SG_vs_k141_8731||lt2gene Low-quality
SG_vs_k141_8736||lt2gene Low-quality
SG_vs_k141_41674||lt2gene Not-determined
SG_vs_k141_13665||lt2gene Low-quality
SG_vs_k141_29937||lt2gene Low-quality
SG_vs_k141_41691||lt2gene Not-determined
SG_vs_k141_41697||lt2gene Low-quality
SG_vs_k141_19869||lt2gene Low-quality
SG_vs_k141_46404||lt2gene Low-quality
SG_vs_k141_53481||lt2gene Low-quality
SG_vs_k141_22438||lt2gene Low-quality
SG_vs_k141_50786||lt2gene Low-quality
SG_vs_k141_29988||lt2gene Not-determined
SG_vs_k141_41728||lt2gene Low-quality
SG_vs_k141_53499||lt2gene Low-quality
SG_vs_k141_8776||lt2gene Low-quality
SG_vs_k141_11015||lt2gene Low-quality
SG_vs_k141_53527||lt2gene Not-determined
SG_vs_k141_18352||lt2gene Low-quality
SG_vs_k141_4382||lt2gene Not-determined
SG_vs_k141_18356||lt2gene Low-quality
SG_vs_k141_15871||lt2gene Low-quality
SG_vs_k141_4388||lt2gene Low-quality
SG_vs_k141_11033||lt2gene Not-determined
SG_vs_k141_55241||lt2gene Not-determined
SG_vs_k141_15891||lt2gene Low-quality
SG_vs_k141_55258||lt2gene Low-quality
SG_vs_k141_11067||lt2gene Low-quality
SG_vs_k141_36780||lt2gene Low-quality
SG_vs_k141_22516||lt2gene Not-determined
SG_vs_k141_34918||lt2gene Low-quality
SG_vs_k141_32174||lt2gene Low-quality
SG_vs_k141_46496||lt2gene Low-quality
SG_vs_k141_55272||lt2gene Low-quality
SG_vs_k141_36794||lt2gene Low-quality
SG_vs_k141_36799||lt2gene Not-determined
SG_vs_k141_41817||lt2gene Not-determined
SG_vs_k141_43810||lt2gene Low-quality
SG_vs_k141_41825||lt2gene Low-quality
SG_vs_k141_13723||lt2gene Low-quality
SG_vs_k141_1506||lt2gene Not-determined
SG_vs_k141_11091||lt2gene Not-determined
SG_vs_k141_39137||lt2gene Low-quality
SG_vs_k141_50849||lt2gene Low-quality
SG_vs_k141_32238||lt2gene Not-determined
SG_vs_k141_34963||lt2gene Low-quality
SG_vs_k141_15953||lt2gene Low-quality
SG_vs_k141_55404||lt2gene Low-quality
SG_vs_k141_50882||lt2gene Low-quality
SG_vs_k141_53643||lt2gene Not-determined
SG_vs_k141_35007||lt2gene Low-quality
SG_vs_k141_15984||lt2gene Low-quality
SG_vs_k141_18437||lt2gene Low-quality
SG_vs_k141_25424||lt2gene Low-quality
SG_vs_k141_53691||lt2gene Low-quality
SG_vs_k141_50908||lt2gene Not-determined
SG_vs_k141_46609||lt2gene Low-quality
SG_vs_k141_46613||lt2gene Low-quality
SG_vs_k141_41917||lt2gene Low-quality
SG_vs_k141_36868||lt2gene Low-quality
SG_vs_k141_16017||lt2gene Not-determined
SG_vs_k141_55473||lt2gene Low-quality
SG_vs_k141_55476||lt2gene Low-quality
SG_vs_k141_18503||lt2gene Not-determined
SG_vs_k141_55500||lt2gene Low-quality
SG_vs_k141_13808||lt2gene Low-quality
SG_vs_k141_39194||lt2gene Low-quality
SG_vs_k141_11134||lt2gene Low-quality
SG_vs_k141_22720||lt2gene Not-determined
SG_vs_k141_22722||lt2gene Low-quality
SG_vs_k141_22726||lt2gene Low-quality
SG_vs_k141_32347||lt2gene Not-determined
SG_vs_k141_39226||lt2gene Low-quality
SG_vs_k141_18541||lt2gene Not-determined
SG_vs_k141_11184||lt2gene Low-quality
SG_vs_k141_42046||lt2gene Low-quality
SG_vs_k141_4593||lt2gene Low-quality
SG_vs_k141_22763||lt2gene Low-quality
SG_vs_k141_18584||lt2gene Not-determined
SG_vs_k141_48467||lt2gene Low-quality
SG_vs_k141_50988||lt2gene Low-quality
SG_vs_k141_8807||lt2gene Low-quality
SG_vs_k141_13938||lt2gene Low-quality
SG_vs_k141_8816||lt2gene Low-quality
SG_vs_k141_6533||lt2gene Not-determined
SG_vs_k141_6541||lt2gene Low-quality
SG_vs_k141_43854||lt2gene Low-quality
SG_vs_k141_46701||lt2gene Low-quality
SG_vs_k141_30262||lt2gene Not-determined
SG_vs_k141_30271||lt2gene Low-quality
SG_vs_k141_32499||lt2gene Low-quality
SG_vs_k141_22830||lt2gene Low-quality
SG_vs_k141_48539||lt2gene Low-quality
SG_vs_k141_6555||lt2gene Not-determined
SG_vs_k141_27535||lt2gene Low-quality
SG_vs_k141_53773||lt2gene Low-quality
SG_vs_k141_20015||lt2gene Low-quality
SG_vs_k141_30284||lt2gene Not-determined
SG_vs_k141_51002||lt2gene Low-quality
SG_vs_k141_48550||lt2gene Low-quality
SG_vs_k141_42163||lt2gene Low-quality
SG_vs_k141_30296||lt2gene Low-quality
SG_vs_k141_32557||lt2gene Low-quality
SG_vs_k141_39305||lt2gene Low-quality
SG_vs_k141_42196||lt2gene Low-quality
SG_vs_k141_1626||lt2gene Low-quality
SG_vs_k141_42222||lt2gene Low-quality
SG_vs_k141_53809||lt2gene Low-quality
SG_vs_k141_22888||lt2gene Low-quality
SG_vs_k141_6601||lt2gene Low-quality
SG_vs_k141_6602||lt2gene Low-quality
SG_vs_k141_42269||lt2gene Low-quality
SG_vs_k141_22909||lt2gene Low-quality
SG_vs_k141_39360||lt2gene Not-determined
SG_vs_k141_22919||lt2gene Low-quality
SG_vs_k141_1650||lt2gene Low-quality
SG_vs_k141_42282||lt2gene Low-quality
SG_vs_k141_18628||lt2gene Low-quality
SG_vs_k141_42303||lt2gene Not-determined
SG_vs_k141_14020||lt2gene Low-quality
SG_vs_k141_32622||lt2gene Low-quality
SG_vs_k141_39397||lt2gene Low-quality
SG_vs_k141_11255||lt2gene Low-quality
SG_vs_k141_22961||lt2gene Not-determined
SG_vs_k141_20139||lt2gene Not-determined
SG_vs_k141_32652||lt2gene Low-quality
SG_vs_k141_53870||lt2gene Low-quality
SG_vs_k141_18676||lt2gene Low-quality
SG_vs_k141_32692||lt2gene Not-determined
SG_vs_k141_1774||lt2gene Low-quality
SG_vs_k141_48597||lt2gene Low-quality
SG_vs_k141_39477||lt2gene Not-determined
SG_vs_k141_39478||lt2gene Low-quality
SG_vs_k141_43988||lt2gene Not-determined
SG_vs_k141_1788||lt2gene Low-quality
SG_vs_k141_16218||lt2gene Not-determined
SG_vs_k141_32743||lt2gene Low-quality
SG_vs_k141_44014||lt2gene Not-determined
SG_vs_k141_16234||lt2gene Low-quality
SG_vs_k141_16235||lt2gene Low-quality
SG_vs_k141_11330||lt2gene Not-determined
SG_vs_k141_16250||lt2gene Low-quality
SG_vs_k141_11336||lt2gene Low-quality
SG_vs_k141_44038||lt2gene Low-quality
SG_vs_k141_23090||lt2gene Low-quality
SG_vs_k141_55543||lt2gene Low-quality
SG_vs_k141_32789||lt2gene Not-determined
SG_vs_k141_6730||lt2gene Not-determined
SG_vs_k141_6731||lt2gene Low-quality
SG_vs_k141_16280||lt2gene Low-quality
SG_vs_k141_20252||lt2gene Low-quality
SG_vs_k141_51033||lt2gene Low-quality
SG_vs_k141_18740||lt2gene Low-quality
SG_vs_k141_53946||lt2gene Not-determined
SG_vs_k141_46787||lt2gene Not-determined
SG_vs_k141_42402||lt2gene Not-determined
SG_vs_k141_27568||lt2gene Low-quality
SG_vs_k141_16289||lt2gene Low-quality
SG_vs_k141_20264||lt2gene Low-quality
SG_vs_k141_44071||lt2gene Low-quality
SG_vs_k141_32836||lt2gene Low-quality
SG_vs_k141_11355||lt2gene Low-quality
SG_vs_k141_53968||lt2gene Low-quality
SG_vs_k141_23164||lt2gene Low-quality
SG_vs_k141_6764||lt2gene Not-determined
SG_vs_k141_44146||lt2gene Low-quality
SG_vs_k141_18775||lt2gene Low-quality
SG_vs_k141_11407||lt2gene Not-determined
SG_vs_k141_32869||lt2gene Low-quality
SG_vs_k141_6774||lt2gene Low-quality
SG_vs_k141_8925||lt2gene Low-quality
SG_vs_k141_18779||lt2gene Not-determined
SG_vs_k141_23169||lt2gene Low-quality
SG_vs_k141_23175||lt2gene Low-quality
SG_vs_k141_44171||lt2gene Not-determined
SG_vs_k141_44175||lt2gene Not-determined
SG_vs_k141_6816||lt2gene Low-quality
SG_vs_k141_6824||lt2gene Low-quality
SG_vs_k141_39574||lt2gene Low-quality
SG_vs_k141_30455||lt2gene Not-determined
SG_vs_k141_39594||lt2gene Low-quality
SG_vs_k141_23242||lt2gene Low-quality
SG_vs_k141_44219||lt2gene Low-quality
SG_vs_k141_27648||lt2gene Low-quality
SG_vs_k141_11468||lt2gene Low-quality
SG_vs_k141_32944||lt2gene Not-determined
SG_vs_k141_32953||lt2gene Not-determined
SG_vs_k141_35341||lt2gene Not-determined
SG_vs_k141_6855||lt2gene Low-quality
SG_vs_k141_20395||lt2gene Low-quality
SG_vs_k141_20401||lt2gene Low-quality
SG_vs_k141_11518||lt2gene Low-quality
SG_vs_k141_6869||lt2gene Low-quality
SG_vs_k141_27688||lt2gene Low-quality
SG_vs_k141_27689||lt2gene Not-determined
SG_vs_k141_11526||lt2gene Low-quality
SG_vs_k141_46894||lt2gene Low-quality
SG_vs_k141_23361||lt2gene Not-determined
SG_vs_k141_6909||lt2gene Low-quality
SG_vs_k141_30489||lt2gene Low-quality
SG_vs_k141_46925||lt2gene Not-determined
SG_vs_k141_25731||lt2gene Low-quality
SG_vs_k141_16340||lt2gene Not-determined
SG_vs_k141_46930||lt2gene Low-quality
SG_vs_k141_39638||lt2gene Low-quality
SG_vs_k141_16357||lt2gene Low-quality
SG_vs_k141_30507||lt2gene Low-quality
SG_vs_k141_23413||lt2gene Low-quality
SG_vs_k141_44230||lt2gene Not-determined
SG_vs_k141_55608||lt2gene Low-quality
SG_vs_k141_30533||lt2gene Low-quality
SG_vs_k141_6942||lt2gene Low-quality
SG_vs_k141_39707||lt2gene Low-quality
SG_vs_k141_39710||lt2gene Not-determined
SG_vs_k141_30558||lt2gene Not-determined
SG_vs_k141_23458||lt2gene Low-quality
SG_vs_k141_37053||lt2gene Low-quality
SG_vs_k141_7009||lt2gene Low-quality
SG_vs_k141_44270||lt2gene Low-quality
SG_vs_k141_25833||lt2gene Low-quality
SG_vs_k141_16398||lt2gene Low-quality
SG_vs_k141_25840||lt2gene Low-quality
SG_vs_k141_11697||lt2gene Low-quality
SG_vs_k141_55724||lt2gene Low-quality
SG_vs_k141_30606||lt2gene Low-quality
SG_vs_k141_49026||lt2gene Low-quality
SG_vs_k141_23516||lt2gene Not-determined
SG_vs_k141_39808||lt2gene Low-quality
SG_vs_k141_11724||lt2gene Low-quality
SG_vs_k141_49046||lt2gene Low-quality
SG_vs_k141_55752||lt2gene Low-quality
SG_vs_k141_47008||lt2gene Low-quality
SG_vs_k141_39833||lt2gene Low-quality
SG_vs_k141_47010||lt2gene Low-quality
SG_vs_k141_44375||lt2gene Low-quality
SG_vs_k141_20484||lt2gene Low-quality
SG_vs_k141_37181||lt2gene Not-determined
SG_vs_k141_9037||lt2gene Low-quality
SG_vs_k141_49095||lt2gene Low-quality
SG_vs_k141_49102||lt2gene Low-quality
SG_vs_k141_9081||lt2gene Not-determined
SG_vs_k141_37242||lt2gene Low-quality
SG_vs_k141_44456||lt2gene Not-determined
SG_vs_k141_37257||lt2gene Not-determined
SG_vs_k141_40002||lt2gene Not-determined
SG_vs_k141_40003||lt2gene Not-determined
SG_vs_k141_37273||lt2gene Not-determined
SG_vs_k141_9141||lt2gene Not-determined
SG_vs_k141_9146||lt2gene Low-quality
SG_vs_k141_37288||lt2gene Low-quality
SG_vs_k141_2020||lt2gene Low-quality
SG_vs_k141_44497||lt2gene Low-quality
SG_vs_k141_20593||lt2gene Low-quality
SG_vs_k141_9184||lt2gene Low-quality
SG_vs_k141_55842||lt2gene Not-determined
SG_vs_k141_20615||lt2gene Low-quality
SG_vs_k141_51127||lt2gene Low-quality
SG_vs_k141_51139||lt2gene Low-quality
SG_vs_k141_2096||lt2gene Not-determined
SG_vs_k141_27732||lt2gene Not-determined
SG_vs_k141_9263||lt2gene Low-quality
SG_vs_k141_9275||lt2gene Not-determined
SG_vs_k141_44537||lt2gene Low-quality
SG_vs_k141_49326||lt2gene Low-quality
SG_vs_k141_56015||lt2gene Low-quality
SG_vs_k141_44582||lt2gene Not-determined
SG_vs_k141_20635||lt2gene Low-quality
SG_vs_k141_49362||lt2gene Low-quality
SG_vs_k141_56027||lt2gene Low-quality
SG_vs_k141_20669||lt2gene Not-determined
SG_vs_k141_51284||lt2gene Low-quality
SG_vs_k141_47106||lt2gene Low-quality
SG_vs_k141_37454||lt2gene Low-quality
SG_vs_k141_37473||lt2gene Not-determined
SG_vs_k141_44647||lt2gene Low-quality
SG_vs_k141_37478||lt2gene Low-quality
SG_vs_k141_56043||lt2gene Low-quality
SG_vs_k141_44707||lt2gene Low-quality
SG_vs_k141_56118||lt2gene Not-determined
SG_vs_k141_44738||lt2gene Not-determined
SG_vs_k141_56165||lt2gene Low-quality
SG_vs_k141_56168||lt2gene Low-quality
SG_vs_k141_56179||lt2gene Low-quality
SG_vs_k141_56190||lt2gene Low-quality
SG_vs_k141_20750||lt2gene Low-quality
SG_vs_k141_37617||lt2gene Low-quality
SG_vs_k141_56226||lt2gene Low-quality
SG_vs_k141_20787||lt2gene Low-quality
SG_vs_k141_37653||lt2gene Low-quality
SG_vs_k141_37666||lt2gene Low-quality
SG_vs_k141_37681||lt2gene Low-quality
SG_vs_k141_51545||lt2gene Low-quality
SG_vs_k141_20906||lt2gene Low-quality
SG_vs_k141_2355||lt2gene Not-determined
SG_vs_k141_51606||lt2gene Not-determined
SG_vs_k141_20934||lt2gene Low-quality
SG_vs_k141_56270||lt2gene Not-determined
SG_vs_k141_56283||lt2gene Low-quality
SG_vs_k141_51640||lt2gene Not-determined
SG_vs_k141_20965||lt2gene Not-determined
SG_vs_k141_20997||lt2gene Not-determined
SG_vs_k141_51684||lt2gene Low-quality
SG_vs_k141_51745||lt2gene Low-quality
SG_vs_k141_21028||lt2gene Low-quality
SG_vs_k141_21035||lt2gene Not-determined
SG_vs_k141_28053||lt2gene Low-quality
SG_vs_k141_51796||lt2gene Low-quality
SG_vs_k141_28145||lt2gene Low-quality
SG_vs_k141_28153||lt2gene Not-determined
SG_vs_k141_28199||lt2gene Low-quality
SG_vs_k141_21116||lt2gene Low-quality
SG_vs_k141_21177||lt2gene Low-quality
SG_vs_k141_28249||lt2gene Not-determined
SB_extracted_k141_182226 Not-determined
SB_extracted_k141_43391 Not-determined
SB_extracted_k141_69424 Low-quality
SB_extracted_k141_43393 Not-determined
SB_extracted_k141_104133 Not-determined
SB_extracted_k141_8679 Not-determined
SB_extracted_k141_138842 Low-quality
SB_extracted_k141_112812 Not-determined
SB_extracted_k141_164873 Not-determined
SB_extracted_k141_138843 Not-determined
SB_extracted_k141_190907 Not-determined
SB_extracted_k141_95460 Not-determined
SB_extracted_k141_2 Not-determined
SB_extracted_k141_17358 Not-determined
SB_extracted_k141_60748 Not-determined
SB_extracted_k141_5 Not-determined
SB_extracted_k141_182228 Not-determined
SB_extracted_k141_7 Not-determined
SB_extracted_k141_147525 Not-determined
SB_extracted_k141_182231 Not-determined
SB_extracted_k141_78109 Not-determined
SB_extracted_k141_112821 Not-determined
SB_extracted_k141_190914 Not-determined
SB_extracted_k141_190915 Not-determined
SB_extracted_k141_26040 Not-determined
SB_extracted_k141_147527 Not-determined
SB_extracted_k141_190916 Not-determined
SB_extracted_k141_199590 Not-determined
SB_extracted_k141_156204 Not-determined
SB_extracted_k141_69435 Not-determined
SB_extracted_k141_104142 Low-quality
SB_extracted_k141_112826 Low-quality
SB_extracted_k141_130171 Not-determined
SB_extracted_k141_112828 Not-determined
SB_extracted_k141_17366 Not-determined
SB_extracted_k141_104147 Not-determined
SB_extracted_k141_60756 Not-determined
SB_extracted_k141_130175 Not-determined
SB_extracted_k141_173554 Not-determined
SB_extracted_k141_199598 Not-determined
SB_extracted_k141_164879 Not-determined
SB_extracted_k141_190921 Not-determined
SB_extracted_k141_138849 Not-determined
SB_extracted_k141_164883 Not-determined
SB_extracted_k141_190923 Not-determined
SB_extracted_k141_112838 Not-determined
SB_extracted_k141_8689 Not-determined
SB_extracted_k141_86789 Low-quality
SB_extracted_k141_26055 Not-determined
SB_extracted_k141_86790 Not-determined
SB_extracted_k141_95476 Not-determined
SB_extracted_k141_52084 Not-determined
SB_extracted_k141_52085 Not-determined
SB_extracted_k141_52086 Not-determined
SB_extracted_k141_182241 Not-determined
SB_extracted_k141_52088 Not-determined
SB_extracted_k141_173561 Not-determined
SB_extracted_k141_26060 Not-determined
SB_extracted_k141_173564 Not-determined
SB_extracted_k141_156216 Not-determined
SB_extracted_k141_164890 Not-determined
SB_extracted_k141_121506 Not-determined
SB_extracted_k141_190931 Not-determined
SB_extracted_k141_52093 Not-determined
SB_extracted_k141_104151 Not-determined
SB_extracted_k141_86801 Not-determined
SB_extracted_k141_104152 Not-determined
SB_extracted_k141_156227 Not-determined
SB_extracted_k141_112846 Not-determined
SB_extracted_k141_60782 Not-determined
SB_extracted_k141_69444 Low-quality
SB_extracted_k141_138860 Not-determined
SB_extracted_k141_190940 Not-determined
SB_extracted_k141_156232 Not-determined
SB_extracted_k141_69447 Not-determined
SB_extracted_k141_26074 Not-determined
SB_extracted_k141_190944 Not-determined
SB_extracted_k141_86808 Not-determined
SB_extracted_k141_69448 Not-determined
SB_extracted_k141_199611 Not-determined
SB_extracted_k141_182253 Not-determined
SB_extracted_k141_104163 Not-determined
SB_extracted_k141_138864 Not-determined
SB_extracted_k141_37 Not-determined
SB_extracted_k141_60794 Low-quality
SB_extracted_k141_182256 Not-determined
SB_extracted_k141_121522 Not-determined
SB_extracted_k141_156234 Low-quality
SB_extracted_k141_8697 Not-determined
SB_extracted_k141_130185 Not-determined
SB_extracted_k141_104174 Not-determined
SB_extracted_k141_43422 Not-determined
SB_extracted_k141_112851 Not-determined
SB_extracted_k141_138872 Not-determined
SB_extracted_k141_147561 Not-determined
SB_extracted_k141_121529 Not-determined
SB_extracted_k141_52117 Not-determined
SB_extracted_k141_182262 Not-determined
SB_extracted_k141_78115 Not-determined
SB_extracted_k141_182263 Not-determined
SB_extracted_k141_78117 Not-determined
SB_extracted_k141_26079 Not-determined
SB_extracted_k141_112853 Not-determined
SB_extracted_k141_112854 Not-determined
SB_extracted_k141_78119 Not-determined
SB_extracted_k141_156244 Not-determined
SB_extracted_k141_34762 Not-determined
SB_extracted_k141_95510 Not-determined
SB_extracted_k141_50 Not-determined
SB_extracted_k141_34763 Low-quality
SB_extracted_k141_104186 Not-determined
SB_extracted_k141_156248 Low-quality
SB_extracted_k141_17388 Low-quality
SB_extracted_k141_182271 Not-determined
SB_extracted_k141_43430 Not-determined
SB_extracted_k141_182272 Not-determined
SB_extracted_k141_69462 Not-determined
SB_extracted_k141_43431 Not-determined
SB_extracted_k141_86825 Not-determined
SB_extracted_k141_147564 Not-determined
SB_extracted_k141_199616 Not-determined
SB_extracted_k141_104191 Not-determined
SB_extracted_k141_78130 Not-determined
SB_extracted_k141_199617 Low-quality
SB_extracted_k141_43436 Low-quality
SB_extracted_k141_60799 Not-determined
SB_extracted_k141_78136 Low-quality
SB_extracted_k141_60801 Not-determined
SB_extracted_k141_60802 Not-determined
SB_extracted_k141_78141 Not-determined
SB_extracted_k141_60808 Not-determined
SB_extracted_k141_199621 Not-determined
SB_extracted_k141_95516 Not-determined
SB_extracted_k141_43440 Not-determined
SB_extracted_k141_43441 Not-determined
SB_extracted_k141_86842 Not-determined
SB_extracted_k141_17395 Not-determined
SB_extracted_k141_78156 Not-determined
SB_extracted_k141_199626 Not-determined
SB_extracted_k141_60809 Not-determined
SB_extracted_k141_86843 Not-determined
SB_extracted_k141_69470 Not-determined
SB_extracted_k141_17397 Not-determined
SB_extracted_k141_26096 Not-determined
SB_extracted_k141_138895 Low-quality
SB_extracted_k141_17403 Not-determined
SB_extracted_k141_69475 Not-determined
SB_extracted_k141_17404 Low-quality
SB_extracted_k141_112881 Low-quality
SB_extracted_k141_43450 Not-determined
SB_extracted_k141_147584 Not-determined
SB_extracted_k141_17406 Not-determined
SB_extracted_k141_156266 Not-determined
SB_extracted_k141_190964 Not-determined
SB_extracted_k141_52120 Not-determined
SB_extracted_k141_69477 Not-determined
SB_extracted_k141_17407 Not-determined
SB_extracted_k141_52121 Low-quality

| | |
|---|---|
| S8_extracted_k141_17410 | Not-determined |
| S8_extracted_k141_190967 | Not-determined |
| S8_extracted_k141_69479 | Not-determined |
| S8_extracted_k141_17411 | Not-determined |
| S8_extracted_k141_86856 | Not-determined |
| S8_extracted_k141_78162 | Low-quality |
| S8_extracted_k141_156271 | Not-determined |
| S8_extracted_k141_156273 | Not-determined |
| S8_extracted_k141_17416 | Low-quality |
| S8_extracted_k141_70 | Low-quality |
| S8_extracted_k141_156277 | Not-determined |
| S8_extracted_k141_17419 | Low-quality |
| S8_extracted_k141_199635 | Low-quality |
| S8_extracted_k141_17420 | Not-determined |
| S8_extracted_k141_164916 | Not-determined |
| S8_extracted_k141_52126 | Not-determined |
| S8_extracted_k141_147590 | Not-determined |
| S8_extracted_k141_130196 | Not-determined |
| S8_extracted_k141_121567 | Not-determined |
| S8_extracted_k141_147591 | Not-determined |
| S8_extracted_k141_86868 | Not-determined |
| S8_extracted_k141_147594 | Low-quality |
| S8_extracted_k141_84 | Not-determined |
| S8_extracted_k141_190982 | Not-determined |
| S8_extracted_k141_156288 | Not-determined |
| S8_extracted_k141_190985 | Not-determined |
| S8_extracted_k141_190986 | Not-determined |
| S8_extracted_k141_121570 | Not-determined |
| S8_extracted_k141_88 | Not-determined |
| S8_extracted_k141_112893 | Not-determined |
| S8_extracted_k141_86872 | Not-determined |
| S8_extracted_k141_69484 | Not-determined |
| S8_extracted_k141_138911 | Low-quality |
| S8_extracted_k141_190988 | Not-determined |
| S8_extracted_k141_60826 | Not-determined |
| S8_extracted_k141_164924 | Not-determined |
| S8_extracted_k141_60828 | Not-determined |
| S8_extracted_k141_147598 | Not-determined |
| S8_extracted_k141_121575 | Low-quality |
| S8_extracted_k141_69486 | Not-determined |
| S8_extracted_k141_164928 | Not-determined |
| S8_extracted_k141_17427 | Not-determined |
| S8_extracted_k141_121578 | Not-determined |
| S8_extracted_k141_164931 | Not-determined |
| S8_extracted_k141_121582 | Low-quality |
| S8_extracted_k141_78168 | Low-quality |
| S8_extracted_k141_26122 | Not-determined |
| S8_extracted_k141_69491 | Not-determined |
| S8_extracted_k141_78169 | Not-determined |
| S8_extracted_k141_130215 | Not-determined |
| S8_extracted_k141_78170 | Not-determined |
| S8_extracted_k141_60842 | Low-quality |
| S8_extracted_k141_52139 | Low-quality |
| S8_extracted_k141_78176 | Not-determined |
| S8_extracted_k141_26129 | Not-determined |
| S8_extracted_k141_26130 | Not-determined |
| S8_extracted_k141_8760 | Low-quality |
| S8_extracted_k141_43474 | Not-determined |
| S8_extracted_k141_164940 | Not-determined |
| S8_extracted_k141_104208 | Low-quality |
| S8_extracted_k141_8763 | Not-determined |
| S8_extracted_k141_69499 | Not-determined |
| S8_extracted_k141_43480 | Not-determined |
| S8_extracted_k141_164946 | Low-quality |
| S8_extracted_k141_52146 | Not-determined |
| S8_extracted_k141_105 | Not-determined |
| S8_extracted_k141_130223 | Not-determined |
| S8_extracted_k141_43481 | Low-quality |
| S8_extracted_k141_8768 | Not-determined |
| S8_extracted_k141_104216 | Not-determined |
| S8_extracted_k141_130224 | Not-determined |
| S8_extracted_k141_69508 | Not-determined |
| S8_extracted_k141_34774 | Not-determined |
| S8_extracted_k141_130226 | Not-determined |
| S8_extracted_k141_86893 | Not-determined |
| S8_extracted_k141_113 | Not-determined |
| S8_extracted_k141_130231 | Not-determined |
| S8_extracted_k141_34777 | Not-determined |
| S8_extracted_k141_86897 | Not-determined |
| S8_extracted_k141_191003 | Not-determined |
| S8_extracted_k141_78186 | Not-determined |
| S8_extracted_k141_130234 | Not-determined |
| S8_extracted_k141_8772 | Not-determined |
| S7_vs_66177\|\|full | Medium-quality |
| S8_extracted_k141_199661 | Not-determined |
| S8_extracted_k141_43487 | Not-determined |
| S8_extracted_k141_130241 | Not-determined |
| S8_extracted_k141_86905 | Not-determined |
| S8_extracted_k141_130242 | Not-determined |
| S8_extracted_k141_173601 | Low-quality |
| S8_extracted_k141_147623 | Not-determined |
| S8_extracted_k141_191013 | Not-determined |
| S8_extracted_k141_119 | Not-determined |
| S8_extracted_k141_173604 | Not-determined |
| S8_extracted_k141_104224 | Low-quality |
| S8_extracted_k141_52155 | Not-determined |
| S8_extracted_k141_138931 | Not-determined |
| S8_extracted_k141_123 | Not-determined |
| S8_extracted_k141_191017 | Not-determined |
| S8_extracted_k141_95536 | Low-quality |
| S8_extracted_k141_138934 | Not-determined |
| S8_extracted_k141_43492 | Not-determined |
| S8_extracted_k141_8785 | Low-quality |
| S8_extracted_k141_52157 | Not-determined |
| S8_extracted_k141_173607 | Not-determined |
| S8_extracted_k141_95539 | Not-determined |
| S8_extracted_k141_138940 | Not-determined |
| S8_extracted_k141_164966 | Not-determined |
| S8_extracted_k141_26157 | Low-quality |
| S8_extracted_k141_86920 | Not-determined |
| S8_extracted_k141_130245 | Not-determined |
| S8_extracted_k141_156313 | Low-quality |
| S8_extracted_k141_60851 | Not-determined |
| S8_extracted_k141_199664 | Not-determined |
| S8_extracted_k141_95544 | Not-determined |
| S8_extracted_k141_52163 | Not-determined |
| S8_extracted_k141_138946 | Not-determined |
| S8_extracted_k141_199665 | Not-determined |
| S8_extracted_k141_147640 | Not-determined |
| S8_extracted_k141_78193 | Not-determined |
| S8_extracted_k141_95545 | Not-determined |
| S8_extracted_k141_17464 | Not-determined |
| S8_extracted_k141_138948 | Not-determined |
| S8_extracted_k141_17465 | Not-determined |
| S8_extracted_k141_138949 | Not-determined |
| S8_extracted_k141_78194 | Not-determined |
| S8_extracted_k141_78195 | Not-determined |
| S8_extracted_k141_191027 | Low-quality |
| S8_extracted_k141_17468 | Not-determined |
| S8_extracted_k141_95548 | Not-determined |
| S8_extracted_k141_191028 | Not-determined |
| S8_extracted_k141_156319 | Low-quality |
| S8_extracted_k141_95550 | Not-determined |
| S8_extracted_k141_112921 | Not-determined |
| S8_extracted_k141_112926 | Not-determined |
| S8_extracted_k141_95554 | Not-determined |
| S8_extracted_k141_34790 | Low-quality |
| S8_extracted_k141_121601 | Not-determined |
| S8_extracted_k141_43504 | Not-determined |
| S8_extracted_k141_147647 | Not-determined |
| S8_extracted_k141_164981 | Not-determined |
| S8_extracted_k141_156322 | Low-quality |
| S8_extracted_k141_140 | Not-determined |
| S8_extracted_k141_34793 | Not-determined |
| S8_extracted_k141_43508 | Not-determined |
| S8_extracted_k141_138962 | Not-determined |
| S8_extracted_k141_156326 | Not-determined |
| S8_extracted_k141_95558 | Low-quality |
| S8_extracted_k141_78202 | Not-determined |
| S8_extracted_k141_156328 | Low-quality |
| S8_extracted_k141_78203 | Not-determined |
| S8_extracted_k141_95562 | Not-determined |
| S8_extracted_k141_156332 | Not-determined |
| S8_extracted_k141_78206 | Not-determined |
| S8_extracted_k141_26179 | Not-determined |
| S8_extracted_k141_121604 | Not-determined |
| S8_extracted_k141_130253 | Low-quality |
| S8_extracted_k141_144 | Not-determined |
| S8_extracted_k141_147664 | Low-quality |
| S8_extracted_k141_130257 | Not-determined |
| S8_extracted_k141_104239 | Not-determined |
| S8_extracted_k141_130258 | Not-determined |
| S8_extracted_k141_34802 | Not-determined |
| S8_extracted_k141_52172 | Not-determined |
| S8_extracted_k141_34804 | Low-quality |
| S8_extracted_k141_86941 | Not-determined |
| S8_extracted_k141_130260 | Not-determined |
| S8_extracted_k141_164990 | Not-determined |
| S8_extracted_k141_151 | Not-determined |
| S8_extracted_k141_199686 | Not-determined |
| S8_extracted_k141_173626 | Not-determined |
| S8_extracted_k141_147666 | Not-determined |
| S8_extracted_k141_8805 | Not-determined |
| S8_extracted_k141_34810 | Not-determined |
| S8_extracted_k141_104242 | Not-determined |
| S8_extracted_k141_191052 | Not-determined |
| S8_extracted_k141_60869 | Not-determined |
| S8_extracted_k141_173632 | Not-determined |
| S8_extracted_k141_173634 | Not-determined |
| S8_extracted_k141_164997 | Not-determined |
| S8_extracted_k141_156341 | Not-determined |
| S8_extracted_k141_112944 | Low-quality |
| S8_extracted_k141_104244 | Not-determined |
| S8_extracted_k141_104245 | Low-quality |
| S8_extracted_k141_8813 | Not-determined |
| S8_extracted_k141_199696 | Not-determined |
| S8_extracted_k141_130267 | Not-determined |
| S8_extracted_k141_165 | Not-determined |
| S8_extracted_k141_121615 | Not-determined |
| S8_extracted_k141_138975 | Not-determined |
| S8_extracted_k141_95579 | Not-determined |
| S8_extracted_k141_104251 | Not-determined |
| S8_extracted_k141_112948 | Not-determined |
| S8_extracted_k141_86960 | Not-determined |
| S8_extracted_k141_156347 | Not-determined |
| S8_extracted_k141_168 | Not-determined |
| S8_extracted_k141_69515 | Low-quality |
| S8_extracted_k141_69516 | Not-determined |
| S8_extracted_k141_26200 | Not-determined |
| S8_extracted_k141_121622 | Not-determined |
| S8_extracted_k141_138976 | Not-determined |
| S8_extracted_k141_121623 | Low-quality |
| S8_extracted_k141_34817 | Not-determined |
| S8_extracted_k141_138978 | Not-determined |
| S8_extracted_k141_112952 | Not-determined |
| S8_extracted_k141_121629 | Not-determined |
| S8_extracted_k141_173647 | Not-determined |
| S8_extracted_k141_173648 | Not-determined |
| S8_extracted_k141_78242 | Not-determined |
| S8_extracted_k141_173649 | Not-determined |
| S8_extracted_k141_8821 | Not-determined |
| S8_extracted_k141_60887 | Not-determined |
| S8_extracted_k141_191076 | Low-quality |
| S8_extracted_k141_121633 | Not-determined |
| S8_extracted_k141_52198 | Not-determined |
| S8_extracted_k141_8827 | Not-determined |
| S8_extracted_k141_8828 | Not-determined |
| S8_extracted_k141_69524 | Not-determined |
| S8_extracted_k141_121638 | Not-determined |
| S8_extracted_k141_121640 | Low-quality |
| S8_extracted_k141_86970 | Not-determined |
| S8_extracted_k141_173660 | Not-determined |
| S8_extracted_k141_173661 | Not-determined |
| S8_extracted_k141_199707 | Not-determined |
| S8_extracted_k141_60893 | Low-quality |
| S8_extracted_k141_86972 | Not-determined |
| S8_extracted_k141_112967 | Not-determined |
| S8_extracted_k141_121645 | Not-determined |
| S8_extracted_k141_52203 | Not-determined |
| S8_extracted_k141_8834 | Low-quality |
| S8_extracted_k141_180 | Not-determined |
| S8_extracted_k141_165020 | Low-quality |
| S8_extracted_k141_60897 | Not-determined |
| S8_extracted_k141_182 | Not-determined |
| S8_extracted_k141_86981 | Not-determined |
| S8_extracted_k141_52207 | Not-determined |
| S8_extracted_k141_165023 | Not-determined |
| S8_extracted_k141_26225 | Not-determined |
| S8_extracted_k141_199710 | Not-determined |
| S8_extracted_k141_112975 | Not-determined |
| S8_extracted_k141_86987 | Not-determined |
| S8_extracted_k141_191085 | Not-determined |
| S8_extracted_k141_189 | Not-determined |
| S8_extracted_k141_34837 | Not-determined |
| S8_extracted_k141_34838 | Not-determined |
| S8_extracted_k141_34839 | Not-determined |
| S8_extracted_k141_8840 | Not-determined |
| S8_extracted_k141_86991 | Not-determined |
| S8_extracted_k141_26230 | Not-determined |
| S8_extracted_k141_86993 | Low-quality |
| S8_extracted_k141_86994 | Not-determined |
| S8_extracted_k141_191089 | Not-determined |
| S8_extracted_k141_173667 | Not-determined |
| S8_extracted_k141_86995 | Not-determined |
| S8_extracted_k141_8846 | Not-determined |
| S8_extracted_k141_156355 | Low-quality |
| S8_extracted_k141_78258 | Not-determined |
| S8_extracted_k141_191092 | Not-determined |
| S8_extracted_k141_17490 | Not-determined |
| S8_extracted_k141_43522 | Not-determined |
| S8_extracted_k141_130275 | Not-determined |
| S8_extracted_k141_26231 | Not-determined |
| S8_extracted_k141_138985 | Not-determined |
| S8_extracted_k141_86997 | Not-determined |
| S8_extracted_k141_86998 | Not-determined |
| S8_extracted_k141_8849 | Not-determined |
| S8_extracted_k141_199725 | Not-determined |
| S8_extracted_k141_138991 | Not-determined |
| S8_extracted_k141_198 | Not-determined |
| S8_extracted_k141_87003 | Not-determined |
| S8_extracted_k141_199727 | Not-determined |
| S8_extracted_k141_34857 | Not-determined |
| S8_extracted_k141_130277 | Not-determined |
| S8_extracted_k141_60911 | Not-determined |
| S8_extracted_k141_60912 | Not-determined |
| S8_extracted_k141_34862 | Not-determined |
| S8_extracted_k141_138998 | Low-quality |
| S8_extracted_k141_17502 | Not-determined |
| S8_extracted_k141_165040 | Low-quality |
| S8_extracted_k141_112993 | Not-determined |
| S8_extracted_k141_112994 | Low-quality |
| S8_extracted_k141_87017 | Not-determined |
| S8_extracted_k141_112999 | Not-determined |
| S8_extracted_k141_43538 | Not-determined |
| S8_extracted_k141_139000 | Not-determined |
| S8_extracted_k141_216 | Not-determined |
| S8_extracted_k141_8862 | Not-determined |
| S8_extracted_k141_113004 | Not-determined |
| S8_extracted_k141_52222 | Not-determined |
| S8_extracted_k141_113006 | Not-determined |
| S8_extracted_k141_60924 | Not-determined |
| S8_extracted_k141_191114 | Not-determined |
| S8_extracted_k141_43546 | Not-determined |
| S8_extracted_k141_191116 | Not-determined |
| S8_extracted_k141_52229 | Not-determined |
| S8_extracted_k141_191118 | Not-determined |
| S8_extracted_k141_223 | Not-determined |
| S8_extracted_k141_139006 | Not-determined |
| S8_extracted_k141_8869 | Not-determined |
| S8_extracted_k141_121671 | Not-determined |
| S8_extracted_k141_173687 | Not-determined |
| S8_extracted_k141_139012 | Not-determined |
| S8_extracted_k141_34879 | Not-determined |
| S8_extracted_k141_139013 | Not-determined |
| S8_extracted_k141_34880 | Not-determined |
| S8_extracted_k141_8873 | Not-determined |
| S8_extracted_k141_69537 | Not-determined |
| S8_extracted_k141_130297 | Not-determined |
| S8_extracted_k141_229 | Low-quality |
| S8_extracted_k141_191126 | Not-determined |
| S8_extracted_k141_52241 | Not-determined |
| S8_extracted_k141_173695 | Not-determined |
| S8_extracted_k141_52243 | Not-determined |
| S8_extracted_k141_87033 | Not-determined |
| S8_extracted_k141_43551 | Not-determined |
| S8_extracted_k141_34882 | Not-determined |
| S8_extracted_k141_17521 | Not-determined |
| S8_extracted_k141_231 | Not-determined |
| S8_extracted_k141_121676 | Not-determined |
| S8_extracted_k141_173700 | Not-determined |
| S8_extracted_k141_87040 | Not-determined |
| S8_extracted_k141_173703 | Not-determined |
| S8_extracted_k141_8885 | Not-determined |
| S8_extracted_k141_191142 | Not-determined |
| S8_extracted_k141_8887 | Not-determined |
| S8_extracted_k141_191143 | Not-determined |
| S8_extracted_k141_8889 | Low-quality |
| S8_extracted_k141_121689 | Not-determined |
| S8_extracted_k141_78283 | Low-quality |
| S8_extracted_k141_87046 | Not-determined |
| S8_extracted_k141_191147 | Not-determined |
| S8_extracted_k141_191149 | Not-determined |
| S8_extracted_k141_69550 | Not-determined |
| S8_extracted_k141_130316 | Not-determined |
| S8_extracted_k141_69552 | Not-determined |
| S8_extracted_k141_139040 | Low-quality |
| S8_extracted_k141_34912 | Not-determined |
| S8_extracted_k141_244 | Not-determined |
| S8_extracted_k141_87059 | Not-determined |
| S8_extracted_k141_52253 | Not-determined |
| S8_extracted_k141_87060 | Not-determined |
| S8_extracted_k141_156367 | Low-quality |
| S8_extracted_k141_52255 | Not-determined |
| S8_extracted_k141_248 | Not-determined |
| S8_extracted_k141_130324 | Not-determined |
| S8_extracted_k141_130326 | Not-determined |
| S8_extracted_k141_60962 | Not-determined |
| S8_extracted_k141_87065 | Not-determined |
| S8_extracted_k141_69564 | Not-determined |
| S8_extracted_k141_139043 | Not-determined |
| S8_extracted_k141_173714 | Not-determined |
| S8_extracted_k141_43576 | Low-quality |
| S8_extracted_k141_256 | Not-determined |
| S8_extracted_k141_34930 | Not-determined |
| S8_extracted_k141_78298 | Low-quality |
| S8_extracted_k141_8908 | Not-determined |
| S8_extracted_k141_43578 | Not-determined |
| S8_extracted_k141_156374 | Not-determined |
| S8_extracted_k141_87073 | Not-determined |
| S8_extracted_k141_43581 | Not-determined |
| S8_extracted_k141_121700 | Not-determined |
| S8_extracted_k141_78303 | Not-determined |
| S8_extracted_k141_173718 | Low-quality |
| S8_extracted_k141_60969 | Not-determined |
| S8_extracted_k141_139052 | Not-determined |
| S8_extracted_k141_139054 | Low-quality |
| S8_extracted_k141_156382 | Low-quality |
| S8_extracted_k141_60971 | Not-determined |
| S8_extracted_k141_78306 | Not-determined |
| S8_extracted_k141_173725 | Not-determined |
| S8_extracted_k141_130339 | Not-determined |
| S8_extracted_k141_78309 | Not-determined |
| S8_extracted_k141_60974 | Low-quality |
| S8_extracted_k141_139057 | Not-determined |
| S8_extracted_k141_113032 | Not-determined |
| S8_extracted_k141_78314 | Not-determined |
| S8_extracted_k141_43589 | Not-determined |
| S8_extracted_k141_87080 | Not-determined |
| S8_extracted_k141_113035 | Not-determined |
| S8_extracted_k141_139063 | Not-determined |
| S8_extracted_k141_139064 | Not-determined |
| S8_extracted_k141_69583 | Not-determined |
| S8_extracted_k141_182290 | Not-determined |
| S8_extracted_k141_279 | Not-determined |
| S8_extracted_k141_87086 | Not-determined |
| S8_extracted_k141_139068 | Not-determined |
| S8_extracted_k141_113042 | Not-determined |
| S8_extracted_k141_8919 | Not-determined |
| S8_extracted_k141_165094 | Not-determined |
| S8_extracted_k141_69588 | Not-determined |
| S8_extracted_k141_52274 | Not-determined |
| S8_extracted_k141_284 | Not-determined |
| S8_extracted_k141_130349 | Low-quality |
| S8_extracted_k141_139074 | Not-determined |
| S8_extracted_k141_139075 | Not-determined |
| S8_extracted_k141_87089 | Low-quality |
| S8_extracted_k141_113047 | Not-determined |
| S8_extracted_k141_43599 | Low-quality |
| S8_extracted_k141_26255 | Not-determined |
| S8_extracted_k141_113048 | Not-determined |
| S8_extracted_k141_52278 | Not-determined |
| S8_extracted_k141_43604 | Not-determined |
| S8_extracted_k141_191158 | Not-determined |
| S8_extracted_k141_52280 | Not-determined |
| S8_extracted_k141_87095 | Not-determined |
| S8_extracted_k141_182305 | Not-determined |
| S8_extracted_k141_130360 | Not-determined |
| S8_extracted_k141_8929 | Not-determined |
| S8_extracted_k141_104269 | Low-quality |
| S8_extracted_k141_78325 | Low-quality |
| S8_extracted_k141_8932 | Not-determined |
| S8_extracted_k141_182310 | Not-determined |
| S8_extracted_k141_121723 | Not-determined |
| S8_extracted_k141_43614 | Not-determined |
| S8_extracted_k141_52283 | Not-determined |
| S8_extracted_k141_8935 | Low-quality |
| S8_extracted_k141_52284 | Not-determined |
| S8_extracted_k141_52285 | Not-determined |
| S8_extracted_k141_165103 | Not-determined |
| S8_extracted_k141_52287 | Not-determined |
| S8_extracted_k141_8939 | Low-quality |
| S8_extracted_k141_130369 | Not-determined |
| S8_extracted_k141_121727 | Not-determined |
| S8_extracted_k141_130370 | Not-determined |
| S8_extracted_k141_147682 | Not-determined |
| S8_extracted_k141_156405 | Not-determined |
| S8_extracted_k141_87109 | Not-determined |
| S8_extracted_k141_182319 | Low-quality |
| S8_extracted_k141_87111 | Not-determined |
| S8_extracted_k141_87113 | Not-determined |
| S8_extracted_k141_34972 | Not-determined |
| S8_extracted_k141_147688 | Low-quality |
| S8_extracted_k141_87116 | Not-determined |
| S8_extracted_k141_8946 | Not-determined |
| S8_extracted_k141_34974 | Not-determined |
| S8_extracted_k141_302 | Not-determined |
| S8_extracted_k141_26273 | Not-determined |
| S8_extracted_k141_121730 | Not-determined |
| S8_extracted_k141_121731 | Low-quality |
| S8_extracted_k141_306 | Not-determined |
| S8_extracted_k141_87127 | Not-determined |
| S8_extracted_k141_121733 | Not-determined |
| S8_extracted_k141_104289 | Not-determined |
| S8_extracted_k141_113066 | Not-determined |
| S8_extracted_k141_113067 | Low-quality |
| S8_extracted_k141_121738 | Not-determined |
| S8_extracted_k141_69607 | Not-determined |
| S8_extracted_k141_104291 | Not-determined |
| S8_extracted_k141_34980 | Low-quality |
| S8_extracted_k141_69609 | Not-determined |
| S8_extracted_k141_309 | Not-determined |
| S8_extracted_k141_52304 | Not-determined |
| S8_extracted_k141_78339 | Not-determined |
| S8_extracted_k141_104299 | Low-quality |
| S8_extracted_k141_8965 | Not-determined |
| S8_extracted_k141_191189 | Not-determined |
| S8_extracted_k141_78344 | Not-determined |
| S8_extracted_k141_43629 | Not-determined |
| S8_extracted_k141_104308 | Not-determined |
| S8_extracted_k141_43632 | Not-determined |
| S8_extracted_k141_156431 | Not-determined |
| S8_extracted_k141_43638 | Not-determined |
| S8_extracted_k141_78349 | Not-determined |
| S8_extracted_k141_78350 | Not-determined |
| S8_extracted_k141_156433 | Not-determined |
| S8_extracted_k141_139088 | Low-quality |
| S8_extracted_k141_43641 | Not-determined |
| S8_extracted_k141_43643 | Not-determined |
| S8_extracted_k141_322 | Not-determined |
| S8_extracted_k141_52320 | Not-determined |
| S8_extracted_k141_121754 | Not-determined |
| S8_extracted_k141_130405 | Not-determined |
| S8_extracted_k141_43646 | Not-determined |
| S8_extracted_k141_113079 | Not-determined |
| S8_extracted_k141_87136 | Not-determined |
| S8_extracted_k141_191198 | Low-quality |
| S8_extracted_k141_130408 | Not-determined |
| S8_extracted_k141_69614 | Not-determined |
| S8_extracted_k141_130411 | Not-determined |
| S8_extracted_k141_95600 | Not-determined |
| S8_extracted_k141_199754 | Not-determined |
| S8_extracted_k141_8982 | Not-determined |
| S8_extracted_k141_113085 | Not-determined |
| S8_extracted_k141_69615 | Not-determined |
| S8_extracted_k141_121759 | Not-determined |
| S8_extracted_k141_43653 | Not-determined |
| S8_extracted_k141_35000 | Not-determined |
| S8_extracted_k141_130414 | Not-determined |
| S8_extracted_k141_8985 | Not-determined |
| S8_extracted_k141_69618 | Not-determined |
| S8_extracted_k141_43654 | Not-determined |
| S8_extracted_k141_199760 | Not-determined |
| S8_extracted_k141_121763 | Not-determined |
| S8_extracted_k141_87145 | Not-determined |
| S8_extracted_k141_199762 | Not-determined |
| S8_extracted_k141_60983 | Not-determined |
| S8_extracted_k141_95608 | Not-determined |
| S8_extracted_k141_95609 | Not-determined |
| S8_extracted_k141_130422 | Not-determined |
| S8_extracted_k141_328 | Not-determined |
| S8_extracted_k141_199764 | Not-determined |
| S8_extracted_k141_139097 | Not-determined |
| S8_extracted_k141_121766 | Not-determined |
| S8_extracted_k141_69628 | Not-determined |
| S8_extracted_k141_121767 | Not-determined |
| S8_extracted_k141_165129 | Low-quality |
| S8_extracted_k141_165130 | Not-determined |
| S8_extracted_k141_78360 | Not-determined |
| S8_extracted_k141_121770 | Not-determined |
| S8_extracted_k141_69631 | Not-determined |
| S8_extracted_k141_78362 | Not-determined |
| S8_extracted_k141_191217 | Not-determined |
| S8_extracted_k141_182342 | Not-determined |
| S8_extracted_k141_35005 | Low-quality |
| S8_extracted_k141_191219 | Not-determined |
| S8_extracted_k141_156445 | Not-determined |
| S8_extracted_k141_182345 | Low-quality |
| S8_extracted_k141_26288 | Not-determined |
| S8_extracted_k141_182346 | Not-determined |
| S8_extracted_k141_199773 | Not-determined |
| S8_extracted_k141_121778 | Not-determined |
| S8_extracted_k141_130429 | Not-determined |
| S8_extracted_k141_9005 | Not-determined |
| S8_extracted_k141_199775 | Not-determined |
| S8_extracted_k141_104328 | Not-determined |
| S8_extracted_k141_130434 | Not-determined |
| S8_extracted_k141_130435 | Not-determined |
| S8_extracted_k141_165135 | Not-determined |
| S8_extracted_k141_69633 | Not-determined |
| S8_extracted_k141_9011 | Not-determined |
| S8_extracted_k141_104332 | Not-determined |
| S8_extracted_k141_182355 | Not-determined |
| S8_extracted_k141_69637 | Not-determined |
| S8_extracted_k141_113100 | Not-determined |
| S8_extracted_k141_17527 | Not-determined |
| S8_extracted_k141_349 | Not-determined |
| S8_extracted_k141_350 | Low-quality |
| S8_extracted_k141_78375 | Low-quality |
| S8_extracted_k141_130440 | Not-determined |
| S8_extracted_k141_69638 | Not-determined |
| S8_extracted_k141_113103 | Not-determined |
| S8_extracted_k141_87151 | Not-determined |
| S8_extracted_k141_35008 | Not-determined |
| S8_extracted_k141_52331 | Not-determined |
| S8_extracted_k141_87152 | Not-determined |
| S8_extracted_k141_35010 | Not-determined |
| S8_extracted_k141_26304 | Low-quality |
| S8_extracted_k141_78385 | Not-determined |
| S8_extracted_k141_87158 | Not-determined |
| S8_extracted_k141_78387 | Not-determined |
| S8_extracted_k141_156460 | Not-determined |
| S8_extracted_k141_61014 | Not-determined |
| S8_extracted_k141_61017 | Not-determined |
| S8_extracted_k141_182359 | Not-determined |
| S8_extracted_k141_78394 | Not-determined |
| S8_extracted_k141_191223 | Not-determined |
| S8_extracted_k141_52340 | Not-determined |
| S8_extracted_k141_139112 | Low-quality |
| S8_extracted_k141_61021 | Low-quality |
| S8_extracted_k141_139114 | Not-determined |
| S8_extracted_k141_165163 | Not-determined |
| S8_extracted_k141_130459 | Not-determined |
| S8_extracted_k141_52343 | Not-determined |
| S8_extracted_k141_35019 | Not-determined |
| S8_extracted_k141_130461 | Not-determined |
| S8_extracted_k141_182367 | Not-determined |
| S8_extracted_k141_130463 | Not-determined |
| S8_extracted_k141_130465 | Not-determined |
| S8_extracted_k141_61030 | Not-determined |
| S8_extracted_k141_130468 | Not-determined |
| S8_extracted_k141_69645 | Low-quality |
| S8_extracted_k141_61040 | Not-determined |
| S8_extracted_k141_130471 | Not-determined |
| S8_extracted_k141_61042 | Not-determined |
| S8_extracted_k141_87162 | Not-determined |
| S8_extracted_k141_113116 | Not-determined |
| S8_extracted_k141_182370 | Not-determined |
| S8_extracted_k141_87165 | Not-determined |
| S8_extracted_k141_113118 | Low-quality |
| S8_extracted_k141_61045 | Not-determined |
| S8_extracted_k141_130473 | Not-determined |
| S8_extracted_k141_26311 | Not-determined |
| S8_extracted_k141_87168 | Not-determined |
| S8_extracted_k141_69653 | Not-determined |
| S8_extracted_k141_113125 | Low-quality |
| S8_extracted_k141_130477 | Low-quality |
| S8_extracted_k141_61050 | Not-determined |
| S8_extracted_k141_139122 | Low-quality |
| S8_extracted_k141_113128 | Not-determined |

| | |
|---|---|
| S8_extracted_k141_87174 | Not-determined |
| S8_extracted_k141_87175 | Not-determined |
| S8_extracted_k141_173742 | Not-determined |
| S8_extracted_k141_182381 | Not-determined |
| S8_extracted_k141_173744 | Not-determined |
| S8_extracted_k141_9035 | Not-determined |
| S8_extracted_k141_130482 | Not-determined |
| S8_extracted_k141_139129 | Not-determined |
| S8_extracted_k141_9040 | Not-determined |
| S8_extracted_k141_52359 | Not-determined |
| S8_extracted_k141_9041 | Not-determined |
| S8_extracted_k141_182387 | Not-determined |
| S8_extracted_k141_61064 | Not-determined |
| S8_extracted_k141_182393 | Not-determined |
| S8_extracted_k141_156470 | Not-determined |
| S8_extracted_k141_95636 | Not-determined |
| S8_extracted_k141_173746 | Low-quality |
| S8_extracted_k141_199805 | Not-determined |
| S8_extracted_k141_173747 | Not-determined |
| S8_extracted_k141_95638 | Not-determined |
| S8_extracted_k141_95639 | Not-determined |
| S8_extracted_k141_61069 | Not-determined |
| S8_extracted_k141_26325 | Not-determined |
| S8_extracted_k141_156476 | Not-determined |
| S8_extracted_k141_173751 | Not-determined |
| S8_extracted_k141_199811 | Not-determined |
| S8_extracted_k141_156479 | Not-determined |
| S8_extracted_k141_156480 | Not-determined |
| S8_extracted_k141_173753 | Not-determined |
| S8_extracted_k141_173754 | Not-determined |
| S8_extracted_k141_69669 | Not-determined |
| S8_extracted_k141_113147 | Not-determined |
| S8_extracted_k141_199814 | Not-determined |
| S8_extracted_k141_130486 | Not-determined |
| S8_extracted_k141_199815 | Not-determined |
| S8_extracted_k141_199816 | Not-determined |
| S8_extracted_k141_199817 | Not-determined |
| S8_extracted_k141_69673 | Not-determined |
| S8_extracted_k141_199819 | Not-determined |
| S8_extracted_k141_156485 | Not-determined |
| S8_extracted_k141_139140 | Not-determined |
| S8_extracted_k141_61086 | Not-determined |
| S8_extracted_k141_78422 | Not-determined |
| S8_extracted_k141_199825 | Not-determined |
| S8_extracted_k141_43664 | Not-determined |
| S8_extracted_k141_182414 | Not-determined |
| S8_extracted_k141_17539 | Low-quality |
| S8_extracted_k141_173770 | Not-determined |
| S8_extracted_k141_121781 | Low-quality |
| S8_extracted_k141_87189 | Not-determined |
| S8_extracted_k141_87190 | Not-determined |
| S13_vs_97702\|\|full | High-quality |
| S8_extracted_k141_61093 | Not-determined |
| S8_extracted_k141_78428 | Low-quality |
| S8_extracted_k141_357 | Low-quality |
| S8_extracted_k141_61094 | Not-determined |
| S8_extracted_k141_130504 | Not-determined |
| S8_extracted_k141_191237 | Low-quality |
| S8_extracted_k141_156490 | Not-determined |
| S8_extracted_k141_104372 | Not-determined |
| S8_extracted_k141_87196 | Not-determined |
| S8_extracted_k141_52399 | Not-determined |
| S8_extracted_k141_61100 | Not-determined |
| S8_extracted_k141_61101 | Not-determined |
| S8_extracted_k141_130510 | Low-quality |
| S8_extracted_k141_156494 | Low-quality |
| S8_extracted_k141_78432 | Low-quality |
| S8_extracted_k141_199832 | Not-determined |
| S8_extracted_k141_113150 | Not-determined |
| S8_extracted_k141_191243 | Not-determined |
| S8_extracted_k141_113151 | Not-determined |
| S8_extracted_k141_191245 | Not-determined |
| S8_extracted_k141_199836 | Not-determined |
| S8_extracted_k141_43673 | Not-determined |
| S8_extracted_k141_121797 | Not-determined |
| S8_extracted_k141_78437 | Not-determined |
| S8_extracted_k141_35053 | Not-determined |
| S8_extracted_k141_26347 | Not-determined |
| S8_extracted_k141_199844 | Not-determined |
| S8_extracted_k141_104383 | Not-determined |
| S8_extracted_k141_199846 | Not-determined |
| S8_extracted_k141_182421 | Not-determined |
| S8_extracted_k141_87212 | Not-determined |
| S8_extracted_k141_173782 | Not-determined |
| S8_extracted_k141_191251 | Low-quality |
| S8_extracted_k141_87213 | Not-determined |
| S8_extracted_k141_156510 | Not-determined |
| S8_extracted_k141_87214 | Not-determined |
| S8_extracted_k141_113156 | Not-determined |
| S8_extracted_k141_121809 | Not-determined |
| S8_extracted_k141_173786 | Not-determined |
| S8_extracted_k141_121812 | Not-determined |
| S8_extracted_k141_191254 | Not-determined |
| S8_extracted_k141_156517 | Not-determined |
| S8_extracted_k141_199849 | Not-determined |
| S8_extracted_k141_87218 | Low-quality |
| S8_extracted_k141_78444 | Not-determined |
| S8_extracted_k141_173789 | Not-determined |
| S8_extracted_k141_43681 | Not-determined |
| S8_extracted_k141_113160 | Not-determined |
| S8_extracted_k141_17541 | Low-quality |
| S8_extracted_k141_113162 | Not-determined |
| S8_extracted_k141_191260 | Not-determined |
| S8_extracted_k141_139155 | Not-determined |
| S8_extracted_k141_35068 | Not-determined |
| S8_extracted_k141_139157 | Not-determined |
| S8_extracted_k141_78448 | Low-quality |
| S8_extracted_k141_191265 | Low-quality |
| S8_extracted_k141_26350 | Not-determined |
| S8_extracted_k141_121826 | Low-quality |
| S8_extracted_k141_191267 | Not-determined |
| S8_extracted_k141_95650 | Not-determined |
| S8_extracted_k141_378 | Not-determined |
| S8_extracted_k141_130529 | Not-determined |
| S8_extracted_k141_121828 | Not-determined |
| S8_extracted_k141_139166 | Low-quality |
| S8_extracted_k141_69683 | Not-determined |
| S8_extracted_k141_113171 | Not-determined |
| S8_extracted_k141_380 | Low-quality |
| S8_extracted_k141_182436 | Not-determined |
| S8_extracted_k141_173802 | Not-determined |
| S8_extracted_k141_26360 | Not-determined |
| S8_extracted_k141_173803 | Not-determined |
| S8_extracted_k141_87230 | Not-determined |
| S8_extracted_k141_61109 | Not-determined |
| S8_extracted_k141_130535 | Not-determined |
| S8_extracted_k141_139177 | Not-determined |
| S8_extracted_k141_199858 | Not-determined |
| S8_extracted_k141_17555 | Not-determined |
| S8_extracted_k141_87236 | Not-determined |
| S8_extracted_k141_35071 | Not-determined |
| S8_extracted_k141_35072 | Not-determined |
| S8_extracted_k141_182440 | Low-quality |
| S8_extracted_k141_199864 | Not-determined |
| S8_extracted_k141_17558 | Not-determined |
| S8_extracted_k141_52410 | Not-determined |
| S8_extracted_k141_191271 | Not-determined |
| S8_extracted_k141_78458 | Not-determined |
| S8_extracted_k141_130543 | Not-determined |
| S8_extracted_k141_17562 | Not-determined |
| S8_extracted_k141_182446 | Low-quality |
| S8_extracted_k141_95662 | Not-determined |
| S8_extracted_k141_182447 | Not-determined |
| S8_extracted_k141_139178 | Not-determined |
| S8_extracted_k141_104417 | Not-determined |
| S8_extracted_k141_191273 | Not-determined |
| S8_extracted_k141_43692 | Not-determined |
| S8_extracted_k141_121839 | Not-determined |
| S8_extracted_k141_35085 | Not-determined |
| S8_extracted_k141_395 | Not-determined |
| S8_extracted_k141_95665 | Not-determined |
| S8_extracted_k141_191277 | Not-determined |
| S8_extracted_k141_95667 | Not-determined |
| S8_extracted_k141_199875 | Low-quality |
| S8_extracted_k141_9081 | Not-determined |
| S8_extracted_k141_173813 | Not-determined |
| S8_extracted_k141_52417 | Low-quality |
| S8_extracted_k141_191287 | Not-determined |
| S8_extracted_k141_173816 | Not-determined |
| S8_extracted_k141_87259 | Not-determined |
| S8_extracted_k141_95672 | Not-determined |
| S8_extracted_k141_95675 | Not-determined |
| S8_extracted_k141_104422 | Not-determined |
| S8_extracted_k141_191292 | Not-determined |
| S8_extracted_k141_173819 | Low-quality |
| S8_extracted_k141_191293 | Not-determined |
| S8_extracted_k141_78479 | Not-determined |
| S8_extracted_k141_399 | Low-quality |
| S8_extracted_k141_191295 | Not-determined |
| S8_extracted_k141_87266 | Not-determined |
| S8_extracted_k141_191296 | Not-determined |
| S8_extracted_k141_26374 | Not-determined |
| S8_extracted_k141_173823 | Not-determined |
| S8_extracted_k141_104431 | Not-determined |
| S8_extracted_k141_87271 | Not-determined |
| S8_extracted_k141_121852 | Not-determined |
| S8_extracted_k141_43709 | Not-determined |
| S8_extracted_k141_121853 | Not-determined |
| S8_extracted_k141_9095 | Not-determined |
| S8_extracted_k141_121856 | Not-determined |
| S8_extracted_k141_156568 | Not-determined |
| S8_extracted_k141_191302 | Not-determined |
| S8_extracted_k141_409 | Not-determined |
| S8_extracted_k141_139196 | Not-determined |
| S8_extracted_k141_61111 | Not-determined |
| S8_extracted_k141_199887 | Not-determined |
| S8_extracted_k141_61114 | Not-determined |
| S8_extracted_k141_26386 | Not-determined |
| S8_extracted_k141_191308 | Not-determined |
| S8_extracted_k141_173828 | Not-determined |
| S8_extracted_k141_411 | Low-quality |
| S8_extracted_k141_95686 | Low-quality |
| S8_extracted_k141_78480 | Low-quality |
| S8_extracted_k141_173829 | Not-determined |
| S8_extracted_k141_191314 | Not-determined |
| S8_extracted_k141_156576 | Not-determined |
| S8_extracted_k141_156579 | Not-determined |
| S8_extracted_k141_147709 | Not-determined |
| S8_extracted_k141_52433 | Not-determined |
| S8_extracted_k141_61120 | Not-determined |
| S8_extracted_k141_147710 | Low-quality |
| S8_extracted_k141_69693 | Not-determined |
| S8_extracted_k141_147711 | Not-determined |
| S8_extracted_k141_78488 | Not-determined |
| S8_extracted_k141_26398 | Not-determined |
| S8_extracted_k141_26399 | Not-determined |
| S8_extracted_k141_191325 | Not-determined |
| S8_extracted_k141_419 | Not-determined |
| S8_extracted_k141_121863 | Not-determined |
| S8_extracted_k141_87279 | Not-determined |
| S8_extracted_k141_52440 | Not-determined |
| S8_extracted_k141_156581 | Not-determined |
| S8_extracted_k141_104439 | Low-quality |
| S8_extracted_k141_147715 | Not-determined |
| S8_extracted_k141_421 | Not-determined |
| S8_extracted_k141_191332 | Not-determined |
| S8_extracted_k141_87284 | Not-determined |
| S8_extracted_k141_425 | Not-determined |
| S8_extracted_k141_426 | Not-determined |
| S8_extracted_k141_87285 | Low-quality |
| S8_extracted_k141_52443 | Not-determined |
| S8_extracted_k141_26407 | Not-determined |
| S8_extracted_k141_35097 | Not-determined |
| S8_extracted_k141_95708 | Not-determined |
| S8_extracted_k141_61129 | Not-determined |
| S8_extracted_k141_182468 | Not-determined |
| S8_extracted_k141_182469 | Not-determined |
| S8_extracted_k141_182470 | Low-quality |
| S8_extracted_k141_35101 | Not-determined |
| S8_extracted_k141_182472 | Not-determined |
| S8_extracted_k141_147720 | Not-determined |
| S8_extracted_k141_191341 | Low-quality |
| S8_extracted_k141_199901 | Not-determined |
| S8_extracted_k141_191342 | Not-determined |
| S8_extracted_k141_121875 | Not-determined |
| S8_extracted_k141_113197 | Not-determined |
| S8_extracted_k141_17576 | Not-determined |
| S8_extracted_k141_165267 | Low-quality |
| S8_extracted_k141_78504 | Not-determined |
| S8_extracted_k141_165269 | Not-determined |
| S8_extracted_k141_130565 | Not-determined |
| S8_extracted_k141_78510 | Not-determined |
| S8_extracted_k141_182488 | Low-quality |
| S8_extracted_k141_78511 | Not-determined |
| S8_extracted_k141_43725 | Not-determined |
| S8_extracted_k141_182490 | Not-determined |
| S8_extracted_k141_35113 | Not-determined |
| S8_extracted_k141_191350 | Not-determined |
| S8_extracted_k141_87293 | Not-determined |
| S8_extracted_k141_165281 | Not-determined |
| S8_extracted_k141_52460 | Not-determined |
| S8_extracted_k141_443 | Not-determined |
| S8_extracted_k141_444 | Not-determined |
| S8_extracted_k141_130577 | Not-determined |
| S8_extracted_k141_52461 | Not-determined |
| S8_extracted_k141_130579 | Not-determined |
| S8_extracted_k141_17589 | Not-determined |
| S8_extracted_k141_52463 | Not-determined |
| S8_extracted_k141_35125 | Not-determined |
| S8_extracted_k141_130582 | Low-quality |
| S8_extracted_k141_17594 | Not-determined |
| S8_extracted_k141_87299 | Not-determined |
| S8_extracted_k141_191362 | Not-determined |
| S8_extracted_k141_35129 | Not-determined |
| S8_extracted_k141_191363 | Not-determined |
| S8_extracted_k141_26427 | Not-determined |
| S8_extracted_k141_199921 | Not-determined |
| S8_extracted_k141_130589 | Not-determined |
| S8_extracted_k141_182510 | Not-determined |
| S8_extracted_k141_87301 | Not-determined |
| S8_extracted_k141_139200 | Not-determined |
| S8_extracted_k141_182511 | Not-determined |
| S8_extracted_k141_182513 | Not-determined |
| S8_extracted_k141_191372 | Not-determined |
| S8_extracted_k141_182514 | Not-determined |
| S8_extracted_k141_191374 | Not-determined |
| S8_extracted_k141_35137 | Not-determined |
| S8_extracted_k141_199923 | Not-determined |
| S8_extracted_k141_17601 | Not-determined |
| S8_extracted_k141_182516 | Not-determined |
| S8_extracted_k141_130592 | Not-determined |
| S8_extracted_k141_43743 | Not-determined |
| S8_extracted_k141_43744 | Not-determined |
| S8_extracted_k141_139202 | Low-quality |
| S8_extracted_k141_130594 | Not-determined |
| S8_extracted_k141_52475 | Not-determined |
| S8_extracted_k141_191378 | Not-determined |
| S8_extracted_k141_43750 | Not-determined |
| S8_extracted_k141_139205 | Low-quality |
| S8_extracted_k141_147750 | Not-determined |
| S8_extracted_k141_130597 | Not-determined |
| S8_extracted_k141_52478 | Not-determined |
| S8_extracted_k141_52479 | Not-determined |
| S8_extracted_k141_139208 | Not-determined |
| S8_extracted_k141_121908 | Not-determined |
| S8_extracted_k141_121909 | Not-determined |
| S8_extracted_k141_78548 | Not-determined |
| S8_extracted_k141_182524 | Not-determined |
| S8_extracted_k141_130601 | Not-determined |
| S8_extracted_k141_139212 | Low-quality |
| S8_extracted_k141_191387 | Not-determined |
| S8_extracted_k141_173851 | Not-determined |
| S8_extracted_k141_35151 | Not-determined |
| S8_extracted_k141_43754 | Not-determined |
| S8_extracted_k141_35154 | Not-determined |
| S8_extracted_k141_139219 | Not-determined |
| S8_extracted_k141_9117 | Not-determined |
| S8_extracted_k141_69712 | Not-determined |
| S8_extracted_k141_139222 | Not-determined |
| S8_extracted_k141_182535 | Not-determined |
| S8_extracted_k141_26439 | Not-determined |
| S8_extracted_k141_191391 | Not-determined |
| S8_extracted_k141_165302 | Not-determined |
| S8_extracted_k141_182536 | Not-determined |
| S8_extracted_k141_191392 | Not-determined |
| S8_extracted_k141_130608 | Not-determined |
| S8_extracted_k141_69715 | Not-determined |
| S8_extracted_k141_191395 | Not-determined |
| S8_extracted_k141_173859 | Not-determined |
| S8_extracted_k141_87323 | Not-determined |
| S8_extracted_k141_147765 | Not-determined |
| S8_extracted_k141_78556 | Not-determined |
| S8_extracted_k141_199950 | Not-determined |
| S8_extracted_k141_121923 | Not-determined |
| S8_extracted_k141_78557 | Not-determined |
| S8_extracted_k141_87330 | Not-determined |
| S8_extracted_k141_9128 | Not-determined |
| S8_extracted_k141_130614 | Not-determined |
| S8_extracted_k141_165311 | Not-determined |
| S8_extracted_k141_130615 | Not-determined |
| S8_extracted_k141_95743 | Low-quality |
| S8_extracted_k141_182540 | Low-quality |
| S8_extracted_k141_9132 | Not-determined |
| S8_extracted_k141_182541 | Not-determined |
| S8_extracted_k141_43769 | Low-quality |
| S8_extracted_k141_35161 | Not-determined |
| S8_extracted_k141_165314 | Not-determined |
| S8_extracted_k141_173864 | Not-determined |
| S8_extracted_k141_9137 | Low-quality |
| S8_extracted_k141_9138 | Not-determined |
| S8_extracted_k141_87331 | Not-determined |
| S8_extracted_k141_139224 | Low-quality |
| S8_extracted_k141_9139 | Not-determined |
| S8_extracted_k141_35164 | Not-determined |
| S8_extracted_k141_95752 | Not-determined |
| S8_extracted_k141_95753 | Not-determined |
| S8_extracted_k141_139227 | Not-determined |
| S8_extracted_k141_35166 | Not-determined |
| S8_extracted_k141_139228 | Not-determined |
| S8_extracted_k141_173875 | Not-determined |
| S8_extracted_k141_9146 | Not-determined |
| S8_extracted_k141_165320 | Low-quality |
| S8_extracted_k141_147766 | Low-quality |
| S8_extracted_k141_147767 | Not-determined |
| S8_extracted_k141_104483 | Not-determined |
| S8_extracted_k141_191400 | Not-determined |
| S8_extracted_k141_95761 | Not-determined |
| S8_extracted_k141_165324 | Not-determined |
| S8_extracted_k141_165325 | Not-determined |
| S8_extracted_k141_173877 | Low-quality |
| S8_extracted_k141_104486 | Not-determined |
| S8_extracted_k141_191407 | Not-determined |
| S8_extracted_k141_139234 | Low-quality |
| S8_extracted_k141_199958 | Not-determined |
| S8_extracted_k141_460 | Low-quality |
| S8_extracted_k141_165329 | Not-determined |
| S8_extracted_k141_199961 | Not-determined |
| S8_extracted_k141_69722 | Not-determined |
| S8_extracted_k141_78575 | Not-determined |
| S8_extracted_k141_139237 | Not-determined |
| S8_extracted_k141_61146 | Not-determined |
| S8_extracted_k141_43779 | Not-determined |
| S8_extracted_k141_173884 | Not-determined |
| S8_extracted_k141_69723 | Not-determined |
| S8_extracted_k141_52486 | Not-determined |
| S8_extracted_k141_173885 | Not-determined |
| S8_extracted_k141_95768 | Not-determined |
| S8_extracted_k141_87339 | Not-determined |
| S8_extracted_k141_470 | Not-determined |
| S8_extracted_k141_43782 | Not-determined |
| S8_extracted_k141_474 | Not-determined |
| S8_extracted_k141_199968 | Not-determined |
| S8_extracted_k141_87342 | Not-determined |
| S8_extracted_k141_147778 | Not-determined |
| S8_extracted_k141_61152 | Not-determined |
| S8_extracted_k141_69728 | Not-determined |
| S8_extracted_k141_61153 | Not-determined |
| S8_extracted_k141_69730 | Low-quality |
| S8_extracted_k141_78580 | Not-determined |
| S8_extracted_k141_35181 | Low-quality |
| S8_extracted_k141_191419 | Not-determined |
| S8_extracted_k141_69731 | Low-quality |
| S8_extracted_k141_52494 | Not-determined |
| S8_extracted_k141_104509 | Not-determined |
| S8_extracted_k141_199970 | Not-determined |
| S8_extracted_k141_69732 | Not-determined |
| S8_extracted_k141_479 | Not-determined |
| S8_extracted_k141_191425 | Not-determined |
| S8_extracted_k141_173900 | Not-determined |
| S8_extracted_k141_130630 | Not-determined |
| S8_extracted_k141_481 | Not-determined |
| S8_extracted_k141_69735 | Not-determined |
| S8_extracted_k141_483 | Not-determined |
| S8_extracted_k141_130632 | Not-determined |
| S8_extracted_k141_147791 | Not-determined |
| S8_extracted_k141_78590 | Not-determined |
| S8_extracted_k141_173904 | Not-determined |
| S8_extracted_k141_35187 | Not-determined |
| S8_extracted_k141_191429 | Not-determined |
| S8_extracted_k141_121931 | Not-determined |
| S8_extracted_k141_52506 | Not-determined |
| S8_extracted_k141_173908 | Not-determined |
| S8_extracted_k141_165348 | Not-determined |
| S8_extracted_k141_173909 | Not-determined |
| S8_extracted_k141_121933 | Not-determined |
| S8_extracted_k141_165349 | Not-determined |
| S8_extracted_k141_78598 | Not-determined |
| S8_extracted_k141_173910 | Not-determined |
| S8_extracted_k141_78599 | Not-determined |
| S8_extracted_k141_87354 | Not-determined |
| S8_extracted_k141_43803 | Not-determined |
| S8_extracted_k141_121934 | Not-determined |
| S8_extracted_k141_191440 | Not-determined |
| S8_extracted_k141_130635 | Not-determined |
| S8_extracted_k141_35188 | Not-determined |
| S8_extracted_k141_69746 | Not-determined |
| S8_extracted_k141_165354 | Not-determined |
| S8_extracted_k141_78600 | Not-determined |
| S8_extracted_k141_87363 | Low-quality |
| S8_extracted_k141_191441 | Not-determined |
| S8_extracted_k141_35197 | Low-quality |
| S8_extracted_k141_43811 | Not-determined |
| S8_extracted_k141_87365 | Not-determined |
| S8_extracted_k141_130640 | Not-determined |
| S8_extracted_k141_104512 | Not-determined |
| S8_extracted_k141_9169 | Not-determined |
| S8_extracted_k141_104514 | Not-determined |
| S8_extracted_k141_78609 | Not-determined |
| S8_extracted_k141_87370 | Not-determined |
| S8_extracted_k141_498 | Not-determined |
| S8_extracted_k141_69754 | Low-quality |
| S8_extracted_k141_87375 | Not-determined |
| S8_extracted_k141_500 | Low-quality |
| S8_extracted_k141_139250 | Not-determined |
| S8_extracted_k141_95775 | Low-quality |
| S8_extracted_k141_95777 | Not-determined |
| S8_extracted_k141_165370 | Not-determined |
| S8_extracted_k141_121946 | Not-determined |
| S8_extracted_k141_35198 | Low-quality |
| S8_extracted_k141_95779 | Not-determined |
| S8_extracted_k141_35199 | Not-determined |
| S8_extracted_k141_52529 | Not-determined |
| S8_extracted_k141_17611 | Not-determined |
| S8_extracted_k141_113217 | Not-determined |
| S8_extracted_k141_165376 | Low-quality |
| S8_extracted_k141_173916 | Low-quality |
| S8_extracted_k141_9181 | Not-determined |
| S8_extracted_k141_43815 | Not-determined |
| S8_extracted_k141_95785 | Low-quality |
| S8_extracted_k141_78622 | Not-determined |
| S8_extracted_k141_191454 | Not-determined |
| S8_extracted_k141_121954 | Not-determined |
| S8_extracted_k141_191457 | Not-determined |
| S8_extracted_k141_35204 | Low-quality |
| S8_extracted_k141_78626 | Not-determined |
| S8_extracted_k141_165383 | Not-determined |
| S8_extracted_k141_191459 | Not-determined |
| S8_extracted_k141_104530 | Not-determined |
| S8_extracted_k141_61169 | Not-determined |
| S8_extracted_k141_121957 | Not-determined |
| S8_extracted_k141_104531 | Not-determined |
| S8_extracted_k141_26477 | Not-determined |
| S8_extracted_k141_165389 | Not-determined |
| S8_extracted_k141_78630 | Not-determined |
| S8_extracted_k141_130662 | Not-determined |
| S8_extracted_k141_104538 | Not-determined |
| S8_extracted_k141_9197 | Not-determined |
| S8_extracted_k141_121962 | Not-determined |
| S8_extracted_k141_9199 | Not-determined |
| S8_extracted_k141_35214 | Not-determined |
| S8_extracted_k141_130667 | Not-determined |
| S8_extracted_k141_78637 | Not-determined |
| S8_extracted_k141_528 | Not-determined |
| S8_extracted_k141_9204 | Not-determined |
| S8_extracted_k141_52545 | Not-determined |
| S8_extracted_k141_531 | Not-determined |
| S8_extracted_k141_130669 | Not-determined |
| S8_extracted_k141_139275 | Not-determined |
| S8_extracted_k141_139278 | Not-determined |
| S8_extracted_k141_43821 | Not-determined |
| S8_extracted_k141_139282 | Not-determined |
| S8_extracted_k141_130675 | Not-determined |
| S8_extracted_k141_17634 | Not-determined |
| S8_extracted_k141_78638 | Not-determined |
| S8_extracted_k141_165399 | Not-determined |
| S8_extracted_k141_61194 | Not-determined |
| S8_extracted_k141_139286 | Not-determined |
| S8_extracted_k141_130678 | Not-determined |
| S8_extracted_k141_121973 | Not-determined |
| S8_extracted_k141_104552 | Not-determined |
| S8_extracted_k141_121975 | Not-determined |
| S8_extracted_k141_541 | Not-determined |
| S8_extracted_k141_165403 | Not-determined |
| S8_extracted_k141_191478 | Not-determined |
| S8_extracted_k141_130682 | Not-determined |
| S8_extracted_k141_87394 | Not-determined |
| S8_extracted_k141_43823 | Low-quality |
| S8_extracted_k141_61206 | Not-determined |
| S8_extracted_k141_191484 | Not-determined |
| S8_extracted_k141_61208 | Not-determined |
| S8_extracted_k141_121985 | Not-determined |
| S8_extracted_k141_191489 | Not-determined |
| S8_extracted_k141_43827 | Low-quality |
| S8_extracted_k141_17646 | Low-quality |
| S8_extracted_k141_43828 | Not-determined |
| S8_extracted_k141_52564 | Not-determined |
| S8_extracted_k141_165413 | Not-determined |
| S8_extracted_k141_139290 | Not-determined |
| S8_extracted_k141_17648 | Not-determined |
| S8_extracted_k141_43832 | Not-determined |
| S8_extracted_k141_165415 | Not-determined |
| S8_extracted_k141_87399 | Low-quality |
| S8_extracted_k141_52568 | Not-determined |
| S8_extracted_k141_43836 | Not-determined |
| S8_extracted_k141_545 | Not-determined |
| S8_extracted_k141_35222 | Not-determined |
| S8_extracted_k141_17656 | Not-determined |
| S8_extracted_k141_121992 | Low-quality |
| S8_extracted_k141_52569 | Not-determined |
| S8_extracted_k141_61215 | Not-determined |
| S8_extracted_k141_121993 | Not-determined |
| S8_extracted_k141_165422 | Not-determined |
| S8_extracted_k141_173928 | Not-determined |
| S8_extracted_k141_147849 | Not-determined |
| S8_extracted_k141_130690 | Not-determined |
| S8_extracted_k141_52577 | Not-determined |
| S8_extracted_k141_139303 | Not-determined |
| S8_extracted_k141_147857 | Not-determined |
| S8_extracted_k141_35230 | Not-determined |
| S8_extracted_k141_78654 | Low-quality |
| S8_extracted_k141_104579 | Not-determined |
| S8_extracted_k141_559 | Not-determined |
| S8_extracted_k141_78656 | Not-determined |
| S8_extracted_k141_26512 | Not-determined |
| S8_extracted_k141_139308 | Not-determined |
| S8_extracted_k141_130695 | Not-determined |
| S8_extracted_k141_52585 | Not-determined |
| S8_extracted_k141_35240 | Not-determined |
| S8_extracted_k141_139312 | Not-determined |
| S8_extracted_k141_87411 | Not-determined |
| S8_extracted_k141_104583 | Not-determined |
| S8_extracted_k141_130700 | Not-determined |
| S8_extracted_k141_104585 | Not-determined |
| S8_extracted_k141_52590 | Not-determined |
| S8_extracted_k141_182550 | Not-determined |
| S8_extracted_k141_122002 | Low-quality |
| S8_extracted_k141_52594 | Not-determined |
| S8_extracted_k141_61236 | Not-determined |
| S8_extracted_k141_139314 | Not-determined |
| S8_extracted_k141_173938 | Not-determined |
| S8_extracted_k141_61237 | Low-quality |
| S8_extracted_k141_104588 | Not-determined |
| S8_extracted_k141_165437 | Not-determined |
| S8_extracted_k141_35247 | Not-determined |
| S8_extracted_k141_104589 | Not-determined |
| S8_extracted_k141_104590 | Not-determined |
| S8_extracted_k141_104591 | Not-determined |
| S8_extracted_k141_26516 | Low-quality |
| S8_extracted_k141_147871 | Not-determined |
| S8_extracted_k141_104595 | Not-determined |
| S8_extracted_k141_173942 | Not-determined |
| S8_extracted_k141_43865 | Not-determined |
| S8_extracted_k141_52599 | Not-determined |
| S8_extracted_k141_17671 | Not-determined |
| S8_extracted_k141_104599 | Low-quality |
| S8_extracted_k141_61243 | Not-determined |
| S8_extracted_k141_139315 | Low-quality |
| S8_extracted_k141_17673 | Not-determined |
| S8_extracted_k141_165443 | Not-determined |
| S8_extracted_k141_104603 | Not-determined |
| S8_extracted_k141_173947 | Not-determined |
| S8_extracted_k141_35258 | Not-determined |
| S8_extracted_k141_104606 | Not-determined |
| S8_extracted_k141_165448 | Not-determined |
| S8_extracted_k141_139317 | Not-determined |
| S8_extracted_k141_165451 | Not-determined |
| S8_extracted_k141_139319 | Not-determined |
| S8_extracted_k141_113233 | Low-quality |
| S8_extracted_k141_52607 | Low-quality |

| | |
|---|---|
| S8_extracted_k141_26520 | Not-determined |
| S8_extracted_k141_122020 | Not-determined |
| S8_extracted_k141_104607 | Low-quality |
| S8_extracted_k141_61252 | Not-determined |
| S8_extracted_k141_191531 | Not-determined |
| S8_extracted_k141_17678 | Not-determined |
| S8_extracted_k141_95841 | Not-determined |
| S8_extracted_k141_35266 | Low-quality |
| S8_extracted_k141_17680 | Not-determined |
| S8_extracted_k141_139325 | Not-determined |
| S8_extracted_k141_17681 | Not-determined |
| S8_extracted_k141_191533 | Not-determined |
| S8_extracted_k141_35269 | Not-determined |
| S8_extracted_k141_139329 | Not-determined |
| S8_extracted_k141_122022 | Low-quality |
| S8_extracted_k141_191537 | Not-determined |
| S8_extracted_k141_122024 | Not-determined |
| S8_extracted_k141_95848 | Not-determined |
| S8_extracted_k141_139331 | Not-determined |
| S8_extracted_k141_182555 | Not-determined |
| S8_extracted_k141_26530 | Not-determined |
| S8_extracted_k141_182557 | Not-determined |
| S8_extracted_k141_578 | Not-determined |
| S8_extracted_k141_165457 | Not-determined |
| S8_extracted_k141_69773 | Not-determined |
| S8_extracted_k141_191541 | Not-determined |
| S8_extracted_k141_182564 | Not-determined |
| S8_extracted_k141_17690 | Not-determined |
| S8_extracted_k141_182566 | Not-determined |
| S8_extracted_k141_69778 | Not-determined |
| S8_extracted_k141_95855 | Not-determined |
| S8_extracted_k141_104623 | Not-determined |
| S8_extracted_k141_182567 | Not-determined |
| S8_extracted_k141_104624 | Not-determined |
| S8_extracted_k141_191546 | Not-determined |
| S8_extracted_k141_139332 | Low-quality |
| S8_extracted_k141_61264 | Not-determined |
| S8_extracted_k141_191550 | Not-determined |
| S8_extracted_k141_139335 | Low-quality |
| S8_extracted_k141_26535 | Not-determined |
| S8_extracted_k141_191551 | Not-determined |
| S8_extracted_k141_165463 | Not-determined |
| S8_extracted_k141_61271 | Not-determined |
| S8_extracted_k141_156594 | Not-determined |
| S8_extracted_k141_35281 | Not-determined |
| S8_extracted_k141_165468 | Low-quality |
| S8_extracted_k141_9221 | Not-determined |
| S8_extracted_k141_95866 | Not-determined |
| S8_extracted_k141_182582 | Not-determined |
| S8_extracted_k141_95867 | Not-determined |
| S8_extracted_k141_17701 | Not-determined |
| S8_extracted_k141_147897 | Not-determined |
| S8_extracted_k141_165471 | Low-quality |
| S8_extracted_k141_17704 | Low-quality |
| S8_extracted_k141_52617 | Not-determined |
| S8_extracted_k141_165475 | Not-determined |
| S8_extracted_k141_595 | Not-determined |
| S8_extracted_k141_130714 | Not-determined |
| S8_extracted_k141_122035 | Not-determined |
| S8_extracted_k141_596 | Not-determined |
| S8_extracted_k141_69785 | Not-determined |
| S8_extracted_k141_69786 | Not-determined |
| S8_extracted_k141_26545 | Not-determined |
| S8_extracted_k141_61288 | Not-determined |
| S8_extracted_k141_104632 | Not-determined |
| S8_extracted_k141_182587 | Not-determined |
| S8_extracted_k141_52619 | Not-determined |
| S8_extracted_k141_139353 | Low-quality |
| S8_extracted_k141_191554 | Low-quality |
| S8_extracted_k141_95883 | Not-determined |
| S8_extracted_k141_52626 | Not-determined |
| S8_extracted_k141_603 | Not-determined |
| S8_extracted_k141_87435 | Not-determined |
| S8_extracted_k141_191556 | Low-quality |
| S8_extracted_k141_52628 | Not-determined |
| S8_extracted_k141_165481 | Not-determined |
| S8_extracted_k141_607 | Not-determined |
| S8_extracted_k141_52631 | Not-determined |
| S8_extracted_k141_182593 | Not-determined |
| S8_extracted_k141_122047 | Not-determined |
| S8_extracted_k141_122048 | Not-determined |
| S8_extracted_k141_130723 | Not-determined |
| S8_extracted_k141_61297 | Not-determined |
| S8_extracted_k141_130724 | Not-determined |
| S8_extracted_k141_609 | Not-determined |
| S8_extracted_k141_191560 | Not-determined |
| S8_extracted_k141_130726 | Not-determined |
| S8_extracted_k141_191561 | Not-determined |
| S8_extracted_k141_191563 | Not-determined |
| S8_extracted_k141_52637 | Not-determined |
| S8_extracted_k141_52638 | Not-determined |
| S8_extracted_k141_147919 | Not-determined |
| S8_extracted_k141_52640 | Not-determined |
| S8_extracted_k141_191566 | Not-determined |
| S8_extracted_k141_104651 | Low-quality |
| S8_extracted_k141_182599 | Not-determined |
| S8_extracted_k141_191574 | Not-determined |
| S8_extracted_k141_52648 | Not-determined |
| S8_extracted_k141_35303 | Not-determined |
| S8_extracted_k141_35304 | Not-determined |
| S8_extracted_k141_122057 | Low-quality |
| S8_extracted_k141_52650 | Low-quality |
| S8_extracted_k141_182605 | Not-determined |
| S8_extracted_k141_52653 | Not-determined |
| S8_extracted_k141_87457 | Not-determined |
| S8_extracted_k141_104659 | Not-determined |
| S8_extracted_k141_9246 | Not-determined |
| S8_extracted_k141_61317 | Not-determined |
| S8_extracted_k141_122063 | Not-determined |
| S8_extracted_k141_139364 | Not-determined |
| S8_extracted_k141_69816 | Not-determined |
| S8_extracted_k141_61321 | Not-determined |
| S8_extracted_k141_61322 | Not-determined |
| S8_extracted_k141_35307 | Not-determined |
| S8_extracted_k141_139369 | Not-determined |
| S8_extracted_k141_199989 | Not-determined |
| S8_extracted_k141_122069 | Not-determined |
| S8_extracted_k141_52674 | Not-determined |
| S8_extracted_k141_95901 | Not-determined |
| S8_extracted_k141_621 | Low-quality |
| S8_extracted_k141_165511 | Not-determined |
| S8_extracted_k141_17728 | Not-determined |
| S8_extracted_k141_139374 | Not-determined |
| S8_extracted_k141_147938 | Not-determined |
| S8_extracted_k141_626 | Not-determined |
| S8_extracted_k141_61328 | Low-quality |
| S8_extracted_k141_87464 | Low-quality |
| S8_extracted_k141_200004 | Not-determined |
| S8_extracted_k141_139378 | Not-determined |
| S8_extracted_k141_17731 | Not-determined |
| S8_extracted_k141_130732 | Not-determined |
| S8_extracted_k141_165518 | Not-determined |
| S8_extracted_k141_173971 | Not-determined |
| S8_extracted_k141_147946 | Not-determined |
| S8_extracted_k141_139382 | Not-determined |
| S8_extracted_k141_130734 | Not-determined |
| S8_extracted_k141_182612 | Not-determined |
| S8_extracted_k141_165520 | Not-determined |
| S8_extracted_k141_130735 | Not-determined |
| S8_extracted_k141_182613 | Not-determined |
| S8_extracted_k141_165522 | Not-determined |
| S8_extracted_k141_17737 | Not-determined |
| S8_extracted_k141_9252 | Not-determined |
| S8_extracted_k141_173978 | Not-determined |
| S8_extracted_k141_165526 | Not-determined |
| S8_extracted_k141_200020 | Not-determined |
| S8_extracted_k141_87478 | Not-determined |
| S8_extracted_k141_69835 | Not-determined |
| S8_extracted_k141_87483 | Not-determined |
| S8_extracted_k141_147957 | Not-determined |
| S8_extracted_k141_147960 | Not-determined |
| S8_extracted_k141_52683 | Not-determined |
| S8_extracted_k141_122088 | Not-determined |
| S8_extracted_k141_147967 | Not-determined |
| S8_extracted_k141_147971 | Not-determined |
| S8_extracted_k141_182624 | Not-determined |
| S8_extracted_k141_147972 | Not-determined |
| S8_extracted_k141_200034 | Low-quality |
| S8_extracted_k141_147974 | Not-determined |
| S8_extracted_k141_69838 | Not-determined |
| S8_extracted_k141_104682 | Not-determined |
| S8_extracted_k141_122100 | Low-quality |
| S8_extracted_k141_69839 | Not-determined |
| S8_extracted_k141_87494 | Not-determined |
| S8_extracted_k141_104685 | Not-determined |
| S8_extracted_k141_78666 | Not-determined |
| S8_extracted_k141_52694 | Not-determined |
| S8_extracted_k141_122102 | Not-determined |
| S8_extracted_k141_173999 | Not-determined |
| S8_extracted_k141_17755 | Not-determined |
| S8_extracted_k141_69843 | Not-determined |
| S8_extracted_k141_664 | Not-determined |
| S8_extracted_k141_147986 | Low-quality |
| S8_extracted_k141_52696 | Not-determined |
| S8_extracted_k141_87502 | Not-determined |
| S8_extracted_k141_665 | Not-determined |
| S8_extracted_k141_113273 | Low-quality |
| S8_extracted_k141_113274 | Not-determined |
| S8_extracted_k141_122106 | Not-determined |
| S8_extracted_k141_104696 | Not-determined |
| S8_extracted_k141_104698 | Not-determined |
| S8_extracted_k141_113281 | Not-determined |
| S8_extracted_k141_69853 | Not-determined |
| S8_extracted_k141_130768 | Not-determined |
| S8_extracted_k141_191603 | Not-determined |
| S8_extracted_k141_113284 | Not-determined |
| S8_extracted_k141_174010 | Low-quality |
| S8_extracted_k141_130769 | Low-quality |
| S8_extracted_k141_113288 | Not-determined |
| S8_extracted_k141_191605 | Not-determined |
| S8_extracted_k141_130771 | Not-determined |
| S8_extracted_k141_17759 | Not-determined |
| S8_extracted_k141_139392 | Not-determined |
| S8_extracted_k141_69864 | Not-determined |
| S8_extracted_k141_104706 | Not-determined |
| S8_extracted_k141_95932 | Not-determined |
| S8_extracted_k141_61347 | Low-quality |
| S8_extracted_k141_17763 | Not-determined |
| S8_extracted_k141_69871 | Not-determined |
| S8_extracted_k141_95934 | Low-quality |
| S8_extracted_k141_95935 | Not-determined |
| S8_extracted_k141_95936 | Not-determined |
| S8_extracted_k141_61352 | Not-determined |
| S8_extracted_k141_95937 | Not-determined |
| S8_extracted_k141_148006 | Not-determined |
| S8_extracted_k141_95940 | Not-determined |
| S8_extracted_k141_174017 | Not-determined |
| S8_extracted_k141_174018 | Not-determined |
| S8_extracted_k141_26550 | Not-determined |
| S8_extracted_k141_200043 | Not-determined |
| S8_extracted_k141_52716 | Not-determined |
| S8_extracted_k141_174028 | Low-quality |
| S8_extracted_k141_35310 | Not-determined |
| S8_extracted_k141_26552 | Not-determined |
| S8_extracted_k141_165547 | Not-determined |
| S8_extracted_k141_35311 | Not-determined |
| S8_extracted_k141_35312 | Not-determined |
| S8_extracted_k141_95951 | Not-determined |
| S8_extracted_k141_122125 | Not-determined |
| S8_extracted_k141_78679 | Not-determined |
| S8_extracted_k141_95954 | Not-determined |
| S8_extracted_k141_52728 | Not-determined |
| S8_extracted_k141_679 | Low-quality |
| S8_extracted_k141_52729 | Not-determined |
| S8_extracted_k141_122130 | Not-determined |
| S8_extracted_k141_139405 | Not-determined |
| S8_extracted_k141_95957 | Not-determined |
| S8_extracted_k141_165556 | Not-determined |
| S8_extracted_k141_104723 | Not-determined |
| S8_extracted_k141_113309 | Not-determined |
| S8_extracted_k141_95960 | Not-determined |
| S8_extracted_k141_148023 | Not-determined |
| S8_extracted_k141_182636 | Low-quality |
| S8_extracted_k141_69882 | Not-determined |
| S8_extracted_k141_681 | Not-determined |
| S8_extracted_k141_95963 | Not-determined |
| S8_extracted_k141_26560 | Not-determined |
| S8_extracted_k141_122138 | Not-determined |
| S8_extracted_k141_156620 | Not-determined |
| S8_extracted_k141_191624 | Not-determined |
| S8_extracted_k141_35326 | Not-determined |
| S8_extracted_k141_61356 | Not-determined |
| S8_extracted_k141_17782 | Not-determined |
| S8_extracted_k141_182642 | Not-determined |
| S8_extracted_k141_148028 | Not-determined |
| S8_extracted_k141_113314 | Not-determined |
| S8_extracted_k141_17783 | Not-determined |
| S8_extracted_k141_191626 | Low-quality |
| S8_extracted_k141_113317 | Not-determined |
| S8_extracted_k141_191630 | Not-determined |
| S8_extracted_k141_113322 | Low-quality |
| S8_extracted_k141_174036 | Not-determined |
| S8_extracted_k141_174038 | Not-determined |
| S8_extracted_k141_687 | Not-determined |
| S8_extracted_k141_43872 | Not-determined |
| S8_extracted_k141_87514 | Not-determined |
| S8_extracted_k141_43873 | Not-determined |
| S8_extracted_k141_200057 | Not-determined |
| S8_extracted_k141_139421 | Not-determined |
| S8_extracted_k141_78698 | Not-determined |
| S8_extracted_k141_165584 | Not-determined |
| S8_extracted_k141_52733 | Low-quality |
| S8_extracted_k141_165585 | Not-determined |
| S8_extracted_k141_95976 | Not-determined |
| S8_extracted_k141_165586 | Low-quality |
| S8_extracted_k141_52735 | Low-quality |
| S8_extracted_k141_130805 | Not-determined |
| S8_extracted_k141_87519 | Not-determined |
| S8_extracted_k141_191644 | Not-determined |
| S8_extracted_k141_17789 | Not-determined |
| S8_extracted_k141_95981 | Not-determined |
| S8_extracted_k141_200063 | Not-determined |
| S8_extracted_k141_165592 | Low-quality |
| S8_extracted_k141_200064 | Not-determined |
| S8_extracted_k141_87522 | Not-determined |
| S8_extracted_k141_113334 | Not-determined |
| S8_extracted_k141_148039 | Not-determined |
| S8_extracted_k141_156629 | Not-determined |
| S8_extracted_k141_139432 | Not-determined |
| S8_extracted_k141_122149 | Not-determined |
| S8_extracted_k141_69899 | Not-determined |
| S8_extracted_k141_69900 | Not-determined |
| S8_extracted_k141_26575 | Low-quality |
| S8_extracted_k141_165597 | Not-determined |
| S8_extracted_k141_61370 | Low-quality |
| S8_extracted_k141_156634 | Not-determined |
| S8_extracted_k141_156635 | Not-determined |
| S8_extracted_k141_61373 | Not-determined |
| S8_extracted_k141_200069 | Not-determined |
| S8_extracted_k141_17798 | Not-determined |
| S8_extracted_k141_200072 | Not-determined |
| S8_extracted_k141_52744 | Not-determined |
| S8_extracted_k141_52745 | Not-determined |
| S8_extracted_k141_200075 | Not-determined |
| S8_extracted_k141_61379 | Not-determined |
| S8_extracted_k141_95983 | Not-determined |
| S8_extracted_k141_148055 | Not-determined |
| S8_extracted_k141_17802 | Not-determined |
| S8_extracted_k141_148061 | Not-determined |
| S8_extracted_k141_69908 | Not-determined |
| S8_extracted_k141_26587 | Not-determined |
| S8_extracted_k141_61381 | Not-determined |
| S8_extracted_k141_78711 | Not-determined |
| S8_extracted_k141_35333 | Low-quality |
| S8_extracted_k141_78712 | Not-determined |
| S8_extracted_k141_156647 | Low-quality |
| S8_extracted_k141_165603 | Not-determined |
| S8_extracted_k141_69911 | Not-determined |
| S8_extracted_k141_697 | Not-determined |
| S8_extracted_k141_122159 | Not-determined |
| S8_extracted_k141_156653 | Low-quality |
| S8_extracted_k141_35338 | Not-determined |
| S8_extracted_k141_61391 | Low-quality |
| S8_extracted_k141_182650 | Low-quality |
| S8_extracted_k141_139446 | Not-determined |
| S8_extracted_k141_61393 | Not-determined |
| S8_extracted_k141_139448 | Not-determined |
| S8_extracted_k141_156657 | Low-quality |
| S8_extracted_k141_122165 | Not-determined |
| S8_extracted_k141_702 | Not-determined |
| S8_extracted_k141_17814 | Not-determined |
| S8_extracted_k141_165604 | Not-determined |
| S8_extracted_k141_61399 | Not-determined |
| S8_extracted_k141_130838 | Not-determined |
| S8_extracted_k141_130839 | Low-quality |
| S8_extracted_k141_182657 | Not-determined |
| S8_extracted_k141_122170 | Not-determined |
| S8_extracted_k141_122172 | Not-determined |
| S8_extracted_k141_709 | Not-determined |
| S8_extracted_k141_156665 | Not-determined |
| S8_extracted_k141_61402 | Not-determined |
| S8_extracted_k141_156666 | Not-determined |
| S8_extracted_k141_122174 | Not-determined |
| S8_extracted_k141_200083 | Not-determined |
| S8_extracted_k141_182659 | Not-determined |
| S8_extracted_k141_113343 | Low-quality |
| S8_extracted_k141_191654 | Not-determined |
| S8_extracted_k141_200087 | Not-determined |
| S8_extracted_k141_69921 | Not-determined |
| S8_extracted_k141_17820 | Not-determined |
| S8_extracted_k141_95993 | Not-determined |
| S8_extracted_k141_17821 | Not-determined |
| S8_extracted_k141_35349 | Not-determined |
| S8_extracted_k141_156675 | Not-determined |
| S8_extracted_k141_165612 | Not-determined |
| S8_extracted_k141_87553 | Not-determined |
| S8_extracted_k141_35350 | Low-quality |
| S8_extracted_k141_17825 | Low-quality |
| S8_extracted_k141_122186 | Not-determined |
| S8_extracted_k141_715 | Not-determined |
| S8_extracted_k141_43906 | Not-determined |
| S8_extracted_k141_165613 | Low-quality |
| S8_extracted_k141_96002 | Not-determined |
| S8_extracted_k141_61417 | Low-quality |
| S8_extracted_k141_130843 | Not-determined |
| S8_extracted_k141_17830 | Low-quality |
| S8_extracted_k141_87562 | Not-determined |
| S8_extracted_k141_17831 | Not-determined |
| S8_extracted_k141_113355 | Not-determined |
| S8_extracted_k141_9270 | Not-determined |
| S8_extracted_k141_17834 | Low-quality |
| S8_extracted_k141_139468 | Not-determined |
| S8_extracted_k141_130846 | Low-quality |
| S8_extracted_k141_113358 | Not-determined |
| S8_extracted_k141_9275 | Not-determined |
| S8_extracted_k141_130847 | Not-determined |
| S8_extracted_k141_200089 | Not-determined |
| S8_extracted_k141_726 | Not-determined |
| S8_extracted_k141_87566 | Not-determined |
| S8_extracted_k141_96009 | Not-determined |
| S8_extracted_k141_156694 | Not-determined |
| S8_extracted_k141_139472 | Not-determined |
| S8_extracted_k141_148093 | Not-determined |
| S8_extracted_k141_43918 | Not-determined |
| S8_extracted_k141_130856 | Not-determined |
| S8_extracted_k141_78752 | Not-determined |
| S8_extracted_k141_35358 | Not-determined |
| S8_extracted_k141_730 | Not-determined |
| S8_extracted_k141_69935 | Not-determined |
| S8_extracted_k141_26606 | Not-determined |
| S8_extracted_k141_122198 | Not-determined |
| S8_extracted_k141_9281 | Not-determined |
| S8_extracted_k141_69936 | Not-determined |
| S8_extracted_k141_191665 | Not-determined |
| S8_extracted_k141_139477 | Not-determined |
| S8_extracted_k141_122202 | Not-determined |
| S8_extracted_k141_35361 | Low-quality |
| S8_extracted_k141_35362 | Not-determined |
| S8_extracted_k141_139479 | Not-determined |
| S8_extracted_k141_113372 | Not-determined |
| S8_extracted_k141_113375 | Not-determined |
| S8_extracted_k141_139486 | Low-quality |
| S8_extracted_k141_165629 | Low-quality |
| S8_extracted_k141_139488 | Not-determined |
| S8_extracted_k141_739 | Low-quality |
| S8_extracted_k141_139490 | Not-determined |
| S8_extracted_k141_156705 | Low-quality |
| S8_extracted_k141_78758 | Not-determined |
| S8_extracted_k141_174071 | Not-determined |
| S8_extracted_k141_165632 | Not-determined |
| S8_extracted_k141_96012 | Not-determined |
| S8_extracted_k141_113378 | Low-quality |
| S8_extracted_k141_174077 | Not-determined |
| S8_extracted_k141_148112 | Not-determined |
| S8_extracted_k141_87578 | Not-determined |
| S8_extracted_k141_156711 | Not-determined |
| S8_extracted_k141_43933 | Not-determined |
| S8_extracted_k141_174079 | Not-determined |
| S8_extracted_k141_174080 | Not-determined |
| S8_extracted_k141_96017 | Not-determined |
| S8_extracted_k141_78760 | Not-determined |
| S8_extracted_k141_174081 | Not-determined |
| S8_extracted_k141_96019 | Not-determined |
| S8_extracted_k141_61437 | Not-determined |
| S8_extracted_k141_26613 | Not-determined |
| S8_extracted_k141_26614 | Not-determined |
| S8_extracted_k141_113388 | Not-determined |
| S8_extracted_k141_130878 | Not-determined |
| S8_extracted_k141_104733 | Low-quality |
| S8_extracted_k141_35380 | Not-determined |
| S8_extracted_k141_17837 | Not-determined |
| S8_extracted_k141_69959 | Not-determined |
| S8_extracted_k141_96027 | Not-determined |
| S8_extracted_k141_9310 | Not-determined |
| S8_extracted_k141_96028 | Not-determined |
| S8_extracted_k141_165644 | Not-determined |
| S8_extracted_k141_104738 | Not-determined |
| S8_extracted_k141_174092 | Not-determined |
| S8_extracted_k141_17846 | Not-determined |
| S8_extracted_k141_96031 | Low-quality |
| S8_extracted_k141_760 | Not-determined |
| S8_extracted_k141_69964 | Not-determined |
| S8_extracted_k141_165646 | Not-determined |
| S8_extracted_k141_17847 | Not-determined |
| S8_extracted_k141_9321 | Not-determined |
| S8_extracted_k141_96037 | Not-determined |
| S8_extracted_k141_174098 | Not-determined |
| S8_extracted_k141_139498 | Low-quality |
| S8_extracted_k141_9323 | Low-quality |
| S8_extracted_k141_96039 | Not-determined |
| S8_extracted_k141_78769 | Low-quality |
| S8_extracted_k141_43949 | Not-determined |
| S8_extracted_k141_43951 | Not-determined |
| S8_extracted_k141_182697 | Not-determined |
| S8_extracted_k141_26629 | Not-determined |
| S8_extracted_k141_96043 | Not-determined |
| S8_extracted_k141_52793 | Not-determined |
| S8_extracted_k141_174111 | Low-quality |
| S8_extracted_k141_17854 | Not-determined |
| S8_extracted_k141_139509 | Not-determined |
| S8_extracted_k141_200106 | Not-determined |
| S8_extracted_k141_9333 | Not-determined |
| S8_extracted_k141_87606 | Not-determined |
| S8_extracted_k141_52798 | Not-determined |
| S8_extracted_k141_17860 | Not-determined |
| S8_extracted_k141_26636 | Not-determined |
| S8_extracted_k141_87607 | Low-quality |
| S8_extracted_k141_17861 | Not-determined |
| S8_extracted_k141_69989 | Not-determined |
| S8_extracted_k141_174115 | Not-determined |
| S8_extracted_k141_78783 | Not-determined |
| S8_extracted_k141_104757 | Not-determined |
| S8_extracted_k141_773 | Not-determined |
| S8_extracted_k141_61453 | Not-determined |
| S8_extracted_k141_52805 | Not-determined |
| S8_extracted_k141_69991 | Low-quality |
| S8_extracted_k141_17864 | Low-quality |
| S8_extracted_k141_26641 | Not-determined |
| S8_extracted_k141_43971 | Not-determined |
| S8_extracted_k141_174123 | Not-determined |
| S8_extracted_k141_61458 | Not-determined |
| S8_extracted_k141_17870 | Not-determined |
| S8_extracted_k141_43973 | Low-quality |
| S8_extracted_k141_26643 | Not-determined |
| S8_extracted_k141_43977 | Not-determined |
| S8_extracted_k141_26645 | Not-determined |
| S8_extracted_k141_191690 | Not-determined |
| S8_extracted_k141_43978 | Not-determined |
| S8_extracted_k141_156736 | Not-determined |
| S8_extracted_k141_96054 | Low-quality |
| S8_extracted_k141_9345 | Low-quality |
| S8_extracted_k141_52814 | Not-determined |
| S8_extracted_k141_43981 | Low-quality |
| S8_extracted_k141_148114 | Not-determined |
| S8_extracted_k141_191694 | Not-determined |
| S8_extracted_k141_139525 | Low-quality |
| S8_extracted_k141_156740 | Not-determined |
| S8_extracted_k141_52820 | Not-determined |
| S8_extracted_k141_174131 | Not-determined |
| S8_extracted_k141_182710 | Low-quality |
| S8_extracted_k141_156742 | Not-determined |
| S8_extracted_k141_17885 | Not-determined |
| S8_extracted_k141_191701 | Not-determined |
| S8_extracted_k141_43987 | Not-determined |
| S8_extracted_k141_156746 | Not-determined |
| S8_extracted_k141_17892 | Not-determined |
| S8_extracted_k141_17893 | Not-determined |
| S8_extracted_k141_191703 | Not-determined |
| S8_extracted_k141_26652 | Low-quality |
| S8_extracted_k141_52821 | Not-determined |
| S8_extracted_k141_78800 | Not-determined |
| S8_extracted_k141_43992 | Not-determined |
| S8_extracted_k141_17899 | Not-determined |
| S8_extracted_k141_61476 | Low-quality |
| S8_extracted_k141_52823 | Not-determined |
| S8_extracted_k141_87632 | Not-determined |
| S8_extracted_k141_148121 | Low-quality |
| S8_extracted_k141_87633 | Not-determined |
| S8_extracted_k141_44000 | Not-determined |
| S8_extracted_k141_87634 | Not-determined |
| S8_extracted_k141_104768 | Not-determined |
| S8_extracted_k141_182718 | Not-determined |
| S8_extracted_k141_148125 | Not-determined |
| S8_extracted_k141_104769 | Not-determined |
| S8_extracted_k141_191714 | Not-determined |
| S8_extracted_k141_87639 | Not-determined |
| S8_extracted_k141_26660 | Not-determined |
| S8_extracted_k141_191719 | Not-determined |
| S8_extracted_k141_191720 | Not-determined |
| S8_extracted_k141_52829 | Not-determined |
| S8_extracted_k141_78809 | Not-determined |
| S8_extracted_k141_156764 | Not-determined |
| S8_extracted_k141_26664 | Not-determined |
| S8_extracted_k141_52832 | Not-determined |
| S8_extracted_k141_17902 | Not-determined |
| S8_extracted_k141_26666 | Not-determined |
| S8_extracted_k141_78810 | Not-determined |
| S8_extracted_k141_78812 | Not-determined |
| S8_extracted_k141_165666 | Not-determined |
| S17_vs_k141_159527\|\|full | High-quality |
| S8_extracted_k141_191729 | Not-determined |
| S8_extracted_k141_182726 | Low-quality |
| S8_extracted_k141_191731 | Not-determined |
| S8_extracted_k141_200107 | Low-quality |
| S8_extracted_k141_26673 | Not-determined |
| S8_extracted_k141_139544 | Not-determined |
| S8_extracted_k141_130929 | Not-determined |
| S8_extracted_k141_26677 | Not-determined |
| S8_extracted_k141_122210 | Not-determined |
| S8_extracted_k141_87650 | Not-determined |
| S8_extracted_k141_182733 | Not-determined |
| S8_extracted_k141_87651 | Not-determined |
| S8_extracted_k141_44022 | Not-determined |
| S8_extracted_k141_78818 | Not-determined |
| S8_extracted_k141_104782 | Not-determined |
| S8_extracted_k141_156773 | Not-determined |
| S8_extracted_k141_808 | Not-determined |
| S8_extracted_k141_104784 | Not-determined |
| S8_extracted_k141_130939 | Low-quality |
| S8_extracted_k141_122220 | Not-determined |
| S8_extracted_k141_165678 | Not-determined |
| S8_extracted_k141_182739 | Not-determined |
| S8_extracted_k141_174144 | Not-determined |
| S8_extracted_k141_26687 | Not-determined |
| S8_extracted_k141_78825 | Not-determined |
| S8_extracted_k141_17914 | Not-determined |
| S8_extracted_k141_9372 | Low-quality |
| S8_extracted_k141_816 | Not-determined |
| S8_extracted_k141_26689 | Not-determined |
| S8_extracted_k141_70002 | Not-determined |
| S8_extracted_k141_61503 | Not-determined |
| S8_extracted_k141_200114 | Not-determined |
| S8_extracted_k141_26691 | Not-determined |
| S8_extracted_k141_819 | Not-determined |
| S8_extracted_k141_52845 | Not-determined |
| S8_extracted_k141_52846 | Not-determined |
| S8_extracted_k141_182741 | Not-determined |
| S8_extracted_k141_165682 | Not-determined |
| S8_extracted_k141_104795 | Not-determined |
| S8_extracted_k141_104797 | Not-determined |
| S8_extracted_k141_70005 | Not-determined |
| S8_extracted_k141_191743 | Not-determined |
| S8_extracted_k141_96062 | Not-determined |
| S8_extracted_k141_17923 | Not-determined |
| S8_extracted_k141_165687 | Not-determined |
| S8_extracted_k141_96064 | Not-determined |
| S8_extracted_k141_165688 | Not-determined |
| S8_extracted_k141_96065 | Not-determined |
| S8_extracted_k141_52855 | Not-determined |
| S8_extracted_k141_44033 | Not-determined |
| S8_extracted_k141_52856 | Not-determined |
| S8_extracted_k141_96067 | Not-determined |
| S8_extracted_k141_113427 | Not-determined |
| S8_extracted_k141_96070 | Not-determined |
| S8_extracted_k141_156785 | Low-quality |
| S8_extracted_k141_113430 | Not-determined |
| S8_extracted_k141_44042 | Not-determined |
| S8_extracted_k141_26699 | Not-determined |
| S8_extracted_k141_156789 | Not-determined |

| | |
|---|---|
| S8_extracted_k141_96073 | Not-determined |
| S8_extracted_k141_78842 | Not-determined |
| S8_extracted_k141_35394 | Low-quality |
| S8_extracted_k141_9392 | Not-determined |
| S8_extracted_k141_35395 | Low-quality |
| S8_extracted_k141_96076 | Not-determined |
| S8_extracted_k141_96078 | Not-determined |
| S8_extracted_k141_44044 | Not-determined |
| S8_extracted_k141_70011 | Not-determined |
| S8_extracted_k141_35398 | Not-determined |
| S8_extracted_k141_104803 | Not-determined |
| S8_extracted_k141_130963 | Not-determined |
| S8_extracted_k141_9401 | Not-determined |
| S8_extracted_k141_130964 | Not-determined |
| S8_extracted_k141_130966 | Not-determined |
| S8_extracted_k141_35406 | Not-determined |
| S8_extracted_k141_9404 | Not-determined |
| S8_extracted_k141_122248 | Not-determined |
| S8_extracted_k141_70021 | Low-quality |
| S8_extracted_k141_130970 | Not-determined |
| S8_extracted_k141_35408 | Not-determined |
| S8_extracted_k141_182745 | Not-determined |
| S8_extracted_k141_122254 | Not-determined |
| S8_extracted_k141_9411 | Not-determined |
| S8_extracted_k141_122261 | Not-determined |
| S8_extracted_k141_35416 | Not-determined |
| S8_extracted_k141_87664 | Not-determined |
| S8_extracted_k141_44060 | Not-determined |
| S8_extracted_k141_26715 | Not-determined |
| S8_extracted_k141_87666 | Not-determined |
| S8_extracted_k141_96091 | Not-determined |
| S8_extracted_k141_78855 | Not-determined |
| S8_extracted_k141_96092 | Not-determined |
| S8_extracted_k141_78856 | Not-determined |
| S8_extracted_k141_87669 | Not-determined |
| S8_extracted_k141_35421 | Not-determined |
| S8_extracted_k141_44065 | Not-determined |
| S8_extracted_k141_70033 | Low-quality |
| S8_extracted_k141_44066 | Not-determined |
| S8_extracted_k141_87672 | Not-determined |
| S8_extracted_k141_139588 | Not-determined |
| S8_extracted_k141_139589 | Not-determined |
| S8_extracted_k141_35424 | Not-determined |
| S8_extracted_k141_148145 | Not-determined |
| S8_extracted_k141_17945 | Not-determined |
| S8_extracted_k141_182754 | Not-determined |
| S8_extracted_k141_26724 | Not-determined |
| S8_extracted_k141_17946 | Not-determined |
| S8_extracted_k141_148149 | Low-quality |
| S8_extracted_k141_165721 | Not-determined |
| S8_extracted_k141_26731 | Not-determined |
| S8_extracted_k141_139596 | Not-determined |
| S8_extracted_k141_856 | Not-determined |
| S8_extracted_k141_70042 | Not-determined |
| S8_extracted_k141_17950 | Not-determined |
| S8_extracted_k141_9430 | Not-determined |
| S8_extracted_k141_35432 | Not-determined |
| S8_extracted_k141_860 | Not-determined |
| S8_extracted_k141_26737 | Not-determined |
| S8_extracted_k141_26739 | Not-determined |
| S8_extracted_k141_87686 | Not-determined |
| S8_extracted_k141_148155 | Not-determined |
| S8_extracted_k141_122281 | Not-determined |
| S8_extracted_k141_867 | Not-determined |
| S8_extracted_k141_868 | Not-determined |
| S8_extracted_k141_122283 | Not-determined |
| S8_extracted_k141_182768 | Not-determined |
| S8_extracted_k141_870 | Not-determined |
| S8_extracted_k141_87693 | Not-determined |
| S8_extracted_k141_871 | Not-determined |
| S8_extracted_k141_96101 | Not-determined |
| S8_extracted_k141_200124 | Not-determined |
| S8_extracted_k141_874 | Not-determined |
| S8_extracted_k141_87694 | Not-determined |
| S8_extracted_k141_26750 | Not-determined |
| S8_extracted_k141_139599 | Not-determined |
| S8_extracted_k141_182773 | Not-determined |
| S8_extracted_k141_139602 | Not-determined |
| S8_extracted_k141_96109 | Not-determined |
| S8_extracted_k141_35445 | Not-determined |
| S8_extracted_k141_139605 | Not-determined |
| S8_extracted_k141_87698 | Not-determined |
| S8_extracted_k141_61508 | Not-determined |
| S8_extracted_k141_148166 | Not-determined |
| S8_extracted_k141_131003 | Not-determined |
| S8_extracted_k141_9439 | Not-determined |
| S8_extracted_k141_191762 | Low-quality |
| S8_extracted_k141_87700 | Not-determined |
| S8_extracted_k141_122294 | Not-determined |
| S8_extracted_k141_182775 | Not-determined |
| S8_extracted_k141_200135 | Not-determined |
| S8_extracted_k141_17973 | Low-quality |
| S8_extracted_k141_35451 | Not-determined |
| S8_extracted_k141_61511 | Not-determined |
| S8_extracted_k141_87704 | Not-determined |
| S8_extracted_k141_96117 | Not-determined |
| S8_extracted_k141_70055 | Not-determined |
| S8_extracted_k141_148175 | Not-determined |
| S8_extracted_k141_35459 | Not-determined |
| S8_extracted_k141_139610 | Not-determined |
| S8_extracted_k141_165740 | Low-quality |
| S8_extracted_k141_78890 | Not-determined |
| S8_extracted_k141_9447 | Not-determined |
| S8_extracted_k141_165741 | Not-determined |
| S8_extracted_k141_165742 | Not-determined |
| S8_extracted_k141_35461 | Low-quality |
| S8_extracted_k141_156804 | Not-determined |
| S8_extracted_k141_61520 | Low-quality |
| S8_extracted_k141_96128 | Low-quality |
| S8_extracted_k141_26766 | Not-determined |
| S8_extracted_k141_61521 | Not-determined |
| S8_extracted_k141_113441 | Low-quality |
| S8_extracted_k141_139615 | Not-determined |
| S8_extracted_k141_139618 | Not-determined |
| S8_extracted_k141_122305 | Not-determined |
| S8_extracted_k141_191774 | Not-determined |
| S8_extracted_k141_191776 | Not-determined |
| S8_extracted_k141_87714 | Not-determined |
| S8_extracted_k141_148180 | Not-determined |
| S8_extracted_k141_113447 | Not-determined |
| S8_extracted_k141_165750 | Not-determined |
| S8_extracted_k141_52858 | Not-determined |
| S8_extracted_k141_9458 | Not-determined |
| S8_extracted_k141_104849 | Not-determined |
| S8_extracted_k141_44118 | Not-determined |
| S8_extracted_k141_9459 | Not-determined |
| S8_extracted_k141_191782 | Not-determined |
| S8_extracted_k141_44120 | Low-quality |
| S8_extracted_k141_96137 | Not-determined |
| S8_extracted_k141_26776 | Low-quality |
| S8_extracted_k141_96139 | Not-determined |
| S8_extracted_k141_191787 | Not-determined |
| S8_extracted_k141_200148 | Not-determined |
| S8_extracted_k141_191789 | Not-determined |
| S8_extracted_k141_52862 | Not-determined |
| S8_extracted_k141_104851 | Not-determined |
| S8_extracted_k141_96144 | Not-determined |
| S8_extracted_k141_139633 | Not-determined |
| S8_extracted_k141_78912 | Low-quality |
| S8_extracted_k141_191794 | Not-determined |
| S8_extracted_k141_122320 | Not-determined |
| S8_extracted_k141_78916 | Not-determined |
| S8_extracted_k141_52869 | Not-determined |
| S8_extracted_k141_52870 | Not-determined |
| S8_extracted_k141_26782 | Not-determined |
| S8_extracted_k141_191796 | Not-determined |
| S8_extracted_k141_87727 | Not-determined |
| S8_extracted_k141_113457 | Low-quality |
| S8_extracted_k141_156817 | Not-determined |
| S8_extracted_k141_191798 | Not-determined |
| S8_extracted_k141_200159 | Not-determined |
| S8_extracted_k141_78924 | Not-determined |
| S8_extracted_k141_61538 | Not-determined |
| S8_extracted_k141_78925 | Not-determined |
| S8_extracted_k141_61540 | Not-determined |
| S8_extracted_k141_191802 | Not-determined |
| S8_extracted_k141_897 | Not-determined |
| S8_extracted_k141_899 | Not-determined |
| S8_extracted_k141_148201 | Not-determined |
| S8_extracted_k141_26791 | Not-determined |
| S8_extracted_k141_26794 | Not-determined |
| S8_extracted_k141_165770 | Not-determined |
| S8_extracted_k141_96159 | Not-determined |
| S8_extracted_k141_139641 | Not-determined |
| S8_extracted_k141_104865 | Not-determined |
| S8_extracted_k141_182796 | Not-determined |
| S8_extracted_k141_156830 | Not-determined |
| S8_extracted_k141_78934 | Not-determined |
| S8_extracted_k141_139644 | Not-determined |
| S8_extracted_k141_44151 | Not-determined |
| S8_extracted_k141_139647 | Not-determined |
| S8_extracted_k141_70074 | Not-determined |
| S8_extracted_k141_52877 | Not-determined |
| S8_extracted_k141_52878 | Not-determined |
| S8_extracted_k141_139649 | Not-determined |
| S8_extracted_k141_52880 | Not-determined |
| S8_extracted_k141_52881 | Not-determined |
| S8_extracted_k141_113468 | Low-quality |
| S8_extracted_k141_26802 | Not-determined |
| S8_extracted_k141_191824 | Not-determined |
| S8_extracted_k141_113469 | Not-determined |
| S8_extracted_k141_156839 | Not-determined |
| S8_extracted_k141_916 | Not-determined |
| S8_extracted_k141_156840 | Low-quality |
| S8_extracted_k141_87749 | Not-determined |
| S8_extracted_k141_61560 | Not-determined |
| S8_extracted_k141_191827 | Not-determined |
| S8_extracted_k141_44157 | Not-determined |
| S8_extracted_k141_191829 | Not-determined |
| S8_extracted_k141_52888 | Not-determined |
| S8_extracted_k141_104870 | Not-determined |
| S8_extracted_k141_52889 | Not-determined |
| S8_extracted_k141_191831 | Not-determined |
| S8_extracted_k141_921 | Not-determined |
| S8_extracted_k141_922 | Not-determined |
| S8_extracted_k141_156845 | Not-determined |
| S8_extracted_k141_35490 | Low-quality |
| S8_extracted_k141_26805 | Not-determined |
| S8_extracted_k141_26806 | Not-determined |
| S8_extracted_k141_52890 | Not-determined |
| S8_extracted_k141_96169 | Not-determined |
| S8_extracted_k141_182813 | Not-determined |
| S8_extracted_k141_26808 | Not-determined |
| S8_extracted_k141_182814 | Not-determined |
| S8_extracted_k141_182815 | Low-quality |
| S8_extracted_k141_44173 | Not-determined |
| S8_extracted_k141_156851 | Not-determined |
| S8_extracted_k141_78951 | Not-determined |
| S8_extracted_k141_9484 | Not-determined |
| S8_extracted_k141_35500 | Not-determined |
| S8_extracted_k141_9485 | Not-determined |
| S8_extracted_k141_78954 | Not-determined |
| S8_extracted_k141_26814 | Not-determined |
| S8_extracted_k141_35502 | Not-determined |
| S8_extracted_k141_61569 | Not-determined |
| S8_extracted_k141_44174 | Not-determined |
| S8_extracted_k141_96177 | Low-quality |
| S8_extracted_k141_148220 | Not-determined |
| S8_extracted_k141_17991 | Low-quality |
| S8_extracted_k141_17992 | Not-determined |
| S8_extracted_k141_61572 | Not-determined |
| S8_extracted_k141_927 | Not-determined |
| S8_extracted_k141_122362 | Not-determined |
| S8_extracted_k141_35506 | Not-determined |
| S8_extracted_k141_70106 | Not-determined |
| S8_extracted_k141_9489 | Not-determined |
| S8_extracted_k141_44185 | Not-determined |
| S8_extracted_k141_78965 | Not-determined |
| S8_extracted_k141_26823 | Not-determined |
| S8_extracted_k141_9494 | Not-determined |
| S8_extracted_k141_191856 | Not-determined |
| S8_extracted_k141_139681 | Low-quality |
| S8_extracted_k141_61580 | Not-determined |
| S8_extracted_k141_148228 | Not-determined |
| S8_extracted_k141_191861 | Not-determined |
| S8_extracted_k141_96188 | Not-determined |
| S8_extracted_k141_148232 | Not-determined |
| S8_extracted_k141_78968 | Not-determined |
| S8_extracted_k141_26831 | Not-determined |
| S8_extracted_k141_122371 | Not-determined |
| S8_extracted_k141_148237 | Not-determined |
| S8_extracted_k141_148238 | Not-determined |
| S16_vs_8493\|\|full | High-quality |
| S8_extracted_k141_96194 | Not-determined |
| S8_extracted_k141_191869 | Not-determined |
| S8_extracted_k141_139685 | Not-determined |
| S8_extracted_k141_113490 | Not-determined |
| S8_extracted_k141_948 | Not-determined |
| S8_extracted_k141_131014 | Not-determined |
| S8_extracted_k141_70115 | Low-quality |
| S8_extracted_k141_949 | Not-determined |
| S8_extracted_k141_156855 | Not-determined |
| S8_extracted_k141_104881 | Not-determined |
| S8_extracted_k141_131016 | Low-quality |
| S8_extracted_k141_139687 | Not-determined |
| S8_extracted_k141_78977 | Not-determined |
| S8_extracted_k141_191874 | Not-determined |
| S8_extracted_k141_96198 | Not-determined |
| S8_extracted_k141_131019 | Not-determined |
| S8_extracted_k141_70121 | Not-determined |
| S8_extracted_k141_78982 | Low-quality |
| S8_extracted_k141_156866 | Not-determined |
| S8_extracted_k141_113500 | Not-determined |
| S8_extracted_k141_96202 | Not-determined |
| S8_extracted_k141_26843 | Not-determined |
| S8_extracted_k141_44203 | Not-determined |
| S8_extracted_k141_96208 | Not-determined |
| S8_extracted_k141_52898 | Not-determined |
| S8_extracted_k141_174162 | Not-determined |
| S8_extracted_k141_35540 | Not-determined |
| S8_extracted_k141_61592 | Not-determined |
| S8_extracted_k141_962 | Not-determined |
| S8_extracted_k141_78991 | Not-determined |
| S8_extracted_k141_44208 | Not-determined |
| S8_extracted_k141_964 | Not-determined |
| S8_extracted_k141_78994 | Not-determined |
| S8_extracted_k141_156881 | Not-determined |
| S8_extracted_k141_9518 | Not-determined |
| S8_extracted_k141_148244 | Not-determined |
| S8_extracted_k141_148245 | Not-determined |
| S8_extracted_k141_131043 | Not-determined |
| S8_extracted_k141_61603 | Not-determined |
| S8_extracted_k141_139698 | Not-determined |
| S8_extracted_k141_971 | Not-determined |
| S8_extracted_k141_96227 | Not-determined |
| S8_extracted_k141_70134 | Not-determined |
| S8_extracted_k141_148248 | Low-quality |
| S8_extracted_k141_104893 | Not-determined |
| S8_extracted_k141_139703 | Not-determined |
| S8_extracted_k141_61610 | Not-determined |
| S8_extracted_k141_104898 | Not-determined |
| S8_extracted_k141_122401 | Not-determined |
| S8_extracted_k141_61615 | Low-quality |
| S8_extracted_k141_52902 | Not-determined |
| S8_extracted_k141_148252 | Not-determined |
| S8_extracted_k141_104902 | Not-determined |
| S8_extracted_k141_191888 | Not-determined |
| S8_extracted_k141_191892 | Not-determined |
| S8_extracted_k141_191893 | Not-determined |
| S8_extracted_k141_156882 | Not-determined |
| S8_extracted_k141_52912 | Not-determined |
| S8_extracted_k141_182852 | Not-determined |
| S8_extracted_k141_131057 | Low-quality |
| S8_extracted_k141_191898 | Not-determined |
| S8_extracted_k141_52914 | Not-determined |
| S8_extracted_k141_182853 | Not-determined |
| S8_extracted_k141_52917 | Not-determined |
| S8_extracted_k141_131059 | Not-determined |
| S8_extracted_k141_174188 | Not-determined |
| S8_extracted_k141_18023 | Low-quality |
| S8_extracted_k141_18025 | Not-determined |
| S8_extracted_k141_182858 | Not-determined |
| S8_extracted_k141_174190 | Not-determined |
| S8_extracted_k141_139713 | Not-determined |
| S8_extracted_k141_104908 | Not-determined |
| S8_extracted_k141_113534 | Low-quality |
| S8_extracted_k141_973 | Not-determined |
| S8_extracted_k141_9533 | Not-determined |
| S8_extracted_k141_122408 | Not-determined |
| S8_extracted_k141_148269 | Not-determined |
| S8_extracted_k141_131065 | Not-determined |
| S8_extracted_k141_174197 | Not-determined |
| S8_extracted_k141_9537 | Not-determined |
| S8_extracted_k141_191903 | Not-determined |
| S8_extracted_k141_191904 | Not-determined |
| S8_extracted_k141_79004 | Not-determined |
| S4_extracted_30103 | Medium-quality |
| S8_extracted_k141_79005 | Not-determined |
| S8_extracted_k141_35559 | Not-determined |
| S8_extracted_k141_104915 | Not-determined |
| S8_extracted_k141_156897 | Not-determined |
| S8_extracted_k141_122416 | Not-determined |
| S8_extracted_k141_96241 | Not-determined |
| S8_extracted_k141_148277 | Not-determined |
| S8_extracted_k141_122419 | Not-determined |
| S8_extracted_k141_182864 | Not-determined |
| S8_extracted_k141_61629 | Not-determined |
| S8_extracted_k141_52925 | Not-determined |
| S8_extracted_k141_96245 | Not-determined |
| S8_extracted_k141_104920 | Not-determined |
| S8_extracted_k141_200166 | Not-determined |
| S8_extracted_k141_191916 | Not-determined |
| S8_extracted_k141_191917 | Not-determined |
| S8_extracted_k141_182869 | Not-determined |
| S8_extracted_k141_148290 | Not-determined |
| S8_extracted_k141_182872 | Not-determined |
| S8_extracted_k141_18039 | Low-quality |
| S8_extracted_k141_131076 | Not-determined |
| S8_extracted_k141_113551 | Not-determined |
| S8_extracted_k141_96253 | Not-determined |
| S8_extracted_k141_986 | Low-quality |
| S8_extracted_k141_200170 | Not-determined |
| S8_extracted_k141_131082 | Not-determined |
| S8_extracted_k141_104931 | Not-determined |
| S8_extracted_k141_139738 | Not-determined |
| S8_extracted_k141_104935 | Not-determined |
| S8_extracted_k141_104936 | Not-determined |
| S8_extracted_k141_122427 | Not-determined |
| S8_extracted_k141_9552 | Not-determined |
| S8_extracted_k141_191924 | Not-determined |
| S8_extracted_k141_191925 | Not-determined |
| S8_extracted_k141_79017 | Not-determined |
| S8_extracted_k141_18046 | Not-determined |
| S8_extracted_k141_9553 | Not-determined |
| S8_extracted_k141_26898 | Not-determined |
| S8_extracted_k141_26899 | Not-determined |
| S8_extracted_k141_96258 | Not-determined |
| S8_extracted_k141_139741 | Not-determined |
| S8_extracted_k141_998 | Not-determined |
| S8_extracted_k141_122434 | Not-determined |
| S8_extracted_k141_156904 | Not-determined |
| S8_extracted_k141_131094 | Not-determined |
| S8_extracted_k141_191931 | Not-determined |
| S8_extracted_k141_139743 | Not-determined |
| S8_extracted_k141_26903 | Not-determined |
| S8_extracted_k141_96263 | Not-determined |
| S8_extracted_k141_104947 | Not-determined |
| S8_extracted_k141_26905 | Not-determined |
| S8_extracted_k141_18048 | Not-determined |
| S8_extracted_k141_18049 | Not-determined |
| S8_extracted_k141_104951 | Not-determined |
| S8_extracted_k141_148298 | Not-determined |
| S8_extracted_k141_148299 | Not-determined |
| S8_extracted_k141_148300 | Not-determined |
| S8_extracted_k141_52946 | Not-determined |
| S8_extracted_k141_61641 | Not-determined |
| S8_extracted_k141_26910 | Not-determined |
| S8_extracted_k141_156916 | Not-determined |
| S8_extracted_k141_148304 | Not-determined |
| S8_extracted_k141_18053 | Not-determined |
| S8_extracted_k141_87764 | Not-determined |
| S8_extracted_k141_139747 | Not-determined |
| S8_extracted_k141_9562 | Not-determined |
| S8_extracted_k141_18057 | Not-determined |
| S8_extracted_k141_18058 | Not-determined |
| S8_extracted_k141_61648 | Not-determined |
| S8_extracted_k141_9566 | Not-determined |
| S8_extracted_k141_139752 | Not-determined |
| S8_extracted_k141_9567 | Not-determined |
| S8_extracted_k141_113566 | Not-determined |
| S8_extracted_k141_26921 | Not-determined |
| S8_extracted_k141_156925 | Not-determined |
| S8_extracted_k141_131114 | Not-determined |
| S8_extracted_k141_200183 | Not-determined |
| S8_extracted_k141_174212 | Low-quality |
| S8_extracted_k141_200184 | Not-determined |
| S8_extracted_k141_35564 | Not-determined |
| S8_extracted_k141_200186 | Not-determined |
| S8_extracted_k141_191946 | Not-determined |
| S8_extracted_k141_79027 | Not-determined |
| S8_extracted_k141_35567 | Not-determined |
| S8_extracted_k141_26925 | Not-determined |
| S8_extracted_k141_113573 | Not-determined |
| S8_extracted_k141_113574 | Not-determined |
| S8_extracted_k141_113575 | Not-determined |
| S8_extracted_k141_191952 | Not-determined |
| S8_extracted_k141_113577 | Not-determined |
| S8_extracted_k141_200190 | Not-determined |
| S8_extracted_k141_52949 | Low-quality |
| S8_extracted_k141_122443 | Not-determined |
| S8_extracted_k141_174216 | Low-quality |
| S8_extracted_k141_113578 | Not-determined |
| S8_extracted_k141_1008 | Not-determined |
| S8_extracted_k141_191958 | Low-quality |
| S8_extracted_k141_1009 | Not-determined |
| S8_extracted_k141_113581 | Not-determined |
| S8_extracted_k141_200194 | Not-determined |
| S8_extracted_k141_35576 | Not-determined |
| S8_extracted_k141_148317 | Not-determined |
| S8_extracted_k141_61656 | Low-quality |
| S8_extracted_k141_156936 | Low-quality |
| S8_extracted_k141_61657 | Not-determined |
| S8_extracted_k141_156937 | Not-determined |
| S8_extracted_k141_191962 | Not-determined |
| S8_extracted_k141_139758 | Not-determined |
| S8_extracted_k141_35581 | Not-determined |
| S8_extracted_k141_44225 | Not-determined |
| S8_extracted_k141_87776 | Not-determined |
| S8_extracted_k141_61663 | Not-determined |
| S8_extracted_k141_61664 | Not-determined |
| S8_extracted_k141_61666 | Low-quality |
| S8_extracted_k141_174222 | Not-determined |
| S8_extracted_k141_44231 | Not-determined |
| S8_extracted_k141_156938 | Not-determined |
| S8_extracted_k141_113592 | Not-determined |
| S8_extracted_k141_52962 | Low-quality |
| S8_extracted_k141_139765 | Not-determined |
| S8_extracted_k141_96289 | Not-determined |
| S8_extracted_k141_174226 | Not-determined |
| S8_extracted_k141_131118 | Not-determined |
| S8_extracted_k141_1025 | Not-determined |
| S8_extracted_k141_18075 | Low-quality |
| S8_extracted_k141_96290 | Not-determined |
| S8_extracted_k141_148332 | Not-determined |
| S8_extracted_k141_156941 | Not-determined |
| S8_extracted_k141_79042 | Not-determined |
| S8_extracted_k141_165784 | Not-determined |
| S8_extracted_k141_79043 | Not-determined |
| S8_extracted_k141_139771 | Not-determined |
| S8_extracted_k141_35593 | Not-determined |
| S8_extracted_k141_96301 | Not-determined |
| S8_extracted_k141_35598 | Not-determined |
| S8_extracted_k141_79054 | Not-determined |
| S8_extracted_k141_44237 | Not-determined |
| S4_vs_VIRSorter_30103-cat_2 | Medium-quality |
| S8_extracted_k141_165792 | Not-determined |
| S8_extracted_k141_191973 | Not-determined |
| S8_extracted_k141_79057 | Not-determined |
| S8_extracted_k141_79058 | Not-determined |
| S8_extracted_k141_156955 | Not-determined |
| S8_extracted_k141_104960 | Not-determined |
| S8_extracted_k141_26944 | Not-determined |
| S8_extracted_k141_87789 | Low-quality |
| S8_extracted_k141_35603 | Not-determined |
| S8_extracted_k141_156956 | Not-determined |
| S8_extracted_k141_156958 | Not-determined |
| S8_extracted_k141_131124 | Low-quality |
| S8_extracted_k141_44238 | Not-determined |
| S8_extracted_k141_87794 | Not-determined |
| S8_extracted_k141_182887 | Not-determined |
| S8_extracted_k141_131126 | Low-quality |
| S8_extracted_k141_104969 | Not-determined |
| S8_extracted_k141_174241 | Not-determined |
| S8_extracted_k141_18092 | Not-determined |
| S8_extracted_k141_35610 | Low-quality |
| S8_extracted_k141_122463 | Not-determined |
| S8_extracted_k141_174244 | Not-determined |
| S8_extracted_k141_104973 | Not-determined |
| S8_extracted_k141_113608 | Not-determined |
| S8_extracted_k141_104975 | Not-determined |
| S8_extracted_k141_174245 | Not-determined |
| S8_extracted_k141_131129 | Not-determined |
| S8_extracted_k141_87802 | Not-determined |
| S8_extracted_k141_26952 | Not-determined |
| S8_extracted_k141_113611 | Not-determined |
| S8_extracted_k141_113612 | Not-determined |
| S8_extracted_k141_96317 | Not-determined |
| S8_extracted_k141_165797 | Not-determined |
| S8_extracted_k141_113613 | Not-determined |
| S8_extracted_k141_156973 | Not-determined |
| S8_extracted_k141_96319 | Not-determined |
| S8_extracted_k141_191983 | Not-determined |
| S8_extracted_k141_122468 | Not-determined |
| S8_extracted_k141_35628 | Not-determined |
| S8_extracted_k141_131140 | Not-determined |
| S8_extracted_k141_131141 | Low-quality |
| S8_extracted_k141_131142 | Not-determined |
| S8_extracted_k141_26959 | Not-determined |
| S8_extracted_k141_44250 | Not-determined |
| S8_extracted_k141_104981 | Not-determined |
| S8_extracted_k141_26961 | Not-determined |
| S8_extracted_k141_148352 | Not-determined |
| S8_extracted_k141_156980 | Not-determined |
| S8_extracted_k141_122476 | Not-determined |
| S8_extracted_k141_191987 | Not-determined |
| S8_extracted_k141_139793 | Not-determined |
| S8_extracted_k141_139795 | Not-determined |
| S8_extracted_k141_61697 | Low-quality |
| S8_extracted_k141_200200 | Not-determined |
| S8_extracted_k141_26968 | Low-quality |
| S8_extracted_k141_35630 | Not-determined |
| S8_extracted_k141_61698 | Low-quality |
| S8_extracted_k141_79072 | Not-determined |
| S8_extracted_k141_26970 | Not-determined |
| S8_extracted_k141_191994 | Not-determined |
| S8_extracted_k141_191995 | Not-determined |
| S8_extracted_k141_35633 | Not-determined |
| S8_extracted_k141_113616 | Not-determined |
| S8_extracted_k141_96329 | Not-determined |
| S8_extracted_k141_191998 | Not-determined |
| S8_extracted_k141_35641 | Not-determined |
| S8_extracted_k141_122488 | Not-determined |
| S8_extracted_k141_148354 | Not-determined |
| S8_extracted_k141_35643 | Low-quality |
| S8_extracted_k141_44262 | Not-determined |
| S8_extracted_k141_87811 | Not-determined |
| S8_extracted_k141_52989 | Not-determined |
| S8_extracted_k141_156991 | Not-determined |
| S8_extracted_k141_70177 | Not-determined |
| S8_extracted_k141_182909 | Not-determined |
| S8_extracted_k141_131153 | Not-determined |
| S8_extracted_k141_52991 | Not-determined |
| S8_extracted_k141_192003 | Not-determined |
| S8_extracted_k141_122496 | Not-determined |
| S8_extracted_k141_61705 | Not-determined |
| S8_extracted_k141_122498 | Not-determined |
| S8_extracted_k141_156998 | Not-determined |
| S8_extracted_k141_61707 | Not-determined |
| S8_extracted_k141_35648 | Low-quality |
| S8_extracted_k141_157000 | Not-determined |
| S8_extracted_k141_104998 | Not-determined |
| S8_extracted_k141_131156 | Not-determined |
| S8_extracted_k141_174254 | Low-quality |
| S8_extracted_k141_182917 | Not-determined |
| S8_extracted_k141_113627 | Not-determined |
| S8_extracted_k141_96340 | Not-determined |
| S8_extracted_k141_79082 | Not-determined |
| S8_extracted_k141_26984 | Not-determined |
| S8_extracted_k141_174258 | Not-determined |
| S8_extracted_k141_96342 | Not-determined |
| S8_extracted_k141_139803 | Not-determined |
| S8_extracted_k141_139804 | Not-determined |
| S8_extracted_k141_87820 | Low-quality |
| S8_extracted_k141_61716 | Not-determined |
| S8_extracted_k141_87822 | Not-determined |
| S8_extracted_k141_122510 | Not-determined |
| S8_extracted_k141_122513 | Not-determined |
| S8_extracted_k141_131161 | Low-quality |
| S8_extracted_k141_70185 | Not-determined |
| S8_extracted_k141_26991 | Low-quality |
| S8_extracted_k141_26992 | Not-determined |
| S8_extracted_k141_200210 | Not-determined |
| S8_extracted_k141_165814 | Not-determined |
| S8_extracted_k141_131163 | Not-determined |
| S8_extracted_k141_113636 | Not-determined |
| S8_extracted_k141_26994 | Not-determined |
| S8_extracted_k141_44269 | Low-quality |
| S8_extracted_k141_87827 | Not-determined |
| S8_extracted_k141_113639 | Not-determined |
| S8_extracted_k141_122520 | Not-determined |
| S8_extracted_k141_157014 | Not-determined |
| S8_extracted_k141_70189 | Not-determined |
| S8_extracted_k141_139810 | Low-quality |
| S8_extracted_k141_79091 | Not-determined |
| S8_extracted_k141_182925 | Not-determined |
| S8_extracted_k141_44274 | Not-determined |
| S8_extracted_k141_165819 | Low-quality |
| S8_extracted_k141_165820 | Not-determined |
| S8_extracted_k141_139812 | Low-quality |

| | |
|---|---|
| SB_extracted_k141_182929 | Not-determined |
| SB_extracted_k141_27003 | Not-determined |
| SB_extracted_k141_27005 | Not-determined |
| SB_extracted_k141_79100 | Not-determined |
| SB_extracted_k141_79101 | Not-determined |
| SB_extracted_k141_96356 | Not-determined |
| SB_extracted_k141_27009 | Not-determined |
| SB_extracted_k141_79102 | Not-determined |
| SB_extracted_k141_9577 | Not-determined |
| SB_extracted_k141_113649 | Low-quality |
| SB_extracted_k141_192008 | Low-quality |
| SB_extracted_k141_200217 | Not-determined |
| SB_extracted_k141_53012 | Not-determined |
| SB_extracted_k141_192011 | Not-determined |
| SB_extracted_k141_200221 | Not-determined |
| SB_extracted_k141_87841 | Not-determined |
| SB_extracted_k141_35679 | Not-determined |
| SB_extracted_k141_200222 | Not-determined |
| SB_extracted_k141_61733 | Not-determined |
| SB_extracted_k141_79108 | Not-determined |
| SB_extracted_k141_79109 | Not-determined |
| SB_extracted_k141_192016 | Not-determined |
| SB_extracted_k141_87845 | Not-determined |
| SB_extracted_k141_35680 | Not-determined |
| SB_extracted_k141_9587 | Not-determined |
| SB_extracted_k141_79114 | Not-determined |
| SB_extracted_k141_53015 | Low-quality |
| SB_extracted_k141_200228 | Not-determined |
| SB_extracted_k141_174279 | Not-determined |
| SB_extracted_k141_122534 | Not-determined |
| SB_extracted_k141_87849 | Not-determined |
| SB_extracted_k141_192019 | Not-determined |
| SB_extracted_k141_27021 | Not-determined |
| SB_extracted_k141_87852 | Not-determined |
| SB_extracted_k141_139825 | Not-determined |
| SB_extracted_k141_113654 | Low-quality |
| SB_extracted_k141_139826 | Not-determined |
| SB_extracted_k141_87854 | Not-determined |
| SB_extracted_k141_165835 | Not-determined |
| SB_extracted_k141_174281 | Not-determined |
| SB_extracted_k141_53022 | Not-determined |
| SB_extracted_k141_53023 | Not-determined |
| SB_extracted_k141_9592 | Not-determined |
| SB_extracted_k141_174286 | Not-determined |
| SB_extracted_k141_87862 | Not-determined |
| SB_extracted_k141_9595 | Not-determined |
| SB_extracted_k141_105027 | Not-determined |
| SB_extracted_k141_139833 | Not-determined |
| SB_extracted_k141_148374 | Not-determined |
| SB_extracted_k141_35699 | Not-determined |
| SB_extracted_k141_87866 | Not-determined |
| SB_extracted_k141_148380 | Not-determined |
| SB_extracted_k141_148381 | Not-determined |
| SB_extracted_k141_165851 | Not-determined |
| SB_extracted_k141_113679 | Not-determined |
| SB_extracted_k141_139843 | Low-quality |
| SB_extracted_k141_192034 | Low-quality |
| SB_extracted_k141_96369 | Not-determined |
| SB_extracted_k141_157032 | Not-determined |
| SB_extracted_k141_113684 | Low-quality |
| SB_extracted_k141_113685 | Not-determined |
| SB_extracted_k141_70197 | Not-determined |
| SB_extracted_k141_113686 | Not-determined |
| SB_extracted_k141_148387 | Low-quality |
| SB_extracted_k141_192038 | Low-quality |
| SB_extracted_k141_96374 | Not-determined |
| SB_extracted_k141_174292 | Low-quality |
| SB_extracted_k141_174295 | Not-determined |
| SB_extracted_k141_105038 | Not-determined |
| SB_extracted_k141_174298 | Not-determined |
| SB_extracted_k141_44309 | Not-determined |
| SB_extracted_k141_96380 | Not-determined |
| SB_extracted_k141_96381 | Not-determined |
| SB_extracted_k141_139853 | Not-determined |
| SB_extracted_k141_96384 | Not-determined |
| SB_extracted_k141_105042 | Not-determined |
| SB_extracted_k141_113692 | Not-determined |
| SB_extracted_k141_200257 | Low-quality |
| SB_extracted_k141_192044 | Not-determined |
| SB_extracted_k141_174300 | Not-determined |
| SB_extracted_k141_192045 | Low-quality |
| SB_extracted_k141_139858 | Not-determined |
| SB_extracted_k141_9634 | Low-quality |
| SB_extracted_k141_35719 | Not-determined |
| SB_extracted_k141_9637 | Not-determined |
| SB_extracted_k141_113698 | Not-determined |
| SB_extracted_k141_157046 | Not-determined |
| SB_extracted_k141_79138 | Not-determined |
| SB_extracted_k141_79140 | Not-determined |
| SB_extracted_k141_79142 | Not-determined |
| SB_extracted_k141_174306 | Low-quality |
| SB_extracted_k141_113705 | Not-determined |
| SB_extracted_k141_174307 | Not-determined |
| SB_extracted_k141_9641 | Not-determined |
| SB_extracted_k141_139866 | Not-determined |
| SB_extracted_k141_96398 | Low-quality |
| SB_extracted_k141_131202 | Not-determined |
| SB_extracted_k141_79143 | Not-determined |
| SB_extracted_k141_131204 | Not-determined |
| SB_extracted_k141_174313 | Not-determined |
| SB_extracted_k141_87880 | Not-determined |
| SB_extracted_k141_70206 | Not-determined |
| SB_extracted_k141_139869 | Not-determined |
| SB_extracted_k141_87886 | Not-determined |
| SB_extracted_k141_27029 | Not-determined |
| SB_extracted_k141_9648 | Low-quality |
| SB_extracted_k141_61756 | Not-determined |
| SB_extracted_k141_96408 | Not-determined |
| SB_extracted_k141_131211 | Not-determined |
| SB_extracted_k141_35739 | Not-determined |
| SB_extracted_k141_131212 | Not-determined |
| SB_extracted_k141_113723 | Not-determined |
| SB_extracted_k141_35741 | Not-determined |
| SB_extracted_k141_53038 | Not-determined |
| SB_extracted_k141_61757 | Low-quality |
| SB_extracted_k141_18107 | Not-determined |
| SB_extracted_k141_53042 | Not-determined |
| SB_extracted_k141_200279 | Not-determined |
| SB_extracted_k141_18108 | Not-determined |
| SB_extracted_k141_27037 | Not-determined |
| SB_extracted_k141_174322 | Not-determined |
| SB_extracted_k141_139872 | Not-determined |
| SB_extracted_k141_18112 | Not-determined |
| SB_extracted_k141_18113 | Not-determined |
| SB_extracted_k141_87907 | Low-quality |
| SB_extracted_k141_113735 | Not-determined |
| SB_extracted_k141_18118 | Not-determined |
| SB_extracted_k141_113736 | Not-determined |
| SB_extracted_k141_165867 | Not-determined |
| SB_extracted_k141_70216 | Not-determined |
| SB_extracted_k141_174330 | Not-determined |
| SB_extracted_k141_200280 | Low-quality |
| SB_extracted_k141_18121 | Not-determined |
| SB_extracted_k141_139877 | Not-determined |
| SB_extracted_k141_200281 | Not-determined |
| SB_extracted_k141_157050 | Low-quality |
| SB_extracted_k141_79174 | Not-determined |
| SB_extracted_k141_18122 | Not-determined |
| SB_extracted_k141_105059 | Not-determined |
| SB_extracted_k141_87914 | Not-determined |
| SB_extracted_k141_113741 | Not-determined |
| SB_extracted_k141_113743 | Not-determined |
| SB_extracted_k141_79180 | Not-determined |
| SB_extracted_k141_165878 | Not-determined |
| SB_extracted_k141_105061 | Not-determined |
| SB_extracted_k141_192066 | Not-determined |
| SB_extracted_k141_131217 | Not-determined |
| SB_extracted_k141_9659 | Not-determined |
| SB_extracted_k141_113750 | Not-determined |
| SB_extracted_k141_131220 | Not-determined |
| SB_extracted_k141_165881 | Not-determined |
| SB_extracted_k141_96429 | Not-determined |
| SB_extracted_k141_131221 | Not-determined |
| SB_extracted_k141_87924 | Not-determined |
| SB_extracted_k141_131223 | Not-determined |
| SB_extracted_k141_18128 | Not-determined |
| SB_extracted_k141_53059 | Not-determined |
| SB_extracted_k141_174350 | Not-determined |
| SB_extracted_k141_27063 | Not-determined |
| SB_extracted_k141_79187 | Not-determined |
| SB_extracted_k141_96431 | Low-quality |
| SB_extracted_k141_27065 | Not-determined |
| SB_extracted_k141_192074 | Low-quality |
| SB_extracted_k141_174351 | Not-determined |
| SB_extracted_k141_35759 | Not-determined |
| SB_extracted_k141_35760 | Not-determined |
| SB_extracted_k141_27068 | Low-quality |
| SB_extracted_k141_18129 | Not-determined |
| SB_extracted_k141_18130 | Not-determined |
| SB_extracted_k141_27070 | Not-determined |
| SB_extracted_k141_53062 | Not-determined |
| SB_extracted_k141_61777 | Low-quality |
| SB_extracted_k141_61779 | Not-determined |
| SB_extracted_k141_96435 | Not-determined |
| SB_extracted_k141_87931 | Not-determined |
| SB_extracted_k141_105071 | Not-determined |
| SB_extracted_k141_105072 | Not-determined |
| SB_extracted_k141_165895 | Not-determined |
| SB_extracted_k141_18131 | Not-determined |
| SB_extracted_k141_139894 | Not-determined |
| SB_extracted_k141_87935 | Low-quality |
| SB_extracted_k141_113751 | Low-quality |
| SB_extracted_k141_18133 | Not-determined |
| SB_extracted_k141_200305 | Not-determined |
| SB_extracted_k141_105081 | Not-determined |
| SB_extracted_k141_9675 | Not-determined |
| SB_extracted_k141_96438 | Low-quality |
| SB_extracted_k141_148396 | Not-determined |
| SB_extracted_k141_139900 | Not-determined |
| SB_extracted_k141_105083 | Not-determined |
| SB_extracted_k141_35770 | Not-determined |
| SB_extracted_k141_96439 | Not-determined |
| SB_extracted_k141_35771 | Not-determined |
| SB_extracted_k141_18139 | Not-determined |
| SB_extracted_k141_53078 | Not-determined |
| SB_extracted_k141_122562 | Not-determined |
| SB_extracted_k141_79202 | Not-determined |
| SB_extracted_k141_113756 | Not-determined |
| SB_extracted_k141_35778 | Not-determined |
| SB_extracted_k141_35779 | Not-determined |
| SB_extracted_k141_148403 | Not-determined |
| SB_extracted_k141_1062 | Not-determined |
| SB_extracted_k141_148404 | Low-quality |
| SB_extracted_k141_122568 | Not-determined |
| SB_extracted_k141_174361 | Not-determined |
| SB_extracted_k141_87941 | Not-determined |
| SB_extracted_k141_165914 | Not-determined |
| SB_extracted_k141_1068 | Not-determined |
| SB_extracted_k141_157065 | Low-quality |
| SB_extracted_k141_139917 | Not-determined |
| SB_extracted_k141_131236 | Not-determined |
| SB_extracted_k141_105104 | Not-determined |
| SB_extracted_k141_113760 | Not-determined |
| SB_extracted_k141_9684 | Not-determined |
| SB_extracted_k141_113761 | Not-determined |
| SB_extracted_k141_96449 | Not-determined |
| SB_extracted_k141_148416 | Not-determined |
| SB_extracted_k141_131238 | Not-determined |
| SB_extracted_k141_148417 | Not-determined |
| SB_extracted_k141_200315 | Not-determined |
| SB_extracted_k141_148418 | Not-determined |
| SB_extracted_k141_148419 | Not-determined |
| SB_extracted_k141_96452 | Not-determined |
| SB_extracted_k141_131240 | Not-determined |
| SB_extracted_k141_27106 | Not-determined |
| SB_extracted_k141_192104 | Not-determined |
| SB_extracted_k141_148421 | Not-determined |
| SB_extracted_k141_1081 | Not-determined |
| SB_extracted_k141_35793 | Not-determined |
| SB_extracted_k141_192106 | Not-determined |
| SB_extracted_k141_1082 | Not-determined |
| SB_extracted_k141_105111 | Not-determined |
| SB_extracted_k141_105112 | Not-determined |
| SB_extracted_k141_139925 | Not-determined |
| SB_extracted_k141_9691 | Not-determined |
| SB_extracted_k141_70222 | Not-determined |
| SB_extracted_k141_105114 | Not-determined |
| SB_extracted_k141_157074 | Not-determined |
| SB_extracted_k141_157075 | Not-determined |
| SB_extracted_k141_70224 | Not-determined |
| SB_extracted_k141_79215 | Not-determined |
| SB_extracted_k141_18156 | Not-determined |
| SB_extracted_k141_174375 | Not-determined |
| SB_extracted_k141_18158 | Low-quality |
| SB_extracted_k141_53087 | Not-determined |
| SB_extracted_k141_174377 | Not-determined |
| SB_extracted_k141_148433 | Low-quality |
| SB_extracted_k141_70229 | Not-determined |
| SB_extracted_k141_131252 | Not-determined |
| SB_extracted_k141_9697 | Not-determined |
| SB_extracted_k141_105127 | Not-determined |
| SB_extracted_k141_148439 | Not-determined |
| SB_extracted_k141_79223 | Low-quality |
| SB_extracted_k141_139933 | Not-determined |
| SB_extracted_k141_61789 | Not-determined |
| SB_extracted_k141_18168 | Not-determined |
| SB_extracted_k141_53095 | Not-determined |
| SB_extracted_k141_148443 | Not-determined |
| SB_extracted_k141_96478 | Not-determined |
| SB_extracted_k141_165924 | Low-quality |
| SB_extracted_k141_139938 | Not-determined |
| SB_extracted_k141_165925 | Not-determined |
| SB_extracted_k141_87950 | Not-determined |
| SB_extracted_k141_131257 | Not-determined |
| SB_extracted_k141_70237 | Not-determined |
| SB_extracted_k141_87954 | Not-determined |
| SB_extracted_k141_87956 | Not-determined |
| SB_extracted_k141_79229 | Low-quality |
| SB_extracted_k141_70240 | Not-determined |
| SB_extracted_k141_87957 | Not-determined |
| SB_extracted_k141_35812 | Not-determined |
| SB_extracted_k141_27130 | Not-determined |
| SB_extracted_k141_61802 | Not-determined |
| SB_extracted_k141_35814 | Not-determined |
| SB_extracted_k141_27131 | Not-determined |
| SB_extracted_k141_9711 | Not-determined |
| SB_extracted_k141_27134 | Not-determined |
| SB_extracted_k141_131263 | Not-determined |
| SB_extracted_k141_148448 | Not-determined |
| SB_extracted_k141_113769 | Not-determined |
| SB_extracted_k141_53111 | Not-determined |
| SB_extracted_k141_113770 | Not-determined |
| SB_extracted_k141_18178 | Not-determined |
| SB_extracted_k141_200329 | Low-quality |
| SB_extracted_k141_96488 | Not-determined |
| SB_extracted_k141_200330 | Not-determined |
| SB_extracted_k141_96490 | Not-determined |
| SB_extracted_k141_35827 | Not-determined |
| SB_extracted_k141_35828 | Not-determined |
| SB_extracted_k141_148455 | Not-determined |
| SB_extracted_k141_174399 | Not-determined |
| SB_extracted_k141_165933 | Not-determined |
| SB_extracted_k141_131272 | Not-determined |
| SB_extracted_k141_174404 | Not-determined |
| SB_extracted_k141_174405 | Not-determined |
| SB_extracted_k141_192138 | Not-determined |
| SB_extracted_k141_131275 | Not-determined |
| SB_extracted_k141_9732 | Not-determined |
| SB_extracted_k141_18188 | Not-determined |
| SB_extracted_k141_1120 | Not-determined |
| SB_extracted_k141_165943 | Not-determined |
| SB_extracted_k141_192139 | Low-quality |
| SB_extracted_k141_35836 | Not-determined |
| SB_extracted_k141_35838 | Not-determined |
| SB_extracted_k141_61827 | Not-determined |
| SB_extracted_k141_96497 | Not-determined |
| SB_extracted_k141_165945 | Not-determined |
| SB_extracted_k141_192144 | Not-determined |
| SB_extracted_k141_87969 | Not-determined |
| SB_extracted_k141_157110 | Not-determined |
| SB_extracted_k141_61831 | Not-determined |
| SB_extracted_k141_61833 | Not-determined |
| SB_extracted_k141_61834 | Not-determined |
| SB_extracted_k141_61835 | Not-determined |
| SB_extracted_k141_87977 | Low-quality |
| SB_extracted_k141_61837 | Not-determined |
| SB_extracted_k141_105143 | Not-determined |
| SB_extracted_k141_27146 | Not-determined |
| SB_extracted_k141_113783 | Not-determined |
| SB_extracted_k141_70277 | Not-determined |
| SB_extracted_k141_61839 | Not-determined |
| SB_extracted_k141_174417 | Not-determined |
| SB_extracted_k141_96501 | Not-determined |
| SB_extracted_k141_1124 | Not-determined |
| SB_extracted_k141_192151 | Low-quality |
| SB_extracted_k141_139968 | Not-determined |
| SB_extracted_k141_131296 | Not-determined |
| SB_extracted_k141_192153 | Not-determined |
| SB_extracted_k141_139969 | Not-determined |
| SB_extracted_k141_18206 | Low-quality |
| SB_extracted_k141_96504 | Not-determined |
| SB_extracted_k141_87989 | Not-determined |
| SB_extracted_k141_18209 | Not-determined |
| SB_extracted_k141_53134 | Not-determined |
| SB_extracted_k141_122599 | Low-quality |
| SB_extracted_k141_35854 | Not-determined |
| SB_extracted_k141_18212 | Not-determined |
| SB_extracted_k141_113798 | Not-determined |
| SB_extracted_k141_35855 | Not-determined |
| SB_extracted_k141_192159 | Not-determined |
| SB_extracted_k141_35857 | Not-determined |
| SB_extracted_k141_122605 | Not-determined |
| SB_extracted_k141_131301 | Not-determined |
| SB_extracted_k141_18216 | Not-determined |
| SB_extracted_k141_79272 | Not-determined |
| SB_extracted_k141_139976 | Not-determined |
| SB_extracted_k141_35859 | Not-determined |
| SB_extracted_k141_113802 | Not-determined |
| SB_extracted_k141_1134 | Not-determined |
| SB_extracted_k141_113803 | Not-determined |
| SB_extracted_k141_79274 | Not-determined |
| SB_extracted_k141_122610 | Not-determined |
| SB_extracted_k141_27165 | Not-determined |
| SB_extracted_k141_139979 | Low-quality |
| SB_extracted_k141_122611 | Not-determined |
| SB_extracted_k141_96509 | Not-determined |
| SB_extracted_k141_174420 | Not-determined |
| SB_extracted_k141_131309 | Not-determined |
| SB_extracted_k141_18224 | Not-determined |
| SB_extracted_k141_27171 | Not-determined |
| SB_extracted_k141_192163 | Not-determined |
| SB_extracted_k141_1145 | Not-determined |
| SB_extracted_k141_53154 | Not-determined |
| SB_extracted_k141_79287 | Not-determined |
| SB_extracted_k141_87995 | Not-determined |
| SB_extracted_k141_96520 | Not-determined |
| SB_extracted_k141_79288 | Not-determined |
| SB_extracted_k141_96523 | Not-determined |
| SB_extracted_k141_105162 | Not-determined |
| SB_extracted_k141_79289 | Not-determined |
| SB_extracted_k141_79291 | Not-determined |
| SB_extracted_k141_35877 | Not-determined |
| SB_extracted_k141_105163 | Not-determined |
| SB_extracted_k141_192172 | Not-determined |
| SB_extracted_k141_96529 | Not-determined |
| SB_extracted_k141_9746 | Not-determined |
| SB_extracted_k141_79298 | Not-determined |
| SB_extracted_k141_148519 | Not-determined |
| SB_extracted_k141_87999 | Not-determined |
| SB_extracted_k141_35890 | Not-determined |
| SB_extracted_k141_70304 | Low-quality |
| SB_extracted_k141_79303 | Not-determined |
| SB_extracted_k141_61851 | Not-determined |
| SB_extracted_k141_148528 | Not-determined |
| SB_extracted_k141_70310 | Not-determined |
| SB_extracted_k141_79306 | Not-determined |
| SB_extracted_k141_96539 | Not-determined |
| SB_extracted_k141_148532 | Not-determined |
| SB_extracted_k141_148534 | Not-determined |
| SB_extracted_k141_61855 | Not-determined |
| SB_extracted_k141_70313 | Not-determined |
| SB_extracted_k141_157126 | Not-determined |
| SB_extracted_k141_70315 | Low-quality |
| SB_extracted_k141_70316 | Low-quality |
| SB_extracted_k141_35901 | Not-determined |
| SB_extracted_k141_96549 | Not-determined |
| SB_extracted_k141_88005 | Not-determined |
| SB_extracted_k141_79316 | Not-determined |
| SB_extracted_k141_131327 | Not-determined |
| SB_extracted_k141_157133 | Not-determined |
| SB_extracted_k141_140002 | Not-determined |
| SB_extracted_k141_18236 | Not-determined |
| SB_extracted_k141_35904 | Not-determined |
| SB_extracted_k141_148538 | Not-determined |
| SB_extracted_k141_148540 | Low-quality |
| SB_extracted_k141_131332 | Not-determined |
| SB_extracted_k141_1164 | Low-quality |
| SB_extracted_k141_122642 | Not-determined |
| SB_extracted_k141_131333 | Low-quality |
| SB_extracted_k141_105167 | Not-determined |
| SB_extracted_k141_131335 | Not-determined |
| SB_extracted_k141_1166 | Not-determined |
| SB_extracted_k141_27211 | Not-determined |
| SB_extracted_k141_53182 | Not-determined |
| SB_extracted_k141_44324 | Not-determined |
| SB_extracted_k141_157140 | Not-determined |
| SB_extracted_k141_27213 | Not-determined |
| SB_extracted_k141_157141 | Not-determined |
| SB_extracted_k141_1168 | Low-quality |
| SB_extracted_k141_79323 | Not-determined |
| SB_extracted_k141_27214 | Not-determined |
| SB_extracted_k141_131340 | Not-determined |
| SB_extracted_k141_9757 | Not-determined |
| SB_extracted_k141_105174 | Not-determined |
| SB_extracted_k141_192184 | Not-determined |
| SB_extracted_k141_122648 | Not-determined |
| SB_extracted_k141_157147 | Not-determined |
| SB_extracted_k141_1171 | Not-determined |
| SB_extracted_k141_9761 | Not-determined |
| SB_extracted_k141_122651 | Not-determined |
| SB_extracted_k141_9762 | Low-quality |
| SB_extracted_k141_131345 | Not-determined |
| SB_extracted_k141_27220 | Not-determined |
| SB_extracted_k141_192185 | Not-determined |
| SB_extracted_k141_131348 | Not-determined |
| SB_extracted_k141_18243 | Not-determined |
| SB_extracted_k141_131349 | Low-quality |
| SB_extracted_k141_192189 | Not-determined |
| SB_extracted_k141_1177 | Not-determined |
| SB_extracted_k141_140005 | Low-quality |
| SB_extracted_k141_148554 | Not-determined |
| SB_extracted_k141_79337 | Not-determined |
| SB_extracted_k141_131351 | Low-quality |
| SB_extracted_k141_148555 | Low-quality |
| SB_extracted_k141_165963 | Not-determined |
| SB_extracted_k141_105184 | Not-determined |
| SB_extracted_k141_131353 | Not-determined |
| SB_extracted_k141_27227 | Not-determined |
| SB_extracted_k141_174446 | Not-determined |
| SB_extracted_k141_18254 | Not-determined |
| SB_extracted_k141_9770 | Not-determined |
| SB_extracted_k141_165967 | Not-determined |
| SB_extracted_k141_148560 | Not-determined |
| SB_extracted_k141_18256 | Not-determined |
| SB_extracted_k141_1186 | Not-determined |
| SB_extracted_k141_70340 | Not-determined |
| SB_extracted_k141_122661 | Not-determined |
| SB_extracted_k141_157169 | Not-determined |
| SB_extracted_k141_200341 | Not-determined |
| SB_extracted_k141_53203 | Not-determined |
| SB_extracted_k141_131357 | Not-determined |
| SB_extracted_k141_165971 | Not-determined |
| SB_extracted_k141_131360 | Not-determined |
| SB_extracted_k141_131362 | Not-determined |
| SB_extracted_k141_122669 | Not-determined |
| SB_extracted_k141_27237 | Low-quality |
| SB_extracted_k141_79349 | Not-determined |
| SB_extracted_k141_1198 | Not-determined |
| SB_extracted_k141_1200 | Not-determined |
| SB_extracted_k141_9779 | Not-determined |
| SB_extracted_k141_148570 | Not-determined |
| SB_extracted_k141_53213 | Not-determined |
| SB_extracted_k141_148571 | Not-determined |
| SB_extracted_k141_200353 | Not-determined |
| SB_extracted_k141_148573 | Not-determined |
| SB_extracted_k141_148576 | Not-determined |
| SB_extracted_k141_27246 | Not-determined |
| SB_extracted_k141_1205 | Not-determined |
| SB_extracted_k141_96556 | Not-determined |
| SB_extracted_k141_9783 | Not-determined |
| SB_extracted_k141_70363 | Not-determined |
| SB_extracted_k141_96557 | Not-determined |
| SB_extracted_k141_53224 | Not-determined |
| SB_extracted_k141_1213 | Not-determined |
| SB_extracted_k141_1214 | Not-determined |
| SB_extracted_k141_200359 | Not-determined |
| SB_extracted_k141_148592 | Not-determined |
| SB_extracted_k141_122671 | Not-determined |
| SB_extracted_k141_192214 | Not-determined |
| SB_extracted_k141_53225 | Low-quality |
| SB_extracted_k141_9796 | Not-determined |
| SB_extracted_k141_27258 | Low-quality |
| SB_extracted_k141_1215 | Not-determined |
| SB_extracted_k141_148598 | Low-quality |
| SB_extracted_k141_148600 | Low-quality |
| SB_extracted_k141_1219 | Not-determined |
| SB_extracted_k141_61907 | Not-determined |
| SB_extracted_k141_70378 | Not-determined |
| SB_extracted_k141_35956 | Not-determined |
| SB_extracted_k141_79363 | Low-quality |
| SB_extracted_k141_79364 | Low-quality |
| SB_extracted_k141_165985 | Not-determined |
| SB_extracted_k141_70381 | Not-determined |
| SB_extracted_k141_140042 | Not-determined |
| SB_extracted_k141_35961 | Not-determined |
| SB_extracted_k141_9806 | Low-quality |
| SB_extracted_k141_140043 | Not-determined |
| SB_extracted_k141_79368 | Not-determined |
| SB_extracted_k141_182933 | Not-determined |
| SB_extracted_k141_131386 | Not-determined |
| SB_extracted_k141_61910 | Not-determined |
| SB_extracted_k141_131387 | Not-determined |
| SB_extracted_k141_35963 | Not-determined |
| SB_extracted_k141_192232 | Not-determined |
| SB_extracted_k141_1230 | Not-determined |
| SB_extracted_k141_140044 | Low-quality |
| SB_extracted_k141_174465 | Not-determined |
| SB_extracted_k141_174467 | Not-determined |
| SB_extracted_k141_165990 | Not-determined |
| SB_extracted_k141_200367 | Not-determined |
| SB_extracted_k141_165991 | Not-determined |
| SB_extracted_k141_122689 | Not-determined |
| SB_extracted_k141_182937 | Not-determined |
| SB_extracted_k141_165995 | Not-determined |
| SB_extracted_k141_70396 | Not-determined |
| SB_extracted_k141_157191 | Not-determined |
| SB_extracted_k141_148612 | Not-determined |
| SB_extracted_k141_35968 | Not-determined |
| SB_extracted_k141_200373 | Not-determined |
| SB_extracted_k141_192241 | Not-determined |
| SB_extracted_k141_200376 | Not-determined |
| SB_vs_k141_174559\|\|full | Medium-quality |
| SB_extracted_k141_1245 | Not-determined |
| SB_extracted_k141_96559 | Low-quality |
| SB_extracted_k141_192243 | Not-determined |
| SB_extracted_k141_192244 | Not-determined |
| SB_extracted_k141_61927 | Not-determined |
| SB_extracted_k141_88015 | Not-determined |
| SB_extracted_k141_18269 | Not-determined |
| SB_extracted_k141_35972 | Not-determined |
| SB_extracted_k141_70402 | Not-determined |
| SB_extracted_k141_113811 | Not-determined |
| SB_extracted_k141_35973 | Not-determined |
| SB_extracted_k141_96561 | Not-determined |
| SB_extracted_k141_166003 | Not-determined |
| SB_extracted_k141_35974 | Not-determined |
| SB_extracted_k141_1251 | Not-determined |
| SB_extracted_k141_174476 | Not-determined |
| SB_extracted_k141_200383 | Not-determined |
| SB_extracted_k141_1253 | Not-determined |
| SB_extracted_k141_157201 | Not-determined |
| SB_extracted_k141_1254 | Not-determined |
| SB_extracted_k141_192252 | Not-determined |
| SB_extracted_k141_148622 | Not-determined |
| SB_extracted_k141_88018 | Not-determined |
| SB_extracted_k141_148626 | Not-determined |
| SB_extracted_k141_148629 | Not-determined |
| SB_extracted_k141_131413 | Low-quality |
| SB_extracted_k141_53232 | Low-quality |
| SB_extracted_k141_70409 | Not-determined |
| SB_extracted_k141_35977 | Not-determined |
| SB_extracted_k141_88022 | Not-determined |
| SB_extracted_k141_148632 | Not-determined |
| SB_extracted_k141_157212 | Low-quality |
| SB_extracted_k141_166016 | Not-determined |
| SB_extracted_k141_27285 | Not-determined |
| SB_extracted_k141_157213 | Not-determined |
| SB_extracted_k141_200394 | Not-determined |
| SB_extracted_k141_88024 | Not-determined |
| SB_extracted_k141_79374 | Not-determined |
| SB_extracted_k141_113827 | Not-determined |
| SB_extracted_k141_148635 | Not-determined |
| SB_extracted_k141_44376 | Not-determined |
| SB_extracted_k141_148636 | Not-determined |
| SB_extracted_k141_122703 | Not-determined |
| SB_extracted_k141_88026 | Not-determined |
| SB_extracted_k141_44377 | Not-determined |
| SB_extracted_k141_200400 | Not-determined |
| SB_extracted_k141_53243 | Low-quality |
| SB_extracted_k141_61937 | Not-determined |
| SB_extracted_k141_157215 | Not-determined |
| SB_extracted_k141_53245 | Not-determined |
| SB_extracted_k141_96580 | Not-determined |
| SB_extracted_k141_122709 | Not-determined |
| SB_extracted_k141_174489 | Not-determined |
| SB_extracted_k141_88032 | Low-quality |
| SB_extracted_k141_192269 | Not-determined |
| SB_extracted_k141_200404 | Low-quality |
| SB_extracted_k141_113836 | Not-determined |

| | |
|---|---|
| S8_extracted_k141_131419 | Low-quality |
| S8_extracted_k141_166023 | Not-determined |
| S8_extracted_k141_157217 | Not-determined |
| S8_extracted_k141_174492 | Not-determined |
| S8_extracted_k141_148644 | Not-determined |
| S8_extracted_k141_96584 | Not-determined |
| S8_extracted_k141_79391 | Not-determined |
| S8_extracted_k141_122720 | Not-determined |
| S8_extracted_k141_192272 | Not-determined |
| S8_extracted_k141_113843 | Not-determined |
| S8_extracted_k141_18282 | Low-quality |
| S8_extracted_k141_174499 | Not-determined |
| S8_extracted_k141_174500 | Not-determined |
| S8_extracted_k141_157220 | Not-determined |
| S8_extracted_k141_44389 | Not-determined |
| S8_extracted_k141_131427 | Not-determined |
| S8_extracted_k141_157224 | Low-quality |
| S8_extracted_k141_18288 | Not-determined |
| S8_extracted_k141_27298 | Not-determined |
| S8_extracted_k141_131428 | Not-determined |
| S8_extracted_k141_105219 | Not-determined |
| S8_extracted_k141_79392 | Not-determined |
| S8_extracted_k141_131430 | Not-determined |
| S8_extracted_k141_53254 | Not-determined |
| S8_extracted_k141_200423 | Not-determined |
| S8_extracted_k141_18293 | Not-determined |
| S8_extracted_k141_61944 | Not-determined |
| S8_extracted_k141_53258 | Not-determined |
| S8_extracted_k141_157226 | Not-determined |
| S8_extracted_k141_122732 | Not-determined |
| S8_extracted_k141_18297 | Not-determined |
| S8_extracted_k141_157232 | Not-determined |
| S8_extracted_k141_27304 | Not-determined |
| S8_extracted_k141_61950 | Not-determined |
| S8_extracted_k141_79399 | Low-quality |
| S8_extracted_k141_44395 | Not-determined |
| S8_extracted_k141_96596 | Low-quality |
| S8_extracted_k141_53264 | Not-determined |
| S8_extracted_k141_61952 | Not-determined |
| S8_extracted_k141_96597 | Not-determined |
| S8_extracted_k141_27306 | Not-determined |
| S8_extracted_k141_105233 | Low-quality |
| S8_extracted_k141_79403 | Not-determined |
| S8_extracted_k141_157240 | Not-determined |
| S8_extracted_k141_200435 | Not-determined |
| S8_extracted_k141_18304 | Not-determined |
| S8_extracted_k141_27312 | Not-determined |
| S8_extracted_k141_192288 | Not-determined |
| S8_extracted_k141_61956 | Not-determined |
| S8_extracted_k141_182973 | Not-determined |
| S8_extracted_k141_157251 | Low-quality |
| S8_extracted_k141_61959 | Not-determined |
| S8_extracted_k141_192298 | Low-quality |
| S8_extracted_k141_157252 | Not-determined |
| S8_extracted_k141_44410 | Not-determined |
| S8_extracted_k141_79410 | Not-determined |
| S8_extracted_k141_18313 | Low-quality |
| S8_extracted_k141_200444 | Not-determined |
| S8_extracted_k141_182975 | Not-determined |
| S8_extracted_k141_200447 | Not-determined |
| S8_extracted_k141_44412 | Not-determined |
| S8_extracted_k141_105246 | Not-determined |
| S8_extracted_k141_148665 | Not-determined |
| S8_extracted_k141_182980 | Not-determined |
| S8_extracted_k141_113878 | Not-determined |
| S8_extracted_k141_35986 | Not-determined |
| S8_extracted_k141_61967 | Not-determined |
| S8_extracted_k141_27332 | Not-determined |
| S8_extracted_k141_166033 | Not-determined |
| S8_extracted_k141_61970 | Not-determined |
| S8_extracted_k141_148671 | Low-quality |
| S8_extracted_k141_18326 | Not-determined |
| S8_extracted_k141_44414 | Not-determined |
| S8_extracted_k141_35996 | Not-determined |
| S8_extracted_k141_192324 | Not-determined |
| S8_extracted_k141_105266 | Not-determined |
| S8_extracted_k141_113880 | Not-determined |
| S8_extracted_k141_192326 | Not-determined |
| S8_extracted_k141_96621 | Not-determined |
| S8_extracted_k141_182995 | Low-quality |
| S8_extracted_k141_192327 | Not-determined |
| S8_extracted_k141_36001 | Not-determined |
| S8_extracted_k141_166046 | Not-determined |
| S8_extracted_k141_44417 | Low-quality |
| S8_extracted_k141_166049 | Not-determined |
| S8_extracted_k141_36008 | Not-determined |
| S8_extracted_k141_18333 | Not-determined |
| S8_extracted_k141_122756 | Not-determined |
| S8_extracted_k141_183005 | Not-determined |
| S8_extracted_k141_174512 | Not-determined |
| S8_extracted_k141_44418 | Not-determined |
| S8_extracted_k141_192328 | Not-determined |
| S8_extracted_k141_105271 | Not-determined |
| S8_extracted_k141_79435 | Not-determined |
| S8_extracted_k141_113884 | Not-determined |
| S8_extracted_k141_61974 | Not-determined |
| S8_extracted_k141_79437 | Not-determined |
| S8_extracted_k141_1292 | Low-quality |
| S8_extracted_k141_61976 | Not-determined |
| S8_extracted_k141_79441 | Not-determined |
| S8_extracted_k141_96636 | Not-determined |
| S8_extracted_k141_1296 | Not-determined |
| S8_extracted_k141_61981 | Not-determined |
| S8_extracted_k141_96637 | Low-quality |
| S8_extracted_k141_27353 | Not-determined |
| S8_extracted_k141_79445 | Not-determined |
| S8_extracted_k141_166063 | Not-determined |
| S8_extracted_k141_44428 | Not-determined |
| S8_extracted_k141_61984 | Not-determined |
| S8_extracted_k141_105277 | Not-determined |
| S8_extracted_k141_122765 | Not-determined |
| S8_extracted_k141_113892 | Not-determined |
| S8_extracted_k141_1299 | Not-determined |
| S8_extracted_k141_105281 | Not-determined |
| S8_extracted_k141_113895 | Not-determined |
| S8_extracted_k141_122766 | Low-quality |
| S8_extracted_k141_166068 | Not-determined |
| S8_extracted_k141_1300 | Not-determined |
| S8_extracted_k141_61996 | Not-determined |
| S8_extracted_k141_27363 | Not-determined |
| S8_extracted_k141_1302 | Low-quality |
| S8_extracted_k141_27365 | Not-determined |
| S8_extracted_k141_122770 | Low-quality |
| S8_extracted_k141_157291 | Low-quality |
| S8_extracted_k141_88060 | Not-determined |
| S8_extracted_k141_88061 | Not-determined |
| S8_extracted_k141_174518 | Low-quality |
| S8_extracted_k141_122776 | Not-determined |
| S8_extracted_k141_96650 | Not-determined |
| S8_extracted_k141_36016 | Not-determined |
| S8_extracted_k141_192344 | Not-determined |
| S8_extracted_k141_36017 | Not-determined |
| S8_extracted_k141_105295 | Not-determined |
| S8_extracted_k141_36019 | Not-determined |
| S8_extracted_k141_53291 | Not-determined |
| S8_extracted_k141_157296 | Not-determined |
| S8_extracted_k141_174524 | Not-determined |
| S8_extracted_k141_122785 | Not-determined |
| S8_extracted_k141_113901 | Not-determined |
| S8_extracted_k141_122788 | Not-determined |
| S8_extracted_k141_166078 | Not-determined |
| S8_extracted_k141_53295 | Not-determined |
| S8_extracted_k141_192352 | Not-determined |
| S8_extracted_k141_157300 | Not-determined |
| S8_extracted_k141_166081 | Not-determined |
| S8_extracted_k141_96654 | Not-determined |
| S8_extracted_k141_88076 | Not-determined |
| S8_extracted_k141_62000 | Not-determined |
| S8_extracted_k141_79467 | Not-determined |
| S8_extracted_k141_157303 | Not-determined |
| S8_extracted_k141_166086 | Not-determined |
| S8_extracted_k141_140065 | Low-quality |
| S8_extracted_k141_157304 | Not-determined |
| S8_extracted_k141_36029 | Not-determined |
| S8_extracted_k141_192355 | Not-determined |
| S8_extracted_k141_27400 | Not-determined |
| S8_extracted_k141_200455 | Not-determined |
| S8_extracted_k141_62002 | Not-determined |
| S8_extracted_k141_192357 | Low-quality |
| S8_extracted_k141_113910 | Not-determined |
| S8_extracted_k141_62006 | Low-quality |
| S8_extracted_k141_183035 | Not-determined |
| S8_extracted_k141_140067 | Not-determined |
| S8_extracted_k141_18351 | Not-determined |
| S8_extracted_k141_174535 | Not-determined |
| S8_extracted_k141_157307 | Not-determined |
| S8_extracted_k141_96660 | Not-determined |
| S8_extracted_k141_122793 | Not-determined |
| S8_extracted_k141_183038 | Not-determined |
| S8_extracted_k141_183039 | Not-determined |
| S8_extracted_k141_140072 | Not-determined |
| S8_extracted_k141_62011 | Not-determined |
| S8_extracted_k141_27410 | Not-determined |
| S8_extracted_k141_113916 | Low-quality |
| S8_extracted_k141_131447 | Low-quality |
| S8_extracted_k141_1333 | Not-determined |
| S8_extracted_k141_36043 | Low-quality |
| S8_extracted_k141_140076 | Not-determined |
| S8_extracted_k141_105301 | Not-determined |
| S8_extracted_k141_1335 | Low-quality |
| S8_extracted_k141_53312 | Not-determined |
| S8_extracted_k141_27412 | Not-determined |
| S8_extracted_k141_53315 | Not-determined |
| S8_extracted_k141_88088 | Not-determined |
| S8_extracted_k141_113923 | Not-determined |
| S8_extracted_k141_131450 | Not-determined |
| S8_extracted_k141_105305 | Not-determined |
| S8_extracted_k141_96671 | Not-determined |
| S8_extracted_k141_192370 | Not-determined |
| S8_extracted_k141_131454 | Not-determined |
| S8_extracted_k141_18359 | Not-determined |
| S8_extracted_k141_1338 | Not-determined |
| S8_extracted_k141_131456 | Not-determined |
| S8_extracted_k141_27415 | Not-determined |
| S8_extracted_k141_166097 | Not-determined |
| S8_extracted_k141_122797 | Not-determined |
| S8_extracted_k141_105312 | Not-determined |
| S8_extracted_k141_88095 | Low-quality |
| S8_extracted_k141_96673 | Not-determined |
| S8_extracted_k141_157312 | Not-determined |
| S8_extracted_k141_192380 | Not-determined |
| S8_extracted_k141_96677 | Not-determined |
| S8_extracted_k141_122799 | Not-determined |
| S8_extracted_k141_27419 | Not-determined |
| S8_extracted_k141_18363 | Not-determined |
| S8_extracted_k141_88100 | Not-determined |
| S8_extracted_k141_27420 | Not-determined |
| S8_extracted_k141_200468 | Not-determined |
| S8_extracted_k141_27422 | Not-determined |
| S8_extracted_k141_174552 | Not-determined |
| S8_extracted_k141_192381 | Not-determined |
| S8_extracted_k141_148718 | Not-determined |
| S8_extracted_k141_122801 | Low-quality |
| S8_extracted_k141_200473 | Not-determined |
| S8_extracted_k141_96679 | Not-determined |
| S8_extracted_k141_148720 | Not-determined |
| S8_extracted_k141_79476 | Not-determined |
| S8_extracted_k141_27428 | Not-determined |
| S8_extracted_k141_27430 | Not-determined |
| S8_extracted_k141_27431 | Not-determined |
| S8_extracted_k141_18365 | Not-determined |
| S8_extracted_k141_200477 | Not-determined |
| S8_extracted_k141_1339 | Not-determined |
| S8_extracted_k141_27432 | Not-determined |
| S8_extracted_k141_79479 | Not-determined |
| S8_extracted_k141_88110 | Not-determined |
| S8_extracted_k141_1344 | Not-determined |
| S8_extracted_k141_148724 | Not-determined |
| S8_extracted_k141_79484 | Not-determined |
| S8_extracted_k141_148727 | Not-determined |
| S8_extracted_k141_105323 | Low-quality |
| S8_extracted_k141_113937 | Not-determined |
| S8_extracted_k141_200484 | Low-quality |
| S8_extracted_k141_105327 | Not-determined |
| S8_extracted_k141_105328 | Not-determined |
| S8_extracted_k141_113940 | Not-determined |
| S8_extracted_k141_88113 | Not-determined |
| S8_extracted_k141_148731 | Not-determined |
| S8_extracted_k141_53333 | Not-determined |
| S8_extracted_k141_131464 | Low-quality |
| S8_extracted_k141_62031 | Low-quality |
| S8_extracted_k141_53334 | Not-determined |
| S8_extracted_k141_200490 | Not-determined |
| S8_extracted_k141_131466 | Not-determined |
| S8_extracted_k141_27444 | Not-determined |
| S8_extracted_k141_53336 | Not-determined |
| S8_extracted_k141_200492 | Not-determined |
| S8_extracted_k141_53338 | Not-determined |
| S8_extracted_k141_27448 | Not-determined |
| S8_extracted_k141_79490 | Not-determined |
| S8_extracted_k141_44462 | Not-determined |
| S8_extracted_k141_27457 | Not-determined |
| S8_extracted_k141_113951 | Not-determined |
| S8_extracted_k141_148735 | Low-quality |
| S8_extracted_k141_122824 | Not-determined |
| S8_extracted_k141_79498 | Not-determined |
| S8_extracted_k141_62038 | Low-quality |
| S8_extracted_k141_192407 | Not-determined |
| S8_extracted_k141_79500 | Not-determined |
| S8_extracted_k141_105345 | Not-determined |
| S8_extracted_k141_131476 | Not-determined |
| S8_extracted_k141_105346 | Not-determined |
| S16_vs_100473\|\|full | Medium-quality |
| S8_extracted_k141_53349 | Not-determined |
| S8_extracted_k141_96698 | Not-determined |
| S8_extracted_k141_36060 | Not-determined |
| S8_extracted_k141_36061 | Not-determined |
| S8_extracted_k141_79505 | Not-determined |
| S8_extracted_k141_166128 | Not-determined |
| S8_extracted_k141_36063 | Not-determined |
| S8_extracted_k141_96702 | Not-determined |
| S8_extracted_k141_62042 | Not-determined |
| S8_extracted_k141_18386 | Not-determined |
| S8_extracted_k141_44466 | Low-quality |
| S8_extracted_k141_79509 | Low-quality |
| S8_extracted_k141_96707 | Not-determined |
| S8_extracted_k141_36069 | Not-determined |
| S8_extracted_k141_105355 | Not-determined |
| S8_extracted_k141_200512 | Low-quality |
| S8_extracted_k141_1367 | Not-determined |
| S8_extracted_k141_62043 | Not-determined |
| S8_extracted_k141_200514 | Not-determined |
| S8_extracted_k141_36076 | Not-determined |
| S8_extracted_k141_148761 | Not-determined |
| S8_extracted_k141_1372 | Not-determined |
| S8_extracted_k141_53367 | Not-determined |
| S8_extracted_k141_88146 | Not-determined |
| S8_extracted_k141_88147 | Not-determined |
| S8_extracted_k141_1374 | Not-determined |
| S8_extracted_k141_174584 | Not-determined |
| S8_extracted_k141_53368 | Low-quality |
| S8_extracted_k141_1375 | Not-determined |
| S8_extracted_k141_1376 | Not-determined |
| S8_extracted_k141_96724 | Not-determined |
| S8_extracted_k141_174585 | Low-quality |
| S8_extracted_k141_122849 | Not-determined |
| S8_extracted_k141_44479 | Not-determined |
| S8_extracted_k141_27473 | Not-determined |
| S8_extracted_k141_174590 | Low-quality |
| S8_extracted_k141_1380 | Not-determined |
| S8_extracted_k141_27475 | Not-determined |
| S8_extracted_k141_62058 | Not-determined |
| S8_extracted_k141_200531 | Not-determined |
| S8_extracted_k141_157333 | Not-determined |
| S8_extracted_k141_96727 | Not-determined |
| S8_extracted_k141_131491 | Not-determined |
| S8_extracted_k141_148770 | Not-determined |
| S3_vs_k141_57422\|\|full | Medium-quality |
| S8_extracted_k141_9808 | Not-determined |
| S8_extracted_k141_53374 | Not-determined |
| S8_extracted_k141_9809 | Not-determined |
| S8_extracted_k141_27481 | Not-determined |
| S8_extracted_k141_79512 | Not-determined |
| S8_extracted_k141_44486 | Not-determined |
| S8_extracted_k141_27485 | Not-determined |
| S8_extracted_k141_44487 | Not-determined |
| S8_extracted_k141_96732 | Not-determined |
| S8_extracted_k141_200533 | Not-determined |
| S8_extracted_k141_200535 | Not-determined |
| S8_extracted_k141_192408 | Low-quality |
| S8_extracted_k141_27492 | Not-determined |
| S8_extracted_k141_53382 | Low-quality |
| S8_extracted_k141_96734 | Not-determined |
| S8_extracted_k141_18413 | Not-determined |
| S8_extracted_k141_1396 | Not-determined |
| S8_extracted_k141_192412 | Not-determined |
| S8_extracted_k141_192413 | Not-determined |
| S8_extracted_k141_174605 | Not-determined |
| S8_extracted_k141_1399 | Not-determined |
| S8_extracted_k141_122859 | Not-determined |
| S8_extracted_k141_9829 | Low-quality |
| S8_extracted_k141_44501 | Not-determined |
| S8_extracted_k141_122860 | Not-determined |
| S8_extracted_k141_140124 | Not-determined |
| S8_extracted_k141_192417 | Not-determined |
| S8_extracted_k141_131508 | Not-determined |
| S8_extracted_k141_105398 | Not-determined |
| S8_extracted_k141_44506 | Not-determined |
| S8_extracted_k141_53391 | Not-determined |
| S8_extracted_k141_166154 | Not-determined |
| S8_extracted_k141_122863 | Not-determined |
| S8_extracted_k141_131517 | Not-determined |
| S8_extracted_k141_192425 | Not-determined |
| S8_extracted_k141_113998 | Low-quality |
| S8_extracted_k141_18417 | Not-determined |
| S8_extracted_k141_166160 | Not-determined |
| S8_extracted_k141_131519 | Low-quality |
| S8_extracted_k141_192427 | Not-determined |
| S8_extracted_k141_53402 | Not-determined |
| S8_extracted_k141_114000 | Not-determined |
| S8_extracted_k141_62087 | Not-determined |
| S8_extracted_k141_157352 | Not-determined |
| S8_extracted_k141_1414 | Not-determined |
| S8_extracted_k141_44517 | Not-determined |
| S8_extracted_k141_44518 | Not-determined |
| S8_extracted_k141_105403 | Not-determined |
| S8_extracted_k141_1420 | Not-determined |
| S8_extracted_k141_192435 | Not-determined |
| S8_extracted_k141_36119 | Not-determined |
| S8_extracted_k141_200556 | Low-quality |
| S8_extracted_k141_200557 | Not-determined |
| S8_extracted_k141_131524 | Not-determined |
| S8_extracted_k141_27513 | Not-determined |
| S8_extracted_k141_18425 | Not-determined |
| S8_extracted_k141_9834 | Not-determined |
| S8_extracted_k141_27514 | Not-determined |
| S8_extracted_k141_18427 | Not-determined |
| S8_extracted_k141_27515 | Not-determined |
| S8_extracted_k141_27516 | Not-determined |
| S8_extracted_k141_157363 | Not-determined |
| S8_extracted_k141_18430 | Not-determined |
| S8_extracted_k141_96746 | Not-determined |
| S8_extracted_k141_9837 | Low-quality |
| S8_extracted_k141_200562 | Not-determined |
| S8_extracted_k141_140128 | Low-quality |
| S8_extracted_k141_200566 | Not-determined |
| S8_extracted_k141_27522 | Not-determined |
| S8_extracted_k141_183074 | Not-determined |
| S8_extracted_k141_200567 | Not-determined |
| S8_extracted_k141_114008 | Not-determined |
| S8_extracted_k141_114009 | Not-determined |
| S8_extracted_k141_27523 | Not-determined |
| S8_extracted_k141_53406 | Low-quality |
| S8_extracted_k141_122877 | Low-quality |
| S8_extracted_k141_200570 | Not-determined |
| S8_extracted_k141_9840 | Low-quality |
| S8_extracted_k141_174616 | Not-determined |
| S8_extracted_k141_88168 | Not-determined |
| S8_extracted_k141_9841 | Not-determined |
| S8_extracted_k141_157373 | Not-determined |
| S8_extracted_k141_157374 | Not-determined |
| S8_extracted_k141_140143 | Not-determined |
| S8_extracted_k141_18439 | Not-determined |
| S8_extracted_k141_140144 | Not-determined |
| S8_extracted_k141_192445 | Not-determined |
| S8_extracted_k141_9844 | Not-determined |
| S8_extracted_k141_27528 | Not-determined |
| S8_extracted_k141_18442 | Not-determined |
| S8_extracted_k141_62112 | Not-determined |
| S8_extracted_k141_88170 | Low-quality |
| S8_extracted_k141_18445 | Not-determined |
| S8_extracted_k141_53419 | Low-quality |
| S8_extracted_k141_148779 | Not-determined |
| S8_extracted_k141_192448 | Not-determined |
| S8_extracted_k141_157381 | Not-determined |
| S8_extracted_k141_174629 | Not-determined |
| S8_extracted_k141_79532 | Not-determined |
| S8_extracted_k141_166170 | Not-determined |
| S8_extracted_k141_148785 | Not-determined |
| S8_extracted_k141_88174 | Not-determined |
| S8_extracted_k141_148786 | Not-determined |
| S8_extracted_k141_105432 | Low-quality |
| S8_extracted_k141_70468 | Low-quality |
| S8_extracted_k141_88177 | Not-determined |
| S8_extracted_k141_166173 | Not-determined |
| S8_extracted_k141_18453 | Not-determined |
| S8_extracted_k141_192460 | Not-determined |
| S8_extracted_k141_96769 | Not-determined |
| S8_extracted_k141_114014 | Low-quality |
| S8_extracted_k141_105434 | Low-quality |
| S8_extracted_k141_53432 | Not-determined |
| S8_extracted_k141_140161 | Not-determined |
| S8_extracted_k141_96772 | Not-determined |
| S8_extracted_k141_18456 | Not-determined |
| S8_extracted_k141_96776 | Not-determined |
| S8_extracted_k141_96779 | Not-determined |
| S8_extracted_k141_96781 | Not-determined |
| S8_extracted_k141_44527 | Low-quality |
| S8_extracted_k141_96784 | Not-determined |
| S8_extracted_k141_44530 | Not-determined |
| S8_extracted_k141_96785 | Not-determined |
| S8_extracted_k141_122894 | Not-determined |
| S8_extracted_k141_148800 | Not-determined |
| S8_extracted_k141_157399 | Not-determined |
| S8_extracted_k141_44539 | Not-determined |
| S8_extracted_k141_1445 | Not-determined |
| S8_extracted_k141_1446 | Not-determined |
| S8_extracted_k141_1448 | Not-determined |
| S8_extracted_k141_79546 | Not-determined |
| S8_extracted_k141_122896 | Low-quality |
| S8_extracted_k141_1450 | Not-determined |
| S8_extracted_k141_157404 | Not-determined |
| S8_extracted_k141_70490 | Not-determined |
| S8_extracted_k141_183093 | Low-quality |
| S8_extracted_k141_79552 | Not-determined |
| S8_extracted_k141_174639 | Not-determined |
| S8_extracted_k141_166187 | Not-determined |
| S8_extracted_k141_9864 | Low-quality |
| S8_extracted_k141_166188 | Low-quality |
| S8_extracted_k141_1457 | Not-determined |
| S8_extracted_k141_114036 | Not-determined |
| S8_extracted_k141_96789 | Not-determined |
| S8_extracted_k141_70496 | Not-determined |
| S8_extracted_k141_174642 | Not-determined |
| S8_extracted_k141_148807 | Not-determined |
| S8_extracted_k141_192470 | Low-quality |
| S8_extracted_k141_70499 | Not-determined |
| S8_extracted_k141_157410 | Not-determined |
| S8_extracted_k141_88181 | Not-determined |
| S8_extracted_k141_96794 | Low-quality |
| S8_extracted_k141_9875 | Not-determined |
| S8_extracted_k141_131532 | Not-determined |
| S8_extracted_k141_36159 | Not-determined |
| S8_extracted_k141_174647 | Not-determined |
| S8_extracted_k141_192474 | Not-determined |
| S8_extracted_k141_140182 | Not-determined |
| S8_extracted_k141_140183 | Not-determined |
| S8_extracted_k141_114043 | Not-determined |
| S8_extracted_k141_1467 | Not-determined |
| S8_extracted_k141_27558 | Not-determined |
| S8_extracted_k141_183095 | Not-determined |
| S8_extracted_k141_166192 | Not-determined |
| S8_extracted_k141_174653 | Not-determined |
| S8_extracted_k141_157419 | Low-quality |
| S8_extracted_k141_96803 | Not-determined |
| S8_extracted_k141_96805 | Not-determined |
| S8_extracted_k141_174655 | Not-determined |
| S8_extracted_k141_192478 | Not-determined |
| S8_extracted_k141_166196 | Not-determined |
| S8_extracted_k141_174657 | Not-determined |
| S8_extracted_k141_131536 | Not-determined |
| S8_extracted_k141_148820 | Not-determined |
| S8_extracted_k141_18488 | Not-determined |
| S8_extracted_k141_131538 | Not-determined |
| S8_extracted_k141_131539 | Not-determined |
| S8_extracted_k141_53473 | Not-determined |
| S8_extracted_k141_18490 | Not-determined |
| S8_extracted_k141_18491 | Not-determined |
| S8_extracted_k141_88188 | Not-determined |
| S8_extracted_k141_36164 | Low-quality |
| S8_extracted_k141_140196 | Not-determined |
| S8_extracted_k141_140197 | Not-determined |
| S8_extracted_k141_192484 | Not-determined |
| S8_extracted_k141_44548 | Not-determined |
| S8_extracted_k141_114046 | Low-quality |
| S8_extracted_k141_53480 | Not-determined |
| S8_extracted_k141_53481 | Not-determined |
| S8_extracted_k141_9903 | Not-determined |
| S8_extracted_k141_36173 | Not-determined |
| S8_extracted_k141_114052 | Not-determined |
| S8_extracted_k141_157431 | Not-determined |
| S8_extracted_k141_44550 | Low-quality |
| S8_extracted_k141_148832 | Not-determined |
| S8_extracted_k141_36176 | Not-determined |
| S8_extracted_k141_70520 | Not-determined |
| S8_extracted_k141_148833 | Not-determined |
| S8_extracted_k141_44553 | Not-determined |
| S8_extracted_k141_44556 | Not-determined |
| S8_extracted_k141_148838 | Not-determined |
| S8_extracted_k141_122919 | Not-determined |
| S8_extracted_k141_53487 | Not-determined |
| S8_extracted_k141_53490 | Not-determined |
| S8_extracted_k141_140215 | Not-determined |
| S8_extracted_k141_1477 | Not-determined |
| S8_extracted_k141_36182 | Not-determined |
| S8_extracted_k141_88197 | Not-determined |
| S8_extracted_k141_114062 | Low-quality |
| S8_extracted_k141_122920 | Low-quality |
| S8_extracted_k141_140221 | Not-determined |
| S8_extracted_k141_44564 | Not-determined |
| S8_extracted_k141_96833 | Not-determined |
| S8_extracted_k141_44566 | Not-determined |
| S8_extracted_k141_88201 | Not-determined |
| S8_extracted_k141_105464 | Not-determined |
| S8_extracted_k141_114070 | Not-determined |
| S8_extracted_k141_96841 | Not-determined |
| S8_extracted_k141_44573 | Not-determined |
| S8_extracted_k141_192512 | Not-determined |
| S8_extracted_k141_105477 | Not-determined |
| S8_extracted_k141_131548 | Not-determined |
| S8_extracted_k141_105478 | Not-determined |
| S8_extracted_k141_140232 | Not-determined |
| S8_extracted_k141_88211 | Not-determined |
| S8_extracted_k141_9929 | Not-determined |
| S8_extracted_k141_1486 | Not-determined |
| S8_extracted_k141_157445 | Not-determined |
| S8_extracted_k141_122921 | Low-quality |
| S8_extracted_k141_174664 | Not-determined |
| S8_extracted_k141_96843 | Low-quality |
| S8_extracted_k141_88214 | Not-determined |
| S8_extracted_k141_36197 | Not-determined |
| S8_extracted_k141_44577 | Not-determined |
| S8_extracted_k141_96850 | Not-determined |
| S8_extracted_k141_192519 | Not-determined |
| S8_extracted_k141_157450 | Not-determined |
| S8_extracted_k141_36199 | Not-determined |
| S8_extracted_k141_9935 | Not-determined |
| S8_extracted_k141_1495 | Low-quality |
| S8_extracted_k141_9938 | Not-determined |
| S8_extracted_k141_27591 | Not-determined |
| S8_extracted_k141_183125 | Not-determined |
| S8_extracted_k141_192523 | Not-determined |
| S8_extracted_k141_192526 | Not-determined |
| S8_extracted_k141_148853 | Not-determined |
| S8_extracted_k141_70548 | Not-determined |
| S8_extracted_k141_183129 | Not-determined |
| S8_extracted_k141_18522 | Not-determined |
| S8_extracted_k141_200578 | Not-determined |
| S8_extracted_k141_148856 | Not-determined |
| S8_extracted_k141_148858 | Not-determined |
| S8_extracted_k141_174675 | Not-determined |
| S8_extracted_k141_44582 | Not-determined |
| S8_extracted_k141_9943 | Not-determined |
| S8_extracted_k141_122935 | Not-determined |
| S8_extracted_k141_79561 | Not-determined |
| S8_extracted_k141_114089 | Low-quality |
| S8_extracted_k141_1502 | Not-determined |
| S8_extracted_k141_200584 | Not-determined |
| S8_extracted_k141_183137 | Not-determined |
| S8_extracted_k141_36211 | Not-determined |
| S8_extracted_k141_157460 | Not-determined |
| S8_extracted_k141_18532 | Not-determined |
| S8_extracted_k141_183139 | Not-determined |
| S8_extracted_k141_122937 | Not-determined |
| S8_extracted_k141_174685 | Not-determined |
| S8_extracted_k141_9949 | Not-determined |
| S8_extracted_k141_114096 | Not-determined |
| S8_extracted_k141_36217 | Not-determined |
| S8_extracted_k141_70557 | Not-determined |
| S8_extracted_k141_200590 | Not-determined |
| S8_extracted_k141_183145 | Not-determined |
| S8_extracted_k141_44592 | Not-determined |
| S8_extracted_k141_183147 | Not-determined |
| S8_extracted_k141_114100 | Not-determined |
| S8_extracted_k141_114101 | Not-determined |
| S8_extracted_k141_200593 | Not-determined |
| S8_extracted_k141_122941 | Low-quality |
| S8_extracted_k141_1514 | Not-determined |
| S8_extracted_k141_148861 | Low-quality |

| | |
|---|---|
| S8_extracted_k141_166238 | Not-determined |
| S8_extracted_k141_36224 | Not-determined |
| S8_extracted_k141_36226 | Not-determined |
| S8_extracted_k141_27607 | Not-determined |
| S8_extracted_k141_18550 | Not-determined |
| S8_extracted_k141_140253 | Not-determined |
| S8_extracted_k141_36229 | Not-determined |
| S8_extracted_k141_1518 | Not-determined |
| S8_extracted_k141_140254 | Not-determined |
| S8_extracted_k141_1520 | Not-determined |
| S8_extracted_k141_27610 | Not-determined |
| S8_extracted_k141_36232 | Not-determined |
| S8_extracted_k141_148868 | Not-determined |
| S8_extracted_k141_174694 | Not-determined |
| S8_extracted_k141_122950 | Not-determined |
| S8_extracted_k141_114107 | Not-determined |
| S8_extracted_k141_27614 | Not-determined |
| S8_extracted_k141_183152 | Low-quality |
| S8_extracted_k141_96875 | Not-determined |
| S8_extracted_k141_96876 | Not-determined |
| S8_extracted_k141_9967 | Not-determined |
| S8_extracted_k141_18560 | Not-determined |
| S8_extracted_k141_174695 | Low-quality |
| S8_extracted_k141_174696 | Low-quality |
| S8_extracted_k141_70575 | Not-determined |
| S8_extracted_k141_79572 | Not-determined |
| S8_extracted_k141_114118 | Not-determined |
| S8_extracted_k141_27622 | Not-determined |
| S8_extracted_k141_183157 | Not-determined |
| S8_extracted_k141_27624 | Not-determined |
| S8_extracted_k141_183159 | Not-determined |
| S8_extracted_k141_96879 | Not-determined |
| S8_extracted_k141_114125 | Not-determined |
| S8_extracted_k141_36246 | Not-determined |
| S8_extracted_k141_79580 | Not-determined |
| S8_extracted_k141_79583 | Not-determined |
| S8_extracted_k141_96880 | Not-determined |
| S8_extracted_k141_174705 | Not-determined |
| S8_extracted_k141_192541 | Not-determined |
| S8_extracted_k141_79584 | Not-determined |
| S8_extracted_k141_44601 | Not-determined |
| S8_extracted_k141_44603 | Not-determined |
| S8_extracted_k141_174707 | Not-determined |
| S8_extracted_k141_1530 | Not-determined |
| S8_extracted_k141_53518 | Not-determined |
| S8_extracted_k141_140270 | Not-determined |
| S8_extracted_k141_166251 | Not-determined |
| S8_extracted_k141_79589 | Not-determined |
| S8_extracted_k141_96888 | Not-determined |
| S8_extracted_k141_1534 | Not-determined |
| S8_extracted_k141_174710 | Not-determined |
| S8_extracted_k141_1536 | Not-determined |
| S8_extracted_k141_1537 | Not-determined |
| S8_extracted_k141_53523 | Not-determined |
| S8_extracted_k141_105501 | Low-quality |
| S8_extracted_k141_1539 | Low-quality |
| S8_extracted_k141_27639 | Not-determined |
| S8_extracted_k141_105503 | Not-determined |
| S8_extracted_k141_36263 | Not-determined |
| S8_extracted_k141_1542 | Not-determined |
| S8_extracted_k141_1543 | Not-determined |
| S8_extracted_k141_122961 | Not-determined |
| S8_extracted_k141_79597 | Not-determined |
| S8_extracted_k141_140280 | Not-determined |
| S8_extracted_k141_183165 | Low-quality |
| S8_extracted_k141_105510 | Not-determined |
| S8_extracted_k141_122967 | Not-determined |
| S8_extracted_k141_192558 | Not-determined |
| S8_extracted_k141_131551 | Low-quality |
| S8_extracted_k141_70616 | Low-quality |
| S8_extracted_k141_27643 | Not-determined |
| S8_extracted_k141_122969 | Not-determined |
| S8_extracted_k141_27647 | Not-determined |
| S8_extracted_k141_140295 | Not-determined |
| S8_extracted_k141_105517 | Not-determined |
| S8_extracted_k141_183170 | Not-determined |
| S8_extracted_k141_200611 | Low-quality |
| S8_extracted_k141_70621 | Not-determined |
| S8_extracted_k141_140299 | Low-quality |
| S8_extracted_k141_200613 | Not-determined |
| S8_extracted_k141_166265 | Not-determined |
| S8_extracted_k141_36266 | Not-determined |
| S8_extracted_k141_70623 | Not-determined |
| S8_extracted_k141_114143 | Low-quality |
| S8_extracted_k141_9979 | Low-quality |
| S8_extracted_k141_157523 | Low-quality |
| S8_extracted_k141_140306 | Not-determined |
| S8_extracted_k141_9984 | Low-quality |
| S8_extracted_k141_70627 | Low-quality |
| S8_extracted_k141_131561 | Low-quality |
| S8_extracted_k141_131562 | Not-determined |
| S8_extracted_k141_70629 | Not-determined |
| S8_extracted_k141_96899 | Not-determined |
| S8_extracted_k141_96901 | Not-determined |
| S8_extracted_k141_53545 | Not-determined |
| S8_extracted_k141_183183 | Not-determined |
| S8_extracted_k141_140312 | Not-determined |
| S8_extracted_k141_114150 | Low-quality |
| S8_extracted_k141_183184 | Not-determined |
| S8_extracted_k141_53548 | Not-determined |
| S8_extracted_k141_70640 | Not-determined |
| S8_extracted_k141_96906 | Not-determined |
| S8_extracted_k141_53551 | Not-determined |
| S8_extracted_k141_140321 | Not-determined |
| S8_extracted_k141_18601 | Not-determined |
| S8_extracted_k141_1564 | Low-quality |
| S8_extracted_k141_79621 | Not-determined |
| S8_extracted_k141_200624 | Low-quality |
| S8_extracted_k141_166289 | Not-determined |
| S8_extracted_k141_53555 | Not-determined |
| S8_extracted_k141_70642 | Not-determined |
| S8_extracted_k141_79625 | Not-determined |
| S8_extracted_k141_174748 | Not-determined |
| S8_extracted_k141_183191 | Not-determined |
| S8_extracted_k141_200627 | Not-determined |
| S8_extracted_k141_140329 | Not-determined |
| S8_extracted_k141_200629 | Not-determined |
| S8_extracted_k141_18605 | Not-determined |
| S8_extracted_k141_200630 | Not-determined |
| S8_extracted_k141_148886 | Low-quality |
| S8_extracted_k141_183193 | Not-determined |
| S8_extracted_k141_131569 | Not-determined |
| S8_extracted_k141_79631 | Not-determined |
| S8_extracted_k141_18608 | Not-determined |
| S8_extracted_k141_114170 | Not-determined |
| S8_extracted_k141_70649 | Not-determined |
| S8_extracted_k141_200636 | Low-quality |
| S8_extracted_k141_18610 | Not-determined |
| S8_extracted_k141_53564 | Not-determined |
| S8_extracted_k141_174753 | Not-determined |
| S8_extracted_k141_200640 | Not-determined |
| S8_extracted_k141_53566 | Not-determined |
| S8_extracted_k141_157535 | Not-determined |
| S8_extracted_k141_105531 | Not-determined |
| S8_extracted_k141_70651 | Not-determined |
| S8_extracted_k141_114174 | Not-determined |
| S8_extracted_k141_18611 | Low-quality |
| S8_extracted_k141_96922 | Low-quality |
| S8_extracted_k141_18612 | Low-quality |
| S8_extracted_k141_157536 | Not-determined |
| S8_extracted_k141_18613 | Not-determined |
| S8_extracted_k141_79643 | Not-determined |
| S8_extracted_k141_140336 | Not-determined |
| S8_extracted_k141_200653 | Not-determined |
| S8_extracted_k141_96929 | Not-determined |
| S8_extracted_k141_96930 | Not-determined |
| S8_extracted_k141_1576 | Not-determined |
| S8_extracted_k141_27663 | Not-determined |
| S8_extracted_k141_44619 | Not-determined |
| S8_extracted_k141_200657 | Not-determined |
| S8_extracted_k141_53582 | Not-determined |
| S8_extracted_k141_27666 | Not-determined |
| S8_extracted_k141_200659 | Not-determined |
| S8_extracted_k141_1578 | Not-determined |
| S8_extracted_k141_200662 | Not-determined |
| S8_extracted_k141_79655 | Not-determined |
| S8_extracted_k141_70660 | Not-determined |
| S8_extracted_k141_79664 | Not-determined |
| S8_extracted_k141_131591 | Not-determined |
| S8_extracted_k141_44629 | Not-determined |
| S8_extracted_k141_27674 | Not-determined |
| S8_extracted_k141_44631 | Not-determined |
| S8_extracted_k141_140349 | Not-determined |
| S8_extracted_k141_27678 | Not-determined |
| S8_extracted_k141_140351 | Not-determined |
| S8_extracted_k141_27680 | Not-determined |
| S8_extracted_k141_53589 | Not-determined |
| S8_extracted_k141_140353 | Not-determined |
| S8_extracted_k141_1595 | Not-determined |
| S8_extracted_k141_1596 | Not-determined |
| S8_extracted_k141_53592 | Not-determined |
| S8_extracted_k141_200678 | Not-determined |
| S8_extracted_k141_96949 | Not-determined |
| S8_extracted_k141_122972 | Low-quality |
| S8_extracted_k141_27685 | Not-determined |
| S8_extracted_k141_70673 | Not-determined |
| S8_extracted_k141_79686 | Low-quality |
| S8_extracted_k141_96954 | Not-determined |
| S8_extracted_k141_131601 | Not-determined |
| S8_extracted_k141_140363 | Low-quality |
| S8_extracted_k141_96955 | Not-determined |
| S8_extracted_k141_140365 | Not-determined |
| S8_extracted_k141_18630 | Not-determined |
| S8_extracted_k141_131603 | Not-determined |
| S8_extracted_k141_148914 | Not-determined |
| S8_extracted_k141_131605 | Not-determined |
| S8_extracted_k141_174785 | Not-determined |
| S8_extracted_k141_122981 | Not-determined |
| S8_extracted_k141_44644 | Not-determined |
| S8_extracted_k141_148915 | Not-determined |
| S8_extracted_k141_53612 | Not-determined |
| S8_extracted_k141_70681 | Not-determined |
| S8_extracted_k141_148917 | Not-determined |
| S8_extracted_k141_79695 | Not-determined |
| S8_extracted_k141_131609 | Not-determined |
| S8_extracted_k141_148919 | Not-determined |
| S8_extracted_k141_166302 | Not-determined |
| S8_extracted_k141_166303 | Not-determined |
| S8_extracted_k141_122987 | Not-determined |
| S8_extracted_k141_140370 | Not-determined |
| S8_extracted_k141_44652 | Low-quality |
| S8_extracted_k141_79700 | Low-quality |
| S8_extracted_k141_140373 | Not-determined |
| S8_extracted_k141_183220 | Not-determined |
| S8_extracted_k141_62142 | Not-determined |
| S8_extracted_k141_44660 | Low-quality |
| S8_extracted_k141_36284 | Low-quality |
| S8_extracted_k141_183225 | Not-determined |
| S8_extracted_k141_174793 | Not-determined |
| S8_extracted_k141_79710 | Not-determined |
| S8_extracted_k141_96979 | Not-determined |
| S8_extracted_k141_105576 | Not-determined |
| S8_extracted_k141_131618 | Not-determined |
| S8_extracted_k141_131619 | Not-determined |
| S8_extracted_k141_44666 | Not-determined |
| S8_extracted_k141_27712 | Not-determined |
| S8_extracted_k141_183229 | Not-determined |
| S8_extracted_k141_105580 | Not-determined |
| S8_extracted_k141_70694 | Not-determined |
| S8_extracted_k141_148932 | Not-determined |
| S8_extracted_k141_148933 | Not-determined |
| S8_extracted_k141_62159 | Not-determined |
| S8_extracted_k141_131624 | Low-quality |
| S8_extracted_k141_62160 | Not-determined |
| S8_extracted_k141_140379 | Not-determined |
| S8_extracted_k141_44675 | Not-determined |
| S8_extracted_k141_62162 | Not-determined |
| S8_extracted_k141_140380 | Not-determined |
| S8_extracted_k141_62164 | Not-determined |
| S8_extracted_k141_123004 | Not-determined |
| S8_extracted_k141_36301 | Not-determined |
| S8_extracted_k141_105588 | Not-determined |
| S8_extracted_k141_62165 | Not-determined |
| S8_extracted_k141_96988 | Low-quality |
| S8_extracted_k141_44681 | Not-determined |
| S8_extracted_k141_70697 | Not-determined |
| S8_extracted_k141_18650 | Not-determined |
| S8_extracted_k141_62169 | Not-determined |
| S8_extracted_k141_123013 | Not-determined |
| S8_extracted_k141_105593 | Not-determined |
| S8_extracted_k141_44684 | Not-determined |
| S8_extracted_k141_148945 | Not-determined |
| S8_extracted_k141_131630 | Not-determined |
| S8_extracted_k141_200691 | Not-determined |
| S8_extracted_k141_9996 | Low-quality |
| S8_extracted_k141_123016 | Low-quality |
| S8_extracted_k141_105601 | Not-determined |
| S8_extracted_k141_70708 | Not-determined |
| S8_extracted_k141_36318 | Not-determined |
| S8_extracted_k141_62181 | Not-determined |
| S8_extracted_k141_114192 | Low-quality |
| S8_extracted_k141_131637 | Not-determined |
| S8_extracted_k141_27720 | Not-determined |
| S8_extracted_k141_9999 | Not-determined |
| S8_extracted_k141_131640 | Not-determined |
| S8_extracted_k141_114196 | Not-determined |
| S8_extracted_k141_70715 | Not-determined |
| S8_extracted_k141_123017 | Not-determined |
| S8_extracted_k141_131644 | Not-determined |
| S8_extracted_k141_27726 | Not-determined |
| S8_extracted_k141_166331 | Low-quality |
| S8_extracted_k141_131646 | Not-determined |
| S8_extracted_k141_114200 | Not-determined |
| S8_extracted_k141_131647 | Not-determined |
| S8_extracted_k141_131648 | Not-determined |
| S8_extracted_k141_131649 | Not-determined |
| S8_extracted_k141_62188 | Low-quality |
| S8_extracted_k141_18664 | Not-determined |
| S8_extracted_k141_200704 | Not-determined |
| S8_extracted_k141_131651 | Not-determined |
| S8_extracted_k141_105610 | Not-determined |
| S8_extracted_k141_18666 | Not-determined |
| S8_extracted_k141_174823 | Not-determined |
| S8_extracted_k141_36326 | Not-determined |
| S8_extracted_k141_44705 | Not-determined |
| S8_extracted_k141_36329 | Not-determined |
| S8_extracted_k141_18673 | Not-determined |
| S8_extracted_k141_62195 | Not-determined |
| S8_extracted_k141_105615 | Not-determined |
| S8_extracted_k141_36331 | Not-determined |
| S8_extracted_k141_166340 | Low-quality |
| S8_extracted_k141_27727 | Not-determined |
| S8_extracted_k141_166341 | Not-determined |
| S8_extracted_k141_62202 | Not-determined |
| S8_extracted_k141_200717 | Not-determined |
| S8_extracted_k141_200718 | Not-determined |
| S8_extracted_k141_140390 | Not-determined |
| S8_extracted_k141_62207 | Not-determined |
| S8_extracted_k141_105626 | Not-determined |
| S8_extracted_k141_44711 | Not-determined |
| S8_extracted_k141_166353 | Not-determined |
| S8_extracted_k141_36336 | Not-determined |
| S8_extracted_k141_166354 | Not-determined |
| S8_extracted_k141_36338 | Not-determined |
| S8_extracted_k141_123031 | Not-determined |
| S8_extracted_k141_79728 | Not-determined |
| S8_extracted_k141_70741 | Not-determined |
| S8_extracted_k141_62209 | Not-determined |
| S8_extracted_k141_174850 | Not-determined |
| S8_extracted_k141_192562 | Low-quality |
| S8_extracted_k141_174851 | Not-determined |
| S8_extracted_k141_10003 | Not-determined |
| S8_extracted_k141_174852 | Not-determined |
| S8_extracted_k141_10004 | Not-determined |
| S8_extracted_k141_105634 | Not-determined |
| S8_extracted_k141_1602 | Not-determined |
| S8_extracted_k141_105638 | Not-determined |
| S8_extracted_k141_140405 | Not-determined |
| S8_extracted_k141_131664 | Not-determined |
| S8_extracted_k141_79736 | Not-determined |
| S8_extracted_k141_44725 | Not-determined |
| S8_extracted_k141_123038 | Low-quality |
| S8_extracted_k141_200735 | Not-determined |
| S8_extracted_k141_174863 | Not-determined |
| S8_extracted_k141_123039 | Not-determined |
| S8_extracted_k141_27741 | Not-determined |
| S8_extracted_k141_62215 | Low-quality |
| S8_extracted_k141_200736 | Not-determined |
| S8_extracted_k141_36352 | Not-determined |
| S8_extracted_k141_131668 | Not-determined |
| S8_extracted_k141_27745 | Not-determined |
| S8_extracted_k141_1612 | Not-determined |
| S8_extracted_k141_27747 | Not-determined |
| S8_extracted_k141_1613 | Not-determined |
| S8_extracted_k141_62217 | Low-quality |
| S8_extracted_k141_62218 | Not-determined |
| S8_extracted_k141_10020 | Not-determined |
| S8_extracted_k141_62219 | Not-determined |
| S8_extracted_k141_174867 | Not-determined |
| S8_extracted_k141_131674 | Not-determined |
| S8_extracted_k141_10021 | Not-determined |
| S8_extracted_k141_114212 | Not-determined |
| S8_extracted_k141_114215 | Not-determined |
| S8_extracted_k141_192577 | Not-determined |
| S8_extracted_k141_36353 | Not-determined |
| S8_extracted_k141_166376 | Not-determined |
| S8_extracted_k141_166377 | Not-determined |
| S8_extracted_k141_166378 | Not-determined |
| S8_extracted_k141_131681 | Not-determined |
| S8_extracted_k141_114216 | Not-determined |
| S8_extracted_k141_62231 | Not-determined |
| S8_extracted_k141_131682 | Not-determined |
| S8_extracted_k141_88219 | Not-determined |
| S8_extracted_k141_18687 | Not-determined |
| S8_extracted_k141_183259 | Not-determined |
| S8_extracted_k141_166383 | Low-quality |
| S8_extracted_k141_88221 | Not-determined |
| S8_extracted_k141_105647 | Not-determined |
| S8_extracted_k141_140416 | Not-determined |
| S8_extracted_k141_105648 | Low-quality |
| S8_extracted_k141_166387 | Not-determined |
| S8_extracted_k141_183264 | Not-determined |
| S14_vs_k141_114491\|\|full | Medium-quality |
| S8_extracted_k141_1631 | Not-determined |
| S8_extracted_k141_183266 | Not-determined |
| S8_extracted_k141_70748 | Not-determined |
| S8_extracted_k141_192586 | Not-determined |
| S8_extracted_k141_1634 | Low-quality |
| S8_extracted_k141_1636 | Not-determined |
| S8_extracted_k141_183271 | Not-determined |
| S8_extracted_k141_192590 | Not-determined |
| S8_extracted_k141_18701 | Not-determined |
| S8_extracted_k141_114231 | Not-determined |
| S8_extracted_k141_70753 | Not-determined |
| S8_extracted_k141_62245 | Not-determined |
| S8_extracted_k141_79762 | Not-determined |
| S8_extracted_k141_1643 | Not-determined |
| S8_extracted_k141_1644 | Not-determined |
| S8_extracted_k141_123066 | Not-determined |
| S8_extracted_k141_1646 | Not-determined |
| S8_extracted_k141_174877 | Not-determined |
| S8_extracted_k141_62250 | Not-determined |
| S8_extracted_k141_157548 | Not-determined |
| S8_extracted_k141_183281 | Not-determined |
| S8_extracted_k141_1651 | Not-determined |
| S8_extracted_k141_27760 | Low-quality |
| S8_extracted_k141_166394 | Not-determined |
| S8_extracted_k141_36383 | Not-determined |
| S8_extracted_k141_183285 | Not-determined |
| S8_extracted_k141_36385 | Not-determined |
| S8_extracted_k141_174886 | Not-determined |
| S8_extracted_k141_114247 | Not-determined |
| S8_extracted_k141_174887 | Not-determined |
| S8_extracted_k141_183289 | Not-determined |
| S8_extracted_k141_166398 | Not-determined |
| S8_extracted_k141_183292 | Not-determined |
| S8_extracted_k141_183293 | Not-determined |
| S8_extracted_k141_183295 | Not-determined |
| S8_extracted_k141_131696 | Not-determined |
| S8_extracted_k141_53624 | Not-determined |
| S8_extracted_k141_36395 | Not-determined |
| S8_extracted_k141_97030 | Not-determined |
| S8_extracted_k141_53629 | Not-determined |
| S8_extracted_k141_53632 | Not-determined |
| S8_extracted_k141_1668 | Not-determined |
| S8_extracted_k141_174894 | Not-determined |
| S8_extracted_k141_200765 | Not-determined |
| S8_extracted_k141_183299 | Not-determined |
| S8_extracted_k141_157561 | Low-quality |
| S8_extracted_k141_192614 | Not-determined |
| S8_extracted_k141_157562 | Not-determined |
| S8_extracted_k141_53642 | Not-determined |
| S8_extracted_k141_105664 | Not-determined |
| S8_extracted_k141_36402 | Not-determined |
| S8_extracted_k141_200770 | Not-determined |
| S8_extracted_k141_140432 | Not-determined |
| S8_extracted_k141_105667 | Not-determined |
| S8_extracted_k141_36406 | Not-determined |
| S8_extracted_k141_200776 | Not-determined |
| S8_extracted_k141_105670 | Not-determined |
| S8_extracted_k141_62265 | Not-determined |
| S8_extracted_k141_123093 | Not-determined |
| S8_extracted_k141_53652 | Not-determined |
| S8_extracted_k141_27784 | Not-determined |
| S8_extracted_k141_36408 | Not-determined |
| S8_extracted_k141_27785 | Not-determined |
| S8_extracted_k141_174909 | Not-determined |
| S8_extracted_k141_36409 | Not-determined |
| S8_extracted_k141_97044 | Not-determined |
| S8_extracted_k141_148982 | Not-determined |
| S8_extracted_k141_10059 | Low-quality |
| S8_extracted_k141_1673 | Low-quality |
| S8_extracted_k141_18735 | Not-determined |
| S8_extracted_k141_114270 | Low-quality |
| S8_extracted_k141_18737 | Not-determined |
| S8_extracted_k141_183311 | Low-quality |
| S8_extracted_k141_88262 | Not-determined |
| S8_extracted_k141_10065 | Not-determined |
| S8_extracted_k141_88263 | Not-determined |
| S8_extracted_k141_97050 | Low-quality |
| S8_extracted_k141_53657 | Not-determined |
| S8_extracted_k141_97051 | Low-quality |
| S8_extracted_k141_157575 | Not-determined |
| S8_extracted_k141_1679 | Not-determined |
| S8_extracted_k141_200788 | Not-determined |
| S8_extracted_k141_114275 | Not-determined |
| S8_extracted_k141_200789 | Not-determined |
| S8_extracted_k141_97057 | Not-determined |
| S8_extracted_k141_114278 | Not-determined |
| S8_extracted_k141_36421 | Low-quality |
| S8_extracted_k141_62274 | Not-determined |
| S8_extracted_k141_97059 | Low-quality |
| S8_extracted_k141_157580 | Not-determined |
| S8_extracted_k141_157583 | Not-determined |
| S8_extracted_k141_62277 | Not-determined |
| S8_extracted_k141_36424 | Not-determined |
| S8_extracted_k141_200795 | Not-determined |
| S8_extracted_k141_157584 | Not-determined |
| S8_extracted_k141_53666 | Not-determined |
| S8_extracted_k141_200796 | Not-determined |
| S8_extracted_k141_183322 | Not-determined |
| S8_extracted_k141_53672 | Not-determined |
| S8_extracted_k141_140454 | Low-quality |
| S8_extracted_k141_62283 | Not-determined |
| S8_extracted_k141_157593 | Not-determined |
| S8_extracted_k141_88278 | Not-determined |
| S8_extracted_k141_157595 | Not-determined |
| S8_extracted_k141_1690 | Not-determined |
| S8_extracted_k141_53677 | Not-determined |
| S8_extracted_k141_53678 | Not-determined |
| S8_extracted_k141_131706 | Not-determined |
| S8_extracted_k141_97070 | Not-determined |
| S8_extracted_k141_131707 | Not-determined |
| S2_vs_k141_661\|\|full | Medium-quality |
| S8_extracted_k141_18750 | Low-quality |
| S8_extracted_k141_123120 | Not-determined |
| S8_extracted_k141_166404 | Not-determined |
| S8_extracted_k141_53679 | Not-determined |
| S8_extracted_k141_192649 | Not-determined |
| S8_extracted_k141_123121 | Not-determined |
| S8_extracted_k141_1694 | Not-determined |
| S8_extracted_k141_53680 | Not-determined |
| S8_extracted_k141_192650 | Not-determined |
| S8_extracted_k141_183329 | Low-quality |
| S8_extracted_k141_123126 | Low-quality |
| S8_extracted_k141_53683 | Not-determined |
| S8_extracted_k141_140461 | Not-determined |
| S8_extracted_k141_166410 | Not-determined |
| S8_extracted_k141_183330 | Not-determined |
| S8_extracted_k141_140463 | Not-determined |
| S8_extracted_k141_166412 | Not-determined |
| S8_extracted_k141_192655 | Not-determined |
| S8_extracted_k141_1700 | Not-determined |
| S8_extracted_k141_62292 | Not-determined |
| S8_extracted_k141_166416 | Not-determined |
| S8_extracted_k141_174938 | Not-determined |
| S8_extracted_k141_53689 | Not-determined |
| S8_extracted_k141_192659 | Not-determined |
| S8_extracted_k141_140467 | Not-determined |
| S8_extracted_k141_123131 | Not-determined |
| S8_extracted_k141_114306 | Not-determined |
| S8_extracted_k141_36438 | Not-determined |
| S8_extracted_k141_10076 | Not-determined |
| S8_extracted_k141_166420 | Low-quality |
| S8_extracted_k141_114309 | Not-determined |
| S8_extracted_k141_157611 | Not-determined |
| S8_extracted_k141_200820 | Low-quality |
| S8_extracted_k141_166422 | Not-determined |
| S8_extracted_k141_18764 | Not-determined |
| S8_extracted_k141_114312 | Not-determined |
| S8_extracted_k141_105714 | Not-determined |
| S8_extracted_k141_114314 | Not-determined |
| S8_extracted_k141_140476 | Not-determined |
| S8_extracted_k141_140478 | Not-determined |
| S8_extracted_k141_105716 | Not-determined |
| S8_extracted_k141_53703 | Low-quality |
| S8_extracted_k141_131712 | Not-determined |
| S8_extracted_k141_44753 | Low-quality |
| S8_extracted_k141_114321 | Not-determined |
| S8_extracted_k141_192671 | Not-determined |
| S8_extracted_k141_62310 | Not-determined |
| S8_extracted_k141_192674 | Not-determined |
| S8_extracted_k141_192675 | Not-determined |
| S8_extracted_k141_53712 | Not-determined |
| S8_extracted_k141_192676 | Not-determined |
| S8_extracted_k141_157625 | Not-determined |
| S8_extracted_k141_88306 | Not-determined |
| S8_extracted_k141_88307 | Low-quality |
| S8_extracted_k141_18773 | Low-quality |
| S8_extracted_k141_166437 | Not-determined |
| S8_extracted_k141_10100 | Not-determined |
| S8_extracted_k141_105724 | Not-determined |
| S8_extracted_k141_157627 | Low-quality |
| S8_extracted_k141_97105 | Not-determined |
| S8_extracted_k141_157628 | Low-quality |
| S8_extracted_k141_18778 | Not-determined |
| S8_extracted_k141_157630 | Not-determined |
| S8_extracted_k141_105725 | Low-quality |
| S8_extracted_k141_157632 | Not-determined |
| S16_vs_119834\|\|full | Medium-quality |
| S8_extracted_k141_53725 | Low-quality |
| S8_extracted_k141_18783 | Not-determined |
| S8_extracted_k141_79771 | Not-determined |
| S8_extracted_k141_18784 | Not-determined |
| S8_extracted_k141_18785 | Not-determined |
| S8_extracted_k141_62323 | Not-determined |
| S8_extracted_k141_192683 | Not-determined |
| S8_extracted_k141_166447 | Not-determined |
| S8_extracted_k141_123147 | Not-determined |
| S8_extracted_k141_166448 | Not-determined |
| S8_extracted_k141_36483 | Not-determined |
| S8_extracted_k141_131732 | Not-determined |
| S8_extracted_k141_166450 | Not-determined |
| S8_extracted_k141_62333 | Not-determined |
| S8_extracted_k141_44772 | Not-determined |
| S8_extracted_k141_88329 | Not-determined |
| S8_extracted_k141_123155 | Not-determined |
| S8_extracted_k141_114335 | Not-determined |
| S8_extracted_k141_27804 | Not-determined |
| S8_extracted_k141_131737 | Not-determined |
| S8_extracted_k141_114336 | Not-determined |
| S8_extracted_k141_131740 | Not-determined |
| S8_extracted_k141_79782 | Not-determined |
| S8_extracted_k141_157649 | Not-determined |
| S8_extracted_k141_10126 | Low-quality |
| S8_extracted_k141_192697 | Low-quality |
| S8_extracted_k141_123162 | Not-determined |
| S8_extracted_k141_18806 | Not-determined |
| S8_extracted_k141_123164 | Not-determined |
| S8_extracted_k141_88333 | Not-determined |
| S8_extracted_k141_157652 | Not-determined |
| S8_extracted_k141_114342 | Not-determined |
| S8_extracted_k141_131748 | Not-determined |
| S8_extracted_k141_114344 | Low-quality |
| S8_extracted_k141_105741 | Not-determined |
| S8_extracted_k141_166470 | Not-determined |
| S8_extracted_k141_183339 | Low-quality |
| S8_extracted_k141_131750 | Low-quality |
| S8_extracted_k141_157657 | Not-determined |
| S8_extracted_k141_53746 | Not-determined |
| S8_extracted_k141_88339 | Not-determined |
| S8_extracted_k141_123166 | Low-quality |
| S8_extracted_k141_62345 | Not-determined |
| S8_extracted_k141_123167 | Not-determined |
| S8_extracted_k141_123168 | Not-determined |
| S8_extracted_k141_36500 | Not-determined |
| S8_extracted_k141_105749 | Low-quality |
| S8_extracted_k141_114345 | Not-determined |
| S8_extracted_k141_79789 | Not-determined |
| S8_extracted_k141_140514 | Not-determined |
| S8_extracted_k141_174964 | Not-determined |
| S8_extracted_k141_183349 | Not-determined |
| S8_extracted_k141_18818 | Not-determined |
| S8_extracted_k141_140516 | Not-determined |
| S8_extracted_k141_88348 | Not-determined |
| S8_extracted_k141_174969 | Not-determined |
| S8_extracted_k141_166482 | Not-determined |
| S8_extracted_k141_97129 | Not-determined |
| S8_extracted_k141_183352 | Not-determined |

| | |
|---|---|
| S8_extracted_k141_18819 | Low-quality |
| S8_extracted_k141_200828 | Not-determined |
| S8_extracted_k141_1728 | Not-determined |
| S8_extracted_k141_200832 | Not-determined |
| S8_extracted_k141_200833 | Not-determined |
| S8_extracted_k141_174975 | Not-determined |
| S8_extracted_k141_97137 | Not-determined |
| S8_extracted_k141_131763 | Not-determined |
| S8_extracted_k141_10138 | Not-determined |
| S8_extracted_k141_166488 | Low-quality |
| S8_extracted_k141_192711 | Not-determined |
| S8_extracted_k141_192712 | Not-determined |
| S8_extracted_k141_18827 | Not-determined |
| S8_extracted_k141_140519 | Low-quality |
| S8_extracted_k141_200839 | Not-determined |
| S8_extracted_k141_200841 | Not-determined |
| S8_extracted_k141_123183 | Not-determined |
| S8_extracted_k141_123184 | Not-determined |
| S8_extracted_k141_53758 | Low-quality |
| S8_extracted_k141_200843 | Not-determined |
| S8_extracted_k141_174988 | Low-quality |
| S8_extracted_k141_36514 | Low-quality |
| S8_extracted_k141_44774 | Not-determined |
| S8_extracted_k141_18837 | Not-determined |
| S8_extracted_k141_53765 | Not-determined |
| S8_extracted_k141_88360 | Not-determined |
| S8_extracted_k141_192719 | Not-determined |
| S8_extracted_k141_44776 | Not-determined |
| S8_extracted_k141_27826 | Low-quality |
| S8_extracted_k141_10148 | Low-quality |
| S8_extracted_k141_10149 | Not-determined |
| S8_extracted_k141_105766 | Not-determined |
| S8_extracted_k141_27831 | Not-determined |
| S8_extracted_k141_27832 | Not-determined |
| S8_extracted_k141_27833 | Not-determined |
| S8_extracted_k141_200846 | Not-determined |
| S8_extracted_k141_10157 | Not-determined |
| S8_extracted_k141_97152 | Not-determined |
| S8_extracted_k141_200852 | Not-determined |
| S8_extracted_k141_88373 | Not-determined |
| S8_extracted_k141_175002 | Not-determined |
| S8_extracted_k141_88374 | Not-determined |
| S8_extracted_k141_1745 | Not-determined |
| S8_extracted_k141_166500 | Not-determined |
| S8_extracted_k141_131789 | Not-determined |
| S8_extracted_k141_44793 | Low-quality |
| S8_extracted_k141_131790 | Not-determined |
| S8_extracted_k141_36519 | Not-determined |
| S8_extracted_k141_105784 | Not-determined |
| S8_extracted_k141_88378 | Not-determined |
| S8_extracted_k141_175007 | Not-determined |
| S8_extracted_k141_157665 | Not-determined |
| S8_extracted_k141_53777 | Low-quality |
| S8_extracted_k141_140529 | Not-determined |
| S8_extracted_k141_114353 | Not-determined |
| S8_extracted_k141_27846 | Not-determined |
| S8_extracted_k141_10169 | Not-determined |
| S8_extracted_k141_27848 | Not-determined |
| S8_extracted_k141_105792 | Not-determined |
| S8_extracted_k141_114355 | Not-determined |
| S8_extracted_k141_114357 | Not-determined |
| S8_extracted_k141_157667 | Not-determined |
| S8_extracted_k141_166508 | Low-quality |
| S8_extracted_k141_131800 | Not-determined |
| S8_extracted_k141_166510 | Not-determined |
| S17_extracted_k141_67042 | Medium-quality |
| S8_extracted_k141_36534 | Low-quality |
| S8_extracted_k141_200870 | Low-quality |
| S8_extracted_k141_140539 | Not-determined |
| S8_extracted_k141_192727 | Not-determined |
| S8_extracted_k141_140543 | Not-determined |
| S8_extracted_k141_105805 | Not-determined |
| S8_extracted_k141_166514 | Not-determined |
| S8_extracted_k141_44808 | Not-determined |
| S8_extracted_k141_114360 | Not-determined |
| S8_extracted_k141_157670 | Low-quality |
| S8_extracted_k141_88392 | Not-determined |
| S8_extracted_k141_36535 | Not-determined |
| S8_extracted_k141_192734 | Not-determined |
| S8_extracted_k141_157676 | Not-determined |
| S8_extracted_k141_166518 | Not-determined |
| S8_extracted_k141_18857 | Not-determined |
| S8_extracted_k141_88398 | Not-determined |
| S8_extracted_k141_114363 | Not-determined |
| S8_extracted_k141_53806 | Low-quality |
| S8_extracted_k141_114367 | Not-determined |
| S8_extracted_k141_36547 | Not-determined |
| S8_extracted_k141_53808 | Not-determined |
| S8_extracted_k141_157690 | Not-determined |
| S8_extracted_k141_114373 | Not-determined |
| S8_extracted_k141_166528 | Not-determined |
| S8_extracted_k141_105820 | Low-quality |
| S8_extracted_k141_36559 | Not-determined |
| S8_extracted_k141_166531 | Not-determined |
| S8_extracted_k141_157697 | Not-determined |
| S8_extracted_k141_1760 | Not-determined |
| S8_extracted_k141_166532 | Low-quality |
| S8_extracted_k141_18871 | Not-determined |
| S8_extracted_k141_114377 | Not-determined |
| S8_extracted_k141_1764 | Not-determined |
| S8_extracted_k141_175014 | Low-quality |
| S8_extracted_k141_53816 | Not-determined |
| S8_extracted_k141_53819 | Not-determined |
| S8_extracted_k141_157700 | Not-determined |
| S8_extracted_k141_166536 | Not-determined |
| S8_extracted_k141_140547 | Not-determined |
| S8_extracted_k141_175025 | Not-determined |
| S8_extracted_k141_140548 | Not-determined |
| S8_extracted_k141_97168 | Not-determined |
| S8_extracted_k141_175026 | Not-determined |
| S8_extracted_k141_44820 | Not-determined |
| S8_extracted_k141_1767 | Not-determined |
| S8_extracted_k141_62362 | Not-determined |
| S8_extracted_k141_44821 | Not-determined |
| S8_extracted_k141_166542 | Not-determined |
| S8_extracted_k141_192758 | Not-determined |
| S8_extracted_k141_200883 | Not-determined |
| S8_extracted_k141_27853 | Low-quality |
| S8_extracted_k141_18874 | Not-determined |
| S8_extracted_k141_166550 | Low-quality |
| S8_extracted_k141_53834 | Low-quality |
| S8_extracted_k141_88412 | Not-determined |
| S8_extracted_k141_53835 | Not-determined |
| S8_extracted_k141_79800 | Low-quality |
| S8_extracted_k141_114391 | Not-determined |
| S8_extracted_k141_88414 | Not-determined |
| S8_extracted_k141_10184 | Not-determined |
| S8_extracted_k141_79803 | Low-quality |
| S8_extracted_k141_140558 | Not-determined |
| S8_extracted_k141_88416 | Not-determined |
| S8_extracted_k141_183370 | Not-determined |
| S8_extracted_k141_140559 | Not-determined |
| S8_extracted_k141_192761 | Not-determined |
| S8_extracted_k141_157707 | Not-determined |
| S8_extracted_k141_123208 | Not-determined |
| S8_extracted_k141_97181 | Not-determined |
| S8_extracted_k141_140561 | Low-quality |
| S8_extracted_k141_1773 | Not-determined |
| S8_extracted_k141_200889 | Low-quality |
| S8_extracted_k141_175039 | Not-determined |
| S8_extracted_k141_123212 | Low-quality |
| S8_extracted_k141_79807 | Low-quality |
| S8_extracted_k141_10191 | Not-determined |
| S8_extracted_k141_175042 | Not-determined |
| S8_extracted_k141_97188 | Not-determined |
| S8_extracted_k141_166558 | Not-determined |
| S8_extracted_k141_166559 | Not-determined |
| S8_extracted_k141_53846 | Not-determined |
| S8_extracted_k141_200893 | Not-determined |
| S8_extracted_k141_79810 | Not-determined |
| S8_extracted_k141_175048 | Not-determined |
| S8_extracted_k141_1779 | Not-determined |
| S8_extracted_k141_44843 | Not-determined |
| S8_extracted_k141_97196 | Not-determined |
| S8_extracted_k141_10193 | Low-quality |
| S8_extracted_k141_192770 | Low-quality |
| S8_extracted_k141_114399 | Not-determined |
| S8_extracted_k141_114400 | Not-determined |
| S8_extracted_k141_1782 | Not-determined |
| S8_extracted_k141_114401 | Not-determined |
| S8_extracted_k141_27858 | Low-quality |
| S8_extracted_k141_79817 | Not-determined |
| S8_extracted_k141_114406 | Not-determined |
| S8_extracted_k141_53853 | Not-determined |
| S8_extracted_k141_36583 | Not-determined |
| S8_extracted_k141_140573 | Not-determined |
| S8_extracted_k141_18885 | Not-determined |
| S8_extracted_k141_123222 | Low-quality |
| S8_extracted_k141_105835 | Not-determined |
| S8_extracted_k141_175053 | Not-determined |
| S8_extracted_k141_53855 | Not-determined |
| S8_extracted_k141_114414 | Not-determined |
| S8_extracted_k141_123226 | Not-determined |
| S8_extracted_k141_123228 | Not-determined |
| S8_extracted_k141_1787 | Low-quality |
| S8_extracted_k141_114415 | Not-determined |
| S8_extracted_k141_97201 | Not-determined |
| S8_extracted_k141_27871 | Not-determined |
| S8_extracted_k141_1790 | Not-determined |
| S8_extracted_k141_114418 | Not-determined |
| S8_extracted_k141_97202 | Not-determined |
| S8_extracted_k141_123234 | Not-determined |
| S8_extracted_k141_36589 | Not-determined |
| S8_extracted_k141_53858 | Not-determined |
| S8_extracted_k141_123240 | Not-determined |
| S8_extracted_k141_123241 | Not-determined |
| S8_extracted_k141_1799 | Not-determined |
| S8_extracted_k141_18904 | Not-determined |
| S8_extracted_k141_18905 | Not-determined |
| S8_extracted_k141_149001 | Not-determined |
| S8_extracted_k141_183382 | Not-determined |
| S8_extracted_k141_1800 | Not-determined |
| S8_extracted_k141_175062 | Not-determined |
| S8_extracted_k141_18908 | Not-determined |
| S8_extracted_k141_149006 | Low-quality |
| S8_extracted_k141_53864 | Not-determined |
| S8_extracted_k141_1802 | Not-determined |
| S8_extracted_k141_114430 | Not-determined |
| S8_extracted_k141_192795 | Not-determined |
| S8_extracted_k141_114434 | Not-determined |
| S8_extracted_k141_18911 | Not-determined |
| S8_extracted_k141_18912 | Not-determined |
| S8_extracted_k141_166588 | Not-determined |
| S8_extracted_k141_88442 | Not-determined |
| S8_extracted_k141_175070 | Not-determined |
| S8_extracted_k141_200905 | Not-determined |
| S8_extracted_k141_123246 | Not-determined |
| S8_extracted_k141_200909 | Not-determined |
| S8_extracted_k141_123247 | Not-determined |
| S8_extracted_k141_97216 | Not-determined |
| S8_extracted_k141_175074 | Not-determined |
| S8_extracted_k141_149012 | Not-determined |
| S8_extracted_k141_149013 | Not-determined |
| S8_extracted_k141_97218 | Not-determined |
| S8_extracted_k141_149016 | Not-determined |
| S8_extracted_k141_97220 | Low-quality |
| S8_extracted_k141_97221 | Not-determined |
| S8_extracted_k141_53873 | Not-determined |
| S8_extracted_k141_157716 | Not-determined |
| S8_extracted_k141_114444 | Not-determined |
| S8_extracted_k141_149022 | Not-determined |
| S8_extracted_k141_157719 | Not-determined |
| S8_extracted_k141_183402 | Not-determined |
| S8_extracted_k141_175091 | Not-determined |
| S8_extracted_k141_149026 | Not-determined |
| S8_extracted_k141_97229 | Low-quality |
| S8_extracted_k141_175093 | Not-determined |
| S8_extracted_k141_166597 | Not-determined |
| S8_extracted_k141_149030 | Not-determined |
| S8_extracted_k141_183411 | Not-determined |
| S8_extracted_k141_114453 | Not-determined |
| S8_extracted_k141_149032 | Not-determined |
| S8_extracted_k141_166602 | Not-determined |
| S8_extracted_k141_53878 | Not-determined |
| S8_extracted_k141_53879 | Not-determined |
| S8_extracted_k141_114456 | Low-quality |
| S8_extracted_k141_114457 | Not-determined |
| S8_extracted_k141_166603 | Not-determined |
| S8_extracted_k141_157732 | Not-determined |
| S8_extracted_k141_200932 | Not-determined |
| S8_extracted_k141_149041 | Not-determined |
| S8_extracted_k141_183416 | Not-determined |
| S8_extracted_k141_88459 | Not-determined |
| S8_extracted_k141_44891 | Not-determined |
| S8_extracted_k141_114462 | Low-quality |
| S8_extracted_k141_44892 | Not-determined |
| S8_extracted_k141_200938 | Not-determined |
| S8_extracted_k141_149045 | Not-determined |
| S8_extracted_k141_149049 | Not-determined |
| S8_extracted_k141_149050 | Not-determined |
| S8_extracted_k141_1821 | Not-determined |
| S8_extracted_k141_44901 | Not-determined |
| S8_extracted_k141_183424 | Not-determined |
| S8_extracted_k141_44904 | Not-determined |
| S8_extracted_k141_123269 | Not-determined |
| S8_extracted_k141_183425 | Not-determined |
| S8_extracted_k141_200945 | Not-determined |
| S8_extracted_k141_88467 | Not-determined |
| S8_extracted_k141_10203 | Low-quality |
| S8_extracted_k141_200947 | Not-determined |
| S8_extracted_k141_62370 | Not-determined |
| S8_extracted_k141_62372 | Not-determined |
| S8_extracted_k141_88470 | Not-determined |
| S8_extracted_k141_79848 | Not-determined |
| S8_extracted_k141_123272 | Not-determined |
| S8_extracted_k141_140621 | Low-quality |
| S8_extracted_k141_183433 | Not-determined |
| S8_extracted_k141_88473 | Not-determined |
| S8_extracted_k141_175122 | Low-quality |
| S8_extracted_k141_114487 | Not-determined |
| S8_extracted_k141_36598 | Low-quality |
| S8_extracted_k141_192828 | Not-determined |
| S8_extracted_k141_79853 | Not-determined |
| S8_extracted_k141_1829 | Low-quality |
| S8_extracted_k141_175125 | Low-quality |
| S8_extracted_k141_114497 | Not-determined |
| S8_extracted_k141_166616 | Not-determined |
| S8_extracted_k141_175127 | Not-determined |
| S8_extracted_k141_62380 | Not-determined |
| S8_extracted_k141_44911 | Low-quality |
| S8_extracted_k141_10216 | Not-determined |
| S8_extracted_k141_79859 | Low-quality |
| S8_extracted_k141_1839 | Not-determined |
| S8_extracted_k141_79862 | Not-determined |
| S8_extracted_k141_114504 | Not-determined |
| S8_extracted_k141_149068 | Not-determined |
| S8_extracted_k141_97264 | Not-determined |
| S8_extracted_k141_149070 | Not-determined |
| S8_extracted_k141_175132 | Low-quality |
| S8_extracted_k141_18931 | Low-quality |
| S8_extracted_k141_140633 | Not-determined |
| S8_extracted_k141_183442 | Not-determined |
| S8_extracted_k141_123283 | Not-determined |
| S3_vs_k141_127411\|\|full | High-quality |
| S8_extracted_k141_97271 | Low-quality |
| S8_extracted_k141_97272 | Not-determined |
| S8_extracted_k141_18939 | Not-determined |
| S8_extracted_k141_36604 | Low-quality |
| S8_extracted_k141_183447 | Not-determined |
| S8_extracted_k141_97278 | Not-determined |
| S8_extracted_k141_97280 | Low-quality |
| S8_extracted_k141_140640 | Low-quality |
| S8_extracted_k141_105857 | Not-determined |
| S8_extracted_k141_183453 | Low-quality |
| S8_extracted_k141_70763 | Not-determined |
| S8_extracted_k141_131818 | Not-determined |
| S8_extracted_k141_131819 | Not-determined |
| S8_extracted_k141_149087 | Not-determined |
| S8_extracted_k141_149088 | Not-determined |
| S8_extracted_k141_123292 | Not-determined |
| S8_extracted_k141_36612 | Not-determined |
| S8_extracted_k141_131820 | Not-determined |
| S8_extracted_k141_97293 | Not-determined |
| S8_extracted_k141_175158 | Not-determined |
| S8_extracted_k141_105866 | Low-quality |
| S8_extracted_k141_192846 | Not-determined |
| S8_extracted_k141_53919 | Not-determined |
| S8_extracted_k141_70772 | Not-determined |
| S8_extracted_k141_1844 | Low-quality |
| S8_extracted_k141_10245 | Not-determined |
| S8_extracted_k141_79882 | Low-quality |
| S8_extracted_k141_175161 | Not-determined |
| S8_extracted_k141_88505 | Not-determined |
| S8_extracted_k141_166632 | Not-determined |
| S8_extracted_k141_36616 | Not-determined |
| S8_extracted_k141_88509 | Not-determined |
| S8_extracted_k141_166634 | Not-determined |
| S8_extracted_k141_140651 | Not-determined |
| S8_extracted_k141_18960 | Not-determined |
| S8_extracted_k141_123305 | Not-determined |
| S8_extracted_k141_36618 | Low-quality |
| S8_extracted_k141_79891 | Not-determined |
| S8_extracted_k141_123306 | Low-quality |
| S8_extracted_k141_97299 | Not-determined |
| S8_extracted_k141_140654 | Not-determined |
| S8_extracted_k141_53931 | Not-determined |
| S8_extracted_k141_10250 | Not-determined |
| S8_extracted_k141_88513 | Not-determined |
| S8_extracted_k141_97300 | Not-determined |
| S8_extracted_k141_131834 | Low-quality |
| S8_extracted_k141_18964 | Not-determined |
| S8_extracted_k141_131835 | Not-determined |
| S8_extracted_k141_131837 | Not-determined |
| S8_extracted_k141_79892 | Not-determined |
| S8_extracted_k141_70779 | Not-determined |
| S8_extracted_k141_36630 | Not-determined |
| S8_extracted_k141_97308 | Not-determined |
| S8_extracted_k141_123312 | Low-quality |
| S8_extracted_k141_36632 | Not-determined |
| S8_extracted_k141_105875 | Not-determined |
| S8_extracted_k141_88523 | Not-determined |
| S8_extracted_k141_105876 | Not-determined |
| S8_extracted_k141_10253 | Not-determined |
| S8_extracted_k141_140664 | Not-determined |
| S8_extracted_k141_123317 | Not-determined |
| S8_extracted_k141_53934 | Not-determined |
| S8_extracted_k141_70784 | Not-determined |
| S8_extracted_k141_44923 | Not-determined |
| S8_extracted_k141_62389 | Not-determined |
| S8_extracted_k141_131845 | Not-determined |
| S8_extracted_k141_140672 | Not-determined |
| S8_extracted_k141_131847 | Not-determined |
| S8_extracted_k141_183483 | Not-determined |
| S8_extracted_k141_53943 | Not-determined |
| S8_extracted_k141_131852 | Not-determined |
| S8_extracted_k141_27923 | Low-quality |
| S8_extracted_k141_183486 | Not-determined |
| S8_extracted_k141_70788 | Not-determined |
| S8_extracted_k141_166643 | Not-determined |
| S8_extracted_k141_183491 | Not-determined |
| S8_extracted_k141_123324 | Low-quality |
| S8_extracted_k141_62393 | Not-determined |
| S8_extracted_k141_88526 | Not-determined |
| S8_extracted_k141_88529 | Not-determined |
| S8_extracted_k141_70794 | Not-determined |
| S8_extracted_k141_140679 | Not-determined |
| S8_extracted_k141_200971 | Not-determined |
| S8_extracted_k141_10259 | Not-determined |
| S8_extracted_k141_88531 | Not-determined |
| S8_extracted_k141_44944 | Not-determined |
| S8_extracted_k141_62397 | Not-determined |
| S8_extracted_k141_149111 | Not-determined |
| S8_extracted_k141_79909 | Not-determined |
| S8_extracted_k141_200975 | Not-determined |
| S8_extracted_k141_114515 | Not-determined |
| S8_extracted_k141_166648 | Not-determined |
| S8_extracted_k141_53947 | Not-determined |
| S8_extracted_k141_166650 | Low-quality |
| S8_extracted_k141_149113 | Not-determined |
| S8_extracted_k141_131854 | Low-quality |
| S8_extracted_k141_62400 | Not-determined |
| S8_extracted_k141_27934 | Not-determined |
| S8_extracted_k141_62401 | Not-determined |
| S8_extracted_k141_79915 | Not-determined |
| S8_extracted_k141_79916 | Not-determined |
| S8_extracted_k141_62403 | Not-determined |
| S8_extracted_k141_114516 | Not-determined |
| S8_extracted_k141_53950 | Not-determined |
| S8_extracted_k141_18979 | Not-determined |
| S8_extracted_k141_27939 | Not-determined |
| S8_extracted_k141_123340 | Not-determined |
| S8_extracted_k141_131864 | Not-determined |
| S8_extracted_k141_79919 | Not-determined |
| S8_extracted_k141_27941 | Low-quality |
| S8_extracted_k141_27944 | Not-determined |
| S8_extracted_k141_149123 | Not-determined |
| S8_extracted_k141_192876 | Low-quality |
| S8_extracted_k141_149125 | Low-quality |
| S8_extracted_k141_10277 | Low-quality |
| S8_extracted_k141_79924 | Not-determined |
| S8_extracted_k141_131872 | Not-determined |
| S8_extracted_k141_192880 | Not-determined |
| S8_extracted_k141_44948 | Not-determined |
| S8_extracted_k141_123344 | Not-determined |
| S8_extracted_k141_131874 | Not-determined |
| S8_extracted_k141_44949 | Not-determined |
| S8_extracted_k141_1857 | Low-quality |
| S8_extracted_k141_140693 | Not-determined |
| S8_extracted_k141_166666 | Not-determined |
| S8_extracted_k141_36681 | Not-determined |
| S8_extracted_k141_140695 | Not-determined |
| S8_extracted_k141_18982 | Not-determined |
| S8_extracted_k141_27949 | Low-quality |
| S8_extracted_k141_53961 | Not-determined |
| S8_extracted_k141_131877 | Not-determined |
| S8_extracted_k141_70807 | Not-determined |
| S8_extracted_k141_123347 | Low-quality |
| S8_extracted_k141_192886 | Not-determined |
| S8_extracted_k141_88547 | Not-determined |
| S8_extracted_k141_149128 | Low-quality |
| S8_extracted_k141_70810 | Not-determined |
| S8_extracted_k141_79929 | Not-determined |
| S8_extracted_k141_10280 | Not-determined |
| S8_extracted_k141_70812 | Not-determined |
| S8_extracted_k141_44956 | Not-determined |
| S8_extracted_k141_88550 | Not-determined |
| S8_extracted_k141_79933 | Not-determined |
| S8_extracted_k141_131885 | Not-determined |
| S8_extracted_k141_140697 | Not-determined |
| S8_extracted_k141_123351 | Not-determined |
| S8_extracted_k141_10283 | Not-determined |
| S8_extracted_k141_88551 | Low-quality |
| S8_extracted_k141_79934 | Not-determined |
| S8_extracted_k141_36690 | Not-determined |
| S8_extracted_k141_27955 | Not-determined |
| S8_extracted_k141_123353 | Not-determined |
| S8_extracted_k141_53969 | Not-determined |
| S8_extracted_k141_10284 | Not-determined |
| S8_extracted_k141_53971 | Not-determined |
| S8_extracted_k141_105900 | Not-determined |
| S8_extracted_k141_140704 | Not-determined |
| S8_extracted_k141_140705 | Not-determined |
| S8_extracted_k141_27961 | Not-determined |
| S8_extracted_k141_183502 | Not-determined |
| S8_extracted_k141_27963 | Not-determined |
| S8_extracted_k141_200996 | Not-determined |
| S8_extracted_k141_27967 | Low-quality |
| S8_extracted_k141_10295 | Not-determined |
| S8_extracted_k141_1876 | Not-determined |
| S8_extracted_k141_157749 | Not-determined |
| S8_extracted_k141_10299 | Not-determined |
| S8_extracted_k141_149142 | Not-determined |
| S8_extracted_k141_62421 | Not-determined |
| S8_extracted_k141_149144 | Not-determined |
| S8_extracted_k141_53982 | Not-determined |
| S8_extracted_k141_183507 | Not-determined |
| S8_extracted_k141_53984 | Not-determined |
| S8_extracted_k141_105910 | Not-determined |
| S8_extracted_k141_62425 | Not-determined |
| S8_extracted_k141_62426 | Not-determined |
| S8_extracted_k141_105912 | Low-quality |
| S8_extracted_k141_88565 | Not-determined |
| S8_extracted_k141_131898 | Not-determined |
| S8_extracted_k141_88567 | Not-determined |
| S8_extracted_k141_166673 | Not-determined |
| S8_extracted_k141_183517 | Not-determined |
| S8_extracted_k141_183519 | Not-determined |
| S8_extracted_k141_114537 | Low-quality |
| S8_extracted_k141_157764 | Not-determined |
| S8_extracted_k141_157765 | Not-determined |
| S8_extracted_k141_201009 | Not-determined |
| S8_extracted_k141_36702 | Low-quality |
| S8_extracted_k141_183526 | Not-determined |
| S8_extracted_k141_149155 | Not-determined |
| S8_extracted_k141_131906 | Not-determined |
| S8_extracted_k141_62434 | Not-determined |
| S8_extracted_k141_157773 | Not-determined |
| S8_extracted_k141_192891 | Not-determined |
| S8_extracted_k141_114544 | Not-determined |
| S8_extracted_k141_183533 | Not-determined |
| S8_extracted_k141_105927 | Not-determined |
| S8_extracted_k141_79958 | Not-determined |
| S8_extracted_k141_44991 | Not-determined |
| S8_extracted_k141_97318 | Not-determined |
| S8_extracted_k141_1897 | Not-determined |
| S8_extracted_k141_44994 | Not-determined |
| S8_extracted_k141_1899 | Not-determined |
| S8_extracted_k141_97319 | Low-quality |
| S8_extracted_k141_201021 | Not-determined |
| S8_extracted_k141_97323 | Not-determined |
| S8_extracted_k141_36708 | Not-determined |
| S8_extracted_k141_157784 | Not-determined |
| S8_extracted_k141_201026 | Not-determined |
| S8_extracted_k141_54006 | Not-determined |
| S8_extracted_k141_183537 | Low-quality |
| S8_extracted_k141_79967 | Not-determined |
| S8_extracted_k141_54008 | Not-determined |
| S8_extracted_k141_149167 | Not-determined |
| S8_extracted_k141_62447 | Not-determined |
| S8_extracted_k141_201028 | Low-quality |
| S8_extracted_k141_97328 | Not-determined |
| S8_extracted_k141_36713 | Not-determined |
| S8_extracted_k141_10324 | Not-determined |
| S8_extracted_k141_36716 | Not-determined |
| S8_extracted_k141_44998 | Not-determined |
| S8_extracted_k141_201034 | Not-determined |
| S8_extracted_k141_36717 | Not-determined |
| S8_extracted_k141_10327 | Not-determined |
| S8_extracted_k141_70819 | Not-determined |
| S8_extracted_k141_10328 | Low-quality |
| S8_extracted_k141_36720 | Not-determined |
| S8_extracted_k141_10331 | Not-determined |
| S8_extracted_k141_45005 | Not-determined |
| S8_extracted_k141_131924 | Not-determined |
| S8_extracted_k141_36723 | Not-determined |
| S8_extracted_k141_54020 | Not-determined |
| S8_extracted_k141_201040 | Not-determined |
| S8_extracted_k141_54022 | Not-determined |
| S8_extracted_k141_79977 | Not-determined |
| S8_extracted_k141_70822 | Not-determined |
| S8_extracted_k141_54025 | Not-determined |
| S8_extracted_k141_123366 | Not-determined |
| S8_extracted_k141_70825 | Low-quality |
| S8_extracted_k141_54028 | Not-determined |
| S8_extracted_k141_183544 | Not-determined |
| S8_extracted_k141_131933 | Low-quality |
| S8_extracted_k141_175174 | Not-determined |
| S8_extracted_k141_36725 | Not-determined |
| S8_extracted_k141_105944 | Not-determined |
| S8_extracted_k141_10342 | Not-determined |
| S8_extracted_k141_79984 | Low-quality |
| S8_extracted_k141_201041 | Not-determined |
| S8_extracted_k141_36729 | Not-determined |
| S8_extracted_k141_149183 | Low-quality |
| S8_extracted_k141_105949 | Not-determined |
| S8_extracted_k141_70832 | Low-quality |
| S8_extracted_k141_157799 | Low-quality |
| S8_extracted_k141_79987 | Not-determined |
| S8_extracted_k141_131936 | Low-quality |
| S8_extracted_k141_79988 | Not-determined |
| S8_extracted_k141_54035 | Not-determined |
| S8_extracted_k141_123377 | Not-determined |
| S8_extracted_k141_70836 | Not-determined |
| S8_extracted_k141_54036 | Not-determined |
| S8_extracted_k141_175184 | Not-determined |
| S8_extracted_k141_123381 | Not-determined |
| S8_extracted_k141_45021 | Not-determined |
| S8_extracted_k141_88575 | Not-determined |
| S8_extracted_k141_131940 | Not-determined |
| S8_extracted_k141_149193 | Not-determined |
| S8_extracted_k141_70837 | Not-determined |
| S8_extracted_k141_70838 | Not-determined |
| S8_extracted_k141_70841 | Low-quality |
| S8_extracted_k141_79991 | Not-determined |
| S8_extracted_k141_36742 | Low-quality |
| S8_extracted_k141_131949 | Not-determined |
| S8_extracted_k141_45025 | Low-quality |
| S8_extracted_k141_45026 | Not-determined |
| S8_extracted_k141_10354 | Not-determined |
| S8_extracted_k141_28026 | Not-determined |
| S8_extracted_k141_123383 | Not-determined |
| S8_extracted_k141_192905 | Not-determined |
| S8_extracted_k141_62470 | Low-quality |
| S8_extracted_k141_54042 | Not-determined |
| S8_extracted_k141_79996 | Not-determined |
| S8_extracted_k141_54043 | Not-determined |
| S8_extracted_k141_28033 | Not-determined |
| S8_extracted_k141_36751 | Not-determined |
| S8_extracted_k141_123387 | Not-determined |
| S8_extracted_k141_192909 | Low-quality |
| S8_extracted_k141_28036 | Not-determined |
| S8_extracted_k141_88589 | Not-determined |
| S8_extracted_k141_45027 | Not-determined |
| S8_extracted_k141_70847 | Low-quality |
| S8_extracted_k141_149203 | Not-determined |
| S8_extracted_k141_192914 | Not-determined |
| S8_extracted_k141_183565 | Not-determined |
| S8_extracted_k141_149205 | Not-determined |
| S8_extracted_k141_131958 | Not-determined |

| | |
|---|---|
| SB_extracted_k141_157808 | Not-determined |
| SB_extracted_k141_70851 | Not-determined |
| SB_extracted_k141_149210 | Not-determined |
| SB_extracted_k141_10363 | Not-determined |
| SB_extracted_k141_201047 | Not-determined |
| SB_extracted_k141_88601 | Not-determined |
| SB_extracted_k141_183571 | Not-determined |
| SB_extracted_k141_54044 | Not-determined |
| SB_extracted_k141_10365 | Not-determined |
| SB_extracted_k141_28045 | Not-determined |
| SB_extracted_k141_175200 | Not-determined |
| SB_extracted_k141_54046 | Low-quality |
| SB_extracted_k141_80007 | Not-determined |
| SB_extracted_k141_36760 | Not-determined |
| SB_extracted_k141_88605 | Not-determined |
| SB_extracted_k141_88606 | Low-quality |
| SB_extracted_k141_28048 | Not-determined |
| SB_extracted_k141_36762 | Not-determined |
| SB_extracted_k141_175204 | Low-quality |
| SB_extracted_k141_19050 | Not-determined |
| SB_extracted_k141_183574 | Not-determined |
| SB_extracted_k141_10369 | Not-determined |
| SB_extracted_k141_175207 | Not-determined |
| SB_extracted_k141_80011 | Not-determined |
| SB_extracted_k141_157817 | Low-quality |
| SB_extracted_k141_19053 | Not-determined |
| SB_extracted_k141_114566 | Not-determined |
| SB_extracted_k141_201056 | Not-determined |
| SB_extracted_k141_123396 | Not-determined |
| SB_extracted_k141_88613 | Low-quality |
| SB_extracted_k141_45032 | Not-determined |
| SB_extracted_k141_140713 | Low-quality |
| SB_extracted_k141_192932 | Not-determined |
| SB_extracted_k141_114568 | Not-determined |
| SB_extracted_k141_175211 | Not-determined |
| SB_extracted_k141_149219 | Not-determined |
| SB_extracted_k141_175215 | Not-determined |
| SB_extracted_k141_62507 | Not-determined |
| SB_extracted_k141_10376 | Not-determined |
| SB_extracted_k141_28065 | Not-determined |
| SB_extracted_k141_157825 | Not-determined |
| SB_extracted_k141_62508 | Low-quality |
| SB_extracted_k141_157826 | Not-determined |
| SB_extracted_k141_114577 | Low-quality |
| SB_extracted_k141_80022 | Not-determined |
| SB_extracted_k141_36769 | Not-determined |
| SB_extracted_k141_105963 | Not-determined |
| SB_extracted_k141_54063 | Not-determined |
| SB_extracted_k141_80026 | Not-determined |
| SB_extracted_k141_28071 | Not-determined |
| SB_extracted_k141_123405 | Not-determined |
| SB_extracted_k141_80029 | Not-determined |
| SB_extracted_k141_54066 | Not-determined |
| SB_extracted_k141_80030 | Not-determined |
| SB_extracted_k141_54067 | Not-determined |
| SB_extracted_k141_157831 | Not-determined |
| SB_extracted_k141_183583 | Not-determined |
| SB_extracted_k141_54068 | Not-determined |
| SB_extracted_k141_80032 | Not-determined |
| SB_extracted_k141_80033 | Not-determined |
| SB_extracted_k141_19059 | Not-determined |
| SB_extracted_k141_123408 | Not-determined |
| SB_extracted_k141_149241 | Not-determined |
| SB_extracted_k141_62519 | Not-determined |
| SB_extracted_k141_19061 | Not-determined |
| SB_extracted_k141_192943 | Not-determined |
| SB_extracted_k141_10379 | Not-determined |
| SB_extracted_k141_123415 | Low-quality |
| SB_extracted_k141_114588 | Not-determined |
| SB_extracted_k141_45034 | Not-determined |
| SB_extracted_k141_19065 | Not-determined |
| SB_extracted_k141_183589 | Not-determined |
| SB_extracted_k141_140730 | Not-determined |
| SB_extracted_k141_19066 | Not-determined |
| SB_extracted_k141_105975 | Not-determined |
| SB_extracted_k141_10385 | Not-determined |
| SB_extracted_k141_105978 | Not-determined |
| SB_extracted_k141_80044 | Not-determined |
| SB_extracted_k141_36779 | Not-determined |
| SB_extracted_k141_28083 | Not-determined |
| SB_extracted_k141_105980 | Not-determined |
| SB_extracted_k141_157838 | Not-determined |
| SB_extracted_k141_80046 | Not-determined |
| SB_extracted_k141_192953 | Not-determined |
| SB_extracted_k141_123425 | Not-determined |
| SB_extracted_k141_149261 | Not-determined |
| SB_extracted_k141_80052 | Not-determined |
| SB_extracted_k141_28085 | Not-determined |
| SB_extracted_k141_10400 | Low-quality |
| SB_extracted_k141_201065 | Not-determined |
| SB_extracted_k141_183599 | Not-determined |
| SB_extracted_k141_183600 | Not-determined |
| SB_extracted_k141_149268 | Not-determined |
| SB_extracted_k141_105993 | Not-determined |
| SB_extracted_k141_105994 | Low-quality |
| SB_extracted_k141_45047 | Not-determined |
| SB_extracted_k141_10407 | Not-determined |
| SB_extracted_k141_149271 | Not-determined |
| SB_extracted_k141_45048 | Not-determined |
| SB_extracted_k141_140733 | Not-determined |
| SB_extracted_k141_28091 | Low-quality |
| SB_extracted_k141_105996 | Not-determined |
| SB_extracted_k141_45051 | Not-determined |
| SB_extracted_k141_140737 | Not-determined |
| SB_extracted_k141_149273 | Not-determined |
| SB_extracted_k141_114603 | Not-determined |
| SB_extracted_k141_105998 | Not-determined |
| SB_extracted_k141_157849 | Not-determined |
| SB_extracted_k141_36792 | Low-quality |
| SB_extracted_k141_131970 | Not-determined |
| SB_extracted_k141_201069 | Not-determined |
| SB_extracted_k141_106000 | Not-determined |
| SB_extracted_k141_62537 | Not-determined |
| SB_extracted_k141_28099 | Not-determined |
| SB_extracted_k141_19081 | Low-quality |
| SB_extracted_k141_88637 | Not-determined |
| SB_extracted_k141_19082 | Not-determined |
| SB_extracted_k141_88641 | Not-determined |
| SB_extracted_k141_106003 | Not-determined |
| SB_extracted_k141_201076 | Not-determined |
| SB_extracted_k141_54087 | Not-determined |
| SB_extracted_k141_62543 | Not-determined |
| SB_extracted_k141_62544 | Not-determined |
| SB_extracted_k141_1902 | Low-quality |
| SB_extracted_k141_28105 | Not-determined |
| SB_extracted_k141_123440 | Not-determined |
| SB_extracted_k141_28108 | Not-determined |
| SB_extracted_k141_140756 | Not-determined |
| SB_extracted_k141_28110 | Not-determined |
| SB_extracted_k141_54093 | Not-determined |
| SB_extracted_k141_131984 | Not-determined |
| SB_extracted_k141_140759 | Not-determined |
| SB_extracted_k141_140760 | Not-determined |
| SB_extracted_k141_201080 | Not-determined |
| SB_extracted_k141_140761 | Not-determined |
| SB_extracted_k141_88652 | Not-determined |
| SB_extracted_k141_140762 | Not-determined |
| SB_extracted_k141_36802 | Not-determined |
| SB_extracted_k141_19095 | Not-determined |
| SB_extracted_k141_123444 | Not-determined |
| SB_extracted_k141_106019 | Not-determined |
| SB_extracted_k141_28113 | Not-determined |
| SB_extracted_k141_36803 | Not-determined |
| SB_extracted_k141_19096 | Not-determined |
| SB_extracted_k141_1915 | Not-determined |
| SB_extracted_k141_80069 | Not-determined |
| SB_extracted_k141_140767 | Not-determined |
| SB_extracted_k141_1916 | Not-determined |
| SB_extracted_k141_123445 | Not-determined |
| SB_extracted_k141_88655 | Not-determined |
| SB_extracted_k141_62550 | Not-determined |
| SB_extracted_k141_140769 | Not-determined |
| SB_extracted_k141_88656 | Low-quality |
| SB_extracted_k141_201086 | Not-determined |
| SB_extracted_k141_157865 | Not-determined |
| SB_extracted_k141_36811 | Not-determined |
| SB_extracted_k141_123448 | Not-determined |
| SG_extracted_k141_45228 | Medium-quality |
| SB_extracted_k141_183617 | Not-determined |
| SB_extracted_k141_149296 | Not-determined |
| SB_extracted_k141_166678 | Not-determined |
| SB_extracted_k141_80076 | Not-determined |
| SB_extracted_k141_149297 | Not-determined |
| SB_extracted_k141_114631 | Not-determined |
| SB_extracted_k141_157867 | Not-determined |
| SB_extracted_k141_183621 | Low-quality |
| SB_extracted_k141_140775 | Not-determined |
| SB_extracted_k141_114636 | Not-determined |
| SB_extracted_k141_166686 | Not-determined |
| SB_extracted_k141_140776 | Not-determined |
| SB_extracted_k141_157872 | Not-determined |
| SB_extracted_k141_157873 | Not-determined |
| SB_extracted_k141_140778 | Not-determined |
| SB_extracted_k141_54101 | Not-determined |
| SB_extracted_k141_201091 | Not-determined |
| SB_extracted_k141_88663 | Not-determined |
| SB_extracted_k141_36822 | Not-determined |
| SB_extracted_k141_54103 | Not-determined |
| SB_extracted_k141_157875 | Not-determined |
| SB_extracted_k141_132004 | Not-determined |
| SB_extracted_k141_149303 | Not-determined |
| SB_extracted_k141_80083 | Not-determined |
| SB_extracted_k141_123463 | Not-determined |
| SB_extracted_k141_54104 | Low-quality |
| SB_extracted_k141_28131 | Not-determined |
| SB_extracted_k141_175237 | Not-determined |
| SB_extracted_k141_201096 | Not-determined |
| SB_extracted_k141_19115 | Not-determined |
| SB_extracted_k141_114647 | Not-determined |
| SB_extracted_k141_175239 | Not-determined |
| SB_extracted_k141_183625 | Low-quality |
| SB_extracted_k141_10428 | Not-determined |
| SB_extracted_k141_140786 | Not-determined |
| SB_extracted_k141_175241 | Not-determined |
| SB_extracted_k141_19121 | Not-determined |
| SB_extracted_k141_19122 | Not-determined |
| SB_extracted_k141_54110 | Not-determined |
| SB_extracted_k141_114652 | Not-determined |
| SB_extracted_k141_54111 | Not-determined |
| SB_extracted_k141_88673 | Not-determined |
| SB_extracted_k141_149308 | Not-determined |
| SB_extracted_k141_166702 | Not-determined |
| SB_extracted_k141_175243 | Low-quality |
| SB_extracted_k141_166703 | Not-determined |
| SB_extracted_k141_201099 | Not-determined |
| SB_extracted_k141_149309 | Not-determined |
| SB_extracted_k141_166705 | Not-determined |
| SB_extracted_k141_157887 | Not-determined |
| SB_extracted_k141_149310 | Not-determined |
| SB_extracted_k141_19131 | Not-determined |
| SB_extracted_k141_45066 | Low-quality |
| SB_extracted_k141_192981 | Not-determined |
| SB_extracted_k141_132012 | Low-quality |
| SB_extracted_k141_88680 | Not-determined |
| SB_extracted_k141_19133 | Low-quality |
| SB_extracted_k141_45071 | Not-determined |
| SB_extracted_k141_28138 | Not-determined |
| SB_extracted_k141_183630 | Not-determined |
| SB_extracted_k141_36830 | Not-determined |
| SB_extracted_k141_149317 | Not-determined |
| SB_extracted_k141_88684 | Low-quality |
| SB_extracted_k141_36833 | Not-determined |
| SB_extracted_k141_192989 | Not-determined |
| SB_extracted_k141_157898 | Not-determined |
| SB_extracted_k141_149318 | Low-quality |
| SB_extracted_k141_88686 | Low-quality |
| SB_extracted_k141_183632 | Not-determined |
| SB_extracted_k141_28147 | Not-determined |
| SB_extracted_k141_10446 | Not-determined |
| SB_extracted_k141_28149 | Not-determined |
| SB_extracted_k141_1944 | Not-determined |
| SB_extracted_k141_166722 | Not-determined |
| SB_extracted_k141_166723 | Not-determined |
| SB_extracted_k141_106023 | Not-determined |
| SB_extracted_k141_45082 | Not-determined |
| SB_extracted_k141_1947 | Not-determined |
| SB_extracted_k141_166726 | Not-determined |
| SB_extracted_k141_1950 | Not-determined |
| SB_extracted_k141_193002 | Not-determined |
| SB_extracted_k141_193003 | Low-quality |
| SB_extracted_k141_1952 | Not-determined |
| SB_extracted_k141_193004 | Not-determined |
| SB_extracted_k141_80107 | Not-determined |
| SB_extracted_k141_1955 | Not-determined |
| SB_extracted_k141_36842 | Not-determined |
| SB_extracted_k141_157908 | Not-determined |
| SB_extracted_k141_201108 | Low-quality |
| SB_extracted_k141_97362 | Not-determined |
| SB_extracted_k141_88696 | Not-determined |
| SB_extracted_k141_62579 | Not-determined |
| SB_extracted_k141_183649 | Not-determined |
| SB_extracted_k141_1960 | Not-determined |
| SB_extracted_k141_106031 | Not-determined |
| SB_extracted_k141_132019 | Not-determined |
| SB_extracted_k141_97365 | Not-determined |
| SB_extracted_k141_132020 | Not-determined |
| SB_extracted_k141_97366 | Not-determined |
| SB_extracted_k141_183654 | Not-determined |
| SB_extracted_k141_19166 | Not-determined |
| SB_extracted_k141_19170 | Not-determined |
| SB_extracted_k141_183663 | Low-quality |
| SB_extracted_k141_193020 | Not-determined |
| SB_extracted_k141_132028 | Not-determined |
| SB_extracted_k141_114672 | Not-determined |
| SB_extracted_k141_45106 | Not-determined |
| SB_extracted_k141_114673 | Not-determined |
| SB_extracted_k141_62586 | Not-determined |
| SB_extracted_k141_114674 | Not-determined |
| SB_extracted_k141_80126 | Not-determined |
| SB_extracted_k141_201120 | Low-quality |
| SB_extracted_k141_36866 | Not-determined |
| SB_extracted_k141_183669 | Not-determined |
| SB_extracted_k141_106039 | Not-determined |
| SB_extracted_k141_157931 | Not-determined |
| SB_extracted_k141_19178 | Not-determined |
| SB_extracted_k141_140811 | Not-determined |
| SB_extracted_k141_1974 | Not-determined |
| SB_extracted_k141_183674 | Not-determined |
| SB_extracted_k141_106042 | Not-determined |
| SB_extracted_k141_19182 | Not-determined |
| SB_extracted_k141_193021 | Not-determined |
| SB_extracted_k141_36873 | Not-determined |
| SB_extracted_k141_201129 | Not-determined |
| SB_extracted_k141_54139 | Not-determined |
| SB_extracted_k141_19186 | Low-quality |
| SB_extracted_k141_10452 | Low-quality |
| SB_extracted_k141_132036 | Not-determined |
| SB_extracted_k141_54141 | Not-determined |
| SB_extracted_k141_88715 | Not-determined |
| SB_extracted_k141_193028 | Not-determined |
| SB_extracted_k141_132038 | Low-quality |
| SB_extracted_k141_1979 | Not-determined |
| SB_extracted_k141_80129 | Not-determined |
| SB_extracted_k141_175253 | Not-determined |
| SB_extracted_k141_140817 | Not-determined |
| SB_extracted_k141_157941 | Low-quality |
| SB_extracted_k141_62598 | Not-determined |
| SB_extracted_k141_45113 | Not-determined |
| SB_extracted_k141_88722 | Not-determined |
| SB_extracted_k141_36877 | Not-determined |
| SB_extracted_k141_201134 | Low-quality |
| SB_extracted_k141_166748 | Not-determined |
| SB_extracted_k141_175261 | Not-determined |
| SB_extracted_k141_28176 | Low-quality |
| SB_extracted_k141_88728 | Not-determined |
| SB_extracted_k141_80142 | Not-determined |
| SB_extracted_k141_123496 | Not-determined |
| SB_extracted_k141_106055 | Not-determined |
| SB_extracted_k141_10460 | Not-determined |
| SB_extracted_k141_54153 | Not-determined |
| SB_extracted_k141_166752 | Not-determined |
| SB_extracted_k141_123498 | Not-determined |
| SB_extracted_k141_193041 | Not-determined |
| SB_extracted_k141_132046 | Not-determined |
| SB_extracted_k141_123500 | Not-determined |
| SB_extracted_k141_175263 | Not-determined |
| SB_extracted_k141_114690 | Not-determined |
| SB_extracted_k141_193042 | Not-determined |
| SB_extracted_k141_114692 | Not-determined |
| SB_extracted_k141_123506 | Not-determined |
| SB_extracted_k141_106060 | Not-determined |
| SB_extracted_k141_157948 | Not-determined |
| SB_extracted_k141_54157 | Not-determined |
| SB_extracted_k141_28182 | Not-determined |
| SB_extracted_k141_149348 | Low-quality |
| SB_extracted_k141_36885 | Not-determined |
| SB_extracted_k141_106064 | Not-determined |
| SB_extracted_k141_36888 | Not-determined |
| SB_extracted_k141_123509 | Not-determined |
| SB_extracted_k141_62613 | Not-determined |
| SB_extracted_k141_123511 | Low-quality |
| SB_extracted_k141_70868 | Low-quality |
| SB_extracted_k141_54161 | Not-determined |
| SB_extracted_k141_114695 | Not-determined |
| SB_extracted_k141_88739 | Not-determined |
| SB_extracted_k141_1995 | Not-determined |
| SB_extracted_k141_88740 | Not-determined |
| SB_extracted_k141_62615 | Not-determined |
| SB_extracted_k141_166764 | Low-quality |
| SB_extracted_k141_132059 | Not-determined |
| SB_extracted_k141_175279 | Not-determined |
| SB_extracted_k141_132060 | Not-determined |
| SB_extracted_k141_45127 | Not-determined |
| SB_extracted_k141_157950 | Not-determined |
| SB_extracted_k141_157951 | Not-determined |
| SB_extracted_k141_132065 | Low-quality |
| SB_extracted_k141_166767 | Not-determined |
| SB_extracted_k141_175283 | Not-determined |
| SB_extracted_k141_149354 | Not-determined |
| SB_extracted_k141_123520 | Not-determined |
| SB_extracted_k141_2002 | Not-determined |
| SB_extracted_k141_106068 | Not-determined |
| SB_extracted_k141_36900 | Not-determined |
| SB_extracted_k141_2003 | Not-determined |
| SB_extracted_k141_97401 | Not-determined |
| SB_extracted_k141_97403 | Not-determined |
| SB_extracted_k141_201162 | Not-determined |
| SB_extracted_k141_123522 | Not-determined |
| SB_extracted_k141_123523 | Not-determined |
| SB_extracted_k141_201163 | Low-quality |
| SB_extracted_k141_175289 | Not-determined |
| SB_extracted_k141_193047 | Low-quality |
| SB_extracted_k141_183692 | Not-determined |
| SB_extracted_k141_45131 | Not-determined |
| SB_extracted_k141_157958 | Not-determined |
| SB_extracted_k141_123532 | Low-quality |
| SB_extracted_k141_132074 | Not-determined |
| SB_extracted_k141_157960 | Not-determined |
| SB_extracted_k141_80163 | Not-determined |
| SB_extracted_k141_193055 | Not-determined |
| SB_extracted_k141_88757 | Low-quality |
| SB_extracted_k141_28203 | Not-determined |
| SB_extracted_k141_70877 | Not-determined |
| SB_extracted_k141_183701 | Not-determined |
| SB_extracted_k141_36918 | Not-determined |
| SB_extracted_k141_106089 | Not-determined |
| SB_extracted_k141_2010 | Not-determined |
| SB_extracted_k141_183706 | Not-determined |
| SB_extracted_k141_36924 | Not-determined |
| SB_extracted_k141_45138 | Low-quality |
| SB_extracted_k141_2016 | Not-determined |
| SB_extracted_k141_80172 | Not-determined |
| SB_extracted_k141_36929 | Not-determined |
| SB_extracted_k141_62646 | Low-quality |
| SB_extracted_k141_114713 | Not-determined |
| SB_extracted_k141_62647 | Low-quality |
| SB_extracted_k141_132078 | Low-quality |
| SB_extracted_k141_106094 | Not-determined |
| SB_extracted_k141_114716 | Not-determined |
| SB_extracted_k141_54182 | Not-determined |
| SB_extracted_k141_157977 | Not-determined |
| SB_extracted_k141_201179 | Not-determined |
| SB_extracted_k141_106099 | Low-quality |
| SB_extracted_k141_36938 | Not-determined |
| SB_extracted_k141_106101 | Not-determined |
| SB_extracted_k141_97434 | Not-determined |
| SB_extracted_k141_106102 | Low-quality |
| SB_extracted_k141_70901 | Not-determined |
| SB_extracted_k141_70902 | Not-determined |
| SB_extracted_k141_132091 | Not-determined |
| SB_extracted_k141_149376 | Not-determined |
| SB_extracted_k141_166778 | Not-determined |
| SB_extracted_k141_175306 | Not-determined |
| SB_extracted_k141_183718 | Not-determined |
| SB_extracted_k141_62660 | Not-determined |
| SB_extracted_k141_201185 | Not-determined |
| SB_extracted_k141_201186 | Not-determined |
| SB_extracted_k141_123555 | Not-determined |
| SB_extracted_k141_62661 | Not-determined |
| SB_extracted_k141_36939 | Not-determined |
| SB_extracted_k141_149377 | Low-quality |
| SB_extracted_k141_123556 | Not-determined |
| SB_extracted_k141_175309 | Not-determined |
| SB_extracted_k141_80179 | Not-determined |
| SB_extracted_k141_36940 | Low-quality |
| SB_extracted_k141_157988 | Not-determined |
| SB_extracted_k141_175313 | Not-determined |
| SB_extracted_k141_97447 | Not-determined |
| SB_extracted_k141_166782 | Not-determined |
| SB_extracted_k141_166784 | Low-quality |
| SB_extracted_k141_132104 | Not-determined |
| SB_extracted_k141_62672 | Not-determined |
| SB_extracted_k141_166787 | Not-determined |
| SB_extracted_k141_175326 | Not-determined |
| SB_extracted_k141_62676 | Not-determined |
| SB_extracted_k141_201191 | Not-determined |
| SB_extracted_k141_157997 | Not-determined |
| SB_extracted_k141_70919 | Not-determined |
| SB_extracted_k141_45157 | Not-determined |
| SB_extracted_k141_149394 | Low-quality |
| SB_extracted_k141_106121 | Not-determined |
| SB_extracted_k141_114727 | Not-determined |
| SB_extracted_k141_175331 | Not-determined |
| SB_extracted_k141_201194 | Not-determined |
| SB_extracted_k141_166790 | Not-determined |
| SB_extracted_k141_106124 | Low-quality |
| SB_extracted_k141_70924 | Low-quality |
| SB_extracted_k141_70925 | Not-determined |
| SB_extracted_k141_114731 | Not-determined |
| SB_extracted_k141_80194 | Not-determined |
| SB_extracted_k141_114733 | Not-determined |
| SB_extracted_k141_183724 | Low-quality |
| SB_extracted_k141_45164 | Not-determined |
| SB_extracted_k141_70930 | Not-determined |
| SB_extracted_k141_158006 | Low-quality |
| SB_extracted_k141_2037 | Low-quality |
| SB_extracted_k141_123569 | Not-determined |
| SB_extracted_k141_183733 | Not-determined |
| SB_extracted_k141_183734 | Not-determined |
| SB_extracted_k141_149403 | Low-quality |
| SB_extracted_k141_183736 | Not-determined |
| SB_extracted_k141_175337 | Low-quality |
| SB_extracted_k141_166802 | Not-determined |
| SB_extracted_k141_166803 | Not-determined |
| SB_extracted_k141_88793 | Not-determined |
| SB_extracted_k141_123570 | Low-quality |
| SB_extracted_k141_201209 | Not-determined |
| SB_extracted_k141_158015 | Not-determined |
| SB_extracted_k141_2048 | Not-determined |
| SB_extracted_k141_62691 | Not-determined |
| SB_extracted_k141_158016 | Not-determined |
| SB_extracted_k141_88799 | Not-determined |
| SB_extracted_k141_166808 | Not-determined |
| SB_extracted_k141_193059 | Not-determined |
| SB_extracted_k141_193061 | Not-determined |
| SB_extracted_k141_70942 | Low-quality |
| SB_extracted_k141_201212 | Not-determined |
| SB_extracted_k141_193065 | Not-determined |
| SB_extracted_k141_140860 | Not-determined |
| SB_extracted_k141_70947 | Not-determined |
| SB_extracted_k141_70948 | Not-determined |
| SB_extracted_k141_106151 | Not-determined |
| SB_extracted_k141_201214 | Not-determined |
| SB_extracted_k141_45170 | Low-quality |
| SB_extracted_k141_2055 | Not-determined |
| SB_extracted_k141_70953 | Low-quality |
| SB_extracted_k141_140868 | Not-determined |
| SB_extracted_k141_140869 | Not-determined |
| SB_extracted_k141_149410 | Not-determined |
| SB_extracted_k141_114752 | Not-determined |
| SB_extracted_k141_114754 | Not-determined |
| SB_extracted_k141_88814 | Not-determined |
| SB_extracted_k141_80204 | Low-quality |
| SB_extracted_k141_62705 | Low-quality |
| SB_extracted_k141_158032 | Not-determined |
| SB_extracted_k141_10494 | Not-determined |
| SB_extracted_k141_19199 | Not-determined |
| SB_extracted_k141_62707 | Not-determined |
| SB_extracted_k141_166822 | Not-determined |
| SB_extracted_k141_45187 | Not-determined |
| SB_extracted_k141_19202 | Not-determined |
| SB_extracted_k141_201225 | Not-determined |
| SB_extracted_k141_114763 | Not-determined |
| SB_extracted_k141_158038 | Low-quality |
| SB_extracted_k141_123587 | Not-determined |
| SB_extracted_k141_2065 | Not-determined |
| SB_extracted_k141_88815 | Low-quality |
| SB_extracted_k141_2066 | Not-determined |
| SB_extracted_k141_10500 | Low-quality |
| SB_extracted_k141_123590 | Not-determined |
| SB_extracted_k141_80211 | Low-quality |
| SB_extracted_k141_175355 | Not-determined |
| SB_extracted_k141_80212 | Not-determined |
| SB_extracted_k141_114764 | Not-determined |
| SB_extracted_k141_10501 | Not-determined |
| SB_extracted_k141_80213 | Not-determined |
| SB_extracted_k141_193090 | Not-determined |
| SB_extracted_k141_2070 | Not-determined |
| SB_extracted_k141_88818 | Not-determined |
| SB_extracted_k141_149418 | Not-determined |
| SB_extracted_k141_62715 | Not-determined |
| SB_extracted_k141_54186 | Not-determined |
| SB_extracted_k141_2072 | Not-determined |
| SB_extracted_k141_166827 | Not-determined |
| SB_extracted_k141_88824 | Low-quality |
| SB_extracted_k141_19210 | Not-determined |
| SB_extracted_k141_158044 | Not-determined |
| SB_extracted_k141_158045 | Low-quality |
| SB_extracted_k141_140884 | Not-determined |
| SB_extracted_k141_140885 | Not-determined |
| SB_extracted_k141_166830 | Not-determined |
| SB_extracted_k141_2074 | Not-determined |
| SB_extracted_k141_2075 | Not-determined |
| SB_extracted_k141_19217 | Not-determined |
| SB_extracted_k141_19218 | Not-determined |
| SB_extracted_k141_28240 | Not-determined |
| SB_extracted_k141_175364 | Not-determined |
| SB_extracted_k141_114777 | Not-determined |
| SB_extracted_k141_28245 | Not-determined |
| SB_extracted_k141_106165 | Low-quality |
| SB_extracted_k141_149421 | Not-determined |
| SB_extracted_k141_114781 | Not-determined |
| SB_extracted_k141_149425 | Not-determined |
| SB_extracted_k141_175372 | Not-determined |
| SB_extracted_k141_158063 | Not-determined |
| SB_extracted_k141_19224 | Low-quality |
| SB_extracted_k141_45197 | Not-determined |
| SB_extracted_k141_123606 | Not-determined |
| SB_extracted_k141_183754 | Not-determined |
| SB_extracted_k141_140900 | Not-determined |
| SB_extracted_k141_70965 | Low-quality |
| SB_extracted_k141_19231 | Not-determined |
| SB_extracted_k141_132118 | Not-determined |
| SB_extracted_k141_88836 | Not-determined |
| SB_extracted_k141_175382 | Not-determined |
| SB_extracted_k141_106177 | Not-determined |
| SB_extracted_k141_193117 | Not-determined |
| SB_extracted_k141_88838 | Not-determined |
| SB_extracted_k141_201258 | Not-determined |
| SB_extracted_k141_70970 | Not-determined |
| SB_extracted_k141_123611 | Not-determined |
| SB_extracted_k141_175385 | Low-quality |
| SB_extracted_k141_201261 | Not-determined |
| SB_extracted_k141_88839 | Not-determined |
| SB_extracted_k141_158074 | Not-determined |
| SB_extracted_k141_54211 | Not-determined |
| SB_extracted_k141_114795 | Not-determined |
| SB_extracted_k141_106183 | Not-determined |
| SB_extracted_k141_2095 | Not-determined |
| SB_extracted_k141_183763 | Not-determined |
| SB_extracted_k141_70977 | Not-determined |
| SB_extracted_k141_62738 | Not-determined |
| SB_extracted_k141_106188 | Not-determined |
| SB_extracted_k141_54220 | Not-determined |
| SB_extracted_k141_2101 | Not-determined |
| SB_extracted_k141_193126 | Not-determined |
| SB_extracted_k141_2102 | Not-determined |
| SB_extracted_k141_45217 | Not-determined |
| SB_extracted_k141_132140 | Not-determined |
| SB_extracted_k141_114803 | Not-determined |
| SB_extracted_k141_201272 | Not-determined |
| SB_extracted_k141_36966 | Not-determined |
| SB_extracted_k141_45221 | Low-quality |
| SB_extracted_k141_45223 | Not-determined |
| SB_extracted_k141_193131 | Not-determined |
| SB_extracted_k141_140910 | Not-determined |
| SB_extracted_k141_62752 | Not-determined |
| SB_extracted_k141_201277 | Not-determined |
| SB_extracted_k141_175402 | Low-quality |
| SB_extracted_k141_193132 | Not-determined |
| SB_extracted_k141_54230 | Not-determined |
| SB_extracted_k141_45225 | Not-determined |
| SB_extracted_k141_62754 | Low-quality |
| SB_extracted_k141_149448 | Not-determined |
| SB_extracted_k141_123619 | Not-determined |
| SB_extracted_k141_45227 | Not-determined |
| SB_extracted_k141_149450 | Not-determined |
| SB_extracted_k141_62760 | Not-determined |
| SB_extracted_k141_149452 | Not-determined |

| | |
|---|---|
| SB_extracted_k141_54236 | Not-determined |
| SB_extracted_k141_166833 | Not-determined |
| SB_extracted_k141_175408 | Low-quality |
| SB_extracted_k141_28252 | Low-quality |
| SB_extracted_k141_88867 | Not-determined |
| SB_extracted_k141_193139 | Not-determined |
| SB_extracted_k141_62768 | Not-determined |
| SB_extracted_k141_54238 | Not-determined |
| SB_extracted_k141_88870 | Not-determined |
| SB_extracted_k141_19257 | Not-determined |
| SB_extracted_k141_193140 | Low-quality |
| SB_extracted_k141_201286 | Not-determined |
| SB_extracted_k141_88874 | Not-determined |
| SB_extracted_k141_114824 | Not-determined |
| SB_extracted_k141_166842 | Not-determined |
| SB_extracted_k141_28259 | Not-determined |
| SB_extracted_k141_71001 | Not-determined |
| SB_extracted_k141_166844 | Not-determined |
| SB_extracted_k141_193142 | Low-quality |
| SB_extracted_k141_28260 | Not-determined |
| SB_extracted_k141_2120 | Not-determined |
| SB_extracted_k141_45241 | Not-determined |
| SB_extracted_k141_183780 | Not-determined |
| SB_extracted_k141_201291 | Not-determined |
| SB_extracted_k141_54252 | Not-determined |
| SB_extracted_k141_54254 | Not-determined |
| SB_extracted_k141_2127 | Not-determined |
| SB_extracted_k141_123633 | Not-determined |
| SB_extracted_k141_45245 | Not-determined |
| SB_extracted_k141_106203 | Not-determined |
| SB_extracted_k141_183787 | Low-quality |
| SB_extracted_k141_183788 | Not-determined |
| SB_extracted_k141_28271 | Not-determined |
| SB_extracted_k141_123637 | Not-determined |
| SB_extracted_k141_183790 | Not-determined |
| SB_extracted_k141_123638 | Not-determined |
| SB_extracted_k141_175421 | Low-quality |
| SB_extracted_k141_45249 | Low-quality |
| SB_extracted_k141_45252 | Not-determined |
| SB_extracted_k141_19270 | Not-determined |
| SB_extracted_k141_183794 | Not-determined |
| SB_extracted_k141_201300 | Not-determined |
| SB_extracted_k141_140934 | Not-determined |
| SB_extracted_k141_54268 | Not-determined |
| SB_extracted_k141_106210 | Not-determined |
| SB_extracted_k141_88894 | Not-determined |
| SB_extracted_k141_175428 | Not-determined |
| SB_extracted_k141_175429 | Not-determined |
| SB_extracted_k141_193156 | Not-determined |
| SB_extracted_k141_123646 | Not-determined |
| SB_extracted_k141_71016 | Not-determined |
| SB_extracted_k141_71018 | Not-determined |
| SB_extracted_k141_201307 | Not-determined |
| SB_extracted_k141_2146 | Not-determined |
| SB_extracted_k141_114843 | Not-determined |
| SB_extracted_k141_193158 | Not-determined |
| SB_extracted_k141_193159 | Not-determined |
| SB_extracted_k141_175438 | Not-determined |
| SB_extracted_k141_193160 | Not-determined |
| SB_extracted_k141_166860 | Not-determined |
| SB_extracted_k141_71023 | Not-determined |
| SB_extracted_k141_140949 | Not-determined |
| SB_extracted_k141_114847 | Not-determined |
| SB_extracted_k141_54283 | Low-quality |
| SB_extracted_k141_140953 | Not-determined |
| SB_extracted_k141_201313 | Not-determined |
| SB_extracted_k141_114849 | Not-determined |
| SB_extracted_k141_201315 | Not-determined |
| SB_extracted_k141_45269 | Not-determined |
| SB_extracted_k141_166867 | Not-determined |
| SB_extracted_k141_193169 | Not-determined |
| SB_extracted_k141_62796 | Not-determined |
| SB_extracted_k141_88910 | Not-determined |
| SB_extracted_k141_28290 | Not-determined |
| SB_extracted_k141_28292 | Not-determined |
| SB_extracted_k141_106223 | Not-determined |
| SB_extracted_k141_62802 | Not-determined |
| SB_extracted_k141_114857 | Not-determined |
| SB_extracted_k141_28296 | Not-determined |
| SB_extracted_k141_2154 | Low-quality |
| SB_extracted_k141_19285 | Not-determined |
| SB_extracted_k141_114859 | Not-determined |
| SB_extracted_k141_97465 | Low-quality |
| SB_extracted_k141_149474 | Low-quality |
| SB_extracted_k141_149475 | Not-determined |
| SB_extracted_k141_106234 | Not-determined |
| SB_extracted_k141_175450 | Not-determined |
| SB_extracted_k141_193177 | Not-determined |
| SB_extracted_k141_97468 | Not-determined |
| SB_extracted_k141_201321 | Not-determined |
| SB_extracted_k141_88921 | Not-determined |
| SB_extracted_k141_2159 | Not-determined |
| SB_extracted_k141_114865 | Not-determined |
| SB_extracted_k141_166881 | Not-determined |
| SB_extracted_k141_193183 | Not-determined |
| SB_extracted_k141_19287 | Low-quality |
| SB_extracted_k141_114867 | Not-determined |
| SB_extracted_k141_175453 | Not-determined |
| SB_extracted_k141_54293 | Not-determined |
| SB_extracted_k141_201326 | Not-determined |
| SB_extracted_k141_71039 | Not-determined |
| SB_extracted_k141_158085 | Not-determined |
| SB_extracted_k141_201328 | Low-quality |
| SB_extracted_k141_193187 | Not-determined |
| SB_extracted_k141_123665 | Not-determined |
| SB_extracted_k141_201330 | Low-quality |
| SB_extracted_k141_166886 | Not-determined |
| SB_extracted_k141_19294 | Not-determined |
| SB_extracted_k141_71050 | Not-determined |
| SB_extracted_k141_71051 | Not-determined |
| SB_extracted_k141_88935 | Not-determined |
| SB_extracted_k141_19299 | Not-determined |
| SB_extracted_k141_183815 | Not-determined |
| SB_extracted_k141_166893 | Not-determined |
| SB_extracted_k141_166894 | Low-quality |
| SB_extracted_k141_10509 | Not-determined |
| SB_extracted_k141_106245 | Not-determined |
| SB_extracted_k141_71061 | Not-determined |
| SB_extracted_k141_114884 | Not-determined |
| SB_extracted_k141_10513 | Not-determined |
| SB_extracted_k141_193197 | Not-determined |
| SB_extracted_k141_45277 | Not-determined |
| SB_extracted_k141_193198 | Not-determined |
| SB_extracted_k141_19308 | Not-determined |
| SB_extracted_k141_114886 | Not-determined |
| SB_extracted_k141_19310 | Low-quality |
| SB_extracted_k141_140975 | Not-determined |
| SB_extracted_k141_45283 | Not-determined |
| SB_extracted_k141_71066 | Not-determined |
| SB_extracted_k141_45285 | Not-determined |
| SB_extracted_k141_71067 | Low-quality |
| SB_extracted_k141_45286 | Low-quality |
| SB_extracted_k141_114889 | Not-determined |
| SB_extracted_k141_45288 | Not-determined |
| SB_extracted_k141_36997 | Not-determined |
| SB_extracted_k141_140978 | Not-determined |
| SB_extracted_k141_166897 | Not-determined |
| SB_extracted_k141_54322 | Not-determined |
| SB_extracted_k141_114891 | Not-determined |
| SB_extracted_k141_193203 | Not-determined |
| SB_extracted_k141_10522 | Not-determined |
| SB_extracted_k141_45289 | Not-determined |
| SB_extracted_k141_201340 | Not-determined |
| SB_extracted_k141_97487 | Not-determined |
| SB_extracted_k141_62819 | Not-determined |
| SB_extracted_k141_193207 | Not-determined |
| SB_extracted_k141_193208 | Not-determined |
| SB_extracted_k141_97489 | Not-determined |
| SB_extracted_k141_10529 | Not-determined |
| SB_extracted_k141_88952 | Low-quality |
| SB_extracted_k141_37006 | Not-determined |
| SB_extracted_k141_45295 | Not-determined |
| SB_extracted_k141_140985 | Not-determined |
| SB_extracted_k141_10531 | Not-determined |
| SB_extracted_k141_106267 | Low-quality |
| SB_extracted_k141_10532 | Not-determined |
| SB_extracted_k141_158113 | Low-quality |
| SB_extracted_k141_114907 | Not-determined |
| SB_extracted_k141_54328 | Not-determined |
| SB_extracted_k141_106269 | Not-determined |
| SB_extracted_k141_132146 | Low-quality |
| SB_extracted_k141_114909 | Not-determined |
| SB_extracted_k141_132147 | Not-determined |
| SB_extracted_k141_88959 | Not-determined |
| SB_extracted_k141_114912 | Not-determined |
| SB_extracted_k141_62828 | Not-determined |
| SB_extracted_k141_140989 | Not-determined |
| SB_extracted_k141_37016 | Not-determined |
| SB_extracted_k141_2171 | Not-determined |
| SB_extracted_k141_37017 | Not-determined |
| SB_extracted_k141_201345 | Not-determined |
| SB_extracted_k141_175486 | Not-determined |
| SB_extracted_k141_123691 | Not-determined |
| SB_extracted_k141_97495 | Not-determined |
| SB_extracted_k141_132155 | Not-determined |
| SB_extracted_k141_166906 | Not-determined |
| SB_extracted_k141_71088 | Not-determined |
| SB_extracted_k141_71090 | Not-determined |
| SB_extracted_k141_97497 | Low-quality |
| SB_extracted_k141_97498 | Not-determined |
| SB_extracted_k141_45307 | Not-determined |
| SB_extracted_k141_132158 | Not-determined |
| SB_extracted_k141_71092 | Not-determined |
| SB_extracted_k141_149524 | Not-determined |
| SB_extracted_k141_45311 | Low-quality |
| SB_extracted_k141_106280 | Not-determined |
| SB_extracted_k141_183824 | Not-determined |
| SB_extracted_k141_37025 | Not-determined |
| SB_extracted_k141_2180 | Not-determined |
| SB_extracted_k141_54343 | Low-quality |
| SB_extracted_k141_141002 | Low-quality |
| SB_extracted_k141_149533 | Not-determined |
| SB_extracted_k141_62838 | Not-determined |
| SB_extracted_k141_71094 | Not-determined |
| SB_extracted_k141_97511 | Not-determined |
| SB_extracted_k141_45318 | Not-determined |
| SB_extracted_k141_97513 | Not-determined |
| SB_extracted_k141_2185 | Not-determined |
| SB_extracted_k141_54349 | Not-determined |
| SB_extracted_k141_37034 | Not-determined |
| SB_extracted_k141_88986 | Not-determined |
| SB_extracted_k141_149543 | Not-determined |
| SB_extracted_k141_132173 | Not-determined |
| SB_extracted_k141_54353 | Not-determined |
| SB_extracted_k141_97525 | Not-determined |
| SB_extracted_k141_201366 | Not-determined |
| SB_extracted_k141_149549 | Not-determined |
| SB_extracted_k141_19358 | Not-determined |
| SB_extracted_k141_45329 | Not-determined |
| SB_extracted_k141_193219 | Not-determined |
| SB_extracted_k141_2195 | Not-determined |
| SB_extracted_k141_149552 | Not-determined |
| SB_extracted_k141_201374 | Not-determined |
| SB_extracted_k141_2198 | Not-determined |
| SB_extracted_k141_62853 | Not-determined |
| SB_extracted_k141_54356 | Not-determined |
| SB_extracted_k141_19361 | Not-determined |
| SB_extracted_k141_166921 | Not-determined |
| SB_extracted_k141_114941 | Not-determined |
| SB_extracted_k141_193220 | Not-determined |
| SB_extracted_k141_54358 | Not-determined |
| SB_extracted_k141_193222 | Not-determined |
| SB_extracted_k141_193223 | Not-determined |
| SB_extracted_k141_62856 | Not-determined |
| SB_extracted_k141_132178 | Not-determined |
| SB_extracted_k141_54362 | Not-determined |
| SB_extracted_k141_37057 | Not-determined |
| SB_extracted_k141_158132 | Not-determined |
| SB_extracted_k141_183842 | Not-determined |
| SB_extracted_k141_19367 | Low-quality |
| SB_extracted_k141_123705 | Not-determined |
| SB_extracted_k141_123706 | Not-determined |
| SB_extracted_k141_97539 | Not-determined |
| SB_extracted_k141_37061 | Low-quality |
| SB_extracted_k141_62862 | Not-determined |
| SB_extracted_k141_88996 | Not-determined |
| SB_extracted_k141_54369 | Not-determined |
| SB_extracted_k141_54371 | Not-determined |
| SB_extracted_k141_106305 | Not-determined |
| SB_extracted_k141_62864 | Not-determined |
| SB_extracted_k141_123714 | Not-determined |
| SB_extracted_k141_62867 | Not-determined |
| SB_extracted_k141_37069 | Not-determined |
| SB_extracted_k141_45342 | Low-quality |
| SB_extracted_k141_166933 | Not-determined |
| SB_extracted_k141_123718 | Low-quality |
| SB_extracted_k141_132194 | Low-quality |
| SB_extracted_k141_175520 | Not-determined |
| SB_extracted_k141_166934 | Not-determined |
| SB_extracted_k141_123719 | Low-quality |
| SB_extracted_k141_183853 | Low-quality |
| SB_extracted_k141_149569 | Not-determined |
| SB_extracted_k141_80242 | Not-determined |
| SB_extracted_k141_54374 | Low-quality |
| SB_extracted_k141_54382 | Not-determined |
| SB_extracted_k141_2216 | Not-determined |
| SB_extracted_k141_45351 | Not-determined |
| SB_extracted_k141_106313 | Not-determined |
| SB_extracted_k141_2219 | Not-determined |
| SB_extracted_k141_2220 | Not-determined |
| SB_extracted_k141_123728 | Low-quality |
| SB_extracted_k141_149584 | Not-determined |
| SB_extracted_k141_28332 | Not-determined |
| SB_extracted_k141_19380 | Low-quality |
| SB_extracted_k141_149587 | Not-determined |
| SB_extracted_k141_2225 | Not-determined |
| SB_extracted_k141_10565 | Not-determined |
| SB_extracted_k141_10566 | Not-determined |
| SB_extracted_k141_97558 | Not-determined |
| SB_extracted_k141_37087 | Not-determined |
| SB_extracted_k141_62881 | Not-determined |
| SB_extracted_k141_193250 | Not-determined |
| SB_extracted_k141_97563 | Not-determined |
| SB_extracted_k141_106320 | Not-determined |
| SB_extracted_k141_123741 | Not-determined |
| SB_extracted_k141_97566 | Not-determined |
| SB_extracted_k141_62886 | Not-determined |
| SB_extracted_k141_28342 | Low-quality |
| SB_extracted_k141_183861 | Not-determined |
| SB_extracted_k141_28343 | Not-determined |
| SB_extracted_k141_106322 | Not-determined |
| SB_extracted_k141_123745 | Not-determined |
| SB_extracted_k141_201389 | Not-determined |
| SB_extracted_k141_97573 | Not-determined |
| SB_extracted_k141_28344 | Not-determined |
| SB_extracted_k141_106329 | Low-quality |
| SB_extracted_k141_54389 | Low-quality |
| SB_extracted_k141_166947 | Not-determined |
| SB_extracted_k141_54390 | Not-determined |
| SB_extracted_k141_175542 | Not-determined |
| SB_extracted_k141_71134 | Not-determined |
| SB_extracted_k141_45367 | Low-quality |
| SB_extracted_k141_97578 | Not-determined |
| SB_extracted_k141_28346 | Not-determined |
| SB_extracted_k141_123751 | Not-determined |
| SB_extracted_k141_106333 | Low-quality |
| SB_extracted_k141_183872 | Not-determined |
| SB_extracted_k141_62895 | Not-determined |
| SB_extracted_k141_132203 | Not-determined |
| SB_extracted_k141_62897 | Not-determined |
| SB_extracted_k141_71140 | Not-determined |
| SB_extracted_k141_80275 | Low-quality |
| SB_extracted_k141_97583 | Not-determined |
| SB_extracted_k141_132205 | Not-determined |
| SB_extracted_k141_71143 | Not-determined |
| SB_extracted_k141_10570 | Not-determined |
| SB_extracted_k141_2232 | Low-quality |
| SB_extracted_k141_183878 | Low-quality |
| SB_extracted_k141_10573 | Not-determined |
| SB_extracted_k141_37108 | Not-determined |
| SB_extracted_k141_183880 | Not-determined |
| SB_extracted_k141_123758 | Not-determined |
| SB_extracted_k141_89010 | Not-determined |
| SB_extracted_k141_201398 | Not-determined |
| SB_extracted_k141_80285 | Not-determined |
| SB_extracted_k141_19413 | Not-determined |
| SB_extracted_k141_132216 | Not-determined |
| SB_extracted_k141_123762 | Not-determined |
| SB_extracted_k141_201408 | Not-determined |
| SB_extracted_k141_54410 | Not-determined |
| SB_extracted_k141_97588 | Low-quality |
| SB_extracted_k141_2241 | Not-determined |
| SB_extracted_k141_28357 | Low-quality |
| SB_extracted_k141_62905 | Not-determined |
| SB_extracted_k141_149627 | Not-determined |
| SB_extracted_k141_28360 | Not-determined |
| SB_extracted_k141_54418 | Not-determined |
| SB_extracted_k141_175569 | Not-determined |
| SB_extracted_k141_28363 | Not-determined |
| SB_extracted_k141_123766 | Not-determined |
| SB_extracted_k141_201414 | Low-quality |
| SB_extracted_k141_97597 | Not-determined |
| SB_extracted_k141_132227 | Not-determined |
| SB_extracted_k141_183896 | Not-determined |
| SB_extracted_k141_54427 | Not-determined |
| SB_extracted_k141_80306 | Not-determined |
| SB_extracted_k141_71153 | Low-quality |
| SB_extracted_k141_201417 | Not-determined |
| SB_extracted_k141_54432 | Not-determined |
| SB_extracted_k141_175575 | Not-determined |
| SB_extracted_k141_28367 | Not-determined |
| SB_extracted_k141_158155 | Not-determined |
| SB_extracted_k141_158156 | Not-determined |
| SB_extracted_k141_54434 | Not-determined |
| SB_extracted_k141_166962 | Low-quality |
| SB_extracted_k141_175578 | Not-determined |
| SB_extracted_k141_106354 | Not-determined |
| SB_extracted_k141_158159 | Not-determined |
| SB_extracted_k141_158160 | Not-determined |
| SB_extracted_k141_106358 | Not-determined |
| SB_extracted_k141_19421 | Not-determined |
| SB_extracted_k141_183906 | Not-determined |
| SB_extracted_k141_28377 | Not-determined |
| SB_extracted_k141_149644 | Not-determined |
| SB_extracted_k141_71156 | Not-determined |
| SB_extracted_k141_183910 | Not-determined |
| SB_extracted_k141_2265 | Not-determined |
| SB_extracted_k141_71157 | Not-determined |
| SB_extracted_k141_183913 | Low-quality |
| SB_extracted_k141_19425 | Not-determined |
| SB_extracted_k141_193281 | Not-determined |
| SB_extracted_k141_19426 | Not-determined |
| SB_extracted_k141_166975 | Not-determined |
| SB_extracted_k141_28384 | Not-determined |
| SB_extracted_k141_141009 | Not-determined |
| SB_extracted_k141_28385 | Not-determined |
| SB_extracted_k141_183915 | Low-quality |
| SB_extracted_k141_2267 | Not-determined |
| SB_extracted_k141_28386 | Not-determined |
| SB_extracted_k141_62923 | Low-quality |
| SB_extracted_k141_28387 | Not-determined |
| SB_extracted_k141_89032 | Low-quality |
| SB_extracted_k141_141013 | Not-determined |
| SB_extracted_k141_28388 | Not-determined |
| SB_extracted_k141_175591 | Not-determined |
| SB_extracted_k141_106369 | Not-determined |
| SB_extracted_k141_71164 | Not-determined |
| SB_extracted_k141_201428 | Low-quality |
| SB_extracted_k141_80325 | Not-determined |
| SB_extracted_k141_166982 | Not-determined |
| SB_extracted_k141_2275 | Not-determined |
| SB_extracted_k141_80327 | Not-determined |
| SB_extracted_k141_193290 | Low-quality |
| SB_extracted_k141_201432 | Not-determined |
| SB_extracted_k141_71171 | Not-determined |
| SB_extracted_k141_201434 | Not-determined |
| SB_extracted_k141_123788 | Not-determined |
| SB_extracted_k141_158176 | Not-determined |
| SB_extracted_k141_106383 | Not-determined |
| SB_extracted_k141_175602 | Not-determined |
| SB_extracted_k141_89039 | Not-determined |
| SB_extracted_k141_62937 | Low-quality |
| SB_extracted_k141_132245 | Not-determined |
| SB_extracted_k141_193300 | Not-determined |
| SB_extracted_k141_166994 | Not-determined |
| SB_extracted_k141_114950 | Low-quality |
| SB_extracted_k141_62940 | Not-determined |
| SB_extracted_k141_28397 | Not-determined |
| SB_extracted_k141_132247 | Low-quality |
| SB_extracted_k141_132248 | Not-determined |
| SB_extracted_k141_166999 | Not-determined |
| SB_extracted_k141_71181 | Not-determined |
| SB_extracted_k141_54445 | Not-determined |
| SB_extracted_k141_2292 | Not-determined |
| SB_extracted_k141_141023 | Not-determined |
| SB_extracted_k141_71184 | Not-determined |
| SB_extracted_k141_149666 | Not-determined |
| SB_extracted_k141_167006 | Not-determined |
| SB_extracted_k141_71187 | Not-determined |
| SB_extracted_k141_149667 | Not-determined |
| SB_extracted_k141_114964 | Low-quality |
| SB_extracted_k141_167011 | Not-determined |
| SB_extracted_k141_28408 | Not-determined |
| SB_extracted_k141_19436 | Not-determined |
| SB_extracted_k141_54447 | Not-determined |
| SB_extracted_k141_158193 | Not-determined |
| SB_extracted_k141_193317 | Not-determined |
| SB_extracted_k141_10597 | Not-determined |
| SB_extracted_k141_97609 | Low-quality |
| SB_extracted_k141_28411 | Not-determined |
| SB_extracted_k141_158198 | Not-determined |
| SB_extracted_k141_158199 | Not-determined |
| SB_extracted_k141_158200 | Not-determined |
| SB_extracted_k141_167025 | Not-determined |
| SB_extracted_k141_141034 | Low-quality |
| SB_extracted_k141_71201 | Not-determined |
| SB_extracted_k141_89051 | Not-determined |
| SB_extracted_k141_149680 | Not-determined |
| SB_extracted_k141_89054 | Not-determined |
| SB_extracted_k141_97621 | Low-quality |
| SB_extracted_k141_2303 | Not-determined |
| SB_extracted_k141_97623 | Not-determined |
| SB_extracted_k141_106406 | Not-determined |
| SB_extracted_k141_141039 | Not-determined |
| SB_extracted_k141_80353 | Not-determined |
| SB_extracted_k141_123795 | Not-determined |
| SB_extracted_k141_97626 | Low-quality |
| SB_extracted_k141_106409 | Not-determined |
| SB_extracted_k141_193320 | Not-determined |
| SB_extracted_k141_71207 | Not-determined |
| SB_extracted_k141_62951 | Low-quality |
| SB_extracted_k141_2305 | Not-determined |
| SB_extracted_k141_114974 | Not-determined |
| SB_extracted_k141_10613 | Not-determined |
| SB_extracted_k141_141045 | Not-determined |
| SB_extracted_k141_114975 | Not-determined |
| SB_extracted_k141_54454 | Low-quality |
| SB_extracted_k141_10614 | Not-determined |
| SB_extracted_k141_132269 | Not-determined |
| SB_extracted_k141_2306 | Not-determined |
| SB_extracted_k141_158210 | Not-determined |
| SB_extracted_k141_158211 | Not-determined |
| SB_extracted_k141_19451 | Not-determined |
| SB_extracted_k141_167048 | Low-quality |
| SB_extracted_k141_106420 | Not-determined |
| SB_extracted_k141_28432 | Low-quality |
| SB_extracted_k141_54463 | Not-determined |
| SB_extracted_k141_114980 | Not-determined |
| SB_extracted_k141_2314 | Not-determined |
| SB_extracted_k141_114981 | Not-determined |
| SB_extracted_k141_54464 | Not-determined |
| SB_extracted_k141_97635 | Not-determined |
| SB_extracted_k141_158214 | Not-determined |
| SB_extracted_k141_28436 | Not-determined |
| SB_extracted_k141_201447 | Low-quality |
| SB_extracted_k141_97639 | Low-quality |
| SB_extracted_k141_175628 | Not-determined |
| SB_extracted_k141_175629 | Not-determined |
| SB_extracted_k141_158220 | Not-determined |
| SB_extracted_k141_193335 | Not-determined |
| SB_extracted_k141_2320 | Not-determined |
| SB_extracted_k141_114989 | Low-quality |
| SB_extracted_k141_175633 | Not-determined |
| SB_extracted_k141_132275 | Not-determined |
| SB_extracted_k141_132276 | Not-determined |
| SB_extracted_k141_106434 | Not-determined |
| SB_extracted_k141_28443 | Not-determined |
| SB_extracted_k141_89075 | Not-determined |
| SB_extracted_k141_80365 | Low-quality |
| SB_extracted_k141_28444 | Not-determined |
| SB_extracted_k141_114993 | Not-determined |
| SB_extracted_k141_89076 | Not-determined |
| SB_extracted_k141_80367 | Not-determined |
| SB_extracted_k141_2326 | Not-determined |
| SB_extracted_k141_132280 | Not-determined |
| SB_extracted_k141_80371 | Not-determined |
| SB_extracted_k141_37115 | Not-determined |
| SB_extracted_k141_114997 | Not-determined |
| SB_extracted_k141_37118 | Not-determined |
| SB_extracted_k141_106439 | Not-determined |
| SB_extracted_k141_62970 | Not-determined |
| SB_extracted_k141_193349 | Not-determined |
| SB_extracted_k141_80376 | Not-determined |
| SB_extracted_k141_114999 | Low-quality |
| SB_extracted_k141_97647 | Not-determined |
| SB_extracted_k141_167056 | Not-determined |
| SB_extracted_k141_132287 | Not-determined |
| SB_extracted_k141_54469 | Not-determined |
| SB_extracted_k141_97649 | Low-quality |
| SB_extracted_k141_37127 | Not-determined |
| SB_extracted_k141_193354 | Not-determined |
| SB_extracted_k141_37129 | Not-determined |
| SB_extracted_k141_89088 | Not-determined |
| SB_extracted_k141_106447 | Not-determined |
| SB_extracted_k141_2331 | Not-determined |
| SB_extracted_k141_193356 | Not-determined |
| SB_extracted_k141_10629 | Low-quality |
| SB_extracted_k141_97656 | Not-determined |
| SB_extracted_k141_10631 | Low-quality |
| SB_extracted_k141_123802 | Not-determined |
| SB_extracted_k141_10639 | Not-determined |
| SB_extracted_k141_193366 | Not-determined |
| SB_extracted_k141_141055 | Not-determined |
| SB_extracted_k141_80396 | Not-determined |
| SB_extracted_k141_141057 | Not-determined |
| SB_extracted_k141_97661 | Not-determined |
| SB_extracted_k141_19464 | Not-determined |
| SB_extracted_k141_193368 | Not-determined |
| SB_extracted_k141_167063 | Not-determined |
| SB_extracted_k141_10643 | Not-determined |
| SB_extracted_k141_28466 | Not-determined |
| SB_extracted_k141_62995 | Not-determined |
| SB_extracted_k141_28471 | Not-determined |
| SB_extracted_k141_45402 | Not-determined |
| SB_extracted_k141_2341 | Not-determined |
| SB_extracted_k141_193375 | Not-determined |
| SB_extracted_k141_19468 | Not-determined |
| SB_extracted_k141_28479 | Not-determined |
| SB_extracted_k141_123812 | Not-determined |
| SB_extracted_k141_141065 | Not-determined |
| SB_extracted_k141_10651 | Not-determined |
| SB_extracted_k141_167069 | Not-determined |
| SB_extracted_k141_54488 | Not-determined |
| SB_extracted_k141_10654 | Not-determined |
| SB_extracted_k141_2346 | Not-determined |
| SB_extracted_k141_37138 | Not-determined |
| SB_extracted_k141_63001 | Not-determined |
| SB_extracted_k141_10656 | Low-quality |
| SB_extracted_k141_10658 | Not-determined |
| SB_extracted_k141_97667 | Not-determined |
| SB_extracted_k141_201475 | Low-quality |
| SB_extracted_k141_10660 | Low-quality |
| SB_extracted_k141_80413 | Not-determined |
| SB_extracted_k141_115009 | Not-determined |
| SB_extracted_k141_45408 | Not-determined |
| SB_extracted_k141_37144 | Low-quality |
| SB_extracted_k141_28487 | Not-determined |
| SB_extracted_k141_141074 | Not-determined |
| SB_extracted_k141_10670 | Not-determined |
| SB_extracted_k141_28489 | Low-quality |
| SB_extracted_k141_80420 | Not-determined |
| SB_extracted_k141_141077 | Not-determined |
| SB_extracted_k141_19483 | Not-determined |
| SB_extracted_k141_123828 | Low-quality |
| SB_extracted_k141_201483 | Not-determined |
| SB_extracted_k141_149696 | Not-determined |
| SB_extracted_k141_123831 | Not-determined |
| SB_extracted_k141_37157 | Low-quality |
| SB_extracted_k141_149698 | Not-determined |
| SB_extracted_k141_37159 | Not-determined |
| SB_extracted_k141_89103 | Not-determined |
| SB_extracted_k141_106479 | Not-determined |
| SB_extracted_k141_167076 | Not-determined |
| SB_extracted_k141_28501 | Not-determined |
| SB_extracted_k141_141085 | Not-determined |
| SB_extracted_k141_132324 | Not-determined |
| SB_extracted_k141_97687 | Not-determined |
| SB_extracted_k141_193411 | Low-quality |
| SB_extracted_k141_80429 | Not-determined |
| SB_extracted_k141_123842 | Not-determined |
| SB_extracted_k141_97691 | Not-determined |
| SB_extracted_k141_123844 | Not-determined |
| SB_extracted_k141_141091 | Not-determined |
| SB_extracted_k141_141093 | Not-determined |
| SB_extracted_k141_115042 | Not-determined |
| SB_extracted_k141_183924 | Not-determined |
| SB_extracted_k141_28510 | Not-determined |
| SB_extracted_k141_19499 | Not-determined |
| SB_extracted_k141_106486 | Not-determined |
| SB_extracted_k141_28513 | Not-determined |
| SB_extracted_k141_183928 | Low-quality |
| SB_extracted_k141_54500 | Not-determined |
| SB_extracted_k141_183931 | Not-determined |
| SB_extracted_k141_132336 | Not-determined |
| SB_extracted_k141_37168 | Not-determined |
| SB_extracted_k141_37169 | Not-determined |
| SB_extracted_k141_10689 | Low-quality |
| SB_extracted_k141_2360 | Not-determined |
| SB_extracted_k141_28519 | Low-quality |

| | |
|---|---|
| S8_extracted_k141_201493 | Not-determined |
| S8_extracted_k141_19506 | Not-determined |
| S8_extracted_k141_63023 | Not-determined |
| S8_extracted_k141_89123 | Not-determined |
| S8_extracted_k141_80445 | Not-determined |
| S8_extracted_k141_106494 | Not-determined |
| S8_extracted_k141_141104 | Not-determined |
| S8_extracted_k141_106497 | Not-determined |
| S8_extracted_k141_141106 | Not-determined |
| S8_extracted_k141_10699 | Not-determined |
| S8_extracted_k141_123850 | Low-quality |
| S8_extracted_k141_141111 | Not-determined |
| S8_extracted_k141_106505 | Not-determined |
| S8_extracted_k141_10701 | Not-determined |
| S13_vs_12957\|\|full | Medium-quality |
| S8_extracted_k141_97707 | Not-determined |
| S8_extracted_k141_167093 | Not-determined |
| S8_extracted_k141_89136 | Not-determined |
| S8_extracted_k141_201507 | Not-determined |
| S8_extracted_k141_89137 | Not-determined |
| S8_extracted_k141_28537 | Not-determined |
| S8_extracted_k141_45417 | Not-determined |
| S8_extracted_k141_97712 | Low-quality |
| S8_extracted_k141_28539 | Not-determined |
| S8_extracted_k141_37176 | Not-determined |
| S8_extracted_k141_37177 | Not-determined |
| S8_extracted_k141_167098 | Not-determined |
| S8_extracted_k141_19526 | Not-determined |
| S8_extracted_k141_141128 | Low-quality |
| S8_extracted_k141_123855 | Not-determined |
| S8_extracted_k141_45420 | Not-determined |
| S8_extracted_k141_63039 | Not-determined |
| S8_extracted_k141_106509 | Not-determined |
| S8_extracted_k141_19530 | Not-determined |
| S8_extracted_k141_106511 | Not-determined |
| S8_extracted_k141_63040 | Not-determined |
| S8_extracted_k141_97728 | Not-determined |
| S8_extracted_k141_19533 | Not-determined |
| S8_extracted_k141_175676 | Not-determined |
| S8_extracted_k141_175677 | Low-quality |
| S8_extracted_k141_63041 | Not-determined |
| S8_extracted_k141_97731 | Not-determined |
| S8_extracted_k141_63045 | Not-determined |
| S8_extracted_k141_123861 | Not-determined |
| S8_extracted_k141_141138 | Not-determined |
| S8_extracted_k141_28549 | Not-determined |
| S8_extracted_k141_97734 | Not-determined |
| S8_extracted_k141_97736 | Not-determined |
| S8_extracted_k141_10726 | Not-determined |
| S8_extracted_k141_167110 | Not-determined |
| S8_extracted_k141_37193 | Not-determined |
| S8_extracted_k141_63052 | Not-determined |
| S8_extracted_k141_80457 | Not-determined |
| S8_extracted_k141_28553 | Not-determined |
| S8_extracted_k141_141141 | Not-determined |
| S8_extracted_k141_167113 | Not-determined |
| S8_extracted_k141_106518 | Not-determined |
| S8_extracted_k141_63057 | Not-determined |
| S8_extracted_k141_80461 | Not-determined |
| S8_extracted_k141_193420 | Not-determined |
| S8_extracted_k141_106521 | Not-determined |
| S8_extracted_k141_106522 | Not-determined |
| S8_extracted_k141_28558 | Not-determined |
| S8_extracted_k141_175692 | Not-determined |
| S8_extracted_k141_183950 | Not-determined |
| S8_extracted_k141_167117 | Not-determined |
| S8_extracted_k141_80470 | Not-determined |
| S8_extracted_k141_123875 | Low-quality |
| S8_extracted_k141_141152 | Not-determined |
| S8_extracted_k141_167121 | Not-determined |
| S8_extracted_k141_10742 | Not-determined |
| S8_extracted_k141_141156 | Not-determined |
| S8_extracted_k141_80480 | Not-determined |
| S8_extracted_k141_37210 | Not-determined |
| S8_extracted_k141_71241 | Not-determined |
| S8_extracted_k141_106534 | Not-determined |
| S8_extracted_k141_71244 | Not-determined |
| S8_extracted_k141_183954 | Low-quality |
| S8_extracted_k141_37219 | Not-determined |
| S8_extracted_k141_183956 | Low-quality |
| S8_extracted_k141_123889 | Low-quality |
| S8_extracted_k141_106538 | Low-quality |
| S8_extracted_k141_123890 | Not-determined |
| S8_extracted_k141_80483 | Not-determined |
| S8_extracted_k141_37222 | Not-determined |
| S8_extracted_k141_106542 | Not-determined |
| S3_vs_k141_90640\|\|full | Medium-quality |
| S8_extracted_k141_19568 | Not-determined |
| S8_extracted_k141_167135 | Not-determined |
| S8_extracted_k141_193432 | Not-determined |
| S8_extracted_k141_71257 | Not-determined |
| S8_extracted_k141_183962 | Not-determined |
| S8_extracted_k141_10767 | Not-determined |
| S8_extracted_k141_201531 | Not-determined |
| S8_extracted_k141_141158 | Low-quality |
| S8_extracted_k141_167140 | Not-determined |
| S8_extracted_k141_37227 | Not-determined |
| S8_extracted_k141_106550 | Not-determined |
| S8_extracted_k141_158231 | Low-quality |
| S8_extracted_k141_183966 | Not-determined |
| S8_extracted_k141_175707 | Not-determined |
| S8_extracted_k141_63090 | Not-determined |
| S8_extracted_k141_123901 | Not-determined |
| S8_extracted_k141_97767 | Low-quality |
| S8_extracted_k141_10774 | Not-determined |
| S8_extracted_k141_19585 | Not-determined |
| S8_extracted_k141_106554 | Not-determined |
| S8_extracted_k141_2391 | Not-determined |
| S8_extracted_k141_97768 | Not-determined |
| S8_extracted_k141_106556 | Not-determined |
| S8_extracted_k141_106557 | Low-quality |
| S8_extracted_k141_2393 | Not-determined |
| S8_extracted_k141_193440 | Low-quality |
| S8_extracted_k141_175715 | Not-determined |
| S8_extracted_k141_106559 | Low-quality |
| S8_extracted_k141_2394 | Not-determined |
| S8_extracted_k141_175716 | Not-determined |
| S8_extracted_k141_19589 | Not-determined |
| S8_extracted_k141_45449 | Not-determined |
| S8_extracted_k141_19592 | Not-determined |
| S8_extracted_k141_106563 | Not-determined |
| S8_extracted_k141_123904 | Not-determined |
| S8_extracted_k141_28575 | Not-determined |
| S8_extracted_k141_97773 | Not-determined |
| S8_extracted_k141_28576 | Not-determined |
| S8_extracted_k141_193446 | Not-determined |
| S8_extracted_k141_28578 | Not-determined |
| S8_extracted_k141_71271 | Not-determined |
| S8_extracted_k141_71272 | Not-determined |
| S14_extracted_k141_93134 | Medium-quality |
| S8_extracted_k141_97776 | Low-quality |
| S8_extracted_k141_183974 | Not-determined |
| S8_extracted_k141_54515 | Low-quality |
| S8_extracted_k141_123907 | Low-quality |
| S8_extracted_k141_183975 | Not-determined |
| S8_extracted_k141_193450 | Not-determined |
| S8_extracted_k141_37236 | Low-quality |
| S8_extracted_k141_201549 | Not-determined |
| S8_extracted_k141_19603 | Not-determined |
| S8_extracted_k141_71278 | Not-determined |
| S8_extracted_k141_28584 | Not-determined |
| S8_extracted_k141_28585 | Low-quality |
| S8_extracted_k141_19605 | Not-determined |
| S8_extracted_k141_2406 | Not-determined |
| S8_extracted_k141_80491 | Not-determined |
| S8_extracted_k141_45450 | Not-determined |
| S8_extracted_k141_106566 | Not-determined |
| S8_extracted_k141_123914 | Low-quality |
| S8_extracted_k141_45451 | Not-determined |
| S8_extracted_k141_123915 | Not-determined |
| S8_extracted_k141_106567 | Not-determined |
| S8_extracted_k141_106568 | Not-determined |
| S8_extracted_k141_106569 | Not-determined |
| S8_extracted_k141_63102 | Low-quality |
| S8_extracted_k141_19616 | Not-determined |
| S8_extracted_k141_37249 | Low-quality |
| S8_extracted_k141_201557 | Not-determined |
| S8_extracted_k141_201559 | Not-determined |
| S8_extracted_k141_19620 | Not-determined |
| S8_extracted_k141_71290 | Not-determined |
| S8_extracted_k141_106579 | Not-determined |
| S8_extracted_k141_63108 | Low-quality |
| S8_extracted_k141_28604 | Not-determined |
| S8_extracted_k141_2416 | Not-determined |
| S8_extracted_k141_115065 | Not-determined |
| S8_extracted_k141_45455 | Not-determined |
| S8_extracted_k141_149715 | Not-determined |
| S8_extracted_k141_115066 | Not-determined |
| S8_extracted_k141_63110 | Not-determined |
| S8_extracted_k141_80507 | Not-determined |
| S8_extracted_k141_45463 | Low-quality |
| S8_extracted_k141_28606 | Not-determined |
| S8_extracted_k141_201572 | Not-determined |
| S8_extracted_k141_28607 | Not-determined |
| S8_extracted_k141_63115 | Not-determined |
| S8_extracted_k141_193466 | Not-determined |
| S8_extracted_k141_37256 | Not-determined |
| S8_extracted_k141_193467 | Not-determined |
| S8_extracted_k141_2419 | Not-determined |
| S8_extracted_k141_37257 | Not-determined |
| S8_extracted_k141_63119 | Not-determined |
| S8_extracted_k141_193469 | Not-determined |
| S8_extracted_k141_149716 | Not-determined |
| S8_extracted_k141_167166 | Low-quality |
| S8_extracted_k141_193471 | Not-determined |
| S8_extracted_k141_183999 | Not-determined |
| S8_extracted_k141_28615 | Not-determined |
| S8_extracted_k141_201579 | Not-determined |
| S8_extracted_k141_45475 | Not-determined |
| S8_extracted_k141_175755 | Not-determined |
| S8_extracted_k141_28622 | Low-quality |
| S8_extracted_k141_37260 | Not-determined |
| S8_extracted_k141_132363 | Not-determined |
| S8_extracted_k141_132364 | Not-determined |
| S8_extracted_k141_54523 | Not-determined |
| S8_extracted_k141_28624 | Not-determined |
| S8_extracted_k141_115071 | Not-determined |
| S8_extracted_k141_63130 | Not-determined |
| S8_extracted_k141_115075 | Not-determined |
| S8_extracted_k141_2432 | Not-determined |
| S8_extracted_k141_184005 | Not-determined |
| S8_extracted_k141_201580 | Not-determined |
| S8_extracted_k141_115078 | Not-determined |
| S8_extracted_k141_132375 | Low-quality |
| S8_extracted_k141_2436 | Not-determined |
| S8_extracted_k141_193486 | Low-quality |
| S8_extracted_k141_193489 | Not-determined |
| S8_extracted_k141_132380 | Not-determined |
| S8_extracted_k141_2440 | Not-determined |
| S8_extracted_k141_115088 | Not-determined |
| S8_extracted_k141_37282 | Not-determined |
| S8_extracted_k141_201592 | Not-determined |
| S8_extracted_k141_123946 | Low-quality |
| S8_extracted_k141_115089 | Not-determined |
| S8_extracted_k141_28636 | Not-determined |
| S8_extracted_k141_115092 | Not-determined |
| S8_extracted_k141_123947 | Not-determined |
| S8_extracted_k141_19644 | Not-determined |
| S8_extracted_k141_193495 | Not-determined |
| S8_extracted_k141_63157 | Not-determined |
| S8_extracted_k141_115099 | Not-determined |
| S8_extracted_k141_132388 | Low-quality |
| S8_extracted_k141_158248 | Not-determined |
| S8_extracted_k141_10790 | Low-quality |
| S8_extracted_k141_63164 | Not-determined |
| S8_extracted_k141_123958 | Not-determined |
| S8_extracted_k141_63165 | Not-determined |
| S8_extracted_k141_28647 | Not-determined |
| S8_extracted_k141_184034 | Not-determined |
| S8_extracted_k141_80515 | Not-determined |
| S8_extracted_k141_54536 | Low-quality |
| S8_extracted_k141_63167 | Not-determined |
| S8_extracted_k141_123960 | Not-determined |
| S8_extracted_k141_19655 | Not-determined |
| S8_extracted_k141_123961 | Low-quality |
| S8_extracted_k141_2446 | Not-determined |
| S8_extracted_k141_184035 | Not-determined |
| S8_extracted_k141_28651 | Not-determined |
| S8_extracted_k141_123963 | Not-determined |
| S8_extracted_k141_19658 | Not-determined |
| S8_extracted_k141_123965 | Not-determined |
| S8_extracted_k141_123966 | Not-determined |
| S8_extracted_k141_28652 | Not-determined |
| S8_extracted_k141_123967 | Not-determined |
| S8_extracted_k141_80523 | Not-determined |
| S8_extracted_k141_80525 | Not-determined |
| S8_extracted_k141_184041 | Not-determined |
| S8_extracted_k141_201624 | Not-determined |
| S8_extracted_k141_45483 | Not-determined |
| S8_extracted_k141_2454 | Not-determined |
| S8_extracted_k141_28656 | Low-quality |
| S8_extracted_k141_19665 | Not-determined |
| S8_extracted_k141_167189 | Not-determined |
| S8_extracted_k141_19668 | Not-determined |
| S8_extracted_k141_167191 | Not-determined |
| S8_extracted_k141_2456 | Not-determined |
| S8_extracted_k141_123978 | Not-determined |
| S8_extracted_k141_167193 | Not-determined |
| S8_extracted_k141_132410 | Not-determined |
| S8_extracted_k141_201635 | Not-determined |
| S8_extracted_k141_54540 | Low-quality |
| S8_extracted_k141_80541 | Not-determined |
| S8_extracted_k141_63183 | Not-determined |
| S8_extracted_k141_28667 | Not-determined |
| S8_extracted_k141_2466 | Not-determined |
| S8_extracted_k141_80545 | Not-determined |
| S8_extracted_k141_19678 | Not-determined |
| S8_extracted_k141_28669 | Not-determined |
| S8_extracted_k141_167197 | Not-determined |
| S8_extracted_k141_28670 | Not-determined |
| S8_extracted_k141_19681 | Not-determined |
| S8_extracted_k141_63189 | Not-determined |
| S8_extracted_k141_10803 | Not-determined |
| S8_extracted_k141_63192 | Not-determined |
| S8_extracted_k141_201649 | Not-determined |
| S8_extracted_k141_123994 | Not-determined |
| S8_extracted_k141_80552 | Low-quality |
| S8_extracted_k141_167199 | Not-determined |
| S8_extracted_k141_201650 | Not-determined |
| S8_extracted_k141_19685 | Low-quality |
| S8_extracted_k141_201660 | Not-determined |
| S8_extracted_k141_28683 | Not-determined |
| S8_extracted_k141_175780 | Not-determined |
| S8_extracted_k141_10811 | Low-quality |
| S8_extracted_k141_124001 | Not-determined |
| S8_extracted_k141_141229 | Not-determined |
| S8_extracted_k141_132430 | Not-determined |
| S8_extracted_k141_184061 | Not-determined |
| S8_extracted_k141_37337 | Not-determined |
| S8_extracted_k141_28691 | Low-quality |
| S8_extracted_k141_28693 | Not-determined |
| S8_extracted_k141_141238 | Not-determined |
| S8_extracted_k141_54549 | Not-determined |
| S8_extracted_k141_10819 | Not-determined |
| S8_extracted_k141_132433 | Not-determined |
| S8_extracted_k141_132434 | Not-determined |
| S8_extracted_k141_63206 | Not-determined |
| S8_extracted_k141_28698 | Not-determined |
| S8_extracted_k141_80556 | Low-quality |
| S8_extracted_k141_201673 | Not-determined |
| S8_extracted_k141_45486 | Low-quality |
| S8_extracted_k141_141243 | Not-determined |
| S8_extracted_k141_201675 | Not-determined |
| S8_extracted_k141_175793 | Not-determined |
| S8_extracted_k141_124010 | Not-determined |
| S8_extracted_k141_149769 | Not-determined |
| S8_extracted_k141_124011 | Not-determined |
| S8_extracted_k141_80559 | Not-determined |
| S8_extracted_k141_193511 | Not-determined |
| S8_extracted_k141_28702 | Not-determined |
| S8_extracted_k141_124012 | Not-determined |
| S8_extracted_k141_45490 | Not-determined |
| S8_extracted_k141_149772 | Low-quality |
| S8_extracted_k141_193512 | Not-determined |
| S8_extracted_k141_28704 | Not-determined |
| S8_extracted_k141_175797 | Not-determined |
| S8_extracted_k141_2478 | Low-quality |
| S8_extracted_k141_80565 | Not-determined |
| S8_extracted_k141_149775 | Not-determined |
| S8_extracted_k141_45494 | Not-determined |
| S8_extracted_k141_2480 | Not-determined |
| S8_extracted_k141_132438 | Not-determined |
| S8_extracted_k141_10828 | Not-determined |
| S8_extracted_k141_45495 | Low-quality |
| S8_extracted_k141_149779 | Not-determined |
| S8_extracted_k141_141248 | Not-determined |
| S8_extracted_k141_45497 | Not-determined |
| S8_extracted_k141_37349 | Not-determined |
| S8_extracted_k141_28711 | Low-quality |
| S8_extracted_k141_80571 | Not-determined |
| S8_extracted_k141_80574 | Not-determined |
| S8_extracted_k141_63219 | Low-quality |
| S8_extracted_k141_10833 | Not-determined |
| S8_extracted_k141_124022 | Not-determined |
| S8_extracted_k141_141257 | Not-determined |
| S8_extracted_k141_63226 | Not-determined |
| S8_extracted_k141_115110 | Not-determined |
| S8_extracted_k141_124026 | Not-determined |
| S8_extracted_k141_54561 | Low-quality |
| S8_extracted_k141_10837 | Not-determined |
| S8_extracted_k141_141259 | Not-determined |
| S8_extracted_k141_193520 | Not-determined |
| S8_extracted_k141_19709 | Not-determined |
| S8_extracted_k141_193523 | Not-determined |
| S8_extracted_k141_149792 | Not-determined |
| S8_extracted_k141_141260 | Not-determined |
| S8_extracted_k141_28727 | Not-determined |
| S8_extracted_k141_201691 | Not-determined |
| S8_extracted_k141_71323 | Not-determined |
| S8_extracted_k141_63233 | Not-determined |
| S8_extracted_k141_54576 | Not-determined |
| S8_extracted_k141_175814 | Low-quality |
| S8_extracted_k141_63236 | Low-quality |
| S8_extracted_k141_149797 | Not-determined |
| S8_extracted_k141_149798 | Not-determined |
| S8_extracted_k141_10845 | Not-determined |
| S8_extracted_k141_28732 | Not-determined |
| S8_extracted_k141_193532 | Not-determined |
| S8_extracted_k141_132448 | Not-determined |
| S8_extracted_k141_71328 | Not-determined |
| S8_extracted_k141_175819 | Not-determined |
| S8_extracted_k141_124038 | Not-determined |
| S8_extracted_k141_10848 | Not-determined |
| S8_extracted_k141_80589 | Not-determined |
| S8_extracted_k141_10849 | Low-quality |
| S8_extracted_k141_115118 | Not-determined |
| S8_extracted_k141_193535 | Not-determined |
| S8_extracted_k141_132452 | Not-determined |
| S8_extracted_k141_175822 | Not-determined |
| S8_extracted_k141_132453 | Not-determined |
| S8_extracted_k141_80592 | Not-determined |
| S8_extracted_k141_124042 | Not-determined |
| S8_extracted_k141_124043 | Not-determined |
| S8_extracted_k141_71336 | Not-determined |
| S8_extracted_k141_141274 | Not-determined |
| S8_extracted_k141_175824 | Not-determined |
| S8_extracted_k141_175825 | Not-determined |
| S8_extracted_k141_124046 | Low-quality |
| S8_extracted_k141_115122 | Not-determined |
| S8_extracted_k141_28740 | Not-determined |
| S8_extracted_k141_124055 | Low-quality |
| S8_extracted_k141_80601 | Not-determined |
| S8_extracted_k141_80602 | Not-determined |
| S8_extracted_k141_175835 | Not-determined |
| S8_extracted_k141_71344 | Not-determined |
| S8_extracted_k141_37355 | Not-determined |
| S8_extracted_k141_175837 | Low-quality |
| S8_extracted_k141_80603 | Not-determined |
| S8_extracted_k141_193546 | Not-determined |
| S8_extracted_k141_71348 | Not-determined |
| S8_extracted_k141_19720 | Not-determined |
| S8_extracted_k141_10856 | Not-determined |
| S8_extracted_k141_19722 | Not-determined |
| S8_extracted_k141_28759 | Not-determined |
| S8_extracted_k141_80609 | Low-quality |
| S8_extracted_k141_184069 | Not-determined |
| S8_extracted_k141_19724 | Not-determined |
| S8_extracted_k141_71355 | Not-determined |
| S8_extracted_k141_71359 | Not-determined |
| S8_extracted_k141_167216 | Not-determined |
| S8_extracted_k141_175848 | Not-determined |
| S8_extracted_k141_167219 | Not-determined |
| S8_extracted_k141_28772 | Not-determined |
| S8_extracted_k141_63253 | Not-determined |
| S8_extracted_k141_167223 | Not-determined |
| S8_extracted_k141_141298 | Not-determined |
| S8_extracted_k141_89170 | Not-determined |
| S8_extracted_k141_115128 | Not-determined |
| S8_extracted_k141_63257 | Not-determined |
| S8_extracted_k141_45529 | Not-determined |
| S8_extracted_k141_149815 | Low-quality |
| S8_extracted_k141_10872 | Not-determined |
| S8_extracted_k141_115130 | Not-determined |
| S8_extracted_k141_54591 | Not-determined |
| S8_extracted_k141_80618 | Low-quality |
| S8_extracted_k141_80619 | Not-determined |
| S8_extracted_k141_89176 | Not-determined |
| S8_extracted_k141_193572 | Not-determined |
| S8_extracted_k141_71363 | Not-determined |
| S8_extracted_k141_89177 | Not-determined |
| S8_extracted_k141_193575 | Not-determined |
| S8_extracted_k141_175853 | Not-determined |
| S8_extracted_k141_167230 | Not-determined |
| S8_extracted_k141_71365 | Not-determined |
| S8_extracted_k141_167232 | Not-determined |
| S8_extracted_k141_10881 | Not-determined |
| S8_extracted_k141_167233 | Not-determined |
| S8_extracted_k141_184099 | Not-determined |
| S8_extracted_k141_167235 | Not-determined |
| S8_extracted_k141_158258 | Not-determined |
| S8_extracted_k141_158259 | Not-determined |
| S8_extracted_k141_115133 | Not-determined |
| S8_extracted_k141_106596 | Not-determined |
| S8_extracted_k141_63269 | Not-determined |
| S8_extracted_k141_19748 | Low-quality |
| S8_extracted_k141_54607 | Not-determined |
| S8_extracted_k141_115140 | Not-determined |
| S8_extracted_k141_167243 | Not-determined |
| S8_extracted_k141_149827 | Low-quality |
| S8_extracted_k141_63270 | Low-quality |
| S8_extracted_k141_19751 | Not-determined |
| S8_extracted_k141_124063 | Not-determined |
| S8_extracted_k141_175870 | Not-determined |
| S8_extracted_k141_54609 | Not-determined |
| S8_extracted_k141_54612 | Not-determined |
| S8_extracted_k141_115147 | Not-determined |
| S8_extracted_k141_2508 | Not-determined |
| S8_extracted_k141_132479 | Not-determined |
| S8_extracted_k141_80645 | Not-determined |
| S8_extracted_k141_89180 | Not-determined |
| S8_extracted_k141_71372 | Not-determined |
| S8_extracted_k141_54614 | Not-determined |
| S8_extracted_k141_175877 | Not-determined |
| S8_extracted_k141_28805 | Not-determined |
| S8_extracted_k141_10899 | Not-determined |
| S8_extracted_k141_175879 | Not-determined |
| S8_extracted_k141_149841 | Not-determined |
| S8_extracted_k141_28807 | Not-determined |
| S8_extracted_k141_124077 | Not-determined |
| S8_extracted_k141_37371 | Not-determined |
| S8_extracted_k141_2510 | Not-determined |
| S8_extracted_k141_124080 | Not-determined |
| S8_extracted_k141_80650 | Not-determined |
| S8_extracted_k141_45539 | Not-determined |
| S8_extracted_k141_124082 | Not-determined |
| S8_extracted_k141_167257 | Not-determined |
| S8_extracted_k141_132485 | Not-determined |
| S14_vs_k141_99344\|\|full | Medium-quality |
| S8_extracted_k141_54621 | Not-determined |
| S8_extracted_k141_167259 | Not-determined |
| S8_extracted_k141_71376 | Not-determined |
| S8_extracted_k141_37375 | Not-determined |
| S8_extracted_k141_115157 | Not-determined |
| S8_extracted_k141_80655 | Not-determined |
| S8_extracted_k141_54622 | Not-determined |
| S8_extracted_k141_115159 | Not-determined |
| S8_extracted_k141_10908 | Not-determined |
| S8_extracted_k141_149849 | Not-determined |
| S8_extracted_k141_124087 | Not-determined |
| S8_extracted_k141_10911 | Not-determined |
| S8_extracted_k141_80657 | Not-determined |
| S8_extracted_k141_80658 | Not-determined |
| S8_extracted_k141_80661 | Not-determined |
| S8_extracted_k141_141310 | Not-determined |
| S8_extracted_k141_10914 | Not-determined |
| S8_extracted_k141_89187 | Not-determined |
| S8_extracted_k141_193592 | Not-determined |
| S8_extracted_k141_71380 | Not-determined |
| S8_extracted_k141_124095 | Not-determined |
| S8_extracted_k141_45555 | Low-quality |
| S8_extracted_k141_19753 | Not-determined |
| S8_extracted_k141_115163 | Not-determined |
| S8_extracted_k141_71383 | Not-determined |
| S8_extracted_k141_132497 | Not-determined |
| S8_extracted_k141_10917 | Not-determined |
| S8_extracted_k141_2520 | Not-determined |
| S8_extracted_k141_10918 | Not-determined |
| S8_extracted_k141_2521 | Not-determined |
| S8_extracted_k141_193595 | Not-determined |
| S8_extracted_k141_19756 | Not-determined |
| S8_extracted_k141_89193 | Not-determined |
| S8_extracted_k141_19758 | Not-determined |
| S8_extracted_k141_19759 | Not-determined |
| S8_extracted_k141_124102 | Not-determined |
| S8_extracted_k141_175901 | Not-determined |
| S8_extracted_k141_28813 | Low-quality |
| S8_extracted_k141_184115 | Not-determined |
| S8_extracted_k141_45559 | Not-determined |
| S8_extracted_k141_19762 | Not-determined |
| S8_extracted_k141_54628 | Not-determined |
| S8_extracted_k141_193604 | Not-determined |
| S8_extracted_k141_28816 | Not-determined |
| S8_extracted_k141_37392 | Not-determined |
| S8_extracted_k141_193606 | Not-determined |
| S8_extracted_k141_115172 | Low-quality |
| S8_extracted_k141_167270 | Low-quality |
| S8_extracted_k141_115174 | Not-determined |
| S8_extracted_k141_63278 | Not-determined |
| S8_extracted_k141_37395 | Not-determined |
| S8_extracted_k141_19765 | Not-determined |
| S8_extracted_k141_184126 | Not-determined |
| S8_extracted_k141_63279 | Not-determined |
| S8_extracted_k141_193611 | Not-determined |
| S8_extracted_k141_149871 | Low-quality |
| S8_extracted_k141_19766 | Not-determined |
| S8_extracted_k141_2531 | Not-determined |
| S8_extracted_k141_184132 | Not-determined |
| S8_extracted_k141_19769 | Not-determined |
| S8_extracted_k141_28830 | Not-determined |
| S8_extracted_k141_193612 | Not-determined |
| S8_extracted_k141_80674 | Not-determined |
| S8_extracted_k141_28832 | Not-determined |
| S8_extracted_k141_63287 | Not-determined |
| S8_extracted_k141_89210 | Not-determined |
| S8_extracted_k141_71394 | Not-determined |
| S8_extracted_k141_149880 | Not-determined |
| S8_extracted_k141_63292 | Not-determined |
| S8_extracted_k141_45573 | Not-determined |
| S8_extracted_k141_149882 | Not-determined |
| S8_extracted_k141_28840 | Not-determined |
| S8_extracted_k141_141335 | Not-determined |
| S8_extracted_k141_45576 | Low-quality |
| S8_extracted_k141_149883 | Not-determined |
| S8_extracted_k141_89218 | Not-determined |
| S8_extracted_k141_71404 | Not-determined |
| S8_extracted_k141_2544 | Not-determined |
| S8_extracted_k141_89219 | Not-determined |
| S8_extracted_k141_184138 | Not-determined |
| S8_extracted_k141_201711 | Not-determined |
| S8_extracted_k141_89222 | Not-determined |
| S8_extracted_k141_141342 | Not-determined |
| S8_extracted_k141_193614 | Not-determined |
| S8_extracted_k141_10935 | Not-determined |
| S8_extracted_k141_132521 | Not-determined |
| S8_extracted_k141_19781 | Not-determined |
| S8_extracted_k141_45586 | Low-quality |
| S8_extracted_k141_158268 | Not-determined |
| S8_extracted_k141_19782 | Not-determined |
| S8_extracted_k141_80686 | Not-determined |
| S8_extracted_k141_132523 | Not-determined |
| S8_extracted_k141_193618 | Not-determined |
| S8_extracted_k141_124121 | Low-quality |
| S8_extracted_k141_149888 | Not-determined |
| S8_extracted_k141_193619 | Not-determined |
| S8_extracted_k141_71414 | Low-quality |
| S8_extracted_k141_2558 | Not-determined |
| S8_extracted_k141_158271 | Not-determined |
| S8_extracted_k141_71416 | Not-determined |
| S8_extracted_k141_54634 | Not-determined |
| S8_extracted_k141_201712 | Not-determined |
| S8_extracted_k141_71419 | Not-determined |
| S8_extracted_k141_10940 | Not-determined |
| S8_extracted_k141_184149 | Not-determined |
| S8_extracted_k141_124122 | Not-determined |
| S8_extracted_k141_132527 | Not-determined |
| S8_extracted_k141_167282 | Not-determined |
| S8_extracted_k141_184153 | Not-determined |
| S8_extracted_k141_2562 | Not-determined |
| S8_extracted_k141_149897 | Low-quality |
| S8_extracted_k141_19794 | Not-determined |
| S8_extracted_k141_71424 | Not-determined |
| S8_extracted_k141_124129 | Not-determined |
| S8_extracted_k141_175931 | Not-determined |
| S8_extracted_k141_97792 | Not-determined |
| S8_extracted_k141_71433 | Not-determined |
| S8_extracted_k141_45588 | Low-quality |
| S8_extracted_k141_141351 | Not-determined |
| S8_extracted_k141_45592 | Not-determined |
| S8_extracted_k141_19801 | Not-determined |

| | |
|---|---|
| S8_extracted_k141_141353 | Low-quality |
| S8_extracted_k141_28847 | Not-determined |
| S8_extracted_k141_158289 | Not-determined |
| S8_extracted_k141_158290 | Not-determined |
| S8_extracted_k141_2581 | Not-determined |
| S8_extracted_k141_115197 | Not-determined |
| S8_extracted_k141_141356 | Not-determined |
| S8_extracted_k141_158292 | Not-determined |
| S8_extracted_k141_193636 | Not-determined |
| S8_extracted_k141_158293 | Not-determined |
| S8_extracted_k141_97806 | Not-determined |
| S8_extracted_k141_106638 | Not-determined |
| S8_extracted_k141_175942 | Not-determined |
| S8_extracted_k141_2588 | Not-determined |
| S8_extracted_k141_141364 | Not-determined |
| S8_extracted_k141_149910 | Not-determined |
| S8_extracted_k141_106643 | Not-determined |
| S8_extracted_k141_167301 | Not-determined |
| S8_extracted_k141_106645 | Not-determined |
| S8_extracted_k141_97819 | Not-determined |
| S8_extracted_k141_2592 | Not-determined |
| S8_extracted_k141_149913 | Low-quality |
| S8_extracted_k141_175947 | Not-determined |
| S8_extracted_k141_28851 | Not-determined |
| S8_extracted_k141_167303 | Low-quality |
| S8_extracted_k141_45609 | Not-determined |
| S8_extracted_k141_132551 | Not-determined |
| S8_extracted_k141_167305 | Not-determined |
| S8_extracted_k141_115199 | Low-quality |
| S8_extracted_k141_141378 | Not-determined |
| S8_extracted_k141_45614 | Not-determined |
| S8_extracted_k141_167309 | Not-determined |
| S8_extracted_k141_10945 | Not-determined |
| S8_extracted_k141_19819 | Not-determined |
| S8_extracted_k141_115204 | Not-determined |
| S8_extracted_k141_54670 | Not-determined |
| S8_extracted_k141_19821 | Not-determined |
| S8_extracted_k141_63331 | Not-determined |
| S8_extracted_k141_80716 | Not-determined |
| S8_extracted_k141_132564 | Not-determined |
| S8_extracted_k141_54675 | Not-determined |
| S8_extracted_k141_124139 | Not-determined |
| S8_extracted_k141_115209 | Not-determined |
| S8_extracted_k141_19824 | Not-determined |
| S8_extracted_k141_89236 | Not-determined |
| S8_extracted_k141_45622 | Not-determined |
| S8_extracted_k141_193643 | Not-determined |
| S8_extracted_k141_10952 | Not-determined |
| S8_extracted_k141_19826 | Not-determined |
| S8_extracted_k141_106650 | Not-determined |
| S8_extracted_k141_201724 | Not-determined |
| S8_extracted_k141_193647 | Low-quality |
| S8_extracted_k141_106651 | Not-determined |
| S8_extracted_k141_10955 | Not-determined |
| S8_extracted_k141_193648 | Not-determined |
| S8_extracted_k141_175966 | Not-determined |
| S8_extracted_k141_89239 | Not-determined |
| S8_extracted_k141_201727 | Not-determined |
| S8_extracted_k141_10960 | Not-determined |
| S8_extracted_k141_97833 | Not-determined |
| S8_extracted_k141_2609 | Not-determined |
| S8_extracted_k141_106656 | Not-determined |
| S8_extracted_k141_175972 | Not-determined |
| S8_extracted_k141_193655 | Not-determined |
| S8_extracted_k141_124153 | Low-quality |
| S8_extracted_k141_132575 | Not-determined |
| S8_extracted_k141_63341 | Not-determined |
| S8_extracted_k141_175975 | Not-determined |
| S8_extracted_k141_80730 | Not-determined |
| S8_extracted_k141_106663 | Not-determined |
| S8_extracted_k141_158316 | Not-determined |
| S8_extracted_k141_184179 | Not-determined |
| S8_extracted_k141_201733 | Not-determined |
| S8_extracted_k141_54679 | Not-determined |
| S8_extracted_k141_132578 | Not-determined |
| S8_extracted_k141_28871 | Not-determined |
| S8_extracted_k141_175979 | Not-determined |
| S8_extracted_k141_63346 | Not-determined |
| S8_extracted_k141_89250 | Not-determined |
| S8_extracted_k141_106669 | Not-determined |
| S8_extracted_k141_141400 | Not-determined |
| S8_extracted_k141_19842 | Not-determined |
| S8_extracted_k141_54682 | Low-quality |
| S8_extracted_k141_201737 | Not-determined |
| S8_extracted_k141_45634 | Not-determined |
| S8_extracted_k141_2620 | Not-determined |
| S8_extracted_k141_54684 | Low-quality |
| S8_extracted_k141_193659 | Not-determined |
| S8_extracted_k141_28875 | Not-determined |
| S8_extracted_k141_184188 | Not-determined |
| S8_extracted_k141_132581 | Low-quality |
| S8_extracted_k141_193664 | Not-determined |
| S8_extracted_k141_149921 | Not-determined |
| S8_extracted_k141_141409 | Low-quality |
| S8_extracted_k141_141410 | Not-determined |
| S8_extracted_k141_45637 | Not-determined |
| S8_extracted_k141_28879 | Not-determined |
| S8_extracted_k141_158319 | Low-quality |
| S8_extracted_k141_28881 | Not-determined |
| S8_extracted_k141_141414 | Low-quality |
| S8_extracted_k141_10983 | Not-determined |
| S8_extracted_k141_45642 | Not-determined |
| S8_extracted_k141_175995 | Not-determined |
| S8_extracted_k141_158325 | Low-quality |
| S8_extracted_k141_106682 | Not-determined |
| S8_extracted_k141_141420 | Not-determined |
| S8_extracted_k141_175997 | Not-determined |
| S8_extracted_k141_141421 | Not-determined |
| S8_extracted_k141_28884 | Not-determined |
| S8_extracted_k141_184193 | Not-determined |
| S8_extracted_k141_141424 | Not-determined |
| S8_extracted_k141_115231 | Not-determined |
| S8_extracted_k141_184198 | Not-determined |
| S8_extracted_k141_175998 | Low-quality |
| S8_extracted_k141_19851 | Not-determined |
| S8_extracted_k141_45650 | Not-determined |
| S8_extracted_k141_106684 | Not-determined |
| S8_extracted_k141_193677 | Not-determined |
| S8_extracted_k141_106685 | Not-determined |
| S8_extracted_k141_71478 | Not-determined |
| S8_extracted_k141_97848 | Not-determined |
| S8_extracted_k141_45653 | Low-quality |
| S8_extracted_k141_45654 | Not-determined |
| S8_extracted_k141_19858 | Not-determined |
| S8_extracted_k141_45660 | Not-determined |
| S8_extracted_k141_106688 | Not-determined |
| S8_extracted_k141_45661 | Not-determined |
| S8_extracted_k141_63358 | Low-quality |
| S8_extracted_k141_97855 | Not-determined |
| S8_extracted_k141_141440 | Not-determined |
| S8_extracted_k141_45665 | Not-determined |
| S8_extracted_k141_28894 | Not-determined |
| S8_extracted_k141_141442 | Not-determined |
| S8_extracted_k141_176011 | Not-determined |
| S8_extracted_k141_97858 | Not-determined |
| S8_extracted_k141_176012 | Low-quality |
| S8_extracted_k141_141444 | Not-determined |
| S8_extracted_k141_19870 | Not-determined |
| S8_extracted_k141_193688 | Not-determined |
| S8_extracted_k141_28896 | Not-determined |
| S8_extracted_k141_193689 | Not-determined |
| S8_extracted_k141_71491 | Not-determined |
| S8_extracted_k141_158332 | Not-determined |
| S8_extracted_k141_115249 | Not-determined |
| S8_extracted_k141_19872 | Not-determined |
| S8_extracted_k141_124171 | Not-determined |
| S8_extracted_k141_184209 | Low-quality |
| S8_extracted_k141_149931 | Not-determined |
| S8_extracted_k141_184212 | Not-determined |
| S8_extracted_k141_71496 | Not-determined |
| S8_extracted_k141_45673 | Not-determined |
| S8_extracted_k141_106700 | Not-determined |
| S8_extracted_k141_71500 | Not-determined |
| S8_extracted_k141_63367 | Not-determined |
| S8_extracted_k141_19879 | Low-quality |
| S8_extracted_k141_19880 | Not-determined |
| S8_extracted_k141_167351 | Not-determined |
| S8_extracted_k141_141460 | Not-determined |
| S8_extracted_k141_71503 | Not-determined |
| S8_extracted_k141_158341 | Not-determined |
| S8_extracted_k141_28907 | Not-determined |
| S8_extracted_k141_97870 | Not-determined |
| S8_extracted_k141_45685 | Not-determined |
| S8_extracted_k141_124185 | Not-determined |
| S8_extracted_k141_167357 | Not-determined |
| S8_extracted_k141_184230 | Low-quality |
| S8_extracted_k141_124188 | Low-quality |
| S8_extracted_k141_124189 | Not-determined |
| S8_extracted_k141_141474 | Not-determined |
| S8_extracted_k141_2629 | Not-determined |
| S8_extracted_k141_45693 | Not-determined |
| S8_extracted_k141_63379 | Not-determined |
| S8_extracted_k141_115259 | Not-determined |
| S8_extracted_k141_124191 | Low-quality |
| S8_extracted_k141_19888 | Not-determined |
| S8_extracted_k141_158346 | Not-determined |
| S8_extracted_k141_184236 | Not-determined |
| S8_extracted_k141_89287 | Not-determined |
| S8_extracted_k141_19889 | Not-determined |
| S8_extracted_k141_184238 | Not-determined |
| S8_extracted_k141_2635 | Not-determined |
| S8_extracted_k141_141483 | Not-determined |
| S8_extracted_k141_184241 | Not-determined |
| S8_extracted_k141_167367 | Not-determined |
| S8_extracted_k141_193719 | Not-determined |
| S8_extracted_k141_124198 | Not-determined |
| S8_extracted_k141_158353 | Not-determined |
| S8_extracted_k141_63393 | Not-determined |
| S8_extracted_k141_115261 | Not-determined |
| S8_extracted_k141_89297 | Not-determined |
| S8_extracted_k141_141486 | Not-determined |
| S8_extracted_k141_193723 | Not-determined |
| S8_extracted_k141_63401 | Not-determined |
| S8_extracted_k141_45701 | Not-determined |
| S8_extracted_k141_89298 | Not-determined |
| S8_extracted_k141_89302 | Not-determined |
| S8_extracted_k141_54695 | Not-determined |
| S8_extracted_k141_71512 | Not-determined |
| S8_extracted_k141_158360 | Not-determined |
| S8_extracted_k141_2640 | Not-determined |
| S8_extracted_k141_106723 | Not-determined |
| S8_extracted_k141_193732 | Not-determined |
| S8_extracted_k141_132613 | Not-determined |
| S8_extracted_k141_97888 | Not-determined |
| S8_extracted_k141_63408 | Not-determined |
| S8_extracted_k141_167373 | Not-determined |
| S8_extracted_k141_115264 | Not-determined |
| S8_extracted_k141_89304 | Not-determined |
| S8_extracted_k141_54702 | Low-quality |
| S8_extracted_k141_106727 | Not-determined |
| S8_extracted_k141_2643 | Not-determined |
| S8_extracted_k141_124209 | Not-determined |
| S8_extracted_k141_167377 | Not-determined |
| S8_extracted_k141_193736 | Not-determined |
| S8_extracted_k141_115270 | Not-determined |
| S8_extracted_k141_37412 | Not-determined |
| S8_extracted_k141_176070 | Not-determined |
| S8_extracted_k141_115273 | Not-determined |
| S8_extracted_k141_158370 | Low-quality |
| S8_extracted_k141_2651 | Not-determined |
| S8_extracted_k141_132617 | Not-determined |
| S8_extracted_k141_124213 | Not-determined |
| S8_extracted_k141_141498 | Low-quality |
| S8_extracted_k141_141499 | Not-determined |
| S8_extracted_k141_37422 | Not-determined |
| S8_extracted_k141_124217 | Not-determined |
| S8_extracted_k141_167389 | Not-determined |
| S8_extracted_k141_97893 | Not-determined |
| S8_extracted_k141_149966 | Low-quality |
| S8_extracted_k141_124221 | Not-determined |
| S8_extracted_k141_71522 | Not-determined |
| S8_extracted_k141_141505 | Not-determined |
| S8_extracted_k141_2659 | Not-determined |
| S8_extracted_k141_115282 | Not-determined |
| S8_extracted_k141_45720 | Not-determined |
| S8_extracted_k141_176084 | Not-determined |
| S8_extracted_k141_80757 | Low-quality |
| S8_extracted_k141_45721 | Not-determined |
| S8_extracted_k141_193752 | Not-determined |
| S8_extracted_k141_2662 | Not-determined |
| S8_extracted_k141_115285 | Not-determined |
| S8_extracted_k141_167395 | Not-determined |
| S8_extracted_k141_63430 | Not-determined |
| S8_extracted_k141_115286 | Low-quality |
| S8_extracted_k141_45725 | Not-determined |
| S8_extracted_k141_37440 | Not-determined |
| S8_extracted_k141_132624 | Not-determined |
| S8_extracted_k141_2664 | Not-determined |
| S8_extracted_k141_176093 | Not-determined |
| S8_extracted_k141_45726 | Not-determined |
| S8_extracted_k141_45728 | Not-determined |
| S8_extracted_k141_89329 | Not-determined |
| S8_extracted_k141_71536 | Low-quality |
| S8_extracted_k141_10994 | Not-determined |
| S8_extracted_k141_193757 | Low-quality |
| S8_extracted_k141_106742 | Not-determined |
| S8_extracted_k141_106743 | Not-determined |
| S8_extracted_k141_176098 | Not-determined |
| S8_extracted_k141_63442 | Not-determined |
| S8_extracted_k141_141512 | Not-determined |
| S8_extracted_k141_45730 | Low-quality |
| S8_extracted_k141_63443 | Low-quality |
| S8_extracted_k141_132635 | Not-determined |
| S8_extracted_k141_124235 | Not-determined |
| S8_extracted_k141_63444 | Not-determined |
| S8_extracted_k141_124238 | Not-determined |
| S8_extracted_k141_63445 | Not-determined |
| S8_extracted_k141_97911 | Not-determined |
| S8_extracted_k141_63446 | Not-determined |
| S8_extracted_k141_193765 | Low-quality |
| S8_extracted_k141_124240 | Not-determined |
| S8_extracted_k141_97913 | Not-determined |
| S8_extracted_k141_193766 | Low-quality |
| S8_extracted_k141_132644 | Not-determined |
| S8_extracted_k141_149983 | Not-determined |
| S8_extracted_k141_176108 | Not-determined |
| S8_extracted_k141_11002 | Not-determined |
| S8_extracted_k141_201763 | Not-determined |
| S8_extracted_k141_97921 | Not-determined |
| S8_extracted_k141_106759 | Low-quality |
| S8_extracted_k141_19903 | Not-determined |
| S8_extracted_k141_106761 | Not-determined |
| S8_extracted_k141_176117 | Not-determined |
| S8_extracted_k141_141530 | Not-determined |
| S8_extracted_k141_45748 | Not-determined |
| S8_extracted_k141_124251 | Low-quality |
| S8_extracted_k141_45749 | Not-determined |
| S8_extracted_k141_2671 | Not-determined |
| S8_extracted_k141_141533 | Not-determined |
| S8_extracted_k141_89344 | Low-quality |
| S8_extracted_k141_80784 | Not-determined |
| S8_extracted_k141_124256 | Not-determined |
| S8_extracted_k141_149988 | Not-determined |
| S8_extracted_k141_176121 | Not-determined |
| S8_extracted_k141_71554 | Not-determined |
| S8_extracted_k141_2677 | Not-determined |
| S8_extracted_k141_97923 | Low-quality |
| S8_extracted_k141_28921 | Not-determined |
| S8_extracted_k141_132663 | Not-determined |
| S8_extracted_k141_141535 | Not-determined |
| S8_extracted_k141_176126 | Not-determined |
| S8_extracted_k141_97925 | Not-determined |
| S8_extracted_k141_71561 | Not-determined |
| S8_extracted_k141_2682 | Low-quality |
| S8_extracted_k141_106768 | Not-determined |
| S8_extracted_k141_124261 | Not-determined |
| S8_extracted_k141_89351 | Not-determined |
| S8_extracted_k141_149999 | Not-determined |
| S8_extracted_k141_184265 | Not-determined |
| S8_extracted_k141_28929 | Not-determined |
| S8_extracted_k141_71562 | Not-determined |
| S8_extracted_k141_176132 | Not-determined |
| S8_extracted_k141_28931 | Not-determined |
| S8_extracted_k141_63473 | Not-determined |
| S8_extracted_k141_2688 | Not-determined |
| S8_extracted_k141_132668 | Not-determined |
| S8_extracted_k141_176136 | Not-determined |
| S8_extracted_k141_45773 | Not-determined |
| S8_extracted_k141_184271 | Not-determined |
| S8_extracted_k141_167431 | Not-determined |
| S8_extracted_k141_11019 | Not-determined |
| S8_extracted_k141_54732 | Not-determined |
| S8_extracted_k141_158411 | Not-determined |
| S8_extracted_k141_158412 | Not-determined |
| S8_extracted_k141_106774 | Not-determined |
| S8_extracted_k141_28937 | Not-determined |
| S8_extracted_k141_201786 | Not-determined |
| S8_extracted_k141_2696 | Not-determined |
| S8_extracted_k141_63486 | Not-determined |
| S8_extracted_k141_106778 | Low-quality |
| S8_extracted_k141_167433 | Not-determined |
| S8_extracted_k141_201792 | Not-determined |
| S8_extracted_k141_201793 | Not-determined |
| S8_extracted_k141_19932 | Not-determined |
| S8_extracted_k141_97941 | Low-quality |
| S8_extracted_k141_184275 | Not-determined |
| S8_extracted_k141_184276 | Not-determined |
| S8_extracted_k141_158421 | Not-determined |
| S8_extracted_k141_89358 | Not-determined |
| S8_extracted_k141_115319 | Not-determined |
| S8_extracted_k141_80811 | Not-determined |
| S8_extracted_k141_201797 | Low-quality |
| S8_extracted_k141_176148 | Not-determined |
| S8_extracted_k141_11027 | Not-determined |
| S8_extracted_k141_63496 | Not-determined |
| S8_extracted_k141_124279 | Not-determined |
| S8_extracted_k141_176151 | Not-determined |
| S8_extracted_k141_28949 | Not-determined |
| S8_extracted_k141_176156 | Not-determined |
| S8_extracted_k141_89361 | Not-determined |
| S8_extracted_k141_150013 | Not-determined |
| S8_extracted_k141_193799 | Not-determined |
| S8_extracted_k141_201805 | Not-determined |
| S8_extracted_k141_150015 | Not-determined |
| S8_extracted_k141_2710 | Not-determined |
| S8_extracted_k141_167452 | Low-quality |
| S8_extracted_k141_150020 | Not-determined |
| S8_extracted_k141_150021 | Not-determined |
| S8_extracted_k141_71564 | Not-determined |
| S8_extracted_k141_167459 | Not-determined |
| S8_extracted_k141_37469 | Not-determined |
| S8_extracted_k141_201815 | Low-quality |
| S8_extracted_k141_193807 | Not-determined |
| S8_extracted_k141_132682 | Not-determined |
| S8_extracted_k141_124291 | Not-determined |
| S8_extracted_k141_158436 | Not-determined |
| S8_extracted_k141_11046 | Not-determined |
| S8_extracted_k141_106792 | Not-determined |
| S8_extracted_k141_141541 | Not-determined |
| S8_extracted_k141_89371 | Not-determined |
| S8_extracted_k141_124295 | Not-determined |
| S8_extracted_k141_89373 | Not-determined |
| S8_extracted_k141_167465 | Not-determined |
| S8_extracted_k141_19954 | Not-determined |
| S8_extracted_k141_132685 | Not-determined |
| S8_extracted_k141_37476 | Not-determined |
| S8_extracted_k141_71577 | Not-determined |
| S8_extracted_k141_19955 | Not-determined |
| S8_extracted_k141_132686 | Not-determined |
| S8_extracted_k141_63502 | Not-determined |
| S8_extracted_k141_71579 | Not-determined |
| S8_extracted_k141_63503 | Not-determined |
| S8_extracted_k141_2721 | Not-determined |
| S8_extracted_k141_132690 | Not-determined |
| S8_extracted_k141_63504 | Not-determined |
| S8_extracted_k141_150031 | Not-determined |
| S8_extracted_k141_167473 | Not-determined |
| S8_extracted_k141_106799 | Not-determined |
| S8_extracted_k141_158448 | Not-determined |
| S8_extracted_k141_158449 | Not-determined |
| S8_extracted_k141_19963 | Not-determined |
| S8_extracted_k141_54769 | Not-determined |
| S8_extracted_k141_37484 | Not-determined |
| S8_extracted_k141_54771 | Not-determined |
| S8_extracted_k141_167477 | Not-determined |
| S8_extracted_k141_115323 | Not-determined |
| S8_extracted_k141_2726 | Not-determined |
| S8_extracted_k141_19968 | Not-determined |
| S8_extracted_k141_115324 | Not-determined |
| S8_extracted_k141_19969 | Not-determined |
| S8_extracted_k141_115325 | Not-determined |
| S8_extracted_k141_141554 | Not-determined |
| S8_extracted_k141_97952 | Low-quality |
| S8_extracted_k141_80833 | Not-determined |
| S8_extracted_k141_124313 | Not-determined |
| S8_extracted_k141_97953 | Not-determined |
| S8_extracted_k141_124314 | Not-determined |
| S8_extracted_k141_97955 | Not-determined |
| S8_extracted_k141_124316 | Not-determined |
| S8_extracted_k141_11062 | Not-determined |
| S8_extracted_k141_167493 | Not-determined |
| S8_extracted_k141_37490 | Not-determined |
| S8_extracted_k141_37493 | Not-determined |
| S8_extracted_k141_176165 | Not-determined |
| S8_extracted_k141_2738 | Not-determined |
| S8_extracted_k141_124318 | Not-determined |
| S8_extracted_k141_176168 | Not-determined |
| S8_extracted_k141_176169 | Not-determined |
| S8_extracted_k141_11067 | Not-determined |
| S13_extracted_28309 | Medium-quality |
| S8_extracted_k141_89393 | Not-determined |
| S8_extracted_k141_2744 | Not-determined |
| S8_extracted_k141_63516 | Not-determined |
| S8_extracted_k141_54785 | Not-determined |
| S8_extracted_k141_97974 | Not-determined |
| S8_extracted_k141_124321 | Not-determined |
| S8_extracted_k141_201848 | Not-determined |
| S8_extracted_k141_2747 | Not-determined |
| S8_extracted_k141_54787 | Low-quality |
| S8_extracted_k141_19995 | Not-determined |
| S8_extracted_k141_89397 | Not-determined |
| S8_extracted_k141_11071 | Not-determined |
| S8_extracted_k141_54789 | Not-determined |
| S8_extracted_k141_54790 | Not-determined |
| S8_extracted_k141_97981 | Not-determined |
| S8_extracted_k141_19999 | Not-determined |
| S8_extracted_k141_176175 | Not-determined |
| S8_extracted_k141_80840 | Not-determined |
| S8_extracted_k141_37508 | Not-determined |
| S8_extracted_k141_89407 | Not-determined |
| S8_extracted_k141_37511 | Not-determined |
| S8_extracted_k141_115340 | Not-determined |
| S8_extracted_k141_2761 | Low-quality |
| S8_extracted_k141_115341 | Not-determined |
| S8_extracted_k141_80852 | Not-determined |
| S8_extracted_k141_80853 | Low-quality |
| S8_extracted_k141_124336 | Not-determined |
| S8_extracted_k141_201857 | Not-determined |
| S8_extracted_k141_106818 | Low-quality |
| S8_extracted_k141_201858 | Not-determined |
| S8_extracted_k141_63528 | Not-determined |
| S8_extracted_k141_115345 | Not-determined |
| S8_extracted_k141_63529 | Not-determined |
| S8_extracted_k141_37515 | Not-determined |
| S8_extracted_k141_115347 | Not-determined |
| S8_extracted_k141_2769 | Not-determined |
| S8_extracted_k141_124339 | Low-quality |
| S8_extracted_k141_63533 | Not-determined |
| S8_extracted_k141_89416 | Not-determined |
| S8_extracted_k141_124341 | Low-quality |
| S8_extracted_k141_80858 | Not-determined |
| S8_extracted_k141_193822 | Not-determined |
| S8_extracted_k141_124342 | Not-determined |
| S8_extracted_k141_115348 | Not-determined |
| S8_extracted_k141_124345 | Not-determined |
| S8_extracted_k141_193826 | Not-determined |
| S8_extracted_k141_45783 | Not-determined |
| S12_vs_k141_42408\|\|full | Medium-quality |
| S8_extracted_k141_184295 | Low-quality |
| S8_extracted_k141_45787 | Not-determined |
| S8_extracted_k141_28958 | Not-determined |
| S8_extracted_k141_115352 | Not-determined |
| S8_extracted_k141_45790 | Not-determined |
| S8_extracted_k141_115353 | Not-determined |
| S8_extracted_k141_89429 | Not-determined |
| S8_extracted_k141_20008 | Not-determined |
| S8_extracted_k141_89431 | Not-determined |
| S8_extracted_k141_28963 | Not-determined |
| S8_extracted_k141_80860 | Low-quality |
| S8_extracted_k141_184303 | Not-determined |
| S8_extracted_k141_11097 | Not-determined |
| S8_extracted_k141_124351 | Not-determined |
| S8_extracted_k141_63546 | Not-determined |
| S8_extracted_k141_45797 | Not-determined |
| S8_extracted_k141_184307 | Not-determined |
| S8_extracted_k141_89438 | Not-determined |
| S8_extracted_k141_28969 | Not-determined |
| S8_extracted_k141_106831 | Not-determined |
| S8_extracted_k141_89440 | Not-determined |
| S8_extracted_k141_106832 | Not-determined |
| S8_extracted_k141_167520 | Not-determined |
| S8_extracted_k141_89444 | Low-quality |
| S8_extracted_k141_176190 | Not-determined |
| S8_extracted_k141_115364 | Not-determined |
| S8_extracted_k141_124354 | Low-quality |
| S8_extracted_k141_167522 | Not-determined |
| S8_extracted_k141_124355 | Not-determined |
| S8_extracted_k141_89447 | Low-quality |
| S8_extracted_k141_20015 | Not-determined |
| S8_extracted_k141_20016 | Not-determined |
| S8_extracted_k141_176195 | Not-determined |
| S8_extracted_k141_115368 | Not-determined |
| S8_extracted_k141_45803 | Not-determined |
| S8_extracted_k141_158471 | Not-determined |
| S8_extracted_k141_115371 | Not-determined |
| S8_extracted_k141_124361 | Not-determined |
| S8_extracted_k141_106846 | Not-determined |
| S8_extracted_k141_193840 | Not-determined |
| S8_extracted_k141_11113 | Not-determined |
| S8_extracted_k141_158473 | Not-determined |
| S8_extracted_k141_124362 | Not-determined |
| S8_extracted_k141_98016 | Not-determined |
| S8_extracted_k141_193841 | Not-determined |
| S8_extracted_k141_11115 | Not-determined |
| S8_extracted_k141_201888 | Not-determined |
| S8_extracted_k141_167526 | Not-determined |
| S8_extracted_k141_89457 | Not-determined |
| S8_extracted_k141_63558 | Not-determined |
| S8_extracted_k141_89458 | Not-determined |
| S8_extracted_k141_124370 | Not-determined |
| S8_extracted_k141_28985 | Not-determined |
| S8_extracted_k141_184325 | Not-determined |
| S8_extracted_k141_11123 | Not-determined |
| S8_extracted_k141_28987 | Low-quality |
| S8_extracted_k141_80892 | Not-determined |
| S8_extracted_k141_20027 | Not-determined |
| S8_extracted_k141_176207 | Not-determined |
| S8_extracted_k141_150048 | Not-determined |
| S8_extracted_k141_167532 | Low-quality |
| S8_extracted_k141_11126 | Not-determined |
| S8_extracted_k141_28989 | Not-determined |
| S8_extracted_k141_176210 | Not-determined |
| S8_extracted_k141_176213 | Not-determined |
| S8_extracted_k141_184332 | Not-determined |
| S8_extracted_k141_98030 | Not-determined |
| S8_extracted_k141_176216 | Not-determined |
| S8_extracted_k141_106867 | Not-determined |
| S8_extracted_k141_89466 | Not-determined |
| S8_extracted_k141_176217 | Not-determined |
| S8_extracted_k141_89468 | Low-quality |
| S8_extracted_k141_28996 | Not-determined |
| S8_extracted_k141_158490 | Not-determined |
| S8_extracted_k141_11137 | Not-determined |
| S8_extracted_k141_80902 | Not-determined |
| S8_extracted_k141_80903 | Not-determined |
| S8_extracted_k141_29001 | Not-determined |
| S8_extracted_k141_37518 | Low-quality |
| S8_extracted_k141_184344 | Not-determined |
| S8_extracted_k141_98036 | Not-determined |
| S8_extracted_k141_201904 | Not-determined |
| S8_extracted_k141_80910 | Not-determined |
| S8_extracted_k141_115388 | Not-determined |
| S8_extracted_k141_201905 | Not-determined |
| S8_extracted_k141_29004 | Not-determined |
| S8_extracted_k141_29005 | Not-determined |
| S8_extracted_k141_29011 | Not-determined |
| S8_extracted_k141_150060 | Not-determined |
| S8_extracted_k141_63570 | Not-determined |
| S8_extracted_k141_80918 | Low-quality |
| S8_extracted_k141_167547 | Not-determined |
| S8_extracted_k141_124374 | Not-determined |
| S9_vs_k141_70236\|\|full | Medium-quality |
| S8_extracted_k141_158500 | Not-determined |
| S8_extracted_k141_167548 | Not-determined |
| S8_extracted_k141_54823 | Low-quality |
| S8_extracted_k141_124376 | Not-determined |
| S8_extracted_k141_54824 | Not-determined |
| S8_extracted_k141_80921 | Not-determined |
| S8_extracted_k141_71591 | Not-determined |
| S8_extracted_k141_176232 | Not-determined |
| S8_extracted_k141_150071 | Low-quality |
| S8_extracted_k141_106881 | Not-determined |
| S8_extracted_k141_150072 | Not-determined |
| S8_extracted_k141_201911 | Not-determined |
| S8_extracted_k141_37527 | Not-determined |
| S8_extracted_k141_80924 | Not-determined |
| S8_extracted_k141_80925 | Not-determined |
| S8_extracted_k141_89497 | Not-determined |
| S8_extracted_k141_167555 | Not-determined |
| S8_extracted_k141_89500 | Not-determined |
| S8_extracted_k141_115407 | Not-determined |
| S8_extracted_k141_184359 | Not-determined |
| S8_extracted_k141_201917 | Low-quality |
| S8_extracted_k141_115410 | Not-determined |
| S8_extracted_k141_54829 | Not-determined |
| S8_extracted_k141_11153 | Not-determined |
| S8_extracted_k141_98049 | Not-determined |
| S8_extracted_k141_115412 | Not-determined |
| S8_extracted_k141_45825 | Not-determined |

| | |
|---|---|
| S8_extracted_k141_106889 | Not-determined |
| S8_extracted_k141_45826 | Not-determined |
| S8_extracted_k141_184369 | Not-determined |
| S8_extracted_k141_2786 | Not-determined |
| S8_extracted_k141_63596 | Low-quality |
| S8_extracted_k141_45830 | Low-quality |
| S8_extracted_k141_54840 | Not-determined |
| S8_extracted_k141_124391 | Not-determined |
| S8_extracted_k141_158510 | Not-determined |
| S8_extracted_k141_11162 | Not-determined |
| S8_extracted_k141_71610 | Low-quality |
| S8_extracted_k141_184377 | Not-determined |
| S8_extracted_k141_201929 | Not-determined |
| S8_extracted_k141_158513 | Not-determined |
| S8_extracted_k141_63599 | Low-quality |
| S8_extracted_k141_45838 | Not-determined |
| S8_extracted_k141_193844 | Not-determined |
| S8_extracted_k141_89512 | Not-determined |
| S8_extracted_k141_167580 | Not-determined |
| S8_extracted_k141_63604 | Not-determined |
| S8_extracted_k141_124403 | Not-determined |
| S8_extracted_k141_71618 | Not-determined |
| S8_extracted_k141_158516 | Not-determined |
| S8_extracted_k141_45843 | Not-determined |
| S8_extracted_k141_167583 | Not-determined |
| S8_extracted_k141_106899 | Not-determined |
| S8_extracted_k141_37555 | Not-determined |
| S8_extracted_k141_158525 | Not-determined |
| S8_extracted_k141_80953 | Not-determined |
| S8_extracted_k141_167590 | Not-determined |
| S8_extracted_k141_54850 | Not-determined |
| S8_extracted_k141_98065 | Not-determined |
| S8_extracted_k141_141563 | Not-determined |
| S8_extracted_k141_141565 | Not-determined |
| S8_extracted_k141_80954 | Not-determined |
| S8_extracted_k141_141567 | Not-determined |
| S8_extracted_k141_80955 | Not-determined |
| S8_extracted_k141_132720 | Not-determined |
| S8_extracted_k141_20076 | Not-determined |
| S8_extracted_k141_106907 | Not-determined |
| S8_extracted_k141_45848 | Low-quality |
| S8_extracted_k141_89520 | Not-determined |
| S8_extracted_k141_71630 | Not-determined |
| S8_extracted_k141_158530 | Not-determined |
| S8_extracted_k141_63618 | Low-quality |
| S8_extracted_k141_158531 | Not-determined |
| S8_extracted_k141_184390 | Not-determined |
| S8_extracted_k141_167606 | Not-determined |
| S8_extracted_k141_158532 | Not-determined |
| S8_extracted_k141_115438 | Not-determined |
| S8_extracted_k141_141575 | Not-determined |
| S8_extracted_k141_98076 | Not-determined |
| S8_extracted_k141_106917 | Not-determined |
| S8_extracted_k141_89527 | Not-determined |
| S8_extracted_k141_106918 | Not-determined |
| S8_extracted_k141_20088 | Not-determined |
| S8_extracted_k141_115439 | Not-determined |
| S8_extracted_k141_71636 | Low-quality |
| S8_extracted_k141_20091 | Not-determined |
| S8_extracted_k141_54862 | Not-determined |
| S8_extracted_k141_176274 | Not-determined |
| S8_extracted_k141_115441 | Low-quality |
| S8_extracted_k141_63621 | Low-quality |
| S8_extracted_k141_141585 | Not-determined |
| S8_extracted_k141_54865 | Not-determined |
| S8_extracted_k141_141588 | Low-quality |
| S8_extracted_k141_54866 | Low-quality |
| S8_extracted_k141_89528 | Not-determined |
| S8_extracted_k141_106925 | Not-determined |
| S8_extracted_k141_115452 | Not-determined |
| S8_extracted_k141_11178 | Not-determined |
| S8_extracted_k141_45868 | Not-determined |
| S8_extracted_k141_167623 | Low-quality |
| S8_extracted_k141_45869 | Not-determined |
| S8_extracted_k141_98089 | Not-determined |
| S8_extracted_k141_132736 | Low-quality |
| S8_extracted_k141_158552 | Not-determined |
| S8_extracted_k141_11180 | Not-determined |
| S8_extracted_k141_98091 | Not-determined |
| S8_extracted_k141_158554 | Low-quality |
| S8_extracted_k141_98093 | Not-determined |
| S8_extracted_k141_106930 | Low-quality |
| S8_extracted_k141_45873 | Not-determined |
| S8_extracted_k141_63628 | Low-quality |
| S8_extracted_k141_184399 | Not-determined |
| S8_extracted_k141_98099 | Not-determined |
| S8_extracted_k141_106938 | Not-determined |
| S8_extracted_k141_158563 | Not-determined |
| S8_extracted_k141_63637 | Not-determined |
| S8_extracted_k141_124407 | Not-determined |
| S8_extracted_k141_54876 | Not-determined |
| S8_extracted_k141_201948 | Low-quality |
| S8_extracted_k141_176289 | Not-determined |
| S8_extracted_k141_124410 | Not-determined |
| S8_extracted_k141_54877 | Low-quality |
| S8_extracted_k141_54878 | Not-determined |
| S8_extracted_k141_141608 | Not-determined |
| S8_extracted_k141_132749 | Not-determined |
| S8_extracted_k141_98113 | Not-determined |
| S8_extracted_k141_124414 | Not-determined |
| S8_extracted_k141_201950 | Not-determined |
| S8_extracted_k141_20111 | Not-determined |
| S8_extracted_k141_132758 | Low-quality |
| S8_extracted_k141_98119 | Not-determined |
| S8_extracted_k141_115473 | Not-determined |
| S8_extracted_k141_184416 | Low-quality |
| S8_extracted_k141_54880 | Not-determined |
| S8_extracted_k141_37580 | Not-determined |
| S8_extracted_k141_80969 | Not-determined |
| S8_extracted_k141_54881 | Not-determined |
| S8_extracted_k141_115476 | Not-determined |
| S8_extracted_k141_63642 | Not-determined |
| S8_extracted_k141_115478 | Not-determined |
| S8_extracted_k141_98125 | Not-determined |
| S8_extracted_k141_201960 | Not-determined |
| S8_extracted_k141_158572 | Not-determined |
| S8_extracted_k141_20118 | Not-determined |
| S8_extracted_k141_45890 | Not-determined |
| S8_extracted_k141_54888 | Not-determined |
| S8_extracted_k141_141619 | Not-determined |
| S8_extracted_k141_132766 | Low-quality |
| S8_extracted_k141_54889 | Not-determined |
| S8_extracted_k141_54890 | Not-determined |
| S8_extracted_k141_124422 | Not-determined |
| S8_extracted_k141_45892 | Not-determined |
| S8_extracted_k141_141622 | Not-determined |
| S8_extracted_k141_54891 | Low-quality |
| S8_extracted_k141_54892 | Not-determined |
| S8_extracted_k141_115480 | Low-quality |
| S8_extracted_k141_141624 | Not-determined |
| S8_extracted_k141_80976 | Not-determined |
| S8_extracted_k141_184431 | Not-determined |
| S8_extracted_k141_54896 | Not-determined |
| S8_extracted_k141_158586 | Not-determined |
| S8_extracted_k141_20128 | Not-determined |
| S8_extracted_k141_71643 | Not-determined |
| S8_extracted_k141_141628 | Not-determined |
| S8_extracted_k141_176317 | Not-determined |
| S8_extracted_k141_80979 | Not-determined |
| S8_extracted_k141_158587 | Not-determined |
| S8_extracted_k141_176320 | Not-determined |
| S8_extracted_k141_89533 | Not-determined |
| S8_extracted_k141_54898 | Not-determined |
| S8_extracted_k141_37586 | Not-determined |
| S8_extracted_k141_98137 | Not-determined |
| S8_extracted_k141_45899 | Not-determined |
| S8_extracted_k141_20132 | Low-quality |
| S8_extracted_k141_71651 | Not-determined |
| S8_extracted_k141_63659 | Not-determined |
| S8_extracted_k141_115495 | Not-determined |
| S8_extracted_k141_141631 | Not-determined |
| S8_extracted_k141_37591 | Not-determined |
| S8_extracted_k141_176331 | Not-determined |
| S8_extracted_k141_184437 | Not-determined |
| S8_extracted_k141_150145 | Not-determined |
| S8_extracted_k141_80987 | Not-determined |
| S8_extracted_k141_37595 | Not-determined |
| S8_extracted_k141_150148 | Not-determined |
| S8_extracted_k141_115502 | Not-determined |
| S8_extracted_k141_80991 | Not-determined |
| S8_extracted_k141_37598 | Not-determined |
| S8_extracted_k141_54907 | Low-quality |
| S8_extracted_k141_63663 | Not-determined |
| S8_extracted_k141_124428 | Not-determined |
| S8_extracted_k141_54909 | Low-quality |
| S8_extracted_k141_176338 | Low-quality |
| S8_extracted_k141_89545 | Not-determined |
| S8_extracted_k141_115510 | Not-determined |
| S8_extracted_k141_115512 | Low-quality |
| S8_extracted_k141_45920 | Not-determined |
| S8_extracted_k141_20136 | Not-determined |
| S8_extracted_k141_98147 | Not-determined |
| S8_extracted_k141_89549 | Not-determined |
| S8_extracted_k141_115514 | Not-determined |
| S8_extracted_k141_98151 | Not-determined |
| S8_extracted_k141_193854 | Not-determined |
| S8_extracted_k141_201967 | Low-quality |
| S8_extracted_k141_158603 | Not-determined |
| S8_extracted_k141_167643 | Not-determined |
| S8_extracted_k141_124435 | Low-quality |
| S8_extracted_k141_115521 | Not-determined |
| S8_extracted_k141_71674 | Not-determined |
| S8_extracted_k141_158607 | Low-quality |
| S8_extracted_k141_81003 | Low-quality |
| S8_extracted_k141_63677 | Not-determined |
| S8_extracted_k141_98156 | Not-determined |
| S8_extracted_k141_45927 | Not-determined |
| S8_extracted_k141_63679 | Not-determined |
| S8_extracted_k141_63680 | Not-determined |
| S8_extracted_k141_71679 | Not-determined |
| S8_extracted_k141_81008 | Not-determined |
| S8_extracted_k141_45932 | Not-determined |
| S8_extracted_k141_167654 | Not-determined |
| S8_extracted_k141_158617 | Not-determined |
| S8_extracted_k141_20148 | Not-determined |
| S8_extracted_k141_81016 | Not-determined |
| S8_extracted_k141_184456 | Not-determined |
| S8_extracted_k141_63685 | Not-determined |
| S8_extracted_k141_71690 | Not-determined |
| S8_extracted_k141_115536 | Not-determined |
| S8_extracted_k141_201984 | Not-determined |
| S8_extracted_k141_63687 | Not-determined |
| S8_extracted_k141_45938 | Not-determined |
| S8_extracted_k141_63690 | Not-determined |
| S12_vs_k141_176437\|\|full | Medium-quality |
| S8_extracted_k141_37609 | Not-determined |
| S8_extracted_k141_11192 | Not-determined |
| S8_extracted_k141_201987 | Not-determined |
| S8_extracted_k141_124447 | Not-determined |
| S8_extracted_k141_98170 | Not-determined |
| S8_extracted_k141_20155 | Not-determined |
| S8_extracted_k141_54917 | Not-determined |
| S8_extracted_k141_81027 | Not-determined |
| S8_extracted_k141_81028 | Not-determined |
| S8_extracted_k141_89552 | Low-quality |
| S8_extracted_k141_71699 | Low-quality |
| S8_extracted_k141_54919 | Not-determined |
| S8_extracted_k141_115545 | Not-determined |
| S8_extracted_k141_45941 | Not-determined |
| S8_extracted_k141_54921 | Not-determined |
| S8_extracted_k141_193874 | Not-determined |
| S8_extracted_k141_193875 | Not-determined |
| S8_extracted_k141_54927 | Not-determined |
| S8_extracted_k141_63696 | Not-determined |
| S8_extracted_k141_167666 | Not-determined |
| S8_extracted_k141_63698 | Not-determined |
| S8_extracted_k141_202002 | Low-quality |
| S8_extracted_k141_124460 | Not-determined |
| S8_extracted_k141_37625 | Not-determined |
| S8_extracted_k141_176370 | Not-determined |
| S8_extracted_k141_98175 | Not-determined |
| S8_extracted_k141_54936 | Not-determined |
| S8_extracted_k141_202008 | Not-determined |
| S8_extracted_k141_98178 | Not-determined |
| S8_extracted_k141_124467 | Not-determined |
| S8_extracted_k141_81038 | Not-determined |
| S8_extracted_k141_45954 | Not-determined |
| S8_extracted_k141_176373 | Not-determined |
| S8_extracted_k141_202011 | Not-determined |
| S8_extracted_k141_193887 | Not-determined |
| S8_extracted_k141_63704 | Not-determined |
| S8_extracted_k141_193888 | Low-quality |
| S8_extracted_k141_98186 | Not-determined |
| S8_extracted_k141_45958 | Not-determined |
| S8_extracted_k141_89560 | Not-determined |
| S8_extracted_k141_115563 | Not-determined |
| S8_extracted_k141_98188 | Not-determined |
| S8_extracted_k141_45961 | Not-determined |
| S8_extracted_k141_11219 | Not-determined |
| S8_extracted_k141_54940 | Not-determined |
| S8_extracted_k141_63708 | Not-determined |
| S8_extracted_k141_98192 | Not-determined |
| S8_extracted_k141_132787 | Not-determined |
| S8_extracted_k141_2807 | Not-determined |
| S8_extracted_k141_98194 | Not-determined |
| S8_extracted_k141_193896 | Not-determined |
| S8_extracted_k141_37641 | Not-determined |
| S8_extracted_k141_202016 | Not-determined |
| S8_extracted_k141_176386 | Not-determined |
| S8_extracted_k141_106972 | Not-determined |
| S8_extracted_k141_63713 | Not-determined |
| S8_extracted_k141_54941 | Not-determined |
| S8_extracted_k141_141653 | Low-quality |
| S8_extracted_k141_124481 | Not-determined |
| S8_extracted_k141_141655 | Not-determined |
| S8_extracted_k141_81046 | Not-determined |
| S8_extracted_k141_202022 | Not-determined |
| S8_extracted_k141_81050 | Not-determined |
| S8_extracted_k141_158656 | Not-determined |
| S8_extracted_k141_115570 | Not-determined |
| S8_extracted_k141_20174 | Not-determined |
| S8_extracted_k141_98205 | Not-determined |
| S8_extracted_k141_81054 | Not-determined |
| S8_extracted_k141_11231 | Low-quality |
| S8_extracted_k141_45980 | Not-determined |
| S8_extracted_k141_81055 | Low-quality |
| S8_extracted_k141_20180 | Not-determined |
| S8_extracted_k141_11234 | Not-determined |
| S8_extracted_k141_106984 | Not-determined |
| S8_extracted_k141_106986 | Not-determined |
| S8_extracted_k141_20183 | Not-determined |
| S8_extracted_k141_106987 | Not-determined |
| S8_extracted_k141_98212 | Not-determined |
| S8_extracted_k141_176390 | Not-determined |
| S8_extracted_k141_141660 | Not-determined |
| S8_extracted_k141_193903 | Not-determined |
| S8_extracted_k141_106990 | Not-determined |
| S8_extracted_k141_141661 | Not-determined |
| S8_extracted_k141_98215 | Not-determined |
| S8_extracted_k141_115580 | Not-determined |
| S8_extracted_k141_124491 | Not-determined |
| S8_extracted_k141_132805 | Not-determined |
| S8_extracted_k141_89577 | Not-determined |
| S8_extracted_k141_89578 | Not-determined |
| S8_extracted_k141_98220 | Not-determined |
| S8_extracted_k141_54951 | Not-determined |
| S8_extracted_k141_98223 | Not-determined |
| S8_extracted_k141_132807 | Not-determined |
| S8_extracted_k141_81059 | Not-determined |
| S8_extracted_k141_176394 | Not-determined |
| S8_extracted_k141_141668 | Not-determined |
| S8_extracted_k141_106997 | Not-determined |
| S8_extracted_k141_150162 | Low-quality |
| S8_extracted_k141_54956 | Not-determined |
| S8_extracted_k141_11251 | Not-determined |
| S8_extracted_k141_107005 | Not-determined |
| S8_extracted_k141_98228 | Low-quality |
| S8_extracted_k141_98229 | Not-determined |
| S8_extracted_k141_37655 | Not-determined |
| S8_extracted_k141_98233 | Not-determined |
| S8_extracted_k141_176397 | Not-determined |
| S8_extracted_k141_115592 | Not-determined |
| S8_extracted_k141_81074 | Not-determined |
| S8_extracted_k141_141674 | Not-determined |
| S8_extracted_k141_98235 | Not-determined |
| S8_extracted_k141_132813 | Not-determined |
| S8_extracted_k141_11260 | Not-determined |
| S8_extracted_k141_81075 | Low-quality |
| S8_extracted_k141_150163 | Not-determined |
| S8_extracted_k141_176400 | Not-determined |
| S8_extracted_k141_81076 | Not-determined |
| S8_extracted_k141_11261 | Not-determined |
| S8_extracted_k141_89592 | Low-quality |
| S8_extracted_k141_132816 | Not-determined |
| S8_extracted_k141_124503 | Not-determined |
| S8_extracted_k141_158681 | Not-determined |
| S8_extracted_k141_115593 | Not-determined |
| S8_extracted_k141_132822 | Not-determined |
| S8_extracted_k141_202042 | Not-determined |
| S8_extracted_k141_81087 | Not-determined |
| S8_extracted_k141_115596 | Not-determined |
| S8_extracted_k141_20196 | Not-determined |
| S8_extracted_k141_193915 | Not-determined |
| S8_extracted_k141_45994 | Not-determined |
| S8_extracted_k141_167677 | Not-determined |
| S8_extracted_k141_193917 | Not-determined |
| S8_extracted_k141_124509 | Not-determined |
| S8_extracted_k141_11272 | Not-determined |
| S8_extracted_k141_45996 | Not-determined |
| S8_extracted_k141_115602 | Not-determined |
| S8_extracted_k141_202049 | Not-determined |
| S8_extracted_k141_107024 | Not-determined |
| S8_extracted_k141_193921 | Not-determined |
| S8_extracted_k141_20198 | Not-determined |
| S8_extracted_k141_132825 | Not-determined |
| S8_extracted_k141_2838 | Not-determined |
| S8_extracted_k141_20200 | Not-determined |
| S8_extracted_k141_132829 | Not-determined |
| S8_extracted_k141_150177 | Not-determined |
| S8_extracted_k141_11279 | Not-determined |
| S8_extracted_k141_54981 | Low-quality |
| S8_extracted_k141_176416 | Low-quality |
| S8_extracted_k141_20203 | Not-determined |
| S8_extracted_k141_115608 | Not-determined |
| S8_extracted_k141_54985 | Not-determined |
| S8_extracted_k141_71707 | Not-determined |
| S8_extracted_k141_115610 | Not-determined |
| S8_extracted_k141_20208 | Not-determined |
| S8_extracted_k141_115611 | Not-determined |
| S8_extracted_k141_115613 | Not-determined |
| S8_extracted_k141_54988 | Not-determined |
| S8_extracted_k141_89609 | Not-determined |
| S8_extracted_k141_176425 | Not-determined |
| S8_extracted_k141_37663 | Not-determined |
| S8_extracted_k141_202055 | Not-determined |
| S8_extracted_k141_150186 | Not-determined |
| S8_extracted_k141_167682 | Not-determined |
| S8_extracted_k141_202062 | Not-determined |
| S8_extracted_k141_37669 | Not-determined |
| S8_extracted_k141_81105 | Low-quality |
| S8_extracted_k141_63734 | Low-quality |
| S8_extracted_k141_20212 | Not-determined |
| S8_extracted_k141_37670 | Not-determined |
| S8_extracted_k141_124523 | Not-determined |
| S8_extracted_k141_2854 | Not-determined |
| S8_extracted_k141_29041 | Low-quality |
| S8_extracted_k141_29043 | Low-quality |
| S8_extracted_k141_167686 | Not-determined |
| S8_extracted_k141_46016 | Low-quality |
| S8_extracted_k141_63738 | Not-determined |
| S8_extracted_k141_132837 | Not-determined |
| S8_extracted_k141_20215 | Not-determined |
| S8_extracted_k141_54999 | Not-determined |
| S8_extracted_k141_107047 | Not-determined |
| S8_extracted_k141_2856 | Not-determined |
| S8_extracted_k141_20219 | Not-determined |
| S8_extracted_k141_107050 | Not-determined |
| S8_extracted_k141_202072 | Not-determined |
| S8_extracted_k141_37677 | Not-determined |
| S8_extracted_k141_20220 | Not-determined |
| S8_extracted_k141_98265 | Not-determined |
| S8_extracted_k141_29048 | Not-determined |
| S8_extracted_k141_55003 | Not-determined |
| S8_extracted_k141_167688 | Not-determined |
| S8_extracted_k141_98267 | Not-determined |
| S8_extracted_k141_63750 | Low-quality |
| S8_extracted_k141_202075 | Not-determined |
| S8_extracted_k141_167690 | Low-quality |
| S8_extracted_k141_167692 | Not-determined |
| S8_extracted_k141_20228 | Not-determined |
| S8_extracted_k141_29054 | Not-determined |
| S8_extracted_k141_98275 | Not-determined |
| S8_extracted_k141_29055 | Low-quality |
| S8_extracted_k141_132851 | Not-determined |
| S8_extracted_k141_2859 | Not-determined |
| S8_extracted_k141_132854 | Not-determined |
| S8_extracted_k141_20231 | Not-determined |
| S8_extracted_k141_63752 | Not-determined |
| S8_extracted_k141_167698 | Not-determined |
| S8_extracted_k141_202083 | Not-determined |
| S8_extracted_k141_11290 | Not-determined |
| S8_extracted_k141_37683 | Low-quality |
| S8_extracted_k141_11291 | Not-determined |
| S8_extracted_k141_55013 | Not-determined |
| S8_extracted_k141_124529 | Low-quality |
| S8_extracted_k141_29064 | Not-determined |
| S8_extracted_k141_11295 | Not-determined |
| S8_extracted_k141_132862 | Not-determined |
| S8_extracted_k141_46020 | Not-determined |
| S8_extracted_k141_20241 | Not-determined |
| S8_extracted_k141_63758 | Not-determined |
| S8_extracted_k141_132864 | Low-quality |
| S8_extracted_k141_46022 | Not-determined |
| S8_extracted_k141_107056 | Not-determined |
| S8_extracted_k141_158687 | Not-determined |
| S8_extracted_k141_2873 | Not-determined |
| S8_extracted_k141_89613 | Not-determined |
| S8_extracted_k141_46027 | Not-determined |
| S8_extracted_k141_132868 | Not-determined |
| S8_extracted_k141_81116 | Not-determined |
| S8_extracted_k141_46029 | Not-determined |
| S8_extracted_k141_55021 | Not-determined |
| S8_extracted_k141_184460 | Not-determined |
| S8_extracted_k141_141702 | Not-determined |
| S8_extracted_k141_184462 | Not-determined |
| S8_extracted_k141_89621 | Not-determined |
| S8_extracted_k141_20248 | Not-determined |
| S8_extracted_k141_184464 | Not-determined |
| S8_extracted_k141_46034 | Not-determined |
| S8_extracted_k141_37692 | Not-determined |
| S8_extracted_k141_55022 | Not-determined |
| S8_extracted_k141_37693 | Not-determined |
| S8_extracted_k141_63764 | Low-quality |
| S8_extracted_k141_107060 | Not-determined |
| S8_extracted_k141_55026 | Not-determined |
| S8_extracted_k141_184469 | Not-determined |
| S8_extracted_k141_115634 | Not-determined |
| S8_extracted_k141_20257 | Not-determined |
| S8_extracted_k141_20258 | Not-determined |
| S8_extracted_k141_55029 | Not-determined |
| S8_extracted_k141_115635 | Not-determined |
| S8_extracted_k141_55030 | Not-determined |
| S8_extracted_k141_124539 | Not-determined |
| S8_extracted_k141_115636 | Not-determined |
| S8_extracted_k141_55031 | Low-quality |
| S8_extracted_k141_20262 | Not-determined |
| S8_extracted_k141_202103 | Not-determined |
| S8_extracted_k141_55035 | Not-determined |
| S8_extracted_k141_37697 | Not-determined |
| S8_extracted_k141_98303 | Not-determined |
| S8_extracted_k141_89634 | Not-determined |
| S8_extracted_k141_29077 | Not-determined |
| S8_extracted_k141_81131 | Not-determined |
| S8_extracted_k141_46042 | Not-determined |
| S8_extracted_k141_89635 | Not-determined |
| S8_extracted_k141_158700 | Not-determined |
| S8_extracted_k141_29081 | Not-determined |
| S8_extracted_k141_141723 | Not-determined |
| S8_extracted_k141_158703 | Low-quality |
| S8_extracted_k141_37699 | Not-determined |
| S8_extracted_k141_46044 | Not-determined |
| S8_extracted_k141_63771 | Not-determined |
| S8_extracted_k141_107073 | Not-determined |
| S8_extracted_k141_11315 | Not-determined |
| S8_extracted_k141_176455 | Not-determined |
| S8_extracted_k141_29086 | Not-determined |
| S8_extracted_k141_46049 | Not-determined |
| S8_extracted_k141_29088 | Not-determined |
| S8_extracted_k141_107078 | Not-determined |
| S8_extracted_k141_115649 | Not-determined |
| S8_extracted_k141_20275 | Not-determined |
| S8_extracted_k141_71739 | Low-quality |
| S8_extracted_k141_115652 | Not-determined |
| S8_extracted_k141_37710 | Not-determined |
| S8_extracted_k141_158713 | Not-determined |
| S8_extracted_k141_115655 | Not-determined |
| S8_extracted_k141_150214 | Not-determined |
| S8_extracted_k141_46056 | Not-determined |
| S8_extracted_k141_176464 | Not-determined |
| S8_extracted_k141_55047 | Not-determined |
| S8_extracted_k141_46057 | Not-determined |
| S8_extracted_k141_176465 | Not-determined |
| S8_extracted_k141_98307 | Low-quality |
| S8_extracted_k141_29096 | Not-determined |
| S8_extracted_k141_98308 | Not-determined |
| S8_extracted_k141_132878 | Not-determined |
| S8_extracted_k141_124557 | Not-determined |
| S8_extracted_k141_63779 | Not-determined |
| S8_extracted_k141_150224 | Not-determined |
| S8_extracted_k141_107081 | Not-determined |
| S8_extracted_k141_115661 | Not-determined |
| S8_extracted_k141_176467 | Not-determined |
| S8_extracted_k141_20286 | Not-determined |
| S8_extracted_k141_141736 | Not-determined |
| S8_extracted_k141_124561 | Not-determined |
| S8_extracted_k141_107084 | Not-determined |
| S8_extracted_k141_89658 | Not-determined |
| S8_extracted_k141_176470 | Low-quality |
| S8_extracted_k141_71751 | Not-determined |
| S8_extracted_k141_98318 | Not-determined |
| S8_extracted_k141_98320 | Not-determined |
| S8_extracted_k141_98321 | Not-determined |
| S8_extracted_k141_20287 | Not-determined |
| S8_extracted_k141_107090 | Not-determined |
| S8_extracted_k141_29105 | Low-quality |
| S8_extracted_k141_71753 | Low-quality |
| S8_extracted_k141_202118 | Not-determined |
| S8_extracted_k141_46070 | Not-determined |
| S8_extracted_k141_37723 | Not-determined |
| S8_extracted_k141_107097 | Not-determined |
| S8_extracted_k141_98324 | Low-quality |
| S8_extracted_k141_107098 | Not-determined |
| S8_extracted_k141_89666 | Not-determined |
| S8_extracted_k141_46073 | Not-determined |
| S8_extracted_k141_37727 | Not-determined |
| S8_extracted_k141_89671 | Not-determined |
| S8_extracted_k141_158723 | Not-determined |
| S8_extracted_k141_29113 | Not-determined |
| S8_extracted_k141_71757 | Not-determined |
| S8_extracted_k141_37729 | Not-determined |
| S8_extracted_k141_107099 | Low-quality |
| S8_extracted_k141_46075 | Not-determined |
| S8_extracted_k141_46078 | Not-determined |
| S8_extracted_k141_98327 | Not-determined |
| S8_extracted_k141_98329 | Not-determined |
| S8_extracted_k141_37733 | Not-determined |
| S8_extracted_k141_150232 | Low-quality |
| S8_extracted_k141_46081 | Not-determined |
| S8_extracted_k141_37734 | Not-determined |
| S8_extracted_k141_141760 | Not-determined |
| S8_extracted_k141_158728 | Not-determined |
| S8_extracted_k141_71765 | Not-determined |
| S8_extracted_k141_132898 | Low-quality |
| S8_extracted_k141_132899 | Not-determined |
| S8_extracted_k141_124576 | Not-determined |
| S8_extracted_k141_132902 | Not-determined |
| S8_extracted_k141_2900 | Not-determined |
| S8_extracted_k141_141767 | Not-determined |
| S8_extracted_k141_132907 | Low-quality |
| S8_extracted_k141_115677 | Not-determined |
| S8_extracted_k141_89682 | Not-determined |
| S8_extracted_k141_176479 | Not-determined |
| S8_extracted_k141_115680 | Not-determined |
| S8_extracted_k141_37745 | Not-determined |
| S8_extracted_k141_107108 | Not-determined |
| S8_extracted_k141_176480 | Not-determined |
| S8_extracted_k141_184476 | Not-determined |
| S8_extracted_k141_176482 | Not-determined |
| S8_extracted_k141_132917 | Not-determined |
| S8_extracted_k141_202128 | Not-determined |
| S8_extracted_k141_20319 | Not-determined |
| S8_extracted_k141_132919 | Not-determined |
| S8_extracted_k141_2911 | Not-determined |
| S8_extracted_k141_202131 | Not-determined |
| S8_extracted_k141_71785 | Not-determined |
| S8_extracted_k141_132921 | Not-determined |
| S8_extracted_k141_2914 | Low-quality |
| S8_extracted_k141_202132 | Low-quality |
| S8_extracted_k141_29118 | Not-determined |
| S8_extracted_k141_55067 | Not-determined |
| S8_extracted_k141_46090 | Not-determined |
| S8_extracted_k141_89693 | Not-determined |
| S8_extracted_k141_132927 | Not-determined |
| S8_extracted_k141_158743 | Not-determined |
| S8_extracted_k141_2920 | Not-determined |
| S8_extracted_k141_46093 | Not-determined |
| S8_extracted_k141_158746 | Not-determined |
| S8_extracted_k141_158749 | Not-determined |
| S8_extracted_k141_150254 | Not-determined |
| S8_extracted_k141_132934 | Not-determined |
| S8_extracted_k141_98343 | Not-determined |
| S8_extracted_k141_184491 | Not-determined |
| S8_extracted_k141_29127 | Not-determined |
| S8_extracted_k141_46096 | Not-determined |
| S8_extracted_k141_2925 | Low-quality |
| S8_extracted_k141_37753 | Not-determined |
| S8_extracted_k141_29129 | Not-determined |
| S8_extracted_k141_132936 | Low-quality |
| S8_extracted_k141_2928 | Not-determined |

| | |
|---|---|
| SB_extracted_k141_141772 | Not-determined |
| SB_extracted_k141_184494 | Not-determined |
| SB_extracted_k141_176498 | Not-determined |
| SB_extracted_k141_55084 | Not-determined |
| SB_extracted_k141_98347 | Not-determined |
| SB_extracted_k141_98349 | Not-determined |
| SB_extracted_k141_202142 | Not-determined |
| SB_extracted_k141_158760 | Low-quality |
| SB_extracted_k141_55091 | Not-determined |
| SB_extracted_k141_132946 | Not-determined |
| SB_extracted_k141_20332 | Not-determined |
| SB_extracted_k141_176504 | Not-determined |
| SB_extracted_k141_98356 | Not-determined |
| SB_extracted_k141_124605 | Low-quality |
| SB_extracted_k141_2933 | Not-determined |
| SB_extracted_k141_124606 | Not-determined |
| SB_extracted_k141_184507 | Not-determined |
| SB_extracted_k141_193953 | Not-determined |
| SB_extracted_k141_98358 | Not-determined |
| SB_extracted_k141_202145 | Not-determined |
| SB_extracted_k141_29139 | Not-determined |
| SB_extracted_k141_71812 | Low-quality |
| SB_extracted_k141_202146 | Not-determined |
| SB_extracted_k141_176510 | Not-determined |
| SB_extracted_k141_202147 | Not-determined |
| SB_extracted_k141_55094 | Low-quality |
| SB_extracted_k141_107139 | Not-determined |
| SB_extracted_k141_124611 | Not-determined |
| SB_extracted_k141_184510 | Not-determined |
| SB_extracted_k141_193957 | Not-determined |
| SB_extracted_k141_29145 | Not-determined |
| SB_extracted_k141_124614 | Not-determined |
| SB_extracted_k141_29146 | Low-quality |
| SB_extracted_k141_89722 | Not-determined |
| SB_extracted_k141_150268 | Not-determined |
| SB_extracted_k141_2940 | Low-quality |
| SB_extracted_k141_20345 | Low-quality |
| SB_extracted_k141_98362 | Not-determined |
| SB_extracted_k141_2941 | Not-determined |
| SB_extracted_k141_29149 | Low-quality |
| SB_extracted_k141_89726 | Not-determined |
| SB_extracted_k141_150270 | Not-determined |
| SB_extracted_k141_63808 | Not-determined |
| SB_extracted_k141_37770 | Not-determined |
| SB_extracted_k141_63811 | Not-determined |
| SB_extracted_k141_63813 | Not-determined |
| SB_extracted_k141_63814 | Not-determined |
| SB_extracted_k141_176524 | Not-determined |
| SB_extracted_k141_150279 | Not-determined |
| SB_extracted_k141_98368 | Not-determined |
| SB_extracted_k141_71831 | Not-determined |
| SB_extracted_k141_81141 | Not-determined |
| SB_extracted_k141_55101 | Not-determined |
| SB_extracted_k141_176525 | Not-determined |
| SB_extracted_k141_132971 | Not-determined |
| SB_extracted_k141_55102 | Not-determined |
| SB_extracted_k141_89738 | Low-quality |
| SB_extracted_k141_150287 | Not-determined |
| SB_extracted_k141_81148 | Not-determined |
| SB_extracted_k141_150291 | Not-determined |
| SB_extracted_k141_89745 | Not-determined |
| SB_extracted_k141_55109 | Not-determined |
| SB_extracted_k141_2961 | Not-determined |
| SB_extracted_k141_81154 | Not-determined |
| SB_extracted_k141_202155 | Not-determined |
| SB_extracted_k141_202156 | Not-determined |
| SB_extracted_k141_115701 | Low-quality |
| SB_extracted_k141_81158 | Not-determined |
| SB_extracted_k141_71840 | Not-determined |
| SB_extracted_k141_98380 | Not-determined |
| SB_extracted_k141_2962 | Not-determined |
| SB_extracted_k141_29185 | Not-determined |
| SB_extracted_k141_29186 | Not-determined |
| SB_extracted_k141_132984 | Not-determined |
| SB_extracted_k141_37786 | Not-determined |
| SB_extracted_k141_141795 | Not-determined |
| SB_extracted_k141_29190 | Low-quality |
| SB_extracted_k141_193966 | Not-determined |
| SB_extracted_k141_46112 | Not-determined |
| SB_extracted_k141_81168 | Not-determined |
| SB_extracted_k141_89768 | Not-determined |
| SB_extracted_k141_89769 | Not-determined |
| SB_extracted_k141_81170 | Not-determined |
| SB_extracted_k141_37793 | Not-determined |
| SB_extracted_k141_158776 | Not-determined |
| SB_extracted_k141_89774 | Not-determined |
| SB_extracted_k141_46115 | Not-determined |
| SB_extracted_k141_81174 | Not-determined |
| SB_extracted_k141_115717 | Not-determined |
| SB_extracted_k141_184532 | Not-determined |
| SB_extracted_k141_81176 | Not-determined |
| SB_extracted_k141_184533 | Not-determined |
| SB_extracted_k141_2976 | Not-determined |
| SB_extracted_k141_55128 | Not-determined |
| SB_extracted_k141_202168 | Not-determined |
| SB_extracted_k141_20361 | Low-quality |
| SB_extracted_k141_141806 | Not-determined |
| SB_extracted_k141_158783 | Not-determined |
| SB_extracted_k141_46125 | Not-determined |
| SB_extracted_k141_37801 | Not-determined |
| SB_extracted_k141_124623 | Not-determined |
| SB_extracted_k141_29202 | Not-determined |
| SB_extracted_k141_193984 | Not-determined |
| SB_extracted_k141_55134 | Not-determined |
| SB_extracted_k141_29204 | Not-determined |
| SB_extracted_k141_176538 | Not-determined |
| SB_extracted_k141_46131 | Not-determined |
| SB_extracted_k141_2983 | Not-determined |
| SB_extracted_k141_202173 | Not-determined |
| SB_extracted_k141_98392 | Low-quality |
| SB_extracted_k141_81187 | Not-determined |
| SB_extracted_k141_46135 | Not-determined |
| SB_extracted_k141_141814 | Not-determined |
| SB_extracted_k141_55139 | Not-determined |
| SB_extracted_k141_124634 | Not-determined |
| SB_extracted_k141_2984 | Not-determined |
| SB_extracted_k141_55141 | Not-determined |
| SB_extracted_k141_11384 | Low-quality |
| SB_extracted_k141_124636 | Not-determined |
| SB_extracted_k141_176547 | Not-determined |
| SB_extracted_k141_11386 | Not-determined |
| SB_extracted_k141_202178 | Not-determined |
| SB_extracted_k141_89782 | Not-determined |
| SB_extracted_k141_11387 | Not-determined |
| SB_extracted_k141_124639 | Low-quality |
| SB_extracted_k141_133021 | Not-determined |
| SB_extracted_k141_176548 | Low-quality |
| SB_extracted_k141_202183 | Not-determined |
| SB_extracted_k141_107178 | Not-determined |
| SB_extracted_k141_193988 | Not-determined |
| SB_extracted_k141_202186 | Not-determined |
| SB_extracted_k141_81195 | Not-determined |
| SB_extracted_k141_29208 | Not-determined |
| SB_extracted_k141_158799 | Not-determined |
| SB_extracted_k141_2992 | Low-quality |
| SB_extracted_k141_55147 | Not-determined |
| [illegible] | [illegible] |
| SB_extracted_k141_158967 | Not-determined |
| SB_extracted_k141_107346 | Not-determined |
| SB_extracted_k141_158969 | Not-determined |
| SB_extracted_k141_3154 | Not-determined |
| SB_extracted_k141_72007 | Not-determined |
| SB_extracted_k141_46268 | Not-determined |
| SB_extracted_k141_29391 | Not-determined |
| SB_extracted_k141_142000 | Low-quality |
| SB_extracted_k141_55247 | Not-determined |
| SB_extracted_k141_3159 | Low-quality |
| SB_extracted_k141_133210 | Not-determined |
| SB_extracted_k141_184706 | Not-determined |
| SB_extracted_k141_133213 | Low-quality |
| SB_extracted_k141_133214 | Not-determined |
| SB_extracted_k141_72014 | Not-determined |
| SB_extracted_k141_11553 | Not-determined |
| SB_extracted_k141_37944 | Not-determined |
| SB_extracted_k141_37945 | Not-determined |
| SB_extracted_k141_158981 | Not-determined |
| SB_extracted_k141_63987 | Not-determined |
| SB_extracted_k141_194181 | Not-determined |
| SB_extracted_k141_158982 | Low-quality |
| SB_extracted_k141_202366 | Not-determined |
| SB_extracted_k141_98577 | Not-determined |
| SB_extracted_k141_150474 | Not-determined |
| SB_extracted_k141_150475 | Not-determined |
| SB_extracted_k141_20516 | Low-quality |
| SB_extracted_k141_133225 | Not-determined |
| SB_extracted_k141_176709 | Not-determined |
| SB_extracted_k141_107352 | Not-determined |
| SB_extracted_k141_150477 | Not-determined |
| SB_extracted_k141_63991 | Not-determined |
| SB_extracted_k141_55259 | Not-determined |
| SB_extracted_k141_89899 | Not-determined |
| SB_extracted_k141_98580 | Not-determined |
| SB_extracted_k141_184720 | Not-determined |
| SB_extracted_k141_11566 | Not-determined |
| SB_extracted_k141_107355 | Not-determined |
| SB_extracted_k141_63992 | Not-determined |
| SB_extracted_k141_158988 | Not-determined |
| SB_extracted_k141_63993 | Not-determined |
| SB_extracted_k141_176712 | Not-determined |
| SB_extracted_k141_167884 | Not-determined |
| SB_extracted_k141_29413 | Not-determined |
| SB_extracted_k141_184723 | Not-determined |
| SB_extracted_k141_115909 | Not-determined |
| SB_extracted_k141_98584 | Not-determined |
| SB_extracted_k141_150481 | Not-determined |
| SB_extracted_k141_3177 | Not-determined |

| | |
|---|---|
| SB_extracted_k141_167891 | Not-determined |
| SB_extracted_k141_37955 | Not-determined |
| SB_extracted_k141_142013 | Not-determined |
| SB_extracted_k141_37956 | Not-determined |
| SB_extracted_k141_98587 | Not-determined |
| SB_extracted_k141_29419 | Not-determined |
| SB_extracted_k141_124791 | Not-determined |
| SB_extracted_k141_72036 | Not-determined |
| SB_extracted_k141_29420 | Not-determined |
| SB_extracted_k141_167894 | Low-quality |
| SB_extracted_k141_107365 | Not-determined |
| SB_extracted_k141_167895 | Not-determined |
| SB_extracted_k141_107368 | Not-determined |
| SB_extracted_k141_3179 | Not-determined |
| SB_extracted_k141_150488 | Not-determined |
| SB_extracted_k141_107371 | Not-determined |
| SB_extracted_k141_81263 | Not-determined |
| SB_extracted_k141_98595 | Not-determined |
| SB_extracted_k141_37964 | Not-determined |
| SB_extracted_k141_20533 | Not-determined |
| SB_extracted_k141_3183 | Not-determined |
| SB_extracted_k141_167905 | Not-determined |
| SB_extracted_k141_89901 | Low-quality |
| SB_extracted_k141_37971 | Not-determined |
| SB_extracted_k141_20538 | Not-determined |
| SB_extracted_k141_158999 | Not-determined |
| SB_extracted_k141_37973 | Not-determined |
| SB_extracted_k141_159001 | Not-determined |
| SB_extracted_k141_29436 | Not-determined |
| SB_extracted_k141_29437 | Not-determined |
| SB_extracted_k141_3186 | Low-quality |
| SB_extracted_k141_89906 | Not-determined |
| SB_extracted_k141_81271 | Low-quality |
| SB_extracted_k141_150501 | Not-determined |
| SB_extracted_k141_159005 | Low-quality |
| SB_extracted_k141_29441 | Not-determined |
| SB_extracted_k141_81273 | Not-determined |
| SB_extracted_k141_89907 | Low-quality |
| SB_extracted_k141_81274 | Not-determined |
| SB_extracted_k141_159006 | Not-determined |
| SB_extracted_k141_81275 | Not-determined |
| SB_extracted_k141_20547 | Not-determined |
| SB_extracted_k141_64007 | Not-determined |
| SB_extracted_k141_194197 | Not-determined |
| SB_extracted_k141_29447 | Not-determined |
| SB_extracted_k141_202374 | Not-determined |
| SB_extracted_k141_29449 | Low-quality |
| SB_extracted_k141_89908 | Not-determined |
| SB_extracted_k141_98597 | Low-quality |
| SB_extracted_k141_81283 | Low-quality |
| SB_extracted_k141_72045 | Not-determined |
| SB_extracted_k141_184745 | Not-determined |
| SB_extracted_k141_81288 | Not-determined |
| SB_extracted_k141_167919 | Not-determined |
| SB_extracted_k141_29453 | Not-determined |
| SB_extracted_k141_98606 | Not-determined |
| SB_extracted_k141_124814 | Not-determined |
| SB_extracted_k141_115944 | Low-quality |
| SB_extracted_k141_167924 | Not-determined |
| SB_extracted_k141_11598 | Not-determined |
| SB_extracted_k141_81290 | Low-quality |
| SB_extracted_k141_81295 | Not-determined |
| SB_extracted_k141_167928 | Not-determined |
| SB_extracted_k141_194212 | Not-determined |
| SB_extracted_k141_124821 | Not-determined |
| SB_extracted_k141_98617 | Not-determined |
| SB_extracted_k141_159021 | Not-determined |
| SB_extracted_k141_150516 | Not-determined |
| SB_extracted_k141_64023 | Not-determined |
| SB_extracted_k141_167941 | Not-determined |
| SB_extracted_k141_38003 | Not-determined |
| SB_extracted_k141_115951 | Not-determined |
| SB_extracted_k141_202388 | Not-determined |
| SB_extracted_k141_64024 | Not-determined |
| SB_extracted_k141_167942 | Not-determined |
| SB_extracted_k141_38007 | Low-quality |
| SB_extracted_k141_133252 | Low-quality |
| SB_extracted_k141_64027 | Not-determined |
| SB_extracted_k141_202391 | Not-determined |
| SB_extracted_k141_159027 | Not-determined |
| SB_extracted_k141_64030 | Not-determined |
| SB_extracted_k141_167944 | Not-determined |
| SB_extracted_k141_64031 | Low-quality |
| SB_extracted_k141_38009 | Not-determined |
| SB_extracted_k141_98630 | Not-determined |
| SB_extracted_k141_11608 | Not-determined |
| SB_extracted_k141_184760 | Not-determined |
| SB_extracted_k141_124829 | Not-determined |
| SB_extracted_k141_107390 | Not-determined |
| SB_extracted_k141_167946 | Not-determined |
| SB_extracted_k141_115956 | Not-determined |
| SB_extracted_k141_72070 | Low-quality |
| SB_extracted_k141_124830 | Not-determined |
| SB_extracted_k141_81300 | Not-determined |
| SB_extracted_k141_167948 | Not-determined |
| SB_extracted_k141_133255 | Not-determined |
| SB_extracted_k141_55265 | Not-determined |
| SB_extracted_k141_11613 | Not-determined |
| SB_extracted_k141_20572 | Not-determined |
| SB_extracted_k141_20573 | Not-determined |
| SB_extracted_k141_202398 | Not-determined |
| SB_extracted_k141_81303 | Not-determined |
| SB_extracted_k141_167950 | Not-determined |
| SB_extracted_k141_202400 | Low-quality |
| SB_extracted_k141_133261 | Not-determined |
| SB_extracted_k141_107398 | Not-determined |
| SB_extracted_k141_142042 | Not-determined |
| SB_extracted_k141_64042 | Not-determined |
| SB_extracted_k141_107402 | Not-determined |
| SB_extracted_k141_133264 | Not-determined |
| SB_extracted_k141_202403 | Low-quality |
| SB_extracted_k141_55273 | Not-determined |
| SB_extracted_k141_81311 | Not-determined |
| SB_extracted_k141_38019 | Not-determined |
| SB_extracted_k141_150525 | Low-quality |
| SB_extracted_k141_55275 | Not-determined |
| SB_extracted_k141_55277 | Not-determined |
| SB_extracted_k141_29465 | Not-determined |
| SB_extracted_k141_38024 | Not-determined |
| SB_extracted_k141_124832 | Low-quality |
| SB_extracted_k141_202407 | Not-determined |
| SB_extracted_k141_133271 | Not-determined |
| SB_extracted_k141_159048 | Low-quality |
| SB_extracted_k141_159051 | Not-determined |
| SB_extracted_k141_167967 | Not-determined |
| SB_extracted_k141_142057 | Not-determined |
| SB_extracted_k141_167970 | Not-determined |
| SB_extracted_k141_167972 | Not-determined |
| SB_extracted_k141_72093 | Not-determined |
| SB_extracted_k141_107413 | Not-determined |
| SB_extracted_k141_20588 | Not-determined |
| SB_extracted_k141_89939 | Not-determined |
| SB_extracted_k141_142064 | Not-determined |
| SB_extracted_k141_150533 | Not-determined |
| SB_extracted_k141_133291 | Not-determined |
| SB_extracted_k141_38034 | Not-determined |
| SB_extracted_k141_150540 | Not-determined |
| SB_extracted_k141_184778 | Not-determined |
| SB_extracted_k141_150543 | Not-determined |
| SB_extracted_k141_38039 | Not-determined |
| SB_extracted_k141_107424 | Not-determined |
| SB_extracted_k141_72105 | Not-determined |
| SB_extracted_k141_81321 | Not-determined |
| SB_extracted_k141_176728 | Not-determined |
| SB_extracted_k141_107428 | Not-determined |
| SB_extracted_k141_72108 | Not-determined |
| SB_extracted_k141_167989 | Not-determined |
| SB_extracted_k141_133304 | Not-determined |
| SB_extracted_k141_55302 | Not-determined |
| SB_extracted_k141_107429 | Not-determined |
| SB_extracted_k141_142067 | Low-quality |
| SB_extracted_k141_150551 | Low-quality |
| SB_extracted_k141_81323 | Not-determined |
| SB_extracted_k141_29486 | Not-determined |
| SB_extracted_k141_11637 | Low-quality |
| SB_extracted_k141_64057 | Not-determined |
| SB_extracted_k141_72115 | Not-determined |
| SB_extracted_k141_64062 | Not-determined |
| SB_extracted_k141_29490 | Low-quality |
| SB_extracted_k141_159076 | Not-determined |
| SB_extracted_k141_159077 | Not-determined |
| SB_extracted_k141_142071 | Not-determined |
| SB_extracted_k141_176734 | Not-determined |
| SB_extracted_k141_168001 | Not-determined |
| SB_extracted_k141_176735 | Not-determined |
| SB_extracted_k141_55308 | Not-determined |
| SB_extracted_k141_168003 | Not-determined |
| SB_extracted_k141_168005 | Not-determined |
| SB_extracted_k141_55312 | Not-determined |
| SB_extracted_k141_202434 | Not-determined |
| SB_extracted_k141_159086 | Not-determined |
| SB_extracted_k141_159088 | Not-determined |
| SB_extracted_k141_98633 | Not-determined |
| SB_extracted_k141_168010 | Not-determined |
| SB_extracted_k141_142077 | Not-determined |
| SB_extracted_k141_159091 | Not-determined |
| SB_extracted_k141_64067 | Not-determined |
| SB_extracted_k141_64068 | Not-determined |
| SB_extracted_k141_38049 | Not-determined |
| SB_extracted_k141_89959 | Not-determined |
| SB_extracted_k141_142081 | Not-determined |
| SB_extracted_k141_55313 | Not-determined |
| SB_extracted_k141_168018 | Not-determined |
| SB_extracted_k141_64072 | Low-quality |
| SB_extracted_k141_89961 | Not-determined |
| SB_extracted_k141_184791 | Not-determined |
| SB_extracted_k141_11651 | Not-determined |
| SB_extracted_k141_29494 | Not-determined |
| SB_extracted_k141_168021 | Not-determined |
| SB_extracted_k141_11653 | Not-determined |
| SB_extracted_k141_81333 | Low-quality |
| SB_extracted_k141_64075 | Not-determined |
| SB_extracted_k141_64076 | Not-determined |
| SB_extracted_k141_81336 | Not-determined |
| SB_extracted_k141_98637 | Not-determined |
| SB_extracted_k141_55319 | Not-determined |
| SB_extracted_k141_168025 | Not-determined |
| SB_extracted_k141_202448 | Not-determined |
| SB_extracted_k141_11654 | Low-quality |
| SB_extracted_k141_202450 | Low-quality |
| SB_extracted_k141_202451 | Not-determined |
| SB_extracted_k141_142087 | Not-determined |
| SB_extracted_k141_29499 | Not-determined |
| SB_extracted_k141_38061 | Not-determined |
| SB_extracted_k141_55326 | Not-determined |
| SB_extracted_k141_11656 | Not-determined |
| SB_extracted_k141_46273 | Not-determined |
| SB_extracted_k141_98648 | Not-determined |
| SB_extracted_k141_168031 | Low-quality |
| SB_extracted_k141_202454 | Not-determined |
| SB_extracted_k141_202455 | Not-determined |
| SB_extracted_k141_159099 | Not-determined |
| SB_extracted_k141_107464 | Not-determined |
| SB_extracted_k141_64086 | Not-determined |
| SB_extracted_k141_107466 | Not-determined |
| SB_extracted_k141_159103 | Not-determined |
| SB_extracted_k141_168032 | Low-quality |
| SB_extracted_k141_107467 | Not-determined |
| SB_extracted_k141_202459 | Not-determined |
| SB_extracted_k141_184806 | Low-quality |
| SB_extracted_k141_159105 | Not-determined |
| SB_extracted_k141_55338 | Not-determined |
| SB_extracted_k141_168036 | Not-determined |
| SB_extracted_k141_107470 | Not-determined |
| SB_extracted_k141_29508 | Low-quality |
| SB_extracted_k141_159112 | Not-determined |
| SB_extracted_k141_142095 | Not-determined |
| SB_extracted_k141_184813 | Not-determined |
| SB_extracted_k141_202463 | Not-determined |
| SB_extracted_k141_64093 | Not-determined |
| SB_extracted_k141_159116 | Not-determined |
| SB_extracted_k141_176755 | Not-determined |
| SB_extracted_k141_55349 | Not-determined |
| SB_extracted_k141_159120 | Not-determined |
| SB_extracted_k141_168044 | Not-determined |
| SB_extracted_k141_168045 | Low-quality |
| SB_extracted_k141_176760 | Not-determined |
| SB_extracted_k141_159124 | Not-determined |
| SB_extracted_k141_176761 | Not-determined |
| SB_extracted_k141_194247 | Not-determined |
| SB_extracted_k141_89969 | Not-determined |
| SB_extracted_k141_176764 | Not-determined |
| SB_extracted_k141_89972 | Not-determined |
| SB_extracted_k141_194248 | Not-determined |
| SB_extracted_k141_89973 | Not-determined |
| SB_extracted_k141_194249 | Not-determined |
| SB_extracted_k141_194250 | Not-determined |
| SB_extracted_k141_184825 | Not-determined |
| SB_extracted_k141_202479 | Not-determined |
| SB_extracted_k141_55353 | Not-determined |
| SB_extracted_k141_176776 | Low-quality |
| SB_extracted_k141_29524 | Not-determined |
| SB_extracted_k141_159135 | Not-determined |
| SB_extracted_k141_46303 | Not-determined |
| SB_extracted_k141_55355 | Not-determined |
| SB_extracted_k141_98665 | Not-determined |
| SB_extracted_k141_81365 | Not-determined |
| SB_extracted_k141_64110 | Not-determined |
| SB_extracted_k141_11689 | Not-determined |
| SB_extracted_k141_55360 | Not-determined |
| SB_extracted_k141_176785 | Not-determined |
| SB_extracted_k141_46307 | Not-determined |
| SB_extracted_k141_81371 | Not-determined |
| SB_extracted_k141_176787 | Not-determined |
| SB_extracted_k141_98671 | Not-determined |
| SB_extracted_k141_64115 | Not-determined |
| SB_extracted_k141_150585 | Not-determined |
| SB_extracted_k141_81376 | Not-determined |
| SB_extracted_k141_64118 | Not-determined |
| SB_extracted_k141_55364 | Low-quality |
| SB_extracted_k141_168056 | Low-quality |
| SB_extracted_k141_38089 | Not-determined |
| SB_extracted_k141_184833 | Not-determined |
| SB_extracted_k141_159149 | Low-quality |
| SB_extracted_k141_72119 | Not-determined |
| SB_extracted_k141_184839 | Not-determined |
| SB_extracted_k141_202492 | Not-determined |
| SB_extracted_k141_124878 | Not-determined |
| SB_extracted_k141_11704 | Not-determined |
| SB_extracted_k141_72122 | Not-determined |
| SB_extracted_k141_81380 | Not-determined |
| SB_extracted_k141_20623 | Not-determined |
| SB_extracted_k141_38106 | Not-determined |
| SB_extracted_k141_81388 | Not-determined |
| SB_extracted_k141_38109 | Not-determined |
| SB_extracted_k141_20634 | Not-determined |
| SB_extracted_k141_124897 | Not-determined |
| SB_extracted_k141_142108 | Not-determined |
| SB_extracted_k141_142109 | Not-determined |
| SB_extracted_k141_81395 | Not-determined |
| SB_extracted_k141_168074 | Not-determined |
| SB_extracted_k141_81397 | Not-determined |
| SB_extracted_k141_202511 | Not-determined |
| SB_extracted_k141_81399 | Not-determined |
| SB_extracted_k141_184846 | Not-determined |
| SB_extracted_k141_64145 | Not-determined |
| SB_extracted_k141_20641 | Not-determined |
| SB_extracted_k141_194272 | Not-determined |
| SB_extracted_k141_124903 | Low-quality |
| SB_extracted_k141_194273 | Not-determined |
| SB_extracted_k141_184849 | Not-determined |
| SB_extracted_k141_64148 | Low-quality |
| SB_extracted_k141_20643 | Not-determined |
| SB_extracted_k141_184854 | Not-determined |
| SB_extracted_k141_29534 | Not-determined |
| SB_extracted_k141_3228 | Not-determined |
| SB_extracted_k141_202522 | Not-determined |
| SB_extracted_k141_98684 | Low-quality |
| SB_extracted_k141_55371 | Not-determined |
| SB_extracted_k141_194279 | Low-quality |
| SB_extracted_k141_124914 | Not-determined |
| SB_extracted_k141_194280 | Not-determined |
| SB_extracted_k141_64152 | Not-determined |
| SB_extracted_k141_168089 | Not-determined |
| SB_extracted_k141_55374 | Low-quality |
| SB_extracted_k141_142118 | Not-determined |
| SB_extracted_k141_159160 | Not-determined |
| SB_extracted_k141_107490 | Not-determined |
| SB_extracted_k141_184865 | Not-determined |
| SB_extracted_k141_55379 | Not-determined |
| SB_extracted_k141_55380 | Not-determined |
| SB_extracted_k141_89981 | Low-quality |
| SB_extracted_k141_81405 | Not-determined |
| SB_extracted_k141_168095 | Not-determined |
| SB_extracted_k141_20652 | Not-determined |
| SB_extracted_k141_20653 | Not-determined |
| SB_extracted_k141_107498 | Not-determined |
| SB_extracted_k141_168097 | Not-determined |
| SB_extracted_k141_184869 | Not-determined |
| SB_extracted_k141_194288 | Not-determined |
| SB_extracted_k141_3244 | Not-determined |
| SB_extracted_k141_107499 | Not-determined |
| SB_extracted_k141_107500 | Not-determined |
| SB_extracted_k141_20659 | Not-determined |
| SB_extracted_k141_159168 | Not-determined |
| SB_extracted_k141_20660 | Not-determined |
| SB_extracted_k141_81408 | Not-determined |
| SB_extracted_k141_176802 | Low-quality |
| SB_extracted_k141_142139 | Not-determined |
| SB_extracted_k141_89992 | Not-determined |
| SB_extracted_k141_98700 | Not-determined |
| SB_extracted_k141_89993 | Not-determined |
| SB_extracted_k141_55389 | Not-determined |
| SB_extracted_k141_89994 | Not-determined |
| SB_extracted_k141_107507 | Not-determined |
| SB_extracted_k141_81412 | Not-determined |
| SB_extracted_k141_176808 | Not-determined |
| SB_extracted_k141_124920 | Not-determined |
| SB_extracted_k141_38156 | Not-determined |
| SB_extracted_k141_202540 | Not-determined |
| SB_extracted_k141_107508 | Not-determined |
| SB_extracted_k141_11750 | Not-determined |
| SB_extracted_k141_184887 | Not-determined |
| SB_extracted_k141_194300 | Not-determined |
| SB_extracted_k141_194301 | Not-determined |
| SB_extracted_k141_107514 | Not-determined |
| SB_extracted_k141_29561 | Not-determined |
| SB_extracted_k141_20672 | Not-determined |
| SB_extracted_k141_72161 | Low-quality |
| SB_extracted_k141_20673 | Not-determined |
| SB_extracted_k141_81428 | Not-determined |
| SB_extracted_k141_107519 | Low-quality |
| SB_extracted_k141_55403 | Not-determined |
| SB_extracted_k141_176825 | Not-determined |
| SB_extracted_k141_194311 | Not-determined |
| SB_extracted_k141_90004 | Not-determined |
| SB_extracted_k141_184890 | Not-determined |
| SB_extracted_k141_184891 | Not-determined |
| SB_extracted_k141_176830 | Not-determined |
| SB_extracted_k141_81437 | Low-quality |
| SB_extracted_k141_72173 | Low-quality |
| SB_extracted_k141_176832 | Not-determined |
| SB_extracted_k141_38168 | Not-determined |
| SB_extracted_k141_20679 | Not-determined |
| SB_extracted_k141_176833 | Not-determined |
| SB_extracted_k141_81441 | Not-determined |
| SB_extracted_k141_124943 | Not-determined |
| SB_extracted_k141_72178 | Not-determined |
| SB_extracted_k141_55412 | Not-determined |
| SB_extracted_k141_107527 | Not-determined |
| SB_extracted_k141_55413 | Low-quality |
| S12_vs_k141_170614\|\|full | Medium-quality |
| SB_extracted_k141_38171 | Not-determined |
| SB_extracted_k141_184898 | Not-determined |
| SB_extracted_k141_107531 | Not-determined |
| SB_extracted_k141_107532 | Not-determined |
| SB_extracted_k141_184902 | Low-quality |
| SB_extracted_k141_3262 | Low-quality |
| SB_extracted_k141_176837 | Not-determined |
| SB_extracted_k141_38178 | Not-determined |
| SB_extracted_k141_81453 | Not-determined |
| SB_extracted_k141_202545 | Not-determined |
| SB_extracted_k141_3267 | Not-determined |
| SB_extracted_k141_107537 | Not-determined |
| SB_extracted_k141_55424 | Not-determined |
| SB_extracted_k141_11771 | Not-determined |
| SB_extracted_k141_133311 | Not-determined |
| SB_extracted_k141_46352 | Not-determined |
| SB_extracted_k141_124959 | Not-determined |
| SB_extracted_k141_46353 | Not-determined |
| SB_extracted_k141_90005 | Not-determined |
| SB_extracted_k141_38181 | Not-determined |
| SB_extracted_k141_29573 | Not-determined |
| SB_extracted_k141_194336 | Not-determined |
| SB_extracted_k141_176850 | Not-determined |
| SB_extracted_k141_194339 | Not-determined |
| SB_extracted_k141_159184 | Not-determined |
| SB_extracted_k141_29576 | Not-determined |
| SB_extracted_k141_20698 | Not-determined |
| SB_extracted_k141_72191 | Not-determined |
| SB_extracted_k141_142151 | Low-quality |
| SB_extracted_k141_46362 | Not-determined |
| SB_extracted_k141_194343 | Not-determined |
| SB_extracted_k141_176855 | Not-determined |
| SB_extracted_k141_90009 | Not-determined |
| SB_extracted_k141_159187 | Not-determined |
| SB_extracted_k141_3278 | Not-determined |
| SB_extracted_k141_72193 | Not-determined |
| SB_extracted_k141_159189 | Not-determined |
| SB_extracted_k141_72194 | Not-determined |
| SB_extracted_k141_3281 | Not-determined |
| SB_extracted_k141_107547 | Not-determined |
| SB_extracted_k141_29588 | Not-determined |
| SB_extracted_k141_107549 | Not-determined |
| SB_extracted_k141_184918 | Not-determined |
| SB_extracted_k141_81462 | Not-determined |
| SB_extracted_k141_11775 | Not-determined |
| SB_extracted_k141_3290 | Not-determined |
| SB_extracted_k141_72205 | Not-determined |
| SB_extracted_k141_133325 | Not-determined |
| SB_extracted_k141_38192 | Not-determined |
| SB_extracted_k141_133327 | Not-determined |
| SB_extracted_k141_107552 | Not-determined |
| SB_extracted_k141_81472 | Not-determined |
| SB_extracted_k141_38194 | Not-determined |
| SB_extracted_k141_72206 | Not-determined |
| SB_extracted_k141_194361 | Not-determined |
| SB_extracted_k141_107555 | Not-determined |
| SB_extracted_k141_72207 | Not-determined |
| SB_extracted_k141_29603 | Not-determined |
| SB_extracted_k141_176879 | Not-determined |
| SB_extracted_k141_176880 | Not-determined |
| SB_extracted_k141_194364 | Not-determined |
| SB_extracted_k141_20713 | Not-determined |
| SB_extracted_k141_3295 | Not-determined |
| SB_extracted_k141_115974 | Not-determined |
| SB_extracted_k141_159208 | Not-determined |
| SB_extracted_k141_90028 | Not-determined |
| SB_extracted_k141_184928 | Not-determined |
| SB_extracted_k141_20715 | Not-determined |
| SB_extracted_k141_90031 | Not-determined |
| SB_extracted_k141_11784 | Not-determined |
| SB_extracted_k141_11785 | Not-determined |
| SB_extracted_k141_55430 | Not-determined |
| SB_extracted_k141_115975 | Not-determined |
| SB_extracted_k141_194369 | Not-determined |
| SB_extracted_k141_20717 | Not-determined |
| SB_extracted_k141_46380 | Not-determined |
| SB_extracted_k141_90033 | Low-quality |
| SB_extracted_k141_55435 | Not-determined |
| SB_extracted_k141_72219 | Not-determined |
| SB_extracted_k141_72221 | Not-determined |
| SB_extracted_k141_115986 | Not-determined |
| SB_extracted_k141_107559 | Not-determined |
| SB_extracted_k141_72223 | Not-determined |
| SB_extracted_k141_115990 | Not-determined |
| SB_extracted_k141_184939 | Not-determined |
| SB_extracted_k141_159212 | Not-determined |
| SB_extracted_k141_38213 | Not-determined |
| SB_extracted_k141_159213 | Not-determined |
| SB_extracted_k141_90041 | Not-determined |
| SB_extracted_k141_55443 | Not-determined |
| SB_extracted_k141_125003 | Not-determined |
| SB_extracted_k141_176887 | Not-determined |
| SB_extracted_k141_125004 | Not-determined |
| SB_extracted_k141_184944 | Not-determined |
| SB_extracted_k141_55446 | Not-determined |
| SB_extracted_k141_46383 | Not-determined |
| SB_extracted_k141_184946 | Not-determined |
| SB_extracted_k141_72231 | Not-determined |
| SB_extracted_k141_194375 | Not-determined |
| SB_extracted_k141_55447 | Not-determined |
| SB_extracted_k141_159220 | Not-determined |
| SB_extracted_k141_55448 | Not-determined |
| SB_extracted_k141_90045 | Not-determined |
| SB_extracted_k141_55449 | Not-determined |
| SB_extracted_k141_20733 | Not-determined |
| SB_extracted_k141_3309 | Low-quality |
| SB_extracted_k141_176895 | Low-quality |
| SB_extracted_k141_20735 | Not-determined |
| SB_extracted_k141_55451 | Not-determined |
| SB_extracted_k141_90050 | Not-determined |
| SB_extracted_k141_116002 | Not-determined |
| SB_extracted_k141_90055 | Not-determined |
| SB_extracted_k141_133343 | Not-determined |
| SB_extracted_k141_133344 | Not-determined |
| SB_extracted_k141_20745 | Not-determined |
| SB_extracted_k141_159230 | Not-determined |
| SB_extracted_k141_194383 | Not-determined |
| SB_extracted_k141_38223 | Not-determined |
| SB_extracted_k141_184956 | Not-determined |
| SB_extracted_k141_194386 | Not-determined |
| SB_extracted_k141_3320 | Not-determined |
| SB_extracted_k141_20748 | Not-determined |
| SB_extracted_k141_194388 | Not-determined |
| SB_extracted_k141_20749 | Not-determined |
| SB_extracted_k141_125009 | Not-determined |
| SB_extracted_k141_90065 | Not-determined |
| SB_extracted_k141_194390 | Not-determined |
| SB_extracted_k141_81485 | Not-determined |
| SB_extracted_k141_38226 | Not-determined |
| SB_extracted_k141_38227 | Not-determined |
| SB_extracted_k141_184960 | Not-determined |
| SB_extracted_k141_133357 | Not-determined |
| SB_extracted_k141_133359 | Not-determined |
| SB_extracted_k141_20751 | Not-determined |
| SB_extracted_k141_133361 | Not-determined |
| SB_extracted_k141_142166 | Not-determined |
| SB_extracted_k141_194396 | Not-determined |
| SB_extracted_k141_72249 | Not-determined |
| SB_extracted_k141_98749 | Not-determined |
| SB_extracted_k141_20756 | Not-determined |
| SB_extracted_k141_98753 | Not-determined |
| SB_extracted_k141_72252 | Not-determined |
| SB_extracted_k141_72253 | Low-quality |
| SB_extracted_k141_55470 | Not-determined |
| SB_extracted_k141_3325 | Not-determined |
| SB_extracted_k141_107569 | Low-quality |
| SB_extracted_k141_81490 | Low-quality |
| SB_extracted_k141_81494 | Not-determined |
| SB_extracted_k141_194400 | Not-determined |
| SB_extracted_k141_176914 | Not-determined |
| SB_extracted_k141_133373 | Not-determined |
| SB_extracted_k141_81499 | Not-determined |
| SB_extracted_k141_194403 | Low-quality |
| SB_extracted_k141_81500 | Not-determined |
| SB_extracted_k141_176916 | Low-quality |
| SB_extracted_k141_81502 | Not-determined |
| SB_extracted_k141_107578 | Not-determined |
| SB_extracted_k141_98766 | Not-determined |
| SB_extracted_k141_133380 | Not-determined |
| SB_extracted_k141_107582 | Not-determined |
| SB_extracted_k141_29641 | Not-determined |
| SB_extracted_k141_3346 | Not-determined |
| SB_extracted_k141_125014 | Not-determined |
| SB_extracted_k141_20770 | Not-determined |
| SB_extracted_k141_20771 | Not-determined |
| SB_extracted_k141_150596 | Not-determined |
| SB_extracted_k141_3349 | Not-determined |
| SB_extracted_k141_150601 | Not-determined |
| SB_extracted_k141_3358 | Not-determined |
| SB_extracted_k141_150604 | Not-determined |
| SB_extracted_k141_142197 | Not-determined |
| SB_extracted_k141_194418 | Not-determined |
| SB_extracted_k141_150608 | Low-quality |
| SB_extracted_k141_3360 | Low-quality |
| SB_extracted_k141_81517 | Not-determined |
| SB_extracted_k141_72276 | Not-determined |
| SB_extracted_k141_116017 | Not-determined |
| SB_extracted_k141_150611 | Not-determined |
| SB_extracted_k141_38243 | Not-determined |
| SB_extracted_k141_116024 | Not-determined |
| SB_extracted_k141_81521 | Low-quality |
| SB_extracted_k141_194424 | Not-determined |
| SB_extracted_k141_133386 | Not-determined |
| SB_extracted_k141_159260 | Not-determined |
| SB_extracted_k141_125018 | Not-determined |
| SB_extracted_k141_150620 | Not-determined |
| SB_extracted_k141_142202 | Not-determined |
| SB_extracted_k141_3377 | Not-determined |
| SB_extracted_k141_29668 | Not-determined |
| SB_extracted_k141_90076 | Not-determined |
| SB_extracted_k141_29672 | Not-determined |
| SB_extracted_k141_90077 | Not-determined |
| SB_extracted_k141_29673 | Not-determined |
| SB_extracted_k141_72290 | Not-determined |
| SB_extracted_k141_90079 | Not-determined |
| SB_extracted_k141_20791 | Not-determined |
| SB_extracted_k141_29676 | Not-determined |
| SB_extracted_k141_81536 | Not-determined |
| SB_extracted_k141_20792 | Not-determined |
| SB_extracted_k141_98772 | Not-determined |
| SB_extracted_k141_98774 | Not-determined |
| SB_extracted_k141_20794 | Not-determined |
| SB_extracted_k141_46386 | Not-determined |
| SB_extracted_k141_194437 | Not-determined |
| SB_extracted_k141_20797 | Low-quality |
| SB_extracted_k141_90086 | Not-determined |
| SB_extracted_k141_107593 | Not-determined |

| | |
|---|---|
| S8_extracted_k141_3386 | Not-determined |
| S8_extracted_k141_142205 | Not-determined |
| S8_extracted_k141_125022 | Low-quality |
| S8_extracted_k141_72294 | Not-determined |
| S8_extracted_k141_133404 | Not-determined |
| S8_extracted_k141_81545 | Not-determined |
| S8_extracted_k141_159275 | Not-determined |
| S8_extracted_k141_194444 | Not-determined |
| S8_extracted_k141_168122 | Not-determined |
| S8_extracted_k141_116038 | Not-determined |
| S8_extracted_k141_98777 | Not-determined |
| S8_extracted_k141_107598 | Not-determined |
| S8_extracted_k141_98778 | Not-determined |
| S8_extracted_k141_142212 | Low-quality |
| S8_extracted_k141_133414 | Not-determined |
| S8_extracted_k141_46397 | Not-determined |
| S8_extracted_k141_81549 | Not-determined |
| S8_extracted_k141_98780 | Low-quality |
| S8_extracted_k141_194449 | Not-determined |
| S8_extracted_k141_142216 | Not-determined |
| S8_extracted_k141_168130 | Not-determined |
| S8_extracted_k141_202572 | Not-determined |
| S8_extracted_k141_159285 | Not-determined |
| S8_extracted_k141_202573 | Not-determined |
| S8_extracted_k141_38258 | Not-determined |
| S8_extracted_k141_133420 | Not-determined |
| S8_extracted_k141_46402 | Not-determined |
| S8_extracted_k141_29689 | Not-determined |
| S8_extracted_k141_72307 | Not-determined |
| S8_extracted_k141_29690 | Not-determined |
| S8_extracted_k141_168135 | Not-determined |
| S8_extracted_k141_3404 | Not-determined |
| S8_extracted_k141_46406 | Not-determined |
| S8_extracted_k141_98787 | Not-determined |
| S8_extracted_k141_176937 | Not-determined |
| S8_extracted_k141_72312 | Not-determined |
| S8_vs_k141_162053\|\|full | Medium-quality |
| S8_extracted_k141_20810 | Not-determined |
| S8_extracted_k141_55477 | Not-determined |
| S8_extracted_k141_168143 | Low-quality |
| S8_extracted_k141_107618 | Not-determined |
| S8_extracted_k141_98789 | Not-determined |
| S8_extracted_k141_90097 | Not-determined |
| S8_extracted_k141_90098 | Not-determined |
| S8_extracted_k141_202581 | Not-determined |
| S8_extracted_k141_29703 | Low-quality |
| S8_extracted_k141_38273 | Low-quality |
| S8_extracted_k141_29704 | Not-determined |
| S8_extracted_k141_98802 | Not-determined |
| S8_extracted_k141_159307 | Not-determined |
| S8_extracted_k141_81554 | Low-quality |
| S8_extracted_k141_55478 | Low-quality |
| S8_extracted_k141_159311 | Not-determined |
| S8_extracted_k141_55484 | Not-determined |
| S8_extracted_k141_98805 | Not-determined |
| S8_extracted_k141_98806 | Not-determined |
| S8_extracted_k141_133434 | Not-determined |
| S8_extracted_k141_90112 | Not-determined |
| S8_extracted_k141_107625 | Not-determined |
| S8_extracted_k141_176950 | Not-determined |
| S8_extracted_k141_176952 | Not-determined |
| S8_extracted_k141_38279 | Not-determined |
| S8_extracted_k141_29717 | Not-determined |
| S8_extracted_k141_90119 | Not-determined |
| S8_extracted_k141_168153 | Not-determined |
| S8_extracted_k141_38281 | Not-determined |
| S1_vs_k141_37646\|\|full | Medium-quality |
| S8_extracted_k141_107633 | Low-quality |
| S8_extracted_k141_20818 | Low-quality |
| S8_extracted_k141_38287 | Not-determined |
| S8_extracted_k141_20819 | Low-quality |
| S8_extracted_k141_133439 | Not-determined |
| S8_extracted_k141_159325 | Not-determined |
| S8_extracted_k141_38288 | Not-determined |
| S8_extracted_k141_46430 | Not-determined |
| S8_extracted_k141_55496 | Low-quality |
| S8_extracted_k141_168161 | Not-determined |
| S8_extracted_k141_55498 | Not-determined |
| S8_extracted_k141_3416 | Not-determined |
| S8_extracted_k141_194457 | Not-determined |
| S8_extracted_k141_194458 | Not-determined |
| S8_extracted_k141_90131 | Not-determined |
| S8_extracted_k141_133445 | Not-determined |
| S8_extracted_k141_46436 | Low-quality |
| S8_extracted_k141_202595 | Not-determined |
| S8_extracted_k141_194462 | Not-determined |
| S8_extracted_k141_202596 | Not-determined |
| S8_extracted_k141_133448 | Not-determined |
| S8_extracted_k141_133454 | Low-quality |
| S8_extracted_k141_150642 | Not-determined |
| S8_extracted_k141_202598 | Not-determined |
| S8_extracted_k141_38291 | Not-determined |
| S8_extracted_k141_46445 | Not-determined |
| S8_extracted_k141_116082 | Low-quality |
| S8_extracted_k141_142245 | Not-determined |
| S8_extracted_k141_55505 | Low-quality |
| S8_extracted_k141_133459 | Not-determined |
| S8_extracted_k141_29731 | Low-quality |
| S8_extracted_k141_202602 | Low-quality |
| S8_extracted_k141_159336 | Not-determined |
| S8_extracted_k141_133463 | Low-quality |
| S8_extracted_k141_107641 | Not-determined |
| S8_extracted_k141_90143 | Not-determined |
| S8_extracted_k141_116090 | Not-determined |
| S8_extracted_k141_202604 | Not-determined |
| S8_extracted_k141_20833 | Not-determined |
| S8_extracted_k141_176974 | Not-determined |
| S8_extracted_k141_133468 | Not-determined |
| S8_extracted_k141_55514 | Not-determined |
| S8_extracted_k141_29739 | Not-determined |
| S8_extracted_k141_20837 | Not-determined |
| S8_extracted_k141_55516 | Not-determined |
| S8_extracted_k141_90147 | Not-determined |
| S8_extracted_k141_29742 | Not-determined |
| S8_extracted_k141_159344 | Not-determined |
| S8_extracted_k141_55518 | Not-determined |
| S8_extracted_k141_46458 | Not-determined |
| S8_extracted_k141_185035 | Not-determined |
| S8_extracted_k141_194464 | Low-quality |
| S8_extracted_k141_81581 | Low-quality |
| S8_extracted_k141_90151 | Not-determined |
| S8_extracted_k141_133478 | Not-determined |
| S8_extracted_k141_107648 | Not-determined |
| S8_extracted_k141_29747 | Not-determined |
| S8_extracted_k141_90153 | Not-determined |
| S8_extracted_k141_20845 | Not-determined |
| S8_extracted_k141_125043 | Not-determined |
| S8_extracted_k141_29750 | Not-determined |
| S8_extracted_k141_194470 | Not-determined |
| S8_extracted_k141_142257 | Not-determined |
| S8_extracted_k141_20850 | Not-determined |
| S8_extracted_k141_133485 | Not-determined |
| S8_extracted_k141_3434 | Not-determined |
| S8_extracted_k141_29754 | Not-determined |
| S8_extracted_k141_116094 | Not-determined |
| S8_extracted_k141_133488 | Not-determined |
| S8_extracted_k141_3436 | Not-determined |
| S8_extracted_k141_81582 | Low-quality |
| S8_extracted_k141_159352 | Not-determined |
| S8_extracted_k141_29758 | Not-determined |
| S8_extracted_k141_81584 | Not-determined |
| S8_extracted_k141_176991 | Not-determined |
| S8_extracted_k141_159353 | Not-determined |
| S8_extracted_k141_107652 | Not-determined |
| S8_extracted_k141_176992 | Not-determined |
| S8_extracted_k141_20860 | Not-determined |
| S8_extracted_k141_98827 | Not-determined |
| S8_extracted_k141_150655 | Not-determined |
| S8_extracted_k141_98830 | Not-determined |
| S8_extracted_k141_159360 | Not-determined |
| S8_extracted_k141_3438 | Not-determined |
| S8_extracted_k141_20864 | Not-determined |
| S8_extracted_k141_3440 | Not-determined |
| S8_extracted_k141_194479 | Not-determined |
| S8_extracted_k141_168201 | Not-determined |
| S8_extracted_k141_159366 | Not-determined |
| S8_extracted_k141_194481 | Low-quality |
| S8_extracted_k141_29769 | Not-determined |
| S3_extracted_k141_83536 | Medium-quality |
| S8_extracted_k141_177002 | Not-determined |
| S8_extracted_k141_150659 | Not-determined |
| S8_extracted_k141_168204 | Not-determined |
| S8_extracted_k141_55543 | Not-determined |
| S8_extracted_k141_90162 | Not-determined |
| S8_extracted_k141_38302 | Not-determined |
| S8_extracted_k141_20868 | Not-determined |
| S8_extracted_k141_90167 | Low-quality |
| S8_extracted_k141_81590 | Not-determined |
| S8_extracted_k141_168214 | Not-determined |
| S8_extracted_k141_159377 | Not-determined |
| S8_extracted_k141_90172 | Not-determined |
| S8_extracted_k141_133523 | Not-determined |
| S8_extracted_k141_98846 | Not-determined |
| S8_extracted_k141_90174 | Not-determined |
| S8_extracted_k141_72337 | Low-quality |
| S8_extracted_k141_81596 | Not-determined |
| S8_extracted_k141_202626 | Not-determined |
| S8_extracted_k141_142297 | Not-determined |
| S8_extracted_k141_133528 | Not-determined |
| S8_extracted_k141_125062 | Low-quality |
| S8_extracted_k141_125063 | Not-determined |
| S8_extracted_k141_72341 | Not-determined |
| S8_extracted_k141_46495 | Not-determined |
| S8_extracted_k141_133532 | Not-determined |
| S8_extracted_k141_72343 | Not-determined |
| S8_extracted_k141_194493 | Not-determined |
| S8_extracted_k141_81599 | Not-determined |
| S7_extracted_69348 | Medium-quality |
| S8_extracted_k141_29791 | Not-determined |
| S8_extracted_k141_107675 | Not-determined |
| S8_extracted_k141_81603 | Not-determined |
| S8_extracted_k141_194495 | Not-determined |
| S8_extracted_k141_107678 | Not-determined |
| S8_extracted_k141_202632 | Not-determined |
| S8_extracted_k141_20890 | Not-determined |
| S8_extracted_k141_125070 | Low-quality |
| S8_extracted_k141_150676 | Not-determined |
| S8_extracted_k141_46502 | Not-determined |
| S8_extracted_k141_55559 | Not-determined |
| S8_extracted_k141_116117 | Not-determined |
| S8_extracted_k141_20894 | Not-determined |
| S8_extracted_k141_81608 | Not-determined |
| S8_extracted_k141_168226 | Not-determined |
| S8_extracted_k141_46506 | Not-determined |
| S8_extracted_k141_159386 | Not-determined |
| S8_extracted_k141_142308 | Low-quality |
| S8_extracted_k141_81612 | Not-determined |
| S8_extracted_k141_159389 | Not-determined |
| S8_extracted_k141_168230 | Not-determined |
| S8_extracted_k141_202639 | Not-determined |
| S8_extracted_k141_72355 | Not-determined |
| S8_extracted_k141_72357 | Not-determined |
| S8_extracted_k141_72358 | Not-determined |
| S8_extracted_k141_3486 | Low-quality |
| S8_extracted_k141_90183 | Low-quality |
| S8_extracted_k141_159395 | Not-determined |
| S8_extracted_k141_72360 | Not-determined |
| S8_extracted_k141_98865 | Not-determined |
| S8_extracted_k141_20897 | Low-quality |
| S8_extracted_k141_98868 | Not-determined |
| S8_extracted_k141_55565 | Not-determined |
| S8_extracted_k141_46526 | Not-determined |
| S8_extracted_k141_11814 | Not-determined |
| S8_extracted_k141_72367 | Not-determined |
| S8_extracted_k141_72368 | Not-determined |
| S8_extracted_k141_159403 | Not-determined |
| S8_extracted_k141_55566 | Not-determined |
| S8_extracted_k141_202644 | Not-determined |
| S8_extracted_k141_194507 | Low-quality |
| S8_extracted_k141_98874 | Not-determined |
| S8_extracted_k141_29803 | Not-determined |
| S8_extracted_k141_55573 | Not-determined |
| S8_extracted_k141_11820 | Not-determined |
| S8_extracted_k141_29810 | Not-determined |
| S8_extracted_k141_20903 | Not-determined |
| S8_extracted_k141_29813 | Not-determined |
| S8_extracted_k141_159406 | Low-quality |
| S8_extracted_k141_142315 | Not-determined |
| S8_extracted_k141_20906 | Low-quality |
| S8_extracted_k141_107706 | Not-determined |
| S8_extracted_k141_194519 | Not-determined |
| S8_extracted_k141_55577 | Not-determined |
| S8_extracted_k141_159413 | Not-determined |
| S8_extracted_k141_81620 | Not-determined |
| S8_extracted_k141_202657 | Not-determined |
| S8_extracted_k141_185043 | Not-determined |
| S8_extracted_k141_29821 | Not-determined |
| S8_extracted_k141_116130 | Not-determined |
| S8_extracted_k141_11826 | Not-determined |
| S8_extracted_k141_150709 | Not-determined |
| S8_extracted_k141_46536 | Not-determined |
| S8_extracted_k141_107714 | Not-determined |
| S8_extracted_k141_168249 | Low-quality |
| S8_extracted_k141_202661 | Not-determined |
| S8_extracted_k141_11831 | Not-determined |
| S8_extracted_k141_194525 | Low-quality |
| S8_extracted_k141_46540 | Not-determined |
| S8_extracted_k141_185047 | Not-determined |
| S8_extracted_k141_107721 | Not-determined |
| S8_extracted_k141_11837 | Low-quality |
| S8_extracted_k141_194528 | Low-quality |
| S8_extracted_k141_3520 | Not-determined |
| S8_extracted_k141_46542 | Not-determined |
| S8_extracted_k141_107724 | Not-determined |
| S8_extracted_k141_46543 | Not-determined |
| S8_extracted_k141_55590 | Low-quality |
| S8_extracted_k141_11842 | Not-determined |
| S8_extracted_k141_11845 | Not-determined |
| S8_extracted_k141_202671 | Low-quality |
| S8_extracted_k141_98884 | Low-quality |
| S8_extracted_k141_168260 | Not-determined |
| S8_extracted_k141_194533 | Low-quality |
| S8_extracted_k141_185054 | Not-determined |
| S8_extracted_k141_202674 | Low-quality |
| S8_extracted_k141_150724 | Not-determined |
| S8_extracted_k141_98886 | Not-determined |
| S8_extracted_k141_185060 | Not-determined |
| S8_extracted_k141_185062 | Not-determined |
| S8_extracted_k141_90200 | Not-determined |
| S8_extracted_k141_98887 | Not-determined |
| S8_extracted_k141_20912 | Not-determined |
| S8_extracted_k141_90201 | Not-determined |
| S8_extracted_k141_90204 | Not-determined |
| S8_extracted_k141_107737 | Not-determined |
| S8_extracted_k141_20916 | Not-determined |
| S8_extracted_k141_81635 | Low-quality |
| S8_extracted_k141_90206 | Not-determined |
| S8_extracted_k141_159424 | Not-determined |
| S8_extracted_k141_20919 | Not-determined |
| S8_extracted_k141_168273 | Not-determined |
| S8_extracted_k141_168274 | Not-determined |
| S8_extracted_k141_81638 | Not-determined |
| S8_extracted_k141_194543 | Not-determined |
| S8_extracted_k141_159427 | Not-determined |
| S8_extracted_k141_202691 | Not-determined |
| S8_extracted_k141_20921 | Not-determined |
| S8_extracted_k141_159429 | Not-determined |
| S8_extracted_k141_46555 | Not-determined |
| S8_extracted_k141_98903 | Not-determined |
| S8_extracted_k141_159433 | Not-determined |
| S8_extracted_k141_46560 | Not-determined |
| S8_extracted_k141_202696 | Not-determined |
| S8_extracted_k141_159434 | Not-determined |
| S8_extracted_k141_116148 | Not-determined |
| S8_extracted_k141_3554 | Not-determined |
| S8_extracted_k141_90215 | Not-determined |
| S8_extracted_k141_185068 | Not-determined |
| S8_extracted_k141_116152 | Not-determined |
| S8_extracted_k141_194562 | Not-determined |
| S8_extracted_k141_107755 | Not-determined |
| S8_extracted_k141_72402 | Not-determined |
| S8_extracted_k141_98910 | Not-determined |
| S8_extracted_k141_20930 | Not-determined |
| S8_extracted_k141_150738 | Low-quality |
| S8_extracted_k141_125073 | Not-determined |
| S8_extracted_k141_116161 | Not-determined |
| S8_extracted_k141_125074 | Low-quality |
| S8_extracted_k141_116165 | Not-determined |
| S8_extracted_k141_81648 | Not-determined |
| S8_extracted_k141_107758 | Low-quality |
| S8_extracted_k141_46567 | Not-determined |
| S8_extracted_k141_125080 | Not-determined |
| S8_extracted_k141_107761 | Not-determined |
| S8_extracted_k141_185081 | Low-quality |
| S8_extracted_k141_46572 | Not-determined |
| S8_extracted_k141_185083 | Not-determined |
| S8_extracted_k141_98922 | Not-determined |
| S8_extracted_k141_185084 | Not-determined |
| S8_extracted_k141_185085 | Not-determined |
| S8_extracted_k141_20935 | Not-determined |
| S8_extracted_k141_98924 | Low-quality |
| S8_extracted_k141_142359 | Not-determined |
| S8_extracted_k141_90227 | Not-determined |
| S8_extracted_k141_125085 | Not-determined |
| S8_extracted_k141_125088 | Not-determined |
| S8_extracted_k141_194590 | Not-determined |
| S8_extracted_k141_185093 | Not-determined |
| S8_extracted_k141_55621 | Low-quality |
| S8_extracted_k141_90230 | Not-determined |
| S8_extracted_k141_90232 | Low-quality |
| S8_extracted_k141_142365 | Low-quality |
| S8_extracted_k141_142366 | Not-determined |
| S8_extracted_k141_185097 | Not-determined |
| S8_extracted_k141_142368 | Not-determined |
| S8_extracted_k141_150753 | Not-determined |
| S8_extracted_k141_159446 | Not-determined |
| S8_extracted_k141_202735 | Low-quality |
| S8_extracted_k141_107773 | Not-determined |
| S8_extracted_k141_107775 | Not-determined |
| S8_extracted_k141_107776 | Not-determined |
| S8_extracted_k141_194594 | Not-determined |
| S8_extracted_k141_142370 | Not-determined |
| S8_extracted_k141_185102 | Not-determined |
| S8_extracted_k141_116183 | Low-quality |
| S8_extracted_k141_142373 | Not-determined |
| S8_extracted_k141_107782 | Not-determined |
| S8_extracted_k141_185106 | Not-determined |
| S8_extracted_k141_142375 | Low-quality |
| S8_extracted_k141_29830 | Not-determined |
| S8_extracted_k141_81656 | Not-determined |
| S8_extracted_k141_46584 | Not-determined |
| S8_extracted_k141_168308 | Not-determined |
| S8_extracted_k141_81659 | Not-determined |
| S8_extracted_k141_142378 | Not-determined |
| S8_extracted_k141_46588 | Not-determined |
| S8_extracted_k141_72423 | Not-determined |
| S8_extracted_k141_116190 | Not-determined |
| S8_extracted_k141_72424 | Not-determined |
| S8_extracted_k141_125107 | Not-determined |
| S8_extracted_k141_177060 | Not-determined |
| S8_extracted_k141_90237 | Not-determined |
| S8_extracted_k141_46592 | Not-determined |
| S8_extracted_k141_98943 | Not-determined |
| S8_extracted_k141_202743 | Not-determined |
| S8_extracted_k141_29841 | Not-determined |
| S8_extracted_k141_3577 | Not-determined |
| S8_extracted_k141_98944 | Not-determined |
| S8_extracted_k141_20957 | Not-determined |
| S8_extracted_k141_159464 | Not-determined |
| S8_extracted_k141_55637 | Not-determined |
| S8_extracted_k141_168311 | Low-quality |
| S8_extracted_k141_81671 | Not-determined |
| S8_extracted_k141_159469 | Not-determined |
| S8_extracted_k141_177073 | Not-determined |
| S8_extracted_k141_202747 | Not-determined |
| S8_extracted_k141_168314 | Not-determined |
| S8_extracted_k141_177074 | Low-quality |
| S8_extracted_k141_125117 | Not-determined |
| S8_extracted_k141_3585 | Not-determined |
| S8_extracted_k141_116202 | Not-determined |
| S8_extracted_k141_177077 | Low-quality |
| S8_extracted_k141_150759 | Low-quality |
| S8_extracted_k141_11858 | Not-determined |
| S8_extracted_k141_3591 | Not-determined |
| S8_extracted_k141_98951 | Not-determined |
| S8_extracted_k141_107799 | Not-determined |
| S8_extracted_k141_20961 | Not-determined |
| S8_extracted_k141_20962 | Not-determined |
| S8_extracted_k141_81675 | Not-determined |
| S8_extracted_k141_168321 | Not-determined |
| S8_extracted_k141_20963 | Not-determined |
| S8_extracted_k141_107802 | Low-quality |
| S8_extracted_k141_46602 | Not-determined |
| S8_extracted_k141_107803 | Not-determined |
| S8_extracted_k141_81678 | Not-determined |
| S8_extracted_k141_46603 | Low-quality |
| S8_extracted_k141_125127 | Not-determined |
| S8_extracted_k141_46606 | Not-determined |
| S8_extracted_k141_55648 | Not-determined |
| S8_extracted_k141_90245 | Not-determined |
| S8_extracted_k141_20967 | Not-determined |
| S8_extracted_k141_159483 | Not-determined |
| S8_extracted_k141_159485 | Not-determined |
| S8_extracted_k141_125134 | Low-quality |
| S8_extracted_k141_81683 | Not-determined |
| S8_extracted_k141_46609 | Not-determined |
| S8_extracted_k141_29861 | Not-determined |
| S8_extracted_k141_125137 | Not-determined |
| S8_extracted_k141_177092 | Not-determined |
| S8_extracted_k141_55652 | Low-quality |
| S8_extracted_k141_177093 | Not-determined |
| S8_extracted_k141_168329 | Not-determined |
| S8_extracted_k141_125139 | Not-determined |
| S8_extracted_k141_177097 | Not-determined |
| S8_extracted_k141_81685 | Not-determined |
| S8_extracted_k141_159493 | Not-determined |
| S8_extracted_k141_159494 | Not-determined |
| S8_extracted_k141_98959 | Not-determined |
| S8_extracted_k141_202758 | Not-determined |
| S8_extracted_k141_142395 | Low-quality |
| S8_extracted_k141_116218 | Not-determined |
| S8_extracted_k141_177098 | Not-determined |
| S8_extracted_k141_150771 | Not-determined |
| S8_extracted_k141_90251 | Not-determined |
| S8_extracted_k141_46615 | Not-determined |
| S8_extracted_k141_3608 | Not-determined |
| S8_extracted_k141_46616 | Not-determined |
| S8_extracted_k141_29866 | Not-determined |
| S8_extracted_k141_29867 | Not-determined |
| S8_extracted_k141_168335 | Low-quality |
| S8_extracted_k141_116220 | Low-quality |
| S8_extracted_k141_185124 | Not-determined |
| S8_extracted_k141_90257 | Not-determined |
| S8_extracted_k141_150777 | Not-determined |
| S8_extracted_k141_125144 | Not-determined |
| S8_extracted_k141_3615 | Not-determined |
| S8_extracted_k141_168341 | Not-determined |
| S8_extracted_k141_98962 | Not-determined |
| S8_extracted_k141_3620 | Not-determined |
| S8_extracted_k141_98965 | Not-determined |
| S8_extracted_k141_98968 | Not-determined |
| S8_extracted_k141_3624 | Not-determined |
| S8_extracted_k141_107815 | Not-determined |
| S8_extracted_k141_3626 | Not-determined |
| S8_extracted_k141_98977 | Not-determined |
| S8_extracted_k141_3627 | Low-quality |
| S8_extracted_k141_177112 | Not-determined |
| S8_extracted_k141_107819 | Not-determined |
| S8_extracted_k141_3629 | Not-determined |
| S8_extracted_k141_90275 | Low-quality |
| S8_extracted_k141_125157 | Not-determined |
| S8_extracted_k141_177117 | Low-quality |
| S8_extracted_k141_168359 | Low-quality |
| S8_extracted_k141_90283 | Not-determined |
| S8_extracted_k141_38305 | Not-determined |
| S8_extracted_k141_98985 | Not-determined |
| S8_extracted_k141_38310 | Not-determined |
| S3_vs_k141_7137\|\|full | Medium-quality |
| S8_extracted_k141_185129 | Not-determined |
| S8_extracted_k141_3637 | Not-determined |
| S8_extracted_k141_185131 | Not-determined |
| S8_extracted_k141_159501 | Not-determined |
| S8_extracted_k141_98990 | Low-quality |
| S8_extracted_k141_3642 | Not-determined |
| S8_extracted_k141_159504 | Not-determined |
| S8_extracted_k141_20984 | Not-determined |
| S8_extracted_k141_202764 | Not-determined |
| S8_extracted_k141_150789 | Not-determined |
| S8_extracted_k141_38313 | Not-determined |
| S8_extracted_k141_168370 | Low-quality |
| S8_extracted_k141_72462 | Not-determined |
| S8_extracted_k141_98991 | Not-determined |
| S8_extracted_k141_116241 | Not-determined |
| S8_extracted_k141_116242 | Not-determined |
| S8_extracted_k141_81698 | Not-determined |
| S8_extracted_k141_177129 | Not-determined |
| S8_extracted_k141_81701 | Not-determined |
| S8_extracted_k141_3653 | Low-quality |
| S8_extracted_k141_107836 | Not-determined |
| S8_extracted_k141_29890 | Low-quality |
| S8_extracted_k141_11893 | Not-determined |
| S8_extracted_k141_55672 | Not-determined |
| S8_extracted_k141_46630 | Not-determined |
| S8_extracted_k141_159511 | Low-quality |
| S8_extracted_k141_81704 | Not-determined |
| S8_extracted_k141_3656 | Low-quality |
| S8_extracted_k141_81706 | Not-determined |
| S8_extracted_k141_168373 | Not-determined |
| S8_extracted_k141_116248 | Not-determined |
| S8_extracted_k141_159515 | Not-determined |
| S8_extracted_k141_72474 | Not-determined |
| S8_extracted_k141_159516 | Not-determined |
| S8_extracted_k141_90301 | Not-determined |
| S8_extracted_k141_72476 | Not-determined |
| S8_extracted_k141_90305 | Not-determined |
| S8_extracted_k141_194631 | Not-determined |
| S8_extracted_k141_90310 | Low-quality |
| S8_extracted_k141_11902 | Not-determined |
| S8_extracted_k141_72484 | Not-determined |
| S8_extracted_k141_46639 | Not-determined |
| S8_extracted_k141_142416 | Not-determined |
| S8_extracted_k141_11905 | Not-determined |
| S8_extracted_k141_38318 | Not-determined |
| S8_extracted_k141_20992 | Not-determined |
| S8_extracted_k141_72486 | Low-quality |
| S8_extracted_k141_72487 | Not-determined |
| S8_extracted_k141_159523 | Not-determined |
| S8_extracted_k141_125189 | Not-determined |
| S8_extracted_k141_11912 | Low-quality |
| S8_extracted_k141_150800 | Not-determined |
| S8_extracted_k141_72489 | Not-determined |
| S8_extracted_k141_177135 | Low-quality |
| S8_extracted_k141_125190 | Not-determined |
| S8_extracted_k141_46640 | Low-quality |
| S8_extracted_k141_46641 | Not-determined |
| S8_extracted_k141_46644 | Not-determined |
| S8_extracted_k141_90314 | Not-determined |
| S8_extracted_k141_125192 | Not-determined |
| S8_extracted_k141_99018 | Not-determined |
| S8_extracted_k141_194642 | Not-determined |
| S8_extracted_k141_81711 | Low-quality |
| S8_extracted_k141_55689 | Not-determined |
| S8_extracted_k141_107861 | Not-determined |
| S8_extracted_k141_168397 | Not-determined |
| S8_extracted_k141_168398 | Not-determined |
| S8_extracted_k141_107864 | Not-determined |
| S8_extracted_k141_142432 | Not-determined |
| S8_extracted_k141_150815 | Not-determined |
| S8_extracted_k141_38329 | Not-determined |
| S8_extracted_k141_150817 | Low-quality |
| S8_extracted_k141_72503 | Not-determined |
| S8_extracted_k141_99026 | Not-determined |
| S8_extracted_k141_159535 | Not-determined |
| S8_extracted_k141_185156 | Not-determined |
| S8_extracted_k141_116272 | Not-determined |
| S8_extracted_k141_55699 | Not-determined |
| S8_extracted_k141_3674 | Not-determined |
| S8_extracted_k141_194645 | Not-determined |
| S8_extracted_k141_99032 | Not-determined |
| S8_extracted_k141_55701 | Not-determined |
| S8_extracted_k141_29913 | Not-determined |
| S8_extracted_k141_202789 | Not-determined |
| S8_extracted_k141_159542 | Not-determined |
| S8_extracted_k141_3676 | Not-determined |
| S8_extracted_k141_150823 | Not-determined |
| S8_extracted_k141_185160 | Not-determined |
| S8_extracted_k141_168405 | Low-quality |
| S8_extracted_k141_177139 | Not-determined |
| S8_extracted_k141_46657 | Low-quality |
| S8_extracted_k141_116279 | Not-determined |
| S8_extracted_k141_194653 | Not-determined |
| S8_extracted_k141_3682 | Not-determined |
| S8_extracted_k141_55704 | Not-determined |
| S8_extracted_k141_3684 | Not-determined |
| S8_extracted_k141_116281 | Not-determined |
| S8_extracted_k141_116283 | Not-determined |
| S8_extracted_k141_3689 | Not-determined |
| S8_extracted_k141_90331 | Not-determined |
| S8_extracted_k141_150833 | Not-determined |
| S8_extracted_k141_142440 | Not-determined |
| S8_extracted_k141_125202 | Not-determined |
| S8_extracted_k141_125203 | Not-determined |
| S8_extracted_k141_159545 | Not-determined |
| S8_extracted_k141_116289 | Not-determined |
| S8_extracted_k141_168410 | Not-determined |
| S8_extracted_k141_99049 | Not-determined |
| S8_extracted_k141_29926 | Not-determined |
| S8_extracted_k141_99050 | Not-determined |
| S8_extracted_k141_29929 | Not-determined |
| S8_extracted_k141_90338 | Not-determined |
| S8_extracted_k141_159546 | Not-determined |
| S8_extracted_k141_202808 | Not-determined |
| S8_extracted_k141_125207 | Not-determined |
| S8_extracted_k141_90341 | Not-determined |
| S8_extracted_k141_29931 | Not-determined |
| S8_extracted_k141_99053 | Not-determined |
| S8_extracted_k141_159550 | Not-determined |
| S8_extracted_k141_168415 | Not-determined |
| S8_extracted_k141_90342 | Not-determined |

| | |
|---|---|
| S8_extracted_k141_55712 | Not-determined |
| S8_extracted_k141_194658 | Not-determined |
| S8_extracted_k141_177146 | Not-determined |
| S8_extracted_k141_116290 | Low-quality |
| S8_extracted_k141_99054 | Not-determined |
| S8_extracted_k141_46674 | Not-determined |
| S8_extracted_k141_46675 | Not-determined |
| S8_extracted_k141_81716 | Not-determined |
| S8_extracted_k141_125211 | Not-determined |
| S8_extracted_k141_90349 | Not-determined |
| S8_extracted_k141_116298 | Not-determined |
| S8_extracted_k141_46680 | Not-determined |
| S8_extracted_k141_90350 | Not-determined |
| S8_extracted_k141_168418 | Not-determined |
| S8_extracted_k141_55717 | Not-determined |
| S8_extracted_k141_29935 | Not-determined |
| S8_extracted_k141_168420 | Not-determined |
| S8_extracted_k141_116300 | Low-quality |
| S8_extracted_k141_168421 | Not-determined |
| S8_extracted_k141_3697 | Not-determined |
| S8_extracted_k141_177155 | Not-determined |
| S8_extracted_k141_177156 | Not-determined |
| S8_extracted_k141_185186 | Not-determined |
| S8_extracted_k141_194663 | Low-quality |
| S8_extracted_k141_3700 | Not-determined |
| S8_extracted_k141_72526 | Not-determined |
| S8_extracted_k141_168429 | Not-determined |
| S8_extracted_k141_177159 | Not-determined |
| S8_extracted_k141_29945 | Not-determined |
| S8_extracted_k141_185187 | Not-determined |
| S8_extracted_k141_159556 | Not-determined |
| S8_extracted_k141_90358 | Not-determined |
| S8_extracted_k141_72531 | Not-determined |
| S8_extracted_k141_107885 | Not-determined |
| S8_extracted_k141_177160 | Low-quality |
| S8_extracted_k141_159558 | Not-determined |
| S8_extracted_k141_116313 | Not-determined |
| S8_extracted_k141_159561 | Not-determined |
| S8_extracted_k141_194674 | Not-determined |
| S8_extracted_k141_168438 | Not-determined |
| S8_extracted_k141_159565 | Not-determined |
| S8_extracted_k141_159566 | Not-determined |
| S8_extracted_k141_55734 | Not-determined |
| S8_extracted_k141_72534 | Not-determined |
| S8_extracted_k141_90367 | Not-determined |
| S8_extracted_k141_168440 | Not-determined |
| S8_extracted_k141_116316 | Not-determined |
| S8_extracted_k141_177168 | Not-determined |
| S8_extracted_k141_116318 | Not-determined |
| S8_extracted_k141_194681 | Not-determined |
| S8_extracted_k141_194682 | Not-determined |
| S8_extracted_k141_125233 | Low-quality |
| S8_extracted_k141_55742 | Not-determined |
| S8_extracted_k141_72546 | Not-determined |
| S8_extracted_k141_116324 | Low-quality |
| S8_extracted_k141_142472 | Not-determined |
| S8_extracted_k141_72548 | Not-determined |
| S8_extracted_k141_81734 | Not-determined |
| S8_extracted_k141_55743 | Not-determined |
| S8_extracted_k141_159582 | Not-determined |
| S8_extracted_k141_202831 | Not-determined |
| S8_extracted_k141_81738 | Not-determined |
| S8_extracted_k141_3720 | Not-determined |
| S8_extracted_k141_29963 | Not-determined |
| S8_extracted_k141_159586 | Not-determined |
| S8_extracted_k141_55751 | Not-determined |
| S8_extracted_k141_81741 | Not-determined |
| S8_extracted_k141_99080 | Not-determined |
| S8_extracted_k141_55753 | Not-determined |
| S8_extracted_k141_3725 | Not-determined |
| S8_extracted_k141_99082 | Low-quality |
| S8_extracted_k141_116330 | Not-determined |
| S8_extracted_k141_99083 | Not-determined |
| S8_extracted_k141_116334 | Not-determined |
| S8_extracted_k141_99087 | Not-determined |
| S8_extracted_k141_168465 | Not-determined |
| S8_extracted_k141_90379 | Not-determined |
| S8_extracted_k141_125245 | Not-determined |
| S8_extracted_k141_29974 | Not-determined |
| S8_extracted_k141_90382 | Not-determined |
| S8_extracted_k141_81743 | Low-quality |
| S8_extracted_k141_125248 | Low-quality |
| S8_extracted_k141_81744 | Not-determined |
| S8_extracted_k141_142487 | Not-determined |
| S8_extracted_k141_90385 | Low-quality |
| S8_extracted_k141_125251 | Not-determined |
| S8_extracted_k141_116339 | Not-determined |
| S8_extracted_k141_202838 | Not-determined |
| S8_extracted_k141_90386 | Not-determined |
| S8_extracted_k141_55757 | Not-determined |
| S8_extracted_k141_99091 | Not-determined |
| S8_extracted_k141_72563 | Not-determined |
| S8_extracted_k141_150858 | Not-determined |
| S8_extracted_k141_99094 | Not-determined |
| S8_extracted_k141_64158 | Not-determined |
| S8_extracted_k141_46705 | Not-determined |
| S8_extracted_k141_159595 | Not-determined |
| S8_extracted_k141_81747 | Low-quality |
| S8_extracted_k141_64160 | Low-quality |
| S8_extracted_k141_3727 | Low-quality |
| S8_extracted_k141_150863 | Not-determined |
| S8_extracted_k141_142493 | Not-determined |
| S8_extracted_k141_64163 | Not-determined |
| S8_extracted_k141_81749 | Not-determined |
| S8_extracted_k141_29984 | Not-determined |
| S8_extracted_k141_81751 | Not-determined |
| S8_extracted_k141_99102 | Not-determined |
| S8_extracted_k141_90393 | Not-determined |
| S8_extracted_k141_64167 | Not-determined |
| S8_extracted_k141_29988 | Not-determined |
| S8_extracted_k141_55769 | Low-quality |
| S8_extracted_k141_168468 | Not-determined |
| S8_extracted_k141_29989 | Not-determined |
| S8_extracted_k141_29990 | Not-determined |
| S8_extracted_k141_55771 | Not-determined |
| S8_extracted_k141_72571 | Not-determined |
| S8_extracted_k141_72573 | Not-determined |
| S8_extracted_k141_99108 | Not-determined |
| S8_extracted_k141_202854 | Not-determined |
| S8_extracted_k141_55774 | Not-determined |
| S8_extracted_k141_55776 | Not-determined |
| S8_extracted_k141_72578 | Low-quality |
| S8_extracted_k141_116347 | Not-determined |
| S8_extracted_k141_55779 | Not-determined |
| S8_extracted_k141_55780 | Not-determined |
| S8_extracted_k141_29991 | Not-determined |
| S8_extracted_k141_72580 | Not-determined |
| S8_extracted_k141_116350 | Not-determined |
| S8_extracted_k141_142503 | Not-determined |
| S8_extracted_k141_81762 | Not-determined |
| S8_extracted_k141_150875 | Not-determined |
| S8_extracted_k141_29994 | Not-determined |
| S8_extracted_k141_3733 | Not-determined |
| S8_extracted_k141_116354 | Low-quality |
| S8_extracted_k141_125262 | Not-determined |
| S8_extracted_k141_116355 | Not-determined |
| S8_extracted_k141_185210 | Not-determined |
| S8_extracted_k141_72586 | Not-determined |
| S8_extracted_k141_55787 | Not-determined |
| S8_extracted_k141_72587 | Not-determined |
| S8_extracted_k141_202866 | Not-determined |
| S8_extracted_k141_81765 | Not-determined |
| S8_extracted_k141_99113 | Not-determined |
| S8_extracted_k141_30004 | Low-quality |
| S8_extracted_k141_3736 | Low-quality |
| S8_extracted_k141_72588 | Not-determined |
| S8_extracted_k141_21006 | Not-determined |
| S8_extracted_k141_107925 | Low-quality |
| S8_extracted_k141_125267 | Not-determined |
| S8_extracted_k141_81768 | Not-determined |
| S8_extracted_k141_185214 | Low-quality |
| S8_extracted_k141_168480 | Not-determined |
| S8_extracted_k141_133563 | Not-determined |
| S8_extracted_k141_168482 | Not-determined |
| S8_extracted_k141_202874 | Not-determined |
| S8_extracted_k141_46723 | Not-determined |
| S8_extracted_k141_21015 | Not-determined |
| S8_extracted_k141_107926 | Not-determined |
| S8_extracted_k141_116370 | Not-determined |
| S8_extracted_k141_30008 | Not-determined |
| S8_extracted_k141_125275 | Not-determined |
| S8_extracted_k141_116371 | Not-determined |
| S8_extracted_k141_116373 | Not-determined |
| S8_extracted_k141_46728 | Low-quality |
| S8_extracted_k141_116376 | Not-determined |
| S8_extracted_k141_21020 | Not-determined |
| S8_extracted_k141_21021 | Not-determined |
| S8_extracted_k141_30014 | Not-determined |
| S8_extracted_k141_72605 | Not-determined |
| S8_extracted_k141_168487 | Not-determined |
| S8_extracted_k141_46732 | Not-determined |
| S8_extracted_k141_46733 | Not-determined |
| S8_extracted_k141_107939 | Not-determined |
| S8_extracted_k141_72609 | Not-determined |
| S8_extracted_k141_72610 | Not-determined |
| S8_extracted_k141_185219 | Low-quality |
| S8_extracted_k141_30020 | Not-determined |
| S8_extracted_k141_30021 | Not-determined |
| S8_extracted_k141_72612 | Low-quality |
| S8_extracted_k141_90408 | Not-determined |
| S8_extracted_k141_142516 | Not-determined |
| S8_extracted_k141_46735 | Not-determined |
| S8_extracted_k141_21032 | Low-quality |
| S8_extracted_k141_142517 | Not-determined |
| S8_extracted_k141_30025 | Not-determined |
| S8_extracted_k141_3758 | Not-determined |
| S8_extracted_k141_90414 | Not-determined |
| S8_extracted_k141_168494 | Not-determined |
| S8_extracted_k141_72617 | Not-determined |
| S8_extracted_k141_46741 | Not-determined |
| S8_extracted_k141_72619 | Not-determined |
| S8_extracted_k141_159633 | Not-determined |
| S8_extracted_k141_116383 | Not-determined |
| S8_extracted_k141_185229 | Not-determined |
| S8_extracted_k141_159634 | Not-determined |
| S8_extracted_k141_30027 | Not-determined |
| S8_extracted_k141_202892 | Not-determined |
| S8_extracted_k141_64196 | Not-determined |
| S8_extracted_k141_202894 | Not-determined |
| S8_extracted_k141_177221 | Not-determined |
| S8_extracted_k141_168504 | Not-determined |
| S8_extracted_k141_116390 | Not-determined |
| S8_extracted_k141_194737 | Not-determined |
| S8_extracted_k141_159643 | Not-determined |
| S8_extracted_k141_168505 | Not-determined |
| S8_extracted_k141_150888 | Not-determined |
| S8_extracted_k141_21040 | Not-determined |
| S8_extracted_k141_3774 | Not-determined |
| S8_extracted_k141_3776 | Not-determined |
| S8_extracted_k141_159645 | Low-quality |
| S8_extracted_k141_64202 | Not-determined |
| S8_extracted_k141_150892 | Not-determined |
| S8_extracted_k141_116395 | Not-determined |
| S8_extracted_k141_142532 | Not-determined |
| S8_extracted_k141_3781 | Not-determined |
| S8_extracted_k141_142533 | Low-quality |
| S8_extracted_k141_64205 | Not-determined |
| S8_extracted_k141_99138 | Not-determined |
| S8_extracted_k141_142534 | Not-determined |
| S8_extracted_k141_168513 | Low-quality |
| S8_extracted_k141_107961 | Not-determined |
| S8_extracted_k141_142535 | Not-determined |
| S8_extracted_k141_142536 | Not-determined |
| S8_extracted_k141_64208 | Not-determined |
| S8_extracted_k141_116399 | Not-determined |
| S8_extracted_k141_133577 | Not-determined |
| S8_extracted_k141_99140 | Not-determined |
| S8_extracted_k141_46752 | Not-determined |
| S8_extracted_k141_107965 | Not-determined |
| S8_extracted_k141_116401 | Not-determined |
| S8_extracted_k141_64214 | Not-determined |
| S8_extracted_k141_107969 | Not-determined |
| S8_extracted_k141_72643 | Not-determined |
| S8_extracted_k141_150903 | Not-determined |
| S8_extracted_k141_64217 | Not-determined |
| S8_extracted_k141_72645 | Not-determined |
| S8_extracted_k141_55808 | Low-quality |
| S8_extracted_k141_72646 | Not-determined |
| S8_extracted_k141_3788 | Not-determined |
| S8_extracted_k141_21046 | Not-determined |
| S8_extracted_k141_194756 | Not-determined |
| S8_extracted_k141_21049 | Not-determined |
| S8_extracted_k141_99153 | Not-determined |
| S8_extracted_k141_133580 | Not-determined |
| S8_extracted_k141_72652 | Not-determined |
| S8_extracted_k141_64223 | Not-determined |
| S8_extracted_k141_30053 | Not-determined |
| S8_extracted_k141_21052 | Not-determined |
| S8_extracted_k141_38340 | Not-determined |
| S8_extracted_k141_55811 | Not-determined |
| S8_extracted_k141_150909 | Not-determined |
| S8_extracted_k141_21053 | Not-determined |
| S8_extracted_k141_177238 | Not-determined |
| S8_extracted_k141_150911 | Not-determined |
| S8_extracted_k141_46762 | Low-quality |
| S8_extracted_k141_46763 | Not-determined |
| S8_extracted_k141_55814 | Not-determined |
| S8_extracted_k141_142550 | Not-determined |
| S8_extracted_k141_99160 | Not-determined |
| S8_extracted_k141_30056 | Low-quality |
| S8_extracted_k141_150913 | Not-determined |
| S8_extracted_k141_177241 | Not-determined |
| S8_extracted_k141_64230 | Not-determined |
| S8_extracted_k141_55815 | Not-determined |
| S8_extracted_k141_72656 | Not-determined |
| S8_extracted_k141_3798 | Not-determined |
| S8_extracted_k141_159652 | Not-determined |
| S8_extracted_k141_55821 | Not-determined |
| S8_extracted_k141_99162 | Not-determined |
| S8_extracted_k141_30064 | Not-determined |
| S8_extracted_k141_55822 | Not-determined |
| S8_extracted_k141_21062 | Not-determined |
| S8_extracted_k141_38344 | Not-determined |
| S8_extracted_k141_107973 | Not-determined |
| S8_extracted_k141_168529 | Not-determined |
| S8_extracted_k141_142559 | Not-determined |
| S8_extracted_k141_3805 | Low-quality |
| S8_extracted_k141_116420 | Not-determined |
| S8_extracted_k141_168535 | Not-determined |
| S8_extracted_k141_142561 | Not-determined |
| S8_extracted_k141_38352 | Not-determined |
| S8_extracted_k141_3810 | Not-determined |
| S8_extracted_k141_38354 | Not-determined |
| S8_extracted_k141_142565 | Not-determined |
| S8_extracted_k141_168539 | Not-determined |
| S8_extracted_k141_194766 | Not-determined |
| S8_extracted_k141_64236 | Not-determined |
| S8_extracted_k141_177246 | Not-determined |
| S8_extracted_k141_194767 | Not-determined |
| S8_extracted_k141_46770 | Not-determined |
| S8_extracted_k141_194768 | Not-determined |
| S8_extracted_k141_55836 | Not-determined |
| S8_extracted_k141_21080 | Not-determined |
| S8_extracted_k141_107980 | Not-determined |
| S8_extracted_k141_202914 | Low-quality |
| S8_extracted_k141_72666 | Not-determined |
| S8_extracted_k141_64239 | Not-determined |
| S8_extracted_k141_3812 | Low-quality |
| S8_extracted_k141_46772 | Not-determined |
| S8_extracted_k141_64240 | Not-determined |
| S8_extracted_k141_21082 | Not-determined |
| S8_extracted_k141_90434 | Not-determined |
| S8_extracted_k141_21083 | Not-determined |
| S8_extracted_k141_168542 | Low-quality |
| S8_extracted_k141_21084 | Not-determined |
| S8_extracted_k141_72668 | Not-determined |
| S8_extracted_k141_21085 | Not-determined |
| S8_extracted_k141_21086 | Not-determined |
| S8_extracted_k141_202919 | Not-determined |
| S8_extracted_k141_177254 | Not-determined |
| S8_extracted_k141_133612 | Not-determined |
| S8_extracted_k141_90436 | Not-determined |
| S8_extracted_k141_150919 | Not-determined |
| S8_extracted_k141_125301 | Not-determined |
| S8_extracted_k141_125302 | Not-determined |
| S8_extracted_k141_38370 | Low-quality |
| S8_extracted_k141_159657 | Not-determined |
| S8_extracted_k141_142577 | Not-determined |
| S8_extracted_k141_133616 | Not-determined |
| S8_extracted_k141_21091 | Not-determined |
| S8_extracted_k141_177261 | Not-determined |
| S8_extracted_k141_133617 | Not-determined |
| S8_extracted_k141_168545 | Low-quality |
| S8_extracted_k141_3825 | Not-determined |
| S8_extracted_k141_159659 | Not-determined |
| S8_extracted_k141_46782 | Not-determined |
| S8_extracted_k141_177265 | Not-determined |
| S8_extracted_k141_142581 | Not-determined |
| S8_extracted_k141_150928 | Low-quality |
| S8_extracted_k141_142582 | Not-determined |
| S8_extracted_k141_142583 | Not-determined |
| S8_extracted_k141_55844 | Not-determined |
| S8_extracted_k141_55845 | Not-determined |
| S8_extracted_k141_55846 | Not-determined |
| S8_extracted_k141_38376 | Not-determined |
| S8_extracted_k141_142585 | Not-determined |
| S8_extracted_k141_177272 | Not-determined |
| S8_extracted_k141_142586 | Not-determined |
| S8_extracted_k141_72687 | Not-determined |
| S8_extracted_k141_3837 | Not-determined |
| S8_extracted_k141_11939 | Not-determined |
| S8_extracted_k141_72690 | Not-determined |
| S8_extracted_k141_90452 | Low-quality |
| S8_extracted_k141_168553 | Not-determined |
| S8_extracted_k141_168555 | Low-quality |
| S8_extracted_k141_168556 | Low-quality |
| S8_extracted_k141_202935 | Not-determined |
| S8_extracted_k141_99187 | Not-determined |
| S8_extracted_k141_116432 | Not-determined |
| S8_extracted_k141_133630 | Not-determined |
| S8_extracted_k141_133631 | Not-determined |
| S8_extracted_k141_3844 | Not-determined |
| S8_extracted_k141_3845 | Not-determined |
| S8_extracted_k141_30077 | Low-quality |
| S8_extracted_k141_99188 | Not-determined |
| S8_extracted_k141_133634 | Low-quality |
| S8_extracted_k141_150946 | Not-determined |
| S8_extracted_k141_30078 | Low-quality |
| S8_extracted_k141_150948 | Not-determined |
| S8_extracted_k141_116439 | Low-quality |
| S8_extracted_k141_30079 | Not-determined |
| S8_extracted_k141_64274 | Low-quality |
| S8_extracted_k141_30082 | Not-determined |
| S8_extracted_k141_55865 | Not-determined |
| S8_extracted_k141_150952 | Not-determined |
| S8_extracted_k141_90456 | Not-determined |
| S8_extracted_k141_99197 | Low-quality |
| S8_extracted_k141_125326 | Not-determined |
| S8_extracted_k141_38387 | Not-determined |
| S8_extracted_k141_202942 | Not-determined |
| S8_extracted_k141_38389 | Not-determined |
| S8_extracted_k141_90458 | Not-determined |
| S8_extracted_k141_177290 | Not-determined |
| S8_extracted_k141_46813 | Not-determined |
| S8_extracted_k141_11944 | Not-determined |
| S8_extracted_k141_38393 | Not-determined |
| S8_extracted_k141_38394 | Not-determined |
| S8_extracted_k141_11945 | Not-determined |
| S8_extracted_k141_168562 | Low-quality |
| S8_extracted_k141_133647 | Not-determined |
| S8_extracted_k141_108010 | Not-determined |
| S8_extracted_k141_125330 | Not-determined |
| S8_extracted_k141_116452 | Not-determined |
| S8_extracted_k141_55867 | Not-determined |
| S8_extracted_k141_3859 | Low-quality |
| S8_extracted_k141_72700 | Not-determined |
| S8_extracted_k141_125331 | Not-determined |
| S8_extracted_k141_142608 | Low-quality |
| S8_extracted_k141_133651 | Not-determined |
| S8_extracted_k141_30094 | Not-determined |
| S8_extracted_k141_142610 | Not-determined |
| S8_extracted_k141_116457 | Not-determined |
| S8_extracted_k141_72703 | Not-determined |
| S8_extracted_k141_11951 | Low-quality |
| S8_extracted_k141_30098 | Not-determined |
| S8_extracted_k141_177300 | Not-determined |
| S8_extracted_k141_72706 | Not-determined |
| S8_extracted_k141_3869 | Not-determined |
| S8_extracted_k141_168568 | Not-determined |
| S8_extracted_k141_55871 | Not-determined |
| S8_extracted_k141_99211 | Not-determined |
| S8_extracted_k141_21119 | Low-quality |
| S8_extracted_k141_90470 | Not-determined |
| S8_extracted_k141_72709 | Not-determined |
| S8_extracted_k141_108019 | Not-determined |
| S8_extracted_k141_30106 | Not-determined |
| S8_extracted_k141_108020 | Low-quality |
| S8_extracted_k141_64299 | Not-determined |
| S8_extracted_k141_150971 | Not-determined |
| S8_extracted_k141_30108 | Not-determined |
| S8_extracted_k141_72710 | Not-determined |
| S8_extracted_k141_108023 | Not-determined |
| S8_extracted_k141_150973 | Not-determined |
| S8_extracted_k141_177307 | Not-determined |
| S8_extracted_k141_150977 | Not-determined |
| S8_extracted_k141_142617 | Not-determined |
| S8_extracted_k141_142618 | Not-determined |
| S8_extracted_k141_185241 | Not-determined |
| S8_extracted_k141_177309 | Not-determined |
| S8_extracted_k141_177310 | Not-determined |
| S8_extracted_k141_177313 | Not-determined |
| S8_extracted_k141_21128 | Not-determined |
| S8_extracted_k141_21130 | Not-determined |
| S8_extracted_k141_90478 | Not-determined |
| S8_extracted_k141_11963 | Not-determined |
| S8_extracted_k141_108027 | Not-determined |
| S8_extracted_k141_21136 | Not-determined |
| S8_extracted_k141_108030 | Not-determined |
| S8_extracted_k141_90482 | Not-determined |
| S8_extracted_k141_150985 | Not-determined |
| S8_extracted_k141_202959 | Not-determined |
| S8_extracted_k141_90484 | Not-determined |
| S8_extracted_k141_125348 | Not-determined |
| S8_extracted_k141_168579 | Low-quality |
| S8_extracted_k141_90487 | Not-determined |
| S8_extracted_k141_21139 | Not-determined |
| S8_extracted_k141_108035 | Not-determined |
| S8_extracted_k141_202965 | Not-determined |
| S8_extracted_k141_55892 | Not-determined |
| S8_extracted_k141_116474 | Not-determined |
| S8_extracted_k141_55893 | Not-determined |
| S8_extracted_k141_90491 | Not-determined |
| S8_extracted_k141_177316 | Low-quality |
| S8_extracted_k141_202971 | Not-determined |
| S8_extracted_k141_90494 | Not-determined |
| S8_extracted_k141_38408 | Not-determined |
| S8_extracted_k141_72733 | Low-quality |
| S8_extracted_k141_11974 | Low-quality |
| S8_extracted_k141_108042 | Not-determined |
| S8_extracted_k141_72734 | Low-quality |
| S8_extracted_k141_142639 | Not-determined |
| S8_extracted_k141_185268 | Not-determined |
| S8_extracted_k141_142640 | Not-determined |
| S8_extracted_k141_11978 | Not-determined |
| S8_extracted_k141_46823 | Not-determined |
| S8_extracted_k141_177318 | Not-determined |
| S8_extracted_k141_194790 | Not-determined |
| S8_extracted_k141_168588 | Low-quality |
| S8_extracted_k141_185273 | Not-determined |
| S8_extracted_k141_185276 | Not-determined |
| S8_extracted_k141_38414 | Not-determined |
| S8_extracted_k141_55908 | Not-determined |
| S8_extracted_k141_125369 | Not-determined |
| S8_extracted_k141_99239 | Not-determined |
| S8_extracted_k141_99240 | Not-determined |
| S8_extracted_k141_194799 | Not-determined |
| S8_extracted_k141_168593 | Not-determined |
| S8_extracted_k141_55911 | Not-determined |
| S8_extracted_k141_11984 | Not-determined |
| S8_extracted_k141_72742 | Not-determined |
| S8_extracted_k141_185279 | Not-determined |
| S8_extracted_k141_116484 | Not-determined |
| S8_extracted_k141_55915 | Not-determined |
| S8_extracted_k141_30140 | Not-determined |
| S8_extracted_k141_38419 | Not-determined |
| S8_extracted_k141_90495 | Not-determined |
| S8_extracted_k141_55918 | Not-determined |
| S8_extracted_k141_81776 | Not-determined |
| S8_extracted_k141_194804 | Not-determined |
| S8_extracted_k141_81777 | Not-determined |
| S8_extracted_k141_116488 | Not-determined |
| S8_extracted_k141_30144 | Not-determined |
| S8_extracted_k141_30145 | Not-determined |
| S8_extracted_k141_55923 | Not-determined |
| S8_extracted_k141_3883 | Low-quality |
| S8_extracted_k141_3884 | Not-determined |
| S8_extracted_k141_21156 | Not-determined |
| S8_extracted_k141_108049 | Low-quality |
| S8_extracted_k141_30148 | Not-determined |
| S8_extracted_k141_142653 | Not-determined |
| S8_extracted_k141_168607 | Not-determined |
| S8_extracted_k141_133676 | Not-determined |
| S8_extracted_k141_21159 | Not-determined |
| S8_extracted_k141_151015 | Not-determined |
| S8_extracted_k141_99247 | Not-determined |
| S8_extracted_k141_38433 | Not-determined |
| S8_extracted_k141_151018 | Not-determined |
| S8_extracted_k141_11999 | Not-determined |
| S8_extracted_k141_168608 | Not-determined |
| S8_extracted_k141_133689 | Not-determined |
| S8_extracted_k141_202975 | Not-determined |
| S8_extracted_k141_125379 | Not-determined |
| S8_extracted_k141_81792 | Not-determined |
| S8_extracted_k141_30163 | Not-determined |
| S8_extracted_k141_12002 | Not-determined |
| S8_extracted_k141_133696 | Not-determined |
| S8_extracted_k141_72756 | Not-determined |
| S8_extracted_k141_81797 | Not-determined |
| S8_extracted_k141_151034 | Not-determined |
| S8_extracted_k141_38447 | Not-determined |
| S8_extracted_k141_12004 | Not-determined |
| S8_extracted_k141_30165 | Not-determined |
| S8_extracted_k141_81800 | Low-quality |
| S8_extracted_k141_133700 | Not-determined |
| S8_extracted_k141_55937 | Not-determined |
| S8_extracted_k141_72759 | Low-quality |
| S8_extracted_k141_55938 | Not-determined |
| S8_extracted_k141_168617 | Not-determined |
| S8_extracted_k141_168620 | Not-determined |
| S8_extracted_k141_151038 | Not-determined |
| S8_extracted_k141_168621 | Not-determined |
| S8_extracted_k141_12008 | Not-determined |
| S8_extracted_k141_81806 | Not-determined |
| S8_extracted_k141_125388 | Not-determined |
| S8_extracted_k141_90507 | Not-determined |
| S8_extracted_k141_90509 | Not-determined |
| S8_extracted_k141_90510 | Low-quality |
| S8_extracted_k141_202981 | Not-determined |
| S8_extracted_k141_194813 | Not-determined |
| S8_extracted_k141_21170 | Not-determined |
| S8_extracted_k141_125398 | Not-determined |
| S8_extracted_k141_12018 | Not-determined |
| S8_extracted_k141_99270 | Not-determined |
| S8_extracted_k141_38461 | Not-determined |
| S8_extracted_k141_64313 | Low-quality |
| S8_extracted_k141_133716 | Not-determined |
| S8_extracted_k141_194820 | Not-determined |
| S8_extracted_k141_168626 | Not-determined |
| S8_extracted_k141_108073 | Not-determined |
| S8_extracted_k141_168628 | Not-determined |
| S8_extracted_k141_38465 | Not-determined |
| S8_extracted_k141_151046 | Low-quality |
| S8_extracted_k141_125407 | Not-determined |
| S8_extracted_k141_90523 | Not-determined |
| S8_extracted_k141_99280 | Not-determined |
| S8_extracted_k141_202997 | Not-determined |
| S8_extracted_k141_12023 | Not-determined |
| S8_extracted_k141_116517 | Not-determined |
| S8_extracted_k141_12027 | Not-determined |
| S8_extracted_k141_12030 | Not-determined |
| S8_extracted_k141_12033 | Not-determined |
| S8_extracted_k141_125413 | Not-determined |
| S8_extracted_k141_72766 | Not-determined |
| S8_extracted_k141_64324 | Not-determined |
| S8_extracted_k141_64325 | Not-determined |
| S8_extracted_k141_151055 | Not-determined |
| S8_extracted_k141_21189 | Not-determined |
| S8_extracted_k141_108076 | Not-determined |
| S8_extracted_k141_21190 | Not-determined |
| S8_extracted_k141_30198 | Not-determined |
| S8_extracted_k141_177337 | Not-determined |
| S8_extracted_k141_30200 | Not-determined |
| S8_extracted_k141_99288 | Not-determined |
| S8_extracted_k141_151059 | Not-determined |
| S8_extracted_k141_151061 | Not-determined |
| S8_extracted_k141_72769 | Not-determined |
| S8_extracted_k141_30206 | Not-determined |
| S8_extracted_k141_90538 | Not-determined |
| S8_extracted_k141_108083 | Low-quality |
| S8_extracted_k141_116532 | Not-determined |
| S8_extracted_k141_108085 | Not-determined |
| S8_extracted_k141_12045 | Not-determined |
| S8_extracted_k141_142699 | Not-determined |
| S8_extracted_k141_194836 | Not-determined |
| S8_extracted_k141_168648 | Not-determined |
| S8_extracted_k141_55979 | Not-determined |
| S8_extracted_k141_151069 | Not-determined |
| S8_extracted_k141_99301 | Not-determined |
| S8_extracted_k141_168653 | Not-determined |
| S8_extracted_k141_3895 | Not-determined |
| S8_extracted_k141_30217 | Not-determined |
| S8_extracted_k141_38471 | Not-determined |
| S8_extracted_k141_168656 | Not-determined |
| S8_extracted_k141_38474 | Not-determined |
| S8_extracted_k141_108095 | Not-determined |
| S8_extracted_k141_38475 | Not-determined |
| S8_extracted_k141_133749 | Not-determined |
| S8_extracted_k141_168659 | Low-quality |
| S8_extracted_k141_185307 | Not-determined |

| | |
|---|---|
| S8_extracted_k141_116550 | Not-determined |
| S8_extracted_k141_90553 | Not-determined |
| S8_extracted_k141_168662 | Not-determined |
| S8_extracted_k141_125442 | Not-determined |
| S8_extracted_k141_194844 | Not-determined |
| S8_extracted_k141_125443 | Not-determined |
| S8_extracted_k141_99315 | Low-quality |
| S8_extracted_k141_12056 | Low-quality |
| S8_extracted_k141_194847 | Not-determined |
| S8_extracted_k141_81829 | Not-determined |
| S8_extracted_k141_55993 | Not-determined |
| S8_extracted_k141_125447 | Not-determined |
| S8_extracted_k141_90555 | Low-quality |
| S8_extracted_k141_159699 | Not-determined |
| S8_extracted_k141_3898 | Not-determined |
| S8_extracted_k141_30226 | Not-determined |
| S8_extracted_k141_185313 | Low-quality |
| S8_extracted_k141_125451 | Not-determined |
| S8_extracted_k141_64354 | Low-quality |
| S8_extracted_k141_38480 | Low-quality |
| S8_extracted_k141_185316 | Not-determined |
| S5_vs_k141_86016\|\|full | Medium-quality |
| S8_extracted_k141_125454 | Not-determined |
| S8_extracted_k141_133757 | Not-determined |
| S8_extracted_k141_90561 | Not-determined |
| S8_extracted_k141_21206 | Not-determined |
| S8_extracted_k141_99327 | Low-quality |
| S8_extracted_k141_30234 | Not-determined |
| S8_extracted_k141_12069 | Low-quality |
| S8_extracted_k141_133762 | Not-determined |
| S8_extracted_k141_142707 | Not-determined |
| S8_extracted_k141_81836 | Not-determined |
| S8_extracted_k141_30238 | Not-determined |
| S8_extracted_k141_151074 | Not-determined |
| S8_extracted_k141_151075 | Not-determined |
| S8_extracted_k141_116554 | Not-determined |
| S8_extracted_k141_81838 | Not-determined |
| S8_extracted_k141_168668 | Not-determined |
| S8_extracted_k141_116556 | Not-determined |
| S8_extracted_k141_133767 | Not-determined |
| S8_extracted_k141_133768 | Not-determined |
| S8_extracted_k141_116558 | Not-determined |
| S8_extracted_k141_185325 | Not-determined |
| S8_extracted_k141_3911 | Not-determined |
| S8_extracted_k141_177366 | Not-determined |
| S8_extracted_k141_30242 | Not-determined |
| S8_extracted_k141_159713 | Not-determined |
| S8_extracted_k141_185331 | Low-quality |
| S8_extracted_k141_185333 | Not-determined |
| S8_extracted_k141_168676 | Not-determined |
| S8_extracted_k141_133780 | Not-determined |
| S8_extracted_k141_38489 | Not-determined |
| S8_extracted_k141_133781 | Not-determined |
| S8_extracted_k141_185338 | Not-determined |
| S8_extracted_k141_168681 | Not-determined |
| S8_extracted_k141_90579 | Not-determined |
| S8_extracted_k141_142726 | Not-determined |
| S8_extracted_k141_159721 | Not-determined |
| S8_extracted_k141_90580 | Not-determined |
| S8_extracted_k141_3929 | Not-determined |
| S8_extracted_k141_133792 | Low-quality |
| S8_extracted_k141_159723 | Not-determined |
| S8_extracted_k141_99347 | Not-determined |
| S8_extracted_k141_177379 | Not-determined |
| S8_extracted_k141_81849 | Not-determined |
| S8_extracted_k141_12088 | Low-quality |
| S8_extracted_k141_142730 | Not-determined |
| S8_extracted_k141_38496 | Not-determined |
| S8_extracted_k141_3934 | Not-determined |
| S8_extracted_k141_185350 | Not-determined |
| S8_extracted_k141_142735 | Not-determined |
| S8_extracted_k141_3940 | Not-determined |
| S8_extracted_k141_64361 | Not-determined |
| S8_extracted_k141_38501 | Not-determined |
| S8_extracted_k141_38503 | Not-determined |
| S8_extracted_k141_203043 | Low-quality |
| S8_extracted_k141_133801 | Not-determined |
| S8_extracted_k141_90589 | Not-determined |
| S8_extracted_k141_81852 | Not-determined |
| S8_extracted_k141_194856 | Not-determined |
| S8_extracted_k141_159737 | Not-determined |
| S8_extracted_k141_151085 | Not-determined |
| S8_extracted_k141_168690 | Not-determined |
| S8_extracted_k141_12097 | Not-determined |
| S8_extracted_k141_194859 | Not-determined |
| S8_extracted_k141_64364 | Not-determined |
| S8_extracted_k141_3952 | Not-determined |
| S7_vs_52002\|\|full | Medium-quality |
| S8_extracted_k141_12101 | Not-determined |
| S8_extracted_k141_90600 | Not-determined |
| S8_extracted_k141_151091 | Not-determined |
| S8_extracted_k141_30248 | Not-determined |
| S8_extracted_k141_177398 | Low-quality |
| S8_extracted_k141_30252 | Not-determined |
| S8_extracted_k141_72777 | Not-determined |
| S8_extracted_k141_12103 | Low-quality |
| S8_extracted_k141_64369 | Not-determined |
| S8_extracted_k141_151094 | Not-determined |
| S8_extracted_k141_38510 | Not-determined |
| S8_extracted_k141_81860 | Not-determined |
| S8_extracted_k141_12107 | Not-determined |
| S8_extracted_k141_133809 | Low-quality |
| S8_extracted_k141_177403 | Not-determined |
| S8_extracted_k141_194867 | Not-determined |
| S8_extracted_k141_81862 | Not-determined |
| S8_extracted_k141_30257 | Not-determined |
| S8_extracted_k141_64375 | Not-determined |
| S8_extracted_k141_159755 | Not-determined |
| S8_extracted_k141_12111 | Not-determined |
| S8_extracted_k141_12112 | Not-determined |
| S8_extracted_k141_142761 | Low-quality |
| S8_extracted_k141_90609 | Not-determined |
| S8_extracted_k141_185368 | Not-determined |
| S8_extracted_k141_21211 | Low-quality |
| S8_extracted_k141_12114 | Low-quality |
| S8_extracted_k141_38514 | Not-determined |
| S8_extracted_k141_142767 | Not-determined |
| S8_extracted_k141_30264 | Not-determined |
| S8_extracted_k141_194872 | Not-determined |
| S8_extracted_k141_3967 | Not-determined |
| S8_extracted_k141_30269 | Low-quality |
| S8_extracted_k141_90617 | Not-determined |
| S8_extracted_k141_177413 | Not-determined |
| S8_extracted_k141_3969 | Low-quality |
| S8_extracted_k141_168710 | Not-determined |
| S12_vs_k141_38725\|\|full | Medium-quality |
| S8_extracted_k141_194873 | Not-determined |
| S8_extracted_k141_185375 | Low-quality |
| S8_extracted_k141_168714 | Not-determined |
| S8_extracted_k141_38521 | Not-determined |
| S8_extracted_k141_64383 | Not-determined |
| S8_extracted_k141_151102 | Not-determined |
| S8_extracted_k141_185378 | Not-determined |
| S8_extracted_k141_3973 | Not-determined |
| S8_extracted_k141_12121 | Not-determined |
| S8_extracted_k141_12123 | Not-determined |
| S8_extracted_k141_30276 | Not-determined |
| S8_extracted_k141_151106 | Not-determined |
| S8_extracted_k141_108119 | Not-determined |
| S8_extracted_k141_159773 | Not-determined |
| S8_extracted_k141_108121 | Not-determined |
| S8_extracted_k141_177426 | Not-determined |
| S8_extracted_k141_185384 | Not-determined |
| S8_extracted_k141_12125 | Low-quality |
| S8_extracted_k141_12127 | Low-quality |
| S8_extracted_k141_108122 | Not-determined |
| S8_extracted_k141_194886 | Not-determined |
| S8_extracted_k141_133830 | Not-determined |
| S8_extracted_k141_125482 | Not-determined |
| S8_extracted_k141_203048 | Not-determined |
| S8_extracted_k141_12129 | Not-determined |
| S8_extracted_k141_21214 | Not-determined |
| S8_extracted_k141_142778 | Not-determined |
| S8_extracted_k141_30286 | Low-quality |
| S8_extracted_k141_133834 | Not-determined |
| S8_extracted_k141_3984 | Not-determined |
| S8_extracted_k141_21219 | Not-determined |
| S8_extracted_k141_3986 | Not-determined |
| S8_extracted_k141_30289 | Not-determined |
| S8_extracted_k141_125491 | Not-determined |
| S8_extracted_k141_177434 | Not-determined |
| S8_extracted_k141_125494 | Not-determined |
| S8_extracted_k141_177437 | Not-determined |
| S8_extracted_k141_46846 | Low-quality |
| S8_extracted_k141_90634 | Not-determined |
| S8_extracted_k141_38529 | Not-determined |
| S8_extracted_k141_90635 | Not-determined |
| S8_extracted_k141_108135 | Not-determined |
| S8_extracted_k141_38532 | Not-determined |
| S8_extracted_k141_203062 | Not-determined |
| S8_extracted_k141_159788 | Not-determined |
| S8_extracted_k141_90638 | Not-determined |
| S8_extracted_k141_30303 | Not-determined |
| S8_extracted_k141_12148 | Not-determined |
| S8_extracted_k141_203066 | Not-determined |
| S8_extracted_k141_151126 | Not-determined |
| S8_extracted_k141_203067 | Not-determined |
| S8_extracted_k141_90641 | Not-determined |
| S8_extracted_k141_177445 | Not-determined |
| S8_extracted_k141_133840 | Not-determined |
| S8_extracted_k141_203069 | Not-determined |
| S8_extracted_k141_194902 | Not-determined |
| S8_extracted_k141_151129 | Not-determined |
| S8_extracted_k141_90644 | Low-quality |
| S8_extracted_k141_46860 | Not-determined |
| S8_extracted_k141_108138 | Not-determined |
| S8_extracted_k141_90646 | Not-determined |
| S8_extracted_k141_151132 | Not-determined |
| S8_extracted_k141_90648 | Not-determined |
| S8_extracted_k141_46864 | Low-quality |
| S8_extracted_k141_185408 | Not-determined |
| S8_extracted_k141_108143 | Not-determined |
| S8_extracted_k141_56023 | Not-determined |
| S8_extracted_k141_194907 | Not-determined |
| S8_extracted_k141_168730 | Not-determined |
| S8_extracted_k141_21236 | Not-determined |
| S8_extracted_k141_72812 | Not-determined |
| S8_extracted_k141_21240 | Not-determined |
| S8_extracted_k141_38540 | Not-determined |
| S8_extracted_k141_21242 | Not-determined |
| S8_extracted_k141_99358 | Low-quality |
| S8_extracted_k141_12164 | Not-determined |
| S8_extracted_k141_194914 | Not-determined |
| S8_extracted_k141_21245 | Low-quality |
| S8_extracted_k141_168737 | Not-determined |
| S8_extracted_k141_72824 | Not-determined |
| S8_extracted_k141_203073 | Not-determined |
| S8_extracted_k141_3995 | Not-determined |
| S8_extracted_k141_185411 | Low-quality |
| S8_extracted_k141_168744 | Not-determined |
| S8_extracted_k141_90660 | Not-determined |
| S8_extracted_k141_64409 | Low-quality |
| S8_extracted_k141_46872 | Not-determined |
| S8_extracted_k141_72829 | Not-determined |
| S8_extracted_k141_194920 | Not-determined |
| S8_extracted_k141_81871 | Not-determined |
| S8_extracted_k141_4008 | Low-quality |
| S8_extracted_k141_177449 | Not-determined |
| S8_extracted_k141_81873 | Not-determined |
| S8_extracted_k141_46880 | Not-determined |
| S8_extracted_k141_125536 | Not-determined |
| S8_extracted_k141_177454 | Not-determined |
| S8_extracted_k141_177455 | Not-determined |
| S8_extracted_k141_133882 | Not-determined |
| S8_extracted_k141_30335 | Not-determined |
| S8_extracted_k141_4018 | Low-quality |
| S8_extracted_k141_64425 | Not-determined |
| S8_extracted_k141_81878 | Not-determined |
| S8_extracted_k141_168753 | Low-quality |
| S8_extracted_k141_108162 | Not-determined |
| S8_extracted_k141_72839 | Not-determined |
| S8_extracted_k141_177459 | Not-determined |
| S8_extracted_k141_177460 | Not-determined |
| S8_extracted_k141_30341 | Not-determined |
| S8_extracted_k141_64429 | Not-determined |
| S8_extracted_k141_108166 | Not-determined |
| S8_extracted_k141_12174 | Not-determined |
| S8_extracted_k141_90668 | Not-determined |
| S8_extracted_k141_30343 | Not-determined |
| S8_extracted_k141_64432 | Not-determined |
| S8_extracted_k141_56041 | Not-determined |
| S8_extracted_k141_56043 | Low-quality |
| S8_extracted_k141_4024 | Low-quality |
| S8_extracted_k141_64435 | Not-determined |
| S8_extracted_k141_185425 | Not-determined |
| S8_extracted_k141_108172 | Not-determined |
| S8_extracted_k141_90672 | Not-determined |
| S8_extracted_k141_21260 | Not-determined |
| S8_extracted_k141_12180 | Not-determined |
| S8_extracted_k141_116605 | Not-determined |
| S8_extracted_k141_12182 | Not-determined |
| S8_extracted_k141_185428 | Not-determined |
| S8_extracted_k141_185430 | Not-determined |
| S8_extracted_k141_46897 | Not-determined |
| S8_extracted_k141_203081 | Not-determined |
| S8_extracted_k141_203082 | Not-determined |
| S8_extracted_k141_203083 | Not-determined |
| S8_extracted_k141_12189 | Not-determined |
| S8_extracted_k141_38558 | Not-determined |
| S8_extracted_k141_168767 | Not-determined |
| S8_extracted_k141_168768 | Not-determined |
| S8_extracted_k141_203085 | Not-determined |
| S8_extracted_k141_90678 | Not-determined |
| S8_extracted_k141_203087 | Not-determined |
| S8_extracted_k141_72843 | Not-determined |
| S8_extracted_k141_177465 | Not-determined |
| S8_extracted_k141_203089 | Not-determined |
| S8_extracted_k141_56056 | Not-determined |
| S8_extracted_k141_72846 | Not-determined |
| S8_extracted_k141_142799 | Not-determined |
| S8_extracted_k141_125546 | Not-determined |
| S8_extracted_k141_4033 | Not-determined |
| S8_extracted_k141_72848 | Low-quality |
| S8_extracted_k141_90688 | Not-determined |
| S8_extracted_k141_4036 | Not-determined |
| S8_extracted_k141_194944 | Not-determined |
| S8_extracted_k141_4038 | Not-determined |
| S8_extracted_k141_185435 | Not-determined |
| S8_extracted_k141_30360 | Not-determined |
| S8_extracted_k141_4042 | Not-determined |
| S8_extracted_k141_81883 | Not-determined |
| S8_extracted_k141_108182 | Not-determined |
| S8_extracted_k141_194946 | Low-quality |
| S8_extracted_k141_38575 | Not-determined |
| S8_extracted_k141_177469 | Not-determined |
| S8_extracted_k141_72854 | Not-determined |
| S8_extracted_k141_151149 | Not-determined |
| S8_extracted_k141_108186 | Low-quality |
| S8_extracted_k141_108187 | Not-determined |
| S8_extracted_k141_116619 | Not-determined |
| S8_extracted_k141_116620 | Not-determined |
| S8_extracted_k141_90706 | Not-determined |
| S8_extracted_k141_108190 | Not-determined |
| S8_extracted_k141_194955 | Not-determined |
| S8_extracted_k141_12196 | Not-determined |
| S8_extracted_k141_159811 | Not-determined |
| S8_extracted_k141_81891 | Not-determined |
| S8_extracted_k141_12197 | Not-determined |
| S8_extracted_k141_72861 | Not-determined |
| S8_extracted_k141_194956 | Not-determined |
| S8_extracted_k141_159814 | Low-quality |
| S8_extracted_k141_46901 | Not-determined |
| S8_extracted_k141_177477 | Not-determined |
| S8_extracted_k141_64455 | Not-determined |
| S8_extracted_k141_30368 | Not-determined |
| S8_extracted_k141_108192 | Not-determined |
| S8_extracted_k141_72865 | Not-determined |
| S8_extracted_k141_21288 | Not-determined |
| S8_extracted_k141_46904 | Not-determined |
| S8_extracted_k141_12201 | Not-determined |
| S8_extracted_k141_203092 | Low-quality |
| S8_extracted_k141_108193 | Not-determined |
| S8_extracted_k141_116628 | Not-determined |
| S8_extracted_k141_108194 | Not-determined |
| S8_extracted_k141_81896 | Not-determined |
| S8_extracted_k141_177482 | Not-determined |
| S8_extracted_k141_185458 | Not-determined |
| S8_extracted_k141_203097 | Not-determined |
| S8_extracted_k141_142819 | Low-quality |
| S8_extracted_k141_194965 | Not-determined |
| S8_extracted_k141_151163 | Not-determined |
| S8_extracted_k141_12203 | Not-determined |
| S8_extracted_k141_46908 | Not-determined |
| S8_extracted_k141_203099 | Not-determined |
| S8_extracted_k141_90719 | Low-quality |
| S8_extracted_k141_56079 | Not-determined |
| S8_extracted_k141_12205 | Low-quality |
| S8_extracted_k141_72866 | Not-determined |
| S8_extracted_k141_194967 | Not-determined |
| S8_extracted_k141_56081 | Not-determined |
| S8_extracted_k141_125570 | Not-determined |
| S8_extracted_k141_46915 | Not-determined |
| S8_extracted_k141_125571 | Not-determined |
| S8_extracted_k141_203106 | Not-determined |
| S8_extracted_k141_99365 | Not-determined |
| S8_extracted_k141_125573 | Not-determined |
| S8_extracted_k141_194973 | Not-determined |
| S8_extracted_k141_99368 | Not-determined |
| S8_extracted_k141_56084 | Not-determined |
| S8_extracted_k141_90728 | Not-determined |
| S8_extracted_k141_203109 | Not-determined |
| S8_extracted_k141_142826 | Not-determined |
| S8_extracted_k141_99374 | Not-determined |
| S8_extracted_k141_30380 | Not-determined |
| S8_extracted_k141_185474 | Low-quality |
| S8_extracted_k141_203118 | Not-determined |
| S8_extracted_k141_151176 | Not-determined |
| S8_extracted_k141_81903 | Low-quality |
| S8_extracted_k141_185478 | Not-determined |
| S8_extracted_k141_125579 | Not-determined |
| S8_extracted_k141_81905 | Low-quality |
| S8_extracted_k141_194981 | Not-determined |
| S8_extracted_k141_177495 | Not-determined |
| S8_extracted_k141_56091 | Not-determined |
| S8_extracted_k141_56093 | Not-determined |
| S8_extracted_k141_185482 | Not-determined |
| S8_extracted_k141_116636 | Low-quality |
| S8_extracted_k141_203122 | Not-determined |
| S8_extracted_k141_151178 | Not-determined |
| S8_extracted_k141_142835 | Low-quality |
| S8_extracted_k141_46929 | Not-determined |
| S8_extracted_k141_21295 | Not-determined |
| S8_extracted_k141_99390 | Not-determined |
| S8_extracted_k141_21296 | Not-determined |
| S8_extracted_k141_133926 | Not-determined |
| S8_extracted_k141_194988 | Not-determined |
| S8_extracted_k141_133928 | Not-determined |
| S8_extracted_k141_30394 | Not-determined |
| S8_extracted_k141_99395 | Not-determined |
| S8_extracted_k141_46933 | Low-quality |
| S8_extracted_k141_30396 | Not-determined |
| S8_extracted_k141_203132 | Not-determined |
| S8_extracted_k141_72880 | Low-quality |
| S8_extracted_k141_142847 | Not-determined |
| S8_extracted_k141_194996 | Not-determined |
| S8_extracted_k141_203135 | Not-determined |
| S8_extracted_k141_142852 | Not-determined |
| S8_extracted_k141_81910 | Not-determined |
| S8_extracted_k141_99403 | Not-determined |
| S8_extracted_k141_46935 | Not-determined |
| S8_extracted_k141_46937 | Low-quality |
| S8_extracted_k141_90751 | Not-determined |
| S8_extracted_k141_81915 | Not-determined |
| S8_extracted_k141_30407 | Not-determined |
| S8_extracted_k141_81916 | Not-determined |
| S8_extracted_k141_125591 | Not-determined |
| S8_extracted_k141_195003 | Not-determined |
| S8_extracted_k141_177512 | Not-determined |
| S8_extracted_k141_81919 | Not-determined |
| S8_extracted_k141_108214 | Not-determined |
| S8_extracted_k141_30411 | Low-quality |
| S8_extracted_k141_72883 | Low-quality |
| S8_extracted_k141_81923 | Not-determined |
| S8_extracted_k141_133936 | Low-quality |
| S8_extracted_k141_90762 | Not-determined |
| S8_extracted_k141_133937 | Low-quality |
| S8_extracted_k141_72886 | Not-determined |
| S8_extracted_k141_81925 | Not-determined |
| S8_extracted_k141_151189 | Not-determined |
| S8_extracted_k141_159835 | Not-determined |
| S8_extracted_k141_125596 | Not-determined |
| S8_extracted_k141_159836 | Not-determined |
| S8_extracted_k141_203147 | Not-determined |
| S8_extracted_k141_195016 | Not-determined |
| S8_extracted_k141_38611 | Not-determined |
| S8_extracted_k141_133941 | Not-determined |
| S8_extracted_k141_159841 | Not-determined |
| S8_extracted_k141_64469 | Not-determined |
| S8_extracted_k141_159842 | Not-determined |
| S8_extracted_k141_38614 | Not-determined |
| S8_extracted_k141_46956 | Not-determined |
| S8_extracted_k141_56106 | Not-determined |
| S8_extracted_k141_38622 | Low-quality |
| S8_extracted_k141_21303 | Not-determined |
| S8_extracted_k141_99414 | Low-quality |
| S8_extracted_k141_159860 | Low-quality |
| S8_extracted_k141_72898 | Not-determined |
| S8_extracted_k141_72899 | Not-determined |
| S8_extracted_k141_90780 | Not-determined |
| S8_extracted_k141_4051 | Not-determined |
| S8_extracted_k141_4052 | Not-determined |
| S8_extracted_k141_203163 | Not-determined |
| S8_extracted_k141_38634 | Not-determined |
| S8_extracted_k141_151206 | Not-determined |
| S8_extracted_k141_99424 | Not-determined |
| S8_extracted_k141_46976 | Not-determined |
| S8_extracted_k141_151210 | Low-quality |
| S8_extracted_k141_177530 | Low-quality |
| S8_extracted_k141_195026 | Not-determined |
| S8_extracted_k141_21316 | Not-determined |
| S8_extracted_k141_21319 | Not-determined |
| S8_extracted_k141_195029 | Not-determined |
| S8_extracted_k141_108229 | Not-determined |
| S8_extracted_k141_177534 | Low-quality |
| S8_extracted_k141_46984 | Low-quality |
| S8_extracted_k141_125612 | Not-determined |
| S8_extracted_k141_99427 | Not-determined |
| S8_extracted_k141_177542 | Not-determined |
| S8_extracted_k141_177543 | Not-determined |
| S8_extracted_k141_177544 | Not-determined |
| S8_extracted_k141_159877 | Not-determined |
| S8_extracted_k141_21330 | Not-determined |
| S8_extracted_k141_21332 | Not-determined |
| S8_extracted_k141_46989 | Not-determined |
| S8_extracted_k141_177548 | Not-determined |
| S8_extracted_k141_142896 | Not-determined |
| S8_extracted_k141_21337 | Not-determined |
| S8_extracted_k141_159879 | Low-quality |
| S8_extracted_k141_142897 | Not-determined |
| S8_extracted_k141_133956 | Not-determined |
| S8_extracted_k141_159881 | Not-determined |
| S8_extracted_k141_125620 | Not-determined |
| S8_extracted_k141_177551 | Not-determined |
| S8_extracted_k141_151228 | Not-determined |
| S8_extracted_k141_81949 | Not-determined |
| S8_vs_k141_81691\|\|full | Medium-quality |
| S8_extracted_k141_81952 | Not-determined |
| S8_extracted_k141_56134 | Not-determined |
| S8_extracted_k141_142904 | Not-determined |
| S8_extracted_k141_159888 | Not-determined |
| S8_extracted_k141_90795 | Not-determined |
| S8_extracted_k141_177554 | Low-quality |
| S8_extracted_k141_38656 | Not-determined |
| S8_extracted_k141_38657 | Not-determined |
| S8_extracted_k141_46998 | Not-determined |
| S8_extracted_k141_108249 | Not-determined |
| S8_extracted_k141_72918 | Not-determined |
| S8_extracted_k141_56141 | Not-determined |
| S8_extracted_k141_151233 | Not-determined |
| S8_extracted_k141_90798 | Not-determined |
| S8_extracted_k141_46999 | Not-determined |
| S8_extracted_k141_90800 | Not-determined |
| S8_extracted_k141_151236 | Not-determined |
| S8_extracted_k141_56145 | Not-determined |
| S8_extracted_k141_12220 | Not-determined |
| S8_extracted_k141_203194 | Not-determined |
| S8_extracted_k141_30429 | Low-quality |
| S8_extracted_k141_64490 | Not-determined |
| S8_extracted_k141_64491 | Not-determined |
| S8_extracted_k141_47005 | Not-determined |
| S8_extracted_k141_159899 | Not-determined |
| S8_extracted_k141_108254 | Not-determined |
| S8_extracted_k141_30432 | Not-determined |
| S8_extracted_k141_185490 | Not-determined |
| S8_extracted_k141_203197 | Not-determined |
| S8_extracted_k141_108255 | Not-determined |
| S8_extracted_k141_38665 | Not-determined |
| S8_extracted_k141_81963 | Not-determined |
| S8_extracted_k141_185492 | Not-determined |
| S8_extracted_k141_64495 | Low-quality |
| S8_extracted_k141_151244 | Not-determined |
| S8_extracted_k141_133966 | Not-determined |
| S8_extracted_k141_177566 | Not-determined |
| S8_extracted_k141_21355 | Not-determined |
| S8_extracted_k141_56154 | Not-determined |
| S8_extracted_k141_64496 | Not-determined |
| S8_extracted_k141_64497 | Not-determined |
| S8_extracted_k141_203204 | Not-determined |
| S8_extracted_k141_159901 | Not-determined |
| S8_extracted_k141_12226 | Not-determined |
| S8_extracted_k141_38672 | Not-determined |
| S8_extracted_k141_185496 | Not-determined |
| S2_extracted_k141_17850 | Medium-quality |
| S8_extracted_k141_81970 | Not-determined |
| S8_extracted_k141_21361 | Not-determined |
| S8_extracted_k141_12229 | Not-determined |
| S8_extracted_k141_203207 | Not-determined |
| S8_extracted_k141_99448 | Not-determined |
| S8_extracted_k141_133972 | Not-determined |
| S8_extracted_k141_203211 | Not-determined |
| S8_extracted_k141_12234 | Not-determined |
| S8_extracted_k141_99452 | Low-quality |
| S8_extracted_k141_99453 | Not-determined |
| S8_extracted_k141_185499 | Low-quality |
| S8_extracted_k141_203213 | Not-determined |
| S8_extracted_k141_195042 | Not-determined |
| S8_extracted_k141_151249 | Not-determined |
| S8_extracted_k141_168809 | Not-determined |
| S8_extracted_k141_142919 | Low-quality |
| S8_extracted_k141_151253 | Not-determined |
| S8_extracted_k141_108272 | Not-determined |
| S8_extracted_k141_12239 | Not-determined |
| S8_extracted_k141_108273 | Not-determined |
| S8_extracted_k141_12242 | Not-determined |
| S8_extracted_k141_108276 | Not-determined |
| S8_extracted_k141_21370 | Not-determined |
| S8_extracted_k141_133979 | Not-determined |
| S8_extracted_k141_203217 | Not-determined |
| S8_extracted_k141_108280 | Not-determined |
| S8_extracted_k141_203219 | Not-determined |
| S8_extracted_k141_125633 | Not-determined |
| S8_extracted_k141_108281 | Low-quality |
| S8_extracted_k141_177579 | Not-determined |
| S8_extracted_k141_72935 | Not-determined |
| S8_extracted_k141_142927 | Not-determined |
| S8_extracted_k141_142928 | Not-determined |
| S8_extracted_k141_21375 | Not-determined |
| S8_extracted_k141_142932 | Not-determined |
| S8_extracted_k141_81977 | Low-quality |
| S8_extracted_k141_133984 | Low-quality |
| S8_extracted_k141_90834 | Not-determined |
| S8_extracted_k141_108288 | Low-quality |
| S8_extracted_k141_81978 | Not-determined |
| S8_extracted_k141_195062 | Not-determined |
| S8_extracted_k141_159919 | Not-determined |
| S8_extracted_k141_90835 | Not-determined |
| S8_extracted_k141_12255 | Low-quality |
| S8_extracted_k141_12256 | Not-determined |
| S8_extracted_k141_81982 | Not-determined |
| S8_extracted_k141_81983 | Low-quality |
| S8_extracted_k141_125646 | Not-determined |
| S8_extracted_k141_12258 | Not-determined |
| S8_extracted_k141_30450 | Low-quality |
| S8_extracted_k141_177593 | Not-determined |
| S8_extracted_k141_90844 | Not-determined |
| S8_extracted_k141_99476 | Not-determined |
| S8_extracted_k141_195071 | Not-determined |
| S8_extracted_k141_108292 | Low-quality |
| S8_extracted_k141_81987 | Low-quality |
| S8_extracted_k141_125652 | Not-determined |
| S8_extracted_k141_108294 | Not-determined |
| S8_extracted_k141_21384 | Not-determined |
| S8_extracted_k141_21386 | Not-determined |
| S8_extracted_k141_116639 | Low-quality |
| S8_extracted_k141_30456 | Not-determined |
| S8_extracted_k141_30458 | Not-determined |
| S8_extracted_k141_203223 | Low-quality |
| S8_extracted_k141_81991 | Not-determined |
| S8_extracted_k141_12268 | Not-determined |
| S8_extracted_k141_116642 | Not-determined |
| S8_extracted_k141_90847 | Not-determined |
| S8_extracted_k141_90848 | Not-determined |
| S8_extracted_k141_38685 | Not-determined |
| S8_extracted_k141_81992 | Not-determined |
| S8_extracted_k141_108298 | Not-determined |
| S8_extracted_k141_168827 | Not-determined |
| S8_extracted_k141_99482 | Not-determined |
| S8_extracted_k141_125660 | Not-determined |
| S8_extracted_k141_125662 | Not-determined |
| S8_extracted_k141_116648 | Not-determined |
| S8_extracted_k141_90853 | Not-determined |
| S8_extracted_k141_125663 | Not-determined |
| S8_extracted_k141_90854 | Not-determined |
| S8_extracted_k141_108302 | Not-determined |
| S8_extracted_k141_108304 | Not-determined |
| S8_extracted_k141_21392 | Not-determined |
| S8_extracted_k141_99487 | Not-determined |
| S8_extracted_k141_108307 | Not-determined |
| S8_extracted_k141_125664 | Low-quality |

| | |
|---|---|
| S8_extracted_k141_12277 | Not-determined |
| S8_extracted_k141_90863 | Not-determined |
| S8_extracted_k141_90864 | Not-determined |
| S8_extracted_k141_151279 | Not-determined |
| S8_extracted_k141_177613 | Not-determined |
| S8_extracted_k141_90865 | Not-determined |
| S8_extracted_k141_116655 | Not-determined |
| S8_extracted_k141_81997 | Not-determined |
| S8_extracted_k141_47016 | Not-determined |
| S8_extracted_k141_151282 | Not-determined |
| S8_extracted_k141_151283 | Not-determined |
| S8_extracted_k141_125667 | Not-determined |
| S8_extracted_k141_12283 | Not-determined |
| S8_extracted_k141_82003 | Not-determined |
| S8_extracted_k141_12284 | Not-determined |
| S8_extracted_k141_38697 | Not-determined |
| S8_extracted_k141_47023 | Low-quality |
| S8_extracted_k141_82006 | Not-determined |
| S8_extracted_k141_142955 | Not-determined |
| S8_extracted_k141_47024 | Not-determined |
| S8_extracted_k141_99499 | Not-determined |
| S8_extracted_k141_116660 | Not-determined |
| S8_extracted_k141_151290 | Not-determined |
| S8_extracted_k141_30482 | Not-determined |
| S8_extracted_k141_99500 | Not-determined |
| S8_extracted_k141_185521 | Not-determined |
| S8_extracted_k141_56184 | Not-determined |
| S8_extracted_k141_177617 | Not-determined |
| S8_extracted_k141_90868 | Low-quality |
| S8_extracted_k141_185522 | Not-determined |
| S8_extracted_k141_90869 | Not-determined |
| S8_extracted_k141_12296 | Not-determined |
| S8_extracted_k141_151296 | Not-determined |
| S8_extracted_k141_56186 | Not-determined |
| S8_extracted_k141_125678 | Not-determined |
| S8_extracted_k141_151299 | Not-determined |
| S8_extracted_k141_30486 | Not-determined |
| S8_extracted_k141_99511 | Not-determined |
| S8_extracted_k141_38700 | Low-quality |
| S8_extracted_k141_12300 | Not-determined |
| S8_extracted_k141_116664 | Low-quality |
| S8_extracted_k141_30492 | Not-determined |
| S8_extracted_k141_151306 | Not-determined |
| S8_extracted_k141_12303 | Not-determined |
| S8_extracted_k141_203234 | Not-determined |
| S8_extracted_k141_151312 | Not-determined |
| S8_extracted_k141_134016 | Not-determined |
| S8_extracted_k141_12309 | Low-quality |
| S8_extracted_k141_168832 | Not-determined |
| S8_extracted_k141_82031 | Low-quality |
| S8_extracted_k141_72978 | Not-determined |
| S8_extracted_k141_56194 | Not-determined |
| S8_extracted_k141_72979 | Not-determined |
| S8_extracted_k141_90881 | Not-determined |
| S8_extracted_k141_177630 | Low-quality |
| S8_extracted_k141_195082 | Not-determined |
| S8_extracted_k141_38718 | Not-determined |
| S8_extracted_k141_47052 | Not-determined |
| S8_extracted_k141_168844 | Not-determined |
| S8_extracted_k141_12317 | Not-determined |
| S8_extracted_k141_38720 | Not-determined |
| S8_extracted_k141_168845 | Not-determined |
| S8_extracted_k141_72986 | Not-determined |
| S8_extracted_k141_134028 | Not-determined |
| S8_extracted_k141_203249 | Not-determined |
| S8_extracted_k141_64522 | Not-determined |
| S8_extracted_k141_90883 | Not-determined |
| S8_extracted_k141_72988 | Not-determined |
| S8_extracted_k141_12322 | Not-determined |
| S8_extracted_k141_177636 | Not-determined |
| S8_extracted_k141_99519 | Not-determined |
| S8_extracted_k141_82048 | Not-determined |
| S8_extracted_k141_203252 | Not-determined |
| S8_extracted_k141_21411 | Not-determined |
| S8_extracted_k141_99522 | Not-determined |
| S8_extracted_k141_90886 | Not-determined |
| S8_extracted_k141_90887 | Not-determined |
| S8_extracted_k141_90890 | Not-determined |
| S8_extracted_k141_195091 | Not-determined |
| S8_extracted_k141_125681 | Not-determined |
| S8_extracted_k141_64526 | Not-determined |
| S8_extracted_k141_125682 | Not-determined |
| S8_extracted_k141_30506 | Not-determined |
| S8_extracted_k141_64529 | Not-determined |
| S8_extracted_k141_12328 | Not-determined |
| S8_extracted_k141_38730 | Not-determined |
| S8_extracted_k141_125684 | Not-determined |
| S8_extracted_k141_12329 | Not-determined |
| S8_extracted_k141_125685 | Not-determined |
| S8_extracted_k141_12333 | Not-determined |
| S8_extracted_k141_177642 | Low-quality |
| S8_extracted_k141_21419 | Not-determined |
| S8_extracted_k141_203260 | Not-determined |
| S8_extracted_k141_30511 | Not-determined |
| S8_extracted_k141_168859 | Not-determined |
| S8_extracted_k141_125690 | Not-determined |
| S8_extracted_k141_125691 | Not-determined |
| S8_extracted_k141_90904 | Not-determined |
| S8_extracted_k141_90905 | Not-determined |
| S8_extracted_k141_177648 | Not-determined |
| S8_extracted_k141_12337 | Not-determined |
| S8_extracted_k141_177649 | Not-determined |
| S8_extracted_k141_12338 | Not-determined |
| S8_extracted_k141_47058 | Not-determined |
| S8_extracted_k141_177652 | Not-determined |
| S8_extracted_k141_203267 | Not-determined |
| S8_extracted_k141_108310 | Low-quality |
| S8_extracted_k141_203268 | Not-determined |
| S8_extracted_k141_125696 | Not-determined |
| S8_extracted_k141_47062 | Not-determined |
| S8_extracted_k141_108311 | Not-determined |
| S8_extracted_k141_38738 | Not-determined |
| S8_extracted_k141_12344 | Not-determined |
| S8_extracted_k141_30518 | Not-determined |
| S8_extracted_k141_47067 | Not-determined |
| S8_extracted_k141_82058 | Not-determined |
| S8_extracted_k141_30520 | Low-quality |
| S8_extracted_k141_4082 | Not-determined |
| S8_extracted_k141_185533 | Not-determined |
| S8_extracted_k141_168869 | Not-determined |
| S8_extracted_k141_21433 | Not-determined |
| S8_extracted_k141_47072 | Not-determined |
| S8_extracted_k141_90915 | Not-determined |
| S8_extracted_k141_21435 | Low-quality |
| S8_extracted_k141_177662 | Not-determined |
| S8_extracted_k141_203274 | Not-determined |
| S8_extracted_k141_90917 | Not-determined |
| S8_extracted_k141_64536 | Not-determined |
| S8_extracted_k141_38747 | Low-quality |
| S8_extracted_k141_12358 | Not-determined |
| S8_extracted_k141_99548 | Not-determined |
| S8_extracted_k141_177669 | Not-determined |
| S8_extracted_k141_203280 | Not-determined |
| S8_extracted_k141_134054 | Not-determined |
| S8_extracted_k141_72996 | Not-determined |
| S8_extracted_k141_12362 | Not-determined |
| S8_extracted_k141_47080 | Not-determined |
| S8_extracted_k141_108327 | Not-determined |
| S8_extracted_k141_168875 | Not-determined |
| S8_extracted_k141_99557 | Not-determined |
| S8_extracted_k141_125715 | Not-determined |
| S8_extracted_k141_108332 | Not-determined |
| S8_extracted_k141_12368 | Not-determined |
| S8_extracted_k141_99558 | Not-determined |
| S8_extracted_k141_134060 | Not-determined |
| S8_extracted_k141_12371 | Not-determined |
| S8_extracted_k141_12374 | Not-determined |
| S8_extracted_k141_38768 | Not-determined |
| S8_extracted_k141_90929 | Not-determined |
| S8_extracted_k141_21438 | Low-quality |
| S8_extracted_k141_177689 | Not-determined |
| S8_extracted_k141_134066 | Not-determined |
| S8_extracted_k141_116689 | Not-determined |
| S8_extracted_k141_12382 | Not-determined |
| S8_extracted_k141_12384 | Not-determined |
| S8_extracted_k141_108334 | Low-quality |
| S8_extracted_k141_159958 | Not-determined |
| S8_extracted_k141_30538 | Not-determined |
| S8_extracted_k141_134073 | Low-quality |
| S8_extracted_k141_21444 | Not-determined |
| S8_extracted_k141_30539 | Not-determined |
| S8_extracted_k141_108336 | Not-determined |
| S8_extracted_k141_151355 | Not-determined |
| S8_extracted_k141_185550 | Not-determined |
| S8_extracted_k141_12388 | Not-determined |
| S8_extracted_k141_21445 | Not-determined |
| S8_extracted_k141_116690 | Not-determined |
| S8_extracted_k141_134078 | Not-determined |
| S8_extracted_k141_47099 | Not-determined |
| S8_extracted_k141_82063 | Not-determined |
| S8_extracted_k141_82064 | Not-determined |
| S8_extracted_k141_64538 | Not-determined |
| S8_extracted_k141_21448 | Low-quality |
| S8_extracted_k141_99563 | Not-determined |
| S8_extracted_k141_108341 | Not-determined |
| S8_extracted_k141_90939 | Low-quality |
| S8_extracted_k141_203291 | Not-determined |
| S8_extracted_k141_203292 | Not-determined |
| S8_extracted_k141_116693 | Not-determined |
| S8_extracted_k141_99565 | Not-determined |
| S8_extracted_k141_151366 | Not-determined |
| S8_extracted_k141_185559 | Low-quality |
| S8_extracted_k141_134082 | Not-determined |
| S8_extracted_k141_159971 | Not-determined |
| S8_extracted_k141_195112 | Not-determined |
| S8_extracted_k141_195114 | Not-determined |
| S8_extracted_k141_30550 | Not-determined |
| S8_extracted_k141_21457 | Low-quality |
| S8_extracted_k141_38784 | Not-determined |
| S8_extracted_k141_4125 | Not-determined |
| S8_extracted_k141_82079 | Not-determined |
| S8_extracted_k141_151374 | Not-determined |
| S8_extracted_k141_99574 | Not-determined |
| S8_extracted_k141_151376 | Not-determined |
| S8_extracted_k141_159977 | Not-determined |
| S8_extracted_k141_56254 | Not-determined |
| S8_extracted_k141_177717 | Not-determined |
| S8_extracted_k141_168901 | Not-determined |
| S8_extracted_k141_99576 | Not-determined |
| S8_extracted_k141_116705 | Not-determined |
| S8_extracted_k141_99578 | Low-quality |
| S8_extracted_k141_30557 | Not-determined |
| S8_extracted_k141_99584 | Not-determined |
| S8_extracted_k141_73004 | Not-determined |
| S8_extracted_k141_168907 | Not-determined |
| S8_extracted_k141_177726 | Not-determined |
| S8_extracted_k141_177727 | Not-determined |
| S8_extracted_k141_12410 | Not-determined |
| S8_extracted_k141_30558 | Low-quality |
| S8_extracted_k141_4131 | Not-determined |
| S8_extracted_k141_30561 | Not-determined |
| S8_extracted_k141_168910 | Not-determined |
| S8_extracted_k141_116715 | Not-determined |
| S8_extracted_k141_185573 | Not-determined |
| S8_extracted_k141_56263 | Not-determined |
| S8_extracted_k141_82097 | Not-determined |
| S8_extracted_k141_12419 | Not-determined |
| S8_extracted_k141_21464 | Not-determined |
| S8_extracted_k141_159984 | Low-quality |
| S8_extracted_k141_90962 | Not-determined |
| S8_extracted_k141_82101 | Low-quality |
| S8_extracted_k141_116717 | Low-quality |
| S8_extracted_k141_21469 | Not-determined |
| S8_extracted_k141_82106 | Not-determined |
| S8_extracted_k141_82107 | Not-determined |
| S8_extracted_k141_99600 | Not-determined |
| S8_extracted_k141_185582 | Not-determined |
| S8_extracted_k141_82110 | Not-determined |
| S8_extracted_k141_177736 | Not-determined |
| S8_extracted_k141_142983 | Not-determined |
| S8_extracted_k141_177739 | Not-determined |
| S8_extracted_k141_4147 | Not-determined |
| S8_extracted_k141_195140 | Not-determined |
| S8_extracted_k141_108357 | Not-determined |
| S8_extracted_k141_90975 | Not-determined |
| S8_extracted_k141_56273 | Not-determined |
| S8_extracted_k141_195144 | Not-determined |
| S8_extracted_k141_108359 | Not-determined |
| S8_extracted_k141_56274 | Not-determined |
| S8_extracted_k141_82122 | Low-quality |
| S8_extracted_k141_64560 | Not-determined |
| S8_extracted_k141_12437 | Not-determined |
| S8_extracted_k141_108361 | Not-determined |
| S8_extracted_k141_12438 | Not-determined |
| S8_extracted_k141_12440 | Not-determined |
| S8_extracted_k141_108366 | Low-quality |
| S8_extracted_k141_142994 | Low-quality |
| S8_extracted_k141_108370 | Not-determined |
| S8_extracted_k141_82132 | Not-determined |
| S8_extracted_k141_21479 | Not-determined |
| S8_extracted_k141_195149 | Not-determined |
| S8_extracted_k141_90985 | Not-determined |
| S8_extracted_k141_195152 | Not-determined |
| S8_extracted_k141_12449 | Not-determined |
| S8_extracted_k141_185585 | Low-quality |
| S8_extracted_k141_64572 | Not-determined |
| S8_extracted_k141_12451 | Not-determined |
| S8_extracted_k141_21483 | Not-determined |
| S8_extracted_k141_90990 | Not-determined |
| S8_extracted_k141_116736 | Not-determined |
| S8_extracted_k141_168926 | Not-determined |
| S8_extracted_k141_185589 | Not-determined |
| S8_extracted_k141_90991 | Not-determined |
| S8_extracted_k141_12455 | Not-determined |
| S8_extracted_k141_185593 | Not-determined |
| S8_extracted_k141_185594 | Not-determined |
| S8_extracted_k141_143003 | Not-determined |
| S8_extracted_k141_56296 | Not-determined |
| S8_extracted_k141_116744 | Not-determined |
| S8_extracted_k141_168937 | Not-determined |
| S8_extracted_k141_160002 | Not-determined |
| S8_extracted_k141_12463 | Not-determined |
| S8_extracted_k141_108380 | Not-determined |
| S8_extracted_k141_21493 | Not-determined |
| S8_extracted_k141_56301 | Not-determined |
| S8_extracted_k141_195157 | Not-determined |
| S8_extracted_k141_12466 | Not-determined |
| S8_extracted_k141_90994 | Low-quality |
| S8_extracted_k141_177766 | Not-determined |
| S8_extracted_k141_160006 | Not-determined |
| S8_extracted_k141_160009 | Not-determined |
| S8_extracted_k141_73038 | Not-determined |
| S8_extracted_k141_64583 | Not-determined |
| S8_extracted_k141_195166 | Not-determined |
| S8_extracted_k141_64584 | Not-determined |
| S8_extracted_k141_90997 | Not-determined |
| S8_extracted_k141_125728 | Not-determined |
| S8_extracted_k141_116755 | Not-determined |
| S8_extracted_k141_12472 | Not-determined |
| S16_extracted_147326 | Medium-quality |
| S8_extracted_k141_185598 | Not-determined |
| S8_extracted_k141_160013 | Not-determined |
| S8_extracted_k141_134118 | Not-determined |
| S8_extracted_k141_21504 | Not-determined |
| S8_extracted_k141_195173 | Not-determined |
| S8_extracted_k141_143014 | Not-determined |
| S8_extracted_k141_38789 | Not-determined |
| S8_extracted_k141_91000 | Not-determined |
| S8_extracted_k141_38792 | Not-determined |
| S8_extracted_k141_160018 | Not-determined |
| S8_extracted_k141_73048 | Not-determined |
| S8_extracted_k141_21505 | Not-determined |
| S8_extracted_k141_47119 | Not-determined |
| S8_extracted_k141_160019 | Not-determined |
| S8_extracted_k141_143015 | Low-quality |
| S8_extracted_k141_21508 | Not-determined |
| S8_extracted_k141_185609 | Not-determined |
| S8_extracted_k141_108398 | Not-determined |
| S8_extracted_k141_134125 | Not-determined |
| S8_extracted_k141_73055 | Not-determined |
| S8_extracted_k141_108400 | Not-determined |
| S8_extracted_k141_12477 | Not-determined |
| S8_extracted_k141_160023 | Not-determined |
| S8_extracted_k141_143020 | Not-determined |
| S8_extracted_k141_38802 | Not-determined |
| S8_extracted_k141_125742 | Not-determined |
| S8_extracted_k141_47128 | Not-determined |
| S8_extracted_k141_195178 | Not-determined |
| S8_extracted_k141_195179 | Not-determined |
| S8_extracted_k141_91009 | Not-determined |
| S8_extracted_k141_177771 | Low-quality |
| S8_extracted_k141_134131 | Low-quality |
| S8_extracted_k141_125749 | Not-determined |
| S8_extracted_k141_12485 | Not-determined |
| S8_extracted_k141_73064 | Not-determined |
| S8_extracted_k141_91019 | Not-determined |
| S8_extracted_k141_177777 | Not-determined |
| S8_extracted_k141_91020 | Not-determined |
| S8_extracted_k141_160038 | Low-quality |
| S8_extracted_k141_21517 | Not-determined |
| S4_extracted_5021 | Medium-quality |
| S8_extracted_k141_21518 | Not-determined |
| S8_extracted_k141_73074 | Not-determined |
| S8_extracted_k141_195191 | Low-quality |
| S8_extracted_k141_21519 | Not-determined |
| S8_extracted_k141_73075 | Not-determined |
| S8_extracted_k141_21521 | Not-determined |
| S8_extracted_k141_38819 | Not-determined |
| S8_extracted_k141_108413 | Not-determined |
| S8_extracted_k141_160047 | Not-determined |
| S8_extracted_k141_125759 | Not-determined |
| S8_extracted_k141_108414 | Not-determined |
| S8_extracted_k141_56341 | Not-determined |
| S8_extracted_k141_195198 | Not-determined |
| S8_extracted_k141_82186 | Not-determined |
| S8_extracted_k141_151385 | Not-determined |
| S8_extracted_k141_195204 | Not-determined |
| S8_extracted_k141_203315 | Not-determined |
| S8_extracted_k141_177789 | Not-determined |
| S8_extracted_k141_4157 | Not-determined |
| S8_extracted_k141_4159 | Not-determined |
| S8_extracted_k141_21533 | Not-determined |
| S8_extracted_k141_56345 | Not-determined |
| S8_extracted_k141_47150 | Not-determined |
| S8_extracted_k141_73088 | Low-quality |
| S8_extracted_k141_160064 | Not-determined |
| S8_extracted_k141_108423 | Not-determined |
| S8_extracted_k141_177791 | Not-determined |
| S8_extracted_k141_73091 | Not-determined |
| S8_extracted_k141_82190 | Not-determined |
| S8_extracted_k141_21537 | Low-quality |
| S8_extracted_k141_64619 | Low-quality |
| S8_extracted_k141_21538 | Not-determined |
| S8_extracted_k141_116772 | Not-determined |
| S8_extracted_k141_38820 | Not-determined |
| S8_extracted_k141_56351 | Not-determined |
| S8_extracted_k141_134153 | Not-determined |
| S8_extracted_k141_12512 | Not-determined |
| S8_extracted_k141_4162 | Not-determined |
| S8_extracted_k141_151403 | Not-determined |
| S8_extracted_k141_99616 | Not-determined |
| S8_extracted_k141_82200 | Not-determined |
| S8_extracted_k141_151404 | Not-determined |
| S8_extracted_k141_151405 | Not-determined |
| S8_extracted_k141_160070 | Not-determined |
| S8_extracted_k141_108432 | Not-determined |
| S8_extracted_k141_143054 | Not-determined |
| S8_extracted_k141_12517 | Not-determined |
| S8_extracted_k141_134158 | Not-determined |
| S8_extracted_k141_56358 | Not-determined |
| S8_extracted_k141_12520 | Not-determined |
| S8_extracted_k141_99623 | Not-determined |
| S8_extracted_k141_203318 | Not-determined |
| S8_extracted_k141_4172 | Not-determined |
| S8_extracted_k141_38829 | Low-quality |
| S8_extracted_k141_4173 | Low-quality |
| S8_extracted_k141_99630 | Not-determined |
| S8_extracted_k141_143061 | Not-determined |
| S8_extracted_k141_177800 | Low-quality |
| S8_extracted_k141_4175 | Low-quality |
| S8_extracted_k141_143062 | Not-determined |
| S8_extracted_k141_38834 | Not-determined |
| S8_extracted_k141_99634 | Not-determined |
| S8_extracted_k141_99637 | Not-determined |
| S8_extracted_k141_185644 | Not-determined |
| S8_extracted_k141_21548 | Not-determined |
| S8_extracted_k141_56368 | Not-determined |
| S8_extracted_k141_108443 | Not-determined |
| S8_extracted_k141_116789 | Low-quality |
| S8_extracted_k141_160089 | Not-determined |
| S8_extracted_k141_82206 | Not-determined |
| S8_extracted_k141_56371 | Not-determined |
| S8_extracted_k141_64636 | Not-determined |
| S8_extracted_k141_177811 | Not-determined |
| S8_extracted_k141_203322 | Low-quality |
| S8_extracted_k141_203325 | Not-determined |
| S8_extracted_k141_4192 | Not-determined |
| S8_extracted_k141_143074 | Not-determined |
| S8_extracted_k141_203326 | Low-quality |
| S8_extracted_k141_73100 | Not-determined |
| S8_extracted_k141_168965 | Not-determined |
| S8_extracted_k141_168966 | Not-determined |
| S8_extracted_k141_185651 | Not-determined |
| S8_extracted_k141_185652 | Not-determined |
| S8_extracted_k141_151420 | Not-determined |
| S8_extracted_k141_195241 | Not-determined |
| S8_extracted_k141_38842 | Low-quality |
| S8_extracted_k141_151422 | Not-determined |
| S8_extracted_k141_73105 | Not-determined |
| S8_extracted_k141_56383 | Not-determined |
| S8_extracted_k141_56384 | Not-determined |
| S8_extracted_k141_177823 | Not-determined |
| S8_extracted_k141_73110 | Not-determined |
| S8_extracted_k141_203331 | Not-determined |
| S8_extracted_k141_185656 | Not-determined |
| S8_extracted_k141_168975 | Not-determined |
| S8_extracted_k141_108453 | Not-determined |
| S8_extracted_k141_195248 | Low-quality |
| S8_extracted_k141_12549 | Not-determined |
| S8_extracted_k141_151430 | Not-determined |
| S8_extracted_k141_185660 | Not-determined |
| S4_vs_VIRSorter_5021-cat_2 | Medium-quality |
| S8_extracted_k141_64645 | Not-determined |
| S8_extracted_k141_4206 | Not-determined |
| S8_extracted_k141_160095 | Low-quality |
| S8_extracted_k141_91035 | Not-determined |
| S8_extracted_k141_73119 | Not-determined |
| S8_extracted_k141_177838 | Not-determined |
| S8_extracted_k141_73120 | Not-determined |
| S8_extracted_k141_64648 | Not-determined |
| S8_extracted_k141_91036 | Not-determined |
| S8_extracted_k141_125783 | Not-determined |
| S8_extracted_k141_195249 | Not-determined |
| S8_extracted_k141_91038 | Not-determined |
| S8_extracted_k141_185665 | Not-determined |
| S8_extracted_k141_168988 | Not-determined |
| S8_extracted_k141_91040 | Low-quality |
| S8_extracted_k141_116796 | Not-determined |
| S8_extracted_k141_47189 | Not-determined |
| S8_extracted_k141_91041 | Not-determined |
| S8_extracted_k141_151439 | Not-determined |
| S8_extracted_k141_151441 | Not-determined |
| S8_extracted_k141_185670 | Low-quality |
| S8_extracted_k141_177839 | Not-determined |
| S8_extracted_k141_21578 | Not-determined |
| S8_extracted_k141_195257 | Not-determined |
| S8_extracted_k141_4226 | Not-determined |
| S8_extracted_k141_168998 | Not-determined |
| S8_extracted_k141_47195 | Not-determined |
| S8_extracted_k141_91056 | Not-determined |
| S8_extracted_k141_73138 | Not-determined |
| S8_extracted_k141_56407 | Not-determined |
| S8_extracted_k141_56408 | Not-determined |
| S8_extracted_k141_143081 | Low-quality |
| S8_extracted_k141_125802 | Not-determined |
| S8_extracted_k141_177841 | Not-determined |
| S8_extracted_k141_38866 | Not-determined |
| S8_extracted_k141_64672 | Not-determined |
| S8_extracted_k141_4233 | Low-quality |
| S8_extracted_k141_64673 | Not-determined |
| S8_extracted_k141_185683 | Not-determined |
| S8_extracted_k141_99667 | Not-determined |
| S8_extracted_k141_195268 | Not-determined |
| S8_extracted_k141_99668 | Not-determined |
| S8_extracted_k141_125806 | Not-determined |
| S8_extracted_k141_125807 | Not-determined |
| S8_extracted_k141_82225 | Not-determined |
| S8_extracted_k141_185686 | Not-determined |
| S8_extracted_k141_91062 | Not-determined |
| S8_extracted_k141_151453 | Not-determined |
| S8_extracted_k141_151454 | Not-determined |
| S8_extracted_k141_177843 | Not-determined |
| S8_extracted_k141_73146 | Not-determined |
| S8_extracted_k141_91068 | Not-determined |
| S8_extracted_k141_99671 | Not-determined |
| S8_extracted_k141_91069 | Not-determined |
| S8_extracted_k141_177849 | Not-determined |
| S8_extracted_k141_82232 | Not-determined |
| S8_extracted_k141_21597 | Low-quality |
| S8_extracted_k141_73151 | Not-determined |
| S8_extracted_k141_73155 | Not-determined |
| S8_extracted_k141_82237 | Not-determined |
| S8_extracted_k141_47214 | Not-determined |
| S8_extracted_k141_143091 | Not-determined |
| S8_extracted_k141_134190 | Not-determined |
| S8_extracted_k141_99681 | Not-determined |
| S8_extracted_k141_82244 | Not-determined |
| S8_extracted_k141_38878 | Not-determined |
| S8_extracted_k141_151466 | Not-determined |
| S8_extracted_k141_99684 | Low-quality |
| S8_extracted_k141_125823 | Not-determined |
| S8_extracted_k141_108495 | Low-quality |
| S8_extracted_k141_143099 | Not-determined |
| S8_extracted_k141_91077 | Not-determined |
| S8_extracted_k141_99689 | Not-determined |
| S8_extracted_k141_116823 | Low-quality |
| S8_extracted_k141_82250 | Not-determined |
| S8_extracted_k141_99693 | Not-determined |
| S8_extracted_k141_169029 | Not-determined |
| S8_extracted_k141_73169 | Low-quality |
| S8_extracted_k141_108501 | Not-determined |
| S8_extracted_k141_203378 | Low-quality |
| S8_extracted_k141_99694 | Not-determined |
| S8_extracted_k141_21607 | Not-determined |
| S8_extracted_k141_99696 | Low-quality |
| S8_extracted_k141_169031 | Not-determined |
| S8_extracted_k141_195283 | Not-determined |
| S8_extracted_k141_203380 | Not-determined |
| S8_extracted_k141_82261 | Not-determined |
| S8_extracted_k141_203383 | Not-determined |
| S8_extracted_k141_56418 | Not-determined |
| S8_extracted_k141_169035 | Not-determined |
| S8_extracted_k141_4252 | Not-determined |
| S8_extracted_k141_125830 | Low-quality |
| S8_extracted_k141_169037 | Not-determined |
| S8_extracted_k141_82266 | Not-determined |
| S8_extracted_k141_108510 | Not-determined |
| S8_extracted_k141_56424 | Not-determined |
| S8_extracted_k141_185694 | Not-determined |
| S8_extracted_k141_12553 | Not-determined |
| S8_extracted_k141_108511 | Low-quality |
| S8_extracted_k141_195293 | Not-determined |
| S8_extracted_k141_108512 | Not-determined |
| S8_extracted_k141_195295 | Not-determined |
| S8_extracted_k141_91091 | Not-determined |
| S8_extracted_k141_21625 | Not-determined |
| S8_extracted_k141_47223 | Low-quality |
| S8_extracted_k141_143126 | Not-determined |
| S8_extracted_k141_169040 | Low-quality |
| S8_extracted_k141_143127 | Not-determined |
| S8_extracted_k141_12558 | Not-determined |
| S8_extracted_k141_195296 | Not-determined |
| S8_extracted_k141_169045 | Not-determined |
| S8_extracted_k141_195297 | Not-determined |
| S8_extracted_k141_151485 | Not-determined |
| S8_extracted_k141_21628 | Not-determined |
| S8_extracted_k141_169046 | Not-determined |
| S8_extracted_k141_203392 | Not-determined |
| S8_extracted_k141_151488 | Not-determined |
| S8_extracted_k141_143131 | Not-determined |
| S8_extracted_k141_47224 | Low-quality |
| S8_extracted_k141_21629 | Not-determined |
| S8_extracted_k141_195303 | Not-determined |
| S8_extracted_k141_108521 | Low-quality |
| S8_extracted_k141_21633 | Not-determined |
| S8_extracted_k141_64698 | Not-determined |
| S8_extracted_k141_12564 | Not-determined |
| S8_extracted_k141_73198 | Not-determined |
| S8_extracted_k141_195304 | Low-quality |
| S8_extracted_k141_195307 | Not-determined |
| S8_extracted_k141_99704 | Low-quality |
| S8_extracted_k141_64702 | Not-determined |
| S8_extracted_k141_195309 | Not-determined |
| S8_extracted_k141_203399 | Low-quality |
| S8_extracted_k141_99705 | Not-determined |
| S8_extracted_k141_64704 | Low-quality |
| S8_extracted_k141_82286 | Not-determined |
| S8_extracted_k141_73201 | Low-quality |
| S8_extracted_k141_56443 | Not-determined |
| S8_extracted_k141_99706 | Not-determined |
| S8_extracted_k141_47230 | Not-determined |
| S8_extracted_k141_4270 | Not-determined |
| S8_extracted_k141_91106 | Not-determined |
| S8_extracted_k141_108528 | Not-determined |
| S8_extracted_k141_82291 | Not-determined |
| S8_extracted_k141_4273 | Not-determined |
| S8_extracted_k141_169050 | Not-determined |
| S8_extracted_k141_99712 | Not-determined |
| S8_extracted_k141_177884 | Not-determined |
| S8_extracted_k141_108531 | Low-quality |
| S8_extracted_k141_125844 | Not-determined |
| S8_extracted_k141_38910 | Not-determined |
| S8_extracted_k141_125846 | Not-determined |
| S8_extracted_k141_21642 | Not-determined |
| S8_extracted_k141_125848 | Not-determined |
| S8_extracted_k141_108532 | Not-determined |
| S8_extracted_k141_21643 | Not-determined |
| S8_extracted_k141_82293 | Not-determined |
| S8_extracted_k141_4283 | Not-determined |

| | |
|---|---|
| SB_extracted_k141_134216 | Not-determined |
| SB_extracted_k141_38913 | Not-determined |
| SB_extracted_k141_47246 | Not-determined |
| SB_extracted_k141_47249 | Not-determined |
| SB_extracted_k141_143140 | Low-quality |
| SB_extracted_k141_169059 | Not-determined |
| SB_extracted_k141_4288 | Not-determined |
| SB_extracted_k141_185712 | Not-determined |
| SB_extracted_k141_56456 | Not-determined |
| SB_extracted_k141_4293 | Not-determined |
| SB_extracted_k141_108545 | Not-determined |
| SB_extracted_k141_38923 | Not-determined |
| SB_extracted_k141_47256 | Not-determined |
| SB_extracted_k141_108548 | Not-determined |
| SB_extracted_k141_91114 | Not-determined |
| SB_extracted_k141_108549 | Not-determined |
| SB_extracted_k141_177896 | Low-quality |
| SB_extracted_k141_134227 | Not-determined |
| SB_extracted_k141_185719 | Low-quality |
| SB_extracted_k141_134229 | Not-determined |
| SB_extracted_k141_73211 | Not-determined |
| SB_extracted_k141_56459 | Not-determined |
| SB_extracted_k141_116850 | Not-determined |
| SB_extracted_k141_169064 | Not-determined |
| SB_extracted_k141_169066 | Not-determined |
| SB_extracted_k141_73215 | Not-determined |
| SB_extracted_k141_125870 | Low-quality |
| SB_extracted_k141_151510 | Not-determined |
| SB_extracted_k141_203422 | Not-determined |
| SB_extracted_k141_73220 | Not-determined |
| SB_extracted_k141_12590 | Not-determined |
| SB_extracted_k141_143159 | Not-determined |
| SB_extracted_k141_82304 | Not-determined |
| SB_extracted_k141_125875 | Not-determined |
| SB_extracted_k141_125876 | Not-determined |
| SB_extracted_k141_82307 | Not-determined |
| SB_extracted_k141_169075 | Low-quality |
| SB_extracted_k141_82308 | Not-determined |
| SB_extracted_k141_143163 | Not-determined |
| SB_extracted_k141_82312 | Not-determined |
| SB_extracted_k141_151516 | Not-determined |
| SB_extracted_k141_82313 | Not-determined |
| SB_extracted_k141_91132 | Not-determined |
| SB_extracted_k141_73228 | Not-determined |
| SB_extracted_k141_203432 | Not-determined |
| SB_extracted_k141_125883 | Not-determined |
| SB_extracted_k141_73229 | Not-determined |
| SB_extracted_k141_125884 | Not-determined |
| SB_extracted_k141_12595 | Not-determined |
| SB_extracted_k141_195322 | Low-quality |
| SB_extracted_k141_21653 | Not-determined |
| SB_extracted_k141_4318 | Not-determined |
| SB_extracted_k141_177909 | Low-quality |
| SB_extracted_k141_38942 | Low-quality |
| SB_extracted_k141_73235 | Not-determined |
| SB_extracted_k141_195328 | Not-determined |
| SB_extracted_k141_134262 | Not-determined |
| SB_extracted_k141_116859 | Not-determined |
| SB_extracted_k141_151529 | Not-determined |
| SB_extracted_k141_195331 | Not-determined |
| SB_extracted_k141_4322 | Not-determined |
| SB_extracted_k141_116862 | Low-quality |
| SB_extracted_k141_134265 | Not-determined |
| SB_extracted_k141_56487 | Not-determined |
| SB_extracted_k141_82321 | Not-determined |
| SB_extracted_k141_143177 | Not-determined |
| SB_extracted_k141_12601 | Not-determined |
| SB_extracted_k141_56495 | Not-determined |
| SB_extracted_k141_56496 | Not-determined |
| SB_extracted_k141_30590 | Not-determined |
| SB_extracted_k141_73240 | Not-determined |
| SB_extracted_k141_12604 | Not-determined |
| SB_extracted_k141_64725 | Not-determined |
| SB_extracted_k141_64726 | Low-quality |
| SB_extracted_k141_195344 | Not-determined |
| SB_extracted_k141_125905 | Not-determined |
| SB_extracted_k141_30597 | Not-determined |
| SB_extracted_k141_195347 | Low-quality |
| SB_extracted_k141_169093 | Not-determined |
| SB_extracted_k141_203450 | Not-determined |
| SB_extracted_k141_143184 | Not-determined |
| SB_extracted_k141_64731 | Not-determined |
| SB_extracted_k141_82333 | Not-determined |
| SB_extracted_k141_185741 | Not-determined |
| SB_extracted_k141_151538 | Low-quality |
| SB_extracted_k141_125912 | Not-determined |
| SB_extracted_k141_82336 | Not-determined |
| SB_extracted_k141_151541 | Not-determined |
| SB_extracted_k141_12609 | Low-quality |
| SB_extracted_k141_38957 | Not-determined |
| SB_extracted_k141_169101 | Not-determined |
| SB_extracted_k141_125915 | Not-determined |
| SB_extracted_k141_56505 | Low-quality |
| SB_extracted_k141_116866 | Low-quality |
| SB_extracted_k141_203459 | Low-quality |
| SB_extracted_k141_185746 | Not-determined |
| SB_extracted_k141_169107 | Not-determined |
| SB_extracted_k141_56509 | Not-determined |
| SB_extracted_k141_195361 | Not-determined |
| SB_extracted_k141_82342 | Not-determined |
| SB_extracted_k141_4334 | Not-determined |
| SB_extracted_k141_195362 | Low-quality |
| SB_extracted_k141_143194 | Low-quality |
| SB_extracted_k141_116871 | Not-determined |
| SB_extracted_k141_116874 | Not-determined |
| SB_extracted_k141_125922 | Not-determined |
| SB_extracted_k141_185754 | Not-determined |
| SB_extracted_k141_64743 | Not-determined |
| SB_extracted_k141_134285 | Not-determined |
| SB_extracted_k141_177933 | Not-determined |
| SB_extracted_k141_203466 | Not-determined |
| SB_extracted_k141_64746 | Not-determined |
| SB_extracted_k141_116879 | Not-determined |
| SB_extracted_k141_134291 | Not-determined |
| SB_extracted_k141_21686 | Not-determined |
| SB_extracted_k141_4338 | Low-quality |
| SB_extracted_k141_169122 | Not-determined |
| SB_extracted_k141_99730 | Not-determined |
| SB_extracted_k141_143207 | Not-determined |
| SB_extracted_k141_64750 | Low-quality |
| SB_extracted_k141_203479 | Not-determined |
| SB_extracted_k141_169132 | Low-quality |
| SB_extracted_k141_143212 | Not-determined |
| SB_extracted_k141_12626 | Not-determined |
| SB_extracted_k141_169135 | Not-determined |
| SB_extracted_k141_125933 | Not-determined |
| SB_extracted_k141_99735 | Not-determined |
| SB_extracted_k141_203482 | Low-quality |
| SB_extracted_k141_73270 | Not-determined |
| SB_extracted_k141_169138 | Not-determined |
| SB_extracted_k141_21688 | Not-determined |
| SB_extracted_k141_38982 | Not-determined |
| SB_extracted_k141_64757 | Not-determined |
| SB_extracted_k141_64758 | Not-determined |
| SB_extracted_k141_134300 | Not-determined |
| SB_extracted_k141_73272 | Not-determined |
| SB_extracted_k141_38985 | Not-determined |
| SB_extracted_k141_38988 | Not-determined |
| SB_extracted_k141_143221 | Low-quality |
| SB_extracted_k141_151555 | Low-quality |
| SB_extracted_k141_21691 | Not-determined |
| SB_extracted_k141_4341 | Not-determined |
| SB_extracted_k141_73275 | Not-determined |
| SB_extracted_k141_151558 | Not-determined |
| SB_extracted_k141_12632 | Not-determined |
| SB_extracted_k141_169147 | Not-determined |
| SB_extracted_k141_125943 | Low-quality |
| SB_extracted_k141_143226 | Not-determined |
| SB_extracted_k141_82358 | Not-determined |
| SB_extracted_k141_116907 | Low-quality |
| SB_extracted_k141_82359 | Not-determined |
| SB_extracted_k141_12637 | Not-determined |
| SB_extracted_k141_99749 | Low-quality |
| SB_extracted_k141_12639 | Not-determined |
| SB_extracted_k141_64763 | Not-determined |
| SB_extracted_k141_4354 | Low-quality |
| SB_extracted_k141_125947 | Not-determined |
| SB_extracted_k141_108574 | Not-determined |
| SB_extracted_k141_160113 | Not-determined |
| SB_extracted_k141_203487 | Not-determined |
| SB_extracted_k141_21698 | Not-determined |
| SB_extracted_k141_4355 | Not-determined |
| SB_extracted_k141_203488 | Not-determined |
| SB_extracted_k141_30615 | Low-quality |
| SB_extracted_k141_4358 | Not-determined |
| SB_extracted_k141_108576 | Low-quality |
| SB_extracted_k141_4359 | Not-determined |
| SB_extracted_k141_99755 | Not-determined |
| SB_extracted_k141_143230 | Not-determined |
| SB_extracted_k141_30618 | Not-determined |
| SB_extracted_k141_73282 | Not-determined |
| SB_extracted_k141_143231 | Not-determined |
| SB_extracted_k141_64768 | Not-determined |
| SB_extracted_k141_99760 | Not-determined |
| SB_extracted_k141_185772 | Not-determined |
| SB_extracted_k141_21704 | Not-determined |
| SB_extracted_k141_185773 | Not-determined |
| SB_extracted_k141_185774 | Not-determined |
| SB_extracted_k141_4362 | Not-determined |
| SB_extracted_k141_160116 | Not-determined |
| SB_extracted_k141_203499 | Not-determined |
| SB_extracted_k141_160117 | Not-determined |
| SB_extracted_k141_21708 | Not-determined |
| SB_extracted_k141_82367 | Not-determined |
| SB_extracted_k141_30622 | Not-determined |
| SB_extracted_k141_116924 | Not-determined |
| SB_extracted_k141_134310 | Not-determined |
| SB_extracted_k141_99771 | Not-determined |
| SB_extracted_k141_30624 | Not-determined |
| SB_extracted_k141_73289 | Low-quality |
| SB_extracted_k141_151582 | Not-determined |
| SB_extracted_k141_143248 | Not-determined |
| SB_extracted_k141_56540 | Not-determined |
| SB_extracted_k141_203511 | Not-determined |
| SB_extracted_k141_56541 | Not-determined |
| SB_extracted_k141_116935 | Not-determined |
| SB_extracted_k141_134318 | Not-determined |
| SB_extracted_k141_195370 | Low-quality |
| SB_extracted_k141_185786 | Not-determined |
| SB_extracted_k141_151587 | Not-determined |
| SB_extracted_k141_177965 | Not-determined |
| SB_extracted_k141_30633 | Not-determined |
| SB_extracted_k141_160130 | Not-determined |
| SB_extracted_k141_21727 | Not-determined |
| SB_extracted_k141_151592 | Low-quality |
| SB_extracted_k141_195374 | Not-determined |
| SB_extracted_k141_108600 | Not-determined |
| SB_extracted_k141_21729 | Not-determined |
| SB_extracted_k141_108601 | Low-quality |
| SB_extracted_k141_169163 | Low-quality |
| SB_extracted_k141_177969 | Not-determined |
| SB_extracted_k141_177971 | Low-quality |
| SB_extracted_k141_134328 | Low-quality |
| SB_extracted_k141_108603 | Not-determined |
| SB_extracted_k141_134329 | Not-determined |
| SB_extracted_k141_56550 | Not-determined |
| SB_extracted_k141_116944 | Not-determined |
| SB_extracted_k141_203522 | Low-quality |
| SB_extracted_k141_99780 | Low-quality |
| SB_extracted_k141_30643 | Not-determined |
| SB_extracted_k141_4380 | Not-determined |
| SB_extracted_k141_56555 | Not-determined |
| SB_extracted_k141_169168 | Not-determined |
| SB_extracted_k141_125972 | Not-determined |
| SB_extracted_k141_134334 | Not-determined |
| SB_extracted_k141_21738 | Not-determined |
| SB_extracted_k141_4384 | Not-determined |
| SB_extracted_k141_125974 | Low-quality |
| SB_extracted_k141_4386 | Not-determined |
| SB_extracted_k141_143257 | Not-determined |
| SB_extracted_k141_195376 | Not-determined |
| SB_extracted_k141_169172 | Not-determined |
| SB_extracted_k141_21744 | Not-determined |
| SB_extracted_k141_125980 | Not-determined |
| SB_extracted_k141_169174 | Not-determined |
| SB_extracted_k141_64786 | Not-determined |
| SB_extracted_k141_108616 | Not-determined |
| SB_extracted_k141_64788 | Not-determined |
| SB_extracted_k141_12668 | Not-determined |
| SB_extracted_k141_169181 | Not-determined |
| SB_extracted_k141_134348 | Not-determined |
| SB_extracted_k141_4394 | Not-determined |
| SB_extracted_k141_169184 | Low-quality |
| SB_extracted_k141_64791 | Not-determined |
| SB_extracted_k141_4397 | Not-determined |
| SB_extracted_k141_195379 | Low-quality |
| SB_extracted_k141_143261 | Not-determined |
| SB_extracted_k141_134354 | Not-determined |
| SB_extracted_k141_56566 | Not-determined |
| SB_extracted_k141_39007 | Not-determined |
| SB_extracted_k141_151602 | Not-determined |
| SB_extracted_k141_151603 | Not-determined |
| SB_extracted_k141_30650 | Not-determined |
| SB_extracted_k141_64792 | Low-quality |
| SB_extracted_k141_4400 | Not-determined |
| SB_extracted_k141_151607 | Not-determined |
| SB_extracted_k141_169190 | Not-determined |
| SB_extracted_k141_169191 | Not-determined |
| SB_extracted_k141_21756 | Not-determined |
| SB_extracted_k141_12680 | Low-quality |
| SB_extracted_k141_64796 | Not-determined |
| SB_extracted_k141_160148 | Not-determined |
| SB_extracted_k141_116955 | Not-determined |
| SB_extracted_k141_64798 | Low-quality |
| SB_extracted_k141_116957 | Not-determined |
| SB_extracted_k141_4410 | Not-determined |
| SB_extracted_k141_47273 | Not-determined |
| SB_extracted_k141_177992 | Not-determined |
| SB_extracted_k141_160149 | Low-quality |
| SB_extracted_k141_30657 | Not-determined |
| SB_extracted_k141_160151 | Not-determined |
| SB_extracted_k141_21765 | Not-determined |
| SB_extracted_k141_143272 | Not-determined |
| SB_extracted_k141_12684 | Not-determined |
| SB_extracted_k141_30663 | Low-quality |
| SB_extracted_k141_125996 | Not-determined |
| SB_extracted_k141_195394 | Not-determined |
| SB_extracted_k141_39020 | Not-determined |
| SB_extracted_k141_134358 | Not-determined |
| SB_extracted_k141_143281 | Not-determined |
| SB_extracted_k141_108632 | Not-determined |
| SB_extracted_k141_4421 | Not-determined |
| SB_extracted_k141_4422 | Not-determined |
| SB_extracted_k141_160158 | Not-determined |
| SB_extracted_k141_30668 | Not-determined |
| SB_extracted_k141_108635 | Not-determined |
| SB_extracted_k141_56571 | Not-determined |
| SB_extracted_k141_73317 | Not-determined |
| SB_extracted_k141_30670 | Not-determined |
| SB_extracted_k141_151612 | Not-determined |
| SB_extracted_k141_108637 | Not-determined |
| SB_extracted_k141_108639 | Not-determined |
| SB_extracted_k141_185792 | Not-determined |
| SB_extracted_k141_203545 | Not-determined |
| SB_extracted_k141_143284 | Not-determined |
| SB_extracted_k141_160165 | Not-determined |
| SB_extracted_k141_126005 | Not-determined |
| SB_extracted_k141_195404 | Not-determined |
| SB_extracted_k141_160166 | Not-determined |
| SB_extracted_k141_185795 | Not-determined |
| SB_extracted_k141_116973 | Not-determined |
| SB_extracted_k141_185796 | Not-determined |
| SB_extracted_k141_143286 | Low-quality |
| SB_extracted_k141_108642 | Not-determined |
| SB_extracted_k141_134361 | Not-determined |
| SB_extracted_k141_126010 | Low-quality |
| SB_extracted_k141_12697 | Not-determined |
| SB_extracted_k141_30679 | Not-determined |
| SB_extracted_k141_195414 | Not-determined |
| SB_extracted_k141_12699 | Not-determined |
| SB_extracted_k141_21779 | Low-quality |
| SB_extracted_k141_47284 | Low-quality |
| SB_extracted_k141_30682 | Not-determined |
| SB_extracted_k141_64819 | Not-determined |
| SB_extracted_k141_169207 | Not-determined |
| SB_extracted_k141_195418 | Low-quality |
| SB_extracted_k141_39031 | Not-determined |
| SB_extracted_k141_47290 | Not-determined |
| SB_extracted_k141_12707 | Not-determined |
| SB_extracted_k141_47292 | Not-determined |
| SB_extracted_k141_195422 | Low-quality |
| SB_extracted_k141_56581 | Not-determined |
| SB_extracted_k141_203565 | Not-determined |
| SB_extracted_k141_12711 | Not-determined |
| SB_extracted_k141_203566 | Not-determined |
| SB_extracted_k141_21787 | Not-determined |
| SB_extracted_k141_4442 | Not-determined |
| SB_extracted_k141_151621 | Not-determined |
| SB_extracted_k141_4446 | Not-determined |
| SB_extracted_k141_21790 | Not-determined |
| SB_extracted_k141_47302 | Not-determined |
| SB_extracted_k141_108659 | Not-determined |
| SB_extracted_k141_39042 | Not-determined |
| SB_extracted_k141_143293 | Low-quality |
| SB_extracted_k141_117000 | Not-determined |
| SB_extracted_k141_56590 | Not-determined |
| SB_extracted_k141_99798 | Not-determined |
| SB_extracted_k141_195440 | Low-quality |
| SB_extracted_k141_21800 | Not-determined |
| SB_extracted_k141_82377 | Not-determined |
| SB_extracted_k141_160197 | Not-determined |
| SB_extracted_k141_169228 | Not-determined |
| SB_extracted_k141_117007 | Not-determined |
| SB_extracted_k141_21804 | Not-determined |
| SB_extracted_k141_169231 | Not-determined |
| SB_extracted_k141_39055 | Not-determined |
| SB_extracted_k141_117011 | Not-determined |
| SB_extracted_k141_82384 | Not-determined |
| SB_extracted_k141_4455 | Not-determined |
| SB_extracted_k141_56601 | Not-determined |
| SB_extracted_k141_21810 | Not-determined |
| SB_extracted_k141_169236 | Not-determined |
| SB_extracted_k141_82386 | Not-determined |
| SB_extracted_k141_99807 | Not-determined |
| SB_extracted_k141_143298 | Not-determined |
| SB_extracted_k141_169238 | Not-determined |
| SB_extracted_k141_73339 | Low-quality |
| SB_extracted_k141_169239 | Not-determined |
| SB_extracted_k141_64840 | Low-quality |
| SB_extracted_k141_4460 | Not-determined |
| SB_extracted_k141_117018 | Not-determined |
| SB_extracted_k141_73343 | Not-determined |
| SB_extracted_k141_99812 | Not-determined |
| SB_extracted_k141_126039 | Not-determined |
| SB_extracted_k141_151639 | Not-determined |
| SB_extracted_k141_99815 | Not-determined |
| SB_extracted_k141_126041 | Not-determined |
| SB_extracted_k141_169242 | Low-quality |
| SB_extracted_k141_56606 | Not-determined |
| SB_extracted_k141_178029 | Not-determined |
| SB_extracted_k141_21816 | Not-determined |
| SB_extracted_k141_73348 | Not-determined |
| SB_extracted_k141_30735 | Not-determined |
| SB_extracted_k141_82392 | Not-determined |
| SB_extracted_k141_4464 | Not-determined |
| SB_extracted_k141_30737 | Not-determined |
| SB_extracted_k141_39066 | Low-quality |
| SB_extracted_k141_4465 | Not-determined |
| SB_extracted_k141_99822 | Not-determined |
| SB_extracted_k141_47318 | Not-determined |
| SB_extracted_k141_195455 | Not-determined |
| SB_extracted_k141_56610 | Not-determined |
| SB_extracted_k141_56612 | Not-determined |
| SB_extracted_k141_160226 | Low-quality |
| SB_extracted_k141_160227 | Not-determined |
| SB_extracted_k141_56613 | Not-determined |
| SB_extracted_k141_143309 | Not-determined |
| SB_extracted_k141_160230 | Not-determined |
| SB_extracted_k141_108683 | Not-determined |
| SB_extracted_k141_82399 | Not-determined |
| SB_extracted_k141_39075 | Not-determined |
| SB_extracted_k141_4476 | Not-determined |
| SB_extracted_k141_73361 | Not-determined |
| SB_extracted_k141_195464 | Not-determined |
| SB_extracted_k141_73365 | Not-determined |
| SB_extracted_k141_64853 | Not-determined |
| SB_extracted_k141_47331 | Low-quality |
| SB_extracted_k141_117036 | Not-determined |
| SB_extracted_k141_73369 | Not-determined |
| SB_extracted_k141_143320 | Not-determined |
| SB_extracted_k141_160239 | Low-quality |
| SB_extracted_k141_47336 | Not-determined |
| SB_extracted_k141_39077 | Not-determined |
| SB_extracted_k141_47339 | Not-determined |
| SB_extracted_k141_117041 | Not-determined |
| SB_extracted_k141_73376 | Not-determined |
| SB_extracted_k141_134394 | Not-determined |
| SB_extracted_k141_195470 | Not-determined |
| SB_extracted_k141_12723 | Not-determined |
| SB_extracted_k141_134397 | Not-determined |
| SB_extracted_k141_39078 | Low-quality |
| SB_extracted_k141_64862 | Not-determined |
| SB_extracted_k141_134398 | Not-determined |
| SB_extracted_k141_99832 | Not-determined |
| SB_extracted_k141_73383 | Not-determined |
| SB_extracted_k141_108706 | Not-determined |
| SB_extracted_k141_151652 | Not-determined |
| SB_extracted_k141_73384 | Not-determined |
| SB_extracted_k141_99834 | Not-determined |
| SB_extracted_k141_108708 | Not-determined |
| SB_extracted_k141_73387 | Low-quality |
| SB_extracted_k141_82415 | Not-determined |
| SB_extracted_k141_73391 | Not-determined |
| SB_extracted_k141_56621 | Low-quality |
| SB_extracted_k141_203608 | Not-determined |
| SB_extracted_k141_185802 | Not-determined |
| SB_extracted_k141_47347 | Not-determined |
| SB_extracted_k141_203610 | Not-determined |
| SB_extracted_k141_82423 | Not-determined |
| SB_extracted_k141_39085 | Not-determined |
| SB_extracted_k141_151662 | Not-determined |
| SB_extracted_k141_39087 | Not-determined |
| SB_extracted_k141_47351 | Not-determined |
| SB_extracted_k141_99847 | Not-determined |
| SB_extracted_k141_21847 | Low-quality |
| SB_extracted_k141_56629 | Low-quality |
| SB_extracted_k141_39089 | Not-determined |
| SB_extracted_k141_91141 | Not-determined |
| SB_extracted_k141_64889 | Not-determined |
| SB_extracted_k141_82433 | Not-determined |
| SB_extracted_k141_203620 | Not-determined |
| SB_extracted_k141_4488 | Low-quality |
| SB_extracted_k141_73410 | Not-determined |
| SB_extracted_k141_169268 | Not-determined |
| SB_extracted_k141_73413 | Not-determined |
| SB_extracted_k141_30759 | Not-determined |
| SB_extracted_k141_21855 | Not-determined |
| SB_extracted_k141_30760 | Not-determined |
| SB_extracted_k141_64895 | Not-determined |
| SB_extracted_k141_99851 | Low-quality |
| SB_extracted_k141_47363 | Not-determined |
| SB_extracted_k141_203625 | Not-determined |
| SB_extracted_k141_91149 | Not-determined |
| SB_extracted_k141_47365 | Not-determined |
| SB_extracted_k141_134420 | Not-determined |
| SB_extracted_k141_4498 | Not-determined |
| SB_extracted_k141_4501 | Not-determined |
| SB_extracted_k141_30766 | Not-determined |
| SB_extracted_k141_178057 | Low-quality |
| SB_extracted_k141_151673 | Not-determined |
| SB_extracted_k141_134425 | Not-determined |
| SB_extracted_k141_178059 | Not-determined |
| SB_extracted_k141_30767 | Not-determined |
| SB_extracted_k141_169276 | Not-determined |
| SB_extracted_k141_195488 | Not-determined |
| SB_extracted_k141_12759 | Not-determined |
| SB_extracted_k141_73426 | Not-determined |
| SB_extracted_k141_82446 | Low-quality |
| SB_extracted_k141_185807 | Low-quality |
| SB_extracted_k141_134427 | Not-determined |
| SB_extracted_k141_185809 | Not-determined |
| SB_extracted_k141_178060 | Not-determined |
| SB_extracted_k141_151679 | Not-determined |
| SB_extracted_k141_160271 | Not-determined |
| SB_extracted_k141_169280 | Not-determined |
| SB_extracted_k141_4509 | Low-quality |
| SB_extracted_k141_178063 | Not-determined |
| SB_extracted_k141_4510 | Not-determined |
| SB_extracted_k141_73432 | Not-determined |
| SB_extracted_k141_21871 | Not-determined |
| SB_extracted_k141_56647 | Not-determined |
| SB_extracted_k141_117058 | Not-determined |
| SB_extracted_k141_169286 | Not-determined |
| SB_extracted_k141_91160 | Not-determined |
| SB_extracted_k141_151691 | Not-determined |
| SB_extracted_k141_160283 | Low-quality |
| SB_extracted_k141_185816 | Not-determined |
| SB_extracted_k141_195499 | Not-determined |
| SB_extracted_k141_91163 | Not-determined |
| SB_extracted_k141_4520 | Not-determined |
| SB_extracted_k141_151695 | Not-determined |
| SB_extracted_k141_82452 | Not-determined |
| SB_extracted_k141_151696 | Not-determined |
| SB_extracted_k141_39118 | Not-determined |
| SB_extracted_k141_99879 | Not-determined |
| SB_extracted_k141_99881 | Not-determined |
| SB_extracted_k141_195504 | Not-determined |
| SB_extracted_k141_117070 | Not-determined |
| SB_extracted_k141_151702 | Not-determined |
| SB_extracted_k141_82456 | Not-determined |
| SB_extracted_k141_160291 | Not-determined |
| SB_extracted_k141_82458 | Not-determined |
| SB_extracted_k141_185824 | Not-determined |
| SB_extracted_k141_185825 | Not-determined |
| SB_extracted_k141_160293 | Not-determined |
| SB_extracted_k141_99883 | Not-determined |
| SB_extracted_k141_47385 | Not-determined |
| SB_extracted_k141_99884 | Not-determined |
| SB_extracted_k141_117073 | Not-determined |
| SB_extracted_k141_160294 | Not-determined |
| SB_extracted_k141_39126 | Not-determined |
| SB_extracted_k141_185829 | Not-determined |
| SB_extracted_k141_73453 | Not-determined |
| SB_extracted_k141_4533 | Low-quality |
| SB_extracted_k141_82465 | Not-determined |
| SB_extracted_k141_30798 | Not-determined |
| SB_extracted_k141_99889 | Not-determined |
| SB_extracted_k141_117078 | Not-determined |
| SB_extracted_k141_47386 | Not-determined |
| SB_extracted_k141_99890 | Not-determined |
| SB_extracted_k141_30799 | Not-determined |
| SB_extracted_k141_73455 | Not-determined |
| SB_extracted_k141_134451 | Low-quality |
| SB_extracted_k141_12785 | Not-determined |
| SB_extracted_k141_178081 | Not-determined |
| SB_extracted_k141_160299 | Not-determined |
| SB_extracted_k141_160300 | Not-determined |
| SB_extracted_k141_160301 | Not-determined |
| SB_extracted_k141_160303 | Not-determined |
| SB_extracted_k141_151721 | Not-determined |
| SB_extracted_k141_56657 | Low-quality |
| SB_extracted_k141_117085 | Not-determined |
| SB_extracted_k141_73463 | Not-determined |
| SB_extracted_k141_203657 | Not-determined |
| SB_extracted_k141_64910 | Low-quality |
| SB_extracted_k141_64911 | Not-determined |
| SB_extracted_k141_143330 | Low-quality |
| SB_extracted_k141_30803 | Not-determined |
| SB_extracted_k141_73466 | Not-determined |
| SB_extracted_k141_30806 | Not-determined |
| SB_extracted_k141_4550 | Not-determined |
| SB_extracted_k141_4551 | Not-determined |
| SB_extracted_k141_64914 | Not-determined |
| SB_extracted_k141_151732 | Not-determined |
| SB_extracted_k141_47404 | Not-determined |
| SB_extracted_k141_73468 | Not-determined |
| SB_extracted_k141_64917 | Not-determined |
| SB_extracted_k141_178087 | Not-determined |
| SB_extracted_k141_73469 | Not-determined |
| SB_extracted_k141_185843 | Not-determined |
| SB_extracted_k141_143338 | Not-determined |
| SB_extracted_k141_203667 | Not-determined |
| SB_extracted_k141_151736 | Low-quality |
| SB_extracted_k141_12792 | Low-quality |
| SB_extracted_k141_195518 | Not-determined |
| SB_extracted_k141_185845 | Not-determined |
| SB_extracted_k141_12793 | Not-determined |
| SB_extracted_k141_73472 | Low-quality |
| SB_extracted_k141_91193 | Not-determined |
| SB_extracted_k141_151738 | Not-determined |
| SB_extracted_k141_169312 | Not-determined |
| SB_extracted_k141_64923 | Not-determined |
| SB_extracted_k141_169314 | Not-determined |
| SB_extracted_k141_195520 | Not-determined |
| SB_extracted_k141_169317 | Not-determined |
| SB_extracted_k141_12796 | Not-determined |
| SB_extracted_k141_21892 | Not-determined |
| SB_extracted_k141_195522 | Not-determined |
| SB_extracted_k141_12798 | Low-quality |
| SB_extracted_k141_178104 | Not-determined |
| SB_extracted_k141_185847 | Not-determined |
| SB_extracted_k141_21897 | Not-determined |
| SB_extracted_k141_21898 | Not-determined |
| SB_extracted_k141_4563 | Not-determined |
| SB_extracted_k141_4564 | Not-determined |
| SB_extracted_k141_82494 | Not-determined |
| SB_extracted_k141_56673 | Not-determined |
| SB_extracted_k141_47414 | Not-determined |
| SB_extracted_k141_64929 | Not-determined |
| SB_extracted_k141_185855 | Not-determined |
| SB_extracted_k141_82500 | Not-determined |
| SB_extracted_k141_56675 | Not-determined |
| SB_extracted_k141_82504 | Not-determined |
| SB_extracted_k141_82505 | Not-determined |
| SB_extracted_k141_99913 | Low-quality |
| SB_extracted_k141_64938 | Not-determined |
| SB_extracted_k141_30816 | Not-determined |
| SB_extracted_k141_203688 | Not-determined |
| SB_extracted_k141_203690 | Not-determined |

| | |
|---|---|
| SB_extracted_k141_30819 | Not-determined |
| SB_extracted_k141_30820 | Low-quality |
| SB_extracted_k141_117103 | Not-determined |
| SB_extracted_k141_160346 | Not-determined |
| SB_extracted_k141_99917 | Low-quality |
| SB_extracted_k141_4579 | Not-determined |
| SB_extracted_k141_82512 | Not-determined |
| SB_extracted_k141_82514 | Not-determined |
| SB_extracted_k141_151757 | Not-determined |
| SB_extracted_k141_169339 | Not-determined |
| SB_extracted_k141_151759 | Not-determined |
| SB_extracted_k141_82518 | Not-determined |
| SB_extracted_k141_4586 | Not-determined |
| SB_extracted_k141_178123 | Not-determined |
| SB_extracted_k141_4589 | Not-determined |
| SB_vs_k141_136235\|\|full | Medium-quality |
| SB_extracted_k141_151767 | Not-determined |
| SB_extracted_k141_73486 | Not-determined |
| SB_extracted_k141_151768 | Low-quality |
| SB_extracted_k141_151770 | Not-determined |
| SB_extracted_k141_64955 | Low-quality |
| SB_extracted_k141_91198 | Not-determined |
| SB_extracted_k141_99926 | Not-determined |
| SB_extracted_k141_99927 | Not-determined |
| SB_extracted_k141_203694 | Not-determined |
| SB_extracted_k141_160365 | Low-quality |
| SB_extracted_k141_203695 | Not-determined |
| SB_extracted_k141_151775 | Not-determined |
| SB_extracted_k141_203696 | Not-determined |
| SB_extracted_k141_151776 | Not-determined |
| SB_extracted_k141_82531 | Not-determined |
| SB_extracted_k141_99929 | Not-determined |
| SB_extracted_k141_195536 | Not-determined |
| SB_extracted_k141_169350 | Not-determined |
| SB_extracted_k141_108716 | Not-determined |
| SB_extracted_k141_99934 | Not-determined |
| SB_extracted_k141_203699 | Low-quality |
| SB_extracted_k141_64967 | Low-quality |
| SB_extracted_k141_169355 | Not-determined |
| SB_extracted_k141_82542 | Not-determined |
| SB_extracted_k141_169356 | Not-determined |
| SB_extracted_k141_195544 | Not-determined |
| SB_extracted_k141_21910 | Not-determined |
| SB_extracted_k141_82544 | Not-determined |
| SB_extracted_k141_30833 | Not-determined |
| SB_extracted_k141_91206 | Not-determined |
| SB_extracted_k141_30835 | Not-determined |
| SB_extracted_k141_56682 | Not-determined |
| SB_extracted_k141_160377 | Not-determined |
| SB_extracted_k141_108730 | Not-determined |
| SB_extracted_k141_169359 | Not-determined |
| SB_extracted_k141_108731 | Not-determined |
| SB_extracted_k141_82552 | Not-determined |
| SB_extracted_k141_30841 | Not-determined |
| SB_extracted_k141_39173 | Not-determined |
| SB_extracted_k141_30842 | Not-determined |
| SB_extracted_k141_30843 | Not-determined |
| SB_extracted_k141_21914 | Not-determined |
| SB_extracted_k141_169364 | Low-quality |
| SB_extracted_k141_108735 | Not-determined |
| SB_extracted_k141_108737 | Not-determined |
| SB_extracted_k141_39178 | Not-determined |
| SB_extracted_k141_108738 | Not-determined |
| SB_extracted_k141_203713 | Not-determined |
| SB_extracted_k141_143361 | Not-determined |
| SB_extracted_k141_185895 | Not-determined |
| SB_extracted_k141_21919 | Not-determined |
| SB_extracted_k141_151792 | Not-determined |
| SB_extracted_k141_64979 | Not-determined |
| SB_extracted_k141_169369 | Not-determined |
| SB_extracted_k141_185896 | Not-determined |
| SB_extracted_k141_169370 | Not-determined |
| SB_extracted_k141_143362 | Not-determined |
| SB_extracted_k141_39182 | Not-determined |
| SB_extracted_k141_178148 | Low-quality |
| SB_extracted_k141_178152 | Not-determined |
| SB_extracted_k141_64986 | Not-determined |
| SB_extracted_k141_12827 | Not-determined |
| SB_extracted_k141_30845 | Not-determined |
| SB_extracted_k141_117131 | Not-determined |
| SB_extracted_k141_39191 | Not-determined |
| SB_extracted_k141_203718 | Not-determined |
| SB_extracted_k141_117135 | Low-quality |
| SB_extracted_k141_91225 | Not-determined |
| SB_extracted_k141_169378 | Not-determined |
| SB_extracted_k141_178156 | Low-quality |
| SB_extracted_k141_91227 | Not-determined |
| SB_extracted_k141_64988 | Not-determined |
| SB_extracted_k141_143367 | Low-quality |
| SB_extracted_k141_185901 | Not-determined |
| SB_extracted_k141_151799 | Not-determined |
| SB_extracted_k141_143368 | Not-determined |
| SB_extracted_k141_151801 | Not-determined |
| SB_extracted_k141_185903 | Low-quality |
| SB_extracted_k141_64990 | Not-determined |
| SB_extracted_k141_185904 | Not-determined |
| SB_extracted_k141_151804 | Not-determined |
| SB_extracted_k141_185905 | Not-determined |
| SB_extracted_k141_151808 | Not-determined |
| SB_extracted_k141_21930 | Low-quality |
| SB_extracted_k141_117144 | Not-determined |
| SB_extracted_k141_143373 | Not-determined |
| SB_extracted_k141_21933 | Not-determined |
| SB_extracted_k141_151809 | Not-determined |
| SB_extracted_k141_73503 | Low-quality |
| SB_extracted_k141_73504 | Not-determined |
| SB_extracted_k141_134465 | Not-determined |
| SB_extracted_k141_134466 | Not-determined |
| SB_extracted_k141_178176 | Not-determined |
| SB_extracted_k141_56704 | Low-quality |
| SB_extracted_k141_169381 | Low-quality |
| SB_extracted_k141_185910 | Not-determined |
| SB_extracted_k141_203731 | Not-determined |
| SB_extracted_k141_73514 | Not-determined |
| SB_extracted_k141_21950 | Not-determined |
| SB_extracted_k141_169383 | Not-determined |
| SB_extracted_k141_117154 | Low-quality |
| SB_extracted_k141_203733 | Not-determined |
| SB_extracted_k141_99972 | Not-determined |
| SB_extracted_k141_178184 | Not-determined |
| SB_extracted_k141_178186 | Not-determined |
| SB_extracted_k141_47474 | Low-quality |
| SB_extracted_k141_178187 | Not-determined |
| SB_extracted_k141_169389 | Not-determined |
| SB_extracted_k141_108757 | Not-determined |
| SB_extracted_k141_47475 | Not-determined |
| SB_extracted_k141_21955 | Not-determined |
| SB_extracted_k141_151823 | Low-quality |
| SB_extracted_k141_134472 | Not-determined |
| SB_extracted_k141_99980 | Not-determined |
| SB_extracted_k141_73521 | Not-determined |
| SB_extracted_k141_134474 | Not-determined |
| SB_extracted_k141_21959 | Not-determined |
| SB_extracted_k141_178194 | Not-determined |
| SB_extracted_k141_151826 | Not-determined |
| SB_extracted_k141_108765 | Not-determined |
| SB_extracted_k141_91251 | Not-determined |
| SB_extracted_k141_65003 | Not-determined |
| SB_extracted_k141_185920 | Not-determined |
| SB_extracted_k141_108770 | Not-determined |
| SB_extracted_k141_91252 | Not-determined |
| SB_extracted_k141_99987 | Not-determined |
| SB_extracted_k141_21963 | Not-determined |
| SB_extracted_k141_203736 | Not-determined |
| SB_extracted_k141_108774 | Not-determined |
| SB_extracted_k141_178204 | Not-determined |
| SB_extracted_k141_21965 | Low-quality |
| SB_extracted_k141_117167 | Low-quality |
| SB_extracted_k141_21967 | Low-quality |
| SB_extracted_k141_65015 | Not-determined |
| SB_extracted_k141_143406 | Not-determined |
| SB_extracted_k141_65018 | Not-determined |
| SB_extracted_k141_143408 | Not-determined |
| SB_extracted_k141_203745 | Not-determined |
| SB_extracted_k141_47489 | Not-determined |
| SB_extracted_k141_108782 | Not-determined |
| SB_extracted_k141_203749 | Not-determined |
| SB_extracted_k141_39205 | Not-determined |
| SB_extracted_k141_134486 | Not-determined |
| SB_extracted_k141_178217 | Not-determined |
| SB_extracted_k141_134490 | Not-determined |
| SB_extracted_k141_21983 | Not-determined |
| SB_extracted_k141_151847 | Low-quality |
| SB_extracted_k141_65022 | Not-determined |
| SB_extracted_k141_21987 | Not-determined |
| SB_extracted_k141_117175 | Not-determined |
| SB_extracted_k141_151854 | Low-quality |
| SB_extracted_k141_203754 | Not-determined |
| SB_extracted_k141_108787 | Not-determined |
| SB_extracted_k141_47493 | Low-quality |
| SB_extracted_k141_47494 | Not-determined |
| SB_extracted_k141_203759 | Not-determined |
| SB_extracted_k141_108792 | Not-determined |
| SB_extracted_k141_185960 | Not-determined |
| SB_extracted_k141_47500 | Not-determined |
| SB_extracted_k141_203762 | Not-determined |
| SB_extracted_k141_65031 | Not-determined |
| SB_extracted_k141_39219 | Not-determined |
| SB_extracted_k141_21996 | Not-determined |
| SB_extracted_k141_203765 | Not-determined |
| SB_extracted_k141_203766 | Not-determined |
| SB_extracted_k141_91266 | Not-determined |
| SB_extracted_k141_195598 | Not-determined |
| SB_extracted_k141_151866 | Not-determined |
| SB_extracted_k141_47508 | Not-determined |
| SB_extracted_k141_91269 | Not-determined |
| SB_extracted_k141_178229 | Not-determined |
| SB_extracted_k141_117188 | Not-determined |
| SB_extracted_k141_91271 | Not-determined |
| SB_extracted_k141_178230 | Not-determined |
| SB_extracted_k141_169403 | Not-determined |
| SB_extracted_k141_195604 | Not-determined |
| SB_extracted_k141_185974 | Not-determined |
| SB_extracted_k141_117194 | Not-determined |
| SB_extracted_k141_39231 | Not-determined |
| SB_extracted_k141_195608 | Not-determined |
| SB_extracted_k141_39232 | Not-determined |
| SB_extracted_k141_117197 | Not-determined |
| SB_extracted_k141_203772 | Not-determined |
| SB_extracted_k141_151870 | Not-determined |
| SB_extracted_k141_151872 | Not-determined |
| SB_extracted_k141_126065 | Not-determined |
| SB_extracted_k141_91276 | Not-determined |
| SB_extracted_k141_126068 | Not-determined |
| SB_extracted_k141_65044 | Not-determined |
| SB_extracted_k141_56731 | Not-determined |
| SB_extracted_k141_12893 | Not-determined |
| SB_extracted_k141_91279 | Not-determined |
| SB_extracted_k141_151878 | Not-determined |
| SB_extracted_k141_126071 | Not-determined |
| SB_extracted_k141_39241 | Not-determined |
| SB_extracted_k141_12895 | Not-determined |
| SB_extracted_k141_47532 | Not-determined |
| SB_extracted_k141_56737 | Not-determined |
| SB_extracted_k141_12901 | Low-quality |
| SB_extracted_k141_56743 | Not-determined |
| SB_extracted_k141_56744 | Not-determined |
| SB_extracted_k141_12902 | Not-determined |
| SB_extracted_k141_108825 | Not-determined |
| SB_extracted_k141_100021 | Not-determined |
| SB_extracted_k141_178253 | Low-quality |
| SB_extracted_k141_134524 | Not-determined |
| SB_extracted_k141_56750 | Not-determined |
| SB_extracted_k141_47543 | Not-determined |
| SB_extracted_k141_108826 | Not-determined |
| SB_extracted_k141_117229 | Not-determined |
| SB_extracted_k141_12921 | Not-determined |
| SB_extracted_k141_126086 | Not-determined |
| SB_extracted_k141_178257 | Not-determined |
| SB_extracted_k141_22000 | Not-determined |
| SB_extracted_k141_126088 | Not-determined |
| SB_extracted_k141_47552 | Not-determined |
| SB_extracted_k141_126091 | Not-determined |
| SB_extracted_k141_117235 | Low-quality |
| SB_extracted_k141_203789 | Not-determined |
| SB_extracted_k141_91296 | Not-determined |
| SB_extracted_k141_195629 | Not-determined |
| SB_extracted_k141_22009 | Not-determined |
| SB_extracted_k141_100039 | Not-determined |
| SB_extracted_k141_134535 | Low-quality |
| SB_extracted_k141_22014 | Not-determined |
| SB_extracted_k141_39265 | Low-quality |
| SB_extracted_k141_30927 | Not-determined |
| SB_extracted_k141_134538 | Not-determined |
| SB_extracted_k141_30928 | Not-determined |
| SG_vs_k141_45228\|\|full | Medium-quality |
| SB_extracted_k141_91308 | Low-quality |
| SB_extracted_k141_30932 | Not-determined |
| SB_extracted_k141_22020 | Not-determined |
| SB_extracted_k141_73544 | Not-determined |
| SB_extracted_k141_22021 | Not-determined |
| SB_extracted_k141_126105 | Not-determined |
| SB_extracted_k141_22024 | Not-determined |
| SB_extracted_k141_91313 | Not-determined |
| SB_extracted_k141_195643 | Not-determined |
| SB_extracted_k141_56769 | Not-determined |
| SB_extracted_k141_126114 | Not-determined |
| SB_extracted_k141_143422 | Not-determined |
| SB_extracted_k141_30939 | Not-determined |
| SB_extracted_k141_134545 | Low-quality |
| SB_extracted_k141_100045 | Not-determined |
| SB_extracted_k141_47564 | Not-determined |
| SB_extracted_k141_117256 | Not-determined |
| SB_extracted_k141_143423 | Not-determined |
| SB_extracted_k141_117258 | Not-determined |
| SB_extracted_k141_178275 | Not-determined |
| SB_extracted_k141_91318 | Not-determined |
| SB_extracted_k141_203807 | Not-determined |
| SB_extracted_k141_203809 | Not-determined |
| SB_extracted_k141_169423 | Not-determined |
| SB_extracted_k141_126120 | Low-quality |
| SB_extracted_k141_91323 | Not-determined |
| SB_extracted_k141_30945 | Not-determined |
| SB_extracted_k141_108855 | Not-determined |
| SB_extracted_k141_108857 | Not-determined |
| SB_extracted_k141_185989 | Not-determined |
| SB_extracted_k141_22029 | Not-determined |
| SB_extracted_k141_22031 | Not-determined |
| SB_extracted_k141_203813 | Not-determined |
| SB_extracted_k141_47580 | Not-determined |
| SB_extracted_k141_143435 | Not-determined |
| SB_extracted_k141_30951 | Not-determined |
| SB_extracted_k141_169434 | Not-determined |
| SB_extracted_k141_143440 | Not-determined |
| SB_extracted_k141_39277 | Not-determined |
| SB_extracted_k141_169435 | Not-determined |
| SB_extracted_k141_117265 | Not-determined |
| SB_extracted_k141_195660 | Not-determined |
| SB_extracted_k141_56773 | Not-determined |
| SB_extracted_k141_65095 | Not-determined |
| SB_extracted_k141_22041 | Not-determined |
| SB_extracted_k141_117269 | Not-determined |
| SB_extracted_k141_73568 | Not-determined |
| SB_extracted_k141_203821 | Not-determined |
| SB_extracted_k141_73570 | Not-determined |
| SB_extracted_k141_47590 | Low-quality |
| SB_extracted_k141_22046 | Not-determined |
| SB_extracted_k141_56778 | Low-quality |
| SB_extracted_k141_47592 | Not-determined |
| SB_extracted_k141_185997 | Not-determined |
| SB_extracted_k141_39288 | Not-determined |
| SB_extracted_k141_195668 | Not-determined |
| SB_extracted_k141_117276 | Not-determined |
| SB_extracted_k141_185998 | Not-determined |
| SB_extracted_k141_91340 | Not-determined |
| SB_extracted_k141_91342 | Not-determined |
| SB_extracted_k141_56784 | Not-determined |
| SB_extracted_k141_65107 | Not-determined |
| SB_extracted_k141_22053 | Not-determined |
| SB_extracted_k141_65108 | Not-determined |
| SB_extracted_k141_39293 | Low-quality |
| SB_extracted_k141_65110 | Low-quality |
| SB_extracted_k141_22054 | Not-determined |
| SB_extracted_k141_143458 | Not-determined |
| SB_extracted_k141_203834 | Not-determined |
| SB_extracted_k141_73584 | Not-determined |
| SB_extracted_k141_82593 | Not-determined |
| SB_extracted_k141_126134 | Not-determined |
| SB_extracted_k141_117281 | Low-quality |
| SB_extracted_k141_143462 | Not-determined |
| SB_extracted_k141_203837 | Not-determined |
| SB_extracted_k141_73589 | Not-determined |
| SB_extracted_k141_100058 | Not-determined |
| SB_extracted_k141_203839 | Not-determined |
| SB_extracted_k141_100059 | Not-determined |
| SB_extracted_k141_143463 | Not-determined |
| SB_extracted_k141_203840 | Not-determined |
| SB_extracted_k141_22063 | Not-determined |
| SB_extracted_k141_22064 | Not-determined |
| SB_extracted_k141_117287 | Not-determined |
| SB_extracted_k141_91351 | Not-determined |
| SB_extracted_k141_203842 | Not-determined |
| SB_extracted_k141_117289 | Not-determined |
| SB_extracted_k141_143467 | Not-determined |
| SB_extracted_k141_169442 | Not-determined |
| SB_extracted_k141_186013 | Not-determined |
| SB_extracted_k141_65124 | Not-determined |
| SB_extracted_k141_143469 | Not-determined |
| SB_extracted_k141_195678 | Not-determined |
| SB_extracted_k141_169445 | Not-determined |
| SB_extracted_k141_47603 | Not-determined |
| SB_extracted_k141_100068 | Not-determined |
| SB_extracted_k141_126143 | Not-determined |
| SB_extracted_k141_100069 | Not-determined |
| SB_extracted_k141_39308 | Not-determined |
| SB_extracted_k141_100070 | Not-determined |
| SB_extracted_k141_186020 | Not-determined |
| SB_extracted_k141_143472 | Not-determined |
| SB_extracted_k141_203853 | Not-determined |
| SB_extracted_k141_169451 | Not-determined |
| SB_extracted_k141_203860 | Not-determined |
| SB_extracted_k141_126147 | Not-determined |
| SB_extracted_k141_169456 | Not-determined |
| SB_extracted_k141_186028 | Low-quality |
| SB_extracted_k141_73598 | Not-determined |
| SB_extracted_k141_82611 | Not-determined |
| SB_extracted_k141_91365 | Not-determined |
| SB_extracted_k141_39314 | Not-determined |
| SB_extracted_k141_12938 | Not-determined |
| SB_extracted_k141_39316 | Not-determined |
| SB_extracted_k141_22077 | Not-determined |
| SB_extracted_k141_82613 | Not-determined |
| SB_extracted_k141_203869 | Not-determined |
| SB_extracted_k141_186035 | Not-determined |
| SB_extracted_k141_117302 | Not-determined |
| SB_extracted_k141_56819 | Not-determined |
| SB_extracted_k141_203875 | Not-determined |
| SB_extracted_k141_22085 | Not-determined |
| SB_extracted_k141_151888 | Not-determined |
| SB_extracted_k141_169476 | Not-determined |
| SB_extracted_k141_143491 | Not-determined |
| SB_extracted_k141_178280 | Not-determined |
| SB_extracted_k141_151892 | Not-determined |
| SB_extracted_k141_82618 | Not-determined |
| SB_extracted_k141_22092 | Not-determined |
| SB_extracted_k141_151893 | Low-quality |
| SB_extracted_k141_56825 | Not-determined |
| SB_extracted_k141_82621 | Not-determined |
| SB_extracted_k141_186042 | Not-determined |
| SB_extracted_k141_151900 | Not-determined |
| SB_extracted_k141_22094 | Not-determined |
| SB_extracted_k141_30965 | Not-determined |
| SB_extracted_k141_186049 | Not-determined |
| SB_extracted_k141_143502 | Not-determined |
| SB_extracted_k141_143505 | Not-determined |
| SB_extracted_k141_151906 | Not-determined |
| SB_extracted_k141_126169 | Low-quality |
| SB_extracted_k141_82631 | Not-determined |
| SB_extracted_k141_186050 | Low-quality |
| SB_extracted_k141_126171 | Not-determined |
| SB_extracted_k141_186051 | Not-determined |
| SB_extracted_k141_22107 | Low-quality |
| SB_extracted_k141_143509 | Low-quality |
| SB_extracted_k141_143511 | Not-determined |
| SB_extracted_k141_134582 | Not-determined |
| SB_extracted_k141_117329 | Not-determined |
| SB_extracted_k141_151910 | Not-determined |
| SB_extracted_k141_151911 | Not-determined |
| SB_extracted_k141_117332 | Not-determined |
| SB_extracted_k141_151914 | Not-determined |
| SB_extracted_k141_73611 | Not-determined |
| SB_extracted_k141_73612 | Not-determined |
| SB_extracted_k141_143516 | Not-determined |
| SB_extracted_k141_56831 | Not-determined |
| SB_extracted_k141_73613 | Low-quality |
| SB_extracted_k141_203895 | Not-determined |
| SB_extracted_k141_143520 | Not-determined |
| SB_extracted_k141_143521 | Not-determined |
| SB_extracted_k141_151915 | Not-determined |
| SB_extracted_k141_134590 | Not-determined |
| SB_extracted_k141_12947 | Low-quality |
| SB_extracted_k141_151917 | Not-determined |
| SB_extracted_k141_160390 | Not-determined |
| SB_extracted_k141_100091 | Not-determined |
| SB_extracted_k141_151919 | Low-quality |
| SB_extracted_k141_12953 | Not-determined |
| SB_extracted_k141_22127 | Not-determined |
| SB_extracted_k141_178299 | Not-determined |
| SB_extracted_k141_160395 | Not-determined |
| SB_extracted_k141_73625 | Low-quality |
| SB_extracted_k141_22130 | Not-determined |
| SB_extracted_k141_73628 | Not-determined |
| SB_extracted_k141_91385 | Not-determined |
| SB_extracted_k141_22134 | Not-determined |
| SB_extracted_k141_160400 | Not-determined |
| SB_extracted_k141_143528 | Not-determined |
| SB_extracted_k141_178303 | Not-determined |
| SB_extracted_k141_65134 | Low-quality |
| SB_extracted_k141_151927 | Not-determined |
| SB_extracted_k141_22137 | Not-determined |
| SB_extracted_k141_160406 | Not-determined |
| SB_extracted_k141_91391 | Not-determined |
| SB_extracted_k141_169502 | Not-determined |
| SB_extracted_k141_22141 | Not-determined |
| SB_extracted_k141_151935 | Not-determined |
| SB_extracted_k141_203903 | Not-determined |
| SB_extracted_k141_22147 | Low-quality |
| SB_extracted_k141_178309 | Low-quality |
| SB_extracted_k141_100102 | Not-determined |
| SB_extracted_k141_39330 | Not-determined |
| SB_extracted_k141_91398 | Not-determined |
| SB_extracted_k141_100103 | Not-determined |
| SB_extracted_k141_143534 | Not-determined |
| SB_extracted_k141_169507 | Not-determined |
| SB_extracted_k141_143541 | Not-determined |
| SB_extracted_k141_30980 | Not-determined |
| SB_extracted_k141_151941 | Not-determined |
| SB_extracted_k141_178316 | Not-determined |
| SB_extracted_k141_12964 | Not-determined |
| SB_extracted_k141_169511 | Not-determined |
| SB_extracted_k141_151945 | Not-determined |
| SB_extracted_k141_73638 | Not-determined |
| SB_extracted_k141_100116 | Not-determined |
| SB_extracted_k141_143544 | Not-determined |
| SB_extracted_k141_195686 | Not-determined |
| SB_extracted_k141_56860 | Low-quality |
| SB_extracted_k141_22157 | Low-quality |
| SB_extracted_k141_100120 | Not-determined |
| SB_extracted_k141_151948 | Low-quality |
| SB_extracted_k141_117344 | Not-determined |
| SB_extracted_k141_134606 | Not-determined |
| SB_extracted_k141_73642 | Not-determined |
| SB_extracted_k141_178321 | Not-determined |
| SB_extracted_k141_91414 | Not-determined |
| SB_extracted_k141_195695 | Not-determined |
| SB_extracted_k141_65156 | Not-determined |
| SB_extracted_k141_30988 | Not-determined |
| SB_extracted_k141_151955 | Not-determined |
| SB_extracted_k141_12970 | Not-determined |
| SB_extracted_k141_117354 | Low-quality |
| SB_extracted_k141_56872 | Not-determined |
| SB_extracted_k141_65162 | Not-determined |
| SB_extracted_k141_178324 | Not-determined |
| SB_extracted_k141_56874 | Not-determined |
| SB_extracted_k141_65164 | Not-determined |
| SB_extracted_k141_91420 | Not-determined |
| SB_extracted_k141_65167 | Not-determined |
| SB_extracted_k141_12979 | Not-determined |
| SB_extracted_k141_65170 | Not-determined |
| SB_extracted_k141_195717 | Not-determined |
| SB_extracted_k141_203919 | Not-determined |
| SB_extracted_k141_31001 | Low-quality |
| SB_extracted_k141_126199 | Not-determined |
| SB_extracted_k141_160417 | Not-determined |
| SB_extracted_k141_31005 | Not-determined |
| SB_extracted_k141_134620 | Not-determined |
| SB_extracted_k141_117359 | Not-determined |
| SB_extracted_k141_143566 | Not-determined |
| SB_extracted_k141_126202 | Not-determined |
| SB_extracted_k141_100139 | Not-determined |
| SB_extracted_k141_91431 | Not-determined |
| SB_extracted_k141_151976 | Not-determined |
| SB_extracted_k141_160419 | Not-determined |
| SB_extracted_k141_31011 | Low-quality |
| SB_extracted_k141_39340 | Not-determined |
| SB_extracted_k141_117368 | Not-determined |
| SB_extracted_k141_134629 | Low-quality |
| SB_extracted_k141_12994 | Not-determined |
| SB_extracted_k141_195731 | Not-determined |
| SB_extracted_k141_160428 | Not-determined |
| SB_extracted_k141_169533 | Low-quality |
| SB_extracted_k141_160429 | Not-determined |
| SB_extracted_k141_126215 | Not-determined |
| SB_extracted_k141_134631 | Not-determined |
| SB_extracted_k141_22180 | Low-quality |
| SB_extracted_k141_203935 | Low-quality |
| SB_extracted_k141_12999 | Not-determined |
| SB_extracted_k141_117375 | Low-quality |
| SB_extracted_k141_39350 | Not-determined |
| SB_extracted_k141_117379 | Not-determined |
| SB_extracted_k141_134634 | Not-determined |
| SB_extracted_k141_169542 | Not-determined |
| SB_extracted_k141_143587 | Not-determined |
| SB_extracted_k141_13002 | Not-determined |
| SB_extracted_k141_151993 | Not-determined |
| SB_extracted_k141_126226 | Low-quality |
| SB_extracted_k141_73663 | Low-quality |
| SB_extracted_k141_13003 | Not-determined |
| SB_extracted_k141_82645 | Not-determined |
| SB_extracted_k141_31019 | Not-determined |
| SB_extracted_k141_39356 | Not-determined |
| SB_extracted_k141_151997 | Not-determined |
| SB_extracted_k141_160432 | Not-determined |
| SB_extracted_k141_117387 | Not-determined |
| SB_extracted_k141_178349 | Not-determined |
| SB_extracted_k141_82650 | Not-determined |
| SB_extracted_k141_117388 | Not-determined |
| SB_extracted_k141_143594 | Not-determined |
| SB_extracted_k141_91438 | Not-determined |
| SB_extracted_k141_31024 | Not-determined |
| SB_extracted_k141_152002 | Not-determined |
| SB_extracted_k141_117392 | Not-determined |
| SB_extracted_k141_100158 | Low-quality |
| SB_extracted_k141_100161 | Not-determined |
| SB_extracted_k141_143598 | Not-determined |
| SB_extracted_k141_160439 | Not-determined |
| SB_extracted_k141_186069 | Not-determined |
| SB_extracted_k141_169547 | Not-determined |
| SB_extracted_k141_13019 | Not-determined |
| SB_extracted_k141_169548 | Not-determined |
| SB_extracted_k141_169551 | Not-determined |
| SB_extracted_k141_22200 | Not-determined |
| SB_extracted_k141_203949 | Not-determined |
| SB_extracted_k141_13022 | Not-determined |
| SB_extracted_k141_47664 | Not-determined |
| SB_extracted_k141_152014 | Not-determined |
| SB_extracted_k141_47665 | Not-determined |
| SB_extracted_k141_203951 | Not-determined |
| SB_extracted_k141_143606 | Not-determined |
| SB_extracted_k141_56895 | Not-determined |
| SB_extracted_k141_134649 | Not-determined |
| SB_extracted_k141_100172 | Not-determined |
| SB_extracted_k141_195747 | Not-determined |
| SB_extracted_k141_22207 | Not-determined |
| SB_extracted_k141_22209 | Not-determined |
| SB_extracted_k141_65198 | Not-determined |
| SB_extracted_k141_22210 | Not-determined |
| SB_extracted_k141_169561 | Not-determined |
| SB_extracted_k141_13034 | Not-determined |
| SB_extracted_k141_178362 | Low-quality |
| SB_extracted_k141_178363 | Not-determined |
| SB_extracted_k141_152031 | Not-determined |
| SB_extracted_k141_108893 | Not-determined |
| SB_extracted_k141_22211 | Not-determined |
| SB_extracted_k141_100179 | Not-determined |
| SB_extracted_k141_169567 | Not-determined |
| SB_extracted_k141_134657 | Not-determined |
| SB_extracted_k141_178366 | Not-determined |
| SB_extracted_k141_195755 | Not-determined |
| SB_extracted_k141_169568 | Not-determined |
| SB_extracted_k141_134659 | Not-determined |
| SB_extracted_k141_195757 | Not-determined |
| SB_extracted_k141_13038 | Low-quality |
| SB_extracted_k141_56901 | Not-determined |
| SB_extracted_k141_39364 | Not-determined |
| SB_extracted_k141_108898 | Low-quality |
| SB_extracted_k141_4602 | Not-determined |
| SB_extracted_k141_22222 | Not-determined |
| SB_extracted_k141_4604 | Not-determined |
| SB_extracted_k141_169572 | Not-determined |
| SB_extracted_k141_39368 | Not-determined |
| SB_extracted_k141_22229 | Not-determined |
| SB_extracted_k141_31063 | Not-determined |
| SB_extracted_k141_13040 | Not-determined |
| SB_extracted_k141_178376 | Not-determined |
| SB_extracted_k141_13041 | Not-determined |
| SB_extracted_k141_169576 | Not-determined |
| SB_extracted_k141_65216 | Not-determined |
| SB_extracted_k141_117400 | Not-determined |

| | |
|---|---|
| SB_extracted_k141_31065 | Low-quality |
| SB_extracted_k141_169579 | Not-determined |
| SB_extracted_k141_4614 | Not-determined |
| SB_extracted_k141_56907 | Not-determined |
| SB_extracted_k141_152045 | Low-quality |
| SB_extracted_k141_13048 | Not-determined |
| SB_extracted_k141_4617 | Not-determined |
| SB_extracted_k141_56910 | Not-determined |
| SB_extracted_k141_100187 | Low-quality |
| SB_extracted_k141_31071 | Not-determined |
| SB_extracted_k141_65218 | Not-determined |
| SB_extracted_k141_65220 | Not-determined |
| SB_extracted_k141_65221 | Not-determined |
| SB_extracted_k141_4623 | Not-determined |
| SB_extracted_k141_13052 | Low-quality |
| SB_extracted_k141_108907 | Low-quality |
| SB_extracted_k141_203968 | Not-determined |
| SB_extracted_k141_31075 | Not-determined |
| SB_extracted_k141_186092 | Not-determined |
| SB_extracted_k141_4628 | Not-determined |
| SB_extracted_k141_100195 | Not-determined |
| SB_extracted_k141_100196 | Not-determined |
| SB_extracted_k141_47694 | Low-quality |
| SB_extracted_k141_4632 | Not-determined |
| SB_extracted_k141_203973 | Not-determined |
| SB_extracted_k141_73699 | Not-determined |
| SB_extracted_k141_31078 | Not-determined |
| SB_extracted_k141_65229 | Not-determined |
| SB_extracted_k141_203975 | Not-determined |
| SB_extracted_k141_31079 | Low-quality |
| SB_extracted_k141_4638 | Not-determined |
| SB_extracted_k141_22233 | Not-determined |
| SB_extracted_k141_203981 | Not-determined |
| SB_extracted_k141_56917 | Not-determined |
| SB_extracted_k141_117416 | Not-determined |
| SB_extracted_k141_143609 | Not-determined |
| SB_extracted_k141_65237 | Not-determined |
| SB_extracted_k141_186098 | Not-determined |
| SB_extracted_k141_39382 | Not-determined |
| SB_extracted_k141_169593 | Not-determined |
| SB_extracted_k141_203987 | Not-determined |
| SB_extracted_k141_4646 | Not-determined |
| SB_extracted_k141_73708 | Not-determined |
| SB_extracted_k141_31086 | Not-determined |
| SB_extracted_k141_186099 | Not-determined |
| SB_extracted_k141_31089 | Not-determined |
| SB_extracted_k141_73712 | Not-determined |
| SB_extracted_k141_47706 | Not-determined |
| SB_extracted_k141_13058 | Low-quality |
| SB_extracted_k141_56925 | Not-determined |
| SB_extracted_k141_160477 | Low-quality |
| SB_extracted_k141_13062 | Not-determined |
| SB_extracted_k141_39395 | Not-determined |
| SB_extracted_k141_13063 | Not-determined |
| SB_extracted_k141_56927 | Not-determined |
| SB_extracted_k141_13065 | Not-determined |
| SB_extracted_k141_203993 | Not-determined |
| SB_extracted_k141_13066 | Not-determined |
| SB_extracted_k141_13068 | Not-determined |
| SB_extracted_k141_13069 | Not-determined |
| SB_extracted_k141_47712 | Not-determined |
| SB_extracted_k141_100210 | Not-determined |
| SB_extracted_k141_22256 | Low-quality |
| SB_extracted_k141_56930 | Not-determined |
| SB_extracted_k141_152070 | Not-determined |
| SB_extracted_k141_203995 | Not-determined |
| SB_extracted_k141_134705 | Not-determined |
| SB_extracted_k141_169605 | Not-determined |
| SB_extracted_k141_117428 | Low-quality |
| SB_extracted_k141_169606 | Not-determined |
| SB_extracted_k141_47716 | Not-determined |
| SB_extracted_k141_108921 | Low-quality |
| SB_extracted_k141_134707 | Low-quality |
| SB_extracted_k141_143630 | Not-determined |
| SB_extracted_k141_13076 | Not-determined |
| SB_extracted_k141_73725 | Low-quality |
| SB_extracted_k141_186108 | Not-determined |
| SB_extracted_k141_13078 | Not-determined |
| SB_extracted_k141_56934 | Not-determined |
| SB_extracted_k141_108927 | Not-determined |
| SB_extracted_k141_73727 | Not-determined |
| SB_extracted_k141_204009 | Low-quality |
| SB_extracted_k141_108930 | Low-quality |
| SB_extracted_k141_152072 | Not-determined |
| SB_extracted_k141_13082 | Not-determined |
| SB_extracted_k141_108932 | Not-determined |
| SB_extracted_k141_4665 | Not-determined |
| SB_extracted_k141_73732 | Not-determined |
| SB_extracted_k141_91454 | Not-determined |
| SB_extracted_k141_13084 | Not-determined |
| SB_extracted_k141_73735 | Not-determined |
| SB_extracted_k141_160490 | Not-determined |
| SB_extracted_k141_134719 | Not-determined |
| SB_extracted_k141_143635 | Not-determined |
| SB_extracted_k141_73744 | Not-determined |
| SB_extracted_k141_134722 | Not-determined |
| SB_extracted_k141_82675 | Not-determined |
| SB_extracted_k141_117446 | Not-determined |
| SB_extracted_k141_108941 | Not-determined |
| SB_extracted_k141_65249 | Not-determined |
| SB_extracted_k141_91459 | Not-determined |
| SB_extracted_k141_204016 | Not-determined |
| SB_extracted_k141_31104 | Not-determined |
| SB_extracted_k141_65252 | Low-quality |
| SB_extracted_k141_65253 | Not-determined |
| SB_extracted_k141_65254 | Not-determined |
| SB_extracted_k141_22272 | Not-determined |
| SB_extracted_k141_82683 | Not-determined |
| SB_extracted_k141_204019 | Not-determined |
| SB_extracted_k141_47735 | Not-determined |
| SB_extracted_k141_31106 | Not-determined |
| SB_extracted_k141_152082 | Not-determined |
| SB_extracted_k141_152083 | Not-determined |
| SB_extracted_k141_22275 | Not-determined |
| SB_extracted_k141_22276 | Not-determined |
| SB_extracted_k141_13095 | Not-determined |
| SB_extracted_k141_47738 | Not-determined |
| SB_extracted_k141_108945 | Not-determined |
| SB_extracted_k141_117449 | Not-determined |
| SB_extracted_k141_65261 | Not-determined |
| SB_extracted_k141_143654 | Not-determined |
| SB_extracted_k141_134728 | Not-determined |
| SB_extracted_k141_117451 | Not-determined |
| SB_extracted_k141_47742 | Low-quality |
| SB_extracted_k141_31115 | Low-quality |
| SB_extracted_k141_100237 | Not-determined |
| SB_extracted_k141_22285 | Not-determined |
| SB_extracted_k141_195775 | Not-determined |
| SB_extracted_k141_100239 | Not-determined |
| SB_extracted_k141_13100 | Not-determined |
| SB_extracted_k141_4675 | Not-determined |
| SB_extracted_k141_22288 | Low-quality |
| SB_extracted_k141_195776 | Low-quality |
| SB_extracted_k141_100241 | Not-determined |
| SB_extracted_k141_100243 | Not-determined |
| SB_extracted_k141_204038 | Not-determined |
| SB_extracted_k141_195780 | Not-determined |
| SB_extracted_k141_204041 | Not-determined |
| SB_extracted_k141_143665 | Not-determined |
| SB_extracted_k141_100246 | Not-determined |
| SB_extracted_k141_117463 | Not-determined |
| SB_extracted_k141_100247 | Not-determined |
| SB_extracted_k141_65270 | Low-quality |
| S17_vs_k141_77057\|\|full | Medium-quality |
| SB_extracted_k141_204045 | Not-determined |
| SB_extracted_k141_31123 | Not-determined |
| SB_extracted_k141_47752 | Low-quality |
| SB_extracted_k141_126242 | Not-determined |
| SB_extracted_k141_82699 | Not-determined |
| SB_extracted_k141_160517 | Not-determined |
| SB_extracted_k141_195786 | Not-determined |
| SB_extracted_k141_4689 | Not-determined |
| SB_extracted_k141_47757 | Not-determined |
| SB_extracted_k141_108965 | Not-determined |
| SB_extracted_k141_65280 | Not-determined |
| SB_extracted_k141_47759 | Not-determined |
| SB_extracted_k141_91479 | Not-determined |
| SB_extracted_k141_65283 | Not-determined |
| SB_extracted_k141_22311 | Not-determined |
| SB_extracted_k141_4699 | Not-determined |
| SB_extracted_k141_73776 | Not-determined |
| SB_extracted_k141_65291 | Not-determined |
| SB_extracted_k141_39424 | Low-quality |
| SB_extracted_k141_108971 | Not-determined |
| SB_extracted_k141_22321 | Not-determined |
| SB_extracted_k141_204063 | Not-determined |
| SB_extracted_k141_108973 | Not-determined |
| SB_extracted_k141_73784 | Not-determined |
| SB_extracted_k141_39435 | Not-determined |
| SB_extracted_k141_126251 | Not-determined |
| SB_extracted_k141_195807 | Not-determined |
| SB_extracted_k141_108975 | Not-determined |
| SB_extracted_k141_65298 | Not-determined |
| SB_extracted_k141_22330 | Not-determined |
| SB_extracted_k141_126254 | Not-determined |
| SB_extracted_k141_65300 | Not-determined |
| SB_extracted_k141_56972 | Not-determined |
| SB_extracted_k141_4711 | Low-quality |
| SB_extracted_k141_47767 | Not-determined |
| SB_extracted_k141_65303 | Not-determined |
| SB_extracted_k141_73786 | Low-quality |
| SB_extracted_k141_4715 | Not-determined |
| SB_extracted_k141_4716 | Not-determined |
| SB_extracted_k141_47774 | Not-determined |
| SB_extracted_k141_152104 | Not-determined |
| SB_extracted_k141_143694 | Not-determined |
| SB_extracted_k141_73793 | Low-quality |
| SB_extracted_k141_204080 | Not-determined |
| SB_extracted_k141_56981 | Not-determined |
| SB_extracted_k141_160530 | Not-determined |
| SB_extracted_k141_100271 | Not-determined |
| SB_extracted_k141_169636 | Not-determined |
| SB_extracted_k141_31158 | Not-determined |
| SB_extracted_k141_22334 | Not-determined |
| SB_extracted_k141_56988 | Low-quality |
| SB_extracted_k141_117495 | Not-determined |
| SB_extracted_k141_204084 | Not-determined |
| SB_extracted_k141_204085 | Not-determined |
| SB_extracted_k141_39447 | Not-determined |
| SB_extracted_k141_117496 | Not-determined |
| SB_extracted_k141_56994 | Not-determined |
| SB_extracted_k141_47785 | Not-determined |
| SB_extracted_k141_56995 | Not-determined |
| SB_extracted_k141_134748 | Not-determined |
| SB_extracted_k141_47787 | Not-determined |
| SB_extracted_k141_22337 | Not-determined |
| SB_extracted_k141_47788 | Not-determined |
| SB_extracted_k141_134750 | Low-quality |
| SB_extracted_k141_169638 | Low-quality |
| SB_extracted_k141_73808 | Not-determined |
| SB_extracted_k141_65312 | Not-determined |
| SB_extracted_k141_73810 | Not-determined |
| SB_extracted_k141_39455 | Low-quality |
| SB_extracted_k141_134757 | Not-determined |
| SB_extracted_k141_13112 | Low-quality |
| SB_extracted_k141_143711 | Low-quality |
| SB_extracted_k141_73812 | Not-determined |
| SB_extracted_k141_178383 | Low-quality |
| SB_extracted_k141_195817 | Not-determined |
| SB_extracted_k141_82729 | Not-determined |
| SB_extracted_k141_73818 | Not-determined |
| SB_extracted_k141_57005 | Not-determined |
| SB_extracted_k141_22344 | Not-determined |
| SB_extracted_k141_57007 | Not-determined |
| SB_extracted_k141_31167 | Not-determined |
| SB_extracted_k141_22346 | Not-determined |
| SB_extracted_k141_47797 | Low-quality |
| SB_extracted_k141_126290 | Not-determined |
| SB_extracted_k141_178387 | Not-determined |
| SB_extracted_k141_100282 | Not-determined |
| SB_extracted_k141_31169 | Not-determined |
| SB_extracted_k141_22351 | Not-determined |
| SB_extracted_k141_178393 | Not-determined |
| SB_extracted_k141_152119 | Not-determined |
| SB_extracted_k141_143715 | Not-determined |
| SB_extracted_k141_47804 | Not-determined |
| SB_extracted_k141_143716 | Not-determined |
| SB_extracted_k141_57021 | Not-determined |
| SB_extracted_k141_73830 | Not-determined |
| SB_extracted_k141_152121 | Not-determined |
| SB_extracted_k141_169643 | Not-determined |
| SB_extracted_k141_152122 | Low-quality |
| SB_extracted_k141_117510 | Not-determined |
| SB_extracted_k141_22356 | Not-determined |
| SB_extracted_k141_204095 | Not-determined |
| SB_extracted_k141_143719 | Not-determined |
| SB_extracted_k141_22358 | Not-determined |
| SB_extracted_k141_169647 | Not-determined |
| SB_extracted_k141_100296 | Not-determined |
| SB_extracted_k141_195839 | Not-determined |
| SB_extracted_k141_195840 | Not-determined |
| SB_extracted_k141_13131 | Not-determined |
| SB_extracted_k141_13132 | Not-determined |
| SB_extracted_k141_47812 | Not-determined |
| SB_extracted_k141_195845 | Not-determined |
| SB_extracted_k141_109002 | Not-determined |
| SB_extracted_k141_109003 | Not-determined |
| SB_extracted_k141_73841 | Not-determined |
| SB_extracted_k141_117528 | Not-determined |
| SB_extracted_k141_31183 | Not-determined |
| SB_extracted_k141_134796 | Not-determined |
| SB_extracted_k141_143734 | Not-determined |
| SB_extracted_k141_22365 | Not-determined |
| SB_extracted_k141_109004 | Not-determined |
| SB_extracted_k141_134801 | Not-determined |
| SB_extracted_k141_47816 | Low-quality |
| SB_extracted_k141_152134 | Not-determined |
| SB_extracted_k141_117540 | Not-determined |
| SB_extracted_k141_47819 | Not-determined |
| SB_extracted_k141_39485 | Not-determined |
| SB_extracted_k141_65345 | Not-determined |
| SB_extracted_k141_82752 | Not-determined |
| SB_extracted_k141_143741 | Not-determined |
| SB_extracted_k141_73853 | Not-determined |
| SB_extracted_k141_13157 | Not-determined |
| SB_extracted_k141_117542 | Not-determined |
| SB_extracted_k141_22373 | Not-determined |
| SB_extracted_k141_109016 | Not-determined |
| SB_extracted_k141_152142 | Not-determined |
| SB_extracted_k141_169662 | Not-determined |
| SB_extracted_k141_143743 | Low-quality |
| SB_extracted_k141_31193 | Not-determined |
| SB_extracted_k141_57052 | Not-determined |
| SB_extracted_k141_204103 | Not-determined |
| SB_extracted_k141_195861 | Not-determined |
| SB_extracted_k141_39493 | Not-determined |
| SB_extracted_k141_22381 | Not-determined |
| SB_extracted_k141_91513 | Not-determined |
| SB_extracted_k141_100333 | Not-determined |
| SB_extracted_k141_31199 | Low-quality |
| SB_extracted_k141_91514 | Low-quality |
| SB_extracted_k141_152148 | Not-determined |
| SB_extracted_k141_13166 | Not-determined |
| SB_extracted_k141_169671 | Not-determined |
| SB_extracted_k141_22388 | Not-determined |
| SB_extracted_k141_204111 | Not-determined |
| SB_extracted_k141_204112 | Not-determined |
| SB_extracted_k141_47835 | Not-determined |
| SB_extracted_k141_82754 | Low-quality |
| SB_extracted_k141_204113 | Not-determined |
| SB_extracted_k141_117554 | Low-quality |
| SB_extracted_k141_73865 | Not-determined |
| SB_extracted_k141_117555 | Not-determined |
| SB_extracted_k141_160543 | Not-determined |
| SB_extracted_k141_47836 | Not-determined |
| SB_extracted_k141_22390 | Not-determined |
| SB_extracted_k141_100338 | Not-determined |
| SB_extracted_k141_169679 | Not-determined |
| SB_extracted_k141_143761 | Not-determined |
| SB_extracted_k141_143762 | Not-determined |
| SB_extracted_k141_91523 | Not-determined |
| SB_extracted_k141_109027 | Not-determined |
| SB_extracted_k141_100339 | Not-determined |
| SB_extracted_k141_57069 | Not-determined |
| SB_extracted_k141_126318 | Not-determined |
| SB_extracted_k141_91524 | Not-determined |
| SB_extracted_k141_65365 | Not-determined |
| SB_extracted_k141_22397 | Not-determined |
| SB_extracted_k141_204118 | Not-determined |
| SB_extracted_k141_117564 | Not-determined |
| SB_extracted_k141_204119 | Low-quality |
| SB_extracted_k141_152161 | Not-determined |
| SB_extracted_k141_126321 | Not-determined |
| SB_extracted_k141_47842 | Not-determined |
| SB_extracted_k141_47843 | Not-determined |
| SB_extracted_k141_143768 | Not-determined |
| SB_extracted_k141_126324 | Not-determined |
| SB_extracted_k141_186131 | Not-determined |
| SB_extracted_k141_195877 | Not-determined |
| SB_extracted_k141_152165 | Not-determined |
| SB_extracted_k141_22409 | Not-determined |
| SB_extracted_k141_39505 | Not-determined |
| SB_extracted_k141_109044 | Not-determined |
| SB_extracted_k141_109048 | Not-determined |
| SB_extracted_k141_169697 | Low-quality |
| SB_extracted_k141_195892 | Not-determined |
| SB_extracted_k141_152166 | Not-determined |
| SB_extracted_k141_22419 | Not-determined |
| SB_extracted_k141_22425 | Not-determined |
| SB_extracted_k141_117584 | Not-determined |
| SB_extracted_k141_65393 | Not-determined |
| SB_extracted_k141_195899 | Not-determined |
| SB_extracted_k141_204141 | Not-determined |
| SB_extracted_k141_117587 | Not-determined |
| SB_extracted_k141_204143 | Not-determined |
| SB_extracted_k141_109062 | Not-determined |
| SB_extracted_k141_178427 | Not-determined |
| SB_extracted_k141_31215 | Not-determined |
| SB_extracted_k141_143797 | Not-determined |
| SB_extracted_k141_195903 | Low-quality |
| SB_extracted_k141_109067 | Low-quality |
| SB_extracted_k141_152184 | Not-determined |
| SB_extracted_k141_22430 | Not-determined |
| SB_extracted_k141_178429 | Not-determined |
| SB_extracted_k141_152187 | Not-determined |
| SB_extracted_k141_13190 | Low-quality |
| SB_extracted_k141_13191 | Not-determined |
| SB_extracted_k141_178430 | Not-determined |
| SB_extracted_k141_31219 | Not-determined |
| SB_extracted_k141_178432 | Not-determined |
| SB_extracted_k141_65403 | Not-determined |
| SB_extracted_k141_143805 | Not-determined |
| SB_extracted_k141_152192 | Not-determined |
| SB_extracted_k141_91554 | Not-determined |
| SB_extracted_k141_204153 | Not-determined |
| SB_extracted_k141_117602 | Not-determined |
| SB_extracted_k141_39524 | Not-determined |
| SB_extracted_k141_169718 | Low-quality |
| SB_extracted_k141_39525 | Low-quality |
| SB_extracted_k141_117606 | Not-determined |
| SB_extracted_k141_169720 | Not-determined |
| SB_extracted_k141_13199 | Low-quality |
| SB_extracted_k141_82788 | Not-determined |
| SB_extracted_k141_143820 | Not-determined |
| SB_extracted_k141_134807 | Not-determined |
| SB_extracted_k141_82789 | Not-determined |
| SB_extracted_k141_134808 | Not-determined |
| SB_extracted_k141_91563 | Not-determined |
| SB_extracted_k141_109084 | Not-determined |
| SB_extracted_k141_22456 | Not-determined |
| SB_extracted_k141_134810 | Low-quality |
| SB_extracted_k141_195916 | Not-determined |
| SB_extracted_k141_126362 | Not-determined |
| SB_extracted_k141_169725 | Not-determined |
| SB_extracted_k141_65406 | Low-quality |
| SB_extracted_k141_82792 | Not-determined |
| SB_extracted_k141_169726 | Low-quality |
| SB_extracted_k141_109086 | Not-determined |
| SB_extracted_k141_57111 | Not-determined |
| SB_extracted_k141_195923 | Not-determined |
| SB_extracted_k141_65411 | Not-determined |
| SB_extracted_k141_47858 | Not-determined |
| SB_extracted_k141_39538 | Not-determined |
| SB_extracted_k141_143829 | Not-determined |
| SB_extracted_k141_160577 | Not-determined |
| SB_extracted_k141_204165 | Not-determined |
| SB_extracted_k141_39546 | Not-determined |
| SB_extracted_k141_47860 | Low-quality |
| SB_extracted_k141_39547 | Not-determined |
| SB_extracted_k141_31236 | Low-quality |
| SB_extracted_k141_47864 | Not-determined |
| SB_extracted_k141_57118 | Not-determined |
| SB_extracted_k141_47865 | Not-determined |
| SB_extracted_k141_47866 | Not-determined |
| SB_extracted_k141_91577 | Not-determined |
| SB_extracted_k141_4770 | Not-determined |
| SB_extracted_k141_47867 | Not-determined |
| SB_extracted_k141_47870 | Not-determined |
| SB_extracted_k141_22463 | Not-determined |
| SB_extracted_k141_117619 | Not-determined |
| SB_extracted_k141_13206 | Low-quality |
| SB_extracted_k141_39553 | Not-determined |
| SB_extracted_k141_65422 | Not-determined |
| SB_extracted_k141_39555 | Not-determined |
| SB_extracted_k141_169737 | Low-quality |
| SB_extracted_k141_91586 | Not-determined |
| SB_extracted_k141_22466 | Not-determined |
| SB_extracted_k141_13210 | Low-quality |
| SB_extracted_k141_91589 | Not-determined |
| SB_extracted_k141_152208 | Not-determined |
| SB_extracted_k141_65426 | Not-determined |
| SB_extracted_k141_204176 | Not-determined |
| SB_extracted_k141_47876 | Not-determined |
| SB_extracted_k141_169744 | Not-determined |
| SB_extracted_k141_13213 | Low-quality |
| SB_extracted_k141_31246 | Not-determined |
| SB_extracted_k141_160589 | Not-determined |
| SB_extracted_k141_195938 | Not-determined |
| SB_extracted_k141_204178 | Not-determined |
| SB_extracted_k141_4782 | Not-determined |
| SB_extracted_k141_22471 | Not-determined |
| SB_extracted_k141_204180 | Not-determined |
| SB_extracted_k141_186147 | Not-determined |
| SB_extracted_k141_109098 | Not-determined |
| SB_extracted_k141_31251 | Not-determined |
| SB_extracted_k141_22475 | Not-determined |
| SB_extracted_k141_39570 | Not-determined |
| SB_extracted_k141_22476 | Not-determined |
| SB_extracted_k141_117624 | Not-determined |
| SB_extracted_k141_117625 | Not-determined |
| SB_extracted_k141_143841 | Not-determined |
| SB_extracted_k141_65436 | Not-determined |
| SB_extracted_k141_195944 | Not-determined |
| SB_extracted_k141_160596 | Not-determined |
| SB_extracted_k141_65438 | Not-determined |
| SB_extracted_k141_91603 | Not-determined |
| SB_extracted_k141_143845 | Not-determined |
| SB_extracted_k141_178446 | Not-determined |
| SB_extracted_k141_152221 | Not-determined |
| SB_extracted_k141_160601 | Not-determined |
| SB_extracted_k141_195949 | Not-determined |
| SB_extracted_k141_178449 | Not-determined |
| SB_extracted_k141_169751 | Low-quality |
| SB_extracted_k141_31256 | Low-quality |
| SB_extracted_k141_195951 | Not-determined |
| SB_extracted_k141_195952 | Low-quality |
| SB_extracted_k141_152225 | Not-determined |
| SB_extracted_k141_160606 | Not-determined |
| SB_extracted_k141_160607 | Not-determined |
| SB_extracted_k141_195955 | Not-determined |
| SB_extracted_k141_65448 | Not-determined |
| SB_extracted_k141_143848 | Not-determined |
| SB_extracted_k141_57135 | Not-determined |
| SB_extracted_k141_22478 | Low-quality |
| SB_extracted_k141_169762 | Not-determined |
| SB_extracted_k141_39580 | Not-determined |
| SB_extracted_k141_109113 | Not-determined |
| SB_extracted_k141_169765 | Low-quality |
| SB_extracted_k141_178459 | Low-quality |
| SB_extracted_k141_160609 | Low-quality |
| SB_extracted_k141_39585 | Not-determined |
| SB_extracted_k141_126396 | Not-determined |
| SB_extracted_k141_109118 | Not-determined |
| SB_extracted_k141_109119 | Not-determined |
| SB_extracted_k141_57139 | Not-determined |
| SB_extracted_k141_91618 | Not-determined |
| SB_extracted_k141_169768 | Not-determined |
| SB_extracted_k141_186148 | Not-determined |
| SB_extracted_k141_169769 | Not-determined |
| SB_extracted_k141_195971 | Not-determined |
| SB_extracted_k141_82824 | Not-determined |
| SB_extracted_k141_39587 | Not-determined |
| SB_extracted_k141_65456 | Not-determined |
| SB_extracted_k141_178465 | Not-determined |
| SB_extracted_k141_22494 | Not-determined |
| SB_extracted_k141_39589 | Not-determined |
| SB_extracted_k141_22495 | Not-determined |
| SB_extracted_k141_169773 | Not-determined |
| SB_extracted_k141_65460 | Not-determined |
| SB_extracted_k141_82828 | Not-determined |
| SB_extracted_k141_117637 | Not-determined |
| SB_extracted_k141_57148 | Not-determined |
| SB_extracted_k141_152229 | Not-determined |
| SB_extracted_k141_57150 | Not-determined |
| SB_extracted_k141_22499 | Not-determined |
| SB_extracted_k141_169778 | Not-determined |
| SB_extracted_k141_152231 | Low-quality |
| SB_extracted_k141_152232 | Not-determined |
| SB_extracted_k141_117643 | Not-determined |
| SB_extracted_k141_152233 | Not-determined |
| SB_extracted_k141_195977 | Not-determined |
| SB_extracted_k141_13218 | Not-determined |
| SB_extracted_k141_143869 | Not-determined |
| SB_extracted_k141_126407 | Not-determined |
| SB_extracted_k141_152237 | Not-determined |
| SB_extracted_k141_126409 | Not-determined |
| SB_extracted_k141_57155 | Not-determined |
| SB_extracted_k141_39602 | Not-determined |
| SB_extracted_k141_169787 | Not-determined |
| SB_extracted_k141_178471 | Not-determined |
| SB_extracted_k141_22508 | Not-determined |
| SB_extracted_k141_204200 | Not-determined |
| SB_extracted_k141_169791 | Not-determined |
| SB_extracted_k141_152243 | Not-determined |
| SB_extracted_k141_91631 | Not-determined |
| SB_extracted_k141_73895 | Not-determined |
| SB_extracted_k141_126421 | Not-determined |
| SB_extracted_k141_126422 | Low-quality |
| SB_extracted_k141_195984 | Not-determined |
| SB_extracted_k141_126423 | Low-quality |
| SB_extracted_k141_204205 | Not-determined |
| SB_extracted_k141_31270 | Not-determined |
| SB_extracted_k141_22513 | Not-determined |
| SB_extracted_k141_39611 | Not-determined |
| SB_extracted_k141_134853 | Not-determined |
| SB_extracted_k141_169803 | Low-quality |
| SB_extracted_k141_91636 | Not-determined |
| SB_extracted_k141_13223 | Not-determined |
| SB_extracted_k141_126431 | Not-determined |
| SB_extracted_k141_13224 | Not-determined |
| SB_extracted_k141_4820 | Not-determined |
| SB_extracted_k141_169807 | Not-determined |
| SB_extracted_k141_73901 | Not-determined |
| SB_extracted_k141_152249 | Low-quality |
| SB_extracted_k141_91645 | Not-determined |
| SB_extracted_k141_134858 | Not-determined |
| SB_extracted_k141_195997 | Not-determined |
| SB_extracted_k141_65472 | Not-determined |
| SB_extracted_k141_204225 | Not-determined |
| SB_extracted_k141_4824 | Not-determined |
| SB_extracted_k141_22524 | Low-quality |
| SB_extracted_k141_91649 | Not-determined |
| SB_extracted_k141_4830 | Not-determined |
| SB_extracted_k141_134863 | Not-determined |
| SB_extracted_k141_31280 | Not-determined |
| SB_extracted_k141_117648 | Not-determined |
| SB_extracted_k141_39634 | Not-determined |
| SB_extracted_k141_13241 | Not-determined |
| SB_extracted_k141_39637 | Not-determined |
| SB_extracted_k141_178493 | Not-determined |
| SB_extracted_k141_169815 | Not-determined |
| SB_extracted_k141_39639 | Not-determined |
| SB_extracted_k141_73917 | Not-determined |
| SB_extracted_k141_109128 | Not-determined |
| SB_extracted_k141_57177 | Not-determined |
| SB_extracted_k141_4839 | Not-determined |
| SB_extracted_k141_91654 | Not-determined |
| SB_extracted_k141_196011 | Not-determined |
| SB_extracted_k141_126443 | Not-determined |
| SB_extracted_k141_13244 | Not-determined |
| SB_extracted_k141_186172 | Not-determined |
| SB_extracted_k141_73923 | Not-determined |
| SB_extracted_k141_196013 | Not-determined |
| SB_extracted_k141_196014 | Not-determined |
| SB_extracted_k141_117658 | Not-determined |
| SB_extracted_k141_169819 | Not-determined |
| SB_extracted_k141_134875 | Not-determined |
| SB_extracted_k141_39644 | Low-quality |
| SB_extracted_k141_109142 | Not-determined |
| SB_extracted_k141_178502 | Not-determined |
| SB_extracted_k141_109144 | Low-quality |
| SB_extracted_k141_196018 | Not-determined |
| SB_extracted_k141_117665 | Low-quality |
| SB_extracted_k141_31301 | Not-determined |
| SB_extracted_k141_143904 | Not-determined |
| SB_extracted_k141_169829 | Low-quality |
| SB_extracted_k141_178507 | Low-quality |
| SB_extracted_k141_47882 | Not-determined |
| SB_extracted_k141_39646 | Low-quality |
| SB_extracted_k141_73933 | Low-quality |
| SB_extracted_k141_178512 | Not-determined |
| SB_extracted_k141_178513 | Not-determined |
| SB_extracted_k141_169831 | Not-determined |
| SB_extracted_k141_204246 | Not-determined |
| SB_extracted_k141_109154 | Not-determined |
| SB_extracted_k141_57193 | Not-determined |
| SB_extracted_k141_39655 | Not-determined |
| SB_extracted_k141_13252 | Not-determined |
| SB_extracted_k141_109155 | Low-quality |

| | |
|---|---|
| S8_extracted_k141_196023 | Not-determined |
| S8_extracted_k141_31310 | Not-determined |
| S8_extracted_k141_22533 | Not-determined |
| S8_extracted_k141_4859 | Not-determined |
| S8_extracted_k141_13258 | Not-determined |
| S8_extracted_k141_134893 | Not-determined |
| S8_extracted_k141_13261 | Not-determined |
| S8_extracted_k141_22536 | Not-determined |
| S8_extracted_k141_22537 | Not-determined |
| S8_extracted_k141_82863 | Not-determined |
| S8_extracted_k141_82864 | Low-quality |
| S8_extracted_k141_178522 | Low-quality |
| S8_extracted_k141_13264 | Not-determined |
| S8_extracted_k141_169837 | Not-determined |
| S8_extracted_k141_13265 | Not-determined |
| S8_extracted_k141_91673 | Not-determined |
| S8_extracted_k141_73945 | Not-determined |
| S8_extracted_k141_82868 | Not-determined |
| S8_extracted_k141_186181 | Low-quality |
| S8_extracted_k141_186182 | Not-determined |
| S8_extracted_k141_117685 | Not-determined |
| S8_extracted_k141_82870 | Low-quality |
| S8_extracted_k141_117687 | Not-determined |
| S8_extracted_k141_152274 | Not-determined |
| S8_extracted_k141_169845 | Not-determined |
| S8_extracted_k141_169846 | Not-determined |
| S8_extracted_k141_22542 | Not-determined |
| S8_extracted_k141_22543 | Not-determined |
| S8_extracted_k141_4870 | Not-determined |
| S8_extracted_k141_196036 | Not-determined |
| S8_extracted_k141_186190 | Not-determined |
| S8_extracted_k141_169850 | Not-determined |
| S8_extracted_k141_13272 | Low-quality |
| S8_extracted_k141_31326 | Not-determined |
| S8_extracted_k141_196041 | Not-determined |
| S8_extracted_k141_109168 | Not-determined |
| S8_extracted_k141_47893 | Not-determined |
| S8_extracted_k141_169855 | Not-determined |
| S8_extracted_k141_169857 | Not-determined |
| S8_extracted_k141_109171 | Not-determined |
| S8_extracted_k141_109172 | Not-determined |
| S8_extracted_k141_196049 | Not-determined |
| S8_extracted_k141_134905 | Not-determined |
| S8_extracted_k141_4882 | Not-determined |
| S8_extracted_k141_13285 | Not-determined |
| S8_extracted_k141_73962 | Not-determined |
| S8_extracted_k141_186205 | Low-quality |
| S8_extracted_k141_126466 | Low-quality |
| S8_extracted_k141_152285 | Low-quality |
| S8_extracted_k141_57199 | Not-determined |
| S8_extracted_k141_47905 | Not-determined |
| S8_extracted_k141_13288 | Low-quality |
| S8_extracted_k141_57204 | Not-determined |
| S8_extracted_k141_152289 | Not-determined |
| S8_extracted_k141_169864 | Low-quality |
| S8_extracted_k141_160642 | Not-determined |
| S8_extracted_k141_169867 | Not-determined |
| S8_extracted_k141_47908 | Not-determined |
| S8_extracted_k141_31342 | Not-determined |
| S8_extracted_k141_39682 | Not-determined |
| S8_extracted_k141_31343 | Not-determined |
| S8_extracted_k141_57213 | Not-determined |
| S8_extracted_k141_91698 | Not-determined |
| S8_extracted_k141_82889 | Not-determined |
| S8_extracted_k141_143936 | Not-determined |
| S8_extracted_k141_109189 | Not-determined |
| S8_extracted_k141_143938 | Not-determined |
| S8_extracted_k141_13297 | Not-determined |
| S8_extracted_k141_82896 | Not-determined |
| S8_extracted_k141_169876 | Not-determined |
| S8_extracted_k141_31352 | Not-determined |
| S8_extracted_k141_126473 | Not-determined |
| S8_extracted_k141_57222 | Low-quality |
| S8_extracted_k141_91707 | Not-determined |
| S8_extracted_k141_65497 | Not-determined |
| S8_extracted_k141_126478 | Not-determined |
| S8_extracted_k141_160652 | Not-determined |
| S8_extracted_k141_57226 | Not-determined |
| S8_extracted_k141_73983 | Not-determined |
| S8_extracted_k141_126479 | Not-determined |
| S8_extracted_k141_143942 | Not-determined |
| S8_extracted_k141_13303 | Not-determined |
| S8_extracted_k141_82899 | Not-determined |
| S8_extracted_k141_22560 | Not-determined |
| S8_extracted_k141_65505 | Low-quality |
| S8_extracted_k141_73986 | Not-determined |
| S8_extracted_k141_13306 | Not-determined |
| S8_extracted_k141_73988 | Not-determined |
| S8_extracted_k141_39688 | Not-determined |
| S8_extracted_k141_169886 | Not-determined |
| S8_extracted_k141_82904 | Not-determined |
| S8_extracted_k141_31360 | Low-quality |
| S8_extracted_k141_186210 | Not-determined |
| S8_extracted_k141_134918 | Not-determined |
| S8_extracted_k141_31361 | Not-determined |
| S8_extracted_k141_169889 | Not-determined |
| S8_extracted_k141_82906 | Not-determined |
| S8_extracted_k141_178556 | Low-quality |
| S8_extracted_k141_31363 | Not-determined |
| S8_extracted_k141_31365 | Not-determined |
| S8_extracted_k141_91722 | Low-quality |
| S8_extracted_k141_82909 | Not-determined |
| S8_extracted_k141_91723 | Not-determined |
| S8_extracted_k141_117707 | Low-quality |
| S8_extracted_k141_143952 | Not-determined |
| S8_extracted_k141_152313 | Not-determined |
| S8_extracted_k141_143956 | Not-determined |
| S8_extracted_k141_82914 | Not-determined |
| S8_extracted_k141_109202 | Not-determined |
| S8_extracted_k141_91728 | Not-determined |
| S8_extracted_k141_126490 | Not-determined |
| S8_extracted_k141_31371 | Not-determined |
| S8_extracted_k141_152321 | Not-determined |
| S8_extracted_k141_152322 | Not-determined |
| S8_extracted_k141_126495 | Low-quality |
| S8_extracted_k141_13316 | Not-determined |
| S8_extracted_k141_4884 | Not-determined |
| S8_extracted_k141_31374 | Not-determined |
| S8_extracted_k141_143964 | Not-determined |
| S8_extracted_k141_178572 | Not-determined |
| S8_extracted_k141_109209 | Not-determined |
| S8_extracted_k141_143967 | Low-quality |
| S8_extracted_k141_186222 | Not-determined |
| S8_extracted_k141_91736 | Not-determined |
| S8_extracted_k141_117718 | Low-quality |
| S8_extracted_k141_178578 | Not-determined |
| S8_extracted_k141_31378 | Not-determined |
| S8_extracted_k141_178582 | Not-determined |
| S8_extracted_k141_134940 | Not-determined |
| S8_extracted_k141_57254 | Not-determined |
| S8_extracted_k141_82921 | Not-determined |
| S8_extracted_k141_39719 | Not-determined |
| S8_extracted_k141_13329 | Not-determined |
| S8_extracted_k141_152338 | Not-determined |
| S8_extracted_k141_169925 | Not-determined |
| S8_extracted_k141_143978 | Not-determined |
| S8_extracted_k141_169931 | Not-determined |
| S8_extracted_k141_169933 | Not-determined |
| S8_extracted_k141_143980 | Not-determined |
| S8_extracted_k141_13335 | Not-determined |
| S8_extracted_k141_91748 | Not-determined |
| S8_extracted_k141_82928 | Not-determined |
| S8_extracted_k141_13336 | Low-quality |
| S8_extracted_k141_13337 | Not-determined |
| S8_extracted_k141_117733 | Not-determined |
| S8_extracted_k141_178594 | Not-determined |
| S8_extracted_k141_152349 | Not-determined |
| S8_extracted_k141_117737 | Not-determined |
| S8_extracted_k141_39726 | Low-quality |
| S8_extracted_k141_57269 | Not-determined |
| S8_extracted_k141_91749 | Not-determined |
| S8_extracted_k141_178596 | Not-determined |
| S8_extracted_k141_13341 | Not-determined |
| S8_extracted_k141_117741 | Not-determined |
| S8_extracted_k141_186231 | Not-determined |
| S8_extracted_k141_31386 | Not-determined |
| S8_extracted_k141_117744 | Not-determined |
| S8_extracted_k141_39729 | Not-determined |
| S8_extracted_k141_57272 | Not-determined |
| S8_extracted_k141_82936 | Not-determined |
| S8_extracted_k141_169943 | Not-determined |
| S8_extracted_k141_178603 | Not-determined |
| S8_extracted_k141_13347 | Not-determined |
| S8_extracted_k141_204275 | Not-determined |
| S8_extracted_k141_4910 | Not-determined |
| S8_extracted_k141_91753 | Not-determined |
| S8_extracted_k141_4912 | Low-quality |
| S8_extracted_k141_39732 | Not-determined |
| S8_extracted_k141_160669 | Not-determined |
| S8_extracted_k141_186237 | Not-determined |
| S8_extracted_k141_13352 | Not-determined |
| S8_extracted_k141_178607 | Not-determined |
| S8_extracted_k141_109234 | Not-determined |
| S8_extracted_k141_160671 | Low-quality |
| S8_extracted_k141_186239 | Low-quality |
| S8_extracted_k141_57276 | Not-determined |
| S8_extracted_k141_22567 | Not-determined |
| S8_extracted_k141_31391 | Not-determined |
| S8_extracted_k141_39737 | Not-determined |
| S8_extracted_k141_186241 | Not-determined |
| S8_extracted_k141_47914 | Low-quality |
| S8_extracted_k141_47916 | Not-determined |
| S8_extracted_k141_143988 | Not-determined |
| S8_extracted_k141_204282 | Not-determined |
| S8_extracted_k141_57280 | Not-determined |
| S8_extracted_k141_4919 | Not-determined |
| S7_vs_93097\|\|full | Medium-quality |
| S8_extracted_k141_47921 | Not-determined |
| S8_extracted_k141_204286 | Low-quality |
| S8_extracted_k141_186248 | Not-determined |
| S8_extracted_k141_31398 | Not-determined |
| S8_extracted_k141_143998 | Not-determined |
| S8_extracted_k141_117764 | Low-quality |
| S8_extracted_k141_126503 | Low-quality |
| S8_extracted_k141_109239 | Low-quality |
| S8_extracted_k141_91769 | Not-determined |
| S8_extracted_k141_178611 | Not-determined |
| S8_extracted_k141_117768 | Not-determined |
| S8_extracted_k141_39747 | Not-determined |
| S8_extracted_k141_186258 | Not-determined |
| S8_extracted_k141_65533 | Not-determined |
| S8_extracted_k141_204292 | Low-quality |
| S8_extracted_k141_31406 | Not-determined |
| S8_extracted_k141_91779 | Not-determined |
| S8_extracted_k141_178618 | Not-determined |
| S8_extracted_k141_152372 | Not-determined |
| S8_extracted_k141_91782 | Not-determined |
| S8_extracted_k141_109241 | Not-determined |
| S8_extracted_k141_186264 | Not-determined |
| S8_extracted_k141_91783 | Low-quality |
| S8_extracted_k141_152375 | Low-quality |
| S8_extracted_k141_186265 | Not-determined |
| S8_extracted_k141_82948 | Not-determined |
| S8_extracted_k141_117776 | Not-determined |
| S8_extracted_k141_47936 | Low-quality |
| S8_extracted_k141_109246 | Not-determined |
| S8_extracted_k141_39762 | Not-determined |
| S8_extracted_k141_109247 | Not-determined |
| S8_extracted_k141_160683 | Not-determined |
| S8_extracted_k141_160687 | Not-determined |
| S8_extracted_k141_47948 | Low-quality |
| S8_extracted_k141_178629 | Not-determined |
| S8_extracted_k141_91793 | Not-determined |
| S8_extracted_k141_186279 | Low-quality |
| S8_extracted_k141_22611 | Not-determined |
| S8_extracted_k141_57309 | Low-quality |
| S8_extracted_k141_91794 | Not-determined |
| S8_extracted_k141_57311 | Not-determined |
| S8_extracted_k141_160691 | Not-determined |
| S8_extracted_k141_91797 | Low-quality |
| S8_extracted_k141_82965 | Low-quality |
| S8_extracted_k141_82967 | Low-quality |
| S8_extracted_k141_109251 | Low-quality |
| S8_extracted_k141_82971 | Not-determined |
| S8_extracted_k141_178637 | Not-determined |
| S8_extracted_k141_47950 | Not-determined |
| S8_extracted_k141_73991 | Low-quality |
| S8_extracted_k141_91803 | Not-determined |
| S8_extracted_k141_144027 | Not-determined |
| S8_extracted_k141_22626 | Not-determined |
| S8_extracted_k141_196068 | Low-quality |
| S8_extracted_k141_47956 | Not-determined |
| S8_extracted_k141_204311 | Low-quality |
| S8_extracted_k141_91809 | Not-determined |
| S8_extracted_k141_204315 | Not-determined |
| S8_extracted_k141_160697 | Not-determined |
| S8_extracted_k141_204316 | Not-determined |
| S8_extracted_k141_152403 | Not-determined |
| S8_extracted_k141_204317 | Not-determined |
| S8_extracted_k141_65542 | Low-quality |
| S8_extracted_k141_22630 | Not-determined |
| S8_extracted_k141_22631 | Not-determined |
| S8_extracted_k141_47960 | Not-determined |
| S8_extracted_k141_144038 | Not-determined |
| S8_extracted_k141_73992 | Not-determined |
| S8_extracted_k141_91814 | Not-determined |
| S8_extracted_k141_22635 | Not-determined |
| S8_extracted_k141_117790 | Not-determined |
| S8_extracted_k141_4940 | Not-determined |
| S8_extracted_k141_178643 | Not-determined |
| S8_extracted_k141_47964 | Not-determined |
| S8_extracted_k141_57327 | Not-determined |
| S8_extracted_k141_178645 | Not-determined |
| S8_extracted_k141_126527 | Not-determined |
| S8_extracted_k141_22641 | Not-determined |
| S8_extracted_k141_144046 | Not-determined |
| S8_extracted_k141_65553 | Not-determined |
| S8_extracted_k141_91818 | Not-determined |
| S8_extracted_k141_196087 | Not-determined |
| S8_extracted_k141_91820 | Not-determined |
| S8_extracted_k141_204333 | Not-determined |
| S8_extracted_k141_65555 | Not-determined |
| S8_extracted_k141_65557 | Not-determined |
| S8_extracted_k141_152418 | Not-determined |
| S8_extracted_k141_160702 | Not-determined |
| S8_extracted_k141_65559 | Not-determined |
| S8_extracted_k141_82992 | Not-determined |
| S8_extracted_k141_65560 | Not-determined |
| S8_extracted_k141_91825 | Not-determined |
| S8_extracted_k141_186310 | Not-determined |
| S8_extracted_k141_65565 | Not-determined |
| S8_extracted_k141_47977 | Not-determined |
| S8_extracted_k141_186314 | Not-determined |
| S8_extracted_k141_82995 | Not-determined |
| S8_extracted_k141_196096 | Not-determined |
| S8_extracted_k141_13392 | Low-quality |
| S8_extracted_k141_178655 | Not-determined |
| S8_extracted_k141_169963 | Not-determined |
| S8_extracted_k141_91831 | Not-determined |
| S8_extracted_k141_73994 | Low-quality |
| S8_extracted_k141_65571 | Not-determined |
| S8_extracted_k141_186323 | Not-determined |
| S8_extracted_k141_47985 | Not-determined |
| S8_extracted_k141_47986 | Not-determined |
| S17_vs_k141_67042\|\|full | Medium-quality |
| S8_extracted_k141_47987 | Not-determined |
| S8_extracted_k141_22645 | Not-determined |
| S8_extracted_k141_65574 | Not-determined |
| S8_extracted_k141_22647 | Not-determined |
| S8_extracted_k141_22649 | Not-determined |
| S8_extracted_k141_134946 | Not-determined |
| S8_extracted_k141_196106 | Low-quality |
| S8_extracted_k141_57334 | Low-quality |
| S8_extracted_k141_134948 | Not-determined |
| S8_extracted_k141_22650 | Not-determined |
| S8_extracted_k141_109282 | Not-determined |
| S8_extracted_k141_57335 | Not-determined |
| S8_extracted_k141_109284 | Not-determined |
| S8_extracted_k141_47992 | Not-determined |
| S8_extracted_k141_57336 | Not-determined |
| S8_extracted_k141_178668 | Low-quality |
| S8_extracted_k141_47994 | Not-determined |
| S8_extracted_k141_196111 | Not-determined |
| S8_extracted_k141_126538 | Not-determined |
| S8_extracted_k141_47995 | Not-determined |
| S8_extracted_k141_83009 | Not-determined |
| S8_extracted_k141_178673 | Not-determined |
| S8_extracted_k141_134949 | Not-determined |
| S8_extracted_k141_22654 | Not-determined |
| S8_extracted_k141_186339 | Not-determined |
| S8_extracted_k141_57339 | Not-determined |
| S8_extracted_k141_169981 | Low-quality |
| S8_extracted_k141_186341 | Not-determined |
| S8_extracted_k141_57342 | Not-determined |
| S8_extracted_k141_186342 | Not-determined |
| S8_extracted_k141_57343 | Not-determined |
| S8_extracted_k141_57344 | Not-determined |
| S8_extracted_k141_169989 | Not-determined |
| S8_extracted_k141_169990 | Not-determined |
| S8_extracted_k141_83024 | Not-determined |
| S8_extracted_k141_39772 | Not-determined |
| S8_extracted_k141_169994 | Not-determined |
| S8_extracted_k141_13417 | Not-determined |
| S8_extracted_k141_39780 | Not-determined |
| S8_extracted_k141_152432 | Not-determined |
| S8_extracted_k141_65591 | Not-determined |
| S8_extracted_k141_134958 | Not-determined |
| S8_extracted_k141_13425 | Not-determined |
| S8_extracted_k141_126551 | Not-determined |
| S8_extracted_k141_57351 | Not-determined |
| S8_extracted_k141_13426 | Not-determined |
| S8_extracted_k141_4947 | Not-determined |
| S8_extracted_k141_22670 | Not-determined |
| S8_extracted_k141_4948 | Not-determined |
| S8_extracted_k141_4949 | Not-determined |
| S8_extracted_k141_109299 | Not-determined |
| S8_extracted_k141_83030 | Not-determined |
| S8_extracted_k141_109300 | Not-determined |
| S8_extracted_k141_83031 | Not-determined |
| S8_extracted_k141_109301 | Not-determined |
| S8_extracted_k141_152436 | Not-determined |
| S8_extracted_k141_186356 | Not-determined |
| S8_extracted_k141_134965 | Not-determined |
| S8_extracted_k141_109303 | Not-determined |
| S8_extracted_k141_152439 | Not-determined |
| S8_extracted_k141_186361 | Not-determined |
| S8_extracted_k141_134967 | Not-determined |
| S8_extracted_k141_4957 | Not-determined |
| S8_extracted_k141_39792 | Not-determined |
| S8_extracted_k141_22686 | Not-determined |
| S8_extracted_k141_83035 | Low-quality |
| S8_extracted_k141_13431 | Not-determined |
| S8_extracted_k141_170000 | Not-determined |
| S8_extracted_k141_170001 | Not-determined |
| S8_extracted_k141_4960 | Not-determined |
| S8_extracted_k141_22690 | Not-determined |
| S8_extracted_k141_170002 | Low-quality |
| S8_extracted_k141_186367 | Low-quality |
| S8_extracted_k141_57364 | Not-determined |
| S8_extracted_k141_83039 | Low-quality |
| S8_extracted_k141_170004 | Not-determined |
| S8_extracted_k141_65608 | Not-determined |
| S8_extracted_k141_57367 | Low-quality |
| S8_extracted_k141_126564 | Low-quality |
| S8_extracted_k141_57371 | Not-determined |
| S8_extracted_k141_39806 | Low-quality |
| S8_extracted_k141_186376 | Not-determined |
| S8_extracted_k141_160724 | Not-determined |
| S8_extracted_k141_186378 | Not-determined |
| S8_extracted_k141_152455 | Not-determined |
| S8_extracted_k141_57374 | Not-determined |
| S14_vs_k141_93134\|\|full | Medium-quality |
| S8_extracted_k141_74027 | Not-determined |
| S8_extracted_k141_39814 | Not-determined |
| S8_extracted_k141_65615 | Not-determined |
| S8_extracted_k141_152459 | Not-determined |
| S8_extracted_k141_160732 | Not-determined |
| S8_extracted_k141_74036 | Not-determined |
| S8_extracted_k141_186384 | Not-determined |
| S8_extracted_k141_144099 | Low-quality |
| S8_extracted_k141_204348 | Not-determined |
| S8_extracted_k141_152466 | Not-determined |
| S8_extracted_k141_152470 | Not-determined |
| S8_extracted_k141_160734 | Not-determined |
| S8_extracted_k141_39823 | Not-determined |
| S8_extracted_k141_83045 | Not-determined |
| S8_extracted_k141_117804 | Not-determined |
| S8_extracted_k141_48023 | Not-determined |
| S8_extracted_k141_22698 | Not-determined |
| S8_extracted_k141_39831 | Not-determined |
| S8_extracted_k141_196124 | Low-quality |
| S8_extracted_k141_22707 | Not-determined |
| S8_extracted_k141_74039 | Not-determined |
| S8_extracted_k141_39833 | Not-determined |
| S8_extracted_k141_57384 | Not-determined |
| S8_extracted_k141_117813 | Not-determined |
| S8_extracted_k141_83047 | Not-determined |
| S8_extracted_k141_65626 | Low-quality |
| S8_extracted_k141_65627 | Not-determined |
| S8_extracted_k141_39837 | Not-determined |
| S8_extracted_k141_117816 | Not-determined |
| S8_extracted_k141_39838 | Not-determined |
| S8_extracted_k141_152480 | Not-determined |
| S8_extracted_k141_204364 | Not-determined |
| S8_extracted_k141_152481 | Not-determined |
| S8_extracted_k141_39839 | Not-determined |
| S8_extracted_k141_144116 | Not-determined |
| S8_extracted_k141_196133 | Not-determined |
| S8_extracted_k141_186399 | Not-determined |
| S8_extracted_k141_74048 | Not-determined |
| S8_extracted_k141_186403 | Low-quality |
| S8_extracted_k141_13467 | Not-determined |
| S8_extracted_k141_57388 | Not-determined |
| S8_extracted_k141_48034 | Not-determined |
| S8_extracted_k141_109328 | Not-determined |
| S8_extracted_k141_178691 | Not-determined |
| S8_extracted_k141_196145 | Not-determined |
| S8_extracted_k141_39843 | Low-quality |
| S8_extracted_k141_39844 | Not-determined |
| S8_extracted_k141_65633 | Not-determined |
| S8_extracted_k141_144137 | Not-determined |
| S8_extracted_k141_22720 | Not-determined |
| S8_extracted_k141_83061 | Low-quality |
| S8_extracted_k141_196151 | Not-determined |
| S8_extracted_k141_83062 | Not-determined |
| S8_extracted_k141_170030 | Not-determined |
| S8_extracted_k141_83063 | Not-determined |
| S8_extracted_k141_13471 | Not-determined |
| S8_extracted_k141_83065 | Not-determined |
| S8_extracted_k141_178697 | Not-determined |
| S8_extracted_k141_178698 | Not-determined |
| S8_extracted_k141_13474 | Low-quality |
| S8_extracted_k141_22726 | Low-quality |
| S8_extracted_k141_186409 | Not-determined |
| S8_extracted_k141_22728 | Not-determined |
| S8_extracted_k141_13476 | Not-determined |
| S8_extracted_k141_74063 | Not-determined |
| S8_extracted_k141_186410 | Not-determined |
| S8_extracted_k141_65635 | Not-determined |
| S8_extracted_k141_117824 | Not-determined |
| S8_extracted_k141_74064 | Not-determined |
| S8_extracted_k141_117826 | Low-quality |
| S8_extracted_k141_160752 | Not-determined |
| S8_extracted_k141_196156 | Not-determined |
| S8_extracted_k141_117828 | Not-determined |
| S8_extracted_k141_22732 | Low-quality |
| S8_extracted_k141_170039 | Not-determined |
| S8_extracted_k141_22734 | Not-determined |
| S8_extracted_k141_22735 | Not-determined |
| S8_extracted_k141_152491 | Not-determined |
| S8_extracted_k141_4981 | Not-determined |
| S8_extracted_k141_22738 | Not-determined |
| S8_extracted_k141_48043 | Not-determined |
| S8_extracted_k141_48044 | Not-determined |
| S8_extracted_k141_13486 | Not-determined |
| S8_extracted_k141_196158 | Not-determined |
| S8_extracted_k141_160762 | Not-determined |
| S8_extracted_k141_152494 | Not-determined |
| S8_extracted_k141_39864 | Not-determined |
| S8_extracted_k141_48048 | Not-determined |
| S8_extracted_k141_152496 | Not-determined |
| S8_extracted_k141_170047 | Not-determined |
| S8_extracted_k141_39868 | Low-quality |
| S8_extracted_k141_83083 | Not-determined |
| S8_extracted_k141_83084 | Not-determined |
| S8_extracted_k141_48051 | Low-quality |
| S8_extracted_k141_13498 | Not-determined |
| S8_extracted_k141_117835 | Not-determined |
| S8_extracted_k141_39872 | Not-determined |
| S8_extracted_k141_196164 | Not-determined |
| S8_extracted_k141_186426 | Low-quality |
| S8_extracted_k141_160772 | Low-quality |
| S8_extracted_k141_178706 | Not-determined |
| S8_extracted_k141_74072 | Not-determined |
| S8_extracted_k141_196167 | Not-determined |
| S8_extracted_k141_22756 | Not-determined |
| S8_extracted_k141_74075 | Not-determined |
| S8_extracted_k141_196169 | Not-determined |
| S8_extracted_k141_196170 | Not-determined |
| S8_extracted_k141_48059 | Low-quality |
| S8_extracted_k141_152498 | Not-determined |
| S8_extracted_k141_48060 | Not-determined |
| S8_extracted_k141_186431 | Not-determined |
| S8_extracted_k141_57408 | Not-determined |
| S8_extracted_k141_170059 | Not-determined |
| S8_extracted_k141_83095 | Not-determined |
| S8_extracted_k141_160778 | Not-determined |
| S8_extracted_k141_152501 | Not-determined |
| S8_extracted_k141_186435 | Not-determined |
| S8_extracted_k141_57411 | Not-determined |
| S8_extracted_k141_13508 | Not-determined |
| S8_extracted_k141_83099 | Not-determined |
| S8_extracted_k141_57412 | Not-determined |
| S8_extracted_k141_109352 | Not-determined |
| S8_extracted_k141_13511 | Not-determined |
| S8_extracted_k141_144159 | Not-determined |
| S8_extracted_k141_152504 | Not-determined |
| S8_extracted_k141_74090 | Not-determined |
| S8_extracted_k141_160786 | Not-determined |
| S8_extracted_k141_152505 | Not-determined |
| S8_extracted_k141_170071 | Not-determined |
| S8_extracted_k141_144161 | Not-determined |
| S8_extracted_k141_83106 | Not-determined |
| S8_extracted_k141_144162 | Low-quality |
| S8_extracted_k141_39893 | Low-quality |
| S8_extracted_k141_160790 | Not-determined |
| S8_extracted_k141_144165 | Not-determined |
| S8_extracted_k141_152507 | Not-determined |
| S8_extracted_k141_144167 | Not-determined |
| S8_extracted_k141_178727 | Not-determined |
| S8_extracted_k141_48073 | Not-determined |
| S8_extracted_k141_13519 | Not-determined |
| S8_extracted_k141_65660 | Low-quality |
| S8_extracted_k141_152508 | Not-determined |
| S8_extracted_k141_65661 | Not-determined |
| S8_extracted_k141_5007 | Low-quality |
| S8_extracted_k141_22763 | Not-determined |
| S8_extracted_k141_57423 | Not-determined |
| S8_extracted_k141_152512 | Not-determined |
| S8_extracted_k141_39904 | Not-determined |
| S8_extracted_k141_196183 | Not-determined |
| S8_extracted_k141_186457 | Not-determined |
| S8_extracted_k141_22766 | Not-determined |
| S8_extracted_k141_48079 | Not-determined |
| S8_extracted_k141_144173 | Not-determined |
| S8_extracted_k141_22767 | Not-determined |
| S8_extracted_k141_22769 | Not-determined |
| S8_extracted_k141_48081 | Not-determined |
| S8_extracted_k141_117846 | Not-determined |
| S8_extracted_k141_39909 | Not-determined |
| S8_extracted_k141_48083 | Not-determined |
| S8_extracted_k141_134996 | Not-determined |
| S8_extracted_k141_117849 | Not-determined |
| S8_extracted_k141_152525 | Not-determined |
| S8_extracted_k141_83116 | Not-determined |
| S8_extracted_k141_160798 | Low-quality |
| S8_extracted_k141_65668 | Low-quality |
| S8_extracted_k141_160800 | Not-determined |
| S8_extracted_k141_186469 | Not-determined |
| S8_extracted_k141_109361 | Not-determined |
| S8_extracted_k141_65671 | Not-determined |
| S8_extracted_k141_22774 | Low-quality |
| S8_extracted_k141_186473 | Not-determined |
| S8_extracted_k141_57436 | Not-determined |
| S8_extracted_k141_160809 | Not-determined |
| S8_extracted_k141_57440 | Not-determined |
| S8_extracted_k141_22782 | Not-determined |
| S8_extracted_k141_204381 | Not-determined |
| S8_extracted_k141_48094 | Not-determined |
| S8_extracted_k141_83124 | Not-determined |
| S8_extracted_k141_5026 | Low-quality |
| S8_extracted_k141_39928 | Not-determined |
| S8_extracted_k141_22788 | Not-determined |
| S8_extracted_k141_22791 | Not-determined |
| S8_extracted_k141_13543 | Not-determined |
| S8_extracted_k141_186484 | Not-determined |
| S8_extracted_k141_152542 | Not-determined |
| S8_extracted_k141_152543 | Low-quality |
| S8_extracted_k141_196200 | Not-determined |
| S8_extracted_k141_109380 | Not-determined |
| S8_extracted_k141_186486 | Low-quality |
| S8_extracted_k141_160827 | Not-determined |
| S8_extracted_k141_160828 | Not-determined |
| S8_extracted_k141_170098 | Not-determined |
| S8_extracted_k141_22796 | Not-determined |
| S8_extracted_k141_83131 | Not-determined |
| S8_extracted_k141_39941 | Not-determined |
| S8_extracted_k141_65675 | Not-determined |
| S8_extracted_k141_117870 | Not-determined |
| S8_extracted_k141_39943 | Not-determined |
| S8_extracted_k141_65681 | Not-determined |
| S8_extracted_k141_91840 | Not-determined |
| S8_extracted_k141_204389 | Not-determined |
| S8_extracted_k141_39945 | Not-determined |
| S8_extracted_k141_39947 | Not-determined |
| S8_extracted_k141_109388 | Not-determined |
| S8_extracted_k141_204393 | Low-quality |
| S8_extracted_k141_5040 | Not-determined |
| S8_extracted_k141_135015 | Not-determined |

| | |
|---|---|
| S8_extracted_k141_170108 | Not-determined |
| S8_extracted_k141_170109 | Not-determined |
| S8_extracted_k141_13550 | Not-determined |
| S8_extracted_k141_109392 | Not-determined |
| S8_extracted_k141_48115 | Not-determined |
| S8_extracted_k141_57453 | Not-determined |
| S8_extracted_k141_13553 | Not-determined |
| S8_extracted_k141_22806 | Not-determined |
| S8_extracted_k141_109398 | Not-determined |
| S8_extracted_k141_39954 | Not-determined |
| S8_extracted_k141_117872 | Not-determined |
| S8_extracted_k141_83141 | Low-quality |
| S8_extracted_k141_117875 | Not-determined |
| S8_extracted_k141_117876 | Not-determined |
| S8_extracted_k141_65704 | Not-determined |
| S8_extracted_k141_91858 | Not-determined |
| S8_extracted_k141_22809 | Not-determined |
| S8_extracted_k141_135027 | Not-determined |
| S8_extracted_k141_117880 | Not-determined |
| S8_extracted_k141_117882 | Not-determined |
| S8_extracted_k141_65708 | Not-determined |
| S8_extracted_k141_83148 | Low-quality |
| S8_extracted_k141_83150 | Not-determined |
| S8_extracted_k141_117884 | Not-determined |
| S8_extracted_k141_48130 | Not-determined |
| S8_extracted_k141_39966 | Not-determined |
| S8_extracted_k141_83151 | Not-determined |
| S8_extracted_k141_13564 | Not-determined |
| S8_extracted_k141_91867 | Not-determined |
| S8_extracted_k141_83153 | Not-determined |
| S8_extracted_k141_186498 | Not-determined |
| S8_extracted_k141_117889 | Not-determined |
| S8_extracted_k141_39975 | Not-determined |
| S8_extracted_k141_117892 | Not-determined |
| S8_extracted_k141_48134 | Not-determined |
| S8_extracted_k141_117893 | Not-determined |
| S8_extracted_k141_91872 | Not-determined |
| S8_extracted_k141_186503 | Not-determined |
| S8_extracted_k141_91873 | Not-determined |
| S8_extracted_k141_65709 | Low-quality |
| S8_extracted_k141_178755 | Not-determined |
| S8_extracted_k141_178756 | Low-quality |
| S8_extracted_k141_204421 | Not-determined |
| S8_extracted_k141_48136 | Not-determined |
| S8_extracted_k141_39981 | Not-determined |
| S8_extracted_k141_65712 | Not-determined |
| S8_extracted_k141_5062 | Not-determined |
| S8_extracted_k141_178760 | Not-determined |
| S8_extracted_k141_22818 | Not-determined |
| S8_extracted_k141_160857 | Not-determined |
| S8_extracted_k141_48141 | Low-quality |
| S8_extracted_k141_170143 | Not-determined |
| S8_extracted_k141_48142 | Not-determined |
| S8_extracted_k141_135048 | Not-determined |
| S8_extracted_k141_204427 | Not-determined |
| S8_extracted_k141_22827 | Not-determined |
| S8_extracted_k141_186517 | Low-quality |
| S8_extracted_k141_91895 | Not-determined |
| S8_extracted_k141_196209 | Not-determined |
| S8_extracted_k141_160859 | Not-determined |
| S8_extracted_k141_170154 | Not-determined |
| S8_extracted_k141_196215 | Not-determined |
| S8_extracted_k141_135060 | Not-determined |
| S8_extracted_k141_40008 | Not-determined |
| S8_extracted_k141_109426 | Not-determined |
| S8_extracted_k141_109427 | Not-determined |
| S8_extracted_k141_204440 | Not-determined |
| S8_extracted_k141_109430 | Not-determined |
| S8_extracted_k141_109432 | Not-determined |
| S8_extracted_k141_13598 | Not-determined |
| S8_extracted_k141_22830 | Not-determined |
| S8_extracted_k141_117896 | Low-quality |
| S8_extracted_k141_13600 | Not-determined |
| S8_extracted_k141_178778 | Not-determined |
| S8_extracted_k141_13602 | Not-determined |
| S8_extracted_k141_57487 | Low-quality |
| S8_extracted_k141_196223 | Not-determined |
| S8_extracted_k141_170163 | Not-determined |
| S8_extracted_k141_135071 | Not-determined |
| S8_extracted_k141_65725 | Not-determined |
| S8_extracted_k141_170167 | Not-determined |
| S8_extracted_k141_83192 | Not-determined |
| S8_extracted_k141_196235 | Not-determined |
| S8_extracted_k141_178788 | Not-determined |
| S8_extracted_k141_117909 | Not-determined |
| S8_extracted_k141_109442 | Not-determined |
| S8_extracted_k141_5083 | Not-determined |
| S8_extracted_k141_13609 | Not-determined |
| S8_extracted_k141_65734 | Not-determined |
| S8_extracted_k141_91912 | Not-determined |
| S8_extracted_k141_65735 | Not-determined |
| S8_extracted_k141_204445 | Not-determined |
| S8_extracted_k141_152570 | Low-quality |
| S8_extracted_k141_109444 | Not-determined |
| S8_extracted_k141_135076 | Not-determined |
| S8_extracted_k141_5086 | Not-determined |
| S8_extracted_k141_91913 | Not-determined |
| S8_extracted_k141_196238 | Not-determined |
| S8_extracted_k141_204447 | Not-determined |
| S8_extracted_k141_65736 | Not-determined |
| S8_extracted_k141_48173 | Not-determined |
| S8_extracted_k141_48174 | Not-determined |
| S8_extracted_k141_170171 | Low-quality |
| S8_extracted_k141_196241 | Not-determined |
| S8_extracted_k141_5090 | Not-determined |
| S8_extracted_k141_48176 | Not-determined |
| S8_extracted_k141_135082 | Not-determined |
| S8_extracted_k141_65743 | Not-determined |
| S8_extracted_k141_204456 | Not-determined |
| S8_extracted_k141_204457 | Not-determined |
| S8_extracted_k141_135085 | Not-determined |
| S8_extracted_k141_57496 | Not-determined |
| S8_extracted_k141_91923 | Low-quality |
| S8_extracted_k141_135087 | Not-determined |
| S8_extracted_k141_170178 | Not-determined |
| S8_extracted_k141_170179 | Not-determined |
| S8_extracted_k141_160882 | Not-determined |
| S8_extracted_k141_57500 | Not-determined |
| S8_extracted_k141_204461 | Not-determined |
| S8_extracted_k141_204462 | Not-determined |
| S8_extracted_k141_186542 | Low-quality |
| S8_extracted_k141_170183 | Not-determined |
| S8_extracted_k141_126597 | Not-determined |
| S8_extracted_k141_178798 | Not-determined |
| S8_extracted_k141_91930 | Not-determined |
| S8_extracted_k141_135089 | Not-determined |
| S8_extracted_k141_152583 | Not-determined |
| S8_extracted_k141_186546 | Not-determined |
| S8_extracted_k141_13629 | Not-determined |
| S8_extracted_k141_65754 | Not-determined |
| S8_extracted_k141_91932 | Not-determined |
| S8_extracted_k141_186549 | Not-determined |
| S8_extracted_k141_22856 | Not-determined |
| S8_extracted_k141_22858 | Not-determined |
| S8_extracted_k141_91937 | Not-determined |
| S8_extracted_k141_186553 | Not-determined |
| S8_extracted_k141_135092 | Not-determined |
| S8_extracted_k141_65760 | Low-quality |
| S8_extracted_k141_83230 | Not-determined |
| S8_extracted_k141_117927 | Not-determined |
| S8_extracted_k141_83233 | Not-determined |
| S8_extracted_k141_57511 | Not-determined |
| S8_extracted_k141_196248 | Not-determined |
| S8_extracted_k141_152598 | Not-determined |
| S8_extracted_k141_160903 | Not-determined |
| S8_extracted_k141_91949 | Not-determined |
| S8_extracted_k141_83241 | Low-quality |
| S8_extracted_k141_170193 | Not-determined |
| S8_extracted_k141_160906 | Not-determined |
| S8_extracted_k141_135103 | Not-determined |
| S8_extracted_k141_204481 | Low-quality |
| S8_extracted_k141_135104 | Not-determined |
| S8_extracted_k141_152607 | Not-determined |
| S8_extracted_k141_178805 | Not-determined |
| S8_extracted_k141_152608 | Not-determined |
| S8_extracted_k141_65769 | Not-determined |
| S8_extracted_k141_57513 | Low-quality |
| S8_extracted_k141_160909 | Not-determined |
| S8_extracted_k141_83245 | Not-determined |
| S8_extracted_k141_135110 | Not-determined |
| S8_extracted_k141_152611 | Not-determined |
| S8_extracted_k141_22887 | Not-determined |
| S8_extracted_k141_31435 | Low-quality |
| S7_vs_112914\|\|full | Medium-quality |
| S8_extracted_k141_22890 | Not-determined |
| S8_extracted_k141_5094 | Not-determined |
| S8_extracted_k141_48201 | Not-determined |
| S8_extracted_k141_144182 | Low-quality |
| S8_extracted_k141_83256 | Low-quality |
| S8_extracted_k141_196263 | Not-determined |
| S8_extracted_k141_117946 | Not-determined |
| S8_extracted_k141_57531 | Not-determined |
| S8_extracted_k141_170209 | Not-determined |
| S8_extracted_k141_160918 | Not-determined |
| S8_extracted_k141_117949 | Not-determined |
| S8_extracted_k141_91962 | Low-quality |
| S8_extracted_k141_152626 | Not-determined |
| S8_extracted_k141_91964 | Not-determined |
| S8_extracted_k141_117951 | Not-determined |
| S8_extracted_k141_22897 | Not-determined |
| S8_extracted_k141_74118 | Not-determined |
| S8_extracted_k141_160923 | Not-determined |
| S8_extracted_k141_40033 | Not-determined |
| S8_extracted_k141_83261 | Not-determined |
| S8_extracted_k141_196278 | Not-determined |
| S8_extracted_k141_22902 | Not-determined |
| S8_extracted_k141_57536 | Not-determined |
| S8_extracted_k141_178839 | Not-determined |
| S8_extracted_k141_48217 | Low-quality |
| S8_extracted_k141_13660 | Not-determined |
| S8_extracted_k141_83262 | Not-determined |
| S8_extracted_k141_57537 | Not-determined |
| S8_extracted_k141_74119 | Low-quality |
| S8_extracted_k141_196281 | Not-determined |
| S8_extracted_k141_40039 | Not-determined |
| S8_extracted_k141_40040 | Not-determined |
| S8_extracted_k141_65779 | Low-quality |
| S8_extracted_k141_5109 | Not-determined |
| S8_extracted_k141_135139 | Low-quality |
| S8_extracted_k141_186576 | Not-determined |
| S8_extracted_k141_109488 | Not-determined |
| S8_extracted_k141_48223 | Low-quality |
| S8_extracted_k141_178848 | Not-determined |
| S8_extracted_k141_74125 | Not-determined |
| S8_extracted_k141_13670 | Not-determined |
| S8_extracted_k141_126602 | Not-determined |
| S8_extracted_k141_126603 | Not-determined |
| S8_extracted_k141_22913 | Not-determined |
| S8_extracted_k141_83271 | Not-determined |
| S8_extracted_k141_13673 | Not-determined |
| S8_extracted_k141_117961 | Not-determined |
| S8_extracted_k141_152644 | Not-determined |
| S8_extracted_k141_22917 | Low-quality |
| S8_extracted_k141_31457 | Low-quality |
| S8_extracted_k141_74133 | Not-determined |
| S8_extracted_k141_178850 | Not-determined |
| S8_extracted_k141_117963 | Low-quality |
| S8_extracted_k141_117964 | Not-determined |
| S8_extracted_k141_117965 | Not-determined |
| S8_extracted_k141_186580 | Not-determined |
| S8_extracted_k141_196299 | Not-determined |
| S8_extracted_k141_74136 | Not-determined |
| S8_extracted_k141_40052 | Not-determined |
| S8_extracted_k141_22921 | Not-determined |
| S8_extracted_k141_160940 | Low-quality |
| S8_extracted_k141_170229 | Not-determined |
| S8_extracted_k141_109494 | Not-determined |
| S8_extracted_k141_117970 | Not-determined |
| S8_extracted_k141_152646 | Not-determined |
| S8_extracted_k141_152647 | Low-quality |
| S8_extracted_k141_160945 | Not-determined |
| S8_extracted_k141_5125 | Not-determined |
| S8_extracted_k141_5127 | Not-determined |
| S8_extracted_k141_65798 | Not-determined |
| S8_extracted_k141_40056 | Not-determined |
| S8_extracted_k141_126625 | Not-determined |
| S8_extracted_k141_170233 | Not-determined |
| S8_extracted_k141_22931 | Not-determined |
| S8_extracted_k141_74143 | Not-determined |
| S8_extracted_k141_152652 | Not-determined |
| S8_extracted_k141_196311 | Low-quality |
| S8_extracted_k141_5130 | Not-determined |
| S8_extracted_k141_74145 | Not-determined |
| S8_extracted_k141_31469 | Low-quality |
| S8_extracted_k141_31472 | Not-determined |
| S8_extracted_k141_57553 | Not-determined |
| S8_extracted_k141_74150 | Not-determined |
| S8_extracted_k141_57554 | Not-determined |
| S8_extracted_k141_186587 | Not-determined |
| S8_extracted_k141_204499 | Low-quality |
| S8_extracted_k141_91984 | Not-determined |
| S8_extracted_k141_178862 | Not-determined |
| S8_extracted_k141_31476 | Not-determined |
| S8_extracted_k141_40059 | Low-quality |
| S8_extracted_k141_160958 | Not-determined |
| S8_extracted_k141_126637 | Not-determined |
| S8_extracted_k141_31480 | Not-determined |
| S8_extracted_k141_170249 | Not-determined |
| S8_extracted_k141_126638 | Not-determined |
| S8_extracted_k141_91993 | Not-determined |
| S8_extracted_k141_74156 | Not-determined |
| S8_extracted_k141_83295 | Not-determined |
| S8_extracted_k141_83296 | Not-determined |
| S8_extracted_k141_135163 | Low-quality |
| S8_extracted_k141_178869 | Not-determined |
| S8_extracted_k141_13695 | Not-determined |
| S8_extracted_k141_5140 | Not-determined |
| S8_extracted_k141_5141 | Not-determined |
| S8_extracted_k141_170262 | Not-determined |
| S8_extracted_k141_170263 | Not-determined |
| S8_extracted_k141_22957 | Not-determined |
| S8_extracted_k141_109502 | Low-quality |
| S8_extracted_k141_65804 | Not-determined |
| S8_extracted_k141_31489 | Not-determined |
| S8_extracted_k141_5145 | Not-determined |
| S8_extracted_k141_117997 | Not-determined |
| S8_extracted_k141_152670 | Not-determined |
| S8_extracted_k141_196332 | Not-determined |
| S8_extracted_k141_13701 | Not-determined |
| S8_extracted_k141_170266 | Not-determined |
| S8_extracted_k141_83304 | Not-determined |
| S8_extracted_k141_152674 | Not-determined |
| S8_extracted_k141_31492 | Low-quality |
| S8_extracted_k141_126647 | Low-quality |
| S8_extracted_k141_160972 | Not-determined |
| S8_extracted_k141_92006 | Not-determined |
| S8_extracted_k141_178881 | Not-determined |
| S8_extracted_k141_178882 | Low-quality |
| S8_extracted_k141_74170 | Not-determined |
| S8_extracted_k141_74171 | Not-determined |
| S8_extracted_k141_135175 | Not-determined |
| S8_extracted_k141_126651 | Not-determined |
| S8_extracted_k141_152679 | Not-determined |
| S8_extracted_k141_22968 | Low-quality |
| S8_extracted_k141_92007 | Not-determined |
| S8_extracted_k141_40074 | Not-determined |
| S8_extracted_k141_13708 | Not-determined |
| S8_extracted_k141_74174 | Not-determined |
| S8_extracted_k141_5156 | Not-determined |
| S8_extracted_k141_22970 | Not-determined |
| S8_extracted_k141_152683 | Not-determined |
| S8_extracted_k141_170277 | Not-determined |
| S8_extracted_k141_118006 | Not-determined |
| S8_extracted_k141_118008 | Not-determined |
| S8_extracted_k141_83312 | Not-determined |
| S8_extracted_k141_152689 | Not-determined |
| S8_extracted_k141_109510 | Not-determined |
| S8_extracted_k141_118010 | Not-determined |
| S8_extracted_k141_83314 | Not-determined |
| S8_extracted_k141_5161 | Not-determined |
| S8_extracted_k141_178890 | Not-determined |
| S8_extracted_k141_109513 | Not-determined |
| S8_extracted_k141_83317 | Low-quality |
| S8_extracted_k141_126662 | Low-quality |
| S8_extracted_k141_92013 | Not-determined |
| S8_extracted_k141_204524 | Not-determined |
| S8_extracted_k141_170280 | Not-determined |
| S8_extracted_k141_109515 | Not-determined |
| S8_extracted_k141_135179 | Not-determined |
| S8_extracted_k141_83319 | Not-determined |
| S8_extracted_k141_5165 | Not-determined |
| S8_extracted_k141_40085 | Not-determined |
| S8_extracted_k141_92017 | Not-determined |
| S8_extracted_k141_74185 | Not-determined |
| S8_extracted_k141_5169 | Not-determined |
| S8_extracted_k141_204525 | Not-determined |
| S8_extracted_k141_31493 | Low-quality |
| S8_extracted_k141_65817 | Not-determined |
| S8_extracted_k141_178900 | Not-determined |
| S8_extracted_k141_57588 | Not-determined |
| S8_extracted_k141_65818 | Not-determined |
| S8_extracted_k141_83324 | Not-determined |
| S8_extracted_k141_109523 | Not-determined |
| S8_extracted_k141_65820 | Low-quality |
| S8_extracted_k141_57592 | Not-determined |
| S8_extracted_k141_109524 | Not-determined |
| S8_extracted_k141_196351 | Not-determined |
| S8_extracted_k141_5177 | Not-determined |
| S8_extracted_k141_160980 | Not-determined |
| S8_extracted_k141_5181 | Not-determined |
| S8_extracted_k141_74196 | Low-quality |
| S8_extracted_k141_160983 | Not-determined |
| S8_extracted_k141_144205 | Not-determined |
| S8_extracted_k141_160987 | Not-determined |
| S8_extracted_k141_83334 | Not-determined |
| S8_extracted_k141_178913 | Not-determined |
| S8_extracted_k141_126681 | Not-determined |
| S8_extracted_k141_178916 | Not-determined |
| S8_extracted_k141_204539 | Not-determined |
| S8_extracted_k141_178917 | Low-quality |
| S8_extracted_k141_5187 | Not-determined |
| S8_extracted_k141_178918 | Not-determined |
| S8_extracted_k141_65832 | Not-determined |
| S8_extracted_k141_92027 | Not-determined |
| S8_extracted_k141_178921 | Not-determined |
| S8_extracted_k141_74202 | Not-determined |
| S8_extracted_k141_74203 | Not-determined |
| S8_extracted_k141_160997 | Not-determined |
| S8_extracted_k141_48229 | Not-determined |
| S8_extracted_k141_161000 | Not-determined |
| S8_extracted_k141_204545 | Not-determined |
| S8_extracted_k141_48231 | Not-determined |
| S8_extracted_k141_74208 | Not-determined |
| S8_extracted_k141_204549 | Not-determined |
| S8_extracted_k141_144212 | Not-determined |
| S8_extracted_k141_170291 | Not-determined |
| S8_extracted_k141_57599 | Not-determined |
| S8_extracted_k141_65840 | Not-determined |
| S8_extracted_k141_92035 | Not-determined |
| S8_extracted_k141_74213 | Low-quality |
| S8_extracted_k141_204551 | Not-determined |
| S8_extracted_k141_126689 | Not-determined |
| S8_extracted_k141_109546 | Not-determined |
| S8_extracted_k141_109554 | Not-determined |
| S8_extracted_k141_204560 | Not-determined |
| S8_extracted_k141_161012 | Not-determined |
| S8_extracted_k141_22986 | Not-determined |
| S8_extracted_k141_178926 | Not-determined |
| S8_extracted_k141_161014 | Not-determined |
| S8_extracted_k141_109561 | Not-determined |
| S8_extracted_k141_161017 | Not-determined |
| S8_extracted_k141_74227 | Not-determined |
| S8_extracted_k141_57615 | Not-determined |
| S8_extracted_k141_65851 | Not-determined |
| S8_extracted_k141_135188 | Not-determined |
| S8_extracted_k141_48237 | Not-determined |
| S8_extracted_k141_170301 | Not-determined |
| S8_extracted_k141_48238 | Not-determined |
| S8_extracted_k141_135192 | Low-quality |
| S8_extracted_k141_31516 | Not-determined |
| S8_extracted_k141_83348 | Not-determined |
| S8_extracted_k141_204565 | Not-determined |
| S8_extracted_k141_65855 | Not-determined |
| S8_extracted_k141_5207 | Not-determined |
| S8_extracted_k141_178935 | Low-quality |
| S8_extracted_k141_178936 | Not-determined |
| S8_extracted_k141_74233 | Not-determined |
| S8_extracted_k141_126709 | Not-determined |
| S8_extracted_k141_92052 | Not-determined |
| S8_extracted_k141_23000 | Not-determined |
| S8_extracted_k141_170307 | Low-quality |
| S8_extracted_k141_161028 | Not-determined |
| S13_vs_28309\|\|full | Medium-quality |
| S8_extracted_k141_204570 | Low-quality |
| S8_extracted_k141_144224 | Not-determined |
| S8_extracted_k141_57629 | Not-determined |
| S8_extracted_k141_48244 | Not-determined |
| S8_extracted_k141_92060 | Low-quality |
| S8_extracted_k141_48246 | Not-determined |
| S8_extracted_k141_92061 | Not-determined |
| S8_extracted_k141_178941 | Not-determined |
| S8_extracted_k141_178942 | Not-determined |
| S8_extracted_k141_23003 | Low-quality |
| S8_extracted_k141_92065 | Not-determined |
| S8_extracted_k141_126719 | Not-determined |
| S8_extracted_k141_83363 | Low-quality |
| S8_extracted_k141_144232 | Not-determined |
| S8_extracted_k141_161037 | Not-determined |
| S8_extracted_k141_109570 | Not-determined |
| S8_extracted_k141_178952 | Not-determined |
| S8_extracted_k141_144234 | Not-determined |
| S8_extracted_k141_152719 | Not-determined |
| S8_extracted_k141_31534 | Not-determined |
| S8_extracted_k141_178955 | Low-quality |
| S8_extracted_k141_48259 | Not-determined |
| S8_extracted_k141_83369 | Not-determined |
| S8_extracted_k141_178960 | Not-determined |
| S8_extracted_k141_170320 | Not-determined |
| S8_extracted_k141_74252 | Not-determined |
| S8_extracted_k141_92072 | Not-determined |
| S8_extracted_k141_57635 | Low-quality |
| S8_extracted_k141_23008 | Not-determined |
| S8_extracted_k141_161044 | Not-determined |
| S8_extracted_k141_178962 | Low-quality |
| S8_extracted_k141_178963 | Not-determined |
| S8_extracted_k141_178964 | Not-determined |
| S8_extracted_k141_178965 | Not-determined |
| S8_extracted_k141_13716 | Not-determined |
| S8_extracted_k141_152731 | Not-determined |
| S8_extracted_k141_57636 | Not-determined |
| S8_extracted_k141_13720 | Not-determined |
| S8_extracted_k141_178967 | Not-determined |
| S8_extracted_k141_135201 | Not-determined |
| S8_extracted_k141_178969 | Not-determined |
| S8_extracted_k141_31543 | Not-determined |
| S8_extracted_k141_126726 | Not-determined |
| S8_extracted_k141_118057 | Not-determined |
| S8_extracted_k141_48266 | Not-determined |
| S8_extracted_k141_57643 | Not-determined |
| S8_extracted_k141_48267 | Not-determined |
| S8_extracted_k141_135204 | Not-determined |
| S8_extracted_k141_144246 | Not-determined |
| S8_extracted_k141_118062 | Not-determined |
| S8_extracted_k141_135207 | Not-determined |
| S8_extracted_k141_57648 | Not-determined |
| S8_extracted_k141_196357 | Not-determined |
| S8_extracted_k141_31553 | Not-determined |
| S8_extracted_k141_83372 | Not-determined |
| S8_extracted_k141_135209 | Not-determined |
| S8_extracted_k141_161063 | Low-quality |
| S8_extracted_k141_161064 | Not-determined |
| S8_extracted_k141_74269 | Not-determined |
| S8_extracted_k141_161066 | Not-determined |
| S8_extracted_k141_92087 | Not-determined |
| S8_extracted_k141_178981 | Not-determined |
| S8_extracted_k141_118067 | Not-determined |
| S8_extracted_k141_92091 | Not-determined |
| S8_extracted_k141_126743 | Not-determined |
| S8_extracted_k141_126744 | Not-determined |
| S8_extracted_k141_152745 | Not-determined |
| S8_extracted_k141_161070 | Not-determined |
| S8_extracted_k141_178987 | Not-determined |
| S8_extracted_k141_178988 | Not-determined |
| S8_extracted_k141_57658 | Low-quality |
| S8_extracted_k141_135212 | Not-determined |
| S8_extracted_k141_204588 | Not-determined |
| S8_extracted_k141_178989 | Not-determined |
| S8_extracted_k141_126746 | Not-determined |
| S8_extracted_k141_178990 | Not-determined |
| S8_extracted_k141_109586 | Not-determined |
| S8_extracted_k141_57659 | Low-quality |
| S8_extracted_k141_196369 | Low-quality |
| S8_extracted_k141_170341 | Not-determined |
| S8_extracted_k141_5212 | Not-determined |
| S8_extracted_k141_135214 | Not-determined |
| S8_extracted_k141_144252 | Not-determined |
| S8_extracted_k141_5214 | Not-determined |
| S8_extracted_k141_170347 | Not-determined |
| S8_extracted_k141_161076 | Not-determined |
| S8_extracted_k141_170349 | Not-determined |
| S8_extracted_k141_118078 | Not-determined |
| S8_extracted_k141_152757 | Not-determined |
| S8_extracted_k141_161080 | Not-determined |
| S8_extracted_k141_152758 | Not-determined |
| S8_extracted_k141_170353 | Not-determined |
| S8_extracted_k141_144258 | Not-determined |
| S8_extracted_k141_126752 | Not-determined |
| S8_extracted_k141_83387 | Not-determined |
| S8_extracted_k141_57669 | Not-determined |
| S8_extracted_k141_179001 | Not-determined |
| S8_extracted_k141_135220 | Not-determined |
| S8_extracted_k141_31561 | Not-determined |
| S8_extracted_k141_135221 | Not-determined |
| S8_extracted_k141_83390 | Not-determined |
| S8_extracted_k141_179006 | Not-determined |
| S8_extracted_k141_126755 | Not-determined |
| S8_extracted_k141_170358 | Not-determined |
| S8_extracted_k141_31564 | Not-determined |
| S8_extracted_k141_57676 | Not-determined |
| S8_extracted_k141_13740 | Not-determined |
| S8_extracted_k141_126764 | Not-determined |
| S8_extracted_k141_31565 | Low-quality |
| S8_extracted_k141_74292 | Not-determined |
| S8_extracted_k141_57678 | Low-quality |
| S8_extracted_k141_144268 | Low-quality |
| S8_extracted_k141_196393 | Not-determined |
| S8_extracted_k141_118092 | Not-determined |
| S8_extracted_k141_144269 | Not-determined |
| S8_extracted_k141_126766 | Not-determined |
| S8_extracted_k141_23021 | Not-determined |
| S8_extracted_k141_179017 | Not-determined |
| S8_extracted_k141_144272 | Not-determined |
| S8_extracted_k141_57683 | Not-determined |
| S8_extracted_k141_179018 | Low-quality |
| S8_extracted_k141_83405 | Not-determined |
| S8_extracted_k141_118100 | Low-quality |
| S8_extracted_k141_31572 | Not-determined |
| S8_extracted_k141_144282 | Not-determined |
| S8_extracted_k141_118104 | Not-determined |
| S8_extracted_k141_48306 | Not-determined |
| S8_extracted_k141_186625 | Not-determined |
| S8_extracted_k141_196401 | Not-determined |
| S8_extracted_k141_152779 | Not-determined |
| S8_extracted_k141_135243 | Low-quality |
| S8_extracted_k141_74301 | Not-determined |
| S8_extracted_k141_74302 | Not-determined |
| S8_extracted_k141_57687 | Not-determined |
| S8_extracted_k141_186629 | Low-quality |
| S8_extracted_k141_152782 | Not-determined |
| S8_extracted_k141_23030 | Not-determined |
| S8_extracted_k141_144293 | Not-determined |
| S8_extracted_k141_5234 | Low-quality |
| S8_extracted_k141_152786 | Not-determined |
| S8_extracted_k141_126776 | Not-determined |
| S8_extracted_k141_161109 | Low-quality |
| S8_extracted_k141_170380 | Not-determined |
| S8_extracted_k141_196416 | Not-determined |
| S8_extracted_k141_83426 | Not-determined |
| S8_extracted_k141_135254 | Not-determined |
| S8_extracted_k141_109616 | Not-determined |
| S8_extracted_k141_31597 | Not-determined |
| S8_extracted_k141_74316 | Not-determined |
| S8_extracted_k141_179042 | Not-determined |
| S8_extracted_k141_92131 | Not-determined |
| S8_extracted_k141_31599 | Not-determined |
| S8_extracted_k141_5246 | Not-determined |
| S8_extracted_k141_57693 | Low-quality |
| S8_extracted_k141_48320 | Not-determined |
| S8_extracted_k141_83436 | Not-determined |
| S8_extracted_k141_92137 | Low-quality |
| S8_extracted_k141_196428 | Not-determined |
| S8_extracted_k141_31603 | Not-determined |
| S8_extracted_k141_31604 | Not-determined |
| S8_extracted_k141_31605 | Low-quality |
| S8_extracted_k141_48325 | Not-determined |
| S8_extracted_k141_109620 | Low-quality |
| S8_extracted_k141_109621 | Not-determined |
| S8_extracted_k141_48326 | Not-determined |
| S8_extracted_k141_170390 | Not-determined |
| S8_extracted_k141_109622 | Not-determined |
| S8_extracted_k141_144311 | Not-determined |
| S8_extracted_k141_48330 | Low-quality |
| S8_extracted_k141_65859 | Not-determined |
| S8_extracted_k141_65860 | Not-determined |
| S8_extracted_k141_74328 | Not-determined |
| S8_extracted_k141_186639 | Not-determined |
| S8_extracted_k141_57701 | Not-determined |
| S8_extracted_k141_5255 | Not-determined |
| S8_extracted_k141_31616 | Not-determined |
| S8_extracted_k141_31617 | Not-determined |
| S8_extracted_k141_135271 | Not-determined |
| S8_extracted_k141_170397 | Not-determined |
| S8_extracted_k141_161124 | Not-determined |
| S8_extracted_k141_204598 | Not-determined |
| S8_extracted_k141_31619 | Not-determined |
| S8_extracted_k141_126792 | Not-determined |
| S17_vs_k141_84456\|\|full | Medium-quality |
| S8_extracted_k141_92144 | Not-determined |
| S8_extracted_k141_144324 | Not-determined |

| | |
|---|---|
| S8_extracted_k141_135278 | Not-determined |
| S8_extracted_k141_196435 | Not-determined |
| S8_extracted_k141_5263 | Not-determined |
| S8_extracted_k141_196436 | Not-determined |
| S8_extracted_k141_65868 | Not-determined |
| S8_extracted_k141_57707 | Not-determined |
| S8_extracted_k141_204602 | Not-determined |
| S8_extracted_k141_13763 | Not-determined |
| S8_extracted_k141_92146 | Not-determined |
| S8_extracted_k141_5266 | Not-determined |
| S8_extracted_k141_92149 | Low-quality |
| S8_extracted_k141_135281 | Not-determined |
| S8_extracted_k141_31626 | Not-determined |
| S8_extracted_k141_126795 | Not-determined |
| S8_extracted_k141_126796 | Not-determined |
| S8_extracted_k141_135285 | Low-quality |
| S8_extracted_k141_135286 | Not-determined |
| S8_extracted_k141_5269 | Not-determined |
| S8_extracted_k141_196445 | Not-determined |
| S8_extracted_k141_5271 | Not-determined |
| S8_extracted_k141_204603 | Not-determined |
| S8_extracted_k141_118153 | Not-determined |
| S8_extracted_k141_144334 | Not-determined |
| S8_extracted_k141_196447 | Not-determined |
| S8_extracted_k141_196448 | Not-determined |
| S8_extracted_k141_126801 | Low-quality |
| S8_extracted_k141_57715 | Not-determined |
| S8_extracted_k141_179057 | Not-determined |
| S8_extracted_k141_186659 | Not-determined |
| S8_extracted_k141_126802 | Not-determined |
| S8_extracted_k141_135293 | Not-determined |
| S8_extracted_k141_57717 | Not-determined |
| S8_extracted_k141_144339 | Not-determined |
| S8_extracted_k141_126806 | Not-determined |
| S8_extracted_k141_57719 | Not-determined |
| S8_extracted_k141_83459 | Not-determined |
| S8_extracted_k141_135295 | Not-determined |
| S8_extracted_k141_109635 | Not-determined |
| S8_extracted_k141_57721 | Not-determined |
| S8_extracted_k141_83460 | Not-determined |
| S8_extracted_k141_100347 | Not-determined |
| S8_extracted_k141_48352 | Not-determined |
| S8_extracted_k141_100348 | Not-determined |
| S8_extracted_k141_5275 | Low-quality |
| S8_extracted_k141_196462 | Not-determined |
| S8_extracted_k141_204607 | Not-determined |
| S8_extracted_k141_144343 | Not-determined |
| S8_extracted_k141_48356 | Not-determined |
| S8_extracted_k141_48358 | Not-determined |
| S8_extracted_k141_161144 | Not-determined |
| S8_extracted_k141_57727 | Not-determined |
| S8_extracted_k141_83465 | Not-determined |
| S8_extracted_k141_92169 | Not-determined |
| S8_extracted_k141_170423 | Not-determined |
| S8_extracted_k141_126814 | Not-determined |
| S8_extracted_k141_144345 | Not-determined |
| S8_extracted_k141_92170 | Not-determined |
| S8_extracted_k141_74340 | Not-determined |
| S8_extracted_k141_5285 | Not-determined |
| S8_extracted_k141_57732 | Not-determined |
| S8_extracted_k141_92171 | Not-determined |
| S8_extracted_k141_109646 | Not-determined |
| S8_extracted_k141_204611 | Not-determined |
| S8_extracted_k141_74344 | Not-determined |
| S8_extracted_k141_196469 | Not-determined |
| S8_extracted_k141_48361 | Not-determined |
| S8_extracted_k141_48362 | Low-quality |
| S8_extracted_k141_144350 | Not-determined |
| S8_extracted_k141_144351 | Low-quality |
| S8_extracted_k141_135308 | Not-determined |
| S8_extracted_k141_152821 | Not-determined |
| S8_extracted_k141_100364 | Low-quality |
| S8_extracted_k141_144353 | Not-determined |
| S8_extracted_k141_204615 | Not-determined |
| S8_extracted_k141_135310 | Not-determined |
| S8_extracted_k141_83469 | Not-determined |
| S8_extracted_k141_135311 | Not-determined |
| S8_extracted_k141_144356 | Not-determined |
| S8_extracted_k141_74348 | Not-determined |
| S8_extracted_k141_92179 | Not-determined |
| S8_extracted_k141_204620 | Not-determined |
| S8_extracted_k141_83473 | Not-determined |
| S8_extracted_k141_126821 | Not-determined |
| S8_extracted_k141_48368 | Not-determined |
| S8_extracted_k141_144360 | Not-determined |
| S8_extracted_k141_204624 | Not-determined |
| S8_extracted_k141_57745 | Not-determined |
| S8_extracted_k141_204627 | Not-determined |
| S8_extracted_k141_92184 | Not-determined |
| S8_extracted_k141_74354 | Not-determined |
| S8_extracted_k141_100379 | Not-determined |
| S8_extracted_k141_135316 | Not-determined |
| S8_extracted_k141_5297 | Not-determined |
| S8_extracted_k141_31645 | Low-quality |
| S8_extracted_k141_100382 | Not-determined |
| S8_extracted_k141_83476 | Not-determined |
| S8_extracted_k141_23062 | Not-determined |
| S8_extracted_k141_31647 | Not-determined |
| S8_extracted_k141_152833 | Not-determined |
| S8_extracted_k141_40099 | Low-quality |
| S8_extracted_k141_161148 | Not-determined |
| S8_extracted_k141_161149 | Not-determined |
| S8_extracted_k141_118181 | Not-determined |
| S8_extracted_k141_23064 | Not-determined |
| S8_extracted_k141_57751 | Not-determined |
| S8_extracted_k141_92192 | Not-determined |
| S8_extracted_k141_57752 | Not-determined |
| S8_extracted_k141_83480 | Not-determined |
| S8_extracted_k141_23066 | Not-determined |
| S8_extracted_k141_92196 | Not-determined |
| S8_extracted_k141_204632 | Not-determined |
| S8_extracted_k141_83484 | Not-determined |
| S8_extracted_k141_83485 | Not-determined |
| S8_extracted_k141_23070 | Not-determined |
| S8_extracted_k141_74363 | Not-determined |
| S8_extracted_k141_161154 | Not-determined |
| S8_extracted_k141_40106 | Not-determined |
| S8_extracted_k141_23071 | Not-determined |
| S8_extracted_k141_92198 | Not-determined |
| S8_extracted_k141_83487 | Not-determined |
| S8_extracted_k141_144367 | Not-determined |
| S8_extracted_k141_65902 | Not-determined |
| S8_extracted_k141_161158 | Not-determined |
| S8_extracted_k141_135324 | Not-determined |
| S8_extracted_k141_126823 | Not-determined |
| S8_extracted_k141_118189 | Not-determined |
| S8_extracted_k141_23074 | Not-determined |
| S8_extracted_k141_118191 | Not-determined |
| S8_extracted_k141_161162 | Not-determined |
| S8_extracted_k141_126825 | Not-determined |
| S8_extracted_k141_161163 | Not-determined |
| S8_extracted_k141_126826 | Not-determined |
| S8_extracted_k141_196492 | Low-quality |
| S8_extracted_k141_31660 | Low-quality |
| S8_extracted_k141_100395 | Not-determined |
| S8_extracted_k141_118195 | Not-determined |
| S8_extracted_k141_144376 | Not-determined |
| S8_extracted_k141_5317 | Not-determined |
| S8_extracted_k141_48373 | Low-quality |
| S8_extracted_k141_100398 | Not-determined |
| S8_extracted_k141_57771 | Not-determined |
| S8_extracted_k141_13772 | Not-determined |
| S8_extracted_k141_40114 | Not-determined |
| S8_extracted_k141_100400 | Not-determined |
| S8_extracted_k141_152849 | Not-determined |
| S8_extracted_k141_65914 | Not-determined |
| S8_extracted_k141_13774 | Not-determined |
| S8_extracted_k141_23082 | Not-determined |
| S8_extracted_k141_13775 | Not-determined |
| S8_extracted_k141_31670 | Not-determined |
| S8_extracted_k141_100403 | Not-determined |
| S8_extracted_k141_23084 | Not-determined |
| S8_extracted_k141_48384 | Not-determined |
| S8_extracted_k141_135330 | Not-determined |
| S8_extracted_k141_100404 | Not-determined |
| S8_extracted_k141_100405 | Low-quality |
| S8_extracted_k141_57777 | Not-determined |
| S8_extracted_k141_100407 | Low-quality |
| S8_extracted_k141_48385 | Not-determined |
| S8_extracted_k141_135333 | Not-determined |
| S8_extracted_k141_144381 | Not-determined |
| S8_extracted_k141_179092 | Not-determined |
| S8_extracted_k141_144382 | Not-determined |
| S8_extracted_k141_31679 | Not-determined |
| S8_extracted_k141_5337 | Not-determined |
| S8_extracted_k141_5338 | Not-determined |
| S8_extracted_k141_100418 | Not-determined |
| S8_extracted_k141_196503 | Not-determined |
| S8_extracted_k141_48396 | Not-determined |
| S8_extracted_k141_196504 | Not-determined |
| S8_extracted_k141_161172 | Not-determined |
| S8_extracted_k141_179095 | Not-determined |
| S8_extracted_k141_23093 | Not-determined |
| S8_extracted_k141_179098 | Not-determined |
| S8_extracted_k141_179099 | Not-determined |
| S8_extracted_k141_48405 | Not-determined |
| S8_extracted_k141_83503 | Not-determined |
| S8_extracted_k141_196511 | Not-determined |
| S8_extracted_k141_31696 | Not-determined |
| S8_extracted_k141_161181 | Not-determined |
| S8_extracted_k141_31697 | Not-determined |
| S8_extracted_k141_83507 | Not-determined |
| S8_extracted_k141_135348 | Not-determined |
| S8_extracted_k141_83508 | Not-determined |
| S8_extracted_k141_74400 | Not-determined |
| S8_extracted_k141_31700 | Low-quality |
| S8_extracted_k141_152863 | Low-quality |
| S8_extracted_k141_135353 | Not-determined |
| S8_extracted_k141_48411 | Not-determined |
| S8_extracted_k141_204651 | Not-determined |
| S8_extracted_k141_179106 | Low-quality |
| S8_extracted_k141_74408 | Not-determined |
| S8_extracted_k141_186694 | Not-determined |
| S8_extracted_k141_204654 | Not-determined |
| S8_extracted_k141_57782 | Not-determined |
| S8_extracted_k141_179109 | Not-determined |
| S8_extracted_k141_161187 | Not-determined |
| S8_extracted_k141_186697 | Not-determined |
| S8_extracted_k141_161188 | Not-determined |
| S8_extracted_k141_152873 | Not-determined |
| S8_extracted_k141_31711 | Not-determined |
| S8_extracted_k141_57787 | Not-determined |
| S8_extracted_k141_48422 | Not-determined |
| S8_extracted_k141_5343 | Low-quality |
| S8_extracted_k141_204664 | Not-determined |
| S8_extracted_k141_92205 | Not-determined |
| S8_extracted_k141_186709 | Not-determined |
| S8_extracted_k141_152878 | Not-determined |
| S8_extracted_k141_135364 | Not-determined |
| S8_extracted_k141_204667 | Not-determined |
| S8_extracted_k141_92210 | Not-determined |
| S8_extracted_k141_186712 | Not-determined |
| S8_extracted_k141_118218 | Not-determined |
| S8_extracted_k141_48430 | Not-determined |
| S8_extracted_k141_100423 | Not-determined |
| S8_extracted_k141_109667 | Not-determined |
| S8_extracted_k141_100424 | Not-determined |
| S8_extracted_k141_179121 | Low-quality |
| S8_extracted_k141_31716 | Not-determined |
| S8_extracted_k141_109669 | Not-determined |
| S8_extracted_k141_204672 | Low-quality |
| S8_extracted_k141_31719 | Not-determined |
| S8_extracted_k141_144392 | Not-determined |
| S8_extracted_k141_100426 | Not-determined |
| S8_extracted_k141_31722 | Not-determined |
| S8_extracted_k141_186721 | Not-determined |
| S8_extracted_k141_144393 | Low-quality |
| S8_extracted_k141_109673 | Not-determined |
| S8_extracted_k141_5348 | Not-determined |
| S8_extracted_k141_161200 | Not-determined |
| S8_extracted_k141_100427 | Not-determined |
| S8_extracted_k141_40153 | Not-determined |
| S8_extracted_k141_135367 | Not-determined |
| S8_extracted_k141_57799 | Not-determined |
| S8_extracted_k141_196534 | Not-determined |
| S8_extracted_k141_31727 | Not-determined |
| S8_extracted_k141_144396 | Not-determined |
| S8_extracted_k141_5353 | Not-determined |
| S8_extracted_k141_161204 | Not-determined |
| S2_vs_k141_17850\|\|full | Medium-quality |
| S8_extracted_k141_144397 | Not-determined |
| S8_extracted_k141_161206 | Not-determined |
| S8_extracted_k141_170431 | Not-determined |
| S8_extracted_k141_144398 | Not-determined |
| S8_extracted_k141_74430 | Not-determined |
| S8_extracted_k141_57802 | Not-determined |
| S8_extracted_k141_31730 | Not-determined |
| S8_extracted_k141_5357 | Not-determined |
| S8_extracted_k141_179125 | Not-determined |
| S8_extracted_k141_100435 | Not-determined |
| S8_extracted_k141_161211 | Not-determined |
| S8_extracted_k141_196542 | Not-determined |
| S8_extracted_k141_31734 | Not-determined |
| S8_extracted_k141_144405 | Not-determined |
| S8_extracted_k141_57807 | Not-determined |
| S8_extracted_k141_204674 | Not-determined |
| S8_extracted_k141_48452 | Not-determined |
| S8_extracted_k141_5361 | Not-determined |
| S8_extracted_k141_100440 | Not-determined |
| S8_extracted_k141_92218 | Not-determined |
| S8_extracted_k141_135370 | Not-determined |
| S8_extracted_k141_57808 | Low-quality |
| S8_extracted_k141_135371 | Not-determined |
| S8_extracted_k141_109685 | Not-determined |
| S8_extracted_k141_48455 | Not-determined |
| S8_extracted_k141_40162 | Not-determined |
| S8_extracted_k141_48457 | Not-determined |
| S8_extracted_k141_196548 | Not-determined |
| S8_extracted_k141_161219 | Not-determined |
| S8_extracted_k141_179138 | Not-determined |
| S8_extracted_k141_40165 | Not-determined |
| S8_extracted_k141_13806 | Not-determined |
| S8_extracted_k141_135376 | Not-determined |
| S8_extracted_k141_186739 | Not-determined |
| S8_extracted_k141_135379 | Not-determined |
| S8_extracted_k141_196555 | Not-determined |
| S8_extracted_k141_31740 | Not-determined |
| S8_extracted_k141_152899 | Not-determined |
| S8_extracted_k141_152900 | Not-determined |
| S8_extracted_k141_65945 | Not-determined |
| S8_extracted_k141_5380 | Low-quality |
| S8_extracted_k141_100457 | Not-determined |
| S8_extracted_k141_100458 | Low-quality |
| S8_extracted_k141_5381 | Not-determined |
| S8_extracted_k141_144419 | Not-determined |
| S8_extracted_k141_126853 | Not-determined |
| S8_extracted_k141_109699 | Not-determined |
| S8_extracted_k141_31748 | Not-determined |
| S8_extracted_k141_204695 | Not-determined |
| S8_extracted_k141_161225 | Not-determined |
| S8_extracted_k141_118240 | Not-determined |
| S8_extracted_k141_161227 | Not-determined |
| S8_extracted_k141_92229 | Not-determined |
| S8_extracted_k141_40177 | Not-determined |
| S8_extracted_k141_170438 | Not-determined |
| S8_extracted_k141_196571 | Not-determined |
| S8_extracted_k141_161231 | Not-determined |
| S8_extracted_k141_57824 | Not-determined |
| S8_extracted_k141_196573 | Low-quality |
| S8_extracted_k141_152902 | Low-quality |
| S8_extracted_k141_100471 | Not-determined |
| S8_extracted_k141_186756 | Not-determined |
| S8_extracted_k141_135391 | Not-determined |
| S8_extracted_k141_186757 | Not-determined |
| S8_extracted_k141_57832 | Not-determined |
| S8_extracted_k141_65964 | Not-determined |
| S8_extracted_k141_100476 | Not-determined |
| S8_extracted_k141_204707 | Not-determined |
| S8_extracted_k141_40182 | Not-determined |
| S8_extracted_k141_57833 | Not-determined |
| S8_extracted_k141_152908 | Not-determined |
| S8_extracted_k141_100478 | Not-determined |
| S8_extracted_k141_118251 | Not-determined |
| S8_extracted_k141_74453 | Not-determined |
| S8_extracted_k141_92240 | Low-quality |
| S8_extracted_k141_186763 | Not-determined |
| S8_extracted_k141_186765 | Not-determined |
| S8_extracted_k141_57838 | Not-determined |
| S8_extracted_k141_204712 | Not-determined |
| S8_extracted_k141_152914 | Not-determined |
| S8_extracted_k141_118253 | Not-determined |
| S8_extracted_k141_186767 | Not-determined |
| S8_extracted_k141_65974 | Low-quality |
| S8_extracted_k141_170449 | Not-determined |
| S8_extracted_k141_179175 | Not-determined |
| S8_extracted_k141_118255 | Not-determined |
| S8_extracted_k141_152919 | Not-determined |
| S8_extracted_k141_196581 | Not-determined |
| S8_extracted_k141_152920 | Not-determined |
| S8_extracted_k141_118257 | Not-determined |
| S8_extracted_k141_100490 | Not-determined |
| S8_extracted_k141_204713 | Not-determined |
| S8_extracted_k141_179179 | Low-quality |
| S8_extracted_k141_170455 | Not-determined |
| S8_extracted_k141_186771 | Not-determined |
| S8_extracted_k141_204714 | Not-determined |
| S8_extracted_k141_179182 | Not-determined |
| S8_extracted_k141_135413 | Not-determined |
| S8_extracted_k141_40203 | Not-determined |
| S8_extracted_k141_13812 | Not-determined |
| S8_extracted_k141_65986 | Not-determined |
| S8_extracted_k141_100503 | Not-determined |
| S8_extracted_k141_186777 | Not-determined |
| S8_extracted_k141_109701 | Not-determined |
| S8_extracted_k141_135419 | Not-determined |
| S8_extracted_k141_5393 | Not-determined |
| S8_extracted_k141_204720 | Not-determined |
| S8_extracted_k141_100509 | Not-determined |
| S8_extracted_k141_83528 | Not-determined |
| S8_extracted_k141_13816 | Not-determined |
| S8_extracted_k141_48468 | Not-determined |
| S8_extracted_k141_40219 | Not-determined |
| S8_extracted_k141_13818 | Not-determined |
| S8_extracted_k141_65990 | Low-quality |
| S8_extracted_k141_179187 | Not-determined |
| S8_extracted_k141_40222 | Not-determined |
| S8_extracted_k141_83535 | Not-determined |
| S8_extracted_k141_31758 | Low-quality |
| S8_extracted_k141_179190 | Not-determined |
| S8_extracted_k141_196599 | Not-determined |
| S8_extracted_k141_74464 | Not-determined |
| S8_extracted_k141_57858 | Not-determined |
| S8_extracted_k141_152934 | Not-determined |
| S8_extracted_k141_186786 | Not-determined |
| S8_extracted_k141_74467 | Not-determined |
| S8_extracted_k141_186787 | Not-determined |
| S8_extracted_k141_74468 | Not-determined |
| S8_extracted_k141_31761 | Low-quality |
| S8_extracted_k141_152936 | Not-determined |
| S8_extracted_k141_48473 | Not-determined |
| S8_extracted_k141_126857 | Not-determined |
| S8_extracted_k141_92270 | Not-determined |
| S8_extracted_k141_100513 | Low-quality |
| S8_extracted_k141_48476 | Not-determined |
| S8_extracted_k141_40228 | Low-quality |
| S8_extracted_k141_186789 | Not-determined |
| S8_extracted_k141_100516 | Not-determined |
| S8_extracted_k141_57867 | Not-determined |
| S8_extracted_k141_118279 | Not-determined |
| S8_extracted_k141_100518 | Not-determined |
| S8_extracted_k141_204730 | Not-determined |
| S8_extracted_k141_92274 | Not-determined |
| S8_extracted_k141_118281 | Not-determined |
| S8_extracted_k141_57868 | Low-quality |
| S8_extracted_k141_135437 | Not-determined |
| S8_extracted_k141_13821 | Not-determined |
| S8_extracted_k141_109709 | Not-determined |
| S8_extracted_k141_57870 | Not-determined |
| S8_extracted_k141_40235 | Not-determined |
| S8_extracted_k141_109711 | Not-determined |
| S8_extracted_k141_170462 | Not-determined |
| S8_extracted_k141_74475 | Low-quality |
| S8_extracted_k141_66001 | Not-determined |
| S8_extracted_k141_170463 | Not-determined |
| S8_extracted_k141_152942 | Not-determined |
| S8_extracted_k141_83552 | Not-determined |
| S8_extracted_k141_152943 | Not-determined |
| S8_extracted_k141_74476 | Not-determined |
| S8_extracted_k141_83553 | Not-determined |
| S8_extracted_k141_92282 | Low-quality |
| S8_extracted_k141_57871 | Not-determined |
| S8_extracted_k141_135443 | Not-determined |
| S8_extracted_k141_92285 | Not-determined |
| S8_extracted_k141_152947 | Low-quality |
| S8_extracted_k141_23151 | Not-determined |
| S8_extracted_k141_152948 | Not-determined |
| S8_extracted_k141_135449 | Not-determined |
| S8_extracted_k141_13825 | Not-determined |
| S8_extracted_k141_48483 | Not-determined |
| S8_extracted_k141_186800 | Not-determined |
| S8_extracted_k141_23154 | Not-determined |
| S8_extracted_k141_186801 | Not-determined |
| S8_extracted_k141_83560 | Not-determined |
| S8_extracted_k141_179199 | Not-determined |
| S8_extracted_k141_92290 | Not-determined |
| S8_extracted_k141_179201 | Not-determined |
| S8_extracted_k141_161268 | Not-determined |
| S8_extracted_k141_161269 | Not-determined |
| S8_extracted_k141_57882 | Not-determined |
| S8_extracted_k141_100526 | Not-determined |
| S8_extracted_k141_92293 | Not-determined |
| S8_extracted_k141_5411 | Not-determined |
| S8_extracted_k141_100529 | Not-determined |
| S8_extracted_k141_5412 | Not-determined |
| S8_extracted_k141_48488 | Not-determined |
| S8_extracted_k141_109714 | Not-determined |
| S8_extracted_k141_40244 | Not-determined |
| S8_extracted_k141_40245 | Not-determined |
| S8_extracted_k141_40246 | Not-determined |
| S8_extracted_k141_57890 | Not-determined |
| S8_extracted_k141_23166 | Not-determined |
| S8_extracted_k141_23167 | Not-determined |
| S8_extracted_k141_126880 | Not-determined |
| S8_extracted_k141_196611 | Not-determined |
| S8_extracted_k141_100533 | Low-quality |
| S8_extracted_k141_13841 | Not-determined |
| S8_extracted_k141_40250 | Not-determined |
| S8_extracted_k141_48493 | Not-determined |
| S8_extracted_k141_92298 | Not-determined |
| S8_extracted_k141_5421 | Not-determined |
| S8_extracted_k141_13842 | Not-determined |
| S8_extracted_k141_23171 | Not-determined |
| S8_extracted_k141_31795 | Not-determined |
| S8_extracted_k141_118305 | Not-determined |
| S8_extracted_k141_23174 | Not-determined |
| S8_extracted_k141_31797 | Not-determined |
| S8_extracted_k141_74497 | Not-determined |
| S8_extracted_k141_170480 | Not-determined |
| S8_extracted_k141_196619 | Not-determined |
| S8_extracted_k141_13849 | Not-determined |
| S8_extracted_k141_5428 | Not-determined |
| S8_extracted_k141_100543 | Not-determined |
| S8_extracted_k141_23178 | Not-determined |
| S8_extracted_k141_100545 | Not-determined |
| S8_extracted_k141_170483 | Not-determined |
| S8_extracted_k141_48502 | Not-determined |
| S8_extracted_k141_196623 | Not-determined |
| S8_extracted_k141_40258 | Not-determined |
| S8_extracted_k141_31803 | Not-determined |
| S8_extracted_k141_161279 | Not-determined |
| S8_extracted_k141_196626 | Not-determined |
| S8_extracted_k141_5433 | Not-determined |
| S8_extracted_k141_5434 | Not-determined |
| S8_extracted_k141_100550 | Not-determined |
| S8_extracted_k141_118313 | Not-determined |
| S8_extracted_k141_100551 | Not-determined |
| S8_extracted_k141_126886 | Not-determined |
| S8_extracted_k141_66005 | Not-determined |
| S8_extracted_k141_5435 | Not-determined |
| S8_extracted_k141_83577 | Not-determined |
| S8_extracted_k141_23182 | Not-determined |
| S8_extracted_k141_13855 | Not-determined |
| S8_extracted_k141_100556 | Not-determined |
| S8_extracted_k141_152979 | Low-quality |
| S8_extracted_k141_66007 | Low-quality |
| S8_extracted_k141_31807 | Low-quality |
| S8_extracted_k141_83582 | Not-determined |
| S8_extracted_k141_126891 | Not-determined |
| S8_extracted_k141_5437 | Not-determined |
| S8_extracted_k141_74507 | Not-determined |
| S8_extracted_k141_196629 | Not-determined |
| S8_extracted_k141_23190 | Not-determined |
| S8_extracted_k141_66009 | Low-quality |
| S8_extracted_k141_170495 | Not-determined |
| S8_extracted_k141_100563 | Not-determined |
| S8_extracted_k141_186821 | Not-determined |
| S8_extracted_k141_83587 | Low-quality |
| S8_extracted_k141_5445 | Not-determined |
| S8_extracted_k141_126900 | Low-quality |
| S8_extracted_k141_83589 | Not-determined |
| S8_extracted_k141_100567 | Low-quality |
| S8_extracted_k141_135491 | Not-determined |
| S8_extracted_k141_126903 | Low-quality |
| S8_extracted_k141_74516 | Not-determined |
| S8_extracted_k141_13871 | Not-determined |
| S8_extracted_k141_135493 | Not-determined |
| S8_extracted_k141_152997 | Not-determined |
| S8_extracted_k141_135495 | Not-determined |
| S8_extracted_k141_196646 | Not-determined |
| S8_extracted_k141_118328 | Not-determined |
| S8_extracted_k141_126910 | Not-determined |
| S8_extracted_k141_13880 | Not-determined |
| S8_extracted_k141_153004 | Not-determined |
| S8_extracted_k141_48530 | Not-determined |
| S8_extracted_k141_40271 | Not-determined |
| S8_extracted_k141_118329 | Not-determined |
| S8_extracted_k141_186832 | Not-determined |
| S8_extracted_k141_109733 | Not-determined |
| S8_extracted_k141_144432 | Not-determined |
| S8_extracted_k141_126915 | Not-determined |
| S8_extracted_k141_40274 | Not-determined |
| S8_extracted_k141_118333 | Not-determined |
| S8_extracted_k141_196647 | Not-determined |
| S8_extracted_k141_118335 | Not-determined |
| S8_extracted_k141_126918 | Not-determined |
| S8_extracted_k141_153010 | Not-determined |
| S8_extracted_k141_186834 | Not-determined |
| S8_extracted_k141_31836 | Not-determined |
| S8_extracted_k141_144435 | Not-determined |
| S8_extracted_k141_118338 | Not-determined |
| S8_extracted_k141_135503 | Not-determined |
| S8_extracted_k141_186837 | Not-determined |
| S8_extracted_k141_92341 | Not-determined |
| S8_extracted_k141_109743 | Not-determined |
| S8_extracted_k141_31841 | Low-quality |
| S8_extracted_k141_161304 | Not-determined |
| S8_extracted_k141_170513 | Not-determined |
| S8_extracted_k141_13891 | Not-determined |
| S8_extracted_k141_92344 | Not-determined |
| S8_extracted_k141_31846 | Not-determined |
| S8_extracted_k141_153019 | Not-determined |
| S8_extracted_k141_170517 | Not-determined |
| S8_extracted_k141_135512 | Low-quality |
| S8_extracted_k141_83606 | Not-determined |
| S8_extracted_k141_66029 | Not-determined |
| S8_extracted_k141_135515 | Not-determined |
| S8_extracted_k141_161313 | Not-determined |
| S8_extracted_k141_170519 | Not-determined |
| S8_extracted_k141_153027 | Not-determined |
| S8_extracted_k141_109753 | Not-determined |
| S8_extracted_k141_153028 | Not-determined |
| S8_extracted_k141_118355 | Not-determined |
| S8_extracted_k141_48559 | Not-determined |
| S8_extracted_k141_66035 | Not-determined |
| S8_extracted_k141_83618 | Not-determined |
| S8_extracted_k141_31853 | Not-determined |
| S8_extracted_k141_66037 | Low-quality |
| S8_extracted_k141_100587 | Not-determined |
| S8_extracted_k141_186858 | Not-determined |
| S8_extracted_k141_74534 | Not-determined |
| S8_extracted_k141_118364 | Not-determined |
| S8_extracted_k141_161328 | Not-determined |
| S8_extracted_k141_74539 | Not-determined |
| S8_extracted_k141_100593 | Low-quality |
| S8_extracted_k141_135521 | Not-determined |
| S8_extracted_k141_40284 | Not-determined |
| S8_extracted_k141_92352 | Not-determined |
| S8_extracted_k141_66043 | Not-determined |
| S8_extracted_k141_57951 | Low-quality |
| S8_extracted_k141_31861 | Not-determined |
| S8_extracted_k141_126932 | Not-determined |
| S8_extracted_k141_66048 | Low-quality |
| S8_extracted_k141_5483 | Not-determined |
| S8_extracted_k141_170539 | Not-determined |
| S8_extracted_k141_40296 | Not-determined |
| S8_extracted_k141_161333 | Not-determined |
| S8_extracted_k141_74550 | Not-determined |
| S8_extracted_k141_153046 | Not-determined |
| S8_extracted_k141_5485 | Not-determined |
| S8_extracted_k141_109763 | Not-determined |
| S8_extracted_k141_126939 | Low-quality |
| S8_extracted_k141_170545 | Not-determined |
| S8_extracted_k141_126940 | Not-determined |
| S8_extracted_k141_92361 | Not-determined |
| S8_extracted_k141_170549 | Not-determined |
| S8_extracted_k141_126941 | Not-determined |
| S8_extracted_k141_170551 | Not-determined |
| S8_extracted_k141_100605 | Not-determined |
| S8_extracted_k141_40307 | Not-determined |
| S8_extracted_k141_40308 | Not-determined |
| S8_extracted_k141_57964 | Not-determined |
| S8_extracted_k141_5495 | Not-determined |
| S8_extracted_k141_135532 | Low-quality |
| S8_extracted_k141_92366 | Not-determined |
| S8_extracted_k141_57969 | Not-determined |
| S8_extracted_k141_5500 | Not-determined |
| S8_extracted_k141_5502 | Not-determined |
| S8_extracted_k141_135536 | Not-determined |
| S8_extracted_k141_92367 | Low-quality |

| | |
|---|---|
| S8_extracted_k141_161336 | Low-quality |
| S8_extracted_k141_118374 | Not-determined |
| S8_extracted_k141_153061 | Not-determined |
| S8_extracted_k141_66054 | Low-quality |
| S8_extracted_k141_144466 | Not-determined |
| S8_extracted_k141_92370 | Not-determined |
| S8_extracted_k141_144468 | Not-determined |
| S8_extracted_k141_186869 | Not-determined |
| S8_extracted_k141_144469 | Not-determined |
| S8_extracted_k141_92374 | Not-determined |
| S8_extracted_k141_170561 | Not-determined |
| S8_extracted_k141_100620 | Not-determined |
| S8_extracted_k141_31874 | Not-determined |
| S8_extracted_k141_74566 | Not-determined |
| S8_extracted_k141_135549 | Not-determined |
| S8_extracted_k141_31877 | Low-quality |
| S7_vs_69348\|\|full | Medium-quality |
| S8_extracted_k141_5506 | Not-determined |
| S8_extracted_k141_196672 | Not-determined |
| S8_extracted_k141_186879 | Not-determined |
| S8_extracted_k141_40315 | Not-determined |
| S8_extracted_k141_135555 | Not-determined |
| S8_extracted_k141_109780 | Not-determined |
| S8_extracted_k141_100624 | Low-quality |
| S8_extracted_k141_5509 | Low-quality |
| S8_extracted_k141_5510 | Not-determined |
| S8_extracted_k141_161339 | Low-quality |
| S8_extracted_k141_5513 | Low-quality |
| S8_extracted_k141_118387 | Not-determined |
| S8_extracted_k141_92380 | Not-determined |
| S8_extracted_k141_100626 | Low-quality |
| S8_extracted_k141_118388 | Not-determined |
| S8_extracted_k141_92382 | Not-determined |
| S8_extracted_k141_161345 | Not-determined |
| S8_extracted_k141_66068 | Not-determined |
| S8_extracted_k141_170569 | Not-determined |
| S8_extracted_k141_40320 | Not-determined |
| S8_extracted_k141_13910 | Low-quality |
| S8_extracted_k141_100631 | Not-determined |
| S8_extracted_k141_135563 | Not-determined |
| S8_extracted_k141_109794 | Not-determined |
| S8_extracted_k141_153084 | Not-determined |
| S8_extracted_k141_66078 | Not-determined |
| S8_extracted_k141_66079 | Not-determined |
| S8_extracted_k141_40327 | Not-determined |
| S8_extracted_k141_40328 | Not-determined |
| S8_extracted_k141_186886 | Low-quality |
| S8_extracted_k141_57979 | Not-determined |
| S8_extracted_k141_109799 | Not-determined |
| S8_extracted_k141_161359 | Not-determined |
| S8_extracted_k141_126965 | Not-determined |
| S8_extracted_k141_204777 | Not-determined |
| S8_extracted_k141_153097 | Low-quality |
| S8_extracted_k141_109802 | Not-determined |
| S8_extracted_k141_92394 | Not-determined |
| S8_extracted_k141_100636 | Not-determined |
| S8_extracted_k141_126969 | Not-determined |
| S8_extracted_k141_153099 | Low-quality |
| S8_extracted_k141_161364 | Not-determined |
| S8_extracted_k141_74590 | Not-determined |
| S8_extracted_k141_161365 | Not-determined |
| S8_extracted_k141_144485 | Not-determined |
| S8_extracted_k141_40335 | Not-determined |
| S8_extracted_k141_126971 | Not-determined |
| S8_extracted_k141_144486 | Not-determined |
| S8_extracted_k141_5524 | Not-determined |
| S8_extracted_k141_170582 | Not-determined |
| S8_extracted_k141_204780 | Not-determined |
| S8_extracted_k141_126975 | Not-determined |
| S8_extracted_k141_57986 | Not-determined |
| S8_extracted_k141_109814 | Not-determined |
| S8_extracted_k141_118413 | Not-determined |
| S8_extracted_k141_126977 | Not-determined |
| S8_extracted_k141_153106 | Not-determined |
| S8_extracted_k141_126979 | Not-determined |
| S8_extracted_k141_57990 | Not-determined |
| S8_extracted_k141_170586 | Not-determined |
| S8_extracted_k141_109817 | Not-determined |
| S8_extracted_k141_126982 | Not-determined |
| S8_extracted_k141_100648 | Not-determined |
| S8_extracted_k141_186896 | Not-determined |
| S8_extracted_k141_100650 | Not-determined |
| S8_extracted_k141_74601 | Not-determined |
| S8_extracted_k141_186898 | Not-determined |
| S8_extracted_k141_100652 | Not-determined |
| S8_extracted_k141_186900 | Not-determined |
| S8_extracted_k141_118417 | Low-quality |
| S8_extracted_k141_135581 | Not-determined |
| S8_extracted_k141_161373 | Low-quality |
| S8_extracted_k141_40343 | Not-determined |
| S8_extracted_k141_204789 | Not-determined |
| S8_extracted_k141_118420 | Not-determined |
| S8_extracted_k141_57995 | Not-determined |
| S8_extracted_k141_153109 | Not-determined |
| S8_extracted_k141_204791 | Not-determined |
| S8_extracted_k141_144499 | Not-determined |
| S8_extracted_k141_135583 | Not-determined |
| S8_extracted_k141_40348 | Not-determined |
| S8_extracted_k141_153113 | Low-quality |
| S8_extracted_k141_13928 | Not-determined |
| S8_extracted_k141_170593 | Low-quality |
| S8_extracted_k141_92413 | Not-determined |
| S8_extracted_k141_48620 | Not-determined |
| S8_extracted_k141_161378 | Low-quality |
| S8_extracted_k141_144510 | Not-determined |
| S8_extracted_k141_109833 | Not-determined |
| S8_extracted_k141_118429 | Not-determined |
| S8_extracted_k141_48622 | Not-determined |
| S8_extracted_k141_144512 | Not-determined |
| S8_extracted_k141_161380 | Not-determined |
| S8_extracted_k141_13932 | Not-determined |
| S8_extracted_k141_118432 | Not-determined |
| S8_extracted_k141_144515 | Not-determined |
| S8_extracted_k141_31904 | Not-determined |
| S8_extracted_k141_170601 | Not-determined |
| S8_extracted_k141_92421 | Not-determined |
| S8_extracted_k141_196697 | Not-determined |
| S8_extracted_k141_31906 | Not-determined |
| S8_extracted_k141_13937 | Not-determined |
| S8_extracted_k141_58003 | Not-determined |
| S8_extracted_k141_31908 | Not-determined |
| S8_extracted_k141_118439 | Not-determined |
| S8_extracted_k141_92424 | Not-determined |
| S8_extracted_k141_31909 | Not-determined |
| S8_extracted_k141_170609 | Not-determined |
| S8_extracted_k141_196704 | Not-determined |
| S8_extracted_k141_40363 | Low-quality |
| S8_extracted_k141_204804 | Not-determined |
| S8_extracted_k141_48625 | Not-determined |
| S8_extracted_k141_40365 | Not-determined |
| S8_extracted_k141_196708 | Not-determined |
| S8_extracted_k141_170614 | Not-determined |
| S8_extracted_k141_109836 | Not-determined |
| S8_extracted_k141_118448 | Low-quality |
| S8_extracted_k141_196710 | Not-determined |
| S8_extracted_k141_40367 | Not-determined |
| S8_extracted_k141_31913 | Not-determined |
| S8_extracted_k141_144520 | Not-determined |
| S8_extracted_k141_5545 | Not-determined |
| S8_extracted_k141_92436 | Not-determined |
| S8_extracted_k141_170620 | Not-determined |
| S8_extracted_k141_153122 | Not-determined |
| S8_extracted_k141_92440 | Not-determined |
| S8_extracted_k141_144527 | Not-determined |
| S8_extracted_k141_40370 | Low-quality |
| S8_extracted_k141_135604 | Low-quality |
| S8_extracted_k141_13952 | Not-determined |
| S8_extracted_k141_58016 | Not-determined |
| S8_extracted_k141_40372 | Not-determined |
| S8_extracted_k141_135606 | Not-determined |
| S8_extracted_k141_118456 | Not-determined |
| S8_extracted_k141_109849 | Not-determined |
| S8_extracted_k141_127009 | Low-quality |
| S8_extracted_k141_179212 | Not-determined |
| S8_extracted_k141_135607 | Not-determined |
| S8_extracted_k141_161412 | Not-determined |
| S8_extracted_k141_13953 | Not-determined |
| S8_extracted_k141_161414 | Not-determined |
| S8_extracted_k141_13954 | Not-determined |
| S8_extracted_k141_144534 | Not-determined |
| S8_extracted_k141_179214 | Not-determined |
| S8_extracted_k141_23205 | Low-quality |
| S8_extracted_k141_23206 | Not-determined |
| S8_extracted_k141_135608 | Not-determined |
| S8_extracted_k141_204816 | Not-determined |
| S8_extracted_k141_161416 | Not-determined |
| S8_extracted_k141_161418 | Not-determined |
| S8_extracted_k141_118463 | Not-determined |
| S8_extracted_k141_109855 | Not-determined |
| S8_extracted_k141_58023 | Not-determined |
| S8_extracted_k141_170629 | Not-determined |
| S8_extracted_k141_92451 | Not-determined |
| S8_extracted_k141_109857 | Not-determined |
| S8_extracted_k141_179215 | Not-determined |
| S8_extracted_k141_161422 | Not-determined |
| S8_extracted_k141_144537 | Not-determined |
| S8_extracted_k141_118466 | Not-determined |
| S8_extracted_k141_179218 | Not-determined |
| S8_extracted_k141_118468 | Low-quality |
| S8_extracted_k141_109860 | Not-determined |
| S8_extracted_k141_196734 | Not-determined |
| S8_extracted_k141_144540 | Not-determined |
| S8_extracted_k141_196735 | Not-determined |
| S8_extracted_k141_74637 | Not-determined |
| S8_extracted_k141_118473 | Not-determined |
| S8_extracted_k141_23221 | Not-determined |
| S8_extracted_k141_186937 | Not-determined |
| S8_extracted_k141_127017 | Not-determined |
| S8_extracted_k141_118475 | Not-determined |
| S8_extracted_k141_40396 | Not-determined |
| S8_extracted_k141_74640 | Not-determined |
| S8_extracted_k141_118476 | Not-determined |
| S8_extracted_k141_5565 | Not-determined |
| S8_extracted_k141_5566 | Not-determined |
| S8_extracted_k141_144545 | Low-quality |
| S8_extracted_k141_144546 | Low-quality |
| S8_extracted_k141_5569 | Not-determined |
| S8_extracted_k141_92463 | Not-determined |
| S8_extracted_k141_31922 | Not-determined |
| S8_extracted_k141_179224 | Not-determined |
| S8_extracted_k141_127019 | Not-determined |
| S8_extracted_k141_186942 | Not-determined |
| S8_extracted_k141_92465 | Not-determined |
| S8_extracted_k141_196738 | Not-determined |
| S8_extracted_k141_5570 | Not-determined |
| S8_extracted_k141_13971 | Not-determined |
| S8_extracted_k141_48639 | Not-determined |
| S8_extracted_k141_58032 | Not-determined |
| S8_extracted_k141_13972 | Not-determined |
| S8_extracted_k141_179230 | Not-determined |
| S8_extracted_k141_13978 | Not-determined |
| S8_extracted_k141_161438 | Not-determined |
| S8_extracted_k141_31929 | Not-determined |
| S8_extracted_k141_23234 | Not-determined |
| S8_extracted_k141_74646 | Not-determined |
| S8_extracted_k141_58038 | Not-determined |
| S8_extracted_k141_109875 | Not-determined |
| S8_extracted_k141_153127 | Not-determined |
| S8_extracted_k141_23236 | Not-determined |
| S8_extracted_k141_23237 | Not-determined |
| S8_extracted_k141_13981 | Low-quality |
| S8_extracted_k141_74649 | Low-quality |
| S8_extracted_k141_161444 | Not-determined |
| S8_extracted_k141_13982 | Not-determined |
| S8_extracted_k141_127026 | Not-determined |
| S8_extracted_k141_40404 | Not-determined |
| S8_extracted_k141_179242 | Not-determined |
| S8_extracted_k141_186956 | Not-determined |
| S8_extracted_k141_92477 | Not-determined |
| S8_extracted_k141_204835 | Not-determined |
| S8_extracted_k141_153132 | Low-quality |
| S8_extracted_k141_40408 | Not-determined |
| S8_extracted_k141_204838 | Not-determined |
| S8_extracted_k141_66102 | Not-determined |
| S8_extracted_k141_144560 | Low-quality |
| S8_extracted_k141_48649 | Not-determined |
| S8_extracted_k141_196749 | Not-determined |
| S8_extracted_k141_48650 | Not-determined |
| S8_extracted_k141_31946 | Not-determined |
| S8_extracted_k141_48651 | Low-quality |
| S8_extracted_k141_58049 | Low-quality |
| S8_extracted_k141_196752 | Not-determined |
| S8_extracted_k141_186965 | Not-determined |
| S8_extracted_k141_23246 | Low-quality |
| S8_extracted_k141_13992 | Not-determined |
| S8_extracted_k141_13996 | Not-determined |
| S8_extracted_k141_161446 | Not-determined |
| S8_extracted_k141_144561 | Low-quality |
| S8_extracted_k141_109893 | Not-determined |
| S8_extracted_k141_144562 | Not-determined |
| S8_extracted_k141_92484 | Not-determined |
| S8_extracted_k141_40412 | Not-determined |
| S8_extracted_k141_135629 | Low-quality |
| S8_extracted_k141_66112 | Not-determined |
| S8_extracted_k141_40416 | Low-quality |
| S8_extracted_k141_153148 | Not-determined |
| S8_extracted_k141_153149 | Not-determined |
| S8_extracted_k141_170633 | Not-determined |
| S8_extracted_k141_14000 | Low-quality |
| S8_extracted_k141_118512 | Low-quality |
| S8_extracted_k141_58059 | Low-quality |
| S8_extracted_k141_135636 | Not-determined |
| S8_extracted_k141_66114 | Low-quality |
| S8_extracted_k141_196765 | Not-determined |
| S8_extracted_k141_23257 | Low-quality |
| S8_extracted_k141_135637 | Not-determined |
| S8_extracted_k141_135638 | Low-quality |
| S8_extracted_k141_100663 | Not-determined |
| S8_extracted_k141_58061 | Not-determined |
| S8_extracted_k141_179271 | Not-determined |
| S8_extracted_k141_14006 | Low-quality |
| S8_extracted_k141_135645 | Low-quality |
| S8_extracted_k141_135646 | Not-determined |
| S8_extracted_k141_170647 | Not-determined |
| S8_extracted_k141_127051 | Not-determined |
| S8_extracted_k141_153153 | Low-quality |
| S8_extracted_k141_161467 | Not-determined |
| S8_extracted_k141_186976 | Not-determined |
| S8_extracted_k141_127053 | Not-determined |
| S8_extracted_k141_14010 | Not-determined |
| S8_extracted_k141_179275 | Not-determined |
| S8_extracted_k141_135650 | Not-determined |
| S8_extracted_k141_118523 | Not-determined |
| S8_extracted_k141_40430 | Not-determined |
| S8_extracted_k141_170652 | Not-determined |
| S8_extracted_k141_14013 | Not-determined |
| S8_extracted_k141_23261 | Not-determined |
| S8_extracted_k141_40435 | Not-determined |
| S8_extracted_k141_179277 | Not-determined |
| S8_extracted_k141_179278 | Not-determined |
| S8_extracted_k141_58066 | Not-determined |
| S8_extracted_k141_40436 | Not-determined |
| S8_extracted_k141_204852 | Not-determined |
| S8_extracted_k141_14018 | Not-determined |
| S8_extracted_k141_14020 | Low-quality |
| S8_extracted_k141_48676 | Not-determined |
| S8_extracted_k141_161474 | Not-determined |
| S8_extracted_k141_100681 | Not-determined |
| S8_extracted_k141_31975 | Not-determined |
| S8_extracted_k141_14025 | Low-quality |
| S8_extracted_k141_92500 | Not-determined |
| S8_extracted_k141_135659 | Not-determined |
| S8_extracted_k141_100684 | Not-determined |
| S8_extracted_k141_186992 | Not-determined |
| S8_extracted_k141_196773 | Not-determined |
| S8_extracted_k141_92503 | Not-determined |
| S8_extracted_k141_135665 | Low-quality |
| S8_extracted_k141_196775 | Low-quality |
| S8_extracted_k141_153165 | Low-quality |
| S8_extracted_k141_58068 | Not-determined |
| S8_extracted_k141_100690 | Low-quality |
| S8_extracted_k141_135669 | Not-determined |
| S8_extracted_k141_100691 | Not-determined |
| S8_extracted_k141_186996 | Not-determined |
| S8_extracted_k141_179286 | Not-determined |
| S8_extracted_k141_153172 | Not-determined |
| S8_extracted_k141_153173 | Not-determined |
| S8_extracted_k141_58073 | Not-determined |
| S8_extracted_k141_196782 | Not-determined |
| S8_extracted_k141_48691 | Not-determined |
| S8_extracted_k141_14035 | Not-determined |
| S8_extracted_k141_109903 | Not-determined |
| S8_extracted_k141_161486 | Not-determined |
| S8_extracted_k141_48693 | Not-determined |
| S8_extracted_k141_144582 | Not-determined |
| S8_extracted_k141_144583 | Not-determined |
| S8_extracted_k141_161488 | Low-quality |
| S8_extracted_k141_66146 | Not-determined |
| S8_extracted_k141_170665 | Not-determined |
| S8_extracted_k141_31979 | Not-determined |
| S8_extracted_k141_161490 | Not-determined |
| S8_extracted_k141_14039 | Not-determined |
| S8_extracted_k141_170669 | Not-determined |
| S8_extracted_k141_204863 | Not-determined |
| S8_extracted_k141_14044 | Not-determined |
| S8_extracted_k141_135680 | Low-quality |
| S8_extracted_k141_204865 | Not-determined |
| S8_extracted_k141_144591 | Not-determined |
| S8_extracted_k141_31987 | Not-determined |
| S8_extracted_k141_40455 | Not-determined |
| S8_extracted_k141_118540 | Not-determined |
| S8_extracted_k141_118543 | Not-determined |
| S8_extracted_k141_48709 | Not-determined |
| S8_extracted_k141_204873 | Not-determined |
| S8_extracted_k141_118544 | Not-determined |
| S8_extracted_k141_187007 | Not-determined |
| S8_extracted_k141_31996 | Not-determined |
| S8_extracted_k141_23286 | Not-determined |
| S8_extracted_k141_135685 | Low-quality |
| S8_extracted_k141_48714 | Not-determined |
| S8_extracted_k141_14055 | Not-determined |
| S8_extracted_k141_31999 | Not-determined |
| S8_extracted_k141_179313 | Not-determined |
| S8_extracted_k141_135693 | Not-determined |
| S8_extracted_k141_135694 | Not-determined |
| S8_extracted_k141_66155 | Not-determined |
| S8_extracted_k141_170675 | Not-determined |
| S8_extracted_k141_32004 | Not-determined |
| S8_extracted_k141_23293 | Not-determined |
| S8_extracted_k141_40465 | Not-determined |
| S8_extracted_k141_48723 | Not-determined |
| S8_extracted_k141_66158 | Low-quality |
| S8_extracted_k141_74678 | Not-determined |
| S8_extracted_k141_179322 | Not-determined |
| S8_extracted_k141_74680 | Not-determined |
| S8_extracted_k141_170681 | Not-determined |
| S8_extracted_k141_161512 | Not-determined |
| S8_extracted_k141_5622 | Not-determined |
| S8_extracted_k141_109927 | Not-determined |
| S8_extracted_k141_66162 | Not-determined |
| S8_extracted_k141_196796 | Not-determined |
| S8_extracted_k141_135704 | Not-determined |
| S8_extracted_k141_92524 | Not-determined |
| S8_extracted_k141_100725 | Not-determined |
| S8_extracted_k141_66163 | Not-determined |
| S8_extracted_k141_92525 | Not-determined |
| S8_extracted_k141_66164 | Low-quality |
| S8_extracted_k141_100726 | Not-determined |
| S8_extracted_k141_127085 | Not-determined |
| S8_extracted_k141_127086 | Not-determined |
| S8_extracted_k141_14072 | Not-determined |
| S8_extracted_k141_100728 | Not-determined |
| S8_extracted_k141_23308 | Not-determined |
| S8_extracted_k141_118554 | Not-determined |
| S8_extracted_k141_66169 | Not-determined |
| S8_extracted_k141_161522 | Not-determined |
| S8_extracted_k141_74695 | Not-determined |
| S8_extracted_k141_100733 | Not-determined |
| S8_extracted_k141_66174 | Not-determined |
| S8_extracted_k141_40477 | Not-determined |
| S8_extracted_k141_92529 | Not-determined |
| S8_extracted_k141_66176 | Not-determined |
| S8_extracted_k141_40481 | Not-determined |
| S8_extracted_k141_196811 | Not-determined |
| S8_extracted_k141_135714 | Not-determined |
| S8_extracted_k141_135715 | Not-determined |
| S8_extracted_k141_118566 | Not-determined |
| S8_extracted_k141_23319 | Not-determined |
| S8_extracted_k141_170692 | Not-determined |
| S8_extracted_k141_109936 | Not-determined |
| S8_extracted_k141_5639 | Not-determined |
| S8_extracted_k141_144614 | Not-determined |
| S8_extracted_k141_32007 | Not-determined |
| S8_extracted_k141_58110 | Not-determined |
| S8_extracted_k141_144621 | Not-determined |
| S8_extracted_k141_74710 | Not-determined |
| S8_extracted_k141_204911 | Not-determined |
| S8_extracted_k141_100749 | Not-determined |
| S8_extracted_k141_170705 | Not-determined |
| S8_extracted_k141_161528 | Not-determined |
| S8_extracted_k141_144628 | Not-determined |
| S8_extracted_k141_179347 | Not-determined |
| S8_extracted_k141_204921 | Not-determined |
| S8_extracted_k141_40491 | Not-determined |
| S8_extracted_k141_32014 | Not-determined |
| S8_extracted_k141_32015 | Not-determined |
| S8_extracted_k141_109946 | Not-determined |
| S8_extracted_k141_58118 | Not-determined |
| S8_extracted_k141_100752 | Not-determined |
| S8_extracted_k141_5658 | Not-determined |
| S8_extracted_k141_48746 | Not-determined |
| S8_extracted_k141_204926 | Not-determined |
| S8_extracted_k141_109947 | Not-determined |
| S8_extracted_k141_109948 | Not-determined |
| S8_extracted_k141_144635 | Low-quality |
| S8_extracted_k141_74720 | Not-determined |
| S8_extracted_k141_161536 | Not-determined |
| S8_extracted_k141_5664 | Not-determined |
| S8_extracted_k141_196821 | Not-determined |
| S8_extracted_k141_161537 | Not-determined |
| S8_extracted_k141_161538 | Not-determined |
| S8_extracted_k141_109954 | Not-determined |
| S8_extracted_k141_179359 | Not-determined |
| S8_extracted_k141_58121 | Not-determined |
| S8_extracted_k141_118581 | Not-determined |
| S8_extracted_k141_74725 | Not-determined |
| S8_extracted_k141_118583 | Not-determined |
| S8_extracted_k141_153190 | Not-determined |
| S8_extracted_k141_153192 | Low-quality |
| S8_extracted_k141_144643 | Not-determined |
| S8_extracted_k141_48750 | Not-determined |
| S8_extracted_k141_161541 | Not-determined |
| S8_extracted_k141_144646 | Not-determined |
| S8_extracted_k141_23343 | Not-determined |
| S8_extracted_k141_153197 | Low-quality |
| S8_extracted_k141_23344 | Not-determined |
| S8_extracted_k141_5674 | Not-determined |
| S8_extracted_k141_204942 | Not-determined |
| S8_extracted_k141_5675 | Low-quality |
| S8_extracted_k141_66213 | Not-determined |
| S8_extracted_k141_14095 | Not-determined |
| S8_extracted_k141_161544 | Low-quality |
| S8_extracted_k141_109966 | Not-determined |
| S8_extracted_k141_74733 | Not-determined |
| S8_extracted_k141_153199 | Not-determined |
| S8_extracted_k141_153202 | Not-determined |
| S8_extracted_k141_100771 | Not-determined |
| S8_extracted_k141_161549 | Not-determined |
| S8_extracted_k141_118597 | Not-determined |
| S8_extracted_k141_196840 | Not-determined |
| S8_extracted_k141_153208 | Not-determined |
| S8_extracted_k141_40516 | Low-quality |
| S8_extracted_k141_161555 | Not-determined |
| S8_extracted_k141_32024 | Not-determined |
| S8_extracted_k141_100778 | Not-determined |
| S8_extracted_k141_83642 | Low-quality |
| S8_extracted_k141_153219 | Not-determined |
| S8_extracted_k141_5691 | Not-determined |
| S8_extracted_k141_83645 | Low-quality |
| S8_extracted_k141_161563 | Not-determined |
| S8_extracted_k141_153225 | Not-determined |
| S8_extracted_k141_23361 | Not-determined |
| S8_extracted_k141_179376 | Not-determined |
| S8_extracted_k141_118610 | Not-determined |
| S8_extracted_k141_74749 | Not-determined |
| S8_extracted_k141_14107 | Low-quality |
| S8_extracted_k141_40529 | Not-determined |
| S8_extracted_k141_153227 | Not-determined |
| S8_extracted_k141_118613 | Low-quality |
| S8_extracted_k141_153229 | Low-quality |
| S8_extracted_k141_144657 | Not-determined |
| S8_extracted_k141_118618 | Not-determined |
| S8_extracted_k141_40532 | Not-determined |
| S8_extracted_k141_118619 | Not-determined |
| S8_extracted_k141_32036 | Not-determined |
| S8_extracted_k141_32039 | Not-determined |
| S8_extracted_k141_74753 | Not-determined |
| S8_extracted_k141_196845 | Not-determined |
| S8_extracted_k141_100791 | Not-determined |
| S8_extracted_k141_196846 | Not-determined |
| S8_extracted_k141_153233 | Not-determined |
| S8_extracted_k141_179382 | Not-determined |
| S8_extracted_k141_161572 | Not-determined |
| S8_extracted_k141_196847 | Not-determined |
| S8_extracted_k141_23367 | Not-determined |
| S8_extracted_k141_109996 | Low-quality |
| S8_extracted_k141_48768 | Not-determined |
| S8_extracted_k141_196852 | Low-quality |
| S8_extracted_k141_179385 | Not-determined |
| S8_extracted_k141_196853 | Not-determined |
| S8_extracted_k141_196854 | Not-determined |
| S8_extracted_k141_196855 | Not-determined |
| S8_extracted_k141_14122 | Low-quality |
| S8_extracted_k141_66230 | Low-quality |
| S8_extracted_k141_110003 | Not-determined |
| S8_extracted_k141_204983 | Not-determined |
| S8_extracted_k141_204984 | Not-determined |
| S8_extracted_k141_118627 | Not-determined |
| S8_extracted_k141_204985 | Not-determined |
| S8_extracted_k141_161584 | Not-determined |
| S8_extracted_k141_40550 | Not-determined |
| S8_extracted_k141_66235 | Low-quality |
| S8_extracted_k141_66238 | Not-determined |
| S8_extracted_k141_100805 | Not-determined |
| S8_extracted_k141_196860 | Not-determined |
| S8_extracted_k141_100806 | Not-determined |
| S8_extracted_k141_66240 | Not-determined |
| S8_extracted_k141_100808 | Not-determined |
| S8_extracted_k141_110009 | Low-quality |
| S8_extracted_k141_32047 | Not-determined |
| S8_extracted_k141_48783 | Not-determined |
| S8_extracted_k141_32049 | Not-determined |
| S8_extracted_k141_23383 | Not-determined |
| S8_extracted_k141_153243 | Not-determined |
| S8_extracted_k141_204991 | Not-determined |
| S8_extracted_k141_144671 | Not-determined |
| S8_extracted_k141_187032 | Not-determined |
| S8_extracted_k141_179391 | Not-determined |
| S8_extracted_k141_58141 | Low-quality |
| S8_extracted_k141_204995 | Not-determined |
| S8_extracted_k141_161586 | Not-determined |
| S8_extracted_k141_40562 | Not-determined |
| S8_extracted_k141_83672 | Not-determined |
| S8_extracted_k141_196866 | Not-determined |
| S8_extracted_k141_187037 | Not-determined |
| S8_extracted_k141_23393 | Not-determined |
| S8_extracted_k141_40566 | Not-determined |
| S8_extracted_k141_48790 | Low-quality |
| S8_extracted_k141_110021 | Not-determined |
| S8_extracted_k141_48793 | Not-determined |
| S8_extracted_k141_196876 | Low-quality |
| S8_extracted_k141_48797 | Not-determined |
| S8_extracted_k141_58152 | Low-quality |
| S8_extracted_k141_161595 | Not-determined |
| S8_extracted_k141_196878 | Not-determined |
| S8_extracted_k141_48799 | Not-determined |
| S8_extracted_k141_153249 | Not-determined |
| S8_extracted_k141_48802 | Not-determined |
| S8_extracted_k141_5715 | Low-quality |
| S8_extracted_k141_83684 | Not-determined |
| S8_extracted_k141_100819 | Not-determined |
| S8_extracted_k141_100820 | Not-determined |
| S8_extracted_k141_66256 | Low-quality |
| S8_extracted_k141_187053 | Not-determined |
| S8_extracted_k141_83687 | Not-determined |
| S8_extracted_k141_196887 | Low-quality |
| S8_extracted_k141_100823 | Not-determined |
| S8_extracted_k141_66262 | Not-determined |
| S8_extracted_k141_40581 | Not-determined |
| S8_extracted_k141_48808 | Not-determined |
| S8_extracted_k141_5719 | Not-determined |
| S8_extracted_k141_74799 | Not-determined |
| S8_extracted_k141_196892 | Low-quality |
| S8_extracted_k141_83689 | Not-determined |
| S8_extracted_k141_187059 | Not-determined |
| S8_extracted_k141_48817 | Not-determined |
| S8_extracted_k141_58159 | Not-determined |
| S8_extracted_k141_48818 | Not-determined |
| S8_extracted_k141_196896 | Not-determined |
| S8_extracted_k141_196897 | Not-determined |
| S8_extracted_k141_127108 | Not-determined |
| S8_extracted_k141_48824 | Not-determined |
| S8_extracted_k141_58166 | Not-determined |
| S8_extracted_k141_144693 | Not-determined |
| S8_extracted_k141_179395 | Not-determined |
| S8_extracted_k141_23410 | Low-quality |
| S8_extracted_k141_100831 | Not-determined |
| S8_extracted_k141_48826 | Not-determined |
| S8_extracted_k141_58167 | Not-determined |
| S8_extracted_k141_23411 | Not-determined |
| S8_extracted_k141_48827 | Not-determined |
| S8_extracted_k141_205016 | Not-determined |
| S8_extracted_k141_100833 | Not-determined |
| S8_extracted_k141_23412 | Low-quality |
| S8_extracted_k141_153259 | Not-determined |
| S8_extracted_k141_74811 | Not-determined |
| S8_extracted_k141_205017 | Not-determined |
| S8_extracted_k141_48829 | Not-determined |
| S8_extracted_k141_74812 | Not-determined |

| | |
|---|---|
| S8_extracted_k141_100834 | Low-quality |
| S8_extracted_k141_179399 | Not-determined |
| S8_extracted_k141_179401 | Not-determined |
| S8_extracted_k141_100837 | Low-quality |
| S8_extracted_k141_66272 | Not-determined |
| S8_extracted_k141_14136 | Not-determined |
| S8_extracted_k141_179403 | Not-determined |
| S8_extracted_k141_5737 | Not-determined |
| S8_extracted_k141_74815 | Not-determined |
| S8_extracted_k141_144704 | Low-quality |
| S8_extracted_k141_74817 | Not-determined |
| S8_extracted_k141_205024 | Not-determined |
| S8_extracted_k141_14142 | Not-determined |
| S8_extracted_k141_127117 | Not-determined |
| S8_extracted_k141_187073 | Not-determined |
| S8_extracted_k141_83706 | Not-determined |
| S8_extracted_k141_187074 | Not-determined |
| S8_extracted_k141_5745 | Not-determined |
| S8_extracted_k141_127119 | Not-determined |
| S8_extracted_k141_14145 | Not-determined |
| S8_extracted_k141_48835 | Not-determined |
| S8_extracted_k141_179414 | Not-determined |
| S8_extracted_k141_179415 | Not-determined |
| S8_extracted_k141_92536 | Low-quality |
| S8_extracted_k141_48837 | Not-determined |
| S8_extracted_k141_144717 | Low-quality |
| S8_extracted_k141_187082 | Not-determined |
| S8_extracted_k141_205036 | Not-determined |
| S8_extracted_k141_161622 | Not-determined |
| S8_extracted_k141_205039 | Not-determined |
| S8_extracted_k141_110040 | Not-determined |
| S8_extracted_k141_127130 | Not-determined |
| S8_extracted_k141_110041 | Not-determined |
| S8_extracted_k141_205042 | Low-quality |
| S8_extracted_k141_74826 | Not-determined |
| S8_extracted_k141_66284 | Not-determined |
| S8_extracted_k141_205044 | Not-determined |
| S8_extracted_k141_14157 | Not-determined |
| S8_extracted_k141_5758 | Not-determined |
| S8_extracted_k141_144724 | Not-determined |
| S8_extracted_k141_14158 | Not-determined |
| S8_extracted_k141_127137 | Low-quality |
| S8_extracted_k141_205048 | Not-determined |
| S8_extracted_k141_161635 | Not-determined |
| S8_extracted_k141_100862 | Not-determined |
| S8_extracted_k141_127138 | Not-determined |
| S8_extracted_k141_153266 | Not-determined |
| S8_extracted_k141_144727 | Not-determined |
| S8_extracted_k141_74835 | Not-determined |
| S8_extracted_k141_179417 | Not-determined |
| S8_extracted_k141_32087 | Not-determined |
| S8_extracted_k141_127148 | Not-determined |
| S8_extracted_k141_92549 | Low-quality |
| S8_extracted_k141_100866 | Not-determined |
| S8_extracted_k141_92550 | Not-determined |
| S8_extracted_k141_92552 | Not-determined |
| S8_extracted_k141_153273 | Not-determined |
| S8_extracted_k141_196926 | Low-quality |
| S8_extracted_k141_179419 | Not-determined |
| S8_extracted_k141_23429 | Not-determined |
| S8_extracted_k141_83720 | Low-quality |
| S8_extracted_k141_187100 | Not-determined |
| S8_extracted_k141_161649 | Not-determined |
| S8_extracted_k141_187102 | Not-determined |
| S8_extracted_k141_83723 | Not-determined |
| S8_extracted_k141_23434 | Not-determined |
| S8_extracted_k141_92558 | Not-determined |
| S8_extracted_k141_144746 | Not-determined |
| S8_extracted_k141_187107 | Not-determined |
| S8_extracted_k141_100874 | Not-determined |
| S8_extracted_k141_144748 | Not-determined |
| S8_extracted_k141_23439 | Not-determined |
| S8_extracted_k141_32106 | Not-determined |
| S8_extracted_k141_5780 | Not-determined |
| S8_extracted_k141_5781 | Not-determined |
| S8_extracted_k141_144754 | Not-determined |
| S8_extracted_k141_5782 | Not-determined |
| S8_extracted_k141_205074 | Not-determined |
| S8_extracted_k141_144756 | Low-quality |
| S8_extracted_k141_92568 | Not-determined |
| S8_extracted_k141_110079 | Not-determined |
| S8_extracted_k141_5788 | Not-determined |
| S8_extracted_k141_5789 | Not-determined |
| S8_extracted_k141_92574 | Not-determined |
| S8_extracted_k141_92575 | Not-determined |
| S8_extracted_k141_144760 | Not-determined |
| S8_extracted_k141_5791 | Not-determined |
| S8_extracted_k141_127171 | Not-determined |
| S8_extracted_k141_92580 | Not-determined |
| S8_extracted_k141_32114 | Not-determined |
| S8_extracted_k141_110085 | Not-determined |
| S8_extracted_k141_32115 | Low-quality |
| S8_extracted_k141_40608 | Not-determined |
| S8_extracted_k141_74861 | Not-determined |
| S8_extracted_k141_187115 | Not-determined |
| S8_extracted_k141_144764 | Not-determined |
| S8_extracted_k141_118689 | Not-determined |
| S8_extracted_k141_58196 | Not-determined |
| S8_extracted_k141_58197 | Not-determined |
| S8_extracted_k141_187116 | Low-quality |
| S8_extracted_k141_170712 | Not-determined |
| S8_extracted_k141_32120 | Not-determined |
| S8_extracted_k141_110089 | Not-determined |
| S8_extracted_k141_74868 | Not-determined |
| S8_extracted_k141_66310 | Not-determined |
| S8_extracted_k141_205095 | Not-determined |
| S8_extracted_k141_187122 | Not-determined |
| S8_extracted_k141_58204 | Not-determined |
| S8_extracted_k141_40621 | Not-determined |
| S8_extracted_k141_179434 | Not-determined |
| S8_extracted_k141_161660 | Not-determined |
| S8_extracted_k141_187124 | Not-determined |
| S8_extracted_k141_32127 | Not-determined |
| S8_extracted_k141_161662 | Not-determined |
| S8_extracted_k141_127183 | Not-determined |
| S8_extracted_k141_100902 | Not-determined |
| S8_extracted_k141_66315 | Not-determined |
| S8_extracted_k141_127186 | Low-quality |
| S8_extracted_k141_153294 | Not-determined |
| S8_extracted_k141_118705 | Not-determined |
| S8_extracted_k141_118708 | Low-quality |
| S8_extracted_k141_153300 | Low-quality |
| S8_extracted_k141_118710 | Not-determined |
| S8_extracted_k141_110105 | Not-determined |
| S8_extracted_k141_161671 | Low-quality |
| S8_extracted_k141_66318 | Not-determined |
| S8_extracted_k141_32138 | Not-determined |
| S8_extracted_k141_5807 | Not-determined |
| S8_extracted_k141_74888 | Not-determined |
| S8_extracted_k141_196935 | Not-determined |
| S8_extracted_k141_161675 | Not-determined |
| S8_extracted_k141_196939 | Not-determined |
| S8_extracted_k141_48870 | Not-determined |
| S8_extracted_k141_66321 | Not-determined |
| S8_extracted_k141_40628 | Not-determined |
| S8_extracted_k141_40629 | Not-determined |
| S8_extracted_k141_5812 | Not-determined |
| S8_extracted_k141_48872 | Not-determined |
| S8_extracted_k141_66322 | Not-determined |
| S8_extracted_k141_196944 | Not-determined |
| S8_extracted_k141_170732 | Not-determined |
| S8_extracted_k141_92606 | Not-determined |
| S8_extracted_k141_32148 | Low-quality |
| S8_extracted_k141_187142 | Not-determined |
| S8_extracted_k141_179461 | Not-determined |
| S8_extracted_k141_144793 | Not-determined |
| S8_extracted_k141_187144 | Not-determined |
| S8_extracted_k141_144794 | Not-determined |
| S8_extracted_k141_74900 | Not-determined |
| S8_extracted_k141_170738 | Not-determined |
| S8_extracted_k141_66328 | Not-determined |
| S8_extracted_k141_187145 | Not-determined |
| S8_extracted_k141_32149 | Not-determined |
| S8_extracted_k141_170740 | Not-determined |
| S8_extracted_k141_110117 | Not-determined |
| S8_extracted_k141_74904 | Not-determined |
| S8_extracted_k141_118727 | Not-determined |
| S8_extracted_k141_144796 | Not-determined |
| S8_extracted_k141_32156 | Not-determined |
| S8_extracted_k141_153309 | Not-determined |
| S8_extracted_k141_153310 | Not-determined |
| S8_extracted_k141_127213 | Not-determined |
| S8_extracted_k141_58211 | Not-determined |
| S8_extracted_k141_66334 | Not-determined |
| S8_extracted_k141_205108 | Low-quality |
| S8_extracted_k141_48883 | Not-determined |
| S8_extracted_k141_153315 | Not-determined |
| S8_extracted_k141_5825 | Not-determined |
| S8_extracted_k141_5827 | Not-determined |
| S8_extracted_k141_23450 | Not-determined |
| S8_extracted_k141_5830 | Not-determined |
| S8_extracted_k141_23452 | Not-determined |
| S8_extracted_k141_58218 | Not-determined |
| S8_extracted_k141_23453 | Low-quality |
| S8_extracted_k141_58219 | Not-determined |
| S8_extracted_k141_127223 | Not-determined |
| S8_extracted_k141_196955 | Not-determined |
| S8_extracted_k141_187157 | Not-determined |
| S8_extracted_k141_48892 | Not-determined |
| S8_extracted_k141_110125 | Not-determined |
| S8_extracted_k141_187160 | Not-determined |
| S8_extracted_k141_110126 | Not-determined |
| S8_extracted_k141_196959 | Not-determined |
| S8_extracted_k141_14176 | Not-determined |
| S8_extracted_k141_74907 | Not-determined |
| S8_extracted_k141_144806 | Not-determined |
| S8_extracted_k141_179473 | Not-determined |
| S8_extracted_k141_110130 | Not-determined |
| S8_extracted_k141_127229 | Not-determined |
| S8_extracted_k141_179476 | Not-determined |
| S8_extracted_k141_5839 | Not-determined |
| S8_extracted_k141_127231 | Not-determined |
| S8_extracted_k141_74913 | Not-determined |
| S8_extracted_k141_179479 | Not-determined |
| S8_extracted_k141_32177 | Not-determined |
| S8_extracted_k141_179480 | Not-determined |
| S8_extracted_k141_32178 | Not-determined |
| S8_extracted_k141_161700 | Not-determined |
| S8_extracted_k141_92627 | Not-determined |
| S8_extracted_k141_179482 | Not-determined |
| S8_extracted_k141_179483 | Not-determined |
| S8_extracted_k141_32182 | Not-determined |
| S8_extracted_k141_196968 | Not-determined |
| S8_extracted_k141_144820 | Not-determined |
| S8_extracted_k141_196969 | Low-quality |
| S8_extracted_k141_187176 | Not-determined |
| S8_extracted_k141_14195 | Not-determined |
| S8_extracted_k141_5845 | Not-determined |
| S8_extracted_k141_100935 | Not-determined |
| S8_extracted_k141_48907 | Not-determined |
| S8_extracted_k141_48909 | Not-determined |
| S8_extracted_k141_118746 | Not-determined |
| S8_extracted_k141_205123 | Low-quality |
| S8_extracted_k141_23468 | Not-determined |
| S8_extracted_k141_170743 | Not-determined |
| S8_extracted_k141_5851 | Low-quality |
| S8_extracted_k141_118749 | Not-determined |
| S8_extracted_k141_5853 | Not-determined |
| S8_extracted_k141_144828 | Not-determined |
| S8_extracted_k141_170744 | Not-determined |
| S8_extracted_k141_170745 | Not-determined |
| S8_extracted_k141_14206 | Not-determined |
| S8_extracted_k141_5857 | Not-determined |
| S8_extracted_k141_196979 | Not-determined |
| S8_extracted_k141_144832 | Not-determined |
| S8_extracted_k141_23474 | Not-determined |
| S8_extracted_k141_170749 | Not-determined |
| S8_extracted_k141_40656 | Low-quality |
| S8_extracted_k141_92636 | Not-determined |
| S8_extracted_k141_48916 | Not-determined |
| S8_extracted_k141_100943 | Not-determined |
| S8_extracted_k141_5860 | Low-quality |
| S8_extracted_k141_100946 | Not-determined |
| S8_extracted_k141_14211 | Not-determined |
| S8_extracted_k141_187191 | Not-determined |
| S8_extracted_k141_205133 | Not-determined |
| S8_extracted_k141_66357 | Not-determined |
| S8_extracted_k141_40664 | Not-determined |
| S8_extracted_k141_196984 | Low-quality |
| S8_extracted_k141_58242 | Not-determined |
| S8_extracted_k141_196985 | Low-quality |
| S8_extracted_k141_144840 | Not-determined |
| S8_extracted_k141_66360 | Not-determined |
| S8_extracted_k141_58245 | Not-determined |
| S8_extracted_k141_205140 | Not-determined |
| S8_extracted_k141_187194 | Low-quality |
| S8_extracted_k141_5867 | Low-quality |
| S8_extracted_k141_58247 | Not-determined |
| S8_extracted_k141_127256 | Not-determined |
| S8_extracted_k141_205141 | Not-determined |
| S8_extracted_k141_144845 | Low-quality |
| S8_extracted_k141_127260 | Not-determined |
| S8_extracted_k141_58252 | Not-determined |
| S8_extracted_k141_100952 | Not-determined |
| S8_extracted_k141_23479 | Not-determined |
| S8_extracted_k141_110148 | Not-determined |
| S8_extracted_k141_92649 | Not-determined |
| S8_extracted_k141_14224 | Not-determined |
| S8_extracted_k141_179508 | Not-determined |
| S8_extracted_k141_40676 | Not-determined |
| S8_extracted_k141_48936 | Not-determined |
| S8_extracted_k141_170758 | Not-determined |
| S8_extracted_k141_66374 | Not-determined |
| S8_extracted_k141_92651 | Low-quality |
| S8_extracted_k141_170759 | Not-determined |
| S8_extracted_k141_74944 | Not-determined |
| S8_extracted_k141_14229 | Not-determined |
| S8_extracted_k141_5882 | Not-determined |
| S8_extracted_k141_100959 | Not-determined |
| S8_extracted_k141_5885 | Not-determined |
| S8_extracted_k141_92655 | Not-determined |
| S8_extracted_k141_92657 | Not-determined |
| S8_extracted_k141_92659 | Not-determined |
| S8_extracted_k141_144869 | Not-determined |
| S8_extracted_k141_66377 | Not-determined |
| S8_extracted_k141_127267 | Not-determined |
| S8_extracted_k141_187203 | Not-determined |
| S8_extracted_k141_66379 | Not-determined |
| S8_extracted_k141_40685 | Low-quality |
| S8_extracted_k141_48944 | Not-determined |
| S8_extracted_k141_14237 | Not-determined |
| S8_extracted_k141_66382 | Not-determined |
| S8_extracted_k141_170769 | Low-quality |
| S8_extracted_k141_14239 | Not-determined |
| S8_extracted_k141_110160 | Not-determined |
| S8_extracted_k141_170770 | Not-determined |
| S8_extracted_k141_127270 | Not-determined |
| S8_extracted_k141_74956 | Not-determined |
| S8_extracted_k141_118755 | Not-determined |
| S8_extracted_k141_40692 | Not-determined |
| S8_extracted_k141_74958 | Not-determined |
| S8_extracted_k141_205167 | Low-quality |
| S8_extracted_k141_23493 | Not-determined |
| S8_extracted_k141_74959 | Not-determined |
| S8_extracted_k141_179522 | Not-determined |
| S8_extracted_k141_110164 | Low-quality |
| S8_extracted_k141_170776 | Not-determined |
| S8_extracted_k141_66395 | Not-determined |
| S8_extracted_k141_23499 | Low-quality |
| S8_extracted_k141_127271 | Not-determined |
| S8_extracted_k141_205175 | Not-determined |
| S8_extracted_k141_205176 | Not-determined |
| S8_extracted_k141_205177 | Not-determined |
| S8_extracted_k141_100977 | Not-determined |
| S8_extracted_k141_144875 | Not-determined |
| S8_extracted_k141_74961 | Not-determined |
| S8_extracted_k141_205181 | Not-determined |
| S8_extracted_k141_48959 | Low-quality |
| S8_extracted_k141_5904 | Not-determined |
| S8_extracted_k141_205183 | Not-determined |
| S8_extracted_k141_92673 | Not-determined |
| S8_extracted_k141_58260 | Low-quality |
| S8_extracted_k141_58261 | Not-determined |
| S8_extracted_k141_127281 | Not-determined |
| S8_extracted_k141_205190 | Not-determined |
| S8_extracted_k141_127283 | Not-determined |
| S8_extracted_k141_40712 | Not-determined |
| S8_extracted_k141_170791 | Not-determined |
| S8_extracted_k141_23514 | Not-determined |
| S8_extracted_k141_23517 | Low-quality |
| S8_extracted_k141_23518 | Not-determined |
| S8_extracted_k141_74980 | Not-determined |
| S8_extracted_k141_40717 | Not-determined |
| S8_extracted_k141_58271 | Not-determined |
| S8_extracted_k141_23521 | Not-determined |
| S8_extracted_k141_66406 | Not-determined |
| S8_extracted_k141_14254 | Not-determined |
| S8_extracted_k141_23523 | Not-determined |
| S8_extracted_k141_110186 | Not-determined |
| S8_extracted_k141_23524 | Low-quality |
| S8_extracted_k141_127301 | Not-determined |
| S8_extracted_k141_170802 | Not-determined |
| S8_extracted_k141_23525 | Low-quality |
| S8_extracted_k141_48977 | Not-determined |
| S8_extracted_k141_23526 | Not-determined |
| S8_extracted_k141_205201 | Not-determined |
| S8_extracted_k141_58275 | Not-determined |
| S8_extracted_k141_48980 | Not-determined |
| S8_extracted_k141_14260 | Low-quality |
| S8_extracted_k141_144884 | Not-determined |
| S8_extracted_k141_48982 | Not-determined |
| S8_extracted_k141_110193 | Not-determined |
| S8_extracted_k141_187242 | Not-determined |
| S8_extracted_k141_23529 | Low-quality |
| S8_extracted_k141_110195 | Not-determined |
| S8_extracted_k141_58281 | Not-determined |
| S8_extracted_k141_58282 | Not-determined |
| S8_extracted_k141_144888 | Not-determined |
| S8_extracted_k141_170809 | Not-determined |
| S8_extracted_k141_118779 | Not-determined |
| S8_extracted_k141_23533 | Not-determined |
| S8_extracted_k141_205206 | Not-determined |
| S8_extracted_k141_66421 | Not-determined |
| S8_extracted_k141_170811 | Not-determined |
| S8_extracted_k141_205208 | Not-determined |
| S8_extracted_k141_127308 | Low-quality |
| S8_extracted_k141_66425 | Low-quality |
| S8_extracted_k141_144891 | Not-determined |
| S8_extracted_k141_83753 | Low-quality |
| S8_extracted_k141_100986 | Low-quality |
| S8_extracted_k141_48994 | Not-determined |
| S8_extracted_k141_144894 | Not-determined |
| S8_extracted_k141_83757 | Not-determined |
| S8_extracted_k141_92689 | Not-determined |
| S8_extracted_k141_135740 | Not-determined |
| S8_extracted_k141_92692 | Not-determined |
| S8_extracted_k141_5916 | Not-determined |
| S8_extracted_k141_5917 | Not-determined |
| S8_extracted_k141_66427 | Not-determined |
| S8_extracted_k141_83760 | Not-determined |
| S8_extracted_k141_187260 | Not-determined |
| S8_extracted_k141_92693 | Not-determined |
| S8_extracted_k141_40744 | Not-determined |
| S8_extracted_k141_14267 | Not-determined |
| S8_extracted_k141_153321 | Not-determined |
| S8_extracted_k141_66431 | Low-quality |
| S8_extracted_k141_5922 | Not-determined |
| S8_extracted_k141_144901 | Not-determined |
| S8_extracted_k141_144902 | Not-determined |
| S8_extracted_k141_58293 | Not-determined |
| S8_extracted_k141_127314 | Not-determined |
| S8_extracted_k141_187265 | Not-determined |
| S8_extracted_k141_153327 | Not-determined |
| S8_extracted_k141_40746 | Low-quality |
| S8_extracted_k141_153329 | Not-determined |
| S8_extracted_k141_205215 | Not-determined |
| S8_extracted_k141_187267 | Not-determined |
| S8_extracted_k141_135748 | Not-determined |
| S8_extracted_k141_144909 | Not-determined |
| S8_extracted_k141_187269 | Low-quality |
| S8_extracted_k141_23541 | Not-determined |
| S8_extracted_k141_40748 | Not-determined |
| S8_extracted_k141_23544 | Not-determined |
| S8_extracted_k141_110208 | Not-determined |
| S8_extracted_k141_197009 | Not-determined |
| S8_extracted_k141_58298 | Not-determined |
| S8_extracted_k141_135754 | Not-determined |
| S8_extracted_k141_40754 | Low-quality |
| S8_extracted_k141_127320 | Not-determined |
| S8_extracted_k141_58303 | Not-determined |
| S8_extracted_k141_40756 | Not-determined |
| S8_extracted_k141_110212 | Not-determined |
| S8_extracted_k141_170825 | Not-determined |
| S8_extracted_k141_23550 | Not-determined |
| S8_extracted_k141_58309 | Not-determined |
| S8_extracted_k141_14282 | Not-determined |
| S8_extracted_k141_153340 | Low-quality |
| S8_extracted_k141_23554 | Not-determined |
| S8_extracted_k141_144922 | Low-quality |
| S8_extracted_k141_144923 | Not-determined |
| S8_extracted_k141_187283 | Not-determined |
| S8_extracted_k141_66440 | Not-determined |
| S8_extracted_k141_170835 | Not-determined |
| S8_extracted_k141_101008 | Not-determined |
| S8_extracted_k141_14285 | Not-determined |
| S8_extracted_k141_40768 | Not-determined |
| S8_extracted_k141_205242 | Not-determined |
| S8_extracted_k141_40769 | Not-determined |
| S8_extracted_k141_170844 | Not-determined |
| S8_extracted_k141_66444 | Not-determined |
| S8_extracted_k141_197030 | Not-determined |
| S8_extracted_k141_101018 | Not-determined |
| S8_extracted_k141_127330 | Not-determined |
| S8_extracted_k141_153355 | Not-determined |
| S8_extracted_k141_5947 | Not-determined |
| S8_extracted_k141_197032 | Low-quality |
| S8_extracted_k141_127335 | Not-determined |
| S8_extracted_k141_127336 | Not-determined |
| S8_extracted_k141_144940 | Low-quality |
| S8_extracted_k141_5950 | Not-determined |
| S8_extracted_k141_40776 | Not-determined |
| S8_extracted_k141_58320 | Not-determined |
| S8_extracted_k141_5951 | Low-quality |
| S8_extracted_k141_205250 | Not-determined |
| S8_extracted_k141_197033 | Not-determined |
| S8_extracted_k141_58322 | Not-determined |
| S8_extracted_k141_110216 | Not-determined |
| S8_extracted_k141_58323 | Not-determined |
| S8_extracted_k141_205254 | Low-quality |
| S8_extracted_k141_170852 | Not-determined |
| S8_extracted_k141_135777 | Not-determined |
| S8_extracted_k141_101026 | Low-quality |
| S8_extracted_k141_5961 | Not-determined |
| S8_extracted_k141_187292 | Not-determined |
| S8_extracted_k141_5965 | Not-determined |
| S8_extracted_k141_14295 | Not-determined |
| S8_extracted_k141_101031 | Not-determined |
| S8_extracted_k141_179530 | Not-determined |
| S8_extracted_k141_75004 | Low-quality |
| S8_extracted_k141_144946 | Not-determined |
| S8_extracted_k141_14298 | Not-determined |
| S8_extracted_k141_144949 | Not-determined |
| S8_extracted_k141_23583 | Low-quality |
| S8_extracted_k141_75007 | Not-determined |
| S8_extracted_k141_179533 | Not-determined |
| S8_extracted_k141_153366 | Not-determined |
| S8_extracted_k141_170857 | Not-determined |
| S8_extracted_k141_197044 | Not-determined |
| S8_extracted_k141_40789 | Not-determined |
| S8_extracted_k141_101039 | Not-determined |
| S8_extracted_k141_23592 | Not-determined |
| S8_extracted_k141_179538 | Not-determined |
| S8_extracted_k141_144955 | Not-determined |
| S8_extracted_k141_58330 | Not-determined |
| S8_extracted_k141_144956 | Not-determined |
| S8_extracted_k141_14310 | Not-determined |
| S8_extracted_k141_92699 | Not-determined |
| S8_extracted_k141_23595 | Not-determined |
| S8_extracted_k141_153378 | Not-determined |
| S8_extracted_k141_197055 | Not-determined |
| S8_extracted_k141_170867 | Not-determined |
| S8_extracted_k141_58335 | Not-determined |
| S8_extracted_k141_110233 | Not-determined |
| S8_extracted_k141_14313 | Low-quality |
| S8_extracted_k141_92706 | Low-quality |
| S8_extracted_k141_14316 | Low-quality |
| S8_extracted_k141_83802 | Not-determined |
| S8_extracted_k141_196670 | Medium-quality |
| S8_extracted_k141_153383 | Not-determined |
| S8_extracted_k141_179543 | Not-determined |
| S8_extracted_k141_187298 | Not-determined |
| S8_extracted_k141_153385 | Not-determined |
| S8_extracted_k141_118814 | Not-determined |
| S8_extracted_k141_14325 | Not-determined |
| S8_extracted_k141_110245 | Not-determined |
| S8_extracted_k141_92717 | Not-determined |
| S8_extracted_k141_161711 | Not-determined |
| S8_extracted_k141_118815 | Not-determined |
| S8_extracted_k141_127364 | Not-determined |
| S8_extracted_k141_161717 | Not-determined |
| S8_extracted_k141_179554 | Low-quality |
| S8_extracted_k141_23618 | Low-quality |
| S8_extracted_k141_118819 | Not-determined |
| S8_extracted_k141_23623 | Not-determined |
| S8_extracted_k141_23627 | Not-determined |
| S8_extracted_k141_118824 | Not-determined |
| S8_extracted_k141_144972 | Not-determined |
| S8_extracted_k141_135809 | Not-determined |
| S8_extracted_k141_58348 | Not-determined |
| S8_extracted_k141_161727 | Not-determined |
| S8_extracted_k141_144977 | Low-quality |
| S8_extracted_k141_127369 | Not-determined |
| S8_extracted_k141_144981 | Not-determined |
| S8_extracted_k141_14336 | Not-determined |
| S8_extracted_k141_144982 | Not-determined |
| S8_extracted_k141_118827 | Not-determined |
| S8_extracted_k141_179573 | Not-determined |
| S8_extracted_k141_6002 | Not-determined |
| S8_extracted_k141_58354 | Not-determined |
| S8_extracted_k141_135815 | Not-determined |
| S8_extracted_k141_58355 | Not-determined |
| S8_extracted_k141_153407 | Not-determined |
| S8_extracted_k141_14341 | Not-determined |
| S8_extracted_k141_161734 | Not-determined |
| S8_extracted_k141_92733 | Not-determined |
| S8_extracted_k141_14342 | Not-determined |
| S8_extracted_k141_23640 | Not-determined |
| S8_extracted_k141_135822 | Low-quality |
| S8_extracted_k141_6007 | Not-determined |
| S8_extracted_k141_118834 | Low-quality |
| S8_extracted_k141_110265 | Not-determined |
| S8_extracted_k141_127378 | Low-quality |
| S8_extracted_k141_161739 | Not-determined |
| S8_extracted_k141_135828 | Not-determined |
| S8_extracted_k141_153417 | Low-quality |
| S8_extracted_k141_144992 | Low-quality |
| S8_extracted_k141_58364 | Low-quality |
| S8_extracted_k141_153419 | Not-determined |
| S8_extracted_k141_23650 | Not-determined |
| S8_extracted_k141_40815 | Not-determined |
| S8_extracted_k141_66450 | Not-determined |
| S8_extracted_k141_161744 | Not-determined |
| S8_extracted_k141_66451 | Not-determined |
| S8_extracted_k141_161745 | Not-determined |
| S8_extracted_k141_92746 | Not-determined |
| S8_extracted_k141_40818 | Not-determined |
| S8_extracted_k141_6010 | Not-determined |
| S8_extracted_k141_179587 | Not-determined |
| S8_extracted_k141_101068 | Not-determined |
| S8_extracted_k141_161752 | Not-determined |
| S8_extracted_k141_58371 | Low-quality |
| S8_extracted_k141_179588 | Low-quality |
| S8_extracted_k141_110285 | Not-determined |
| S8_extracted_k141_6015 | Not-determined |
| S8_extracted_k141_187341 | Not-determined |
| S8_extracted_k141_101075 | Not-determined |
| S8_extracted_k141_66457 | Not-determined |
| S8_extracted_k141_75044 | Not-determined |
| S8_extracted_k141_135841 | Not-determined |
| S8_extracted_k141_6022 | Not-determined |
| S8_extracted_k141_127394 | Not-determined |
| S8_extracted_k141_101076 | Low-quality |
| S8_extracted_k141_110292 | Not-determined |
| S8_extracted_k141_135844 | Not-determined |
| S8_extracted_k141_58380 | Low-quality |
| S8_extracted_k141_40833 | Not-determined |
| S8_extracted_k141_161758 | Not-determined |
| S8_extracted_k141_75048 | Not-determined |
| S8_extracted_k141_23657 | Low-quality |
| S8_extracted_k141_187348 | Not-determined |
| S8_extracted_k141_75055 | Not-determined |
| S8_extracted_k141_66468 | Not-determined |
| S8_extracted_k141_92753 | Not-determined |
| S8_extracted_k141_187352 | Not-determined |
| S8_extracted_k141_23663 | Not-determined |
| S8_extracted_k141_66473 | Low-quality |
| S8_extracted_k141_127401 | Low-quality |
| S8_extracted_k141_127405 | Not-determined |
| S8_extracted_k141_14355 | Not-determined |
| S8_extracted_k141_83846 | Low-quality |
| S8_extracted_k141_135857 | Not-determined |
| S8_extracted_k141_110307 | Not-determined |
| S8_extracted_k141_75065 | Not-determined |
| S8_extracted_k141_127407 | Low-quality |
| S8_extracted_k141_83848 | Low-quality |
| S8_extracted_k141_23667 | Not-determined |
| S8_extracted_k141_14359 | Not-determined |
| S8_extracted_k141_192724 | Medium-quality |
| S8_extracted_k141_118869 | Not-determined |
| S8_extracted_k141_14362 | Not-determined |
| S8_extracted_k141_187361 | Not-determined |
| S8_extracted_k141_118870 | Not-determined |
| S8_extracted_k141_6026 | Low-quality |
| S8_extracted_k141_179606 | Not-determined |
| S8_extracted_k141_40842 | Not-determined |
| S8_extracted_k141_187363 | Not-determined |
| S8_extracted_k141_135863 | Not-determined |
| S8_extracted_k141_83856 | Not-determined |
| S8_extracted_k141_40846 | Not-determined |

| | |
|---|---|
| S8_extracted_k141_197063 | Not-determined |
| S8_extracted_k141_153465 | Not-determined |
| S8_extracted_k141_101102 | Not-determined |
| S8_extracted_k141_153467 | Low-quality |
| S8_extracted_k141_75072 | Not-determined |
| S8_extracted_k141_83861 | Not-determined |
| S8_extracted_k141_14368 | Not-determined |
| S8_extracted_k141_23680 | Not-determined |
| S8_extracted_k141_110323 | Not-determined |
| S8_extracted_k141_205265 | Not-determined |
| S8_extracted_k141_23683 | Not-determined |
| S8_extracted_k141_187374 | Not-determined |
| S8_extracted_k141_197072 | Low-quality |
| S8_extracted_k141_127427 | Not-determined |
| S8_extracted_k141_197074 | Not-determined |
| S8_extracted_k141_205268 | Low-quality |
| S7_extracted_36627 | Medium-quality |
| S8_extracted_k141_118883 | Not-determined |
| S8_extracted_k141_127434 | Not-determined |
| S8_extracted_k141_135867 | Low-quality |
| S8_extracted_k141_135868 | Not-determined |
| S8_extracted_k141_92767 | Low-quality |
| S8_extracted_k141_135870 | Not-determined |
| S8_extracted_k141_197081 | Not-determined |
| S8_extracted_k141_66484 | Not-determined |
| S8_extracted_k141_32196 | Not-determined |
| S8_extracted_k141_205276 | Not-determined |
| S8_extracted_k141_32199 | Not-determined |
| S8_extracted_k141_187386 | Low-quality |
| S8_extracted_k141_66487 | Not-determined |
| S5_vs_k141_20268\|\|full | Medium-quality |
| S8_extracted_k141_127442 | Not-determined |
| S8_extracted_k141_153483 | Not-determined |
| S8_extracted_k141_58423 | Not-determined |
| S8_extracted_k141_161788 | Not-determined |
| S8_extracted_k141_110331 | Not-determined |
| S8_extracted_k141_187392 | Not-determined |
| S8_extracted_k141_66488 | Low-quality |
| S8_extracted_k141_32207 | Not-determined |
| S8_extracted_k141_14379 | Not-determined |
| S8_extracted_k141_6057 | Not-determined |
| S8_extracted_k141_92775 | Not-determined |
| S8_extracted_k141_197100 | Not-determined |
| S8_extracted_k141_92776 | Not-determined |
| S8_extracted_k141_14381 | Not-determined |
| S8_extracted_k141_197101 | Not-determined |
| S8_extracted_k141_127447 | Low-quality |
| S8_extracted_k141_197103 | Not-determined |
| S8_extracted_k141_153489 | Low-quality |
| S8_extracted_k141_153490 | Not-determined |
| S8_extracted_k141_101117 | Not-determined |
| S8_extracted_k141_153491 | Not-determined |
| S8_extracted_k141_197107 | Not-determined |
| S8_extracted_k141_49019 | Not-determined |
| S8_extracted_k141_135873 | Not-determined |
| S8_extracted_k141_118900 | Not-determined |
| S8_extracted_k141_161792 | Not-determined |
| S8_extracted_k141_92785 | Not-determined |
| S8_extracted_k141_187400 | Not-determined |
| S8_extracted_k141_58427 | Not-determined |
| S8_extracted_k141_118901 | Not-determined |
| S8_extracted_k141_92786 | Low-quality |
| S8_extracted_k141_40870 | Not-determined |
| S8_extracted_k141_118902 | Not-determined |
| S8_extracted_k141_153498 | Not-determined |
| S8_extracted_k141_153499 | Not-determined |
| S8_extracted_k141_153500 | Not-determined |
| S8_extracted_k141_58429 | Not-determined |
| S8_extracted_k141_179633 | Not-determined |
| S8_extracted_k141_187404 | Not-determined |
| S8_extracted_k141_66494 | Not-determined |
| S8_extracted_k141_187407 | Not-determined |
| S8_extracted_k141_153503 | Not-determined |
| S8_extracted_k141_75097 | Not-determined |
| S8_extracted_k141_187410 | Not-determined |
| S8_extracted_k141_23714 | Not-determined |
| S8_extracted_k141_153505 | Not-determined |
| S8_extracted_k141_161800 | Not-determined |
| S8_extracted_k141_187411 | Not-determined |
| S8_extracted_k141_135875 | Not-determined |
| S8_extracted_k141_14392 | Not-determined |
| S8_extracted_k141_75100 | Not-determined |
| S8_extracted_k141_49035 | Not-determined |
| S8_extracted_k141_40882 | Not-determined |
| S8_extracted_k141_170890 | Not-determined |
| S8_extracted_k141_179640 | Not-determined |
| S8_extracted_k141_75103 | Low-quality |
| S8_extracted_k141_23718 | Not-determined |
| S8_extracted_k141_153509 | Not-determined |
| S8_extracted_k141_83890 | Not-determined |
| S8_extracted_k141_153513 | Low-quality |
| S8_extracted_k141_101124 | Not-determined |
| S8_extracted_k141_32223 | Not-determined |
| S8_extracted_k141_32224 | Not-determined |
| S8_extracted_k141_170897 | Not-determined |
| S8_extracted_k141_101126 | Not-determined |
| S8_extracted_k141_118913 | Not-determined |
| S8_extracted_k141_75111 | Not-determined |
| S8_extracted_k141_153523 | Not-determined |
| S8_extracted_k141_6076 | Not-determined |
| S8_extracted_k141_110358 | Not-determined |
| S8_extracted_k141_135882 | Not-determined |
| S8_extracted_k141_58440 | Not-determined |
| S8_extracted_k141_6078 | Low-quality |
| S8_extracted_k141_49045 | Not-determined |
| S8_extracted_k141_14409 | Not-determined |
| S8_extracted_k141_6079 | Not-determined |
| S8_extracted_k141_127452 | Low-quality |
| S8_extracted_k141_66505 | Not-determined |
| S8_extracted_k141_58442 | Low-quality |
| S8_extracted_k141_118915 | Not-determined |
| S8_extracted_k141_153529 | Not-determined |
| S8_extracted_k141_170905 | Not-determined |
| S8_extracted_k141_205298 | Not-determined |
| S8_extracted_k141_66509 | Not-determined |
| S8_extracted_k141_49052 | Not-determined |
| S8_extracted_k141_153534 | Not-determined |
| S8_extracted_k141_187425 | Not-determined |
| S8_extracted_k141_83901 | Not-determined |
| S8_extracted_k141_127456 | Not-determined |
| S8_extracted_k141_118917 | Low-quality |
| S8_extracted_k141_14414 | Not-determined |
| S8_extracted_k141_127458 | Not-determined |
| S8_extracted_k141_66514 | Not-determined |
| S8_extracted_k141_135887 | Not-determined |
| S8_extracted_k141_6090 | Not-determined |
| S8_extracted_k141_66518 | Low-quality |
| S8_extracted_k141_66519 | Not-determined |
| S8_extracted_k141_66520 | Not-determined |
| S8_extracted_k141_205308 | Not-determined |
| S8_extracted_k141_118924 | Not-determined |
| S8_extracted_k141_66526 | Not-determined |
| S8_extracted_k141_66527 | Not-determined |
| S8_extracted_k141_135897 | Not-determined |
| S8_extracted_k141_32246 | Not-determined |
| S8_extracted_k141_118926 | Not-determined |
| S8_extracted_k141_6096 | Not-determined |
| S8_extracted_k141_153548 | Not-determined |
| S8_extracted_k141_23753 | Not-determined |
| S8_extracted_k141_101140 | Low-quality |
| S8_extracted_k141_187436 | Not-determined |
| S8_extracted_k141_187437 | Not-determined |
| S8_extracted_k141_153555 | Not-determined |
| S8_extracted_k141_153558 | Not-determined |
| S8_extracted_k141_153559 | Not-determined |
| S8_extracted_k141_32252 | Low-quality |
| S8_extracted_k141_23756 | Not-determined |
| S8_extracted_k141_14434 | Not-determined |
| S8_extracted_k141_153563 | Not-determined |
| S8_extracted_k141_83922 | Not-determined |
| S8_extracted_k141_110381 | Not-determined |
| S8_extracted_k141_14435 | Not-determined |
| S8_extracted_k141_32255 | Not-determined |
| S8_extracted_k141_75130 | Not-determined |
| S8_extracted_k141_23757 | Not-determined |
| S8_extracted_k141_32256 | Not-determined |
| S8_extracted_k141_83924 | Not-determined |
| S8_extracted_k141_161842 | Low-quality |
| S8_extracted_k141_127479 | Not-determined |
| S8_extracted_k141_83925 | Not-determined |
| S8_extracted_k141_161844 | Not-determined |
| S8_extracted_k141_161845 | Not-determined |
| S8_extracted_k141_101156 | Not-determined |
| S8_extracted_k141_135909 | Not-determined |
| S8_extracted_k141_83928 | Not-determined |
| S8_extracted_k141_75136 | Not-determined |
| S8_extracted_k141_135910 | Not-determined |
| S8_extracted_k141_14442 | Low-quality |
| S8_extracted_k141_14443 | Not-determined |
| S8_extracted_k141_75137 | Not-determined |
| S8_extracted_k141_40907 | Not-determined |
| S8_extracted_k141_14444 | Not-determined |
| S8_extracted_k141_144998 | Not-determined |
| S8_extracted_k141_49081 | Not-determined |
| S8_extracted_k141_40910 | Not-determined |
| S8_extracted_k141_127489 | Not-determined |
| S8_extracted_k141_49084 | Not-determined |
| S8_extracted_k141_153568 | Not-determined |
| S8_extracted_k141_127490 | Not-determined |
| S8_extracted_k141_110391 | Not-determined |
| S8_extracted_k141_40917 | Not-determined |
| S8_extracted_k141_127497 | Not-determined |
| S8_extracted_k141_145006 | Not-determined |
| S8_extracted_k141_40920 | Not-determined |
| S8_extracted_k141_32266 | Not-determined |
| S8_extracted_k141_110397 | Not-determined |
| S8_extracted_k141_187443 | Not-determined |
| S8_extracted_k141_75151 | Low-quality |
| S8_extracted_k141_179659 | Not-determined |
| S8_extracted_k141_32271 | Not-determined |
| S8_extracted_k141_101166 | Low-quality |
| S8_extracted_k141_179665 | Not-determined |
| S8_extracted_k141_205329 | Not-determined |
| S8_extracted_k141_23772 | Not-determined |
| S7_extracted_32559 | Medium-quality |
| S8_extracted_k141_75154 | Not-determined |
| S8_extracted_k141_66529 | Not-determined |
| S8_extracted_k141_127511 | Not-determined |
| S8_extracted_k141_75155 | Not-determined |
| S8_extracted_k141_40930 | Not-determined |
| S8_extracted_k141_40931 | Not-determined |
| S8_extracted_k141_14453 | Not-determined |
| S8_extracted_k141_161873 | Not-determined |
| S8_extracted_k141_118949 | Not-determined |
| S8_extracted_k141_187457 | Low-quality |
| S8_extracted_k141_161875 | Not-determined |
| S8_extracted_k141_127516 | Low-quality |
| S8_extracted_k141_179674 | Not-determined |
| S8_extracted_k141_110409 | Not-determined |
| S8_extracted_k141_58487 | Not-determined |
| S8_extracted_k141_101176 | Not-determined |
| S8_extracted_k141_205337 | Not-determined |
| S8_extracted_k141_127519 | Low-quality |
| S8_extracted_k141_14463 | Not-determined |
| S8_extracted_k141_49094 | Not-determined |
| S8_extracted_k141_58489 | Not-determined |
| S8_extracted_k141_179680 | Not-determined |
| S8_extracted_k141_179683 | Not-determined |
| S8_extracted_k141_58490 | Low-quality |
| S8_extracted_k141_153580 | Not-determined |
| S8_extracted_k141_161880 | Not-determined |
| S8_extracted_k141_110417 | Not-determined |
| S8_extracted_k141_58491 | Not-determined |
| S8_extracted_k141_83938 | Not-determined |
| S8_extracted_k141_127532 | Not-determined |
| S8_extracted_k141_135942 | Low-quality |
| S8_extracted_k141_179689 | Not-determined |
| S8_extracted_k141_32285 | Not-determined |
| S8_extracted_k141_170936 | Not-determined |
| S8_extracted_k141_153588 | Not-determined |
| S8_extracted_k141_153589 | Not-determined |
| S8_extracted_k141_40940 | Not-determined |
| S8_extracted_k141_153590 | Not-determined |
| S8_extracted_k141_40941 | Not-determined |
| S8_extracted_k141_127539 | Low-quality |
| S8_extracted_k141_40944 | Not-determined |
| S8_extracted_k141_40946 | Low-quality |
| S8_extracted_k141_40947 | Not-determined |
| S8_extracted_k141_23790 | Not-determined |
| S8_extracted_k141_187485 | Low-quality |
| S8_extracted_k141_153595 | Not-determined |
| S8_extracted_k141_14473 | Low-quality |
| S8_extracted_k141_127542 | Not-determined |
| S8_extracted_k141_197122 | Low-quality |
| S8_extracted_k141_49104 | Low-quality |
| S8_extracted_k141_66550 | Not-determined |
| S8_extracted_k141_14475 | Not-determined |
| S8_extracted_k141_66551 | Not-determined |
| S8_extracted_k141_197123 | Not-determined |
| S8_extracted_k141_127546 | Not-determined |
| S8_extracted_k141_110426 | Low-quality |
| S8_extracted_k141_49105 | Low-quality |
| S8_extracted_k141_23791 | Not-determined |
| S8_extracted_k141_135952 | Not-determined |
| S8_extracted_k141_23792 | Not-determined |
| S8_extracted_k141_127548 | Not-determined |
| S8_extracted_k141_179698 | Not-determined |
| S8_extracted_k141_49109 | Not-determined |
| S8_extracted_k141_127553 | Not-determined |
| S8_extracted_k141_110431 | Low-quality |
| S8_extracted_k141_49111 | Low-quality |
| S8_extracted_k141_49112 | Not-determined |
| S8_extracted_k141_127554 | Not-determined |
| S8_extracted_k141_49114 | Not-determined |
| S8_extracted_k141_32298 | Not-determined |
| S8_extracted_k141_23794 | Not-determined |
| S8_extracted_k141_49116 | Not-determined |
| S8_extracted_k141_58503 | Not-determined |
| S8_extracted_k141_66557 | Low-quality |
| S8_extracted_k141_110440 | Not-determined |
| S8_extracted_k141_197133 | Not-determined |
| S8_extracted_k141_6161 | Not-determined |
| S8_extracted_k141_118969 | Not-determined |
| S8_extracted_k141_75191 | Not-determined |
| S8_extracted_k141_118970 | Not-determined |
| S8_extracted_k141_170956 | Not-determined |
| S8_extracted_k141_145034 | Not-determined |
| S8_extracted_k141_197139 | Low-quality |
| S8_extracted_k141_75195 | Not-determined |
| S8_extracted_k141_58510 | Not-determined |
| S8_extracted_k141_170964 | Low-quality |
| S8_extracted_k141_23805 | Not-determined |
| S8_extracted_k141_101202 | Not-determined |
| S8_extracted_k141_145036 | Not-determined |
| S8_extracted_k141_110454 | Not-determined |
| S8_extracted_k141_23808 | Low-quality |
| S8_extracted_k141_66566 | Not-determined |
| S8_extracted_k141_118977 | Not-determined |
| S8_extracted_k141_101204 | Not-determined |
| S8_extracted_k141_101205 | Not-determined |
| S8_extracted_k141_135969 | Not-determined |
| S8_extracted_k141_6168 | Not-determined |
| S8_extracted_k141_127575 | Not-determined |
| S8_extracted_k141_6171 | Not-determined |
| S8_extracted_k141_110462 | Low-quality |
| S8_extracted_k141_197147 | Low-quality |
| S8_extracted_k141_110463 | Not-determined |
| S8_extracted_k141_118980 | Not-determined |
| S8_extracted_k141_83955 | Not-determined |
| S8_extracted_k141_145045 | Low-quality |
| S8_extracted_k141_118983 | Not-determined |
| S8_extracted_k141_153610 | Not-determined |
| S8_extracted_k141_170980 | Not-determined |
| S8_extracted_k141_145046 | Not-determined |
| S8_extracted_k141_40966 | Low-quality |
| S8_extracted_k141_110467 | Not-determined |
| S8_extracted_k141_161908 | Low-quality |
| S8_extracted_k141_161909 | Not-determined |
| S8_extracted_k141_40969 | Not-determined |
| S8_extracted_k141_66569 | Low-quality |
| S8_extracted_k141_153612 | Not-determined |
| S8_extracted_k141_127585 | Not-determined |
| S8_extracted_k141_127586 | Not-determined |
| S8_extracted_k141_101213 | Low-quality |
| S8_extracted_k141_14498 | Not-determined |
| S8_extracted_k141_32306 | Not-determined |
| S8_extracted_k141_118988 | Low-quality |
| S8_extracted_k141_6174 | Not-determined |
| S8_extracted_k141_83963 | Not-determined |
| S8_extracted_k141_58527 | Not-determined |
| S8_extracted_k141_6176 | Not-determined |
| S8_extracted_k141_118989 | Not-determined |
| S8_extracted_k141_101217 | Not-determined |
| S8_extracted_k141_6178 | Not-determined |
| S8_extracted_k141_153618 | Not-determined |
| S8_extracted_k141_197160 | Not-determined |
| S8_extracted_k141_32312 | Not-determined |
| S8_extracted_k141_197162 | Not-determined |
| S8_extracted_k141_58530 | Not-determined |
| S8_extracted_k141_32313 | Not-determined |
| S8_extracted_k141_135989 | Not-determined |
| S8_extracted_k141_6183 | Not-determined |
| S8_extracted_k141_32315 | Not-determined |
| S8_extracted_k141_118993 | Not-determined |
| S8_extracted_k141_145059 | Low-quality |
| S8_extracted_k141_197167 | Not-determined |
| S8_extracted_k141_135990 | Not-determined |
| S8_extracted_k141_6186 | Not-determined |
| S8_extracted_k141_197168 | Not-determined |
| S8_extracted_k141_40982 | Not-determined |
| S8_extracted_k141_127597 | Not-determined |
| S8_extracted_k141_197169 | Not-determined |
| S8_extracted_k141_58539 | Not-determined |
| S8_extracted_k141_23829 | Low-quality |
| S8_extracted_k141_83971 | Not-determined |
| S8_extracted_k141_127601 | Not-determined |
| S8_extracted_k141_135995 | Not-determined |
| S8_extracted_k141_161914 | Not-determined |
| S8_extracted_k141_75218 | Low-quality |
| S8_extracted_k141_171003 | Not-determined |
| S8_extracted_k141_161915 | Not-determined |
| S8_extracted_k141_32327 | Not-determined |
| S8_extracted_k141_58547 | Not-determined |
| S8_extracted_k141_101229 | Not-determined |
| S8_extracted_k141_14509 | Not-determined |
| S8_extracted_k141_205362 | Not-determined |
| S8_extracted_k141_58548 | Not-determined |
| S8_extracted_k141_171007 | Not-determined |
| S8_extracted_k141_145070 | Not-determined |
| S8_extracted_k141_197177 | Not-determined |
| S8_extracted_k141_23834 | Low-quality |
| S8_extracted_k141_171009 | Not-determined |
| S8_extracted_k141_49121 | Not-determined |
| S8_extracted_k141_75225 | Not-determined |
| S8_extracted_k141_75226 | Not-determined |
| S8_extracted_k141_101235 | Not-determined |
| S8_extracted_k141_41001 | Not-determined |
| S8_extracted_k141_205368 | Not-determined |
| S8_extracted_k141_135996 | Low-quality |
| S8_extracted_k141_205370 | Not-determined |
| S8_extracted_k141_83981 | Not-determined |
| S8_extracted_k141_161921 | Not-determined |
| S8_extracted_k141_23839 | Not-determined |
| S8_extracted_k141_205373 | Not-determined |
| S8_extracted_k141_153639 | Not-determined |
| S8_extracted_k141_161924 | Not-determined |
| S8_extracted_k141_49128 | Not-determined |
| S8_extracted_k141_179708 | Not-determined |
| S8_extracted_k141_101242 | Not-determined |
| S8_extracted_k141_161926 | Not-determined |
| S8_extracted_k141_23841 | Not-determined |
| S8_extracted_k141_187509 | Not-determined |
| S8_extracted_k141_66583 | Not-determined |
| S8_extracted_k141_14520 | Not-determined |
| S8_extracted_k141_83986 | Not-determined |
| S8_extracted_k141_179710 | Low-quality |
| S8_extracted_k141_171020 | Not-determined |
| S8_extracted_k141_58558 | Not-determined |
| S8_extracted_k141_136007 | Not-determined |
| S8_extracted_k141_92789 | Not-determined |
| S8_extracted_k141_6190 | Not-determined |
| S8_extracted_k141_101254 | Low-quality |
| S8_extracted_k141_66592 | Not-determined |
| S8_extracted_k141_187518 | Not-determined |
| S8_extracted_k141_145090 | Not-determined |
| S8_extracted_k141_153650 | Not-determined |
| S8_extracted_k141_83990 | Not-determined |
| S8_extracted_k141_119021 | Not-determined |
| S8_extracted_k141_66598 | Not-determined |
| S8_extracted_k141_41019 | Not-determined |
| S8_extracted_k141_92802 | Not-determined |
| S8_extracted_k141_6197 | Not-determined |
| S8_extracted_k141_110474 | Not-determined |
| S8_extracted_k141_92803 | Not-determined |
| S8_extracted_k141_145093 | Low-quality |
| S8_extracted_k141_6199 | Not-determined |
| S8_extracted_k141_14532 | Not-determined |
| S8_extracted_k141_171027 | Not-determined |
| S8_extracted_k141_101260 | Not-determined |
| S8_extracted_k141_92806 | Not-determined |
| S8_extracted_k141_75248 | Low-quality |
| S8_extracted_k141_110478 | Not-determined |
| S8_extracted_k141_66601 | Not-determined |
| S8_extracted_k141_197198 | Not-determined |
| S8_extracted_k141_14534 | Not-determined |
| S8_extracted_k141_14535 | Not-determined |
| S8_extracted_k141_14536 | Not-determined |
| S8_extracted_k141_161937 | Not-determined |
| S8_extracted_k141_127615 | Not-determined |
| S8_extracted_k141_179728 | Low-quality |
| S8_extracted_k141_161939 | Not-determined |
| S8_extracted_k141_179729 | Not-determined |
| S8_extracted_k141_41030 | Low-quality |
| S8_extracted_k141_153659 | Not-determined |
| S8_extracted_k141_110483 | Low-quality |
| S8_extracted_k141_14539 | Not-determined |
| S8_extracted_k141_23855 | Not-determined |
| S8_extracted_k141_58574 | Not-determined |
| S8_extracted_k141_187528 | Not-determined |
| S8_extracted_k141_58576 | Not-determined |
| S8_extracted_k141_49145 | Not-determined |
| S8_extracted_k141_171040 | Not-determined |
| S8_extracted_k141_161942 | Not-determined |
| S8_extracted_k141_84001 | Not-determined |
| S8_extracted_k141_75261 | Not-determined |
| S8_extracted_k141_179737 | Not-determined |
| S8_extracted_k141_58581 | Not-determined |
| S8_extracted_k141_92819 | Not-determined |
| S8_extracted_k141_49147 | Not-determined |
| S8_extracted_k141_119032 | Not-determined |
| S8_extracted_k141_127624 | Low-quality |
| S8_extracted_k141_187538 | Not-determined |
| S8_extracted_k141_49152 | Not-determined |
| S8_extracted_k141_49153 | Not-determined |
| S8_extracted_k141_153668 | Not-determined |
| S8_extracted_k141_58590 | Not-determined |
| S8_extracted_k141_110484 | Low-quality |
| S8_extracted_k141_110485 | Not-determined |
| S8_extracted_k141_161948 | Not-determined |
| S8_extracted_k141_75271 | Not-determined |
| S8_extracted_k141_92828 | Not-determined |
| S8_extracted_k141_75272 | Not-determined |
| S8_extracted_k141_14547 | Low-quality |
| S8_extracted_k141_75273 | Not-determined |
| S8_extracted_k141_110487 | Not-determined |
| S8_extracted_k141_6215 | Not-determined |
| S8_extracted_k141_127628 | Not-determined |
| S8_extracted_k141_197222 | Not-determined |
| S8_extracted_k141_127631 | Not-determined |
| S8_extracted_k141_58594 | Not-determined |
| S8_extracted_k141_187547 | Not-determined |
| S8_extracted_k141_197223 | Not-determined |
| S8_extracted_k141_14553 | Not-determined |
| S8_extracted_k141_110491 | Not-determined |
| S8_extracted_k141_187548 | Not-determined |
| S8_extracted_k141_110493 | Not-determined |
| S8_extracted_k141_197225 | Low-quality |
| S8_extracted_k141_58603 | Not-determined |
| S8_extracted_k141_171051 | Not-determined |
| S8_extracted_k141_187552 | Not-determined |
| S8_extracted_k141_153678 | Not-determined |
| S8_extracted_k141_110497 | Not-determined |
| S8_extracted_k141_153680 | Low-quality |
| S8_extracted_k141_153681 | Not-determined |
| S8_extracted_k141_153682 | Not-determined |
| S8_extracted_k141_66630 | Not-determined |
| S8_extracted_k141_136041 | Not-determined |
| S8_extracted_k141_179754 | Not-determined |
| S8_extracted_k141_6227 | Low-quality |
| S8_extracted_k141_75290 | Not-determined |
| S8_extracted_k141_197237 | Not-determined |
| S8_extracted_k141_119056 | Not-determined |
| S8_extracted_k141_127637 | Not-determined |
| S8_extracted_k141_66634 | Not-determined |
| S8_extracted_k141_92847 | Not-determined |
| S8_extracted_k141_58610 | Not-determined |
| S8_extracted_k141_14559 | Not-determined |
| S8_extracted_k141_66638 | Not-determined |
| S8_extracted_k141_92854 | Not-determined |
| S8_extracted_k141_145119 | Not-determined |
| S8_extracted_k141_75293 | Low-quality |
| S8_extracted_k141_84025 | Not-determined |
| S8_extracted_k141_153694 | Not-determined |
| S8_extracted_k141_127641 | Not-determined |
| S8_extracted_k141_187557 | Not-determined |
| S8_extracted_k141_171057 | Not-determined |
| S8_extracted_k141_161954 | Not-determined |
| S8_extracted_k141_66645 | Not-determined |
| S8_extracted_k141_187559 | Not-determined |
| S8_extracted_k141_161956 | Not-determined |
| S8_extracted_k141_66646 | Not-determined |
| S8_extracted_k141_32345 | Not-determined |
| S8_extracted_k141_75299 | Not-determined |
| S8_extracted_k141_75300 | Not-determined |
| S8_extracted_k141_136054 | Not-determined |
| S8_extracted_k141_92863 | Not-determined |
| S8_extracted_k141_136056 | Not-determined |
| S8_extracted_k141_171067 | Not-determined |
| S8_extracted_k141_179769 | Not-determined |
| S8_extracted_k141_23867 | Not-determined |
| S8_extracted_k141_14569 | Not-determined |
| S8_extracted_k141_179773 | Not-determined |
| S8_extracted_k141_136062 | Not-determined |
| S8_extracted_k141_58618 | Low-quality |
| S8_extracted_k141_92870 | Not-determined |
| S8_extracted_k141_14572 | Not-determined |
| S8_extracted_k141_6232 | Not-determined |
| S8_extracted_k141_136066 | Low-quality |
| S8_extracted_k141_136067 | Not-determined |
| S8_extracted_k141_171073 | Not-determined |
| S8_extracted_k141_58620 | Not-determined |
| S8_extracted_k141_14575 | Low-quality |
| S8_extracted_k141_84032 | Low-quality |
| S8_extracted_k141_110515 | Not-determined |
| S8_extracted_k141_171075 | Not-determined |
| S8_extracted_k141_110518 | Not-determined |
| S8_extracted_k141_32353 | Not-determined |
| S8_extracted_k141_161961 | Not-determined |
| S8_extracted_k141_161962 | Not-determined |
| S8_extracted_k141_187577 | Low-quality |
| S8_extracted_k141_6237 | Not-determined |
| S8_extracted_k141_119087 | Not-determined |
| S8_extracted_k141_92876 | Not-determined |
| S8_extracted_k141_23872 | Not-determined |
| S8_extracted_k141_75315 | Low-quality |
| S8_extracted_k141_119091 | Not-determined |
| S8_extracted_k141_187580 | Not-determined |
| S8_extracted_k141_127654 | Not-determined |
| S8_extracted_k141_171091 | Not-determined |
| S8_extracted_k141_187582 | Not-determined |
| S8_extracted_k141_66652 | Low-quality |
| S8_extracted_k141_161965 | Not-determined |
| S7_vs_36627\|\|full | Medium-quality |
| S8_extracted_k141_92878 | Not-determined |
| S8_extracted_k141_127657 | Not-determined |
| S8_extracted_k141_6245 | Not-determined |
| S8_extracted_k141_171098 | Not-determined |
| S8_extracted_k141_187585 | Not-determined |
| S8_extracted_k141_92882 | Not-determined |
| S8_extracted_k141_136082 | Not-determined |
| S8_extracted_k141_92885 | Not-determined |
| S8_extracted_k141_179781 | Not-determined |
| S8_extracted_k141_23881 | Not-determined |
| S8_extracted_k141_197247 | Not-determined |
| S8_extracted_k141_58642 | Not-determined |
| S8_extracted_k141_110529 | Not-determined |
| S8_extracted_k141_153710 | Not-determined |
| S8_extracted_k141_136087 | Not-determined |
| S8_extracted_k141_32364 | Low-quality |
| S8_extracted_k141_32365 | Not-determined |
| S8_extracted_k141_127663 | Not-determined |
| S8_extracted_k141_32366 | Not-determined |
| S8_extracted_k141_32368 | Not-determined |
| S8_extracted_k141_119107 | Low-quality |
| S8_extracted_k141_161980 | Not-determined |
| S8_extracted_k141_110533 | Low-quality |
| S8_extracted_k141_32369 | Not-determined |
| S8_vs_k141_196670\|\|full | Medium-quality |
| S8_extracted_k141_92893 | Not-determined |
| S8_extracted_k141_101285 | Not-determined |
| S8_extracted_k141_161983 | Low-quality |
| S8_extracted_k141_84047 | Not-determined |
| S8_extracted_k141_197258 | Not-determined |
| S8_extracted_k141_23888 | Not-determined |
| S8_extracted_k141_14586 | Not-determined |
| S8_extracted_k141_197261 | Not-determined |
| S8_extracted_k141_32375 | Not-determined |
| S8_extracted_k141_161985 | Not-determined |
| S8_extracted_k141_136099 | Not-determined |
| S8_extracted_k141_161987 | Not-determined |
| S8_extracted_k141_127664 | Not-determined |
| S8_extracted_k141_75336 | Not-determined |
| S8_extracted_k141_75338 | Not-determined |
| S8_extracted_k141_205386 | Not-determined |
| S8_extracted_k141_84049 | Low-quality |
| S8_extracted_k141_14593 | Not-determined |
| S8_extracted_k141_84050 | Not-determined |
| S8_extracted_k141_75341 | Not-determined |
| S8_extracted_k141_110542 | Not-determined |

| | |
|---|---|
| SB_extracted_k141_205388 | Low-quality |
| SB_extracted_k141_6256 | Not-determined |
| SB_extracted_k141_205391 | Not-determined |
| SB_extracted_k141_205392 | Not-determined |
| SB_extracted_k141_32383 | Not-determined |
| SB_extracted_k141_162000 | Not-determined |
| SB_extracted_k141_171115 | Not-determined |
| SB_extracted_k141_75349 | Not-determined |
| SB_extracted_k141_197273 | Not-determined |
| SB_extracted_k141_179812 | Low-quality |
| SB_extracted_k141_58661 | Low-quality |
| SB_extracted_k141_162002 | Not-determined |
| SB_extracted_k141_6265 | Not-determined |
| SB_extracted_k141_162006 | Not-determined |
| SB_extracted_k141_23903 | Low-quality |
| SB_extracted_k141_153716 | Not-determined |
| SB_extracted_k141_162009 | Not-determined |
| SB_extracted_k141_110553 | Not-determined |
| SB_extracted_k141_153718 | Not-determined |
| SB_extracted_k141_58671 | Not-determined |
| SB_extracted_k141_162011 | Low-quality |
| SB_extracted_k141_153720 | Not-determined |
| SB_extracted_k141_101310 | Low-quality |
| SB_extracted_k141_14616 | Not-determined |
| SB_extracted_k141_92898 | Not-determined |
| SB_extracted_k141_23907 | Not-determined |
| SB_extracted_k141_205393 | Not-determined |
| SB_extracted_k141_75364 | Not-determined |
| SB_extracted_k141_197275 | Not-determined |
| SB_extracted_k141_205394 | Not-determined |
| SB_extracted_k141_187595 | Not-determined |
| SB_extracted_k141_92901 | Not-determined |
| SB_extracted_k141_205395 | Not-determined |
| SB_extracted_k141_6280 | Not-determined |
| SB_extracted_k141_84073 | Not-determined |
| SB_extracted_k141_84074 | Not-determined |
| SB_extracted_k141_92904 | Not-determined |
| SB_extracted_k141_14622 | Not-determined |
| SB_extracted_k141_32398 | Low-quality |
| SB_extracted_k141_23914 | Not-determined |
| SB_extracted_k141_136117 | Not-determined |
| SB_extracted_k141_6287 | Not-determined |
| SB_extracted_k141_153731 | Not-determined |
| SB_extracted_k141_187603 | Not-determined |
| SB_extracted_k141_197286 | Not-determined |
| SB_extracted_k141_6290 | Low-quality |
| SB_extracted_k141_197287 | Not-determined |
| SB_extracted_k141_127688 | Not-determined |
| SB_extracted_k141_179822 | Low-quality |
| SB_extracted_k141_162029 | Not-determined |
| SB_extracted_k141_197288 | Not-determined |
| SB_extracted_k141_23923 | Not-determined |
| SB_extracted_k141_179827 | Not-determined |
| SB_extracted_k141_145144 | Not-determined |
| SB_extracted_k141_127690 | Not-determined |
| SB_extracted_k141_75372 | Not-determined |
| SB_extracted_k141_14627 | Not-determined |
| SB_extracted_k141_14628 | Not-determined |
| SB_extracted_k141_32412 | Not-determined |
| SB_extracted_k141_179834 | Not-determined |
| SB_extracted_k141_197297 | Not-determined |
| SB_extracted_k141_162040 | Not-determined |
| SB_extracted_k141_136125 | Not-determined |
| SB_extracted_k141_92923 | Not-determined |
| SB_extracted_k141_127702 | Not-determined |
| SB_extracted_k141_197300 | Not-determined |
| SB_extracted_k141_32417 | Not-determined |
| SB_extracted_k141_14634 | Not-determined |
| SB_extracted_k141_153751 | Not-determined |
| SB_extracted_k141_6298 | Low-quality |
| SB_extracted_k141_14638 | Not-determined |
| SB_extracted_k141_153753 | Not-determined |
| SB_extracted_k141_153755 | Not-determined |
| SB_extracted_k141_84105 | Not-determined |
| SB_extracted_k141_14641 | Not-determined |
| SB_extracted_k141_32422 | Not-determined |
| SB_extracted_k141_162047 | Not-determined |
| SB_extracted_k141_110589 | Not-determined |
| SB_extracted_k141_127714 | Low-quality |
| SB_extracted_k141_6312 | Not-determined |
| SB_extracted_k141_75391 | Not-determined |
| SB_extracted_k141_179845 | Not-determined |
| SB_extracted_k141_171149 | Not-determined |
| SB_extracted_k141_110591 | Not-determined |
| SB_extracted_k141_75394 | Not-determined |
| SB_extracted_k141_75395 | Not-determined |
| SB_extracted_k141_75399 | Not-determined |
| SB_extracted_k141_84114 | Not-determined |
| SB_extracted_k141_75400 | Not-determined |
| SB_extracted_k141_92929 | Low-quality |
| SB_extracted_k141_84116 | Not-determined |
| SB_extracted_k141_14651 | Low-quality |
| SB_extracted_k141_66699 | Not-determined |
| SB_extracted_k141_75402 | Low-quality |
| SB_extracted_k141_92934 | Not-determined |
| SB_extracted_k141_32436 | Not-determined |
| SB_extracted_k141_14655 | Not-determined |
| SB_extracted_k141_136131 | Not-determined |
| SB_extracted_k141_110601 | Low-quality |
| SB_extracted_k141_92940 | Not-determined |
| SB_extracted_k141_197325 | Low-quality |
| SB_extracted_k141_110602 | Not-determined |
| SB_extracted_k141_127731 | Not-determined |
| SB_extracted_k141_179846 | Low-quality |
| SB_extracted_k141_101329 | Not-determined |
| SB_extracted_k141_92944 | Not-determined |
| SB_extracted_k141_187614 | Not-determined |
| SB_extracted_k141_101335 | Low-quality |
| SB_extracted_k141_187615 | Not-determined |
| SB_extracted_k141_58693 | Not-determined |
| SB_extracted_k141_205414 | Not-determined |
| SB_extracted_k141_14667 | Not-determined |
| SB_extracted_k141_187621 | Low-quality |
| SB_extracted_k141_110611 | Not-determined |
| SB_extracted_k141_145162 | Not-determined |
| SB_extracted_k141_101339 | Not-determined |
| SB_extracted_k141_171170 | Not-determined |
| SB_extracted_k141_136136 | Not-determined |
| SB_extracted_k141_187629 | Not-determined |
| SB_extracted_k141_127739 | Not-determined |
| SB_extracted_k141_101340 | Not-determined |
| SB_extracted_k141_75405 | Not-determined |
| SB_extracted_k141_145168 | Not-determined |
| SB_extracted_k141_127741 | Not-determined |
| SB_extracted_k141_179858 | Not-determined |
| SB_extracted_k141_197334 | Not-determined |
| SB_extracted_k141_32441 | Not-determined |
| SB_extracted_k141_6342 | Not-determined |
| SB_extracted_k141_6344 | Not-determined |
| SB_extracted_k141_153791 | Not-determined |
| SB_extracted_k141_58711 | Not-determined |
| SB_extracted_k141_136141 | Not-determined |
| SB_extracted_k141_6346 | Not-determined |
| SB_extracted_k141_205430 | Not-determined |
| SB_extracted_k141_205431 | Not-determined |
| SB_extracted_k141_23932 | Not-determined |
| SB_extracted_k141_197340 | Not-determined |
| SB_extracted_k141_187643 | Not-determined |
| SB_extracted_k141_75409 | Low-quality |
| SB_extracted_k141_58720 | Not-determined |
| SB_extracted_k141_162063 | Not-determined |
| SB_extracted_k141_101363 | Not-determined |
| SB_extracted_k141_153801 | Not-determined |
| SB_extracted_k141_171178 | Not-determined |
| SB_extracted_k141_187644 | Not-determined |
| SB_extracted_k141_119117 | Not-determined |
| SB_extracted_k141_197342 | Low-quality |
| SB_extracted_k141_110624 | Not-determined |
| SB_extracted_k141_101367 | Low-quality |
| SB_extracted_k141_23938 | Not-determined |
| SB_extracted_k141_101369 | Not-determined |
| SB_extracted_k141_66717 | Not-determined |
| SB_extracted_k141_136148 | Not-determined |
| SB_extracted_k141_66718 | Not-determined |
| SB_extracted_k141_101371 | Not-determined |
| SB_extracted_k141_187653 | Not-determined |
| SB_extracted_k141_179876 | Not-determined |
| SB_extracted_k141_187654 | Not-determined |
| SB_extracted_k141_197347 | Not-determined |
| SB_extracted_k141_23942 | Not-determined |
| SB_extracted_k141_75422 | Not-determined |
| SB_extracted_k141_66722 | Low-quality |
| SB_extracted_k141_32456 | Low-quality |
| SB_extracted_k141_75423 | Not-determined |
| SB_extracted_k141_75424 | Not-determined |
| SB_extracted_k141_197349 | Not-determined |
| SB_extracted_k141_32458 | Not-determined |
| SB_extracted_k141_58734 | Not-determined |
| SB_extracted_k141_127744 | Not-determined |
| SB_extracted_k141_205447 | Low-quality |
| SB_extracted_k141_14690 | Not-determined |
| SB_extracted_k141_58738 | Low-quality |
| SB_extracted_k141_179878 | Low-quality |
| SB_extracted_k141_153817 | Not-determined |
| SB_extracted_k141_127747 | Not-determined |
| SB_extracted_k141_23944 | Not-determined |
| SB_extracted_k141_153820 | Not-determined |
| SB_extracted_k141_92984 | Not-determined |
| SB_extracted_k141_14693 | Low-quality |
| SB_extracted_k141_6361 | Not-determined |
| SB_extracted_k141_58742 | Not-determined |
| SB_extracted_k141_23946 | Not-determined |
| SB_extracted_k141_127752 | Not-determined |
| SB_extracted_k141_32470 | Not-determined |
| SB_extracted_k141_23948 | Not-determined |
| SB_extracted_k141_75437 | Not-determined |
| SB_extracted_k141_119129 | Not-determined |
| SB_extracted_k141_110628 | Not-determined |
| SB_extracted_k141_6367 | Not-determined |
| SB_extracted_k141_110631 | Not-determined |
| SB_extracted_k141_32473 | Not-determined |
| SB_extracted_k141_110632 | Not-determined |
| SB_extracted_k141_119133 | Not-determined |
| SB_extracted_k141_136150 | Not-determined |
| SB_extracted_k141_75440 | Not-determined |
| SB_extracted_k141_119135 | Not-determined |
| SB_extracted_k141_119136 | Not-determined |
| SB_extracted_k141_110635 | Not-determined |
| SB_extracted_k141_179883 | Not-determined |
| SB_extracted_k141_58749 | Low-quality |
| SB_extracted_k141_75445 | Not-determined |
| SB_extracted_k141_32476 | Not-determined |
| SB_extracted_k141_110637 | Not-determined |
| SB_extracted_k141_92994 | Not-determined |
| SB_extracted_k141_119142 | Not-determined |
| SB_extracted_k141_179888 | Not-determined |
| SB_extracted_k141_92998 | Not-determined |
| SB_extracted_k141_127758 | Not-determined |
| SB_extracted_k141_110644 | Not-determined |
| SB_extracted_k141_119148 | Not-determined |
| SB_extracted_k141_93000 | Low-quality |
| SB_extracted_k141_205464 | Not-determined |
| SB_extracted_k141_153838 | Not-determined |
| SB_extracted_k141_187681 | Not-determined |
| SB_extracted_k141_205466 | Low-quality |
| SB_extracted_k141_205467 | Not-determined |
| SB_extracted_k141_187683 | Low-quality |
| SB_extracted_k141_41047 | Low-quality |
| SB_extracted_k141_41048 | Low-quality |
| SB_extracted_k141_58758 | Not-determined |
| SB_extracted_k141_119156 | Not-determined |
| SB_extracted_k141_93002 | Not-determined |
| SB_extracted_k141_101387 | Low-quality |
| SB_extracted_k141_93003 | Not-determined |
| SB_extracted_k141_6389 | Not-determined |
| SB_extracted_k141_110651 | Not-determined |
| SB_extracted_k141_119158 | Not-determined |
| SB_extracted_k141_179913 | Not-determined |
| SB_extracted_k141_153848 | Low-quality |
| SB_extracted_k141_110653 | Not-determined |
| SB_extracted_k141_127768 | Not-determined |
| SB_extracted_k141_110656 | Not-determined |
| SB_extracted_k141_58768 | Not-determined |
| SB_extracted_k141_136163 | Not-determined |
| SB_extracted_k141_179922 | Not-determined |
| SB_extracted_k141_179923 | Not-determined |
| SB_extracted_k141_101390 | Not-determined |
| SB_extracted_k141_84152 | Not-determined |
| SB_extracted_k141_93014 | Not-determined |
| SB_extracted_k141_6395 | Not-determined |
| SB_extracted_k141_93015 | Not-determined |
| SB_extracted_k141_14707 | Not-determined |
| SB_extracted_k141_162087 | Not-determined |
| SB_extracted_k141_127780 | Not-determined |
| SB_extracted_k141_179931 | Not-determined |
| SB_extracted_k141_179933 | Not-determined |
| SB_extracted_k141_119172 | Not-determined |
| SB_extracted_k141_14708 | Not-determined |
| SB_extracted_k141_14709 | Not-determined |
| SB_extracted_k141_66747 | Not-determined |
| SB_extracted_k141_6405 | Not-determined |
| SB_extracted_k141_41065 | Not-determined |
| SB_extracted_k141_110665 | Not-determined |
| SB_extracted_k141_93022 | Not-determined |
| SB_extracted_k141_145179 | Not-determined |
| SB_extracted_k141_119179 | Not-determined |
| SB_extracted_k141_101405 | Not-determined |
| SB_extracted_k141_41070 | Low-quality |
| SB_extracted_k141_41071 | Not-determined |
| SB_extracted_k141_197405 | Not-determined |
| SB_extracted_k141_66753 | Not-determined |
| SB_extracted_k141_197408 | Low-quality |
| SB_extracted_k141_84168 | Low-quality |
| SB_extracted_k141_84169 | Not-determined |
| SB_extracted_k141_136184 | Not-determined |
| SB_extracted_k141_162100 | Not-determined |
| SB_extracted_k141_136185 | Not-determined |
| SB_extracted_k141_32514 | Not-determined |
| SB_extracted_k141_153861 | Not-determined |
| SB_extracted_k141_187697 | Not-determined |
| SB_extracted_k141_187698 | Not-determined |
| SB_extracted_k141_119188 | Not-determined |
| SB_extracted_k141_93033 | Not-determined |
| SB_extracted_k141_58796 | Not-determined |
| SB_extracted_k141_153865 | Not-determined |
| SB_extracted_k141_171184 | Low-quality |
| SB_extracted_k141_14720 | Not-determined |
| SB_extracted_k141_66762 | Not-determined |
| SB_extracted_k141_32519 | Not-determined |
| SB_extracted_k141_187702 | Low-quality |
| SB_extracted_k141_41089 | Not-determined |
| SB_extracted_k141_101413 | Not-determined |
| SB_extracted_k141_119192 | Not-determined |
| SB_extracted_k141_127797 | Not-determined |
| SB_extracted_k141_84181 | Not-determined |
| SB_extracted_k141_187705 | Not-determined |
| SB_extracted_k141_153870 | Not-determined |
| SB_extracted_k141_179951 | Not-determined |
| SB_extracted_k141_205493 | Not-determined |
| SB_extracted_k141_162103 | Low-quality |
| SB_extracted_k141_179953 | Not-determined |
| SB_extracted_k141_32529 | Not-determined |
| SB_extracted_k141_58798 | Not-determined |
| SB_extracted_k141_119196 | Not-determined |
| SB_extracted_k141_162108 | Not-determined |
| SB_extracted_k141_58801 | Not-determined |
| SB_extracted_k141_153871 | Not-determined |
| SB_extracted_k141_58804 | Not-determined |
| SB_extracted_k141_32532 | Not-determined |
| SB_extracted_k141_187708 | Low-quality |
| SB_extracted_k141_58805 | Not-determined |
| SB_extracted_k141_197418 | Low-quality |
| SB_extracted_k141_14732 | Not-determined |
| SB_extracted_k141_101426 | Not-determined |
| SB_extracted_k141_66776 | Not-determined |
| SB_extracted_k141_84187 | Not-determined |
| SB_extracted_k141_119197 | Low-quality |
| SB_extracted_k141_179959 | Not-determined |
| SB_extracted_k141_119198 | Not-determined |
| SB_extracted_k141_58810 | Not-determined |
| SB_extracted_k141_153873 | Not-determined |
| SB_extracted_k141_119201 | Not-determined |
| SB_extracted_k141_32539 | Not-determined |
| SB_extracted_k141_205500 | Not-determined |
| SB_extracted_k141_41100 | Not-determined |
| SB_extracted_k141_197423 | Not-determined |
| SB_extracted_k141_32543 | Not-determined |
| SB_extracted_k141_162123 | Not-determined |
| SB_extracted_k141_179966 | Not-determined |
| SB_extracted_k141_197427 | Not-determined |
| SB_extracted_k141_171216 | Not-determined |
| SB_extracted_k141_153882 | Not-determined |
| SB_extracted_k141_93052 | Low-quality |
| SB_extracted_k141_14743 | Not-determined |
| SB_extracted_k141_110681 | Not-determined |
| SB_extracted_k141_75457 | Not-determined |
| SB_extracted_k141_75459 | Not-determined |
| SB_extracted_k141_153886 | Not-determined |
| SB_extracted_k141_41109 | Not-determined |
| SB_extracted_k141_153888 | Not-determined |
| SB_extracted_k141_187712 | Not-determined |
| SB_extracted_k141_32551 | Not-determined |
| SB_extracted_k141_197440 | Not-determined |
| SB_extracted_k141_6435 | Not-determined |
| SB_extracted_k141_205513 | Not-determined |
| SB_extracted_k141_75469 | Not-determined |
| SB_extracted_k141_6437 | Not-determined |
| SB_extracted_k141_93056 | Not-determined |
| SB_extracted_k141_162127 | Not-determined |
| SB_extracted_k141_145208 | Not-determined |
| SB_extracted_k141_153895 | Not-determined |
| SB_extracted_k141_179979 | Not-determined |
| SB_extracted_k141_75473 | Not-determined |
| SB_extracted_k141_187723 | Low-quality |
| SB_extracted_k141_110686 | Not-determined |
| SB_extracted_k141_14746 | Not-determined |
| SB_extracted_k141_49181 | Not-determined |
| SB_extracted_k141_41118 | Not-determined |
| SB_extracted_k141_6443 | Not-determined |
| SB_extracted_k141_162135 | Not-determined |
| SB_extracted_k141_6445 | Not-determined |
| SB_extracted_k141_6449 | Low-quality |
| SB_extracted_k141_14750 | Not-determined |
| SB_extracted_k141_101460 | Not-determined |
| SB_extracted_k141_14751 | Not-determined |
| SB_extracted_k141_32558 | Not-determined |
| SB_extracted_k141_179982 | Not-determined |
| SB_extracted_k141_145216 | Not-determined |
| SB_extracted_k141_58832 | Not-determined |
| SB_extracted_k141_66803 | Low-quality |
| SB_extracted_k141_49191 | Not-determined |
| SB_extracted_k141_119221 | Not-determined |
| SB_extracted_k141_66805 | Not-determined |
| SB_extracted_k141_179985 | Not-determined |
| SB_extracted_k141_171236 | Not-determined |
| SB_extracted_k141_127829 | Not-determined |
| SB_extracted_k141_136214 | Not-determined |
| SB_extracted_k141_179987 | Not-determined |
| SB_extracted_k141_6454 | Not-determined |
| SB_extracted_k141_93078 | Not-determined |
| SB_extracted_k141_153905 | Not-determined |
| SB_extracted_k141_84208 | Not-determined |
| SB_extracted_k141_127840 | Not-determined |
| SB_extracted_k141_58838 | Not-determined |
| SB_extracted_k141_14753 | Not-determined |
| SB_extracted_k141_145230 | Not-determined |
| SB_extracted_k141_32571 | Not-determined |
| SB_extracted_k141_58839 | Low-quality |
| SB_extracted_k141_6455 | Not-determined |
| SB_extracted_k141_153910 | Not-determined |
| SB_extracted_k141_41129 | Not-determined |
| SB_extracted_k141_14758 | Not-determined |
| SB_extracted_k141_171248 | Not-determined |
| SB_extracted_k141_127845 | Low-quality |
| SB_extracted_k141_180001 | Not-determined |
| SB_extracted_k141_93090 | Not-determined |
| SB_extracted_k141_6466 | Not-determined |
| SB_extracted_k141_101467 | Not-determined |
| SB_extracted_k141_153916 | Not-determined |
| SB_extracted_k141_153918 | Not-determined |
| SB_extracted_k141_171254 | Not-determined |
| SB_extracted_k141_136221 | Not-determined |
| SB_extracted_k141_180003 | Not-determined |
| SB_extracted_k141_41135 | Not-determined |
| SB_extracted_k141_180004 | Not-determined |
| SB_extracted_k141_136222 | Not-determined |
| SB_extracted_k141_153922 | Low-quality |
| SB_extracted_k141_84220 | Low-quality |
| SB_extracted_k141_162163 | Low-quality |
| SB_extracted_k141_84222 | Not-determined |
| SB_extracted_k141_136224 | Not-determined |
| SB_extracted_k141_145249 | Not-determined |
| SB_extracted_k141_145250 | Not-determined |
| SB_extracted_k141_101473 | Not-determined |
| SB_extracted_k141_41139 | Low-quality |
| SB_extracted_k141_75489 | Not-determined |
| SB_extracted_k141_58851 | Not-determined |
| SB_extracted_k141_41140 | Not-determined |
| SB_extracted_k141_101475 | Not-determined |
| SB_extracted_k141_197446 | Not-determined |
| SB_extracted_k141_41142 | Not-determined |
| SB_extracted_k141_41143 | Not-determined |
| SB_extracted_k141_101476 | Not-determined |
| SB_extracted_k141_49207 | Not-determined |
| SB_extracted_k141_162167 | Not-determined |
| SB_extracted_k141_32581 | Not-determined |
| SB_extracted_k141_58856 | Not-determined |
| SB_extracted_k141_58857 | Not-determined |
| SB_extracted_k141_49210 | Low-quality |
| SB_extracted_k141_84226 | Low-quality |
| SB_extracted_k141_197451 | Not-determined |
| SB_extracted_k141_101481 | Not-determined |
| SB_extracted_k141_14771 | Not-determined |
| SB_extracted_k141_180014 | Not-determined |
| SB_extracted_k141_58861 | Not-determined |
| SB_extracted_k141_41144 | Not-determined |
| SB_extracted_k141_49216 | Not-determined |
| SB_extracted_k141_58863 | Not-determined |
| SB_extracted_k141_32585 | Not-determined |
| SB_extracted_k141_145263 | Not-determined |
| SB_extracted_k141_153936 | Not-determined |
| SB_extracted_k141_145264 | Not-determined |
| SB_extracted_k141_75495 | Not-determined |
| SB_extracted_k141_75496 | Not-determined |
| SB_extracted_k141_197456 | Not-determined |
| SB_extracted_k141_162178 | Not-determined |
| SB_extracted_k141_110702 | Not-determined |
| SB_extracted_k141_145267 | Not-determined |
| SB_extracted_k141_153939 | Not-determined |
| SB_extracted_k141_162180 | Not-determined |
| SB_extracted_k141_75500 | Not-determined |
| SB_extracted_k141_49226 | Not-determined |
| SB_extracted_k141_84236 | Not-determined |
| SB_extracted_k141_49230 | Not-determined |
| SB_extracted_k141_6473 | Not-determined |
| SB_extracted_k141_110715 | Not-determined |
| SB_extracted_k141_180027 | Not-determined |
| SB_extracted_k141_49234 | Not-determined |
| SB_extracted_k141_32598 | Not-determined |
| SB_extracted_k141_58871 | Not-determined |
| SB_extracted_k141_171269 | Not-determined |
| SB_extracted_k141_49235 | Not-determined |
| SB_extracted_k141_110719 | Not-determined |
| SB_extracted_k141_171270 | Not-determined |
| SB_extracted_k141_41168 | Not-determined |
| SB_extracted_k141_32599 | Low-quality |
| SB_extracted_k141_162186 | Low-quality |
| SB_extracted_k141_127864 | Not-determined |
| SB_extracted_k141_75505 | Not-determined |
| SB_extracted_k141_205542 | Not-determined |
| SB_extracted_k141_101493 | Not-determined |
| S5_vs_k141_145605\|\|full | Medium-quality |
| SB_extracted_k141_110724 | Not-determined |
| SB_extracted_k141_49242 | Not-determined |
| SB_extracted_k141_171278 | Not-determined |
| SB_extracted_k141_205548 | Not-determined |
| SB_extracted_k141_197466 | Not-determined |
| SB_extracted_k141_153947 | Not-determined |
| SB_extracted_k141_145277 | Not-determined |
| S7_vs_32559\|\|full | Medium-quality |
| SB_extracted_k141_101496 | Not-determined |
| SB_extracted_k141_205551 | Not-determined |
| SB_extracted_k141_153948 | Low-quality |
| SB_extracted_k141_171289 | Not-determined |
| SB_extracted_k141_32610 | Not-determined |
| SB_extracted_k141_119254 | Not-determined |
| SB_extracted_k141_32612 | Not-determined |
| SB_extracted_k141_58885 | Low-quality |
| SB_extracted_k141_110737 | Not-determined |
| SB_extracted_k141_32613 | Not-determined |
| SB_extracted_k141_23960 | Low-quality |
| SB_extracted_k141_136241 | Low-quality |
| SB_extracted_k141_171294 | Not-determined |
| SB_extracted_k141_101502 | Not-determined |
| SB_extracted_k141_136242 | Not-determined |
| SB_extracted_k141_93103 | Not-determined |
| SB_extracted_k141_23964 | Not-determined |
| SB_extracted_k141_145284 | Not-determined |
| SB_extracted_k141_101505 | Not-determined |
| SB_extracted_k141_171300 | Not-determined |
| SB_extracted_k141_23966 | Not-determined |
| SB_extracted_k141_145288 | Not-determined |
| SB_extracted_k141_180036 | Not-determined |
| SB_extracted_k141_205556 | Not-determined |
| SB_extracted_k141_145289 | Not-determined |
| SB_extracted_k141_66833 | Not-determined |
| SB_extracted_k141_197480 | Not-determined |
| SB_extracted_k141_101508 | Not-determined |
| SB_extracted_k141_197481 | Not-determined |
| SB_extracted_k141_23969 | Not-determined |
| SB_extracted_k141_66837 | Not-determined |
| SB_extracted_k141_171305 | Not-determined |
| SB_extracted_k141_75525 | Not-determined |
| SB_extracted_k141_32624 | Low-quality |
| SB_extracted_k141_49254 | Not-determined |
| SB_extracted_k141_93116 | Not-determined |
| SB_extracted_k141_49255 | Not-determined |
| SB_extracted_k141_197491 | Not-determined |
| SB_extracted_k141_110741 | Not-determined |
| SB_extracted_k141_110742 | Not-determined |
| SB_extracted_k141_93120 | Not-determined |
| SB_extracted_k141_145291 | Low-quality |
| SB_extracted_k141_58905 | Not-determined |
| SB_extracted_k141_171319 | Not-determined |
| SB_extracted_k141_171320 | Not-determined |
| SB_extracted_k141_14802 | Not-determined |
| SB_extracted_k141_180049 | Not-determined |
| SB_extracted_k141_205573 | Not-determined |
| SB_extracted_k141_180050 | Not-determined |
| SB_extracted_k141_58907 | Not-determined |
| SB_extracted_k141_41193 | Not-determined |
| SB_extracted_k141_136250 | Not-determined |
| SB_extracted_k141_119261 | Not-determined |
| SB_extracted_k141_58908 | Not-determined |
| SB_extracted_k141_14804 | Low-quality |
| SB_extracted_k141_205577 | Not-determined |
| SB_extracted_k141_180053 | Low-quality |
| SB_extracted_k141_205578 | Not-determined |
| SB_extracted_k141_180054 | Not-determined |
| SB_extracted_k141_197498 | Not-determined |
| SB_extracted_k141_171330 | Not-determined |
| SB_extracted_k141_41198 | Low-quality |
| SB_extracted_k141_49259 | Not-determined |
| SB_extracted_k141_84263 | Not-determined |
| SB_extracted_k141_84264 | Not-determined |
| SB_extracted_k141_41200 | Not-determined |
| SB_extracted_k141_41201 | Not-determined |
| SB_extracted_k141_23980 | Not-determined |
| SB_extracted_k141_101526 | Not-determined |
| SB_extracted_k141_93128 | Not-determined |
| SB_extracted_k141_14809 | Not-determined |
| SB_extracted_k141_75542 | Low-quality |
| SB_extracted_k141_14810 | Not-determined |
| SB_extracted_k141_205584 | Not-determined |
| SB_extracted_k141_145300 | Low-quality |
| SB_extracted_k141_14814 | Not-determined |
| SB_extracted_k141_205589 | Not-determined |
| SB_extracted_k141_127881 | Low-quality |
| SB_extracted_k141_101533 | Not-determined |
| SB_extracted_k141_6509 | Not-determined |
| SB_extracted_k141_58918 | Not-determined |
| SB_extracted_k141_171333 | Not-determined |
| SB_extracted_k141_6511 | Not-determined |
| SB_extracted_k141_180070 | Not-determined |
| SB_extracted_k141_101535 | Not-determined |
| SB_extracted_k141_84270 | Not-determined |
| SB_extracted_k141_75549 | Not-determined |
| SB_extracted_k141_41212 | Not-determined |
| SB_extracted_k141_6515 | Not-determined |
| SB_extracted_k141_75552 | Not-determined |
| SB_extracted_k141_49279 | Low-quality |
| SB_extracted_k141_58927 | Low-quality |
| SB_extracted_k141_75554 | Low-quality |
| SB_extracted_k141_127887 | Not-determined |
| SB_extracted_k141_41218 | Not-determined |
| SB_extracted_k141_101541 | Not-determined |
| SB_extracted_k141_110767 | Not-determined |
| SB_extracted_k141_145312 | Not-determined |
| SB_extracted_k141_84280 | Not-determined |
| SB_extracted_k141_84281 | Not-determined |
| SB_extracted_k141_66859 | Not-determined |
| SB_extracted_k141_127891 | Not-determined |
| SB_extracted_k141_58933 | Not-determined |
| SB_extracted_k141_197521 | Low-quality |
| SB_extracted_k141_101548 | Not-determined |
| SB_extracted_k141_119284 | Not-determined |
| SB_extracted_k141_153962 | Not-determined |
| SB_extracted_k141_101549 | Not-determined |
| SB_extracted_k141_6522 | Low-quality |
| SB_extracted_k141_197524 | Not-determined |
| SB_extracted_k141_171351 | Not-determined |
| SB_extracted_k141_136274 | Not-determined |
| SB_extracted_k141_14834 | Not-determined |
| SB_extracted_k141_75565 | Low-quality |
| SB_extracted_k141_110778 | Not-determined |
| SB_extracted_k141_205605 | Not-determined |
| SB_extracted_k141_93144 | Not-determined |
| SB_extracted_k141_75566 | Not-determined |
| SB_extracted_k141_180086 | Not-determined |

| | |
|---|---|
| S8_extracted_k141_84290 | Not-determined |
| S8_extracted_k141_75568 | Not-determined |
| S8_extracted_k141_171360 | Not-determined |
| S8_extracted_k141_205607 | Not-determined |
| S8_extracted_k141_110785 | Not-determined |
| S8_extracted_k141_66861 | Not-determined |
| S8_extracted_k141_205609 | Not-determined |
| S8_extracted_k141_197527 | Not-determined |
| S8_extracted_k141_119298 | Not-determined |
| S8_extracted_k141_119299 | Not-determined |
| S8_extracted_k141_110788 | Not-determined |
| S8_extracted_k141_171365 | Not-determined |
| S8_extracted_k141_14838 | Not-determined |
| S8_extracted_k141_66868 | Not-determined |
| S8_extracted_k141_110792 | Not-determined |
| S8_extracted_k141_14840 | Not-determined |
| S8_extracted_k141_110794 | Not-determined |
| S8_extracted_k141_66871 | Not-determined |
| S8_extracted_k141_101559 | Not-determined |
| S8_extracted_k141_24000 | Not-determined |
| S8_extracted_k141_93164 | Not-determined |
| S8_extracted_k141_119302 | Not-determined |
| S8_extracted_k141_14844 | Not-determined |
| S8_extracted_k141_205616 | Not-determined |
| S8_extracted_k141_101564 | Not-determined |
| S8_extracted_k141_58943 | Not-determined |
| S8_extracted_k141_101565 | Not-determined |
| S8_extracted_k141_205618 | Not-determined |
| S8_extracted_k141_127910 | Not-determined |
| S8_extracted_k141_197534 | Not-determined |
| S8_extracted_k141_127912 | Not-determined |
| S8_extracted_k141_171377 | Not-determined |
| S8_extracted_k141_14854 | Not-determined |
| S8_extracted_k141_14855 | Not-determined |
| S8_extracted_k141_119313 | Low-quality |
| S8_extracted_k141_24006 | Not-determined |
| S8_extracted_k141_205621 | Not-determined |
| S8_extracted_k141_119317 | Not-determined |
| S8_extracted_k141_205623 | Not-determined |
| S8_extracted_k141_93174 | Not-determined |
| S8_extracted_k141_110808 | Not-determined |
| S8_extracted_k141_14859 | Not-determined |
| S8_extracted_k141_101577 | Not-determined |
| S8_extracted_k141_6531 | Not-determined |
| S8_extracted_k141_145333 | Not-determined |
| S8_extracted_k141_119319 | Not-determined |
| S8_extracted_k141_24012 | Not-determined |
| S8_extracted_k141_171385 | Not-determined |
| S8_extracted_k141_75581 | Not-determined |
| S8_extracted_k141_110814 | Not-determined |
| S8_extracted_k141_171388 | Not-determined |
| S8_extracted_k141_171389 | Not-determined |
| S8_extracted_k141_136288 | Not-determined |
| S8_extracted_k141_24017 | Not-determined |
| S8_extracted_k141_101582 | Not-determined |
| S8_extracted_k141_6537 | Not-determined |
| S8_extracted_k141_205627 | Not-determined |
| S8_extracted_k141_84308 | Not-determined |
| S8_extracted_k141_24022 | Not-determined |
| S8_extracted_k141_75585 | Low-quality |
| S8_extracted_k141_110817 | Low-quality |
| S8_extracted_k141_84314 | Not-determined |
| S8_extracted_k141_205629 | Not-determined |
| S8_extracted_k141_180104 | Not-determined |
| S8_extracted_k141_66896 | Not-determined |
| S8_extracted_k141_205633 | Low-quality |
| S8_extracted_k141_205634 | Not-determined |
| S8_extracted_k141_136298 | Not-determined |
| S8_extracted_k141_127927 | Not-determined |
| S8_extracted_k141_14882 | Low-quality |
| S8_extracted_k141_101594 | Not-determined |
| S8_extracted_k141_171400 | Not-determined |
| S8_extracted_k141_32648 | Low-quality |
| S8_extracted_k141_24027 | Not-determined |
| S8_extracted_k141_14885 | Not-determined |
| S8_extracted_k141_101596 | Not-determined |
| S8_extracted_k141_41224 | Not-determined |
| S8_extracted_k141_75601 | Not-determined |
| S8_extracted_k141_205641 | Not-determined |
| S8_extracted_k141_41225 | Not-determined |
| S8_extracted_k141_14889 | Not-determined |
| S8_extracted_k141_145350 | Not-determined |
| S8_extracted_k141_136302 | Not-determined |
| S8_extracted_k141_66909 | Low-quality |
| S8_extracted_k141_197555 | Not-determined |
| S8_extracted_k141_153993 | Not-determined |
| S8_extracted_k141_24034 | Not-determined |
| S8_extracted_k141_32656 | Not-determined |
| S8_extracted_k141_75606 | Not-determined |
| S8_extracted_k141_127934 | Not-determined |
| S8_extracted_k141_32658 | Not-determined |
| S8_extracted_k141_153995 | Not-determined |
| S8_extracted_k141_32660 | Not-determined |
| S8_extracted_k141_110831 | Not-determined |
| S8_extracted_k141_162201 | Not-determined |
| S8_extracted_k141_24038 | Not-determined |
| S8_extracted_k141_171413 | Not-determined |
| S8_extracted_k141_145355 | Low-quality |
| S8_extracted_k141_127943 | Not-determined |
| S8_extracted_k141_154001 | Not-determined |
| S8_extracted_k141_101605 | Not-determined |
| S8_extracted_k141_162205 | Low-quality |
| S8_extracted_k141_110841 | Not-determined |
| S8_extracted_k141_14901 | Not-determined |
| S8_extracted_k141_110842 | Not-determined |
| S8_extracted_k141_171416 | Not-determined |
| S8_extracted_k141_127949 | Not-determined |
| S8_extracted_k141_197564 | Not-determined |
| S8_extracted_k141_6551 | Not-determined |
| S8_extracted_k141_205658 | Not-determined |
| S8_extracted_k141_145361 | Not-determined |
| S8_extracted_k141_6553 | Not-determined |
| S8_extracted_k141_162207 | Not-determined |
| S8_extracted_k141_93182 | Not-determined |
| S8_extracted_k141_197568 | Not-determined |
| S8_extracted_k141_93184 | Low-quality |
| S8_extracted_k141_127951 | Not-determined |
| S8_extracted_k141_58955 | Not-determined |
| S8_extracted_k141_75613 | Not-determined |
| S8_extracted_k141_205669 | Not-determined |
| S8_extracted_k141_197573 | Not-determined |
| S8_extracted_k141_205672 | Not-determined |
| S8_extracted_k141_93193 | Not-determined |
| S8_extracted_k141_14911 | Not-determined |
| S8_extracted_k141_58961 | Not-determined |
| S8_extracted_k141_93197 | Not-determined |
| S8_extracted_k141_197584 | Not-determined |
| S8_extracted_k141_101626 | Not-determined |
| S8_extracted_k141_32670 | Not-determined |
| S8_extracted_k141_14914 | Not-determined |
| S8_extracted_k141_6562 | Low-quality |
| S8_extracted_k141_136338 | Not-determined |
| S8_extracted_k141_101627 | Not-determined |
| S8_extracted_k141_197588 | Not-determined |
| S8_extracted_k141_101629 | Not-determined |
| S8_extracted_k141_197590 | Not-determined |
| S8_extracted_k141_93203 | Not-determined |
| S8_extracted_k141_197591 | Not-determined |
| S8_extracted_k141_197593 | Not-determined |
| S8_extracted_k141_66929 | Low-quality |
| S8_extracted_k141_41236 | Not-determined |
| S8_extracted_k141_180124 | Low-quality |
| S8_extracted_k141_58974 | Not-determined |
| S8_extracted_k141_136342 | Not-determined |
| S8_extracted_k141_136343 | Not-determined |
| S8_extracted_k141_93209 | Not-determined |
| S8_extracted_k141_32679 | Not-determined |
| S8_extracted_k141_6567 | Low-quality |
| S8_extracted_k141_136345 | Not-determined |
| S8_extracted_k141_6568 | Not-determined |
| S8_extracted_k141_84350 | Low-quality |
| S8_extracted_k141_84351 | Not-determined |
| S8_extracted_k141_6571 | Not-determined |
| S8_extracted_k141_93211 | Not-determined |
| S8_extracted_k141_32684 | Not-determined |
| S8_extracted_k141_145393 | Not-determined |
| S8_extracted_k141_32689 | Not-determined |
| S8_extracted_k141_6574 | Not-determined |
| S8_extracted_k141_93214 | Not-determined |
| S8_extracted_k141_197599 | Not-determined |
| S8_extracted_k141_32693 | Not-determined |
| S8_extracted_k141_24074 | Not-determined |
| S8_extracted_k141_6577 | Not-determined |
| S8_extracted_k141_171444 | Not-determined |
| S8_extracted_k141_197601 | Not-determined |
| S8_extracted_k141_75632 | Not-determined |
| S8_extracted_k141_41252 | Not-determined |
| S8_extracted_k141_180138 | Not-determined |
| S8_extracted_k141_180139 | Not-determined |
| S8_extracted_k141_58987 | Low-quality |
| S8_extracted_k141_180141 | Not-determined |
| S8_extracted_k141_14932 | Not-determined |
| S8_extracted_k141_58990 | Not-determined |
| S8_extracted_k141_24080 | Not-determined |
| S8_extracted_k141_6591 | Not-determined |
| S8_extracted_k141_154030 | Not-determined |
| S8_extracted_k141_24081 | Not-determined |
| S8_extracted_k141_24084 | Not-determined |
| S3_vs_k141_1031\|\|full | Medium-quality |
| S8_extracted_k141_154034 | Not-determined |
| S8_extracted_k141_162238 | Not-determined |
| S8_extracted_k141_93222 | Not-determined |
| S8_extracted_k141_93223 | Not-determined |
| S8_extracted_k141_154036 | Not-determined |
| S8_extracted_k141_171453 | Not-determined |
| S8_extracted_k141_6594 | Low-quality |
| S8_extracted_k141_93227 | Not-determined |
| S8_extracted_k141_197616 | Not-determined |
| S8_extracted_k141_162240 | Not-determined |
| S8_extracted_k141_154040 | Not-determined |
| S8_extracted_k141_93231 | Not-determined |
| S8_extracted_k141_180149 | Not-determined |
| S8_extracted_k141_58995 | Low-quality |
| S8_extracted_k141_24088 | Not-determined |
| S8_extracted_k141_180150 | Not-determined |
| S8_extracted_k141_75648 | Not-determined |
| S8_extracted_k141_84373 | Not-determined |
| S8_extracted_k141_6596 | Not-determined |
| S8_extracted_k141_101650 | Not-determined |
| S8_extracted_k141_101651 | Low-quality |
| S8_extracted_k141_162247 | Not-determined |
| S8_extracted_k141_75655 | Low-quality |
| S8_extracted_k141_180154 | Not-determined |
| S8_extracted_k141_154051 | Not-determined |
| S8_extracted_k141_24098 | Not-determined |
| S8_extracted_k141_205696 | Not-determined |
| S8_extracted_k141_75662 | Low-quality |
| S8_extracted_k141_136354 | Not-determined |
| S8_extracted_k141_101653 | Not-determined |
| S8_extracted_k141_14944 | Not-determined |
| S8_extracted_k141_6608 | Not-determined |
| S8_extracted_k141_6611 | Low-quality |
| S8_extracted_k141_14946 | Not-determined |
| S8_extracted_k141_75667 | Not-determined |
| S8_extracted_k141_84385 | Not-determined |
| S8_extracted_k141_136362 | Not-determined |
| S8_extracted_k141_187756 | Not-determined |
| S8_extracted_k141_84389 | Low-quality |
| S8_extracted_k141_59009 | Not-determined |
| S8_extracted_k141_187758 | Not-determined |
| S8_extracted_k141_75674 | Not-determined |
| S8_extracted_k141_66971 | Not-determined |
| S8_extracted_k141_136365 | Not-determined |
| S8_extracted_k141_197626 | Not-determined |
| S8_extracted_k141_180170 | Not-determined |
| S8_extracted_k141_41270 | Not-determined |
| S8_extracted_k141_84394 | Not-determined |
| S8_extracted_k141_187761 | Not-determined |
| S8_extracted_k141_154063 | Not-determined |
| S8_extracted_k141_187763 | Not-determined |
| S8_extracted_k141_136369 | Low-quality |
| S8_extracted_k141_101666 | Not-determined |
| S8_extracted_k141_136372 | Not-determined |
| S8_extracted_k141_180180 | Not-determined |
| S8_extracted_k141_127964 | Low-quality |
| S8_extracted_k141_101670 | Not-determined |
| S8_extracted_k141_101672 | Not-determined |
| S8_extracted_k141_66983 | Low-quality |
| S8_extracted_k141_205712 | Low-quality |
| S8_extracted_k141_197632 | Not-determined |
| S8_extracted_k141_180187 | Not-determined |
| S5_vs_k141_145593\|\|full | High-quality |
| S8_extracted_k141_127971 | Not-determined |
| S8_extracted_k141_59019 | Low-quality |
| S8_extracted_k141_101677 | Not-determined |
| S8_extracted_k141_205722 | Low-quality |
| S8_extracted_k141_136386 | Not-determined |
| S8_extracted_k141_119344 | Not-determined |
| S8_extracted_k141_24110 | Low-quality |
| S8_extracted_k141_32714 | Not-determined |
| S8_extracted_k141_187777 | Low-quality |
| S8_extracted_k141_162252 | Not-determined |
| S8_extracted_k141_136390 | Not-determined |
| S8_extracted_k141_101685 | Not-determined |
| S8_extracted_k141_101686 | Low-quality |
| S8_extracted_k141_197640 | Not-determined |
| S8_extracted_k141_145425 | Not-determined |
| S8_extracted_k141_101689 | Not-determined |
| S8_extracted_k141_145427 | Not-determined |
| S8_extracted_k141_24118 | Not-determined |
| S8_extracted_k141_154072 | Not-determined |
| S8_extracted_k141_187788 | Not-determined |
| S8_extracted_k141_101692 | Not-determined |
| S8_extracted_k141_180202 | Not-determined |
| S8_extracted_k141_24119 | Low-quality |
| S8_extracted_k141_162253 | Not-determined |
| S8_extracted_k141_24120 | Not-determined |
| S8_extracted_k141_180205 | Not-determined |
| S8_extracted_k141_6632 | Low-quality |
| S8_extracted_k141_84413 | Low-quality |
| S8_extracted_k141_154078 | Not-determined |
| S8_extracted_k141_205730 | Not-determined |
| S8_extracted_k141_145433 | Not-determined |
| S8_extracted_k141_119356 | Not-determined |
| S8_extracted_k141_127981 | Low-quality |
| S8_extracted_k141_6635 | Not-determined |
| S8_extracted_k141_145436 | Not-determined |
| S8_extracted_k141_84421 | Not-determined |
| S8_extracted_k141_84422 | Not-determined |
| S8_extracted_k141_154086 | Not-determined |
| S8_extracted_k141_32727 | Low-quality |
| S8_extracted_k141_32728 | Not-determined |
| S8_extracted_k141_101696 | Not-determined |
| S8_extracted_k141_136404 | Not-determined |
| S8_extracted_k141_205735 | Not-determined |
| S8_extracted_k141_154089 | Not-determined |
| S8_extracted_k141_24124 | Not-determined |
| S8_extracted_k141_180213 | Not-determined |
| S8_extracted_k141_14984 | Not-determined |
| S8_extracted_k141_101698 | Not-determined |
| S8_extracted_k141_32732 | Not-determined |
| S8_extracted_k141_101699 | Not-determined |
| S8_extracted_k141_145441 | Not-determined |
| S8_extracted_k141_32733 | Low-quality |
| S8_extracted_k141_162269 | Not-determined |
| S8_extracted_k141_32736 | Not-determined |
| S8_extracted_k141_145443 | Not-determined |
| S8_extracted_k141_32737 | Not-determined |
| S8_extracted_k141_119371 | Not-determined |
| S8_extracted_k141_162271 | Low-quality |
| S8_extracted_k141_180218 | Not-determined |
| S8_extracted_k141_145448 | Not-determined |
| S8_extracted_k141_162274 | Not-determined |
| S8_extracted_k141_49311 | Not-determined |
| S8_extracted_k141_145449 | Not-determined |
| S8_extracted_k141_24133 | Not-determined |
| S8_extracted_k141_127991 | Not-determined |
| S8_extracted_k141_127992 | Not-determined |
| S8_extracted_k141_171484 | Not-determined |
| S8_extracted_k141_136414 | Low-quality |
| S8_extracted_k141_171485 | Not-determined |
| S8_extracted_k141_136417 | Not-determined |
| S8_extracted_k141_24138 | Not-determined |
| S8_extracted_k141_75712 | Not-determined |
| S8_extracted_k141_171488 | Not-determined |
| S8_extracted_k141_14998 | Low-quality |
| S8_extracted_k141_75714 | Not-determined |
| S8_extracted_k141_171489 | Not-determined |
| S8_extracted_k141_136418 | Not-determined |
| S8_extracted_k141_154102 | Not-determined |
| S8_extracted_k141_84441 | Not-determined |
| S8_extracted_k141_84442 | Not-determined |
| S8_extracted_k141_205754 | Not-determined |
| S8_extracted_k141_59050 | Low-quality |
| S8_extracted_k141_162287 | Not-determined |
| S8_extracted_k141_171497 | Not-determined |
| S8_extracted_k141_197655 | Not-determined |
| S8_extracted_k141_128000 | Not-determined |
| S8_extracted_k141_205758 | Not-determined |
| S8_extracted_k141_136424 | Not-determined |
| S8_extracted_k141_205761 | Not-determined |
| S8_extracted_k141_145462 | Not-determined |
| S8_extracted_k141_197662 | Not-determined |
| S8_extracted_k141_145463 | Not-determined |
| S8_extracted_k141_101715 | Not-determined |
| S8_extracted_k141_180234 | Not-determined |
| S8_extracted_k141_180235 | Low-quality |
| S8_extracted_k141_75727 | Not-determined |
| S8_extracted_k141_84444 | Not-determined |
| S8_extracted_k141_128003 | Low-quality |
| S8_extracted_k141_162292 | Not-determined |
| S8_extracted_k141_67029 | Not-determined |
| S8_extracted_k141_162294 | Not-determined |
| S8_extracted_k141_154115 | Low-quality |
| S8_extracted_k141_59061 | Not-determined |
| S8_extracted_k141_24147 | Not-determined |
| S8_extracted_k141_101724 | Not-determined |
| S8_extracted_k141_32756 | Not-determined |
| S8_extracted_k141_171509 | Not-determined |
| S8_extracted_k141_128008 | Not-determined |
| S8_extracted_k141_197671 | Low-quality |
| S8_extracted_k141_32758 | Not-determined |
| S8_extracted_k141_101730 | Not-determined |
| S8_extracted_k141_119391 | Not-determined |
| S8_extracted_k141_180243 | Not-determined |
| S8_extracted_k141_67038 | Not-determined |
| S8_extracted_k141_101731 | Low-quality |
| S8_extracted_k141_128017 | Not-determined |
| S8_extracted_k141_187809 | Not-determined |
| S8_extracted_k141_180246 | Not-determined |
| S8_extracted_k141_187810 | Not-determined |
| S8_extracted_k141_6664 | Not-determined |
| S8_extracted_k141_187813 | Not-determined |
| S8_extracted_k141_145478 | Not-determined |
| S8_extracted_k141_205772 | Not-determined |
| S8_extracted_k141_101735 | Not-determined |
| S8_extracted_k141_84450 | Not-determined |
| S8_extracted_k141_136433 | Not-determined |
| S8_extracted_k141_180247 | Not-determined |
| S8_extracted_k141_84451 | Not-determined |
| S8_extracted_k141_154123 | Low-quality |
| S8_extracted_k141_67043 | Not-determined |
| S8_extracted_k141_93251 | Not-determined |
| S8_extracted_k141_197679 | Low-quality |
| S8_extracted_k141_93252 | Not-determined |
| S8_extracted_k141_49317 | Not-determined |
| S8_extracted_k141_180250 | Not-determined |
| S8_extracted_k141_24162 | Not-determined |
| S8_extracted_k141_180251 | Not-determined |
| S8_extracted_k141_197680 | Not-determined |
| S8_extracted_k141_180252 | Not-determined |
| S8_extracted_k141_101737 | Not-determined |
| S8_extracted_k141_154127 | Not-determined |
| S8_extracted_k141_197682 | Not-determined |
| S8_extracted_k141_49319 | Not-determined |
| S8_extracted_k141_59072 | Not-determined |
| S8_extracted_k141_6668 | Not-determined |
| S8_extracted_k141_154129 | Not-determined |
| S8_extracted_k141_32768 | Not-determined |
| S8_extracted_k141_75745 | Not-determined |
| S8_extracted_k141_171521 | Low-quality |
| S8_extracted_k141_205785 | Low-quality |
| S8_extracted_k141_145494 | Not-determined |
| S8_extracted_k141_154132 | Not-determined |
| S8_extracted_k141_154133 | Not-determined |
| S8_extracted_k141_101744 | Not-determined |
| S8_extracted_k141_49331 | Not-determined |
| S8_extracted_k141_67052 | Not-determined |
| S8_extracted_k141_59080 | Not-determined |
| S8_extracted_k141_128039 | Not-determined |
| S8_extracted_k141_49333 | Not-determined |
| S8_extracted_k141_145501 | Not-determined |
| S8_extracted_k141_101747 | Not-determined |
| S8_extracted_k141_49338 | Not-determined |
| S8_extracted_k141_205790 | Not-determined |
| S8_extracted_k141_32775 | Not-determined |
| S8_extracted_k141_75755 | Not-determined |
| S8_extracted_k141_154136 | Not-determined |
| S8_extracted_k141_75756 | Not-determined |
| S8_extracted_k141_162320 | Not-determined |
| S8_extracted_k141_180263 | Not-determined |
| S8_extracted_k141_32779 | Not-determined |
| S8_extracted_k141_59087 | Not-determined |
| S8_extracted_k141_101754 | Not-determined |
| S8_extracted_k141_93271 | Not-determined |
| S8_extracted_k141_75760 | Not-determined |
| S8_extracted_k141_75761 | Not-determined |
| S8_extracted_k141_197700 | Not-determined |
| S8_extracted_k141_32781 | Not-determined |
| S8_extracted_k141_67066 | Not-determined |
| S8_extracted_k141_75762 | Not-determined |
| S8_extracted_k141_41279 | Not-determined |
| S8_extracted_k141_59091 | Not-determined |
| S8_extracted_k141_49342 | Low-quality |
| S8_extracted_k141_49343 | Not-determined |
| S8_extracted_k141_6686 | Not-determined |
| S8_extracted_k141_180271 | Not-determined |
| S8_extracted_k141_67069 | Not-determined |
| S8_extracted_k141_180274 | Not-determined |
| S8_extracted_k141_49346 | Not-determined |
| S8_extracted_k141_119409 | Not-determined |
| S8_extracted_k141_84481 | Not-determined |
| S8_extracted_k141_119410 | Not-determined |
| S8_extracted_k141_24177 | Not-determined |
| S8_extracted_k141_67082 | Not-determined |
| S8_extracted_k141_32792 | Low-quality |
| S8_extracted_k141_180284 | Not-determined |
| S8_extracted_k141_75773 | Not-determined |
| S8_extracted_k141_162329 | Not-determined |
| S8_extracted_k141_197718 | Not-determined |
| S8_extracted_k141_128060 | Not-determined |
| S8_extracted_k141_84487 | Not-determined |
| S8_extracted_k141_24182 | Not-determined |
| S8_extracted_k141_93279 | Not-determined |
| S8_extracted_k141_24183 | Not-determined |
| S8_extracted_k141_145521 | Not-determined |
| S8_extracted_k141_84489 | Not-determined |
| S8_extracted_k141_162337 | Not-determined |
| S8_extracted_k141_75785 | Not-determined |
| S8_extracted_k141_128072 | Not-determined |
| S8_extracted_k141_84491 | Not-determined |
| S8_extracted_k141_171555 | Not-determined |
| S8_extracted_k141_41297 | Low-quality |
| S8_extracted_k141_101778 | Not-determined |
| S8_extracted_k141_84495 | Not-determined |
| S8_extracted_k141_154163 | Not-determined |
| S8_extracted_k141_197721 | Not-determined |
| S8_extracted_k141_32806 | Not-determined |
| S8_extracted_k141_41302 | Not-determined |
| S8_extracted_k141_171560 | Not-determined |
| S8_extracted_k141_171562 | Not-determined |
| S8_extracted_k141_180295 | Not-determined |
| S8_extracted_k141_93287 | Not-determined |
| S8_extracted_k141_24192 | Low-quality |
| S8_extracted_k141_197726 | Not-determined |
| S8_extracted_k141_136466 | Not-determined |
| S8_extracted_k141_136468 | Not-determined |
| S8_extracted_k141_145531 | Low-quality |
| S8_extracted_k141_128083 | Not-determined |
| S8_extracted_k141_6706 | Not-determined |
| S8_extracted_k141_162343 | Not-determined |
| S8_extracted_k141_24200 | Not-determined |
| S8_extracted_k141_67084 | Not-determined |
| S8_extracted_k141_162344 | Not-determined |
| S8_extracted_k141_180301 | Not-determined |
| S8_extracted_k141_171573 | Not-determined |
| S8_extracted_k141_59109 | Not-determined |
| S8_extracted_k141_6710 | Not-determined |
| S8_extracted_k141_59110 | Not-determined |
| S8_extracted_k141_59112 | Not-determined |
| S8_extracted_k141_136475 | Not-determined |
| S8_extracted_k141_24201 | Not-determined |
| S8_extracted_k141_171574 | Not-determined |
| S8_extracted_k141_110854 | Not-determined |
| S8_extracted_k141_187830 | Not-determined |
| S8_extracted_k141_145541 | Not-determined |
| S8_extracted_k141_67093 | Low-quality |
| S8_extracted_k141_75801 | Not-determined |
| S8_extracted_k141_101801 | Not-determined |
| S8_extracted_k141_171581 | Not-determined |
| S8_extracted_k141_49367 | Not-determined |
| S8_extracted_k141_171583 | Not-determined |
| S8_extracted_k141_24205 | Not-determined |
| S8_extracted_k141_171584 | Not-determined |
| S8_extracted_k141_75806 | Not-determined |
| S8_extracted_k141_84516 | Not-determined |
| S8_extracted_k141_84517 | Not-determined |
| S8_extracted_k141_162360 | Not-determined |
| S8_extracted_k141_180312 | Not-determined |
| S8_extracted_k141_171587 | Not-determined |
| S8_extracted_k141_145545 | Not-determined |
| S8_extracted_k141_136487 | Low-quality |
| S8_extracted_k141_180315 | Not-determined |
| S8_extracted_k141_101808 | Not-determined |
| S8_extracted_k141_6722 | Not-determined |
| S8_extracted_k141_197737 | Not-determined |
| S8_extracted_k141_119440 | Not-determined |
| S8_extracted_k141_162368 | Not-determined |
| S8_extracted_k141_75813 | Not-determined |
| S8_extracted_k141_187836 | Not-determined |
| S8_extracted_k141_128091 | Not-determined |
| S8_extracted_k141_84522 | Not-determined |
| S8_extracted_k141_101816 | Not-determined |
| S8_extracted_k141_128094 | Not-determined |
| S8_extracted_k141_110870 | Low-quality |
| S8_extracted_k141_136496 | Low-quality |
| S8_extracted_k141_162375 | Not-determined |
| S8_extracted_k141_187838 | Not-determined |
| S8_extracted_k141_162376 | Not-determined |
| S8_extracted_k141_49375 | Not-determined |
| S8_extracted_k141_84525 | Not-determined |
| S8_extracted_k141_75819 | Not-determined |
| S8_extracted_k141_171607 | Not-determined |
| S8_extracted_k141_145554 | Not-determined |
| S8_extracted_k141_101825 | Low-quality |
| S8_extracted_k141_59132 | Not-determined |
| S8_extracted_k141_84532 | Not-determined |
| S8_extracted_k141_101828 | Not-determined |
| S8_extracted_k141_93311 | Not-determined |
| S8_extracted_k141_75831 | Low-quality |
| S8_extracted_k141_59136 | Not-determined |
| S8_extracted_k141_187846 | Not-determined |
| S8_extracted_k141_75832 | Low-quality |
| S8_extracted_k141_24218 | Low-quality |
| S8_extracted_k141_187847 | Not-determined |
| S8_extracted_k141_197756 | Low-quality |
| S8_extracted_k141_24221 | Not-determined |
| S8_extracted_k141_59140 | Not-determined |
| S8_extracted_k141_128105 | Low-quality |
| S8_extracted_k141_162387 | Not-determined |
| S8_extracted_k141_59141 | Not-determined |
| S8_extracted_k141_145564 | Low-quality |
| S8_extracted_k141_154189 | Not-determined |
| S8_extracted_k141_84542 | Not-determined |
| S8_extracted_k141_6732 | Not-determined |
| S8_extracted_k141_145565 | Not-determined |
| S8_extracted_k141_128110 | Not-determined |
| S8_extracted_k141_59144 | Not-determined |
| S8_extracted_k141_205798 | Not-determined |
| S8_extracted_k141_205800 | Not-determined |
| S8_extracted_k141_24226 | Not-determined |
| S8_extracted_k141_6739 | Not-determined |
| S8_extracted_k141_6741 | Low-quality |
| S8_extracted_k141_145566 | Not-determined |
| S8_extracted_k141_110886 | Not-determined |
| S8_extracted_k141_24234 | Not-determined |
| S8_extracted_k141_197763 | Not-determined |
| S8_extracted_k141_6745 | Not-determined |
| S8_extracted_k141_110892 | Not-determined |
| S8_extracted_k141_84557 | Not-determined |
| S8_extracted_k141_32821 | Not-determined |
| S8_extracted_k141_6747 | Not-determined |
| S8_extracted_k141_154199 | Low-quality |
| S8_extracted_k141_59154 | Not-determined |
| S8_extracted_k141_162400 | Low-quality |
| S8_extracted_k141_136508 | Not-determined |
| S8_extracted_k141_6749 | Not-determined |
| S8_extracted_k141_205809 | Low-quality |
| S8_extracted_k141_145575 | Not-determined |
| S8_extracted_k141_32823 | Not-determined |
| S8_extracted_k141_187859 | Not-determined |
| S8_extracted_k141_24243 | Not-determined |
| S8_extracted_k141_67120 | Not-determined |
| S8_extracted_k141_24244 | Not-determined |
| S8_extracted_k141_171622 | Low-quality |
| S8_extracted_k141_24246 | Not-determined |
| S8_extracted_k141_93342 | Not-determined |
| S8_extracted_k141_67125 | Not-determined |
| S8_extracted_k141_101849 | Not-determined |
| S8_extracted_k141_101850 | Not-determined |
| S8_extracted_k141_154205 | Not-determined |
| S8_extracted_k141_49408 | Not-determined |
| S8_extracted_k141_67127 | Not-determined |
| S8_extracted_k141_101853 | Not-determined |
| S8_extracted_k141_187870 | Not-determined |
| S8_extracted_k141_187871 | Not-determined |
| S8_extracted_k141_67128 | Not-determined |
| S8_extracted_k141_145581 | Not-determined |
| S8_extracted_k141_171629 | Not-determined |

| | |
|---|---|
| S8_extracted_k141_6754 | Low-quality |
| S8_extracted_k141_197777 | Not-determined |
| S8_extracted_k141_145583 | Not-determined |
| S8_extracted_k141_75866 | Not-determined |
| S8_extracted_k141_197778 | Not-determined |
| S8_extracted_k141_171631 | Not-determined |
| S8_extracted_k141_32833 | Not-determined |
| S8_extracted_k141_6758 | Not-determined |
| S8_extracted_k141_154211 | Not-determined |
| S8_extracted_k141_32835 | Not-determined |
| S8_extracted_k141_59166 | Not-determined |
| S8_extracted_k141_93349 | Not-determined |
| S8_extracted_k141_197784 | Not-determined |
| S8_extracted_k141_84579 | Not-determined |
| S8_extracted_k141_101861 | Not-determined |
| S8_extracted_k141_24254 | Not-determined |
| S8_extracted_k141_84580 | Not-determined |
| S8_extracted_k141_49410 | Not-determined |
| S8_extracted_k141_128119 | Not-determined |
| S8_extracted_k141_15030 | Not-determined |
| S8_extracted_k141_187882 | Not-determined |
| S8_extracted_k141_162422 | Not-determined |
| S8_extracted_k141_197791 | Not-determined |
| S8_extracted_k141_162423 | Not-determined |
| S8_extracted_k141_187884 | Not-determined |
| S8_extracted_k141_145597 | Not-determined |
| S8_extracted_k141_15034 | Low-quality |
| S8_extracted_k141_84588 | Not-determined |
| S8_extracted_k141_119454 | Not-determined |
| S8_extracted_k141_49411 | Not-determined |
| S8_extracted_k141_154224 | Low-quality |
| S8_extracted_k141_101871 | Not-determined |
| S8_extracted_k141_171646 | Not-determined |
| S8_extracted_k141_101874 | Not-determined |
| S8_extracted_k141_187888 | Not-determined |
| S8_extracted_k141_101875 | Low-quality |
| S8_extracted_k141_24262 | Not-determined |
| S8_extracted_k141_119457 | Not-determined |
| S8_extracted_k141_162429 | Not-determined |
| S8_extracted_k141_24263 | Not-determined |
| S8_extracted_k141_101877 | Not-determined |
| S8_extracted_k141_136524 | Not-determined |
| S8_extracted_k141_15042 | Not-determined |
| S8_extracted_k141_145601 | Low-quality |
| S8_extracted_k141_145602 | Not-determined |
| S8_extracted_k141_162434 | Not-determined |
| S8_extracted_k141_187893 | Low-quality |
| S8_extracted_k141_59184 | Not-determined |
| S8_extracted_k141_180359 | Not-determined |
| S8_extracted_k141_187896 | Not-determined |
| S8_extracted_k141_24265 | Low-quality |
| S8_extracted_k141_67135 | Not-determined |
| S8_extracted_k141_32850 | Not-determined |
| S8_extracted_k141_171651 | Not-determined |
| S8_extracted_k141_24267 | Not-determined |
| S8_extracted_k141_67136 | Not-determined |
| S8_extracted_k141_32853 | Not-determined |
| S8_extracted_k141_136530 | Not-determined |
| S8_extracted_k141_6770 | Not-determined |
| S8_extracted_k141_197795 | Not-determined |
| S8_extracted_k141_145610 | Not-determined |
| S8_extracted_k141_67140 | Not-determined |
| S8_extracted_k141_171654 | Not-determined |
| S8_extracted_k141_24272 | Not-determined |
| S8_extracted_k141_128144 | Not-determined |
| S8_extracted_k141_6773 | Not-determined |
| S8_extracted_k141_162442 | Not-determined |
| S8_extracted_k141_197796 | Not-determined |
| S8_extracted_k141_180371 | Not-determined |
| S8_extracted_k141_101890 | Not-determined |
| S8_extracted_k141_75887 | Low-quality |
| S8_extracted_k141_101892 | Not-determined |
| S8_extracted_k141_15054 | Not-determined |
| S8_extracted_k141_136536 | Not-determined |
| S8_extracted_k141_136537 | Not-determined |
| S8_extracted_k141_24283 | Not-determined |
| S8_extracted_k141_136538 | Not-determined |
| S8_extracted_k141_136539 | Not-determined |
| S8_extracted_k141_187911 | Not-determined |
| S8_extracted_k141_162453 | Not-determined |
| S8_extracted_k141_67151 | Not-determined |
| S8_extracted_k141_187914 | Not-determined |
| S8_extracted_k141_59207 | Not-determined |
| S8_extracted_k141_84608 | Not-determined |
| S8_extracted_k141_171662 | Not-determined |
| S8_extracted_k141_6781 | Low-quality |
| S8_extracted_k141_136545 | Not-determined |
| S8_extracted_k141_67154 | Not-determined |
| S8_extracted_k141_119467 | Low-quality |
| S8_extracted_k141_24290 | Not-determined |
| S8_extracted_k141_119472 | Not-determined |
| S8_extracted_k141_197806 | Not-determined |
| S8_extracted_k141_101900 | Not-determined |
| S8_extracted_k141_6793 | Not-determined |
| S8_extracted_k141_101903 | Not-determined |
| S8_extracted_k141_59220 | Not-determined |
| S8_extracted_k141_128168 | Not-determined |
| S8_extracted_k141_15073 | Not-determined |
| S8_extracted_k141_93383 | Low-quality |
| S8_extracted_k141_24296 | Not-determined |
| S8_extracted_k141_110897 | Not-determined |
| S8_extracted_k141_49428 | Not-determined |
| S8_extracted_k141_67158 | Not-determined |
| S8_extracted_k141_84629 | Not-determined |
| S8_extracted_k141_15075 | Not-determined |
| S8_extracted_k141_197817 | Low-quality |
| S8_extracted_k141_32870 | Not-determined |
| S8_extracted_k141_171676 | Not-determined |
| S8_extracted_k141_101917 | Not-determined |
| S8_extracted_k141_171677 | Not-determined |
| S8_extracted_k141_162456 | Not-determined |
| S8_extracted_k141_162457 | Not-determined |
| S8_extracted_k141_32873 | Not-determined |
| S8_extracted_k141_101921 | Not-determined |
| S8_extracted_k141_101922 | Not-determined |
| S8_extracted_k141_162460 | Not-determined |
| S8_extracted_k141_101923 | Low-quality |
| S8_extracted_k141_15080 | Not-determined |
| S8_extracted_k141_197828 | Not-determined |
| S8_extracted_k141_59230 | Not-determined |
| S8_extracted_k141_171689 | Not-determined |
| S8_extracted_k141_101927 | Not-determined |
| S8_extracted_k141_6813 | Not-determined |
| S8_extracted_k141_101928 | Not-determined |
| S8_extracted_k141_67169 | Low-quality |
| S8_extracted_k141_145637 | Not-determined |
| S8_extracted_k141_171692 | Not-determined |
| S8_extracted_k141_128183 | Not-determined |
| S8_extracted_k141_180380 | Low-quality |
| S8_extracted_k141_67172 | Not-determined |
| S8_extracted_k141_187918 | Low-quality |
| S8_extracted_k141_171695 | Not-determined |
| S8_extracted_k141_101930 | Not-determined |
| S8_extracted_k141_101931 | Not-determined |
| S8_extracted_k141_49435 | Not-determined |
| S8_extracted_k141_119486 | Not-determined |
| S8_extracted_k141_145642 | Not-determined |
| S8_extracted_k141_67177 | Low-quality |
| S8_extracted_k141_145643 | Not-determined |
| S8_extracted_k141_180386 | Not-determined |
| S8_extracted_k141_171699 | Not-determined |
| S8_extracted_k141_49438 | Not-determined |
| S8_extracted_k141_162468 | Not-determined |
| S8_extracted_k141_67182 | Not-determined |
| S8_extracted_k141_24313 | Not-determined |
| S8_extracted_k141_119495 | Not-determined |
| S8_extracted_k141_145651 | Not-determined |
| S8_extracted_k141_171701 | Low-quality |
| S8_extracted_k141_110908 | Low-quality |
| S8_extracted_k141_49450 | Low-quality |
| S8_extracted_k141_67191 | Not-determined |
| S8_extracted_k141_171705 | Not-determined |
| S8_extracted_k141_162479 | Not-determined |
| S8_extracted_k141_84649 | Low-quality |
| S8_extracted_k141_101936 | Not-determined |
| S8_extracted_k141_84652 | Not-determined |
| S8_extracted_k141_101937 | Not-determined |
| S8_extracted_k141_6822 | Not-determined |
| S8_extracted_k141_119505 | Not-determined |
| S8_extracted_k141_180403 | Not-determined |
| S8_extracted_k141_84654 | Not-determined |
| S8_extracted_k141_171711 | Not-determined |
| S8_extracted_k141_154233 | Not-determined |
| S8_extracted_k141_6823 | Not-determined |
| S8_extracted_k141_171714 | Not-determined |
| S8_extracted_k141_110914 | Not-determined |
| S8_extracted_k141_59249 | Low-quality |
| S8_extracted_k141_128212 | Not-determined |
| S8_extracted_k141_110915 | Low-quality |
| S8_extracted_k141_136561 | Not-determined |
| S8_extracted_k141_145656 | Low-quality |
| S8_extracted_k141_24326 | Not-determined |
| S8_extracted_k141_49458 | Not-determined |
| S8_extracted_k141_110919 | Not-determined |
| S8_extracted_k141_145660 | Not-determined |
| S8_extracted_k141_154239 | Not-determined |
| S8_extracted_k141_75907 | Not-determined |
| S8_extracted_k141_145661 | Not-determined |
| S8_extracted_k141_24330 | Not-determined |
| S8_extracted_k141_41327 | Low-quality |
| S8_extracted_k141_67207 | Not-determined |
| S8_extracted_k141_24332 | Not-determined |
| S8_extracted_k141_171724 | Not-determined |
| S8_extracted_k141_24333 | Not-determined |
| S8_extracted_k141_67210 | Not-determined |
| S8_extracted_k141_145663 | Low-quality |
| S8_extracted_k141_67212 | Not-determined |
| S8_extracted_k141_49468 | Not-determined |
| S8_extracted_k141_84664 | Not-determined |
| S8_extracted_k141_171726 | Not-determined |
| S8_extracted_k141_41335 | Not-determined |
| S8_extracted_k141_67215 | Not-determined |
| S8_extracted_k141_41338 | Not-determined |
| S8_extracted_k141_110930 | Not-determined |
| S8_extracted_k141_75914 | Not-determined |
| S8_extracted_k141_75916 | Low-quality |
| S8_extracted_k141_171736 | Not-determined |
| S8_extracted_k141_84671 | Not-determined |
| S8_extracted_k141_136583 | Low-quality |
| S8_extracted_k141_205827 | Not-determined |
| S8_extracted_k141_15129 | Not-determined |
| S8_extracted_k141_136585 | Not-determined |
| S8_extracted_k141_6844 | Not-determined |
| S8_extracted_k141_197851 | Low-quality |
| S8_extracted_k141_110935 | Not-determined |
| S8_extracted_k141_101957 | Not-determined |
| S8_extracted_k141_15133 | Not-determined |
| S8_extracted_k141_162491 | Not-determined |
| S8_extracted_k141_162495 | Not-determined |
| S8_extracted_k141_180420 | Not-determined |
| S8_extracted_k141_75927 | Not-determined |
| S8_extracted_k141_180421 | Not-determined |
| S8_extracted_k141_84683 | Not-determined |
| S8_extracted_k141_84684 | Not-determined |
| S8_extracted_k141_187955 | Not-determined |
| S8_extracted_k141_187956 | Not-determined |
| S8_extracted_k141_6849 | Not-determined |
| S8_extracted_k141_128222 | Low-quality |
| S8_extracted_k141_197863 | Low-quality |
| S8_extracted_k141_180427 | Not-determined |
| S8_extracted_k141_59284 | Not-determined |
| S8_extracted_k141_128225 | Not-determined |
| S8_extracted_k141_15140 | Not-determined |
| S8_extracted_k141_24345 | Not-determined |
| S8_extracted_k141_49493 | Not-determined |
| S8_extracted_k141_15142 | Not-determined |
| S8_extracted_k141_128229 | Not-determined |
| S8_extracted_k141_49496 | Not-determined |
| S8_extracted_k141_15146 | Not-determined |
| S8_extracted_k141_110942 | Not-determined |
| S8_extracted_k141_171752 | Not-determined |
| S8_extracted_k141_136599 | Not-determined |
| S8_extracted_k141_197865 | Not-determined |
| S8_extracted_k141_15149 | Not-determined |
| S8_extracted_k141_162510 | Low-quality |
| S8_extracted_k141_49499 | Not-determined |
| S8_extracted_k141_59292 | Not-determined |
| S8_extracted_k141_59293 | Not-determined |
| S8_extracted_k141_162512 | Not-determined |
| S8_extracted_k141_128232 | Low-quality |
| S8_extracted_k141_136604 | Not-determined |
| S8_extracted_k141_6866 | Not-determined |
| S8_extracted_k141_101965 | Low-quality |
| S8_extracted_k141_84702 | Not-determined |
| S8_extracted_k141_197869 | Not-determined |
| S8_extracted_k141_197873 | Not-determined |
| S8_extracted_k141_180440 | Not-determined |
| S8_extracted_k141_136612 | Not-determined |
| S8_extracted_k141_59303 | Not-determined |
| S8_extracted_k141_136613 | Not-determined |
| S8_extracted_k141_59304 | Not-determined |
| S8_extracted_k141_6876 | Not-determined |
| S8_extracted_k141_15155 | Not-determined |
| S8_extracted_k141_15158 | Not-determined |
| S8_extracted_k141_15160 | Not-determined |
| S8_extracted_k141_75949 | Not-determined |
| S8_extracted_k141_180447 | Not-determined |
| S8_extracted_k141_180449 | Not-determined |
| S8_extracted_k141_110960 | Not-determined |
| S8_extracted_k141_49501 | Not-determined |
| S8_extracted_k141_59311 | Not-determined |
| S8_extracted_k141_93388 | Not-determined |
| S8_extracted_k141_171773 | Not-determined |
| S8_extracted_k141_197886 | Not-determined |
| S8_extracted_k141_171774 | Not-determined |
| S8_extracted_k141_145678 | Not-determined |
| S8_extracted_k141_93389 | Not-determined |
| S8_extracted_k141_187977 | Not-determined |
| S8_extracted_k141_49507 | Not-determined |
| S8_extracted_k141_187978 | Not-determined |
| S8_extracted_k141_145682 | Not-determined |
| S8_extracted_k141_205835 | Low-quality |
| S8_extracted_k141_162518 | Not-determined |
| S8_extracted_k141_136616 | Not-determined |
| S8_extracted_k141_180454 | Not-determined |
| S8_extracted_k141_180455 | Not-determined |
| S8_extracted_k141_15167 | Not-determined |
| S8_extracted_k141_6882 | Not-determined |
| S8_extracted_k141_93399 | Not-determined |
| S8_extracted_k141_24350 | Not-determined |
| S8_extracted_k141_59324 | Not-determined |
| S8_extracted_k141_136623 | Not-determined |
| S8_extracted_k141_110973 | Not-determined |
| S8_extracted_k141_187991 | Not-determined |
| S8_extracted_k141_145697 | Not-determined |
| S8_extracted_k141_119530 | Low-quality |
| S8_extracted_k141_171785 | Not-determined |
| S8_extracted_k141_145699 | Not-determined |
| S8_extracted_k141_110980 | Low-quality |
| S8_extracted_k141_136635 | Not-determined |
| S8_extracted_k141_171788 | Not-determined |
| S8_extracted_k141_49522 | Not-determined |
| S8_extracted_k141_110984 | Not-determined |
| S8_extracted_k141_145704 | Not-determined |
| S8_extracted_k141_136637 | Not-determined |
| S8_extracted_k141_110987 | Not-determined |
| S8_extracted_k141_162523 | Not-determined |
| S8_extracted_k141_84730 | Not-determined |
| S8_extracted_k141_49529 | Not-determined |
| S8_extracted_k141_128257 | Not-determined |
| S8_extracted_k141_197900 | Low-quality |
| S8_extracted_k141_145706 | Not-determined |
| S8_extracted_k141_59333 | Not-determined |
| S8_extracted_k141_128263 | Not-determined |
| S8_extracted_k141_145708 | Not-determined |
| S8_extracted_k141_136645 | Not-determined |
| S8_extracted_k141_171793 | Low-quality |
| S8_extracted_k141_84736 | Not-determined |
| S8_extracted_k141_93421 | Low-quality |
| S8_extracted_k141_128265 | Low-quality |
| S8_extracted_k141_162535 | Not-determined |
| S8_extracted_k141_187999 | Not-determined |
| S8_extracted_k141_128269 | Not-determined |
| S8_extracted_k141_205855 | Not-determined |
| S8_extracted_k141_110993 | Not-determined |
| S8_extracted_k141_171801 | Not-determined |
| S8_extracted_k141_119552 | Low-quality |
| S8_extracted_k141_15183 | Not-determined |
| S8_extracted_k141_110995 | Not-determined |
| S8_extracted_k141_197902 | Not-determined |
| S8_extracted_k141_136658 | Not-determined |
| S8_extracted_k141_180479 | Not-determined |
| S8_extracted_k141_110998 | Not-determined |
| S8_extracted_k141_205861 | Not-determined |
| S8_extracted_k141_205862 | Not-determined |
| S8_extracted_k141_24371 | Not-determined |
| S8_extracted_k141_171805 | Not-determined |
| S8_extracted_k141_180480 | Low-quality |
| S8_extracted_k141_24373 | Not-determined |
| S8_extracted_k141_128279 | Not-determined |
| S8_extracted_k141_171806 | Low-quality |
| S8_extracted_k141_136663 | Not-determined |
| S8_extracted_k141_49552 | Not-determined |
| S8_extracted_k141_136664 | Not-determined |
| S8_extracted_k141_136666 | Not-determined |
| S8_extracted_k141_188013 | Not-determined |
| S8_extracted_k141_24383 | Not-determined |
| S8_extracted_k141_136668 | Not-determined |
| S8_extracted_k141_205868 | Not-determined |
| S8_extracted_k141_59357 | Not-determined |
| S8_extracted_k141_32917 | Not-determined |
| S8_extracted_k141_75989 | Not-determined |
| S8_extracted_k141_111015 | Not-determined |
| S8_extracted_k141_205877 | Not-determined |
| S8_extracted_k141_188021 | Not-determined |
| S8_extracted_k141_6902 | Not-determined |
| S8_extracted_k141_188024 | Not-determined |
| S8_extracted_k141_32920 | Not-determined |
| S8_extracted_k141_75995 | Not-determined |
| S8_extracted_k141_84764 | Not-determined |
| S8_extracted_k141_136675 | Low-quality |
| S8_extracted_k141_93444 | Not-determined |
| S8_extracted_k141_102006 | Low-quality |
| S8_extracted_k141_15206 | Not-determined |
| S8_extracted_k141_171819 | Not-determined |
| S8_extracted_k141_32926 | Not-determined |
| S8_extracted_k141_32927 | Low-quality |
| S8_extracted_k141_93446 | Not-determined |
| S8_extracted_k141_24401 | Not-determined |
| S8_extracted_k141_75999 | Not-determined |
| S8_extracted_k141_76000 | Not-determined |
| S8_extracted_k141_76003 | Not-determined |
| S8_extracted_k141_102010 | Not-determined |
| S8_extracted_k141_145745 | Not-determined |
| S8_extracted_k141_145747 | Not-determined |
| S8_extracted_k141_15215 | Not-determined |
| S8_extracted_k141_102014 | Not-determined |
| S8_extracted_k141_119570 | Not-determined |
| S8_extracted_k141_84782 | Not-determined |
| S8_extracted_k141_145755 | Not-determined |
| S8_extracted_k141_93447 | Not-determined |
| S8_extracted_k141_111032 | Not-determined |
| S8_extracted_k141_93449 | Low-quality |
| S8_extracted_k141_6932 | Low-quality |
| S8_extracted_k141_197913 | Not-determined |
| S8_extracted_k141_188031 | Not-determined |
| S8_extracted_k141_128287 | Not-determined |
| S8_extracted_k141_145761 | Not-determined |
| S8_extracted_k141_93451 | Low-quality |
| S8_extracted_k141_162553 | Not-determined |
| S8_extracted_k141_128288 | Not-determined |
| S8_extracted_k141_205895 | Not-determined |
| S8_extracted_k141_128289 | Not-determined |
| S8_extracted_k141_24424 | Not-determined |
| S8_extracted_k141_59390 | Not-determined |
| S8_extracted_k141_188034 | Not-determined |
| S8_extracted_k141_128290 | Not-determined |
| S8_extracted_k141_67233 | Not-determined |
| S8_extracted_k141_76017 | Not-determined |
| S8_extracted_k141_15218 | Not-determined |
| S8_extracted_k141_49575 | Low-quality |
| S8_extracted_k141_59393 | Not-determined |
| S8_extracted_k141_128291 | Low-quality |
| S8_extracted_k141_76019 | Not-determined |
| S8_extracted_k141_84791 | Not-determined |
| S8_extracted_k141_188037 | Not-determined |
| S8_extracted_k141_102027 | Not-determined |
| S8_extracted_k141_188039 | Not-determined |
| S7_extracted_5843 | Medium-quality |
| S8_extracted_k141_67235 | Not-determined |
| S8_extracted_k141_24428 | Low-quality |
| S8_extracted_k141_136687 | Not-determined |
| S8_extracted_k141_93454 | Not-determined |
| S8_extracted_k141_128293 | Not-determined |
| S8_extracted_k141_197922 | Not-determined |
| S8_extracted_k141_188042 | Not-determined |
| S8_extracted_k141_24430 | Not-determined |
| S8_extracted_k141_59398 | Not-determined |
| S8_extracted_k141_6941 | Low-quality |
| S8_extracted_k141_93458 | Not-determined |
| S8_extracted_k141_49577 | Low-quality |
| S8_extracted_k141_67244 | Not-determined |
| S8_extracted_k141_136692 | Not-determined |
| S8_extracted_k141_188049 | Not-determined |
| S8_extracted_k141_171833 | Not-determined |
| S8_extracted_k141_171834 | Not-determined |
| S8_extracted_k141_67246 | Not-determined |
| S8_extracted_k141_41392 | Not-determined |
| S8_extracted_k141_15229 | Not-determined |
| S8_extracted_k141_84798 | Not-determined |
| S8_extracted_k141_59409 | Not-determined |
| S8_extracted_k141_188050 | Not-determined |
| S8_extracted_k141_24435 | Low-quality |
| S8_extracted_k141_102030 | Not-determined |
| S8_extracted_k141_171837 | Not-determined |
| S8_extracted_k141_188052 | Not-determined |
| S8_extracted_k141_84803 | Not-determined |
| S8_extracted_k141_84804 | Not-determined |
| S8_extracted_k141_93468 | Not-determined |
| S8_extracted_k141_128305 | Low-quality |
| S8_extracted_k141_188054 | Not-determined |
| S8_extracted_k141_49585 | Not-determined |
| S8_extracted_k141_24438 | Not-determined |
| S8_extracted_k141_67254 | Low-quality |
| S8_extracted_k141_41400 | Low-quality |
| S8_extracted_k141_93470 | Not-determined |
| S8_extracted_k141_49589 | Not-determined |
| S8_extracted_k141_162572 | Not-determined |
| S8_extracted_k141_188056 | Not-determined |
| S8_extracted_k141_205918 | Not-determined |
| S8_extracted_k141_145787 | Not-determined |
| S8_extracted_k141_59422 | Not-determined |
| S8_extracted_k141_67260 | Not-determined |
| S8_extracted_k141_32957 | Not-determined |
| S8_extracted_k141_111057 | Not-determined |
| S8_extracted_k141_102037 | Not-determined |
| S8_extracted_k141_162580 | Not-determined |
| S8_extracted_k141_102041 | Not-determined |
| S8_extracted_k141_111063 | Not-determined |
| S8_extracted_k141_6952 | Not-determined |
| S8_extracted_k141_49597 | Not-determined |
| S8_extracted_k141_205925 | Not-determined |
| S8_extracted_k141_128310 | Not-determined |
| S8_extracted_k141_41409 | Not-determined |
| S8_extracted_k141_76045 | Not-determined |
| S8_extracted_k141_136698 | Not-determined |
| S8_extracted_k141_111071 | Not-determined |
| S8_extracted_k141_93485 | Not-determined |
| S8_extracted_k141_111072 | Not-determined |
| S8_extracted_k141_32966 | Not-determined |
| S8_extracted_k141_197947 | Not-determined |
| S8_extracted_k141_102048 | Not-determined |
| S8_extracted_k141_41410 | Not-determined |
| S8_extracted_k141_24460 | Not-determined |
| S8_extracted_k141_197950 | Low-quality |
| S8_extracted_k141_41413 | Not-determined |
| S8_extracted_k141_162595 | Low-quality |
| S8_extracted_k141_128318 | Low-quality |
| S8_extracted_k141_197957 | Not-determined |
| S8_extracted_k141_59446 | Not-determined |
| S8_extracted_k141_102067 | Not-determined |
| S8_extracted_k141_188081 | Not-determined |
| S8_extracted_k141_102068 | Not-determined |
| S8_extracted_k141_102069 | Not-determined |
| S8_extracted_k141_41426 | Not-determined |
| S8_extracted_k141_162605 | Low-quality |
| S8_extracted_k141_197961 | Not-determined |
| S8_extracted_k141_102073 | Not-determined |
| S8_extracted_k141_136709 | Low-quality |
| S8_extracted_k141_171853 | Not-determined |
| S8_extracted_k141_188089 | Not-determined |
| S8_extracted_k141_59452 | Not-determined |
| S8_extracted_k141_41432 | Low-quality |
| S8_extracted_k141_171856 | Not-determined |
| S8_extracted_k141_197967 | Not-determined |
| S8_extracted_k141_32973 | Not-determined |
| S8_extracted_k141_59456 | Low-quality |
| S8_extracted_k141_197973 | Not-determined |
| S8_extracted_k141_93503 | Not-determined |
| S8_extracted_k141_59459 | Not-determined |
| S8_extracted_k141_205946 | Not-determined |
| S8_extracted_k141_128337 | Low-quality |
| S8_extracted_k141_180537 | Not-determined |
| S8_extracted_k141_128338 | Not-determined |
| S8_extracted_k141_180538 | Not-determined |
| S8_extracted_k141_67298 | Not-determined |
| S8_extracted_k141_111089 | Not-determined |
| S8_extracted_k141_41439 | Not-determined |
| S8_extracted_k141_111090 | Not-determined |
| S8_extracted_k141_171864 | Not-determined |
| S8_extracted_k141_162610 | Not-determined |
| S8_extracted_k141_41441 | Not-determined |
| S8_extracted_k141_102090 | Not-determined |
| S8_extracted_k141_162611 | Not-determined |
| S8_extracted_k141_180547 | Not-determined |
| S8_extracted_k141_76077 | Not-determined |
| S8_extracted_k141_171866 | Not-determined |
| S8_extracted_k141_84825 | Low-quality |
| S8_extracted_k141_41446 | Low-quality |
| S8_extracted_k141_111097 | Not-determined |
| S8_extracted_k141_76079 | Not-determined |
| S8_extracted_k141_93515 | Not-determined |
| S8_extracted_k141_171869 | Not-determined |
| S8_extracted_k141_41450 | Not-determined |
| S8_extracted_k141_171870 | Not-determined |
| S8_extracted_k141_128348 | Not-determined |
| S8_extracted_k141_84830 | Not-determined |
| S8_extracted_k141_32987 | Not-determined |
| S8_extracted_k141_49624 | Not-determined |
| S8_extracted_k141_205954 | Not-determined |
| S8_extracted_k141_197988 | Not-determined |
| S8_extracted_k141_84832 | Not-determined |
| S8_extracted_k141_93520 | Not-determined |
| S8_extracted_k141_128353 | Not-determined |
| S8_extracted_k141_49626 | Not-determined |
| S8_extracted_k141_76083 | Not-determined |
| S8_extracted_k141_171873 | Not-determined |
| S8_extracted_k141_76085 | Not-determined |
| S8_extracted_k141_41456 | Not-determined |
| S8_extracted_k141_128363 | Low-quality |
| S8_extracted_k141_180549 | Not-determined |
| S8_extracted_k141_197991 | Not-determined |
| S8_extracted_k141_145819 | Not-determined |
| S8_extracted_k141_32994 | Not-determined |
| S8_extracted_k141_119594 | Not-determined |
| S8_extracted_k141_93525 | Not-determined |
| S8_extracted_k141_145821 | Not-determined |
| S8_extracted_k141_128365 | Not-determined |
| S8_extracted_k141_49638 | Not-determined |
| S8_extracted_k141_93527 | Not-determined |
| S8_extracted_k141_171887 | Low-quality |
| S8_extracted_k141_180552 | Not-determined |
| S8_extracted_k141_119599 | Not-determined |
| S8_extracted_k141_145824 | Low-quality |
| S8_extracted_k141_197996 | Not-determined |
| S8_extracted_k141_119601 | Low-quality |
| S8_extracted_k141_59482 | Not-determined |
| S8_extracted_k141_119604 | Not-determined |
| S8_extracted_k141_93535 | Not-determined |
| S8_extracted_k141_59484 | Not-determined |
| S8_extracted_k141_67304 | Not-determined |
| S8_extracted_k141_49650 | Not-determined |
| S8_extracted_k141_145837 | Not-determined |
| S8_extracted_k141_171897 | Not-determined |
| S8_extracted_k141_180559 | Not-determined |
| S8_extracted_k141_205977 | Not-determined |
| S8_extracted_k141_171899 | Not-determined |
| S8_extracted_k141_33001 | Not-determined |
| S8_extracted_k141_67310 | Not-determined |
| S8_extracted_k141_67314 | Not-determined |
| S8_extracted_k141_162630 | Not-determined |
| S8_extracted_k141_128378 | Not-determined |
| S8_extracted_k141_162631 | Low-quality |
| S8_extracted_k141_145840 | Not-determined |
| S8_extracted_k141_162632 | Not-determined |
| S8_extracted_k141_205980 | Not-determined |
| S8_extracted_k141_49662 | Not-determined |
| S8_extracted_k141_188121 | Not-determined |
| S8_extracted_k141_136766 | Not-determined |
| S8_extracted_k141_49665 | Not-determined |
| S8_extracted_k141_111107 | Not-determined |
| S8_extracted_k141_188127 | Not-determined |
| S8_extracted_k141_33009 | Not-determined |
| S8_extracted_k141_145847 | Not-determined |
| S8_extracted_k141_111110 | Not-determined |
| S8_extracted_k141_76122 | Not-determined |
| S8_extracted_k141_205988 | Not-determined |
| S8_extracted_k141_136771 | Low-quality |
| S8_extracted_k141_198012 | Not-determined |
| S8_extracted_k141_67322 | Not-determined |
| S8_extracted_k141_198014 | Not-determined |
| S8_extracted_k141_33017 | Not-determined |
| S8_extracted_k141_145851 | Not-determined |
| S8_extracted_k141_49668 | Low-quality |
| S8_extracted_k141_188132 | Low-quality |
| S8_extracted_k141_198018 | Not-determined |
| S8_extracted_k141_198019 | Not-determined |
| S8_extracted_k141_136775 | Not-determined |
| S8_extracted_k141_15247 | Not-determined |

| | |
|---|---|
| SB_extracted_k141_67328 | Not-determined |
| SB_extracted_k141_136777 | Not-determined |
| SB_extracted_k141_136778 | Not-determined |
| SB_extracted_k141_59500 | Not-determined |
| SB_extracted_k141_162645 | Not-determined |
| SB_extracted_k141_180583 | Not-determined |
| SB_extracted_k141_162648 | Not-determined |
| SB_extracted_k141_93563 | Not-determined |
| SB_extracted_k141_15251 | Not-determined |
| SB_extracted_k141_67334 | Not-determined |
| SB_extracted_k141_171928 | Not-determined |
| SB_extracted_k141_102106 | Not-determined |
| SB_extracted_k141_67336 | Not-determined |
| SB_extracted_k141_93565 | Not-determined |
| SB_extracted_k141_15254 | Not-determined |
| SB_extracted_k141_145867 | Not-determined |
| SB_extracted_k141_67339 | Not-determined |
| SB_extracted_k141_171932 | Low-quality |
| SB_extracted_k141_33031 | Not-determined |
| SB_extracted_k141_206000 | Not-determined |
| SB_extracted_k141_102114 | Not-determined |
| SB_extracted_k141_162659 | Not-determined |
| SB_extracted_k141_206002 | Not-determined |
| SB_extracted_k141_198038 | Low-quality |
| SB_extracted_k141_206004 | Not-determined |
| SB_extracted_k141_49685 | Not-determined |
| SB_extracted_k141_206008 | Not-determined |
| SB_extracted_k141_145871 | Low-quality |
| SB_extracted_k141_119617 | Not-determined |
| SB_extracted_k141_206009 | Not-determined |
| SB_extracted_k141_198048 | Not-determined |
| SB_extracted_k141_76145 | Not-determined |
| SB_extracted_k141_198049 | Not-determined |
| SB_extracted_k141_33047 | Low-quality |
| SB_extracted_k141_180589 | Not-determined |
| SB_extracted_k141_198053 | Low-quality |
| SB_extracted_k141_188153 | Not-determined |
| SB_extracted_k141_49695 | Not-determined |
| SB_extracted_k141_111113 | Not-determined |
| SB_extracted_k141_206014 | Not-determined |
| SB_extracted_k141_119624 | Low-quality |
| SB_extracted_k141_162672 | Not-determined |
| SB_extracted_k141_41461 | Not-determined |
| SB_extracted_k141_171948 | Not-determined |
| SB_extracted_k141_76149 | Not-determined |
| SB_extracted_k141_145878 | Not-determined |
| SB_extracted_k141_49700 | Low-quality |
| SB_extracted_k141_136820 | Not-determined |
| SB_extracted_k141_111116 | Not-determined |
| SB_extracted_k141_102121 | Not-determined |
| SB_extracted_k141_111118 | Not-determined |
| SB_extracted_k141_136822 | Not-determined |
| SB_extracted_k141_180606 | Not-determined |
| SB_extracted_k141_59520 | Not-determined |
| SB_extracted_k141_111121 | Not-determined |
| SB_extracted_k141_119636 | Not-determined |
| SB_extracted_k141_59523 | Low-quality |
| SB_extracted_k141_162686 | Not-determined |
| SB_extracted_k141_76153 | Not-determined |
| SB_extracted_k141_171951 | Not-determined |
| SB_extracted_k141_162689 | Not-determined |
| SB_extracted_k141_171953 | Not-determined |
| SB_extracted_k141_41463 | Not-determined |
| SB_extracted_k141_198060 | Not-determined |
| SB_extracted_k141_119639 | Not-determined |
| SB_extracted_k141_76155 | Not-determined |
| SB_extracted_k141_24478 | Not-determined |
| SB_extracted_k141_180613 | Low-quality |
| SB_extracted_k141_102132 | Not-determined |
| SB_extracted_k141_198062 | Not-determined |
| SB_extracted_k141_171962 | Not-determined |
| SB_extracted_k141_59529 | Not-determined |
| SB_extracted_k141_41477 | Not-determined |
| SB_extracted_k141_206020 | Low-quality |
| SB_extracted_k141_76159 | Not-determined |
| SB_extracted_k141_102138 | Low-quality |
| SB_extracted_k141_6972 | Not-determined |
| SB_extracted_k141_136836 | Not-determined |
| SB_extracted_k141_76162 | Not-determined |
| SB_extracted_k141_15278 | Not-determined |
| SB_extracted_k141_119649 | Not-determined |
| SB_extracted_k141_162699 | Not-determined |
| SB_extracted_k141_111139 | Not-determined |
| SB_extracted_k141_49722 | Not-determined |
| SB_extracted_k141_136844 | Not-determined |
| SB_extracted_k141_15286 | Not-determined |
| SB_extracted_k141_162706 | Not-determined |
| SB_extracted_k141_84851 | Not-determined |
| SB_extracted_k141_136848 | Not-determined |
| SB_extracted_k141_49728 | Not-determined |
| SB_extracted_k141_111146 | Not-determined |
| SB_extracted_k141_49730 | Not-determined |
| SB_extracted_k141_111148 | Not-determined |
| SB_extracted_k141_111149 | Not-determined |
| SB_extracted_k141_59537 | Not-determined |
| SB_extracted_k141_111152 | Not-determined |
| SB_extracted_k141_84852 | Low-quality |
| SB_extracted_k141_136851 | Not-determined |
| SB_extracted_k141_84856 | Not-determined |
| SB_extracted_k141_6981 | Not-determined |
| SB_extracted_k141_15293 | Low-quality |
| SB_extracted_k141_119657 | Not-determined |
| SB_extracted_k141_119658 | Not-determined |
| SB_extracted_k141_93584 | Not-determined |
| SB_extracted_k141_102166 | Not-determined |
| SB_extracted_k141_180638 | Not-determined |
| SB_extracted_k141_76178 | Not-determined |
| SB_extracted_k141_24502 | Not-determined |
| SB_extracted_k141_111162 | Not-determined |
| SB_extracted_k141_162713 | Not-determined |
| SB_extracted_k141_49734 | Not-determined |
| SB_extracted_k141_136858 | Low-quality |
| SB_extracted_k141_15298 | Not-determined |
| SB_extracted_k141_180641 | Not-determined |
| SB_extracted_k141_136860 | Not-determined |
| SB_extracted_k141_206027 | Not-determined |
| SB_extracted_k141_84863 | Not-determined |
| SB_extracted_k141_111166 | Not-determined |
| SB_extracted_k141_84864 | Not-determined |
| SB_extracted_k141_188180 | Not-determined |
| SB_extracted_k141_102169 | Low-quality |
| SB_extracted_k141_136865 | Not-determined |
| SB_extracted_k141_6987 | Not-determined |
| SB_extracted_k141_111171 | Not-determined |
| SB_extracted_k141_24517 | Not-determined |
| SB_extracted_k141_49740 | Not-determined |
| SB_extracted_k141_162719 | Not-determined |
| SB_extracted_k141_41502 | Not-determined |
| SB_extracted_k141_145914 | Not-determined |
| SB_extracted_k141_15309 | Not-determined |
| SB_extracted_k141_172005 | Not-determined |
| SB_extracted_k141_188184 | Not-determined |
| SB_extracted_k141_84877 | Not-determined |
| SB_extracted_k141_162723 | Not-determined |
| SB_extracted_k141_180647 | Not-determined |
| SB_extracted_k141_102173 | Not-determined |
| SB_extracted_k141_180648 | Low-quality |
| SB_extracted_k141_162725 | Low-quality |
| SB_extracted_k141_24524 | Not-determined |
| SB_extracted_k141_180650 | Not-determined |
| SB_extracted_k141_119663 | Low-quality |
| SB_extracted_k141_93589 | Not-determined |
| SB_extracted_k141_6989 | Not-determined |
| SB_extracted_k141_180652 | Not-determined |
| SB_extracted_k141_111182 | Not-determined |
| SB_extracted_k141_162729 | Not-determined |
| SB_extracted_k141_24532 | Not-determined |
| SB_extracted_k141_84887 | Not-determined |
| SB_extracted_k141_172013 | Not-determined |
| SB_extracted_k141_49746 | Low-quality |
| SB_extracted_k141_162731 | Low-quality |
| SB_extracted_k141_111184 | Low-quality |
| SB_extracted_k141_93598 | Not-determined |
| SB_extracted_k141_41518 | Not-determined |
| SB_extracted_k141_180656 | Low-quality |
| SB_extracted_k141_76203 | Low-quality |
| SB_extracted_k141_93603 | Not-determined |
| SB_extracted_k141_180659 | Not-determined |
| SB_extracted_k141_15328 | Not-determined |
| SB_extracted_k141_84893 | Not-determined |
| SB_extracted_k141_76205 | Not-determined |
| SB_extracted_k141_49753 | Not-determined |
| SB_extracted_k141_111191 | Not-determined |
| SB_extracted_k141_33068 | Not-determined |
| SB_extracted_k141_188192 | Not-determined |
| SB_extracted_k141_93608 | Not-determined |
| SB_extracted_k141_136885 | Not-determined |
| SB_extracted_k141_15332 | Not-determined |
| SB_extracted_k141_41526 | Not-determined |
| SB_extracted_k141_119673 | Not-determined |
| SB_extracted_k141_102194 | Not-determined |
| SB_extracted_k141_172026 | Not-determined |
| SB_extracted_k141_180665 | Not-determined |
| SB_extracted_k141_145937 | Low-quality |
| SB_extracted_k141_180666 | Low-quality |
| SB_extracted_k141_93614 | Not-determined |
| SB_extracted_k141_136890 | Not-determined |
| SB_extracted_k141_172030 | Not-determined |
| SB_extracted_k141_93619 | Low-quality |
| SB_extracted_k141_84906 | Not-determined |
| SB_extracted_k141_49757 | Low-quality |
| SB_extracted_k141_84907 | Not-determined |
| SB_extracted_k141_111207 | Not-determined |
| SB_extracted_k141_102199 | Not-determined |
| SB_extracted_k141_111210 | Not-determined |
| SB_extracted_k141_145945 | Not-determined |
| SB_extracted_k141_93630 | Not-determined |
| SB_extracted_k141_102202 | Not-determined |
| SB_extracted_k141_206039 | Not-determined |
| SB_extracted_k141_49762 | Not-determined |
| SB_extracted_k141_206040 | Not-determined |
| SB_extracted_k141_162758 | Not-determined |
| SB_extracted_k141_84913 | Not-determined |
| SB_extracted_k141_41534 | Not-determined |
| SB_extracted_k141_49766 | Not-determined |
| SB_extracted_k141_15348 | Low-quality |
| SB_extracted_k141_93633 | Not-determined |
| SB_extracted_k141_162763 | Not-determined |
| SB_extracted_k141_102209 | Not-determined |
| SB_extracted_k141_119697 | Not-determined |
| SB_extracted_k141_7016 | Not-determined |
| SB_extracted_k141_172034 | Not-determined |
| SB_extracted_k141_102211 | Not-determined |
| SB_extracted_k141_172035 | Not-determined |
| SB_extracted_k141_7017 | Not-determined |
| SB_extracted_k141_49769 | Not-determined |
| SB_extracted_k141_111223 | Not-determined |
| SB_extracted_k141_33090 | Not-determined |
| SB_extracted_k141_15356 | Not-determined |
| SB_extracted_k141_49771 | Low-quality |
| SB_extracted_k141_15357 | Not-determined |
| SB_extracted_k141_102217 | Not-determined |
| SB_extracted_k141_41547 | Not-determined |
| SB_extracted_k141_162777 | Not-determined |
| SB_extracted_k141_206056 | Not-determined |
| SB_extracted_k141_93644 | Not-determined |
| SB_extracted_k141_172041 | Low-quality |
| SB_extracted_k141_162779 | Not-determined |
| SB_extracted_k141_15360 | Not-determined |
| SB_extracted_k141_128393 | Not-determined |
| SB_extracted_k141_15362 | Not-determined |
| SB_extracted_k141_162780 | Not-determined |
| SB_extracted_k141_15363 | Not-determined |
| SB_extracted_k141_162782 | Low-quality |
| SB_extracted_k141_49775 | Not-determined |
| SB_extracted_k141_67367 | Low-quality |
| SB_extracted_k141_24547 | Not-determined |
| SB_extracted_k141_67370 | Low-quality |
| SB_extracted_k141_188234 | Low-quality |
| SB_extracted_k141_206062 | Not-determined |
| SB_extracted_k141_49778 | Not-determined |
| SB_extracted_k141_180687 | Not-determined |
| SB_extracted_k141_188235 | Low-quality |
| SB_extracted_k141_180688 | Not-determined |
| SB_extracted_k141_206065 | Not-determined |
| SB_extracted_k141_24551 | Not-determined |
| SB_extracted_k141_206067 | Not-determined |
| SB_extracted_k141_102229 | Not-determined |
| SB_extracted_k141_111247 | Low-quality |
| SB_extracted_k141_102230 | Not-determined |
| SB_extracted_k141_41560 | Not-determined |
| SB_extracted_k141_172048 | Not-determined |
| SB_extracted_k141_93655 | Not-determined |
| SB_extracted_k141_172052 | Not-determined |
| SB_extracted_k141_145975 | Not-determined |
| SB_extracted_k141_172055 | Not-determined |
| SB_extracted_k141_180696 | Not-determined |
| SB_extracted_k141_41561 | Not-determined |
| SB_extracted_k141_162797 | Low-quality |
| SB_extracted_k141_172057 | Not-determined |
| SB_extracted_k141_172058 | Not-determined |
| SB_extracted_k141_93660 | Not-determined |
| [illegible] | [illegible] |

| | |
|---|---|
| S8_extracted_k141_128533 | Not-determined |
| S8_extracted_k141_128534 | Not-determined |
| S8_extracted_k141_102376 | Not-determined |
| S8_extracted_k141_15485 | Not-determined |
| S8_extracted_k141_85095 | Not-determined |
| S8_extracted_k141_180839 | Not-determined |
| S8_extracted_k141_198233 | Not-determined |
| S8_extracted_k141_102379 | Not-determined |
| S8_extracted_k141_111399 | Not-determined |
| S8_extracted_k141_85097 | Not-determined |
| S8_extracted_k141_111401 | Not-determined |
| S8_extracted_k141_7149 | Not-determined |
| S8_extracted_k141_15489 | Not-determined |
| S8_extracted_k141_24695 | Not-determined |
| S8_extracted_k141_111402 | Not-determined |
| S8_extracted_k141_154255 | Not-determined |
| S8_extracted_k141_93780 | Low-quality |
| S8_extracted_k141_154256 | Not-determined |
| S8_extracted_k141_206189 | Not-determined |
| S8_extracted_k141_119797 | Not-determined |
| S8_extracted_k141_146097 | Not-determined |
| S8_extracted_k141_137015 | Not-determined |
| S8_extracted_k141_180844 | Not-determined |
| S8_extracted_k141_188374 | Not-determined |
| S8_extracted_k141_67476 | Not-determined |
| S8_extracted_k141_162964 | Not-determined |
| S8_extracted_k141_128548 | Not-determined |
| S8_extracted_k141_180852 | Not-determined |
| S8_extracted_k141_15501 | Not-determined |
| S8_extracted_k141_162969 | Not-determined |
| S8_extracted_k141_198244 | Not-determined |
| S8_extracted_k141_188380 | Not-determined |
| S8_extracted_k141_67484 | Not-determined |
| S8_extracted_k141_188382 | Low-quality |
| S8_extracted_k141_15505 | Not-determined |
| S8_extracted_k141_188384 | Not-determined |
| S8_extracted_k141_102384 | Not-determined |
| S8_extracted_k141_41727 | Not-determined |
| S8_extracted_k141_7165 | Not-determined |
| S8_extracted_k141_41728 | Not-determined |
| S8_extracted_k141_154269 | Not-determined |
| S8_extracted_k141_162972 | Not-determined |
| S8_extracted_k141_154270 | Not-determined |
| S8_extracted_k141_198253 | Not-determined |
| S8_extracted_k141_41732 | Not-determined |
| S8_extracted_k141_24713 | Not-determined |
| S8_extracted_k141_102390 | Not-determined |
| S8_extracted_k141_198258 | Not-determined |
| S8_extracted_k141_162977 | Not-determined |
| S8_extracted_k141_119809 | Not-determined |
| S8_extracted_k141_24719 | Not-determined |
| S8_extracted_k141_93797 | Not-determined |
| S8_extracted_k141_206215 | Not-determined |
| S8_extracted_k141_146109 | Not-determined |
| S8_extracted_k141_24723 | Not-determined |
| S8_extracted_k141_162980 | Low-quality |
| S8_extracted_k141_15514 | Not-determined |
| S8_extracted_k141_188391 | Not-determined |
| S8_extracted_k141_102395 | Not-determined |
| S8_extracted_k141_119817 | Not-determined |
| S8_extracted_k141_102396 | Not-determined |
| S8_extracted_k141_146114 | Not-determined |
| S8_extracted_k141_33266 | Not-determined |
| S8_extracted_k141_111426 | Not-determined |
| S8_extracted_k141_24732 | Not-determined |
| S8_extracted_k141_180870 | Not-determined |
| S8_extracted_k141_7179 | Not-determined |
| S8_extracted_k141_146115 | Not-determined |
| S8_extracted_k141_119818 | Low-quality |
| S8_extracted_k141_24733 | Not-determined |
| S8_extracted_k141_102404 | Not-determined |
| S8_extracted_k141_198269 | Not-determined |
| S8_extracted_k141_198270 | Not-determined |
| S8_extracted_k141_137035 | Not-determined |
| S8_extracted_k141_146118 | Not-determined |
| S8_extracted_k141_188402 | Not-determined |
| S8_extracted_k141_111434 | Not-determined |
| S8_extracted_k141_24738 | Not-determined |
| S8_extracted_k141_198275 | Not-determined |
| S8_extracted_k141_93814 | Not-determined |
| S8_extracted_k141_137042 | Not-determined |
| S8_extracted_k141_33270 | Not-determined |
| S8_extracted_k141_33271 | Not-determined |
| S8_extracted_k141_128556 | Low-quality |
| S8_extracted_k141_137043 | Not-determined |
| S8_extracted_k141_49890 | Low-quality |
| S8_extracted_k141_198276 | Not-determined |
| S8_extracted_k141_119820 | Low-quality |
| S8_extracted_k141_41750 | Not-determined |
| S8_extracted_k141_188412 | Not-determined |
| S8_extracted_k141_188413 | Low-quality |
| S8_extracted_k141_7196 | Not-determined |
| S8_extracted_k141_41753 | Not-determined |
| S8_extracted_k141_111440 | Not-determined |
| S8_extracted_k141_111441 | Not-determined |
| S8_extracted_k141_188416 | Not-determined |
| S8_extracted_k141_15524 | Not-determined |
| S8_extracted_k141_146127 | Not-determined |
| S8_extracted_k141_111446 | Not-determined |
| S8_extracted_k141_180875 | Low-quality |
| S8_extracted_k141_198282 | Low-quality |
| S8_extracted_k141_49902 | Not-determined |
| S8_extracted_k141_119825 | Low-quality |
| S8_extracted_k141_49903 | Not-determined |
| S8_extracted_k141_67506 | Not-determined |
| S8_extracted_k141_41754 | Not-determined |
| S8_extracted_k141_67507 | Not-determined |
| S8_extracted_k141_111449 | Not-determined |
| S8_extracted_k141_119826 | Not-determined |
| S8_extracted_k141_67510 | Not-determined |
| S8_extracted_k141_24743 | Not-determined |
| S8_extracted_k141_85134 | Not-determined |
| S8_extracted_k141_41757 | Not-determined |
| S8_extracted_k141_85135 | Not-determined |
| S8_extracted_k141_119832 | Not-determined |
| S8_extracted_k141_85138 | Not-determined |
| S8_extracted_k141_198292 | Low-quality |
| S8_extracted_k141_146134 | Not-determined |
| S8_extracted_k141_119835 | Not-determined |
| S8_extracted_k141_146136 | Not-determined |
| S8_extracted_k141_162995 | Not-determined |
| S8_extracted_k141_85140 | Not-determined |
| S8_extracted_k141_188425 | Not-determined |
| S8_extracted_k141_41764 | Not-determined |
| S8_extracted_k141_59649 | Low-quality |
| S8_extracted_k141_188427 | Not-determined |
| S8_extracted_k141_24750 | Not-determined |
| S8_extracted_k141_154282 | Not-determined |
| S8_extracted_k141_24753 | Not-determined |
| S8_extracted_k141_180893 | Not-determined |
| S8_extracted_k141_146139 | Not-determined |
| S8_extracted_k141_49922 | Not-determined |
| S8_extracted_k141_163002 | Low-quality |
| S8_extracted_k141_102411 | Low-quality |
| S8_extracted_k141_15542 | Not-determined |
| S8_extracted_k141_85146 | Not-determined |
| S8_extracted_k141_119843 | Not-determined |
| S8_extracted_k141_172186 | Not-determined |
| S8_extracted_k141_33285 | Not-determined |
| S8_extracted_k141_33286 | Not-determined |
| S8_extracted_k141_198297 | Not-determined |
| S8_extracted_k141_111474 | Not-determined |
| S8_extracted_k141_119850 | Low-quality |
| S8_extracted_k141_102417 | Not-determined |
| S8_extracted_k141_93836 | Not-determined |
| S8_extracted_k141_146145 | Low-quality |
| S8_extracted_k141_172194 | Not-determined |
| S8_extracted_k141_137051 | Not-determined |
| S8_extracted_k141_111477 | Low-quality |
| S8_extracted_k141_206226 | Not-determined |
| S8_extracted_k141_137052 | Not-determined |
| S8_extracted_k141_24754 | Not-determined |
| S8_extracted_k141_146148 | Not-determined |
| S8_extracted_k141_85155 | Not-determined |
| S8_extracted_k141_102420 | Not-determined |
| S8_extracted_k141_85159 | Not-determined |
| S8_extracted_k141_93840 | Not-determined |
| S8_extracted_k141_146151 | Not-determined |
| S8_extracted_k141_102421 | Low-quality |
| S8_extracted_k141_85162 | Not-determined |
| S8_extracted_k141_24761 | Low-quality |
| S8_extracted_k141_33292 | Not-determined |
| S8_extracted_k141_198313 | Not-determined |
| S8_extracted_k141_33294 | Not-determined |
| S8_extracted_k141_67533 | Low-quality |
| S8_extracted_k141_137056 | Not-determined |
| S8_extracted_k141_67537 | Low-quality |
| S8_extracted_k141_172207 | Low-quality |
| S8_extracted_k141_119867 | Low-quality |
| S8_extracted_k141_119868 | Not-determined |
| S8_extracted_k141_119870 | Not-determined |
| S8_extracted_k141_85166 | Not-determined |
| S8_extracted_k141_41782 | Not-determined |
| S8_extracted_k141_93850 | Not-determined |
| S8_extracted_k141_119871 | Not-determined |
| S8_extracted_k141_41783 | Not-determined |
| S8_extracted_k141_15563 | Not-determined |
| S8_extracted_k141_206231 | Not-determined |
| S8_extracted_k141_146167 | Not-determined |
| S8_extracted_k141_15565 | Not-determined |
| S8_extracted_k141_119875 | Not-determined |
| S8_extracted_k141_163033 | Not-determined |
| S8_extracted_k141_7234 | Not-determined |
| S8_extracted_k141_146168 | Not-determined |
| S8_extracted_k141_188458 | Not-determined |
| S8_extracted_k141_206237 | Not-determined |
| S8_extracted_k141_33309 | Not-determined |
| S8_extracted_k141_85167 | Low-quality |
| S8_extracted_k141_41790 | Not-determined |
| S8_extracted_k141_206240 | Not-determined |
| S8_extracted_k141_180906 | Not-determined |
| S8_extracted_k141_128580 | Not-determined |
| S8_extracted_k141_206244 | Not-determined |
| S8_extracted_k141_172223 | Not-determined |
| S8_extracted_k141_172224 | Not-determined |
| S8_extracted_k141_146178 | Low-quality |
| S8_extracted_k141_198320 | Not-determined |
| S8_extracted_k141_24774 | Not-determined |
| S8_extracted_k141_15576 | Not-determined |
| S8_extracted_k141_24777 | Not-determined |
| S8_extracted_k141_198326 | Not-determined |
| S8_extracted_k141_24779 | Not-determined |
| S8_extracted_k141_33317 | Not-determined |
| S8_extracted_k141_172229 | Not-determined |
| S8_extracted_k141_85179 | Not-determined |
| S8_extracted_k141_24782 | Not-determined |
| S8_extracted_k141_163039 | Not-determined |
| S8_extracted_k141_172231 | Not-determined |
| S8_extracted_k141_198330 | Not-determined |
| S8_extracted_k141_172232 | Not-determined |
| S8_extracted_k141_33319 | Low-quality |
| S8_extracted_k141_15581 | Low-quality |
| S8_extracted_k141_111491 | Not-determined |
| S8_extracted_k141_188459 | Not-determined |
| S8_extracted_k141_15584 | Not-determined |
| S8_extracted_k141_41801 | Not-determined |
| S8_extracted_k141_188461 | Not-determined |
| S8_extracted_k141_137077 | Not-determined |
| S8_extracted_k141_206254 | Not-determined |
| S8_extracted_k141_111498 | Not-determined |
| S8_extracted_k141_24792 | Not-determined |
| S8_extracted_k141_154292 | Not-determined |
| S8_extracted_k141_41807 | Not-determined |
| S8_extracted_k141_102429 | Low-quality |
| S8_extracted_k141_198344 | Not-determined |
| S8_extracted_k141_49953 | Not-determined |
| S8_extracted_k141_111508 | Not-determined |
| S8_extracted_k141_188473 | Not-determined |
| S8_extracted_k141_206264 | Not-determined |
| S8_extracted_k141_24802 | Not-determined |
| S8_extracted_k141_172245 | Not-determined |
| S8_extracted_k141_41812 | Not-determined |
| S8_extracted_k141_198348 | Not-determined |
| S8_extracted_k141_172247 | Low-quality |
| S8_extracted_k141_85192 | Not-determined |
| S8_extracted_k141_119877 | Not-determined |
| S8_extracted_k141_15593 | Not-determined |
| S8_extracted_k141_137089 | Not-determined |
| S8_extracted_k141_128597 | Low-quality |
| S8_extracted_k141_15597 | Not-determined |
| S8_extracted_k141_24805 | Not-determined |
| S8_extracted_k141_163064 | Not-determined |
| S8_extracted_k141_15598 | Not-determined |
| S8_extracted_k141_146192 | Not-determined |
| S8_extracted_k141_15599 | Not-determined |
| S8_extracted_k141_146193 | Low-quality |
| S8_extracted_k141_188480 | Not-determined |
| S8_extracted_k141_154303 | Not-determined |
| S8_extracted_k141_119881 | Not-determined |
| S8_extracted_k141_137092 | Not-determined |
| S8_extracted_k141_198360 | Low-quality |
| S8_extracted_k141_163069 | Not-determined |
| S8_extracted_k141_59711 | Not-determined |
| S8_extracted_k141_163070 | Not-determined |
| S8_extracted_k141_59712 | Not-determined |
| S8_extracted_k141_85202 | Not-determined |
| S8_extracted_k141_85203 | Low-quality |
| S8_extracted_k141_163072 | Not-determined |
| S8_extracted_k141_49970 | Not-determined |
| S8_extracted_k141_163073 | Not-determined |
| S8_extracted_k141_33350 | Not-determined |
| S8_extracted_k141_128603 | Not-determined |
| S8_extracted_k141_154306 | Low-quality |
| S8_extracted_k141_180924 | Not-determined |
| S8_extracted_k141_163075 | Not-determined |
| S8_extracted_k141_180926 | Not-determined |
| S8_extracted_k141_111519 | Not-determined |
| S8_extracted_k141_137101 | Not-determined |
| S8_extracted_k141_102436 | Not-determined |
| S8_extracted_k141_172259 | Not-determined |
| S8_extracted_k141_15605 | Not-determined |
| S8_extracted_k141_93890 | Low-quality |
| S8_extracted_k141_59724 | Not-determined |
| S8_extracted_k141_67547 | Not-determined |
| S8_extracted_k141_180930 | Not-determined |
| S8_extracted_k141_119889 | Not-determined |
| S8_extracted_k141_119890 | Low-quality |
| S8_extracted_k141_49980 | Not-determined |
| S8_extracted_k141_119891 | Not-determined |
| S8_extracted_k141_172268 | Not-determined |
| S8_extracted_k141_172269 | Low-quality |
| S8_extracted_k141_49984 | Not-determined |
| S8_extracted_k141_128606 | Not-determined |
| S8_extracted_k141_24816 | Not-determined |
| S8_extracted_k141_85217 | Not-determined |
| S8_extracted_k141_15612 | Not-determined |
| S8_extracted_k141_188485 | Not-determined |
| S8_extracted_k141_67560 | Not-determined |
| S8_extracted_k141_33362 | Not-determined |
| S8_extracted_k141_128610 | Not-determined |
| S8_extracted_k141_188486 | Low-quality |
| S8_extracted_k141_93905 | Low-quality |
| S8_extracted_k141_146212 | Low-quality |
| S8_extracted_k141_172276 | Not-determined |
| S8_extracted_k141_128612 | Not-determined |
| S8_extracted_k141_137111 | Not-determined |
| S8_extracted_k141_41848 | Not-determined |
| S8_extracted_k141_180948 | Not-determined |
| S8_extracted_k141_33367 | Not-determined |
| S8_extracted_k141_180949 | Not-determined |
| S8_extracted_k141_198365 | Not-determined |
| S8_extracted_k141_198366 | Not-determined |
| S8_extracted_k141_93913 | Not-determined |
| S8_extracted_k141_76363 | Not-determined |
| S8_extracted_k141_137116 | Not-determined |
| S8_extracted_k141_128617 | Not-determined |
| S7_vs_5843\|\|full | Medium-quality |
| S8_extracted_k141_41855 | Not-determined |
| S8_extracted_k141_67567 | Not-determined |
| S8_extracted_k141_41857 | Not-determined |
| S8_extracted_k141_67568 | Not-determined |
| S8_extracted_k141_93916 | Not-determined |
| S8_extracted_k141_146217 | Not-determined |
| S8_extracted_k141_50000 | Low-quality |
| S8_extracted_k141_7244 | Low-quality |
| S8_extracted_k141_33373 | Not-determined |
| S8_extracted_k141_33374 | Not-determined |
| S8_extracted_k141_41860 | Not-determined |
| S8_extracted_k141_163100 | Not-determined |
| S8_extracted_k141_102461 | Not-determined |
| S8_extracted_k141_172292 | Not-determined |
| S8_extracted_k141_41863 | Not-determined |
| S8_extracted_k141_163102 | Not-determined |
| S8_extracted_k141_163103 | Not-determined |
| S8_extracted_k141_137129 | Low-quality |
| S8_extracted_k141_180957 | Not-determined |
| S8_extracted_k141_128621 | Low-quality |
| S8_extracted_k141_15626 | Not-determined |
| S8_extracted_k141_76375 | Not-determined |
| S8_extracted_k141_163106 | Low-quality |
| S8_extracted_k141_206303 | Not-determined |
| S8_extracted_k141_50011 | Not-determined |
| S8_extracted_k141_137134 | Not-determined |
| S8_extracted_k141_188503 | Not-determined |
| S8_extracted_k141_7255 | Not-determined |
| S8_extracted_k141_111546 | Not-determined |
| S8_extracted_k141_15631 | Not-determined |
| S8_extracted_k141_172300 | Not-determined |
| S8_extracted_k141_93923 | Not-determined |
| S8_extracted_k141_85224 | Not-determined |
| S8_extracted_k141_119913 | Not-determined |
| S8_extracted_k141_102474 | Not-determined |
| S8_extracted_k141_67582 | Not-determined |
| S8_extracted_k141_119915 | Not-determined |
| S8_extracted_k141_146230 | Not-determined |
| S8_extracted_k141_119916 | Not-determined |
| S8_extracted_k141_7261 | Not-determined |
| S8_extracted_k141_50017 | Not-determined |
| S8_extracted_k141_85230 | Not-determined |
| S8_extracted_k141_163115 | Not-determined |
| S8_extracted_k141_85231 | Not-determined |
| S8_extracted_k141_15634 | Not-determined |
| S8_extracted_k141_119923 | Not-determined |
| S8_extracted_k141_180962 | Not-determined |
| S8_extracted_k141_188511 | Not-determined |
| S8_extracted_k141_67586 | Not-determined |
| S8_extracted_k141_128629 | Not-determined |
| S8_extracted_k141_33399 | Not-determined |
| S8_extracted_k141_198386 | Not-determined |
| S8_extracted_k141_172317 | Low-quality |
| S8_extracted_k141_7270 | Not-determined |
| S8_extracted_k141_163120 | Low-quality |
| S8_extracted_k141_146240 | Not-determined |
| S8_extracted_k141_119932 | Not-determined |
| S8_extracted_k141_146241 | Not-determined |
| S8_extracted_k141_15640 | Not-determined |
| S8_extracted_k141_33402 | Not-determined |
| S8_extracted_k141_7272 | Not-determined |
| S8_extracted_k141_206321 | Not-determined |
| S8_extracted_k141_188520 | Not-determined |
| S8_extracted_k141_59729 | Not-determined |
| S8_extracted_k141_111564 | Low-quality |
| S8_extracted_k141_50036 | Not-determined |
| S8_extracted_k141_15642 | Not-determined |
| S8_extracted_k141_119944 | Not-determined |
| S8_extracted_k141_59732 | Not-determined |
| S8_extracted_k141_206325 | Not-determined |
| S8_extracted_k141_7277 | Not-determined |
| S8_extracted_k141_137141 | Not-determined |
| S8_extracted_k141_198391 | Not-determined |
| S8_extracted_k141_67595 | Not-determined |
| S8_extracted_k141_137142 | Not-determined |
| S8_extracted_k141_119946 | Not-determined |
| S8_extracted_k141_67596 | Low-quality |
| S8_extracted_k141_93946 | Not-determined |
| S8_extracted_k141_198393 | Low-quality |
| S8_extracted_k141_7279 | Not-determined |
| S8_extracted_k141_137144 | Low-quality |
| S8_extracted_k141_50040 | Not-determined |
| S8_extracted_k141_137145 | Low-quality |
| S8_extracted_k141_119948 | Not-determined |
| S8_extracted_k141_146249 | Not-determined |
| S8_extracted_k141_15646 | Not-determined |
| S8_extracted_k141_128641 | Not-determined |
| S8_extracted_k141_111572 | Not-determined |
| S8_extracted_k141_111573 | Not-determined |
| S8_extracted_k141_102494 | Not-determined |
| S8_extracted_k141_172333 | Not-determined |
| S8_extracted_k141_50045 | Not-determined |
| S8_extracted_k141_119952 | Not-determined |
| S8_extracted_k141_180977 | Not-determined |
| S8_extracted_k141_128643 | Not-determined |
| S8_extracted_k141_102496 | Not-determined |
| S8_extracted_k141_50047 | Not-determined |
| S8_extracted_k141_111577 | Not-determined |
| S8_extracted_k141_41878 | Not-determined |
| S8_extracted_k141_7281 | Not-determined |
| S8_extracted_k141_163134 | Not-determined |
| S8_extracted_k141_198401 | Not-determined |
| S8_extracted_k141_76398 | Not-determined |
| S8_extracted_k141_111579 | Not-determined |
| S8_extracted_k141_102502 | Not-determined |
| S8_extracted_k141_172337 | Not-determined |
| S8_extracted_k141_15658 | Not-determined |
| S8_extracted_k141_33417 | Not-determined |
| S8_extracted_k141_41884 | Not-determined |
| S8_extracted_k141_198407 | Not-determined |
| S8_extracted_k141_128651 | Low-quality |
| S8_extracted_k141_93956 | Not-determined |
| S8_extracted_k141_154322 | Not-determined |
| S8_extracted_k141_15661 | Not-determined |
| S8_extracted_k141_102505 | Not-determined |
| S8_extracted_k141_172338 | Not-determined |
| S8_extracted_k141_180990 | Not-determined |
| S8_extracted_k141_85250 | Low-quality |
| S8_extracted_k141_85251 | Not-determined |
| S8_extracted_k141_172341 | Not-determined |
| S8_extracted_k141_180992 | Not-determined |
| S8_extracted_k141_128656 | Not-determined |
| S8_extracted_k141_128658 | Not-determined |
| S8_extracted_k141_172344 | Not-determined |
| S8_extracted_k141_7293 | Not-determined |
| S8_extracted_k141_67620 | Not-determined |
| S8_extracted_k141_85254 | Low-quality |
| S8_extracted_k141_111591 | Not-determined |
| S8_extracted_k141_50056 | Not-determined |
| S8_extracted_k141_76405 | Not-determined |
| S8_extracted_k141_50057 | Not-determined |
| S8_extracted_k141_163146 | Not-determined |
| S8_extracted_k141_67623 | Not-determined |
| S8_extracted_k141_154326 | Not-determined |
| S8_extracted_k141_85263 | Not-determined |
| S8_extracted_k141_154328 | Not-determined |
| S8_extracted_k141_206347 | Not-determined |
| S8_extracted_k141_93970 | Not-determined |
| S8_extracted_k141_111603 | Not-determined |
| S8_extracted_k141_119971 | Not-determined |
| S8_extracted_k141_15668 | Not-determined |
| S8_extracted_k141_85266 | Not-determined |
| S8_extracted_k141_119977 | Not-determined |
| S8_extracted_k141_206356 | Not-determined |
| S8_extracted_k141_181005 | Not-determined |
| S8_extracted_k141_128672 | Not-determined |
| S8_extracted_k141_181006 | Not-determined |
| S8_extracted_k141_50060 | Not-determined |
| S8_extracted_k141_163166 | Not-determined |
| S8_extracted_k141_119979 | Not-determined |
| S8_extracted_k141_7310 | Not-determined |
| S8_extracted_k141_33445 | Not-determined |
| S8_extracted_k141_76419 | Not-determined |
| S8_extracted_k141_33450 | Not-determined |
| S8_extracted_k141_67657 | Not-determined |
| S8_extracted_k141_111619 | Not-determined |
| S8_extracted_k141_67658 | Not-determined |
| S8_extracted_k141_119984 | Not-determined |
| S8_extracted_k141_154346 | Not-determined |
| S8_extracted_k141_128682 | Not-determined |
| S8_extracted_k141_50069 | Not-determined |
| S8_extracted_k141_15680 | Not-determined |
| S8_extracted_k141_7312 | Not-determined |
| S8_extracted_k141_67661 | Not-determined |
| S8_extracted_k141_119990 | Not-determined |
| S8_extracted_k141_111624 | Low-quality |
| S8_extracted_k141_137197 | Low-quality |
| S8_extracted_k141_137198 | Low-quality |
| S8_extracted_k141_206368 | Low-quality |
| S8_extracted_k141_137200 | Not-determined |
| S8_extracted_k141_163174 | Not-determined |
| S8_extracted_k141_154350 | Not-determined |
| S8_extracted_k141_67669 | Not-determined |
| S8_extracted_k141_163175 | Not-determined |
| S8_extracted_k141_206372 | Not-determined |
| S8_extracted_k141_67671 | Not-determined |
| S8_extracted_k141_181026 | Low-quality |
| S8_extracted_k141_7318 | Not-determined |
| S8_extracted_k141_128692 | Not-determined |
| S8_extracted_k141_119995 | Not-determined |
| S8_extracted_k141_15685 | Not-determined |
| S8_extracted_k141_154355 | Low-quality |
| S8_extracted_k141_206375 | Not-determined |
| S8_extracted_k141_154357 | Not-determined |
| S8_extracted_k141_181033 | Not-determined |
| S8_extracted_k141_181036 | Not-determined |
| S8_extracted_k141_76440 | Low-quality |
| S8_extracted_k141_33456 | Not-determined |
| S8_extracted_k141_41938 | Not-determined |
| S8_extracted_k141_41939 | Not-determined |
| S8_extracted_k141_172375 | Not-determined |
| S8_extracted_k141_76442 | Not-determined |
| S8_extracted_k141_188527 | Not-determined |
| S8_extracted_k141_33460 | Not-determined |
| S8_extracted_k141_50083 | Low-quality |
| S8_extracted_k141_33461 | Not-determined |
| S8_extracted_k141_120004 | Not-determined |
| S8_extracted_k141_50085 | Not-determined |
| S8_extracted_k141_198435 | Not-determined |
| S8_extracted_k141_50087 | Not-determined |
| S8_extracted_k141_120007 | Not-determined |
| S8_extracted_k141_206385 | Not-determined |
| S8_extracted_k141_50090 | Not-determined |
| S5_extracted_k141_55687 | Medium-quality |
| S8_extracted_k141_128697 | Not-determined |
| S8_extracted_k141_120014 | Not-determined |
| S8_extracted_k141_128698 | Not-determined |
| S8_extracted_k141_188531 | Not-determined |
| S8_extracted_k141_67688 | Not-determined |
| S8_extracted_k141_172382 | Not-determined |
| S8_extracted_k141_15708 | Low-quality |
| S8_extracted_k141_172383 | Not-determined |
| S8_extracted_k141_41942 | Not-determined |
| S8_extracted_k141_41944 | Not-determined |
| S8_extracted_k141_15711 | Not-determined |
| S8_extracted_k141_172387 | Not-determined |
| S8_extracted_k141_198439 | Not-determined |
| S8_extracted_k141_85291 | Not-determined |
| S8_extracted_k141_172388 | Not-determined |
| S8_extracted_k141_128701 | Not-determined |
| S8_extracted_k141_33468 | Not-determined |
| S8_extracted_k141_15715 | Low-quality |
| S8_extracted_k141_7344 | Not-determined |
| S8_extracted_k141_154366 | Not-determined |
| S8_extracted_k141_188535 | Not-determined |
| S8_extracted_k141_198445 | Not-determined |
| S8_extracted_k141_111636 | Not-determined |
| S8_extracted_k141_198448 | Not-determined |
| S8_extracted_k141_181056 | Not-determined |
| S8_extracted_k141_128711 | Not-determined |
| S8_extracted_k141_181058 | Not-determined |
| S8_extracted_k141_128714 | Not-determined |
| S8_extracted_k141_76451 | Not-determined |
| S8_extracted_k141_102520 | Not-determined |
| S8_extracted_k141_128715 | Not-determined |
| S8_extracted_k141_76452 | Not-determined |
| S8_extracted_k141_33476 | Not-determined |
| S8_extracted_k141_111642 | Not-determined |
| S8_extracted_k141_128718 | Not-determined |
| S8_extracted_k141_50105 | Not-determined |
| S8_extracted_k141_137220 | Low-quality |
| S8_extracted_k141_128720 | Not-determined |
| S8_extracted_k141_50107 | Not-determined |
| S8_extracted_k141_120024 | Not-determined |
| S8_extracted_k141_128722 | Not-determined |
| S8_extracted_k141_50109 | Not-determined |
| S8_extracted_k141_154380 | Not-determined |
| S8_extracted_k141_128725 | Not-determined |
| S8_extracted_k141_41956 | Not-determined |
| S8_extracted_k141_33488 | Not-determined |
| S8_extracted_k141_50113 | Not-determined |
| S8_extracted_k141_67696 | Not-determined |
| S8_extracted_k141_172409 | Low-quality |
| S8_extracted_k141_76471 | Not-determined |
| S8_extracted_k141_188541 | Not-determined |
| S8_extracted_k141_188542 | Not-determined |
| S8_extracted_k141_67700 | Not-determined |
| S8_extracted_k141_188546 | Low-quality |
| S8_extracted_k141_198464 | Not-determined |
| S8_extracted_k141_128733 | Not-determined |
| S8_extracted_k141_206403 | Not-determined |
| S8_extracted_k141_76478 | Not-determined |
| S8_extracted_k141_198468 | Not-determined |
| S8_extracted_k141_206409 | Not-determined |
| S12_vs_k141_183752\|\|full | Medium-quality |
| S8_extracted_k141_154397 | Not-determined |
| S8_extracted_k141_172417 | Not-determined |
| S8_extracted_k141_93978 | Not-determined |
| S8_extracted_k141_41973 | Not-determined |
| S8_extracted_k141_93981 | Low-quality |
| S8_extracted_k141_206412 | Not-determined |
| S8_extracted_k141_188554 | Not-determined |
| S8_extracted_k141_172420 | Low-quality |
| S8_extracted_k141_188557 | Not-determined |
| S8_extracted_k141_41977 | Low-quality |
| S8_extracted_k141_24822 | Not-determined |
| S8_extracted_k141_41978 | Not-determined |
| S8_extracted_k141_67716 | Not-determined |
| S8_extracted_k141_120043 | Low-quality |
| S8_extracted_k141_206418 | Not-determined |
| S8_extracted_k141_93989 | Low-quality |
| S8_extracted_k141_172428 | Not-determined |

| | |
|---|---|
| S8_extracted_k141_24831 | Not-determined |
| S8_extracted_k141_154413 | Not-determined |
| S8_extracted_k141_24836 | Not-determined |
| S8_extracted_k141_67724 | Not-determined |
| S8_extracted_k141_50140 | Not-determined |
| S8_extracted_k141_93994 | Not-determined |
| S8_extracted_k141_93995 | Low-quality |
| S8_extracted_k141_154417 | Not-determined |
| S8_extracted_k141_24837 | Not-determined |
| S8_extracted_k141_102554 | Not-determined |
| S8_extracted_k141_50148 | Not-determined |
| S8_extracted_k141_33513 | Not-determined |
| S8_extracted_k141_137240 | Not-determined |
| S8_extracted_k141_120054 | Low-quality |
| S8_extracted_k141_206432 | Not-determined |
| S8_extracted_k141_94002 | Not-determined |
| S8_extracted_k141_41989 | Not-determined |
| S8_extracted_k141_120058 | Not-determined |
| S8_extracted_k141_111652 | Low-quality |
| S8_extracted_k141_94005 | Low-quality |
| S8_extracted_k141_120061 | Not-determined |
| S8_extracted_k141_206439 | Not-determined |
| S8_extracted_k141_50158 | Not-determined |
| S8_extracted_k141_111657 | Not-determined |
| S8_extracted_k141_120064 | Not-determined |
| S8_extracted_k141_102571 | Not-determined |
| S8_extracted_k141_102572 | Not-determined |
| S8_extracted_k141_94016 | Not-determined |
| S8_extracted_k141_42000 | Not-determined |
| S8_extracted_k141_102575 | Low-quality |
| S8_extracted_k141_94018 | Not-determined |
| S8_extracted_k141_50160 | Not-determined |
| S8_extracted_k141_111665 | Not-determined |
| S8_extracted_k141_42005 | Not-determined |
| S8_extracted_k141_181079 | Not-determined |
| S8_extracted_k141_137245 | Not-determined |
| S8_extracted_k141_163181 | Not-determined |
| S8_extracted_k141_137247 | Not-determined |
| S8_extracted_k141_154434 | Not-determined |
| S8_extracted_k141_128775 | Low-quality |
| S8_extracted_k141_206452 | Not-determined |
| S8_extracted_k141_206454 | Not-determined |
| S8_extracted_k141_206457 | Not-determined |
| S8_extracted_k141_102582 | Not-determined |
| S8_extracted_k141_198506 | Not-determined |
| S8_extracted_k141_128776 | Not-determined |
| S8_extracted_k141_102583 | Not-determined |
| S8_extracted_k141_206459 | Not-determined |
| S8_extracted_k141_128779 | Not-determined |
| S8_extracted_k141_206461 | Low-quality |
| S8_extracted_k141_111672 | Not-determined |
| S8_extracted_k141_102588 | Low-quality |
| S8_extracted_k141_111674 | Low-quality |
| S8_extracted_k141_85318 | Not-determined |
| S8_extracted_k141_33535 | Low-quality |
| S8_extracted_k141_102592 | Not-determined |
| S8_extracted_k141_42015 | Not-determined |
| S8_extracted_k141_94025 | Not-determined |
| S8_extracted_k141_128782 | Not-determined |
| S8_extracted_k141_7357 | Not-determined |
| S8_extracted_k141_172454 | Not-determined |
| S8_extracted_k141_128784 | Not-determined |
| S8_extracted_k141_33538 | Not-determined |
| S8_extracted_k141_85328 | Not-determined |
| S8_extracted_k141_85329 | Not-determined |
| S8_extracted_k141_172457 | Not-determined |
| S8_extracted_k141_85330 | Not-determined |
| S8_extracted_k141_154447 | Not-determined |
| S8_extracted_k141_163188 | Not-determined |
| S8_extracted_k141_42023 | Low-quality |
| S8_extracted_k141_102600 | Not-determined |
| S8_extracted_k141_85332 | Not-determined |
| S8_extracted_k141_42025 | Not-determined |
| S8_extracted_k141_67734 | Not-determined |
| S8_extracted_k141_111682 | Not-determined |
| S8_extracted_k141_50175 | Not-determined |
| S8_extracted_k141_42027 | Not-determined |
| S8_extracted_k141_198516 | Not-determined |
| S8_extracted_k141_154451 | Not-determined |
| S8_extracted_k141_206476 | Not-determined |
| S8_extracted_k141_102607 | Not-determined |
| S8_extracted_k141_154454 | Not-determined |
| S8_extracted_k141_111689 | Not-determined |
| S8_extracted_k141_128794 | Not-determined |
| S8_extracted_k141_206481 | Not-determined |
| S8_extracted_k141_120068 | Not-determined |
| S8_extracted_k141_154457 | Not-determined |
| S8_extracted_k141_128797 | Not-determined |
| S8_extracted_k141_154460 | Not-determined |
| S8_extracted_k141_120074 | Not-determined |
| S8_extracted_k141_67756 | Not-determined |
| S8_extracted_k141_206488 | Not-determined |
| S8_extracted_k141_181103 | Not-determined |
| S8_extracted_k141_67758 | Not-determined |
| S8_extracted_k141_76551 | Not-determined |
| S8_extracted_k141_120077 | Not-determined |
| S8_extracted_k141_85344 | Not-determined |
| S8_extracted_k141_85345 | Not-determined |
| S8_extracted_k141_76554 | Not-determined |
| S8_extracted_k141_198535 | Not-determined |
| S8_extracted_k141_76556 | Not-determined |
| S8_extracted_k141_67762 | Not-determined |
| S8_extracted_k141_154467 | Not-determined |
| S8_extracted_k141_154468 | Not-determined |
| S8_extracted_k141_163205 | Not-determined |
| S8_extracted_k141_154470 | Not-determined |
| S8_extracted_k141_181111 | Not-determined |
| S8_extracted_k141_198542 | Not-determined |
| S8_extracted_k141_120086 | Not-determined |
| S8_extracted_k141_154473 | Not-determined |
| S8_extracted_k141_50202 | Low-quality |
| S8_extracted_k141_198544 | Not-determined |
| S8_extracted_k141_163212 | Not-determined |
| S8_extracted_k141_33561 | Not-determined |
| S8_extracted_k141_198550 | Not-determined |
| S8_extracted_k141_172474 | Not-determined |
| S8_extracted_k141_42046 | Not-determined |
| S8_extracted_k141_206496 | Not-determined |
| S8_extracted_k141_172476 | Not-determined |
| S8_extracted_k141_198553 | Not-determined |
| S8_extracted_k141_94043 | Not-determined |
| S8_extracted_k141_188604 | Not-determined |
| S8_extracted_k141_67763 | Not-determined |
| S8_extracted_k141_76569 | Not-determined |
| S8_extracted_k141_137311 | Not-determined |
| S8_extracted_k141_198557 | Not-determined |
| S8_extracted_k141_206500 | Not-determined |
| S8_extracted_k141_76577 | Not-determined |
| S8_extracted_k141_137312 | Not-determined |
| S8_extracted_k141_33564 | Not-determined |
| S8_extracted_k141_198562 | Low-quality |
| S8_extracted_k141_154490 | Not-determined |
| S8_extracted_k141_76584 | Not-determined |
| S8_extracted_k141_50218 | Not-determined |
| S8_extracted_k141_198564 | Low-quality |
| S8_extracted_k141_198565 | Not-determined |
| S8_extracted_k141_50219 | Not-determined |
| S8_extracted_k141_181123 | Not-determined |
| S8_extracted_k141_188623 | Low-quality |
| S8_extracted_k141_137317 | Not-determined |
| S8_extracted_k141_33570 | Not-determined |
| S8_extracted_k141_24842 | Not-determined |
| S8_extracted_k141_7397 | Not-determined |
| S8_extracted_k141_137321 | Not-determined |
| S8_extracted_k141_188626 | Not-determined |
| S8_extracted_k141_137323 | Not-determined |
| S8_extracted_k141_24844 | Not-determined |
| S8_extracted_k141_7401 | Not-determined |
| S8_extracted_k141_137325 | Not-determined |
| S8_extracted_k141_154503 | Not-determined |
| S8_extracted_k141_188629 | Not-determined |
| S8_extracted_k141_94054 | Not-determined |
| S8_extracted_k141_181139 | Not-determined |
| S8_extracted_k141_33579 | Not-determined |
| S8_extracted_k141_24851 | Not-determined |
| S8_extracted_k141_111739 | Not-determined |
| S8_extracted_k141_172490 | Not-determined |
| S8_extracted_k141_188634 | Not-determined |
| S8_extracted_k141_120109 | Not-determined |
| S8_extracted_k141_188637 | Not-determined |
| S8_extracted_k141_188638 | Low-quality |
| S8_extracted_k141_163228 | Not-determined |
| S8_extracted_k141_120111 | Not-determined |
| S8_extracted_k141_67785 | Not-determined |
| S8_extracted_k141_102609 | Not-determined |
| S8_extracted_k141_172495 | Not-determined |
| S8_extracted_k141_198587 | Not-determined |
| S8_extracted_k141_181143 | Not-determined |
| S8_extracted_k141_76607 | Not-determined |
| S8_extracted_k141_181144 | Not-determined |
| S8_extracted_k141_120114 | Not-determined |
| S8_extracted_k141_154517 | Not-determined |
| S8_extracted_k141_76608 | Not-determined |
| S8_extracted_k141_206513 | Not-determined |
| S8_extracted_k141_111741 | Not-determined |
| S8_extracted_k141_24856 | Not-determined |
| S8_extracted_k141_172500 | Low-quality |
| S8_extracted_k141_146268 | Not-determined |
| S8_extracted_k141_33590 | Not-determined |
| S8_extracted_k141_67798 | Not-determined |
| S8_extracted_k141_188654 | Not-determined |
| S8_extracted_k141_154525 | Not-determined |
| S8_extracted_k141_67800 | Low-quality |
| S8_extracted_k141_172505 | Not-determined |
| S8_extracted_k141_206520 | Not-determined |
| S8_extracted_k141_24859 | Low-quality |
| S8_extracted_k141_94070 | Low-quality |
| S8_extracted_k141_181149 | Not-determined |
| S8_extracted_k141_7407 | Not-determined |
| S8_extracted_k141_181151 | Not-determined |
| S8_extracted_k141_120121 | Not-determined |
| S8_extracted_k141_128846 | Not-determined |
| S8_extracted_k141_102624 | Not-determined |
| S8_extracted_k141_33594 | Not-determined |
| S8_extracted_k141_50247 | Low-quality |
| S8_extracted_k141_24865 | Not-determined |
| S8_extracted_k141_206522 | Not-determined |
| S8_extracted_k141_111755 | Not-determined |
| S8_extracted_k141_163238 | Not-determined |
| S8_extracted_k141_137350 | Not-determined |
| S8_extracted_k141_172509 | Not-determined |
| S8_extracted_k141_163242 | Not-determined |
| S8_extracted_k141_154539 | Not-determined |
| S8_extracted_k141_137354 | Not-determined |
| S8_extracted_k141_33600 | Not-determined |
| S8_extracted_k141_76619 | Not-determined |
| S8_extracted_k141_33603 | Not-determined |
| S8_extracted_k141_102639 | Not-determined |
| S8_extracted_k141_172522 | Not-determined |
| S8_extracted_k141_154543 | Not-determined |
| S8_extracted_k141_188665 | Not-determined |
| S8_extracted_k141_76623 | Not-determined |
| S8_extracted_k141_111765 | Not-determined |
| S8_extracted_k141_188666 | Not-determined |
| S8_extracted_k141_33615 | Not-determined |
| S8_extracted_k141_163246 | Not-determined |
| S8_extracted_k141_128854 | Not-determined |
| S8_extracted_k141_102649 | Not-determined |
| S8_extracted_k141_102650 | Not-determined |
| S8_extracted_k141_111770 | Not-determined |
| S8_extracted_k141_128859 | Not-determined |
| S8_extracted_k141_111771 | Not-determined |
| S8_extracted_k141_76632 | Not-determined |
| S8_extracted_k141_7431 | Not-determined |
| S8_extracted_k141_111774 | Not-determined |
| S8_extracted_k141_128860 | Not-determined |
| S8_extracted_k141_94099 | Not-determined |
| S8_extracted_k141_188671 | Not-determined |
| S8_extracted_k141_198599 | Not-determined |
| S8_extracted_k141_198601 | Not-determined |
| S8_extracted_k141_111775 | Low-quality |
| S8_extracted_k141_146291 | Not-determined |
| S8_extracted_k141_137374 | Not-determined |
| S8_extracted_k141_33627 | Not-determined |
| S8_extracted_k141_154547 | Not-determined |
| S8_extracted_k141_111780 | Low-quality |
| S8_extracted_k141_50266 | Not-determined |
| S8_extracted_k141_111781 | Low-quality |
| S8_extracted_k141_137381 | Not-determined |
| S8_extracted_k141_188678 | Not-determined |
| S8_extracted_k141_198610 | Not-determined |
| S8_extracted_k141_33636 | Not-determined |
| S8_extracted_k141_42056 | Not-determined |
| S8_extracted_k141_24884 | Not-determined |
| S8_extracted_k141_42060 | Not-determined |
| S8_extracted_k141_198614 | Not-determined |
| S8_extracted_k141_7442 | Not-determined |
| S8_extracted_k141_24886 | Not-determined |
| S8_extracted_k141_76645 | Not-determined |
| S8_extracted_k141_154551 | Not-determined |
| S8_extracted_k141_146302 | Not-determined |
| S8_extracted_k141_154555 | Not-determined |
| S8_extracted_k141_7447 | Not-determined |
| S8_extracted_k141_94110 | Not-determined |
| S8_extracted_k141_67829 | Not-determined |
| S8_extracted_k141_146306 | Low-quality |
| S8_extracted_k141_137386 | Not-determined |
| S8_extracted_k141_76650 | Not-determined |
| S8_extracted_k141_7451 | Not-determined |
| S8_extracted_k141_181191 | Not-determined |
| S8_extracted_k141_7453 | Low-quality |
| S8_extracted_k141_85374 | Not-determined |
| S8_extracted_k141_7454 | Not-determined |
| S8_extracted_k141_128870 | Not-determined |
| S8_extracted_k141_85376 | Not-determined |
| S8_extracted_k141_76653 | Not-determined |
| S8_extracted_k141_111789 | Not-determined |
| S8_extracted_k141_24901 | Not-determined |
| S8_extracted_k141_154558 | Not-determined |
| S8_extracted_k141_181197 | Low-quality |
| S8_extracted_k141_94121 | Not-determined |
| S8_extracted_k141_181199 | Not-determined |
| S8_extracted_k141_7460 | Not-determined |
| S8_extracted_k141_128872 | Low-quality |
| S8_extracted_k141_76661 | Not-determined |
| S8_extracted_k141_33655 | Not-determined |
| S8_extracted_k141_198635 | Not-determined |
| S8_extracted_k141_198636 | Not-determined |
| S8_extracted_k141_128875 | Not-determined |
| S8_extracted_k141_94125 | Not-determined |
| S8_extracted_k141_154574 | Not-determined |
| S8_extracted_k141_163283 | Not-determined |
| S8_extracted_k141_24909 | Not-determined |
| S8_extracted_k141_24911 | Not-determined |
| S8_extracted_k141_181212 | Not-determined |
| S8_extracted_k141_33659 | Not-determined |
| S8_extracted_k141_163287 | Not-determined |
| S8_extracted_k141_85387 | Low-quality |
| S8_extracted_k141_128884 | Not-determined |
| S8_extracted_k141_198640 | Low-quality |
| S8_extracted_k141_146324 | Not-determined |
| S8_extracted_k141_154577 | Low-quality |
| S8_extracted_k141_146326 | Not-determined |
| S8_extracted_k141_94130 | Not-determined |
| S8_extracted_k141_94131 | Not-determined |
| S8_extracted_k141_7476 | Not-determined |
| S8_extracted_k141_120153 | Not-determined |
| S8_extracted_k141_146330 | Not-determined |
| S8_extracted_k141_172550 | Not-determined |
| S8_extracted_k141_59743 | Not-determined |
| S8_extracted_k141_137406 | Not-determined |
| S8_extracted_k141_128888 | Not-determined |
| S8_extracted_k141_50287 | Not-determined |
| S8_extracted_k141_137408 | Not-determined |
| S8_extracted_k141_24921 | Not-determined |
| S8_extracted_k141_76670 | Not-determined |
| S8_extracted_k141_137409 | Not-determined |
| S8_extracted_k141_76671 | Not-determined |
| S8_extracted_k141_85396 | Not-determined |
| S8_extracted_k141_137410 | Not-determined |
| S8_extracted_k141_188699 | Not-determined |
| S8_extracted_k141_181225 | Not-determined |
| S8_extracted_k141_198648 | Not-determined |
| S8_extracted_k141_7484 | Not-determined |
| S8_extracted_k141_42082 | Not-determined |
| S8_extracted_k141_94136 | Not-determined |
| S8_extracted_k141_42085 | Not-determined |
| S8_extracted_k141_102693 | Not-determined |
| S8_extracted_k141_172558 | Not-determined |
| S8_extracted_k141_85403 | Low-quality |
| S8_extracted_k141_188702 | Not-determined |
| S8_extracted_k141_128897 | Not-determined |
| S8_extracted_k141_146339 | Not-determined |
| S8_extracted_k141_42090 | Low-quality |
| S8_extracted_k141_50300 | Not-determined |
| S8_extracted_k141_102695 | Not-determined |
| S8_extracted_k141_146341 | Not-determined |
| S8_extracted_k141_120161 | Not-determined |
| S8_extracted_k141_172564 | Not-determined |
| S8_extracted_k141_94138 | Low-quality |
| S8_extracted_k141_137418 | Not-determined |
| S8_extracted_k141_94139 | Low-quality |
| S8_extracted_k141_7488 | Low-quality |
| S8_extracted_k141_24932 | Not-determined |
| S8_extracted_k141_59758 | Not-determined |
| S8_extracted_k141_181231 | Low-quality |
| S8_extracted_k141_42096 | Low-quality |
| S8_extracted_k141_198653 | Not-determined |
| S8_extracted_k141_33675 | Not-determined |
| S8_extracted_k141_181233 | Not-determined |
| S8_extracted_k141_188709 | Not-determined |
| S8_extracted_k141_24935 | Not-determined |
| S8_extracted_k141_137423 | Not-determined |
| S8_extracted_k141_50308 | Not-determined |
| S8_extracted_k141_154603 | Not-determined |
| S8_extracted_k141_50309 | Not-determined |
| S8_extracted_k141_50310 | Not-determined |
| S8_extracted_k141_146350 | Not-determined |
| S8_extracted_k141_33682 | Not-determined |
| S8_extracted_k141_198659 | Not-determined |
| S8_extracted_k141_85415 | Not-determined |
| S8_extracted_k141_206536 | Not-determined |
| S8_extracted_k141_24944 | Not-determined |
| S14_extracted_k141_117164 | Medium-quality |
| S8_extracted_k141_111791 | Low-quality |
| S8_extracted_k141_85420 | Low-quality |
| S8_extracted_k141_33694 | Not-determined |
| S8_extracted_k141_102704 | Not-determined |
| S8_extracted_k141_111793 | Not-determined |
| S8_extracted_k141_120169 | Not-determined |
| S8_extracted_k141_111795 | Not-determined |
| S8_extracted_k141_206545 | Not-determined |
| S8_extracted_k141_188715 | Not-determined |
| S8_extracted_k141_102711 | Not-determined |
| S8_extracted_k141_128910 | Low-quality |
| S8_extracted_k141_50321 | Not-determined |
| S8_extracted_k141_94156 | Not-determined |
| S8_extracted_k141_59771 | Not-determined |
| S8_extracted_k141_94159 | Not-determined |
| S8_extracted_k141_198664 | Not-determined |
| S8_extracted_k141_42110 | Not-determined |
| S8_extracted_k141_94161 | Not-determined |
| S8_extracted_k141_59775 | Low-quality |
| S8_extracted_k141_137442 | Not-determined |
| S8_extracted_k141_50324 | Not-determined |
| S8_extracted_k141_120178 | Not-determined |
| S8_extracted_k141_7510 | Not-determined |
| S8_extracted_k141_154614 | Not-determined |
| S8_extracted_k141_67860 | Not-determined |
| S8_extracted_k141_76683 | Not-determined |
| S8_extracted_k141_33700 | Not-determined |
| S8_extracted_k141_76684 | Not-determined |
| S8_extracted_k141_42111 | Not-determined |
| S8_extracted_k141_154615 | Not-determined |
| S8_extracted_k141_146370 | Not-determined |
| S8_extracted_k141_7513 | Not-determined |
| S8_extracted_k141_15752 | Not-determined |
| S8_extracted_k141_24962 | Not-determined |
| S8_extracted_k141_146372 | Not-determined |
| S8_extracted_k141_111803 | Not-determined |
| S8_extracted_k141_163323 | Low-quality |
| S8_extracted_k141_128916 | Not-determined |
| S8_extracted_k141_15754 | Low-quality |
| S8_extracted_k141_24965 | Not-determined |
| S8_extracted_k141_163325 | Not-determined |
| S8_extracted_k141_154618 | Not-determined |
| S8_extracted_k141_120187 | Not-determined |
| S8_extracted_k141_198672 | Not-determined |
| S8_extracted_k141_85439 | Not-determined |
| S8_extracted_k141_181251 | Not-determined |
| S8_extracted_k141_15758 | Not-determined |
| S8_extracted_k141_67864 | Not-determined |
| S8_extracted_k141_198677 | Not-determined |
| S8_extracted_k141_137450 | Not-determined |
| S8_extracted_k141_154621 | Not-determined |
| S8_extracted_k141_33715 | Not-determined |
| S8_extracted_k141_7524 | Not-determined |
| S8_extracted_k141_42114 | Not-determined |
| S8_extracted_k141_163332 | Not-determined |
| S8_extracted_k141_181257 | Not-determined |
| S8_extracted_k141_76694 | Low-quality |
| S8_extracted_k141_137453 | Not-determined |
| S8_extracted_k141_154627 | Not-determined |
| S8_extracted_k141_76696 | Not-determined |
| S8_extracted_k141_137455 | Not-determined |
| S8_extracted_k141_33721 | Not-determined |
| S8_extracted_k141_198687 | Not-determined |
| S8_extracted_k141_94177 | Not-determined |
| S8_extracted_k141_42124 | Not-determined |
| S8_extracted_k141_94178 | Not-determined |
| S8_extracted_k141_67872 | Not-determined |
| S8_extracted_k141_67873 | Not-determined |
| S8_extracted_k141_7536 | Not-determined |
| S8_extracted_k141_128935 | Low-quality |
| S8_extracted_k141_163345 | Not-determined |
| S8_extracted_k141_7539 | Not-determined |
| S8_extracted_k141_146387 | Not-determined |
| S8_extracted_k141_7541 | Not-determined |
| S8_extracted_k141_76701 | Not-determined |
| S8_extracted_k141_67877 | Not-determined |
| S8_extracted_k141_33731 | Not-determined |
| S8_extracted_k141_85451 | Not-determined |
| S8_extracted_k141_67881 | Not-determined |
| S8_extracted_k141_154631 | Low-quality |
| S8_extracted_k141_94182 | Not-determined |
| S8_extracted_k141_163351 | Not-determined |
| S8_extracted_k141_94183 | Not-determined |
| S8_extracted_k141_128947 | Not-determined |
| S8_extracted_k141_42131 | Low-quality |
| S8_extracted_k141_33733 | Low-quality |
| S8_extracted_k141_76708 | Low-quality |
| S8_extracted_k141_7550 | Not-determined |
| S8_extracted_k141_76710 | Not-determined |
| S8_extracted_k141_7551 | Not-determined |
| S8_extracted_k141_137465 | Not-determined |
| S8_extracted_k141_163357 | Not-determined |
| S8_extracted_k141_198696 | Not-determined |
| S8_extracted_k141_7557 | Not-determined |
| S8_extracted_k141_128957 | Not-determined |
| S8_extracted_k141_154641 | Not-determined |
| S8_extracted_k141_146395 | Not-determined |
| S8_extracted_k141_76721 | Not-determined |
| S8_extracted_k141_198698 | Not-determined |
| S8_extracted_k141_24990 | Not-determined |
| S8_extracted_k141_146397 | Not-determined |
| S8_extracted_k141_128961 | Not-determined |
| S8_extracted_k141_154642 | Low-quality |
| S8_extracted_k141_42136 | Not-determined |
| S8_extracted_k141_50341 | Not-determined |
| S8_extracted_k141_137471 | Not-determined |
| S8_extracted_k141_137472 | Not-determined |
| S8_extracted_k141_120210 | Not-determined |
| S8_extracted_k141_50343 | Low-quality |
| S8_extracted_k141_163363 | Not-determined |
| S8_extracted_k141_206569 | Low-quality |
| S8_extracted_k141_163366 | Not-determined |
| S8_extracted_k141_67895 | Not-determined |
| S8_extracted_k141_76728 | Not-determined |
| S8_extracted_k141_120215 | Not-determined |
| S8_extracted_k141_42139 | Not-determined |
| S8_extracted_k141_67897 | Not-determined |
| S8_extracted_k141_85457 | Not-determined |
| S8_extracted_k141_120218 | Not-determined |
| S8_extracted_k141_50348 | Not-determined |
| S8_extracted_k141_163370 | Not-determined |
| S8_extracted_k141_137477 | Low-quality |
| S8_extracted_k141_94200 | Not-determined |
| S8_extracted_k141_67903 | Not-determined |
| S8_extracted_k141_76735 | Not-determined |
| S8_extracted_k141_15765 | Not-determined |
| S8_extracted_k141_59803 | Low-quality |
| S8_extracted_k141_128976 | Not-determined |
| S8_extracted_k141_172609 | Not-determined |
| S8_extracted_k141_163380 | Not-determined |
| S8_extracted_k141_7576 | Not-determined |
| S8_extracted_k141_15770 | Not-determined |
| S8_extracted_k141_76741 | Not-determined |
| S8_extracted_k141_59806 | Not-determined |
| S8_extracted_k141_181271 | Not-determined |
| S8_extracted_k141_198717 | Not-determined |
| S8_extracted_k141_67909 | Not-determined |
| S8_extracted_k141_163385 | Not-determined |
| S8_extracted_k141_67911 | Not-determined |
| S8_extracted_k141_33756 | Not-determined |
| S8_extracted_k141_198725 | Not-determined |
| S8_extracted_k141_76753 | Not-determined |
| S8_extracted_k141_128990 | Not-determined |
| S8_extracted_k141_59810 | Not-determined |
| S8_extracted_k141_120241 | Not-determined |
| S8_extracted_k141_33761 | Low-quality |
| S8_extracted_k141_50362 | Not-determined |
| S8_extracted_k141_198729 | Not-determined |
| S8_extracted_k141_67916 | Low-quality |
| S8_extracted_k141_7582 | Not-determined |
| S8_extracted_k141_198732 | Not-determined |
| S8_extracted_k141_7584 | Not-determined |
| S8_extracted_k141_128999 | Not-determined |
| S8_extracted_k141_24993 | Low-quality |
| S8_extracted_k141_67918 | Not-determined |
| S8_extracted_k141_50365 | Not-determined |
| S8_extracted_k141_67920 | Not-determined |
| S8_extracted_k141_85468 | Not-determined |
| S8_extracted_k141_59819 | Not-determined |
| S8_extracted_k141_15784 | Low-quality |
| S8_extracted_k141_163396 | Not-determined |
| S8_extracted_k141_154655 | Not-determined |
| S8_extracted_k141_76766 | Low-quality |
| S8_extracted_k141_146432 | Not-determined |
| S8_extracted_k141_7593 | Not-determined |
| S8_extracted_k141_154660 | Not-determined |
| S8_extracted_k141_154661 | Not-determined |
| S8_extracted_k141_50371 | Not-determined |
| S8_extracted_k141_50373 | Not-determined |
| S8_extracted_k141_206579 | Low-quality |
| S8_extracted_k141_85475 | Not-determined |
| S8_extracted_k141_146440 | Low-quality |
| S8_extracted_k141_15797 | Not-determined |
| S8_extracted_k141_154668 | Not-determined |
| S8_extracted_k141_25008 | Not-determined |
| S8_extracted_k141_146442 | Low-quality |
| S8_extracted_k141_50378 | Low-quality |
| S8_extracted_k141_146446 | Not-determined |
| S8_extracted_k141_198751 | Not-determined |
| S8_extracted_k141_146448 | Not-determined |
| S8_extracted_k141_154673 | Not-determined |
| S8_extracted_k141_198754 | Not-determined |
| S8_extracted_k141_94227 | Not-determined |
| S8_extracted_k141_25014 | Not-determined |
| S8_extracted_k141_172639 | Not-determined |
| S8_extracted_k141_76790 | Not-determined |
| S8_extracted_k141_198756 | Not-determined |
| S8_extracted_k141_172643 | Not-determined |
| S8_extracted_k141_120253 | Not-determined |
| S8_extracted_k141_59828 | Not-determined |
| S8_extracted_k141_59831 | Not-determined |
| S8_extracted_k141_50381 | Not-determined |
| S8_extracted_k141_76796 | Not-determined |
| S8_extracted_k141_85477 | Not-determined |
| S8_extracted_k141_59833 | Not-determined |
| S8_extracted_k141_94231 | Not-determined |
| S8_extracted_k141_198762 | Not-determined |
| S8_extracted_k141_172648 | Not-determined |
| S8_extracted_k141_120257 | Low-quality |
| S8_extracted_k141_129035 | Not-determined |
| S8_extracted_k141_85480 | Not-determined |
| S8_extracted_k141_67940 | Not-determined |
| S8_extracted_k141_85483 | Not-determined |
| S8_extracted_k141_42184 | Not-determined |
| S8_extracted_k141_85484 | Not-determined |
| S8_extracted_k141_67942 | Not-determined |
| S8_extracted_k141_50390 | Not-determined |
| S8_extracted_k141_146459 | Not-determined |
| S8_extracted_k141_94235 | Low-quality |
| S8_extracted_k141_15818 | Not-determined |
| S8_extracted_k141_7609 | Not-determined |
| S8_extracted_k141_206603 | Not-determined |
| S8_extracted_k141_120265 | Low-quality |
| S8_extracted_k141_59850 | Not-determined |
| S8_extracted_k141_85487 | Not-determined |
| S8_extracted_k141_198771 | Not-determined |
| S8_extracted_k141_50396 | Not-determined |
| S8_extracted_k141_206608 | Not-determined |
| S8_extracted_k141_172668 | Not-determined |
| S8_extracted_k141_129049 | Not-determined |
| S8_extracted_k141_198778 | Not-determined |
| S8_extracted_k141_33807 | Low-quality |
| S8_extracted_k141_129052 | Not-determined |
| S8_extracted_k141_67955 | Not-determined |
| S8_extracted_k141_163411 | Not-determined |
| S8_extracted_k141_154679 | Not-determined |
| S8_extracted_k141_33810 | Not-determined |
| S8_extracted_k141_181278 | Not-determined |
| S8_extracted_k141_198780 | Not-determined |
| S8_extracted_k141_15825 | Not-determined |
| S8_extracted_k141_76823 | Not-determined |
| S8_extracted_k141_163417 | Not-determined |
| S8_extracted_k141_120276 | Not-determined |
| S8_extracted_k141_120277 | Not-determined |
| S8_extracted_k141_206613 | Not-determined |

| | |
|---|---|
| S8_extracted_k141_120279 | Not-determined |
| S8_extracted_k141_206616 | Not-determined |
| S8_extracted_k141_154686 | Not-determined |
| S8_extracted_k141_154687 | Not-determined |
| S8_extracted_k141_76827 | Not-determined |
| S8_extracted_k141_120282 | Not-determined |
| S8_extracted_k141_42193 | Not-determined |
| S8_extracted_k141_120283 | Not-determined |
| S8_extracted_k141_198787 | Not-determined |
| S8_extracted_k141_154692 | Not-determined |
| S8_extracted_k141_129056 | Not-determined |
| S8_extracted_k141_42195 | Not-determined |
| S8_extracted_k141_146461 | Not-determined |
| S8_extracted_k141_154694 | Not-determined |
| S8_extracted_k141_172675 | Not-determined |
| S8_extracted_k141_129057 | Not-determined |
| S8_extracted_k141_188732 | Not-determined |
| S8_extracted_k141_67957 | Not-determined |
| S8_extracted_k141_146462 | Not-determined |
| S8_extracted_k141_42199 | Low-quality |
| S8_extracted_k141_206618 | Not-determined |
| S8_extracted_k141_129062 | Low-quality |
| S8_extracted_k141_76829 | Not-determined |
| S8_extracted_k141_7625 | Low-quality |
| S8_extracted_k141_172677 | Not-determined |
| S8_extracted_k141_67958 | Not-determined |
| S8_extracted_k141_172679 | Low-quality |
| S8_extracted_k141_206625 | Not-determined |
| S8_extracted_k141_206628 | Not-determined |
| S8_extracted_k141_129066 | Not-determined |
| S8_extracted_k141_42206 | Not-determined |
| S8_extracted_k141_206630 | Not-determined |
| S8_extracted_k141_206631 | Not-determined |
| S8_extracted_k141_94243 | Not-determined |
| S8_extracted_k141_59875 | Not-determined |
| S8_extracted_k141_7626 | Low-quality |
| S8_extracted_k141_198802 | Not-determined |
| S8_extracted_k141_67960 | Not-determined |
| S8_extracted_k141_7627 | Not-determined |
| S8_extracted_k141_163433 | Not-determined |
| S8_extracted_k141_198803 | Not-determined |
| S8_extracted_k141_154703 | Low-quality |
| S8_extracted_k141_198805 | Low-quality |
| S8_extracted_k141_181302 | Not-determined |
| S8_extracted_k141_198807 | Not-determined |
| S8_extracted_k141_7631 | Not-determined |
| S8_extracted_k141_59884 | Not-determined |
| S8_extracted_k141_85510 | Not-determined |
| S8_extracted_k141_25057 | Not-determined |
| S8_extracted_k141_146477 | Not-determined |
| S8_extracted_k141_33834 | Not-determined |
| S8_extracted_k141_33836 | Not-determined |
| S8_extracted_k141_85519 | Not-determined |
| S8_extracted_k141_50420 | Not-determined |
| S8_extracted_k141_76843 | Not-determined |
| S8_extracted_k141_85523 | Not-determined |
| S8_extracted_k141_198818 | Not-determined |
| S8_extracted_k141_188752 | Not-determined |
| S8_extracted_k141_94249 | Not-determined |
| S8_extracted_k141_154714 | Not-determined |
| S8_extracted_k141_33843 | Not-determined |
| S8_extracted_k141_67975 | Not-determined |
| S8_extracted_k141_181318 | Not-determined |
| S8_extracted_k141_15832 | Not-determined |
| S8_extracted_k141_7642 | Not-determined |
| S8_extracted_k141_76849 | Not-determined |
| S8_extracted_k141_7643 | Not-determined |
| S8_extracted_k141_188760 | Not-determined |
| S8_extracted_k141_188761 | Not-determined |
| S8_extracted_k141_146479 | Not-determined |
| S8_extracted_k141_120332 | Not-determined |
| S8_extracted_k141_42223 | Not-determined |
| S8_extracted_k141_67978 | Not-determined |
| S8_extracted_k141_94259 | Not-determined |
| S8_extracted_k141_67979 | Not-determined |
| S8_extracted_k141_120336 | Not-determined |
| S8_extracted_k141_42227 | Not-determined |
| S8_extracted_k141_129090 | Not-determined |
| S8_extracted_k141_120338 | Not-determined |
| S8_extracted_k141_94262 | Not-determined |
| S8_extracted_k141_94263 | Not-determined |
| S8_extracted_k141_198830 | Low-quality |
| S8_extracted_k141_67984 | Not-determined |
| S8_extracted_k141_15835 | Low-quality |
| S8_extracted_k141_25084 | Low-quality |
| S8_extracted_k141_154720 | Not-determined |
| S8_extracted_k141_198835 | Not-determined |
| S8_extracted_k141_154727 | Not-determined |
| S8_extracted_k141_154729 | Not-determined |
| S8_extracted_k141_102718 | Not-determined |
| S8_extracted_k141_25092 | Not-determined |
| S8_extracted_k141_129100 | Not-determined |
| S8_extracted_k141_120351 | Not-determined |
| S8_extracted_k141_129101 | Not-determined |
| S8_extracted_k141_120352 | Not-determined |
| S8_extracted_k141_59904 | Not-determined |
| S8_extracted_k141_146491 | Not-determined |
| S8_extracted_k141_181326 | Not-determined |
| S8_extracted_k141_137506 | Not-determined |
| S8_extracted_k141_120362 | Not-determined |
| S8_extracted_k141_25097 | Not-determined |
| S8_extracted_k141_129107 | Low-quality |
| S8_extracted_k141_188770 | Not-determined |
| S8_extracted_k141_42246 | Not-determined |
| S8_extracted_k141_102723 | Not-determined |
| S8_extracted_k141_206659 | Not-determined |
| S8_extracted_k141_172720 | Low-quality |
| S8_extracted_k141_206660 | Not-determined |
| S8_extracted_k141_42249 | Not-determined |
| S8_extracted_k141_206662 | Not-determined |
| S8_extracted_k141_137512 | Not-determined |
| S8_extracted_k141_102729 | Not-determined |
| S8_extracted_k141_129110 | Low-quality |
| S8_extracted_k141_181330 | Not-determined |
| S8_extracted_k141_59914 | Not-determined |
| S8_extracted_k141_85550 | Not-determined |
| S8_extracted_k141_102730 | Not-determined |
| S8_extracted_k141_33855 | Low-quality |
| S8_extracted_k141_59918 | Not-determined |
| S8_extracted_k141_85554 | Not-determined |
| S8_extracted_k141_198848 | Low-quality |
| S8_extracted_k141_188775 | Not-determined |
| S8_extracted_k141_59923 | Not-determined |
| S8_extracted_k141_172735 | Not-determined |
| S8_extracted_k141_146501 | Low-quality |
| S8_extracted_k141_198850 | Low-quality |
| S8_extracted_k141_15854 | Not-determined |
| S8_extracted_k141_172739 | Not-determined |
| S8_extracted_k141_42263 | Not-determined |
| S8_extracted_k141_146506 | Not-determined |
| S8_extracted_k141_146507 | Not-determined |
| S8_extracted_k141_120376 | Not-determined |
| S8_extracted_k141_172743 | Not-determined |
| S8_extracted_k141_33863 | Not-determined |
| S8_extracted_k141_146511 | Not-determined |
| S8_extracted_k141_181342 | Not-determined |
| S8_extracted_k141_94277 | Not-determined |
| S8_extracted_k141_76858 | Not-determined |
| S8_extracted_k141_15863 | Not-determined |
| S8_extracted_k141_68003 | Not-determined |
| S8_extracted_k141_120381 | Not-determined |
| S8_extracted_k141_68004 | Low-quality |
| S8_extracted_k141_68005 | Not-determined |
| S8_extracted_k141_68006 | Not-determined |
| S8_extracted_k141_94280 | Not-determined |
| S8_extracted_k141_137520 | Not-determined |
| S8_extracted_k141_7656 | Not-determined |
| S8_extracted_k141_94283 | Low-quality |
| S8_extracted_k141_42271 | Not-determined |
| S8_extracted_k141_102756 | Not-determined |
| S8_extracted_k141_181350 | Not-determined |
| S8_extracted_k141_188794 | Not-determined |
| S8_extracted_k141_154734 | Not-determined |
| S8_extracted_k141_111822 | Not-determined |
| S8_extracted_k141_102758 | Not-determined |
| S8_extracted_k141_188797 | Not-determined |
| S8_extracted_k141_76869 | Not-determined |
| S8_extracted_k141_172748 | Not-determined |
| S8_extracted_k141_111828 | Not-determined |
| S8_extracted_k141_85562 | Not-determined |
| S8_extracted_k141_85563 | Not-determined |
| S8_extracted_k141_181356 | Low-quality |
| S8_extracted_k141_111832 | Low-quality |
| S8_extracted_k141_172754 | Not-determined |
| S8_extracted_k141_172756 | Not-determined |
| S8_extracted_k141_206688 | Not-determined |
| S8_extracted_k141_68014 | Not-determined |
| S8_extracted_k141_172757 | Not-determined |
| S8_extracted_k141_76872 | Not-determined |
| S8_extracted_k141_94292 | Not-determined |
| S8_extracted_k141_154742 | Not-determined |
| S8_extracted_k141_146522 | Not-determined |
| S8_extracted_k141_120393 | Not-determined |
| S8_extracted_k141_102764 | Not-determined |
| S8_extracted_k141_206692 | Not-determined |
| S8_extracted_k141_76878 | Not-determined |
| S8_extracted_k141_146527 | Not-determined |
| S8_extracted_k141_137526 | Not-determined |
| S8_extracted_k141_59940 | Not-determined |
| S8_extracted_k141_154749 | Not-determined |
| S8_extracted_k141_94299 | Not-determined |
| S8_extracted_k141_137533 | Not-determined |
| S8_extracted_k141_120403 | Not-determined |
| S8_extracted_k141_85568 | Not-determined |
| S8_extracted_k141_15881 | Not-determined |
| S8_extracted_k141_181367 | Not-determined |
| S8_extracted_k141_120406 | Not-determined |
| S8_extracted_k141_120407 | Not-determined |
| S8_extracted_k141_172759 | Not-determined |
| S8_extracted_k141_7660 | Not-determined |
| S8_extracted_k141_188799 | Not-determined |
| S8_extracted_k141_94304 | Not-determined |
| S8_extracted_k141_188803 | Not-determined |
| S8_extracted_k141_15892 | Not-determined |
| S8_extracted_k141_15894 | Low-quality |
| S8_extracted_k141_50449 | Not-determined |
| S8_extracted_k141_76891 | Low-quality |
| S8_extracted_k141_50451 | Not-determined |
| S8_extracted_k141_15897 | Not-determined |
| S8_extracted_k141_129124 | Not-determined |
| S8_extracted_k141_68027 | Not-determined |
| S8_extracted_k141_188806 | Not-determined |
| S8_extracted_k141_172768 | Not-determined |
| S8_extracted_k141_76894 | Low-quality |
| S8_extracted_k141_7667 | Not-determined |
| S8_extracted_k141_188812 | Not-determined |
| S8_extracted_k141_206703 | Not-determined |
| S8_extracted_k141_137548 | Not-determined |
| S8_extracted_k141_7669 | Not-determined |
| S8_extracted_k141_163438 | Not-determined |
| S8_extracted_k141_33894 | Not-determined |
| S8_extracted_k141_129129 | Not-determined |
| S8_extracted_k141_85576 | Not-determined |
| S8_extracted_k141_206710 | Not-determined |
| S8_extracted_k141_59951 | Not-determined |
| S8_extracted_k141_102793 | Not-determined |
| S8_extracted_k141_102794 | Not-determined |
| S8_extracted_k141_7675 | Not-determined |
| S8_extracted_k141_68035 | Not-determined |
| S8_extracted_k141_85580 | Low-quality |
| S8_extracted_k141_68036 | Not-determined |
| S8_extracted_k141_59954 | Not-determined |
| S8_extracted_k141_154763 | Not-determined |
| S8_extracted_k141_59955 | Not-determined |
| S8_extracted_k141_59957 | Not-determined |
| S8_extracted_k141_33904 | Not-determined |
| S8_extracted_k141_7678 | Not-determined |
| S8_extracted_k141_146541 | Not-determined |
| S8_extracted_k141_33906 | Low-quality |
| S8_extracted_k141_50469 | Not-determined |
| S8_extracted_k141_111872 | Not-determined |
| S8_extracted_k141_198879 | Not-determined |
| S8_extracted_k141_137555 | Not-determined |
| S8_extracted_k141_188822 | Not-determined |
| S8_extracted_k141_68039 | Not-determined |
| S8_extracted_k141_59966 | Not-determined |
| S8_extracted_k141_59967 | Not-determined |
| S8_extracted_k141_94319 | Low-quality |
| S8_extracted_k141_85599 | Not-determined |
| S8_extracted_k141_76909 | Not-determined |
| S8_extracted_k141_129154 | Not-determined |
| S8_extracted_k141_94323 | Not-determined |
| S8_extracted_k141_33914 | Not-determined |
| S8_extracted_k141_163447 | Not-determined |
| S8_extracted_k141_137562 | Low-quality |
| S8_extracted_k141_163448 | Not-determined |
| S8_extracted_k141_129156 | Not-determined |
| S8_extracted_k141_146555 | Not-determined |
| S8_extracted_k141_163449 | Low-quality |
| S8_extracted_k141_7689 | Not-determined |
| S8_extracted_k141_33915 | Low-quality |
| S8_extracted_k141_181384 | Not-determined |
| S8_extracted_k141_111887 | Not-determined |
| S8_extracted_k141_146561 | Not-determined |
| S8_extracted_k141_33918 | Not-determined |
| S8_extracted_k141_68052 | Not-determined |
| S8_extracted_k141_68053 | Not-determined |
| S8_extracted_k141_146563 | Not-determined |
| S8_extracted_k141_181393 | Not-determined |
| S8_extracted_k141_94332 | Not-determined |
| S8_extracted_k141_68058 | Not-determined |
| S8_extracted_k141_76928 | Not-determined |
| S8_extracted_k141_7701 | Not-determined |
| S8_extracted_k141_85613 | Not-determined |
| S8_extracted_k141_68063 | Not-determined |
| S8_extracted_k141_206737 | Not-determined |
| S8_extracted_k141_85615 | Not-determined |
| S8_extracted_k141_154791 | Not-determined |
| S8_extracted_k141_76932 | Not-determined |
| S8_extracted_k141_172808 | Not-determined |
| S8_extracted_k141_137581 | Low-quality |
| S8_extracted_k141_7707 | Not-determined |
| S8_extracted_k141_172812 | Not-determined |
| S8_extracted_k141_111904 | Low-quality |
| S8_extracted_k141_129167 | Not-determined |
| S8_extracted_k141_94339 | Not-determined |
| S8_extracted_k141_198902 | Not-determined |
| S8_extracted_k141_154796 | Not-determined |
| S8_extracted_k141_181406 | Not-determined |
| S8_extracted_k141_42278 | Not-determined |
| S8_extracted_k141_154799 | Not-determined |
| S8_extracted_k141_181408 | Not-determined |
| S8_extracted_k141_25110 | Not-determined |
| S8_extracted_k141_198905 | Not-determined |
| S8_extracted_k141_181411 | Not-determined |
| S8_extracted_k141_172819 | Not-determined |
| S8_extracted_k141_172820 | Not-determined |
| S8_extracted_k141_172821 | Low-quality |
| S8_extracted_k141_163460 | Low-quality |
| S8_extracted_k141_146588 | Not-determined |
| S8_extracted_k141_42286 | Not-determined |
| S8_extracted_k141_137599 | Not-determined |
| S8_extracted_k141_42288 | Not-determined |
| S8_extracted_k141_76949 | Not-determined |
| S8_extracted_k141_146590 | Not-determined |
| S8_extracted_k141_7728 | Not-determined |
| S8_extracted_k141_85633 | Not-determined |
| S8_extracted_k141_102847 | Not-determined |
| S8_extracted_k141_146592 | Not-determined |
| S8_extracted_k141_94351 | Not-determined |
| S8_extracted_k141_76954 | Not-determined |
| S8_extracted_k141_94353 | Low-quality |
| S8_extracted_k141_137605 | Not-determined |
| S8_extracted_k141_33955 | Low-quality |
| S8_extracted_k141_120440 | Not-determined |
| S8_extracted_k141_7736 | Not-determined |
| S8_extracted_k141_172830 | Not-determined |
| S8_extracted_k141_188843 | Not-determined |
| S8_extracted_k141_7738 | Low-quality |
| S8_extracted_k141_76960 | Not-determined |
| S8_extracted_k141_111919 | Not-determined |
| S8_extracted_k141_206760 | Not-determined |
| S8_extracted_k141_76962 | Not-determined |
| S8_extracted_k141_111920 | Not-determined |
| S8_extracted_k141_206763 | Not-determined |
| S8_extracted_k141_172833 | Not-determined |
| S8_extracted_k141_42298 | Not-determined |
| S8_extracted_k141_76966 | Not-determined |
| S8_extracted_k141_60015 | Not-determined |
| S8_extracted_k141_146605 | Not-determined |
| S8_extracted_k141_60016 | Low-quality |
| S8_extracted_k141_25119 | Not-determined |
| S8_extracted_k141_76968 | Not-determined |
| S8_extracted_k141_146608 | Not-determined |
| S8_extracted_k141_188850 | Not-determined |
| S8_extracted_k141_111930 | Not-determined |
| S8_extracted_k141_76974 | Not-determined |
| S8_extracted_k141_15928 | Not-determined |
| S8_extracted_k141_188852 | Not-determined |
| S8_extracted_k141_76976 | Not-determined |
| S8_extracted_k141_111932 | Not-determined |
| S8_extracted_k141_111933 | Not-determined |
| S8_extracted_k141_7749 | Not-determined |
| S8_extracted_k141_146616 | Not-determined |
| S8_extracted_k141_85656 | Not-determined |
| S8_extracted_k141_102866 | Not-determined |
| S8_extracted_k141_181430 | Not-determined |
| S8_extracted_k141_102867 | Not-determined |
| S8_extracted_k141_7754 | Low-quality |
| S8_extracted_k141_198927 | Not-determined |
| S8_extracted_k141_85659 | Not-determined |
| S8_extracted_k141_198929 | Not-determined |
| S8_extracted_k141_85660 | Not-determined |
| S8_extracted_k141_33977 | Not-determined |
| S8_extracted_k141_25128 | Not-determined |
| S8_extracted_k141_85661 | Not-determined |
| S8_vs_k141_196247\|\|full | Medium-quality |
| S8_extracted_k141_60028 | Not-determined |
| S8_extracted_k141_137614 | Not-determined |
| S8_extracted_k141_181434 | Not-determined |
| S8_extracted_k141_76977 | Not-determined |
| S8_extracted_k141_137616 | Not-determined |
| S8_extracted_k141_68089 | Not-determined |
| S8_extracted_k141_111941 | Not-determined |
| S8_extracted_k141_94364 | Not-determined |
| S8_extracted_k141_33982 | Not-determined |
| S8_extracted_k141_60036 | Not-determined |
| S8_extracted_k141_68093 | Not-determined |
| S8_extracted_k141_7764 | Not-determined |
| S8_extracted_k141_42301 | Not-determined |
| S8_extracted_k141_146623 | Not-determined |
| S8_extracted_k141_42304 | Not-determined |
| S8_extracted_k141_137623 | Not-determined |
| S8_extracted_k141_76984 | Not-determined |
| S8_extracted_k141_76985 | Not-determined |
| S8_extracted_k141_146625 | Not-determined |
| S8_extracted_k141_198932 | Not-determined |
| S8_extracted_k141_102880 | Not-determined |
| S8_extracted_k141_68096 | Not-determined |
| S8_extracted_k141_129219 | Low-quality |
| S8_extracted_k141_85672 | Not-determined |
| S8_extracted_k141_42308 | Low-quality |
| S8_extracted_k141_15935 | Not-determined |
| S8_extracted_k141_188870 | Not-determined |
| S8_extracted_k141_42311 | Not-determined |
| S8_extracted_k141_94372 | Not-determined |
| S8_extracted_k141_85673 | Not-determined |
| S8_extracted_k141_76991 | Not-determined |
| S8_extracted_k141_33994 | Not-determined |
| S8_extracted_k141_188874 | Not-determined |
| S8_extracted_k141_33995 | Low-quality |
| S8_extracted_k141_129226 | Not-determined |
| S8_extracted_k141_76994 | Not-determined |
| S8_extracted_k141_154826 | Not-determined |
| S8_extracted_k141_154827 | Not-determined |
| S8_extracted_k141_7780 | Not-determined |
| S8_extracted_k141_7781 | Not-determined |
| S8_extracted_k141_85682 | Not-determined |
| S8_extracted_k141_137632 | Low-quality |
| S8_extracted_k141_76999 | Not-determined |
| S8_extracted_k141_181465 | Not-determined |
| S8_extracted_k141_129237 | Not-determined |
| S8_extracted_k141_68114 | Not-determined |
| S8_extracted_k141_198951 | Not-determined |
| S8_extracted_k141_42330 | Not-determined |
| S8_extracted_k141_102890 | Not-determined |
| S8_extracted_k141_34002 | Not-determined |
| S8_extracted_k141_102891 | Not-determined |
| S8_extracted_k141_60048 | Not-determined |
| S8_extracted_k141_77008 | Not-determined |
| S8_extracted_k141_137651 | Not-determined |
| S8_extracted_k141_181470 | Not-determined |
| S8_extracted_k141_146634 | Not-determined |
| S8_extracted_k141_137653 | Not-determined |
| S8_extracted_k141_102894 | Not-determined |
| S8_extracted_k141_146637 | Not-determined |
| S8_extracted_k141_68122 | Low-quality |
| S8_extracted_k141_154830 | Not-determined |
| S8_extracted_k141_85688 | Not-determined |
| S8_extracted_k141_68126 | Low-quality |
| S8_extracted_k141_102896 | Not-determined |
| S8_extracted_k141_15958 | Not-determined |
| S8_extracted_k141_94393 | Not-determined |
| S8_extracted_k141_206798 | Low-quality |
| S8_extracted_k141_146640 | Not-determined |
| S8_extracted_k141_146641 | Not-determined |
| S8_extracted_k141_206802 | Not-determined |
| S8_extracted_k141_7798 | Not-determined |
| S8_extracted_k141_198961 | Low-quality |
| S8_extracted_k141_25157 | Low-quality |
| S8_extracted_k141_77019 | Not-determined |
| S8_extracted_k141_188896 | Not-determined |
| S8_extracted_k141_181477 | Not-determined |
| S8_extracted_k141_77024 | Not-determined |
| S8_extracted_k141_102904 | Not-determined |
| S8_extracted_k141_42333 | Not-determined |
| S8_extracted_k141_102905 | Not-determined |
| S8_extracted_k141_163478 | Not-determined |
| S8_extracted_k141_85696 | Not-determined |
| S8_extracted_k141_34014 | Not-determined |
| S8_extracted_k141_25165 | Not-determined |
| S8_extracted_k141_188900 | Not-determined |
| S8_extracted_k141_181481 | Low-quality |
| S8_extracted_k141_188903 | Not-determined |
| S8_extracted_k141_85701 | Not-determined |
| S8_extracted_k141_102911 | Not-determined |
| S8_extracted_k141_111975 | Not-determined |
| S8_extracted_k141_181483 | Not-determined |
| S8_extracted_k141_85703 | Not-determined |
| S8_extracted_k141_181484 | Not-determined |
| S8_extracted_k141_34021 | Not-determined |
| S8_extracted_k141_85705 | Low-quality |
| S8_extracted_k141_198975 | Not-determined |
| S8_extracted_k141_181491 | Not-determined |
| S8_extracted_k141_198976 | Not-determined |
| S8_extracted_k141_34026 | Not-determined |
| S8_extracted_k141_154837 | Not-determined |
| S8_extracted_k141_111981 | Not-determined |
| S8_extracted_k141_137673 | Not-determined |
| S8_extracted_k141_34029 | Not-determined |
| S8_extracted_k141_94414 | Not-determined |
| S8_extracted_k141_68140 | Low-quality |
| S8_extracted_k141_34031 | Not-determined |
| S8_extracted_k141_129240 | Not-determined |
| S8_extracted_k141_129241 | Low-quality |
| S8_extracted_k141_34032 | Low-quality |
| S8_extracted_k141_206819 | Not-determined |
| S8_extracted_k141_163497 | Not-determined |
| S8_extracted_k141_94424 | Not-determined |
| S8_extracted_k141_102920 | Not-determined |
| S8_extracted_k141_198987 | Low-quality |
| S8_extracted_k141_188919 | Low-quality |
| S8_extracted_k141_42349 | Not-determined |
| S8_extracted_k141_94428 | Not-determined |
| S8_extracted_k141_129244 | Not-determined |
| S8_extracted_k141_60091 | Not-determined |
| S8_extracted_k141_77044 | Not-determined |
| S8_extracted_k141_42350 | Not-determined |
| S8_extracted_k141_198994 | Not-determined |
| S8_extracted_k141_146660 | Not-determined |
| S8_extracted_k141_120452 | Not-determined |
| S8_extracted_k141_60098 | Not-determined |
| S8_extracted_k141_77050 | Not-determined |
| S8_extracted_k141_146664 | Not-determined |
| S8_extracted_k141_94438 | Not-determined |
| S8_extracted_k141_206825 | Not-determined |
| S8_extracted_k141_154852 | Not-determined |
| S8_extracted_k141_206827 | Not-determined |
| S8_extracted_k141_7817 | Low-quality |
| S8_extracted_k141_42357 | Not-determined |
| S8_extracted_k141_25190 | Not-determined |
| S8_extracted_k141_68152 | Not-determined |
| S8_extracted_k141_206833 | Not-determined |
| S8_extracted_k141_42359 | Not-determined |
| S8_extracted_k141_77060 | Not-determined |
| S8_extracted_k141_111995 | Not-determined |
| S8_extracted_k141_25195 | Not-determined |
| S8_extracted_k141_7823 | Not-determined |
| S8_extracted_k141_68157 | Not-determined |
| S8_extracted_k141_60111 | Not-determined |
| S8_extracted_k141_102937 | Not-determined |
| S8_extracted_k141_7828 | Not-determined |
| S8_extracted_k141_25199 | Not-determined |
| S8_extracted_k141_102940 | Not-determined |
| S8_extracted_k141_102944 | Not-determined |
| S8_extracted_k141_25202 | Not-determined |
| S8_extracted_k141_199007 | Not-determined |
| S8_extracted_k141_199009 | Not-determined |
| S8_extracted_k141_188923 | Not-determined |
| S8_extracted_k141_199010 | Not-determined |
| S8_extracted_k141_42375 | Not-determined |
| S8_extracted_k141_7831 | Not-determined |
| S8_extracted_k141_34056 | Not-determined |
| S8_extracted_k141_199014 | Not-determined |
| S8_extracted_k141_7835 | Not-determined |
| S8_extracted_k141_60120 | Not-determined |
| S8_extracted_k141_34057 | Not-determined |
| S8_extracted_k141_146679 | Not-determined |
| S8_extracted_k141_163521 | Low-quality |
| S8_extracted_k141_137706 | Not-determined |
| S8_extracted_k141_112009 | Not-determined |
| S8_extracted_k141_206845 | Low-quality |
| S8_extracted_k141_181506 | Not-determined |
| S8_extracted_k141_137707 | Not-determined |
| S8_extracted_k141_199019 | Not-determined |
| S8_extracted_k141_206849 | Not-determined |
| S8_extracted_k141_206851 | Not-determined |
| S8_extracted_k141_112012 | Low-quality |
| S8_extracted_k141_188936 | Not-determined |
| S8_extracted_k141_112014 | Not-determined |
| S8_extracted_k141_7838 | Low-quality |
| S8_extracted_k141_85737 | Low-quality |
| S8_extracted_k141_60124 | Not-determined |
| S8_extracted_k141_15994 | Not-determined |
| S8_extracted_k141_94448 | Not-determined |
| S8_extracted_k141_15997 | Not-determined |
| S8_extracted_k141_94449 | Not-determined |
| S8_extracted_k141_112017 | Not-determined |
| S8_extracted_k141_206853 | Not-determined |
| S8_extracted_k141_34069 | Not-determined |
| S8_extracted_k141_85744 | Not-determined |
| S8_extracted_k141_7839 | Not-determined |
| S8_extracted_k141_102958 | Not-determined |
| S8_extracted_k141_199027 | Low-quality |
| S8_extracted_k141_85746 | Not-determined |
| S8_extracted_k141_94454 | Not-determined |
| S8_extracted_k141_112022 | Not-determined |
| S8_extracted_k141_102961 | Low-quality |
| S8_extracted_k141_181516 | Not-determined |
| S8_extracted_k141_163533 | Not-determined |
| S8_extracted_k141_85748 | Not-determined |
| S8_extracted_k141_199031 | Not-determined |
| S8_extracted_k141_112025 | Not-determined |
| S8_extracted_k141_199033 | Not-determined |
| S8_extracted_k141_85750 | Not-determined |
| S8_extracted_k141_16004 | Not-determined |
| S8_extracted_k141_60131 | Not-determined |
| S8_extracted_k141_42383 | Low-quality |
| S8_extracted_k141_199035 | Not-determined |
| S8_extracted_k141_154868 | Not-determined |
| S8_extracted_k141_154869 | Not-determined |
| S8_extracted_k141_77088 | Not-determined |
| S8_extracted_k141_163540 | Not-determined |
| S8_extracted_k141_34074 | Not-determined |
| S8_extracted_k141_7855 | Not-determined |
| S8_extracted_k141_137735 | Not-determined |
| S8_extracted_k141_129260 | Not-determined |
| S8_extracted_k141_146697 | Not-determined |
| S8_extracted_k141_34081 | Not-determined |
| S8_extracted_k141_60144 | Not-determined |
| S8_extracted_k141_163543 | Not-determined |
| S8_extracted_k141_137738 | Not-determined |
| S8_extracted_k141_94468 | Not-determined |
| S8_extracted_k141_172837 | Not-determined |
| S8_extracted_k141_146702 | Not-determined |
| S8_extracted_k141_146703 | Not-determined |
| S8_extracted_k141_172838 | Not-determined |
| S8_extracted_k141_206863 | Not-determined |
| S8_extracted_k141_25214 | Not-determined |
| S8_extracted_k141_199043 | Not-determined |
| S8_extracted_k141_206865 | Not-determined |
| S8_extracted_k141_137744 | Not-determined |
| S8_extracted_k141_137745 | Not-determined |
| S8_extracted_k141_7870 | Not-determined |
| S8_extracted_k141_112038 | Not-determined |
| S8_extracted_k141_163547 | Low-quality |
| S8_extracted_k141_94472 | Not-determined |
| S8_extracted_k141_172846 | Not-determined |
| S8_extracted_k141_188947 | Not-determined |
| S8_extracted_k141_199047 | Not-determined |
| S8_extracted_k141_42398 | Not-determined |
| S8_extracted_k141_206874 | Not-determined |
| S8_extracted_k141_172850 | Not-determined |
| S8_extracted_k141_154884 | Not-determined |
| S8_extracted_k141_146709 | Not-determined |
| S8_extracted_k141_85757 | Not-determined |
| S8_extracted_k141_146710 | Not-determined |
| S8_extracted_k141_137752 | Not-determined |
| S8_extracted_k141_163553 | Not-determined |

| | |
|---|---|
| S8_extracted_k141_188951 | Not-determined |
| S8_extracted_k141_60156 | Not-determined |
| S8_extracted_k141_188952 | Not-determined |
| S8_extracted_k141_102993 | Low-quality |
| S8_extracted_k141_112042 | Not-determined |
| S8_extracted_k141_188955 | Not-determined |
| S8_extracted_k141_154893 | Not-determined |
| S8_extracted_k141_154894 | Not-determined |
| S8_extracted_k141_120505 | Not-determined |
| S8_extracted_k141_34090 | Not-determined |
| S8_extracted_k141_181541 | Low-quality |
| S8_extracted_k141_154899 | Not-determined |
| S8_extracted_k141_85760 | Low-quality |
| S8_extracted_k141_60162 | Not-determined |
| S8_extracted_k141_94485 | Not-determined |
| S8_extracted_k141_68169 | Low-quality |
| S8_extracted_k141_154902 | Not-determined |
| S8_extracted_k141_163560 | Not-determined |
| S8_extracted_k141_181542 | Not-determined |
| S8_extracted_k141_199050 | Not-determined |
| S8_extracted_k141_34093 | Not-determined |
| S8_extracted_k141_146723 | Low-quality |
| S8_extracted_k141_7880 | Not-determined |
| S8_extracted_k141_34097 | Not-determined |
| S8_extracted_k141_146727 | Not-determined |
| S8_extracted_k141_146728 | Not-determined |
| S8_extracted_k141_120518 | Not-determined |
| S8_extracted_k141_137768 | Not-determined |
| S8_extracted_k141_85767 | Not-determined |
| S8_extracted_k141_16039 | Not-determined |
| S8_extracted_k141_188968 | Not-determined |
| S8_extracted_k141_85768 | Low-quality |
| S8_extracted_k141_154914 | Not-determined |
| S8_extracted_k141_16041 | Not-determined |
| S8_extracted_k141_137772 | Not-determined |
| S8_extracted_k141_94494 | Not-determined |
| S8_extracted_k141_103002 | Not-determined |
| S8_extracted_k141_188970 | Low-quality |
| S8_extracted_k141_206890 | Not-determined |
| S8_extracted_k141_68178 | Not-determined |
| S8_extracted_k141_199052 | Not-determined |
| S8_extracted_k141_112062 | Low-quality |
| S8_extracted_k141_146734 | Not-determined |
| S8_extracted_k141_25246 | Not-determined |
| S8_extracted_k141_137778 | Not-determined |
| S8_extracted_k141_103008 | Not-determined |
| S8_extracted_k141_120524 | Not-determined |
| S8_extracted_k141_154920 | Not-determined |
| S8_extracted_k141_188974 | Not-determined |
| S8_extracted_k141_85775 | Not-determined |
| S8_extracted_k141_7886 | Not-determined |
| S8_extracted_k141_112067 | Low-quality |
| S8_extracted_k141_181559 | Not-determined |
| S8_extracted_k141_94505 | Not-determined |
| S8_extracted_k141_7888 | Not-determined |
| S8_extracted_k141_137784 | Not-determined |
| S8_extracted_k141_60177 | Not-determined |
| S8_extracted_k141_85777 | Not-determined |
| S8_extracted_k141_137785 | Not-determined |
| S8_extracted_k141_154923 | Not-determined |
| S8_extracted_k141_68182 | Not-determined |
| S8_extracted_k141_188978 | Low-quality |
| S8_extracted_k141_25249 | Not-determined |
| S8_extracted_k141_188979 | Not-determined |
| S8_extracted_k141_199061 | Not-determined |
| S8_extracted_k141_137789 | Not-determined |
| S8_extracted_k141_199062 | Not-determined |
| S8_extracted_k141_77093 | Not-determined |
| S8_extracted_k141_146741 | Low-quality |
| S8_extracted_k141_120527 | Not-determined |
| S8_extracted_k141_16052 | Not-determined |
| S8_extracted_k141_60183 | Not-determined |
| S8_extracted_k141_42401 | Not-determined |
| S8_extracted_k141_42403 | Not-determined |
| S8_extracted_k141_120531 | Low-quality |
| S8_extracted_k141_120533 | Not-determined |
| S8_extracted_k141_112081 | Not-determined |
| S8_extracted_k141_34118 | Low-quality |
| S8_extracted_k141_34121 | Not-determined |
| S8_extracted_k141_85788 | Not-determined |
| S8_extracted_k141_34126 | Not-determined |
| S8_extracted_k141_146747 | Not-determined |
| S8_extracted_k141_94520 | Not-determined |
| S8_extracted_k141_94521 | Not-determined |
| S8_extracted_k141_94523 | Not-determined |
| S8_extracted_k141_199076 | Not-determined |
| S8_extracted_k141_112084 | Not-determined |
| S8_extracted_k141_120540 | Low-quality |
| S8_extracted_k141_137800 | Not-determined |
| S8_extracted_k141_77103 | Not-determined |
| S8_extracted_k141_112086 | Not-determined |
| S8_extracted_k141_120543 | Not-determined |
| S8_extracted_k141_154939 | Not-determined |
| S8_extracted_k141_120545 | Not-determined |
| S8_extracted_k141_206903 | Not-determined |
| S8_extracted_k141_25265 | Low-quality |
| S8_extracted_k141_68193 | Not-determined |
| S8_extracted_k141_34136 | Not-determined |
| S8_extracted_k141_137804 | Low-quality |
| S8_extracted_k141_199079 | Not-determined |
| S8_extracted_k141_25266 | Not-determined |
| S8_extracted_k141_34138 | Not-determined |
| S8_extracted_k141_7908 | Low-quality |
| S8_extracted_k141_146761 | Low-quality |
| S8_extracted_k141_103028 | Not-determined |
| S8_extracted_k141_146762 | Not-determined |
| S8_extracted_k141_85806 | Not-determined |
| S8_extracted_k141_68201 | Not-determined |
| S8_extracted_k141_85810 | Not-determined |
| S8_extracted_k141_77113 | Not-determined |
| S8_extracted_k141_60204 | Low-quality |
| S8_extracted_k141_199089 | Not-determined |
| S8_extracted_k141_25276 | Low-quality |
| S8_extracted_k141_103033 | Not-determined |
| S8_extracted_k141_25278 | Not-determined |
| S8_extracted_k141_60209 | Not-determined |
| S8_extracted_k141_85820 | Not-determined |
| S8_extracted_k141_206914 | Not-determined |
| S8_extracted_k141_25283 | Not-determined |
| S8_extracted_k141_189012 | Not-determined |
| S8_extracted_k141_85823 | Not-determined |
| S8_extracted_k141_112092 | Low-quality |
| S8_extracted_k141_77123 | Not-determined |
| S8_extracted_k141_94546 | Not-determined |
| S8_extracted_k141_103042 | Not-determined |
| S8_extracted_k141_68202 | Not-determined |
| S8_extracted_k141_85826 | Not-determined |
| S8_extracted_k141_172882 | Not-determined |
| S8_extracted_k141_172883 | Not-determined |
| S8_extracted_k141_85827 | Low-quality |
| S8_extracted_k141_68204 | Not-determined |
| S8_extracted_k141_25287 | Low-quality |
| S8_extracted_k141_181589 | Not-determined |
| S8_extracted_k141_85830 | Not-determined |
| S8_extracted_k141_16087 | Not-determined |
| S8_extracted_k141_25291 | Not-determined |
| S8_extracted_k141_103044 | Not-determined |
| S8_extracted_k141_120579 | Low-quality |
| S8_extracted_k141_42424 | Low-quality |
| S8_extracted_k141_103049 | Not-determined |
| S8_extracted_k141_7920 | Not-determined |
| S8_extracted_k141_206922 | Low-quality |
| S8_extracted_k141_16095 | Not-determined |
| S8_extracted_k141_25299 | Not-determined |
| S8_extracted_k141_129295 | Not-determined |
| S8_extracted_k141_77134 | Not-determined |
| S8_extracted_k141_77139 | Low-quality |
| S8_extracted_k141_206930 | Low-quality |
| S8_extracted_k141_16103 | Not-determined |
| S8_extracted_k141_42433 | Not-determined |
| S8_extracted_k141_68223 | Not-determined |
| S8_extracted_k141_120587 | Not-determined |
| S8_extracted_k141_16108 | Not-determined |
| S8_extracted_k141_7931 | Not-determined |
| S8_extracted_k141_154976 | Not-determined |
| S8_extracted_k141_16111 | Not-determined |
| S8_extracted_k141_34171 | Not-determined |
| S8_extracted_k141_103059 | Not-determined |
| S8_extracted_k141_34174 | Not-determined |
| S8_extracted_k141_85852 | Not-determined |
| S8_extracted_k141_77143 | Not-determined |
| S8_extracted_k141_85853 | Not-determined |
| S8_extracted_k141_146791 | Not-determined |
| S8_extracted_k141_181608 | Not-determined |
| S8_extracted_k141_112117 | Not-determined |
| S8_extracted_k141_146793 | Not-determined |
| S8_extracted_k141_16116 | Not-determined |
| S8_extracted_k141_154980 | Not-determined |
| S8_extracted_k141_34180 | Not-determined |
| S8_extracted_k141_77147 | Not-determined |
| S8_extracted_k141_199111 | Not-determined |
| S8_extracted_k141_137831 | Low-quality |
| S8_extracted_k141_146797 | Not-determined |
| S8_extracted_k141_112123 | Not-determined |
| S8_extracted_k141_7941 | Not-determined |
| S8_extracted_k141_163569 | Not-determined |
| S8_extracted_k141_16122 | Not-determined |
| S8_extracted_k141_189029 | Not-determined |
| S8_extracted_k141_77150 | Not-determined |
| S8_extracted_k141_181615 | Not-determined |
| S8_extracted_k141_154983 | Not-determined |
| S8_extracted_k141_163571 | Not-determined |
| S8_extracted_k141_7945 | Not-determined |
| S8_extracted_k141_163573 | Not-determined |
| S8_extracted_k141_163574 | Not-determined |
| S8_extracted_k141_181621 | Not-determined |
| S8_extracted_k141_199119 | Not-determined |
| S8_extracted_k141_189032 | Not-determined |
| S8_extracted_k141_199122 | Not-determined |
| S8_extracted_k141_34190 | Not-determined |
| S8_extracted_k141_103079 | Not-determined |
| S8_extracted_k141_94585 | Not-determined |
| S8_extracted_k141_25325 | Not-determined |
| S8_extracted_k141_129317 | Not-determined |
| S8_extracted_k141_137834 | Not-determined |
| S8_extracted_k141_34193 | Not-determined |
| S8_extracted_k141_103083 | Not-determined |
| S8_extracted_k141_42446 | Not-determined |
| S8_extracted_k141_154985 | Not-determined |
| S8_extracted_k141_163581 | Not-determined |
| S8_extracted_k141_154987 | Not-determined |
| S8_extracted_k141_94590 | Not-determined |
| S8_extracted_k141_120604 | Not-determined |
| S8_extracted_k141_199134 | Low-quality |
| S8_extracted_k141_181629 | Not-determined |
| S8_extracted_k141_199137 | Not-determined |
| S8_extracted_k141_112137 | Not-determined |
| S8_extracted_k141_77159 | Not-determined |
| S8_extracted_k141_68245 | Not-determined |
| S8_extracted_k141_103084 | Low-quality |
| S8_extracted_k141_34208 | Not-determined |
| S8_extracted_k141_42463 | Not-determined |
| S8_extracted_k141_25339 | Not-determined |
| S8_extracted_k141_199141 | Not-determined |
| S8_extracted_k141_60265 | Not-determined |
| S8_extracted_k141_199142 | Not-determined |
| S8_extracted_k141_112141 | Not-determined |
| S8_extracted_k141_25340 | Not-determined |
| S8_extracted_k141_25341 | Not-determined |
| S8_extracted_k141_199145 | Not-determined |
| S8_extracted_k141_77165 | Not-determined |
| S8_extracted_k141_42467 | Not-determined |
| S8_extracted_k141_120616 | Not-determined |
| S8_extracted_k141_77166 | Not-determined |
| S8_extracted_k141_60272 | Low-quality |
| S8_extracted_k141_206949 | Not-determined |
| S8_extracted_k141_172898 | Not-determined |
| S8_extracted_k141_94597 | Low-quality |
| S8_extracted_k141_146806 | Not-determined |
| S8_extracted_k141_103093 | Not-determined |
| S8_extracted_k141_199150 | Low-quality |
| S8_extracted_k141_103094 | Low-quality |
| S8_extracted_k141_129331 | Not-determined |
| S8_extracted_k141_146809 | Not-determined |
| S8_extracted_k141_181646 | Low-quality |
| S8_extracted_k141_120625 | Not-determined |
| S8_extracted_k141_94602 | Not-determined |
| S8_extracted_k141_206955 | Not-determined |
| S8_extracted_k141_120626 | Not-determined |
| S8_extracted_k141_154996 | Low-quality |
| S8_extracted_k141_25357 | Not-determined |
| S8_extracted_k141_16157 | Not-determined |
| S8_extracted_k141_60273 | Not-determined |
| S8_extracted_k141_189054 | Not-determined |
| S8_extracted_k141_25359 | Not-determined |
| S8_extracted_k141_154999 | Not-determined |
| S8_extracted_k141_16160 | Not-determined |
| S8_extracted_k141_68252 | Not-determined |
| S8_extracted_k141_42473 | Not-determined |
| S8_extracted_k141_172910 | Not-determined |
| S8_extracted_k141_129337 | Not-determined |
| S8_extracted_k141_85889 | Not-determined |
| S8_extracted_k141_206958 | Not-determined |
| S8_extracted_k141_42478 | Not-determined |
| S8_extracted_k141_112159 | Low-quality |
| S8_extracted_k141_199159 | Not-determined |
| S8_extracted_k141_181658 | Not-determined |
| S8_extracted_k141_181659 | Not-determined |
| S8_extracted_k141_129341 | Not-determined |
| S8_extracted_k141_163610 | Not-determined |
| S8_extracted_k141_77176 | Not-determined |
| S8_extracted_k141_77177 | Not-determined |
| S8_extracted_k141_16163 | Not-determined |
| S8_extracted_k141_112163 | Not-determined |
| S8_extracted_k141_25367 | Not-determined |
| S8_extracted_k141_181661 | Not-determined |
| S8_extracted_k141_42480 | Not-determined |
| S8_extracted_k141_60279 | Not-determined |
| S8_extracted_k141_163615 | Low-quality |
| S8_extracted_k141_181663 | Not-determined |
| S8_extracted_k141_85894 | Not-determined |
| S8_extracted_k141_25369 | Not-determined |
| S8_extracted_k141_172914 | Not-determined |
| S8_extracted_k141_172916 | Not-determined |
| S8_extracted_k141_199160 | Not-determined |
| SS_vs_k141_55687\|\|full | Medium-quality |
| S8_extracted_k141_163617 | Not-determined |
| S8_extracted_k141_181666 | Not-determined |
| S8_extracted_k141_103111 | Not-determined |
| S8_extracted_k141_77184 | Not-determined |
| S8_extracted_k141_112164 | Low-quality |
| S8_extracted_k141_60285 | Not-determined |
| S8_extracted_k141_16168 | Not-determined |
| S8_extracted_k141_42487 | Not-determined |
| S8_extracted_k141_68269 | Not-determined |
| S8_extracted_k141_181674 | Not-determined |
| S8_extracted_k141_103117 | Not-determined |
| S8_extracted_k141_103118 | Low-quality |
| S8_extracted_k141_137843 | Not-determined |
| S8_extracted_k141_60297 | Low-quality |
| S8_extracted_k141_85910 | Not-determined |
| S8_extracted_k141_77193 | Not-determined |
| S8_extracted_k141_25381 | Not-determined |
| S8_extracted_k141_77194 | Not-determined |
| S8_extracted_k141_34228 | Not-determined |
| S8_extracted_k141_112169 | Not-determined |
| S8_extracted_k141_77197 | Not-determined |
| S8_extracted_k141_34230 | Not-determined |
| S8_extracted_k141_137847 | Not-determined |
| S8_extracted_k141_103123 | Not-determined |
| S8_extracted_k141_137853 | Not-determined |
| S8_extracted_k141_112175 | Not-determined |
| S8_extracted_k141_112176 | Not-determined |
| S8_extracted_k141_189078 | Not-determined |
| S8_extracted_k141_146834 | Not-determined |
| S8_extracted_k141_137856 | Not-determined |
| S8_extracted_k141_77203 | Not-determined |
| S8_extracted_k141_129360 | Not-determined |
| S8_extracted_k141_137858 | Not-determined |
| S8_extracted_k141_85918 | Not-determined |
| S8_extracted_k141_199169 | Not-determined |
| S8_extracted_k141_129366 | Not-determined |
| S8_extracted_k141_112184 | Not-determined |
| S8_extracted_k141_85921 | Not-determined |
| S8_extracted_k141_103133 | Not-determined |
| S8_extracted_k141_77207 | Not-determined |
| S8_extracted_k141_172921 | Low-quality |
| S8_extracted_k141_129369 | Not-determined |
| S8_extracted_k141_181690 | Low-quality |
| S8_extracted_k141_34245 | Not-determined |
| S8_extracted_k141_34246 | Low-quality |
| S8_extracted_k141_199176 | Not-determined |
| S8_extracted_k141_129371 | Not-determined |
| S8_extracted_k141_129372 | Not-determined |
| S8_extracted_k141_68282 | Not-determined |
| S8_extracted_k141_68284 | Not-determined |
| S8_extracted_k141_68286 | Not-determined |
| S8_extracted_k141_85923 | Not-determined |
| S8_extracted_k141_129376 | Not-determined |
| S8_extracted_k141_146846 | Not-determined |
| S8_extracted_k141_85926 | Not-determined |
| S8_extracted_k141_206971 | Not-determined |
| S8_extracted_k141_77215 | Low-quality |
| S8_extracted_k141_137868 | Not-determined |
| S8_extracted_k141_85931 | Not-determined |
| S8_extracted_k141_181696 | Low-quality |
| S8_extracted_k141_25404 | Not-determined |
| S8_extracted_k141_199189 | Not-determined |
| S8_extracted_k141_68298 | Not-determined |
| S8_extracted_k141_137871 | Not-determined |
| S8_extracted_k141_163663 | Not-determined |
| S8_extracted_k141_199192 | Not-determined |
| S8_extracted_k141_25409 | Not-determined |
| S8_extracted_k141_129387 | Low-quality |
| S8_extracted_k141_199194 | Not-determined |
| S8_extracted_k141_146860 | Not-determined |
| S8_extracted_k141_181699 | Not-determined |
| S8_extracted_k141_60328 | Not-determined |
| S8_extracted_k141_189091 | Not-determined |
| S8_extracted_k141_68304 | Not-determined |
| S8_extracted_k141_60331 | Low-quality |
| S8_extracted_k141_146866 | Not-determined |
| S8_extracted_k141_146867 | Not-determined |
| S8_extracted_k141_77226 | Not-determined |
| S8_extracted_k141_189101 | Not-determined |
| S8_extracted_k141_25416 | Not-determined |
| S8_extracted_k141_189102 | Not-determined |
| S8_extracted_k141_103142 | Not-determined |
| S8_extracted_k141_120629 | Not-determined |
| S8_extracted_k141_7980 | Not-determined |
| S8_extracted_k141_181706 | Not-determined |
| S8_extracted_k141_172940 | Not-determined |
| S8_extracted_k141_68311 | Not-determined |
| S8_extracted_k141_7982 | Not-determined |
| S8_extracted_k141_68312 | Not-determined |
| S8_extracted_k141_68313 | Not-determined |
| S8_extracted_k141_85952 | Not-determined |
| S8_extracted_k141_7985 | Low-quality |
| S8_extracted_k141_172943 | Not-determined |
| S8_extracted_k141_68317 | Not-determined |
| S8_extracted_k141_172947 | Not-determined |
| S8_extracted_k141_146879 | Not-determined |
| S8_extracted_k141_68318 | Not-determined |
| S8_extracted_k141_120640 | Not-determined |
| S8_extracted_k141_77240 | Not-determined |
| S8_extracted_k141_181713 | Low-quality |
| S8_extracted_k141_112198 | Not-determined |
| S8_extracted_k141_85958 | Not-determined |
| S8_extracted_k141_85959 | Not-determined |
| S8_extracted_k141_181714 | Not-determined |
| S8_extracted_k141_77245 | Not-determined |
| S8_extracted_k141_85963 | Not-determined |
| S3_extracted_k141_87896 | Medium-quality |
| S8_extracted_k141_68319 | Low-quality |
| S8_extracted_k141_137900 | Not-determined |
| S8_extracted_k141_137901 | Not-determined |
| S8_extracted_k141_16176 | Low-quality |
| S8_extracted_k141_189106 | Not-determined |
| S8_extracted_k141_16177 | Not-determined |
| S8_extracted_k141_68324 | Low-quality |
| S8_extracted_k141_181716 | Not-determined |
| S8_extracted_k141_25435 | Not-determined |
| S8_extracted_k141_163673 | Not-determined |
| S8_extracted_k141_112205 | Not-determined |
| S8_extracted_k141_206996 | Not-determined |
| S8_extracted_k141_16186 | Low-quality |
| S8_extracted_k141_8004 | Low-quality |
| S8_extracted_k141_163679 | Not-determined |
| S8_extracted_k141_146894 | Low-quality |
| S8_extracted_k141_129412 | Not-determined |
| S8_extracted_k141_112208 | Not-determined |
| S8_extracted_k141_60350 | Not-determined |
| S8_extracted_k141_172956 | Not-determined |
| S8_extracted_k141_68337 | Not-determined |
| S8_extracted_k141_172960 | Not-determined |
| S8_extracted_k141_129416 | Not-determined |
| S8_extracted_k141_181734 | Not-determined |
| S8_extracted_k141_16197 | Not-determined |
| S8_extracted_k141_8013 | Not-determined |
| S8_extracted_k141_181736 | Not-determined |
| S8_extracted_k141_181738 | Low-quality |
| S8_extracted_k141_25450 | Not-determined |
| S8_extracted_k141_112216 | Not-determined |
| S8_extracted_k141_16204 | Not-determined |
| S8_extracted_k141_42498 | Not-determined |
| S8_extracted_k141_8018 | Not-determined |
| S8_extracted_k141_68343 | Low-quality |
| S8_extracted_k141_68344 | Not-determined |
| S8_extracted_k141_189127 | Low-quality |
| S8_extracted_k141_120649 | Not-determined |
| S8_extracted_k141_85982 | Not-determined |
| S8_extracted_k141_189130 | Not-determined |
| S8_extracted_k141_163690 | Not-determined |
| S8_extracted_k141_77268 | Not-determined |
| S8_extracted_k141_163691 | Not-determined |
| S8_extracted_k141_112226 | Not-determined |
| S8_extracted_k141_181744 | Not-determined |
| S8_extracted_k141_207016 | Not-determined |
| S8_extracted_k141_77269 | Not-determined |
| S8_extracted_k141_163693 | Not-determined |
| S8_extracted_k141_189135 | Not-determined |
| S8_extracted_k141_172970 | Low-quality |
| S8_extracted_k141_112229 | Not-determined |
| S8_extracted_k141_42508 | Not-determined |
| S8_extracted_k141_207020 | Not-determined |
| S8_extracted_k141_77276 | Low-quality |
| S8_extracted_k141_137919 | Not-determined |
| S8_extracted_k141_172973 | Not-determined |
| S8_extracted_k141_163697 | Not-determined |
| S8_extracted_k141_207021 | Low-quality |
| S8_extracted_k141_42510 | Not-determined |
| S8_extracted_k141_112231 | Not-determined |
| S8_extracted_k141_207023 | Not-determined |
| S8_extracted_k141_120656 | Not-determined |
| S8_extracted_k141_77281 | Not-determined |
| S8_extracted_k141_137924 | Not-determined |
| S8_extracted_k141_155024 | Not-determined |
| S8_extracted_k141_42513 | Not-determined |
| S8_extracted_k141_112232 | Not-determined |
| S8_extracted_k141_199232 | Not-determined |
| S8_extracted_k141_120657 | Low-quality |
| S8_extracted_k141_8029 | Low-quality |
| S8_extracted_k141_25465 | Not-determined |
| S8_extracted_k141_112234 | Not-determined |
| S8_extracted_k141_199233 | Not-determined |
| S8_extracted_k141_137929 | Not-determined |
| S8_extracted_k141_42515 | Not-determined |
| S8_extracted_k141_146909 | Not-determined |
| S8_extracted_k141_42517 | Not-determined |
| S8_extracted_k141_112236 | Not-determined |
| S8_extracted_k141_85999 | Not-determined |
| S8_extracted_k141_172980 | Not-determined |
| S8_extracted_k141_146912 | Not-determined |
| S8_extracted_k141_137933 | Not-determined |
| S8_extracted_k141_189147 | Not-determined |
| S8_extracted_k141_189148 | Not-determined |
| S8_extracted_k141_163710 | Not-determined |
| S8_extracted_k141_181760 | Not-determined |
| S8_extracted_k141_120666 | Low-quality |
| S8_extracted_k141_146915 | Low-quality |
| S8_extracted_k141_120668 | Not-determined |
| S8_extracted_k141_172984 | Not-determined |
| S8_extracted_k141_86004 | Not-determined |
| S8_extracted_k141_137938 | Not-determined |
| S8_extracted_k141_181763 | Not-determined |
| S8_extracted_k141_146918 | Not-determined |
| S8_extracted_k141_120671 | Not-determined |
| S8_extracted_k141_86007 | Not-determined |
| S8_extracted_k141_189156 | Not-determined |
| S8_extracted_k141_25481 | Not-determined |
| S8_extracted_k141_77299 | Not-determined |
| S8_extracted_k141_60382 | Not-determined |
| S8_extracted_k141_163716 | Not-determined |
| S8_extracted_k141_146921 | Not-determined |
| S8_extracted_k141_50502 | Not-determined |
| S8_extracted_k141_189157 | Not-determined |
| S8_extracted_k141_163721 | Not-determined |
| S8_extracted_k141_86008 | Low-quality |
| S8_extracted_k141_77302 | Low-quality |
| S8_extracted_k141_16227 | Not-determined |
| S8_extracted_k141_189161 | Not-determined |
| S8_extracted_k141_50509 | Not-determined |
| S8_extracted_k141_173001 | Not-determined |
| S8_extracted_k141_146931 | Not-determined |
| S8_extracted_k141_155054 | Not-determined |
| S8_extracted_k141_68365 | Not-determined |
| S8_extracted_k141_120683 | Not-determined |
| S8_extracted_k141_120684 | Not-determined |
| S8_extracted_k141_60393 | Not-determined |
| S8_extracted_k141_189162 | Not-determined |
| S8_extracted_k141_137956 | Not-determined |
| S8_extracted_k141_199248 | Not-determined |
| S8_extracted_k141_155062 | Not-determined |
| S8_extracted_k141_137960 | Not-determined |
| S8_extracted_k141_199253 | Low-quality |
| S8_extracted_k141_181777 | Not-determined |
| S8_extracted_k141_8053 | Not-determined |
| S8_extracted_k141_77314 | Not-determined |
| S8_extracted_k141_181781 | Not-determined |
| S8_extracted_k141_86023 | Not-determined |
| S8_extracted_k141_42530 | Not-determined |
| S8_extracted_k141_199255 | Not-determined |
| S8_extracted_k141_42531 | Not-determined |
| S8_extracted_k141_181783 | Not-determined |
| S8_extracted_k141_120697 | Not-determined |
| S8_extracted_k141_8060 | Not-determined |
| S8_extracted_k141_181786 | Not-determined |
| S8_extracted_k141_42536 | Not-determined |
| S8_extracted_k141_112274 | Not-determined |
| S8_extracted_k141_86027 | Not-determined |
| S8_extracted_k141_199262 | Not-determined |
| S8_extracted_k141_42540 | Not-determined |
| S8_extracted_k141_155079 | Low-quality |
| S8_extracted_k141_42541 | Not-determined |
| S8_extracted_k141_181790 | Not-determined |
| S8_extracted_k141_50512 | Not-determined |
| S8_extracted_k141_155083 | Not-determined |
| S8_extracted_k141_50513 | Not-determined |
| S8_extracted_k141_86032 | Not-determined |
| S8_extracted_k141_155085 | Not-determined |
| S8_extracted_k141_16237 | Low-quality |
| S8_extracted_k141_155086 | Not-determined |
| S8_extracted_k141_146950 | Not-determined |
| S8_extracted_k141_173017 | Not-determined |
| S8_extracted_k141_120705 | Not-determined |
| S8_extracted_k141_16239 | Low-quality |
| S7_extracted_135778 | Complete |
| S8_extracted_k141_120707 | Not-determined |
| S8_extracted_k141_68389 | Not-determined |
| S8_extracted_k141_68391 | Not-determined |
| S8_extracted_k141_25494 | Not-determined |
| S8_extracted_k141_189188 | Low-quality |
| S8_extracted_k141_129426 | Not-determined |
| S8_extracted_k141_68393 | Not-determined |
| S8_extracted_k141_163745 | Not-determined |
| S8_extracted_k141_129428 | Not-determined |
| S8_extracted_k141_8066 | Not-determined |
| S8_extracted_k141_146955 | Low-quality |
| S8_extracted_k141_146958 | Not-determined |
| S8_extracted_k141_77338 | Not-determined |
| S8_extracted_k141_77339 | Not-determined |
| S8_extracted_k141_77340 | Not-determined |
| S8_extracted_k141_60406 | Not-determined |
| S8_extracted_k141_8075 | Not-determined |
| S8_extracted_k141_60414 | Low-quality |
| S8_extracted_k141_8078 | Not-determined |
| S8_extracted_k141_163748 | Low-quality |
| S8_extracted_k141_8079 | Not-determined |
| S8_extracted_k141_8080 | Not-determined |
| S8_extracted_k141_77347 | Not-determined |
| S8_extracted_k141_16243 | Not-determined |
| S8_extracted_k141_42565 | Not-determined |
| S8_extracted_k141_68395 | Not-determined |
| S8_extracted_k141_16245 | Low-quality |
| S8_extracted_k141_129434 | Not-determined |
| S8_extracted_k141_155100 | Low-quality |
| S8_extracted_k141_60417 | Low-quality |
| S8_extracted_k141_155101 | Not-determined |
| S8_extracted_k141_60419 | Not-determined |
| S8_extracted_k141_16250 | Not-determined |
| S8_extracted_k141_189196 | Not-determined |
| S8_extracted_k141_163751 | Low-quality |
| S8_extracted_k141_42569 | Not-determined |
| S8_extracted_k141_163753 | Not-determined |
| S8_extracted_k141_181806 | Low-quality |
| S8_extracted_k141_16254 | Not-determined |
| S8_extracted_k141_207051 | Not-determined |
| S8_extracted_k141_129442 | Not-determined |
| S8_extracted_k141_173031 | Not-determined |
| S8_extracted_k141_112286 | Not-determined |
| S8_extracted_k141_16255 | Not-determined |
| S8_extracted_k141_129444 | Not-determined |
| S8_extracted_k141_129445 | Not-determined |
| S8_extracted_k141_16256 | Low-quality |
| S8_extracted_k141_16257 | Not-determined |
| S8_extracted_k141_77353 | Not-determined |
| S8_extracted_k141_77354 | Not-determined |
| S8_extracted_k141_112290 | Not-determined |
| S8_extracted_k141_207057 | Low-quality |

| | |
|---|---|
| SB_extracted_k141_120723 | Not-determined |
| SB_extracted_k141_189199 | Not-determined |
| SB_extracted_k141_146975 | Not-determined |
| SB_extracted_k141_8092 | Not-determined |
| SB_extracted_k141_137979 | Not-determined |
| SB_extracted_k141_50530 | Not-determined |
| SB_extracted_k141_189202 | Not-determined |
| SB_extracted_k141_137981 | Not-determined |
| SB_extracted_k141_112291 | Not-determined |
| SB_extracted_k141_137982 | Not-determined |
| SB_extracted_k141_77359 | Not-determined |
| SB_extracted_k141_207061 | Low-quality |
| SB_extracted_k141_16265 | Not-determined |
| SB_extracted_k141_60430 | Low-quality |
| SB_extracted_k141_8098 | Not-determined |
| SB_extracted_k141_25515 | Not-determined |
| SB_extracted_k141_103151 | Not-determined |
| SB_extracted_k141_112299 | Not-determined |
| SB_extracted_k141_16270 | Not-determined |
| SB_extracted_k141_16272 | Not-determined |
| SB_extracted_k141_112303 | Not-determined |
| SB_extracted_k141_60435 | Not-determined |
| SB_extracted_k141_207066 | Not-determined |
| SB_extracted_k141_25523 | Not-determined |
| SB_extracted_k141_94651 | Not-determined |
| SB_extracted_k141_77369 | Not-determined |
| SB_extracted_k141_50542 | Not-determined |
| SB_extracted_k141_42590 | Not-determined |
| SB_extracted_k141_112314 | Not-determined |
| SB_extracted_k141_77374 | Not-determined |
| SB_extracted_k141_173050 | Not-determined |
| SB_extracted_k141_181828 | Not-determined |
| SB_extracted_k141_112318 | Not-determined |
| SB_extracted_k141_112321 | Low-quality |
| SB_extracted_k141_42598 | Not-determined |
| SB_extracted_k141_112322 | Not-determined |
| SB_extracted_k141_25527 | Not-determined |
| SB_extracted_k141_68403 | Not-determined |
| SB_extracted_k141_50550 | Not-determined |
| SB_extracted_k141_94661 | Not-determined |
| SB_extracted_k141_120731 | Not-determined |
| SB_extracted_k141_94662 | Not-determined |
| SB_extracted_k141_155123 | Not-determined |
| SB_extracted_k141_34286 | Low-quality |
| SB_extracted_k141_103161 | Not-determined |
| SB_extracted_k141_103162 | Not-determined |
| SB_extracted_k141_120733 | Not-determined |
| SB_extracted_k141_8107 | Not-determined |
| SB_extracted_k141_146995 | Not-determined |
| SB_extracted_k141_181834 | Low-quality |
| SB_extracted_k141_189208 | Low-quality |
| SB_extracted_k141_86050 | Low-quality |
| SB_extracted_k141_16282 | Low-quality |
| SB_extracted_k141_60454 | Not-determined |
| SB_extracted_k141_94668 | Not-determined |
| SB_extracted_k141_77386 | Low-quality |
| SB_extracted_k141_120740 | Not-determined |
| SB_extracted_k141_68408 | Not-determined |
| SB_extracted_k141_112330 | Not-determined |
| SB_extracted_k141_138003 | Not-determined |
| SB_extracted_k141_8111 | Not-determined |
| SB_extracted_k141_155132 | Not-determined |
| SB_extracted_k141_112333 | Not-determined |
| SB_extracted_k141_60457 | Not-determined |
| SB_extracted_k141_42607 | Not-determined |
| SB_extracted_k141_68412 | Not-determined |
| SB_extracted_k141_155134 | Not-determined |
| SB_extracted_k141_181837 | Not-determined |
| SB_extracted_k141_68418 | Not-determined |
| SB_extracted_k141_8117 | Low-quality |
| SB_extracted_k141_16296 | Not-determined |
| SB_extracted_k141_25535 | Not-determined |
| SB_extracted_k141_25536 | Not-determined |
| SB_extracted_k141_34294 | Not-determined |
| SB_extracted_k141_42608 | Not-determined |
| SB_extracted_k141_112345 | Not-determined |
| SB_extracted_k141_181843 | Not-determined |
| SB_extracted_k141_42612 | Not-determined |
| SB_extracted_k141_138013 | Low-quality |
| SB_extracted_k141_50558 | Not-determined |
| SB_extracted_k141_60465 | Not-determined |
| SB_extracted_k141_207095 | Low-quality |
| SB_extracted_k141_189224 | Low-quality |
| SB_extracted_k141_103176 | Not-determined |
| SB_extracted_k141_147006 | Not-determined |
| SB_extracted_k141_60470 | Not-determined |
| SB_extracted_k141_112354 | Not-determined |
| SB_extracted_k141_42619 | Not-determined |
| SB_extracted_k141_77399 | Not-determined |
| SB_extracted_k141_42621 | Not-determined |
| SB_extracted_k141_207103 | Not-determined |
| SB_extracted_k141_199277 | Not-determined |
| SB_extracted_k141_138026 | Not-determined |
| SB_extracted_k141_147008 | Not-determined |
| SB_extracted_k141_68432 | Not-determined |
| SB_extracted_k141_155146 | Not-determined |
| SB_extracted_k141_199278 | Not-determined |
| SB_extracted_k141_138029 | Not-determined |
| SB_extracted_k141_94686 | Not-determined |
| SB_extracted_k141_112362 | Not-determined |
| SB_extracted_k141_50564 | Not-determined |
| SB_extracted_k141_147011 | Not-determined |
| SB_extracted_k141_77403 | Not-determined |
| SB_extracted_k141_207108 | Low-quality |
| SB_extracted_k141_50568 | Not-determined |
| SB_extracted_k141_181852 | Not-determined |
| SB_extracted_k141_199286 | Not-determined |
| SB_extracted_k141_199288 | Not-determined |
| SB_extracted_k141_199290 | Not-determined |
| SB_extracted_k141_199291 | Not-determined |
| SB_extracted_k141_155156 | Not-determined |
| SB_extracted_k141_189244 | Not-determined |
| SB_extracted_k141_16322 | Not-determined |
| SB_extracted_k141_42631 | Not-determined |
| SB_extracted_k141_86067 | Not-determined |
| SB_extracted_k141_103193 | Low-quality |
| SB_extracted_k141_120750 | Not-determined |
| SB_extracted_k141_120753 | Not-determined |
| [illegible] | [illegible] |
| SB_extracted_k141_138078 | Not-determined |
| SB_extracted_k141_16382 | Not-determined |
| SB_extracted_k141_25570 | Not-determined |
| SB_extracted_k141_25571 | Not-determined |
| SB_extracted_k141_112407 | Not-determined |
| SB_extracted_k141_207149 | Not-determined |
| SB_extracted_k141_8171 | Not-determined |
| SB_extracted_k141_50631 | Not-determined |
| SB_extracted_k141_189309 | Not-determined |
| SB_extracted_k141_138082 | Not-determined |
| SB_extracted_k141_129522 | Not-determined |
| SB_extracted_k141_112411 | Not-determined |
| SB_extracted_k141_16385 | Not-determined |
| SB_extracted_k141_129524 | Not-determined |
| SB_extracted_k141_16388 | Low-quality |
| SB_extracted_k141_34352 | Not-determined |
| SB_extracted_k141_50637 | Not-determined |
| SB_extracted_k141_207153 | Not-determined |
| SB_extracted_k141_207154 | Not-determined |
| SB_extracted_k141_189313 | Not-determined |
| SB_extracted_k141_50638 | Low-quality |
| SB_extracted_k141_112416 | Not-determined |
| SB_extracted_k141_189315 | Not-determined |
| SB_extracted_k141_207157 | Not-determined |
| SB_extracted_k141_50640 | Not-determined |
| SB_extracted_k141_25576 | Not-determined |
| SB_extracted_k141_86097 | Not-determined |
| SB_extracted_k141_25580 | Not-determined |
| SB_extracted_k141_173142 | Not-determined |
| SB_extracted_k141_163776 | Not-determined |
| SB_extracted_k141_86100 | Not-determined |
| SB_extracted_k141_77468 | Not-determined |
| SB_extracted_k141_199340 | Low-quality |
| SB_extracted_k141_163777 | Low-quality |
| SB_extracted_k141_60522 | Not-determined |
| SB_extracted_k141_16394 | Not-determined |
| SB_extracted_k141_86104 | Not-determined |
| SB_extracted_k141_86105 | Not-determined |
| SB_extracted_k141_155233 | Not-determined |
| SB_extracted_k141_199341 | Not-determined |
| SB_extracted_k141_16398 | Not-determined |
| SB_extracted_k141_181904 | Not-determined |
| SB_extracted_k141_199343 | Not-determined |
| SB_extracted_k141_138101 | Not-determined |
| SB_extracted_k141_173148 | Not-determined |
| SB_extracted_k141_94767 | Not-determined |
| SB_extracted_k141_129536 | Low-quality |
| SB_extracted_k141_16403 | Not-determined |
| SB_extracted_k141_207172 | Not-determined |
| SB_extracted_k141_60530 | Not-determined |
| SB_extracted_k141_207174 | Not-determined |
| SB_extracted_k141_147066 | Not-determined |
| SB_extracted_k141_25586 | Not-determined |
| SB_extracted_k141_173151 | Not-determined |
| SB_extracted_k141_199353 | Not-determined |
| SB_extracted_k141_42689 | Not-determined |
| SB_extracted_k141_60532 | Not-determined |
| SB_extracted_k141_50652 | Not-determined |
| SB_extracted_k141_199356 | Not-determined |
| SB_extracted_k141_25590 | Not-determined |
| SB_extracted_k141_163781 | Not-determined |
| SB_extracted_k141_60534 | Not-determined |
| SB_extracted_k141_77480 | Not-determined |
| SB_extracted_k141_77481 | Not-determined |
| SB_extracted_k141_207175 | Not-determined |
| SB_extracted_k141_25594 | Not-determined |
| SB_extracted_k141_189328 | Low-quality |
| SB_extracted_k141_155244 | Not-determined |
| SB_extracted_k141_77484 | Not-determined |
| SB_extracted_k141_25598 | Not-determined |
| SB_extracted_k141_189331 | Low-quality |
| SB_extracted_k141_103260 | Not-determined |
| SB_extracted_k141_77487 | Not-determined |
| SB_extracted_k141_34367 | Not-determined |
| SB_extracted_k141_8190 | Not-determined |
| SB_extracted_k141_94776 | Not-determined |
| SB_extracted_k141_60542 | Not-determined |
| SB_extracted_k141_155248 | Not-determined |
| SB_extracted_k141_34368 | Not-determined |
| SB_extracted_k141_155250 | Not-determined |
| SB_extracted_k141_77491 | Not-determined |
| SB_extracted_k141_25603 | Not-determined |
| SB_extracted_k141_77492 | Not-determined |
| SB_extracted_k141_147085 | Not-determined |
| SB_extracted_k141_181914 | Not-determined |
| SB_extracted_k141_163792 | Not-determined |
| SB_extracted_k141_42705 | Not-determined |
| SB_extracted_k141_68529 | Not-determined |
| SB_extracted_k141_207183 | Not-determined |
| SB_extracted_k141_25611 | Not-determined |
| SB_extracted_k141_34376 | Not-determined |
| SB_extracted_k141_147092 | Not-determined |
| SB_extracted_k141_68537 | Not-determined |
| SB_extracted_k141_16413 | Not-determined |
| SB_extracted_k141_94782 | Not-determined |
| SB_extracted_k141_103268 | Not-determined |
| SB_extracted_k141_60554 | Not-determined |
| SB_extracted_k141_50665 | Low-quality |
| SB_extracted_k141_207191 | Not-determined |
| SB_extracted_k141_50666 | Low-quality |
| SB_extracted_k141_60557 | Not-determined |
| SB_extracted_k141_16419 | Not-determined |
| SB_extracted_k141_86115 | Not-determined |
| SB_extracted_k141_181923 | Low-quality |
| SB_extracted_k141_138120 | Not-determined |
| SB_extracted_k141_68538 | Not-determined |
| SB_extracted_k141_163798 | Not-determined |
| SB_extracted_k141_34382 | Not-determined |
| SB_extracted_k141_155258 | Not-determined |
| SB_extracted_k141_155259 | Not-determined |
| SB_extracted_k141_50669 | Not-determined |
| SB_extracted_k141_147102 | Not-determined |
| SB_extracted_k141_50670 | Not-determined |
| SB_extracted_k141_138122 | Not-determined |
| SB_extracted_k141_207193 | Not-determined |
| SB_extracted_k141_50671 | Not-determined |
| SB_extracted_k141_189349 | Not-determined |
| SB_extracted_k141_68544 | Not-determined |
| SB_extracted_k141_207199 | Not-determined |
| SB_extracted_k141_112428 | Not-determined |
| SB_extracted_k141_129568 | Not-determined |
| SB_extracted_k141_103281 | Not-determined |
| SB_extracted_k141_189356 | Not-determined |
| [illegible] | [illegible] |

| | |
|---|---|
| S8_extracted_k141_16515 | Not-determined |
| S8_extracted_k141_182006 | Not-determined |
| S8_extracted_k141_155348 | Not-determined |
| S8_extracted_k141_182007 | Not-determined |
| S8_extracted_k141_103358 | Low-quality |
| S8_extracted_k141_94815 | Not-determined |
| S8_extracted_k141_103361 | Low-quality |
| S8_extracted_k141_25706 | Low-quality |
| S8_extracted_k141_77629 | Not-determined |
| S8_extracted_k141_207269 | Not-determined |
| S8_extracted_k141_182012 | Not-determined |
| S8_extracted_k141_163914 | Not-determined |
| S8_extracted_k141_129650 | Not-determined |
| S8_extracted_k141_16525 | Not-determined |
| S8_extracted_k141_129651 | Not-determined |
| S8_extracted_k141_138225 | Not-determined |
| S8_extracted_k141_25708 | Not-determined |
| S8_extracted_k141_207273 | Not-determined |
| S8_extracted_k141_86205 | Low-quality |
| S8_extracted_k141_182017 | Low-quality |
| S8_extracted_k141_103366 | Not-determined |
| S8_extracted_k141_34515 | Not-determined |
| S8_extracted_k141_112528 | Not-determined |
| S8_extracted_k141_129660 | Low-quality |
| S8_extracted_k141_112530 | Not-determined |
| S8_extracted_k141_120829 | Not-determined |
| S8_extracted_k141_120830 | Not-determined |
| S8_extracted_k141_103368 | Low-quality |
| S8_extracted_k141_199454 | Not-determined |
| S8_extracted_k141_163922 | Not-determined |
| S8_extracted_k141_163925 | Not-determined |
| S8_extracted_k141_120836 | Not-determined |
| S8_extracted_k141_182023 | Not-determined |
| S8_extracted_k141_182025 | Not-determined |
| S8_extracted_k141_155358 | Not-determined |
| S8_extracted_k141_182026 | Not-determined |
| S8_extracted_k141_86217 | Not-determined |
| S8_extracted_k141_94832 | Not-determined |
| S8_extracted_k141_163930 | Not-determined |
| S8_extracted_k141_103377 | Not-determined |
| S8_extracted_k141_103378 | Not-determined |
| S8_extracted_k141_199461 | Not-determined |
| S8_extracted_k141_138234 | Not-determined |
| S8_extracted_k141_25717 | Not-determined |
| S8_extracted_k141_199463 | Low-quality |
| S8_extracted_k141_129673 | Not-determined |
| S8_extracted_k141_86223 | Not-determined |
| S8_extracted_k141_16541 | Not-determined |
| S8_extracted_k141_138237 | Not-determined |
| S8_extracted_k141_8292 | Not-determined |
| S8_extracted_k141_182033 | Not-determined |
| S8_extracted_k141_182035 | Not-determined |
| S8_extracted_k141_112544 | Not-determined |
| S8_extracted_k141_25723 | Not-determined |
| S8_extracted_k141_182038 | Not-determined |
| S8_extracted_k141_207293 | Low-quality |
| S8_extracted_k141_94836 | Low-quality |
| S8_extracted_k141_42797 | Not-determined |
| S8_extracted_k141_120849 | Not-determined |
| S8_extracted_k141_103389 | Not-determined |
| S8_extracted_k141_120851 | Low-quality |
| S8_extracted_k141_16549 | Not-determined |
| S8_extracted_k141_199469 | Not-determined |
| S8_extracted_k141_25726 | Low-quality |
| S8_extracted_k141_138246 | Not-determined |
| S8_extracted_k141_199470 | Not-determined |
| S8_extracted_k141_94842 | Not-determined |
| S8_extracted_k141_120852 | Not-determined |
| S8_extracted_k141_112549 | Not-determined |
| S8_extracted_k141_50734 | Not-determined |
| S8_extracted_k141_207300 | Not-determined |
| S8_extracted_k141_120853 | Not-determined |
| S8_extracted_k141_112550 | Not-determined |
| S8_extracted_k141_207306 | Not-determined |
| S8_extracted_k141_120857 | Not-determined |
| S8_extracted_k141_112554 | Not-determined |
| S8_extracted_k141_138250 | Not-determined |
| S8_extracted_k141_120858 | Not-determined |
| S8_extracted_k141_182050 | Low-quality |
| S8_extracted_k141_103398 | Not-determined |
| S8_extracted_k141_86231 | Not-determined |
| S8_extracted_k141_182051 | Low-quality |
| S8_extracted_k141_112558 | Low-quality |
| S8_extracted_k141_77661 | Low-quality |
| S8_extracted_k141_182052 | Not-determined |
| S8_extracted_k141_189492 | Not-determined |
| S8_extracted_k141_94852 | Not-determined |
| S8_extracted_k141_182054 | Not-determined |
| S8_extracted_k141_155371 | Not-determined |
| S8_extracted_k141_94853 | Not-determined |
| S8_extracted_k141_86234 | Not-determined |
| S8_extracted_k141_42809 | Not-determined |
| S8_extracted_k141_94854 | Not-determined |
| S8_extracted_k141_163950 | Not-determined |
| S8_extracted_k141_138256 | Not-determined |
| S8_extracted_k141_138257 | Not-determined |
| S12_vs_k141_74280\|\|full | Medium-quality |
| S8_extracted_k141_207308 | Not-determined |
| S8_extracted_k141_120863 | Not-determined |
| S8_extracted_k141_112562 | Not-determined |
| S8_extracted_k141_16563 | Not-determined |
| S8_extracted_k141_199482 | Not-determined |
| S8_extracted_k141_163953 | Not-determined |
| S8_extracted_k141_86240 | Not-determined |
| S8_extracted_k141_155377 | Not-determined |
| S8_extracted_k141_120867 | Not-determined |
| S8_extracted_k141_112565 | Low-quality |
| S8_extracted_k141_207313 | Low-quality |
| S8_extracted_k141_163956 | Low-quality |
| S8_extracted_k141_163957 | Not-determined |
| S8_extracted_k141_86248 | Not-determined |
| S8_extracted_k141_147133 | Low-quality |
| S8_extracted_k141_8310 | Not-determined |
| S8_extracted_k141_163959 | Not-determined |
| S8_extracted_k141_147134 | Not-determined |
| S8_extracted_k141_155379 | Not-determined |
| S8_extracted_k141_207316 | Low-quality |
| S8_extracted_k141_155381 | Low-quality |
| S8_extracted_k141_147137 | Low-quality |
| S8_extracted_k141_147138 | Not-determined |
| S8_extracted_k141_77674 | Not-determined |
| S8_extracted_k141_199490 | Low-quality |
| S8_extracted_k141_138266 | Not-determined |
| S8_extracted_k141_25749 | Not-determined |
| S8_extracted_k141_199492 | Not-determined |
| S8_extracted_k141_120883 | Not-determined |
| S8_extracted_k141_129696 | Not-determined |
| S8_extracted_k141_77684 | Not-determined |
| S8_extracted_k141_182078 | Low-quality |
| S8_extracted_k141_207323 | Not-determined |
| S8_extracted_k141_163965 | Not-determined |
| S8_extracted_k141_86257 | Not-determined |
| S8_extracted_k141_189511 | Low-quality |
| S8_extracted_k141_129698 | Not-determined |
| S8_extracted_k141_8325 | Not-determined |
| S8_extracted_k141_68677 | Not-determined |
| S8_extracted_k141_199499 | Not-determined |
| S8_extracted_k141_199501 | Not-determined |
| S8_extracted_k141_120890 | Not-determined |
| S8_extracted_k141_182088 | Not-determined |
| S8_extracted_k141_120892 | Not-determined |
| S8_extracted_k141_189518 | Not-determined |
| S8_extracted_k141_68681 | Not-determined |
| S8_extracted_k141_163969 | Not-determined |
| S8_extracted_k141_77694 | Not-determined |
| S8_extracted_k141_68685 | Not-determined |
| S8_extracted_k141_94881 | Not-determined |
| S8_extracted_k141_182094 | Not-determined |
| S8_extracted_k141_182095 | Not-determined |
| S8_extracted_k141_94882 | Not-determined |
| S8_extracted_k141_94883 | Not-determined |
| S8_extracted_k141_94885 | Not-determined |
| S8_extracted_k141_8333 | Not-determined |
| S8_extracted_k141_8334 | Not-determined |
| S8_extracted_k141_155395 | Not-determined |
| S8_extracted_k141_16601 | Low-quality |
| S8_extracted_k141_207336 | Not-determined |
| S8_extracted_k141_94892 | Not-determined |
| S8_extracted_k141_155399 | Not-determined |
| S8_extracted_k141_147152 | Not-determined |
| S8_extracted_k141_25763 | Low-quality |
| S8_extracted_k141_86268 | Not-determined |
| S8_extracted_k141_189529 | Not-determined |
| S8_extracted_k141_129701 | Not-determined |
| S8_extracted_k141_182096 | Low-quality |
| S8_extracted_k141_25764 | Not-determined |
| S8_extracted_k141_16606 | Not-determined |
| S8_extracted_k141_163974 | Not-determined |
| S8_extracted_k141_16608 | Not-determined |
| S8_extracted_k141_77698 | Not-determined |
| S8_extracted_k141_163975 | Low-quality |
| S8_extracted_k141_147156 | Not-determined |
| S8_extracted_k141_68690 | Not-determined |
| S8_extracted_k141_199511 | Not-determined |
| S8_extracted_k141_16615 | Not-determined |
| S8_extracted_k141_25772 | Not-determined |
| S8_extracted_k141_120904 | Not-determined |
| S8_extracted_k141_42830 | Not-determined |
| S8_extracted_k141_60614 | Not-determined |
| S8_extracted_k141_8338 | Low-quality |
| S8_extracted_k141_68695 | Not-determined |
| S8_extracted_k141_86280 | Not-determined |
| S8_extracted_k141_163978 | Not-determined |
| S8_extracted_k141_189543 | Low-quality |
| S8_extracted_k141_120906 | Not-determined |
| S8_extracted_k141_138276 | Low-quality |
| S8_extracted_k141_199518 | Not-determined |
| S8_extracted_k141_199519 | Not-determined |
| S8_extracted_k141_182098 | Low-quality |
| S8_extracted_k141_138278 | Not-determined |
| S8_extracted_k141_163980 | Not-determined |
| S8_extracted_k141_138281 | Not-determined |
| S8_extracted_k141_8347 | Not-determined |
| S8_extracted_k141_120909 | Not-determined |
| S8_extracted_k141_129713 | Not-determined |
| S8_extracted_k141_86284 | Not-determined |
| S8_extracted_k141_189549 | Not-determined |
| S8_extracted_k141_86286 | Not-determined |
| S8_extracted_k141_207344 | Not-determined |
| S8_extracted_k141_199528 | Not-determined |
| S8_extracted_k141_86288 | Not-determined |
| S8_extracted_k141_68703 | Low-quality |
| S8_extracted_k141_60624 | Not-determined |
| S8_extracted_k141_94897 | Not-determined |
| S8_extracted_k141_189555 | Not-determined |
| S8_extracted_k141_25791 | Not-determined |
| S8_extracted_k141_94904 | Low-quality |
| S8_extracted_k141_34543 | Not-determined |
| S8_extracted_k141_199533 | Not-determined |
| S8_extracted_k141_8355 | Not-determined |
| S8_extracted_k141_60628 | Not-determined |
| S8_extracted_k141_8357 | Not-determined |
| S8_extracted_k141_77721 | Not-determined |
| S8_extracted_k141_120911 | Not-determined |
| S8_extracted_k141_42844 | Not-determined |
| S8_extracted_k141_8359 | Not-determined |
| S8_extracted_k141_8361 | Low-quality |
| S8_extracted_k141_182108 | Low-quality |
| S8_extracted_k141_86299 | Not-determined |
| S8_extracted_k141_86300 | Not-determined |
| S8_extracted_k141_182109 | Not-determined |
| S8_extracted_k141_163993 | Not-determined |
| S7_vs_135778\|\|full | High-quality |
| S8_extracted_k141_138294 | Not-determined |
| S8_extracted_k141_182116 | Not-determined |
| S8_extracted_k141_42850 | Not-determined |
| S8_extracted_k141_60640 | Not-determined |
| S8_extracted_k141_34557 | Low-quality |
| S8_extracted_k141_207357 | Not-determined |
| S8_extracted_k141_34558 | Not-determined |
| S8_extracted_k141_182121 | Not-determined |
| S8_extracted_k141_103457 | Not-determined |
| S8_extracted_k141_8375 | Not-determined |
| S8_extracted_k141_42856 | Low-quality |
| S8_extracted_k141_34563 | Not-determined |
| S8_extracted_k141_147174 | Not-determined |
| S8_extracted_k141_155416 | Not-determined |
| S8_extracted_k141_155417 | Not-determined |
| S8_extracted_k141_207363 | Not-determined |
| S8_extracted_k141_50764 | Low-quality |
| S8_extracted_k141_163998 | Not-determined |
| S8_extracted_k141_120923 | Not-determined |
| S8_extracted_k141_42860 | Not-determined |
| S8_extracted_k141_68713 | Not-determined |
| S8_extracted_k141_42863 | Not-determined |
| S8_extracted_k141_173236 | Not-determined |
| S8_extracted_k141_8377 | Not-determined |
| S8_extracted_k141_155419 | Not-determined |
| S8_extracted_k141_182128 | Low-quality |
| S8_extracted_k141_68719 | Not-determined |
| S8_extracted_k141_199558 | Not-determined |
| S8_extracted_k141_199560 | Not-determined |
| S8_extracted_k141_199561 | Not-determined |
| S8_extracted_k141_155424 | Not-determined |
| S8_extracted_k141_129740 | Not-determined |
| S8_extracted_k141_42870 | Not-determined |
| S8_extracted_k141_8379 | Not-determined |
| S8_extracted_k141_129742 | Not-determined |
| S8_extracted_k141_16628 | Not-determined |
| S8_extracted_k141_129744 | Not-determined |
| S8_extracted_k141_120933 | Not-determined |
| S8_extracted_k141_8380 | Not-determined |
| S8_extracted_k141_164004 | Not-determined |
| S8_extracted_k141_86321 | Not-determined |
| S8_extracted_k141_138299 | Not-determined |
| S8_extracted_k141_86322 | Not-determined |
| S8_extracted_k141_60656 | Not-determined |
| S8_extracted_k141_138303 | Not-determined |
| S8_extracted_k141_60658 | Not-determined |
| S8_extracted_k141_16634 | Not-determined |
| S8_extracted_k141_8387 | Not-determined |
| S8_extracted_k141_147185 | Not-determined |
| S8_extracted_k141_16636 | Low-quality |
| S8_extracted_k141_50778 | Not-determined |
| S8_extracted_k141_129756 | Not-determined |
| S8_extracted_k141_155438 | Not-determined |
| S8_extracted_k141_155439 | Not-determined |
| S8_extracted_k141_8393 | Not-determined |
| S8_extracted_k141_120940 | Not-determined |
| S8_extracted_k141_16642 | Not-determined |
| S8_extracted_k141_207389 | Not-determined |
| S8_extracted_k141_129761 | Not-determined |
| S8_extracted_k141_50783 | Not-determined |
| S8_extracted_k141_103472 | Not-determined |
| S8_extracted_k141_8397 | Not-determined |
| S8_extracted_k141_50784 | Not-determined |
| S8_extracted_k141_34570 | Not-determined |
| S8_extracted_k141_164020 | Not-determined |
| S8_extracted_k141_77757 | Not-determined |
| S8_extracted_k141_16646 | Not-determined |
| S8_extracted_k141_34572 | Not-determined |
| S8_extracted_k141_103475 | Not-determined |
| S8_extracted_k141_68741 | Low-quality |
| S8_extracted_k141_103478 | Not-determined |
| S8_extracted_k141_60673 | Not-determined |
| S8_extracted_k141_68744 | Not-determined |
| S8_extracted_k141_182150 | Not-determined |
| S8_extracted_k141_189583 | Not-determined |
| S8_extracted_k141_50792 | Not-determined |
| S8_extracted_k141_42883 | Not-determined |
| S8_extracted_k141_94914 | Not-determined |
| S8_extracted_k141_155448 | Low-quality |
| S8_extracted_k141_155451 | Not-determined |
| S8_extracted_k141_103482 | Not-determined |
| S8_extracted_k141_16660 | Low-quality |
| S8_extracted_k141_164022 | Not-determined |
| S8_extracted_k141_189591 | Not-determined |
| S8_extracted_k141_129764 | Not-determined |
| S8_extracted_k141_164024 | Not-determined |
| S8_extracted_k141_182155 | Not-determined |
| S8_extracted_k141_173255 | Not-determined |
| S8_extracted_k141_103488 | Not-determined |
| S8_extracted_k141_155455 | Not-determined |
| S8_extracted_k141_129767 | Not-determined |
| S8_extracted_k141_77771 | Not-determined |
| S8_extracted_k141_182156 | Not-determined |
| S8_extracted_k141_120946 | Not-determined |
| S8_extracted_k141_94918 | Not-determined |
| S8_extracted_k141_8408 | Not-determined |
| S8_extracted_k141_77773 | Not-determined |
| S8_extracted_k141_147203 | Not-determined |
| S8_extracted_k141_94919 | Not-determined |
| S8_extracted_k141_103492 | Not-determined |
| S8_extracted_k141_189594 | Not-determined |
| S8_extracted_k141_182158 | Not-determined |
| S8_extracted_k141_155460 | Not-determined |
| S8_extracted_k141_60688 | Not-determined |
| S8_extracted_k141_189598 | Not-determined |
| S8_extracted_k141_173260 | Not-determined |
| S8_extracted_k141_147210 | Not-determined |
| S8_extracted_k141_94924 | Not-determined |
| S8_extracted_k141_189600 | Not-determined |
| S8_extracted_k141_16665 | Not-determined |
| S8_extracted_k141_103498 | Not-determined |
| S8_extracted_k141_103499 | Not-determined |
| S8_extracted_k141_34587 | Not-determined |
| S8_extracted_k141_103501 | Not-determined |
| S8_extracted_k141_120956 | Not-determined |
| S8_extracted_k141_147218 | Not-determined |
| S8_extracted_k141_94930 | Not-determined |
| S8_extracted_k141_86357 | Not-determined |
| S8_extracted_k141_164029 | Not-determined |
| S8_extracted_k141_164030 | Not-determined |
| S8_extracted_k141_103505 | Not-determined |
| S8_extracted_k141_68758 | Not-determined |
| S8_extracted_k141_120960 | Not-determined |
| S8_extracted_k141_60695 | Not-determined |
| S8_extracted_k141_173269 | Not-determined |
| S8_extracted_k141_189609 | Not-determined |
| S8_extracted_k141_94934 | Not-determined |
| S8_extracted_k141_60697 | Not-determined |
| S8_extracted_k141_155464 | Not-determined |
| S8_extracted_k141_155466 | Not-determined |
| S8_extracted_k141_86360 | Not-determined |
| S8_extracted_k141_16674 | Not-determined |
| S8_extracted_k141_129788 | Not-determined |
| S8_extracted_k141_120961 | Not-determined |
| S8_extracted_k141_86365 | Not-determined |
| S8_extracted_k141_86366 | Not-determined |
| S8_extracted_k141_189614 | Not-determined |
| S8_extracted_k141_16675 | Not-determined |
| S8_extracted_k141_25820 | Not-determined |
| S8_extracted_k141_129794 | Not-determined |
| S8_extracted_k141_94945 | Not-determined |
| S8_extracted_k141_147234 | Not-determined |
| S8_extracted_k141_189621 | Low-quality |
| S8_extracted_k141_120971 | Not-determined |
| S8_extracted_k141_138347 | Not-determined |
| S8_extracted_k141_103518 | Low-quality |
| S8_extracted_k141_155479 | Not-determined |
| S8_extracted_k141_68775 | Not-determined |
| S8_extracted_k141_94951 | Not-determined |
| S8_extracted_k141_103520 | Not-determined |
| S8_extracted_k141_77797 | Not-determined |
| S8_extracted_k141_60715 | Not-determined |
| S8_extracted_k141_189627 | Not-determined |
| S8_extracted_k141_207429 | Not-determined |
| S8_extracted_k141_86377 | Not-determined |
| S8_extracted_k141_129805 | Not-determined |
| S8_extracted_k141_8439 | Not-determined |
| S8_extracted_k141_155485 | Not-determined |
| S8_extracted_k141_94963 | Not-determined |
| S8_extracted_k141_25832 | Low-quality |
| S8_extracted_k141_34610 | Not-determined |
| S8_extracted_k141_207435 | Not-determined |
| S8_extracted_k141_155487 | Not-determined |
| S8_extracted_k141_164045 | Not-determined |
| S8_extracted_k141_120986 | Not-determined |
| S8_extracted_k141_120987 | Not-determined |
| S8_extracted_k141_112595 | Not-determined |
| S8_extracted_k141_112597 | Not-determined |
| S8_extracted_k141_8448 | Not-determined |
| S8_extracted_k141_94970 | Not-determined |
| S8_extracted_k141_8449 | Not-determined |
| S8_extracted_k141_129813 | Not-determined |
| S8_extracted_k141_129814 | Low-quality |
| S8_extracted_k141_77815 | Not-determined |
| S8_extracted_k141_155496 | Not-determined |
| S8_extracted_k141_147247 | Not-determined |
| S8_extracted_k141_155498 | Not-determined |
| S8_extracted_k141_129822 | Not-determined |
| S8_extracted_k141_173273 | Low-quality |
| S8_extracted_k141_77819 | Not-determined |
| S8_extracted_k141_68788 | Not-determined |
| S8_extracted_k141_68789 | Not-determined |
| S8_extracted_k141_182198 | Not-determined |
| S8_extracted_k141_138364 | Not-determined |
| S8_extracted_k141_103544 | Not-determined |
| S8_extracted_k141_173275 | Not-determined |
| S8_extracted_k141_8457 | Not-determined |
| S8_extracted_k141_86405 | Not-determined |
| S8_extracted_k141_138365 | Not-determined |
| S8_extracted_k141_129826 | Not-determined |
| S8_extracted_k141_68794 | Not-determined |
| S8_extracted_k141_8458 | Not-determined |
| S8_extracted_k141_138369 | Not-determined |
| S8_extracted_k141_16695 | Not-determined |
| S8_extracted_k141_164062 | Not-determined |
| S8_extracted_k141_42896 | Not-determined |
| S8_extracted_k141_155506 | Not-determined |
| S8_extracted_k141_68798 | Not-determined |
| S8_extracted_k141_164066 | Not-determined |
| S8_extracted_k141_94990 | Not-determined |
| S8_extracted_k141_86414 | Low-quality |
| S8_extracted_k141_121004 | Not-determined |
| S8_extracted_k141_147256 | Not-determined |
| S8_extracted_k141_16699 | Not-determined |
| S8_extracted_k141_94991 | Not-determined |
| S8_extracted_k141_138374 | Not-determined |
| S8_extracted_k141_94992 | Not-determined |
| S8_extracted_k141_182202 | Not-determined |
| S8_extracted_k141_173282 | Not-determined |
| S8_extracted_k141_121008 | Not-determined |
| S8_extracted_k141_164067 | Not-determined |
| S8_extracted_k141_42902 | Low-quality |
| S8_extracted_k141_164068 | Low-quality |
| S8_extracted_k141_138383 | Not-determined |
| S8_extracted_k141_8462 | Not-determined |
| S8_extracted_k141_121013 | Not-determined |
| S8_extracted_k141_94999 | Not-determined |
| S8_extracted_k141_147269 | Not-determined |
| S8_extracted_k141_155511 | Not-determined |
| S8_extracted_k141_129840 | Not-determined |
| S8_extracted_k141_129841 | Not-determined |
| S8_extracted_k141_16709 | Not-determined |
| S8_extracted_k141_207442 | Not-determined |
| S8_extracted_k141_86423 | Not-determined |
| S8_extracted_k141_16711 | Not-determined |
| S8_extracted_k141_121020 | Not-determined |
| S8_extracted_k141_182216 | Not-determined |
| S8_extracted_k141_182218 | Not-determined |
| S8_extracted_k141_147271 | Not-determined |
| S8_extracted_k141_155517 | Not-determined |
| S8_extracted_k141_207446 | Not-determined |
| S8_extracted_k141_189668 | Low-quality |
| S8_extracted_k141_173294 | Not-determined |
| S8_extracted_k141_8469 | Not-determined |
| S8_extracted_k141_138395 | Not-determined |
| S8_extracted_k141_103568 | Not-determined |
| S8_extracted_k141_207452 | Not-determined |
| S8_extracted_k141_68825 | Not-determined |
| S8_extracted_k141_207454 | Not-determined |
| S8_extracted_k141_121035 | Low-quality |
| S8_extracted_k141_164078 | Not-determined |
| S8_extracted_k141_42914 | Not-determined |
| S8_extracted_k141_207457 | Not-determined |
| S8_extracted_k141_147278 | Not-determined |
| S8_extracted_k141_147279 | Not-determined |
| S8_extracted_k141_173296 | Not-determined |
| S8_extracted_k141_60737 | Not-determined |
| S8_extracted_k141_147284 | Not-determined |
| S8_extracted_k141_95006 | Not-determined |
| S8_extracted_k141_129849 | Not-determined |
| S8_extracted_k141_155525 | Not-determined |
| S8_extracted_k141_207462 | Not-determined |
| S8_extracted_k141_189685 | Not-determined |
| S8_extracted_k141_77843 | Not-determined |
| S8_extracted_k141_155528 | Not-determined |
| S8_extracted_k141_121041 | Low-quality |
| S8_extracted_k141_189687 | Not-determined |
| S8_extracted_k141_189688 | Not-determined |
| S8_extracted_k141_189689 | Not-determined |
| S8_extracted_k141_189690 | Not-determined |
| S8_extracted_k141_95013 | Not-determined |
| S8_extracted_k141_121044 | Not-determined |
| S8_extracted_k141_16725 | Not-determined |
| S8_extracted_k141_95015 | Not-determined |
| S8_extracted_k141_8494 | Not-determined |
| S8_extracted_k141_95017 | Not-determined |
| S8_extracted_k141_147302 | Not-determined |
| S8_extracted_k141_129862 | Not-determined |
| S8_extracted_k141_86439 | Not-determined |
| S8_extracted_k141_189695 | Not-determined |
| S8_extracted_k141_8495 | Not-determined |
| S8_extracted_k141_147307 | Not-determined |
| S8_extracted_k141_189698 | Low-quality |
| S8_extracted_k141_121051 | Not-determined |
| S8_extracted_k141_207472 | Not-determined |
| S8_extracted_k141_164093 | Not-determined |
| S8_extracted_k141_95019 | Not-determined |
| S8_extracted_k141_8498 | Not-determined |
| S8_extracted_k141_164097 | Not-determined |
| S8_extracted_k141_155541 | Not-determined |
| S8_extracted_k141_16730 | Not-determined |
| S8_extracted_k141_138417 | Low-quality |
| S8_extracted_k141_68844 | Not-determined |
| S8_extracted_k141_34663 | Not-determined |
| S8_extracted_k141_103583 | Low-quality |
| S8_extracted_k141_164099 | Not-determined |
| S8_extracted_k141_164102 | Not-determined |
| S8_extracted_k141_138423 | Not-determined |
| S8_extracted_k141_86455 | Not-determined |
| S8_extracted_k141_164104 | Not-determined |
| S8_extracted_k141_207482 | Not-determined |
| S8_extracted_k141_207484 | Not-determined |
| S8_extracted_k141_164107 | Not-determined |
| S8_extracted_k141_173300 | Not-determined |
| S8_extracted_k141_95030 | Not-determined |
| S8_extracted_k141_173301 | Not-determined |
| S8_extracted_k141_189713 | Not-determined |
| S8_extracted_k141_68847 | Low-quality |
| S8_extracted_k141_129869 | Not-determined |
| S8_extracted_k141_155548 | Not-determined |
| S8_extracted_k141_173304 | Low-quality |
| S8_extracted_k141_42939 | Not-determined |
| S8_extracted_k141_42940 | Low-quality |
| S8_extracted_k141_121067 | Not-determined |
| S8_extracted_k141_77854 | Not-determined |
| S8_extracted_k141_77855 | Not-determined |
| S8_extracted_k141_121069 | Not-determined |
| S8_extracted_k141_129877 | Not-determined |
| S8_extracted_k141_16742 | Not-determined |
| S8_extracted_k141_129879 | Not-determined |
| S8_extracted_k141_103593 | Not-determined |
| S8_extracted_k141_86470 | Not-determined |
| S8_extracted_k141_129883 | Not-determined |
| S8_extracted_k141_34685 | Not-determined |
| S8_extracted_k141_34687 | Not-determined |
| S8_extracted_k141_42948 | Not-determined |
| S8_extracted_k141_155562 | Not-determined |
| S8_extracted_k141_8508 | Not-determined |
| S8_extracted_k141_129886 | Not-determined |
| S8_extracted_k141_129888 | Not-determined |
| S8_extracted_k141_34690 | Not-determined |
| S8_extracted_k141_68853 | Not-determined |
| S8_extracted_k141_121080 | Not-determined |
| S8_extracted_k141_77869 | Not-determined |
| S8_extracted_k141_42955 | Not-determined |
| S8_extracted_k141_207491 | Not-determined |
| S8_extracted_k141_189731 | Not-determined |
| S8_extracted_k141_173315 | Not-determined |
| S8_extracted_k141_95041 | Low-quality |
| S8_extracted_k141_42958 | Not-determined |
| S8_extracted_k141_68863 | Not-determined |
| S8_extracted_k141_68865 | Not-determined |
| S8_extracted_k141_207500 | Not-determined |
| S8_extracted_k141_77876 | Not-determined |
| S8_extracted_k141_155568 | Not-determined |
| S8_extracted_k141_86482 | Not-determined |
| S8_extracted_k141_103604 | Low-quality |
| S8_extracted_k141_16756 | Not-determined |
| S8_extracted_k141_50818 | Low-quality |
| S8_extracted_k141_77880 | Not-determined |
| S8_extracted_k141_16760 | Not-determined |
| S8_extracted_k141_42965 | Not-determined |
| S8_extracted_k141_95053 | Low-quality |
| S8_extracted_k141_50822 | Not-determined |
| S8_extracted_k141_155573 | Not-determined |
| S8_extracted_k141_121087 | Not-determined |
| S8_extracted_k141_129901 | Not-determined |
| S8_extracted_k141_103610 | Not-determined |
| S8_extracted_k141_77884 | Not-determined |
| S8_extracted_k141_103612 | Not-determined |
| S8_extracted_k141_173325 | Not-determined |
| S8_extracted_k141_42972 | Low-quality |
| S8_extracted_k141_173326 | Low-quality |
| S8_extracted_k141_189741 | Low-quality |
| S8_extracted_k141_77888 | Not-determined |
| S8_extracted_k141_95063 | Not-determined |
| S8_extracted_k141_103616 | Not-determined |
| S8_extracted_k141_8534 | Not-determined |
| S8_extracted_k141_86496 | Not-determined |
| S8_extracted_k141_207505 | Not-determined |
| S8_extracted_k141_95066 | Not-determined |
| S8_extracted_k141_68877 | Not-determined |
| S8_extracted_k141_164116 | Not-determined |
| S8_extracted_k141_25861 | Not-determined |
| S8_extracted_k141_86499 | Not-determined |
| S8_extracted_k141_8539 | Not-determined |

| | |
|---|---|
| S8_extracted_k141_8542 | Not-determined |
| S8_extracted_k141_25863 | Not-determined |
| S8_extracted_k141_207510 | Low-quality |
| S8_extracted_k141_103621 | Not-determined |
| S8_extracted_k141_8544 | Not-determined |
| S8_extracted_k141_86501 | Not-determined |
| S8_extracted_k141_68880 | Not-determined |
| S8_extracted_k141_189742 | Not-determined |
| S8_extracted_k141_50830 | Not-determined |
| S8_extracted_k141_103622 | Not-determined |
| S8_extracted_k141_173336 | Not-determined |
| S8_extracted_k141_34709 | Not-determined |
| S8_extracted_k141_155594 | Not-determined |
| S8_extracted_k141_103624 | Not-determined |
| S8_extracted_k141_42980 | Low-quality |
| S8_extracted_k141_95072 | Not-determined |
| S8_extracted_k141_50835 | Not-determined |
| S8_extracted_k141_189748 | Not-determined |
| S8_extracted_k141_50839 | Not-determined |
| S8_extracted_k141_16776 | Not-determined |
| S8_extracted_k141_164130 | Not-determined |
| S8_extracted_k141_68888 | Not-determined |
| S8_extracted_k141_50842 | Low-quality |
| S8_extracted_k141_189754 | Low-quality |
| S8_extracted_k141_50845 | Not-determined |
| S8_extracted_k141_189755 | Not-determined |
| S8_extracted_k141_8555 | Not-determined |
| S8_extracted_k141_8556 | Not-determined |
| S8_extracted_k141_8557 | Low-quality |
| S8_extracted_k141_121106 | Not-determined |
| S8_extracted_k141_189759 | Not-determined |
| S8_extracted_k141_164138 | Low-quality |
| S8_extracted_k141_25873 | Not-determined |
| S8_extracted_k141_164139 | Low-quality |
| S8_extracted_k141_68897 | Not-determined |
| S8_extracted_k141_121112 | Not-determined |
| S8_extracted_k141_86527 | Not-determined |
| S8_extracted_k141_103635 | Not-determined |
| S8_extracted_k141_138469 | Not-determined |
| S8_extracted_k141_16791 | Not-determined |
| S8_extracted_k141_138470 | Not-determined |
| S8_extracted_k141_43003 | Not-determined |
| S8_extracted_k141_138471 | Not-determined |
| S8_extracted_k141_129929 | Not-determined |
| S8_extracted_k141_121117 | Not-determined |
| S8_extracted_k141_103642 | Not-determined |
| S8_extracted_k141_16798 | Not-determined |
| S8_extracted_k141_207516 | Low-quality |
| S8_extracted_k141_16799 | Not-determined |
| S8_extracted_k141_68913 | Not-determined |
| S8_extracted_k141_164148 | Not-determined |
| S8_extracted_k141_103645 | Not-determined |
| S8_extracted_k141_129937 | Not-determined |
| S8_extracted_k141_95093 | Low-quality |
| S8_extracted_k141_164150 | Low-quality |
| S8_extracted_k141_8561 | Not-determined |
| S8_extracted_k141_138483 | Not-determined |
| S8_extracted_k141_86536 | Not-determined |
| S8_extracted_k141_8565 | Not-determined |
| S8_extracted_k141_207523 | Low-quality |
| S8_extracted_k141_207524 | Not-determined |
| S8_extracted_k141_8568 | Not-determined |
| S8_extracted_k141_68918 | Not-determined |
| S8_extracted_k141_95101 | Not-determined |
| S8_extracted_k141_207525 | Not-determined |
| S8_extracted_k141_155608 | Low-quality |
| S8_extracted_k141_25900 | Not-determined |
| S8_extracted_k141_43015 | Not-determined |
| S8_extracted_k141_164157 | Not-determined |
| S8_extracted_k141_16803 | Not-determined |
| S8_extracted_k141_8571 | Not-determined |
| S8_extracted_k141_129945 | Low-quality |
| S8_extracted_k141_50858 | Not-determined |
| S8_extracted_k141_77924 | Not-determined |
| S8_extracted_k141_155615 | Low-quality |
| S8_extracted_k141_86544 | Not-determined |
| S8_extracted_k141_138492 | Not-determined |
| S8_extracted_k141_121126 | Low-quality |
| S8_extracted_k141_95105 | Not-determined |
| S10_extracted_57705 | Medium-quality |
| S8_extracted_k141_147339 | Not-determined |
| S8_extracted_k141_207527 | Not-determined |
| S8_extracted_k141_68930 | Not-determined |
| S8_extracted_k141_50868 | Not-determined |
| S8_extracted_k141_43027 | Not-determined |
| S8_extracted_k141_155617 | Not-determined |
| S8_extracted_k141_147345 | Not-determined |
| S8_extracted_k141_50871 | Not-determined |
| S8_extracted_k141_50872 | Not-determined |
| S8_extracted_k141_43028 | Not-determined |
| S8_extracted_k141_103660 | Not-determined |
| S8_extracted_k141_95109 | Not-determined |
| S8_extracted_k141_86552 | Not-determined |
| S8_extracted_k141_147348 | Not-determined |
| S8_extracted_k141_25912 | Not-determined |
| S8_extracted_k141_147350 | Not-determined |
| S8_extracted_k141_68934 | Not-determined |
| S8_extracted_k141_86557 | Not-determined |
| S8_extracted_k141_121144 | Not-determined |
| S8_extracted_k141_103667 | Not-determined |
| S8_extracted_k141_86560 | Not-determined |
| S8_extracted_k141_86562 | Low-quality |
| S8_extracted_k141_77939 | Not-determined |
| S8_extracted_k141_173350 | Low-quality |
| S8_extracted_k141_103670 | Low-quality |
| S8_extracted_k141_43036 | Low-quality |
| S8_extracted_k141_16817 | Not-determined |
| S8_extracted_k141_138517 | Low-quality |
| S8_extracted_k141_77943 | Not-determined |
| S8_extracted_k141_86566 | Low-quality |
| S8_extracted_k141_95126 | Not-determined |
| S8_extracted_k141_50897 | Not-determined |
| S8_extracted_k141_8591 | Not-determined |
| S8_extracted_k141_86568 | Not-determined |
| S8_extracted_k141_155630 | Low-quality |
| S8_extracted_k141_103678 | Not-determined |
| S8_extracted_k141_16822 | Not-determined |
| S8_extracted_k141_86571 | Not-determined |
| S8_extracted_k141_103684 | Not-determined |
| S8_extracted_k141_43046 | Not-determined |
| S8_extracted_k141_25926 | Not-determined |
| S8_extracted_k141_43048 | Not-determined |
| S8_extracted_k141_207544 | Not-determined |
| S8_extracted_k141_16825 | Not-determined |
| S8_extracted_k141_43052 | Not-determined |
| S8_extracted_k141_86575 | Not-determined |
| S8_extracted_k141_16828 | Not-determined |
| S8_extracted_k141_173363 | Not-determined |
| S8_extracted_k141_8602 | Not-determined |
| S8_extracted_k141_25932 | Not-determined |
| S8_extracted_k141_16830 | Not-determined |
| S8_extracted_k141_207546 | Not-determined |
| S8_extracted_k141_147367 | Not-determined |
| S8_extracted_k141_121163 | Not-determined |
| S8_extracted_k141_86582 | Not-determined |
| S8_extracted_k141_138531 | Low-quality |
| S8_extracted_k141_50908 | Not-determined |
| S8_extracted_k141_207551 | Not-determined |
| S8_extracted_k141_103689 | Not-determined |
| S8_extracted_k141_129993 | Not-determined |
| S8_extracted_k141_95131 | Low-quality |
| S8_extracted_k141_50910 | Not-determined |
| S8_extracted_k141_50912 | Not-determined |
| S8_extracted_k141_25935 | Not-determined |
| S8_extracted_k141_86585 | Not-determined |
| S8_extracted_k141_86586 | Not-determined |
| S8_extracted_k141_8610 | Not-determined |
| S8_extracted_k141_138539 | Not-determined |
| S8_extracted_k141_207553 | Not-determined |
| S8_extracted_k141_50916 | Not-determined |
| S8_extracted_k141_103696 | Not-determined |
| S8_extracted_k141_16839 | Not-determined |
| S8_extracted_k141_121177 | Not-determined |
| S8_extracted_k141_138541 | Not-determined |
| S8_extracted_k141_77954 | Not-determined |
| S8_extracted_k141_130001 | Not-determined |
| S8_extracted_k141_8614 | Not-determined |
| S8_extracted_k141_138545 | Not-determined |
| S8_extracted_k141_8616 | Not-determined |
| S8_extracted_k141_112613 | Not-determined |
| S8_extracted_k141_8617 | Not-determined |
| S8_extracted_k141_130005 | Not-determined |
| S8_extracted_k141_112615 | Not-determined |
| S8_extracted_k141_121187 | Not-determined |
| S8_extracted_k141_112616 | Not-determined |
| S8_extracted_k141_8619 | Not-determined |
| S8_extracted_k141_68951 | Not-determined |
| S8_extracted_k141_121191 | Not-determined |
| S8_extracted_k141_121192 | Not-determined |
| S8_extracted_k141_25943 | Not-determined |
| S8_extracted_k141_155643 | Not-determined |
| S8_extracted_k141_16851 | Not-determined |
| S8_extracted_k141_103706 | Not-determined |
| S8_extracted_k141_77969 | Not-determined |
| S8_extracted_k141_50936 | Not-determined |
| S8_extracted_k141_189764 | Not-determined |
| S8_extracted_k141_112622 | Low-quality |
| S8_extracted_k141_103712 | Not-determined |
| S8_extracted_k141_95136 | Not-determined |
| S8_extracted_k141_103714 | Not-determined |
| S8_extracted_k141_8638 | Low-quality |
| S8_extracted_k141_121205 | Not-determined |
| S8_extracted_k141_112630 | Not-determined |
| S8_extracted_k141_189771 | Not-determined |
| S8_extracted_k141_207572 | Not-determined |
| S8_extracted_k141_77980 | Not-determined |
| S8_extracted_k141_8646 | Not-determined |
| S8_extracted_k141_112634 | Not-determined |
| S8_extracted_k141_8648 | Not-determined |
| S8_extracted_k141_95144 | Low-quality |
| S8_extracted_k141_112636 | Not-determined |
| S8_extracted_k141_130025 | Not-determined |
| S8_extracted_k141_112637 | Not-determined |
| S8_extracted_k141_130026 | Not-determined |
| S8_extracted_k141_16865 | Not-determined |
| S8_extracted_k141_86604 | Not-determined |
| S8_extracted_k141_147384 | Not-determined |
| S8_extracted_k141_207576 | Not-determined |
| S8_extracted_k141_50947 | Not-determined |
| S8_extracted_k141_121221 | Not-determined |
| S8_extracted_k141_130029 | Not-determined |
| S8_extracted_k141_95154 | Not-determined |
| S8_extracted_k141_147390 | Not-determined |
| S8_extracted_k141_77998 | Not-determined |
| S8_extracted_k141_207583 | Not-determined |
| S8_extracted_k141_112647 | Low-quality |
| S8_extracted_k141_138553 | Not-determined |
| S8_extracted_k141_86606 | Not-determined |
| S8_extracted_k141_8658 | Not-determined |
| S8_extracted_k141_138556 | Not-determined |
| S8_extracted_k141_103729 | Not-determined |
| S8_extracted_k141_68980 | Not-determined |
| S8_extracted_k141_138560 | Not-determined |
| S8_extracted_k141_121230 | Low-quality |
| S8_extracted_k141_147398 | Low-quality |
| S8_extracted_k141_173380 | Not-determined |
| S8_extracted_k141_95168 | Not-determined |
| S8_extracted_k141_138563 | Low-quality |
| S8_extracted_k141_155660 | Not-determined |
| S8_extracted_k141_95169 | Not-determined |
| S8_extracted_k141_130038 | Not-determined |
| S8_extracted_k141_78005 | Not-determined |
| S8_extracted_k141_173384 | Not-determined |
| S8_extracted_k141_138565 | Not-determined |
| S8_extracted_k141_130039 | Not-determined |
| S8_extracted_k141_95171 | Not-determined |
| S8_extracted_k141_112654 | Not-determined |
| S8_extracted_k141_16872 | Not-determined |
| S8_extracted_k141_86609 | Not-determined |
| S8_extracted_k141_155662 | Not-determined |
| S8_extracted_k141_50962 | Not-determined |
| S8_extracted_k141_68982 | Not-determined |
| S8_extracted_k141_68983 | Not-determined |
| S8_extracted_k141_138567 | Not-determined |
| S8_extracted_k141_189781 | Low-quality |
| S8_extracted_k141_95175 | Not-determined |
| S8_extracted_k141_173390 | Not-determined |
| S8_extracted_k141_155670 | Low-quality |
| S8_extracted_k141_25969 | Not-determined |
| S8_extracted_k141_95180 | Low-quality |
| S8_extracted_k141_68992 | Not-determined |
| S8_extracted_k141_155672 | Not-determined |
| S8_extracted_k141_95181 | Not-determined |
| S8_extracted_k141_147407 | Not-determined |
| S8_extracted_k141_138574 | Not-determined |
| S8_extracted_k141_147409 | Not-determined |
| S8_extracted_k141_43077 | Not-determined |
| S8_extracted_k141_130048 | Not-determined |
| S8_extracted_k141_189791 | Not-determined |
| S8_extracted_k141_173398 | Low-quality |
| S8_extracted_k141_43080 | Not-determined |
| S8_extracted_k141_138578 | Low-quality |
| S8_extracted_k141_69000 | Not-determined |
| S8_extracted_k141_130053 | Not-determined |
| S8_extracted_k141_95188 | Not-determined |
| S8_extracted_k141_138581 | Not-determined |
| S8_extracted_k141_173401 | Low-quality |
| S8_extracted_k141_95190 | Not-determined |
| S8_extracted_k141_25978 | Not-determined |
| S8_extracted_k141_138585 | Not-determined |
| S8_extracted_k141_155679 | Low-quality |
| S8_extracted_k141_103749 | Not-determined |
| S8_extracted_k141_155681 | Not-determined |
| S8_extracted_k141_138588 | Not-determined |
| S8_extracted_k141_147417 | Not-determined |
| S8_extracted_k141_189801 | Not-determined |
| S8_extracted_k141_43087 | Not-determined |
| S8_extracted_k141_207598 | Low-quality |
| S8_extracted_k141_43089 | Not-determined |
| S8_extracted_k141_95201 | Not-determined |
| S8_extracted_k141_69005 | Not-determined |
| S8_extracted_k141_138591 | Not-determined |
| S8_extracted_k141_43091 | Not-determined |
| S8_extracted_k141_78012 | Not-determined |
| S8_extracted_k141_16895 | Not-determined |
| S8_extracted_k141_164231 | Not-determined |
| S8_extracted_k141_78014 | Not-determined |
| S8_extracted_k141_103757 | Not-determined |
| S8_extracted_k141_207603 | Not-determined |
| S8_extracted_k141_147427 | Not-determined |
| S8_extracted_k141_112681 | Not-determined |
| S8_extracted_k141_155688 | Not-determined |
| S8_extracted_k141_207606 | Not-determined |
| S8_extracted_k141_69015 | Not-determined |
| S8_extracted_k141_138600 | Low-quality |
| S8_extracted_k141_69016 | Not-determined |
| S8_extracted_k141_78024 | Not-determined |
| S8_extracted_k141_130076 | Not-determined |
| S8_extracted_k141_103759 | Not-determined |
| S8_extracted_k141_164239 | Not-determined |
| S8_extracted_k141_69017 | Not-determined |
| S8_extracted_k141_130077 | Not-determined |
| S8_extracted_k141_147433 | Not-determined |
| S8_extracted_k141_155697 | Not-determined |
| S8_extracted_k141_164243 | Not-determined |
| S8_extracted_k141_173407 | Not-determined |
| S8_extracted_k141_78028 | Not-determined |
| S8_extracted_k141_43102 | Not-determined |
| S8_extracted_k141_78029 | Not-determined |
| S8_extracted_k141_173409 | Not-determined |
| S8_extracted_k141_95209 | Low-quality |
| S8_extracted_k141_25989 | Not-determined |
| S8_extracted_k141_130080 | Not-determined |
| S8_extracted_k141_189809 | Not-determined |
| S8_extracted_k141_16902 | Low-quality |
| S8_extracted_k141_155699 | Not-determined |
| S8_extracted_k141_138603 | Not-determined |
| S8_extracted_k141_130083 | Not-determined |
| S8_extracted_k141_138605 | Not-determined |
| S8_extracted_k141_95216 | Not-determined |
| S8_extracted_k141_16903 | Not-determined |
| S8_extracted_k141_25991 | Not-determined |
| S8_extracted_k141_50999 | Low-quality |
| S8_extracted_k141_25993 | Not-determined |
| S8_extracted_k141_112695 | Low-quality |
| S8_extracted_k141_207614 | Not-determined |
| S8_extracted_k141_16908 | Not-determined |
| S8_extracted_k141_207617 | Not-determined |
| S8_extracted_k141_189815 | Not-determined |
| S8_extracted_k141_51001 | Not-determined |
| S8_extracted_k141_69033 | Low-quality |
| S8_extracted_k141_130089 | Not-determined |
| S8_extracted_k141_207618 | Not-determined |
| S8_extracted_k141_95229 | Not-determined |
| S8_extracted_k141_78037 | Not-determined |
| S8_extracted_k141_138613 | Not-determined |
| S8_extracted_k141_78039 | Not-determined |
| S8_extracted_k141_112705 | Not-determined |
| S8_extracted_k141_130092 | Not-determined |
| S8_extracted_k141_43112 | Not-determined |
| S8_extracted_k141_207622 | Not-determined |
| S8_extracted_k141_112707 | Not-determined |
| S8_extracted_k141_189816 | Not-determined |
| S8_extracted_k141_69049 | Not-determined |
| S8_extracted_k141_95237 | Not-determined |
| S8_extracted_k141_147462 | Not-determined |
| S8_extracted_k141_155713 | Not-determined |
| S8_extracted_k141_130101 | Not-determined |
| S8_extracted_k141_173425 | Not-determined |
| S8_extracted_k141_112717 | Not-determined |
| S8_extracted_k141_103777 | Not-determined |
| S8_extracted_k141_112718 | Not-determined |
| S8_extracted_k141_155716 | Not-determined |
| S8_extracted_k141_103779 | Not-determined |
| S8_extracted_k141_189825 | Low-quality |
| S8_extracted_k141_189826 | Not-determined |
| S8_extracted_k141_26005 | Not-determined |
| S8_extracted_k141_189827 | Not-determined |
| S8_extracted_k141_189828 | Not-determined |
| S8_extracted_k141_189829 | Not-determined |
| S8_extracted_k141_16920 | Not-determined |
| S8_extracted_k141_189830 | Not-determined |
| S8_extracted_k141_173427 | Not-determined |
| S8_extracted_k141_16921 | Not-determined |
| S8_extracted_k141_155722 | Not-determined |
| S8_extracted_k141_16923 | Not-determined |
| S8_extracted_k141_95249 | Not-determined |
| S8_extracted_k141_26013 | Not-determined |
| S8_extracted_k141_78056 | Not-determined |
| S8_extracted_k141_103783 | Low-quality |
| S8_extracted_k141_69067 | Not-determined |
| S8_extracted_k141_16928 | Not-determined |
| S8_extracted_k141_78058 | Not-determined |
| S8_extracted_k141_8672 | Not-determined |
| S8_extracted_k141_69071 | Not-determined |
| S8_extracted_k141_103786 | Not-determined |
| S8_extracted_k141_112728 | Not-determined |
| S8_extracted_k141_78063 | Not-determined |
| S8_extracted_k141_155731 | Not-determined |
| S8_extracted_k141_173438 | Low-quality |
| S8_extracted_k141_16930 | Not-determined |
| S8_extracted_k141_51014 | Not-determined |
| S8_extracted_k141_189840 | Not-determined |
| S8_extracted_k141_95264 | Not-determined |
| S8_extracted_k141_112733 | Low-quality |
| S8_extracted_k141_189842 | Not-determined |
| S8_extracted_k141_43137 | Not-determined |
| S8_extracted_k141_164292 | Not-determined |
| S8_extracted_k141_8677 | Not-determined |
| S8_extracted_k141_189850 | Not-determined |
| S8_extracted_k141_86644 | Not-determined |
| S8_extracted_k141_164298 | Not-determined |
| S8_extracted_k141_86646 | Not-determined |
| S8_extracted_k141_78084 | Not-determined |
| S8_extracted_k141_147481 | Low-quality |
| S8_extracted_k141_51033 | Not-determined |
| S8_extracted_k141_78090 | Not-determined |
| S8_extracted_k141_86652 | Not-determined |
| S8_extracted_k141_207623 | Low-quality |
| S8_extracted_k141_173449 | Low-quality |
| S8_extracted_k141_189858 | Not-determined |
| S8_extracted_k141_173451 | Not-determined |
| S8_extracted_k141_207627 | Low-quality |
| S8_extracted_k141_130106 | Low-quality |
| S8_extracted_k141_26029 | Not-determined |
| S8_extracted_k141_173454 | Not-determined |
| S8_extracted_k141_51034 | Not-determined |
| S8_extracted_k141_26032 | Not-determined |
| S8_extracted_k141_189862 | Low-quality |
| S8_extracted_k141_207631 | Not-determined |
| S8_extracted_k141_121292 | Low-quality |
| S8_extracted_k141_43147 | Not-determined |
| S8_extracted_k141_207632 | Not-determined |
| S8_extracted_k141_43148 | Not-determined |
| S8_extracted_k141_16946 | Low-quality |
| S8_extracted_k141_130115 | Not-determined |
| S8_extracted_k141_164308 | Not-determined |
| S8_extracted_k141_78095 | Not-determined |
| S8_extracted_k141_164310 | Not-determined |
| S8_extracted_k141_130116 | Not-determined |
| S8_extracted_k141_164311 | Not-determined |
| S8_extracted_k141_130117 | Not-determined |
| S8_extracted_k141_164312 | Not-determined |
| S8_extracted_k141_69086 | Not-determined |
| S8_extracted_k141_189872 | Not-determined |
| S8_extracted_k141_130121 | Not-determined |
| S8_extracted_k141_164315 | Low-quality |
| S8_extracted_k141_155744 | Not-determined |
| S8_extracted_k141_16949 | Not-determined |
| S8_extracted_k141_173472 | Not-determined |
| S8_extracted_k141_189875 | Not-determined |
| S8_extracted_k141_155747 | Not-determined |
| S8_extracted_k141_207639 | Not-determined |
| S8_extracted_k141_207640 | Not-determined |
| S8_extracted_k141_173473 | Not-determined |
| S8_extracted_k141_147502 | Not-determined |
| S8_extracted_k141_130128 | Not-determined |
| S8_extracted_k141_155748 | Not-determined |
| S8_extracted_k141_112744 | Not-determined |
| S8_extracted_k141_164322 | Not-determined |
| S8_extracted_k141_112746 | Low-quality |
| S8_extracted_k141_155749 | Not-determined |
| S8_extracted_k141_16956 | Not-determined |
| S8_extracted_k141_164325 | Not-determined |
| S8_extracted_k141_173479 | Not-determined |
| S8_extracted_k141_155752 | Not-determined |
| S8_extracted_k141_147511 | Not-determined |
| S8_extracted_k141_155755 | Not-determined |
| S8_extracted_k141_43163 | Not-determined |
| S8_extracted_k141_121305 | Not-determined |
| S8_extracted_k141_147517 | Not-determined |
| S8_extracted_k141_95291 | Not-determined |
| S8_extracted_k141_16966 | Not-determined |
| S8_extracted_k141_164327 | Not-determined |
| S8_extracted_k141_43172 | Not-determined |
| S8_extracted_k141_86680 | Not-determined |
| S8_extracted_k141_51040 | Not-determined |
| S8_extracted_k141_164335 | Not-determined |
| S8_extracted_k141_155763 | Not-determined |
| S8_extracted_k141_155765 | Not-determined |
| S8_extracted_k141_164340 | Low-quality |
| S8_extracted_k141_51044 | Not-determined |
| S8_extracted_k141_207659 | Low-quality |
| S8_extracted_k141_164343 | Not-determined |
| S8_extracted_k141_121319 | Not-determined |
| S8_extracted_k141_103803 | Not-determined |
| S8_extracted_k141_103805 | Not-determined |
| S8_extracted_k141_43186 | Not-determined |
| S8_extracted_k141_207661 | Not-determined |
| S8_extracted_k141_130154 | Not-determined |
| S8_extracted_k141_51056 | Not-determined |
| S8_extracted_k141_51059 | Not-determined |
| S8_extracted_k141_16985 | Not-determined |
| S8_extracted_k141_69112 | Not-determined |
| S8_extracted_k141_112763 | Not-determined |
| S8_extracted_k141_95304 | Not-determined |
| S8_extracted_k141_95306 | Not-determined |
| S8_extracted_k141_86692 | Not-determined |
| S8_extracted_k141_51062 | Not-determined |
| S8_extracted_k141_95310 | Low-quality |
| S8_extracted_k141_189898 | Not-determined |
| S8_extracted_k141_43192 | Low-quality |
| S8_extracted_k141_173489 | Not-determined |
| S8_extracted_k141_51067 | Not-determined |
| S8_extracted_k141_112778 | Not-determined |
| S8_extracted_k141_173491 | Not-determined |
| S8_extracted_k141_43195 | Not-determined |
| S8_extracted_k141_51070 | Not-determined |
| S8_extracted_k141_43198 | Not-determined |
| S8_extracted_k141_16994 | Low-quality |
| S8_extracted_k141_155794 | Low-quality |
| S8_extracted_k141_103815 | Not-determined |
| S8_extracted_k141_95323 | Not-determined |
| S8_extracted_k141_112786 | Not-determined |
| S8_extracted_k141_95324 | Low-quality |
| S8_extracted_k141_69124 | Not-determined |
| S8_extracted_k141_121342 | Not-determined |
| S8_extracted_k141_207673 | Not-determined |
| S8_extracted_k141_103818 | Not-determined |
| S8_extracted_k141_95329 | Not-determined |
| S8_extracted_k141_103822 | Not-determined |
| S8_extracted_k141_112793 | Not-determined |
| S8_extracted_k141_189900 | Not-determined |
| S8_extracted_k141_51078 | Not-determined |
| S8_extracted_k141_112794 | Not-determined |
| S8_extracted_k141_95333 | Not-determined |
| S8_extracted_k141_103824 | Not-determined |
| S8_extracted_k141_51079 | Not-determined |
| S8_extracted_k141_189902 | Not-determined |
| S8_extracted_k141_86707 | Not-determined |
| S8_extracted_k141_112797 | Not-determined |
| S8_extracted_k141_103828 | Not-determined |
| S8_extracted_k141_121350 | Not-determined |
| S8_extracted_k141_51084 | Not-determined |
| S8_extracted_k141_103831 | Not-determined |
| S8_extracted_k141_173508 | Not-determined |
| [illegible] | [illegible] |
| S8_extracted_k141_69134 | Not-determined |
| S8_extracted_k141_112806 | Not-determined |
| S8_extracted_k141_69136 | Not-determined |
| S8_extracted_k141_51097 | Not-determined |
| S8_extracted_k141_86720 | Not-determined |
| S8_extracted_k141_173525 | Not-determined |
| S8_extracted_k141_95350 | Not-determined |
| S8_extracted_k141_51102 | Not-determined |
| S8_extracted_k141_207692 | Not-determined |
| S8_extracted_k141_86723 | Not-determined |
| S8_extracted_k141_17001 | Not-determined |
| S8_extracted_k141_189919 | Not-determined |
| S8_extracted_k141_103839 | Not-determined |
| S8_extracted_k141_95353 | Not-determined |
| S8_extracted_k141_69142 | Not-determined |
| S8_extracted_k141_86729 | Not-determined |
| S8_extracted_k141_69143 | Not-determined |
| S8_extracted_k141_207698 | Not-determined |
| S8_extracted_k141_164373 | Not-determined |
| S8_extracted_k141_189930 | Not-determined |
| S8_extracted_k141_189933 | Not-determined |
| S8_extracted_k141_51117 | Not-determined |
| S8_extracted_k141_95360 | Not-determined |
| S8_extracted_k141_207701 | Not-determined |
| S8_extracted_k141_189939 | Low-quality |
| S8_extracted_k141_69155 | Not-determined |
| S8_extracted_k141_121391 | Not-determined |
| S8_extracted_k141_51125 | Not-determined |
| S8_extracted_k141_164378 | Not-determined |
| S8_extracted_k141_173533 | Not-determined |
| S8_extracted_k141_69157 | Not-determined |
| S8_extracted_k141_69161 | Not-determined |
| S8_extracted_k141_173537 | Not-determined |
| S8_extracted_k141_121397 | Not-determined |
| S8_extracted_k141_121398 | Not-determined |
| S8_extracted_k141_95371 | Not-determined |
| S8_extracted_k141_95372 | Not-determined |
| S8_extracted_k141_103864 | Not-determined |
| S8_extracted_k141_51136 | Not-determined |
| S8_extracted_k141_103866 | Not-determined |
| S8_extracted_k141_164385 | Not-determined |
| S8_extracted_k141_189961 | Not-determined |
| S8_extracted_k141_86760 | Not-determined |
| S8_extracted_k141_86761 | Not-determined |
| S8_extracted_k141_86764 | Not-determined |
| S8_extracted_k141_138617 | Not-determined |
| S8_extracted_k141_95377 | Not-determined |
| S8_extracted_k141_86766 | Not-determined |
| S8_extracted_k141_17013 | Low-quality |
| S8_extracted_k141_69182 | Not-determined |
| S8_extracted_k141_69183 | Not-determined |
| S8_extracted_k141_138621 | Not-determined |
| S8_extracted_k141_155829 | Not-determined |
| S8_extracted_k141_51149 | Not-determined |
| S8_extracted_k141_95381 | Low-quality |
| S8_extracted_k141_51153 | Low-quality |
| S8_extracted_k141_138624 | Not-determined |
| S8_extracted_k141_121420 | Not-determined |
| S8_extracted_k141_121422 | Not-determined |
| S8_extracted_k141_155838 | Low-quality |
| S8_extracted_k141_164391 | Not-determined |
| S8_extracted_k141_155839 | Not-determined |
| S8_extracted_k141_164394 | Not-determined |
| S8_extracted_k141_86777 | Low-quality |
| S8_extracted_k141_17032 | Not-determined |
| S8_extracted_k141_17033 | Not-determined |
| S8_extracted_k141_95391 | Not-determined |
| S8_extracted_k141_121428 | Low-quality |
| S8_extracted_k141_69198 | Not-determined |
| S8_extracted_k141_17038 | Low-quality |
| S8_extracted_k141_95395 | Not-determined |
| S8_extracted_k141_51182 | Not-determined |
| S8_extracted_k141_155842 | Not-determined |
| S8_extracted_k141_69204 | Not-determined |
| S8_extracted_k141_69206 | Not-determined |
| S8_extracted_k141_69210 | Not-determined |
| S8_extracted_k141_189979 | Not-determined |
| S8_extracted_k141_17044 | Low-quality |

| | |
|---|---|
| S8_extracted_k141_103889 | Not-determined |
| S8_extracted_k141_138647 | Not-determined |
| S8_extracted_k141_69223 | Not-determined |
| S8_extracted_k141_138648 | Not-determined |
| S8_extracted_k141_95412 | Not-determined |
| S8_extracted_k141_95413 | Not-determined |
| S8_extracted_k141_17055 | Not-determined |
| S8_extracted_k141_103892 | Not-determined |
| S8_extracted_k141_207740 | Not-determined |
| S8_extracted_k141_51206 | Not-determined |
| S8_extracted_k141_69229 | Not-determined |
| S8_extracted_k141_95416 | Not-determined |
| S8_extracted_k141_138650 | Not-determined |
| S8_extracted_k141_69233 | Not-determined |
| S8_extracted_k141_121457 | Not-determined |
| S8_extracted_k141_51214 | Not-determined |
| S8_extracted_k141_138657 | Not-determined |
| S8_extracted_k141_103893 | Not-determined |
| S8_extracted_k141_95419 | Not-determined |
| S8_extracted_k141_164416 | Not-determined |
| S8_extracted_k141_164417 | Not-determined |
| S8_extracted_k141_121460 | Not-determined |
| S8_extracted_k141_121464 | Not-determined |
| S8_extracted_k141_121467 | Not-determined |
| S8_extracted_k141_190011 | Not-determined |
| S8_extracted_k141_190012 | Not-determined |
| S8_extracted_k141_190013 | Not-determined |
| S8_extracted_k141_69241 | Not-determined |
| S8_extracted_k141_43217 | Not-determined |
| S8_extracted_k141_121471 | Not-determined |
| S8_extracted_k141_95429 | Not-determined |
| S8_extracted_k141_190023 | Not-determined |
| S8_extracted_k141_190026 | Not-determined |
| S8_extracted_k141_121479 | Not-determined |
| S8_extracted_k141_69255 | Not-determined |
| S8_extracted_k141_164435 | Not-determined |
| S8_extracted_k141_121482 | Low-quality |
| S8_extracted_k141_121484 | Low-quality |
| S8_extracted_k141_43233 | Low-quality |
| S8_extracted_k141_155859 | Not-determined |
| S8_extracted_k141_190038 | Not-determined |
| S8_extracted_k141_51233 | Not-determined |
| S8_extracted_k141_103915 | Not-determined |
| S8_extracted_k141_103917 | Not-determined |
| S8_extracted_k141_138702 | Not-determined |
| S8_extracted_k141_51238 | Not-determined |
| S8_extracted_k141_103935 | Not-determined |
| S8_extracted_k141_51243 | Not-determined |
| S8_extracted_k141_69284 | Low-quality |
| S8_extracted_k141_69285 | Not-determined |
| S8_extracted_k141_69293 | Not-determined |
| S8_extracted_k141_164452 | Not-determined |
| S8_extracted_k141_155879 | Low-quality |
| S8_extracted_k141_164457 | Not-determined |
| S8_extracted_k141_155880 | Low-quality |
| S8_extracted_k141_51254 | Low-quality |
| S8_extracted_k141_164458 | Not-determined |
| S8_extracted_k141_43268 | Not-determined |
| S8_extracted_k141_164462 | Not-determined |
| S8_extracted_k141_155887 | Low-quality |
| S8_extracted_k141_51266 | Not-determined |
| S8_extracted_k141_17063 | Not-determined |
| S8_extracted_k141_69299 | Not-determined |
| S8_extracted_k141_51275 | Not-determined |
| S8_extracted_k141_138720 | Not-determined |
| S8_extracted_k141_51278 | Not-determined |
| S8_extracted_k141_51279 | Not-determined |
| S8_extracted_k141_17068 | Low-quality |
| S8_extracted_k141_164474 | Not-determined |
| S8_extracted_k141_43277 | Low-quality |
| S8_extracted_k141_95451 | Not-determined |
| S8_extracted_k141_164475 | Not-determined |
| S8_extracted_k141_155900 | Low-quality |
| S8_extracted_k141_17076 | Not-determined |
| S8_extracted_k141_43281 | Not-determined |
| S8_extracted_k141_155903 | Not-determined |
| S8_extracted_k141_164483 | Not-determined |
| S8_extracted_k141_138732 | Not-determined |
| S8_extracted_k141_43286 | Not-determined |
| S8_extracted_k141_51295 | Not-determined |
| S8_extracted_k141_51296 | Not-determined |
| S8_extracted_k141_155912 | Not-determined |
| S8_extracted_k141_155913 | Not-determined |
| S8_extracted_k141_69324 | Not-determined |
| S8_extracted_k141_17086 | Not-determined |
| S8_extracted_k141_190090 | Not-determined |
| S8_extracted_k141_17090 | Not-determined |
| S8_extracted_k141_190098 | Not-determined |
| S8_extracted_k141_155920 | Not-determined |
| S8_extracted_k141_164496 | Not-determined |
| S8_extracted_k141_17093 | Not-determined |
| S8_extracted_k141_190102 | Not-determined |
| S8_extracted_k141_155923 | Not-determined |
| S8_extracted_k141_51317 | Low-quality |
| S8_extracted_k141_17102 | Not-determined |
| S8_extracted_k141_43298 | Not-determined |
| S8_extracted_k141_155935 | Not-determined |
| S8_extracted_k141_43301 | Low-quality |
| S8_extracted_k141_17107 | Low-quality |
| S8_extracted_k141_190107 | Not-determined |
| S8_extracted_k141_155938 | Not-determined |
| S8_extracted_k141_138755 | Not-determined |
| S8_extracted_k141_155942 | Not-determined |
| S8_extracted_k141_155944 | Not-determined |
| S8_extracted_k141_69363 | Not-determined |
| S8_extracted_k141_164514 | Not-determined |
| S8_extracted_k141_155948 | Low-quality |
| S8_extracted_k141_164519 | Not-determined |
| S8_extracted_k141_164520 | Not-determined |
| S8_extracted_k141_164521 | Not-determined |
| S8_extracted_k141_43315 | Not-determined |
| S8_extracted_k141_69369 | Not-determined |
| S8_extracted_k141_190134 | Not-determined |
| S8_extracted_k141_190137 | Not-determined |
| S8_extracted_k141_43322 | Low-quality |
| S8_extracted_k141_155952 | Not-determined |
| S8_extracted_k141_69381 | Not-determined |
| S8_extracted_k141_138765 | Not-determined |
| S8_extracted_k141_69383 | Not-determined |
| S8_extracted_k141_43332 | Not-determined |
| S8_extracted_k141_69392 | Not-determined |
| S8_extracted_k141_164535 | Not-determined |
| S8_extracted_k141_164540 | Not-determined |
| S8_extracted_k141_43338 | Not-determined |
| S8_extracted_k141_69399 | Not-determined |
| S8_extracted_k141_43342 | Not-determined |
| S8_extracted_k141_155964 | Low-quality |
| S8_extracted_k141_43346 | Not-determined |
| S8_extracted_k141_155965 | Not-determined |
| S8_extracted_k141_43349 | Not-determined |
| S8_extracted_k141_43351 | Not-determined |
| S8_extracted_k141_138784 | Not-determined |
| S8_extracted_k141_207755 | Not-determined |
| S8_extracted_k141_164548 | Low-quality |
| S8_extracted_k141_190160 | Not-determined |
| S8_extracted_k141_43357 | Not-determined |
| S8_extracted_k141_43358 | Not-determined |
| S8_extracted_k141_43359 | Not-determined |
| S8_extracted_k141_164552 | Not-determined |
| S8_extracted_k141_190164 | Not-determined |
| S8_extracted_k141_43362 | Not-determined |
| S8_extracted_k141_190167 | Not-determined |
| S8_extracted_k141_43363 | Not-determined |
| S8_extracted_k141_164556 | Not-determined |
| S8_extracted_k141_207760 | Not-determined |
| S8_extracted_k141_43367 | Not-determined |
| S8_extracted_k141_207763 | Not-determined |
| S8_extracted_k141_69420 | Low-quality |
| S8_extracted_k141_69422 | Not-determined |
| S8_extracted_k141_43370 | Low-quality |
| S8_extracted_k141_164567 | Not-determined |
| S8_extracted_k141_190177 | Not-determined |
| S8_extracted_k141_138799 | Not-determined |
| S8_extracted_k141_155988 | Low-quality |
| S8_extracted_k141_190184 | Low-quality |
| S8_extracted_k141_164573 | Not-determined |
| S8_extracted_k141_207777 | Not-determined |
| S8_extracted_k141_43381 | Not-determined |
| S8_extracted_k141_190191 | Not-determined |
| S8_extracted_k141_207783 | Not-determined |
| S8_extracted_k141_43383 | Not-determined |
| S8_extracted_k141_190196 | Low-quality |
| S8_extracted_k141_43386 | Not-determined |
| S8_extracted_k141_164590 | Not-determined |
| S8_extracted_k141_190207 | Not-determined |
| S8_extracted_k141_156009 | Not-determined |
| S8_extracted_k141_207792 | Not-determined |
| S8_extracted_k141_156012 | Low-quality |
| S8_extracted_k141_156013 | Not-determined |
| S8_extracted_k141_207795 | Not-determined |
| S8_extracted_k141_103950 | Not-determined |
| S8_extracted_k141_138801 | Low-quality |
| S8_extracted_k141_51324 | Not-determined |
| S8_extracted_k141_138805 | Not-determined |
| S8_extracted_k141_51330 | Low-quality |
| S8_extracted_k141_156022 | Not-determined |
| S8_extracted_k141_51334 | Not-determined |
| S8_extracted_k141_103962 | Not-determined |
| S8_extracted_k141_207806 | Not-determined |
| S8_extracted_k141_190216 | Not-determined |
| S8_extracted_k141_51337 | Not-determined |
| S8_extracted_k141_164607 | Not-determined |
| S8_extracted_k141_51347 | Not-determined |
| S8_extracted_k141_51348 | Not-determined |
| S8_extracted_k141_190230 | Low-quality |
| S8_extracted_k141_164611 | Low-quality |
| S8_extracted_k141_51355 | Low-quality |
| S8_extracted_k141_51356 | Not-determined |
| S8_extracted_k141_51357 | Not-determined |
| S8_extracted_k141_190241 | Not-determined |
| S8_extracted_k141_156038 | Not-determined |
| S8_extracted_k141_51358 | Not-determined |
| S8_extracted_k141_156042 | Not-determined |
| S8_extracted_k141_156043 | Not-determined |
| S8_extracted_k141_156047 | Not-determined |
| S8_extracted_k141_103975 | Not-determined |
| S8_extracted_k141_103976 | Not-determined |
| S8_extracted_k141_103978 | Not-determined |
| S8_extracted_k141_190254 | Not-determined |
| S8_extracted_k141_190258 | Not-determined |
| S8_extracted_k141_138837 | Not-determined |
| S8_extracted_k141_103985 | Not-determined |
| S8_extracted_k141_17115 | Not-determined |
| S8_extracted_k141_190265 | Not-determined |
| S8_extracted_k141_190269 | Not-determined |
| S8_extracted_k141_103988 | Not-determined |
| S8_extracted_k141_207812 | Not-determined |
| S8_extracted_k141_190271 | Not-determined |
| S8_extracted_k141_207815 | Not-determined |
| S8_extracted_k141_190272 | Not-determined |
| S8_extracted_k141_17122 | Not-determined |
| S8_extracted_k141_164652 | Not-determined |
| S8_extracted_k141_17126 | Not-determined |
| S8_extracted_k141_207817 | Not-determined |
| S8_extracted_k141_190276 | Not-determined |
| S8_extracted_k141_156057 | Not-determined |
| S8_extracted_k141_207827 | Not-determined |
| S8_extracted_k141_164667 | Not-determined |
| S8_extracted_k141_104001 | Not-determined |
| S8_extracted_k141_156067 | Not-determined |
| S8_extracted_k141_104005 | Not-determined |
| S8_extracted_k141_156069 | Not-determined |
| S8_extracted_k141_207837 | Not-determined |
| S8_extracted_k141_164672 | Not-determined |
| S8_extracted_k141_164673 | Not-determined |
| S8_extracted_k141_207842 | Not-determined |
| S8_extracted_k141_17152 | Not-determined |
| S8_extracted_k141_207846 | Not-determined |
| S8_extracted_k141_190290 | Low-quality |
| S8_extracted_k141_156078 | Not-determined |
| S8_extracted_k141_190294 | Not-determined |
| S8_extracted_k141_190305 | Not-determined |
| S8_extracted_k141_207867 | Not-determined |
| S8_extracted_k141_207871 | Low-quality |
| S8_extracted_k141_164697 | Not-determined |
| S8_extracted_k141_51371 | Not-determined |
| S8_extracted_k141_17168 | Not-determined |
| S8_extracted_k141_156086 | Not-determined |
| S8_extracted_k141_51375 | Not-determined |
| S8_extracted_k141_156089 | Low-quality |
| S8_extracted_k141_104045 | Not-determined |
| S8_extracted_k141_17169 | Not-determined |
| S8_extracted_k141_207880 | Not-determined |
| S8_extracted_k141_190321 | Low-quality |
| S8_extracted_k141_104047 | Not-determined |
| S8_extracted_k141_207888 | Not-determined |
| S8_extracted_k141_190332 | Not-determined |
| S8_extracted_k141_190339 | Not-determined |
| S8_extracted_k141_156097 | Not-determined |
| S8_extracted_k141_104051 | Not-determined |
| S8_extracted_k141_51381 | Not-determined |
| S8_extracted_k141_207892 | Not-determined |
| S8_extracted_k141_156101 | Not-determined |
| S8_extracted_k141_51385 | Not-determined |
| S8_extracted_k141_207894 | Not-determined |
| S8_extracted_k141_207895 | Not-determined |
| S8_extracted_k141_207896 | Not-determined |
| S8_extracted_k141_17177 | Not-determined |
| S8_extracted_k141_207897 | Not-determined |
| S8_extracted_k141_164725 | Not-determined |
| S8_extracted_k141_207901 | Not-determined |
| S8_extracted_k141_17185 | Not-determined |
| S8_extracted_k141_156105 | Not-determined |
| S8_extracted_k141_207910 | Not-determined |
| S8_extracted_k141_156113 | Not-determined |
| S8_extracted_k141_207914 | Not-determined |
| S8_extracted_k141_164733 | Not-determined |
| S8_extracted_k141_156122 | Not-determined |
| S8_extracted_k141_207921 | Not-determined |
| S8_extracted_k141_104059 | Not-determined |
| S8_extracted_k141_104060 | Not-determined |
| S8_extracted_k141_207930 | Not-determined |
| S8_extracted_k141_207934 | Not-determined |
| S8_extracted_k141_104065 | Not-determined |
| S8_extracted_k141_207939 | Low-quality |
| S8_extracted_k141_156131 | Not-determined |
| S8_extracted_k141_156132 | Not-determined |
| S8_extracted_k141_207946 | Not-determined |
| S8_extracted_k141_207950 | Not-determined |
| S8_extracted_k141_17199 | Not-determined |
| S8_extracted_k141_17205 | Not-determined |
| S8_extracted_k141_17206 | Low-quality |
| S8_extracted_k141_164745 | Not-determined |
| S8_extracted_k141_51425 | Not-determined |
| S8_extracted_k141_51427 | Not-determined |
| S8_extracted_k141_51428 | Not-determined |
| S8_extracted_k141_51430 | Not-determined |
| S8_extracted_k141_17212 | Not-determined |
| S8_extracted_k141_156143 | Not-determined |
| S8_extracted_k141_17217 | Low-quality |
| S8_extracted_k141_104091 | Not-determined |
| S8_extracted_k141_104093 | Not-determined |
| S8_extracted_k141_104096 | Not-determined |
| S8_extracted_k141_104098 | Not-determined |
| S8_extracted_k141_51446 | Not-determined |
| S8_extracted_k141_190362 | Not-determined |
| S8_extracted_k141_156154 | Not-determined |
| S8_extracted_k141_164763 | Not-determined |
| S8_extracted_k141_207972 | Not-determined |
| S8_extracted_k141_17235 | Not-determined |
| S8_extracted_k141_17237 | Not-determined |
| S8_extracted_k141_17239 | Not-determined |
| S8_extracted_k141_156161 | Low-quality |
| S8_extracted_k141_104130 | Not-determined |
| S8_extracted_k141_51463 | Not-determined |
| S8_extracted_k141_104131 | Not-determined |
| S8_extracted_k141_156168 | Not-determined |
| S8_extracted_k141_51466 | Not-determined |
| S8_extracted_k141_190379 | Not-determined |
| S8_extracted_k141_51469 | Not-determined |
| S8_extracted_k141_190382 | Not-determined |
| S8_extracted_k141_51473 | Not-determined |
| S8_extracted_k141_190387 | Not-determined |
| S8_extracted_k141_51474 | Not-determined |
| S8_extracted_k141_207997 | Not-determined |
| S8_extracted_k141_190391 | Not-determined |
| S8_extracted_k141_164791 | Not-determined |
| S8_extracted_k141_208002 | Not-determined |
| S8_extracted_k141_17264 | Not-determined |
| S8_extracted_k141_156173 | Not-determined |
| S8_extracted_k141_17271 | Not-determined |
| S8_extracted_k141_51490 | Not-determined |
| S8_extracted_k141_190402 | Not-determined |
| S8_extracted_k141_208012 | Not-determined |
| S8_extracted_k141_17281 | Not-determined |
| S8_extracted_k141_17282 | Not-determined |
| S8_extracted_k141_208018 | Not-determined |
| S8_extracted_k141_51501 | Not-determined |
| S8_extracted_k141_51502 | Not-determined |
| S8_extracted_k141_208020 | Not-determined |
| S8_extracted_k141_164806 | Not-determined |
| S8_extracted_k141_17288 | Not-determined |
| S8_extracted_k141_208025 | Not-determined |
| S8_extracted_k141_208035 | Not-determined |
| S8_extracted_k141_51509 | Not-determined |
| S8_extracted_k141_208043 | Not-determined |
| S8_extracted_k141_208046 | Not-determined |
| S8_extracted_k141_17294 | Low-quality |
| S8_extracted_k141_17302 | Not-determined |
| S8_extracted_k141_17305 | Not-determined |
| S8_extracted_k141_190426 | Not-determined |
| S8_extracted_k141_17308 | Not-determined |
| S8_extracted_k141_164809 | Low-quality |
| S8_extracted_k141_208064 | Not-determined |
| S8_extracted_k141_208065 | Not-determined |
| S8_extracted_k141_17316 | Not-determined |
| S8_extracted_k141_208070 | Not-determined |
| S8_extracted_k141_208071 | Not-determined |
| S8_extracted_k141_190450 | Not-determined |
| S8_extracted_k141_190452 | Not-determined |
| S8_extracted_k141_208078 | Not-determined |
| S8_extracted_k141_190455 | Not-determined |
| S8_extracted_k141_164817 | Not-determined |
| S8_extracted_k141_164818 | Not-determined |
| S8_extracted_k141_190458 | Not-determined |
| S8_extracted_k141_208082 | Not-determined |
| S8_extracted_k141_164823 | Not-determined |
| S8_extracted_k141_51512 | Not-determined |
| S8_extracted_k141_51515 | Not-determined |
| S8_extracted_k141_208083 | Low-quality |
| S8_extracted_k141_51517 | Not-determined |
| S8_extracted_k141_164831 | Not-determined |
| S8_extracted_k141_51521 | Not-determined |
| S8_extracted_k141_190476 | Low-quality |
| S8_extracted_k141_190477 | Not-determined |
| S8_extracted_k141_190480 | Not-determined |
| S8_extracted_k141_51525 | Not-determined |
| S8_extracted_k141_190481 | Not-determined |
| S8_extracted_k141_190483 | Not-determined |
| S8_extracted_k141_190489 | Not-determined |
| S8_extracted_k141_164849 | Not-determined |
| S8_extracted_k141_190491 | Not-determined |
| S8_extracted_k141_190493 | Not-determined |
| S8_extracted_k141_190502 | Not-determined |
| S8_extracted_k141_164860 | Not-determined |
| S8_extracted_k141_51542 | Low-quality |
| S8_extracted_k141_51545 | Not-determined |
| S8_extracted_k141_164865 | Not-determined |
| S8_extracted_k141_164866 | Not-determined |
| S8_extracted_k141_17325 | Not-determined |
| S8_extracted_k141_17326 | Not-determined |
| S8_extracted_k141_51548 | Low-quality |
| S8_extracted_k141_17331 | Not-determined |
| S8_extracted_k141_17342 | Not-determined |
| S8_extracted_k141_190537 | Not-determined |
| S8_extracted_k141_51572 | Not-determined |
| S8_extracted_k141_190538 | Not-determined |
| S8_extracted_k141_51576 | Not-determined |
| S8_extracted_k141_17346 | Low-quality |
| S8_extracted_k141_51578 | Not-determined |
| S8_extracted_k141_190543 | Not-determined |
| S8_extracted_k141_17350 | Not-determined |
| S3_extracted_k141_127168 | Complete |
| S8_extracted_k141_51585 | Not-determined |
| S8_extracted_k141_51589 | Not-determined |
| S8_extracted_k141_51594 | Not-determined |
| S8_extracted_k141_51598 | Not-determined |
| S8_extracted_k141_190558 | Not-determined |
| S8_extracted_k141_51609 | Not-determined |
| S8_extracted_k141_51611 | Not-determined |
| S8_extracted_k141_190562 | Not-determined |
| S8_extracted_k141_51621 | Not-determined |
| S8_extracted_k141_51625 | Not-determined |
| S8_extracted_k141_51626 | Not-determined |
| S8_extracted_k141_51630 | Low-quality |
| S8_extracted_k141_190567 | Low-quality |
| S8_extracted_k141_190570 | Not-determined |
| S8_extracted_k141_51633 | Not-determined |
| S8_extracted_k141_190591 | Not-determined |
| S8_extracted_k141_51641 | Low-quality |
| S8_extracted_k141_190597 | Not-determined |
| S8_extracted_k141_51644 | Not-determined |
| S8_extracted_k141_51646 | Not-determined |
| S8_extracted_k141_190603 | Not-determined |
| S8_extracted_k141_51650 | Not-determined |
| S8_extracted_k141_190608 | Not-determined |
| S8_extracted_k141_190613 | Not-determined |
| S8_extracted_k141_51652 | Low-quality |
| S8_extracted_k141_51656 | Not-determined |
| S8_extracted_k141_51671 | Not-determined |
| S8_extracted_k141_51672 | Not-determined |
| S8_extracted_k141_190633 | Not-determined |
| S8_extracted_k141_190635 | Not-determined |
| S8_extracted_k141_51679 | Not-determined |
| S8_extracted_k141_190642 | Not-determined |
| S8_extracted_k141_51683 | Low-quality |
| S8_extracted_k141_51689 | Not-determined |
| S8_extracted_k141_51692 | Not-determined |
| S8_extracted_k141_51693 | Not-determined |
| S8_extracted_k141_51694 | Not-determined |
| S8_extracted_k141_51700 | Not-determined |
| S8_extracted_k141_51703 | Not-determined |
| S8_extracted_k141_51707 | Not-determined |
| S8_extracted_k141_51708 | Not-determined |
| S8_extracted_k141_190695 | Low-quality |
| S8_extracted_k141_51736 | Low-quality |
| S8_extracted_k141_51743 | Not-determined |
| S8_extracted_k141_190712 | Low-quality |
| S8_extracted_k141_51755 | Not-determined |
| S8_extracted_k141_51766 | Not-determined |
| S8_extracted_k141_51768 | Not-determined |
| S8_extracted_k141_51781 | Not-determined |
| S8_extracted_k141_51797 | Not-determined |
| S8_extracted_k141_190739 | Not-determined |
| S8_extracted_k141_190744 | Not-determined |
| S8_extracted_k141_190746 | Not-determined |
| S8_extracted_k141_190748 | Not-determined |
| S8_extracted_k141_190749 | Not-determined |
| S8_extracted_k141_190752 | Not-determined |
| S8_extracted_k141_51807 | Not-determined |
| S8_extracted_k141_190762 | Not-determined |
| S8_extracted_k141_51811 | Not-determined |
| S8_extracted_k141_190766 | Not-determined |
| S8_extracted_k141_190776 | Not-determined |
| S8_extracted_k141_51829 | Not-determined |
| S8_extracted_k141_190781 | Not-determined |
| S8_extracted_k141_190784 | Low-quality |
| S8_extracted_k141_190787 | Not-determined |
| S8_extracted_k141_51838 | Low-quality |
| S8_extracted_k141_190802 | Not-determined |
| S8_extracted_k141_190810 | Not-determined |
| S8_extracted_k141_190819 | Not-determined |
| S8_extracted_k141_190821 | Not-determined |
| S8_extracted_k141_51853 | Low-quality |
| S8_extracted_k141_51871 | Not-determined |
| S8_extracted_k141_190843 | Not-determined |
| S8_extracted_k141_190867 | Not-determined |
| S12_extracted_k141_8173 | Medium-quality |
| S8_extracted_k141_190869 | Not-determined |
| S8_extracted_k141_190871 | Not-determined |
| S8_extracted_k141_190872 | Not-determined |
| S8_extracted_k141_190874 | Low-quality |
| S8_extracted_k141_190877 | Not-determined |
| S8_extracted_k141_51903 | Not-determined |
| S8_extracted_k141_190895 | Low-quality |
| S15_extracted_71616 | Medium-quality |
| S8_extracted_k141_51947 | Low-quality |
| S8_extracted_k141_51955 | Not-determined |
| S8_extracted_k141_51966 | Not-determined |
| S8_extracted_k141_51972 | Not-determined |
| S8_extracted_k141_51994 | Not-determined |
| S8_extracted_k141_52001 | Not-determined |
| S8_extracted_k141_52002 | Not-determined |
| S8_extracted_k141_52007 | Not-determined |
| S8_extracted_k141_52008 | Not-determined |
| S8_extracted_k141_52014 | Not-determined |
| S8_extracted_k141_52020 | Not-determined |
| S8_extracted_k141_52042 | Not-determined |
| S9_extracted_k141_34736 | Medium-quality |
| S8_extracted_k141_52061 | Not-determined |
| S11_vs_k141_37666\|\|full | Medium-quality |
| S15_vs_71616\|\|full | Medium-quality |
| S9_vs_k141_34736\|\|full | Medium-quality |
| S8_extracted_k141_208116 | Not-determined |
| S5_vs_k141_142450\|\|full | Medium-quality |
| S6_vs_k141_3\|\|full | Medium-quality |
| S2_extracted_k141_3356 | Complete |
| S8_extracted_k141_208143 | Not-determined |
| S8_extracted_k141_208144 | Not-determined |
| S8_extracted_k141_208145 | Not-determined |
| S10_vs_57705\|\|full | Medium-quality |
| S8_extracted_k141_208151 | Not-determined |
| S8_extracted_k141_208152 | Not-determined |
| S8_extracted_k141_208153 | Not-determined |
| S8_extracted_k141_208158 | Not-determined |
| S8_extracted_k141_208161 | Not-determined |
| S8_extracted_k141_208163 | Not-determined |
| S8_extracted_k141_208173 | Not-determined |
| S8_extracted_k141_208177 | Not-determined |
| S8_extracted_k141_208178 | Low-quality |
| S8_extracted_k141_208179 | Not-determined |
| S8_extracted_k141_208183 | Not-determined |
| S8_extracted_k141_208187 | Not-determined |
| S8_extracted_k141_208188 | Not-determined |
| S8_extracted_k141_208193 | Not-determined |
| S8_extracted_k141_208195 | Not-determined |
| S8_extracted_k141_208196 | Not-determined |
| S8_extracted_k141_208210 | Not-determined |
| S8_extracted_k141_208211 | Not-determined |
| S8_extracted_k141_208212 | Low-quality |
| S8_extracted_k141_208219 | Low-quality |
| S8_extracted_k141_208225 | Not-determined |
| S8_extracted_k141_208227 | Not-determined |
| S8_extracted_k141_208229 | Not-determined |
| S8_extracted_k141_208230 | Not-determined |
| S8_extracted_k141_208233 | Not-determined |
| S8_extracted_k141_208236 | Not-determined |
| S8_extracted_k141_208241 | Not-determined |
| S8_extracted_k141_208242 | Not-determined |
| S8_extracted_k141_208248 | Not-determined |
| S8_extracted_k141_208253 | Not-determined |
| S8_vs_k141_156354\|\|full | Not-determined |
| S7_extracted_27437 | Complete |
| S15_extracted_55639 | High-quality |
| S12_vs_k141_8173\|\|full | Medium-quality |
| S11_vs_k141_104053\|\|full | High-quality |
| S7_vs_138188\|\|full | High-quality |
| S8_vs_k141_156882\|\|full | Not-determined |
| S8_vs_k141_156883\|\|full | Not-determined |
| S14_extracted_k141_45290 | High-quality |
| S8_vs_k141_87809\|\|full | Not-determined |
| S3_extracted_k141_83074 | Medium-quality |
| S14_extracted_k141_109356 | Complete |
| S3_vs_k141_127168\|\|full | High-quality |
| S6_vs_k141_56493\|\|full | High-quality |
| S8_vs_k141_182422\|\|full | Not-determined |
| S2_vs_k141_3356\|\|full | High-quality |
| S13_extracted_131208 | Medium-quality |
| S7_vs_27437\|\|full | High-quality |
| S15_vs_55639\|\|full | High-quality |
| S7_vs_119492\|\|full | Medium-quality |
| S4_extracted_2524 | High-quality |
| S4_vs_VIRSorter_2524-cat_2 | High-quality |
| S8_vs_k141_115089\|\|full | Not-determined |
| S8_vs_k141_176212\|\|full | Low-quality |
| S14_vs_k141_109356\|\|full | High-quality |
| S3_vs_k141_83074\|\|full | Medium-quality |
| S2_extracted_k141_12860 | Medium-quality |
| S8_vs_k141_81164\|\|full | Not-determined |
| S13_extracted_91189 | High-quality |
| S15_vs_1407\|\|full | High-quality |
| S14_vs_k141_45290\|\|full | High-quality |
| S4_extracted_16485 | High-quality |
| S8_vs_k141_3621\|\|full | Not-determined |
| S16_extracted_173133 | Complete |
| S8_vs_k141_4221\|\|full | Not-determined |
| S17_vs_k141_58025\|\|full | High-quality |
| S8_vs_k141_39683\|\|full | Not-determined |
| S2_vs_k141_12860\|\|full | Medium-quality |
| S8_vs_k141_68398\|\|full | High-quality |
| S8_vs_k141_196354\|\|full | Low-quality |
| S13_vs_131208\|\|full | Medium-quality |
| S8_vs_k141_31758\|\|full | Low-quality |
| S7_extracted_71527 | High-quality |
| S16_vs_173133\|\|full | Medium-quality |
| S3_extracted_k141_84832 | Medium-quality |
| S3_vs_k141_87896\|\|full | Medium-quality |
| S7_vs_71527\|\|full | High-quality |
| S8_vs_k141_147256\|\|full | Not-determined |
| S13_vs_91189\|\|full | High-quality |
| S9_vs_k141_75290\|\|full | High-quality |
| S13_extracted_1770 | High-quality |
| S9_extracted_k141_75363 | Medium-quality |
| S3_vs_k141_84832\|\|full | Medium-quality |
| S8_vs_k141_208108\|\|full | Not-determined |
| S6_extracted_k141_47238 | High-quality |
| S5_extracted_k141_63825 | Complete |
| S9_vs_k141_75363\|\|full | Medium-quality |
| S13_vs_1770\|\|full | High-quality |

S5_vs_k141_124035||full High-quality
S17_extracted_k141_172132 Complete
S7_extracted_47343 Complete
S17_vs_k141_172132||full Complete
S7_vs_47343||full Complete
S6_extracted_k141_3435 Medium-quality
S6_vs_k141_3435||full Medium-quality
S5_vs_k141_63825||full High-quality
S8_vs_k141_208135||full Low-quality
S8_vs_k141_208142||full Not-determined
S8_vs_k141_208144||full Not-determined
S6_vs_k141_47238||full High-quality
S8_vs_k141_208147||full Not-determined
S8_vs_k141_208148||full Low-quality
S8_vs_k141_208152||full Not-determined
S8_vs_k141_208157||full Not-determined
S8_vs_k141_112815||full Low-quality
S8_vs_k141_26037||full Low-quality
S8_vs_k141_43399||full Low-quality
S8_vs_k141_104140||full Low-quality
S8_vs_k141_34726||full Low-quality
S8_vs_k141_121492||full Low-quality
S8_vs_k141_95469||full Not-determined
S8_vs_k141_156209||full Low-quality
S8_vs_k141_182234||full Not-determined
S8_vs_k141_86789||full Low-quality
S8_vs_k141_17371||full Not-determined
S8_vs_k141_121505||full Not-determined
S8_vs_k141_190930||full Not-determined
S8_vs_k141_121509||full Not-determined
S8_vs_k141_104154||full Low-quality
S8_vs_k141_147564||full Low-quality
S8_vs_k141_34||full Not-determined
S8_vs_k141_69448||full Not-determined
S8_vs_k141_26077||full Low-quality
S8_vs_k141_130187||full Not-determined
S8_vs_k141_78120||full Not-determined
S8_vs_k141_26081||full Not-determined
S8_vs_k141_199613||full Low-quality
S8_vs_k141_34765||full Not-determined
S8_vs_k141_56||full Not-determined
S8_vs_k141_43432||full Not-determined
S8_vs_k141_156254||full Low-quality
S8_vs_k141_199617||full Low-quality
S8_vs_k141_199618||full Not-determined
S8_vs_k141_182281||full Not-determined
S8_vs_k141_26094||full Low-quality
S8_vs_k141_138892||full Not-determined
S8_vs_k141_147582||full Low-quality
S8_vs_k141_26097||full Not-determined
S8_vs_k141_78161||full Low-quality
S8_vs_k141_164910||full Not-determined
S8_vs_k141_26100||full Low-quality
S8_vs_k141_60814||full Low-quality
S8_vs_k141_70||full Low-quality
S8_vs_k141_130194||full Not-determined
S8_vs_k141_130196||full Not-determined
S8_vs_k141_69482||full Low-quality
S8_vs_k141_17421||full Not-determined
S8_vs_k141_199641||full Not-determined
S8_vs_k141_78174||full Low-quality
S8_vs_k141_69494||full Low-quality
S8_vs_k141_104208||full Low-quality
S8_vs_k141_52143||full Not-determined
S8_vs_k141_52145||full Low-quality
S8_vs_k141_190996||full Low-quality
S8_vs_k141_43481||full Low-quality
S8_vs_k141_121586||full Low-quality
S8_vs_k141_78181||full Low-quality
S8_vs_k141_111||full Not-determined
S8_vs_k141_78187||full Low-quality
S8_vs_k141_69509||full Low-quality
S8_vs_k141_8772||full Low-quality
S9_extracted_k141_6033 Medium-quality
S8_vs_k141_147621||full Not-determined
S8_vs_k141_104224||full Low-quality
S8_vs_k141_95536||full Low-quality
S8_vs_k141_130243||full Low-quality
S8_vs_k141_60850||full Not-determined
S8_vs_k141_156313||full Low-quality
S8_vs_k141_86922||full Low-quality
S8_vs_k141_112915||full Low-quality
S8_vs_k141_34783||full Not-determined
S8_vs_k141_8794||full Low-quality
S8_vs_k141_60861||full Low-quality
S8_vs_k141_156322||full Low-quality
S8_vs_k141_199669||full Low-quality
S8_vs_k141_78199||full Low-quality
S8_vs_k141_34798||full Low-quality
S8_vs_k141_86933||full Not-determined
S8_vs_k141_52167||full Not-determined
S8_vs_k141_130253||full Low-quality
S8_vs_k141_130255||full Not-determined
S8_vs_k141_95567||full Low-quality
S8_vs_k141_60866||full Not-determined
S8_vs_k141_78226||full Not-determined
S8_vs_k141_52186||full Not-determined
S8_vs_k141_69515||full Low-quality
S8_vs_k141_199697||full Not-determined
S8_vs_k141_121623||full Low-quality
S8_vs_k141_199698||full Low-quality
S8_vs_k141_8820||full Not-determined
S8_vs_k141_52200||full Low-quality
S8_vs_k141_199706||full Low-quality
S8_vs_k141_86972||full Not-determined
S8_vs_k141_60894||full Low-quality
S8_vs_k141_8834||full Low-quality
S8_vs_k141_112971||full Not-determined
S8_vs_k141_60896||full Low-quality
S9_vs_k141_6033||full Medium-quality
S8_vs_k141_26228||full Low-quality
S8_vs_k141_34842||full Low-quality
S8_vs_k141_86993||full Low-quality
S8_vs_k141_138980||full Low-quality
S8_vs_k141_199717||full Low-quality
S8_vs_k141_130274||full Low-quality
S8_vs_k141_173668||full Not-determined
S8_vs_k141_86996||full Not-determined
S8_vs_k141_191094||full Not-determined
S8_vs_k141_138998||full Low-quality
S8_vs_k141_165040||full Low-quality
S8_vs_k141_112993||full Not-determined
S8_vs_k141_17503||full Not-determined
S8_vs_k141_130282||full Low-quality
S8_vs_k141_78262||full Low-quality
S8_vs_k141_138999||full Not-determined
S8_vs_k141_17505||full Not-determined
S8_vs_k141_130283||full Not-determined
S8_vs_k141_121667||full Not-determined
S8_vs_k141_87021||full Low-quality
S8_vs_k141_121670||full Low-quality
S8_vs_k141_191124||full Not-determined
S8_vs_k141_165070||full Not-determined
S8_vs_k141_60933||full Not-determined
S8_vs_k141_199735||full Not-determined
S8_vs_k141_121678||full Low-quality
S8_vs_k141_43558||full Low-quality
S8_vs_k141_173702||full Low-quality
S8_vs_k141_43560||full Low-quality
S8_vs_k141_60944||full Not-determined
S8_vs_k141_78283||full Low-quality
S8_vs_k141_60955||full Not-determined
S8_vs_k141_243||full Low-quality
S8_vs_k141_246||full Low-quality
S8_vs_k141_156367||full Low-quality
S8_vs_k141_130323||full Low-quality
S8_vs_k141_60961||full Not-determined
S8_vs_k141_8904||full Low-quality
S8_vs_k141_139043||full Not-determined
S8_vs_k141_253||full Low-quality
S8_vs_k141_173717||full Low-quality
S8_vs_k141_130331||full Low-quality
S8_vs_k141_256||full Low-quality
S8_vs_k141_156368||full Low-quality
S8_vs_k141_78298||full Low-quality
S8_vs_k141_139052||full Not-determined
S8_vs_k141_87074||full Not-determined
S8_vs_k141_43584||full Low-quality
S8_vs_k141_78314||full Not-determined
S8_vs_k141_52269||full Low-quality
S8_vs_k141_52270||full Not-determined
S8_vs_k141_113044||full Not-determined
S8_vs_k141_69592||full Not-determined
S8_vs_k141_43599||full Low-quality
S8_vs_k141_165095||full Not-determined
S8_vs_k141_104269||full Low-quality
S8_vs_k141_78325||full Low-quality
S8_vs_k141_26259||full Not-determined
S8_vs_k141_43615||full Not-determined
S8_vs_k141_87106||full Low-quality
S8_vs_k141_26264||full Not-determined
S11_extracted_k141_28645 Complete
S8_vs_k141_52292||full Low-quality
S8_vs_k141_104282||full Not-determined
S8_vs_k141_121731||full Low-quality
S8_vs_k141_182327||full Low-quality
S8_vs_k141_113064||full Low-quality
S8_vs_k141_52295||full Low-quality
S8_vs_k141_34979||full Low-quality
S8_vs_k141_147689||full Not-determined
S8_vs_k141_78334||full Low-quality
S8_vs_k141_8970||full Not-determined
S8_vs_k141_52319||full Not-determined
S8_vs_k141_326||full Not-determined
S8_vs_k141_69621||full Not-determined
S13_extracted_147589 Complete
S8_vs_k141_60984||full Low-quality
S8_vs_k141_26284||full Not-determined
S8_vs_k141_35004||full Low-quality
S8_vs_k141_8998||full Not-determined
S8_vs_k141_43659||full Low-quality
S8_vs_k141_199772||full Low-quality
S8_vs_k141_156447||full Not-determined
S8_vs_k141_60997||full Low-quality
S8_vs_k141_26297||full Not-determined
S8_vs_k141_95613||full Low-quality
S8_vs_k141_182354||full Low-quality
S8_vs_k141_156449||full Not-determined
S8_vs_k141_113104||full Low-quality
S8_vs_k141_52334||full Not-determined
S8_vs_k141_156459||full Low-quality
S8_vs_k141_78396||full Not-determined
S8_vs_k141_139112||full Low-quality
S8_vs_k141_147694||full Low-quality
S8_vs_k141_139117||full Low-quality
S8_vs_k141_139120||full Low-quality
S8_vs_k141_113123||full Not-determined
S8_vs_k141_139122||full Low-quality
S8_vs_k141_173741||full Low-quality
S8_vs_k141_165190||full Low-quality
S8_vs_k141_35031||full Not-determined
S8_vs_k141_35037||full Low-quality
S8_vs_k141_182386||full Not-determined
S8_vs_k141_26319||full Low-quality
S8_vs_k141_156469||full Low-quality
S8_vs_k141_182392||full Not-determined
S8_vs_k141_78409||full Low-quality
S8_vs_k141_52374||full Not-determined
S8_vs_k141_199809||full Low-quality
S8_vs_k141_69664||full Low-quality
S8_vs_k141_17539||full Low-quality
S8_vs_k141_61091||full Low-quality
S8_vs_k141_121781||full Low-quality
S11_extracted_k141_41696 Medium-quality
S8_vs_k141_78428||full Low-quality
S8_vs_k141_357||full Low-quality
S8_vs_k141_199828||full Not-determined
S8_vs_k141_191237||full Low-quality
S8_vs_k141_130510||full Low-quality
S8_vs_k141_78432||full Low-quality
S8_vs_k141_87211||full Not-determined
S8_vs_k141_191251||full Low-quality
S8_vs_k141_9046||full Low-quality
S8_vs_k141_17541||full Low-quality
S8_vs_k141_43683||full Not-determined
S8_vs_k141_374||full Low-quality
S8_vs_k141_139165||full Low-quality
S8_vs_k141_113169||full Low-quality
S8_vs_k141_139166||full Low-quality
S8_vs_k141_199856||full Low-quality
S8_vs_k141_182436||full Not-determined
S8_vs_k141_17549||full Not-determined
S8_vs_k141_139171||full Low-quality
S8_vs_k141_43686||full Not-determined
S13_vs_147589||full High-quality
S8_vs_k141_69684||full Low-quality
S8_vs_k141_130538||full Not-determined
S8_vs_k141_130541||full Not-determined
S8_vs_k141_26362||full Low-quality
S8_vs_k141_394||full Not-determined
S8_vs_k141_9077||full Not-determined
S8_vs_k141_182456||full Low-quality
S8_vs_k141_78469||full Not-determined
S8_vs_k141_26370||full Not-determined
S8_vs_k141_173818||full Low-quality
S8_vs_k141_399||full Low-quality
S8_vs_k141_104431||full Not-determined
S8_vs_k141_139195||full Not-determined
S8_vs_k141_165250||full Not-determined
S8_vs_k141_95685||full Not-determined
S8_vs_k141_78480||full Low-quality
S8_vs_k141_9105||full Not-determined
S8_vs_k141_413||full Low-quality
S8_vs_k141_52435||full Low-quality
S8_vs_k141_104439||full Low-quality
S8_vs_k141_17567||full Not-determined
S8_vs_k141_121864||full Not-determined
S8_vs_k141_26406||full Not-determined
S8_vs_k141_104442||full Not-determined
S8_vs_k141_17573||full Not-determined
S8_vs_k141_182476||full Low-quality
S8_vs_k141_104446||full Low-quality
S8_vs_k141_165267||full Low-quality
S8_vs_k141_113199||full Not-determined
S8_vs_k141_52456||full Low-quality
S8_vs_k141_432||full Not-determined
S8_vs_k141_191356||full Low-quality
S8_vs_k141_87293||full Not-determined
S8_vs_k141_441||full Low-quality
S8_vs_k141_43731||full Low-quality
S8_vs_k141_147736||full Low-quality
S8_vs_k141_87297||full Not-determined
S8_vs_k141_130582||full Low-quality
S8_vs_k141_199922||full Low-quality
S8_vs_k141_78539||full Low-quality
S8_vs_k141_104452||full Low-quality
S8_vs_k141_139202||full Low-quality
S8_vs_k141_121904||full Not-determined
S8_vs_k141_130600||full Not-determined
S8_vs_k141_121910||full Low-quality
S8_vs_k141_35156||full Not-determined
S8_vs_k141_165304||full Not-determined
S8_vs_k141_182540||full Low-quality
S8_vs_k141_43769||full Low-quality
S8_vs_k141_26447||full Low-quality
S8_vs_k141_191398||full Low-quality
S8_vs_k141_199951||full Low-quality
S8_vs_k141_95765||full Low-quality
S8_vs_k141_460||full Low-quality
S8_vs_k141_78574||full Low-quality
S8_vs_k141_35176||full Low-quality
S8_vs_k141_61148||full Low-quality
S8_vs_k141_130624||full Low-quality
S8_vs_k141_78578||full Low-quality
S8_vs_k141_35181||full Low-quality
S8_vs_k141_69731||full Low-quality
S8_vs_k141_61154||full Low-quality
S8_vs_k141_78583||full Low-quality
S8_vs_k141_104511||full Not-determined
S8_vs_k141_9150||full Not-determined
S8_vs_k141_487||full Not-determined
S8_vs_k141_95769||full Not-determined
S8_vs_k141_147799||full Low-quality
S8_vs_k141_87363||full Low-quality
S8_vs_k141_52513||full Not-determined
S8_vs_k141_104520||full Low-quality
S8_vs_k141_95775||full Low-quality
S8_vs_k141_121946||full Not-determined
S8_vs_k141_35198||full Low-quality
S8_vs_k141_17608||full Low-quality
S8_vs_k141_509||full Low-quality
S8_vs_k141_191448||full Not-determined
S8_vs_k141_130657||full Low-quality
S8_vs_k141_87388||full Not-determined
S8_vs_k141_9191||full Not-determined
S8_vs_k141_17636||full Not-determined
S8_vs_k141_121968||full Not-determined
S8_vs_k141_9210||full Not-determined
S8_vs_k141_61199||full Low-quality
S8_vs_k141_26498||full Low-quality
S8_vs_k141_43823||full Low-quality
S8_vs_k141_9215||full Not-determined
S8_vs_k141_130683||full Not-determined
S8_vs_k141_26501||full Low-quality
S8_vs_k141_61209||full Not-determined
S8_vs_k141_43827||full Low-quality
S8_vs_k141_17646||full Low-quality
S8_vs_k141_35220||full Not-determined
S8_vs_k141_95801||full Low-quality
S8_vs_k141_61221||full Not-determined
S8_vs_k141_191510||full Not-determined
S8_vs_k141_43853||full Not-determined
S8_vs_k141_52581||full Not-determined
S8_vs_k141_121996||full Low-quality
S8_vs_k141_191513||full Not-determined
S8_vs_k141_165428||full Low-quality
S8_vs_k141_130693||full Low-quality
S8_vs_k141_560||full Low-quality
S8_vs_k141_78660||full Not-determined
S8_vs_k141_35242||full Not-determined
S8_vs_k141_122001||full Low-quality
S8_vs_k141_87414||full Low-quality
S8_vs_k141_52595||full Low-quality
S8_vs_k141_139315||full Low-quality
S8_vs_k141_173946||full Not-determined
S8_vs_k141_104605||full Not-determined
S8_vs_k141_173948||full Not-determined
S8_vs_k141_113233||full Low-quality
S8_vs_k141_104607||full Low-quality
S8_vs_k141_165454||full Not-determined
S8_vs_k141_139322||full Low-quality
S8_vs_k141_35268||full Not-determined
S8_vs_k141_87426||full Low-quality
S8_vs_k141_122022||full Low-quality
S8_vs_k141_61253||full Low-quality
S8_vs_k141_95849||full Not-determined
S8_vs_k141_104617||full Low-quality
S8_vs_k141_122025||full Low-quality
S8_vs_k141_122027||full Low-quality
S8_vs_k141_26532||full Not-determined
S8_vs_k141_139332||full Low-quality
S8_vs_k141_61264||full Not-determined
S8_vs_k141_95861||full Low-quality
S8_vs_k141_147893||full Low-quality
S8_vs_k141_589||full Low-quality
S8_vs_k141_139342||full Not-determined
S8_vs_k141_95869||full Not-determined
S8_vs_k141_139353||full Low-quality
S8_vs_k141_191556||full Low-quality
S8_vs_k141_35294||full Low-quality
S8_vs_k141_52631||full Not-determined
S8_vs_k141_69793||full Low-quality
S8_vs_k141_9232||full Low-quality
S8_vs_k141_61296||full Low-quality
S8_vs_k141_52634||full Not-determined
S8_vs_k141_147919||full Not-determined
S8_vs_k141_95896||full Not-determined
S8_vs_k141_35296||full Not-determined
S8_vs_k141_61307||full Low-quality
S8_vs_k141_165501||full Low-quality
S8_vs_k141_147922||full Not-determined
S8_vs_k141_9251||full Low-quality
S11_vs_k141_41696||full Medium-quality
S8_vs_k141_87460||full Not-determined
S8_vs_k141_165513||full Not-determined
S8_vs_k141_147942||full Not-determined
S8_vs_k141_104667||full Low-quality
S8_vs_k141_17739||full Not-determined
S8_vs_k141_104668||full Low-quality
S8_vs_k141_9254||full Not-determined
S8_vs_k141_173991||full Not-determined
S8_vs_k141_200034||full Low-quality
S8_vs_k141_191594||full Low-quality
S8_vs_k141_113277||full Not-determined
S8_vs_k141_130769||full Low-quality
S8_vs_k141_147994||full Not-determined
S8_vs_k141_147995||full Low-quality
S8_vs_k141_95927||full Low-quality
S13_extracted_101804 Medium-quality
S8_vs_k141_122122||full Not-determined
S8_vs_k141_148006||full Not-determined
S8_vs_k141_191608||full Not-determined
S8_vs_k141_191609||full Not-determined
S8_vs_k141_95946||full Not-determined
S8_vs_k141_35310||full Not-determined
S8_vs_k141_26552||full Not-determined
S8_vs_k141_26559||full Low-quality
S8_vs_k141_139407||full Low-quality
S8_vs_k141_156613||full Low-quality
S8_vs_k141_95962||full Low-quality
S8_vs_k141_26560||full Not-determined
S8_vs_k141_113313||full Not-determined
S8_vs_k141_35325||full Not-determined
S8_vs_k141_87506||full Low-quality
S8_vs_k141_130798||full Not-determined
S8_vs_k141_122141||full Not-determined
S8_vs_k141_43870||full Not-determined
S8_vs_k141_191632||full Low-quality
S8_vs_k141_139418||full Not-determined
S8_vs_k141_165591||full Low-quality
S8_vs_k141_174042||full Not-determined
S8_vs_k141_61370||full Low-quality
S8_vs_k141_78705||full Low-quality
S8_vs_k141_139433||full Not-determined
S8_vs_k141_122152||full Not-determined
S8_vs_k141_130821||full Low-quality
S8_vs_k141_87531||full Low-quality
S8_vs_k141_122158||full Low-quality
S8_vs_k141_17803||full Low-quality
S8_vs_k141_35333||full Low-quality
S8_vs_k141_182650||full Low-quality
S8_vs_k141_148070||full Not-determined
S8_vs_k141_113343||full Low-quality
S8_vs_k141_52769||full Low-quality
S8_vs_k141_61411||full Not-determined
S8_vs_k141_174056||full Low-quality
S8_vs_k141_96004||full Low-quality
S8_vs_k141_78737||full Low-quality
S8_vs_k141_43909||full Not-determined
S8_vs_k141_148088||full Low-quality
S8_vs_k141_130846||full Low-quality
S8_vs_k141_61421||full Not-determined
S8_vs_k141_182685||full Low-quality
S8_vs_k141_43919||full Low-quality
S8_vs_k141_78756||full Not-determined
S8_vs_k141_9283||full Not-determined
S8_vs_k141_148098||full Low-quality
S8_vs_k141_174067||full Not-determined
S8_vs_k141_736||full Low-quality
S8_vs_k141_165630||full Not-determined
S8_vs_k141_156705||full Low-quality
S8_vs_k141_174069||full Low-quality
S8_vs_k141_165634||full Low-quality
S8_vs_k141_745||full Not-determined
S8_vs_k141_61437||full Not-determined
S8_vs_k141_156712||full Low-quality
S8_vs_k141_174085||full Low-quality
S8_vs_k141_104733||full Low-quality
S8_vs_k141_104737||full Not-determined
S8_vs_k141_130889||full Low-quality
S8_vs_k141_17848||full Low-quality
S8_vs_k141_52784||full Not-determined
S8_vs_k141_139497||full Not-determined
S8_vs_k141_104748||full Not-determined
S8_vs_k141_174112||full Not-determined
S8_vs_k141_156723||full Low-quality
S8_vs_k141_17864||full Low-quality
S8_vs_k141_78784||full Not-determined
S8_vs_k141_165660||full Low-quality
S8_vs_k141_191688||full Not-determined
S8_vs_k141_43973||full Low-quality
S8_vs_k141_87621||full Low-quality
S8_vs_k141_139525||full Low-quality
S8_vs_k141_43985||full Low-quality
S8_vs_k141_9347||full Low-quality
S8_vs_k141_26652||full Low-quality
S8_vs_k141_87627||full Not-determined
S8_vs_k141_139541||full Not-determined
S8_vs_k141_61476||full Low-quality
S8_vs_k141_104764||full Low-quality
S8_vs_k141_26661||full Low-quality
S8_vs_k141_165666||full Not-determined
S13_vs_101804||full Medium-quality
S8_vs_k141_182726||full Low-quality
S8_vs_k141_200107||full Low-quality
S8_vs_k141_17906||full Low-quality
S8_vs_k141_9367||full Low-quality
S8_vs_k141_17907||full Low-quality
S8_vs_k141_156773||full Low-quality
S8_vs_k141_139551||full Low-quality
S8_vs_k141_44024||full Not-determined
S8_vs_k141_139556||full Not-determined
S8_vs_k141_26692||full Low-quality
S8_vs_k141_139557||full Not-determined
S8_vs_k141_156783||full Not-determined
S8_vs_k141_104800||full Not-determined
S8_vs_k141_96073||full Not-determined
S8_vs_k141_824||full Not-determined
S8_vs_k141_78846||full Not-determined
S8_vs_k141_165698||full Low-quality
S8_vs_k141_26712||full Low-quality
S8_vs_k141_130973||full Not-determined
S8_vs_k141_9408||full Low-quality
S8_vs_k141_139579||full Not-determined
S8_vs_k141_104812||full Low-quality
S8_vs_k141_130978||full Not-determined
S8_vs_k141_35417||full Low-quality
S8_vs_k141_44061||full Not-determined
S8_vs_k141_9415||full Not-determined
S8_vs_k141_35423||full Not-determined
S8_vs_k141_848||full Not-determined
S8_vs_k141_70035||full Not-determined
S8_vs_k141_148146||full Low-quality
S8_vs_k141_139594||full Low-quality
S8_vs_k141_853||full Not-determined
S8_vs_k141_148151||full Not-determined
S8_vs_k141_122270||full Low-quality
S8_vs_k141_87683||full Low-quality
S8_vs_k141_78870||full Not-determined
S8_vs_k141_96104||full Low-quality
S8_vs_k141_131000||full Not-determined
S8_vs_k141_122290||full Low-quality
S8_vs_k141_191762||full Low-quality
S8_vs_k141_165738||full Low-quality
S8_vs_k141_26758||full Not-determined
S8_vs_k141_61511||full Not-determined
S8_vs_k141_122296||full Not-determined
S8_vs_k141_17976||full Low-quality
S8_vs_k141_165740||full Low-quality
S8_vs_k141_104834||full Low-quality
S8_vs_k141_104836||full Low-quality
S8_vs_k141_61520||full Low-quality
S8_vs_k141_148178||full Low-quality
S8_vs_k141_888||full Low-quality
S8_vs_k141_9455||full Low-quality
S8_vs_k141_113446||full Low-quality
S8_vs_k141_890||full Low-quality
S8_vs_k141_26775||full Low-quality
S8_vs_k141_35469||full Not-determined
S8_vs_k141_200146||full Low-quality
S8_vs_k141_26776||full Low-quality
S8_vs_k141_165752||full Low-quality
S8_vs_k141_78906||full Low-quality
S8_vs_k141_893||full Not-determined
S8_vs_k141_44125||full Low-quality
S8_vs_k141_200156||full Low-quality
S8_vs_k141_61533||full Low-quality
S8_vs_k141_9460||full Not-determined
S8_vs_k141_70063||full Low-quality
S8_vs_k141_44132||full Not-determined
S8_vs_k141_200158||full Not-determined
S8_vs_k141_35481||full Low-quality
S8_vs_k141_9463||full Low-quality
S8_vs_k141_182797||full Low-quality
S8_vs_k141_35483||full Not-determined
S8_vs_k141_61558||full Not-determined
S8_vs_k141_61559||full Low-quality
S8_vs_k141_148209||full Not-determined
S8_vs_k141_78944||full Low-quality
S8_vs_k141_156843||full Not-determined
S8_vs_k141_35490||full Low-quality
S8_vs_k141_113473||full Not-determined
S8_vs_k141_61568||full Not-determined
S8_vs_k141_44174||full Not-determined
S8_vs_k141_927||full Not-determined
S8_vs_k141_17999||full Not-determined
S8_vs_k141_26823||full Not-determined
S8_vs_k141_9496||full Not-determined
S8_vs_k141_70115||full Low-quality
S8_vs_k141_35522||full Not-determined
S8_vs_k141_18013||full Not-determined
S8_vs_k141_191878||full Low-quality
S8_vs_k141_104888||full Low-quality
S8_vs_k141_104889||full Not-determined
S8_vs_k141_122387||full Low-quality
S8_vs_k141_131032||full Low-quality
S8_vs_k141_35539||full Not-determined
S8_vs_k141_44206||full Not-determined
S8_vs_k141_78990||full Not-determined

S8_vs_k141_131037||full Not-determined
S8_vs_k141_174166||full Not-determined
S8_vs_k141_182847||full Low-quality
S8_vs_k141_26863||full Not-determined
S8_vs_k141_35545||full Low-quality
S8_vs_k141_35546||full Low-quality
S8_vs_k141_18023||full Low-quality
S8_vs_k141_104908||full Not-determined
S8_vs_k141_139715||full Low-quality
S12_vs_k141_137069||full Medium-quality
S8_vs_k141_52925||full Not-determined
S8_vs_k141_122420||full Not-determined
S8_vs_k141_9547||full Low-quality
S8_vs_k141_131088||full Not-determined
S8_vs_k141_191926||full Low-quality
S8_vs_k141_156903||full Not-determined
S8_vs_k141_26904||full Not-determined
S8_vs_k141_1001||full Low-quality
S8_vs_k141_52945||full Low-quality
S8_vs_k141_191942||full Low-quality
S8_vs_k141_18060||full Low-quality
S8_vs_k141_113578||full Not-determined
S8_vs_k141_191958||full Low-quality
S8_vs_k141_35576||full Not-determined
S8_vs_k141_87771||full Not-determined
S8_vs_k141_148323||full Low-quality
S8_vs_k141_174221||full Low-quality
S8_vs_k141_1021||full Low-quality
S8_vs_k141_96289||full Not-determined
S8_vs_k141_18075||full Low-quality
S8_vs_k141_122457||full Low-quality
S8_vs_k141_44236||full Not-determined
S8_vs_k141_61681||full Low-quality
S8_vs_k141_52971||full Low-quality
S8_vs_k141_156952||full Not-determined
S8_vs_k141_18083||full Not-determined
S8_vs_k141_26947||full Not-determined
S8_vs_k141_18091||full Not-determined
S8_vs_k141_131136||full Low-quality
S8_vs_k141_52980||full Not-determined
S8_vs_k141_44246||full Not-determined
S8_vs_k141_52983||full Low-quality
S8_vs_k141_61696||full Low-quality
S8_vs_k141_35630||full Not-determined
S8_vs_k141_200203||full Low-quality
S8_vs_k141_52985||full Not-determined
S8_vs_k141_200204||full Low-quality
S8_vs_k141_44261||full Not-determined
S8_vs_k141_44264||full Low-quality
S8_vs_k141_26975||full Low-quality
S8_vs_k141_35648||full Low-quality
S8_vs_k141_157000||full Not-determined
S8_vs_k141_122503||full Low-quality
S8_vs_k141_87820||full Low-quality
S8_vs_k141_157007||full Low-quality
S8_vs_k141_87822||full Not-determined
S8_vs_k141_70179||full Not-determined
S8_vs_k141_35657||full Not-determined
S8_vs_k141_148361||full Low-quality
S8_vs_k141_96350||full Low-quality
S8_vs_k141_44267||full Not-determined
S8_vs_k141_165814||full Not-determined
S8_vs_k141_44269||full Low-quality
S8_vs_k141_157016||full Low-quality
S8_vs_k141_157017||full Low-quality
S8_vs_k141_182928||full Low-quality
S8_vs_k141_139812||full Low-quality
S8_vs_k141_200213||full Not-determined
S8_vs_k141_44277||full Low-quality
S8_vs_k141_139817||full Low-quality
S8_vs_k141_53007||full Low-quality
S8_vs_k141_113649||full Low-quality
S8_vs_k141_192008||full Low-quality
S8_vs_k141_192014||full Not-determined
S8_vs_k141_18095||full Not-determined
S8_vs_k141_165832||full Low-quality
S8_vs_k141_35688||full Not-determined
S8_vs_k141_139828||full Not-determined
S8_vs_k141_165839||full Not-determined
S8_vs_k141_87865||full Low-quality
S8_vs_k141_113666||full Not-determined
S8_vs_k141_192026||full Not-determined
S8_vs_k141_131184||full Not-determined
S8_vs_k141_9599||full Not-determined
S8_vs_k141_79128||full Not-determined
S8_vs_k141_139840||full Not-determined
S8_vs_k141_200242||full Low-quality
S8_vs_k141_148387||full Low-quality
S8_vs_k141_192038||full Low-quality
S8_vs_k141_157041||full Low-quality
S8_vs_k141_87870||full Not-determined
S8_vs_k141_139857||full Low-quality
S8_vs_k141_200258||full Not-determined
S8_vs_k141_79136||full Not-determined
S8_vs_k141_113698||full Not-determined
S8_vs_k141_165854||full Low-quality
S8_vs_k141_131207||full Low-quality
S8_vs_k141_53035||full Low-quality
S8_vs_k141_96408||full Not-determined
S8_vs_k141_61757||full Low-quality
S8_vs_k141_35747||full Not-determined
S8_vs_k141_27043||full Not-determined
S8_vs_k141_113739||full Low-quality
S8_vs_k141_53054||full Low-quality
S8_vs_k141_96427||full Not-determined
S8_vs_k141_192074||full Low-quality
S8_vs_k141_139888||full Not-determined
S8_vs_k141_61776||full Low-quality
S11_vs_k141_28645||full Medium-quality
S8_vs_k141_79191||full Low-quality
S8_vs_k141_79193||full Not-determined
S8_vs_k141_148394||full Not-determined
S8_vs_k141_113751||full Low-quality
S8_vs_k141_18137||full Not-determined
S8_vs_k141_9683||full Not-determined
S8_vs_k141_53077||full Not-determined
S8_vs_k141_122567||full Low-quality
S8_vs_k141_113758||full Not-determined
S8_vs_k141_165916||full Low-quality
S8_vs_k141_165920||full Not-determined
S8_vs_k141_192112||full Low-quality
S8_vs_k141_1088||full Low-quality
S8_vs_k141_79215||full Not-determined
S8_vs_k141_157078||full Not-determined
S8_vs_k141_27119||full Not-determined
S8_vs_k141_192120||full Not-determined
S8_vs_k141_79223||full Low-quality
S8_vs_k141_165924||full Low-quality
S8_vs_k141_131255||full Low-quality
S8_vs_k141_174389||full Low-quality
S8_vs_k141_1105||full Low-quality
S8_vs_k141_192127||full Not-determined
S8_vs_k141_70241||full Low-quality
S8_vs_k141_87961||full Not-determined
S8_vs_k141_27136||full Not-determined
S8_vs_k141_61806||full Not-determined
S8_vs_k141_148449||full Low-quality
S8_vs_k141_96485||full Not-determined
S8_vs_k141_200329||full Low-quality
S8_vs_k141_61814||full Low-quality
S8_vs_k141_61816||full Low-quality
S8_vs_k141_192139||full Low-quality
S8_vs_k141_157109||full Low-quality
S8_vs_k141_174412||full Low-quality
S8_vs_k141_53123||full Not-determined
S8_vs_k141_192151||full Low-quality
S8_vs_k141_157115||full Not-determined
S8_vs_k141_113793||full Low-quality
S8_vs_k141_122599||full Low-quality
S8_vs_k141_139977||full Not-determined
S8_vs_k141_131305||full Low-quality
S8_vs_k141_122614||full Not-determined
S8_vs_k141_122625||full Not-determined
S8_vs_k141_174426||full Not-determined
S8_vs_k141_192175||full Not-determined
S8_vs_k141_70304||full Low-quality
S8_vs_k141_27191||full Not-determined
S8_vs_k141_79314||full Not-determined
S8_vs_k141_9747||full Not-determined
S8_vs_k141_1164||full Low-quality
S8_vs_k141_53177||full Low-quality
S8_vs_k141_140003||full Low-quality
S8_vs_k141_44320||full Not-determined
S8_vs_k141_174438||full Low-quality
S8_vs_k141_79328||full Low-quality
S8_vs_k141_131349||full Low-quality
S8_vs_k141_148553||full Low-quality
S8_vs_k141_18244||full Low-quality
S8_vs_k141_131351||full Low-quality
S8_vs_k141_148555||full Low-quality
S8_vs_k141_27226||full Low-quality
S8_vs_k141_200340||full Not-determined
S8_vs_k141_174451||full Not-determined
S8_vs_k141_27236||full Low-quality
S8_vs_k141_27237||full Low-quality
S8_vs_k141_79349||full Not-determined
S8_vs_k141_140021||full Low-quality
S8_vs_k141_131367||full Low-quality
S8_vs_k141_192203||full Not-determined
S8_vs_k141_27256||full Not-determined
S8_vs_k141_27257||full Not-determined
S8_vs_k141_131380||full Low-quality
S8_vs_k141_53225||full Low-quality
S8_vs_k141_148598||full Low-quality
S8_vs_k141_140044||full Low-quality
S8_vs_k141_148611||full Low-quality
S8_vs_k141_1238||full Low-quality
S8_vs_k141_1243||full Low-quality
S13_vs_87108||full Medium-quality
S8_vs_k141_18272||full Low-quality
S8_vs_k141_35975||full Not-determined
S8_vs_k141_131413||full Low-quality
S8_vs_k141_166011||full Not-determined
S8_vs_k141_18277||full Low-quality
S8_vs_k141_113832||full Low-quality
S8_vs_k141_79383||full Not-determined
S8_vs_k141_44380||full Not-determined
S8_vs_k141_1271||full Low-quality
S8_vs_k141_53247||full Low-quality
S8_vs_k141_105210||full Not-determined
S8_vs_k141_174498||full Low-quality
S8_vs_k141_18282||full Low-quality
S8_vs_k141_148647||full Not-determined
S8_vs_k141_27303||full Not-determined
S8_vs_k141_105227||full Low-quality
S8_vs_k141_79399||full Low-quality
S8_vs_k141_88052||full Low-quality
S8_vs_k141_105236||full Not-determined
S8_vs_k141_113862||full Low-quality
S8_vs_k141_27313||full Not-determined
S8_vs_k141_192298||full Low-quality
S8_vs_k141_200440||full Low-quality
S8_vs_k141_148658||full Not-determined
S8_vs_k141_27322||full Low-quality
S8_vs_k141_61964||full Low-quality
S8_vs_k141_79415||full Not-determined
S8_vs_k141_122751||full Low-quality
S8_vs_k141_1278||full Low-quality
S8_vs_k141_148671||full Low-quality
S8_vs_k141_18325||full Low-quality
S8_vs_k141_36000||full Low-quality
S8_vs_k141_157271||full Low-quality
S8_vs_k141_18333||full Not-determined
S8_vs_k141_105269||full Not-determined
S8_vs_k141_1292||full Low-quality
S8_vs_k141_166057||full Not-determined
S8_vs_k141_27355||full Low-quality
S8_vs_k141_166070||full Low-quality
S8_vs_k141_183022||full Not-determined
S8_vs_k141_192341||full Not-determined
S8_vs_k141_44439||full Low-quality
S8_vs_k141_79459||full Not-determined
S8_vs_k141_105291||full Low-quality
S8_vs_k141_166077||full Low-quality
S8_vs_k141_79466||full Not-determined
S8_vs_k141_148699||full Low-quality
S8_vs_k141_140065||full Low-quality
S8_vs_k141_1327||full Low-quality
S8_vs_k141_174530||full Not-determined
S8_vs_k141_27401||full Low-quality
S8_vs_k141_183035||full Not-determined
S8_vs_k141_157306||full Low-quality
S8_vs_k141_122793||full Not-determined
S8_vs_k141_192364||full Not-determined
S8_vs_k141_113916||full Low-quality
S8_vs_k141_131447||full Low-quality
S8_vs_k141_88084||full Not-determined
S8_vs_k141_174536||full Not-determined
S8_vs_k141_88093||full Low-quality
S8_vs_k141_62016||full Low-quality
S8_vs_k141_131459||full Low-quality
S8_vs_k141_122802||full Not-determined
S8_vs_k141_18364||full Low-quality
S8_vs_k141_166104||full Low-quality
S8_vs_k141_44451||full Not-determined
S8_vs_k141_131469||full Low-quality
S8_vs_k141_27452||full Low-quality
S8_vs_k141_53341||full Not-determined
S8_vs_k141_96692||full Low-quality
S7_vs_138116||full Medium-quality
S8_vs_k141_1361||full Low-quality
S8_vs_k141_88124||full Not-determined
S8_vs_k141_200508||full Not-determined
S8_vs_k141_96708||full Low-quality
S8_vs_k141_96713||full Not-determined
S8_vs_k141_96715||full Low-quality
S8_vs_k141_62050||full Low-quality
S8_vs_k141_131482||full Not-determined
S8_vs_k141_36081||full Not-determined
S8_vs_k141_96722||full Not-determined
S8_vs_k141_122848||full Not-determined
S8_vs_k141_174590||full Low-quality
S8_vs_k141_44483||full Low-quality
S8_vs_k141_148768||full Not-determined
S8_vs_k141_18408||full Low-quality
S8_vs_k141_174591||full Not-determined
S8_vs_k141_131492||full Low-quality
S8_vs_k141_53377||full Low-quality
S8_vs_k141_174595||full Not-determined
S8_vs_k141_183053||full Not-determined
S8_vs_k141_62067||full Not-determined
S8_vs_k141_192408||full Low-quality
S8_vs_k141_1391||full Not-determined
S8_vs_k141_96734||full Not-determined
S8_vs_k141_9829||full Low-quality
S8_vs_k141_96735||full Not-determined
S8_vs_k141_113991||full Not-determined
S8_vs_k141_166158||full Low-quality
S8_vs_k141_113998||full Low-quality
S8_vs_k141_18419||full Low-quality
S8_vs_k141_114003||full Not-determined
S8_vs_k141_27511||full Low-quality
S8_vs_k141_96745||full Not-determined
S8_vs_k141_131525||full Low-quality
S8_vs_k141_27521||full Low-quality
S8_vs_k141_53406||full Low-quality
S8_vs_k141_183075||full Low-quality
S8_vs_k141_174615||full Not-determined
S8_vs_k141_53407||full Not-determined
S8_vs_k141_105412||full Low-quality
S8_vs_k141_192444||full Not-determined
S8_vs_k141_157378||full Not-determined
S8_vs_k141_140149||full Low-quality
S8_vs_k141_148782||full Low-quality
S8_vs_k141_70468||full Low-quality
S8_vs_k141_27531||full Not-determined
S8_vs_k141_114014||full Low-quality
S8_vs_k141_18454||full Low-quality
S8_vs_k141_1434||full Not-determined
S8_vs_k141_166183||full Not-determined
S8_vs_k141_140175||full Not-determined
S8_vs_k141_157407||full Low-quality
S8_vs_k141_9864||full Low-quality
S8_vs_k141_148809||full Not-determined
S8_vs_k141_122904||full Not-determined
S8_vs_k141_166191||full Low-quality
S8_vs_k141_53468||full Not-determined
S8_vs_k141_36164||full Low-quality
S8_vs_k141_70513||full Low-quality
S8_vs_k141_1471||full Not-determined
S8_vs_k141_36172||full Not-determined
S8_vs_k141_53482||full Not-determined
S8_vs_k141_122920||full Low-quality
S8_vs_k141_36189||full Low-quality
S8_vs_k141_166212||full Low-quality
S8_vs_k141_114073||full Low-quality
S8_vs_k141_140231||full Not-determined
S8_vs_k141_122921||full Low-quality
S8_vs_k141_148843||full Not-determined
S8_vs_k141_96843||full Low-quality
S8_vs_k141_140235||full Low-quality
S8_vs_k141_122922||full Low-quality
S8_vs_k141_183119||full Low-quality
S8_vs_k141_1491||full Low-quality
S8_vs_k141_148855||full Not-determined
S8_vs_k141_166224||full Low-quality
S8_vs_k141_44581||full Not-determined
S8_vs_k141_44583||full Not-determined
S8_vs_k141_122933||full Not-determined
S8_vs_k141_44585||full Not-determined
S8_vs_k141_183135||full Not-determined
S8_vs_k141_53501||full Not-determined
S8_vs_k141_183136||full Low-quality
S8_vs_k141_44589||full Low-quality
S8_vs_k141_27602||full Not-determined
S8_vs_k141_122941||full Low-quality
S8_vs_k141_174691||full Low-quality
S8_vs_k141_148865||full Not-determined
S8_vs_k141_9959||full Not-determined
S8_vs_k141_148868||full Not-determined
S8_vs_k141_183152||full Low-quality
S8_vs_k141_44594||full Low-quality
S8_vs_k141_79568||full Low-quality
S8_vs_k141_27627||full Not-determined
S8_vs_k141_192537||full Low-quality
S8_vs_k141_183160||full Low-quality
S8_vs_k141_174703||full Low-quality
S8_vs_k141_1528||full Low-quality
S8_vs_k141_140275||full Not-determined
S8_vs_k141_70599||full Not-determined
S8_vs_k141_105501||full Low-quality
S8_vs_k141_1539||full Low-quality
S8_vs_k141_44610||full Low-quality
S8_vs_k141_183165||full Low-quality
S8_vs_k141_27641||full Not-determined
S8_vs_k141_131551||full Low-quality
S8_vs_k141_27643||full Not-determined
S8_vs_k141_1550||full Not-determined
S8_vs_k141_140296||full Not-determined
S8_vs_k141_200611||full Low-quality
S8_vs_k141_114143||full Low-quality
S8_vs_k141_9979||full Low-quality
S8_vs_k141_53537||full Not-determined
S8_vs_k141_96895||full Low-quality
S8_vs_k141_114150||full Low-quality
S8_vs_k141_157532||full Low-quality
S8_vs_k141_1564||full Low-quality
S8_vs_k141_18605||full Not-determined
S8_vs_k141_70644||full Low-quality
S8_vs_k141_183208||full Low-quality
S8_vs_k141_27669||full Not-determined
S8_vs_k141_105544||full Not-determined
S8_vs_k141_157543||full Not-determined
S8_vs_k141_44633||full Not-determined
S8_vs_k141_1596||full Not-determined
S8_vs_k141_148909||full Not-determined
S8_vs_k141_105554||full Low-quality
S8_vs_k141_122972||full Low-quality
S8_vs_k141_27683||full Not-determined
S8_vs_k141_79686||full Low-quality
S8_vs_k141_18629||full Low-quality
S8_vs_k141_27690||full Low-quality
S8_vs_k141_166299||full Not-determined
S8_vs_k141_44652||full Low-quality
S8_vs_k141_148924||full Low-quality
S8_vs_k141_62145||full Low-quality
S8_vs_k141_148927||full Low-quality
S8_vs_k141_174800||full Low-quality
S8_vs_k141_183232||full Low-quality
S8_vs_k141_62165||full Not-determined
S8_vs_k141_131628||full Low-quality
S8_vs_k141_62168||full Low-quality
S8_vs_k141_140381||full Low-quality
S8_vs_k141_44686||full Low-quality
S8_vs_k141_166324||full Not-determined
S8_vs_k141_9996||full Low-quality
S8_vs_k141_114186||full Low-quality
S8_vs_k141_70713||full Low-quality
S8_vs_k141_166333||full Not-determined
S8_vs_k141_105613||full Low-quality
S8_vs_k141_166340||full Low-quality
S8_vs_k141_62199||full Not-determined
S8_vs_k141_123028||full Low-quality
S8_vs_k141_166350||full Not-determined
S8_vs_k141_192562||full Low-quality
S8_vs_k141_79731||full Not-determined
S8_vs_k141_27736||full Not-determined
S8_vs_k141_79735||full Low-quality
S8_vs_k141_123038||full Low-quality
S8_vs_k141_114216||full Not-determined
S8_vs_k141_36357||full Low-quality
S8_vs_k141_140415||full Low-quality
S8_vs_k141_70746||full Low-quality
S7_extracted_138019 Medium-quality
S8_vs_k141_1630||full Not-determined
S8_vs_k141_18695||full Not-determined
S8_vs_k141_27753||full Not-determined
S8_vs_k141_88226||full Not-determined
S8_vs_k141_157545||full Low-quality
S8_vs_k141_88230||full Low-quality
S8_vs_k141_1645||full Low-quality
S8_vs_k141_174877||full Not-determined
S1_extracted_k141_35633 Medium-quality
S8_vs_k141_200755||full Not-determined
S8_vs_k141_27760||full Low-quality
S8_vs_k141_36382||full Not-determined
S8_vs_k141_18714||full Not-determined
S8_vs_k141_200759||full Not-determined
S8_vs_k141_174894||full Not-determined
S8_vs_k141_123086||full Not-determined
S8_vs_k141_27776||full Low-quality
S8_vs_k141_183307||full Low-quality
S8_vs_k141_36403||full Not-determined
S8_vs_k141_18723||full Low-quality
S8_vs_k141_62264||full Not-determined
S8_vs_k141_157571||full Not-determined
S8_vs_k141_10062||full Low-quality
S8_vs_k141_174913||full Low-quality
S8_vs_k141_97048||full Not-determined
S8_vs_k141_157573||full Low-quality
S8_vs_k141_53658||full Low-quality
S8_vs_k141_10068||full Not-determined
S8_vs_k141_88268||full Low-quality
S8_vs_k141_53663||full Not-determined
S8_vs_k141_183315||full Not-determined
S8_vs_k141_53664||full Low-quality
S8_vs_k141_114282||full Low-quality
S8_vs_k141_114286||full Low-quality
S8_vs_k141_97064||full Low-quality
S8_vs_k141_114287||full Not-determined
S1_vs_k141_35633||full Medium-quality
S8_vs_k141_18750||full Low-quality
S8_vs_k141_192648||full Low-quality
S8_vs_k141_105702||full Low-quality
S8_vs_k141_174928||full Low-quality
S8_vs_k141_123126||full Low-quality
S8_vs_k141_157598||full Not-determined
S8_vs_k141_166420||full Low-quality
S8_vs_k141_166421||full Not-determined
S8_vs_k141_62300||full Not-determined
S8_vs_k141_97081||full Not-determined
S8_vs_k141_88290||full Low-quality
S8_vs_k141_36448||full Not-determined
S8_vs_k141_18768||full Low-quality
S8_vs_k141_36453||full Low-quality
S8_vs_k141_131710||full Low-quality
S8_vs_k141_53703||full Low-quality
S8_vs_k141_44750||full Not-determined
S8_vs_k141_88301||full Not-determined
S8_vs_k141_97090||full Not-determined
S8_vs_k141_36466||full Low-quality
S8_vs_k141_157627||full Low-quality
S8_vs_k141_166439||full Not-determined
S8_vs_k141_53729||full Not-determined
S8_vs_k141_44769||full Low-quality
S8_vs_k141_131730||full Low-quality
S8_vs_k141_44771||full Not-determined
S8_vs_k141_62333||full Not-determined
S8_vs_k141_166455||full Low-quality
S8_vs_k141_53739||full Not-determined
S8_vs_k141_114335||full Not-determined
S8_vs_k141_157650||full Not-determined
S8_vs_k141_166468||full Low-quality
S8_vs_k141_183339||full Low-quality
S8_vs_k141_123166||full Low-quality
S8_vs_k141_105749||full Low-quality
S8_vs_k141_183347||full Not-determined
S8_vs_k141_18819||full Low-quality
S8_vs_k141_140518||full Low-quality
S8_vs_k141_192711||full Not-determined
S8_vs_k141_123185||full Low-quality
S8_vs_k141_36512||full Not-determined
S8_vs_k141_174989||full Low-quality
S8_vs_k141_27826||full Low-quality
S8_vs_k141_166494||full Low-quality
S8_vs_k141_27828||full Low-quality
S8_vs_k141_1742||full Not-determined
S8_vs_k141_44793||full Low-quality
S8_vs_k141_10166||full Not-determined
S8_vs_k141_53777||full Low-quality
S8_vs_k141_105792||full Not-determined
S8_vs_k141_105795||full Low-quality
S8_vs_k141_192724||full Low-quality
S8_vs_k141_200870||full Low-quality
S8_vs_k141_192730||full Not-determined
S8_vs_k141_18852||full Not-determined
S8_vs_k141_166516||full Low-quality
S8_vs_k141_53806||full Low-quality
S8_vs_k141_123203||full Not-determined
S8_vs_k141_97156||full Not-determined
S8_vs_k141_105820||full Low-quality
S8_vs_k141_44813||full Low-quality
S8_vs_k141_175014||full Low-quality
S8_vs_k141_157700||full Low-quality
S8_vs_k141_166538||full Low-quality
S8_vs_k141_1766||full Not-determined
S8_vs_k141_27853||full Low-quality
S8_vs_k141_53834||full Low-quality
S8_vs_k141_79800||full Low-quality
S8_vs_k141_140557||full Low-quality
S8_vs_k141_183369||full Low-quality
S8_vs_k141_140561||full Low-quality
S8_vs_k141_27855||full Low-quality
S8_vs_k141_166566||full Not-determined
S8_vs_k141_114407||full Not-determined
S8_vs_k141_166568||full Low-quality
S8_vs_k141_36585||full Not-determined
S8_vs_k141_18899||full Low-quality
S8_vs_k141_53857||full Low-quality
S8_vs_k141_1800||full Not-determined
S8_vs_k141_88440||full Not-determined
S8_vs_k141_192795||full Not-determined
S8_vs_k141_123244||full Not-determined
S8_vs_k141_149011||full Not-determined
S8_vs_k141_97215||full Not-determined
S8_vs_k141_149017||full Not-determined
S8_vs_k141_200915||full Not-determined
S8_vs_k141_140600||full Low-quality
S8_vs_k141_97229||full Low-quality
S8_vs_k141_192810||full Low-quality
S8_vs_k141_88453||full Not-determined
S8_vs_k141_166601||full Not-determined
S8_vs_k141_114454||full Not-determined
S8_vs_k141_1811||full Not-determined
S8_vs_k141_157730||full Not-determined
S8_vs_k141_183412||full Low-quality
S8_vs_k141_140607||full Not-determined
S8_vs_k141_183414||full Not-determined
S8_vs_k141_200938||full Not-determined
S8_vs_k141_53888||full Not-determined
S8_vs_k141_1818||full Low-quality
S8_vs_k141_192815||full Low-quality
S8_vs_k141_140617||full Not-determined
S8_vs_k141_10204||full Not-determined
S8_vs_k141_183427||full Low-quality
S8_vs_k141_62372||full Not-determined
S8_vs_k141_114486||full Low-quality
S8_vs_k141_149060||full Low-quality
S8_vs_k141_200951||full Low-quality
S8_vs_k141_44911||full Low-quality
S8_vs_k141_79859||full Low-quality
S8_vs_k141_10221||full Low-quality
S8_vs_k141_18937||full Low-quality
S8_vs_k141_97271||full Low-quality
S8_vs_k141_123284||full Not-determined
S8_vs_k141_10227||full Low-quality
S8_vs_k141_36604||full Low-quality
S8_vs_k141_200964||full Not-determined
S8_vs_k141_105856||full Not-determined
S8_vs_k141_131817||full Not-determined
S8_vs_k141_105865||full Not-determined
S8_vs_k141_183460||full Not-determined
S8_vs_k141_1844||full Low-quality
S8_vs_k141_53921||full Low-quality
S8_vs_k141_183466||full Low-quality
S8_vs_k141_10246||full Not-determined
S8_vs_k141_183471||full Low-quality
S8_vs_k141_62384||full Not-determined
S8_vs_k141_53932||full Not-determined

| | |
|---|---|
| S8_vs_k141_123323\|\|full | Not-determined |
| S8_vs_k141_27923\|\|full | Low-quality |
| S8_vs_k141_149108\|\|full | Not-determined |
| S8_vs_k141_140677\|\|full | Not-determined |
| S8_vs_k141_183493\|\|full | Not-determined |
| S8_vs_k141_166645\|\|full | Not-determined |
| S8_vs_k141_183495\|\|full | Low-quality |
| S8_vs_k141_131854\|\|full | Low-quality |
| S8_vs_k141_27934\|\|full | Not-determined |
| S8_vs_k141_192876\|\|full | Low-quality |
| S8_vs_k141_200984\|\|full | Low-quality |
| S8_vs_k141_70805\|\|full | Low-quality |
| S8_vs_k141_10279\|\|full | Low-quality |
| S8_vs_k141_123347\|\|full | Low-quality |
| S8_vs_k141_53964\|\|full | Not-determined |
| S8_vs_k141_44951\|\|full | Not-determined |
| S8_vs_k141_149128\|\|full | Low-quality |
| S8_vs_k141_114520\|\|full | Not-determined |
| S8_vs_k141_10281\|\|full | Not-determined |
| S8_vs_k141_53973\|\|full | Not-determined |
| S8_vs_k141_88552\|\|full | Low-quality |
| S8_vs_k141_27970\|\|full | Low-quality |
| S8_vs_k141_157748\|\|full | Low-quality |
| S8_vs_k141_105907\|\|full | Not-determined |
| S8_vs_k141_44975\|\|full | Not-determined |
| S8_vs_k141_105921\|\|full | Not-determined |
| S8_vs_k141_105928\|\|full | Low-quality |
| S8_vs_k141_97317\|\|full | Low-quality |
| S8_vs_k141_97321\|\|full | Low-quality |
| S8_vs_k141_114548\|\|full | Not-determined |
| S8_vs_k141_36708\|\|full | Not-determined |
| S8_vs_k141_183537\|\|full | Low-quality |
| S8_vs_k141_123360\|\|full | Low-quality |
| S8_vs_k141_62450\|\|full | Low-quality |
| S8_vs_k141_149176\|\|full | Not-determined |
| S8_vs_k141_201035\|\|full | Low-quality |
| S8_vs_k141_149182\|\|full | Not-determined |
| S8_vs_k141_19011\|\|full | Low-quality |
| S8_vs_k141_79984\|\|full | Low-quality |
| S8_vs_k141_123370\|\|full | Low-quality |
| S8_vs_k141_70832\|\|full | Low-quality |
| S8_vs_k141_36730\|\|full | Not-determined |
| S8_vs_k141_131936\|\|full | Low-quality |
| S8_vs_k141_123376\|\|full | Low-quality |
| S8_vs_k141_79989\|\|full | Low-quality |
| S8_vs_k141_183547\|\|full | Low-quality |
| S8_vs_k141_45019\|\|full | Not-determined |
| S8_vs_k141_88573\|\|full | Low-quality |
| S8_vs_k141_105957\|\|full | Not-determined |
| S8_vs_k141_192909\|\|full | Low-quality |
| S8_vs_k141_10359\|\|full | Low-quality |
| S8_vs_k141_131956\|\|full | Not-determined |
| S8_vs_k141_157809\|\|full | Not-determined |
| S8_vs_k141_10368\|\|full | Not-determined |
| S8_vs_k141_28053\|\|full | Not-determined |
| S8_vs_k141_140713\|\|full | Low-quality |
| S8_vs_k141_54056\|\|full | Not-determined |
| S8_vs_k141_131963\|\|full | Not-determined |
| S8_vs_k141_80019\|\|full | Not-determined |
| S8_vs_k141_62508\|\|full | Low-quality |
| S8_vs_k141_157830\|\|full | Low-quality |
| S8_vs_k141_62518\|\|full | Not-determined |
| S8_vs_k141_149239\|\|full | Not-determined |
| S8_vs_k141_157834\|\|full | Low-quality |
| S8_vs_k141_28078\|\|full | Low-quality |
| S8_vs_k141_54076\|\|full | Low-quality |
| S8_vs_k141_157835\|\|full | Low-quality |
| S8_vs_k141_36777\|\|full | Not-determined |
| S8_vs_k141_123424\|\|full | Low-quality |
| S8_vs_k141_36785\|\|full | Not-determined |
| S8_vs_k141_149272\|\|full | Low-quality |
| S8_vs_k141_192967\|\|full | Low-quality |
| S8_vs_k141_140740\|\|full | Low-quality |
| S8_vs_k141_201069\|\|full | Not-determined |
| S8_vs_k141_62536\|\|full | Not-determined |
| S8_vs_k141_140747\|\|full | Low-quality |
| S8_vs_k141_106001\|\|full | Not-determined |
| S8_vs_k141_192969\|\|full | Not-determined |
| S8_vs_k141_97333\|\|full | Low-quality |
| S8_vs_k141_1903\|\|full | Low-quality |
| S8_vs_k141_62547\|\|full | Low-quality |
| S8_vs_k141_80060\|\|full | Low-quality |
| S8_vs_k141_149279\|\|full | Not-determined |
| S8_vs_k141_183608\|\|full | Not-determined |
| S8_vs_k141_28112\|\|full | Not-determined |
| S8_vs_k141_54094\|\|full | Not-determined |
| S8_vs_k141_131988\|\|full | Not-determined |
| S7_vs_138019\|\|full | Medium-quality |
| S8_vs_k141_10416\|\|full | Not-determined |
| S8_vs_k141_157866\|\|full | Low-quality |
| S8_vs_k141_88658\|\|full | Not-determined |
| S8_vs_k141_149299\|\|full | Not-determined |
| S8_vs_k141_36820\|\|full | Not-determined |
| S5_vs_k141_145633\|\|full | Medium-quality |
| S8_vs_k141_123460\|\|full | Not-determined |
| S8_vs_k141_54104\|\|full | Low-quality |
| S8_vs_k141_19114\|\|full | Not-determined |
| S8_vs_k141_10426\|\|full | Low-quality |
| S8_vs_k141_132007\|\|full | Low-quality |
| S8_vs_k141_10427\|\|full | Low-quality |
| S8_vs_k141_80084\|\|full | Low-quality |
| S8_vs_k141_175243\|\|full | Low-quality |
| S8_vs_k141_175248\|\|full | Low-quality |
| S8_vs_k141_88686\|\|full | Low-quality |
| S8_vs_k141_1942\|\|full | Low-quality |
| S8_vs_k141_1943\|\|full | Low-quality |
| S8_vs_k141_123470\|\|full | Not-determined |
| S8_vs_k141_183637\|\|full | Not-determined |
| S8_vs_k141_28154\|\|full | Not-determined |
| S8_vs_k141_1950\|\|full | Not-determined |
| S8_vs_k141_62577\|\|full | Low-quality |
| S8_vs_k141_132016\|\|full | Low-quality |
| S8_vs_k141_123481\|\|full | Low-quality |
| S8_vs_k141_36851\|\|full | Low-quality |
| S8_vs_k141_114668\|\|full | Not-determined |
| S8_vs_k141_54121\|\|full | Not-determined |
| S8_vs_k141_157919\|\|full | Not-determined |
| S8_vs_k141_140805\|\|full | Low-quality |
| S8_vs_k141_80126\|\|full | Not-determined |
| S8_vs_k141_54136\|\|full | Not-determined |
| S8_vs_k141_1976\|\|full | Not-determined |
| S8_vs_k141_10451\|\|full | Low-quality |
| S8_vs_k141_10452\|\|full | Low-quality |
| S8_vs_k141_175252\|\|full | Not-determined |
| S8_vs_k141_123492\|\|full | Not-determined |
| S8_vs_k141_193029\|\|full | Not-determined |
| S8_vs_k141_97372\|\|full | Low-quality |
| S8_vs_k141_28173\|\|full | Low-quality |
| S8_vs_k141_36876\|\|full | Low-quality |
| S8_vs_k141_201134\|\|full | Low-quality |
| S8_vs_k141_88725\|\|full | Low-quality |
| S8_vs_k141_28176\|\|full | Low-quality |
| S8_vs_k141_54152\|\|full | Low-quality |
| S8_vs_k141_28182\|\|full | Not-determined |
| S8_vs_k141_88731\|\|full | Not-determined |
| S8_vs_k141_1987\|\|full | Not-determined |
| S8_vs_k141_149348\|\|full | Low-quality |
| S8_vs_k141_193044\|\|full | Not-determined |
| S8_vs_k141_132053\|\|full | Not-determined |
| S8_vs_k141_149352\|\|full | Low-quality |
| S8_vs_k141_70868\|\|full | Low-quality |
| S8_vs_k141_80155\|\|full | Not-determined |
| S8_vs_k141_193046\|\|full | Not-determined |
| S8_vs_k141_201149\|\|full | Low-quality |
| S8_vs_k141_88752\|\|full | Low-quality |
| S8_vs_k141_193047\|\|full | Low-quality |
| S8_vs_k141_201164\|\|full | Low-quality |
| S8_vs_k141_88757\|\|full | Low-quality |
| S8_vs_k141_97417\|\|full | Low-quality |
| S8_vs_k141_157962\|\|full | Not-determined |
| S8_vs_k141_132076\|\|full | Low-quality |
| S8_vs_k141_157968\|\|full | Not-determined |
| S8_vs_k141_62647\|\|full | Low-quality |
| S8_vs_k141_132078\|\|full | Low-quality |
| S8_vs_k141_166769\|\|full | Low-quality |
| S8_vs_k141_157974\|\|full | Not-determined |
| S8_vs_k141_114719\|\|full | Not-determined |
| S8_vs_k141_132085\|\|full | Low-quality |
| S8_vs_k141_106100\|\|full | Low-quality |
| S8_vs_k141_157982\|\|full | Low-quality |
| S8_vs_k141_149377\|\|full | Low-quality |
| S8_vs_k141_45151\|\|full | Low-quality |
| S8_vs_k141_80185\|\|full | Not-determined |
| S8_vs_k141_166791\|\|full | Low-quality |
| S8_vs_k141_166792\|\|full | Not-determined |
| S8_vs_k141_201202\|\|full | Not-determined |
| S8_vs_k141_2037\|\|full | Low-quality |
| S8_vs_k141_183732\|\|full | Low-quality |
| S8_vs_k141_175337\|\|full | Low-quality |
| S8_vs_k141_123570\|\|full | Low-quality |
| S8_vs_k141_166807\|\|full | Low-quality |
| S8_vs_k141_201210\|\|full | Low-quality |
| S8_vs_k141_106145\|\|full | Not-determined |
| S8_vs_k141_123577\|\|full | Low-quality |
| S8_vs_k141_2050\|\|full | Low-quality |
| S8_vs_k141_201211\|\|full | Low-quality |
| S8_vs_k141_149408\|\|full | Low-quality |
| S8_vs_k141_19188\|\|full | Low-quality |
| S8_vs_k141_45170\|\|full | Low-quality |
| S8_vs_k141_193067\|\|full | Low-quality |
| S8_vs_k141_80204\|\|full | Low-quality |
| S8_vs_k141_62705\|\|full | Low-quality |
| S8_vs_k141_10500\|\|full | Low-quality |
| S9_extracted_k141_934 | Medium-quality |
| S8_vs_k141_193098\|\|full | Not-determined |
| S8_vs_k141_80222\|\|full | Not-determined |
| S8_vs_k141_45192\|\|full | Low-quality |
| S8_vs_k141_132109\|\|full | Low-quality |
| S8_vs_k141_106165\|\|full | Low-quality |
| S8_vs_k141_149421\|\|full | Not-determined |
| S8_vs_k141_123603\|\|full | Not-determined |
| S8_vs_k141_106171\|\|full | Not-determined |
| S8_vs_k141_62731\|\|full | Not-determined |
| S8_vs_k141_114783\|\|full | Low-quality |
| S8_vs_k141_36953\|\|full | Not-determined |
| S8_vs_k141_19236\|\|full | Not-determined |
| S8_vs_k141_54212\|\|full | Not-determined |
| S8_vs_k141_62736\|\|full | Not-determined |
| S8_vs_k141_158078\|\|full | Low-quality |
| S8_vs_k141_2098\|\|full | Not-determined |
| S8_vs_k141_36961\|\|full | Not-determined |
| S8_vs_k141_45213\|\|full | Low-quality |
| S8_vs_k141_36962\|\|full | Not-determined |
| S8_vs_k141_19242\|\|full | Low-quality |
| S8_vs_k141_175395\|\|full | Low-quality |
| S8_vs_k141_2104\|\|full | Low-quality |
| S8_vs_k141_70989\|\|full | Low-quality |
| S8_vs_k141_201277\|\|full | Not-determined |
| S8_vs_k141_114809\|\|full | Not-determined |
| S8_vs_k141_36970\|\|full | Low-quality |
| S8_vs_k141_106194\|\|full | Low-quality |
| S8_vs_k141_2113\|\|full | Low-quality |
| S8_vs_k141_62769\|\|full | Low-quality |
| S8_vs_k141_123624\|\|full | Low-quality |
| S8_vs_k141_149463\|\|full | Low-quality |
| S8_vs_k141_149464\|\|full | Low-quality |
| S8_vs_k141_193142\|\|full | Low-quality |
| S8_vs_k141_88879\|\|full | Low-quality |
| S8_vs_k141_183787\|\|full | Low-quality |
| S8_vs_k141_175417\|\|full | Low-quality |
| S8_vs_k141_106204\|\|full | Low-quality |
| S8_vs_k141_140930\|\|full | Low-quality |
| S8_vs_k141_62783\|\|full | Not-determined |
| S8_vs_k141_123644\|\|full | Not-determined |
| S8_vs_k141_28288\|\|full | Not-determined |
| S8_vs_k141_106220\|\|full | Not-determined |
| S8_vs_k141_71034\|\|full | Low-quality |
| S8_vs_k141_123657\|\|full | Low-quality |
| S8_vs_k141_2154\|\|full | Low-quality |
| S8_vs_k141_97465\|\|full | Low-quality |
| S8_vs_k141_36995\|\|full | Not-determined |
| S8_vs_k141_2158\|\|full | Low-quality |
| S8_vs_k141_201322\|\|full | Low-quality |
| S8_vs_k141_106236\|\|full | Not-determined |
| S8_vs_k141_19291\|\|full | Low-quality |
| S8_vs_k141_123664\|\|full | Low-quality |
| S8_vs_k141_71044\|\|full | Low-quality |
| S8_vs_k141_201330\|\|full | Low-quality |
| S8_vs_k141_201333\|\|full | Low-quality |
| S8_vs_k141_193189\|\|full | Not-determined |
| S8_vs_k141_166891\|\|full | Low-quality |
| S8_vs_k141_149503\|\|full | Not-determined |
| S8_vs_k141_88938\|\|full | Low-quality |
| S8_vs_k141_106246\|\|full | Not-determined |
| S8_vs_k141_10511\|\|full | Not-determined |
| S8_vs_k141_19307\|\|full | Not-determined |
| S8_vs_k141_114885\|\|full | Not-determined |
| S8_vs_k141_54315\|\|full | Not-determined |
| S8_vs_k141_114888\|\|full | Not-determined |
| S8_vs_k141_97484\|\|full | Low-quality |
| S8_vs_k141_54323\|\|full | Not-determined |
| S8_vs_k141_10527\|\|full | Low-quality |
| S8_vs_k141_19318\|\|full | Not-determined |
| S8_vs_k141_158113\|\|full | Low-quality |
| S8_vs_k141_175483\|\|full | Low-quality |
| S8_vs_k141_123684\|\|full | Low-quality |
| S8_vs_k141_132149\|\|full | Not-determined |
| S8_vs_k141_45301\|\|full | Not-determined |
| S8_vs_k141_54335\|\|full | Low-quality |
| S8_vs_k141_166913\|\|full | Low-quality |
| S8_vs_k141_88976\|\|full | Not-determined |
| S8_vs_k141_166915\|\|full | Low-quality |
| S8_vs_k141_54343\|\|full | Low-quality |
| S8_vs_k141_183827\|\|full | Not-determined |
| S8_vs_k141_123696\|\|full | Not-determined |
| S8_vs_k141_158129\|\|full | Low-quality |
| S8_vs_k141_19354\|\|full | Not-determined |
| S8_vs_k141_19355\|\|full | Low-quality |
| S8_vs_k141_54350\|\|full | Low-quality |
| S8_vs_k141_45323\|\|full | Low-quality |
| S8_vs_k141_175506\|\|full | Not-determined |
| S8_vs_k141_106295\|\|full | Not-determined |
| S8_vs_k141_10545\|\|full | Low-quality |
| S8_vs_k141_132178\|\|full | Not-determined |
| S8_vs_k141_166923\|\|full | Low-quality |
| S8_vs_k141_62858\|\|full | Not-determined |
| S8_vs_k141_62859\|\|full | Low-quality |
| S7_vs_30665\|\|full | Medium-quality |
| S8_vs_k141_193229\|\|full | Low-quality |
| S8_vs_k141_62861\|\|full | Low-quality |
| S8_vs_k141_106302\|\|full | Not-determined |
| S8_vs_k141_45338\|\|full | Not-determined |
| S8_vs_k141_80237\|\|full | Low-quality |
| S8_vs_k141_123720\|\|full | Low-quality |
| S8_vs_k141_80247\|\|full | Not-determined |
| S8_vs_k141_28326\|\|full | Not-determined |
| S8_vs_k141_149578\|\|full | Low-quality |
| S8_vs_k141_166944\|\|full | Low-quality |
| S8_vs_k141_45352\|\|full | Low-quality |
| S8_vs_k141_123728\|\|full | Low-quality |
| S8_vs_k141_19380\|\|full | Low-quality |
| S8_vs_k141_123737\|\|full | Not-determined |
| S8_vs_k141_183857\|\|full | Not-determined |
| S8_vs_k141_62883\|\|full | Not-determined |
| S8_vs_k141_54388\|\|full | Low-quality |
| S8_vs_k141_37098\|\|full | Low-quality |
| S8_vs_k141_54393\|\|full | Not-determined |
| S8_vs_k141_201394\|\|full | Low-quality |
| S8_vs_k141_2232\|\|full | Low-quality |
| S8_vs_k141_183878\|\|full | Low-quality |
| S8_vs_k141_89009\|\|full | Low-quality |
| S8_vs_k141_149617\|\|full | Not-determined |
| S8_vs_k141_183888\|\|full | Not-determined |
| S8_vs_k141_132218\|\|full | Not-determined |
| S8_vs_k141_10585\|\|full | Not-determined |
| S8_vs_k141_158146\|\|full | Low-quality |
| S8_vs_k141_106344\|\|full | Low-quality |
| S8_vs_k141_183894\|\|full | Not-determined |
| S8_vs_k141_158151\|\|full | Low-quality |
| S8_vs_k141_71153\|\|full | Low-quality |
| S8_vs_k141_166959\|\|full | Low-quality |
| S8_vs_k141_106353\|\|full | Not-determined |
| S8_vs_k141_149633\|\|full | Low-quality |
| S8_vs_k141_158162\|\|full | Low-quality |
| S8_vs_k141_106357\|\|full | Low-quality |
| S8_vs_k141_132230\|\|full | Not-determined |
| S8_vs_k141_123776\|\|full | Low-quality |
| S8_vs_k141_175585\|\|full | Not-determined |
| S8_vs_k141_158171\|\|full | Low-quality |
| S8_vs_k141_193282\|\|full | Low-quality |
| S8_vs_k141_175590\|\|full | Low-quality |
| S8_vs_k141_149646\|\|full | Low-quality |
| S8_vs_k141_166982\|\|full | Not-determined |
| S8_vs_k141_175593\|\|full | Not-determined |
| S8_vs_k141_201431\|\|full | Not-determined |
| S8_vs_k141_158174\|\|full | Low-quality |
| S8_vs_k141_175601\|\|full | Low-quality |
| S8_vs_k141_114950\|\|full | Low-quality |
| S8_vs_k141_2288\|\|full | Not-determined |
| S8_vs_k141_2292\|\|full | Not-determined |
| S8_vs_k141_114965\|\|full | Not-determined |
| S8_vs_k141_114966\|\|full | Not-determined |
| S8_vs_k141_141032\|\|full | Low-quality |
| S8_vs_k141_175613\|\|full | Not-determined |
| S8_vs_k141_114968\|\|full | Not-determined |
| S8_vs_k141_193318\|\|full | Not-determined |
| S8_vs_k141_62951\|\|full | Low-quality |
| S8_vs_k141_10616\|\|full | Not-determined |
| S8_vs_k141_97633\|\|full | Not-determined |
| S8_vs_k141_28432\|\|full | Low-quality |
| S8_vs_k141_106427\|\|full | Low-quality |
| S8_vs_k141_80365\|\|full | Low-quality |
| S8_vs_k141_80375\|\|full | Low-quality |
| S8_vs_k141_97649\|\|full | Low-quality |
| S8_vs_k141_37125\|\|full | Not-determined |
| S8_vs_k141_175646\|\|full | Not-determined |
| S8_vs_k141_10628\|\|full | Low-quality |
| S8_vs_k141_132290\|\|full | Low-quality |
| S8_vs_k141_62984\|\|full | Not-determined |
| S8_vs_k141_10631\|\|full | Low-quality |
| S8_vs_k141_149685\|\|full | Not-determined |
| S8_vs_k141_28451\|\|full | Low-quality |
| S8_vs_k141_45396\|\|full | Low-quality |
| S8_vs_k141_2336\|\|full | Not-determined |
| S8_vs_k141_28474\|\|full | Low-quality |
| S8_vs_k141_37134\|\|full | Low-quality |
| S8_vs_k141_97663\|\|full | Not-determined |
| S8_vs_k141_97665\|\|full | Low-quality |
| S8_vs_k141_19472\|\|full | Not-determined |
| S8_vs_k141_201475\|\|full | Low-quality |
| S8_vs_k141_28489\|\|full | Low-quality |
| S8_vs_k141_201482\|\|full | Low-quality |
| S8_vs_k141_80426\|\|full | Low-quality |
| S8_vs_k141_115031\|\|full | Not-determined |
| S8_vs_k141_28505\|\|full | Low-quality |
| S8_vs_k141_37162\|\|full | Low-quality |
| S8_vs_k141_167080\|\|full | Low-quality |
| S8_vs_k141_28519\|\|full | Low-quality |
| S8_vs_k141_183936\|\|full | Low-quality |
| S8_vs_k141_141103\|\|full | Low-quality |
| S9_vs_k141_934\|\|full | Medium-quality |
| S8_vs_k141_80453\|\|full | Not-determined |
| S8_vs_k141_175661\|\|full | Not-determined |
| S8_vs_k141_89138\|\|full | Low-quality |
| S8_vs_k141_141128\|\|full | Low-quality |
| S8_vs_k141_28546\|\|full | Not-determined |
| S8_vs_k141_19529\|\|full | Not-determined |
| S8_vs_k141_132346\|\|full | Low-quality |
| S8_vs_k141_45421\|\|full | Low-quality |
| S8_vs_k141_37191\|\|full | Low-quality |
| S8_vs_k141_175680\|\|full | Low-quality |
| S8_vs_k141_80457\|\|full | Not-determined |
| S8_vs_k141_10727\|\|full | Low-quality |
| S8_vs_k141_123865\|\|full | Low-quality |
| S8_vs_k141_37198\|\|full | Not-determined |
| S8_vs_k141_63058\|\|full | Not-determined |
| S8_vs_k141_106520\|\|full | Low-quality |
| S8_vs_k141_45424\|\|full | Low-quality |
| S8_vs_k141_28556\|\|full | Low-quality |
| S8_vs_k141_123871\|\|full | Low-quality |
| S8_vs_k141_80468\|\|full | Not-determined |
| S8_vs_k141_97745\|\|full | Low-quality |
| S8_vs_k141_193428\|\|full | Not-determined |
| S8_vs_k141_106543\|\|full | Not-determined |
| S8_vs_k141_201531\|\|full | Not-determined |
| S8_vs_k141_141158\|\|full | Low-quality |
| S8_vs_k141_2380\|\|full | Not-determined |
| S8_vs_k141_106549\|\|full | Low-quality |
| S8_vs_k141_123900\|\|full | Not-determined |
| S8_vs_k141_71262\|\|full | Not-determined |
| S8_vs_k141_175709\|\|full | Not-determined |
| S8_vs_k141_97768\|\|full | Not-determined |
| S8_vs_k141_193451\|\|full | Not-determined |
| S8_vs_k141_175725\|\|full | Low-quality |
| S8_vs_k141_63102\|\|full | Low-quality |
| S8_vs_k141_201561\|\|full | Low-quality |
| S8_vs_k141_183987\|\|full | Low-quality |
| S8_vs_k141_115062\|\|full | Not-determined |
| S8_vs_k141_63108\|\|full | Low-quality |
| S8_vs_k141_123928\|\|full | Low-quality |
| S8_vs_k141_183990\|\|full | Not-determined |
| S8_vs_k141_28605\|\|full | Low-quality |
| S8_vs_k141_175746\|\|full | Not-determined |
| S8_vs_k141_2417\|\|full | Not-determined |
| S8_vs_k141_54518\|\|full | Low-quality |
| S8_vs_k141_149716\|\|full | Not-determined |
| S8_vs_k141_2422\|\|full | Low-quality |
| S8_vs_k141_193476\|\|full | Low-quality |
| S8_vs_k141_141170\|\|full | Not-determined |
| S8_vs_k141_193486\|\|full | Low-quality |
| S8_vs_k141_19653\|\|full | Low-quality |
| S8_vs_k141_28651\|\|full | Not-determined |
| S8_vs_k141_167184\|\|full | Not-determined |
| S8_vs_k141_37309\|\|full | Not-determined |
| S8_vs_k141_28656\|\|full | Low-quality |
| S8_vs_k141_63175\|\|full | Low-quality |
| S8_vs_k141_19676\|\|full | Not-determined |
| S8_vs_k141_141219\|\|full | Low-quality |
| S8_vs_k141_28675\|\|full | Not-determined |
| S8_vs_k141_141226\|\|full | Low-quality |
| S8_vs_k141_184059\|\|full | Not-determined |
| S8_vs_k141_201666\|\|full | Low-quality |
| S8_vs_k141_10815\|\|full | Not-determined |
| S8_vs_k141_184064\|\|full | Not-determined |
| S8_vs_k141_149765\|\|full | Not-determined |
| S8_vs_k141_45486\|\|full | Low-quality |
| S8_vs_k141_141244\|\|full | Not-determined |
| S8_vs_k141_193515\|\|full | Not-determined |
| S8_vs_k141_28711\|\|full | Low-quality |
| S8_vs_k141_10832\|\|full | Low-quality |
| S8_vs_k141_149786\|\|full | Low-quality |
| S8_vs_k141_54561\|\|full | Low-quality |
| S8_vs_k141_193530\|\|full | Not-determined |
| S8_vs_k141_201693\|\|full | Low-quality |
| S8_vs_k141_10849\|\|full | Low-quality |
| S8_vs_k141_28737\|\|full | Not-determined |
| S8_vs_k141_149802\|\|full | Low-quality |
| S8_vs_k141_124057\|\|full | Not-determined |
| S8_vs_k141_80607\|\|full | Not-determined |
| S8_vs_k141_149813\|\|full | Low-quality |
| S8_vs_k141_141299\|\|full | Low-quality |
| S8_vs_k141_175850\|\|full | Not-determined |
| S8_vs_k141_167226\|\|full | Low-quality |
| S9_extracted_k141_90866 | Complete |
| S8_vs_k141_132476\|\|full | Low-quality |
| S8_vs_k141_167247\|\|full | Not-determined |
| S8_vs_k141_63272\|\|full | Not-determined |
| S8_vs_k141_89178\|\|full | Low-quality |
| S8_vs_k141_167253\|\|full | Low-quality |
| S8_vs_k141_19752\|\|full | Not-determined |
| S9_vs_k141_90866\|\|full | High-quality |
| S8_vs_k141_2518\|\|full | Not-determined |
| S8_vs_k141_45555\|\|full | Low-quality |
| S8_vs_k141_149857\|\|full | Low-quality |
| S8_vs_k141_141318\|\|full | Not-determined |
| S8_vs_k141_167266\|\|full | Not-determined |
| S8_vs_k141_80668\|\|full | Low-quality |
| S8_vs_k141_28813\|\|full | Low-quality |
| S8_vs_k141_184122\|\|full | Not-determined |
| S17_vs_k141_133231\|\|full | High-quality |
| S8_vs_k141_158265\|\|full | Not-determined |
| S8_vs_k141_89209\|\|full | Low-quality |
| S8_vs_k141_80676\|\|full | Low-quality |
| S8_vs_k141_184135\|\|full | Not-determined |
| S8_vs_k141_19777\|\|full | Low-quality |
| S8_vs_k141_132514\|\|full | Not-determined |
| S8_vs_k141_19778\|\|full | Low-quality |
| S8_vs_k141_45586\|\|full | Low-quality |
| S8_vs_k141_45587\|\|full | Low-quality |
| S8_vs_k141_63301\|\|full | Not-determined |
| S8_vs_k141_80687\|\|full | Not-determined |
| S8_vs_k141_141347\|\|full | Not-determined |
| S8_vs_k141_175931\|\|full | Not-determined |
| S8_vs_k141_149899\|\|full | Not-determined |
| S8_vs_k141_45588\|\|full | Low-quality |
| S8_vs_k141_97800\|\|full | Not-determined |
| S8_vs_k141_2579\|\|full | Not-determined |
| S8_vs_k141_175938\|\|full | Not-determined |
| S8_vs_k141_97809\|\|full | Not-determined |
| S8_vs_k141_124130\|\|full | Low-quality |
| S8_vs_k141_106638\|\|full | Not-determined |
| S8_vs_k141_19805\|\|full | Not-determined |
| S8_vs_k141_106645\|\|full | Not-determined |
| S8_vs_k141_175946\|\|full | Not-determined |
| S8_vs_k141_10942\|\|full | Not-determined |
| S8_vs_k141_141373\|\|full | Low-quality |
| S8_vs_k141_10944\|\|full | Not-determined |
| S8_vs_k141_115199\|\|full | Low-quality |
| S8_vs_k141_2597\|\|full | Low-quality |
| S8_vs_k141_80713\|\|full | Not-determined |
| S8_vs_k141_97823\|\|full | Not-determined |
| S8_vs_k141_201721\|\|full | Low-quality |
| S8_vs_k141_89236\|\|full | Not-determined |
| S8_vs_k141_132569\|\|full | Not-determined |
| S8_vs_k141_158306\|\|full | Low-quality |
| S8_vs_k141_115213\|\|full | Not-determined |
| S8_vs_k141_132572\|\|full | Low-quality |
| S8_vs_k141_124153\|\|full | Low-quality |
| S8_vs_k141_19836\|\|full | Not-determined |
| S8_vs_k141_45624\|\|full | Low-quality |
| S8_vs_k141_10964\|\|full | Low-quality |
| S8_vs_k141_141400\|\|full | Not-determined |
| S8_vs_k141_175982\|\|full | Not-determined |
| S8_vs_k141_28872\|\|full | Low-quality |
| S8_vs_k141_115225\|\|full | Low-quality |
| S8_vs_k141_158319\|\|full | Low-quality |
| S8_vs_k141_89258\|\|full | Not-determined |
| S8_vs_k141_97846\|\|full | Not-determined |
| S8_vs_k141_115232\|\|full | Low-quality |
| S8_vs_k141_175998\|\|full | Low-quality |
| S8_vs_k141_19850\|\|full | Low-quality |
| S8_vs_k141_106684\|\|full | Not-determined |
| S8_vs_k141_97848\|\|full | Not-determined |
| S8_vs_k141_63358\|\|full | Low-quality |
| S8_vs_k141_149927\|\|full | Not-determined |
| S8_vs_k141_45670\|\|full | Low-quality |
| S8_vs_k141_45675\|\|full | Low-quality |
| S8_vs_k141_141474\|\|full | Not-determined |
| S8_vs_k141_176045\|\|full | Low-quality |
| S8_vs_k141_124191\|\|full | Low-quality |
| S8_vs_k141_97878\|\|full | Not-determined |
| S8_vs_k141_63388\|\|full | Low-quality |
| S8_vs_k141_184244\|\|full | Not-determined |
| S8_vs_k141_63402\|\|full | Low-quality |
| S8_vs_k141_80750\|\|full | Low-quality |
| S8_vs_k141_106724\|\|full | Low-quality |
| S8_vs_k141_193735\|\|full | Not-determined |
| S8_vs_k141_63411\|\|full | Not-determined |
| S8_vs_k141_124210\|\|full | Low-quality |
| S8_vs_k141_158370\|\|full | Low-quality |
| S8_vs_k141_71513\|\|full | Low-quality |
| S8_vs_k141_176074\|\|full | Low-quality |
| S8_vs_k141_158381\|\|full | Low-quality |
| S8_vs_k141_80759\|\|full | Low-quality |
| S8_vs_k141_37439\|\|full | Low-quality |
| S8_vs_k141_124229\|\|full | Low-quality |
| S1_extracted_k141_125650 | Medium-quality |
| S8_vs_k141_193757\|\|full | Low-quality |
| S8_vs_k141_37441\|\|full | Not-determined |
| S8_vs_k141_193765\|\|full | Low-quality |
| S8_vs_k141_158393\|\|full | Not-determined |
| S8_vs_k141_89339\|\|full | Not-determined |
| S8_vs_k141_63452\|\|full | Not-determined |
| S8_vs_k141_80771\|\|full | Not-determined |
| S8_vs_k141_184253\|\|full | Low-quality |
| S8_vs_k141_71550\|\|full | Low-quality |
| S8_vs_k141_124251\|\|full | Low-quality |
| S8_vs_k141_54709\|\|full | Not-determined |
| S8_vs_k141_167413\|\|full | Low-quality |
| S8_vs_k141_141534\|\|full | Low-quality |
| S8_vs_k141_97923\|\|full | Low-quality |
| S8_vs_k141_167418\|\|full | Low-quality |
| S8_vs_k141_2678\|\|full | Low-quality |
| S8_vs_k141_54718\|\|full | Not-determined |
| S8_vs_k141_124259\|\|full | Low-quality |
| S8_vs_k141_45763\|\|full | Low-quality |
| S8_vs_k141_11013\|\|full | Not-determined |
| S8_vs_k141_2682\|\|full | Low-quality |
| S8_vs_k141_115311\|\|full | Not-determined |
| S8_vs_k141_193788\|\|full | Low-quality |
| S8_vs_k141_132669\|\|full | Low-quality |
| S8_vs_k141_2693\|\|full | Low-quality |
| S8_vs_k141_106773\|\|full | Low-quality |
| S8_vs_k141_80806\|\|full | Not-determined |
| S8_vs_k141_115318\|\|full | Not-determined |
| S8_vs_k141_167435\|\|full | Low-quality |
| S8_vs_k141_201797\|\|full | Low-quality |
| S8_vs_k141_19939\|\|full | Not-determined |
| S8_vs_k141_28948\|\|full | Low-quality |
| S8_vs_k141_184279\|\|full | Low-quality |
| S8_vs_k141_80816\|\|full | Not-determined |
| S8_vs_k141_19940\|\|full | Low-quality |
| S8_vs_k141_11037\|\|full | Not-determined |
| S8_vs_k141_184285\|\|full | Not-determined |
| S8_vs_k141_167463\|\|full | Low-quality |
| S8_vs_k141_71575\|\|full | Low-quality |
| S8_vs_k141_37476\|\|full | Not-determined |
| S8_vs_k141_167468\|\|full | Low-quality |
| S8_vs_k141_2718\|\|full | Not-determined |

| | |
|---|---|
| S8_vs_k141_89377\|\|full | Low-quality |
| S8_vs_k141_158446\|\|full | Low-quality |
| S8_vs_k141_201819\|\|full | Not-determined |
| S8_vs_k141_2730\|\|full | Not-determined |
| S8_vs_k141_176165\|\|full | Not-determined |
| S8_vs_k141_201838\|\|full | Low-quality |
| S8_vs_k141_80835\|\|full | Not-determined |
| S8_vs_k141_201841\|\|full | Low-quality |
| S1_vs_k141_125650\|\|full | Medium-quality |
| S8_vs_k141_80836\|\|full | Low-quality |
| S8_vs_k141_97983\|\|full | Not-determined |
| S8_vs_k141_54793\|\|full | Low-quality |
| S8_vs_k141_97985\|\|full | Low-quality |
| S8_vs_k141_115339\|\|full | Not-determined |
| S8_vs_k141_45780\|\|full | Low-quality |
| S8_vs_k141_124336\|\|full | Not-determined |
| S8_vs_k141_106818\|\|full | Low-quality |
| S8_vs_k141_89414\|\|full | Low-quality |
| S8_vs_k141_176184\|\|full | Low-quality |
| S8_vs_k141_158459\|\|full | Not-determined |
| S6_extracted_k141_6314 | Medium-quality |
| S8_vs_k141_80860\|\|full | Low-quality |
| S8_vs_k141_97995\|\|full | Not-determined |
| S8_vs_k141_54804\|\|full | Low-quality |
| S8_vs_k141_124354\|\|full | Low-quality |
| S8_vs_k141_28976\|\|full | Low-quality |
| S8_vs_k141_63552\|\|full | Low-quality |
| S8_vs_k141_80876\|\|full | Not-determined |
| S8_vs_k141_184321\|\|full | Not-determined |
| S8_vs_k141_176200\|\|full | Low-quality |
| S8_vs_k141_201897\|\|full | Not-determined |
| S8_vs_k141_28992\|\|full | Low-quality |
| S8_vs_k141_20037\|\|full | Low-quality |
| S8_vs_k141_63567\|\|full | Low-quality |
| S8_vs_k141_176225\|\|full | Low-quality |
| S8_vs_k141_20039\|\|full | Not-determined |
| S8_vs_k141_54823\|\|full | Low-quality |
| S8_vs_k141_37526\|\|full | Not-determined |
| S8_vs_k141_54828\|\|full | Low-quality |
| S8_vs_k141_98046\|\|full | Not-determined |
| S8_vs_k141_71601\|\|full | Not-determined |
| S8_vs_k141_167558\|\|full | Low-quality |
| S8_vs_k141_132698\|\|full | Low-quality |
| S8_vs_k141_132700\|\|full | Not-determined |
| S8_vs_k141_167559\|\|full | Low-quality |
| S8_vs_k141_20066\|\|full | Not-determined |
| S8_vs_k141_201919\|\|full | Not-determined |
| S8_vs_k141_80937\|\|full | Not-determined |
| S8_vs_k141_11160\|\|full | Low-quality |
| S8_vs_k141_29025\|\|full | Not-determined |
| S8_vs_k141_124396\|\|full | Not-determined |
| S8_vs_k141_132707\|\|full | Low-quality |
| S8_vs_k141_71613\|\|full | Low-quality |
| S8_vs_k141_158514\|\|full | Not-determined |
| S8_vs_k141_184380\|\|full | Low-quality |
| S8_vs_k141_176263\|\|full | Not-determined |
| S8_vs_k141_37556\|\|full | Not-determined |
| S8_vs_k141_45848\|\|full | Low-quality |
| S8_vs_k141_141571\|\|full | Not-determined |
| S8_vs_k141_98070\|\|full | Not-determined |
| S8_vs_k141_132724\|\|full | Not-determined |
| S8_vs_k141_115441\|\|full | Low-quality |
| S8_vs_k141_63621\|\|full | Low-quality |
| S8_vs_k141_54864\|\|full | Low-quality |
| S8_vs_k141_106924\|\|full | Low-quality |
| S8_vs_k141_45863\|\|full | Not-determined |
| S8_vs_k141_167624\|\|full | Low-quality |
| S8_vs_k141_63628\|\|full | Low-quality |
| S8_vs_k141_184402\|\|full | Not-determined |
| S8_vs_k141_158566\|\|full | Low-quality |
| S8_vs_k141_54877\|\|full | Low-quality |
| S8_vs_k141_115469\|\|full | Not-determined |
| S8_vs_k141_176298\|\|full | Low-quality |
| S8_vs_k141_124411\|\|full | Not-determined |
| S8_vs_k141_150122\|\|full | Low-quality |
| S8_vs_k141_132762\|\|full | Not-determined |
| S8_vs_k141_150125\|\|full | Not-determined |
| S8_vs_k141_150128\|\|full | Low-quality |
| S8_vs_k141_124419\|\|full | Not-determined |
| S8_vs_k141_20119\|\|full | Low-quality |
| S8_vs_k141_63650\|\|full | Low-quality |
| S8_vs_k141_176313\|\|full | Low-quality |
| S8_vs_k141_141620\|\|full | Low-quality |
| S8_vs_k141_80975\|\|full | Not-determined |
| S8_vs_k141_20132\|\|full | Low-quality |
| S8_vs_k141_54902\|\|full | Low-quality |
| S8_vs_k141_98141\|\|full | Not-determined |
| S8_vs_k141_158598\|\|full | Not-determined |
| S8_vs_k141_37600\|\|full | Not-determined |
| S8_vs_k141_201967\|\|full | Low-quality |
| S8_vs_k141_184453\|\|full | Low-quality |
| S8_vs_k141_115531\|\|full | Low-quality |
| S8_vs_k141_11189\|\|full | Low-quality |
| S8_vs_k141_124445\|\|full | Not-determined |
| S8_vs_k141_158626\|\|full | Low-quality |
| S8_vs_k141_81032\|\|full | Not-determined |
| S8_vs_k141_141645\|\|full | Low-quality |
| S8_vs_k141_106955\|\|full | Not-determined |
| S8_vs_k141_45946\|\|full | Low-quality |
| S8_vs_k141_63700\|\|full | Not-determined |
| S8_vs_k141_98180\|\|full | Low-quality |
| S8_vs_k141_37634\|\|full | Not-determined |
| S8_vs_k141_158646\|\|full | Not-determined |
| S8_vs_k141_124470\|\|full | Low-quality |
| S8_vs_k141_106964\|\|full | Not-determined |
| S8_vs_k141_89563\|\|full | Not-determined |
| S8_vs_k141_81043\|\|full | Low-quality |
| S8_vs_k141_124473\|\|full | Not-determined |
| S8_vs_k141_29036\|\|full | Not-determined |
| S8_vs_k141_45969\|\|full | Not-determined |
| S8_vs_k141_202020\|\|full | Low-quality |
| S8_vs_k141_89569\|\|full | Low-quality |
| S8_vs_k141_132802\|\|full | Not-determined |
| S8_vs_k141_106988\|\|full | Not-determined |
| S8_vs_k141_54945\|\|full | Low-quality |
| S8_vs_k141_158661\|\|full | Not-determined |
| S8_vs_k141_81071\|\|full | Low-quality |
| S8_vs_k141_11254\|\|full | Not-determined |
| S8_vs_k141_2835\|\|full | Low-quality |
| S8_vs_k141_81086\|\|full | Low-quality |
| S8_vs_k141_132824\|\|full | Low-quality |
| S8_vs_k141_176413\|\|full | Not-determined |
| S8_vs_k141_46002\|\|full | Not-determined |
| S8_vs_k141_167681\|\|full | Low-quality |
| S8_vs_k141_98253\|\|full | Low-quality |
| S8_vs_k141_37674\|\|full | Not-determined |
| S8_vs_k141_141680\|\|full | Not-determined |
| S8_vs_k141_202073\|\|full | Not-determined |
| S8_vs_k141_167690\|\|full | Low-quality |
| S8_vs_k141_132848\|\|full | Low-quality |
| S8_vs_k141_2857\|\|full | Not-determined |
| S8_vs_k141_11286\|\|full | Not-determined |
| S8_vs_k141_63753\|\|full | Not-determined |
| S8_vs_k141_124526\|\|full | Low-quality |
| S8_vs_k141_193927\|\|full | Not-determined |
| S8_vs_k141_202089\|\|full | Low-quality |
| S8_vs_k141_150197\|\|full | Low-quality |
| S8_vs_k141_71727\|\|full | Not-determined |
| S8_vs_k141_150200\|\|full | Not-determined |
| S8_vs_k141_89628\|\|full | Not-determined |
| S8_vs_k141_202099\|\|full | Not-determined |
| S8_vs_k141_29071\|\|full | Not-determined |
| S8_vs_k141_71730\|\|full | Low-quality |
| S8_vs_k141_37694\|\|full | Low-quality |
| S8_vs_k141_107070\|\|full | Low-quality |
| S8_vs_k141_20267\|\|full | Low-quality |
| S8_vs_k141_158704\|\|full | Not-determined |
| S8_vs_k141_141728\|\|full | Not-determined |
| S8_vs_k141_89646\|\|full | Low-quality |
| S8_vs_k141_124552\|\|full | Low-quality |
| S8_vs_k141_71739\|\|full | Low-quality |
| S8_vs_k141_20282\|\|full | Not-determined |
| S8_vs_k141_89659\|\|full | Not-determined |
| S8_vs_k141_29105\|\|full | Low-quality |
| S8_vs_k141_150226\|\|full | Low-quality |
| S8_vs_k141_202119\|\|full | Not-determined |
| S8_vs_k141_98324\|\|full | Low-quality |
| S8_vs_k141_46073\|\|full | Not-determined |
| S8_vs_k141_89673\|\|full | Low-quality |
| S8_vs_k141_46080\|\|full | Low-quality |
| S8_vs_k141_124572\|\|full | Low-quality |
| S8_vs_k141_158727\|\|full | Low-quality |
| S8_vs_k141_115671\|\|full | Not-determined |
| S8_vs_k141_89679\|\|full | Not-determined |
| S8_vs_k141_141770\|\|full | Low-quality |
| S8_vs_k141_37751\|\|full | Not-determined |
| S8_vs_k141_98336\|\|full | Low-quality |
| S8_vs_k141_89696\|\|full | Not-determined |
| S8_vs_k141_46095\|\|full | Low-quality |
| S8_vs_k141_98343\|\|full | Not-determined |
| S8_vs_k141_89701\|\|full | Not-determined |
| S8_vs_k141_184493\|\|full | Not-determined |
| S8_vs_k141_98346\|\|full | Low-quality |
| S8_vs_k141_11347\|\|full | Low-quality |
| S8_vs_k141_132941\|\|full | Not-determined |
| S8_vs_k141_55088\|\|full | Low-quality |
| S8_vs_k141_71802\|\|full | Not-determined |
| S8_vs_k141_55092\|\|full | Low-quality |
| S8_vs_k141_2931\|\|full | Low-quality |
| S8_vs_k141_55094\|\|full | Low-quality |
| S8_vs_k141_150263\|\|full | Not-determined |
| S8_vs_k141_29149\|\|full | Low-quality |
| S8_vs_k141_71824\|\|full | Low-quality |
| S8_vs_k141_202153\|\|full | Not-determined |
| S8_vs_k141_98370\|\|full | Not-determined |
| S8_vs_k141_141787\|\|full | Low-quality |
| S8_vs_k141_81150\|\|full | Low-quality |
| S8_vs_k141_89758\|\|full | Low-quality |
| S8_vs_k141_55115\|\|full | Not-determined |
| S8_vs_k141_150309\|\|full | Low-quality |
| S8_vs_k141_107162\|\|full | Low-quality |
| S8_vs_k141_193963\|\|full | Not-determined |
| S8_vs_k141_141796\|\|full | Not-determined |
| S8_vs_k141_20357\|\|full | Not-determined |
| S8_vs_k141_150322\|\|full | Low-quality |
| S8_vs_k141_141802\|\|full | Not-determined |
| S8_vs_k141_184531\|\|full | Low-quality |
| S8_vs_k141_71863\|\|full | Not-determined |
| S8_vs_k141_202171\|\|full | Not-determined |
| S8_vs_k141_29205\|\|full | Not-determined |
| S8_vs_k141_176548\|\|full | Low-quality |
| S8_vs_k141_37805\|\|full | Low-quality |
| S8_vs_k141_184542\|\|full | Low-quality |
| S8_vs_k141_184543\|\|full | Low-quality |
| S8_vs_k141_71871\|\|full | Not-determined |
| S8_vs_k141_55149\|\|full | Not-determined |
| S8_vs_k141_141827\|\|full | Not-determined |
| S8_vs_k141_107189\|\|full | Low-quality |
| S8_vs_k141_193998\|\|full | Not-determined |
| S8_vs_k141_98413\|\|full | Not-determined |
| S8_vs_k141_98414\|\|full | Low-quality |
| S8_vs_k141_81205\|\|full | Not-determined |
| S8_vs_k141_98420\|\|full | Low-quality |
| S8_vs_k141_202206\|\|full | Low-quality |
| S8_vs_k141_63840\|\|full | Low-quality |
| S8_vs_k141_167709\|\|full | Low-quality |
| S8_vs_k141_81208\|\|full | Low-quality |
| S8_vs_k141_55168\|\|full | Low-quality |
| S8_vs_k141_29236\|\|full | Not-determined |
| S8_vs_k141_107209\|\|full | Low-quality |
| S8_vs_k141_98425\|\|full | Low-quality |
| S8_vs_k141_98426\|\|full | Low-quality |
| S8_vs_k141_107211\|\|full | Not-determined |
| S8_vs_k141_63851\|\|full | Low-quality |
| S8_vs_k141_176602\|\|full | Not-determined |
| S8_vs_k141_46157\|\|full | Low-quality |
| S8_vs_k141_107224\|\|full | Not-determined |
| S8_vs_k141_194025\|\|full | Low-quality |
| S8_vs_k141_81221\|\|full | Not-determined |
| S8_vs_k141_71932\|\|full | Not-determined |
| S8_vs_k141_11444\|\|full | Low-quality |
| S8_vs_k141_124677\|\|full | Not-determined |
| S8_vs_k141_202223\|\|full | Not-determined |
| S8_vs_k141_98450\|\|full | Not-determined |
| S8_vs_k141_11451\|\|full | Low-quality |
| S8_vs_k141_20421\|\|full | Low-quality |
| S8_vs_k141_194056\|\|full | Not-determined |
| S8_vs_k141_150338\|\|full | Low-quality |
| S8_vs_k141_89805\|\|full | Low-quality |
| S8_vs_k141_158879\|\|full | Low-quality |
| S8_vs_k141_202258\|\|full | Low-quality |
| S8_vs_k141_3059\|\|full | Not-determined |
| S8_vs_k141_3062\|\|full | Low-quality |
| S8_vs_k141_11474\|\|full | Low-quality |
| S8_vs_k141_158895\|\|full | Not-determined |
| S8_vs_k141_133093\|\|full | Not-determined |
| S5_extracted_k141_84047 | Medium-quality |
| S8_vs_k141_167765\|\|full | Low-quality |
| S8_vs_k141_46180\|\|full | Not-determined |
| S8_vs_k141_202274\|\|full | Low-quality |
| S8_vs_k141_20447\|\|full | Low-quality |
| S8_vs_k141_167776\|\|full | Low-quality |
| S8_vs_k141_20454\|\|full | Low-quality |
| S8_vs_k141_167777\|\|full | Not-determined |
| S8_vs_k141_98505\|\|full | Not-determined |
| S8_vs_k141_176613\|\|full | Not-determined |
| S8_vs_k141_3070\|\|full | Low-quality |
| S8_vs_k141_167779\|\|full | Not-determined |
| S8_vs_k141_107270\|\|full | Not-determined |
| S8_vs_k141_115812\|\|full | Low-quality |
| S8_vs_k141_63897\|\|full | Low-quality |
| S8_vs_k141_98513\|\|full | Not-determined |
| S8_vs_k141_158910\|\|full | Not-determined |
| S8_vs_k141_167797\|\|full | Low-quality |
| S8_vs_k141_71935\|\|full | Low-quality |
| S8_vs_k141_71936\|\|full | Low-quality |
| S8_vs_k141_167801\|\|full | Low-quality |
| S8_vs_k141_115829\|\|full | Not-determined |
| S8_vs_k141_158913\|\|full | Not-determined |
| S8_vs_k141_176634\|\|full | Low-quality |
| S8_vs_k141_71950\|\|full | Low-quality |
| S8_vs_k141_46211\|\|full | Low-quality |
| S8_vs_k141_158924\|\|full | Not-determined |
| S8_vs_k141_55185\|\|full | Not-determined |
| S8_vs_k141_158926\|\|full | Low-quality |
| S8_vs_k141_141949\|\|full | Low-quality |
| S8_vs_k141_20461\|\|full | Low-quality |
| S8_vs_k141_176661\|\|full | Low-quality |
| S8_vs_k141_71972\|\|full | Not-determined |
| S8_vs_k141_184663\|\|full | Not-determined |
| S8_vs_k141_98546\|\|full | Not-determined |
| S8_vs_k141_63944\|\|full | Not-determined |
| S8_vs_k141_158946\|\|full | Low-quality |
| S8_vs_k141_98551\|\|full | Not-determined |
| S8_vs_k141_29339\|\|full | Not-determined |
| S8_vs_k141_167828\|\|full | Low-quality |
| S8_vs_k141_167832\|\|full | Low-quality |
| S8_vs_k141_63960\|\|full | Low-quality |
| S8_vs_k141_37909\|\|full | Low-quality |
| S8_vs_k141_98560\|\|full | Not-determined |
| S8_vs_k141_184679\|\|full | Not-determined |
| S8_vs_k141_55217\|\|full | Not-determined |
| S8_vs_k141_29356\|\|full | Not-determined |
| S8_vs_k141_107334\|\|full | Not-determined |
| S8_vs_k141_71981\|\|full | Not-determined |
| S8_vs_k141_202320\|\|full | Low-quality |
| S8_vs_k141_194138\|\|full | Low-quality |
| S8_vs_k141_20490\|\|full | Low-quality |
| S8_vs_k141_11530\|\|full | Not-determined |
| S8_vs_k141_55221\|\|full | Low-quality |
| S8_vs_k141_71987\|\|full | Low-quality |
| S8_vs_k141_20493\|\|full | Low-quality |
| S8_vs_k141_133191\|\|full | Not-determined |
| S8_vs_k141_176691\|\|full | Not-determined |
| S8_vs_k141_158951\|\|full | Not-determined |
| S8_vs_k141_46253\|\|full | Low-quality |
| S8_vs_k141_115896\|\|full | Not-determined |
| S8_vs_k141_29377\|\|full | Low-quality |
| S8_vs_k141_37932\|\|full | Not-determined |
| S8_vs_k141_167854\|\|full | Not-determined |
| S8_vs_k141_71991\|\|full | Low-quality |
| S8_vs_k141_167855\|\|full | Low-quality |
| S8_vs_k141_107341\|\|full | Not-determined |
| S8_vs_k141_115899\|\|full | Not-determined |
| S8_vs_k141_107342\|\|full | Low-quality |
| S8_vs_k141_55239\|\|full | Low-quality |
| S8_vs_k141_194173\|\|full | Low-quality |
| S8_vs_k141_167876\|\|full | Low-quality |
| S8_vs_k141_158982\|\|full | Low-quality |
| S8_vs_k141_202367\|\|full | Low-quality |
| S8_vs_k141_63988\|\|full | Not-determined |
| S8_vs_k141_11565\|\|full | Low-quality |
| S8_vs_k141_55263\|\|full | Not-determined |
| S8_vs_k141_194186\|\|full | Not-determined |
| S8_vs_k141_194188\|\|full | Low-quality |
| S8_vs_k141_184728\|\|full | Not-determined |
| S8_vs_k141_150484\|\|full | Not-determined |
| S8_vs_k141_37958\|\|full | Not-determined |
| S8_vs_k141_3181\|\|full | Low-quality |
| S8_vs_k141_81265\|\|full | Low-quality |
| S8_vs_k141_89901\|\|full | Low-quality |
| S8_vs_k141_133228\|\|full | Low-quality |
| S8_vs_k141_11577\|\|full | Not-determined |
| S8_vs_k141_107373\|\|full | Not-determined |
| S8_vs_k141_81267\|\|full | Low-quality |
| S8_vs_k141_89907\|\|full | Low-quality |
| S8_vs_k141_202372\|\|full | Low-quality |
| S8_vs_k141_202373\|\|full | Low-quality |
| S8_vs_k141_159007\|\|full | Not-determined |
| S8_vs_k141_98597\|\|full | Low-quality |
| S8_vs_k141_167912\|\|full | Low-quality |
| S8_vs_k141_194200\|\|full | Low-quality |
| S8_vs_k141_37990\|\|full | Low-quality |
| S8_vs_k141_98613\|\|full | Low-quality |
| S8_vs_k141_11601\|\|full | Low-quality |
| S8_vs_k141_194213\|\|full | Not-determined |
| S8_vs_k141_115946\|\|full | Not-determined |
| S8_vs_k141_176717\|\|full | Low-quality |
| S8_vs_k141_38002\|\|full | Low-quality |
| S8_vs_k141_194217\|\|full | Low-quality |
| S8_vs_k141_159023\|\|full | Not-determined |
| S8_vs_k141_20567\|\|full | Not-determined |
| S8_vs_k141_150525\|\|full | Low-quality |
| S8_vs_k141_124832\|\|full | Low-quality |
| S8_vs_k141_124837\|\|full | Low-quality |
| S8_vs_k141_29476\|\|full | Not-determined |
| S17_extracted_k141_128147 | High-quality |
| S8_vs_k141_20591\|\|full | Low-quality |
| S8_vs_k141_29482\|\|full | Not-determined |
| S8_vs_k141_176726\|\|full | Not-determined |
| S8_vs_k141_142067\|\|full | Low-quality |
| S8_vs_k141_29490\|\|full | Low-quality |
| S8_vs_k141_202432\|\|full | Low-quality |
| S8_vs_k141_202433\|\|full | Not-determined |
| S8_vs_k141_142076\|\|full | Low-quality |
| S8_vs_k141_3218\|\|full | Not-determined |
| S8_vs_k141_29492\|\|full | Not-determined |
| S8_vs_k141_142084\|\|full | Low-quality |
| S8_vs_k141_11654\|\|full | Low-quality |
| S8_vs_k141_107454\|\|full | Low-quality |
| S8_vs_k141_176746\|\|full | Low-quality |
| S8_vs_k141_38068\|\|full | Low-quality |
| S8_vs_k141_168032\|\|full | Low-quality |
| S8_vs_k141_38072\|\|full | Low-quality |
| S8_vs_k141_184810\|\|full | Not-determined |
| S8_vs_k141_202469\|\|full | Low-quality |
| S8_vs_k141_194246\|\|full | Not-determined |
| S8_vs_k141_20598\|\|full | Low-quality |
| S8_vs_k141_38081\|\|full | Not-determined |
| S8_vs_k141_55354\|\|full | Not-determined |
| S8_vs_k141_38085\|\|full | Low-quality |
| S8_vs_k141_202487\|\|full | Not-determined |
| S8_vs_k141_64113\|\|full | Low-quality |
| S8_vs_k141_194259\|\|full | Low-quality |
| S8_vs_k141_81377\|\|full | Not-determined |
| S17_vs_k141_128147\|\|full | High-quality |
| S8_vs_k141_202492\|\|full | Low-quality |
| S8_vs_k141_11704\|\|full | Not-determined |
| S8_vs_k141_11705\|\|full | Low-quality |
| S5_vs_k141_84047\|\|full | Medium-quality |
| S8_vs_k141_184841\|\|full | Low-quality |
| S8_vs_k141_3225\|\|full | Low-quality |
| S8_vs_k141_64148\|\|full | Low-quality |
| S8_vs_k141_20642\|\|full | Not-determined |
| S8_vs_k141_29531\|\|full | Not-determined |
| S8_vs_k141_98681\|\|full | Low-quality |
| S8_vs_k141_194276\|\|full | Not-determined |
| S8_vs_k141_168086\|\|full | Low-quality |
| S8_vs_k141_98684\|\|full | Low-quality |
| S8_vs_k141_46317\|\|full | Not-determined |
| S8_vs_k141_202526\|\|full | Not-determined |
| S8_vs_k141_11728\|\|full | Low-quality |
| S8_vs_k141_168091\|\|full | Not-determined |
| S8_vs_k141_107492\|\|full | Low-quality |
| S8_vs_k141_81403\|\|full | Not-determined |
| S8_vs_k141_194285\|\|full | Not-determined |
| S8_vs_k141_159165\|\|full | Not-determined |
| S8_vs_k141_176800\|\|full | Not-determined |
| S8_vs_k141_81408\|\|full | Not-determined |
| S8_vs_k141_20661\|\|full | Not-determined |
| S8_vs_k141_176802\|\|full | Low-quality |
| S8_vs_k141_11741\|\|full | Low-quality |
| S8_vs_k141_72158\|\|full | Not-determined |
| S8_vs_k141_202535\|\|full | Low-quality |
| S8_vs_k141_142143\|\|full | Not-determined |
| S8_vs_k141_29550\|\|full | Low-quality |
| S8_vs_k141_81418\|\|full | Low-quality |
| S8_vs_k141_46333\|\|full | Low-quality |
| S8_vs_k141_20670\|\|full | Not-determined |
| S8_vs_k141_194304\|\|full | Low-quality |
| S8_vs_k141_168114\|\|full | Low-quality |
| S8_vs_k141_107521\|\|full | Not-determined |
| S8_vs_k141_176830\|\|full | Not-determined |
| S8_vs_k141_81437\|\|full | Low-quality |
| S6_vs_k141_6314\|\|full | Medium-quality |
| S8_vs_k141_194320\|\|full | Not-determined |
| S8_vs_k141_107531\|\|full | Not-determined |
| S8_vs_k141_3262\|\|full | Low-quality |
| S8_vs_k141_176836\|\|full | Low-quality |
| S8_vs_k141_107535\|\|full | Not-determined |
| S8_vs_k141_55421\|\|full | Low-quality |
| S8_vs_k141_133309\|\|full | Not-determined |
| S8_vs_k141_159177\|\|full | Not-determined |
| S8_vs_k141_90005\|\|full | Not-determined |
| S8_vs_k141_81454\|\|full | Low-quality |
| S8_vs_k141_142151\|\|full | Low-quality |
| S8_vs_k141_107547\|\|full | Not-determined |
| S8_extracted_k141_43483 | Medium-quality |
| S8_vs_k141_159191\|\|full | Not-determined |
| S8_vs_k141_159193\|\|full | Not-determined |
| S8_vs_k141_194349\|\|full | Low-quality |
| S8_vs_k141_159195\|\|full | Not-determined |
| S8_vs_k141_38195\|\|full | Low-quality |
| S8_vs_k141_20710\|\|full | Not-determined |
| S8_vs_k141_72213\|\|full | Low-quality |
| S8_vs_k141_29608\|\|full | Not-determined |
| S8_vs_k141_72217\|\|full | Low-quality |
| S8_vs_k141_124988\|\|full | Not-determined |
| S8_vs_k141_29611\|\|full | Low-quality |
| S8_vs_k141_142153\|\|full | Not-determined |
| S8_vs_k141_3305\|\|full | Low-quality |
| S8_vs_k141_3312\|\|full | Low-quality |
| S8_vs_k141_194381\|\|full | Not-determined |
| S8_vs_k141_116005\|\|full | Not-determined |
| S8_vs_k141_55453\|\|full | Low-quality |
| S8_vs_k141_72243\|\|full | Not-determined |
| S8_vs_k141_159237\|\|full | Not-determined |
| S8_vs_k141_194397\|\|full | Not-determined |
| S8_vs_k141_107569\|\|full | Low-quality |
| S8_vs_k141_125011\|\|full | Low-quality |
| S8_vs_k141_184965\|\|full | Low-quality |
| S8_vs_k141_142173\|\|full | Low-quality |
| S8_vs_k141_159248\|\|full | Low-quality |
| S8_vs_k141_194403\|\|full | Low-quality |
| S8_vs_k141_176916\|\|full | Low-quality |
| S8_vs_k141_3336\|\|full | Low-quality |
| S8_vs_k141_142192\|\|full | Not-determined |
| S8_vs_k141_3348\|\|full | Not-determined |
| S8_vs_k141_142195\|\|full | Not-determined |
| S8_vs_k141_150608\|\|full | Low-quality |
| S8_vs_k141_184984\|\|full | Not-determined |
| S8_vs_k141_159258\|\|full | Not-determined |
| S8_vs_k141_38244\|\|full | Not-determined |
| S8_vs_k141_125022\|\|full | Low-quality |
| S8_vs_k141_38254\|\|full | Not-determined |
| S8_vs_k141_125029\|\|full | Not-determined |
| S8_vs_k141_107598\|\|full | Not-determined |
| S8_vs_k141_142212\|\|full | Low-quality |
| S8_vs_k141_20799\|\|full | Not-determined |
| S8_vs_k141_116045\|\|full | Low-quality |
| S8_vs_k141_20803\|\|full | Low-quality |
| S8_vs_k141_133422\|\|full | Not-determined |
| S8_vs_k141_20810\|\|full | Not-determined |
| S8_vs_k141_185018\|\|full | Low-quality |
| S8_vs_k141_98789\|\|full | Not-determined |
| S8_vs_k141_72319\|\|full | Not-determined |
| S8_vs_k141_116048\|\|full | Not-determined |
| S8_vs_k141_107623\|\|full | Low-quality |
| S8_vs_k141_29703\|\|full | Low-quality |
| S8_vs_k141_81554\|\|full | Low-quality |
| S8_vs_k141_168147\|\|full | Low-quality |
| S8_vs_k141_46417\|\|full | Low-quality |
| S8_vs_k141_55487\|\|full | Not-determined |
| S8_vs_k141_150625\|\|full | Low-quality |
| S8_vs_k141_38280\|\|full | Low-quality |
| S8_vs_k141_55490\|\|full | Low-quality |
| S8_vs_k141_46426\|\|full | Not-determined |
| S10_extracted_25241 | Medium-quality |
| S8_vs_k141_20818\|\|full | Low-quality |
| S8_vs_k141_194463\|\|full | Low-quality |
| S8_vs_k141_116082\|\|full | Low-quality |
| S8_vs_k141_90144\|\|full | Not-determined |
| S8_vs_k141_125031\|\|full | Low-quality |
| S8_vs_k141_107642\|\|full | Low-quality |
| S8_vs_k141_133481\|\|full | Low-quality |
| S8_vs_k141_90154\|\|full | Not-determined |
| S8_vs_k141_125044\|\|full | Not-determined |
| S8_vs_k141_194470\|\|full | Not-determined |
| S8_vs_k141_55525\|\|full | Low-quality |
| S8_vs_k141_46469\|\|full | Low-quality |
| S8_vs_k141_176986\|\|full | Not-determined |
| S8_vs_k141_81582\|\|full | Low-quality |
| S8_vs_k141_142267\|\|full | Not-determined |
| S8_vs_k141_150652\|\|full | Low-quality |
| S8_vs_k141_176995\|\|full | Low-quality |
| S8_vs_k141_159361\|\|full | Not-determined |
| S8_vs_k141_29769\|\|full | Not-determined |
| S8_vs_k141_125050\|\|full | Not-determined |
| S8_vs_k141_90167\|\|full | Low-quality |
| S8_vs_k141_194483\|\|full | Not-determined |
| S8_vs_k141_55555\|\|full | Low-quality |
| S8_vs_k141_3472\|\|full | Not-determined |
| S8_vs_k141_177015\|\|full | Low-quality |
| S8_vs_k141_125070\|\|full | Low-quality |
| S8_vs_k141_168224\|\|full | Low-quality |
| S8_vs_k141_3475\|\|full | Low-quality |
| S8_vs_k141_150677\|\|full | Low-quality |
| S8_vs_k141_72346\|\|full | Low-quality |
| S8_vs_k141_150678\|\|full | Low-quality |
| S8_vs_k141_150679\|\|full | Low-quality |
| S8_vs_k141_20897\|\|full | Low-quality |
| S8_vs_k141_29801\|\|full | Not-determined |
| S8_vs_k141_202645\|\|full | Low-quality |
| S8_vs_k141_55574\|\|full | Low-quality |
| S8_vs_k141_11821\|\|full | Not-determined |
| S8_vs_k141_159407\|\|full | Low-quality |
| S8_vs_k141_177021\|\|full | Low-quality |
| S10_vs_25241\|\|full | Medium-quality |
| S8_vs_k141_194521\|\|full | Low-quality |
| S8_vs_k141_177024\|\|full | Low-quality |
| S8_vs_k141_194525\|\|full | Low-quality |
| S8_vs_k141_150715\|\|full | Low-quality |
| S8_vs_k141_177029\|\|full | Not-determined |
| S8_vs_k141_194532\|\|full | Not-determined |
| S8_vs_k141_55593\|\|full | Low-quality |
| S8_vs_k141_98884\|\|full | Low-quality |
| S8_vs_k141_168260\|\|full | Not-determined |
| S8_vs_k141_107727\|\|full | Not-determined |
| S8_vs_k141_90195\|\|full | Low-quality |
| S8_vs_k141_98890\|\|full | Low-quality |
| S8_vs_k141_90202\|\|full | Low-quality |
| S8_vs_k141_194537\|\|full | Not-determined |
| S8_vs_k141_185063\|\|full | Low-quality |
| S8_vs_k141_81634\|\|full | Not-determined |
| S8_vs_k141_159424\|\|full | Not-determined |
| S8_vs_k141_20920\|\|full | Low-quality |
| S8_vs_k141_3544\|\|full | Low-quality |
| S8_vs_k141_72393\|\|full | Not-determined |
| S8_vs_k141_168282\|\|full | Not-determined |
| S8_vs_k141_202694\|\|full | Not-determined |
| S8_vs_k141_72398\|\|full | Low-quality |
| S8_vs_k141_150733\|\|full | Not-determined |
| S8_vs_k141_150734\|\|full | Not-determined |
| S8_vs_k141_125074\|\|full | Low-quality |
| S8_vs_k141_46566\|\|full | Not-determined |
| S8_vs_k141_81650\|\|full | Low-quality |
| S8_vs_k141_98924\|\|full | Low-quality |
| S8_vs_k141_72412\|\|full | Not-determined |
| S8_vs_k141_98928\|\|full | Low-quality |
| S8_vs_k141_177046\|\|full | Not-determined |
| S8_vs_k141_55621\|\|full | Low-quality |
| S8_vs_k141_168306\|\|full | Not-determined |
| S8_vs_k141_159447\|\|full | Not-determined |
| S8_vs_k141_3564\|\|full | Low-quality |
| S8_vs_k141_116185\|\|full | Low-quality |
| S8_vs_k141_72420\|\|full | Not-determined |
| S8_vs_k141_194601\|\|full | Not-determined |
| S8_vs_k141_150756\|\|full | Not-determined |
| S8_vs_k141_72426\|\|full | Low-quality |
| S8_vs_k141_194604\|\|full | Not-determined |
| S8_vs_k141_168311\|\|full | Low-quality |
| S8_vs_k141_72428\|\|full | Not-determined |
| S8_vs_k141_168312\|\|full | Low-quality |
| S8_vs_k141_125115\|\|full | Low-quality |
| S8_vs_k141_159473\|\|full | Not-determined |
| S8_vs_k141_11854\|\|full | Not-determined |
| S8_vs_k141_194610\|\|full | Low-quality |
| S8_vs_k141_125116\|\|full | Not-determined |
| S8_vs_k141_98949\|\|full | Low-quality |

| | |
|---|---|
| S8_vs_k141_150760\|\|full | Not-determined |
| S8_vs_k141_98955\|\|full | Low-quality |
| S8_vs_k141_142388\|\|full | Low-quality |
| S8_vs_k141_3597\|\|full | Low-quality |
| S8_vs_k141_177090\|\|full | Not-determined |
| S8_vs_k141_90248\|\|full | Not-determined |
| S8_vs_k141_3601\|\|full | Low-quality |
| S8_vs_k141_177098\|\|full | Low-quality |
| S8_vs_k141_142396\|\|full | Low-quality |
| S8_vs_k141_116220\|\|full | Low-quality |
| S8_vs_k141_20968\|\|full | Low-quality |
| S8_vs_k141_168341\|\|full | Not-determined |
| S8_vs_k141_29871\|\|full | Not-determined |
| S8_vs_k141_90264\|\|full | Not-determined |
| S8_vs_k141_98968\|\|full | Not-determined |
| S8_vs_k141_55659\|\|full | Not-determined |
| S8_vs_k141_150783\|\|full | Not-determined |
| S3_extracted_k141_109277 | Medium-quality |
| S8_vs_k141_81692\|\|full | Not-determined |
| S8_vs_k141_150788\|\|full | Not-determined |
| S8_vs_k141_177123\|\|full | Low-quality |
| S8_vs_k141_81697\|\|full | Not-determined |
| S8_vs_k141_3651\|\|full | Not-determined |
| S8_vs_k141_29890\|\|full | Low-quality |
| S8_vs_k141_3656\|\|full | Low-quality |
| S8_vs_k141_72471\|\|full | Not-determined |
| S8_vs_k141_202772\|\|full | Not-determined |
| S8_vs_k141_20991\|\|full | Low-quality |
| S8_vs_k141_125182\|\|full | Low-quality |
| S8_vs_k141_72486\|\|full | Low-quality |
| S8_vs_k141_11914\|\|full | Low-quality |
| S8_vs_k141_177135\|\|full | Low-quality |
| S8_vs_k141_72490\|\|full | Low-quality |
| S8_vs_k141_46640\|\|full | Low-quality |
| S8_vs_k141_116256\|\|full | Low-quality |
| S8_vs_k141_125191\|\|full | Low-quality |
| S8_vs_k141_81711\|\|full | Low-quality |
| S8_vs_k141_150814\|\|full | Low-quality |
| S8_vs_k141_99025\|\|full | Not-determined |
| S8_vs_k141_202785\|\|full | Not-determined |
| S8_vs_k141_194644\|\|full | Low-quality |
| S8_vs_k141_38334\|\|full | Low-quality |
| S8_vs_k141_168405\|\|full | Low-quality |
| S8_vs_k141_185161\|\|full | Not-determined |
| S8_vs_k141_194655\|\|full | Not-determined |
| S8_vs_k141_72508\|\|full | Not-determined |
| S8_vs_k141_55708\|\|full | Low-quality |
| S8_vs_k141_168414\|\|full | Not-determined |
| S8_vs_k141_107869\|\|full | Low-quality |
| S8_vs_k141_116290\|\|full | Low-quality |
| S8_vs_k141_168416\|\|full | Not-determined |
| S8_vs_k141_116293\|\|full | Not-determined |
| S8_vs_k141_72513\|\|full | Low-quality |
| S8_vs_k141_202814\|\|full | Low-quality |
| S8_vs_k141_168425\|\|full | Not-determined |
| S8_vs_k141_194663\|\|full | Low-quality |
| S8_vs_k141_3699\|\|full | Not-determined |
| S8_vs_k141_177160\|\|full | Low-quality |
| S8_vs_k141_29947\|\|full | Not-determined |
| S8_vs_k141_194681\|\|full | Not-determined |
| S8_vs_k141_159570\|\|full | Not-determined |
| S8_vs_k141_3713\|\|full | Low-quality |
| S8_vs_k141_125233\|\|full | Low-quality |
| S8_vs_k141_3714\|\|full | Not-determined |
| S8_vs_k141_116324\|\|full | Low-quality |
| S8_vs_k141_29958\|\|full | Not-determined |
| S8_vs_k141_116325\|\|full | Low-quality |
| S8_vs_k141_142480\|\|full | Low-quality |
| S8_vs_k141_159590\|\|full | Not-determined |
| S8_vs_k141_81743\|\|full | Low-quality |
| S8_vs_k141_90385\|\|full | Low-quality |
| S8_vs_k141_99090\|\|full | Not-determined |
| S8_vs_k141_125252\|\|full | Low-quality |
| S8_vs_k141_202839\|\|full | Low-quality |
| S8_vs_k141_142489\|\|full | Low-quality |
| S8_vs_k141_107910\|\|full | Low-quality |
| S8_vs_k141_3727\|\|full | Low-quality |
| S8_vs_k141_133540\|\|full | Low-quality |
| S8_vs_k141_116343\|\|full | Low-quality |
| S8_vs_k141_64170\|\|full | Low-quality |
| S8_vs_k141_150872\|\|full | Low-quality |
| S8_vs_k141_64176\|\|full | Low-quality |
| S8_vs_k141_177178\|\|full | Not-determined |
| S8_vs_k141_168475\|\|full | Not-determined |
| S8_vs_k141_3736\|\|full | Low-quality |
| S8_vs_k141_107925\|\|full | Low-quality |
| S8_vs_k141_81773\|\|full | Low-quality |
| S8_vs_k141_125276\|\|full | Not-determined |
| S8_vs_k141_177198\|\|full | Low-quality |
| S8_vs_k141_168487\|\|full | Not-determined |
| S8_vs_k141_202882\|\|full | Not-determined |
| S8_vs_k141_168508\|\|full | Low-quality |
| S8_vs_k141_142542\|\|full | Low-quality |
| S8_vs_k141_30046\|\|full | Low-quality |
| S8_vs_k141_55811\|\|full | Not-determined |
| S8_vs_k141_142547\|\|full | Not-determined |
| S8_vs_k141_202906\|\|full | Low-quality |
| S8_vs_k141_116411\|\|full | Not-determined |
| S8_vs_k141_125293\|\|full | Low-quality |
| S8_vs_k141_142561\|\|full | Not-determined |
| S8_vs_k141_11928\|\|full | Low-quality |
| S8_vs_k141_3811\|\|full | Not-determined |
| S8_vs_k141_202914\|\|full | Low-quality |
| S8_vs_k141_168542\|\|full | Low-quality |
| S8_vs_k141_168545\|\|full | Low-quality |
| S8_vs_k141_159658\|\|full | Not-determined |
| S8_vs_k141_99175\|\|full | Not-determined |
| S8_vs_k141_168555\|\|full | Low-quality |
| S8_vs_k141_30077\|\|full | Low-quality |
| S8_vs_k141_99188\|\|full | Not-determined |
| S8_vs_k141_90455\|\|full | Low-quality |
| S8_vs_k141_55866\|\|full | Not-determined |
| S8_vs_k141_90468\|\|full | Low-quality |
| S8_vs_k141_11953\|\|full | Low-quality |
| S8_vs_k141_55870\|\|full | Low-quality |
| S8_vs_k141_168575\|\|full | Low-quality |
| S8_vs_k141_30117\|\|full | Not-determined |
| S8_vs_k141_168580\|\|full | Not-determined |
| S8_vs_k141_108034\|\|full | Not-determined |
| S8_vs_k141_168581\|\|full | Not-determined |
| S8_vs_k141_194779\|\|full | Not-determined |
| S8_vs_k141_168582\|\|full | Not-determined |
| S8_vs_k141_177316\|\|full | Low-quality |
| S8_vs_k141_168584\|\|full | Not-determined |
| S8_vs_k141_21140\|\|full | Not-determined |
| S8_vs_k141_38410\|\|full | Low-quality |
| S8_vs_k141_150993\|\|full | Low-quality |
| S8_vs_k141_108046\|\|full | Low-quality |
| S8_vs_k141_177333\|\|full | Low-quality |
| S8_vs_k141_99243\|\|full | Not-determined |
| S8_vs_k141_3882\|\|full | Low-quality |
| S8_vs_k141_11992\|\|full | Low-quality |
| S8_vs_k141_133679\|\|full | Not-determined |
| S8_vs_k141_55927\|\|full | Not-determined |
| S8_vs_k141_125372\|\|full | Low-quality |
| S8_vs_k141_38435\|\|full | Low-quality |
| S8_vs_k141_30156\|\|full | Low-quality |
| S8_vs_k141_38438\|\|full | Low-quality |
| S8_vs_k141_142673\|\|full | Low-quality |
| S8_vs_k141_202979\|\|full | Not-determined |
| S8_vs_k141_81801\|\|full | Low-quality |
| S8_vs_k141_72759\|\|full | Low-quality |
| S8_vs_k141_99267\|\|full | Low-quality |
| S8_vs_k141_12011\|\|full | Not-determined |
| S8_vs_k141_133710\|\|full | Not-determined |
| S8_vs_k141_151041\|\|full | Not-determined |
| S8_vs_k141_64313\|\|full | Low-quality |
| S8_vs_k141_90518\|\|full | Not-determined |
| S8_vs_k141_125404\|\|full | Not-determined |
| S8_vs_k141_30194\|\|full | Low-quality |
| S8_vs_k141_203002\|\|full | Not-determined |
| S8_vs_k141_116519\|\|full | Low-quality |
| S8_vs_k141_72768\|\|full | Low-quality |
| S8_vs_k141_90532\|\|full | Not-determined |
| S8_vs_k141_133730\|\|full | Not-determined |
| S8_vs_k141_168643\|\|full | Low-quality |
| S8_vs_k141_142696\|\|full | Low-quality |
| S8_vs_k141_151063\|\|full | Not-determined |
| S8_vs_k141_99296\|\|full | Low-quality |
| S8_vs_k141_12047\|\|full | Low-quality |
| S8_vs_k141_30210\|\|full | Not-determined |
| S8_vs_k141_3894\|\|full | Low-quality |
| S8_vs_k141_12049\|\|full | Low-quality |
| S8_vs_k141_194841\|\|full | Low-quality |
| S8_vs_k141_64350\|\|full | Low-quality |
| S8_vs_k141_99315\|\|full | Low-quality |
| S8_vs_k141_90555\|\|full | Low-quality |
| S8_vs_k141_159697\|\|full | Not-determined |
| S8_vs_k141_99319\|\|full | Low-quality |
| S8_vs_k141_30224\|\|full | Not-determined |
| S8_vs_k141_38480\|\|full | Low-quality |
| S8_vs_k141_43483\|\|full | Medium-quality |
| S8_vs_k141_185322\|\|full | Low-quality |
| S8_vs_k141_168668\|\|full | Not-determined |
| S8_vs_k141_81839\|\|full | Low-quality |
| S8_vs_k141_185324\|\|full | Not-determined |
| S8_vs_k141_159710\|\|full | Low-quality |
| S8_vs_k141_99337\|\|full | Not-determined |
| S8_vs_k141_3918\|\|full | Not-determined |
| S8_vs_k141_81846\|\|full | Low-quality |
| S8_vs_k141_12081\|\|full | Not-determined |
| S8_vs_k141_159720\|\|full | Not-determined |
| S8_vs_k141_133792\|\|full | Low-quality |
| S8_vs_k141_38496\|\|full | Not-determined |
| S8_vs_k141_142732\|\|full | Low-quality |
| S8_vs_k141_125471\|\|full | Not-determined |
| S8_vs_k141_151081\|\|full | Not-determined |
| S8_vs_k141_38507\|\|full | Low-quality |
| S8_vs_k141_81854\|\|full | Low-quality |
| S8_vs_k141_168689\|\|full | Low-quality |
| S8_vs_k141_3951\|\|full | Low-quality |
| S8_vs_k141_168691\|\|full | Not-determined |
| S8_vs_k141_64366\|\|full | Low-quality |
| S8_vs_k141_133807\|\|full | Low-quality |
| S8_vs_k141_185363\|\|full | Not-determined |
| S8_vs_k141_38509\|\|full | Low-quality |
| S8_vs_k141_133809\|\|full | Low-quality |
| S8_vs_k141_21210\|\|full | Low-quality |
| S8_vs_k141_30263\|\|full | Low-quality |
| S8_vs_k141_194871\|\|full | Not-determined |
| S8_vs_k141_116584\|\|full | Not-determined |
| S8_vs_k141_3969\|\|full | Low-quality |
| S12_vs_k141_164056\|\|full | High-quality |
| S8_vs_k141_30274\|\|full | Low-quality |
| S8_vs_k141_56011\|\|full | Not-determined |
| S8_vs_k141_12125\|\|full | Low-quality |
| S8_vs_k141_151110\|\|full | Not-determined |
| S8_vs_k141_38524\|\|full | Not-determined |
| S8_vs_k141_90628\|\|full | Low-quality |
| S8_vs_k141_38527\|\|full | Low-quality |
| S8_vs_k141_46846\|\|full | Low-quality |
| S8_vs_k141_21220\|\|full | Low-quality |
| S8_vs_k141_159788\|\|full | Not-determined |
| S8_vs_k141_46856\|\|full | Low-quality |
| S8_vs_k141_30310\|\|full | Low-quality |
| S8_vs_k141_194906\|\|full | Not-determined |
| S8_vs_k141_151134\|\|full | Low-quality |
| S8_vs_k141_99358\|\|full | Low-quality |
| S8_vs_k141_3992\|\|full | Not-determined |
| S8_vs_k141_90659\|\|full | Not-determined |
| S8_vs_k141_185411\|\|full | Low-quality |
| S8_vs_k141_108155\|\|full | Not-determined |
| S8_vs_k141_90660\|\|full | Not-determined |
| S8_vs_k141_108160\|\|full | Low-quality |
| S8_vs_k141_185418\|\|full | Low-quality |
| S8_vs_k141_56036\|\|full | Low-quality |
| S8_vs_k141_168753\|\|full | Low-quality |
| S8_vs_k141_116596\|\|full | Not-determined |
| S8_vs_k141_168757\|\|full | Low-quality |
| S8_vs_k141_64433\|\|full | Low-quality |
| S8_vs_k141_142787\|\|full | Not-determined |
| S8_vs_k141_108173\|\|full | Low-quality |
| S8_vs_k141_12184\|\|full | Low-quality |
| S8_vs_k141_38557\|\|full | Not-determined |
| S8_vs_k141_168766\|\|full | Not-determined |
| S8_vs_k141_4029\|\|full | Low-quality |
| S8_vs_k141_125544\|\|full | Low-quality |
| S8_vs_k141_125546\|\|full | Not-determined |
| S8_vs_k141_90694\|\|full | Low-quality |
| S8_vs_k141_194946\|\|full | Low-quality |
| S8_vs_k141_177468\|\|full | Not-determined |
| S8_vs_k141_108189\|\|full | Low-quality |
| S8_vs_k141_185449\|\|full | Not-determined |
| S8_vs_k141_90709\|\|full | Low-quality |
| S8_vs_k141_12200\|\|full | Low-quality |
| S8_vs_k141_203092\|\|full | Low-quality |
| S8_vs_k141_159819\|\|full | Not-determined |
| S8_vs_k141_46906\|\|full | Not-determined |
| S8_vs_k141_90719\|\|full | Low-quality |
| S8_vs_k141_12205\|\|full | Low-quality |
| S4_extracted_17948 | High-quality |
| S8_vs_k141_90726\|\|full | Low-quality |
| S8_vs_k141_108206\|\|full | Low-quality |
| S8_vs_k141_12207\|\|full | Low-quality |
| S8_vs_k141_81897\|\|full | Low-quality |
| S8_vs_k141_151174\|\|full | Not-determined |
| S8_vs_k141_81903\|\|full | Low-quality |
| S8_vs_k141_90730\|\|full | Not-determined |
| S8_vs_k141_194986\|\|full | Not-determined |
| S8_vs_k141_203129\|\|full | Not-determined |
| S8_vs_k141_46933\|\|full | Low-quality |
| S8_vs_k141_159825\|\|full | Low-quality |
| S8_vs_k141_38596\|\|full | Not-determined |
| S8_vs_k141_133932\|\|full | Low-quality |
| S8_vs_k141_159826\|\|full | Not-determined |
| S8_vs_k141_168784\|\|full | Not-determined |
| S8_vs_k141_203154\|\|full | Low-quality |
| S8_vs_k141_81928\|\|full | Not-determined |
| S8_vs_k141_99414\|\|full | Low-quality |
| S8_vs_k141_203160\|\|full | Not-determined |
| S8_vs_k141_203161\|\|full | Low-quality |
| S8_vs_k141_56118\|\|full | Not-determined |
| S8_vs_k141_177529\|\|full | Low-quality |
| S8_vs_k141_195023\|\|full | Not-determined |
| S8_vs_k141_177530\|\|full | Low-quality |
| S8_vs_k141_133942\|\|full | Low-quality |
| S8_vs_k141_108227\|\|full | Low-quality |
| S8_vs_k141_46982\|\|full | Low-quality |
| S8_vs_k141_108232\|\|full | Not-determined |
| S8_vs_k141_125614\|\|full | Low-quality |
| S8_vs_k141_159879\|\|full | Low-quality |
| S8_vs_k141_46997\|\|full | Low-quality |
| S10_extracted_47170 | Medium-quality |
| S8_vs_k141_38652\|\|full | Not-determined |
| S8_vs_k141_142902\|\|full | Not-determined |
| S8_vs_k141_21342\|\|full | Not-determined |
| S8_vs_k141_99437\|\|full | Low-quality |
| S8_vs_k141_21349\|\|full | Low-quality |
| S8_vs_k141_12223\|\|full | Low-quality |
| S3_extracted_k141_67828 | Complete |
| S8_vs_k141_56158\|\|full | Low-quality |
| S8_vs_k141_142919\|\|full | Low-quality |
| S8_vs_k141_90820\|\|full | Low-quality |
| S8_vs_k141_12244\|\|full | Low-quality |
| S8_vs_k141_108275\|\|full | Low-quality |
| S8_vs_k141_142926\|\|full | Low-quality |
| S8_vs_k141_30441\|\|full | Low-quality |
| S8_vs_k141_12253\|\|full | Not-determined |
| S8_vs_k141_38676\|\|full | Low-quality |
| S8_vs_k141_133984\|\|full | Low-quality |
| S8_vs_k141_38678\|\|full | Not-determined |
| S8_vs_k141_64502\|\|full | Low-quality |
| S8_vs_k141_81983\|\|full | Low-quality |
| S8_vs_k141_30450\|\|full | Low-quality |
| S8_vs_k141_90843\|\|full | Not-determined |
| S8_vs_k141_203221\|\|full | Not-determined |
| S8_vs_k141_81987\|\|full | Low-quality |
| S8_vs_k141_38680\|\|full | Not-determined |
| S8_vs_k141_203223\|\|full | Low-quality |
| S8_vs_k141_185503\|\|full | Not-determined |
| S8_vs_k141_168828\|\|full | Not-determined |
| S8_vs_k141_177609\|\|full | Not-determined |
| S8_vs_k141_81993\|\|full | Not-determined |
| S8_vs_k141_108305\|\|full | Not-determined |
| S8_vs_k141_99487\|\|full | Not-determined |
| S8_vs_k141_56168\|\|full | Low-quality |
| S8_vs_k141_125664\|\|full | Low-quality |
| S8_vs_k141_47018\|\|full | Low-quality |
| S8_vs_k141_56176\|\|full | Not-determined |
| S8_vs_k141_82000\|\|full | Low-quality |
| S8_vs_k141_185512\|\|full | Not-determined |
| S8_vs_k141_12281\|\|full | Not-determined |
| S8_vs_k141_72949\|\|full | Not-determined |
| S8_vs_k141_133999\|\|full | Low-quality |
| S8_vs_k141_90868\|\|full | Low-quality |
| S8_vs_k141_195076\|\|full | Low-quality |
| S8_vs_k141_72959\|\|full | Low-quality |
| S8_vs_k141_195081\|\|full | Not-determined |
| S8_vs_k141_134017\|\|full | Low-quality |
| S8_vs_k141_116674\|\|full | Not-determined |
| S8_vs_k141_38708\|\|full | Low-quality |
| S8_vs_k141_168833\|\|full | Low-quality |
| S8_vs_k141_151320\|\|full | Low-quality |
| S8_vs_k141_12316\|\|full | Low-quality |
| S8_vs_k141_82041\|\|full | Low-quality |
| S8_vs_k141_99519\|\|full | Not-determined |
| S8_vs_k141_64523\|\|full | Not-determined |
| S8_vs_k141_47053\|\|full | Not-determined |
| S8_vs_k141_47060\|\|full | Not-determined |
| S8_vs_k141_30514\|\|full | Low-quality |
| S8_vs_k141_108310\|\|full | Low-quality |
| S8_vs_k141_185530\|\|full | Low-quality |
| S8_vs_k141_203269\|\|full | Not-determined |
| S8_vs_k141_38740\|\|full | Not-determined |
| S8_vs_k141_125699\|\|full | Low-quality |
| S8_vs_k141_185532\|\|full | Low-quality |
| S8_vs_k141_168869\|\|full | Not-determined |
| S8_vs_k141_125704\|\|full | Not-determined |
| S8_vs_k141_185536\|\|full | Not-determined |
| S8_vs_k141_177669\|\|full | Not-determined |
| S8_vs_k141_108324\|\|full | Not-determined |
| S8_vs_k141_168874\|\|full | Not-determined |
| S8_vs_k141_21438\|\|full | Low-quality |
| S8_vs_k141_108334\|\|full | Low-quality |
| S8_vs_k141_134073\|\|full | Low-quality |
| S8_vs_k141_185547\|\|full | Not-determined |
| S8_vs_k141_47095\|\|full | Low-quality |
| S8_vs_k141_159963\|\|full | Low-quality |
| S8_vs_k141_30542\|\|full | Not-determined |
| S8_vs_k141_38780\|\|full | Not-determined |
| S8_vs_k141_195108\|\|full | Not-determined |
| S8_vs_k141_185559\|\|full | Low-quality |
| S8_vs_k141_21451\|\|full | Not-determined |
| S8_vs_k141_159974\|\|full | Low-quality |
| S8_vs_k141_134084\|\|full | Low-quality |
| S8_vs_k141_12407\|\|full | Low-quality |
| S8_vs_k141_73002\|\|full | Not-determined |
| S8_vs_k141_64549\|\|full | Not-determined |
| S8_vs_k141_99587\|\|full | Low-quality |
| S8_vs_k141_21460\|\|full | Low-quality |
| S8_vs_k141_30558\|\|full | Low-quality |
| S8_vs_k141_168908\|\|full | Not-determined |
| S8_vs_k141_4137\|\|full | Low-quality |
| S8_vs_k141_116717\|\|full | Low-quality |
| S8_vs_k141_99604\|\|full | Not-determined |
| S8_vs_k141_168919\|\|full | Low-quality |
| S8_vs_k141_177745\|\|full | Low-quality |
| S8_vs_k141_56281\|\|full | Low-quality |
| S8_vs_k141_116731\|\|full | Low-quality |
| S8_vs_k141_56282\|\|full | Not-determined |
| S8_vs_k141_56283\|\|full | Not-determined |
| S8_vs_k141_177750\|\|full | Not-determined |
| S8_vs_k141_12447\|\|full | Not-determined |
| S8_vs_k141_185585\|\|full | Low-quality |
| S8_vs_k141_56289\|\|full | Low-quality |
| S8_vs_k141_185587\|\|full | Not-determined |
| S8_vs_k141_56292\|\|full | Low-quality |
| S8_vs_k141_64575\|\|full | Not-determined |
| S8_vs_k141_73023\|\|full | Low-quality |
| S8_vs_k141_56295\|\|full | Low-quality |
| S9_extracted_k141_26805 | Complete |
| S8_vs_k141_73029\|\|full | Low-quality |
| S8_vs_k141_90994\|\|full | Low-quality |
| S8_vs_k141_108383\|\|full | Not-determined |
| S8_vs_k141_143008\|\|full | Low-quality |
| S8_vs_k141_21496\|\|full | Not-determined |
| S8_vs_k141_203303\|\|full | Not-determined |
| S8_vs_k141_56309\|\|full | Low-quality |
| S14_extracted_k141_43340 | High-quality |
| S8_vs_k141_143013\|\|full | Low-quality |
| S8_vs_k141_143014\|\|full | Not-determined |
| S8_vs_k141_64587\|\|full | Low-quality |
| S8_vs_k141_143015\|\|full | Low-quality |
| S8_vs_k141_125743\|\|full | Low-quality |
| S8_vs_k141_134128\|\|full | Not-determined |
| S8_vs_k141_56326\|\|full | Not-determined |
| S8_vs_k141_12482\|\|full | Low-quality |
| S8_vs_k141_134131\|\|full | Low-quality |
| S8_vs_k141_47132\|\|full | Not-determined |
| S8_vs_k141_64598\|\|full | Low-quality |
| S3_vs_k141_67828\|\|full | High-quality |
| S8_vs_k141_143030\|\|full | Not-determined |
| S14_vs_k141_67094\|\|0_partial | High-quality |
| S8_vs_k141_21518\|\|full | Not-determined |
| S8_vs_k141_38817\|\|full | Low-quality |
| S8_vs_k141_82183\|\|full | Low-quality |
| S8_vs_k141_73080\|\|full | Low-quality |
| S8_vs_k141_134140\|\|full | Low-quality |
| S8_vs_k141_82190\|\|full | Not-determined |
| S8_vs_k141_38820\|\|full | Not-determined |
| S8_vs_k141_4161\|\|full | Low-quality |
| S8_vs_k141_108427\|\|full | Not-determined |
| S8_vs_k141_195216\|\|full | Not-determined |
| S8_vs_k141_82196\|\|full | Not-determined |
| S8_vs_k141_12516\|\|full | Low-quality |
| S8_vs_k141_4171\|\|full | Low-quality |
| S8_vs_k141_185629\|\|full | Low-quality |
| S8_vs_k141_195219\|\|full | Not-determined |
| S8_vs_k141_4173\|\|full | Low-quality |
| S8_vs_k141_21547\|\|full | Low-quality |
| S8_vs_k141_64627\|\|full | Not-determined |
| S8_vs_k141_116789\|\|full | Low-quality |
| S8_vs_k141_160091\|\|full | Low-quality |
| S8_vs_k141_108446\|\|full | Not-determined |
| S8_vs_k141_177812\|\|full | Low-quality |
| S8_vs_k141_177821\|\|full | Not-determined |
| S14_vs_k141_43340\|\|full | High-quality |
| S8_vs_k141_160095\|\|full | Low-quality |
| S8_vs_k141_168991\|\|full | Not-determined |
| S8_vs_k141_160102\|\|full | Low-quality |
| S8_vs_k141_195258\|\|full | Not-determined |
| S8_vs_k141_134178\|\|full | Low-quality |
| S8_vs_k141_91058\|\|full | Low-quality |
| S8_vs_k141_64671\|\|full | Low-quality |
| S8_vs_k141_160108\|\|full | Not-determined |
| S8_vs_k141_73142\|\|full | Low-quality |
| S8_vs_k141_91060\|\|full | Low-quality |
| S8_vs_k141_73145\|\|full | Low-quality |
| S8_vs_k141_203355\|\|full | Not-determined |
| S8_vs_k141_99670\|\|full | Not-determined |
| S8_vs_k141_203364\|\|full | Not-determined |
| S8_vs_k141_134188\|\|full | Low-quality |
| S8_vs_k141_143090\|\|full | Not-determined |
| S8_vs_k141_151466\|\|full | Not-determined |
| S8_vs_k141_177854\|\|full | Low-quality |
| S8_vs_k141_91074\|\|full | Not-determined |
| S8_vs_k141_47215\|\|full | Low-quality |
| S8_vs_k141_38881\|\|full | Low-quality |
| S8_vs_k141_47220\|\|full | Not-determined |
| S8_vs_k141_116828\|\|full | Not-determined |
| S8_vs_k141_21607\|\|full | Low-quality |
| S8_vs_k141_143118\|\|full | Not-determined |
| S8_vs_k141_195288\|\|full | Not-determined |
| S8_vs_k141_108511\|\|full | Low-quality |
| S8_vs_k141_47223\|\|full | Low-quality |
| S8_vs_k141_91092\|\|full | Not-determined |
| S8_vs_k141_21628\|\|full | Low-quality |
| S8_vs_k141_47224\|\|full | Low-quality |
| S8_vs_k141_169047\|\|full | Low-quality |
| S8_vs_k141_12563\|\|full | Low-quality |
| S8_vs_k141_151490\|\|full | Not-determined |
| S8_vs_k141_169048\|\|full | Not-determined |
| S8_vs_k141_99704\|\|full | Low-quality |
| S8_vs_k141_64704\|\|full | Low-quality |
| S8_vs_k141_73201\|\|full | Low-quality |
| S8_vs_k141_4270\|\|full | Not-determined |
| S8_vs_k141_125838\|\|full | Low-quality |
| S8_vs_k141_177881\|\|full | Low-quality |
| S8_vs_k141_134213\|\|full | Not-determined |
| S8_vs_k141_195319\|\|full | Not-determined |
| S8_vs_k141_185701\|\|full | Low-quality |
| S8_vs_k141_12579\|\|full | Low-quality |
| S8_vs_k141_73209\|\|full | Not-determined |
| S8_vs_k141_47261\|\|full | Not-determined |
| S8_vs_k141_82298\|\|full | Not-determined |
| S8_vs_k141_12586\|\|full | Low-quality |
| S8_vs_k141_125870\|\|full | Low-quality |
| S8_vs_k141_91122\|\|full | Not-determined |
| S8_vs_k141_73221\|\|full | Not-determined |
| S8_vs_k141_134240\|\|full | Low-quality |
| S8_vs_k141_185724\|\|full | Not-determined |
| S8_vs_k141_151516\|\|full | Not-determined |
| S8_vs_k141_195322\|\|full | Low-quality |
| S8_vs_k141_38942\|\|full | Low-quality |
| S8_vs_k141_91136\|\|full | Low-quality |
| S8_vs_k141_143174\|\|full | Low-quality |
| S8_vs_k141_56498\|\|full | Low-quality |
| S8_vs_k141_185739\|\|full | Low-quality |
| S8_vs_k141_4328\|\|full | Low-quality |
| S8_vs_k141_82335\|\|full | Not-determined |
| S8_vs_k141_21670\|\|full | Low-quality |
| S8_vs_k141_116866\|\|full | Low-quality |
| S8_vs_k141_82341\|\|full | Not-determined |
| S8_vs_k141_12612\|\|full | Not-determined |
| S8_vs_k141_185750\|\|full | Not-determined |
| S8_vs_k141_116871\|\|full | Not-determined |
| S8_vs_k141_12619\|\|full | Low-quality |
| S8_vs_k141_125930\|\|full | Low-quality |
| S8_vs_k141_38978\|\|full | Not-determined |
| S8_vs_k141_108554\|\|full | Low-quality |
| S8_vs_k141_125939\|\|full | Low-quality |
| S8_vs_k141_73274\|\|full | Low-quality |
| S8_vs_k141_134301\|\|full | Not-determined |
| S8_vs_k141_56521\|\|full | Not-determined |
| S8_vs_k141_4344\|\|full | Low-quality |
| S8_vs_k141_116906\|\|full | Not-determined |
| S8_vs_k141_125944\|\|full | Not-determined |
| S8_vs_k141_203486\|\|full | Not-determined |
| S8_vs_k141_125948\|\|full | Low-quality |
| S8_vs_k141_12648\|\|full | Low-quality |
| S8_vs_k141_56527\|\|full | Not-determined |
| S8_vs_k141_56528\|\|full | Low-quality |
| S8_vs_k141_125954\|\|full | Not-determined |
| S8_vs_k141_160116\|\|full | Not-determined |
| S8_vs_k141_21708\|\|full | Not-determined |
| S8_vs_k141_73291\|\|full | Low-quality |
| S8_vs_k141_169154\|\|full | Low-quality |
| S8_vs_k141_195370\|\|full | Low-quality |
| S8_vs_k141_108596\|\|full | Low-quality |
| S8_vs_k141_21729\|\|full | Not-determined |
| S8_vs_k141_195375\|\|full | Low-quality |
| S8_vs_k141_99780\|\|full | Low-quality |
| S8_vs_k141_169169\|\|full | Low-quality |
| S8_vs_k141_125976\|\|full | Low-quality |
| S8_vs_k141_21745\|\|full | Low-quality |
| S8_vs_k141_195379\|\|full | Low-quality |
| S8_vs_k141_21749\|\|full | Not-determined |
| S7_vs_135828\|\|full | High-quality |
| S8_vs_k141_73300\|\|full | Low-quality |
| S8_vs_k141_160149\|\|full | Low-quality |
| S8_vs_k141_47275\|\|full | Low-quality |
| S8_vs_k141_12683\|\|full | Not-determined |
| S8_vs_k141_177996\|\|full | Not-determined |
| S8_vs_k141_30666\|\|full | Low-quality |
| S8_vs_k141_108631\|\|full | Not-determined |
| S8_vs_k141_160159\|\|full | Low-quality |
| S8_vs_k141_160162\|\|full | Not-determined |
| S8_vs_k141_169202\|\|full | Low-quality |
| S8_vs_k141_12691\|\|full | Low-quality |
| S8_vs_k141_39030\|\|full | Low-quality |
| S8_vs_k141_178016\|\|full | Not-determined |
| S8_vs_k141_126017\|\|full | Low-quality |
| S8_vs_k141_73332\|\|full | Low-quality |
| S8_vs_k141_143289\|\|full | Not-determined |
| S8_vs_k141_47297\|\|full | Not-determined |
| S8_vs_k141_4447\|\|full | Not-determined |
| S8_vs_k141_64831\|\|full | Low-quality |
| S8_vs_k141_12715\|\|full | Low-quality |
| S8_vs_k141_203573\|\|full | Not-determined |
| S8_vs_k141_117017\|\|full | Not-determined |
| S8_vs_k141_4458\|\|full | Not-determined |
| S8_vs_k141_203581\|\|full | Low-quality |
| S8_vs_k141_169242\|\|full | Low-quality |
| S8_vs_k141_73347\|\|full | Not-determined |
| S8_vs_k141_47315\|\|full | Low-quality |
| S8_vs_k141_39066\|\|full | Low-quality |
| S8_vs_k141_143305\|\|full | Not-determined |
| S8_vs_k141_151643\|\|full | Low-quality |
| S8_vs_k141_73355\|\|full | Not-determined |
| S8_vs_k141_64849\|\|full | Not-determined |
| S8_vs_k141_82408\|\|full | Low-quality |
| S8_vs_k141_143322\|\|full | Low-quality |
| S8_vs_k141_4483\|\|full | Low-quality |
| S8_vs_k141_47346\|\|full | Low-quality |
| S8_vs_k141_39082\|\|full | Low-quality |
| S8_vs_k141_12732\|\|full | Low-quality |
| S8_vs_k141_203613\|\|full | Not-determined |
| S8_vs_k141_178054\|\|full | Low-quality |
| S8_vs_k141_56629\|\|full | Low-quality |
| S8_vs_k141_47352\|\|full | Not-determined |
| S8_vs_k141_134409\|\|full | Low-quality |
| S8_vs_k141_21850\|\|full | Low-quality |
| S8_vs_k141_21851\|\|full | Low-quality |
| S8_vs_k141_4488\|\|full | Low-quality |
| S8_vs_k141_30757\|\|full | Not-determined |
| S8_vs_k141_151663\|\|full | Not-determined |
| S8_vs_k141_99851\|\|full | Low-quality |
| S8_vs_k141_4495\|\|full | Low-quality |
| S8_vs_k141_21857\|\|full | Low-quality |

| | |
|---|---|
| S8_vs_k141_73421\|\|full | Low-quality |
| S8_vs_k141_56637\|\|full | Not-determined |
| S8_vs_k141_178057\|\|full | Low-quality |
| S8_vs_k141_185807\|\|full | Low-quality |
| S8_vs_k141_21864\|\|full | Not-determined |
| S8_vs_k141_30776\|\|full | Not-determined |
| S8_vs_k141_91157\|\|full | Not-determined |
| S8_vs_k141_91160\|\|full | Not-determined |
| S8_vs_k141_21875\|\|full | Low-quality |
| S8_vs_k141_99877\|\|full | Low-quality |
| S8_vs_k141_117068\|\|full | Not-determined |
| S8_vs_k141_134442\|\|full | Low-quality |
| S8_vs_k141_151704\|\|full | Low-quality |
| S8_vs_k141_4529\|\|full | Low-quality |
| S8_vs_k141_178072\|\|full | Not-determined |
| S8_vs_k141_4533\|\|full | Low-quality |
| S8_vs_k141_169298\|\|full | Not-determined |
| S8_vs_k141_30798\|\|full | Not-determined |
| S8_vs_k141_47387\|\|full | Not-determined |
| S8_vs_k141_160296\|\|full | Low-quality |
| S8_vs_k141_151713\|\|full | Low-quality |
| S8_vs_k141_39132\|\|full | Not-determined |
| S8_vs_k141_160298\|\|full | Not-determined |
| S8_vs_k141_47394\|\|full | Low-quality |
| S8_vs_k141_64910\|\|full | Low-quality |
| S8_vs_k141_203659\|\|full | Low-quality |
| S8_vs_k141_185842\|\|full | Not-determined |
| S8_vs_k141_91188\|\|full | Low-quality |
| S8_vs_k141_117088\|\|full | Not-determined |
| S8_vs_k141_73467\|\|full | Not-determined |
| S8_vs_k141_4557\|\|full | Low-quality |
| S8_vs_k141_82485\|\|full | Not-determined |
| S8_vs_k141_21896\|\|full | Not-determined |
| S8_vs_k141_185848\|\|full | Not-determined |
| S8_vs_k141_178110\|\|full | Low-quality |
| S8_vs_k141_203685\|\|full | Low-quality |
| S8_vs_k141_99913\|\|full | Low-quality |
| S8_vs_k141_30817\|\|full | Not-determined |
| S8_vs_k141_151751\|\|full | Low-quality |
| S8_vs_k141_160343\|\|full | Low-quality |
| S8_vs_k141_99915\|\|full | Not-determined |
| S8_vs_k141_99917\|\|full | Low-quality |
| S8_vs_k141_151754\|\|full | Low-quality |
| S8_vs_k141_195525\|\|full | Low-quality |
| S8_vs_k141_169338\|\|full | Not-determined |
| S8_vs_k141_47429\|\|full | Low-quality |
| S8_vs_k141_178119\|\|full | Not-determined |
| S10_vs_43473\|\|full | High-quality |
| S8_vs_k141_64952\|\|full | Not-determined |
| S8_vs_k141_178125\|\|full | Low-quality |
| S8_vs_k141_160362\|\|full | Low-quality |
| S8_vs_k141_108713\|\|full | Not-determined |
| S8_vs_k141_82536\|\|full | Low-quality |
| S8_vs_k141_117112\|\|full | Not-determined |
| S8_vs_k141_39162\|\|full | Not-determined |
| S8_vs_k141_99936\|\|full | Low-quality |
| S8_vs_k141_99937\|\|full | Not-determined |
| S8_vs_k141_47450\|\|full | Low-quality |
| S8_vs_k141_21913\|\|full | Not-determined |
| S8_vs_k141_39175\|\|full | Not-determined |
| S8_vs_k141_178148\|\|full | Low-quality |
| S8_vs_k141_151793\|\|full | Not-determined |
| S8_vs_k141_151794\|\|full | Not-determined |
| S8_vs_k141_117135\|\|full | Low-quality |
| S8_vs_k141_56700\|\|full | Low-quality |
| S8_vs_k141_82570\|\|full | Not-determined |
| S8_vs_k141_73503\|\|full | Low-quality |
| S8_vs_k141_21939\|\|full | Low-quality |
| S8_vs_k141_143383\|\|full | Not-determined |
| S8_vs_k141_56704\|\|full | Low-quality |
| S8_vs_k141_169381\|\|full | Low-quality |
| S8_vs_k141_117154\|\|full | Low-quality |
| S8_vs_k141_151823\|\|full | Low-quality |
| S8_vs_k141_195581\|\|full | Low-quality |
| S8_vs_k141_143395\|\|full | Not-determined |
| S8_vs_k141_169390\|\|full | Not-determined |
| S8_vs_k141_56714\|\|full | Low-quality |
| S8_vs_k141_143403\|\|full | Not-determined |
| S8_vs_k141_203742\|\|full | Not-determined |
| S8_vs_k141_108783\|\|full | Low-quality |
| S8_vs_k141_47492\|\|full | Not-determined |
| S8_vs_k141_91261\|\|full | Not-determined |
| S8_vs_k141_21984\|\|full | Low-quality |
| S8_vs_k141_99997\|\|full | Low-quality |
| S8_vs_k141_65024\|\|full | Not-determined |
| S8_vs_k141_151863\|\|full | Low-quality |
| S8_vs_k141_126053\|\|full | Not-determined |
| S8_vs_k141_39230\|\|full | Not-determined |
| S8_vs_k141_91273\|\|full | Low-quality |
| S8_vs_k141_203774\|\|full | Not-determined |
| S8_vs_k141_151875\|\|full | Low-quality |
| S8_vs_k141_12894\|\|full | Not-determined |
| S8_vs_k141_39249\|\|full | Not-determined |
| S8_vs_k141_12901\|\|full | Low-quality |
| S8_vs_k141_108821\|\|full | Low-quality |
| S8_vs_k141_195615\|\|full | Low-quality |
| S8_vs_k141_178256\|\|full | Not-determined |
| S8_vs_k141_47550\|\|full | Low-quality |
| S8_vs_k141_117235\|\|full | Low-quality |
| S8_vs_k141_134529\|\|full | Low-quality |
| S8_vs_k141_195627\|\|full | Low-quality |
| S8_vs_k141_117242\|\|full | Low-quality |
| S8_vs_k141_195635\|\|full | Low-quality |
| S8_vs_k141_134535\|\|full | Low-quality |
| S8_vs_k141_108840\|\|full | Not-determined |
| S8_vs_k141_117243\|\|full | Low-quality |
| S8_vs_k141_65078\|\|full | Low-quality |
| S8_vs_k141_47554\|\|full | Low-quality |
| S8_vs_k141_30934\|\|full | Not-determined |
| S8_vs_k141_169411\|\|full | Not-determined |
| S8_vs_k141_203801\|\|full | Low-quality |
| S8_vs_k141_91315\|\|full | Not-determined |
| S8_vs_k141_134545\|\|full | Low-quality |
| S8_vs_k141_108852\|\|full | Low-quality |
| S8_vs_k141_73562\|\|full | Not-determined |
| S8_vs_k141_169428\|\|full | Low-quality |
| S8_vs_k141_100049\|\|full | Low-quality |
| S8_vs_k141_117262\|\|full | Low-quality |
| S8_vs_k141_169431\|\|full | Not-determined |
| S8_vs_k141_56772\|\|full | Low-quality |
| S8_vs_k141_91332\|\|full | Low-quality |
| S8_vs_k141_82581\|\|full | Not-determined |
| S8_vs_k141_22049\|\|full | Low-quality |
| S8_vs_k141_126131\|\|full | Low-quality |
| S8_vs_k141_143458\|\|full | Not-determined |
| S8_vs_k141_134557\|\|full | Low-quality |
| S8_vs_k141_82596\|\|full | Not-determined |
| S8_vs_k141_195677\|\|full | Not-determined |
| S8_vs_k141_47601\|\|full | Not-determined |
| S8_vs_k141_100066\|\|full | Low-quality |
| S8_vs_k141_186021\|\|full | Not-determined |
| S8_vs_k141_186025\|\|full | Not-determined |
| S8_vs_k141_186029\|\|full | Not-determined |
| S8_vs_k141_143483\|\|full | Low-quality |
| S8_vs_k141_108870\|\|full | Not-determined |
| S8_vs_k141_91367\|\|full | Not-determined |
| S8_vs_k141_82625\|\|full | Low-quality |
| S8_vs_k141_203882\|\|full | Not-determined |
| S8_vs_k141_30967\|\|full | Not-determined |
| S8_vs_k141_117325\|\|full | Not-determined |
| S8_vs_k141_108885\|\|full | Not-determined |
| S8_vs_k141_178291\|\|full | Low-quality |
| S8_vs_k141_151919\|\|full | Low-quality |
| S8_vs_k141_12954\|\|full | Not-determined |
| S8_vs_k141_178300\|\|full | Low-quality |
| S8_vs_k141_73629\|\|full | Low-quality |
| S8_vs_k141_100096\|\|full | Low-quality |
| S8_vs_k141_178309\|\|full | Low-quality |
| S8_vs_k141_100101\|\|full | Not-determined |
| S8_vs_k141_134591\|\|full | Low-quality |
| S8_vs_k141_91403\|\|full | Not-determined |
| S8_vs_k141_134601\|\|full | Low-quality |
| S8_vs_k141_56860\|\|full | Low-quality |
| S8_vs_k141_169514\|\|full | Low-quality |
| S8_vs_k141_100124\|\|full | Low-quality |
| S8_vs_k141_30994\|\|full | Not-determined |
| S8_vs_k141_56877\|\|full | Low-quality |
| S8_vs_k141_73644\|\|full | Not-determined |
| S8_vs_k141_151973\|\|full | Low-quality |
| S8_vs_k141_56888\|\|full | Not-determined |
| S8_vs_k141_31011\|\|full | Low-quality |
| S8_vs_k141_195732\|\|full | Not-determined |
| S8_vs_k141_169536\|\|full | Low-quality |
| S8_vs_k141_73663\|\|full | Not-determined |
| S8_vs_k141_134637\|\|full | Not-determined |
| S8_vs_k141_143605\|\|full | Not-determined |
| S8_vs_k141_31036\|\|full | Not-determined |
| S8_vs_k141_31038\|\|full | Not-determined |
| S8_vs_k141_178362\|\|full | Low-quality |
| S8_vs_k141_134657\|\|full | Not-determined |
| S8_vs_k141_134666\|\|full | Low-quality |
| S8_vs_k141_178377\|\|full | Low-quality |
| S8_vs_k141_65217\|\|full | Low-quality |
| S8_vs_k141_203962\|\|full | Not-determined |
| S8_vs_k141_100187\|\|full | Low-quality |
| S8_vs_k141_134671\|\|full | Not-determined |
| S8_vs_k141_152046\|\|full | Low-quality |
| S8_vs_k141_47687\|\|full | Low-quality |
| S8_vs_k141_108907\|\|full | Low-quality |
| S8_vs_k141_186088\|\|full | Low-quality |
| S8_vs_k141_47694\|\|full | Low-quality |
| S8_vs_k141_4632\|\|full | Not-determined |
| S8_vs_k141_31079\|\|full | Low-quality |
| S8_vs_k141_134684\|\|full | Low-quality |
| S8_vs_k141_4639\|\|full | Low-quality |
| S9_extracted_k141_23901 | Complete |
| S8_vs_k141_73706\|\|full | Not-determined |
| S8_vs_k141_186099\|\|full | Not-determined |
| S8_vs_k141_160477\|\|full | Low-quality |
| S8_vs_k141_82671\|\|full | Not-determined |
| S8_vs_k141_143623\|\|full | Low-quality |
| S8_vs_k141_169601\|\|full | Not-determined |
| S8_vs_k141_100211\|\|full | Not-determined |
| S8_vs_k141_100214\|\|full | Not-determined |
| S8_vs_k141_160485\|\|full | Not-determined |
| S8_vs_k141_204011\|\|full | Not-determined |
| S8_vs_k141_31098\|\|full | Low-quality |
| S8_vs_k141_134720\|\|full | Low-quality |
| S8_vs_k141_22275\|\|full | Not-determined |
| S8_vs_k141_39408\|\|full | Not-determined |
| S8_vs_k141_47742\|\|full | Low-quality |
| S8_vs_k141_31115\|\|full | Low-quality |
| S8_vs_k141_143659\|\|full | Not-determined |
| S8_vs_k141_22288\|\|full | Low-quality |
| S8_vs_k141_117453\|\|full | Not-determined |
| S8_vs_k141_82688\|\|full | Not-determined |
| S9_vs_k141_26805\|\|full | High-quality |
| S8_vs_k141_39418\|\|full | Not-determined |
| S8_vs_k141_160512\|\|full | Low-quality |
| S8_vs_k141_108969\|\|full | Not-determined |
| S8_vs_k141_31134\|\|full | Low-quality |
| S8_vs_k141_39424\|\|full | Low-quality |
| S8_vs_k141_204055\|\|full | Low-quality |
| S8_vs_k141_65293\|\|full | Not-determined |
| S8_vs_k141_143687\|\|full | Not-determined |
| S8_vs_k141_126249\|\|full | Not-determined |
| S8_vs_k141_4704\|\|full | Low-quality |
| S8_vs_k141_47765\|\|full | Not-determined |
| S8_vs_k141_169620\|\|full | Not-determined |
| S8_vs_k141_4714\|\|full | Low-quality |
| S8_vs_k141_195809\|\|full | Not-determined |
| S8_vs_k141_91499\|\|full | Low-quality |
| S8_vs_k141_91500\|\|full | Low-quality |
| S8_vs_k141_169627\|\|full | Not-determined |
| S8_vs_k141_82717\|\|full | Not-determined |
| S8_vs_k141_117496\|\|full | Not-determined |
| S8_vs_k141_22335\|\|full | Not-determined |
| S8_vs_k141_152109\|\|full | Low-quality |
| S8_vs_k141_169638\|\|full | Low-quality |
| S8_vs_k141_143707\|\|full | Not-determined |
| S8_vs_k141_126282\|\|full | Low-quality |
| S8_vs_k141_82722\|\|full | Not-determined |
| S8_vs_k141_13112\|\|full | Low-quality |
| S8_vs_k141_13113\|\|full | Not-determined |
| S8_vs_k141_134771\|\|full | Low-quality |
| S8_vs_k141_134776\|\|full | Low-quality |
| S8_vs_k141_82738\|\|full | Not-determined |
| S8_vs_k141_31178\|\|full | Low-quality |
| S8_vs_k141_65336\|\|full | Not-determined |
| S8_vs_k141_126302\|\|full | Low-quality |
| S8_vs_k141_100307\|\|full | Not-determined |
| S8_vs_k141_109004\|\|full | Not-determined |
| S8_vs_k141_143740\|\|full | Low-quality |
| S8_vs_k141_65343\|\|full | Not-determined |
| S8_vs_k141_100325\|\|full | Low-quality |
| S8_vs_k141_143743\|\|full | Low-quality |
| S8_vs_k141_47822\|\|full | Not-determined |
| S8_vs_k141_117543\|\|full | Not-determined |
| S8_vs_k141_100330\|\|full | Low-quality |
| S8_vs_k141_169671\|\|full | Not-determined |
| S8_vs_k141_73866\|\|full | Low-quality |
| S8_vs_k141_47836\|\|full | Not-determined |
| S8_vs_k141_204119\|\|full | Low-quality |
| S8_vs_k141_13178\|\|full | Low-quality |
| S8_vs_k141_47839\|\|full | Not-determined |
| S8_vs_k141_13179\|\|full | Low-quality |
| S8_vs_k141_82769\|\|full | Not-determined |
| S8_vs_k141_152162\|\|full | Not-determined |
| S8_vs_k141_117571\|\|full | Not-determined |
| S8_vs_k141_143775\|\|full | Low-quality |
| S8_vs_k141_73879\|\|full | Not-determined |
| S8_vs_k141_91532\|\|full | Low-quality |
| S8_vs_k141_169700\|\|full | Not-determined |
| S8_vs_k141_117583\|\|full | Low-quality |
| S8_vs_k141_13184\|\|full | Low-quality |
| S8_vs_k141_169708\|\|full | Low-quality |
| S8_vs_k141_195909\|\|full | Low-quality |
| S8_vs_k141_152193\|\|full | Low-quality |
| S8_vs_k141_117599\|\|full | Low-quality |
| S14_extracted_k141_37500 | High-quality |
| S8_vs_k141_160565\|\|full | Not-determined |
| S8_vs_k141_39525\|\|full | Low-quality |
| S8_vs_k141_4756\|\|full | Low-quality |
| S8_vs_k141_13199\|\|full | Low-quality |
| S8_vs_k141_13200\|\|full | Low-quality |
| S8_vs_k141_117607\|\|full | Low-quality |
| S8_vs_k141_57108\|\|full | Not-determined |
| S8_vs_k141_65406\|\|full | Low-quality |
| S8_vs_k141_169726\|\|full | Low-quality |
| S8_vs_k141_195924\|\|full | Not-determined |
| S8_vs_k141_47860\|\|full | Low-quality |
| S8_vs_k141_160578\|\|full | Not-determined |
| S8_vs_k141_204171\|\|full | Low-quality |
| S8_vs_k141_117618\|\|full | Not-determined |
| S8_vs_k141_169733\|\|full | Not-determined |
| S8_vs_k141_117621\|\|full | Not-determined |
| S8_vs_k141_13213\|\|full | Low-quality |
| S8_vs_k141_195935\|\|full | Not-determined |
| S8_vs_k141_195944\|\|full | Not-determined |
| S8_vs_k141_152219\|\|full | Low-quality |
| S8_vs_k141_169751\|\|full | Low-quality |
| S8_vs_k141_65440\|\|full | Not-determined |
| S8_vs_k141_31256\|\|full | Low-quality |
| S8_vs_k141_126384\|\|full | Not-determined |
| S8_vs_k141_152225\|\|full | Not-determined |
| S8_vs_k141_22482\|\|full | Low-quality |
| S8_vs_k141_65451\|\|full | Not-determined |
| S8_vs_k141_178459\|\|full | Low-quality |
| S8_vs_k141_160609\|\|full | Low-quality |
| S8_vs_k141_91621\|\|full | Low-quality |
| S8_vs_k141_152228\|\|full | Low-quality |
| S8_vs_k141_31266\|\|full | Low-quality |
| S8_vs_k141_204192\|\|full | Low-quality |
| S8_vs_k141_91626\|\|full | Low-quality |
| S8_vs_k141_143868\|\|full | Not-determined |
| S8_vs_k141_169783\|\|full | Not-determined |
| S8_vs_k141_178474\|\|full | Low-quality |
| S8_vs_k141_126428\|\|full | Not-determined |
| S8_vs_k141_91636\|\|full | Not-determined |
| S8_vs_k141_204214\|\|full | Low-quality |
| S8_vs_k141_13223\|\|full | Not-determined |
| S8_vs_k141_134855\|\|full | Low-quality |
| S8_vs_k141_57160\|\|full | Not-determined |
| S8_vs_k141_143875\|\|full | Not-determined |
| S8_vs_k141_126433\|\|full | Not-determined |
| S8_vs_k141_169810\|\|full | Low-quality |
| S8_vs_k141_65472\|\|full | Not-determined |
| S8_vs_k141_22524\|\|full | Low-quality |
| S8_vs_k141_91647\|\|full | Low-quality |
| S8_vs_k141_22525\|\|full | Low-quality |
| S8_vs_k141_178493\|\|full | Not-determined |
| S8_vs_k141_178496\|\|full | Not-determined |
| S8_vs_k141_31287\|\|full | Not-determined |
| S8_vs_k141_186173\|\|full | Not-determined |
| S8_vs_k141_22526\|\|full | Low-quality |
| S8_vs_k141_82853\|\|full | Not-determined |
| S8_vs_k141_204233\|\|full | Low-quality |
| S8_vs_k141_39644\|\|full | Low-quality |
| S8_vs_k141_178507\|\|full | Low-quality |
| S8_vs_k141_152270\|\|full | Low-quality |
| S8_vs_k141_143907\|\|full | Low-quality |
| S8_vs_k141_73935\|\|full | Not-determined |
| S8_vs_k141_57195\|\|full | Low-quality |
| S8_vs_k141_31317\|\|full | Low-quality |
| S8_vs_k141_39660\|\|full | Not-determined |
| S8_vs_k141_186181\|\|full | Low-quality |
| S8_vs_k141_117694\|\|full | Low-quality |
| S8_vs_k141_31327\|\|full | Low-quality |
| S8_vs_k141_109170\|\|full | Not-determined |
| S8_vs_k141_47900\|\|full | Low-quality |
| S8_vs_k141_22551\|\|full | Low-quality |
| S8_vs_k141_47901\|\|full | Not-determined |
| S8_vs_k141_152285\|\|full | Low-quality |
| S8_vs_k141_169864\|\|full | Low-quality |
| S8_vs_k141_39678\|\|full | Not-determined |
| S8_vs_k141_13290\|\|full | Not-determined |
| S8_vs_k141_31342\|\|full | Not-determined |
| S8_vs_k141_91698\|\|full | Not-determined |
| S8_vs_k141_178532\|\|full | Not-determined |
| S8_vs_k141_169870\|\|full | Not-determined |
| S8_vs_k141_143939\|\|full | Low-quality |
| S8_vs_k141_160651\|\|full | Not-determined |
| S8_vs_k141_82897\|\|full | Low-quality |
| S8_vs_k141_57222\|\|full | Low-quality |
| S8_vs_k141_186207\|\|full | Low-quality |
| S8_vs_k141_143942\|\|full | Not-determined |
| S8_vs_k141_186208\|\|full | Not-determined |
| S8_vs_k141_143946\|\|full | Low-quality |
| S8_vs_k141_152311\|\|full | Low-quality |
| S8_vs_k141_117707\|\|full | Low-quality |
| S8_vs_k141_109196\|\|full | Not-determined |
| S14_vs_k141_37500\|\|full | High-quality |
| S8_vs_k141_186215\|\|full | Low-quality |
| S8_vs_k141_13314\|\|full | Not-determined |
| S8_vs_k141_57231\|\|full | Not-determined |
| S8_vs_k141_39704\|\|full | Low-quality |
| S3_vs_k141_77156\|\|full | High-quality |
| S8_vs_k141_186219\|\|full | Not-determined |
| S8_vs_k141_57242\|\|full | Low-quality |
| S8_vs_k141_117718\|\|full | Low-quality |
| S8_vs_k141_39724\|\|full | Low-quality |
| S8_vs_k141_13336\|\|full | Low-quality |
| S8_vs_k141_109230\|\|full | Not-determined |
| S8_vs_k141_117733\|\|full | Not-determined |
| S8_vs_k141_91749\|\|full | Not-determined |
| S8_vs_k141_186230\|\|full | Not-determined |
| S8_vs_k141_178597\|\|full | Low-quality |
| S8_vs_k141_82932\|\|full | Not-determined |
| S8_vs_k141_57275\|\|full | Low-quality |
| S8_vs_k141_160671\|\|full | Low-quality |
| S8_vs_k141_47914\|\|full | Low-quality |
| S3_extracted_k141_59078 | High-quality |
| S8_vs_k141_22573\|\|full | Not-determined |
| S8_vs_k141_126503\|\|full | Low-quality |
| S8_vs_k141_109239\|\|full | Low-quality |
| S8_vs_k141_13360\|\|full | Not-determined |
| S8_vs_k141_178610\|\|full | Low-quality |
| S8_vs_k141_13361\|\|full | Low-quality |
| S8_vs_k141_144005\|\|full | Low-quality |
| S8_vs_k141_126511\|\|full | Low-quality |
| S8_vs_k141_39759\|\|full | Low-quality |
| S8_vs_k141_152380\|\|full | Low-quality |
| S8_vs_k141_31425\|\|full | Not-determined |
| S8_vs_k141_160690\|\|full | Not-determined |
| S8_vs_k141_91797\|\|full | Low-quality |
| S8_vs_k141_82965\|\|full | Low-quality |
| S8_vs_k141_109250\|\|full | Not-determined |
| S9_vs_k141_23901\|\|full | High-quality |
| S8_vs_k141_178635\|\|full | Low-quality |
| S8_vs_k141_186293\|\|full | Not-determined |
| S8_vs_k141_57316\|\|full | Low-quality |
| S8_vs_k141_204311\|\|full | Low-quality |
| S8_vs_k141_65542\|\|full | Low-quality |
| S8_vs_k141_196075\|\|full | Low-quality |
| S8_vs_k141_82978\|\|full | Low-quality |
| S8_vs_k141_186308\|\|full | Low-quality |
| S8_vs_k141_204338\|\|full | Low-quality |
| S8_vs_k141_144063\|\|full | Low-quality |
| S8_vs_k141_73994\|\|full | Low-quality |
| S8_extracted_k141_208119 | Complete |
| S8_vs_k141_73998\|\|full | Low-quality |
| S8_vs_k141_204340\|\|full | Low-quality |
| S8_vs_k141_178665\|\|full | Not-determined |
| S8_vs_k141_83006\|\|full | Not-determined |
| S8_vs_k141_169978\|\|full | Low-quality |
| S8_vs_k141_169981\|\|full | Low-quality |
| S8_vs_k141_57340\|\|full | Low-quality |
| S8_vs_k141_22660\|\|full | Low-quality |
| S8_vs_k141_109294\|\|full | Low-quality |
| S8_vs_k141_126546\|\|full | Not-determined |
| S8_vs_k141_109296\|\|full | Low-quality |
| S8_vs_k141_169996\|\|full | Not-determined |
| S8_vs_k141_169998\|\|full | Low-quality |
| S8_vs_k141_83035\|\|full | Low-quality |
| S8_vs_k141_152443\|\|full | Not-determined |
| S8_vs_k141_65603\|\|full | Not-determined |
| S8_vs_k141_144085\|\|full | Not-determined |
| S8_vs_k141_13439\|\|full | Low-quality |
| S8_vs_k141_57369\|\|full | Low-quality |
| S8_vs_k141_74021\|\|full | Low-quality |
| S8_vs_k141_39804\|\|full | Low-quality |
| S8_vs_k141_170008\|\|full | Low-quality |
| S8_vs_k141_4974\|\|full | Not-determined |
| S3_vs_k141_59078\|\|full | High-quality |
| S8_vs_k141_144099\|\|full | Low-quality |
| S8_vs_k141_160734\|\|full | Not-determined |
| S8_vs_k141_39824\|\|full | Not-determined |
| S8_vs_k141_57381\|\|full | Low-quality |
| S8_vs_k141_196122\|\|full | Not-determined |
| S8_vs_k141_170017\|\|full | Not-determined |
| S8_vs_k141_57385\|\|full | Not-determined |
| S8_vs_k141_178690\|\|full | Low-quality |
| S8_vs_k141_22718\|\|full | Low-quality |
| S8_vs_k141_160745\|\|full | Low-quality |
| S8_vs_k141_117819\|\|full | Low-quality |
| S8_vs_k141_48044\|\|full | Not-determined |
| S8_vs_k141_4984\|\|full | Not-determined |
| S8_vs_k141_196163\|\|full | Not-determined |
| S8_vs_k141_144151\|\|full | Low-quality |
| S8_vs_k141_186426\|\|full | Low-quality |
| S8_vs_k141_109346\|\|full | Low-quality |
| S8_vs_k141_109348\|\|full | Not-determined |
| S8_vs_k141_39880\|\|full | Low-quality |
| S8_vs_k141_74082\|\|full | Not-determined |
| S8_vs_k141_152503\|\|full | Low-quality |
| S8_vs_k141_109352\|\|full | Not-determined |
| S8_vs_k141_196174\|\|full | Not-determined |
| S8_vs_k141_109353\|\|full | Low-quality |
| S8_vs_k141_22757\|\|full | Not-determined |
| S8_vs_k141_39896\|\|full | Not-determined |
| S8_vs_k141_22761\|\|full | Low-quality |
| S8_vs_k141_5007\|\|full | Low-quality |
| S8_vs_k141_13523\|\|full | Low-quality |
| S8_vs_k141_144172\|\|full | Not-determined |
| S8_vs_k141_126577\|\|full | Low-quality |
| S8_vs_k141_57429\|\|full | Low-quality |
| S8_vs_k141_13532\|\|full | Low-quality |
| S8_vs_k141_22770\|\|full | Not-determined |
| S8_vs_k141_39914\|\|full | Low-quality |
| S8_vs_k141_83122\|\|full | Not-determined |
| S8_vs_k141_196192\|\|full | Not-determined |
| S8_vs_k141_5025\|\|full | Not-determined |
| S8_vs_k141_152541\|\|full | Not-determined |
| S8_vs_k141_117859\|\|full | Not-determined |
| S8_vs_k141_48105\|\|full | Low-quality |
| S8_vs_k141_204392\|\|full | Not-determined |
| S8_vs_k141_13544\|\|full | Not-determined |
| S8_vs_k141_91841\|\|full | Low-quality |
| S8_vs_k141_22803\|\|full | Not-determined |
| S8_vs_k141_117871\|\|full | Not-determined |
| S8_vs_k141_5047\|\|full | Not-determined |
| S8_vs_k141_109406\|\|full | Low-quality |
| S8_vs_k141_135041\|\|full | Not-determined |
| S8_vs_k141_5069\|\|full | Not-determined |
| S8_vs_k141_186515\|\|full | Not-determined |
| S8_vs_k141_135056\|\|full | Low-quality |
| S8_vs_k141_170149\|\|full | Not-determined |
| S3_vs_k141_127442\|\|full | High-quality |
| S8_vs_k141_160859\|\|full | Not-determined |
| S8_vs_k141_204432\|\|full | Not-determined |
| S8_vs_k141_160863\|\|full | Low-quality |
| S8_vs_k141_109433\|\|full | Low-quality |
| S8_vs_k141_135073\|\|full | Not-determined |
| S8_vs_k141_83192\|\|full | Not-determined |
| S5_vs_k141_7444\|\|0_partial | Medium-quality |
| S8_vs_k141_204444\|\|full | Low-quality |
| S8_vs_k141_178789\|\|full | Low-quality |
| S8_vs_k141_152575\|\|full | Not-determined |
| S8_vs_k141_109451\|\|full | Not-determined |
| S8_vs_k141_91923\|\|full | Low-quality |
| S8_vs_k141_22847\|\|full | Not-determined |
| S8_vs_k141_91925\|\|full | Low-quality |
| S8_vs_k141_186541\|\|full | Not-determined |
| S8_vs_k141_186545\|\|full | Low-quality |
| S8_vs_k141_204464\|\|full | Not-determined |
| S8_vs_k141_117923\|\|full | Not-determined |
| S8_vs_k141_117927\|\|full | Not-determined |
| S8_vs_k141_208119\|\|full | High-quality |
| S8_vs_k141_91949\|\|full | Not-determined |
| S8_vs_k141_117929\|\|full | Not-determined |
| S8_vs_k141_196254\|\|full | Not-determined |
| S8_vs_k141_170196\|\|full | Not-determined |
| S8_vs_k141_170197\|\|full | Low-quality |
| S8_vs_k141_57520\|\|full | Not-determined |
| S8_vs_k141_65775\|\|full | Low-quality |
| S8_vs_k141_170202\|\|full | Low-quality |
| S8_extracted_k141_208115 | Complete |
| S8_vs_k141_91951\|\|full | Not-determined |
| S8_vs_k141_5092\|\|full | Low-quality |
| S8_vs_k141_31441\|\|full | Low-quality |
| S8_vs_k141_152620\|\|full | Low-quality |
| S8_vs_k141_109473\|\|full | Not-determined |
| S8_vs_k141_186560\|\|full | Low-quality |
| S8_vs_k141_204495\|\|full | Not-determined |
| S8_vs_k141_91962\|\|full | Low-quality |
| S8_vs_k141_152625\|\|full | Low-quality |
| S8_vs_k141_109477\|\|full | Low-quality |
| S8_vs_k141_13656\|\|full | Low-quality |
| S8_vs_k141_13659\|\|full | Low-quality |
| S8_vs_k141_74119\|\|full | Low-quality |
| S8_vs_k141_186572\|\|full | Not-determined |
| S8_vs_k141_109494\|\|full | Not-determined |
| S8_vs_k141_152645\|\|full | Not-determined |
| S8_vs_k141_109497\|\|full | Not-determined |
| S8_vs_k141_40056\|\|full | Not-determined |
| S8_vs_k141_5128\|\|full | Not-determined |
| S8_vs_k141_31469\|\|full | Low-quality |
| S8_vs_k141_204497\|\|full | Not-determined |
| S8_vs_k141_22945\|\|full | Low-quality |
| S8_vs_k141_91989\|\|full | Low-quality |
| S8_vs_k141_170250\|\|full | Low-quality |
| S8_vs_k141_74161\|\|full | Low-quality |
| S8_vs_k141_92001\|\|full | Not-determined |
| S12_extracted_k141_188846 | Complete |
| S8_vs_k141_144187\|\|full | Low-quality |
| S8_vs_k141_196345\|\|full | Low-quality |
| S8_vs_k141_74180\|\|full | Low-quality |
| S8_vs_k141_135179\|\|full | Not-determined |
| S8_vs_k141_31493\|\|full | Low-quality |
| S8_vs_k141_178901\|\|full | Not-determined |
| S8_vs_k141_160978\|\|full | Low-quality |
| S8_vs_k141_5180\|\|full | Not-determined |
| S8_vs_k141_83328\|\|full | Low-quality |
| S8_vs_k141_160993\|\|full | Not-determined |
| S8_vs_k141_65832\|\|full | Not-determined |
| S8_vs_k141_118035\|\|full | Not-determined |
| S8_vs_k141_48231\|\|full | Not-determined |
| S8_vs_k141_109543\|\|full | Not-determined |
| S8_vs_k141_118036\|\|full | Low-quality |
| S8_vs_k141_204553\|\|full | Not-determined |
| S8_vs_k141_170298\|\|full | Low-quality |
| S8_vs_k141_57604\|\|full | Not-determined |
| S8_vs_k141_135181\|\|full | Low-quality |
| S8_vs_k141_5201\|\|full | Low-quality |
| S8_vs_k141_48241\|\|full | Not-determined |
| S8_vs_k141_204570\|\|full | Low-quality |
| S8_vs_k141_178938\|\|full | Low-quality |
| S8_vs_k141_161034\|\|full | Low-quality |
| S8_vs_k141_83363\|\|full | Low-quality |
| S8_vs_k141_92066\|\|full | Low-quality |
| S8_vs_k141_31538\|\|full | Not-determined |
| S8_vs_k141_118049\|\|full | Low-quality |
| S8_vs_k141_13713\|\|full | Not-determined |
| S8_vs_k141_170325\|\|full | Not-determined |
| S8_vs_k141_31546\|\|full | Not-determined |
| S8_vs_k141_178979\|\|full | Not-determined |
| S8_vs_k141_208115\|\|full | High-quality |
| S8_vs_k141_144251\|\|full | Low-quality |
| S8_vs_k141_13723\|\|full | Not-determined |
| S8_vs_k141_135215\|\|full | Not-determined |
| S8_vs_k141_13728\|\|full | Not-determined |
| S8_vs_k141_152764\|\|full | Not-determined |
| S8_vs_k141_5222\|\|full | Low-quality |
| S8_vs_k141_5223\|\|full | Low-quality |
| S8_vs_k141_161091\|\|full | Not-determined |
| S8_vs_k141_179011\|\|full | Not-determined |
| S8_vs_k141_135234\|\|full | Not-determined |
| S8_vs_k141_48294\|\|full | Low-quality |
| S8_vs_k141_179018\|\|full | Low-quality |

| | |
|---|---|
| S8_vs_k141_170372\|\|full | Low-quality |
| S8_vs_k141_196400\|\|full | Low-quality |
| S8_vs_k141_186629\|\|full | Low-quality |
| S8_vs_k141_5234\|\|full | Low-quality |
| S8_vs_k141_161102\|\|full | Not-determined |
| S8_vs_k141_83419\|\|full | Not-determined |
| S8_vs_k141_161109\|\|full | Low-quality |
| S8_vs_k141_83422\|\|full | Low-quality |
| S8_vs_k141_135251\|\|full | Low-quality |
| S8_vs_k141_179044\|\|full | Not-determined |
| S8_vs_k141_23043\|\|full | Low-quality |
| S8_vs_k141_109620\|\|full | Low-quality |
| S8_vs_k141_196429\|\|full | Not-determined |
| S8_vs_k141_126790\|\|full | Low-quality |
| S7_vs_26757\|\|full | Medium-quality |
| S8_vs_k141_196435\|\|full | Not-determined |
| S8_vs_k141_126799\|\|full | Not-determined |
| S8_vs_k141_126801\|\|full | Low-quality |
| S8_vs_k141_83453\|\|full | Low-quality |
| S8_vs_k141_74331\|\|full | Low-quality |
| S8_vs_k141_109649\|\|full | Low-quality |
| S8_vs_k141_83469\|\|full | Not-determined |
| S8_vs_k141_74347\|\|full | Low-quality |
| S8_vs_k141_65894\|\|full | Not-determined |
| S8_vs_k141_152829\|\|full | Not-determined |
| S8_vs_k141_92188\|\|full | Low-quality |
| S8_vs_k141_31645\|\|full | Low-quality |
| S8_vs_k141_40099\|\|full | Low-quality |
| S8_vs_k141_196484\|\|full | Low-quality |
| S8_vs_k141_74364\|\|full | Low-quality |
| S8_vs_k141_126825\|\|full | Not-determined |
| S8_vs_k141_31660\|\|full | Low-quality |
| S8_vs_k141_48373\|\|full | Low-quality |
| S8_vs_k141_186672\|\|full | Low-quality |
| S8_vs_k141_65907\|\|full | Low-quality |
| S8_vs_k141_196497\|\|full | Low-quality |
| S8_vs_k141_196500\|\|full | Low-quality |
| S8_vs_k141_74390\|\|full | Low-quality |
| S8_vs_k141_152861\|\|full | Low-quality |
| S8_vs_k141_5342\|\|full | Not-determined |
| S8_vs_k141_23104\|\|full | Not-determined |
| S8_vs_k141_118210\|\|full | Low-quality |
| S8_vs_k141_57783\|\|full | Low-quality |
| S8_vs_k141_118211\|\|full | Low-quality |
| S8_vs_k141_100420\|\|full | Low-quality |
| S8_vs_k141_5343\|\|full | Low-quality |
| S8_vs_k141_31715\|\|full | Not-determined |
| S8_vs_k141_152878\|\|full | Not-determined |
| S8_vs_k141_40150\|\|full | Not-determined |
| S8_vs_k141_196531\|\|full | Not-determined |
| S8_vs_k141_40157\|\|full | Not-determined |
| S8_vs_k141_92211\|\|full | Low-quality |
| S8_vs_k141_48449\|\|full | Not-determined |
| S8_vs_k141_196544\|\|full | Not-determined |
| S8_vs_k141_57809\|\|full | Not-determined |
| S8_vs_k141_161216\|\|full | Not-determined |
| S8_vs_k141_161222\|\|full | Not-determined |
| S8_vs_k141_109691\|\|full | Not-determined |
| S8_vs_k141_40173\|\|full | Low-quality |
| S8_vs_k141_118244\|\|full | Low-quality |
| S8_vs_k141_170439\|\|full | Not-determined |
| S8_vs_k141_204706\|\|full | Low-quality |
| S8_vs_k141_135400\|\|full | Low-quality |
| S8_vs_k141_100484\|\|full | Not-determined |
| S8_vs_k141_65976\|\|full | Low-quality |
| S8_vs_k141_83520\|\|full | Not-determined |
| S8_vs_k141_186774\|\|full | Not-determined |
| S8_vs_k141_74454\|\|full | Low-quality |
| S8_vs_k141_40205\|\|full | Low-quality |
| S8_vs_k141_65990\|\|full | Low-quality |
| S8_vs_k141_152932\|\|full | Low-quality |
| S8_vs_k141_196598\|\|full | Not-determined |
| S8_vs_k141_118275\|\|full | Not-determined |
| S8_vs_k141_100513\|\|full | Low-quality |
| S8_vs_k141_74470\|\|full | Not-determined |
| S8_vs_k141_170462\|\|full | Not-determined |
| S8_vs_k141_204735\|\|full | Not-determined |
| S8_vs_k141_13822\|\|full | Not-determined |
| S8_vs_k141_204738\|\|full | Not-determined |
| S8_vs_k141_152947\|\|full | Low-quality |
| S8_vs_k141_74480\|\|full | Low-quality |
| S8_vs_k141_109713\|\|full | Not-determined |
| S8_vs_k141_74490\|\|full | Not-determined |
| S8_vs_k141_83570\|\|full | Low-quality |
| S8_vs_k141_48494\|\|full | Low-quality |
| S8_vs_k141_109718\|\|full | Not-determined |
| S8_vs_k141_196615\|\|full | Low-quality |
| S8_vs_k141_83574\|\|full | Low-quality |
| S8_vs_k141_31798\|\|full | Low-quality |
| S8_vs_k141_83575\|\|full | Not-determined |
| S8_vs_k141_74503\|\|full | Low-quality |
| S8_vs_k141_100559\|\|full | Low-quality |
| S8_vs_k141_170490\|\|full | Not-determined |
| S8_vs_k141_13862\|\|full | Not-determined |
| S8_vs_k141_100567\|\|full | Low-quality |
| S8_vs_k141_57917\|\|full | Low-quality |
| S8_vs_k141_153002\|\|full | Not-determined |
| S8_vs_k141_92331\|\|full | Low-quality |
| S8_vs_k141_135497\|\|full | Not-determined |
| S8_vs_k141_153014\|\|full | Low-quality |
| S8_vs_k141_31841\|\|full | Low-quality |
| S8_vs_k141_135513\|\|full | Low-quality |
| S8_vs_k141_109752\|\|full | Not-determined |
| S8_vs_k141_153029\|\|full | Low-quality |
| S8_vs_k141_40278\|\|full | Low-quality |
| S8_vs_k141_48567\|\|full | Low-quality |
| S8_vs_k141_135520\|\|full | Not-determined |
| S8_vs_k141_109759\|\|full | Low-quality |
| S8_vs_k141_100595\|\|full | Not-determined |
| S8_vs_k141_40288\|\|full | Not-determined |
| S8_vs_k141_100597\|\|full | Low-quality |
| S8_vs_k141_5481\|\|full | Low-quality |
| S8_vs_k141_135526\|\|full | Low-quality |
| S8_vs_k141_170536\|\|full | Low-quality |
| S8_vs_k141_161333\|\|full | Not-determined |
| S8_vs_k141_153050\|\|full | Not-determined |
| S8_vs_k141_57964\|\|full | Not-determined |
| S8_vs_k141_135532\|\|full | Low-quality |
| S8_vs_k141_92365\|\|full | Low-quality |
| S8_vs_k141_92367\|\|full | Low-quality |
| S8_vs_k141_109774\|\|full | Low-quality |
| S8_vs_k141_40312\|\|full | Low-quality |
| S8_vs_k141_31877\|\|full | Low-quality |
| S3_extracted_k141_127431 | Complete |
| S8_vs_k141_5507\|\|full | Not-determined |
| S8_vs_k141_196674\|\|full | Low-quality |
| S8_vs_k141_118383\|\|full | Not-determined |
| S8_vs_k141_204761\|\|full | Not-determined |
| S8_vs_k141_100625\|\|full | Not-determined |
| S8_vs_k141_31882\|\|full | Low-quality |
| S8_vs_k141_204766\|\|full | Not-determined |
| S8_vs_k141_13910\|\|full | Low-quality |
| S8_vs_k141_170575\|\|full | Not-determined |
| S8_vs_k141_186886\|\|full | Low-quality |
| S8_vs_k141_118399\|\|full | Low-quality |
| S8_vs_k141_48599\|\|full | Not-determined |
| S8_vs_k141_40337\|\|full | Not-determined |
| S8_vs_k141_204780\|\|full | Not-determined |
| S8_vs_k141_170585\|\|full | Not-determined |
| S8_vs_k141_74595\|\|full | Low-quality |
| S8_vs_k141_135582\|\|full | Low-quality |
| S8_vs_k141_57997\|\|full | Not-determined |
| S8_vs_k141_170593\|\|full | Low-quality |
| S8_vs_k141_109825\|\|full | Not-determined |
| S8_vs_k141_40352\|\|full | Low-quality |
| S8_vs_k141_186911\|\|full | Not-determined |
| S8_vs_k141_40356\|\|full | Not-determined |
| S8_vs_k141_204793\|\|full | Low-quality |
| S8_vs_k141_170603\|\|full | Not-determined |
| S8_vs_k141_186920\|\|full | Low-quality |
| S8_vs_k141_144516\|\|full | Not-determined |
| S8_vs_k141_40366\|\|full | Not-determined |
| S8_vs_k141_58007\|\|full | Low-quality |
| S8_vs_k141_204806\|\|full | Not-determined |
| S8_vs_k141_186924\|\|full | Not-determined |
| S9_extracted_k141_90852 | Complete |
| S8_vs_k141_23202\|\|full | Low-quality |
| S8_vs_k141_186925\|\|full | Low-quality |
| S8_vs_k141_204812\|\|full | Not-determined |
| S8_vs_k141_23205\|\|full | Low-quality |
| S8_vs_k141_58021\|\|full | Low-quality |
| S8_vs_k141_170629\|\|full | Not-determined |
| S8_vs_k141_13959\|\|full | Not-determined |
| S8_vs_k141_127015\|\|full | Low-quality |
| S8_vs_k141_127016\|\|full | Not-determined |
| S8_vs_k141_135617\|\|full | Low-quality |
| S8_vs_k141_196738\|\|full | Not-determined |
| S8_vs_k141_13979\|\|full | Low-quality |
| S8_vs_k141_5582\|\|full | Not-determined |
| S8_vs_k141_13980\|\|full | Low-quality |
| S8_vs_k141_13981\|\|full | Low-quality |
| S8_vs_k141_153132\|\|full | Low-quality |
| S8_vs_k141_109882\|\|full | Not-determined |
| S8_vs_k141_196753\|\|full | Not-determined |
| S8_vs_k141_13988\|\|full | Low-quality |
| S8_vs_k141_48652\|\|full | Low-quality |
| S8_vs_k141_58050\|\|full | Not-determined |
| S8_vs_k141_58053\|\|full | Low-quality |
| S8_vs_k141_135629\|\|full | Low-quality |
| S8_vs_k141_196761\|\|full | Low-quality |
| S8_vs_k141_92486\|\|full | Not-determined |
| S8_vs_k141_40416\|\|full | Low-quality |
| S8_vs_k141_5592\|\|full | Low-quality |
| S8_vs_k141_196763\|\|full | Not-determined |
| S8_vs_k141_127048\|\|full | Low-quality |
| S8_vs_k141_118515\|\|full | Not-determined |
| S8_vs_k141_23259\|\|full | Not-determined |
| S8_vs_k141_40428\|\|full | Low-quality |
| S8_vs_k141_66125\|\|full | Not-determined |
| S8_vs_k141_66129\|\|full | Not-determined |
| S8_vs_k141_14027\|\|full | Not-determined |
| S8_vs_k141_153165\|\|full | Low-quality |
| S8_vs_k141_40442\|\|full | Low-quality |
| S8_vs_k141_144577\|\|full | Low-quality |
| S8_vs_k141_161480\|\|full | Low-quality |
| S8_vs_k141_179290\|\|full | Low-quality |
| S8_vs_k141_135673\|\|full | Low-quality |
| S8_vs_k141_31977\|\|full | Not-determined |
| S8_vs_k141_14035\|\|full | Not-determined |
| S8_vs_k141_144583\|\|full | Not-determined |
| S8_vs_k141_187001\|\|full | Low-quality |
| S8_vs_k141_187004\|\|full | Low-quality |
| S8_vs_k141_196789\|\|full | Not-determined |
| S8_vs_k141_135685\|\|full | Low-quality |
| S8_vs_k141_23290\|\|full | Low-quality |
| S8_vs_k141_118547\|\|full | Low-quality |
| S8_vs_k141_153182\|\|full | Not-determined |
| S8_vs_k141_135694\|\|full | Not-determined |
| S8_vs_k141_66151\|\|full | Not-determined |
| S8_vs_k141_109919\|\|full | Not-determined |
| S8_vs_k141_204880\|\|full | Low-quality |
| S8_vs_k141_66158\|\|full | Low-quality |
| S8_vs_k141_144598\|\|full | Not-determined |
| S8_vs_k141_144599\|\|full | Low-quality |
| S8_vs_k141_144602\|\|full | Not-determined |
| S8_vs_k141_14071\|\|full | Low-quality |
| S8_vs_k141_66167\|\|full | Low-quality |
| S8_vs_k141_161523\|\|full | Not-determined |
| S8_vs_k141_196805\|\|full | Not-determined |
| S8_vs_k141_127092\|\|full | Low-quality |
| S8_vs_k141_40477\|\|full | Not-determined |
| S8_vs_k141_40481\|\|full | Not-determined |
| S8_vs_k141_179337\|\|full | Low-quality |
| S8_vs_k141_92532\|\|full | Not-determined |
| S8_vs_k141_14078\|\|full | Not-determined |
| S8_vs_k141_118572\|\|full | Not-determined |
| S8_vs_k141_74711\|\|full | Not-determined |
| S8_vs_k141_196815\|\|full | Not-determined |
| S8_vs_k141_48741\|\|full | Low-quality |
| S8_vs_k141_32013\|\|full | Not-determined |
| S8_vs_k141_14082\|\|full | Not-determined |
| S8_vs_k141_74713\|\|full | Low-quality |
| S8_vs_k141_23333\|\|full | Not-determined |
| S8_vs_k141_23334\|\|full | Low-quality |
| S8_vs_k141_5668\|\|full | Low-quality |
| S8_vs_k141_48748\|\|full | Low-quality |
| S8_vs_k141_48749\|\|full | Low-quality |
| S8_vs_k141_204937\|\|full | Low-quality |
| S8_vs_k141_153197\|\|full | Low-quality |
| S8_vs_k141_161544\|\|full | Low-quality |
| S8_vs_k141_170710\|\|full | Not-determined |
| S8_vs_k141_40516\|\|full | Low-quality |
| S8_vs_k141_179367\|\|full | Not-determined |
| S8_vs_k141_66219\|\|full | Not-determined |
| S8_vs_k141_40527\|\|full | Low-quality |
| S8_vs_k141_48759\|\|full | Low-quality |
| S8_vs_k141_161570\|\|full | Not-determined |
| S8_vs_k141_179381\|\|full | Not-determined |
| S8_vs_k141_109996\|\|full | Low-quality |
| S8_vs_k141_179384\|\|full | Not-determined |
| S8_vs_k141_204980\|\|full | Not-determined |
| S8_vs_k141_179386\|\|full | Low-quality |
| S8_vs_k141_74773\|\|full | Low-quality |
| S8_vs_k141_32045\|\|full | Low-quality |
| S8_vs_k141_110009\|\|full | Low-quality |
| S8_vs_k141_74774\|\|full | Low-quality |
| S8_vs_k141_58141\|\|full | Low-quality |
| S8_vs_k141_187036\|\|full | Low-quality |
| S8_vs_k141_83680\|\|full | Not-determined |
| S8_vs_k141_58154\|\|full | Not-determined |
| S8_vs_k141_5716\|\|full | Low-quality |
| S8_vs_k141_205008\|\|full | Not-determined |
| S8_vs_k141_83686\|\|full | Not-determined |
| S8_vs_k141_74798\|\|full | Low-quality |
| S8_vs_k141_153251\|\|full | Low-quality |
| S8_vs_k141_196890\|\|full | Low-quality |
| S8_vs_k141_83689\|\|full | Not-determined |
| S8_vs_k141_32070\|\|full | Not-determined |
| S8_vs_k141_58168\|\|full | Low-quality |
| S8_vs_k141_127109\|\|full | Low-quality |
| S8_vs_k141_83698\|\|full | Not-determined |
| S8_vs_k141_74813\|\|full | Low-quality |
| S8_vs_k141_32077\|\|full | Not-determined |
| S8_vs_k141_74819\|\|full | Low-quality |
| S8_vs_k141_58181\|\|full | Low-quality |
| S8_vs_k141_110039\|\|full | Low-quality |
| S8_vs_k141_205042\|\|full | Low-quality |
| S8_vs_k141_92539\|\|full | Not-determined |
| S8_vs_k141_144722\|\|full | Not-determined |
| S8_vs_k141_110047\|\|full | Low-quality |
| S8_vs_k141_161638\|\|full | Not-determined |
| S8_vs_k141_179418\|\|full | Low-quality |
| S8_vs_k141_92549\|\|full | Low-quality |
| S8_vs_k141_161647\|\|full | Low-quality |
| S8_vs_k141_83720\|\|full | Low-quality |
| S8_vs_k141_196927\|\|full | Not-determined |
| S8_vs_k141_5778\|\|full | Low-quality |
| S8_vs_k141_161654\|\|full | Not-determined |
| S8_vs_k141_23444\|\|full | Not-determined |
| S8_vs_k141_205085\|\|full | Not-determined |
| S8_vs_k141_92583\|\|full | Low-quality |
| S8_vs_k141_66307\|\|full | Not-determined |
| S8_vs_k141_66314\|\|full | Low-quality |
| S8_vs_k141_92596\|\|full | Not-determined |
| S8_vs_k141_179441\|\|full | Not-determined |
| S8_vs_k141_127191\|\|full | Low-quality |
| S8_vs_k141_5817\|\|full | Low-quality |
| S8_vs_k141_58209\|\|full | Not-determined |
| S8_vs_k141_153309\|\|full | Not-determined |
| S8_vs_k141_5822\|\|full | Not-determined |
| S8_vs_k141_187155\|\|full | Low-quality |
| S8_vs_k141_205122\|\|full | Low-quality |
| S8_vs_k141_179470\|\|full | Not-determined |
| S8_vs_k141_14176\|\|full | Not-determined |
| S8_vs_k141_74906\|\|full | Low-quality |
| S8_vs_k141_5836\|\|full | Low-quality |
| S8_vs_k141_100922\|\|full | Low-quality |
| S8_vs_k141_161694\|\|full | Not-determined |
| S8_vs_k141_100924\|\|full | Not-determined |
| S8_vs_k141_161696\|\|full | Low-quality |
| S8_vs_k141_23458\|\|full | Low-quality |
| S8_vs_k141_74921\|\|full | Low-quality |
| S8_vs_k141_100933\|\|full | Low-quality |
| S8_vs_k141_66349\|\|full | Not-determined |
| S8_vs_k141_74927\|\|full | Low-quality |
| S8_vs_k141_74928\|\|full | Low-quality |
| S8_vs_k141_205123\|\|full | Low-quality |
| S8_vs_k141_66351\|\|full | Low-quality |
| S8_vs_k141_66352\|\|full | Low-quality |
| S8_vs_k141_187193\|\|full | Low-quality |
| S8_vs_k141_118752\|\|full | Not-determined |
| S8_vs_k141_187194\|\|full | Low-quality |
| S8_vs_k141_5867\|\|full | Low-quality |
| S8_vs_k141_196993\|\|full | Low-quality |
| S8_vs_k141_100952\|\|full | Not-determined |
| S8_vs_k141_179505\|\|full | Not-determined |
| S8_vs_k141_92651\|\|full | Low-quality |
| S8_vs_k141_48938\|\|full | Low-quality |
| S8_vs_k141_74946\|\|full | Low-quality |
| S8_vs_k141_66376\|\|full | Not-determined |
| S8_vs_k141_179516\|\|full | Not-determined |
| S8_vs_k141_127268\|\|full | Low-quality |
| S8_vs_k141_66379\|\|full | Not-determined |
| S8_vs_k141_179521\|\|full | Low-quality |
| S8_vs_k141_118756\|\|full | Low-quality |
| S8_vs_k141_14246\|\|full | Not-determined |
| S8_vs_k141_48959\|\|full | Low-quality |
| S8_vs_k141_100981\|\|full | Low-quality |
| S8_vs_k141_14252\|\|full | Not-determined |
| S8_vs_k141_187229\|\|full | Not-determined |
| S8_vs_k141_74976\|\|full | Low-quality |
| S8_vs_k141_23517\|\|full | Low-quality |
| S8_vs_k141_127295\|\|full | Low-quality |
| S8_vs_k141_110187\|\|full | Not-determined |
| S8_vs_k141_100986\|\|full | Low-quality |
| S8_vs_k141_40743\|\|full | Low-quality |
| S8_vs_k141_135743\|\|full | Low-quality |
| S8_vs_k141_40746\|\|full | Low-quality |
| S8_vs_k141_187274\|\|full | Not-determined |
| S8_vs_k141_40754\|\|full | Low-quality |
| S8_vs_k141_110211\|\|full | Not-determined |
| S8_vs_k141_5935\|\|full | Not-determined |
| S8_vs_k141_58302\|\|full | Not-determined |
| S8_vs_k141_153335\|\|full | Low-quality |
| S8_vs_k141_74992\|\|full | Not-determined |
| S8_vs_k141_197015\|\|full | Not-determined |
| S8_vs_k141_197019\|\|full | Low-quality |
| S8_vs_k141_101009\|\|full | Low-quality |
| S8_vs_k141_144927\|\|full | Not-determined |
| S8_vs_k141_75000\|\|full | Low-quality |
| S8_vs_k141_153356\|\|full | Not-determined |
| S8_vs_k141_101020\|\|full | Not-determined |
| S8_vs_k141_197034\|\|full | Low-quality |
| S8_vs_k141_5959\|\|full | Not-determined |
| S8_vs_k141_40783\|\|full | Not-determined |
| S3_extracted_k141_54498 | High-quality |
| S8_vs_k141_153361\|\|full | Low-quality |
| S8_vs_k141_187294\|\|full | Low-quality |
| S8_vs_k141_101038\|\|full | Low-quality |
| S8_vs_k141_5973\|\|full | Not-determined |
| S8_vs_k141_58328\|\|full | Not-determined |
| S8_vs_k141_14309\|\|full | Not-determined |
| S8_vs_k141_23594\|\|full | Low-quality |
| S8_vs_k141_127350\|\|full | Not-determined |
| S8_vs_k141_118803\|\|full | Not-determined |
| S8_vs_k141_101048\|\|full | Not-determined |
| S8_vs_k141_5982\|\|full | Low-quality |
| S13_extracted_150407 | Complete |
| S8_vs_k141_170873\|\|full | Low-quality |
| S8_vs_k141_40800\|\|full | Not-determined |
| S8_vs_k141_118811\|\|full | Not-determined |
| S8_vs_k141_58336\|\|full | Low-quality |
| S8_vs_k141_110253\|\|full | Low-quality |
| S8_vs_k141_144968\|\|full | Not-determined |
| S8_vs_k141_187314\|\|full | Not-determined |
| S8_vs_k141_14341\|\|full | Not-determined |
| S8_vs_k141_179584\|\|full | Not-determined |
| S8_vs_k141_153417\|\|full | Low-quality |
| S8_vs_k141_92741\|\|full | Not-determined |
| S8_vs_k141_40815\|\|full | Not-determined |
| S8_vs_k141_118838\|\|full | Low-quality |
| S8_vs_k141_179588\|\|full | Low-quality |
| S8_vs_k141_101073\|\|full | Low-quality |
| S8_vs_k141_101076\|\|full | Low-quality |
| S8_vs_k141_58380\|\|full | Low-quality |
| S8_vs_k141_66458\|\|full | Low-quality |
| S8_vs_k141_127395\|\|full | Not-determined |
| S8_vs_k141_23657\|\|full | Low-quality |
| S8_vs_k141_66462\|\|full | Not-determined |
| S8_vs_k141_23663\|\|full | Not-determined |
| S8_vs_k141_135851\|\|full | Low-quality |
| S8_vs_k141_161771\|\|full | Low-quality |
| S8_vs_k141_75063\|\|full | Not-determined |
| S12_vs_k141_188846\|\|full | High-quality |
| S3_vs_k141_54498\|\|full | High-quality |
| S8_vs_k141_179605\|\|full | Not-determined |
| S8_vs_k141_6026\|\|full | Low-quality |
| S8_vs_k141_118875\|\|full | Low-quality |
| S8_vs_k141_40847\|\|full | Not-determined |
| S8_vs_k141_153467\|\|full | Low-quality |
| S8_vs_k141_83859\|\|full | Low-quality |
| S8_vs_k141_23678\|\|full | Not-determined |
| S8_vs_k141_161774\|\|full | Low-quality |
| S8_vs_k141_92766\|\|full | Low-quality |
| S8_vs_k141_14369\|\|full | Not-determined |
| S8_vs_k141_197072\|\|full | Low-quality |
| S8_vs_k141_66480\|\|full | Low-quality |
| S8_vs_k141_118881\|\|full | Not-determined |
| S8_vs_k141_161777\|\|full | Low-quality |
| S8_vs_k141_179623\|\|full | Low-quality |
| S8_vs_k141_135867\|\|full | Low-quality |
| S8_vs_k141_92767\|\|full | Low-quality |
| S8_vs_k141_127437\|\|full | Low-quality |
| S13_vs_150407\|\|full | High-quality |
| S8_vs_k141_153482\|\|full | Low-quality |
| S8_vs_k141_6049\|\|full | Low-quality |
| S8_vs_k141_40861\|\|full | Low-quality |
| S8_vs_k141_66488\|\|full | Low-quality |
| S8_vs_k141_153487\|\|full | Not-determined |
| S8_vs_k141_127445\|\|full | Low-quality |
| S8_vs_k141_110338\|\|full | Not-determined |
| S8_vs_k141_110339\|\|full | Not-determined |
| S8_vs_k141_179635\|\|full | Low-quality |
| S8_vs_k141_75103\|\|full | Low-quality |
| S8_vs_k141_49038\|\|full | Not-determined |
| S8_vs_k141_40889\|\|full | Low-quality |
| S8_vs_k141_49040\|\|full | Low-quality |
| S8_vs_k141_205288\|\|full | Not-determined |
| S8_vs_k141_32227\|\|full | Low-quality |
| S8_vs_k141_101130\|\|full | Not-determined |
| S8_vs_k141_118914\|\|full | Not-determined |
| S8_vs_k141_58442\|\|full | Low-quality |
| S8_vs_k141_187422\|\|full | Not-determined |
| S8_vs_k141_161816\|\|full | Low-quality |
| S8_vs_k141_118917\|\|full | Low-quality |
| S8_vs_k141_127458\|\|full | Not-determined |
| S8_vs_k141_6090\|\|full | Not-determined |
| S8_vs_k141_49063\|\|full | Low-quality |
| S8_vs_k141_153546\|\|full | Low-quality |
| S8_vs_k141_101140\|\|full | Low-quality |
| S8_vs_k141_153550\|\|full | Low-quality |
| S8_vs_k141_83916\|\|full | Not-determined |
| S8_vs_k141_6107\|\|full | Not-determined |
| S8_vs_k141_6109\|\|full | Low-quality |
| S8_vs_k141_23759\|\|full | Not-determined |
| S8_vs_k141_58463\|\|full | Low-quality |
| S8_vs_k141_40912\|\|full | Not-determined |
| S8_vs_k141_127490\|\|full | Not-determined |
| S8_vs_k141_145001\|\|full | Not-determined |
| S8_vs_k141_32265\|\|full | Low-quality |
| S8_vs_k141_161860\|\|full | Not-determined |
| S8_vs_k141_118945\|\|full | Low-quality |
| S8_vs_k141_179654\|\|full | Not-determined |
| S8_vs_k141_145009\|\|full | Not-determined |
| S8_vs_k141_75151\|\|full | Low-quality |
| S8_vs_k141_161862\|\|full | Low-quality |
| S8_vs_k141_127498\|\|full | Low-quality |
| S8_vs_k141_14445\|\|full | Not-determined |
| S8_vs_k141_49092\|\|full | Low-quality |
| S8_vs_k141_110400\|\|full | Not-determined |
| S12_extracted_k141_188838 | Complete |
| S8_vs_k141_187457\|\|full | Low-quality |
| S8_vs_k141_127516\|\|full | Low-quality |
| S8_vs_k141_179674\|\|full | Not-determined |
| S8_vs_k141_32277\|\|full | Not-determined |
| S8_vs_k141_127517\|\|full | Low-quality |
| S8_vs_k141_110412\|\|full | Low-quality |
| S8_vs_k141_75163\|\|full | Not-determined |
| S8_vs_k141_135936\|\|full | Low-quality |
| S8_vs_k141_58490\|\|full | Low-quality |
| S8_vs_k141_118957\|\|full | Not-determined |
| S8_vs_k141_40937\|\|full | Not-determined |
| S8_vs_k141_14467\|\|full | Low-quality |
| S8_vs_k141_14469\|\|full | Not-determined |
| S8_vs_k141_101183\|\|full | Low-quality |
| S8_vs_k141_127539\|\|full | Low-quality |
| S8_vs_k141_14473\|\|full | Low-quality |
| S8_vs_k141_197122\|\|full | Low-quality |
| S8_vs_k141_110425\|\|full | Not-determined |
| S8_vs_k141_49105\|\|full | Low-quality |
| S8_vs_k141_6147\|\|full | Low-quality |
| S8_vs_k141_58498\|\|full | Low-quality |
| S8_vs_k141_161891\|\|full | Not-determined |
| S8_vs_k141_32296\|\|full | Not-determined |
| S8_vs_k141_6156\|\|full | Low-quality |
| S8_vs_k141_66557\|\|full | Low-quality |
| S8_vs_k141_40961\|\|full | Low-quality |
| S8_vs_k141_153598\|\|full | Not-determined |
| S8_vs_k141_197139\|\|full | Low-quality |
| S8_vs_k141_75201\|\|full | Not-determined |
| S8_vs_k141_40966\|\|full | Low-quality |
| S8_vs_k141_101210\|\|full | Not-determined |
| S8_vs_k141_66569\|\|full | Low-quality |
| S8_vs_k141_145052\|\|full | Not-determined |
| S8_vs_k141_118988\|\|full | Low-quality |
| S8_vs_k141_135980\|\|full | Low-quality |
| S8_vs_k141_135990\|\|full | Not-determined |
| S8_vs_k141_83967\|\|full | Not-determined |
| S8_vs_k141_58540\|\|full | Low-quality |
| S8_vs_k141_23833\|\|full | Not-determined |
| S8_vs_k141_49120\|\|full | Low-quality |
| S8_vs_k141_197182\|\|full | Not-determined |
| S8_vs_k141_101240\|\|full | Low-quality |
| S8_vs_k141_75230\|\|full | Low-quality |
| S8_vs_k141_41004\|\|full | Not-determined |
| S8_vs_k141_58555\|\|full | Low-quality |
| S8_vs_k141_197189\|\|full | Low-quality |
| S8_vs_k141_58560\|\|full | Low-quality |
| S8_vs_k141_110472\|\|full | Not-determined |
| S8_vs_k141_153648\|\|full | Low-quality |
| S8_vs_k141_161931\|\|full | Not-determined |
| S8_vs_k141_145093\|\|full | Low-quality |
| S8_vs_k141_110479\|\|full | Not-determined |
| S8_vs_k141_179727\|\|full | Not-determined |
| S8_vs_k141_58570\|\|full | Low-quality |
| S8_vs_k141_145098\|\|full | Not-determined |
| S8_vs_k141_153664\|\|full | Low-quality |
| S8_vs_k141_127624\|\|full | Low-quality |
| S8_vs_k141_101268\|\|full | Not-determined |
| S8_vs_k141_110484\|\|full | Low-quality |
| S8_vs_k141_41036\|\|full | Low-quality |
| S8_vs_k141_119038\|\|full | Low-quality |
| S8_vs_k141_127625\|\|full | Low-quality |
| S8_vs_k141_41037\|\|full | Not-determined |
| S8_vs_k141_14551\|\|full | Not-determined |
| S8_vs_k141_110490\|\|full | Low-quality |
| S8_vs_k141_14556\|\|full | Not-determined |
| S8_vs_k141_171051\|\|full | Not-determined |
| S8_vs_k141_101276\|\|full | Low-quality |
| S8_vs_k141_179751\|\|full | Not-determined |
| S8_vs_k141_32340\|\|full | Not-determined |
| S8_vs_k141_66631\|\|full | Low-quality |
| S8_vs_k141_14558\|\|full | Not-determined |
| S8_vs_k141_197241\|\|full | Not-determined |
| S8_vs_k141_153689\|\|full | Low-quality |
| S8_vs_k141_110507\|\|full | Not-determined |
| S8_vs_k141_84024\|\|full | Low-quality |
| S8_vs_k141_153695\|\|full | Not-determined |
| S8_vs_k141_179765\|\|full | Not-determined |
| S8_vs_k141_84028\|\|full | Not-determined |
| S8_vs_k141_110513\|\|full | Low-quality |
| S8_vs_k141_32353\|\|full | Not-determined |
| S8_vs_k141_92877\|\|full | Not-determined |
| S9_vs_k141_90863\|\|full | High-quality |
| S8_vs_k141_23876\|\|full | Low-quality |
| S8_vs_k141_119107\|\|full | Low-quality |
| S8_vs_k141_6249\|\|full | Not-determined |
| S8_vs_k141_110533\|\|full | Low-quality |
| S8_extracted_k141_208146 | Complete |
| S8_vs_k141_179794\|\|full | Low-quality |
| S8_vs_k141_119112\|\|full | Low-quality |
| S8_vs_k141_171107\|\|full | Low-quality |
| S8_vs_k141_187589\|\|full | Low-quality |
| S8_vs_k141_179801\|\|full | Not-determined |
| S8_vs_k141_197267\|\|full | Low-quality |
| S8_vs_k141_205388\|\|full | Low-quality |
| S8_vs_k141_58661\|\|full | Not-determined |
| S8_vs_k141_110545\|\|full | Not-determined |
| S8_vs_k141_171120\|\|full | Not-determined |
| S8_vs_k141_32388\|\|full | Low-quality |
| S8_vs_k141_23914\|\|full | Not-determined |
| S8_vs_k141_197285\|\|full | Low-quality |
| S8_vs_k141_6290\|\|full | Low-quality |
| S8_vs_k141_23924\|\|full | Not-determined |
| S8_vs_k141_127689\|\|full | Low-quality |
| S8_vs_k141_127691\|\|full | Low-quality |
| S8_vs_k141_127699\|\|full | Low-quality |
| S8_vs_k141_153750\|\|full | Not-determined |
| S8_vs_k141_6298\|\|full | Low-quality |
| S8_vs_k141_75379\|\|full | Low-quality |
| S8_vs_k141_197305\|\|full | Not-determined |
| S8_vs_k141_84106\|\|full | Low-quality |
| S8_vs_k141_171148\|\|full | Not-determined |
| S8_vs_k141_32425\|\|full | Low-quality |
| S8_vs_k141_92929\|\|full | Low-quality |
| S8_vs_k141_32430\|\|full | Low-quality |
| S8_vs_k141_153772\|\|full | Not-determined |
| S8_vs_k141_14651\|\|full | Low-quality |
| S8_vs_k141_110601\|\|full | Low-quality |

| | |
|---|---|
| S8_vs_k141_197325\|\|full | Low-quality |
| S8_vs_k141_179846\|\|full | Low-quality |
| S8_vs_k141_145158\|\|full | Low-quality |
| S8_vs_k141_14669\|\|full | Low-quality |
| S8_vs_k141_92948\|\|full | Low-quality |
| S8_vs_k141_205417\|\|full | Low-quality |
| S8_vs_k141_205418\|\|full | Low-quality |
| S8_vs_k141_187632\|\|full | Low-quality |
| S8_vs_k141_110614\|\|full | Low-quality |
| S8_vs_k141_75408\|\|full | Not-determined |
| S8_vs_k141_197340\|\|full | Not-determined |
| S8_vs_k141_32444\|\|full | Not-determined |
| S8_vs_k141_179868\|\|full | Low-quality |
| S8_vs_k141_6349\|\|full | Not-determined |
| S8_vs_k141_179870\|\|full | Not-determined |
| S8_vs_k141_66719\|\|full | Low-quality |
| S8_vs_k141_32454\|\|full | Not-determined |
| S8_vs_k141_14685\|\|full | Not-determined |
| S8_vs_k141_162069\|\|full | Not-determined |
| S8_vs_k141_179878\|\|full | Low-quality |
| S8_vs_k141_119126\|\|full | Not-determined |
| S8_vs_k141_119131\|\|full | Low-quality |
| S8_vs_k141_205460\|\|full | Low-quality |
| S8_vs_k141_92990\|\|full | Not-determined |
| S8_vs_k141_153827\|\|full | Not-determined |
| S8_vs_k141_92992\|\|full | Low-quality |
| S8_vs_k141_153832\|\|full | Not-determined |
| S8_vs_k141_14696\|\|full | Not-determined |
| S8_vs_k141_197371\|\|full | Low-quality |
| S8_vs_k141_32478\|\|full | Not-determined |
| S8_vs_k141_93000\|\|full | Low-quality |
| S8_vs_k141_41047\|\|full | Low-quality |
| S8_vs_k141_32485\|\|full | Not-determined |
| S8_vs_k141_101387\|\|full | Low-quality |
| S8_vs_k141_41060\|\|full | Low-quality |
| S12_vs_k141_183891\|\|full | High-quality |
| S8_vs_k141_197401\|\|full | Low-quality |
| S8_vs_k141_41070\|\|full | Low-quality |
| S8_vs_k141_58795\|\|full | Not-determined |
| S8_vs_k141_14718\|\|full | Not-determined |
| S8_vs_k141_66755\|\|full | Low-quality |
| S8_vs_k141_127791\|\|full | Low-quality |
| S8_vs_k141_187702\|\|full | Low-quality |
| S8_vs_k141_119193\|\|full | Not-determined |
| S8_vs_k141_162103\|\|full | Low-quality |
| S8_vs_k141_187706\|\|full | Not-determined |
| S8_vs_k141_197418\|\|full | Low-quality |
| S8_vs_k141_84186\|\|full | Not-determined |
| S8_vs_k141_145196\|\|full | Not-determined |
| S8_vs_k141_84191\|\|full | Low-quality |
| S8_vs_k141_6422\|\|full | Low-quality |
| S8_vs_k141_75456\|\|full | Low-quality |
| S8_vs_k141_93052\|\|full | Low-quality |
| S8_vs_k141_32546\|\|full | Low-quality |
| S8_vs_k141_205506\|\|full | Low-quality |
| S8_vs_k141_205508\|\|full | Low-quality |
| S8_vs_k141_179975\|\|full | Not-determined |
| S8_vs_k141_58822\|\|full | Not-determined |
| S8_vs_k141_32554\|\|full | Low-quality |
| S8_vs_k141_136212\|\|full | Not-determined |
| S8_vs_k141_187725\|\|full | Low-quality |
| S8_vs_k141_84203\|\|full | Low-quality |
| S8_vs_k141_93064\|\|full | Low-quality |
| S8_vs_k141_41122\|\|full | Not-determined |
| S8_vs_k141_171234\|\|full | Not-determined |
| S8_vs_k141_6451\|\|full | Not-determined |
| S8_vs_k141_205520\|\|full | Low-quality |
| S8_vs_k141_162153\|\|full | Low-quality |
| S8_vs_k141_127845\|\|full | Low-quality |
| S8_vs_k141_127849\|\|full | Not-determined |
| S8_vs_k141_162160\|\|full | Not-determined |
| S8_vs_k141_75484\|\|full | Low-quality |
| S8_vs_k141_136223\|\|full | Not-determined |
| S8_vs_k141_41138\|\|full | Not-determined |
| S8_vs_k141_205527\|\|full | Low-quality |
| S8_vs_k141_119238\|\|full | Not-determined |
| S8_vs_k141_153930\|\|full | Not-determined |
| S8_vs_k141_119241\|\|full | Not-determined |
| S12_vs_k141_188838\|\|full | High-quality |
| S8_vs_k141_205529\|\|full | Not-determined |
| S8_vs_k141_66816\|\|full | Not-determined |
| S8_vs_k141_127852\|\|full | Not-determined |
| S8_vs_k141_180023\|\|full | Low-quality |
| S8_vs_k141_32596\|\|full | Low-quality |
| S8_vs_k141_171267\|\|full | Low-quality |
| S8_vs_k141_127858\|\|full | Low-quality |
| S8_vs_k141_93093\|\|full | Low-quality |
| S8_vs_k141_32599\|\|full | Low-quality |
| S8_vs_k141_171271\|\|full | Not-determined |
| S8_vs_k141_180032\|\|full | Low-quality |
| S8_vs_k141_75504\|\|full | Low-quality |
| S8_vs_k141_84241\|\|full | Low-quality |
| S8_vs_k141_110723\|\|full | Low-quality |
| S8_vs_k141_208146\|\|full | High-quality |
| S8_vs_k141_49241\|\|full | Low-quality |
| S1_vs_k141_3267\|\|full | High-quality |
| S8_vs_k141_75512\|\|full | Not-determined |
| S8_vs_k141_153948\|\|full | Low-quality |
| S8_vs_k141_23956\|\|full | Not-determined |
| S8_vs_k141_136241\|\|full | Low-quality |
| S8_vs_k141_110739\|\|full | Not-determined |
| S8_vs_k141_41180\|\|full | Not-determined |
| S8_vs_k141_101502\|\|full | Not-determined |
| S8_vs_k141_6495\|\|full | Low-quality |
| S8_vs_k141_136245\|\|full | Low-quality |
| S8_vs_k141_41188\|\|full | Not-determined |
| S8_vs_k141_180044\|\|full | Low-quality |
| S8_vs_k141_14802\|\|full | Not-determined |
| S8_vs_k141_180049\|\|full | Not-determined |
| S8_vs_k141_145294\|\|full | Not-determined |
| S8_vs_k141_153950\|\|full | Not-determined |
| S8_vs_k141_41199\|\|full | Low-quality |
| S8_vs_k141_58913\|\|full | Not-determined |
| S8_vs_k141_41204\|\|full | Not-determined |
| S8_vs_k141_197509\|\|full | Low-quality |
| S8_vs_k141_93135\|\|full | Low-quality |
| S8_vs_k141_197510\|\|full | Not-determined |
| S8_vs_k141_119273\|\|full | Low-quality |
| S8_vs_k141_14823\|\|full | Low-quality |
| S8_vs_k141_84272\|\|full | Low-quality |
| S8_vs_k141_49279\|\|full | Low-quality |
| S8_vs_k141_197513\|\|full | Not-determined |
| S8_vs_k141_75554\|\|full | Low-quality |
| S8_vs_k141_171342\|\|full | Not-determined |
| S8_vs_k141_32627\|\|full | Not-determined |
| S8_vs_k141_6522\|\|full | Low-quality |
| S8_vs_k141_136274\|\|full | Not-determined |
| S8_vs_k141_119293\|\|full | Low-quality |
| S8_vs_k141_205610\|\|full | Low-quality |
| S8_vs_k141_205612\|\|full | Low-quality |
| S8_vs_k141_119303\|\|full | Not-determined |
| S8_vs_k141_153967\|\|full | Not-determined |
| S8_vs_k141_58943\|\|full | Not-determined |
| S8_vs_k141_93169\|\|full | Low-quality |
| S8_vs_k141_24002\|\|full | Not-determined |
| S8_vs_k141_58944\|\|full | Not-determined |
| S8_vs_k141_84300\|\|full | Low-quality |
| S8_vs_k141_205624\|\|full | Not-determined |
| S8_vs_k141_197539\|\|full | Not-determined |
| S8_vs_k141_180095\|\|full | Low-quality |
| S8_vs_k141_171391\|\|full | Not-determined |
| S8_vs_k141_145342\|\|full | Not-determined |
| S8_vs_k141_110817\|\|full | Low-quality |
| S8_vs_k141_145344\|\|full | Not-determined |
| S8_vs_k141_136293\|\|full | Low-quality |
| S8_vs_k141_145348\|\|full | Not-determined |
| S8_vs_k141_119336\|\|full | Not-determined |
| S8_vs_k141_66903\|\|full | Low-quality |
| S8_vs_k141_14886\|\|full | Not-determined |
| S8_vs_k141_162198\|\|full | Low-quality |
| S8_vs_k141_180109\|\|full | Low-quality |
| S8_vs_k141_136303\|\|full | Not-determined |
| S8_vs_k141_162203\|\|full | Not-determined |
| S8_vs_k141_162206\|\|full | Not-determined |
| S8_vs_k141_197571\|\|full | Low-quality |
| S8_vs_k141_14906\|\|full | Low-quality |
| S8_vs_k141_93202\|\|full | Not-determined |
| S8_vs_k141_154017\|\|full | Low-quality |
| S8_vs_k141_75622\|\|full | Low-quality |
| S8_vs_k141_84350\|\|full | Low-quality |
| S8_vs_k141_171432\|\|full | Not-determined |
| S8_vs_k141_145392\|\|full | Not-determined |
| S8_vs_k141_180127\|\|full | Low-quality |
| S8_vs_k141_75628\|\|full | Not-determined |
| S8_vs_k141_154026\|\|full | Not-determined |
| S8_vs_k141_93216\|\|full | Not-determined |
| S8_vs_k141_58987\|\|full | Low-quality |
| S1_vs_k141_100019\|\|full | Medium-quality |
| S8_vs_k141_171449\|\|full | Low-quality |
| S8_vs_k141_101640\|\|full | Low-quality |
| S8_vs_k141_66944\|\|full | Low-quality |
| S8_vs_k141_6593\|\|full | Low-quality |
| S8_vs_k141_197610\|\|full | Not-determined |
| S8_vs_k141_6594\|\|full | Low-quality |
| S8_vs_k141_162241\|\|full | Not-determined |
| S8_vs_k141_154050\|\|full | Not-determined |
| S8_vs_k141_75655\|\|full | Low-quality |
| S8_vs_k141_66952\|\|full | Low-quality |
| S8_vs_k141_101652\|\|full | Not-determined |
| S8_vs_k141_59002\|\|full | Low-quality |
| S8_vs_k141_93238\|\|full | Low-quality |
| S8_vs_k141_84389\|\|full | Low-quality |
| S8_vs_k141_14958\|\|full | Low-quality |
| S8_vs_k141_66979\|\|full | Low-quality |
| S8_vs_k141_127965\|\|full | Low-quality |
| S3_extracted_k141_101591 | Medium-quality |
| S8_vs_k141_59021\|\|full | Not-determined |
| S8_vs_k141_119346\|\|full | Low-quality |
| S8_vs_k141_197639\|\|full | Not-determined |
| S8_vs_k141_162254\|\|full | Not-determined |
| S8_vs_k141_171475\|\|full | Not-determined |
| S8_vs_k141_32727\|\|full | Low-quality |
| S8_vs_k141_24124\|\|full | Not-determined |
| S8_vs_k141_84428\|\|full | Not-determined |
| S8_vs_k141_119372\|\|full | Not-determined |
| S8_vs_k141_6653\|\|full | Not-determined |
| S8_vs_k141_119377\|\|full | Not-determined |
| S8_vs_k141_162286\|\|full | Low-quality |
| S8_vs_k141_101710\|\|full | Not-determined |
| S8_vs_k141_101713\|\|full | Not-determined |
| S8_vs_k141_187798\|\|full | Low-quality |
| S8_vs_k141_145459\|\|full | Not-determined |
| S8_vs_k141_59060\|\|full | Low-quality |
| S8_vs_k141_24146\|\|full | Not-determined |
| S8_vs_k141_197671\|\|full | Low-quality |
| S8_vs_k141_187803\|\|full | Low-quality |
| S8_vs_k141_75737\|\|full | Low-quality |
| S8_vs_k141_154123\|\|full | Low-quality |
| S8_vs_k141_32766\|\|full | Low-quality |
| S8_vs_k141_59077\|\|full | Low-quality |
| S8_vs_k141_145498\|\|full | Not-determined |
| S8_vs_k141_75749\|\|full | Low-quality |
| S8_vs_k141_101750\|\|full | Low-quality |
| S8_vs_k141_145505\|\|full | Not-determined |
| S8_vs_k141_93268\|\|full | Not-determined |
| S8_vs_k141_171538\|\|full | Low-quality |
| S8_vs_k141_101756\|\|full | Low-quality |
| S8_vs_k141_101757\|\|full | Not-determined |
| S8_vs_k141_49342\|\|full | Low-quality |
| S8_vs_k141_145507\|\|full | Not-determined |
| S8_vs_k141_75770\|\|full | Low-quality |
| S8_vs_k141_32797\|\|full | Low-quality |
| S8_vs_k141_119412\|\|full | Not-determined |
| S8_vs_k141_41297\|\|full | Low-quality |
| S8_vs_k141_154160\|\|full | Not-determined |
| S8_vs_k141_119417\|\|full | Not-determined |
| S8_vs_k141_128076\|\|full | Low-quality |
| S8_vs_k141_84503\|\|full | Low-quality |
| S8_vs_k141_24192\|\|full | Low-quality |
| S8_vs_k141_41303\|\|full | Low-quality |
| S8_vs_k141_162341\|\|full | Low-quality |
| S8_vs_k141_171570\|\|full | Not-determined |
| S8_vs_k141_101797\|\|full | Low-quality |
| S8_vs_k141_154170\|\|full | Low-quality |
| S8_vs_k141_24203\|\|full | Low-quality |
| S8_vs_k141_180306\|\|full | Low-quality |
| S8_vs_k141_136486\|\|full | Low-quality |
| S8_vs_k141_75810\|\|full | Low-quality |
| S8_vs_k141_49370\|\|full | Not-determined |
| S8_vs_k141_197736\|\|full | Not-determined |
| S8_vs_k141_136490\|\|full | Low-quality |
| S8_vs_k141_101814\|\|full | Not-determined |
| S8_vs_k141_154178\|\|full | Low-quality |
| S8_vs_k141_84526\|\|full | Not-determined |
| S8_vs_k141_187841\|\|full | Low-quality |
| S8_vs_k141_101825\|\|full | Low-quality |
| S8_vs_k141_180324\|\|full | Low-quality |
| S8_vs_k141_101829\|\|full | Low-quality |
| S8_vs_k141_49395\|\|full | Low-quality |
| S8_vs_k141_154199\|\|full | Low-quality |
| S8_vs_k141_162400\|\|full | Low-quality |
| S8_vs_k141_32822\|\|full | Not-determined |
| S8_vs_k141_110895\|\|full | Not-determined |
| S8_vs_k141_171622\|\|full | Low-quality |
| S8_vs_k141_162405\|\|full | Not-determined |
| S8_vs_k141_49407\|\|full | Low-quality |
| S8_vs_k141_197776\|\|full | Not-determined |
| S8_vs_k141_128116\|\|full | Low-quality |
| S8_vs_k141_24254\|\|full | Not-determined |
| S8_vs_k141_59170\|\|full | Low-quality |
| S8_vs_k141_6765\|\|full | Not-determined |
| S8_vs_k141_75874\|\|full | Low-quality |
| S8_vs_k141_101867\|\|full | Not-determined |
| S8_vs_k141_49411\|\|full | Not-determined |
| S8_vs_k141_180354\|\|full | Low-quality |
| S8_vs_k141_15044\|\|full | Not-determined |
| S8_vs_k141_32847\|\|full | Not-determined |
| S8_vs_k141_24265\|\|full | Low-quality |
| S8_vs_k141_93367\|\|full | Low-quality |
| S8_vs_k141_75887\|\|full | Low-quality |
| S8_vs_k141_67144\|\|full | Not-determined |
| S8_vs_k141_49423\|\|full | Not-determined |
| S8_vs_k141_119467\|\|full | Low-quality |
| S8_vs_k141_197809\|\|full | Not-determined |
| S17_vs_k141_177210\|\|full | High-quality |
| S8_vs_k141_128171\|\|full | Not-determined |
| S8_vs_k141_32868\|\|full | Low-quality |
| S8_vs_k141_128173\|\|full | Low-quality |
| S8_vs_k141_24301\|\|full | Low-quality |
| S8_vs_k141_101923\|\|full | Low-quality |
| S8_vs_k141_128179\|\|full | Low-quality |
| S8_vs_k141_67169\|\|full | Low-quality |
| S8_vs_k141_59234\|\|full | Not-determined |
| S8_vs_k141_32889\|\|full | Not-determined |
| S8_vs_k141_59235\|\|full | Not-determined |
| S8_vs_k141_171701\|\|full | Low-quality |
| S8_vs_k141_180392\|\|full | Low-quality |
| S8_vs_k141_128199\|\|full | Low-quality |
| S8_vs_k141_84645\|\|full | Low-quality |
| S8_vs_k141_24317\|\|full | Not-determined |
| S8_vs_k141_41320\|\|full | Low-quality |
| S8_vs_k141_145654\|\|full | Not-determined |
| S12_vs_k141_188832\|\|full | High-quality |
| S8_vs_k141_67198\|\|full | Low-quality |
| S8_vs_k141_84657\|\|full | Not-determined |
| S8_vs_k141_187927\|\|full | Low-quality |
| S8_vs_k141_6824\|\|full | Low-quality |
| S8_vs_k141_24325\|\|full | Low-quality |
| S8_vs_k141_75902\|\|full | Not-determined |
| S8_vs_k141_136567\|\|full | Low-quality |
| S8_vs_k141_110920\|\|full | Not-determined |
| S8_vs_k141_15121\|\|full | Not-determined |
| S8_vs_k141_110927\|\|full | Low-quality |
| S8_vs_k141_15123\|\|full | Not-determined |
| S8_vs_k141_110932\|\|full | Low-quality |
| S8_vs_k141_49480\|\|full | Low-quality |
| S8_vs_k141_49483\|\|full | Low-quality |
| S8_vs_k141_197851\|\|full | Not-determined |
| S8_vs_k141_187946\|\|full | Low-quality |
| S8_vs_k141_49484\|\|full | Not-determined |
| S8_vs_k141_49487\|\|full | Not-determined |
| S8_vs_k141_197865\|\|full | Not-determined |
| S8_vs_k141_180432\|\|full | Not-determined |
| S8_vs_k141_41375\|\|full | Not-determined |
| S8_vs_k141_136601\|\|full | Not-determined |
| S8_vs_k141_197867\|\|full | Low-quality |
| S8_vs_k141_128232\|\|full | Low-quality |
| S8_vs_k141_59297\|\|full | Low-quality |
| S8_vs_k141_180437\|\|full | Not-determined |
| S8_vs_k141_171761\|\|full | Low-quality |
| S8_vs_k141_162516\|\|full | Not-determined |
| S8_vs_k141_128239\|\|full | Not-determined |
| S8_vs_k141_145673\|\|full | Not-determined |
| S8_vs_k141_49508\|\|full | Low-quality |
| S8_vs_k141_205835\|\|full | Low-quality |
| S8_vs_k141_110963\|\|full | Not-determined |
| S8_vs_k141_128244\|\|full | Low-quality |
| S8_vs_k141_145686\|\|full | Low-quality |
| S8_vs_k141_59320\|\|full | Not-determined |
| S8_vs_k141_128255\|\|full | Low-quality |
| S8_vs_k141_24351\|\|full | Not-determined |
| S8_vs_k141_6887\|\|full | Not-determined |
| S8_vs_k141_197901\|\|full | Low-quality |
| S8_vs_k141_24360\|\|full | Low-quality |
| S8_vs_k141_171793\|\|full | Low-quality |
| S8_vs_k141_128265\|\|full | Low-quality |
| S8_vs_k141_162534\|\|full | Low-quality |
| S8_vs_k141_93424\|\|full | Not-determined |
| S8_vs_k141_180480\|\|full | Low-quality |
| S8_vs_k141_171806\|\|full | Low-quality |
| S8_vs_k141_171807\|\|full | Not-determined |
| S8_vs_k141_6897\|\|full | Not-determined |
| S8_vs_k141_6898\|\|full | Low-quality |
| S8_vs_k141_101994\|\|full | Low-quality |
| S8_vs_k141_6900\|\|full | Not-determined |
| S8_vs_k141_119558\|\|full | Low-quality |
| S8_vs_k141_136675\|\|full | Low-quality |
| S8_vs_k141_145741\|\|full | Not-determined |
| S8_vs_k141_32925\|\|full | Low-quality |
| S8_vs_k141_111021\|\|full | Low-quality |
| S8_vs_k141_162539\|\|full | Low-quality |
| S8_vs_k141_15214\|\|full | Not-determined |
| S8_vs_k141_171823\|\|full | Not-determined |
| S8_vs_k141_145753\|\|full | Low-quality |
| S8_vs_k141_49574\|\|full | Not-determined |
| S8_vs_k141_93448\|\|full | Low-quality |
| S8_vs_k141_32934\|\|full | Not-determined |
| S8_vs_k141_67230\|\|full | Low-quality |
| S8_vs_k141_15218\|\|full | Not-determined |
| S8_vs_k141_49575\|\|full | Low-quality |
| S8_vs_k141_111037\|\|full | Low-quality |
| S8_vs_k141_59391\|\|full | Not-determined |
| S8_vs_k141_128291\|\|full | Low-quality |
| S11_vs_k141_60245\|\|full | Medium-quality |
| S8_vs_k141_205905\|\|full | Low-quality |
| S8_vs_k141_84792\|\|full | Not-determined |
| S8_vs_k141_145772\|\|full | Not-determined |
| S8_vs_k141_49577\|\|full | Low-quality |
| S8_vs_k141_197925\|\|full | Not-determined |
| S8_vs_k141_128297\|\|full | Not-determined |
| S8_vs_k141_162564\|\|full | Not-determined |
| S8_vs_k141_24435\|\|full | Low-quality |
| S8_vs_k141_6943\|\|full | Low-quality |
| S8_vs_k141_67254\|\|full | Low-quality |
| S8_vs_k141_32955\|\|full | Not-determined |
| S8_vs_k141_32958\|\|full | Low-quality |
| S8_vs_k141_76038\|\|full | Low-quality |
| S8_vs_k141_84809\|\|full | Low-quality |
| S8_vs_k141_32959\|\|full | Not-determined |
| S8_vs_k141_41406\|\|full | Not-determined |
| S8_vs_k141_205927\|\|full | Low-quality |
| S8_vs_k141_59433\|\|full | Low-quality |
| S8_vs_k141_197950\|\|full | Low-quality |
| S8_vs_k141_205935\|\|full | Not-determined |
| S8_vs_k141_93490\|\|full | Not-determined |
| S8_vs_k141_128318\|\|full | Low-quality |
| S8_vs_k141_41416\|\|full | Low-quality |
| S8_vs_k141_59447\|\|full | Low-quality |
| S8_vs_k141_197959\|\|full | Low-quality |
| S8_vs_k141_76054\|\|full | Not-determined |
| S8_vs_k141_162605\|\|full | Low-quality |
| S8_vs_k141_67288\|\|full | Not-determined |
| S8_vs_k141_136709\|\|full | Low-quality |
| S8_vs_k141_162609\|\|full | Low-quality |
| S8_vs_k141_136727\|\|full | Low-quality |
| S8_vs_k141_111094\|\|full | Low-quality |
| S8_vs_k141_128343\|\|full | Not-determined |
| S8_vs_k141_67301\|\|full | Not-determined |
| S8_vs_k141_145802\|\|full | Low-quality |
| S8_vs_k141_145803\|\|full | Low-quality |
| S8_vs_k141_93520\|\|full | Not-determined |
| S8_vs_k141_93523\|\|full | Not-determined |
| S8_vs_k141_171887\|\|full | Low-quality |
| S8_vs_k141_93530\|\|full | Low-quality |
| S8_vs_k141_145824\|\|full | Low-quality |
| S8_vs_k141_205967\|\|full | Not-determined |
| S8_vs_k141_205969\|\|full | Low-quality |
| S8_vs_k141_93536\|\|full | Not-determined |
| S8_vs_k141_49651\|\|full | Not-determined |
| S8_vs_k141_145838\|\|full | Not-determined |
| S8_vs_k141_33002\|\|full | Low-quality |
| S8_vs_k141_67318\|\|full | Low-quality |
| S8_vs_k141_111111\|\|full | Low-quality |
| S8_vs_k141_15240\|\|full | Low-quality |
| S8_vs_k141_15244\|\|full | Low-quality |
| S8_vs_k141_59497\|\|full | Low-quality |
| S8_vs_k141_67328\|\|full | Not-determined |
| S8_vs_k141_205993\|\|full | Not-determined |
| S8_vs_k141_33025\|\|full | Low-quality |
| S8_vs_k141_136782\|\|full | Low-quality |
| S8_vs_k141_198038\|\|full | Low-quality |
| S8_vs_k141_171934\|\|full | Not-determined |
| S8_vs_k141_145874\|\|full | Low-quality |
| S8_vs_k141_198053\|\|full | Low-quality |
| S8_vs_k141_59509\|\|full | Not-determined |
| S8_vs_k141_119622\|\|full | Not-determined |
| S8_vs_k141_180595\|\|full | Low-quality |
| S8_vs_k141_119627\|\|full | Low-quality |
| S8_vs_k141_111114\|\|full | Low-quality |
| S8_vs_k141_111116\|\|full | Not-determined |
| S8_vs_k141_76152\|\|full | Low-quality |
| S8_vs_k141_171951\|\|full | Not-determined |
| S8_vs_k141_15271\|\|full | Low-quality |
| S8_vs_k141_162691\|\|full | Not-determined |
| S8_vs_k141_24481\|\|full | Low-quality |
| S8_vs_k141_102137\|\|full | Not-determined |
| S8_vs_k141_188167\|\|full | Not-determined |
| S8_vs_k141_136842\|\|full | Low-quality |
| S8_vs_k141_111140\|\|full | Low-quality |
| S8_vs_k141_188174\|\|full | Not-determined |
| S8_vs_k141_171979\|\|full | Not-determined |
| S8_vs_k141_41485\|\|full | Low-quality |
| S8_vs_k141_59536\|\|full | Low-quality |
| S8_vs_k141_84852\|\|full | Low-quality |
| S8_vs_k141_145905\|\|full | Low-quality |
| S8_vs_k141_188175\|\|full | Low-quality |
| S8_vs_k141_49734\|\|full | Not-determined |
| S8_vs_k141_119660\|\|full | Not-determined |
| S8_vs_k141_136862\|\|full | Low-quality |
| S8_vs_k141_24511\|\|full | Low-quality |
| S8_vs_k141_15306\|\|full | Not-determined |
| S8_vs_k141_119663\|\|full | Low-quality |
| S8_vs_k141_93589\|\|full | Not-determined |
| S8_vs_k141_41513\|\|full | Not-determined |
| S8_vs_k141_6991\|\|full | Not-determined |
| S8_vs_k141_76203\|\|full | Low-quality |
| S8_vs_k141_6994\|\|full | Low-quality |
| S8_vs_k141_172021\|\|full | Low-quality |
| S8_vs_k141_102191\|\|full | Not-determined |
| S8_vs_k141_145941\|\|full | Not-determined |
| S8_vs_k141_162753\|\|full | Not-determined |
| S8_vs_k141_41531\|\|full | Low-quality |
| S8_vs_k141_49764\|\|full | Not-determined |
| S8_vs_k141_15351\|\|full | Low-quality |
| S8_vs_k141_111217\|\|full | Not-determined |
| S8_vs_k141_128388\|\|full | Not-determined |
| S8_vs_k141_41551\|\|full | Low-quality |
| S8_vs_k141_84928\|\|full | Low-quality |
| S8_vs_k141_145962\|\|full | Not-determined |
| S8_vs_k141_188234\|\|full | Low-quality |
| S8_vs_k141_188235\|\|full | Low-quality |
| S8_vs_k141_41555\|\|full | Not-determined |
| S8_vs_k141_172050\|\|full | Low-quality |
| S8_vs_k141_102235\|\|full | Low-quality |
| S8_vs_k141_7030\|\|full | Low-quality |
| S8_vs_k141_15382\|\|full | Low-quality |
| S8_vs_k141_162794\|\|full | Low-quality |
| S8_vs_k141_128411\|\|full | Not-determined |
| S8_vs_k141_102241\|\|full | Low-quality |
| S8_vs_k141_93667\|\|full | Low-quality |
| S8_vs_k141_76221\|\|full | Not-determined |
| S8_vs_k141_162801\|\|full | Not-determined |
| S8_vs_k141_145988\|\|full | Low-quality |
| S8_vs_k141_67381\|\|full | Not-determined |
| S8_vs_k141_49793\|\|full | Low-quality |
| S8_vs_k141_102254\|\|full | Low-quality |
| S8_vs_k141_93671\|\|full | Not-determined |
| S8_vs_k141_198105\|\|full | Not-determined |
| S8_vs_k141_67392\|\|full | Low-quality |
| S8_vs_k141_188273\|\|full | Low-quality |
| S8_vs_k141_206087\|\|full | Low-quality |
| S8_vs_k141_198115\|\|full | Not-determined |
| S8_vs_k141_7053\|\|full | Not-determined |
| S8_vs_k141_146015\|\|full | Low-quality |
| S8_vs_k141_24580\|\|full | Not-determined |
| S8_vs_k141_49803\|\|full | Not-determined |
| S8_vs_k141_41588\|\|full | Not-determined |
| S8_vs_k141_188276\|\|full | Low-quality |
| S8_vs_k141_33134\|\|full | Low-quality |
| S8_vs_k141_188277\|\|full | Not-determined |
| S8_vs_k141_102285\|\|full | Not-determined |
| S8_vs_k141_146016\|\|full | Low-quality |
| S8_vs_k141_180734\|\|full | Not-determined |
| S8_vs_k141_206099\|\|full | Not-determined |
| S8_vs_k141_188284\|\|full | Not-determined |
| S8_vs_k141_24602\|\|full | Not-determined |
| S8_vs_k141_84996\|\|full | Low-quality |
| S8_vs_k141_24603\|\|full | Low-quality |
| S8_vs_k141_180752\|\|full | Low-quality |
| S3_vs_k141_101591\|\|full | Medium-quality |
| S8_vs_k141_162862\|\|full | Not-determined |
| S8_vs_k141_85003\|\|full | Low-quality |
| S8_vs_k141_59562\|\|full | Low-quality |
| S8_vs_k141_206117\|\|full | Not-determined |
| S8_vs_k141_162866\|\|full | Low-quality |
| S8_vs_k141_67451\|\|full | Not-determined |
| S8_vs_k141_128436\|\|full | Not-determined |
| S8_vs_k141_76277\|\|full | Low-quality |
| S8_vs_k141_111330\|\|full | Low-quality |
| S8_vs_k141_7083\|\|full | Low-quality |
| S8_vs_k141_111337\|\|full | Low-quality |
| S8_vs_k141_188309\|\|full | Not-determined |
| S13_vs_90967\|\|full | Medium-quality |
| S8_vs_k141_162884\|\|full | Low-quality |
| S8_vs_k141_49838\|\|full | Low-quality |
| S8_vs_k141_206121\|\|full | Low-quality |
| S8_vs_k141_146044\|\|full | Low-quality |
| S8_vs_k141_15435\|\|full | Low-quality |
| S8_vs_k141_7097\|\|full | Low-quality |
| S8_vs_k141_128462\|\|full | Not-determined |
| S8_vs_k141_198163\|\|full | Not-determined |
| S8_vs_k141_136935\|\|full | Low-quality |
| S8_vs_k141_67469\|\|full | Low-quality |
| S8_vs_k141_85031\|\|full | Low-quality |
| S8_vs_k141_24647\|\|full | Not-determined |
| S8_vs_k141_7113\|\|full | Low-quality |
| S8_vs_k141_162903\|\|full | Not-determined |
| S8_vs_k141_76299\|\|full | Low-quality |
| S8_vs_k141_172133\|\|full | Not-determined |
| S8_vs_k141_7116\|\|full | Not-determined |
| S8_vs_k141_188326\|\|full | Low-quality |
| S8_vs_k141_206146\|\|full | Not-determined |
| S8_vs_k141_172148\|\|full | Not-determined |
| S8_vs_k141_85045\|\|full | Low-quality |
| S8_vs_k141_198181\|\|full | Low-quality |
| S8_vs_k141_59604\|\|full | Not-determined |
| S8_vs_k141_206149\|\|full | Not-determined |
| S8_vs_k141_59610\|\|full | Not-determined |
| S8_vs_k141_128493\|\|full | Low-quality |
| S8_vs_k141_76322\|\|full | Not-determined |
| S8_vs_k141_162933\|\|full | Not-determined |
| S8_vs_k141_162934\|\|full | Low-quality |
| S8_vs_k141_111369\|\|full | Not-determined |
| S8_vs_k141_119786\|\|full | Low-quality |
| S8_vs_k141_111370\|\|full | Low-quality |
| S8_vs_k141_206155\|\|full | Not-determined |
| S8_vs_k141_7135\|\|full | Not-determined |
| S8_vs_k141_15471\|\|full | Not-determined |
| S8_vs_k141_85073\|\|full | Low-quality |
| S8_vs_k141_119795\|\|full | Low-quality |
| S8_vs_k141_136981\|\|full | Low-quality |
| S8_vs_k141_180823\|\|full | Low-quality |
| S8_vs_k141_85083\|\|full | Low-quality |
| S8_vs_k141_102364\|\|full | Not-determined |
| S8_vs_k141_172171\|\|full | Not-determined |
| S8_vs_k141_15480\|\|full | Low-quality |
| S8_vs_k141_7144\|\|full | Low-quality |
| S8_vs_k141_41715\|\|full | Low-quality |
| S8_vs_k141_7147\|\|full | Not-determined |
| S8_vs_k141_154252\|\|full | Not-determined |
| S8_vs_k141_24695\|\|full | Not-determined |
| S8_vs_k141_154264\|\|full | Low-quality |
| S8_vs_k141_111418\|\|full | Low-quality |
| S8_vs_k141_137023\|\|full | Low-quality |
| S8_vs_k141_198261\|\|full | Low-quality |
| S8_vs_k141_137028\|\|full | Not-determined |
| S8_vs_k141_119818\|\|full | Low-quality |
| S8_vs_k141_188400\|\|full | Low-quality |
| S8_vs_k141_146117\|\|full | Not-determined |
| S8_vs_k141_128556\|\|full | Low-quality |
| S8_vs_k141_49890\|\|full | Low-quality |
| S8_vs_k141_119820\|\|full | Low-quality |
| S8_vs_k141_67498\|\|full | Low-quality |
| S8_vs_k141_137048\|\|full | Low-quality |
| S8_vs_k141_180875\|\|full | Low-quality |
| S8_vs_k141_180878\|\|full | Low-quality |
| S8_vs_k141_59642\|\|full | Low-quality |
| S8_vs_k141_33282\|\|full | Not-determined |
| S8_vs_k141_146133\|\|full | Low-quality |

| | |
|---|---|
| S8_vs_k141_59649\|\|full | Low-quality |
| S8_vs_k141_154279\|\|full | Low-quality |
| S8_vs_k141_49919\|\|full | Low-quality |
| S8_vs_k141_163002\|\|full | Low-quality |
| S8_vs_k141_102411\|\|full | Low-quality |
| S8_vs_k141_93828\|\|full | Not-determined |
| S8_vs_k141_41767\|\|full | Low-quality |
| S8_vs_k141_85147\|\|full | Low-quality |
| S8_vs_k141_41771\|\|full | Low-quality |
| S8_vs_k141_172194\|\|full | Not-determined |
| S8_vs_k141_102421\|\|full | Low-quality |
| S8_vs_k141_24761\|\|full | Low-quality |
| S8_vs_k141_206229\|\|full | Low-quality |
| S8_vs_k141_163020\|\|full | Not-determined |
| S8_vs_k141_67533\|\|full | Low-quality |
| S8_vs_k141_172207\|\|full | Low-quality |
| S8_vs_k141_119867\|\|full | Low-quality |
| S8_vs_k141_41785\|\|full | Low-quality |
| S8_vs_k141_33306\|\|full | Low-quality |
| S8_vs_k141_85167\|\|full | Low-quality |
| S8_vs_k141_15570\|\|full | Not-determined |
| S8_vs_k141_59677\|\|full | Not-determined |
| S8_vs_k141_180910\|\|full | Low-quality |
| S8_vs_k141_111490\|\|full | Low-quality |
| S8_vs_k141_41800\|\|full | Not-determined |
| S8_vs_k141_137076\|\|full | Low-quality |
| S8_vs_k141_111510\|\|full | Low-quality |
| S8_vs_k141_172247\|\|full | Low-quality |
| S8_vs_k141_188476\|\|full | Low-quality |
| S8_vs_k141_59700\|\|full | Not-determined |
| S8_vs_k141_137089\|\|full | Not-determined |
| S8_vs_k141_128597\|\|full | Low-quality |
| S8_vs_k141_111511\|\|full | Low-quality |
| S8_vs_k141_49967\|\|full | Low-quality |
| S8_vs_k141_24812\|\|full | Low-quality |
| S8_vs_k141_102438\|\|full | Not-determined |
| S8_vs_k141_180928\|\|full | Low-quality |
| S8_vs_k141_33361\|\|full | Low-quality |
| S8_vs_k141_24817\|\|full | Not-determined |
| S8_vs_k141_188486\|\|full | Low-quality |
| S3_vs_k141_51931\|\|full | Medium-quality |
| S8_vs_k141_67567\|\|full | Not-determined |
| S8_vs_k141_7244\|\|full | Low-quality |
| S8_vs_k141_198373\|\|full | Low-quality |
| S8_vs_k141_41862\|\|full | Low-quality |
| S8_vs_k141_111544\|\|full | Low-quality |
| S8_vs_k141_180957\|\|full | Not-determined |
| S8_vs_k141_67578\|\|full | Not-determined |
| S8_vs_k141_33387\|\|full | Not-determined |
| S8_vs_k141_128629\|\|full | Not-determined |
| S8_vs_k141_172317\|\|full | Low-quality |
| S8_vs_k141_163120\|\|full | Low-quality |
| S8_vs_k141_128635\|\|full | Low-quality |
| S8_vs_k141_206317\|\|full | Not-determined |
| S8_vs_k141_67588\|\|full | Low-quality |
| S8_vs_k141_206327\|\|full | Low-quality |
| S8_vs_k141_137145\|\|full | Low-quality |
| S8_vs_k141_206332\|\|full | Not-determined |
| S8_vs_k141_146253\|\|full | Low-quality |
| S8_vs_k141_76399\|\|full | Not-determined |
| S8_vs_k141_41885\|\|full | Low-quality |
| S8_vs_k141_154322\|\|full | Not-determined |
| S8_vs_k141_85248\|\|full | Not-determined |
| S8_vs_k141_85250\|\|full | Low-quality |
| S8_vs_k141_128654\|\|full | Not-determined |
| S8_vs_k141_76401\|\|full | Low-quality |
| S8_vs_k141_85254\|\|full | Low-quality |
| S8_vs_k141_198411\|\|full | Not-determined |
| S8_vs_k141_93968\|\|full | Not-determined |
| S8_vs_k141_76406\|\|full | Low-quality |
| S8_vs_k141_119978\|\|full | Low-quality |
| S8_vs_k141_7310\|\|full | Not-determined |
| S8_vs_k141_181010\|\|full | Not-determined |
| S8_vs_k141_15675\|\|full | Not-determined |
| S8_vs_k141_198416\|\|full | Low-quality |
| S8_vs_k141_41920\|\|full | Low-quality |
| S8_vs_k141_67670\|\|full | Low-quality |
| S8_vs_k141_7318\|\|full | Not-determined |
| S8_vs_k141_50077\|\|full | Not-determined |
| S8_vs_k141_33451\|\|full | Low-quality |
| S8_vs_k141_137205\|\|full | Low-quality |
| S8_vs_k141_33454\|\|full | Low-quality |
| S8_vs_k141_15688\|\|full | Low-quality |
| S8_vs_k141_15696\|\|full | Low-quality |
| S8_vs_k141_85286\|\|full | Not-determined |
| S8_vs_k141_172378\|\|full | Not-determined |
| S8_vs_k141_50089\|\|full | Low-quality |
| S1_vs_k141_3435\|\|full | Medium-quality |
| S8_vs_k141_128696\|\|full | Not-determined |
| S8_vs_k141_181049\|\|full | Low-quality |
| S8_vs_k141_154360\|\|full | Not-determined |
| S8_vs_k141_102517\|\|full | Low-quality |
| S8_vs_k141_120018\|\|full | Not-determined |
| S8_vs_k141_111634\|\|full | Low-quality |
| S8_vs_k141_120020\|\|full | Not-determined |
| S8_vs_k141_41954\|\|full | Not-determined |
| S8_vs_k141_33485\|\|full | Not-determined |
| S8_vs_k141_154381\|\|full | Low-quality |
| S8_vs_k141_172409\|\|full | Low-quality |
| S8_vs_k141_154388\|\|full | Not-determined |
| S8_vs_k141_41969\|\|full | Low-quality |
| S8_vs_k141_33497\|\|full | Low-quality |
| S8_vs_k141_128737\|\|full | Low-quality |
| S16_vs_173141\|\|full | Medium-quality |
| S8_vs_k141_188555\|\|full | Not-determined |
| S8_vs_k141_120041\|\|full | Low-quality |
| S8_vs_k141_206416\|\|full | Low-quality |
| S8_vs_k141_120043\|\|full | Low-quality |
| S8_vs_k141_33505\|\|full | Not-determined |
| S8_vs_k141_24837\|\|full | Not-determined |
| S8_vs_k141_120054\|\|full | Low-quality |
| S8_vs_k141_172437\|\|full | Not-determined |
| S8_vs_k141_76508\|\|full | Not-determined |
| S8_vs_k141_33514\|\|full | Not-determined |
| S8_vs_k141_206451\|\|full | Not-determined |
| S8_vs_k141_198506\|\|full | Not-determined |
| S8_vs_k141_33535\|\|full | Low-quality |
| S8_vs_k141_181085\|\|full | Not-determined |
| S8_vs_k141_42023\|\|full | Low-quality |
| S8_vs_k141_128795\|\|full | Not-determined |
| S8_vs_k141_50180\|\|full | Not-determined |
| S8_vs_k141_137281\|\|full | Low-quality |
| S8_vs_k141_94033\|\|full | Low-quality |
| S8_vs_k141_7374\|\|full | Low-quality |
| S8_vs_k141_137288\|\|full | Not-determined |
| S8_vs_k141_33554\|\|full | Not-determined |
| S8_vs_k141_94034\|\|full | Low-quality |
| S8_vs_k141_181108\|\|full | Not-determined |
| S8_vs_k141_137303\|\|full | Not-determined |
| S8_vs_k141_154471\|\|full | Not-determined |
| S8_vs_k141_137305\|\|full | Not-determined |
| S8_vs_k141_33559\|\|full | Not-determined |
| S8_vs_k141_42045\|\|full | Not-determined |
| S8_vs_k141_206496\|\|full | Not-determined |
| S8_vs_k141_67763\|\|full | Not-determined |
| S8_vs_k141_42047\|\|full | Not-determined |
| S8_vs_k141_198556\|\|full | Low-quality |
| S8_vs_k141_120094\|\|full | Not-determined |
| S8_vs_k141_111707\|\|full | Low-quality |
| S8_vs_k141_198563\|\|full | Low-quality |
| S8_vs_k141_188622\|\|full | Low-quality |
| S8_vs_k141_50223\|\|full | Low-quality |
| S8_vs_k141_50225\|\|full | Not-determined |
| S8_vs_k141_206508\|\|full | Not-determined |
| S8_vs_k141_163225\|\|full | Low-quality |
| S8_vs_k141_163226\|\|full | Not-determined |
| S8_vs_k141_113059\|\|full | Medium-quality |
| S8_vs_k141_154512\|\|full | Low-quality |
| S8_vs_k141_128833\|\|full | Not-determined |
| S8_vs_k141_154522\|\|full | Low-quality |
| S8_vs_k141_24859\|\|full | Low-quality |
| S8_vs_k141_33591\|\|full | Not-determined |
| S8_vs_k141_172507\|\|full | Not-determined |
| S8_vs_k141_154530\|\|full | Not-determined |
| S8_vs_k141_7411\|\|full | Low-quality |
| S8_vs_k141_76611\|\|full | Low-quality |
| S8_vs_k141_146282\|\|full | Not-determined |
| S8_vs_k141_111761\|\|full | Not-determined |
| S8_vs_k141_76631\|\|full | Not-determined |
| S8_vs_k141_128860\|\|full | Not-determined |
| S8_vs_k141_198598\|\|full | Low-quality |
| S8_vs_k141_172530\|\|full | Not-determined |
| S8_vs_k141_85350\|\|full | Not-determined |
| S8_vs_k141_146296\|\|full | Low-quality |
| S8_vs_k141_120135\|\|full | Low-quality |
| S8_vs_k141_67820\|\|full | Low-quality |
| S8_vs_k141_198617\|\|full | Not-determined |
| S8_vs_k141_154554\|\|full | Not-determined |
| S8_vs_k141_7447\|\|full | Not-determined |
| S8_vs_k141_67828\|\|full | Not-determined |
| S8_vs_k141_33646\|\|full | Low-quality |
| S8_vs_k141_111784\|\|full | Not-determined |
| S8_vs_k141_24897\|\|full | Low-quality |
| S8_vs_k141_154561\|\|full | Low-quality |
| S8_vs_k141_181197\|\|full | Low-quality |
| S8_vs_k141_137394\|\|full | Not-determined |
| S8_vs_k141_50273\|\|full | Low-quality |
| S8_vs_k141_128872\|\|full | Low-quality |
| S8_vs_k141_188693\|\|full | Not-determined |
| S8_vs_k141_128886\|\|full | Low-quality |
| S8_vs_k141_42077\|\|full | Not-determined |
| S8_vs_k141_181221\|\|full | Not-determined |
| S8_vs_k141_163292\|\|full | Not-determined |
| S8_vs_k141_198643\|\|full | Not-determined |
| S8_vs_k141_33666\|\|full | Not-determined |
| S8_vs_k141_128894\|\|full | Low-quality |
| S8_vs_k141_24927\|\|full | Not-determined |
| S16_vs_120605\|\|full | Medium-quality |
| S8_vs_k141_146339\|\|full | Not-determined |
| S8_vs_k141_67849\|\|full | Low-quality |
| S8_vs_k141_7488\|\|full | Low-quality |
| S8_vs_k141_206528\|\|full | Low-quality |
| S8_vs_k141_85406\|\|full | Low-quality |
| S8_vs_k141_181231\|\|full | Low-quality |
| S8_vs_k141_42096\|\|full | Low-quality |
| S8_vs_k141_94148\|\|full | Low-quality |
| S8_vs_k141_120163\|\|full | Low-quality |
| S8_vs_k141_181240\|\|full | Not-determined |
| S8_extracted_k141_94361 | Medium-quality |
| S8_vs_k141_120169\|\|full | Not-determined |
| S8_vs_k141_128910\|\|full | Low-quality |
| S8_vs_k141_154614\|\|full | Not-determined |
| S8_vs_k141_137443\|\|full | Low-quality |
| S8_vs_k141_128915\|\|full | Not-determined |
| S8_vs_k141_15754\|\|full | Low-quality |
| S8_vs_k141_94168\|\|full | Low-quality |
| S8_vs_k141_137451\|\|full | Low-quality |
| S8_vs_k141_7527\|\|full | Low-quality |
| S8_vs_k141_76694\|\|full | Low-quality |
| S8_vs_k141_85448\|\|full | Low-quality |
| S8_vs_k141_146388\|\|full | Not-determined |
| S8_vs_k141_111805\|\|full | Not-determined |
| S8_vs_k141_146390\|\|full | Not-determined |
| S8_vs_k141_154629\|\|full | Not-determined |
| S8_vs_k141_42131\|\|full | Low-quality |
| S8_vs_k141_172596\|\|full | Low-quality |
| S8_vs_k141_33733\|\|full | Low-quality |
| S8_vs_k141_163352\|\|full | Not-determined |
| S8_vs_k141_94189\|\|full | Low-quality |
| S8_vs_k141_76722\|\|full | Not-determined |
| S8_vs_k141_206568\|\|full | Not-determined |
| S8_vs_k141_120214\|\|full | Not-determined |
| S8_vs_k141_85455\|\|full | Not-determined |
| S8_vs_k141_42139\|\|full | Not-determined |
| S8_vs_k141_137477\|\|full | Low-quality |
| S8_vs_k141_128987\|\|full | Low-quality |
| S8_vs_k141_59810\|\|full | Not-determined |
| S8_vs_k141_15778\|\|full | Not-determined |
| S8_vs_k141_24993\|\|full | Low-quality |
| S8_vs_k141_24995\|\|full | Not-determined |
| S8_vs_k141_50368\|\|full | Low-quality |
| S8_vs_k141_24999\|\|full | Not-determined |
| S8_vs_k141_25002\|\|full | Low-quality |
| S8_vs_k141_198740\|\|full | Low-quality |
| S8_vs_k141_206579\|\|full | Low-quality |
| S8_vs_k141_206580\|\|full | Not-determined |
| S8_vs_k141_50376\|\|full | Not-determined |
| S8_vs_k141_206587\|\|full | Low-quality |
| S8_vs_k141_120257\|\|full | Low-quality |
| S8_vs_k141_120259\|\|full | Low-quality |
| S8_vs_k141_120265\|\|full | Low-quality |
| S8_vs_k141_129053\|\|full | Not-determined |
| S8_vs_k141_188733\|\|full | Not-determined |
| S8_vs_k141_42205\|\|full | Low-quality |
| S8_vs_k141_181300\|\|full | Not-determined |
| S8_vs_k141_172683\|\|full | Not-determined |
| S8_vs_k141_172685\|\|full | Not-determined |
| S8_vs_k141_76836\|\|full | Not-determined |
| S8_vs_k141_94249\|\|full | Not-determined |
| S8_vs_k141_7640\|\|full | Not-determined |
| S8_vs_k141_25080\|\|full | Low-quality |
| S8_vs_k141_85539\|\|full | Not-determined |
| S8_vs_k141_15835\|\|full | Low-quality |
| S8_vs_k141_137495\|\|full | Low-quality |
| S8_vs_k141_25084\|\|full | Low-quality |
| S8_vs_k141_33845\|\|full | Not-determined |
| S8_vs_k141_188764\|\|full | Low-quality |
| S8_vs_k141_206651\|\|full | Not-determined |
| S8_vs_k141_25092\|\|full | Not-determined |
| S8_vs_k141_137504\|\|full | Not-determined |
| S8_vs_k141_172715\|\|full | Low-quality |
| S8_vs_k141_15843\|\|full | Not-determined |
| S8_vs_k141_172720\|\|full | Low-quality |
| S8_vs_k141_15847\|\|full | Not-determined |
| S8_vs_k141_120372\|\|full | Not-determined |
| S8_vs_k141_94361\|\|full | Medium-quality |
| S8_vs_k141_76858\|\|full | Not-determined |
| S8_vs_k141_33869\|\|full | Not-determined |
| S8_vs_k141_76867\|\|full | Low-quality |
| S8_vs_k141_102757\|\|full | Not-determined |
| S8_vs_k141_172746\|\|full | Low-quality |
| S8_vs_k141_15873\|\|full | Not-determined |
| S8_vs_k141_76872\|\|full | Not-determined |
| S8_vs_k141_102763\|\|full | Not-determined |
| S3_extracted_k141_127426 | Medium-quality |
| S8_vs_k141_146524\|\|full | Low-quality |
| S8_vs_k141_94294\|\|full | Low-quality |
| S8_vs_k141_198861\|\|full | Low-quality |
| S8_vs_k141_15874\|\|full | Not-determined |
| S8_vs_k141_50440\|\|full | Not-determined |
| S8_vs_k141_181366\|\|full | Low-quality |
| S8_vs_k141_76884\|\|full | Low-quality |
| S8_vs_k141_33879\|\|full | Low-quality |
| S8_vs_k141_68025\|\|full | Low-quality |
| S8_vs_k141_111854\|\|full | Low-quality |
| S8_vs_k141_76894\|\|full | Low-quality |
| S8_vs_k141_111855\|\|full | Low-quality |
| S8_vs_k141_129134\|\|full | Not-determined |
| S8_vs_k141_163441\|\|full | Low-quality |
| S8_vs_k141_146534\|\|full | Low-quality |
| S8_vs_k141_85580\|\|full | Low-quality |
| S8_vs_k141_15914\|\|full | Not-determined |
| S8_vs_k141_50467\|\|full | Not-determined |
| S8_vs_k141_206718\|\|full | Not-determined |
| S8_vs_k141_137549\|\|full | Not-determined |
| S8_vs_k141_33907\|\|full | Low-quality |
| S8_vs_k141_33910\|\|full | Not-determined |
| S8_vs_k141_15925\|\|full | Low-quality |
| S8_vs_k141_102805\|\|full | Low-quality |
| S8_vs_k141_137562\|\|full | Low-quality |
| S8_vs_k141_33915\|\|full | Low-quality |
| S8_vs_k141_111886\|\|full | Low-quality |
| S8_vs_k141_94331\|\|full | Not-determined |
| S8_vs_k141_188824\|\|full | Not-determined |
| S8_vs_k141_146565\|\|full | Not-determined |
| S8_vs_k141_188827\|\|full | Not-determined |
| S8_vs_k141_172806\|\|full | Low-quality |
| S8_vs_k141_206735\|\|full | Low-quality |
| S8_vs_k141_85617\|\|full | Not-determined |
| S8_vs_k141_137581\|\|full | Low-quality |
| S8_vs_k141_7711\|\|full | Low-quality |
| S8_vs_k141_42278\|\|full | Not-determined |
| S8_vs_k141_163459\|\|full | Low-quality |
| S8_vs_k141_181408\|\|full | Not-determined |
| S8_vs_k141_129174\|\|full | Not-determined |
| S8_vs_k141_102841\|\|full | Not-determined |
| S8_vs_k141_163460\|\|full | Low-quality |
| S8_vs_k141_172825\|\|full | Low-quality |
| S8_vs_k141_198909\|\|full | Not-determined |
| S8_vs_k141_188839\|\|full | Not-determined |
| S8_vs_k141_102849\|\|full | Low-quality |
| S8_vs_k141_146601\|\|full | Low-quality |
| S8_vs_k141_181415\|\|full | Not-determined |
| S8_vs_k141_198916\|\|full | Low-quality |
| S8_vs_k141_60016\|\|full | Low-quality |
| S8_vs_k141_188848\|\|full | Low-quality |
| S8_vs_k141_181424\|\|full | Low-quality |
| S8_vs_k141_198918\|\|full | Not-determined |
| S8_vs_k141_111934\|\|full | Low-quality |
| S8_vs_k141_137609\|\|full | Low-quality |
| S8_vs_k141_102865\|\|full | Low-quality |
| S8_vs_k141_33975\|\|full | Not-determined |
| S8_vs_k141_137612\|\|full | Low-quality |
| S3_vs_k141_127426\|\|full | Medium-quality |
| S8_vs_k141_129219\|\|full | Low-quality |
| S8_vs_k141_137626\|\|full | Low-quality |
| S8_vs_k141_76988\|\|full | Low-quality |
| S8_vs_k141_94371\|\|full | Low-quality |
| S8_vs_k141_206786\|\|full | Not-determined |
| S8_vs_k141_25143\|\|full | Not-determined |
| S8_vs_k141_137632\|\|full | Low-quality |
| S8_vs_k141_102882\|\|full | Not-determined |
| S8_vs_k141_33998\|\|full | Low-quality |
| S8_vs_k141_129232\|\|full | Low-quality |
| S8_vs_k141_42327\|\|full | Low-quality |
| S8_vs_k141_15954\|\|full | Low-quality |
| S8_vs_k141_206798\|\|full | Low-quality |
| S8_vs_k141_25151\|\|full | Not-determined |
| S8_vs_k141_60058\|\|full | Low-quality |
| S8_vs_k141_102898\|\|full | Not-determined |
| S8_vs_k141_206805\|\|full | Low-quality |
| S8_vs_k141_137655\|\|full | Low-quality |
| S8_vs_k141_198961\|\|full | Low-quality |
| S8_vs_k141_25157\|\|full | Low-quality |
| S8_vs_k141_137656\|\|full | Low-quality |
| S8_vs_k141_198966\|\|full | Low-quality |
| S8_vs_k141_15971\|\|full | Not-determined |
| S8_vs_k141_60079\|\|full | Low-quality |
| S8_vs_k141_137667\|\|full | Not-determined |
| S8_vs_k141_111980\|\|full | Low-quality |
| S8_vs_k141_85715\|\|full | Not-determined |
| S8_vs_k141_25173\|\|full | Not-determined |
| S8_vs_k141_34032\|\|full | Low-quality |
| S8_vs_k141_77038\|\|full | Low-quality |
| S8_vs_k141_181497\|\|full | Low-quality |
| S8_vs_k141_94431\|\|full | Not-determined |
| S8_vs_k141_60094\|\|full | Low-quality |
| S8_vs_k141_102931\|\|full | Low-quality |
| S8_vs_k141_34042\|\|full | Not-determined |
| S8_vs_k141_154853\|\|full | Low-quality |
| S8_vs_k141_163506\|\|full | Low-quality |
| S8_vs_k141_77053\|\|full | Low-quality |
| S8_vs_k141_60101\|\|full | Low-quality |
| S8_vs_k141_163513\|\|full | Low-quality |
| S8_vs_k141_206835\|\|full | Not-determined |
| S8_vs_k141_77064\|\|full | Not-determined |
| S8_vs_k141_68160\|\|full | Not-determined |
| S8_vs_k141_163518\|\|full | Low-quality |
| S8_vs_k141_137703\|\|full | Not-determined |
| S8_vs_k141_206844\|\|full | Not-determined |
| S8_vs_k141_206845\|\|full | Low-quality |
| S8_vs_k141_163522\|\|full | Low-quality |
| S8_vs_k141_7837\|\|full | Not-determined |
| S8_vs_k141_163528\|\|full | Low-quality |
| S8_vs_k141_199022\|\|full | Not-determined |
| S8_vs_k141_102957\|\|full | Low-quality |
| S8_vs_k141_77083\|\|full | Not-determined |
| S8_vs_k141_120471\|\|full | Low-quality |
| S8_vs_k141_42383\|\|full | Low-quality |
| S8_vs_k141_16012\|\|full | Low-quality |
| S8_vs_k141_199039\|\|full | Not-determined |
| S8_vs_k141_7870\|\|full | Not-determined |
| S8_vs_k141_7871\|\|full | Not-determined |
| S8_vs_k141_16021\|\|full | Low-quality |
| S8_vs_k141_137751\|\|full | Not-determined |
| S8_vs_k141_94479\|\|full | Low-quality |
| S8_vs_k141_25231\|\|full | Low-quality |
| S8_vs_k141_85758\|\|full | Not-determined |
| S8_vs_k141_154898\|\|full | Not-determined |
| S8_vs_k141_206878\|\|full | Not-determined |
| S8_vs_k141_206885\|\|full | Low-quality |
| S8_vs_k141_154920\|\|full | Not-determined |
| S8_vs_k141_103010\|\|full | Low-quality |
| S8_vs_k141_146740\|\|full | Not-determined |
| S8_vs_k141_112073\|\|full | Low-quality |
| S8_vs_k141_146743\|\|full | Low-quality |
| S8_vs_k141_42405\|\|full | Not-determined |
| S8_vs_k141_94519\|\|full | Low-quality |
| S8_vs_k141_68184\|\|full | Low-quality |
| S8_vs_k141_25259\|\|full | Not-determined |
| S8_vs_k141_34130\|\|full | Not-determined |
| S8_vs_k141_154938\|\|full | Low-quality |
| S8_vs_k141_137802\|\|full | Not-determined |
| S8_vs_k141_103027\|\|full | Not-determined |
| S8_vs_k141_7908\|\|full | Low-quality |
| S8_vs_k141_146761\|\|full | Low-quality |
| S8_vs_k141_206904\|\|full | Low-quality |
| S8_vs_k141_77117\|\|full | Not-determined |
| S8_vs_k141_85817\|\|full | Low-quality |
| S8_vs_k141_103035\|\|full | Low-quality |
| S8_vs_k141_137807\|\|full | Low-quality |
| S8_vs_k141_85827\|\|full | Low-quality |
| S8_vs_k141_60219\|\|full | Not-determined |
| S8_vs_k141_146774\|\|full | Not-determined |
| S8_vs_k141_120579\|\|full | Low-quality |
| S8_vs_k141_42424\|\|full | Low-quality |
| S8_vs_k141_154962\|\|full | Not-determined |
| S8_vs_k141_103052\|\|full | Not-determined |
| S8_vs_k141_120582\|\|full | Low-quality |
| S8_vs_k141_146783\|\|full | Not-determined |
| S8_vs_k141_129294\|\|full | Not-determined |
| S8_vs_k141_206927\|\|full | Not-determined |
| S8_vs_k141_112104\|\|full | Low-quality |
| S8_vs_k141_206930\|\|full | Low-quality |
| S8_vs_k141_68223\|\|full | Not-determined |
| S8_vs_k141_42436\|\|full | Not-determined |
| S8_vs_k141_7930\|\|full | Not-determined |
| S8_vs_k141_129301\|\|full | Low-quality |
| S8_vs_k141_68225\|\|full | Not-determined |
| S8_vs_k141_137822\|\|full | Low-quality |
| S8_vs_k141_206939\|\|full | Not-determined |
| S8_vs_k141_154979\|\|full | Low-quality |
| S8_vs_k141_154981\|\|full | Low-quality |
| S8_vs_k141_7937\|\|full | Not-determined |
| S8_vs_k141_112130\|\|full | Low-quality |
| S8_vs_k141_154984\|\|full | Not-determined |
| S8_vs_k141_206944\|\|full | Low-quality |
| S8_vs_k141_16140\|\|full | Low-quality |
| S8_vs_k141_199134\|\|full | Low-quality |
| S8_vs_k141_206945\|\|full | Not-determined |
| S8_vs_k141_85880\|\|full | Low-quality |
| S8_vs_k141_103084\|\|full | Low-quality |
| S8_vs_k141_189040\|\|full | Low-quality |
| S8_vs_k141_42466\|\|full | Low-quality |
| S8_vs_k141_7964\|\|full | Low-quality |
| S8_vs_k141_163595\|\|full | Not-determined |
| S8_vs_k141_129334\|\|full | Not-determined |
| S8_vs_k141_42472\|\|full | Not-determined |
| S8_vs_k141_163601\|\|full | Low-quality |
| S8_vs_k141_112159\|\|full | Low-quality |
| S8_vs_k141_77181\|\|full | Not-determined |
| S16_vs_101671\|\|full | Medium-quality |
| S8_vs_k141_112164\|\|full | Low-quality |
| S8_vs_k141_103118\|\|full | Low-quality |
| S8_vs_k141_181680\|\|full | Low-quality |
| S8_vs_k141_77194\|\|full | Not-determined |
| S8_vs_k141_163630\|\|full | Not-determined |
| S8_vs_k141_112174\|\|full | Low-quality |
| S8_vs_k141_137855\|\|full | Low-quality |
| S8_vs_k141_172919\|\|full | Low-quality |
| S8_vs_k141_77205\|\|full | Not-determined |
| S8_vs_k141_181690\|\|full | Low-quality |
| S8_vs_k141_206967\|\|full | Not-determined |
| S8_vs_k141_60316\|\|full | Low-quality |
| S8_vs_k141_7975\|\|full | Low-quality |
| S8_vs_k141_103136\|\|full | Low-quality |
| S8_vs_k141_68303\|\|full | Low-quality |
| S8_vs_k141_112189\|\|full | Not-determined |
| S8_vs_k141_120627\|\|full | Low-quality |
| S8_vs_k141_189103\|\|full | Low-quality |
| S8_vs_k141_189104\|\|full | Low-quality |
| S8_vs_k141_199204\|\|full | Not-determined |
| S8_vs_k141_172945\|\|full | Low-quality |
| S8_vs_k141_60337\|\|full | Low-quality |
| S8_vs_k141_181710\|\|full | Low-quality |
| S8_vs_k141_85958\|\|full | Not-determined |
| S8_vs_k141_137896\|\|full | Low-quality |
| S8_vs_k141_25427\|\|full | Not-determined |
| S3_extracted_k141_22968 | Medium-quality |
| S8_vs_k141_68219\|\|full | Low-quality |
| S8_vs_k141_25430\|\|full | Not-determined |
| S8_vs_k141_129400\|\|full | Low-quality |
| S8_vs_k141_68328\|\|full | Not-determined |
| S8_vs_k141_77250\|\|full | Not-determined |
| S8_vs_k141_172950\|\|full | Low-quality |
| S8_vs_k141_16186\|\|full | Low-quality |
| S8_vs_k141_68331\|\|full | Not-determined |
| S8_vs_k141_16189\|\|full | Not-determined |
| S8_vs_k141_207007\|\|full | Low-quality |
| S8_vs_k141_181739\|\|full | Not-determined |
| S8_vs_k141_60357\|\|full | Not-determined |
| S8_vs_k141_172970\|\|full | Low-quality |
| S8_vs_k141_163694\|\|full | Not-determined |
| S8_vs_k141_112230\|\|full | Low-quality |
| S8_vs_k141_120655\|\|full | Low-quality |
| S8_vs_k141_172974\|\|full | Not-determined |
| S8_vs_k141_207030\|\|full | Low-quality |
| S8_vs_k141_77293\|\|full | Low-quality |
| S8_vs_k141_60376\|\|full | Not-determined |
| S8_vs_k141_146915\|\|full | Low-quality |
| S8_vs_k141_86008\|\|full | Low-quality |
| S8_vs_k141_120682\|\|full | Low-quality |
| S8_vs_k141_60392\|\|full | Low-quality |
| S8_vs_k141_16228\|\|full | Low-quality |
| S8_vs_k141_181777\|\|full | Not-determined |
| S8_vs_k141_60397\|\|full | Not-determined |
| S8_vs_k141_8054\|\|full | Low-quality |
| S8_vs_k141_86029\|\|full | Low-quality |
| S8_vs_k141_86033\|\|full | Not-determined |
| S12_vs_k141_70241\|\|full | Medium-quality |
| S8_vs_k141_146953\|\|full | Low-quality |
| S8_vs_k141_189191\|\|full | Low-quality |
| S8_vs_k141_173024\|\|full | Not-determined |
| S8_vs_k141_94622\|\|full | Low-quality |
| S8_vs_k141_120717\|\|full | Not-determined |
| S8_vs_k141_60414\|\|full | Low-quality |
| S8_vs_k141_163748\|\|full | Low-quality |
| S8_vs_k141_68395\|\|full | Not-determined |
| S8_vs_k141_60417\|\|full | Low-quality |
| S8_vs_k141_137974\|\|full | Low-quality |
| S8_vs_k141_25504\|\|full | Low-quality |
| S8_vs_k141_94644\|\|full | Low-quality |
| S8_vs_k141_94645\|\|full | Low-quality |
| S3_vs_k141_51005\|\|full | Medium-quality |
| S8_vs_k141_207068\|\|full | Not-determined |
| S8_vs_k141_60440\|\|full | Low-quality |
| S8_vs_k141_146981\|\|full | Not-determined |
| S8_vs_k141_120725\|\|full | Low-quality |
| S8_vs_k141_34279\|\|full | Low-quality |
| S8_vs_k141_155119\|\|full | Not-determined |
| S8_vs_k141_173057\|\|full | Not-determined |
| S8_vs_k141_120728\|\|full | Low-quality |
| S8_vs_k141_34286\|\|full | Low-quality |
| S8_vs_k141_207072\|\|full | Low-quality |
| S8_vs_k141_86050\|\|full | Low-quality |
| S8_vs_k141_16282\|\|full | Low-quality |
| S8_vs_k141_86052\|\|full | Low-quality |
| S8_vs_k141_60463\|\|full | Not-determined |
| S8_vs_k141_189224\|\|full | Low-quality |
| S8_vs_k141_207098\|\|full | Low-quality |
| S8_vs_k141_138019\|\|full | Not-determined |
| S8_vs_k141_147008\|\|full | Not-determined |
| S8_vs_k141_207108\|\|full | Low-quality |
| S8_vs_k141_189239\|\|full | Not-determined |
| S8_vs_k141_16324\|\|full | Not-determined |
| S8_vs_k141_147016\|\|full | Low-quality |
| S8_vs_k141_155163\|\|full | Not-determined |
| S8_vs_k141_42647\|\|full | Low-quality |
| S8_vs_k141_129488\|\|full | Low-quality |
| S8_vs_k141_50589\|\|full | Not-determined |
| S8_vs_k141_50591\|\|full | Low-quality |
| S8_vs_k141_147037\|\|full | Low-quality |
| S8_vs_k141_147038\|\|full | Low-quality |
| S8_vs_k141_112392\|\|full | Not-determined |
| S8_vs_k141_34334\|\|full | Low-quality |
| S8_vs_k141_8155\|\|full | Not-determined |
| S8_vs_k141_207139\|\|full | Not-determined |
| S8_vs_k141_34345\|\|full | Not-determined |
| S8_vs_k141_42672\|\|full | Not-determined |
| S8_vs_k141_50616\|\|full | Low-quality |
| S8_vs_k141_8167\|\|full | Not-determined |
| S8_vs_k141_147049\|\|full | Low-quality |
| S8_vs_k141_112399\|\|full | Low-quality |
| S8_vs_k141_34347\|\|full | Low-quality |
| S8_vs_k141_147052\|\|full | Low-quality |
| S8_vs_k141_68510\|\|full | Low-quality |
| S8_vs_k141_173126\|\|full | Low-quality |
| S8_vs_k141_16384\|\|full | Low-quality |
| S8_vs_k141_25573\|\|full | Low-quality |
| S8_vs_k141_147056\|\|full | Low-quality |
| S8_vs_k141_147057\|\|full | Low-quality |
| S8_vs_k141_173136\|\|full | Low-quality |
| S8_vs_k141_163777\|\|full | Low-quality |
| S8_vs_k141_181901\|\|full | Low-quality |
| S8_vs_k141_68516\|\|full | Low-quality |
| S8_vs_k141_181905\|\|full | Not-determined |
| S8_vs_k141_189320\|\|full | Not-determined |
| S8_vs_k141_207170\|\|full | Low-quality |
| S8_vs_k141_129536\|\|full | Low-quality |

| | |
|---|---|
| S8_vs_k141_94779\|\|full | Low-quality |
| S8_vs_k141_207190\|\|full | Not-determined |
| S8_vs_k141_181923\|\|full | Low-quality |
| S8_vs_k141_207193\|\|full | Not-determined |
| S8_vs_k141_189347\|\|full | Not-determined |
| S8_vs_k141_129561\|\|full | Low-quality |
| S8_vs_k141_207198\|\|full | Low-quality |
| S8_vs_k141_8208\|\|full | Not-determined |
| S8_vs_k141_138125\|\|full | Low-quality |
| S8_vs_k141_25614\|\|full | Low-quality |
| S8_vs_k141_60565\|\|full | Not-determined |
| S8_vs_k141_42719\|\|full | Not-determined |
| S8_vs_k141_103283\|\|full | Not-determined |
| S8_vs_k141_8216\|\|full | Low-quality |
| S8_vs_k141_138136\|\|full | Not-determined |
| S8_vs_k141_16450\|\|full | Low-quality |
| S8_vs_k141_163820\|\|full | Not-determined |
| S8_vs_k141_50685\|\|full | Low-quality |
| S8_vs_k141_8231\|\|full | Low-quality |
| S8_vs_k141_147124\|\|full | Low-quality |
| S8_vs_k141_138148\|\|full | Low-quality |
| S8_vs_k141_103305\|\|full | Low-quality |
| S8_vs_k141_68578\|\|full | Low-quality |
| S8_vs_k141_163821\|\|full | Low-quality |
| S8_vs_k141_60589\|\|full | Not-determined |
| S8_vs_k141_155297\|\|full | Low-quality |
| S8_vs_k141_181943\|\|full | Low-quality |
| S8_vs_k141_138162\|\|full | Not-determined |
| S8_vs_k141_86152\|\|full | Not-determined |
| S8_vs_k141_189388\|\|full | Not-determined |
| S8_vs_k141_34420\|\|full | Low-quality |
| S8_vs_k141_181950\|\|full | Not-determined |
| S8_vs_k141_42738\|\|full | Low-quality |
| S8_vs_k141_68593\|\|full | Low-quality |
| S8_vs_k141_50697\|\|full | Not-determined |
| S8_vs_k141_155312\|\|full | Not-determined |
| S8_vs_k141_199399\|\|full | Not-determined |
| S8_vs_k141_77561\|\|full | Not-determined |
| S8_vs_k141_189410\|\|full | Low-quality |
| S8_vs_k141_8262\|\|full | Low-quality |
| S8_vs_k141_8266\|\|full | Not-determined |
| S8_vs_k141_50708\|\|full | Low-quality |
| S8_vs_k141_155325\|\|full | Not-determined |
| S8_vs_k141_138198\|\|full | Not-determined |
| S8_vs_k141_34465\|\|full | Not-determined |
| S8_vs_k141_77586\|\|full | Low-quality |
| S8_vs_k141_8278\|\|full | Low-quality |
| S8_vs_k141_68617\|\|full | Low-quality |
| S8_vs_k141_34475\|\|full | Not-determined |
| S8_vs_k141_77596\|\|full | Not-determined |
| S8_vs_k141_34484\|\|full | Not-determined |
| S8_vs_k141_25687\|\|full | Low-quality |
| S8_vs_k141_189448\|\|full | Not-determined |
| S8_vs_k141_207254\|\|full | Low-quality |
| S8_vs_k141_86192\|\|full | Low-quality |
| S8_vs_k141_189454\|\|full | Not-determined |
| S8_vs_k141_103356\|\|full | Not-determined |
| S8_vs_k141_8283\|\|full | Low-quality |
| S8_vs_k141_207268\|\|full | Low-quality |
| S8_vs_k141_34506\|\|full | Not-determined |
| S8_vs_k141_155356\|\|full | Low-quality |
| S8_vs_k141_25708\|\|full | Not-determined |
| S8_vs_k141_163915\|\|full | Not-determined |
| S8_vs_k141_86205\|\|full | Low-quality |
| S8_vs_k141_182017\|\|full | Low-quality |
| S8_vs_k141_94823\|\|full | Low-quality |
| S8_vs_k141_25709\|\|full | Low-quality |
| S8_vs_k141_155357\|\|full | Not-determined |
| S8_vs_k141_120839\|\|full | Low-quality |
| S8_vs_k141_207282\|\|full | Low-quality |
| S8_vs_k141_42789\|\|full | Not-determined |
| S8_vs_k141_199463\|\|full | Low-quality |
| S8_vs_k141_103381\|\|full | Not-determined |
| S8_vs_k141_34530\|\|full | Low-quality |
| S8_vs_k141_94836\|\|full | Low-quality |
| S8_vs_k141_112546\|\|full | Not-determined |
| S8_vs_k141_50731\|\|full | Not-determined |
| S8_vs_k141_182043\|\|full | Not-determined |
| S8_vs_k141_189485\|\|full | Not-determined |
| S8_vs_k141_16552\|\|full | Not-determined |
| S12_extracted_k141_188827 | Complete |
| S8_vs_k141_112563\|\|full | Low-quality |
| S8_vs_k141_16566\|\|full | Low-quality |
| S8_vs_k141_182064\|\|full | Low-quality |
| S8_vs_k141_207314\|\|full | Low-quality |
| S8_vs_k141_25743\|\|full | Low-quality |
| S8_vs_k141_147133\|\|full | Low-quality |
| S8_vs_k141_77672\|\|full | Low-quality |
| S8_vs_k141_207316\|\|full | Low-quality |
| S8_vs_k141_147141\|\|full | Low-quality |
| S8_vs_k141_163962\|\|full | Not-determined |
| S8_vs_k141_86258\|\|full | Low-quality |
| S8_vs_k141_182093\|\|full | Not-determined |
| S8_vs_k141_207334\|\|full | Not-determined |
| S8_vs_k141_120896\|\|full | Not-determined |
| S8_vs_k141_155394\|\|full | Low-quality |
| S8_vs_k141_25758\|\|full | Low-quality |
| S8_vs_k141_155397\|\|full | Not-determined |
| S8_vs_k141_182096\|\|full | Low-quality |
| S8_vs_k141_147154\|\|full | Low-quality |
| S8_vs_k141_68687\|\|full | Not-determined |
| S8_vs_k141_103423\|\|full | Low-quality |
| S8_vs_k141_8337\|\|full | Low-quality |
| S8_vs_k141_25770\|\|full | Low-quality |
| S8_vs_k141_138275\|\|full | Low-quality |
| S8_vs_k141_60613\|\|full | Low-quality |
| S8_vs_k141_8338\|\|full | Low-quality |
| S8_vs_k141_182098\|\|full | Low-quality |
| S8_vs_k141_86284\|\|full | Not-determined |
| S8_vs_k141_103428\|\|full | Low-quality |
| S8_vs_k141_50749\|\|full | Not-determined |
| S8_vs_k141_138282\|\|full | Low-quality |
| S8_vs_k141_34539\|\|full | Not-determined |
| S8_vs_k141_25791\|\|full | Not-determined |
| S8_vs_k141_50751\|\|full | Low-quality |
| S8_vs_k141_86292\|\|full | Not-determined |
| S8_vs_k141_199538\|\|full | Low-quality |
| S8_vs_k141_86297\|\|full | Low-quality |
| S8_vs_k141_182108\|\|full | Low-quality |
| S8_vs_k141_155407\|\|full | Not-determined |
| S1_extracted_k141_140259 | Medium-quality |
| S8_vs_k141_86302\|\|full | Low-quality |
| S8_vs_k141_147166\|\|full | Low-quality |
| S8_vs_k141_163994\|\|full | Low-quality |
| S8_vs_k141_77731\|\|full | Not-determined |
| S8_vs_k141_68710\|\|full | Not-determined |
| S8_vs_k141_147173\|\|full | Low-quality |
| S8_vs_k141_182122\|\|full | Not-determined |
| S8_vs_k141_77739\|\|full | Low-quality |
| S8_vs_k141_155418\|\|full | Not-determined |
| S8_vs_k141_147178\|\|full | Not-determined |
| S8_vs_k141_25802\|\|full | Not-determined |
| S8_vs_k141_42866\|\|full | Not-determined |
| S8_vs_k141_129737\|\|full | Low-quality |
| S8_vs_k141_189578\|\|full | Not-determined |
| S8_vs_k141_155430\|\|full | Low-quality |
| S8_vs_k141_182136\|\|full | Not-determined |
| S8_vs_k141_60654\|\|full | Low-quality |
| S8_vs_k141_129749\|\|full | Not-determined |
| S8_vs_k141_50777\|\|full | Low-quality |
| S8_vs_k141_182137\|\|full | Low-quality |
| S8_vs_k141_77748\|\|full | Low-quality |
| S8_vs_k141_68736\|\|full | Not-determined |
| S8_vs_k141_199576\|\|full | Not-determined |
| S8_vs_k141_129759\|\|full | Not-determined |
| S8_vs_k141_199578\|\|full | Low-quality |
| S8_vs_k141_16643\|\|full | Not-determined |
| S8_vs_k141_60671\|\|full | Not-determined |
| S8_vs_k141_68741\|\|full | Low-quality |
| S8_vs_k141_42881\|\|full | Low-quality |
| S8_vs_k141_77761\|\|full | Not-determined |
| S8_vs_k141_129764\|\|full | Not-determined |
| S8_vs_k141_120944\|\|full | Low-quality |
| S8_vs_k141_94916\|\|full | Not-determined |
| S8_vs_k141_8407\|\|full | Not-determined |
| S8_vs_k141_68753\|\|full | Low-quality |
| S8_vs_k141_34580\|\|full | Not-determined |
| S8_vs_k141_34585\|\|full | Low-quality |
| S8_vs_k141_16667\|\|full | Not-determined |
| S8_vs_k141_34597\|\|full | Low-quality |
| S8_vs_k141_16674\|\|full | Not-determined |
| S8_vs_k141_34598\|\|full | Not-determined |
| S8_vs_k141_129790\|\|full | Low-quality |
| S8_vs_k141_182178\|\|full | Low-quality |
| S8_vs_k141_164035\|\|full | Not-determined |
| S8_vs_k141_103518\|\|full | Low-quality |
| S8_vs_k141_34604\|\|full | Low-quality |
| S8_vs_k141_86373\|\|full | Low-quality |
| S8_vs_k141_68777\|\|full | Low-quality |
| S8_vs_k141_207428\|\|full | Not-determined |
| S8_vs_k141_8441\|\|full | Low-quality |
| S8_vs_k141_34617\|\|full | Not-determined |
| S8_vs_k141_25840\|\|full | Low-quality |
| S8_vs_k141_86395\|\|full | Low-quality |
| S8_vs_k141_103539\|\|full | Low-quality |
| S8_vs_k141_42887\|\|full | Low-quality |
| S8_vs_k141_173273\|\|full | Low-quality |
| S8_vs_k141_16690\|\|full | Not-determined |
| S8_vs_k141_173276\|\|full | Low-quality |
| S8_vs_k141_77827\|\|full | Not-determined |
| S8_vs_k141_94990\|\|full | Not-determined |
| S8_vs_k141_155507\|\|full | Not-determined |
| S8_vs_k141_121011\|\|full | Low-quality |
| S8_vs_k141_86419\|\|full | Not-determined |
| S8_vs_k141_129837\|\|full | Low-quality |
| S8_vs_k141_173289\|\|full | Low-quality |
| S8_vs_k141_207441\|\|full | Not-determined |
| S8_vs_k141_121021\|\|full | Not-determined |
| S8_vs_k141_173292\|\|full | Low-quality |
| S8_vs_k141_34640\|\|full | Low-quality |
| S8_vs_k141_155516\|\|full | Low-quality |
| S8_vs_k141_42907\|\|full | Low-quality |
| S8_vs_k141_129843\|\|full | Not-determined |
| S8_vs_k141_121035\|\|full | Low-quality |
| S8_vs_k141_16717\|\|full | Low-quality |
| S8_vs_k141_182222\|\|full | Not-determined |
| S8_vs_k141_77838\|\|full | Low-quality |
| S8_vs_k141_164079\|\|full | Low-quality |
| S8_vs_k141_8477\|\|full | Not-determined |
| S8_vs_k141_60736\|\|full | Not-determined |
| S8_vs_k141_86425\|\|full | Not-determined |
| S8_vs_k141_155523\|\|full | Not-determined |
| S8_vs_k141_147289\|\|full | Low-quality |
| S8_vs_k141_207467\|\|full | Not-determined |
| S8_vs_k141_164087\|\|full | Low-quality |
| S8_vs_k141_34655\|\|full | Low-quality |
| S8_vs_k141_189698\|\|full | Low-quality |
| S8_vs_k141_95018\|\|full | Not-determined |
| S8_vs_k141_77846\|\|full | Low-quality |
| S8_vs_k141_103583\|\|full | Low-quality |
| S8_vs_k141_164104\|\|full | Not-determined |
| S8_vs_k141_68847\|\|full | Low-quality |
| S8_vs_k141_207489\|\|full | Not-determined |
| S8_vs_k141_95041\|\|full | Low-quality |
| S8_vs_k141_103604\|\|full | Low-quality |
| S8_vs_k141_34691\|\|full | Low-quality |
| S8_vs_k141_155582\|\|full | Low-quality |
| S8_vs_k141_25862\|\|full | Low-quality |
| S8_vs_k141_34708\|\|full | Low-quality |
| S8_vs_k141_164129\|\|full | Low-quality |
| S8_vs_k141_8553\|\|full | Not-determined |
| S8_vs_k141_95076\|\|full | Low-quality |
| S8_vs_k141_68890\|\|full | Not-determined |
| S8_vs_k141_25872\|\|full | Low-quality |
| S8_vs_k141_164138\|\|full | Low-quality |
| S8_vs_k141_42990\|\|full | Low-quality |
| S8_vs_k141_164141\|\|full | Low-quality |
| S8_vs_k141_68909\|\|full | Low-quality |
| S8_vs_k141_25891\|\|full | Low-quality |
| S8_vs_k141_68916\|\|full | Not-determined |
| S8_vs_k141_155608\|\|full | Low-quality |
| S8_vs_k141_25899\|\|full | Not-determined |
| S8_vs_k141_50857\|\|full | Not-determined |
| S1_vs_k141_140259\|\|full | Medium-quality |
| S8_vs_k141_68931\|\|full | Low-quality |
| S8_vs_k141_155618\|\|full | Not-determined |
| S8_vs_k141_50877\|\|full | Low-quality |
| S8_vs_k141_95113\|\|full | Not-determined |
| S8_vs_k141_121149\|\|full | Not-determined |
| S8_vs_k141_103670\|\|full | Low-quality |
| S8_vs_k141_43036\|\|full | Low-quality |
| S8_vs_k141_173352\|\|full | Low-quality |
| S8_vs_k141_50893\|\|full | Low-quality |
| S8_vs_k141_43044\|\|full | Not-determined |
| S8_vs_k141_147359\|\|full | Low-quality |
| S8_vs_k141_129979\|\|full | Low-quality |
| S8_vs_k141_138523\|\|full | Not-determined |
| S8_vs_k141_50904\|\|full | Low-quality |
| S8_vs_k141_138531\|\|full | Low-quality |
| S8_vs_k141_8607\|\|full | Not-determined |
| S8_vs_k141_129996\|\|full | Not-determined |
| S8_vs_k141_129999\|\|full | Low-quality |
| S8_vs_k141_25938\|\|full | Low-quality |
| S8_vs_k141_103703\|\|full | Not-determined |
| S8_vs_k141_207557\|\|full | Not-determined |
| S8_vs_k141_68953\|\|full | Not-determined |
| S8_vs_k141_112621\|\|full | Low-quality |
| S8_vs_k141_112622\|\|full | Low-quality |
| S8_vs_k141_16858\|\|full | Low-quality |
| S8_vs_k141_77983\|\|full | Low-quality |
| S8_vs_k141_86603\|\|full | Low-quality |
| S8_vs_k141_147384\|\|full | Not-determined |
| S8_vs_k141_121223\|\|full | Low-quality |
| S8_vs_k141_112647\|\|full | Low-quality |
| S8_vs_k141_121234\|\|full | Low-quality |
| S8_vs_k141_95180\|\|full | Low-quality |
| S8_vs_k141_155671\|\|full | Low-quality |
| S8_vs_k141_164218\|\|full | Not-determined |
| S8_vs_k141_103742\|\|full | Low-quality |
| S8_vs_k141_189795\|\|full | Low-quality |
| S8_vs_k141_155679\|\|full | Low-quality |
| S8_vs_k141_103748\|\|full | Not-determined |
| S8_vs_k141_207597\|\|full | Low-quality |
| S8_vs_k141_103756\|\|full | Low-quality |
| S8_vs_k141_112678\|\|full | Low-quality |
| S8_vs_k141_189803\|\|full | Low-quality |
| S8_vs_k141_121248\|\|full | Low-quality |
| S8_vs_k141_121250\|\|full | Not-determined |
| S8_vs_k141_25990\|\|full | Not-determined |
| S8_vs_k141_189811\|\|full | Not-determined |
| S8_vs_k141_25991\|\|full | Not-determined |
| S8_vs_k141_78033\|\|full | Low-quality |
| S8_vs_k141_147449\|\|full | Not-determined |
| S8_vs_k141_43109\|\|full | Low-quality |
| S8_vs_k141_103768\|\|full | Low-quality |
| S8_vs_k141_43113\|\|full | Low-quality |
| S8_vs_k141_26002\|\|full | Not-determined |
| S8_vs_k141_189821\|\|full | Not-determined |
| S8_vs_k141_130105\|\|full | Not-determined |
| S8_vs_k141_95243\|\|full | Not-determined |
| S8_vs_k141_121270\|\|full | Low-quality |
| S8_vs_k141_173430\|\|full | Not-determined |
| S8_vs_k141_155728\|\|full | Low-quality |
| S8_vs_k141_43134\|\|full | Not-determined |
| S8_vs_k141_51012\|\|full | Not-determined |
| S8_vs_k141_51016\|\|full | Low-quality |
| S8_vs_k141_69077\|\|full | Low-quality |
| S8_vs_k141_189845\|\|full | Not-determined |
| S8_vs_k141_8676\|\|full | Low-quality |
| S8_vs_k141_164299\|\|full | Not-determined |
| S8_vs_k141_207623\|\|full | Low-quality |
| S8_vs_k141_155732\|\|full | Not-determined |
| S8_vs_k141_207627\|\|full | Low-quality |
| S8_vs_k141_155736\|\|full | Low-quality |
| S8_vs_k141_43147\|\|full | Not-determined |
| S8_vs_k141_130113\|\|full | Not-determined |
| S8_vs_k141_16946\|\|full | Low-quality |
| S8_vs_k141_147494\|\|full | Not-determined |
| S8_vs_k141_69087\|\|full | Not-determined |
| S8_vs_k141_112746\|\|full | Low-quality |
| S8_vs_k141_173478\|\|full | Low-quality |
| S8_vs_k141_147506\|\|full | Low-quality |
| S8_vs_k141_147514\|\|full | Low-quality |
| S8_vs_k141_103797\|\|full | Low-quality |
| S8_vs_k141_43180\|\|full | Low-quality |
| S8_vs_k141_103802\|\|full | Low-quality |
| S8_vs_k141_207662\|\|full | Low-quality |
| S8_vs_k141_121325\|\|full | Low-quality |
| S8_vs_k141_86696\|\|full | Low-quality |
| S8_vs_k141_16989\|\|full | Not-determined |
| S8_vs_k141_207669\|\|full | Low-quality |
| S8_vs_k141_16991\|\|full | Low-quality |
| S8_vs_k141_86699\|\|full | Not-determined |
| S8_vs_k141_207674\|\|full | Not-determined |
| S8_vs_k141_51083\|\|full | Low-quality |
| S8_vs_k141_121351\|\|full | Not-determined |
| S8_vs_k141_207685\|\|full | Low-quality |
| S8_vs_k141_103832\|\|full | Not-determined |
| S8_vs_k141_43208\|\|full | Not-determined |
| S8_vs_k141_95343\|\|full | Not-determined |
| S8_vs_k141_155805\|\|full | Not-determined |
| S8_vs_k141_121376\|\|full | Low-quality |
| S8_vs_k141_51109\|\|full | Low-quality |
| S8_vs_k141_69146\|\|full | Not-determined |
| S8_vs_k141_95365\|\|full | Low-quality |
| S8_vs_k141_173543\|\|full | Not-determined |
| S8_vs_k141_51129\|\|full | Not-determined |
| S8_vs_k141_69165\|\|full | Not-determined |
| S8_vs_k141_17007\|\|full | Not-determined |
| S8_vs_k141_86750\|\|full | Low-quality |
| S8_vs_k141_207711\|\|full | Low-quality |
| S8_vs_k141_164385\|\|full | Not-determined |
| S8_vs_k141_86756\|\|full | Low-quality |
| S8_vs_k141_69179\|\|full | Low-quality |
| S8_vs_k141_17013\|\|full | Low-quality |
| S8_vs_k141_51147\|\|full | Not-determined |
| S8_vs_k141_17023\|\|full | Not-determined |
| S8_vs_k141_86777\|\|full | Low-quality |
| S8_vs_k141_17037\|\|full | Low-quality |
| S8_vs_k141_95392\|\|full | Not-determined |
| S8_vs_k141_155840\|\|full | Not-determined |
| S8_vs_k141_17038\|\|full | Low-quality |
| S8_vs_k141_207729\|\|full | Low-quality |
| S8_vs_k141_17041\|\|full | Low-quality |
| S8_vs_k141_17044\|\|full | Low-quality |
| S8_vs_k141_51200\|\|full | Not-determined |
| S8_vs_k141_69226\|\|full | Not-determined |
| S8_vs_k141_69230\|\|full | Not-determined |
| S8_vs_k141_164413\|\|full | Low-quality |
| S12_vs_k141_188827\|\|full | High-quality |
| S8_vs_k141_164422\|\|full | Low-quality |
| S8_vs_k141_164424\|\|full | Low-quality |
| S8_vs_k141_121472\|\|full | Not-determined |
| S8_vs_k141_138678\|\|full | Low-quality |
| S8_vs_k141_43234\|\|full | Not-determined |
| S8_vs_k141_164439\|\|full | Low-quality |
| S8_vs_k141_164443\|\|full | Not-determined |
| S8_vs_k141_190049\|\|full | Not-determined |
| S8_vs_k141_69284\|\|full | Low-quality |
| S8_vs_k141_43257\|\|full | Not-determined |
| S8_vs_k141_190072\|\|full | Low-quality |
| S8_vs_k141_190074\|\|full | Low-quality |
| S8_vs_k141_138713\|\|full | Low-quality |
| S8_vs_k141_155891\|\|full | Low-quality |
| S8_vs_k141_17091\|\|full | Low-quality |
| S8_vs_k141_138749\|\|full | Low-quality |
| S8_vs_k141_164500\|\|full | Low-quality |
| S8_vs_k141_207749\|\|full | Low-quality |
| S8_vs_k141_138755\|\|full | Not-determined |
| S8_vs_k141_69370\|\|full | Not-determined |
| S8_vs_k141_164524\|\|full | Low-quality |
| S8_vs_k141_164534\|\|full | Low-quality |
| S8_vs_k141_69387\|\|full | Not-determined |
| S8_vs_k141_69420\|\|full | Low-quality |
| S8_vs_k141_155980\|\|full | Not-determined |
| S8_vs_k141_207778\|\|full | Low-quality |
| S8_vs_k141_103944\|\|full | Low-quality |
| S8_vs_k141_164598\|\|full | Not-determined |
| S8_vs_k141_138807\|\|full | Low-quality |
| S8_vs_k141_138809\|\|full | Low-quality |
| S8_vs_k141_190219\|\|full | Not-determined |
| S8_vs_k141_164611\|\|full | Low-quality |
| S8_vs_k141_190236\|\|full | Low-quality |
| S8_vs_k141_156036\|\|full | Low-quality |
| S8_vs_k141_156045\|\|full | Not-determined |
| S8_vs_k141_138835\|\|full | Not-determined |
| S8_vs_k141_138838\|\|full | Not-determined |
| S8_vs_k141_164644\|\|full | Low-quality |
| S8_vs_k141_103988\|\|full | Not-determined |
| S8_vs_k141_103993\|\|full | Low-quality |
| S8_vs_k141_104000\|\|full | Low-quality |
| S8_vs_k141_156073\|\|full | Low-quality |
| S8_vs_k141_164683\|\|full | Low-quality |
| S8_vs_k141_207862\|\|full | Not-determined |
| S8_vs_k141_190302\|\|full | Not-determined |
| S8_vs_k141_207868\|\|full | Not-determined |
| S8_vs_k141_207871\|\|full | Low-quality |
| S8_vs_k141_104032\|\|full | Not-determined |
| S8_vs_k141_164700\|\|full | Not-determined |
| S8_vs_k141_156087\|\|full | Not-determined |
| S8_vs_k141_17169\|\|full | Not-determined |
| S8_vs_k141_190323\|\|full | Not-determined |
| S8_vs_k141_207881\|\|full | Low-quality |
| S8_vs_k141_190331\|\|full | Not-determined |
| S8_vs_k141_190343\|\|full | Low-quality |
| S8_vs_k141_207900\|\|full | Low-quality |
| S8_vs_k141_17188\|\|full | Low-quality |
| S8_vs_k141_164738\|\|full | Not-determined |
| S8_vs_k141_104070\|\|full | Not-determined |
| S8_vs_k141_51416\|\|full | Not-determined |
| S8_vs_k141_207951\|\|full | Low-quality |
| S8_vs_k141_17211\|\|full | Low-quality |
| S8_vs_k141_156141\|\|full | Not-determined |
| S8_vs_k141_17217\|\|full | Low-quality |
| S8_vs_k141_164754\|\|full | Not-determined |
| S8_vs_k141_51449\|\|full | Low-quality |
| S8_vs_k141_207971\|\|full | Not-determined |
| S8_vs_k141_104114\|\|full | Not-determined |
| S8_vs_k141_17231\|\|full | Not-determined |
| S8_vs_k141_190387\|\|full | Not-determined |
| S8_vs_k141_190388\|\|full | Low-quality |
| S8_vs_k141_17278\|\|full | Not-determined |
| S8_vs_k141_51506\|\|full | Low-quality |
| S8_vs_k141_17294\|\|full | Low-quality |
| S8_vs_k141_164809\|\|full | Low-quality |
| S8_vs_k141_208072\|\|full | Not-determined |
| S8_vs_k141_51512\|\|full | Not-determined |
| S8_vs_k141_208083\|\|full | Low-quality |
| S8_vs_k141_208097\|\|full | Not-determined |
| S8_vs_k141_164834\|\|full | Low-quality |
| S8_vs_k141_164836\|\|full | Low-quality |
| S8_vs_k141_51542\|\|full | Low-quality |
| S8_vs_k141_190519\|\|full | Not-determined |
| S8_vs_k141_190521\|\|full | Low-quality |
| S8_vs_k141_51548\|\|full | Low-quality |
| S8_vs_k141_17338\|\|full | Low-quality |
| S8_vs_k141_51566\|\|full | Not-determined |
| S8_vs_k141_17355\|\|full | Low-quality |
| S8_vs_k141_51589\|\|full | Not-determined |
| S8_vs_k141_51593\|\|full | Low-quality |
| S8_vs_k141_51620\|\|full | Not-determined |
| S8_vs_k141_190567\|\|full | Not-determined |
| S8_vs_k141_190617\|\|full | Low-quality |
| S8_vs_k141_190619\|\|full | Not-determined |
| S8_vs_k141_190645\|\|full | Low-quality |
| S8_vs_k141_190678\|\|full | Not-determined |
| S8_vs_k141_190695\|\|full | Low-quality |
| S8_vs_k141_190759\|\|full | Low-quality |
| S8_vs_k141_51819\|\|full | Not-determined |
| S8_vs_k141_51824\|\|full | Not-determined |
| S8_vs_k141_190779\|\|full | Low-quality |
| S8_vs_k141_190836\|\|full | Not-determined |
| S9_vs_k141_42609\|\|lt2gene | Medium-quality |
| S8_vs_k141_51928\|\|full | Not-determined |
| S8_vs_k141_51954\|\|full | Low-quality |
| S8_vs_k141_52025\|\|full | Low-quality |
| S14_vs_k141_97835\|\|full | Medium-quality |
| S8_vs_k141_52054\|\|full | Not-determined |
| S8_vs_k141_78664\|\|0_partial | Low-quality |
| S8_vs_k141_35390\|\|0_partial | Not-determined |
| S8_vs_k141_35561\|\|0_partial | Low-quality |
| S8_vs_k141_79829\|\|0_partial | Low-quality |
| S8_vs_k141_62364\|\|0_partial | Not-determined |
| S8_vs_k141_70754\|\|0_partial | Low-quality |
| S8_vs_k141_114505\|\|0_partial | Low-quality |
| S8_vs_k141_175167\|\|0_partial | Low-quality |
| S8_vs_k141_149336\|\|0_partial | Low-quality |
| S8_vs_k141_2668\|\|0_partial | Not-determined |
| S8_vs_k141_2792\|\|0_partial | Low-quality |
| S8_vs_k141_184630\|\|0_partial | Low-quality |
| S8_vs_k141_124782\|\|0_partial | Low-quality |
| S8_vs_k141_29825\|\|0_partial | Not-determined |
| S8_vs_k141_133535\|\|0_partial | Low-quality |
| S8_vs_k141_64156\|\|0_partial | Low-quality |
| S8_vs_k141_116675\|\|0_partial | Low-quality |
| S8_vs_k141_185981\|\|0_partial | Not-determined |
| S8_vs_k141_100346\|\|0_partial | Low-quality |
| S8_vs_k141_153320\|\|0_partial | Low-quality |
| S8_vs_k141_23926\|\|0_partial | Low-quality |
| S8_vs_k141_24820\|\|0_partial | Low-quality |
| S8_vs_k141_103148\|\|0_partial | Low-quality |
| S8_vs_k141_138842\|\|lt2gene | Low-quality |
| S8_vs_k141_104142\|\|lt2gene | Low-quality |
| S8_vs_k141_147532\|\|lt2gene | Low-quality |
| S8_vs_k141_34730\|\|lt2gene | Not-determined |
| S8_vs_k141_164877\|\|lt2gene | Not-determined |
| S8_vs_k141_190926\|\|lt2gene | Low-quality |
| S8_vs_k141_52087\|\|lt2gene | Low-quality |
| S8_vs_k141_86798\|\|lt2gene | Low-quality |
| S8_vs_k141_60794\|\|lt2gene | Low-quality |
| S8_vs_k141_43424\|\|lt2gene | Low-quality |
| S8_vs_k141_156246\|\|lt2gene | Low-quality |
| S8_vs_k141_34763\|\|lt2gene | Low-quality |
| S8_vs_k141_104187\|\|lt2gene | Not-determined |
| S8_vs_k141_156251\|\|lt2gene | Low-quality |
| S8_vs_k141_138890\|\|lt2gene | Low-quality |
| S8_vs_k141_43443\|\|lt2gene | Low-quality |
| S8_vs_k141_17419\|\|lt2gene | Low-quality |
| S8_vs_k141_112897\|\|lt2gene | Not-determined |
| S8_vs_k141_130210\|\|lt2gene | Low-quality |
| S8_vs_k141_121582\|\|lt2gene | Low-quality |
| S8_vs_k141_26135\|\|lt2gene | Low-quality |
| S8_vs_k141_173592\|\|lt2gene | Low-quality |
| S8_vs_k141_199660\|\|lt2gene | Low-quality |
| S8_vs_k141_95538\|\|lt2gene | Low-quality |
| S8_vs_k141_52164\|\|lt2gene | Low-quality |
| S8_vs_k141_147650\|\|lt2gene | Low-quality |
| S8_vs_k141_26185\|\|lt2gene | Low-quality |
| S8_vs_k141_112944\|\|lt2gene | Low-quality |
| S8_vs_k141_173643\|\|lt2gene | Low-quality |
| S8_vs_k141_60876\|\|lt2gene | Low-quality |
| S8_vs_k141_69529\|\|lt2gene | Low-quality |
| S8_vs_k141_165020\|\|lt2gene | Low-quality |
| S8_vs_k141_26225\|\|lt2gene | Not-determined |
| S8_vs_k141_69535\|\|lt2gene | Low-quality |
| S8_vs_k141_34863\|\|lt2gene | Not-determined |
| S8_vs_k141_112994\|\|lt2gene | Low-quality |
| S8_vs_k141_191134\|\|lt2gene | Not-determined |
| S8_vs_k141_173715\|\|lt2gene | Not-determined |
| S8_vs_k141_139054\|\|lt2gene | Low-quality |
| S8_vs_k141_156382\|\|lt2gene | Low-quality |
| S8_vs_k141_139058\|\|lt2gene | Low-quality |
| S8_vs_k141_139073\|\|lt2gene | Low-quality |
| S8_vs_k141_182307\|\|lt2gene | Low-quality |
| S8_vs_k141_34955\|\|lt2gene | Low-quality |
| S8_vs_k141_34958\|\|lt2gene | Low-quality |
| S8_vs_k141_182310\|\|lt2gene | Not-determined |
| S8_vs_k141_34963\|\|lt2gene | Low-quality |
| S8_vs_k141_301\|\|lt2gene | Not-determined |
| S8_vs_k141_104299\|\|lt2gene | Low-quality |
| S8_vs_k141_314\|\|lt2gene | Not-determined |
| S8_vs_k141_139088\|\|lt2gene | Low-quality |
| S8_vs_k141_69612\|\|lt2gene | Low-quality |
| S8_vs_k141_130421\|\|lt2gene | Low-quality |
| S8_vs_k141_104323\|\|lt2gene | Low-quality |
| S8_vs_k141_69625\|\|lt2gene | Low-quality |
| S8_vs_k141_95615\|\|lt2gene | Low-quality |
| S8_vs_k141_165147\|\|lt2gene | Low-quality |
| S8_vs_k141_104345\|\|lt2gene | Not-determined |
| S8_vs_k141_165156\|\|lt2gene | Low-quality |
| S8_vs_k141_104349\|\|lt2gene | Not-determined |
| S8_vs_k141_182366\|\|lt2gene | Low-quality |
| S8_vs_k141_165177\|\|lt2gene | Low-quality |
| S8_vs_k141_87173\|\|lt2gene | Low-quality |
| S8_vs_k141_35046\|\|lt2gene | Low-quality |
| S8_vs_k141_199813\|\|lt2gene | Not-determined |
| S8_vs_k141_17532\|\|lt2gene | Low-quality |
| S8_vs_k141_78424\|\|lt2gene | Not-determined |
| S8_vs_k141_43666\|\|lt2gene | Low-quality |
| S8_vs_k141_121783\|\|lt2gene | Low-quality |
| S8_vs_k141_26338\|\|lt2gene | Not-determined |
| S8_vs_k141_61103\|\|lt2gene | Low-quality |
| S8_vs_k141_182423\|\|lt2gene | Low-quality |
| S8_vs_k141_78443\|\|lt2gene | Low-quality |
| S8_vs_k141_87218\|\|lt2gene | Low-quality |
| S8_vs_k141_156523\|\|lt2gene | Not-determined |
| S8_vs_k141_78448\|\|lt2gene | Low-quality |
| S8_vs_k141_17545\|\|lt2gene | Not-determined |
| S8_vs_k141_95659\|\|lt2gene | Not-determined |
| S8_vs_k141_182440\|\|lt2gene | Low-quality |
| S8_vs_k141_388\|\|lt2gene | Low-quality |
| S8_vs_k141_182444\|\|lt2gene | Not-determined |
| S8_vs_k141_17562\|\|lt2gene | Not-determined |
| S8_vs_k141_9093\|\|lt2gene | Low-quality |
| S8_vs_k141_26384\|\|lt2gene | Not-determined |
| S8_vs_k141_121862\|\|lt2gene | Low-quality |
| S8_vs_k141_43713\|\|lt2gene | Low-quality |
| S8_vs_k141_95693\|\|lt2gene | Low-quality |
| S8_vs_k141_147710\|\|lt2gene | Low-quality |
| S8_vs_k141_182470\|\|lt2gene | Low-quality |
| S8_vs_k141_35104\|\|lt2gene | Low-quality |
| S8_vs_k141_17576\|\|lt2gene | Not-determined |
| S8_vs_k141_182486\|\|lt2gene | Low-quality |
| S8_vs_k141_52461\|\|lt2gene | Not-determined |
| S8_vs_k141_17599\|\|lt2gene | Not-determined |
| S8_vs_k141_182519\|\|lt2gene | Low-quality |
| S8_vs_k141_26441\|\|lt2gene | Low-quality |
| S8_vs_k141_9147\|\|lt2gene | Low-quality |
| S8_vs_k141_199953\|\|lt2gene | Low-quality |

| | |
|---|---|
| S8_vs_k141_139234\|\|lt2gene | Low-quality |
| S8_vs_k141_69730\|\|lt2gene | Low-quality |
| S8_vs_k141_147782\|\|lt2gene | Not-determined |
| S8_vs_k141_52495\|\|lt2gene | Not-determined |
| S8_vs_k141_78591\|\|lt2gene | Low-quality |
| S8_vs_k141_35191\|\|lt2gene | Not-determined |
| S8_vs_k141_35192\|\|lt2gene | Low-quality |
| S8_vs_k141_43807\|\|lt2gene | Not-determined |
| S8_vs_k141_121944\|\|lt2gene | Low-quality |
| S8_vs_k141_95787\|\|lt2gene | Low-quality |
| S8_vs_k141_191464\|\|lt2gene | Low-quality |
| S8_vs_k141_525\|\|lt2gene | Not-determined |
| S8_vs_k141_87411\|\|lt2gene | Not-determined |
| S8_vs_k141_130700\|\|lt2gene | Not-determined |
| S8_vs_k141_165434\|\|lt2gene | Low-quality |
| S8_vs_k141_122003\|\|lt2gene | Low-quality |
| S8_vs_k141_61237\|\|lt2gene | Low-quality |
| S8_vs_k141_104589\|\|lt2gene | Not-determined |
| S8_vs_k141_35250\|\|lt2gene | Low-quality |
| S8_vs_k141_104599\|\|lt2gene | Low-quality |
| S8_vs_k141_139321\|\|lt2gene | Low-quality |
| S8_vs_k141_52607\|\|lt2gene | Low-quality |
| S8_vs_k141_139324\|\|lt2gene | Low-quality |
| S8_vs_k141_17681\|\|lt2gene | Not-determined |
| S8_vs_k141_69762\|\|lt2gene | Low-quality |
| S8_vs_k141_95855\|\|lt2gene | Not-determined |
| S8_vs_k141_35279\|\|lt2gene | Not-determined |
| S8_vs_k141_165468\|\|lt2gene | Low-quality |
| S8_vs_k141_17704\|\|lt2gene | Low-quality |
| S8_vs_k141_17711\|\|lt2gene | Low-quality |
| S8_vs_k141_9235\|\|lt2gene | Not-determined |
| S8_vs_k141_165496\|\|lt2gene | Low-quality |
| S8_vs_k141_182606\|\|lt2gene | Low-quality |
| S8_vs_k141_17721\|\|lt2gene | Low-quality |
| S8_vs_k141_87464\|\|lt2gene | Low-quality |
| S8_vs_k141_87484\|\|lt2gene | Not-determined |
| S8_vs_k141_122121\|\|lt2gene | Low-quality |
| S8_vs_k141_130787\|\|lt2gene | Low-quality |
| S8_vs_k141_174028\|\|lt2gene | Low-quality |
| S8_vs_k141_95948\|\|lt2gene | Low-quality |
| S8_vs_k141_677\|\|lt2gene | Low-quality |
| S8_vs_k141_182634\|\|lt2gene | Not-determined |
| S8_vs_k141_52740\|\|lt2gene | Low-quality |
| S8_vs_k141_122153\|\|lt2gene | Not-determined |
| S8_vs_k141_61375\|\|lt2gene | Low-quality |
| S8_vs_k141_113341\|\|lt2gene | Not-determined |
| S8_vs_k141_61377\|\|lt2gene | Low-quality |
| S8_vs_k141_61379\|\|lt2gene | Not-determined |
| S8_vs_k141_139437\|\|lt2gene | Low-quality |
| S8_vs_k141_156653\|\|lt2gene | Low-quality |
| S8_vs_k141_708\|\|lt2gene | Not-determined |
| S8_vs_k141_95995\|\|lt2gene | Low-quality |
| S8_vs_k141_17825\|\|lt2gene | Low-quality |
| S8_vs_k141_17830\|\|lt2gene | Low-quality |
| S8_vs_k141_17834\|\|lt2gene | Low-quality |
| S8_vs_k141_43913\|\|lt2gene | Low-quality |
| S8_vs_k141_139486\|\|lt2gene | Low-quality |
| S8_vs_k141_174083\|\|lt2gene | Low-quality |
| S8_vs_k141_69955\|\|lt2gene | Low-quality |
| S8_vs_k141_130885\|\|lt2gene | Low-quality |
| S8_vs_k141_61462\|\|lt2gene | Low-quality |
| S8_vs_k141_96054\|\|lt2gene | Low-quality |
| S8_vs_k141_156741\|\|lt2gene | Not-determined |
| S8_vs_k141_61472\|\|lt2gene | Low-quality |
| S8_vs_k141_9351\|\|lt2gene | Low-quality |
| S8_vs_k141_139550\|\|lt2gene | Not-determined |
| S8_vs_k141_9370\|\|lt2gene | Low-quality |
| S8_vs_k141_52844\|\|lt2gene | Low-quality |
| S8_vs_k141_122235\|\|lt2gene | Low-quality |
| S8_vs_k141_26707\|\|lt2gene | Low-quality |
| S8_vs_k141_70015\|\|lt2gene | Low-quality |
| S8_vs_k141_70016\|\|lt2gene | Low-quality |
| S8_vs_k141_70021\|\|lt2gene | Low-quality |
| S8_vs_k141_70024\|\|lt2gene | Low-quality |
| S8_vs_k141_35412\|\|lt2gene | Low-quality |
| S8_vs_k141_130977\|\|lt2gene | Low-quality |
| S8_vs_k141_78859\|\|lt2gene | Low-quality |
| S8_vs_k141_867\|\|lt2gene | Not-determined |
| S8_vs_k141_70048\|\|lt2gene | Low-quality |
| S8_vs_k141_26753\|\|lt2gene | Low-quality |
| S8_vs_k141_96128\|\|lt2gene | Low-quality |
| S8_vs_k141_191785\|\|lt2gene | Low-quality |
| S8_vs_k141_44123\|\|lt2gene | Low-quality |
| S8_vs_k141_104852\|\|lt2gene | Low-quality |
| S8_vs_k141_87734\|\|lt2gene | Low-quality |
| S8_vs_k141_156828\|\|lt2gene | Low-quality |
| S8_vs_k141_26796\|\|lt2gene | Low-quality |
| S8_vs_k141_191824\|\|lt2gene | Not-determined |
| S8_vs_k141_920\|\|lt2gene | Not-determined |
| S8_vs_k141_70093\|\|lt2gene | Low-quality |
| S8_vs_k141_17995\|\|lt2gene | Low-quality |
| S8_vs_k141_18011\|\|lt2gene | Low-quality |
| S8_vs_k141_131013\|\|lt2gene | Not-determined |
| S8_vs_k141_96197\|\|lt2gene | Low-quality |
| S8_vs_k141_131021\|\|lt2gene | Not-determined |
| S8_vs_k141_96202\|\|lt2gene | Not-determined |
| S8_vs_k141_96204\|\|lt2gene | Low-quality |
| S8_vs_k141_44204\|\|lt2gene | Not-determined |
| S8_vs_k141_122391\|\|lt2gene | Low-quality |
| S8_vs_k141_113508\|\|lt2gene | Low-quality |
| S8_vs_k141_61600\|\|lt2gene | Not-determined |
| S8_vs_k141_104898\|\|lt2gene | Not-determined |
| S8_vs_k141_191888\|\|lt2gene | Not-determined |
| S8_vs_k141_9532\|\|lt2gene | Not-determined |
| S8_vs_k141_139716\|\|lt2gene | Low-quality |
| S8_vs_k141_131070\|\|lt2gene | Not-determined |
| S8_vs_k141_52929\|\|lt2gene | Low-quality |
| S8_vs_k141_986\|\|lt2gene | Low-quality |
| S8_vs_k141_96273\|\|lt2gene | Not-determined |
| S8_vs_k141_9567\|\|lt2gene | Not-determined |
| S8_vs_k141_113567\|\|lt2gene | Low-quality |
| S8_vs_k141_122447\|\|lt2gene | Low-quality |
| S8_vs_k141_113584\|\|lt2gene | Low-quality |
| S8_vs_k141_61656\|\|lt2gene | Low-quality |
| S8_vs_k141_26937\|\|lt2gene | Low-quality |
| S8_vs_k141_61659\|\|lt2gene | Not-determined |
| S8_vs_k141_52962\|\|lt2gene | Low-quality |
| S8_vs_k141_79051\|\|lt2gene | Low-quality |
| S8_vs_k141_174233\|\|lt2gene | Low-quality |
| S8_vs_k141_70147\|\|lt2gene | Low-quality |
| S8_vs_k141_131142\|\|lt2gene | Not-determined |
| S8_vs_k141_156979\|\|lt2gene | Not-determined |
| S8_vs_k141_26968\|\|lt2gene | Low-quality |
| S8_vs_k141_61698\|\|lt2gene | Low-quality |
| S8_vs_k141_131152\|\|lt2gene | Low-quality |
| S8_vs_k141_70175\|\|lt2gene | Low-quality |
| S8_vs_k141_52995\|\|lt2gene | Low-quality |
| S8_vs_k141_157006\|\|lt2gene | Low-quality |
| S8_vs_k141_87858\|\|lt2gene | Low-quality |
| S8_vs_k141_44302\|\|lt2gene | Low-quality |
| S8_vs_k141_96367\|\|lt2gene | Low-quality |
| S8_vs_k141_9621\|\|lt2gene | Low-quality |
| S8_vs_k141_139855\|\|lt2gene | Low-quality |
| S8_vs_k141_200257\|\|lt2gene | Low-quality |
| S8_vs_k141_122548\|\|lt2gene | Low-quality |
| S8_vs_k141_27068\|\|lt2gene | Low-quality |
| S8_vs_k141_61777\|\|lt2gene | Low-quality |
| S8_vs_k141_53071\|\|lt2gene | Not-determined |
| S8_vs_k141_96438\|\|lt2gene | Low-quality |
| S8_vs_k141_35773\|\|lt2gene | Low-quality |
| S8_vs_k141_105089\|\|lt2gene | Low-quality |
| S8_vs_k141_18141\|\|lt2gene | Not-determined |
| S8_vs_k141_148402\|\|lt2gene | Low-quality |
| S8_vs_k141_174361\|\|lt2gene | Not-determined |
| S8_vs_k141_157065\|\|lt2gene | Low-quality |
| S8_vs_k141_139917\|\|lt2gene | Not-determined |
| S8_vs_k141_139926\|\|lt2gene | Low-quality |
| S8_vs_k141_96459\|\|lt2gene | Low-quality |
| S8_vs_k141_96480\|\|lt2gene | Not-determined |
| S8_vs_k141_87962\|\|lt2gene | Low-quality |
| S8_vs_k141_70250\|\|lt2gene | Not-determined |
| S8_vs_k141_96512\|\|lt2gene | Low-quality |
| S8_vs_k141_122620\|\|lt2gene | Low-quality |
| S8_vs_k141_122623\|\|lt2gene | Low-quality |
| S8_vs_k141_148509\|\|lt2gene | Low-quality |
| S8_vs_k141_148512\|\|lt2gene | Not-determined |
| S8_vs_k141_122638\|\|lt2gene | Low-quality |
| S8_vs_k141_139996\|\|lt2gene | Low-quality |
| S8_vs_k141_148540\|\|lt2gene | Low-quality |
| S8_vs_k141_44323\|\|lt2gene | Not-determined |
| S8_vs_k141_9762\|\|lt2gene | Low-quality |
| S8_vs_k141_122653\|\|lt2gene | Low-quality |
| S8_vs_k141_27234\|\|lt2gene | Not-determined |
| S8_vs_k141_192202\|\|lt2gene | Not-determined |
| S8_vs_k141_148600\|\|lt2gene | Low-quality |
| S8_vs_k141_165986\|\|lt2gene | Not-determined |
| S8_vs_k141_131389\|\|lt2gene | Low-quality |
| S8_vs_k141_1235\|\|lt2gene | Not-determined |
| S8_vs_k141_53234\|\|lt2gene | Low-quality |
| S8_vs_k141_157212\|\|lt2gene | Low-quality |
| S8_vs_k141_140054\|\|lt2gene | Low-quality |
| S8_vs_k141_53243\|\|lt2gene | Low-quality |
| S8_vs_k141_79384\|\|lt2gene | Low-quality |
| S8_vs_k141_88032\|\|lt2gene | Low-quality |
| S8_vs_k141_113837\|\|lt2gene | Not-determined |
| S8_vs_k141_113838\|\|lt2gene | Low-quality |
| S8_vs_k141_174494\|\|lt2gene | Low-quality |
| S8_vs_k141_174496\|\|lt2gene | Low-quality |
| S8_vs_k141_88040\|\|lt2gene | Low-quality |
| S8_vs_k141_18283\|\|lt2gene | Not-determined |
| S8_vs_k141_157224\|\|lt2gene | Low-quality |
| S8_vs_k141_157251\|\|lt2gene | Low-quality |
| S8_vs_k141_113875\|\|lt2gene | Not-determined |
| S8_vs_k141_166047\|\|lt2gene | Not-determined |
| S8_vs_k141_183004\|\|lt2gene | Low-quality |
| S8_vs_k141_122758\|\|lt2gene | Low-quality |
| S8_vs_k141_166064\|\|lt2gene | Not-determined |
| S8_vs_k141_122766\|\|lt2gene | Low-quality |
| S8_vs_k141_192350\|\|lt2gene | Low-quality |
| S8_vs_k141_27394\|\|lt2gene | Low-quality |
| S8_vs_k141_200460\|\|lt2gene | Low-quality |
| S8_vs_k141_1334\|\|lt2gene | Not-determined |
| S8_vs_k141_200484\|\|lt2gene | Low-quality |
| S8_vs_k141_105339\|\|lt2gene | Low-quality |
| S8_vs_k141_166125\|\|lt2gene | Low-quality |
| S8_vs_k141_105357\|\|lt2gene | Low-quality |
| S8_vs_k141_36082\|\|lt2gene | Low-quality |
| S8_vs_k141_105371\|\|lt2gene | Low-quality |
| S8_vs_k141_192413\|\|lt2gene | Not-determined |
| S8_vs_k141_131507\|\|lt2gene | Low-quality |
| S8_vs_k141_192440\|\|lt2gene | Low-quality |
| S8_vs_k141_96747\|\|lt2gene | Low-quality |
| S8_vs_k141_62101\|\|lt2gene | Low-quality |
| S8_vs_k141_122877\|\|lt2gene | Low-quality |
| S8_vs_k141_62124\|\|lt2gene | Low-quality |
| S8_vs_k141_105431\|\|lt2gene | Low-quality |
| S8_vs_k141_166174\|\|lt2gene | Low-quality |
| S8_vs_k141_157388\|\|lt2gene | Not-determined |
| S8_vs_k141_62132\|\|lt2gene | Not-determined |
| S8_vs_k141_114022\|\|lt2gene | Low-quality |
| S8_vs_k141_44544\|\|lt2gene | Low-quality |
| S8_vs_k141_114032\|\|lt2gene | Low-quality |
| S8_vs_k141_114035\|\|lt2gene | Low-quality |
| S8_vs_k141_1465\|\|lt2gene | Low-quality |
| S8_vs_k141_27550\|\|lt2gene | Low-quality |
| S8_vs_k141_88184\|\|lt2gene | Not-determined |
| S8_vs_k141_9902\|\|lt2gene | Not-determined |
| S8_vs_k141_9906\|\|lt2gene | Low-quality |
| S8_vs_k141_148837\|\|lt2gene | Low-quality |
| S8_vs_k141_148838\|\|lt2gene | Not-determined |
| S8_vs_k141_105471\|\|lt2gene | Low-quality |
| S8_vs_k141_9924\|\|lt2gene | Not-determined |
| S8_vs_k141_105478\|\|lt2gene | Not-determined |
| S8_vs_k141_140233\|\|lt2gene | Low-quality |
| S8_vs_k141_1487\|\|lt2gene | Low-quality |
| S8_vs_k141_140236\|\|lt2gene | Low-quality |
| S8_vs_k141_1492\|\|lt2gene | Low-quality |
| S8_vs_k141_44578\|\|lt2gene | Not-determined |
| S8_vs_k141_174677\|\|lt2gene | Low-quality |
| S8_vs_k141_114089\|\|lt2gene | Low-quality |
| S8_vs_k141_174686\|\|lt2gene | Low-quality |
| S8_vs_k141_96868\|\|lt2gene | Low-quality |
| S8_vs_k141_70575\|\|lt2gene | Not-determined |
| S8_vs_k141_70600\|\|lt2gene | Not-determined |
| S8_vs_k141_96893\|\|lt2gene | Low-quality |
| S8_vs_k141_140299\|\|lt2gene | Low-quality |
| S8_vs_k141_18585\|\|lt2gene | Not-determined |
| S8_vs_k141_131561\|\|lt2gene | Low-quality |
| S8_vs_k141_70645\|\|lt2gene | Not-determined |
| S8_vs_k141_53564\|\|lt2gene | Not-determined |
| S8_vs_k141_53567\|\|lt2gene | Not-determined |
| S8_vs_k141_131576\|\|lt2gene | Low-quality |
| S8_vs_k141_157538\|\|lt2gene | Low-quality |
| S8_vs_k141_200665\|\|lt2gene | Low-quality |
| S8_vs_k141_183209\|\|lt2gene | Low-quality |
| S8_vs_k141_140363\|\|lt2gene | Low-quality |
| S8_vs_k141_44660\|\|lt2gene | Low-quality |
| S8_vs_k141_36284\|\|lt2gene | Low-quality |
| S8_vs_k141_36294\|\|lt2gene | Low-quality |
| S8_vs_k141_123016\|\|lt2gene | Low-quality |
| S8_vs_k141_105605\|\|lt2gene | Low-quality |
| S8_vs_k141_18657\|\|lt2gene | Low-quality |
| S8_vs_k141_140389\|\|lt2gene | Low-quality |
| S8_vs_k141_105652\|\|lt2gene | Not-determined |
| S8_vs_k141_97027\|\|lt2gene | Low-quality |
| S8_vs_k141_97031\|\|lt2gene | Low-quality |
| S8_vs_k141_123094\|\|lt2gene | Not-determined |
| S8_vs_k141_97047\|\|lt2gene | Low-quality |
| S8_vs_k141_183311\|\|lt2gene | Low-quality |
| S8_vs_k141_105688\|\|lt2gene | Low-quality |
| S8_vs_k141_97061\|\|lt2gene | Not-determined |
| S8_vs_k141_18767\|\|lt2gene | Not-determined |
| S8_vs_k141_62302\|\|lt2gene | Not-determined |
| S8_vs_k141_44753\|\|lt2gene | Low-quality |
| S8_vs_k141_36474\|\|lt2gene | Low-quality |
| S8_vs_k141_88312\|\|lt2gene | Low-quality |
| S8_vs_k141_62322\|\|lt2gene | Not-determined |
| S8_vs_k141_192697\|\|lt2gene | Low-quality |
| S8_vs_k141_18810\|\|lt2gene | Not-determined |
| S8_vs_k141_27812\|\|lt2gene | Not-determined |
| S8_vs_k141_123174\|\|lt2gene | Low-quality |
| S8_vs_k141_53754\|\|lt2gene | Low-quality |
| S8_vs_k141_105759\|\|lt2gene | Low-quality |
| S8_vs_k141_192720\|\|lt2gene | Low-quality |
| S8_vs_k141_53768\|\|lt2gene | Low-quality |
| S8_vs_k141_140535\|\|lt2gene | Low-quality |
| S8_vs_k141_18860\|\|lt2gene | Low-quality |
| S8_vs_k141_36550\|\|lt2gene | Not-determined |
| S8_vs_k141_36555\|\|lt2gene | Low-quality |
| S8_vs_k141_175017\|\|lt2gene | Low-quality |
| S8_vs_k141_79803\|\|lt2gene | Low-quality |
| S8_vs_k141_88427\|\|lt2gene | Not-determined |
| S8_vs_k141_123222\|\|lt2gene | Low-quality |
| S8_vs_k141_114418\|\|lt2gene | Not-determined |
| S8_vs_k141_36587\|\|lt2gene | Low-quality |
| S8_vs_k141_105848\|\|lt2gene | Not-determined |
| S8_vs_k141_53871\|\|lt2gene | Low-quality |
| S8_vs_k141_97220\|\|lt2gene | Low-quality |
| S8_vs_k141_27888\|\|lt2gene | Low-quality |
| S8_vs_k141_157721\|\|lt2gene | Low-quality |
| S8_vs_k141_200921\|\|lt2gene | Low-quality |
| S8_vs_k141_114456\|\|lt2gene | Low-quality |
| S8_vs_k141_149038\|\|lt2gene | Low-quality |
| S8_vs_k141_183417\|\|lt2gene | Not-determined |
| S8_vs_k141_114462\|\|lt2gene | Low-quality |
| S8_vs_k141_44899\|\|lt2gene | Low-quality |
| S8_vs_k141_149057\|\|lt2gene | Low-quality |
| S8_vs_k141_10203\|\|lt2gene | Low-quality |
| S8_vs_k141_140621\|\|lt2gene | Low-quality |
| S8_vs_k141_36603\|\|lt2gene | Not-determined |
| S8_vs_k141_140640\|\|lt2gene | Low-quality |
| S8_vs_k141_79883\|\|lt2gene | Not-determined |
| S8_vs_k141_88522\|\|lt2gene | Low-quality |
| S8_vs_k141_27917\|\|lt2gene | Low-quality |
| S8_vs_k141_140675\|\|lt2gene | Low-quality |
| S8_vs_k141_62392\|\|lt2gene | Low-quality |
| S8_vs_k141_166658\|\|lt2gene | Low-quality |
| S8_vs_k141_1857\|\|lt2gene | Low-quality |
| S8_vs_k141_123349\|\|lt2gene | Low-quality |
| S8_vs_k141_27967\|\|lt2gene | Low-quality |
| S8_vs_k141_105910\|\|lt2gene | Not-determined |
| S8_vs_k141_62427\|\|lt2gene | Not-determined |
| S8_vs_k141_36695\|\|lt2gene | Low-quality |
| S8_vs_k141_105917\|\|lt2gene | Not-determined |
| S8_vs_k141_1889\|\|lt2gene | Low-quality |
| S8_vs_k141_114545\|\|lt2gene | Low-quality |
| S8_vs_k141_27990\|\|lt2gene | Low-quality |
| S8_vs_k141_62442\|\|lt2gene | Low-quality |
| S8_vs_k141_131914\|\|lt2gene | Not-determined |
| S8_vs_k141_62447\|\|lt2gene | Not-determined |
| S8_vs_k141_97329\|\|lt2gene | Low-quality |
| S8_vs_k141_28000\|\|lt2gene | Not-determined |
| S8_vs_k141_70825\|\|lt2gene | Low-quality |
| S8_vs_k141_131933\|\|lt2gene | Low-quality |
| S8_vs_k141_10341\|\|lt2gene | Low-quality |
| S8_vs_k141_54046\|\|lt2gene | Low-quality |
| S8_vs_k141_175204\|\|lt2gene | Low-quality |
| S8_vs_k141_54051\|\|lt2gene | Low-quality |
| S8_vs_k141_149236\|\|lt2gene | Low-quality |
| S8_vs_k141_175224\|\|lt2gene | Not-determined |
| S8_vs_k141_36772\|\|lt2gene | Low-quality |
| S8_vs_k141_123415\|\|lt2gene | Low-quality |
| S8_vs_k141_157836\|\|lt2gene | Low-quality |
| S8_vs_k141_192952\|\|lt2gene | Low-quality |
| S8_vs_k141_88624\|\|lt2gene | Low-quality |
| S8_vs_k141_10399\|\|lt2gene | Not-determined |
| S8_vs_k141_10400\|\|lt2gene | Low-quality |
| S8_vs_k141_123438\|\|lt2gene | Low-quality |
| S8_vs_k141_88646\|\|lt2gene | Low-quality |
| S8_vs_k141_131981\|\|lt2gene | Low-quality |
| S8_vs_k141_131983\|\|lt2gene | Not-determined |
| S8_vs_k141_97338\|\|lt2gene | Low-quality |
| S8_vs_k141_140760\|\|lt2gene | Not-determined |
| S8_vs_k141_88656\|\|lt2gene | Low-quality |
| S8_vs_k141_183621\|\|lt2gene | Low-quality |
| S8_vs_k141_183625\|\|lt2gene | Low-quality |
| S8_vs_k141_88684\|\|lt2gene | Low-quality |
| S8_vs_k141_80101\|\|lt2gene | Low-quality |
| S8_vs_k141_149332\|\|lt2gene | Low-quality |
| S8_vs_k141_201108\|\|lt2gene | Low-quality |
| S8_vs_k141_166732\|\|lt2gene | Low-quality |
| S8_vs_k141_193011\|\|lt2gene | Low-quality |
| S8_vs_k141_157915\|\|lt2gene | Low-quality |
| S8_vs_k141_1983\|\|lt2gene | Low-quality |
| S8_vs_k141_166749\|\|lt2gene | Not-determined |
| S8_vs_k141_97379\|\|lt2gene | Not-determined |
| S8_vs_k141_80152\|\|lt2gene | Low-quality |
| S8_vs_k141_123511\|\|lt2gene | Low-quality |
| S8_vs_k141_88739\|\|lt2gene | Not-determined |
| S8_vs_k141_132065\|\|lt2gene | Low-quality |
| S8_vs_k141_62646\|\|lt2gene | Low-quality |
| S8_vs_k141_106102\|\|lt2gene | Low-quality |
| S8_vs_k141_183718\|\|lt2gene | Not-determined |
| S8_vs_k141_62675\|\|lt2gene | Low-quality |
| S8_vs_k141_149394\|\|lt2gene | Low-quality |
| S8_vs_k141_114727\|\|lt2gene | Not-determined |
| S8_vs_k141_158006\|\|lt2gene | Low-quality |
| S8_vs_k141_149403\|\|lt2gene | Low-quality |
| S8_vs_k141_70938\|\|lt2gene | Not-determined |
| S8_vs_k141_70942\|\|lt2gene | Low-quality |
| S8_vs_k141_140870\|\|lt2gene | Low-quality |
| S8_vs_k141_45181\|\|lt2gene | Not-determined |
| S8_vs_k141_193087\|\|lt2gene | Low-quality |
| S8_vs_k141_114770\|\|lt2gene | Not-determined |
| S8_vs_k141_19216\|\|lt2gene | Low-quality |
| S8_vs_k141_114773\|\|lt2gene | Low-quality |
| S8_vs_k141_201251\|\|lt2gene | Not-determined |
| S8_vs_k141_175385\|\|lt2gene | Low-quality |
| S8_vs_k141_88851\|\|lt2gene | Low-quality |
| S8_vs_k141_62754\|\|lt2gene | Low-quality |
| S8_vs_k141_175408\|\|lt2gene | Low-quality |
| S8_vs_k141_149455\|\|lt2gene | Not-determined |
| S8_vs_k141_140924\|\|lt2gene | Low-quality |
| S8_vs_k141_54240\|\|lt2gene | Not-determined |
| S8_vs_k141_193143\|\|lt2gene | Low-quality |
| S8_vs_k141_2129\|\|lt2gene | Not-determined |
| S8_vs_k141_45244\|\|lt2gene | Not-determined |
| S8_vs_k141_19265\|\|lt2gene | Not-determined |
| S8_vs_k141_175421\|\|lt2gene | Low-quality |
| S8_vs_k141_175422\|\|lt2gene | Low-quality |
| S8_vs_k141_175424\|\|lt2gene | Low-quality |
| S8_vs_k141_54283\|\|lt2gene | Low-quality |
| S8_vs_k141_183808\|\|lt2gene | Not-determined |
| S8_vs_k141_45274\|\|lt2gene | Low-quality |
| S8_vs_k141_166871\|\|lt2gene | Low-quality |
| S8_vs_k141_149474\|\|lt2gene | Low-quality |
| S8_vs_k141_71043\|\|lt2gene | Low-quality |
| S8_vs_k141_201331\|\|lt2gene | Low-quality |
| S8_vs_k141_201334\|\|lt2gene | Low-quality |
| S8_vs_k141_114878\|\|lt2gene | Low-quality |
| S8_vs_k141_158107\|\|lt2gene | Not-determined |
| S8_vs_k141_71076\|\|lt2gene | Not-determined |
| S8_vs_k141_88957\|\|lt2gene | Not-determined |
| S8_vs_k141_62832\|\|lt2gene | Low-quality |
| S8_vs_k141_140997\|\|lt2gene | Low-quality |
| S8_vs_k141_54341\|\|lt2gene | Not-determined |
| S8_vs_k141_88983\|\|lt2gene | Low-quality |
| S8_vs_k141_54351\|\|lt2gene | Low-quality |
| S8_vs_k141_37061\|\|lt2gene | Low-quality |
| S8_vs_k141_106304\|\|lt2gene | Low-quality |
| S8_vs_k141_89000\|\|lt2gene | Low-quality |
| S8_vs_k141_45342\|\|lt2gene | Low-quality |
| S8_vs_k141_10557\|\|lt2gene | Low-quality |
| S8_vs_k141_37087\|\|lt2gene | Not-determined |
| S8_vs_k141_201384\|\|lt2gene | Low-quality |
| S8_vs_k141_80260\|\|lt2gene | Low-quality |
| S8_vs_k141_28342\|\|lt2gene | Low-quality |
| S8_vs_k141_106325\|\|lt2gene | Low-quality |
| S8_vs_k141_54411\|\|lt2gene | Low-quality |
| S8_vs_k141_166953\|\|lt2gene | Not-determined |
| S8_vs_k141_2252\|\|lt2gene | Low-quality |
| S8_vs_k141_2259\|\|lt2gene | Low-quality |
| S8_vs_k141_19419\|\|lt2gene | Low-quality |
| S8_vs_k141_183915\|\|lt2gene | Low-quality |
| S8_vs_k141_167002\|\|lt2gene | Not-determined |
| S8_vs_k141_141044\|\|lt2gene | Low-quality |
| S8_vs_k141_80357\|\|lt2gene | Not-determined |
| S8_vs_k141_71210\|\|lt2gene | Not-determined |
| S8_vs_k141_201447\|\|lt2gene | Low-quality |
| S8_vs_k141_106435\|\|lt2gene | Not-determined |
| S8_vs_k141_97654\|\|lt2gene | Low-quality |
| S8_vs_k141_54477\|\|lt2gene | Not-determined |
| S8_vs_k141_106465\|\|lt2gene | Low-quality |
| S8_vs_k141_80412\|\|lt2gene | Low-quality |
| S8_vs_k141_167071\|\|lt2gene | Low-quality |
| S8_vs_k141_37144\|\|lt2gene | Low-quality |
| S8_vs_k141_37157\|\|lt2gene | Low-quality |
| S8_vs_k141_193406\|\|lt2gene | Low-quality |
| S8_vs_k141_201496\|\|lt2gene | Not-determined |
| S8_vs_k141_201509\|\|lt2gene | Low-quality |
| S8_vs_k141_97712\|\|lt2gene | Low-quality |
| S8_vs_k141_175666\|\|lt2gene | Low-quality |
| S8_vs_k141_19545\|\|lt2gene | Low-quality |
| S8_vs_k141_28566\|\|lt2gene | Low-quality |
| S8_vs_k141_97767\|\|lt2gene | Low-quality |
| S8_vs_k141_106557\|\|lt2gene | Low-quality |
| S8_vs_k141_106559\|\|lt2gene | Low-quality |
| S8_vs_k141_71281\|\|lt2gene | Low-quality |
| S8_vs_k141_45463\|\|lt2gene | Low-quality |
| S8_vs_k141_175750\|\|lt2gene | Not-determined |
| S8_vs_k141_201578\|\|lt2gene | Low-quality |
| S8_vs_k141_63150\|\|lt2gene | Low-quality |
| S8_vs_k141_184020\|\|lt2gene | Low-quality |
| S8_vs_k141_132388\|\|lt2gene | Low-quality |
| S8_vs_k141_10797\|\|lt2gene | Low-quality |
| S8_vs_k141_132393\|\|lt2gene | Not-determined |
| S8_vs_k141_175787\|\|lt2gene | Not-determined |
| S8_vs_k141_2478\|\|lt2gene | Low-quality |
| S8_vs_k141_71319\|\|lt2gene | Low-quality |
| S8_vs_k141_141261\|\|lt2gene | Not-determined |
| S8_vs_k141_141276\|\|lt2gene | Low-quality |
| S8_vs_k141_71340\|\|lt2gene | Not-determined |
| S8_vs_k141_19736\|\|lt2gene | Low-quality |
| S8_vs_k141_141295\|\|lt2gene | Low-quality |
| S8_vs_k141_132473\|\|lt2gene | Low-quality |
| S8_vs_k141_115140\|\|lt2gene | Not-determined |
| S8_vs_k141_175874\|\|lt2gene | Low-quality |
| S8_vs_k141_54618\|\|lt2gene | Not-determined |
| S8_vs_k141_37374\|\|lt2gene | Low-quality |
| S8_vs_k141_184118\|\|lt2gene | Low-quality |
| S8_vs_k141_80673\|\|lt2gene | Low-quality |
| S8_vs_k141_89205\|\|lt2gene | Low-quality |
| S8_vs_k141_184142\|\|lt2gene | Low-quality |
| S8_vs_k141_71414\|\|lt2gene | Low-quality |
| S8_vs_k141_193631\|\|lt2gene | Low-quality |
| S8_vs_k141_141353\|\|lt2gene | Low-quality |
| S8_vs_k141_106634\|\|lt2gene | Low-quality |
| S8_vs_k141_71447\|\|lt2gene | Low-quality |
| S8_vs_k141_10953\|\|lt2gene | Not-determined |
| S8_vs_k141_193647\|\|lt2gene | Low-quality |
| S8_vs_k141_97831\|\|lt2gene | Low-quality |
| S8_vs_k141_63338\|\|lt2gene | Low-quality |
| S8_vs_k141_19840\|\|lt2gene | Low-quality |
| S8_vs_k141_97844\|\|lt2gene | Not-determined |
| S8_vs_k141_184197\|\|lt2gene | Not-determined |
| S8_vs_k141_167336\|\|lt2gene | Low-quality |
| S8_vs_k141_19863\|\|lt2gene | Low-quality |
| S8_vs_k141_176009\|\|lt2gene | Low-quality |
| S8_vs_k141_176016\|\|lt2gene | Low-quality |
| S8_vs_k141_184215\|\|lt2gene | Low-quality |
| S8_vs_k141_28902\|\|lt2gene | Low-quality |
| S8_vs_k141_184230\|\|lt2gene | Low-quality |
| S8_vs_k141_2626\|\|lt2gene | Low-quality |
| S8_vs_k141_63378\|\|lt2gene | Low-quality |
| S8_vs_k141_19892\|\|lt2gene | Low-quality |
| S8_vs_k141_80757\|\|lt2gene | Low-quality |
| S8_vs_k141_2665\|\|lt2gene | Low-quality |
| S8_vs_k141_132627\|\|lt2gene | Not-determined |
| S8_vs_k141_176109\|\|lt2gene | Low-quality |
| S8_vs_k141_19918\|\|lt2gene | Low-quality |
| S8_vs_k141_97942\|\|lt2gene | Low-quality |
| S8_vs_k141_167449\|\|lt2gene | Low-quality |
| S8_vs_k141_37465\|\|lt2gene | Low-quality |
| S8_vs_k141_201831\|\|lt2gene | Not-determined |
| S8_vs_k141_167494\|\|lt2gene | Not-determined |
| S8_vs_k141_89389\|\|lt2gene | Not-determined |
| S8_vs_k141_2738\|\|lt2gene | Not-determined |
| S8_vs_k141_89398\|\|lt2gene | Low-quality |
| S8_vs_k141_11085\|\|lt2gene | Not-determined |
| S8_vs_k141_201858\|\|lt2gene | Not-determined |
| S8_vs_k141_124338\|\|lt2gene | Not-determined |
| S8_vs_k141_97994\|\|lt2gene | Low-quality |
| S8_vs_k141_11099\|\|lt2gene | Low-quality |
| S8_vs_k141_89444\|\|lt2gene | Low-quality |
| S8_vs_k141_80878\|\|lt2gene | Low-quality |
| S8_vs_k141_158478\|\|lt2gene | Low-quality |
| S8_vs_k141_20022\|\|lt2gene | Not-determined |
| S8_vs_k141_176213\|\|lt2gene | Not-determined |
| S8_vs_k141_184333\|\|lt2gene | Low-quality |
| S8_vs_k141_89468\|\|lt2gene | Low-quality |
| S8_vs_k141_124373\|\|lt2gene | Low-quality |
| S8_vs_k141_29016\|\|lt2gene | Low-quality |
| S8_vs_k141_124381\|\|lt2gene | Low-quality |
| S8_vs_k141_115406\|\|lt2gene | Not-determined |
| S8_vs_k141_115408\|\|lt2gene | Not-determined |
| S8_vs_k141_201917\|\|lt2gene | Low-quality |
| S8_vs_k141_201921\|\|lt2gene | Low-quality |
| S8_vs_k141_80944\|\|lt2gene | Not-determined |
| S8_vs_k141_132708\|\|lt2gene | Not-determined |
| S8_vs_k141_150094\|\|lt2gene | Not-determined |
| S8_vs_k141_45852\|\|lt2gene | Low-quality |
| S8_vs_k141_106929\|\|lt2gene | Not-determined |
| S8_vs_k141_132743\|\|lt2gene | Not-determined |
| S8_vs_k141_132758\|\|lt2gene | Low-quality |
| S8_vs_k141_184415\|\|lt2gene | Low-quality |
| S8_vs_k141_150129\|\|lt2gene | Low-quality |
| S8_vs_k141_37586\|\|lt2gene | Not-determined |
| S8_vs_k141_71651\|\|lt2gene | Not-determined |
| S8_vs_k141_176326\|\|lt2gene | Low-quality |
| S8_vs_k141_158596\|\|lt2gene | Low-quality |
| S8_vs_k141_193857\|\|lt2gene | Not-determined |
| S8_vs_k141_54914\|\|lt2gene | Low-quality |
| S8_vs_k141_37615\|\|lt2gene | Low-quality |
| S8_vs_k141_81027\|\|lt2gene | Not-determined |
| S8_vs_k141_176358\|\|lt2gene | Low-quality |
| S8_vs_k141_63715\|\|lt2gene | Low-quality |
| S8_vs_k141_89590\|\|lt2gene | Low-quality |
| S8_vs_k141_81075\|\|lt2gene | Low-quality |
| S8_vs_k141_193916\|\|lt2gene | Low-quality |
| S8_vs_k141_20206\|\|lt2gene | Not-determined |
| S8_vs_k141_107036\|\|lt2gene | Low-quality |
| S8_vs_k141_81100\|\|lt2gene | Low-quality |
| S8_vs_k141_202064\|\|lt2gene | Low-quality |
| S8_vs_k141_202068\|\|lt2gene | Low-quality |
| S8_vs_k141_37676\|\|lt2gene | Low-quality |
| S8_vs_k141_141695\|\|lt2gene | Low-quality |
| S8_vs_k141_202095\|\|lt2gene | Low-quality |
| S8_vs_k141_55031\|\|lt2gene | Low-quality |
| S8_vs_k141_37695\|\|lt2gene | Low-quality |
| S8_vs_k141_29099\|\|lt2gene | Low-quality |
| S8_vs_k141_55050\|\|lt2gene | Low-quality |
| S8_vs_k141_46060\|\|lt2gene | Low-quality |
| S8_vs_k141_71747\|\|lt2gene | Low-quality |
| S8_vs_k141_63797\|\|lt2gene | Low-quality |
| S8_vs_k141_71781\|\|lt2gene | Low-quality |
| S8_vs_k141_20324\|\|lt2gene | Low-quality |
| S8_vs_k141_132934\|\|lt2gene | Not-determined |
| S8_vs_k141_71795\|\|lt2gene | Not-determined |
| S8_vs_k141_71799\|\|lt2gene | Low-quality |
| S8_vs_k141_11350\|\|lt2gene | Low-quality |
| S8_vs_k141_158760\|\|lt2gene | Low-quality |
| S8_vs_k141_71806\|\|lt2gene | Low-quality |
| S8_vs_k141_158765\|\|lt2gene | Low-quality |
| S8_vs_k141_29146\|\|lt2gene | Low-quality |
| S8_vs_k141_2946\|\|lt2gene | Low-quality |
| S8_vs_k141_55105\|\|lt2gene | Low-quality |
| S8_vs_k141_107153\|\|lt2gene | Not-determined |
| S8_vs_k141_81155\|\|lt2gene | Low-quality |
| S8_vs_k141_81165\|\|lt2gene | Not-determined |
| S8_vs_k141_29197\|\|lt2gene | Not-determined |
| S8_vs_k141_81173\|\|lt2gene | Low-quality |
| S8_vs_k141_98395\|\|lt2gene | Low-quality |
| S8_vs_k141_11384\|\|lt2gene | Low-quality |
| S8_vs_k141_37816\|\|lt2gene | Low-quality |
| S8_vs_k141_107188\|\|lt2gene | Low-quality |
| S8_vs_k141_107192\|\|lt2gene | Low-quality |
| S8_vs_k141_133036\|\|lt2gene | Low-quality |
| S8_vs_k141_176576\|\|lt2gene | Low-quality |
| S8_vs_k141_37838\|\|lt2gene | Not-determined |

| | |
|---|---|
| SB_vs_k141_71892\|\|lt2gene | Low-quality |
| SB_vs_k141_71894\|\|lt2gene | Low-quality |
| SB_vs_k141_71908\|\|lt2gene | Not-determined |
| SB_vs_k141_133063\|\|lt2gene | Low-quality |
| SB_vs_k141_11439\|\|lt2gene | Low-quality |
| SB_vs_k141_124688\|\|lt2gene | Low-quality |
| SB_vs_k141_63887\|\|lt2gene | Low-quality |
| SB_vs_k141_89801\|\|lt2gene | Low-quality |
| SB_vs_k141_3037\|\|lt2gene | Not-determined |
| SB_vs_k141_3043\|\|lt2gene | Low-quality |
| SB_vs_k141_184594\|\|lt2gene | Low-quality |
| SB_vs_k141_115794\|\|lt2gene | Low-quality |
| SB_vs_k141_3056\|\|lt2gene | Not-determined |
| SB_vs_k141_98496\|\|lt2gene | Not-determined |
| SB_vs_k141_167775\|\|lt2gene | Low-quality |
| SB_vs_k141_150353\|\|lt2gene | Low-quality |
| SB_vs_k141_194082\|\|lt2gene | Low-quality |
| SB_vs_k141_89827\|\|lt2gene | Not-determined |
| SB_vs_k141_29311\|\|lt2gene | Low-quality |
| SB_vs_k141_194089\|\|lt2gene | Low-quality |
| SB_vs_k141_141935\|\|lt2gene | Low-quality |
| SB_vs_k141_115840\|\|lt2gene | Not-determined |
| SB_vs_k141_63950\|\|lt2gene | Low-quality |
| SB_vs_k141_184670\|\|lt2gene | Low-quality |
| SB_vs_k141_20482\|\|lt2gene | Low-quality |
| SB_vs_k141_20485\|\|lt2gene | Not-determined |
| SB_vs_k141_133178\|\|lt2gene | Not-determined |
| SB_vs_k141_133201\|\|lt2gene | Low-quality |
| SB_vs_k141_158962\|\|lt2gene | Low-quality |
| SB_vs_k141_20504\|\|lt2gene | Low-quality |
| SB_vs_k141_158969\|\|lt2gene | Not-determined |
| SB_vs_k141_133209\|\|lt2gene | Low-quality |
| SB_vs_k141_184710\|\|lt2gene | Low-quality |
| SB_vs_k141_202364\|\|lt2gene | Low-quality |
| SB_vs_k141_11570\|\|lt2gene | Not-determined |
| SB_vs_k141_124806\|\|lt2gene | Low-quality |
| SB_vs_k141_107383\|\|lt2gene | Not-determined |
| SB_vs_k141_72053\|\|lt2gene | Low-quality |
| SB_vs_k141_142022\|\|lt2gene | Low-quality |
| SB_vs_k141_133244\|\|lt2gene | Not-determined |
| SB_vs_k141_11603\|\|lt2gene | Low-quality |
| SB_vs_k141_98624\|\|lt2gene | Low-quality |
| SB_vs_k141_202390\|\|lt2gene | Low-quality |
| SB_vs_k141_38007\|\|lt2gene | Low-quality |
| SB_vs_k141_142032\|\|lt2gene | Not-determined |
| SB_vs_k141_64031\|\|lt2gene | Low-quality |
| SB_vs_k141_64036\|\|lt2gene | Low-quality |
| SB_vs_k141_159037\|\|lt2gene | Low-quality |
| SB_vs_k141_142046\|\|lt2gene | Not-determined |
| SB_vs_k141_107404\|\|lt2gene | Low-quality |
| SB_vs_k141_29466\|\|lt2gene | Low-quality |
| SB_vs_k141_20579\|\|lt2gene | Low-quality |
| SB_vs_k141_150551\|\|lt2gene | Low-quality |
| SB_vs_k141_202429\|\|lt2gene | Low-quality |
| SB_vs_k141_11646\|\|lt2gene | Low-quality |
| SB_vs_k141_202435\|\|lt2gene | Low-quality |
| SB_vs_k141_89955\|\|lt2gene | Low-quality |
| SB_vs_k141_11649\|\|lt2gene | Not-determined |
| SB_vs_k141_64072\|\|lt2gene | Low-quality |
| SB_vs_k141_98637\|\|lt2gene | Not-determined |
| SB_vs_k141_202450\|\|lt2gene | Low-quality |
| SB_vs_k141_168035\|\|lt2gene | Low-quality |
| SB_vs_k141_150567\|\|lt2gene | Low-quality |
| SB_vs_k141_124860\|\|lt2gene | Not-determined |
| SB_vs_k141_202474\|\|lt2gene | Low-quality |
| SB_vs_k141_194254\|\|lt2gene | Low-quality |
| SB_vs_k141_20607\|\|lt2gene | Low-quality |
| SB_vs_k141_64124\|\|lt2gene | Not-determined |
| SB_vs_k141_64137\|\|lt2gene | Low-quality |
| SB_vs_k141_64147\|\|lt2gene | Not-determined |
| SB_vs_k141_194279\|\|lt2gene | Low-quality |
| SB_vs_k141_168113\|\|lt2gene | Low-quality |
| SB_vs_k141_29556\|\|lt2gene | Low-quality |
| SB_vs_k141_20673\|\|lt2gene | Not-determined |
| SB_vs_k141_38165\|\|lt2gene | Not-determined |
| SB_vs_k141_55413\|\|lt2gene | Low-quality |
| SB_vs_k141_115959\|\|lt2gene | Low-quality |
| SB_vs_k141_38192\|\|lt2gene | Not-determined |
| SB_vs_k141_159205\|\|lt2gene | Low-quality |
| SB_vs_k141_142154\|\|lt2gene | Low-quality |
| SB_vs_k141_125000\|\|lt2gene | Low-quality |
| SB_vs_k141_3309\|\|lt2gene | Low-quality |
| SB_vs_k141_38227\|\|lt2gene | Not-determined |
| SB_vs_k141_133365\|\|lt2gene | Not-determined |
| SB_vs_k141_29636\|\|lt2gene | Low-quality |
| SB_vs_k141_81508\|\|lt2gene | Low-quality |
| SB_vs_k141_20769\|\|lt2gene | Not-determined |
| SB_vs_k141_116009\|\|lt2gene | Low-quality |
| SB_vs_k141_3355\|\|lt2gene | Not-determined |
| SB_vs_k141_3358\|\|lt2gene | Not-determined |
| SB_vs_k141_133385\|\|lt2gene | Low-quality |
| SB_vs_k141_3371\|\|lt2gene | Low-quality |
| SB_vs_k141_116034\|\|lt2gene | Not-determined |
| SB_vs_k141_185016\|\|lt2gene | Not-determined |
| SB_vs_k141_159291\|\|lt2gene | Low-quality |
| SB_vs_k141_107619\|\|lt2gene | Low-quality |
| SB_vs_k141_168145\|\|lt2gene | Low-quality |
| SB_vs_k141_90102\|\|lt2gene | Low-quality |
| SB_vs_k141_90103\|\|lt2gene | Low-quality |
| SB_vs_k141_38275\|\|lt2gene | Low-quality |
| SB_vs_k141_133435\|\|lt2gene | Not-determined |
| SB_vs_k141_55496\|\|lt2gene | Low-quality |
| SB_vs_k141_116074\|\|lt2gene | Low-quality |
| SB_vs_k141_3418\|\|lt2gene | Low-quality |
| SB_vs_k141_168174\|\|lt2gene | Low-quality |
| SB_vs_k141_133454\|\|lt2gene | Low-quality |
| SB_vs_k141_38290\|\|lt2gene | Not-determined |
| SB_vs_k141_133461\|\|lt2gene | Not-determined |
| SB_vs_k141_90148\|\|lt2gene | Low-quality |
| SB_vs_k141_81581\|\|lt2gene | Low-quality |
| SB_vs_k141_29747\|\|lt2gene | Not-determined |
| SB_vs_k141_159352\|\|lt2gene | Not-determined |
| SB_vs_k141_29760\|\|lt2gene | Low-quality |
| SB_vs_k141_29784\|\|lt2gene | Low-quality |
| SB_vs_k141_72337\|\|lt2gene | Low-quality |
| SB_vs_k141_98847\|\|lt2gene | Not-determined |
| SB_vs_k141_11798\|\|lt2gene | Low-quality |
| SB_vs_k141_20906\|\|lt2gene | Low-quality |
| SB_vs_k141_168250\|\|lt2gene | Not-determined |
| SB_vs_k141_194528\|\|lt2gene | Low-quality |
| SB_vs_k141_150718\|\|lt2gene | Low-quality |
| SB_vs_k141_150726\|\|lt2gene | Not-determined |
| SB_vs_k141_107741\|\|lt2gene | Low-quality |
| SB_vs_k141_20929\|\|lt2gene | Low-quality |
| SB_vs_k141_107759\|\|lt2gene | Not-determined |
| SB_vs_k141_125080\|\|lt2gene | Not-determined |
| SB_vs_k141_98923\|\|lt2gene | Low-quality |
| SB_vs_k141_202735\|\|lt2gene | Low-quality |
| SB_vs_k141_81657\|\|lt2gene | Low-quality |
| SB_vs_k141_116192\|\|lt2gene | Not-determined |
| SB_vs_k141_116200\|\|lt2gene | Low-quality |
| SB_vs_k141_55652\|\|lt2gene | Low-quality |
| SB_vs_k141_202761\|\|lt2gene | Not-determined |
| SB_vs_k141_177117\|\|lt2gene | Low-quality |
| SB_vs_k141_116236\|\|lt2gene | Low-quality |
| SB_vs_k141_98990\|\|lt2gene | Low-quality |
| SB_vs_k141_125169\|\|lt2gene | Low-quality |
| SB_vs_k141_98995\|\|lt2gene | Low-quality |
| SB_vs_k141_116249\|\|lt2gene | Low-quality |
| SB_vs_k141_107844\|\|lt2gene | Low-quality |
| SB_vs_k141_55690\|\|lt2gene | Low-quality |
| SB_vs_k141_150817\|\|lt2gene | Low-quality |
| SB_vs_k141_150818\|\|lt2gene | Low-quality |
| SB_vs_k141_3675\|\|lt2gene | Not-determined |
| SB_vs_k141_90327\|\|lt2gene | Not-determined |
| SB_vs_k141_202804\|\|lt2gene | Not-determined |
| SB_vs_k141_72517\|\|lt2gene | Not-determined |
| SB_vs_k141_116300\|\|lt2gene | Low-quality |
| SB_vs_k141_99062\|\|lt2gene | Not-determined |
| SB_vs_k141_185186\|\|lt2gene | Not-determined |
| SB_vs_k141_3707\|\|lt2gene | Not-determined |
| SB_vs_k141_116334\|\|lt2gene | Not-determined |
| SB_vs_k141_177173\|\|lt2gene | Low-quality |
| SB_vs_k141_72578\|\|lt2gene | Low-quality |
| SB_vs_k141_202857\|\|lt2gene | Low-quality |
| SB_vs_k141_159621\|\|lt2gene | Low-quality |
| SB_vs_k141_46733\|\|lt2gene | Not-determined |
| SB_vs_k141_99128\|\|lt2gene | Low-quality |
| SB_vs_k141_3762\|\|lt2gene | Low-quality |
| SB_vs_k141_168511\|\|lt2gene | Not-determined |
| SB_vs_k141_64218\|\|lt2gene | Not-determined |
| SB_vs_k141_194759\|\|lt2gene | Low-quality |
| SB_vs_k141_55818\|\|lt2gene | Not-determined |
| SB_vs_k141_55822\|\|lt2gene | Not-determined |
| SB_vs_k141_150933\|\|lt2gene | Low-quality |
| SB_vs_k141_11940\|\|lt2gene | Low-quality |
| SB_vs_k141_90452\|\|lt2gene | Low-quality |
| SB_vs_k141_64268\|\|lt2gene | Low-quality |
| SB_vs_k141_21103\|\|lt2gene | Low-quality |
| SB_vs_k141_116446\|\|lt2gene | Low-quality |
| SB_vs_k141_177289\|\|lt2gene | Not-determined |
| SB_vs_k141_30091\|\|lt2gene | Low-quality |
| SB_vs_k141_55867\|\|lt2gene | Not-determined |
| SB_vs_k141_116454\|\|lt2gene | Not-determined |
| SB_vs_k141_11951\|\|lt2gene | Low-quality |
| SB_vs_k141_99209\|\|lt2gene | Low-quality |
| SB_vs_k141_150968\|\|lt2gene | Low-quality |
| SB_vs_k141_125342\|\|lt2gene | Low-quality |
| SB_vs_k141_11967\|\|lt2gene | Low-quality |
| SB_vs_k141_168579\|\|lt2gene | Low-quality |
| SB_vs_k141_55890\|\|lt2gene | Low-quality |
| SB_vs_k141_72734\|\|lt2gene | Low-quality |
| SB_vs_k141_168588\|\|lt2gene | Low-quality |
| SB_vs_k141_194796\|\|lt2gene | Low-quality |
| SB_vs_k141_81775\|\|lt2gene | Low-quality |
| SB_vs_k141_81784\|\|lt2gene | Not-determined |
| SB_vs_k141_30167\|\|lt2gene | Not-determined |
| SB_vs_k141_12009\|\|lt2gene | Low-quality |
| SB_vs_k141_99265\|\|lt2gene | Low-quality |
| SB_vs_k141_108063\|\|lt2gene | Low-quality |
| SB_vs_k141_99273\|\|lt2gene | Not-determined |
| SB_vs_k141_30211\|\|lt2gene | Low-quality |
| SB_vs_k141_99312\|\|lt2gene | Low-quality |
| SB_vs_k141_108107\|\|lt2gene | Low-quality |
| SB_vs_k141_133760\|\|lt2gene | Low-quality |
| SB_vs_k141_125465\|\|lt2gene | Not-determined |
| SB_vs_k141_3914\|\|lt2gene | Not-determined |
| SB_vs_k141_38490\|\|lt2gene | Low-quality |
| SB_vs_k141_159724\|\|lt2gene | Low-quality |
| SB_vs_k141_142729\|\|lt2gene | Low-quality |
| SB_vs_k141_90588\|\|lt2gene | Low-quality |
| SB_vs_k141_142748\|\|lt2gene | Low-quality |
| SB_vs_k141_90602\|\|lt2gene | Low-quality |
| SB_vs_k141_12103\|\|lt2gene | Low-quality |
| SB_vs_k141_185375\|\|lt2gene | Low-quality |
| SB_vs_k141_194877\|\|lt2gene | Not-determined |
| SB_vs_k141_168716\|\|lt2gene | Low-quality |
| SB_vs_k141_159773\|\|lt2gene | Not-determined |
| SB_vs_k141_12137\|\|lt2gene | Low-quality |
| SB_vs_k141_21236\|\|lt2gene | Not-determined |
| SB_vs_k141_38539\|\|lt2gene | Low-quality |
| SB_vs_k141_21245\|\|lt2gene | Low-quality |
| SB_vs_k141_64409\|\|lt2gene | Low-quality |
| SB_vs_k141_21251\|\|lt2gene | Not-determined |
| SB_vs_k141_4018\|\|lt2gene | Low-quality |
| SB_vs_k141_4020\|\|lt2gene | Low-quality |
| SB_vs_k141_133902\|\|lt2gene | Low-quality |
| SB_vs_k141_21274\|\|lt2gene | Low-quality |
| SB_vs_k141_133906\|\|lt2gene | Low-quality |
| SB_vs_k141_142835\|\|lt2gene | Low-quality |
| SB_vs_k141_185487\|\|lt2gene | Low-quality |
| SB_vs_k141_72880\|\|lt2gene | Low-quality |
| SB_vs_k141_142858\|\|lt2gene | Low-quality |
| SB_vs_k141_72883\|\|lt2gene | Low-quality |
| SB_vs_k141_203152\|\|lt2gene | Low-quality |
| SB_vs_k141_72888\|\|lt2gene | Low-quality |
| SB_vs_k141_90775\|\|lt2gene | Low-quality |
| SB_vs_k141_159860\|\|lt2gene | Low-quality |
| SB_vs_k141_21306\|\|lt2gene | Not-determined |
| SB_vs_k141_56122\|\|lt2gene | Low-quality |
| SB_vs_k141_151208\|\|lt2gene | Low-quality |
| SB_vs_k141_195028\|\|lt2gene | Low-quality |
| SB_vs_k141_177545\|\|lt2gene | Not-determined |
| SB_vs_k141_30429\|\|lt2gene | Low-quality |
| SB_vs_k141_168801\|\|lt2gene | Low-quality |
| SB_vs_k141_99452\|\|lt2gene | Low-quality |
| SB_vs_k141_72933\|\|lt2gene | Low-quality |
| SB_vs_k141_81977\|\|lt2gene | Low-quality |
| SB_vs_k141_108288\|\|lt2gene | Low-quality |
| SB_vs_k141_12255\|\|lt2gene | Low-quality |
| SB_vs_k141_47023\|\|lt2gene | Low-quality |
| SB_vs_k141_30484\|\|lt2gene | Not-determined |
| SB_vs_k141_99511\|\|lt2gene | Not-determined |
| SB_vs_k141_116664\|\|lt2gene | Low-quality |
| SB_vs_k141_195087\|\|lt2gene | Low-quality |
| SB_vs_k141_151332\|\|lt2gene | Low-quality |
| SB_vs_k141_30502\|\|lt2gene | Low-quality |
| SB_vs_k141_99543\|\|lt2gene | Low-quality |
| SB_vs_k141_108325\|\|lt2gene | Low-quality |
| SB_vs_k141_38756\|\|lt2gene | Low-quality |
| SB_vs_k141_134057\|\|lt2gene | Low-quality |
| SB_vs_k141_12378\|\|lt2gene | Not-determined |
| SB_vs_k141_47092\|\|lt2gene | Low-quality |
| SB_vs_k141_203287\|\|lt2gene | Not-determined |
| SB_vs_k141_12392\|\|lt2gene | Low-quality |
| SB_vs_k141_30544\|\|lt2gene | Low-quality |
| SB_vs_k141_90939\|\|lt2gene | Low-quality |
| SB_vs_k141_203294\|\|lt2gene | Low-quality |
| SB_vs_k141_90945\|\|lt2gene | Low-quality |
| SB_vs_k141_177722\|\|lt2gene | Not-determined |
| SB_vs_k141_56269\|\|lt2gene | Not-determined |
| SB_vs_k141_108364\|\|lt2gene | Low-quality |
| SB_vs_k141_90985\|\|lt2gene | Not-determined |
| SB_vs_k141_116737\|\|lt2gene | Not-determined |
| SB_vs_k141_73026\|\|lt2gene | Low-quality |
| SB_vs_k141_21491\|\|lt2gene | Low-quality |
| SB_vs_k141_56314\|\|lt2gene | Low-quality |
| SB_vs_k141_168947\|\|lt2gene | Low-quality |
| SB_vs_k141_73056\|\|lt2gene | Low-quality |
| SB_vs_k141_134138\|\|lt2gene | Low-quality |
| SB_vs_k141_12496\|\|lt2gene | Low-quality |
| SB_vs_k141_64619\|\|lt2gene | Low-quality |
| SB_vs_k141_12511\|\|lt2gene | Not-determined |
| SB_vs_k141_160073\|\|lt2gene | Low-quality |
| SB_vs_k141_185633\|\|lt2gene | Low-quality |
| SB_vs_k141_143063\|\|lt2gene | Low-quality |
| SB_vs_k141_177801\|\|lt2gene | Low-quality |
| SB_vs_k141_151412\|\|lt2gene | Low-quality |
| SB_vs_k141_56380\|\|lt2gene | Low-quality |
| SB_vs_k141_38842\|\|lt2gene | Low-quality |
| SB_vs_k141_195248\|\|lt2gene | Low-quality |
| SB_vs_k141_64677\|\|lt2gene | Low-quality |
| SB_vs_k141_47209\|\|lt2gene | Low-quality |
| SB_vs_k141_134191\|\|lt2gene | Not-determined |
| SB_vs_k141_143110\|\|lt2gene | Low-quality |
| SB_vs_k141_99696\|\|lt2gene | Low-quality |
| SB_vs_k141_21627\|\|lt2gene | Low-quality |
| SB_vs_k141_108516\|\|lt2gene | Low-quality |
| SB_vs_k141_203399\|\|lt2gene | Low-quality |
| SB_vs_k141_177879\|\|lt2gene | Not-determined |
| SB_vs_k141_203402\|\|lt2gene | Low-quality |
| SB_vs_k141_30569\|\|lt2gene | Low-quality |
| SB_vs_k141_169055\|\|lt2gene | Low-quality |
| SB_vs_k141_185715\|\|lt2gene | Low-quality |
| SB_vs_k141_125862\|\|lt2gene | Low-quality |
| SB_vs_k141_169075\|\|lt2gene | Low-quality |
| SB_vs_k141_56476\|\|lt2gene | Low-quality |
| SB_vs_k141_203434\|\|lt2gene | Low-quality |
| SB_vs_k141_143173\|\|lt2gene | Not-determined |
| SB_vs_k141_116862\|\|lt2gene | Low-quality |
| SB_vs_k141_30593\|\|lt2gene | Low-quality |
| SB_vs_k141_195347\|\|lt2gene | Low-quality |
| SB_vs_k141_21677\|\|lt2gene | Low-quality |
| SB_vs_k141_143195\|\|lt2gene | Not-determined |
| SB_vs_k141_82346\|\|lt2gene | Low-quality |
| SB_vs_k141_4339\|\|lt2gene | Low-quality |
| SB_vs_k141_143203\|\|lt2gene | Low-quality |
| SB_vs_k141_143221\|\|lt2gene | Low-quality |
| SB_vs_k141_82360\|\|lt2gene | Low-quality |
| SB_vs_k141_4351\|\|lt2gene | Low-quality |
| SB_vs_k141_21700\|\|lt2gene | Low-quality |
| SB_vs_k141_143239\|\|lt2gene | Low-quality |
| SB_vs_k141_4364\|\|lt2gene | Low-quality |
| SB_vs_k141_12656\|\|lt2gene | Low-quality |
| SB_vs_k141_185785\|\|lt2gene | Low-quality |
| SB_vs_k141_151592\|\|lt2gene | Low-quality |
| SB_vs_k141_108601\|\|lt2gene | Low-quality |
| SB_vs_k141_143256\|\|lt2gene | Low-quality |
| SB_vs_k141_73301\|\|lt2gene | Not-determined |
| SB_vs_k141_4413\|\|lt2gene | Low-quality |
| SB_vs_k141_4419\|\|lt2gene | Low-quality |
| SB_vs_k141_143286\|\|lt2gene | Low-quality |
| SB_vs_k141_185798\|\|lt2gene | Not-determined |
| SB_vs_k141_126010\|\|lt2gene | Low-quality |
| SB_vs_k141_21776\|\|lt2gene | Low-quality |
| SB_vs_k141_47284\|\|lt2gene | Low-quality |
| SB_vs_k141_195439\|\|lt2gene | Not-determined |
| SB_vs_k141_4455\|\|lt2gene | Not-determined |
| SB_vs_k141_73339\|\|lt2gene | Low-quality |
| SB_vs_k141_134382\|\|lt2gene | Low-quality |
| SB_vs_k141_39068\|\|lt2gene | Low-quality |
| SB_vs_k141_195462\|\|lt2gene | Low-quality |
| SB_vs_k141_108689\|\|lt2gene | Low-quality |
| SB_vs_k141_160235\|\|lt2gene | Not-determined |
| SB_vs_k141_39091\|\|lt2gene | Low-quality |
| SB_vs_k141_91147\|\|lt2gene | Low-quality |
| SB_vs_k141_73420\|\|lt2gene | Low-quality |
| SB_vs_k141_160283\|\|lt2gene | Low-quality |
| SB_vs_k141_39131\|\|lt2gene | Low-quality |
| SB_vs_k141_185838\|\|lt2gene | Low-quality |
| SB_vs_k141_151723\|\|lt2gene | Low-quality |
| SB_vs_k141_169328\|\|lt2gene | Low-quality |
| SB_vs_k141_185859\|\|lt2gene | Low-quality |
| SB_vs_k141_30820\|\|lt2gene | Low-quality |
| SB_vs_k141_39147\|\|lt2gene | Not-determined |
| SB_vs_k141_82525\|\|lt2gene | Low-quality |
| SB_vs_k141_151772\|\|lt2gene | Low-quality |
| SB_vs_k141_64955\|\|lt2gene | Low-quality |
| SB_vs_k141_178128\|\|lt2gene | Low-quality |
| SB_vs_k141_203699\|\|lt2gene | Low-quality |
| SB_vs_k141_203715\|\|lt2gene | Low-quality |
| SB_vs_k141_178156\|\|lt2gene | Low-quality |
| SB_vs_k141_185903\|\|lt2gene | Low-quality |
| SB_vs_k141_195572\|\|lt2gene | Not-determined |
| SB_vs_k141_30867\|\|lt2gene | Low-quality |
| SB_vs_k141_21974\|\|lt2gene | Low-quality |
| SB_vs_k141_12861\|\|lt2gene | Low-quality |
| SB_vs_k141_39208\|\|lt2gene | Low-quality |
| SB_vs_k141_151847\|\|lt2gene | Low-quality |
| SB_vs_k141_178235\|\|lt2gene | Low-quality |
| SB_vs_k141_117230\|\|lt2gene | Low-quality |
| SB_vs_k141_39265\|\|lt2gene | Low-quality |
| SB_vs_k141_143412\|\|lt2gene | Low-quality |
| SB_vs_k141_108842\|\|lt2gene | Low-quality |
| SB_vs_k141_65110\|\|lt2gene | Low-quality |
| SB_vs_k141_134560\|\|lt2gene | Low-quality |
| SB_vs_k141_169460\|\|lt2gene | Low-quality |
| SB_vs_k141_186044\|\|lt2gene | Low-quality |
| SB_vs_k141_56830\|\|lt2gene | Low-quality |
| SB_vs_k141_22094\|\|lt2gene | Not-determined |
| SB_vs_k141_126169\|\|lt2gene | Low-quality |
| SB_vs_k141_12947\|\|lt2gene | Low-quality |
| SB_vs_k141_100087\|\|lt2gene | Low-quality |
| SB_vs_k141_56848\|\|lt2gene | Low-quality |
| SB_vs_k141_151930\|\|lt2gene | Low-quality |
| SB_vs_k141_143535\|\|lt2gene | Low-quality |
| SB_vs_k141_151948\|\|lt2gene | Low-quality |
| SB_vs_k141_143549\|\|lt2gene | Low-quality |
| SB_vs_k141_195701\|\|lt2gene | Low-quality |
| SB_vs_k141_12991\|\|lt2gene | Not-determined |
| SB_vs_k141_203935\|\|lt2gene | Low-quality |
| SB_vs_k141_31029\|\|lt2gene | Low-quality |
| SB_vs_k141_203946\|\|lt2gene | Low-quality |
| SB_vs_k141_203949\|\|lt2gene | Not-determined |
| SB_vs_k141_22212\|\|lt2gene | Low-quality |
| SB_vs_k141_134660\|\|lt2gene | Not-determined |
| SB_vs_k141_108898\|\|lt2gene | Low-quality |
| SB_vs_k141_65210\|\|lt2gene | Low-quality |
| SB_vs_k141_186087\|\|lt2gene | Not-determined |
| SB_vs_k141_4612\|\|lt2gene | Low-quality |
| SB_vs_k141_4614\|\|lt2gene | Not-determined |
| SB_vs_k141_47693\|\|lt2gene | Low-quality |
| SB_vs_k141_169589\|\|lt2gene | Low-quality |
| SB_vs_k141_73709\|\|lt2gene | Low-quality |
| SB_vs_k141_22247\|\|lt2gene | Low-quality |
| SB_vs_k141_134697\|\|lt2gene | Low-quality |
| SB_vs_k141_152065\|\|lt2gene | Low-quality |
| SB_vs_k141_117428\|\|lt2gene | Low-quality |
| SB_vs_k141_134707\|\|lt2gene | Low-quality |
| SB_vs_k141_169610\|\|lt2gene | Low-quality |
| SB_vs_k141_117432\|\|lt2gene | Not-determined |
| SB_vs_k141_143635\|\|lt2gene | Not-determined |
| SB_vs_k141_143636\|\|lt2gene | Not-determined |
| SB_vs_k141_160495\|\|lt2gene | Low-quality |
| SB_vs_k141_31103\|\|lt2gene | Low-quality |
| SB_vs_k141_22280\|\|lt2gene | Not-determined |
| SB_vs_k141_91463\|\|lt2gene | Low-quality |
| SB_vs_k141_108954\|\|lt2gene | Low-quality |
| SB_vs_k141_195778\|\|lt2gene | Low-quality |
| SB_vs_k141_22291\|\|lt2gene | Low-quality |
| SB_vs_k141_47752\|\|lt2gene | Low-quality |
| SB_vs_k141_152097\|\|lt2gene | Low-quality |
| SB_vs_k141_160528\|\|lt2gene | Not-determined |
| SB_vs_k141_100270\|\|lt2gene | Low-quality |
| SB_vs_k141_56994\|\|lt2gene | Not-determined |
| SB_vs_k141_143710\|\|lt2gene | Not-determined |
| SB_vs_k141_143711\|\|lt2gene | Low-quality |
| SB_vs_k141_57049\|\|lt2gene | Low-quality |
| SB_vs_k141_152169\|\|lt2gene | Low-quality |
| SB_vs_k141_65400\|\|lt2gene | Low-quality |
| SB_vs_k141_13190\|\|lt2gene | Low-quality |
| SB_vs_k141_117597\|\|lt2gene | Low-quality |
| SB_vs_k141_143818\|\|lt2gene | Low-quality |
| SB_vs_k141_91569\|\|lt2gene | Not-determined |
| SB_vs_k141_13208\|\|lt2gene | Low-quality |
| SB_vs_k141_13210\|\|lt2gene | Low-quality |
| SB_vs_k141_195943\|\|lt2gene | Low-quality |
| SB_vs_k141_57136\|\|lt2gene | Low-quality |
| SB_vs_k141_169765\|\|lt2gene | Low-quality |
| SB_vs_k141_117640\|\|lt2gene | Not-determined |
| SB_vs_k141_4810\|\|lt2gene | Low-quality |
| SB_vs_k141_126422\|\|lt2gene | Low-quality |
| SB_vs_k141_152249\|\|lt2gene | Low-quality |
| SB_vs_k141_13245\|\|lt2gene | Low-quality |
| SB_vs_k141_4860\|\|lt2gene | Low-quality |
| SB_vs_k141_152275\|\|lt2gene | Not-determined |
| SB_vs_k141_186205\|\|lt2gene | Low-quality |
| SB_vs_k141_126470\|\|lt2gene | Not-determined |
| SB_vs_k141_169872\|\|lt2gene | Low-quality |
| SB_vs_k141_73988\|\|lt2gene | Not-determined |
| SB_vs_k141_178553\|\|lt2gene | Low-quality |
| SB_vs_k141_178556\|\|lt2gene | Low-quality |
| SB_vs_k141_178560\|\|lt2gene | Low-quality |
| SB_vs_k141_143954\|\|lt2gene | Low-quality |
| SB_vs_k141_126495\|\|lt2gene | Low-quality |
| SB_vs_k141_186222\|\|lt2gene | Not-determined |
| SB_vs_k141_178578\|\|lt2gene | Not-determined |
| SB_vs_k141_178613\|\|lt2gene | Low-quality |
| SB_vs_k141_22593\|\|lt2gene | Low-quality |
| SB_vs_k141_91783\|\|lt2gene | Low-quality |
| SB_vs_k141_47936\|\|lt2gene | Low-quality |
| SB_vs_k141_47948\|\|lt2gene | Low-quality |
| SB_vs_k141_22609\|\|lt2gene | Not-determined |
| SB_vs_k141_178633\|\|lt2gene | Low-quality |
| SB_vs_k141_73991\|\|lt2gene | Low-quality |
| SB_vs_k141_22626\|\|lt2gene | Not-determined |
| SB_vs_k141_22630\|\|lt2gene | Not-determined |
| SB_vs_k141_196076\|\|lt2gene | Low-quality |
| SB_vs_k141_134950\|\|lt2gene | Low-quality |
| SB_vs_k141_65602\|\|lt2gene | Low-quality |
| SB_vs_k141_57361\|\|lt2gene | Low-quality |
| SB_vs_k141_144090\|\|lt2gene | Low-quality |
| SB_vs_k141_109308\|\|lt2gene | Not-determined |
| SB_vs_k141_22695\|\|lt2gene | Low-quality |
| SB_vs_k141_39823\|\|lt2gene | Not-determined |
| SB_vs_k141_48029\|\|lt2gene | Not-determined |
| SB_vs_k141_39843\|\|lt2gene | Low-quality |
| SB_vs_k141_196148\|\|lt2gene | Not-determined |
| SB_vs_k141_196152\|\|lt2gene | Not-determined |
| SB_vs_k141_22732\|\|lt2gene | Low-quality |
| SB_vs_k141_22739\|\|lt2gene | Low-quality |
| SB_vs_k141_160772\|\|lt2gene | Low-quality |
| SB_vs_k141_48059\|\|lt2gene | Low-quality |
| SB_vs_k141_48074\|\|lt2gene | Low-quality |
| SB_vs_k141_186472\|\|lt2gene | Low-quality |
| SB_vs_k141_5038\|\|lt2gene | Low-quality |
| SB_vs_k141_186486\|\|lt2gene | Low-quality |
| SB_vs_k141_170097\|\|lt2gene | Not-determined |
| SB_vs_k141_170104\|\|lt2gene | Low-quality |
| SB_vs_k141_40006\|\|lt2gene | Not-determined |
| SB_vs_k141_48145\|\|lt2gene | Low-quality |
| SB_vs_k141_178776\|\|lt2gene | Not-determined |
| SB_vs_k141_170162\|\|lt2gene | Low-quality |
| SB_vs_k141_135072\|\|lt2gene | Not-determined |
| SB_vs_k141_204452\|\|lt2gene | Not-determined |
| SB_vs_k141_126596\|\|lt2gene | Low-quality |
| SB_vs_k141_135095\|\|lt2gene | Not-determined |
| SB_vs_k141_65760\|\|lt2gene | Low-quality |
| SB_vs_k141_196248\|\|lt2gene | Not-determined |
| SB_vs_k141_178817\|\|lt2gene | Low-quality |
| SB_vs_k141_135123\|\|lt2gene | Not-determined |
| SB_vs_k141_135133\|\|lt2gene | Low-quality |
| SB_vs_k141_135136\|\|lt2gene | Low-quality |
| SB_vs_k141_5106\|\|lt2gene | Low-quality |
| SB_vs_k141_135139\|\|lt2gene | Low-quality |
| SB_vs_k141_22908\|\|lt2gene | Low-quality |
| SB_vs_k141_48223\|\|lt2gene | Low-quality |
| SB_vs_k141_178852\|\|lt2gene | Low-quality |
| SB_vs_k141_40051\|\|lt2gene | Not-determined |
| SB_vs_k141_109492\|\|lt2gene | Low-quality |
| SB_vs_k141_126616\|\|lt2gene | Low-quality |
| SB_vs_k141_5131\|\|lt2gene | Low-quality |
| SB_vs_k141_170257\|\|lt2gene | Low-quality |
| SB_vs_k141_126659\|\|lt2gene | Not-determined |
| SB_vs_k141_118012\|\|lt2gene | Not-determined |
| SB_vs_k141_170281\|\|lt2gene | Not-determined |
| SB_vs_k141_5197\|\|lt2gene | Low-quality |
| SB_vs_k141_109565\|\|lt2gene | Low-quality |
| SB_vs_k141_178935\|\|lt2gene | Low-quality |
| SB_vs_k141_92058\|\|lt2gene | Low-quality |
| SB_vs_k141_92060\|\|lt2gene | Low-quality |
| SB_vs_k141_92065\|\|lt2gene | Not-determined |
| SB_vs_k141_83365\|\|lt2gene | Low-quality |
| SB_vs_k141_178955\|\|lt2gene | Low-quality |
| SB_vs_k141_118051\|\|lt2gene | Low-quality |
| SB_vs_k141_74261\|\|lt2gene | Not-determined |
| SB_vs_k141_13735\|\|lt2gene | Low-quality |
| SB_vs_k141_109604\|\|lt2gene | Low-quality |
| SB_vs_k141_118095\|\|lt2gene | Not-determined |
| SB_vs_k141_118100\|\|lt2gene | Low-quality |
| SB_vs_k141_92116\|\|lt2gene | Not-determined |
| SB_vs_k141_23037\|\|lt2gene | Low-quality |
| SB_vs_k141_57689\|\|lt2gene | Not-determined |
| SB_vs_k141_152805\|\|lt2gene | Low-quality |
| SB_vs_k141_57714\|\|lt2gene | Low-quality |
| SB_vs_k141_152815\|\|lt2gene | Not-determined |
| SB_vs_k141_31629\|\|lt2gene | Low-quality |
| SB_vs_k141_196463\|\|lt2gene | Low-quality |
| SB_vs_k141_5279\|\|lt2gene | Low-quality |
| SB_vs_k141_170421\|\|lt2gene | Low-quality |
| SB_vs_k141_126813\|\|lt2gene | Not-determined |
| SB_vs_k141_5287\|\|lt2gene | Low-quality |
| SB_vs_k141_109648\|\|lt2gene | Low-quality |
| SB_vs_k141_118171\|\|lt2gene | Low-quality |
| SB_vs_k141_100377\|\|lt2gene | Not-determined |
| SB_vs_k141_144369\|\|lt2gene | Low-quality |
| SB_vs_k141_100391\|\|lt2gene | Low-quality |
| SB_vs_k141_100395\|\|lt2gene | Not-determined |
| SB_vs_k141_31662\|\|lt2gene | Low-quality |
| SB_vs_k141_100400\|\|lt2gene | Not-determined |
| SB_vs_k141_100405\|\|lt2gene | Low-quality |
| SB_vs_k141_135338\|\|lt2gene | Not-determined |
| SB_vs_k141_135352\|\|lt2gene | Low-quality |
| SB_vs_k141_57781\|\|lt2gene | Low-quality |
| SB_vs_k141_48424\|\|lt2gene | Low-quality |
| SB_vs_k141_179121\|\|lt2gene | Low-quality |
| SB_vs_k141_48435\|\|lt2gene | Low-quality |
| SB_vs_k141_109689\|\|lt2gene | Not-determined |
| SB_vs_k141_186737\|\|lt2gene | Low-quality |
| SB_vs_k141_5380\|\|lt2gene | Low-quality |
| SB_vs_k141_65979\|\|lt2gene | Not-determined |
| SB_vs_k141_196589\|\|lt2gene | Not-determined |
| SB_vs_k141_204725\|\|lt2gene | Not-determined |
| SB_vs_k141_204731\|\|lt2gene | Not-determined |
| SB_vs_k141_83545\|\|lt2gene | Not-determined |
| SB_vs_k141_83547\|\|lt2gene | Not-determined |
| SB_vs_k141_118286\|\|lt2gene | Not-determined |
| SB_vs_k141_92282\|\|lt2gene | Low-quality |
| SB_vs_k141_83561\|\|lt2gene | Not-determined |
| SB_vs_k141_57880\|\|lt2gene | Not-determined |
| SB_vs_k141_161270\|\|lt2gene | Not-determined |
| SB_vs_k141_126873\|\|lt2gene | Low-quality |
| SB_vs_k141_100533\|\|lt2gene | Low-quality |
| SB_vs_k141_31800\|\|lt2gene | Low-quality |
| SB_vs_k141_135472\|\|lt2gene | Low-quality |
| SB_vs_k141_5433\|\|lt2gene | Not-determined |
| SB_vs_k141_23184\|\|lt2gene | Low-quality |
| SB_vs_k141_100557\|\|lt2gene | Low-quality |
| SB_vs_k141_66007\|\|lt2gene | Low-quality |
| SB_vs_k141_152982\|\|lt2gene | Not-determined |
| SB_vs_k141_170500\|\|lt2gene | Low-quality |
| SB_vs_k141_170511\|\|lt2gene | Low-quality |
| SB_vs_k141_83600\|\|lt2gene | Not-determined |
| SB_vs_k141_100587\|\|lt2gene | Not-determined |
| SB_vs_k141_40285\|\|lt2gene | Not-determined |
| SB_vs_k141_92353\|\|lt2gene | Low-quality |
| SB_vs_k141_74546\|\|lt2gene | Low-quality |
| SB_vs_k141_196673\|\|lt2gene | Not-determined |
| SB_vs_k141_153066\|\|lt2gene | Low-quality |
| SB_vs_k141_5509\|\|lt2gene | Low-quality |
| SB_vs_k141_5513\|\|lt2gene | Low-quality |
| SB_vs_k141_74587\|\|lt2gene | Not-determined |
| SB_vs_k141_40329\|\|lt2gene | Low-quality |
| SB_vs_k141_153099\|\|lt2gene | Low-quality |
| SB_vs_k141_126992\|\|lt2gene | Not-determined |
| SB_vs_k141_48612\|\|lt2gene | Low-quality |
| SB_vs_k141_66092\|\|lt2gene | Low-quality |
| SB_vs_k141_118448\|\|lt2gene | Low-quality |
| SB_vs_k141_109849\|\|lt2gene | Not-determined |

S8_vs_k141_127009||lt2gene Low-quality
S8_vs_k141_40385||lt2gene Low-quality
S8_vs_k141_58024||lt2gene Low-quality
S8_vs_k141_179216||lt2gene Low-quality
S8_vs_k141_118487||lt2gene Low-quality
S8_vs_k141_58047||lt2gene Low-quality
S8_vs_k141_48651||lt2gene Low-quality
S8_vs_k141_58051||lt2gene Not-determined
S8_vs_k141_23246||lt2gene Low-quality
S8_vs_k141_144562||lt2gene Not-determined
S8_vs_k141_144569||lt2gene Low-quality
S8_vs_k141_118512||lt2gene Low-quality
S8_vs_k141_204842||lt2gene Low-quality
S8_vs_k141_161454||lt2gene Low-quality
S8_vs_k141_186969||lt2gene Low-quality
S8_vs_k141_127050||lt2gene Low-quality
S8_vs_k141_58083||lt2gene Low-quality
S8_vs_k141_135680||lt2gene Low-quality
S8_vs_k141_14050||lt2gene Not-determined
S8_vs_k141_23286||lt2gene Not-determined
S8_vs_k141_135695||lt2gene Low-quality
S8_vs_k141_196791||lt2gene Not-determined
S8_vs_k141_66164||lt2gene Low-quality
S8_vs_k141_196810||lt2gene Not-determined
S8_vs_k141_179348||lt2gene Low-quality
S8_vs_k141_179349||lt2gene Low-quality
S8_vs_k141_40493||lt2gene Low-quality
S8_vs_k141_74720||lt2gene Not-determined
S8_vs_k141_144636||lt2gene Not-determined
S8_vs_k141_32026||lt2gene Not-determined
S8_vs_k141_118605||lt2gene Low-quality
S8_vs_k141_32031||lt2gene Low-quality
S8_vs_k141_161567||lt2gene Low-quality
S8_vs_k141_153229||lt2gene Low-quality
S8_vs_k141_204974||lt2gene Not-determined
S8_vs_k141_48763||lt2gene Not-determined
S8_vs_k141_109995||lt2gene Low-quality
S8_vs_k141_196852||lt2gene Low-quality
S8_vs_k141_14122||lt2gene Low-quality
S8_vs_k141_187022||lt2gene Not-determined
S8_vs_k141_48780||lt2gene Not-determined
S8_vs_k141_58152||lt2gene Low-quality
S8_vs_k141_5715||lt2gene Low-quality
S8_vs_k141_196892||lt2gene Low-quality
S8_vs_k141_110035||lt2gene Low-quality
S8_vs_k141_205018||lt2gene Low-quality
S8_vs_k141_66270||lt2gene Low-quality
S8_vs_k141_100837||lt2gene Low-quality
S8_vs_k141_5739||lt2gene Not-determined
S8_vs_k141_58177||lt2gene Low-quality
S8_vs_k141_161628||lt2gene Low-quality
S8_vs_k141_127135||lt2gene Not-determined
S8_vs_k141_127137||lt2gene Low-quality
S8_vs_k141_74836||lt2gene Not-determined
S8_vs_k141_23423||lt2gene Low-quality
S8_vs_k141_196926||lt2gene Low-quality
S8_vs_k141_32115||lt2gene Low-quality
S8_vs_k141_187116||lt2gene Low-quality
S8_vs_k141_153292||lt2gene Low-quality
S8_vs_k141_153300||lt2gene Low-quality
S8_vs_k141_170733||lt2gene Low-quality
S8_vs_k141_127220||lt2gene Not-determined
S8_vs_k141_205119||lt2gene Not-determined
S8_vs_k141_74918||lt2gene Low-quality
S8_vs_k141_187176||lt2gene Not-determined
S8_vs_k141_118747||lt2gene Low-quality
S8_vs_k141_74930||lt2gene Low-quality
S8_vs_k141_196978||lt2gene Low-quality
S8_vs_k141_127247||lt2gene Low-quality
S8_vs_k141_187187||lt2gene Low-quality
S8_vs_k141_196984||lt2gene Low-quality
S8_vs_k141_196985||lt2gene Low-quality
S8_vs_k141_196987||lt2gene Low-quality
S8_vs_k141_5881||lt2gene Low-quality
S8_vs_k141_14240||lt2gene Low-quality
S8_vs_k141_66391||lt2gene Low-quality
S8_vs_k141_205168||lt2gene Low-quality
S8_vs_k141_100975||lt2gene Not-determined
S8_vs_k141_23514||lt2gene Not-determined
S8_vs_k141_23518||lt2gene Not-determined
S8_vs_k141_66405||lt2gene Low-quality
S8_vs_k141_23521||lt2gene Not-determined
S8_vs_k141_23525||lt2gene Low-quality
S8_vs_k141_110192||lt2gene Low-quality
S8_vs_k141_23529||lt2gene Low-quality
S8_vs_k141_66431||lt2gene Low-quality
S8_vs_k141_14273||lt2gene Not-determined
S8_vs_k141_127333||lt2gene Not-determined
S8_vs_k141_205254||lt2gene Low-quality
S8_vs_k141_23583||lt2gene Low-quality
S8_vs_k141_153373||lt2gene Not-determined
S8_vs_k141_135794||lt2gene Low-quality
S8_vs_k141_14313||lt2gene Low-quality
S8_vs_k141_127353||lt2gene Low-quality
S8_vs_k141_161714||lt2gene Low-quality
S8_vs_k141_179554||lt2gene Low-quality
S8_vs_k141_23618||lt2gene Low-quality
S8_vs_k141_144977||lt2gene Low-quality
S8_vs_k141_92732||lt2gene Low-quality
S8_vs_k141_161744||lt2gene Not-determined
S8_vs_k141_135846||lt2gene Low-quality
S8_vs_k141_110300||lt2gene Low-quality
S8_vs_k141_153443||lt2gene Low-quality
S8_vs_k141_118865||lt2gene Low-quality
S8_vs_k141_23673||lt2gene Low-quality
S8_vs_k141_32187||lt2gene Not-determined
S8_vs_k141_205268||lt2gene Low-quality
S8_vs_k141_23693||lt2gene Low-quality
S8_vs_k141_92785||lt2gene Not-determined
S8_vs_k141_75098||lt2gene Low-quality
S8_vs_k141_179650||lt2gene Low-quality
S8_vs_k141_135879||lt2gene Not-determined
S8_vs_k141_23736||lt2gene Low-quality
S8_vs_k141_75120||lt2gene Not-determined
S8_vs_k141_66521||lt2gene Low-quality
S8_vs_k141_110377||lt2gene Not-determined
S8_vs_k141_58458||lt2gene Not-determined
S8_vs_k141_135906||lt2gene Not-determined
S8_vs_k141_14440||lt2gene Low-quality
S8_vs_k141_6120||lt2gene Low-quality
S8_vs_k141_187446||lt2gene Not-determined
S8_vs_k141_75157||lt2gene Not-determined
S8_vs_k141_161877||lt2gene Not-determined
S8_vs_k141_49103||lt2gene Low-quality
S8_vs_k141_6140||lt2gene Low-quality
S8_vs_k141_187478||lt2gene Low-quality
S8_vs_k141_6160||lt2gene Low-quality
S8_vs_k141_170960||lt2gene Low-quality
S8_vs_k141_23808||lt2gene Low-quality
S8_vs_k141_145039||lt2gene Low-quality
S8_vs_k141_197147||lt2gene Low-quality
S8_vs_k141_197148||lt2gene Low-quality
S8_vs_k141_145045||lt2gene Low-quality
S8_vs_k141_145049||lt2gene Not-determined
S8_vs_k141_6185||lt2gene Low-quality
S8_vs_k141_83989||lt2gene Low-quality
S8_vs_k141_171024||lt2gene Low-quality
S8_vs_k141_23851||lt2gene Not-determined
S8_vs_k141_41026||lt2gene Not-determined
S8_vs_k141_66605||lt2gene Low-quality
S8_vs_k141_75255||lt2gene Low-quality
S8_vs_k141_197206||lt2gene Low-quality
S8_vs_k141_179744||lt2gene Not-determined
S8_vs_k141_75274||lt2gene Low-quality
S8_vs_k141_92832||lt2gene Low-quality
S8_vs_k141_6223||lt2gene Not-determined
S8_vs_k141_66624||lt2gene Not-determined
S8_vs_k141_136040||lt2gene Low-quality
S8_vs_k141_101282||lt2gene Not-determined
S8_vs_k141_75293||lt2gene Low-quality
S8_vs_k141_171067||lt2gene Not-determined
S8_vs_k141_161969||lt2gene Low-quality
S8_vs_k141_110529||lt2gene Not-determined
S8_vs_k141_119110||lt2gene Not-determined
S8_vs_k141_66678||lt2gene Low-quality
S8_vs_k141_6270||lt2gene Low-quality
S8_vs_k141_205395||lt2gene Not-determined
S8_vs_k141_32398||lt2gene Low-quality
S8_vs_k141_179822||lt2gene Low-quality
S8_vs_k141_101328||lt2gene Not-determined
S8_vs_k141_187616||lt2gene Not-determined
S8_vs_k141_6333||lt2gene Not-determined
S8_vs_k141_205430||lt2gene Not-determined
S8_vs_k141_119116||lt2gene Low-quality
S8_vs_k141_205439||lt2gene Not-determined
S8_vs_k141_153807||lt2gene Low-quality
S8_vs_k141_75414||lt2gene Low-quality
S8_vs_k141_179872||lt2gene Low-quality
S8_vs_k141_205447||lt2gene Low-quality
S8_vs_k141_14693||lt2gene Low-quality
S8_vs_k141_205455||lt2gene Low-quality
S8_vs_k141_153843||lt2gene Low-quality
S8_vs_k141_153848||lt2gene Low-quality
S8_vs_k141_162082||lt2gene Low-quality
S8_vs_k141_32505||lt2gene Low-quality
S8_vs_k141_41068||lt2gene Low-quality
S8_vs_k141_145181||lt2gene Low-quality
S8_vs_k141_127788||lt2gene Not-determined
S8_vs_k141_101408||lt2gene Low-quality
S8_vs_k141_171184||lt2gene Low-quality
S8_vs_k141_205490||lt2gene Low-quality
S8_vs_k141_162110||lt2gene Not-determined
S8_vs_k141_127813||lt2gene Low-quality
S8_vs_k141_58818||lt2gene Low-quality
S8_vs_k141_110683||lt2gene Low-quality
S8_vs_k141_66794||lt2gene Not-determined
S8_vs_k141_66800||lt2gene Low-quality
S8_vs_k141_180000||lt2gene Low-quality
S8_vs_k141_6474||lt2gene Low-quality
S8_vs_k141_205539||lt2gene Low-quality
S8_vs_k141_162186||lt2gene Low-quality
S8_vs_k141_205546||lt2gene Low-quality
S8_vs_k141_171287||lt2gene Low-quality
S8_vs_k141_136239||lt2gene Low-quality
S8_vs_k141_75516||lt2gene Low-quality
S8_vs_k141_41183||lt2gene Not-determined
S8_vs_k141_84263||lt2gene Not-determined
S8_vs_k141_75542||lt2gene Low-quality
S8_vs_k141_93129||lt2gene Low-quality
S8_vs_k141_6503||lt2gene Low-quality
S8_vs_k141_180066||lt2gene Low-quality
S8_vs_k141_49269||lt2gene Low-quality
S8_vs_k141_136262||lt2gene Not-determined
S8_vs_k141_127883||lt2gene Low-quality
S8_vs_k141_119277||lt2gene Low-quality
S8_vs_k141_32626||lt2gene Not-determined
S8_vs_k141_127910||lt2gene Not-determined
S8_vs_k141_93176||lt2gene Not-determined
S8_vs_k141_14866||lt2gene Low-quality
S8_vs_k141_14869||lt2gene Low-quality
S8_vs_k141_75585||lt2gene Low-quality
S8_vs_k141_66912||lt2gene Low-quality
S8_vs_k141_93185||lt2gene Low-quality
S8_vs_k141_84340||lt2gene Not-determined
S8_vs_k141_58965||lt2gene Not-determined
S8_vs_k141_197599||lt2gene Not-determined
S8_vs_k141_171445||lt2gene Not-determined
S8_vs_k141_171446||lt2gene Low-quality
S8_vs_k141_197611||lt2gene Low-quality
S8_vs_k141_101651||lt2gene Low-quality
S8_vs_k141_6611||lt2gene Low-quality
S8_vs_k141_101657||lt2gene Low-quality
S8_vs_k141_205702||lt2gene Not-determined
S8_vs_k141_127964||lt2gene Low-quality
S8_vs_k141_66983||lt2gene Low-quality
S8_vs_k141_187775||lt2gene Low-quality
S8_vs_k141_24119||lt2gene Low-quality
S8_vs_k141_171477||lt2gene Low-quality
S8_vs_k141_119365||lt2gene Low-quality
S8_vs_k141_171499||lt2gene Low-quality
S8_vs_k141_180235||lt2gene Low-quality
S8_vs_k141_128003||lt2gene Low-quality
S8_vs_k141_154115||lt2gene Low-quality
S8_vs_k141_6669||lt2gene Low-quality
S8_vs_k141_154130||lt2gene Low-quality
S8_vs_k141_205785||lt2gene Low-quality
S8_vs_k141_84465||lt2gene Not-determined
S8_vs_k141_197719||lt2gene Not-determined
S8_vs_k141_75780||lt2gene Low-quality
S8_vs_k141_128069||lt2gene Low-quality
S8_vs_k141_101777||lt2gene Low-quality
S8_vs_k141_197721||lt2gene Not-determined
S8_vs_k141_32807||lt2gene Not-determined
S8_vs_k141_110850||lt2gene Not-determined
S8_vs_k141_145531||lt2gene Low-quality
S8_vs_k141_75803||lt2gene Not-determined
S8_vs_k141_101803||lt2gene Low-quality
S8_vs_k141_180307||lt2gene Low-quality
S8_vs_k141_162363||lt2gene Low-quality
S8_vs_k141_101811||lt2gene Low-quality
S8_vs_k141_136496||lt2gene Low-quality
S8_vs_k141_84530||lt2gene Not-determined
S8_vs_k141_24211||lt2gene Not-determined
S8_vs_k141_154183||lt2gene Low-quality
S8_vs_k141_6740||lt2gene Low-quality
S8_vs_k141_6741||lt2gene Low-quality
S8_vs_k141_24252||lt2gene Low-quality
S8_vs_k141_93349||lt2gene Not-determined
S8_vs_k141_24258||lt2gene Not-determined
S8_vs_k141_180345||lt2gene Not-determined
S8_vs_k141_205818||lt2gene Not-determined
S8_vs_k141_101883||lt2gene Low-quality
S8_vs_k141_136540||lt2gene Low-quality
S8_vs_k141_84610||lt2gene Low-quality
S8_vs_k141_128159||lt2gene Low-quality
S8_vs_k141_15068||lt2gene Low-quality
S8_vs_k141_67157||lt2gene Low-quality
S8_vs_k141_145650||lt2gene Low-quality
S8_vs_k141_59249||lt2gene Low-quality
S8_vs_k141_75916||lt2gene Low-quality
S8_vs_k141_180413||lt2gene Low-quality
S8_vs_k141_15138||lt2gene Low-quality
S8_vs_k141_197863||lt2gene Low-quality
S8_vs_k141_128229||lt2gene Not-determined
S8_vs_k141_162510||lt2gene Low-quality
S8_vs_k141_187967||lt2gene Low-quality
S8_vs_k141_59314||lt2gene Not-determined
S8_vs_k141_49509||lt2gene Low-quality
S8_vs_k141_110978||lt2gene Low-quality
S8_vs_k141_197900||lt2gene Low-quality
S8_vs_k141_119552||lt2gene Low-quality
S8_vs_k141_93449||lt2gene Low-quality
S8_vs_k141_24428||lt2gene Low-quality
S8_vs_k141_197922||lt2gene Not-determined
S8_vs_k141_67257||lt2gene Low-quality
S8_vs_k141_180509||lt2gene Low-quality
S8_vs_k141_188064||lt2gene Low-quality
S8_vs_k141_145800||lt2gene Not-determined
S8_vs_k141_84822||lt2gene Low-quality
S8_vs_k141_111086||lt2gene Low-quality
S8_vs_k141_205942||lt2gene Not-determined
S8_vs_k141_188096||lt2gene Not-determined
S8_vs_k141_136719||lt2gene Low-quality
S8_vs_k141_102083||lt2gene Low-quality
S8_vs_k141_136725||lt2gene Low-quality
S8_vs_k141_93512||lt2gene Low-quality
S8_vs_k141_171885||lt2gene Not-determined
S8_vs_k141_128371||lt2gene Low-quality
S8_vs_k141_171906||lt2gene Low-quality
S8_vs_k141_162635||lt2gene Low-quality
S8_vs_k141_33010||lt2gene Low-quality
S8_vs_k141_76131||lt2gene Not-determined
S8_vs_k141_162646||lt2gene Low-quality
S8_vs_k141_49677||lt2gene Not-determined
S8_vs_k141_33034||lt2gene Low-quality
S8_vs_k141_119624||lt2gene Low-quality
S8_vs_k141_180601||lt2gene Low-quality
S8_vs_k141_162688||lt2gene Low-quality
S8_vs_k141_59525||lt2gene Low-quality
S8_vs_k141_102138||lt2gene Low-quality
S8_vs_k141_15293||lt2gene Low-quality
S8_vs_k141_102169||lt2gene Low-quality
S8_vs_k141_49745||lt2gene Low-quality
S8_vs_k141_188187||lt2gene Low-quality
S8_vs_k141_84892||lt2gene Low-quality
S8_vs_k141_180660||lt2gene Low-quality
S8_vs_k141_41521||lt2gene Low-quality
S8_vs_k141_119674||lt2gene Low-quality
S8_vs_k141_7007||lt2gene Low-quality
S8_vs_k141_15372||lt2gene Low-quality
S8_vs_k141_24566||lt2gene Low-quality
S8_vs_k141_84944||lt2gene Not-determined
S8_vs_k141_162806||lt2gene Low-quality
S8_vs_k141_76229||lt2gene Low-quality
S8_vs_k141_146002||lt2gene Low-quality
S8_vs_k141_119711||lt2gene Low-quality
S8_vs_k141_188272||lt2gene Not-determined
S8_vs_k141_206095||lt2gene Low-quality
S8_vs_k141_162853||lt2gene Low-quality
S8_vs_k141_33139||lt2gene Low-quality
S8_vs_k141_67433||lt2gene Low-quality
S8_vs_k141_41633||lt2gene Low-quality
S8_vs_k141_198153||lt2gene Low-quality
S8_vs_k141_111334||lt2gene Low-quality
S8_vs_k141_146051||lt2gene Low-quality
S8_vs_k141_76302||lt2gene Low-quality
S8_vs_k141_172145||lt2gene Not-determined
S8_vs_k141_59612||lt2gene Low-quality
S8_vs_k141_172163||lt2gene Low-quality
S8_vs_k141_180819||lt2gene Not-determined
S8_vs_k141_7139||lt2gene Low-quality
S8_vs_k141_7155||lt2gene Low-quality
S8_vs_k141_41726||lt2gene Not-determined
S8_vs_k141_15510||lt2gene Low-quality
S8_vs_k141_7175||lt2gene Low-quality
S8_vs_k141_188426||lt2gene Not-determined
S8_vs_k141_188427||lt2gene Not-determined
S8_vs_k141_7217||lt2gene Low-quality
S8_vs_k141_67528||lt2gene Low-quality
S8_vs_k141_33295||lt2gene Low-quality
S8_vs_k141_206246||lt2gene Low-quality
S8_vs_k141_128593||lt2gene Not-determined
S8_vs_k141_172236||lt2gene Low-quality
S8_vs_k141_198341||lt2gene Low-quality
S8_vs_k141_163050||lt2gene Not-determined
S8_vs_k141_154299||lt2gene Low-quality
S8_vs_k141_85193||lt2gene Low-quality
S8_vs_k141_85197||lt2gene Low-quality
S8_vs_k141_137090||lt2gene Low-quality
S8_vs_k141_163075||lt2gene Not-determined
S8_vs_k141_76356||lt2gene Low-quality
S8_vs_k141_102446||lt2gene Not-determined
S8_vs_k141_163089||lt2gene Low-quality
S8_vs_k141_163096||lt2gene Not-determined
S8_vs_k141_50000||lt2gene Low-quality
S8_vs_k141_50001||lt2gene Low-quality
S8_vs_k141_41859||lt2gene Low-quality
S8_vs_k141_137129||lt2gene Low-quality
S8_vs_k141_33386||lt2gene Not-determined
S8_vs_k141_119917||lt2gene Not-determined
S8_vs_k141_188511||lt2gene Not-determined
S8_vs_k141_33398||lt2gene Low-quality
S8_vs_k141_76384||lt2gene Low-quality
S8_vs_k141_33403||lt2gene Low-quality
S8_vs_k141_111564||lt2gene Low-quality
S8_vs_k141_128639||lt2gene Low-quality
S8_vs_k141_172332||lt2gene Low-quality
S8_vs_k141_206340||lt2gene Low-quality
S8_vs_k141_76403||lt2gene Low-quality
S8_vs_k141_111600||lt2gene Low-quality
S8_vs_k141_67633||lt2gene Low-quality
S8_vs_k141_33437||lt2gene Low-quality
S8_vs_k141_128672||lt2gene Not-determined
S8_vs_k141_128679||lt2gene Low-quality
S8_vs_k141_67662||lt2gene Low-quality
S8_vs_k141_50078||lt2gene Low-quality
S8_vs_k141_137209||lt2gene Not-determined
S8_vs_k141_33458||lt2gene Low-quality
S8_vs_k141_206380||lt2gene Low-quality
S8_vs_k141_50083||lt2gene Low-quality
S8_vs_k141_76444||lt2gene Low-quality
S8_vs_k141_7339||lt2gene Low-quality
S8_vs_k141_128710||lt2gene Low-quality
S8_vs_k141_15723||lt2gene Low-quality
S8_vs_k141_85303||lt2gene Low-quality
S8_vs_k141_128736||lt2gene Low-quality
S8_vs_k141_50130||lt2gene Not-determined
S8_vs_k141_172435||lt2gene Low-quality
S8_vs_k141_198485||lt2gene Not-determined
S8_vs_k141_111652||lt2gene Low-quality
S8_vs_k141_102582||lt2gene Not-determined
S8_vs_k141_94020||lt2gene Not-determined
S8_vs_k141_67748||lt2gene Not-determined
S8_vs_k141_137282||lt2gene Low-quality
S8_vs_k141_111698||lt2gene Not-determined
S8_vs_k141_76550||lt2gene Not-determined
S8_vs_k141_50202||lt2gene Low-quality
S8_vs_k141_198562||lt2gene Low-quality
S8_vs_k141_172483||lt2gene Low-quality
S8_vs_k141_67775||lt2gene Low-quality
S8_vs_k141_181129||lt2gene Low-quality
S8_vs_k141_188628||lt2gene Low-quality
S8_vs_k141_137334||lt2gene Low-quality
S8_vs_k141_172500||lt2gene Low-quality
S8_vs_k141_67800||lt2gene Low-quality
S8_vs_k141_120120||lt2gene Low-quality
S8_vs_k141_50247||lt2gene Low-quality
S8_vs_k141_76616||lt2gene Low-quality
S8_vs_k141_137356||lt2gene Low-quality
S8_vs_k141_33614||lt2gene Low-quality
S8_vs_k141_102654||lt2gene Not-determined
S8_vs_k141_33621||lt2gene Low-quality
S8_vs_k141_111781||lt2gene Low-quality
S8_vs_k141_67825||lt2gene Low-quality
S8_vs_k141_172540||lt2gene Low-quality
S8_vs_k141_146306||lt2gene Low-quality
S8_vs_k141_181190||lt2gene Low-quality
S8_vs_k141_50274||lt2gene Low-quality
S8_vs_k141_198636||lt2gene Not-determined
S8_vs_k141_24911||lt2gene Not-determined
S8_vs_k141_24914||lt2gene Not-determined
S8_vs_k141_137413||lt2gene Low-quality
S8_vs_k141_59748||lt2gene Not-determined
S8_vs_k141_102703||lt2gene Low-quality
S8_vs_k141_163308||lt2gene Low-quality
S8_vs_k141_67853||lt2gene Low-quality
S8_vs_k141_24946||lt2gene Low-quality
S8_vs_k141_85420||lt2gene Low-quality
S8_vs_k141_24949||lt2gene Low-quality
S8_vs_k141_59775||lt2gene Low-quality
S8_vs_k141_85435||lt2gene Low-quality
S8_vs_k141_128918||lt2gene Low-quality
S8_vs_k141_154622||lt2gene Low-quality
S8_vs_k141_42126||lt2gene Not-determined
S8_vs_k141_33723||lt2gene Low-quality
S8_vs_k141_7543||lt2gene Not-determined
S8_vs_k141_154630||lt2gene Low-quality
S8_vs_k141_120209||lt2gene Low-quality
S8_vs_k141_7554||lt2gene Low-quality
S8_vs_k141_128965||lt2gene Low-quality
S8_vs_k141_50343||lt2gene Low-quality
S8_vs_k141_206569||lt2gene Low-quality
S8_vs_k141_198706||lt2gene Low-quality
S8_vs_k141_42146||lt2gene Low-quality
S8_vs_k141_163380||lt2gene Not-determined
S8_vs_k141_198718||lt2gene Low-quality
S8_vs_k141_50351||lt2gene Not-determined
S8_vs_k141_94210||lt2gene Low-quality
S8_vs_k141_67916||lt2gene Low-quality
S8_vs_k141_59815||lt2gene Low-quality
S8_vs_k141_163404||lt2gene Not-determined
S8_vs_k141_154665||lt2gene Low-quality
S8_vs_k141_33779||lt2gene Not-determined
S8_vs_k141_25027||lt2gene Low-quality
S8_vs_k141_120261||lt2gene Low-quality
S8_vs_k141_172662||lt2gene Not-determined
S8_vs_k141_154695||lt2gene Not-determined
S8_vs_k141_154696||lt2gene Not-determined
S8_vs_k141_7626||lt2gene Low-quality
S8_vs_k141_154703||lt2gene Low-quality
S8_vs_k141_172687||lt2gene Not-determined
S8_vs_k141_85523||lt2gene Not-determined
S8_vs_k141_85529||lt2gene Low-quality
S8_vs_k141_76851||lt2gene Not-determined
S8_vs_k141_59909||lt2gene Low-quality
S8_vs_k141_172729||lt2gene Not-determined
S8_vs_k141_111817||lt2gene Low-quality
S8_vs_k141_198850||lt2gene Low-quality
S8_vs_k141_188778||lt2gene Low-quality
S8_vs_k141_206676||lt2gene Low-quality
S8_vs_k141_181354||lt2gene Low-quality
S8_vs_k141_181356||lt2gene Low-quality
S8_vs_k141_111832||lt2gene Low-quality
S8_vs_k141_137536||lt2gene Low-quality
S8_vs_k141_15886||lt2gene Low-quality
S8_vs_k141_15889||lt2gene Low-quality
S8_vs_k141_172781||lt2gene Low-quality
S8_vs_k141_33909||lt2gene Low-quality
S8_vs_k141_137556||lt2gene Low-quality
S8_vs_k141_94319||lt2gene Low-quality
S8_vs_k141_102818||lt2gene Low-quality
S8_vs_k141_172821||lt2gene Low-quality
S8_vs_k141_7738||lt2gene Low-quality
S8_vs_k141_102857||lt2gene Low-quality
S8_vs_k141_137620||lt2gene Low-quality
S8_vs_k141_68097||lt2gene Low-quality
S8_vs_k141_94373||lt2gene Low-quality
S8_vs_k141_77000||lt2gene Low-quality
S8_vs_k141_111963||lt2gene Low-quality
S8_vs_k141_68121||lt2gene Low-quality
S8_vs_k141_163486||lt2gene Low-quality
S8_vs_k141_60089||lt2gene Not-determined
S8_vs_k141_154848||lt2gene Not-determined
S8_vs_k141_85729||lt2gene Not-determined
S8_vs_k141_77063||lt2gene Low-quality
S8_vs_k141_34051||lt2gene Low-quality
S8_vs_k141_34063||lt2gene Not-determined
S8_vs_k141_15989||lt2gene Low-quality
S8_vs_k141_60123||lt2gene Not-determined
S8_vs_k141_85742||lt2gene Low-quality
S8_vs_k141_129256||lt2gene Low-quality
S8_vs_k141_199027||lt2gene Low-quality
S8_vs_k141_94463||lt2gene Low-quality
S8_vs_k141_16014||lt2gene Low-quality
S8_vs_k141_137730||lt2gene Not-determined
S8_vs_k141_146696||lt2gene Not-determined
S8_vs_k141_137738||lt2gene Not-determined
S8_vs_k141_163547||lt2gene Low-quality
S8_vs_k141_172851||lt2gene Low-quality
S8_vs_k141_102993||lt2gene Low-quality
S8_vs_k141_34091||lt2gene Low-quality
S8_vs_k141_68169||lt2gene Low-quality
S8_vs_k141_206884||lt2gene Low-quality
S8_vs_k141_25241||lt2gene Low-quality
S8_vs_k141_188970||lt2gene Low-quality
S8_vs_k141_112062||lt2gene Low-quality
S8_vs_k141_34108||lt2gene Low-quality
S8_vs_k141_112080||lt2gene Low-quality
S8_vs_k141_120547||lt2gene Low-quality
S8_vs_k141_60199||lt2gene Low-quality
S8_vs_k141_34145||lt2gene Low-quality
S8_vs_k141_181583||lt2gene Low-quality
S8_vs_k141_94540||lt2gene Not-determined
S8_vs_k141_154950||lt2gene Low-quality
S8_vs_k141_181590||lt2gene Low-quality
S8_vs_k141_60228||lt2gene Low-quality
S8_vs_k141_206922||lt2gene Low-quality
S8_vs_k141_137831||lt2gene Low-quality
S8_vs_k141_163569||lt2gene Not-determined
S8_vs_k141_199114||lt2gene Not-determined
S8_vs_k141_25328||lt2gene Low-quality
S8_vs_k141_25329||lt2gene Low-quality
S8_vs_k141_77161||lt2gene Low-quality
S8_vs_k141_60272||lt2gene Low-quality
S8_vs_k141_94597||lt2gene Low-quality
S8_vs_k141_112150||lt2gene Low-quality
S8_vs_k141_181656||lt2gene Low-quality
S8_vs_k141_60279||lt2gene Not-determined
S8_vs_k141_163615||lt2gene Low-quality
S8_vs_k141_189068||lt2gene Low-quality
S8_vs_k141_25380||lt2gene Low-quality
S8_vs_k141_34229||lt2gene Not-determined
S8_vs_k141_103121||lt2gene Low-quality
S8_vs_k141_181689||lt2gene Low-quality
S8_vs_k141_103126||lt2gene Low-quality
S8_vs_k141_34246||lt2gene Low-quality
S8_vs_k141_68281||lt2gene Not-determined
S8_vs_k141_206969||lt2gene Low-quality
S8_vs_k141_60315||lt2gene Low-quality
S8_vs_k141_68296||lt2gene Low-quality
S8_vs_k141_34253||lt2gene Low-quality
S8_vs_k141_34254||lt2gene Low-quality
S8_vs_k141_60327||lt2gene Low-quality
S8_vs_k141_172930||lt2gene Low-quality
S8_vs_k141_137882||lt2gene Low-quality
S8_vs_k141_7985||lt2gene Low-quality
S8_vs_k141_112202||lt2gene Not-determined
S8_vs_k141_137921||lt2gene Low-quality
S8_vs_k141_60366||lt2gene Low-quality
S8_vs_k141_137924||lt2gene Not-determined
S8_vs_k141_163706||lt2gene Low-quality
S8_vs_k141_112240||lt2gene Not-determined
S8_vs_k141_16220||lt2gene Not-determined
S8_vs_k141_112262||lt2gene Not-determined
S8_vs_k141_120687||lt2gene Low-quality
S8_vs_k141_173009||lt2gene Low-quality
S8_vs_k141_8055||lt2gene Not-determined
S8_vs_k141_86023||lt2gene Not-determined
S8_vs_k141_163744||lt2gene Low-quality
S8_vs_k141_94639||lt2gene Low-quality
S8_vs_k141_34277||lt2gene Low-quality
S8_vs_k141_94654||lt2gene Low-quality
S8_vs_k141_155117||lt2gene Not-determined
S8_vs_k141_8109||lt2gene Not-determined
S8_vs_k141_50555||lt2gene Low-quality
S8_vs_k141_103165||lt2gene Low-quality
S8_vs_k141_189216||lt2gene Low-quality
S8_vs_k141_16320||lt2gene Low-quality
S8_vs_k141_173066||lt2gene Not-determined

| | |
|---|---|
| S8_vs_k141_68453\|\|lt2gene | Low-quality |
| S8_vs_k141_199305\|\|lt2gene | Low-quality |
| S8_vs_k141_68461\|\|lt2gene | Low-quality |
| S8_vs_k141_77420\|\|lt2gene | Not-determined |
| S8_vs_k141_181870\|\|lt2gene | Low-quality |
| S8_vs_k141_50592\|\|lt2gene | Low-quality |
| S8_vs_k141_181872\|\|lt2gene | Low-quality |
| S8_vs_k141_129496\|\|lt2gene | Low-quality |
| S8_vs_k141_103236\|\|lt2gene | Not-determined |
| S8_vs_k141_155217\|\|lt2gene | Low-quality |
| S8_vs_k141_50638\|\|lt2gene | Low-quality |
| S8_vs_k141_163775\|\|lt2gene | Not-determined |
| S8_vs_k141_155229\|\|lt2gene | Low-quality |
| S8_vs_k141_207166\|\|lt2gene | Not-determined |
| S8_vs_k141_94768\|\|lt2gene | Not-determined |
| S8_vs_k141_77484\|\|lt2gene | Not-determined |
| S8_vs_k141_34374\|\|lt2gene | Not-determined |
| S8_vs_k141_189343\|\|lt2gene | Low-quality |
| S8_vs_k141_8199\|\|lt2gene | Not-determined |
| S8_vs_k141_50665\|\|lt2gene | Low-quality |
| S8_vs_k141_50666\|\|lt2gene | Low-quality |
| S8_vs_k141_103273\|\|lt2gene | Low-quality |
| S8_vs_k141_189365\|\|lt2gene | Not-determined |
| S8_vs_k141_68565\|\|lt2gene | Low-quality |
| S8_vs_k141_77530\|\|lt2gene | Low-quality |
| S8_vs_k141_112445\|\|lt2gene | Low-quality |
| S8_vs_k141_207222\|\|lt2gene | Low-quality |
| S8_vs_k141_189375\|\|lt2gene | Low-quality |
| S8_vs_k141_8238\|\|lt2gene | Not-determined |
| S8_vs_k141_34408\|\|lt2gene | Not-determined |
| S8_vs_k141_199378\|\|lt2gene | Low-quality |
| S8_vs_k141_103318\|\|lt2gene | Low-quality |
| S8_vs_k141_163851\|\|lt2gene | Low-quality |
| S8_vs_k141_138183\|\|lt2gene | Not-determined |
| S8_vs_k141_50703\|\|lt2gene | Low-quality |
| S8_vs_k141_86181\|\|lt2gene | Low-quality |
| S8_vs_k141_129629\|\|lt2gene | Not-determined |
| S8_vs_k141_138220\|\|lt2gene | Low-quality |
| S8_vs_k141_16532\|\|lt2gene | Low-quality |
| S8_vs_k141_77632\|\|lt2gene | Low-quality |
| S8_vs_k141_129659\|\|lt2gene | Low-quality |
| S8_vs_k141_163929\|\|lt2gene | Low-quality |
| S8_vs_k141_120848\|\|lt2gene | Low-quality |
| S8_vs_k141_103386\|\|lt2gene | Low-quality |
| S8_vs_k141_16547\|\|lt2gene | Not-determined |
| S8_vs_k141_189486\|\|lt2gene | Not-determined |
| S8_vs_k141_94847\|\|lt2gene | Not-determined |
| S8_vs_k141_112553\|\|lt2gene | Low-quality |
| S8_vs_k141_112557\|\|lt2gene | Low-quality |
| S8_vs_k141_182051\|\|lt2gene | Low-quality |
| S8_vs_k141_25736\|\|lt2gene | Low-quality |
| S8_vs_k141_112565\|\|lt2gene | Low-quality |
| S8_vs_k141_207312\|\|lt2gene | Low-quality |
| S8_vs_k141_163956\|\|lt2gene | Low-quality |
| S8_vs_k141_77673\|\|lt2gene | Low-quality |
| S8_vs_k141_189511\|\|lt2gene | Low-quality |
| S8_vs_k141_103419\|\|lt2gene | Low-quality |
| S8_vs_k141_25763\|\|lt2gene | Low-quality |
| S8_vs_k141_189534\|\|lt2gene | Low-quality |
| S8_vs_k141_25781\|\|lt2gene | Low-quality |
| S8_vs_k141_103429\|\|lt2gene | Low-quality |
| S8_vs_k141_138286\|\|lt2gene | Not-determined |
| S8_vs_k141_155412\|\|lt2gene | Low-quality |
| S8_vs_k141_42858\|\|lt2gene | Not-determined |
| S8_vs_k141_163998\|\|lt2gene | Not-determined |
| S8_vs_k141_68717\|\|lt2gene | Low-quality |
| S8_vs_k141_68726\|\|lt2gene | Low-quality |
| S8_vs_k141_155425\|\|lt2gene | Low-quality |
| S8_vs_k141_16636\|\|lt2gene | Low-quality |
| S8_vs_k141_16642\|\|lt2gene | Not-determined |
| S8_vs_k141_199579\|\|lt2gene | Not-determined |
| S8_vs_k141_16646\|\|lt2gene | Not-determined |
| S8_vs_k141_42883\|\|lt2gene | Not-determined |
| S8_vs_k141_86341\|\|lt2gene | Low-quality |
| S8_vs_k141_16660\|\|lt2gene | Low-quality |
| S8_vs_k141_77786\|\|lt2gene | Low-quality |
| S8_vs_k141_77788\|\|lt2gene | Low-quality |
| S8_vs_k141_112583\|\|lt2gene | Low-quality |
| S8_vs_k141_112588\|\|lt2gene | Low-quality |
| S8_vs_k141_94977\|\|lt2gene | Low-quality |
| S8_vs_k141_77818\|\|lt2gene | Not-determined |
| S8_vs_k141_173280\|\|lt2gene | Low-quality |
| S8_vs_k141_86414\|\|lt2gene | Low-quality |
| S8_vs_k141_42902\|\|lt2gene | Low-quality |
| S8_vs_k141_42905\|\|lt2gene | Low-quality |
| S8_vs_k141_8471\|\|lt2gene | Low-quality |
| S8_vs_k141_42911\|\|lt2gene | Not-determined |
| S8_vs_k141_60741\|\|lt2gene | Not-determined |
| S8_vs_k141_8488\|\|lt2gene | Low-quality |
| S8_vs_k141_155538\|\|lt2gene | Low-quality |
| S8_vs_k141_16732\|\|lt2gene | Low-quality |
| S8_vs_k141_138428\|\|lt2gene | Low-quality |
| S8_vs_k141_42940\|\|lt2gene | Low-quality |
| S8_vs_k141_103595\|\|lt2gene | Not-determined |
| S8_vs_k141_129902\|\|lt2gene | Not-determined |
| S8_vs_k141_173326\|\|lt2gene | Low-quality |
| S8_vs_k141_8534\|\|lt2gene | Not-determined |
| S8_vs_k141_8539\|\|lt2gene | Not-determined |
| S8_vs_k141_86505\|\|lt2gene | Low-quality |
| S8_vs_k141_164128\|\|lt2gene | Low-quality |
| S8_vs_k141_129914\|\|lt2gene | Low-quality |
| S8_vs_k141_189754\|\|lt2gene | Low-quality |
| S8_vs_k141_25878\|\|lt2gene | Not-determined |
| S8_vs_k141_95094\|\|lt2gene | Low-quality |
| S8_vs_k141_207523\|\|lt2gene | Low-quality |
| S8_vs_k141_86538\|\|lt2gene | Low-quality |
| S8_vs_k141_129945\|\|lt2gene | Low-quality |
| S8_vs_k141_68924\|\|lt2gene | Low-quality |
| S8_vs_k141_16809\|\|lt2gene | Low-quality |
| S8_vs_k141_147345\|\|lt2gene | Not-determined |
| S8_vs_k141_16814\|\|lt2gene | Not-determined |
| S8_vs_k141_86559\|\|lt2gene | Not-determined |
| S8_vs_k141_86562\|\|lt2gene | Low-quality |
| S8_vs_k141_138528\|\|lt2gene | Low-quality |
| S8_vs_k141_86579\|\|lt2gene | Low-quality |
| S8_vs_k141_16836\|\|lt2gene | Low-quality |
| S8_vs_k141_121174\|\|lt2gene | Low-quality |
| S8_vs_k141_121175\|\|lt2gene | Not-determined |
| S8_vs_k141_103694\|\|lt2gene | Low-quality |
| S8_vs_k141_147379\|\|lt2gene | Not-determined |
| S8_vs_k141_50919\|\|lt2gene | Low-quality |
| S8_vs_k141_8615\|\|lt2gene | Not-determined |
| S8_vs_k141_68947\|\|lt2gene | Low-quality |
| S8_vs_k141_130007\|\|lt2gene | Low-quality |
| S8_vs_k141_50942\|\|lt2gene | Not-determined |
| S8_vs_k141_77977\|\|lt2gene | Low-quality |
| S8_vs_k141_103721\|\|lt2gene | Low-quality |
| S8_vs_k141_138551\|\|lt2gene | Not-determined |
| S8_vs_k141_68979\|\|lt2gene | Low-quality |
| S8_vs_k141_86607\|\|lt2gene | Low-quality |
| S8_vs_k141_155664\|\|lt2gene | Low-quality |
| S8_vs_k141_86619\|\|lt2gene | Not-determined |
| S8_vs_k141_189796\|\|lt2gene | Low-quality |
| S8_vs_k141_95191\|\|lt2gene | Low-quality |
| S8_vs_k141_147418\|\|lt2gene | Low-quality |
| S8_vs_k141_43092\|\|lt2gene | Not-determined |
| S8_vs_k141_207608\|\|lt2gene | Low-quality |
| S8_vs_k141_69024\|\|lt2gene | Not-determined |
| S8_vs_k141_95209\|\|lt2gene | Low-quality |
| S8_vs_k141_95214\|\|lt2gene | Low-quality |
| S8_vs_k141_164261\|\|lt2gene | Not-determined |
| S8_vs_k141_164274\|\|lt2gene | Low-quality |
| S8_vs_k141_189828\|\|lt2gene | Not-determined |
| S8_vs_k141_69062\|\|lt2gene | Low-quality |
| S8_vs_k141_69074\|\|lt2gene | Low-quality |
| S8_vs_k141_147482\|\|lt2gene | Low-quality |
| S8_vs_k141_43142\|\|lt2gene | Low-quality |
| S8_vs_k141_78089\|\|lt2gene | Low-quality |
| S8_vs_k141_16945\|\|lt2gene | Low-quality |
| S8_vs_k141_78097\|\|lt2gene | Low-quality |
| S8_vs_k141_95291\|\|lt2gene | Not-determined |
| S8_vs_k141_95312\|\|lt2gene | Low-quality |
| S8_vs_k141_95319\|\|lt2gene | Low-quality |
| S8_vs_k141_112797\|\|lt2gene | Not-determined |
| S8_vs_k141_155802\|\|lt2gene | Low-quality |
| S8_vs_k141_17008\|\|lt2gene | Low-quality |
| S8_vs_k141_86762\|\|lt2gene | Low-quality |
| S8_vs_k141_86763\|\|lt2gene | Low-quality |
| S8_vs_k141_51150\|\|lt2gene | Low-quality |
| S8_vs_k141_164394\|\|lt2gene | Not-determined |
| S8_vs_k141_189966\|\|lt2gene | Low-quality |
| S8_vs_k141_138642\|\|lt2gene | Low-quality |
| S8_vs_k141_103887\|\|lt2gene | Not-determined |
| S8_vs_k141_138652\|\|lt2gene | Low-quality |
| S8_vs_k141_103899\|\|lt2gene | Low-quality |
| S8_vs_k141_190017\|\|lt2gene | Low-quality |
| S8_vs_k141_69250\|\|lt2gene | Low-quality |
| S8_vs_k141_190032\|\|lt2gene | Low-quality |
| S8_vs_k141_121484\|\|lt2gene | Low-quality |
| S8_vs_k141_164440\|\|lt2gene | Low-quality |
| S8_vs_k141_103925\|\|lt2gene | Low-quality |
| S8_vs_k141_190069\|\|lt2gene | Low-quality |
| S8_vs_k141_138705\|\|lt2gene | Low-quality |
| S8_vs_k141_51254\|\|lt2gene | Low-quality |
| S8_vs_k141_155894\|\|lt2gene | Low-quality |
| S8_vs_k141_69313\|\|lt2gene | Low-quality |
| S8_vs_k141_43287\|\|lt2gene | Low-quality |
| S8_vs_k141_190100\|\|lt2gene | Low-quality |
| S8_vs_k141_17107\|\|lt2gene | Low-quality |
| S8_vs_k141_69365\|\|lt2gene | Low-quality |
| S8_vs_k141_69369\|\|lt2gene | Not-determined |
| S8_vs_k141_190142\|\|lt2gene | Not-determined |
| S8_vs_k141_164548\|\|lt2gene | Low-quality |
| S8_vs_k141_69413\|\|lt2gene | Low-quality |
| S8_vs_k141_207759\|\|lt2gene | Not-determined |
| S8_vs_k141_156031\|\|lt2gene | Low-quality |
| S8_vs_k141_138821\|\|lt2gene | Low-quality |
| S8_vs_k141_164623\|\|lt2gene | Not-determined |
| S8_vs_k141_164626\|\|lt2gene | Low-quality |
| S8_vs_k141_164658\|\|lt2gene | Low-quality |
| S8_vs_k141_104016\|\|lt2gene | Low-quality |
| S8_vs_k141_17150\|\|lt2gene | Not-determined |
| S8_vs_k141_164678\|\|lt2gene | Low-quality |
| S8_vs_k141_207865\|\|lt2gene | Low-quality |
| S8_vs_k141_156089\|\|lt2gene | Low-quality |
| S8_vs_k141_164706\|\|lt2gene | Low-quality |
| S8_vs_k141_164721\|\|lt2gene | Not-determined |
| S8_vs_k141_207939\|\|lt2gene | Low-quality |
| S8_vs_k141_51459\|\|lt2gene | Low-quality |
| S8_vs_k141_164770\|\|lt2gene | Low-quality |
| S8_vs_k141_190380\|\|lt2gene | Low-quality |
| S8_vs_k141_156181\|\|lt2gene | Not-determined |
| S8_vs_k141_156190\|\|lt2gene | Low-quality |
| S8_vs_k141_190422\|\|lt2gene | Not-determined |
| S8_vs_k141_208060\|\|lt2gene | Low-quality |
| S8_vs_k141_51516\|\|lt2gene | Low-quality |
| S8_vs_k141_190476\|\|lt2gene | Low-quality |
| S8_vs_k141_190486\|\|lt2gene | Not-determined |
| S8_vs_k141_51543\|\|lt2gene | Low-quality |
| S8_vs_k141_51575\|\|lt2gene | Low-quality |
| S8_vs_k141_51742\|\|lt2gene | Low-quality |
| S8_vs_k141_190739\|\|lt2gene | Not-determined |
| S8_vs_k141_190784\|\|lt2gene | Low-quality |
| S8_vs_k141_190810\|\|lt2gene | Not-determined |
| S8_vs_k141_190811\|\|lt2gene | Low-quality |
| S8_vs_k141_51854\|\|lt2gene | Low-quality |
| S8_vs_k141_190859\|\|lt2gene | Low-quality |

| | Count | Mean | Median | SD |
|---|---|---|---|---|
| Complete (100% completeness) | 273 | 65799.9 | 53491 | 42978.7 |
| High quality (90-100% completeness) | 992 | 55083.8 | 44111.5 | 35459.7 |
| Medium quality (50-90% completeness) | 1002 | 33906.4 | 27336.5 | 27394.5 |
| Low quality (< 50% completeness) | 97632 | 1852.3 | 835 | 3746.6 |
| Undetermined | 211309 | 907.5 | 459 | 1570.1 |
| Total UViGs | 311208 | 31509.98 | 27336.5 | 18712.78 |
| Sum of 50-100% genome completeness | 2267 | 51596.7 | 44111.5 | 7793.695 |
